## Supplementary material for "Comprehensive profiling of the human intestinal DNA virome and prediction of disease-associated bacterial hosts in severe Myalgic Encephalomyelitis/Chronic Fatigue Syndrome (ME/CFS)": S Table 4 WMS UViGs

| contig_id | checkv_quality |
|---|---|
| 17-106-final-viral-combined_k141_18\|\|full | Not-determined |
| 17-106-final-viral-combined_k141_29611\|\|full | Complete |
| 17-106-final-viral-combined_k141_118569\|\|full | Not-determined |
| 17-106-final-viral-combined_k141_61921\|\|full | Not-determined |
| 17-106-final-viral-combined_k141_78884\|\|full | Low-quality |
| 18-109-final-viral-combined_k141_62119\|\|full | Medium-quality |
| 17-106-final-viral-combined_k141_128234\|\|full | Not-determined |
| 17-106-final-viral-combined_k141_128240\|\|full | Not-determined |
| 17-106-final-viral-combined_k141_128251\|\|full | Low-quality |
| 17-106-final-viral-combined_k141_80221\|\|full | Not-determined |
| 17-106-final-viral-combined_k141_112305\|\|full | Not-determined |
| 17-106-final-viral-combined_k141_5361\|\|full | Not-determined |
| 17-106-final-viral-combined_k141_48140\|\|full | Low-quality |
| 17-106-final-viral-combined_k141_117660\|\|full | Low-quality |
| 17-106-final-viral-combined_k141_101618\|\|full | Not-determined |
| 17-106-final-viral-combined_k141_101621\|\|full | Low-quality |
| 17-106-final-viral-combined_k141_106974\|\|full | Low-quality |
| 17-106-final-viral-combined_k141_21414\|\|full | Not-determined |
| 17-106-final-viral-combined_k141_37449\|\|full | Not-determined |
| 17-106-final-viral-combined_k141_48156\|\|full | Low-quality |
| 17-106-final-viral-combined_k141_106986\|\|full | Not-determined |
| 17-106-final-viral-combined_k141_10738\|\|full | Low-quality |
| 17-106-final-viral-combined_k141_101645\|\|full | Not-determined |
| 17-106-final-viral-combined_k141_42795\|\|full | Low-quality |
| 17-106-final-viral-combined_k141_107007\|\|full | Not-determined |
| 17-106-final-viral-combined_k141_117704\|\|full | Not-determined |
| 17-106-final-viral-combined_k141_32119\|\|full | Not-determined |
| 17-106-final-viral-combined_k141_117715\|\|full | Not-determined |
| 17-106-final-viral-combined_k141_64239\|\|full | Not-determined |
| 17-106-final-viral-combined_k141_16080\|\|full | Not-determined |
| 17-106-final-viral-combined_k141_107011\|\|full | Low-quality |
| 17-106-final-viral-combined_k141_96317\|\|full | Not-determined |
| 17-106-final-viral-combined_k141_80259\|\|full | Not-determined |
| 17-106-final-viral-combined_k141_47\|\|full | Low-quality |
| 17-106-final-viral-combined_k141_32130\|\|full | Not-determined |
| 17-106-final-viral-combined_k141_16105\|\|full | Low-quality |
| 17-106-final-viral-combined_k141_42858\|\|full | Not-determined |
| 17-106-final-viral-combined_k141_16107\|\|full | Not-determined |
| 17-106-final-viral-combined_k141_101657\|\|full | Not-determined |
| 17-106-final-viral-combined_k141_26779\|\|full | Not-determined |
| 17-106-final-viral-combined_k141_74918\|\|full | Not-determined |
| 17-106-final-viral-combined_k141_48178\|\|full | Not-determined |
| 17-106-final-viral-combined_k141_26784\|\|full | Low-quality |
| 17-106-final-viral-combined_k141_82\|\|full | Low-quality |
| 17-106-final-viral-combined_k141_58878\|\|full | Low-quality |
| 17-106-final-viral-combined_k141_86\|\|full | Not-determined |
| 17-106-final-viral-combined_k141_101673\|\|full | Not-determined |
| 17-106-final-viral-combined_k141_90953\|\|full | Low-quality |
| 17-106-final-viral-combined_k141_101\|\|full | Not-determined |
| 17-106-final-viral-combined_k141_104\|\|full | Low-quality |
| 17-106-final-viral-combined_k141_58897\|\|full | Not-determined |
| 17-106-final-viral-combined_k141_85669\|\|full | Low-quality |
| 17-106-final-viral-combined_k141_90970\|\|full | Low-quality |
| 17-106-final-viral-combined_k141_16127\|\|full | Low-quality |
| 17-106-final-viral-combined_k141_53528\|\|full | Low-quality |
| 17-106-final-viral-combined_k141_16134\|\|full | Not-determined |
| 17-106-final-viral-combined_k141_53535\|\|full | Not-determined |
| 17-106-final-viral-combined_k141_85697\|\|full | Low-quality |
| 17-106-final-viral-combined_k141_26789\|\|full | Not-determined |
| 17-106-final-viral-combined_k141_117781\|\|full | Not-determined |
| 17-106-final-viral-combined_k141_26797\|\|full | Not-determined |
| 17-106-final-viral-combined_k141_21485\|\|full | Low-quality |
| 17-106-final-viral-combined_k141_168\|\|full | Not-determined |
| 17-106-final-viral-combined_k141_117793\|\|full | Not-determined |
| 17-106-final-viral-combined_k141_96345\|\|full | Low-quality |
| 17-106-final-viral-combined_k141_69588\|\|full | Not-determined |
| 17-106-final-viral-combined_k141_96349\|\|full | Low-quality |
| 17-106-final-viral-combined_k141_21512\|\|full | Low-quality |
| 17-106-final-viral-combined_k141_10862\|\|full | Not-determined |
| 17-106-final-viral-combined_k141_112457\|\|full | Not-determined |
| 17-106-final-viral-combined_k141_74987\|\|full | Not-determined |
| 17-106-final-viral-combined_k141_74996\|\|full | Low-quality |
| 17-106-final-viral-combined_k141_37568\|\|full | Not-determined |
| 17-106-final-viral-combined_k141_75008\|\|full | Low-quality |
| 17-106-final-viral-combined_k141_206\|\|full | Low-quality |
| 17-106-final-viral-combined_k141_96377\|\|full | Low-quality |
| 17-106-final-viral-combined_k141_75027\|\|full | Not-determined |
| 17-106-final-viral-combined_k141_75033\|\|full | Not-determined |
| 17-106-final-viral-combined_k141_91050\|\|full | Not-determined |
| 17-106-final-viral-combined_k141_75046\|\|full | Low-quality |
| 17-106-final-viral-combined_k141_26848\|\|full | Low-quality |
| 17-106-final-viral-combined_k141_96403\|\|full | Not-determined |
| 17-106-final-viral-combined_k141_101800\|\|full | Not-determined |
| 17-106-final-viral-combined_k141_48284\|\|full | Low-quality |
| 17-106-final-viral-combined_k141_10887\|\|full | Low-quality |
| 17-106-final-viral-combined_k141_69621\|\|full | Low-quality |
| 17-106-final-viral-combined_k141_48293\|\|full | Low-quality |
| 17-106-final-viral-combined_k141_58912\|\|full | Low-quality |
| 17-106-final-viral-combined_k141_69628\|\|full | Not-determined |
| 17-106-final-viral-combined_k141_75064\|\|full | Not-determined |
| 17-106-final-viral-combined_k141_32206\|\|full | Low-quality |
| 17-106-final-viral-combined_k141_85743\|\|full | Low-quality |
| 17-106-final-viral-combined_k141_123110\|\|full | Low-quality |
| 17-106-final-viral-combined_k141_123111\|\|full | Low-quality |
| 17-106-final-viral-combined_k141_42987\|\|full | Low-quality |
| 17-106-final-viral-combined_k141_21550\|\|full | Low-quality |
| 17-106-final-viral-combined_k141_123115\|\|full | Not-determined |
| 17-106-final-viral-combined_k141_32219\|\|full | Low-quality |
| 17-106-final-viral-combined_k141_85755\|\|full | Not-determined |
| 17-106-final-viral-combined_k141_80327\|\|full | Low-quality |
| 17-106-final-viral-combined_k141_69645\|\|full | Not-determined |
| 17-106-final-viral-combined_k141_58944\|\|full | Low-quality |
| 17-106-final-viral-combined_k141_16218\|\|full | Low-quality |
| 17-106-final-viral-combined_k141_43024\|\|full | Not-determined |
| 17-106-final-viral-combined_k141_287\|\|full | Not-determined |
| 17-106-final-viral-combined_k141_16230\|\|full | Not-determined |
| 17-106-final-viral-combined_k141_85775\|\|full | Low-quality |
| 17-106-final-viral-combined_k141_5476\|\|full | Not-determined |
| 17-106-final-viral-combined_k141_299\|\|full | Not-determined |
| 17-106-final-viral-combined_k141_305\|\|full | Not-determined |
| 17-106-final-viral-combined_k141_85784\|\|full | Not-determined |
| 17-106-final-viral-combined_k141_16240\|\|full | Not-determined |
| 17-106-final-viral-combined_k141_53545\|\|full | Low-quality |
| 17-106-final-viral-combined_k141_112569\|\|full | Not-determined |
| 17-106-final-viral-combined_k141_91116\|\|full | Low-quality |
| 17-106-final-viral-combined_k141_75120\|\|full | Low-quality |
| 17-106-final-viral-combined_k141_112595\|\|full | Low-quality |
| 17-106-final-viral-combined_k141_317\|\|full | Low-quality |
| 17-106-final-viral-combined_k141_32277\|\|full | Low-quality |
| 17-106-final-viral-combined_k141_26934\|\|full | Not-determined |
| 17-106-final-viral-combined_k141_32282\|\|full | Not-determined |
| 17-106-final-viral-combined_k141_123189\|\|full | Low-quality |
| 17-106-final-viral-combined_k141_80384\|\|full | Not-determined |
| 17-106-final-viral-combined_k141_32287\|\|full | Not-determined |
| 17-106-final-viral-combined_k141_32289\|\|full | Not-determined |
| 17-106-final-viral-combined_k141_123194\|\|full | Not-determined |
| 17-106-final-viral-combined_k141_48351\|\|full | Not-determined |
| 17-106-final-viral-combined_k141_48360\|\|full | Low-quality |
| 17-106-final-viral-combined_k141_59008\|\|full | Low-quality |
| 17-106-final-viral-combined_k141_26968\|\|full | Not-determined |
| 17-106-final-viral-combined_k141_43064\|\|full | Low-quality |
| 17-106-final-viral-combined_k141_16253\|\|full | Not-determined |
| 17-106-final-viral-combined_k141_117850\|\|full | Not-determined |
| 18-102-final-viral-combined_k141_140996\|\|full | Complete |
| 17-106-final-viral-combined_k141_16257\|\|full | Low-quality |
| 17-106-final-viral-combined_k141_26972\|\|full | Low-quality |
| 17-106-final-viral-combined_k141_333\|\|full | Low-quality |
| 17-106-final-viral-combined_k141_48364\|\|full | Low-quality |
| 17-106-final-viral-combined_k141_75179\|\|full | Low-quality |
| 17-106-final-viral-combined_k141_16277\|\|full | Low-quality |
| 17-106-final-viral-combined_k141_80423\|\|full | Not-determined |
| 17-106-final-viral-combined_k141_96509\|\|full | Low-quality |
| 17-106-final-viral-combined_k141_64290\|\|full | Low-quality |
| 17-106-final-viral-combined_k141_85819\|\|full | Low-quality |
| 17-106-final-viral-combined_k141_16296\|\|full | Not-determined |
| 17-106-final-viral-combined_k141_27000\|\|full | Not-determined |
| 17-106-final-viral-combined_k141_101848\|\|full | Not-determined |
| 17-106-final-viral-combined_k141_27009\|\|full | Not-determined |
| 17-106-final-viral-combined_k141_80447\|\|full | Low-quality |
| 17-106-final-viral-combined_k141_69697\|\|full | Not-determined |
| 17-106-final-viral-combined_k141_96528\|\|full | Not-determined |
| 17-106-final-viral-combined_k141_64319\|\|full | Low-quality |
| 17-106-final-viral-combined_k141_101861\|\|full | Low-quality |
| 17-106-final-viral-combined_k141_59086\|\|full | Low-quality |
| 17-106-final-viral-combined_k141_59090\|\|full | Low-quality |
| 17-106-final-viral-combined_k141_96561\|\|full | Low-quality |
| 17-106-final-viral-combined_k141_21587\|\|full | Not-determined |
| 17-106-final-viral-combined_k141_75233\|\|full | Low-quality |
| 17-106-final-viral-combined_k141_64367\|\|full | Low-quality |
| 17-106-final-viral-combined_k141_64371\|\|full | Low-quality |
| 17-106-final-viral-combined_k141_80509\|\|full | Not-determined |
| 17-106-final-viral-combined_k141_48407\|\|full | Low-quality |
| 17-106-final-viral-combined_k141_32337\|\|full | Not-determined |
| 17-106-final-viral-combined_k141_117949\|\|full | Not-determined |
| 17-106-final-viral-combined_k141_117952\|\|full | Low-quality |
| 17-106-final-viral-combined_k141_16354\|\|full | Not-determined |
| 17-106-final-viral-combined_k141_48425\|\|full | Not-determined |
| 17-106-final-viral-combined_k141_16357\|\|full | Low-quality |
| 17-106-final-viral-combined_k141_16364\|\|full | Low-quality |
| 17-106-final-viral-combined_k141_441\|\|full | Low-quality |
| 17-106-final-viral-combined_k141_43125\|\|full | Not-determined |
| 17-106-final-viral-combined_k141_37598\|\|full | Not-determined |
| 17-106-final-viral-combined_k141_101956\|\|full | Low-quality |
| 17-106-final-viral-combined_k141_48449\|\|full | Low-quality |
| 17-106-final-viral-combined_k141_32350\|\|full | Low-quality |
| 17-106-final-viral-combined_k141_48451\|\|full | Not-determined |
| 17-106-final-viral-combined_k141_64400\|\|full | Not-determined |
| 17-106-final-viral-combined_k141_32353\|\|full | Low-quality |
| 17-106-final-viral-combined_k141_85928\|\|full | Not-determined |
| 17-106-final-viral-combined_k141_59107\|\|full | Not-determined |
| 17-106-final-viral-combined_k141_112628\|\|full | Not-determined |
| 17-106-final-viral-combined_k141_112634\|\|full | Low-quality |
| 17-106-final-viral-combined_k141_75255\|\|full | Low-quality |
| 17-106-final-viral-combined_k141_117961\|\|full | Low-quality |
| 17-106-final-viral-combined_k141_479\|\|full | Not-determined |
| 17-106-final-viral-combined_k141_485\|\|full | Not-determined |
| 17-106-final-viral-combined_k141_101997\|\|full | Low-quality |
| 17-106-final-viral-combined_k141_117972\|\|full | Not-determined |
| 17-106-final-viral-combined_k141_27116\|\|full | Not-determined |
| 17-106-final-viral-combined_k141_59132\|\|full | Not-determined |
| 17-106-final-viral-combined_k141_80582\|\|full | Low-quality |
| 17-106-final-viral-combined_k141_37634\|\|full | Low-quality |
| 17-106-final-viral-combined_k141_498\|\|full | Low-quality |
| 17-106-final-viral-combined_k141_59146\|\|full | Not-determined |
| 17-106-final-viral-combined_k141_102045\|\|full | Low-quality |
| 17-106-final-viral-combined_k141_59149\|\|full | Low-quality |
| 17-106-final-viral-combined_k141_59156\|\|full | Low-quality |
| 17-106-final-viral-combined_k141_112695\|\|full | Not-determined |
| 17-106-final-viral-combined_k141_80603\|\|full | Not-determined |
| 17-106-final-viral-combined_k141_96609\|\|full | Low-quality |
| 17-106-final-viral-combined_k141_43141\|\|full | Low-quality |
| 17-106-final-viral-combined_k141_53724\|\|full | Low-quality |
| 17-106-final-viral-combined_k141_102061\|\|full | Not-determined |
| 17-106-final-viral-combined_k141_59179\|\|full | Not-determined |
| 17-106-final-viral-combined_k141_53731\|\|full | Not-determined |
| 17-106-final-viral-combined_k141_118003\|\|full | Low-quality |
| 17-106-final-viral-combined_k141_96645\|\|full | Not-determined |
| 17-106-final-viral-combined_k141_48495\|\|full | Not-determined |
| 17-106-final-viral-combined_k141_5552\|\|full | Low-quality |
| 17-106-final-viral-combined_k141_75346\|\|full | Low-quality |
| 17-106-final-viral-combined_k141_53735\|\|full | Low-quality |
| 17-106-final-viral-combined_k141_112718\|\|full | Not-determined |
| 17-106-final-viral-combined_k141_37676\|\|full | Not-determined |
| 17-106-final-viral-combined_k141_16439\|\|full | Not-determined |
| 17-106-final-viral-combined_k141_21601\|\|full | Not-determined |
| 17-106-final-viral-combined_k141_112735\|\|full | Not-determined |
| 17-106-final-viral-combined_k141_27191\|\|full | Low-quality |
| 17-106-final-viral-combined_k141_43179\|\|full | Low-quality |
| 17-106-final-viral-combined_k141_53780\|\|full | Low-quality |
| 17-106-final-viral-combined_k141_96686\|\|full | Not-determined |
| 17-106-final-viral-combined_k141_80653\|\|full | Not-determined |
| 17-106-final-viral-combined_k141_21613\|\|full | Low-quality |
| 17-106-final-viral-combined_k141_5571\|\|full | Low-quality |
| 17-106-final-viral-combined_k141_5572\|\|full | Not-determined |
| 17-106-final-viral-combined_k141_107113\|\|full | Not-determined |
| 17-106-final-viral-combined_k141_118029\|\|full | Low-quality |
| 18-102-final-viral-combined_k141_13246\|\|full | Medium-quality |
| 17-106-final-viral-combined_k141_53788\|\|full | Low-quality |
| 17-106-final-viral-combined_k141_102119\|\|full | Not-determined |
| 17-106-final-viral-combined_k141_107136\|\|full | Low-quality |
| 17-106-final-viral-combined_k141_48570\|\|full | Low-quality |
| 17-106-final-viral-combined_k141_59247\|\|full | Not-determined |
| 17-106-final-viral-combined_k141_69805\|\|full | Low-quality |
| 17-106-final-viral-combined_k141_48592\|\|full | Low-quality |
| 17-106-final-viral-combined_k141_118102\|\|full | Low-quality |
| 17-106-final-viral-combined_k141_102132\|\|full | Not-determined |
| 17-106-final-viral-combined_k141_59250\|\|full | Not-determined |
| 17-106-final-viral-combined_k141_96745\|\|full | Not-determined |
| 17-106-final-viral-combined_k141_43195\|\|full | Not-determined |
| 17-106-final-viral-combined_k141_43198\|\|full | Not-determined |
| 17-106-final-viral-combined_k141_592\|\|full | Low-quality |
| 17-106-final-viral-combined_k141_69835\|\|full | Not-determined |
| 17-106-final-viral-combined_k141_53844\|\|full | Not-determined |
| 17-106-final-viral-combined_k141_48628\|\|full | Not-determined |
| 17-106-final-viral-combined_k141_59260\|\|full | Low-quality |
| 17-106-final-viral-combined_k141_607\|\|full | Low-quality |
| 17-106-final-viral-combined_k141_5657\|\|full | Low-quality |
| 17-106-final-viral-combined_k141_48650\|\|full | Low-quality |
| 17-106-final-viral-combined_k141_610\|\|full | Not-determined |
| 17-106-final-viral-combined_k141_5668\|\|full | Not-determined |
| 17-106-final-viral-combined_k141_21658\|\|full | Low-quality |
| 17-106-final-viral-combined_k141_59282\|\|full | Not-determined |
| 17-106-final-viral-combined_k141_21668\|\|full | Low-quality |
| 17-106-final-viral-combined_k141_53863\|\|full | Not-determined |
| 17-106-final-viral-combined_k141_43228\|\|full | Not-determined |
| 17-106-final-viral-combined_k141_118121\|\|full | Low-quality |
| 17-106-final-viral-combined_k141_43232\|\|full | Low-quality |
| 17-106-final-viral-combined_k141_59296\|\|full | Not-determined |
| 17-106-final-viral-combined_k141_118134\|\|full | Not-determined |
| 17-106-final-viral-combined_k141_118149\|\|full | Not-determined |
| 17-106-final-viral-combined_k141_80701\|\|full | Low-quality |
| 17-106-final-viral-combined_k141_107208\|\|full | Not-determined |
| 17-106-final-viral-combined_k141_37713\|\|full | Low-quality |
| 17-106-final-viral-combined_k141_32382\|\|full | Low-quality |
| 17-106-final-viral-combined_k141_96864\|\|full | Not-determined |
| 17-106-final-viral-combined_k141_32383\|\|full | Not-determined |
| 17-106-final-viral-combined_k141_10970\|\|full | Low-quality |
| 17-106-final-viral-combined_k141_10972\|\|full | Not-determined |
| 17-106-final-viral-combined_k141_59345\|\|full | Not-determined |
| 17-106-final-viral-combined_k141_37742\|\|full | Low-quality |
| 17-106-final-viral-combined_k141_59346\|\|full | Not-determined |
| 17-106-final-viral-combined_k141_123341\|\|full | Low-quality |
| 17-106-final-viral-combined_k141_96881\|\|full | Low-quality |
| 17-106-final-viral-combined_k141_37761\|\|full | Not-determined |
| 17-106-final-viral-combined_k141_32398\|\|full | Not-determined |
| 17-106-final-viral-combined_k141_43252\|\|full | Low-quality |
| 17-106-final-viral-combined_k141_80745\|\|full | Low-quality |
| 17-106-final-viral-combined_k141_59385\|\|full | Low-quality |
| 17-106-final-viral-combined_k141_86103\|\|full | Not-determined |
| 17-106-final-viral-combined_k141_696\|\|full | Low-quality |
| 17-106-final-viral-combined_k141_702\|\|full | Not-determined |
| 17-106-final-viral-combined_k141_86118\|\|full | Not-determined |
| 17-106-final-viral-combined_k141_705\|\|full | Low-quality |
| 17-106-final-viral-combined_k141_37803\|\|full | Not-determined |
| 17-106-final-viral-combined_k141_32442\|\|full | Low-quality |
| 17-106-final-viral-combined_k141_43275\|\|full | Low-quality |
| 17-106-final-viral-combined_k141_102219\|\|full | Not-determined |
| 17-106-final-viral-combined_k141_27245\|\|full | Low-quality |
| 17-106-final-viral-combined_k141_21753\|\|full | Not-determined |
| 17-106-final-viral-combined_k141_48768\|\|full | Not-determined |
| 17-106-final-viral-combined_k141_86168\|\|full | Low-quality |
| 17-106-final-viral-combined_k141_96950\|\|full | Low-quality |
| 17-106-final-viral-combined_k141_5730\|\|full | Low-quality |
| 17-106-final-viral-combined_k141_102224\|\|full | Low-quality |
| 17-106-final-viral-combined_k141_53913\|\|full | Not-determined |
| 17-106-final-viral-combined_k141_102239\|\|full | Not-determined |
| 17-106-final-viral-combined_k141_80780\|\|full | Not-determined |
| 17-106-final-viral-combined_k141_5749\|\|full | Low-quality |
| 17-106-final-viral-combined_k141_118237\|\|full | Not-determined |
| 17-106-final-viral-combined_k141_37848\|\|full | Low-quality |
| 17-106-final-viral-combined_k141_21772\|\|full | Not-determined |
| 17-106-final-viral-combined_k141_107259\|\|full | Low-quality |
| 17-106-final-viral-combined_k141_16513\|\|full | Not-determined |
| 17-106-final-viral-combined_k141_80817\|\|full | Low-quality |
| 17-106-final-viral-combined_k141_5767\|\|full | Not-determined |
| 18-105-final-viral-combined_k141_72410\|\|full | Complete |
| 17-106-final-viral-combined_k141_118243\|\|full | Low-quality |
| 17-106-final-viral-combined_k141_37869\|\|full | Not-determined |
| 17-106-final-viral-combined_k141_5776\|\|full | Not-determined |
| 17-106-final-viral-combined_k141_102270\|\|full | Not-determined |
| 17-106-final-viral-combined_k141_751\|\|full | Low-quality |
| 17-106-final-viral-combined_k141_27269\|\|full | Not-determined |
| 17-106-final-viral-combined_k141_16525\|\|full | Not-determined |
| 17-106-final-viral-combined_k141_102275\|\|full | Low-quality |
| 17-106-final-viral-combined_k141_102276\|\|full | Not-determined |
| 17-106-final-viral-combined_k141_37880\|\|full | Not-determined |
| 17-106-final-viral-combined_k141_102277\|\|full | Low-quality |
| 17-106-final-viral-combined_k141_27278\|\|full | Not-determined |
| 17-106-final-viral-combined_k141_107292\|\|full | Low-quality |
| 17-106-final-viral-combined_k141_768\|\|full | Not-determined |
| 17-106-final-viral-combined_k141_112863\|\|full | Low-quality |
| 17-106-final-viral-combined_k141_80879\|\|full | Low-quality |
| 17-106-final-viral-combined_k141_59476\|\|full | Low-quality |
| 17-106-final-viral-combined_k141_107318\|\|full | Not-determined |
| 17-106-final-viral-combined_k141_16580\|\|full | Not-determined |
| 17-106-final-viral-combined_k141_59480\|\|full | Low-quality |
| 17-106-final-viral-combined_k141_102292\|\|full | Low-quality |
| 17-106-final-viral-combined_k141_118286\|\|full | Low-quality |
| 17-106-final-viral-combined_k141_123414\|\|full | Low-quality |
| 17-106-final-viral-combined_k141_80897\|\|full | Not-determined |
| 17-106-final-viral-combined_k141_102297\|\|full | Not-determined |
| 17-106-final-viral-combined_k141_118292\|\|full | Not-determined |
| 17-106-final-viral-combined_k141_59492\|\|full | Not-determined |
| 17-106-final-viral-combined_k141_112903\|\|full | Not-determined |
| 17-106-final-viral-combined_k141_5808\|\|full | Low-quality |
| 17-106-final-viral-combined_k141_43404\|\|full | Low-quality |
| 17-106-final-viral-combined_k141_822\|\|full | Not-determined |
| 17-106-final-viral-combined_k141_59510\|\|full | Not-determined |
| 17-106-final-viral-combined_k141_112914\|\|full | Low-quality |
| 17-106-final-viral-combined_k141_824\|\|full | Not-determined |
| 17-106-final-viral-combined_k141_112916\|\|full | Not-determined |
| 17-106-final-viral-combined_k141_43418\|\|full | Not-determined |
| 17-106-final-viral-combined_k141_43428\|\|full | Not-determined |
| 18-102-final-viral-combined_k141_185494\|\|full | Medium-quality |
| 17-106-final-viral-combined_k141_43439\|\|full | Low-quality |
| 17-106-final-viral-combined_k141_102344\|\|full | Not-determined |
| 17-106-final-viral-combined_k141_43447\|\|full | Low-quality |
| 17-106-final-viral-combined_k141_86247\|\|full | Not-determined |
| 17-106-final-viral-combined_k141_118366\|\|full | Not-determined |
| 17-106-final-viral-combined_k141_37967\|\|full | Low-quality |
| 17-106-final-viral-combined_k141_53997\|\|full | Low-quality |
| 17-106-final-viral-combined_k141_11063\|\|full | Low-quality |
| 17-106-final-viral-combined_k141_69877\|\|full | Not-determined |
| 17-106-final-viral-combined_k141_80975\|\|full | Not-determined |
| 17-106-final-viral-combined_k141_898\|\|full | Not-determined |
| 17-106-final-viral-combined_k141_54007\|\|full | Not-determined |
| 17-106-final-viral-combined_k141_59595\|\|full | Not-determined |
| 17-106-final-viral-combined_k141_11076\|\|full | Low-quality |
| 17-106-final-viral-combined_k141_5886\|\|full | Low-quality |
| 17-106-final-viral-combined_k141_102361\|\|full | Not-determined |
| 17-106-final-viral-combined_k141_97083\|\|full | Low-quality |
| 17-106-final-viral-combined_k141_5895\|\|full | Low-quality |
| 17-106-final-viral-combined_k141_102368\|\|full | Not-determined |
| 17-106-final-viral-combined_k141_69881\|\|full | Low-quality |
| 17-106-final-viral-combined_k141_16597\|\|full | Not-determined |
| 17-106-final-viral-combined_k141_32630\|\|full | Not-determined |
| 17-106-final-viral-combined_k141_5922\|\|full | Not-determined |
| 17-106-final-viral-combined_k141_916\|\|full | Not-determined |
| 17-106-final-viral-combined_k141_107387\|\|full | Low-quality |
| 17-106-final-viral-combined_k141_48856\|\|full | Not-determined |
| 17-106-final-viral-combined_k141_27348\|\|full | Low-quality |
| 17-106-final-viral-combined_k141_64655\|\|full | Low-quality |
| 17-106-final-viral-combined_k141_48862\|\|full | Not-determined |
| 17-106-final-viral-combined_k141_91353\|\|full | Not-determined |
| 17-106-final-viral-combined_k141_5932\|\|full | Not-determined |
| 17-106-final-viral-combined_k141_64659\|\|full | Not-determined |
| 17-106-final-viral-combined_k141_5937\|\|full | Not-determined |
| 17-106-final-viral-combined_k141_64669\|\|full | Not-determined |
| 17-106-final-viral-combined_k141_38008\|\|full | Not-determined |
| 17-106-final-viral-combined_k141_64676\|\|full | Not-determined |
| 17-106-final-viral-combined_k141_97142\|\|full | Not-determined |
| 17-106-final-viral-combined_k141_5951\|\|full | Not-determined |
| 17-106-final-viral-combined_k141_97146\|\|full | Low-quality |
| 17-106-final-viral-combined_k141_21933\|\|full | Not-determined |
| 17-106-final-viral-combined_k141_32669\|\|full | Not-determined |
| 17-106-final-viral-combined_k141_11091\|\|full | Low-quality |
| 17-106-final-viral-combined_k141_11092\|\|full | Low-quality |
| 17-106-final-viral-combined_k141_64705\|\|full | Low-quality |
| 17-106-final-viral-combined_k141_48891\|\|full | Not-determined |
| 17-106-final-viral-combined_k141_102429\|\|full | Low-quality |
| 17-106-final-viral-combined_k141_64706\|\|full | Not-determined |
| 17-106-final-viral-combined_k141_5977\|\|full | Low-quality |
| 17-106-final-viral-combined_k141_32690\|\|full | Not-determined |
| 17-106-final-viral-combined_k141_81047\|\|full | Low-quality |
| 17-106-final-viral-combined_k141_64718\|\|full | Low-quality |
| 17-106-final-viral-combined_k141_81052\|\|full | Not-determined |
| 17-106-final-viral-combined_k141_64721\|\|full | Not-determined |
| 17-106-final-viral-combined_k141_992\|\|full | Low-quality |
| 17-106-final-viral-combined_k141_59660\|\|full | Low-quality |
| 17-106-final-viral-combined_k141_54037\|\|full | Low-quality |
| 17-106-final-viral-combined_k141_64734\|\|full | Low-quality |
| 17-106-final-viral-combined_k141_11101\|\|full | Not-determined |
| 17-106-final-viral-combined_k141_48911\|\|full | Not-determined |
| 17-106-final-viral-combined_k141_81063\|\|full | Not-determined |
| 17-106-final-viral-combined_k141_69955\|\|full | Not-determined |
| 17-106-final-viral-combined_k141_11112\|\|full | Not-determined |
| 17-106-final-viral-combined_k141_91374\|\|full | Not-determined |
| 17-106-final-viral-combined_k141_102489\|\|full | Low-quality |
| 17-106-final-viral-combined_k141_118456\|\|full | Low-quality |
| 17-106-final-viral-combined_k141_81091\|\|full | Low-quality |
| 17-106-final-viral-combined_k141_97169\|\|full | Low-quality |
| 17-106-final-viral-combined_k141_97179\|\|full | Low-quality |
| 17-106-final-viral-combined_k141_118475\|\|full | Low-quality |
| 17-106-final-viral-combined_k141_32759\|\|full | Low-quality |
| 17-106-final-viral-combined_k141_81104\|\|full | Not-determined |
| 17-106-final-viral-combined_k141_91393\|\|full | Low-quality |
| 17-106-final-viral-combined_k141_11148\|\|full | Low-quality |
| 17-106-final-viral-combined_k141_69999\|\|full | Low-quality |
| 17-106-final-viral-combined_k141_107416\|\|full | Not-determined |
| 17-106-final-viral-combined_k141_107418\|\|full | Low-quality |
| 17-106-final-viral-combined_k141_70007\|\|full | Not-determined |
| 17-106-final-viral-combined_k141_91415\|\|full | Low-quality |
| 17-106-final-viral-combined_k141_38048\|\|full | Not-determined |
| 17-106-final-viral-combined_k141_11167\|\|full | Not-determined |
| 17-106-final-viral-combined_k141_38057\|\|full | Low-quality |
| 17-106-final-viral-combined_k141_86397\|\|full | Not-determined |
| 17-106-final-viral-combined_k141_38061\|\|full | Not-determined |
| 17-106-final-viral-combined_k141_48980\|\|full | Not-determined |
| 17-106-final-viral-combined_k141_123552\|\|full | Not-determined |
| 17-106-final-viral-combined_k141_38065\|\|full | Low-quality |
| 17-106-final-viral-combined_k141_48998\|\|full | Not-determined |
| 17-106-final-viral-combined_k141_11177\|\|full | Low-quality |
| 17-106-final-viral-combined_k141_48999\|\|full | Not-determined |
| 17-106-final-viral-combined_k141_27431\|\|full | Not-determined |
| 17-106-final-viral-combined_k141_59681\|\|full | Not-determined |
| 17-106-final-viral-combined_k141_91439\|\|full | Low-quality |
| 17-106-final-viral-combined_k141_38094\|\|full | Not-determined |
| 17-106-final-viral-combined_k141_1115\|\|full | Not-determined |
| 17-106-final-viral-combined_k141_64752\|\|full | Not-determined |
| 17-106-final-viral-combined_k141_6012\|\|full | Not-determined |
| 17-106-final-viral-combined_k141_70044\|\|full | Low-quality |
| 17-106-final-viral-combined_k141_64765\|\|full | Low-quality |
| 17-106-final-viral-combined_k141_27448\|\|full | Not-determined |
| 17-106-final-viral-combined_k141_81148\|\|full | Low-quality |
| 17-106-final-viral-combined_k141_86447\|\|full | Not-determined |
| 17-106-final-viral-combined_k141_38121\|\|full | Not-determined |
| 17-106-final-viral-combined_k141_70063\|\|full | Not-determined |
| 17-106-final-viral-combined_k141_86463\|\|full | Low-quality |
| 17-106-final-viral-combined_k141_81163\|\|full | Low-quality |
| 17-106-final-viral-combined_k141_27471\|\|full | Not-determined |
| 17-106-final-viral-combined_k141_64784\|\|full | Not-determined |
| 17-106-final-viral-combined_k141_64785\|\|full | Not-determined |
| 17-106-final-viral-combined_k141_70099\|\|full | Low-quality |
| 17-106-final-viral-combined_k141_118531\|\|full | Low-quality |
| 17-106-final-viral-combined_k141_81192\|\|full | Not-determined |
| 17-106-final-viral-combined_k141_54087\|\|full | Not-determined |
| 17-106-final-viral-combined_k141_97219\|\|full | Not-determined |
| 17-106-final-viral-combined_k141_75374\|\|full | Low-quality |
| 17-106-final-viral-combined_k141_16695\|\|full | Not-determined |
| 17-106-final-viral-combined_k141_97234\|\|full | Not-determined |
| 17-106-final-viral-combined_k141_54102\|\|full | Not-determined |
| 17-106-final-viral-combined_k141_38176\|\|full | Not-determined |
| 17-106-final-viral-combined_k141_11239\|\|full | Low-quality |
| 17-106-final-viral-combined_k141_38183\|\|full | Not-determined |
| 17-106-final-viral-combined_k141_81263\|\|full | Not-determined |
| 17-106-final-viral-combined_k141_11255\|\|full | Low-quality |
| 17-106-final-viral-combined_k141_11266\|\|full | Low-quality |
| 17-106-final-viral-combined_k141_54136\|\|full | Not-determined |
| 17-106-final-viral-combined_k141_107449\|\|full | Not-determined |
| 17-106-final-viral-combined_k141_123594\|\|full | Low-quality |
| 17-106-final-viral-combined_k141_1240\|\|full | Low-quality |
| 17-106-final-viral-combined_k141_16719\|\|full | Low-quality |
| 17-106-final-viral-combined_k141_91559\|\|full | Not-determined |
| 17-106-final-viral-combined_k141_27506\|\|full | Not-determined |
| 17-106-final-viral-combined_k141_1255\|\|full | Not-determined |
| 17-106-final-viral-combined_k141_123614\|\|full | Not-determined |
| 17-106-final-viral-combined_k141_118553\|\|full | Not-determined |
| 17-106-final-viral-combined_k141_81286\|\|full | Low-quality |
| 17-106-final-viral-combined_k141_91580\|\|full | Not-determined |
| 17-106-final-viral-combined_k141_113062\|\|full | Not-determined |
| 17-106-final-viral-combined_k141_54162\|\|full | Not-determined |
| 17-106-final-viral-combined_k141_91604\|\|full | Not-determined |
| 17-106-final-viral-combined_k141_1262\|\|full | Not-determined |
| 17-106-final-viral-combined_k141_97292\|\|full | Low-quality |
| 17-106-final-viral-combined_k141_113074\|\|full | Not-determined |
| 17-106-final-viral-combined_k141_11298\|\|full | Low-quality |
| 17-106-final-viral-combined_k141_123664\|\|full | Not-determined |
| 17-106-final-viral-combined_k141_59785\|\|full | Not-determined |
| 17-106-final-viral-combined_k141_123667\|\|full | Not-determined |
| 17-106-final-viral-combined_k141_27514\|\|full | Low-quality |
| 17-106-final-viral-combined_k141_107528\|\|full | Not-determined |
| 17-106-final-viral-combined_k141_113103\|\|full | Not-determined |
| 17-106-final-viral-combined_k141_75458\|\|full | Low-quality |
| 17-106-final-viral-combined_k141_32978\|\|full | Low-quality |
| 17-106-final-viral-combined_k141_11326\|\|full | Low-quality |
| 17-106-final-viral-combined_k141_64838\|\|full | Low-quality |
| 17-106-final-viral-combined_k141_64845\|\|full | Not-determined |
| 17-106-final-viral-combined_k141_27549\|\|full | Low-quality |
| 17-106-final-viral-combined_k141_86572\|\|full | Not-determined |
| 17-106-final-viral-combined_k141_22091\|\|full | Not-determined |
| 17-106-final-viral-combined_k141_43462\|\|full | Low-quality |
| 17-106-final-viral-combined_k141_11355\|\|full | Low-quality |
| 17-106-final-viral-combined_k141_64859\|\|full | Not-determined |
| 17-106-final-viral-combined_k141_86580\|\|full | Not-determined |
| 17-106-final-viral-combined_k141_1324\|\|full | Not-determined |
| 17-106-final-viral-combined_k141_43472\|\|full | Low-quality |
| 17-106-final-viral-combined_k141_43474\|\|full | Not-determined |
| 17-106-final-viral-combined_k141_86584\|\|full | Not-determined |
| 17-106-final-viral-combined_k141_43477\|\|full | Not-determined |
| 17-106-final-viral-combined_k141_102627\|\|full | Low-quality |
| 17-106-final-viral-combined_k141_102631\|\|full | Not-determined |
| 17-106-final-viral-combined_k141_123754\|\|full | Not-determined |
| 17-106-final-viral-combined_k141_91678\|\|full | Not-determined |
| 17-106-final-viral-combined_k141_86593\|\|full | Low-quality |
| 17-106-final-viral-combined_k141_102639\|\|full | Not-determined |
| 17-106-final-viral-combined_k141_38284\|\|full | Not-determined |
| 17-106-final-viral-combined_k141_27618\|\|full | Not-determined |
| 17-106-final-viral-combined_k141_43500\|\|full | Low-quality |
| 17-106-final-viral-combined_k141_6078\|\|full | Low-quality |
| 17-106-final-viral-combined_k141_22099\|\|full | Low-quality |
| 17-106-final-viral-combined_k141_33038\|\|full | Low-quality |
| 17-106-final-viral-combined_k141_22108\|\|full | Low-quality |
| 17-106-final-viral-combined_k141_43519\|\|full | Low-quality |
| 17-106-final-viral-combined_k141_123787\|\|full | Low-quality |
| 17-106-final-viral-combined_k141_64935\|\|full | Not-determined |
| 17-106-final-viral-combined_k141_6108\|\|full | Not-determined |
| 17-106-final-viral-combined_k141_81319\|\|full | Low-quality |
| 17-106-final-viral-combined_k141_27648\|\|full | Low-quality |
| 17-106-final-viral-combined_k141_64944\|\|full | Not-determined |
| 17-106-final-viral-combined_k141_59823\|\|full | Low-quality |
| 17-106-final-viral-combined_k141_81344\|\|full | Not-determined |
| 17-106-final-viral-combined_k141_118657\|\|full | Not-determined |
| 17-106-final-viral-combined_k141_64967\|\|full | Low-quality |
| 17-106-final-viral-combined_k141_11400\|\|full | Not-determined |
| 17-106-final-viral-combined_k141_27687\|\|full | Not-determined |
| 17-106-final-viral-combined_k141_33106\|\|full | Not-determined |
| 17-106-final-viral-combined_k141_91715\|\|full | Not-determined |
| 17-106-final-viral-combined_k141_59889\|\|full | Not-determined |
| 17-106-final-viral-combined_k141_33112\|\|full | Low-quality |
| 17-106-final-viral-combined_k141_64990\|\|full | Not-determined |
| 17-106-final-viral-combined_k141_43537\|\|full | Low-quality |
| 17-106-final-viral-combined_k141_107631\|\|full | Low-quality |
| 17-106-final-viral-combined_k141_22136\|\|full | Not-determined |
| 17-106-final-viral-combined_k141_43540\|\|full | Low-quality |
| 17-106-final-viral-combined_k141_54363\|\|full | Not-determined |
| 17-106-final-viral-combined_k141_22147\|\|full | Low-quality |
| 17-106-final-viral-combined_k141_64998\|\|full | Not-determined |
| 17-106-final-viral-combined_k141_123815\|\|full | Low-quality |
| 17-106-final-viral-combined_k141_33158\|\|full | Not-determined |
| 17-106-final-viral-combined_k141_59929\|\|full | Not-determined |
| 17-106-final-viral-combined_k141_33161\|\|full | Low-quality |
| 17-106-final-viral-combined_k141_65020\|\|full | Not-determined |
| 17-106-final-viral-combined_k141_43575\|\|full | Low-quality |
| 17-106-final-viral-combined_k141_22165\|\|full | Low-quality |
| 17-106-final-viral-combined_k141_11423\|\|full | Not-determined |
| 17-106-final-viral-combined_k141_54400\|\|full | Low-quality |
| 17-106-final-viral-combined_k141_6194\|\|full | Not-determined |
| 17-106-final-viral-combined_k141_81396\|\|full | Low-quality |
| 17-106-final-viral-combined_k141_11424\|\|full | Not-determined |
| 17-106-final-viral-combined_k141_22181\|\|full | Low-quality |
| 17-106-final-viral-combined_k141_102708\|\|full | Not-determined |
| 17-106-final-viral-combined_k141_102709\|\|full | Low-quality |
| 17-106-final-viral-combined_k141_86623\|\|full | Low-quality |
| 17-106-final-viral-combined_k141_11440\|\|full | Not-determined |
| 17-106-final-viral-combined_k141_86625\|\|full | Low-quality |
| 17-106-final-viral-combined_k141_65043\|\|full | Low-quality |
| 17-106-final-viral-combined_k141_107701\|\|full | Not-determined |
| 17-106-final-viral-combined_k141_97394\|\|full | Low-quality |
| 17-106-final-viral-combined_k141_54434\|\|full | Not-determined |
| 17-106-final-viral-combined_k141_65062\|\|full | Not-determined |
| 17-106-final-viral-combined_k141_113207\|\|full | Low-quality |
| 17-106-final-viral-combined_k141_113210\|\|full | Low-quality |
| 17-106-final-viral-combined_k141_43609\|\|full | Not-determined |
| 17-106-final-viral-combined_k141_49194\|\|full | Low-quality |
| 17-106-final-viral-combined_k141_91796\|\|full | Low-quality |
| 17-106-final-viral-combined_k141_49217\|\|full | Low-quality |
| 17-106-final-viral-combined_k141_27794\|\|full | Not-determined |
| 17-106-final-viral-combined_k141_75493\|\|full | Not-determined |
| 17-106-final-viral-combined_k141_27796\|\|full | Not-determined |
| 17-106-final-viral-combined_k141_43624\|\|full | Not-determined |
| 17-106-final-viral-combined_k141_27824\|\|full | Not-determined |
| 17-106-final-viral-combined_k141_43635\|\|full | Not-determined |
| 17-106-final-viral-combined_k141_86696\|\|full | Low-quality |
| 17-106-final-viral-combined_k141_97435\|\|full | Low-quality |
| 17-106-final-viral-combined_k141_81418\|\|full | Low-quality |
| 17-106-final-viral-combined_k141_38336\|\|full | Low-quality |
| 17-106-final-viral-combined_k141_107785\|\|full | Low-quality |
| 17-106-final-viral-combined_k141_75548\|\|full | Not-determined |

| | |
|---|---|
| 17-106-final-viral-combined_k141_6290\|\|full | Low-quality |
| 17-106-final-viral-combined_k141_54444\|\|full | Not-determined |
| 17-106-final-viral-combined_k141_27853\|\|full | Low-quality |
| 17-106-final-viral-combined_k141_1480\|\|full | Not-determined |
| 17-106-final-viral-combined_k141_27859\|\|full | Low-quality |
| 17-106-final-viral-combined_k141_86742\|\|full | Low-quality |
| 17-106-final-viral-combined_k141_16775\|\|full | Not-determined |
| 17-106-final-viral-combined_k141_38350\|\|full | Low-quality |
| 17-106-final-viral-combined_k141_86752\|\|full | Not-determined |
| 17-106-final-viral-combined_k141_91837\|\|full | Not-determined |
| 17-106-final-viral-combined_k141_6327\|\|full | Low-quality |
| 17-106-final-viral-combined_k141_86755\|\|full | Low-quality |
| 17-106-final-viral-combined_k141_22349\|\|full | Low-quality |
| 17-106-final-viral-combined_k141_97492\|\|full | Low-quality |
| 17-106-final-viral-combined_k141_102840\|\|full | Not-determined |
| 17-106-final-viral-combined_k141_86762\|\|full | Low-quality |
| 17-106-final-viral-combined_k141_54503\|\|full | Low-quality |
| 17-106-final-viral-combined_k141_54509\|\|full | Low-quality |
| 17-106-final-viral-combined_k141_97506\|\|full | Not-determined |
| 17-106-final-viral-combined_k141_54511\|\|full | Not-determined |
| 17-106-final-viral-combined_k141_27906\|\|full | Not-determined |
| 17-106-final-viral-combined_k141_86779\|\|full | Low-quality |
| 17-106-final-viral-combined_k141_11519\|\|full | Low-quality |
| 17-106-final-viral-combined_k141_81461\|\|full | Low-quality |
| 17-106-final-viral-combined_k141_43653\|\|full | Not-determined |
| 17-106-final-viral-combined_k141_22387\|\|full | Not-determined |
| 17-106-final-viral-combined_k141_118774\|\|full | Low-quality |
| 17-106-final-viral-combined_k141_27913\|\|full | Low-quality |
| 17-106-final-viral-combined_k141_107862\|\|full | Not-determined |
| 17-106-final-viral-combined_k141_70220\|\|full | Not-determined |
| 17-106-final-viral-combined_k141_70223\|\|full | Low-quality |
| 17-106-final-viral-combined_k141_118782\|\|full | Not-determined |
| 17-106-final-viral-combined_k141_6357\|\|full | Not-determined |
| 17-106-final-viral-combined_k141_97514\|\|full | Low-quality |
| 17-106-final-viral-combined_k141_22427\|\|full | Not-determined |
| 17-106-final-viral-combined_k141_11542\|\|full | Not-determined |
| 17-106-final-viral-combined_k141_70257\|\|full | Low-quality |
| 17-106-final-viral-combined_k141_22437\|\|full | Not-determined |
| 17-106-final-viral-combined_k141_16838\|\|full | Low-quality |
| 17-106-final-viral-combined_k141_6387\|\|full | Not-determined |
| 17-106-final-viral-combined_k141_75612\|\|full | Not-determined |
| 17-106-final-viral-combined_k141_70274\|\|full | Not-determined |
| 17-106-final-viral-combined_k141_123994\|\|full | Low-quality |
| 17-106-final-viral-combined_k141_123995\|\|full | Not-determined |
| 17-106-final-viral-combined_k141_70282\|\|full | Low-quality |
| 17-106-final-viral-combined_k141_65132\|\|full | Not-determined |
| 17-106-final-viral-combined_k141_124013\|\|full | Not-determined |
| 17-106-final-viral-combined_k141_70305\|\|full | Not-determined |
| 17-106-final-viral-combined_k141_118836\|\|full | Not-determined |
| 17-106-final-viral-combined_k141_54609\|\|full | Low-quality |
| 17-106-final-viral-combined_k141_70318\|\|full | Low-quality |
| 17-106-final-viral-combined_k141_65169\|\|full | Not-determined |
| 17-106-final-viral-combined_k141_16859\|\|full | Not-determined |
| 17-106-final-viral-combined_k141_102893\|\|full | Not-determined |
| 17-106-final-viral-combined_k141_81554\|\|full | Low-quality |
| 17-106-final-viral-combined_k141_1627\|\|full | Not-determined |
| 17-106-final-viral-combined_k141_70344\|\|full | Not-determined |
| 17-106-final-viral-combined_k141_86827\|\|full | Not-determined |
| 17-106-final-viral-combined_k141_54629\|\|full | Low-quality |
| 17-106-final-viral-combined_k141_124050\|\|full | Not-determined |
| 17-106-final-viral-combined_k141_22475\|\|full | Not-determined |
| 17-106-final-viral-combined_k141_22476\|\|full | Not-determined |
| 17-106-final-viral-combined_k141_6416\|\|full | Not-determined |
| 17-106-final-viral-combined_k141_70354\|\|full | Low-quality |
| 17-106-final-viral-combined_k141_49282\|\|full | Low-quality |
| 17-106-final-viral-combined_k141_6430\|\|full | Not-determined |
| 17-106-final-viral-combined_k141_65211\|\|full | Low-quality |
| 17-106-final-viral-combined_k141_6432\|\|full | Low-quality |
| 17-106-final-viral-combined_k141_75678\|\|full | Low-quality |
| 17-106-final-viral-combined_k141_6434\|\|full | Not-determined |
| 17-106-final-viral-combined_k141_49286\|\|full | Not-determined |
| 17-106-final-viral-combined_k141_60034\|\|full | Not-determined |
| 17-106-final-viral-combined_k141_91869\|\|full | Low-quality |
| 17-106-final-viral-combined_k141_118884\|\|full | Not-determined |
| 17-106-final-viral-combined_k141_6448\|\|full | Not-determined |
| 17-106-final-viral-combined_k141_6455\|\|full | Low-quality |
| 17-106-final-viral-combined_k141_81619\|\|full | Not-determined |
| 17-106-final-viral-combined_k141_22494\|\|full | Not-determined |
| 17-106-final-viral-combined_k141_75715\|\|full | Not-determined |
| 17-106-final-viral-combined_k141_54658\|\|full | Not-determined |
| 17-106-final-viral-combined_k141_124081\|\|full | Not-determined |
| 17-106-final-viral-combined_k141_60074\|\|full | Low-quality |
| 17-106-final-viral-combined_k141_43712\|\|full | Low-quality |
| 17-106-final-viral-combined_k141_11616\|\|full | Low-quality |
| 17-106-final-viral-combined_k141_22510\|\|full | Not-determined |
| 17-106-final-viral-combined_k141_118909\|\|full | Not-determined |
| 17-106-final-viral-combined_k141_1655\|\|full | Low-quality |
| 17-106-final-viral-combined_k141_49306\|\|full | Low-quality |
| 17-106-final-viral-combined_k141_113286\|\|full | Not-determined |
| 17-106-final-viral-combined_k141_118920\|\|full | Low-quality |
| 17-106-final-viral-combined_k141_118931\|\|full | Low-quality |
| 17-106-final-viral-combined_k141_65250\|\|full | Not-determined |
| 17-106-final-viral-combined_k141_97550\|\|full | Not-determined |
| 17-106-final-viral-combined_k141_49339\|\|full | Not-determined |
| 17-106-final-viral-combined_k141_102925\|\|full | Low-quality |
| 17-106-final-viral-combined_k141_6478\|\|full | Low-quality |
| 17-106-final-viral-combined_k141_124119\|\|full | Not-determined |
| 17-106-final-viral-combined_k141_1700\|\|full | Not-determined |
| 17-106-final-viral-combined_k141_49355\|\|full | Low-quality |
| 17-106-final-viral-combined_k141_91982\|\|full | Not-determined |
| 17-106-final-viral-combined_k141_113325\|\|full | Not-determined |
| 17-106-final-viral-combined_k141_118970\|\|full | Not-determined |
| 17-106-final-viral-combined_k141_1717\|\|full | Not-determined |
| 17-106-final-viral-combined_k141_49367\|\|full | Not-determined |
| 17-106-final-viral-combined_k141_22573\|\|full | Low-quality |
| 17-106-final-viral-combined_k141_81678\|\|full | Low-quality |
| 17-106-final-viral-combined_k141_113334\|\|full | Not-determined |
| 17-106-final-viral-combined_k141_81683\|\|full | Not-determined |
| 17-106-final-viral-combined_k141_75772\|\|full | Low-quality |
| 17-106-final-viral-combined_k141_102958\|\|full | Low-quality |
| 17-106-final-viral-combined_k141_102971\|\|full | Low-quality |
| 17-106-final-viral-combined_k141_54741\|\|full | Not-determined |
| 17-106-final-viral-combined_k141_49407\|\|full | Not-determined |
| 17-106-final-viral-combined_k141_54760\|\|full | Not-determined |
| 17-106-final-viral-combined_k141_60152\|\|full | Not-determined |
| 17-106-final-viral-combined_k141_81767\|\|full | Not-determined |
| 17-106-final-viral-combined_k141_97607\|\|full | Low-quality |
| 17-106-final-viral-combined_k141_43812\|\|full | Not-determined |
| 17-106-final-viral-combined_k141_17003\|\|full | Low-quality |
| 17-106-final-viral-combined_k141_43817\|\|full | Low-quality |
| 17-106-final-viral-combined_k141_49423\|\|full | Low-quality |
| 17-106-final-viral-combined_k141_70472\|\|full | Not-determined |
| 17-106-final-viral-combined_k141_70474\|\|full | Low-quality |
| 17-106-final-viral-combined_k141_70479\|\|full | Not-determined |
| 17-106-final-viral-combined_k141_92090\|\|full | Low-quality |
| 17-106-final-viral-combined_k141_92092\|\|full | Not-determined |
| 17-106-final-viral-combined_k141_1749\|\|full | Not-determined |
| 17-106-final-viral-combined_k141_107976\|\|full | Low-quality |
| 17-106-final-viral-combined_k141_43832\|\|full | Not-determined |
| 17-106-final-viral-combined_k141_107986\|\|full | Not-determined |
| 17-106-final-viral-combined_k141_119017\|\|full | Not-determined |
| 17-106-final-viral-combined_k141_38477\|\|full | Low-quality |
| 17-106-final-viral-combined_k141_119028\|\|full | Not-determined |
| 17-106-final-viral-combined_k141_70537\|\|full | Low-quality |
| 17-106-final-viral-combined_k141_75877\|\|full | Low-quality |
| 17-106-final-viral-combined_k141_97633\|\|full | Not-determined |
| 17-106-final-viral-combined_k141_22701\|\|full | Not-determined |
| 17-106-final-viral-combined_k141_22706\|\|full | Not-determined |
| 17-106-final-viral-combined_k141_75888\|\|full | Not-determined |
| 17-106-final-viral-combined_k141_65378\|\|full | Not-determined |
| 17-106-final-viral-combined_k141_60207\|\|full | Low-quality |
| 17-106-final-viral-combined_k141_119043\|\|full | Not-determined |
| 17-106-final-viral-combined_k141_43866\|\|full | Low-quality |
| 17-106-final-viral-combined_k141_38500\|\|full | Low-quality |
| 17-106-final-viral-combined_k141_60228\|\|full | Not-determined |
| 17-106-final-viral-combined_k141_75911\|\|full | Low-quality |
| 17-106-final-viral-combined_k141_86868\|\|full | Low-quality |
| 17-106-final-viral-combined_k141_28041\|\|full | Low-quality |
| 17-106-final-viral-combined_k141_28051\|\|full | Not-determined |
| 17-106-final-viral-combined_k141_124178\|\|full | Not-determined |
| 17-106-final-viral-combined_k141_17102\|\|full | Not-determined |
| 17-106-final-viral-combined_k141_17103\|\|full | Low-quality |
| 17-106-final-viral-combined_k141_49477\|\|full | Low-quality |
| 17-106-final-viral-combined_k141_17104\|\|full | Not-determined |
| 17-106-final-viral-combined_k141_49486\|\|full | Not-determined |
| 17-106-final-viral-combined_k141_65428\|\|full | Not-determined |
| 17-106-final-viral-combined_k141_54866\|\|full | Low-quality |
| 17-106-final-viral-combined_k141_97696\|\|full | Not-determined |
| 17-106-final-viral-combined_k141_22758\|\|full | Not-determined |
| 17-106-final-viral-combined_k141_43917\|\|full | Not-determined |
| 17-106-final-viral-combined_k141_70577\|\|full | Not-determined |
| 17-106-final-viral-combined_k141_54870\|\|full | Not-determined |
| 17-106-final-viral-combined_k141_22762\|\|full | Low-quality |
| 17-106-final-viral-combined_k141_22764\|\|full | Low-quality |
| 17-106-final-viral-combined_k141_124228\|\|full | Not-determined |
| 17-106-final-viral-combined_k141_60288\|\|full | Low-quality |
| 17-106-final-viral-combined_k141_22768\|\|full | Not-determined |
| 17-106-final-viral-combined_k141_54874\|\|full | Not-determined |
| 17-106-final-viral-combined_k141_108094\|\|full | Not-determined |
| 17-106-final-viral-combined_k141_11752\|\|full | Low-quality |
| 17-106-final-viral-combined_k141_6731\|\|full | Low-quality |
| 17-106-final-viral-combined_k141_124242\|\|full | Not-determined |
| 17-106-final-viral-combined_k141_108098\|\|full | Not-determined |
| 17-106-final-viral-combined_k141_60318\|\|full | Not-determined |
| 17-106-final-viral-combined_k141_108099\|\|full | Low-quality |
| 17-106-final-viral-combined_k141_60336\|\|full | Not-determined |
| 17-106-final-viral-combined_k141_28124\|\|full | Low-quality |
| 17-106-final-viral-combined_k141_28127\|\|full | Not-determined |
| 17-106-final-viral-combined_k141_103026\|\|full | Low-quality |
| 17-106-final-viral-combined_k141_6747\|\|full | Low-quality |
| 17-106-final-viral-combined_k141_38556\|\|full | Not-determined |
| 17-106-final-viral-combined_k141_103047\|\|full | Low-quality |
| 17-106-final-viral-combined_k141_103048\|\|full | Not-determined |
| 17-106-final-viral-combined_k141_17150\|\|full | Low-quality |
| 17-106-final-viral-combined_k141_17156\|\|full | Low-quality |
| 17-106-final-viral-combined_k141_28154\|\|full | Not-determined |
| 17-106-final-viral-combined_k141_60361\|\|full | Not-determined |
| 17-106-final-viral-combined_k141_113537\|\|full | Not-determined |
| 17-106-final-viral-combined_k141_6823\|\|full | Low-quality |
| 17-106-final-viral-combined_k141_108121\|\|full | Low-quality |
| 18-109-final-viral-combined_k141_3697\|\|full | Medium-quality |
| 17-106-final-viral-combined_k141_124277\|\|full | Not-determined |
| 17-106-final-viral-combined_k141_124283\|\|full | Not-determined |
| 17-106-final-viral-combined_k141_38572\|\|full | Low-quality |
| 17-106-final-viral-combined_k141_70662\|\|full | Not-determined |
| 17-106-final-viral-combined_k141_22875\|\|full | Not-determined |
| 17-106-final-viral-combined_k141_70667\|\|full | Not-determined |
| 17-106-final-viral-combined_k141_28178\|\|full | Not-determined |
| 17-106-final-viral-combined_k141_124289\|\|full | Not-determined |
| 17-106-final-viral-combined_k141_65537\|\|full | Not-determined |
| 17-106-final-viral-combined_k141_103057\|\|full | Low-quality |
| 17-106-final-viral-combined_k141_6877\|\|full | Not-determined |
| 17-106-final-viral-combined_k141_113567\|\|full | Low-quality |
| 17-106-final-viral-combined_k141_6880\|\|full | Low-quality |
| 17-106-final-viral-combined_k141_22906\|\|full | Not-determined |
| 17-106-final-viral-combined_k141_113575\|\|full | Low-quality |
| 17-106-final-viral-combined_k141_70683\|\|full | Not-determined |
| 17-106-final-viral-combined_k141_44026\|\|full | Not-determined |
| 17-106-final-viral-combined_k141_28228\|\|full | Low-quality |
| 17-106-final-viral-combined_k141_60417\|\|full | Not-determined |
| 17-106-final-viral-combined_k141_28229\|\|full | Low-quality |
| 17-109-final-viral-combined_k141_201125\|\|full | Medium-quality |
| 17-106-final-viral-combined_k141_60431\|\|full | Not-determined |
| 17-106-final-viral-combined_k141_103062\|\|full | Low-quality |
| 17-106-final-viral-combined_k141_17227\|\|full | Not-determined |
| 17-106-final-viral-combined_k141_44063\|\|full | Low-quality |
| 17-106-final-viral-combined_k141_54996\|\|full | Low-quality |
| 17-106-final-viral-combined_k141_49568\|\|full | Not-determined |
| 17-106-final-viral-combined_k141_44066\|\|full | Low-quality |
| 17-106-final-viral-combined_k141_17245\|\|full | Not-determined |
| 17-106-final-viral-combined_k141_49571\|\|full | Not-determined |
| 17-106-final-viral-combined_k141_103096\|\|full | Low-quality |
| 17-106-final-viral-combined_k141_17262\|\|full | Not-determined |
| 17-106-final-viral-combined_k141_28235\|\|full | Not-determined |
| 17-106-final-viral-combined_k141_103107\|\|full | Low-quality |
| 17-106-final-viral-combined_k141_103108\|\|full | Not-determined |
| 17-106-final-viral-combined_k141_17275\|\|full | Not-determined |
| 17-106-final-viral-combined_k141_119052\|\|full | Not-determined |
| 17-106-final-viral-combined_k141_113624\|\|full | Low-quality |
| 17-106-final-viral-combined_k141_22965\|\|full | Low-quality |
| 17-106-final-viral-combined_k141_113626\|\|full | Not-determined |
| 17-106-final-viral-combined_k141_113629\|\|full | Low-quality |
| 17-106-final-viral-combined_k141_17305\|\|full | Not-determined |
| 17-106-final-viral-combined_k141_103150\|\|full | Low-quality |
| 17-106-final-viral-combined_k141_124315\|\|full | Not-determined |
| 17-106-final-viral-combined_k141_113640\|\|full | Low-quality |
| 17-106-final-viral-combined_k141_113641\|\|full | Not-determined |
| 17-106-final-viral-combined_k141_33313\|\|full | Low-quality |
| 17-106-final-viral-combined_k141_81877\|\|full | Not-determined |
| 17-106-final-viral-combined_k141_60466\|\|full | Not-determined |
| 17-106-final-viral-combined_k141_86977\|\|full | Low-quality |
| 17-106-final-viral-combined_k141_70788\|\|full | Not-determined |
| 17-106-final-viral-combined_k141_65634\|\|full | Not-determined |
| 17-106-final-viral-combined_k141_22993\|\|full | Not-determined |
| 17-106-final-viral-combined_k141_113662\|\|full | Not-determined |
| 17-106-final-viral-combined_k141_81905\|\|full | Low-quality |
| 17-106-final-viral-combined_k141_87001\|\|full | Low-quality |
| 17-106-final-viral-combined_k141_23007\|\|full | Not-determined |
| 17-106-final-viral-combined_k141_65661\|\|full | Not-determined |
| 17-106-final-viral-combined_k141_124367\|\|full | Low-quality |
| 17-106-final-viral-combined_k141_49672\|\|full | Not-determined |
| 17-106-final-viral-combined_k141_103194\|\|full | Not-determined |
| 17-106-final-viral-combined_k141_23041\|\|full | Low-quality |
| 17-106-final-viral-combined_k141_103203\|\|full | Not-determined |
| 17-106-final-viral-combined_k141_87023\|\|full | Not-determined |
| 17-106-final-viral-combined_k141_23045\|\|full | Low-quality |
| 17-106-final-viral-combined_k141_65668\|\|full | Low-quality |
| 17-106-final-viral-combined_k141_103223\|\|full | Low-quality |
| 17-106-final-viral-combined_k141_65678\|\|full | Low-quality |
| 17-106-final-viral-combined_k141_65689\|\|full | Not-determined |
| 17-106-final-viral-combined_k141_55042\|\|full | Not-determined |
| 17-106-final-viral-combined_k141_11866\|\|full | Not-determined |
| 17-106-final-viral-combined_k141_87066\|\|full | Not-determined |
| 17-106-final-viral-combined_k141_87106\|\|full | Not-determined |
| 17-106-final-viral-combined_k141_103271\|\|full | Not-determined |
| 17-106-final-viral-combined_k141_49726\|\|full | Not-determined |
| 17-106-final-viral-combined_k141_17376\|\|full | Not-determined |
| 17-106-final-viral-combined_k141_28305\|\|full | Not-determined |
| 17-106-final-viral-combined_k141_55092\|\|full | Low-quality |
| 17-106-final-viral-combined_k141_97820\|\|full | Not-determined |
| 17-106-final-viral-combined_k141_28314\|\|full | Not-determined |
| 17-106-final-viral-combined_k141_1897\|\|full | Not-determined |
| 17-106-final-viral-combined_k141_92358\|\|full | Low-quality |
| 17-106-final-viral-combined_k141_28321\|\|full | Not-determined |
| 17-106-final-viral-combined_k141_38725\|\|full | Not-determined |
| 17-106-final-viral-combined_k141_1917\|\|full | Low-quality |
| 17-106-final-viral-combined_k141_49760\|\|full | Low-quality |
| 17-106-final-viral-combined_k141_70868\|\|full | Not-determined |
| 17-106-final-viral-combined_k141_17427\|\|full | Low-quality |
| 17-106-final-viral-combined_k141_108225\|\|full | Low-quality |
| 17-106-final-viral-combined_k141_11877\|\|full | Low-quality |
| 17-106-final-viral-combined_k141_17436\|\|full | Low-quality |
| 17-106-final-viral-combined_k141_108227\|\|full | Not-determined |
| 17-106-final-viral-combined_k141_75960\|\|full | Not-determined |
| 17-106-final-viral-combined_k141_60515\|\|full | Not-determined |
| 17-106-final-viral-combined_k141_92410\|\|full | Low-quality |
| 17-106-final-viral-combined_k141_108241\|\|full | Low-quality |
| 17-106-final-viral-combined_k141_49774\|\|full | Low-quality |
| 17-106-final-viral-combined_k141_44248\|\|full | Low-quality |
| 17-106-final-viral-combined_k141_11903\|\|full | Low-quality |
| 17-106-final-viral-combined_k141_81967\|\|full | Not-determined |
| 17-106-final-viral-combined_k141_81970\|\|full | Low-quality |
| 17-106-final-viral-combined_k141_55146\|\|full | Low-quality |
| 17-106-final-viral-combined_k141_44278\|\|full | Not-determined |
| 17-106-final-viral-combined_k141_76004\|\|full | Not-determined |
| 17-106-final-viral-combined_k141_97840\|\|full | Not-determined |
| 17-106-final-viral-combined_k141_119138\|\|full | Not-determined |
| 17-106-final-viral-combined_k141_76013\|\|full | Not-determined |
| 17-106-final-viral-combined_k141_70929\|\|full | Not-determined |
| 17-106-final-viral-combined_k141_38821\|\|full | Not-determined |
| 17-106-final-viral-combined_k141_55180\|\|full | Low-quality |
| 17-106-final-viral-combined_k141_108280\|\|full | Not-determined |
| 17-106-final-viral-combined_k141_65852\|\|full | Low-quality |
| 17-106-final-viral-combined_k141_60575\|\|full | Not-determined |
| 17-106-final-viral-combined_k141_60577\|\|full | Low-quality |
| 17-106-final-viral-combined_k141_49846\|\|full | Not-determined |
| 17-106-final-viral-combined_k141_65859\|\|full | Not-determined |
| 17-106-final-viral-combined_k141_108283\|\|full | Low-quality |
| 17-106-final-viral-combined_k141_11919\|\|full | Low-quality |
| 17-106-final-viral-combined_k141_23081\|\|full | Not-determined |
| 17-106-final-viral-combined_k141_92486\|\|full | Low-quality |
| 17-106-final-viral-combined_k141_65877\|\|full | Not-determined |
| 17-106-final-viral-combined_k141_60600\|\|full | Not-determined |
| 17-106-final-viral-combined_k141_70947\|\|full | Low-quality |
| 17-106-final-viral-combined_k141_17525\|\|full | Not-determined |
| 17-106-final-viral-combined_k141_65882\|\|full | Low-quality |
| 17-106-final-viral-combined_k141_60610\|\|full | Not-determined |
| 18-106-final-viral-combined_k141_82597\|\|full | Complete |
| 17-106-final-viral-combined_k141_124520\|\|full | Low-quality |
| 17-106-final-viral-combined_k141_23130\|\|full | Not-determined |
| 17-106-final-viral-combined_k141_76068\|\|full | Not-determined |
| 17-106-final-viral-combined_k141_65921\|\|full | Low-quality |
| 17-106-final-viral-combined_k141_124536\|\|full | Low-quality |
| 17-106-final-viral-combined_k141_11964\|\|full | Low-quality |
| 17-106-final-viral-combined_k141_7129\|\|full | Low-quality |
| 17-106-final-viral-combined_k141_23142\|\|full | Not-determined |
| 17-106-final-viral-combined_k141_49880\|\|full | Not-determined |
| 17-106-final-viral-combined_k141_60651\|\|full | Low-quality |
| 17-106-final-viral-combined_k141_7153\|\|full | Not-determined |
| 17-106-final-viral-combined_k141_33405\|\|full | Not-determined |
| 17-106-final-viral-combined_k141_119177\|\|full | Low-quality |
| 17-106-final-viral-combined_k141_92496\|\|full | Low-quality |
| 17-106-final-viral-combined_k141_65965\|\|full | Low-quality |
| 17-106-final-viral-combined_k141_7173\|\|full | Low-quality |
| 17-106-final-viral-combined_k141_92516\|\|full | Not-determined |
| 17-106-final-viral-combined_k141_49893\|\|full | Not-determined |
| 17-106-final-viral-combined_k141_82087\|\|full | Low-quality |
| 17-106-final-viral-combined_k141_33421\|\|full | Not-determined |
| 17-106-final-viral-combined_k141_2125\|\|full | Low-quality |
| 17-106-final-viral-combined_k141_119203\|\|full | Low-quality |
| 17-106-final-viral-combined_k141_2129\|\|full | Low-quality |
| 17-106-final-viral-combined_k141_44299\|\|full | Low-quality |
| 17-106-final-viral-combined_k141_2130\|\|full | Not-determined |
| 17-106-final-viral-combined_k141_124574\|\|full | Not-determined |
| 17-106-final-viral-combined_k141_17589\|\|full | Not-determined |
| 17-106-final-viral-combined_k141_55305\|\|full | Not-determined |
| 17-106-final-viral-combined_k141_17591\|\|full | Not-determined |
| 17-106-final-viral-combined_k141_82096\|\|full | Low-quality |
| 17-106-final-viral-combined_k141_7196\|\|full | Low-quality |
| 17-106-final-viral-combined_k141_92541\|\|full | Not-determined |
| 17-106-final-viral-combined_k141_49951\|\|full | Not-determined |
| 17-106-final-viral-combined_k141_103358\|\|full | Not-determined |
| 17-106-final-viral-combined_k141_12024\|\|full | Low-quality |
| 17-106-final-viral-combined_k141_71022\|\|full | Not-determined |
| 17-106-final-viral-combined_k141_55339\|\|full | Not-determined |
| 17-106-final-viral-combined_k141_38974\|\|full | Low-quality |
| 17-106-final-viral-combined_k141_92569\|\|full | Low-quality |
| 17-106-final-viral-combined_k141_55349\|\|full | Not-determined |
| 17-106-final-viral-combined_k141_103379\|\|full | Low-quality |
| 17-106-final-viral-combined_k141_82135\|\|full | Not-determined |
| 17-106-final-viral-combined_k141_82136\|\|full | Not-determined |
| 17-106-final-viral-combined_k141_2199\|\|full | Low-quality |
| 17-106-final-viral-combined_k141_97913\|\|full | Not-determined |
| 17-106-final-viral-combined_k141_113720\|\|full | Low-quality |
| 17-106-final-viral-combined_k141_113724\|\|full | Not-determined |
| 17-106-final-viral-combined_k141_50006\|\|full | Low-quality |
| 17-106-final-viral-combined_k141_50017\|\|full | Low-quality |
| 17-106-final-viral-combined_k141_108379\|\|full | Not-determined |
| 17-106-final-viral-combined_k141_55392\|\|full | Low-quality |
| 17-106-final-viral-combined_k141_113740\|\|full | Not-determined |
| 17-106-final-viral-combined_k141_55404\|\|full | Not-determined |
| 17-106-final-viral-combined_k141_7225\|\|full | Not-determined |
| 17-106-final-viral-combined_k141_108405\|\|full | Low-quality |
| 17-106-final-viral-combined_k141_55416\|\|full | Not-determined |
| 17-106-final-viral-combined_k141_97945\|\|full | Not-determined |
| 17-106-final-viral-combined_k141_97946\|\|full | Low-quality |
| 17-106-final-viral-combined_k141_124633\|\|full | Low-quality |
| 17-106-final-viral-combined_k141_97948\|\|full | Not-determined |
| 17-106-final-viral-combined_k141_60754\|\|full | Not-determined |
| 17-106-final-viral-combined_k141_50064\|\|full | Low-quality |
| 17-106-final-viral-combined_k141_55427\|\|full | Low-quality |
| 17-106-final-viral-combined_k141_7232\|\|full | Not-determined |
| 17-106-final-viral-combined_k141_60763\|\|full | Not-determined |
| 17-106-final-viral-combined_k141_12102\|\|full | Low-quality |
| 17-106-final-viral-combined_k141_66079\|\|full | Low-quality |
| 17-106-final-viral-combined_k141_60777\|\|full | Low-quality |
| 17-106-final-viral-combined_k141_44363\|\|full | Low-quality |
| 17-106-final-viral-combined_k141_82228\|\|full | Not-determined |
| 17-106-final-viral-combined_k141_12108\|\|full | Not-determined |
| 17-106-final-viral-combined_k141_23284\|\|full | Low-quality |
| 17-106-final-viral-combined_k141_87284\|\|full | Low-quality |
| 17-106-final-viral-combined_k141_2256\|\|full | Low-quality |
| 17-106-final-viral-combined_k141_82235\|\|full | Low-quality |
| 17-106-final-viral-combined_k141_7269\|\|full | Not-determined |
| 17-106-final-viral-combined_k141_17742\|\|full | Low-quality |
| 17-106-final-viral-combined_k141_17744\|\|full | Not-determined |
| 17-106-final-viral-combined_k141_23303\|\|full | Not-determined |
| 17-106-final-viral-combined_k141_17754\|\|full | Low-quality |
| 17-106-final-viral-combined_k141_23304\|\|full | Low-quality |
| 17-106-final-viral-combined_k141_119332\|\|full | Not-determined |
| 17-106-final-viral-combined_k141_55451\|\|full | Not-determined |
| 17-106-final-viral-combined_k141_23306\|\|full | Not-determined |
| 17-106-final-viral-combined_k141_39000\|\|full | Low-quality |
| 17-106-final-viral-combined_k141_17759\|\|full | Not-determined |
| 17-106-final-viral-combined_k141_60811\|\|full | Not-determined |
| 17-106-final-viral-combined_k141_12127\|\|full | Low-quality |
| 17-106-final-viral-combined_k141_50122\|\|full | Not-determined |
| 17-106-final-viral-combined_k141_12150\|\|full | Not-determined |
| 17-106-final-viral-combined_k141_87304\|\|full | Not-determined |
| 17-106-final-viral-combined_k141_12154\|\|full | Low-quality |
| 17-106-final-viral-combined_k141_71074\|\|full | Low-quality |
| 17-106-final-viral-combined_k141_17788\|\|full | Not-determined |
| 17-106-final-viral-combined_k141_17789\|\|full | Not-determined |
| 17-106-final-viral-combined_k141_28427\|\|full | Not-determined |
| 17-106-final-viral-combined_k141_55492\|\|full | Not-determined |
| 17-106-final-viral-combined_k141_50158\|\|full | Low-quality |
| 17-106-final-viral-combined_k141_28440\|\|full | Low-quality |
| 17-106-final-viral-combined_k141_12184\|\|full | Not-determined |
| 17-106-final-viral-combined_k141_108428\|\|full | Low-quality |
| 17-106-final-viral-combined_k141_2318\|\|full | Low-quality |
| 17-106-final-viral-combined_k141_33471\|\|full | Low-quality |
| 17-106-final-viral-combined_k141_33473\|\|full | Not-determined |
| 17-106-final-viral-combined_k141_55507\|\|full | Not-determined |
| 17-106-final-viral-combined_k141_12205\|\|full | Not-determined |
| 17-106-final-viral-combined_k141_82293\|\|full | Not-determined |
| 17-106-final-viral-combined_k141_66153\|\|full | Low-quality |
| 17-106-final-viral-combined_k141_33481\|\|full | Low-quality |
| 17-106-final-viral-combined_k141_7344\|\|full | Low-quality |
| 17-106-final-viral-combined_k141_108429\|\|full | Low-quality |
| 17-106-final-viral-combined_k141_60838\|\|full | Low-quality |
| 17-106-final-viral-combined_k141_66160\|\|full | Not-determined |
| 17-106-final-viral-combined_k141_50211\|\|full | Not-determined |
| 17-106-final-viral-combined_k141_119377\|\|full | Not-determined |
| 17-106-final-viral-combined_k141_28496\|\|full | Low-quality |
| 17-106-final-viral-combined_k141_17820\|\|full | Low-quality |
| 17-106-final-viral-combined_k141_98042\|\|full | Low-quality |
| 17-106-final-viral-combined_k141_60841\|\|full | Low-quality |
| 17-106-final-viral-combined_k141_60850\|\|full | Low-quality |
| 17-106-final-viral-combined_k141_87381\|\|full | Not-determined |
| 17-106-final-viral-combined_k141_2348\|\|full | Not-determined |
| 17-106-final-viral-combined_k141_55538\|\|full | Low-quality |
| 17-106-final-viral-combined_k141_71118\|\|full | Not-determined |
| 17-106-final-viral-combined_k141_7397\|\|full | Low-quality |
| 17-106-final-viral-combined_k141_7398\|\|full | Low-quality |
| 17-106-final-viral-combined_k141_119403\|\|full | Low-quality |
| 17-106-final-viral-combined_k141_71124\|\|full | Not-determined |
| 17-106-final-viral-combined_k141_55551\|\|full | Not-determined |
| 17-106-final-viral-combined_k141_119413\|\|full | Low-quality |
| 17-106-final-viral-combined_k141_92671\|\|full | Not-determined |
| 17-106-final-viral-combined_k141_50268\|\|full | Low-quality |
| 17-106-final-viral-combined_k141_66239\|\|full | Low-quality |
| 17-106-final-viral-combined_k141_98074\|\|full | Not-determined |
| 17-106-final-viral-combined_k141_87411\|\|full | Not-determined |
| 17-106-final-viral-combined_k141_71154\|\|full | Not-determined |
| 17-106-final-viral-combined_k141_28563\|\|full | Low-quality |
| 17-106-final-viral-combined_k141_28569\|\|full | Not-determined |
| 17-106-final-viral-combined_k141_119428\|\|full | Not-determined |
| 17-106-final-viral-combined_k141_7450\|\|full | Low-quality |
| 17-106-final-viral-combined_k141_50302\|\|full | Low-quality |
| 17-106-final-viral-combined_k141_60878\|\|full | Low-quality |
| 17-106-final-viral-combined_k141_98115\|\|full | Not-determined |
| 17-106-final-viral-combined_k141_28591\|\|full | Low-quality |
| 17-106-final-viral-combined_k141_98127\|\|full | Not-determined |
| 17-106-final-viral-combined_k141_60894\|\|full | Not-determined |
| 17-106-final-viral-combined_k141_92731\|\|full | Not-determined |
| 17-106-final-viral-combined_k141_92736\|\|full | Low-quality |
| 17-106-final-viral-combined_k141_98152\|\|full | Not-determined |
| 17-106-final-viral-combined_k141_71209\|\|full | Not-determined |
| 17-106-final-viral-combined_k141_103556\|\|full | Not-determined |
| 17-106-final-viral-combined_k141_33510\|\|full | Not-determined |
| 17-106-final-viral-combined_k141_103559\|\|full | Not-determined |
| 17-106-final-viral-combined_k141_39169\|\|full | Not-determined |
| 17-106-final-viral-combined_k141_17888\|\|full | Low-quality |
| 17-106-final-viral-combined_k141_66252\|\|full | Low-quality |
| 17-106-final-viral-combined_k141_12352\|\|full | Low-quality |
| 17-106-final-viral-combined_k141_76277\|\|full | Low-quality |
| 17-106-final-viral-combined_k141_39183\|\|full | Not-determined |
| 17-106-final-viral-combined_k141_87483\|\|full | Not-determined |
| 17-106-final-viral-combined_k141_71234\|\|full | Low-quality |
| 17-106-final-viral-combined_k141_7474\|\|full | Low-quality |
| 17-106-final-viral-combined_k141_66268\|\|full | Not-determined |
| 17-106-final-viral-combined_k141_87489\|\|full | Not-determined |
| 17-106-final-viral-combined_k141_28619\|\|full | Low-quality |
| 17-106-final-viral-combined_k141_7492\|\|full | Not-determined |
| 17-106-final-viral-combined_k141_87497\|\|full | Low-quality |
| 17-106-final-viral-combined_k141_87498\|\|full | Low-quality |
| 17-106-final-viral-combined_k141_39210\|\|full | Low-quality |
| 17-106-final-viral-combined_k141_28623\|\|full | Low-quality |
| 17-106-final-viral-combined_k141_71254\|\|full | Not-determined |
| 17-106-final-viral-combined_k141_12363\|\|full | Not-determined |
| 17-106-final-viral-combined_k141_82342\|\|full | Low-quality |
| 17-106-final-viral-combined_k141_113809\|\|full | Low-quality |
| 17-106-final-viral-combined_k141_92842\|\|full | Low-quality |
| 17-106-final-viral-combined_k141_92846\|\|full | Low-quality |
| 17-106-final-viral-combined_k141_76292\|\|full | Low-quality |
| 17-106-final-viral-combined_k141_124698\|\|full | Not-determined |
| 17-106-final-viral-combined_k141_28657\|\|full | Not-determined |
| 17-106-final-viral-combined_k141_87551\|\|full | Not-determined |
| 17-106-final-viral-combined_k141_7511\|\|full | Low-quality |
| 17-106-final-viral-combined_k141_28662\|\|full | Not-determined |
| 17-106-final-viral-combined_k141_39253\|\|full | Not-determined |
| 17-106-final-viral-combined_k141_82389\|\|full | Low-quality |
| 17-106-final-viral-combined_k141_108495\|\|full | Not-determined |
| 17-106-final-viral-combined_k141_82397\|\|full | Not-determined |
| 17-106-final-viral-combined_k141_103620\|\|full | Not-determined |
| 17-106-final-viral-combined_k141_124733\|\|full | Not-determined |
| 17-106-final-viral-combined_k141_108505\|\|full | Not-determined |
| 17-106-final-viral-combined_k141_103626\|\|full | Low-quality |
| 17-106-final-viral-combined_k141_17935\|\|full | Low-quality |
| 17-106-final-viral-combined_k141_108514\|\|full | Not-determined |
| 17-106-final-viral-combined_k141_71289\|\|full | Low-quality |
| 17-106-final-viral-combined_k141_71300\|\|full | Not-determined |
| 17-106-final-viral-combined_k141_12394\|\|full | Not-determined |
| 17-106-final-viral-combined_k141_87581\|\|full | Not-determined |
| 17-106-final-viral-combined_k141_12398\|\|full | Low-quality |
| 17-106-final-viral-combined_k141_17959\|\|full | Not-determined |
| 17-106-final-viral-combined_k141_98184\|\|full | Low-quality |
| 17-106-final-viral-combined_k141_12403\|\|full | Not-determined |
| 17-106-final-viral-combined_k141_98192\|\|full | Not-determined |
| 17-106-final-viral-combined_k141_87596\|\|full | Not-determined |
| 17-106-final-viral-combined_k141_50388\|\|full | Not-determined |
| 17-106-final-viral-combined_k141_2500\|\|full | Low-quality |
| 17-106-final-viral-combined_k141_98205\|\|full | Low-quality |
| 17-106-final-viral-combined_k141_18004\|\|full | Low-quality |
| 17-106-final-viral-combined_k141_87615\|\|full | Not-determined |
| 17-106-final-viral-combined_k141_39301\|\|full | Low-quality |
| 17-106-final-viral-combined_k141_55573\|\|full | Low-quality |
| 17-106-final-viral-combined_k141_124759\|\|full | Low-quality |
| 17-106-final-viral-combined_k141_98211\|\|full | Low-quality |
| 17-106-final-viral-combined_k141_119546\|\|full | Low-quality |
| 17-106-final-viral-combined_k141_55574\|\|full | Low-quality |
| 17-106-final-viral-combined_k141_98215\|\|full | Low-quality |
| 17-106-final-viral-combined_k141_119555\|\|full | Low-quality |
| 17-106-final-viral-combined_k141_12453\|\|full | Low-quality |
| 17-106-final-viral-combined_k141_119563\|\|full | Not-determined |
| 17-106-final-viral-combined_k141_82431\|\|full | Low-quality |
| 17-106-final-viral-combined_k141_82433\|\|full | Low-quality |
| 17-106-final-viral-combined_k141_108578\|\|full | Low-quality |
| 17-106-final-viral-combined_k141_82440\|\|full | Low-quality |
| 17-106-final-viral-combined_k141_113858\|\|full | Low-quality |
| 17-106-final-viral-combined_k141_76387\|\|full | Low-quality |
| 17-106-final-viral-combined_k141_18028\|\|full | Low-quality |
| 17-106-final-viral-combined_k141_124797\|\|full | Low-quality |
| 17-106-final-viral-combined_k141_55609\|\|full | Low-quality |
| 17-106-final-viral-combined_k141_18051\|\|full | Low-quality |
| 17-106-final-viral-combined_k141_7613\|\|full | Low-quality |
| 17-106-final-viral-combined_k141_108599\|\|full | Not-determined |
| 17-106-final-viral-combined_k141_18067\|\|full | Low-quality |
| 17-106-final-viral-combined_k141_124813\|\|full | Not-determined |
| 17-106-final-viral-combined_k141_50415\|\|full | Low-quality |
| 17-106-final-viral-combined_k141_28683\|\|full | Low-quality |
| 17-106-final-viral-combined_k141_92950\|\|full | Not-determined |
| 17-106-final-viral-combined_k141_28690\|\|full | Low-quality |
| 17-106-final-viral-combined_k141_18090\|\|full | Not-determined |
| 17-106-final-viral-combined_k141_82481\|\|full | Not-determined |
| 17-106-final-viral-combined_k141_82483\|\|full | Low-quality |
| 17-106-final-viral-combined_k141_113920\|\|full | Not-determined |
| 17-106-final-viral-combined_k141_82487\|\|full | Not-determined |
| 17-106-final-viral-combined_k141_18095\|\|full | Not-determined |
| 17-106-final-viral-combined_k141_76463\|\|full | Not-determined |
| 17-106-final-viral-combined_k141_18109\|\|full | Low-quality |
| 17-106-final-viral-combined_k141_33673\|\|full | Low-quality |
| 17-106-final-viral-combined_k141_119606\|\|full | Not-determined |
| 17-106-final-viral-combined_k141_103750\|\|full | Low-quality |
| 17-106-final-viral-combined_k141_18144\|\|full | Low-quality |
| 17-106-final-viral-combined_k141_7645\|\|full | Low-quality |
| 17-106-final-viral-combined_k141_71343\|\|full | Low-quality |
| 17-106-final-viral-combined_k141_28718\|\|full | Not-determined |
| 17-106-final-viral-combined_k141_12576\|\|full | Low-quality |
| 17-106-final-viral-combined_k141_50471\|\|full | Low-quality |
| 17-106-final-viral-combined_k141_12584\|\|full | Low-quality |
| 17-106-final-viral-combined_k141_108695\|\|full | Not-determined |
| 17-106-final-viral-combined_k141_76521\|\|full | Low-quality |
| 17-106-final-viral-combined_k141_71354\|\|full | Low-quality |
| 17-106-final-viral-combined_k141_71362\|\|full | Not-determined |
| 17-106-final-viral-combined_k141_82522\|\|full | Not-determined |
| 17-106-final-viral-combined_k141_71364\|\|full | Not-determined |
| 17-106-final-viral-combined_k141_18219\|\|full | Low-quality |
| 17-106-final-viral-combined_k141_71370\|\|full | Not-determined |
| 17-106-final-viral-combined_k141_23411\|\|full | Not-determined |
| 17-106-final-viral-combined_k141_23416\|\|full | Low-quality |
| 17-106-final-viral-combined_k141_12642\|\|full | Low-quality |
| 17-106-final-viral-combined_k141_18256\|\|full | Not-determined |
| 17-106-final-viral-combined_k141_87696\|\|full | Not-determined |
| 17-106-final-viral-combined_k141_76540\|\|full | Low-quality |
| 17-106-final-viral-combined_k141_71378\|\|full | Not-determined |
| 17-106-final-viral-combined_k141_12645\|\|full | Low-quality |
| 17-106-final-viral-combined_k141_23440\|\|full | Low-quality |
| 17-106-final-viral-combined_k141_18268\|\|full | Low-quality |

| | |
|---|---|
| 17-106-final-viral-combined_k141_108699\|\|full | Low-quality |
| 17-106-final-viral-combined_k141_23445\|\|full | Low-quality |
| 17-106-final-viral-combined_k141_18274\|\|full | Not-determined |
| 17-106-final-viral-combined_k141_23447\|\|full | Not-determined |
| 17-106-final-viral-combined_k141_39433\|\|full | Low-quality |
| 17-106-final-viral-combined_k141_18287\|\|full | Not-determined |
| 17-106-final-viral-combined_k141_18290\|\|full | Not-determined |
| 17-106-final-viral-combined_k141_82615\|\|full | Low-quality |
| 17-106-final-viral-combined_k141_71406\|\|full | Not-determined |
| 17-106-final-viral-combined_k141_119617\|\|full | Not-determined |
| 17-106-final-viral-combined_k141_23461\|\|full | Low-quality |
| 17-106-final-viral-combined_k141_23467\|\|full | Not-determined |
| 17-106-final-viral-combined_k141_12694\|\|full | Low-quality |
| 17-106-final-viral-combined_k141_71424\|\|full | Not-determined |
| 17-106-final-viral-combined_k141_113982\|\|full | Not-determined |
| 17-106-final-viral-combined_k141_113988\|\|full | Not-determined |
| 17-106-final-viral-combined_k141_113997\|\|full | Not-determined |
| 17-106-final-viral-combined_k141_114000\|\|full | Not-determined |
| 17-106-final-viral-combined_k141_50533\|\|full | Not-determined |
| 17-106-final-viral-combined_k141_12725\|\|full | Low-quality |
| 17-106-final-viral-combined_k141_82669\|\|full | Low-quality |
| 17-106-final-viral-combined_k141_76595\|\|full | Low-quality |
| 17-106-final-viral-combined_k141_44479\|\|full | Low-quality |
| 17-106-final-viral-combined_k141_12737\|\|full | Not-determined |
| 17-106-final-viral-combined_k141_103855\|\|full | Low-quality |
| 17-106-final-viral-combined_k141_87729\|\|full | Not-determined |
| 17-106-final-viral-combined_k141_50568\|\|full | Not-determined |
| 17-106-final-viral-combined_k141_12746\|\|full | Not-determined |
| 17-106-final-viral-combined_k141_23515\|\|full | Low-quality |
| 17-106-final-viral-combined_k141_87743\|\|full | Low-quality |
| 17-106-final-viral-combined_k141_44505\|\|full | Not-determined |
| 17-106-final-viral-combined_k141_23526\|\|full | Not-determined |
| 17-106-final-viral-combined_k141_82726\|\|full | Low-quality |
| 17-106-final-viral-combined_k141_124857\|\|full | Medium-quality |
| 17-106-final-viral-combined_k141_108725\|\|full | Low-quality |
| 17-106-final-viral-combined_k141_124865\|\|full | Not-determined |
| 17-106-final-viral-combined_k141_23564\|\|full | Low-quality |
| 17-106-final-viral-combined_k141_23567\|\|full | Low-quality |
| 17-106-final-viral-combined_k141_103918\|\|full | Not-determined |
| 17-106-final-viral-combined_k141_55759\|\|full | Not-determined |
| 17-106-final-viral-combined_k141_12802\|\|full | Not-determined |
| 17-106-final-viral-combined_k141_28782\|\|full | Low-quality |
| 17-106-final-viral-combined_k141_124874\|\|full | Low-quality |
| 17-106-final-viral-combined_k141_12809\|\|full | Low-quality |
| 17-106-final-viral-combined_k141_124878\|\|full | Not-determined |
| 17-106-final-viral-combined_k141_12816\|\|full | Not-determined |
| 17-106-final-viral-combined_k141_124894\|\|full | Low-quality |
| 17-106-final-viral-combined_k141_108776\|\|full | Not-determined |
| 17-106-final-viral-combined_k141_87806\|\|full | Low-quality |
| 17-106-final-viral-combined_k141_23618\|\|full | Not-determined |
| 17-106-final-viral-combined_k141_82741\|\|full | Not-determined |
| 17-106-final-viral-combined_k141_108784\|\|full | Not-determined |
| 17-106-final-viral-combined_k141_44547\|\|full | Not-determined |
| 17-106-final-viral-combined_k141_93043\|\|full | Low-quality |
| 17-106-final-viral-combined_k141_87838\|\|full | Not-determined |
| 17-106-final-viral-combined_k141_108807\|\|full | Low-quality |
| 17-106-final-viral-combined_k141_87843\|\|full | Low-quality |
| 17-106-final-viral-combined_k141_23622\|\|full | Not-determined |
| 17-106-final-viral-combined_k141_66392\|\|full | Low-quality |
| 17-106-final-viral-combined_k141_87857\|\|full | Not-determined |
| 17-106-final-viral-combined_k141_98268\|\|full | Low-quality |
| 17-106-final-viral-combined_k141_124935\|\|full | Not-determined |
| 17-106-final-viral-combined_k141_7851\|\|full | Not-determined |
| 17-106-final-viral-combined_k141_71509\|\|full | Not-determined |
| 17-106-final-viral-combined_k141_76730\|\|full | Low-quality |
| 18-102-final-viral-combined_k141_225908\|\|full | Medium-quality |
| 17-106-final-viral-combined_k141_28813\|\|full | Low-quality |
| 17-106-final-viral-combined_k141_7880\|\|full | Not-determined |
| 17-106-final-viral-combined_k141_28817\|\|full | Not-determined |
| 17-106-final-viral-combined_k141_18353\|\|full | Low-quality |
| 17-106-final-viral-combined_k141_33805\|\|full | Not-determined |
| 17-106-final-viral-combined_k141_2716\|\|full | Not-determined |
| 17-106-final-viral-combined_k141_33812\|\|full | Not-determined |
| 17-106-final-viral-combined_k141_98281\|\|full | Not-determined |
| 17-106-final-viral-combined_k141_55803\|\|full | Low-quality |
| 17-106-final-viral-combined_k141_2735\|\|full | Not-determined |
| 17-106-final-viral-combined_k141_66432\|\|full | Not-determined |
| 17-106-final-viral-combined_k141_18377\|\|full | Low-quality |
| 17-106-final-viral-combined_k141_39556\|\|full | Low-quality |
| 17-106-final-viral-combined_k141_55814\|\|full | Not-determined |
| 17-106-final-viral-combined_k141_2748\|\|full | Not-determined |
| 17-106-final-viral-combined_k141_82838\|\|full | Low-quality |
| 17-106-final-viral-combined_k141_82840\|\|full | Not-determined |
| 17-106-final-viral-combined_k141_28846\|\|full | Low-quality |
| 17-106-final-viral-combined_k141_2756\|\|full | Not-determined |
| 17-106-final-viral-combined_k141_66447\|\|full | Not-determined |
| 17-106-final-viral-combined_k141_18395\|\|full | Low-quality |
| 17-106-final-viral-combined_k141_103955\|\|full | Low-quality |
| 17-106-final-viral-combined_k141_66458\|\|full | Not-determined |
| 17-106-final-viral-combined_k141_93068\|\|full | Not-determined |
| 17-106-final-viral-combined_k141_39585\|\|full | Low-quality |
| 17-106-final-viral-combined_k141_2765\|\|full | Not-determined |
| 17-106-final-viral-combined_k141_66468\|\|full | Not-determined |
| 17-106-final-viral-combined_k141_125036\|\|full | Low-quality |
| 17-106-final-viral-combined_k141_71548\|\|full | Low-quality |
| 17-106-final-viral-combined_k141_55853\|\|full | Low-quality |
| 17-106-final-viral-combined_k141_39618\|\|full | Low-quality |
| 17-106-final-viral-combined_k141_66494\|\|full | Not-determined |
| 17-106-final-viral-combined_k141_39619\|\|full | Low-quality |
| 17-106-final-viral-combined_k141_76777\|\|full | Not-determined |
| 17-106-final-viral-combined_k141_87947\|\|full | Not-determined |
| 17-106-final-viral-combined_k141_7934\|\|full | Low-quality |
| 17-106-final-viral-combined_k141_28888\|\|full | Not-determined |
| 17-106-final-viral-combined_k141_55863\|\|full | Low-quality |
| 17-106-final-viral-combined_k141_28891\|\|full | Low-quality |
| 17-106-final-viral-combined_k141_33875\|\|full | Not-determined |
| 17-106-final-viral-combined_k141_28895\|\|full | Not-determined |
| 17-106-final-viral-combined_k141_119682\|\|full | Not-determined |
| 17-106-final-viral-combined_k141_39641\|\|full | Not-determined |
| 17-106-final-viral-combined_k141_93102\|\|full | Low-quality |
| 17-106-final-viral-combined_k141_50754\|\|full | Not-determined |
| 17-106-final-viral-combined_k141_93111\|\|full | Not-determined |
| 17-106-final-viral-combined_k141_50775\|\|full | Not-determined |
| 17-106-final-viral-combined_k141_71579\|\|full | Low-quality |
| 17-106-final-viral-combined_k141_125111\|\|full | Not-determined |
| 17-106-final-viral-combined_k141_125114\|\|full | Low-quality |
| 17-106-final-viral-combined_k141_104032\|\|full | Not-determined |
| 17-106-final-viral-combined_k141_71606\|\|full | Low-quality |
| 17-106-final-viral-combined_k141_88019\|\|full | Low-quality |
| 17-106-final-viral-combined_k141_44647\|\|full | Not-determined |
| 17-106-final-viral-combined_k141_39689\|\|full | Low-quality |
| 17-106-final-viral-combined_k141_7946\|\|full | Not-determined |
| 17-106-final-viral-combined_k141_104056\|\|full | Low-quality |
| 17-106-final-viral-combined_k141_88033\|\|full | Not-determined |
| 17-106-final-viral-combined_k141_50810\|\|full | Low-quality |
| 17-106-final-viral-combined_k141_119695\|\|full | Low-quality |
| 17-106-final-viral-combined_k141_50813\|\|full | Not-determined |
| 17-106-final-viral-combined_k141_104070\|\|full | Low-quality |
| 17-106-final-viral-combined_k141_119698\|\|full | Not-determined |
| 17-106-final-viral-combined_k141_39711\|\|full | Not-determined |
| 17-106-final-viral-combined_k141_23716\|\|full | Not-determined |
| 17-106-final-viral-combined_k141_55983\|\|full | Low-quality |
| 17-106-final-viral-combined_k141_114104\|\|full | Not-determined |
| 17-106-final-viral-combined_k141_50826\|\|full | Not-determined |
| 17-106-final-viral-combined_k141_119729\|\|full | Not-determined |
| 17-106-final-viral-combined_k141_83057\|\|full | Not-determined |
| 17-106-final-viral-combined_k141_7991\|\|full | Not-determined |
| 17-106-final-viral-combined_k141_23741\|\|full | Low-quality |
| 18-105-final-viral-combined_k141_210959\|\|full | Medium-quality |
| 17-106-final-viral-combined_k141_61362\|\|full | Low-quality |
| 17-106-final-viral-combined_k141_88077\|\|full | Low-quality |
| 17-106-final-viral-combined_k141_18470\|\|full | Not-determined |
| 17-106-final-viral-combined_k141_8026\|\|full | Not-determined |
| 17-106-final-viral-combined_k141_50838\|\|full | Not-determined |
| 17-106-final-viral-combined_k141_83112\|\|full | Not-determined |
| 17-106-final-viral-combined_k141_8034\|\|full | Not-determined |
| 17-106-final-viral-combined_k141_88107\|\|full | Not-determined |
| 17-106-final-viral-combined_k141_104094\|\|full | Low-quality |
| 17-106-final-viral-combined_k141_23790\|\|full | Not-determined |
| 17-106-final-viral-combined_k141_88109\|\|full | Low-quality |
| 17-106-final-viral-combined_k141_104097\|\|full | Low-quality |
| 17-106-final-viral-combined_k141_66624\|\|full | Not-determined |
| 17-106-final-viral-combined_k141_18499\|\|full | Not-determined |
| 17-106-final-viral-combined_k141_18502\|\|full | Not-determined |
| 17-106-final-viral-combined_k141_88130\|\|full | Not-determined |
| 17-106-final-viral-combined_k141_18503\|\|full | Not-determined |
| 17-106-final-viral-combined_k141_83138\|\|full | Low-quality |
| 17-106-final-viral-combined_k141_66639\|\|full | Not-determined |
| 17-106-final-viral-combined_k141_28998\|\|full | Low-quality |
| 17-106-final-viral-combined_k141_76842\|\|full | Low-quality |
| 17-106-final-viral-combined_k141_23810\|\|full | Not-determined |
| 17-106-final-viral-combined_k141_29016\|\|full | Not-determined |
| 17-106-final-viral-combined_k141_12856\|\|full | Low-quality |
| 17-106-final-viral-combined_k141_18555\|\|full | Not-determined |
| 17-106-final-viral-combined_k141_29025\|\|full | Not-determined |
| 17-106-final-viral-combined_k141_56119\|\|full | Not-determined |
| 17-106-final-viral-combined_k141_56130\|\|full | Low-quality |
| 17-106-final-viral-combined_k141_39747\|\|full | Low-quality |
| 17-106-final-viral-combined_k141_12886\|\|full | Low-quality |
| 18-105-final-viral-combined_k141_161191\|\|full | Medium-quality |
| 17-106-final-viral-combined_k141_83158\|\|full | Not-determined |
| 17-106-final-viral-combined_k141_18579\|\|full | Not-determined |
| 17-106-final-viral-combined_k141_119864\|\|full | Not-determined |
| 17-106-final-viral-combined_k141_18580\|\|full | Low-quality |
| 17-106-final-viral-combined_k141_76881\|\|full | Not-determined |
| 17-106-final-viral-combined_k141_29035\|\|full | Low-quality |
| 17-106-final-viral-combined_k141_104188\|\|full | Not-determined |
| 17-106-final-viral-combined_k141_119869\|\|full | Not-determined |
| 17-106-final-viral-combined_k141_104199\|\|full | Low-quality |
| 17-106-final-viral-combined_k141_93207\|\|full | Low-quality |
| 17-106-final-viral-combined_k141_119878\|\|full | Low-quality |
| 17-106-final-viral-combined_k141_93220\|\|full | Low-quality |
| 17-106-final-viral-combined_k141_18601\|\|full | Not-determined |
| 17-106-final-viral-combined_k141_39778\|\|full | Low-quality |
| 17-106-final-viral-combined_k141_76914\|\|full | Not-determined |
| 17-106-final-viral-combined_k141_23826\|\|full | Not-determined |
| 17-106-final-viral-combined_k141_23827\|\|full | Not-determined |
| 17-106-final-viral-combined_k141_2960\|\|full | Not-determined |
| 17-106-final-viral-combined_k141_8095\|\|full | Not-determined |
| 17-106-final-viral-combined_k141_119905\|\|full | Not-determined |
| 17-106-final-viral-combined_k141_83225\|\|full | Not-determined |
| 17-106-final-viral-combined_k141_83231\|\|full | Not-determined |
| 17-106-final-viral-combined_k141_23848\|\|full | Low-quality |
| 17-106-final-viral-combined_k141_44815\|\|full | Not-determined |
| 17-106-final-viral-combined_k141_8129\|\|full | Low-quality |
| 17-106-final-viral-combined_k141_108932\|\|full | Not-determined |
| 17-106-final-viral-combined_k141_119941\|\|full | Not-determined |
| 17-106-final-viral-combined_k141_76946\|\|full | Not-determined |
| 17-109-final-viral-combined_k141_320727\|\|full | High-quality |
| 17-106-final-viral-combined_k141_44856\|\|full | Not-determined |
| 17-106-final-viral-combined_k141_29073\|\|full | Not-determined |
| 17-106-final-viral-combined_k141_104257\|\|full | Low-quality |
| 17-106-final-viral-combined_k141_3000\|\|full | Low-quality |
| 17-106-final-viral-combined_k141_29084\|\|full | Not-determined |
| 17-106-final-viral-combined_k141_83270\|\|full | Not-determined |
| 17-106-final-viral-combined_k141_119966\|\|full | Not-determined |
| 17-106-final-viral-combined_k141_29087\|\|full | Not-determined |
| 17-106-final-viral-combined_k141_12969\|\|full | Not-determined |
| 17-106-final-viral-combined_k141_8177\|\|full | Not-determined |
| 17-106-final-viral-combined_k141_12979\|\|full | Low-quality |
| 17-106-final-viral-combined_k141_12989\|\|full | Low-quality |
| 17-106-final-viral-combined_k141_119993\|\|full | Not-determined |
| 17-106-final-viral-combined_k141_83306\|\|full | Not-determined |
| 17-106-final-viral-combined_k141_71786\|\|full | Not-determined |
| 17-106-final-viral-combined_k141_18698\|\|full | Not-determined |
| 17-106-final-viral-combined_k141_98450\|\|full | Not-determined |
| 17-106-final-viral-combined_k141_18704\|\|full | Low-quality |
| 17-106-final-viral-combined_k141_76981\|\|full | Not-determined |
| 17-106-final-viral-combined_k141_104266\|\|full | Low-quality |
| 17-106-final-viral-combined_k141_29136\|\|full | Low-quality |
| 17-106-final-viral-combined_k141_108965\|\|full | Low-quality |
| 17-106-final-viral-combined_k141_18726\|\|full | Not-determined |
| 17-106-final-viral-combined_k141_66679\|\|full | Low-quality |
| 17-106-final-viral-combined_k141_8243\|\|full | Low-quality |
| 17-106-final-viral-combined_k141_50920\|\|full | Low-quality |
| 17-106-final-viral-combined_k141_18730\|\|full | Not-determined |
| 17-106-final-viral-combined_k141_83323\|\|full | Low-quality |
| 17-106-final-viral-combined_k141_13061\|\|full | Low-quality |
| 17-106-final-viral-combined_k141_66697\|\|full | Low-quality |
| 17-106-final-viral-combined_k141_13075\|\|full | Not-determined |
| 17-106-final-viral-combined_k141_18749\|\|full | Not-determined |
| 17-106-final-viral-combined_k141_39845\|\|full | Not-determined |
| 17-106-final-viral-combined_k141_13077\|\|full | Not-determined |
| 17-106-final-viral-combined_k141_50940\|\|full | Not-determined |
| 17-106-final-viral-combined_k141_77032\|\|full | Low-quality |
| 17-106-final-viral-combined_k141_114343\|\|full | Not-determined |
| 17-106-final-viral-combined_k141_83339\|\|full | Low-quality |
| 17-106-final-viral-combined_k141_114356\|\|full | Not-determined |
| 17-106-final-viral-combined_k141_114357\|\|full | Low-quality |
| 17-106-final-viral-combined_k141_108985\|\|full | Not-determined |
| 17-106-final-viral-combined_k141_61619\|\|full | Not-determined |
| 17-106-final-viral-combined_k141_13093\|\|full | Not-determined |
| 17-106-final-viral-combined_k141_93347\|\|full | Not-determined |
| 17-106-final-viral-combined_k141_83368\|\|full | Not-determined |
| 17-106-final-viral-combined_k141_61625\|\|full | Low-quality |
| 17-106-final-viral-combined_k141_93363\|\|full | Low-quality |
| 17-106-final-viral-combined_k141_34197\|\|full | Low-quality |
| 17-106-final-viral-combined_k141_83385\|\|full | Low-quality |
| 17-106-final-viral-combined_k141_34199\|\|full | Not-determined |
| 17-106-final-viral-combined_k141_120030\|\|full | Low-quality |
| 17-106-final-viral-combined_k141_56251\|\|full | Low-quality |
| 17-106-final-viral-combined_k141_77080\|\|full | Not-determined |
| 17-106-final-viral-combined_k141_50977\|\|full | Low-quality |
| 17-106-final-viral-combined_k141_98497\|\|full | Low-quality |
| 17-106-final-viral-combined_k141_44949\|\|full | Not-determined |
| 17-106-final-viral-combined_k141_120040\|\|full | Not-determined |
| 17-106-final-viral-combined_k141_50996\|\|full | Low-quality |
| 17-106-final-viral-combined_k141_24029\|\|full | Low-quality |
| 17-106-final-viral-combined_k141_109040\|\|full | Low-quality |
| 17-106-final-viral-combined_k141_71895\|\|full | Low-quality |
| 17-106-final-viral-combined_k141_39869\|\|full | Not-determined |
| 17-106-final-viral-combined_k141_98520\|\|full | Not-determined |
| 17-106-final-viral-combined_k141_77130\|\|full | Low-quality |
| 17-106-final-viral-combined_k141_114452\|\|full | Low-quality |
| 17-106-final-viral-combined_k141_125241\|\|full | Not-determined |
| 17-106-final-viral-combined_k141_104303\|\|full | Low-quality |
| 17-106-final-viral-combined_k141_24056\|\|full | Not-determined |
| 17-106-final-viral-combined_k141_13151\|\|full | Not-determined |
| 17-106-final-viral-combined_k141_71908\|\|full | Not-determined |
| 17-106-final-viral-combined_k141_8347\|\|full | Not-determined |
| 17-106-final-viral-combined_k141_71918\|\|full | Low-quality |
| 17-106-final-viral-combined_k141_34244\|\|full | Low-quality |
| 17-106-final-viral-combined_k141_18835\|\|full | Low-quality |
| 17-106-final-viral-combined_k141_29187\|\|full | Not-determined |
| 17-106-final-viral-combined_k141_34271\|\|full | Not-determined |
| 17-106-final-viral-combined_k141_45052\|\|full | Not-determined |
| 17-106-final-viral-combined_k141_98560\|\|full | Low-quality |
| 17-106-final-viral-combined_k141_120176\|\|full | Not-determined |
| 17-106-final-viral-combined_k141_83417\|\|full | Low-quality |
| 17-106-final-viral-combined_k141_24153\|\|full | Low-quality |
| 17-106-final-viral-combined_k141_13250\|\|full | Not-determined |
| 17-106-final-viral-combined_k141_18893\|\|full | Not-determined |
| 17-106-final-viral-combined_k141_83431\|\|full | Low-quality |
| 17-106-final-viral-combined_k141_18899\|\|full | Low-quality |
| 17-106-final-viral-combined_k141_18902\|\|full | Not-determined |
| 17-106-final-viral-combined_k141_77191\|\|full | Low-quality |
| 17-106-final-viral-combined_k141_39921\|\|full | Not-determined |
| 17-106-final-viral-combined_k141_104402\|\|full | Not-determined |
| 17-106-final-viral-combined_k141_61670\|\|full | Not-determined |
| 17-106-final-viral-combined_k141_13280\|\|full | Not-determined |
| 17-106-final-viral-combined_k141_77206\|\|full | Not-determined |
| 17-106-final-viral-combined_k141_45088\|\|full | Not-determined |
| 17-106-final-viral-combined_k141_98595\|\|full | Not-determined |
| 17-106-final-viral-combined_k141_98596\|\|full | Low-quality |
| 17-106-final-viral-combined_k141_13294\|\|full | Low-quality |
| 17-106-final-viral-combined_k141_18947\|\|full | Not-determined |
| 17-106-final-viral-combined_k141_109090\|\|full | Not-determined |
| 17-106-final-viral-combined_k141_13308\|\|full | Not-determined |
| 17-106-final-viral-combined_k141_45113\|\|full | Not-determined |
| 17-106-final-viral-combined_k141_83477\|\|full | Low-quality |
| 17-106-final-viral-combined_k141_8440\|\|full | Not-determined |
| 17-106-final-viral-combined_k141_120229\|\|full | Not-determined |
| 17-106-final-viral-combined_k141_77236\|\|full | Not-determined |
| 17-106-final-viral-combined_k141_13325\|\|full | Not-determined |
| 17-106-final-viral-combined_k141_77243\|\|full | Low-quality |
| 17-106-final-viral-combined_k141_125289\|\|full | Low-quality |
| 17-106-final-viral-combined_k141_114626\|\|full | Not-determined |
| 17-106-final-viral-combined_k141_77248\|\|full | Not-determined |
| 17-106-final-viral-combined_k141_24188\|\|full | Low-quality |
| 17-106-final-viral-combined_k141_125295\|\|full | Not-determined |
| 17-106-final-viral-combined_k141_98652\|\|full | Low-quality |
| 17-106-final-viral-combined_k141_109123\|\|full | Not-determined |
| 17-106-final-viral-combined_k141_19005\|\|full | Low-quality |
| 17-106-final-viral-combined_k141_88171\|\|full | Low-quality |
| 17-106-final-viral-combined_k141_93448\|\|full | Low-quality |
| 17-106-final-viral-combined_k141_56382\|\|full | Low-quality |
| 17-106-final-viral-combined_k141_39984\|\|full | Low-quality |
| 17-106-final-viral-combined_k141_83534\|\|full | Low-quality |
| 17-106-final-viral-combined_k141_29274\|\|full | Low-quality |
| 17-106-final-viral-combined_k141_45168\|\|full | Low-quality |
| 17-106-final-viral-combined_k141_34360\|\|full | Low-quality |
| 17-106-final-viral-combined_k141_45169\|\|full | Low-quality |
| 17-106-final-viral-combined_k141_34362\|\|full | Low-quality |
| 17-106-final-viral-combined_k141_51131\|\|full | Not-determined |
| 17-106-final-viral-combined_k141_56403\|\|full | Low-quality |
| 17-106-final-viral-combined_k141_13338\|\|full | Not-determined |
| 17-106-final-viral-combined_k141_13342\|\|full | Not-determined |
| 17-106-final-viral-combined_k141_51145\|\|full | Low-quality |
| 17-106-final-viral-combined_k141_125354\|\|full | Not-determined |
| 17-106-final-viral-combined_k141_45200\|\|full | Low-quality |
| 17-106-final-viral-combined_k141_120265\|\|full | Low-quality |
| 17-106-final-viral-combined_k141_71960\|\|full | Low-quality |
| 17-106-final-viral-combined_k141_77343\|\|full | Not-determined |
| 17-106-final-viral-combined_k141_125371\|\|full | Low-quality |
| 17-106-final-viral-combined_k141_71973\|\|full | Not-determined |
| 17-106-final-viral-combined_k141_114689\|\|full | Low-quality |
| 17-106-final-viral-combined_k141_45238\|\|full | Not-determined |
| 17-106-final-viral-combined_k141_13369\|\|full | Low-quality |
| 17-106-final-viral-combined_k141_24258\|\|full | Low-quality |
| 17-106-final-viral-combined_k141_77355\|\|full | Not-determined |
| 17-106-final-viral-combined_k141_8502\|\|full | Low-quality |
| 17-106-final-viral-combined_k141_8506\|\|full | Not-determined |
| 17-106-final-viral-combined_k141_66732\|\|full | Not-determined |
| 17-106-final-viral-combined_k141_61797\|\|full | Not-determined |
| 17-106-final-viral-combined_k141_88193\|\|full | Low-quality |
| 17-106-final-viral-combined_k141_71998\|\|full | Low-quality |
| 18-109-final-viral-combined_k141_20921\|\|full | Medium-quality |
| 17-106-final-viral-combined_k141_8514\|\|full | Not-determined |
| 17-106-final-viral-combined_k141_66740\|\|full | Low-quality |
| 17-106-final-viral-combined_k141_56467\|\|full | Not-determined |
| 17-106-final-viral-combined_k141_61803\|\|full | Low-quality |
| 17-106-final-viral-combined_k141_66753\|\|full | Low-quality |
| 17-106-final-viral-combined_k141_45266\|\|full | Not-determined |
| 17-106-final-viral-combined_k141_13390\|\|full | Not-determined |
| 17-106-final-viral-combined_k141_66758\|\|full | Not-determined |
| 17-106-final-viral-combined_k141_61812\|\|full | Low-quality |
| 17-106-final-viral-combined_k141_61819\|\|full | Not-determined |
| 17-106-final-viral-combined_k141_61823\|\|full | Low-quality |
| 17-106-final-viral-combined_k141_8547\|\|full | Low-quality |
| 17-106-final-viral-combined_k141_72026\|\|full | Low-quality |
| 17-106-final-viral-combined_k141_77388\|\|full | Low-quality |
| 17-106-final-viral-combined_k141_88220\|\|full | Low-quality |
| 17-106-final-viral-combined_k141_34456\|\|full | Low-quality |
| 17-106-final-viral-combined_k141_40092\|\|full | Low-quality |
| 17-106-final-viral-combined_k141_93513\|\|full | Low-quality |
| 17-106-final-viral-combined_k141_3160\|\|full | Not-determined |
| 17-106-final-viral-combined_k141_93520\|\|full | Not-determined |
| 17-106-final-viral-combined_k141_88245\|\|full | Low-quality |
| 17-106-final-viral-combined_k141_77403\|\|full | Not-determined |
| 17-106-final-viral-combined_k141_77404\|\|full | Not-determined |
| 17-106-final-viral-combined_k141_56512\|\|full | Not-determined |
| 17-106-final-viral-combined_k141_13432\|\|full | Not-determined |
| 17-106-final-viral-combined_k141_93538\|\|full | Not-determined |
| 17-106-final-viral-combined_k141_77412\|\|full | Not-determined |
| 17-106-final-viral-combined_k141_83619\|\|full | Low-quality |
| 17-106-final-viral-combined_k141_29300\|\|full | Low-quality |
| 17-106-final-viral-combined_k141_45333\|\|full | Low-quality |
| 17-106-final-viral-combined_k141_29316\|\|full | Not-determined |
| 17-106-final-viral-combined_k141_19081\|\|full | Not-determined |
| 17-106-final-viral-combined_k141_77448\|\|full | Low-quality |
| 17-106-final-viral-combined_k141_56516\|\|full | Low-quality |
| 17-106-final-viral-combined_k141_56518\|\|full | Low-quality |
| 17-106-final-viral-combined_k141_66822\|\|full | Not-determined |
| 17-106-final-viral-combined_k141_56527\|\|full | Low-quality |
| 17-106-final-viral-combined_k141_29350\|\|full | Not-determined |
| 17-106-final-viral-combined_k141_125455\|\|full | Not-determined |
| 17-106-final-viral-combined_k141_93553\|\|full | Not-determined |
| 17-106-final-viral-combined_k141_66831\|\|full | Low-quality |
| 17-106-final-viral-combined_k141_93556\|\|full | Not-determined |
| 17-106-final-viral-combined_k141_24316\|\|full | Not-determined |
| 17-106-final-viral-combined_k141_125464\|\|full | Low-quality |
| 17-106-final-viral-combined_k141_3210\|\|full | Not-determined |
| 17-106-final-viral-combined_k141_3214\|\|full | Low-quality |
| 17-106-final-viral-combined_k141_40176\|\|full | Low-quality |
| 17-106-final-viral-combined_k141_56542\|\|full | Low-quality |
| 17-106-final-viral-combined_k141_29356\|\|full | Low-quality |
| 17-106-final-viral-combined_k141_29363\|\|full | Not-determined |
| 17-106-final-viral-combined_k141_56567\|\|full | Not-determined |
| 17-106-final-viral-combined_k141_3232\|\|full | Not-determined |
| 17-106-final-viral-combined_k141_34557\|\|full | Low-quality |
| 17-106-final-viral-combined_k141_56575\|\|full | Not-determined |
| 17-106-final-viral-combined_k141_125508\|\|full | Not-determined |
| 17-106-final-viral-combined_k141_45397\|\|full | Low-quality |
| 17-106-final-viral-combined_k141_77472\|\|full | Not-determined |
| 17-106-final-viral-combined_k141_40195\|\|full | Low-quality |
| 17-106-final-viral-combined_k141_3241\|\|full | Not-determined |
| 17-106-final-viral-combined_k141_109158\|\|full | Not-determined |
| 17-106-final-viral-combined_k141_98830\|\|full | Not-determined |
| 17-106-final-viral-combined_k141_19160\|\|full | Low-quality |
| 17-106-final-viral-combined_k141_66886\|\|full | Not-determined |
| 17-106-final-viral-combined_k141_77495\|\|full | Not-determined |
| 17-106-final-viral-combined_k141_13483\|\|full | Low-quality |
| 17-106-final-viral-combined_k141_19165\|\|full | Low-quality |
| 17-106-final-viral-combined_k141_83693\|\|full | Not-determined |
| 17-106-final-viral-combined_k141_13492\|\|full | Low-quality |
| 17-106-final-viral-combined_k141_66896\|\|full | Not-determined |
| 17-106-final-viral-combined_k141_77512\|\|full | Not-determined |
| 17-106-final-viral-combined_k141_34616\|\|full | Low-quality |
| 17-106-final-viral-combined_k141_34618\|\|full | Not-determined |
| 17-106-final-viral-combined_k141_40227\|\|full | Not-determined |
| 17-106-final-viral-combined_k141_114849\|\|full | Not-determined |
| 17-106-final-viral-combined_k141_45423\|\|full | Not-determined |
| 17-106-final-viral-combined_k141_40230\|\|full | Low-quality |
| 17-106-final-viral-combined_k141_45440\|\|full | Low-quality |
| 17-106-final-viral-combined_k141_83714\|\|full | Low-quality |
| 17-106-final-viral-combined_k141_40233\|\|full | Low-quality |
| 17-106-final-viral-combined_k141_40235\|\|full | Not-determined |
| 17-106-final-viral-combined_k141_13519\|\|full | Not-determined |
| 17-106-final-viral-combined_k141_56653\|\|full | Not-determined |
| 17-106-final-viral-combined_k141_40244\|\|full | Not-determined |
| 17-106-final-viral-combined_k141_19185\|\|full | Low-quality |
| 17-106-final-viral-combined_k141_114890\|\|full | Not-determined |
| 17-106-final-viral-combined_k141_56665\|\|full | Not-determined |
| 17-106-final-viral-combined_k141_88317\|\|full | Not-determined |
| 17-106-final-viral-combined_k141_56679\|\|full | Not-determined |
| 17-106-final-viral-combined_k141_83744\|\|full | Not-determined |
| 17-106-final-viral-combined_k141_40274\|\|full | Low-quality |
| 17-106-final-viral-combined_k141_61936\|\|full | Low-quality |
| 17-106-final-viral-combined_k141_72175\|\|full | Not-determined |
| 17-106-final-viral-combined_k141_114902\|\|full | Low-quality |
| 17-106-final-viral-combined_k141_77539\|\|full | Low-quality |
| 17-106-final-viral-combined_k141_93650\|\|full | Low-quality |
| 17-106-final-viral-combined_k141_56691\|\|full | Low-quality |
| 17-106-final-viral-combined_k141_66940\|\|full | Not-determined |
| 17-106-final-viral-combined_k141_40283\|\|full | Low-quality |
| 17-106-final-viral-combined_k141_61954\|\|full | Not-determined |
| 17-106-final-viral-combined_k141_40290\|\|full | Low-quality |
| 17-106-final-viral-combined_k141_56714\|\|full | Not-determined |
| 17-106-final-viral-combined_k141_72200\|\|full | Low-quality |
| 17-106-final-viral-combined_k141_8611\|\|full | Not-determined |
| 17-106-final-viral-combined_k141_56720\|\|full | Not-determined |
| 17-106-final-viral-combined_k141_66953\|\|full | Low-quality |
| 17-106-final-viral-combined_k141_8613\|\|full | Not-determined |
| 17-106-final-viral-combined_k141_3339\|\|full | Low-quality |
| 17-106-final-viral-combined_k141_51266\|\|full | Not-determined |
| 17-106-final-viral-combined_k141_40304\|\|full | Low-quality |
| 17-106-final-viral-combined_k141_83763\|\|full | Low-quality |
| 17-106-final-viral-combined_k141_93691\|\|full | Not-determined |
| 17-106-final-viral-combined_k141_8653\|\|full | Not-determined |
| 17-106-final-viral-combined_k141_98957\|\|full | Low-quality |
| 17-106-final-viral-combined_k141_83776\|\|full | Low-quality |
| 17-106-final-viral-combined_k141_98965\|\|full | Not-determined |
| 17-106-final-viral-combined_k141_51280\|\|full | Low-quality |
| 17-106-final-viral-combined_k141_120578\|\|full | Low-quality |
| 17-106-final-viral-combined_k141_104477\|\|full | Low-quality |
| 17-106-final-viral-combined_k141_51295\|\|full | Not-determined |
| 17-106-final-viral-combined_k141_120591\|\|full | Low-quality |
| 18-105-final-viral-combined_k141_108794\|\|full | Complete |
| 17-106-final-viral-combined_k141_8666\|\|full | Not-determined |
| 17-106-final-viral-combined_k141_83818\|\|full | Low-quality |
| 17-106-final-viral-combined_k141_120604\|\|full | Not-determined |
| 17-106-final-viral-combined_k141_93724\|\|full | Low-quality |
| 17-106-final-viral-combined_k141_114979\|\|full | Not-determined |
| 17-106-final-viral-combined_k141_109220\|\|full | Low-quality |
| 17-106-final-viral-combined_k141_40313\|\|full | Low-quality |
| 17-106-final-viral-combined_k141_93726\|\|full | Low-quality |
| 17-106-final-viral-combined_k141_83831\|\|full | Not-determined |
| 17-106-final-viral-combined_k141_62019\|\|full | Not-determined |
| 17-106-final-viral-combined_k141_93735\|\|full | Low-quality |
| 17-106-final-viral-combined_k141_34793\|\|full | Not-determined |
| 17-106-final-viral-combined_k141_88423\|\|full | Not-determined |
| 17-106-final-viral-combined_k141_83865\|\|full | Low-quality |
| 17-106-final-viral-combined_k141_125608\|\|full | Not-determined |
| 17-106-final-viral-combined_k141_19236\|\|full | Low-quality |
| 17-106-final-viral-combined_k141_88439\|\|full | Low-quality |
| 17-106-final-viral-combined_k141_34810\|\|full | Not-determined |
| 17-106-final-viral-combined_k141_88450\|\|full | Not-determined |
| 17-106-final-viral-combined_k141_125616\|\|full | Not-determined |
| 17-106-final-viral-combined_k141_56781\|\|full | Low-quality |
| 17-106-final-viral-combined_k141_34821\|\|full | Not-determined |
| 17-106-final-viral-combined_k141_120630\|\|full | Not-determined |
| 17-106-final-viral-combined_k141_13607\|\|full | Not-determined |
| 17-106-final-viral-combined_k141_29550\|\|full | Low-quality |
| 17-106-final-viral-combined_k141_51366\|\|full | Low-quality |
| 17-106-final-viral-combined_k141_77640\|\|full | Not-determined |
| 17-106-final-viral-combined_k141_24458\|\|full | Low-quality |
| 17-106-final-viral-combined_k141_99042\|\|full | Low-quality |
| 17-106-final-viral-combined_k141_83909\|\|full | Not-determined |
| 17-106-final-viral-combined_k141_34877\|\|full | Low-quality |
| 17-106-final-viral-combined_k141_45520\|\|full | Low-quality |
| 17-106-final-viral-combined_k141_29598\|\|full | Low-quality |
| 17-106-final-viral-combined_k141_88497\|\|full | Not-determined |
| 17-106-final-viral-combined_k141_120676\|\|full | Not-determined |
| 17-106-final-viral-combined_k141_109282\|\|full | Low-quality |
| 17-106-final-viral-combined_k141_51407\|\|full | Low-quality |
| 17-106-final-viral-combined_k141_125650\|\|full | Low-quality |
| 17-106-final-viral-combined_k141_104589\|\|full | Not-determined |
| 17-106-final-viral-combined_k141_125651\|\|full | Not-determined |
| 17-106-final-viral-combined_k141_109300\|\|full | Not-determined |
| 17-106-final-viral-combined_k141_93772\|\|full | Not-determined |
| 17-106-final-viral-combined_k141_77661\|\|full | Low-quality |
| 17-106-final-viral-combined_k141_13693\|\|full | Low-quality |
| 17-106-final-viral-combined_k141_99059\|\|full | Not-determined |
| 17-106-final-viral-combined_k141_99070\|\|full | Not-determined |
| 17-106-final-viral-combined_k141_51419\|\|full | Low-quality |
| 17-106-final-viral-combined_k141_99107\|\|full | Not-determined |
| 17-106-final-viral-combined_k141_88518\|\|full | Not-determined |
| 17-106-final-viral-combined_k141_77714\|\|full | Low-quality |
| 17-106-final-viral-combined_k141_19273\|\|full | Not-determined |
| 17-106-final-viral-combined_k141_34916\|\|full | Low-quality |
| 17-106-final-viral-combined_k141_56803\|\|full | Not-determined |
| 17-106-final-viral-combined_k141_24557\|\|full | Not-determined |
| 17-106-final-viral-combined_k141_24562\|\|full | Low-quality |
| 17-106-final-viral-combined_k141_99145\|\|full | Low-quality |
| 17-106-final-viral-combined_k141_115022\|\|full | Not-determined |
| 17-106-final-viral-combined_k141_56813\|\|full | Low-quality |
| 17-106-final-viral-combined_k141_120727\|\|full | Not-determined |
| 17-106-final-viral-combined_k141_45534\|\|full | Low-quality |
| 17-106-final-viral-combined_k141_40359\|\|full | Not-determined |
| 17-106-final-viral-combined_k141_34945\|\|full | Not-determined |
| 17-106-final-viral-combined_k141_67111\|\|full | Low-quality |
| 17-106-final-viral-combined_k141_56822\|\|full | Not-determined |
| 17-106-final-viral-combined_k141_72230\|\|full | Not-determined |
| 17-106-final-viral-combined_k141_3381\|\|full | Low-quality |
| 17-106-final-viral-combined_k141_34957\|\|full | Low-quality |
| 17-106-final-viral-combined_k141_45544\|\|full | Low-quality |
| 17-106-final-viral-combined_k141_88583\|\|full | Not-determined |
| 17-106-final-viral-combined_k141_51484\|\|full | Not-determined |
| 17-106-final-viral-combined_k141_88598\|\|full | Not-determined |
| 17-106-final-viral-combined_k141_56855\|\|full | Not-determined |
| 17-106-final-viral-combined_k141_77796\|\|full | Not-determined |
| 17-106-final-viral-combined_k141_67153\|\|full | Low-quality |
| 17-106-final-viral-combined_k141_83936\|\|full | Low-quality |
| 17-106-final-viral-combined_k141_67159\|\|full | Low-quality |
| 17-106-final-viral-combined_k141_34974\|\|full | Low-quality |
| 17-106-final-viral-combined_k141_67177\|\|full | Low-quality |
| 17-106-final-viral-combined_k141_51489\|\|full | Low-quality |
| 17-106-final-viral-combined_k141_88632\|\|full | Not-determined |
| 18-106-final-viral-combined_k141_118427\|\|full | Complete |
| 17-106-final-viral-combined_k141_34993\|\|full | Low-quality |
| 17-106-final-viral-combined_k141_51498\|\|full | Not-determined |
| 17-106-final-viral-combined_k141_125712\|\|full | Not-determined |
| 17-106-final-viral-combined_k141_13737\|\|full | Low-quality |
| 17-106-final-viral-combined_k141_67190\|\|full | Low-quality |
| 17-106-final-viral-combined_k141_19361\|\|full | Low-quality |
| 17-106-final-viral-combined_k141_8853\|\|full | Low-quality |
| 17-106-final-viral-combined_k141_40392\|\|full | Not-determined |
| 17-106-final-viral-combined_k141_120791\|\|full | Not-determined |
| 17-106-final-viral-combined_k141_51524\|\|full | Low-quality |
| 17-106-final-viral-combined_k141_56932\|\|full | Low-quality |
| 17-106-final-viral-combined_k141_88662\|\|full | Low-quality |
| 17-106-final-viral-combined_k141_115069\|\|full | Low-quality |
| 17-106-final-viral-combined_k141_35014\|\|full | Low-quality |
| 17-106-final-viral-combined_k141_120803\|\|full | Not-determined |
| 17-106-final-viral-combined_k141_13758\|\|full | Low-quality |
| 17-106-final-viral-combined_k141_125750\|\|full | Low-quality |
| 17-106-final-viral-combined_k141_62129\|\|full | Not-determined |
| 17-106-final-viral-combined_k141_99246\|\|full | Low-quality |
| 17-106-final-viral-combined_k141_115085\|\|full | Low-quality |
| 17-106-final-viral-combined_k141_115088\|\|full | Not-determined |
| 17-106-final-viral-combined_k141_88664\|\|full | Low-quality |
| 17-106-final-viral-combined_k141_72314\|\|full | Low-quality |
| 17-106-final-viral-combined_k141_104675\|\|full | Not-determined |
| 17-106-final-viral-combined_k141_35045\|\|full | Not-determined |
| 17-106-final-viral-combined_k141_120812\|\|full | Low-quality |
| 17-106-final-viral-combined_k141_72319\|\|full | Low-quality |
| 17-106-final-viral-combined_k141_125766\|\|full | Low-quality |
| 17-106-final-viral-combined_k141_35052\|\|full | Low-quality |
| 17-106-final-viral-combined_k141_109387\|\|full | Low-quality |
| 17-106-final-viral-combined_k141_120824\|\|full | Not-determined |
| 17-106-final-viral-combined_k141_125769\|\|full | Low-quality |
| 17-106-final-viral-combined_k141_67232\|\|full | Not-determined |
| 17-106-final-viral-combined_k141_19452\|\|full | Low-quality |
| 17-106-final-viral-combined_k141_56989\|\|full | Low-quality |
| 17-106-final-viral-combined_k141_51604\|\|full | Not-determined |
| 17-106-final-viral-combined_k141_40449\|\|full | Low-quality |
| 17-106-final-viral-combined_k141_77844\|\|full | Not-determined |
| 17-106-final-viral-combined_k141_72336\|\|full | Low-quality |
| 17-106-final-viral-combined_k141_62213\|\|full | Low-quality |

| | |
|---|---|
| 17-106-final-viral-combined_k141_77851\|\|full | Not-determined |
| 17-106-final-viral-combined_k141_57010\|\|full | Not-determined |
| 17-106-final-viral-combined_k141_13841\|\|full | Not-determined |
| 17-106-final-viral-combined_k141_19464\|\|full | Not-determined |
| 17-106-final-viral-combined_k141_8876\|\|full | Low-quality |
| 17-106-final-viral-combined_k141_24651\|\|full | Not-determined |
| 17-106-final-viral-combined_k141_99255\|\|full | Not-determined |
| 17-106-final-viral-combined_k141_24657\|\|full | Not-determined |
| 17-106-final-viral-combined_k141_57026\|\|full | Not-determined |
| 17-106-final-viral-combined_k141_77868\|\|full | Not-determined |
| 17-106-final-viral-combined_k141_24661\|\|full | Not-determined |
| 17-106-final-viral-combined_k141_99263\|\|full | Low-quality |
| 17-106-final-viral-combined_k141_99266\|\|full | Low-quality |
| 17-106-final-viral-combined_k141_19514\|\|full | Not-determined |
| 17-106-final-viral-combined_k141_88706\|\|full | Low-quality |
| 17-106-final-viral-combined_k141_77907\|\|full | Low-quality |
| 17-106-final-viral-combined_k141_88708\|\|full | Low-quality |
| 17-106-final-viral-combined_k141_35112\|\|full | Low-quality |
| 17-106-final-viral-combined_k141_35116\|\|full | Not-determined |
| 17-106-final-viral-combined_k141_35129\|\|full | Low-quality |
| 17-106-final-viral-combined_k141_45764\|\|full | Not-determined |
| 17-106-final-viral-combined_k141_83971\|\|full | Low-quality |
| 17-106-final-viral-combined_k141_125847\|\|full | Low-quality |
| 17-106-final-viral-combined_k141_62260\|\|full | Not-determined |
| 18-104-final-viral-combined_k141_57963\|\|full | Medium-quality |
| 17-106-final-viral-combined_k141_19546\|\|full | Not-determined |
| 17-106-final-viral-combined_k141_120925\|\|full | Low-quality |
| 17-106-final-viral-combined_k141_8891\|\|full | Not-determined |
| 17-106-final-viral-combined_k141_3468\|\|full | Low-quality |
| 17-106-final-viral-combined_k141_120932\|\|full | Low-quality |
| 17-106-final-viral-combined_k141_88759\|\|full | Low-quality |
| 17-106-final-viral-combined_k141_115184\|\|full | Low-quality |
| 17-106-final-viral-combined_k141_8919\|\|full | Low-quality |
| 17-106-final-viral-combined_k141_125873\|\|full | Low-quality |
| 17-106-final-viral-combined_k141_57141\|\|full | Low-quality |
| 17-106-final-viral-combined_k141_35190\|\|full | Low-quality |
| 17-106-final-viral-combined_k141_45785\|\|full | Not-determined |
| 17-106-final-viral-combined_k141_57148\|\|full | Low-quality |
| 17-106-final-viral-combined_k141_40517\|\|full | Not-determined |
| 17-106-final-viral-combined_k141_45801\|\|full | Low-quality |
| 17-106-final-viral-combined_k141_57170\|\|full | Low-quality |
| 17-106-final-viral-combined_k141_45807\|\|full | Low-quality |
| 17-106-final-viral-combined_k141_88795\|\|full | Low-quality |
| 17-106-final-viral-combined_k141_3539\|\|full | Not-determined |
| 17-106-final-viral-combined_k141_99329\|\|full | Not-determined |
| 17-106-final-viral-combined_k141_3543\|\|full | Not-determined |
| 17-106-final-viral-combined_k141_109446\|\|full | Low-quality |
| 17-106-final-viral-combined_k141_62292\|\|full | Low-quality |
| 17-106-final-viral-combined_k141_120976\|\|full | Not-determined |
| 17-106-final-viral-combined_k141_120991\|\|full | Low-quality |
| 17-106-final-viral-combined_k141_57208\|\|full | Not-determined |
| 17-106-final-viral-combined_k141_62310\|\|full | Not-determined |
| 17-106-final-viral-combined_k141_19595\|\|full | Low-quality |
| 17-106-final-viral-combined_k141_35248\|\|full | Not-determined |
| 17-106-final-viral-combined_k141_62319\|\|full | Low-quality |
| 17-106-final-viral-combined_k141_35256\|\|full | Low-quality |
| 17-106-final-viral-combined_k141_24800\|\|full | Low-quality |
| 17-106-final-viral-combined_k141_125941\|\|full | Not-determined |
| 17-106-final-viral-combined_k141_115223\|\|full | Not-determined |
| 17-106-final-viral-combined_k141_78006\|\|full | Not-determined |
| 17-106-final-viral-combined_k141_9034\|\|full | Not-determined |
| 17-106-final-viral-combined_k141_51769\|\|full | Low-quality |
| 17-106-final-viral-combined_k141_13937\|\|full | Low-quality |
| 17-106-final-viral-combined_k141_99376\|\|full | Low-quality |
| 17-106-final-viral-combined_k141_78014\|\|full | Low-quality |
| 17-106-final-viral-combined_k141_121035\|\|full | Low-quality |
| 17-106-final-viral-combined_k141_88888\|\|full | Low-quality |
| 17-106-final-viral-combined_k141_78021\|\|full | Not-determined |
| 17-106-final-viral-combined_k141_121043\|\|full | Low-quality |
| 17-106-final-viral-combined_k141_121049\|\|full | Not-determined |
| 17-106-final-viral-combined_k141_67276\|\|full | Not-determined |
| 17-106-final-viral-combined_k141_109503\|\|full | Low-quality |
| 17-106-final-viral-combined_k141_57284\|\|full | Low-quality |
| 17-106-final-viral-combined_k141_62372\|\|full | Low-quality |
| 17-106-final-viral-combined_k141_24856\|\|full | Not-determined |
| 17-106-final-viral-combined_k141_125989\|\|full | Not-determined |
| 17-106-final-viral-combined_k141_121091\|\|full | Low-quality |
| 17-106-final-viral-combined_k141_57293\|\|full | Not-determined |
| 17-106-final-viral-combined_k141_40544\|\|full | Not-determined |
| 17-106-final-viral-combined_k141_109521\|\|full | Low-quality |
| 17-106-final-viral-combined_k141_57305\|\|full | Not-determined |
| 17-106-final-viral-combined_k141_45840\|\|full | Low-quality |
| 18-102-final-viral-combined_k141_147065\|\|full | High-quality |
| 17-106-final-viral-combined_k141_62401\|\|full | Not-determined |
| 17-106-final-viral-combined_k141_3609\|\|full | Not-determined |
| 17-106-final-viral-combined_k141_62410\|\|full | Not-determined |
| 17-106-final-viral-combined_k141_78067\|\|full | Low-quality |
| 17-106-final-viral-combined_k141_35389\|\|full | Not-determined |
| 17-106-final-viral-combined_k141_126033\|\|full | Low-quality |
| 17-106-final-viral-combined_k141_62427\|\|full | Not-determined |
| 17-106-final-viral-combined_k141_88980\|\|full | Not-determined |
| 17-106-final-viral-combined_k141_88995\|\|full | Not-determined |
| 17-106-final-viral-combined_k141_3650\|\|full | Low-quality |
| 17-106-final-viral-combined_k141_72397\|\|full | Low-quality |
| 17-106-final-viral-combined_k141_45870\|\|full | Low-quality |
| 17-106-final-viral-combined_k141_35406\|\|full | Low-quality |
| 17-106-final-viral-combined_k141_29645\|\|full | Not-determined |
| 17-106-final-viral-combined_k141_99466\|\|full | Low-quality |
| 17-106-final-viral-combined_k141_9114\|\|full | Low-quality |
| 17-106-final-viral-combined_k141_19698\|\|full | Low-quality |
| 17-106-final-viral-combined_k141_57341\|\|full | Not-determined |
| 17-106-final-viral-combined_k141_19706\|\|full | Low-quality |
| 17-106-final-viral-combined_k141_29664\|\|full | Not-determined |
| 17-106-final-viral-combined_k141_109552\|\|full | Low-quality |
| 17-106-final-viral-combined_k141_109554\|\|full | Low-quality |
| 17-106-final-viral-combined_k141_89018\|\|full | Low-quality |
| 17-106-final-viral-combined_k141_51949\|\|full | Not-determined |
| 17-106-final-viral-combined_k141_99483\|\|full | Not-determined |
| 17-106-final-viral-combined_k141_72428\|\|full | Low-quality |
| 17-106-final-viral-combined_k141_51956\|\|full | Not-determined |
| 17-106-final-viral-combined_k141_51958\|\|full | Low-quality |
| 17-106-final-viral-combined_k141_109571\|\|full | Low-quality |
| 17-106-final-viral-combined_k141_67383\|\|full | Not-determined |
| 17-106-final-viral-combined_k141_104791\|\|full | Low-quality |
| 17-106-final-viral-combined_k141_3709\|\|full | Not-determined |
| 17-106-final-viral-combined_k141_78141\|\|full | Low-quality |
| 17-106-final-viral-combined_k141_40587\|\|full | Not-determined |
| 17-106-final-viral-combined_k141_104792\|\|full | Low-quality |
| 17-106-final-viral-combined_k141_57377\|\|full | Not-determined |
| 17-106-final-viral-combined_k141_109588\|\|full | Not-determined |
| 17-106-final-viral-combined_k141_19728\|\|full | Not-determined |
| 17-106-final-viral-combined_k141_62472\|\|full | Low-quality |
| 17-106-final-viral-combined_k141_51966\|\|full | Low-quality |
| 17-106-final-viral-combined_k141_51973\|\|full | Low-quality |
| 17-106-final-viral-combined_k141_62477\|\|full | Not-determined |
| 17-106-final-viral-combined_k141_72458\|\|full | Low-quality |
| 17-106-final-viral-combined_k141_78167\|\|full | Not-determined |
| 17-106-final-viral-combined_k141_104815\|\|full | Low-quality |
| 17-106-final-viral-combined_k141_62485\|\|full | Not-determined |
| 17-106-final-viral-combined_k141_57395\|\|full | Not-determined |
| 17-106-final-viral-combined_k141_115394\|\|full | Low-quality |
| 17-106-final-viral-combined_k141_3745\|\|full | Not-determined |
| 17-106-final-viral-combined_k141_24942\|\|full | Not-determined |
| 17-106-final-viral-combined_k141_24946\|\|full | Low-quality |
| 17-106-final-viral-combined_k141_115400\|\|full | Low-quality |
| 17-106-final-viral-combined_k141_115407\|\|full | Low-quality |
| 17-106-final-viral-combined_k141_89046\|\|full | Low-quality |
| 17-106-final-viral-combined_k141_115414\|\|full | Low-quality |
| 17-106-final-viral-combined_k141_51996\|\|full | Low-quality |
| 17-106-final-viral-combined_k141_115419\|\|full | Not-determined |
| 17-106-final-viral-combined_k141_78204\|\|full | Not-determined |
| 17-106-final-viral-combined_k141_89059\|\|full | Low-quality |
| 17-106-final-viral-combined_k141_52005\|\|full | Not-determined |
| 17-106-final-viral-combined_k141_45915\|\|full | Not-determined |
| 17-106-final-viral-combined_k141_104856\|\|full | Not-determined |
| 17-106-final-viral-combined_k141_29708\|\|full | Not-determined |
| 17-106-final-viral-combined_k141_104862\|\|full | Not-determined |
| 17-106-final-viral-combined_k141_62513\|\|full | Low-quality |
| 17-106-final-viral-combined_k141_29711\|\|full | Not-determined |
| 17-106-final-viral-combined_k141_121235\|\|full | Low-quality |
| 17-106-final-viral-combined_k141_62531\|\|full | Low-quality |
| 17-106-final-viral-combined_k141_24997\|\|full | Not-determined |
| 17-106-final-viral-combined_k141_121242\|\|full | Not-determined |
| 17-106-final-viral-combined_k141_93866\|\|full | Low-quality |
| 17-106-final-viral-combined_k141_57429\|\|full | Not-determined |
| 17-106-final-viral-combined_k141_62542\|\|full | Low-quality |
| 17-106-final-viral-combined_k141_62549\|\|full | Low-quality |
| 17-106-final-viral-combined_k141_93892\|\|full | Low-quality |
| 17-106-final-viral-combined_k141_121262\|\|full | Low-quality |
| 17-106-final-viral-combined_k141_3796\|\|full | Not-determined |
| 17-106-final-viral-combined_k141_72491\|\|full | Not-determined |
| 17-106-final-viral-combined_k141_109643\|\|full | Not-determined |
| 17-106-final-viral-combined_k141_62593\|\|full | Low-quality |
| 17-106-final-viral-combined_k141_45935\|\|full | Not-determined |
| 18-101-final-viral-combined_k141_220022\|\|full | High-quality |
| 17-106-final-viral-combined_k141_115484\|\|full | Low-quality |
| 17-106-final-viral-combined_k141_3817\|\|full | Low-quality |
| 17-106-final-viral-combined_k141_89121\|\|full | Not-determined |
| 17-106-final-viral-combined_k141_99590\|\|full | Low-quality |
| 17-106-final-viral-combined_k141_99598\|\|full | Not-determined |
| 17-106-final-viral-combined_k141_99600\|\|full | Low-quality |
| 17-106-final-viral-combined_k141_109662\|\|full | Low-quality |
| 17-106-final-viral-combined_k141_89134\|\|full | Low-quality |
| 17-106-final-viral-combined_k141_93907\|\|full | Low-quality |
| 17-106-final-viral-combined_k141_84194\|\|full | Not-determined |
| 17-106-final-viral-combined_k141_40610\|\|full | Low-quality |
| 17-106-final-viral-combined_k141_67478\|\|full | Not-determined |
| 17-106-final-viral-combined_k141_52087\|\|full | Low-quality |
| 17-106-final-viral-combined_k141_93918\|\|full | Low-quality |
| 17-106-final-viral-combined_k141_67485\|\|full | Not-determined |
| 17-106-final-viral-combined_k141_104976\|\|full | Low-quality |
| 17-106-final-viral-combined_k141_67492\|\|full | Not-determined |
| 17-106-final-viral-combined_k141_9179\|\|full | Not-determined |
| 17-106-final-viral-combined_k141_89155\|\|full | Low-quality |
| 17-106-final-viral-combined_k141_67507\|\|full | Not-determined |
| 17-106-final-viral-combined_k141_115543\|\|full | Not-determined |
| 17-106-final-viral-combined_k141_121350\|\|full | Low-quality |
| 17-106-final-viral-combined_k141_35413\|\|full | Low-quality |
| 17-106-final-viral-combined_k141_104994\|\|full | Low-quality |
| 17-106-final-viral-combined_k141_72581\|\|full | Not-determined |
| 17-106-final-viral-combined_k141_121357\|\|full | Low-quality |
| 17-106-final-viral-combined_k141_126059\|\|full | Not-determined |
| 17-106-final-viral-combined_k141_35415\|\|full | Not-determined |
| 17-106-final-viral-combined_k141_29789\|\|full | Not-determined |
| 17-106-final-viral-combined_k141_14075\|\|full | Not-determined |
| 17-106-final-viral-combined_k141_29806\|\|full | Not-determined |
| 17-106-final-viral-combined_k141_93964\|\|full | Not-determined |
| 17-106-final-viral-combined_k141_115564\|\|full | Not-determined |
| 17-106-final-viral-combined_k141_109697\|\|full | Low-quality |
| 17-106-final-viral-combined_k141_99643\|\|full | Not-determined |
| 17-106-final-viral-combined_k141_93973\|\|full | Not-determined |
| 17-106-final-viral-combined_k141_72598\|\|full | Low-quality |
| 17-106-final-viral-combined_k141_25065\|\|full | Not-determined |
| 17-106-final-viral-combined_k141_72601\|\|full | Not-determined |
| 17-106-final-viral-combined_k141_45959\|\|full | Not-determined |
| 17-106-final-viral-combined_k141_84236\|\|full | Not-determined |
| 17-106-final-viral-combined_k141_9204\|\|full | Low-quality |
| 17-106-final-viral-combined_k141_126119\|\|full | Low-quality |
| 17-106-final-viral-combined_k141_84249\|\|full | Not-determined |
| 17-106-final-viral-combined_k141_84250\|\|full | Not-determined |
| 17-106-final-viral-combined_k141_109714\|\|full | Not-determined |
| 17-108_k141_32691 | Medium-quality |
| 17-106-final-viral-combined_k141_35469\|\|full | Low-quality |
| 17-106-final-viral-combined_k141_78363\|\|full | Low-quality |
| 17-106-final-viral-combined_k141_126139\|\|full | Low-quality |
| 17-106-final-viral-combined_k141_45998\|\|full | Not-determined |
| 17-108-final-viral-combined_k141_32691\|\|full | Medium-quality |
| 17-106-final-viral-combined_k141_46011\|\|full | Not-determined |
| 17-106-final-viral-combined_k141_35503\|\|full | Low-quality |
| 17-106-final-viral-combined_k141_67544\|\|full | Not-determined |
| 17-106-final-viral-combined_k141_99679\|\|full | Not-determined |
| 17-106-final-viral-combined_k141_19922\|\|full | Not-determined |
| 17-106-final-viral-combined_k141_14125\|\|full | Low-quality |
| 17-106-final-viral-combined_k141_14128\|\|full | Not-determined |
| 17-106-final-viral-combined_k141_35522\|\|full | Not-determined |
| 17-106-final-viral-combined_k141_78425\|\|full | Not-determined |
| 17-106-final-viral-combined_k141_67563\|\|full | Not-determined |
| 17-106-final-viral-combined_k141_84281\|\|full | Not-determined |
| 17-106-final-viral-combined_k141_25125\|\|full | Low-quality |
| 17-106-final-viral-combined_k141_14155\|\|full | Not-determined |
| 17-106-final-viral-combined_k141_9230\|\|full | Not-determined |
| 17-106-final-viral-combined_k141_105058\|\|full | Not-determined |
| 17-106-final-viral-combined_k141_62601\|\|full | Low-quality |
| 17-106-final-viral-combined_k141_105067\|\|full | Not-determined |
| 17-106-final-viral-combined_k141_109774\|\|full | Low-quality |
| 17-106-final-viral-combined_k141_121464\|\|full | Not-determined |
| [illegible] | [illegible] |

17-106-final-viral-combined_k141_68467||full Low-quality
17-106-final-viral-combined_k141_84913||full Not-determined
17-106-final-viral-combined_k141_46556||full Low-quality
17-106-final-viral-combined_k141_14887||full Low-quality
17-106-final-viral-combined_k141_57937||full Not-determined
17-106-final-viral-combined_k141_63461||full Low-quality
17-106-final-viral-combined_k141_63464||full Not-determined
17-106-final-viral-combined_k141_9884||full Not-determined
17-106-final-viral-combined_k141_30435||full Not-determined
17-106-final-viral-combined_k141_100267||full Low-quality
17-106-final-viral-combined_k141_4527||full Low-quality
17-106-final-viral-combined_k141_14896||full Low-quality
17-106-final-viral-combined_k141_79115||full Not-determined
17-106-final-viral-combined_k141_122257||full Not-determined
17-106-final-viral-combined_k141_73339||full Not-determined
17-106-final-viral-combined_k141_110324||full Not-determined
17-106-final-viral-combined_k141_36196||full Not-determined
17-106-final-viral-combined_k141_79116||full Low-quality
17-106-final-viral-combined_k141_73357||full Low-quality
17-106-final-viral-combined_k141_14906||full Not-determined
17-106-final-viral-combined_k141_73358||full Low-quality
17-106-final-viral-combined_k141_30461||full Not-determined
17-106-final-viral-combined_k141_52797||full Low-quality
17-106-final-viral-combined_k141_14914||full Low-quality
17-106-final-viral-combined_k141_30468||full Low-quality
17-106-final-viral-combined_k141_57984||full Not-determined
17-106-final-viral-combined_k141_79133||full Not-determined
17-106-final-viral-combined_k141_30472||full Not-determined
17-106-final-viral-combined_k141_126647||full Low-quality
17-106-final-viral-combined_k141_46609||full Low-quality
17-106-final-viral-combined_k141_79144||full Low-quality
17-106-final-viral-combined_k141_41287||full Not-determined
17-106-final-viral-combined_k141_46610||full Not-determined
17-106-final-viral-combined_k141_30483||full Not-determined
17-106-final-viral-combined_k141_41289||full Not-determined
17-106-final-viral-combined_k141_9918||full Not-determined
17-106-final-viral-combined_k141_9929||full Not-determined
17-106-final-viral-combined_k141_126656||full Not-determined
17-106-final-viral-combined_k141_25760||full Low-quality
17-106-final-viral-combined_k141_110343||full Low-quality
17-106-final-viral-combined_k141_63493||full Low-quality
17-106-final-viral-combined_k141_63495||full Low-quality
17-106-final-viral-combined_k141_46634||full Not-determined
17-106-final-viral-combined_k141_25771||full Low-quality
17-106-final-viral-combined_k141_90144||full Low-quality
17-106-final-viral-combined_k141_41324||full Low-quality
17-106-final-viral-combined_k141_63502||full Not-determined
17-106-final-viral-combined_k141_58051||full Not-determined
17-106-final-viral-combined_k141_126669||full Low-quality
17-106-final-viral-combined_k141_90167||full Not-determined
17-106-final-viral-combined_k141_79206||full Not-determined
17-106-final-viral-combined_k141_94690||full Not-determined
17-106-final-viral-combined_k141_36219||full Low-quality
17-106-final-viral-combined_k141_68596||full Not-determined
17-106-final-viral-combined_k141_63519||full Not-determined
17-106-final-viral-combined_k141_36222||full Low-quality
17-106-final-viral-combined_k141_94703||full Low-quality
17-106-final-viral-combined_k141_63538||full Not-determined
17-106-final-viral-combined_k141_110381||full Low-quality
17-106-final-viral-combined_k141_126713||full Not-determined
17-106-final-viral-combined_k141_126714||full Low-quality
17-106-final-viral-combined_k141_79232||full Low-quality
17-106-final-viral-combined_k141_90185||full Not-determined
17-106-final-viral-combined_k141_116329||full Low-quality
17-106-final-viral-combined_k141_100323||full Low-quality
17-106-final-viral-combined_k141_94730||full Not-determined
17-106-final-viral-combined_k141_90196||full Low-quality
17-106-final-viral-combined_k141_30587||full Low-quality
17-106-final-viral-combined_k141_63565||full Low-quality
18-106-final-viral-combined_k141_135223||full Medium-quality
17-106-final-viral-combined_k141_79262||full Not-determined
17-106-final-viral-combined_k141_46680||full Not-determined
17-106-final-viral-combined_k141_105507||full Not-determined
17-106-final-viral-combined_k141_30608||full Not-determined
17-106-final-viral-combined_k141_46690||full Low-quality
17-106-final-viral-combined_k141_122305||full Not-determined
18-105-final-viral-combined_k141_78434||full Medium-quality
17-106-final-viral-combined_k141_105521||full Low-quality
17-106-final-viral-combined_k141_52892||full Low-quality
17-106-final-viral-combined_k141_4589||full Low-quality
17-106-final-viral-combined_k141_100368||full Not-determined
17-106-final-viral-combined_k141_20512||full Low-quality
17-106-final-viral-combined_k141_52901||full Not-determined
17-106-final-viral-combined_k141_46702||full Low-quality
17-106-final-viral-combined_k141_4592||full Not-determined
17-106-final-viral-combined_k141_20525||full Low-quality
17-106-final-viral-combined_k141_79303||full Low-quality
17-106-final-viral-combined_k141_105543||full Low-quality
17-106-final-viral-combined_k141_52911||full Not-determined
17-106-final-viral-combined_k141_68636||full Not-determined
17-106-final-viral-combined_k141_94795||full Not-determined
17-106-final-viral-combined_k141_41476||full Low-quality
17-106-final-viral-combined_k141_68652||full Not-determined
17-106-final-viral-combined_k141_94800||full Not-determined
17-106-final-viral-combined_k141_105561||full Not-determined
17-106-final-viral-combined_k141_90270||full Low-quality
17-106-final-viral-combined_k141_36269||full Not-determined
17-106-final-viral-combined_k141_41506||full Not-determined
17-106-final-viral-combined_k141_36274||full Low-quality
17-106-final-viral-combined_k141_41507||full Not-determined
17-106-final-viral-combined_k141_122348||full Not-determined
17-106-final-viral-combined_k141_36276||full Low-quality
17-106-final-viral-combined_k141_20582||full Low-quality
17-106-final-viral-combined_k141_122359||full Low-quality
17-106-final-viral-combined_k141_30636||full Not-determined
17-106-final-viral-combined_k141_116398||full Not-determined
17-106-final-viral-combined_k141_36299||full Not-determined
17-106-final-viral-combined_k141_68700||full Low-quality
17-106-final-viral-combined_k141_73451||full Low-quality
17-106-final-viral-combined_k141_63666||full Not-determined
17-106-final-viral-combined_k141_58080||full Not-determined
17-106-final-viral-combined_k141_105623||full Low-quality
17-106-final-viral-combined_k141_30666||full Low-quality
17-106-final-viral-combined_k141_20597||full Low-quality
17-106-final-viral-combined_k141_30672||full Not-determined
17-106-final-viral-combined_k141_90325||full Low-quality
17-106-final-viral-combined_k141_20602||full Not-determined
17-106-final-viral-combined_k141_90327||full Not-determined
17-106-final-viral-combined_k141_58105||full Low-quality
17-106-final-viral-combined_k141_122429||full Low-quality
17-106-final-viral-combined_k141_20618||full Not-determined
17-106-final-viral-combined_k141_79414||full Low-quality
17-106-final-viral-combined_k141_58120||full Not-determined
17-106-final-viral-combined_k141_94849||full Low-quality
17-106-final-viral-combined_k141_58125||full Not-determined
17-106-final-viral-combined_k141_20634||full Low-quality
17-106-final-viral-combined_k141_126742||full Low-quality
17-106-final-viral-combined_k141_52999||full Not-determined
17-106-final-viral-combined_k141_122453||full Low-quality
17-106-final-viral-combined_k141_15058||full Not-determined
17-106-final-viral-combined_k141_126758||full Not-determined
17-106-final-viral-combined_k141_100477||full Low-quality
17-106-final-viral-combined_k141_126764||full Low-quality
17-106-final-viral-combined_k141_79459||full Low-quality
17-106-final-viral-combined_k141_94893||full Low-quality
17-106-final-viral-combined_k141_58167||full Low-quality
17-106-final-viral-combined_k141_15065||full Not-determined
17-106-final-viral-combined_k141_126769||full Not-determined
17-106-final-viral-combined_k141_90372||full Not-determined
17-106-final-viral-combined_k141_15080||full Low-quality
17-106-final-viral-combined_k141_36373||full Not-determined
17-106-final-viral-combined_k141_84979||full Not-determined
17-106-final-viral-combined_k141_100514||full Not-determined
17-106-final-viral-combined_k141_122479||full Not-determined
17-106-final-viral-combined_k141_94911||full Low-quality
17-106-final-viral-combined_k141_63772||full Low-quality
17-106-final-viral-combined_k141_36387||full Low-quality
17-106-final-viral-combined_k141_94913||full Low-quality
17-106-final-viral-combined_k141_122484||full Not-determined
17-106-final-viral-combined_k141_4687||full Not-determined
17-106-final-viral-combined_k141_30743||full Not-determined
17-106-final-viral-combined_k141_110439||full Not-determined
17-106-final-viral-combined_k141_73514||full Not-determined
17-106-final-viral-combined_k141_122506||full Not-determined
17-106-final-viral-combined_k141_90415||full Low-quality
17-106-final-viral-combined_k141_85024||full Not-determined
17-106-final-viral-combined_k141_68850||full Not-determined
17-106-final-viral-combined_k141_79515||full Not-determined
17-106-final-viral-combined_k141_122543||full Not-determined
17-106-final-viral-combined_k141_10070||full Not-determined
17-106-final-viral-combined_k141_126804||full Low-quality
17-106-final-viral-combined_k141_68881||full Not-determined
17-106-final-viral-combined_k141_63829||full Not-determined
17-106-final-viral-combined_k141_53079||full Low-quality
17-106-final-viral-combined_k141_68888||full Not-determined
17-106-final-viral-combined_k141_90452||full Not-determined
17-106-final-viral-combined_k141_15132||full Low-quality
17-106-final-viral-combined_k141_110446||full Low-quality
17-106-final-viral-combined_k141_85059||full Not-determined
17-106-final-viral-combined_k141_116440||full Low-quality
17-106-final-viral-combined_k141_68905||full Not-determined
17-106-final-viral-combined_k141_79532||full Not-determined
17-106-final-viral-combined_k141_110452||full Low-quality
17-106-final-viral-combined_k141_79547||full Low-quality
17-106-final-viral-combined_k141_110458||full Not-determined
17-106-final-viral-combined_k141_73568||full Low-quality
17-106-final-viral-combined_k141_53119||full Not-determined
17-106-final-viral-combined_k141_126819||full Low-quality
17-106-final-viral-combined_k141_94997||full Not-determined
17-106-final-viral-combined_k141_90479||full Not-determined
18-102-final-viral-combined_k141_62839||full High-quality
17-106-final-viral-combined_k141_4767||full Low-quality
17-106-final-viral-combined_k141_53137||full Not-determined
17-106-final-viral-combined_k141_95014||full Not-determined
17-106-final-viral-combined_k141_95017||full Low-quality
17-106-final-viral-combined_k141_10141||full Not-determined
17-106-final-viral-combined_k141_46771||full Not-determined
17-106-final-viral-combined_k141_100615||full Not-determined
17-106-final-viral-combined_k141_100623||full Low-quality
17-106-final-viral-combined_k141_85102||full Low-quality
17-106-final-viral-combined_k141_41563||full Low-quality
17-106-final-viral-combined_k141_36470||full Low-quality
17-106-final-viral-combined_k141_53159||full Not-determined
17-106-final-viral-combined_k141_85111||full Low-quality
17-106-final-viral-combined_k141_63904||full Not-determined
17-106-final-viral-combined_k141_36480||full Low-quality
17-106-final-viral-combined_k141_79607||full Not-determined
17-106-final-viral-combined_k141_46804||full Not-determined
17-106-final-viral-combined_k141_26095||full Low-quality
17-106-final-viral-combined_k141_26100||full Low-quality
17-106-final-viral-combined_k141_10161||full Low-quality
17-106-final-viral-combined_k141_36526||full Not-determined
17-106-final-viral-combined_k141_122612||full Not-determined
17-106-final-viral-combined_k141_26126||full Low-quality
17-106-final-viral-combined_k141_20809||full Low-quality
17-106-final-viral-combined_k141_100710||full Not-determined
17-106-final-viral-combined_k141_79640||full Low-quality
17-106-final-viral-combined_k141_41615||full Not-determined
17-106-final-viral-combined_k141_116492||full Not-determined
17-106-final-viral-combined_k141_90591||full Not-determined
17-106-final-viral-combined_k141_122632||full Low-quality
17-106-final-viral-combined_k141_85168||full Low-quality
17-106-final-viral-combined_k141_41644||full Not-determined
17-106-final-viral-combined_k141_85174||full Not-determined
17-106-final-viral-combined_k141_4834||full Not-determined
17-106-final-viral-combined_k141_85188||full Low-quality
17-106-final-viral-combined_k141_110571||full Not-determined
17-106-final-viral-combined_k141_85194||full Low-quality
17-106-final-viral-combined_k141_26173||full Not-determined
17-106-final-viral-combined_k141_15230||full Not-determined
17-106-final-viral-combined_k141_10213||full Not-determined
17-106-final-viral-combined_k141_69051||full Low-quality
17-106-final-viral-combined_k141_53178||full Not-determined
17-106-final-viral-combined_k141_95072||full Not-determined
17-106-final-viral-combined_k141_4872||full Not-determined
17-106-final-viral-combined_k141_79726||full Low-quality
17-106-final-viral-combined_k141_46865||full Low-quality
17-106-final-viral-combined_k141_15267||full Low-quality
17-106-final-viral-combined_k141_79735||full Low-quality
17-106-final-viral-combined_k141_10250||full Low-quality
17-106-final-viral-combined_k141_30842||full Low-quality
17-106-final-viral-combined_k141_85217||full Low-quality
17-106-final-viral-combined_k141_90681||full Not-determined
17-106-final-viral-combined_k141_30868||full Low-quality
17-106-final-viral-combined_k141_90695||full Low-quality
17-106-final-viral-combined_k141_53217||full Not-determined
17-106-final-viral-combined_k141_100772||full Low-quality
17-106-final-viral-combined_k141_63990||full Low-quality
17-106-final-viral-combined_k141_116609||full Not-determined
17-106-final-viral-combined_k141_100783||full Not-determined
17-106-final-viral-combined_k141_100784||full Not-determined
17-106-final-viral-combined_k141_69102||full Not-determined
17-106-final-viral-combined_k141_64007||full Low-quality
17-106-final-viral-combined_k141_69103||full Not-determined
17-106-final-viral-combined_k141_73694||full Low-quality
17-106-final-viral-combined_k141_64016||full Low-quality
17-106-final-viral-combined_k141_64032||full Low-quality
17-106-final-viral-combined_k141_36694||full Not-determined
17-106-final-viral-combined_k141_100813||full Low-quality
17-106-final-viral-combined_k141_126981||full Not-determined
17-106-final-viral-combined_k141_53253||full Low-quality
17-106-final-viral-combined_k141_41747||full Low-quality
17-106-final-viral-combined_k141_20912||full Low-quality
17-106-final-viral-combined_k141_126987||full Low-quality
17-106-final-viral-combined_k141_100825||full Low-quality
17-106-final-viral-combined_k141_85306||full Low-quality
17-106-final-viral-combined_k141_73721||full Low-quality
17-106-final-viral-combined_k141_46890||full Low-quality
17-106-final-viral-combined_k141_73731||full Low-quality
17-106-final-viral-combined_k141_85311||full Not-determined
17-106-final-viral-combined_k141_41787||full Low-quality
17-106-final-viral-combined_k141_26272||full Low-quality
17-106-final-viral-combined_k141_4936||full Not-determined
17-106-final-viral-combined_k141_36747||full Not-determined
17-106-final-viral-combined_k141_116671||full Low-quality
17-106-final-viral-combined_k141_30932||full Low-quality
17-106-final-viral-combined_k141_116677||full Low-quality
17-106-final-viral-combined_k141_53295||full Not-determined
17-106-final-viral-combined_k141_85328||full Not-determined
17-106-final-viral-combined_k141_116694||full Not-determined
17-106-final-viral-combined_k141_36764||full Low-quality
17-106-final-viral-combined_k141_15396||full Low-quality
17-106-final-viral-combined_k141_41802||full Not-determined
17-106-final-viral-combined_k141_26287||full Not-determined
17-106-final-viral-combined_k141_26288||full Low-quality
17-106-final-viral-combined_k141_90790||full Low-quality
17-106-final-viral-combined_k141_30971||full Low-quality
17-106-final-viral-combined_k141_26296||full Low-quality
17-106-final-viral-combined_k141_41818||full Not-determined
17-106-final-viral-combined_k141_36798||full Low-quality
17-106-final-viral-combined_k141_90808||full Not-determined
17-106-final-viral-combined_k141_30979||full Not-determined
17-106-final-viral-combined_k141_73784||full Not-determined
17-106-final-viral-combined_k141_90818||full Not-determined
17-106-final-viral-combined_k141_36810||full Low-quality
17-106-final-viral-combined_k141_69175||full Not-determined
17-106-final-viral-combined_k141_90828||full Low-quality
17-106-final-viral-combined_k141_90833||full Low-quality
17-106-final-viral-combined_k141_64149||full Low-quality
17-106-final-viral-combined_k141_73804||full Not-determined
17-106-final-viral-combined_k141_105817||full Low-quality
17-106-final-viral-combined_k141_58366||full Not-determined
17-106-final-viral-combined_k141_90858||full Not-determined
17-106-final-viral-combined_k141_36873||full Not-determined
17-106-final-viral-combined_k141_73816||full Low-quality
17-106-final-viral-combined_k141_85431||full Not-determined
17-106-final-viral-combined_k141_36904||full Low-quality
17-106-final-viral-combined_k141_31057||full Not-determined
17-106-final-viral-combined_k141_116758||full Not-determined
17-106-final-viral-combined_k141_85438||full Not-determined
17-106-final-viral-combined_k141_116765||full Not-determined
17-106-final-viral-combined_k141_41907||full Not-determined
17-106-final-viral-combined_k141_36940||full Low-quality
17-106-final-viral-combined_k141_36944||full Low-quality
18-102-final-viral-combined_k141_5791||full Medium-quality
17-106-final-viral-combined_k141_85456||full Low-quality
17-106-final-viral-combined_k141_41930||full Low-quality
17-106-final-viral-combined_k141_116787||full Not-determined
17-106-final-viral-combined_k141_90897||full Not-determined
17-106-final-viral-combined_k141_101000||full Low-quality
17-106-final-viral-combined_k141_15463||full Not-determined
17-106-final-viral-combined_k141_53378||full Low-quality
17-106-final-viral-combined_k141_69210||full Not-determined
17-106-final-viral-combined_k141_53384||full Low-quality
17-106-final-viral-combined_k141_85489||full Low-quality
17-106-final-viral-combined_k141_5086||full Not-determined
17-106-final-viral-combined_k141_105840||full Low-quality
17-106-final-viral-combined_k141_15499||full Not-determined
17-106-final-viral-combined_k141_105854||full Not-determined
17-106-final-viral-combined_k141_10324||full Low-quality
17-106-final-viral-combined_k141_53426||full Not-determined
17-106-final-viral-combined_k141_58435||full Low-quality
17-106-final-viral-combined_k141_15532||full Low-quality
17-106-final-viral-combined_k141_79889||full Not-determined
17-106-final-viral-combined_k141_31066||full Not-determined
17-106-final-viral-combined_k141_37040||full Low-quality
17-106-final-viral-combined_k141_79931||full Low-quality
17-106-final-viral-combined_k141_58458||full Not-determined
17-106-final-viral-combined_k141_105901||full Not-determined
18-102-final-viral-combined_k141_181391||full Medium-quality
17-106-final-viral-combined_k141_73918||full Low-quality
17-106-final-viral-combined_k141_5120||full Low-quality
17-106-final-viral-combined_k141_46927||full Low-quality
17-106-final-viral-combined_k141_101109||full Not-determined
17-106-final-viral-combined_k141_122746||full Not-determined
17-106-final-viral-combined_k141_122747||full Not-determined
17-106-final-viral-combined_k141_122749||full Not-determined
17-106-final-viral-combined_k141_101111||full Not-determined
17-106-final-viral-combined_k141_10362||full Not-determined
17-106-final-viral-combined_k141_69305||full Low-quality
17-106-final-viral-combined_k141_26458||full Not-determined
17-106-final-viral-combined_k141_15624||full Not-determined
17-106-final-viral-combined_k141_15629||full Not-determined
17-106-final-viral-combined_k141_21093||full Low-quality
17-106-final-viral-combined_k141_58515||full Not-determined
17-106-final-viral-combined_k141_105955||full Low-quality
17-106-final-viral-combined_k141_37113||full Not-determined
17-106-final-viral-combined_k141_101186||full Low-quality
17-106-final-viral-combined_k141_73968||full Low-quality
17-106-final-viral-combined_k141_37121||full Not-determined
17-106-final-viral-combined_k141_37126||full Not-determined
17-106-final-viral-combined_k141_58532||full Not-determined
17-106-final-viral-combined_k141_26533||full Not-determined
17-106-final-viral-combined_k141_101207||full Not-determined
17-106-final-viral-combined_k141_37141||full Not-determined
17-106-final-viral-combined_k141_47013||full Low-quality
17-106-final-viral-combined_k141_26559||full Not-determined
17-106-final-viral-combined_k141_47018||full Not-determined
17-106-final-viral-combined_k141_15695||full Low-quality
17-106-final-viral-combined_k141_37161||full Low-quality
17-106-final-viral-combined_k141_37162||full Not-determined
17-106-final-viral-combined_k141_10446||full Low-quality
17-106-final-viral-combined_k141_26573||full Low-quality
17-106-final-viral-combined_k141_69382||full Not-determined
17-106-final-viral-combined_k141_21126||full Low-quality
17-106-final-viral-combined_k141_42037||full Low-quality
17-106-final-viral-combined_k141_21140||full Low-quality
17-106-final-viral-combined_k141_26592||full Not-determined
17-106-final-viral-combined_k141_127086||full Not-determined
17-106-final-viral-combined_k141_47048||full Not-determined
17-106-final-viral-combined_k141_5130||full Not-determined
17-106-final-viral-combined_k141_58568||full Low-quality
17-106-final-viral-combined_k141_26615||full Not-determined
17-106-final-viral-combined_k141_127104||full Low-quality
17-106-final-viral-combined_k141_10451||full Low-quality
17-106-final-viral-combined_k141_95312||full Low-quality
17-106-final-viral-combined_k141_15729||full Not-determined
17-106-final-viral-combined_k141_26632||full Low-quality
17-106-final-viral-combined_k141_74040||full Not-determined
17-106-final-viral-combined_k141_69416||full Low-quality
17-106-final-viral-combined_k141_21177||full Low-quality
17-106-final-viral-combined_k141_10460||full Not-determined
17-106-final-viral-combined_k141_74060||full Low-quality
17-106-final-viral-combined_k141_26643||full Low-quality
17-106-final-viral-combined_k141_58581||full Low-quality
17-106-final-viral-combined_k141_74063||full Not-determined
18-102-final-viral-combined_k141_183036||full High-quality
17-106-final-viral-combined_k141_21192||full Low-quality
17-106-final-viral-combined_k141_58588||full Low-quality
17-106-final-viral-combined_k141_21198||full Not-determined
17-106-final-viral-combined_k141_15776||full Not-determined
17-106-final-viral-combined_k141_101251||full Not-determined
17-106-final-viral-combined_k141_26652||full Not-determined
17-106-final-viral-combined_k141_26654||full Not-determined
17-106-final-viral-combined_k141_26656||full Not-determined
17-106-final-viral-combined_k141_37196||full Low-quality
17-106-final-viral-combined_k141_101270||full Not-determined
17-106-final-viral-combined_k141_26666||full Not-determined
17-106-final-viral-combined_k141_15801||full Low-quality
17-106-final-viral-combined_k141_58608||full Not-determined
17-106-final-viral-combined_k141_47096||full Not-determined
17-106-final-viral-combined_k141_80036||full Not-determined
17-106-final-viral-combined_k141_95399||full Not-determined
17-106-final-viral-combined_k141_58621||full Low-quality
17-106-final-viral-combined_k141_74121||full Not-determined
17-106-final-viral-combined_k141_37250||full Low-quality
17-106-final-viral-combined_k141_37252||full Low-quality
17-106-final-viral-combined_k141_10518||full Not-determined
17-106-final-viral-combined_k141_37253||full Low-quality
17-106-final-viral-combined_k141_37257||full Not-determined
17-106-final-viral-combined_k141_15849||full Not-determined
17-106-final-viral-combined_k141_42162||full Not-determined
17-106-final-viral-combined_k141_21289||full Low-quality
17-106-final-viral-combined_k141_37289||full Not-determined
17-106-final-viral-combined_k141_31296||full Not-determined
17-106-final-viral-combined_k141_21301||full Not-determined
17-106-final-viral-combined_k141_122950||full Not-determined
17-106-final-viral-combined_k141_101376||full Not-determined
17-106-final-viral-combined_k141_26687||full Low-quality
17-106-final-viral-combined_k141_74175||full Not-determined
17-106-final-viral-combined_k141_21310||full Not-determined
17-106-final-viral-combined_k141_15894||full Low-quality
17-106-final-viral-combined_k141_42180||full Not-determined
17-106-final-viral-combined_k141_122966||full Low-quality
17-106-final-viral-combined_k141_80110||full Not-determined
17-106-final-viral-combined_k141_116905||full Not-determined
17-106-final-viral-combined_k141_58694||full Low-quality
17-106-final-viral-combined_k141_74179||full Low-quality
17-106-final-viral-combined_k141_42202||full Not-determined
17-106-final-viral-combined_k141_101405||full Low-quality
17-106-final-viral-combined_k141_116938||full Not-determined
17-106-final-viral-combined_k141_10606||full Low-quality
17-106-final-viral-combined_k141_74202||full Low-quality
17-106-final-viral-combined_k141_42214||full Not-determined
17-106-final-viral-combined_k141_58727||full Not-determined
17-106-final-viral-combined_k141_74227||full Not-determined
17-106-final-viral-combined_k141_58747||full Not-determined
17-106-final-viral-combined_k141_127148||full Low-quality
17-106-final-viral-combined_k141_95436||full Low-quality
17-106-final-viral-combined_k141_47111||full Not-determined
17-106-final-viral-combined_k141_47115||full Not-determined
17-106-final-viral-combined_k141_58759||full Low-quality
17-106-final-viral-combined_k141_80158||full Low-quality
17-106-final-viral-combined_k141_47119||full Not-determined
17-106-final-viral-combined_k141_127159||full Not-determined
17-106-final-viral-combined_k141_37400||full Low-quality
17-106-final-viral-combined_k141_15970||full Low-quality
17-106-final-viral-combined_k141_80179||full Low-quality
17-106-final-viral-combined_k141_106135||full Not-determined
17-106-final-viral-combined_k141_42267||full Not-determined
17-106-final-viral-combined_k141_42268||full Low-quality
17-106-final-viral-combined_k141_58788||full Low-quality
17-106-final-viral-combined_k141_31418||full Low-quality
17-106-final-viral-combined_k141_31425||full Not-determined
17-106-final-viral-combined_k141_101441||full Not-determined
17-106-final-viral-combined_k141_58804||full Low-quality
17-106-final-viral-combined_k141_31440||full Not-determined
17-106-final-viral-combined_k141_5150||full Low-quality
17-106-final-viral-combined_k141_127211||full Low-quality
17-106-final-viral-combined_k141_117052||full Not-determined
17-106-final-viral-combined_k141_5164||full Low-quality
17-106-final-viral-combined_k141_127222||full Low-quality
17-106-final-viral-combined_k141_95550||full Not-determined
17-106-final-viral-combined_k141_16017||full Not-determined
17-106-final-viral-combined_k141_5216||full Not-determined
17-106-final-viral-combined_k141_117093||full Not-determined
17-106-final-viral-combined_k141_95563||full Low-quality
17-106-final-viral-combined_k141_5223||full Low-quality
17-106-final-viral-combined_k141_101504||full Low-quality
17-106-final-viral-combined_k141_117094||full Not-determined
17-106-final-viral-combined_k141_5228||full Not-determined
17-106-final-viral-combined_k141_117119||full Low-quality
17-106-final-viral-combined_k141_117124||full Not-determined
17-109-final-viral-combined_k141_297545||full Medium-quality
17-106-final-viral-combined_k141_74315||full Not-determined
17-106-final-viral-combined_k141_101527||full Not-determined
17-106-final-viral-combined_k141_127271||full Low-quality
17-106-final-viral-combined_k141_106161||full Not-determined
17-106-final-viral-combined_k141_127273||full Not-determined
17-106-final-viral-combined_k141_106166||full Not-determined
17-106-final-viral-combined_k141_106167||full Not-determined
17-106-final-viral-combined_k141_117152||full Not-determined
17-106-final-viral-combined_k141_127281||full Not-determined
17-106-final-viral-combined_k141_101551||full Not-determined
17-106-final-viral-combined_k141_74363||full Low-quality
17-106-final-viral-combined_k141_16024||full Not-determined
17-106-final-viral-combined_k141_31484||full Low-quality
17-106-final-viral-combined_k141_16032||full Not-determined
17-106-final-viral-combined_k141_31489||full Low-quality
17-106-final-viral-combined_k141_47196||full Not-determined
17-106-final-viral-combined_k141_16039||full Low-quality
17-106-final-viral-combined_k141_5294||full Low-quality
17-106-final-viral-combined_k141_42364||full Low-quality
17-106-final-viral-combined_k141_95624||full Not-determined
17-106-final-viral-combined_k141_31520||full Low-quality
17-106-final-viral-combined_k141_106242||full Not-determined
17-106-final-viral-combined_k141_74404||full Low-quality
17-106-final-viral-combined_k141_47218||full Low-quality
17-106-final-viral-combined_k141_117186||full Low-quality
17-106-final-viral-combined_k141_106254||full Low-quality
17-106-final-viral-combined_k141_106258||full Not-determined
17-106-final-viral-combined_k141_101602||full Not-determined
17-106-final-viral-combined_k141_31567||full Low-quality
17-106-final-viral-combined_k141_127319||full Not-determined
17-106-final-viral-combined_k141_106302||full Low-quality
17-106-final-viral-combined_k141_74455||full Low-quality
17-106-final-viral-combined_k141_127345||full Low-quality
17-106-final-viral-combined_k141_74458||full Low-quality
17-106-final-viral-combined_k141_31614||full Not-determined
17-106-final-viral-combined_k141_95700||full Not-determined
17-106-final-viral-combined_k141_42439||full Not-determined
17-106-final-viral-combined_k141_106308||full Not-determined
17-106-final-viral-combined_k141_127364||full Low-quality
17-106-final-viral-combined_k141_42455||full Not-determined
17-106-final-viral-combined_k141_31682||full Not-determined
17-106-final-viral-combined_k141_110827||full Not-determined
17-106-final-viral-combined_k141_42484||full Low-quality
17-106-final-viral-combined_k141_74484||full Not-determined
17-106-final-viral-combined_k141_95738||full Not-determined
17-106-final-viral-combined_k141_95743||full Low-quality
17-106-final-viral-combined_k141_110846||full Not-determined
17-106-final-viral-combined_k141_106379||full Not-determined
17-106-final-viral-combined_k141_95758||full Low-quality
17-106-final-viral-combined_k141_95764||full Low-quality
17-106-final-viral-combined_k141_117244||full Low-quality
17-106-final-viral-combined_k141_106397||full Not-determined
17-106-final-viral-combined_k141_110875||full Not-determined
17-106-final-viral-combined_k141_47311||full Not-determined
17-106-final-viral-combined_k141_74507||full Not-determined
17-106-final-viral-combined_k141_47314||full Low-quality
17-106-final-viral-combined_k141_117275||full Not-determined
17-106-final-viral-combined_k141_42565||full Low-quality
17-106-final-viral-combined_k141_42572||full Not-determined
17-106-final-viral-combined_k141_110908||full Not-determined
17-106-final-viral-combined_k141_47363||full Not-determined
17-106-final-viral-combined_k141_95798||full Not-determined
17-106-final-viral-combined_k141_106469||full Not-determined
17-106-final-viral-combined_k141_74532||full Low-quality
17-106-final-viral-combined_k141_95813||full Low-quality
17-106-final-viral-combined_k141_117317||full Not-determined
17-106-final-viral-combined_k141_95825||full Not-determined
17-106-final-viral-combined_k141_74571||full Low-quality
17-106-final-viral-combined_k141_47413||full Low-quality
17-106-final-viral-combined_k141_106504||full Not-determined
17-106-final-viral-combined_k141_95853||full Low-quality
17-106-final-viral-combined_k141_127435||full Low-quality
17-106-final-viral-combined_k141_95869||full Low-quality
17-106-final-viral-combined_k141_117424||full Not-determined
17-106-final-viral-combined_k141_127484||full Low-quality
17-106-final-viral-combined_k141_110937||full Not-determined
17-106-final-viral-combined_k141_117483||full Low-quality
17-106-final-viral-combined_k141_95959||full Not-determined
17-106-final-viral-combined_k141_47494||full Low-quality
17-106-final-viral-combined_k141_106570||full Not-determined
17-106-final-viral-combined_k141_106576||full Low-quality
17-106-final-viral-combined_k141_42606||full Not-determined
17-106-final-viral-combined_k141_96018||full Not-determined
17-106-final-viral-combined_k141_106607||full Not-determined
17-106-final-viral-combined_k141_106612||full Not-determined
17-106-final-viral-combined_k141_106617||full Not-determined
17-106-final-viral-combined_k141_47508||full Low-quality
17-106-final-viral-combined_k141_111003||full Not-determined
17-106-final-viral-combined_k141_74707||full Not-determined
17-106-final-viral-combined_k141_127537||full Not-determined
17-106-final-viral-combined_k141_127540||full Not-determined
17-106-final-viral-combined_k141_127545||full Not-determined
17-106-final-viral-combined_k141_47545||full Not-determined
17-106-final-viral-combined_k141_42663||full Not-determined
17-106-final-viral-combined_k141_47550||full Not-determined
17-106-final-viral-combined_k141_106662||full Not-determined
17-106-final-viral-combined_k141_117641||full Low-quality
17-106-final-viral-combined_k141_42719||full Not-determined
17-106-final-viral-combined_k141_47584||full Low-quality
17-106-final-viral-combined_k141_47595||full Not-determined
17-106-final-viral-combined_k141_47600||full Low-quality
17-106-final-viral-combined_k141_106687||full Low-quality
17-106-final-viral-combined_k141_47603||full Not-determined
17-106-final-viral-combined_k141_96155||full Low-quality
17-106-final-viral-combined_k141_47606||full Low-quality
17-106-final-viral-combined_k141_31827||full Not-determined
17-106-final-viral-combined_k141_127660||full Not-determined
17-106-final-viral-combined_k141_31834||full Not-determined
17-106-final-viral-combined_k141_42761||full Low-quality
17-106-final-viral-combined_k141_96214||full Low-quality
17-106-final-viral-combined_k141_42766||full Low-quality
17-106-final-viral-combined_k141_31854||full Not-determined
17-106-final-viral-combined_k141_127677||full Low-quality
17-106-final-viral-combined_k141_127682||full Low-quality
17-106-final-viral-combined_k141_47677||full Low-quality

17-106-final-viral-combined_k141_127690||full Low-quality
17-106-final-viral-combined_k141_31862||full Low-quality
17-106-final-viral-combined_k141_106733||full Not-determined
17-106-final-viral-combined_k141_106750||full Low-quality
17-106-final-viral-combined_k141_106751||full Not-determined
17-106-final-viral-combined_k141_106800||full Low-quality
17-106-final-viral-combined_k141_106820||full Low-quality
17-106-final-viral-combined_k141_127771||full Not-determined
17-106-final-viral-combined_k141_111151||full Not-determined
17-106-final-viral-combined_k141_31973||full Low-quality
17-106-final-viral-combined_k141_111167||full Low-quality
17-106-final-viral-combined_k141_74790||full Low-quality
17-106-final-viral-combined_k141_106860||full Low-quality
17-106-final-viral-combined_k141_111171||full Not-determined
17-106-final-viral-combined_k141_127812||full Not-determined
17-106-final-viral-combined_k141_106874||full Low-quality
17-106-final-viral-combined_k141_32025||full Not-determined
17-106-final-viral-combined_k141_32027||full Not-determined
17-106-final-viral-combined_k141_74830||full Not-determined
17-106-final-viral-combined_k141_127871||full Low-quality
17-106-final-viral-combined_k141_127891||full Not-determined
17-106-final-viral-combined_k141_47774||full Not-determined
17-106-final-viral-combined_k141_111201||full Not-determined
17-106-final-viral-combined_k141_127903||full Not-determined
17-106-final-viral-combined_k141_47807||full Not-determined
17-106-final-viral-combined_k141_32086||full Not-determined
17-106-final-viral-combined_k141_127926||full Low-quality
17-106-final-viral-combined_k141_111231||full Low-quality
17-106-final-viral-combined_k141_111243||full Not-determined
17-106-final-viral-combined_k141_111265||full Low-quality
17-106-final-viral-combined_k141_47867||full Not-determined
17-106-final-viral-combined_k141_127971||full Not-determined
17-106-final-viral-combined_k141_47875||full Low-quality
17-106-final-viral-combined_k141_111336||full Not-determined
17-106-final-viral-combined_k141_47905||full Low-quality
17-106-final-viral-combined_k141_47925||full Not-determined
17-106-final-viral-combined_k141_47928||full Not-determined
17-106-final-viral-combined_k141_111379||full Not-determined
17-106-final-viral-combined_k141_47976||full Low-quality
17-106-final-viral-combined_k141_128023||full Low-quality
17-106-final-viral-combined_k141_48030||full Not-determined
17-106-final-viral-combined_k141_111515||full Not-determined
17-106-final-viral-combined_k141_48062||full Low-quality
17-106-final-viral-combined_k141_128101||full Not-determined
17-106-final-viral-combined_k141_128110||full Low-quality
17-106-final-viral-combined_k141_48117||full Low-quality
17-106-final-viral-combined_k141_128198||full Not-determined
17-106-final-viral-combined_k141_128208||full Not-determined
17-106-final-viral-combined_k141_128215||full Low-quality
17-106-final-viral-combined_k141_111764||full Low-quality
17-106-final-viral-combined_k141_111791||full Not-determined
17-106-final-viral-combined_k141_111794||full Low-quality
17-106-final-viral-combined_k141_111873||full Not-determined
17-106-final-viral-combined_k141_111884||full Not-determined
17-106-final-viral-combined_k141_111935||full Not-determined
17-106-final-viral-combined_k141_111982||full Low-quality
17-106-final-viral-combined_k141_112023||full Low-quality
17-106-final-viral-combined_k141_112067||full Not-determined
17-106-final-viral-combined_k141_112081||full Low-quality
17-106-final-viral-combined_k141_112112||full Low-quality
17-106-final-viral-combined_k141_112260||full Low-quality
17-106-final-viral-combined_k141_112277||full Low-quality
17-106-final-viral-combined_k141_48174||0_partial Not-determined
18-105-final-viral-combined_k141_56146||full Medium-quality
17-106-final-viral-combined_k141_43110||0_partial Not-determined
18-106-final-viral-combined_k141_50721||full Medium-quality
17-106-final-viral-combined_k141_16462||0_partial Low-quality
17-106-final-viral-combined_k141_32355||0_partial Low-quality
17-106-final-viral-combined_k141_27201||0_partial Low-quality
17-106-final-viral-combined_k141_5696||0_partial Not-determined
17-106-final-viral-combined_k141_102665||0_partial Low-quality
17-106-final-viral-combined_k141_123844||0_partial Low-quality
17-106-final-viral-combined_k141_65073||0_partial Low-quality
17-106-final-viral-combined_k141_59997||0_partial Not-determined
17-106-final-viral-combined_k141_102994||0_partial Low-quality
18-109-final-viral-combined_k141_169375||full High-quality
17-106-final-viral-combined_k141_54941||0_partial Not-determined
17-106-final-viral-combined_k141_44292||0_partial Not-determined
17-106-final-viral-combined_k141_103478||0_partial Low-quality
18-109-final-viral-combined_k141_167867||full Medium-quality
17-106-final-viral-combined_k141_23386||0_partial Not-determined
18-104-final-viral-combined_k141_121978||full Complete
18-102-final-viral-combined_k141_84608||full Medium-quality
17-106-final-viral-combined_k141_66356||0_partial Not-determined
17-106-final-viral-combined_k141_71523||0_partial Low-quality
17-109-final-viral-combined_k141_151843||full High-quality
17-106-final-viral-combined_k141_93117||0_partial Not-determined
17-106-final-viral-combined_k141_39807||0_partial Low-quality
17-106-final-viral-combined_k141_24172||0_partial Low-quality
17-106-final-viral-combined_k141_88140||0_partial Low-quality
17-106-final-viral-combined_k141_24310||0_partial Low-quality
17-106-final-viral-combined_k141_61898||0_partial Not-determined
KM-final-viral-combined_k141_95360||full Complete
17-106-final-viral-combined_k141_72213||0_partial Low-quality
17-106-final-viral-combined_k141_83924||0_partial Low-quality
17-106-final-viral-combined_k141_45779||0_partial Low-quality
17-106-final-viral-combined_k141_93806||0_partial Low-quality
17-106-final-viral-combined_k141_19806||0_partial Not-determined
17-106-final-viral-combined_k141_57441||0_partial Low-quality
17-106-final-viral-combined_k141_126041||0_partial Not-determined
17-106-final-viral-combined_k141_14106||0_partial Low-quality
17-106-final-viral-combined_k141_109742||0_partial Low-quality
17-106-final-viral-combined_k141_57695||0_partial Low-quality
17-106-final-viral-combined_k141_14742||0_partial Not-determined
17-106-final-viral-combined_k141_9647||0_partial Not-determined
17-106-final-viral-combined_k141_14810||0_partial Not-determined
17-106-final-viral-combined_k141_4518||0_partial Not-determined
17-106-final-viral-combined_k141_58078||0_partial Not-determined
17-106-final-viral-combined_k141_63934||0_partial Not-determined
17-106-final-viral-combined_k141_30805||0_partial Low-quality
17-106-final-viral-combined_k141_10274||0_partial Not-determined
KM-final-viral-combined_k141_248321||full High-quality
17-106-final-viral-combined_k141_42136||0_partial Low-quality
17-106-final-viral-combined_k141_10665||0_partial Not-determined
17-106-final-viral-combined_k141_74649||0_partial Low-quality
17-106-final-viral-combined_k141_96070||0_partial Not-determined
17-106-final-viral-combined_k141_106708||0_partial Not-determined
17-106-final-viral-combined_k141_101611||lt2gene Low-quality
17-106-final-viral-combined_k141_80233||lt2gene Not-determined
17-106-final-viral-combined_k141_37456||lt2gene Not-determined
17-106-final-viral-combined_k141_112342||lt2gene Not-determined
17-106-final-viral-combined_k141_10745||lt2gene Low-quality
17-106-final-viral-combined_k141_112349||lt2gene Not-determined
17-106-final-viral-combined_k141_117713||lt2gene Low-quality
17-106-final-viral-combined_k141_112365||lt2gene Low-quality
17-106-final-viral-combined_k141_58894||lt2gene Not-determined
17-106-final-viral-combined_k141_85665||lt2gene Low-quality
17-106-final-viral-combined_k141_37510||lt2gene Not-determined
17-106-final-viral-combined_k141_21471||lt2gene Low-quality
17-106-final-viral-combined_k141_21506||lt2gene Low-quality
17-106-final-viral-combined_k141_74976||lt2gene Low-quality
17-106-final-viral-combined_k141_64263||lt2gene Not-determined
17-106-final-viral-combined_k141_75023||lt2gene Low-quality
17-106-final-viral-combined_k141_91037||lt2gene Low-quality
17-106-final-viral-combined_k141_75058||lt2gene Low-quality
17-106-final-viral-combined_k141_58926||lt2gene Low-quality
17-106-final-viral-combined_k141_91066||lt2gene Low-quality
17-106-final-viral-combined_k141_10916||lt2gene Low-quality
17-106-final-viral-combined_k141_32234||lt2gene Low-quality
17-106-final-viral-combined_k141_42998||lt2gene Low-quality
17-106-final-viral-combined_k141_96431||lt2gene Low-quality
17-106-final-viral-combined_k141_123156||lt2gene Low-quality
17-106-final-viral-combined_k141_123158||lt2gene Low-quality
17-106-final-viral-combined_k141_80340||lt2gene Low-quality
17-106-final-viral-combined_k141_117805||lt2gene Low-quality
17-106-final-viral-combined_k141_80378||lt2gene Low-quality
17-106-final-viral-combined_k141_321||lt2gene Low-quality
17-106-final-viral-combined_k141_96490||lt2gene Low-quality
17-106-final-viral-combined_k141_75177||lt2gene Low-quality
17-106-final-viral-combined_k141_26998||lt2gene Low-quality
17-106-final-viral-combined_k141_386||lt2gene Low-quality
17-106-final-viral-combined_k141_53636||lt2gene Low-quality
17-106-final-viral-combined_k141_53679||lt2gene Low-quality
17-106-final-viral-combined_k141_419||lt2gene Low-quality
17-106-final-viral-combined_k141_439||lt2gene Low-quality
17-106-final-viral-combined_k141_43133||lt2gene Low-quality
17-106-final-viral-combined_k141_85947||lt2gene Not-determined
17-106-final-viral-combined_k141_85979||lt2gene Low-quality
17-106-final-viral-combined_k141_37636||lt2gene Low-quality
17-106-final-viral-combined_k141_16422||lt2gene Low-quality
17-106-final-viral-combined_k141_75345||lt2gene Not-determined
17-106-final-viral-combined_k141_43150||lt2gene Low-quality
17-106-final-viral-combined_k141_43154||lt2gene Low-quality
17-106-final-viral-combined_k141_5566||lt2gene Low-quality
17-106-final-viral-combined_k141_64456||lt2gene Low-quality
17-106-final-viral-combined_k141_69820||lt2gene Low-quality
17-106-final-viral-combined_k141_69827||lt2gene Low-quality
17-106-final-viral-combined_k141_48631||lt2gene Not-determined
17-106-final-viral-combined_k141_53859||lt2gene Not-determined
17-106-final-viral-combined_k141_107198||lt2gene Low-quality
17-106-final-viral-combined_k141_48698||lt2gene Low-quality
17-106-final-viral-combined_k141_107223||lt2gene Not-determined
17-106-final-viral-combined_k141_37760||lt2gene Low-quality
17-106-final-viral-combined_k141_32414||lt2gene Low-quality
17-106-final-viral-combined_k141_32418||lt2gene Low-quality
17-106-final-viral-combined_k141_118173||lt2gene Low-quality
17-106-final-viral-combined_k141_10986||lt2gene Not-determined
17-106-final-viral-combined_k141_102209||lt2gene Low-quality
17-106-final-viral-combined_k141_96946||lt2gene Low-quality
17-106-final-viral-combined_k141_48781||lt2gene Low-quality
17-106-final-viral-combined_k141_37842||lt2gene Low-quality
17-106-final-viral-combined_k141_16500||lt2gene Low-quality
17-106-final-viral-combined_k141_43323||lt2gene Low-quality
17-106-final-viral-combined_k141_107274||lt2gene Low-quality
17-106-final-viral-combined_k141_43328||lt2gene Low-quality
17-106-final-viral-combined_k141_16524||lt2gene Low-quality
17-106-final-viral-combined_k141_107296||lt2gene Low-quality
17-106-final-viral-combined_k141_59437||lt2gene Low-quality
17-106-final-viral-combined_k141_37891||lt2gene Low-quality
17-106-final-viral-combined_k141_112871||lt2gene Low-quality
17-106-final-viral-combined_k141_59477||lt2gene Low-quality
17-106-final-viral-combined_k141_48827||lt2gene Low-quality
17-106-final-viral-combined_k141_102303||lt2gene Low-quality
17-106-final-viral-combined_k141_80908||lt2gene Low-quality
17-106-final-viral-combined_k141_112921||lt2gene Low-quality
17-106-final-viral-combined_k141_32598||lt2gene Low-quality
17-106-final-viral-combined_k141_864||lt2gene Low-quality
17-106-final-viral-combined_k141_69876||lt2gene Low-quality
17-106-final-viral-combined_k141_37990||lt2gene Low-quality
17-106-final-viral-combined_k141_900||lt2gene Low-quality
17-106-final-viral-combined_k141_102372||lt2gene Low-quality
17-106-final-viral-combined_k141_107393||lt2gene Low-quality
17-106-final-viral-combined_k141_69905||lt2gene Low-quality
17-106-final-viral-combined_k141_32667||lt2gene Low-quality
17-106-final-viral-combined_k141_64693||lt2gene Low-quality
17-106-final-viral-combined_k141_91356||lt2gene Low-quality
17-106-final-viral-combined_k141_102444||lt2gene Low-quality
17-106-final-viral-combined_k141_54050||lt2gene Not-determined
17-106-final-viral-combined_k141_1031||lt2gene Not-determined
17-106-final-viral-combined_k141_123509||lt2gene Not-determined
17-106-final-viral-combined_k141_81099||lt2gene Low-quality
17-106-final-viral-combined_k141_118479||lt2gene Low-quality
17-106-final-viral-combined_k141_38052||lt2gene Low-quality
17-106-final-viral-combined_k141_97200||lt2gene Low-quality
17-106-final-viral-combined_k141_16651||lt2gene Low-quality
17-106-final-viral-combined_k141_16652||lt2gene Low-quality
17-106-final-viral-combined_k141_86415||lt2gene Low-quality
17-106-final-viral-combined_k141_1095||lt2gene Low-quality
17-106-final-viral-combined_k141_102564||lt2gene Low-quality
17-106-final-viral-combined_k141_27444||lt2gene Low-quality
17-106-final-viral-combined_k141_81122||lt2gene Not-determined
17-106-final-viral-combined_k141_38102||lt2gene Not-determined
17-106-final-viral-combined_k141_59722||lt2gene Not-determined
17-106-final-viral-combined_k141_16684||lt2gene Not-determined
17-106-final-viral-combined_k141_91493||lt2gene Low-quality
17-106-final-viral-combined_k141_75373||lt2gene Low-quality
17-106-final-viral-combined_k141_97230||lt2gene Not-determined
17-106-final-viral-combined_k141_70120||lt2gene Low-quality
17-106-final-viral-combined_k141_81238||lt2gene Not-determined
17-106-final-viral-combined_k141_32868||lt2gene Low-quality
17-106-final-viral-combined_k141_54109||lt2gene Not-determined
17-106-final-viral-combined_k141_11259||lt2gene Low-quality
17-106-final-viral-combined_k141_54130||lt2gene Not-determined
17-106-final-viral-combined_k141_32903||lt2gene Not-determined
17-106-final-viral-combined_k141_86507||lt2gene Low-quality
17-106-final-viral-combined_k141_75429||lt2gene Low-quality
17-106-final-viral-combined_k141_59774||lt2gene Not-determined
17-106-final-viral-combined_k141_123687||lt2gene Low-quality
17-106-final-viral-combined_k141_1292||lt2gene Low-quality
17-106-final-viral-combined_k141_49049||lt2gene Not-determined
17-106-final-viral-combined_k141_64848||lt2gene Low-quality
17-106-final-viral-combined_k141_75479||lt2gene Low-quality
17-106-final-viral-combined_k141_22088||lt2gene Low-quality
17-106-final-viral-combined_k141_64869||lt2gene Not-determined
17-106-final-viral-combined_k141_27593||lt2gene Low-quality
17-106-final-viral-combined_k141_86603||lt2gene Low-quality
17-106-final-viral-combined_k141_6090||lt2gene Low-quality
17-106-final-viral-combined_k141_38292||lt2gene Not-determined
17-106-final-viral-combined_k141_64950||lt2gene Low-quality
17-106-final-viral-combined_k141_1368||lt2gene Not-determined
17-106-final-viral-combined_k141_27703||lt2gene Low-quality
17-106-final-viral-combined_k141_22155||lt2gene Not-determined
17-106-final-viral-combined_k141_113204||lt2gene Low-quality
17-106-final-viral-combined_k141_107680||lt2gene Not-determined
17-106-final-viral-combined_k141_11452||lt2gene Low-quality
17-106-final-viral-combined_k141_6233||lt2gene Low-quality
17-106-final-viral-combined_k141_118716||lt2gene Not-determined
17-106-final-viral-combined_k141_49214||lt2gene Low-quality
17-106-final-viral-combined_k141_97417||lt2gene Low-quality
17-106-final-viral-combined_k141_38321||lt2gene Low-quality
17-106-final-viral-combined_k141_38328||lt2gene Not-determined
17-106-final-viral-combined_k141_86729||lt2gene Low-quality
17-106-final-viral-combined_k141_75568||lt2gene Low-quality
17-106-final-viral-combined_k141_54470||lt2gene Low-quality
17-106-final-viral-combined_k141_123876||lt2gene Low-quality
17-106-final-viral-combined_k141_16786||lt2gene Not-determined
17-106-final-viral-combined_k141_118771||lt2gene Low-quality
17-106-final-viral-combined_k141_91846||lt2gene Not-determined
17-106-final-viral-combined_k141_118806||lt2gene Low-quality
17-106-final-viral-combined_k141_81508||lt2gene Low-quality
17-106-final-viral-combined_k141_6398||lt2gene Low-quality
17-106-final-viral-combined_k141_75621||lt2gene Low-quality
17-106-final-viral-combined_k141_65175||lt2gene Low-quality
17-106-final-viral-combined_k141_113241||lt2gene Low-quality
17-106-final-viral-combined_k141_86824||lt2gene Low-quality
17-106-final-viral-combined_k141_118867||lt2gene Low-quality
17-106-final-viral-combined_k141_65195||lt2gene Not-determined
17-106-final-viral-combined_k141_22481||lt2gene Low-quality
17-106-final-viral-combined_k141_113251||lt2gene Low-quality
17-106-final-viral-combined_k141_1647||lt2gene Low-quality
17-106-final-viral-combined_k141_81613||lt2gene Not-determined
17-106-final-viral-combined_k141_75718||lt2gene Low-quality
17-106-final-viral-combined_k141_54673||lt2gene Low-quality
17-106-final-viral-combined_k141_91936||lt2gene Low-quality
17-106-final-viral-combined_k141_91942||lt2gene Low-quality
17-106-final-viral-combined_k141_97563||lt2gene Low-quality
17-106-final-viral-combined_k141_43734||lt2gene Low-quality
17-106-final-viral-combined_k141_97572||lt2gene Not-determined
17-106-final-viral-combined_k141_91979||lt2gene Low-quality
17-106-final-viral-combined_k141_43745||lt2gene Low-quality
17-106-final-viral-combined_k141_65295||lt2gene Low-quality
17-106-final-viral-combined_k141_92041||lt2gene Not-determined
17-106-final-viral-combined_k141_70435||lt2gene Low-quality
17-106-final-viral-combined_k141_75807||lt2gene Low-quality
17-106-final-viral-combined_k141_107930||lt2gene Low-quality
17-106-final-viral-combined_k141_1728||lt2gene Not-determined
17-106-final-viral-combined_k141_17010||lt2gene Not-determined
17-106-final-viral-combined_k141_113432||lt2gene Low-quality
17-106-final-viral-combined_k141_70526||lt2gene Low-quality
17-106-final-viral-combined_k141_65372||lt2gene Low-quality
17-106-final-viral-combined_k141_54839||lt2gene Low-quality
17-106-final-viral-combined_k141_119047||lt2gene Low-quality
17-106-final-viral-combined_k141_108038||lt2gene Low-quality
17-106-final-viral-combined_k141_108039||lt2gene Low-quality
17-106-final-viral-combined_k141_92151||lt2gene Low-quality
17-106-final-viral-combined_k141_92172||lt2gene Low-quality
17-106-final-viral-combined_k141_65444||lt2gene Low-quality
17-106-final-viral-combined_k141_65490||lt2gene Low-quality
17-106-final-viral-combined_k141_38555||lt2gene Not-determined
17-106-final-viral-combined_k141_97747||lt2gene Low-quality
17-106-final-viral-combined_k141_92261||lt2gene Not-determined
17-106-final-viral-combined_k141_70652||lt2gene Low-quality
17-106-final-viral-combined_k141_108143||lt2gene Low-quality
17-106-final-viral-combined_k141_70665||lt2gene Low-quality
17-106-final-viral-combined_k141_97766||lt2gene Low-quality
17-106-final-viral-combined_k141_60427||lt2gene Low-quality
17-106-final-viral-combined_k141_44041||lt2gene Not-determined
17-106-final-viral-combined_k141_54978||lt2gene Low-quality
17-106-final-viral-combined_k141_92312||lt2gene Low-quality
17-106-final-viral-combined_k141_60447||lt2gene Not-determined
17-106-final-viral-combined_k141_103127||lt2gene Not-determined
17-106-final-viral-combined_k141_103143||lt2gene Not-determined
17-106-final-viral-combined_k141_49619||lt2gene Low-quality
17-106-final-viral-combined_k141_81883||lt2gene Low-quality
17-106-final-viral-combined_k141_6963||lt2gene Low-quality
17-106-final-viral-combined_k141_6967||lt2gene Not-determined
17-106-final-viral-combined_k141_124360||lt2gene Low-quality
17-106-final-viral-combined_k141_6971||lt2gene Low-quality
17-106-final-viral-combined_k141_113672||lt2gene Not-determined
17-106-final-viral-combined_k141_23006||lt2gene Not-determined
17-106-final-viral-combined_k141_23013||lt2gene Not-determined
17-106-final-viral-combined_k141_44117||lt2gene Low-quality
17-106-final-viral-combined_k141_55045||lt2gene Low-quality
17-106-final-viral-combined_k141_17351||lt2gene Low-quality
17-106-final-viral-combined_k141_49702||lt2gene Low-quality
17-106-final-viral-combined_k141_108200||lt2gene Not-determined
17-106-final-viral-combined_k141_49733||lt2gene Not-determined
17-106-final-viral-combined_k141_103318||lt2gene Not-determined
17-106-final-viral-combined_k141_28318||lt2gene Low-quality
17-106-final-viral-combined_k141_75944||lt2gene Not-determined
17-106-final-viral-combined_k141_38741||lt2gene Low-quality
17-106-final-viral-combined_k141_17421||lt2gene Low-quality
17-106-final-viral-combined_k141_87166||lt2gene Low-quality
17-106-final-viral-combined_k141_119087||lt2gene Low-quality
17-106-final-viral-combined_k141_33370||lt2gene Low-quality
17-106-final-viral-combined_k141_11894||lt2gene Low-quality
17-106-final-viral-combined_k141_1963||lt2gene Low-quality
17-106-final-viral-combined_k141_65778||lt2gene Not-determined
17-106-final-viral-combined_k141_76002||lt2gene Low-quality
17-106-final-viral-combined_k141_44285||lt2gene Low-quality
17-106-final-viral-combined_k141_17465||lt2gene Low-quality
17-106-final-viral-combined_k141_17474||lt2gene Low-quality
17-106-final-viral-combined_k141_23068||lt2gene Low-quality
17-106-final-viral-combined_k141_65861||lt2gene Low-quality
17-106-final-viral-combined_k141_11949||lt2gene Not-determined
17-106-final-viral-combined_k141_55218||lt2gene Low-quality
17-106-final-viral-combined_k141_124522||lt2gene Low-quality
17-106-final-viral-combined_k141_60615||lt2gene Low-quality
17-106-final-viral-combined_k141_60632||lt2gene Low-quality
17-106-final-viral-combined_k141_11972||lt2gene Low-quality
17-106-final-viral-combined_k141_60649||lt2gene Low-quality
17-106-final-viral-combined_k141_119171||lt2gene Low-quality
17-106-final-viral-combined_k141_11980||lt2gene Low-quality
17-106-final-viral-combined_k141_7158||lt2gene Not-determined
17-106-final-viral-combined_k141_92504||lt2gene Low-quality
17-106-final-viral-combined_k141_97874||lt2gene Low-quality
17-106-final-viral-combined_k141_49895||lt2gene Low-quality
17-106-final-viral-combined_k141_33422||lt2gene Not-determined
17-106-final-viral-combined_k141_70998||lt2gene Low-quality
17-106-final-viral-combined_k141_76111||lt2gene Not-determined
17-106-final-viral-combined_k141_33446||lt2gene Low-quality
17-106-final-viral-combined_k141_103332||lt2gene Low-quality
17-106-final-viral-combined_k141_12013||lt2gene Low-quality
17-106-final-viral-combined_k141_17643||lt2gene Low-quality
17-106-final-viral-combined_k141_49983||lt2gene Low-quality
17-106-final-viral-combined_k141_17661||lt2gene Low-quality
17-106-final-viral-combined_k141_55403||lt2gene Low-quality
17-106-final-viral-combined_k141_12063||lt2gene Low-quality
17-106-final-viral-combined_k141_17683||lt2gene Low-quality
17-106-final-viral-combined_k141_23272||lt2gene Low-quality
17-106-final-viral-combined_k141_60766||lt2gene Low-quality
17-106-final-viral-combined_k141_66084||lt2gene Low-quality
17-106-final-viral-combined_k141_50086||lt2gene Not-determined
17-106-final-viral-combined_k141_17757||lt2gene Low-quality
17-106-final-viral-combined_k141_38999||lt2gene Low-quality
17-106-final-viral-combined_k141_44379||lt2gene Low-quality
17-106-final-viral-combined_k141_71066||lt2gene Low-quality
17-106-final-viral-combined_k141_50131||lt2gene Low-quality
17-106-final-viral-combined_k141_50145||lt2gene Low-quality
17-106-final-viral-combined_k141_28465||lt2gene Low-quality
17-106-final-viral-combined_k141_28495||lt2gene Low-quality
17-106-final-viral-combined_k141_87371||lt2gene Not-determined
17-106-final-viral-combined_k141_50230||lt2gene Low-quality
17-106-final-viral-combined_k141_87384||lt2gene Low-quality
17-106-final-viral-combined_k141_119398||lt2gene Low-quality
17-106-final-viral-combined_k141_108464||lt2gene Low-quality
17-106-final-viral-combined_k141_66218||lt2gene Low-quality
17-106-final-viral-combined_k141_2371||lt2gene Low-quality
17-106-final-viral-combined_k141_87404||lt2gene Low-quality
17-106-final-viral-combined_k141_66246||lt2gene Not-determined
17-106-final-viral-combined_k141_28565||lt2gene Low-quality
17-106-final-viral-combined_k141_50295||lt2gene Low-quality
17-106-final-viral-combined_k141_98105||lt2gene Low-quality
17-106-final-viral-combined_k141_60901||lt2gene Low-quality
17-106-final-viral-combined_k141_39143||lt2gene Not-determined
17-106-final-viral-combined_k141_113763||lt2gene Low-quality
17-106-final-viral-combined_k141_12325||lt2gene Low-quality
17-106-final-viral-combined_k141_17897||lt2gene Not-determined
17-106-final-viral-combined_k141_103597||lt2gene Not-determined
17-106-final-viral-combined_k141_33556||lt2gene Low-quality
17-106-final-viral-combined_k141_124690||lt2gene Low-quality
17-106-final-viral-combined_k141_82348||lt2gene Low-quality
17-106-final-viral-combined_k141_98172||lt2gene Low-quality
17-106-final-viral-combined_k141_92864||lt2gene Low-quality
17-106-final-viral-combined_k141_71286||lt2gene Low-quality
17-106-final-viral-combined_k141_119514||lt2gene Low-quality
17-106-final-viral-combined_k141_98193||lt2gene Low-quality
17-106-final-viral-combined_k141_124782||lt2gene Low-quality
17-106-final-viral-combined_k141_12464||lt2gene Low-quality
17-106-final-viral-combined_k141_7587||lt2gene Low-quality
17-106-final-viral-combined_k141_18031||lt2gene Low-quality
17-106-final-viral-combined_k141_82458||lt2gene Low-quality
17-106-final-viral-combined_k141_113874||lt2gene Low-quality
17-106-final-viral-combined_k141_12525||lt2gene Low-quality
17-106-final-viral-combined_k141_50437||lt2gene Low-quality
17-106-final-viral-combined_k141_113917||lt2gene Low-quality
17-106-final-viral-combined_k141_103726||lt2gene Not-determined
17-106-final-viral-combined_k141_108630||lt2gene Low-quality
17-106-final-viral-combined_k141_103754||lt2gene Low-quality
17-106-final-viral-combined_k141_61134||lt2gene Low-quality
17-106-final-viral-combined_k141_18201||lt2gene Low-quality
17-106-final-viral-combined_k141_18209||lt2gene Low-quality
17-106-final-viral-combined_k141_61221||lt2gene Low-quality
17-106-final-viral-combined_k141_82598||lt2gene Low-quality
17-106-final-viral-combined_k141_50503||lt2gene Low-quality
17-106-final-viral-combined_k141_61251||lt2gene Low-quality
17-106-final-viral-combined_k141_82650||lt2gene Not-determined
17-106-final-viral-combined_k141_87730||lt2gene Not-determined
17-106-final-viral-combined_k141_33766||lt2gene Low-quality
17-106-final-viral-combined_k141_44543||lt2gene Low-quality
17-106-final-viral-combined_k141_18314||lt2gene Low-quality
17-106-final-viral-combined_k141_7819||lt2gene Low-quality
17-106-final-viral-combined_k141_108809||lt2gene Low-quality
17-106-final-viral-combined_k141_50642||lt2gene Low-quality
17-106-final-viral-combined_k141_7856||lt2gene Not-determined
17-106-final-viral-combined_k141_108846||lt2gene Low-quality
17-106-final-viral-combined_k141_50683||lt2gene Low-quality
17-106-final-viral-combined_k141_108852||lt2gene Low-quality
17-106-final-viral-combined_k141_33834||lt2gene Low-quality
17-106-final-viral-combined_k141_76754||lt2gene Low-quality
17-106-final-viral-combined_k141_125026||lt2gene Low-quality
17-106-final-viral-combined_k141_2780||lt2gene Low-quality
17-106-final-viral-combined_k141_23660||lt2gene Low-quality
17-106-final-viral-combined_k141_28879||lt2gene Low-quality
17-106-final-viral-combined_k141_33862||lt2gene Low-quality
17-106-final-viral-combined_k141_55847||lt2gene Low-quality
17-106-final-viral-combined_k141_66492||lt2gene Low-quality
17-106-final-viral-combined_k141_2794||lt2gene Low-quality
17-106-final-viral-combined_k141_2800||lt2gene Not-determined
17-106-final-viral-combined_k141_39650||lt2gene Not-determined
17-106-final-viral-combined_k141_50785||lt2gene Low-quality
17-106-final-viral-combined_k141_66539||lt2gene Not-determined
17-106-final-viral-combined_k141_44643||lt2gene Not-determined
17-106-final-viral-combined_k141_88043||lt2gene Low-quality
17-106-final-viral-combined_k141_83036||lt2gene Low-quality
17-106-final-viral-combined_k141_44666||lt2gene Not-determined
17-106-final-viral-combined_k141_33922||lt2gene Low-quality
17-106-final-viral-combined_k141_2851||lt2gene Not-determined
17-106-final-viral-combined_k141_98327||lt2gene Low-quality
17-106-final-viral-combined_k141_23738||lt2gene Low-quality
17-106-final-viral-combined_k141_119750||lt2gene Not-determined
17-106-final-viral-combined_k141_61344||lt2gene Low-quality
17-106-final-viral-combined_k141_119769||lt2gene Not-determined
17-106-final-viral-combined_k141_76805||lt2gene Low-quality
17-106-final-viral-combined_k141_71679||lt2gene Low-quality
17-106-final-viral-combined_k141_93130||lt2gene Low-quality
17-106-final-viral-combined_k141_104124||lt2gene Low-quality
17-106-final-viral-combined_k141_18520||lt2gene Low-quality
17-106-final-viral-combined_k141_34015||lt2gene Not-determined
17-106-final-viral-combined_k141_2884||lt2gene Low-quality
17-106-final-viral-combined_k141_34038||lt2gene Low-quality
17-106-final-viral-combined_k141_2904||lt2gene Not-determined
17-106-final-viral-combined_k141_2911||lt2gene Low-quality
17-106-final-viral-combined_k141_56140||lt2gene Low-quality
17-106-final-viral-combined_k141_93198||lt2gene Not-determined
17-106-final-viral-combined_k141_76899||lt2gene Low-quality
17-106-final-viral-combined_k141_76901||lt2gene Low-quality
17-106-final-viral-combined_k141_12908||lt2gene Low-quality
17-106-final-viral-combined_k141_61440||lt2gene Low-quality
17-106-final-viral-combined_k141_39792||lt2gene Low-quality
17-106-final-viral-combined_k141_61459||lt2gene Low-quality
17-106-final-viral-combined_k141_12928||lt2gene Low-quality
17-106-final-viral-combined_k141_119925||lt2gene Low-quality
17-106-final-viral-combined_k141_44820||lt2gene Not-determined
17-106-final-viral-combined_k141_44837||lt2gene Low-quality
17-106-final-viral-combined_k141_44849||lt2gene Not-determined
17-106-final-viral-combined_k141_50908||lt2gene Not-determined
17-106-final-viral-combined_k141_13003||lt2gene Not-determined
17-106-final-viral-combined_k141_13018||lt2gene Low-quality
17-106-final-viral-combined_k141_18706||lt2gene Low-quality
17-106-final-viral-combined_k141_76971||lt2gene Not-determined
17-106-final-viral-combined_k141_98465||lt2gene Low-quality
17-106-final-viral-combined_k141_34122||lt2gene Low-quality
17-106-final-viral-combined_k141_83336||lt2gene Low-quality
17-106-final-viral-combined_k141_109001||lt2gene Low-quality
17-106-final-viral-combined_k141_3099||lt2gene Low-quality
17-106-final-viral-combined_k141_13121||lt2gene Low-quality
17-106-final-viral-combined_k141_93384||lt2gene Not-determined
17-106-final-viral-combined_k141_13135||lt2gene Low-quality
17-106-final-viral-combined_k141_29157||lt2gene Low-quality
17-106-final-viral-combined_k141_120084||lt2gene Not-determined
17-106-final-viral-combined_k141_39892||lt2gene Low-quality
17-106-final-viral-combined_k141_24094||lt2gene Low-quality
17-106-final-viral-combined_k141_34241||lt2gene Low-quality
17-106-final-viral-combined_k141_104338||lt2gene Not-determined
17-106-final-viral-combined_k141_120140||lt2gene Low-quality
17-106-final-viral-combined_k141_51033||lt2gene Low-quality
17-106-final-viral-combined_k141_29207||lt2gene Low-quality
17-106-final-viral-combined_k141_77165||lt2gene Not-determined
17-106-final-viral-combined_k141_13248||lt2gene Low-quality
17-106-final-viral-combined_k141_98594||lt2gene Low-quality
17-106-final-viral-combined_k141_18948||lt2gene Low-quality
17-106-final-viral-combined_k141_98610||lt2gene Low-quality
17-106-final-viral-combined_k141_109094||lt2gene Low-quality
17-106-final-viral-combined_k141_45128||lt2gene Low-quality
17-106-final-viral-combined_k141_56369||lt2gene Low-quality
17-106-final-viral-combined_k141_88172||lt2gene Low-quality
17-106-final-viral-combined_k141_51099||lt2gene Low-quality
17-106-final-viral-combined_k141_98662||lt2gene Low-quality
17-106-final-viral-combined_k141_120263||lt2gene Low-quality
17-106-final-viral-combined_k141_125367||lt2gene Not-determined
17-106-final-viral-combined_k141_40042||lt2gene Low-quality
17-106-final-viral-combined_k141_125392||lt2gene Low-quality
17-106-final-viral-combined_k141_13385||lt2gene Low-quality
17-106-final-viral-combined_k141_40065||lt2gene Low-quality
17-106-final-viral-combined_k141_125415||lt2gene Not-determined
17-106-final-viral-combined_k141_13426||lt2gene Low-quality
17-106-final-viral-combined_k141_93518||lt2gene Low-quality
17-106-final-viral-combined_k141_19063||lt2gene Low-quality
17-106-final-viral-combined_k141_120384||lt2gene Low-quality
17-106-final-viral-combined_k141_72073||lt2gene Not-determined
17-106-final-viral-combined_k141_114795||lt2gene Low-quality
17-106-final-viral-combined_k141_19102||lt2gene Not-determined
17-106-final-viral-combined_k141_56533||lt2gene Low-quality
17-106-final-viral-combined_k141_40167||lt2gene Low-quality
17-106-final-viral-combined_k141_72119||lt2gene Low-quality
17-106-final-viral-combined_k141_125467||lt2gene Low-quality
17-106-final-viral-combined_k141_19115||lt2gene Low-quality
17-106-final-viral-combined_k141_98816||lt2gene Low-quality
17-106-final-viral-combined_k141_83672||lt2gene Low-quality
17-106-final-viral-combined_k141_114809||lt2gene Low-quality
17-106-final-viral-combined_k141_114819||lt2gene Not-determined
17-106-final-viral-combined_k141_29389||lt2gene Low-quality
17-106-final-viral-combined_k141_13505||lt2gene Low-quality
17-106-final-viral-combined_k141_120499||lt2gene Low-quality
17-106-final-viral-combined_k141_34667||lt2gene Low-quality
17-106-final-viral-combined_k141_3309||lt2gene Low-quality
17-106-final-viral-combined_k141_98896||lt2gene Low-quality
17-106-final-viral-combined_k141_98901||lt2gene Low-quality
17-106-final-viral-combined_k141_114935||lt2gene Low-quality
17-106-final-viral-combined_k141_114936||lt2gene Low-quality
17-106-final-viral-combined_k141_104449||lt2gene Low-quality
17-106-final-viral-combined_k141_19199||lt2gene Low-quality
17-106-final-viral-combined_k141_93717||lt2gene Low-quality
17-106-final-viral-combined_k141_8665||lt2gene Low-quality
17-106-final-viral-combined_k141_88401||lt2gene Not-determined
17-106-final-viral-combined_k141_51330||lt2gene Low-quality
17-106-final-viral-combined_k141_67039||lt2gene Low-quality
17-106-final-viral-combined_k141_120624||lt2gene Low-quality
17-106-final-viral-combined_k141_109249||lt2gene Not-determined
17-106-final-viral-combined_k141_125637||lt2gene Low-quality
17-106-final-viral-combined_k141_104553||lt2gene Not-determined
17-106-final-viral-combined_k141_24515||lt2gene Low-quality
17-106-final-viral-combined_k141_88516||lt2gene Low-quality
17-106-final-viral-combined_k141_34922||lt2gene Low-quality
17-106-final-viral-combined_k141_19302||lt2gene Low-quality
17-106-final-viral-combined_k141_51467||lt2gene Low-quality
17-106-final-viral-combined_k141_115027||lt2gene Low-quality
17-106-final-viral-combined_k141_8791||lt2gene Low-quality
17-106-final-viral-combined_k141_19321||lt2gene Low-quality
17-106-final-viral-combined_k141_72259||lt2gene Not-determined
17-106-final-viral-combined_k141_62085||lt2gene Not-determined
17-106-final-viral-combined_k141_67165||lt2gene Not-determined
17-106-final-viral-combined_k141_99213||lt2gene Low-quality
17-106-final-viral-combined_k141_109323||lt2gene Low-quality
17-106-final-viral-combined_k141_99220||lt2gene Not-determined
17-106-final-viral-combined_k141_19347||lt2gene Not-determined
17-106-final-viral-combined_k141_115064||lt2gene Low-quality
17-106-final-viral-combined_k141_45625||lt2gene Low-quality

17-106-final-viral-combined_k141_8862||lt2gene Low-quality
17-106-final-viral-combined_k141_19372||lt2gene Not-determined
17-106-final-viral-combined_k141_8864||lt2gene Low-quality
17-106-final-viral-combined_k141_35022||lt2gene Low-quality
17-106-final-viral-combined_k141_62139||lt2gene Low-quality
17-106-final-viral-combined_k141_51569||lt2gene Not-determined
17-106-final-viral-combined_k141_24610||lt2gene Low-quality
17-106-final-viral-combined_k141_104677||lt2gene Low-quality
17-106-final-viral-combined_k141_13803||lt2gene Low-quality
17-106-final-viral-combined_k141_45670||lt2gene Low-quality
17-106-final-viral-combined_k141_19494||lt2gene Low-quality
17-106-final-viral-combined_k141_57040||lt2gene Low-quality
17-106-final-viral-combined_k141_104724||lt2gene Low-quality
17-106-final-viral-combined_k141_24718||lt2gene Not-determined
17-106-final-viral-combined_k141_104734||lt2gene Low-quality
17-106-final-viral-combined_k141_57165||lt2gene Not-determined
17-106-final-viral-combined_k141_51712||lt2gene Low-quality
17-106-final-viral-combined_k141_8958||lt2gene Not-determined
17-106-final-viral-combined_k141_83988||lt2gene Not-determined
17-106-final-viral-combined_k141_8976||lt2gene Low-quality
17-106-final-viral-combined_k141_120981||lt2gene Not-determined
17-106-final-viral-combined_k141_125927||lt2gene Low-quality
17-106-final-viral-combined_k141_62329||lt2gene Low-quality
17-106-final-viral-combined_k141_13925||lt2gene Low-quality
17-106-final-viral-combined_k141_115232||lt2gene Low-quality
17-106-final-viral-combined_k141_24825||lt2gene Low-quality
17-106-final-viral-combined_k141_35331||lt2gene Low-quality
17-106-final-viral-combined_k141_78049||lt2gene Not-determined
17-106-final-viral-combined_k141_19647||lt2gene Not-determined
17-106-final-viral-combined_k141_78073||lt2gene Low-quality
17-106-final-viral-combined_k141_35399||lt2gene Low-quality
17-106-final-viral-combined_k141_51886||lt2gene Low-quality
17-106-final-viral-combined_k141_9087||lt2gene Low-quality
17-106-final-viral-combined_k141_40566||lt2gene Not-determined
17-106-final-viral-combined_k141_3681||lt2gene Not-determined
17-106-final-viral-combined_k141_57336||lt2gene Low-quality
17-106-final-viral-combined_k141_29658||lt2gene Low-quality
17-106-final-viral-combined_k141_115347||lt2gene Low-quality
17-106-final-viral-combined_k141_109576||lt2gene Not-determined
17-106-final-viral-combined_k141_84078||lt2gene Not-determined
17-106-final-viral-combined_k141_78144||lt2gene Low-quality
17-106-final-viral-combined_k141_121191||lt2gene Low-quality
17-106-final-viral-combined_k141_57376||lt2gene Low-quality
17-106-final-viral-combined_k141_109589||lt2gene Low-quality
17-106-final-viral-combined_k141_29705||lt2gene Low-quality
17-106-final-viral-combined_k141_62519||lt2gene Not-determined
17-106-final-viral-combined_k141_109618||lt2gene Low-quality
17-106-final-viral-combined_k141_89095||lt2gene Low-quality
17-106-final-viral-combined_k141_19818||lt2gene Not-determined
17-106-final-viral-combined_k141_109649||lt2gene Low-quality
17-106-final-viral-combined_k141_72518||lt2gene Not-determined
17-106-final-viral-combined_k141_45942||lt2gene Low-quality
17-106-final-viral-combined_k141_14018||lt2gene Low-quality
17-106-final-viral-combined_k141_29776||lt2gene Not-determined
17-106-final-viral-combined_k141_14051||lt2gene Low-quality
17-106-final-viral-combined_k141_72565||lt2gene Low-quality
17-106-final-viral-combined_k141_78251||lt2gene Low-quality
17-106-final-viral-combined_k141_67516||lt2gene Low-quality
17-106-final-viral-combined_k141_93950||lt2gene Low-quality
17-106-final-viral-combined_k141_89198||lt2gene Low-quality
17-106-final-viral-combined_k141_115554||lt2gene Low-quality
17-106-final-viral-combined_k141_78316||lt2gene Low-quality
17-106-final-viral-combined_k141_115563||lt2gene Not-determined
17-106-final-viral-combined_k141_89230||lt2gene Low-quality
17-106-final-viral-combined_k141_84247||lt2gene Low-quality
17-106-final-viral-combined_k141_67534||lt2gene Not-determined
17-106-final-viral-combined_k141_89253||lt2gene Low-quality
17-106-final-viral-combined_k141_115599||lt2gene Not-determined
17-106-final-viral-combined_k141_115603||lt2gene Low-quality
17-106-final-viral-combined_k141_3886||lt2gene Low-quality
17-106-final-viral-combined_k141_94049||lt2gene Low-quality
17-106-final-viral-combined_k141_94060||lt2gene Low-quality
17-106-final-viral-combined_k141_121422||lt2gene Low-quality
17-106-final-viral-combined_k141_126168||lt2gene Low-quality
17-106-final-viral-combined_k141_89302||lt2gene Low-quality
17-106-final-viral-combined_k141_67578||lt2gene Not-determined
17-106-final-viral-combined_k141_3934||lt2gene Low-quality
17-106-final-viral-combined_k141_78448||lt2gene Not-determined
17-106-final-viral-combined_k141_126201||lt2gene Low-quality
17-106-final-viral-combined_k141_67614||lt2gene Not-determined
17-106-final-viral-combined_k141_105072||lt2gene Low-quality
17-106-final-viral-combined_k141_62627||lt2gene Not-determined
17-106-final-viral-combined_k141_78482||lt2gene Low-quality
17-106-final-viral-combined_k141_35546||lt2gene Low-quality
17-106-final-viral-combined_k141_9291||lt2gene Not-determined
17-106-final-viral-combined_k141_78578||lt2gene Not-determined
17-106-final-viral-combined_k141_25233||lt2gene Low-quality
17-106-final-viral-combined_k141_9345||lt2gene Low-quality
17-106-final-viral-combined_k141_46147||lt2gene Not-determined
17-106-final-viral-combined_k141_126307||lt2gene Low-quality
17-106-final-viral-combined_k141_9413||lt2gene Low-quality
17-106-final-viral-combined_k141_121629||lt2gene Not-determined
17-106-final-viral-combined_k141_89485||lt2gene Low-quality
17-106-final-viral-combined_k141_57568||lt2gene Low-quality
17-106-final-viral-combined_k141_67851||lt2gene Low-quality
17-106-final-viral-combined_k141_62825||lt2gene Low-quality
17-106-final-viral-combined_k141_14480||lt2gene Low-quality
17-106-final-viral-combined_k141_4132||lt2gene Low-quality
17-106-final-viral-combined_k141_20075||lt2gene Not-determined
17-106-final-viral-combined_k141_20078||lt2gene Not-determined
17-106-final-viral-combined_k141_99805||lt2gene Not-determined
17-106-final-viral-combined_k141_9454||lt2gene Low-quality
17-106-final-viral-combined_k141_4156||lt2gene Low-quality
17-106-final-viral-combined_k141_72790||lt2gene Not-determined
17-106-final-viral-combined_k141_40859||lt2gene Not-determined
17-106-final-viral-combined_k141_9488||lt2gene Low-quality
17-106-final-viral-combined_k141_78768||lt2gene Not-determined
17-106-final-viral-combined_k141_30023||lt2gene Low-quality
17-106-final-viral-combined_k141_121767||lt2gene Low-quality
17-106-final-viral-combined_k141_78811||lt2gene Low-quality
17-106-final-viral-combined_k141_94158||lt2gene Low-quality
17-106-final-viral-combined_k141_72830||lt2gene Low-quality
17-106-final-viral-combined_k141_9577||lt2gene Low-quality
17-106-final-viral-combined_k141_121831||lt2gene Not-determined
17-106-final-viral-combined_k141_68010||lt2gene Low-quality
17-106-final-viral-combined_k141_57682||lt2gene Low-quality
17-106-final-viral-combined_k141_72887||lt2gene Not-determined
17-106-final-viral-combined_k141_105197||lt2gene Low-quality
17-106-final-viral-combined_k141_14622||lt2gene Low-quality
17-106-final-viral-combined_k141_41003||lt2gene Low-quality
17-106-final-viral-combined_k141_126498||lt2gene Low-quality
17-106-final-viral-combined_k141_99949||lt2gene Not-determined
17-106-final-viral-combined_k141_121897||lt2gene Low-quality
17-106-final-viral-combined_k141_25512||lt2gene Low-quality
17-106-final-viral-combined_k141_25530||lt2gene Low-quality
17-106-final-viral-combined_k141_94241||lt2gene Low-quality
17-106-final-viral-combined_k141_41057||lt2gene Low-quality
17-106-final-viral-combined_k141_105312||lt2gene Not-determined
17-106-final-viral-combined_k141_25584||lt2gene Not-determined
17-106-final-viral-combined_k141_4343||lt2gene Low-quality
17-106-final-viral-combined_k141_30255||lt2gene Not-determined
17-106-final-viral-combined_k141_35837||lt2gene Low-quality
17-106-final-viral-combined_k141_63100||lt2gene Low-quality
17-106-final-viral-combined_k141_57735||lt2gene Low-quality
17-106-final-viral-combined_k141_57743||lt2gene Low-quality
17-106-final-viral-combined_k141_20264||lt2gene Low-quality
17-106-final-viral-combined_k141_105401||lt2gene Low-quality
17-106-final-viral-combined_k141_78981||lt2gene Low-quality
17-106-final-viral-combined_k141_35905||lt2gene Low-quality
17-106-final-viral-combined_k141_73098||lt2gene Not-determined
17-106-final-viral-combined_k141_46346||lt2gene Not-determined
17-106-final-viral-combined_k141_100079||lt2gene Not-determined
17-106-final-viral-combined_k141_14783||lt2gene Low-quality
17-106-final-viral-combined_k141_4419||lt2gene Low-quality
17-106-final-viral-combined_k141_57787||lt2gene Low-quality
17-106-final-viral-combined_k141_68297||lt2gene Low-quality
17-106-final-viral-combined_k141_89949||lt2gene Low-quality
17-106-final-viral-combined_k141_105452||lt2gene Low-quality
17-106-final-viral-combined_k141_100133||lt2gene Low-quality
17-106-final-viral-combined_k141_100156||lt2gene Low-quality
17-106-final-viral-combined_k141_9700||lt2gene Low-quality
17-106-final-viral-combined_k141_105470||lt2gene Low-quality
17-106-final-viral-combined_k141_9730||lt2gene Low-quality
17-106-final-viral-combined_k141_73193||lt2gene Low-quality
17-106-final-viral-combined_k141_63356||lt2gene Not-determined
17-106-final-viral-combined_k141_68387||lt2gene Low-quality
17-106-final-viral-combined_k141_57895||lt2gene Low-quality
17-106-final-viral-combined_k141_122148||lt2gene Not-determined
17-106-final-viral-combined_k141_68397||lt2gene Low-quality
17-106-final-viral-combined_k141_94599||lt2gene Low-quality
17-106-final-viral-combined_k141_52728||lt2gene Low-quality
17-106-final-viral-combined_k141_41230||lt2gene Low-quality
17-106-final-viral-combined_k141_90062||lt2gene Low-quality
17-106-final-viral-combined_k141_73225||lt2gene Low-quality
17-106-final-viral-combined_k141_73246||lt2gene Low-quality
17-106-final-viral-combined_k141_14862||lt2gene Low-quality
17-106-final-viral-combined_k141_79082||lt2gene Low-quality
17-106-final-viral-combined_k141_20447||lt2gene Low-quality
17-106-final-viral-combined_k141_46578||lt2gene Low-quality
17-106-final-viral-combined_k141_46588||lt2gene Not-determined
17-106-final-viral-combined_k141_79124||lt2gene Low-quality
17-106-final-viral-combined_k141_52804||lt2gene Low-quality
17-106-final-viral-combined_k141_57978||lt2gene Low-quality
17-106-final-viral-combined_k141_79134||lt2gene Low-quality
17-106-final-viral-combined_k141_14919||lt2gene Low-quality
17-106-final-viral-combined_k141_73374||lt2gene Low-quality
17-106-final-viral-combined_k141_52808||lt2gene Low-quality
17-106-final-viral-combined_k141_52810||lt2gene Not-determined
17-106-final-viral-combined_k141_9930||lt2gene Low-quality
17-106-final-viral-combined_k141_52842||lt2gene Low-quality
17-106-final-viral-combined_k141_30540||lt2gene Low-quality
17-106-final-viral-combined_k141_122277||lt2gene Not-determined
17-106-final-viral-combined_k141_79233||lt2gene Low-quality
17-106-final-viral-combined_k141_41404||lt2gene Low-quality
17-106-final-viral-combined_k141_90213||lt2gene Low-quality
17-106-final-viral-combined_k141_79301||lt2gene Low-quality
17-106-final-viral-combined_k141_79326||lt2gene Low-quality
17-106-final-viral-combined_k141_52941||lt2gene Low-quality
17-106-final-viral-combined_k141_52943||lt2gene Low-quality
17-106-final-viral-combined_k141_79334||lt2gene Low-quality
17-106-final-viral-combined_k141_25856||lt2gene Low-quality
17-106-final-viral-combined_k141_36304||lt2gene Not-determined
17-106-final-viral-combined_k141_36316||lt2gene Low-quality
17-106-final-viral-combined_k141_52967||lt2gene Not-determined
17-106-final-viral-combined_k141_52988||lt2gene Low-quality
17-106-final-viral-combined_k141_52995||lt2gene Not-determined
17-106-final-viral-combined_k141_63726||lt2gene Low-quality
17-106-final-viral-combined_k141_53004||lt2gene Low-quality
17-106-final-viral-combined_k141_30730||lt2gene Low-quality
17-106-final-viral-combined_k141_58158||lt2gene Low-quality
17-106-final-viral-combined_k141_4672||lt2gene Not-determined
17-106-final-viral-combined_k141_36384||lt2gene Low-quality
17-106-final-viral-combined_k141_94914||lt2gene Low-quality
17-106-final-viral-combined_k141_94924||lt2gene Not-determined
17-106-final-viral-combined_k141_85015||lt2gene Not-determined
17-106-final-viral-combined_k141_25991||lt2gene Low-quality
17-106-final-viral-combined_k141_85032||lt2gene Low-quality
17-106-final-viral-combined_k141_63816||lt2gene Low-quality
17-106-final-viral-combined_k141_90442||lt2gene Low-quality
17-106-final-viral-combined_k141_90448||lt2gene Low-quality
17-106-final-viral-combined_k141_85057||lt2gene Low-quality
17-106-final-viral-combined_k141_90451||lt2gene Low-quality
17-106-final-viral-combined_k141_100574||lt2gene Not-determined
17-106-final-viral-combined_k141_20749||lt2gene Low-quality
17-106-final-viral-combined_k141_85093||lt2gene Not-determined
17-106-final-viral-combined_k141_26056||lt2gene Low-quality
17-106-final-viral-combined_k141_63871||lt2gene Low-quality
17-106-final-viral-combined_k141_95032||lt2gene Not-determined
17-106-final-viral-combined_k141_79625||lt2gene Not-determined
17-106-final-viral-combined_k141_100675||lt2gene Low-quality
17-106-final-viral-combined_k141_15174||lt2gene Low-quality
17-106-final-viral-combined_k141_15195||lt2gene Low-quality
17-106-final-viral-combined_k141_41638||lt2gene Not-determined
17-106-final-viral-combined_k141_79681||lt2gene Low-quality
17-106-final-viral-combined_k141_36618||lt2gene Low-quality
17-106-final-viral-combined_k141_20868||lt2gene Low-quality
17-106-final-viral-combined_k141_15274||lt2gene Not-determined
17-106-final-viral-combined_k141_36662||lt2gene Low-quality
17-106-final-viral-combined_k141_110643||lt2gene Low-quality
17-106-final-viral-combined_k141_30879||lt2gene Low-quality
17-106-final-viral-combined_k141_63999||lt2gene Low-quality
17-106-final-viral-combined_k141_90716||lt2gene Low-quality
17-106-final-viral-combined_k141_69101||lt2gene Low-quality
17-106-final-viral-combined_k141_73709||lt2gene Low-quality
17-106-final-viral-combined_k141_20916||lt2gene Low-quality
17-106-final-viral-combined_k141_30925||lt2gene Low-quality
17-106-final-viral-combined_k141_30939||lt2gene Low-quality
17-106-final-viral-combined_k141_64108||lt2gene Low-quality
17-106-final-viral-combined_k141_110670||lt2gene Low-quality
17-106-final-viral-combined_k141_36811||lt2gene Low-quality
17-106-final-viral-combined_k141_69178||lt2gene Not-determined
17-106-final-viral-combined_k141_58343||lt2gene Not-determined
17-106-final-viral-combined_k141_90865||lt2gene Low-quality
17-106-final-viral-combined_k141_5042||lt2gene Not-determined
17-106-final-viral-combined_k141_31042||lt2gene Not-determined
17-106-final-viral-combined_k141_31045||lt2gene Not-determined
17-106-final-viral-combined_k141_73823||lt2gene Not-determined
17-106-final-viral-combined_k141_85429||lt2gene Low-quality
17-106-final-viral-combined_k141_116748||lt2gene Not-determined
17-106-final-viral-combined_k141_110682||lt2gene Low-quality
17-106-final-viral-combined_k141_26371||lt2gene Low-quality
17-106-final-viral-combined_k141_10282||lt2gene Low-quality
17-106-final-viral-combined_k141_36951||lt2gene Not-determined
17-106-final-viral-combined_k141_116792||lt2gene Low-quality
17-106-final-viral-combined_k141_100995||lt2gene Low-quality
17-106-final-viral-combined_k141_53386||lt2gene Low-quality
17-106-final-viral-combined_k141_53399||lt2gene Low-quality
17-106-final-viral-combined_k141_37024||lt2gene Not-determined
17-106-final-viral-combined_k141_53452||lt2gene Low-quality
17-106-final-viral-combined_k141_21042||lt2gene Low-quality
17-106-final-viral-combined_k141_26411||lt2gene Low-quality
17-106-final-viral-combined_k141_26415||lt2gene Not-determined
17-106-final-viral-combined_k141_95228||lt2gene Low-quality
17-106-final-viral-combined_k141_122830||lt2gene Low-quality
17-106-final-viral-combined_k141_21139||lt2gene Low-quality
17-106-final-viral-combined_k141_95278||lt2gene Low-quality
17-106-final-viral-combined_k141_127090||lt2gene Low-quality
17-106-final-viral-combined_k141_69422||lt2gene Not-determined
17-106-final-viral-combined_k141_10485||lt2gene Not-determined
17-106-final-viral-combined_k141_37187||lt2gene Low-quality
17-106-final-viral-combined_k141_74074||lt2gene Low-quality
17-106-final-viral-combined_k141_122892||lt2gene Low-quality
17-106-final-viral-combined_k141_37235||lt2gene Low-quality
17-106-final-viral-combined_k141_69503||lt2gene Low-quality
17-106-final-viral-combined_k141_69506||lt2gene Not-determined
17-106-final-viral-combined_k141_37264||lt2gene Not-determined
17-106-final-viral-combined_k141_10547||lt2gene Low-quality
17-106-final-viral-combined_k141_101372||lt2gene Low-quality
17-106-final-viral-combined_k141_116900||lt2gene Low-quality
17-106-final-viral-combined_k141_80112||lt2gene Low-quality
17-106-final-viral-combined_k141_42188||lt2gene Not-determined
17-106-final-viral-combined_k141_116919||lt2gene Low-quality
17-106-final-viral-combined_k141_122992||lt2gene Low-quality
17-106-final-viral-combined_k141_21337||lt2gene Low-quality
17-106-final-viral-combined_k141_116943||lt2gene Low-quality
17-106-final-viral-combined_k141_37361||lt2gene Low-quality
17-106-final-viral-combined_k141_37421||lt2gene Low-quality
17-106-final-viral-combined_k141_58787||lt2gene Low-quality
17-106-final-viral-combined_k141_31443||lt2gene Low-quality
17-106-final-viral-combined_k141_117078||lt2gene Low-quality
17-106-final-viral-combined_k141_117101||lt2gene Low-quality
17-106-final-viral-combined_k141_101511||lt2gene Low-quality
17-106-final-viral-combined_k141_74331||lt2gene Not-determined
17-106-final-viral-combined_k141_101550||lt2gene Not-determined
17-106-final-viral-combined_k141_101566||lt2gene Low-quality
17-106-final-viral-combined_k141_47198||lt2gene Low-quality
17-106-final-viral-combined_k141_117166||lt2gene Not-determined
17-106-final-viral-combined_k141_31530||lt2gene Not-determined
17-106-final-viral-combined_k141_74423||lt2gene Low-quality
17-106-final-viral-combined_k141_42428||lt2gene Low-quality
17-106-final-viral-combined_k141_31551||lt2gene Low-quality
17-106-final-viral-combined_k141_31553||lt2gene Low-quality
17-106-final-viral-combined_k141_31561||lt2gene Low-quality
17-106-final-viral-combined_k141_95695||lt2gene Not-determined
17-106-final-viral-combined_k141_31651||lt2gene Not-determined
17-106-final-viral-combined_k141_47283||lt2gene Low-quality
17-106-final-viral-combined_k141_95752||lt2gene Not-determined
17-106-final-viral-combined_k141_110869||lt2gene Not-determined
17-106-final-viral-combined_k141_117288||lt2gene Low-quality
17-106-final-viral-combined_k141_117320||lt2gene Not-determined
17-106-final-viral-combined_k141_95977||lt2gene Low-quality
17-106-final-viral-combined_k141_95988||lt2gene Low-quality
17-106-final-viral-combined_k141_96012||lt2gene Low-quality
17-106-final-viral-combined_k141_74690||lt2gene Low-quality
17-106-final-viral-combined_k141_96034||lt2gene Low-quality
17-106-final-viral-combined_k141_96057||lt2gene Not-determined
17-106-final-viral-combined_k141_74730||lt2gene Not-determined
17-106-final-viral-combined_k141_74743||lt2gene Low-quality
17-106-final-viral-combined_k141_42698||lt2gene Low-quality
17-106-final-viral-combined_k141_31786||lt2gene Low-quality
17-106-final-viral-combined_k141_127601||lt2gene Low-quality
17-106-final-viral-combined_k141_31820||lt2gene Low-quality
17-106-final-viral-combined_k141_127715||lt2gene Low-quality
17-106-final-viral-combined_k141_31909||lt2gene Low-quality
17-106-final-viral-combined_k141_111121||lt2gene Low-quality
17-106-final-viral-combined_k141_106822||lt2gene Low-quality
17-106-final-viral-combined_k141_127785||lt2gene Low-quality
17-106-final-viral-combined_k141_74818||lt2gene Low-quality
17-106-final-viral-combined_k141_127969||lt2gene Low-quality
17-106-final-viral-combined_k141_127980||lt2gene Low-quality
17-106-final-viral-combined_k141_47944||lt2gene Low-quality
17-106-final-viral-combined_k141_111435||lt2gene Not-determined
17-106-final-viral-combined_k141_48028||lt2gene Low-quality
17-106-final-viral-combined_k141_111535||lt2gene Not-determined
17-106-final-viral-combined_k141_128156||lt2gene Not-determined
17-106-final-viral-combined_k141_128177||lt2gene Not-determined
17-106-final-viral-combined_k141_128190||lt2gene Low-quality
17-106-final-viral-combined_k141_111669||lt2gene Low-quality
17-106-final-viral-combined_k141_128193||lt2gene Low-quality
17-106-final-viral-combined_k141_111852||lt2gene Low-quality
17-106-final-viral-combined_k141_111867||lt2gene Not-determined
17-106_k141_21392 Not-determined
17-106_k141_117648 Not-determined
17-106_k141_96259 Not-determined
17-106_k141_6 Not-determined
17-106_k141_5350 Not-determined
17-106_k141_48138 Not-determined
17-106_k141_10698 Not-determined
17-106_k141_90918 Not-determined
17-106_k141_64187 Not-determined
17-106_k141_80220 Not-determined
17-106_k141_112305 Not-determined
17-106_k141_106961 Not-determined
17-106_k141_96263 Not-determined
17-106_k141_122993 Not-determined
17-106_k141_48147 Not-determined
17-106_k141_101613 Not-determined
17-106_k141_90931 Not-determined
17-106_k141_28 Not-determined
17-106_k141_117669 Not-determined
17-106_k141_21407 Low-quality
17-106_k141_10714 Not-determined
17-106_k141_42791 Not-determined
17-106_k141_39 Not-determined
17-106_k141_85580 Not-determined
17-106_k141_85583 Not-determined
17-106_k141_101633 Not-determined
17-106_k141_45 Not-determined
17-106_k141_48159 Not-determined
17-106_k141_53485 Not-determined
17-106_k141_106986 Not-determined
17-106_k141_106987 Not-determined
17-106_k141_101639 Not-determined
17-106_k141_85587 Not-determined
17-106_k141_37465 Not-determined
17-106_k141_112338 Not-determined
17-106_k141_10739 Not-determined
17-106_k141_117685 Not-determined
17-106_k141_32105 Not-determined
17-106_k141_96302 Not-determined
17-106_k141_10744 Not-determined
17-106_k141_117695 Not-determined
17-106_k141_112352 Not-determined
17-106_k141_107001 Not-determined
17-106_k141_5393 Not-determined
17-106_k141_58856 Not-determined
17-106_k141_107004 Not-determined
17-106_k141_53492 Low-quality
17-106_k141_117704 Not-determined
17-106_k141_117710 Not-determined
17-106_k141_10758 Not-determined
17-106_k141_21434 Low-quality
17-106_k141_64238 Not-determined
17-106_k141_64239 Not-determined
17-106_k141_21440 Not-determined
17-106_k141_10765 Not-determined
17-106_k141_32124 Not-determined
17-106_k141_69568 Not-determined
17-106_k141_117728 Not-determined
17-106_k141_122999 Not-determined
17-106_k141_5399 Not-determined
17-106_k141_123000 Not-determined
17-106_k141_117733 Low-quality
17-106_k141_21449 Not-determined
17-106_k141_107016 Not-determined
17-106_k141_80264 Not-determined
17-106_k141_21457 Not-determined
17-106_k141_26758 Not-determined
17-106_k141_55 Not-determined
17-106_k141_37476 Not-determined
17-106_k141_42862 Not-determined
17-106_k141_37481 Not-determined
17-106_k141_123015 Not-determined
17-106_k141_85640 Not-determined
17-106_k141_42866 Not-determined
17-106_k141_5414 Low-quality
17-106_k141_85643 Not-determined
17-106_k141_53505 Not-determined
17-106_k141_74904 Not-determined
17-106_k141_74905 Not-determined
17-106_k141_53508 Not-determined
17-106_k141_58861 Low-quality
17-106_k141_96341 Low-quality
17-106_k141_107042 Not-determined
17-106_k141_107047 Not-determined
17-106_k141_10813 Not-determined
17-106_k141_93 Not-determined
17-106_k141_94 Not-determined
17-106_k141_90954 Not-determined
17-106_k141_107053 Not-determined
17-106_k141_107055 Not-determined
17-106_k141_97 Not-determined
17-106_k141_104 Low-quality
17-106_k141_58897 Not-determined
17-106_k141_101688 Not-determined
17-106_k141_101689 Not-determined
17-106_k141_58903 Low-quality
17-106_k141_101704 Not-determined
17-106_k141_48218 Not-determined
17-106_k141_117755 Not-determined
17-106_k141_21462 Not-determined
17-106_k141_90976 Not-determined
17-106_k141_90979 Not-determined
17-106_k141_132 Not-determined
17-106_k141_134 Not-determined
17-106_k141_48234 Not-determined
17-106_k141_21474 Low-quality
17-106_k141_117771 Not-determined
17-106_k141_117773 Low-quality
17-106_k141_101719 Not-determined
17-106_k141_16148 Not-determined
17-106_k141_85704 Low-quality
17-106_k141_157 Not-determined
17-106_k141_21476 Not-determined
17-106_k141_32143 Not-determined
17-106_k141_26796 Not-determined
17-106_k141_32148 Not-determined
17-106_k141_32149 Not-determined
17-106_k141_16158 Not-determined
17-106_k141_16159 Not-determined
17-106_k141_26799 Not-determined
17-106_k141_32152 Not-determined
17-106_k141_85710 Not-determined
17-106_k141_37527 Not-determined
17-106_k141_21494 Not-determined
17-106_k141_37536 Not-determined
17-106_k141_37538 Not-determined
17-106_k141_107070 Not-determined
17-106_k141_112434 Not-determined
17-106_k141_74961 Not-determined
17-106_k141_37551 Low-quality
17-106_k141_74965 Not-determined
17-106_k141_37559 Not-determined
17-106_k141_21509 Not-determined
17-106_k141_69588 Not-determined
17-106_k141_21511 Not-determined
17-106_k141_42922 Not-determined
17-106_k141_69592 Not-determined
17-106_k141_69605 Not-determined
17-106_k141_74988 Not-determined
17-106_k141_183 Not-determined
17-106_k141_74993 Not-determined
17-106_k141_74994 Not-determined
17-106_k141_101765 Low-quality
17-106_k141_37565 Not-determined
17-106_k141_21540 Not-determined
17-106_k141_64267 Not-determined
17-106_k141_10881 Not-determined
17-106_k141_75005 Not-determined
17-106_k141_197 Not-determined
17-106_k141_112475 Not-determined
17-106_k141_96376 Not-determined
17-106_k141_204 Not-determined
17-106_k141_75015 Not-determined
17-106_k141_26817 Not-determined
17-106_k141_26822 Not-determined
17-106_k141_26825 Not-determined
17-106_k141_26831 Not-determined
17-106_k141_96384 Not-determined
17-106_k141_112490 Not-determined
17-106_k141_96388 Not-determined
17-106_k141_85712 Not-determined
17-106_k141_101786 Not-determined
17-106_k141_117800 Not-determined
17-106_k141_96396 Not-determined
17-106_k141_112503 Not-determined
17-106_k141_85718 Not-determined
17-106_k141_85722 Not-determined
17-106_k141_85725 Not-determined
17-106_k141_16204 Not-determined
17-106_k141_42951 Not-determined
17-106_k141_26847 Low-quality
17-106_k141_112515 Not-determined
17-106_k141_42953 Not-determined
17-106_k141_101802 Not-determined
17-106_k141_42961 Not-determined
17-106_k141_69615 Not-determined
17-106_k141_10884 Not-determined
17-106_k141_48288 Not-determined
17-106_k141_48291 Not-determined
17-106_k141_42969 Not-determined
17-106_k141_42972 Not-determined
17-106_k141_10896 Not-determined
17-106_k141_48294 Not-determined
17-106_k141_85732 Not-determined
17-106_k141_42982 Not-determined
17-106_k141_101818 Not-determined
17-106_k141_75065 Not-determined
17-106_k141_69633 Not-determined
17-106_k141_80302 Not-determined
17-106_k141_58932 Not-determined
17-106_k141_32208 Not-determined
17-106_k141_42987 Low-quality
17-106_k141_26862 Not-determined
17-106_k141_10916 Low-quality
17-106_k141_75072 Low-quality
17-106_k141_123113 Not-determined
17-106_k141_26872 Not-determined
17-106_k141_123122 Not-determined
17-106_k141_32223 Not-determined
17-106_k141_123124 Not-determined
17-106_k141_112529 Not-determined
17-106_k141_123128 Not-determined
17-106_k141_80327 Low-quality
17-106_k141_96422 Not-determined
17-106_k141_42998 Low-quality
17-106_k141_267 Not-determined
17-106_k141_21564 Not-determined
17-106_k141_58949 Not-determined
17-106_k141_58953 Not-determined
17-106_k141_277 Not-determined
17-106_k141_96431 Low-quality
17-106_k141_48304 Not-determined
17-106_k141_21575 Not-determined
17-106_k141_21577 Not-determined
17-106_k141_43016 Not-determined
17-106_k141_58962 Not-determined
17-106_k141_16215 Not-determined
17-106_k141_37577 Not-determined
17-106_k141_75110 Not-determined
17-106_k141_123148 Not-determined
17-106_k141_123149 Low-quality
17-106_k141_289 Not-determined
17-106_k141_32240 Not-determined
17-106_k141_32246 Not-determined
17-106_k141_32247 Not-determined
17-106_k141_32249 Not-determined
17-106_k141_85781 Not-determined
17-106_k141_85784 Not-determined
17-106_k141_117805 Low-quality
17-106_k141_85785 Not-determined
17-106_k141_58977 Not-determined
17-106_k141_53545 Low-quality
17-106_k141_85794 Not-determined
17-106_k141_53549 Not-determined
17-106_k141_5494 Not-determined
17-106_k141_112569 Not-determined
17-106_k141_80349 Not-determined
17-106_k141_5504 Not-determined
17-106_k141_5505 Not-determined
17-106_k141_48325 Low-quality
17-106_k141_26931 Not-determined
17-106_k141_48326 Not-determined
17-106_k141_5512 Not-determined
17-106_k141_58992 Not-determined
17-106_k141_123166 Not-determined
17-106_k141_80367 Not-determined
17-106_k141_80371 Not-determined
17-106_k141_58997 Not-determined

| | |
|---|---|
| 17-106_k141_123172 | Not-determined |
| 17-106_k141_91117 | Not-determined |
| 17-106_k141_80378 | Low-quality |
| 17-106_k141_75125 | Not-determined |
| 17-106_k141_32262 | Not-determined |
| 17-106_k141_75128 | Not-determined |
| 17-106_k141_32264 | Not-determined |
| 17-106_k141_32277 | Low-quality |
| 17-106_k141_43047 | Not-determined |
| 17-106_k141_96453 | Low-quality |
| 17-106_k141_43052 | Not-determined |
| 17-106_k141_43055 | Not-determined |
| 17-106_k141_43059 | Not-determined |
| 17-106_k141_80384 | Not-determined |
| 17-106_k141_117818 | Not-determined |
| 17-106_k141_80389 | Not-determined |
| 17-106_k141_117823 | Not-determined |
| 17-106_k141_80395 | Not-determined |
| 17-106_k141_123199 | Not-determined |
| 17-106_k141_80400 | Low-quality |
| 17-106_k141_96468 | Not-determined |
| 17-106_k141_26958 | Not-determined |
| 17-106_k141_101824 | Low-quality |
| 17-106_k141_59016 | Not-determined |
| 17-106_k141_59026 | Not-determined |
| 17-106_k141_75159 | Not-determined |
| 17-106_k141_59035 | Not-determined |
| 17-106_k141_101841 | Not-determined |
| 17-106_k141_53573 | Not-determined |
| 17-106_k141_75168 | Not-determined |
| 17-106_k141_16262 | Not-determined |
| 17-106_k141_333 | Low-quality |
| 17-106_k141_59042 | Not-determined |
| 17-106_k141_48364 | Low-quality |
| 17-106_k141_64282 | Not-determined |
| 17-106_k141_339 | Not-determined |
| 17-106_k141_80418 | Not-determined |
| 17-106_k141_32302 | Not-determined |
| 17-106_k141_75180 | Not-determined |
| 17-106_k141_101845 | Not-determined |
| 17-106_k141_96501 | Not-determined |
| 17-106_k141_75185 | Low-quality |
| 17-106_k141_53597 | Not-determined |
| 17-106_k141_117870 | Not-determined |
| 17-106_k141_26993 | Not-determined |
| 17-106_k141_16287 | Low-quality |
| 17-106_k141_117875 | Not-determined |
| 17-106_k141_80429 | Not-determined |
| 17-106_k141_96506 | Low-quality |
| 17-106_k141_85816 | Not-determined |
| 17-106_k141_80431 | Not-determined |
| 17-106_k141_48379 | Not-determined |
| 17-106_k141_64297 | Not-determined |
| 17-106_k141_85824 | Not-determined |
| 17-106_k141_27001 | Not-determined |
| 17-106_k141_91134 | Not-determined |
| 17-106_k141_64302 | Not-determined |
| 17-106_k141_48380 | Low-quality |
| 17-106_k141_80441 | Not-determined |
| 17-106_k141_16307 | Not-determined |
| 17-106_k141_48384 | Not-determined |
| 17-106_k141_96514 | Not-determined |
| 17-106_k141_117890 | Not-determined |
| 17-106_k141_117891 | Not-determined |
| 17-106_k141_365 | Low-quality |
| 17-106_k141_27016 | Not-determined |
| 17-106_k141_64310 | Not-determined |
| 17-106_k141_69697 | Not-determined |
| 17-106_k141_48392 | Not-determined |
| 17-106_k141_96531 | Not-determined |
| 17-106_k141_85843 | Low-quality |
| 17-106_k141_85847 | Not-determined |
| 17-106_k141_69701 | Not-determined |
| 17-106_k141_69702 | Low-quality |
| 17-106_k141_96539 | Not-determined |
| 17-106_k141_80463 | Not-determined |
| 17-106_k141_27022 | Not-determined |
| 17-106_k141_384 | Not-determined |
| 17-106_k141_27024 | Not-determined |
| 17-106_k141_64323 | Not-determined |
| 17-106_k141_117902 | Not-determined |
| 17-106_k141_80471 | Not-determined |
| 17-106_k141_16322 | Not-determined |
| 17-106_k141_59086 | Low-quality |
| 17-106_k141_101869 | Not-determined |
| 17-106_k141_101873 | Not-determined |
| 17-106_k141_64327 | Not-determined |
| 17-106_k141_27038 | Not-determined |
| 17-106_k141_64334 | Not-determined |
| 17-106_k141_96560 | Not-determined |
| 17-106_k141_59094 | Not-determined |
| 17-106_k141_101876 | Not-determined |
| 17-106_k141_80486 | Not-determined |
| 17-106_k141_75216 | Not-determined |
| 17-106_k141_80488 | Not-determined |
| 17-106_k141_64348 | Not-determined |
| 17-106_k141_75223 | Not-determined |
| 17-106_k141_64355 | Low-quality |
| 17-106_k141_53658 | Not-determined |
| 17-106_k141_399 | Not-determined |
| 17-106_k141_32308 | Low-quality |
| 17-106_k141_117925 | Not-determined |
| 17-106_k141_101907 | Not-determined |
| 17-106_k141_64384 | Not-determined |
| 17-106_k141_64385 | Not-determined |
| 17-106_k141_64387 | Not-determined |
| 17-106_k141_32322 | Not-determined |
| 17-106_k141_117935 | Not-determined |
| 17-106_k141_117944 | Not-determined |
| 17-106_k141_101921 | Not-determined |
| 17-106_k141_413 | Not-determined |
| 17-106_k141_117947 | Not-determined |
| 17-106_k141_421 | Not-determined |
| 17-106_k141_85874 | Not-determined |
| 17-106_k141_429 | Not-determined |
| 17-106_k141_436 | Not-determined |
| 17-106_k141_16360 | Not-determined |
| 17-106_k141_440 | Not-determined |
| 17-106_k141_85894 | Not-determined |
| 17-106_k141_43124 | Not-determined |
| 17-106_k141_37603 | Not-determined |
| 17-106_k141_48448 | Not-determined |
| 17-106_k141_101966 | Not-determined |
| 17-106_k141_80516 | Not-determined |
| 17-106_k141_101971 | Not-determined |
| 17-106_k141_80517 | Not-determined |
| 17-106_k141_16372 | Not-determined |
| 17-106_k141_461 | Not-determined |
| 17-106_k141_85932 | Not-determined |
| 17-106_k141_59105 | Not-determined |
| 17-106_k141_16382 | Not-determined |
| 17-106_k141_96593 | Not-determined |
| 17-106_k141_16387 | Not-determined |
| 17-106_k141_27093 | Low-quality |
| 17-106_k141_85936 | Not-determined |
| 17-106_k141_27094 | Not-determined |
| 17-106_k141_112636 | Not-determined |
| 17-106_k141_48462 | Not-determined |
| 17-106_k141_75249 | Not-determined |
| 17-106_k141_59124 | Not-determined |
| 17-106_k141_476 | Not-determined |
| 17-106_k141_80547 | Not-determined |
| 17-106_k141_112648 | Not-determined |
| 17-106_k141_75259 | Not-determined |
| 17-106_k141_112653 | Not-determined |
| 17-106_k141_117962 | Not-determined |
| 17-106_k141_85954 | Not-determined |
| 17-106_k141_117965 | Not-determined |
| 17-106_k141_117966 | Low-quality |
| 17-106_k141_27108 | Not-determined |
| 17-106_k141_27114 | Not-determined |
| 17-106_k141_75275 | Not-determined |
| 17-106_k141_80552 | Not-determined |
| 17-106_k141_80557 | Not-determined |
| 17-106_k141_75279 | Not-determined |
| 17-106_k141_75283 | Not-determined |
| 17-106_k141_37617 | Not-determined |
| 17-106_k141_80562 | Low-quality |
| 17-106_k141_69771 | Low-quality |
| 17-106_k141_85981 | Not-determined |
| 17-106_k141_102019 | Not-determined |
| 17-106_k141_69773 | Not-determined |
| 17-106_k141_27131 | Not-determined |
| 17-106_k141_80582 | Low-quality |
| 17-106_k141_117980 | Not-determined |
| 17-106_k141_59140 | Not-determined |
| 17-106_k141_102037 | Not-determined |
| 17-106_k141_53692 | Not-determined |
| 17-106_k141_112675 | Not-determined |
| 17-106_k141_496 | Not-determined |
| 17-106_k141_53695 | Not-determined |
| 17-106_k141_37635 | Not-determined |
| 17-106_k141_85996 | Not-determined |
| 17-106_k141_59155 | Not-determined |
| 17-106_k141_53700 | Low-quality |
| 17-106_k141_53703 | Not-determined |
| 17-106_k141_112691 | Not-determined |
| 17-106_k141_75318 | Not-determined |
| 17-106_k141_37659 | Not-determined |
| 17-106_k141_53711 | Not-determined |
| 17-106_k141_59158 | Not-determined |
| 17-106_k141_96609 | Low-quality |
| 17-106_k141_59165 | Not-determined |
| 17-106_k141_64426 | Not-determined |
| 17-106_k141_16418 | Low-quality |
| 17-106_k141_53722 | Not-determined |
| 17-106_k141_123226 | Not-determined |
| 17-106_k141_112700 | Not-determined |
| 17-106_k141_80614 | Low-quality |
| 17-106_k141_96629 | Not-determined |
| 17-106_k141_75342 | Not-determined |
| 17-106_k141_27139 | Not-determined |
| 17-106_k141_123244 | Not-determined |
| 17-106_k141_53727 | Not-determined |
| 17-106_k141_80621 | Not-determined |
| 17-106_k141_123246 | Not-determined |
| 17-106_k141_16429 | Not-determined |
| 17-106_k141_48488 | Low-quality |
| 17-106_k141_118003 | Low-quality |
| 17-106_k141_80628 | Not-determined |
| 17-106_k141_37668 | Not-determined |
| 17-106_k141_27149 | Not-determined |
| 17-106_k141_27152 | Low-quality |
| 17-106_k141_123253 | Not-determined |
| 17-106_k141_43160 | Not-determined |
| 17-106_k141_27154 | Not-determined |
| 17-106_k141_112715 | Not-determined |
| 17-106_k141_53735 | Low-quality |
| 17-106_k141_53736 | Not-determined |
| 17-106_k141_96659 | Not-determined |
| 17-106_k141_80632 | Low-quality |
| 17-106_k141_80635 | Not-determined |
| 17-106_k141_27162 | Not-determined |
| 17-106_k141_43169 | Not-determined |
| 17-106_k141_43171 | Not-determined |
| 17-106_k141_43174 | Not-determined |
| 17-106_k141_80637 | Not-determined |
| 17-106_k141_16444 | Not-determined |
| 17-106_k141_37691 | Not-determined |
| 17-106_k141_37693 | Not-determined |
| 17-106_k141_48530 | Not-determined |
| 17-106_k141_96674 | Not-determined |
| 17-106_k141_112737 | Not-determined |
| 17-106_k141_59200 | Not-determined |
| 17-106_k141_21605 | Not-determined |
| 17-106_k141_75361 | Not-determined |
| 17-106_k141_107098 | Not-determined |
| 17-106_k141_102082 | Not-determined |
| 17-106_k141_118009 | Not-determined |
| 17-106_k141_21613 | Low-quality |
| 17-106_k141_123304 | Not-determined |
| 17-106_k141_102091 | Not-determined |
| 17-106_k141_118014 | Not-determined |
| 17-106_k141_102093 | Low-quality |
| 17-106_k141_102094 | Not-determined |
| 17-106_k141_102095 | Not-determined |
| 17-106_k141_107112 | Low-quality |
| 17-106_k141_123307 | Low-quality |
| 17-106_k141_118024 | Not-determined |
| 17-106_k141_64457 | Not-determined |
| 17-106_k141_59224 | Not-determined |
| 17-106_k141_64460 | Not-determined |
| 17-106_k141_107114 | Not-determined |
| 17-106_k141_59228 | Not-determined |
| 17-106_k141_5593 | Not-determined |
| 17-106_k141_102110 | Not-determined |
| 17-106_k141_118036 | Not-determined |
| 17-106_k141_118040 | Low-quality |
| 17-106_k141_48563 | Not-determined |
| 17-106_k141_107128 | Not-determined |
| 17-106_k141_107129 | Not-determined |
| 17-106_k141_59229 | Low-quality |
| 17-109-final-viral-combined_k141_51076\|\|full | Medium-quality |
| 17-106_k141_118058 | Not-determined |
| 17-106_k141_112753 | Not-determined |
| 17-106_k141_107136 | Low-quality |
| 17-106_k141_53801 | Not-determined |
| 17-106_k141_69795 | Not-determined |
| 17-106_k141_69796 | Not-determined |
| 17-106_k141_107143 | Not-determined |
| 17-106_k141_102127 | Not-determined |
| 17-106_k141_48581 | Not-determined |
| 17-106_k141_576 | Not-determined |
| 17-106_k141_118076 | Not-determined |
| 17-106_k141_5597 | Not-determined |
| 17-106_k141_5600 | Not-determined |
| 17-106_k141_86011 | Not-determined |
| 17-106_k141_107155 | Not-determined |
| 17-106_k141_107157 | Low-quality |
| 17-106_k141_86014 | Not-determined |
| 17-106_k141_21629 | Not-determined |
| 17-106_k141_107160 | Not-determined |
| 17-106_k141_64508 | Not-determined |
| 17-106_k141_21630 | Not-determined |
| 17-106_k141_86024 | Not-determined |
| 17-106_k141_21634 | Not-determined |
| 17-106_k141_96738 | Not-determined |
| 17-106_k141_86029 | Low-quality |
| 17-106_k141_48594 | Not-determined |
| 17-106_k141_16466 | Not-determined |
| 17-106_k141_5621 | Not-determined |
| 17-106_k141_69823 | Not-determined |
| 17-106_k141_69827 | Low-quality |
| 17-106_k141_5633 | Not-determined |
| 17-106_k141_598 | Not-determined |
| 17-106_k141_43202 | Not-determined |
| 17-106_k141_96751 | Not-determined |
| 17-106_k141_107174 | Low-quality |
| 17-106_k141_5645 | Not-determined |
| 17-106_k141_102143 | Not-determined |
| 17-106_k141_48630 | Not-determined |
| 17-106_k141_86059 | Not-determined |
| 17-106_k141_96761 | Not-determined |
| 17-106_k141_107180 | Not-determined |
| 17-106_k141_48640 | Not-determined |
| 17-106_k141_86064 | Not-determined |
| 17-106_k141_48642 | Not-determined |
| 17-106_k141_118110 | Not-determined |
| 17-106_k141_43211 | Not-determined |
| 17-106_k141_53853 | Not-determined |
| 17-106_k141_53854 | Low-quality |
| 17-106_k141_96777 | Not-determined |
| 17-106_k141_59273 | Not-determined |
| 17-106_k141_43217 | Not-determined |
| 17-106_k141_21659 | Not-determined |
| 17-106_k141_48659 | Not-determined |
| 17-106_k141_21661 | Not-determined |
| 17-106_k141_43221 | Low-quality |
| 17-106_k141_59294 | Not-determined |
| 17-106_k141_96793 | Not-determined |
| 17-106_k141_32358 | Not-determined |
| 17-106_k141_59298 | Not-determined |
| 17-106_k141_96807 | Not-determined |
| 17-106_k141_96808 | Not-determined |
| 17-106_k141_629 | Not-determined |
| 17-106_k141_59304 | Not-determined |
| 17-106_k141_86071 | Not-determined |
| 17-106_k141_59307 | Not-determined |
| 17-106_k141_32365 | Not-determined |
| 17-106_k141_59316 | Not-determined |
| 17-106_k141_650 | Not-determined |
| 17-106_k141_59317 | Not-determined |
| 17-106_k141_660 | Not-determined |
| 17-106_k141_53884 | Not-determined |
| 17-106_k141_48678 | Not-determined |
| 17-106_k141_107206 | Not-determined |
| 17-106_k141_96839 | Not-determined |
| 17-106_k141_48688 | Not-determined |
| 17-106_k141_48691 | Not-determined |
| 17-106_k141_48692 | Not-determined |
| 17-106_k141_96844 | Not-determined |
| 17-106_k141_107209 | Not-determined |
| 17-106_k141_37713 | Low-quality |
| 17-106_k141_118170 | Not-determined |
| 17-106_k141_48699 | Not-determined |
| 17-106_k141_21701 | Not-determined |
| 17-106_k141_10963 | Not-determined |
| 17-106_k141_32375 | Not-determined |
| 17-106_k141_59335 | Not-determined |
| 17-106_k141_21705 | Not-determined |
| 17-106_k141_32378 | Not-determined |
| 17-106_k141_21706 | Not-determined |
| 17-106_k141_91149 | Not-determined |
| 17-106_k141_37725 | Low-quality |
| 17-106_k141_91153 | Not-determined |
| 17-106_k141_10971 | Not-determined |
| 17-106_k141_59339 | Low-quality |
| 17-106_k141_37729 | Low-quality |
| 17-106_k141_10974 | Not-determined |
| 17-106_k141_37734 | Not-determined |
| 17-106_k141_91170 | Low-quality |
| 17-106_k141_37749 | Not-determined |
| 17-106_k141_48705 | Not-determined |
| 17-106_k141_102163 | Not-determined |
| 17-106_k141_48708 | Not-determined |
| 17-106_k141_91194 | Not-determined |
| 17-106_k141_48713 | Not-determined |
| 17-106_k141_5688 | Not-determined |
| 17-106_k141_96892 | Not-determined |
| 17-106_k141_32404 | Not-determined |
| 17-106_k141_64548 | Not-determined |
| 17-106_k141_64551 | Not-determined |
| 17-106_k141_59364 | Not-determined |
| 17-106_k141_102167 | Not-determined |
| 17-106_k141_59367 | Not-determined |
| 17-106_k141_80739 | Not-determined |
| 17-106_k141_91215 | Not-determined |
| 17-106_k141_32419 | Not-determined |
| 17-106_k141_48728 | Not-determined |
| 17-106_k141_64558 | Not-determined |
| 17-106_k141_91222 | Not-determined |
| 17-106_k141_64563 | Not-determined |
| 17-106_k141_118173 | Low-quality |
| 17-106_k141_80753 | Low-quality |
| 17-106_k141_118180 | Not-determined |
| 17-106_k141_86102 | Not-determined |
| 17-106_k141_59385 | Low-quality |
| 17-106_k141_59386 | Not-determined |
| 17-106_k141_43269 | Not-determined |
| 17-106_k141_43271 | Not-determined |
| 17-106_k141_80769 | Not-determined |
| 17-106_k141_27205 | Not-determined |
| 17-106_k141_37792 | Not-determined |
| 17-106_k141_91244 | Not-determined |
| 17-106_k141_21731 | Not-determined |
| 17-106_k141_96917 | Not-determined |
| 17-106_k141_86119 | Not-determined |
| 17-106_k141_707 | Not-determined |
| 17-106_k141_712 | Not-determined |
| 17-106_k141_21738 | Not-determined |
| 17-106_k141_27223 | Not-determined |
| 17-106_k141_27224 | Not-determined |
| 17-106_k141_102204 | Not-determined |
| 17-106_k141_102205 | Not-determined |
| 17-106_k141_37805 | Not-determined |
| 17-106_k141_86143 | Not-determined |
| 17-106_k141_37811 | Not-determined |
| 17-106_k141_5700 | Not-determined |
| 17-106_k141_48753 | Not-determined |
| 17-106_k141_86149 | Not-determined |
| 17-106_k141_123378 | Not-determined |
| 17-106_k141_86152 | Not-determined |
| 17-106_k141_43278 | Not-determined |
| 17-106_k141_43281 | Not-determined |
| 17-106_k141_32452 | Not-determined |
| 17-106_k141_86163 | Not-determined |
| 17-106_k141_64574 | Not-determined |
| 17-106_k141_5718 | Not-determined |
| 17-106_k141_21753 | Not-determined |
| 17-106_k141_32459 | Not-determined |
| 17-106_k141_96949 | Not-determined |
| 17-106_k141_102224 | Low-quality |
| 17-106_k141_16489 | Not-determined |
| 17-106_k141_102230 | Low-quality |
| 17-106_k141_37826 | Not-determined |
| 17-106_k141_102234 | Not-determined |
| 17-106_k141_96963 | Low-quality |
| 17-106_k141_80781 | Not-determined |
| 17-106_k141_5749 | Low-quality |
| 17-106_k141_102247 | Not-determined |
| 17-106_k141_43285 | Not-determined |
| 17-106_k141_43286 | Not-determined |
| 17-106_k141_43288 | Low-quality |
| 17-106_k141_43289 | Not-determined |
| 17-106_k141_53933 | Not-determined |
| 17-106_k141_43296 | Low-quality |
| 17-106_k141_86196 | Not-determined |
| 17-106_k141_48793 | Low-quality |
| 17-106_k141_118238 | Not-determined |
| 17-106_k141_48795 | Not-determined |
| 17-106_k141_96978 | Not-determined |
| 17-106_k141_80803 | Not-determined |
| 17-106_k141_112802 | Not-determined |
| 17-106_k141_735 | Not-determined |
| 17-106_k141_5756 | Low-quality |
| 17-106_k141_5759 | Not-determined |
| 17-106_k141_112811 | Not-determined |
| 17-106_k141_112815 | Low-quality |
| 17-106_k141_123385 | Not-determined |
| 17-106_k141_123386 | Not-determined |
| 17-106_k141_5769 | Not-determined |
| 17-106_k141_37862 | Not-determined |
| 17-106_k141_102262 | Not-determined |
| 17-106_k141_112818 | Low-quality |
| 17-106_k141_107272 | Not-determined |
| 17-106_k141_107274 | Low-quality |
| 17-106_k141_64585 | Not-determined |
| 17-106_k141_37869 | Not-determined |
| 17-106_k141_53956 | Not-determined |
| 17-106_k141_27258 | Not-determined |
| 17-106_k141_5775 | Not-determined |
| 17-106_k141_27261 | Not-determined |
| 17-106_k141_80830 | Not-determined |
| 17-106_k141_64595 | Not-determined |
| 17-106_k141_80832 | Not-determined |
| 17-106_k141_123395 | Not-determined |
| 17-106_k141_752 | Not-determined |
| 17-106_k141_64600 | Not-determined |
| 17-106_k141_43333 | Not-determined |
| 17-106_k141_27272 | Low-quality |
| 17-106_k141_112843 | Not-determined |
| 17-106_k141_80843 | Low-quality |
| 17-106_k141_16531 | Not-determined |
| 17-106_k141_11009 | Low-quality |
| 17-106_k141_80851 | Not-determined |
| 17-106_k141_102279 | Low-quality |
| 17-106_k141_765 | Not-determined |
| 17-106_k141_112845 | Not-determined |
| 17-106_k141_80854 | Not-determined |
| 17-106_k141_59437 | Low-quality |
| 17-106_k141_118254 | Not-determined |
| 17-106_k141_118256 | Not-determined |
| 17-106_k141_107302 | Not-determined |
| 17-106_k141_32486 | Not-determined |
| 17-106_k141_27291 | Not-determined |
| 17-106_k141_768 | Not-determined |
| 17-106_k141_27295 | Not-determined |
| 17-106_k141_59449 | Not-determined |
| 17-106_k141_80876 | Not-determined |
| 17-106_k141_48813 | Not-determined |
| 17-106_k141_27305 | Low-quality |
| 17-106_k141_27307 | Not-determined |
| 17-106_k141_59464 | Not-determined |
| 17-106_k141_80884 | Not-determined |
| 17-106_k141_784 | Not-determined |
| 17-106_k141_16557 | Not-determined |
| 17-106_k141_43369 | Low-quality |
| 17-106_k141_789 | Not-determined |
| 17-106_k141_16567 | Not-determined |
| 17-106_k141_59477 | Low-quality |
| 17-106_k141_43380 | Not-determined |
| 17-106_k141_97025 | Not-determined |
| 17-106_k141_16572 | Not-determined |
| 17-106_k141_16573 | Low-quality |
| 17-106_k141_799 | Not-determined |
| 17-106_k141_32515 | Low-quality |
| 17-106_k141_16577 | Low-quality |
| 17-106_k141_112875 | Not-determined |
| 17-106_k141_32524 | Not-determined |
| 17-106_k141_59480 | Low-quality |
| 17-106_k141_80889 | Not-determined |
| 17-106_k141_97037 | Not-determined |
| 17-106_k141_86202 | Not-determined |
| 17-106_k141_80893 | Not-determined |
| 17-106_k141_59484 | Not-determined |
| 17-106_k141_118286 | Low-quality |
| 17-106_k141_91267 | Not-determined |
| 17-106_k141_37935 | Not-determined |
| 17-106_k141_21828 | Not-determined |
| 17-106_k141_80903 | Not-determined |
| 17-106_k141_809 | Low-quality |
| 17-106_k141_5807 | Not-determined |
| 17-106_k141_32539 | Not-determined |
| 17-106_k141_97053 | Not-determined |
| 17-106_k141_43401 | Not-determined |
| 17-106_k141_118296 | Not-determined |
| 17-106_k141_59493 | Not-determined |
| 17-106_k141_80911 | Not-determined |
| 17-106_k141_32550 | Not-determined |
| 17-106_k141_43404 | Low-quality |
| 17-106_k141_32552 | Not-determined |
| 17-106_k141_818 | Not-determined |
| 17-106_k141_91292 | Not-determined |
| 17-106_k141_97062 | Not-determined |
| 17-106_k141_118315 | Low-quality |
| 17-106_k141_5813 | Not-determined |
| 17-106_k141_32567 | Not-determined |
| 17-106_k141_43421 | Not-determined |
| 17-106_k141_43426 | Not-determined |
| 17-106_k141_86228 | Not-determined |
| 17-106_k141_43428 | Not-determined |
| 17-106_k141_102330 | Not-determined |
| 17-106_k141_118323 | Not-determined |
| 17-106_k141_43437 | Not-determined |
| 17-106_k141_37953 | Low-quality |
| 17-106_k141_80923 | Not-determined |
| 17-106_k141_53996 | Not-determined |
| 17-106_k141_32589 | Not-determined |
| 17-106_k141_112927 | Not-determined |
| 17-106_k141_102342 | Not-determined |
| 17-106_k141_59534 | Not-determined |
| 17-106_k141_32597 | Not-determined |
| 17-106_k141_21851 | Not-determined |
| 17-106_k141_118338 | Not-determined |
| 17-106_k141_112939 | Not-determined |
| 17-106_k141_59546 | Not-determined |
| 17-106_k141_86245 | Not-determined |
| 17-106_k141_102356 | Not-determined |
| 17-106_k141_5833 | Not-determined |
| 17-106_k141_37962 | Not-determined |
| 17-106_k141_37964 | Not-determined |
| 17-106_k141_882 | Not-determined |
| 17-106_k141_5853 | Low-quality |
| 17-106_k141_112951 | Not-determined |
| 17-106_k141_112956 | Not-determined |
| 17-106_k141_118358 | Low-quality |
| 17-106_k141_5858 | Not-determined |
| 17-106_k141_21870 | Low-quality |
| 17-106_k141_91316 | Not-determined |
| 17-106_k141_21879 | Not-determined |
| 17-106_k141_64611 | Not-determined |
| 17-106_k141_59591 | Not-determined |
| 17-106_k141_54007 | Not-determined |
| 17-106_k141_86296 | Not-determined |
| 17-106_k141_80983 | Low-quality |
| 17-106_k141_112972 | Not-determined |
| 17-106_k141_5881 | Not-determined |
| 17-106_k141_97072 | Not-determined |
| 17-106_k141_32613 | Low-quality |
| 17-106_k141_11080 | Not-determined |
| 17-106_k141_112982 | Not-determined |
| 17-106_k141_21897 | Not-determined |
| 17-106_k141_11081 | Not-determined |
| 17-106_k141_86317 | Not-determined |
| 17-106_k141_5893 | Not-determined |
| 17-106_k141_91332 | Low-quality |
| 17-106_k141_80996 | Not-determined |
| 17-106_k141_102367 | Not-determined |
| 17-106_k141_27340 | Not-determined |
| 17-106_k141_81001 | Not-determined |
| 17-106_k141_905 | Not-determined |
| 17-106_k141_27343 | Not-determined |
| 17-106_k141_123461 | Not-determined |
| 17-106_k141_107372 | Low-quality |
| 17-106_k141_102372 | Low-quality |
| 17-106_k141_907 | Low-quality |
| 17-106_k141_909 | Low-quality |
| 17-106_k141_911 | Not-determined |
| 17-106_k141_123466 | Not-determined |

| | |
|---|---|
| 17-106_k141_107375 | Not-determined |
| 17-106_k141_11086 | Not-determined |
| 17-106_k141_5915 | Not-determined |
| 17-106_k141_5916 | Not-determined |
| 17-106_k141_123471 | Not-determined |
| 17-106_k141_107384 | Not-determined |
| 17-106_k141_32632 | Not-determined |
| 17-106_k141_922 | Not-determined |
| 17-106_k141_107387 | Low-quality |
| 17-106_k141_5925 | Not-determined |
| 17-106_k141_64654 | Not-determined |
| 17-106_k141_48857 | Not-determined |
| 17-106_k141_32640 | Not-determined |
| 17-106_k141_32641 | Not-determined |
| 17-106_k141_27352 | Not-determined |
| 17-106_k141_27356 | Not-determined |
| 17-106_k141_118405 | Not-determined |
| 17-106_k141_97131 | Not-determined |
| 17-106_k141_64665 | Not-determined |
| 17-106_k141_118410 | Not-determined |
| 17-106_k141_97138 | Not-determined |
| 17-106_k141_21908 | Not-determined |
| 17-106_k141_32652 | Not-determined |
| 17-106_k141_27363 | Not-determined |
| 17-106_k141_21914 | Not-determined |
| 17-106_k141_21920 | Not-determined |
| 17-106_k141_69907 | Not-determined |
| 17-106_k141_118421 | Not-determined |
| 17-106_k141_27377 | Not-determined |
| 17-106_k141_16609 | Not-determined |
| 17-106_k141_5951 | Not-determined |
| 17-106_k141_59618 | Not-determined |
| 17-106_k141_5959 | Not-determined |
| 17-106_k141_27387 | Not-determined |
| 17-106_k141_951 | Not-determined |
| 17-106_k141_64688 | Not-determined |
| 17-106_k141_953 | Not-determined |
| 17-106_k141_16619 | Not-determined |
| 17-106_k141_69923 | Low-quality |
| 17-106_k141_958 | Not-determined |
| 17-106_k141_965 | Not-determined |
| 17-106_k141_5966 | Low-quality |
| 17-106_k141_59647 | Not-determined |
| 17-106_k141_64699 | Not-determined |
| 17-106_k141_32672 | Not-determined |
| 17-106_k141_64702 | Not-determined |
| 17-106_k141_59653 | Low-quality |
| 17-106_k141_971 | Not-determined |
| 17-106_k141_123492 | Not-determined |
| 17-106_k141_5975 | Not-determined |
| 17-106_k141_64706 | Not-determined |
| 17-106_k141_11095 | Not-determined |
| 17-106_k141_980 | Not-determined |
| 17-106_k141_81047 | Low-quality |
| 17-106_k141_102443 | Not-determined |
| 17-106_k141_81050 | Not-determined |
| 17-106_k141_64718 | Low-quality |
| 17-106_k141_987 | Not-determined |
| 17-106_k141_118444 | Not-determined |
| 17-106_k141_102451 | Not-determined |
| 17-106_k141_64731 | Not-determined |
| 17-106_k141_64734 | Low-quality |
| 17-106_k141_59664 | Not-determined |
| 17-106_k141_69948 | Not-determined |
| 17-106_k141_6004 | Not-determined |
| 17-106_k141_69951 | Not-determined |
| 17-106_k141_102467 | Not-determined |
| 17-106_k141_11106 | Not-determined |
| 17-106_k141_81072 | Not-determined |
| 17-106_k141_32731 | Not-determined |
| 17-106_k141_102486 | Not-determined |
| 17-106_k141_21954 | Low-quality |
| 17-106_k141_81087 | Low-quality |
| 17-106_k141_69968 | Not-determined |
| 17-106_k141_69971 | Not-determined |
| 17-106_k141_21968 | Not-determined |
| 17-106_k141_102495 | Not-determined |
| 17-106_k141_102496 | Not-determined |
| 17-106_k141_32755 | Not-determined |
| 17-106_k141_91379 | Not-determined |
| 17-106_k141_69979 | Not-determined |
| 17-106_k141_86372 | Not-determined |
| 17-106_k141_21976 | Not-determined |
| 17-106_k141_38015 | Not-determined |
| 17-106_k141_81099 | Low-quality |
| 17-106_k141_118473 | Not-determined |
| 17-106_k141_81104 | Not-determined |
| 17-106_k141_38029 | Not-determined |
| 17-106_k141_91393 | Low-quality |
| 17-106_k141_102532 | Not-determined |
| 17-106_k141_118479 | Low-quality |
| 17-106_k141_102534 | Low-quality |
| 17-106_k141_118481 | Not-determined |
| 17-106_k141_69999 | Low-quality |
| 17-106_k141_118487 | Not-determined |
| 17-106_k141_27398 | Not-determined |
| 17-106_k141_38038 | Not-determined |
| 17-106_k141_27399 | Not-determined |
| 17-106_k141_38039 | Not-determined |
| 17-106_k141_118490 | Not-determined |
| 17-106_k141_102542 | Not-determined |
| 17-106_k141_38043 | Not-determined |
| 17-106_k141_48958 | Not-determined |
| 17-106_k141_86385 | Not-determined |
| 17-106_k141_86387 | Not-determined |
| 17-106_k141_123545 | Not-determined |
| 17-106_k141_97196 | Not-determined |
| 17-106_k141_97197 | Not-determined |
| 17-106_k141_48979 | Not-determined |
| 17-106_k141_1066 | Not-determined |
| 17-106_k141_81109 | Not-determined |
| 17-106_k141_81112 | Not-determined |
| 17-106_k141_97202 | Not-determined |
| 17-106_k141_32804 | Not-determined |
| 17-106_k141_11176 | Not-determined |
| 17-106_k141_123558 | Low-quality |
| 17-106_k141_48989 | Not-determined |
| 17-106_k141_32810 | Not-determined |
| 17-106_k141_11177 | Low-quality |
| 17-106_k141_86418 | Not-determined |
| 17-106_k141_86419 | Not-determined |
| 17-106_k141_102563 | Not-determined |
| 17-106_k141_64741 | Not-determined |
| 17-106_k141_21990 | Not-determined |
| 17-106_k141_1099 | Not-determined |
| 17-106_k141_86428 | Not-determined |
| 17-106_k141_21997 | Not-determined |
| 17-106_k141_49002 | Low-quality |
| 17-106_k141_27438 | Not-determined |
| 17-106_k141_59688 | Not-determined |
| 17-106_k141_91437 | Not-determined |
| 17-106_k141_49008 | Not-determined |
| 17-106_k141_27444 | Low-quality |
| 17-106_k141_64751 | Not-determined |
| 17-106_k141_81116 | Not-determined |
| 17-106_k141_38091 | Not-determined |
| 17-106_k141_59705 | Low-quality |
| 17-106_k141_1116 | Not-determined |
| 17-106_k141_86438 | Not-determined |
| 17-106_k141_1121 | Not-determined |
| 17-106_k141_38103 | Not-determined |
| 17-106_k141_38106 | Not-determined |
| 17-106_k141_91455 | Not-determined |
| 17-106_k141_91459 | Not-determined |
| 17-106_k141_64765 | Low-quality |
| 17-106_k141_38114 | Not-determined |
| 17-106_k141_27449 | Not-determined |
| 17-106_k141_27452 | Not-determined |
| 17-106_k141_70062 | Not-determined |
| 17-106_k141_81153 | Low-quality |
| 17-106_k141_86454 | Low-quality |
| 17-106_k141_59726 | Not-determined |
| 17-106_k141_81164 | Not-determined |
| 17-106_k141_86465 | Not-determined |
| 17-106_k141_16684 | Not-determined |
| 17-106_k141_1154 | Not-determined |
| 17-106_k141_91492 | Not-determined |
| 17-106_k141_1158 | Not-determined |
| 17-106_k141_91499 | Not-determined |
| 17-106_k141_38135 | Not-determined |
| 17-106_k141_70093 | Not-determined |
| 17-106_k141_6019 | Not-determined |
| 17-106_k141_70099 | Low-quality |
| 17-106_k141_113017 | Low-quality |
| 17-106_k141_81193 | Not-determined |
| 17-106_k141_1187 | Not-determined |
| 17-106_k141_54088 | Not-determined |
| 17-106_k141_64800 | Not-determined |
| 17-106_k141_27476 | Not-determined |
| 17-106_k141_1197 | Not-determined |
| 17-106_k141_75381 | Low-quality |
| 17-106_k141_86476 | Not-determined |
| 17-106_k141_86478 | Not-determined |
| 17-106_k141_32850 | Not-determined |
| 17-106_k141_38162 | Not-determined |
| 17-106_k141_97227 | Not-determined |
| 17-106_k141_91524 | Not-determined |
| 17-106_k141_32855 | Not-determined |
| 17-106_k141_97230 | Not-determined |
| 17-106_k141_11237 | Not-determined |
| 17-106_k141_1205 | Not-determined |
| 17-106_k141_32858 | Not-determined |
| 17-106_k141_1208 | Not-determined |
| 17-106_k141_38166 | Not-determined |
| 17-106_k141_38169 | Not-determined |
| 17-106_k141_81232 | Not-determined |
| 17-106_k141_91532 | Not-determined |
| 17-106_k141_1214 | Not-determined |
| 17-106_k141_32867 | Not-determined |
| 17-106_k141_81239 | Not-determined |
| 17-106_k141_38172 | Not-determined |
| 17-106_k141_81248 | Not-determined |
| 17-106_k141_54108 | Not-determined |
| 17-106_k141_54109 | Not-determined |
| 17-106_k141_11239 | Low-quality |
| 17-106_k141_22024 | Not-determined |
| 17-106_k141_22025 | Not-determined |
| 17-106_k141_97251 | Not-determined |
| 17-106_k141_38191 | Not-determined |
| 17-106_k141_22027 | Not-determined |
| 17-106_k141_32886 | Not-determined |
| 17-106_k141_11262 | Not-determined |
| 17-106_k141_27501 | Not-determined |
| 17-106_k141_54135 | Not-determined |
| 17-106_k141_11270 | Low-quality |
| 17-106_k141_22030 | Not-determined |
| 17-106_k141_123590 | Not-determined |
| 17-106_k141_16711 | Not-determined |
| 17-106_k141_123592 | Low-quality |
| 17-106_k141_123593 | Low-quality |
| 17-106_k141_91556 | Not-determined |
| 17-106_k141_81265 | Not-determined |
| 17-106_k141_86492 | Not-determined |
| 17-106_k141_113038 | Not-determined |
| 17-106_k141_1238 | Not-determined |
| 17-106_k141_81269 | Not-determined |
| 17-106_k141_86495 | Not-determined |
| 17-106_k141_32903 | Not-determined |
| 17-106_k141_123609 | Not-determined |
| 17-106_k141_107459 | Not-determined |
| 17-106_k141_81281 | Not-determined |
| 17-106_k141_81282 | Not-determined |
| 17-106_k141_107463 | Not-determined |
| 17-106_k141_38212 | Not-determined |
| 17-106_k141_107468 | Not-determined |
| 17-106_k141_16726 | Low-quality |
| 17-106_k141_59740 | Not-determined |
| 17-106_k141_86501 | Not-determined |
| 17-106_k141_102591 | Not-determined |
| 17-106_k141_32918 | Not-determined |
| 17-106_k141_86510 | Not-determined |
| 17-106_k141_32922 | Not-determined |
| 17-106_k141_86516 | Not-determined |
| 17-106_k141_16730 | Low-quality |
| 17-106_k141_118562 | Not-determined |
| 17-106_k141_81301 | Not-determined |
| 17-106_k141_81302 | Not-determined |
| 17-106_k141_75432 | Not-determined |
| 17-106_k141_81307 | Not-determined |
| 17-106_k141_59749 | Not-determined |
| 17-106_k141_59751 | Not-determined |
| 17-106_k141_59753 | Not-determined |
| 17-106_k141_107486 | Not-determined |
| 17-106_k141_123633 | Low-quality |
| 17-106_k141_118569 | Not-determined |
| 17-106_k141_118570 | Not-determined |
| 17-106_k141_91595 | Not-determined |
| 17-106_k141_97274 | Low-quality |
| 17-106_k141_123637 | Not-determined |
| 17-106_k141_38246 | Not-determined |
| 17-106_k141_107493 | Not-determined |
| 17-106_k141_54158 | Not-determined |
| 17-106_k141_6034 | Not-determined |
| 17-106_k141_59772 | Not-determined |
| 17-106_k141_107504 | Not-determined |
| 17-106_k141_102607 | Not-determined |
| 17-106_k141_123654 | Not-determined |
| 17-106_k141_11293 | Not-determined |
| 17-106_k141_1263 | Low-quality |
| 17-106_k141_97293 | Not-determined |
| 17-106_k141_11296 | Not-determined |
| 17-106_k141_11297 | Not-determined |
| 17-106_k141_123663 | Not-determined |
| 17-106_k141_107509 | Not-determined |
| 17-106_k141_54171 | Not-determined |
| 17-106_k141_107516 | Not-determined |
| 17-106_k141_32953 | Low-quality |
| 17-106_k141_32954 | Not-determined |
| 17-106_k141_113082 | Not-determined |
| 17-106_k141_123682 | Not-determined |
| 17-106_k141_1282 | Not-determined |
| 17-106_k141_54184 | Not-determined |
| 17-106_k141_123687 | Low-quality |
| 17-106_k141_123688 | Not-determined |
| 17-106_k141_97307 | Not-determined |
| 17-106_k141_123693 | Not-determined |
| 17-106_k141_1294 | Not-determined |
| 17-106_k141_54201 | Not-determined |
| 17-106_k141_123701 | Not-determined |
| 17-106_k141_27523 | Not-determined |
| 17-106_k141_86532 | Low-quality |
| 17-106_k141_86534 | Not-determined |
| 17-106_k141_27526 | Not-determined |
| 17-106_k141_32974 | Not-determined |
| 17-106_k141_86539 | Not-determined |
| 17-106_k141_54211 | Not-determined |
| 17-106_k141_123712 | Not-determined |
| 17-106_k141_118607 | Not-determined |
| 17-106_k141_49027 | Low-quality |
| 17-106_k141_113117 | Not-determined |
| 17-106_k141_64829 | Not-determined |
| 17-106_k141_49029 | Not-determined |
| 17-106_k141_49033 | Not-determined |
| 17-106_k141_91645 | Not-determined |
| 17-106_k141_86543 | Low-quality |
| 17-106_k141_32990 | Not-determined |
| 17-106_k141_86554 | Not-determined |
| 17-106_k141_91656 | Not-determined |
| 17-106_k141_11320 | Not-determined |
| 17-106_k141_11323 | Not-determined |
| 17-106_k141_33001 | Not-determined |
| 17-106_k141_11326 | Low-quality |
| 17-106_k141_64840 | Not-determined |
| 17-106_k141_49049 | Not-determined |
| 17-106_k141_27547 | Not-determined |
| 17-106_k141_11342 | Low-quality |
| 17-106_k141_75480 | Not-determined |
| 17-106_k141_38256 | Not-determined |
| 17-106_k141_64856 | Not-determined |
| 17-106_k141_1323 | Not-determined |
| 17-106_k141_54246 | Low-quality |
| 17-106_k141_118632 | Not-determined |
| 17-106_k141_118633 | Low-quality |
| 17-106_k141_118637 | Not-determined |
| 17-106_k141_6049 | Not-determined |
| 17-106_k141_38267 | Not-determined |
| 17-106_k141_27572 | Not-determined |
| 17-106_k141_27575 | Not-determined |
| 17-106_k141_38271 | Not-determined |
| 17-106_k141_1330 | Not-determined |
| 17-106_k141_38275 | Not-determined |
| 17-106_k141_64864 | Not-determined |
| 17-106_k141_64868 | Not-determined |
| 17-106_k141_1336 | Not-determined |
| 17-106_k141_107586 | Not-determined |
| 17-106_k141_64870 | Not-determined |
| 17-106_k141_33018 | Not-determined |
| 17-106_k141_43482 | Not-determined |
| 17-106_k141_1339 | Not-determined |
| 17-106_k141_64874 | Not-determined |
| 17-106_k141_49085 | Not-determined |
| 17-106_k141_27595 | Not-determined |
| 17-106_k141_27600 | Not-determined |
| 17-106_k141_107592 | Not-determined |
| 17-106_k141_123755 | Low-quality |
| 17-106_k141_64893 | Not-determined |
| 17-106_k141_54271 | Not-determined |
| 17-106_k141_86593 | Low-quality |
| 17-106_k141_54273 | Not-determined |
| 17-106_k141_86598 | Not-determined |
| 17-106_k141_86600 | Not-determined |
| 17-106_k141_54285 | Not-determined |
| 17-106_k141_102647 | Not-determined |
| 17-106_k141_54290 | Low-quality |
| 17-106_k141_54292 | Not-determined |
| 17-106_k141_54294 | Not-determined |
| 17-106_k141_123769 | Not-determined |
| 17-106_k141_107610 | Not-determined |
| 17-106_k141_113130 | Not-determined |
| 17-106_k141_27626 | Not-determined |
| 17-106_k141_113133 | Not-determined |
| 17-106_k141_43507 | Not-determined |
| 17-106_k141_123775 | Not-determined |
| 17-106_k141_54302 | Not-determined |
| 17-106_k141_22108 | Low-quality |
| 17-106_k141_102650 | Not-determined |
| 17-106_k141_64913 | Not-determined |
| 17-106_k141_43513 | Not-determined |
| 17-106_k141_43516 | Not-determined |
| 17-106_k141_33050 | Not-determined |
| 17-106_k141_113139 | Not-determined |
| 17-106_k141_43518 | Not-determined |
| 17-106_k141_27633 | Not-determined |
| 17-106_k141_38291 | Not-determined |
| 17-106_k141_54314 | Not-determined |
| 17-106_k141_123787 | Low-quality |
| 17-106_k141_64937 | Not-determined |
| 17-106_k141_123793 | Not-determined |
| 17-106_k141_107619 | Not-determined |
| 17-106_k141_64942 | Not-determined |
| 17-106_k141_123795 | Not-determined |
| 17-106_k141_123796 | Not-determined |
| 17-106_k141_113177 | Not-determined |
| 17-106_k141_27656 | Not-determined |
| 17-106_k141_118655 | Not-determined |
| 17-106_k141_59838 | Not-determined |
| 17-106_k141_33066 | Not-determined |
| 17-106_k141_6152 | Not-determined |
| 17-106_k141_6153 | Not-determined |
| 17-106_k141_118673 | Not-determined |
| 17-106_k141_118682 | Not-determined |
| 17-106_k141_64969 | Not-determined |
| 17-106_k141_1369 | Not-determined |
| 17-106_k141_1370 | Not-determined |
| 17-106_k141_118687 | Not-determined |
| 17-106_k141_33082 | Not-determined |
| 17-106_k141_33084 | Not-determined |
| 17-106_k141_49109 | Not-determined |
| 17-106_k141_33086 | Not-determined |
| 17-106_k141_33091 | Not-determined |
| 17-106_k141_1382 | Not-determined |
| 17-106_k141_59874 | Not-determined |
| 17-106_k141_59885 | Not-determined |
| 17-106_k141_33103 | Not-determined |
| 17-106_k141_49125 | Not-determined |
| 17-106_k141_91726 | Not-determined |
| 17-106_k141_64988 | Not-determined |
| 17-106_k141_54352 | Low-quality |
| 17-106_k141_33128 | Not-determined |
| 17-106_k141_54356 | Low-quality |
| 17-106_k141_33131 | Not-determined |
| 17-106_k141_43539 | Not-determined |
| 17-106_k141_81371 | Not-determined |
| 17-106_k141_43541 | Not-determined |
| 17-106_k141_54360 | Not-determined |
| 17-106_k141_54366 | Not-determined |
| 17-106_k141_59916 | Not-determined |
| 17-106_k141_102673 | Low-quality |
| 17-106_k141_22144 | Not-determined |
| 17-106_k141_43552 | Not-determined |
| 17-106_k141_107637 | Not-determined |
| 17-106_k141_49137 | Not-determined |
| 17-106_k141_43555 | Not-determined |
| 17-106_k141_27708 | Not-determined |
| 17-106_k141_102677 | Not-determined |
| 17-106_k141_54380 | Not-determined |
| 17-106_k141_59921 | Not-determined |
| 17-106_k141_59925 | Not-determined |
| 17-106_k141_102682 | Not-determined |
| 17-106_k141_27712 | Not-determined |
| 17-106_k141_102686 | Not-determined |
| 17-106_k141_102698 | Not-determined |
| 17-106_k141_6178 | Low-quality |
| 17-106_k141_6183 | Not-determined |
| 17-106_k141_33164 | Not-determined |
| 17-106_k141_43575 | Low-quality |
| 17-106_k141_97338 | Not-determined |
| 17-106_k141_97341 | Not-determined |
| 17-106_k141_97343 | Low-quality |
| 17-106_k141_81382 | Not-determined |
| 17-106_k141_97345 | Not-determined |
| 17-106_k141_6194 | Not-determined |
| 17-106_k141_54404 | Not-determined |
| 17-106_k141_54405 | Not-determined |
| 17-106_k141_59940 | Not-determined |
| 17-106_k141_97360 | Not-determined |
| 17-106_k141_59941 | Not-determined |
| 17-106_k141_22184 | Not-determined |
| 17-106_k141_107680 | Not-determined |
| 17-106_k141_11430 | Not-determined |
| 17-106_k141_6202 | Not-determined |
| 17-106_k141_54415 | Not-determined |
| 17-106_k141_102709 | Low-quality |
| 17-106_k141_33186 | Not-determined |
| 17-106_k141_11439 | Not-determined |
| 17-106_k141_22194 | Not-determined |
| 17-106_k141_102720 | Not-determined |
| 17-106_k141_27755 | Not-determined |
| 17-106_k141_65048 | Not-determined |
| 17-106_k141_107701 | Not-determined |
| 17-106_k141_59975 | Not-determined |
| 17-106_k141_59980 | Not-determined |
| 17-106_k141_6226 | Not-determined |
| 17-106_k141_97398 | Not-determined |
| 17-106_k141_27766 | Not-determined |
| 17-106_k141_54438 | Not-determined |
| 17-106_k141_27770 | Not-determined |
| 17-106_k141_33197 | Low-quality |
| 17-106_k141_91749 | Not-determined |
| 17-106_k141_33199 | Not-determined |
| 17-106_k141_33201 | Not-determined |
| 17-106_k141_107718 | Not-determined |
| 17-106_k141_91753 | Not-determined |
| 17-106_k141_11461 | Not-determined |
| 17-106_k141_97410 | Low-quality |
| 17-106_k141_43604 | Not-determined |
| 17-106_k141_91756 | Not-determined |
| 17-106_k141_33214 | Not-determined |
| 17-106_k141_1418 | Not-determined |
| 17-106_k141_107731 | Not-determined |
| 17-106_k141_65062 | Not-determined |
| 17-106_k141_91763 | Not-determined |
| 17-106_k141_22234 | Not-determined |
| 17-106_k141_22235 | Not-determined |
| 17-106_k141_59990 | Not-determined |
| 17-106_k141_27777 | Not-determined |
| 17-106_k141_118716 | Not-determined |
| 17-106_k141_91786 | Not-determined |
| 17-106_k141_43614 | Not-determined |
| 17-106_k141_1432 | Not-determined |
| 17-106_k141_22255 | Low-quality |
| 17-106_k141_49206 | Not-determined |
| 17-106_k141_1435 | Not-determined |
| 17-106_k141_91796 | Low-quality |
| 17-106_k141_49216 | Low-quality |
| 17-106_k141_49218 | Low-quality |
| 17-106_k141_97416 | Low-quality |
| 17-106_k141_107754 | Not-determined |
| 17-106_k141_1453 | Not-determined |
| 17-106_k141_97422 | Not-determined |
| 17-106_k141_86655 | Not-determined |
| 17-106_k141_86657 | Low-quality |
| 17-106_k141_27795 | Not-determined |
| 17-106_k141_16756 | Not-determined |
| 17-106_k141_43622 | Not-determined |
| 17-106_k141_75509 | Not-determined |
| 17-106_k141_38308 | Not-determined |
| 17-106_k141_38310 | Not-determined |
| 17-106_k141_22289 | Not-determined |
| 17-106_k141_22291 | Not-determined |
| 17-106_k141_75519 | Not-determined |
| 17-106_k141_75521 | Not-determined |
| 17-106_k141_38315 | Not-determined |
| 17-106_k141_86685 | Not-determined |
| 17-106_k141_11467 | Not-determined |
| 17-106_k141_27825 | Not-determined |
| 17-106_k141_75528 | Not-determined |
| 17-106_k141_38320 | Not-determined |
| 17-106_k141_38322 | Not-determined |
| 17-106_k141_107759 | Not-determined |
| 17-106_k141_38323 | Not-determined |
| 17-106_k141_22309 | Not-determined |
| 17-106_k141_43638 | Not-determined |
| 17-106_k141_27837 | Not-determined |
| 17-106_k141_81425 | Low-quality |
| 17-106_k141_102810 | Not-determined |
| 17-106_k141_86721 | Not-determined |
| 17-106_k141_11484 | Not-determined |
| 17-106_k141_27853 | Low-quality |
| 17-106_k141_38346 | Not-determined |
| 17-106_k141_38347 | Not-determined |
| 17-106_k141_1480 | Not-determined |
| 17-106_k141_75556 | Not-determined |
| 17-106_k141_22317 | Not-determined |
| 17-106_k141_27860 | Not-determined |
| 17-106_k141_86738 | Not-determined |
| 17-106_k141_86742 | Low-quality |
| 17-106_k141_54464 | Not-determined |
| 17-106_k141_1496 | Not-determined |
| 17-106_k141_107825 | Not-determined |
| 17-106_k141_38349 | Low-quality |
| 17-106_k141_22339 | Not-determined |
| 17-106_k141_38352 | Not-determined |
| 17-106_k141_54486 | Not-determined |
| 17-106_k141_11488 | Not-determined |
| 17-106_k141_11490 | Not-determined |
| 17-106_k141_11491 | Not-determined |
| 17-106_k141_54489 | Not-determined |
| 17-106_k141_97479 | Not-determined |
| 17-106_k141_81444 | Not-determined |
| 17-106_k141_75575 | Not-determined |
| 17-106_k141_16796 | Not-determined |
| 17-106_k141_27880 | Not-determined |
| 17-106_k141_107838 | Not-determined |
| 17-106_k141_118736 | Not-determined |
| 17-106_k141_118737 | Not-determined |
| 17-106_k141_118738 | Not-determined |
| 17-106_k141_27895 | Not-determined |
| 17-106_k141_97502 | Not-determined |
| 17-106_k141_97503 | Not-determined |
| 17-106_k141_6332 | Not-determined |
| 17-106_k141_107853 | Not-determined |
| 17-106_k141_6338 | Not-determined |
| 17-106_k141_27905 | Low-quality |
| 17-106_k141_11503 | Not-determined |
| 17-106_k141_70197 | Not-determined |
| 17-106_k141_54511 | Not-determined |
| 17-106_k141_70198 | Not-determined |
| 17-106_k141_123892 | Not-determined |
| 17-106_k141_1505 | Not-determined |
| 17-106_k141_123894 | Not-determined |
| 17-106_k141_6347 | Not-determined |
| 17-106_k141_6348 | Not-determined |
| 17-106_k141_123909 | Not-determined |
| 17-106_k141_22382 | Not-determined |
| 17-106_k141_123911 | Low-quality |
| 17-106_k141_1509 | Not-determined |
| 17-106_k141_70214 | Low-quality |
| 17-106_k141_102847 | Not-determined |
| 17-106_k141_54537 | Not-determined |
| 17-106_k141_107864 | Not-determined |
| 17-106_k141_1514 | Not-determined |
| 17-106_k141_1520 | Not-determined |
| 17-106_k141_43665 | Not-determined |
| 17-106_k141_1524 | Not-determined |
| 17-106_k141_123938 | Not-determined |
| 17-106_k141_65074 | Not-determined |
| 17-106_k141_65077 | Not-determined |
| 17-106_k141_81486 | Not-determined |
| 17-106_k141_70231 | Not-determined |
| 17-106_k141_22416 | Not-determined |
| 17-106_k141_54563 | Not-determined |
| 17-106_k141_16829 | Not-determined |
| 17-106_k141_6363 | Not-determined |
| 17-106_k141_107898 | Not-determined |
| 17-106_k141_118798 | Not-determined |
| 17-106_k141_54567 | Not-determined |
| 17-106_k141_54569 | Not-determined |
| 17-106_k141_97518 | Not-determined |

| | |
|---|---|
| 17-106_k141_123953 | Not-determined |
| 17-106_k141_123954 | Not-determined |
| 17-106_k141_123958 | Not-determined |
| 17-106_k141_123961 | Not-determined |
| 17-106_k141_118805 | Not-determined |
| 17-106_k141_43683 | Not-determined |
| 17-106_k141_123967 | Not-determined |
| 17-106_k141_70252 | Not-determined |
| 17-106_k141_11551 | Not-determined |
| 17-106_k141_1544 | Not-determined |
| 17-106_k141_54575 | Not-determined |
| 17-106_k141_123973 | Not-determined |
| 17-106_k141_27947 | Not-determined |
| 17-106_k141_27948 | Not-determined |
| 17-106_k141_65103 | Not-determined |
| 17-106_k141_65118 | Not-determined |
| 17-106_k141_102866 | Not-determined |
| 17-106_k141_102874 | Not-determined |
| 17-106_k141_65124 | Not-determined |
| 17-106_k141_1571 | Not-determined |
| 17-106_k141_124000 | Not-determined |
| 17-106_k141_65129 | Not-determined |
| 17-106_k141_124003 | Not-determined |
| 17-106_k141_81508 | Low-quality |
| 17-106_k141_81515 | Not-determined |
| 17-106_k141_86806 | Not-determined |
| 17-106_k141_86810 | Not-determined |
| 17-106_k141_11562 | Not-determined |
| 17-106_k141_6394 | Not-determined |
| 17-106_k141_16843 | Not-determined |
| 17-106_k141_49253 | Not-determined |
| 17-106_k141_65147 | Not-determined |
| 17-106_k141_16852 | Not-determined |
| 17-106_k141_16855 | Not-determined |
| 17-106_k141_27982 | Low-quality |
| 17-106_k141_65155 | Not-determined |
| 17-106_k141_65157 | Not-determined |
| 17-106_k141_1603 | Not-determined |
| 17-106_k141_113236 | Not-determined |
| 17-106_k141_65162 | Not-determined |
| 17-106_k141_113237 | Not-determined |
| 17-106_k141_65164 | Not-determined |
| 17-106_k141_81552 | Not-determined |
| 17-106_k141_28001 | Low-quality |
| 17-106_k141_49265 | Low-quality |
| 17-106_k141_1613 | Not-determined |
| 17-106_k141_28007 | Not-determined |
| 17-106_k141_22456 | Not-determined |
| 17-106_k141_124043 | Low-quality |
| 17-106_k141_102896 | Not-determined |
| 17-106_k141_124048 | Not-determined |
| 17-106_k141_86814 | Not-determined |
| 17-106_k141_86817 | Not-determined |
| 17-106_k141_28016 | Not-determined |
| 17-106_k141_118854 | Not-determined |
| 17-106_k141_54617 | Not-determined |
| 17-106_k141_49274 | Not-determined |
| 17-106_k141_113242 | Not-determined |
| 17-106_k141_86824 | Low-quality |
| 17-106_k141_11589 | Not-determined |
| 17-106_k141_11590 | Not-determined |
| 17-106_k141_54629 | Low-quality |
| 17-106_k141_11598 | Not-determined |
| 17-106_k141_70348 | Not-determined |
| 17-106_k141_11602 | Not-determined |
| 17-106_k141_75648 | Not-determined |
| 17-106_k141_124059 | Not-determined |
| 17-106_k141_65196 | Low-quality |
| 17-106_k141_6418 | Not-determined |
| 17-106_k141_22484 | Not-determined |
| 17-106_k141_86842 | Not-determined |
| 17-106_k141_49282 | Low-quality |
| 17-106_k141_1644 | Not-determined |
| 17-106_k141_118878 | Not-determined |
| 17-106_k141_113254 | Not-determined |
| 17-106_k141_75667 | Low-quality |
| 17-106_k141_113259 | Not-determined |
| 17-106_k141_54642 | Not-determined |
| 17-106_k141_91867 | Not-determined |
| 17-106_k141_60030 | Not-determined |
| 17-106_k141_75684 | Low-quality |
| 17-106_k141_75688 | Not-determined |
| 17-106_k141_60036 | Not-determined |
| 17-106_k141_6443 | Not-determined |
| 17-106_k141_75699 | Not-determined |
| 17-106_k141_118888 | Not-determined |
| 17-106_k141_6452 | Not-determined |
| 17-106_k141_6453 | Not-determined |
| 17-106_k141_6454 | Not-determined |
| 17-106_k141_81615 | Not-determined |
| 17-106_k141_75706 | Not-determined |
| 17-106_k141_54646 | Not-determined |
| 17-106_k141_38376 | Not-determined |
| 17-106_k141_118900 | Not-determined |
| 17-106_k141_75715 | Not-determined |
| 17-106_k141_54655 | Not-determined |
| 17-106_k141_54656 | Not-determined |
| 17-106_k141_124078 | Not-determined |
| 17-106_k141_75718 | Low-quality |
| 17-106_k141_43705 | Not-determined |
| 17-106_k141_81627 | Not-determined |
| 17-106_k141_124080 | Not-determined |
| 17-106_k141_91892 | Not-determined |
| 17-106_k141_60070 | Not-determined |
| 17-106_k141_75723 | Not-determined |
| 17-106_k141_91898 | Not-determined |
| 17-106_k141_38389 | Not-determined |
| 17-106_k141_91903 | Not-determined |
| 17-106_k141_91915 | Not-determined |
| 17-106_k141_11621 | Not-determined |
| 17-106_k141_16898 | Not-determined |
| 17-106_k141_38401 | Not-determined |
| 17-106_k141_54668 | Not-determined |
| 17-106_k141_16901 | Not-determined |
| 17-106_k141_54673 | Low-quality |
| 17-106_k141_49303 | Not-determined |
| 17-106_k141_60093 | Not-determined |
| 17-106_k141_65235 | Not-determined |
| 17-106_k141_54681 | Not-determined |
| 17-106_k141_11626 | Not-determined |
| 17-106_k141_60096 | Not-determined |
| 17-106_k141_43722 | Not-determined |
| 17-106_k141_91932 | Not-determined |
| 17-106_k141_97542 | Not-determined |
| 17-106_k141_70378 | Not-determined |
| 17-106_k141_16907 | Not-determined |
| 17-106_k141_118919 | Not-determined |
| 17-106_k141_102915 | Not-determined |
| 17-106_k141_16914 | Not-determined |
| 17-106_k141_91940 | Not-determined |
| 17-106_k141_11631 | Not-determined |
| 17-106_k141_113298 | Not-determined |
| 17-106_k141_113299 | Not-determined |
| 17-106_k141_81654 | Not-determined |
| 17-106_k141_113301 | Not-determined |
| 17-106_k141_118931 | Low-quality |
| 17-106_k141_49330 | Not-determined |
| 17-106_k141_22548 | Not-determined |
| 17-106_k141_22549 | Not-determined |
| 17-106_k141_113309 | Not-determined |
| 17-106_k141_1678 | Not-determined |
| 17-106_k141_22553 | Low-quality |
| 17-106_k141_102926 | Not-determined |
| 17-106_k141_1684 | Not-determined |
| 17-106_k141_11662 | Not-determined |
| 17-106_k141_6478 | Low-quality |
| 17-106_k141_6481 | Low-quality |
| 17-106_k141_91967 | Not-determined |
| 17-106_k141_97566 | Not-determined |
| 17-106_k141_124125 | Not-determined |
| 17-106_k141_102944 | Not-determined |
| 17-106_k141_43737 | Not-determined |
| 17-106_k141_75749 | Not-determined |
| 17-106_k141_75751 | Not-determined |
| 17-106_k141_118966 | Not-determined |
| 17-106_k141_118973 | Not-determined |
| 17-106_k141_70413 | Not-determined |
| 17-106_k141_92001 | Not-determined |
| 17-106_k141_92005 | Not-determined |
| 17-106_k141_54706 | Low-quality |
| 17-106_k141_92006 | Not-determined |
| 17-106_k141_75769 | Not-determined |
| 17-106_k141_22582 | Not-determined |
| 17-106_k141_75773 | Not-determined |
| 17-106_k141_6531 | Not-determined |
| 17-106_k141_65295 | Low-quality |
| 17-106_k141_38409 | Not-determined |
| 17-106_k141_16979 | Not-determined |
| 17-106_k141_75781 | Not-determined |
| 17-106_k141_81718 | Not-determined |
| 17-106_k141_118979 | Not-determined |
| 17-106_k141_65314 | Not-determined |
| 17-106_k141_92034 | Not-determined |
| 17-106_k141_38422 | Not-determined |
| 17-106_k141_92039 | Not-determined |
| 17-106_k141_92040 | Not-determined |
| 17-106_k141_102971 | Low-quality |
| 17-106_k141_49387 | Not-determined |
| 17-106_k141_11692 | Not-determined |
| 17-106_k141_118994 | Not-determined |
| 17-106_k141_81731 | Not-determined |
| 17-106_k141_60129 | Low-quality |
| 17-106_k141_81734 | Low-quality |
| 17-106_k141_60130 | Not-determined |
| 17-106_k141_92051 | Not-determined |
| 17-106_k141_92053 | Not-determined |
| 17-106_k141_124129 | Not-determined |
| 17-106_k141_102986 | Not-determined |
| 17-106_k141_11707 | Low-quality |
| 17-106_k141_75803 | Not-determined |
| 17-106_k141_107926 | Not-determined |
| 17-106_k141_49394 | Not-determined |
| 17-106_k141_92066 | Not-determined |
| 17-106_k141_6549 | Not-determined |
| 17-106_k141_107930 | Low-quality |
| 17-106_k141_75810 | Not-determined |
| 17-106_k141_54752 | Low-quality |
| 17-106_k141_54755 | Not-determined |
| 17-106_k141_60148 | Not-determined |
| 17-106_k141_97597 | Not-determined |
| 17-106_k141_22608 | Not-determined |
| 17-106_k141_49415 | Not-determined |
| 17-106_k141_81763 | Not-determined |
| 17-106_k141_38425 | Not-determined |
| 17-106_k141_16989 | Not-determined |
| 17-106_k141_22615 | Not-determined |
| 17-106_k141_43802 | Not-determined |
| 17-106_k141_60164 | Not-determined |
| 17-106_k141_113404 | Not-determined |
| 17-106_k141_60167 | Not-determined |
| 17-106_k141_107941 | Not-determined |
| 17-106_k141_97609 | Not-determined |
| 17-106_k141_1736 | Not-determined |
| 17-106_k141_81778 | Not-determined |
| 17-106_k141_70470 | Not-determined |
| 17-106_k141_1739 | Not-determined |
| 17-106_k141_17017 | Not-determined |
| 17-106_k141_70472 | Not-determined |
| 17-106_k141_22640 | Not-determined |
| 17-106_k141_81791 | Not-determined |
| 17-106_k141_60169 | Not-determined |
| 17-106_k141_54778 | Not-determined |
| 17-106_k141_60174 | Not-determined |
| 17-106_k141_60178 | Not-determined |
| 17-106_k141_97617 | Not-determined |
| 17-106_k141_60183 | Not-determined |
| 17-106_k141_107965 | Not-determined |
| 17-106_k141_6613 | Not-determined |
| 17-106_k141_17035 | Not-determined |
| 17-106_k141_17036 | Not-determined |
| 17-106_k141_124135 | Not-determined |
| 17-106_k141_22666 | Low-quality |
| 17-106_k141_54807 | Not-determined |
| 17-106_k141_92106 | Not-determined |
| 17-106_k141_38466 | Not-determined |
| 17-106_k141_75848 | Not-determined |
| 17-106_k141_70508 | Not-determined |
| 17-106_k141_119018 | Low-quality |
| 17-106_k141_70514 | Not-determined |
| 17-106_k141_75857 | Low-quality |
| 17-106_k141_17050 | Not-determined |
| 17-106_k141_22680 | Not-determined |
| 17-106_k141_75868 | Low-quality |
| 17-106_k141_97622 | Low-quality |
| 17-106_k141_102991 | Low-quality |
| 17-106_k141_22691 | Not-determined |
| 17-106_k141_33220 | Not-determined |
| 17-106_k141_75871 | Low-quality |
| 17-106_k141_38490 | Not-determined |
| 17-106_k141_97629 | Not-determined |
| 17-106_k141_54829 | Not-determined |
| 17-106_k141_75876 | Not-determined |
| 17-106_k141_43843 | Not-determined |
| 17-106_k141_54833 | Low-quality |
| 17-106_k141_43849 | Not-determined |
| 17-106_k141_92124 | Not-determined |
| 17-106_k141_108015 | Not-determined |
| 17-106_k141_113439 | Not-determined |
| 17-106_k141_75881 | Low-quality |
| 17-106_k141_49436 | Not-determined |
| 17-106_k141_49437 | Not-determined |
| 17-106_k141_108020 | Not-determined |
| 17-106_k141_6642 | Not-determined |
| 17-106_k141_54839 | Low-quality |
| 17-106_k141_108024 | Not-determined |
| 17-106_k141_70544 | Not-determined |
| 17-106_k141_6648 | Not-determined |
| 17-106_k141_97644 | Not-determined |
| 17-106_k141_86857 | Not-determined |
| 17-106_k141_113453 | Low-quality |
| 17-106_k141_86863 | Not-determined |
| 17-106_k141_49454 | Low-quality |
| 17-106_k141_6659 | Low-quality |
| 17-106_k141_17075 | Not-determined |
| 17-106_k141_54857 | Not-determined |
| 17-106_k141_60222 | Low-quality |
| 17-106_k141_108040 | Low-quality |
| 17-106_k141_75909 | Not-determined |
| 17-106_k141_75911 | Low-quality |
| 17-106_k141_113462 | Low-quality |
| 17-106_k141_92144 | Not-determined |
| 17-106_k141_65395 | Not-determined |
| 17-106_k141_65397 | Not-determined |
| 17-106_k141_28051 | Not-determined |
| 17-106_k141_60247 | Not-determined |
| 17-106_k141_28053 | Low-quality |
| 17-106_k141_49467 | Low-quality |
| 17-106_k141_6698 | Not-determined |
| 17-106_k141_43886 | Low-quality |
| 17-106_k141_6704 | Not-determined |
| 17-106_k141_92160 | Not-determined |
| 17-106_k141_6708 | Not-determined |
| 17-106_k141_75933 | Low-quality |
| 17-106_k141_60258 | Not-determined |
| 17-106_k141_22726 | Not-determined |
| 17-106_k141_124189 | Not-determined |
| 17-106_k141_49477 | Low-quality |
| 17-106_k141_22734 | Not-determined |
| 17-106_k141_49478 | Not-determined |
| 17-106_k141_28071 | Low-quality |
| 17-106_k141_65420 | Not-determined |
| 17-106_k141_43896 | Not-determined |
| 17-106_k141_49485 | Not-determined |
| 17-106_k141_11726 | Not-determined |
| 17-106_k141_43902 | Not-determined |
| 17-106_k141_65423 | Not-determined |
| 17-106_k141_97687 | Not-determined |
| 17-106_k141_60277 | Not-determined |
| 17-106_k141_54863 | Not-determined |
| 17-106_k141_54864 | Not-determined |
| 17-106_k141_70566 | Not-determined |
| 17-106_k141_86878 | Not-determined |
| 17-106_k141_43910 | Not-determined |
| 17-106_k141_113498 | Not-determined |
| 17-106_k141_49490 | Not-determined |
| 17-106_k141_43914 | Not-determined |
| 17-106_k141_70570 | Not-determined |
| 17-106_k141_28089 | Not-determined |
| 17-106_k141_124222 | Not-determined |
| 17-106_k141_70578 | Not-determined |
| 17-106_k141_65446 | Low-quality |
| 17-106_k141_22764 | Low-quality |
| 17-106_k141_108089 | Not-determined |
| 17-106_k141_70589 | Not-determined |
| 17-106_k141_28102 | Low-quality |
| 17-106_k141_6723 | Not-determined |
| 17-106_k141_22771 | Not-determined |
| 17-106_k141_70594 | Not-determined |
| 17-106_k141_28111 | Not-determined |
| 17-106_k141_70600 | Not-determined |
| 17-106_k141_65474 | Not-determined |
| 17-106_k141_65476 | Not-determined |
| 17-106_k141_65477 | Not-determined |
| 17-106_k141_65478 | Not-determined |
| 17-106_k141_54887 | Not-determined |
| 17-106_k141_81797 | Not-determined |
| 17-106_k141_65481 | Not-determined |
| 17-106_k141_86927 | Not-determined |
| 17-106_k141_65485 | Not-determined |
| 17-106_k141_43925 | Not-determined |
| 17-106_k141_54891 | Not-determined |
| 17-106_k141_108101 | Not-determined |
| 17-106_k141_81799 | Not-determined |
| 17-106_k141_103009 | Not-determined |
| 17-106_k141_65499 | Low-quality |
| 17-106_k141_6738 | Not-determined |
| 17-106_k141_38535 | Not-determined |
| 17-106_k141_103017 | Low-quality |
| 17-106_k141_54897 | Low-quality |
| 17-106_k141_38540 | Not-determined |
| 17-106_k141_60344 | Not-determined |
| 17-106_k141_54903 | Not-determined |
| 17-106_k141_54909 | Not-determined |
| 17-106_k141_54911 | Not-determined |
| 17-106_k141_1774 | Not-determined |
| 17-106_k141_103036 | Not-determined |
| 17-106_k141_54918 | Not-determined |
| 17-106_k141_17144 | Not-determined |
| 17-106_k141_22796 | Not-determined |
| 17-106_k141_54925 | Not-determined |
| 17-106_k141_6764 | Not-determined |
| 17-106_k141_103047 | Low-quality |
| 17-106_k141_124259 | Not-determined |
| 17-106_k141_124260 | Not-determined |
| 17-106_k141_22805 | Not-determined |
| 17-106_k141_22812 | Not-determined |
| 17-106_k141_17157 | Not-determined |
| 17-106_k141_49521 | Not-determined |
| 17-106_k141_38562 | Not-determined |
| 17-106_k141_124268 | Not-determined |
| 17-106_k141_124269 | Not-determined |
| 17-106_k141_70626 | Low-quality |
| 17-106_k141_60386 | Not-determined |
| 17-106_k141_60393 | Not-determined |
| 17-106_k141_70634 | Not-determined |
| 17-106_k141_54932 | Low-quality |
| 17-106_k141_92277 | Not-determined |
| 17-106_k141_6813 | Not-determined |
| 17-106_k141_6814 | Not-determined |
| 17-106_k141_22843 | Not-determined |
| 17-106_k141_113532 | Not-determined |
| 17-106_k141_113534 | Not-determined |
| 17-106_k141_6819 | Not-determined |
| 17-106_k141_22853 | Not-determined |
| 17-106_k141_113540 | Not-determined |
| 17-106_k141_113543 | Not-determined |
| 17-106_k141_113546 | Not-determined |
| 17-106_k141_70659 | Not-determined |
| 17-106_k141_6834 | Not-determined |
| 17-106_k141_49540 | Not-determined |
| 17-106_k141_22868 | Not-determined |
| 17-106_k141_108139 | Not-determined |
| 17-106_k141_65523 | Not-determined |
| 17-106_k141_28174 | Not-determined |
| 17-106_k141_6855 | Not-determined |
| 17-106_k141_70669 | Not-determined |
| 17-106_k141_49554 | Not-determined |
| 17-106_k141_6860 | Not-determined |
| 17-106_k141_113559 | Not-determined |
| 17-106_k141_70677 | Not-determined |
| 17-106_k141_28191 | Not-determined |
| 17-106_k141_28193 | Not-determined |
| 17-106_k141_86950 | Not-determined |
| 17-106_k141_22890 | Not-determined |
| 17-106_k141_17195 | Not-determined |
| 17-106_k141_6879 | Not-determined |
| 17-106_k141_22897 | Not-determined |
| 17-106_k141_113569 | Not-determined |
| 17-106_k141_97766 | Low-quality |
| 17-106_k141_22911 | Not-determined |
| 17-106_k141_22912 | Not-determined |
| 17-106_k141_70683 | Not-determined |
| 17-106_k141_113584 | Not-determined |
| 17-106_k141_70689 | Not-determined |
| 17-106_k141_60419 | Not-determined |
| 17-106_k141_44030 | Not-determined |
| 17-106_k141_33231 | Low-quality |
| 17-106_k141_60422 | Not-determined |
| 17-106_k141_33236 | Not-determined |
| 17-106_k141_22925 | Not-determined |
| 17-106_k141_70707 | Not-determined |
| 17-106_k141_6907 | Not-determined |
| 17-106_k141_6908 | Not-determined |
| 17-106_k141_6919 | Not-determined |
| 17-106_k141_54978 | Low-quality |
| 17-106_k141_17228 | Not-determined |
| 17-106_k141_70723 | Not-determined |
| 17-106_k141_70724 | Not-determined |
| 17-106_k141_103076 | Not-determined |
| 17-106_k141_54992 | Not-determined |
| 17-106_k141_54994 | Not-determined |
| 17-106_k141_44066 | Low-quality |
| 17-106_k141_70734 | Not-determined |
| 17-106_k141_17249 | Not-determined |
| 17-106_k141_113615 | Not-determined |
| 17-106_k141_103088 | Not-determined |
| 17-106_k141_70743 | Not-determined |
| 17-106_k141_60450 | Not-determined |
| 17-106_k141_17259 | Not-determined |
| 17-106_k141_65581 | Not-determined |
| 17-106_k141_33269 | Not-determined |
| 17-106_k141_81841 | Not-determined |
| 17-106_k141_33270 | Not-determined |
| 17-106_k141_70750 | Not-determined |
| 17-106_k141_22952 | Not-determined |
| 17-106_k141_17272 | Not-determined |
| 17-106_k141_28235 | Not-determined |
| 17-106_k141_70753 | Not-determined |
| 17-106_k141_28236 | Not-determined |
| 17-106_k141_70754 | Not-determined |
| 17-106_k141_49591 | Not-determined |
| 17-106_k141_44091 | Not-determined |
| 17-106_k141_81852 | Low-quality |
| 17-106_k141_113618 | Not-determined |
| 17-106_k141_49596 | Not-determined |
| 17-106_k141_113620 | Not-determined |
| 17-106_k141_38578 | Not-determined |
| 17-106_k141_38579 | Not-determined |
| 17-106_k141_60459 | Not-determined |
| 17-106_k141_103122 | Not-determined |
| 17-106_k141_103129 | Not-determined |
| 17-106_k141_119062 | Not-determined |
| 17-106_k141_22972 | Not-determined |
| 17-106_k141_113632 | Not-determined |
| 17-106_k141_6938 | Not-determined |
| 17-106_k141_81870 | Not-determined |
| 17-106_k141_81872 | Not-determined |
| 17-106_k141_1793 | Not-determined |
| 17-106_k141_70776 | Not-determined |
| 17-106_k141_55012 | Not-determined |
| 17-106_k141_103140 | Not-determined |
| 17-106_k141_22982 | Not-determined |
| 17-106_k141_92326 | Not-determined |
| 17-106_k141_49616 | Low-quality |
| 17-106_k141_92333 | Not-determined |
| 17-106_k141_38591 | Not-determined |
| 17-106_k141_1809 | Not-determined |
| 17-106_k141_38593 | Not-determined |
| 17-106_k141_33297 | Not-determined |
| 17-106_k141_108186 | Not-determined |
| 17-106_k141_103156 | Low-quality |
| 17-106_k141_1816 | Not-determined |
| 17-106_k141_17329 | Not-determined |
| 17-106_k141_38602 | Not-determined |
| 17-106_k141_124323 | Not-determined |
| 17-106_k141_124325 | Not-determined |
| 17-106_k141_86970 | Not-determined |
| 17-106_k141_65624 | Not-determined |
| 17-106_k141_28262 | Not-determined |
| 17-106_k141_70781 | Not-determined |
| 17-106_k141_124335 | Not-determined |
| 17-106_k141_70784 | Not-determined |
| 17-106_k141_124336 | Low-quality |
| 17-106_k141_86977 | Low-quality |
| 17-106_k141_38614 | Not-determined |
| 17-106_k141_33333 | Not-determined |
| 17-106_k141_38618 | Not-determined |
| 17-106_k141_33334 | Not-determined |
| 17-106_k141_49656 | Not-determined |
| 17-106_k141_49659 | Not-determined |
| 17-106_k141_97782 | Not-determined |
| 17-106_k141_113655 | Not-determined |
| 17-106_k141_124342 | Not-determined |
| 17-106_k141_124348 | Not-determined |
| 17-106_k141_113663 | Not-determined |
| 17-106_k141_6963 | Low-quality |
| 17-106_k141_113665 | Not-determined |
| 17-106_k141_124363 | Not-determined |
| 17-106_k141_6974 | Not-determined |
| 17-106_k141_33355 | Not-determined |
| 17-106_k141_87007 | Not-determined |
| 17-106_k141_6983 | Not-determined |
| 17-106_k141_28275 | Not-determined |
| 17-106_k141_81922 | Not-determined |
| 17-106_k141_23018 | Not-determined |
| 17-106_k141_28281 | Not-determined |
| 17-106_k141_65661 | Not-determined |
| 17-106_k141_38637 | Not-determined |
| 17-106_k141_87013 | Not-determined |
| 17-106_k141_97797 | Low-quality |
| 17-106_k141_11815 | Not-determined |
| 17-106_k141_49680 | Not-determined |
| 17-106_k141_103197 | Not-determined |
| 17-106_k141_38647 | Not-determined |
| 17-106_k141_103202 | Low-quality |
| 17-106_k141_44110 | Not-determined |
| 17-106_k141_55025 | Not-determined |
| 17-106_k141_49691 | Not-determined |
| 17-106_k141_38657 | Not-determined |
| 17-106_k141_81948 | Not-determined |
| 17-106_k141_87035 | Not-determined |
| 17-106_k141_38663 | Low-quality |
| 17-106_k141_6999 | Not-determined |
| 17-106_k141_103221 | Not-determined |
| 17-106_k141_103226 | Not-determined |
| 17-106_k141_81953 | Not-determined |
| 17-106_k141_124410 | Not-determined |
| 17-106_k141_65688 | Not-determined |
| 17-106_k141_70794 | Not-determined |
| 17-106_k141_44132 | Not-determined |
| 17-106_k141_70806 | Not-determined |
| 17-106_k141_11869 | Not-determined |
| 17-106_k141_55046 | Not-determined |
| 17-106_k141_65694 | Not-determined |
| 17-106_k141_103240 | Low-quality |
| 17-106_k141_103242 | Not-determined |
| 17-106_k141_87061 | Not-determined |
| 17-106_k141_103243 | Not-determined |
| 17-106_k141_70808 | Not-determined |
| 17-106_k141_44150 | Not-determined |
| 17-106_k141_103249 | Not-determined |
| 17-106_k141_103251 | Not-determined |
| 17-106_k141_17342 | Not-determined |
| 17-106_k141_55058 | Not-determined |
| 17-106_k141_70813 | Not-determined |
| 17-106_k141_70818 | Not-determined |
| 17-106_k141_49706 | Not-determined |
| 17-106_k141_17365 | Not-determined |
| 17-106_k141_7019 | Low-quality |
| 17-106_k141_124422 | Not-determined |
| 17-106_k141_97803 | Not-determined |
| 17-106_k141_44177 | Not-determined |
| 17-106_k141_87100 | Not-determined |
| 17-106_k141_108196 | Not-determined |
| 17-106_k141_38678 | Not-determined |
| 17-106_k141_124442 | Not-determined |
| 17-106_k141_38683 | Not-determined |
| 17-106_k141_38686 | Not-determined |
| 17-106_k141_103286 | Not-determined |
| 17-106_k141_49731 | Not-determined |
| 17-106_k141_65735 | Not-determined |
| 17-106_k141_124449 | Low-quality |
| 17-106_k141_108214 | Not-determined |
| 17-106_k141_108215 | Not-determined |
| 17-106_k141_124461 | Not-determined |
| 17-106_k141_1883 | Not-determined |
| 17-106_k141_103296 | Not-determined |
| 17-106_k141_108221 | Not-determined |
| 17-106_k141_108223 | Not-determined |
| 17-106_k141_55086 | Not-determined |
| 17-106_k141_103303 | Not-determined |
| 17-106_k141_44188 | Not-determined |
| 17-106_k141_44189 | Not-determined |
| 17-106_k141_49739 | Low-quality |
| 17-106_k141_97816 | Not-determined |
| 17-106_k141_17380 | Not-determined |
| 17-106_k141_124471 | Not-determined |
| 17-106_k141_124473 | Not-determined |
| 17-106_k141_119076 | Not-determined |

| | |
|---|---|
| 17-106_k141_28311 | Not-determined |
| 17-106_k141_55093 | Not-determined |
| 17-106_k141_97821 | Not-determined |
| 17-106_k141_97824 | Not-determined |
| 17-106_k141_44199 | Not-determined |
| 17-106_k141_1889 | Not-determined |
| 17-106_k141_49741 | Not-determined |
| 17-106_k141_44207 | Not-determined |
| 17-106_k141_49742 | Not-determined |
| 17-106_k141_44215 | Not-determined |
| 17-106_k141_38718 | Not-determined |
| 17-106_k141_92360 | Not-determined |
| 17-106_k141_70848 | Not-determined |
| 17-106_k141_28325 | Not-determined |
| 17-106_k141_38727 | Not-determined |
| 17-106_k141_49755 | Not-determined |
| 17-106_k141_38736 | Not-determined |
| 17-106_k141_17417 | Not-determined |
| 17-106_k141_38740 | Not-determined |
| 17-106_k141_60484 | Not-determined |
| 17-106_k141_60487 | Not-determined |
| 17-106_k141_87158 | Not-determined |
| 17-106_k141_108228 | Not-determined |
| 17-106_k141_1931 | Not-determined |
| 17-106_k141_119079 | Not-determined |
| 17-106_k141_87165 | Low-quality |
| 17-106_k141_11884 | Low-quality |
| 17-106_k141_38768 | Not-determined |
| 17-106_k141_17454 | Not-determined |
| 17-106_k141_1948 | Not-determined |
| 17-106_k141_92414 | Not-determined |
| 17-106_k141_38780 | Not-determined |
| 17-106_k141_11894 | Low-quality |
| 17-106_k141_11895 | Not-determined |
| 17-106_k141_33372 | Not-determined |
| 17-106_k141_92420 | Not-determined |
| 17-106_k141_87184 | Not-determined |
| 17-106_k141_7056 | Not-determined |
| 17-106_k141_65754 | Not-determined |
| 17-106_k141_11900 | Not-determined |
| 17-106_k141_33383 | Not-determined |
| 17-106_k141_38783 | Not-determined |
| 17-106_k141_49779 | Not-determined |
| 17-106_k141_33386 | Not-determined |
| 17-106_k141_7070 | Not-determined |
| 17-106_k141_38791 | Not-determined |
| 17-106_k141_119113 | Not-determined |
| 17-106_k141_75989 | Not-determined |
| 17-106_k141_1969 | Not-determined |
| 17-106_k141_65763 | Not-determined |
| 17-106_k141_1974 | Not-determined |
| 17-106_k141_70898 | Not-determined |
| 17-106_k141_92429 | Not-determined |
| 17-106_k141_65768 | Not-determined |
| 17-106_k141_75990 | Not-determined |
| 17-106_k141_92439 | Not-determined |
| 17-106_k141_92440 | Not-determined |
| 17-106_k141_87187 | Not-determined |
| 17-106_k141_49816 | Not-determined |
| 17-106_k141_49819 | Not-determined |
| 17-106_k141_1996 | Not-determined |
| 17-106_k141_49821 | Low-quality |
| 17-106_k141_70919 | Not-determined |
| 17-106_k141_81999 | Not-determined |
| 17-106_k141_44282 | Not-determined |
| 17-106_k141_65817 | Not-determined |
| 17-106_k141_82001 | Not-determined |
| 17-106_k141_82003 | Not-determined |
| 17-106_k141_55160 | Not-determined |
| 17-106_k141_82009 | Not-determined |
| 17-106_k141_65831 | Not-determined |
| 17-106_k141_49831 | Not-determined |
| 17-106_k141_7097 | Not-determined |
| 17-106_k141_33392 | Not-determined |
| 17-106_k141_2018 | Not-determined |
| 17-106_k141_49836 | Not-determined |
| 17-106_k141_65840 | Low-quality |
| 17-106_k141_55167 | Not-determined |
| 17-106_k141_49844 | Not-determined |
| 17-106_k141_60561 | Not-determined |
| 17-106_k141_2032 | Not-determined |
| 17-106_k141_70927 | Low-quality |
| 17-106_k141_70929 | Not-determined |
| 17-106_k141_97851 | Not-determined |
| 17-106_k141_38823 | Not-determined |
| 17-106_k141_60572 | Not-determined |
| 17-106_k141_108278 | Not-determined |
| 17-106_k141_17501 | Not-determined |
| 17-106_k141_65858 | Not-determined |
| 17-106_k141_38837 | Not-determined |
| 17-106_k141_76018 | Not-determined |
| 17-106_k141_11921 | Not-determined |
| 17-106_k141_11926 | Not-determined |
| 17-106_k141_92485 | Not-determined |
| 17-106_k141_23082 | Not-determined |
| 17-106_k141_60582 | Not-determined |
| 17-106_k141_108294 | Not-determined |
| 17-106_k141_60586 | Not-determined |
| 17-106_k141_55198 | Not-determined |
| 17-106_k141_23092 | Not-determined |
| 17-106_k141_70941 | Not-determined |
| 17-106_k141_11946 | Not-determined |
| 17-106_k141_11947 | Low-quality |
| 17-106_k141_23097 | Not-determined |
| 17-106_k141_17517 | Not-determined |
| 17-106_k141_70945 | Low-quality |
| 17-106_k141_60598 | Not-determined |
| 17-106_k141_60601 | Not-determined |
| 17-106_k141_108312 | Not-determined |
| 17-106_k141_11956 | Not-determined |
| 17-106_k141_23107 | Not-determined |
| 17-106_k141_76051 | Not-determined |
| 17-106_k141_124517 | Not-determined |
| 17-106_k141_23127 | Not-determined |
| 17-106_k141_124524 | Not-determined |
| 17-106_k141_23134 | Not-determined |
| 17-106_k141_17542 | Not-determined |
| 17-106_k141_82070 | Not-determined |
| 17-106_k141_33397 | Not-determined |
| 17-106_k141_124533 | Not-determined |
| 17-106_k141_76073 | Not-determined |
| 17-106_k141_23139 | Not-determined |
| 17-106_k141_65923 | Not-determined |
| 17-106_k141_55234 | Not-determined |
| 17-106_k141_17551 | Not-determined |
| 17-106_k141_55239 | Not-determined |
| 17-106_k141_55240 | Not-determined |
| 17-106_k141_55243 | Not-determined |
| 17-106_k141_60622 | Not-determined |
| 17-106_k141_11964 | Low-quality |
| 17-106_k141_55248 | Not-determined |
| 17-106_k141_55251 | Not-determined |
| 17-106_k141_49873 | Not-determined |
| 17-106_k141_49874 | Not-determined |
| 17-106_k141_65938 | Not-determined |
| 17-106_k141_17562 | Not-determined |
| 17-106_k141_60639 | Not-determined |
| 17-106_k141_23150 | Not-determined |
| 17-106_k141_70965 | Low-quality |
| 17-106_k141_119166 | Not-determined |
| 17-106_k141_55274 | Not-determined |
| 17-106_k141_119171 | Low-quality |
| 17-106_k141_92492 | Low-quality |
| 17-106_k141_65959 | Not-determined |
| 17-106_k141_2097 | Not-determined |
| 17-106_k141_33404 | Not-determined |
| 17-106_k141_55284 | Low-quality |
| 17-106_k141_11979 | Not-determined |
| 17-106_k141_11980 | Low-quality |
| 17-106_k141_119177 | Low-quality |
| 17-106_k141_11983 | Not-determined |
| 17-106_k141_33407 | Low-quality |
| 17-106_k141_2108 | Not-determined |
| 17-106_k141_87227 | Not-determined |
| 17-106_k141_7171 | Not-determined |
| 17-106_k141_55289 | Low-quality |
| 17-106_k141_55291 | Not-determined |
| 17-106_k141_97875 | Low-quality |
| 17-106_k141_87233 | Not-determined |
| 17-106_k141_87234 | Not-determined |
| 17-106_k141_49893 | Not-determined |
| 17-106_k141_2112 | Not-determined |
| 17-106_k141_82087 | Low-quality |
| 17-106_k141_87238 | Not-determined |
| 17-106_k141_49899 | Not-determined |
| 17-106_k141_23181 | Not-determined |
| 17-106_k141_49901 | Not-determined |
| 17-106_k141_65976 | Low-quality |
| 17-106_k141_119189 | Not-determined |
| 17-106_k141_65978 | Not-determined |
| 17-106_k141_119191 | Not-determined |
| 17-106_k141_33422 | Not-determined |
| 17-106_k141_87243 | Not-determined |
| 17-106_k141_124553 | Not-determined |
| 17-106_k141_124556 | Not-determined |
| 17-106_k141_49909 | Not-determined |
| 17-106_k141_65987 | Not-determined |
| 17-106_k141_65988 | Not-determined |
| 17-106_k141_17575 | Not-determined |
| 17-106_k141_33426 | Not-determined |
| 17-106_k141_119214 | Not-determined |
| 17-106_k141_17584 | Not-determined |
| 17-106_k141_44300 | Not-determined |
| 17-106_k141_124572 | Not-determined |
| 17-106_k141_33430 | Not-determined |
| 17-106_k141_49926 | Low-quality |
| 17-106_k141_33433 | Not-determined |
| 17-106_k141_49929 | Not-determined |
| 17-106_k141_124578 | Not-determined |
| 17-106_k141_119224 | Not-determined |
| 17-106_k141_2138 | Not-determined |
| 17-106_k141_23197 | Not-determined |
| 17-106_k141_82095 | Not-determined |
| 17-106_k141_33437 | Not-determined |
| 17-106_k141_44308 | Not-determined |
| 17-106_k141_38907 | Not-determined |
| 17-106_k141_119234 | Not-determined |
| 17-106_k141_44312 | Not-determined |
| 17-106_k141_33443 | Not-determined |
| 17-106_k141_33446 | Low-quality |
| 17-106_k141_55314 | Not-determined |
| 17-106_k141_60679 | Not-determined |
| 17-106_k141_92533 | Not-determined |
| 17-106_k141_76128 | Not-determined |
| 17-106_k141_82102 | Not-determined |
| 17-106_k141_2153 | Not-determined |
| 17-106_k141_2155 | Not-determined |
| 17-106_k141_7195 | Not-determined |
| 17-106_k141_76135 | Low-quality |
| 17-106_k141_7196 | Low-quality |
| 17-106_k141_7201 | Not-determined |
| 17-106_k141_49947 | Low-quality |
| 17-106_k141_7207 | Low-quality |
| 17-106_k141_92547 | Not-determined |
| 17-106_k141_12005 | Not-determined |
| 17-106_k141_103355 | Not-determined |
| 17-106_k141_38951 | Not-determined |
| 17-106_k141_12013 | Low-quality |
| 17-106_k141_38953 | Not-determined |
| 17-106_k141_2164 | Not-determined |
| 17-106_k141_103358 | Not-determined |
| 17-106_k141_71020 | Low-quality |
| 17-106_k141_108335 | Not-determined |
| 17-106_k141_23223 | Not-determined |
| 17-106_k141_103363 | Not-determined |
| 17-106_k141_38973 | Not-determined |
| 17-106_k141_23227 | Not-determined |
| 17-106_k141_23232 | Not-determined |
| 17-106_k141_55340 | Not-determined |
| 17-106_k141_108349 | Not-determined |
| 17-106_k141_92567 | Not-determined |
| 17-106_k141_119277 | Not-determined |
| 17-106_k141_66008 | Not-determined |
| 17-106_k141_66010 | Not-determined |
| 17-106_k141_2183 | Not-determined |
| 17-106_k141_103380 | Not-determined |
| 17-106_k141_113708 | Not-determined |
| 17-106_k141_82124 | Not-determined |
| 17-106_k141_38996 | Not-determined |
| 17-106_k141_108356 | Not-determined |
| 17-106_k141_66021 | Not-determined |
| 17-106_k141_60694 | Not-determined |
| 17-106_k141_23266 | Not-determined |
| 17-106_k141_60703 | Not-determined |
| 17-106_k141_60704 | Not-determined |
| 17-106_k141_17656 | Not-determined |
| 17-106_k141_66026 | Not-determined |
| 17-106_k141_108358 | Not-determined |
| 17-106_k141_92589 | Not-determined |
| 17-106_k141_60713 | Not-determined |
| 17-106_k141_7215 | Not-determined |
| 17-106_k141_76180 | Not-determined |
| 17-106_k141_76185 | Not-determined |
| 17-106_k141_82143 | Not-determined |
| 17-106_k141_71044 | Not-determined |
| 17-106_k141_17670 | Not-determined |
| 17-106_k141_97916 | Not-determined |
| 17-106_k141_108370 | Not-determined |
| 17-106_k141_66042 | Low-quality |
| 17-106_k141_113728 | Not-determined |
| 17-106_k141_82156 | Not-determined |
| 17-106_k141_2222 | Low-quality |
| 17-106_k141_82167 | Not-determined |
| 17-106_k141_12052 | Not-determined |
| 17-106_k141_76189 | Not-determined |
| 17-106_k141_44346 | Not-determined |
| 17-106_k141_55403 | Low-quality |
| 17-106_k141_12063 | Low-quality |
| 17-106_k141_108397 | Not-determined |
| 17-106_k141_55405 | Not-determined |
| 17-106_k141_66055 | Not-determined |
| 17-106_k141_97936 | Not-determined |
| 17-106_k141_55414 | Not-determined |
| 17-106_k141_60730 | Not-determined |
| 17-106_k141_17693 | Not-determined |
| 17-106_k141_103426 | Not-determined |
| 17-106_k141_97946 | Low-quality |
| 17-106_k141_124635 | Not-determined |
| 17-106_k141_55418 | Not-determined |
| 17-106_k141_12081 | Low-quality |
| 17-106_k141_108409 | Not-determined |
| 17-106_k141_23272 | Low-quality |
| 17-106_k141_108418 | Not-determined |
| 17-106_k141_76220 | Not-determined |
| 17-106_k141_82194 | Not-determined |
| 17-106_k141_76224 | Not-determined |
| 17-106_k141_82198 | Not-determined |
| 17-106_k141_50068 | Not-determined |
| 17-106_k141_76227 | Not-determined |
| 17-106_k141_82208 | Not-determined |
| 17-106_k141_66070 | Not-determined |
| 17-106_k141_50074 | Not-determined |
| 17-106_k141_124638 | Not-determined |
| 17-106_k141_50076 | Low-quality |
| 17-106_k141_60765 | Not-determined |
| 17-106_k141_12102 | Low-quality |
| 17-106_k141_87275 | Not-determined |
| 17-106_k141_2254 | Not-determined |
| 17-106_k141_2259 | Not-determined |
| 17-106_k141_2262 | Not-determined |
| 17-106_k141_7265 | Not-determined |
| 17-106_k141_23289 | Not-determined |
| 17-106_k141_87287 | Not-determined |
| 17-106_k141_66091 | Low-quality |
| 17-106_k141_55446 | Not-determined |
| 17-106_k141_71061 | Not-determined |
| 17-106_k141_66097 | Not-determined |
| 17-106_k141_97956 | Not-determined |
| 17-106_k141_7287 | Not-determined |
| 17-106_k141_76242 | Not-determined |
| 17-106_k141_23304 | Low-quality |
| 17-106_k141_50103 | Not-determined |
| 17-106_k141_82265 | Not-determined |
| 17-106_k141_66110 | Not-determined |
| 17-106_k141_55453 | Not-determined |
| 17-106_k141_23306 | Not-determined |
| 17-106_k141_76253 | Not-determined |
| 17-106_k141_60811 | Not-determined |
| 17-106_k141_66117 | Not-determined |
| 17-106_k141_23313 | Not-determined |
| 17-106_k141_66120 | Not-determined |
| 17-106_k141_28379 | Not-determined |
| 17-106_k141_12127 | Low-quality |
| 17-106_k141_119339 | Not-determined |
| 17-106_k141_2293 | Not-determined |
| 17-106_k141_7296 | Not-determined |
| 17-106_k141_7298 | Not-determined |
| 17-106_k141_7299 | Not-determined |
| 17-106_k141_76266 | Not-determined |
| 17-106_k141_28390 | Not-determined |
| 17-106_k141_12136 | Not-determined |
| 17-106_k141_50113 | Low-quality |
| 17-106_k141_2302 | Not-determined |
| 17-106_k141_17782 | Not-determined |
| 17-106_k141_39008 | Not-determined |
| 17-106_k141_97981 | Not-determined |
| 17-106_k141_28401 | Not-determined |
| 17-106_k141_50122 | Not-determined |
| 17-106_k141_39018 | Not-determined |
| 17-106_k141_39019 | Not-determined |
| 17-106_k141_44388 | Not-determined |
| 17-106_k141_12151 | Not-determined |
| 17-106_k141_82283 | Not-determined |
| 17-106_k141_50133 | Not-determined |
| 17-106_k141_55484 | Not-determined |
| 17-106_k141_12154 | Low-quality |
| 17-106_k141_23341 | Not-determined |
| 17-106_k141_28419 | Not-determined |
| 17-106_k141_23347 | Not-determined |
| 17-106_k141_50139 | Low-quality |
| 17-106_k141_55486 | Not-determined |
| 17-106_k141_66135 | Low-quality |
| 17-106_k141_66136 | Low-quality |
| 17-106_k141_124649 | Not-determined |
| 17-106_k141_66137 | Not-determined |
| 17-106_k141_97999 | Not-determined |
| 17-106_k141_82290 | Not-determined |
| 17-106_k141_55500 | Not-determined |
| 17-106_k141_50160 | Not-determined |
| 17-106_k141_28440 | Low-quality |
| 17-106_k141_28441 | Not-determined |
| 17-106_k141_12186 | Not-determined |
| 17-106_k141_2318 | Low-quality |
| 17-106_k141_2319 | Not-determined |
| 17-106_k141_28449 | Not-determined |
| 17-106_k141_12196 | Not-determined |
| 17-106_k141_12197 | Not-determined |
| 17-106_k141_23356 | Not-determined |
| 17-106_k141_28454 | Not-determined |
| 17-106_k141_7321 | Not-determined |
| 17-106_k141_87330 | Not-determined |
| 17-106_k141_28457 | Not-determined |
| 17-106_k141_12207 | Not-determined |
| 17-106_k141_124656 | Not-determined |
| 17-106_k141_7336 | Not-determined |
| 17-106_k141_60830 | Not-determined |
| 17-106_k141_60832 | Not-determined |
| 17-106_k141_98018 | Not-determined |
| 17-106_k141_82310 | Not-determined |
| 17-106_k141_7344 | Low-quality |
| 17-106_k141_108429 | Low-quality |
| 17-106_k141_124665 | Not-determined |
| 17-106_k141_60838 | Low-quality |
| 17-106_k141_50200 | Not-determined |
| 17-106_k141_12231 | Not-determined |
| 17-106_k141_33492 | Not-determined |
| 17-106_k141_108448 | Not-determined |
| 17-106_k141_28485 | Not-determined |
| 17-106_k141_108452 | Not-determined |
| 17-106_k141_66177 | Not-determined |
| 17-106_k141_92635 | Not-determined |
| 17-106_k141_7370 | Low-quality |
| 17-106_k141_39055 | Not-determined |
| 17-106_k141_12235 | Not-determined |
| 17-106_k141_87371 | Not-determined |
| 17-106_k141_50230 | Low-quality |
| 17-106_k141_28504 | Not-determined |
| 17-106_k141_71102 | Not-determined |
| 17-106_k141_17820 | Low-quality |
| 17-106_k141_71106 | Not-determined |
| 17-106_k141_50234 | Not-determined |
| 17-106_k141_66198 | Not-determined |
| 17-106_k141_60841 | Low-quality |
| 17-106_k141_60847 | Not-determined |
| 17-106_k141_71110 | Not-determined |
| 17-106_k141_50243 | Not-determined |
| 17-106_k141_2349 | Not-determined |
| 17-106_k141_55538 | Low-quality |
| 17-106_k141_66206 | Not-determined |
| 17-106_k141_119401 | Low-quality |
| 17-106_k141_55541 | Not-determined |
| 17-106_k141_71122 | Not-determined |
| 17-106_k141_92658 | Not-determined |
| 17-106_k141_98048 | Not-determined |
| 17-106_k141_17842 | Not-determined |
| 17-106_k141_7401 | Not-determined |
| 17-106_k141_87397 | Low-quality |
| 17-106_k141_119403 | Low-quality |
| 17-106_k141_39073 | Not-determined |
| 17-106_k141_98066 | Not-determined |
| 17-106_k141_98068 | Not-determined |
| 17-106_k141_2362 | Low-quality |
| 17-106_k141_39076 | Not-determined |
| 17-106_k141_119413 | Low-quality |
| 17-106_k141_50261 | Not-determined |
| 17-106_k141_39077 | Low-quality |
| 17-106_k141_39078 | Not-determined |
| 17-106_k141_23372 | Not-determined |
| 17-106_k141_92672 | Not-determined |
| 17-106_k141_23374 | Not-determined |
| 17-106_k141_7418 | Not-determined |
| 17-106_k141_71138 | Not-determined |
| 17-106_k141_87405 | Not-determined |
| 17-106_k141_50272 | Not-determined |
| 17-106_k141_7428 | Not-determined |
| 17-106_k141_50274 | Not-determined |
| 17-106_k141_66250 | Not-determined |
| 17-106_k141_71154 | Not-determined |
| 17-106_k141_103522 | Not-determined |
| 17-106_k141_98095 | Not-determined |
| 17-106_k141_71173 | Not-determined |
| 17-106_k141_119430 | Not-determined |
| 17-106_k141_50296 | Not-determined |
| 17-106_k141_87425 | Not-determined |
| 17-106_k141_98100 | Not-determined |
| 17-106_k141_98102 | Not-determined |
| 17-106_k141_39110 | Not-determined |
| 17-106_k141_39117 | Not-determined |
| 17-106_k141_28574 | Not-determined |
| 17-106_k141_28575 | Not-determined |
| 17-106_k141_17866 | Not-determined |
| 17-106_k141_60873 | Not-determined |
| 17-106_k141_28577 | Not-determined |
| 17-106_k141_28581 | Not-determined |
| 17-106_k141_98109 | Not-determined |
| 17-106_k141_71196 | Not-determined |
| 17-106_k141_92719 | Not-determined |
| 17-106_k141_12304 | Not-determined |
| 17-106_k141_98132 | Not-determined |
| 17-106_k141_98135 | Not-determined |
| 17-106_k141_98142 | Not-determined |
| 17-106_k141_39134 | Not-determined |
| 17-106_k141_92739 | Not-determined |
| 17-106_k141_50318 | Not-determined |
| 17-106_k141_92741 | Not-determined |
| 17-106_k141_39139 | Not-determined |
| 17-106_k141_60917 | Not-determined |
| 17-106_k141_103539 | Not-determined |
| 17-106_k141_92757 | Not-determined |
| 17-106_k141_103545 | Not-determined |
| 17-106_k141_92762 | Not-determined |
| 17-106_k141_113762 | Not-determined |
| 17-106_k141_71212 | Not-determined |
| 17-106_k141_60945 | Not-determined |
| 17-106_k141_60954 | Not-determined |
| 17-106_k141_103560 | Not-determined |
| 17-106_k141_33527 | Not-determined |
| 17-106_k141_113776 | Not-determined |
| 17-106_k141_103568 | Low-quality |
| 17-106_k141_39172 | Not-determined |
| 17-106_k141_71225 | Not-determined |
| 17-106_k141_66253 | Not-determined |
| 17-106_k141_66256 | Not-determined |
| 17-106_k141_87476 | Not-determined |
| 17-106_k141_76277 | Low-quality |
| 17-106_k141_103583 | Not-determined |
| 17-106_k141_76282 | Not-determined |
| 17-106_k141_39190 | Not-determined |
| 17-106_k141_66266 | Low-quality |
| 17-106_k141_66267 | Not-determined |
| 17-106_k141_39194 | Not-determined |
| 17-106_k141_7483 | Not-determined |
| 17-106_k141_39198 | Not-determined |
| 17-106_k141_7489 | Not-determined |
| 17-106_k141_39205 | Not-determined |
| 17-106_k141_66270 | Not-determined |
| 17-106_k141_17916 | Low-quality |
| 17-106_k141_7495 | Low-quality |
| 17-106_k141_60965 | Not-determined |
| 17-106_k141_82324 | Not-determined |
| 17-106_k141_39212 | Not-determined |
| 17-106_k141_60970 | Not-determined |
| 17-106_k141_60971 | Not-determined |
| 17-106_k141_28622 | Low-quality |
| 17-106_k141_60978 | Not-determined |
| 17-106_k141_71252 | Not-determined |
| 17-106_k141_12358 | Not-determined |
| 17-106_k141_87513 | Not-determined |
| 17-106_k141_60982 | Not-determined |
| 17-106_k141_66281 | Not-determined |
| 17-106_k141_28637 | Low-quality |
| 17-106_k141_39226 | Not-determined |
| 17-106_k141_82343 | Not-determined |
| 17-106_k141_39230 | Not-determined |
| 17-106_k141_87527 | Not-determined |
| 17-106_k141_113818 | Not-determined |
| 17-106_k141_108469 | Not-determined |
| 17-106_k141_2438 | Not-determined |
| 17-106_k141_82350 | Low-quality |
| 17-106_k141_119468 | Not-determined |
| 17-106_k141_12372 | Not-determined |
| 17-106_k141_98172 | Low-quality |
| 17-106_k141_92859 | Not-determined |
| 17-106_k141_92861 | Not-determined |
| 17-106_k141_50368 | Not-determined |
| 17-106_k141_82360 | Not-determined |
| 17-106_k141_2448 | Not-determined |
| 17-106_k141_2456 | Not-determined |
| 17-106_k141_39249 | Not-determined |
| 17-106_k141_87561 | Not-determined |
| 17-106_k141_119486 | Not-determined |
| 17-106_k141_7519 | Not-determined |
| 17-106_k141_2471 | Not-determined |
| 17-106_k141_2473 | Not-determined |
| 17-106_k141_7523 | Not-determined |
| 17-106_k141_124721 | Not-determined |
| 17-106_k141_108485 | Not-determined |
| 17-106_k141_87573 | Not-determined |
| 17-106_k141_17925 | Not-determined |
| 17-106_k141_33571 | Not-determined |
| 17-106_k141_108506 | Not-determined |
| 17-106_k141_71289 | Low-quality |
| 17-106_k141_108520 | Not-determined |
| 17-106_k141_23392 | Not-determined |
| 17-106_k141_119511 | Low-quality |
| 17-106_k141_108522 | Not-determined |
| 17-106_k141_119520 | Not-determined |
| 17-106_k141_39264 | Not-determined |
| 17-106_k141_17951 | Not-determined |
| 17-106_k141_119528 | Not-determined |
| 17-106_k141_23401 | Not-determined |
| 17-106_k141_71305 | Not-determined |
| 17-106_k141_17964 | Not-determined |
| 17-106_k141_119531 | Not-determined |
| 17-106_k141_17965 | Not-determined |
| 17-106_k141_119534 | Not-determined |
| 17-106_k141_98186 | Not-determined |
| 17-106_k141_103643 | Not-determined |
| 17-106_k141_87590 | Not-determined |
| 17-106_k141_12403 | Not-determined |
| 17-106_k141_98192 | Not-determined |
| 17-106_k141_39272 | Not-determined |
| 17-106_k141_12422 | Not-determined |
| 17-106_k141_103669 | Not-determined |
| 17-106_k141_17988 | Not-determined |
| 17-106_k141_33606 | Not-determined |
| 17-106_k141_92889 | Not-determined |
| 17-106_k141_17994 | Not-determined |
| 17-106_k141_2488 | Not-determined |
| 17-106_k141_2490 | Not-determined |
| 17-106_k141_18001 | Not-determined |
| 17-106_k141_61018 | Not-determined |
| 17-106_k141_103685 | Not-determined |
| 17-106_k141_103689 | Not-determined |
| 17-106_k141_87612 | Not-determined |
| 17-106_k141_124746 | Not-determined |
| 17-106_k141_39297 | Not-determined |
| 17-106_k141_76344 | Not-determined |
| 17-106_k141_66297 | Not-determined |
| 17-106_k141_92909 | Not-determined |
| 17-106_k141_18004 | Low-quality |
| 17-106_k141_66300 | Low-quality |
| 17-106_k141_71316 | Not-determined |
| 17-106_k141_61028 | Not-determined |
| 17-106_k141_39301 | Low-quality |
| 17-106_k141_92913 | Not-determined |
| 17-106_k141_108555 | Not-determined |
| 17-106_k141_61043 | Not-determined |
| 17-106_k141_12445 | Not-determined |

| | |
|---|---|
| 17-106_k141_12448 | Not-determined |
| 17-106_k141_108559 | Not-determined |
| 17-106_k141_66317 | Not-determined |
| 17-106_k141_55580 | Not-determined |
| 17-106_k141_124769 | Not-determined |
| 17-106_k141_98216 | Not-determined |
| 17-106_k141_124770 | Not-determined |
| 17-106_k141_119557 | Not-determined |
| 17-106_k141_61053 | Not-determined |
| 17-106_k141_61054 | Not-determined |
| 17-106_k141_82421 | Not-determined |
| 17-106_k141_66323 | Not-determined |
| 17-106_k141_66326 | Not-determined |
| 17-106_k141_39306 | Low-quality |
| 17-106_k141_12455 | Not-determined |
| 17-106_k141_12457 | Not-determined |
| 17-106_k141_113850 | Not-determined |
| 17-106_k141_18012 | Not-determined |
| 17-106_k141_12459 | Not-determined |
| 17-106_k141_113854 | Not-determined |
| 17-106_k141_98221 | Low-quality |
| 17-106_k141_76370 | Not-determined |
| 17-106_k141_7563 | Not-determined |
| 17-106_k141_2531 | Not-determined |
| 17-106_k141_12462 | Not-determined |
| 17-106_k141_12463 | Not-determined |
| 17-106_k141_98229 | Not-determined |
| 17-106_k141_12464 | Low-quality |
| 17-106_k141_108576 | Not-determined |
| 17-106_k141_7568 | Not-determined |
| 17-106_k141_7569 | Not-determined |
| 17-106_k141_2541 | Not-determined |
| 17-106_k141_119581 | Not-determined |
| 17-106_k141_2542 | Not-determined |
| 17-106_k141_39316 | Not-determined |
| 17-106_k141_108578 | Low-quality |
| 17-106_k141_98240 | Not-determined |
| 17-106_k141_82440 | Low-quality |
| 17-106_k141_61069 | Not-determined |
| 17-106_k141_98245 | Not-determined |
| 17-106_k141_61073 | Not-determined |
| 17-106_k141_7585 | Not-determined |
| 17-106_k141_12481 | Not-determined |
| 17-106_k141_7588 | Not-determined |
| 17-106_k141_12484 | Low-quality |
| 17-106_k141_113861 | Not-determined |
| 17-106_k141_76389 | Not-determined |
| 17-106_k141_82450 | Not-determined |
| 17-106_k141_103703 | Not-determined |
| 17-106_k141_113866 | Not-determined |
| 17-106_k141_82453 | Not-determined |
| 17-106_k141_18031 | Low-quality |
| 17-106_k141_76396 | Not-determined |
| 17-106_k141_61088 | Not-determined |
| 17-106_k141_76397 | Not-determined |
| 17-106_k141_76403 | Low-quality |
| 17-106_k141_113876 | Not-determined |
| 17-106_k141_12502 | Not-determined |
| 17-106_k141_61101 | Low-quality |
| 17-106_k141_28670 | Not-determined |
| 17-106_k141_28672 | Not-determined |
| 17-106_k141_33630 | Not-determined |
| 17-106_k141_113891 | Not-determined |
| 17-106_k141_108597 | Not-determined |
| 17-106_k141_50404 | Not-determined |
| 17-106_k141_18055 | Not-determined |
| 17-106_k141_108598 | Low-quality |
| 17-106_k141_76419 | Not-determined |
| 17-106_k141_113901 | Not-determined |
| 17-106_k141_76423 | Not-determined |
| 17-106_k141_76425 | Not-determined |
| 17-106_k141_113908 | Not-determined |
| 17-106_k141_28686 | Not-determined |
| 17-106_k141_2580 | Low-quality |
| 17-106_k141_92952 | Low-quality |
| 17-106_k141_103713 | Not-determined |
| 17-106_k141_103715 | Not-determined |
| 17-106_k141_76438 | Not-determined |
| 17-106_k141_50434 | Not-determined |
| 17-106_k141_82475 | Low-quality |
| 17-106_k141_76440 | Not-determined |
| 17-106_k141_2587 | Not-determined |
| 17-106_k141_28690 | Low-quality |
| 17-106_k141_2588 | Not-determined |
| 17-106_k141_124818 | Low-quality |
| 17-106_k141_2593 | Not-determined |
| 17-106_k141_18090 | Not-determined |
| 17-106_k141_108616 | Not-determined |
| 17-106_k141_2596 | Not-determined |
| 17-106_k141_108621 | Low-quality |
| 17-106_k141_82487 | Not-determined |
| 17-106_k141_12534 | Low-quality |
| 17-106_k141_18101 | Not-determined |
| 17-106_k141_33655 | Not-determined |
| 17-106_k141_39335 | Not-determined |
| 17-106_k141_39338 | Not-determined |
| 17-106_k141_18105 | Not-determined |
| 17-106_k141_66352 | Not-determined |
| 17-106_k141_113931 | Low-quality |
| 17-106_k141_113932 | Not-determined |
| 17-106_k141_33666 | Not-determined |
| 17-106_k141_124842 | Not-determined |
| 17-106_k141_39349 | Not-determined |
| 17-106_k141_12545 | Not-determined |
| 17-106_k141_61115 | Low-quality |
| 17-106_k141_124846 | Not-determined |
| 17-106_k141_39354 | Not-determined |
| 17-106_k141_92978 | Not-determined |
| 17-106_k141_108648 | Low-quality |
| 17-106_k141_18129 | Not-determined |
| 17-106_k141_39359 | Not-determined |
| 17-106_k141_18130 | Not-determined |
| 17-106_k141_61124 | Not-determined |
| 17-106_k141_119607 | Low-quality |
| 17-106_k141_61126 | Not-determined |
| 17-106_k141_12552 | Not-determined |
| 17-106_k141_39364 | Not-determined |
| 17-106_k141_18138 | Not-determined |
| 17-106_k141_12556 | Not-determined |
| 17-106_k141_39365 | Low-quality |
| 17-106_k141_103754 | Low-quality |
| 17-106_k141_7649 | Not-determined |
| 17-106_k141_76475 | Not-determined |
| 17-106_k141_18149 | Not-determined |
| 17-106_k141_103761 | Low-quality |
| 17-106_k141_33692 | Not-determined |
| 17-106_k141_18153 | Not-determined |
| 17-106_k141_92996 | Not-determined |
| 17-106_k141_50460 | Not-determined |
| 17-106_k141_71342 | Not-determined |
| 17-106_k141_39386 | Not-determined |
| 17-106_k141_50462 | Not-determined |
| 17-106_k141_76488 | Not-determined |
| 17-106_k141_93003 | Not-determined |
| 17-106_k141_61139 | Not-determined |
| 17-106_k141_61141 | Not-determined |
| 17-106_k141_108677 | Not-determined |
| 17-106_k141_12571 | Not-determined |
| 17-106_k141_2604 | Not-determined |
| 17-106_k141_18172 | Not-determined |
| 17-106_k141_12575 | Not-determined |
| 17-106_k141_33697 | Not-determined |
| 17-106_k141_18176 | Not-determined |
| 17-106_k141_113957 | Not-determined |
| 17-106_k141_33702 | Not-determined |
| 17-106_k141_28727 | Not-determined |
| 17-106_k141_76507 | Not-determined |
| 17-106_k141_18189 | Not-determined |
| 17-106_k141_82496 | Not-determined |
| 17-106_k141_82498 | Low-quality |
| 17-106_k141_76516 | Not-determined |
| 17-106_k141_33718 | Not-determined |
| 17-106_k141_76518 | Low-quality |
| 17-106_k141_28738 | Not-determined |
| 17-106_k141_44416 | Not-determined |
| 17-106_k141_61182 | Not-determined |
| 17-106_k141_87638 | Not-determined |
| 17-106_k141_87639 | Not-determined |
| 17-106_k141_76528 | Not-determined |
| 17-106_k141_33732 | Not-determined |
| 17-106_k141_82514 | Not-determined |
| 17-106_k141_12601 | Not-determined |
| 17-106_k141_12604 | Not-determined |
| 17-106_k141_71360 | Not-determined |
| 17-106_k141_12613 | Not-determined |
| 17-106_k141_39398 | Not-determined |
| 17-106_k141_61203 | Not-determined |
| 17-106_k141_61204 | Not-determined |
| 17-106_k141_71368 | Low-quality |
| 17-106_k141_44428 | Not-determined |
| 17-106_k141_18221 | Not-determined |
| 17-106_k141_44433 | Not-determined |
| 17-106_k141_18228 | Not-determined |
| 17-106_k141_71370 | Not-determined |
| 17-106_k141_12629 | Not-determined |
| 17-106_k141_71374 | Low-quality |
| 17-106_k141_87683 | Not-determined |
| 17-106_k141_87685 | Not-determined |
| 17-106_k141_39417 | Not-determined |
| 17-106_k141_12639 | Not-determined |
| 17-106_k141_18242 | Not-determined |
| 17-106_k141_87692 | Not-determined |
| 17-106_k141_61212 | Not-determined |
| 17-106_k141_76533 | Not-determined |
| 17-106_k141_39420 | Not-determined |
| 17-106_k141_39421 | Not-determined |
| 17-106_k141_76540 | Low-quality |
| 17-106_k141_23436 | Not-determined |
| 17-106_k141_2640 | Not-determined |
| 17-106_k141_2643 | Not-determined |
| 17-106_k141_82589 | Not-determined |
| 17-106_k141_50482 | Not-determined |
| 17-106_k141_18275 | Not-determined |
| 17-106_k141_103773 | Not-determined |
| 17-106_k141_39428 | Not-determined |
| 17-106_k141_103775 | Not-determined |
| 17-106_k141_44445 | Not-determined |
| 17-106_k141_39433 | Low-quality |
| 17-106_k141_12675 | Low-quality |
| 17-106_k141_2667 | Not-determined |
| 17-106_k141_18286 | Not-determined |
| 17-106_k141_2670 | Not-determined |
| 17-106_k141_71400 | Not-determined |
| 17-106_k141_103784 | Not-determined |
| 17-106_k141_103785 | Not-determined |
| 17-106_k141_71403 | Not-determined |
| 17-106_k141_39446 | Not-determined |
| 17-106_k141_55641 | Not-determined |
| 17-106_k141_55644 | Not-determined |
| 17-106_k141_82623 | Not-determined |
| 17-106_k141_119623 | Not-determined |
| 17-106_k141_55658 | Not-determined |
| 17-106_k141_50497 | Not-determined |
| 17-106_k141_55661 | Not-determined |
| 17-106_k141_7677 | Not-determined |
| 17-106_k141_103813 | Not-determined |
| 17-106_k141_23474 | Not-determined |
| 17-106_k141_82637 | Not-determined |
| 17-106_k141_44463 | Low-quality |
| 17-106_k141_50511 | Not-determined |
| 17-106_k141_61253 | Not-determined |
| 17-106_k141_103832 | Not-determined |
| 17-106_k141_23481 | Not-determined |
| 17-106_k141_87713 | Low-quality |
| 17-106_k141_50531 | Not-determined |
| 17-106_k141_114004 | Not-determined |
| 17-106_k141_55681 | Not-determined |
| 17-106_k141_50536 | Not-determined |
| 17-106_k141_7687 | Not-determined |
| 17-106_k141_71442 | Not-determined |
| 17-106_k141_50540 | Not-determined |
| 17-106_k141_108705 | Not-determined |
| 17-106_k141_76589 | Not-determined |
| 17-106_k141_76596 | Not-determined |
| 17-106_k141_55690 | Not-determined |
| 17-106_k141_44483 | Not-determined |
| 17-106_k141_103850 | Not-determined |
| 17-106_k141_76606 | Not-determined |
| 17-106_k141_55705 | Not-determined |
| 17-106_k141_50562 | Not-determined |
| 17-106_k141_82693 | Not-determined |
| 17-106_k141_44490 | Not-determined |
| 17-106_k141_12742 | Low-quality |
| 17-106_k141_103857 | Not-determined |
| 17-106_k141_103863 | Not-determined |
| 17-106_k141_7715 | Not-determined |
| 17-106_k141_103874 | Not-determined |
| 17-106_k141_50576 | Not-determined |
| 17-106_k141_7720 | Low-quality |
| 17-106_k141_7722 | Not-determined |
| 17-106_k141_44500 | Not-determined |
| 17-106_k141_76627 | Not-determined |
| 17-106_k141_23515 | Low-quality |
| 17-106_k141_71449 | Not-determined |
| 17-106_k141_50584 | Not-determined |
| 17-106_k141_7730 | Not-determined |
| 17-106_k141_50588 | Not-determined |
| 17-106_k141_82719 | Not-determined |
| 17-106_k141_103880 | Not-determined |
| 17-106_k141_103882 | Not-determined |
| 17-106_k141_7733 | Not-determined |
| 17-106_k141_82724 | Not-determined |
| 17-106_k141_2685 | Not-determined |
| 17-106_k141_7735 | Not-determined |
| 17-106_k141_50598 | Not-determined |
| 17-106_k141_124863 | Not-determined |
| 17-106_k141_23550 | Not-determined |
| 17-106_k141_23554 | Not-determined |
| 17-106_k141_87751 | Not-determined |
| 17-106_k141_7753 | Not-determined |
| 17-106_k141_87759 | Not-determined |
| 17-106_k141_7758 | Not-determined |
| 17-106_k141_108728 | Low-quality |
| 17-106_k141_71454 | Not-determined |
| 17-106_k141_12774 | Not-determined |
| 17-106_k141_76639 | Not-determined |
| 17-106_k141_12778 | Not-determined |
| 17-106_k141_12784 | Not-determined |
| 17-106_k141_12789 | Not-determined |
| 17-106_k141_23586 | Not-determined |
| 17-106_k141_23588 | Not-determined |
| 17-106_k141_7784 | Low-quality |
| 17-106_k141_23597 | Not-determined |
| 17-106_k141_71481 | Not-determined |
| 17-106_k141_108746 | Not-determined |
| 17-106_k141_12807 | Not-determined |
| 17-106_k141_55771 | Not-determined |
| 17-106_k141_61267 | Not-determined |
| 17-106_k141_12809 | Low-quality |
| 17-106_k141_66370 | Not-determined |
| 17-106_k141_33775 | Not-determined |
| 17-106_k141_108762 | Not-determined |
| 17-106_k141_39484 | Not-determined |
| 17-106_k141_66373 | Not-determined |
| 17-106_k141_66376 | Not-determined |
| 17-106_k141_87792 | Not-determined |
| 17-106_k141_66377 | Not-determined |
| 17-106_k141_12817 | Not-determined |
| 17-106_k141_50627 | Not-determined |
| 17-106_k141_108773 | Not-determined |
| 17-106_k141_44533 | Not-determined |
| 17-106_k141_50631 | Not-determined |
| 17-106_k141_124887 | Not-determined |
| 17-106_k141_50632 | Not-determined |
| 17-106_k141_44537 | Not-determined |
| 17-106_k141_61292 | Not-determined |
| 17-106_k141_61294 | Not-determined |
| 17-106_k141_44541 | Not-determined |
| 17-106_k141_82737 | Not-determined |
| 17-106_k141_82739 | Not-determined |
| 17-106_k141_108779 | Not-determined |
| 17-106_k141_108781 | Not-determined |
| 17-106_k141_108787 | Not-determined |
| 17-106_k141_44546 | Not-determined |
| 17-106_k141_108789 | Not-determined |
| 17-106_k141_61303 | Not-determined |
| 17-106_k141_76690 | Not-determined |
| 17-106_k141_87824 | Not-determined |
| 17-106_k141_76693 | Not-determined |
| 17-106_k141_103943 | Not-determined |
| 17-106_k141_108795 | Low-quality |
| 17-106_k141_7813 | Not-determined |
| 17-106_k141_12839 | Not-determined |
| 17-106_k141_66382 | Not-determined |
| 17-106_k141_76698 | Not-determined |
| 17-106_k141_108798 | Not-determined |
| 17-106_k141_66384 | Not-determined |
| 17-106_k141_87835 | Not-determined |
| 17-106_k141_87842 | Not-determined |
| 17-106_k141_108805 | Not-determined |
| 17-106_k141_23620 | Not-determined |
| 17-106_k141_124914 | Not-determined |
| 17-106_k141_124916 | Not-determined |
| 17-106_k141_124918 | Not-determined |
| 17-106_k141_82756 | Not-determined |
| 17-106_k141_93049 | Not-determined |
| 17-106_k141_108813 | Not-determined |
| 17-106_k141_71491 | Not-determined |
| 17-106_k141_124925 | Low-quality |
| 17-106_k141_98264 | Not-determined |
| 17-106_k141_82766 | Not-determined |
| 17-106_k141_50642 | Low-quality |
| 17-106_k141_124936 | Not-determined |
| 17-106_k141_71499 | Not-determined |
| 17-106_k141_82785 | Not-determined |
| 17-106_k141_87866 | Not-determined |
| 17-106_k141_50650 | Not-determined |
| 17-106_k141_50651 | Not-determined |
| 17-106_k141_124947 | Not-determined |
| 17-106_k141_7853 | Not-determined |
| 17-106_k141_87875 | Not-determined |
| 17-106_k141_7861 | Not-determined |
| 17-106_k141_7862 | Not-determined |
| 17-106_k141_76730 | Low-quality |
| 17-106_k141_87880 | Not-determined |
| 17-106_k141_71518 | Low-quality |
| 17-106_k141_7869 | Not-determined |
| 17-106_k141_33801 | Not-determined |
| 17-106_k141_55788 | Not-determined |
| 17-106_k141_33802 | Low-quality |
| 17-106_k141_44608 | Not-determined |
| 17-106_k141_7879 | Not-determined |
| 17-106_k141_44611 | Not-determined |
| 17-106_k141_50662 | Not-determined |
| 17-106_k141_50672 | Not-determined |
| 17-106_k141_2717 | Not-determined |
| 17-106_k141_124960 | Not-determined |
| 17-106_k141_98279 | Not-determined |
| 17-106_k141_28828 | Not-determined |
| 17-106_k141_28829 | Not-determined |
| 17-106_k141_124967 | Not-determined |
| 17-106_k141_39542 | Not-determined |
| 17-106_k141_39545 | Not-determined |
| 17-106_k141_124975 | Not-determined |
| 17-106_k141_18370 | Not-determined |
| 17-106_k141_2744 | Not-determined |
| 17-106_k141_108851 | Low-quality |
| 17-106_k141_50689 | Low-quality |
| 17-106_k141_55816 | Not-determined |
| 17-106_k141_2748 | Not-determined |
| 17-106_k141_50697 | Not-determined |
| 17-106_k141_33834 | Low-quality |
| 17-106_k141_66442 | Not-determined |
| 17-106_k141_18390 | Not-determined |
| 17-106_k141_50702 | Not-determined |
| 17-106_k141_82844 | Not-determined |
| 17-106_k141_2754 | Not-determined |
| 17-106_k141_33841 | Not-determined |
| 17-106_k141_87896 | Low-quality |
| 17-106_k141_103951 | Not-determined |
| 17-106_k141_33844 | Not-determined |
| 17-106_k141_125000 | Not-determined |
| 17-106_k141_103955 | Low-quality |
| 17-106_k141_125005 | Low-quality |
| 17-106_k141_55841 | Not-determined |
| 17-106_k141_50716 | Not-determined |
| 17-106_k141_103964 | Not-determined |
| 17-106_k141_23645 | Not-determined |
| 17-106_k141_28864 | Not-determined |
| 17-106_k141_93069 | Not-determined |
| 17-106_k141_93070 | Not-determined |
| 17-106_k141_39594 | Not-determined |
| 17-106_k141_2768 | Not-determined |
| 17-106_k141_125019 | Not-determined |
| 17-106_k141_18415 | Not-determined |
| 17-106_k141_82869 | Not-determined |
| 17-106_k141_71530 | Low-quality |
| 17-106_k141_119641 | Not-determined |
| 17-106_k141_2771 | Not-determined |
| 17-106_k141_66472 | Not-determined |
| 17-106_k141_23655 | Not-determined |
| 17-106_k141_125027 | Not-determined |
| 17-106_k141_119654 | Not-determined |
| 17-106_k141_82879 | Not-determined |
| 17-106_k141_71536 | Not-determined |
| 17-106_k141_7913 | Not-determined |
| 17-106_k141_103983 | Not-determined |
| 17-106_k141_87930 | Not-determined |
| 17-106_k141_18424 | Not-determined |
| 17-106_k141_76774 | Not-determined |
| 17-106_k141_39614 | Not-determined |
| 17-106_k141_71545 | Not-determined |
| 17-106_k141_93084 | Not-determined |
| 17-106_k141_55847 | Low-quality |
| 17-106_k141_18431 | Not-determined |
| 17-106_k141_82889 | Not-determined |
| 17-106_k141_55851 | Not-determined |
| 17-106_k141_71550 | Not-determined |
| 17-106_k141_2790 | Not-determined |
| 17-106_k141_7926 | Not-determined |
| 17-106_k141_119668 | Not-determined |
| 17-106_k141_55853 | Low-quality |
| 17-106_k141_28885 | Low-quality |
| 17-106_k141_2793 | Not-determined |
| 17-106_k141_82893 | Not-determined |
| 17-106_k141_39619 | Low-quality |
| 17-106_k141_98298 | Not-determined |
| 17-106_k141_125058 | Low-quality |
| 17-106_k141_71557 | Not-determined |
| 17-106_k141_71558 | Not-determined |
| 17-106_k141_2800 | Not-determined |
| 17-106_k141_76781 | Not-determined |
| 17-106_k141_7934 | Low-quality |
| 17-106_k141_76784 | Not-determined |
| 17-106_k141_33869 | Not-determined |
| 17-106_k141_7938 | Not-determined |
| 17-106_k141_55872 | Not-determined |
| 17-106_k141_98304 | Not-determined |
| 17-106_k141_93095 | Not-determined |
| 17-106_k141_55876 | Low-quality |
| 17-106_k141_33876 | Not-determined |
| 17-106_k141_23666 | Not-determined |
| 17-106_k141_55880 | Not-determined |
| 17-106_k141_87967 | Not-determined |
| 17-106_k141_71568 | Not-determined |
| 17-106_k141_82919 | Low-quality |
| 17-106_k141_93105 | Not-determined |
| 17-106_k141_87975 | Low-quality |
| 17-106_k141_33892 | Not-determined |
| 17-106_k141_82928 | Not-determined |
| 17-106_k141_50762 | Not-determined |
| 17-106_k141_71577 | Not-determined |
| 17-106_k141_114036 | Not-determined |
| 17-106_k141_50774 | Not-determined |
| 17-106_k141_87990 | Low-quality |
| 17-106_k141_71579 | Low-quality |
| 17-106_k141_71581 | Not-determined |
| 17-106_k141_71583 | Not-determined |
| 17-106_k141_114049 | Not-determined |
| 17-106_k141_98311 | Not-determined |
| 17-106_k141_50778 | Not-determined |
| 17-106_k141_125097 | Low-quality |
| 17-106_k141_82953 | Not-determined |
| 17-106_k141_114055 | Low-quality |
| 17-106_k141_71588 | Not-determined |
| 17-106_k141_104019 | Low-quality |
| 17-106_k141_114056 | Not-determined |
| 17-106_k141_23694 | Not-determined |
| 17-106_k141_28908 | Low-quality |
| 17-106_k141_114060 | Not-determined |
| 17-106_k141_66534 | Not-determined |
| 17-106_k141_50797 | Not-determined |
| 17-106_k141_39665 | Not-determined |
| 17-106_k141_28916 | Not-determined |
| 17-106_k141_55909 | Not-determined |
| 17-106_k141_82972 | Not-determined |
| 17-106_k141_50802 | Not-determined |
| 17-106_k141_82990 | Not-determined |
| 17-106_k141_82991 | Not-determined |
| 17-106_k141_82993 | Not-determined |
| 17-106_k141_88027 | Not-determined |
| 17-106_k141_50807 | Not-determined |
| 17-106_k141_28948 | Not-determined |
| 17-106_k141_125129 | Not-determined |
| 17-106_k141_83003 | Not-determined |
| 17-106_k141_7945 | Not-determined |
| 17-106_k141_55954 | Not-determined |
| 17-106_k141_33909 | Not-determined |
| 17-106_k141_104058 | Low-quality |
| 17-106_k141_114070 | Not-determined |
| 17-106_k141_7955 | Low-quality |
| 17-106_k141_83020 | Not-determined |
| 17-106_k141_28963 | Not-determined |
| 17-106_k141_114077 | Low-quality |
| 17-106_k141_83029 | Not-determined |
| 17-106_k141_119691 | Low-quality |
| 17-106_k141_39699 | Not-determined |
| 17-106_k141_119692 | Not-determined |
| 17-106_k141_50811 | Not-determined |
| 17-106_k141_55967 | Low-quality |
| 17-106_k141_71622 | Not-determined |
| 17-106_k141_83039 | Not-determined |
| 17-106_k141_55972 | Not-determined |
| 17-106_k141_55975 | Not-determined |
| 17-106_k141_7971 | Not-determined |
| 17-106_k141_125143 | Not-determined |
| 17-106_k141_39715 | Not-determined |
| 17-106_k141_66557 | Not-determined |
| 17-106_k141_33915 | Not-determined |
| 17-106_k141_83043 | Not-determined |
| 17-106_k141_44665 | Not-determined |
| 17-106_k141_33922 | Low-quality |
| 17-106_k141_39719 | Not-determined |
| 17-106_k141_119708 | Not-determined |
| 17-106_k141_33928 | Not-determined |
| 17-106_k141_33930 | Not-determined |
| 17-106_k141_83049 | Not-determined |
| 17-106_k141_114103 | Not-determined |
| 17-106_k141_125164 | Not-determined |
| 17-106_k141_33946 | Not-determined |
| 17-106_k141_2839 | Not-determined |
| 17-106_k141_61319 | Not-determined |
| 17-106_k141_71643 | Not-determined |
| 17-106_k141_50826 | Not-determined |
| 17-106_k141_23723 | Not-determined |
| 17-106_k141_83055 | Low-quality |
| 17-106_k141_33954 | Not-determined |
| 17-106_k141_98327 | Low-quality |
| 17-106_k141_23731 | Not-determined |
| 17-106_k141_61329 | Not-determined |
| 17-106_k141_119735 | Not-determined |
| 17-106_k141_119737 | Not-determined |
| 17-106_k141_23742 | Low-quality |
| 17-106_k141_83062 | Not-determined |
| 17-106_k141_83063 | Not-determined |
| 17-106_k141_44697 | Low-quality |
| 17-106_k141_61332 | Not-determined |
| 17-106_k141_98335 | Not-determined |
| 17-106_k141_66592 | Not-determined |
| 17-106_k141_8003 | Not-determined |
| 17-106_k141_8005 | Not-determined |
| 17-106_k141_83070 | Not-determined |
| 17-106_k141_56001 | Not-determined |
| 17-106_k141_61344 | Low-quality |
| 17-106_k141_56005 | Not-determined |
| 17-106_k141_8009 | Not-determined |
| 17-106_k141_8010 | Not-determined |
| 17-106_k141_18446 | Not-determined |
| 17-106_k141_18450 | Not-determined |
| 17-106_k141_114147 | Not-determined |
| 17-106_k141_56008 | Not-determined |
| 17-106_k141_114153 | Not-determined |
| 17-106_k141_71648 | Low-quality |
| 17-106_k141_18459 | Not-determined |
| 17-106_k141_61361 | Not-determined |
| 17-106_k141_83098 | Not-determined |
| 17-106_k141_83099 | Not-determined |
| 17-106_k141_119772 | Not-determined |
| 17-106_k141_88092 | Not-determined |
| 17-106_k141_8033 | Not-determined |
| 17-106_k141_23768 | Not-determined |
| 17-106_k141_50839 | Not-determined |
| 17-106_k141_50840 | Not-determined |
| 17-106_k141_56044 | Not-determined |
| 17-106_k141_66613 | Not-determined |
| 17-106_k141_76804 | Not-determined |
| 17-106_k141_56047 | Not-determined |
| 17-106_k141_8038 | Not-determined |
| 17-106_k141_23783 | Not-determined |
| 17-106_k141_104090 | Not-determined |
| 17-106_k141_2862 | Not-determined |
| 17-106_k141_2867 | Not-determined |
| 17-106_k141_76814 | Low-quality |
| 17-106_k141_98372 | Not-determined |
| 17-106_k141_23788 | Not-determined |
| 17-106_k141_8055 | Not-determined |
| 17-106_k141_8062 | Not-determined |
| 17-106_k141_44710 | Not-determined |
| 17-106_k141_88115 | Not-determined |

| | |
|---|---|
| 17-106_k141_88119 | Not-determined |
| 17-106_k141_76822 | Not-determined |
| 17-106_k141_66632 | Not-determined |
| 17-106_k141_98389 | Not-determined |
| 17-106_k141_93130 | Low-quality |
| 17-106_k141_44722 | Not-determined |
| 17-106_k141_119799 | Not-determined |
| 17-106_k141_44726 | Not-determined |
| 17-106_k141_98391 | Not-determined |
| 17-106_k141_104122 | Not-determined |
| 17-106_k141_23805 | Not-determined |
| 17-106_k141_23806 | Not-determined |
| 17-106_k141_104124 | Low-quality |
| 17-106_k141_34006 | Low-quality |
| 17-106_k141_44729 | Not-determined |
| 17-106_k141_18519 | Not-determined |
| 17-106_k141_66646 | Not-determined |
| 17-106_k141_119816 | Not-determined |
| 17-106_k141_56079 | Not-determined |
| 17-106_k141_34021 | Not-determined |
| 17-106_k141_8067 | Not-determined |
| 17-106_k141_56093 | Not-determined |
| 17-106_k141_76845 | Not-determined |
| 17-106_k141_93155 | Not-determined |
| 17-106_k141_93158 | Not-determined |
| 17-106_k141_93159 | Not-determined |
| 17-106_k141_93162 | Not-determined |
| 17-106_k141_12856 | Low-quality |
| 17-106_k141_56108 | Not-determined |
| 17-106_k141_56109 | Not-determined |
| 17-106_k141_104153 | Not-determined |
| 17-106_k141_76859 | Not-determined |
| 17-106_k141_56111 | Not-determined |
| 17-106_k141_39739 | Low-quality |
| 17-106_k141_39740 | Not-determined |
| 17-106_k141_12863 | Low-quality |
| 17-106_k141_2891 | Not-determined |
| 17-106_k141_61391 | Not-determined |
| 17-106_k141_8076 | Not-determined |
| 17-106_k141_2904 | Not-determined |
| 17-106_k141_93181 | Not-determined |
| 17-106_k141_18561 | Not-determined |
| 17-106_k141_18564 | Not-determined |
| 17-106_k141_12885 | Not-determined |
| 18-109-final-viral-combined_k141_33166\|\|full | High-quality |
| 17-106_k141_104172 | Not-determined |
| 17-106_k141_8092 | Not-determined |
| 17-106_k141_76874 | Not-determined |
| 17-106_k141_76875 | Not-determined |
| 17-106_k141_44760 | Not-determined |
| 17-106_k141_44767 | Low-quality |
| 17-106_k141_83170 | Not-determined |
| 17-106_k141_39760 | Not-determined |
| 17-106_k141_83183 | Not-determined |
| 17-106_k141_34044 | Low-quality |
| 17-106_k141_71686 | Not-determined |
| 17-106_k141_104188 | Not-determined |
| 17-106_k141_71690 | Not-determined |
| 17-106_k141_71691 | Not-determined |
| 17-106_k141_34047 | Not-determined |
| 17-106_k141_44778 | Not-determined |
| 17-106_k141_93205 | Not-determined |
| 17-106_k141_83191 | Not-determined |
| 17-106_k141_2929 | Not-determined |
| 17-106_k141_44787 | Not-determined |
| 17-106_k141_34054 | Not-determined |
| 17-106_k141_93207 | Low-quality |
| 17-106_k141_39771 | Not-determined |
| 17-106_k141_76897 | Not-determined |
| 17-106_k141_29045 | Not-determined |
| 17-106_k141_104208 | Low-quality |
| 17-106_k141_114180 | Not-determined |
| 17-106_k141_76908 | Not-determined |
| 17-106_k141_12895 | Not-determined |
| 17-106_k141_34069 | Low-quality |
| 17-106_k141_12896 | Low-quality |
| 17-106_k141_34071 | Not-determined |
| 17-106_k141_76916 | Not-determined |
| 17-106_k141_108878 | Not-determined |
| 17-106_k141_56160 | Not-determined |
| 17-106_k141_12905 | Low-quality |
| 17-106_k141_2959 | Not-determined |
| 17-106_k141_23828 | Not-determined |
| 17-106_k141_71696 | Not-determined |
| 17-106_k141_12908 | Low-quality |
| 17-106_k141_39785 | Not-determined |
| 17-106_k141_56161 | Not-determined |
| 17-106_k141_39790 | Not-determined |
| 17-106_k141_56162 | Not-determined |
| 17-106_k141_83213 | Not-determined |
| 17-106_k141_39792 | Low-quality |
| 17-106_k141_12921 | Not-determined |
| 17-106_k141_8107 | Not-determined |
| 17-106_k141_114197 | Not-determined |
| 17-106_k141_2969 | Not-determined |
| 17-106_k141_61460 | Not-determined |
| 17-106_k141_61461 | Not-determined |
| 17-106_k141_2973 | Not-determined |
| 17-106_k141_71718 | Low-quality |
| 17-106_k141_71720 | Not-determined |
| 17-106_k141_83223 | Not-determined |
| 17-106_k141_83224 | Not-determined |
| 17-106_k141_76937 | Not-determined |
| 17-106_k141_2979 | Low-quality |
| 17-106_k141_61466 | Not-determined |
| 17-106_k141_34081 | Not-determined |
| 17-106_k141_61468 | Not-determined |
| 17-106_k141_12927 | Not-determined |
| 17-106_k141_12928 | Low-quality |
| 17-106_k141_34083 | Not-determined |
| 17-106_k141_108915 | Not-determined |
| 17-106_k141_104230 | Not-determined |
| 17-106_k141_83232 | Not-determined |
| 17-106_k141_12936 | Not-determined |
| 17-106_k141_23848 | Low-quality |
| 17-106_k141_23849 | Low-quality |
| 17-106_k141_12938 | Not-determined |
| 17-106_k141_108921 | Not-determined |
| 17-106_k141_119929 | Not-determined |
| 17-106_k141_8129 | Low-quality |
| 17-106_k141_61486 | Not-determined |
| 17-106_k141_44822 | Not-determined |
| 17-106_k141_12945 | Not-determined |
| 17-106_k141_61507 | Not-determined |
| 17-106_k141_83258 | Not-determined |
| 17-106_k141_114204 | Low-quality |
| 17-106_k141_119951 | Not-determined |
| 17-106_k141_119954 | Not-determined |
| 17-106_k141_83259 | Not-determined |
| 17-106_k141_23855 | Low-quality |
| 17-106_k141_2988 | Not-determined |
| 17-106_k141_50861 | Not-determined |
| 17-106_k141_29066 | Not-determined |
| 17-106_k141_8163 | Not-determined |
| 17-106_k141_93240 | Not-determined |
| 17-106_k141_93241 | Not-determined |
| 17-106_k141_23865 | Not-determined |
| 17-106_k141_29077 | Not-determined |
| 17-106_k141_98409 | Not-determined |
| 17-106_k141_71736 | Low-quality |
| 17-106_k141_98412 | Not-determined |
| 17-106_k141_93258 | Not-determined |
| 17-106_k141_39811 | Not-determined |
| 17-106_k141_29083 | Not-determined |
| 17-106_k141_93263 | Not-determined |
| 17-106_k141_3017 | Not-determined |
| 17-106_k141_114255 | Not-determined |
| 17-106_k141_71754 | Not-determined |
| 17-106_k141_50890 | Not-determined |
| 17-106_k141_8177 | Not-determined |
| 17-106_k141_61541 | Not-determined |
| 17-106_k141_50903 | Not-determined |
| 17-106_k141_12982 | Low-quality |
| 17-106_k141_66655 | Not-determined |
| 17-106_k141_44880 | Not-determined |
| 17-106_k141_114270 | Not-determined |
| 17-106_k141_66657 | Not-determined |
| 17-106_k141_83290 | Not-determined |
| 17-106_k141_44881 | Not-determined |
| 17-106_k141_83295 | Low-quality |
| 17-106_k141_56182 | Not-determined |
| 17-106_k141_3030 | Not-determined |
| 17-106_k141_93276 | Not-determined |
| 17-106_k141_114278 | Not-determined |
| 17-106_k141_12998 | Not-determined |
| 17-106_k141_119993 | Not-determined |
| 17-106_k141_61550 | Low-quality |
| 17-106_k141_8199 | Not-determined |
| 17-106_k141_13003 | Not-determined |
| 17-106_k141_8205 | Not-determined |
| 17-106_k141_44894 | Low-quality |
| 17-106_k141_18699 | Not-determined |
| 17-106_k141_98451 | Not-determined |
| 17-106_k141_13021 | Not-determined |
| 17-106_k141_71790 | Not-determined |
| 17-106_k141_3036 | Low-quality |
| 17-106_k141_13024 | Not-determined |
| 17-106_k141_29120 | Not-determined |
| 17-106_k141_13027 | Not-determined |
| 17-106_k141_71797 | Not-determined |
| 17-106_k141_13032 | Not-determined |
| 17-106_k141_108950 | Not-determined |
| 17-106_k141_18712 | Not-determined |
| 17-106_k141_71801 | Not-determined |
| 17-106_k141_104265 | Not-determined |
| 17-106_k141_98460 | Not-determined |
| 17-106_k141_3045 | Not-determined |
| 17-106_k141_50915 | Not-determined |
| 17-106_k141_104268 | Not-determined |
| 17-106_k141_71808 | Not-determined |
| 17-106_k141_104269 | Not-determined |
| 17-106_k141_34116 | Low-quality |
| 17-106_k141_34120 | Not-determined |
| 17-106_k141_34122 | Low-quality |
| 17-106_k141_3052 | Not-determined |
| 17-106_k141_13047 | Not-determined |
| 17-106_k141_114315 | Not-determined |
| 17-106_k141_76995 | Not-determined |
| 17-106_k141_50916 | Low-quality |
| 17-106_k141_18725 | Not-determined |
| 17-106_k141_34135 | Not-determined |
| 17-106_k141_61585 | Not-determined |
| 17-106_k141_8245 | Not-determined |
| 17-106_k141_8246 | Low-quality |
| 17-106_k141_50922 | Not-determined |
| 17-106_k141_114328 | Not-determined |
| 17-106_k141_23914 | Not-determined |
| 17-106_k141_66684 | Not-determined |
| 17-106_k141_18737 | Not-determined |
| 17-106_k141_61598 | Not-determined |
| 17-106_k141_8258 | Not-determined |
| 17-106_k141_71817 | Not-determined |
| 17-106_k141_23931 | Not-determined |
| 17-106_k141_125207 | Not-determined |
| 17-106_k141_50936 | Not-determined |
| 17-106_k141_50937 | Not-determined |
| 17-106_k141_8269 | Not-determined |
| 17-106_k141_114339 | Not-determined |
| 17-106_k141_93317 | Not-determined |
| 17-106_k141_83333 | Not-determined |
| 17-106_k141_50939 | Not-determined |
| 17-106_k141_114340 | Low-quality |
| 17-106_k141_39845 | Not-determined |
| 17-106_k141_34145 | Not-determined |
| 17-106_k141_93326 | Not-determined |
| 17-106_k141_77038 | Not-determined |
| 17-106_k141_18757 | Not-determined |
| 17-106_k141_83340 | Not-determined |
| 17-106_k141_114350 | Not-determined |
| 17-106_k141_34155 | Not-determined |
| 17-106_k141_18763 | Not-determined |
| 17-106_k141_114355 | Not-determined |
| 17-106_k141_77044 | Not-determined |
| 17-106_k141_77050 | Low-quality |
| 17-106_k141_108992 | Not-determined |
| 17-106_k141_93342 | Not-determined |
| 17-106_k141_23972 | Not-determined |
| 17-106_k141_13091 | Low-quality |
| 17-106_k141_34169 | Not-determined |
| 17-106_k141_3085 | Not-determined |
| 17-106_k141_93348 | Not-determined |
| 17-106_k141_34172 | Not-determined |
| 17-106_k141_34173 | Low-quality |
| 17-106_k141_34174 | Not-determined |
| 17-106_k141_56237 | Not-determined |
| 17-106_k141_71836 | Not-determined |
| 17-106_k141_71837 | Low-quality |
| 17-106_k141_56241 | Low-quality |
| 17-106_k141_56242 | Not-determined |
| 17-106_k141_71842 | Low-quality |
| 17-106_k141_23991 | Not-determined |
| 17-106_k141_3089 | Not-determined |
| 17-106_k141_23993 | Not-determined |
| 17-106_k141_13114 | Not-determined |
| 17-106_k141_34191 | Not-determined |
| 17-106_k141_77073 | Not-determined |
| 17-106_k141_61639 | Not-determined |
| 17-106_k141_56250 | Not-determined |
| 17-106_k141_50960 | Not-determined |
| 17-106_k141_34200 | Not-determined |
| 17-106_k141_50966 | Not-determined |
| 17-106_k141_61647 | Low-quality |
| 17-106_k141_18785 | Not-determined |
| 17-106_k141_56251 | Low-quality |
| 17-106_k141_34205 | Not-determined |
| 17-106_k141_71860 | Not-determined |
| 17-106_k141_61660 | Not-determined |
| 17-106_k141_44931 | Not-determined |
| 17-106_k141_83401 | Not-determined |
| 17-106_k141_3108 | Low-quality |
| 17-106_k141_83407 | Not-determined |
| 17-106_k141_71870 | Not-determined |
| 17-106_k141_83409 | Not-determined |
| 17-106_k141_8285 | Not-determined |
| 17-106_k141_13126 | Not-determined |
| 17-106_k141_93376 | Not-determined |
| 17-106_k141_8287 | Not-determined |
| 17-106_k141_8289 | Low-quality |
| 17-106_k141_120036 | Not-determined |
| 17-106_k141_39851 | Not-determined |
| 17-106_k141_13135 | Low-quality |
| 17-106_k141_24025 | Not-determined |
| 17-106_k141_13138 | Not-determined |
| 17-106_k141_98502 | Not-determined |
| 17-106_k141_120046 | Not-determined |
| 17-106_k141_77109 | Not-determined |
| 17-106_k141_120049 | Not-determined |
| 17-106_k141_114421 | Not-determined |
| 17-106_k141_77114 | Not-determined |
| 17-106_k141_120057 | Not-determined |
| 17-106_k141_8313 | Not-determined |
| 17-106_k141_8314 | Not-determined |
| 17-106_k141_71891 | Not-determined |
| 17-106_k141_77119 | Low-quality |
| 17-106_k141_114434 | Not-determined |
| 17-106_k141_120071 | Not-determined |
| 17-106_k141_98514 | Not-determined |
| 17-106_k141_98518 | Not-determined |
| 17-106_k141_18802 | Not-determined |
| 17-106_k141_44991 | Low-quality |
| 17-106_k141_125234 | Low-quality |
| 17-106_k141_114451 | Low-quality |
| 17-106_k141_45001 | Not-determined |
| 17-106_k141_29154 | Not-determined |
| 17-106_k141_29155 | Not-determined |
| 17-106_k141_125240 | Not-determined |
| 17-106_k141_125241 | Not-determined |
| 17-106_k141_120085 | Not-determined |
| 17-106_k141_56274 | Not-determined |
| 17-106_k141_120088 | Not-determined |
| 17-106_k141_39882 | Not-determined |
| 17-106_k141_45011 | Not-determined |
| 17-106_k141_125251 | Not-determined |
| 17-106_k141_29161 | Not-determined |
| 17-106_k141_93402 | Not-determined |
| 17-106_k141_8337 | Not-determined |
| 17-106_k141_56291 | Not-determined |
| 17-106_k141_18805 | Not-determined |
| 17-106_k141_56293 | Not-determined |
| 17-106_k141_120102 | Low-quality |
| 17-106_k141_104308 | Not-determined |
| 17-106_k141_8344 | Not-determined |
| 17-106_k141_125266 | Not-determined |
| 17-106_k141_13170 | Low-quality |
| 17-106_k141_8346 | Not-determined |
| 17-106_k141_13173 | Low-quality |
| 17-106_k141_13175 | Not-determined |
| 17-106_k141_56312 | Not-determined |
| 17-106_k141_104317 | Not-determined |
| 17-106_k141_13179 | Not-determined |
| 17-106_k141_120115 | Not-determined |
| 17-106_k141_45025 | Not-determined |
| 17-106_k141_45029 | Not-determined |
| 17-106_k141_120117 | Not-determined |
| 17-106_k141_114483 | Not-determined |
| 17-106_k141_120119 | Not-determined |
| 17-106_k141_13182 | Not-determined |
| 17-106_k141_24110 | Not-determined |
| 17-106_k141_24111 | Not-determined |
| 17-106_k141_18829 | Not-determined |
| 17-106_k141_34241 | Low-quality |
| 17-106_k141_18834 | Not-determined |
| 17-106_k141_34249 | Not-determined |
| 17-106_k141_45039 | Not-determined |
| 17-106_k141_29175 | Not-determined |
| 17-106_k141_18843 | Not-determined |
| 17-106_k141_29177 | Not-determined |
| 17-106_k141_34257 | Not-determined |
| 17-106_k141_120142 | Not-determined |
| 17-106_k141_98534 | Not-determined |
| 17-106_k141_18844 | Not-determined |
| 17-106_k141_98541 | Not-determined |
| 17-106_k141_51015 | Not-determined |
| 17-106_k141_51021 | Not-determined |
| 17-106_k141_24126 | Not-determined |
| 17-106_k141_13215 | Not-determined |
| 17-106_k141_114514 | Low-quality |
| 17-106_k141_120150 | Not-determined |
| 17-106_k141_29199 | Not-determined |
| 17-106_k141_8396 | Low-quality |
| 17-106_k141_18868 | Not-determined |
| 17-106_k141_29207 | Low-quality |
| 17-106_k141_120163 | Not-determined |
| 17-106_k141_120165 | Low-quality |
| 17-106_k141_34286 | Not-determined |
| 17-106_k141_120169 | Not-determined |
| 17-106_k141_77153 | Low-quality |
| 17-106_k141_8406 | Low-quality |
| 17-106_k141_77156 | Not-determined |
| 17-106_k141_45059 | Not-determined |
| 17-106_k141_120179 | Not-determined |
| 17-106_k141_120180 | Not-determined |
| 17-106_k141_24152 | Not-determined |
| 17-106_k141_104355 | Not-determined |
| 17-106_k141_98575 | Not-determined |
| 17-106_k141_39903 | Not-determined |
| 17-106_k141_83424 | Not-determined |
| 17-106_k141_39908 | Not-determined |
| 17-106_k141_18896 | Not-determined |
| 17-106_k141_120185 | Not-determined |
| 17-106_k141_13257 | Not-determined |
| 17-106_k141_120190 | Not-determined |
| 17-106_k141_24161 | Not-determined |
| 17-106_k141_13261 | Not-determined |
| 17-106_k141_77185 | Not-determined |
| 17-106_k141_83438 | Not-determined |
| 17-106_k141_39914 | Not-determined |
| 17-106_k141_39915 | Not-determined |
| 17-106_k141_13266 | Not-determined |
| 17-106_k141_83450 | Not-determined |
| 17-106_k141_18911 | Not-determined |
| 17-106_k141_77198 | Low-quality |
| 17-106_k141_104392 | Not-determined |
| 17-106_k141_8417 | Not-determined |
| 17-106_k141_83458 | Not-determined |
| 17-106_k141_45077 | Not-determined |
| 17-106_k141_8419 | Not-determined |
| 17-106_k141_18916 | Not-determined |
| 17-106_k141_34332 | Not-determined |
| 17-106_k141_8424 | Not-determined |
| 17-106_k141_18922 | Low-quality |
| 17-106_k141_120216 | Not-determined |
| 17-106_k141_77211 | Not-determined |
| 17-106_k141_114562 | Not-determined |
| 17-106_k141_39940 | Not-determined |
| 17-106_k141_109065 | Not-determined |
| 17-106_k141_77221 | Not-determined |
| 17-106_k141_45093 | Not-determined |
| 17-106_k141_3131 | Not-determined |
| 17-106_k141_18936 | Not-determined |
| 17-106_k141_109071 | Not-determined |
| 17-106_k141_109072 | Not-determined |
| 17-106_k141_109076 | Not-determined |
| 17-106_k141_29245 | Not-determined |
| 17-106_k141_109081 | Not-determined |
| 17-106_k141_13297 | Not-determined |
| 17-106_k141_98603 | Not-determined |
| 17-106_k141_13302 | Not-determined |
| 17-106_k141_61686 | Not-determined |
| 17-106_k141_18949 | Not-determined |
| 17-106_k141_109092 | Low-quality |
| 17-106_k141_98617 | Not-determined |
| 17-106_k141_61701 | Not-determined |
| 17-106_k141_114600 | Not-determined |
| 17-106_k141_114601 | Low-quality |
| 17-106_k141_13317 | Not-determined |
| 17-106_k141_77235 | Low-quality |
| 17-106_k141_114610 | Not-determined |
| 17-106_k141_13323 | Not-determined |
| 17-106_k141_3137 | Not-determined |
| 17-106_k141_24177 | Low-quality |
| 17-106_k141_56359 | Not-determined |
| 17-106_k141_109102 | Not-determined |
| 17-106_k141_109103 | Not-determined |
| 17-106_k141_51064 | Not-determined |
| 17-106_k141_8450 | Not-determined |
| 17-106_k141_88141 | Not-determined |
| 17-106_k141_39954 | Not-determined |
| 17-106_k141_109114 | Not-determined |
| 17-106_k141_18987 | Not-determined |
| 17-106_k141_51072 | Not-determined |
| 17-106_k141_71933 | Not-determined |
| 17-106_k141_88156 | Not-determined |
| 17-106_k141_51078 | Low-quality |
| 17-106_k141_39962 | Not-determined |
| 17-106_k141_39968 | Not-determined |
| 17-106_k141_83503 | Not-determined |
| 17-106_k141_98652 | Low-quality |
| 17-106_k141_24193 | Not-determined |
| 17-106_k141_125306 | Not-determined |
| 17-106_k141_19002 | Not-determined |
| 17-106_k141_114636 | Low-quality |
| 17-106_k141_51089 | Not-determined |
| 17-106_k141_45143 | Not-determined |
| 17-106_k141_51090 | Not-determined |
| 17-106_k141_88172 | Low-quality |
| 17-106_k141_88175 | Not-determined |
| 17-106_k141_51097 | Not-determined |
| 17-106_k141_83518 | Not-determined |
| 17-106_k141_24351 | Not-determined |
| 17-106_k141_114650 | Not-determined |
| 17-106_k141_56386 | Not-determined |
| 17-106_k141_125318 | Not-determined |
| 17-106_k141_114653 | Not-determined |
| 17-106_k141_125320 | Not-determined |
| 17-106_k141_83525 | Not-determined |
| 17-106_k141_56392 | Not-determined |
| 17-106_k141_29265 | Low-quality |
| 17-106_k141_24208 | Not-determined |
| 17-106_k141_45164 | Low-quality |
| 17-106_k141_40001 | Not-determined |
| 17-106_k141_125329 | Not-determined |
| 17-106_k141_40004 | Not-determined |
| 17-106_k141_125330 | Not-determined |
| 17-106_k141_77290 | Not-determined |
| 17-106_k141_77292 | Low-quality |
| 17-106_k141_61735 | Not-determined |
| 17-106_k141_71957 | Low-quality |
| 17-106_k141_61744 | Not-determined |
| 17-106_k141_114666 | Not-determined |
| 17-106_k141_34362 | Low-quality |
| 17-106_k141_120247 | Not-determined |
| 17-106_k141_45188 | Not-determined |
| 17-106_k141_34368 | Not-determined |
| 17-106_k141_45193 | Not-determined |
| 17-106_k141_45198 | Not-determined |
| 17-106_k141_120262 | Not-determined |
| 17-106_k141_120269 | Not-determined |
| 17-106_k141_125363 | Not-determined |
| 17-106_k141_40013 | Not-determined |
| 17-106_k141_24232 | Not-determined |
| 17-106_k141_125365 | Not-determined |
| 17-106_k141_24233 | Not-determined |
| 17-106_k141_61779 | Not-determined |
| 17-106_k141_51162 | Not-determined |
| 17-106_k141_24244 | Not-determined |
| 17-106_k141_51164 | Not-determined |
| 17-106_k141_88180 | Low-quality |
| 17-106_k141_24246 | Not-determined |
| 17-106_k141_45220 | Not-determined |
| 17-106_k141_66712 | Not-determined |
| 17-106_k141_120272 | Not-determined |
| 17-106_k141_29284 | Low-quality |
| 17-106_k141_29285 | Not-determined |
| 17-106_k141_120274 | Not-determined |
| 17-106_k141_66718 | Not-determined |
| 17-106_k141_45231 | Not-determined |
| 17-106_k141_120278 | Not-determined |
| 17-106_k141_40032 | Not-determined |
| 17-106_k141_114698 | Not-determined |
| 17-106_k141_13367 | Low-quality |
| 17-106_k141_51181 | Not-determined |
| 17-106_k141_3156 | Not-determined |
| 17-106_k141_40039 | Not-determined |
| 17-106_k141_114716 | Not-determined |
| 17-106_k141_71989 | Not-determined |
| 17-106_k141_51189 | Not-determined |
| 17-106_k141_114718 | Not-determined |
| 17-106_k141_24269 | Not-determined |
| 17-106_k141_34404 | Not-determined |
| 17-106_k141_24274 | Not-determined |
| 17-106_k141_66731 | Not-determined |
| 17-106_k141_51199 | Not-determined |
| 17-106_k141_45253 | Not-determined |
| 17-106_k141_34408 | Not-determined |
| 17-106_k141_13379 | Not-determined |
| 17-106_k141_13380 | Low-quality |
| 17-106_k141_125392 | Low-quality |
| 17-106_k141_56459 | Not-determined |
| 17-106_k141_13385 | Low-quality |
| 17-106_k141_125395 | Not-determined |
| 18-105-final-viral-combined_k141_59064\|\|full | Medium-quality |
| 17-106_k141_98680 | Not-determined |
| 17-106_k141_45258 | Not-determined |
| 17-106_k141_98682 | Not-determined |
| 17-106_k141_56467 | Not-determined |
| 17-106_k141_8526 | Not-determined |
| 17-106_k141_45264 | Not-determined |
| 17-106_k141_8529 | Low-quality |
| 17-106_k141_34428 | Not-determined |
| 17-106_k141_77373 | Not-determined |
| 17-106_k141_56474 | Not-determined |
| 17-106_k141_61807 | Not-determined |
| 17-106_k141_93470 | Not-determined |
| 17-106_k141_114728 | Not-determined |
| 17-106_k141_120302 | Not-determined |
| 17-106_k141_120304 | Not-determined |
| 17-106_k141_61816 | Not-determined |
| 17-106_k141_34435 | Not-determined |
| 17-106_k141_66765 | Not-determined |
| 17-106_k141_13396 | Not-determined |
| 17-106_k141_8535 | Not-determined |
| 17-106_k141_24282 | Not-determined |
| 17-106_k141_61823 | Low-quality |
| 17-106_k141_120313 | Not-determined |
| 17-106_k141_125415 | Not-determined |
| 17-106_k141_125418 | Not-determined |
| 17-106_k141_34443 | Not-determined |
| 17-106_k141_13406 | Not-determined |
| 17-106_k141_51228 | Not-determined |
| 17-106_k141_120324 | Low-quality |
| 17-106_k141_77388 | Low-quality |
| 17-106_k141_120333 | Not-determined |
| 17-106_k141_56492 | Not-determined |
| 17-106_k141_40083 | Not-determined |
| 17-106_k141_83593 | Not-determined |
| 17-106_k141_13420 | Not-determined |
| 17-106_k141_93513 | Low-quality |
| 17-106_k141_40095 | Not-determined |
| 17-106_k141_93515 | Not-determined |
| 17-106_k141_56497 | Not-determined |
| 17-106_k141_83600 | Not-determined |
| 17-106_k141_13427 | Not-determined |
| 17-106_k141_93518 | Low-quality |
| 17-106_k141_34463 | Not-determined |
| 17-106_k141_56506 | Not-determined |
| 17-106_k141_88242 | Not-determined |
| 17-106_k141_114760 | Not-determined |
| 17-106_k141_72047 | Not-determined |
| 17-106_k141_88244 | Not-determined |
| 17-106_k141_51238 | Not-determined |
| 17-106_k141_77406 | Not-determined |
| 17-106_k141_98705 | Not-determined |
| 17-106_k141_120365 | Not-determined |
| 17-106_k141_93534 | Not-determined |
| 17-106_k141_77410 | Not-determined |
| 17-106_k141_61855 | Low-quality |
| 17-106_k141_29295 | Not-determined |
| 17-106_k141_72059 | Not-determined |

| | |
|---|---|
| 17-106_k141_125422 | Not-determined |
| 17-106_k141_45309 | Not-determined |
| 17-106_k141_83620 | Not-determined |
| 17-106_k141_83621 | Not-determined |
| 17-106_k141_114785 | Not-determined |
| 17-106_k141_45315 | Not-determined |
| 17-106_k141_40118 | Not-determined |
| 17-106_k141_45318 | Not-determined |
| 17-106_k141_34494 | Not-determined |
| 17-106_k141_88254 | Low-quality |
| 17-106_k141_98735 | Not-determined |
| 17-106_k141_45332 | Not-determined |
| 17-106_k141_98742 | Not-determined |
| 17-106_k141_72086 | Not-determined |
| 17-106_k141_83634 | Not-determined |
| 17-106_k141_45338 | Not-determined |
| 17-106_k141_19075 | Not-determined |
| 17-106_k141_114804 | Not-determined |
| 17-106_k141_19080 | Not-determined |
| 17-106_k141_72095 | Not-determined |
| 17-106_k141_29313 | Not-determined |
| 17-106_k141_34500 | Low-quality |
| 17-106_k141_72098 | Not-determined |
| 17-106_k141_120410 | Not-determined |
| 17-106_k141_125433 | Not-determined |
| 17-106_k141_72104 | Not-determined |
| 17-106_k141_29325 | Not-determined |
| 17-106_k141_72110 | Not-determined |
| 17-106_k141_45348 | Not-determined |
| 17-106_k141_56516 | Low-quality |
| 17-106_k141_120415 | Low-quality |
| 17-106_k141_77455 | Not-determined |
| 17-106_k141_56519 | Not-determined |
| 17-106_k141_56522 | Not-determined |
| 17-106_k141_66823 | Not-determined |
| 17-106_k141_29343 | Not-determined |
| 17-106_k141_120427 | Not-determined |
| 17-106_k141_125444 | Not-determined |
| 17-106_k141_93545 | Not-determined |
| 17-106_k141_93547 | Not-determined |
| 17-106_k141_83647 | Not-determined |
| 17-106_k141_93551 | Not-determined |
| 17-106_k141_8560 | Not-determined |
| 17-106_k141_56531 | Not-determined |
| 17-106_k141_56532 | Not-determined |
| 17-106_k141_56533 | Low-quality |
| 17-106_k141_98768 | Not-determined |
| 17-106_k141_56536 | Not-determined |
| 17-106_k141_40167 | Low-quality |
| 17-106_k141_56538 | Low-quality |
| 17-106_k141_72117 | Low-quality |
| 17-106_k141_3211 | Not-determined |
| 17-106_k141_45366 | Not-determined |
| 17-106_k141_45367 | Not-determined |
| 17-106_k141_66842 | Not-determined |
| 17-106_k141_88263 | Not-determined |
| 17-106_k141_45373 | Not-determined |
| 17-106_k141_34525 | Not-determined |
| 17-106_k141_24330 | Not-determined |
| 17-106_k141_34531 | Not-determined |
| 17-106_k141_56556 | Not-determined |
| 17-106_k141_34536 | Not-determined |
| 17-106_k141_125487 | Not-determined |
| 17-106_k141_93563 | Not-determined |
| 17-106_k141_19122 | Not-determined |
| 17-106_k141_19125 | Not-determined |
| 17-106_k141_125496 | Low-quality |
| 17-106_k141_40182 | Not-determined |
| 17-106_k141_98810 | Not-determined |
| 17-106_k141_24348 | Not-determined |
| 17-106_k141_98818 | Not-determined |
| 17-106_k141_66859 | Not-determined |
| 17-106_k141_120446 | Not-determined |
| 17-106_k141_34557 | Low-quality |
| 17-106_k141_72133 | Not-determined |
| 17-106_k141_45397 | Low-quality |
| 17-106_k141_45398 | Not-determined |
| 17-106_k141_19142 | Not-determined |
| 17-106_k141_40196 | Not-determined |
| 17-106_k141_40199 | Not-determined |
| 17-106_k141_24363 | Not-determined |
| 17-106_k141_83667 | Not-determined |
| 17-106_k141_109157 | Not-determined |
| 17-106_k141_40211 | Not-determined |
| 17-106_k141_83678 | Not-determined |
| 17-106_k141_66874 | Not-determined |
| 17-106_k141_40215 | Not-determined |
| 17-106_k141_34586 | Not-determined |
| 17-106_k141_66883 | Not-determined |
| 17-106_k141_56602 | Not-determined |
| 17-106_k141_120458 | Not-determined |
| 17-106_k141_13479 | Not-determined |
| 17-106_k141_120460 | Low-quality |
| 17-106_k141_56608 | Not-determined |
| 17-106_k141_120464 | Not-determined |
| 17-106_k141_13486 | Not-determined |
| 17-106_k141_19167 | Not-determined |
| 17-106_k141_19168 | Not-determined |
| 17-106_k141_77506 | Low-quality |
| 17-106_k141_29379 | Not-determined |
| 17-106_k141_120471 | Not-determined |
| 17-106_k141_93582 | Not-determined |
| 17-106_k141_83701 | Not-determined |
| 17-106_k141_98846 | Low-quality |
| 17-106_k141_114837 | Not-determined |
| 17-106_k141_34619 | Not-determined |
| 17-106_k141_45422 | Low-quality |
| 17-106_k141_98852 | Not-determined |
| 17-106_k141_3267 | Not-determined |
| 17-106_k141_3274 | Not-determined |
| 17-106_k141_88285 | Low-quality |
| 17-106_k141_61902 | Not-determined |
| 17-106_k141_34630 | Not-determined |
| 17-106_k141_3279 | Not-determined |
| 17-106_k141_45446 | Not-determined |
| 17-106_k141_93590 | Not-determined |
| 17-106_k141_83716 | Not-determined |
| 17-106_k141_125515 | Not-determined |
| 17-106_k141_93599 | Not-determined |
| 17-106_k141_93604 | Not-determined |
| 17-106_k141_40241 | Low-quality |
| 17-106_k141_83723 | Not-determined |
| 17-106_k141_83724 | Not-determined |
| 17-106_k141_56663 | Not-determined |
| 17-106_k141_40255 | Not-determined |
| 17-106_k141_56666 | Not-determined |
| 17-106_k141_45457 | Not-determined |
| 17-106_k141_56669 | Not-determined |
| 17-106_k141_56672 | Not-determined |
| 17-106_k141_93616 | Low-quality |
| 17-106_k141_98890 | Not-determined |
| 17-106_k141_88320 | Not-determined |
| 17-106_k141_114899 | Not-determined |
| 17-106_k141_93621 | Not-determined |
| 17-106_k141_45464 | Low-quality |
| 17-106_k141_45466 | Not-determined |
| 17-106_k141_120509 | Not-determined |
| 17-106_k141_93630 | Not-determined |
| 17-106_k141_125543 | Low-quality |
| 17-106_k141_93632 | Not-determined |
| 17-106_k141_83744 | Not-determined |
| 17-106_k141_93635 | Not-determined |
| 17-106_k141_93637 | Not-determined |
| 17-106_k141_13530 | Not-determined |
| 17-106_k141_83757 | Low-quality |
| 17-106_k141_93646 | Not-determined |
| 17-106_k141_72179 | Not-determined |
| 17-106_k141_77538 | Not-determined |
| 17-106_k141_13537 | Not-determined |
| 17-106_k141_8578 | Not-determined |
| 17-106_k141_3309 | Low-quality |
| 17-106_k141_88341 | Not-determined |
| 17-106_k141_125561 | Not-determined |
| 17-106_k141_56691 | Low-quality |
| 17-106_k141_98897 | Not-determined |
| 17-106_k141_77545 | Not-determined |
| 17-106_k141_114915 | Not-determined |
| 17-106_k141_51256 | Not-determined |
| 17-106_k141_98901 | Low-quality |
| 17-106_k141_72193 | Not-determined |
| 17-106_k141_24380 | Not-determined |
| 17-106_k141_45474 | Not-determined |
| 17-106_k141_34695 | Not-determined |
| 17-106_k141_34696 | Not-determined |
| 17-106_k141_29450 | Not-determined |
| 17-106_k141_45477 | Not-determined |
| 17-106_k141_8599 | Not-determined |
| 17-106_k141_88362 | Not-determined |
| 17-106_k141_8602 | Not-determined |
| 17-106_k141_34706 | Low-quality |
| 17-106_k141_8604 | Not-determined |
| 17-106_k141_114926 | Not-determined |
| 17-106_k141_8608 | Low-quality |
| 17-106_k141_88368 | Not-determined |
| 17-106_k141_114930 | Not-determined |
| 17-106_k141_34715 | Not-determined |
| 17-106_k141_98927 | Not-determined |
| 17-106_k141_114936 | Low-quality |
| 17-106_k141_34720 | Not-determined |
| 17-106_k141_98929 | Not-determined |
| 17-106_k141_61964 | Not-determined |
| 17-106_k141_120524 | Not-determined |
| 17-106_k141_61970 | Not-determined |
| 17-106_k141_51261 | Not-determined |
| 17-106_k141_114942 | Not-determined |
| 17-106_k141_77570 | Not-determined |
| 17-106_k141_66963 | Not-determined |
| 17-106_k141_66964 | Not-determined |
| 17-106_k141_61975 | Not-determined |
| 17-106_k141_93669 | Not-determined |
| 17-106_k141_34726 | Not-determined |
| 17-106_k141_93673 | Not-determined |
| 17-106_k141_34729 | Not-determined |
| 17-106_k141_56727 | Low-quality |
| 17-106_k141_104436 | Not-determined |
| 17-106_k141_114960 | Not-determined |
| 17-106_k141_114965 | Not-determined |
| 17-106_k141_93693 | Not-determined |
| 17-106_k141_77576 | Not-determined |
| 17-106_k141_77577 | Not-determined |
| 17-106_k141_93699 | Not-determined |
| 17-106_k141_114967 | Not-determined |
| 17-106_k141_19199 | Low-quality |
| 17-106_k141_19201 | Not-determined |
| 17-106_k141_56747 | Not-determined |
| 17-106_k141_120574 | Not-determined |
| 17-106_k141_66991 | Not-determined |
| 17-106_k141_61987 | Not-determined |
| 17-106_k141_66994 | Not-determined |
| 17-106_k141_29504 | Not-determined |
| 17-106_k141_66996 | Low-quality |
| 17-106_k141_66999 | Low-quality |
| 17-106_k141_51291 | Not-determined |
| 17-106_k141_51297 | Not-determined |
| 17-106_k141_51306 | Not-determined |
| 17-106_k141_120591 | Low-quality |
| 17-106_k141_120592 | Not-determined |
| 18-109-final-viral-combined_k141_5625\|\|full | Medium-quality |
| 17-106_k141_114972 | Not-determined |
| 17-106_k141_114973 | Not-determined |
| 17-106_k141_114975 | Not-determined |
| 17-106_k141_24388 | Not-determined |
| 17-106_k141_56763 | Low-quality |
| 17-106_k141_8671 | Not-determined |
| 17-106_k141_88387 | Not-determined |
| 17-106_k141_56766 | Low-quality |
| 17-106_k141_62015 | Not-determined |
| 17-106_k141_83822 | Not-determined |
| 17-106_k141_104515 | Not-determined |
| 17-106_k141_8685 | Not-determined |
| 17-106_k141_56772 | Not-determined |
| 17-106_k141_34776 | Not-determined |
| 17-106_k141_125579 | Not-determined |
| 17-106_k141_51321 | Not-determined |
| 17-106_k141_114986 | Not-determined |
| 17-106_k141_109222 | Not-determined |
| 17-106_k141_51329 | Not-determined |
| 17-106_k141_93735 | Low-quality |
| 17-106_k141_125582 | Low-quality |
| 17-106_k141_83843 | Not-determined |
| 17-106_k141_51334 | Not-determined |
| 17-106_k141_8702 | Not-determined |
| 17-106_k141_77620 | Not-determined |
| 17-106_k141_77621 | Not-determined |
| 17-106_k141_13594 | Low-quality |
| 17-106_k141_19221 | Not-determined |
| 17-106_k141_24415 | Not-determined |
| 17-106_k141_98998 | Low-quality |
| 17-106_k141_104529 | Not-determined |
| 17-106_k141_40320 | Not-determined |
| 17-106_k141_125607 | Not-determined |
| 17-106_k141_67061 | Not-determined |
| 17-106_k141_51345 | Not-determined |
| 17-106_k141_109230 | Not-determined |
| 17-106_k141_109231 | Not-determined |
| 17-106_k141_88439 | Low-quality |
| 17-106_k141_88448 | Not-determined |
| 17-106_k141_88449 | Not-determined |
| 17-106_k141_109234 | Not-determined |
| 17-106_k141_109235 | Not-determined |
| 17-106_k141_115011 | Not-determined |
| 17-106_k141_56776 | Not-determined |
| 17-106_k141_125616 | Not-determined |
| 17-106_k141_24425 | Low-quality |
| 17-106_k141_88453 | Not-determined |
| 17-106_k141_34822 | Not-determined |
| 17-106_k141_51359 | Not-determined |
| 17-106_k141_56788 | Not-determined |
| 17-106_k141_88456 | Not-determined |
| 17-106_k141_51361 | Not-determined |
| 17-106_k141_29550 | Low-quality |
| 17-106_k141_88464 | Not-determined |
| 17-106_k141_99021 | Low-quality |
| 17-106_k141_109245 | Not-determined |
| 17-106_k141_83883 | Low-quality |
| 17-106_k141_29562 | Not-determined |
| 17-106_k141_125625 | Not-determined |
| 17-106_k141_83896 | Not-determined |
| 17-106_k141_77638 | Not-determined |
| 17-106_k141_24448 | Not-determined |
| 17-106_k141_62039 | Not-determined |
| 17-106_k141_8725 | Low-quality |
| 17-106_k141_24456 | Not-determined |
| 17-106_k141_51386 | Not-determined |
| 17-106_k141_34843 | Not-determined |
| 17-106_k141_13625 | Not-determined |
| 17-106_k141_99042 | Low-quality |
| 17-106_k141_125637 | Low-quality |
| 17-106_k141_104554 | Not-determined |
| 17-106_k141_45503 | Not-determined |
| 17-106_k141_13635 | Not-determined |
| 17-106_k141_51395 | Not-determined |
| 17-106_k141_51397 | Not-determined |
| 17-106_k141_83916 | Not-determined |
| 17-106_k141_45507 | Not-determined |
| 17-106_k141_24476 | Not-determined |
| 17-106_k141_93742 | Not-determined |
| 17-106_k141_34867 | Not-determined |
| 17-106_k141_104560 | Not-determined |
| 17-106_k141_120669 | Not-determined |
| 17-106_k141_104564 | Low-quality |
| 17-106_k141_8740 | Not-determined |
| 17-106_k141_29593 | Not-determined |
| 17-106_k141_45518 | Not-determined |
| 17-106_k141_29599 | Not-determined |
| 17-106_k141_13658 | Not-determined |
| 17-106_k141_51406 | Not-determined |
| 17-106_k141_29605 | Not-determined |
| 17-106_k141_24488 | Not-determined |
| 17-106_k141_104582 | Not-determined |
| 17-106_k141_24489 | Not-determined |
| 17-106_k141_93758 | Not-determined |
| 17-106_k141_13675 | Not-determined |
| 17-106_k141_104587 | Not-determined |
| 17-106_k141_125647 | Not-determined |
| 17-106_k141_77655 | Not-determined |
| 17-106_k141_51414 | Not-determined |
| 17-106_k141_77660 | Not-determined |
| 17-106_k141_51418 | Not-determined |
| 17-106_k141_13686 | Not-determined |
| 17-106_k141_104592 | Not-determined |
| 17-106_k141_109305 | Not-determined |
| 17-106_k141_120686 | Not-determined |
| 17-106_k141_109311 | Not-determined |
| 17-106_k141_77667 | Not-determined |
| 17-106_k141_13695 | Not-determined |
| 17-106_k141_109314 | Not-determined |
| 17-106_k141_13696 | Not-determined |
| 17-106_k141_77670 | Not-determined |
| 17-106_k141_13700 | Not-determined |
| 17-106_k141_34891 | Not-determined |
| 17-106_k141_13705 | Not-determined |
| 17-106_k141_13707 | Low-quality |
| 17-106_k141_34892 | Not-determined |
| 17-106_k141_24518 | Not-determined |
| 17-106_k141_24524 | Low-quality |
| 17-106_k141_99076 | Not-determined |
| 17-106_k141_77691 | Not-determined |
| 17-106_k141_125662 | Not-determined |
| 17-106_k141_99088 | Not-determined |
| 17-106_k141_67097 | Not-determined |
| 17-106_k141_99090 | Not-determined |
| 17-106_k141_99091 | Not-determined |
| 17-106_k141_125667 | Not-determined |
| 17-106_k141_24543 | Not-determined |
| 17-106_k141_104620 | Not-determined |
| 17-106_k141_88517 | Not-determined |
| 17-106_k141_51424 | Not-determined |
| 17-106_k141_77708 | Not-determined |
| 17-106_k141_77712 | Not-determined |
| 17-106_k141_120695 | Not-determined |
| 17-106_k141_34898 | Not-determined |
| 17-106_k141_77723 | Not-determined |
| 17-106_k141_125680 | Not-determined |
| 17-106_k141_77724 | Not-determined |
| 17-106_k141_88526 | Not-determined |
| 17-106_k141_125684 | Not-determined |
| 17-106_k141_125686 | Low-quality |
| 17-106_k141_19274 | Not-determined |
| 17-106_k141_88529 | Not-determined |
| 17-106_k141_24552 | Not-determined |
| 17-106_k141_88535 | Not-determined |
| 17-106_k141_77741 | Not-determined |
| 17-106_k141_88538 | Not-determined |
| 17-106_k141_115019 | Not-determined |
| 17-106_k141_77749 | Not-determined |
| 17-106_k141_40341 | Not-determined |
| 17-106_k141_34920 | Not-determined |
| 17-106_k141_3362 | Not-determined |
| 17-106_k141_72214 | Not-determined |
| 17-106_k141_72215 | Not-determined |
| 17-106_k141_120716 | Not-determined |
| 17-106_k141_19297 | Not-determined |
| 17-106_k141_19298 | Not-determined |
| 17-106_k141_19300 | Not-determined |
| 17-106_k141_99147 | Not-determined |
| 17-106_k141_120722 | Not-determined |
| 17-106_k141_19302 | Low-quality |
| 17-106_k141_40348 | Not-determined |
| 17-106_k141_93793 | Not-determined |
| 17-106_k141_72217 | Not-determined |
| 17-106_k141_88553 | Not-determined |
| 17-106_k141_24569 | Not-determined |
| 17-106_k141_77756 | Not-determined |
| 17-106_k141_40351 | Not-determined |
| 17-106_k141_51468 | Low-quality |
| 17-106_k141_34941 | Not-determined |
| 17-106_k141_93801 | Not-determined |
| 17-106_k141_8789 | Not-determined |
| 17-106_k141_88562 | Not-determined |
| 17-106_k141_24572 | Not-determined |
| 17-106_k141_24574 | Not-determined |
| 17-106_k141_72226 | Not-determined |
| 17-106_k141_51473 | Not-determined |
| 17-106_k141_19317 | Not-determined |
| 17-106_k141_24581 | Not-determined |
| 17-106_k141_115029 | Low-quality |
| 17-106_k141_115030 | Not-determined |
| 17-106_k141_72232 | Not-determined |
| 17-106_k141_3381 | Low-quality |
| 17-106_k141_88570 | Not-determined |
| 17-106_k141_88574 | Not-determined |
| 17-106_k141_99174 | Not-determined |
| 17-106_k141_77771 | Not-determined |
| 17-106_k141_24599 | Not-determined |
| 17-106_k141_72253 | Not-determined |
| 17-106_k141_51480 | Not-determined |
| 17-106_k141_67133 | Not-determined |
| 17-106_k141_88589 | Not-determined |
| 17-106_k141_72258 | Not-determined |
| 17-106_k141_83927 | Not-determined |
| 17-106_k141_72260 | Not-determined |
| 17-106_k141_56854 | Not-determined |
| 17-106_k141_77790 | Not-determined |
| 17-106_k141_77793 | Not-determined |
| 17-106_k141_8802 | Not-determined |
| 17-106_k141_8804 | Not-determined |
| 17-106_k141_88605 | Not-determined |
| 17-106_k141_56864 | Low-quality |
| 17-106_k141_77799 | Not-determined |
| 17-106_k141_45563 | Not-determined |
| 17-106_k141_62082 | Not-determined |
| 17-106_k141_45565 | Not-determined |
| 17-106_k141_56872 | Not-determined |
| 17-106_k141_77803 | Not-determined |
| 17-106_k141_62087 | Not-determined |
| 17-106_k141_8807 | Low-quality |
| 17-106_k141_56877 | Not-determined |
| 17-106_k141_72268 | Not-determined |
| 17-106_k141_72269 | Not-determined |
| 17-106_k141_62094 | Not-determined |
| 17-106_k141_99207 | Not-determined |
| 17-106_k141_13715 | Not-determined |
| 17-106_k141_3413 | Not-determined |
| 17-106_k141_34984 | Low-quality |
| 17-106_k141_88621 | Not-determined |
| 17-106_k141_56887 | Not-determined |
| 17-106_k141_99213 | Low-quality |
| 17-106_k141_51486 | Low-quality |
| 17-106_k141_67173 | Not-determined |
| 17-106_k141_67176 | Not-determined |
| 17-106_k141_120750 | Not-determined |
| 17-106_k141_67177 | Low-quality |
| 17-106_k141_51489 | Low-quality |
| 17-106_k141_72285 | Not-determined |
| 17-106_k141_8835 | Not-determined |
| 17-106_k141_115050 | Not-determined |
| 17-106_k141_56902 | Not-determined |
| 17-106_k141_45594 | Not-determined |
| 17-106_k141_72291 | Low-quality |
| 17-106_k141_120758 | Not-determined |
| 17-106_k141_120761 | Low-quality |
| 17-106_k141_115057 | Not-determined |
| 17-106_k141_120764 | Not-determined |
| 17-106_k141_120765 | Not-determined |
| 17-106_k141_109331 | Not-determined |
| 17-106_k141_51500 | Not-determined |
| 17-106_k141_45605 | Not-determined |
| 17-106_k141_40377 | Not-determined |
| 17-106_k141_45606 | Not-determined |
| 17-106_k141_13739 | Not-determined |
| 17-106_k141_51506 | Low-quality |
| 17-106_k141_19354 | Not-determined |
| 17-106_k141_72295 | Not-determined |
| 17-106_k141_109348 | Not-determined |
| 17-106_k141_115064 | Low-quality |
| 17-106_k141_19363 | Not-determined |
| 17-106_k141_19368 | Not-determined |
| 17-106_k141_51521 | Not-determined |
| 17-106_k141_3419 | Not-determined |
| 17-106_k141_99238 | Not-determined |
| 17-106_k141_40397 | Not-determined |
| 17-106_k141_8862 | Low-quality |
| 17-106_k141_88659 | Not-determined |
| 17-106_k141_125745 | Not-determined |
| 17-106_k141_45635 | Not-determined |
| 17-106_k141_51539 | Not-determined |
| 17-106_k141_13764 | Not-determined |
| 17-106_k141_13765 | Not-determined |
| 17-106_k141_115079 | Not-determined |
| 17-106_k141_115080 | Not-determined |
| 17-106_k141_3428 | Not-determined |
| 17-106_k141_125760 | Not-determined |
| 17-106_k141_51555 | Not-determined |
| 17-106_k141_62138 | Not-determined |
| 17-106_k141_51556 | Not-determined |
| 17-106_k141_77823 | Not-determined |
| 17-106_k141_51560 | Not-determined |
| 17-106_k141_115093 | Not-determined |
| 17-106_k141_77830 | Not-determined |
| 17-106_k141_19412 | Not-determined |
| 17-106_k141_40418 | Not-determined |
| 17-106_k141_88675 | Not-determined |
| 17-106_k141_19416 | Not-determined |
| 17-106_k141_62158 | Not-determined |
| 17-106_k141_56978 | Not-determined |
| 17-106_k141_35032 | Not-determined |
| 17-106_k141_24613 | Not-determined |
| 17-106_k141_72314 | Low-quality |
| 17-106_k141_104674 | Not-determined |
| 17-106_k141_88687 | Not-determined |
| 17-106_k141_13781 | Not-determined |
| 17-106_k141_19430 | Not-determined |
| 17-106_k141_104677 | Low-quality |
| 17-106_k141_51585 | Not-determined |
| 17-106_k141_13798 | Not-determined |
| 17-106_k141_51593 | Not-determined |
| 17-106_k141_13805 | Not-determined |
| 17-106_k141_109387 | Low-quality |
| 17-106_k141_45649 | Not-determined |
| 17-106_k141_35054 | Not-determined |
| 17-106_k141_19451 | Not-determined |
| 17-106_k141_72332 | Not-determined |
| 17-106_k141_62187 | Not-determined |
| 17-106_k141_109396 | Not-determined |
| 17-106_k141_40448 | Not-determined |
| 17-106_k141_24622 | Not-determined |
| 17-106_k141_35062 | Not-determined |
| 17-106_k141_35065 | Not-determined |
| 17-106_k141_13822 | Not-determined |
| 17-106_k141_24630 | Not-determined |
| 17-106_k141_45675 | Not-determined |
| 17-106_k141_13828 | Not-determined |
| 17-106_k141_125773 | Not-determined |
| 17-106_k141_57009 | Not-determined |
| 17-106_k141_62220 | Not-determined |
| 17-106_k141_77854 | Not-determined |
| 17-106_k141_24647 | Not-determined |
| 17-106_k141_13842 | Not-determined |
| 17-106_k141_40468 | Not-determined |
| 17-106_k141_19465 | Not-determined |
| 17-106_k141_13845 | Not-determined |
| 17-106_k141_77856 | Not-determined |
| 17-106_k141_72356 | Not-determined |
| 17-106_k141_40477 | Not-determined |
| 17-106_k141_77857 | Not-determined |
| 17-106_k141_72363 | Not-determined |
| 17-106_k141_8881 | Not-determined |
| 17-106_k141_72364 | Not-determined |
| 17-106_k141_72365 | Not-determined |
| 17-106_k141_40481 | Not-determined |
| 17-106_k141_72367 | Not-determined |
| 17-106_k141_77863 | Not-determined |
| 17-106_k141_77864 | Not-determined |
| 17-106_k141_13855 | Not-determined |
| 17-106_k141_99259 | Not-determined |
| 17-106_k141_24658 | Not-determined |
| 17-106_k141_120863 | Not-determined |
| 17-106_k141_125804 | Not-determined |
| 17-106_k141_77873 | Not-determined |
| 17-106_k141_45696 | Not-determined |
| 17-106_k141_24664 | Not-determined |
| 17-106_k141_88702 | Not-determined |
| 17-106_k141_57035 | Low-quality |
| 17-106_k141_19492 | Not-determined |
| 17-106_k141_24671 | Not-determined |
| 17-106_k141_35097 | Not-determined |
| 17-106_k141_99266 | Low-quality |
| 17-106_k141_115105 | Not-determined |
| 17-106_k141_35107 | Not-determined |
| 17-106_k141_45724 | Not-determined |
| 17-106_k141_77907 | Low-quality |
| 17-106_k141_57070 | Not-determined |
| 17-106_k141_35112 | Low-quality |
| 17-106_k141_35117 | Not-determined |
| 17-106_k141_62241 | Not-determined |
| 17-106_k141_115125 | Not-determined |
| 17-106_k141_62244 | Not-determined |
| 17-106_k141_125827 | Not-determined |
| 17-106_k141_125828 | Not-determined |
| 17-106_k141_104703 | Not-determined |
| 17-106_k141_88722 | Not-determined |
| 17-106_k141_45749 | Low-quality |
| 17-106_k141_19533 | Not-determined |
| 17-106_k141_13863 | Not-determined |
| 17-106_k141_88725 | Not-determined |
| 17-106_k141_24679 | Not-determined |
| 17-106_k141_57092 | Not-determined |
| 17-106_k141_24681 | Not-determined |
| 17-106_k141_13870 | Not-determined |
| 17-106_k141_120919 | Not-determined |
| 17-106_k141_13878 | Not-determined |
| 17-106_k141_104726 | Low-quality |
| 17-106_k141_83978 | Not-determined |
| 17-106_k141_115146 | Not-determined |
| 17-106_k141_24692 | Not-determined |
| 17-106_k141_120923 | Not-determined |
| 17-106_k141_88753 | Not-determined |
| 17-106_k141_57103 | Not-determined |

| | |
|---|---|
| 17-106_k141_35156 | Not-determined |
| 17-106_k141_120928 | Not-determined |
| 17-106_k141_3466 | Not-determined |
| 17-106_k141_24700 | Not-determined |
| 17-106_k141_3468 | Low-quality |
| 17-106_k141_35162 | Not-determined |
| 17-106_k141_125867 | Not-determined |
| 17-106_k141_35163 | Not-determined |
| 17-106_k141_24702 | Not-determined |
| 17-106_k141_8896 | Not-determined |
| 17-106_k141_115181 | Not-determined |
| 17-106_k141_24712 | Not-determined |
| 17-106_k141_3490 | Not-determined |
| 17-106_k141_24713 | Not-determined |
| 17-106_k141_24714 | Not-determined |
| 17-106_k141_35172 | Not-determined |
| 17-106_k141_115186 | Low-quality |
| 17-106_k141_57133 | Not-determined |
| 17-106_k141_35173 | Not-determined |
| 17-106_k141_77932 | Not-determined |
| 17-106_k141_35179 | Not-determined |
| 17-106_k141_8919 | Low-quality |
| 17-106_k141_104738 | Not-determined |
| 17-106_k141_51686 | Not-determined |
| 17-106_k141_8927 | Not-determined |
| 17-106_k141_8928 | Not-determined |
| 17-106_k141_115200 | Not-determined |
| 17-106_k141_88766 | Not-determined |
| 17-106_k141_45789 | Low-quality |
| 17-106_k141_3512 | Not-determined |
| 17-106_k141_3516 | Not-determined |
| 17-106_k141_125891 | Not-determined |
| 17-106_k141_120958 | Not-determined |
| 17-106_k141_8941 | Not-determined |
| 17-106_k141_77933 | Low-quality |
| 17-106_k141_77934 | Not-determined |
| 17-106_k141_35203 | Not-determined |
| 17-106_k141_24727 | Not-determined |
| 17-106_k141_62279 | Not-determined |
| 17-106_k141_45807 | Low-quality |
| 17-106_k141_40522 | Not-determined |
| 17-106_k141_57173 | Not-determined |
| 17-106_k141_35211 | Not-determined |
| 17-106_k141_88802 | Not-determined |
| 17-106_k141_19576 | Not-determined |
| 17-106_k141_77965 | Not-determined |
| 17-106_k141_125902 | Not-determined |
| 17-106_k141_24745 | Not-determined |
| 17-106_k141_125904 | Not-determined |
| 17-106_k141_19580 | Not-determined |
| 17-106_k141_109437 | Not-determined |
| 17-106_k141_88815 | Not-determined |
| 17-106_k141_3548 | Not-determined |
| 17-106_k141_8971 | Not-determined |
| 17-106_k141_19590 | Not-determined |
| 17-106_k141_109450 | Not-determined |
| 17-106_k141_99342 | Not-determined |
| 17-106_k141_120981 | Not-determined |
| 17-106_k141_99344 | Not-determined |
| 17-106_k141_57204 | Not-determined |
| 17-106_k141_88831 | Not-determined |
| 17-106_k141_8983 | Not-determined |
| 17-106_k141_99351 | Not-determined |
| 17-106_k141_125919 | Not-determined |
| 17-106_k141_8986 | Not-determined |
| 17-106_k141_125922 | Not-determined |
| 17-106_k141_24773 | Not-determined |
| 17-106_k141_24774 | Not-determined |
| 17-106_k141_62312 | Not-determined |
| 17-106_k141_8988 | Low-quality |
| 17-106_k141_125925 | Not-determined |
| 17-106_k141_84016 | Not-determined |
| 17-106_k141_120999 | Not-determined |
| 17-106_k141_51745 | Not-determined |
| 17-106_k141_24784 | Not-determined |
| 17-106_k141_62319 | Low-quality |
| 17-106_k141_121003 | Not-determined |
| 17-106_k141_35252 | Not-determined |
| 17-106_k141_8995 | Not-determined |
| 17-106_k141_121006 | Not-determined |
| 17-106_k141_77993 | Not-determined |
| 17-106_k141_24793 | Not-determined |
| 17-106_k141_62321 | Low-quality |
| 17-106_k141_62322 | Not-determined |
| 17-106_k141_9005 | Not-determined |
| 17-106_k141_35259 | Not-determined |
| 17-106_k141_62327 | Not-determined |
| 17-106_k141_125939 | Not-determined |
| 17-106_k141_3550 | Not-determined |
| 17-106_k141_35265 | Not-determined |
| 17-106_k141_88861 | Not-determined |
| 17-106_k141_115231 | Not-determined |
| 17-106_k141_35279 | Not-determined |
| 17-106_k141_9031 | Not-determined |
| 17-106_k141_67253 | Not-determined |
| 17-106_k141_13934 | Low-quality |
| 17-106_k141_24825 | Low-quality |
| 17-106_k141_99376 | Low-quality |
| 17-106_k141_109490 | Not-determined |
| 17-106_k141_51775 | Not-determined |
| 17-106_k141_62338 | Not-determined |
| 17-106_k141_35280 | Not-determined |
| 17-106_k141_99386 | Not-determined |
| 17-106_k141_57251 | Not-determined |
| 17-106_k141_62348 | Not-determined |
| 17-106_k141_9057 | Low-quality |
| 17-106_k141_24835 | Low-quality |
| 17-106_k141_51791 | Not-determined |
| 17-106_k141_57259 | Not-determined |
| 17-106_k141_51793 | Not-determined |
| 17-106_k141_115256 | Not-determined |
| 17-106_k141_57267 | Not-determined |
| 17-106_k141_57274 | Not-determined |
| 17-106_k141_78030 | Not-determined |
| 17-106_k141_62353 | Not-determined |
| 17-106_k141_78031 | Not-determined |
| 17-106_k141_78032 | Not-determined |
| 17-106_k141_125965 | Not-determined |
| 17-106_k141_67279 | Not-determined |
| 17-106_k141_99405 | Not-determined |
| 17-106_k141_125967 | Not-determined |
| 17-106_k141_121059 | Not-determined |
| 17-106_k141_45813 | Not-determined |
| 17-106_k141_88908 | Not-determined |
| 17-106_k141_62366 | Not-determined |
| 17-106_k141_121070 | Not-determined |
| 17-106_k141_35322 | Not-determined |
| 17-106_k141_19629 | Not-determined |
| 17-106_k141_125980 | Not-determined |
| 17-106_k141_121078 | Not-determined |
| 17-106_k141_121083 | Low-quality |
| 17-106_k141_24861 | Not-determined |
| 17-106_k141_51809 | Not-determined |
| 17-106_k141_121090 | Not-determined |
| 17-106_k141_51818 | Not-determined |
| 17-106_k141_88938 | Not-determined |
| 17-106_k141_35344 | Not-determined |
| 17-106_k141_99422 | Not-determined |
| 17-106_k141_99424 | Not-determined |
| 17-106_k141_51826 | Not-determined |
| 17-106_k141_88945 | Not-determined |
| 17-106_k141_3581 | Not-determined |
| 17-106_k141_109513 | Not-determined |
| 17-106_k141_3583 | Not-determined |
| 17-106_k141_35348 | Low-quality |
| 17-106_k141_121095 | Not-determined |
| 17-106_k141_51838 | Not-determined |
| 17-106_k141_121101 | Not-determined |
| 17-106_k141_88953 | Not-determined |
| 17-106_k141_88957 | Not-determined |
| 17-106_k141_104755 | Not-determined |
| 17-106_k141_126015 | Not-determined |
| 17-106_k141_57306 | Not-determined |
| 17-106_k141_67314 | Not-determined |
| 17-106_k141_126019 | Not-determined |
| 17-106_k141_67317 | Not-determined |
| 17-106_k141_109534 | Not-determined |
| 17-106_k141_35374 | Not-determined |
| 17-106_k141_29618 | Not-determined |
| 17-106_k141_35377 | Not-determined |
| 17-106_k141_126024 | Not-determined |
| 17-106_k141_104774 | Not-determined |
| 17-106_k141_19655 | Not-determined |
| 17-106_k141_121115 | Not-determined |
| 17-106_k141_45850 | Low-quality |
| 17-106_k141_51867 | Not-determined |
| 17-106_k141_57313 | Not-determined |
| 17-106_k141_57314 | Not-determined |
| 17-106_k141_62419 | Not-determined |
| 17-106_k141_88971 | Not-determined |
| 17-106_k141_19661 | Not-determined |
| 17-106_k141_19662 | Not-determined |
| 17-106_k141_88976 | Not-determined |
| 17-106_k141_35390 | Not-determined |
| 17-106_k141_3629 | Not-determined |
| 17-106_k141_121128 | Not-determined |
| 17-106_k141_88979 | Not-determined |
| 17-106_k141_35395 | Not-determined |
| 17-106_k141_35399 | Low-quality |
| 17-106_k141_9076 | Not-determined |
| 17-106_k141_121140 | Not-determined |
| 17-106_k141_29629 | Not-determined |
| 17-106_k141_45865 | Not-determined |
| 17-106_k141_88999 | Low-quality |
| 17-106_k141_99446 | Not-determined |
| 17-106_k141_121153 | Not-determined |
| 17-106_k141_99449 | Not-determined |
| 17-106_k141_45866 | Not-determined |
| 17-106_k141_67339 | Not-determined |
| 17-106_k141_51889 | Not-determined |
| 17-106_k141_29635 | Not-determined |
| 17-106_k141_9088 | Not-determined |
| 17-106_k141_72397 | Low-quality |
| 17-106_k141_3658 | Not-determined |
| 17-106_k141_72399 | Low-quality |
| 17-106_k141_29643 | Not-determined |
| 17-106_k141_51905 | Not-determined |
| 17-106_k141_57323 | Not-determined |
| 17-106_k141_72404 | Not-determined |
| 17-106_k141_67349 | Not-determined |
| 17-106_k141_67352 | Not-determined |
| 17-106_k141_121177 | Not-determined |
| 17-106_k141_24914 | Not-determined |
| 17-106_k141_29652 | Not-determined |
| 17-106_k141_51915 | Not-determined |
| 17-106_k141_3680 | Not-determined |
| 17-106_k141_9112 | Not-determined |
| 17-106_k141_57340 | Not-determined |
| 17-106_k141_78094 | Not-determined |
| 17-106_k141_78096 | Not-determined |
| 17-106_k141_40570 | Not-determined |
| 17-106_k141_72413 | Not-determined |
| 17-106_k141_115347 | Low-quality |
| 17-106_k141_78100 | Not-determined |
| 17-106_k141_19710 | Not-determined |
| 17-106_k141_57343 | Not-determined |
| 17-106_k141_109552 | Low-quality |
| 17-106_k141_72425 | Not-determined |
| 17-106_k141_72426 | Low-quality |
| 17-106_k141_51944 | Not-determined |
| 17-106_k141_99475 | Low-quality |
| 17-106_k141_3686 | Low-quality |
| 17-106_k141_45884 | Not-determined |
| 17-106_k141_62460 | Not-determined |
| 17-106_k141_24934 | Not-determined |
| 17-106_k141_72442 | Not-determined |
| 17-106_k141_99504 | Not-determined |
| 17-106_k141_19717 | Not-determined |
| 17-106_k141_3705 | Not-determined |
| 17-106_k141_19721 | Not-determined |
| 17-106_k141_19724 | Not-determined |
| 17-106_k141_78139 | Not-determined |
| 17-106_k141_84082 | Not-determined |
| 17-106_k141_109579 | Low-quality |
| 17-106_k141_45892 | Not-determined |
| 17-106_k141_99515 | Not-determined |
| 17-106_k141_45893 | Not-determined |
| 17-106_k141_99517 | Not-determined |
| 17-106_k141_109580 | Not-determined |
| 17-106_k141_78144 | Low-quality |
| 17-106_k141_72452 | Not-determined |
| 17-106_k141_84088 | Not-determined |
| 17-106_k141_115368 | Not-determined |
| 17-106_k141_67394 | Not-determined |
| 17-106_k141_62472 | Low-quality |
| 17-106_k141_84098 | Not-determined |
| 17-106_k141_99530 | Not-determined |
| 17-106_k141_51970 | Not-determined |
| 17-106_k141_115377 | Not-determined |
| 17-106_k141_84102 | Low-quality |
| 17-106_k141_104807 | Not-determined |
| 17-106_k141_104810 | Not-determined |
| 17-106_k141_115385 | Not-determined |
| 17-106_k141_62479 | Low-quality |
| 17-106_k141_3725 | Not-determined |
| 17-106_k141_78173 | Not-determined |
| 17-106_k141_19752 | Low-quality |
| 17-106_k141_84117 | Not-determined |
| 17-106_k141_84118 | Not-determined |
| 17-106_k141_62485 | Not-determined |
| 17-106_k141_57396 | Not-determined |
| 17-106_k141_19755 | Not-determined |
| 17-106_k141_62492 | Not-determined |
| 17-106_k141_19762 | Not-determined |
| 17-106_k141_57411 | Not-determined |
| 17-106_k141_121205 | Not-determined |
| 17-106_k141_24945 | Not-determined |
| 17-106_k141_99548 | Not-determined |
| 17-106_k141_72465 | Not-determined |
| 17-106_k141_24948 | Not-determined |
| 17-106_k141_24952 | Not-determined |
| 17-106_k141_84131 | Not-determined |
| 17-106_k141_72471 | Not-determined |
| 17-106_k141_99555 | Not-determined |
| 17-106_k141_115407 | Low-quality |
| 17-106_k141_115408 | Not-determined |
| 17-106_k141_78183 | Not-determined |
| 17-106_k141_24960 | Not-determined |
| 17-106_k141_24961 | Not-determined |
| 17-106_k141_84138 | Not-determined |
| 17-106_k141_78196 | Not-determined |
| 17-106_k141_78198 | Not-determined |
| 17-106_k141_24967 | Not-determined |
| 17-106_k141_115414 | Low-quality |
| 17-106_k141_78201 | Not-determined |
| 17-106_k141_89053 | Not-determined |
| 17-106_k141_109611 | Low-quality |
| 17-106_k141_89059 | Low-quality |
| 17-106_k141_19800 | Not-determined |
| 17-106_k141_89062 | Not-determined |
| 17-106_k141_62506 | Low-quality |
| 17-106_k141_52010 | Not-determined |
| 17-106_k141_121211 | Not-determined |
| 17-106_k141_45915 | Not-determined |
| 17-106_k141_29701 | Not-determined |
| 17-106_k141_115430 | Not-determined |
| 17-106_k141_93842 | Low-quality |
| 17-106_k141_121218 | Not-determined |
| 17-106_k141_93843 | Not-determined |
| 17-106_k141_121222 | Not-determined |
| 17-106_k141_52017 | Not-determined |
| 17-106_k141_52018 | Not-determined |
| 17-106_k141_62514 | Not-determined |
| 17-106_k141_62516 | Not-determined |
| 17-106_k141_62518 | Not-determined |
| 17-106_k141_109620 | Not-determined |
| 17-106_k141_93853 | Not-determined |
| 17-106_k141_57416 | Not-determined |
| 17-106_k141_57417 | Not-determined |
| 17-106_k141_62530 | Low-quality |
| 17-106_k141_29723 | Not-determined |
| 17-106_k141_89080 | Not-determined |
| 17-106_k141_93861 | Not-determined |
| 17-106_k141_78223 | Not-determined |
| 17-106_k141_19807 | Not-determined |
| 17-106_k141_104879 | Not-determined |
| 17-106_k141_29731 | Not-determined |
| 17-106_k141_121245 | Not-determined |
| 17-106_k141_84172 | Not-determined |
| 17-106_k141_13971 | Not-determined |
| 17-106_k141_45919 | Not-determined |
| 17-106_k141_57434 | Not-determined |
| 17-106_k141_121253 | Not-determined |
| 17-106_k141_115443 | Not-determined |
| 17-106_k141_62555 | Not-determined |
| 17-106_k141_13979 | Low-quality |
| 17-106_k141_13980 | Not-determined |
| 17-106_k141_62561 | Not-determined |
| 17-106_k141_13985 | Not-determined |
| 17-106_k141_115451 | Low-quality |
| 17-106_k141_67426 | Not-determined |
| 17-106_k141_67427 | Low-quality |
| 17-106_k141_67428 | Not-determined |
| 17-106_k141_104914 | Low-quality |
| 17-106_k141_72487 | Not-determined |
| 17-106_k141_62577 | Not-determined |
| 17-106_k141_121267 | Not-determined |
| 17-106_k141_67431 | Not-determined |
| 17-106_k141_52035 | Not-determined |
| 17-106_k141_115466 | Not-determined |
| 17-106_k141_121273 | Not-determined |
| 17-106_k141_109641 | Low-quality |
| 17-106_k141_89094 | Not-determined |
| 17-106_k141_121277 | Not-determined |
| 17-106_k141_109644 | Not-determined |
| 17-106_k141_52046 | Not-determined |
| 17-106_k141_121282 | Not-determined |
| 17-106_k141_45935 | Not-determined |
| 17-106_k141_29762 | Not-determined |
| 17-106_k141_3814 | Not-determined |
| 17-106_k141_45936 | Not-determined |
| 17-106_k141_109649 | Low-quality |
| 17-106_k141_109650 | Not-determined |
| 17-106_k141_115483 | Not-determined |
| 17-106_k141_29767 | Not-determined |
| 17-106_k141_104932 | Not-determined |
| 17-106_k141_121289 | Not-determined |
| 17-106_k141_52053 | Not-determined |
| 17-106_k141_52056 | Not-determined |
| 17-106_k141_104937 | Not-determined |
| 17-106_k141_52058 | Not-determined |
| 17-106_k141_45944 | Not-determined |
| 17-106_k141_3822 | Not-determined |
| 17-106_k141_52069 | Low-quality |
| 17-106_k141_99590 | Low-quality |
| 17-106_k141_99595 | Not-determined |
| 17-106_k141_121295 | Not-determined |
| 17-106_k141_3838 | Not-determined |
| 17-106_k141_40597 | Not-determined |
| 17-106_k141_89128 | Not-determined |
| 17-106_k141_99600 | Low-quality |
| 17-106_k141_40601 | Not-determined |
| 17-106_k141_14004 | Not-determined |
| 17-106_k141_89132 | Low-quality |
| 17-106_k141_93905 | Low-quality |
| 17-106_k141_57446 | Not-determined |
| 17-106_k141_9166 | Not-determined |
| 17-106_k141_14016 | Not-determined |
| 17-106_k141_93912 | Not-determined |
| 17-106_k141_14020 | Not-determined |
| 17-106_k141_89145 | Not-determined |
| 17-106_k141_14023 | Not-determined |
| 17-106_k141_72539 | Not-determined |
| 17-106_k141_14025 | Not-determined |
| 17-106_k141_3842 | Not-determined |
| 17-106_k141_29778 | Not-determined |
| 17-106_k141_14046 | Not-determined |
| 17-106_k141_72549 | Not-determined |
| 17-106_k141_72554 | Not-determined |
| 17-106_k141_84210 | Not-determined |
| 17-106_k141_14055 | Not-determined |
| 17-106_k141_93928 | Not-determined |
| 17-106_k141_121326 | Not-determined |
| 17-106_k141_89168 | Not-determined |
| 17-106_k141_78251 | Low-quality |
| 17-106_k141_78264 | Not-determined |
| 17-106_k141_67508 | Not-determined |
| 17-106_k141_29780 | Not-determined |
| 17-106_k141_40644 | Not-determined |
| 17-106_k141_109676 | Not-determined |
| 17-106_k141_67516 | Low-quality |
| 17-106_k141_40649 | Not-determined |
| 17-106_k141_78280 | Low-quality |
| 17-106_k141_52101 | Low-quality |
| 17-106_k141_93943 | Not-determined |
| 17-106_k141_126048 | Not-determined |
| 17-106_k141_29785 | Not-determined |
| 17-106_k141_67519 | Not-determined |
| 17-106_k141_25052 | Low-quality |
| 17-106_k141_126058 | Not-determined |
| 17-106_k141_14057 | Not-determined |
| 17-106_k141_35419 | Not-determined |
| 17-106_k141_40671 | Not-determined |
| 17-106_k141_89207 | Not-determined |
| 17-106_k141_40673 | Not-determined |
| 17-106_k141_29794 | Not-determined |
| 17-106_k141_52114 | Not-determined |
| 17-106_k141_78301 | Not-determined |
| 17-106_k141_35429 | Not-determined |
| 17-106_k141_115550 | Not-determined |
| 17-106_k141_126077 | Not-determined |
| 17-106_k141_52119 | Not-determined |
| 17-106_k141_78314 | Not-determined |
| 17-106_k141_78317 | Not-determined |
| 17-106_k141_78320 | Low-quality |
| 17-106_k141_35448 | Not-determined |
| 17-106_k141_35452 | Not-determined |
| 17-106_k141_78325 | Not-determined |
| 17-106_k141_93972 | Not-determined |
| 17-106_k141_78334 | Low-quality |
| 17-106_k141_121390 | Not-determined |
| 17-106_k141_126104 | Not-determined |
| 17-106_k141_105026 | Not-determined |
| 17-106_k141_93982 | Not-determined |
| 17-106_k141_99648 | Not-determined |
| 17-106_k141_89235 | Not-determined |
| 17-106_k141_89237 | Not-determined |
| 17-106_k141_93989 | Low-quality |
| 17-106_k141_126112 | Not-determined |
| 17-106_k141_84236 | Not-determined |
| 17-106_k141_45964 | Not-determined |
| 17-106_k141_3862 | Not-determined |
| 17-106_k141_3864 | Not-determined |
| 17-106_k141_84245 | Not-determined |
| 17-106_k141_126117 | Not-determined |
| 17-106_k141_3872 | Not-determined |
| 17-106_k141_72611 | Not-determined |
| 17-106_k141_45974 | Not-determined |
| 17-106_k141_3874 | Not-determined |
| 17-106_k141_29838 | Not-determined |
| 17-106_k141_9211 | Not-determined |
| 17-106_k141_115567 | Not-determined |
| 17-106_k141_9215 | Low-quality |
| 17-106_k141_93994 | Not-determined |
| 17-106_k141_45977 | Not-determined |
| 17-106_k141_45980 | Not-determined |
| 17-106_k141_84263 | Not-determined |
| 17-106_k141_121395 | Not-determined |
| 17-106_k141_94010 | Not-determined |
| 17-106_k141_19841 | Not-determined |
| 17-106_k141_35470 | Not-determined |
| 17-106_k141_67521 | Not-determined |
| 17-106_k141_35474 | Not-determined |
| 17-106_k141_126138 | Low-quality |
| 17-106_k141_9222 | Not-determined |
| 17-106_k141_19851 | Low-quality |
| 17-106_k141_35478 | Not-determined |
| 17-106_k141_9225 | Low-quality |
| 17-106_k141_9226 | Not-determined |
| 17-106_k141_67531 | Not-determined |
| 17-106_k141_9228 | Not-determined |
| 17-106_k141_99674 | Not-determined |
| 17-106_k141_78375 | Low-quality |
| 17-106_k141_25087 | Low-quality |
| 17-106_k141_78379 | Not-determined |
| 17-106_k141_19877 | Not-determined |
| 17-106_k141_126149 | Not-determined |
| 17-106_k141_67536 | Not-determined |
| 17-106_k141_67537 | Not-determined |
| 17-106_k141_19886 | Not-determined |
| 17-106_k141_94049 | Low-quality |
| 17-106_k141_35500 | Low-quality |
| 17-106_k141_67542 | Not-determined |
| 17-106_k141_89272 | Not-determined |
| 17-106_k141_121419 | Not-determined |
| 17-106_k141_78394 | Low-quality |
| 17-106_k141_121426 | Not-determined |
| 17-106_k141_89276 | Low-quality |
| 17-106_k141_67555 | Low-quality |
| 17-106_k141_126161 | Not-determined |
| 17-106_k141_89280 | Not-determined |
| 17-106_k141_109740 | Not-determined |
| 17-106_k141_19915 | Not-determined |
| 17-106_k141_3898 | Not-determined |
| 17-106_k141_19922 | Not-determined |
| 17-106_k141_3903 | Not-determined |
| 17-106_k141_89289 | Not-determined |
| 17-106_k141_3911 | Not-determined |
| 17-106_k141_78416 | Not-determined |
| 17-106_k141_19946 | Not-determined |
| 17-106_k141_25104 | Not-determined |
| 17-106_k141_67564 | Not-determined |
| 17-106_k141_25106 | Not-determined |
| 17-106_k141_25109 | Not-determined |
| 17-106_k141_78428 | Not-determined |
| 17-106_k141_25115 | Low-quality |
| 17-106_k141_29848 | Not-determined |
| 17-106_k141_3928 | Not-determined |
| 17-106_k141_19963 | Not-determined |
| 17-106_k141_67584 | Not-determined |
| 17-106_k141_19968 | Not-determined |
| 17-106_k141_78444 | Not-determined |
| 17-106_k141_89325 | Not-determined |
| 17-106_k141_19975 | Not-determined |
| 17-106_k141_14165 | Not-determined |
| 17-106_k141_105060 | Not-determined |
| 17-106_k141_126198 | Not-determined |
| 17-106_k141_29856 | Not-determined |
| 17-106_k141_62604 | Not-determined |
| 17-106_k141_126201 | Low-quality |
| 17-106_k141_46048 | Not-determined |
| 17-106_k141_78456 | Low-quality |
| 17-106_k141_126203 | Not-determined |
| 17-106_k141_35533 | Not-determined |
| 17-106_k141_9237 | Not-determined |
| 17-106_k141_9238 | Not-determined |
| 17-106_k141_14170 | Not-determined |
| 17-106_k141_115625 | Not-determined |
| 17-106_k141_29869 | Not-determined |
| 17-106_k141_115629 | Not-determined |
| 17-106_k141_29874 | Not-determined |
| 17-106_k141_121455 | Not-determined |
| 17-106_k141_121456 | Not-determined |
| 17-106_k141_14177 | Low-quality |
| 17-106_k141_3955 | Not-determined |
| 17-106_k141_3958 | Not-determined |
| 17-106_k141_9246 | Not-determined |
| 17-106_k141_46065 | Not-determined |
| 17-106_k141_67618 | Not-determined |
| 17-106_k141_89355 | Not-determined |
| 17-106_k141_25132 | Not-determined |
| 17-106_k141_62625 | Not-determined |
| 17-106_k141_109789 | Not-determined |
| 17-106_k141_25136 | Not-determined |
| 17-106_k141_109790 | Not-determined |
| 17-106_k141_25137 | Not-determined |
| 17-106_k141_105080 | Not-determined |
| 17-106_k141_109796 | Not-determined |
| 17-106_k141_89363 | Low-quality |
| 17-106_k141_62635 | Not-determined |
| 17-106_k141_78484 | Not-determined |
| 17-106_k141_52169 | Not-determined |
| 17-106_k141_19989 | Not-determined |
| 17-106_k141_126243 | Not-determined |
| 17-106_k141_126247 | Low-quality |
| 17-106_k141_121475 | Not-determined |
| 17-106_k141_3995 | Not-determined |
| 17-106_k141_89392 | Not-determined |
| 17-106_k141_121488 | Not-determined |
| 17-106_k141_67643 | Not-determined |
| 17-106_k141_35541 | Not-determined |
| 17-106_k141_115671 | Not-determined |
| 17-106_k141_35545 | Not-determined |
| 17-106_k141_62642 | Not-determined |
| 17-106_k141_29890 | Not-determined |
| 17-106_k141_35552 | Not-determined |
| 17-106_k141_62649 | Not-determined |
| 17-106_k141_78513 | Not-determined |
| 17-106_k141_115677 | Not-determined |
| 17-106_k141_25158 | Not-determined |
| 17-106_k141_35554 | Low-quality |
| 17-106_k141_78516 | Not-determined |
| 17-106_k141_9280 | Not-determined |
| 17-106_k141_9281 | Not-determined |
| 17-106_k141_62655 | Not-determined |
| 17-106_k141_25162 | Not-determined |
| 17-106_k141_78524 | Not-determined |
| 17-106_k141_115684 | Not-determined |
| 17-106_k141_115685 | Not-determined |
| 17-106_k141_99693 | Not-determined |
| 17-106_k141_121519 | Not-determined |
| 17-106_k141_115693 | Not-determined |
| 17-106_k141_14238 | Not-determined |
| 17-106_k141_25178 | Not-determined |
| 17-106_k141_78535 | Not-determined |
| 17-106_k141_14246 | Not-determined |
| 17-106_k141_57453 | Not-determined |
| 17-106_k141_57455 | Not-determined |
| 17-106_k141_115699 | Low-quality |

| | |
|---|---|
| 17-106_k141_35579 | Not-determined |
| 17-106_k141_89409 | Low-quality |
| 17-106_k141_9309 | Not-determined |
| 17-106_k141_84286 | Not-determined |
| 17-106_k141_57468 | Not-determined |
| 17-106_k141_67683 | Not-determined |
| 17-106_k141_84290 | Not-determined |
| 17-106_k141_14264 | Not-determined |
| 17-106_k141_4010 | Not-determined |
| 17-106_k141_25192 | Not-determined |
| 17-106_k141_52183 | Not-determined |
| 17-106_k141_62700 | Low-quality |
| 17-106_k141_52185 | Not-determined |
| 17-106_k141_14273 | Low-quality |
| 17-106_k141_126267 | Low-quality |
| 17-106_k141_72629 | Not-determined |
| 17-106_k141_115722 | Not-determined |
| 17-106_k141_57479 | Not-determined |
| 17-106_k141_121543 | Not-determined |
| 17-106_k141_72631 | Not-determined |
| 17-106_k141_72632 | Not-determined |
| 17-106_k141_84305 | Not-determined |
| 17-106_k141_121548 | Not-determined |
| 17-106_k141_52195 | Not-determined |
| 17-106_k141_84311 | Not-determined |
| 17-106_k141_14284 | Not-determined |
| 17-106_k141_84314 | Not-determined |
| 17-106_k141_78576 | Low-quality |
| 17-106_k141_14295 | Not-determined |
| 17-106_k141_40687 | Not-determined |
| 17-106_k141_40700 | Not-determined |
| 17-106_k141_40701 | Low-quality |
| 17-106_k141_126276 | Not-determined |
| 17-106_k141_52224 | Not-determined |
| 17-106_k141_121568 | Not-determined |
| 17-106_k141_78589 | Not-determined |
| 17-106_k141_62739 | Not-determined |
| 17-106_k141_52233 | Not-determined |
| 17-106_k141_67692 | Not-determined |
| 17-106_k141_14308 | Not-determined |
| 17-106_k141_52237 | Not-determined |
| 17-106_k141_121579 | Not-determined |
| 17-106_k141_121583 | Not-determined |
| 17-106_k141_57500 | Not-determined |
| 17-106_k141_72652 | Low-quality |
| 17-106_k141_35615 | Not-determined |
| 17-106_k141_89451 | Not-determined |
| 17-106_k141_115774 | Not-determined |
| 17-106_k141_89454 | Not-determined |
| 17-106_k141_89456 | Not-determined |
| 17-106_k141_19998 | Not-determined |
| 17-106_k141_25252 | Not-determined |
| 17-106_k141_35625 | Not-determined |
| 17-106_k141_9362 | Not-determined |
| 17-106_k141_14334 | Not-determined |
| 17-106_k141_14335 | Low-quality |
| 17-106_k141_40736 | Not-determined |
| 17-106_k141_35629 | Low-quality |
| 17-106_k141_89466 | Not-determined |
| 17-106_k141_57524 | Not-determined |
| 17-106_k141_57525 | Not-determined |
| 17-106_k141_46137 | Not-determined |
| 17-106_k141_35633 | Not-determined |
| 17-106_k141_40746 | Not-determined |
| 17-106_k141_67733 | Not-determined |
| 17-106_k141_126280 | Not-determined |
| 17-106_k141_9380 | Low-quality |
| 17-106_k141_89478 | Not-determined |
| 17-106_k141_62759 | Low-quality |
| 17-106_k141_25264 | Not-determined |
| 17-106_k141_14350 | Not-determined |
| 17-106_k141_121599 | Not-determined |
| 17-106_k141_9389 | Not-determined |
| 17-106_k141_78621 | Low-quality |
| 17-106_k141_35641 | Not-determined |
| 17-106_k141_35643 | Not-determined |
| 17-106_k141_35644 | Not-determined |
| 17-106_k141_62772 | Not-determined |
| 17-106_k141_121605 | Not-determined |
| 17-106_k141_20008 | Not-determined |
| 17-106_k141_78628 | Not-determined |
| 17-106_k141_9396 | Not-determined |
| 17-106_k141_78631 | Not-determined |
| 17-106_k141_9398 | Not-determined |
| 17-106_k141_126310 | Low-quality |
| 17-106_k141_126312 | Not-determined |
| 17-106_k141_72697 | Not-determined |
| 17-106_k141_9403 | Not-determined |
| 17-106_k141_14378 | Not-determined |
| 17-106_k141_9409 | Not-determined |
| 17-106_k141_67771 | Low-quality |
| 17-106_k141_126330 | Not-determined |
| 17-106_k141_126332 | Not-determined |
| 17-106_k141_35673 | Not-determined |
| 17-106_k141_67786 | Not-determined |
| 17-106_k141_121630 | Not-determined |
| 17-106_k141_20045 | Not-determined |
| 17-106_k141_4059 | Not-determined |
| 17-106_k141_121635 | Not-determined |
| 17-106_k141_121640 | Not-determined |
| 17-106_k141_67794 | Not-determined |
| 17-106_k141_72704 | Not-determined |
| 17-106_k141_4068 | Not-determined |
| 17-106_k141_14417 | Not-determined |
| 17-106_k141_109864 | Not-determined |
| 17-106_k141_78682 | Not-determined |
| 17-106_k141_72712 | Low-quality |
| 17-106_k141_14422 | Not-determined |
| 17-106_k141_78693 | Low-quality |
| 17-106_k141_109871 | Not-determined |
| 17-106_k141_14433 | Not-determined |
| 17-106_k141_14434 | Not-determined |
| 17-106_k141_25315 | Not-determined |
| 17-106_k141_99734 | Not-determined |
| 17-106_k141_4083 | Low-quality |
| 17-106_k141_72715 | Not-determined |
| 17-106_k141_14443 | Not-determined |
| 17-106_k141_25320 | Not-determined |
| 17-106_k141_14445 | Not-determined |
| 17-106_k141_40764 | Not-determined |
| 17-106_k141_25326 | Not-determined |
| 17-106_k141_4089 | Not-determined |
| 17-106_k141_25328 | Not-determined |
| 17-106_k141_109894 | Not-determined |
| 17-106_k141_99748 | Not-determined |
| 17-106_k141_14447 | Not-determined |
| 17-106_k141_72722 | Not-determined |
| 17-106_k141_72723 | Not-determined |
| 17-106_k141_99754 | Low-quality |
| 17-106_k141_14454 | Not-determined |
| 17-106_k141_14455 | Not-determined |
| 17-106_k141_89502 | Not-determined |
| 17-106_k141_62807 | Not-determined |
| 17-106_k141_62808 | Not-determined |
| 17-106_k141_40780 | Not-determined |
| 17-106_k141_115832 | Not-determined |
| 17-106_k141_57555 | Low-quality |
| 17-106_k141_40790 | Not-determined |
| 17-106_k141_4101 | Not-determined |
| 17-106_k141_72730 | Not-determined |
| 17-106_k141_99775 | Not-determined |
| 17-106_k141_4109 | Not-determined |
| 17-106_k141_29985 | Low-quality |
| 17-106_k141_78715 | Not-determined |
| 17-106_k141_57581 | Not-determined |
| 17-106_k141_57585 | Not-determined |
| 17-106_k141_99780 | Not-determined |
| 17-106_k141_78718 | Not-determined |
| 17-106_k141_72739 | Not-determined |
| 17-106_k141_52326 | Not-determined |
| 17-106_k141_52329 | Not-determined |
| 17-106_k141_62840 | Not-determined |
| 17-106_k141_62841 | Not-determined |
| 17-106_k141_52332 | Not-determined |
| 17-106_k141_109903 | Not-determined |
| 17-106_k141_14481 | Not-determined |
| 17-106_k141_52335 | Not-determined |
| 17-106_k141_72749 | Not-determined |
| 17-106_k141_109907 | Not-determined |
| 17-106_k141_72752 | Low-quality |
| 17-106_k141_109911 | Not-determined |
| 17-106_k141_52338 | Not-determined |
| 17-106_k141_109915 | Not-determined |
| 17-106_k141_78730 | Not-determined |
| 17-106_k141_105111 | Not-determined |
| 17-106_k141_4145 | Not-determined |
| 17-106_k141_84371 | Not-determined |
| 17-106_k141_40822 | Not-determined |
| 17-106_k141_52346 | Not-determined |
| 17-106_k141_57615 | Not-determined |
| 17-106_k141_4160 | Not-determined |
| 17-106_k141_57621 | Not-determined |
| 17-106_k141_52355 | Not-determined |
| 17-106_k141_78754 | Not-determined |
| 17-106_k141_57631 | Not-determined |
| 17-106_k141_9467 | Not-determined |
| 17-106_k141_57634 | Not-determined |
| 17-106_k141_121709 | Not-determined |
| 17-106_k141_9469 | Not-determined |
| 17-106_k141_62868 | Not-determined |
| 17-106_k141_40841 | Not-determined |
| 17-106_k141_99818 | Not-determined |
| 17-106_k141_99819 | Not-determined |
| 17-106_k141_40848 | Low-quality |
| 17-106_k141_52364 | Not-determined |
| 17-106_k141_52368 | Not-determined |
| 17-106_k141_9484 | Not-determined |
| 17-106_k141_109942 | Not-determined |
| 17-106_k141_72798 | Not-determined |
| 17-106_k141_121725 | Not-determined |
| 17-106_k141_105134 | Not-determined |
| 17-106_k141_89527 | Not-determined |
| 17-106_k141_99830 | Not-determined |
| 17-106_k141_40858 | Not-determined |
| 17-106_k141_89528 | Not-determined |
| 17-106_k141_89530 | Not-determined |
| 17-106_k141_99837 | Not-determined |
| 17-106_k141_25337 | Low-quality |
| 17-106_k141_99840 | Not-determined |
| 17-106_k141_40867 | Not-determined |
| 17-106_k141_35687 | Not-determined |
| 17-106_k141_121742 | Low-quality |
| 17-106_k141_78768 | Not-determined |
| 17-106_k141_99847 | Not-determined |
| 17-106_k141_35690 | Not-determined |
| 17-106_k141_94095 | Not-determined |
| 17-106_k141_78773 | Not-determined |
| 17-106_k141_20112 | Not-determined |
| 17-106_k141_20115 | Not-determined |
| 17-106_k141_40873 | Not-determined |
| 17-106_k141_20119 | Not-determined |
| 17-106_k141_84420 | Not-determined |
| 17-106_k141_20124 | Not-determined |
| 17-106_k141_72808 | Not-determined |
| 17-106_k141_40874 | Not-determined |
| 17-106_k141_35702 | Not-determined |
| 17-106_k141_121753 | Not-determined |
| 17-106_k141_9494 | Low-quality |
| 17-106_k141_94110 | Not-determined |
| 17-106_k141_25359 | Not-determined |
| 17-106_k141_14524 | Not-determined |
| 17-106_k141_67916 | Not-determined |
| 17-106_k141_94119 | Not-determined |
| 17-106_k141_126344 | Not-determined |
| 17-106_k141_25367 | Not-determined |
| 17-106_k141_67923 | Not-determined |
| 17-106_k141_9513 | Not-determined |
| 17-106_k141_35720 | Not-determined |
| 17-106_k141_126358 | Not-determined |
| 17-106_k141_94140 | Not-determined |
| 17-106_k141_62891 | Not-determined |
| 17-106_k141_62893 | Not-determined |
| 17-106_k141_121767 | Low-quality |
| 17-106_k141_30037 | Not-determined |
| 17-106_k141_105146 | Not-determined |
| 17-106_k141_9516 | Not-determined |
| 17-106_k141_72815 | Not-determined |
| 17-106_k141_62904 | Low-quality |
| 17-106_k141_94150 | Not-determined |
| 17-106_k141_62907 | Not-determined |
| 17-106_k141_99892 | Not-determined |
| 17-106_k141_25382 | Not-determined |
| 17-106_k141_35729 | Not-determined |
| 17-106_k141_25386 | Not-determined |
| 17-106_k141_62915 | Low-quality |
| 17-106_k141_126366 | Not-determined |
| 17-106_k141_121783 | Not-determined |
| 17-106_k141_25392 | Not-determined |
| 17-106_k141_62922 | Not-determined |
| 17-106_k141_121787 | Not-determined |
| 17-106_k141_126372 | Not-determined |
| 17-106_k141_67948 | Not-determined |
| 17-106_k141_115894 | Not-determined |
| 17-106_k141_99902 | Not-determined |
| 17-106_k141_52421 | Not-determined |
| 17-106_k141_67958 | Not-determined |
| 17-106_k141_115905 | Not-determined |
| 17-106_k141_109996 | Not-determined |
| 17-106_k141_62928 | Low-quality |
| 17-106_k141_25403 | Not-determined |
| 17-106_k141_126384 | Not-determined |
| 17-106_k141_99919 | Not-determined |
| 17-106_k141_46179 | Not-determined |
| 17-106_k141_46180 | Not-determined |
| 17-106_k141_89584 | Not-determined |
| 17-106_k141_72837 | Not-determined |
| 17-106_k141_4242 | Not-determined |
| 17-106_k141_9553 | Not-determined |
| 17-106_k141_30084 | Not-determined |
| 17-106_k141_20129 | Low-quality |
| 17-106_k141_115934 | Not-determined |
| 17-106_k141_46195 | Not-determined |
| 17-106_k141_89597 | Not-determined |
| 17-106_k141_94189 | Not-determined |
| 17-106_k141_9558 | Not-determined |
| 17-106_k141_40916 | Not-determined |
| 17-106_k141_105156 | Not-determined |
| 17-106_k141_126409 | Not-determined |
| 17-106_k141_94199 | Not-determined |
| 17-106_k141_62960 | Not-determined |
| 17-106_k141_94202 | Not-determined |
| 17-106_k141_20159 | Not-determined |
| 17-106_k141_67985 | Not-determined |
| 17-106_k141_9577 | Low-quality |
| 17-106_k141_67987 | Not-determined |
| 17-106_k141_46216 | Not-determined |
| 17-106_k141_9583 | Not-determined |
| 17-106_k141_67991 | Not-determined |
| 17-106_k141_9584 | Not-determined |
| 17-106_k141_46220 | Not-determined |
| 17-106_k141_46222 | Not-determined |
| 17-106_k141_14595 | Not-determined |
| 17-106_k141_121837 | Not-determined |
| 17-106_k141_46228 | Not-determined |
| 17-106_k141_35762 | Not-determined |
| 17-106_k141_72855 | Not-determined |
| 17-106_k141_110037 | Not-determined |
| 17-106_k141_78875 | Not-determined |
| 17-106_k141_72857 | Not-determined |
| 17-106_k141_40957 | Not-determined |
| 17-106_k141_72860 | Low-quality |
| 17-106_k141_25475 | Not-determined |
| 17-106_k141_52439 | Not-determined |
| 17-106_k141_72863 | Not-determined |
| 17-106_k141_35767 | Not-determined |
| 17-106_k141_9603 | Not-determined |
| 17-106_k141_68010 | Low-quality |
| 17-106_k141_30122 | Not-determined |
| 17-106_k141_52450 | Not-determined |
| 17-106_k141_72868 | Not-determined |
| 17-106_k141_30129 | Not-determined |
| 17-106_k141_35773 | Not-determined |
| 17-106_k141_68020 | Not-determined |
| 17-106_k141_68021 | Not-determined |
| 17-106_k141_78889 | Not-determined |
| 17-106_k141_57693 | Not-determined |
| 17-106_k141_25486 | Not-determined |
| 17-106_k141_20176 | Not-determined |
| 17-106_k141_30140 | Low-quality |
| 17-106_k141_20178 | Not-determined |
| 17-106_k141_25498 | Not-determined |
| 17-106_k141_72887 | Not-determined |
| 17-106_k141_25499 | Not-determined |
| 17-106_k141_52463 | Not-determined |
| 17-106_k141_46255 | Not-determined |
| 17-106_k141_46256 | Not-determined |
| 17-106_k141_126468 | Not-determined |
| 17-106_k141_46258 | Not-determined |
| 17-106_k141_105193 | Not-determined |
| 17-106_k141_52470 | Not-determined |
| 17-106_k141_20193 | Not-determined |
| 17-106_k141_105197 | Low-quality |
| 17-106_k141_52477 | Not-determined |
| 17-106_k141_9628 | Not-determined |
| 17-106_k141_121860 | Not-determined |
| 17-106_k141_40997 | Not-determined |
| 17-106_k141_40998 | Not-determined |
| 17-106_k141_72899 | Not-determined |
| 17-106_k141_121866 | Not-determined |
| 17-106_k141_105204 | Not-determined |
| 17-106_k141_57697 | Not-determined |
| 17-106_k141_89662 | Not-determined |
| 17-106_k141_121869 | Not-determined |
| 17-106_k141_62998 | Low-quality |
| 17-106_k141_62999 | Not-determined |
| 17-106_k141_105214 | Not-determined |
| 17-106_k141_68032 | Not-determined |
| 17-106_k141_105217 | Not-determined |
| 17-106_k141_121875 | Not-determined |
| 17-106_k141_126494 | Not-determined |
| 17-106_k141_126500 | Not-determined |
| 17-106_k141_105223 | Not-determined |
| 17-106_k141_105224 | Not-determined |
| 17-106_k141_68040 | Low-quality |
| 17-106_k141_52496 | Low-quality |
| 17-106_k141_121879 | Not-determined |
| 17-106_k141_89673 | Not-determined |
| 17-106_k141_20213 | Not-determined |
| 17-106_k141_30186 | Not-determined |
| 17-106_k141_78897 | Not-determined |
| 17-106_k141_52516 | Not-determined |
| 17-106_k141_63027 | Not-determined |
| 17-106_k141_30195 | Not-determined |
| 17-106_k141_25509 | Not-determined |
| 17-106_k141_78899 | Not-determined |
| 17-106_k141_105244 | Not-determined |
| 17-106_k141_121897 | Low-quality |
| 17-106_k141_63046 | Not-determined |
| 17-106_k141_4274 | Not-determined |
| 17-106_k141_105255 | Not-determined |
| 17-106_k141_30219 | Not-determined |
| 17-106_k141_121900 | Not-determined |
| 17-106_k141_78912 | Not-determined |
| 17-106_k141_89699 | Not-determined |
| 17-106_k141_99966 | Not-determined |
| 17-106_k141_35817 | Not-determined |
| 17-106_k141_25526 | Not-determined |
| 17-106_k141_78918 | Not-determined |
| 17-106_k141_35819 | Not-determined |
| 17-106_k141_35821 | Not-determined |
| 17-106_k141_30232 | Not-determined |
| 17-106_k141_30234 | Low-quality |
| 17-106_k141_72914 | Not-determined |
| 17-106_k141_35826 | Not-determined |
| 17-106_k141_4292 | Not-determined |
| 17-106_k141_110057 | Not-determined |
| 17-106_k141_110058 | Not-determined |
| 17-106_k141_4301 | Low-quality |
| 17-106_k141_20227 | Not-determined |
| 17-106_k141_9639 | Not-determined |
| 17-106_k141_9644 | Not-determined |
| 17-106_k141_89715 | Not-determined |
| 17-106_k141_78926 | Not-determined |
| 17-106_k141_25544 | Not-determined |
| 17-106_k141_63058 | Not-determined |
| 17-106_k141_94266 | Not-determined |
| 17-106_k141_84508 | Not-determined |
| 17-106_k141_25549 | Not-determined |
| 17-106_k141_110072 | Not-determined |
| 17-106_k141_68075 | Not-determined |
| 17-106_k141_25553 | Not-determined |
| 17-106_k141_100003 | Not-determined |
| 17-106_k141_72920 | Not-determined |
| 17-106_k141_105300 | Not-determined |
| 17-106_k141_121911 | Not-determined |
| 17-106_k141_25566 | Not-determined |
| 17-106_k141_72927 | Not-determined |
| 17-106_k141_84517 | Not-determined |
| 17-106_k141_84522 | Not-determined |
| 17-106_k141_25580 | Low-quality |
| 17-106_k141_25585 | Not-determined |
| 17-106_k141_4320 | Not-determined |
| 17-106_k141_105321 | Not-determined |
| 17-106_k141_68090 | Not-determined |
| 17-106_k141_57713 | Not-determined |
| 17-106_k141_105324 | Low-quality |
| 17-106_k141_105325 | Not-determined |
| 17-106_k141_30246 | Not-determined |
| 17-106_k141_100028 | Not-determined |
| 17-106_k141_89742 | Not-determined |
| 17-106_k141_115984 | Not-determined |
| 17-106_k141_63088 | Not-determined |
| 17-106_k141_4332 | Low-quality |
| 17-106_k141_84534 | Not-determined |
| 17-106_k141_46295 | Not-determined |
| 17-106_k141_4340 | Not-determined |
| 17-106_k141_4343 | Low-quality |
| 17-106_k141_121933 | Not-determined |
| 17-106_k141_110087 | Not-determined |
| 17-106_k141_30255 | Not-determined |
| 17-106_k141_105339 | Not-determined |
| 17-106_k141_105340 | Not-determined |
| 17-106_k141_105341 | Not-determined |
| 17-106_k141_25606 | Not-determined |
| 17-106_k141_25609 | Not-determined |
| 17-106_k141_30265 | Not-determined |
| 17-106_k141_30266 | Not-determined |
| 17-106_k141_100042 | Not-determined |
| 17-106_k141_84552 | Not-determined |
| 17-106_k141_35833 | Not-determined |
| 17-106_k141_30274 | Low-quality |
| 17-106_k141_110103 | Not-determined |
| 17-106_k141_14636 | Low-quality |
| 17-106_k141_14637 | Not-determined |
| 17-106_k141_116001 | Not-determined |
| 17-106_k141_84559 | Not-determined |
| 17-106_k141_25619 | Not-determined |
| 17-106_k141_41073 | Not-determined |
| 17-106_k141_110113 | Not-determined |
| 17-106_k141_14643 | Not-determined |
| 17-106_k141_72972 | Not-determined |
| 17-106_k141_121963 | Not-determined |
| 17-106_k141_121965 | Not-determined |
| 17-106_k141_46308 | Not-determined |
| 17-106_k141_57723 | Not-determined |
| 17-106_k141_52562 | Not-determined |
| 17-106_k141_121968 | Not-determined |
| 17-106_k141_25629 | Low-quality |
| 17-106_k141_25634 | Not-determined |
| 17-106_k141_63101 | Not-determined |
| 17-106_k141_94307 | Not-determined |
| 17-106_k141_35853 | Not-determined |
| 17-106_k141_20253 | Not-determined |
| 17-106_k141_25642 | Not-determined |
| 17-106_k141_68132 | Low-quality |
| 17-106_k141_72991 | Not-determined |
| 17-106_k141_25644 | Not-determined |
| 17-106_k141_25645 | Not-determined |
| 17-106_k141_68135 | Not-determined |
| 17-106_k141_14668 | Not-determined |
| 17-106_k141_46321 | Not-determined |
| 17-106_k141_35860 | Not-determined |
| 17-106_k141_105363 | Not-determined |
| 17-106_k141_30296 | Not-determined |
| 17-106_k141_116026 | Not-determined |
| 17-106_k141_57749 | Not-determined |
| 17-106_k141_105367 | Not-determined |
| 17-106_k141_73005 | Not-determined |
| 17-106_k141_105371 | Not-determined |
| 17-106_k141_116030 | Not-determined |
| 17-106_k141_73015 | Not-determined |
| 17-106_k141_14691 | Not-determined |
| 17-106_k141_41086 | Not-determined |
| 17-106_k141_57764 | Not-determined |
| 17-106_k141_73027 | Not-determined |
| 17-106_k141_105383 | Not-determined |
| 17-106_k141_110126 | Not-determined |
| 17-106_k141_122001 | Not-determined |
| 17-106_k141_68158 | Not-determined |
| 17-106_k141_110142 | Not-determined |
| 17-106_k141_110143 | Not-determined |
| 17-106_k141_35870 | Not-determined |
| 17-106_k141_89825 | Not-determined |
| 17-106_k141_89826 | Not-determined |
| 17-106_k141_105401 | Low-quality |
| 17-106_k141_116048 | Not-determined |
| 17-106_k141_41116 | Not-determined |
| 17-106_k141_94368 | Not-determined |
| 17-106_k141_94371 | Not-determined |
| 17-106_k141_110155 | Not-determined |
| 17-106_k141_68192 | Not-determined |
| 17-106_k141_94373 | Not-determined |
| 17-106_k141_84585 | Not-determined |
| 17-106_k141_126533 | Not-determined |
| 17-106_k141_52596 | Low-quality |
| 17-106_k141_52602 | Not-determined |
| 17-106_k141_84588 | Low-quality |
| 17-106_k141_52605 | Low-quality |
| 17-106_k141_94378 | Not-determined |
| 17-106_k141_94381 | Not-determined |
| 17-106_k141_89882 | Not-determined |
| 17-106_k141_4352 | Not-determined |
| 17-106_k141_4355 | Not-determined |
| 17-106_k141_84605 | Not-determined |
| 17-106_k141_63141 | Not-determined |
| 17-106_k141_94396 | Not-determined |
| 17-106_k141_126548 | Not-determined |
| 17-106_k141_126549 | Not-determined |
| 17-106_k141_73072 | Not-determined |
| 17-106_k141_78980 | Not-determined |
| 17-106_k141_63150 | Not-determined |
| 17-106_k141_84614 | Not-determined |
| 17-106_k141_84618 | Not-determined |
| 17-106_k141_63157 | Not-determined |
| 17-106_k141_126556 | Not-determined |
| 17-106_k141_73082 | Not-determined |
| 17-106_k141_63165 | Not-determined |
| 17-106_k141_78987 | Not-determined |
| 17-106_k141_73093 | Not-determined |
| 17-106_k141_68250 | Not-determined |
| 17-106_k141_68254 | Not-determined |
| 17-106_k141_35915 | Not-determined |
| 17-106_k141_20280 | Not-determined |
| 17-106_k141_110160 | Not-determined |
| 17-106_k141_35919 | Not-determined |
| 17-106_k141_100056 | Not-determined |
| 17-106_k141_68263 | Low-quality |
| 17-106_k141_100057 | Not-determined |
| 17-106_k141_110164 | Not-determined |
| 17-106_k141_73103 | Not-determined |
| 17-106_k141_46350 | Not-determined |
| 17-106_k141_4396 | Not-determined |
| 17-106_k141_110166 | Low-quality |
| 17-106_k141_122015 | Not-determined |
| 17-106_k141_63181 | Not-determined |
| 17-106_k141_46359 | Not-determined |
| 17-106_k141_68273 | Not-determined |
| 17-106_k141_110172 | Not-determined |
| 17-106_k141_105412 | Not-determined |
| 17-106_k141_84646 | Not-determined |
| 17-106_k141_84648 | Not-determined |
| 17-106_k141_84653 | Not-determined |
| 17-106_k141_41157 | Low-quality |
| 17-106_k141_94432 | Low-quality |
| 17-106_k141_94433 | Low-quality |
| 17-106_k141_41164 | Not-determined |
| 17-106_k141_100079 | Not-determined |
| 17-106_k141_4400 | Low-quality |
| 17-106_k141_105427 | Not-determined |
| 17-106_k141_68287 | Not-determined |
| 17-106_k141_94438 | Not-determined |
| 17-106_k141_94443 | Not-determined |
| 17-106_k141_25665 | Not-determined |
| 17-106_k141_116108 | Low-quality |
| 17-106_k141_57782 | Not-determined |
| 17-106_k141_100097 | Not-determined |
| 17-106_k141_116116 | Not-determined |
| 17-106_k141_100105 | Not-determined |
| 17-106_k141_35938 | Not-determined |
| 17-106_k141_4421 | Not-determined |
| 17-106_k141_57789 | Not-determined |
| 17-106_k141_35939 | Not-determined |
| 17-106_k141_110191 | Not-determined |
| 17-106_k141_68297 | Low-quality |
| 17-106_k141_30332 | Not-determined |
| 17-106_k141_46384 | Not-determined |
| 17-106_k141_30336 | Not-determined |
| 17-106_k141_68302 | Not-determined |
| 17-106_k141_9651 | Not-determined |
| 17-106_k141_94455 | Not-determined |
| 17-106_k141_63219 | Not-determined |
| 17-106_k141_100116 | Not-determined |
| 17-106_k141_68313 | Not-determined |
| 17-106_k141_30345 | Low-quality |
| 17-106_k141_46393 | Not-determined |
| 17-106_k141_30351 | Not-determined |
| 17-106_k141_110218 | Not-determined |
| 17-106_k141_9672 | Not-determined |
| 17-106_k141_122055 | Not-determined |

| | |
|---|---|
| 17-106_k141_41175 | Not-determined |
| 17-106_k141_84704 | Not-determined |
| 17-106_k141_122057 | Not-determined |
| 17-106_k141_73144 | Not-determined |
| 17-106_k141_20312 | Not-determined |
| 17-106_k141_35952 | Not-determined |
| 17-106_k141_35954 | Not-determined |
| 17-106_k141_100130 | Not-determined |
| 17-106_k141_116150 | Not-determined |
| 17-106_k141_100134 | Not-determined |
| 17-106_k141_94481 | Not-determined |
| 17-106_k141_30359 | Low-quality |
| 17-106_k141_35959 | Not-determined |
| 17-106_k141_20321 | Not-determined |
| 17-106_k141_100139 | Not-determined |
| 17-106_k141_9685 | Not-determined |
| 17-106_k141_46418 | Low-quality |
| 17-106_k141_46419 | Low-quality |
| 17-106_k141_46421 | Not-determined |
| 17-106_k141_41197 | Not-determined |
| 17-106_k141_9690 | Low-quality |
| 17-106_k141_63268 | Not-determined |
| 17-106_k141_100152 | Not-determined |
| 17-106_k141_20338 | Not-determined |
| 17-106_k141_41205 | Not-determined |
| 17-106_k141_126566 | Not-determined |
| 17-106_k141_30372 | Not-determined |
| 17-106_k141_46430 | Not-determined |
| 17-106_k141_9703 | Not-determined |
| 17-106_k141_84727 | Not-determined |
| 17-106_k141_46440 | Not-determined |
| 17-106_k141_9711 | Low-quality |
| 17-106_k141_84728 | Not-determined |
| 17-106_k141_52673 | Not-determined |
| 17-106_k141_126573 | Not-determined |
| 17-106_k141_126574 | Not-determined |
| 17-106_k141_73169 | Not-determined |
| 17-106_k141_9716 | Not-determined |
| 17-106_k141_9717 | Not-determined |
| 17-106_k141_122080 | Not-determined |
| 17-106_k141_122081 | Not-determined |
| 17-106_k141_122082 | Not-determined |
| 17-106_k141_105466 | Not-determined |
| 17-106_k141_63291 | Not-determined |
| 17-106_k141_30378 | Not-determined |
| 17-106_k141_63293 | Not-determined |
| 17-106_k141_105471 | Not-determined |
| 17-106_k141_30382 | Not-determined |
| 17-106_k141_100162 | Not-determined |
| 17-106_k141_57839 | Not-determined |
| 17-106_k141_110237 | Not-determined |
| 17-106_k141_100169 | Low-quality |
| 17-106_k141_122093 | Not-determined |
| 17-106_k141_84741 | Not-determined |
| 17-106_k141_84743 | Not-determined |
| 17-106_k141_84748 | Not-determined |
| 17-106_k141_57848 | Not-determined |
| 17-106_k141_46466 | Not-determined |
| 17-106_k141_46467 | Not-determined |
| 17-106_k141_4442 | Not-determined |
| 17-106_k141_4443 | Not-determined |
| 17-106_k141_46472 | Not-determined |
| 17-106_k141_122107 | Not-determined |
| 17-106_k141_94521 | Not-determined |
| 17-106_k141_36012 | Not-determined |
| 17-106_k141_89964 | Not-determined |
| 17-106_k141_89968 | Not-determined |
| 17-106_k141_4452 | Not-determined |
| 17-106_k141_9756 | Not-determined |
| 17-106_k141_84768 | Not-determined |
| 17-106_k141_84769 | Not-determined |
| 17-106_k141_68347 | Not-determined |
| 17-106_k141_100193 | Not-determined |
| 17-106_k141_110258 | Not-determined |
| 17-106_k141_89984 | Not-determined |
| 17-106_k141_30393 | Not-determined |
| 17-106_k141_100195 | Not-determined |
| 17-106_k141_73186 | Not-determined |
| 17-106_k141_100197 | Not-determined |
| 17-106_k141_94537 | Not-determined |
| 17-106_k141_30398 | Not-determined |
| 17-106_k141_116220 | Not-determined |
| 17-106_k141_122119 | Not-determined |
| 17-106_k141_89996 | Not-determined |
| 17-106_k141_73200 | Not-determined |
| 17-106_k141_126587 | Not-determined |
| 17-106_k141_100201 | Not-determined |
| 17-106_k141_4468 | Not-determined |
| 17-106_k141_90001 | Not-determined |
| 17-106_k141_63331 | Not-determined |
| 17-106_k141_126595 | Low-quality |
| 17-106_k141_90003 | Not-determined |
| 17-106_k141_90006 | Not-determined |
| 17-106_k141_126604 | Not-determined |
| 17-106_k141_122130 | Not-determined |
| 17-106_k141_94563 | Not-determined |
| 17-106_k141_94566 | Not-determined |
| 17-106_k141_63353 | Not-determined |
| 17-106_k141_90013 | Not-determined |
| 17-106_k141_46482 | Not-determined |
| 17-106_k141_4478 | Not-determined |
| 17-106_k141_100212 | Not-determined |
| 17-106_k141_116242 | Not-determined |
| 17-106_k141_84816 | Low-quality |
| 17-106_k141_94586 | Not-determined |
| 17-106_k141_90022 | Not-determined |
| 17-106_k141_20372 | Not-determined |
| 17-106_k141_100221 | Not-determined |
| 17-106_k141_4486 | Not-determined |
| 17-106_k141_36068 | Not-determined |
| 17-106_k141_90026 | Low-quality |
| 17-106_k141_116247 | Not-determined |
| 17-106_k141_46509 | Not-determined |
| 17-106_k141_90027 | Not-determined |
| 17-106_k141_4497 | Not-determined |
| 17-106_k141_20377 | Not-determined |
| 17-106_k141_122156 | Not-determined |
| 17-106_k141_52728 | Low-quality |
| 17-106_k141_52731 | Low-quality |
| 17-106_k141_90039 | Not-determined |
| 17-106_k141_46523 | Not-determined |
| 17-106_k141_116265 | Not-determined |
| 17-106_k141_20379 | Not-determined |
| 17-106_k141_122166 | Not-determined |
| 17-106_k141_9827 | Low-quality |
| 17-106_k141_41217 | Not-determined |
| 17-106_k141_68413 | Not-determined |
| 17-106_k141_68415 | Not-determined |
| 17-106_k141_52742 | Not-determined |
| 17-106_k141_20395 | Low-quality |
| 17-106_k141_4509 | Not-determined |
| 17-106_k141_52748 | Not-determined |
| 17-106_k141_122170 | Not-determined |
| 17-106_k141_36087 | Not-determined |
| 17-106_k141_41222 | Low-quality |
| 17-106_k141_122179 | Not-determined |
| 17-106_k141_30413 | Not-determined |
| 17-106_k141_79038 | Not-determined |
| 17-106_k141_52759 | Not-determined |
| 17-106_k141_90057 | Not-determined |
| 17-106_k141_79042 | Low-quality |
| 17-106_k141_14811 | Not-determined |
| 17-106_k141_63406 | Not-determined |
| 17-106_k141_84864 | Not-determined |
| 17-106_k141_63414 | Not-determined |
| 17-106_k141_73226 | Not-determined |
| 17-106_k141_14831 | Not-determined |
| 17-106_k141_63421 | Not-determined |
| 17-106_k141_25687 | Not-determined |
| 17-106_k141_73232 | Not-determined |
| 17-106_k141_14832 | Not-determined |
| 17-106_k141_90075 | Not-determined |
| 17-106_k141_100242 | Not-determined |
| 17-106_k141_9858 | Not-determined |
| 17-106_k141_25692 | Not-determined |
| 17-106_k141_90077 | Low-quality |
| 17-106_k141_84882 | Not-determined |
| 17-106_k141_41249 | Not-determined |
| 17-106_k141_14842 | Low-quality |
| 17-106_k141_14844 | Not-determined |
| 17-106_k141_63431 | Low-quality |
| 17-106_k141_30421 | Not-determined |
| 17-106_k141_63433 | Not-determined |
| 17-106_k141_84890 | Not-determined |
| 17-106_k141_122205 | Low-quality |
| 17-106_k141_41258 | Not-determined |
| 17-106_k141_84894 | Low-quality |
| 17-106_k141_122207 | Not-determined |
| 17-106_k141_41259 | Low-quality |
| 17-106_k141_20426 | Not-determined |
| 17-106_k141_84895 | Low-quality |
| 17-106_k141_36117 | Low-quality |
| 17-106_k141_84901 | Not-determined |
| 17-106_k141_30432 | Not-determined |
| 17-106_k141_68463 | Not-determined |
| 17-106_k141_73271 | Not-determined |
| 17-106_k141_14878 | Not-determined |
| 17-106_k141_90098 | Not-determined |
| 17-106_k141_57923 | Not-determined |
| 17-106_k141_36139 | Not-determined |
| 17-106_k141_68476 | Not-determined |
| 17-106_k141_68477 | Not-determined |
| 17-106_k141_68480 | Not-determined |
| 17-106_k141_122224 | Not-determined |
| 17-106_k141_73298 | Not-determined |
| 17-106_k141_84937 | Not-determined |
| 17-106_k141_79086 | Not-determined |
| 17-106_k141_79087 | Not-determined |
| 17-106_k141_63460 | Not-determined |
| 17-106_k141_36164 | Not-determined |
| 17-106_k141_122233 | Not-determined |
| 17-106_k141_68490 | Not-determined |
| 17-106_k141_9869 | Low-quality |
| 17-106_k141_36173 | Not-determined |
| 17-106_k141_52773 | Not-determined |
| 17-106_k141_100254 | Not-determined |
| 17-106_k141_36174 | Low-quality |
| 17-106_k141_36176 | Not-determined |
| 17-106_k141_116288 | Not-determined |
| 17-106_k141_100260 | Low-quality |
| 17-106_k141_100261 | Not-determined |
| 17-106_k141_52780 | Not-determined |
| 17-106_k141_79101 | Not-determined |
| 17-106_k141_14890 | Not-determined |
| 17-106_k141_79107 | Not-determined |
| 17-106_k141_100268 | Not-determined |
| 17-106_k141_4527 | Low-quality |
| 17-106_k141_110317 | Not-determined |
| 17-106_k141_73329 | Not-determined |
| 17-106_k141_116299 | Not-determined |
| 17-106_k141_73330 | Not-determined |
| 17-106_k141_79114 | Not-determined |
| 17-106_k141_110319 | Not-determined |
| 17-106_k141_4532 | Not-determined |
| 17-106_k141_73336 | Not-determined |
| 17-106_k141_25711 | Not-determined |
| 17-106_k141_4535 | Not-determined |
| 17-106_k141_36191 | Not-determined |
| 17-106_k141_41272 | Not-determined |
| 17-106_k141_90110 | Not-determined |
| 17-106_k141_30450 | Not-determined |
| 17-106_k141_57964 | Not-determined |
| 17-106_k141_4544 | Not-determined |
| 17-106_k141_20461 | Not-determined |
| 17-106_k141_126636 | Not-determined |
| 17-106_k141_79122 | Not-determined |
| 17-106_k141_30459 | Not-determined |
| 17-106_k141_4550 | Not-determined |
| 17-106_k141_68517 | Not-determined |
| 17-106_k141_9891 | Low-quality |
| 17-106_k141_126640 | Low-quality |
| 17-106_k141_14912 | Not-determined |
| 17-106_k141_4556 | Not-determined |
| 17-106_k141_30464 | Not-determined |
| 17-106_k141_68524 | Not-determined |
| 17-106_k141_30466 | Not-determined |
| 17-106_k141_30467 | Not-determined |
| 17-106_k141_126644 | Not-determined |
| 17-106_k141_36211 | Not-determined |
| 17-106_k141_25724 | Not-determined |
| 17-106_k141_68526 | Not-determined |
| 17-106_k141_4570 | Not-determined |
| 17-106_k141_25731 | Low-quality |
| 17-106_k141_14919 | Low-quality |
| 17-106_k141_9896 | Not-determined |
| 17-106_k141_63471 | Low-quality |
| 17-106_k141_68542 | Not-determined |
| 17-106_k141_30481 | Not-determined |
| 17-106_k141_57997 | Not-determined |
| 17-106_k141_9901 | Not-determined |
| 17-106_k141_9904 | Not-determined |
| 17-106_k141_9906 | Not-determined |
| 17-106_k141_68549 | Not-determined |
| 17-106_k141_30487 | Not-determined |
| 17-106_k141_9915 | Not-determined |
| 17-106_k141_41293 | Not-determined |
| 17-106_k141_46618 | Not-determined |
| 17-106_k141_68563 | Not-determined |
| 17-106_k141_52823 | Not-determined |
| 17-106_k141_9927 | Not-determined |
| 17-106_k141_46627 | Not-determined |
| 17-106_k141_63485 | Low-quality |
| 17-106_k141_46628 | Not-determined |
| 17-106_k141_52830 | Not-determined |
| 17-106_k141_46629 | Not-determined |
| 17-106_k141_9930 | Low-quality |
| 17-106_k141_14929 | Not-determined |
| 17-106_k141_58028 | Not-determined |
| 17-106_k141_52835 | Not-determined |
| 17-106_k141_122266 | Not-determined |
| 17-106_k141_58033 | Not-determined |
| 17-106_k141_100283 | Not-determined |
| 17-106_k141_90137 | Not-determined |
| 17-106_k141_79180 | Not-determined |
| 17-106_k141_14938 | Not-determined |
| 17-106_k141_100287 | Not-determined |
| 17-106_k141_46637 | Low-quality |
| 17-106_k141_58040 | Not-determined |
| 17-106_k141_41319 | Not-determined |
| 17-106_k141_41322 | Not-determined |
| 17-106_k141_73409 | Not-determined |
| 17-106_k141_73411 | Low-quality |
| 17-106_k141_41331 | Not-determined |
| 17-106_k141_41333 | Not-determined |
| 17-106_k141_94685 | Not-determined |
| 17-106_k141_41336 | Not-determined |
| 17-106_k141_126662 | Not-determined |
| 17-106_k141_41341 | Not-determined |
| 17-106_k141_100308 | Not-determined |
| 17-106_k141_90171 | Not-determined |
| 17-106_k141_41348 | Low-quality |
| 17-106_k141_30553 | Not-determined |
| 17-106_k141_30556 | Not-determined |
| 17-106_k141_110372 | Low-quality |
| 17-106_k141_63517 | Not-determined |
| 17-106_k141_122289 | Not-determined |
| 17-106_k141_73431 | Not-determined |
| 17-106_k141_122290 | Not-determined |
| 17-106_k141_122294 | Not-determined |
| 17-106_k141_41361 | Not-determined |
| 17-106_k141_68608 | Not-determined |
| 17-106_k141_68609 | Not-determined |
| 17-106_k141_79224 | Not-determined |
| 17-106_k141_110374 | Not-determined |
| 17-106_k141_94710 | Not-determined |
| 17-106_k141_36230 | Not-determined |
| 17-106_k141_110380 | Not-determined |
| 17-106_k141_58064 | Not-determined |
| 17-106_k141_58066 | Low-quality |
| 17-106_k141_90183 | Not-determined |
| 17-106_k141_68619 | Not-determined |
| 17-106_k141_36238 | Not-determined |
| 17-106_k141_36242 | Not-determined |
| 17-106_k141_41382 | Not-determined |
| 17-106_k141_63541 | Not-determined |
| 17-106_k141_110383 | Not-determined |
| 17-106_k141_116326 | Not-determined |
| 17-106_k141_79232 | Low-quality |
| 17-106_k141_100320 | Not-determined |
| 17-106_k141_79237 | Not-determined |
| 17-106_k141_14993 | Not-determined |
| 17-106_k141_100323 | Low-quality |
| 17-106_k141_36255 | Not-determined |
| 17-106_k141_25785 | Not-determined |
| 17-106_k141_41398 | Not-determined |
| 17-106_k141_116334 | Not-determined |
| 17-106_k141_41401 | Not-determined |
| 17-106_k141_25788 | Not-determined |
| 17-106_k141_25790 | Not-determined |
| 17-106_k141_116337 | Not-determined |
| 17-106_k141_63560 | Not-determined |
| 17-106_k141_105481 | Not-determined |
| 17-106_k141_90190 | Not-determined |
| 17-106_k141_90193 | Not-determined |
| 17-106_k141_41409 | Not-determined |
| 17-106_k141_100337 | Not-determined |
| 17-106_k141_25805 | Low-quality |
| 17-106_k141_41416 | Low-quality |
| 17-106_k141_90201 | Not-determined |
| 17-106_k141_25809 | Not-determined |
| 17-106_k141_79251 | Low-quality |
| 17-106_k141_30586 | Not-determined |
| 17-106_k141_100347 | Not-determined |
| 17-106_k141_90205 | Not-determined |
| 17-106_k141_46677 | Not-determined |
| 17-106_k141_63565 | Low-quality |
| 17-106_k141_63567 | Not-determined |
| 17-106_k141_30596 | Low-quality |
| 17-106_k141_105497 | Not-determined |
| 17-106_k141_4577 | Not-determined |
| 17-106_k141_30598 | Not-determined |
| 17-106_k141_105499 | Low-quality |
| 17-106_k141_20488 | Not-determined |
| 17-106_k141_63583 | Not-determined |
| 17-106_k141_52871 | Not-determined |
| 17-106_k141_30601 | Low-quality |
| 17-106_k141_100349 | Not-determined |
| 17-106_k141_52872 | Not-determined |
| 17-106_k141_30605 | Not-determined |
| 17-106_k141_46685 | Low-quality |
| 17-106_k141_105508 | Low-quality |
| 17-106_k141_105509 | Not-determined |
| 17-106_k141_4580 | Not-determined |
| 17-106_k141_90206 | Low-quality |
| 17-106_k141_94761 | Not-determined |
| 17-106_k141_105511 | Not-determined |
| 17-106_k141_100355 | Not-determined |
| 17-106_k141_52879 | Not-determined |
| 17-106_k141_110403 | Not-determined |
| 17-106_k141_52883 | Not-determined |
| 17-106_k141_20495 | Not-determined |
| 17-106_k141_116367 | Not-determined |
| 17-106_k141_20498 | Not-determined |
| 17-106_k141_79285 | Not-determined |
| 17-106_k141_79286 | Not-determined |
| 17-106_k141_4584 | Not-determined |
| 17-106_k141_9937 | Not-determined |
| 17-106_k141_20505 | Not-determined |
| 17-106_k141_20507 | Not-determined |
| 17-106_k141_20510 | Not-determined |
| 17-106_k141_90214 | Not-determined |
| 17-106_k141_52892 | Low-quality |
| 17-106_k141_110408 | Not-determined |
| 17-106_k141_63616 | Not-determined |
| 17-106_k141_63623 | Not-determined |
| 17-106_k141_20526 | Not-determined |
| 17-106_k141_41466 | Not-determined |
| 17-106_k141_79304 | Not-determined |
| 17-106_k141_52907 | Not-determined |
| 17-106_k141_94793 | Not-determined |
| 17-106_k141_90231 | Not-determined |
| 17-106_k141_122314 | Not-determined |
| 17-106_k141_41475 | Not-determined |
| 17-106_k141_46708 | Not-determined |
| 17-106_k141_100393 | Not-determined |
| 17-106_k141_52918 | Not-determined |
| 17-106_k141_20547 | Not-determined |
| 17-106_k141_79308 | Not-determined |
| 17-106_k141_122317 | Not-determined |
| 17-106_k141_79311 | Not-determined |
| 17-106_k141_52922 | Not-determined |
| 17-106_k141_52927 | Not-determined |
| 17-106_k141_90242 | Not-determined |
| 17-106_k141_110414 | Not-determined |
| 17-106_k141_41490 | Not-determined |
| 17-106_k141_90245 | Not-determined |
| 17-106_k141_46712 | Not-determined |
| 17-106_k141_4618 | Not-determined |
| 17-106_k141_4619 | Not-determined |
| 17-106_k141_52931 | Not-determined |
| 17-106_k141_94805 | Not-determined |
| 17-106_k141_4626 | Not-determined |
| 17-106_k141_25821 | Low-quality |
| 17-106_k141_4628 | Not-determined |
| 17-106_k141_4630 | Not-determined |
| 17-106_k141_52941 | Low-quality |
| 17-106_k141_122332 | Not-determined |
| 17-106_k141_79332 | Not-determined |
| 17-106_k141_52949 | Not-determined |
| 17-106_k141_100404 | Not-determined |
| 17-106_k141_122339 | Not-determined |
| 17-106_k141_105574 | Not-determined |
| 17-106_k141_36269 | Not-determined |
| 17-106_k141_100405 | Not-determined |
| 17-106_k141_122341 | Not-determined |
| 17-106_k141_90279 | Not-determined |
| 17-106_k141_36274 | Low-quality |
| 17-106_k141_90287 | Not-determined |
| 17-106_k141_90291 | Not-determined |
| 17-106_k141_25840 | Not-determined |
| 17-106_k141_25841 | Not-determined |
| 17-106_k141_116390 | Not-determined |
| 17-106_k141_94809 | Not-determined |
| 17-106_k141_20579 | Not-determined |
| 17-106_k141_36284 | Not-determined |
| 17-106_k141_116394 | Not-determined |
| 17-106_k141_79358 | Not-determined |
| 17-106_k141_36288 | Low-quality |
| 17-106_k141_15001 | Not-determined |
| 17-106_k141_90300 | Not-determined |
| 17-106_k141_15004 | Not-determined |
| 17-106_k141_122359 | Low-quality |
| 17-106_k141_90307 | Not-determined |
| 17-106_k141_15018 | Not-determined |
| 17-106_k141_15022 | Not-determined |
| 17-106_k141_73456 | Not-determined |
| 17-106_k141_63667 | Not-determined |
| 17-106_k141_25866 | Low-quality |
| 17-106_k141_63669 | Not-determined |
| 17-106_k141_79373 | Not-determined |
| 17-106_k141_73468 | Not-determined |
| 17-106_k141_36329 | Not-determined |
| 17-106_k141_122399 | Not-determined |
| 17-106_k141_15040 | Not-determined |
| 17-106_k141_58084 | Not-determined |
| 17-106_k141_58085 | Not-determined |
| 17-106_k141_58088 | Not-determined |
| 17-106_k141_105631 | Not-determined |
| 17-106_k141_20599 | Not-determined |
| 17-106_k141_122412 | Not-determined |
| 17-106_k141_122417 | Not-determined |
| 17-106_k141_58101 | Not-determined |
| 17-106_k141_94833 | Not-determined |
| 17-106_k141_94836 | Not-determined |
| 17-106_k141_90330 | Not-determined |
| 17-106_k141_20607 | Not-determined |
| 17-106_k141_25887 | Not-determined |
| 17-106_k141_79410 | Not-determined |
| 17-106_k141_79412 | Not-determined |
| 17-106_k141_30689 | Not-determined |
| 17-106_k141_63691 | Not-determined |
| 17-106_k141_20618 | Not-determined |
| 17-106_k141_10012 | Not-determined |
| 17-106_k141_122441 | Not-determined |
| 17-106_k141_52976 | Not-determined |
| 17-106_k141_63706 | Not-determined |
| 17-106_k141_68764 | Not-determined |
| 17-106_k141_100448 | Not-determined |
| 17-106_k141_63719 | Not-determined |
| 17-106_k141_79426 | Not-determined |
| 17-106_k141_36338 | Not-determined |
| 17-106_k141_30710 | Not-determined |
| 17-106_k141_25911 | Not-determined |
| 17-106_k141_36342 | Not-determined |
| 17-106_k141_52991 | Not-determined |
| 17-106_k141_126740 | Not-determined |
| 17-106_k141_79439 | Not-determined |
| 17-106_k141_63720 | Not-determined |
| 17-106_k141_63722 | Not-determined |
| 17-106_k141_30722 | Not-determined |
| 17-106_k141_126748 | Not-determined |
| 17-106_k141_126749 | Not-determined |
| 17-106_k141_122453 | Low-quality |
| 17-106_k141_20655 | Not-determined |
| 17-106_k141_15050 | Not-determined |
| 17-106_k141_126750 | Not-determined |
| 17-106_k141_53004 | Low-quality |
| 17-106_k141_4649 | Low-quality |
| 17-106_k141_30727 | Not-determined |
| 17-106_k141_122457 | Not-determined |
| 17-106_k141_126756 | Not-determined |
| 17-106_k141_68794 | Not-determined |
| 17-106_k141_30730 | Low-quality |
| 17-106_k141_68803 | Not-determined |
| 17-106_k141_58158 | Low-quality |
| 17-106_k141_126762 | Not-determined |
| 17-106_k141_58162 | Not-determined |
| 17-106_k141_30737 | Low-quality |
| 17-106_k141_68808 | Not-determined |
| 17-106_k141_100489 | Not-determined |
| 17-106_k141_15064 | Not-determined |
| 17-106_k141_79460 | Not-determined |
| 17-106_k141_79461 | Not-determined |
| 17-106_k141_25934 | Not-determined |
| 17-106_k141_79463 | Not-determined |
| 17-106_k141_84968 | Not-determined |
| 17-106_k141_84970 | Not-determined |
| 17-106_k141_94900 | Not-determined |
| 17-106_k141_122462 | Low-quality |
| 17-106_k141_58169 | Not-determined |
| 17-106_k141_58170 | Not-determined |
| 17-106_k141_84972 | Not-determined |
| 17-106_k141_15071 | Not-determined |
| 17-106_k141_4668 | Not-determined |
| 17-106_k141_94905 | Not-determined |
| 17-106_k141_46741 | Not-determined |
| 17-106_k141_46743 | Not-determined |
| 17-106_k141_36371 | Not-determined |
| 17-106_k141_73498 | Not-determined |
| 17-106_k141_36377 | Not-determined |
| 17-106_k141_73501 | Not-determined |
| 17-106_k141_90385 | Not-determined |
| 17-106_k141_46752 | Not-determined |
| 17-106_k141_58187 | Not-determined |
| 17-106_k141_110431 | Not-determined |
| 17-106_k141_122479 | Not-determined |
| 17-106_k141_36382 | Not-determined |
| 17-106_k141_73504 | Not-determined |
| 17-106_k141_46757 | Not-determined |
| 17-106_k141_20664 | Not-determined |
| 17-106_k141_90396 | Not-determined |
| 17-106_k141_63772 | Low-quality |
| 17-106_k141_25981 | Not-determined |
| 17-106_k141_68832 | Not-determined |
| 17-106_k141_58213 | Not-determined |
| 17-106_k141_100541 | Not-determined |
| 17-106_k141_73523 | Not-determined |
| 17-106_k141_4702 | Not-determined |
| 17-106_k141_4705 | Not-determined |
| 17-106_k141_94934 | Not-determined |
| 17-106_k141_46768 | Not-determined |
| 17-106_k141_10040 | Not-determined |
| 17-106_k141_94935 | Not-determined |
| 17-106_k141_122519 | Not-determined |
| 17-106_k141_20687 | Low-quality |
| 17-106_k141_36423 | Not-determined |
| 17-106_k141_94945 | Not-determined |
| 17-106_k141_10054 | Not-determined |
| 17-106_k141_10055 | Not-determined |
| 17-106_k141_126797 | Not-determined |
| 17-106_k141_94951 | Not-determined |
| 17-106_k141_25999 | Not-determined |
| 17-106_k141_94955 | Not-determined |
| 17-106_k141_53056 | Not-determined |
| 17-106_k141_85041 | Not-determined |
| 17-106_k141_10066 | Not-determined |
| 17-106_k141_15112 | Not-determined |
| 17-106_k141_63826 | Not-determined |
| 17-106_k141_63827 | Not-determined |
| 17-106_k141_53058 | Not-determined |
| 17-106_k141_20698 | Not-determined |
| 17-106_k141_30756 | Not-determined |
| 17-106_k141_26018 | Not-determined |
| 17-106_k141_53064 | Not-determined |
| 17-106_k141_53065 | Not-determined |
| 17-106_k141_53066 | Low-quality |
| 17-106_k141_30766 | Not-determined |
| 17-106_k141_90437 | Not-determined |
| 17-106_k141_90438 | Not-determined |
| 17-106_k141_10071 | Not-determined |
| 17-106_k141_4718 | Not-determined |
| 17-106_k141_85049 | Not-determined |
| 17-106_k141_94970 | Not-determined |
| 17-106_k141_90444 | Not-determined |
| 17-106_k141_30778 | Not-determined |
| 17-106_k141_20710 | Not-determined |
| 17-106_k141_85055 | Not-determined |
| 17-106_k141_90447 | Not-determined |
| 17-106_k141_30781 | Not-determined |

| | |
|---|---|
| 17-106_k141_90450 | Not-determined |
| 17-106_k141_36457 | Not-determined |
| 17-106_k141_79530 | Not-determined |
| 17-106_k141_10089 | Not-determined |
| 17-106_k141_90455 | Not-determined |
| 17-106_k141_63835 | Not-determined |
| 17-106_k141_63841 | Not-determined |
| 17-106_k141_122553 | Not-determined |
| 17-106_k141_68902 | Not-determined |
| 17-106_k141_110450 | Not-determined |
| 17-106_k141_10095 | Not-determined |
| 17-106_k141_85069 | Low-quality |
| 17-106_k141_116444 | Not-determined |
| 17-106_k141_94986 | Not-determined |
| 17-106_k141_68908 | Not-determined |
| 17-106_k141_30798 | Low-quality |
| 17-106_k141_79540 | Not-determined |
| 17-106_k141_85077 | Low-quality |
| 17-106_k141_53105 | Not-determined |
| 17-106_k141_79544 | Not-determined |
| 17-106_k141_79547 | Low-quality |
| 17-106_k141_100567 | Low-quality |
| 17-106_k141_10120 | Not-determined |
| 17-106_k141_94995 | Not-determined |
| 17-106_k141_10122 | Not-determined |
| 17-106_k141_79554 | Not-determined |
| 17-106_k141_79555 | Not-determined |
| 17-106_k141_10126 | Not-determined |
| 17-106_k141_53114 | Not-determined |
| 17-106_k141_100575 | Not-determined |
| 17-106_k141_41527 | Not-determined |
| 17-106_k141_79559 | Not-determined |
| 17-106_k141_100579 | Not-determined |
| 17-106_k141_79564 | Low-quality |
| 17-106_k141_20749 | Low-quality |
| 17-106_k141_53118 | Not-determined |
| 17-106_k141_100581 | Not-determined |
| 17-106_k141_94997 | Not-determined |
| 17-106_k141_68922 | Low-quality |
| 17-106_k141_79573 | Not-determined |
| 17-106_k141_90483 | Not-determined |
| 17-106_k141_4765 | Not-determined |
| 17-106_k141_68927 | Not-determined |
| 17-106_k141_53127 | Not-determined |
| 17-106_k141_90490 | Not-determined |
| 17-106_k141_20764 | Not-determined |
| 17-106_k141_105686 | Low-quality |
| 17-106_k141_90492 | Not-determined |
| 17-106_k141_90493 | Not-determined |
| 17-106_k141_126832 | Not-determined |
| 17-106_k141_26056 | Low-quality |
| 17-106_k141_41554 | Not-determined |
| 17-106_k141_100609 | Not-determined |
| 17-106_k141_100614 | Not-determined |
| 17-106_k141_46777 | Not-determined |
| 17-106_k141_10155 | Not-determined |
| 17-106_k141_79582 | Not-determined |
| 17-106_k141_46780 | Low-quality |
| 17-106_k141_10159 | Not-determined |
| 17-106_k141_20773 | Not-determined |
| 17-106_k141_110489 | Not-determined |
| 17-106_k141_100624 | Not-determined |
| 17-106_k141_100627 | Not-determined |
| 17-106_k141_20777 | Not-determined |
| 17-106_k141_90505 | Not-determined |
| 17-106_k141_90512 | Not-determined |
| 17-106_k141_53164 | Low-quality |
| 17-106_k141_100640 | Not-determined |
| 17-106_k141_110502 | Not-determined |
| 17-106_k141_53166 | Not-determined |
| 17-106_k141_79600 | Not-determined |
| 17-106_k141_36482 | Not-determined |
| 17-106_k141_79606 | Not-determined |
| 17-106_k141_79608 | Not-determined |
| 17-106_k141_41575 | Not-determined |
| 17-106_k141_85117 | Not-determined |
| 17-106_k141_110510 | Not-determined |
| 17-106_k141_85127 | Not-determined |
| 17-106_k141_26095 | Low-quality |
| 17-106_k141_100675 | Low-quality |
| 17-106_k141_68997 | Not-determined |
| 17-106_k141_41592 | Not-determined |
| 17-106_k141_26101 | Not-determined |
| 17-106_k141_126859 | Not-determined |
| 17-106_k141_116468 | Not-determined |
| 17-106_k141_26104 | Not-determined |
| 17-106_k141_36511 | Low-quality |
| 17-106_k141_10162 | Not-determined |
| 17-106_k141_46819 | Low-quality |
| 17-106_k141_116472 | Not-determined |
| 17-106_k141_36524 | Not-determined |
| 17-106_k141_116474 | Not-determined |
| 17-106_k141_10170 | Not-determined |
| 17-106_k141_15180 | Not-determined |
| 17-106_k141_20807 | Not-determined |
| 17-106_k141_110531 | Not-determined |
| 17-106_k141_122610 | Not-determined |
| 17-106_k141_15183 | Low-quality |
| 17-106_k141_36531 | Not-determined |
| 17-106_k141_46825 | Not-determined |
| 17-106_k141_69006 | Not-determined |
| 17-106_k141_110536 | Not-determined |
| 17-106_k141_116486 | Not-determined |
| 17-106_k141_69008 | Not-determined |
| 17-106_k141_90566 | Not-determined |
| 17-106_k141_15198 | Not-determined |
| 17-106_k141_20818 | Low-quality |
| 17-106_k141_69023 | Not-determined |
| 17-106_k141_15199 | Not-determined |
| 17-106_k141_15203 | Not-determined |
| 17-106_k141_116493 | Not-determined |
| 17-106_k141_79651 | Not-determined |
| 17-106_k141_46830 | Not-determined |
| 17-106_k141_79654 | Not-determined |
| 17-106_k141_46835 | Not-determined |
| 17-106_k141_10200 | Not-determined |
| 17-106_k141_69033 | Not-determined |
| 17-106_k141_10207 | Low-quality |
| 17-106_k141_95050 | Not-determined |
| 17-106_k141_79665 | Not-determined |
| 17-106_k141_110550 | Not-determined |
| 17-106_k141_85162 | Not-determined |
| 17-106_k141_20833 | Not-determined |
| 17-106_k141_15217 | Not-determined |
| 17-106_k141_41649 | Not-determined |
| 17-106_k141_4826 | Not-determined |
| 17-106_k141_110568 | Not-determined |
| 17-106_k141_110569 | Not-determined |
| 17-106_k141_4834 | Not-determined |
| 17-106_k141_36573 | Not-determined |
| 17-106_k141_85181 | Not-determined |
| 17-106_k141_41663 | Not-determined |
| 17-106_k141_4849 | Not-determined |
| 17-106_k141_110573 | Not-determined |
| 17-106_k141_36583 | Not-determined |
| 17-106_k141_30810 | Not-determined |
| 17-106_k141_36585 | Not-determined |
| 17-106_k141_73597 | Not-determined |
| 17-106_k141_73598 | Not-determined |
| 17-106_k141_15230 | Not-determined |
| 17-106_k141_85198 | Not-determined |
| 17-106_k141_126896 | Not-determined |
| 17-106_k141_63952 | Not-determined |
| 17-106_k141_126900 | Not-determined |
| 17-106_k141_30816 | Not-determined |
| 17-106_k141_110589 | Not-determined |
| 17-106_k141_116563 | Not-determined |
| 17-106_k141_90628 | Not-determined |
| 17-106_k141_53174 | Not-determined |
| 17-106_k141_95068 | Not-determined |
| 17-106_k141_53175 | Not-determined |
| 17-106_k141_53176 | Not-determined |
| 17-106_k141_4860 | Not-determined |
| 17-106_k141_63978 | Low-quality |
| 17-106_k141_15250 | Not-determined |
| 17-106_k141_73608 | Not-determined |
| 17-106_k141_73609 | Not-determined |
| 17-106_k141_15257 | Not-determined |
| 17-106_k141_15259 | Not-determined |
| 17-106_k141_36611 | Not-determined |
| 17-106_k141_105694 | Low-quality |
| 17-106_k141_15265 | Not-determined |
| 17-106_k141_46867 | Not-determined |
| 17-106_k141_73617 | Not-determined |
| 17-106_k141_90655 | Not-determined |
| 17-106_k141_26188 | Not-determined |
| 17-106_k141_20858 | Not-determined |
| 17-106_k141_36619 | Not-determined |
| 17-106_k141_73621 | Not-determined |
| 17-106_k141_79735 | Low-quality |
| 17-106_k141_26192 | Not-determined |
| 17-106_k141_30838 | Not-determined |
| 17-106_k141_95080 | Low-quality |
| 17-106_k141_20862 | Not-determined |
| 17-106_k141_90676 | Not-determined |
| 17-106_k141_126926 | Not-determined |
| 17-106_k141_126932 | Low-quality |
| 17-106_k141_69061 | Not-determined |
| 17-106_k141_53203 | Low-quality |
| 17-106_k141_46878 | Not-determined |
| 17-106_k141_26207 | Not-determined |
| 17-106_k141_126938 | Not-determined |
| 17-106_k141_126940 | Not-determined |
| 17-106_k141_126941 | Not-determined |
| 17-106_k141_116571 | Not-determined |
| 17-106_k141_95089 | Not-determined |
| 17-106_k141_46883 | Not-determined |
| 17-106_k141_20879 | Not-determined |
| 17-106_k141_90693 | Not-determined |
| 17-106_k141_116583 | Not-determined |
| 17-106_k141_15287 | Not-determined |
| 17-106_k141_100762 | Not-determined |
| 17-106_k141_36656 | Not-determined |
| 17-106_k141_69071 | Not-determined |
| 17-106_k141_116591 | Not-determined |
| 17-106_k141_69079 | Not-determined |
| 17-106_k141_20883 | Not-determined |
| 17-106_k141_126967 | Not-determined |
| 17-106_k141_63987 | Not-determined |
| 17-106_k141_100780 | Low-quality |
| 17-106_k141_36680 | Low-quality |
| 17-106_k141_10266 | Not-determined |
| 17-106_k141_69099 | Not-determined |
| 17-106_k141_41721 | Not-determined |
| 17-106_k141_41722 | Not-determined |
| 17-106_k141_10268 | Not-determined |
| 17-106_k141_41723 | Not-determined |
| 17-106_k141_10270 | Not-determined |
| 17-106_k141_53237 | Not-determined |
| 17-106_k141_53238 | Not-determined |
| 17-106_k141_90735 | Low-quality |
| 17-106_k141_53243 | Not-determined |
| 17-106_k141_73694 | Low-quality |
| 17-106_k141_100804 | Not-determined |
| 17-106_k141_64019 | Not-determined |
| 17-106_k141_64021 | Not-determined |
| 17-106_k141_90740 | Not-determined |
| 17-106_k141_30895 | Not-determined |
| 17-106_k141_64027 | Not-determined |
| 17-106_k141_64030 | Not-determined |
| 17-106_k141_90751 | Not-determined |
| 17-106_k141_73702 | Low-quality |
| 17-106_k141_73703 | Not-determined |
| 17-106_k141_73704 | Not-determined |
| 17-106_k141_69109 | Not-determined |
| 17-106_k141_36698 | Not-determined |
| 17-106_k141_30911 | Not-determined |
| 17-106_k141_79786 | Not-determined |
| 17-106_k141_15330 | Not-determined |
| 17-106_k141_15332 | Not-determined |
| 17-106_k141_116648 | Not-determined |
| 17-106_k141_79788 | Not-determined |
| 17-106_k141_53253 | Low-quality |
| 17-106_k141_126987 | Low-quality |
| 17-106_k141_85291 | Not-determined |
| 17-106_k141_15335 | Not-determined |
| 17-106_k141_30925 | Low-quality |
| 17-106_k141_85302 | Not-determined |
| 17-106_k141_4929 | Not-determined |
| 17-106_k141_58241 | Not-determined |
| 17-106_k141_15351 | Low-quality |
| 17-106_k141_41774 | Not-determined |
| 17-106_k141_15355 | Not-determined |
| 17-106_k141_15356 | Not-determined |
| 17-106_k141_36730 | Not-determined |
| 17-106_k141_58258 | Not-determined |
| 17-106_k141_85311 | Not-determined |
| 17-106_k141_15358 | Not-determined |
| 17-106_k141_53271 | Not-determined |
| 17-106_k141_73740 | Not-determined |
| 17-106_k141_58267 | Not-determined |
| 17-106_k141_36739 | Not-determined |
| 17-106_k141_41788 | Not-determined |
| 17-106_k141_36741 | Not-determined |
| 17-106_k141_20929 | Not-determined |
| 17-106_k141_73744 | Not-determined |
| 17-106_k141_15367 | Not-determined |
| 17-106_k141_69133 | Not-determined |
| 17-106_k141_15373 | Not-determined |
| 17-106_k141_58277 | Not-determined |
| 17-106_k141_116666 | Not-determined |
| 17-106_k141_73750 | Not-determined |
| 17-106_k141_73752 | Not-determined |
| 17-106_k141_100878 | Not-determined |
| 17-106_k141_30933 | Not-determined |
| 17-106_k141_4948 | Not-determined |
| 17-106_k141_4949 | Not-determined |
| 17-106_k141_53297 | Low-quality |
| 17-106_k141_30942 | Low-quality |
| 17-106_k141_4958 | Not-determined |
| 17-106_k141_116690 | Not-determined |
| 17-106_k141_30950 | Not-determined |
| 17-106_k141_58298 | Not-determined |
| 17-106_k141_79820 | Not-determined |
| 17-106_k141_58299 | Not-determined |
| 17-106_k141_116696 | Not-determined |
| 17-106_k141_73765 | Not-determined |
| 17-106_k141_58304 | Not-determined |
| 17-106_k141_36770 | Not-determined |
| 17-106_k141_116700 | Not-determined |
| 17-106_k141_30958 | Not-determined |
| 17-106_k141_85345 | Not-determined |
| 17-106_k141_30963 | Not-determined |
| 17-106_k141_58319 | Not-determined |
| 17-106_k141_53324 | Not-determined |
| 17-106_k141_58324 | Not-determined |
| 17-106_k141_36780 | Not-determined |
| 17-106_k141_90794 | Not-determined |
| 17-106_k141_64111 | Not-determined |
| 17-106_k141_64114 | Not-determined |
| 17-106_k141_58331 | Not-determined |
| 17-106_k141_53337 | Not-determined |
| 17-106_k141_100905 | Not-determined |
| 17-106_k141_36795 | Not-determined |
| 17-106_k141_36797 | Not-determined |
| 17-106_k141_90805 | Not-determined |
| 17-106_k141_41822 | Not-determined |
| 17-106_k141_105752 | Low-quality |
| 17-106_k141_127025 | Not-determined |
| 17-106_k141_127026 | Not-determined |
| 17-106_k141_127031 | Not-determined |
| 17-106_k141_105770 | Not-determined |
| 17-106_k141_79844 | Not-determined |
| 17-106_k141_36811 | Low-quality |
| 17-106_k141_36812 | Not-determined |
| 17-106_k141_30995 | Not-determined |
| 17-106_k141_85371 | Not-determined |
| 17-106_k141_36818 | Not-determined |
| 17-106_k141_79853 | Not-determined |
| 17-106_k141_79856 | Not-determined |
| 17-106_k141_90828 | Low-quality |
| 17-106_k141_36827 | Not-determined |
| 17-106_k141_31006 | Not-determined |
| 17-106_k141_31012 | Not-determined |
| 17-106_k141_36832 | Not-determined |
| 17-106_k141_4999 | Not-determined |
| 17-106_k141_36833 | Not-determined |
| 17-106_k141_85384 | Not-determined |
| 17-106_k141_53358 | Not-determined |
| 17-106_k141_100923 | Not-determined |
| 17-106_k141_36840 | Not-determined |
| 17-106_k141_58342 | Not-determined |
| 17-106_k141_69183 | Not-determined |
| 17-106_k141_41847 | Not-determined |
| 17-106_k141_58348 | Not-determined |
| 17-106_k141_69184 | Not-determined |
| 17-106_k141_36851 | Not-determined |
| 17-106_k141_105812 | Low-quality |
| 17-106_k141_41854 | Not-determined |
| 17-106_k141_85401 | Not-determined |
| 17-106_k141_5020 | Not-determined |
| 17-106_k141_58355 | Not-determined |
| 17-106_k141_36863 | Not-determined |
| 17-106_k141_58364 | Not-determined |
| 17-106_k141_85416 | Not-determined |
| 17-106_k141_85417 | Not-determined |
| 17-106_k141_36866 | Not-determined |
| 17-106_k141_95104 | Not-determined |
| 17-106_k141_85423 | Not-determined |
| 17-106_k141_41870 | Not-determined |
| 17-106_k141_90866 | Not-determined |
| 17-106_k141_36880 | Not-determined |
| 17-106_k141_90870 | Not-determined |
| 17-106_k141_5035 | Not-determined |
| 17-106_k141_26349 | Not-determined |
| 17-106_k141_41881 | Not-determined |
| 17-106_k141_41882 | Low-quality |
| 17-106_k141_36890 | Not-determined |
| 17-106_k141_41886 | Not-determined |
| 17-106_k141_5042 | Not-determined |
| 17-106_k141_15432 | Not-determined |
| 17-106_k141_73819 | Not-determined |
| 17-106_k141_31044 | Not-determined |
| 17-106_k141_31045 | Not-determined |
| 17-106_k141_116742 | Not-determined |
| 17-106_k141_31048 | Not-determined |
| 17-106_k141_73822 | Not-determined |
| 17-106_k141_73823 | Not-determined |
| 17-106_k141_116748 | Not-determined |
| 17-106_k141_41898 | Not-determined |
| 17-106_k141_85432 | Not-determined |
| 17-106_k141_122651 | Not-determined |
| 17-106_k141_73834 | Low-quality |
| 17-106_k141_100967 | Not-determined |
| 17-106_k141_26369 | Not-determined |
| 17-106_k141_73837 | Not-determined |
| 17-106_k141_122659 | Low-quality |
| 17-106_k141_122664 | Not-determined |
| 17-106_k141_10278 | Not-determined |
| 17-106_k141_122668 | Not-determined |
| 17-106_k141_110702 | Not-determined |
| 17-106_k141_10281 | Not-determined |
| 17-106_k141_41906 | Not-determined |
| 17-106_k141_122673 | Not-determined |
| 17-106_k141_90886 | Not-determined |
| 17-106_k141_122677 | Not-determined |
| 17-106_k141_53368 | Not-determined |
| 17-106_k141_20939 | Not-determined |
| 17-106_k141_36948 | Not-determined |
| 17-106_k141_73863 | Not-determined |
| 17-106_k141_41927 | Low-quality |
| 17-106_k141_10295 | Not-determined |
| 17-106_k141_95120 | Not-determined |
| 17-106_k141_36954 | Not-determined |
| 17-106_k141_122693 | Not-determined |
| 17-106_k141_58380 | Not-determined |
| 17-106_k141_41937 | Not-determined |
| 17-106_k141_100991 | Not-determined |
| 17-106_k141_95131 | Not-determined |
| 17-106_k141_41947 | Not-determined |
| 17-106_k141_20945 | Not-determined |
| 17-106_k141_41955 | Not-determined |
| 17-106_k141_85471 | Not-determined |
| 17-106_k141_101006 | Not-determined |
| 17-106_k141_101009 | Not-determined |
| 17-106_k141_26385 | Not-determined |
| 17-106_k141_69215 | Not-determined |
| 17-106_k141_53384 | Low-quality |
| 17-106_k141_105831 | Not-determined |
| 17-106_k141_53386 | Low-quality |
| 17-106_k141_90905 | Not-determined |
| 17-106_k141_36986 | Not-determined |
| 17-106_k141_26392 | Not-determined |
| 17-106_k141_58405 | Not-determined |
| 17-106_k141_58406 | Not-determined |
| 17-106_k141_20957 | Not-determined |
| 17-106_k141_69224 | Not-determined |
| 17-106_k141_85489 | Low-quality |
| 17-106_k141_122702 | Not-determined |
| 17-106_k141_53398 | Not-determined |
| 17-106_k141_5083 | Not-determined |
| 17-106_k141_53399 | Low-quality |
| 17-106_k141_105840 | Low-quality |
| 17-106_k141_101028 | Not-determined |
| 17-106_k141_105842 | Not-determined |
| 17-106_k141_53406 | Not-determined |
| 17-106_k141_105843 | Not-determined |
| 17-106_k141_53409 | Not-determined |
| 17-106_k141_10313 | Not-determined |
| 17-106_k141_5092 | Not-determined |
| 17-106_k141_10314 | Not-determined |
| 17-106_k141_15499 | Not-determined |
| 17-106_k141_69254 | Not-determined |
| 17-106_k141_122723 | Not-determined |
| 17-106_k141_41969 | Not-determined |
| 17-106_k141_53418 | Not-determined |
| 17-106_k141_69260 | Not-determined |
| 17-106_k141_122729 | Low-quality |
| 17-106_k141_10326 | Not-determined |
| 17-106_k141_105865 | Not-determined |
| 17-106_k141_15514 | Not-determined |
| 17-106_k141_10338 | Not-determined |
| 17-106_k141_122739 | Not-determined |
| 17-106_k141_20991 | Not-determined |
| 17-106_k141_20996 | Not-determined |
| 17-106_k141_85520 | Not-determined |
| 17-106_k141_36996 | Not-determined |
| 17-106_k141_53434 | Not-determined |
| 17-106_k141_21002 | Not-determined |
| 17-106_k141_79880 | Not-determined |
| 17-106_k141_73874 | Not-determined |
| 17-106_k141_31062 | Not-determined |
| 17-106_k141_101055 | Not-determined |
| 17-106_k141_21017 | Not-determined |
| 17-106_k141_37017 | Not-determined |
| 17-106_k141_15545 | Not-determined |
| 17-106_k141_101064 | Not-determined |
| 17-106_k141_85553 | Low-quality |
| 17-106_k141_79893 | Not-determined |
| 17-106_k141_101066 | Not-determined |
| 17-106_k141_73888 | Low-quality |
| 17-106_k141_37024 | Not-determined |
| 17-106_k141_37025 | Not-determined |
| 17-106_k141_15555 | Not-determined |
| 17-106_k141_95175 | Not-determined |
| 17-106_k141_53441 | Not-determined |
| 17-106_k141_110715 | Not-determined |
| 17-106_k141_101079 | Not-determined |
| 17-106_k141_15574 | Low-quality |
| 17-106_k141_79901 | Not-determined |
| 17-106_k141_79903 | Not-determined |
| 17-106_k141_101083 | Not-determined |
| 17-106_k141_53447 | Not-determined |
| 17-106_k141_53450 | Not-determined |
| 17-106_k141_79915 | Not-determined |
| 17-106_k141_53455 | Not-determined |
| 17-106_k141_41985 | Not-determined |
| 17-106_k141_41991 | Not-determined |
| 17-106_k141_37048 | Not-determined |
| 17-106_k141_101088 | Not-determined |
| 17-106_k141_37049 | Not-determined |
| 17-106_k141_31118 | Not-determined |
| 17-106_k141_79934 | Not-determined |
| 17-106_k141_122743 | Not-determined |
| 17-106_k141_122744 | Not-determined |
| 17-106_k141_31129 | Not-determined |
| 17-106_k141_31130 | Not-determined |
| 17-106_k141_73914 | Low-quality |
| 17-106_k141_105910 | Low-quality |
| 17-106_k141_26415 | Not-determined |
| 17-106_k141_105913 | Not-determined |
| 17-106_k141_21049 | Not-determined |
| 17-106_k141_31136 | Not-determined |
| 17-106_k141_73922 | Not-determined |
| 17-106_k141_105922 | Not-determined |
| 17-106_k141_58465 | Low-quality |
| 17-106_k141_58468 | Low-quality |
| 17-106_k141_46931 | Not-determined |
| 17-106_k141_15602 | Not-determined |
| 17-106_k141_122748 | Not-determined |
| 17-106_k141_26428 | Not-determined |
| 17-106_k141_105933 | Not-determined |
| 17-106_k141_122757 | Not-determined |
| 17-106_k141_73937 | Not-determined |
| 17-106_k141_73939 | Not-determined |
| 17-106_k141_58480 | Not-determined |
| 17-106_k141_101121 | Not-determined |
| 17-106_k141_58484 | Not-determined |
| 17-106_k141_26445 | Not-determined |
| 17-106_k141_21075 | Not-determined |
| 17-106_k141_69296 | Not-determined |
| 17-106_k141_21082 | Not-determined |
| 17-106_k141_53470 | Not-determined |
| 17-106_k141_69304 | Not-determined |
| 17-106_k141_69309 | Not-determined |
| 17-106_k141_69310 | Not-determined |
| 17-106_k141_95212 | Not-determined |
| 17-106_k141_101137 | Not-determined |
| 17-106_k141_26463 | Not-determined |
| 17-106_k141_101142 | Low-quality |
| 17-106_k141_37086 | Not-determined |
| 17-106_k141_26469 | Not-determined |
| 17-106_k141_26474 | Not-determined |
| 17-106_k141_101151 | Not-determined |
| 17-106_k141_58507 | Not-determined |
| 17-106_k141_101160 | Not-determined |
| 17-106_k141_46951 | Not-determined |
| 17-106_k141_101168 | Low-quality |
| 17-106_k141_79953 | Not-determined |
| 17-106_k141_46968 | Low-quality |
| 17-106_k141_79959 | Not-determined |
| 17-106_k141_101185 | Not-determined |
| 17-106_k141_15644 | Not-determined |
| 17-106_k141_10407 | Not-determined |
| 17-106_k141_26506 | Not-determined |
| 17-106_k141_95241 | Not-determined |
| 17-106_k141_122779 | Not-determined |
| 17-106_k141_10410 | Not-determined |
| 17-106_k141_105964 | Not-determined |
| 17-106_k141_26509 | Low-quality |
| 17-106_k141_73970 | Not-determined |
| 17-106_k141_37121 | Not-determined |
| 17-106_k141_15652 | Not-determined |
| 17-106_k141_79970 | Not-determined |
| 17-106_k141_122789 | Not-determined |
| 17-106_k141_69336 | Not-determined |
| 17-106_k141_101194 | Not-determined |
| 17-106_k141_79980 | Not-determined |
| 17-106_k141_122806 | Not-determined |
| 17-106_k141_42021 | Not-determined |
| 17-106_k141_101197 | Not-determined |
| 17-106_k141_122809 | Not-determined |
| 17-106_k141_105985 | Not-determined |
| 17-106_k141_105989 | Not-determined |
| 17-106_k141_47001 | Not-determined |
| 17-106_k141_105994 | Not-determined |
| 17-106_k141_15680 | Not-determined |
| 17-106_k141_10431 | Not-determined |
| 17-106_k141_106001 | Not-determined |
| 17-106_k141_74000 | Not-determined |
| 17-106_k141_95255 | Not-determined |
| 17-106_k141_47010 | Low-quality |
| 17-106_k141_37153 | Not-determined |
| 17-106_k141_106007 | Not-determined |
| 17-106_k141_26556 | Not-determined |
| 17-106_k141_127068 | Not-determined |
| 17-106_k141_47012 | Not-determined |
| 17-106_k141_21109 | Not-determined |
| 17-106_k141_21113 | Not-determined |
| 17-106_k141_122829 | Not-determined |
| 17-106_k141_15688 | Not-determined |
| 17-106_k141_101227 | Not-determined |
| 17-106_k141_37160 | Low-quality |
| 17-106_k141_21119 | Not-determined |
| 17-106_k141_37162 | Not-determined |
| 17-106_k141_42024 | Not-determined |
| 17-106_k141_42029 | Not-determined |
| 17-106_k141_21126 | Low-quality |
| 17-106_k141_15704 | Not-determined |
| 17-106_k141_21133 | Not-determined |
| 17-106_k141_15710 | Not-determined |
| 17-106_k141_21138 | Not-determined |
| 17-106_k141_21139 | Low-quality |
| 17-106_k141_31143 | Not-determined |
| 17-106_k141_95276 | Not-determined |
| 17-106_k141_106039 | Not-determined |
| 17-106_k141_47058 | Not-determined |
| 17-106_k141_26608 | Not-determined |
| 17-106_k141_21155 | Not-determined |
| 17-106_k141_74030 | Not-determined |
| 17-106_k141_15718 | Not-determined |
| 17-106_k141_106044 | Low-quality |
| 17-106_k141_127101 | Not-determined |
| 17-106_k141_26614 | Not-determined |
| 17-106_k141_95293 | Not-determined |
| 17-106_k141_58568 | Low-quality |
| 17-106_k141_21161 | Not-determined |
| 17-106_k141_127108 | Not-determined |
| 17-106_k141_42059 | Not-determined |

| | |
|---|---|
| 17-106_k141_26621 | Not-determined |
| 17-106_k141_127114 | Not-determined |
| 17-106_k141_106062 | Low-quality |
| 17-106_k141_122844 | Not-determined |
| 17-106_k141_47070 | Not-determined |
| 17-106_k141_106066 | Not-determined |
| 17-106_k141_74044 | Not-determined |
| 17-106_k141_74047 | Not-determined |
| 17-106_k141_122852 | Not-determined |
| 17-106_k141_15737 | Not-determined |
| 17-106_k141_106067 | Not-determined |
| 17-106_k141_21177 | Low-quality |
| 17-106_k141_15743 | Not-determined |
| 17-106_k141_95322 | Not-determined |
| 17-106_k141_69427 | Not-determined |
| 17-106_k141_15745 | Not-determined |
| 17-106_k141_31178 | Low-quality |
| 17-106_k141_69430 | Not-determined |
| 17-106_k141_10465 | Not-determined |
| 17-106_k141_10473 | Not-determined |
| 18-109-final-viral-combined_k141_157260\|\|full | Medium-quality |
| 17-106_k141_122859 | Not-determined |
| 17-106_k141_116864 | Not-determined |
| 17-106_k141_69445 | Low-quality |
| 17-106_k141_15770 | Not-determined |
| 17-106_k141_31196 | Not-determined |
| 17-106_k141_21201 | Not-determined |
| 17-106_k141_58594 | Not-determined |
| 17-106_k141_58596 | Not-determined |
| 17-106_k141_37181 | Not-determined |
| 17-106_k141_80008 | Not-determined |
| 17-106_k141_74065 | Not-determined |
| 17-106_k141_37187 | Low-quality |
| 17-106_k141_101257 | Not-determined |
| 17-106_k141_21221 | Not-determined |
| 17-106_k141_80018 | Not-determined |
| 17-106_k141_80019 | Low-quality |
| 17-106_k141_15783 | Not-determined |
| 17-106_k141_42121 | Not-determined |
| 17-106_k141_74077 | Low-quality |
| 17-106_k141_95367 | Not-determined |
| 17-106_k141_69458 | Not-determined |
| 17-106_k141_37197 | Not-determined |
| 17-106_k141_74080 | Low-quality |
| 17-106_k141_74081 | Low-quality |
| 17-106_k141_69463 | Not-determined |
| 17-106_k141_26665 | Not-determined |
| 17-106_k141_69465 | Not-determined |
| 17-106_k141_74087 | Not-determined |
| 17-106_k141_101280 | Not-determined |
| 17-106_k141_74088 | Not-determined |
| 17-106_k141_101281 | Low-quality |
| 17-106_k141_15798 | Not-determined |
| 17-106_k141_95380 | Not-determined |
| 17-106_k141_37213 | Not-determined |
| 17-106_k141_74095 | Not-determined |
| 17-106_k141_95384 | Not-determined |
| 17-106_k141_74096 | Not-determined |
| 17-106_k141_26668 | Not-determined |
| 17-106_k141_80034 | Not-determined |
| 17-106_k141_21248 | Not-determined |
| 17-106_k141_101296 | Not-determined |
| 17-106_k141_21250 | Not-determined |
| 17-106_k141_31221 | Not-determined |
| 17-106_k141_101299 | Not-determined |
| 17-106_k141_15813 | Not-determined |
| 17-106_k141_10496 | Low-quality |
| 17-106_k141_122891 | Not-determined |
| 17-106_k141_10505 | Not-determined |
| 17-106_k141_31232 | Not-determined |
| 17-106_k141_101310 | Not-determined |
| 17-106_k141_31236 | Not-determined |
| 17-106_k141_10512 | Not-determined |
| 17-106_k141_31243 | Not-determined |
| 17-106_k141_122906 | Low-quality |
| 17-106_k141_31251 | Not-determined |
| 17-106_k141_127132 | Not-determined |
| 17-106_k141_37252 | Low-quality |
| 17-106_k141_122919 | Not-determined |
| 17-106_k141_101335 | Not-determined |
| 17-106_k141_127140 | Not-determined |
| 17-106_k141_31264 | Not-determined |
| 17-106_k141_101344 | Not-determined |
| 17-106_k141_21267 | Not-determined |
| 17-106_k141_21270 | Not-determined |
| 17-106_k141_15843 | Not-determined |
| 17-106_k141_101362 | Not-determined |
| 17-106_k141_31278 | Not-determined |
| 17-106_k141_10543 | Not-determined |
| 17-106_k141_42156 | Not-determined |
| 17-106_k141_15849 | Not-determined |
| 17-106_k141_58653 | Not-determined |
| 17-106_k141_58655 | Not-determined |
| 17-106_k141_31293 | Not-determined |
| 17-106_k141_80080 | Not-determined |
| 17-106_k141_42168 | Not-determined |
| 17-106_k141_10556 | Not-determined |
| 17-106_k141_101371 | Not-determined |
| 17-106_k141_10563 | Not-determined |
| 17-106_k141_15880 | Not-determined |
| 17-106_k141_26692 | Not-determined |
| 17-106_k141_58676 | Not-determined |
| 17-106_k141_101377 | Not-determined |
| 17-106_k141_31314 | Not-determined |
| 17-106_k141_122955 | Not-determined |
| 17-106_k141_10577 | Not-determined |
| 17-106_k141_42187 | Not-determined |
| 17-106_k141_10583 | Not-determined |
| 17-106_k141_116910 | Not-determined |
| 17-106_k141_31338 | Not-determined |
| 17-106_k141_95419 | Not-determined |
| 17-106_k141_95421 | Not-determined |
| 17-106_k141_80116 | Not-determined |
| 17-106_k141_42205 | Not-determined |
| 17-106_k141_10599 | Not-determined |
| 17-106_k141_101413 | Not-determined |
| 17-106_k141_10606 | Low-quality |
| 17-106_k141_31358 | Not-determined |
| 17-106_k141_116946 | Not-determined |
| 17-106_k141_116949 | Not-determined |
| 17-106_k141_37339 | Not-determined |
| 17-106_k141_37341 | Not-determined |
| 17-106_k141_47099 | Low-quality |
| 17-106_k141_101425 | Low-quality |
| 17-106_k141_116968 | Not-determined |
| 17-106_k141_31387 | Not-determined |
| 17-106_k141_47105 | Not-determined |
| 17-106_k141_127145 | Not-determined |
| 17-106_k141_47112 | Not-determined |
| 17-106_k141_58754 | Not-determined |
| 17-106_k141_21356 | Not-determined |
| 17-106_k141_127153 | Not-determined |
| 17-106_k141_31398 | Not-determined |
| 17-106_k141_95446 | Low-quality |
| 17-106_k141_74239 | Low-quality |
| 17-106_k141_58763 | Not-determined |
| 17-106_k141_74242 | Not-determined |
| 17-106_k141_31404 | Not-determined |
| 17-106_k141_31405 | Not-determined |
| 17-106_k141_80166 | Not-determined |
| 17-106_k141_74248 | Not-determined |
| 17-106_k141_106125 | Not-determined |
| 17-106_k141_116986 | Not-determined |
| 17-106_k141_37398 | Not-determined |
| 17-106_k141_15968 | Not-determined |
| 17-106_k141_80173 | Not-determined |
| 17-106_k141_42252 | Not-determined |
| 17-106_k141_74270 | Not-determined |
| 17-106_k141_10630 | Not-determined |
| 17-106_k141_106134 | Not-determined |
| 17-106_k141_106136 | Low-quality |
| 17-106_k141_10635 | Not-determined |
| 17-106_k141_116997 | Not-determined |
| 17-106_k141_37420 | Not-determined |
| 17-106_k141_116999 | Low-quality |
| 17-106_k141_127174 | Not-determined |
| 17-106_k141_5138 | Not-determined |
| 17-106_k141_127181 | Not-determined |
| 17-106_k141_117005 | Not-determined |
| 17-106_k141_15993 | Not-determined |
| 17-106_k141_58797 | Not-determined |
| 17-106_k141_127186 | Not-determined |
| 17-106_k141_42283 | Not-determined |
| 17-106_k141_95484 | Not-determined |
| 17-106_k141_101444 | Not-determined |
| 17-106_k141_117023 | Not-determined |
| 17-106_k141_58810 | Not-determined |
| 17-106_k141_31428 | Not-determined |
| 17-106_k141_117027 | Not-determined |
| 17-106_k141_117028 | Not-determined |
| 17-106_k141_95490 | Not-determined |
| 17-106_k141_42301 | Not-determined |
| 17-106_k141_42304 | Not-determined |
| 17-106_k141_127202 | Not-determined |
| 17-106_k141_31434 | Not-determined |
| 17-106_k141_117039 | Not-determined |
| 17-106_k141_31436 | Not-determined |
| 17-106_k141_95509 | Not-determined |
| 17-106_k141_42323 | Not-determined |
| 17-106_k141_5151 | Not-determined |
| 17-106_k141_95515 | Not-determined |
| 17-106_k141_117053 | Not-determined |
| 17-106_k141_95532 | Not-determined |
| 17-106_k141_74303 | Not-determined |
| 17-106_k141_5172 | Not-determined |
| 17-106_k141_127219 | Not-determined |
| 17-106_k141_95543 | Not-determined |
| 17-106_k141_74310 | Not-determined |
| 17-106_k141_5185 | Not-determined |
| 17-106_k141_101465 | Not-determined |
| 17-106_k141_47149 | Not-determined |
| 17-106_k141_101466 | Not-determined |
| 17-106_k141_127222 | Low-quality |
| 17-106_k141_127225 | Not-determined |
| 17-106_k141_47164 | Not-determined |
| 17-106_k141_5208 | Not-determined |
| 17-106_k141_10677 | Not-determined |
| 17-106_k141_127258 | Not-determined |
| 17-106_k141_5228 | Not-determined |
| 17-106_k141_10686 | Not-determined |
| 17-106_k141_42333 | Not-determined |
| 17-106_k141_117117 | Not-determined |
| 17-106_k141_101511 | Low-quality |
| 17-106_k141_101512 | Not-determined |
| 17-106_k141_117125 | Low-quality |
| 17-106_k141_5241 | Not-determined |
| 17-106_k141_101517 | Not-determined |
| 17-106_k141_74318 | Not-determined |
| 17-106_k141_74320 | Not-determined |
| 17-106_k141_74329 | Low-quality |
| 17-106_k141_101532 | Not-determined |
| 17-106_k141_74336 | Not-determined |
| 17-106_k141_117139 | Low-quality |
| 17-106_k141_31477 | Not-determined |
| 17-106_k141_47178 | Not-determined |
| 17-106_k141_106165 | Not-determined |
| 17-106_k141_95597 | Not-determined |
| 17-106_k141_42354 | Not-determined |
| 17-106_k141_5274 | Not-determined |
| 17-106_k141_5280 | Not-determined |
| 17-106_k141_16031 | Not-determined |
| 17-106_k141_16032 | Not-determined |
| 17-106_k141_106195 | Not-determined |
| 17-106_k141_127298 | Not-determined |
| 17-106_k141_106200 | Not-determined |
| 17-106_k141_16036 | Low-quality |
| 17-106_k141_47196 | Not-determined |
| 17-106_k141_5292 | Not-determined |
| 17-106_k141_101559 | Not-determined |
| 17-106_k141_74383 | Not-determined |
| 17-106_k141_101565 | Not-determined |
| 17-106_k141_95621 | Low-quality |
| 17-106_k141_31504 | Not-determined |
| 17-106_k141_74391 | Low-quality |
| 17-106_k141_5307 | Not-determined |
| 17-106_k141_106227 | Not-determined |
| 17-106_k141_106229 | Not-determined |
| 17-106_k141_106232 | Not-determined |
| 17-106_k141_31515 | Not-determined |
| 17-106_k141_106235 | Not-determined |
| 17-106_k141_31522 | Not-determined |
| 17-106_k141_47212 | Not-determined |
| 17-106_k141_117180 | Not-determined |
| 17-106_k141_5328 | Not-determined |
| 17-106_k141_106243 | Not-determined |
| 17-106_k141_5332 | Not-determined |
| 17-106_k141_5333 | Not-determined |
| 17-106_k141_101583 | Not-determined |
| 17-106_k141_95649 | Not-determined |
| 17-106_k141_95650 | Not-determined |
| 17-106_k141_117190 | Not-determined |
| 17-106_k141_95653 | Low-quality |
| 17-106_k141_110743 | Not-determined |
| 17-106_k141_106263 | Not-determined |
| 17-106_k141_106265 | Not-determined |
| 17-106_k141_74423 | Low-quality |
| 17-106_k141_31545 | Low-quality |
| 17-106_k141_42420 | Not-determined |
| 17-106_k141_42421 | Not-determined |
| 17-106_k141_42426 | Not-determined |
| 17-106_k141_42428 | Low-quality |
| 17-106_k141_31551 | Low-quality |
| 17-106_k141_101599 | Not-determined |
| 17-106_k141_101604 | Not-determined |
| 17-106_k141_31564 | Low-quality |
| 17-106_k141_106284 | Not-determined |
| 17-106_k141_106293 | Not-determined |
| 17-106_k141_110753 | Not-determined |
| 17-106_k141_95668 | Not-determined |
| 17-106_k141_110755 | Not-determined |
| 17-106_k141_31574 | Not-determined |
| 17-106_k141_74431 | Low-quality |
| 17-106_k141_74438 | Not-determined |
| 17-106_k141_95684 | Not-determined |
| 17-106_k141_110786 | Not-determined |
| 17-106_k141_74457 | Not-determined |
| 17-106_k141_47243 | Not-determined |
| 17-106_k141_47244 | Not-determined |
| 17-106_k141_42437 | Not-determined |
| 17-106_k141_47249 | Not-determined |
| 17-106_k141_47253 | Not-determined |
| 17-106_k141_106311 | Not-determined |
| 17-106_k141_106315 | Not-determined |
| 17-106_k141_127364 | Low-quality |
| 17-106_k141_47258 | Low-quality |
| 17-106_k141_31630 | Not-determined |
| 17-106_k141_106323 | Not-determined |
| 17-106_k141_31635 | Not-determined |
| 17-106_k141_95725 | Low-quality |
| 17-106_k141_95726 | Not-determined |
| 17-106_k141_31652 | Not-determined |
| 17-106_k141_42452 | Not-determined |
| 17-106_k141_31654 | Not-determined |
| 17-106_k141_31658 | Not-determined |
| 17-106_k141_110802 | Not-determined |
| 17-106_k141_31670 | Not-determined |
| 17-106_k141_31679 | Not-determined |
| 17-106_k141_106331 | Not-determined |
| 17-106_k141_106334 | Not-determined |
| 17-106_k141_47281 | Not-determined |
| 17-106_k141_127388 | Not-determined |
| 17-106_k141_106344 | Not-determined |
| 17-106_k141_42491 | Not-determined |
| 17-106_k141_42493 | Not-determined |
| 17-106_k141_110835 | Not-determined |
| 17-106_k141_31695 | Not-determined |
| 17-106_k141_106361 | Not-determined |
| 17-106_k141_95741 | Not-determined |
| 17-106_k141_74493 | Not-determined |
| 17-106_k141_31703 | Not-determined |
| 17-106_k141_31715 | Not-determined |
| 17-106_k141_95751 | Not-determined |
| 17-106_k141_95756 | Not-determined |
| 17-106_k141_106382 | Not-determined |
| 17-106_k141_127404 | Not-determined |
| 17-106_k141_42527 | Not-determined |
| 17-106_k141_106402 | Not-determined |
| 17-106_k141_127414 | Low-quality |
| 17-106_k141_95781 | Not-determined |
| 17-106_k141_95787 | Not-determined |
| 17-106_k141_117264 | Low-quality |
| 17-106_k141_110885 | Not-determined |
| 17-106_k141_47331 | Not-determined |
| 17-106_k141_117275 | Not-determined |
| 17-106_k141_106418 | Not-determined |
| 17-106_k141_42571 | Not-determined |
| 17-106_k141_47343 | Low-quality |
| 17-106_k141_42572 | Low-quality |
| 17-106_k141_42575 | Low-quality |
| 17-106_k141_106437 | Not-determined |
| 17-106_k141_47355 | Not-determined |
| 17-106_k141_106456 | Not-determined |
| 17-106_k141_106461 | Not-determined |
| 17-106_k141_106464 | Not-determined |
| 17-106_k141_106471 | Not-determined |
| 17-106_k141_31739 | Not-determined |
| 17-106_k141_47396 | Not-determined |
| 17-106_k141_95827 | Not-determined |
| 17-106_k141_47399 | Low-quality |
| 17-106_k141_47400 | Not-determined |
| 17-106_k141_74556 | Not-determined |
| 17-106_k141_117335 | Not-determined |
| 17-106_k141_74563 | Not-determined |
| 17-106_k141_106492 | Not-determined |
| 17-106_k141_74583 | Not-determined |
| 17-106_k141_127425 | Not-determined |
| 17-106_k141_117369 | Not-determined |
| 17-106_k141_95846 | Not-determined |
| 17-106_k141_47421 | Not-determined |
| 17-106_k141_95849 | Not-determined |
| 17-106_k141_117386 | Not-determined |
| 17-106_k141_47438 | Not-determined |
| 17-106_k141_117390 | Not-determined |
| 17-106_k141_127437 | Not-determined |
| 17-106_k141_117396 | Not-determined |
| 17-106_k141_74613 | Not-determined |
| 17-106_k141_117403 | Not-determined |
| 17-106_k141_95867 | Low-quality |
| 17-106_k141_95878 | Not-determined |
| 17-106_k141_117425 | Not-determined |
| 17-106_k141_127461 | Not-determined |
| 17-106_k141_117439 | Not-determined |
| 17-106_k141_106520 | Not-determined |
| 17-106_k141_95916 | Not-determined |
| 17-106_k141_95920 | Not-determined |
| 17-106_k141_117465 | Not-determined |
| 17-106_k141_47462 | Not-determined |
| 17-106_k141_95936 | Not-determined |
| 17-106_k141_117476 | Not-determined |
| 17-106_k141_117477 | Not-determined |
| 17-106_k141_117478 | Not-determined |
| 17-106_k141_117480 | Not-determined |
| 17-106_k141_95948 | Not-determined |
| 17-106_k141_95949 | Not-determined |
| 17-106_k141_95956 | Not-determined |
| 17-106_k141_106559 | Not-determined |
| 17-106_k141_95970 | Not-determined |
| 17-106_k141_106565 | Not-determined |
| 17-106_k141_47496 | Not-determined |
| 17-106_k141_95978 | Not-determined |
| 17-106_k141_95989 | Not-determined |
| 17-106_k141_106576 | Low-quality |
| 17-106_k141_117509 | Not-determined |
| 17-106_k141_74662 | Not-determined |
| 17-106_k141_117520 | Not-determined |
| 17-106_k141_106588 | Not-determined |
| 17-106_k141_42611 | Not-determined |
| 17-106_k141_42612 | Not-determined |
| 17-106_k141_127514 | Not-determined |
| 17-106_k141_31746 | Not-determined |
| 17-106_k141_117529 | Not-determined |
| 17-106_k141_110981 | Not-determined |
| 17-106_k141_42638 | Not-determined |
| 17-106_k141_74696 | Not-determined |
| 17-106_k141_42648 | Not-determined |
| 17-106_k141_47504 | Not-determined |
| 17-106_k141_110997 | Not-determined |
| 17-106_k141_74703 | Not-determined |
| 17-106_k141_106629 | Not-determined |
| 17-106_k141_127532 | Not-determined |
| 17-106_k141_127533 | Not-determined |
| 17-106_k141_127536 | Not-determined |
| 17-106_k141_42656 | Not-determined |
| 17-106_k141_96065 | Not-determined |
| 17-106_k141_74725 | Not-determined |
| 17-106_k141_74726 | Not-determined |
| 17-106_k141_127548 | Not-determined |
| 17-106_k141_42662 | Low-quality |
| 17-106_k141_111040 | Not-determined |
| 17-106_k141_42663 | Not-determined |
| 17-106_k141_74743 | Low-quality |
| 17-106_k141_47550 | Not-determined |
| 17-106_k141_111053 | Not-determined |
| 17-106_k141_127559 | Not-determined |
| 17-106_k141_47563 | Not-determined |
| 17-106_k141_42679 | Not-determined |
| 17-106_k141_127565 | Not-determined |
| 17-106_k141_117556 | Not-determined |
| 17-106_k141_74753 | Not-determined |
| 17-106_k141_117563 | Not-determined |
| 17-106_k141_96071 | Not-determined |
| 17-106_k141_117575 | Not-determined |
| 17-106_k141_117586 | Not-determined |
| 17-106_k141_31754 | Low-quality |
| 17-106_k141_117607 | Not-determined |
| 17-106_k141_117611 | Not-determined |
| 17-106_k141_31786 | Low-quality |
| 17-106_k141_96110 | Low-quality |
| 17-106_k141_31794 | Not-determined |
| 17-106_k141_42706 | Not-determined |
| 17-106_k141_117636 | Not-determined |
| 17-106_k141_96119 | Not-determined |
| 17-106_k141_42707 | Not-determined |
| 17-106_k141_127576 | Low-quality |
| 17-106_k141_127585 | Not-determined |
| 17-106_k141_127587 | Not-determined |
| 17-106_k141_127588 | Not-determined |
| 17-106_k141_31812 | Not-determined |
| 17-106_k141_127601 | Low-quality |
| 17-106_k141_96150 | Not-determined |
| 17-106_k141_47599 | Not-determined |
| 17-106_k141_106683 | Not-determined |
| 17-106_k141_127614 | Not-determined |
| 17-106_k141_47603 | Not-determined |
| 17-106_k141_127625 | Not-determined |
| 17-106_k141_111093 | Not-determined |
| 17-106_k141_127637 | Not-determined |
| 17-106_k141_42725 | Low-quality |
| 17-106_k141_106706 | Not-determined |
| 17-106_k141_106707 | Not-determined |
| 17-106_k141_31829 | Not-determined |
| 17-106_k141_47638 | Not-determined |
| 17-106_k141_47642 | Not-determined |
| 17-106_k141_47645 | Not-determined |
| 17-106_k141_127652 | Not-determined |
| 17-106_k141_127654 | Not-determined |
| 17-106_k141_31832 | Not-determined |
| 17-106_k141_127665 | Low-quality |
| 17-106_k141_42748 | Not-determined |
| 17-106_k141_96204 | Not-determined |
| 17-106_k141_47656 | Low-quality |
| 17-106_k141_31844 | Not-determined |
| 17-106_k141_96224 | Not-determined |
| 17-106_k141_96229 | Not-determined |
| 17-106_k141_96234 | Not-determined |
| 17-106_k141_96236 | Not-determined |
| 17-106_k141_96237 | Not-determined |
| 17-106_k141_47666 | Not-determined |
| 17-106_k141_47667 | Not-determined |
| 17-106_k141_127683 | Not-determined |
| 17-106_k141_47671 | Not-determined |
| 17-106_k141_106728 | Not-determined |
| 17-106_k141_47678 | Not-determined |
| 17-106_k141_127694 | Not-determined |
| 17-106_k141_47686 | Not-determined |
| 17-106_k141_31864 | Not-determined |
| 17-106_k141_31871 | Not-determined |
| 17-106_k141_127701 | Not-determined |
| 17-106_k141_47703 | Not-determined |
| 17-106_k141_31887 | Not-determined |
| 17-106_k141_31890 | Not-determined |
| 17-106_k141_127709 | Not-determined |
| 17-106_k141_106734 | Not-determined |
| 17-106_k141_31898 | Not-determined |
| 17-106_k141_106742 | Not-determined |
| 17-106_k141_127712 | Low-quality |
| 17-106_k141_106748 | Not-determined |
| 17-106_k141_127739 | Not-determined |
| 17-106_k141_106778 | Not-determined |
| 17-106_k141_106779 | Not-determined |
| 17-106_k141_31912 | Not-determined |
| 17-106_k141_31935 | Not-determined |
| 17-106_k141_31938 | Not-determined |
| 17-106_k141_127762 | Not-determined |
| 17-106_k141_47710 | Not-determined |
| 17-106_k141_111150 | Low-quality |
| 17-106_k141_31989 | Not-determined |
| 17-106_k141_127788 | Low-quality |
| 17-106_k141_127789 | Not-determined |
| 17-106_k141_127792 | Not-determined |
| 17-106_k141_32004 | Not-determined |
| 17-106_k141_32005 | Not-determined |
| 17-106_k141_74787 | Not-determined |
| 17-106_k141_106863 | Not-determined |
| 17-106_k141_127806 | Not-determined |
| 17-106_k141_106870 | Not-determined |
| 17-106_k141_106871 | Not-determined |
| 17-106_k141_32018 | Not-determined |
| 17-106_k141_74798 | Not-determined |
| 17-106_k141_32021 | Not-determined |
| 17-106_k141_74809 | Not-determined |
| 17-106_k141_32026 | Not-determined |
| 17-106_k141_127828 | Not-determined |
| 17-106_k141_32029 | Not-determined |
| 17-106_k141_32031 | Not-determined |
| 17-106_k141_106887 | Not-determined |
| 17-106_k141_106890 | Not-determined |
| 17-106_k141_106898 | Not-determined |
| 17-106_k141_111178 | Not-determined |
| 17-106_k141_111192 | Not-determined |
| 17-106_k141_47723 | Not-determined |
| 17-106_k141_47727 | Not-determined |
| 17-106_k141_74854 | Not-determined |
| 17-106_k141_47737 | Low-quality |
| 17-106_k141_74867 | Not-determined |
| 17-106_k141_47745 | Not-determined |
| 17-106_k141_47749 | Not-determined |
| 17-106_k141_32037 | Not-determined |
| 17-106_k141_32038 | Not-determined |
| 17-106_k141_32045 | Not-determined |
| 17-106_k141_127892 | Not-determined |
| 17-106_k141_47775 | Low-quality |
| 17-106_k141_32058 | Not-determined |
| 17-106_k141_111207 | Not-determined |
| 17-106_k141_127904 | Low-quality |
| 17-106_k141_32064 | Not-determined |
| 17-106_k141_47827 | Not-determined |
| 17-106_k141_47830 | Not-determined |
| 17-106_k141_47831 | Not-determined |
| 17-106_k141_47838 | Not-determined |
| 17-106_k141_127918 | Not-determined |
| 17-106_k141_111226 | Not-determined |
| 17-106_k141_111227 | Not-determined |
| 17-106_k141_127938 | Not-determined |
| 17-106_k141_111237 | Not-determined |
| 17-106_k141_47863 | Not-determined |
| 17-106_k141_47867 | Not-determined |
| 17-106_k141_111311 | Not-determined |
| 17-106_k141_47915 | Low-quality |
| 17-106_k141_47920 | Not-determined |
| 17-106_k141_47921 | Not-determined |
| 17-106_k141_47926 | Not-determined |
| 17-106_k141_127999 | Not-determined |
| 17-106_k141_47937 | Not-determined |
| 17-106_k141_47956 | Not-determined |
| 17-106_k141_47958 | Not-determined |
| 17-106_k141_111413 | Not-determined |
| 17-106_k141_111449 | Low-quality |
| 17-106_k141_111454 | Not-determined |
| 17-106_k141_47995 | Not-determined |
| 17-106_k141_111465 | Not-determined |
| 17-106_k141_111466 | Not-determined |
| 17-106_k141_111468 | Not-determined |
| 17-106_k141_111471 | Low-quality |
| 17-106_k141_111474 | Not-determined |
| 17-106_k141_48009 | Not-determined |
| 17-106_k141_48012 | Not-determined |
| 17-106_k141_48013 | Not-determined |
| 17-106_k141_111485 | Not-determined |
| 17-106_k141_111488 | Not-determined |
| 17-106_k141_128041 | Not-determined |
| 17-106_k141_128051 | Not-determined |
| 17-106_k141_48037 | Not-determined |
| 17-106_k141_128060 | Not-determined |
| 17-106_k141_111534 | Not-determined |
| 17-106_k141_111537 | Not-determined |
| 17-106_k141_128086 | Not-determined |
| 17-106_k141_48067 | Not-determined |
| 17-106_k141_128121 | Not-determined |
| 17-106_k141_128122 | Not-determined |
| 17-106_k141_111575 | Not-determined |
| 17-106_k141_128134 | Not-determined |
| 17-106_k141_128135 | Not-determined |
| 17-106_k141_111582 | Not-determined |
| 17-106_k141_111591 | Not-determined |
| 17-106_k141_111592 | Low-quality |
| 17-106_k141_48091 | Not-determined |
| 17-106_k141_48112 | Not-determined |
| 17-106_k141_128149 | Not-determined |

| | |
|---|---|
| 17-106_k141_128151 | Not-determined |
| 17-106_k141_111605 | Not-determined |
| 17-106_k141_111610 | Not-determined |
| 17-106_k141_128177 | Not-determined |
| 17-106_k141_111622 | Not-determined |
| 17-106_k141_111634 | Not-determined |
| 17-106_k141_128181 | Not-determined |
| 17-106_k141_111642 | Not-determined |
| 17-106_k141_128189 | Not-determined |
| 17-106_k141_128193 | Low-quality |
| 17-106_k141_128198 | Not-determined |
| 17-106_k141_128211 | Not-determined |
| 17-106_k141_111685 | Low-quality |
| 17-106_k141_111694 | Not-determined |
| 17-106_k141_128230 | Not-determined |
| 18-102-final-viral-combined_k141_135139\|\|full | Medium-quality |
| 17-106_k141_111714 | Not-determined |
| 17-106_k141_111716 | Not-determined |
| 17-106_k141_128245 | Not-determined |
| 17-106_k141_128248 | Not-determined |
| 17-106_k141_128251 | Low-quality |
| 17-106_k141_128255 | Not-determined |
| 17-106_k141_128261 | Not-determined |
| 17-106_k141_128263 | Not-determined |
| 17-106_k141_128264 | Not-determined |
| 17-106_k141_128265 | Not-determined |
| 17-106_k141_128267 | Not-determined |
| 17-106_k141_128269 | Not-determined |
| 17-106_k141_128271 | Not-determined |
| 17-106_k141_128277 | Not-determined |
| 17-106_k141_128284 | Not-determined |
| 17-106_k141_128288 | Not-determined |
| 17-106_k141_128289 | Not-determined |
| 17-106_k141_128294 | Not-determined |
| 17-106_k141_128295 | Not-determined |
| 17-106_k141_128297 | Not-determined |
| 17-106_k141_128299 | Not-determined |
| 17-106_k141_128303 | Not-determined |
| 17-106_k141_128323 | Not-determined |
| 17-106_k141_128330 | Not-determined |
| 17-106_k141_128331 | Not-determined |
| 17-106_k141_128332 | Not-determined |
| 17-106_k141_128335 | Not-determined |
| 17-106_k141_128336 | Low-quality |
| 17-106_k141_128338 | Not-determined |
| 17-106_k141_128339 | Not-determined |
| 17-106_k141_111739 | Not-determined |
| 17-106_k141_111753 | Low-quality |
| 17-106_k141_111764 | Low-quality |
| 17-106_k141_111769 | Not-determined |
| 17-106_k141_111802 | Not-determined |
| 17-106_k141_111815 | Not-determined |
| 17-106_k141_111860 | Not-determined |
| 17-106_k141_111886 | Not-determined |
| 17-106_k141_111901 | Not-determined |
| 17-106_k141_111919 | Not-determined |
| 17-106_k141_111947 | Not-determined |
| 17-106_k141_111964 | Not-determined |
| 17-106_k141_111981 | Not-determined |
| 17-106_k141_111986 | Not-determined |
| 17-106_k141_111993 | Not-determined |
| 17-106_k141_112020 | Not-determined |
| 17-106_k141_112026 | Not-determined |
| 17-106_k141_112040 | Not-determined |
| 17-106_k141_112072 | Not-determined |
| 17-106_k141_112081 | Low-quality |
| 17-106_k141_112107 | Not-determined |
| 17-106_k141_112109 | Not-determined |
| 17-106_k141_112124 | Not-determined |
| 17-106_k141_112128 | Not-determined |
| 17-106_k141_112171 | Low-quality |
| 17-106_k141_112172 | Not-determined |
| 17-106_k141_112182 | Not-determined |
| 17-106_k141_112192 | Not-determined |
| 17-106_k141_112196 | Not-determined |
| 17-106_k141_112201 | Not-determined |
| 17-106_k141_112251 | Not-determined |
| 17-106_k141_112262 | Not-determined |
| 17-106_k141_112274 | Not-determined |
| 17-106_k141_112288 | Not-determined |
| 17-106_k141_112291 | Not-determined |
| 17-108-final-viral-combined_k141_220682\|\|full | Not-determined |
| 18-105-final-viral-combined_k141_29447\|\|full | Medium-quality |
| 17-108-final-viral-combined_k141_278488\|\|full | Low-quality |
| 17-108-final-viral-combined_k141_31646\|\|full | Low-quality |
| KM-final-viral-combined_k141_56061\|\|full | High-quality |
| 17-108-final-viral-combined_k141_281193\|\|full | Not-determined |
| 17-108-final-viral-combined_k141_74576\|\|full | Not-determined |
| 17-108-final-viral-combined_k141_63142\|\|full | Not-determined |
| 17-108-final-viral-combined_k141_161509\|\|full | Low-quality |
| 17-108-final-viral-combined_k141_190241\|\|full | Not-determined |
| 17-108-final-viral-combined_k141_327634\|\|full | Low-quality |
| 17-108-final-viral-combined_k141_330477\|\|full | Not-determined |
| 18-102-final-viral-combined_k141_229529\|\|full | Complete |
| 17-108-final-viral-combined_k141_330480\|\|full | Low-quality |
| 17-108-final-viral-combined_k141_330484\|\|full | Not-determined |
| 17-108-final-viral-combined_k141_179156\|\|full | Not-determined |
| 17-108-final-viral-combined_k141_275627\|\|full | Low-quality |
| 17-108-final-viral-combined_k141_55135\|\|full | Not-determined |
| 17-108-final-viral-combined_k141_96474\|\|full | Not-determined |
| 17-108-final-viral-combined_k141_33\|\|full | Low-quality |
| 17-108-final-viral-combined_k141_137838\|\|full | Not-determined |
| 17-108-final-viral-combined_k141_275643\|\|full | Low-quality |
| 17-108-final-viral-combined_k141_179179\|\|full | Low-quality |
| 17-108-final-viral-combined_k141_110257\|\|full | Not-determined |
| 17-108-final-viral-combined_k141_27578\|\|full | Not-determined |
| 17-108-final-viral-combined_k141_179187\|\|full | Not-determined |
| 17-108-final-viral-combined_k141_151621\|\|full | Not-determined |
| 17-108-final-viral-combined_k141_110272\|\|full | Not-determined |
| 17-108-final-viral-combined_k141_124077\|\|full | Low-quality |
| 17-108-final-viral-combined_k141_179205\|\|full | Low-quality |
| 17-108-final-viral-combined_k141_303216\|\|full | Low-quality |
| 17-108-final-viral-combined_k141_316987\|\|full | Not-determined |
| 17-108-final-viral-combined_k141_206749\|\|full | Not-determined |
| 17-108-final-viral-combined_k141_124092\|\|full | Low-quality |
| 17-108-final-viral-combined_k141_261863\|\|full | Not-determined |
| 17-108-final-viral-combined_k141_137877\|\|full | Not-determined |
| 17-108-final-viral-combined_k141_55172\|\|full | Low-quality |
| 17-108-final-viral-combined_k141_303235\|\|full | Not-determined |
| 17-108-final-viral-combined_k141_13874\|\|full | Not-determined |
| 17-108-final-viral-combined_k141_317021\|\|full | Low-quality |
| 17-108-final-viral-combined_k141_317024\|\|full | Not-determined |
| 17-108-final-viral-combined_k141_137899\|\|full | Not-determined |
| 17-108-final-viral-combined_k141_206772\|\|full | Not-determined |
| 17-108-final-viral-combined_k141_303240\|\|full | Not-determined |
| 17-108-final-viral-combined_k141_151648\|\|full | Not-determined |
| 17-108-final-viral-combined_k141_248127\|\|full | Not-determined |
| 17-108-final-viral-combined_k141_248131\|\|full | Low-quality |
| 17-108-final-viral-combined_k141_248132\|\|full | Not-determined |
| 17-108-final-viral-combined_k141_317033\|\|full | Not-determined |
| 17-108-final-viral-combined_k141_206779\|\|full | Not-determined |
| 17-108-final-viral-combined_k141_206790\|\|full | Low-quality |
| 17-108-final-viral-combined_k141_248146\|\|full | Low-quality |
| 17-108-final-viral-combined_k141_303262\|\|full | Low-quality |
| 17-109-final-viral-combined_k141_368190\|\|full | Complete |
| 17-108-final-viral-combined_k141_41369\|\|full | Not-determined |
| 17-108-final-viral-combined_k141_317054\|\|full | Not-determined |
| 17-108-final-viral-combined_k141_289503\|\|full | Low-quality |
| 17-108-final-viral-combined_k141_192966\|\|full | Not-determined |
| 17-108-final-viral-combined_k141_69000\|\|full | Not-determined |
| 17-108-final-viral-combined_k141_275698\|\|full | Low-quality |
| 17-108-final-viral-combined_k141_137926\|\|full | Low-quality |
| 17-108-final-viral-combined_k141_137931\|\|full | Low-quality |
| 17-108-final-viral-combined_k141_192971\|\|full | Not-determined |
| 17-108-final-viral-combined_k141_82799\|\|full | Not-determined |
| 17-108-final-viral-combined_k141_142\|\|full | Low-quality |
| 17-108-final-viral-combined_k141_234310\|\|full | Low-quality |
| 17-108-final-viral-combined_k141_110327\|\|full | Not-determined |
| 17-108-final-viral-combined_k141_192994\|\|full | Not-determined |
| 17-108-final-viral-combined_k141_206830\|\|full | Not-determined |
| 17-108-final-viral-combined_k141_124189\|\|full | Low-quality |
| 17-108-final-viral-combined_k141_179297\|\|full | Not-determined |
| 17-108-final-viral-combined_k141_41400\|\|full | Not-determined |
| 17-108-final-viral-combined_k141_220599\|\|full | Low-quality |
| 17-108-final-viral-combined_k141_137937\|\|full | Low-quality |
| 17-108-final-viral-combined_k141_124198\|\|full | Not-determined |
| 17-108-final-viral-combined_k141_261933\|\|full | Low-quality |
| 17-108-final-viral-combined_k141_303288\|\|full | Low-quality |
| 17-108-final-viral-combined_k141_137956\|\|full | Low-quality |
| 17-108-final-viral-combined_k141_289520\|\|full | Not-determined |
| 17-108-final-viral-combined_k141_220623\|\|full | Not-determined |
| 17-108-final-viral-combined_k141_234332\|\|full | Low-quality |
| 17-108-final-viral-combined_k141_82837\|\|full | Not-determined |
| 17-108-final-viral-combined_k141_193050\|\|full | Low-quality |
| 17-108-final-viral-combined_k141_289537\|\|full | Not-determined |
| 17-108-final-viral-combined_k141_55233\|\|full | Low-quality |
| 17-108-final-viral-combined_k141_206880\|\|full | Not-determined |
| 17-108-final-viral-combined_k141_41451\|\|full | Not-determined |
| 17-108-final-viral-combined_k141_124225\|\|full | Low-quality |
| 17-108-final-viral-combined_k141_234349\|\|full | Low-quality |
| 17-108-final-viral-combined_k141_27620\|\|full | Low-quality |
| 17-108-final-viral-combined_k141_27625\|\|full | Not-determined |
| 17-108-final-viral-combined_k141_69074\|\|full | Low-quality |
| 17-108-final-viral-combined_k141_27630\|\|full | Low-quality |
| 17-108-final-viral-combined_k141_69075\|\|full | Not-determined |
| 17-108-final-viral-combined_k141_96575\|\|full | Low-quality |
| 17-108-final-viral-combined_k141_27632\|\|full | Low-quality |
| 17-108-final-viral-combined_k141_13974\|\|full | Not-determined |
| 17-108-final-viral-combined_k141_151801\|\|full | Low-quality |
| 17-108-final-viral-combined_k141_69086\|\|full | Not-determined |
| 17-108-final-viral-combined_k141_206910\|\|full | Low-quality |
| 17-108-final-viral-combined_k141_69090\|\|full | Low-quality |
| 17-108-final-viral-combined_k141_165584\|\|full | Low-quality |
| 17-108-final-viral-combined_k141_234385\|\|full | Not-determined |
| 17-108-final-viral-combined_k141_303353\|\|full | Not-determined |
| 17-108-final-viral-combined_k141_165609\|\|full | Not-determined |
| 17-108-final-viral-combined_k141_303354\|\|full | Low-quality |
| 17-108-final-viral-combined_k141_110341\|\|full | Low-quality |
| 17-108-final-viral-combined_k141_179336\|\|full | Low-quality |
| 17-108-final-viral-combined_k141_179337\|\|full | Low-quality |
| 17-108-final-viral-combined_k141_82907\|\|full | Low-quality |
| 17-108-final-viral-combined_k141_303365\|\|full | Low-quality |
| 17-108-final-viral-combined_k141_41476\|\|full | Low-quality |
| 17-108-final-viral-combined_k141_151854\|\|full | Not-determined |
| 17-108-final-viral-combined_k141_82923\|\|full | Low-quality |
| 17-108-final-viral-combined_k141_110355\|\|full | Low-quality |
| 17-108-final-viral-combined_k141_138007\|\|full | Low-quality |
| 17-108-final-viral-combined_k141_165646\|\|full | Not-determined |
| 17-108-final-viral-combined_k141_165654\|\|full | Low-quality |
| 17-108-final-viral-combined_k141_96640\|\|full | Low-quality |
| 17-108-final-viral-combined_k141_275786\|\|full | Low-quality |
| 18-109-final-viral-combined_k141_26217\|\|full | Complete |
| 17-108-final-viral-combined_k141_234411\|\|full | Low-quality |
| 17-108-final-viral-combined_k141_206965\|\|full | Not-determined |
| 17-108-final-viral-combined_k141_124324\|\|full | Not-determined |
| 17-108-final-viral-combined_k141_165683\|\|full | Low-quality |
| 17-108-final-viral-combined_k141_124348\|\|full | Low-quality |
| 18-102-final-viral-combined_k141_98888\|\|full | High-quality |
| 17-108-final-viral-combined_k141_151871\|\|full | Low-quality |
| 17-108-final-viral-combined_k141_193173\|\|full | Low-quality |
| 17-108-final-viral-combined_k141_289579\|\|full | Low-quality |
| 17-108-final-viral-combined_k141_234426\|\|full | Not-determined |
| 17-108-final-viral-combined_k141_110390\|\|full | Low-quality |
| 17-108-final-viral-combined_k141_14064\|\|full | Not-determined |
| 17-108-final-viral-combined_k141_110399\|\|full | Low-quality |
| 17-108-final-viral-combined_k141_83002\|\|full | Low-quality |
| 17-108-final-viral-combined_k141_151896\|\|full | Low-quality |
| 17-108-final-viral-combined_k141_248259\|\|full | Not-determined |
| 17-108-final-viral-combined_k141_69182\|\|full | Not-determined |
| 17-108-final-viral-combined_k141_124393\|\|full | Not-determined |
| 17-108-final-viral-combined_k141_27722\|\|full | Low-quality |
| 17-108-final-viral-combined_k141_220808\|\|full | Not-determined |
| 17-108-final-viral-combined_k141_193193\|\|full | Low-quality |
| 17-108-final-viral-combined_k141_165697\|\|full | Not-determined |
| 17-108-final-viral-combined_k141_138099\|\|full | Not-determined |
| 17-108-final-viral-combined_k141_138104\|\|full | Low-quality |
| 17-108-final-viral-combined_k141_14092\|\|full | Not-determined |
| 17-108-final-viral-combined_k141_193219\|\|full | Low-quality |
| 17-108-final-viral-combined_k141_165709\|\|full | Not-determined |
| 17-108-final-viral-combined_k141_27757\|\|full | Low-quality |
| 17-108-final-viral-combined_k141_138123\|\|full | Low-quality |
| 17-108-final-viral-combined_k141_27758\|\|full | Not-determined |
| 17-108-final-viral-combined_k141_207007\|\|full | Not-determined |
| 17-108-final-viral-combined_k141_110457\|\|full | Low-quality |
| 17-108-final-viral-combined_k141_96692\|\|full | Not-determined |
| 17-108-final-viral-combined_k141_304\|\|full | Low-quality |
| 17-108-final-viral-combined_k141_110474\|\|full | Low-quality |
| 17-108-final-viral-combined_k141_262077\|\|full | Not-determined |
| 17-108-final-viral-combined_k141_96702\|\|full | Low-quality |
| 17-108-final-viral-combined_k141_14121\|\|full | Not-determined |
| 17-108-final-viral-combined_k141_220825\|\|full | Low-quality |
| 17-108-final-viral-combined_k141_313\|\|full | Low-quality |
| 17-108-final-viral-combined_k141_110485\|\|full | Not-determined |
| 17-108-final-viral-combined_k141_55409\|\|full | Not-determined |
| 17-108-final-viral-combined_k141_317254\|\|full | Not-determined |
| 17-108-final-viral-combined_k141_262087\|\|full | Low-quality |
| 17-108-final-viral-combined_k141_55419\|\|full | Low-quality |
| 17-108-final-viral-combined_k141_262093\|\|full | Not-determined |
| 17-108-final-viral-combined_k141_165745\|\|full | Low-quality |
| 17-108-final-viral-combined_k141_289605\|\|full | Not-determined |
| 17-108-final-viral-combined_k141_41510\|\|full | Not-determined |
| 17-108-final-viral-combined_k141_96729\|\|full | Low-quality |
| 17-108-final-viral-combined_k141_207026\|\|full | Low-quality |
| 17-108-final-viral-combined_k141_303495\|\|full | Low-quality |
| 17-108-final-viral-combined_k141_220854\|\|full | Not-determined |
| 17-108-final-viral-combined_k141_207030\|\|full | Low-quality |
| 17-108-final-viral-combined_k141_69207\|\|full | Not-determined |
| 17-108-final-viral-combined_k141_248292\|\|full | Low-quality |
| 17-108-final-viral-combined_k141_69208\|\|full | Low-quality |
| 17-108-final-viral-combined_k141_69211\|\|full | Not-determined |
| 17-108-final-viral-combined_k141_165785\|\|full | Not-determined |
| 17-108-final-viral-combined_k141_248297\|\|full | Low-quality |
| 17-108-final-viral-combined_k141_303506\|\|full | Low-quality |
| 17-108-final-viral-combined_k141_358\|\|full | Not-determined |
| 17-108-final-viral-combined_k141_220861\|\|full | Low-quality |
| 17-108-final-viral-combined_k141_303518\|\|full | Not-determined |
| 17-108-final-viral-combined_k141_220871\|\|full | Not-determined |
| 17-108-final-viral-combined_k141_96737\|\|full | Not-determined |
| 17-108-final-viral-combined_k141_124446\|\|full | Not-determined |
| 17-108-final-viral-combined_k141_27829\|\|full | Low-quality |
| 17-108-final-viral-combined_k141_193271\|\|full | Not-determined |
| 17-108-final-viral-combined_k141_96752\|\|full | Low-quality |
| 17-108-final-viral-combined_k141_262153\|\|full | Low-quality |
| 17-108-final-viral-combined_k141_96760\|\|full | Low-quality |
| 17-108-final-viral-combined_k141_124461\|\|full | Low-quality |
| 17-108-final-viral-combined_k141_41548\|\|full | Low-quality |
| 17-108-final-viral-combined_k141_262171\|\|full | Not-determined |
| 17-108-final-viral-combined_k141_151959\|\|full | Low-quality |
| 17-108-final-viral-combined_k141_207067\|\|full | Low-quality |
| 17-108-final-viral-combined_k141_234538\|\|full | Low-quality |
| 17-108-final-viral-combined_k141_207084\|\|full | Not-determined |
| 17-108-final-viral-combined_k141_234548\|\|full | Low-quality |
| 17-108-final-viral-combined_k141_96781\|\|full | Not-determined |
| 17-108-final-viral-combined_k141_303534\|\|full | Not-determined |
| 17-108-final-viral-combined_k141_234556\|\|full | Low-quality |
| 17-108-final-viral-combined_k141_399\|\|full | Low-quality |
| 17-108-final-viral-combined_k141_165877\|\|full | Low-quality |
| 17-108-final-viral-combined_k141_248395\|\|full | Not-determined |
| 17-108-final-viral-combined_k141_151974\|\|full | Not-determined |
| 17-108-final-viral-combined_k141_41592\|\|full | Not-determined |
| 17-108-final-viral-combined_k141_96823\|\|full | Low-quality |
| 17-108-final-viral-combined_k141_248401\|\|full | Not-determined |
| 17-108-final-viral-combined_k141_83043\|\|full | Low-quality |
| 17-108-final-viral-combined_k141_289690\|\|full | Low-quality |
| 17-108-final-viral-combined_k141_165891\|\|full | Low-quality |
| 17-108-final-viral-combined_k141_55467\|\|full | Low-quality |
| 17-108-final-viral-combined_k141_303559\|\|full | Low-quality |
| 17-108-final-viral-combined_k141_138238\|\|full | Low-quality |
| 17-108-final-viral-combined_k141_110566\|\|full | Not-determined |
| 17-108-final-viral-combined_k141_138247\|\|full | Not-determined |
| 17-108-final-viral-combined_k141_193356\|\|full | Low-quality |
| 17-108-final-viral-combined_k141_165906\|\|full | Not-determined |
| 17-108-final-viral-combined_k141_289704\|\|full | Not-determined |
| 17-108-final-viral-combined_k141_152012\|\|full | Not-determined |
| 17-108-final-viral-combined_k141_69230\|\|full | Not-determined |
| 17-108-final-viral-combined_k141_179547\|\|full | Low-quality |
| 17-108-final-viral-combined_k141_96859\|\|full | Low-quality |
| 17-108-final-viral-combined_k141_69232\|\|full | Low-quality |
| 17-108-final-viral-combined_k141_83070\|\|full | Not-determined |
| 17-108-final-viral-combined_k141_110577\|\|full | Low-quality |
| 17-108-final-viral-combined_k141_110579\|\|full | Low-quality |
| 17-108-final-viral-combined_k141_165927\|\|full | Low-quality |
| 17-108-final-viral-combined_k141_303584\|\|full | Not-determined |
| 17-108-final-viral-combined_k141_440\|\|full | Low-quality |
| 17-108-final-viral-combined_k141_124507\|\|full | Not-determined |
| 17-108-final-viral-combined_k141_152036\|\|full | Not-determined |
| 17-108-final-viral-combined_k141_289712\|\|full | Not-determined |
| 17-108-final-viral-combined_k141_165940\|\|full | Low-quality |
| 17-108-final-viral-combined_k141_165943\|\|full | Not-determined |
| 17-108-final-viral-combined_k141_234605\|\|full | Low-quality |
| 17-108-final-viral-combined_k141_152053\|\|full | Not-determined |
| 17-108-final-viral-combined_k141_207173\|\|full | Low-quality |
| 17-108-final-viral-combined_k141_303615\|\|full | Low-quality |
| 17-108-final-viral-combined_k141_110603\|\|full | Low-quality |
| 17-108-final-viral-combined_k141_165966\|\|full | Low-quality |
| 17-108-final-viral-combined_k141_165969\|\|full | Low-quality |
| 17-108-final-viral-combined_k141_14262\|\|full | Low-quality |
| 17-108-final-viral-combined_k141_41630\|\|full | Low-quality |
| 17-108-final-viral-combined_k141_28013\|\|full | Not-determined |
| 17-108-final-viral-combined_k141_83101\|\|full | Not-determined |
| 17-108-final-viral-combined_k141_14266\|\|full | Not-determined |
| 17-108-final-viral-combined_k141_165989\|\|full | Low-quality |
| 17-108-final-viral-combined_k141_289730\|\|full | Not-determined |
| 17-108-final-viral-combined_k141_138299\|\|full | Not-determined |
| 17-108-final-viral-combined_k141_41659\|\|full | Not-determined |
| 17-108-final-viral-combined_k141_289732\|\|full | Not-determined |
| 17-108-final-viral-combined_k141_96946\|\|full | Low-quality |
| 17-108-final-viral-combined_k141_14296\|\|full | Low-quality |
| 17-108-final-viral-combined_k141_28067\|\|full | Low-quality |
| 17-108-final-viral-combined_k141_166017\|\|full | Low-quality |
| 17-108-final-viral-combined_k141_96949\|\|full | Low-quality |
| 17-108-final-viral-combined_k141_14315\|\|full | Not-determined |
| 17-108-final-viral-combined_k141_110636\|\|full | Not-determined |
| 17-108-final-viral-combined_k141_69335\|\|full | Not-determined |
| 17-108-final-viral-combined_k141_83107\|\|full | Not-determined |
| 17-108-final-viral-combined_k141_69337\|\|full | Not-determined |
| 17-108-final-viral-combined_k141_96961\|\|full | Low-quality |
| 17-108-final-viral-combined_k141_14318\|\|full | Not-determined |
| 17-108-final-viral-combined_k141_303682\|\|full | Not-determined |
| 17-108-final-viral-combined_k141_69346\|\|full | Low-quality |
| 17-108-final-viral-combined_k141_96965\|\|full | Low-quality |
| 17-108-final-viral-combined_k141_41698\|\|full | Low-quality |
| 17-108-final-viral-combined_k141_166030\|\|full | Not-determined |
| 17-108-final-viral-combined_k141_69357\|\|full | Not-determined |
| 17-108-final-viral-combined_k141_69359\|\|full | Low-quality |
| 17-108-final-viral-combined_k141_220937\|\|full | Not-determined |
| 17-108-final-viral-combined_k141_14332\|\|full | Not-determined |
| 17-108-final-viral-combined_k141_83135\|\|full | Not-determined |
| 17-108-final-viral-combined_k141_41710\|\|full | Not-determined |
| 17-108-final-viral-combined_k141_234703\|\|full | Low-quality |
| 17-108-final-viral-combined_k141_69376\|\|full | Not-determined |
| 17-108-final-viral-combined_k141_14349\|\|full | Not-determined |
| 17-108-final-viral-combined_k141_276062\|\|full | Low-quality |
| 17-108-final-viral-combined_k141_248565\|\|full | Not-determined |
| 17-108-final-viral-combined_k141_276066\|\|full | Not-determined |
| 17-108-final-viral-combined_k141_14356\|\|full | Low-quality |
| 17-108-final-viral-combined_k141_96991\|\|full | Not-determined |
| 18-105-final-viral-combined_k141_67650\|\|full | Medium-quality |
| 17-108-final-viral-combined_k141_234719\|\|full | Not-determined |
| 17-108-final-viral-combined_k141_234728\|\|full | Low-quality |
| 17-108-final-viral-combined_k141_317505\|\|full | Low-quality |
| 17-108-final-viral-combined_k141_478\|\|full | Low-quality |
| 17-108-final-viral-combined_k141_14365\|\|full | Not-determined |
| 17-108-final-viral-combined_k141_41742\|\|full | Not-determined |
| 17-108-final-viral-combined_k141_234757\|\|full | Not-determined |
| 17-108-final-viral-combined_k141_289779\|\|full | Low-quality |
| 17-108-final-viral-combined_k141_193402\|\|full | Low-quality |
| 17-108-final-viral-combined_k141_124629\|\|full | Low-quality |
| 18-106-final-viral-combined_k141_56389\|\|full | Medium-quality |
| 17-108-final-viral-combined_k141_234790\|\|full | Not-determined |
| 17-108-final-viral-combined_k141_110701\|\|full | Low-quality |
| 17-108-final-viral-combined_k141_262315\|\|full | Not-determined |
| 17-108-final-viral-combined_k141_152161\|\|full | Low-quality |
| 17-108-final-viral-combined_k141_152167\|\|full | Not-determined |
| 17-108-final-viral-combined_k141_28104\|\|full | Not-determined |
| 17-108-final-viral-combined_k141_276108\|\|full | Low-quality |
| 17-108-final-viral-combined_k141_289799\|\|full | Not-determined |
| 17-108-final-viral-combined_k141_289800\|\|full | Not-determined |
| 17-108-final-viral-combined_k141_193408\|\|full | Not-determined |
| 17-108-final-viral-combined_k141_166086\|\|full | Low-quality |
| 17-108-final-viral-combined_k141_166087\|\|full | Not-determined |
| 17-108-final-viral-combined_k141_193418\|\|full | Not-determined |
| 17-108-final-viral-combined_k141_317546\|\|full | Low-quality |
| 17-108-final-viral-combined_k141_152188\|\|full | Not-determined |
| 17-108-final-viral-combined_k141_193419\|\|full | Not-determined |
| 17-108-final-viral-combined_k141_83201\|\|full | Low-quality |
| 17-108-final-viral-combined_k141_138346\|\|full | Not-determined |
| 17-108-final-viral-combined_k141_248578\|\|full | Low-quality |
| 17-108-final-viral-combined_k141_289837\|\|full | Not-determined |
| 17-108-final-viral-combined_k141_124687\|\|full | Low-quality |
| 17-108-final-viral-combined_k141_193426\|\|full | Low-quality |
| 17-108-final-viral-combined_k141_55652\|\|full | Low-quality |
| 17-108-final-viral-combined_k141_97034\|\|full | Not-determined |
| 17-108-final-viral-combined_k141_138359\|\|full | Low-quality |
| 17-108-final-viral-combined_k141_41853\|\|full | Not-determined |
| 17-108-final-viral-combined_k141_248582\|\|full | Not-determined |
| 17-108-final-viral-combined_k141_97039\|\|full | Low-quality |
| 17-108-final-viral-combined_k141_248583\|\|full | Low-quality |
| 17-108-final-viral-combined_k141_262352\|\|full | Low-quality |
| 17-108-final-viral-combined_k141_262360\|\|full | Not-determined |
| 17-108-final-viral-combined_k141_193446\|\|full | Low-quality |
| 17-108-final-viral-combined_k141_110756\|\|full | Not-determined |
| 17-108-final-viral-combined_k141_221028\|\|full | Not-determined |
| 17-108-final-viral-combined_k141_289855\|\|full | Not-determined |
| 17-108-final-viral-combined_k141_69483\|\|full | Low-quality |
| 17-108-final-viral-combined_k141_14397\|\|full | Low-quality |
| 17-108-final-viral-combined_k141_28164\|\|full | Not-determined |
| 17-108-final-viral-combined_k141_248624\|\|full | Not-determined |
| 17-108-final-viral-combined_k141_97068\|\|full | Not-determined |
| 17-108-final-viral-combined_k141_221049\|\|full | Low-quality |
| 17-108-final-viral-combined_k141_28176\|\|full | Low-quality |
| 17-108-final-viral-combined_k141_14413\|\|full | Not-determined |
| 17-108-final-viral-combined_k141_317588\|\|full | Low-quality |
| 17-108-final-viral-combined_k141_558\|\|full | Low-quality |
| 17-108-final-viral-combined_k141_262411\|\|full | Low-quality |
| 17-108-final-viral-combined_k141_97086\|\|full | Not-determined |
| 17-108-final-viral-combined_k141_179622\|\|full | Low-quality |
| 17-108-final-viral-combined_k141_55713\|\|full | Low-quality |
| 17-108-final-viral-combined_k141_317598\|\|full | Low-quality |
| 17-108-final-viral-combined_k141_166152\|\|full | Not-determined |
| 17-108-final-viral-combined_k141_179642\|\|full | Low-quality |
| 17-108-final-viral-combined_k141_138475\|\|full | Not-determined |
| 17-108-final-viral-combined_k141_14449\|\|full | Low-quality |
| 17-108-final-viral-combined_k141_138485\|\|full | Not-determined |
| 17-108-final-viral-combined_k141_138489\|\|full | Low-quality |
| 17-108-final-viral-combined_k141_248664\|\|full | Not-determined |
| 17-108-final-viral-combined_k141_14466\|\|full | Not-determined |
| 17-108-final-viral-combined_k141_289941\|\|full | Not-determined |
| 17-108-final-viral-combined_k141_207291\|\|full | Not-determined |
| 17-108-final-viral-combined_k141_207298\|\|full | Low-quality |
| 17-108-final-viral-combined_k141_97132\|\|full | Not-determined |
| 17-108-final-viral-combined_k141_289967\|\|full | Not-determined |
| 17-108-final-viral-combined_k141_289968\|\|full | Low-quality |
| 17-108-final-viral-combined_k141_124799\|\|full | Not-determined |
| 17-108-final-viral-combined_k141_41889\|\|full | Low-quality |
| 17-108-final-viral-combined_k141_234936\|\|full | Low-quality |
| 17-108-final-viral-combined_k141_289981\|\|full | Low-quality |
| 17-108-final-viral-combined_k141_289987\|\|full | Low-quality |
| 17-108-final-viral-combined_k141_83281\|\|full | Not-determined |
| 17-108-final-viral-combined_k141_14500\|\|full | Low-quality |
| 17-108-final-viral-combined_k141_97159\|\|full | Not-determined |
| 17-108-final-viral-combined_k141_110814\|\|full | Not-determined |
| 17-108-final-viral-combined_k141_193615\|\|full | Low-quality |
| 17-108-final-viral-combined_k141_166243\|\|full | Low-quality |
| 17-108-final-viral-combined_k141_28248\|\|full | Low-quality |
| 17-108-final-viral-combined_k141_97191\|\|full | Low-quality |
| 17-108-final-viral-combined_k141_28252\|\|full | Low-quality |
| 17-108-final-viral-combined_k141_248701\|\|full | Low-quality |
| 17-108-final-viral-combined_k141_28254\|\|full | Low-quality |
| 17-108-final-viral-combined_k141_207364\|\|full | Low-quality |
| 17-108-final-viral-combined_k141_235008\|\|full | Not-determined |
| 17-108-final-viral-combined_k141_221159\|\|full | Low-quality |
| 17-108-final-viral-combined_k141_303892\|\|full | Not-determined |
| 17-108-final-viral-combined_k141_290010\|\|full | Not-determined |
| 17-108-final-viral-combined_k141_55838\|\|full | Not-determined |
| 17-108-final-viral-combined_k141_221167\|\|full | Not-determined |
| 17-108-final-viral-combined_k141_97207\|\|full | Low-quality |
| 17-108-final-viral-combined_k141_138530\|\|full | Not-determined |
| 17-108-final-viral-combined_k141_235038\|\|full | Low-quality |
| 17-108-final-viral-combined_k141_14547\|\|full | Not-determined |
| 17-108-final-viral-combined_k141_124887\|\|full | Not-determined |
| 17-108-final-viral-combined_k141_290030\|\|full | Not-determined |
| 17-108-final-viral-combined_k141_124891\|\|full | Low-quality |
| 17-108-final-viral-combined_k141_69619\|\|full | Not-determined |
| 17-108-final-viral-combined_k141_166308\|\|full | Low-quality |
| 17-108-final-viral-combined_k141_28286\|\|full | Low-quality |
| 17-108-final-viral-combined_k141_152339\|\|full | Not-determined |
| 17-108-final-viral-combined_k141_290043\|\|full | Low-quality |
| 17-108-final-viral-combined_k141_14573\|\|full | Low-quality |
| 17-108-final-viral-combined_k141_262567\|\|full | Low-quality |
| 17-108-final-viral-combined_k141_41967\|\|full | Low-quality |
| 17-108-final-viral-combined_k141_124914\|\|full | Not-determined |
| 17-108-final-viral-combined_k141_28316\|\|full | Not-determined |
| 17-108-final-viral-combined_k141_235062\|\|full | Low-quality |
| 17-108-final-viral-combined_k141_290055\|\|full | Low-quality |
| 17-108-final-viral-combined_k141_152356\|\|full | Not-determined |
| 17-108-final-viral-combined_k141_248753\|\|full | Low-quality |
| 17-108-final-viral-combined_k141_276341\|\|full | Not-determined |
| 17-108-final-viral-combined_k141_28317\|\|full | Not-determined |
| 17-108-final-viral-combined_k141_221231\|\|full | Not-determined |
| 17-108-final-viral-combined_k141_276342\|\|full | Not-determined |
| 17-108-final-viral-combined_k141_678\|\|full | Not-determined |
| 17-108-final-viral-combined_k141_290066\|\|full | Not-determined |
| 17-108-final-viral-combined_k141_110891\|\|full | Not-determined |
| 17-108-final-viral-combined_k141_110894\|\|full | Not-determined |
| 17-108-final-viral-combined_k141_193702\|\|full | Not-determined |
| 17-108-final-viral-combined_k141_69659\|\|full | Not-determined |
| 17-108-final-viral-combined_k141_69661\|\|full | Not-determined |
| 17-108-final-viral-combined_k141_317703\|\|full | Low-quality |
| 17-108-final-viral-combined_k141_235106\|\|full | Low-quality |
| 17-108-final-viral-combined_k141_303944\|\|full | Not-determined |
| 17-108-final-viral-combined_k141_290104\|\|full | Not-determined |
| 17-108-final-viral-combined_k141_276370\|\|full | Low-quality |
| 17-108-final-viral-combined_k141_69677\|\|full | Low-quality |
| 17-108-final-viral-combined_k141_97292\|\|full | Not-determined |
| 17-108-final-viral-combined_k141_179785\|\|full | Low-quality |
| 17-108-final-viral-combined_k141_138533\|\|full | Low-quality |
| 17-108-final-viral-combined_k141_138549\|\|full | Low-quality |
| 17-108-final-viral-combined_k141_738\|\|full | Low-quality |
| 17-108-final-viral-combined_k141_235142\|\|full | Not-determined |
| 17-108-final-viral-combined_k141_55961\|\|full | Not-determined |
| 17-108-final-viral-combined_k141_754\|\|full | Not-determined |
| 17-108-final-viral-combined_k141_221281\|\|full | Low-quality |
| 17-108-final-viral-combined_k141_276395\|\|full | Not-determined |
| 17-108-final-viral-combined_k141_235152\|\|full | Low-quality |
| 17-108-final-viral-combined_k141_193734\|\|full | Not-determined |
| 17-108-final-viral-combined_k141_55982\|\|full | Not-determined |
| 17-108-final-viral-combined_k141_55983\|\|full | Low-quality |
| 17-108-final-viral-combined_k141_207475\|\|full | Low-quality |
| 17-108-final-viral-combined_k141_235186\|\|full | Not-determined |
| 17-108-final-viral-combined_k141_166423\|\|full | Not-determined |
| 17-108-final-viral-combined_k141_83459\|\|full | Not-determined |
| 17-108-final-viral-combined_k141_262643\|\|full | Low-quality |
| 17-108-final-viral-combined_k141_110981\|\|full | Not-determined |
| 17-108-final-viral-combined_k141_124957\|\|full | Low-quality |
| 17-108-final-viral-combined_k141_28391\|\|full | Low-quality |
| 17-108-final-viral-combined_k141_207502\|\|full | Low-quality |
| 17-108-final-viral-combined_k141_207506\|\|full | Not-determined |
| 17-108-final-viral-combined_k141_276439\|\|full | Low-quality |
| 17-108-final-viral-combined_k141_166426\|\|full | Low-quality |
| 17-108-final-viral-combined_k141_28397\|\|full | Not-determined |
| 17-109-final-viral-combined_k141_111524\|\|full | Medium-quality |
| 17-108-final-viral-combined_k141_317802\|\|full | Not-determined |
| 17-108-final-viral-combined_k141_248859\|\|full | Not-determined |
| 17-108-final-viral-combined_k141_69706\|\|full | Low-quality |
| 17-108-final-viral-combined_k141_97366\|\|full | Low-quality |
| 17-108-final-viral-combined_k141_235212\|\|full | Not-determined |
| 17-108-final-viral-combined_k141_317806\|\|full | Not-determined |
| 17-108-final-viral-combined_k141_124980\|\|full | Not-determined |
| 17-108-final-viral-combined_k141_235215\|\|full | Low-quality |
| 17-108-final-viral-combined_k141_276481\|\|full | Not-determined |
| 17-108-final-viral-combined_k141_262693\|\|full | Not-determined |
| 17-108-final-viral-combined_k141_235220\|\|full | Low-quality |
| 17-108-final-viral-combined_k141_193822\|\|full | Not-determined |
| 17-108-final-viral-combined_k141_304041\|\|full | Not-determined |
| 17-108-final-viral-combined_k141_69727\|\|full | Not-determined |
| 17-108-final-viral-combined_k141_152492\|\|full | Not-determined |
| 17-108-final-viral-combined_k141_193830\|\|full | Not-determined |
| 17-108-final-viral-combined_k141_83551\|\|full | Not-determined |
| 17-108-final-viral-combined_k141_221357\|\|full | Not-determined |
| 17-108-final-viral-combined_k141_317864\|\|full | Not-determined |
| 17-108-final-viral-combined_k141_166477\|\|full | Not-determined |
| 18-110-final-viral-combined_k141_23583\|\|full | Complete |
| 17-108-final-viral-combined_k141_304103\|\|full | Not-determined |
| 17-108-final-viral-combined_k141_111032\|\|full | Low-quality |
| 17-108-final-viral-combined_k141_28424\|\|full | Not-determined |
| 17-108-final-viral-combined_k141_235270\|\|full | Low-quality |
| 17-108-final-viral-combined_k141_28428\|\|full | Low-quality |
| 17-108-final-viral-combined_k141_290135\|\|full | Not-determined |
| 17-108-final-viral-combined_k141_97429\|\|full | Low-quality |
| 17-108-final-viral-combined_k141_28430\|\|full | Low-quality |
| 17-108-final-viral-combined_k141_262750\|\|full | Low-quality |
| 17-108-final-viral-combined_k141_262752\|\|full | Low-quality |
| 17-108-final-viral-combined_k141_179909\|\|full | Low-quality |
| 18-105-final-viral-combined_k141_134743\|\|full | Medium-quality |
| 17-108-final-viral-combined_k141_14663\|\|full | Not-determined |
| 17-108-final-viral-combined_k141_179918\|\|full | Low-quality |
| 17-108-final-viral-combined_k141_290150\|\|full | Low-quality |
| 17-108-final-viral-combined_k141_304113\|\|full | Low-quality |
| 17-108-final-viral-combined_k141_166490\|\|full | Not-determined |
| 17-108-final-viral-combined_k141_42087\|\|full | Low-quality |
| 17-108-final-viral-combined_k141_891\|\|full | Low-quality |
| 17-108-final-viral-combined_k141_179937\|\|full | Not-determined |
| 17-108-final-viral-combined_k141_125034\|\|full | Not-determined |
| 17-108-final-viral-combined_k141_152533\|\|full | Not-determined |
| 17-108-final-viral-combined_k141_14684\|\|full | Not-determined |
| 17-108-final-viral-combined_k141_28440\|\|full | Not-determined |
| 17-108-final-viral-combined_k141_111082\|\|full | Not-determined |
| 17-108-final-viral-combined_k141_69788\|\|full | Not-determined |
| 17-108-final-viral-combined_k141_69790\|\|full | Not-determined |
| 17-108-final-viral-combined_k141_914\|\|full | Not-determined |
| 17-108-final-viral-combined_k141_262781\|\|full | Low-quality |
| 17-108-final-viral-combined_k141_166522\|\|full | Low-quality |
| 17-108-final-viral-combined_k141_83624\|\|full | Not-determined |
| 17-108-final-viral-combined_k141_97491\|\|full | Low-quality |
| 17-108-final-viral-combined_k141_304139\|\|full | Not-determined |
| 17-108-final-viral-combined_k141_317908\|\|full | Low-quality |
| 17-108-final-viral-combined_k141_235354\|\|full | Low-quality |
| 17-108-final-viral-combined_k141_111104\|\|full | Low-quality |

17-108-final-viral-combined_k141_14713||full Low-quality
17-108-final-viral-combined_k141_207638||full Low-quality
17-108-final-viral-combined_k141_152609||full Low-quality
17-108-final-viral-combined_k141_166576||full Not-determined
17-108-final-viral-combined_k141_97531||full Low-quality
17-108-final-viral-combined_k141_125119||full Not-determined
17-108-final-viral-combined_k141_125126||full Not-determined
17-108-final-viral-combined_k141_248996||full Not-determined
17-108-final-viral-combined_k141_152628||full Not-determined
17-108-final-viral-combined_k141_42176||full Low-quality
17-108-final-viral-combined_k141_977||full Not-determined
17-108-final-viral-combined_k141_276570||full Not-determined
17-108-final-viral-combined_k141_28478||full Low-quality
17-108-final-viral-combined_k141_125135||full Not-determined
17-108-final-viral-combined_k141_983||full Low-quality
17-108-final-viral-combined_k141_83703||full Not-determined
17-108-final-viral-combined_k141_28483||full Not-determined
17-108-final-viral-combined_k141_276581||full Not-determined
17-108-final-viral-combined_k141_207668||full Low-quality
17-108-final-viral-combined_k141_207674||full Low-quality
17-108-final-viral-combined_k141_69815||full Not-determined
17-108-final-viral-combined_k141_166600||full Low-quality
17-108-final-viral-combined_k141_166608||full Not-determined
17-108-final-viral-combined_k141_207691||full Not-determined
17-108-final-viral-combined_k141_180040||full Not-determined
17-108-final-viral-combined_k141_304194||full Not-determined
17-108-final-viral-combined_k141_152665||full Not-determined
17-108-final-viral-combined_k141_56074||full Not-determined
17-108-final-viral-combined_k141_152666||full Not-determined
17-108-final-viral-combined_k141_14777||full Not-determined
17-108-final-viral-combined_k141_276616||full Not-determined
17-108-final-viral-combined_k141_14778||full Not-determined
17-108-final-viral-combined_k141_83735||full Not-determined
17-108-final-viral-combined_k141_97600||full Not-determined
17-108-final-viral-combined_k141_83742||full Not-determined
17-108-final-viral-combined_k141_42211||full Low-quality
17-108-final-viral-combined_k141_97609||full Not-determined
17-108-final-viral-combined_k141_14794||full Low-quality
17-108-final-viral-combined_k141_83754||full Low-quality
17-108-final-viral-combined_k141_97612||full Low-quality
17-108-final-viral-combined_k141_180077||full Not-determined
17-108-final-viral-combined_k141_69852||full Low-quality
17-108-final-viral-combined_k141_97632||full Low-quality
17-108-final-viral-combined_k141_69862||full Low-quality
17-108-final-viral-combined_k141_111189||full Low-quality
17-108-final-viral-combined_k141_290231||full Low-quality
17-108-final-viral-combined_k141_221462||full Not-determined
17-108-final-viral-combined_k141_262831||full Not-determined
17-108-final-viral-combined_k141_290235||full Not-determined
17-108-final-viral-combined_k141_290241||full Low-quality
17-108-final-viral-combined_k141_221480||full Low-quality
17-108-final-viral-combined_k141_221483||full Not-determined
17-108-final-viral-combined_k141_138794||full Low-quality
17-108-final-viral-combined_k141_69880||full Low-quality
17-108-final-viral-combined_k141_166697||full Low-quality
17-108-final-viral-combined_k141_290247||full Low-quality
17-108-final-viral-combined_k141_42215||full Not-determined
17-108-final-viral-combined_k141_276638||full Not-determined
17-108-final-viral-combined_k141_262871||full Low-quality
17-108-final-viral-combined_k141_42219||full Low-quality
17-106-final-viral-combined_k141_109322||full High-quality
18-105-final-viral-combined_k141_81754||full Medium-quality
17-108-final-viral-combined_k141_83805||full Not-determined
17-108-final-viral-combined_k141_14870||full Not-determined
17-108-final-viral-combined_k141_125193||full Not-determined
17-108-final-viral-combined_k141_56123||full Not-determined
17-108-final-viral-combined_k141_152773||full Not-determined
17-108-final-viral-combined_k141_42234||full Not-determined
17-108-final-viral-combined_k141_290278||full Not-determined
17-108-final-viral-combined_k141_235569||full Low-quality
17-108-final-viral-combined_k141_166735||full Low-quality
17-108-final-viral-combined_k141_152788||full Not-determined
17-108-final-viral-combined_k141_125209||full Low-quality
17-108-final-viral-combined_k141_166736||full Low-quality
17-108-final-viral-combined_k141_42262||full Low-quality
17-108-final-viral-combined_k141_276663||full Low-quality
17-108-final-viral-combined_k141_56172||full Low-quality
17-108-final-viral-combined_k141_249117||full Low-quality
17-108-final-viral-combined_k141_304274||full Not-determined
17-108-final-viral-combined_k141_207824||full Not-determined
17-108-final-viral-combined_k141_207825||full Low-quality
17-108-final-viral-combined_k141_42282||full Not-determined
17-108-final-viral-combined_k141_221549||full Not-determined
17-108-final-viral-combined_k141_207831||full Not-determined
17-108-final-viral-combined_k141_304287||full Not-determined
17-108-final-viral-combined_k141_249131||full Not-determined
17-108-final-viral-combined_k141_138826||full Not-determined
17-108-final-viral-combined_k141_193956||full Not-determined
17-108-final-viral-combined_k141_249142||full Low-quality
17-108-final-viral-combined_k141_235594||full Not-determined
17-108-final-viral-combined_k141_152802||full Not-determined
17-108-final-viral-combined_k141_14892||full Not-determined
17-108-final-viral-combined_k141_125234||full Low-quality
17-108-final-viral-combined_k141_221578||full Not-determined
17-108-final-viral-combined_k141_28530||full Low-quality
17-108-final-viral-combined_k141_28535||full Not-determined
17-108-final-viral-combined_k141_235644||full Low-quality
17-108-final-viral-combined_k141_235648||full Low-quality
17-108-final-viral-combined_k141_193989||full Not-determined
17-108-final-viral-combined_k141_14951||full Not-determined
17-108-final-viral-combined_k141_14952||full Low-quality
17-108-final-viral-combined_k141_83882||full Low-quality
17-108-final-viral-combined_k141_1161||full Low-quality
17-108-final-viral-combined_k141_14959||full Low-quality
17-108-final-viral-combined_k141_1162||full Low-quality
17-108-final-viral-combined_k141_207892||full Low-quality
17-108-final-viral-combined_k141_125273||full Low-quality
17-108-final-viral-combined_k141_97810||full Low-quality
17-108-final-viral-combined_k141_235683||full Low-quality
17-108-final-viral-combined_k141_194005||full Not-determined
17-108-final-viral-combined_k141_28579||full Not-determined
17-108-final-viral-combined_k141_194006||full Low-quality
17-108-final-viral-combined_k141_14967||full Low-quality
17-108-final-viral-combined_k141_262998||full Not-determined
17-108-final-viral-combined_k141_1185||full Low-quality
17-108-final-viral-combined_k141_97828||full Low-quality
17-108-final-viral-combined_k141_138886||full Low-quality
17-108-final-viral-combined_k141_14978||full Low-quality
17-108-final-viral-combined_k141_235715||full Not-determined
17-108-final-viral-combined_k141_290433||full Low-quality
17-108-final-viral-combined_k141_138897||full Not-determined
17-108-final-viral-combined_k141_152825||full Not-determined
17-108-final-viral-combined_k141_56201||full Not-determined
17-108-final-viral-combined_k141_111303||full Not-determined
17-108-final-viral-combined_k141_83943||full Low-quality
17-108-final-viral-combined_k141_207949||full Low-quality
17-108-final-viral-combined_k141_263031||full Low-quality
17-108-final-viral-combined_k141_180228||full Low-quality
17-108-final-viral-combined_k141_290456||full Not-determined
17-108-final-viral-combined_k141_42398||full Not-determined
17-108-final-viral-combined_k141_263038||full Low-quality
17-108-final-viral-combined_k141_180240||full Not-determined
17-108-final-viral-combined_k141_194037||full Low-quality
17-108-final-viral-combined_k141_152848||full Low-quality
17-108-final-viral-combined_k141_276829||full Low-quality
17-108-final-viral-combined_k141_125323||full Not-determined
17-108-final-viral-combined_k141_249267||full Not-determined
17-108-final-viral-combined_k141_304378||full Not-determined
17-108-final-viral-combined_k141_208002||full Low-quality
17-108-final-viral-combined_k141_125333||full Not-determined
17-108-final-viral-combined_k141_208008||full Not-determined
17-108-final-viral-combined_k141_208013||full Low-quality
17-108-final-viral-combined_k141_276868||full Not-determined
17-108-final-viral-combined_k141_194047||full Low-quality
17-108-final-viral-combined_k141_180286||full Not-determined
17-108-final-viral-combined_k141_208028||full Not-determined
17-108-final-viral-combined_k141_208029||full Low-quality
17-108-final-viral-combined_k141_235827||full Low-quality
17-108-final-viral-combined_k141_276897||full Low-quality
17-108-final-viral-combined_k141_97926||full Low-quality
17-108-final-viral-combined_k141_304450||full Not-determined
17-108-final-viral-combined_k141_276899||full Low-quality
17-108-final-viral-combined_k141_263100||full Low-quality
17-108-final-viral-combined_k141_290474||full Not-determined
17-108-final-viral-combined_k141_70008||full Low-quality
17-108-final-viral-combined_k141_42454||full Low-quality
17-108-final-viral-combined_k141_290482||full Low-quality
17-108-final-viral-combined_k141_194059||full Low-quality
17-108-final-viral-combined_k141_263112||full Low-quality
17-108-final-viral-combined_k141_84003||full Not-determined
17-108-final-viral-combined_k141_15036||full Low-quality
17-108-final-viral-combined_k141_42477||full Low-quality
17-108-final-viral-combined_k141_249326||full Low-quality
17-108-final-viral-combined_k141_28625||full Low-quality
17-108-final-viral-combined_k141_111359||full Low-quality
17-108-final-viral-combined_k141_111363||full Not-determined
17-108-final-viral-combined_k141_290506||full Not-determined
17-108-final-viral-combined_k141_304497||full Low-quality
17-108-final-viral-combined_k141_263137||full Not-determined
17-108-final-viral-combined_k141_166939||full Low-quality
17-108-final-viral-combined_k141_194075||full Low-quality
17-108-final-viral-combined_k141_28648||full Not-determined
17-108-final-viral-combined_k141_235888||full Low-quality
17-108-final-viral-combined_k141_318046||full Low-quality
17-108-final-viral-combined_k141_263155||full Low-quality
17-108-final-viral-combined_k141_125424||full Not-determined
17-108-final-viral-combined_k141_70034||full Not-determined
17-108-final-viral-combined_k141_42544||full Not-determined
17-108-final-viral-combined_k141_221651||full Not-determined
17-108-final-viral-combined_k141_125431||full Not-determined
17-108-final-viral-combined_k141_180333||full Low-quality
17-108-final-viral-combined_k141_276962||full Low-quality
17-108-final-viral-combined_k141_125438||full Low-quality
17-108-final-viral-combined_k141_304521||full Not-determined
17-108-final-viral-combined_k141_180340||full Not-determined
17-108-final-viral-combined_k141_98023||full Low-quality
17-108-final-viral-combined_k141_208039||full Low-quality
17-108-final-viral-combined_k141_304529||full Low-quality
17-108-final-viral-combined_k141_98028||full Not-determined
17-108-final-viral-combined_k141_276980||full Not-determined
17-108-final-viral-combined_k141_70065||full Not-determined
17-108-final-viral-combined_k141_98034||full Not-determined
17-108-final-viral-combined_k141_208056||full Not-determined
17-108-final-viral-combined_k141_98039||full Not-determined
17-108-final-viral-combined_k141_263184||full Low-quality
17-108-final-viral-combined_k141_98049||full Low-quality
17-108-final-viral-combined_k141_304560||full Low-quality
17-108-final-viral-combined_k141_290523||full Low-quality
17-108-final-viral-combined_k141_15104||full Not-determined
17-108-final-viral-combined_k141_304572||full Not-determined
17-108-final-viral-combined_k141_180367||full Not-determined
17-108-final-viral-combined_k141_263205||full Not-determined
17-108-final-viral-combined_k141_263208||full Not-determined
17-108-final-viral-combined_k141_152916||full Low-quality
17-108-final-viral-combined_k141_1294||full Low-quality
17-108-final-viral-combined_k141_318116||full Not-determined
17-108-final-viral-combined_k141_304586||full Low-quality
17-108-final-viral-combined_k141_111400||full Not-determined
17-108-final-viral-combined_k141_249411||full Low-quality
17-108-final-viral-combined_k141_208094||full Low-quality
17-108-final-viral-combined_k141_208096||full Low-quality
17-108-final-viral-combined_k141_70113||full Low-quality
17-108-final-viral-combined_k141_138943||full Low-quality
17-108-final-viral-combined_k141_98086||full Low-quality
17-108-final-viral-combined_k141_221771||full Not-determined
17-108-final-viral-combined_k141_84050||full Low-quality
17-108-final-viral-combined_k141_194166||full Not-determined
17-108-final-viral-combined_k141_70124||full Low-quality
17-108-final-viral-combined_k141_98096||full Low-quality
17-108-final-viral-combined_k141_194172||full Low-quality
17-108-final-viral-combined_k141_70134||full Low-quality
17-108-final-viral-combined_k141_221792||full Low-quality
17-108-final-viral-combined_k141_1318||full Low-quality
17-108-final-viral-combined_k141_194178||full Not-determined
17-108-final-viral-combined_k141_194179||full Not-determined
17-108-final-viral-combined_k141_304635||full Low-quality
17-108-final-viral-combined_k141_70147||full Not-determined
17-108-final-viral-combined_k141_1325||full Low-quality
17-108-final-viral-combined_k141_28808||full Low-quality
17-108-final-viral-combined_k141_180394||full Low-quality
17-108-final-viral-combined_k141_194195||full Low-quality
17-108-final-viral-combined_k141_1327||full Not-determined
17-108-final-viral-combined_k141_221827||full Low-quality
17-108-final-viral-combined_k141_180409||full Not-determined
17-108-final-viral-combined_k141_70161||full Low-quality
17-108-final-viral-combined_k141_138981||full Low-quality
17-108-final-viral-combined_k141_318155||full Low-quality
17-108-final-viral-combined_k141_84094||full Not-determined
17-108-final-viral-combined_k141_166975||full Low-quality
17-108-final-viral-combined_k141_70175||full Low-quality
17-108-final-viral-combined_k141_98124||full Not-determined
17-108-final-viral-combined_k141_235970||full Not-determined
17-108-final-viral-combined_k141_138983||full Not-determined
17-108-final-viral-combined_k141_1346||full Low-quality
17-108-final-viral-combined_k141_98127||full Low-quality
17-108-final-viral-combined_k141_138987||full Not-determined
17-108-final-viral-combined_k141_235986||full Low-quality
17-108-final-viral-combined_k141_249504||full Not-determined
17-108-final-viral-combined_k141_318161||full Low-quality
18-105-final-viral-combined_k141_90315||full High-quality
17-108-final-viral-combined_k141_166998||full Low-quality
17-108-final-viral-combined_k141_318179||full Not-determined
17-108-final-viral-combined_k141_208165||full Low-quality
17-108-final-viral-combined_k141_208167||full Low-quality
17-108-final-viral-combined_k141_70231||full Low-quality
17-108-final-viral-combined_k141_221847||full Low-quality
17-108-final-viral-combined_k141_167014||full Low-quality
17-108-final-viral-combined_k141_84140||full Low-quality
17-108-final-viral-combined_k141_194243||full Not-determined
17-108-final-viral-combined_k141_28900||full Low-quality
17-108-final-viral-combined_k141_180451||full Not-determined
17-108-final-viral-combined_k141_318209||full Not-determined
17-108-final-viral-combined_k141_249518||full Low-quality
17-108-final-viral-combined_k141_15244||full Low-quality
17-108-final-viral-combined_k141_208195||full Not-determined
17-108-final-viral-combined_k141_180462||full Not-determined
17-108-final-viral-combined_k141_15261||full Low-quality
17-108-final-viral-combined_k141_263316||full Low-quality
17-108-final-viral-combined_k141_304728||full Not-determined
17-108-final-viral-combined_k141_290674||full Not-determined
17-108-final-viral-combined_k141_180486||full Low-quality
17-108-final-viral-combined_k141_194298||full Low-quality
17-108-final-viral-combined_k141_277120||full Not-determined
17-108-final-viral-combined_k141_290683||full Low-quality
17-108-final-viral-combined_k141_111441||full Not-determined
17-108-final-viral-combined_k141_180500||full Low-quality
17-108-final-viral-combined_k141_304737||full Not-determined
17-108-final-viral-combined_k141_167046||full Low-quality
17-108-final-viral-combined_k141_249534||full Low-quality
17-108-final-viral-combined_k141_221925||full Low-quality
17-108-final-viral-combined_k141_290705||full Low-quality
17-108-final-viral-combined_k141_318242||full Not-determined
17-108-final-viral-combined_k141_290708||full Low-quality
17-108-final-viral-combined_k141_167054||full Not-determined
17-108-final-viral-combined_k141_277171||full Not-determined
17-108-final-viral-combined_k141_84223||full Low-quality
17-108-final-viral-combined_k141_180519||full Not-determined
17-108-final-viral-combined_k141_318255||full Not-determined
17-108-final-viral-combined_k141_180522||full Low-quality
17-108-final-viral-combined_k141_42751||full Low-quality
17-108-final-viral-combined_k141_70307||full Low-quality
17-108-final-viral-combined_k141_153026||full Low-quality
17-108-final-viral-combined_k141_42755||full Low-quality
17-108-final-viral-combined_k141_290741||full Not-determined
17-108-final-viral-combined_k141_249558||full Not-determined
17-108-final-viral-combined_k141_180527||full Low-quality
17-108-final-viral-combined_k141_153033||full Low-quality
17-108-final-viral-combined_k141_249564||full Not-determined
17-108-final-viral-combined_k141_221958||full Not-determined
17-108-final-viral-combined_k141_221962||full Low-quality
17-108-final-viral-combined_k141_70328||full Low-quality
17-108-final-viral-combined_k141_180550||full Low-quality
17-108-final-viral-combined_k141_42787||full Low-quality
17-108-final-viral-combined_k141_208250||full Not-determined
17-108-final-viral-combined_k141_249571||full Not-determined
17-108-final-viral-combined_k141_125676||full Low-quality
17-108-final-viral-combined_k141_208262||full Not-determined
17-108-final-viral-combined_k141_249591||full Low-quality
17-108-final-viral-combined_k141_28934||full Low-quality
17-108-final-viral-combined_k141_249597||full Not-determined
17-108-final-viral-combined_k141_304791||full Not-determined
17-108-final-viral-combined_k141_111490||full Low-quality
17-108-final-viral-combined_k141_236090||full Low-quality
17-108-final-viral-combined_k141_153081||full Low-quality
17-108-final-viral-combined_k141_42825||full Not-determined
17-108-final-viral-combined_k141_277246||full Low-quality
17-108-final-viral-combined_k141_249624||full Not-determined
17-108-final-viral-combined_k141_290804||full Not-determined
17-108-final-viral-combined_k141_249629||full Not-determined
17-108-final-viral-combined_k141_263427||full Not-determined
17-108-final-viral-combined_k141_1461||full Not-determined
17-108-final-viral-combined_k141_125721||full Low-quality
17-108-final-viral-combined_k141_167194||full Not-determined
17-108-final-viral-combined_k141_111503||full Not-determined
17-108-final-viral-combined_k141_1469||full Not-determined
17-108-final-viral-combined_k141_56589||full Not-determined
17-108-final-viral-combined_k141_111506||full Low-quality
17-108-final-viral-combined_k141_290825||full Not-determined
17-108-final-viral-combined_k141_318352||full Not-determined
17-108-final-viral-combined_k141_153110||full Not-determined
17-108-final-viral-combined_k141_153111||full Low-quality
17-108-final-viral-combined_k141_236146||full Not-determined
17-108-final-viral-combined_k141_236165||full Not-determined
17-108-final-viral-combined_k141_125755||full Not-determined
17-108-final-viral-combined_k141_318371||full Low-quality
17-108-final-viral-combined_k141_180631||full Not-determined
17-108-final-viral-combined_k141_318374||full Not-determined
17-108-final-viral-combined_k141_208315||full Not-determined
17-108-final-viral-combined_k141_56625||full Not-determined
17-108-final-viral-combined_k141_1510||full Low-quality
17-108-final-viral-combined_k141_290886||full Not-determined
17-108-final-viral-combined_k141_29048||full Not-determined
17-108-final-viral-combined_k141_180646||full Not-determined
17-108-final-viral-combined_k141_98309||full Low-quality
17-108-final-viral-combined_k141_222067||full Not-determined
17-108-final-viral-combined_k141_56638||full Not-determined
17-108-final-viral-combined_k141_98321||full Not-determined
17-108-final-viral-combined_k141_236191||full Low-quality
17-108-final-viral-combined_k141_139179||full Low-quality
17-108-final-viral-combined_k141_236201||full Not-determined
17-108-final-viral-combined_k141_194475||full Low-quality
17-108-final-viral-combined_k141_194482||full Not-determined
17-108-final-viral-combined_k141_277321||full Not-determined
17-108-final-viral-combined_k141_277325||full Not-determined
17-108-final-viral-combined_k141_1594||full Not-determined
KM-final-viral-combined_k141_186314||full Complete
17-108-final-viral-combined_k141_263485||full Not-determined
17-108-final-viral-combined_k141_139196||full Not-determined
17-108-final-viral-combined_k141_180682||full Low-quality
17-108-final-viral-combined_k141_194509||full Low-quality
17-108-final-viral-combined_k141_290937||full Not-determined
17-108-final-viral-combined_k141_304863||full Not-determined
17-108-final-viral-combined_k141_208367||full Not-determined
17-108-final-viral-combined_k141_180685||full Not-determined
17-108-final-viral-combined_k141_15480||full Not-determined
17-108-final-viral-combined_k141_194525||full Not-determined
17-108-final-viral-combined_k141_208378||full Low-quality
17-108-final-viral-combined_k141_84363||full Not-determined
17-108-final-viral-combined_k141_153187||full Not-determined
17-108-final-viral-combined_k141_98369||full Low-quality
17-108-final-viral-combined_k141_208394||full Not-determined
17-108-final-viral-combined_k141_263516||full Not-determined
17-108-final-viral-combined_k141_84378||full Not-determined
17-108-final-viral-combined_k141_304880||full Not-determined
17-108-final-viral-combined_k141_290980||full Not-determined
17-108-final-viral-combined_k141_15522||full Low-quality
17-108-final-viral-combined_k141_222198||full Not-determined
17-108-final-viral-combined_k141_153243||full Low-quality
17-108-final-viral-combined_k141_15525||full Low-quality
17-108-final-viral-combined_k141_277383||full Not-determined
17-108-final-viral-combined_k141_180726||full Low-quality
17-108-final-viral-combined_k141_291014||full Not-determined
17-108-final-viral-combined_k141_194580||full Low-quality
17-108-final-viral-combined_k141_125856||full Low-quality
17-108-final-viral-combined_k141_277392||full Low-quality
17-108-final-viral-combined_k141_208423||full Not-determined
17-108-final-viral-combined_k141_291031||full Not-determined
17-108-final-viral-combined_k141_304922||full Low-quality
17-108-final-viral-combined_k141_304924||full Low-quality
17-108-final-viral-combined_k141_180743||full Low-quality
18-101_k141_394571 Complete
17-108-final-viral-combined_k141_291042||full Not-determined
17-108-final-viral-combined_k141_194587||full Low-quality
17-108-final-viral-combined_k141_15573||full Low-quality
17-108-final-viral-combined_k141_29134||full Low-quality
17-108-final-viral-combined_k141_236234||full Low-quality
17-108-final-viral-combined_k141_208452||full Not-determined
17-108-final-viral-combined_k141_98394||full Low-quality
17-108-final-viral-combined_k141_84430||full Low-quality
17-108-final-viral-combined_k141_111620||full Low-quality
17-108-final-viral-combined_k141_236236||full Not-determined
17-108-final-viral-combined_k141_291053||full Low-quality
17-108-final-viral-combined_k141_15586||full Not-determined
17-108-final-viral-combined_k141_84433||full Not-determined
17-108-final-viral-combined_k141_56753||full Low-quality
17-108-final-viral-combined_k141_236246||full Low-quality
17-108-final-viral-combined_k141_180782||full Low-quality
17-108-final-viral-combined_k141_56759||full Not-determined
17-108-final-viral-combined_k141_304966||full Low-quality
17-108-final-viral-combined_k141_42928||full Not-determined
17-108-final-viral-combined_k141_98398||full Low-quality
17-108-final-viral-combined_k141_291057||full Low-quality
17-108-final-viral-combined_k141_111652||full Low-quality
17-108-final-viral-combined_k141_111656||full Low-quality
17-108-final-viral-combined_k141_180806||full Low-quality
17-108-final-viral-combined_k141_180807||full Low-quality
17-108-final-viral-combined_k141_222263||full Low-quality
17-108-final-viral-combined_k141_180808||full Low-quality
17-108-final-viral-combined_k141_194597||full Not-determined
17-108-final-viral-combined_k141_236254||full Not-determined
17-108-final-viral-combined_k141_70349||full Not-determined
17-108-final-viral-combined_k141_180824||full Low-quality
17-108-final-viral-combined_k141_139335||full Not-determined
17-108-final-viral-combined_k141_1702||full Not-determined
17-108-final-viral-combined_k141_167421||full Not-determined
17-108-final-viral-combined_k141_42956||full Low-quality
17-108-final-viral-combined_k141_222276||full Low-quality
17-108-final-viral-combined_k141_180827||full Low-quality
17-108-final-viral-combined_k141_15629||full Low-quality
17-108-final-viral-combined_k141_277460||full Not-determined
17-108-final-viral-combined_k141_84449||full Not-determined
17-108-final-viral-combined_k141_304985||full Not-determined
17-108-final-viral-combined_k141_153340||full Low-quality
17-108-final-viral-combined_k141_222289||full Not-determined
17-108-final-viral-combined_k141_15646||full Low-quality
17-108-final-viral-combined_k141_194620||full Not-determined
17-108-final-viral-combined_k141_15652||full Not-determined
17-108-final-viral-combined_k141_208527||full Low-quality
17-108-final-viral-combined_k141_153350||full Not-determined
17-108-final-viral-combined_k141_167448||full Not-determined
17-108-final-viral-combined_k141_263587||full Not-determined
17-108-final-viral-combined_k141_111704||full Low-quality
17-108-final-viral-combined_k141_98435||full Not-determined
17-108-final-viral-combined_k141_43002||full Not-determined
17-108-final-viral-combined_k141_318520||full Low-quality
17-108-final-viral-combined_k141_180858||full Low-quality
17-108-final-viral-combined_k141_43020||full Not-determined
17-108-final-viral-combined_k141_167462||full Low-quality
17-108-final-viral-combined_k141_1715||full Not-determined
17-108-final-viral-combined_k141_98447||full Not-determined
17-108-final-viral-combined_k141_56857||full Not-determined
17-108-final-viral-combined_k141_291100||full Not-determined
17-108-final-viral-combined_k141_56859||full Not-determined
17-108-final-viral-combined_k141_153365||full Low-quality
17-108-final-viral-combined_k141_111715||full Not-determined
17-108-final-viral-combined_k141_208545||full Low-quality
17-108-final-viral-combined_k141_84504||full Low-quality
17-108-final-viral-combined_k141_277499||full Not-determined
17-108-final-viral-combined_k141_15676||full Not-determined
17-108-final-viral-combined_k141_180865||full Not-determined
17-108-final-viral-combined_k141_98465||full Not-determined
17-108-final-viral-combined_k141_56874||full Low-quality
17-108-final-viral-combined_k141_111736||full Low-quality
17-108-final-viral-combined_k141_249813||full Low-quality
17-108-final-viral-combined_k141_222322||full Low-quality
17-108-final-viral-combined_k141_236343||full Not-determined
17-108-final-viral-combined_k141_84536||full Not-determined
17-108-final-viral-combined_k141_43090||full Not-determined
17-108-final-viral-combined_k141_167524||full Low-quality
17-108-final-viral-combined_k141_111758||full Low-quality
17-108-final-viral-combined_k141_167526||full Not-determined
17-108-final-viral-combined_k141_84554||full Low-quality
17-108-final-viral-combined_k141_180894||full Not-determined
17-108-final-viral-combined_k141_180895||full Low-quality
17-108-final-viral-combined_k141_125887||full Not-determined
17-108-final-viral-combined_k141_84580||full Low-quality
17-108-final-viral-combined_k141_56923||full Low-quality
17-108-final-viral-combined_k141_98528||full Low-quality
17-108-final-viral-combined_k141_84587||full Not-determined
17-108-final-viral-combined_k141_84593||full Not-determined
17-108-final-viral-combined_k141_125908||full Low-quality
17-108-final-viral-combined_k141_98540||full Not-determined
17-108-final-viral-combined_k141_305018||full Not-determined
17-108-final-viral-combined_k141_153401||full Low-quality
17-108-final-viral-combined_k141_305022||full Low-quality
17-108-final-viral-combined_k141_222372||full Not-determined
17-108-final-viral-combined_k141_291171||full Low-quality
17-108-final-viral-combined_k141_222378||full Not-determined
17-108-final-viral-combined_k141_29236||full Not-determined
17-108-final-viral-combined_k141_139484||full Low-quality
17-108-final-viral-combined_k141_139498||full Not-determined
17-108-final-viral-combined_k141_236390||full Not-determined
17-108-final-viral-combined_k141_84624||full Low-quality
17-108-final-viral-combined_k141_153428||full Not-determined
17-108-final-viral-combined_k141_56944||full Low-quality
17-108-final-viral-combined_k141_277598||full Not-determined
17-108-final-viral-combined_k141_208690||full Not-determined
17-108-final-viral-combined_k141_291200||full Not-determined
17-108-final-viral-combined_k141_305081||full Not-determined
17-108-final-viral-combined_k141_318628||full Low-quality
17-108-final-viral-combined_k141_111833||full Low-quality
17-108-final-viral-combined_k141_153451||full Not-determined
17-108-final-viral-combined_k141_15710||full Low-quality
17-108-final-viral-combined_k141_291227||full Not-determined
17-108-final-viral-combined_k141_15712||full Not-determined
17-108-final-viral-combined_k141_84665||full Low-quality
17-108-final-viral-combined_k141_139530||full Not-determined
17-108-final-viral-combined_k141_318643||full Low-quality
17-108-final-viral-combined_k141_139532||full Low-quality
17-108-final-viral-combined_k141_194717||full Low-quality
17-108-final-viral-combined_k141_318656||full Low-quality
17-108-final-viral-combined_k141_194736||full Not-determined
17-108-final-viral-combined_k141_98600||full Not-determined
17-108-final-viral-combined_k141_318675||full Not-determined
17-108-final-viral-combined_k141_98603||full Not-determined
17-108-final-viral-combined_k141_194753||full Low-quality
17-108-final-viral-combined_k141_15754||full Not-determined
17-108-final-viral-combined_k141_29300||full Low-quality
17-108-final-viral-combined_k141_194761||full Low-quality
17-108-final-viral-combined_k141_194784||full Low-quality
17-108-final-viral-combined_k141_222501||full Not-determined
17-108-final-viral-combined_k141_57035||full Low-quality
17-108-final-viral-combined_k141_167569||full Not-determined
17-108-final-viral-combined_k141_249860||full Not-determined
17-108-final-viral-combined_k141_222521||full Not-determined
17-108-final-viral-combined_k141_111844||full Not-determined
17-108-final-viral-combined_k141_126019||full Low-quality
17-108-final-viral-combined_k141_180981||full Low-quality
17-108-final-viral-combined_k141_126023||full Not-determined
17-108-final-viral-combined_k141_43184||full Not-determined
17-108-final-viral-combined_k141_43189||full Not-determined
17-108-final-viral-combined_k141_291272||full Not-determined
17-108-final-viral-combined_k141_236477||full Low-quality
17-108-final-viral-combined_k141_249891||full Not-determined
17-108-final-viral-combined_k141_57082||full Not-determined
17-108-final-viral-combined_k141_291275||full Low-quality
17-108-final-viral-combined_k141_167608||full Not-determined
17-108-final-viral-combined_k141_153490||full Low-quality
17-108-final-viral-combined_k141_15775||full Not-determined
17-108-final-viral-combined_k141_57091||full Low-quality
17-108-final-viral-combined_k141_277658||full Not-determined
17-108-final-viral-combined_k141_84739||full Low-quality
17-108-final-viral-combined_k141_1859||full Low-quality
17-108-final-viral-combined_k141_1860||full Low-quality
17-108-final-viral-combined_k141_263794||full Low-quality
17-108-final-viral-combined_k141_194825||full Low-quality
17-108-final-viral-combined_k141_111868||full Not-determined
17-108-final-viral-combined_k141_15782||full Low-quality
17-108-final-viral-combined_k141_43229||full Low-quality
17-108-final-viral-combined_k141_1884||full Low-quality
17-108-final-viral-combined_k141_181029||full Not-determined
17-108-final-viral-combined_k141_249929||full Not-determined
17-108-final-viral-combined_k141_84762||full Low-quality
17-108-final-viral-combined_k141_305209||full Not-determined
17-108-final-viral-combined_k141_291313||full Not-determined
17-108-final-viral-combined_k141_98673||full Not-determined
17-108-final-viral-combined_k141_126059||full Low-quality
17-108-final-viral-combined_k141_111894||full Low-quality
17-108-final-viral-combined_k141_1906||full Not-determined
17-108-final-viral-combined_k141_263833||full Low-quality
17-108-final-viral-combined_k141_1909||full Low-quality
17-108-final-viral-combined_k141_249937||full Not-determined
17-108-final-viral-combined_k141_305230||full Low-quality
17-108-final-viral-combined_k141_43251||full Low-quality
17-108-final-viral-combined_k141_236532||full Low-quality
17-108-final-viral-combined_k141_318741||full Not-determined
17-108-final-viral-combined_k141_139681||full Low-quality
17-108-final-viral-combined_k141_236538||full Not-determined
17-108-final-viral-combined_k141_318748||full Not-determined
17-108-final-viral-combined_k141_318749||full Not-determined
17-108-final-viral-combined_k141_249989||full Not-determined
17-108-final-viral-combined_k141_208877||full Low-quality
17-108-final-viral-combined_k141_194888||full Not-determined
17-108-final-viral-combined_k141_318761||full Not-determined
17-108-final-viral-combined_k141_84807||full Not-determined
17-108-final-viral-combined_k141_291333||full Not-determined
17-108-final-viral-combined_k141_167712||full Not-determined
17-108-final-viral-combined_k141_29384||full Low-quality
17-108-final-viral-combined_k141_139698||full Not-determined
17-108-final-viral-combined_k141_167715||full Low-quality
17-108-final-viral-combined_k141_126093||full Low-quality
17-108-final-viral-combined_k141_98730||full Not-determined
17-108-final-viral-combined_k141_98731||full Low-quality
17-108-final-viral-combined_k141_84828||full Not-determined
17-108-final-viral-combined_k141_98738||full Low-quality
17-108-final-viral-combined_k141_29406||full Low-quality
17-108-final-viral-combined_k141_126100||full Not-determined
17-108-final-viral-combined_k141_222578||full Not-determined
17-108-final-viral-combined_k141_263865||full Not-determined

| | |
|---|---|
| 17-108-final-viral-combined_k141_181101\|\|full | Not-determined |
| 17-108-final-viral-combined_k141_29428\|\|full | Not-determined |
| 17-108-final-viral-combined_k141_43329\|\|full | Not-determined |
| 17-108-final-viral-combined_k141_126114\|\|full | Low-quality |
| 17-108-final-viral-combined_k141_15924\|\|full | Not-determined |
| 17-108-final-viral-combined_k141_263877\|\|full | Low-quality |
| 17-108-final-viral-combined_k141_208956\|\|full | Not-determined |
| 17-108-final-viral-combined_k141_291410\|\|full | Not-determined |
| 17-108-final-viral-combined_k141_167741\|\|full | Low-quality |
| 17-108-final-viral-combined_k141_43337\|\|full | Not-determined |
| 17-108-final-viral-combined_k141_126132\|\|full | Low-quality |
| 17-108-final-viral-combined_k141_263889\|\|full | Not-determined |
| 17-108-final-viral-combined_k141_277850\|\|full | Low-quality |
| 17-108-final-viral-combined_k141_98795\|\|full | Not-determined |
| 17-108-final-viral-combined_k141_29467\|\|full | Low-quality |
| 17-108-final-viral-combined_k141_236617\|\|full | Not-determined |
| 17-108-final-viral-combined_k141_153539\|\|full | Low-quality |
| 17-108-final-viral-combined_k141_70432\|\|full | Low-quality |
| 17-108-final-viral-combined_k141_43361\|\|full | Not-determined |
| 17-108-final-viral-combined_k141_263921\|\|full | Not-determined |
| 17-108-final-viral-combined_k141_153566\|\|full | Not-determined |
| 17-108-final-viral-combined_k141_318847\|\|full | Low-quality |
| 17-108-final-viral-combined_k141_305298\|\|full | Not-determined |
| 17-108-final-viral-combined_k141_2050\|\|full | Low-quality |
| 17-108-final-viral-combined_k141_222621\|\|full | Low-quality |
| 17-108-final-viral-combined_k141_153579\|\|full | Not-determined |
| 17-108-final-viral-combined_k141_209006\|\|full | Not-determined |
| 17-108-final-viral-combined_k141_153584\|\|full | Low-quality |
| 17-108-final-viral-combined_k141_305321\|\|full | Not-determined |
| 17-108-final-viral-combined_k141_43400\|\|full | Not-determined |
| 17-108-final-viral-combined_k141_70453\|\|full | Not-determined |
| 17-108-final-viral-combined_k141_222625\|\|full | Low-quality |
| 17-108-final-viral-combined_k141_57239\|\|full | Not-determined |
| 17-108-final-viral-combined_k141_277905\|\|full | Not-determined |
| 17-108-final-viral-combined_k141_263962\|\|full | Not-determined |
| 17-108-final-viral-combined_k141_167763\|\|full | Not-determined |
| 17-108-final-viral-combined_k141_291465\|\|full | Not-determined |
| 17-108-final-viral-combined_k141_84859\|\|full | Low-quality |
| 17-108-final-viral-combined_k141_209025\|\|full | Not-determined |
| 17-108-final-viral-combined_k141_43402\|\|full | Not-determined |
| 17-108-final-viral-combined_k141_318885\|\|full | Low-quality |
| 17-108-final-viral-combined_k141_57283\|\|full | Low-quality |
| 17-108-final-viral-combined_k141_236712\|\|full | Not-determined |
| 17-108-final-viral-combined_k141_43404\|\|full | Low-quality |
| 17-108-final-viral-combined_k141_236721\|\|full | Not-determined |
| 17-108-final-viral-combined_k141_305367\|\|full | Not-determined |
| 17-108-final-viral-combined_k141_43411\|\|full | Low-quality |
| 17-108-final-viral-combined_k141_318920\|\|full | Not-determined |
| 17-108-final-viral-combined_k141_195011\|\|full | Not-determined |
| 17-108-final-viral-combined_k141_209033\|\|full | Low-quality |
| 17-108-final-viral-combined_k141_167797\|\|full | Low-quality |
| 17-108-final-viral-combined_k141_277969\|\|full | Not-determined |
| 17-108-final-viral-combined_k141_29580\|\|full | Not-determined |
| 17-108-final-viral-combined_k141_126204\|\|full | Not-determined |
| 17-108-final-viral-combined_k141_167800\|\|full | Low-quality |
| 17-108-final-viral-combined_k141_70468\|\|full | Not-determined |
| 17-108-final-viral-combined_k141_236731\|\|full | Low-quality |
| 17-108-final-viral-combined_k141_84904\|\|full | Not-determined |
| 17-108-final-viral-combined_k141_153635\|\|full | Low-quality |
| 17-108-final-viral-combined_k141_305411\|\|full | Not-determined |
| 17-108-final-viral-combined_k141_291493\|\|full | Not-determined |
| 17-108-final-viral-combined_k141_2119\|\|full | Not-determined |
| 17-108-final-viral-combined_k141_153661\|\|full | Low-quality |
| 17-108-final-viral-combined_k141_126235\|\|full | Not-determined |
| 17-108-final-viral-combined_k141_209070\|\|full | Low-quality |
| 17-108-final-viral-combined_k141_139879\|\|full | Not-determined |
| 17-108-final-viral-combined_k141_57355\|\|full | Not-determined |
| 17-108-final-viral-combined_k141_195033\|\|full | Not-determined |
| 17-108-final-viral-combined_k141_318965\|\|full | Not-determined |
| 17-108-final-viral-combined_k141_84925\|\|full | Not-determined |
| 17-108-final-viral-combined_k141_181217\|\|full | Not-determined |
| 17-108-final-viral-combined_k141_153678\|\|full | Not-determined |
| 17-108-final-viral-combined_k141_318971\|\|full | Low-quality |
| 17-108-final-viral-combined_k141_209091\|\|full | Not-determined |
| 17-108-final-viral-combined_k141_16103\|\|full | Not-determined |
| 17-108-final-viral-combined_k141_181225\|\|full | Low-quality |
| 17-108-final-viral-combined_k141_2139\|\|full | Low-quality |
| 17-108-final-viral-combined_k141_209104\|\|full | Not-determined |
| 17-108-final-viral-combined_k141_84956\|\|full | Not-determined |
| 17-108-final-viral-combined_k141_195070\|\|full | Low-quality |
| 17-108-final-viral-combined_k141_209119\|\|full | Not-determined |
| 17-108-final-viral-combined_k141_112100\|\|full | Not-determined |
| 17-108-final-viral-combined_k141_222729\|\|full | Not-determined |
| 17-108-final-viral-combined_k141_167828\|\|full | Not-determined |
| 17-108-final-viral-combined_k141_209135\|\|full | Low-quality |
| 17-108-final-viral-combined_k141_126259\|\|full | Not-determined |
| 17-108-final-viral-combined_k141_278092\|\|full | Not-determined |
| 17-108-final-viral-combined_k141_98900\|\|full | Low-quality |
| 17-108-final-viral-combined_k141_57407\|\|full | Not-determined |
| 17-108-final-viral-combined_k141_139912\|\|full | Not-determined |
| 17-108-final-viral-combined_k141_319018\|\|full | Not-determined |
| 17-108-final-viral-combined_k141_57412\|\|full | Not-determined |
| 17-108-final-viral-combined_k141_70570\|\|full | Not-determined |
| 17-108-final-viral-combined_k141_291530\|\|full | Low-quality |
| 17-108-final-viral-combined_k141_236884\|\|full | Not-determined |
| 17-108-final-viral-combined_k141_236885\|\|full | Low-quality |
| 17-108-final-viral-combined_k141_236889\|\|full | Not-determined |
| 17-108-final-viral-combined_k141_236894\|\|full | Not-determined |
| 18-102_k141_124992 | High-quality |
| 17-108-final-viral-combined_k141_250295\|\|full | Low-quality |
| 17-108-final-viral-combined_k141_209182\|\|full | Not-determined |
| 17-108-final-viral-combined_k141_85010\|\|full | Low-quality |
| 17-108-final-viral-combined_k141_139935\|\|full | Not-determined |
| 17-108-final-viral-combined_k141_98951\|\|full | Not-determined |
| 17-108-final-viral-combined_k141_250315\|\|full | Low-quality |
| 17-108-final-viral-combined_k141_16121\|\|full | Low-quality |
| 17-108-final-viral-combined_k141_43503\|\|full | Low-quality |
| 17-108-final-viral-combined_k141_153732\|\|full | Low-quality |
| 17-108-final-viral-combined_k141_264066\|\|full | Not-determined |
| 17-108-final-viral-combined_k141_2197\|\|full | Not-determined |
| 17-108-final-viral-combined_k141_153743\|\|full | Low-quality |
| 17-108-final-viral-combined_k141_139941\|\|full | Not-determined |
| 17-108-final-viral-combined_k141_305507\|\|full | Not-determined |
| 17-108-final-viral-combined_k141_70616\|\|full | Low-quality |
| 17-108-final-viral-combined_k141_70633\|\|full | Low-quality |
| 17-108-final-viral-combined_k141_57518\|\|full | Low-quality |
| 17-108-final-viral-combined_k141_139958\|\|full | Not-determined |
| 17-108-final-viral-combined_k141_70645\|\|full | Not-determined |
| 17-108-final-viral-combined_k141_236934\|\|full | Low-quality |
| 17-108-final-viral-combined_k141_291579\|\|full | Not-determined |
| 17-108-final-viral-combined_k141_209287\|\|full | Not-determined |
| 17-108-final-viral-combined_k141_195175\|\|full | Low-quality |
| 17-108-final-viral-combined_k141_29735\|\|full | Low-quality |
| 17-108-final-viral-combined_k141_291594\|\|full | Not-determined |
| 17-108-final-viral-combined_k141_250339\|\|full | Low-quality |
| 17-108-final-viral-combined_k141_209295\|\|full | Not-determined |
| 17-108-final-viral-combined_k141_126308\|\|full | Not-determined |
| 17-108-final-viral-combined_k141_278168\|\|full | Low-quality |
| 17-108-final-viral-combined_k141_264131\|\|full | Low-quality |
| 17-108-final-viral-combined_k141_126314\|\|full | Low-quality |
| 18-101-final-viral-combined_k141_394571\|\|full | High-quality |
| 17-108-final-viral-combined_k141_305577\|\|full | Low-quality |
| 17-108-final-viral-combined_k141_250379\|\|full | Low-quality |
| 17-108-final-viral-combined_k141_153796\|\|full | Low-quality |
| 17-108-final-viral-combined_k141_319036\|\|full | Low-quality |
| 17-108-final-viral-combined_k141_167923\|\|full | Not-determined |
| 17-108-final-viral-combined_k141_70697\|\|full | Not-determined |
| 17-108-final-viral-combined_k141_16155\|\|full | Not-determined |
| 17-108-final-viral-combined_k141_126328\|\|full | Low-quality |
| 17-108-final-viral-combined_k141_236944\|\|full | Low-quality |
| 17-108-final-viral-combined_k141_305593\|\|full | Not-determined |
| 17-108-final-viral-combined_k141_209317\|\|full | Low-quality |
| 17-108-final-viral-combined_k141_126341\|\|full | Not-determined |
| 17-108-final-viral-combined_k141_209321\|\|full | Low-quality |
| 17-108-final-viral-combined_k141_126352\|\|full | Not-determined |
| 17-108-final-viral-combined_k141_319045\|\|full | Not-determined |
| 17-108-final-viral-combined_k141_2297\|\|full | Not-determined |
| 17-108-final-viral-combined_k141_264168\|\|full | Not-determined |
| 17-108-final-viral-combined_k141_305611\|\|full | Not-determined |
| 17-108-final-viral-combined_k141_43549\|\|full | Not-determined |
| 17-108-final-viral-combined_k141_29788\|\|full | Not-determined |
| 17-108-final-viral-combined_k141_126391\|\|full | Low-quality |
| 17-108-final-viral-combined_k141_70726\|\|full | Not-determined |
| 17-108-final-viral-combined_k141_319060\|\|full | Low-quality |
| 17-108-final-viral-combined_k141_319065\|\|full | Not-determined |
| 18-102-final-viral-combined_k141_124992\|\|full | High-quality |
| 17-108-final-viral-combined_k141_43578\|\|full | Low-quality |
| 17-108-final-viral-combined_k141_126404\|\|full | Low-quality |
| 17-108-final-viral-combined_k141_278232\|\|full | Not-determined |
| 17-108-final-viral-combined_k141_2336\|\|full | Not-determined |
| 17-108-final-viral-combined_k141_153833\|\|full | Low-quality |
| 17-108-final-viral-combined_k141_29819\|\|full | Not-determined |
| 17-108-final-viral-combined_k141_209357\|\|full | Not-determined |
| 17-108-final-viral-combined_k141_140040\|\|full | Not-determined |
| 17-108-final-viral-combined_k141_222766\|\|full | Not-determined |
| 17-108-final-viral-combined_k141_167967\|\|full | Low-quality |
| 17-108-final-viral-combined_k141_43609\|\|full | Low-quality |
| 17-108-final-viral-combined_k141_70768\|\|full | Low-quality |
| 17-108-final-viral-combined_k141_57615\|\|full | Low-quality |
| 17-108-final-viral-combined_k141_236973\|\|full | Low-quality |
| 17-108-final-viral-combined_k141_167982\|\|full | Low-quality |
| 17-108-final-viral-combined_k141_250437\|\|full | Low-quality |
| 17-108-final-viral-combined_k141_319123\|\|full | Low-quality |
| 17-108-final-viral-combined_k141_222785\|\|full | Not-determined |
| 17-108-final-viral-combined_k141_319127\|\|full | Low-quality |
| 17-108-final-viral-combined_k141_250445\|\|full | Not-determined |
| 17-108-final-viral-combined_k141_222789\|\|full | Not-determined |
| 17-108-final-viral-combined_k141_209410\|\|full | Not-determined |
| 17-108-final-viral-combined_k141_70789\|\|full | Not-determined |
| 17-108-final-viral-combined_k141_99064\|\|full | Low-quality |
| 17-108-final-viral-combined_k141_319130\|\|full | Low-quality |
| 17-108-final-viral-combined_k141_126460\|\|full | Low-quality |
| 17-108-final-viral-combined_k141_319133\|\|full | Low-quality |
| 17-108-final-viral-combined_k141_140080\|\|full | Low-quality |
| 17-108-final-viral-combined_k141_43621\|\|full | Not-determined |
| 17-108-final-viral-combined_k141_195273\|\|full | Low-quality |
| 17-108-final-viral-combined_k141_305704\|\|full | Low-quality |
| 17-108-final-viral-combined_k141_195284\|\|full | Low-quality |
| 17-108-final-viral-combined_k141_181353\|\|full | Low-quality |
| 17-108-final-viral-combined_k141_29840\|\|full | Low-quality |
| 17-108-final-viral-combined_k141_250459\|\|full | Low-quality |
| 17-108-final-viral-combined_k141_29841\|\|full | Not-determined |
| 17-108-final-viral-combined_k141_250460\|\|full | Not-determined |
| 17-108-final-viral-combined_k141_153899\|\|full | Low-quality |
| 17-108-final-viral-combined_k141_43679\|\|full | Not-determined |
| 17-108-final-viral-combined_k141_57682\|\|full | Low-quality |
| 17-108-final-viral-combined_k141_112228\|\|full | Low-quality |
| 17-108-final-viral-combined_k141_250492\|\|full | Not-determined |
| 17-108-final-viral-combined_k141_2432\|\|full | Not-determined |
| 17-108-final-viral-combined_k141_29856\|\|full | Not-determined |
| 17-108-final-viral-combined_k141_43692\|\|full | Not-determined |
| 17-108-final-viral-combined_k141_222847\|\|full | Low-quality |
| 17-108-final-viral-combined_k141_57694\|\|full | Not-determined |
| 17-108-final-viral-combined_k141_29863\|\|full | Low-quality |
| 17-108-final-viral-combined_k141_70840\|\|full | Not-determined |
| 17-108-final-viral-combined_k141_209477\|\|full | Low-quality |
| 17-108-final-viral-combined_k141_70845\|\|full | Not-determined |
| 17-108-final-viral-combined_k141_112259\|\|full | Low-quality |
| 17-108-final-viral-combined_k141_209501\|\|full | Low-quality |
| 17-108-final-viral-combined_k141_305809\|\|full | Not-determined |
| 17-108-final-viral-combined_k141_85099\|\|full | Not-determined |
| 17-108-final-viral-combined_k141_278316\|\|full | Not-determined |
| 17-108-final-viral-combined_k141_70863\|\|full | Not-determined |
| 17-108-final-viral-combined_k141_250516\|\|full | Not-determined |
| 17-108-final-viral-combined_k141_319223\|\|full | Low-quality |
| 17-108-final-viral-combined_k141_209521\|\|full | Not-determined |
| 17-108-final-viral-combined_k141_70875\|\|full | Low-quality |
| 17-108-final-viral-combined_k141_181393\|\|full | Low-quality |
| 17-108-final-viral-combined_k141_209524\|\|full | Not-determined |
| 17-108-final-viral-combined_k141_126580\|\|full | Not-determined |
| 18-106-final-viral-combined_k141_88414\|\|full | Complete |
| 17-108-final-viral-combined_k141_278337\|\|full | Not-determined |
| 17-108-final-viral-combined_k141_181405\|\|full | Low-quality |
| 17-108-final-viral-combined_k141_140224\|\|full | Low-quality |
| 17-108-final-viral-combined_k141_168065\|\|full | Low-quality |
| 17-108-final-viral-combined_k141_57757\|\|full | Low-quality |
| 17-108-final-viral-combined_k141_2536\|\|full | Low-quality |
| 17-108-final-viral-combined_k141_209542\|\|full | Low-quality |
| 17-108-final-viral-combined_k141_222957\|\|full | Not-determined |
| 17-108-final-viral-combined_k141_2552\|\|full | Low-quality |
| 17-108-final-viral-combined_k141_57787\|\|full | Not-determined |
| 17-108-final-viral-combined_k141_168091\|\|full | Not-determined |
| 17-108-final-viral-combined_k141_222972\|\|full | Not-determined |
| 17-108-final-viral-combined_k141_140237\|\|full | Not-determined |
| 17-108-final-viral-combined_k141_57791\|\|full | Low-quality |
| 17-108-final-viral-combined_k141_43763\|\|full | Not-determined |
| 17-108-final-viral-combined_k141_2567\|\|full | Low-quality |
| 17-108-final-viral-combined_k141_57802\|\|full | Not-determined |
| 17-108-final-viral-combined_k141_140250\|\|full | Not-determined |
| 17-108-final-viral-combined_k141_278411\|\|full | Low-quality |
| 17-108-final-viral-combined_k141_57807\|\|full | Low-quality |
| 17-108-final-viral-combined_k141_2587\|\|full | Not-determined |
| 17-108-final-viral-combined_k141_99183\|\|full | Not-determined |
| 17-108-final-viral-combined_k141_99186\|\|full | Not-determined |
| 17-108-final-viral-combined_k141_70962\|\|full | Low-quality |
| 17-108-final-viral-combined_k141_181414\|\|full | Not-determined |
| 17-108-final-viral-combined_k141_140272\|\|full | Not-determined |
| 17-108-final-viral-combined_k141_85122\|\|full | Low-quality |
| 17-108-final-viral-combined_k141_154062\|\|full | Not-determined |
| 17-108-final-viral-combined_k141_209597\|\|full | Not-determined |
| 17-108-final-viral-combined_k141_140288\|\|full | Low-quality |
| 17-108-final-viral-combined_k141_43818\|\|full | Not-determined |
| 17-108-final-viral-combined_k141_154076\|\|full | Not-determined |
| 17-108-final-viral-combined_k141_30034\|\|full | Not-determined |
| 17-108-final-viral-combined_k141_99216\|\|full | Not-determined |
| 17-108-final-viral-combined_k141_237088\|\|full | Not-determined |
| 17-108-final-viral-combined_k141_154085\|\|full | Low-quality |
| 17-108-final-viral-combined_k141_70983\|\|full | Not-determined |
| 17-108-final-viral-combined_k141_319317\|\|full | Not-determined |
| 17-108-final-viral-combined_k141_112328\|\|full | Not-determined |
| 17-108-final-viral-combined_k141_319318\|\|full | Not-determined |
| 17-108-final-viral-combined_k141_250618\|\|full | Low-quality |
| 17-108-final-viral-combined_k141_291744\|\|full | Not-determined |
| 17-108-final-viral-combined_k141_57868\|\|full | Not-determined |
| 17-108-final-viral-combined_k141_2619\|\|full | Not-determined |
| 17-108-final-viral-combined_k141_278457\|\|full | Low-quality |
| 17-108-final-viral-combined_k141_43862\|\|full | Low-quality |
| 17-108-final-viral-combined_k141_168179\|\|full | Not-determined |
| 17-108-final-viral-combined_k141_71008\|\|full | Not-determined |
| 17-108-final-viral-combined_k141_237108\|\|full | Low-quality |
| 17-108-final-viral-combined_k141_112357\|\|full | Low-quality |
| 17-108-final-viral-combined_k141_278473\|\|full | Low-quality |
| 17-108-final-viral-combined_k141_195395\|\|full | Not-determined |
| 17-108-final-viral-combined_k141_112367\|\|full | Not-determined |
| 17-108-final-viral-combined_k141_16242\|\|full | Not-determined |
| 17-108-final-viral-combined_k141_85182\|\|full | Low-quality |
| 17-108-final-viral-combined_k141_223102\|\|full | Low-quality |
| 17-108-final-viral-combined_k141_291762\|\|full | Low-quality |
| 17-108-final-viral-combined_k141_305945\|\|full | Not-determined |
| 17-108-final-viral-combined_k141_264325\|\|full | Not-determined |
| 17-108-final-viral-combined_k141_305957\|\|full | Low-quality |
| 17-108-final-viral-combined_k141_278498\|\|full | Not-determined |
| 17-108-final-viral-combined_k141_278504\|\|full | Not-determined |
| 17-108-final-viral-combined_k141_71040\|\|full | Not-determined |
| 17-108-final-viral-combined_k141_43904\|\|full | Not-determined |
| 17-108-final-viral-combined_k141_57942\|\|full | Low-quality |
| 17-108-final-viral-combined_k141_126619\|\|full | Not-determined |
| 17-108-final-viral-combined_k141_126622\|\|full | Not-determined |
| 17-108-final-viral-combined_k141_43918\|\|full | Low-quality |
| 17-108-final-viral-combined_k141_43924\|\|full | Low-quality |
| 17-108-final-viral-combined_k141_264353\|\|full | Low-quality |
| 17-108-final-viral-combined_k141_71056\|\|full | Not-determined |
| 17-108-final-viral-combined_k141_112409\|\|full | Low-quality |
| 17-108-final-viral-combined_k141_305980\|\|full | Not-determined |
| 17-108-final-viral-combined_k141_71073\|\|full | Low-quality |
| 17-108-final-viral-combined_k141_264378\|\|full | Not-determined |
| 17-108-final-viral-combined_k141_264382\|\|full | Not-determined |
| 17-108-final-viral-combined_k141_43960\|\|full | Low-quality |
| 17-108-final-viral-combined_k141_278558\|\|full | Not-determined |
| 17-108-final-viral-combined_k141_278563\|\|full | Low-quality |
| 17-108-final-viral-combined_k141_99353\|\|full | Low-quality |
| 17-108-final-viral-combined_k141_223162\|\|full | Not-determined |
| 17-108-final-viral-combined_k141_319373\|\|full | Low-quality |
| 17-108-final-viral-combined_k141_209746\|\|full | Low-quality |
| 17-108-final-viral-combined_k141_30153\|\|full | Low-quality |
| 17-108-final-viral-combined_k141_140338\|\|full | Not-determined |
| 17-108-final-viral-combined_k141_291809\|\|full | Not-determined |
| 17-108-final-viral-combined_k141_264405\|\|full | Not-determined |
| 17-108-final-viral-combined_k141_112434\|\|full | Not-determined |
| 17-108-final-viral-combined_k141_237241\|\|full | Low-quality |
| 17-108-final-viral-combined_k141_181538\|\|full | Not-determined |
| 17-108-final-viral-combined_k141_2743\|\|full | Not-determined |
| 17-108-final-viral-combined_k141_58014\|\|full | Low-quality |
| 17-108-final-viral-combined_k141_305996\|\|full | Not-determined |
| 17-108-final-viral-combined_k141_140363\|\|full | Low-quality |
| 17-108-final-viral-combined_k141_250701\|\|full | Not-determined |
| 17-108-final-viral-combined_k141_319381\|\|full | Not-determined |
| 17-108-final-viral-combined_k141_112475\|\|full | Not-determined |
| 17-108-final-viral-combined_k141_58024\|\|full | Low-quality |
| 17-108-final-viral-combined_k141_291830\|\|full | Low-quality |
| KM-final-viral-combined_k141_193958\|\|full | High-quality |
| 17-108-final-viral-combined_k141_319394\|\|full | Low-quality |
| 17-108-final-viral-combined_k141_209769\|\|full | Not-determined |
| 17-108-final-viral-combined_k141_58031\|\|full | Low-quality |
| 17-108-final-viral-combined_k141_223216\|\|full | Not-determined |
| 17-108-final-viral-combined_k141_99386\|\|full | Low-quality |
| 17-108-final-viral-combined_k141_168293\|\|full | Low-quality |
| 17-108-final-viral-combined_k141_99387\|\|full | Low-quality |
| 17-108-final-viral-combined_k141_30178\|\|full | Not-determined |
| 17-108-final-viral-combined_k141_112515\|\|full | Low-quality |
| 17-108-final-viral-combined_k141_264456\|\|full | Low-quality |
| 17-108-final-viral-combined_k141_319432\|\|full | Not-determined |
| 17-108-final-viral-combined_k141_195456\|\|full | Low-quality |
| 17-108-final-viral-combined_k141_71161\|\|full | Not-determined |
| 17-108-final-viral-combined_k141_223232\|\|full | Low-quality |
| 17-108-final-viral-combined_k141_71162\|\|full | Low-quality |
| 17-108-final-viral-combined_k141_154312\|\|full | Low-quality |
| 17-108-final-viral-combined_k141_112535\|\|full | Not-determined |
| 17-108-final-viral-combined_k141_58063\|\|full | Not-determined |
| 17-108-final-viral-combined_k141_209823\|\|full | Not-determined |
| 17-108-final-viral-combined_k141_58070\|\|full | Low-quality |
| 17-108-final-viral-combined_k141_99429\|\|full | Low-quality |
| 17-108-final-viral-combined_k141_278616\|\|full | Not-determined |
| 17-108-final-viral-combined_k141_140453\|\|full | Low-quality |
| 17-108-final-viral-combined_k141_278622\|\|full | Not-determined |
| 17-108-final-viral-combined_k141_209846\|\|full | Low-quality |
| 17-108-final-viral-combined_k141_85305\|\|full | Low-quality |
| 17-108-final-viral-combined_k141_181591\|\|full | Low-quality |
| 17-108-final-viral-combined_k141_85317\|\|full | Not-determined |
| 17-108-final-viral-combined_k141_168339\|\|full | Not-determined |
| 17-108-final-viral-combined_k141_209862\|\|full | Low-quality |
| 17-108-final-viral-combined_k141_112546\|\|full | Not-determined |
| 17-108-final-viral-combined_k141_154333\|\|full | Not-determined |
| 17-108-final-viral-combined_k141_71208\|\|full | Not-determined |
| 17-108-final-viral-combined_k141_209868\|\|full | Low-quality |
| 17-108-final-viral-combined_k141_250800\|\|full | Not-determined |
| 17-108-final-viral-combined_k141_250803\|\|full | Low-quality |
| 17-108-final-viral-combined_k141_250804\|\|full | Low-quality |
| 17-108-final-viral-combined_k141_140481\|\|full | Low-quality |
| 17-108-final-viral-combined_k141_99455\|\|full | Not-determined |
| 17-108-final-viral-combined_k141_168375\|\|full | Low-quality |
| 17-108-final-viral-combined_k141_250813\|\|full | Not-determined |
| 17-108-final-viral-combined_k141_140493\|\|full | Low-quality |
| 17-108-final-viral-combined_k141_209889\|\|full | Low-quality |
| 17-108-final-viral-combined_k141_209893\|\|full | Low-quality |
| 17-108-final-viral-combined_k141_126689\|\|full | Not-determined |
| 17-108-final-viral-combined_k141_2868\|\|full | Low-quality |
| 17-108-final-viral-combined_k141_44034\|\|full | Not-determined |
| 17-108-final-viral-combined_k141_278649\|\|full | Low-quality |
| 17-108-final-viral-combined_k141_181614\|\|full | Low-quality |
| 17-108-final-viral-combined_k141_140508\|\|full | Low-quality |
| 17-108-final-viral-combined_k141_2876\|\|full | Low-quality |
| 17-108-final-viral-combined_k141_16366\|\|full | Not-determined |
| 17-108-final-viral-combined_k141_168389\|\|full | Not-determined |
| 17-108-final-viral-combined_k141_99466\|\|full | Not-determined |
| 17-108-final-viral-combined_k141_168395\|\|full | Low-quality |
| 17-108-final-viral-combined_k141_85371\|\|full | Low-quality |
| 17-108-final-viral-combined_k141_264551\|\|full | Not-determined |
| 17-108-final-viral-combined_k141_223325\|\|full | Low-quality |
| 17-108-final-viral-combined_k141_237438\|\|full | Not-determined |
| 17-108-final-viral-combined_k141_195523\|\|full | Low-quality |
| 17-108-final-viral-combined_k141_44071\|\|full | Not-determined |
| 17-108-final-viral-combined_k141_16399\|\|full | Low-quality |
| 17-108-final-viral-combined_k141_126736\|\|full | Low-quality |
| 17-108-final-viral-combined_k141_306166\|\|full | Not-determined |
| 17-108-final-viral-combined_k141_71279\|\|full | Low-quality |
| 17-108-final-viral-combined_k141_58193\|\|full | Not-determined |
| 17-108-final-viral-combined_k141_71281\|\|full | Low-quality |
| 17-108-final-viral-combined_k141_126760\|\|full | Low-quality |
| 17-108-final-viral-combined_k141_58204\|\|full | Low-quality |
| 17-108-final-viral-combined_k141_306176\|\|full | Low-quality |
| 17-108-final-viral-combined_k141_223346\|\|full | Low-quality |
| 17-108-final-viral-combined_k141_168454\|\|full | Not-determined |
| 17-108-final-viral-combined_k141_44125\|\|full | Not-determined |
| 17-108-final-viral-combined_k141_16424\|\|full | Not-determined |
| 17-108-final-viral-combined_k141_126781\|\|full | Not-determined |
| 17-108-final-viral-combined_k141_71288\|\|full | Low-quality |
| 17-108-final-viral-combined_k141_195557\|\|full | Not-determined |
| 17-108-final-viral-combined_k141_140585\|\|full | Low-quality |
| 17-108-final-viral-combined_k141_264598\|\|full | Not-determined |
| 17-108-final-viral-combined_k141_250860\|\|full | Not-determined |
| 17-108-final-viral-combined_k141_264601\|\|full | Low-quality |
| 17-108-final-viral-combined_k141_291989\|\|full | Not-determined |
| 17-108-final-viral-combined_k141_71310\|\|full | Low-quality |
| 17-108-final-viral-combined_k141_30297\|\|full | Low-quality |
| 17-108-final-viral-combined_k141_2955\|\|full | Low-quality |
| 17-108-final-viral-combined_k141_85447\|\|full | Not-determined |
| 17-108-final-viral-combined_k141_140616\|\|full | Not-determined |
| 17-108-final-viral-combined_k141_154394\|\|full | Not-determined |
| 17-108-final-viral-combined_k141_195610\|\|full | Low-quality |
| 17-108-final-viral-combined_k141_223391\|\|full | Not-determined |
| 17-108-final-viral-combined_k141_292026\|\|full | Not-determined |
| 17-108-final-viral-combined_k141_168515\|\|full | Not-determined |
| 17-108-final-viral-combined_k141_16494\|\|full | Not-determined |
| 17-108-final-viral-combined_k141_292043\|\|full | Not-determined |
| 17-108-final-viral-combined_k141_292048\|\|full | Not-determined |
| 17-108-final-viral-combined_k141_58228\|\|full | Not-determined |
| 17-108-final-viral-combined_k141_168518\|\|full | Low-quality |
| 17-108-final-viral-combined_k141_209942\|\|full | Low-quality |
| 17-108-final-viral-combined_k141_306262\|\|full | Low-quality |
| 17-108-final-viral-combined_k141_99537\|\|full | Low-quality |
| 17-108-final-viral-combined_k141_209946\|\|full | Low-quality |
| 17-108-final-viral-combined_k141_85484\|\|full | Not-determined |
| 17-108-final-viral-combined_k141_154415\|\|full | Low-quality |
| 17-108-final-viral-combined_k141_209947\|\|full | Low-quality |
| 17-108-final-viral-combined_k141_71374\|\|full | Low-quality |
| 17-108-final-viral-combined_k141_168537\|\|full | Low-quality |
| 17-108-final-viral-combined_k141_195634\|\|full | Not-determined |
| 17-108-final-viral-combined_k141_223431\|\|full | Not-determined |
| 17-108-final-viral-combined_k141_112709\|\|full | Not-determined |
| 17-108-final-viral-combined_k141_223435\|\|full | Not-determined |
| 17-108-final-viral-combined_k141_306286\|\|full | Low-quality |
| 17-108-final-viral-combined_k141_278738\|\|full | Not-determined |
| 17-108-final-viral-combined_k141_306288\|\|full | Not-determined |
| 17-108-final-viral-combined_k141_126840\|\|full | Not-determined |
| 17-108-final-viral-combined_k141_181717\|\|full | Not-determined |
| 17-108-final-viral-combined_k141_99553\|\|full | Low-quality |
| 17-108-final-viral-combined_k141_223445\|\|full | Low-quality |
| 17-108-final-viral-combined_k141_278745\|\|full | Not-determined |
| 17-108-final-viral-combined_k141_237557\|\|full | Not-determined |
| 17-108-final-viral-combined_k141_181745\|\|full | Not-determined |
| 17-108-final-viral-combined_k141_181749\|\|full | Low-quality |
| 17-108-final-viral-combined_k141_30383\|\|full | Not-determined |
| 17-108-final-viral-combined_k141_126850\|\|full | Not-determined |
| 17-108-final-viral-combined_k141_223459\|\|full | Not-determined |
| 17-108-final-viral-combined_k141_210009\|\|full | Low-quality |
| 17-108-final-viral-combined_k141_181763\|\|full | Low-quality |
| 17-108-final-viral-combined_k141_168615\|\|full | Not-determined |
| 17-108-final-viral-combined_k141_58310\|\|full | Low-quality |
| 17-108-final-viral-combined_k141_126855\|\|full | Not-determined |
| 17-108-final-viral-combined_k141_181770\|\|full | Not-determined |
| 17-108-final-viral-combined_k141_278786\|\|full | Low-quality |
| 17-108-final-viral-combined_k141_154485\|\|full | Not-determined |
| 17-108-final-viral-combined_k141_58323\|\|full | Not-determined |
| 17-108-final-viral-combined_k141_126864\|\|full | Not-determined |
| 17-108-final-viral-combined_k141_30396\|\|full | Not-determined |
| 17-108-final-viral-combined_k141_63818\|\|full | Complete |
| 17-108-final-viral-combined_k141_195693\|\|full | Low-quality |
| 17-108-final-viral-combined_k141_58327\|\|full | Not-determined |
| 17-108-final-viral-combined_k141_168629\|\|full | Low-quality |
| 17-108-final-viral-combined_k141_168631\|\|full | Low-quality |
| 17-108-final-viral-combined_k141_154493\|\|full | Low-quality |
| 17-108-final-viral-combined_k141_16603\|\|full | Not-determined |
| 17-108-final-viral-combined_k141_319543\|\|full | Low-quality |
| 17-108-final-viral-combined_k141_210043\|\|full | Not-determined |
| 17-108-final-viral-combined_k141_126907\|\|full | Low-quality |
| 17-108-final-viral-combined_k141_168654\|\|full | Not-determined |
| 17-108-final-viral-combined_k141_58350\|\|full | Low-quality |
| 17-108-final-viral-combined_k141_154504\|\|full | Low-quality |
| 17-108-final-viral-combined_k141_126913\|\|full | Not-determined |
| 17-108-final-viral-combined_k141_154505\|\|full | Not-determined |
| 17-108-final-viral-combined_k141_30458\|\|full | Low-quality |
| 17-108-final-viral-combined_k141_99632\|\|full | Low-quality |
| 17-108-final-viral-combined_k141_3041\|\|full | Not-determined |
| 17-108-final-viral-combined_k141_126914\|\|full | Low-quality |
| 17-108-final-viral-combined_k141_278828\|\|full | Low-quality |
| 17-108-final-viral-combined_k141_237626\|\|full | Low-quality |
| 17-108-final-viral-combined_k141_140706\|\|full | Not-determined |
| 17-108-final-viral-combined_k141_181809\|\|full | Low-quality |
| 17-108-final-viral-combined_k141_278834\|\|full | Low-quality |
| 17-108-final-viral-combined_k141_306430\|\|full | Low-quality |
| 17-108-final-viral-combined_k141_250961\|\|full | Low-quality |
| 17-108-final-viral-combined_k141_58388\|\|full | Not-determined |
| 17-108-final-viral-combined_k141_306439\|\|full | Low-quality |
| 17-108-final-viral-combined_k141_99648\|\|full | Low-quality |
| 17-108-final-viral-combined_k141_112765\|\|full | Not-determined |
| 17-108-final-viral-combined_k141_306442\|\|full | Low-quality |
| 17-108-final-viral-combined_k141_154535\|\|full | Low-quality |
| 17-108-final-viral-combined_k141_154536\|\|full | Not-determined |
| 17-108-final-viral-combined_k141_278859\|\|full | Not-determined |
| 17-108-final-viral-combined_k141_126946\|\|full | Low-quality |
| 17-108-final-viral-combined_k141_306454\|\|full | Not-determined |
| 17-108-final-viral-combined_k141_250998\|\|full | Not-determined |
| 17-108-final-viral-combined_k141_210116\|\|full | Not-determined |
| 17-108-final-viral-combined_k141_278866\|\|full | Low-quality |
| 17-108-final-viral-combined_k141_223530\|\|full | Not-determined |
| 17-108-final-viral-combined_k141_251017\|\|full | Low-quality |
| 17-108-final-viral-combined_k141_278870\|\|full | Low-quality |
| 17-108-final-viral-combined_k141_292061\|\|full | Not-determined |
| 17-108-final-viral-combined_k141_126984\|\|full | Not-determined |
| 17-108-final-viral-combined_k141_58405\|\|full | Not-determined |
| 17-108-final-viral-combined_k141_71514\|\|full | Low-quality |
| 17-108-final-viral-combined_k141_181880\|\|full | Low-quality |
| 17-108-final-viral-combined_k141_44294\|\|full | Not-determined |
| 17-108-final-viral-combined_k141_99722\|\|full | Low-quality |
| 17-108-final-viral-combined_k141_71526\|\|full | Low-quality |
| 17-108-final-viral-combined_k141_58425\|\|full | Not-determined |
| 17-108-final-viral-combined_k141_306486\|\|full | Low-quality |
| 17-108-final-viral-combined_k141_278912\|\|full | Low-quality |
| 17-108-final-viral-combined_k141_140743\|\|full | Not-determined |
| 17-108-final-viral-combined_k141_112847\|\|full | Not-determined |
| 17-108-final-viral-combined_k141_168697\|\|full | Not-determined |
| 17-108-final-viral-combined_k141_99747\|\|full | Low-quality |
| 17-108-final-viral-combined_k141_71551\|\|full | Low-quality |
| 17-108-final-viral-combined_k141_264834\|\|full | Low-quality |
| 17-108-final-viral-combined_k141_195758\|\|full | Low-quality |
| 17-108-final-viral-combined_k141_264839\|\|full | Low-quality |
| 17-108-final-viral-combined_k141_223596\|\|full | Low-quality |
| 17-108-final-viral-combined_k141_154659\|\|full | Not-determined |
| 17-108-final-viral-combined_k141_30529\|\|full | Not-determined |
| 17-108-final-viral-combined_k141_181947\|\|full | Low-quality |
| 17-108-final-viral-combined_k141_306508\|\|full | Not-determined |
| 17-108-final-viral-combined_k141_140788\|\|full | Low-quality |
| 17-108-final-viral-combined_k141_140791\|\|full | Not-determined |
| 17-108-final-viral-combined_k141_30534\|\|full | Not-determined |
| 17-108-final-viral-combined_k141_30538\|\|full | Not-determined |
| 17-108-final-viral-combined_k141_140792\|\|full | Low-quality |
| 17-108-final-viral-combined_k141_85609\|\|full | Low-quality |
| 17-108-final-viral-combined_k141_58489\|\|full | Low-quality |
| 17-108-final-viral-combined_k141_237709\|\|full | Not-determined |
| 17-108-final-viral-combined_k141_210126\|\|full | Low-quality |
| 17-108-final-viral-combined_k141_181988\|\|full | Not-determined |
| 17-108-final-viral-combined_k141_181991\|\|full | Low-quality |
| 17-108-final-viral-combined_k141_99757\|\|full | Not-determined |
| 17-108-final-viral-combined_k141_99758\|\|full | Not-determined |
| 17-108-final-viral-combined_k141_30549\|\|full | Low-quality |
| 17-108-final-viral-combined_k141_85645\|\|full | Low-quality |
| 17-108-final-viral-combined_k141_210143\|\|full | Low-quality |
| 17-108-final-viral-combined_k141_30556\|\|full | Low-quality |
| 17-108-final-viral-combined_k141_16717\|\|full | Low-quality |
| 17-108-final-viral-combined_k141_30567\|\|full | Not-determined |
| 17-108-final-viral-combined_k141_319632\|\|full | Low-quality |
| 17-108-final-viral-combined_k141_58498\|\|full | Not-determined |
| 17-108-final-viral-combined_k141_154730\|\|full | Low-quality |
| 17-108-final-viral-combined_k141_140835\|\|full | Not-determined |
| 17-108-final-viral-combined_k141_99763\|\|full | Not-determined |
| 17-108-final-viral-combined_k141_306546\|\|full | Low-quality |
| 17-108-final-viral-combined_k141_210173\|\|full | Not-determined |
| 17-108-final-viral-combined_k141_127048\|\|full | Low-quality |
| 17-108-final-viral-combined_k141_195803\|\|full | Low-quality |
| 17-108-final-viral-combined_k141_210183\|\|full | Not-determined |
| 17-108-final-viral-combined_k141_140846\|\|full | Low-quality |
| 17-108-final-viral-combined_k141_30592\|\|full | Low-quality |
| 17-108-final-viral-combined_k141_140849\|\|full | Not-determined |
| 18-106_k141_138342 | Complete |
| 17-108-final-viral-combined_k141_251082\|\|full | Not-determined |
| 17-108-final-viral-combined_k141_182056\|\|full | Low-quality |
| 17-108-final-viral-combined_k141_237759\|\|full | Not-determined |
| 17-108-final-viral-combined_k141_99776\|\|full | Low-quality |
| 17-108-final-viral-combined_k141_278976\|\|full | Low-quality |
| 17-108-final-viral-combined_k141_251090\|\|full | Low-quality |
| 17-108-final-viral-combined_k141_264927\|\|full | Low-quality |
| 17-108-final-viral-combined_k141_99784\|\|full | Low-quality |
| 17-108-final-viral-combined_k141_292215\|\|full | Not-determined |
| 17-108-final-viral-combined_k141_85677\|\|full | Low-quality |
| 17-108-final-viral-combined_k141_237785\|\|full | Not-determined |
| 17-108-final-viral-combined_k141_168794\|\|full | Not-determined |
| 17-108-final-viral-combined_k141_154800\|\|full | Not-determined |
| 17-108-final-viral-combined_k141_154801\|\|full | Not-determined |
| 17-108-final-viral-combined_k141_44357\|\|full | Low-quality |
| 18-104_k141_50475 | Complete |
| 17-108-final-viral-combined_k141_237807\|\|full | Low-quality |
| 17-108-final-viral-combined_k141_319665\|\|full | Low-quality |
| 17-108-final-viral-combined_k141_251130\|\|full | Low-quality |
| 17-108-final-viral-combined_k141_264949\|\|full | Low-quality |
| 17-108-final-viral-combined_k141_251133\|\|full | Not-determined |
| 17-108-final-viral-combined_k141_279015\|\|full | Not-determined |
| 17-108-final-viral-combined_k141_182114\|\|full | Low-quality |
| 17-108-final-viral-combined_k141_264959\|\|full | Not-determined |
| 17-108-final-viral-combined_k141_264960\|\|full | Not-determined |
| 17-108-final-viral-combined_k141_168839\|\|full | Low-quality |
| 17-108-final-viral-combined_k141_30653\|\|full | Not-determined |
| 17-108-final-viral-combined_k141_306658\|\|full | Not-determined |
| 17-108-final-viral-combined_k141_85728\|\|full | Low-quality |
| 17-108-final-viral-combined_k141_3171\|\|full | Not-determined |
| 17-108-final-viral-combined_k141_127130\|\|full | Low-quality |
| 17-108-final-viral-combined_k141_264978\|\|full | Low-quality |
| 17-108-final-viral-combined_k141_223715\|\|full | Not-determined |
| 17-108-final-viral-combined_k141_71617\|\|full | Not-determined |

| | |
|---|---|
| 17-108-final-viral-combined_k141_16852\|\|full | Not-determined |
| 17-108-final-viral-combined_k141_195895\|\|full | Not-determined |
| 17-108-final-viral-combined_k141_127152\|\|full | Not-determined |
| 17-108-final-viral-combined_k141_71632\|\|full | Low-quality |
| 17-108-final-viral-combined_k141_223739\|\|full | Low-quality |
| 17-108-final-viral-combined_k141_319717\|\|full | Not-determined |
| 17-108-final-viral-combined_k141_168886\|\|full | Not-determined |
| 17-108-final-viral-combined_k141_30701\|\|full | Low-quality |
| 17-108-final-viral-combined_k141_195906\|\|full | Low-quality |
| 17-108-final-viral-combined_k141_16866\|\|full | Low-quality |
| 17-108-final-viral-combined_k141_292248\|\|full | Low-quality |
| 17-108-final-viral-combined_k141_168894\|\|full | Not-determined |
| 17-108-final-viral-combined_k141_85748\|\|full | Low-quality |
| 17-108-final-viral-combined_k141_279088\|\|full | Not-determined |
| 17-108-final-viral-combined_k141_30726\|\|full | Low-quality |
| 17-108-final-viral-combined_k141_85752\|\|full | Not-determined |
| 17-108-final-viral-combined_k141_99877\|\|full | Low-quality |
| 17-108-final-viral-combined_k141_306671\|\|full | Low-quality |
| 17-108-final-viral-combined_k141_195935\|\|full | Not-determined |
| 17-108-final-viral-combined_k141_319754\|\|full | Low-quality |
| 17-108-final-viral-combined_k141_279098\|\|full | Low-quality |
| 17-108-final-viral-combined_k141_99899\|\|full | Low-quality |
| 17-108-final-viral-combined_k141_319774\|\|full | Not-determined |
| 17-108-final-viral-combined_k141_71662\|\|full | Low-quality |
| 17-108-final-viral-combined_k141_99916\|\|full | Not-determined |
| 17-108-final-viral-combined_k141_237851\|\|full | Low-quality |
| 17-108-final-viral-combined_k141_265014\|\|full | Not-determined |
| 17-108-final-viral-combined_k141_99930\|\|full | Low-quality |
| 17-108-final-viral-combined_k141_195993\|\|full | Low-quality |
| 17-108-final-viral-combined_k141_99949\|\|full | Low-quality |
| 17-108-final-viral-combined_k141_279152\|\|full | Low-quality |
| 17-108-final-viral-combined_k141_58598\|\|full | Low-quality |
| 17-108-final-viral-combined_k141_140918\|\|full | Not-determined |
| 17-108-final-viral-combined_k141_279154\|\|full | Not-determined |
| 17-108-final-viral-combined_k141_58607\|\|full | Not-determined |
| 17-108-final-viral-combined_k141_127228\|\|full | Low-quality |
| 17-108-final-viral-combined_k141_30778\|\|full | Not-determined |
| 17-108-final-viral-combined_k141_279160\|\|full | Not-determined |
| 17-108-final-viral-combined_k141_85831\|\|full | Low-quality |
| 17-108-final-viral-combined_k141_127241\|\|full | Not-determined |
| 17-108-final-viral-combined_k141_279164\|\|full | Low-quality |
| 17-108-final-viral-combined_k141_127245\|\|full | Low-quality |
| 17-108-final-viral-combined_k141_210192\|\|full | Not-determined |
| 17-108-final-viral-combined_k141_237896\|\|full | Not-determined |
| 17-108-final-viral-combined_k141_306746\|\|full | Not-determined |
| 17-108-final-viral-combined_k141_196003\|\|full | Not-determined |
| 17-108-final-viral-combined_k141_237902\|\|full | Not-determined |
| 17-108-final-viral-combined_k141_30808\|\|full | Not-determined |
| 17-108-final-viral-combined_k141_265060\|\|full | Not-determined |
| 17-108-final-viral-combined_k141_140958\|\|full | Not-determined |
| 17-108-final-viral-combined_k141_210213\|\|full | Not-determined |
| 17-108-final-viral-combined_k141_58622\|\|full | Not-determined |
| 17-108-final-viral-combined_k141_237909\|\|full | Low-quality |
| 17-108-final-viral-combined_k141_140968\|\|full | Low-quality |
| 18-104-final-viral-combined_k141_50475\|\|full | Complete |
| 17-108-final-viral-combined_k141_237931\|\|full | Not-determined |
| 17-108-final-viral-combined_k141_100009\|\|full | Low-quality |
| 17-108-final-viral-combined_k141_168978\|\|full | Low-quality |
| 17-108-final-viral-combined_k141_319842\|\|full | Low-quality |
| 17-108-final-viral-combined_k141_279215\|\|full | Not-determined |
| 17-108-final-viral-combined_k141_237940\|\|full | Not-determined |
| 17-108-final-viral-combined_k141_44507\|\|full | Low-quality |
| 17-108-final-viral-combined_k141_154875\|\|full | Not-determined |
| 17-108-final-viral-combined_k141_319860\|\|full | Low-quality |
| 17-108-final-viral-combined_k141_169000\|\|full | Not-determined |
| 17-108-final-viral-combined_k141_265104\|\|full | Not-determined |
| 17-108-final-viral-combined_k141_3271\|\|full | Not-determined |
| 17-108-final-viral-combined_k141_306804\|\|full | Not-determined |
| 17-108-final-viral-combined_k141_223879\|\|full | Not-determined |
| 17-108-final-viral-combined_k141_85929\|\|full | Not-determined |
| 17-108-final-viral-combined_k141_3278\|\|full | Low-quality |
| 17-108-final-viral-combined_k141_100032\|\|full | Not-determined |
| 17-108-final-viral-combined_k141_85931\|\|full | Not-determined |
| 17-108-final-viral-combined_k141_100035\|\|full | Not-determined |
| 17-108-final-viral-combined_k141_319881\|\|full | Low-quality |
| 17-108-final-viral-combined_k141_182270\|\|full | Low-quality |
| 17-108-final-viral-combined_k141_100037\|\|full | Not-determined |
| 17-108-final-viral-combined_k141_196050\|\|full | Low-quality |
| 17-108-final-viral-combined_k141_71780\|\|full | Low-quality |
| 17-108-final-viral-combined_k141_251286\|\|full | Not-determined |
| 17-108-final-viral-combined_k141_279251\|\|full | Not-determined |
| 17-108-final-viral-combined_k141_44558\|\|full | Low-quality |
| 17-108-final-viral-combined_k141_154907\|\|full | Not-determined |
| 17-108-final-viral-combined_k141_279270\|\|full | Low-quality |
| 17-108-final-viral-combined_k141_182305\|\|full | Low-quality |
| 17-108-final-viral-combined_k141_100053\|\|full | Not-determined |
| 17-108-final-viral-combined_k141_279272\|\|full | Not-determined |
| 17-108-final-viral-combined_k141_319895\|\|full | Not-determined |
| 17-108-final-viral-combined_k141_169042\|\|full | Not-determined |
| 17-108-final-viral-combined_k141_100069\|\|full | Low-quality |
| 17-108-final-viral-combined_k141_279284\|\|full | Low-quality |
| 17-108-final-viral-combined_k141_223921\|\|full | Low-quality |
| 17-108-final-viral-combined_k141_113099\|\|full | Not-determined |
| 17-108-final-viral-combined_k141_182328\|\|full | Not-determined |
| 17-108-final-viral-combined_k141_292363\|\|full | Low-quality |
| 17-108-final-viral-combined_k141_100079\|\|full | Not-determined |
| 17-108-final-viral-combined_k141_319911\|\|full | Low-quality |
| 17-108-final-viral-combined_k141_154941\|\|full | Low-quality |
| 17-108-final-viral-combined_k141_265141\|\|full | Not-determined |
| 17-108-final-viral-combined_k141_127282\|\|full | Low-quality |
| 17-108-final-viral-combined_k141_100091\|\|full | Not-determined |
| 17-108-final-viral-combined_k141_319913\|\|full | Low-quality |
| 17-108-final-viral-combined_k141_182363\|\|full | Low-quality |
| 17-108-final-viral-combined_k141_210329\|\|full | Not-determined |
| 17-108-final-viral-combined_k141_3330\|\|full | Low-quality |
| 17-108-final-viral-combined_k141_182373\|\|full | Low-quality |
| 17-108-final-viral-combined_k141_127292\|\|full | Low-quality |
| 17-108-final-viral-combined_k141_44657\|\|full | Low-quality |
| 17-108-final-viral-combined_k141_141082\|\|full | Not-determined |
| 17-108-final-viral-combined_k141_30888\|\|full | Not-determined |
| 17-108-final-viral-combined_k141_210337\|\|full | Low-quality |
| 17-108-final-viral-combined_k141_3369\|\|full | Low-quality |
| 17-108-final-viral-combined_k141_154976\|\|full | Not-determined |
| 17-108-final-viral-combined_k141_100124\|\|full | Not-determined |
| 17-108-final-viral-combined_k141_319967\|\|full | Low-quality |
| 17-108-final-viral-combined_k141_182408\|\|full | Low-quality |
| 17-108-final-viral-combined_k141_306873\|\|full | Low-quality |
| 17-108-final-viral-combined_k141_210341\|\|full | Low-quality |
| 17-108-final-viral-combined_k141_3381\|\|full | Not-determined |
| 17-108-final-viral-combined_k141_292412\|\|full | Low-quality |
| 17-108-final-viral-combined_k141_238024\|\|full | Low-quality |
| 18-106-final-viral-combined_k141_138342\|\|full | High-quality |
| 17-108-final-viral-combined_k141_292418\|\|full | Low-quality |
| 17-108-final-viral-combined_k141_127315\|\|full | Low-quality |
| 17-108-final-viral-combined_k141_30936\|\|full | Not-determined |
| 17-108-final-viral-combined_k141_251360\|\|full | Not-determined |
| 17-108-final-viral-combined_k141_127320\|\|full | Low-quality |
| 17-108-final-viral-combined_k141_17033\|\|full | Low-quality |
| 17-108-final-viral-combined_k141_223977\|\|full | Low-quality |
| 17-108-final-viral-combined_k141_251363\|\|full | Not-determined |
| 17-108-final-viral-combined_k141_127327\|\|full | Not-determined |
| 17-108-final-viral-combined_k141_251369\|\|full | Low-quality |
| 17-108-final-viral-combined_k141_86087\|\|full | Low-quality |
| 17-108-final-viral-combined_k141_182451\|\|full | Low-quality |
| 17-108-final-viral-combined_k141_320004\|\|full | Not-determined |
| 17-108-final-viral-combined_k141_320005\|\|full | Low-quality |
| 17-108-final-viral-combined_k141_3417\|\|full | Not-determined |
| 17-108-final-viral-combined_k141_292437\|\|full | Not-determined |
| 17-108-final-viral-combined_k141_320010\|\|full | Not-determined |
| 17-108-final-viral-combined_k141_3421\|\|full | Not-determined |
| 17-108-final-viral-combined_k141_44721\|\|full | Not-determined |
| 17-108-final-viral-combined_k141_3425\|\|full | Low-quality |
| 17-108-final-viral-combined_k141_210369\|\|full | Not-determined |
| 17-108-final-viral-combined_k141_58716\|\|full | Not-determined |
| 17-108-final-viral-combined_k141_100171\|\|full | Not-determined |
| 17-108-final-viral-combined_k141_265191\|\|full | Low-quality |
| 17-108-final-viral-combined_k141_224003\|\|full | Low-quality |
| 17-108-final-viral-combined_k141_100191\|\|full | Low-quality |
| 17-108-final-viral-combined_k141_306968\|\|full | Low-quality |
| 17-108-final-viral-combined_k141_292452\|\|full | Low-quality |
| 17-108-final-viral-combined_k141_306971\|\|full | Not-determined |
| 17-108-final-viral-combined_k141_58743\|\|full | Not-determined |
| 17-108-final-viral-combined_k141_58745\|\|full | Not-determined |
| 17-108-final-viral-combined_k141_58746\|\|full | Low-quality |
| 17-108-final-viral-combined_k141_182489\|\|full | Low-quality |
| 17-108-final-viral-combined_k141_210383\|\|full | Not-determined |
| 17-108-final-viral-combined_k141_58764\|\|full | Low-quality |
| 17-108-final-viral-combined_k141_210386\|\|full | Low-quality |
| 17-108-final-viral-combined_k141_224052\|\|full | Not-determined |
| 17-108-final-viral-combined_k141_44751\|\|full | Not-determined |
| 17-108-final-viral-combined_k141_58788\|\|full | Low-quality |
| 17-108-final-viral-combined_k141_265236\|\|full | Low-quality |
| 17-108-final-viral-combined_k141_169244\|\|full | Not-determined |
| 17-108-final-viral-combined_k141_113224\|\|full | Low-quality |
| 17-108-final-viral-combined_k141_58790\|\|full | Low-quality |
| 17-108-final-viral-combined_k141_155110\|\|full | Low-quality |
| 17-108-final-viral-combined_k141_169247\|\|full | Not-determined |
| 17-108-final-viral-combined_k141_86154\|\|full | Low-quality |
| 17-108-final-viral-combined_k141_86155\|\|full | Not-determined |
| 17-108-final-viral-combined_k141_292514\|\|full | Not-determined |
| 17-108-final-viral-combined_k141_113239\|\|full | Not-determined |
| 17-108-final-viral-combined_k141_265252\|\|full | Low-quality |
| 17-108-final-viral-combined_k141_210426\|\|full | Low-quality |
| 17-108-final-viral-combined_k141_210431\|\|full | Not-determined |
| 17-108-final-viral-combined_k141_3466\|\|full | Not-determined |
| 17-108-final-viral-combined_k141_224085\|\|full | Low-quality |
| 17-108-final-viral-combined_k141_210447\|\|full | Low-quality |
| 17-108-final-viral-combined_k141_100255\|\|full | Low-quality |
| 17-108-final-viral-combined_k141_265268\|\|full | Low-quality |
| 17-108-final-viral-combined_k141_141197\|\|full | Not-determined |
| 17-108-final-viral-combined_k141_127411\|\|full | Not-determined |
| 17-108-final-viral-combined_k141_238131\|\|full | Not-determined |
| 17-108-final-viral-combined_k141_58837\|\|full | Low-quality |
| 17-108-final-viral-combined_k141_306978\|\|full | Low-quality |
| 17-108-final-viral-combined_k141_86223\|\|full | Low-quality |
| 17-108-final-viral-combined_k141_224105\|\|full | Low-quality |
| 17-108-final-viral-combined_k141_169317\|\|full | Not-determined |
| 17-108-final-viral-combined_k141_306989\|\|full | Low-quality |
| 17-108-final-viral-combined_k141_31000\|\|full | Low-quality |
| 17-108-final-viral-combined_k141_3505\|\|full | Not-determined |
| 17-108-final-viral-combined_k141_127436\|\|full | Not-determined |
| 17-108-final-viral-combined_k141_86251\|\|full | Low-quality |
| 17-108-final-viral-combined_k141_86255\|\|full | Low-quality |
| 17-108-final-viral-combined_k141_31018\|\|full | Not-determined |
| 17-108-final-viral-combined_k141_86257\|\|full | Not-determined |
| 17-108-final-viral-combined_k141_113291\|\|full | Not-determined |
| 17-108-final-viral-combined_k141_292585\|\|full | Low-quality |
| 17-108-final-viral-combined_k141_71915\|\|full | Not-determined |
| 17-108-final-viral-combined_k141_17108\|\|full | Low-quality |
| 17-108-final-viral-combined_k141_292602\|\|full | Low-quality |
| 17-108-final-viral-combined_k141_292605\|\|full | Not-determined |
| 17-108-final-viral-combined_k141_86273\|\|full | Low-quality |
| 17-108-final-viral-combined_k141_182617\|\|full | Not-determined |
| 17-108-final-viral-combined_k141_86276\|\|full | Low-quality |
| 17-108-final-viral-combined_k141_182618\|\|full | Low-quality |
| 17-108-final-viral-combined_k141_307036\|\|full | Low-quality |
| 17-108-final-viral-combined_k141_292614\|\|full | Low-quality |
| 17-108-final-viral-combined_k141_100323\|\|full | Not-determined |
| 17-108-final-viral-combined_k141_155163\|\|full | Not-determined |
| 17-108-final-viral-combined_k141_320140\|\|full | Not-determined |
| 17-108-final-viral-combined_k141_127488\|\|full | Not-determined |
| 17-108-final-viral-combined_k141_100327\|\|full | Low-quality |
| 17-108-final-viral-combined_k141_17136\|\|full | Low-quality |
| 17-108-final-viral-combined_k141_141265\|\|full | Low-quality |
| 17-108-final-viral-combined_k141_71947\|\|full | Not-determined |
| 17-108-final-viral-combined_k141_279469\|\|full | Not-determined |
| 17-108-final-viral-combined_k141_127511\|\|full | Not-determined |
| 17-108-final-viral-combined_k141_113315\|\|full | Not-determined |
| 18-105-final-viral-combined_k141_66662\|\|full | High-quality |
| 17-108-final-viral-combined_k141_224181\|\|full | Low-quality |
| 17-108-final-viral-combined_k141_71977\|\|full | Not-determined |
| 17-108-final-viral-combined_k141_224183\|\|full | Not-determined |
| 17-108-final-viral-combined_k141_86336\|\|full | Not-determined |
| 17-108-final-viral-combined_k141_210582\|\|full | Not-determined |
| 17-108-final-viral-combined_k141_292678\|\|full | Low-quality |
| 17-108-final-viral-combined_k141_224185\|\|full | Low-quality |
| 17-108-final-viral-combined_k141_3516\|\|full | Not-determined |
| 17-108-final-viral-combined_k141_320176\|\|full | Not-determined |
| 17-108-final-viral-combined_k141_86343\|\|full | Low-quality |
| 17-108-final-viral-combined_k141_251437\|\|full | Not-determined |
| 17-108-final-viral-combined_k141_44934\|\|full | Not-determined |
| 17-108-final-viral-combined_k141_320191\|\|full | Not-determined |
| 17-108-final-viral-combined_k141_210602\|\|full | Low-quality |
| 17-108-final-viral-combined_k141_44944\|\|full | Not-determined |
| 17-108-final-viral-combined_k141_224213\|\|full | Low-quality |
| 17-108-final-viral-combined_k141_113335\|\|full | Low-quality |
| 17-108-final-viral-combined_k141_141301\|\|full | Low-quality |
| 17-108-final-viral-combined_k141_292697\|\|full | Low-quality |
| 17-108-final-viral-combined_k141_238230\|\|full | Low-quality |
| 17-108-final-viral-combined_k141_113351\|\|full | Low-quality |
| 17-108-final-viral-combined_k141_3536\|\|full | Low-quality |
| 17-108-final-viral-combined_k141_210623\|\|full | Not-determined |
| 17-108-final-viral-combined_k141_72021\|\|full | Low-quality |
| 17-108-final-viral-combined_k141_31075\|\|full | Not-determined |
| 17-108-final-viral-combined_k141_141316\|\|full | Low-quality |
| 17-108-final-viral-combined_k141_279510\|\|full | Low-quality |
| 17-108-final-viral-combined_k141_224240\|\|full | Not-determined |
| 17-108-final-viral-combined_k141_279517\|\|full | Not-determined |
| 17-108-final-viral-combined_k141_196187\|\|full | Not-determined |
| 17-108-final-viral-combined_k141_292739\|\|full | Low-quality |
| 17-108-final-viral-combined_k141_127570\|\|full | Low-quality |
| 17-108-final-viral-combined_k141_44977\|\|full | Not-determined |
| 17-108-final-viral-combined_k141_44982\|\|full | Low-quality |
| 17-108-final-viral-combined_k141_86440\|\|full | Not-determined |
| 17-108-final-viral-combined_k141_279539\|\|full | Not-determined |
| 17-108-final-viral-combined_k141_141349\|\|full | Not-determined |
| 17-108-final-viral-combined_k141_210655\|\|full | Not-determined |
| 17-108-final-viral-combined_k141_141351\|\|full | Not-determined |
| 17-108-final-viral-combined_k141_320260\|\|full | Low-quality |
| 17-108-final-viral-combined_k141_86451\|\|full | Low-quality |
| 17-108-final-viral-combined_k141_320266\|\|full | Not-determined |
| 17-108-final-viral-combined_k141_279556\|\|full | Low-quality |
| 17-108-final-viral-combined_k141_210661\|\|full | Not-determined |
| 17-108-final-viral-combined_k141_307155\|\|full | Low-quality |
| 17-108-final-viral-combined_k141_265290\|\|full | Low-quality |
| 17-108-final-viral-combined_k141_279565\|\|full | Low-quality |
| 17-108-final-viral-combined_k141_210668\|\|full | Not-determined |
| 17-108-final-viral-combined_k141_279572\|\|full | Low-quality |
| 17-108-final-viral-combined_k141_182763\|\|full | Low-quality |
| 17-108-final-viral-combined_k141_45051\|\|full | Low-quality |
| 17-108-final-viral-combined_k141_155290\|\|full | Low-quality |
| 17-108-final-viral-combined_k141_100495\|\|full | Not-determined |
| 17-108-final-viral-combined_k141_45066\|\|full | Low-quality |
| 17-108-final-viral-combined_k141_58996\|\|full | Low-quality |
| 17-108-final-viral-combined_k141_182776\|\|full | Low-quality |
| 17-108-final-viral-combined_k141_320299\|\|full | Low-quality |
| 17-108-final-viral-combined_k141_86490\|\|full | Not-determined |
| 17-108-final-viral-combined_k141_196225\|\|full | Not-determined |
| 17-108-final-viral-combined_k141_196227\|\|full | Low-quality |
| 17-108-final-viral-combined_k141_182785\|\|full | Not-determined |
| 17-108-final-viral-combined_k141_59005\|\|full | Low-quality |
| 17-108-final-viral-combined_k141_17221\|\|full | Low-quality |
| 17-108-final-viral-combined_k141_279617\|\|full | Not-determined |
| 17-108-final-viral-combined_k141_141366\|\|full | Not-determined |
| 17-108-final-viral-combined_k141_141367\|\|full | Not-determined |
| 17-108-final-viral-combined_k141_127635\|\|full | Not-determined |
| 17-108-final-viral-combined_k141_3589\|\|full | Not-determined |
| 17-108-final-viral-combined_k141_320320\|\|full | Low-quality |
| 17-108-final-viral-combined_k141_196243\|\|full | Not-determined |
| 17-108-final-viral-combined_k141_45081\|\|full | Low-quality |
| 17-108-final-viral-combined_k141_210725\|\|full | Not-determined |
| 17-108-final-viral-combined_k141_45093\|\|full | Low-quality |
| 17-108-final-viral-combined_k141_100548\|\|full | Low-quality |
| 17-108-final-viral-combined_k141_86516\|\|full | Low-quality |
| 17-108-final-viral-combined_k141_127675\|\|full | Low-quality |
| 17-108-final-viral-combined_k141_141402\|\|full | Not-determined |
| 17-108-final-viral-combined_k141_292874\|\|full | Not-determined |
| 17-108-final-viral-combined_k141_141417\|\|full | Not-determined |
| 17-108-final-viral-combined_k141_279651\|\|full | Not-determined |
| 17-108-final-viral-combined_k141_307157\|\|full | Low-quality |
| 17-108-final-viral-combined_k141_265404\|\|full | Not-determined |
| 17-108-final-viral-combined_k141_279682\|\|full | Low-quality |
| 17-108-final-viral-combined_k141_182855\|\|full | Not-determined |
| 17-108-final-viral-combined_k141_210782\|\|full | Not-determined |
| 17-108-final-viral-combined_k141_59049\|\|full | Low-quality |
| 17-108-final-viral-combined_k141_113444\|\|full | Not-determined |
| 17-108-final-viral-combined_k141_210789\|\|full | Low-quality |
| 17-108-final-viral-combined_k141_141452\|\|full | Low-quality |
| 17-108-final-viral-combined_k141_279702\|\|full | Not-determined |
| 17-108-final-viral-combined_k141_113448\|\|full | Not-determined |
| 17-108-final-viral-combined_k141_224318\|\|full | Low-quality |
| 17-108-final-viral-combined_k141_86587\|\|full | Not-determined |
| 17-108-final-viral-combined_k141_59061\|\|full | Low-quality |
| 17-108-final-viral-combined_k141_155350\|\|full | Not-determined |
| 17-108-final-viral-combined_k141_182874\|\|full | Low-quality |
| 17-108-final-viral-combined_k141_320393\|\|full | Low-quality |
| 17-108-final-viral-combined_k141_127706\|\|full | Not-determined |
| 17-108-final-viral-combined_k141_251620\|\|full | Low-quality |
| 17-108-final-viral-combined_k141_59073\|\|full | Low-quality |
| 17-109-final-viral-combined_k141_18287\|\|full | Medium-quality |
| 17-108-final-viral-combined_k141_72103\|\|full | Low-quality |
| 17-108-final-viral-combined_k141_307176\|\|full | Low-quality |
| 17-108-final-viral-combined_k141_72106\|\|full | Not-determined |
| 17-108-final-viral-combined_k141_320409\|\|full | Not-determined |
| 17-108-final-viral-combined_k141_141474\|\|full | Not-determined |
| 17-108-final-viral-combined_k141_155378\|\|full | Low-quality |
| 17-108-final-viral-combined_k141_292947\|\|full | Low-quality |
| 17-108-final-viral-combined_k141_155379\|\|full | Not-determined |
| 17-108-final-viral-combined_k141_86619\|\|full | Low-quality |
| 17-108-final-viral-combined_k141_155389\|\|full | Low-quality |
| 17-108-final-viral-combined_k141_251661\|\|full | Low-quality |
| 17-108-final-viral-combined_k141_292971\|\|full | Low-quality |
| 17-108-final-viral-combined_k141_210884\|\|full | Low-quality |
| 17-108-final-viral-combined_k141_100664\|\|full | Low-quality |
| 17-108-final-viral-combined_k141_210890\|\|full | Low-quality |
| 17-108-final-viral-combined_k141_72132\|\|full | Low-quality |
| 17-108-final-viral-combined_k141_210912\|\|full | Low-quality |
| 17-108-final-viral-combined_k141_3686\|\|full | Low-quality |
| 17-108-final-viral-combined_k141_265511\|\|full | Not-determined |
| 17-108-final-viral-combined_k141_155429\|\|full | Not-determined |
| 17-108-final-viral-combined_k141_3692\|\|full | Not-determined |
| 17-108-final-viral-combined_k141_265514\|\|full | Low-quality |
| 17-108-final-viral-combined_k141_155437\|\|full | Low-quality |
| 17-108-final-viral-combined_k141_141525\|\|full | Not-determined |
| 17-108-final-viral-combined_k141_251695\|\|full | Low-quality |
| 17-108-final-viral-combined_k141_155444\|\|full | Low-quality |
| 17-108-final-viral-combined_k141_169566\|\|full | Low-quality |
| 17-108-final-viral-combined_k141_155445\|\|full | Low-quality |
| 17-108-final-viral-combined_k141_86699\|\|full | Not-determined |
| 17-108-final-viral-combined_k141_210945\|\|full | Low-quality |
| 17-108-final-viral-combined_k141_59176\|\|full | Low-quality |
| 17-108-final-viral-combined_k141_141550\|\|full | Not-determined |
| 17-108-final-viral-combined_k141_238317\|\|full | Not-determined |
| 17-108-final-viral-combined_k141_127785\|\|full | Low-quality |
| 17-108-final-viral-combined_k141_141553\|\|full | Not-determined |
| 17-108-final-viral-combined_k141_196346\|\|full | Low-quality |
| 17-108-final-viral-combined_k141_238332\|\|full | Low-quality |
| 17-108-final-viral-combined_k141_320503\|\|full | Low-quality |
| 17-108-final-viral-combined_k141_72198\|\|full | Low-quality |
| 17-108-final-viral-combined_k141_210964\|\|full | Low-quality |
| 17-108-final-viral-combined_k141_72204\|\|full | Low-quality |
| 17-108-final-viral-combined_k141_141573\|\|full | Not-determined |
| 17-108-final-viral-combined_k141_100692\|\|full | Low-quality |
| 17-108-final-viral-combined_k141_238357\|\|full | Low-quality |
| 17-108-final-viral-combined_k141_169599\|\|full | Not-determined |
| 17-108-final-viral-combined_k141_224444\|\|full | Low-quality |
| 17-108-final-viral-combined_k141_17346\|\|full | Low-quality |
| 17-108-final-viral-combined_k141_141581\|\|full | Low-quality |
| 17-108-final-viral-combined_k141_59213\|\|full | Low-quality |
| 17-108-final-viral-combined_k141_17348\|\|full | Low-quality |
| 17-108-final-viral-combined_k141_307293\|\|full | Low-quality |
| 17-108-final-viral-combined_k141_127826\|\|full | Low-quality |
| 17-108-final-viral-combined_k141_72266\|\|full | Low-quality |
| 17-108-final-viral-combined_k141_196395\|\|full | Low-quality |
| 17-108-final-viral-combined_k141_45197\|\|full | Low-quality |
| 17-108-final-viral-combined_k141_86725\|\|full | Not-determined |
| 17-108-final-viral-combined_k141_293040\|\|full | Not-determined |
| 17-108-final-viral-combined_k141_86726\|\|full | Low-quality |
| 17-108-final-viral-combined_k141_86731\|\|full | Low-quality |
| 17-108-final-viral-combined_k141_100718\|\|full | Low-quality |
| 17-108-final-viral-combined_k141_293050\|\|full | Not-determined |
| 17-108-final-viral-combined_k141_279848\|\|full | Not-determined |
| 17-108-final-viral-combined_k141_196421\|\|full | Not-determined |
| 17-108-final-viral-combined_k141_45221\|\|full | Low-quality |
| 17-108-final-viral-combined_k141_155521\|\|full | Not-determined |
| 17-108-final-viral-combined_k141_169652\|\|full | Low-quality |
| 17-108-final-viral-combined_k141_169660\|\|full | Low-quality |
| 17-108-final-viral-combined_k141_31377\|\|full | Not-determined |
| 17-108-final-viral-combined_k141_279870\|\|full | Not-determined |
| 17-108-final-viral-combined_k141_211016\|\|full | Low-quality |
| 17-108-final-viral-combined_k141_279872\|\|full | Low-quality |
| 17-108-final-viral-combined_k141_196445\|\|full | Low-quality |
| 17-108-final-viral-combined_k141_3813\|\|full | Low-quality |
| 17-108-final-viral-combined_k141_155555\|\|full | Not-determined |
| 17-108-final-viral-combined_k141_224464\|\|full | Not-determined |
| 17-108-final-viral-combined_k141_59289\|\|full | Not-determined |
| 17-108-final-viral-combined_k141_224472\|\|full | Low-quality |
| 17-108-final-viral-combined_k141_238403\|\|full | Low-quality |
| 17-108-final-viral-combined_k141_238404\|\|full | Low-quality |
| 17-108-final-viral-combined_k141_17428\|\|full | Not-determined |
| 17-108-final-viral-combined_k141_293129\|\|full | Low-quality |
| 17-108-final-viral-combined_k141_196476\|\|full | Not-determined |
| 17-108-final-viral-combined_k141_141696\|\|full | Low-quality |
| 17-108-final-viral-combined_k141_307386\|\|full | Not-determined |
| 17-108-final-viral-combined_k141_293133\|\|full | Not-determined |
| 17-108-final-viral-combined_k141_196483\|\|full | Not-determined |
| 18-102_k141_128962 | Complete |
| 17-108-final-viral-combined_k141_59315\|\|full | Not-determined |
| 17-108-final-viral-combined_k141_265617\|\|full | Low-quality |
| 17-108-final-viral-combined_k141_100734\|\|full | Not-determined |
| 17-108-final-viral-combined_k141_238424\|\|full | Not-determined |
| 17-108-final-viral-combined_k141_320569\|\|full | Not-determined |
| 17-108-final-viral-combined_k141_72318\|\|full | Not-determined |
| 17-108-final-viral-combined_k141_17449\|\|full | Not-determined |
| 17-108-final-viral-combined_k141_31387\|\|full | Low-quality |
| 17-108-final-viral-combined_k141_279909\|\|full | Not-determined |
| 17-108-final-viral-combined_k141_238438\|\|full | Low-quality |
| 17-108-final-viral-combined_k141_196505\|\|full | Low-quality |
| 17-108-final-viral-combined_k141_113569\|\|full | Not-determined |
| 17-108-final-viral-combined_k141_31399\|\|full | Not-determined |
| 17-108-final-viral-combined_k141_279913\|\|full | Low-quality |
| 17-108-final-viral-combined_k141_86834\|\|full | Not-determined |
| 17-108-final-viral-combined_k141_293157\|\|full | Not-determined |
| 17-108-final-viral-combined_k141_238461\|\|full | Low-quality |
| 17-108-final-viral-combined_k141_251855\|\|full | Low-quality |
| 17-108-final-viral-combined_k141_183034\|\|full | Not-determined |
| 17-108-final-viral-combined_k141_127870\|\|full | Not-determined |
| 17-108-final-viral-combined_k141_127871\|\|full | Low-quality |
| 17-108-final-viral-combined_k141_100749\|\|full | Not-determined |
| 17-108-final-viral-combined_k141_45309\|\|full | Not-determined |
| 17-108-final-viral-combined_k141_86856\|\|full | Low-quality |
| 17-108-final-viral-combined_k141_251870\|\|full | Not-determined |
| 17-108-final-viral-combined_k141_100767\|\|full | Not-determined |
| 17-108-final-viral-combined_k141_279938\|\|full | Not-determined |
| 17-108-final-viral-combined_k141_320601\|\|full | Not-determined |
| 17-108-final-viral-combined_k141_45324\|\|full | Low-quality |
| 17-108-final-viral-combined_k141_251886\|\|full | Not-determined |
| 17-108-final-viral-combined_k141_211141\|\|full | Low-quality |
| 17-108-final-viral-combined_k141_224496\|\|full | Not-determined |
| 17-108-final-viral-combined_k141_169750\|\|full | Low-quality |
| 17-108-final-viral-combined_k141_169752\|\|full | Low-quality |
| 17-108-final-viral-combined_k141_3889\|\|full | Not-determined |
| 17-108-final-viral-combined_k141_113605\|\|full | Not-determined |
| 17-108-final-viral-combined_k141_45353\|\|full | Not-determined |
| 17-108-final-viral-combined_k141_196526\|\|full | Not-determined |
| 17-108-final-viral-combined_k141_320611\|\|full | Not-determined |
| 17-108-final-viral-combined_k141_224508\|\|full | Low-quality |
| 17-108-final-viral-combined_k141_196535\|\|full | Low-quality |
| 17-108-final-viral-combined_k141_279980\|\|full | Not-determined |
| 17-108-final-viral-combined_k141_72366\|\|full | Not-determined |
| 17-108-final-viral-combined_k141_183090\|\|full | Low-quality |
| 17-108-final-viral-combined_k141_196549\|\|full | Low-quality |
| 17-108-final-viral-combined_k141_113627\|\|full | Not-determined |
| 17-108-final-viral-combined_k141_196556\|\|full | Low-quality |
| 17-108-final-viral-combined_k141_155645\|\|full | Low-quality |
| 17-108-final-viral-combined_k141_251922\|\|full | Low-quality |
| 17-108-final-viral-combined_k141_224538\|\|full | Not-determined |
| 17-108-final-viral-combined_k141_280023\|\|full | Low-quality |
| 17-108-final-viral-combined_k141_196565\|\|full | Low-quality |
| 17-108-final-viral-combined_k141_127965\|\|full | Not-determined |
| 17-108-final-viral-combined_k141_238561\|\|full | Low-quality |
| 17-108-final-viral-combined_k141_45402\|\|full | Low-quality |
| 17-108-final-viral-combined_k141_307476\|\|full | Low-quality |
| 17-108-final-viral-combined_k141_183111\|\|full | Not-determined |
| 17-108-final-viral-combined_k141_141818\|\|full | Not-determined |
| 17-108-final-viral-combined_k141_72401\|\|full | Low-quality |
| 17-108-final-viral-combined_k141_224566\|\|full | Low-quality |
| 17-108-final-viral-combined_k141_307488\|\|full | Low-quality |
| 17-108-final-viral-combined_k141_100782\|\|full | Not-determined |
| 17-108-final-viral-combined_k141_280054\|\|full | Not-determined |
| 17-108-final-viral-combined_k141_307510\|\|full | Not-determined |
| 17-108-final-viral-combined_k141_86917\|\|full | Low-quality |
| 17-108-final-viral-combined_k141_183124\|\|full | Low-quality |
| 17-108-final-viral-combined_k141_265795\|\|full | Not-determined |
| 17-108-final-viral-combined_k141_307518\|\|full | Not-determined |
| 17-108-final-viral-combined_k141_72418\|\|full | Not-determined |
| 17-108-final-viral-combined_k141_211211\|\|full | Low-quality |
| 17-108-final-viral-combined_k141_251953\|\|full | Low-quality |
| 17-108-final-viral-combined_k141_265804\|\|full | Low-quality |
| 17-108-final-viral-combined_k141_113702\|\|full | Low-quality |
| 17-108-final-viral-combined_k141_251971\|\|full | Low-quality |
| 17-108-final-viral-combined_k141_280074\|\|full | Low-quality |
| 17-108-final-viral-combined_k141_183131\|\|full | Low-quality |
| 17-108-final-viral-combined_k141_293197\|\|full | Not-determined |
| 17-108-final-viral-combined_k141_196584\|\|full | Low-quality |
| 17-108-final-viral-combined_k141_169829\|\|full | Not-determined |
| 17-108-final-viral-combined_k141_211245\|\|full | Low-quality |
| 17-108-final-viral-combined_k141_31495\|\|full | Low-quality |
| 17-108-final-viral-combined_k141_45457\|\|full | Not-determined |
| 17-108-final-viral-combined_k141_45458\|\|full | Low-quality |
| 17-108-final-viral-combined_k141_141868\|\|full | Not-determined |
| 17-108-final-viral-combined_k141_211311\|\|full | Not-determined |
| 17-108-final-viral-combined_k141_141880\|\|full | Not-determined |
| 17-108-final-viral-combined_k141_196625\|\|full | Not-determined |
| 17-108-final-viral-combined_k141_238717\|\|full | Low-quality |
| 17-108-final-viral-combined_k141_211334\|\|full | Not-determined |
| 17-108-final-viral-combined_k141_211342\|\|full | Not-determined |
| 17-108-final-viral-combined_k141_59408\|\|full | Not-determined |
| 17-108-final-viral-combined_k141_183174\|\|full | Low-quality |
| 17-108-final-viral-combined_k141_141901\|\|full | Not-determined |
| 17-108-final-viral-combined_k141_31568\|\|full | Not-determined |
| 17-108-final-viral-combined_k141_293279\|\|full | Low-quality |
| 17-108-final-viral-combined_k141_59410\|\|full | Low-quality |
| 17-108-final-viral-combined_k141_31572\|\|full | Low-quality |
| 17-108-final-viral-combined_k141_238736\|\|full | Not-determined |
| 17-108-final-viral-combined_k141_169903\|\|full | Low-quality |
| 17-108-final-viral-combined_k141_252044\|\|full | Not-determined |
| 17-108-final-viral-combined_k141_128007\|\|full | Low-quality |
| 17-108-final-viral-combined_k141_307549\|\|full | Not-determined |
| 17-108-final-viral-combined_k141_265892\|\|full | Not-determined |
| 17-108-final-viral-combined_k141_87025\|\|full | Not-determined |
| 17-108-final-viral-combined_k141_141931\|\|full | Not-determined |
| 17-108-final-viral-combined_k141_293302\|\|full | Not-determined |
| 17-108-final-viral-combined_k141_59424\|\|full | Not-determined |
| 17-108-final-viral-combined_k141_72444\|\|full | Not-determined |
| 17-108-final-viral-combined_k141_59431\|\|full | Low-quality |
| 17-108-final-viral-combined_k141_293323\|\|full | Not-determined |
| 17-108-final-viral-combined_k141_87047\|\|full | Low-quality |
| 17-108-final-viral-combined_k141_224695\|\|full | Not-determined |
| 17-108-final-viral-combined_k141_183195\|\|full | Low-quality |
| 17-108-final-viral-combined_k141_224703\|\|full | Not-determined |
| 17-108-final-viral-combined_k141_45524\|\|full | Low-quality |
| 17-108-final-viral-combined_k141_196664\|\|full | Low-quality |
| 17-108-final-viral-combined_k141_238790\|\|full | Low-quality |
| 17-108-final-viral-combined_k141_265942\|\|full | Not-determined |
| 17-108-final-viral-combined_k141_252072\|\|full | Low-quality |
| 17-108-final-viral-combined_k141_196667\|\|full | Low-quality |
| 17-108-final-viral-combined_k141_293337\|\|full | Low-quality |
| 17-108-final-viral-combined_k141_45538\|\|full | Low-quality |
| 17-108-final-viral-combined_k141_169940\|\|full | Low-quality |
| 17-108-final-viral-combined_k141_320679\|\|full | Low-quality |
| 17-108-final-viral-combined_k141_252077\|\|full | Not-determined |
| 17-108-final-viral-combined_k141_4014\|\|full | Low-quality |
| 17-108-final-viral-combined_k141_196679\|\|full | Low-quality |
| 17-108-final-viral-combined_k141_4022\|\|full | Low-quality |
| 17-108-final-viral-combined_k141_196688\|\|full | Low-quality |
| 17-108-final-viral-combined_k141_4024\|\|full | Not-determined |
| 17-108-final-viral-combined_k141_211449\|\|full | Not-determined |
| 17-108-final-viral-combined_k141_141980\|\|full | Not-determined |
| 17-108-final-viral-combined_k141_196708\|\|full | Not-determined |
| 17-108-final-viral-combined_k141_183264\|\|full | Low-quality |
| 17-108-final-viral-combined_k141_238840\|\|full | Not-determined |
| 17-108-final-viral-combined_k141_100979\|\|full | Low-quality |
| 17-108-final-viral-combined_k141_45598\|\|full | Low-quality |
| 17-108-final-viral-combined_k141_196723\|\|full | Not-determined |
| 17-108-final-viral-combined_k141_183284\|\|full | Low-quality |
| 17-108-final-viral-combined_k141_155744\|\|full | Not-determined |
| 17-108-final-viral-combined_k141_142011\|\|full | Not-determined |
| 17-108-final-viral-combined_k141_169982\|\|full | Low-quality |
| 17-108-final-viral-combined_k141_31632\|\|full | Low-quality |
| 17-108-final-viral-combined_k141_142024\|\|full | Not-determined |
| 17-108-final-viral-combined_k141_128089\|\|full | Not-determined |
| 17-108-final-viral-combined_k141_128090\|\|full | Not-determined |
| 17-108-final-viral-combined_k141_170001\|\|full | Low-quality |
| 17-108-final-viral-combined_k141_170008\|\|full | Not-determined |
| 17-108-final-viral-combined_k141_264050\|\|full | Complete |
| 17-108-final-viral-combined_k141_252148\|\|full | Not-determined |
| 17-108-final-viral-combined_k141_31639\|\|full | Low-quality |
| 17-108-final-viral-combined_k141_224752\|\|full | Low-quality |
| 17-108-final-viral-combined_k141_183339\|\|full | Low-quality |
| 17-108-final-viral-combined_k141_155774\|\|full | Low-quality |
| 17-108-final-viral-combined_k141_307624\|\|full | Not-determined |
| 17-108-final-viral-combined_k141_59465\|\|full | Not-determined |
| 17-108-final-viral-combined_k141_87174\|\|full | Not-determined |
| 17-108-final-viral-combined_k141_155786\|\|full | Low-quality |
| 17-108-final-viral-combined_k141_31670\|\|full | Low-quality |
| 17-108-final-viral-combined_k141_170018\|\|full | Not-determined |
| 17-108-final-viral-combined_k141_59468\|\|full | Not-determined |
| 17-108-final-viral-combined_k141_224774\|\|full | Low-quality |
| 17-108-final-viral-combined_k141_293463\|\|full | Not-determined |
| 17-108-final-viral-combined_k141_224781\|\|full | Not-determined |
| 17-108-final-viral-combined_k141_196765\|\|full | Not-determined |
| 17-108-final-viral-combined_k141_155798\|\|full | Low-quality |
| 17-108-final-viral-combined_k141_266058\|\|full | Low-quality |
| 17-108-final-viral-combined_k141_4071\|\|full | Not-determined |
| 17-108-final-viral-combined_k141_238873\|\|full | Low-quality |
| 17-108-final-viral-combined_k141_196784\|\|full | Low-quality |
| 17-108-final-viral-combined_k141_252197\|\|full | Low-quality |
| 17-108-final-viral-combined_k141_196790\|\|full | Low-quality |
| 17-108-final-viral-combined_k141_170065\|\|full | Low-quality |
| 17-108-final-viral-combined_k141_87209\|\|full | Not-determined |
| 17-108-final-viral-combined_k141_142069\|\|full | Low-quality |
| 17-108-final-viral-combined_k141_183386\|\|full | Not-determined |
| 17-108-final-viral-combined_k141_266065\|\|full | Low-quality |
| 17-108-final-viral-combined_k141_293485\|\|full | Low-quality |
| 17-108-final-viral-combined_k141_59514\|\|full | Low-quality |
| 17-108-final-viral-combined_k141_280322\|\|full | Low-quality |
| 17-108-final-viral-combined_k141_87215\|\|full | Low-quality |
| 17-108-final-viral-combined_k141_59520\|\|full | Not-determined |

17-108-final-viral-combined_k141_87218||full Low-quality
17-108-final-viral-combined_k141_252222||full Not-determined
17-108-final-viral-combined_k141_211534||full Not-determined
17-108-final-viral-combined_k141_45722||full Not-determined
17-108-final-viral-combined_k141_142081||full Low-quality
17-108-final-viral-combined_k141_320767||full Low-quality
17-108-final-viral-combined_k141_142090||full Low-quality
17-108-final-viral-combined_k141_224831||full Not-determined
17-108-final-viral-combined_k141_183427||full Not-determined
17-108-final-viral-combined_k141_87230||full Low-quality
17-108-final-viral-combined_k141_211569||full Not-determined
17-108-final-viral-combined_k141_211570||full Not-determined
17-108-final-viral-combined_k141_238964||full Not-determined
17-108-final-viral-combined_k141_211571||full Low-quality
17-108-final-viral-combined_k141_128153||full Low-quality
17-108-final-viral-combined_k141_293530||full Not-determined
17-108-final-viral-combined_k141_87242||full Not-determined
17-108-final-viral-combined_k141_320814||full Low-quality
17-108-final-viral-combined_k141_128177||full Low-quality
17-108-final-viral-combined_k141_113869||full Low-quality
17-108-final-viral-combined_k141_45766||full Not-determined
17-108-final-viral-combined_k141_113875||full Low-quality
17-108-final-viral-combined_k141_31735||full Not-determined
17-108-final-viral-combined_k141_128193||full Not-determined
17-108-final-viral-combined_k141_224870||full Low-quality
17-108-final-viral-combined_k141_320815||full Low-quality
17-108-final-viral-combined_k141_293567||full Low-quality
17-108-final-viral-combined_k141_196849||full Not-determined
17-108-final-viral-combined_k141_128209||full Not-determined
17-108-final-viral-combined_k141_211605||full Low-quality
17-108-final-viral-combined_k141_4155||full Low-quality
17-108-final-viral-combined_k141_307747||full Low-quality
17-108-final-viral-combined_k141_239018||full Low-quality
17-108-final-viral-combined_k141_128218||full Low-quality
17-108-final-viral-combined_k141_239024||full Low-quality
17-108-final-viral-combined_k141_293583||full Not-determined
17-108-final-viral-combined_k141_87287||full Low-quality
17-108-final-viral-combined_k141_113904||full Low-quality
17-108-final-viral-combined_k141_266156||full Low-quality
17-108-final-viral-combined_k141_307756||full Low-quality
17-108-final-viral-combined_k141_128238||full Not-determined
17-108-final-viral-combined_k141_101116||full Not-determined
17-108-final-viral-combined_k141_224892||full Not-determined
17-108-final-viral-combined_k141_211615||full Low-quality
17-108-final-viral-combined_k141_101124||full Not-determined
17-108-final-viral-combined_k141_280409||full Not-determined
17-108-final-viral-combined_k141_252337||full Not-determined
17-108-final-viral-combined_k141_4199||full Not-determined
17-108-final-viral-combined_k141_45812||full Low-quality
17-108-final-viral-combined_k141_266182||full Not-determined
17-108-final-viral-combined_k141_4202||full Not-determined
17-108-final-viral-combined_k141_293636||full Low-quality
17-108-final-viral-combined_k141_293637||full Not-determined
17-108-final-viral-combined_k141_183471||full Not-determined
17-108-final-viral-combined_k141_170146||full Low-quality
17-108-final-viral-combined_k141_224908||full Low-quality
17-108-final-viral-combined_k141_224909||full Not-determined
17-108-final-viral-combined_k141_307791||full Not-determined
17-108-final-viral-combined_k141_239067||full Not-determined
17-108-final-viral-combined_k141_101154||full Low-quality
17-108-final-viral-combined_k141_307793||full Low-quality
17-108-final-viral-combined_k141_59649||full Not-determined
17-108-final-viral-combined_k141_31777||full Low-quality
17-108-final-viral-combined_k141_31782||full Low-quality
17-108-final-viral-combined_k141_183510||full Low-quality
17-108-final-viral-combined_k141_307816||full Low-quality
17-108-final-viral-combined_k141_87301||full Low-quality
17-108-final-viral-combined_k141_252363||full Low-quality
17-108-final-viral-combined_k141_266226||full Low-quality
17-108-final-viral-combined_k141_280469||full Low-quality
17-108-final-viral-combined_k141_224945||full Not-determined
17-108-final-viral-combined_k141_266239||full Not-determined
17-108-final-viral-combined_k141_45866||full Low-quality
17-108-final-viral-combined_k141_239117||full Not-determined
17-108-final-viral-combined_k141_17685||full Low-quality
17-108-final-viral-combined_k141_266244||full Not-determined
17-108-final-viral-combined_k141_239122||full Low-quality
17-108-final-viral-combined_k141_183534||full Not-determined
17-108-final-viral-combined_k141_155959||full Not-determined
17-108-final-viral-combined_k141_4269||full Not-determined
17-108-final-viral-combined_k141_239134||full Low-quality
17-108-final-viral-combined_k141_45876||full Low-quality
17-108-final-viral-combined_k141_307861||full Not-determined
17-108-final-viral-combined_k141_307865||full Not-determined
17-108-final-viral-combined_k141_17694||full Not-determined
17-108-final-viral-combined_k141_72737||full Not-determined
17-108-final-viral-combined_k141_307876||full Not-determined
17-108-final-viral-combined_k141_293682||full Not-determined
17-108-final-viral-combined_k141_17700||full Low-quality
17-108-final-viral-combined_k141_4274||full Not-determined
17-108-final-viral-combined_k141_17703||full Not-determined
17-108-final-viral-combined_k141_211646||full Low-quality
17-108-final-viral-combined_k141_307896||full Not-determined
17-108-final-viral-combined_k141_170229||full Low-quality
17-108-final-viral-combined_k141_4285||full Not-determined
17-108-final-viral-combined_k141_17713||full Low-quality
17-108-final-viral-combined_k141_142244||full Low-quality
17-108-final-viral-combined_k141_128335||full Low-quality
17-108-final-viral-combined_k141_128336||full Low-quality
17-108-final-viral-combined_k141_196991||full Not-determined
17-108-final-viral-combined_k141_266341||full Not-determined
17-108-final-viral-combined_k141_142257||full Low-quality
17-108-final-viral-combined_k141_128342||full Not-determined
17-108-final-viral-combined_k141_101291||full Low-quality
17-108-final-viral-combined_k141_293752||full Not-determined
17-108-final-viral-combined_k141_31913||full Low-quality
17-108-final-viral-combined_k141_293762||full Low-quality
17-108-final-viral-combined_k141_293763||full Not-determined
17-108-final-viral-combined_k141_293765||full Low-quality
17-108-final-viral-combined_k141_59700||full Not-determined
17-108-final-viral-combined_k141_128366||full Low-quality
17-108-final-viral-combined_k141_101322||full Not-determined
17-108-final-viral-combined_k141_320937||full Not-determined
17-108-final-viral-combined_k141_142281||full Low-quality
17-108-final-viral-combined_k141_266398||full Low-quality
17-108-final-viral-combined_k141_128374||full Not-determined
17-108-final-viral-combined_k141_45977||full Not-determined
17-108-final-viral-combined_k141_59725||full Not-determined
17-108-final-viral-combined_k141_239191||full Not-determined
17-108-final-viral-combined_k141_239194||full Not-determined
17-108-final-viral-combined_k141_45984||full Not-determined
17-108-final-viral-combined_k141_320964||full Low-quality
17-108-final-viral-combined_k141_156020||full Not-determined
17-108-final-viral-combined_k141_31957||full Not-determined
17-108-final-viral-combined_k141_45993||full Low-quality
17-108-final-viral-combined_k141_128396||full Not-determined
17-108-final-viral-combined_k141_72833||full Not-determined
17-108-final-viral-combined_k141_128407||full Low-quality
17-108-final-viral-combined_k141_156038||full Low-quality
17-108-final-viral-combined_k141_31959||full Not-determined
18-102-final-viral-combined_k141_128962||full High-quality
17-108-final-viral-combined_k141_72837||full Not-determined
17-108-final-viral-combined_k141_211685||full Low-quality
17-108-final-viral-combined_k141_142327||full Low-quality
17-108-final-viral-combined_k141_252507||full Low-quality
17-108-final-viral-combined_k141_114057||full Low-quality
17-108-final-viral-combined_k141_280643||full Not-determined
17-108-final-viral-combined_k141_320990||full Low-quality
17-108-final-viral-combined_k141_252514||full Low-quality
17-108-final-viral-combined_k141_225086||full Low-quality
17-108-final-viral-combined_k141_225097||full Not-determined
17-108-final-viral-combined_k141_211701||full Low-quality
17-108-final-viral-combined_k141_225099||full Low-quality
17-108-final-viral-combined_k141_211704||full Not-determined
17-108-final-viral-combined_k141_17823||full Not-determined
17-108-final-viral-combined_k141_17836||full Low-quality
17-108-final-viral-combined_k141_183679||full Low-quality
17-108-final-viral-combined_k141_72881||full Not-determined
17-108-final-viral-combined_k141_4417||full Not-determined
17-108-final-viral-combined_k141_183693||full Low-quality
18-109-final-viral-combined_k141_106253||full High-quality
17-108-final-viral-combined_k141_128445||full Low-quality
17-108-final-viral-combined_k141_4424||full Low-quality
17-108-final-viral-combined_k141_197076||full Low-quality
17-108-final-viral-combined_k141_59771||full Not-determined
17-108-final-viral-combined_k141_17850||full Low-quality
17-108-final-viral-combined_k141_211748||full Not-determined
17-108-final-viral-combined_k141_31984||full Low-quality
17-108-final-viral-combined_k141_239251||full Not-determined
17-108-final-viral-combined_k141_225160||full Not-determined
17-108-final-viral-combined_k141_142382||full Not-determined
17-108-final-viral-combined_k141_17856||full Low-quality
17-108-final-viral-combined_k141_17859||full Not-determined
17-108-final-viral-combined_k141_321043||full Not-determined
17-108-final-viral-combined_k141_72904||full Not-determined
17-108-final-viral-combined_k141_252583||full Low-quality
17-108-final-viral-combined_k141_170361||full Not-determined
17-108-final-viral-combined_k141_266452||full Low-quality
17-108-final-viral-combined_k141_46076||full Low-quality
17-108-final-viral-combined_k141_17875||full Low-quality
17-108-final-viral-combined_k141_114169||full Not-determined
17-108-final-viral-combined_k141_252598||full Not-determined
17-108-final-viral-combined_k141_321057||full Low-quality
17-108-final-viral-combined_k141_4451||full Low-quality
17-108-final-viral-combined_k141_101386||full Not-determined
17-108-final-viral-combined_k141_114180||full Low-quality
17-108-final-viral-combined_k141_183772||full Low-quality
17-108-final-viral-combined_k141_114181||full Not-determined
17-108-final-viral-combined_k141_142436||full Not-determined
17-108-final-viral-combined_k141_87425||full Not-determined
17-108-final-viral-combined_k141_101394||full Low-quality
17-108-final-viral-combined_k141_128480||full Not-determined
17-108-final-viral-combined_k141_266464||full Low-quality
17-108-final-viral-combined_k141_72970||full Not-determined
17-108-final-viral-combined_k141_183782||full Not-determined
17-108-final-viral-combined_k141_197126||full Not-determined
17-108-final-viral-combined_k141_46118||full Low-quality
17-108-final-viral-combined_k141_46121||full Not-determined
17-108-final-viral-combined_k141_142453||full Low-quality
17-108-final-viral-combined_k141_321069||full Not-determined
17-108-final-viral-combined_k141_32064||full Not-determined
17-108-final-viral-combined_k141_59814||full Low-quality
17-108-final-viral-combined_k141_4523||full Not-determined
17-108-final-viral-combined_k141_239278||full Not-determined
17-108-final-viral-combined_k141_59819||full Not-determined
17-108-final-viral-combined_k141_225258||full Not-determined
17-108-final-viral-combined_k141_211889||full Not-determined
17-108-final-viral-combined_k141_293833||full Not-determined
17-108-final-viral-combined_k141_239286||full Not-determined
17-108-final-viral-combined_k141_225273||full Not-determined
17-108-final-viral-combined_k141_225274||full Low-quality
17-108-final-viral-combined_k141_4556||full Not-determined
17-108-final-viral-combined_k141_128540||full Low-quality
17-108-final-viral-combined_k141_101443||full Low-quality
17-108-final-viral-combined_k141_46168||full Low-quality
17-108-final-viral-combined_k141_4559||full Low-quality
17-108-final-viral-combined_k141_321116||full Low-quality
17-108-final-viral-combined_k141_128542||full Not-determined
17-108-final-viral-combined_k141_170427||full Low-quality
17-108-final-viral-combined_k141_59850||full Not-determined
17-108-final-viral-combined_k141_73050||full Low-quality
17-108-final-viral-combined_k141_59852||full Low-quality
17-108-final-viral-combined_k141_308080||full Not-determined
17-108-final-viral-combined_k141_87518||full Low-quality
17-108-final-viral-combined_k141_32126||full Not-determined
17-108-final-viral-combined_k141_17913||full Not-determined
17-108-final-viral-combined_k141_197192||full Not-determined
17-108-final-viral-combined_k141_239323||full Not-determined
17-108-final-viral-combined_k141_128548||full Not-determined
17-108-final-viral-combined_k141_170434||full Not-determined
17-108-final-viral-combined_k141_239327||full Low-quality
17-108-final-viral-combined_k141_225302||full Low-quality
17-108-final-viral-combined_k141_321139||full Not-determined
17-108-final-viral-combined_k141_59862||full Not-determined
17-108-final-viral-combined_k141_128554||full Not-determined
17-108-final-viral-combined_k141_211898||full Low-quality
17-108-final-viral-combined_k141_211905||full Not-determined
17-108-final-viral-combined_k141_59868||full Low-quality
17-108-final-viral-combined_k141_170464||full Not-determined
17-108-final-viral-combined_k141_142548||full Not-determined
17-108-final-viral-combined_k141_308134||full Low-quality
17-108-final-viral-combined_k141_142552||full Low-quality
17-108-final-viral-combined_k141_32178||full Not-determined
17-108-final-viral-combined_k141_225376||full Low-quality
17-108-final-viral-combined_k141_266557||full Low-quality
17-108-final-viral-combined_k141_128594||full Low-quality
17-108-final-viral-combined_k141_225377||full Not-determined
17-108-final-viral-combined_k141_239344||full Low-quality
17-108-final-viral-combined_k141_128596||full Not-determined
17-108-final-viral-combined_k141_225378||full Low-quality
17-108-final-viral-combined_k141_17943||full Not-determined
17-108-final-viral-combined_k141_197265||full Low-quality
17-108-final-viral-combined_k141_170495||full Low-quality
17-108-final-viral-combined_k141_101516||full Low-quality
17-108-final-viral-combined_k141_170502||full Not-determined
17-108-final-viral-combined_k141_156259||full Low-quality
17-108-final-viral-combined_k141_197279||full Low-quality
17-108-final-viral-combined_k141_266567||full Not-determined
17-108-final-viral-combined_k141_183871||full Low-quality
17-108-final-viral-combined_k141_59909||full Low-quality
17-108-final-viral-combined_k141_266568||full Low-quality
17-108-final-viral-combined_k141_225408||full Not-determined
17-108-final-viral-combined_k141_46244||full Low-quality
17-108-final-viral-combined_k141_266569||full Not-determined
17-108-final-viral-combined_k141_252701||full Not-determined
17-108-final-viral-combined_k141_114256||full Not-determined
17-108-final-viral-combined_k141_308166||full Low-quality
17-108-final-viral-combined_k141_59920||full Low-quality
17-108-final-viral-combined_k141_308167||full Not-determined
17-108-final-viral-combined_k141_266586||full Not-determined
17-108-final-viral-combined_k141_321203||full Not-determined
17-108-final-viral-combined_k141_239416||full Low-quality
17-108-final-viral-combined_k141_293937||full Not-determined
17-108-final-viral-combined_k141_32211||full Not-determined
17-108-final-viral-combined_k141_280838||full Not-determined
17-108-final-viral-combined_k141_46292||full Low-quality
17-108-final-viral-combined_k141_308175||full Low-quality
17-108-final-viral-combined_k141_170557||full Not-determined
17-108-final-viral-combined_k141_239446||full Low-quality
17-108-final-viral-combined_k141_59978||full Low-quality
17-108-final-viral-combined_k141_293955||full Not-determined
17-108-final-viral-combined_k141_280851||full Not-determined
17-108-final-viral-combined_k141_252765||full Not-determined
17-108-final-viral-combined_k141_114309||full Low-quality
17-108-final-viral-combined_k141_156296||full Not-determined
17-108-final-viral-combined_k141_170575||full Low-quality
17-108-final-viral-combined_k141_59983||full Not-determined
17-108-final-viral-combined_k141_46309||full Low-quality
17-108-final-viral-combined_k141_321266||full Low-quality
17-108-final-viral-combined_k141_32237||full Not-determined
17-108-final-viral-combined_k141_212048||full Low-quality
17-108-final-viral-combined_k141_18011||full Not-determined
17-108-final-viral-combined_k141_293992||full Low-quality
17-108-final-viral-combined_k141_128686||full Low-quality
17-108-final-viral-combined_k141_212063||full Low-quality
17-108-final-viral-combined_k141_212064||full Low-quality
17-108-final-viral-combined_k141_239471||full Not-determined
17-108-final-viral-combined_k141_46317||full Not-determined
17-108-final-viral-combined_k141_212068||full Low-quality
17-108-final-viral-combined_k141_73113||full Not-determined
17-108-final-viral-combined_k141_252803||full Not-determined
17-108-final-viral-combined_k141_46319||full Not-determined
17-108-final-viral-combined_k141_18042||full Not-determined
17-108-final-viral-combined_k141_46321||full Not-determined
17-108-final-viral-combined_k141_156361||full Low-quality
17-108-final-viral-combined_k141_170635||full Low-quality
17-108-final-viral-combined_k141_73125||full Low-quality
17-108-final-viral-combined_k141_46337||full Low-quality
17-108-final-viral-combined_k141_128724||full Low-quality
17-108-final-viral-combined_k141_212074||full Low-quality
17-108-final-viral-combined_k141_18045||full Not-determined
17-108-final-viral-combined_k141_197365||full Not-determined
17-108-final-viral-combined_k141_73139||full Not-determined
17-108-final-viral-combined_k141_156384||full Not-determined
17-108-final-viral-combined_k141_266631||full Not-determined
17-108-final-viral-combined_k141_321320||full Not-determined
17-108-final-viral-combined_k141_18057||full Not-determined
17-108-final-viral-combined_k141_280946||full Not-determined
17-108-final-viral-combined_k141_183928||full Low-quality
17-108-final-viral-combined_k141_294053||full Low-quality
17-108-final-viral-combined_k141_114373||full Not-determined
17-108-final-viral-combined_k141_73153||full Not-determined
17-108-final-viral-combined_k141_4744||full Low-quality
17-108-final-viral-combined_k141_308195||full Low-quality
17-108-final-viral-combined_k141_170673||full Low-quality
17-108-final-viral-combined_k141_212093||full Low-quality
17-108-final-viral-combined_k141_73170||full Not-determined
17-108-final-viral-combined_k141_32294||full Not-determined
17-108-final-viral-combined_k141_18077||full Not-determined
17-108-final-viral-combined_k141_170679||full Not-determined
17-108-final-viral-combined_k141_294077||full Low-quality
17-108-final-viral-combined_k141_128780||full Low-quality
17-108-final-viral-combined_k141_197402||full Low-quality
17-108-final-viral-combined_k141_128792||full Not-determined
17-108-final-viral-combined_k141_114414||full Low-quality
17-108-final-viral-combined_k141_239545||full Not-determined
17-108-final-viral-combined_k141_308218||full Low-quality
17-108-final-viral-combined_k141_308219||full Low-quality
17-108-final-viral-combined_k141_114429||full Not-determined
17-108-final-viral-combined_k141_225585||full Low-quality
17-108-final-viral-combined_k141_114446||full Low-quality
17-108-final-viral-combined_k141_225587||full Low-quality
17-108-final-viral-combined_k141_225589||full Not-determined
17-108-final-viral-combined_k141_225613||full Not-determined
17-108-final-viral-combined_k141_114490||full Low-quality
17-108-final-viral-combined_k141_114491||full Low-quality
17-108-final-viral-combined_k141_101738||full Not-determined
17-108-final-viral-combined_k141_156475||full Low-quality
17-108-final-viral-combined_k141_266758||full Low-quality
17-108-final-viral-combined_k141_170732||full Low-quality
17-108-final-viral-combined_k141_212210||full Low-quality
17-108-final-viral-combined_k141_32374||full Low-quality
17-108-final-viral-combined_k141_4781||full Not-determined
17-108-final-viral-combined_k141_197439||full Low-quality
17-108-final-viral-combined_k141_184008||full Not-determined
17-108-final-viral-combined_k141_239628||full Low-quality
17-108-final-viral-combined_k141_184021||full Low-quality
17-108-final-viral-combined_k141_308316||full Low-quality
17-108-final-viral-combined_k141_128842||full Low-quality
18-102-final-viral-combined_k141_157934||full Complete
17-108-final-viral-combined_k141_128843||full Low-quality
17-108-final-viral-combined_k141_87721||full Low-quality
17-108-final-viral-combined_k141_128850||full Not-determined
17-108-final-viral-combined_k141_87734||full Not-determined
17-108-final-viral-combined_k141_87737||full Low-quality
17-108-final-viral-combined_k141_46419||full Not-determined
17-108-final-viral-combined_k141_60098||full Low-quality
17-108-final-viral-combined_k141_281152||full Low-quality
17-108-final-viral-combined_k141_46431||full Not-determined
17-108-final-viral-combined_k141_281156||full Not-determined
17-108-final-viral-combined_k141_60101||full Not-determined
17-108-final-viral-combined_k141_60104||full Not-determined
17-108-final-viral-combined_k141_321430||full Low-quality
17-108-final-viral-combined_k141_308351||full Not-determined
17-108-final-viral-combined_k141_308353||full Low-quality
17-108-final-viral-combined_k141_197498||full Low-quality
17-108-final-viral-combined_k141_156536||full Low-quality
17-108-final-viral-combined_k141_32453||full Low-quality
17-108-final-viral-combined_k141_32456||full Low-quality
17-108-final-viral-combined_k141_18184||full Not-determined
17-108-final-viral-combined_k141_294128||full Low-quality
17-108-final-viral-combined_k141_212285||full Not-determined
17-108-final-viral-combined_k141_142849||full Not-determined
17-108-final-viral-combined_k141_197534||full Not-determined
17-108-final-viral-combined_k141_114605||full Not-determined
17-108-final-viral-combined_k141_60166||full Not-determined
17-108-final-viral-combined_k141_60168||full Low-quality
17-108-final-viral-combined_k141_252977||full Low-quality
17-108-final-viral-combined_k141_60173||full Low-quality
17-108-final-viral-combined_k141_46462||full Not-determined
17-108-final-viral-combined_k141_239689||full Not-determined
17-108-final-viral-combined_k141_184103||full Not-determined
17-108-final-viral-combined_k141_252991||full Low-quality
17-108-final-viral-combined_k141_32505||full Low-quality
17-108-final-viral-combined_k141_142885||full Low-quality
17-108-final-viral-combined_k141_184106||full Not-determined
17-108-final-viral-combined_k141_197562||full Low-quality
17-108-final-viral-combined_k141_308395||full Low-quality
17-108-final-viral-combined_k141_101916||full Low-quality
17-108-final-viral-combined_k141_60207||full Not-determined
17-108-final-viral-combined_k141_18237||full Not-determined
17-108-final-viral-combined_k141_308412||full Not-determined
17-108-final-viral-combined_k141_142891||full Low-quality
17-108-final-viral-combined_k141_128957||full Not-determined
17-108-final-viral-combined_k141_73299||full Not-determined
17-108-final-viral-combined_k141_184123||full Low-quality
17-108-final-viral-combined_k141_170816||full Low-quality
17-108-final-viral-combined_k141_184124||full Low-quality
17-108-final-viral-combined_k141_156562||full Low-quality
17-108-final-viral-combined_k141_239757||full Not-determined
17-108-final-viral-combined_k141_101935||full Not-determined
17-108-final-viral-combined_k141_156563||full Low-quality
17-108-final-viral-combined_k141_128971||full Not-determined
17-108-final-viral-combined_k141_4872||full Low-quality
17-108-final-viral-combined_k141_87844||full Not-determined
17-108-final-viral-combined_k141_266881||full Not-determined
17-108-final-viral-combined_k141_212302||full Not-determined
17-108-final-viral-combined_k141_32546||full Not-determined
17-108-final-viral-combined_k141_18264||full Not-determined
17-108-final-viral-combined_k141_294209||full Not-determined
17-108-final-viral-combined_k141_128982||full Not-determined
17-108-final-viral-combined_k141_101945||full Not-determined
17-108-final-viral-combined_k141_239774||full Not-determined
17-108-final-viral-combined_k141_308448||full Low-quality
17-108-final-viral-combined_k141_73331||full Low-quality
17-108-final-viral-combined_k141_184155||full Not-determined
17-108-final-viral-combined_k141_281295||full Not-determined
17-108-final-viral-combined_k141_239790||full Low-quality
17-108-final-viral-combined_k141_101947||full Low-quality
17-108-final-viral-combined_k141_170863||full Not-determined
17-108-final-viral-combined_k141_32566||full Low-quality
17-108-final-viral-combined_k141_46519||full Not-determined
17-108-final-viral-combined_k141_308486||full Not-determined
17-108-final-viral-combined_k141_101985||full Not-determined
17-108-final-viral-combined_k141_4908||full Low-quality
17-108-final-viral-combined_k141_321507||full Not-determined
17-108-final-viral-combined_k141_170906||full Low-quality
17-108-final-viral-combined_k141_60295||full Low-quality
17-108-final-viral-combined_k141_32620||full Not-determined
17-108-final-viral-combined_k141_102009||full Low-quality
17-108-final-viral-combined_k141_129008||full Low-quality
17-108-final-viral-combined_k141_32628||full Low-quality
17-108-final-viral-combined_k141_225740||full Low-quality
17-108-final-viral-combined_k141_18315||full Low-quality
17-108-final-viral-combined_k141_197635||full Not-determined
17-108-final-viral-combined_k141_239878||full Low-quality
17-108-final-viral-combined_k141_321533||full Not-determined
17-108-final-viral-combined_k141_60310||full Low-quality
17-108-final-viral-combined_k141_102040||full Low-quality
17-108-final-viral-combined_k141_143000||full Low-quality
17-108-final-viral-combined_k141_46553||full Low-quality
17-108-final-viral-combined_k141_87925||full Low-quality
17-108-final-viral-combined_k141_4957||full Low-quality
17-108-final-viral-combined_k141_143006||full Not-determined
17-108-final-viral-combined_k141_46556||full Low-quality
17-108-final-viral-combined_k141_253144||full Low-quality
17-108-final-viral-combined_k141_170909||full Low-quality
17-108-final-viral-combined_k141_225789||full Not-determined
17-108-final-viral-combined_k141_197672||full Low-quality
17-108-final-viral-combined_k141_308541||full Low-quality
17-108-final-viral-combined_k141_197673||full Low-quality
17-108-final-viral-combined_k141_266925||full Low-quality
17-108-final-viral-combined_k141_281391||full Not-determined
17-108-final-viral-combined_k141_321561||full Low-quality
17-108-final-viral-combined_k141_308554||full Not-determined
17-108-final-viral-combined_k141_114741||full Low-quality
17-108-final-viral-combined_k141_239905||full Not-determined
17-108-final-viral-combined_k141_281396||full Not-determined
17-108-final-viral-combined_k141_225802||full Not-determined
17-108-final-viral-combined_k141_281397||full Not-determined
17-108-final-viral-combined_k141_266936||full Not-determined
17-108-final-viral-combined_k141_266938||full Low-quality
17-108-final-viral-combined_k141_225812||full Not-determined
17-108-final-viral-combined_k141_253181||full Not-determined
17-108-final-viral-combined_k141_87960||full Not-determined
17-108-final-viral-combined_k141_4989||full Not-determined
17-108-final-viral-combined_k141_281419||full Low-quality
17-108-final-viral-combined_k141_239930||full Low-quality
17-108-final-viral-combined_k141_239934||full Not-determined
17-108-final-viral-combined_k141_60321||full Not-determined
17-108-final-viral-combined_k141_170949||full Not-determined
17-108-final-viral-combined_k141_321598||full Not-determined
17-108-final-viral-combined_k141_253202||full Low-quality
17-108-final-viral-combined_k141_321605||full Low-quality
17-108-final-viral-combined_k141_5035||full Low-quality
17-108-final-viral-combined_k141_184189||full Low-quality
17-108-final-viral-combined_k141_212400||full Not-determined
17-108-final-viral-combined_k141_266982||full Not-determined
17-108-final-viral-combined_k141_114764||full Low-quality
17-108-final-viral-combined_k141_102128||full Not-determined
17-108-final-viral-combined_k141_281450||full Low-quality
17-108-final-viral-combined_k141_102132||full Low-quality
17-108-final-viral-combined_k141_266990||full Low-quality
17-108-final-viral-combined_k141_88002||full Low-quality
17-108-final-viral-combined_k141_102149||full Not-determined
17-108-final-viral-combined_k141_88009||full Not-determined
17-108-final-viral-combined_k141_143062||full Not-determined
17-108-final-viral-combined_k141_88011||full Low-quality
17-108-final-viral-combined_k141_102156||full Not-determined
17-108-final-viral-combined_k141_197710||full Low-quality
17-108-final-viral-combined_k141_321652||full Low-quality
17-108-final-viral-combined_k141_5075||full Low-quality
17-108-final-viral-combined_k141_73398||full Low-quality
17-108-final-viral-combined_k141_294408||full Low-quality
17-108-final-viral-combined_k141_184209||full Not-determined
17-108-final-viral-combined_k141_5089||full Not-determined
17-108-final-viral-combined_k141_18413||full Low-quality
17-108-final-viral-combined_k141_267016||full Not-determined
17-108-final-viral-combined_k141_88042||full Low-quality
17-108-final-viral-combined_k141_143098||full Low-quality
17-108-final-viral-combined_k141_18423||full Low-quality
17-108-final-viral-combined_k141_197763||full Low-quality
17-108-final-viral-combined_k141_294429||full Low-quality
17-108-final-viral-combined_k141_114811||full Low-quality
17-108-final-viral-combined_k141_308661||full Low-quality
17-108-final-viral-combined_k141_102187||full Low-quality
17-108-final-viral-combined_k141_129136||full Low-quality
17-108-final-viral-combined_k141_197808||full Not-determined
17-108-final-viral-combined_k141_321666||full Not-determined
17-108-final-viral-combined_k141_212474||full Not-determined
17-108-final-viral-combined_k141_308699||full Not-determined
17-108-final-viral-combined_k141_156734||full Not-determined
17-108-final-viral-combined_k141_267089||full Low-quality
17-108-final-viral-combined_k141_60478||full Low-quality
17-108-final-viral-combined_k141_46666||full Low-quality
17-108-final-viral-combined_k141_18482||full Low-quality
17-108-final-viral-combined_k141_281555||full Not-determined
18-101-final-viral-combined_k141_251778||full Medium-quality
17-108-final-viral-combined_k141_73441||full Low-quality
17-108-final-viral-combined_k141_88084||full Not-determined
17-108-final-viral-combined_k141_225930||full Low-quality
17-108-final-viral-combined_k141_156746||full Not-determined
17-108-final-viral-combined_k141_73450||full Low-quality
17-108-final-viral-combined_k141_240025||full Low-quality
17-108-final-viral-combined_k141_212492||full Low-quality
17-108-final-viral-combined_k141_281579||full Low-quality
17-108-final-viral-combined_k141_114879||full Not-determined
17-108-final-viral-combined_k141_267118||full Not-determined
17-108-final-viral-combined_k141_294458||full Not-determined
17-108-final-viral-combined_k141_60490||full Not-determined
17-108-final-viral-combined_k141_267120||full Not-determined
17-108-final-viral-combined_k141_32704||full Not-determined
17-108-final-viral-combined_k141_143164||full Not-determined
17-108-final-viral-combined_k141_156752||full Not-determined
17-108-final-viral-combined_k141_294469||full Not-determined
17-108-final-viral-combined_k141_308726||full Not-determined
17-108-final-viral-combined_k141_281599||full Not-determined
17-108-final-viral-combined_k141_184317||full Not-determined
17-108-final-viral-combined_k141_32724||full Low-quality
17-108-final-viral-combined_k141_32725||full Not-determined
17-108-final-viral-combined_k141_143184||full Not-determined
17-108-final-viral-combined_k141_143192||full Low-quality
17-108-final-viral-combined_k141_308741||full Not-determined
17-108-final-viral-combined_k141_143193||full Not-determined
17-108-final-viral-combined_k141_281622||full Low-quality
17-108-final-viral-combined_k141_102257||full Not-determined
17-108-final-viral-combined_k141_143200||full Low-quality
17-108-final-viral-combined_k141_114936||full Low-quality
17-108-final-viral-combined_k141_253245||full Low-quality
17-108-final-viral-combined_k141_18533||full Not-determined
17-108-final-viral-combined_k141_281637||full Not-determined
17-108-final-viral-combined_k141_143204||full Not-determined
17-108-final-viral-combined_k141_212556||full Not-determined
17-108-final-viral-combined_k141_308750||full Low-quality
17-108-final-viral-combined_k141_88102||full Low-quality
17-108-final-viral-combined_k141_102266||full Not-determined
17-108-final-viral-combined_k141_129203||full Low-quality
17-108-final-viral-combined_k141_114954||full Not-determined
17-108-final-viral-combined_k141_253255||full Not-determined
17-108-final-viral-combined_k141_281649||full Not-determined
18-102-final-viral-combined_k141_137119||full High-quality
17-108-final-viral-combined_k141_197938||full Not-determined
17-108-final-viral-combined_k141_88124||full Low-quality
17-108-final-viral-combined_k141_129237||full Low-quality
17-108-final-viral-combined_k141_281672||full Not-determined
17-108-final-viral-combined_k141_308804||full Low-quality
17-108-final-viral-combined_k141_88132||full Low-quality
17-108-final-viral-combined_k141_281679||full Low-quality
17-108-final-viral-combined_k141_60520||full Not-determined
17-108-final-viral-combined_k141_18562||full Not-determined
17-108-final-viral-combined_k141_225975||full Low-quality
17-108-final-viral-combined_k141_5246||full Not-determined
17-108-final-viral-combined_k141_225976||full Low-quality
17-108-final-viral-combined_k141_225980||full Low-quality
17-108-final-viral-combined_k141_46779||full Low-quality
17-108-final-viral-combined_k141_184379||full Not-determined
17-108-final-viral-combined_k141_321695||full Low-quality
17-108-final-viral-combined_k141_184382||full Not-determined
17-108-final-viral-combined_k141_197964||full Low-quality
17-108-final-viral-combined_k141_240139||full Not-determined
17-108-final-viral-combined_k141_308832||full Low-quality
17-108-final-viral-combined_k141_225999||full Low-quality
17-108-final-viral-combined_k141_32810||full Not-determined
17-108-final-viral-combined_k141_156843||full Not-determined
17-108-final-viral-combined_k141_321706||full Not-determined
17-108-final-viral-combined_k141_73534||full Low-quality
17-108-final-viral-combined_k141_102326||full Low-quality
17-108-final-viral-combined_k141_115052||full Low-quality
17-108-final-viral-combined_k141_73562||full Not-determined

| | |
|---|---|
| 17-108-final-viral-combined_k141_32838\|\|full | Low-quality |
| 17-108-final-viral-combined_k141_60548\|\|full | Not-determined |
| 17-108-final-viral-combined_k141_115061\|\|full | Low-quality |
| 17-108-final-viral-combined_k141_294528\|\|full | Low-quality |
| 17-108-final-viral-combined_k141_212683\|\|full | Not-determined |
| 17-108-final-viral-combined_k141_129337\|\|full | Not-determined |
| 17-108-final-viral-combined_k141_267154\|\|full | Not-determined |
| 17-108-final-viral-combined_k141_102373\|\|full | Low-quality |
| 17-108-final-viral-combined_k141_184420\|\|full | Not-determined |
| 17-108-final-viral-combined_k141_115095\|\|full | Low-quality |
| 17-108-final-viral-combined_k141_226056\|\|full | Not-determined |
| 17-108-final-viral-combined_k141_129352\|\|full | Low-quality |
| 17-108-final-viral-combined_k141_73611\|\|full | Not-determined |
| 17-108-final-viral-combined_k141_102388\|\|full | Low-quality |
| 17-108-final-viral-combined_k141_198010\|\|full | Low-quality |
| 17-108-final-viral-combined_k141_253417\|\|full | Not-determined |
| 17-108-final-viral-combined_k141_294568\|\|full | Not-determined |
| 17-108-final-viral-combined_k141_212727\|\|full | Low-quality |
| 17-108-final-viral-combined_k141_102396\|\|full | Low-quality |
| 17-108-final-viral-combined_k141_253422\|\|full | Low-quality |
| 18-109-final-viral-combined_k141_218379\|\|0_partial | Complete |
| 17-108-final-viral-combined_k141_73639\|\|full | Low-quality |
| 17-108-final-viral-combined_k141_171066\|\|full | Not-determined |
| 17-108-final-viral-combined_k141_32902\|\|full | Not-determined |
| 17-108-final-viral-combined_k141_294583\|\|full | Low-quality |
| 17-108-final-viral-combined_k141_115148\|\|full | Not-determined |
| 17-108-final-viral-combined_k141_212762\|\|full | Low-quality |
| 17-108-final-viral-combined_k141_73663\|\|full | Not-determined |
| 17-108-final-viral-combined_k141_46838\|\|full | Not-determined |
| 17-108-final-viral-combined_k141_115152\|\|full | Not-determined |
| 17-108-final-viral-combined_k141_171084\|\|full | Not-determined |
| 17-108-final-viral-combined_k141_32929\|\|full | Low-quality |
| 17-108-final-viral-combined_k141_156954\|\|full | Not-determined |
| 17-108-final-viral-combined_k141_294612\|\|full | Not-determined |
| 17-108-final-viral-combined_k141_240277\|\|full | Not-determined |
| 17-108-final-viral-combined_k141_198050\|\|full | Not-determined |
| 17-108-final-viral-combined_k141_294617\|\|full | Low-quality |
| 17-108-final-viral-combined_k141_253447\|\|full | Low-quality |
| 17-108-final-viral-combined_k141_102437\|\|full | Not-determined |
| 17-108-final-viral-combined_k141_46844\|\|full | Not-determined |
| 17-108-final-viral-combined_k141_143285\|\|full | Not-determined |
| 17-108-final-viral-combined_k141_156974\|\|full | Not-determined |
| 17-108-final-viral-combined_k141_115172\|\|full | Not-determined |
| 17-108-final-viral-combined_k141_294627\|\|full | Not-determined |
| 17-108-final-viral-combined_k141_5402\|\|full | Not-determined |
| 17-108-final-viral-combined_k141_88259\|\|full | Not-determined |
| 17-108-final-viral-combined_k141_294629\|\|full | Low-quality |
| 17-108-final-viral-combined_k141_267205\|\|full | Low-quality |
| 17-108-final-viral-combined_k141_294648\|\|full | Low-quality |
| 17-108-final-viral-combined_k141_294658\|\|full | Low-quality |
| 17-108-final-viral-combined_k141_198080\|\|full | Not-determined |
| 17-108-final-viral-combined_k141_184528\|\|full | Not-determined |
| 17-108-final-viral-combined_k141_294666\|\|full | Not-determined |
| 17-108-final-viral-combined_k141_156986\|\|full | Not-determined |
| 17-108-final-viral-combined_k141_184536\|\|full | Low-quality |
| 17-108-final-viral-combined_k141_240336\|\|full | Not-determined |
| 17-108-final-viral-combined_k141_184544\|\|full | Low-quality |
| 17-108-final-viral-combined_k141_5431\|\|full | Not-determined |
| 17-108-final-viral-combined_k141_198084\|\|full | Low-quality |
| 17-108-final-viral-combined_k141_294673\|\|full | Not-determined |
| 17-108-final-viral-combined_k141_156992\|\|full | Low-quality |
| 17-108-final-viral-combined_k141_240340\|\|full | Not-determined |
| 17-108-final-viral-combined_k141_321782\|\|full | Low-quality |
| 17-108-final-viral-combined_k141_184557\|\|full | Low-quality |
| 17-108-final-viral-combined_k141_321787\|\|full | Low-quality |
| 17-108-final-viral-combined_k141_321788\|\|full | Low-quality |
| 17-108-final-viral-combined_k141_281740\|\|full | Low-quality |
| 17-108-final-viral-combined_k141_294692\|\|full | Not-determined |
| 17-108-final-viral-combined_k141_308958\|\|full | Not-determined |
| 17-108-final-viral-combined_k141_281748\|\|full | Not-determined |
| 17-108-final-viral-combined_k141_18600\|\|full | Low-quality |
| 17-108-final-viral-combined_k141_294704\|\|full | Not-determined |
| 17-108-final-viral-combined_k141_88295\|\|full | Not-determined |
| 17-108-final-viral-combined_k141_212804\|\|full | Low-quality |
| 17-108-final-viral-combined_k141_212811\|\|full | Low-quality |
| 17-108-final-viral-combined_k141_18606\|\|full | Not-determined |
| 17-108-final-viral-combined_k141_184588\|\|full | Not-determined |
| 17-108-final-viral-combined_k141_33022\|\|full | Low-quality |
| 17-108-final-viral-combined_k141_157020\|\|full | Low-quality |
| 17-108-final-viral-combined_k141_294738\|\|full | Not-determined |
| 17-108-final-viral-combined_k141_184606\|\|full | Low-quality |
| 17-108-final-viral-combined_k141_46913\|\|full | Not-determined |
| 17-108-final-viral-combined_k141_184610\|\|full | Not-determined |
| 17-108-final-viral-combined_k141_309011\|\|full | Low-quality |
| 17-108-final-viral-combined_k141_60673\|\|full | Not-determined |
| 17-108-final-viral-combined_k141_240405\|\|full | Not-determined |
| 17-108-final-viral-combined_k141_171175\|\|full | Not-determined |
| 17-108-final-viral-combined_k141_294754\|\|full | Low-quality |
| 17-108-final-viral-combined_k141_46932\|\|full | Low-quality |
| 17-108-final-viral-combined_k141_321834\|\|full | Not-determined |
| 17-108-final-viral-combined_k141_88329\|\|full | Low-quality |
| 17-108-final-viral-combined_k141_253539\|\|full | Low-quality |
| 17-108-final-viral-combined_k141_88332\|\|full | Not-determined |
| 17-108-final-viral-combined_k141_212829\|\|full | Not-determined |
| 17-108-final-viral-combined_k141_267282\|\|full | Not-determined |
| 17-108-final-viral-combined_k141_129456\|\|full | Low-quality |
| 17-108-final-viral-combined_k141_267283\|\|full | Low-quality |
| 17-108-final-viral-combined_k141_184637\|\|full | Low-quality |
| 17-108-final-viral-combined_k141_115242\|\|full | Not-determined |
| 17-108-final-viral-combined_k141_157054\|\|full | Low-quality |
| 17-108-final-viral-combined_k141_18662\|\|full | Low-quality |
| 17-108-final-viral-combined_k141_46965\|\|full | Low-quality |
| 17-108-final-viral-combined_k141_88348\|\|full | Low-quality |
| 17-108-final-viral-combined_k141_253582\|\|full | Not-determined |
| 17-108-final-viral-combined_k141_198187\|\|full | Low-quality |
| 17-108-final-viral-combined_k141_143472\|\|full | Low-quality |
| 17-108-final-viral-combined_k141_171219\|\|full | Low-quality |
| 17-108-final-viral-combined_k141_294766\|\|full | Low-quality |
| 17-108-final-viral-combined_k141_226113\|\|full | Low-quality |
| 17-108-final-viral-combined_k141_157092\|\|full | Not-determined |
| 17-108-final-viral-combined_k141_226135\|\|full | Not-determined |
| 17-108-final-viral-combined_k141_184668\|\|full | Not-determined |
| 17-108-final-viral-combined_k141_267316\|\|full | Low-quality |
| 17-108-final-viral-combined_k141_294784\|\|full | Not-determined |
| 17-108-final-viral-combined_k141_267318\|\|full | Not-determined |
| 17-108-final-viral-combined_k141_102640\|\|full | Not-determined |
| 17-108-final-viral-combined_k141_212869\|\|full | Low-quality |
| 17-108-final-viral-combined_k141_321860\|\|full | Not-determined |
| 17-108-final-viral-combined_k141_267341\|\|full | Low-quality |
| 17-108-final-viral-combined_k141_73883\|\|full | Not-determined |
| 17-108-final-viral-combined_k141_18700\|\|full | Not-determined |
| 17-108-final-viral-combined_k141_226159\|\|full | Low-quality |
| 17-108-final-viral-combined_k141_73889\|\|full | Not-determined |
| 17-108-final-viral-combined_k141_115309\|\|full | Not-determined |
| 17-108-final-viral-combined_k141_129506\|\|full | Not-determined |
| 17-108-final-viral-combined_k141_73891\|\|full | Not-determined |
| 17-108-final-viral-combined_k141_102671\|\|full | Low-quality |
| 17-108-final-viral-combined_k141_184681\|\|full | Low-quality |
| 17-108-final-viral-combined_k141_267351\|\|full | Low-quality |
| 17-108-final-viral-combined_k141_73906\|\|full | Low-quality |
| 17-108-final-viral-combined_k141_73908\|\|full | Not-determined |
| 17-108-final-viral-combined_k141_198244\|\|full | Not-determined |
| 17-108-final-viral-combined_k141_88395\|\|full | Not-determined |
| 17-108-final-viral-combined_k141_294832\|\|full | Low-quality |
| 17-108-final-viral-combined_k141_294839\|\|full | Low-quality |
| 17-108-final-viral-combined_k141_129543\|\|full | Not-determined |
| 17-108-final-viral-combined_k141_321873\|\|full | Low-quality |
| 17-108-final-viral-combined_k141_33110\|\|full | Not-determined |
| 17-108-final-viral-combined_k141_198269\|\|full | Low-quality |
| 17-108-final-viral-combined_k141_47099\|\|full | Not-determined |
| KM-final-viral-combined_k141_56006\|\|full | High-quality |
| 17-108-final-viral-combined_k141_33113\|\|full | Not-determined |
| 17-108-final-viral-combined_k141_184695\|\|full | Low-quality |
| 17-108-final-viral-combined_k141_5563\|\|full | Not-determined |
| 17-108-final-viral-combined_k141_88435\|\|full | Low-quality |
| 17-108-final-viral-combined_k141_309192\|\|full | Low-quality |
| 17-108-final-viral-combined_k141_240535\|\|full | Not-determined |
| 17-108-final-viral-combined_k141_47141\|\|full | Not-determined |
| 17-108-final-viral-combined_k141_157151\|\|full | Low-quality |
| 17-108-final-viral-combined_k141_253699\|\|full | Not-determined |
| 17-108-final-viral-combined_k141_321918\|\|full | Low-quality |
| 17-108-final-viral-combined_k141_267363\|\|full | Not-determined |
| 17-108-final-viral-combined_k141_129591\|\|full | Low-quality |
| 17-108-final-viral-combined_k141_33153\|\|full | Not-determined |
| 17-108-final-viral-combined_k141_33155\|\|full | Low-quality |
| 17-108-final-viral-combined_k141_184760\|\|full | Low-quality |
| 17-108-final-viral-combined_k141_157172\|\|full | Low-quality |
| 17-108-final-viral-combined_k141_102712\|\|full | Low-quality |
| 17-108-final-viral-combined_k141_212964\|\|full | Low-quality |
| 17-108-final-viral-combined_k141_129593\|\|full | Not-determined |
| 17-108-final-viral-combined_k141_171340\|\|full | Not-determined |
| 17-108-final-viral-combined_k141_102717\|\|full | Low-quality |
| 17-108-final-viral-combined_k141_184770\|\|full | Not-determined |
| 17-108-final-viral-combined_k141_240585\|\|full | Low-quality |
| 17-108-final-viral-combined_k141_212975\|\|full | Low-quality |
| 17-108-final-viral-combined_k141_5581\|\|full | Low-quality |
| 17-108-final-viral-combined_k141_102742\|\|full | Low-quality |
| 17-108-final-viral-combined_k141_240616\|\|full | Not-determined |
| 17-108-final-viral-combined_k141_212990\|\|full | Low-quality |
| 17-108-final-viral-combined_k141_184791\|\|full | Not-determined |
| 17-108-final-viral-combined_k141_294913\|\|full | Not-determined |
| 17-108-final-viral-combined_k141_33172\|\|full | Low-quality |
| 17-108-final-viral-combined_k141_5604\|\|full | Low-quality |
| 17-108-final-viral-combined_k141_102759\|\|full | Low-quality |
| 17-108-final-viral-combined_k141_253743\|\|full | Low-quality |
| 17-108-final-viral-combined_k141_33179\|\|full | Not-determined |
| 17-108-final-viral-combined_k141_47188\|\|full | Low-quality |
| 17-108-final-viral-combined_k141_294915\|\|full | Not-determined |
| 17-108-final-viral-combined_k141_33181\|\|full | Not-determined |
| 17-108-final-viral-combined_k141_309270\|\|full | Not-determined |
| 18-101-final-viral-combined_k141_304377\|\|full | Complete |
| 17-108-final-viral-combined_k141_33192\|\|full | Low-quality |
| 17-108-final-viral-combined_k141_226236\|\|full | Not-determined |
| 17-108-final-viral-combined_k141_321949\|\|full | Low-quality |
| 17-108-final-viral-combined_k141_74017\|\|full | Low-quality |
| 17-108-final-viral-combined_k141_33212\|\|full | Not-determined |
| 17-108-final-viral-combined_k141_60929\|\|full | Low-quality |
| 17-108-final-viral-combined_k141_294946\|\|full | Low-quality |
| 17-108-final-viral-combined_k141_129642\|\|full | Not-determined |
| 17-108-final-viral-combined_k141_267475\|\|full | Not-determined |
| 17-108-final-viral-combined_k141_157224\|\|full | Not-determined |
| 17-108-final-viral-combined_k141_226248\|\|full | Not-determined |
| 17-108-final-viral-combined_k141_33239\|\|full | Not-determined |
| 17-108-final-viral-combined_k141_240647\|\|full | Low-quality |
| 17-108-final-viral-combined_k141_226260\|\|full | Not-determined |
| 17-108-final-viral-combined_k141_47214\|\|full | Low-quality |
| 17-108-final-viral-combined_k141_157235\|\|full | Low-quality |
| 17-108-final-viral-combined_k141_213034\|\|full | Low-quality |
| 17-108-final-viral-combined_k141_309288\|\|full | Low-quality |
| 17-108-final-viral-combined_k141_143621\|\|full | Low-quality |
| 17-108-final-viral-combined_k141_294999\|\|full | Low-quality |
| 17-108-final-viral-combined_k141_322012\|\|full | Low-quality |
| 17-108-final-viral-combined_k141_102820\|\|full | Not-determined |
| 17-108-final-viral-combined_k141_143622\|\|full | Not-determined |
| 17-108-final-viral-combined_k141_157265\|\|full | Low-quality |
| 17-108-final-viral-combined_k141_253838\|\|full | Low-quality |
| 17-108-final-viral-combined_k141_253857\|\|full | Low-quality |
| 17-108-final-viral-combined_k141_60999\|\|full | Low-quality |
| 17-108-final-viral-combined_k141_253869\|\|full | Not-determined |
| 17-108-final-viral-combined_k141_295020\|\|full | Not-determined |
| 17-108-final-viral-combined_k141_213079\|\|full | Not-determined |
| 17-108-final-viral-combined_k141_295029\|\|full | Low-quality |
| 17-108-final-viral-combined_k141_61014\|\|full | Low-quality |
| 17-108-final-viral-combined_k141_295033\|\|full | Not-determined |
| 17-108-final-viral-combined_k141_295038\|\|full | Not-determined |
| 17-108-final-viral-combined_k141_143674\|\|full | Low-quality |
| 17-108-final-viral-combined_k141_240699\|\|full | Not-determined |
| 17-108-final-viral-combined_k141_213099\|\|full | Not-determined |
| 17-108-final-viral-combined_k141_309356\|\|full | Not-determined |
| 17-108-final-viral-combined_k141_5677\|\|full | Not-determined |
| 17-108-final-viral-combined_k141_47300\|\|full | Low-quality |
| 17-108-final-viral-combined_k141_253889\|\|full | Low-quality |
| 17-108-final-viral-combined_k141_253894\|\|full | Not-determined |
| 17-108-final-viral-combined_k141_157333\|\|full | Low-quality |
| 17-108-final-viral-combined_k141_5688\|\|full | Not-determined |
| 17-108-final-viral-combined_k141_143699\|\|full | Low-quality |
| 17-108-final-viral-combined_k141_88560\|\|full | Not-determined |
| 17-108-final-viral-combined_k141_309384\|\|full | Not-determined |
| 17-108-final-viral-combined_k141_5699\|\|full | Not-determined |
| 17-108-final-viral-combined_k141_322046\|\|full | Low-quality |
| 17-108-final-viral-combined_k141_282052\|\|full | Not-determined |
| 17-108-final-viral-combined_k141_226380\|\|full | Not-determined |
| 17-108-final-viral-combined_k141_33341\|\|full | Low-quality |
| 17-108-final-viral-combined_k141_253934\|\|full | Not-determined |
| 17-108-final-viral-combined_k141_322062\|\|full | Not-determined |
| 17-108-final-viral-combined_k141_240758\|\|full | Low-quality |
| 17-108-final-viral-combined_k141_88579\|\|full | Not-determined |
| 17-108-final-viral-combined_k141_129740\|\|full | Low-quality |
| 17-108-final-viral-combined_k141_253948\|\|full | Not-determined |
| 17-108-final-viral-combined_k141_171462\|\|full | Not-determined |
| 17-108-final-viral-combined_k141_61033\|\|full | Not-determined |
| 17-108-final-viral-combined_k141_213161\|\|full | Not-determined |
| 17-108-final-viral-combined_k141_282079\|\|full | Low-quality |
| 17-108-final-viral-combined_k141_129751\|\|full | Not-determined |
| 17-108-final-viral-combined_k141_253960\|\|full | Not-determined |
| 17-108-final-viral-combined_k141_33379\|\|full | Low-quality |
| 17-108-final-viral-combined_k141_240783\|\|full | Not-determined |
| 17-108-final-viral-combined_k141_47377\|\|full | Not-determined |
| 17-108-final-viral-combined_k141_102922\|\|full | Low-quality |
| 17-108-final-viral-combined_k141_18915\|\|full | Not-determined |
| 17-108-final-viral-combined_k141_198511\|\|full | Low-quality |
| 17-108-final-viral-combined_k141_240798\|\|full | Not-determined |
| 17-108-final-viral-combined_k141_115425\|\|full | Not-determined |
| 17-108-final-viral-combined_k141_143743\|\|full | Not-determined |
| 17-108-final-viral-combined_k141_267667\|\|full | Not-determined |
| 17-108-final-viral-combined_k141_61071\|\|full | Low-quality |
| 17-108-final-viral-combined_k141_295119\|\|full | Low-quality |
| 17-108-final-viral-combined_k141_213229\|\|full | Low-quality |
| 17-108-final-viral-combined_k141_295126\|\|full | Not-determined |
| 17-108-final-viral-combined_k141_115440\|\|full | Low-quality |
| 17-108-final-viral-combined_k141_184869\|\|full | Low-quality |
| 17-108-final-viral-combined_k141_171499\|\|full | Not-determined |
| 17-108-final-viral-combined_k141_129834\|\|full | Low-quality |
| 17-108-final-viral-combined_k141_254030\|\|full | Not-determined |
| 17-108-final-viral-combined_k141_143768\|\|full | Not-determined |
| 17-106-final-viral-combined_k141_64277\|\|full | High-quality |
| 17-108-final-viral-combined_k141_171514\|\|full | Not-determined |
| 17-108-final-viral-combined_k141_157470\|\|full | Low-quality |
| 17-108-final-viral-combined_k141_267707\|\|full | Low-quality |
| 17-108-final-viral-combined_k141_198518\|\|full | Low-quality |
| 17-108-final-viral-combined_k141_115461\|\|full | Not-determined |
| 17-108-final-viral-combined_k141_198523\|\|full | Low-quality |
| 17-108-final-viral-combined_k141_226492\|\|full | Not-determined |
| 17-108-final-viral-combined_k141_157489\|\|full | Low-quality |
| 17-108-final-viral-combined_k141_33476\|\|full | Not-determined |
| 17-108-final-viral-combined_k141_282155\|\|full | Not-determined |
| 17-108-final-viral-combined_k141_171529\|\|full | Not-determined |
| 17-108-final-viral-combined_k141_184917\|\|full | Not-determined |
| 17-108-final-viral-combined_k141_322136\|\|full | Not-determined |
| 17-108-final-viral-combined_k141_282164\|\|full | Low-quality |
| 17-108-final-viral-combined_k141_322149\|\|full | Not-determined |
| 17-108-final-viral-combined_k141_157537\|\|full | Not-determined |
| 17-108-final-viral-combined_k141_19002\|\|full | Low-quality |
| 17-108-final-viral-combined_k141_61147\|\|full | Not-determined |
| 17-108-final-viral-combined_k141_198559\|\|full | Not-determined |
| 17-108-final-viral-combined_k141_226519\|\|full | Not-determined |
| 17-108-final-viral-combined_k141_157565\|\|full | Low-quality |
| 17-108-final-viral-combined_k141_102994\|\|full | Low-quality |
| 17-108-final-viral-combined_k141_309542\|\|full | Low-quality |
| 17-108-final-viral-combined_k141_115496\|\|full | Low-quality |
| 17-108-final-viral-combined_k141_267814\|\|full | Not-determined |
| 17-108-final-viral-combined_k141_157572\|\|full | Not-determined |
| 17-108-final-viral-combined_k141_322197\|\|full | Low-quality |
| 17-108-final-viral-combined_k141_267819\|\|full | Not-determined |
| 17-108-final-viral-combined_k141_226549\|\|full | Low-quality |
| 17-108-final-viral-combined_k141_171577\|\|full | Low-quality |
| 17-108-final-viral-combined_k141_157590\|\|full | Not-determined |
| 17-108-final-viral-combined_k141_240883\|\|full | Low-quality |
| 17-108-final-viral-combined_k141_322218\|\|full | Not-determined |
| 17-108-final-viral-combined_k141_19057\|\|full | Not-determined |
| 17-108-final-viral-combined_k141_157611\|\|full | Low-quality |
| 17-108-final-viral-combined_k141_184984\|\|full | Not-determined |
| 17-108-final-viral-combined_k141_240890\|\|full | Not-determined |
| 17-109-final-viral-combined_k141_282594\|\|full | Medium-quality |
| 17-108-final-viral-combined_k141_103072\|\|full | Not-determined |
| 17-108-final-viral-combined_k141_103073\|\|full | Not-determined |
| 17-108-final-viral-combined_k141_226563\|\|full | Low-quality |
| 17-108-final-viral-combined_k141_240901\|\|full | Low-quality |
| 17-108-final-viral-combined_k141_103076\|\|full | Low-quality |
| 17-108-final-viral-combined_k141_33574\|\|full | Low-quality |
| 17-108-final-viral-combined_k141_33580\|\|full | Low-quality |
| 17-108-final-viral-combined_k141_19099\|\|full | Not-determined |
| 17-108-final-viral-combined_k141_61211\|\|full | Not-determined |
| 17-108-final-viral-combined_k141_115611\|\|full | Not-determined |
| 17-108-final-viral-combined_k141_157668\|\|full | Low-quality |
| 17-108-final-viral-combined_k141_198641\|\|full | Not-determined |
| 17-108-final-viral-combined_k141_213368\|\|full | Not-determined |
| 17-108-final-viral-combined_k141_198658\|\|full | Low-quality |
| 17-108-final-viral-combined_k141_115650\|\|full | Not-determined |
| 17-108-final-viral-combined_k141_322286\|\|full | Not-determined |
| 17-108-final-viral-combined_k141_5920\|\|full | Not-determined |
| 17-108-final-viral-combined_k141_103167\|\|full | Low-quality |
| 17-108-final-viral-combined_k141_267875\|\|full | Low-quality |
| 17-108-final-viral-combined_k141_267883\|\|full | Low-quality |
| 17-108-final-viral-combined_k141_103177\|\|full | Low-quality |
| 17-108-final-viral-combined_k141_157704\|\|full | Not-determined |
| 17-108-final-viral-combined_k141_282291\|\|full | Not-determined |
| 17-108-final-viral-combined_k141_226604\|\|full | Low-quality |
| 17-108-final-viral-combined_k141_143907\|\|full | Low-quality |
| 17-108-final-viral-combined_k141_282297\|\|full | Not-determined |
| 17-108-final-viral-combined_k141_61240\|\|full | Low-quality |
| 17-108-final-viral-combined_k141_295242\|\|full | Low-quality |
| 17-108-final-viral-combined_k141_226613\|\|full | Low-quality |
| 17-108-final-viral-combined_k141_61241\|\|full | Not-determined |
| 17-108-final-viral-combined_k141_295252\|\|full | Not-determined |
| 17-108-final-viral-combined_k141_19156\|\|full | Not-determined |
| 17-108-final-viral-combined_k141_282320\|\|full | Not-determined |
| 17-108-final-viral-combined_k141_226624\|\|full | Not-determined |
| 17-108-final-viral-combined_k141_198700\|\|full | Not-determined |
| 17-108-final-viral-combined_k141_322337\|\|full | Not-determined |
| 17-108-final-viral-combined_k141_103190\|\|full | Not-determined |
| 17-108-final-viral-combined_k141_213381\|\|full | Not-determined |
| 17-108-final-viral-combined_k141_254271\|\|full | Low-quality |
| 17-108-final-viral-combined_k141_322338\|\|full | Not-determined |
| 17-108-final-viral-combined_k141_254282\|\|full | Not-determined |
| 17-108-final-viral-combined_k141_47604\|\|full | Low-quality |
| 17-108-final-viral-combined_k141_226639\|\|full | Low-quality |
| 17-108-final-viral-combined_k141_322354\|\|full | Not-determined |
| 17-108-final-viral-combined_k141_157762\|\|full | Not-determined |
| 17-108-final-viral-combined_k141_47614\|\|full | Low-quality |
| 17-108-final-viral-combined_k141_282358\|\|full | Low-quality |
| 17-108-final-viral-combined_k141_115696\|\|full | Not-determined |
| 17-108-final-viral-combined_k141_61312\|\|full | Low-quality |
| 17-108-final-viral-combined_k141_33645\|\|full | Not-determined |
| 17-108-final-viral-combined_k141_198731\|\|full | Low-quality |
| 17-108-final-viral-combined_k141_171699\|\|full | Low-quality |
| 17-108-final-viral-combined_k141_322392\|\|full | Not-determined |
| 17-108-final-viral-combined_k141_47638\|\|full | Low-quality |
| 17-108-final-viral-combined_k141_309726\|\|full | Low-quality |
| 17-108-final-viral-combined_k141_143975\|\|full | Not-determined |
| 17-108-final-viral-combined_k141_254333\|\|full | Not-determined |
| 17-108-final-viral-combined_k141_74464\|\|full | Not-determined |
| 17-108-final-viral-combined_k141_61339\|\|full | Not-determined |
| 17-108-final-viral-combined_k141_103219\|\|full | Not-determined |
| 17-108-final-viral-combined_k141_185079\|\|full | Low-quality |
| 17-108-final-viral-combined_k141_61361\|\|full | Not-determined |
| 17-108-final-viral-combined_k141_130105\|\|full | Low-quality |
| 17-108-final-viral-combined_k141_47668\|\|full | Not-determined |
| 17-108-final-viral-combined_k141_295337\|\|full | Low-quality |
| 17-108-final-viral-combined_k141_19223\|\|full | Low-quality |
| 17-108-final-viral-combined_k141_185094\|\|full | Not-determined |
| 17-108-final-viral-combined_k141_295353\|\|full | Low-quality |
| 17-108-final-viral-combined_k141_295357\|\|full | Not-determined |
| 17-108-final-viral-combined_k141_322449\|\|full | Low-quality |
| 17-108-final-viral-combined_k141_74498\|\|full | Low-quality |
| 17-108-final-viral-combined_k141_88838\|\|full | Low-quality |
| 17-108-final-viral-combined_k141_61406\|\|full | Low-quality |
| 17-108-final-viral-combined_k141_322466\|\|full | Not-determined |
| 17-108-final-viral-combined_k141_144014\|\|full | Not-determined |
| 17-108-final-viral-combined_k141_33716\|\|full | Not-determined |
| 17-108-final-viral-combined_k141_157870\|\|full | Low-quality |
| 17-108-final-viral-combined_k141_213459\|\|full | Not-determined |
| 17-108-final-viral-combined_k141_254390\|\|full | Low-quality |
| 17-108-final-viral-combined_k141_254393\|\|full | Low-quality |
| 17-108-final-viral-combined_k141_309845\|\|full | Not-determined |
| 17-108-final-viral-combined_k141_295386\|\|full | Low-quality |
| 17-108-final-viral-combined_k141_213474\|\|full | Not-determined |
| 17-108-final-viral-combined_k141_185140\|\|full | Not-determined |
| 17-108-final-viral-combined_k141_171803\|\|full | Not-determined |
| 17-108-final-viral-combined_k141_33758\|\|full | Not-determined |
| 17-108-final-viral-combined_k141_241158\|\|full | Not-determined |
| 17-108-final-viral-combined_k141_103249\|\|full | Low-quality |
| 17-108-final-viral-combined_k141_47698\|\|full | Not-determined |
| 17-108-final-viral-combined_k141_144071\|\|full | Low-quality |
| 17-108-final-viral-combined_k141_309883\|\|full | Low-quality |
| 17-108-final-viral-combined_k141_144079\|\|full | Not-determined |
| 17-108-final-viral-combined_k141_213524\|\|full | Low-quality |
| 17-108-final-viral-combined_k141_295430\|\|full | Low-quality |
| 17-108-final-viral-combined_k141_33773\|\|full | Low-quality |
| 17-108-final-viral-combined_k141_115886\|\|full | Not-determined |
| 17-108-final-viral-combined_k141_236214\|\|full | Medium-quality |
| 17-108-final-viral-combined_k141_322573\|\|full | Low-quality |
| 17-108-final-viral-combined_k141_61507\|\|full | Low-quality |
| 17-108-final-viral-combined_k141_171838\|\|full | Not-determined |
| 17-108-final-viral-combined_k141_254435\|\|full | Low-quality |
| 17-108-final-viral-combined_k141_198863\|\|full | Not-determined |
| 17-108-final-viral-combined_k141_241192\|\|full | Not-determined |
| 17-108-final-viral-combined_k141_322596\|\|full | Not-determined |
| 17-108-final-viral-combined_k141_157977\|\|full | Not-determined |
| 17-108-final-viral-combined_k141_88966\|\|full | Not-determined |
| 17-108-final-viral-combined_k141_61543\|\|full | Low-quality |
| 17-108-final-viral-combined_k141_295490\|\|full | Not-determined |
| 17-108-final-viral-combined_k141_254445\|\|full | Low-quality |
| 17-108-final-viral-combined_k141_157987\|\|full | Low-quality |
| 17-108-final-viral-combined_k141_144137\|\|full | Not-determined |
| 17-108-final-viral-combined_k141_241219\|\|full | Low-quality |
| 17-108-final-viral-combined_k141_282560\|\|full | Low-quality |
| 17-108-final-viral-combined_k141_103307\|\|full | Low-quality |
| 17-108-final-viral-combined_k141_185169\|\|full | Not-determined |
| 17-108-final-viral-combined_k141_103309\|\|full | Not-determined |
| 17-108-final-viral-combined_k141_254464\|\|full | Not-determined |
| 17-108-final-viral-combined_k141_115978\|\|full | Low-quality |
| 17-108-final-viral-combined_k141_171863\|\|full | Not-determined |
| 17-108-final-viral-combined_k141_158015\|\|full | Not-determined |
| 17-108-final-viral-combined_k141_309920\|\|full | Not-determined |
| 17-108-final-viral-combined_k141_171865\|\|full | Not-determined |
| 17-108-final-viral-combined_k141_144163\|\|full | Not-determined |
| 17-108-final-viral-combined_k141_171872\|\|full | Low-quality |
| 17-108-final-viral-combined_k141_241274\|\|full | Low-quality |
| 17-108-final-viral-combined_k141_268061\|\|full | Not-determined |
| 17-108-final-viral-combined_k141_282615\|\|full | Not-determined |
| 17-108-final-viral-combined_k141_254492\|\|full | Low-quality |
| 17-108-final-viral-combined_k141_171883\|\|full | Low-quality |
| 17-108-final-viral-combined_k141_103350\|\|full | Low-quality |
| 17-108-final-viral-combined_k141_213591\|\|full | Low-quality |
| 17-108-final-viral-combined_k141_116018\|\|full | Not-determined |
| 17-108-final-viral-combined_k141_268086\|\|full | Not-determined |
| 17-108-final-viral-combined_k141_282630\|\|full | Not-determined |
| 17-108-final-viral-combined_k141_74648\|\|full | Not-determined |
| 17-108-final-viral-combined_k141_19371\|\|full | Not-determined |
| 17-108-final-viral-combined_k141_282639\|\|full | Low-quality |
| 17-108-final-viral-combined_k141_268099\|\|full | Low-quality |
| 17-108-final-viral-combined_k141_254541\|\|full | Low-quality |
| 17-108-final-viral-combined_k141_322692\|\|full | Low-quality |
| 17-108-final-viral-combined_k141_198918\|\|full | Low-quality |
| 17-108-final-viral-combined_k141_268108\|\|full | Not-determined |
| 17-108-final-viral-combined_k141_103382\|\|full | Not-determined |
| 17-108-final-viral-combined_k141_74658\|\|full | Not-determined |
| 17-108-final-viral-combined_k141_116051\|\|full | Low-quality |
| 17-108-final-viral-combined_k141_47739\|\|full | Not-determined |
| 17-108-final-viral-combined_k141_6271\|\|full | Not-determined |
| 17-108-final-viral-combined_k141_171933\|\|full | Low-quality |
| 17-108-final-viral-combined_k141_116053\|\|full | Not-determined |
| 17-108-final-viral-combined_k141_74670\|\|full | Not-determined |
| 17-108-final-viral-combined_k141_144214\|\|full | Low-quality |
| 17-108-final-viral-combined_k141_322719\|\|full | Low-quality |
| 17-108-final-viral-combined_k141_309974\|\|full | Not-determined |
| 17-108-final-viral-combined_k141_241328\|\|full | Low-quality |
| 17-108-final-viral-combined_k141_309976\|\|full | Low-quality |
| 17-108-final-viral-combined_k141_171950\|\|full | Low-quality |
| 17-108-final-viral-combined_k141_130147\|\|full | Not-determined |
| 17-108-final-viral-combined_k141_171951\|\|full | Not-determined |
| 17-108-final-viral-combined_k141_74678\|\|full | Not-determined |
| 17-108-final-viral-combined_k141_185280\|\|full | Not-determined |
| 17-108-final-viral-combined_k141_198933\|\|full | Low-quality |
| 17-108-final-viral-combined_k141_198960\|\|full | Not-determined |
| 17-108-final-viral-combined_k141_185297\|\|full | Low-quality |
| 17-108-final-viral-combined_k141_144273\|\|full | Low-quality |
| 17-108-final-viral-combined_k141_144276\|\|full | Low-quality |
| 17-108-final-viral-combined_k141_130192\|\|full | Low-quality |
| 17-108-final-viral-combined_k141_61692\|\|full | Not-determined |
| 17-108-final-viral-combined_k141_185310\|\|full | Low-quality |
| 17-108-final-viral-combined_k141_241396\|\|full | Low-quality |
| 17-108-final-viral-combined_k141_310011\|\|full | Not-determined |
| 17-108-final-viral-combined_k141_144291\|\|full | Not-determined |
| 17-108-final-viral-combined_k141_61709\|\|full | Not-determined |
| 17-108-final-viral-combined_k141_241434\|\|full | Not-determined |
| 17-108-final-viral-combined_k141_310043\|\|full | Low-quality |
| 17-108-final-viral-combined_k141_213710\|\|full | Not-determined |
| 17-108-final-viral-combined_k141_310045\|\|full | Not-determined |
| 17-108-final-viral-combined_k141_227013\|\|full | Not-determined |
| 17-108-final-viral-combined_k141_130231\|\|full | Low-quality |
| 17-108-final-viral-combined_k141_6372\|\|full | Not-determined |
| 17-108-final-viral-combined_k141_322787\|\|full | Not-determined |
| 17-108-final-viral-combined_k141_89079\|\|full | Not-determined |
| 17-108-final-viral-combined_k141_268238\|\|full | Not-determined |
| 17-108-final-viral-combined_k141_198995\|\|full | Not-determined |
| 17-108-final-viral-combined_k141_19470\|\|full | Not-determined |
| 17-108-final-viral-combined_k141_89094\|\|full | Low-quality |
| 17-108-final-viral-combined_k141_241481\|\|full | Not-determined |
| 17-108-final-viral-combined_k141_172081\|\|full | Low-quality |
| 17-108-final-viral-combined_k141_6394\|\|full | Not-determined |
| 17-108-final-viral-combined_k141_172085\|\|full | Low-quality |
| 17-108-final-viral-combined_k141_213736\|\|full | Not-determined |
| 17-108-final-viral-combined_k141_130239\|\|full | Low-quality |
| 17-108-final-viral-combined_k141_322809\|\|full | Not-determined |
| 17-108-final-viral-combined_k141_130247\|\|full | Not-determined |
| 17-108-final-viral-combined_k141_254677\|\|full | Low-quality |
| 17-108-final-viral-combined_k141_268286\|\|full | Not-determined |
| 17-108-final-viral-combined_k141_199028\|\|full | Low-quality |
| 17-108-final-viral-combined_k141_295581\|\|full | Not-determined |
| 17-108-final-viral-combined_k141_227046\|\|full | Not-determined |
| 17-108-final-viral-combined_k141_130265\|\|full | Low-quality |
| 17-108-final-viral-combined_k141_61811\|\|full | Not-determined |
| 17-108-final-viral-combined_k141_74766\|\|full | Low-quality |
| 17-108-final-viral-combined_k141_322835\|\|full | Low-quality |
| 17-108-final-viral-combined_k141_103483\|\|full | Not-determined |
| 17-108-final-viral-combined_k141_227054\|\|full | Low-quality |
| 17-108-final-viral-combined_k141_47820\|\|full | Low-quality |
| 17-108-final-viral-combined_k141_130299\|\|full | Not-determined |
| 17-108-final-viral-combined_k141_322838\|\|full | Low-quality |
| 17-108-final-viral-combined_k141_227079\|\|full | Low-quality |
| 17-108-final-viral-combined_k141_213774\|\|full | Not-determined |
| 17-108-final-viral-combined_k141_213778\|\|full | Low-quality |
| 17-108-final-viral-combined_k141_34002\|\|full | Not-determined |
| 17-108-final-viral-combined_k141_254703\|\|full | Low-quality |
| 17-108-final-viral-combined_k141_89170\|\|full | Low-quality |
| 17-108-final-viral-combined_k141_34010\|\|full | Low-quality |
| 17-108-final-viral-combined_k141_322857\|\|full | Not-determined |
| 17-108-final-viral-combined_k141_74807\|\|full | Not-determined |
| 17-108-final-viral-combined_k141_89187\|\|full | Not-determined |
| 17-108-final-viral-combined_k141_310162\|\|full | Low-quality |
| 17-108-final-viral-combined_k141_241532\|\|full | Low-quality |
| 17-108-final-viral-combined_k141_199098\|\|full | Low-quality |
| 17-108-final-viral-combined_k141_310165\|\|full | Low-quality |
| 17-108-final-viral-combined_k141_185437\|\|full | Not-determined |
| 17-108-final-viral-combined_k141_103559\|\|full | Not-determined |
| 17-108-final-viral-combined_k141_322872\|\|full | Not-determined |
| 17-108-final-viral-combined_k141_74837\|\|full | Low-quality |
| 17-108-final-viral-combined_k141_158306\|\|full | Not-determined |
| 17-108-final-viral-combined_k141_322881\|\|full | Not-determined |
| 17-108-final-viral-combined_k141_185466\|\|full | Low-quality |
| 17-108-final-viral-combined_k141_61885\|\|full | Not-determined |
| 17-108-final-viral-combined_k141_185467\|\|full | Low-quality |
| 17-108-final-viral-combined_k141_282748\|\|full | Low-quality |
| 17-108-final-viral-combined_k141_116141\|\|full | Low-quality |
| 17-108-final-viral-combined_k141_268332\|\|full | Not-determined |
| 17-108-final-viral-combined_k141_254765\|\|full | Not-determined |
| 17-108-final-viral-combined_k141_268335\|\|full | Low-quality |
| 17-108-final-viral-combined_k141_74851\|\|full | Not-determined |
| 17-108-final-viral-combined_k141_213849\|\|full | Low-quality |
| 17-108-final-viral-combined_k141_310217\|\|full | Low-quality |
| 17-108-final-viral-combined_k141_61897\|\|full | Not-determined |
| 17-108-final-viral-combined_k141_6560\|\|full | Low-quality |
| 17-108-final-viral-combined_k141_61901\|\|full | Low-quality |
| 17-108-final-viral-combined_k141_254779\|\|full | Not-determined |
| 17-108-final-viral-combined_k141_213863\|\|full | Not-determined |
| 17-108-final-viral-combined_k141_61911\|\|full | Low-quality |
| 17-108-final-viral-combined_k141_172205\|\|full | Not-determined |
| 17-108-final-viral-combined_k141_310229\|\|full | Not-determined |
| 17-108-final-viral-combined_k141_172206\|\|full | Low-quality |
| 17-108-final-viral-combined_k141_322908\|\|full | Not-determined |
| 17-108-final-viral-combined_k141_89231\|\|full | Low-quality |
| 17-108-final-viral-combined_k141_116180\|\|full | Low-quality |
| 17-108-final-viral-combined_k141_103605\|\|full | Not-determined |
| 17-108-final-viral-combined_k141_282779\|\|full | Low-quality |
| 17-108-final-viral-combined_k141_158378\|\|full | Not-determined |
| 17-108-final-viral-combined_k141_241600\|\|full | Low-quality |
| 17-108-final-viral-combined_k141_158379\|\|full | Not-determined |
| 17-108-final-viral-combined_k141_19588\|\|full | Low-quality |
| 17-108-final-viral-combined_k141_241601\|\|full | Low-quality |
| 17-108-final-viral-combined_k141_47927\|\|full | Low-quality |
| 17-108-final-viral-combined_k141_268379\|\|full | Not-determined |
| 17-108-final-viral-combined_k141_158405\|\|full | Low-quality |
| 17-108-final-viral-combined_k141_116212\|\|full | Low-quality |
| 17-108-final-viral-combined_k141_74912\|\|full | Low-quality |
| 17-108-final-viral-combined_k141_6578\|\|full | Not-determined |
| 17-108-final-viral-combined_k141_172266\|\|full | Low-quality |
| 17-108-final-viral-combined_k141_282825\|\|full | Low-quality |
| 17-108-final-viral-combined_k141_310317\|\|full | Not-determined |
| 17-108-final-viral-combined_k141_295693\|\|full | Low-quality |
| 17-108-final-viral-combined_k141_89268\|\|full | Not-determined |
| 17-108-final-viral-combined_k141_116240\|\|full | Low-quality |
| 17-108-final-viral-combined_k141_74947\|\|full | Not-determined |
| 17-108-final-viral-combined_k141_268393\|\|full | Not-determined |
| 17-108-final-viral-combined_k141_310323\|\|full | Not-determined |
| 17-108-final-viral-combined_k141_199136\|\|full | Low-quality |
| 17-108-final-viral-combined_k141_185532\|\|full | Low-quality |
| 17-108-final-viral-combined_k141_295719\|\|full | Low-quality |
| 17-108-final-viral-combined_k141_89287\|\|full | Not-determined |
| 17-108-final-viral-combined_k141_19658\|\|full | Low-quality |
| 17-108-final-viral-combined_k141_241674\|\|full | Not-determined |
| 17-108-final-viral-combined_k141_19660\|\|full | Not-determined |
| 17-108-final-viral-combined_k141_61989\|\|full | Not-determined |
| 17-108-final-viral-combined_k141_158468\|\|full | Low-quality |
| 17-108-final-viral-combined_k141_268422\|\|full | Low-quality |
| 17-108-final-viral-combined_k141_116251\|\|full | Not-determined |
| 17-108-final-viral-combined_k141_172338\|\|full | Not-determined |
| 17-108-final-viral-combined_k141_89315\|\|full | Not-determined |

| | |
|---|---|
| 17-108-final-viral-combined_k141_268434\|\|full | Low-quality |
| 17-108-final-viral-combined_k141_199172\|\|full | Low-quality |
| 17-108-final-viral-combined_k141_19678\|\|full | Not-determined |
| 17-108-final-viral-combined_k141_116261\|\|full | Low-quality |
| 17-108-final-viral-combined_k141_172346\|\|full | Not-determined |
| 17-108-final-viral-combined_k141_130366\|\|full | Low-quality |
| 17-108-final-viral-combined_k141_47958\|\|full | Low-quality |
| 17-108-final-viral-combined_k141_172357\|\|full | Low-quality |
| 17-108-final-viral-combined_k141_47965\|\|full | Not-determined |
| 17-108-final-viral-combined_k141_89341\|\|full | Low-quality |
| 17-108-final-viral-combined_k141_282865\|\|full | Not-determined |
| 17-108-final-viral-combined_k141_47974\|\|full | Not-determined |
| 17-108-final-viral-combined_k141_6644\|\|full | Not-determined |
| 17-108-final-viral-combined_k141_213924\|\|full | Low-quality |
| 17-108-final-viral-combined_k141_199195\|\|full | Low-quality |
| 17-108-final-viral-combined_k141_241698\|\|full | Low-quality |
| 17-108-final-viral-combined_k141_6649\|\|full | Low-quality |
| 17-108-final-viral-combined_k141_47980\|\|full | Not-determined |
| 17-108-final-viral-combined_k141_254913\|\|full | Not-determined |
| 17-108-final-viral-combined_k141_144533\|\|full | Not-determined |
| 17-108-final-viral-combined_k141_62015\|\|full | Not-determined |
| 18-106-final-viral-combined_k141_91039\|\|0_partial | Complete |
| 17-108-final-viral-combined_k141_34202\|\|full | Not-determined |
| 17-108-final-viral-combined_k141_62023\|\|full | Low-quality |
| 17-108-final-viral-combined_k141_103695\|\|full | Not-determined |
| 17-108-final-viral-combined_k141_144559\|\|full | Low-quality |
| 17-108-final-viral-combined_k141_254940\|\|full | Not-determined |
| 17-108-final-viral-combined_k141_172363\|\|full | Low-quality |
| 17-108-final-viral-combined_k141_144563\|\|full | Not-determined |
| 17-108-final-viral-combined_k141_213952\|\|full | Not-determined |
| 17-108-final-viral-combined_k141_199235\|\|full | Low-quality |
| 17-108-final-viral-combined_k141_282903\|\|full | Not-determined |
| 17-108-final-viral-combined_k141_116333\|\|full | Not-determined |
| 17-108-final-viral-combined_k141_310363\|\|full | Not-determined |
| 17-108-final-viral-combined_k141_254970\|\|full | Low-quality |
| 17-108-final-viral-combined_k141_241759\|\|full | Not-determined |
| 17-108-final-viral-combined_k141_254981\|\|full | Low-quality |
| 17-108-final-viral-combined_k141_268487\|\|full | Low-quality |
| 17-108-final-viral-combined_k141_103731\|\|full | Low-quality |
| 17-108-final-viral-combined_k141_322964\|\|full | Low-quality |
| 17-108-final-viral-combined_k141_213973\|\|full | Low-quality |
| 17-108-final-viral-combined_k141_89377\|\|full | Low-quality |
| 17-108-final-viral-combined_k141_227368\|\|full | Not-determined |
| 17-108-final-viral-combined_k141_254995\|\|full | Low-quality |
| 17-108-final-viral-combined_k141_89382\|\|full | Not-determined |
| 17-108-final-viral-combined_k141_295827\|\|full | Not-determined |
| 17-108-final-viral-combined_k141_62071\|\|full | Low-quality |
| 17-108-final-viral-combined_k141_6725\|\|full | Not-determined |
| 17-108-final-viral-combined_k141_213977\|\|full | Not-determined |
| 17-108-final-viral-combined_k141_268505\|\|full | Low-quality |
| 17-108-final-viral-combined_k141_130410\|\|full | Low-quality |
| 17-108-final-viral-combined_k141_199259\|\|full | Not-determined |
| 17-108-final-viral-combined_k141_89384\|\|full | Not-determined |
| 17-108-final-viral-combined_k141_227385\|\|full | Low-quality |
| 17-108-final-viral-combined_k141_19784\|\|full | Low-quality |
| 17-108-final-viral-combined_k141_89401\|\|full | Low-quality |
| 17-108-final-viral-combined_k141_34236\|\|full | Not-determined |
| 17-108-final-viral-combined_k141_62092\|\|full | Low-quality |
| 17-108-final-viral-combined_k141_144596\|\|full | Not-determined |
| 17-108-final-viral-combined_k141_227391\|\|full | Not-determined |
| 17-108-final-viral-combined_k141_282958\|\|full | Low-quality |
| 17-108-final-viral-combined_k141_144598\|\|full | Not-determined |
| 17-108-final-viral-combined_k141_103749\|\|full | Low-quality |
| 17-108-final-viral-combined_k141_89428\|\|full | Not-determined |
| 17-108-final-viral-combined_k141_48013\|\|full | Low-quality |
| 17-108-final-viral-combined_k141_144607\|\|full | Low-quality |
| 17-108-final-viral-combined_k141_199280\|\|full | Low-quality |
| 17-108-final-viral-combined_k141_268548\|\|full | Low-quality |
| 17-108-final-viral-combined_k141_255017\|\|full | Low-quality |
| 17-108-final-viral-combined_k141_158633\|\|full | Not-determined |
| 17-108-final-viral-combined_k141_19818\|\|full | Not-determined |
| 17-108-final-viral-combined_k141_322973\|\|full | Not-determined |
| 17-108-final-viral-combined_k141_62134\|\|full | Not-determined |
| 17-108-final-viral-combined_k141_130456\|\|full | Low-quality |
| 17-108-final-viral-combined_k141_185644\|\|full | Low-quality |
| 17-108-final-viral-combined_k141_62139\|\|full | Not-determined |
| 17-108-final-viral-combined_k141_144619\|\|full | Low-quality |
| 17-108-final-viral-combined_k141_103767\|\|full | Not-determined |
| 17-108-final-viral-combined_k141_322981\|\|full | Not-determined |
| 17-108-final-viral-combined_k141_75058\|\|full | Low-quality |
| 17-108-final-viral-combined_k141_322986\|\|full | Low-quality |
| 17-108-final-viral-combined_k141_295868\|\|full | Not-determined |
| 17-108-final-viral-combined_k141_227427\|\|full | Low-quality |
| 17-108-final-viral-combined_k141_34302\|\|full | Not-determined |
| 17-108-final-viral-combined_k141_185655\|\|full | Low-quality |
| 17-108-final-viral-combined_k141_75066\|\|full | Not-determined |
| 17-108-final-viral-combined_k141_34306\|\|full | Not-determined |
| 17-108-final-viral-combined_k141_62168\|\|full | Low-quality |
| 17-108-final-viral-combined_k141_255061\|\|full | Not-determined |
| 17-108-final-viral-combined_k141_268581\|\|full | Not-determined |
| 17-108-final-viral-combined_k141_144658\|\|full | Low-quality |
| 17-108-final-viral-combined_k141_6801\|\|full | Not-determined |
| 17-108-final-viral-combined_k141_89471\|\|full | Low-quality |
| 17-108-final-viral-combined_k141_322993\|\|full | Low-quality |
| 17-108-final-viral-combined_k141_227447\|\|full | Low-quality |
| 17-108-final-viral-combined_k141_89478\|\|full | Not-determined |
| 17-108-final-viral-combined_k141_322994\|\|full | Low-quality |
| 17-108-final-viral-combined_k141_268596\|\|full | Low-quality |
| 17-108-final-viral-combined_k141_144671\|\|full | Low-quality |
| 17-108-final-viral-combined_k141_89482\|\|full | Not-determined |
| 17-108-final-viral-combined_k141_322998\|\|full | Low-quality |
| 17-108-final-viral-combined_k141_227464\|\|full | Low-quality |
| 17-108-final-viral-combined_k141_323003\|\|full | Not-determined |
| 17-108-final-viral-combined_k141_34367\|\|full | Low-quality |
| 17-108-final-viral-combined_k141_310499\|\|full | Not-determined |
| 17-108-final-viral-combined_k141_144701\|\|full | Low-quality |
| 17-108-final-viral-combined_k141_310508\|\|full | Low-quality |
| 17-108-final-viral-combined_k141_255075\|\|full | Low-quality |
| 17-108-final-viral-combined_k141_310510\|\|full | Not-determined |
| 17-108-final-viral-combined_k141_158697\|\|full | Not-determined |
| 17-108-final-viral-combined_k141_214071\|\|full | Low-quality |
| 17-108-final-viral-combined_k141_310515\|\|full | Not-determined |
| 17-108-final-viral-combined_k141_227504\|\|full | Low-quality |
| 17-108-final-viral-combined_k141_310527\|\|full | Low-quality |
| 17-108-final-viral-combined_k141_103856\|\|full | Low-quality |
| 17-108-final-viral-combined_k141_158709\|\|full | Not-determined |
| 17-108-final-viral-combined_k141_34382\|\|full | Low-quality |
| 17-108-final-viral-combined_k141_172444\|\|full | Not-determined |
| 17-108-final-viral-combined_k141_144725\|\|full | Low-quality |
| 17-108-final-viral-combined_k141_158713\|\|full | Not-determined |
| 17-108-final-viral-combined_k141_116484\|\|full | Not-determined |
| 17-108-final-viral-combined_k141_75185\|\|full | Not-determined |
| 17-108-final-viral-combined_k141_116486\|\|full | Low-quality |
| 17-108-final-viral-combined_k141_48117\|\|full | Low-quality |
| 17-108-final-viral-combined_k141_116489\|\|full | Not-determined |
| 17-108-final-viral-combined_k141_199320\|\|full | Low-quality |
| 17-108-final-viral-combined_k141_268631\|\|full | Low-quality |
| 17-108-final-viral-combined_k141_158727\|\|full | Low-quality |
| 17-108-final-viral-combined_k141_310564\|\|full | Low-quality |
| 17-108-final-viral-combined_k141_34421\|\|full | Not-determined |
| 17-108-final-viral-combined_k141_214104\|\|full | Low-quality |
| 17-108-final-viral-combined_k141_48130\|\|full | Not-determined |
| 17-108-final-viral-combined_k141_199347\|\|full | Low-quality |
| 17-108-final-viral-combined_k141_116497\|\|full | Low-quality |
| 17-108-final-viral-combined_k141_268653\|\|full | Not-determined |
| 17-108-final-viral-combined_k141_48135\|\|full | Not-determined |
| 17-108-final-viral-combined_k141_103892\|\|full | Low-quality |
| 17-108-final-viral-combined_k141_323046\|\|full | Not-determined |
| 17-108-final-viral-combined_k141_144734\|\|full | Not-determined |
| 17-108-final-viral-combined_k141_255129\|\|full | Low-quality |
| 17-108-final-viral-combined_k141_323051\|\|full | Not-determined |
| 17-108-final-viral-combined_k141_295932\|\|full | Low-quality |
| 17-108-final-viral-combined_k141_310594\|\|full | Not-determined |
| 17-108-final-viral-combined_k141_34424\|\|full | Low-quality |
| 17-108-final-viral-combined_k141_214134\|\|full | Not-determined |
| 17-108-final-viral-combined_k141_323056\|\|full | Low-quality |
| 17-108-final-viral-combined_k141_130518\|\|full | Low-quality |
| 17-108-final-viral-combined_k141_310605\|\|full | Low-quality |
| 17-108-final-viral-combined_k141_19943\|\|full | Not-determined |
| 17-108-final-viral-combined_k141_130522\|\|full | Not-determined |
| 17-108-final-viral-combined_k141_48162\|\|full | Low-quality |
| 17-108-final-viral-combined_k141_34449\|\|full | Low-quality |
| 17-108-final-viral-combined_k141_185724\|\|full | Low-quality |
| 17-108-final-viral-combined_k141_199387\|\|full | Not-determined |
| 17-108-final-viral-combined_k141_6909\|\|full | Not-determined |
| 17-108-final-viral-combined_k141_185731\|\|full | Not-determined |
| 17-108-final-viral-combined_k141_227575\|\|full | Not-determined |
| 17-108-final-viral-combined_k141_130546\|\|full | Not-determined |
| 17-108-final-viral-combined_k141_323077\|\|full | Low-quality |
| 17-108-final-viral-combined_k141_255170\|\|full | Low-quality |
| 17-108-final-viral-combined_k141_323078\|\|full | Not-determined |
| 17-108-final-viral-combined_k141_19988\|\|full | Low-quality |
| 17-108-final-viral-combined_k141_130565\|\|full | Not-determined |
| 17-108-final-viral-combined_k141_75233\|\|full | Low-quality |
| 17-108-final-viral-combined_k141_323081\|\|full | Not-determined |
| 17-108-final-viral-combined_k141_62334\|\|full | Low-quality |
| 17-108-final-viral-combined_k141_89595\|\|full | Not-determined |
| 17-108-final-viral-combined_k141_144785\|\|full | Low-quality |
| 17-108-final-viral-combined_k141_48209\|\|full | Low-quality |
| 17-108-final-viral-combined_k141_62349\|\|full | Not-determined |
| 17-108-final-viral-combined_k141_241917\|\|full | Not-determined |
| 17-108-final-viral-combined_k141_227607\|\|full | Low-quality |
| 17-108-final-viral-combined_k141_20009\|\|full | Low-quality |
| 17-108-final-viral-combined_k141_6937\|\|full | Not-determined |
| 17-108-final-viral-combined_k141_20013\|\|full | Not-determined |
| 17-108-final-viral-combined_k141_144803\|\|full | Low-quality |
| 17-108-final-viral-combined_k141_227616\|\|full | Low-quality |
| 17-108-final-viral-combined_k141_62375\|\|full | Low-quality |
| 17-108-final-viral-combined_k141_89616\|\|full | Not-determined |
| 17-108-final-viral-combined_k141_241934\|\|full | Not-determined |
| 17-108-final-viral-combined_k141_20016\|\|full | Low-quality |
| 17-108-final-viral-combined_k141_310684\|\|full | Not-determined |
| 17-108-final-viral-combined_k141_214222\|\|full | Not-determined |
| 17-108-final-viral-combined_k141_227630\|\|full | Not-determined |
| 17-108-final-viral-combined_k141_199450\|\|full | Low-quality |
| 17-108-final-viral-combined_k141_241944\|\|full | Not-determined |
| 17-108-final-viral-combined_k141_75266\|\|full | Not-determined |
| 17-108-final-viral-combined_k141_214232\|\|full | Low-quality |
| 17-108-final-viral-combined_k141_241950\|\|full | Low-quality |
| 17-108-final-viral-combined_k141_20038\|\|full | Not-determined |
| 17-108-final-viral-combined_k141_172559\|\|full | Not-determined |
| 17-108-final-viral-combined_k141_20045\|\|full | Not-determined |
| 17-108-final-viral-combined_k141_75278\|\|full | Low-quality |
| 17-108-final-viral-combined_k141_89625\|\|full | Not-determined |
| 17-108-final-viral-combined_k141_310736\|\|full | Not-determined |
| 17-108-final-viral-combined_k141_6974\|\|full | Not-determined |
| 17-108-final-viral-combined_k141_283081\|\|full | Not-determined |
| 17-108-final-viral-combined_k141_310738\|\|full | Not-determined |
| 17-108-final-viral-combined_k141_295997\|\|full | Not-determined |
| 18-102-final-viral-combined_k141_210822\|\|full | High-quality |
| 17-108-final-viral-combined_k141_323154\|\|full | Low-quality |
| 17-108-final-viral-combined_k141_20063\|\|full | Not-determined |
| 17-108-final-viral-combined_k141_283098\|\|full | Not-determined |
| 17-108-final-viral-combined_k141_310765\|\|full | Low-quality |
| 17-108-final-viral-combined_k141_268767\|\|full | Not-determined |
| 17-108-final-viral-combined_k141_104023\|\|full | Low-quality |
| 17-108-final-viral-combined_k141_48227\|\|full | Low-quality |
| 17-108-final-viral-combined_k141_185767\|\|full | Low-quality |
| 17-108-final-viral-combined_k141_241991\|\|full | Low-quality |
| 17-108-final-viral-combined_k141_255255\|\|full | Not-determined |
| 17-108-final-viral-combined_k141_116619\|\|full | Not-determined |
| 17-108-final-viral-combined_k141_296044\|\|full | Low-quality |
| 17-108-final-viral-combined_k141_116621\|\|full | Low-quality |
| 17-108-final-viral-combined_k141_296048\|\|full | Not-determined |
| 17-108-final-viral-combined_k141_214256\|\|full | Not-determined |
| 17-108-final-viral-combined_k141_130609\|\|full | Not-determined |
| 17-108-final-viral-combined_k141_172648\|\|full | Not-determined |
| 17-108-final-viral-combined_k141_89676\|\|full | Not-determined |
| 17-108-final-viral-combined_k141_75319\|\|full | Low-quality |
| 17-108-final-viral-combined_k141_34573\|\|full | Not-determined |
| 17-108-final-viral-combined_k141_185782\|\|full | Low-quality |
| 17-108-final-viral-combined_k141_34581\|\|full | Not-determined |
| 17-108-final-viral-combined_k141_283148\|\|full | Not-determined |
| 17-108-final-viral-combined_k141_227700\|\|full | Low-quality |
| 17-108-final-viral-combined_k141_296078\|\|full | Low-quality |
| 17-108-final-viral-combined_k141_144918\|\|full | Not-determined |
| 17-108-final-viral-combined_k141_20096\|\|full | Low-quality |
| 17-108-final-viral-combined_k141_199534\|\|full | Low-quality |
| 17-108-final-viral-combined_k141_62468\|\|full | Not-determined |
| 17-108-final-viral-combined_k141_75338\|\|full | Low-quality |
| 17-108-final-viral-combined_k141_34596\|\|full | Low-quality |
| 17-108-final-viral-combined_k141_130640\|\|full | Low-quality |
| 17-108-final-viral-combined_k141_323234\|\|full | Low-quality |
| 17-108-final-viral-combined_k141_7054\|\|full | Not-determined |
| 17-108-final-viral-combined_k141_283178\|\|full | Not-determined |
| 17-108-final-viral-combined_k141_62481\|\|full | Not-determined |
| 17-108-final-viral-combined_k141_75358\|\|full | Not-determined |
| 17-108-final-viral-combined_k141_323259\|\|full | Low-quality |
| 17-108-final-viral-combined_k141_310841\|\|full | Not-determined |
| 18-106-final-viral-combined_k141_72228\|\|full | Medium-quality |
| 17-108-final-viral-combined_k141_310844\|\|full | Low-quality |
| 17-108-final-viral-combined_k141_130664\|\|full | Low-quality |
| 17-108-final-viral-combined_k141_34647\|\|full | Low-quality |
| 17-108-final-viral-combined_k141_283206\|\|full | Not-determined |
| 17-108-final-viral-combined_k141_296134\|\|full | Low-quality |
| 17-108-final-viral-combined_k141_310853\|\|full | Low-quality |
| 17-108-final-viral-combined_k141_296138\|\|full | Not-determined |
| 17-108-final-viral-combined_k141_268830\|\|full | Low-quality |
| 17-108-final-viral-combined_k141_144949\|\|full | Not-determined |
| 17-108-final-viral-combined_k141_130676\|\|full | Not-determined |
| 17-108-final-viral-combined_k141_20160\|\|full | Not-determined |
| 17-108-final-viral-combined_k141_255314\|\|full | Not-determined |
| 17-108-final-viral-combined_k141_34666\|\|full | Low-quality |
| 17-108-final-viral-combined_k141_89728\|\|full | Not-determined |
| 17-108-final-viral-combined_k141_89732\|\|full | Not-determined |
| 17-108-final-viral-combined_k141_227737\|\|full | Low-quality |
| 17-108-final-viral-combined_k141_75384\|\|full | Not-determined |
| 17-108-final-viral-combined_k141_34681\|\|full | Low-quality |
| 17-108-final-viral-combined_k141_48303\|\|full | Not-determined |
| 17-108-final-viral-combined_k141_199584\|\|full | Not-determined |
| 17-108-final-viral-combined_k141_89746\|\|full | Not-determined |
| 17-108-final-viral-combined_k141_34695\|\|full | Low-quality |
| 17-108-final-viral-combined_k141_62523\|\|full | Low-quality |
| 17-108-final-viral-combined_k141_34700\|\|full | Low-quality |
| 17-108-final-viral-combined_k141_20181\|\|full | Low-quality |
| 17-108-final-viral-combined_k141_310877\|\|full | Low-quality |
| 17-108-final-viral-combined_k141_255341\|\|full | Low-quality |
| 18-101-final-viral-combined_k141_131371\|\|full | Complete |
| 17-108-final-viral-combined_k141_255351\|\|full | Low-quality |
| 17-108-final-viral-combined_k141_283223\|\|full | Low-quality |
| 17-108-final-viral-combined_k141_7143\|\|full | Not-determined |
| 17-108-final-viral-combined_k141_310904\|\|full | Low-quality |
| 17-108-final-viral-combined_k141_172802\|\|full | Not-determined |
| 17-108-final-viral-combined_k141_75399\|\|full | Not-determined |
| 17-108-final-viral-combined_k141_104162\|\|full | Low-quality |
| 17-108-final-viral-combined_k141_255378\|\|full | Low-quality |
| 17-108-final-viral-combined_k141_214365\|\|full | Not-determined |
| 17-108-final-viral-combined_k141_116654\|\|full | Low-quality |
| 17-108-final-viral-combined_k141_62588\|\|full | Not-determined |
| 17-108-final-viral-combined_k141_75404\|\|full | Low-quality |
| 17-108-final-viral-combined_k141_145064\|\|full | Not-determined |
| 17-108-final-viral-combined_k141_34753\|\|full | Not-determined |
| 17-108-final-viral-combined_k141_214378\|\|full | Not-determined |
| 17-108-final-viral-combined_k141_172855\|\|full | Low-quality |
| 17-108-final-viral-combined_k141_104191\|\|full | Not-determined |
| 17-108-final-viral-combined_k141_75425\|\|full | Not-determined |
| 17-108-final-viral-combined_k141_242159\|\|full | Not-determined |
| 17-108-final-viral-combined_k141_89807\|\|full | Low-quality |
| 17-108-final-viral-combined_k141_255436\|\|full | Low-quality |
| 17-108-final-viral-combined_k141_145072\|\|full | Low-quality |
| 17-108-final-viral-combined_k141_199640\|\|full | Low-quality |
| 17-108-final-viral-combined_k141_296174\|\|full | Low-quality |
| 17-108-final-viral-combined_k141_255440\|\|full | Low-quality |
| 17-108-final-viral-combined_k141_7201\|\|full | Not-determined |
| 17-108-final-viral-combined_k141_199648\|\|full | Low-quality |
| 17-108-final-viral-combined_k141_255447\|\|full | Low-quality |
| 17-108-final-viral-combined_k141_296183\|\|full | Low-quality |
| 17-108-final-viral-combined_k141_268922\|\|full | Low-quality |
| 17-108-final-viral-combined_k141_75453\|\|full | Not-determined |
| 17-108-final-viral-combined_k141_130763\|\|full | Not-determined |
| 17-108-final-viral-combined_k141_255453\|\|full | Low-quality |
| 17-108-final-viral-combined_k141_130764\|\|full | Not-determined |
| 17-108-final-viral-combined_k141_296193\|\|full | Not-determined |
| 17-108-final-viral-combined_k141_227859\|\|full | Low-quality |
| 17-108-final-viral-combined_k141_242198\|\|full | Low-quality |
| 17-108-final-viral-combined_k141_62642\|\|full | Not-determined |
| 17-108-final-viral-combined_k141_227860\|\|full | Low-quality |
| 17-108-final-viral-combined_k141_62647\|\|full | Low-quality |
| 17-108-final-viral-combined_k141_242200\|\|full | Not-determined |
| 17-108-final-viral-combined_k141_116728\|\|full | Low-quality |
| 17-108-final-viral-combined_k141_116729\|\|full | Not-determined |
| 17-108-final-viral-combined_k141_34814\|\|full | Not-determined |
| 17-108-final-viral-combined_k141_145114\|\|full | Not-determined |
| 17-108-final-viral-combined_k141_130790\|\|full | Not-determined |
| 17-108-final-viral-combined_k141_283286\|\|full | Not-determined |
| 17-108-final-viral-combined_k141_242226\|\|full | Low-quality |
| 17-106-final-viral-combined_k141_9931\|\|full | Medium-quality |
| 17-108-final-viral-combined_k141_242236\|\|full | Low-quality |
| 18-102-final-viral-combined_k141_116221\|\|full | High-quality |
| 17-108-final-viral-combined_k141_268957\|\|full | Low-quality |
| 17-108-final-viral-combined_k141_185932\|\|full | Low-quality |
| 17-108-final-viral-combined_k141_158914\|\|full | Low-quality |
| 17-108-final-viral-combined_k141_214450\|\|full | Not-determined |
| 17-108-final-viral-combined_k141_62692\|\|full | Low-quality |
| 17-108-final-viral-combined_k141_158919\|\|full | Low-quality |
| 17-108-final-viral-combined_k141_242248\|\|full | Low-quality |
| 17-108-final-viral-combined_k141_310949\|\|full | Low-quality |
| 17-108-final-viral-combined_k141_130829\|\|full | Not-determined |
| 17-108-final-viral-combined_k141_20293\|\|full | Low-quality |
| 17-108-final-viral-combined_k141_62708\|\|full | Low-quality |
| 17-108-final-viral-combined_k141_158929\|\|full | Low-quality |
| 17-108-final-viral-combined_k141_283306\|\|full | Low-quality |
| 17-108-final-viral-combined_k141_185959\|\|full | Not-determined |
| 17-108-final-viral-combined_k141_34861\|\|full | Not-determined |
| 17-108-final-viral-combined_k141_214464\|\|full | Low-quality |
| 17-108-final-viral-combined_k141_185963\|\|full | Not-determined |
| 17-108-final-viral-combined_k141_130846\|\|full | Not-determined |
| 17-108-final-viral-combined_k141_62713\|\|full | Not-determined |
| 17-108-final-viral-combined_k141_255543\|\|full | Low-quality |
| 17-108-final-viral-combined_k141_62717\|\|full | Low-quality |
| 17-108-final-viral-combined_k141_20306\|\|full | Not-determined |
| 17-108-final-viral-combined_k141_255547\|\|full | Not-determined |
| 17-108-final-viral-combined_k141_130848\|\|full | Not-determined |
| 17-108-final-viral-combined_k141_34871\|\|full | Not-determined |
| 17-108-final-viral-combined_k141_158946\|\|full | Not-determined |
| 17-108-final-viral-combined_k141_145178\|\|full | Not-determined |
| 17-108-final-viral-combined_k141_89888\|\|full | Not-determined |
| 17-108-final-viral-combined_k141_296287\|\|full | Not-determined |
| 17-108-final-viral-combined_k141_172940\|\|full | Not-determined |
| 17-108-final-viral-combined_k141_145188\|\|full | Not-determined |
| 17-108-final-viral-combined_k141_116814\|\|full | Not-determined |
| 17-108-final-viral-combined_k141_172942\|\|full | Not-determined |
| 17-108-final-viral-combined_k141_145195\|\|full | Not-determined |
| 17-108-final-viral-combined_k141_323424\|\|full | Not-determined |
| 17-108-final-viral-combined_k141_62740\|\|full | Not-determined |
| 17-108-final-viral-combined_k141_172953\|\|full | Not-determined |
| 17-106-final-viral-combined_k141_20485\|\|full | Complete |
| 17-108-final-viral-combined_k141_104266\|\|full | Low-quality |
| 17-108-final-viral-combined_k141_7294\|\|full | Low-quality |
| 17-108-final-viral-combined_k141_214514\|\|full | Low-quality |
| 17-108-final-viral-combined_k141_199715\|\|full | Not-determined |
| 17-108-final-viral-combined_k141_130858\|\|full | Not-determined |
| 17-108-final-viral-combined_k141_268979\|\|full | Low-quality |
| 17-108-final-viral-combined_k141_34911\|\|full | Not-determined |
| 17-108-final-viral-combined_k141_172968\|\|full | Not-determined |
| 17-108-final-viral-combined_k141_75613\|\|full | Not-determined |
| 17-108-final-viral-combined_k141_62752\|\|full | Not-determined |
| 17-108-final-viral-combined_k141_242275\|\|full | Low-quality |
| 17-108-final-viral-combined_k141_159002\|\|full | Not-determined |
| 17-108-final-viral-combined_k141_172975\|\|full | Not-determined |
| 17-108-final-viral-combined_k141_214526\|\|full | Low-quality |
| 17-108-final-viral-combined_k141_186009\|\|full | Not-determined |
| 17-108-final-viral-combined_k141_310985\|\|full | Not-determined |
| 17-108-final-viral-combined_k141_130862\|\|full | Low-quality |
| 17-108-final-viral-combined_k141_116860\|\|full | Not-determined |
| 17-108-final-viral-combined_k141_186039\|\|full | Not-determined |
| 17-108-final-viral-combined_k141_269008\|\|full | Low-quality |
| 17-108-final-viral-combined_k141_255630\|\|full | Not-determined |
| 17-108-final-viral-combined_k141_7321\|\|full | Low-quality |
| 17-108-final-viral-combined_k141_130878\|\|full | Low-quality |
| 17-108-final-viral-combined_k141_228021\|\|full | Not-determined |
| 17-108-final-viral-combined_k141_75662\|\|full | Not-determined |
| 17-108-final-viral-combined_k141_48556\|\|full | Low-quality |
| 17-108-final-viral-combined_k141_173006\|\|full | Not-determined |
| 17-108-final-viral-combined_k141_48561\|\|full | Not-determined |
| 17-108-final-viral-combined_k141_283395\|\|full | Not-determined |
| 17-108-final-viral-combined_k141_75663\|\|full | Low-quality |
| 17-108-final-viral-combined_k141_311018\|\|full | Low-quality |
| 17-108-final-viral-combined_k141_62818\|\|full | Low-quality |
| 17-108-final-viral-combined_k141_255672\|\|full | Not-determined |
| 17-108-final-viral-combined_k141_311022\|\|full | Low-quality |
| 17-108-final-viral-combined_k141_34975\|\|full | Not-determined |
| 17-108-final-viral-combined_k141_199781\|\|full | Not-determined |
| 17-108-final-viral-combined_k141_296376\|\|full | Not-determined |
| 17-108-final-viral-combined_k141_48581\|\|full | Not-determined |
| 17-108-final-viral-combined_k141_116899\|\|full | Low-quality |
| 17-108-final-viral-combined_k141_48582\|\|full | Low-quality |
| 17-108-final-viral-combined_k141_130913\|\|full | Not-determined |
| 17-108-final-viral-combined_k141_199792\|\|full | Not-determined |
| 17-108-final-viral-combined_k141_130914\|\|full | Low-quality |
| 17-108-final-viral-combined_k141_34987\|\|full | Low-quality |
| 17-108-final-viral-combined_k141_186079\|\|full | Not-determined |
| 17-108-final-viral-combined_k141_104323\|\|full | Not-determined |
| 17-108-final-viral-combined_k141_186099\|\|full | Low-quality |
| 17-108-final-viral-combined_k141_35001\|\|full | Not-determined |
| 17-108-final-viral-combined_k141_255685\|\|full | Not-determined |
| 17-108-final-viral-combined_k141_104339\|\|full | Not-determined |
| 17-108-final-viral-combined_k141_255696\|\|full | Not-determined |
| 17-108-final-viral-combined_k141_283452\|\|full | Not-determined |
| 17-108-final-viral-combined_k141_283456\|\|full | Not-determined |
| 17-108-final-viral-combined_k141_173092\|\|full | Low-quality |
| 17-108-final-viral-combined_k141_130959\|\|full | Not-determined |
| 17-108-final-viral-combined_k141_242389\|\|full | Not-determined |
| 17-108-final-viral-combined_k141_186120\|\|full | Low-quality |
| 17-108-final-viral-combined_k141_75697\|\|full | Not-determined |
| 17-108-final-viral-combined_k141_35033\|\|full | Not-determined |
| 17-108-final-viral-combined_k141_35040\|\|full | Not-determined |
| 17-108-final-viral-combined_k141_116954\|\|full | Not-determined |
| 17-108-final-viral-combined_k141_199834\|\|full | Not-determined |
| 17-108-final-viral-combined_k141_228090\|\|full | Low-quality |
| 17-108-final-viral-combined_k141_311102\|\|full | Not-determined |
| 17-108-final-viral-combined_k141_186127\|\|full | Not-determined |
| 17-108-final-viral-combined_k141_35054\|\|full | Not-determined |
| 17-108-final-viral-combined_k141_255749\|\|full | Not-determined |
| 17-108-final-viral-combined_k141_311132\|\|full | Not-determined |
| 17-108-final-viral-combined_k141_228117\|\|full | Not-determined |
| 17-108-final-viral-combined_k141_104371\|\|full | Not-determined |
| 17-108-final-viral-combined_k141_296448\|\|full | Low-quality |
| 17-108-final-viral-combined_k141_131016\|\|full | Not-determined |
| 17-108-final-viral-combined_k141_296465\|\|full | Low-quality |
| 17-108-final-viral-combined_k141_75737\|\|full | Not-determined |
| 17-108-final-viral-combined_k141_228138\|\|full | Not-determined |
| 17-108-final-viral-combined_k141_131037\|\|full | Low-quality |
| 17-108-final-viral-combined_k141_48676\|\|full | Low-quality |
| 17-108-final-viral-combined_k141_7430\|\|full | Not-determined |
| 17-108-final-viral-combined_k141_75740\|\|full | Low-quality |
| 17-108-final-viral-combined_k141_323536\|\|full | Not-determined |
| 17-108-final-viral-combined_k141_75755\|\|full | Low-quality |
| 17-108-final-viral-combined_k141_20479\|\|full | Low-quality |
| 17-108-final-viral-combined_k141_90076\|\|full | Not-determined |
| 17-108-final-viral-combined_k141_7443\|\|full | Not-determined |
| 17-108-final-viral-combined_k141_35105\|\|full | Low-quality |
| 17-108-final-viral-combined_k141_311182\|\|full | Not-determined |
| 17-108-final-viral-combined_k141_255796\|\|full | Low-quality |
| 17-108-final-viral-combined_k141_35124\|\|full | Not-determined |
| 17-108-final-viral-combined_k141_173181\|\|full | Low-quality |
| 17-108-final-viral-combined_k141_90103\|\|full | Low-quality |
| 17-108-final-viral-combined_k141_20498\|\|full | Low-quality |
| 17-108-final-viral-combined_k141_35138\|\|full | Not-determined |
| 17-108-final-viral-combined_k141_296536\|\|full | Not-determined |
| 17-108-final-viral-combined_k141_75797\|\|full | Not-determined |
| 17-108-final-viral-combined_k141_48716\|\|full | Low-quality |
| 17-108-final-viral-combined_k141_186170\|\|full | Low-quality |
| 17-108-final-viral-combined_k141_48726\|\|full | Low-quality |
| 17-108-final-viral-combined_k141_145336\|\|full | Low-quality |
| 17-108-final-viral-combined_k141_35160\|\|full | Low-quality |
| 17-108-final-viral-combined_k141_214665\|\|full | Low-quality |
| 17-108-final-viral-combined_k141_186196\|\|full | Not-determined |
| 17-108-final-viral-combined_k141_117044\|\|full | Low-quality |
| 17-108-final-viral-combined_k141_323586\|\|full | Low-quality |
| 17-108-final-viral-combined_k141_323594\|\|full | Low-quality |
| 17-108-final-viral-combined_k141_35175\|\|full | Not-determined |
| 17-108-final-viral-combined_k141_296588\|\|full | Low-quality |
| 17-108-final-viral-combined_k141_104404\|\|full | Low-quality |
| 17-108-final-viral-combined_k141_323609\|\|full | Low-quality |
| 17-108-final-viral-combined_k141_159163\|\|full | Not-determined |
| 17-108-final-viral-combined_k141_173219\|\|full | Not-determined |
| 17-108-final-viral-combined_k141_20542\|\|full | Not-determined |
| 17-108-final-viral-combined_k141_242526\|\|full | Not-determined |
| 17-108-final-viral-combined_k141_323612\|\|full | Not-determined |
| 17-108-final-viral-combined_k141_173230\|\|full | Not-determined |
| 17-108-final-viral-combined_k141_173231\|\|full | Low-quality |
| 17-108-final-viral-combined_k141_173235\|\|full | Not-determined |
| 17-108-final-viral-combined_k141_269216\|\|full | Low-quality |
| 17-108-final-viral-combined_k141_311238\|\|full | Low-quality |
| 17-108-final-viral-combined_k141_186241\|\|full | Low-quality |
| 17-108-final-viral-combined_k141_35226\|\|full | Not-determined |
| 17-108-final-viral-combined_k141_117121\|\|full | Not-determined |
| 17-108-final-viral-combined_k141_117128\|\|full | Low-quality |
| 17-108-final-viral-combined_k141_311261\|\|full | Not-determined |
| 17-108-final-viral-combined_k141_159198\|\|full | Not-determined |
| 17-108-final-viral-combined_k141_214692\|\|full | Not-determined |
| 17-108-final-viral-combined_k141_35256\|\|full | Low-quality |
| 17-108-final-viral-combined_k141_159201\|\|full | Not-determined |
| 17-108-final-viral-combined_k141_90223\|\|full | Not-determined |
| 17-108-final-viral-combined_k141_283587\|\|full | Low-quality |
| 17-108-final-viral-combined_k141_145412\|\|full | Not-determined |
| 17-108-final-viral-combined_k141_283590\|\|full | Low-quality |
| 17-108-final-viral-combined_k141_117161\|\|full | Low-quality |
| 17-108-final-viral-combined_k141_35275\|\|full | Not-determined |
| 17-108-final-viral-combined_k141_173305\|\|full | Low-quality |
| 17-108-final-viral-combined_k141_242624\|\|full | Not-determined |
| 17-108-final-viral-combined_k141_186300\|\|full | Low-quality |
| 17-108-final-viral-combined_k141_228258\|\|full | Low-quality |
| 17-108-final-viral-combined_k141_159229\|\|full | Low-quality |
| 17-108-final-viral-combined_k141_20633\|\|full | Low-quality |
| 17-108-final-viral-combined_k141_228262\|\|full | Not-determined |
| 17-108-final-viral-combined_k141_311294\|\|full | Not-determined |
| 17-108-final-viral-combined_k141_242641\|\|full | Low-quality |
| 17-108-final-viral-combined_k141_75879\|\|full | Low-quality |
| 17-108-final-viral-combined_k141_173341\|\|full | Not-determined |
| 17-108-final-viral-combined_k141_186329\|\|full | Low-quality |
| 17-108-final-viral-combined_k141_159244\|\|full | Not-determined |
| 17-108-final-viral-combined_k141_20658\|\|full | Low-quality |
| 17-108-final-viral-combined_k141_159258\|\|full | Not-determined |
| 17-108-final-viral-combined_k141_35338\|\|full | Low-quality |
| 17-108-final-viral-combined_k141_104444\|\|full | Low-quality |
| 17-108-final-viral-combined_k141_75907\|\|full | Low-quality |
| 17-108-final-viral-combined_k141_159271\|\|full | Not-determined |
| 17-108-final-viral-combined_k141_159272\|\|full | Low-quality |
| 17-108-final-viral-combined_k141_20675\|\|full | Low-quality |
| 18-102-final-viral-combined_k141_200390\|\|full | Complete |
| 17-108-final-viral-combined_k141_200093\|\|full | Low-quality |
| 17-108-final-viral-combined_k141_228326\|\|full | Low-quality |
| 17-108-final-viral-combined_k141_35354\|\|full | Low-quality |
| 17-108-final-viral-combined_k141_255988\|\|full | Low-quality |
| 17-108-final-viral-combined_k141_200108\|\|full | Not-determined |
| 17-108-final-viral-combined_k141_186370\|\|full | Low-quality |
| 17-108-final-viral-combined_k141_242686\|\|full | Low-quality |
| 17-108-final-viral-combined_k141_7556\|\|full | Low-quality |
| 17-108-final-viral-combined_k141_131247\|\|full | Low-quality |
| 17-108-final-viral-combined_k141_117207\|\|full | Low-quality |
| 17-108-final-viral-combined_k141_311331\|\|full | Not-determined |
| 17-108-final-viral-combined_k141_200117\|\|full | Not-determined |
| 17-108-final-viral-combined_k141_214754\|\|full | Low-quality |
| 17-108-final-viral-combined_k141_269326\|\|full | Low-quality |
| 17-108-final-viral-combined_k141_311345\|\|full | Not-determined |
| 17-108-final-viral-combined_k141_200126\|\|full | Not-determined |
| 17-108-final-viral-combined_k141_214759\|\|full | Low-quality |
| 17-108-final-viral-combined_k141_255992\|\|full | Low-quality |
| 17-108-final-viral-combined_k141_104521\|\|full | Not-determined |
| 17-108-final-viral-combined_k141_90352\|\|full | Not-determined |
| 17-108-final-viral-combined_k141_296702\|\|full | Low-quality |
| 17-108-final-viral-combined_k141_145459\|\|full | Low-quality |
| 17-108-final-viral-combined_k141_159321\|\|full | Not-determined |
| 17-108-final-viral-combined_k141_90365\|\|full | Low-quality |
| 17-108-final-viral-combined_k141_186395\|\|full | Low-quality |
| 17-108-final-viral-combined_k141_159342\|\|full | Low-quality |
| 17-108-final-viral-combined_k141_214804\|\|full | Low-quality |
| 17-108-final-viral-combined_k141_186413\|\|full | Not-determined |
| 17-108-final-viral-combined_k141_214808\|\|full | Low-quality |
| 17-108-final-viral-combined_k141_296729\|\|full | Low-quality |
| 17-108-final-viral-combined_k141_63110\|\|full | Not-determined |
| 17-108-final-viral-combined_k141_104555\|\|full | Not-determined |
| 17-108-final-viral-combined_k141_173471\|\|full | Not-determined |
| 17-108-final-viral-combined_k141_311370\|\|full | Not-determined |
| 17-108-final-viral-combined_k141_256045\|\|full | Low-quality |
| 17-108-final-viral-combined_k141_63128\|\|full | Low-quality |
| 17-108-final-viral-combined_k141_311381\|\|full | Low-quality |
| 17-108-final-viral-combined_k141_296760\|\|full | Not-determined |
| 17-108-final-viral-combined_k141_186455\|\|full | Not-determined |
| 17-108-final-viral-combined_k141_7641\|\|full | Not-determined |
| 17-108-final-viral-combined_k141_256055\|\|full | Not-determined |
| 17-108-final-viral-combined_k141_269433\|\|full | Not-determined |
| 17-108-final-viral-combined_k141_35460\|\|full | Low-quality |
| 17-108-final-viral-combined_k141_323782\|\|full | Low-quality |
| 17-108-final-viral-combined_k141_7647\|\|full | Low-quality |
| 17-108-final-viral-combined_k141_214847\|\|full | Not-determined |
| 17-108-final-viral-combined_k141_228397\|\|full | Low-quality |
| 17-108-final-viral-combined_k141_256064\|\|full | Low-quality |
| 17-108-final-viral-combined_k141_311436\|\|full | Not-determined |
| 17-108-final-viral-combined_k141_214866\|\|full | Not-determined |
| 17-108-final-viral-combined_k141_145490\|\|full | Not-determined |
| 17-108-final-viral-combined_k141_228403\|\|full | Not-determined |
| 17-108-final-viral-combined_k141_35492\|\|full | Not-determined |
| 17-108-final-viral-combined_k141_173507\|\|full | Not-determined |
| 17-108-final-viral-combined_k141_145494\|\|full | Not-determined |
| 17-108-final-viral-combined_k141_159400\|\|full | Low-quality |
| 17-108-final-viral-combined_k141_173511\|\|full | Not-determined |
| 17-108-final-viral-combined_k141_48987\|\|full | Low-quality |
| 17-108-final-viral-combined_k141_7686\|\|full | Not-determined |
| 17-108-final-viral-combined_k141_63171\|\|full | Not-determined |
| 17-108-final-viral-combined_k141_200193\|\|full | Not-determined |
| 17-108-final-viral-combined_k141_200198\|\|full | Low-quality |
| 17-108-final-viral-combined_k141_7696\|\|full | Not-determined |
| 17-108-final-viral-combined_k141_104656\|\|full | Low-quality |
| 17-108-final-viral-combined_k141_117286\|\|full | Low-quality |
| 17-108-final-viral-combined_k141_214891\|\|full | Not-determined |
| 17-108-final-viral-combined_k141_311464\|\|full | Low-quality |
| 17-108-final-viral-combined_k141_63199\|\|full | Not-determined |
| 17-108-final-viral-combined_k141_214892\|\|full | Low-quality |
| 17-108-final-viral-combined_k141_49018\|\|full | Low-quality |
| 17-108-final-viral-combined_k141_173519\|\|full | Low-quality |
| 17-108-final-viral-combined_k141_104672\|\|full | Not-determined |
| 17-108-final-viral-combined_k141_20701\|\|full | Not-determined |
| 17-108-final-viral-combined_k141_214913\|\|full | Low-quality |
| 17-108-final-viral-combined_k141_228417\|\|full | Not-determined |
| 17-108-final-viral-combined_k141_256148\|\|full | Not-determined |
| 17-108-final-viral-combined_k141_283660\|\|full | Not-determined |

| | |
|---|---|
| 17-108-final-viral-combined_k141_214942\|\|full | Low-quality |
| 17-108-final-viral-combined_k141_214944\|\|full | Low-quality |
| 17-108-final-viral-combined_k141_256167\|\|full | Low-quality |
| 17-108-final-viral-combined_k141_186546\|\|full | Low-quality |
| 17-108-final-viral-combined_k141_49050\|\|full | Not-determined |
| 17-108-final-viral-combined_k141_283673\|\|full | Low-quality |
| 17-108-final-viral-combined_k141_35521\|\|full | Not-determined |
| 17-108-final-viral-combined_k141_323823\|\|full | Low-quality |
| 17-108-final-viral-combined_k141_104730\|\|full | Not-determined |
| 17-108-final-viral-combined_k141_145585\|\|full | Low-quality |
| 17-108-final-viral-combined_k141_296844\|\|full | Not-determined |
| 17-108-final-viral-combined_k141_173580\|\|full | Not-determined |
| 17-108-final-viral-combined_k141_35532\|\|full | Not-determined |
| 17-108-final-viral-combined_k141_117370\|\|full | Not-determined |
| 17-108-final-viral-combined_k141_159502\|\|full | Not-determined |
| 17-108-final-viral-combined_k141_145609\|\|full | Low-quality |
| 17-108-final-viral-combined_k141_283716\|\|full | Low-quality |
| 17-108-final-viral-combined_k141_159514\|\|full | Low-quality |
| 17-108-final-viral-combined_k141_296871\|\|full | Not-determined |
| 17-108-final-viral-combined_k141_173590\|\|full | Not-determined |
| 17-108-final-viral-combined_k141_228476\|\|full | Low-quality |
| 17-108-final-viral-combined_k141_117394\|\|full | Not-determined |
| 17-108-final-viral-combined_k141_173594\|\|full | Not-determined |
| 17-108-final-viral-combined_k141_200285\|\|full | Low-quality |
| 17-108-final-viral-combined_k141_35569\|\|full | Low-quality |
| 17-108-final-viral-combined_k141_200288\|\|full | Low-quality |
| 17-108-final-viral-combined_k141_242895\|\|full | Low-quality |
| 17-108-final-viral-combined_k141_256216\|\|full | Low-quality |
| 17-108-final-viral-combined_k141_228485\|\|full | Not-determined |
| 17-108-final-viral-combined_k141_145644\|\|full | Not-determined |
| 17-108-final-viral-combined_k141_186579\|\|full | Low-quality |
| 17-108-final-viral-combined_k141_76075\|\|full | Not-determined |
| 17-108-final-viral-combined_k141_200302\|\|full | Low-quality |
| 17-108-final-viral-combined_k141_296893\|\|full | Not-determined |
| 17-108-final-viral-combined_k141_228486\|\|full | Low-quality |
| 17-108-final-viral-combined_k141_49122\|\|full | Not-determined |
| 17-108-final-viral-combined_k141_186606\|\|full | Low-quality |
| 17-108-final-viral-combined_k141_173605\|\|full | Not-determined |
| 17-108-final-viral-combined_k141_63289\|\|full | Not-determined |
| 17-108-final-viral-combined_k141_7892\|\|full | Low-quality |
| 17-108-final-viral-combined_k141_49134\|\|full | Low-quality |
| 17-108-final-viral-combined_k141_323889\|\|full | Low-quality |
| 17-108-final-viral-combined_k141_311513\|\|full | Low-quality |
| 17-108-final-viral-combined_k141_145671\|\|full | Low-quality |
| 17-108-final-viral-combined_k141_7913\|\|full | Not-determined |
| 17-108-final-viral-combined_k141_35621\|\|full | Low-quality |
| 17-108-final-viral-combined_k141_323901\|\|full | Not-determined |
| 17-108-final-viral-combined_k141_117418\|\|full | Low-quality |
| 17-108-final-viral-combined_k141_311542\|\|full | Low-quality |
| 17-108-final-viral-combined_k141_159576\|\|full | Not-determined |
| 17-108-final-viral-combined_k141_159578\|\|full | Not-determined |
| 17-108-final-viral-combined_k141_311546\|\|full | Not-determined |
| 17-108-final-viral-combined_k141_228557\|\|full | Low-quality |
| 17-108-final-viral-combined_k141_117439\|\|full | Low-quality |
| 17-108-final-viral-combined_k141_7949\|\|full | Low-quality |
| 17-108-final-viral-combined_k141_90574\|\|full | Not-determined |
| 17-108-final-viral-combined_k141_228564\|\|full | Not-determined |
| 17-108-final-viral-combined_k141_200398\|\|full | Low-quality |
| 17-108-final-viral-combined_k141_7955\|\|full | Not-determined |
| 17-108-final-viral-combined_k141_159604\|\|full | Not-determined |
| 17-108-final-viral-combined_k141_323943\|\|full | Low-quality |
| 17-108-final-viral-combined_k141_296954\|\|full | Not-determined |
| 17-108-final-viral-combined_k141_269656\|\|full | Not-determined |
| 17-108-final-viral-combined_k141_256240\|\|full | Low-quality |
| 17-108-final-viral-combined_k141_242964\|\|full | Not-determined |
| 17-108-final-viral-combined_k141_242970\|\|full | Low-quality |
| 17-108-final-viral-combined_k141_20769\|\|full | Low-quality |
| 17-108-final-viral-combined_k141_7989\|\|full | Not-determined |
| 17-108-final-viral-combined_k141_117501\|\|full | Low-quality |
| 17-108-final-viral-combined_k141_173658\|\|full | Low-quality |
| 17-108-final-viral-combined_k141_173661\|\|full | Low-quality |
| 17-108-final-viral-combined_k141_269679\|\|full | Not-determined |
| 17-108-final-viral-combined_k141_269680\|\|full | Not-determined |
| 17-108-final-viral-combined_k141_90610\|\|full | Not-determined |
| 17-108-final-viral-combined_k141_35677\|\|full | Low-quality |
| 17-108-final-viral-combined_k141_131398\|\|full | Not-determined |
| 17-108-final-viral-combined_k141_131399\|\|full | Not-determined |
| 17-108-final-viral-combined_k141_323955\|\|full | Low-quality |
| 17-108-final-viral-combined_k141_63359\|\|full | Not-determined |
| 17-108-final-viral-combined_k141_49198\|\|full | Not-determined |
| 17-108-final-viral-combined_k141_117535\|\|full | Not-determined |
| 17-108-final-viral-combined_k141_242986\|\|full | Not-determined |
| 17-108-final-viral-combined_k141_269687\|\|full | Low-quality |
| 17-108-final-viral-combined_k141_49206\|\|full | Not-determined |
| 17-108-final-viral-combined_k141_131414\|\|full | Not-determined |
| 17-108-final-viral-combined_k141_20818\|\|full | Not-determined |
| 17-108-final-viral-combined_k141_20828\|\|full | Low-quality |
| 17-108-final-viral-combined_k141_90645\|\|full | Low-quality |
| 17-108-final-viral-combined_k141_186687\|\|full | Low-quality |
| 17-108-final-viral-combined_k141_269704\|\|full | Low-quality |
| 17-108-final-viral-combined_k141_76187\|\|full | Low-quality |
| 17-108-final-viral-combined_k141_117573\|\|full | Low-quality |
| 17-108-final-viral-combined_k141_186698\|\|full | Low-quality |
| 17-108-final-viral-combined_k141_296992\|\|full | Not-determined |
| 17-108-final-viral-combined_k141_323976\|\|full | Low-quality |
| 17-108-final-viral-combined_k141_228673\|\|full | Low-quality |
| 17-108-final-viral-combined_k141_145751\|\|full | Low-quality |
| 17-108-final-viral-combined_k141_215046\|\|full | Not-determined |
| 17-108-final-viral-combined_k141_145757\|\|full | Not-determined |
| 17-108-final-viral-combined_k141_117611\|\|full | Low-quality |
| 17-108-final-viral-combined_k141_131470\|\|full | Not-determined |
| 17-108-final-viral-combined_k141_200511\|\|full | Not-determined |
| 17-108-final-viral-combined_k141_228706\|\|full | Not-determined |
| 17-108-final-viral-combined_k141_117619\|\|full | Low-quality |
| 17-108-final-viral-combined_k141_186710\|\|full | Low-quality |
| 17-108-final-viral-combined_k141_243052\|\|full | Not-determined |
| 17-108-final-viral-combined_k141_63394\|\|full | Low-quality |
| 17-108-final-viral-combined_k141_269759\|\|full | Low-quality |
| 17-108-final-viral-combined_k141_8091\|\|full | Low-quality |
| 17-108-final-viral-combined_k141_215083\|\|full | Not-determined |
| 17-108-final-viral-combined_k141_8117\|\|full | Not-determined |
| 17-108-final-viral-combined_k141_186739\|\|full | Not-determined |
| 17-108-final-viral-combined_k141_117637\|\|full | Not-determined |
| 17-108-final-viral-combined_k141_104971\|\|full | Not-determined |
| 17-108-final-viral-combined_k141_186745\|\|full | Low-quality |
| 17-108-final-viral-combined_k141_90738\|\|full | Not-determined |
| 17-108-final-viral-combined_k141_200553\|\|full | Not-determined |
| 17-108-final-viral-combined_k141_186756\|\|full | Not-determined |
| 17-108-final-viral-combined_k141_90741\|\|full | Not-determined |
| 17-108-final-viral-combined_k141_215112\|\|full | Not-determined |
| 17-108-final-viral-combined_k141_20887\|\|full | Not-determined |
| 17-108-final-viral-combined_k141_186763\|\|full | Low-quality |
| 17-108-final-viral-combined_k141_173719\|\|full | Low-quality |
| 17-108-final-viral-combined_k141_283895\|\|full | Low-quality |
| 17-108-final-viral-combined_k141_283896\|\|full | Not-determined |
| 17-108-final-viral-combined_k141_117647\|\|full | Low-quality |
| 17-108-final-viral-combined_k141_76291\|\|full | Low-quality |
| 17-108-final-viral-combined_k141_200572\|\|full | Low-quality |
| 17-108-final-viral-combined_k141_173731\|\|full | Low-quality |
| 17-108-final-viral-combined_k141_131534\|\|full | Not-determined |
| 17-108-final-viral-combined_k141_20918\|\|full | Not-determined |
| 17-108-final-viral-combined_k141_105004\|\|full | Not-determined |
| 17-108-final-viral-combined_k141_228743\|\|full | Low-quality |
| 17-108-final-viral-combined_k141_283917\|\|full | Low-quality |
| 17-108-final-viral-combined_k141_90765\|\|full | Not-determined |
| 17-108-final-viral-combined_k141_200580\|\|full | Not-determined |
| 17-108-final-viral-combined_k141_228746\|\|full | Not-determined |
| 17-108-final-viral-combined_k141_200582\|\|full | Not-determined |
| 17-108-final-viral-combined_k141_173759\|\|full | Low-quality |
| 17-108-final-viral-combined_k141_105008\|\|full | Low-quality |
| 17-108-final-viral-combined_k141_8177\|\|full | Not-determined |
| 17-108-final-viral-combined_k141_131561\|\|full | Not-determined |
| 17-108-final-viral-combined_k141_76332\|\|full | Low-quality |
| 17-108-final-viral-combined_k141_228753\|\|full | Low-quality |
| 17-108-final-viral-combined_k141_173766\|\|full | Low-quality |
| 17-108-final-viral-combined_k141_145831\|\|full | Not-determined |
| 17-108-final-viral-combined_k141_228759\|\|full | Low-quality |
| 17-108-final-viral-combined_k141_324032\|\|full | Low-quality |
| 17-108-final-viral-combined_k141_311776\|\|full | Not-determined |
| 17-108-final-viral-combined_k141_49290\|\|full | Low-quality |
| 17-108-final-viral-combined_k141_145846\|\|full | Low-quality |
| 17-108-final-viral-combined_k141_76364\|\|full | Low-quality |
| 17-108-final-viral-combined_k141_35811\|\|full | Low-quality |
| 17-108-final-viral-combined_k141_76369\|\|full | Not-determined |
| 17-108-final-viral-combined_k141_215154\|\|full | Not-determined |
| 17-108-final-viral-combined_k141_269897\|\|full | Low-quality |
| 17-108-final-viral-combined_k141_145880\|\|full | Not-determined |
| 17-108-final-viral-combined_k141_311801\|\|full | Not-determined |
| 17-108-final-viral-combined_k141_186860\|\|full | Not-determined |
| 17-108-final-viral-combined_k141_76403\|\|full | Low-quality |
| 17-108-final-viral-combined_k141_159741\|\|full | Not-determined |
| 17-108-final-viral-combined_k141_35854\|\|full | Not-determined |
| 17-108-final-viral-combined_k141_311803\|\|full | Low-quality |
| 17-108-final-viral-combined_k141_35869\|\|full | Low-quality |
| 17-108-final-viral-combined_k141_76407\|\|full | Not-determined |
| 17-108-final-viral-combined_k141_105072\|\|full | Low-quality |
| 17-108-final-viral-combined_k141_311830\|\|full | Low-quality |
| 17-108-final-viral-combined_k141_186866\|\|full | Low-quality |
| 17-108-final-viral-combined_k141_311831\|\|full | Low-quality |
| 17-108-final-viral-combined_k141_76408\|\|full | Not-determined |
| 17-108-final-viral-combined_k141_269903\|\|full | Not-determined |
| 17-108-final-viral-combined_k141_90818\|\|full | Low-quality |
| 17-108-final-viral-combined_k141_324075\|\|full | Low-quality |
| 17-108-final-viral-combined_k141_228781\|\|full | Low-quality |
| 17-108-final-viral-combined_k141_117746\|\|full | Low-quality |
| 17-108-final-viral-combined_k141_200627\|\|full | Low-quality |
| 17-108-final-viral-combined_k141_243159\|\|full | Low-quality |
| 17-108-final-viral-combined_k141_76437\|\|full | Not-determined |
| 17-108-final-viral-combined_k141_324092\|\|full | Low-quality |
| 17-108-final-viral-combined_k141_105085\|\|full | Low-quality |
| 17-108-final-viral-combined_k141_186907\|\|full | Not-determined |
| 17-109-final-viral-combined_k141_98046\|\|full | Medium-quality |
| 17-108-final-viral-combined_k141_117749\|\|full | Not-determined |
| 17-108-final-viral-combined_k141_311881\|\|full | Low-quality |
| 17-108-final-viral-combined_k141_145957\|\|full | Not-determined |
| 17-108-final-viral-combined_k141_311888\|\|full | Not-determined |
| 17-108-final-viral-combined_k141_324115\|\|full | Low-quality |
| 17-108-final-viral-combined_k141_76466\|\|full | Not-determined |
| 17-108-final-viral-combined_k141_284068\|\|full | Low-quality |
| 17-108-final-viral-combined_k141_311898\|\|full | Not-determined |
| 17-108-final-viral-combined_k141_243184\|\|full | Low-quality |
| 17-108-final-viral-combined_k141_269955\|\|full | Not-determined |
| 17-108-final-viral-combined_k141_256491\|\|full | Low-quality |
| 17-108-final-viral-combined_k141_76479\|\|full | Not-determined |
| 17-108-final-viral-combined_k141_200670\|\|full | Low-quality |
| 17-108-final-viral-combined_k141_35947\|\|full | Not-determined |
| 17-108-final-viral-combined_k141_186929\|\|full | Low-quality |
| 17-108-final-viral-combined_k141_269961\|\|full | Not-determined |
| 17-108-final-viral-combined_k141_269969\|\|full | Low-quality |
| 17-108-final-viral-combined_k141_297168\|\|full | Low-quality |
| 17-108-final-viral-combined_k141_243218\|\|full | Low-quality |
| 17-108-final-viral-combined_k141_146002\|\|full | Low-quality |
| 17-108-final-viral-combined_k141_21024\|\|full | Low-quality |
| 17-109_k141_401372 | Complete |
| 17-108-final-viral-combined_k141_105111\|\|full | Not-determined |
| 17-108-final-viral-combined_k141_8356\|\|full | Low-quality |
| 17-108-final-viral-combined_k141_200693\|\|full | Low-quality |
| 17-108-final-viral-combined_k141_131697\|\|full | Low-quality |
| 17-108-final-viral-combined_k141_256513\|\|full | Not-determined |
| 17-108-final-viral-combined_k141_243232\|\|full | Low-quality |
| 17-108-final-viral-combined_k141_269994\|\|full | Not-determined |
| 17-108-final-viral-combined_k141_256517\|\|full | Low-quality |
| 17-108-final-viral-combined_k141_76510\|\|full | Low-quality |
| 17-108-final-viral-combined_k141_90905\|\|full | Not-determined |
| 17-108-final-viral-combined_k141_159829\|\|full | Not-determined |
| 17-108-final-viral-combined_k141_8377\|\|full | Low-quality |
| 17-108-final-viral-combined_k141_146039\|\|full | Not-determined |
| 17-108-final-viral-combined_k141_63554\|\|full | Low-quality |
| 17-108-final-viral-combined_k141_284113\|\|full | Not-determined |
| 17-108-final-viral-combined_k141_146044\|\|full | Not-determined |
| 17-108-final-viral-combined_k141_324180\|\|full | Not-determined |
| 17-108-final-viral-combined_k141_270022\|\|full | Low-quality |
| 17-108-final-viral-combined_k141_146057\|\|full | Low-quality |
| 17-108-final-viral-combined_k141_8396\|\|full | Not-determined |
| 17-108-final-viral-combined_k141_21073\|\|full | Low-quality |
| 17-108-final-viral-combined_k141_284142\|\|full | Not-determined |
| 17-108-final-viral-combined_k141_49444\|\|full | Low-quality |
| 17-108-final-viral-combined_k141_90925\|\|full | Not-determined |
| 17-108-final-viral-combined_k141_146066\|\|full | Not-determined |
| 17-108-final-viral-combined_k141_324181\|\|full | Low-quality |
| 17-108-final-viral-combined_k141_49449\|\|full | Low-quality |
| 17-108-final-viral-combined_k141_284157\|\|full | Not-determined |
| 17-108-final-viral-combined_k141_159862\|\|full | Not-determined |
| 17-108-final-viral-combined_k141_8415\|\|full | Not-determined |
| 17-108-final-viral-combined_k141_117825\|\|full | Not-determined |
| 17-108-final-viral-combined_k141_146085\|\|full | Low-quality |
| 17-108-final-viral-combined_k141_146086\|\|full | Not-determined |
| 17-108-final-viral-combined_k141_200768\|\|full | Low-quality |
| 17-108-final-viral-combined_k141_256556\|\|full | Not-determined |
| 17-108-final-viral-combined_k141_159881\|\|full | Low-quality |
| 17-108-final-viral-combined_k141_228835\|\|full | Not-determined |
| 17-108-final-viral-combined_k141_256566\|\|full | Low-quality |
| 17-108-final-viral-combined_k141_297261\|\|full | Not-determined |
| 17-108-final-viral-combined_k141_187000\|\|full | Low-quality |
| 17-108-final-viral-combined_k141_297262\|\|full | Low-quality |
| 17-108-final-viral-combined_k141_90974\|\|full | Not-determined |
| 17-108-final-viral-combined_k141_146117\|\|full | Low-quality |
| 17-108-final-viral-combined_k141_76580\|\|full | Low-quality |
| 17-108-final-viral-combined_k141_117861\|\|full | Low-quality |
| 17-108-final-viral-combined_k141_105243\|\|full | Low-quality |
| 17-108-final-viral-combined_k141_256605\|\|full | Not-determined |
| 17-108-final-viral-combined_k141_8478\|\|full | Not-determined |
| 17-108-final-viral-combined_k141_63652\|\|full | Not-determined |
| 17-108-final-viral-combined_k141_200776\|\|full | Not-determined |
| 17-108-final-viral-combined_k141_324193\|\|full | Low-quality |
| 17-108-final-viral-combined_k141_324201\|\|full | Not-determined |
| 17-108-final-viral-combined_k141_297297\|\|full | Low-quality |
| 17-108-final-viral-combined_k141_215336\|\|full | Low-quality |
| 17-108-final-viral-combined_k141_159936\|\|full | Not-determined |
| 17-108-final-viral-combined_k141_187046\|\|full | Not-determined |
| 17-108-final-viral-combined_k141_284224\|\|full | Not-determined |
| 17-108-final-viral-combined_k141_200797\|\|full | Not-determined |
| 17-108-final-viral-combined_k141_159952\|\|full | Not-determined |
| 17-108-final-viral-combined_k141_311910\|\|full | Not-determined |
| 17-108-final-viral-combined_k141_187058\|\|full | Low-quality |
| 17-108-final-viral-combined_k141_8513\|\|full | Low-quality |
| 17-108-final-viral-combined_k141_36015\|\|full | Low-quality |
| 17-108-final-viral-combined_k141_187068\|\|full | Low-quality |
| 17-108-final-viral-combined_k141_215378\|\|full | Low-quality |
| 17-108-final-viral-combined_k141_324239\|\|full | Not-determined |
| 17-108-final-viral-combined_k141_173989\|\|full | Low-quality |
| 17-108-final-viral-combined_k141_36030\|\|full | Low-quality |
| 17-108-final-viral-combined_k141_297347\|\|full | Low-quality |
| 17-108-final-viral-combined_k141_21145\|\|full | Not-determined |
| 17-108-final-viral-combined_k141_215388\|\|full | Not-determined |
| 17-108-final-viral-combined_k141_324248\|\|full | Low-quality |
| 17-108-final-viral-combined_k141_8530\|\|full | Not-determined |
| 17-108-final-viral-combined_k141_187110\|\|full | Not-determined |
| 17-108-final-viral-combined_k141_117912\|\|full | Low-quality |
| 17-108-final-viral-combined_k141_63698\|\|full | Not-determined |
| 17-108-final-viral-combined_k141_105307\|\|full | Not-determined |
| 17-108-final-viral-combined_k141_228976\|\|full | Low-quality |
| 17-108-final-viral-combined_k141_49548\|\|full | Low-quality |
| 17-108-final-viral-combined_k141_146151\|\|full | Low-quality |
| 17-108-final-viral-combined_k141_76660\|\|full | Not-determined |
| 17-108-final-viral-combined_k141_160007\|\|full | Low-quality |
| 17-108-final-viral-combined_k141_174021\|\|full | Not-determined |
| 17-108-final-viral-combined_k141_146153\|\|full | Low-quality |
| 17-108-final-viral-combined_k141_146157\|\|full | Low-quality |
| 17-108-final-viral-combined_k141_63724\|\|full | Low-quality |
| 17-108-final-viral-combined_k141_131795\|\|full | Low-quality |
| 17-108-final-viral-combined_k141_311954\|\|full | Not-determined |
| 17-108-final-viral-combined_k141_8585\|\|full | Low-quality |
| 17-108-final-viral-combined_k141_117959\|\|full | Not-determined |
| 17-108-final-viral-combined_k141_36082\|\|full | Low-quality |
| 17-108-final-viral-combined_k141_160022\|\|full | Not-determined |
| 17-108-final-viral-combined_k141_49562\|\|full | Low-quality |
| 17-108-final-viral-combined_k141_311965\|\|full | Not-determined |
| 17-108-final-viral-combined_k141_36093\|\|full | Not-determined |
| 17-108-final-viral-combined_k141_160032\|\|full | Not-determined |
| 17-108-final-viral-combined_k141_243372\|\|full | Not-determined |
| 17-108-final-viral-combined_k141_8606\|\|full | Not-determined |
| 17-108-final-viral-combined_k141_229003\|\|full | Not-determined |
| 17-108-final-viral-combined_k141_105349\|\|full | Not-determined |
| 17-108-final-viral-combined_k141_243397\|\|full | Not-determined |
| 17-108-final-viral-combined_k141_284340\|\|full | Low-quality |
| 17-108-final-viral-combined_k141_36117\|\|full | Low-quality |
| 17-108-final-viral-combined_k141_187163\|\|full | Low-quality |
| 17-108-final-viral-combined_k141_76714\|\|full | Low-quality |
| 17-108-final-viral-combined_k141_174057\|\|full | Low-quality |
| 17-108-final-viral-combined_k141_146178\|\|full | Low-quality |
| 17-108-final-viral-combined_k141_160049\|\|full | Low-quality |
| 17-108-final-viral-combined_k141_131836\|\|full | Low-quality |
| 17-108-final-viral-combined_k141_36139\|\|full | Low-quality |
| 17-108-final-viral-combined_k141_21270\|\|full | Low-quality |
| 17-108-final-viral-combined_k141_146191\|\|full | Low-quality |
| 17-108-final-viral-combined_k141_284360\|\|full | Not-determined |
| 17-108-final-viral-combined_k141_200895\|\|full | Not-determined |
| 17-108-final-viral-combined_k141_8639\|\|full | Not-determined |
| 17-108-final-viral-combined_k141_174067\|\|full | Low-quality |
| 17-108-final-viral-combined_k141_270096\|\|full | Low-quality |
| 17-108-final-viral-combined_k141_131843\|\|full | Low-quality |
| 17-108-final-viral-combined_k141_229040\|\|full | Low-quality |
| 17-108-final-viral-combined_k141_146208\|\|full | Low-quality |
| 17-108-final-viral-combined_k141_131845\|\|full | Not-determined |
| 17-109-final-viral-combined_k141_401372\|\|full | High-quality |
| 17-108-final-viral-combined_k141_297437\|\|full | Not-determined |
| 17-108-final-viral-combined_k141_270097\|\|full | Low-quality |
| 17-108-final-viral-combined_k141_131846\|\|full | Not-determined |
| 17-108-final-viral-combined_k141_324336\|\|full | Not-determined |
| 17-108-final-viral-combined_k141_312020\|\|full | Low-quality |
| 17-108-final-viral-combined_k141_36175\|\|full | Not-determined |
| 17-108-final-viral-combined_k141_229055\|\|full | Not-determined |
| 17-108-final-viral-combined_k141_174092\|\|full | Not-determined |
| 17-108-final-viral-combined_k141_21287\|\|full | Not-determined |
| 17-108-final-viral-combined_k141_229059\|\|full | Low-quality |
| 17-108-final-viral-combined_k141_243426\|\|full | Not-determined |
| 17-108-final-viral-combined_k141_174097\|\|full | Not-determined |
| 17-108-final-viral-combined_k141_63770\|\|full | Not-determined |
| 17-108-final-viral-combined_k141_105378\|\|full | Low-quality |
| 17-108-final-viral-combined_k141_312042\|\|full | Low-quality |
| 17-108-final-viral-combined_k141_76736\|\|full | Low-quality |
| 17-108-final-viral-combined_k141_200950\|\|full | Not-determined |
| 17-108-final-viral-combined_k141_21302\|\|full | Not-determined |
| 17-108-final-viral-combined_k141_76743\|\|full | Low-quality |
| 17-108-final-viral-combined_k141_131865\|\|full | Not-determined |
| 17-108-final-viral-combined_k141_200962\|\|full | Not-determined |
| 17-108-final-viral-combined_k141_76759\|\|full | Not-determined |
| 17-108-final-viral-combined_k141_8718\|\|full | Low-quality |
| 17-108-final-viral-combined_k141_160146\|\|full | Not-determined |
| 17-108-final-viral-combined_k141_49703\|\|full | Not-determined |
| 17-108-final-viral-combined_k141_91102\|\|full | Not-determined |
| 17-108-final-viral-combined_k141_297456\|\|full | Not-determined |
| 17-108-final-viral-combined_k141_118083\|\|full | Low-quality |
| 17-108-final-viral-combined_k141_36233\|\|full | Not-determined |
| 17-108-final-viral-combined_k141_243481\|\|full | Low-quality |
| 17-108-final-viral-combined_k141_324364\|\|full | Not-determined |
| 17-108-final-viral-combined_k141_215508\|\|full | Low-quality |
| 17-108-final-viral-combined_k141_284436\|\|full | Not-determined |
| 17-108-final-viral-combined_k141_76789\|\|full | Not-determined |
| 17-108-final-viral-combined_k141_270129\|\|full | Low-quality |
| 17-108-final-viral-combined_k141_270130\|\|full | Low-quality |
| 17-108-final-viral-combined_k141_324371\|\|full | Low-quality |
| 17-108-final-viral-combined_k141_297475\|\|full | Low-quality |
| 17-108-final-viral-combined_k141_201025\|\|full | Not-determined |
| 17-108-final-viral-combined_k141_91146\|\|full | Not-determined |
| 17-108-final-viral-combined_k141_91150\|\|full | Not-determined |
| 17-108-final-viral-combined_k141_118107\|\|full | Not-determined |
| 17-108-final-viral-combined_k141_91159\|\|full | Low-quality |
| 17-108-final-viral-combined_k141_118119\|\|full | Not-determined |
| 17-108-final-viral-combined_k141_324393\|\|full | Not-determined |
| 17-108-final-viral-combined_k141_284491\|\|full | Not-determined |
| 17-108-final-viral-combined_k141_215549\|\|full | Low-quality |
| 17-108-final-viral-combined_k141_160226\|\|full | Low-quality |
| 17-108-final-viral-combined_k141_229184\|\|full | Not-determined |
| 17-108-final-viral-combined_k141_187263\|\|full | Not-determined |
| 17-108-final-viral-combined_k141_256928\|\|full | Low-quality |
| 17-108-final-viral-combined_k141_174223\|\|full | Not-determined |
| 17-108-final-viral-combined_k141_256929\|\|full | Low-quality |
| 17-108-final-viral-combined_k141_118142\|\|full | Not-determined |
| 17-108-final-viral-combined_k141_76869\|\|full | Low-quality |
| 17-108-final-viral-combined_k141_21382\|\|full | Not-determined |
| 17-108-final-viral-combined_k141_215562\|\|full | Low-quality |
| 17-108-final-viral-combined_k141_284514\|\|full | Low-quality |
| 17-108-final-viral-combined_k141_36324\|\|full | Low-quality |
| 17-108-final-viral-combined_k141_105531\|\|full | Not-determined |
| 17-108-final-viral-combined_k141_49839\|\|full | Low-quality |
| 17-108-final-viral-combined_k141_8815\|\|full | Low-quality |
| 17-108-final-viral-combined_k141_174246\|\|full | Low-quality |
| 17-108-final-viral-combined_k141_215584\|\|full | Not-determined |
| 17-108-final-viral-combined_k141_243513\|\|full | Low-quality |
| 17-108-final-viral-combined_k141_76887\|\|full | Not-determined |
| 17-108-final-viral-combined_k141_297500\|\|full | Low-quality |
| 17-108-final-viral-combined_k141_105546\|\|full | Not-determined |
| 17-108-final-viral-combined_k141_229238\|\|full | Low-quality |
| 17-108-final-viral-combined_k141_174266\|\|full | Low-quality |
| 17-108-final-viral-combined_k141_8841\|\|full | Low-quality |
| 17-108-final-viral-combined_k141_256976\|\|full | Low-quality |
| 17-108-final-viral-combined_k141_174277\|\|full | Low-quality |
| 17-108-final-viral-combined_k141_146408\|\|full | Low-quality |
| 17-108-final-viral-combined_k141_91228\|\|full | Low-quality |
| 17-108-final-viral-combined_k141_76897\|\|full | Not-determined |
| 17-108-final-viral-combined_k141_105554\|\|full | Low-quality |
| 18-104-final-viral-combined_k141_41490\|\|full | Medium-quality |
| 17-108-final-viral-combined_k141_105558\|\|full | Low-quality |
| 17-108-final-viral-combined_k141_105559\|\|full | Not-determined |
| 17-108-final-viral-combined_k141_243544\|\|full | Low-quality |
| 17-108-final-viral-combined_k141_174285\|\|full | Low-quality |
| 17-108-final-viral-combined_k141_63822\|\|full | Low-quality |
| 17-108-final-viral-combined_k141_201090\|\|full | Not-determined |
| 17-108-final-viral-combined_k141_21419\|\|full | Low-quality |
| 17-108-final-viral-combined_k141_243555\|\|full | Low-quality |
| 17-108-final-viral-combined_k141_63829\|\|full | Low-quality |
| 17-108-final-viral-combined_k141_76912\|\|full | Low-quality |
| 17-108-final-viral-combined_k141_215631\|\|full | Low-quality |
| 17-108-final-viral-combined_k141_256995\|\|full | Low-quality |
| 17-108-final-viral-combined_k141_105573\|\|full | Low-quality |
| 17-108-final-viral-combined_k141_312160\|\|full | Low-quality |
| 17-108-final-viral-combined_k141_118185\|\|full | Not-determined |
| 17-108-final-viral-combined_k141_160311\|\|full | Not-determined |
| 17-108-final-viral-combined_k141_243586\|\|full | Low-quality |
| 17-108-final-viral-combined_k141_118190\|\|full | Low-quality |
| 17-108-final-viral-combined_k141_229280\|\|full | Low-quality |
| 17-108-final-viral-combined_k141_63854\|\|full | Not-determined |
| 17-108-final-viral-combined_k141_243591\|\|full | Not-determined |
| 17-108-final-viral-combined_k141_270194\|\|full | Not-determined |
| 17-108-final-viral-combined_k141_105597\|\|full | Low-quality |
| 17-108-final-viral-combined_k141_243594\|\|full | Not-determined |
| 17-108-final-viral-combined_k141_146463\|\|full | Not-determined |
| 17-108-final-viral-combined_k141_229283\|\|full | Low-quality |
| 17-108-final-viral-combined_k141_297544\|\|full | Not-determined |
| 17-108-final-viral-combined_k141_76936\|\|full | Low-quality |
| 17-108-final-viral-combined_k141_49926\|\|full | Low-quality |
| 17-108-final-viral-combined_k141_257023\|\|full | Not-determined |
| 17-108-final-viral-combined_k141_49928\|\|full | Not-determined |
| 17-108-final-viral-combined_k141_146470\|\|full | Not-determined |
| 17-108-final-viral-combined_k141_215662\|\|full | Low-quality |
| 17-108-final-viral-combined_k141_160329\|\|full | Low-quality |
| 17-108-final-viral-combined_k141_8931\|\|full | Not-determined |
| 17-108-final-viral-combined_k141_49937\|\|full | Low-quality |
| 17-108-final-viral-combined_k141_243624\|\|full | Low-quality |
| 17-108-final-viral-combined_k141_324496\|\|full | Low-quality |
| 17-108-final-viral-combined_k141_8942\|\|full | Not-determined |
| 17-108-final-viral-combined_k141_146478\|\|full | Low-quality |
| 17-108-final-viral-combined_k141_118235\|\|full | Low-quality |
| 17-108-final-viral-combined_k141_63891\|\|full | Low-quality |
| 17-108-final-viral-combined_k141_243649\|\|full | Not-determined |
| 17-108-final-viral-combined_k141_215700\|\|full | Low-quality |
| 17-108-final-viral-combined_k141_160341\|\|full | Low-quality |
| 17-108-final-viral-combined_k141_146494\|\|full | Low-quality |
| 17-108-final-viral-combined_k141_187345\|\|full | Low-quality |
| 17-108-final-viral-combined_k141_312215\|\|full | Not-determined |
| 17-108-final-viral-combined_k141_160367\|\|full | Low-quality |
| 17-108-final-viral-combined_k141_187358\|\|full | Low-quality |
| 17-108-final-viral-combined_k141_49973\|\|full | Not-determined |
| 17-108-final-viral-combined_k141_91314\|\|full | Low-quality |
| 17-108-final-viral-combined_k141_257061\|\|full | Low-quality |
| 17-108-final-viral-combined_k141_91317\|\|full | Not-determined |
| 17-108-final-viral-combined_k141_257065\|\|full | Low-quality |
| 17-108-final-viral-combined_k141_9009\|\|full | Low-quality |
| 17-108-final-viral-combined_k141_187372\|\|full | Not-determined |
| 17-108-final-viral-combined_k141_284550\|\|full | Low-quality |
| 17-108-final-viral-combined_k141_187383\|\|full | Low-quality |
| 17-108-final-viral-combined_k141_284552\|\|full | Not-determined |
| 17-108-final-viral-combined_k141_174383\|\|full | Not-determined |
| 17-108-final-viral-combined_k141_63951\|\|full | Low-quality |
| 17-108-final-viral-combined_k141_63956\|\|full | Low-quality |
| 17-108-final-viral-combined_k141_9043\|\|full | Not-determined |
| 17-108-final-viral-combined_k141_77004\|\|full | Not-determined |
| 17-108-final-viral-combined_k141_312279\|\|full | Low-quality |
| 17-108-final-viral-combined_k141_312286\|\|full | Not-determined |
| 17-108-final-viral-combined_k141_105710\|\|full | Not-determined |
| 17-108-final-viral-combined_k141_9046\|\|full | Not-determined |
| 17-108-final-viral-combined_k141_50045\|\|full | Low-quality |
| 17-108-final-viral-combined_k141_36434\|\|full | Not-determined |
| 18-106-final-viral-combined_k141_8508\|\|full | Medium-quality |
| 17-108-final-viral-combined_k141_297660\|\|full | Not-determined |
| 17-108-final-viral-combined_k141_297662\|\|full | Low-quality |
| 17-108-final-viral-combined_k141_324584\|\|full | Low-quality |
| 17-108-final-viral-combined_k141_284599\|\|full | Not-determined |
| 17-108-final-viral-combined_k141_9081\|\|full | Low-quality |
| 17-108-final-viral-combined_k141_91352\|\|full | Low-quality |
| 17-108-final-viral-combined_k141_284601\|\|full | Low-quality |
| 17-108-final-viral-combined_k141_270331\|\|full | Not-determined |
| 17-108-final-viral-combined_k141_50081\|\|full | Low-quality |
| 17-108-final-viral-combined_k141_160426\|\|full | Not-determined |
| 17-108-final-viral-combined_k141_64040\|\|full | Not-determined |
| 17-108-final-viral-combined_k141_36437\|\|full | Low-quality |
| 17-108-final-viral-combined_k141_324622\|\|full | Low-quality |
| 17-108-final-viral-combined_k141_105758\|\|full | Not-determined |
| 17-108-final-viral-combined_k141_36438\|\|full | Not-determined |
| 17-108-final-viral-combined_k141_146527\|\|full | Not-determined |
| 17-108-final-viral-combined_k141_257093\|\|full | Low-quality |
| 17-108-final-viral-combined_k141_36447\|\|full | Low-quality |
| 17-108-final-viral-combined_k141_270355\|\|full | Not-determined |
| 17-108-final-viral-combined_k141_105769\|\|full | Not-determined |
| 17-108-final-viral-combined_k141_297698\|\|full | Not-determined |
| 17-108-final-viral-combined_k141_50117\|\|full | Not-determined |
| 17-108-final-viral-combined_k141_270371\|\|full | Not-determined |
| 17-108-final-viral-combined_k141_146538\|\|full | Low-quality |
| 17-108-final-viral-combined_k141_229431\|\|full | Low-quality |
| 17-108-final-viral-combined_k141_118346\|\|full | Low-quality |
| 17-108-final-viral-combined_k141_9128\|\|full | Not-determined |
| 17-108-final-viral-combined_k141_160470\|\|full | Not-determined |
| 17-108-final-viral-combined_k141_284655\|\|full | Low-quality |
| 17-108-final-viral-combined_k141_215781\|\|full | Not-determined |
| 17-108-final-viral-combined_k141_201355\|\|full | Not-determined |
| 17-108-final-viral-combined_k141_284666\|\|full | Not-determined |
| 17-108-final-viral-combined_k141_105795\|\|full | Low-quality |
| 17-108-final-viral-combined_k141_187496\|\|full | Low-quality |
| 17-108-final-viral-combined_k141_50186\|\|full | Not-determined |
| 17-108-final-viral-combined_k141_146564\|\|full | Not-determined |
| 17-108-final-viral-combined_k141_215820\|\|full | Low-quality |
| 17-108-final-viral-combined_k141_77086\|\|full | Low-quality |
| 17-108-final-viral-combined_k141_174486\|\|full | Low-quality |
| 17-108-final-viral-combined_k141_201386\|\|full | Low-quality |
| 17-108-final-viral-combined_k141_160511\|\|full | Low-quality |
| 17-108-final-viral-combined_k141_312423\|\|full | Low-quality |
| 17-108-final-viral-combined_k141_9190\|\|full | Low-quality |
| 17-108-final-viral-combined_k141_21603\|\|full | Low-quality |
| 17-108-final-viral-combined_k141_284700\|\|full | Low-quality |
| 17-108-final-viral-combined_k141_50209\|\|full | Not-determined |
| 17-108-final-viral-combined_k141_257180\|\|full | Not-determined |
| 17-108-final-viral-combined_k141_105807\|\|full | Low-quality |
| 17-108-final-viral-combined_k141_118403\|\|full | Not-determined |
| 17-108-final-viral-combined_k141_284716\|\|full | Low-quality |
| 17-108-final-viral-combined_k141_215863\|\|full | Low-quality |
| 17-108-final-viral-combined_k141_324658\|\|full | Low-quality |
| 17-108-final-viral-combined_k141_270395\|\|full | Low-quality |
| 17-108-final-viral-combined_k141_297775\|\|full | Not-determined |
| 17-108-final-viral-combined_k141_284728\|\|full | Low-quality |
| 17-108-final-viral-combined_k141_174530\|\|full | Low-quality |
| 17-108-final-viral-combined_k141_284731\|\|full | Not-determined |
| 17-108-final-viral-combined_k141_187552\|\|full | Not-determined |
| 17-108-final-viral-combined_k141_201422\|\|full | Low-quality |
| 17-108-final-viral-combined_k141_91495\|\|full | Low-quality |
| 17-108-final-viral-combined_k141_174540\|\|full | Not-determined |
| 17-108-final-viral-combined_k141_257238\|\|full | Not-determined |
| 17-108-final-viral-combined_k141_9258\|\|full | Low-quality |
| 17-108-final-viral-combined_k141_257254\|\|full | Low-quality |
| 17-108-final-viral-combined_k141_270427\|\|full | Not-determined |
| 17-108-final-viral-combined_k141_312468\|\|full | Low-quality |
| 17-108-final-viral-combined_k141_187585\|\|full | Low-quality |
| 17-108-final-viral-combined_k141_270445\|\|full | Low-quality |
| 17-108-final-viral-combined_k141_118442\|\|full | Not-determined |
| 17-108-final-viral-combined_k141_64157\|\|full | Low-quality |
| 17-108-final-viral-combined_k141_229554\|\|full | Low-quality |
| 17-108-final-viral-combined_k141_270484\|\|full | Low-quality |
| 17-108-final-viral-combined_k141_64174\|\|full | Not-determined |
| 17-108-final-viral-combined_k141_50335\|\|full | Not-determined |
| 17-108-final-viral-combined_k141_50336\|\|full | Not-determined |
| 17-108-final-viral-combined_k141_187620\|\|full | Low-quality |
| 17-108-final-viral-combined_k141_284787\|\|full | Low-quality |
| 17-108-final-viral-combined_k141_50348\|\|full | Low-quality |
| 17-108-final-viral-combined_k141_105933\|\|full | Not-determined |
| 17-108-final-viral-combined_k141_160638\|\|full | Low-quality |
| 17-108-final-viral-combined_k141_64190\|\|full | Not-determined |
| 17-108-final-viral-combined_k141_229592\|\|full | Not-determined |
| 17-108-final-viral-combined_k141_270514\|\|full | Not-determined |
| 17-108-final-viral-combined_k141_160646\|\|full | Low-quality |
| 17-108-final-viral-combined_k141_9313\|\|full | Low-quality |
| 17-108-final-viral-combined_k141_174588\|\|full | Not-determined |
| 17-108-final-viral-combined_k141_297865\|\|full | Not-determined |
| 17-108-final-viral-combined_k141_297866\|\|full | Not-determined |
| 17-108-final-viral-combined_k141_284817\|\|full | Not-determined |
| 17-108-final-viral-combined_k141_105940\|\|full | Low-quality |
| 17-108-final-viral-combined_k141_324824\|\|full | Low-quality |
| 17-108-final-viral-combined_k141_160656\|\|full | Low-quality |
| 17-108-final-viral-combined_k141_146660\|\|full | Low-quality |
| 17-108-final-viral-combined_k141_64222\|\|full | Not-determined |
| 17-108-final-viral-combined_k141_297891\|\|full | Not-determined |
| 17-108-final-viral-combined_k141_201510\|\|full | Low-quality |
| 17-108-final-viral-combined_k141_257354\|\|full | Not-determined |
| 17-108-final-viral-combined_k141_312536\|\|full | Low-quality |
| 17-108-final-viral-combined_k141_160694\|\|full | Not-determined |
| 17-108-final-viral-combined_k141_146694\|\|full | Not-determined |
| 17-108-final-viral-combined_k141_105993\|\|full | Not-determined |
| 17-108-final-viral-combined_k141_160703\|\|full | Low-quality |
| 17-108-final-viral-combined_k141_132032\|\|full | Low-quality |
| 17-108-final-viral-combined_k141_118595\|\|full | Not-determined |
| 17-108-final-viral-combined_k141_118596\|\|full | Not-determined |
| 17-108-final-viral-combined_k141_243932\|\|full | Low-quality |
| 17-108-final-viral-combined_k141_284882\|\|full | Low-quality |
| 17-108-final-viral-combined_k141_201545\|\|full | Not-determined |
| 17-108-final-viral-combined_k141_284900\|\|full | Not-determined |
| 17-108-final-viral-combined_k141_270553\|\|full | Not-determined |
| 17-108-final-viral-combined_k141_257374\|\|full | Not-determined |
| 17-108-final-viral-combined_k141_50388\|\|full | Not-determined |
| 17-108-final-viral-combined_k141_118629\|\|full | Not-determined |

| | |
|---|---|
| 17-108-final-viral-combined_k141_284919\|\|full | Not-determined |
| 17-108-final-viral-combined_k141_36590\|\|full | Not-determined |
| 17-108-final-viral-combined_k141_257401\|\|full | Low-quality |
| 17-108-final-viral-combined_k141_132055\|\|full | Not-determined |
| 17-108-final-viral-combined_k141_270585\|\|full | Low-quality |
| 17-108-final-viral-combined_k141_324889\|\|full | Low-quality |
| 17-108-final-viral-combined_k141_284928\|\|full | Low-quality |
| 17-108-final-viral-combined_k141_187733\|\|full | Low-quality |
| 17-108-final-viral-combined_k141_106093\|\|full | Not-determined |
| 17-108-final-viral-combined_k141_284937\|\|full | Not-determined |
| 17-108-final-viral-combined_k141_187736\|\|full | Low-quality |
| 17-108-final-viral-combined_k141_77262\|\|full | Low-quality |
| 17-108-final-viral-combined_k141_284955\|\|full | Not-determined |
| 17-108-final-viral-combined_k141_201606\|\|full | Low-quality |
| 17-108-final-viral-combined_k141_243951\|\|full | Not-determined |
| 17-108-final-viral-combined_k141_91705\|\|full | Low-quality |
| 17-108-final-viral-combined_k141_132123\|\|full | Low-quality |
| 17-108-final-viral-combined_k141_106106\|\|full | Low-quality |
| 17-108-final-viral-combined_k141_77308\|\|full | Not-determined |
| 17-108-final-viral-combined_k141_201636\|\|full | Low-quality |
| 17-108-final-viral-combined_k141_50431\|\|full | Low-quality |
| 17-108-final-viral-combined_k141_215961\|\|full | Low-quality |
| 17-108-final-viral-combined_k141_201640\|\|full | Low-quality |
| 17-108-final-viral-combined_k141_106112\|\|full | Low-quality |
| 17-108-final-viral-combined_k141_91725\|\|full | Low-quality |
| 17-108-final-viral-combined_k141_229768\|\|full | Not-determined |
| 17-108-final-viral-combined_k141_146837\|\|full | Not-determined |
| 17-108-final-viral-combined_k141_257474\|\|full | Low-quality |
| 17-108-final-viral-combined_k141_187827\|\|full | Not-determined |
| 17-108-final-viral-combined_k141_297944\|\|full | Not-determined |
| 17-108-final-viral-combined_k141_229781\|\|full | Low-quality |
| 17-108-final-viral-combined_k141_257486\|\|full | Low-quality |
| 17-108-final-viral-combined_k141_324950\|\|full | Not-determined |
| 17-108-final-viral-combined_k141_132150\|\|full | Not-determined |
| 17-108-final-viral-combined_k141_106152\|\|full | Low-quality |
| 17-108-final-viral-combined_k141_187844\|\|full | Low-quality |
| 17-108-final-viral-combined_k141_229794\|\|full | Low-quality |
| 17-108-final-viral-combined_k141_146851\|\|full | Not-determined |
| 17-108-final-viral-combined_k141_187855\|\|full | Not-determined |
| 17-108-final-viral-combined_k141_201703\|\|full | Low-quality |
| 17-108-final-viral-combined_k141_297962\|\|full | Not-determined |
| 17-108-final-viral-combined_k141_270656\|\|full | Not-determined |
| 17-108-final-viral-combined_k141_257508\|\|full | Not-determined |
| 17-108-final-viral-combined_k141_174717\|\|full | Not-determined |
| 17-108-final-viral-combined_k141_187869\|\|full | Low-quality |
| 17-108-final-viral-combined_k141_270671\|\|full | Not-determined |
| 17-108-final-viral-combined_k141_118722\|\|full | Not-determined |
| 17-108-final-viral-combined_k141_106209\|\|full | Not-determined |
| 17-108-final-viral-combined_k141_270678\|\|full | Low-quality |
| 17-108-final-viral-combined_k141_187877\|\|full | Not-determined |
| 17-108-final-viral-combined_k141_244042\|\|full | Not-determined |
| 17-108-final-viral-combined_k141_270681\|\|full | Not-determined |
| 17-108-final-viral-combined_k141_187891\|\|full | Not-determined |
| 17-108-final-viral-combined_k141_106222\|\|full | Not-determined |
| 17-108-final-viral-combined_k141_325005\|\|full | Low-quality |
| 17-108-final-viral-combined_k141_160832\|\|full | Not-determined |
| 17-108-final-viral-combined_k141_257550\|\|full | Low-quality |
| 17-108-final-viral-combined_k141_91798\|\|full | Not-determined |
| 17-108-final-viral-combined_k141_229852\|\|full | Not-determined |
| 17-108-final-viral-combined_k141_325025\|\|full | Low-quality |
| 17-108-final-viral-combined_k141_146928\|\|full | Not-determined |
| 17-108-final-viral-combined_k141_132216\|\|full | Low-quality |
| 17-108-final-viral-combined_k141_187926\|\|full | Not-determined |
| 17-108-final-viral-combined_k141_215987\|\|full | Not-determined |
| 17-108-final-viral-combined_k141_312698\|\|full | Not-determined |
| 17-108-final-viral-combined_k141_215995\|\|full | Not-determined |
| 17-108-final-viral-combined_k141_312701\|\|full | Low-quality |
| 17-108-final-viral-combined_k141_21794\|\|full | Not-determined |
| 17-108-final-viral-combined_k141_244102\|\|full | Low-quality |
| 17-108-final-viral-combined_k141_257559\|\|full | Not-determined |
| 17-108-final-viral-combined_k141_312706\|\|full | Not-determined |
| 17-108-final-viral-combined_k141_298037\|\|full | Not-determined |
| 17-108-final-viral-combined_k141_146939\|\|full | Low-quality |
| 17-108-final-viral-combined_k141_174757\|\|full | Low-quality |
| 17-108-final-viral-combined_k141_132230\|\|full | Low-quality |
| 17-108-final-viral-combined_k141_187966\|\|full | Not-determined |
| 17-108-final-viral-combined_k141_9590\|\|full | Low-quality |
| 17-108-final-viral-combined_k141_216014\|\|full | Not-determined |
| 17-108-final-viral-combined_k141_132236\|\|full | Not-determined |
| 17-108-final-viral-combined_k141_257596\|\|full | Low-quality |
| 17-108-final-viral-combined_k141_285099\|\|full | Not-determined |
| 17-108-final-viral-combined_k141_77459\|\|full | Low-quality |
| 17-108-final-viral-combined_k141_174775\|\|full | Not-determined |
| 17-108-final-viral-combined_k141_216036\|\|full | Low-quality |
| 17-108-final-viral-combined_k141_325095\|\|full | Not-determined |
| 17-108-final-viral-combined_k141_187992\|\|full | Not-determined |
| 17-108-final-viral-combined_k141_174777\|\|full | Low-quality |
| 17-108-final-viral-combined_k141_187998\|\|full | Not-determined |
| 17-108-final-viral-combined_k141_325104\|\|full | Low-quality |
| 17-108-final-viral-combined_k141_244136\|\|full | Not-determined |
| 17-108-final-viral-combined_k141_325131\|\|full | Not-determined |
| 17-108-final-viral-combined_k141_174806\|\|full | Not-determined |
| 17-108-final-viral-combined_k141_216094\|\|full | Not-determined |
| 17-108-final-viral-combined_k141_188008\|\|full | Low-quality |
| 17-108-final-viral-combined_k141_285164\|\|full | Low-quality |
| 17-108-final-viral-combined_k141_147010\|\|full | Not-determined |
| 17-108-final-viral-combined_k141_312775\|\|full | Not-determined |
| 17-108-final-viral-combined_k141_216099\|\|full | Low-quality |
| 17-108-final-viral-combined_k141_147022\|\|full | Not-determined |
| 17-108-final-viral-combined_k141_285195\|\|full | Not-determined |
| 17-108-final-viral-combined_k141_21893\|\|full | Low-quality |
| 17-108-final-viral-combined_k141_216106\|\|full | Low-quality |
| 17-108-final-viral-combined_k141_257621\|\|full | Not-determined |
| 17-108-final-viral-combined_k141_298126\|\|full | Not-determined |
| 17-108-final-viral-combined_k141_36901\|\|full | Low-quality |
| 17-108-final-viral-combined_k141_229921\|\|full | Low-quality |
| 17-108-final-viral-combined_k141_188033\|\|full | Not-determined |
| 17-108-final-viral-combined_k141_147034\|\|full | Not-determined |
| 17-108-final-viral-combined_k141_216113\|\|full | Low-quality |
| 17-108-final-viral-combined_k141_118831\|\|full | Low-quality |
| 17-108-final-viral-combined_k141_285215\|\|full | Not-determined |
| 18-109-final-viral-combined_k141_217226\|\|full | High-quality |
| 17-108-final-viral-combined_k141_312794\|\|full | Low-quality |
| 17-108-final-viral-combined_k141_161005\|\|full | Not-determined |
| 17-108-final-viral-combined_k141_298146\|\|full | Low-quality |
| 17-108-final-viral-combined_k141_118852\|\|full | Not-determined |
| 17-108-final-viral-combined_k141_50716\|\|full | Low-quality |
| 17-108-final-viral-combined_k141_77549\|\|full | Not-determined |
| 17-108-final-viral-combined_k141_174869\|\|full | Not-determined |
| 17-108-final-viral-combined_k141_118857\|\|full | Low-quality |
| 17-108-final-viral-combined_k141_161037\|\|full | Not-determined |
| 17-108-final-viral-combined_k141_77553\|\|full | Not-determined |
| 17-108-final-viral-combined_k141_50742\|\|full | Low-quality |
| 17-108-final-viral-combined_k141_216136\|\|full | Not-determined |
| 17-108-final-viral-combined_k141_161046\|\|full | Low-quality |
| 17-108-final-viral-combined_k141_161056\|\|full | Low-quality |
| 17-108-final-viral-combined_k141_312811\|\|full | Not-determined |
| 17-108-final-viral-combined_k141_9724\|\|full | Low-quality |
| 17-108-final-viral-combined_k141_244239\|\|full | Not-determined |
| 17-108-final-viral-combined_k141_118899\|\|full | Low-quality |
| 17-108-final-viral-combined_k141_9732\|\|full | Not-determined |
| 17-108-final-viral-combined_k141_132281\|\|full | Low-quality |
| 17-108-final-viral-combined_k141_50768\|\|full | Not-determined |
| 17-108-final-viral-combined_k141_64448\|\|full | Not-determined |
| 17-108-final-viral-combined_k141_36933\|\|full | Low-quality |
| 17-108-final-viral-combined_k141_270847\|\|full | Not-determined |
| 17-108-final-viral-combined_k141_325164\|\|full | Low-quality |
| 17-108-final-viral-combined_k141_132290\|\|full | Not-determined |
| 17-108-final-viral-combined_k141_50780\|\|full | Not-determined |
| 17-108-final-viral-combined_k141_106393\|\|full | Low-quality |
| 17-108-final-viral-combined_k141_91959\|\|full | Not-determined |
| 17-108-final-viral-combined_k141_312855\|\|full | Low-quality |
| 17-108-final-viral-combined_k141_244273\|\|full | Not-determined |
| 17-108-final-viral-combined_k141_285303\|\|full | Low-quality |
| 17-108-final-viral-combined_k141_216193\|\|full | Not-determined |
| 17-108-final-viral-combined_k141_202018\|\|full | Low-quality |
| 17-108-final-viral-combined_k141_50800\|\|full | Not-determined |
| 17-108-final-viral-combined_k141_118936\|\|full | Low-quality |
| 17-108-final-viral-combined_k141_325187\|\|full | Not-determined |
| 17-108-final-viral-combined_k141_244293\|\|full | Low-quality |
| 17-108-final-viral-combined_k141_91999\|\|full | Low-quality |
| 17-108-final-viral-combined_k141_216220\|\|full | Low-quality |
| 17-108-final-viral-combined_k141_298223\|\|full | Low-quality |
| 17-108-final-viral-combined_k141_77620\|\|full | Not-determined |
| 17-108-final-viral-combined_k141_202057\|\|full | Low-quality |
| 17-108-final-viral-combined_k141_147160\|\|full | Low-quality |
| 17-108-final-viral-combined_k141_106428\|\|full | Not-determined |
| 17-108-final-viral-combined_k141_325209\|\|full | Low-quality |
| 17-108-final-viral-combined_k141_9778\|\|full | Not-determined |
| 17-108-final-viral-combined_k141_312911\|\|full | Not-determined |
| 17-108-final-viral-combined_k141_229981\|\|full | Not-determined |
| 17-108-final-viral-combined_k141_312913\|\|full | Low-quality |
| 17-108-final-viral-combined_k141_132363\|\|full | Not-determined |
| 17-108-final-viral-combined_k141_298241\|\|full | Not-determined |
| 17-108-final-viral-combined_k141_257757\|\|full | Not-determined |
| 17-108-final-viral-combined_k141_325227\|\|full | Not-determined |
| 17-108-final-viral-combined_k141_216254\|\|full | Not-determined |
| 17-108-final-viral-combined_k141_132371\|\|full | Low-quality |
| 17-108-final-viral-combined_k141_92019\|\|full | Not-determined |
| 17-108-final-viral-combined_k141_174976\|\|full | Not-determined |
| 17-108-final-viral-combined_k141_147193\|\|full | Low-quality |
| 17-108-final-viral-combined_k141_50841\|\|full | Not-determined |
| 17-108-final-viral-combined_k141_161111\|\|full | Not-determined |
| 17-108-final-viral-combined_k141_77639\|\|full | Not-determined |
| 17-108-final-viral-combined_k141_106448\|\|full | Not-determined |
| 17-108-final-viral-combined_k141_92027\|\|full | Low-quality |
| 17-108-final-viral-combined_k141_50846\|\|full | Low-quality |
| 17-108-final-viral-combined_k141_161121\|\|full | Low-quality |
| 17-108-final-viral-combined_k141_118982\|\|full | Low-quality |
| 17-108-final-viral-combined_k141_132383\|\|full | Not-determined |
| 17-108-final-viral-combined_k141_244323\|\|full | Low-quality |
| 17-108-final-viral-combined_k141_118987\|\|full | Not-determined |
| 17-108-final-viral-combined_k141_188165\|\|full | Low-quality |
| 17-108-final-viral-combined_k141_118989\|\|full | Low-quality |
| 17-108-final-viral-combined_k141_257791\|\|full | Not-determined |
| 17-108-final-viral-combined_k141_230019\|\|full | Not-determined |
| 17-108-final-viral-combined_k141_175014\|\|full | Not-determined |
| 17-108-final-viral-combined_k141_244333\|\|full | Not-determined |
| 17-108-final-viral-combined_k141_77680\|\|full | Not-determined |
| 17-108-final-viral-combined_k141_64569\|\|full | Not-determined |
| 17-108-final-viral-combined_k141_230028\|\|full | Not-determined |
| 17-108-final-viral-combined_k141_77692\|\|full | Low-quality |
| 17-108-final-viral-combined_k141_175039\|\|full | Low-quality |
| 17-108-final-viral-combined_k141_50886\|\|full | Low-quality |
| 17-108-final-viral-combined_k141_37043\|\|full | Low-quality |
| 17-108-final-viral-combined_k141_77717\|\|full | Not-determined |
| 17-108-final-viral-combined_k141_77720\|\|full | Not-determined |
| 17-108-final-viral-combined_k141_257839\|\|full | Not-determined |
| 17-108-final-viral-combined_k141_244352\|\|full | Not-determined |
| 17-108-final-viral-combined_k141_64588\|\|full | Low-quality |
| 17-108-final-viral-combined_k141_64592\|\|full | Not-determined |
| 17-108-final-viral-combined_k141_37059\|\|full | Low-quality |
| 17-108-final-viral-combined_k141_77740\|\|full | Not-determined |
| 17-108-final-viral-combined_k141_92058\|\|full | Not-determined |
| 17-108-final-viral-combined_k141_257859\|\|full | Low-quality |
| 17-108-final-viral-combined_k141_37075\|\|full | Not-determined |
| 17-108-final-viral-combined_k141_21950\|\|full | Not-determined |
| 17-108-final-viral-combined_k141_132442\|\|full | Low-quality |
| 17-108-final-viral-combined_k141_298370\|\|full | Not-determined |
| 17-108-final-viral-combined_k141_325295\|\|full | Not-determined |
| 17-108-final-viral-combined_k141_270928\|\|full | Low-quality |
| 17-108-final-viral-combined_k141_298381\|\|full | Not-determined |
| 17-108-final-viral-combined_k141_188218\|\|full | Not-determined |
| 17-108-final-viral-combined_k141_119087\|\|full | Low-quality |
| 17-108-final-viral-combined_k141_313046\|\|full | Low-quality |
| 17-108-final-viral-combined_k141_202175\|\|full | Low-quality |
| 17-108-final-viral-combined_k141_21967\|\|full | Low-quality |
| 17-108-final-viral-combined_k141_161198\|\|full | Low-quality |
| 17-108-final-viral-combined_k141_244409\|\|full | Low-quality |
| 17-108-final-viral-combined_k141_147268\|\|full | Not-determined |
| 17-108-final-viral-combined_k141_257887\|\|full | Low-quality |
| 17-108-final-viral-combined_k141_147287\|\|full | Not-determined |
| 17-108-final-viral-combined_k141_230077\|\|full | Low-quality |
| 17-108-final-viral-combined_k141_119113\|\|full | Low-quality |
| 17-108-final-viral-combined_k141_202201\|\|full | Not-determined |
| 17-108-final-viral-combined_k141_119117\|\|full | Low-quality |
| 17-108-final-viral-combined_k141_147304\|\|full | Not-determined |
| 17-108-final-viral-combined_k141_202207\|\|full | Not-determined |
| 17-108-final-viral-combined_k141_9897\|\|full | Not-determined |
| 17-108-final-viral-combined_k141_257911\|\|full | Not-determined |
| 17-108-final-viral-combined_k141_50993\|\|full | Not-determined |
| 17-108-final-viral-combined_k141_147312\|\|full | Not-determined |
| 17-108-final-viral-combined_k141_202212\|\|full | Low-quality |
| 17-108-final-viral-combined_k141_270995\|\|full | Not-determined |
| 17-108-final-viral-combined_k141_77765\|\|full | Not-determined |
| 17-108-final-viral-combined_k141_325353\|\|full | Low-quality |
| 17-108-final-viral-combined_k141_51007\|\|full | Low-quality |
| 17-108-final-viral-combined_k141_257931\|\|full | Not-determined |
| 17-108-final-viral-combined_k141_188255\|\|full | Low-quality |
| 17-108-final-viral-combined_k141_313073\|\|full | Low-quality |
| 17-108-final-viral-combined_k141_244481\|\|full | Low-quality |
| 17-108-final-viral-combined_k141_119142\|\|full | Not-determined |
| 17-108-final-viral-combined_k141_119150\|\|full | Not-determined |
| 17-108-final-viral-combined_k141_298387\|\|full | Not-determined |
| 17-108-final-viral-combined_k141_161250\|\|full | Low-quality |
| 17-108-final-viral-combined_k141_51036\|\|full | Not-determined |
| 17-108-final-viral-combined_k141_298394\|\|full | Low-quality |
| 17-108-final-viral-combined_k141_230110\|\|full | Low-quality |
| 17-108-final-viral-combined_k141_51040\|\|full | Not-determined |
| 18-102_k141_128831 | Complete |
| 17-108-final-viral-combined_k141_119176\|\|full | Low-quality |
| 17-108-final-viral-combined_k141_313109\|\|full | Low-quality |
| 17-108-final-viral-combined_k141_202257\|\|full | Not-determined |
| 17-108-final-viral-combined_k141_313114\|\|full | Not-determined |
| 17-108-final-viral-combined_k141_161278\|\|full | Low-quality |
| 17-108-final-viral-combined_k141_202266\|\|full | Low-quality |
| 17-108-final-viral-combined_k141_161292\|\|full | Not-determined |
| 17-108-final-viral-combined_k141_51073\|\|full | Low-quality |
| 17-108-final-viral-combined_k141_298437\|\|full | Not-determined |
| 17-108-final-viral-combined_k141_230167\|\|full | Not-determined |
| 17-108-final-viral-combined_k141_51087\|\|full | Low-quality |
| 17-108-final-viral-combined_k141_325427\|\|full | Low-quality |
| 17-108-final-viral-combined_k141_37151\|\|full | Low-quality |
| 17-108-final-viral-combined_k141_313153\|\|full | Low-quality |
| 17-108-final-viral-combined_k141_64665\|\|full | Low-quality |
| 17-108-final-viral-combined_k141_175192\|\|full | Low-quality |
| 17-108-final-viral-combined_k141_230176\|\|full | Low-quality |
| 17-108-final-viral-combined_k141_77799\|\|full | Not-determined |
| 17-108-final-viral-combined_k141_92172\|\|full | Not-determined |
| 17-108-final-viral-combined_k141_258048\|\|full | Not-determined |
| 17-108-final-viral-combined_k141_147404\|\|full | Not-determined |
| 18-102-final-viral-combined_k141_128831\|\|full | High-quality |
| 17-108-final-viral-combined_k141_106589\|\|full | Not-determined |
| 17-108-final-viral-combined_k141_106590\|\|full | Not-determined |
| 17-108-final-viral-combined_k141_119233\|\|full | Not-determined |
| 17-108-final-viral-combined_k141_216451\|\|full | Low-quality |
| 17-108-final-viral-combined_k141_92202\|\|full | Not-determined |
| 17-108-final-viral-combined_k141_188358\|\|full | Not-determined |
| 17-108-final-viral-combined_k141_216453\|\|full | Not-determined |
| 17-108-final-viral-combined_k141_64714\|\|full | Low-quality |
| 17-108-final-viral-combined_k141_106610\|\|full | Not-determined |
| 17-108-final-viral-combined_k141_258059\|\|full | Low-quality |
| 17-108-final-viral-combined_k141_230211\|\|full | Not-determined |
| 17-108-final-viral-combined_k141_147471\|\|full | Not-determined |
| 17-108-final-viral-combined_k141_51182\|\|full | Low-quality |
| 17-108-final-viral-combined_k141_298516\|\|full | Not-determined |
| 17-108-final-viral-combined_k141_175235\|\|full | Not-determined |
| 17-108-final-viral-combined_k141_92234\|\|full | Not-determined |
| 17-108-final-viral-combined_k141_132575\|\|full | Low-quality |
| 17-108-final-viral-combined_k141_147480\|\|full | Low-quality |
| 17-108-final-viral-combined_k141_119250\|\|full | Not-determined |
| 17-108-final-viral-combined_k141_106627\|\|full | Low-quality |
| 17-108-final-viral-combined_k141_285583\|\|full | Low-quality |
| 17-108-final-viral-combined_k141_92245\|\|full | Low-quality |
| 17-108-final-viral-combined_k141_202349\|\|full | Not-determined |
| 17-108-final-viral-combined_k141_64760\|\|full | Low-quality |
| 17-108-final-viral-combined_k141_132591\|\|full | Not-determined |
| 17-108-final-viral-combined_k141_244557\|\|full | Low-quality |
| 17-108-final-viral-combined_k141_92250\|\|full | Low-quality |
| 17-108-final-viral-combined_k141_106647\|\|full | Not-determined |
| 17-108-final-viral-combined_k141_285595\|\|full | Low-quality |
| 17-108-final-viral-combined_k141_147510\|\|full | Low-quality |
| 17-108-final-viral-combined_k141_230239\|\|full | Not-determined |
| 17-108-final-viral-combined_k141_51214\|\|full | Not-determined |
| 17-108-final-viral-combined_k141_313222\|\|full | Low-quality |
| 17-108-final-viral-combined_k141_132604\|\|full | Not-determined |
| 17-108-final-viral-combined_k141_64774\|\|full | Not-determined |
| 17-108-final-viral-combined_k141_325514\|\|full | Not-determined |
| 17-108-final-viral-combined_k141_298567\|\|full | Low-quality |
| 17-108-final-viral-combined_k141_202383\|\|full | Not-determined |
| 17-108-final-viral-combined_k141_92279\|\|full | Low-quality |
| 17-108-final-viral-combined_k141_271085\|\|full | Not-determined |
| 17-108-final-viral-combined_k141_51234\|\|full | Not-determined |
| 17-108-final-viral-combined_k141_271088\|\|full | Not-determined |
| 17-108-final-viral-combined_k141_92282\|\|full | Not-determined |
| 17-108-final-viral-combined_k141_64780\|\|full | Low-quality |
| 17-108-final-viral-combined_k141_202396\|\|full | Not-determined |
| 17-108-final-viral-combined_k141_77871\|\|full | Low-quality |
| 17-108-final-viral-combined_k141_132632\|\|full | Low-quality |
| 17-108-final-viral-combined_k141_202401\|\|full | Not-determined |
| 17-108-final-viral-combined_k141_175283\|\|full | Low-quality |
| 17-108-final-viral-combined_k141_271097\|\|full | Not-determined |
| 17-108-final-viral-combined_k141_92308\|\|full | Not-determined |
| 17-108-final-viral-combined_k141_92313\|\|full | Not-determined |
| 17-108-final-viral-combined_k141_132649\|\|full | Not-determined |
| 17-108-final-viral-combined_k141_132651\|\|full | Not-determined |
| 17-108-final-viral-combined_k141_230287\|\|full | Low-quality |
| 17-108-final-viral-combined_k141_64820\|\|full | Not-determined |
| 17-108-final-viral-combined_k141_313267\|\|full | Not-determined |
| 17-108-final-viral-combined_k141_271124\|\|full | Low-quality |
| 17-108-final-viral-combined_k141_51270\|\|full | Not-determined |
| 17-108-final-viral-combined_k141_64832\|\|full | Not-determined |
| 17-108-final-viral-combined_k141_106721\|\|full | Not-determined |
| 17-108-final-viral-combined_k141_119328\|\|full | Not-determined |
| 17-108-final-viral-combined_k141_22146\|\|full | Low-quality |
| 17-108-final-viral-combined_k141_285651\|\|full | Not-determined |
| 17-108-final-viral-combined_k141_77906\|\|full | Not-determined |
| 17-108-final-viral-combined_k141_64878\|\|full | Not-determined |
| 17-108-final-viral-combined_k141_298642\|\|full | Low-quality |
| 17-108-final-viral-combined_k141_285677\|\|full | Not-determined |
| 17-108-final-viral-combined_k141_313284\|\|full | Low-quality |
| 17-108-final-viral-combined_k141_92381\|\|full | Low-quality |
| 17-108-final-viral-combined_k141_77915\|\|full | Low-quality |
| 17-108-final-viral-combined_k141_119381\|\|full | Not-determined |
| 17-108-final-viral-combined_k141_161396\|\|full | Not-determined |
| 17-108-final-viral-combined_k141_37365\|\|full | Not-determined |
| 17-108-final-viral-combined_k141_258190\|\|full | Not-determined |
| 17-108-final-viral-combined_k141_216637\|\|full | Low-quality |
| 17-108-final-viral-combined_k141_147625\|\|full | Low-quality |
| 17-108-final-viral-combined_k141_119398\|\|full | Not-determined |
| 17-108-final-viral-combined_k141_106772\|\|full | Not-determined |
| 17-108-final-viral-combined_k141_147637\|\|full | Not-determined |
| 17-108-final-viral-combined_k141_244773\|\|full | Low-quality |
| 17-108-final-viral-combined_k141_132714\|\|full | Not-determined |
| 17-108-final-viral-combined_k141_244775\|\|full | Low-quality |
| 17-108-final-viral-combined_k141_147642\|\|full | Not-determined |
| 17-108-final-viral-combined_k141_77953\|\|full | Low-quality |
| 17-108-final-viral-combined_k141_132731\|\|full | Not-determined |
| 17-108-final-viral-combined_k141_258242\|\|full | Low-quality |
| 17-108-final-viral-combined_k141_77973\|\|full | Low-quality |
| 17-108-final-viral-combined_k141_37438\|\|full | Low-quality |
| 17-108-final-viral-combined_k141_161427\|\|full | Not-determined |
| 17-108-final-viral-combined_k141_161431\|\|full | Low-quality |
| 17-108-final-viral-combined_k141_37447\|\|full | Low-quality |
| 17-108-final-viral-combined_k141_22249\|\|full | Not-determined |
| 17-108-final-viral-combined_k141_258251\|\|full | Not-determined |
| 17-108-final-viral-combined_k141_37466\|\|full | Low-quality |
| 17-108-final-viral-combined_k141_64945\|\|full | Low-quality |
| 17-108-final-viral-combined_k141_271215\|\|full | Low-quality |
| 17-108-final-viral-combined_k141_132765\|\|full | Not-determined |
| 17-108-final-viral-combined_k141_230377\|\|full | Low-quality |
| 17-108-final-viral-combined_k141_64962\|\|full | Not-determined |
| 17-108-final-viral-combined_k141_22292\|\|full | Low-quality |
| 17-108-final-viral-combined_k141_37477\|\|full | Low-quality |
| 17-108-final-viral-combined_k141_298764\|\|full | Not-determined |
| 17-108-final-viral-combined_k141_132773\|\|full | Low-quality |
| 17-108-final-viral-combined_k141_92437\|\|full | Not-determined |
| 17-108-final-viral-combined_k141_313344\|\|full | Not-determined |
| 17-108-final-viral-combined_k141_175474\|\|full | Not-determined |
| 17-108-final-viral-combined_k141_258291\|\|full | Not-determined |
| 17-108-final-viral-combined_k141_106846\|\|full | Not-determined |
| 17-108-final-viral-combined_k141_188519\|\|full | Low-quality |
| 17-108-final-viral-combined_k141_244799\|\|full | Low-quality |
| 17-108-final-viral-combined_k141_313362\|\|full | Low-quality |
| 17-108-final-viral-combined_k141_78053\|\|full | Low-quality |
| 17-108-final-viral-combined_k141_37493\|\|full | Not-determined |
| 17-108-final-viral-combined_k141_188526\|\|full | Low-quality |
| 17-108-final-viral-combined_k141_92462\|\|full | Low-quality |
| 17-108-final-viral-combined_k141_78056\|\|full | Not-determined |
| 17-108-final-viral-combined_k141_64994\|\|full | Not-determined |
| 17-108-final-viral-combined_k141_22328\|\|full | Low-quality |
| 17-108-final-viral-combined_k141_161520\|\|full | Not-determined |
| 17-108-final-viral-combined_k141_106863\|\|full | Low-quality |
| 17-108-final-viral-combined_k141_202544\|\|full | Not-determined |
| 17-108-final-viral-combined_k141_325629\|\|full | Low-quality |
| 17-108-final-viral-combined_k141_65004\|\|full | Not-determined |
| 17-108-final-viral-combined_k141_325632\|\|full | Not-determined |
| 17-108-final-viral-combined_k141_175523\|\|full | Not-determined |
| 17-108-final-viral-combined_k141_216757\|\|full | Low-quality |
| 17-108-final-viral-combined_k141_132826\|\|full | Not-determined |
| 17-108-final-viral-combined_k141_285779\|\|full | Low-quality |
| 17-108-final-viral-combined_k141_78101\|\|full | Not-determined |
| 17-108-final-viral-combined_k141_175543\|\|full | Not-determined |
| 17-108-final-viral-combined_k141_285785\|\|full | Low-quality |
| 17-108-final-viral-combined_k141_22350\|\|full | Not-determined |
| 17-108-final-viral-combined_k141_119495\|\|full | Not-determined |
| 17-108-final-viral-combined_k141_298795\|\|full | Not-determined |
| 17-108-final-viral-combined_k141_285800\|\|full | Low-quality |
| 17-108-final-viral-combined_k141_147757\|\|full | Low-quality |
| 17-108-final-viral-combined_k141_132854\|\|full | Not-determined |
| 17-108-final-viral-combined_k141_313402\|\|full | Not-determined |
| 17-108-final-viral-combined_k141_230473\|\|full | Not-determined |
| 17-108-final-viral-combined_k141_65053\|\|full | Low-quality |
| 17-108-final-viral-combined_k141_216811\|\|full | Not-determined |
| 17-108-final-viral-combined_k141_271306\|\|full | Low-quality |
| 17-108-final-viral-combined_k141_37534\|\|full | Not-determined |
| 17-108-final-viral-combined_k141_285822\|\|full | Low-quality |
| 17-108-final-viral-combined_k141_147790\|\|full | Low-quality |
| 17-108-final-viral-combined_k141_37546\|\|full | Not-determined |
| 17-108-final-viral-combined_k141_51477\|\|full | Low-quality |
| 17-108-final-viral-combined_k141_313437\|\|full | Not-determined |
| 17-108-final-viral-combined_k141_37556\|\|full | Low-quality |
| 17-108-final-viral-combined_k141_22425\|\|full | Low-quality |
| 17-108-final-viral-combined_k141_216831\|\|full | Low-quality |
| 17-108-final-viral-combined_k141_132895\|\|full | Not-determined |
| 17-108-final-viral-combined_k141_51499\|\|full | Not-determined |
| 17-108-final-viral-combined_k141_92532\|\|full | Not-determined |
| 17-108-final-viral-combined_k141_132896\|\|full | Low-quality |
| 17-108-final-viral-combined_k141_161582\|\|full | Low-quality |
| 17-108-final-viral-combined_k141_119559\|\|full | Not-determined |
| 17-108-final-viral-combined_k141_325662\|\|full | Not-determined |
| 17-108-final-viral-combined_k141_325666\|\|full | Low-quality |
| 17-108-final-viral-combined_k141_147812\|\|full | Low-quality |
| 17-108-final-viral-combined_k141_161588\|\|full | Low-quality |
| 17-108-final-viral-combined_k141_271349\|\|full | Not-determined |
| 17-108-final-viral-combined_k141_202630\|\|full | Not-determined |
| 17-108-final-viral-combined_k141_188609\|\|full | Not-determined |
| 17-108-final-viral-combined_k141_51536\|\|full | Low-quality |
| 17-108-final-viral-combined_k141_78177\|\|full | Not-determined |
| 17-108-final-viral-combined_k141_271373\|\|full | Not-determined |
| 17-108-final-viral-combined_k141_92581\|\|full | Not-determined |
| 17-108-final-viral-combined_k141_119596\|\|full | Low-quality |
| 17-108-final-viral-combined_k141_22448\|\|full | Not-determined |
| 17-108-final-viral-combined_k141_313487\|\|full | Low-quality |
| 17-108-final-viral-combined_k141_244878\|\|full | Not-determined |
| 17-108-final-viral-combined_k141_325689\|\|full | Not-determined |
| 17-108-final-viral-combined_k141_92607\|\|full | Low-quality |
| 17-108-final-viral-combined_k141_244885\|\|full | Not-determined |
| 17-108-final-viral-combined_k141_78203\|\|full | Not-determined |
| 17-108-final-viral-combined_k141_65137\|\|full | Not-determined |
| 17-108-final-viral-combined_k141_119623\|\|full | Low-quality |
| 17-108-final-viral-combined_k141_92612\|\|full | Not-determined |
| 17-108-final-viral-combined_k141_106993\|\|full | Not-determined |
| 17-108-final-viral-combined_k141_92616\|\|full | Low-quality |
| 17-108-final-viral-combined_k141_258431\|\|full | Low-quality |
| 17-108-final-viral-combined_k141_161650\|\|full | Not-determined |
| 17-108-final-viral-combined_k141_78228\|\|full | Not-determined |
| 17-108-final-viral-combined_k141_271415\|\|full | Not-determined |
| 17-108-final-viral-combined_k141_147842\|\|full | Low-quality |
| 17-108-final-viral-combined_k141_298862\|\|full | Not-determined |
| 17-108-final-viral-combined_k141_10220\|\|full | Low-quality |
| 17-108-final-viral-combined_k141_313520\|\|full | Not-determined |
| 17-108-final-viral-combined_k141_188652\|\|full | Not-determined |
| 17-108-final-viral-combined_k141_10225\|\|full | Not-determined |
| 17-108-final-viral-combined_k141_285875\|\|full | Low-quality |
| 17-108-final-viral-combined_k141_175625\|\|full | Low-quality |
| 17-108-final-viral-combined_k141_285876\|\|full | Low-quality |
| 17-108-final-viral-combined_k141_202714\|\|full | Low-quality |
| 17-108-final-viral-combined_k141_147861\|\|full | Low-quality |
| 17-108-final-viral-combined_k141_10237\|\|full | Low-quality |
| 17-108-final-viral-combined_k141_161661\|\|full | Low-quality |
| 17-108-final-viral-combined_k141_37652\|\|full | Not-determined |
| 17-108-final-viral-combined_k141_119641\|\|full | Low-quality |
| 17-108-final-viral-combined_k141_202748\|\|full | Not-determined |
| 17-108-final-viral-combined_k141_161675\|\|full | Not-determined |
| 17-108-final-viral-combined_k141_78273\|\|full | Not-determined |
| 17-108-final-viral-combined_k141_271466\|\|full | Low-quality |
| 17-108-final-viral-combined_k141_285909\|\|full | Not-determined |
| 17-108-final-viral-combined_k141_119680\|\|full | Low-quality |
| 17-108-final-viral-combined_k141_188689\|\|full | Low-quality |
| 17-108-final-viral-combined_k141_230617\|\|full | Low-quality |
| 17-108-final-viral-combined_k141_313553\|\|full | Not-determined |
| 17-108-final-viral-combined_k141_119686\|\|full | Low-quality |
| 17-108-final-viral-combined_k141_37656\|\|full | Not-determined |
| 17-108-final-viral-combined_k141_147887\|\|full | Low-quality |
| 17-108-final-viral-combined_k141_188703\|\|full | Not-determined |
| 17-108-final-viral-combined_k141_119687\|\|full | Low-quality |
| 17-108-final-viral-combined_k141_216898\|\|full | Not-determined |
| 17-108-final-viral-combined_k141_313573\|\|full | Low-quality |
| 17-108-final-viral-combined_k141_271483\|\|full | Low-quality |
| 17-108-final-viral-combined_k141_10324\|\|full | Low-quality |
| 17-108-final-viral-combined_k141_161719\|\|full | Not-determined |
| 17-108-final-viral-combined_k141_65241\|\|full | Not-determined |
| 17-108-final-viral-combined_k141_271489\|\|full | Low-quality |
| 17-108-final-viral-combined_k141_258497\|\|full | Low-quality |
| 17-108-final-viral-combined_k141_51704\|\|full | Low-quality |
| 17-108-final-viral-combined_k141_147905\|\|full | Low-quality |
| 17-108-final-viral-combined_k141_188737\|\|full | Not-determined |
| 17-108-final-viral-combined_k141_298946\|\|full | Low-quality |
| 17-108-final-viral-combined_k141_65258\|\|full | Not-determined |
| 17-108-final-viral-combined_k141_37681\|\|full | Low-quality |
| 17-108-final-viral-combined_k141_202796\|\|full | Not-determined |
| 17-108-final-viral-combined_k141_92677\|\|full | Low-quality |
| 17-108-final-viral-combined_k141_107078\|\|full | Not-determined |
| 17-108-final-viral-combined_k141_313586\|\|full | Low-quality |
| 17-108-final-viral-combined_k141_147914\|\|full | Not-determined |
| 17-108-final-viral-combined_k141_65264\|\|full | Not-determined |
| 17-108-final-viral-combined_k141_78315\|\|full | Not-determined |
| 17-108-final-viral-combined_k141_133055\|\|full | Not-determined |
| 17-108-final-viral-combined_k141_147921\|\|full | Not-determined |
| 17-108-final-viral-combined_k141_188752\|\|full | Low-quality |
| 17-108-final-viral-combined_k141_230669\|\|full | Not-determined |
| 17-108-final-viral-combined_k141_313604\|\|full | Low-quality |
| 17-108-final-viral-combined_k141_51749\|\|full | Low-quality |
| 17-108-final-viral-combined_k141_313610\|\|full | Low-quality |
| 17-108-final-viral-combined_k141_10360\|\|full | Low-quality |
| 17-108-final-viral-combined_k141_37714\|\|full | Not-determined |
| 17-108-final-viral-combined_k141_313614\|\|full | Low-quality |
| 17-108-final-viral-combined_k141_298969\|\|full | Low-quality |
| 17-108-final-viral-combined_k141_258552\|\|full | Low-quality |
| 17-108-final-viral-combined_k141_78354\|\|full | Not-determined |
| 17-108-final-viral-combined_k141_37720\|\|full | Low-quality |
| 17-108-final-viral-combined_k141_258558\|\|full | Low-quality |
| 17-108-final-viral-combined_k141_325817\|\|full | Not-determined |
| 17-108-final-viral-combined_k141_298984\|\|full | Low-quality |
| 17-108-final-viral-combined_k141_216975\|\|full | Low-quality |
| 17-108-final-viral-combined_k141_51768\|\|full | Not-determined |
| 17-108-final-viral-combined_k141_325821\|\|full | Not-determined |
| 17-108-final-viral-combined_k141_22631\|\|full | Low-quality |
| 17-108-final-viral-combined_k141_175735\|\|full | Low-quality |
| 17-108-final-viral-combined_k141_78380\|\|full | Not-determined |
| 17-108-final-viral-combined_k141_22646\|\|full | Not-determined |
| 17-108-final-viral-combined_k141_37750\|\|full | Low-quality |
| 17-108-final-viral-combined_k141_285933\|\|full | Not-determined |
| 17-108-final-viral-combined_k141_202859\|\|full | Not-determined |
| 17-108-final-viral-combined_k141_37759\|\|full | Low-quality |
| 17-108-final-viral-combined_k141_175749\|\|full | Low-quality |
| 17-108-final-viral-combined_k141_188809\|\|full | Not-determined |
| 17-108-final-viral-combined_k141_147972\|\|full | Not-determined |
| 17-108-final-viral-combined_k141_51791\|\|full | Not-determined |
| 17-108-final-viral-combined_k141_299014\|\|full | Not-determined |
| 17-108-final-viral-combined_k141_271586\|\|full | Low-quality |
| 17-108-final-viral-combined_k141_202882\|\|full | Low-quality |
| 17-108-final-viral-combined_k141_10419\|\|full | Low-quality |
| 17-108-final-viral-combined_k141_51813\|\|full | Low-quality |
| 17-108-final-viral-combined_k141_299036\|\|full | Not-determined |
| 17-108-final-viral-combined_k141_51826\|\|full | Not-determined |
| 17-108-final-viral-combined_k141_37809\|\|full | Not-determined |
| 17-108-final-viral-combined_k141_10435\|\|full | Low-quality |
| 17-108-final-viral-combined_k141_107181\|\|full | Not-determined |
| 17-108-final-viral-combined_k141_10440\|\|full | Low-quality |
| 17-108-final-viral-combined_k141_65374\|\|full | Low-quality |
| 17-108-final-viral-combined_k141_133135\|\|full | Low-quality |
| 17-108-final-viral-combined_k141_188836\|\|full | Not-determined |
| 17-108-final-viral-combined_k141_78453\|\|full | Not-determined |
| 17-108-final-viral-combined_k141_325889\|\|full | Not-determined |
| 17-108-final-viral-combined_k141_107201\|\|full | Low-quality |
| 17-108-final-viral-combined_k141_202958\|\|full | Not-determined |
| 17-108-final-viral-combined_k141_258635\|\|full | Low-quality |
| 17-108-final-viral-combined_k141_161825\|\|full | Low-quality |
| 17-108-final-viral-combined_k141_271652\|\|full | Low-quality |
| 17-108-final-viral-combined_k141_245111\|\|full | Low-quality |
| 17-108-final-viral-combined_k141_161836\|\|full | Not-determined |
| 17-108-final-viral-combined_k141_175822\|\|full | Not-determined |
| 17-108-final-viral-combined_k141_271676\|\|full | Low-quality |
| 17-108-final-viral-combined_k141_313742\|\|full | Not-determined |
| 17-108-final-viral-combined_k141_202996\|\|full | Low-quality |
| 17-108-final-viral-combined_k141_119824\|\|full | Low-quality |
| 17-108-final-viral-combined_k141_107223\|\|full | Not-determined |
| 17-108-final-viral-combined_k141_325920\|\|full | Low-quality |
| 17-108-final-viral-combined_k141_313750\|\|full | Not-determined |
| 17-108-final-viral-combined_k141_325922\|\|full | Low-quality |
| 17-108-final-viral-combined_k141_51875\|\|full | Not-determined |
| 17-108-final-viral-combined_k141_148085\|\|full | Low-quality |
| 17-108-final-viral-combined_k141_51877\|\|full | Not-determined |
| 17-108-final-viral-combined_k141_51882\|\|full | Not-determined |
| 17-108-final-viral-combined_k141_245152\|\|full | Low-quality |
| 17-108-final-viral-combined_k141_51893\|\|full | Not-determined |
| 17-108-final-viral-combined_k141_65465\|\|full | Low-quality |
| 17-108-final-viral-combined_k141_107248\|\|full | Not-determined |
| 17-108-final-viral-combined_k141_230836\|\|full | Not-determined |
| 17-108-final-viral-combined_k141_188903\|\|full | Low-quality |
| 17-108-final-viral-combined_k141_107251\|\|full | Low-quality |
| 17-108-final-viral-combined_k141_65472\|\|full | Low-quality |
| 17-108-final-viral-combined_k141_325931\|\|full | Low-quality |
| 17-108-final-viral-combined_k141_217110\|\|full | Low-quality |
| 17-108-final-viral-combined_k141_161890\|\|full | Low-quality |
| 17-108-final-viral-combined_k141_92821\|\|full | Low-quality |
| 17-108-final-viral-combined_k141_286049\|\|full | Low-quality |

| | |
|---|---|
| 17-108-final-viral-combined_k141_22783\|\|full | Low-quality |
| 17-108-final-viral-combined_k141_188923\|\|full | Low-quality |
| 17-108-final-viral-combined_k141_51919\|\|full | Not-determined |
| 17-108-final-viral-combined_k141_133185\|\|full | Low-quality |
| 17-108-final-viral-combined_k141_51920\|\|full | Not-determined |
| 17-108-final-viral-combined_k141_65480\|\|full | Low-quality |
| 17-108-final-viral-combined_k141_92856\|\|full | Low-quality |
| 17-108-final-viral-combined_k141_65485\|\|full | Low-quality |
| 17-108-final-viral-combined_k141_188942\|\|full | Not-determined |
| 17-108-final-viral-combined_k141_107280\|\|full | Not-determined |
| 17-108-final-viral-combined_k141_313768\|\|full | Low-quality |
| 17-108-final-viral-combined_k141_299201\|\|full | Not-determined |
| 17-108-final-viral-combined_k141_161925\|\|full | Not-determined |
| 17-108-final-viral-combined_k141_92885\|\|full | Low-quality |
| 17-108-final-viral-combined_k141_133201\|\|full | Not-determined |
| 17-108-final-viral-combined_k141_133206\|\|full | Low-quality |
| 17-108-final-viral-combined_k141_161935\|\|full | Not-determined |
| 17-108-final-viral-combined_k141_65524\|\|full | Low-quality |
| 17-108-final-viral-combined_k141_325950\|\|full | Low-quality |
| 17-108-final-viral-combined_k141_51946\|\|full | Not-determined |
| 17-108-final-viral-combined_k141_230874\|\|full | Not-determined |
| 17-108-final-viral-combined_k141_188983\|\|full | Not-determined |
| 17-108-final-viral-combined_k141_51959\|\|full | Not-determined |
| 17-108-final-viral-combined_k141_119910\|\|full | Low-quality |
| 17-108-final-viral-combined_k141_299220\|\|full | Low-quality |
| 17-108-final-viral-combined_k141_65547\|\|full | Not-determined |
| 17-108-final-viral-combined_k141_313814\|\|full | Not-determined |
| 17-108-final-viral-combined_k141_189007\|\|full | Low-quality |
| 17-108-final-viral-combined_k141_161971\|\|full | Low-quality |
| 17-108-final-viral-combined_k141_175926\|\|full | Low-quality |
| 17-108-final-viral-combined_k141_92934\|\|full | Not-determined |
| 17-108-final-viral-combined_k141_22855\|\|full | Not-determined |
| 17-108-final-viral-combined_k141_299255\|\|full | Not-determined |
| 17-108-final-viral-combined_k141_38038\|\|full | Low-quality |
| 17-108-final-viral-combined_k141_92938\|\|full | Low-quality |
| 17-108-final-viral-combined_k141_38048\|\|full | Not-determined |
| 17-108-final-viral-combined_k141_78557\|\|full | Low-quality |
| 17-108-final-viral-combined_k141_325982\|\|full | Low-quality |
| 17-108-final-viral-combined_k141_78559\|\|full | Low-quality |
| 17-108-final-viral-combined_k141_299286\|\|full | Low-quality |
| 17-108-final-viral-combined_k141_299292\|\|full | Not-determined |
| 17-108-final-viral-combined_k141_119944\|\|full | Not-determined |
| 17-108-final-viral-combined_k141_258785\|\|full | Not-determined |
| 17-108-final-viral-combined_k141_92953\|\|full | Low-quality |
| 17-108-final-viral-combined_k141_258788\|\|full | Low-quality |
| 17-108-final-viral-combined_k141_286134\|\|full | Low-quality |
| 17-108-final-viral-combined_k141_286138\|\|full | Not-determined |
| 17-108-final-viral-combined_k141_299304\|\|full | Not-determined |
| 17-108-final-viral-combined_k141_230929\|\|full | Low-quality |
| 17-108-final-viral-combined_k141_189030\|\|full | Low-quality |
| 17-108-final-viral-combined_k141_203151\|\|full | Low-quality |
| 17-108-final-viral-combined_k141_245271\|\|full | Low-quality |
| 17-108-final-viral-combined_k141_22874\|\|full | Not-determined |
| 17-108-final-viral-combined_k141_162029\|\|full | Not-determined |
| 17-108-final-viral-combined_k141_78599\|\|full | Low-quality |
| 17-108-final-viral-combined_k141_230946\|\|full | Not-determined |
| 17-108-final-viral-combined_k141_299318\|\|full | Low-quality |
| 17-109-final-viral-combined_k141_129799\|\|full | Complete |
| 17-108-final-viral-combined_k141_286154\|\|full | Low-quality |
| 17-108-final-viral-combined_k141_245289\|\|full | Low-quality |
| 17-108-final-viral-combined_k141_10579\|\|full | Low-quality |
| 17-108-final-viral-combined_k141_271772\|\|full | Not-determined |
| 17-108-final-viral-combined_k141_78613\|\|full | Not-determined |
| 17-108-final-viral-combined_k141_299328\|\|full | Low-quality |
| 17-108-final-viral-combined_k141_271782\|\|full | Low-quality |
| 17-108-final-viral-combined_k141_22904\|\|full | Low-quality |
| 17-108-final-viral-combined_k141_258830\|\|full | Low-quality |
| 17-108-final-viral-combined_k141_230957\|\|full | Low-quality |
| 17-108-final-viral-combined_k141_22908\|\|full | Not-determined |
| 17-108-final-viral-combined_k141_148177\|\|full | Not-determined |
| 17-108-final-viral-combined_k141_78633\|\|full | Not-determined |
| 17-108-final-viral-combined_k141_286185\|\|full | Not-determined |
| 17-108-final-viral-combined_k141_313889\|\|full | Low-quality |
| 17-108-final-viral-combined_k141_258844\|\|full | Not-determined |
| 17-108-final-viral-combined_k141_230970\|\|full | Low-quality |
| 17-108-final-viral-combined_k141_107400\|\|full | Not-determined |
| 17-108-final-viral-combined_k141_245309\|\|full | Not-determined |
| 17-108-final-viral-combined_k141_299362\|\|full | Low-quality |
| 17-108-final-viral-combined_k141_176044\|\|full | Low-quality |
| 17-108-final-viral-combined_k141_148191\|\|full | Not-determined |
| 17-108-final-viral-combined_k141_120030\|\|full | Not-determined |
| 17-108-final-viral-combined_k141_38151\|\|full | Not-determined |
| 17-108-final-viral-combined_k141_245323\|\|full | Low-quality |
| 17-108-final-viral-combined_k141_38161\|\|full | Not-determined |
| 17-108-final-viral-combined_k141_107440\|\|full | Low-quality |
| 17-108-final-viral-combined_k141_286234\|\|full | Not-determined |
| 17-108-final-viral-combined_k141_65706\|\|full | Not-determined |
| 17-108-final-viral-combined_k141_107461\|\|full | Not-determined |
| 17-108-final-viral-combined_k141_133361\|\|full | Low-quality |
| 17-108-final-viral-combined_k141_52134\|\|full | Not-determined |
| 17-108-final-viral-combined_k141_93082\|\|full | Low-quality |
| 17-108-final-viral-combined_k141_10605\|\|full | Not-determined |
| 17-108-final-viral-combined_k141_10606\|\|full | Low-quality |
| 17-108-final-viral-combined_k141_326048\|\|full | Low-quality |
| 17-108-final-viral-combined_k141_22952\|\|full | Not-determined |
| 17-108-final-viral-combined_k141_10611\|\|full | Low-quality |
| 17-108-final-viral-combined_k141_271831\|\|full | Low-quality |
| 17-108-final-viral-combined_k141_133385\|\|full | Not-determined |
| 17-108-final-viral-combined_k141_38222\|\|full | Low-quality |
| 17-108-final-viral-combined_k141_231071\|\|full | Not-determined |
| 17-108-final-viral-combined_k141_52162\|\|full | Not-determined |
| 17-108-final-viral-combined_k141_133410\|\|full | Not-determined |
| 17-108-final-viral-combined_k141_10625\|\|full | Low-quality |
| 17-108-final-viral-combined_k141_38224\|\|full | Not-determined |
| 17-108-final-viral-combined_k141_286267\|\|full | Low-quality |
| 17-108-final-viral-combined_k141_107524\|\|full | Low-quality |
| 17-108-final-viral-combined_k141_107526\|\|full | Low-quality |
| 17-108-final-viral-combined_k141_245413\|\|full | Low-quality |
| 17-108-final-viral-combined_k141_231084\|\|full | Low-quality |
| 17-108-final-viral-combined_k141_217300\|\|full | Not-determined |
| 17-108-final-viral-combined_k141_52209\|\|full | Not-determined |
| 17-108-final-viral-combined_k141_299497\|\|full | Low-quality |
| 17-108-final-viral-combined_k141_258961\|\|full | Not-determined |
| 17-108-final-viral-combined_k141_78703\|\|full | Low-quality |
| 17-108-final-viral-combined_k141_52230\|\|full | Not-determined |
| 17-108-final-viral-combined_k141_23003\|\|full | Not-determined |
| 17-108-final-viral-combined_k141_299510\|\|full | Low-quality |
| 17-108-final-viral-combined_k141_148271\|\|full | Low-quality |
| 17-108-final-viral-combined_k141_299519\|\|full | Not-determined |
| 17-108-final-viral-combined_k141_176128\|\|full | Low-quality |
| 17-108-final-viral-combined_k141_52253\|\|full | Low-quality |
| 17-108-final-viral-combined_k141_258998\|\|full | Not-determined |
| 17-108-final-viral-combined_k141_52256\|\|full | Not-determined |
| 17-108-final-viral-combined_k141_52257\|\|full | Not-determined |
| 17-108-final-viral-combined_k141_93167\|\|full | Low-quality |
| 17-108-final-viral-combined_k141_65801\|\|full | Not-determined |
| 17-108-final-viral-combined_k141_245452\|\|full | Low-quality |
| 17-108-final-viral-combined_k141_271908\|\|full | Low-quality |
| 17-108-final-viral-combined_k141_23019\|\|full | Not-determined |
| 17-108-final-viral-combined_k141_189167\|\|full | Low-quality |
| 17-108-final-viral-combined_k141_93196\|\|full | Not-determined |
| 17-108-final-viral-combined_k141_217307\|\|full | Low-quality |
| 17-108-final-viral-combined_k141_203323\|\|full | Not-determined |
| 17-108-final-viral-combined_k141_148305\|\|full | Low-quality |
| 17-108-final-viral-combined_k141_65829\|\|full | Not-determined |
| 17-108-final-viral-combined_k141_65833\|\|full | Low-quality |
| 17-108-final-viral-combined_k141_326164\|\|full | Not-determined |
| 17-108-final-viral-combined_k141_326167\|\|full | Low-quality |
| 17-108-final-viral-combined_k141_65836\|\|full | Not-determined |
| 17-108-final-viral-combined_k141_65840\|\|full | Low-quality |
| 17-108-final-viral-combined_k141_176182\|\|full | Not-determined |
| 17-108-final-viral-combined_k141_38308\|\|full | Not-determined |
| 17-108-final-viral-combined_k141_133539\|\|full | Not-determined |
| 17-108-final-viral-combined_k141_93237\|\|full | Low-quality |
| 17-108-final-viral-combined_k141_10708\|\|full | Low-quality |
| 17-108-final-viral-combined_k141_203351\|\|full | Low-quality |
| 17-108-final-viral-combined_k141_286378\|\|full | Not-determined |
| 17-108-final-viral-combined_k141_38314\|\|full | Low-quality |
| 17-108-final-viral-combined_k141_245522\|\|full | Low-quality |
| 17-108-final-viral-combined_k141_148345\|\|full | Not-determined |
| 17-108-final-viral-combined_k141_326184\|\|full | Not-determined |
| 17-108-final-viral-combined_k141_78790\|\|full | Not-determined |
| 17-108-final-viral-combined_k141_313961\|\|full | Low-quality |
| 17-108-final-viral-combined_k141_271973\|\|full | Low-quality |
| 17-108-final-viral-combined_k141_93262\|\|full | Not-determined |
| 17-108-final-viral-combined_k141_148351\|\|full | Not-determined |
| 17-108-final-viral-combined_k141_162310\|\|full | Low-quality |
| 17-108-final-viral-combined_k141_217372\|\|full | Not-determined |
| 17-108-final-viral-combined_k141_65883\|\|full | Low-quality |
| 17-108-final-viral-combined_k141_176210\|\|full | Not-determined |
| 17-108-final-viral-combined_k141_162328\|\|full | Not-determined |
| 17-108-final-viral-combined_k141_65899\|\|full | Not-determined |
| 17-108-final-viral-combined_k141_65907\|\|full | Low-quality |
| 17-108-final-viral-combined_k141_38329\|\|full | Low-quality |
| 17-108-final-viral-combined_k141_162352\|\|full | Not-determined |
| 17-108-final-viral-combined_k141_176248\|\|full | Low-quality |
| 17-108-final-viral-combined_k141_23122\|\|full | Low-quality |
| 17-108-final-viral-combined_k141_217434\|\|full | Not-determined |
| 17-108-final-viral-combined_k141_162360\|\|full | Not-determined |
| 17-108-final-viral-combined_k141_93325\|\|full | Not-determined |
| 17-108-final-viral-combined_k141_148412\|\|full | Not-determined |
| 17-108-final-viral-combined_k141_245608\|\|full | Not-determined |
| 17-108-final-viral-combined_k141_120059\|\|full | Not-determined |
| 17-108-final-viral-combined_k141_326255\|\|full | Not-determined |
| 17-108-final-viral-combined_k141_162382\|\|full | Low-quality |
| 17-108-final-viral-combined_k141_299603\|\|full | Not-determined |
| 17-108-final-viral-combined_k141_93354\|\|full | Not-determined |
| 17-108-final-viral-combined_k141_38405\|\|full | Low-quality |
| 17-108-final-viral-combined_k141_314041\|\|full | Low-quality |
| 17-108-final-viral-combined_k141_65965\|\|full | Low-quality |
| 17-108-final-viral-combined_k141_326268\|\|full | Low-quality |
| 17-108-final-viral-combined_k141_314054\|\|full | Not-determined |
| 17-108-final-viral-combined_k141_231269\|\|full | Low-quality |
| 17-108-final-viral-combined_k141_107744\|\|full | Low-quality |
| 17-108-final-viral-combined_k141_176303\|\|full | Low-quality |
| 17-108-final-viral-combined_k141_65983\|\|full | Not-determined |
| 17-108-final-viral-combined_k141_299635\|\|full | Not-determined |
| 17-108-final-viral-combined_k141_314066\|\|full | Not-determined |
| 17-108-final-viral-combined_k141_162437\|\|full | Low-quality |
| 17-108-final-viral-combined_k141_299637\|\|full | Low-quality |
| 17-108-final-viral-combined_k141_93388\|\|full | Not-determined |
| 17-108-final-viral-combined_k141_326277\|\|full | Low-quality |
| 17-108-final-viral-combined_k141_23173\|\|full | Low-quality |
| 17-108-final-viral-combined_k141_272030\|\|full | Low-quality |
| 17-108-final-viral-combined_k141_314082\|\|full | Low-quality |
| 17-108-final-viral-combined_k141_176322\|\|full | Not-determined |
| 17-108-final-viral-combined_k141_217499\|\|full | Not-determined |
| 17-108-final-viral-combined_k141_176324\|\|full | Not-determined |
| 17-108-final-viral-combined_k141_120114\|\|full | Low-quality |
| 17-108-final-viral-combined_k141_176329\|\|full | Low-quality |
| 17-108-final-viral-combined_k141_10780\|\|full | Not-determined |
| 17-108-final-viral-combined_k141_107756\|\|full | Low-quality |
| 17-108-final-viral-combined_k141_120122\|\|full | Low-quality |
| 17-108-final-viral-combined_k141_217522\|\|full | Not-determined |
| 17-108-final-viral-combined_k141_162465\|\|full | Low-quality |
| 17-108-final-viral-combined_k141_217532\|\|full | Low-quality |
| 17-108-final-viral-combined_k141_10792\|\|full | Not-determined |
| 17-108-final-viral-combined_k141_286527\|\|full | Not-determined |
| 17-108-final-viral-combined_k141_148488\|\|full | Low-quality |
| 17-108-final-viral-combined_k141_162474\|\|full | Not-determined |
| 17-108-final-viral-combined_k141_10801\|\|full | Not-determined |
| 17-108-final-viral-combined_k141_66052\|\|full | Not-determined |
| 17-108-final-viral-combined_k141_217552\|\|full | Low-quality |
| 17-108-final-viral-combined_k141_107797\|\|full | Low-quality |
| 17-108-final-viral-combined_k141_10837\|\|full | Not-determined |
| 17-108-final-viral-combined_k141_217555\|\|full | Not-determined |
| 17-108-final-viral-combined_k141_176381\|\|full | Low-quality |
| 17-108-final-viral-combined_k141_38483\|\|full | Low-quality |
| 17-108-final-viral-combined_k141_217557\|\|full | Not-determined |
| 18-102-final-viral-combined_k141_62519\|\|full | Medium-quality |
| 17-108-final-viral-combined_k141_107810\|\|full | Low-quality |
| 17-108-final-viral-combined_k141_23251\|\|full | Low-quality |
| 17-108-final-viral-combined_k141_217568\|\|full | Not-determined |
| 17-108-final-viral-combined_k141_93469\|\|full | Low-quality |
| 17-108-final-viral-combined_k141_259209\|\|full | Not-determined |
| 17-108-final-viral-combined_k141_286549\|\|full | Not-determined |
| 17-108-final-viral-combined_k141_314146\|\|full | Low-quality |
| 17-108-final-viral-combined_k141_52430\|\|full | Low-quality |
| 17-108-final-viral-combined_k141_52431\|\|full | Low-quality |
| 17-108-final-viral-combined_k141_176392\|\|full | Not-determined |
| 17-108-final-viral-combined_k141_78874\|\|full | Low-quality |
| 17-108-final-viral-combined_k141_286564\|\|full | Low-quality |
| 17-108-final-viral-combined_k141_148535\|\|full | Not-determined |
| 17-108-final-viral-combined_k141_66105\|\|full | Low-quality |
| 17-108-final-viral-combined_k141_52442\|\|full | Low-quality |
| 17-108-final-viral-combined_k141_299741\|\|full | Low-quality |
| 17-108-final-viral-combined_k141_133808\|\|full | Low-quality |
| 17-108-final-viral-combined_k141_245690\|\|full | Low-quality |
| 17-108-final-viral-combined_k141_314161\|\|full | Not-determined |
| 17-108-final-viral-combined_k141_245701\|\|full | Not-determined |
| 17-108-final-viral-combined_k141_133819\|\|full | Low-quality |
| 17-108-final-viral-combined_k141_259223\|\|full | Low-quality |
| 17-108-final-viral-combined_k141_272097\|\|full | Low-quality |
| 17-108-final-viral-combined_k141_148553\|\|full | Not-determined |
| 17-108-final-viral-combined_k141_272098\|\|full | Not-determined |
| 17-108-final-viral-combined_k141_133825\|\|full | Low-quality |
| 17-108-final-viral-combined_k141_326343\|\|full | Low-quality |
| 17-108-final-viral-combined_k141_272099\|\|full | Low-quality |
| 17-108-final-viral-combined_k141_52476\|\|full | Not-determined |
| 17-108-final-viral-combined_k141_314165\|\|full | Low-quality |
| 17-108-final-viral-combined_k141_299763\|\|full | Low-quality |
| 17-108-final-viral-combined_k141_299767\|\|full | Low-quality |
| 17-108-final-viral-combined_k141_245733\|\|full | Low-quality |
| 17-108-final-viral-combined_k141_245739\|\|full | Low-quality |
| 17-108-final-viral-combined_k141_189352\|\|full | Low-quality |
| 17-108-final-viral-combined_k141_326367\|\|full | Not-determined |
| 17-108-final-viral-combined_k141_314190\|\|full | Low-quality |
| 17-108-final-viral-combined_k141_259263\|\|full | Not-determined |
| 17-108-final-viral-combined_k141_78924\|\|full | Low-quality |
| 17-108-final-viral-combined_k141_314201\|\|full | Low-quality |
| 17-108-final-viral-combined_k141_133857\|\|full | Low-quality |
| 17-108-final-viral-combined_k141_10923\|\|full | Not-determined |
| 17-108-final-viral-combined_k141_10925\|\|full | Low-quality |
| 17-108-final-viral-combined_k141_189365\|\|full | Low-quality |
| 17-108-final-viral-combined_k141_272134\|\|full | Low-quality |
| 17-108-final-viral-combined_k141_107873\|\|full | Not-determined |
| 17-108-final-viral-combined_k141_120212\|\|full | Not-determined |
| 17-108-final-viral-combined_k141_23321\|\|full | Low-quality |
| 17-108-final-viral-combined_k141_299804\|\|full | Not-determined |
| 17-108-final-viral-combined_k141_107876\|\|full | Low-quality |
| 17-108-final-viral-combined_k141_299807\|\|full | Not-determined |
| 17-108-final-viral-combined_k141_272148\|\|full | Low-quality |
| 17-108-final-viral-combined_k141_299810\|\|full | Not-determined |
| 17-108-final-viral-combined_k141_107884\|\|full | Low-quality |
| 17-108-final-viral-combined_k141_162604\|\|full | Low-quality |
| 17-108-final-viral-combined_k141_93568\|\|full | Low-quality |
| 17-108-final-viral-combined_k141_38541\|\|full | Low-quality |
| 17-108-final-viral-combined_k141_299818\|\|full | Low-quality |
| 17-108-final-viral-combined_k141_148610\|\|full | Low-quality |
| 17-108-final-viral-combined_k141_176417\|\|full | Low-quality |
| 17-108-final-viral-combined_k141_52564\|\|full | Low-quality |
| 17-108-final-viral-combined_k141_326427\|\|full | Low-quality |
| 17-108-final-viral-combined_k141_299828\|\|full | Not-determined |
| 17-108-final-viral-combined_k141_52569\|\|full | Low-quality |
| 17-108-final-viral-combined_k141_299830\|\|full | Not-determined |
| 17-108-final-viral-combined_k141_259323\|\|full | Not-determined |
| 17-108-final-viral-combined_k141_120215\|\|full | Low-quality |
| 17-108-final-viral-combined_k141_148618\|\|full | Not-determined |
| 17-108-final-viral-combined_k141_272177\|\|full | Not-determined |
| 17-108-final-viral-combined_k141_38556\|\|full | Not-determined |
| 17-108-final-viral-combined_k141_107913\|\|full | Low-quality |
| 17-108-final-viral-combined_k141_286663\|\|full | Low-quality |
| 17-108-final-viral-combined_k141_259341\|\|full | Low-quality |
| 17-108-final-viral-combined_k141_133942\|\|full | Not-determined |
| 17-108-final-viral-combined_k141_79002\|\|full | Not-determined |
| 17-108-final-viral-combined_k141_11005\|\|full | Low-quality |
| 17-108-final-viral-combined_k141_38576\|\|full | Not-determined |
| 17-108-final-viral-combined_k141_120242\|\|full | Not-determined |
| 17-108-final-viral-combined_k141_52586\|\|full | Not-determined |
| 17-108-final-viral-combined_k141_107940\|\|full | Low-quality |
| 17-108-final-viral-combined_k141_162624\|\|full | Not-determined |
| 17-108-final-viral-combined_k141_286692\|\|full | Not-determined |
| 17-108-final-viral-combined_k141_299858\|\|full | Not-determined |
| 17-108-final-viral-combined_k141_231430\|\|full | Not-determined |
| 17-108-final-viral-combined_k141_23375\|\|full | Low-quality |
| 17-108-final-viral-combined_k141_326483\|\|full | Not-determined |
| 17-108-final-viral-combined_k141_38609\|\|full | Low-quality |
| 17-108-final-viral-combined_k141_314214\|\|full | Not-determined |
| 17-108-final-viral-combined_k141_314220\|\|full | Not-determined |
| 17-108-final-viral-combined_k141_314223\|\|full | Low-quality |
| 17-108-final-viral-combined_k141_52634\|\|full | Low-quality |
| 17-108-final-viral-combined_k141_11048\|\|full | Low-quality |
| 17-108-final-viral-combined_k141_52635\|\|full | Low-quality |
| 17-108-final-viral-combined_k141_259371\|\|full | Low-quality |
| 17-108-final-viral-combined_k141_38625\|\|full | Not-determined |
| 17-108-final-viral-combined_k141_217681\|\|full | Not-determined |
| 18-105-final-viral-combined_k141_26971\|\|0_partial | Complete |
| 17-108-final-viral-combined_k141_272238\|\|full | Not-determined |
| 17-108-final-viral-combined_k141_176502\|\|full | Low-quality |
| 17-108-final-viral-combined_k141_23388\|\|full | Low-quality |
| 17-108-final-viral-combined_k141_93654\|\|full | Low-quality |
| 17-108-final-viral-combined_k141_314239\|\|full | Low-quality |
| 17-108-final-viral-combined_k141_79075\|\|full | Not-determined |
| 17-108-final-viral-combined_k141_231498\|\|full | Low-quality |
| 17-108-final-viral-combined_k141_314244\|\|full | Not-determined |
| 17-108-final-viral-combined_k141_203640\|\|full | Low-quality |
| 17-108-final-viral-combined_k141_162742\|\|full | Not-determined |
| 17-108-final-viral-combined_k141_52713\|\|full | Not-determined |
| 17-108-final-viral-combined_k141_286729\|\|full | Low-quality |
| 17-108-final-viral-combined_k141_52717\|\|full | Low-quality |
| 17-108-final-viral-combined_k141_52722\|\|full | Not-determined |
| 17-108-final-viral-combined_k141_231533\|\|full | Not-determined |
| 17-108-final-viral-combined_k141_162772\|\|full | Low-quality |
| 17-108-final-viral-combined_k141_23419\|\|full | Low-quality |
| 17-108-final-viral-combined_k141_120303\|\|full | Low-quality |
| 17-108-final-viral-combined_k141_231550\|\|full | Low-quality |
| 17-108-final-viral-combined_k141_11116\|\|full | Not-determined |
| 17-108-final-viral-combined_k141_176555\|\|full | Low-quality |
| 17-109-final-viral-combined_k141_189323\|\|full | Medium-quality |
| 17-108-final-viral-combined_k141_314271\|\|full | Not-determined |
| 17-108-final-viral-combined_k141_286765\|\|full | Not-determined |
| 17-108-final-viral-combined_k141_203705\|\|full | Low-quality |
| 17-108-final-viral-combined_k141_286769\|\|full | Low-quality |
| 17-108-final-viral-combined_k141_217715\|\|full | Low-quality |
| 17-108-final-viral-combined_k141_286772\|\|full | Low-quality |
| 17-108-final-viral-combined_k141_176572\|\|full | Not-determined |
| 17-108-final-viral-combined_k141_176586\|\|full | Not-determined |
| 17-108-final-viral-combined_k141_162800\|\|full | Not-determined |
| 17-108-final-viral-combined_k141_314292\|\|full | Not-determined |
| 17-108-final-viral-combined_k141_52786\|\|full | Low-quality |
| 17-108-final-viral-combined_k141_148684\|\|full | Low-quality |
| 17-108-final-viral-combined_k141_23474\|\|full | Not-determined |
| 17-108-final-viral-combined_k141_38740\|\|full | Low-quality |
| 17-108-final-viral-combined_k141_231594\|\|full | Low-quality |
| 17-108-final-viral-combined_k141_79145\|\|full | Low-quality |
| 17-108-final-viral-combined_k141_272304\|\|full | Low-quality |
| 17-108-final-viral-combined_k141_245933\|\|full | Low-quality |
| 17-108-final-viral-combined_k141_66244\|\|full | Low-quality |
| 17-108-final-viral-combined_k141_108043\|\|full | Low-quality |
| 17-108-final-viral-combined_k141_272317\|\|full | Low-quality |
| 17-108-final-viral-combined_k141_23487\|\|full | Not-determined |
| 17-108-final-viral-combined_k141_259459\|\|full | Low-quality |
| 17-108-final-viral-combined_k141_272320\|\|full | Not-determined |
| 17-108-final-viral-combined_k141_162850\|\|full | Not-determined |
| 17-108-final-viral-combined_k141_300027\|\|full | Low-quality |
| 17-108-final-viral-combined_k141_38790\|\|full | Low-quality |
| 17-108-final-viral-combined_k141_231640\|\|full | Not-determined |
| 17-108-final-viral-combined_k141_272332\|\|full | Low-quality |
| 17-108-final-viral-combined_k141_176636\|\|full | Not-determined |
| 17-108-final-viral-combined_k141_162851\|\|full | Low-quality |
| 17-108-final-viral-combined_k141_314324\|\|full | Low-quality |
| 17-108-final-viral-combined_k141_326547\|\|full | Low-quality |
| 17-108-final-viral-combined_k141_23496\|\|full | Not-determined |
| 17-108-final-viral-combined_k141_134140\|\|full | Low-quality |
| 17-108-final-viral-combined_k141_245984\|\|full | Low-quality |
| 17-108-final-viral-combined_k141_120395\|\|full | Not-determined |
| 17-108-final-viral-combined_k141_108088\|\|full | Not-determined |
| 17-108-final-viral-combined_k141_108090\|\|full | Low-quality |
| 17-108-final-viral-combined_k141_120406\|\|full | Low-quality |
| 17-108-final-viral-combined_k141_108095\|\|full | Low-quality |
| 17-108-final-viral-combined_k141_286832\|\|full | Low-quality |
| 17-108-final-viral-combined_k141_259498\|\|full | Low-quality |
| 17-108-final-viral-combined_k141_231681\|\|full | Not-determined |
| 17-108-final-viral-combined_k141_52861\|\|full | Not-determined |
| 17-108-final-viral-combined_k141_246009\|\|full | Low-quality |
| 17-108-final-viral-combined_k141_11200\|\|full | Not-determined |
| 17-108-final-viral-combined_k141_217756\|\|full | Low-quality |
| 17-108-final-viral-combined_k141_326590\|\|full | Not-determined |
| 17-108-final-viral-combined_k141_203779\|\|full | Not-determined |
| 17-108-final-viral-combined_k141_272356\|\|full | Low-quality |
| 17-108-final-viral-combined_k141_231712\|\|full | Not-determined |
| 17-108-final-viral-combined_k141_259525\|\|full | Low-quality |
| 17-108-final-viral-combined_k141_259526\|\|full | Not-determined |
| 17-108-final-viral-combined_k141_272374\|\|full | Low-quality |
| 17-108-final-viral-combined_k141_23546\|\|full | Not-determined |
| 17-108-final-viral-combined_k141_79238\|\|full | Not-determined |
| 17-108-final-viral-combined_k141_162916\|\|full | Low-quality |
| 17-108-final-viral-combined_k141_38858\|\|full | Low-quality |
| 17-108-final-viral-combined_k141_108122\|\|full | Low-quality |
| 17-108-final-viral-combined_k141_203828\|\|full | Low-quality |
| 17-108-final-viral-combined_k141_120477\|\|full | Low-quality |
| 17-108-final-viral-combined_k141_326648\|\|full | Not-determined |
| 17-108-final-viral-combined_k141_203843\|\|full | Low-quality |
| 17-108-final-viral-combined_k141_231733\|\|full | Low-quality |
| 17-108-final-viral-combined_k141_326657\|\|full | Low-quality |
| 17-108-final-viral-combined_k141_259598\|\|full | Low-quality |
| 17-108-final-viral-combined_k141_272392\|\|full | Not-determined |
| 17-108-final-viral-combined_k141_108155\|\|full | Not-determined |
| 17-108-final-viral-combined_k141_300134\|\|full | Low-quality |
| 17-108-final-viral-combined_k141_231740\|\|full | Low-quality |
| 17-108-final-viral-combined_k141_189634\|\|full | Low-quality |
| 17-108-final-viral-combined_k141_79302\|\|full | Not-determined |
| 17-108-final-viral-combined_k141_108180\|\|full | Not-determined |
| 17-108-final-viral-combined_k141_38888\|\|full | Low-quality |
| 18-104-final-viral-combined_k141_10758\|\|full | Medium-quality |
| 17-108-final-viral-combined_k141_79313\|\|full | Low-quality |
| 17-108-final-viral-combined_k141_314386\|\|full | Low-quality |
| 17-108-final-viral-combined_k141_38900\|\|full | Low-quality |
| 17-108-final-viral-combined_k141_66376\|\|full | Low-quality |
| 17-108-final-viral-combined_k141_326684\|\|full | Low-quality |
| 17-108-final-viral-combined_k141_79315\|\|full | Low-quality |
| 17-108-final-viral-combined_k141_120510\|\|full | Low-quality |
| 17-108-final-viral-combined_k141_148887\|\|full | Not-determined |
| 17-108-final-viral-combined_k141_300173\|\|full | Not-determined |
| 17-108-final-viral-combined_k141_120513\|\|full | Low-quality |
| 17-108-final-viral-combined_k141_189673\|\|full | Low-quality |
| 17-108-final-viral-combined_k141_38922\|\|full | Low-quality |
| 17-108-final-viral-combined_k141_79340\|\|full | Low-quality |
| 17-108-final-viral-combined_k141_38926\|\|full | Not-determined |
| 17-108-final-viral-combined_k141_11293\|\|full | Not-determined |
| 17-108-final-viral-combined_k141_203930\|\|full | Low-quality |
| 17-108-final-viral-combined_k141_79354\|\|full | Low-quality |
| 17-108-final-viral-combined_k141_66410\|\|full | Not-determined |
| 17-108-final-viral-combined_k141_259619\|\|full | Not-determined |
| 17-108-final-viral-combined_k141_300220\|\|full | Not-determined |
| 17-108-final-viral-combined_k141_259629\|\|full | Low-quality |
| 17-108-final-viral-combined_k141_93844\|\|full | Not-determined |
| 17-108-final-viral-combined_k141_300241\|\|full | Low-quality |
| 17-108-final-viral-combined_k141_231847\|\|full | Not-determined |
| 17-108-final-viral-combined_k141_52991\|\|full | Not-determined |
| 17-108-final-viral-combined_k141_326714\|\|full | Low-quality |
| 17-108-final-viral-combined_k141_79383\|\|full | Low-quality |
| 17-108-final-viral-combined_k141_66463\|\|full | Low-quality |
| 17-108-final-viral-combined_k141_134225\|\|full | Not-determined |
| 17-108-final-viral-combined_k141_246160\|\|full | Not-determined |
| 17-108-final-viral-combined_k141_246168\|\|full | Low-quality |
| 17-108-final-viral-combined_k141_148947\|\|full | Low-quality |
| 17-108-final-viral-combined_k141_53015\|\|full | Low-quality |
| 17-108-final-viral-combined_k141_66488\|\|full | Not-determined |
| 17-108-final-viral-combined_k141_148959\|\|full | Low-quality |
| 17-108-final-viral-combined_k141_287069\|\|full | Not-determined |
| 17-108-final-viral-combined_k141_272572\|\|full | Not-determined |
| 17-108-final-viral-combined_k141_189701\|\|full | Not-determined |
| 17-108-final-viral-combined_k141_326764\|\|full | Low-quality |
| 17-108-final-viral-combined_k141_246210\|\|full | Not-determined |
| 17-108-final-viral-combined_k141_134265\|\|full | Not-determined |
| 17-108-final-viral-combined_k141_163036\|\|full | Not-determined |
| 17-108-final-viral-combined_k141_259701\|\|full | Low-quality |
| 17-108-final-viral-combined_k141_148982\|\|full | Low-quality |
| 17-108-final-viral-combined_k141_93898\|\|full | Not-determined |
| 17-108-final-viral-combined_k141_189723\|\|full | Not-determined |
| 17-108-final-viral-combined_k141_66553\|\|full | Low-quality |
| 17-108-final-viral-combined_k141_66558\|\|full | Not-determined |
| 17-108-final-viral-combined_k141_23676\|\|full | Not-determined |
| 17-108-final-viral-combined_k141_163068\|\|full | Low-quality |
| 17-108-final-viral-combined_k141_300270\|\|full | Not-determined |
| 17-108-final-viral-combined_k141_120632\|\|full | Not-determined |
| 17-108-final-viral-combined_k141_300272\|\|full | Low-quality |
| 17-108-final-viral-combined_k141_300273\|\|full | Low-quality |
| 17-108-final-viral-combined_k141_189731\|\|full | Not-determined |
| 17-108-final-viral-combined_k141_149002\|\|full | Low-quality |
| 17-108-final-viral-combined_k141_300283\|\|full | Low-quality |
| 17-108-final-viral-combined_k141_66568\|\|full | Low-quality |
| 17-108-final-viral-combined_k141_259765\|\|full | Not-determined |
| 17-108-final-viral-combined_k141_259776\|\|full | Not-determined |
| 17-108-final-viral-combined_k141_38992\|\|full | Not-determined |
| 17-108-final-viral-combined_k141_39002\|\|full | Low-quality |
| 17-108-final-viral-combined_k141_93918\|\|full | Low-quality |
| 17-108-final-viral-combined_k141_53164\|\|full | Low-quality |
| 17-108-final-viral-combined_k141_39009\|\|full | Not-determined |
| 17-108-final-viral-combined_k141_314436\|\|full | Not-determined |
| 17-108-final-viral-combined_k141_93928\|\|full | Not-determined |
| 17-108-final-viral-combined_k141_23772\|\|full | Low-quality |
| 17-108-final-viral-combined_k141_287144\|\|full | Low-quality |
| 17-108-final-viral-combined_k141_287145\|\|full | Low-quality |
| 17-108-final-viral-combined_k141_189779\|\|full | Not-determined |
| 17-108-final-viral-combined_k141_326853\|\|full | Low-quality |
| 17-108-final-viral-combined_k141_11390\|\|full | Low-quality |
| 17-108-final-viral-combined_k141_120710\|\|full | Not-determined |
| 17-108-final-viral-combined_k141_11400\|\|full | Low-quality |
| 17-108-final-viral-combined_k141_53197\|\|full | Low-quality |
| 17-108-final-viral-combined_k141_287161\|\|full | Low-quality |
| 17-108-final-viral-combined_k141_300385\|\|full | Low-quality |
| 17-108-final-viral-combined_k141_272702\|\|full | Not-determined |
| 17-108-final-viral-combined_k141_53200\|\|full | Not-determined |
| 17-108-final-viral-combined_k141_120734\|\|full | Not-determined |
| 17-108-final-viral-combined_k141_149038\|\|full | Not-determined |
| 17-108-final-viral-combined_k141_272710\|\|full | Low-quality |
| 17-108-final-viral-combined_k141_231969\|\|full | Low-quality |
| 17-108-final-viral-combined_k141_39068\|\|full | Not-determined |
| 17-108-final-viral-combined_k141_314483\|\|full | Low-quality |
| 17-108-final-viral-combined_k141_217901\|\|full | Low-quality |
| 17-108-final-viral-combined_k141_217907\|\|full | Low-quality |
| 17-108-final-viral-combined_k141_246385\|\|full | Not-determined |
| 17-108-final-viral-combined_k141_23817\|\|full | Low-quality |
| 17-108-final-viral-combined_k141_11421\|\|full | Not-determined |
| 17-108-final-viral-combined_k141_326904\|\|full | Not-determined |
| 17-108-final-viral-combined_k141_11424\|\|full | Not-determined |
| 17-108-final-viral-combined_k141_134424\|\|full | Not-determined |
| 17-108-final-viral-combined_k141_189844\|\|full | Not-determined |
| 17-108-final-viral-combined_k141_326909\|\|full | Not-determined |
| 17-108-final-viral-combined_k141_11434\|\|full | Low-quality |
| 17-108-final-viral-combined_k141_39098\|\|full | Not-determined |
| 17-108-final-viral-combined_k141_149131\|\|full | Low-quality |
| 17-108-final-viral-combined_k141_176980\|\|full | Low-quality |
| 17-108-final-viral-combined_k141_287220\|\|full | Not-determined |
| 17-108-final-viral-combined_k141_149135\|\|full | Low-quality |
| 17-108-final-viral-combined_k141_134462\|\|full | Not-determined |
| 17-108-final-viral-combined_k141_300466\|\|full | Not-determined |
| 17-108-final-viral-combined_k141_259849\|\|full | Low-quality |
| 17-108-final-viral-combined_k141_176995\|\|full | Not-determined |
| 17-108-final-viral-combined_k141_120779\|\|full | Low-quality |
| 17-108-final-viral-combined_k141_66735\|\|full | Not-determined |
| 17-108-final-viral-combined_k141_314528\|\|full | Not-determined |
| 17-108-final-viral-combined_k141_189869\|\|full | Not-determined |
| 17-108-final-viral-combined_k141_232051\|\|full | Not-determined |
| 17-108-final-viral-combined_k141_108513\|\|full | Not-determined |
| 17-108-final-viral-combined_k141_120785\|\|full | Not-determined |
| 17-108-final-viral-combined_k141_108514\|\|full | Not-determined |
| 17-108-final-viral-combined_k141_134484\|\|full | Low-quality |
| 17-108-final-viral-combined_k141_232059\|\|full | Low-quality |
| 17-108-final-viral-combined_k141_177022\|\|full | Not-determined |
| 17-108-final-viral-combined_k141_39149\|\|full | Not-determined |
| 17-108-final-viral-combined_k141_287258\|\|full | Low-quality |
| 17-108-final-viral-combined_k141_66745\|\|full | Low-quality |
| 17-108-final-viral-combined_k141_23862\|\|full | Not-determined |
| 17-108-final-viral-combined_k141_326963\|\|full | Low-quality |
| 17-108-final-viral-combined_k141_79622\|\|full | Low-quality |
| 17-108-final-viral-combined_k141_246462\|\|full | Low-quality |
| 17-108-final-viral-combined_k141_287277\|\|full | Low-quality |
| 17-108-final-viral-combined_k141_23872\|\|full | Low-quality |
| 17-108-final-viral-combined_k141_53260\|\|full | Low-quality |
| 17-108-final-viral-combined_k141_287278\|\|full | Low-quality |
| 17-108-final-viral-combined_k141_66763\|\|full | Low-quality |
| 17-108-final-viral-combined_k141_149186\|\|full | Not-determined |
| 17-108-final-viral-combined_k141_134527\|\|full | Low-quality |
| 17-108-final-viral-combined_k141_149188\|\|full | Not-determined |
| 17-108-final-viral-combined_k141_177049\|\|full | Low-quality |
| 17-108-final-viral-combined_k141_326980\|\|full | Low-quality |
| 17-108-final-viral-combined_k141_134535\|\|full | Low-quality |
| 17-108-final-viral-combined_k141_79664\|\|full | Low-quality |
| 17-108-final-viral-combined_k141_134545\|\|full | Not-determined |
| 17-108-final-viral-combined_k141_39208\|\|full | Low-quality |
| 17-108-final-viral-combined_k141_272856\|\|full | Not-determined |
| 17-108-final-viral-combined_k141_232097\|\|full | Low-quality |
| 17-108-final-viral-combined_k141_163285\|\|full | Low-quality |
| 17-108-final-viral-combined_k141_300528\|\|full | Not-determined |
| 17-108-final-viral-combined_k141_163296\|\|full | Low-quality |
| 17-108-final-viral-combined_k141_11547\|\|full | Low-quality |
| 17-108-final-viral-combined_k141_246530\|\|full | Low-quality |
| 17-108-final-viral-combined_k141_300551\|\|full | Low-quality |
| 17-108-final-viral-combined_k141_134599\|\|full | Low-quality |
| 17-108-final-viral-combined_k141_11572\|\|full | Not-determined |
| 17-108-final-viral-combined_k141_79734\|\|full | Low-quality |
| 17-108-final-viral-combined_k141_149267\|\|full | Not-determined |
| 17-108-final-viral-combined_k141_177114\|\|full | Not-determined |
| 17-108-final-viral-combined_k141_189912\|\|full | Low-quality |
| 17-108-final-viral-combined_k141_163336\|\|full | Not-determined |
| 17-108-final-viral-combined_k141_327041\|\|full | Low-quality |
| 17-108-final-viral-combined_k141_23942\|\|full | Not-determined |
| 17-108-final-viral-combined_k141_327045\|\|full | Low-quality |
| 17-108-final-viral-combined_k141_163338\|\|full | Not-determined |
| 17-108-final-viral-combined_k141_177123\|\|full | Not-determined |
| 17-108-final-viral-combined_k141_149277\|\|full | Low-quality |
| 17-108-final-viral-combined_k141_23949\|\|full | Not-determined |
| 17-108-final-viral-combined_k141_39279\|\|full | Not-determined |
| 17-108-final-viral-combined_k141_246599\|\|full | Low-quality |
| 17-108-final-viral-combined_k141_53292\|\|full | Not-determined |
| 17-108-final-viral-combined_k141_79765\|\|full | Not-determined |
| 17-108-final-viral-combined_k141_149294\|\|full | Low-quality |
| 17-108-final-viral-combined_k141_189938\|\|full | Not-determined |
| 17-108-final-viral-combined_k141_120946\|\|full | Not-determined |
| 17-108-final-viral-combined_k141_287347\|\|full | Not-determined |
| 17-108-final-viral-combined_k141_246608\|\|full | Not-determined |
| 17-108-final-viral-combined_k141_287352\|\|full | Low-quality |
| 17-108-final-viral-combined_k141_39310\|\|full | Not-determined |
| 17-108-final-viral-combined_k141_246626\|\|full | Low-quality |
| 17-108-final-viral-combined_k141_79794\|\|full | Not-determined |
| 17-108-final-viral-combined_k141_189953\|\|full | Low-quality |

17-108-final-viral-combined_k141_149316||full Low-quality
17-108-final-viral-combined_k141_246629||full Low-quality
17-108-final-viral-combined_k141_327073||full Low-quality
17-108-final-viral-combined_k141_11617||full Not-determined
17-108-final-viral-combined_k141_189966||full Not-determined
17-108-final-viral-combined_k141_287373||full Low-quality
17-108-final-viral-combined_k141_218090||full Not-determined
17-108-final-viral-combined_k141_314598||full Low-quality
17-108-final-viral-combined_k141_177167||full Not-determined
17-108-final-viral-combined_k141_204202||full Not-determined
17-108-final-viral-combined_k141_314613||full Not-determined
17-108-final-viral-combined_k141_190000||full Not-determined
17-108-final-viral-combined_k141_204235||full Low-quality
17-108-final-viral-combined_k141_314635||full Not-determined
17-108-final-viral-combined_k141_66873||full Not-determined
17-108-final-viral-combined_k141_108744||full Not-determined
17-108-final-viral-combined_k141_287393||full Not-determined
17-108-final-viral-combined_k141_314653||full Low-quality
17-108-final-viral-combined_k141_108761||full Low-quality
17-108-final-viral-combined_k141_24088||full Not-determined
17-108-final-viral-combined_k141_11724||full Low-quality
17-108-final-viral-combined_k141_163455||full Low-quality
17-108-final-viral-combined_k141_177232||full Not-determined
17-108-final-viral-combined_k141_11747||full Not-determined
17-108-final-viral-combined_k141_121054||full Low-quality
17-108-final-viral-combined_k141_163470||full Not-determined
17-108-final-viral-combined_k141_218194||full Low-quality
17-108-final-viral-combined_k141_287418||full Low-quality
17-108-final-viral-combined_k141_246741||full Low-quality
17-108-final-viral-combined_k141_39438||full Low-quality
17-108-final-viral-combined_k141_53434||full Not-determined
17-108-final-viral-combined_k141_190052||full Not-determined
17-108-final-viral-combined_k141_11761||full Low-quality
17-108-final-viral-combined_k141_272980||full Low-quality
17-108-final-viral-combined_k141_204299||full Low-quality
17-108-final-viral-combined_k141_11769||full Not-determined
17-108-final-viral-combined_k141_94051||full Low-quality
17-108-final-viral-combined_k141_287438||full Not-determined
17-108-final-viral-combined_k141_177264||full Low-quality
17-108-final-viral-combined_k141_204339||full Low-quality
17-108-final-viral-combined_k141_149463||full Not-determined
17-108-final-viral-combined_k141_300632||full Not-determined
17-108-final-viral-combined_k141_190096||full Not-determined
17-108-final-viral-combined_k141_218204||full Low-quality
17-108-final-viral-combined_k141_163520||full Not-determined
17-108-final-viral-combined_k141_53487||full Not-determined
17-108-final-viral-combined_k141_134772||full Not-determined
17-108-final-viral-combined_k141_11796||full Low-quality
17-108-final-viral-combined_k141_24156||full Not-determined
17-108-final-viral-combined_k141_204350||full Low-quality
17-108-final-viral-combined_k141_287476||full Not-determined
17-108-final-viral-combined_k141_232300||full Low-quality
17-108-final-viral-combined_k141_11820||full Low-quality
17-108-final-viral-combined_k141_232305||full Low-quality
17-108-final-viral-combined_k141_79898||full Low-quality
17-108-final-viral-combined_k141_204362||full Not-determined
17-108-final-viral-combined_k141_177314||full Low-quality
17-108-final-viral-combined_k141_94135||full Not-determined
17-108-final-viral-combined_k141_108853||full Low-quality
17-108-final-viral-combined_k141_287511||full Not-determined
17-108-final-viral-combined_k141_232342||full Not-determined
17-108-final-viral-combined_k141_260133||full Low-quality
17-108-final-viral-combined_k141_300689||full Low-quality
17-108-final-viral-combined_k141_287534||full Low-quality
17-108-final-viral-combined_k141_94158||full Low-quality
17-108-final-viral-combined_k141_273084||full Not-determined
17-108-final-viral-combined_k141_149549||full Not-determined
17-108-final-viral-combined_k141_300700||full Low-quality
17-108-final-viral-combined_k141_327235||full Not-determined
17-108-final-viral-combined_k141_24236||full Low-quality
17-108-final-viral-combined_k141_79978||full Not-determined
17-108-final-viral-combined_k141_67020||full Not-determined
17-108-final-viral-combined_k141_218247||full Not-determined
17-108-final-viral-combined_k141_177347||full Not-determined
17-108-final-viral-combined_k141_287555||full Not-determined
17-108-final-viral-combined_k141_190155||full Low-quality
17-108-final-viral-combined_k141_149563||full Not-determined
17-108-final-viral-combined_k141_273104||full Low-quality
17-108-final-viral-combined_k141_11859||full Not-determined
17-108-final-viral-combined_k141_24238||full Low-quality
17-108-final-viral-combined_k141_218279||full Not-determined
17-108-final-viral-combined_k141_218283||full Not-determined
17-108-final-viral-combined_k141_149569||full Not-determined
17-108-final-viral-combined_k141_300761||full Not-determined
17-108-final-viral-combined_k141_232358||full Not-determined
17-108-final-viral-combined_k141_300762||full Not-determined
17-108-final-viral-combined_k141_67083||full Not-determined
17-108-final-viral-combined_k141_246810||full Low-quality
17-108-final-viral-combined_k141_149573||full Not-determined
17-108-final-viral-combined_k141_190199||full Low-quality
17-108-final-viral-combined_k141_94206||full Not-determined
17-108-final-viral-combined_k141_24261||full Low-quality
17-108-final-viral-combined_k141_218294||full Not-determined
17-108-final-viral-combined_k141_232362||full Not-determined
17-108-final-viral-combined_k141_53561||full Low-quality
17-108-final-viral-combined_k141_218301||full Not-determined
17-108-final-viral-combined_k141_300777||full Not-determined
17-108-final-viral-combined_k141_24265||full Low-quality
17-108-final-viral-combined_k141_80014||full Low-quality
17-108-final-viral-combined_k141_218310||full Low-quality
17-108-final-viral-combined_k141_300783||full Low-quality
17-108-final-viral-combined_k141_300785||full Low-quality
17-108-final-viral-combined_k141_232381||full Not-determined
17-108-final-viral-combined_k141_204467||full Low-quality
17-108-final-viral-combined_k141_39616||full Not-determined
17-108-final-viral-combined_k141_300795||full Not-determined
17-108-final-viral-combined_k141_218314||full Not-determined
17-108-final-viral-combined_k141_67111||full Not-determined
17-108-final-viral-combined_k141_218317||full Not-determined
17-108-final-viral-combined_k141_246860||full Not-determined
17-108-final-viral-combined_k141_11918||full Not-determined
17-108-final-viral-combined_k141_39638||full Low-quality
17-108-final-viral-combined_k141_134925||full Low-quality
17-108-final-viral-combined_k141_327330||full Low-quality
17-108-final-viral-combined_k141_11938||full Low-quality
17-108-final-viral-combined_k141_39652||full Not-determined
17-108-final-viral-combined_k141_149626||full Not-determined
17-108-final-viral-combined_k141_273141||full Not-determined
17-108-final-viral-combined_k141_314841||full Not-determined
17-108-final-viral-combined_k141_327343||full Low-quality
17-108-final-viral-combined_k141_287650||full Low-quality
17-108-final-viral-combined_k141_53646||full Low-quality
17-108-final-viral-combined_k141_11976||full Low-quality
17-108-final-viral-combined_k141_53654||full Not-determined
17-108-final-viral-combined_k141_80131||full Low-quality
17-108-final-viral-combined_k141_260241||full Not-determined
17-108-final-viral-combined_k141_260242||full Low-quality
17-108-final-viral-combined_k141_53659||full Not-determined
17-108-final-viral-combined_k141_260243||full Not-determined
17-108-final-viral-combined_k141_327383||full Low-quality
17-108-final-viral-combined_k141_232412||full Low-quality
17-108-final-viral-combined_k141_287690||full Low-quality
17-108-final-viral-combined_k141_163688||full Not-determined
17-108-final-viral-combined_k141_53678||full Not-determined
17-108-final-viral-combined_k141_327401||full Not-determined
17-108-final-viral-combined_k141_135006||full Low-quality
17-108-final-viral-combined_k141_177536||full Not-determined
17-108-final-viral-combined_k141_246962||full Low-quality
17-108-final-viral-combined_k141_12010||full Low-quality
17-108-final-viral-combined_k141_287706||full Low-quality
17-108-final-viral-combined_k141_24371||full Low-quality
17-108-final-viral-combined_k141_94346||full Not-determined
17-108-final-viral-combined_k141_163716||full Not-determined
17-108-final-viral-combined_k141_314908||full Low-quality
17-108-final-viral-combined_k141_149675||full Low-quality
17-108-final-viral-combined_k141_177546||full Low-quality
17-108-final-viral-combined_k141_67193||full Not-determined
17-108-final-viral-combined_k141_67204||full Not-determined
17-108-final-viral-combined_k141_108987||full Low-quality
17-108-final-viral-combined_k141_149692||full Low-quality
17-108-final-viral-combined_k141_163738||full Low-quality
17-108-final-viral-combined_k141_53705||full Low-quality
17-108-final-viral-combined_k141_232473||full Low-quality
17-108-final-viral-combined_k141_135052||full Not-determined
17-108-final-viral-combined_k141_190288||full Low-quality
17-108-final-viral-combined_k141_287770||full Low-quality
17-108-final-viral-combined_k141_109025||full Not-determined
17-108-final-viral-combined_k141_300840||full Low-quality
17-108-final-viral-combined_k141_39768||full Low-quality
17-108-final-viral-combined_k141_53738||full Low-quality
17-108-final-viral-combined_k141_135076||full Not-determined
17-108-final-viral-combined_k141_327488||full Low-quality
17-108-final-viral-combined_k141_232505||full Low-quality
17-108-final-viral-combined_k141_135080||full Low-quality
17-108-final-viral-combined_k141_287787||full Not-determined
17-108-final-viral-combined_k141_247023||full Low-quality
17-108-final-viral-combined_k141_327506||full Not-determined
17-108-final-viral-combined_k141_135113||full Low-quality
17-108-final-viral-combined_k141_327508||full Low-quality
17-108-final-viral-combined_k141_273241||full Low-quality
17-108-final-viral-combined_k141_218427||full Not-determined
17-108-final-viral-combined_k141_327515||full Not-determined
17-108-final-viral-combined_k141_163822||full Low-quality
17-108-final-viral-combined_k141_135121||full Not-determined
17-108-final-viral-combined_k141_300884||full Not-determined
17-108-final-viral-combined_k141_121354||full Low-quality
17-108-final-viral-combined_k141_190355||full Low-quality
17-108-final-viral-combined_k141_109068||full Not-determined
17-108-final-viral-combined_k141_177619||full Not-determined
17-108-final-viral-combined_k141_247052||full Not-determined
17-108-final-viral-combined_k141_204497||full Not-determined
17-108-final-viral-combined_k141_190359||full Not-determined
17-108-final-viral-combined_k141_149789||full Low-quality
17-108-final-viral-combined_k141_135145||full Low-quality
17-108-final-viral-combined_k141_327520||full Low-quality
17-108-final-viral-combined_k141_177635||full Low-quality
17-108-final-viral-combined_k141_163859||full Low-quality
17-108-final-viral-combined_k141_67256||full Low-quality
17-108-final-viral-combined_k141_109071||full Not-determined
17-108-final-viral-combined_k141_300930||full Low-quality
17-108-final-viral-combined_k141_273292||full Not-determined
17-108-final-viral-combined_k141_109096||full Low-quality
17-108-final-viral-combined_k141_53770||full Not-determined
17-108-final-viral-combined_k141_149835||full Not-determined
17-108-final-viral-combined_k141_190420||full Not-determined
17-108-final-viral-combined_k141_53786||full Not-determined
17-108-final-viral-combined_k141_327565||full Not-determined
17-108-final-viral-combined_k141_163914||full Low-quality
17-108-final-viral-combined_k141_190424||full Not-determined
17-108-final-viral-combined_k141_163916||full Not-determined
17-108-final-viral-combined_k141_190441||full Not-determined
17-108-final-viral-combined_k141_121447||full Low-quality
17-108-final-viral-combined_k141_218535||full Low-quality
17-108-final-viral-combined_k141_53794||full Low-quality
17-108-final-viral-combined_k141_53795||full Low-quality
17-108-final-viral-combined_k141_300977||full Not-determined
17-108-final-viral-combined_k141_218548||full Low-quality
17-108-final-viral-combined_k141_39912||full Not-determined
17-108-final-viral-combined_k141_315038||full Low-quality
17-108-final-viral-combined_k141_121465||full Low-quality
17-108-final-viral-combined_k141_247113||full Low-quality
17-108-final-viral-combined_k141_39918||full Not-determined
17-108-final-viral-combined_k141_80249||full Low-quality
17-108-final-viral-combined_k141_94483||full Not-determined
17-108-final-viral-combined_k141_315046||full Not-determined
17-108-final-viral-combined_k141_163945||full Low-quality
17-108-final-viral-combined_k141_190477||full Low-quality
17-108-final-viral-combined_k141_218573||full Low-quality
17-108-final-viral-combined_k141_94497||full Not-determined
17-108-final-viral-combined_k141_39930||full Not-determined
17-108-final-viral-combined_k141_12082||full Low-quality
17-108-final-viral-combined_k141_53827||full Low-quality
17-108-final-viral-combined_k141_94498||full Not-determined
17-108-final-viral-combined_k141_260412||full Low-quality
17-108-final-viral-combined_k141_109163||full Low-quality
17-108-final-viral-combined_k141_94507||full Not-determined
18-109-final-viral-combined_k141_238231||full Medium-quality
17-108-final-viral-combined_k141_67279||full Not-determined
17-108-final-viral-combined_k141_24585||full Low-quality
17-108-final-viral-combined_k141_260430||full Low-quality
17-108-final-viral-combined_k141_80280||full Low-quality
17-108-final-viral-combined_k141_204555||full Low-quality
17-108-final-viral-combined_k141_80281||full Low-quality
17-108-final-viral-combined_k141_53848||full Low-quality
17-108-final-viral-combined_k141_121552||full Not-determined
17-108-final-viral-combined_k141_163996||full Low-quality
17-108-final-viral-combined_k141_232643||full Low-quality
17-108-final-viral-combined_k141_135184||full Not-determined
17-108-final-viral-combined_k141_164006||full Not-determined
17-108-final-viral-combined_k141_247174||full Low-quality
17-108-final-viral-combined_k141_232649||full Not-determined
17-108-final-viral-combined_k141_12124||full Not-determined
17-108-final-viral-combined_k141_288009||full Low-quality
17-108-final-viral-combined_k141_301029||full Low-quality
17-108-final-viral-combined_k141_190522||full Not-determined
17-108-final-viral-combined_k141_288014||full Low-quality
17-108-final-viral-combined_k141_247182||full Not-determined
17-108-final-viral-combined_k141_190526||full Not-determined
17-108-final-viral-combined_k141_135196||full Low-quality
17-108-final-viral-combined_k141_288028||full Not-determined
17-108-final-viral-combined_k141_121561||full Low-quality
17-108-final-viral-combined_k141_204587||full Low-quality
17-108-final-viral-combined_k141_204588||full Low-quality
17-108-final-viral-combined_k141_80324||full Not-determined
17-108-final-viral-combined_k141_301057||full Low-quality
17-108-final-viral-combined_k141_288046||full Not-determined
17-108-final-viral-combined_k141_135223||full Not-determined
17-108-final-viral-combined_k141_53880||full Not-determined
17-108-final-viral-combined_k141_190543||full Low-quality
17-108-final-viral-combined_k141_94598||full Not-determined
17-108-final-viral-combined_k141_164043||full Not-determined
17-108-final-viral-combined_k141_218652||full Low-quality
17-108-final-viral-combined_k141_247220||full Not-determined
17-108-final-viral-combined_k141_149928||full Not-determined
17-108-final-viral-combined_k141_121616||full Not-determined
17-108-final-viral-combined_k141_121622||full Not-determined
17-108-final-viral-combined_k141_273453||full Low-quality
17-108-final-viral-combined_k141_315152||full Low-quality
17-108-final-viral-combined_k141_247246||full Low-quality
17-108-final-viral-combined_k141_288106||full Low-quality
17-108-final-viral-combined_k141_190594||full Not-determined
17-108-final-viral-combined_k141_149961||full Not-determined
18-101-final-viral-combined_k141_23248||full Complete
17-108-final-viral-combined_k141_288119||full Low-quality
17-108-final-viral-combined_k141_121631||full Low-quality
17-108-final-viral-combined_k141_121633||full Low-quality
17-108-final-viral-combined_k141_40047||full Low-quality
17-108-final-viral-combined_k141_260540||full Low-quality
17-108-final-viral-combined_k141_190624||full Not-determined
17-108-final-viral-combined_k141_327708||full Not-determined
17-108-final-viral-combined_k141_24683||full Low-quality
17-108-final-viral-combined_k141_80385||full Not-determined
17-108-final-viral-combined_k141_288137||full Not-determined
17-108-final-viral-combined_k141_121634||full Not-determined
17-108-final-viral-combined_k141_232767||full Not-determined
17-108-final-viral-combined_k141_190634||full Low-quality
17-108-final-viral-combined_k141_53919||full Low-quality
17-108-final-viral-combined_k141_24695||full Low-quality
17-108-final-viral-combined_k141_232774||full Not-determined
17-108-final-viral-combined_k141_80393||full Low-quality
17-108-final-viral-combined_k141_12240||full Not-determined
17-108-final-viral-combined_k141_135309||full Low-quality
17-108-final-viral-combined_k141_40087||full Low-quality
17-108-final-viral-combined_k141_80419||full Not-determined
17-108-final-viral-combined_k141_53954||full Not-determined
17-108-final-viral-combined_k141_177894||full Not-determined
17-108-final-viral-combined_k141_315196||full Low-quality
17-108-final-viral-combined_k141_315198||full Not-determined
17-108-final-viral-combined_k141_247298||full Low-quality
17-108-final-viral-combined_k141_247301||full Low-quality
17-108-final-viral-combined_k141_273475||full Low-quality
17-108-final-viral-combined_k141_109231||full Not-determined
17-108-final-viral-combined_k141_121710||full Low-quality
17-108-final-viral-combined_k141_260603||full Not-determined
17-108-final-viral-combined_k141_177925||full Low-quality
17-108-final-viral-combined_k141_94679||full Not-determined
17-108-final-viral-combined_k141_247323||full Not-determined
17-108-final-viral-combined_k141_288176||full Low-quality
17-108-final-viral-combined_k141_327765||full Not-determined
17-108-final-viral-combined_k141_247333||full Not-determined
17-108-final-viral-combined_k141_315230||full Not-determined
17-108-final-viral-combined_k141_204694||full Low-quality
17-108-final-viral-combined_k141_190675||full Low-quality
17-108-final-viral-combined_k141_273513||full Not-determined
17-108-final-viral-combined_k141_232855||full Low-quality
17-108-final-viral-combined_k141_273526||full Not-determined
17-108-final-viral-combined_k141_260656||full Low-quality
17-108-final-viral-combined_k141_24737||full Low-quality
17-108-final-viral-combined_k141_12349||full Low-quality
17-108-final-viral-combined_k141_273529||full Not-determined
17-108-final-viral-combined_k141_218752||full Not-determined
17-108-final-viral-combined_k141_260674||full Low-quality
17-108-final-viral-combined_k141_218769||full Not-determined
17-108-final-viral-combined_k141_273541||full Low-quality
17-108-final-viral-combined_k141_204743||full Low-quality
17-108-final-viral-combined_k141_260692||full Low-quality
17-108-final-viral-combined_k141_12389||full Not-determined
17-108-final-viral-combined_k141_24779||full Not-determined
17-108-final-viral-combined_k141_150125||full Low-quality
17-108-final-viral-combined_k141_301122||full Not-determined
17-108-final-viral-combined_k141_109360||full Low-quality
17-108-final-viral-combined_k141_273577||full Low-quality
17-108-final-viral-combined_k141_178041||full Not-determined
17-108-final-viral-combined_k141_232933||full Low-quality
17-108-final-viral-combined_k141_190777||full Low-quality
17-108-final-viral-combined_k141_190789||full Not-determined
17-108-final-viral-combined_k141_67564||full Not-determined
17-108-final-viral-combined_k141_40252||full Not-determined
17-108-final-viral-combined_k141_12454||full Low-quality
17-108-final-viral-combined_k141_40256||full Not-determined
17-108-final-viral-combined_k141_150160||full Low-quality
17-108-final-viral-combined_k141_218827||full Low-quality
17-108-final-viral-combined_k141_40274||full Not-determined
17-108-final-viral-combined_k141_54027||full Not-determined
17-108-final-viral-combined_k141_150170||full Low-quality
17-108-final-viral-combined_k141_150173||full Low-quality
17-108-final-viral-combined_k141_218830||full Low-quality
17-108-final-viral-combined_k141_12486||full Low-quality
17-108-final-viral-combined_k141_273619||full Not-determined
17-108-final-viral-combined_k141_40306||full Not-determined
17-108-final-viral-combined_k141_247367||full Not-determined
17-108-final-viral-combined_k141_315404||full Low-quality
17-108-final-viral-combined_k141_247376||full Not-determined
17-108-final-viral-combined_k141_24864||full Low-quality
17-108-final-viral-combined_k141_150192||full Not-determined
17-108-final-viral-combined_k141_260795||full Not-determined
17-108-final-viral-combined_k141_327820||full Not-determined
17-108-final-viral-combined_k141_273646||full Not-determined
17-108-final-viral-combined_k141_80519||full Low-quality
17-108-final-viral-combined_k141_150210||full Not-determined
17-108-final-viral-combined_k141_67643||full Not-determined
17-108-final-viral-combined_k141_150213||full Low-quality
17-108-final-viral-combined_k141_150214||full Not-determined
17-108-final-viral-combined_k141_247407||full Low-quality
17-108-final-viral-combined_k141_164081||full Low-quality
17-108-final-viral-combined_k141_54116||full Low-quality
17-108-final-viral-combined_k141_247422||full Low-quality
17-108-final-viral-combined_k141_94817||full Low-quality
17-108-final-viral-combined_k141_260837||full Not-determined
17-108-final-viral-combined_k141_327854||full Low-quality
17-108-final-viral-combined_k141_178090||full Not-determined
17-108-final-viral-combined_k141_135581||full Low-quality
17-108-final-viral-combined_k141_301156||full Low-quality
17-108-final-viral-combined_k141_94824||full Low-quality
17-108-final-viral-combined_k141_109524||full Low-quality
17-108-final-viral-combined_k141_301161||full Not-determined
17-108-final-viral-combined_k141_121821||full Not-determined
17-108-final-viral-combined_k141_301165||full Low-quality
17-108-final-viral-combined_k141_94829||full Not-determined
17-108-final-viral-combined_k141_54148||full Low-quality
17-108-final-viral-combined_k141_135601||full Low-quality
17-108-final-viral-combined_k141_12531||full Low-quality
17-108-final-viral-combined_k141_190905||full Low-quality
17-108-final-viral-combined_k141_204859||full Low-quality
17-108-final-viral-combined_k141_178107||full Not-determined
17-108-final-viral-combined_k141_301189||full Low-quality
17-108-final-viral-combined_k141_40383||full Not-determined
17-108-final-viral-combined_k141_204875||full Low-quality
17-108-final-viral-combined_k141_247477||full Low-quality
17-108-final-viral-combined_k141_12564||full Not-determined
17-108-final-viral-combined_k141_135631||full Not-determined
17-108-final-viral-combined_k141_327900||full Not-determined
17-108-final-viral-combined_k141_327903||full Not-determined
17-108-final-viral-combined_k141_301234||full Not-determined
17-108-final-viral-combined_k141_150319||full Not-determined
17-108-final-viral-combined_k141_247486||full Low-quality
17-108-final-viral-combined_k141_204910||full Low-quality
17-108-final-viral-combined_k141_12583||full Not-determined
17-108-final-viral-combined_k141_288277||full Not-determined
17-108-final-viral-combined_k141_301237||full Low-quality
17-108-final-viral-combined_k141_218910||full Low-quality
17-108-final-viral-combined_k141_67748||full Low-quality
17-108-final-viral-combined_k141_247509||full Low-quality
17-108-final-viral-combined_k141_301257||full Not-determined
17-108-final-viral-combined_k141_135679||full Low-quality
17-108-final-viral-combined_k141_260911||full Low-quality
17-108-final-viral-combined_k141_135685||full Not-determined
17-108-final-viral-combined_k141_190972||full Not-determined
17-108-final-viral-combined_k141_327931||full Not-determined
17-108-final-viral-combined_k141_190975||full Not-determined
17-108-final-viral-combined_k141_121916||full Not-determined
17-108-final-viral-combined_k141_327936||full Low-quality
17-108-final-viral-combined_k141_24936||full Low-quality
17-108-final-viral-combined_k141_233178||full Low-quality
17-108-final-viral-combined_k141_80551||full Low-quality
17-108-final-viral-combined_k141_109637||full Not-determined
17-108-final-viral-combined_k141_150384||full Not-determined
17-108-final-viral-combined_k141_150388||full Not-determined
17-108-final-viral-combined_k141_150389||full Low-quality
17-108-final-viral-combined_k141_260954||full Low-quality
17-108-final-viral-combined_k141_12621||full Not-determined
17-108-final-viral-combined_k141_233180||full Low-quality
17-108-final-viral-combined_k141_94935||full Low-quality
17-108-final-viral-combined_k141_273849||full Low-quality
17-108-final-viral-combined_k141_40503||full Low-quality
17-108-final-viral-combined_k141_67824||full Low-quality
17-108-final-viral-combined_k141_164266||full Not-determined
17-108-final-viral-combined_k141_260978||full Low-quality
17-108-final-viral-combined_k141_24955||full Not-determined
17-108-final-viral-combined_k141_260987||full Not-determined
17-108-final-viral-combined_k141_164284||full Not-determined
17-108-final-viral-combined_k141_67843||full Not-determined
17-108-final-viral-combined_k141_247574||full Low-quality
17-108-final-viral-combined_k141_12637||full Not-determined
17-108-final-viral-combined_k141_67845||full Not-determined
17-108-final-viral-combined_k141_233195||full Not-determined
17-108-final-viral-combined_k141_12648||full Low-quality
17-108-final-viral-combined_k141_301313||full Low-quality
17-108-final-viral-combined_k141_80600||full Not-determined
17-108-final-viral-combined_k141_191059||full Not-determined
17-108-final-viral-combined_k141_273868||full Not-determined
17-108-final-viral-combined_k141_109695||full Low-quality
17-108-final-viral-combined_k141_109700||full Not-determined
17-108-final-viral-combined_k141_327990||full Low-quality
17-108-final-viral-combined_k141_204970||full Not-determined
17-108-final-viral-combined_k141_67873||full Low-quality
17-108-final-viral-combined_k141_247594||full Not-determined
17-108-final-viral-combined_k141_40545||full Low-quality
17-108-final-viral-combined_k141_80629||full Not-determined
17-108-final-viral-combined_k141_247602||full Low-quality
17-108-final-viral-combined_k141_109719||full Not-determined
17-108-final-viral-combined_k141_67879||full Low-quality
17-108-final-viral-combined_k141_54270||full Low-quality
17-108-final-viral-combined_k141_122010||full Not-determined
17-108-final-viral-combined_k141_261042||full Not-determined
17-108-final-viral-combined_k141_328018||full Not-determined
17-108-final-viral-combined_k141_67891||full Not-determined
17-108-final-viral-combined_k141_219034||full Low-quality
17-108-final-viral-combined_k141_54280||full Not-determined
17-108-final-viral-combined_k141_94972||full Low-quality
17-108-final-viral-combined_k141_328025||full Not-determined
17-108-final-viral-combined_k141_25022||full Not-determined
17-108-final-viral-combined_k141_273920||full Low-quality
17-108-final-viral-combined_k141_25025||full Low-quality
17-108-final-viral-combined_k141_150453||full Low-quality
17-108-final-viral-combined_k141_247639||full Not-determined
17-108-final-viral-combined_k141_288422||full Not-determined
17-108-final-viral-combined_k141_150458||full Not-determined
17-108-final-viral-combined_k141_54312||full Not-determined
17-108-final-viral-combined_k141_301354||full Low-quality
17-108-final-viral-combined_k141_67915||full Not-determined
17-108-final-viral-combined_k141_135763||full Low-quality
17-108-final-viral-combined_k141_40575||full Not-determined
17-108-final-viral-combined_k141_135778||full Not-determined
17-108-final-viral-combined_k141_219059||full Low-quality
17-108-final-viral-combined_k141_80708||full Not-determined
17-108-final-viral-combined_k141_12748||full Low-quality
17-108-final-viral-combined_k141_109798||full Not-determined
17-108-final-viral-combined_k141_205054||full Low-quality
17-108-final-viral-combined_k141_191101||full Not-determined
17-108-final-viral-combined_k141_315601||full Not-determined
17-108-final-viral-combined_k141_233266||full Low-quality
17-108-final-viral-combined_k141_219078||full Not-determined
17-108-final-viral-combined_k141_109810||full Low-quality
17-108-final-viral-combined_k141_164417||full Low-quality
17-108-final-viral-combined_k141_40616||full Not-determined
17-108-final-viral-combined_k141_219089||full Low-quality
17-108-final-viral-combined_k141_205073||full Not-determined
17-108-final-viral-combined_k141_274009||full Not-determined
17-108-final-viral-combined_k141_205083||full Low-quality
17-108-final-viral-combined_k141_25083||full Low-quality
17-108-final-viral-combined_k141_135832||full Not-determined
17-108-final-viral-combined_k141_301389||full Not-determined
17-108-final-viral-combined_k141_274039||full Not-determined
17-108-final-viral-combined_k141_95055||full Not-determined
17-108-final-viral-combined_k141_40657||full Low-quality
17-108-final-viral-combined_k141_67981||full Low-quality
17-108-final-viral-combined_k141_178362||full Low-quality
17-108-final-viral-combined_k141_122075||full Not-determined
17-108-final-viral-combined_k141_315651||full Not-determined
17-108-final-viral-combined_k141_54406||full Low-quality
17-108-final-viral-combined_k141_12804||full Not-determined
17-108-final-viral-combined_k141_233322||full Not-determined
17-108-final-viral-combined_k141_301407||full Low-quality
17-108-final-viral-combined_k141_67999||full Low-quality
17-108-final-viral-combined_k141_191172||full Low-quality
17-108-final-viral-combined_k141_315674||full Not-determined
17-108-final-viral-combined_k141_288572||full Low-quality
17-108-final-viral-combined_k141_261150||full Low-quality
17-108-final-viral-combined_k141_301444||full Not-determined
17-108-final-viral-combined_k141_54454||full Not-determined
17-108-final-viral-combined_k141_150605||full Not-determined
17-108-final-viral-combined_k141_109892||full Not-determined
17-108-final-viral-combined_k141_288604||full Low-quality
17-108-final-viral-combined_k141_135934||full Not-determined
17-108-final-viral-combined_k141_288610||full Not-determined
17-108-final-viral-combined_k141_315694||full Low-quality
17-108-final-viral-combined_k141_95147||full Low-quality
17-108-final-viral-combined_k141_205111||full Low-quality
17-108-final-viral-combined_k141_233370||full Not-determined
17-108-final-viral-combined_k141_205114||full Low-quality
17-108-final-viral-combined_k141_80833||full Low-quality
17-108-final-viral-combined_k141_247835||full Not-determined
17-108-final-viral-combined_k141_135946||full Not-determined
17-108-final-viral-combined_k141_164493||full Low-quality
17-108-final-viral-combined_k141_328113||full Low-quality
17-108-final-viral-combined_k141_205135||full Low-quality
17-108-final-viral-combined_k141_95173||full Low-quality
17-108-final-viral-combined_k141_301482||full Not-determined
17-108-final-viral-combined_k141_219172||full Not-determined
17-108-final-viral-combined_k141_315728||full Not-determined
17-108-final-viral-combined_k141_261182||full Not-determined
17-108-final-viral-combined_k141_219179||full Not-determined
17-108-final-viral-combined_k141_301502||full Low-quality
17-108-final-viral-combined_k141_109936||full Low-quality
17-108-final-viral-combined_k141_191224||full Low-quality
17-108-final-viral-combined_k141_205155||full Not-determined
17-108-final-viral-combined_k141_219184||full Not-determined
17-108-final-viral-combined_k141_109937||full Low-quality
17-108-final-viral-combined_k141_233418||full Not-determined
17-108-final-viral-combined_k141_219192||full Low-quality
17-108-final-viral-combined_k141_109952||full Low-quality
17-108-final-viral-combined_k141_164514||full Low-quality
17-108-final-viral-combined_k141_164515||full Not-determined
17-108-final-viral-combined_k141_40799||full Not-determined
17-108-final-viral-combined_k141_150675||full Not-determined
17-108-final-viral-combined_k141_205196||full Not-determined
17-108-final-viral-combined_k141_25222||full Not-determined
17-108-final-viral-combined_k141_219200||full Not-determined
17-108-final-viral-combined_k141_274122||full Not-determined
17-108-final-viral-combined_k141_301515||full Low-quality
17-108-final-viral-combined_k141_68126||full Not-determined
17-108-final-viral-combined_k141_219208||full Not-determined
17-108-final-viral-combined_k141_219216||full Not-determined
17-108-final-viral-combined_k141_135994||full Low-quality
17-108-final-viral-combined_k141_328156||full Low-quality
17-108-final-viral-combined_k141_164548||full Low-quality
17-108-final-viral-combined_k141_95220||full Not-determined
17-108-final-viral-combined_k141_328161||full Low-quality
17-108-final-viral-combined_k141_25239||full Low-quality
17-108-final-viral-combined_k141_191295||full Low-quality
17-108-final-viral-combined_k141_122230||full Low-quality
17-108-final-viral-combined_k141_178525||full Not-determined
17-108-final-viral-combined_k141_315770||full Not-determined
17-108-final-viral-combined_k141_150725||full Not-determined
17-108-final-viral-combined_k141_95225||full Not-determined
17-108-final-viral-combined_k141_25252||full Not-determined
17-108-final-viral-combined_k141_191312||full Low-quality
17-108-final-viral-combined_k141_68174||full Not-determined
17-108-final-viral-combined_k141_122262||full Low-quality
17-108-final-viral-combined_k141_191337||full Not-determined
17-108-final-viral-combined_k141_219238||full Not-determined
17-108-final-viral-combined_k141_150758||full Not-determined
17-108-final-viral-combined_k141_95253||full Low-quality
17-108-final-viral-combined_k141_122282||full Low-quality
17-108-final-viral-combined_k141_80893||full Low-quality
17-108-final-viral-combined_k141_219258||full Not-determined
17-108-final-viral-combined_k141_274161||full Not-determined
17-108-final-viral-combined_k141_315820||full Not-determined
17-108-final-viral-combined_k141_219267||full Low-quality
17-108-final-viral-combined_k141_261246||full Not-determined
17-108-final-viral-combined_k141_136036||full Not-determined
17-108-final-viral-combined_k141_219269||full Low-quality
17-108-final-viral-combined_k141_95263||full Not-determined
17-108-final-viral-combined_k141_54592||full Low-quality
17-108-final-viral-combined_k141_219271||full Not-determined
17-108-final-viral-combined_k141_191365||full Not-determined
17-108-final-viral-combined_k141_164615||full Not-determined
17-108-final-viral-combined_k141_328240||full Low-quality

17-108-final-viral-combined_k141_315822||full Not-determined
17-108-final-viral-combined_k141_261270||full Not-determined
17-108-final-viral-combined_k141_261274||full Not-determined
17-108-final-viral-combined_k141_288677||full Not-determined
17-108-final-viral-combined_k141_150816||full Not-determined
17-108-final-viral-combined_k141_288693||full Not-determined
17-108-final-viral-combined_k141_80939||full Low-quality
17-108-final-viral-combined_k141_328271||full Not-determined
17-108-final-viral-combined_k141_328280||full Low-quality
17-108-final-viral-combined_k141_288702||full Low-quality
17-108-final-viral-combined_k141_150836||full Low-quality
17-108-final-viral-combined_k141_13040||full Not-determined
17-108-final-viral-combined_k141_288706||full Low-quality
17-108-final-viral-combined_k141_150851||full Not-determined
17-108-final-viral-combined_k141_261302||full Not-determined
17-108-final-viral-combined_k141_13063||full Low-quality
17-108-final-viral-combined_k141_13067||full Not-determined
17-108-final-viral-combined_k141_95355||full Low-quality
17-108-final-viral-combined_k141_164701||full Low-quality
17-108-final-viral-combined_k141_110117||full Low-quality
17-108-final-viral-combined_k141_178619||full Not-determined
17-108-final-viral-combined_k141_95372||full Not-determined
17-108-final-viral-combined_k141_261318||full Low-quality
17-108-final-viral-combined_k141_315844||full Not-determined
17-108-final-viral-combined_k141_274269||full Low-quality
17-108-final-viral-combined_k141_68205||full Not-determined
17-108-final-viral-combined_k141_54681||full Low-quality
17-108-final-viral-combined_k141_68215||full Low-quality
17-108-final-viral-combined_k141_68217||full Not-determined
17-108-final-viral-combined_k141_178652||full Low-quality
17-108-final-viral-combined_k141_315863||full Low-quality
17-108-final-viral-combined_k141_122369||full Low-quality
17-108-final-viral-combined_k141_68243||full Low-quality
17-108-final-viral-combined_k141_248034||full Not-determined
17-108-final-viral-combined_k141_136137||full Low-quality
17-108-final-viral-combined_k141_81006||full Low-quality
17-108-final-viral-combined_k141_288732||full Low-quality
17-108-final-viral-combined_k141_110185||full Not-determined
17-108-final-viral-combined_k141_301630||full Not-determined
17-108-final-viral-combined_k141_150908||full Not-determined
17-108-final-viral-combined_k141_261397||full Not-determined
17-108-final-viral-combined_k141_13159||full Low-quality
17-108-final-viral-combined_k141_219410||full Low-quality
17-108-final-viral-combined_k141_13162||full Low-quality
17-108-final-viral-combined_k141_191463||full Not-determined
17-108-final-viral-combined_k141_68271||full Low-quality
17-108-final-viral-combined_k141_178671||full Low-quality
17-108-final-viral-combined_k141_122375||full Low-quality
17-108-final-viral-combined_k141_110205||full Low-quality
17-108-final-viral-combined_k141_13173||full Low-quality
17-108-final-viral-combined_k141_54761||full Low-quality
17-108-final-viral-combined_k141_288772||full Not-determined
17-108-final-viral-combined_k141_191472||full Not-determined
17-108-final-viral-combined_k141_25450||full Not-determined
17-108-final-viral-combined_k141_301655||full Low-quality
17-108-final-viral-combined_k141_95439||full Not-determined
17-108-final-viral-combined_k141_301658||full Low-quality
17-108-final-viral-combined_k141_191476||full Low-quality
17-108-final-viral-combined_k141_274323||full Low-quality
17-108-final-viral-combined_k141_25467||full Not-determined
17-108-final-viral-combined_k141_301669||full Low-quality
17-108-final-viral-combined_k141_261454||full Low-quality
17-108-final-viral-combined_k141_41025||full Low-quality
17-108-final-viral-combined_k141_13220||full Low-quality
17-108-final-viral-combined_k141_328345||full Low-quality
17-108-final-viral-combined_k141_41031||full Low-quality
17-108-final-viral-combined_k141_274349||full Not-determined
17-108-final-viral-combined_k141_205397||full Low-quality
17-108-final-viral-combined_k141_95476||full Low-quality
17-108-final-viral-combined_k141_136226||full Low-quality
17-108-final-viral-combined_k141_81083||full Not-determined
17-108-final-viral-combined_k141_95483||full Low-quality
17-108-final-viral-combined_k141_219485||full Low-quality
17-108-final-viral-combined_k141_41053||full Low-quality
17-108-final-viral-combined_k141_288843||full Not-determined
17-108-final-viral-combined_k141_136252||full Not-determined
17-108-final-viral-combined_k141_68330||full Not-determined
17-108-final-viral-combined_k141_301728||full Low-quality
17-108-final-viral-combined_k141_13267||full Low-quality
17-108-final-viral-combined_k141_328403||full Low-quality
17-108-final-viral-combined_k141_25552||full Not-determined
17-108-final-viral-combined_k141_54843||full Not-determined
17-108-final-viral-combined_k141_178755||full Not-determined
17-108-final-viral-combined_k141_41093||full Not-determined
17-108-final-viral-combined_k141_151010||full Low-quality
17-108-final-viral-combined_k141_301769||full Low-quality
17-108-final-viral-combined_k141_136302||full Low-quality
17-108-final-viral-combined_k141_233787||full Low-quality
17-108-final-viral-combined_k141_288889||full Low-quality
17-108-final-viral-combined_k141_219535||full Low-quality
17-108-final-viral-combined_k141_81129||full Not-determined
17-108-final-viral-combined_k141_274366||full Low-quality
17-108-final-viral-combined_k141_41114||full Not-determined
17-108-final-viral-combined_k141_288914||full Low-quality
17-108-final-viral-combined_k141_178787||full Low-quality
17-108-final-viral-combined_k141_178794||full Not-determined
17-108-final-viral-combined_k141_191632||full Low-quality
17-108-final-viral-combined_k141_151050||full Low-quality
17-108-final-viral-combined_k141_219584||full Not-determined
17-108-final-viral-combined_k141_95582||full Not-determined
17-108-final-viral-combined_k141_191648||full Not-determined
17-108-final-viral-combined_k141_274396||full Not-determined
17-108-final-viral-combined_k141_274404||full Low-quality
17-108-final-viral-combined_k141_95589||full Low-quality
17-108-final-viral-combined_k141_136356||full Low-quality
17-108-final-viral-combined_k141_233842||full Not-determined
17-108-final-viral-combined_k141_219612||full Low-quality
17-108-final-viral-combined_k141_274426||full Low-quality
17-108-final-viral-combined_k141_316012||full Low-quality
17-108-final-viral-combined_k141_301929||full Not-determined
17-108-final-viral-combined_k141_95621||full Low-quality
17-108-final-viral-combined_k141_301938||full Not-determined
17-108-final-viral-combined_k141_301944||full Low-quality
17-108-final-viral-combined_k141_316033||full Not-determined
17-108-final-viral-combined_k141_274459||full Low-quality
17-108-final-viral-combined_k141_41128||full Low-quality
17-108-final-viral-combined_k141_191723||full Not-determined
17-108-final-viral-combined_k141_13398||full Not-determined
17-108-final-viral-combined_k141_233903||full Low-quality
17-108-final-viral-combined_k141_274479||full Not-determined
17-108-final-viral-combined_k141_316051||full Low-quality
17-108-final-viral-combined_k141_164967||full Low-quality
17-108-final-viral-combined_k141_41155||full Low-quality
17-108-final-viral-combined_k141_54942||full Low-quality
17-108-final-viral-combined_k141_328443||full Not-determined
17-108-final-viral-combined_k141_233923||full Not-determined
17-108-final-viral-combined_k141_54951||full Low-quality
17-108-final-viral-combined_k141_288981||full Not-determined
17-108-final-viral-combined_k141_41163||full Not-determined
17-108-final-viral-combined_k141_151072||full Low-quality
17-108-final-viral-combined_k141_68492||full Not-determined
17-108-final-viral-combined_k141_205553||full Not-determined
17-108-final-viral-combined_k141_233930||full Not-determined
17-108-final-viral-combined_k141_328454||full Low-quality
17-108-final-viral-combined_k141_205555||full Low-quality
17-108-final-viral-combined_k141_81233||full Low-quality
17-108-final-viral-combined_k141_122659||full Low-quality
17-108-final-viral-combined_k141_68502||full Not-determined
17-108-final-viral-combined_k141_219686||full Not-determined
17-108-final-viral-combined_k141_41193||full Not-determined
17-108-final-viral-combined_k141_289006||full Not-determined
17-108-final-viral-combined_k141_191787||full Not-determined
17-108-final-viral-combined_k141_81271||full Not-determined
17-108-final-viral-combined_k141_289023||full Not-determined
17-108-final-viral-combined_k141_219704||full Not-determined
17-108-final-viral-combined_k141_41204||full Not-determined
17-108-final-viral-combined_k141_233965||full Not-determined
17-108-final-viral-combined_k141_122693||full Not-determined
17-108-final-viral-combined_k141_136444||full Not-determined
17-108-final-viral-combined_k141_122695||full Not-determined
17-108-final-viral-combined_k141_122696||full Low-quality
17-108-final-viral-combined_k141_328498||full Not-determined
17-108-final-viral-combined_k141_274507||full Not-determined
17-108-final-viral-combined_k141_81293||full Low-quality
17-108-final-viral-combined_k141_289044||full Low-quality
17-108-final-viral-combined_k141_191820||full Low-quality
17-108-final-viral-combined_k141_316135||full Not-determined
17-108-final-viral-combined_k141_302027||full Not-determined
17-108-final-viral-combined_k141_219733||full Not-determined
17-108-final-viral-combined_k141_55003||full Not-determined
17-108-final-viral-combined_k141_165049||full Low-quality
17-108-final-viral-combined_k141_191843||full Low-quality
17-108-final-viral-combined_k141_178885||full Not-determined
17-108-final-viral-combined_k141_41243||full Low-quality
17-108-final-viral-combined_k141_81322||full Low-quality
17-108-final-viral-combined_k141_328530||full Low-quality
17-108-final-viral-combined_k141_136490||full Low-quality
17-108-final-viral-combined_k141_261713||full Low-quality
17-108-final-viral-combined_k141_165072||full Low-quality
17-108-final-viral-combined_k141_316162||full Low-quality
17-108-final-viral-combined_k141_234017||full Low-quality
17-108-final-viral-combined_k141_122752||full Low-quality
17-108-final-viral-combined_k141_302054||full Low-quality
17-108-final-viral-combined_k141_68527||full Not-determined
17-108-final-viral-combined_k141_55037||full Low-quality
17-108-final-viral-combined_k141_302058||full Low-quality
17-108-final-viral-combined_k141_178909||full Low-quality
17-108-final-viral-combined_k141_95751||full Low-quality
17-108-final-viral-combined_k141_41279||full Low-quality
17-108-final-viral-combined_k141_219770||full Low-quality
17-108-final-viral-combined_k141_274560||full Low-quality
17-108-final-viral-combined_k141_261762||full Not-determined
17-108-final-viral-combined_k141_151165||full Not-determined
17-108-final-viral-combined_k141_274574||full Not-determined
17-108-final-viral-combined_k141_81386||full Low-quality
17-108-final-viral-combined_k141_234057||full Low-quality
17-108-final-viral-combined_k141_316191||full Low-quality
17-108-final-viral-combined_k141_328565||full Not-determined
17-108-final-viral-combined_k141_55077||full Not-determined
17-108-final-viral-combined_k141_68567||full Low-quality
17-108-final-viral-combined_k141_302109||full Not-determined
17-108-final-viral-combined_k141_165124||full Not-determined
17-108-final-viral-combined_k141_151187||full Low-quality
17-108-final-viral-combined_k141_261789||full Not-determined
17-108-final-viral-combined_k141_81406||full Low-quality
17-108-final-viral-combined_k141_274602||full Not-determined
17-108-final-viral-combined_k141_302117||full Not-determined
17-108-final-viral-combined_k141_274604||full Not-determined
17-108-final-viral-combined_k141_234091||full Low-quality
17-108-final-viral-combined_k141_205673||full Low-quality
17-108-final-viral-combined_k141_151211||full Low-quality
17-108-final-viral-combined_k141_151217||full Low-quality
17-108-final-viral-combined_k141_178953||full Not-determined
17-108-final-viral-combined_k141_289132||full Low-quality
17-108-final-viral-combined_k141_165161||full Low-quality
17-108-final-viral-combined_k141_68602||full Not-determined
17-108-final-viral-combined_k141_136555||full Not-determined
17-108-final-viral-combined_k141_13546||full Low-quality
17-108-final-viral-combined_k141_219811||full Not-determined
17-108-final-viral-combined_k141_136561||full Not-determined
17-108-final-viral-combined_k141_205702||full Not-determined
17-108-final-viral-combined_k141_41342||full Low-quality
17-108-final-viral-combined_k141_95848||full Low-quality
17-108-final-viral-combined_k141_328632||full Low-quality
17-108-final-viral-combined_k141_68627||full Low-quality
17-108-final-viral-combined_k141_81434||full Low-quality
17-108-final-viral-combined_k141_261821||full Low-quality
17-108-final-viral-combined_k141_25886||full Not-determined
17-108-final-viral-combined_k141_328638||full Low-quality
17-108-final-viral-combined_k141_151242||full Not-determined
17-108-final-viral-combined_k141_95855||full Not-determined
17-108-final-viral-combined_k141_13584||full Low-quality
17-108-final-viral-combined_k141_95858||full Low-quality
17-108-final-viral-combined_k141_179010||full Not-determined
17-108-final-viral-combined_k141_13592||full Low-quality
17-108-final-viral-combined_k141_234153||full Low-quality
17-108-final-viral-combined_k141_289182||full Not-determined
17-108-final-viral-combined_k141_136596||full Not-determined
17-108-final-viral-combined_k141_219851||full Low-quality
17-108-final-viral-combined_k141_302217||full Low-quality
17-108-final-viral-combined_k141_289191||full Not-determined
17-108-final-viral-combined_k141_55096||full Low-quality
17-108-final-viral-combined_k141_55099||full Low-quality
17-108-final-viral-combined_k141_179046||full Low-quality
17-108-final-viral-combined_k141_55103||full Low-quality
17-108-final-viral-combined_k141_165226||full Not-determined
17-108-final-viral-combined_k141_274703||full Low-quality
17-108-final-viral-combined_k141_151274||full Not-determined
17-108-final-viral-combined_k141_25941||full Not-determined
17-108-final-viral-combined_k141_302243||full Low-quality
17-108-final-viral-combined_k141_136615||full Not-determined
17-108-final-viral-combined_k141_165248||full Low-quality
17-108-final-viral-combined_k141_302258||full Low-quality
17-108-final-viral-combined_k141_136618||full Not-determined
17-108-final-viral-combined_k141_328682||full Not-determined
17-108-final-viral-combined_k141_165265||full Low-quality
17-108-final-viral-combined_k141_328693||full Not-determined
17-108-final-viral-combined_k141_13660||full Low-quality
17-108-final-viral-combined_k141_219903||full Not-determined
17-108-final-viral-combined_k141_179090||full Not-determined
17-108-final-viral-combined_k141_205796||full Low-quality
17-108-final-viral-combined_k141_95918||full Low-quality
17-108-final-viral-combined_k141_136643||full Low-quality
17-108-final-viral-combined_k141_95919||full Low-quality
17-108-final-viral-combined_k141_289254||full Low-quality
17-108-final-viral-combined_k141_165293||full Low-quality
17-108-final-viral-combined_k141_95920||full Low-quality
17-108-final-viral-combined_k141_122963||full Low-quality
17-108-final-viral-combined_k141_219928||full Low-quality
17-108-final-viral-combined_k141_81558||full Low-quality
17-108-final-viral-combined_k141_151358||full Low-quality
17-108-final-viral-combined_k141_219931||full Not-determined
17-108-final-viral-combined_k141_26014||full Not-determined
17-108-final-viral-combined_k141_26019||full Low-quality
17-108-final-viral-combined_k141_191955||full Not-determined
17-108-final-viral-combined_k141_151375||full Not-determined
17-108-final-viral-combined_k141_81598||full Not-determined
17-108-final-viral-combined_k141_289325||full Low-quality
17-108-final-viral-combined_k141_191990||full Low-quality
17-108-final-viral-combined_k141_179133||full Low-quality
17-108-final-viral-combined_k141_165353||full Low-quality
17-108-final-viral-combined_k141_179142||full Low-quality
17-108-final-viral-combined_k141_179146||full Low-quality
17-108-final-viral-combined_k141_289343||full Not-determined
17-108-final-viral-combined_k141_302379||full Low-quality
17-108-final-viral-combined_k141_136744||full Not-determined
17-108-final-viral-combined_k141_302394||full Low-quality
17-108-final-viral-combined_k141_136747||full Low-quality
17-108-final-viral-combined_k141_26097||full Not-determined
17-108-final-viral-combined_k141_136758||full Not-determined
17-108-final-viral-combined_k141_13761||full Not-determined
17-108-final-viral-combined_k141_192004||full Low-quality
17-108-final-viral-combined_k141_274844||full Low-quality
17-108-final-viral-combined_k141_81637||full Low-quality
17-108-final-viral-combined_k141_81646||full Not-determined
17-108-final-viral-combined_k141_316386||full Low-quality
17-108-final-viral-combined_k141_136796||full Not-determined
17-108-final-viral-combined_k141_289399||full Low-quality
17-108-final-viral-combined_k141_302483||full Low-quality
17-108-final-viral-combined_k141_302484||full Not-determined
17-108-final-viral-combined_k141_192075||full Low-quality
17-108-final-viral-combined_k141_302505||full Low-quality
17-108-final-viral-combined_k141_316436||full Low-quality
17-108-final-viral-combined_k141_81710||full Not-determined
17-108-final-viral-combined_k141_328798||full Not-determined
17-108-final-viral-combined_k141_96107||full Not-determined
17-108-final-viral-combined_k141_68756||full Low-quality
17-108-final-viral-combined_k141_81721||full Low-quality
17-108-final-viral-combined_k141_192109||full Low-quality
17-108-final-viral-combined_k141_302522||full Low-quality
17-108-final-viral-combined_k141_274935||full Low-quality
17-108-final-viral-combined_k141_316459||full Not-determined
17-108-final-viral-combined_k141_328826||full Not-determined
17-108-final-viral-combined_k141_328835||full Low-quality
17-108-final-viral-combined_k141_328840||full Not-determined
17-108-final-viral-combined_k141_123165||full Low-quality
17-108-final-viral-combined_k141_151446||full Not-determined
17-108-final-viral-combined_k141_302584||full Not-determined
17-108-final-viral-combined_k141_220141||full Low-quality
17-108-final-viral-combined_k141_68804||full Low-quality
17-108-final-viral-combined_k141_96210||full Low-quality
17-108-final-viral-combined_k141_96218||full Low-quality
17-108-final-viral-combined_k141_206049||full Low-quality
17-108-final-viral-combined_k141_302618||full Low-quality
17-108-final-viral-combined_k141_151488||full Not-determined
17-108-final-viral-combined_k141_136951||full Not-determined
17-108-final-viral-combined_k141_316516||full Low-quality
17-108-final-viral-combined_k141_81805||full Not-determined
17-108-final-viral-combined_k141_206084||full Low-quality
17-108-final-viral-combined_k141_192190||full Low-quality
17-108-final-viral-combined_k141_192210||full Low-quality
17-108-final-viral-combined_k141_302662||full Not-determined
17-108-final-viral-combined_k141_275068||full Not-determined
17-108-final-viral-combined_k141_192211||full Low-quality
17-108-final-viral-combined_k141_328877||full Low-quality
17-108-final-viral-combined_k141_26341||full Low-quality
17-108-final-viral-combined_k141_68864||full Low-quality
17-108-final-viral-combined_k141_96285||full Low-quality
17-108-final-viral-combined_k141_96287||full Low-quality
17-108-final-viral-combined_k141_81877||full Not-determined
17-108-final-viral-combined_k141_220207||full Not-determined
17-108-final-viral-combined_k141_68874||full Not-determined
17-108-final-viral-combined_k141_123287||full Not-determined
17-108-final-viral-combined_k141_137033||full Not-determined
17-108-final-viral-combined_k141_302736||full Low-quality
17-108-final-viral-combined_k141_137044||full Not-determined
17-108-final-viral-combined_k141_328919||full Low-quality
17-108-final-viral-combined_k141_81916||full Not-determined
17-108-final-viral-combined_k141_192286||full Low-quality
17-108-final-viral-combined_k141_96324||full Low-quality
17-108-final-viral-combined_k141_81919||full Not-determined
17-108-final-viral-combined_k141_123337||full Not-determined
17-108-final-viral-combined_k141_137063||full Low-quality
17-108-final-viral-combined_k141_137073||full Not-determined
17-108-final-viral-combined_k141_26402||full Not-determined
17-108-final-viral-combined_k141_192319||full Low-quality
17-108-final-viral-combined_k141_96357||full Not-determined
17-108-final-viral-combined_k141_81950||full Low-quality
17-108-final-viral-combined_k141_316573||full Low-quality
17-108-final-viral-combined_k141_137128||full Not-determined
17-108-final-viral-combined_k141_96375||full Low-quality
17-108-final-viral-combined_k141_192361||full Not-determined
17-108-final-viral-combined_k141_192363||full Not-determined
17-108-final-viral-combined_k141_123409||full Not-determined
17-108-final-viral-combined_k141_206158||full Low-quality
17-108-final-viral-combined_k141_96396||full Low-quality
17-108-final-viral-combined_k141_275207||full Not-determined
17-108-final-viral-combined_k141_316580||full Low-quality
17-108-final-viral-combined_k141_123458||full Low-quality
17-108-final-viral-combined_k141_220336||full Low-quality
17-108-final-viral-combined_k141_316608||full Not-determined
17-108-final-viral-combined_k141_302866||full Low-quality
17-108-final-viral-combined_k141_206179||full Low-quality
17-108-final-viral-combined_k141_137228||full Low-quality
17-108-final-viral-combined_k141_302875||full Low-quality
17-108-final-viral-combined_k141_3507||full Complete
17-108-final-viral-combined_k141_137242||full Low-quality
17-108-final-viral-combined_k141_275277||full Low-quality
17-108-final-viral-combined_k141_26585||full Not-determined
17-108-final-viral-combined_k141_275289||full Not-determined
17-108-final-viral-combined_k141_328997||full Low-quality
17-108-final-viral-combined_k141_192444||full Low-quality
17-108-final-viral-combined_k141_329005||full Not-determined
17-108-final-viral-combined_k141_329008||full Not-determined
17-108-final-viral-combined_k141_192453||full Not-determined
17-108-final-viral-combined_k141_275309||full Not-determined
17-108-final-viral-combined_k141_137277||full Low-quality
17-108-final-viral-combined_k141_220386||full Low-quality
17-108-final-viral-combined_k141_275323||full Not-determined
17-108-final-viral-combined_k141_26628||full Not-determined
17-108-final-viral-combined_k141_137334||full Low-quality
17-108-final-viral-combined_k141_302966||full Low-quality
17-108-final-viral-combined_k141_275358||full Low-quality
17-108-final-viral-combined_k141_302986||full Low-quality
17-108-final-viral-combined_k141_26670||full Not-determined
17-108-final-viral-combined_k141_220436||full Not-determined
17-108-final-viral-combined_k141_329080||full Low-quality
17-108-final-viral-combined_k141_275376||full Not-determined
17-108-final-viral-combined_k141_329087||full Low-quality
17-108-final-viral-combined_k141_82137||full Not-determined
17-108-final-viral-combined_k141_220464||full Low-quality
17-108-final-viral-combined_k141_26686||full Low-quality
17-108-final-viral-combined_k141_123643||full Not-determined
17-108-final-viral-combined_k141_220472||full Not-determined
17-108-final-viral-combined_k141_82177||full Not-determined
17-108-final-viral-combined_k141_316731||full Low-quality
17-108-final-viral-combined_k141_26706||full Not-determined
17-108-final-viral-combined_k141_123658||full Not-determined
17-108-final-viral-combined_k141_275408||full Low-quality
17-108-final-viral-combined_k141_82198||full Low-quality
17-108-final-viral-combined_k141_303054||full Not-determined
17-108-final-viral-combined_k141_82209||full Not-determined
17-108-final-viral-combined_k141_137426||full Low-quality
17-108-final-viral-combined_k141_137427||full Not-determined
17-108-final-viral-combined_k141_82237||full Low-quality
17-108-final-viral-combined_k141_303066||full Not-determined
17-108-final-viral-combined_k141_303071||full Low-quality
17-108-final-viral-combined_k141_316770||full Low-quality
17-108-final-viral-combined_k141_329164||full Not-determined
17-108-final-viral-combined_k141_137498||full Not-determined
17-108-final-viral-combined_k141_137502||full Low-quality
17-108-final-viral-combined_k141_137509||full Not-determined
17-108-final-viral-combined_k141_316841||full Low-quality
17-108-final-viral-combined_k141_192708||full Not-determined
17-108-final-viral-combined_k141_137547||full Not-determined
17-108-final-viral-combined_k141_137548||full Not-determined
17-108-final-viral-combined_k141_303143||full Low-quality
17-108-final-viral-combined_k141_26833||full Low-quality
17-108-final-viral-combined_k141_303148||full Not-determined
17-108-final-viral-combined_k141_192728||full Low-quality
17-108-final-viral-combined_k141_329193||full Not-determined
17-108-final-viral-combined_k141_82361||full Not-determined
17-108-final-viral-combined_k141_137566||full Not-determined
17-108-final-viral-combined_k141_316880||full Not-determined
17-108-final-viral-combined_k141_316883||full Low-quality
17-108-final-viral-combined_k141_192759||full Low-quality
17-108-final-viral-combined_k141_192761||full Low-quality
17-108-final-viral-combined_k141_192772||full Not-determined
17-108-final-viral-combined_k141_192773||full Not-determined
17-108-final-viral-combined_k141_192796||full Not-determined
17-108-final-viral-combined_k141_137623||full Low-quality
17-108-final-viral-combined_k141_206350||full Not-determined
17-108-final-viral-combined_k141_123858||full Low-quality
17-108-final-viral-combined_k141_275525||full Not-determined
17-108-final-viral-combined_k141_329260||full Not-determined
17-108-final-viral-combined_k141_123879||full Not-determined
17-108-final-viral-combined_k141_329272||full Low-quality
17-108-final-viral-combined_k141_82457||full Low-quality
17-108-final-viral-combined_k141_26933||full Not-determined
17-108-final-viral-combined_k141_275551||full Low-quality
17-108-final-viral-combined_k141_82477||full Low-quality
17-108-final-viral-combined_k141_26956||full Not-determined
17-108-final-viral-combined_k141_123925||full Low-quality
17-108-final-viral-combined_k141_137703||full Low-quality
17-108-final-viral-combined_k141_206462||full Not-determined
17-108-final-viral-combined_k141_329343||full Not-determined
17-108-final-viral-combined_k141_275614||full Not-determined
17-108-final-viral-combined_k141_206481||full Low-quality
17-108-final-viral-combined_k141_206511||full Low-quality
17-108-final-viral-combined_k141_329390||full Low-quality
17-108-final-viral-combined_k141_27010||full Not-determined
17-108-final-viral-combined_k141_27024||full Not-determined
17-108-final-viral-combined_k141_329420||full Low-quality
17-108-final-viral-combined_k141_329421||full Not-determined
17-108-final-viral-combined_k141_124013||full Low-quality
17-108-final-viral-combined_k141_124017||full Not-determined
17-108-final-viral-combined_k141_137806||full Low-quality
17-108-final-viral-combined_k141_206634||full Low-quality
17-108-final-viral-combined_k141_329483||full Low-quality
17-108-final-viral-combined_k141_27115||full Not-determined
17-108-final-viral-combined_k141_206655||full Not-determined
17-108-final-viral-combined_k141_27141||full Not-determined
17-108-final-viral-combined_k141_206692||full Low-quality
17-109-final-viral-combined_k141_334806||full High-quality
17-108-final-viral-combined_k141_27173||full Not-determined
17-108-final-viral-combined_k141_329507||full Low-quality
17-108-final-viral-combined_k141_329512||full Low-quality
17-108-final-viral-combined_k141_329522||full Not-determined
17-108-final-viral-combined_k141_27202||full Not-determined
17-108-final-viral-combined_k141_27219||full Not-determined
17-108-final-viral-combined_k141_329570||full Low-quality
17-108-final-viral-combined_k141_27282||full Not-determined
17-108-final-viral-combined_k141_329661||full Low-quality
17-108-final-viral-combined_k141_329672||full Low-quality
17-108-final-viral-combined_k141_27318||full Low-quality
17-108-final-viral-combined_k141_329708||full Not-determined
17-108-final-viral-combined_k141_329792||full Not-determined
17-108-final-viral-combined_k141_329809||full Not-determined
17-108-final-viral-combined_k141_329840||full Not-determined
17-108-final-viral-combined_k141_27472||full Low-quality
17-108-final-viral-combined_k141_329864||full Not-determined
17-108-final-viral-combined_k141_329871||full Low-quality
17-108-final-viral-combined_k141_330012||full Not-determined
17-108-final-viral-combined_k141_330096||full Low-quality
17-108-final-viral-combined_k141_330119||full Not-determined
17-108-final-viral-combined_k141_330128||full Low-quality
17-108-final-viral-combined_k141_330252||full Not-determined
17-108-final-viral-combined_k141_330253||full Low-quality
17-108-final-viral-combined_k141_330345||full Not-determined
17-108-final-viral-combined_k141_330368||full Not-determined
17-108-final-viral-combined_k141_330398||full Not-determined
17-108-final-viral-combined_k141_165692||0_partial Low-quality
17-108-final-viral-combined_k141_124528||0_partial Not-determined
17-108-final-viral-combined_k141_96978||0_partial Not-determined
17-108-final-viral-combined_k141_248571||0_partial Low-quality
17-108-final-viral-combined_k141_248668||0_partial Not-determined
17-108-final-viral-combined_k141_207561||0_partial Low-quality
17-108-final-viral-combined_k141_180429||0_partial Not-determined
17-108-final-viral-combined_k141_236046||0_partial Not-determined
17-108-final-viral-combined_k141_70339||0_partial Low-quality
17-108-final-viral-combined_k141_153467||0_partial Not-determined
17-108-final-viral-combined_k141_70424||0_partial Low-quality
17-108-final-viral-combined_k141_291525||0_partial Low-quality
17-108-final-viral-combined_k141_98981||0_partial Low-quality
17-108-final-viral-combined_k141_181500||0_partial Low-quality
17-108-final-viral-combined_k141_319509||0_partial Low-quality
17-108-final-viral-combined_k141_113055||0_partial Low-quality
17-108-final-viral-combined_k141_306975||0_partial Low-quality
17-108-final-viral-combined_k141_224490||0_partial Not-determined
17-108-final-viral-combined_k141_293182||0_partial Low-quality
17-108-final-viral-combined_k141_212078||0_partial Low-quality
17-108-final-viral-combined_k141_170745||0_partial Low-quality
17-108-final-viral-combined_k141_266918||0_partial Low-quality
17-108-final-viral-combined_k141_18338||0_partial Not-determined
17-108-final-viral-combined_k141_197700||0_partial Low-quality
17-108-final-viral-combined_k141_143125||0_partial Low-quality
17-108-final-viral-combined_k141_129364||0_partial Low-quality
17-108-final-viral-combined_k141_129668||0_partial Not-determined
17-108-final-viral-combined_k141_102842||0_partial Low-quality
17-108-final-viral-combined_k141_295134||0_partial Low-quality
17-108-final-viral-combined_k141_47427||0_partial Not-determined
17-108-final-viral-combined_k141_6203||0_partial Low-quality
17-108-final-viral-combined_k141_199102||0_partial Not-determined
17-108-final-viral-combined_k141_241798||0_partial Low-quality
17-108-final-viral-combined_k141_172428||0_partial Not-determined
17-108-final-viral-combined_k141_283224||0_partial Not-determined
17-108-final-viral-combined_k141_186153||0_partial Low-quality
17-108-final-viral-combined_k141_200131||0_partial Not-determined
17-108-final-viral-combined_k141_242954||0_partial Not-determined
17-108-final-viral-combined_k141_324356||0_partial Low-quality
17-108-final-viral-combined_k141_146499||0_partial Not-determined
17-108-final-viral-combined_k141_174593||0_partial Low-quality
17-108-final-viral-combined_k141_21942||0_partial Low-quality
17-108-final-viral-combined_k141_9954||0_partial Low-quality
17-108-final-viral-combined_k141_244786||0_partial Low-quality
17-108-final-viral-combined_k141_119802||0_partial Low-quality
17-108-final-viral-combined_k141_299568||0_partial Low-quality
17-108-final-viral-combined_k141_23572||0_partial Low-quality
17-108-final-viral-combined_k141_53204||0_partial Low-quality
17-108-final-viral-combined_k141_315528||0_partial Low-quality
17-108-final-viral-combined_k141_150924||0_partial Low-quality
17-108-final-viral-combined_k141_275441||0_partial Not-determined
17-108-final-viral-combined_k141_27564||lt2gene Not-determined
17-108-final-viral-combined_k141_68915||lt2gene Low-quality
17-108-final-viral-combined_k141_165379||lt2gene Low-quality
17-108-final-viral-combined_k141_82704||lt2gene Not-determined
17-108-final-viral-combined_k141_289438||lt2gene Low-quality
17-108-final-viral-combined_k141_137871||lt2gene Low-quality
17-108-final-viral-combined_k141_317007||lt2gene Low-quality
17-108-final-viral-combined_k141_13866||lt2gene Not-determined
17-108-final-viral-combined_k141_317014||lt2gene Low-quality
17-108-final-viral-combined_k141_124113||lt2gene Low-quality
17-108-final-viral-combined_k141_248108||lt2gene Low-quality
17-108-final-viral-combined_k141_234283||lt2gene Not-determined
17-108-final-viral-combined_k141_68995||lt2gene Not-determined
17-108-final-viral-combined_k141_123||lt2gene Low-quality
17-108-final-viral-combined_k141_69002||lt2gene Not-determined
17-108-final-viral-combined_k141_192977||lt2gene Low-quality
17-108-final-viral-combined_k141_110324||lt2gene Low-quality
17-108-final-viral-combined_k141_124178||lt2gene Low-quality
17-108-final-viral-combined_k141_193018||lt2gene Not-determined
17-108-final-viral-combined_k141_303305||lt2gene Low-quality
17-108-final-viral-combined_k141_248173||lt2gene Low-quality
17-108-final-viral-combined_k141_206886||lt2gene Not-determined
17-108-final-viral-combined_k141_303335||lt2gene Low-quality
17-108-final-viral-combined_k141_55266||lt2gene Low-quality
17-108-final-viral-combined_k141_151791||lt2gene Low-quality
17-108-final-viral-combined_k141_27639||lt2gene Low-quality
17-108-final-viral-combined_k141_193113||lt2gene Low-quality
17-108-final-viral-combined_k141_41468||lt2gene Not-determined
17-108-final-viral-combined_k141_234401||lt2gene Not-determined
17-108-final-viral-combined_k141_151859||lt2gene Low-quality
17-108-final-viral-combined_k141_303383||lt2gene Not-determined
17-108-final-viral-combined_k141_206943||lt2gene Not-determined
17-108-final-viral-combined_k141_303399||lt2gene Not-determined
17-108-final-viral-combined_k141_55343||lt2gene Not-determined
17-108-final-viral-combined_k141_138055||lt2gene Low-quality
17-108-final-viral-combined_k141_14058||lt2gene Not-determined
17-108-final-viral-combined_k141_55359||lt2gene Not-determined
17-108-final-viral-combined_k141_234443||lt2gene Low-quality
17-108-final-viral-combined_k141_110429||lt2gene Low-quality
17-108-final-viral-combined_k141_165721||lt2gene Low-quality
17-108-final-viral-combined_k141_55412||lt2gene Low-quality
17-108-final-viral-combined_k141_220835||lt2gene Low-quality
17-108-final-viral-combined_k141_317271||lt2gene Low-quality
17-108-final-viral-combined_k141_165782||lt2gene Low-quality
17-108-final-viral-combined_k141_303499||lt2gene Low-quality
17-108-final-viral-combined_k141_96787||lt2gene Low-quality
17-108-final-viral-combined_k141_151971||lt2gene Low-quality

| | |
|---|---|
| 17-108-final-viral-combined_k141_419||lt2gene | Not-determined |
| 17-108-final-viral-combined_k141_289698||lt2gene | Low-quality |
| 17-108-final-viral-combined_k141_317354||lt2gene | Not-determined |
| 17-108-final-viral-combined_k141_262214||lt2gene | Not-determined |
| 17-108-final-viral-combined_k141_83066||lt2gene | Not-determined |
| 17-108-final-viral-combined_k141_83072||lt2gene | Low-quality |
| 17-108-final-viral-combined_k141_303576||lt2gene | Low-quality |
| 17-108-final-viral-combined_k141_124504||lt2gene | Low-quality |
| 17-108-final-viral-combined_k141_110580||lt2gene | Low-quality |
| 17-108-final-viral-combined_k141_234587||lt2gene | Low-quality |
| 17-108-final-viral-combined_k141_41627||lt2gene | Low-quality |
| 17-108-final-viral-combined_k141_14248||lt2gene | Low-quality |
| 17-108-final-viral-combined_k141_207200||lt2gene | Low-quality |
| 17-108-final-viral-combined_k141_28017||lt2gene | Not-determined |
| 17-108-final-viral-combined_k141_41637||lt2gene | Low-quality |
| 17-108-final-viral-combined_k141_289745||lt2gene | Not-determined |
| 17-108-final-viral-combined_k141_14294||lt2gene | Low-quality |
| 17-108-final-viral-combined_k141_166008||lt2gene | Not-determined |
| 17-108-final-viral-combined_k141_14303||lt2gene | Not-determined |
| 17-108-final-viral-combined_k141_69340||lt2gene | Low-quality |
| 17-108-final-viral-combined_k141_220933||lt2gene | Low-quality |
| 17-108-final-viral-combined_k141_152129||lt2gene | Not-determined |
| 17-108-final-viral-combined_k141_14357||lt2gene | Low-quality |
| 17-108-final-viral-combined_k141_179607||lt2gene | Not-determined |
| 17-108-final-viral-combined_k141_97012||lt2gene | Low-quality |
| 17-108-final-viral-combined_k141_124612||lt2gene | Low-quality |
| 17-108-final-viral-combined_k141_234816||lt2gene | Low-quality |
| 17-108-final-viral-combined_k141_193435||lt2gene | Low-quality |
| 17-108-final-viral-combined_k141_248614||lt2gene | Low-quality |
| 17-108-final-viral-combined_k141_221043||lt2gene | Low-quality |
| 17-108-final-viral-combined_k141_124722||lt2gene | Not-determined |
| 17-108-final-viral-combined_k141_41873||lt2gene | Low-quality |
| 17-108-final-viral-combined_k141_41876||lt2gene | Low-quality |
| 17-108-final-viral-combined_k141_166141||lt2gene | Not-determined |
| 17-108-final-viral-combined_k141_97092||lt2gene | Low-quality |
| 17-108-final-viral-combined_k141_69506||lt2gene | Not-determined |
| 17-108-final-viral-combined_k141_55729||lt2gene | Low-quality |
| 17-108-final-viral-combined_k141_207281||lt2gene | Low-quality |
| 17-108-final-viral-combined_k141_69557||lt2gene | Low-quality |
| 17-108-final-viral-combined_k141_14482||lt2gene | Not-determined |
| 17-108-final-viral-combined_k141_276224||lt2gene | Low-quality |
| 17-108-final-viral-combined_k141_276251||lt2gene | Low-quality |
| 17-108-final-viral-combined_k141_97173||lt2gene | Low-quality |
| 17-108-final-viral-combined_k141_110829||lt2gene | Not-determined |
| 17-108-final-viral-combined_k141_166256||lt2gene | Not-determined |
| 17-108-final-viral-combined_k141_303897||lt2gene | Low-quality |
| 17-108-final-viral-combined_k141_193663||lt2gene | Low-quality |
| 17-108-final-viral-combined_k141_14557||lt2gene | Low-quality |
| 17-108-final-viral-combined_k141_124900||lt2gene | Low-quality |
| 17-108-final-viral-combined_k141_179748||lt2gene | Not-determined |
| 17-108-final-viral-combined_k141_179757||lt2gene | Low-quality |
| 17-108-final-viral-combined_k141_317680||lt2gene | Low-quality |
| 17-108-final-viral-combined_k141_684||lt2gene | Low-quality |
| 17-108-final-viral-combined_k141_248767||lt2gene | Low-quality |
| 17-108-final-viral-combined_k141_193709||lt2gene | Not-determined |
| 17-108-final-viral-combined_k141_290097||lt2gene | Low-quality |
| 17-108-final-viral-combined_k141_703||lt2gene | Low-quality |
| 17-108-final-viral-combined_k141_166357||lt2gene | Not-determined |
| 17-108-final-viral-combined_k141_83414||lt2gene | Low-quality |
| 17-108-final-viral-combined_k141_28343||lt2gene | Not-determined |
| 17-108-final-viral-combined_k141_276405||lt2gene | Low-quality |
| 17-108-final-viral-combined_k141_179840||lt2gene | Not-determined |
| 17-108-final-viral-combined_k141_207486||lt2gene | Low-quality |
| 17-108-final-viral-combined_k141_276435||lt2gene | Low-quality |
| 17-108-final-viral-combined_k141_221312||lt2gene | Low-quality |
| 17-108-final-viral-combined_k141_124967||lt2gene | Not-determined |
| 17-108-final-viral-combined_k141_262673||lt2gene | Low-quality |
| 17-108-final-viral-combined_k141_166443||lt2gene | Not-determined |
| 17-108-final-viral-combined_k141_248895||lt2gene | Low-quality |
| 17-108-final-viral-combined_k141_235225||lt2gene | Low-quality |
| 17-108-final-viral-combined_k141_28427||lt2gene | Low-quality |
| 17-108-final-viral-combined_k141_276496||lt2gene | Low-quality |
| 17-108-final-viral-combined_k141_111045||lt2gene | Low-quality |
| 17-108-final-viral-combined_k141_262751||lt2gene | Not-determined |
| 17-108-final-viral-combined_k141_97443||lt2gene | Not-determined |
| 17-108-final-viral-combined_k141_235286||lt2gene | Not-determined |
| 17-108-final-viral-combined_k141_896||lt2gene | Low-quality |
| 17-108-final-viral-combined_k141_42095||lt2gene | Low-quality |
| 17-108-final-viral-combined_k141_276508||lt2gene | Low-quality |
| 17-108-final-viral-combined_k141_125055||lt2gene | Low-quality |
| 17-108-final-viral-combined_k141_235338||lt2gene | Low-quality |
| 17-108-final-viral-combined_k141_28457||lt2gene | Low-quality |
| 17-108-final-viral-combined_k141_944||lt2gene | Low-quality |
| 17-108-final-viral-combined_k141_83653||lt2gene | Not-determined |
| 17-108-final-viral-combined_k141_97508||lt2gene | Not-determined |
| 17-108-final-viral-combined_k141_42167||lt2gene | Not-determined |
| 17-108-final-viral-combined_k141_235398||lt2gene | Low-quality |
| 17-108-final-viral-combined_k141_14772||lt2gene | Low-quality |
| 17-108-final-viral-combined_k141_262790||lt2gene | Not-determined |
| 17-108-final-viral-combined_k141_207726||lt2gene | Low-quality |
| 17-108-final-viral-combined_k141_207733||lt2gene | Low-quality |
| 17-108-final-viral-combined_k141_262805||lt2gene | Not-determined |
| 17-108-final-viral-combined_k141_166632||lt2gene | Low-quality |
| 17-108-final-viral-combined_k141_152690||lt2gene | Low-quality |
| 17-108-final-viral-combined_k141_276620||lt2gene | Low-quality |
| 17-108-final-viral-combined_k141_207744||lt2gene | Not-determined |
| 17-108-final-viral-combined_k141_138732||lt2gene | Low-quality |
| 17-108-final-viral-combined_k141_56102||lt2gene | Low-quality |
| 17-108-final-viral-combined_k141_56103||lt2gene | Low-quality |
| 17-108-final-viral-combined_k141_83782||lt2gene | Not-determined |
| 17-108-final-viral-combined_k141_166680||lt2gene | Low-quality |
| 17-108-final-viral-combined_k141_14831||lt2gene | Low-quality |
| 17-108-final-viral-combined_k141_69870||lt2gene | Not-determined |
| 17-108-final-viral-combined_k141_97671||lt2gene | Low-quality |
| 17-108-final-viral-combined_k141_56131||lt2gene | Low-quality |
| 17-108-final-viral-combined_k141_221505||lt2gene | Low-quality |
| 17-108-final-viral-combined_k141_276648||lt2gene | Low-quality |
| 17-108-final-viral-combined_k141_262904||lt2gene | Low-quality |
| 17-108-final-viral-combined_k141_304278||lt2gene | Low-quality |
| 17-108-final-viral-combined_k141_249129||lt2gene | Not-determined |
| 17-108-final-viral-combined_k141_69912||lt2gene | Low-quality |
| 17-108-final-viral-combined_k141_180160||lt2gene | Not-determined |
| 17-108-final-viral-combined_k141_180161||lt2gene | Low-quality |
| 17-108-final-viral-combined_k141_304310||lt2gene | Not-determined |
| 17-108-final-viral-combined_k141_304332||lt2gene | Not-determined |
| 17-108-final-viral-combined_k141_304333||lt2gene | Low-quality |
| 17-108-final-viral-combined_k141_235658||lt2gene | Low-quality |
| 17-108-final-viral-combined_k141_276719||lt2gene | Not-determined |
| 17-108-final-viral-combined_k141_138876||lt2gene | Low-quality |
| 17-108-final-viral-combined_k141_14974||lt2gene | Not-determined |
| 17-108-final-viral-combined_k141_276779||lt2gene | Not-determined |
| 17-108-final-viral-combined_k141_152826||lt2gene | Low-quality |
| 17-108-final-viral-combined_k141_235746||lt2gene | Low-quality |
| 17-108-final-viral-combined_k141_276826||lt2gene | Low-quality |
| 17-108-final-viral-combined_k141_207983||lt2gene | Not-determined |
| 17-108-final-viral-combined_k141_69936||lt2gene | Low-quality |
| 17-108-final-viral-combined_k141_97889||lt2gene | Not-determined |
| 17-108-final-viral-combined_k141_69955||lt2gene | Low-quality |
| 17-108-final-viral-combined_k141_125350||lt2gene | Low-quality |
| 17-108-final-viral-combined_k141_111345||lt2gene | Not-determined |
| 17-108-final-viral-combined_k141_42488||lt2gene | Low-quality |
| 17-108-final-viral-combined_k141_125399||lt2gene | Low-quality |
| 17-108-final-viral-combined_k141_70041||lt2gene | Low-quality |
| 17-108-final-viral-combined_k141_125456||lt2gene | Not-determined |
| 17-108-final-viral-combined_k141_276987||lt2gene | Low-quality |
| 17-108-final-viral-combined_k141_276989||lt2gene | Low-quality |
| 17-108-final-viral-combined_k141_98040||lt2gene | Low-quality |
| 17-108-final-viral-combined_k141_28736||lt2gene | Low-quality |
| 17-108-final-viral-combined_k141_263188||lt2gene | Not-determined |
| 17-108-final-viral-combined_k141_263191||lt2gene | Low-quality |
| 17-108-final-viral-combined_k141_125490||lt2gene | Low-quality |
| 17-108-final-viral-combined_k141_138953||lt2gene | Low-quality |
| 17-108-final-viral-combined_k141_70145||lt2gene | Not-determined |
| 17-108-final-viral-combined_k141_304646||lt2gene | Low-quality |
| 17-108-final-viral-combined_k141_28853||lt2gene | Not-determined |
| 17-108-final-viral-combined_k141_1345||lt2gene | Low-quality |
| 17-108-final-viral-combined_k141_28856||lt2gene | Low-quality |
| 17-108-final-viral-combined_k141_15192||lt2gene | Low-quality |
| 17-108-final-viral-combined_k141_208147||lt2gene | Low-quality |
| 17-108-final-viral-combined_k141_84132||lt2gene | Low-quality |
| 17-108-final-viral-combined_k141_84141||lt2gene | Low-quality |
| 17-108-final-viral-combined_k141_208219||lt2gene | Low-quality |
| 17-108-final-viral-combined_k141_208226||lt2gene | Low-quality |
| 17-108-final-viral-combined_k141_84180||lt2gene | Low-quality |
| 17-108-final-viral-combined_k141_290712||lt2gene | Not-determined |
| 17-108-final-viral-combined_k141_84215||lt2gene | Low-quality |
| 17-108-final-viral-combined_k141_290767||lt2gene | Not-determined |
| 17-108-final-viral-combined_k141_221967||lt2gene | Low-quality |
| 17-108-final-viral-combined_k141_221971||lt2gene | Low-quality |
| 17-108-final-viral-combined_k141_125657||lt2gene | Not-determined |
| 17-108-final-viral-combined_k141_125664||lt2gene | Not-determined |
| 17-108-final-viral-combined_k141_42792||lt2gene | Low-quality |
| 17-108-final-viral-combined_k141_153047||lt2gene | Low-quality |
| 17-108-final-viral-combined_k141_180554||lt2gene | Low-quality |
| 17-108-final-viral-combined_k141_304776||lt2gene | Not-determined |
| 17-108-final-viral-combined_k141_194355||lt2gene | Not-determined |
| 17-108-final-viral-combined_k141_249608||lt2gene | Not-determined |
| 17-108-final-viral-combined_k141_249617||lt2gene | Low-quality |
| 17-108-final-viral-combined_k141_42850||lt2gene | Not-determined |
| 17-108-final-viral-combined_k141_111515||lt2gene | Low-quality |
| 17-108-final-viral-combined_k141_180613||lt2gene | Low-quality |
| 17-108-final-viral-combined_k141_194409||lt2gene | Low-quality |
| 17-108-final-viral-combined_k141_318360||lt2gene | Not-determined |
| 17-108-final-viral-combined_k141_194413||lt2gene | Not-determined |
| 17-108-final-viral-combined_k141_222022||lt2gene | Not-determined |
| 17-108-final-viral-combined_k141_42872||lt2gene | Low-quality |
| 17-108-final-viral-combined_k141_98297||lt2gene | Low-quality |
| 17-108-final-viral-combined_k141_167245||lt2gene | Not-determined |
| 17-108-final-viral-combined_k141_167259||lt2gene | Low-quality |
| 17-108-final-viral-combined_k141_98316||lt2gene | Low-quality |
| 17-108-final-viral-combined_k141_125796||lt2gene | Not-determined |
| 17-108-final-viral-combined_k141_1565||lt2gene | Low-quality |
| 17-108-final-viral-combined_k141_222132||lt2gene | Low-quality |
| 17-108-final-viral-combined_k141_318419||lt2gene | Low-quality |
| 17-108-final-viral-combined_k141_111573||lt2gene | Low-quality |
| 17-108-final-viral-combined_k141_304859||lt2gene | Low-quality |
| 17-108-final-viral-combined_k141_15503||lt2gene | Not-determined |
| 17-108-final-viral-combined_k141_125827||lt2gene | Low-quality |
| 17-108-final-viral-combined_k141_290971||lt2gene | Not-determined |
| 17-108-final-viral-combined_k141_139262||lt2gene | Low-quality |
| 17-108-final-viral-combined_k141_304895||lt2gene | Not-determined |
| 17-108-final-viral-combined_k141_291012||lt2gene | Low-quality |
| 17-108-final-viral-combined_k141_180736||lt2gene | Not-determined |
| 17-108-final-viral-combined_k141_291035||lt2gene | Low-quality |
| 17-108-final-viral-combined_k141_153270||lt2gene | Not-determined |
| 17-108-final-viral-combined_k141_194590||lt2gene | Low-quality |
| 17-108-final-viral-combined_k141_15584||lt2gene | Low-quality |
| 17-108-final-viral-combined_k141_111625||lt2gene | Low-quality |
| 17-108-final-viral-combined_k141_42920||lt2gene | Low-quality |
| 17-108-final-viral-combined_k141_180789||lt2gene | Low-quality |
| 17-108-final-viral-combined_k141_304972||lt2gene | Low-quality |
| 17-108-final-viral-combined_k141_167400||lt2gene | Not-determined |
| 17-108-final-viral-combined_k141_56781||lt2gene | Low-quality |
| 17-108-final-viral-combined_k141_249768||lt2gene | Low-quality |
| 17-108-final-viral-combined_k141_194621||lt2gene | Not-determined |
| 17-108-final-viral-combined_k141_139370||lt2gene | Not-determined |
| 17-108-final-viral-combined_k141_236302||lt2gene | Low-quality |
| 17-108-final-viral-combined_k141_222305||lt2gene | Low-quality |
| 17-108-final-viral-combined_k141_208542||lt2gene | Low-quality |
| 17-108-final-viral-combined_k141_249815||lt2gene | Not-determined |
| 17-108-final-viral-combined_k141_291116||lt2gene | Low-quality |
| 17-108-final-viral-combined_k141_43096||lt2gene | Not-determined |
| 17-108-final-viral-combined_k141_111766||lt2gene | Low-quality |
| 17-108-final-viral-combined_k141_180883||lt2gene | Low-quality |
| 17-108-final-viral-combined_k141_208659||lt2gene | Not-determined |
| 17-108-final-viral-combined_k141_98536||lt2gene | Not-determined |
| 17-108-final-viral-combined_k141_125916||lt2gene | Not-determined |
| 17-108-final-viral-combined_k141_139474||lt2gene | Low-quality |
| 17-108-final-viral-combined_k141_139482||lt2gene | Not-determined |
| 17-108-final-viral-combined_k141_222386||lt2gene | Low-quality |
| 17-108-final-viral-combined_k141_222437||lt2gene | Low-quality |
| 17-108-final-viral-combined_k141_15717||lt2gene | Low-quality |
| 17-108-final-viral-combined_k141_318646||lt2gene | Low-quality |
| 17-108-final-viral-combined_k141_263698||lt2gene | Not-determined |
| 17-108-final-viral-combined_k141_236432||lt2gene | Low-quality |
| 17-108-final-viral-combined_k141_139564||lt2gene | Low-quality |
| 17-108-final-viral-combined_k141_305103||lt2gene | Not-determined |
| 17-108-final-viral-combined_k141_222497||lt2gene | Low-quality |
| 17-108-final-viral-combined_k141_305113||lt2gene | Not-determined |
| 17-108-final-viral-combined_k141_181025||lt2gene | Not-determined |
| 17-108-final-viral-combined_k141_305212||lt2gene | Low-quality |
| 17-108-final-viral-combined_k141_1899||lt2gene | Low-quality |
| 17-108-final-viral-combined_k141_277725||lt2gene | Low-quality |
| 17-108-final-viral-combined_k141_1959||lt2gene | Not-determined |
| 17-108-final-viral-combined_k141_208933||lt2gene | Low-quality |
| 17-108-final-viral-combined_k141_318822||lt2gene | Low-quality |
| 17-108-final-viral-combined_k141_291416||lt2gene | Low-quality |
| 17-108-final-viral-combined_k141_208973||lt2gene | Not-determined |
| 17-108-final-viral-combined_k141_126164||lt2gene | Low-quality |
| 17-108-final-viral-combined_k141_194960||lt2gene | Not-determined |
| 17-108-final-viral-combined_k141_29513||lt2gene | Not-determined |
| 17-108-final-viral-combined_k141_318858||lt2gene | Low-quality |
| 17-108-final-viral-combined_k141_15975||lt2gene | Not-determined |
| 17-108-final-viral-combined_k141_277910||lt2gene | Not-determined |
| 17-108-final-viral-combined_k141_318882||lt2gene | Not-determined |
| 17-108-final-viral-combined_k141_277918||lt2gene | Low-quality |
| 17-108-final-viral-combined_k141_57276||lt2gene | Low-quality |
| 17-108-final-viral-combined_k141_57279||lt2gene | Low-quality |
| 17-108-final-viral-combined_k141_236699||lt2gene | Not-determined |
| 17-108-final-viral-combined_k141_112016||lt2gene | Low-quality |
| 17-108-final-viral-combined_k141_250124||lt2gene | Not-determined |
| 17-108-final-viral-combined_k141_43427||lt2gene | Low-quality |
| 17-108-final-viral-combined_k141_126200||lt2gene | Low-quality |
| 17-108-final-viral-combined_k141_139860||lt2gene | Low-quality |
| 17-108-final-viral-combined_k141_236762||lt2gene | Low-quality |
| 17-108-final-viral-combined_k141_278018||lt2gene | Low-quality |
| 17-108-final-viral-combined_k141_236764||lt2gene | Not-determined |
| 17-108-final-viral-combined_k141_250162||lt2gene | Low-quality |
| 17-108-final-viral-combined_k141_250181||lt2gene | Low-quality |
| 17-108-final-viral-combined_k141_236843||lt2gene | Low-quality |
| 17-108-final-viral-combined_k141_250216||lt2gene | Low-quality |
| 17-108-final-viral-combined_k141_98890||lt2gene | Low-quality |
| 17-108-final-viral-combined_k141_236865||lt2gene | Low-quality |
| 17-108-final-viral-combined_k141_167851||lt2gene | Low-quality |
| 17-108-final-viral-combined_k141_250256||lt2gene | Not-determined |
| 17-108-final-viral-combined_k141_167870||lt2gene | Not-determined |
| 17-108-final-viral-combined_k141_98919||lt2gene | Low-quality |
| 17-108-final-viral-combined_k141_236888||lt2gene | Low-quality |
| 17-108-final-viral-combined_k141_181300||lt2gene | Not-determined |
| 17-108-final-viral-combined_k141_209207||lt2gene | Low-quality |
| 17-108-final-viral-combined_k141_126277||lt2gene | Not-determined |
| 17-108-final-viral-combined_k141_153747||lt2gene | Not-determined |
| 17-108-final-viral-combined_k141_126292||lt2gene | Low-quality |
| 17-108-final-viral-combined_k141_126302||lt2gene | Low-quality |
| 17-108-final-viral-combined_k141_278173||lt2gene | Low-quality |
| 17-108-final-viral-combined_k141_250367||lt2gene | Low-quality |
| 17-108-final-viral-combined_k141_57536||lt2gene | Low-quality |
| 17-108-final-viral-combined_k141_250378||lt2gene | Low-quality |
| 17-108-final-viral-combined_k141_305586||lt2gene | Low-quality |
| 17-108-final-viral-combined_k141_278190||lt2gene | Low-quality |
| 17-108-final-viral-combined_k141_126368||lt2gene | Low-quality |
| 17-108-final-viral-combined_k141_126426||lt2gene | Not-determined |
| 17-108-final-viral-combined_k141_99049||lt2gene | Low-quality |
| 17-108-final-viral-combined_k141_167981||lt2gene | Low-quality |
| 17-108-final-viral-combined_k141_319113||lt2gene | Low-quality |
| 17-108-final-viral-combined_k141_2377||lt2gene | Low-quality |
| 17-108-final-viral-combined_k141_209406||lt2gene | Not-determined |
| 17-108-final-viral-combined_k141_209408||lt2gene | Not-determined |
| 17-108-final-viral-combined_k141_291627||lt2gene | Not-determined |
| 17-108-final-viral-combined_k141_126464||lt2gene | Low-quality |
| 17-108-final-viral-combined_k141_319142||lt2gene | Low-quality |
| 17-108-final-viral-combined_k141_168030||lt2gene | Low-quality |
| 17-108-final-viral-combined_k141_291663||lt2gene | Low-quality |
| 17-108-final-viral-combined_k141_43676||lt2gene | Low-quality |
| 17-108-final-viral-combined_k141_126519||lt2gene | Low-quality |
| 17-108-final-viral-combined_k141_264263||lt2gene | Not-determined |
| 17-108-final-viral-combined_k141_264272||lt2gene | Low-quality |
| 17-108-final-viral-combined_k141_29894||lt2gene | Low-quality |
| 17-108-final-viral-combined_k141_2502||lt2gene | Low-quality |
| 17-108-final-viral-combined_k141_291691||lt2gene | Low-quality |
| 17-108-final-viral-combined_k141_140219||lt2gene | Low-quality |
| 17-108-final-viral-combined_k141_222967||lt2gene | Not-determined |
| 17-108-final-viral-combined_k141_222996||lt2gene | Low-quality |
| 17-108-final-viral-combined_k141_223017||lt2gene | Low-quality |
| 17-108-final-viral-combined_k141_57829||lt2gene | Low-quality |
| 17-108-final-viral-combined_k141_126603||lt2gene | Low-quality |
| 17-108-final-viral-combined_k141_168165||lt2gene | Not-determined |
| 17-108-final-viral-combined_k141_154096||lt2gene | Low-quality |
| 17-108-final-viral-combined_k141_70996||lt2gene | Low-quality |
| 17-108-final-viral-combined_k141_57899||lt2gene | Low-quality |
| 17-108-final-viral-combined_k141_181488||lt2gene | Low-quality |
| 17-108-final-viral-combined_k141_278476||lt2gene | Low-quality |
| 17-108-final-viral-combined_k141_43876||lt2gene | Low-quality |
| 17-108-final-viral-combined_k141_237124||lt2gene | Low-quality |
| 17-108-final-viral-combined_k141_278486||lt2gene | Low-quality |
| 17-108-final-viral-combined_k141_209667||lt2gene | Low-quality |
| 17-108-final-viral-combined_k141_209715||lt2gene | Not-determined |
| 17-108-final-viral-combined_k141_43966||lt2gene | Low-quality |
| 17-108-final-viral-combined_k141_154233||lt2gene | Low-quality |
| 17-108-final-viral-combined_k141_250679||lt2gene | Low-quality |
| 17-108-final-viral-combined_k141_181534||lt2gene | Low-quality |
| 17-108-final-viral-combined_k141_58015||lt2gene | Low-quality |
| 17-108-final-viral-combined_k141_291827||lt2gene | Not-determined |
| 17-108-final-viral-combined_k141_223212||lt2gene | Low-quality |
| 17-108-final-viral-combined_k141_264447||lt2gene | Low-quality |
| 17-108-final-viral-combined_k141_112527||lt2gene | Low-quality |
| 17-108-final-viral-combined_k141_237316||lt2gene | Low-quality |
| 17-108-final-viral-combined_k141_306043||lt2gene | Not-determined |
| 17-108-final-viral-combined_k141_85302||lt2gene | Low-quality |
| 17-108-final-viral-combined_k141_291867||lt2gene | Low-quality |
| 17-108-final-viral-combined_k141_140461||lt2gene | Not-determined |
| 17-108-final-viral-combined_k141_278640||lt2gene | Low-quality |
| 17-108-final-viral-combined_k141_223258||lt2gene | Low-quality |
| 17-108-final-viral-combined_k141_71209||lt2gene | Not-determined |
| 17-108-final-viral-combined_k141_264513||lt2gene | Low-quality |
| 17-108-final-viral-combined_k141_154341||lt2gene | Low-quality |
| 17-108-final-viral-combined_k141_154342||lt2gene | Not-determined |
| 17-108-final-viral-combined_k141_99448||lt2gene | Not-determined |
| 17-108-final-viral-combined_k141_85345||lt2gene | Not-determined |
| 17-108-final-viral-combined_k141_306114||lt2gene | Not-determined |
| 17-108-final-viral-combined_k141_223299||lt2gene | Not-determined |
| 17-108-final-viral-combined_k141_30231||lt2gene | Low-quality |
| 17-108-final-viral-combined_k141_223315||lt2gene | Not-determined |
| 17-108-final-viral-combined_k141_223336||lt2gene | Not-determined |
| 17-108-final-viral-combined_k141_250823||lt2gene | Not-determined |
| 17-108-final-viral-combined_k141_85398||lt2gene | Low-quality |
| 17-108-final-viral-combined_k141_112618||lt2gene | Not-determined |
| 17-108-final-viral-combined_k141_112648||lt2gene | Low-quality |
| 17-108-final-viral-combined_k141_44149||lt2gene | Low-quality |
| 17-108-final-viral-combined_k141_154369||lt2gene | Low-quality |
| 17-108-final-viral-combined_k141_195588||lt2gene | Low-quality |
| 17-108-final-viral-combined_k141_278672||lt2gene | Low-quality |
| 17-108-final-viral-combined_k141_112689||lt2gene | Low-quality |
| 17-108-final-viral-combined_k141_58256||lt2gene | Low-quality |
| 17-108-final-viral-combined_k141_154438||lt2gene | Not-determined |
| 17-108-final-viral-combined_k141_30348||lt2gene | Low-quality |
| 17-108-final-viral-combined_k141_112713||lt2gene | Low-quality |
| 17-108-final-viral-combined_k141_16518||lt2gene | Not-determined |
| 17-108-final-viral-combined_k141_154470||lt2gene | Not-determined |
| 17-108-final-viral-combined_k141_71412||lt2gene | Low-quality |
| 17-108-final-viral-combined_k141_181764||lt2gene | Low-quality |
| 17-108-final-viral-combined_k141_181773||lt2gene | Low-quality |
| 17-108-final-viral-combined_k141_319516||lt2gene | Low-quality |
| 17-108-final-viral-combined_k141_140663||lt2gene | Not-determined |
| 17-108-final-viral-combined_k141_85563||lt2gene | Not-determined |
| 17-108-final-viral-combined_k141_168639||lt2gene | Low-quality |
| 17-108-final-viral-combined_k141_181797||lt2gene | Low-quality |
| 17-108-final-viral-combined_k141_237592||lt2gene | Low-quality |
| 17-108-final-viral-combined_k141_278826||lt2gene | Low-quality |
| 17-108-final-viral-combined_k141_237620||lt2gene | Low-quality |
| 17-108-final-viral-combined_k141_154522||lt2gene | Low-quality |
| 17-108-final-viral-combined_k141_237634||lt2gene | Low-quality |
| 17-108-final-viral-combined_k141_278848||lt2gene | Low-quality |
| 17-108-final-viral-combined_k141_237672||lt2gene | Low-quality |
| 17-108-final-viral-combined_k141_126963||lt2gene | Low-quality |
| 17-108-final-viral-combined_k141_99670||lt2gene | Low-quality |
| 17-108-final-viral-combined_k141_99686||lt2gene | Not-determined |
| 17-108-final-viral-combined_k141_71493||lt2gene | Low-quality |
| 17-108-final-viral-combined_k141_181868||lt2gene | Not-determined |
| 17-108-final-viral-combined_k141_292081||lt2gene | Not-determined |
| 17-108-final-viral-combined_k141_154611||lt2gene | Not-determined |
| 17-108-final-viral-combined_k141_99740||lt2gene | Low-quality |
| 17-108-final-viral-combined_k141_140745||lt2gene | Low-quality |
| 17-108-final-viral-combined_k141_154624||lt2gene | Not-determined |
| 17-108-final-viral-combined_k141_16645||lt2gene | Low-quality |
| 17-108-final-viral-combined_k141_264811||lt2gene | Low-quality |
| 17-108-final-viral-combined_k141_168705||lt2gene | Not-determined |
| 17-108-final-viral-combined_k141_168706||lt2gene | Not-determined |
| 17-108-final-viral-combined_k141_140780||lt2gene | Low-quality |
| 17-108-final-viral-combined_k141_195775||lt2gene | Low-quality |
| 17-108-final-viral-combined_k141_251052||lt2gene | Low-quality |
| 17-108-final-viral-combined_k141_292142||lt2gene | Low-quality |
| 17-108-final-viral-combined_k141_210144||lt2gene | Not-determined |
| 17-108-final-viral-combined_k141_264898||lt2gene | Low-quality |
| 17-108-final-viral-combined_k141_210155||lt2gene | Low-quality |
| 17-108-final-viral-combined_k141_292197||lt2gene | Low-quality |
| 17-108-final-viral-combined_k141_237744||lt2gene | Low-quality |
| 17-108-final-viral-combined_k141_237751||lt2gene | Not-determined |
| 17-108-final-viral-combined_k141_223641||lt2gene | Not-determined |
| 17-108-final-viral-combined_k141_292210||lt2gene | Low-quality |
| 17-108-final-viral-combined_k141_237772||lt2gene | Not-determined |
| 17-108-final-viral-combined_k141_306602||lt2gene | Not-determined |
| 17-108-final-viral-combined_k141_223664||lt2gene | Not-determined |
| 17-108-final-viral-combined_k141_306612||lt2gene | Not-determined |
| 17-108-final-viral-combined_k141_306614||lt2gene | Low-quality |
| 17-108-final-viral-combined_k141_195861||lt2gene | Low-quality |
| 17-108-final-viral-combined_k141_44380||lt2gene | Low-quality |
| 17-108-final-viral-combined_k141_168850||lt2gene | Low-quality |
| 17-108-final-viral-combined_k141_237835||lt2gene | Low-quality |
| 17-108-final-viral-combined_k141_319707||lt2gene | Low-quality |
| 17-108-final-viral-combined_k141_223754||lt2gene | Not-determined |
| 17-108-final-viral-combined_k141_223765||lt2gene | Low-quality |
| 17-108-final-viral-combined_k141_195922||lt2gene | Low-quality |
| 17-108-final-viral-combined_k141_30732||lt2gene | Low-quality |
| 17-108-final-viral-combined_k141_223786||lt2gene | Low-quality |
| 17-108-final-viral-combined_k141_44462||lt2gene | Low-quality |
| 17-108-final-viral-combined_k141_195975||lt2gene | Low-quality |
| 17-108-final-viral-combined_k141_127210||lt2gene | Low-quality |
| 17-108-final-viral-combined_k141_85814||lt2gene | Not-determined |
| 17-108-final-viral-combined_k141_127225||lt2gene | Low-quality |
| 17-108-final-viral-combined_k141_196000||lt2gene | Not-determined |
| 17-108-final-viral-combined_k141_140943||lt2gene | Low-quality |
| 17-108-final-viral-combined_k141_251199||lt2gene | Not-determined |
| 17-108-final-viral-combined_k141_223847||lt2gene | Low-quality |
| 17-108-final-viral-combined_k141_237921||lt2gene | Low-quality |
| 17-108-final-viral-combined_k141_140993||lt2gene | Not-determined |
| 17-108-final-viral-combined_k141_71734||lt2gene | Not-determined |
| 17-108-final-viral-combined_k141_223853||lt2gene | Not-determined |
| 17-108-final-viral-combined_k141_210235||lt2gene | Low-quality |
| 17-108-final-viral-combined_k141_182260||lt2gene | Not-determined |
| 17-108-final-viral-combined_k141_71774||lt2gene | Low-quality |
| 17-108-final-viral-combined_k141_85941||lt2gene | Low-quality |
| 17-108-final-viral-combined_k141_16957||lt2gene | Low-quality |
| 17-108-final-viral-combined_k141_30839||lt2gene | Low-quality |
| 17-108-final-viral-combined_k141_251317||lt2gene | Not-determined |
| 17-108-final-viral-combined_k141_100074||lt2gene | Low-quality |
| 17-108-final-viral-combined_k141_182330||lt2gene | Low-quality |
| 17-108-final-viral-combined_k141_169058||lt2gene | Low-quality |
| 17-108-final-viral-combined_k141_182361||lt2gene | Low-quality |
| 17-108-final-viral-combined_k141_182362||lt2gene | Low-quality |
| 17-108-final-viral-combined_k141_71846||lt2gene | Not-determined |
| 17-108-final-viral-combined_k141_3346||lt2gene | Low-quality |
| 17-108-final-viral-combined_k141_71863||lt2gene | Not-determined |
| 17-108-final-viral-combined_k141_44664||lt2gene | Low-quality |
| 17-108-final-viral-combined_k141_141086||lt2gene | Low-quality |
| 17-108-final-viral-combined_k141_3364||lt2gene | Low-quality |
| 17-108-final-viral-combined_k141_141105||lt2gene | Low-quality |
| 17-108-final-viral-combined_k141_141116||lt2gene | Low-quality |
| 17-108-final-viral-combined_k141_3382||lt2gene | Not-determined |
| 17-108-final-viral-combined_k141_182427||lt2gene | Low-quality |
| 17-108-final-viral-combined_k141_182457||lt2gene | Low-quality |
| 17-108-final-viral-combined_k141_3424||lt2gene | Low-quality |
| 17-108-final-viral-combined_k141_306937||lt2gene | Low-quality |
| 17-108-final-viral-combined_k141_58718||lt2gene | Not-determined |
| 17-108-final-viral-combined_k141_251395||lt2gene | Low-quality |
| 17-108-final-viral-combined_k141_210380||lt2gene | Low-quality |
| 17-108-final-viral-combined_k141_182480||lt2gene | Not-determined |
| 17-108-final-viral-combined_k141_224021||lt2gene | Low-quality |
| 17-108-final-viral-combined_k141_320078||lt2gene | Low-quality |
| 17-108-final-viral-combined_k141_100229||lt2gene | Low-quality |
| 17-108-final-viral-combined_k141_155111||lt2gene | Not-determined |
| 17-108-final-viral-combined_k141_44788||lt2gene | Low-quality |
| 17-108-final-viral-combined_k141_155125||lt2gene | Low-quality |
| 17-108-final-viral-combined_k141_44814||lt2gene | Not-determined |
| 17-108-final-viral-combined_k141_210446||lt2gene | Not-determined |
| 17-108-final-viral-combined_k141_86187||lt2gene | Not-determined |
| 17-108-final-viral-combined_k141_44821||lt2gene | Not-determined |
| 17-108-final-viral-combined_k141_265271||lt2gene | Not-determined |
| 17-108-final-viral-combined_k141_58841||lt2gene | Low-quality |
| 17-108-final-viral-combined_k141_58846||lt2gene | Not-determined |
| 17-108-final-viral-combined_k141_58857||lt2gene | Low-quality |
| 17-108-final-viral-combined_k141_224123||lt2gene | Low-quality |
| 17-108-final-viral-combined_k141_17104||lt2gene | Not-determined |
| 17-108-final-viral-combined_k141_182589||lt2gene | Low-quality |
| 17-108-final-viral-combined_k141_307000||lt2gene | Low-quality |
| 17-108-final-viral-combined_k141_307001||lt2gene | Low-quality |
| 17-108-final-viral-combined_k141_31015||lt2gene | Not-determined |
| 17-108-final-viral-combined_k141_31019||lt2gene | Low-quality |
| 17-108-final-viral-combined_k141_182608||lt2gene | Low-quality |
| 17-108-final-viral-combined_k141_17111||lt2gene | Low-quality |
| 17-108-final-viral-combined_k141_127467||lt2gene | Not-determined |
| 17-108-final-viral-combined_k141_169352||lt2gene | Not-determined |
| 17-108-final-viral-combined_k141_210512||lt2gene | Not-determined |
| 17-108-final-viral-combined_k141_238196||lt2gene | Low-quality |
| 17-108-final-viral-combined_k141_44882||lt2gene | Low-quality |
| 17-108-final-viral-combined_k141_71941||lt2gene | Low-quality |
| 17-108-final-viral-combined_k141_169376||lt2gene | Low-quality |
| 17-108-final-viral-combined_k141_100341||lt2gene | Low-quality |
| 17-108-final-viral-combined_k141_210569||lt2gene | Low-quality |
| 17-108-final-viral-combined_k141_100411||lt2gene | Low-quality |
| 17-108-final-viral-combined_k141_100421||lt2gene | Low-quality |
| 17-108-final-viral-combined_k141_210607||lt2gene | Not-determined |
| 17-108-final-viral-combined_k141_44965||lt2gene | Low-quality |
| 17-108-final-viral-combined_k141_44971||lt2gene | Low-quality |
| 17-108-final-viral-combined_k141_279544||lt2gene | Low-quality |
| 17-108-final-viral-combined_k141_58937||lt2gene | Not-determined |
| 17-108-final-viral-combined_k141_58971||lt2gene | Low-quality |
| 17-108-final-viral-combined_k141_251534||lt2gene | Low-quality |
| 17-108-final-viral-combined_k141_292848||lt2gene | Not-determined |
| 17-108-final-viral-combined_k141_292853||lt2gene | Low-quality |
| 17-108-final-viral-combined_k141_45072||lt2gene | Low-quality |
| 17-108-final-viral-combined_k141_141413||lt2gene | Low-quality |
| 17-108-final-viral-combined_k141_169438||lt2gene | Low-quality |
| 17-108-final-viral-combined_k141_169446||lt2gene | Low-quality |
| 17-108-final-viral-combined_k141_169453||lt2gene | Low-quality |
| 17-108-final-viral-combined_k141_292903||lt2gene | Low-quality |
| 17-108-final-viral-combined_k141_238258||lt2gene | Low-quality |
| 17-108-final-viral-combined_k141_292904||lt2gene | Low-quality |
| 17-108-final-viral-combined_k141_307166||lt2gene | Not-determined |
| 17-108-final-viral-combined_k141_182862||lt2gene | Low-quality |
| 17-108-final-viral-combined_k141_113459||lt2gene | Low-quality |
| 17-108-final-viral-combined_k141_155354||lt2gene | Low-quality |
| 17-108-final-viral-combined_k141_155373||lt2gene | Low-quality |
| 17-108-final-viral-combined_k141_155374||lt2gene | Low-quality |
| 17-108-final-viral-combined_k141_265470||lt2gene | Low-quality |
| 17-108-final-viral-combined_k141_86615||lt2gene | Not-determined |
| 17-108-final-viral-combined_k141_31224||lt2gene | Not-determined |
| 17-108-final-viral-combined_k141_31276||lt2gene | Low-quality |
| 17-108-final-viral-combined_k141_127773||lt2gene | Low-quality |
| 17-108-final-viral-combined_k141_238311||lt2gene | Low-quality |
| 17-108-final-viral-combined_k141_169590||lt2gene | Low-quality |
| 17-108-final-viral-combined_k141_3715||lt2gene | Not-determined |
| 17-108-final-viral-combined_k141_141565||lt2gene | Low-quality |
| 17-108-final-viral-combined_k141_100700||lt2gene | Not-determined |
| 17-108-final-viral-combined_k141_155499||lt2gene | Not-determined |
| 17-108-final-viral-combined_k141_3767||lt2gene | Low-quality |
| 17-108-final-viral-combined_k141_31339||lt2gene | Low-quality |
| 17-108-final-viral-combined_k141_59262||lt2gene | Low-quality |
| 17-108-final-viral-combined_k141_113538||lt2gene | Low-quality |
| 17-108-final-viral-combined_k141_86762||lt2gene | Low-quality |
| 17-108-final-viral-combined_k141_141659||lt2gene | Low-quality |
| 17-108-final-viral-combined_k141_155562||lt2gene | Low-quality |
| 17-108-final-viral-combined_k141_293118||lt2gene | Low-quality |
| 17-108-final-viral-combined_k141_141694||lt2gene | Low-quality |
| 17-108-final-viral-combined_k141_45252||lt2gene | Low-quality |
| 17-108-final-viral-combined_k141_307415||lt2gene | Low-quality |
| 17-108-final-viral-combined_k141_211109||lt2gene | Low-quality |
| 17-108-final-viral-combined_k141_251882||lt2gene | Not-determined |
| 17-108-final-viral-combined_k141_17484||lt2gene | Not-determined |
| 17-108-final-viral-combined_k141_183067||lt2gene | Low-quality |
| 17-108-final-viral-combined_k141_72347||lt2gene | Not-determined |
| 17-108-final-viral-combined_k141_251912||lt2gene | Low-quality |
| 17-108-final-viral-combined_k141_141794||lt2gene | Not-determined |
| 17-108-final-viral-combined_k141_251924||lt2gene | Low-quality |
| 17-108-final-viral-combined_k141_155656||lt2gene | Low-quality |
| 17-108-final-viral-combined_k141_238567||lt2gene | Not-determined |
| 17-108-final-viral-combined_k141_211201||lt2gene | Low-quality |
| 17-108-final-viral-combined_k141_251957||lt2gene | Low-quality |
| 17-108-final-viral-combined_k141_238626||lt2gene | Low-quality |
| 17-108-final-viral-combined_k141_86951||lt2gene | Low-quality |
| 17-108-final-viral-combined_k141_293222||lt2gene | Low-quality |
| 17-108-final-viral-combined_k141_252008||lt2gene | Not-determined |
| 17-108-final-viral-combined_k141_183164||lt2gene | Low-quality |
| 17-108-final-viral-combined_k141_293259||lt2gene | Low-quality |
| 17-108-final-viral-combined_k141_31557||lt2gene | Low-quality |
| 17-108-final-viral-combined_k141_238729||lt2gene | Not-determined |
| 17-108-final-viral-combined_k141_31583||lt2gene | Low-quality |
| 17-108-final-viral-combined_k141_196648||lt2gene | Not-determined |
| 17-108-final-viral-combined_k141_238818||lt2gene | Low-quality |
| 17-108-final-viral-combined_k141_280189||lt2gene | Not-determined |
| 17-108-final-viral-combined_k141_155711||lt2gene | Low-quality |
| 17-108-final-viral-combined_k141_72561||lt2gene | Low-quality |
| 17-108-final-viral-combined_k141_183283||lt2gene | Not-determined |
| 17-108-final-viral-combined_k141_87148||lt2gene | Low-quality |
| 17-108-final-viral-combined_k141_31623||lt2gene | Low-quality |
| 17-108-final-viral-combined_k141_87157||lt2gene | Low-quality |
| 17-108-final-viral-combined_k141_142021||lt2gene | Low-quality |
| 17-108-final-viral-combined_k141_252143||lt2gene | Not-determined |
| 17-108-final-viral-combined_k141_45637||lt2gene | Low-quality |
| 17-108-final-viral-combined_k141_252182||lt2gene | Low-quality |
| 17-108-final-viral-combined_k141_307642||lt2gene | Not-determined |
| 17-108-final-viral-combined_k141_170025||lt2gene | Low-quality |
| 17-108-final-viral-combined_k141_266073||lt2gene | Not-determined |
| 17-108-final-viral-combined_k141_101025||lt2gene | Not-determined |
| 17-108-final-viral-combined_k141_196829||lt2gene | Low-quality |
| 17-108-final-viral-combined_k141_320779||lt2gene | Low-quality |
| 17-108-final-viral-combined_k141_101044||lt2gene | Low-quality |
| 17-108-final-viral-combined_k141_320805||lt2gene | Low-quality |
| 17-108-final-viral-combined_k141_320810||lt2gene | Low-quality |
| 17-108-final-viral-combined_k141_113867||lt2gene | Not-determined |
| 17-108-final-viral-combined_k141_320817||lt2gene | Low-quality |
| 17-108-final-viral-combined_k141_113889||lt2gene | Not-determined |
| 17-108-final-viral-combined_k141_113892||lt2gene | Not-determined |
| 17-108-final-viral-combined_k141_252296||lt2gene | Not-determined |
| 17-108-final-viral-combined_k141_252329||lt2gene | Not-determined |
| 17-108-final-viral-combined_k141_307789||lt2gene | Not-determined |
| 17-108-final-viral-combined_k141_72668||lt2gene | Low-quality |
| 17-108-final-viral-combined_k141_72679||lt2gene | Not-determined |
| 17-108-final-viral-combined_k141_72707||lt2gene | Low-quality |

17-108-final-viral-combined_k141_170216||lt2gene Low-quality
17-108-final-viral-combined_k141_293691||lt2gene Low-quality
17-108-final-viral-combined_k141_113973||lt2gene Low-quality
17-108-final-viral-combined_k141_31879||lt2gene Not-determined
17-108-final-viral-combined_k141_293758||lt2gene Not-determined
17-108-final-viral-combined_k141_170256||lt2gene Not-determined
17-108-final-viral-combined_k141_252458||lt2gene Not-determined
17-108-final-viral-combined_k141_59730||lt2gene Not-determined
17-108-final-viral-combined_k141_72809||lt2gene Not-determined
17-108-final-viral-combined_k141_183603||lt2gene Low-quality
17-108-final-viral-combined_k141_46002||lt2gene Low-quality
17-108-final-viral-combined_k141_4395||lt2gene Low-quality
17-108-final-viral-combined_k141_156054||lt2gene Low-quality
17-108-final-viral-combined_k141_72856||lt2gene Not-determined
17-108-final-viral-combined_k141_156073||lt2gene Low-quality
17-108-final-viral-combined_k141_114092||lt2gene Not-determined
17-108-final-viral-combined_k141_170329||lt2gene Low-quality
17-108-final-viral-combined_k141_170331||lt2gene Not-determined
17-108-final-viral-combined_k141_211724||lt2gene Not-determined
17-108-final-viral-combined_k141_128429||lt2gene Not-determined
17-108-final-viral-combined_k141_101341||lt2gene Low-quality
17-108-final-viral-combined_k141_101345||lt2gene Low-quality
17-108-final-viral-combined_k141_4429||lt2gene Not-determined
17-108-final-viral-combined_k141_321053||lt2gene Low-quality
17-108-final-viral-combined_k141_225200||lt2gene Not-determined
17-108-final-viral-combined_k141_156107||lt2gene Low-quality
17-108-final-viral-combined_k141_211794||lt2gene Low-quality
17-108-final-viral-combined_k141_72934||lt2gene Not-determined
17-108-final-viral-combined_k141_32024||lt2gene Low-quality
17-108-final-viral-combined_k141_183775||lt2gene Not-determined
17-108-final-viral-combined_k141_114191||lt2gene Not-determined
17-108-final-viral-combined_k141_156142||lt2gene Low-quality
17-108-final-viral-combined_k141_87480||lt2gene Low-quality
17-108-final-viral-combined_k141_142471||lt2gene Low-quality
17-108-final-viral-combined_k141_293804||lt2gene Low-quality
17-108-final-viral-combined_k141_252625||lt2gene Low-quality
17-108-final-viral-combined_k141_225270||lt2gene Not-determined
17-108-final-viral-combined_k141_239299||lt2gene Not-determined
17-108-final-viral-combined_k141_4563||lt2gene Low-quality
17-108-final-viral-combined_k141_46180||lt2gene Low-quality
17-108-final-viral-combined_k141_225301||lt2gene Not-determined
17-108-final-viral-combined_k141_239331||lt2gene Low-quality
17-108-final-viral-combined_k141_87536||lt2gene Low-quality
17-108-final-viral-combined_k141_197257||lt2gene Low-quality
17-108-final-viral-combined_k141_4617||lt2gene Not-determined
17-108-final-viral-combined_k141_114239||lt2gene Not-determined
17-108-final-viral-combined_k141_211973||lt2gene Low-quality
17-108-final-viral-combined_k141_156268||lt2gene Low-quality
17-108-final-viral-combined_k141_280800||lt2gene Low-quality
17-108-final-viral-combined_k141_212001||lt2gene Not-determined
17-108-final-viral-combined_k141_280837||lt2gene Low-quality
17-108-final-viral-combined_k141_266603||lt2gene Not-determined
17-108-final-viral-combined_k141_212019||lt2gene Low-quality
17-108-final-viral-combined_k141_59967||lt2gene Not-determined
17-108-final-viral-combined_k141_252755||lt2gene Low-quality
17-108-final-viral-combined_k141_321242||lt2gene Low-quality
17-108-final-viral-combined_k141_183898||lt2gene Low-quality
17-108-final-viral-combined_k141_87617||lt2gene Not-determined
17-108-final-viral-combined_k141_183902||lt2gene Not-determined
17-108-final-viral-combined_k141_321270||lt2gene Low-quality
17-108-final-viral-combined_k141_321272||lt2gene Low-quality
17-108-final-viral-combined_k141_170621||lt2gene Low-quality
17-108-final-viral-combined_k141_197350||lt2gene Not-determined
17-108-final-viral-combined_k141_294012||lt2gene Not-determined
17-108-final-viral-combined_k141_294015||lt2gene Low-quality
17-108-final-viral-combined_k141_294021||lt2gene Low-quality
17-108-final-viral-combined_k141_156374||lt2gene Low-quality
17-108-final-viral-combined_k141_73132||lt2gene Not-determined
17-108-final-viral-combined_k141_280931||lt2gene Not-determined
17-108-final-viral-combined_k141_142702||lt2gene Low-quality
17-108-final-viral-combined_k141_156423||lt2gene Not-determined
17-108-final-viral-combined_k141_197395||lt2gene Low-quality
17-108-final-viral-combined_k141_60031||lt2gene Low-quality
17-108-final-viral-combined_k141_73181||lt2gene Low-quality
17-108-final-viral-combined_k141_252874||lt2gene Low-quality
17-108-final-viral-combined_k141_212142||lt2gene Not-determined
17-108-final-viral-combined_k141_225579||lt2gene Low-quality
17-108-final-viral-combined_k141_252883||lt2gene Low-quality
17-108-final-viral-combined_k141_60047||lt2gene Low-quality
17-108-final-viral-combined_k141_114472||lt2gene Not-determined
17-108-final-viral-combined_k141_183991||lt2gene Not-determined
17-108-final-viral-combined_k141_101725||lt2gene Low-quality
17-108-final-viral-combined_k141_4766||lt2gene Low-quality
17-108-final-viral-combined_k141_281094||lt2gene Not-determined
17-108-final-viral-combined_k141_212214||lt2gene Low-quality
17-108-final-viral-combined_k141_266763||lt2gene Not-determined
17-108-final-viral-combined_k141_212232||lt2gene Not-determined
17-108-final-viral-combined_k141_184028||lt2gene Low-quality
17-108-final-viral-combined_k141_308323||lt2gene Not-determined
17-108-final-viral-combined_k141_114579||lt2gene Low-quality
17-108-final-viral-combined_k141_225680||lt2gene Not-determined
17-108-final-viral-combined_k141_18159||lt2gene Low-quality
17-108-final-viral-combined_k141_87749||lt2gene Low-quality
17-108-final-viral-combined_k141_101819||lt2gene Low-quality
17-108-final-viral-combined_k141_308359||lt2gene Low-quality
17-108-final-viral-combined_k141_156545||lt2gene Not-determined
17-108-final-viral-combined_k141_197532||lt2gene Low-quality
17-108-final-viral-combined_k141_266811||lt2gene Not-determined
17-108-final-viral-combined_k141_266838||lt2gene Not-determined
17-108-final-viral-combined_k141_101929||lt2gene Not-determined
17-108-final-viral-combined_k141_308434||lt2gene Low-quality
17-108-final-viral-combined_k141_60225||lt2gene Low-quality
17-108-final-viral-combined_k141_212311||lt2gene Low-quality
17-108-final-viral-combined_k141_87852||lt2gene Not-determined
17-108-final-viral-combined_k141_266910||lt2gene Low-quality
17-108-final-viral-combined_k141_60244||lt2gene Low-quality
17-108-final-viral-combined_k141_294233||lt2gene Low-quality
17-108-final-viral-combined_k141_60261||lt2gene Low-quality
17-108-final-viral-combined_k141_197626||lt2gene Low-quality
17-108-final-viral-combined_k141_87893||lt2gene Low-quality
17-108-final-viral-combined_k141_142982||lt2gene Low-quality
17-108-final-viral-combined_k141_294269||lt2gene Low-quality
17-108-final-viral-combined_k141_129026||lt2gene Not-determined
17-108-final-viral-combined_k141_321536||lt2gene Low-quality
17-108-final-viral-combined_k141_281366||lt2gene Not-determined
17-108-final-viral-combined_k141_294304||lt2gene Low-quality
17-108-final-viral-combined_k141_129040||lt2gene Not-determined
17-108-final-viral-combined_k141_225794||lt2gene Low-quality
17-108-final-viral-combined_k141_321583||lt2gene Low-quality
17-108-final-viral-combined_k141_321597||lt2gene Low-quality
17-108-final-viral-combined_k141_281458||lt2gene Low-quality
17-108-final-viral-combined_k141_143045||lt2gene Low-quality
17-108-final-viral-combined_k141_308616||lt2gene Low-quality
17-108-final-viral-combined_k141_239956||lt2gene Not-determined
17-108-final-viral-combined_k141_197738||lt2gene Low-quality
17-108-final-viral-combined_k141_143108||lt2gene Low-quality
17-108-final-viral-combined_k141_184249||lt2gene Low-quality
17-108-final-viral-combined_k141_102198||lt2gene Low-quality
17-108-final-viral-combined_k141_73465||lt2gene Low-quality
17-108-final-viral-combined_k141_267134||lt2gene Not-determined
17-108-final-viral-combined_k141_73484||lt2gene Low-quality
17-108-final-viral-combined_k141_129193||lt2gene Not-determined
17-108-final-viral-combined_k141_308775||lt2gene Low-quality
17-108-final-viral-combined_k141_46745||lt2gene Not-determined
17-108-final-viral-combined_k141_253283||lt2gene Low-quality
17-108-final-viral-combined_k141_32766||lt2gene Not-determined
17-108-final-viral-combined_k141_102289||lt2gene Low-quality
17-108-final-viral-combined_k141_5239||lt2gene Not-determined
17-108-final-viral-combined_k141_240137||lt2gene Not-determined
17-108-final-viral-combined_k141_184374||lt2gene Low-quality
17-108-final-viral-combined_k141_60540||lt2gene Low-quality
17-108-final-viral-combined_k141_240147||lt2gene Low-quality
17-108-final-viral-combined_k141_253375||lt2gene Not-determined
17-108-final-viral-combined_k141_73539||lt2gene Low-quality
17-108-final-viral-combined_k141_156854||lt2gene Low-quality
17-108-final-viral-combined_k141_32825||lt2gene Low-quality
17-108-final-viral-combined_k141_115043||lt2gene Low-quality
17-108-final-viral-combined_k141_73568||lt2gene Low-quality
17-108-final-viral-combined_k141_184408||lt2gene Not-determined
17-108-final-viral-combined_k141_46804||lt2gene Low-quality
17-108-final-viral-combined_k141_226044||lt2gene Low-quality
17-108-final-viral-combined_k141_197988||lt2gene Not-determined
17-108-final-viral-combined_k141_88207||lt2gene Low-quality
17-108-final-viral-combined_k141_294576||lt2gene Low-quality
17-108-final-viral-combined_k141_240234||lt2gene Low-quality
17-108-final-viral-combined_k141_60596||lt2gene Not-determined
17-108-final-viral-combined_k141_73673||lt2gene Not-determined
17-108-final-viral-combined_k141_115156||lt2gene Not-determined
17-108-final-viral-combined_k141_32934||lt2gene Low-quality
17-108-final-viral-combined_k141_102436||lt2gene Low-quality
17-108-final-viral-combined_k141_46860||lt2gene Not-determined
17-108-final-viral-combined_k141_32982||lt2gene Low-quality
17-108-final-viral-combined_k141_5432||lt2gene Low-quality
17-108-final-viral-combined_k141_294678||lt2gene Low-quality
17-108-final-viral-combined_k141_294688||lt2gene Low-quality
17-108-final-viral-combined_k141_184590||lt2gene Low-quality
17-108-final-viral-combined_k141_60641||lt2gene Not-determined
17-108-final-viral-combined_k141_184615||lt2gene Not-determined
17-108-final-viral-combined_k141_46922||lt2gene Low-quality
17-108-final-viral-combined_k141_18643||lt2gene Low-quality
17-108-final-viral-combined_k141_309045||lt2gene Low-quality
17-108-final-viral-combined_k141_73798||lt2gene Low-quality
17-108-final-viral-combined_k141_129466||lt2gene Low-quality
17-108-final-viral-combined_k141_102591||lt2gene Not-determined
17-108-final-viral-combined_k141_212841||lt2gene Not-determined
17-108-final-viral-combined_k141_5504||lt2gene Low-quality
17-108-final-viral-combined_k141_253583||lt2gene Low-quality
17-108-final-viral-combined_k141_198197||lt2gene Not-determined
17-108-final-viral-combined_k141_46983||lt2gene Low-quality
17-108-final-viral-combined_k141_102636||lt2gene Low-quality
17-108-final-viral-combined_k141_73894||lt2gene Not-determined
17-108-final-viral-combined_k141_33103||lt2gene Not-determined
17-108-final-viral-combined_k141_88398||lt2gene Low-quality
17-108-final-viral-combined_k141_18734||lt2gene Not-determined
17-108-final-viral-combined_k141_18737||lt2gene Not-determined
17-108-final-viral-combined_k141_18742||lt2gene Low-quality
17-108-final-viral-combined_k141_309161||lt2gene Not-determined
17-108-final-viral-combined_k141_281834||lt2gene Low-quality
17-108-final-viral-combined_k141_60840||lt2gene Low-quality
17-108-final-viral-combined_k141_309244||lt2gene Low-quality
17-108-final-viral-combined_k141_129612||lt2gene Low-quality
17-108-final-viral-combined_k141_240612||lt2gene Low-quality
17-108-final-viral-combined_k141_171377||lt2gene Not-determined
17-108-final-viral-combined_k141_267444||lt2gene Low-quality
17-108-final-viral-combined_k141_226238||lt2gene Low-quality
17-108-final-viral-combined_k141_240644||lt2gene Low-quality
17-108-final-viral-combined_k141_157222||lt2gene Low-quality
17-108-final-viral-combined_k141_198380||lt2gene Low-quality
17-108-final-viral-combined_k141_294981||lt2gene Low-quality
17-108-final-viral-combined_k141_129652||lt2gene Low-quality
17-108-final-viral-combined_k141_33259||lt2gene Not-determined
17-108-final-viral-combined_k141_322016||lt2gene Not-determined
17-108-final-viral-combined_k141_281977||lt2gene Not-determined
17-108-final-viral-combined_k141_33275||lt2gene Low-quality
17-108-final-viral-combined_k141_102840||lt2gene Low-quality
17-108-final-viral-combined_k141_282019||lt2gene Not-determined
17-108-final-viral-combined_k141_267520||lt2gene Not-determined
17-108-final-viral-combined_k141_253881||lt2gene Not-determined
17-108-final-viral-combined_k141_253893||lt2gene Low-quality
17-108-final-viral-combined_k141_88551||lt2gene Not-determined
17-108-final-viral-combined_k141_322037||lt2gene Low-quality
17-108-final-viral-combined_k141_74132||lt2gene Not-determined
17-108-final-viral-combined_k141_157344||lt2gene Not-determined
17-108-final-viral-combined_k141_282047||lt2gene Low-quality
17-108-final-viral-combined_k141_74143||lt2gene Low-quality
17-108-final-viral-combined_k141_253945||lt2gene Low-quality
17-108-final-viral-combined_k141_322085||lt2gene Not-determined
17-108-final-viral-combined_k141_309448||lt2gene Low-quality
17-108-final-viral-combined_k141_47382||lt2gene Low-quality
17-108-final-viral-combined_k141_47386||lt2gene Low-quality
17-108-final-viral-combined_k141_198514||lt2gene Not-determined
17-108-final-viral-combined_k141_322119||lt2gene Low-quality
17-108-final-viral-combined_k141_61103||lt2gene Low-quality
17-108-final-viral-combined_k141_157514||lt2gene Low-quality
17-108-final-viral-combined_k141_18982||lt2gene Low-quality
17-108-final-viral-combined_k141_74327||lt2gene Low-quality
17-108-final-viral-combined_k141_47430||lt2gene Low-quality
17-108-final-viral-combined_k141_309559||lt2gene Not-determined
17-108-final-viral-combined_k141_240885||lt2gene Low-quality
17-108-final-viral-combined_k141_103036||lt2gene Low-quality
17-108-final-viral-combined_k141_254196||lt2gene Low-quality
17-108-final-viral-combined_k141_61202||lt2gene Not-determined
17-108-final-viral-combined_k141_240894||lt2gene Low-quality
17-108-final-viral-combined_k141_47488||lt2gene Low-quality
17-108-final-viral-combined_k141_157656||lt2gene Low-quality
17-108-final-viral-combined_k141_267868||lt2gene Low-quality
17-108-final-viral-combined_k141_115652||lt2gene Low-quality
17-108-final-viral-combined_k141_5943||lt2gene Low-quality
17-108-final-viral-combined_k141_74444||lt2gene Not-determined
17-108-final-viral-combined_k141_309675||lt2gene Low-quality
17-108-final-viral-combined_k141_254301||lt2gene Low-quality
17-108-final-viral-combined_k141_309701||lt2gene Low-quality
17-108-final-viral-combined_k141_254312||lt2gene Low-quality
17-108-final-viral-combined_k141_33655||lt2gene Low-quality
17-108-final-viral-combined_k141_115723||lt2gene Low-quality
17-108-final-viral-combined_k141_88797||lt2gene Low-quality
17-108-final-viral-combined_k141_226681||lt2gene Low-quality
17-108-final-viral-combined_k141_267964||lt2gene Low-quality
17-108-final-viral-combined_k141_241027||lt2gene Low-quality
17-108-final-viral-combined_k141_241044||lt2gene Not-determined
17-108-final-viral-combined_k141_295345||lt2gene Not-determined
17-108-final-viral-combined_k141_171775||lt2gene Low-quality
17-108-final-viral-combined_k141_295372||lt2gene Low-quality
17-108-final-viral-combined_k141_295376||lt2gene Low-quality
17-108-final-viral-combined_k141_213473||lt2gene Low-quality
17-108-final-viral-combined_k141_157901||lt2gene Low-quality
17-108-final-viral-combined_k141_309866||lt2gene Low-quality
17-108-final-viral-combined_k141_309868||lt2gene Not-determined
17-108-final-viral-combined_k141_6136||lt2gene Low-quality
17-108-final-viral-combined_k141_198838||lt2gene Low-quality
17-108-final-viral-combined_k141_115861||lt2gene Low-quality
17-108-final-viral-combined_k141_103266||lt2gene Not-determined
17-108-final-viral-combined_k141_226798||lt2gene Not-determined
17-108-final-viral-combined_k141_322564||lt2gene Low-quality
17-108-final-viral-combined_k141_254438||lt2gene Not-determined
17-108-final-viral-combined_k141_33808||lt2gene Low-quality
17-108-final-viral-combined_k141_322614||lt2gene Low-quality
17-108-final-viral-combined_k141_185171||lt2gene Low-quality
17-108-final-viral-combined_k141_282582||lt2gene Low-quality
17-108-final-viral-combined_k141_88983||lt2gene Low-quality
17-108-final-viral-combined_k141_213568||lt2gene Not-determined
17-108-final-viral-combined_k141_268063||lt2gene Not-determined
17-108-final-viral-combined_k141_268070||lt2gene Low-quality
17-108-final-viral-combined_k141_268071||lt2gene Low-quality
17-108-final-viral-combined_k141_61600||lt2gene Low-quality
17-108-final-viral-combined_k141_226923||lt2gene Low-quality
17-108-final-viral-combined_k141_185243||lt2gene Not-determined
17-108-final-viral-combined_k141_171917||lt2gene Not-determined
17-108-final-viral-combined_k141_171924||lt2gene Low-quality
17-108-final-viral-combined_k141_6265||lt2gene Low-quality
17-108-final-viral-combined_k141_74663||lt2gene Not-determined
17-108-final-viral-combined_k141_213649||lt2gene Not-determined
17-108-final-viral-combined_k141_19418||lt2gene Low-quality
17-108-final-viral-combined_k141_213662||lt2gene Low-quality
17-108-final-viral-combined_k141_158100||lt2gene Low-quality
17-108-final-viral-combined_k141_268133||lt2gene Low-quality
17-108-final-viral-combined_k141_226977||lt2gene Low-quality
17-108-final-viral-combined_k141_241367||lt2gene Low-quality
17-108-final-viral-combined_k141_33892||lt2gene Low-quality
17-108-final-viral-combined_k141_33898||lt2gene Low-quality
17-108-final-viral-combined_k141_254607||lt2gene Not-determined
17-108-final-viral-combined_k141_185300||lt2gene Not-determined
17-108-final-viral-combined_k141_254626||lt2gene Low-quality
17-108-final-viral-combined_k141_241412||lt2gene Not-determined
17-108-final-viral-combined_k141_172009||lt2gene Not-determined
17-108-final-viral-combined_k141_241426||lt2gene Low-quality
17-108-final-viral-combined_k141_130208||lt2gene Not-determined
17-108-final-viral-combined_k141_172027||lt2gene Low-quality
17-108-final-viral-combined_k141_172029||lt2gene Low-quality
17-108-final-viral-combined_k141_268213||lt2gene Low-quality
17-108-final-viral-combined_k141_310054||lt2gene Low-quality
17-108-final-viral-combined_k141_241459||lt2gene Low-quality
17-108-final-viral-combined_k141_241477||lt2gene Low-quality
17-108-final-viral-combined_k141_254678||lt2gene Low-quality
17-108-final-viral-combined_k141_61795||lt2gene Low-quality
17-108-final-viral-combined_k141_227038||lt2gene Not-determined
17-108-final-viral-combined_k141_130266||lt2gene Not-determined
17-108-final-viral-combined_k141_268295||lt2gene Low-quality
17-108-final-viral-combined_k141_47823||lt2gene Low-quality
17-108-final-viral-combined_k141_116114||lt2gene Low-quality
17-108-final-viral-combined_k141_89150||lt2gene Not-determined
17-108-final-viral-combined_k141_172134||lt2gene Not-determined
17-108-final-viral-combined_k141_241522||lt2gene Low-quality
17-108-final-viral-combined_k141_227101||lt2gene Not-determined
17-108-final-viral-combined_k141_34016||lt2gene Low-quality
17-108-final-viral-combined_k141_158289||lt2gene Low-quality
17-108-final-viral-combined_k141_6518||lt2gene Not-determined
17-108-final-viral-combined_k141_227123||lt2gene Low-quality
17-108-final-viral-combined_k141_213854||lt2gene Low-quality
17-108-final-viral-combined_k141_185488||lt2gene Low-quality
17-108-final-viral-combined_k141_185495||lt2gene Low-quality
17-108-final-viral-combined_k141_241576||lt2gene Low-quality
17-108-final-viral-combined_k141_34062||lt2gene Not-determined
17-108-final-viral-combined_k141_172219||lt2gene Low-quality
17-108-final-viral-combined_k141_158374||lt2gene Low-quality
17-108-final-viral-combined_k141_268370||lt2gene Not-determined
17-108-final-viral-combined_k141_158382||lt2gene Not-determined
17-108-final-viral-combined_k141_254822||lt2gene Not-determined
17-108-final-viral-combined_k141_282832||lt2gene Low-quality
17-108-final-viral-combined_k141_172285||lt2gene Not-determined
17-108-final-viral-combined_k141_227210||lt2gene Not-determined
17-108-final-viral-combined_k141_34148||lt2gene Low-quality
17-108-final-viral-combined_k141_254878||lt2gene Low-quality
17-108-final-viral-combined_k141_268411||lt2gene Low-quality
17-108-final-viral-combined_k141_241672||lt2gene Low-quality
17-108-final-viral-combined_k141_34166||lt2gene Low-quality
17-108-final-viral-combined_k141_199188||lt2gene Low-quality
17-108-final-viral-combined_k141_213920||lt2gene Low-quality
17-108-final-viral-combined_k141_158537||lt2gene Not-determined
17-108-final-viral-combined_k141_6645||lt2gene Low-quality
17-108-final-viral-combined_k141_295760||lt2gene Not-determined
17-108-final-viral-combined_k141_295761||lt2gene Not-determined
17-108-final-viral-combined_k141_89353||lt2gene Low-quality
17-108-final-viral-combined_k141_254937||lt2gene Low-quality
17-108-final-viral-combined_k141_144553||lt2gene Low-quality
17-108-final-viral-combined_k141_199219||lt2gene Low-quality
17-108-final-viral-combined_k141_199222||lt2gene Not-determined
17-108-final-viral-combined_k141_19747||lt2gene Low-quality
17-108-final-viral-combined_k141_48007||lt2gene Not-determined
17-108-final-viral-combined_k141_185615||lt2gene Low-quality
17-108-final-viral-combined_k141_268530||lt2gene Low-quality
17-108-final-viral-combined_k141_62145||lt2gene Low-quality
17-108-final-viral-combined_k141_268574||lt2gene Not-determined
17-108-final-viral-combined_k141_282968||lt2gene Low-quality
17-108-final-viral-combined_k141_48053||lt2gene Not-determined
17-108-final-viral-combined_k141_62165||lt2gene Low-quality
17-108-final-viral-combined_k141_34320||lt2gene Low-quality
17-108-final-viral-combined_k141_6802||lt2gene Low-quality
17-108-final-viral-combined_k141_116447||lt2gene Not-determined
17-108-final-viral-combined_k141_19879||lt2gene Not-determined
17-108-final-viral-combined_k141_158659||lt2gene Not-determined
17-108-final-viral-combined_k141_62205||lt2gene Low-quality
17-108-final-viral-combined_k141_75120||lt2gene Low-quality
17-108-final-viral-combined_k141_6830||lt2gene Not-determined
17-108-final-viral-combined_k141_241855||lt2gene Not-determined
17-108-final-viral-combined_k141_310579||lt2gene Not-determined
17-108-final-viral-combined_k141_103902||lt2gene Low-quality
17-108-final-viral-combined_k141_268658||lt2gene Not-determined
17-108-final-viral-combined_k141_48146||lt2gene Low-quality
17-108-final-viral-combined_k141_130528||lt2gene Not-determined
17-108-final-viral-combined_k141_295944||lt2gene Low-quality
17-108-final-viral-combined_k141_323076||lt2gene Not-determined
17-108-final-viral-combined_k141_116551||lt2gene Not-determined
17-108-final-viral-combined_k141_199404||lt2gene Low-quality
17-108-final-viral-combined_k141_144777||lt2gene Low-quality
17-108-final-viral-combined_k141_199423||lt2gene Not-determined
17-108-final-viral-combined_k141_283032||lt2gene Low-quality
17-108-final-viral-combined_k141_89611||lt2gene Low-quality
17-108-final-viral-combined_k141_214220||lt2gene Low-quality
17-108-final-viral-combined_k141_323102||lt2gene Not-determined
17-108-final-viral-combined_k141_75283||lt2gene Not-determined
17-108-final-viral-combined_k141_104005||lt2gene Low-quality
17-108-final-viral-combined_k141_20062||lt2gene Low-quality
17-108-final-viral-combined_k141_323165||lt2gene Low-quality
17-108-final-viral-combined_k141_116606||lt2gene Low-quality
17-108-final-viral-combined_k141_227682||lt2gene Low-quality
17-108-final-viral-combined_k141_104076||lt2gene Low-quality
17-108-final-viral-combined_k141_75341||lt2gene Not-determined
17-108-final-viral-combined_k141_34599||lt2gene Low-quality
17-108-final-viral-combined_k141_75348||lt2gene Low-quality
17-108-final-viral-combined_k141_158767||lt2gene Not-determined
17-108-final-viral-combined_k141_227724||lt2gene Low-quality
17-108-final-viral-combined_k141_89701||lt2gene Low-quality
17-108-final-viral-combined_k141_20106||lt2gene Not-determined
17-108-final-viral-combined_k141_172685||lt2gene Not-determined
17-108-final-viral-combined_k141_227731||lt2gene Low-quality
17-108-final-viral-combined_k141_296111||lt2gene Low-quality
17-108-final-viral-combined_k141_7057||lt2gene Not-determined
17-108-final-viral-combined_k141_20143||lt2gene Not-determined
17-108-final-viral-combined_k141_62487||lt2gene Not-determined
17-108-final-viral-combined_k141_172730||lt2gene Low-quality
17-108-final-viral-combined_k141_242066||lt2gene Low-quality
17-108-final-viral-combined_k141_255298||lt2gene Low-quality
17-108-final-viral-combined_k141_283216||lt2gene Low-quality
17-108-final-viral-combined_k141_48295||lt2gene Low-quality
17-108-final-viral-combined_k141_255333||lt2gene Low-quality
17-108-final-viral-combined_k141_34680||lt2gene Low-quality
17-108-final-viral-combined_k141_199596||lt2gene Low-quality
17-108-final-viral-combined_k141_227756||lt2gene Low-quality
17-108-final-viral-combined_k141_268892||lt2gene Low-quality
17-108-final-viral-combined_k141_214346||lt2gene Low-quality
17-108-final-viral-combined_k141_62553||lt2gene Not-determined
17-108-final-viral-combined_k141_242111||lt2gene Not-determined
17-108-final-viral-combined_k141_104158||lt2gene Low-quality
17-108-final-viral-combined_k141_255393||lt2gene Not-determined
17-108-final-viral-combined_k141_48381||lt2gene Not-determined
17-108-final-viral-combined_k141_214383||lt2gene Low-quality
17-108-final-viral-combined_k141_296156||lt2gene Low-quality
17-108-final-viral-combined_k141_255435||lt2gene Low-quality
17-108-final-viral-combined_k141_62611||lt2gene Low-quality
17-108-final-viral-combined_k141_34781||lt2gene Low-quality
17-108-final-viral-combined_k141_20262||lt2gene Not-determined
17-108-final-viral-combined_k141_296186||lt2gene Low-quality
17-108-final-viral-combined_k141_7235||lt2gene Low-quality
17-108-final-viral-combined_k141_227896||lt2gene Low-quality
17-108-final-viral-combined_k141_34842||lt2gene Low-quality
17-108-final-viral-combined_k141_185930||lt2gene Low-quality
17-108-final-viral-combined_k141_214441||lt2gene Not-determined
17-108-final-viral-combined_k141_130825||lt2gene Not-determined
17-108-final-viral-combined_k141_172901||lt2gene Low-quality
17-108-final-viral-combined_k141_323379||lt2gene Not-determined
17-108-final-viral-combined_k141_283307||lt2gene Not-determined
17-108-final-viral-combined_k141_62724||lt2gene Not-determined
17-108-final-viral-combined_k141_255566||lt2gene Low-quality
17-108-final-viral-combined_k141_62736||lt2gene Low-quality
17-108-final-viral-combined_k141_283371||lt2gene Not-determined
17-108-final-viral-combined_k141_48520||lt2gene Low-quality
17-108-final-viral-combined_k141_242294||lt2gene Not-determined
17-108-final-viral-combined_k141_199745||lt2gene Low-quality
17-108-final-viral-combined_k141_20381||lt2gene Low-quality
17-108-final-viral-combined_k141_269019||lt2gene Not-determined
17-108-final-viral-combined_k141_283414||lt2gene Low-quality
17-108-final-viral-combined_k141_62823||lt2gene Low-quality
17-108-final-viral-combined_k141_228038||lt2gene Low-quality
17-108-final-viral-combined_k141_7363||lt2gene Low-quality
17-108-final-viral-combined_k141_199797||lt2gene Low-quality
17-108-final-viral-combined_k141_242353||lt2gene Low-quality
17-108-final-viral-combined_k141_242373||lt2gene Low-quality
17-108-final-viral-combined_k141_186121||lt2gene Low-quality
17-108-final-viral-combined_k141_116959||lt2gene Low-quality
17-108-final-viral-combined_k141_48640||lt2gene Not-determined
17-108-final-viral-combined_k141_255770||lt2gene Low-quality
17-108-final-viral-combined_k141_20467||lt2gene Not-determined
17-108-final-viral-combined_k141_283501||lt2gene Not-determined
17-108-final-viral-combined_k141_159113||lt2gene Low-quality
17-108-final-viral-combined_k141_90081||lt2gene Low-quality
17-108-final-viral-combined_k141_159116||lt2gene Low-quality
17-108-final-viral-combined_k141_90087||lt2gene Not-determined
17-108-final-viral-combined_k141_75773||lt2gene Low-quality
17-108-final-viral-combined_k141_186160||lt2gene Low-quality
17-108-final-viral-combined_k141_186177||lt2gene Low-quality
17-108-final-viral-combined_k141_214660||lt2gene Not-determined
17-108-final-viral-combined_k141_20539||lt2gene Not-determined
17-108-final-viral-combined_k141_20558||lt2gene Low-quality
17-108-final-viral-combined_k141_20560||lt2gene Low-quality
17-108-final-viral-combined_k141_131108||lt2gene Not-determined
17-108-final-viral-combined_k141_214678||lt2gene Low-quality
17-108-final-viral-combined_k141_35232||lt2gene Low-quality
17-108-final-viral-combined_k141_48804||lt2gene Low-quality
17-108-final-viral-combined_k141_242585||lt2gene Not-determined
17-108-final-viral-combined_k141_200031||lt2gene Low-quality
17-108-final-viral-combined_k141_269272||lt2gene Low-quality
17-108-final-viral-combined_k141_75862||lt2gene Low-quality
17-108-final-viral-combined_k141_311299||lt2gene Not-determined
17-108-final-viral-combined_k141_255954||lt2gene Low-quality
17-108-final-viral-combined_k141_311319||lt2gene Low-quality
17-108-final-viral-combined_k141_104437||lt2gene Not-determined
17-108-final-viral-combined_k141_323683||lt2gene Low-quality
17-108-final-viral-combined_k141_242676||lt2gene Low-quality
17-108-final-viral-combined_k141_104470||lt2gene Not-determined
17-108-final-viral-combined_k141_186365||lt2gene Low-quality
17-108-final-viral-combined_k141_269313||lt2gene Not-determined
17-108-final-viral-combined_k141_35372||lt2gene Low-quality
17-108-final-viral-combined_k141_296698||lt2gene Low-quality
17-108-final-viral-combined_k141_104517||lt2gene Low-quality
17-108-final-viral-combined_k141_242734||lt2gene Low-quality
17-108-final-viral-combined_k141_214791||lt2gene Low-quality
17-108-final-viral-combined_k141_186396||lt2gene Low-quality
17-108-final-viral-combined_k141_242827||lt2gene Low-quality
17-108-final-viral-combined_k141_186472||lt2gene Low-quality
17-108-final-viral-combined_k141_256089||lt2gene Low-quality
17-108-final-viral-combined_k141_104657||lt2gene Not-determined
17-108-final-viral-combined_k141_214890||lt2gene Not-determined
17-108-final-viral-combined_k141_200234||lt2gene Low-quality
17-108-final-viral-combined_k141_200242||lt2gene Low-quality
17-108-final-viral-combined_k141_173550||lt2gene Low-quality
17-108-final-viral-combined_k141_117338||lt2gene Low-quality
17-108-final-viral-combined_k141_186562||lt2gene Low-quality
17-108-final-viral-combined_k141_311487||lt2gene Not-determined
17-108-final-viral-combined_k141_256210||lt2gene Low-quality
17-108-final-viral-combined_k141_35575||lt2gene Not-determined
17-108-final-viral-combined_k141_7861||lt2gene Low-quality
17-108-final-viral-combined_k141_242916||lt2gene Low-quality
17-108-final-viral-combined_k141_7904||lt2gene Low-quality
17-108-final-viral-combined_k141_311520||lt2gene Low-quality
17-108-final-viral-combined_k141_63319||lt2gene Low-quality
17-108-final-viral-combined_k141_49172||lt2gene Low-quality
17-108-final-viral-combined_k141_311560||lt2gene Low-quality
17-108-final-viral-combined_k141_76125||lt2gene Not-determined
17-108-final-viral-combined_k141_296934||lt2gene Low-quality
17-108-final-viral-combined_k141_49177||lt2gene Low-quality
17-108-final-viral-combined_k141_323947||lt2gene Low-quality
17-108-final-viral-combined_k141_159608||lt2gene Low-quality
17-108-final-viral-combined_k141_311583||lt2gene Low-quality
17-108-final-viral-combined_k141_20765||lt2gene Low-quality
17-108-final-viral-combined_k141_200437||lt2gene Not-determined
17-108-final-viral-combined_k141_256255||lt2gene Low-quality
17-108-final-viral-combined_k141_90608||lt2gene Low-quality
17-108-final-viral-combined_k141_35680||lt2gene Low-quality
17-108-final-viral-combined_k141_131400||lt2gene Low-quality
17-108-final-viral-combined_k141_242977||lt2gene Not-determined
17-108-final-viral-combined_k141_117525||lt2gene Low-quality
17-108-final-viral-combined_k141_90640||lt2gene Not-determined
17-108-final-viral-combined_k141_269725||lt2gene Not-determined
17-108-final-viral-combined_k141_104925||lt2gene Not-determined
17-108-final-viral-combined_k141_76242||lt2gene Low-quality
17-108-final-viral-combined_k141_243049||lt2gene Not-determined
17-108-final-viral-combined_k141_145770||lt2gene Low-quality
17-108-final-viral-combined_k141_131492||lt2gene Not-determined
17-108-final-viral-combined_k141_145797||lt2gene Low-quality
17-108-final-viral-combined_k141_8124||lt2gene Not-determined
17-108-final-viral-combined_k141_8135||lt2gene Low-quality
17-108-final-viral-combined_k141_35737||lt2gene Low-quality
17-108-final-viral-combined_k141_173741||lt2gene Low-quality
17-108-final-viral-combined_k141_35758||lt2gene Low-quality
17-108-final-viral-combined_k141_159708||lt2gene Low-quality
17-108-final-viral-combined_k141_173797||lt2gene Low-quality
17-108-final-viral-combined_k141_49350||lt2gene Low-quality
17-108-final-viral-combined_k141_186863||lt2gene Low-quality
17-108-final-viral-combined_k141_35893||lt2gene Low-quality
17-108-final-viral-combined_k141_186891||lt2gene Not-determined
17-108-final-viral-combined_k141_173806||lt2gene Not-determined
17-108-final-viral-combined_k141_117743||lt2gene Not-determined
17-108-final-viral-combined_k141_173818||lt2gene Not-determined
17-108-final-viral-combined_k141_90848||lt2gene Not-determined
17-108-final-viral-combined_k141_76453||lt2gene Low-quality
17-108-final-viral-combined_k141_131644||lt2gene Low-quality
17-108-final-viral-combined_k141_256498||lt2gene Low-quality
17-108-final-viral-combined_k141_145990||lt2gene Low-quality
17-108-final-viral-combined_k141_243212||lt2gene Not-determined
17-108-final-viral-combined_k141_35958||lt2gene Not-determined
17-108-final-viral-combined_k141_49450||lt2gene Low-quality
17-108-final-viral-combined_k141_76549||lt2gene Low-quality
17-108-final-viral-combined_k141_186984||lt2gene Low-quality
17-108-final-viral-combined_k141_117860||lt2gene Low-quality
17-108-final-viral-combined_k141_297278||lt2gene Low-quality
17-108-final-viral-combined_k141_8491||lt2gene Low-quality
17-108-final-viral-combined_k141_311912||lt2gene Low-quality
17-108-final-viral-combined_k141_159959||lt2gene Low-quality
17-108-final-viral-combined_k141_215374||lt2gene Low-quality
17-108-final-viral-combined_k141_200830||lt2gene Low-quality
17-108-final-viral-combined_k141_311928||lt2gene Low-quality
17-108-final-viral-combined_k141_76634||lt2gene Low-quality
17-108-final-viral-combined_k141_8524||lt2gene Not-determined
17-108-final-viral-combined_k141_270068||lt2gene Low-quality
17-108-final-viral-combined_k141_105300||lt2gene Not-determined
17-108-final-viral-combined_k141_228961||lt2gene Not-determined
17-108-final-viral-combined_k141_160001||lt2gene Low-quality
17-108-final-viral-combined_k141_187127||lt2gene Not-determined
17-108-final-viral-combined_k141_76661||lt2gene Not-determined
17-108-final-viral-combined_k141_21218||lt2gene Low-quality
17-108-final-viral-combined_k141_174026||lt2gene Low-quality
17-108-final-viral-combined_k141_76677||lt2gene Not-determined
17-108-final-viral-combined_k141_256709||lt2gene Low-quality
17-108-final-viral-combined_k141_228997||lt2gene Low-quality
17-108-final-viral-combined_k141_243393||lt2gene Low-quality
17-108-final-viral-combined_k141_160037||lt2gene Low-quality
17-108-final-viral-combined_k141_8630||lt2gene Not-determined
17-108-final-viral-combined_k141_117993||lt2gene Not-determined
17-108-final-viral-combined_k141_229042||lt2gene Not-determined
17-108-final-viral-combined_k141_215456||lt2gene Low-quality
17-108-final-viral-combined_k141_270104||lt2gene Not-determined
17-108-final-viral-combined_k141_215481||lt2gene Low-quality
17-108-final-viral-combined_k141_229063||lt2gene Low-quality
17-108-final-viral-combined_k141_49678||lt2gene Low-quality
17-108-final-viral-combined_k141_8722||lt2gene Low-quality
17-108-final-viral-combined_k141_21327||lt2gene Low-quality
17-108-final-viral-combined_k141_270124||lt2gene Low-quality
17-108-final-viral-combined_k141_312060||lt2gene Low-quality

| | |
|---|---|
| 17-108-final-viral-combined_k141_63802\|\|lt2gene | Not-determined |
| 17-108-final-viral-combined_k141_256849\|\|lt2gene | Not-determined |
| 17-108-final-viral-combined_k141_49729\|\|lt2gene | Not-determined |
| 17-108-final-viral-combined_k141_284447\|\|lt2gene | Not-determined |
| 17-108-final-viral-combined_k141_91128\|\|lt2gene | Low-quality |
| 17-108-final-viral-combined_k141_284452\|\|lt2gene | Not-determined |
| 17-108-final-viral-combined_k141_187239\|\|lt2gene | Not-determined |
| 17-108-final-viral-combined_k141_312094\|\|lt2gene | Not-determined |
| 17-108-final-viral-combined_k141_174205\|\|lt2gene | Low-quality |
| 17-108-final-viral-combined_k141_229143\|\|lt2gene | Not-determined |
| 17-108-final-viral-combined_k141_256897\|\|lt2gene | Not-determined |
| 17-108-final-viral-combined_k141_21354\|\|lt2gene | Low-quality |
| 17-108-final-viral-combined_k141_284490\|\|lt2gene | Low-quality |
| 17-108-final-viral-combined_k141_76839\|\|lt2gene | Low-quality |
| 17-108-final-viral-combined_k141_91181\|\|lt2gene | Low-quality |
| 17-108-final-viral-combined_k141_312127\|\|lt2gene | Low-quality |
| 17-108-final-viral-combined_k141_284513\|\|lt2gene | Low-quality |
| 17-108-final-viral-combined_k141_256947\|\|lt2gene | Not-determined |
| 17-108-final-viral-combined_k141_49820\|\|lt2gene | Not-determined |
| 17-108-final-viral-combined_k141_36329\|\|lt2gene | Low-quality |
| 17-108-final-viral-combined_k141_324435\|\|lt2gene | Low-quality |
| 17-108-final-viral-combined_k141_91205\|\|lt2gene | Not-determined |
| 17-108-final-viral-combined_k141_49851\|\|lt2gene | Not-determined |
| 17-108-final-viral-combined_k141_201065\|\|lt2gene | Not-determined |
| 17-108-final-viral-combined_k141_270157\|\|lt2gene | Low-quality |
| 17-108-final-viral-combined_k141_297514\|\|lt2gene | Not-determined |
| 17-108-final-viral-combined_k141_76930\|\|lt2gene | Low-quality |
| 17-108-final-viral-combined_k141_243627\|\|lt2gene | Not-determined |
| 17-108-final-viral-combined_k141_105633\|\|lt2gene | Not-determined |
| 17-108-final-viral-combined_k141_201146\|\|lt2gene | Not-determined |
| 17-108-final-viral-combined_k141_229317\|\|lt2gene | Low-quality |
| 17-108-final-viral-combined_k141_160345\|\|lt2gene | Low-quality |
| 17-108-final-viral-combined_k141_49961\|\|lt2gene | Low-quality |
| 17-108-final-viral-combined_k141_201164\|\|lt2gene | Low-quality |
| 17-108-final-viral-combined_k141_63912\|\|lt2gene | Low-quality |
| 17-108-final-viral-combined_k141_63915\|\|lt2gene | Low-quality |
| 17-108-final-viral-combined_k141_131934\|\|lt2gene | Not-determined |
| 17-108-final-viral-combined_k141_312237\|\|lt2gene | Not-determined |
| 17-108-final-viral-combined_k141_187390\|\|lt2gene | Not-determined |
| 17-108-final-viral-combined_k141_131967\|\|lt2gene | Low-quality |
| 17-108-final-viral-combined_k141_297626\|\|lt2gene | Low-quality |
| 17-108-final-viral-combined_k141_243746\|\|lt2gene | Low-quality |
| 17-108-final-viral-combined_k141_270298\|\|lt2gene | Not-determined |
| 17-108-final-viral-combined_k141_297651\|\|lt2gene | Low-quality |
| 17-108-final-viral-combined_k141_91339\|\|lt2gene | Not-determined |
| [illegible] | [illegible] |

| | |
|---|---|
| 17-108-final-viral-combined_k141_218930\|\|lt2gene | Low-quality |
| 17-108-final-viral-combined_k141_54186\|\|lt2gene | Not-determined |
| 17-108-final-viral-combined_k141_150366\|\|lt2gene | Low-quality |
| 17-108-final-viral-combined_k141_260936\|\|lt2gene | Not-determined |
| 17-108-final-viral-combined_k141_135705\|\|lt2gene | Not-determined |
| 17-108-final-viral-combined_k141_191031\|\|lt2gene | Not-determined |
| 17-108-final-viral-combined_k141_327968\|\|lt2gene | Low-quality |
| 17-108-final-viral-combined_k141_327972\|\|lt2gene | Low-quality |
| 17-108-final-viral-combined_k141_219003\|\|lt2gene | Not-determined |
| 17-108-final-viral-combined_k141_54254\|\|lt2gene | Low-quality |
| 17-108-final-viral-combined_k141_261016\|\|lt2gene | Not-determined |
| 17-108-final-viral-combined_k141_109710\|\|lt2gene | Low-quality |
| 17-108-final-viral-combined_k141_25001\|\|lt2gene | Not-determined |
| 17-108-final-viral-combined_k141_150437\|\|lt2gene | Low-quality |
| 17-108-final-viral-combined_k141_273914\|\|lt2gene | Low-quality |
| 17-108-final-viral-combined_k141_315551\|\|lt2gene | Low-quality |
| 17-108-final-viral-combined_k141_67906\|\|lt2gene | Low-quality |
| 17-108-final-viral-combined_k141_178272\|\|lt2gene | Low-quality |
| 17-108-final-viral-combined_k141_109764\|\|lt2gene | Not-determined |
| 17-108-final-viral-combined_k141_80694\|\|lt2gene | Not-determined |
| 17-108-final-viral-combined_k141_247683\|\|lt2gene | Not-determined |
| 17-108-final-viral-combined_k141_54338\|\|lt2gene | Low-quality |
| 17-108-final-viral-combined_k141_205047\|\|lt2gene | Low-quality |
| 17-108-final-viral-combined_k141_54359\|\|lt2gene | Low-quality |
| 17-108-final-viral-combined_k141_40631\|\|lt2gene | Not-determined |
| 17-108-final-viral-combined_k141_178342\|\|lt2gene | Not-determined |
| 17-108-final-viral-combined_k141_54378\|\|lt2gene | Not-determined |
| 17-108-final-viral-combined_k141_288515\|\|lt2gene | Low-quality |
| 17-108-final-viral-combined_k141_67975\|\|lt2gene | Low-quality |
| 17-108-final-viral-combined_k141_247735\|\|lt2gene | Low-quality |
| 17-108-final-viral-combined_k141_247740\|\|lt2gene | Not-determined |
| 17-108-final-viral-combined_k141_274064\|\|lt2gene | Not-determined |
| 17-108-final-viral-combined_k141_301420\|\|lt2gene | Not-determined |
| 17-108-final-viral-combined_k141_301425\|\|lt2gene | Low-quality |
| 17-108-final-viral-combined_k141_54431\|\|lt2gene | Low-quality |
| 17-108-final-viral-combined_k141_191180\|\|lt2gene | Not-determined |
| 17-108-final-viral-combined_k141_150592\|\|lt2gene | Low-quality |
| 17-108-final-viral-combined_k141_12841\|\|lt2gene | Not-determined |
| 17-108-final-viral-combined_k141_150600\|\|lt2gene | Low-quality |
| 17-108-final-viral-combined_k141_122087\|\|lt2gene | Not-determined |
| 17-108-final-viral-combined_k141_25122\|\|lt2gene | Not-determined |
| 17-108-final-viral-combined_k141_178431\|\|lt2gene | Low-quality |
| 17-108-final-viral-combined_k141_95151\|\|lt2gene | Low-quality |
| 17-108-final-viral-combined_k141_178437\|\|lt2gene | Low-quality |
| 17-108-final-viral-combined_k141_315711\|\|lt2gene | Low-quality |
| 17-108-final-viral-combined_k141_109910\|\|lt2gene | Low-quality |
| 17-108-final-viral-combined_k141_12893\|\|lt2gene | Low-quality |
| 17-108-final-viral-combined_k141_109926\|\|lt2gene | Low-quality |
| 17-108-final-viral-combined_k141_80863\|\|lt2gene | Low-quality |
| 17-108-final-viral-combined_k141_68098\|\|lt2gene | Low-quality |
| 17-108-final-viral-combined_k141_191228\|\|lt2gene | Not-determined |
| 17-108-final-viral-combined_k141_191249\|\|lt2gene | Not-determined |
| 17-108-final-viral-combined_k141_135978\|\|lt2gene | Low-quality |
| 17-108-final-viral-combined_k141_54543\|\|lt2gene | Low-quality |
| 17-108-final-viral-combined_k141_191293\|\|lt2gene | Not-determined |
| 17-108-final-viral-combined_k141_205231\|\|lt2gene | Low-quality |
| 17-108-final-viral-combined_k141_25254\|\|lt2gene | Low-quality |
| 17-108-final-viral-combined_k141_219235\|\|lt2gene | Not-determined |
| 17-108-final-viral-combined_k141_40876\|\|lt2gene | Not-determined |
| 17-108-final-viral-combined_k141_191338\|\|lt2gene | Not-determined |
| 17-108-final-viral-combined_k141_261233\|\|lt2gene | Low-quality |
| 17-108-final-viral-combined_k141_178540\|\|lt2gene | Low-quality |
| 17-108-final-viral-combined_k141_80892\|\|lt2gene | Low-quality |
| 17-108-final-viral-combined_k141_274180\|\|lt2gene | Low-quality |
| 17-108-final-viral-combined_k141_219276\|\|lt2gene | Not-determined |
| 17-108-final-viral-combined_k141_191369\|\|lt2gene | Low-quality |
| 17-108-final-viral-combined_k141_95279\|\|lt2gene | Low-quality |
| 17-108-final-viral-combined_k141_80923\|\|lt2gene | Not-determined |
| 17-108-final-viral-combined_k141_164629\|\|lt2gene | Low-quality |
| 17-108-final-viral-combined_k141_150815\|\|lt2gene | Not-determined |
| 17-108-final-viral-combined_k141_95305\|\|lt2gene | Not-determined |
| 17-108-final-viral-combined_k141_233562\|\|lt2gene | Low-quality |
| 17-108-final-viral-combined_k141_110112\|\|lt2gene | Not-determined |
| 17-108-final-viral-combined_k141_13071\|\|lt2gene | Not-determined |
| 17-108-final-viral-combined_k141_301602\|\|lt2gene | Low-quality |
| 17-108-final-viral-combined_k141_261370\|\|lt2gene | Low-quality |
| 17-108-final-viral-combined_k141_68233\|\|lt2gene | Low-quality |
| 17-108-final-viral-combined_k141_274311\|\|lt2gene | Low-quality |
| 17-108-final-viral-combined_k141_205358\|\|lt2gene | Low-quality |
| 17-108-final-viral-combined_k141_288760\|\|lt2gene | Low-quality |
| 17-108-final-viral-combined_k141_288762\|\|lt2gene | Not-determined |
| 17-108-final-viral-combined_k141_315906\|\|lt2gene | Low-quality |
| 17-108-final-viral-combined_k141_178701\|\|lt2gene | Low-quality |
| 17-108-final-viral-combined_k141_136204\|\|lt2gene | Not-determined |
| 17-108-final-viral-combined_k141_25476\|\|lt2gene | Low-quality |
| 17-108-final-viral-combined_k141_191509\|\|lt2gene | Low-quality |
| 17-108-final-viral-combined_k141_301696\|\|lt2gene | Low-quality |
| 17-108-final-viral-combined_k141_178729\|\|lt2gene | Not-determined |
| 17-108-final-viral-combined_k141_13254\|\|lt2gene | Low-quality |
| 17-108-final-viral-combined_k141_219494\|\|lt2gene | Not-determined |
| 17-108-final-viral-combined_k141_41060\|\|lt2gene | Not-determined |
| 17-108-final-viral-combined_k141_164863\|\|lt2gene | Low-quality |
| 17-108-final-viral-combined_k141_288878\|\|lt2gene | Low-quality |
| 17-108-final-viral-combined_k141_288912\|\|lt2gene | Low-quality |
| 17-108-final-viral-combined_k141_219574\|\|lt2gene | Low-quality |
| 17-108-final-viral-combined_k141_178795\|\|lt2gene | Low-quality |
| 17-108-final-viral-combined_k141_151049\|\|lt2gene | Not-determined |
| 17-108-final-viral-combined_k141_13327\|\|lt2gene | Low-quality |
| 17-108-final-viral-combined_k141_13338\|\|lt2gene | Not-determined |
| 17-108-final-viral-combined_k141_164919\|\|lt2gene | Not-determined |
| 17-108-final-viral-combined_k141_122589\|\|lt2gene | Low-quality |
| 17-108-final-viral-combined_k141_25653\|\|lt2gene | Low-quality |
| 17-108-final-viral-combined_k141_301935\|\|lt2gene | Low-quality |
| 17-108-final-viral-combined_k141_136402\|\|lt2gene | Low-quality |
| 17-108-final-viral-combined_k141_316053\|\|lt2gene | Low-quality |
| 17-108-final-viral-combined_k141_13435\|\|lt2gene | Low-quality |
| 17-108-final-viral-combined_k141_205578\|\|lt2gene | Low-quality |
| 17-108-final-viral-combined_k141_54986\|\|lt2gene | Low-quality |
| 17-108-final-viral-combined_k141_165021\|\|lt2gene | Low-quality |
| 17-108-final-viral-combined_k141_81303\|\|lt2gene | Not-determined |
| 17-108-final-viral-combined_k141_233997\|\|lt2gene | Low-quality |
| 17-108-final-viral-combined_k141_81314\|\|lt2gene | Low-quality |
| 17-108-final-viral-combined_k141_95692\|\|lt2gene | Low-quality |
| 17-108-final-viral-combined_k141_302043\|\|lt2gene | Not-determined |
| 17-108-final-viral-combined_k141_178890\|\|lt2gene | Low-quality |
| 17-108-final-viral-combined_k141_41246\|\|lt2gene | Low-quality |
| 17-108-final-viral-combined_k141_151135\|\|lt2gene | Not-determined |
| 17-108-final-viral-combined_k141_328539\|\|lt2gene | Low-quality |
| 17-108-final-viral-combined_k141_316167\|\|lt2gene | Low-quality |
| 17-108-final-viral-combined_k141_289062\|\|lt2gene | Low-quality |
| 17-108-final-viral-combined_k141_81343\|\|lt2gene | Low-quality |
| 17-108-final-viral-combined_k141_178905\|\|lt2gene | Not-determined |
| 17-108-final-viral-combined_k141_95743\|\|lt2gene | Low-quality |
| 17-108-final-viral-combined_k141_81374\|\|lt2gene | Low-quality |
| 17-108-final-viral-combined_k141_289080\|\|lt2gene | Low-quality |
| 17-108-final-viral-combined_k141_234058\|\|lt2gene | Low-quality |
| 17-108-final-viral-combined_k141_151200\|\|lt2gene | Low-quality |
| 17-108-final-viral-combined_k141_25850\|\|lt2gene | Low-quality |
| 17-108-final-viral-combined_k141_205677\|\|lt2gene | Low-quality |
| 17-108-final-viral-combined_k141_178980\|\|lt2gene | Low-quality |
| 17-108-final-viral-combined_k141_302161\|\|lt2gene | Low-quality |
| 17-108-final-viral-combined_k141_302183\|\|lt2gene | Not-determined |
| 17-108-final-viral-combined_k141_191865\|\|lt2gene | Low-quality |
| 17-108-final-viral-combined_k141_289196\|\|lt2gene | Not-determined |
| 17-108-final-viral-combined_k141_302220\|\|lt2gene | Low-quality |
| 17-108-final-viral-combined_k141_81484\|\|lt2gene | Low-quality |
| 17-108-final-viral-combined_k141_302271\|\|lt2gene | Not-determined |
| 17-108-final-viral-combined_k141_328687\|\|lt2gene | Low-quality |
| 17-108-final-viral-combined_k141_191902\|\|lt2gene | Not-determined |
| 17-108-final-viral-combined_k141_205793\|\|lt2gene | Low-quality |
| 17-108-final-viral-combined_k141_274764\|\|lt2gene | Low-quality |
| 17-108-final-viral-combined_k141_289261\|\|lt2gene | Low-quality |
| 17-108-final-viral-combined_k141_151362\|\|lt2gene | Low-quality |
| 17-108-final-viral-combined_k141_316340\|\|lt2gene | Low-quality |
| 17-108-final-viral-combined_k141_165338\|\|lt2gene | Low-quality |
| 17-108-final-viral-combined_k141_95953\|\|lt2gene | Low-quality |
| 17-108-final-viral-combined_k141_13726\|\|lt2gene | Low-quality |
| 17-108-final-viral-combined_k141_274808\|\|lt2gene | Low-quality |
| 17-108-final-viral-combined_k141_26063\|\|lt2gene | Low-quality |
| 17-108-final-viral-combined_k141_165355\|\|lt2gene | Low-quality |
| 17-108-final-viral-combined_k141_191996\|\|lt2gene | Not-determined |
| 17-108-final-viral-combined_k141_289352\|\|lt2gene | Low-quality |
| 17-108-final-viral-combined_k141_26089\|\|lt2gene | Low-quality |
| 17-108-final-viral-combined_k141_205886\|\|lt2gene | Not-determined |
| 17-108-final-viral-combined_k141_26141\|\|lt2gene | Not-determined |
| 17-108-final-viral-combined_k141_328827\|\|lt2gene | Not-determined |
| 17-108-final-viral-combined_k141_123145\|\|lt2gene | Low-quality |
| 17-108-final-viral-combined_k141_96181\|\|lt2gene | Low-quality |
| 17-108-final-viral-combined_k141_151474\|\|lt2gene | Low-quality |
| 17-108-final-viral-combined_k141_136958\|\|lt2gene | Not-determined |
| 17-108-final-viral-combined_k141_220179\|\|lt2gene | Not-determined |
| 17-108-final-viral-combined_k141_68813\|\|lt2gene | Low-quality |
| 17-108-final-viral-combined_k141_81831\|\|lt2gene | Not-determined |
| 17-108-final-viral-combined_k141_192197\|\|lt2gene | Not-determined |
| 17-108-final-viral-combined_k141_68871\|\|lt2gene | Not-determined |
| 17-108-final-viral-combined_k141_96294\|\|lt2gene | Low-quality |
| 17-108-final-viral-combined_k141_123296\|\|lt2gene | Low-quality |
| 17-108-final-viral-combined_k141_81952\|\|lt2gene | Not-determined |
| 17-108-final-viral-combined_k141_206125\|\|lt2gene | Low-quality |
| 17-108-final-viral-combined_k141_123404\|\|lt2gene | Low-quality |
| 17-108-final-viral-combined_k141_123417\|\|lt2gene | Not-determined |
| 17-108-final-viral-combined_k141_192386\|\|lt2gene | Low-quality |
| 17-108-final-viral-combined_k141_26484\|\|lt2gene | Low-quality |
| 17-108-final-viral-combined_k141_123429\|\|lt2gene | Low-quality |
| 17-108-final-viral-combined_k141_81998\|\|lt2gene | Low-quality |
| 17-108-final-viral-combined_k141_123473\|\|lt2gene | Low-quality |
| 17-108-final-viral-combined_k141_137198\|\|lt2gene | Not-determined |
| 17-108-final-viral-combined_k141_137209\|\|lt2gene | Not-determined |
| 17-108-final-viral-combined_k141_82038\|\|lt2gene | Low-quality |
| 17-108-final-viral-combined_k141_302872\|\|lt2gene | Low-quality |
| 17-108-final-viral-combined_k141_192434\|\|lt2gene | Low-quality |
| 17-108-final-viral-combined_k141_275303\|\|lt2gene | Low-quality |
| 17-108-final-viral-combined_k141_275315\|\|lt2gene | Low-quality |
| 17-108-final-viral-combined_k141_123565\|\|lt2gene | Low-quality |
| 17-108-final-viral-combined_k141_302932\|\|lt2gene | Not-determined |
| 17-108-final-viral-combined_k141_26614\|\|lt2gene | Low-quality |
| 17-108-final-viral-combined_k141_220387\|\|lt2gene | Low-quality |
| 17-108-final-viral-combined_k141_220391\|\|lt2gene | Not-determined |
| 17-108-final-viral-combined_k141_302957\|\|lt2gene | Low-quality |
| 17-108-final-viral-combined_k141_275352\|\|lt2gene | Low-quality |
| 17-108-final-viral-combined_k141_82111\|\|lt2gene | Low-quality |
| 17-108-final-viral-combined_k141_137342\|\|lt2gene | Low-quality |
| 17-108-final-viral-combined_k141_137390\|\|lt2gene | Low-quality |
| 17-108-final-viral-combined_k141_82138\|\|lt2gene | Low-quality |
| 17-108-final-viral-combined_k141_192609\|\|lt2gene | Not-determined |
| 17-108-final-viral-combined_k141_26712\|\|lt2gene | Not-determined |
| 17-108-final-viral-combined_k141_82211\|\|lt2gene | Low-quality |
| 17-108-final-viral-combined_k141_275422\|\|lt2gene | Low-quality |
| 17-108-final-viral-combined_k141_123685\|\|lt2gene | Low-quality |
| 17-108-final-viral-combined_k141_206307\|\|lt2gene | Not-determined |
| 17-108-final-viral-combined_k141_192752\|\|lt2gene | Not-determined |
| 17-108-final-viral-combined_k141_329211\|\|lt2gene | Low-quality |
| 17-108-final-viral-combined_k141_26866\|\|lt2gene | Low-quality |
| 17-108-final-viral-combined_k141_82386\|\|lt2gene | Low-quality |
| 17-108-final-viral-combined_k141_82402\|\|lt2gene | Not-determined |
| 17-108-final-viral-combined_k141_275530\|\|lt2gene | Low-quality |
| 17-108-final-viral-combined_k141_192831\|\|lt2gene | Low-quality |
| 17-108-final-viral-combined_k141_123878\|\|lt2gene | Low-quality |
| 17-108-final-viral-combined_k141_329265\|\|lt2gene | Not-determined |
| 17-108-final-viral-combined_k141_123914\|\|lt2gene | Low-quality |
| 17-108-final-viral-combined_k141_123917\|\|lt2gene | Not-determined |
| 17-108-final-viral-combined_k141_275612\|\|lt2gene | Low-quality |
| 17-108-final-viral-combined_k141_329364\|\|lt2gene | Low-quality |
| 17-108-final-viral-combined_k141_206558\|\|lt2gene | Not-determined |
| 17-108-final-viral-combined_k141_192907\|\|lt2gene | Not-determined |
| 17-108-final-viral-combined_k141_82602\|\|lt2gene | Not-determined |
| 17-108-final-viral-combined_k141_27112\|\|lt2gene | Low-quality |
| 17-108-final-viral-combined_k141_27120\|\|lt2gene | Low-quality |
| 17-108-final-viral-combined_k141_329581\|\|lt2gene | Low-quality |
| 17-108-final-viral-combined_k141_329627\|\|lt2gene | Low-quality |
| 17-108-final-viral-combined_k141_27367\|\|lt2gene | Not-determined |
| 17-108-final-viral-combined_k141_329953\|\|lt2gene | Low-quality |
| 17-108-final-viral-combined_k141_330162\|\|lt2gene | Low-quality |
| 17-108-final-viral-combined_k141_330234\|\|lt2gene | Low-quality |
| 17-108_k141_137810 | Not-determined |
| 17-108_k141_165372 | Not-determined |
| 17-108_k141_165373 | Not-determined |
| 17-108_k141_124032 | Not-determined |
| 17-108_k141_68911 | Not-determined |
| 17-108_k141_220502 | Not-determined |
| 17-108_k141_275627 | Low-quality |
| 17-108_k141_13793 | Not-determined |
| 17-108_k141_27567 | Not-determined |
| 17-108_k141_303204 | Not-determined |
| 17-108_k141_151604 | Not-determined |
| 17-108_k141_179170 | Not-determined |
| 17-108_k141_27 | Not-determined |
| 17-108_k141_275633 | Not-determined |
| 17-108_k141_289419 | Not-determined |
| 17-108_k141_179176 | Not-determined |
| 17-108_k141_137834 | Not-determined |
| 17-108_k141_151619 | Not-determined |
| 17-108_k141_316965 | Not-determined |
| 17-108_k141_124051 | Not-determined |
| 17-108_k141_220522 | Not-determined |
| 17-108_k141_96479 | Not-determined |
| 17-108_k141_13813 | Not-determined |
| 17-108_k141_275649 | Not-determined |
| 17-108_k141_220527 | Low-quality |
| 17-108_k141_42 | Not-determined |
| 17-108_k141_275660 | Not-determined |
| 17-108_k141_46 | Not-determined |
| 17-108_k141_165381 | Not-determined |
| 17-108_k141_13825 | Not-determined |
| 17-108_k141_82715 | Not-determined |
| 17-108_k141_68937 | Not-determined |
| 17-108_k141_124075 | Not-determined |
| 17-108_k141_54 | Not-determined |
| 17-108_k141_275662 | Not-determined |
| 17-108_k141_56 | Not-determined |
| 17-108_k141_275665 | Not-determined |
| 17-108_k141_275666 | Not-determined |
| 17-108_k141_165391 | Not-determined |
| 17-108_k141_63 | Not-determined |
| 17-108_k141_124077 | Low-quality |
| 17-108_k141_179202 | Not-determined |
| 17-108_k141_41347 | Not-determined |
| 17-108_k141_96490 | Not-determined |
| 17-108_k141_55138 | Not-determined |
| 17-108_k141_110277 | Not-determined |
| 17-108_k141_55140 | Not-determined |
| 17-108_k141_13844 | Not-determined |
| 17-108_k141_316984 | Not-determined |
| 17-108_k141_55144 | Not-determined |
| 17-108_k141_55146 | Not-determined |
| 17-108_k141_303216 | Low-quality |
| 17-108_k141_68952 | Not-determined |
| 17-108_k141_124091 | Low-quality |
| 17-108_k141_165399 | Not-determined |
| 17-108_k141_206750 | Not-determined |
| 17-108_k141_137873 | Not-determined |
| 17-108_k141_179222 | Not-determined |
| 17-108_k141_179223 | Not-determined |
| 17-108_k141_124104 | Not-determined |
| 17-108_k141_165418 | Not-determined |
| 17-108_k141_179241 | Not-determined |
| 17-108_k141_13868 | Not-determined |
| 17-108_k141_124110 | Not-determined |
| 17-108_k141_165430 | Not-determined |
| 17-108_k141_261872 | Not-determined |
| 17-108_k141_55182 | Low-quality |
| 17-108_k141_261878 | Not-determined |
| 17-108_k141_317021 | Low-quality |
| 17-108_k141_248115 | Not-determined |
| 17-108_k141_55187 | Low-quality |
| 17-108_k141_55192 | Not-determined |
| 17-108_k141_69 | Not-determined |
| 17-108_k141_73 | Not-determined |
| 17-108_k141_289466 | Not-determined |
| 17-108_k141_13882 | Not-determined |
| 17-108_k141_124125 | Not-determined |
| 17-108_k141_206777 | Not-determined |
| 17-108_k141_165447 | Not-determined |
| 17-108_k141_289478 | Not-determined |
| 17-108_k141_68981 | Not-determined |
| 17-108_k141_13890 | Not-determined |
| 17-108_k141_248134 | Not-determined |
| 17-108_k141_96511 | Low-quality |
| 17-108_k141_206785 | Not-determined |
| 17-108_k141_206789 | Not-determined |
| 17-108_k141_275674 | Not-determined |
| 17-108_k141_289490 | Not-determined |
| 17-108_k141_165463 | Not-determined |
| 17-108_k141_275681 | Not-determined |
| 17-108_k141_275686 | Not-determined |
| 17-108_k141_275687 | Not-determined |
| 17-108_k141_303249 | Not-determined |
| 17-108_k141_82761 | Not-determined |
| 17-108_k141_137918 | Low-quality |
| 17-108_k141_103 | Not-determined |
| 17-108_k141_192952 | Not-determined |
| 17-108_k141_303257 | Not-determined |
| 17-108_k141_220577 | Not-determined |
| 17-108_k141_151665 | Not-determined |
| 17-108_k141_192954 | Not-determined |
| 17-108_k141_137920 | Not-determined |
| 17-108_k141_275697 | Not-determined |
| 17-108_k141_289494 | Not-determined |
| 17-108_k141_289497 | Not-determined |
| 17-108_k141_289498 | Low-quality |
| 17-108_k141_110317 | Not-determined |
| 17-108_k141_317056 | Not-determined |
| 17-108_k141_179276 | Not-determined |
| 17-108_k141_165483 | Not-determined |
| 17-108_k141_41375 | Not-determined |
| 17-108_k141_165486 | Low-quality |
| 17-108_k141_69000 | Not-determined |
| 17-108_k141_317062 | Not-determined |
| 17-108_k141_275701 | Not-determined |
| 17-108_k141_165493 | Not-determined |
| 17-108_k141_151683 | Not-determined |
| 17-108_k141_151684 | Not-determined |
| 17-108_k141_317069 | Not-determined |
| 17-108_k141_220589 | Not-determined |
| 17-108_k141_317075 | Not-determined |
| 17-108_k141_192979 | Not-determined |
| 17-108_k141_165507 | Not-determined |
| 17-108_k141_141 | Not-determined |
| 17-108_k141_27585 | Not-determined |
| 17-108_k141_13913 | Not-determined |
| 17-108_k141_179292 | Not-determined |
| 17-108_k141_275717 | Not-determined |
| 17-108_k141_261908 | Low-quality |
| 17-108_k141_110332 | Not-determined |
| 17-108_k141_317086 | Not-determined |
| 17-108_k141_41394 | Not-determined |
| 17-108_k141_192993 | Low-quality |
| 17-108_k141_165523 | Not-determined |
| 17-108_k141_192995 | Not-determined |
| 17-108_k141_192996 | Not-determined |
| 17-108_k141_206834 | Not-determined |
| 17-108_k141_41399 | Not-determined |
| 17-108_k141_13928 | Not-determined |
| 17-108_k141_220598 | Not-determined |
| 17-108_k141_317092 | Not-determined |
| 17-108_k141_275742 | Not-determined |
| 17-108_k141_303288 | Low-quality |
| 17-108_k141_41410 | Not-determined |
| 17-108_k141_275746 | Not-determined |
| 17-108_k141_193012 | Not-determined |
| 17-108_k141_206851 | Not-determined |
| 17-108_k141_206852 | Not-determined |
| 17-108_k141_179313 | Not-determined |
| 17-108_k141_96541 | Not-determined |
| 17-108_k141_289518 | Not-determined |
| 17-108_k141_234315 | Not-determined |
| 17-108_k141_41422 | Not-determined |
| 17-108_k141_137951 | Not-determined |
| 17-108_k141_234321 | Not-determined |
| 17-108_k141_166 | Not-determined |
| 17-108_k141_167 | Not-determined |
| 17-108_k141_137957 | Not-determined |
| 17-108_k141_206872 | Not-determined |
| 17-108_k141_69057 | Not-determined |
| 17-108_k141_179328 | Not-determined |
| 17-108_k141_220623 | Not-determined |
| 17-108_k141_289529 | Not-determined |
| 17-108_k141_179330 | Not-determined |
| 17-108_k141_179331 | Not-determined |
| 17-108_k141_82830 | Not-determined |
| 17-108_k141_220632 | Not-determined |
| 17-108_k141_317111 | Not-determined |
| 17-108_k141_151732 | Low-quality |
| 17-108_k141_193046 | Not-determined |
| 17-108_k141_234332 | Low-quality |
| 17-108_k141_124216 | Not-determined |
| 17-108_k141_234334 | Not-determined |
| 17-108_k141_179 | Not-determined |
| 17-108_k141_41435 | Not-determined |
| 17-108_k141_151743 | Not-determined |
| 17-108_k141_289536 | Not-determined |
| 17-108_k141_317116 | Not-determined |
| 17-108_k141_27600 | Not-determined |
| 17-108_k141_27602 | Not-determined |
| 17-108_k141_220641 | Not-determined |
| 17-108_k141_41444 | Not-determined |
| 17-108_k141_303315 | Not-determined |
| 17-108_k141_151760 | Not-determined |
| 17-108_k141_96543 | Not-determined |
| 17-108_k141_124222 | Low-quality |
| 17-108_k141_41452 | Not-determined |
| 17-108_k141_55249 | Not-determined |
| 17-108_k141_124225 | Low-quality |
| 17-108_k141_234348 | Not-determined |
| 17-108_k141_165569 | Low-quality |
| 17-108_k141_234353 | Not-determined |
| 17-108_k141_234354 | Not-determined |
| 17-108_k141_124232 | Not-determined |
| 17-108_k141_27621 | Not-determined |
| 17-108_k141_220665 | Low-quality |
| 17-108_k141_220667 | Not-determined |
| 17-108_k141_193074 | Not-determined |
| 17-108_k141_55259 | Not-determined |
| 17-108_k141_234358 | Not-determined |
| 17-108_k141_234360 | Not-determined |
| 17-108_k141_124242 | Not-determined |
| 17-108_k141_69070 | Not-determined |
| 17-108_k141_55262 | Not-determined |
| 17-108_k141_55263 | Not-determined |
| 17-108_k141_193090 | Not-determined |
| 17-108_k141_206894 | Not-determined |
| 17-108_k141_248199 | Not-determined |
| 17-108_k141_193095 | Not-determined |
| 17-108_k141_124258 | Not-determined |
| 17-108_k141_124260 | Not-determined |
| 17-108_k141_96577 | Low-quality |
| 17-108_k141_82874 | Not-determined |
| 17-108_k141_165572 | Not-determined |
| 17-108_k141_82875 | Not-determined |
| 17-108_k141_220690 | Not-determined |
| 17-108_k141_69086 | Not-determined |
| 17-108_k141_69089 | Not-determined |
| 17-108_k141_317144 | Not-determined |
| 17-108_k141_13984 | Not-determined |
| 17-108_k141_151809 | Not-determined |
| 17-108_k141_151811 | Not-determined |
| 17-108_k141_234378 | Not-determined |
| 17-108_k141_206914 | Not-determined |
| 17-108_k141_165587 | Not-determined |
| 17-108_k141_234383 | Not-determined |
| 17-108_k141_13994 | Not-determined |
| 17-108_k141_96593 | Not-determined |
| 17-108_k141_317161 | Not-determined |
| 17-108_k141_96598 | Not-determined |
| 17-108_k141_193130 | Not-determined |
| 17-108_k141_165603 | Not-determined |
| 17-108_k141_124266 | Not-determined |
| 17-108_k141_96606 | Not-determined |
| 17-108_k141_151830 | Not-determined |
| 17-108_k141_317170 | Not-determined |
| 17-108_k141_69106 | Not-determined |
| 17-108_k141_179336 | Low-quality |
| 17-108_k141_41456 | Not-determined |
| 17-108_k141_193147 | Not-determined |
| 17-108_k141_165619 | Not-determined |
| 17-108_k141_179345 | Not-determined |
| 17-108_k141_124283 | Not-determined |
| 17-108_k141_234402 | Not-determined |
| 17-108_k141_165631 | Low-quality |
| 17-108_k141_96617 | Not-determined |
| 17-108_k141_317180 | Not-determined |
| 17-108_k141_317181 | Not-determined |
| 17-108_k141_69131 | Not-determined |
| 17-108_k141_124294 | Not-determined |
| 17-108_k141_138001 | Not-determined |
| 17-108_k141_303377 | Not-determined |
| 17-108_k141_82933 | Not-determined |
| 17-108_k141_82939 | Not-determined |
| 17-108_k141_124300 | Not-determined |
| 17-108_k141_165648 | Not-determined |
| 17-108_k141_69137 | Not-determined |
| 17-108_k141_165653 | Not-determined |
| 17-108_k141_165656 | Not-determined |
| 17-108_k141_138023 | Not-determined |
| 17-108_k141_96643 | Not-determined |
| 17-108_k141_96644 | Not-determined |
| 17-108_k141_206955 | Low-quality |
| 17-108_k141_179376 | Not-determined |
| 17-108_k141_206963 | Not-determined |
| 17-108_k141_96651 | Not-determined |
| 17-108_k141_192 | Not-determined |
| 17-108_k141_55319 | Not-determined |
| 17-108_k141_234420 | Not-determined |
| 17-108_k141_193150 | Not-determined |
| 17-108_k141_248237 | Not-determined |
| 17-108_k141_206971 | Not-determined |
| 17-108_k141_193155 | Not-determined |
| 17-108_k141_193157 | Not-determined |
| 17-108_k141_193159 | Not-determined |
| 17-108_k141_124336 | Not-determined |
| 17-108_k141_55327 | Not-determined |
| 17-108_k141_69147 | Not-determined |
| 17-108_k141_193165 | Not-determined |
| 17-108_k141_179381 | Not-determined |
| 17-108_k141_82987 | Not-determined |
| 17-108_k141_212 | Not-determined |
| 17-108_k141_213 | Not-determined |
| 17-108_k141_96656 | Not-determined |
| 17-108_k141_193172 | Not-determined |
| 17-108_k141_55336 | Not-determined |
| 17-108_k141_55340 | Not-determined |
| 17-106-final-viral-combined_k141_37587\|\|0_partial | Medium-quality |
| 17-108_k141_124358 | Not-determined |
| 17-108_k141_138032 | Not-determined |
| 17-108_k141_317187 | Not-determined |
| 17-108_k141_275824 | Not-determined |
| 17-108_k141_317188 | Not-determined |
| 17-108_k141_124366 | Not-determined |
| 17-108_k141_275829 | Not-determined |
| 17-108_k141_207001 | Not-determined |
| 17-108_k141_110380 | Not-determined |
| 17-108_k141_138044 | Not-determined |
| 17-108_k141_317193 | Not-determined |
| 17-108_k141_193176 | Not-determined |
| 17-108_k141_317195 | Not-determined |
| 17-108_k141_275839 | Not-determined |
| 17-108_k141_220772 | Not-determined |
| 17-108_k141_262007 | Not-determined |
| 17-108_k141_289586 | Not-determined |
| 17-108_k141_220785 | Not-determined |
| 17-108_k141_179396 | Not-determined |
| 17-108_k141_222 | Not-determined |
| 17-108_k141_179401 | Not-determined |
| 17-108_k141_248254 | Not-determined |
| 17-108_k141_275859 | Not-determined |
| 17-108_k141_69159 | Not-determined |
| 17-108_k141_69160 | Not-determined |
| 17-108_k141_220799 | Not-determined |
| 17-108_k141_275868 | Low-quality |
| 17-108_k141_193182 | Not-determined |
| 17-108_k141_151892 | Not-determined |
| 17-108_k141_303428 | Not-determined |
| 17-108_k141_14066 | Low-quality |
| 17-108_k141_83002 | Low-quality |
| 17-108_k141_55353 | Low-quality |
| 17-108_k141_193185 | Not-determined |
| 17-108_k141_55354 | Not-determined |
| 17-108_k141_151896 | Low-quality |
| 17-108_k141_151897 | Not-determined |
| 17-108_k141_262024 | Not-determined |
| 17-108_k141_138080 | Not-determined |
| 17-108_k141_248261 | Not-determined |
| 17-108_k141_257 | Not-determined |
| 17-108_k141_165694 | Low-quality |
| 17-108_k141_193203 | Not-determined |
| 17-108_k141_234434 | Not-determined |
| 17-108_k141_55369 | Not-determined |
| 17-108_k141_248266 | Not-determined |
| 17-108_k141_138102 | Not-determined |
| 17-108_k141_248275 | Not-determined |
| 17-108_k141_234449 | Not-determined |
| 17-108_k141_110429 | Low-quality |
| 17-108_k141_270 | Low-quality |
| 17-108_k141_14095 | Not-determined |
| 17-108_k141_248277 | Not-determined |
| 17-108_k141_179404 | Not-determined |
| 17-108_k141_262040 | Not-determined |
| 17-108_k141_165703 | Not-determined |
| 17-108_k141_55379 | Not-determined |
| 17-108_k141_110450 | Not-determined |
| 17-108_k141_262050 | Not-determined |
| 17-108_k141_234460 | Not-determined |
| 17-108_k141_262053 | Not-determined |
| 17-108_k141_138123 | Low-quality |
| 17-108_k141_124410 | Not-determined |
| 17-108_k141_138134 | Not-determined |
| 17-108_k141_138136 | Not-determined |
| 17-108_k141_234470 | Not-determined |
| 17-108_k141_303461 | Not-determined |
| 17-108_k141_179422 | Not-determined |
| 17-108_k141_124419 | Low-quality |
| 17-108_k141_124422 | Not-determined |
| 17-108_k141_207035 | Not-determined |
| 17-108_k141_110470 | Not-determined |
| 17-108_k141_83021 | Not-determined |
| 17-108_k141_83023 | Not-determined |
| 17-108_k141_55399 | Not-determined |
| 17-108_k141_96702 | Low-quality |
| 17-108_k141_179439 | Not-determined |

| | |
|---|---|
| 17-108_k141_311 | Not-determined |
| 17-108_k141_138151 | Not-determined |
| 17-108_k141_165734 | Not-determined |
| 17-108_k141_83034 | Low-quality |
| 17-108_k141_14125 | Not-determined |
| 17-108_k141_41490 | Low-quality |
| 17-108_k141_69198 | Not-determined |
| 17-108_k141_69199 | Not-determined |
| 17-108_k141_220831 | Not-determined |
| 17-108_k141_220832 | Not-determined |
| 17-108_k141_220834 | Not-determined |
| 17-108_k141_220835 | Low-quality |
| 17-108_k141_320 | Not-determined |
| 17-108_k141_179444 | Not-determined |
| 17-108_k141_14133 | Not-determined |
| 17-108_k141_110493 | Not-determined |
| 17-108_k141_262092 | Not-determined |
| 17-108_k141_303474 | Not-determined |
| 17-108_k141_289601 | Not-determined |
| 17-108_k141_110500 | Not-determined |
| 17-108_k141_96726 | Not-determined |
| 17-108_k141_110502 | Not-determined |
| 17-108_k141_69205 | Not-determined |
| 17-108_k141_262097 | Not-determined |
| 17-108_k141_334 | Not-determined |
| 17-108_k141_303477 | Low-quality |
| 17-108_k141_193248 | Not-determined |
| 17-108_k141_110508 | Not-determined |
| 17-108_k141_248286 | Low-quality |
| 17-108_k141_165752 | Not-determined |
| 17-108_k141_317261 | Not-determined |
| 17-108_k141_220850 | Not-determined |
| 17-108_k141_165776 | Not-determined |
| 17-108_k141_207028 | Not-determined |
| 17-108_k141_234495 | Not-determined |
| 17-108_k141_207029 | Not-determined |
| 17-108_k141_165778 | Not-determined |
| 17-108_k141_234498 | Not-determined |
| 17-108_k141_193250 | Not-determined |
| 17-108_k141_165782 | Low-quality |
| 17-108_k141_289636 | Not-determined |
| 17-108_k141_165790 | Not-determined |
| 17-108_k141_303512 | Not-determined |
| 17-108_k141_27808 | Not-determined |
| 17-108_k141_262125 | Not-determined |
| 17-108_k141_220873 | Not-determined |
| 17-108_k141_262134 | Not-determined |
| 17-108_k141_96744 | Not-determined |
| 17-108_k141_220877 | Low-quality |
| 17-108_k141_234526 | Not-determined |
| 17-108_k141_165794 | Low-quality |
| 17-108_k141_248317 | Not-determined |
| 17-108_k141_262148 | Not-determined |
| 17-108_k141_262149 | Not-determined |
| 17-108_k141_41531 | Not-determined |
| 17-108_k141_41532 | Not-determined |
| 17-108_k141_262153 | Low-quality |
| 17-108_k141_151945 | Not-determined |
| 17-108_k141_41535 | Not-determined |
| 17-108_k141_27835 | Not-determined |
| 17-108_k141_96756 | Not-determined |
| 17-108_k141_27836 | Not-determined |
| 17-108_k141_165801 | Not-determined |
| 17-108_k141_179453 | Not-determined |
| 17-108_k141_193282 | Not-determined |
| 17-108_k141_27840 | Not-determined |
| 17-108_k141_124458 | Not-determined |
| 17-108_k141_124459 | Not-determined |
| 17-108_k141_138173 | Not-determined |
| 17-108_k141_165807 | Not-determined |
| 17-108_k141_124461 | Low-quality |
| 17-108_k141_317302 | Not-determined |
| 17-108_k141_248337 | Not-determined |
| 17-108_k141_151956 | Not-determined |
| 17-108_k141_165812 | Not-determined |
| 17-108_k141_207067 | Low-quality |
| 17-108_k141_165816 | Not-determined |
| 17-108_k141_207070 | Not-determined |
| 17-108_k141_110531 | Low-quality |
| 17-108_k141_27853 | Not-determined |
| 17-108_k141_27855 | Not-determined |
| 17-108_k141_207078 | Not-determined |
| 17-108_k141_165825 | Not-determined |
| 17-108_k141_41564 | Not-determined |
| 17-108_k141_14147 | Not-determined |
| 17-108_k141_41567 | Not-determined |
| 17-108_k141_165831 | Not-determined |
| 17-108_k141_110539 | Not-determined |
| 17-108_k141_138177 | Not-determined |
| 17-108_k141_193295 | Not-determined |
| 17-108_k141_138185 | Not-determined |
| 17-108_k141_14154 | Not-determined |
| 17-108_k141_27893 | Not-determined |
| 17-108_k141_14159 | Low-quality |
| 17-108_k141_138193 | Not-determined |
| 17-108_k141_248378 | Not-determined |
| 17-108_k141_179494 | Not-determined |
| 17-108_k141_151971 | Low-quality |
| 17-108_k141_193318 | Not-determined |
| 17-108_k141_69213 | Low-quality |
| 17-108_k141_165869 | Not-determined |
| 17-108_k141_124469 | Not-determined |
| 17-108_k141_96813 | Low-quality |
| 17-108_k141_234574 | Low-quality |
| 17-108_k141_110548 | Not-determined |
| 17-108_k141_234575 | Not-determined |
| 17-108_k141_96819 | Not-determined |
| 17-108_k141_289671 | Not-determined |
| 17-108_k141_415 | Not-determined |
| 17-108_k141_41593 | Not-determined |
| 17-108_k141_419 | Not-determined |
| 17-108_k141_138212 | Not-determined |
| 17-108_k141_110560 | Not-determined |
| 17-108_k141_138217 | Not-determined |
| 17-108_k141_248409 | Not-determined |
| 17-108_k141_303547 | Not-determined |
| 17-108_k141_248412 | Not-determined |
| 17-108_k141_96838 | Not-determined |
| 17-108_k141_138225 | Not-determined |
| 17-108_k141_27944 | Not-determined |
| 17-108_k141_83048 | Not-determined |
| 17-108_k141_303559 | Low-quality |
| 17-108_k141_96843 | Not-determined |
| 17-108_k141_275932 | Low-quality |
| 17-108_k141_248431 | Not-determined |
| 17-108_k141_14172 | Not-determined |
| 17-108_k141_207141 | Not-determined |
| 17-108_k141_248437 | Not-determined |
| 17-108_k141_275940 | Not-determined |
| 17-108_k141_152004 | Not-determined |
| 17-108_k141_124473 | Not-determined |
| 17-108_k141_262209 | Low-quality |
| 17-108_k141_14183 | Not-determined |
| 17-108_k141_165906 | Not-determined |
| 17-108_k141_262214 | Not-determined |
| 17-108_k141_317374 | Not-determined |
| 17-108_k141_193357 | Not-determined |
| 17-108_k141_165916 | Not-determined |
| 17-108_k141_41607 | Not-determined |
| 17-108_k141_96853 | Not-determined |
| 17-108_k141_275952 | Not-determined |
| 17-108_k141_317379 | Not-determined |
| 17-108_k141_124480 | Not-determined |
| 17-108_k141_41610 | Not-determined |
| 17-108_k141_69232 | Low-quality |
| 17-108_k141_317387 | Not-determined |
| 17-108_k141_138269 | Not-determined |
| 17-108_k141_83072 | Low-quality |
| 17-108_k141_303576 | Low-quality |
| 17-108_k141_303577 | Not-determined |
| 17-108_k141_124498 | Not-determined |
| 17-108_k141_303578 | Not-determined |
| 17-108_k141_165923 | Not-determined |
| 17-108_k141_27973 | Not-determined |
| 17-108_k141_303582 | Not-determined |
| 17-108_k141_69249 | Not-determined |
| 17-108_k141_234583 | Not-determined |
| 17-108_k141_152031 | Not-determined |
| 17-108_k141_124505 | Not-determined |
| 17-108_k141_27978 | Not-determined |
| 17-108_k141_165937 | Not-determined |
| 17-108_k141_110585 | Not-determined |
| 17-108_k141_124508 | Not-determined |
| 17-108_k141_69264 | Not-determined |
| 17-108_k141_248468 | Not-determined |
| 17-108_k141_96879 | Not-determined |
| 17-108_k141_41627 | Low-quality |
| 17-108_k141_234592 | Not-determined |
| 17-108_k141_303595 | Not-determined |
| 17-108_k141_124514 | Not-determined |
| 17-108_k141_452 | Not-determined |
| 17-108_k141_28007 | Not-determined |
| 17-108_k141_28008 | Not-determined |
| 17-108_k141_234602 | Not-determined |
| 17-108_k141_179587 | Not-determined |
| 17-108_k141_179588 | Low-quality |
| 17-108_k141_110599 | Not-determined |
| 17-108_k141_179590 | Not-determined |
| 17-108_k141_275970 | Low-quality |
| 17-108_k141_459 | Not-determined |
| 17-108_k141_124518 | Not-determined |
| 17-108_k141_248476 | Not-determined |
| 17-108_k141_303604 | Not-determined |
| 17-108_k141_303606 | Not-determined |
| 17-108_k141_207175 | Not-determined |
| 17-108_k141_275977 | Not-determined |
| 17-108_k141_303612 | Not-determined |
| 17-108_k141_152058 | Not-determined |
| 17-108_k141_96896 | Not-determined |
| 17-108_k141_14246 | Not-determined |
| 17-108_k141_165969 | Low-quality |
| 17-108_k141_303624 | Not-determined |
| 17-108_k141_165970 | Not-determined |
| 17-108_k141_234631 | Not-determined |
| 17-108_k141_207191 | Not-determined |
| 17-108_k141_207192 | Not-determined |
| 17-108_k141_207199 | Not-determined |
| 17-108_k141_207200 | Low-quality |
| 17-108_k141_248496 | Not-determined |
| 17-108_k141_207210 | Not-determined |
| 17-108_k141_28015 | Not-determined |
| 17-108_k141_41633 | Not-determined |
| 17-108_k141_28024 | Not-determined |
| 17-108_k141_83099 | Not-determined |
| 17-108_k141_234643 | Not-determined |
| 17-108_k141_69295 | Not-determined |
| 17-108_k141_289718 | Not-determined |
| 17-108_k141_96925 | Not-determined |
| 17-108_k141_152076 | Not-determined |
| 17-108_k141_124536 | Not-determined |
| 17-108_k141_317429 | Not-determined |
| 17-108_k141_165987 | Not-determined |
| 17-108_k141_124544 | Not-determined |
| 17-108_k141_276013 | Not-determined |
| 17-108_k141_207232 | Not-determined |
| 17-108_k141_317445 | Not-determined |
| 17-108_k141_96939 | Not-determined |
| 17-108_k141_124546 | Not-determined |
| 17-108_k141_14277 | Not-determined |
| 17-108_k141_207238 | Not-determined |
| 17-108_k141_69308 | Low-quality |
| 17-108_k141_166009 | Not-determined |
| 17-108_k141_166013 | Not-determined |
| 17-108_k141_55551 | Not-determined |
| 17-108_k141_289753 | Not-determined |
| 17-108_k141_276036 | Not-determined |
| 17-108_k141_303673 | Low-quality |
| 17-108_k141_166019 | Not-determined |
| 17-108_k141_234668 | Not-determined |
| 17-108_k141_96956 | Not-determined |
| 17-108_k141_138317 | Not-determined |
| 17-108_k141_69338 | Not-determined |
| 17-108_k141_207245 | Not-determined |
| 17-108_k141_69347 | Not-determined |
| 17-108_k141_303684 | Not-determined |
| 17-108_k141_124582 | Not-determined |
| 17-108_k141_317478 | Not-determined |
| 17-108_k141_110660 | Not-determined |
| 17-108_k141_207251 | Not-determined |
| 17-108_k141_83134 | Not-determined |
| 17-108_k141_207256 | Not-determined |
| 17-108_k141_152116 | Not-determined |
| 17-108_k141_220939 | Not-determined |
| 17-108_k141_303702 | Not-determined |
| 17-108_k141_276045 | Not-determined |
| 17-108_k141_276046 | Not-determined |
| 17-108_k141_14338 | Not-determined |
| 17-108_k141_110671 | Not-determined |
| 17-108_k141_41710 | Not-determined |
| 17-108_k141_276054 | Not-determined |
| 17-108_k141_276055 | Not-determined |
| 17-108_k141_152132 | Low-quality |
| 17-108_k141_83140 | Low-quality |
| 17-108_k141_83142 | Not-determined |
| 17-108_k141_41716 | Not-determined |
| 17-108_k141_276059 | Not-determined |
| 17-108_k141_69380 | Not-determined |
| 17-108_k141_303714 | Not-determined |
| 17-108_k141_276070 | Not-determined |
| 17-108_k141_124601 | Not-determined |
| 17-108_k141_276080 | Not-determined |
| 17-108_k141_234724 | Not-determined |
| 17-108_k141_193376 | Not-determined |
| 17-108_k141_234728 | Low-quality |
| 17-108_k141_193392 | Not-determined |
| 17-108_k141_234738 | Not-determined |
| 17-108_k141_193395 | Not-determined |
| 17-108_k141_166069 | Not-determined |
| 17-108_k141_276092 | Not-determined |
| 17-108_k141_474 | Not-determined |
| 17-108_k141_289765 | Not-determined |
| 17-108_k141_152143 | Not-determined |
| 17-108_k141_317508 | Not-determined |
| 17-108_k141_317516 | Not-determined |
| 17-108_k141_14372 | Not-determined |
| 17-108_k141_110675 | Not-determined |
| 17-108_k141_289779 | Low-quality |
| 17-108_k141_152145 | Not-determined |
| 17-108_k141_41768 | Not-determined |
| 17-108_k141_124613 | Not-determined |
| 17-108_k141_124617 | Not-determined |
| 17-108_k141_152155 | Not-determined |
| 17-108_k141_14384 | Not-determined |
| 17-108_k141_110683 | Not-determined |
| 17-108_k141_14388 | Not-determined |
| 17-108_k141_289783 | Not-determined |
| 17-108_k141_124626 | Not-determined |
| 17-108_k141_124630 | Not-determined |
| 17-109-final-viral-combined_k141_2450\|\|full | Medium-quality |
| 17-108_k141_317528 | Not-determined |
| 17-108_k141_124638 | Not-determined |
| 17-108_k141_28096 | Not-determined |
| 17-108_k141_262314 | Not-determined |
| 17-108_k141_234795 | Not-determined |
| 17-108_k141_152163 | Not-determined |
| 17-108_k141_289795 | Not-determined |
| 17-108_k141_69397 | Not-determined |
| 17-108_k141_41792 | Not-determined |
| 17-108_k141_166076 | Not-determined |
| 17-108_k141_83175 | Not-determined |
| 17-108_k141_83179 | Not-determined |
| 17-108_k141_289801 | Not-determined |
| 17-108_k141_110716 | Not-determined |
| 17-108_k141_317537 | Not-determined |
| 17-108_k141_152175 | Not-determined |
| 17-108_k141_276112 | Not-determined |
| 17-108_k141_317542 | Not-determined |
| 17-108_k141_110721 | Not-determined |
| 17-108_k141_124652 | Low-quality |
| 17-108_k141_138330 | Not-determined |
| 17-108_k141_83186 | Not-determined |
| 17-108_k141_166086 | Low-quality |
| 17-108_k141_69410 | Not-determined |
| 17-108_k141_234816 | Low-quality |
| 17-108_k141_138344 | Not-determined |
| 17-108_k141_41812 | Not-determined |
| 17-108_k141_166090 | Not-determined |
| 17-108_k141_28128 | Not-determined |
| 17-108_k141_234821 | Not-determined |
| 17-108_k141_166097 | Not-determined |
| 17-108_k141_41819 | Not-determined |
| 17-108_k141_69416 | Low-quality |
| 17-108_k141_124675 | Not-determined |
| 17-108_k141_220983 | Not-determined |
| 17-108_k141_124679 | Not-determined |
| 17-108_k141_41825 | Not-determined |
| 17-108_k141_317563 | Not-determined |
| 17-108_k141_234827 | Not-determined |
| 17-108_k141_317567 | Not-determined |
| 17-108_k141_276143 | Not-determined |
| 17-108_k141_289837 | Not-determined |
| 17-108_k141_317572 | Not-determined |
| 17-108_k141_41842 | Not-determined |
| 17-108_k141_276145 | Not-determined |
| 17-108_k141_220991 | Not-determined |
| 17-108_k141_193430 | Not-determined |
| 17-108_k141_166115 | Not-determined |
| 17-108_k141_28137 | Not-determined |
| 17-108_k141_28138 | Not-determined |
| 17-108_k141_69440 | Not-determined |
| 17-108_k141_166118 | Not-determined |
| 17-108_k141_55660 | Not-determined |
| 17-108_k141_69444 | Not-determined |
| 17-108_k141_110737 | Not-determined |
| 17-108_k141_234833 | Not-determined |
| 17-108_k141_28144 | Low-quality |
| 17-108_k141_289846 | Not-determined |
| 17-108_k141_507 | Not-determined |
| 17-108_k141_221008 | Not-determined |
| 18-109_k141_76437 | High-quality |
| 17-108_k141_138366 | Not-determined |
| 17-108_k141_234837 | Not-determined |
| 17-108_k141_55671 | Not-determined |
| 17-108_k141_221018 | Not-determined |
| 17-108_k141_69458 | Not-determined |
| 17-108_k141_69464 | Not-determined |
| 17-108_k141_248586 | Low-quality |
| 17-108_k141_289849 | Not-determined |
| 17-108_k141_69469 | Not-determined |
| 17-108_k141_41858 | Not-determined |
| 17-108_k141_41859 | Not-determined |
| 17-108_k141_221026 | Not-determined |
| 17-108_k141_524 | Not-determined |
| 17-108_k141_83233 | Not-determined |
| 17-108_k141_83235 | Not-determined |
| 17-108_k141_248609 | Not-determined |
| 17-108_k141_531 | Not-determined |
| 17-108_k141_533 | Not-determined |
| 17-108_k141_124697 | Not-determined |
| 17-108_k141_83239 | Not-determined |
| 17-108_k141_535 | Low-quality |
| 17-108_k141_539 | Not-determined |
| 17-108_k141_138399 | Not-determined |
| 17-108_k141_545 | Low-quality |
| 17-108_k141_221046 | Not-determined |
| 17-108_k141_124708 | Not-determined |
| 17-108_k141_14402 | Not-determined |
| 17-108_k141_262385 | Not-determined |
| 17-108_k141_262391 | Not-determined |
| 17-108_k141_152221 | Not-determined |
| 17-108_k141_289877 | Not-determined |
| 17-108_k141_193467 | Not-determined |
| 17-108_k141_193476 | Not-determined |
| 17-108_k141_14416 | Not-determined |
| 17-108_k141_28191 | Not-determined |
| 17-108_k141_28196 | Not-determined |
| 17-108_k141_55706 | Not-determined |
| 17-108_k141_262409 | Not-determined |
| 17-108_k141_193489 | Not-determined |
| 17-108_k141_166141 | Not-determined |
| 17-108_k141_303751 | Low-quality |
| 17-108_k141_234871 | Not-determined |
| 17-108_k141_276175 | Not-determined |
| 17-108_k141_193500 | Not-determined |
| 17-108_k141_97090 | Not-determined |
| 17-108_k141_262421 | Not-determined |
| 17-108_k141_262427 | Not-determined |
| 17-108_k141_568 | Not-determined |
| 17-108_k141_234884 | Not-determined |
| 17-108_k141_152253 | Low-quality |
| 17-108_k141_124762 | Not-determined |
| 17-108_k141_124767 | Not-determined |
| 17-108_k141_152260 | Not-determined |
| 17-108_k141_69526 | Not-determined |
| 17-108_k141_179640 | Not-determined |
| 17-108_k141_152263 | Not-determined |
| 17-108_k141_179642 | Low-quality |
| 17-108_k141_166171 | Not-determined |
| 17-108_k141_14449 | Low-quality |
| 17-108_k141_55741 | Not-determined |
| 17-108_k141_578 | Low-quality |
| 17-108_k141_248653 | Not-determined |
| 17-108_k141_124778 | Not-determined |
| 17-108_k141_179657 | Not-determined |
| 17-108_k141_14457 | Not-determined |
| 17-108_k141_166178 | Not-determined |
| 17-108_k141_138495 | Not-determined |
| 17-108_k141_207279 | Not-determined |
| 17-108_k141_193549 | Not-determined |
| 17-108_k141_166190 | Not-determined |
| 17-108_k141_193551 | Not-determined |
| 17-108_k141_193552 | Not-determined |
| 17-108_k141_276199 | Not-determined |
| 17-108_k141_276200 | Not-determined |
| 17-108_k141_591 | Not-determined |
| 17-108_k141_289947 | Not-determined |
| 17-108_k141_289950 | Not-determined |
| 17-108_k141_55780 | Not-determined |
| 17-108_k141_262456 | Not-determined |
| 17-108_k141_604 | Not-determined |
| 17-108_k141_221130 | Not-determined |
| 17-108_k141_193566 | Not-determined |
| 17-108_k141_289966 | Not-determined |
| 17-108_k141_262462 | Not-determined |
| 17-108_k141_83262 | Not-determined |
| 17-108_k141_83266 | Not-determined |
| 17-108_k141_303833 | Not-determined |
| 17-108_k141_262478 | Not-determined |
| 17-108_k141_289981 | Low-quality |
| 17-108_k141_41895 | Low-quality |
| 17-108_k141_152284 | Not-determined |
| 17-108_k141_124809 | Not-determined |
| 17-108_k141_625 | Not-determined |
| 17-108_k141_234950 | Not-determined |
| 17-108_k141_289990 | Not-determined |
| 17-108_k141_193592 | Not-determined |
| 17-108_k141_289993 | Not-determined |
| 17-108_k141_234953 | Not-determined |
| 17-108_k141_166217 | Low-quality |
| 17-108_k141_262493 | Not-determined |
| 17-108_k141_41901 | Low-quality |
| 17-108_k141_637 | Low-quality |
| 17-108_k141_193603 | Not-determined |
| 17-108_k141_166225 | Not-determined |
| 17-108_k141_152288 | Not-determined |
| 17-108_k141_166229 | Not-determined |
| 17-108_k141_289996 | Not-determined |
| 17-108_k141_234978 | Low-quality |
| 17-108_k141_234979 | Not-determined |
| 17-108_k141_262517 | Not-determined |
| 17-108_k141_207333 | Not-determined |
| 17-108_k141_317608 | Not-determined |
| 17-108_k141_207338 | Not-determined |
| 17-108_k141_262524 | Not-determined |
| 17-108_k141_152299 | Not-determined |
| 17-108_k141_97176 | Not-determined |
| 17-108_k141_276263 | Not-determined |
| 17-108_k141_41912 | Not-determined |
| 17-108_k141_276266 | Not-determined |
| 17-108_k141_69564 | Not-determined |
| 17-108_k141_166255 | Not-determined |
| 17-108_k141_83302 | Not-determined |
| 17-108_k141_262538 | Not-determined |
| 17-108_k141_317626 | Not-determined |
| 17-108_k141_317627 | Not-determined |
| 17-108_k141_110844 | Low-quality |
| 17-108_k141_193628 | Not-determined |
| 17-108_k141_83308 | Not-determined |
| 17-108_k141_276281 | Not-determined |
| 17-108_k141_124844 | Not-determined |
| 17-108_k141_124847 | Not-determined |
| 17-108_k141_69569 | Not-determined |
| 17-108_k141_193635 | Not-determined |
| 17-108_k141_207364 | Low-quality |
| 17-108_k141_303881 | Not-determined |
| 17-108_k141_166270 | Not-determined |
| 17-108_k141_152310 | Not-determined |
| 17-108_k141_235017 | Not-determined |
| 17-108_k141_290002 | Not-determined |
| 17-108_k141_166281 | Low-quality |
| 17-108_k141_290007 | Not-determined |
| 17-108_k141_69582 | Not-determined |
| 17-108_k141_69583 | Not-determined |
| 17-108_k141_69586 | Not-determined |
| 17-108_k141_69590 | Not-determined |
| 17-108_k141_124875 | Not-determined |
| 17-108_k141_124877 | Not-determined |
| 17-108_k141_124879 | Not-determined |
| 17-108_k141_69597 | Low-quality |
| 17-108_k141_166296 | Not-determined |
| 17-108_k141_303900 | Not-determined |
| 17-108_k141_193655 | Not-determined |
| 17-108_k141_28277 | Not-determined |
| 17-108_k141_193657 | Not-determined |
| 17-108_k141_290021 | Not-determined |
| 17-108_k141_28278 | Not-determined |
| 17-108_k141_69605 | Not-determined |
| 17-108_k141_152331 | Not-determined |
| 17-108_k141_69610 | Not-determined |
| 17-108_k141_124890 | Low-quality |
| 17-108_k141_152338 | Not-determined |
| 17-108_k141_179731 | Not-determined |
| 17-108_k141_55855 | Not-determined |
| 17-108_k141_166310 | Not-determined |
| 17-108_k141_97221 | Not-determined |
| 17-108_k141_97223 | Not-determined |
| 17-108_k141_290046 | Not-determined |
| 17-108_k141_317653 | Not-determined |
| 17-108_k141_276330 | Not-determined |
| 17-108_k141_276331 | Not-determined |
| 17-108_k141_28300 | Not-determined |
| 17-108_k141_69633 | Not-determined |
| 17-108_k141_83354 | Not-determined |
| 17-108_k141_110873 | Not-determined |
| 17-108_k141_110874 | Not-determined |
| 17-108_k141_666 | Not-determined |
| 17-108_k141_276339 | Not-determined |
| 17-108_k141_69644 | Not-determined |
| 17-108_k141_221217 | Not-determined |
| 17-108_k141_262577 | Not-determined |
| 17-108_k141_262579 | Not-determined |
| 17-108_k141_124918 | Not-determined |
| 17-108_k141_262582 | Not-determined |
| 17-108_k141_235063 | Not-determined |
| 17-108_k141_675 | Not-determined |
| 17-108_k141_152361 | Not-determined |
| 17-108_k141_110886 | Not-determined |
| 17-108_k141_221232 | Not-determined |
| 17-108_k141_55906 | Not-determined |
| 17-108_k141_55912 | Not-determined |
| 17-108_k141_193688 | Not-determined |
| 17-108_k141_69649 | Not-determined |
| 17-108_k141_83379 | Not-determined |
| 17-108_k141_83380 | Not-determined |
| 17-108_k141_317680 | Low-quality |
| 17-108_k141_110891 | Not-determined |
| 17-108_k141_221238 | Not-determined |
| 17-108_k141_193697 | Not-determined |
| 17-108_k141_317684 | Not-determined |
| 17-108_k141_166333 | Not-determined |
| 17-108_k141_179762 | Low-quality |
| 17-108_k141_207380 | Not-determined |
| 17-108_k141_221243 | Low-quality |
| 17-108_k141_290078 | Not-determined |
| 17-108_k141_235086 | Not-determined |
| 17-108_k141_69651 | Not-determined |
| 17-108_k141_207389 | Not-determined |
| 17-108_k141_193708 | Not-determined |
| 17-108_k141_83395 | Not-determined |
| 17-108_k141_193714 | Not-determined |
| 17-108_k141_317699 | Not-determined |
| 17-108_k141_69661 | Not-determined |
| 17-108_k141_69663 | Not-determined |
| 17-108_k141_124922 | Not-determined |
| 17-108_k141_303942 | Not-determined |
| 17-108_k141_55927 | Not-determined |
| 17-108_k141_69674 | Not-determined |
| 17-108_k141_207410 | Not-determined |
| 17-108_k141_193716 | Not-determined |
| 17-108_k141_110912 | Not-determined |
| 17-108_k141_55937 | Low-quality |
| 17-108_k141_221252 | Low-quality |
| 17-108_k141_41975 | Not-determined |
| 17-108_k141_110923 | Low-quality |
| 17-108_k141_138540 | Not-determined |
| 17-108_k141_166355 | Not-determined |
| 17-108_k141_166356 | Not-determined |
| 17-108_k141_138544 | Not-determined |
| 17-108_k141_317733 | Not-determined |
| 17-108_k141_262600 | Not-determined |
| 17-108_k141_138549 | Low-quality |
| 17-108_k141_152415 | Not-determined |
| 17-108_k141_152419 | Not-determined |
| 17-108_k141_221265 | Not-determined |
| 17-108_k141_262604 | Not-determined |
| 17-108_k141_317742 | Low-quality |
| 17-108_k141_152423 | Not-determined |
| 17-108_k141_55949 | Not-determined |
| 17-108_k141_235135 | Not-determined |

| | |
|---|---|
| 17-108_k141_221269 | Not-determined |
| 17-108_k141_110948 | Not-determined |
| 17-108_k141_124924 | Not-determined |
| 17-108_k141_262611 | Not-determined |
| 17-108_k141_303976 | Not-determined |
| 17-108_k141_317752 | Not-determined |
| 17-108_k141_152433 | Not-determined |
| 17-108_k141_235144 | Not-determined |
| 17-108_k141_303977 | Not-determined |
| 17-108_k141_207447 | Not-determined |
| 17-108_k141_248792 | Not-determined |
| 17-108_k141_752 | Not-determined |
| 17-108_k141_152445 | Not-determined |
| 17-108_k141_28333 | Not-determined |
| 17-108_k141_276392 | Low-quality |
| 17-108_k141_303980 | Not-determined |
| 17-108_k141_138554 | Not-determined |
| 17-108_k141_152450 | Not-determined |
| 17-108_k141_221284 | Not-determined |
| 17-108_k141_166385 | Not-determined |
| 17-108_k141_758 | Not-determined |
| 17-108_k141_83438 | Not-determined |
| 17-108_k141_276405 | Low-quality |
| 17-108_k141_138566 | Not-determined |
| 17-108_k141_221293 | Not-determined |
| 17-108_k141_193744 | Not-determined |
| 17-108_k141_83446 | Not-determined |
| 17-108_k141_166404 | Not-determined |
| 17-108_k141_166412 | Low-quality |
| 17-108_k141_193753 | Not-determined |
| 17-108_k141_28362 | Not-determined |
| 17-108_k141_28364 | Not-determined |
| 17-108_k141_28365 | Not-determined |
| 17-108_k141_28368 | Not-determined |
| 17-108_k141_166419 | Not-determined |
| 17-108_k141_28370 | Not-determined |
| 17-108_k141_179832 | Not-determined |
| 17-108_k141_166422 | Not-determined |
| 17-108_k141_97317 | Not-determined |
| 17-108_k141_28380 | Not-determined |
| 17-108_k141_138591 | Not-determined |
| 17-108_k141_124950 | Not-determined |
| 17-108_k141_14587 | Not-determined |
| 17-108_k141_235189 | Not-determined |
| 17-108_k141_110980 | Not-determined |
| 17-108_k141_248831 | Low-quality |
| 17-108_k141_193780 | Not-determined |
| 17-108_k141_28391 | Low-quality |
| 17-108_k141_138609 | Not-determined |
| 17-108_k141_317792 | Not-determined |
| 17-108_k141_207499 | Not-determined |
| 17-108_k141_97342 | Not-determined |
| 17-108_k141_790 | Not-determined |
| 17-108_k141_97344 | Not-determined |
| 17-108_k141_179853 | Not-determined |
| 17-108_k141_207523 | Not-determined |
| 17-108_k141_262664 | Not-determined |
| 17-108_k141_262665 | Not-determined |
| 17-108_k141_262669 | Not-determined |
| 17-108_k141_207528 | Not-determined |
| 17-108_k141_28400 | Not-determined |
| 17-108_k141_262672 | Not-determined |
| 17-108_k141_138617 | Not-determined |
| 17-108_k141_304031 | Not-determined |
| 17-108_k141_179856 | Not-determined |
| 17-108_k141_179858 | Not-determined |
| 17-108_k141_276461 | Not-determined |
| 17-108_k141_14609 | Not-determined |
| 17-108_k141_262681 | Not-determined |
| 17-108_k141_193799 | Not-determined |
| 17-108_k141_193803 | Not-determined |
| 17-108_k141_97366 | Low-quality |
| 17-108_k141_193808 | Not-determined |
| 17-108_k141_820 | Not-determined |
| 17-108_k141_138629 | Not-determined |
| 17-108_k141_14621 | Not-determined |
| 17-108_k141_207542 | Not-determined |
| 17-108_k141_248872 | Not-determined |
| 17-108_k141_207543 | Not-determined |
| 17-108_k141_97370 | Not-determined |
| 17-108_k141_42007 | Not-determined |
| 17-108_k141_276471 | Not-determined |
| 17-108_k141_276472 | Not-determined |
| 17-108_k141_42010 | Not-determined |
| 17-108_k141_207547 | Not-determined |
| 17-108_k141_138641 | Not-determined |
| 17-108_k141_235215 | Low-quality |
| 17-108_k141_42020 | Not-determined |
| 17-108_k141_28409 | Not-determined |
| 17-108_k141_14632 | Not-determined |
| 17-108_k141_276482 | Not-determined |
| 17-108_k141_235218 | Not-determined |
| 17-108_k141_83512 | Not-determined |
| 17-108_k141_124982 | Not-determined |
| 17-108_k141_166443 | Not-determined |
| 17-108_k141_152487 | Not-determined |
| 17-108_k141_166446 | Not-determined |
| 17-108_k141_28415 | Not-determined |
| 17-108_k141_97384 | Low-quality |
| 17-108_k141_304043 | Not-determined |
| 17-108_k141_83526 | Not-determined |
| 17-108_k141_207558 | Not-determined |
| 17-108_k141_42028 | Low-quality |
| 17-108_k141_304053 | Not-determined |
| 17-108_k141_111002 | Not-determined |
| 17-108_k141_111004 | Not-determined |
| 17-108_k141_111007 | Not-determined |
| 17-108_k141_83541 | Not-determined |
| 17-108_k141_179873 | Not-determined |
| 17-108_k141_235246 | Not-determined |
| 17-108_k141_69739 | Not-determined |
| 17-108_k141_111010 | Not-determined |
| 17-108_k141_42046 | Not-determined |
| 17-108_k141_235257 | Not-determined |
| 17-108_k141_304087 | Not-determined |
| 17-108_k141_304088 | Not-determined |
| 17-108_k141_111019 | Not-determined |
| 17-108_k141_111024 | Not-determined |
| 17-108_k141_304102 | Not-determined |
| 17-108_k141_28421 | Not-determined |
| 17-108_k141_276490 | Not-determined |
| 17-108_k141_97423 | Not-determined |
| 17-108_k141_276495 | Not-determined |
| 17-108_k141_235279 | Not-determined |
| 17-108_k141_152516 | Low-quality |
| 17-108_k141_125010 | Low-quality |
| 17-108_k141_193847 | Not-determined |
| 17-108_k141_179904 | Not-determined |
| 17-108_k141_125015 | Not-determined |
| 17-108_k141_235285 | Not-determined |
| 17-108_k141_235286 | Not-determined |
| 17-108_k141_111051 | Not-determined |
| 17-108_k141_138674 | Not-determined |
| 17-108_k141_97450 | Not-determined |
| 17-108_k141_14666 | Not-determined |
| 17-108_k141_14670 | Not-determined |
| 17-108_k141_69755 | Low-quality |
| 17-108_k141_290154 | Not-determined |
| 17-108_k141_166486 | Not-determined |
| 17-108_k141_83574 | Not-determined |
| 17-108_k141_179931 | Not-determined |
| 17-108_k141_166495 | Not-determined |
| 17-108_k141_179933 | Not-determined |
| 17-108_k141_111068 | Not-determined |
| 17-108_k141_111070 | Not-determined |
| 17-108_k141_83582 | Not-determined |
| 17-108_k141_42090 | Not-determined |
| 17-108_k141_152530 | Not-determined |
| 17-108_k141_317885 | Not-determined |
| 17-108_k141_152534 | Not-determined |
| 17-108_k141_69772 | Not-determined |
| 17-108_k141_152537 | Not-determined |
| 17-108_k141_902 | Not-determined |
| 17-108_k141_166508 | Low-quality |
| 17-108_k141_207588 | Not-determined |
| 17-108_k141_152549 | Not-determined |
| 17-108_k141_235313 | Not-determined |
| 17-108_k141_83598 | Not-determined |
| 17-108_k141_14690 | Not-determined |
| 17-108_k141_28446 | Not-determined |
| 17-108_k141_69781 | Not-determined |
| 17-108_k141_235318 | Not-determined |
| 17-108_k141_42109 | Not-determined |
| 17-108_k141_235323 | Not-determined |
| 17-108_k141_125055 | Low-quality |
| 17-108_k141_125060 | Not-determined |
| 17-108_k141_42117 | Low-quality |
| 17-108_k141_304120 | Not-determined |
| 17-108_k141_83610 | Not-determined |
| 17-108_k141_42126 | Not-determined |
| 17-108_k141_923 | Not-determined |
| 17-108_k141_248931 | Not-determined |
| 17-108_k141_276530 | Low-quality |
| 17-108_k141_152577 | Not-determined |
| 17-108_k141_125074 | Low-quality |
| 17-108_k141_83626 | Not-determined |
| 17-108_k141_69809 | Not-determined |
| 17-108_k141_166532 | Not-determined |
| 17-108_k141_83632 | Not-determined |
| 17-108_k141_235345 | Not-determined |
| 17-108_k141_152584 | Not-determined |
| 17-108_k141_207625 | Not-determined |
| 17-108_k141_207628 | Not-determined |
| 17-108_k141_111097 | Not-determined |
| 17-108_k141_940 | Not-determined |
| 17-108_k141_944 | Low-quality |
| 17-108_k141_248942 | Not-determined |
| 17-108_k141_193885 | Not-determined |
| 17-108_k141_125085 | Not-determined |
| 17-108_k141_179965 | Not-determined |
| 17-108_k141_83653 | Not-determined |
| 17-108_k141_125096 | Not-determined |
| 17-108_k141_125098 | Not-determined |
| 17-108_k141_83666 | Not-determined |
| 17-108_k141_235377 | Not-determined |
| 17-108_k141_304168 | Not-determined |
| 17-108_k141_166577 | Low-quality |
| 17-108_k141_193911 | Not-determined |
| 17-108_k141_235398 | Low-quality |
| 17-108_k141_111135 | Not-determined |
| 17-108_k141_111136 | Not-determined |
| 17-108_k141_111138 | Not-determined |
| 17-108_k141_193918 | Not-determined |
| 17-108_k141_193924 | Not-determined |
| 17-108_k141_179994 | Not-determined |
| 17-108_k141_276576 | Not-determined |
| 17-108_k141_97547 | Not-determined |
| 17-108_k141_193929 | Not-determined |
| 17-108_k141_235422 | Not-determined |
| 17-108_k141_249005 | Not-determined |
| 17-108_k141_249006 | Low-quality |
| 17-108_k141_249009 | Not-determined |
| 17-108_k141_304171 | Not-determined |
| 17-108_k141_14754 | Not-determined |
| 17-108_k141_180011 | Low-quality |
| 17-108_k141_14759 | Not-determined |
| 17-108_k141_207673 | Not-determined |
| 17-108_k141_56056 | Not-determined |
| 17-108_k141_14763 | Not-determined |
| 17-108_k141_249022 | Not-determined |
| 17-108_k141_166599 | Not-determined |
| 17-108_k141_42181 | Not-determined |
| 17-108_k141_180024 | Not-determined |
| 17-108_k141_28492 | Not-determined |
| 17-108_k141_249034 | Not-determined |
| 17-108_k141_69815 | Not-determined |
| 17-108_k141_276593 | Not-determined |
| 17-108_k141_317921 | Not-determined |
| 17-108_k141_276598 | Not-determined |
| 17-108_k141_152643 | Not-determined |
| 17-108_k141_152646 | Not-determined |
| 17-108_k141_97574 | Not-determined |
| 17-108_k141_138701 | Not-determined |
| 17-108_k141_207692 | Not-determined |
| 17-108_k141_28507 | Not-determined |
| 17-108_k141_138705 | Not-determined |
| 17-108_k141_166613 | Not-determined |
| 17-108_k141_180049 | Not-determined |
| 17-108_k141_304203 | Not-determined |
| 17-108_k141_317934 | Not-determined |
| 17-108_k141_180053 | Not-determined |
| 17-108_k141_125152 | Not-determined |
| 17-108_k141_207716 | Not-determined |
| 17-108_k141_83726 | Not-determined |
| 17-108_k141_42194 | Not-determined |
| 17-108_k141_207719 | Not-determined |
| 17-108_k141_207721 | Not-determined |
| 17-108_k141_207722 | Not-determined |
| 17-108_k141_304208 | Not-determined |
| 17-108_k141_152671 | Not-determined |
| 17-108_k141_249049 | Not-determined |
| 17-108_k141_166629 | Not-determined |
| 17-108_k141_83741 | Not-determined |
| 17-108_k141_42206 | Not-determined |
| 17-108_k141_304220 | Not-determined |
| 17-108_k141_69822 | Not-determined |
| 17-108_k141_262805 | Not-determined |
| 17-108_k141_166632 | Low-quality |
| 17-108_k141_249062 | Not-determined |
| 17-108_k141_83752 | Not-determined |
| 17-108_k141_276624 | Not-determined |
| 17-108_k141_221444 | Not-determined |
| 17-108_k141_166640 | Not-determined |
| 17-108_k141_276628 | Not-determined |
| 17-108_k141_14801 | Not-determined |
| 17-108_k141_69838 | Low-quality |
| 17-108_k141_97622 | Not-determined |
| 17-108_k141_138731 | Not-determined |
| 17-108_k141_69841 | Not-determined |
| 17-108_k141_235490 | Not-determined |
| 17-108_k141_207761 | Low-quality |
| 17-108_k141_138736 | Not-determined |
| 17-108_k141_111181 | Not-determined |
| 17-108_k141_207768 | Not-determined |
| 17-108_k141_138740 | Not-determined |
| 17-108_k141_138741 | Not-determined |
| 17-108_k141_14813 | Not-determined |
| 17-108_k141_111186 | Low-quality |
| 17-108_k141_290228 | Not-determined |
| 17-108_k141_249071 | Low-quality |
| 17-108_k141_97635 | Not-determined |
| 17-108_k141_56102 | Low-quality |
| 17-108_k141_235502 | Not-determined |
| 17-108_k141_97640 | Not-determined |
| 17-108_k141_14816 | Low-quality |
| 17-108_k141_166666 | Not-determined |
| 17-108_k141_207779 | Not-determined |
| 17-108_k141_207782 | Not-determined |
| 17-108_k141_14819 | Not-determined |
| 17-108_k141_207783 | Not-determined |
| 17-108_k141_138761 | Not-determined |
| 17-108_k141_166679 | Not-determined |
| 17-108_k141_1030 | Not-determined |
| 17-108_k141_152723 | Not-determined |
| 17-108_k141_1035 | Low-quality |
| 17-108_k141_14831 | Low-quality |
| 17-108_k141_138779 | Not-determined |
| 17-108_k141_221467 | Not-determined |
| 17-108_k141_83787 | Not-determined |
| 17-108_k141_262838 | Not-determined |
| 17-108_k141_14838 | Low-quality |
| 17-108_k141_97652 | Not-determined |
| 17-108_k141_56115 | Not-determined |
| 17-108_k141_83791 | Not-determined |
| 17-108_k141_235505 | Not-determined |
| 17-108_k141_69875 | Not-determined |
| 17-108_k141_152736 | Not-determined |
| 17-108_k141_152737 | Not-determined |
| 17-108_k141_69876 | Not-determined |
| 17-108_k141_69878 | Low-quality |
| 17-108_k141_1063 | Not-determined |
| 17-108_k141_138797 | Not-determined |
| 17-108_k141_276637 | Not-determined |
| 17-108_k141_152748 | Not-determined |
| 17-108_k141_1081 | Not-determined |
| 17-108_k141_276641 | Not-determined |
| 17-108_k141_235514 | Not-determined |
| 17-108_k141_235515 | Not-determined |
| 17-108_k141_317998 | Not-determined |
| 17-108_k141_97658 | Not-determined |
| 17-108_k141_166708 | Not-determined |
| 17-108_k141_180107 | Low-quality |
| 17-108_k141_42222 | Not-determined |
| 17-108_k141_14851 | Not-determined |
| 17-108_k141_235528 | Not-determined |
| 17-108_k141_207796 | Not-determined |
| 17-108_k141_235531 | Not-determined |
| 17-108_k141_152762 | Not-determined |
| 17-108_k141_221490 | Not-determined |
| 17-108_k141_152767 | Not-determined |
| 17-108_k141_69890 | Not-determined |
| 17-108_k141_290264 | Not-determined |
| 17-108_k141_221491 | Not-determined |
| 17-108_k141_83802 | Not-determined |
| 17-108_k141_83803 | Not-determined |
| 17-108_k141_262887 | Not-determined |
| 17-108_k141_249097 | Not-determined |
| 17-108_k141_262891 | Not-determined |
| 17-108_k141_262894 | Not-determined |
| 17-108_k141_125195 | Not-determined |
| 17-108_k141_221498 | Not-determined |
| 17-108_k141_83811 | Not-determined |
| 17-108_k141_152777 | Not-determined |
| 17-108_k141_235556 | Not-determined |
| 17-108_k141_83813 | Not-determined |
| 17-108_k141_42235 | Not-determined |
| 17-108_k141_14874 | Not-determined |
| 17-108_k141_235558 | Not-determined |
| 17-108_k141_14886 | Not-determined |
| 17-108_k141_304251 | Not-determined |
| 17-108_k141_152783 | Not-determined |
| 17-108_k141_290283 | Not-determined |
| 17-108_k141_1120 | Not-determined |
| 17-108_k141_276652 | Not-determined |
| 17-108_k141_262907 | Not-determined |
| 17-108_k141_56164 | Not-determined |
| 17-108_k141_290291 | Not-determined |
| 17-108_k141_28522 | Not-determined |
| 17-108_k141_276666 | Not-determined |
| 17-108_k141_42277 | Not-determined |
| 17-108_k141_56172 | Low-quality |
| 17-108_k141_249119 | Not-determined |
| 17-108_k141_304284 | Not-determined |
| 17-108_k141_221549 | Not-determined |
| 17-108_k141_249127 | Not-determined |
| 17-108_k141_97711 | Not-determined |
| 17-108_k141_125229 | Not-determined |
| 17-108_k141_97719 | Not-determined |
| 17-108_k141_56193 | Not-determined |
| 17-108_k141_193961 | Not-determined |
| 17-108_k141_262917 | Not-determined |
| 17-108_k141_69915 | Not-determined |
| 17-108_k141_193962 | Not-determined |
| 17-108_k141_1133 | Not-determined |
| 17-108_k141_69921 | Low-quality |
| 17-108_k141_97727 | Low-quality |
| 17-108_k141_180149 | Not-determined |
| 17-108_k141_193970 | Not-determined |
| 17-108_k141_111213 | Not-determined |
| 17-108_k141_290345 | Not-determined |
| 17-108_k141_166800 | Not-determined |
| 17-108_k141_318035 | Not-determined |
| 17-108_k141_125234 | Low-quality |
| 17-108_k141_83845 | Not-determined |
| 17-108_k141_14911 | Not-determined |
| 17-108_k141_97743 | Not-determined |
| 17-108_k141_152807 | Not-determined |
| 17-108_k141_111223 | Not-determined |
| 17-108_k141_207867 | Not-determined |
| 17-108_k141_249164 | Not-determined |
| 17-108_k141_28539 | Not-determined |
| 17-108_k141_166805 | Not-determined |
| 17-108_k141_221587 | Not-determined |
| 17-108_k141_97760 | Not-determined |
| 17-108_k141_262957 | Not-determined |
| 17-108_k141_14929 | Not-determined |
| 17-108_k141_180181 | Not-determined |
| 17-108_k141_28549 | Not-determined |
| 17-108_k141_235644 | Low-quality |
| 17-108_k141_262965 | Not-determined |
| 17-108_k141_290365 | Not-determined |
| 17-108_k141_125247 | Not-determined |
| 17-108_k141_166813 | Not-determined |
| 17-108_k141_262976 | Not-determined |
| 17-108_k141_14940 | Not-determined |
| 17-108_k141_235656 | Not-determined |
| 17-108_k141_304333 | Low-quality |
| 17-108_k141_97786 | Not-determined |
| 17-108_k141_235665 | Not-determined |
| 17-108_k141_180198 | Not-determined |
| 17-108_k141_125252 | Low-quality |
| 17-108_k141_235671 | Not-determined |
| 17-108_k141_14952 | Low-quality |
| 17-108_k141_97795 | Not-determined |
| 17-108_k141_125258 | Not-determined |
| 17-108_k141_14955 | Not-determined |
| 17-108_k141_42346 | Not-determined |
| 17-108_k141_235674 | Low-quality |
| 17-108_k141_249173 | Not-determined |
| 17-108_k141_28565 | Not-determined |
| 17-108_k141_111264 | Low-quality |
| 17-108_k141_14959 | Low-quality |
| 17-108_k141_28570 | Not-determined |
| 17-108_k141_249189 | Not-determined |
| 17-108_k141_111273 | Not-determined |
| 17-108_k141_125284 | Not-determined |
| 17-108_k141_235679 | Not-determined |
| 17-108_k141_1167 | Not-determined |
| 17-108_k141_207909 | Not-determined |
| 17-108_k141_207917 | Not-determined |
| 17-108_k141_262997 | Not-determined |
| 17-108_k141_290404 | Not-determined |
| 17-108_k141_249212 | Not-determined |
| 17-108_k141_207929 | Not-determined |
| 17-108_k141_276738 | Not-determined |
| 17-108_k141_276742 | Not-determined |
| 17-108_k141_83903 | Not-determined |
| 17-108_k141_83908 | Not-determined |
| 17-108_k141_276747 | Not-determined |
| 17-108_k141_221594 | Not-determined |
| 17-108_k141_28606 | Not-determined |
| 17-108_k141_276761 | Not-determined |
| 17-108_k141_276766 | Not-determined |
| 17-108_k141_194015 | Not-determined |
| 17-108_k141_276778 | Not-determined |
| 17-108_k141_290429 | Not-determined |
| 17-108_k141_263017 | Not-determined |
| 17-108_k141_235735 | Not-determined |
| 17-108_k141_263019 | Not-determined |
| 17-108_k141_263022 | Not-determined |
| 17-108_k141_263023 | Not-determined |
| 17-108_k141_207945 | Not-determined |
| 17-108_k141_83942 | Not-determined |
| 17-108_k141_290441 | Not-determined |
| 17-108_k141_83946 | Not-determined |
| 17-108_k141_263031 | Low-quality |
| 17-108_k141_15006 | Not-determined |
| 17-108_k141_152835 | Not-determined |
| 17-108_k141_15009 | Not-determined |
| 17-108_k141_290449 | Not-determined |
| 17-108_k141_56204 | Not-determined |
| 17-108_k141_97845 | Not-determined |
| 17-108_k141_249236 | Not-determined |
| 17-108_k141_221618 | Low-quality |
| 17-108_k141_276809 | Not-determined |
| 17-108_k141_221619 | Not-determined |
| 17-108_k141_97854 | Not-determined |
| 17-108_k141_97855 | Not-determined |
| 17-108_k141_180229 | Not-determined |
| 17-108_k141_249241 | Not-determined |
| 17-108_k141_249243 | Not-determined |
| 17-108_k141_42399 | Not-determined |
| 17-108_k141_56214 | Not-determined |
| 17-108_k141_207974 | Not-determined |
| 17-108_k141_1194 | Not-determined |
| 17-108_k141_125308 | Not-determined |
| 17-108_k141_166861 | Not-determined |
| 17-108_k141_152849 | Not-determined |
| 17-108_k141_152852 | Not-determined |
| 17-108_k141_97865 | Not-determined |
| 17-108_k141_235774 | Not-determined |
| 17-108_k141_207992 | Not-determined |
| 17-108_k141_97873 | Not-determined |
| 17-108_k141_276831 | Not-determined |
| 17-108_k141_276833 | Not-determined |
| 17-108_k141_304373 | Not-determined |
| 17-108_k141_180264 | Not-determined |
| 17-108_k141_69928 | Not-determined |
| 17-108_k141_125327 | Not-determined |
| 17-108_k141_249269 | Not-determined |
| 17-108_k141_42412 | Not-determined |
| 17-108_k141_166878 | Not-determined |
| 17-108_k141_56226 | Low-quality |
| 17-108_k141_97887 | Not-determined |
| 17-108_k141_304380 | Not-determined |
| 17-108_k141_208004 | Not-determined |
| 17-108_k141_235795 | Not-determined |
| 17-108_k141_125336 | Not-determined |
| 17-108_k141_235799 | Not-determined |
| 17-108_k141_304394 | Not-determined |
| 17-108_k141_263076 | Not-determined |
| 17-108_k141_304399 | Not-determined |
| 17-108_k141_1231 | Not-determined |
| 17-108_k141_249284 | Not-determined |
| 17-108_k141_235809 | Not-determined |
| 17-108_k141_111309 | Not-determined |
| 17-108_k141_221628 | Not-determined |
| 17-108_k141_56255 | Not-determined |
| 17-108_k141_276873 | Low-quality |
| 17-108_k141_125349 | Not-determined |
| 17-108_k141_166886 | Not-determined |
| 17-108_k141_56256 | Not-determined |
| 17-108_k141_97911 | Not-determined |
| 17-108_k141_180277 | Not-determined |
| 17-108_k141_166890 | Not-determined |
| 17-108_k141_249298 | Not-determined |
| 17-108_k141_166892 | Low-quality |
| 17-108_k141_235817 | Not-determined |
| 17-108_k141_276880 | Not-determined |
| 17-108_k141_235822 | Not-determined |
| 17-108_k141_125365 | Not-determined |
| 17-108_k141_249305 | Not-determined |
| 17-108_k141_56271 | Not-determined |
| 17-108_k141_249312 | Not-determined |
| 17-108_k141_276897 | Low-quality |
| 17-108_k141_249315 | Not-determined |
| 17-108_k141_83982 | Not-determined |
| 17-108_k141_276899 | Low-quality |
| 17-108_k141_249319 | Not-determined |
| 17-108_k141_56292 | Low-quality |
| 17-108_k141_56295 | Not-determined |
| 17-108_k141_304460 | Not-determined |
| 17-108_k141_263103 | Not-determined |
| 17-108_k141_304463 | Low-quality |
| 17-108_k141_111332 | Not-determined |
| 17-108_k141_42454 | Low-quality |
| 17-108_k141_42455 | Not-determined |
| 17-108_k141_56310 | Not-determined |
| 17-108_k141_180296 | Not-determined |
| 17-108_k141_1250 | Not-determined |
| 17-108_k141_84013 | Not-determined |
| 17-108_k141_111348 | Not-determined |
| 17-108_k141_15040 | Not-determined |
| 17-108_k141_84016 | Not-determined |
| 17-108_k141_166921 | Not-determined |
| 17-108_k141_1256 | Not-determined |
| 17-108_k141_249332 | Not-determined |
| 17-108_k141_1261 | Not-determined |
| 17-108_k141_249337 | Not-determined |
| 17-108_k141_125403 | Not-determined |
| 17-108_k141_290495 | Not-determined |
| 17-108_k141_249339 | Not-determined |
| 17-108_k141_235861 | Not-determined |
| 17-108_k141_180304 | Not-determined |
| 17-108_k141_42504 | Not-determined |
| 17-108_k141_15062 | Not-determined |
| 17-108_k141_15063 | Not-determined |
| 17-108_k141_84029 | Not-determined |
| 17-108_k141_235867 | Not-determined |
| 17-108_k141_125408 | Not-determined |
| 17-108_k141_290503 | Not-determined |
| 17-108_k141_15069 | Low-quality |
| 17-108_k141_28647 | Low-quality |
| 17-108_k141_42515 | Not-determined |
| 17-108_k141_166939 | Low-quality |
| 17-108_k141_276921 | Not-determined |
| 17-108_k141_263141 | Not-determined |
| 17-108_k141_235888 | Low-quality |
| 17-108_k141_318043 | Not-determined |
| 17-108_k141_125419 | Not-determined |
| 17-108_k141_42527 | Not-determined |
| 17-108_k141_263154 | Not-determined |
| 17-108_k141_111383 | Not-determined |
| 17-108_k141_276938 | Not-determined |
| 17-108_k141_304510 | Not-determined |
| 17-108_k141_70036 | Not-determined |
| 17-108_k141_42552 | Not-determined |
| 17-108_k141_42553 | Not-determined |
| 17-108_k141_221652 | Not-determined |
| 17-108_k141_97997 | Not-determined |
| 17-108_k141_194095 | Not-determined |
| 17-108_k141_152881 | Not-determined |
| 17-108_k141_276961 | Not-determined |
| 17-108_k141_221670 | Not-determined |
| 17-108_k141_304527 | Not-determined |
| 17-108_k141_152899 | Not-determined |
| 17-108_k141_208042 | Not-determined |
| 17-108_k141_221684 | Not-determined |
| 17-108_k141_208046 | Not-determined |

| | |
|---|---|
| 17-108_k141_208050 | Not-determined |
| 17-108_k141_276983 | Not-determined |
| 17-108_k141_28712 | Not-determined |
| 17-108_k141_98033 | Not-determined |
| 17-108_k141_208055 | Not-determined |
| 17-108_k141_125463 | Not-determined |
| 17-108_k141_235918 | Not-determined |
| 17-108_k141_28722 | Not-determined |
| 17-108_k141_28728 | Not-determined |
| 17-108_k141_263175 | Not-determined |
| 17-108_k141_318087 | Not-determined |
| 17-108_k141_208060 | Not-determined |
| 17-108_k141_263178 | Not-determined |
| 17-108_k141_125472 | Low-quality |
| 17-108_k141_70078 | Not-determined |
| 17-108_k141_98044 | Not-determined |
| 17-108_k141_70080 | Not-determined |
| 17-108_k141_15098 | Not-determined |
| 17-108_k141_221703 | Not-determined |
| 17-108_k141_318101 | Not-determined |
| 17-108_k141_28735 | Not-determined |
| 17-108_k141_98049 | Low-quality |
| 17-108_k141_249377 | Not-determined |
| 17-108_k141_221706 | Not-determined |
| 17-108_k141_15102 | Not-determined |
| 17-108_k141_221708 | Not-determined |
| 17-108_k141_318103 | Not-determined |
| 17-108_k141_221710 | Not-determined |
| 17-108_k141_208066 | Not-determined |
| 17-108_k141_235920 | Not-determined |
| 17-108_k141_70086 | Not-determined |
| 17-108_k141_290520 | Not-determined |
| 17-108_k141_263195 | Not-determined |
| 17-108_k141_152908 | Not-determined |
| 17-108_k141_304563 | Not-determined |
| 17-108_k141_180363 | Not-determined |
| 17-108_k141_290525 | Not-determined |
| 17-108_k141_304570 | Not-determined |
| 17-108_k141_125484 | Not-determined |
| 17-108_k141_221721 | Not-determined |
| 17-108_k141_249394 | Not-determined |
| 17-108_k141_208083 | Not-determined |
| 17-108_k141_221729 | Not-determined |
| 17-108_k141_221734 | Not-determined |
| 17-108_k141_28760 | Not-determined |
| 17-108_k141_180371 | Not-determined |
| 17-108_k141_263211 | Not-determined |
| 17-108_k141_221738 | Not-determined |
| 17-108_k141_277013 | Not-determined |
| 17-108_k141_277019 | Not-determined |
| 17-108_k141_221751 | Not-determined |
| 17-108_k141_304590 | Not-determined |
| 17-108_k141_208089 | Not-determined |
| 17-108_k141_263223 | Not-determined |
| 17-108_k141_15120 | Not-determined |
| 17-108_k141_304600 | Not-determined |
| 17-108_k141_152924 | Not-determined |
| 17-108_k141_125487 | Not-determined |
| 17-108_k141_166959 | Not-determined |
| 17-108_k141_28774 | Not-determined |
| 17-108_k141_208096 | Low-quality |
| 17-108_k141_166962 | Not-determined |
| 17-108_k141_263240 | Not-determined |
| 17-108_k141_152933 | Not-determined |
| 17-108_k141_125497 | Not-determined |
| 17-108_k141_28786 | Not-determined |
| 17-108_k141_28787 | Not-determined |
| 17-108_k141_70123 | Not-determined |
| 17-108_k141_304615 | Not-determined |
| 17-108_k141_1309 | Not-determined |
| 17-108_k141_1315 | Not-determined |
| 17-108_k141_70134 | Low-quality |
| 17-108_k141_290586 | Not-determined |
| 17-108_k141_84063 | Not-determined |
| 17-108_k141_249433 | Low-quality |
| 17-108_k141_235942 | Not-determined |
| 17-108_k141_235943 | Not-determined |
| 17-108_k141_70145 | Not-determined |
| 17-108_k141_235946 | Not-determined |
| 17-108_k141_180387 | Not-determined |
| 17-108_k141_180388 | Not-determined |
| 17-108_k141_42608 | Not-determined |
| 17-108_k141_70149 | Not-determined |
| 17-108_k141_194192 | Not-determined |
| 17-108_k141_194193 | Not-determined |
| 17-108_k141_42614 | Not-determined |
| 17-108_k141_180395 | Not-determined |
| 17-108_k141_290594 | Not-determined |
| 17-108_k141_249454 | Not-determined |
| 17-108_k141_290598 | Not-determined |
| 17-108_k141_15146 | Not-determined |
| 17-108_k141_70153 | Low-quality |
| 17-108_k141_15151 | Not-determined |
| 17-108_k141_42633 | Not-determined |
| 17-108_k141_56390 | Not-determined |
| 17-108_k141_290613 | Not-determined |
| 17-108_k141_1334 | Not-determined |
| 17-108_k141_277047 | Not-determined |
| 17-108_k141_180419 | Not-determined |
| 17-108_k141_249478 | Low-quality |
| 17-108_k141_84090 | Not-determined |
| 17-108_k141_15160 | Low-quality |
| 17-108_k141_56410 | Not-determined |
| 17-108_k141_98119 | Not-determined |
| 17-108_k141_180423 | Not-determined |
| 17-108_k141_28843 | Low-quality |
| 17-108_k141_15162 | Not-determined |
| 17-108_k141_235968 | Not-determined |
| 17-108_k141_166981 | Not-determined |
| 17-108_k141_290624 | Not-determined |
| 17-108_k141_70180 | Not-determined |
| 17-108_k141_249494 | Not-determined |
| 17-108_k141_235974 | Not-determined |
| 17-108_k141_208137 | Not-determined |
| 17-108_k141_208139 | Not-determined |
| 17-108_k141_28856 | Low-quality |
| 17-108_k141_290634 | Not-determined |
| 17-108_k141_290639 | Not-determined |
| 17-108_k141_235986 | Low-quality |
| 17-108_k141_208147 | Low-quality |
| 17-108_k141_125504 | Not-determined |
| 17-108_k141_208148 | Not-determined |
| 17-108_k141_249504 | Not-determined |
| 17-108_k141_15196 | Not-determined |
| 17-108_k141_235994 | Not-determined |
| 17-108_k141_111408 | Low-quality |
| 17-108_k141_125512 | Low-quality |
| 17-108_k141_125513 | Not-determined |
| 17-108_k141_125515 | Not-determined |
| 17-108_k141_318169 | Not-determined |
| 17-108_k141_249511 | Not-determined |
| 17-108_k141_56455 | Not-determined |
| 17-108_k141_304685 | Not-determined |
| 17-108_k141_111415 | Not-determined |
| 17-108_k141_98147 | Not-determined |
| 17-108_k141_236000 | Not-determined |
| 17-108_k141_70211 | Not-determined |
| 17-108_k141_28862 | Not-determined |
| 17-108_k141_139011 | Not-determined |
| 17-108_k141_56460 | Not-determined |
| 17-108_k141_28864 | Not-determined |
| 17-108_k141_139014 | Not-determined |
| 17-108_k141_194224 | Not-determined |
| 17-108_k141_28872 | Not-determined |
| 17-108_k141_208172 | Not-determined |
| 17-108_k141_84132 | Low-quality |
| 17-108_k141_15224 | Not-determined |
| 17-108_k141_15226 | Not-determined |
| 17-108_k141_208177 | Not-determined |
| 17-108_k141_167004 | Not-determined |
| 17-108_k141_277103 | Not-determined |
| 17-108_k141_277105 | Not-determined |
| 17-108_k141_28881 | Not-determined |
| 17-108_k141_28883 | Not-determined |
| 17-108_k141_180437 | Low-quality |
| 17-108_k141_236016 | Not-determined |
| 17-108_k141_236017 | Not-determined |
| 17-108_k141_277110 | Not-determined |
| 17-108_k141_318192 | Not-determined |
| 17-108_k141_167014 | Low-quality |
| 17-108_k141_318198 | Not-determined |
| 17-108_k141_290663 | Not-determined |
| 17-108_k141_221857 | Not-determined |
| 17-108_k141_236021 | Not-determined |
| 17-108_k141_152983 | Not-determined |
| 17-108_k141_304711 | Not-determined |
| 17-108_k141_249518 | Low-quality |
| 17-108_k141_15243 | Not-determined |
| 17-108_k141_70269 | Not-determined |
| 17-108_k141_56484 | Not-determined |
| 17-108_k141_56485 | Not-determined |
| 17-108_k141_318217 | Not-determined |
| 17-108_k141_84156 | Not-determined |
| 17-108_k141_98186 | Not-determined |
| 17-108_k141_290669 | Not-determined |
| 17-108_k141_194285 | Not-determined |
| 17-108_k141_125556 | Not-determined |
| 17-108_k141_263316 | Low-quality |
| 17-108_k141_15264 | Not-determined |
| 17-108_k141_221884 | Not-determined |
| 17-108_k141_56502 | Not-determined |
| 17-108_k141_98191 | Not-determined |
| 17-108_k141_98196 | Not-determined |
| 17-108_k141_98202 | Not-determined |
| 17-108_k141_139042 | Not-determined |
| 17-108_k141_42712 | Not-determined |
| 17-108_k141_56515 | Not-determined |
| 17-108_k141_56519 | Not-determined |
| 17-108_k141_139048 | Not-determined |
| 17-108_k141_15269 | Not-determined |
| 17-108_k141_15272 | Not-determined |
| 17-108_k141_15273 | Not-determined |
| 17-108_k141_194312 | Not-determined |
| 17-108_k141_139051 | Not-determined |
| 17-108_k141_70298 | Not-determined |
| 17-108_k141_180497 | Not-determined |
| 17-108_k141_277146 | Not-determined |
| 17-108_k141_277148 | Not-determined |
| 17-108_k141_318236 | Not-determined |
| 17-108_k141_249533 | Not-determined |
| 17-108_k141_139058 | Not-determined |
| 17-108_k141_249536 | Not-determined |
| 17-108_k141_139063 | Not-determined |
| 17-108_k141_42718 | Not-determined |
| 17-108_k141_56539 | Not-determined |
| 17-108_k141_84199 | Not-determined |
| 17-108_k141_84200 | Not-determined |
| 17-108_k141_304750 | Not-determined |
| 17-108_k141_208242 | Not-determined |
| 17-108_k141_84203 | Not-determined |
| 17-108_k141_125606 | Not-determined |
| 17-108_k141_318251 | Not-determined |
| 17-108_k141_194326 | Not-determined |
| 17-108_k141_290721 | Not-determined |
| 17-108_k141_125614 | Not-determined |
| 17-108_k141_42731 | Not-determined |
| 17-108_k141_125619 | Not-determined |
| 17-108_k141_15298 | Not-determined |
| 17-108_k141_1399 | Not-determined |
| 17-108_k141_42743 | Not-determined |
| 17-108_k141_249548 | Not-determined |
| 17-108_k141_125636 | Not-determined |
| 17-108_k141_194338 | Not-determined |
| 17-108_k141_1413 | Not-determined |
| 17-108_k141_167085 | Not-determined |
| 17-108_k141_42752 | Not-determined |
| 17-108_k141_1414 | Not-determined |
| 17-108_k141_249555 | Not-determined |
| 17-108_k141_153028 | Not-determined |
| 17-108_k141_1421 | Not-determined |
| 17-108_k141_125645 | Not-determined |
| 17-108_k141_290751 | Not-determined |
| 17-108_k141_249562 | Not-determined |
| 17-108_k141_70318 | Low-quality |
| 17-108_k141_180532 | Not-determined |
| 17-108_k141_290763 | Not-determined |
| 17-108_k141_98241 | Not-determined |
| 17-108_k141_249569 | Not-determined |
| 17-108_k141_180538 | Not-determined |
| 17-108_k141_290769 | Not-determined |
| 17-108_k141_84246 | Not-determined |
| 17-108_k141_70328 | Low-quality |
| 17-108_k141_70329 | Low-quality |
| 17-108_k141_318279 | Not-determined |
| 17-108_k141_318283 | Not-determined |
| 17-108_k141_70335 | Low-quality |
| 17-108_k141_1432 | Not-determined |
| 17-108_k141_42793 | Not-determined |
| 17-108_k141_42796 | Not-determined |
| 17-108_k141_153043 | Not-determined |
| 17-108_k141_304766 | Not-determined |
| 17-108_k141_236049 | Not-determined |
| 17-108_k141_318289 | Low-quality |
| 17-108_k141_263346 | Not-determined |
| 17-108_k141_221981 | Not-determined |
| 17-108_k141_263351 | Not-determined |
| 17-108_k141_180553 | Not-determined |
| 17-108_k141_263354 | Not-determined |
| 17-108_k141_208266 | Not-determined |
| 17-108_k141_15344 | Not-determined |
| 17-108_k141_249588 | Not-determined |
| 17-108_k141_221996 | Not-determined |
| 17-108_k141_208278 | Not-determined |
| 17-108_k141_249593 | Low-quality |
| 17-108_k141_139113 | Not-determined |
| 17-108_k141_180565 | Not-determined |
| 17-108_k141_277218 | Not-determined |
| 17-108_k141_304783 | Not-determined |
| 17-108_k141_263381 | Not-determined |
| 17-108_k141_277225 | Not-determined |
| 17-108_k141_98287 | Not-determined |
| 17-108_k141_290792 | Not-determined |
| 17-108_k141_194368 | Not-determined |
| 17-108_k141_167160 | Not-determined |
| 17-108_k141_153073 | Not-determined |
| 17-108_k141_277236 | Not-determined |
| 17-108_k141_236097 | Not-determined |
| 17-108_k141_111496 | Not-determined |
| 17-108_k141_111498 | Not-determined |
| 17-108_k141_15389 | Not-determined |
| 17-108_k141_125713 | Not-determined |
| 17-108_k141_98289 | Not-determined |
| 17-108_k141_277246 | Low-quality |
| 17-108_k141_318324 | Not-determined |
| 17-108_k141_56578 | Not-determined |
| 17-108_k141_208306 | Not-determined |
| 17-108_k141_290806 | Not-determined |
| 17-108_k141_249632 | Not-determined |
| 17-108_k141_194374 | Not-determined |
| 17-108_k141_153087 | Low-quality |
| 17-108_k141_153089 | Not-determined |
| 17-108_k141_1464 | Not-determined |
| 17-108_k141_318337 | Not-determined |
| 17-108_k141_290819 | Low-quality |
| 17-108_k141_15402 | Not-determined |
| 17-108_k141_153097 | Not-determined |
| 17-108_k141_318346 | Not-determined |
| 17-108_k141_1469 | Not-determined |
| 17-108_k141_318351 | Not-determined |
| 17-108_k141_111506 | Low-quality |
| 17-108_k141_111515 | Low-quality |
| 17-108_k141_194393 | Not-determined |
| 17-108_k141_111517 | Low-quality |
| 17-108_k141_194397 | Not-determined |
| 17-108_k141_56597 | Not-determined |
| 17-108_k141_180622 | Not-determined |
| 17-108_k141_139160 | Not-determined |
| 17-108_k141_318361 | Not-determined |
| 17-108_k141_29005 | Not-determined |
| 17-108_k141_125749 | Not-determined |
| 17-108_k141_29009 | Low-quality |
| 17-108_k141_290861 | Not-determined |
| 17-108_k141_277264 | Not-determined |
| 17-108_k141_42889 | Not-determined |
| 17-108_k141_222035 | Not-determined |
| 17-108_k141_153133 | Not-determined |
| 17-108_k141_1496 | Not-determined |
| 17-108_k141_318371 | Low-quality |
| 17-108_k141_290870 | Not-determined |
| 17-108_k141_56623 | Not-determined |
| 17-108_k141_318378 | Not-determined |
| 17-108_k141_222053 | Not-determined |
| 17-108_k141_56631 | Not-determined |
| 17-108_k141_29037 | Not-determined |
| 17-108_k141_263456 | Not-determined |
| 17-108_k141_1521 | Not-determined |
| 17-108_k141_29039 | Not-determined |
| 17-108_k141_139164 | Not-determined |
| 17-108_k141_29043 | Not-determined |
| 17-108_k141_222064 | Not-determined |
| 17-108_k141_318392 | Not-determined |
| 17-108_k141_15428 | Not-determined |
| 17-108_k141_277294 | Not-determined |
| 17-108_k141_180650 | Not-determined |
| 17-108_k141_29057 | Not-determined |
| 17-108_k141_1532 | Low-quality |
| 17-108_k141_15435 | Not-determined |
| 17-108_k141_15436 | Not-determined |
| 17-108_k141_167275 | Not-determined |
| 17-108_k141_290903 | Not-determined |
| 17-108_k141_249688 | Not-determined |
| 17-108_k141_84301 | Not-determined |
| 17-108_k141_1537 | Not-determined |
| 17-108_k141_167277 | Not-determined |
| 17-108_k141_167280 | Low-quality |
| 17-108_k141_139173 | Low-quality |
| 17-108_k141_1545 | Not-determined |
| 17-108_k141_111540 | Not-determined |
| 17-108_k141_263471 | Not-determined |
| 17-108_k141_263472 | Not-determined |
| 17-108_k141_56645 | Not-determined |
| 17-108_k141_56648 | Not-determined |
| 17-108_k141_1560 | Not-determined |
| 17-108_k141_111551 | Not-determined |
| 17-108_k141_125810 | Not-determined |
| 17-108_k141_1586 | Not-determined |
| 17-108_k141_29070 | Not-determined |
| 17-108_k141_1589 | Not-determined |
| 17-108_k141_125815 | Not-determined |
| 17-108_k141_111554 | Not-determined |
| 17-108_k141_194500 | Not-determined |
| 17-108_k141_180664 | Not-determined |
| 17-108_k141_56660 | Not-determined |
| 17-108_k141_263484 | Not-determined |
| 17-108_k141_98358 | Not-determined |
| 17-108_k141_56661 | Not-determined |
| 17-108_k141_139190 | Not-determined |
| 17-108_k141_84332 | Not-determined |
| 17-108_k141_222130 | Not-determined |
| 17-108_k141_318418 | Not-determined |
| 17-108_k141_222133 | Not-determined |
| 17-108_k141_15463 | Low-quality |
| 17-108_k141_194509 | Low-quality |
| 17-108_k141_1608 | Not-determined |
| 17-108_k141_15471 | Not-determined |
| 17-108_k141_194521 | Not-determined |
| 17-108_k141_222147 | Not-determined |
| 17-108_k141_208369 | Not-determined |
| 17-108_k141_222151 | Low-quality |
| 17-108_k141_180688 | Low-quality |
| 17-108_k141_15485 | Not-determined |
| 17-108_k141_56676 | Not-determined |
| 17-108_k141_180696 | Not-determined |
| 17-108_k141_84362 | Not-determined |
| 17-108_k141_1631 | Not-determined |
| 17-108_k141_56681 | Not-determined |
| 17-108_k141_1633 | Not-determined |
| 17-108_k141_222162 | Not-determined |
| 17-108_k141_1635 | Low-quality |
| 17-108_k141_153204 | Not-determined |
| 17-108_k141_56689 | Not-determined |
| 17-108_k141_139227 | Not-determined |
| 17-108_k141_56694 | Not-determined |
| 17-108_k141_139232 | Not-determined |
| 17-108_k141_139238 | Not-determined |
| 17-108_k141_208396 | Not-determined |
| 17-108_k141_153219 | Low-quality |
| 17-108_k141_84373 | Not-determined |
| 17-108_k141_15521 | Not-determined |
| 17-108_k141_318432 | Not-determined |
| 17-108_k141_290979 | Not-determined |
| 17-108_k141_139258 | Not-determined |
| 17-108_k141_318440 | Not-determined |
| 17-108_k141_318447 | Not-determined |
| 17-108_k141_139264 | Not-determined |
| 17-108_k141_290989 | Not-determined |
| 17-108_k141_249727 | Not-determined |
| 17-108_k141_153239 | Low-quality |
| 17-108_k141_304898 | Not-determined |
| 17-108_k141_290994 | Not-determined |
| 17-108_k141_263533 | Not-determined |
| 17-108_k141_277379 | Low-quality |
| 17-108_k141_153247 | Not-determined |
| 17-108_k141_1654 | Not-determined |
| 17-108_k141_153251 | Not-determined |
| 17-108_k141_194576 | Not-determined |
| 17-108_k141_304920 | Not-determined |
| 17-108_k141_125855 | Not-determined |
| 17-108_k141_84404 | Not-determined |
| 17-108_k141_249738 | Not-determined |
| 17-108_k141_318485 | Not-determined |
| 17-108_k141_208423 | Not-determined |
| 17-108_k141_318492 | Not-determined |
| 17-108_k141_291028 | Not-determined |
| 17-108_k141_98373 | Not-determined |
| 17-108_k141_318506 | Not-determined |
| 17-108_k141_304922 | Low-quality |
| 17-108_k141_208431 | Not-determined |
| 17-108_k141_304924 | Low-quality |
| 17-108_k141_180743 | Low-quality |
| 17-108_k141_29122 | Not-determined |
| 17-108_k141_249754 | Low-quality |
| 17-108_k141_84420 | Not-determined |
| 17-108_k141_56732 | Not-determined |
| 17-108_k141_236224 | Not-determined |
| 17-108_k141_15570 | Not-determined |
| 17-108_k141_15572 | Low-quality |
| 17-108_k141_222228 | Not-determined |
| 17-108_k141_15573 | Low-quality |
| 17-108_k141_29134 | Low-quality |
| 17-108_k141_29135 | Not-determined |
| 17-108_k141_56742 | Not-determined |
| 17-108_k141_236234 | Low-quality |
| 17-108_k141_15584 | Low-quality |
| 17-108_k141_222240 | Not-determined |
| 17-108_k141_29141 | Not-determined |
| 17-108_k141_42917 | Not-determined |
| 17-108_k141_42919 | Not-determined |
| 17-108_k141_222246 | Not-determined |
| 17-108_k141_111626 | Not-determined |
| 17-108_k141_222249 | Not-determined |
| 17-108_k141_125861 | Not-determined |
| 17-108_k141_304956 | Not-determined |
| 17-108_k141_236242 | Not-determined |
| 17-108_k141_29155 | Not-determined |
| 17-108_k141_236246 | Low-quality |
| 17-108_k141_180781 | Not-determined |
| 17-108_k141_15612 | Not-determined |
| 17-108_k141_180789 | Low-quality |
| 17-108_k141_304970 | Not-determined |
| 17-108_k141_208472 | Not-determined |
| 17-108_k141_139319 | Not-determined |
| 17-108_k141_98406 | Not-determined |
| 17-108_k141_98409 | Not-determined |
| 17-108_k141_56762 | Not-determined |
| 17-108_k141_180801 | Not-determined |
| 17-108_k141_222262 | Not-determined |
| 17-108_k141_29183 | Not-determined |
| 17-108_k141_42950 | Not-determined |
| 17-108_k141_167402 | Not-determined |
| 17-108_k141_194595 | Not-determined |
| 17-108_k141_194597 | Not-determined |
| 17-108_k141_180813 | Not-determined |
| 17-108_k141_98423 | Not-determined |
| 17-108_k141_222267 | Not-determined |
| 17-108_k141_194605 | Not-determined |
| 17-108_k141_56786 | Not-determined |
| 17-108_k141_194612 | Not-determined |
| 17-108_k141_277445 | Not-determined |
| 17-108_k141_167419 | Not-determined |
| 17-108_k141_194617 | Not-determined |
| 17-108_k141_84445 | Not-determined |
| 17-108_k141_111678 | Not-determined |
| 17-108_k141_70354 | Not-determined |
| 17-108_k141_56808 | Low-quality |
| 17-108_k141_222285 | Not-determined |
| 17-108_k141_277464 | Not-determined |
| 17-108_k141_111691 | Not-determined |
| 17-108_k141_208515 | Not-determined |
| 17-108_k141_153343 | Not-determined |
| 17-108_k141_15649 | Not-determined |
| 17-108_k141_208522 | Not-determined |
| 17-108_k141_277480 | Not-determined |
| 17-108_k141_56819 | Not-determined |
| 17-108_k141_15652 | Not-determined |
| 17-108_k141_15655 | Not-determined |
| 17-108_k141_139373 | Not-determined |
| 17-108_k141_98431 | Not-determined |
| 17-108_k141_208527 | Low-quality |
| 17-108_k141_249794 | Not-determined |
| 17-108_k141_111705 | Not-determined |
| 17-108_k141_236303 | Low-quality |
| 17-108_k141_56835 | Not-determined |
| 17-108_k141_277492 | Not-determined |
| 17-108_k141_84471 | Not-determined |
| 17-108_k141_43012 | Not-determined |
| 17-108_k141_180856 | Not-determined |
| 17-108_k141_15663 | Not-determined |
| 17-108_k141_318532 | Not-determined |
| 17-108_k141_180859 | Not-determined |
| 17-108_k141_208542 | Low-quality |
| 17-108_k141_111712 | Not-determined |
| 17-108_k141_153365 | Low-quality |
| 17-108_k141_84509 | Not-determined |
| 17-108_k141_167476 | Not-determined |
| 17-108_k141_167484 | Not-determined |
| 17-108_k141_277508 | Not-determined |
| 17-108_k141_43058 | Not-determined |
| 17-108_k141_84529 | Not-determined |
| 17-108_k141_236324 | Not-determined |
| 17-108_k141_15683 | Not-determined |
| 17-108_k141_291106 | Not-determined |
| 17-108_k141_249819 | Not-determined |
| 17-108_k141_208571 | Not-determined |
| 17-108_k141_236326 | Not-determined |
| 17-108_k141_222322 | Low-quality |
| 17-108_k141_1732 | Not-determined |
| 17-108_k141_1735 | Not-determined |
| 17-108_k141_291116 | Low-quality |
| 17-108_k141_167509 | Not-determined |
| 17-108_k141_236338 | Not-determined |
| 17-108_k141_84532 | Not-determined |
| 17-108_k141_222330 | Not-determined |
| 17-108_k141_208586 | Not-determined |
| 17-108_k141_98487 | Not-determined |
| 17-108_k141_111752 | Not-determined |
| 17-108_k141_56900 | Not-determined |
| 17-108_k141_1743 | Not-determined |
| 17-108_k141_1744 | Not-determined |
| 17-108_k141_43101 | Low-quality |
| 17-108_k141_1747 | Not-determined |
| 17-108_k141_180879 | Not-determined |
| 17-108_k141_180880 | Not-determined |
| 17-108_k141_180881 | Not-determined |
| 17-108_k141_222349 | Not-determined |
| 17-108_k141_277528 | Low-quality |
| 17-108_k141_98504 | Low-quality |
| 17-108_k141_236369 | Not-determined |
| 17-108_k141_139436 | Not-determined |
| 17-108_k141_208608 | Not-determined |
| 17-108_k141_318562 | Not-determined |
| 17-108_k141_125886 | Not-determined |
| 17-108_k141_84580 | Low-quality |
| 17-108_k141_125890 | Not-determined |
| 17-108_k141_125896 | Not-determined |
| 17-108_k141_125897 | Not-determined |
| 17-108_k141_291144 | Low-quality |
| 17-108_k141_139447 | Not-determined |
| 17-108_k141_111786 | Not-determined |
| 17-108_k141_208646 | Not-determined |
| 17-108_k141_167549 | Not-determined |
| 17-108_k141_277560 | Not-determined |
| 17-108_k141_305017 | Not-determined |
| 17-108_k141_111795 | Not-determined |
| 17-108_k141_153395 | Not-determined |
| 17-108_k141_222364 | Not-determined |
| 17-108_k141_194644 | Not-determined |
| 17-108_k141_291171 | Low-quality |
| 17-108_k141_29232 | Not-determined |
| 17-108_k141_291178 | Not-determined |
| 17-108_k141_222382 | Not-determined |
| 17-108_k141_139481 | Not-determined |
| 17-108_k141_139482 | Not-determined |
| 17-108_k141_98577 | Not-determined |
| 17-108_k141_277573 | Not-determined |
| 17-108_k141_139484 | Low-quality |
| 17-108_k141_111810 | Low-quality |
| 17-108_k141_222396 | Not-determined |
| 17-108_k141_29240 | Not-determined |
| 17-108_k141_139492 | Not-determined |
| 17-108_k141_29243 | Not-determined |
| 17-108_k141_222406 | Not-determined |
| 17-108_k141_84610 | Not-determined |
| 17-108_k141_84612 | Not-determined |
| 17-108_k141_194675 | Not-determined |
| 17-108_k141_194681 | Not-determined |
| 17-108_k141_277593 | Not-determined |
| 17-108_k141_56944 | Low-quality |
| 17-108_k141_180931 | Not-determined |

| | |
|---|---|
| 17-108_k141_180932 | Not-determined |
| 17-108_k141_29263 | Not-determined |
| 17-108_k141_29264 | Not-determined |
| 17-108_k141_180939 | Not-determined |
| 17-108_k141_291203 | Not-determined |
| 17-108_k141_263660 | Not-determined |
| 17-108_k141_153438 | Not-determined |
| 17-108_k141_125938 | Not-determined |
| 17-108_k141_291214 | Not-determined |
| 17-108_k141_222446 | Not-determined |
| 17-108_k141_84637 | Not-determined |
| 17-108_k141_291221 | Not-determined |
| 17-108_k141_84649 | Not-determined |
| 17-108_k141_139520 | Not-determined |
| 17-108_k141_222458 | Low-quality |
| 17-108_k141_222466 | Low-quality |
| 17-108_k141_15710 | Low-quality |
| 17-108_k141_15718 | Not-determined |
| 17-108_k141_194706 | Not-determined |
| 17-108_k141_1762 | Not-determined |
| 17-108_k141_318642 | Not-determined |
| 17-108_k141_194711 | Low-quality |
| 17-108_k141_125945 | Not-determined |
| 17-108_k141_318644 | Not-determined |
| 17-108_k141_194716 | Not-determined |
| 17-108_k141_1773 | Not-determined |
| 17-108_k141_180963 | Not-determined |
| 17-108_k141_84674 | Not-determined |
| 17-108_k141_139536 | Not-determined |
| 17-108_k141_84678 | Not-determined |
| 17-108_k141_1784 | Not-determined |
| 17-108_k141_263701 | Not-determined |
| 17-108_k141_318666 | Not-determined |
| 17-108_k141_56998 | Not-determined |
| 17-108_k141_15741 | Not-determined |
| 17-108_k141_318672 | Not-determined |
| 17-108_k141_139549 | Not-determined |
| 17-108_k141_98601 | Not-determined |
| 17-108_k141_57006 | Not-determined |
| 17-108_k141_180972 | Not-determined |
| 17-108_k141_84695 | Not-determined |
| 17-108_k141_194754 | Not-determined |
| 17-108_k141_29296 | Not-determined |
| 17-108_k141_125961 | Not-determined |
| 17-108_k141_70370 | Not-determined |
| 17-108_k141_29297 | Not-determined |
| 17-108_k141_125962 | Not-determined |
| 17-108_k141_57017 | Not-determined |
| 17-108_k141_305095 | Not-determined |
| 17-108_k141_125964 | Not-determined |
| 17-108_k141_194769 | Not-determined |
| 17-108_k141_208722 | Not-determined |
| 17-108_k141_70377 | Not-determined |
| 17-108_k141_208730 | Not-determined |
| 17-108_k141_249838 | Not-determined |
| 17-108_k141_194790 | Not-determined |
| 17-108_k141_222500 | Not-determined |
| 17-108_k141_1790 | Not-determined |
| 17-108_k141_249842 | Not-determined |
| 17-108_k141_277626 | Not-determined |
| 17-108_k141_70385 | Not-determined |
| 17-108_k141_70387 | Not-determined |
| 17-108_k141_222502 | Not-determined |
| 17-108_k141_194798 | Not-determined |
| 17-108_k141_305120 | Not-determined |
| 17-108_k141_249852 | Not-determined |
| 17-108_k141_208743 | Not-determined |
| 17-108_k141_125990 | Low-quality |
| 17-108_k141_84704 | Not-determined |
| 17-108_k141_167569 | Not-determined |
| 17-108_k141_125994 | Not-determined |
| 17-108_k141_236457 | Not-determined |
| 17-108_k141_84712 | Not-determined |
| 17-108_k141_43168 | Not-determined |
| 17-108_k141_126010 | Not-determined |
| 17-108_k141_167577 | Not-determined |
| 17-108_k141_167578 | Not-determined |
| 17-108_k141_57060 | Not-determined |
| 17-108_k141_126018 | Not-determined |
| 17-108_k141_15765 | Not-determined |
| 17-108_k141_57066 | Not-determined |
| 17-108_k141_126022 | Not-determined |
| 17-108_k141_291263 | Low-quality |
| 17-108_k141_249883 | Not-determined |
| 17-108_k141_305136 | Not-determined |
| 17-108_k141_153487 | Not-determined |
| 17-108_k141_236471 | Not-determined |
| 17-108_k141_139611 | Not-determined |
| 17-108_k141_249890 | Not-determined |
| 17-108_k141_305145 | Low-quality |
| 17-108_k141_84725 | Not-determined |
| 17-108_k141_291279 | Not-determined |
| 17-108_k141_305152 | Not-determined |
| 17-108_k141_263769 | Not-determined |
| 17-108_k141_167611 | Not-determined |
| 17-108_k141_1845 | Not-determined |
| 17-108_k141_43205 | Not-determined |
| 17-108_k141_263772 | Not-determined |
| 17-108_k141_263773 | Not-determined |
| 17-108_k141_236489 | Not-determined |
| 17-108_k141_167617 | Not-determined |
| 17-108_k141_167622 | Not-determined |
| 17-108_k141_305156 | Not-determined |
| 17-108_k141_167625 | Not-determined |
| 17-108_k141_236495 | Not-determined |
| 17-108_k141_263785 | Not-determined |
| 17-108_k141_98644 | Not-determined |
| 17-108_k141_43218 | Not-determined |
| 17-108_k141_139634 | Not-determined |
| 17-108_k141_98648 | Not-determined |
| 17-108_k141_208787 | Not-determined |
| 17-108_k141_277670 | Not-determined |
| 17-108_k141_126037 | Not-determined |
| 17-108_k141_194825 | Low-quality |
| 17-108_k141_98658 | Not-determined |
| 17-108_k141_291294 | Not-determined |
| 17-108_k141_43229 | Low-quality |
| 17-108_k141_208798 | Not-determined |
| 17-108_k141_194830 | Not-determined |
| 17-108_k141_263812 | Low-quality |
| 17-108_k141_305189 | Not-determined |
| 17-108_k141_305191 | Not-determined |
| 17-108_k141_305194 | Not-determined |
| 17-108_k141_181027 | Not-determined |
| 17-108_k141_15793 | Not-determined |
| 17-108_k141_1887 | Not-determined |
| 17-108_k141_263819 | Not-determined |
| 17-108_k141_194847 | Not-determined |
| 17-108_k141_126047 | Not-determined |
| 17-108_k141_181033 | Low-quality |
| 17-108_k141_84763 | Not-determined |
| 17-108_k141_167661 | Not-determined |
| 17-108_k141_277697 | Not-determined |
| 17-108_k141_126055 | Not-determined |
| 17-108_k141_111888 | Not-determined |
| 17-108_k141_84775 | Not-determined |
| 17-108_k141_1904 | Not-determined |
| 17-108_k141_277712 | Not-determined |
| 17-108_k141_249938 | Not-determined |
| 17-108_k141_222534 | Not-determined |
| 17-108_k141_222536 | Not-determined |
| 17-108_k141_167681 | Not-determined |
| 17-108_k141_167684 | Not-determined |
| 17-108_k141_98678 | Not-determined |
| 17-108_k141_84791 | Not-determined |
| 17-108_k141_249943 | Not-determined |
| 17-108_k141_277730 | Not-determined |
| 17-108_k141_98684 | Not-determined |
| 17-108_k141_98687 | Not-determined |
| 17-108_k141_139672 | Low-quality |
| 17-108_k141_277740 | Not-determined |
| 17-108_k141_98695 | Not-determined |
| 17-108_k141_98698 | Not-determined |
| 17-108_k141_167696 | Not-determined |
| 17-108_k141_1921 | Not-determined |
| 17-108_k141_1922 | Not-determined |
| 17-108_k141_222554 | Not-determined |
| 17-108_k141_181044 | Not-determined |
| 17-108_k141_318732 | Low-quality |
| 17-108_k141_318736 | Not-determined |
| 17-108_k141_43251 | Low-quality |
| 17-108_k141_236529 | Not-determined |
| 17-108_k141_126076 | Not-determined |
| 17-108_k141_15848 | Not-determined |
| 17-108_k141_57155 | Not-determined |
| 17-108_k141_277761 | Low-quality |
| 17-108_k141_249982 | Low-quality |
| 17-108_k141_194872 | Low-quality |
| 17-108_k141_1930 | Not-determined |
| 17-108_k141_57166 | Not-determined |
| 17-108_k141_15860 | Not-determined |
| 17-108_k141_305234 | Not-determined |
| 17-108_k141_194881 | Not-determined |
| 17-108_k141_15863 | Not-determined |
| 17-108_k141_29375 | Not-determined |
| 17-108_k141_57168 | Not-determined |
| 17-108_k141_194884 | Not-determined |
| 17-108_k141_318755 | Not-determined |
| 17-108_k141_291326 | Not-determined |
| 17-108_k141_208877 | Low-quality |
| 17-108_k141_98722 | Not-determined |
| 17-108_k141_1945 | Not-determined |
| 17-108_k141_43282 | Not-determined |
| 17-108_k141_181062 | Not-determined |
| 17-108_k141_194894 | Not-determined |
| 17-108_k141_263844 | Not-determined |
| 17-108_k141_98728 | Not-determined |
| 17-108_k141_181067 | Not-determined |
| 17-108_k141_181068 | Not-determined |
| 17-108_k141_139697 | Not-determined |
| 17-108_k141_194902 | Not-determined |
| 17-108_k141_57179 | Not-determined |
| 17-108_k141_236558 | Not-determined |
| 17-108_k141_263848 | Not-determined |
| 17-108_k141_43286 | Not-determined |
| 17-108_k141_194911 | Not-determined |
| 17-108_k141_1954 | Low-quality |
| 17-108_k141_57185 | Not-determined |
| 17-108_k141_318775 | Not-determined |
| 17-108_k141_318777 | Not-determined |
| 17-108_k141_57193 | Not-determined |
| 17-108_k141_57194 | Not-determined |
| 17-108_k141_1959 | Not-determined |
| 17-108_k141_1960 | Not-determined |
| 17-108_k141_43300 | Not-determined |
| 17-108_k141_139709 | Not-determined |
| 17-108_k141_57202 | Low-quality |
| 17-108_k141_139713 | Not-determined |
| 17-108_k141_98745 | Not-determined |
| 17-108_k141_277799 | Not-determined |
| 17-108_k141_15888 | Not-determined |
| 17-108_k141_111909 | Not-determined |
| 17-108_k141_29410 | Not-determined |
| 17-108_k141_277807 | Not-determined |
| 17-108_k141_1971 | Not-determined |
| 17-108_k141_15899 | Not-determined |
| 17-108_k141_250005 | Not-determined |
| 17-108_k141_250007 | Not-determined |
| 17-108_k141_29421 | Not-determined |
| 17-108_k141_98766 | Not-determined |
| 17-108_k141_263867 | Not-determined |
| 17-108_k141_43324 | Not-determined |
| 17-108_k141_291382 | Not-determined |
| 17-108_k141_98774 | Not-determined |
| 17-108_k141_57222 | Not-determined |
| 17-108_k141_98776 | Not-determined |
| 17-108_k141_277819 | Not-determined |
| 17-108_k141_222585 | Not-determined |
| 17-108_k141_57229 | Not-determined |
| 17-108_k141_277826 | Not-determined |
| 17-108_k141_111927 | Not-determined |
| 17-108_k141_181118 | Not-determined |
| 17-108_k141_318811 | Not-determined |
| 17-108_k141_126114 | Low-quality |
| 17-108_k141_291405 | Not-determined |
| 17-108_k141_291406 | Not-determined |
| 17-108_k141_111937 | Not-determined |
| 17-108_k141_29448 | Not-determined |
| 17-108_k141_277834 | Not-determined |
| 17-108_k141_236607 | Not-determined |
| 17-108_k141_111941 | Not-determined |
| 17-108_k141_208957 | Not-determined |
| 17-108_k141_318822 | Low-quality |
| 17-108_k141_277841 | Not-determined |
| 17-108_k141_277842 | Not-determined |
| 17-108_k141_277844 | Not-determined |
| 17-108_k141_15931 | Not-determined |
| 17-108_k141_98783 | Not-determined |
| 17-108_k141_98787 | Not-determined |
| 17-108_k141_318828 | Not-determined |
| 17-108_k141_15940 | Low-quality |
| 17-108_k141_139741 | Not-determined |
| 17-108_k141_318838 | Not-determined |
| 17-108_k141_291438 | Not-determined |
| 17-108_k141_181129 | Not-determined |
| 17-108_k141_153539 | Low-quality |
| 17-108_k141_194952 | Not-determined |
| 17-108_k141_29477 | Not-determined |
| 17-108_k141_208972 | Not-determined |
| 17-108_k141_139747 | Not-determined |
| 17-108_k141_305279 | Not-determined |
| 17-108_k141_181141 | Not-determined |
| 17-108_k141_29486 | Not-determined |
| 17-108_k141_305285 | Not-determined |
| 17-108_k141_305286 | Not-determined |
| 17-108_k141_277865 | Not-determined |
| 17-108_k141_236636 | Low-quality |
| 17-108_k141_153556 | Not-determined |
| 17-108_k141_305291 | Not-determined |
| 17-108_k141_126161 | Not-determined |
| 17-108_k141_181143 | Not-determined |
| 17-108_k141_29496 | Not-determined |
| 17-108_k141_2041 | Not-determined |
| 17-108_k141_236648 | Not-determined |
| 17-108_k141_305294 | Not-determined |
| 17-108_k141_126168 | Not-determined |
| 17-108_k141_181151 | Not-determined |
| 17-108_k141_250069 | Not-determined |
| 17-108_k141_139766 | Not-determined |
| 17-108_k141_29505 | Not-determined |
| 17-108_k141_29506 | Not-determined |
| 17-108_k141_84852 | Not-determined |
| 17-108_k141_277884 | Not-determined |
| 17-108_k141_126172 | Not-determined |
| 17-108_k141_250075 | Not-determined |
| 17-108_k141_29513 | Not-determined |
| 17-108_k141_126176 | Not-determined |
| 17-108_k141_126177 | Not-determined |
| 17-108_k141_2055 | Not-determined |
| 17-108_k141_208997 | Not-determined |
| 17-108_k141_236658 | Not-determined |
| 17-108_k141_126181 | Not-determined |
| 17-108_k141_43386 | Not-determined |
| 17-108_k141_111959 | Not-determined |
| 17-108_k141_153579 | Not-determined |
| 17-108_k141_98802 | Not-determined |
| 17-108_k141_318865 | Not-determined |
| 17-108_k141_236670 | Not-determined |
| 17-108_k141_70445 | Not-determined |
| 17-108_k141_111969 | Low-quality |
| 17-108_k141_15971 | Not-determined |
| 17-108_k141_305328 | Not-determined |
| 17-108_k141_277903 | Not-determined |
| 17-108_k141_209016 | Not-determined |
| 17-108_k141_111973 | Not-determined |
| 17-108_k141_305338 | Not-determined |
| 17-108_k141_15999 | Low-quality |
| 17-108_k141_57255 | Not-determined |
| 17-108_k141_181159 | Low-quality |
| 17-108_k141_263967 | Not-determined |
| 17-108_k141_139795 | Not-determined |
| 17-108_k141_111986 | Not-determined |
| 17-108_k141_139799 | Not-determined |
| 17-108_k141_111991 | Not-determined |
| 17-108_k141_236685 | Not-determined |
| 17-108_k141_111996 | Not-determined |
| 17-108_k141_84864 | Not-determined |
| 17-108_k141_167768 | Not-determined |
| 17-108_k141_16009 | Not-determined |
| 17-108_k141_153595 | Not-determined |
| 17-108_k141_112010 | Not-determined |
| 17-108_k141_277923 | Low-quality |
| 17-108_k141_139811 | Not-determined |
| 17-108_k141_305352 | Not-determined |
| 17-108_k141_112016 | Low-quality |
| 17-108_k141_57286 | Not-determined |
| 17-108_k141_57291 | Not-determined |
| 17-108_k141_305366 | Not-determined |
| 17-108_k141_29549 | Not-determined |
| 17-108_k141_277945 | Not-determined |
| 17-108_k141_222635 | Not-determined |
| 17-108_k141_16026 | Not-determined |
| 17-108_k141_277953 | Not-determined |
| 17-108_k141_84887 | Not-determined |
| 17-108_k141_194997 | Not-determined |
| 17-108_k141_305369 | Not-determined |
| 17-108_k141_264010 | Not-determined |
| 17-108_k141_29574 | Not-determined |
| 17-108_k141_264011 | Not-determined |
| 17-108_k141_264012 | Not-determined |
| 17-108_k141_250123 | Not-determined |
| 17-108_k141_250124 | Not-determined |
| 17-108_k141_2085 | Not-determined |
| 17-108_k141_43421 | Not-determined |
| 17-108_k141_250126 | Not-determined |
| 17-108_k141_43422 | Not-determined |
| 17-108_k141_318926 | Not-determined |
| 17-108_k141_305387 | Not-determined |
| 17-108_k141_305389 | Not-determined |
| 17-108_k141_195021 | Not-determined |
| 17-108_k141_29587 | Not-determined |
| 17-108_k141_126210 | Not-determined |
| 17-108_k141_318938 | Low-quality |
| 17-108_k141_222651 | Not-determined |
| 17-108_k141_126216 | Not-determined |
| 17-108_k141_236731 | Low-quality |
| 17-108_k141_277988 | Not-determined |
| 17-108_k141_29594 | Not-determined |
| 17-108_k141_16052 | Low-quality |
| 17-108_k141_318944 | Not-determined |
| 17-108_k141_305411 | Not-determined |
| 17-108_k141_153644 | Not-determined |
| 17-108_k141_57339 | Not-determined |
| 17-108_k141_153649 | Not-determined |
| 17-108_k141_70482 | Low-quality |
| 17-108_k141_264038 | Not-determined |
| 17-108_k141_16064 | Not-determined |
| 17-108_k141_236759 | Not-determined |
| 17-108_k141_16069 | Not-determined |
| 17-108_k141_264045 | Not-determined |
| 17-108_k141_264047 | Not-determined |
| 17-108_k141_278015 | Not-determined |
| 17-108_k141_2122 | Not-determined |
| 17-108_k141_236764 | Not-determined |
| 17-108_k141_126235 | Not-determined |
| 17-108_k141_209070 | Low-quality |
| 17-108_k141_250151 | Low-quality |
| 17-108_k141_250153 | Not-determined |
| 17-108_k141_139880 | Not-determined |
| 17-108_k141_181214 | Not-determined |
| 17-108_k141_181215 | Not-determined |
| 17-108_k141_318965 | Not-determined |
| 17-108_k141_195044 | Not-determined |
| 17-108_k141_278041 | Not-determined |
| 17-108_k141_16088 | Not-determined |
| 17-108_k141_57366 | Not-determined |
| 17-108_k141_291512 | Not-determined |
| 17-108_k141_195049 | Low-quality |
| 17-108_k141_29631 | Not-determined |
| 17-108_k141_236795 | Not-determined |
| 17-108_k141_29637 | Not-determined |
| 17-108_k141_57382 | Not-determined |
| 17-108_k141_98841 | Not-determined |
| 17-108_k141_195058 | Not-determined |
| 17-108_k141_222692 | Not-determined |
| 17-108_k141_222694 | Not-determined |
| 17-108_k141_70520 | Not-determined |
| 17-108_k141_2139 | Low-quality |
| 17-108_k141_250175 | Not-determined |
| 17-108_k141_278066 | Not-determined |
| 17-108_k141_305457 | Low-quality |
| 17-108_k141_236818 | Not-determined |
| 17-108_k141_236821 | Not-determined |
| 17-108_k141_209111 | Not-determined |
| 17-108_k141_236827 | Not-determined |
| 17-108_k141_250192 | Not-determined |
| 17-108_k141_236835 | Not-determined |
| 17-108_k141_236836 | Not-determined |
| 17-108_k141_250195 | Not-determined |
| 17-108_k141_209117 | Not-determined |
| 17-108_k141_236844 | Not-determined |
| 17-108_k141_98867 | Not-determined |
| 17-108_k141_70536 | Low-quality |
| 17-108_k141_209125 | Not-determined |
| 17-108_k141_2153 | Not-determined |
| 17-108_k141_98879 | Not-determined |
| 17-108_k141_250209 | Not-determined |
| 17-108_k141_222725 | Not-determined |
| 17-108_k141_222726 | Not-determined |
| 17-108_k141_112110 | Not-determined |
| 17-108_k141_236849 | Not-determined |
| 17-108_k141_98891 | Not-determined |
| 17-108_k141_126258 | Not-determined |
| 17-108_k141_250222 | Not-determined |
| 17-108_k141_250224 | Not-determined |
| 17-108_k141_209144 | Not-determined |
| 17-108_k141_167849 | Not-determined |
| 17-108_k141_98901 | Not-determined |
| 17-108_k141_57405 | Not-determined |
| 17-108_k141_139907 | Low-quality |
| 17-108_k141_209154 | Not-determined |
| 17-108_k141_278101 | Not-determined |
| 17-108_k141_319012 | Not-determined |
| 17-108_k141_319014 | Not-determined |
| 17-108_k141_139910 | Low-quality |
| 17-108_k141_70565 | Not-determined |
| 17-108_k141_181276 | Not-determined |
| 17-108_k141_278115 | Not-determined |
| 17-108_k141_278118 | Not-determined |
| 17-108_k141_236881 | Not-determined |
| 17-108_k141_236883 | Low-quality |
| 17-108_k141_278123 | Low-quality |
| 17-108_k141_291528 | Not-determined |
| 17-108_k141_250270 | Not-determined |
| 17-108_k141_291529 | Not-determined |
| 17-108_k141_195106 | Not-determined |
| 17-108_k141_291534 | Not-determined |
| 17-108_k141_84996 | Not-determined |
| 17-108_k141_70580 | Not-determined |
| 17-108_k141_236885 | Low-quality |
| 17-108_k141_98922 | Not-determined |
| 17-108_k141_236888 | Low-quality |
| 17-108_k141_195116 | Not-determined |
| 17-108_k141_195117 | Not-determined |
| 17-108_k141_70591 | Not-determined |
| 17-108_k141_209180 | Not-determined |
| 17-108_k141_195123 | Not-determined |
| 17-108_k141_85007 | Not-determined |
| 17-108_k141_209181 | Not-determined |
| 17-108_k141_250298 | Not-determined |
| 17-108_k141_2170 | Not-determined |
| 17-108_k141_2175 | Not-determined |
| 17-108_k141_305474 | Not-determined |
| 17-108_k141_57458 | Not-determined |
| 17-108_k141_43499 | Not-determined |
| 17-108_k141_43501 | Not-determined |
| 17-108_k141_43504 | Not-determined |
| 17-108_k141_2200 | Low-quality |
| 17-108_k141_305496 | Not-determined |
| 17-108_k141_236906 | Low-quality |
| 17-108_k141_2205 | Not-determined |
| 17-108_k141_209227 | Not-determined |
| 17-108_k141_57486 | Not-determined |
| 17-108_k141_57487 | Not-determined |
| 17-108_k141_209229 | Not-determined |
| 17-108_k141_29680 | Not-determined |
| 17-108_k141_305505 | Not-determined |
| 17-108_k141_43509 | Not-determined |
| 17-108_k141_43510 | Not-determined |
| 17-108_k141_57493 | Not-determined |
| 17-108_k141_305507 | Not-determined |
| 17-108_k141_57497 | Not-determined |
| 17-108_k141_2219 | Not-determined |
| 17-108_k141_2222 | Not-determined |
| 17-108_k141_85034 | Not-determined |
| 17-108_k141_70624 | Not-determined |
| 17-108_k141_57508 | Not-determined |
| 17-108_k141_209253 | Not-determined |
| 17-108_k141_209254 | Not-determined |
| 17-108_k141_57514 | Not-determined |
| 17-108_k141_16141 | Not-determined |
| 17-108_k141_264104 | Not-determined |
| 17-108_k141_209262 | Not-determined |
| 17-108_k141_291575 | Not-determined |
| 17-108_k141_153770 | Not-determined |
| 17-108_k141_153772 | Not-determined |
| 17-108_k141_29715 | Not-determined |
| 17-108_k141_209277 | Not-determined |
| 17-108_k141_236936 | Not-determined |
| 17-108_k141_139968 | Not-determined |
| 17-108_k141_139975 | Not-determined |
| 17-108_k141_250329 | Not-determined |
| 17-108_k141_43515 | Not-determined |
| 17-108_k141_264111 | Not-determined |
| 17-108_k141_264113 | Not-determined |
| 17-108_k141_70662 | Not-determined |
| 17-108_k141_126292 | Low-quality |
| 17-108_k141_195178 | Not-determined |
| 17-108_k141_209292 | Not-determined |
| 17-108_k141_112146 | Not-determined |
| 17-108_k141_278154 | Not-determined |
| 17-108_k141_250342 | Not-determined |
| 17-108_k141_278156 | Low-quality |
| 17-108_k141_126300 | Not-determined |
| 17-108_k141_98978 | Not-determined |
| 17-108_k141_319026 | Not-determined |
| 17-108_k141_2252 | Not-determined |
| 17-108_k141_264129 | Not-determined |
| 17-108_k141_278171 | Not-determined |
| 17-108_k141_264131 | Low-quality |
| 17-108_k141_278180 | Not-determined |
| 17-108_k141_250370 | Not-determined |
| 17-108_k141_29751 | Not-determined |
| 17-108_k141_305578 | Not-determined |
| 17-108_k141_57539 | Low-quality |
| 17-108_k141_195197 | Not-determined |
| 17-108_k141_250379 | Low-quality |
| 17-108_k141_278183 | Not-determined |
| 17-108_k141_250381 | Not-determined |
| 17-108_k141_195208 | Not-determined |
| 17-108_k141_126326 | Not-determined |
| 17-108_k141_195211 | Not-determined |
| 17-108_k141_195214 | Not-determined |
| 17-108_k141_195217 | Not-determined |
| 17-108_k141_195218 | Not-determined |
| 17-108_k141_29766 | Not-determined |
| 17-108_k141_153808 | Not-determined |
| 17-108_k141_236950 | Not-determined |
| 17-108_k141_278198 | Not-determined |
| 17-108_k141_305593 | Not-determined |
| 17-108_k141_264146 | Not-determined |
| 17-108_k141_264149 | Not-determined |
| 17-108_k141_126345 | Not-determined |
| 17-108_k141_278202 | Not-determined |
| 17-108_k141_167934 | Low-quality |
| 17-108_k141_126347 | Not-determined |
| 17-108_k141_264155 | Not-determined |
| 17-108_k141_29778 | Not-determined |
| 17-108_k141_264157 | Not-determined |
| 17-108_k141_43534 | Not-determined |
| 17-108_k141_264162 | Not-determined |
| 17-108_k141_2302 | Not-determined |
| 17-108_k141_57549 | Low-quality |
| 17-108_k141_250388 | Not-determined |
| 17-108_k141_98982 | Not-determined |
| 17-108_k141_29785 | Not-determined |
| 17-108_k141_305612 | Not-determined |
| 17-108_k141_264171 | Not-determined |
| 17-108_k141_250395 | Not-determined |
| 17-108_k141_236959 | Not-determined |
| 17-108_k141_98990 | Not-determined |
| 17-108_k141_264179 | Not-determined |
| 17-108_k141_250406 | Not-determined |
| 17-108_k141_126393 | Not-determined |
| 17-108_k141_305627 | Not-determined |
| 17-108_k141_57568 | Not-determined |
| 17-108_k141_29795 | Not-determined |
| 17-108_k141_57570 | Low-quality |
| 17-108_k141_2328 | Not-determined |
| 17-108_k141_278226 | Not-determined |
| 17-108_k141_29798 | Not-determined |
| 17-108_k141_43573 | Not-determined |
| 17-108_k141_70736 | Not-determined |
| 17-108_k141_126404 | Low-quality |
| 17-108_k141_140017 | Not-determined |
| 17-108_k141_305645 | Not-determined |
| 17-108_k141_43583 | Not-determined |
| 17-108_k141_153822 | Not-determined |
| 17-108_k141_140023 | Not-determined |
| 17-108_k141_112180 | Not-determined |
| 17-108_k141_264215 | Not-determined |
| 17-108_k141_250422 | Not-determined |
| 17-108_k141_29815 | Not-determined |
| 17-108_k141_250423 | Not-determined |
| 17-108_k141_319079 | Not-determined |
| 17-108_k141_167955 | Not-determined |
| 17-108_k141_112187 | Not-determined |
| 17-108_k141_305652 | Not-determined |
| 17-108_k141_167959 | Not-determined |

| | |
|---|---|
| 17-108_k141_16160 | Not-determined |
| 17-108_k141_112195 | Not-determined |
| 17-108_k141_112199 | Not-determined |
| 17-108_k141_99032 | Not-determined |
| 17-108_k141_43598 | Not-determined |
| 17-108_k141_43601 | Not-determined |
| 17-108_k141_140043 | Not-determined |
| 17-108_k141_43608 | Not-determined |
| 17-108_k141_278245 | Not-determined |
| 17-108_k141_319096 | Not-determined |
| 17-108_k141_70777 | Not-determined |
| 17-108_k141_126431 | Not-determined |
| 17-108_k141_167983 | Not-determined |
| 17-108_k141_126434 | Not-determined |
| 17-108_k141_140057 | Not-determined |
| 17-108_k141_195253 | Low-quality |
| 17-108_k141_222784 | Not-determined |
| 17-108_k141_70786 | Not-determined |
| 17-108_k141_319123 | Low-quality |
| 17-108_k141_305685 | Not-determined |
| 17-108_k141_57633 | Not-determined |
| 17-108_k141_29825 | Not-determined |
| 17-108_k141_70787 | Not-determined |
| 17-108_k141_29828 | Not-determined |
| 17-108_k141_319127 | Low-quality |
| 17-108_k141_153867 | Not-determined |
| 17-108_k141_126453 | Not-determined |
| 17-108_k141_236984 | Not-determined |
| 17-108_k141_99069 | Not-determined |
| 17-108_k141_57648 | Not-determined |
| 17-108_k141_43616 | Not-determined |
| 17-108_k141_43618 | Not-determined |
| 17-108_k141_305701 | Not-determined |
| 17-108_k141_140080 | Low-quality |
| 17-108_k141_126467 | Not-determined |
| 17-108_k141_236999 | Not-determined |
| 17-108_k141_195276 | Not-determined |
| 17-108_k141_126471 | Not-determined |
| 17-108_k141_57660 | Not-determined |
| 17-108_k141_29835 | Not-determined |
| 17-108_k141_209432 | Not-determined |
| 17-108_k141_16182 | Not-determined |
| 17-108_k141_29837 | Not-determined |
| 17-108_k141_16183 | Not-determined |
| 17-108_k141_70809 | Low-quality |
| 17-108_k141_291645 | Not-determined |
| 17-108_k141_43638 | Not-determined |
| 17-108_k141_319147 | Not-determined |
| 17-108_k141_168010 | Low-quality |
| 17-108_k141_291653 | Not-determined |
| 17-108_k141_29840 | Low-quality |
| 17-108_k141_43656 | Not-determined |
| 17-108_k141_2406 | Not-determined |
| 17-108_k141_222824 | Not-determined |
| 17-108_k141_57678 | Not-determined |
| 17-108_k141_168030 | Low-quality |
| 17-108_k141_305745 | Not-determined |
| 17-108_k141_99076 | Not-determined |
| 17-108_k141_305746 | Not-determined |
| 17-108_k141_2414 | Not-determined |
| 17-108_k141_2415 | Not-determined |
| 17-108_k141_112212 | Not-determined |
| 17-108_k141_112217 | Not-determined |
| 17-108_k141_168040 | Not-determined |
| 17-108_k141_209443 | Not-determined |
| 17-108_k141_140121 | Not-determined |
| 17-108_k141_126480 | Not-determined |
| 17-108_k141_70822 | Not-determined |
| 17-108_k141_291676 | Not-determined |
| 17-108_k141_43679 | Not-determined |
| 17-108_k141_319165 | Not-determined |
| 17-108_k141_153906 | Not-determined |
| 17-108_k141_153907 | Not-determined |
| 17-108_k141_70830 | Not-determined |
| 17-108_k141_153910 | Not-determined |
| 17-108_k141_140131 | Low-quality |
| 17-108_k141_181373 | Not-determined |
| 17-108_k141_140140 | Not-determined |
| 17-108_k141_99103 | Not-determined |
| 17-108_k141_305773 | Not-determined |
| 17-108_k141_112237 | Not-determined |
| 17-108_k141_2452 | Not-determined |
| 17-108_k141_153918 | Not-determined |
| 17-108_k141_209468 | Not-determined |
| 17-108_k141_126514 | Not-determined |
| 17-108_k141_112253 | Not-determined |
| 17-108_k141_126517 | Not-determined |
| 17-108_k141_2470 | Not-determined |
| 17-108_k141_2472 | Not-determined |
| 17-108_k141_264249 | Not-determined |
| 17-108_k141_29873 | Not-determined |
| 17-108_k141_112259 | Low-quality |
| 17-108_k141_222882 | Not-determined |
| 17-108_k141_70849 | Not-determined |
| 17-108_k141_57722 | Not-determined |
| 17-108_k141_57728 | Not-determined |
| 17-108_k141_209495 | Not-determined |
| 17-108_k141_222895 | Not-determined |
| 17-108_k141_305797 | Not-determined |
| 17-108_k141_264272 | Low-quality |
| 17-108_k141_264275 | Not-determined |
| 17-108_k141_85082 | Not-determined |
| 17-108_k141_85088 | Not-determined |
| 17-108_k141_209508 | Not-determined |
| 17-108_k141_85093 | Low-quality |
| 17-108_k141_278306 | Not-determined |
| 17-108_k141_85095 | Not-determined |
| 17-108_k141_209514 | Not-determined |
| 17-108_k141_112296 | Not-determined |
| 17-108_k141_112298 | Not-determined |
| 17-108_k141_2515 | Not-determined |
| 17-108_k141_278312 | Not-determined |
| 17-108_k141_57745 | Low-quality |
| 17-108_k141_319223 | Low-quality |
| 17-108_k141_2521 | Not-determined |
| 17-108_k141_209520 | Not-determined |
| 17-108_k141_319225 | Not-determined |
| 17-108_k141_278322 | Not-determined |
| 17-108_k141_250521 | Not-determined |
| 17-108_k141_278324 | Not-determined |
| 17-108_k141_57749 | Not-determined |
| 17-108_k141_57752 | Not-determined |
| 17-108_k141_57754 | Not-determined |
| 17-108_k141_57756 | Not-determined |
| 17-108_k141_70880 | Not-determined |
| 17-108_k141_70881 | Not-determined |
| 17-108_k141_126582 | Not-determined |
| 17-108_k141_264310 | Not-determined |
| 17-108_k141_70885 | Not-determined |
| 17-108_k141_70890 | Not-determined |
| 17-108_k141_250536 | Not-determined |
| 17-108_k141_222939 | Not-determined |
| 17-108_k141_70893 | Not-determined |
| 17-108_k141_154001 | Not-determined |
| 17-108_k141_278352 | Not-determined |
| 17-108_k141_154002 | Not-determined |
| 17-108_k141_278355 | Not-determined |
| 17-108_k141_154006 | Not-determined |
| 17-108_k141_291721 | Not-determined |
| 17-108_k141_278358 | Not-determined |
| 17-108_k141_57758 | Not-determined |
| 17-108_k141_70901 | Not-determined |
| 17-108_k141_278370 | Not-determined |
| 17-108_k141_278371 | Not-determined |
| 17-108_k141_168071 | Not-determined |
| 17-108_k141_70903 | Not-determined |
| 17-108_k141_319255 | Not-determined |
| 17-108_k141_278380 | Not-determined |
| 17-108_k141_2540 | Not-determined |
| 17-108_k141_57767 | Not-determined |
| 17-108_k141_57771 | Not-determined |
| 17-108_k141_2551 | Not-determined |
| 17-108_k141_154022 | Not-determined |
| 17-108_k141_99161 | Not-determined |
| 17-108_k141_222967 | Not-determined |
| 17-108_k141_29958 | Not-determined |
| 17-108_k141_222969 | Not-determined |
| 17-108_k141_305866 | Not-determined |
| 17-108_k141_209564 | Not-determined |
| 17-108_k141_57790 | Not-determined |
| 17-108_k141_29969 | Not-determined |
| 17-108_k141_29971 | Not-determined |
| 17-108_k141_70922 | Not-determined |
| 17-108_k141_209574 | Not-determined |
| 17-108_k141_278386 | Not-determined |
| 17-108_k141_278388 | Not-determined |
| 17-108_k141_29974 | Not-determined |
| 17-108_k141_168100 | Low-quality |
| 17-108_k141_209579 | Not-determined |
| 17-108_k141_43767 | Not-determined |
| 17-108_k141_250564 | Not-determined |
| 17-108_k141_223002 | Not-determined |
| 17-108_k141_29983 | Not-determined |
| 17-108_k141_140251 | Not-determined |
| 17-108_k141_140252 | Not-determined |
| 17-108_k141_168107 | Not-determined |
| 17-108_k141_168113 | Not-determined |
| 17-108_k141_2586 | Not-determined |
| 17-108_k141_305889 | Not-determined |
| 17-108_k141_278417 | Not-determined |
| 17-108_k141_250572 | Not-determined |
| 17-108_k141_250573 | Not-determined |
| 17-108_k141_168126 | Not-determined |
| 17-108_k141_2593 | Not-determined |
| 17-108_k141_43783 | Not-determined |
| 17-108_k141_154059 | Not-determined |
| 17-108_k141_250576 | Not-determined |
| 17-108_k141_70961 | Not-determined |
| 17-108_k141_237069 | Not-determined |
| 17-108_k141_126596 | Not-determined |
| 17-108_k141_99189 | Not-determined |
| 17-108_k141_30014 | Not-determined |
| 17-108_k141_237071 | Low-quality |
| 17-108_k141_195311 | Low-quality |
| 17-108_k141_43804 | Low-quality |
| 17-108_k141_250589 | Not-determined |
| 17-108_k141_85119 | Not-determined |
| 17-108_k141_237077 | Not-determined |
| 17-108_k141_181429 | Not-determined |
| 17-108_k141_250599 | Not-determined |
| 17-108_k141_291735 | Not-determined |
| 17-108_k141_195321 | Not-determined |
| 17-108_k141_57846 | Not-determined |
| 17-108_k141_305899 | Not-determined |
| 17-108_k141_250607 | Not-determined |
| 17-108_k141_168149 | Not-determined |
| 17-108_k141_319304 | Not-determined |
| 17-108_k141_278441 | Not-determined |
| 17-108_k141_237082 | Not-determined |
| 17-108_k141_57853 | Not-determined |
| 17-108_k141_319308 | Not-determined |
| 17-108_k141_70978 | Not-determined |
| 17-108_k141_305910 | Not-determined |
| 17-108_k141_70980 | Not-determined |
| 17-108_k141_195338 | Not-determined |
| 17-108_k141_57863 | Not-determined |
| 17-108_k141_291740 | Not-determined |
| 17-108_k141_154086 | Not-determined |
| 17-108_k141_223043 | Not-determined |
| 17-108_k141_223046 | Not-determined |
| 17-108_k141_140297 | Low-quality |
| 17-108_k141_99230 | Low-quality |
| 17-108_k141_43844 | Not-determined |
| 17-108_k141_181450 | Not-determined |
| 17-108_k141_43849 | Not-determined |
| 17-108_k141_43850 | Not-determined |
| 17-108_k141_140308 | Not-determined |
| 17-108_k141_57869 | Not-determined |
| 17-108_k141_57870 | Not-determined |
| 17-108_k141_140311 | Not-determined |
| 17-108_k141_43857 | Not-determined |
| 17-108_k141_181461 | Not-determined |
| 17-108_k141_154099 | Not-determined |
| 17-108_k141_112341 | Not-determined |
| 17-108_k141_112342 | Not-determined |
| 17-108_k141_223054 | Low-quality |
| 17-108_k141_195369 | Not-determined |
| 17-108_k141_112349 | Not-determined |
| 17-108_k141_195377 | Not-determined |
| 17-108_k141_223063 | Low-quality |
| 17-108_k141_2633 | Not-determined |
| 17-108_k141_278469 | Not-determined |
| 17-108_k141_85157 | Not-determined |
| 17-108_k141_2640 | Not-determined |
| 17-108_k141_195388 | Not-determined |
| 17-108_k141_154113 | Low-quality |
| 17-108_k141_57892 | Not-determined |
| 17-108_k141_57893 | Not-determined |
| 17-108_k141_71014 | Not-determined |
| 17-108_k141_30058 | Not-determined |
| 17-108_k141_99255 | Not-determined |
| 17-108_k141_16241 | Not-determined |
| 17-108_k141_278475 | Not-determined |
| 17-108_k141_30064 | Not-determined |
| 17-108_k141_181495 | Not-determined |
| 17-108_k141_319326 | Not-determined |
| 17-108_k141_278482 | Not-determined |
| 17-108_k141_237121 | Not-determined |
| 17-108_k141_195408 | Not-determined |
| 17-108_k141_154141 | Low-quality |
| 17-108_k141_223099 | Not-determined |
| 17-108_k141_154145 | Not-determined |
| 17-108_k141_2656 | Not-determined |
| 17-108_k141_209642 | Not-determined |
| 17-108_k141_57924 | Not-determined |
| 17-108_k141_57925 | Not-determined |
| 17-108_k141_209648 | Not-determined |
| 17-108_k141_57927 | Not-determined |
| 17-108_k141_237135 | Not-determined |
| 17-108_k141_154157 | Not-determined |
| 17-108_k141_319337 | Not-determined |
| 17-108_k141_2671 | Not-determined |
| 17-108_k141_264326 | Not-determined |
| 17-108_k141_2673 | Not-determined |
| 17-108_k141_16262 | Not-determined |
| 17-108_k141_16263 | Not-determined |
| 17-108_k141_237149 | Not-determined |
| 17-108_k141_71032 | Not-determined |
| 17-108_k141_237150 | Not-determined |
| 17-108_k141_209663 | Low-quality |
| 17-108_k141_2681 | Not-determined |
| 17-108_k141_168240 | Not-determined |
| 17-108_k141_305964 | Not-determined |
| 17-108_k141_112376 | Low-quality |
| 17-108_k141_237158 | Not-determined |
| 17-108_k141_57939 | Not-determined |
| 17-108_k141_57940 | Not-determined |
| 17-108_k141_43906 | Not-determined |
| 17-108_k141_264333 | Not-determined |
| 17-108_k141_278510 | Not-determined |
| 17-108_k141_30089 | Not-determined |
| 17-108_k141_99313 | Not-determined |
| 17-108_k141_223124 | Not-determined |
| 17-108_k141_154179 | Low-quality |
| 17-108_k141_264341 | Not-determined |
| 17-108_k141_264346 | Not-determined |
| 17-108_k141_264347 | Not-determined |
| 17-108_k141_278527 | Not-determined |
| 17-108_k141_237173 | Not-determined |
| 17-108_k141_305971 | Not-determined |
| 17-108_k141_112395 | Low-quality |
| 17-108_k141_278530 | Not-determined |
| 17-108_k141_237177 | Not-determined |
| 17-108_k141_43929 | Not-determined |
| 17-108_k141_237181 | Not-determined |
| 17-108_k141_237182 | Not-determined |
| 17-108_k141_154196 | Not-determined |
| 17-108_k141_237186 | Not-determined |
| 17-108_k141_112413 | Not-determined |
| 17-108_k141_223143 | Not-determined |
| 17-108_k141_223145 | Not-determined |
| 17-108_k141_57964 | Not-determined |
| 17-108_k141_43941 | Not-determined |
| 17-108_k141_30114 | Not-determined |
| 17-108_k141_305988 | Low-quality |
| 17-108_k141_71068 | Not-determined |
| 17-108_k141_99323 | Not-determined |
| 17-108_k141_16291 | Not-determined |
| 17-108_k141_112418 | Low-quality |
| 17-108_k141_264377 | Not-determined |
| 17-108_k141_30131 | Not-determined |
| 17-108_k141_250647 | Not-determined |
| 17-108_k141_264378 | Not-determined |
| 17-108_k141_99342 | Not-determined |
| 17-108_k141_16305 | Not-determined |
| 17-108_k141_16307 | Not-determined |
| 17-108_k141_223156 | Not-determined |
| 17-108_k141_223157 | Not-determined |
| 17-108_k141_278558 | Not-determined |
| 17-108_k141_264387 | Not-determined |
| 17-108_k141_278563 | Low-quality |
| 17-108_k141_209743 | Not-determined |
| 17-108_k141_319369 | Not-determined |
| 17-108_k141_2718 | Not-determined |
| 17-108_k141_223165 | Not-determined |
| 17-108_k141_319374 | Not-determined |
| 17-108_k141_181521 | Not-determined |
| 17-108_k141_209746 | Low-quality |
| 17-108_k141_250667 | Not-determined |
| 17-108_k141_319376 | Not-determined |
| 17-108_k141_99360 | Not-determined |
| 17-108_k141_99361 | Not-determined |
| 17-108_k141_250670 | Not-determined |
| 17-108_k141_237237 | Not-determined |
| 17-108_k141_126627 | Not-determined |
| 17-108_k141_126629 | Not-determined |
| 17-108_k141_126631 | Not-determined |
| 17-108_k141_237240 | Not-determined |
| 17-108_k141_291816 | Not-determined |
| 17-108_k141_250679 | Low-quality |
| 17-108_k141_71091 | Not-determined |
| 17-108_k141_57998 | Not-determined |
| 17-108_k141_140347 | Not-determined |
| 17-108_k141_112447 | Not-determined |
| 17-108_k141_140353 | Not-determined |
| 17-108_k141_71103 | Not-determined |
| 17-108_k141_223180 | Not-determined |
| 17-108_k141_112465 | Not-determined |
| 17-108_k141_264416 | Not-determined |
| 17-108_k141_154251 | Not-determined |
| 17-108_k141_306001 | Not-determined |
| 17-108_k141_126650 | Not-determined |
| 17-108_k141_250712 | Not-determined |
| 17-108_k141_112476 | Not-determined |
| 17-108_k141_112477 | Not-determined |
| 17-108_k141_85207 | Not-determined |
| 17-108_k141_291830 | Low-quality |
| 17-108_k141_223204 | Not-determined |
| 17-108_k141_250719 | Low-quality |
| 17-108_k141_319392 | Not-determined |
| 17-108_k141_209771 | Not-determined |
| 17-108_k141_209772 | Not-determined |
| 17-108_k141_168285 | Not-determined |
| 17-108_k141_99381 | Not-determined |
| 17-108_k141_237266 | Not-determined |
| 17-108_k141_264430 | Not-determined |
| 17-108_k141_140383 | Low-quality |
| 17-108_k141_306015 | Not-determined |
| 17-108_k141_306018 | Not-determined |
| 17-108_k141_209784 | Not-determined |
| 17-108_k141_195433 | Not-determined |
| 17-108_k141_154266 | Low-quality |
| 17-108_k141_237273 | Not-determined |
| 17-108_k141_16327 | Not-determined |
| 17-108_k141_209789 | Not-determined |
| 17-108_k141_250745 | Not-determined |
| 17-108_k141_140396 | Not-determined |
| 17-108_k141_154274 | Not-determined |
| 17-108_k141_278583 | Not-determined |
| 17-108_k141_112510 | Not-determined |
| 17-108_k141_291836 | Not-determined |
| 17-108_k141_16333 | Not-determined |
| 17-108_k141_291838 | Not-determined |
| 17-108_k141_264446 | Not-determined |
| 17-108_k141_319423 | Not-determined |
| 17-108_k141_30177 | Not-determined |
| 17-108_k141_264447 | Low-quality |
| 17-108_k141_209800 | Not-determined |
| 17-108_k141_319424 | Not-determined |
| 17-108_k141_264450 | Not-determined |
| 17-108_k141_195443 | Not-determined |
| 17-108_k141_195448 | Not-determined |
| 17-108_k141_71154 | Not-determined |
| 17-108_k141_237297 | Not-determined |
| 17-108_k141_99393 | Not-determined |
| 17-108_k141_209815 | Not-determined |
| 17-108_k141_154299 | Not-determined |
| 17-108_k141_140434 | Not-determined |
| 17-108_k141_99407 | Not-determined |
| 17-108_k141_58054 | Not-determined |
| 17-108_k141_264472 | Not-determined |
| 17-108_k141_237329 | Not-determined |
| 17-108_k141_2785 | Not-determined |
| 17-108_k141_2787 | Not-determined |
| 17-108_k141_264478 | Low-quality |
| 17-108_k141_306034 | Not-determined |
| 17-108_k141_154315 | Low-quality |
| 17-108_k141_264483 | Not-determined |
| 17-108_k141_99426 | Not-determined |
| 17-108_k141_209831 | Low-quality |
| 17-108_k141_85298 | Not-determined |
| 17-108_k141_223247 | Not-determined |
| 17-108_k141_71179 | Not-determined |
| 17-108_k141_181594 | Not-determined |
| 17-108_k141_250785 | Not-determined |
| 17-108_k141_306064 | Not-determined |
| 17-108_k141_2815 | Not-determined |
| 17-108_k141_43981 | Low-quality |
| 17-108_k141_154329 | Not-determined |
| 17-108_k141_154330 | Not-determined |
| 17-108_k141_264507 | Not-determined |
| 17-108_k141_168349 | Not-determined |
| 17-108_k141_306082 | Low-quality |
| 17-108_k141_306092 | Not-determined |
| 17-108_k141_168358 | Not-determined |
| 17-108_k141_99450 | Not-determined |
| 17-108_k141_264522 | Not-determined |
| 17-108_k141_2829 | Not-determined |
| 17-108_k141_85342 | Low-quality |
| 17-108_k141_181610 | Not-determined |
| 17-108_k141_181612 | Not-determined |
| 17-108_k141_223277 | Not-determined |
| 17-108_k141_85351 | Not-determined |
| 17-108_k141_140485 | Not-determined |
| 17-108_k141_195496 | Not-determined |
| 17-108_k141_44011 | Not-determined |
| 17-108_k141_291891 | Not-determined |
| 17-108_k141_223290 | Not-determined |
| 17-108_k141_223291 | Not-determined |
| 17-108_k141_71227 | Not-determined |
| 17-108_k141_168383 | Not-determined |
| 17-108_k141_44020 | Not-determined |
| 17-108_k141_237387 | Not-determined |
| 17-108_k141_16361 | Not-determined |
| 17-108_k141_140502 | Not-determined |
| 17-108_k141_58150 | Not-determined |
| 17-108_k141_58151 | Low-quality |
| 17-108_k141_126695 | Not-determined |
| 17-108_k141_2868 | Low-quality |
| 17-108_k141_2869 | Not-determined |
| 17-108_k141_99461 | Not-determined |
| 17-108_k141_181614 | Low-quality |
| 17-108_k141_264539 | Not-determined |
| 17-108_k141_140509 | Not-determined |
| 17-108_k141_181627 | Not-determined |
| 17-108_k141_237418 | Not-determined |
| 17-108_k141_30236 | Not-determined |
| 17-108_k141_44045 | Not-determined |
| 17-108_k141_195509 | Not-determined |
| 17-108_k141_85371 | Low-quality |
| 17-108_k141_140520 | Not-determined |
| 17-108_k141_168405 | Not-determined |
| 17-108_k141_112556 | Not-determined |
| 17-108_k141_195517 | Not-determined |
| 17-108_k141_16375 | Not-determined |
| 17-108_k141_168417 | Not-determined |
| 17-108_k141_58172 | Not-determined |
| 17-108_k141_58173 | Not-determined |
| 17-108_k141_168422 | Not-determined |
| 17-108_k141_223337 | Not-determined |
| 17-108_k141_306162 | Not-determined |
| 17-108_k141_126730 | Not-determined |
| 17-108_k141_44081 | Not-determined |
| 17-108_k141_223341 | Not-determined |
| 17-108_k141_112587 | Not-determined |
| 17-108_k141_58188 | Not-determined |
| 17-108_k141_112594 | Low-quality |
| 17-108_k141_85399 | Not-determined |
| 17-108_k141_85404 | Low-quality |
| 17-108_k141_291949 | Not-determined |
| 17-108_k141_44095 | Not-determined |
| 17-108_k141_112607 | Not-determined |
| 17-108_k141_44097 | Not-determined |
| 17-108_k141_264583 | Not-determined |
| 17-108_k141_2909 | Not-determined |
| 17-108_k141_250831 | Not-determined |
| 17-108_k141_126760 | Low-quality |
| 17-108_k141_58204 | Low-quality |
| 17-108_k141_195527 | Not-determined |
| 17-108_k141_291960 | Low-quality |
| 17-108_k141_112618 | Not-determined |
| 17-108_k141_58212 | Not-determined |
| 17-108_k141_44116 | Not-determined |
| 17-108_k141_306184 | Not-determined |
| 17-108_k141_168454 | Not-determined |
| 17-108_k141_291961 | Not-determined |
| 17-108_k141_195547 | Not-determined |
| 17-108_k141_223347 | Not-determined |
| 17-108_k141_112636 | Not-determined |
| 17-108_k141_44131 | Not-determined |
| 17-108_k141_126785 | Not-determined |
| 17-108_k141_2934 | Not-determined |
| 17-108_k141_306196 | Not-determined |
| 17-108_k141_195568 | Not-determined |
| 17-108_k141_168472 | Not-determined |
| 17-108_k141_140587 | Not-determined |
| 17-108_k141_264601 | Low-quality |
| 17-108_k141_140588 | Not-determined |
| 17-108_k141_30280 | Not-determined |
| 17-108_k141_112648 | Low-quality |
| 17-108_k141_223365 | Not-determined |
| 17-108_k141_44152 | Not-determined |
| 17-108_k141_168491 | Low-quality |
| 17-108_k141_71311 | Not-determined |
| 17-108_k141_195585 | Not-determined |
| 17-108_k141_291995 | Not-determined |
| 17-108_k141_291997 | Not-determined |
| 17-108_k141_154373 | Not-determined |
| 17-108_k141_2947 | Not-determined |
| 17-108_k141_306219 | Not-determined |
| 17-108_k141_112663 | Not-determined |
| 17-108_k141_71316 | Not-determined |
| 17-108_k141_195592 | Not-determined |
| 17-108_k141_44164 | Not-determined |
| 17-108_k141_292003 | Not-determined |
| 17-108_k141_44165 | Not-determined |
| 17-108_k141_319493 | Not-determined |
| 17-108_k141_126807 | Not-determined |
| 17-108_k141_319494 | Not-determined |
| 17-108_k141_154378 | Not-determined |
| 17-108_k141_2951 | Not-determined |
| 17-108_k141_306224 | Not-determined |
| 17-108_k141_237472 | Not-determined |
| 17-108_k141_292011 | Not-determined |
| 17-108_k141_306226 | Not-determined |
| 17-108_k141_278668 | Not-determined |
| 17-108_k141_44174 | Low-quality |
| 17-108_k141_292018 | Not-determined |
| 17-108_k141_44182 | Not-determined |
| 17-108_k141_154391 | Not-determined |
| 17-108_k141_292022 | Not-determined |
| 17-108_k141_292023 | Not-determined |
| 17-108_k141_30304 | Low-quality |
| 17-108_k141_319503 | Not-determined |
| 17-108_k141_278678 | Not-determined |
| 17-108_k141_195609 | Not-determined |
| 17-108_k141_16485 | Not-determined |
| 17-108_k141_140621 | Not-determined |
| 17-108_k141_85466 | Not-determined |
| 17-108_k141_195615 | Not-determined |
| 17-108_k141_140628 | Not-determined |
| 17-108_k141_44195 | Not-determined |
| 17-108_k141_44196 | Not-determined |
| 17-108_k141_237500 | Not-determined |
| 17-108_k141_71357 | Not-determined |
| 17-108_k141_237505 | Not-determined |
| 17-108_k141_140638 | Not-determined |
| 17-108_k141_140644 | Not-determined |
| 17-108_k141_181668 | Not-determined |
| 17-108_k141_292048 | Not-determined |
| 17-108_k141_85479 | Not-determined |
| 17-108_k141_223413 | Not-determined |
| 17-108_k141_168521 | Not-determined |
| 17-108_k141_58242 | Not-determined |
| 17-108_k141_250915 | Not-determined |
| 17-108_k141_250916 | Not-determined |
| 17-108_k141_99545 | Not-determined |
| 17-108_k141_168522 | Not-determined |
| 17-108_k141_195622 | Not-determined |
| 17-108_k141_223425 | Not-determined |
| 17-108_k141_278708 | Not-determined |
| 17-108_k141_195630 | Low-quality |
| 17-108_k141_58252 | Not-determined |
| 17-108_k141_58256 | Low-quality |
| 17-108_k141_30337 | Not-determined |
| 17-108_k141_168543 | Not-determined |
| 17-108_k141_71382 | Low-quality |
| 17-108_k141_71383 | Not-determined |
| 17-108_k141_181700 | Not-determined |

| | |
|---|---|
| 17-108_k141_250932 | Not-determined |
| 17-108_k141_30346 | Low-quality |
| 17-108_k141_237541 | Not-determined |
| 17-108_k141_154447 | Not-determined |
| 17-108_k141_154450 | Not-determined |
| 17-108_k141_306286 | Low-quality |
| 17-108_k141_16505 | Not-determined |
| 17-108_k141_278737 | Not-determined |
| 17-108_k141_154455 | Not-determined |
| 17-108_k141_181705 | Low-quality |
| 17-108_k141_16517 | Not-determined |
| 17-108_k141_181710 | Not-determined |
| 17-108_k141_278741 | Not-determined |
| 17-108_k141_264629 | Not-determined |
| 17-108_k141_306292 | Not-determined |
| 17-108_k141_112724 | Not-determined |
| 17-108_k141_99552 | Not-determined |
| 17-108_k141_154471 | Not-determined |
| 17-108_k141_44247 | Not-determined |
| 17-108_k141_85518 | Not-determined |
| 17-108_k141_58270 | Not-determined |
| 17-108_k141_58272 | Not-determined |
| 17-108_k141_168572 | Not-determined |
| 17-108_k141_223446 | Not-determined |
| 17-108_k141_30366 | Not-determined |
| 17-108_k141_278747 | Not-determined |
| 17-108_k141_112729 | Not-determined |
| 17-108_k141_112730 | Not-determined |
| 17-108_k141_306304 | Not-determined |
| 17-108_k141_16534 | Not-determined |
| 17-108_k141_30376 | Not-determined |
| 17-108_k141_168593 | Not-determined |
| 17-108_k141_168594 | Not-determined |
| 17-108_k141_30380 | Not-determined |
| 17-108_k141_44260 | Not-determined |
| 17-108_k141_181751 | Not-determined |
| 17-108_k141_223450 | Not-determined |
| 17-108_k141_44268 | Not-determined |
| 17-108_k141_168612 | Not-determined |
| 17-108_k141_99563 | Not-determined |
| 17-108_k141_306326 | Not-determined |
| 17-108_k141_44270 | Not-determined |
| 17-108_k141_16543 | Low-quality |
| 17-108_k141_181762 | Low-quality |
| 17-108_k141_237571 | Not-determined |
| 17-108_k141_181764 | Low-quality |
| 17-108_k141_181767 | Not-determined |
| 17-108_k141_223465 | Not-determined |
| 17-108_k141_16550 | Not-determined |
| 17-108_k141_210013 | Low-quality |
| 17-108_k141_168615 | Not-determined |
| 17-108_k141_16555 | Not-determined |
| 17-108_k141_306340 | Not-determined |
| 17-108_k141_306343 | Not-determined |
| 17-108_k141_195677 | Not-determined |
| 17-108_k141_99589 | Low-quality |
| 17-108_k141_58324 | Not-determined |
| 17-108_k141_195689 | Not-determined |
| 17-108_k141_30398 | Not-determined |
| 17-108_k141_154490 | Not-determined |
| 17-108_k141_99593 | Not-determined |
| 17-108_k141_210028 | Not-determined |
| 17-108_k141_16578 | Not-determined |
| 17-108_k141_58333 | Not-determined |
| 17-108_k141_3023 | Low-quality |
| 17-108_k141_140663 | Not-determined |
| 17-108_k141_16586 | Not-determined |
| 17-108_k141_99600 | Not-determined |
| 17-108_k141_16588 | Not-determined |
| 17-108_k141_16591 | Not-determined |
| 17-108_k141_140666 | Not-determined |
| 17-108_k141_126879 | Not-determined |
| 17-108_k141_168636 | Not-determined |
| 17-108_k141_140675 | Low-quality |
| 17-108_k141_3034 | Not-determined |
| 17-108_k141_126891 | Not-determined |
| 17-108_k141_319535 | Not-determined |
| 17-108_k141_99602 | Not-determined |
| 17-108_k141_319543 | Low-quality |
| 17-108_k141_168651 | Not-determined |
| 17-108_k141_195699 | Not-determined |
| 17-108_k141_195700 | Not-determined |
| 17-108_k141_264682 | Not-determined |
| 17-108_k141_278812 | Not-determined |
| 17-108_k141_195709 | Not-determined |
| 17-108_k141_58355 | Not-determined |
| 17-108_k141_99622 | Not-determined |
| 17-108_k141_99628 | Not-determined |
| 17-108_k141_99629 | Not-determined |
| 17-108_k141_210060 | Not-determined |
| 17-108_k141_30458 | Low-quality |
| 17-108_k141_306397 | Not-determined |
| 17-108_k141_140695 | Low-quality |
| 17-108_k141_195720 | Not-determined |
| 17-108_k141_154513 | Not-determined |
| 17-108_k141_168679 | Not-determined |
| 17-108_k141_140708 | Not-determined |
| 17-108_k141_181812 | Not-determined |
| 17-108_k141_154524 | Not-determined |
| 17-108_k141_58384 | Not-determined |
| 17-108_k141_181814 | Low-quality |
| 17-108_k141_278836 | Not-determined |
| 17-108_k141_237646 | Not-determined |
| 17-108_k141_278841 | Not-determined |
| 17-108_k141_250961 | Low-quality |
| 17-108_k141_250963 | Not-determined |
| 17-108_k141_210083 | Not-determined |
| 17-108_k141_126933 | Not-determined |
| 17-108_k141_112759 | Not-determined |
| 17-108_k141_112763 | Not-determined |
| 17-108_k141_58390 | Not-determined |
| 17-108_k141_264700 | Not-determined |
| 17-108_k141_278850 | Not-determined |
| 17-108_k141_210096 | Not-determined |
| 17-108_k141_99652 | Not-determined |
| 17-108_k141_278858 | Not-determined |
| 17-108_k141_112770 | Not-determined |
| 17-108_k141_264706 | Not-determined |
| 17-108_k141_237672 | Low-quality |
| 17-108_k141_112772 | Not-determined |
| 17-108_k141_154547 | Not-determined |
| 17-108_k141_181844 | Not-determined |
| 17-108_k141_154552 | Not-determined |
| 17-108_k141_126960 | Not-determined |
| 17-108_k141_112786 | Not-determined |
| 17-108_k141_319575 | Low-quality |
| 17-108_k141_264734 | Not-determined |
| 17-108_k141_71489 | Low-quality |
| 17-108_k141_112805 | Not-determined |
| 17-108_k141_99683 | Not-determined |
| 17-108_k141_278866 | Low-quality |
| 17-108_k141_264756 | Not-determined |
| 17-108_k141_223537 | Not-determined |
| 17-108_k141_16616 | Not-determined |
| 17-108_k141_126980 | Not-determined |
| 17-108_k141_58401 | Not-determined |
| 17-108_k141_71506 | Not-determined |
| 17-108_k141_292056 | Not-determined |
| 17-108_k141_3075 | Not-determined |
| 17-108_k141_71512 | Not-determined |
| 17-108_k141_292070 | Not-determined |
| 17-108_k141_112808 | Not-determined |
| 17-108_k141_44289 | Not-determined |
| 17-108_k141_99715 | Not-determined |
| 17-108_k141_264762 | Not-determined |
| 17-108_k141_181876 | Not-determined |
| 17-108_k141_181879 | Not-determined |
| 17-108_k141_112816 | Low-quality |
| 17-108_k141_99720 | Not-determined |
| 17-108_k141_44296 | Not-determined |
| 17-108_k141_168689 | Not-determined |
| 17-108_k141_58419 | Not-determined |
| 17-108_k141_292081 | Not-determined |
| 17-108_k141_264772 | Not-determined |
| 17-108_k141_112832 | Not-determined |
| 17-108_k141_306466 | Not-determined |
| 17-108_k141_58431 | Not-determined |
| 17-108_k141_127012 | Not-determined |
| 17-108_k141_140717 | Not-determined |
| 17-108_k141_16622 | Not-determined |
| 17-108_k141_264787 | Not-determined |
| 17-108_k141_140723 | Not-determined |
| 17-108_k141_306479 | Not-determined |
| 17-108_k141_16629 | Not-determined |
| 17-108_k141_99728 | Not-determined |
| 17-108_k141_154604 | Not-determined |
| 17-108_k141_127030 | Not-determined |
| 17-108_k141_58437 | Not-determined |
| 17-108_k141_264798 | Not-determined |
| 17-108_k141_168694 | Not-determined |
| 17-108_k141_223566 | Not-determined |
| 17-108_k141_154619 | Not-determined |
| 17-108_k141_140745 | Low-quality |
| 17-108_k141_58442 | Not-determined |
| 17-108_k141_99747 | Low-quality |
| 17-108_k141_154628 | Not-determined |
| 17-108_k141_278925 | Not-determined |
| 17-108_k141_30484 | Not-determined |
| 17-108_k141_140759 | Not-determined |
| 17-108_k141_16654 | Not-determined |
| 17-108_k141_264826 | Not-determined |
| 17-108_k141_71545 | Not-determined |
| 17-108_k141_112858 | Not-determined |
| 17-108_k141_44312 | Not-determined |
| 17-108_k141_154640 | Not-determined |
| 17-108_k141_71551 | Low-quality |
| 17-108_k141_16664 | Not-determined |
| 17-108_k141_16665 | Not-determined |
| 17-108_k141_223580 | Not-determined |
| 17-108_k141_223583 | Not-determined |
| 17-108_k141_85587 | Not-determined |
| 17-108_k141_223587 | Not-determined |
| 17-108_k141_292116 | Not-determined |
| 17-108_k141_264841 | Not-determined |
| 17-108_k141_292119 | Not-determined |
| 17-108_k141_30523 | Not-determined |
| 17-108_k141_264845 | Not-determined |
| 17-108_k141_195761 | Low-quality |
| 17-108_k141_264854 | Not-determined |
| 17-108_k141_278950 | Not-determined |
| 17-108_k141_58474 | Not-determined |
| 17-108_k141_140791 | Not-determined |
| 17-108_k141_264864 | Not-determined |
| 17-108_k141_251047 | Not-determined |
| 17-108_k141_85607 | Not-determined |
| 17-108_k141_264872 | Not-determined |
| 17-108_k141_195780 | Not-determined |
| 17-108_k141_168730 | Not-determined |
| 17-108_k141_58482 | Not-determined |
| 17-108_k141_264880 | Low-quality |
| 17-108_k141_85615 | Low-quality |
| 17-108_k141_237696 | Not-determined |
| 17-108_k141_154687 | Not-determined |
| 17-108_k141_168735 | Not-determined |
| 17-108_k141_154690 | Not-determined |
| 17-108_k141_181971 | Not-determined |
| 17-108_k141_319597 | Not-determined |
| 17-108_k141_181977 | Not-determined |
| 17-108_k141_319598 | Not-determined |
| 17-108_k141_237706 | Not-determined |
| 17-108_k141_278964 | Not-determined |
| 17-108_k141_16691 | Not-determined |
| 17-108_k141_237711 | Not-determined |
| 17-108_k141_210130 | Not-determined |
| 17-108_k141_210135 | Not-determined |
| 17-108_k141_3113 | Not-determined |
| 17-108_k141_140814 | Not-determined |
| 17-108_k141_292167 | Low-quality |
| 17-108_k141_16707 | Not-determined |
| 17-108_k141_30556 | Low-quality |
| 17-108_k141_319619 | Not-determined |
| 17-108_k141_182007 | Not-determined |
| 17-108_k141_154721 | Not-determined |
| 17-108_k141_182013 | Not-determined |
| 17-108_k141_319633 | Not-determined |
| 17-108_k141_58502 | Not-determined |
| 17-108_k141_112961 | Not-determined |
| 17-108_k141_127043 | Not-determined |
| 17-108_k141_154736 | Not-determined |
| 17-108_k141_306556 | Not-determined |
| 17-108_k141_292185 | Not-determined |
| 17-108_k141_112967 | Not-determined |
| 17-108_k141_30586 | Not-determined |
| 17-108_k141_182033 | Not-determined |
| 17-108_k141_182037 | Not-determined |
| 17-108_k141_292194 | Not-determined |
| 17-108_k141_237736 | Not-determined |
| 17-108_k141_112976 | Not-determined |
| 17-108_k141_58517 | Not-determined |
| 17-108_k141_16742 | Not-determined |
| 17-108_k141_112985 | Not-determined |
| 17-108_k141_154748 | Not-determined |
| 17-108_k141_112987 | Not-determined |
| 17-108_k141_112989 | Not-determined |
| 17-108_k141_195808 | Not-determined |
| 17-108_k141_210187 | Not-determined |
| 17-108_k141_85658 | Not-determined |
| 17-108_k141_306576 | Not-determined |
| 17-108_k141_99767 | Not-determined |
| 17-108_k141_154766 | Not-determined |
| 18-102-final-viral-combined_k141_6605\|\|full | High-quality |
| 17-108_k141_306582 | Not-determined |
| 17-108_k141_306586 | Not-determined |
| 17-108_k141_3123 | Not-determined |
| 17-108_k141_30609 | Not-determined |
| 17-108_k141_195824 | Not-determined |
| 17-108_k141_127061 | Not-determined |
| 17-108_k141_306590 | Low-quality |
| 17-108_k141_195828 | Not-determined |
| 17-108_k141_292208 | Not-determined |
| 17-108_k141_182060 | Not-determined |
| 17-108_k141_251085 | Not-determined |
| 17-108_k141_251088 | Not-determined |
| 17-108_k141_16777 | Not-determined |
| 17-108_k141_278976 | Low-quality |
| 17-108_k141_278979 | Not-determined |
| 17-108_k141_278980 | Not-determined |
| 17-108_k141_127067 | Not-determined |
| 17-108_k141_71566 | Not-determined |
| 17-108_k141_182070 | Not-determined |
| 17-108_k141_182077 | Low-quality |
| 17-108_k141_16791 | Not-determined |
| 17-108_k141_182086 | Not-determined |
| 17-108_k141_154788 | Not-determined |
| 17-108_k141_251105 | Not-determined |
| 17-108_k141_237780 | Not-determined |
| 17-108_k141_251106 | Not-determined |
| 17-108_k141_264938 | Not-determined |
| 17-108_k141_292227 | Not-determined |
| 17-108_k141_237784 | Not-determined |
| 17-108_k141_168786 | Not-determined |
| 17-108_k141_154792 | Not-determined |
| 17-108_k141_99803 | Not-determined |
| 17-108_k141_127089 | Not-determined |
| 17-108_k141_182093 | Not-determined |
| 17-108_k141_3150 | Not-determined |
| 17-108_k141_127109 | Not-determined |
| 17-108_k141_3154 | Not-determined |
| 17-108_k141_44351 | Not-determined |
| 17-108_k141_182103 | Not-determined |
| 17-108_k141_113019 | Not-determined |
| 17-108_k141_140871 | Not-determined |
| 17-108_k141_30635 | Not-determined |
| 17-108_k141_99828 | Not-determined |
| 17-108_k141_251129 | Not-determined |
| 17-108_k141_58544 | Not-determined |
| 17-108_k141_251137 | Not-determined |
| 17-108_k141_85708 | Not-determined |
| 17-108_k141_279020 | Not-determined |
| 17-108_k141_264958 | Not-determined |
| 17-108_k141_223700 | Not-determined |
| 17-108_k141_279026 | Not-determined |
| 17-108_k141_306644 | Not-determined |
| 17-108_k141_251140 | Not-determined |
| 17-108_k141_16825 | Not-determined |
| 17-108_k141_30653 | Not-determined |
| 17-108_k141_44390 | Not-determined |
| 17-108_k141_182131 | Not-determined |
| 17-108_k141_319686 | Not-determined |
| 17-108_k141_279054 | Not-determined |
| 17-108_k141_127124 | Not-determined |
| 17-108_k141_195886 | Not-determined |
| 17-108_k141_168868 | Not-determined |
| 17-108_k141_44414 | Not-determined |
| 17-108_k141_44415 | Not-determined |
| 17-108_k141_319701 | Not-determined |
| 17-108_k141_99849 | Not-determined |
| 17-108_k141_127146 | Not-determined |
| 17-108_k141_182163 | Not-determined |
| 17-108_k141_237833 | Not-determined |
| 17-108_k141_319710 | Not-determined |
| 17-108_k141_127152 | Not-determined |
| 17-108_k141_195898 | Not-determined |
| 17-108_k141_195899 | Not-determined |
| 17-108_k141_58560 | Not-determined |
| 17-108_k141_264996 | Not-determined |
| 17-108_k141_71633 | Not-determined |
| 17-108_k141_71634 | Not-determined |
| 17-108_k141_71636 | Not-determined |
| 17-108_k141_113038 | Low-quality |
| 17-108_k141_279070 | Not-determined |
| 17-108_k141_195906 | Low-quality |
| 17-108_k141_223747 | Not-determined |
| 17-108_k141_127165 | Not-determined |
| 17-108_k141_223748 | Not-determined |
| 17-108_k141_30705 | Not-determined |
| 17-108_k141_182183 | Not-determined |
| 17-108_k141_30707 | Not-determined |
| 17-108_k141_30708 | Not-determined |
| 17-108_k141_71644 | Not-determined |
| 17-108_k141_30709 | Not-determined |
| 17-108_k141_168891 | Not-determined |
| 17-108_k141_99865 | Not-determined |
| 17-108_k141_292251 | Low-quality |
| 17-108_k141_71652 | Not-determined |
| 17-108_k141_99870 | Not-determined |
| 17-108_k141_237840 | Not-determined |
| 17-108_k141_223762 | Not-determined |
| 17-108_k141_319737 | Not-determined |
| 17-108_k141_223765 | Low-quality |
| 17-108_k141_44428 | Not-determined |
| 17-108_k141_195922 | Low-quality |
| 17-108_k141_319742 | Not-determined |
| 17-108_k141_44433 | Not-determined |
| 17-108_k141_30732 | Low-quality |
| 17-108_k141_113051 | Not-determined |
| 17-108_k141_71657 | Not-determined |
| 17-108_k141_3194 | Not-determined |
| 17-108_k141_30742 | Not-determined |
| 17-108_k141_16881 | Not-determined |
| 17-108_k141_127177 | Not-determined |
| 17-108_k141_306686 | Not-determined |
| 17-108_k141_127183 | Not-determined |
| 17-108_k141_306689 | Not-determined |
| 17-108_k141_319761 | Not-determined |
| 17-108_k141_319763 | Low-quality |
| 17-108_k141_85772 | Not-determined |
| 17-108_k141_306696 | Not-determined |
| 17-108_k141_16904 | Not-determined |
| 17-108_k141_127188 | Not-determined |
| 17-108_k141_44472 | Not-determined |
| 17-108_k141_85777 | Not-determined |
| 17-108_k141_279117 | Not-determined |
| 17-108_k141_85782 | Not-determined |
| 17-108_k141_223800 | Not-determined |
| 17-108_k141_85786 | Not-determined |
| 17-108_k141_279124 | Not-determined |
| 17-108_k141_71663 | Not-determined |
| 17-108_k141_223806 | Not-determined |
| 17-108_k141_127198 | Not-determined |
| 17-108_k141_195975 | Low-quality |
| 17-108_k141_279137 | Not-determined |
| 17-108_k141_182201 | Not-determined |
| 17-108_k141_182203 | Low-quality |
| 17-108_k141_85802 | Not-determined |
| 17-108_k141_306728 | Not-determined |
| 17-108_k141_127207 | Not-determined |
| 17-108_k141_99933 | Not-determined |
| 17-108_k141_279148 | Not-determined |
| 17-108_k141_99937 | Not-determined |
| 17-108_k141_195988 | Not-determined |
| 17-108_k141_3241 | Not-determined |
| 17-108_k141_195993 | Low-quality |
| 17-108_k141_140905 | Not-determined |
| 17-108_k141_168936 | Not-determined |
| 17-108_k141_99950 | Low-quality |
| 17-108_k141_140917 | Not-determined |
| 17-108_k141_3253 | Not-determined |
| 17-108_k141_58605 | Not-determined |
| 17-108_k141_3257 | Not-determined |
| 17-108_k141_251148 | Not-determined |
| 17-108_k141_44489 | Not-determined |
| 17-108_k141_140926 | Low-quality |
| 17-108_k141_265027 | Low-quality |
| 17-108_k141_99954 | Not-determined |
| 17-108_k141_30780 | Not-determined |
| 17-108_k141_251168 | Not-determined |
| 17-108_k141_30785 | Not-determined |
| 17-108_k141_140932 | Not-determined |
| 17-108_k141_99965 | Not-determined |
| 17-108_k141_251175 | Low-quality |
| 17-108_k141_251176 | Not-determined |
| 17-108_k141_265035 | Not-determined |
| 17-108_k141_251182 | Not-determined |
| 17-108_k141_99972 | Not-determined |
| 17-108_k141_99974 | Not-determined |
| 17-108_k141_58614 | Not-determined |
| 17-108_k141_292328 | Not-determined |
| 17-108_k141_30790 | Not-determined |
| 17-108_k141_251186 | Not-determined |
| 17-108_k141_279175 | Not-determined |
| 17-108_k141_127258 | Not-determined |
| 17-108_k141_30796 | Not-determined |
| 17-108_k141_196004 | Not-determined |
| 17-108_k141_16925 | Not-determined |
| 17-108_k141_154844 | Not-determined |
| 17-108_k141_210199 | Not-determined |
| 17-108_k141_85851 | Not-determined |
| 17-108_k141_319826 | Not-determined |
| 17-108_k141_99984 | Not-determined |
| 17-108_k141_30806 | Not-determined |
| 17-108_k141_99990 | Not-determined |
| 17-108_k141_223825 | Not-determined |
| 17-108_k141_223827 | Low-quality |
| 17-108_k141_265067 | Not-determined |
| 17-108_k141_279202 | Not-determined |
| 17-108_k141_237913 | Not-determined |
| 17-108_k141_182219 | Low-quality |
| 17-108_k141_210218 | Not-determined |
| 17-108_k141_251212 | Not-determined |
| 17-108_k141_223846 | Not-determined |
| 17-108_k141_140971 | Not-determined |
| 17-108_k141_85882 | Low-quality |
| 17-108_k141_58634 | Not-determined |
| 17-108_k141_223848 | Not-determined |
| 17-108_k141_237921 | Low-quality |
| 17-108_k141_237924 | Not-determined |
| 17-108_k141_85892 | Not-determined |
| 17-108_k141_58639 | Not-determined |
| 17-108_k141_237928 | Not-determined |
| 17-108_k141_140981 | Not-determined |
| 17-108_k141_182234 | Not-determined |
| 17-108_k141_251229 | Not-determined |
| 17-108_k141_85898 | Not-determined |
| 17-108_k141_251232 | Low-quality |
| 17-108_k141_265092 | Not-determined |
| 17-108_k141_168978 | Low-quality |
| 17-108_k141_306782 | Not-determined |
| 17-108_k141_71740 | Low-quality |
| 17-108_k141_279217 | Not-determined |
| 17-108_k141_306787 | Not-determined |
| 17-108_k141_168991 | Not-determined |
| 17-108_k141_210222 | Not-determined |
| 17-108_k141_85907 | Not-determined |
| 17-108_k141_251248 | Not-determined |
| 17-108_k141_223862 | Not-determined |
| 17-108_k141_182240 | Not-determined |
| 17-108_k141_16929 | Not-determined |
| 17-108_k141_154880 | Not-determined |
| 17-108_k141_306796 | Not-determined |
| 17-108_k141_71754 | Not-determined |
| 17-108_k141_265104 | Not-determined |
| 17-108_k141_210236 | Not-determined |
| 17-108_k141_265110 | Not-determined |
| 17-108_k141_210238 | Not-determined |
| 17-108_k141_265111 | Not-determined |
| 17-108_k141_182252 | Not-determined |
| 17-108_k141_251258 | Not-determined |
| 17-108_k141_319870 | Not-determined |
| 17-108_k141_210243 | Not-determined |
| 17-108_k141_251265 | Not-determined |
| 17-108_k141_210246 | Not-determined |
| 17-108_k141_85925 | Not-determined |
| 17-108_k141_265118 | Not-determined |
| 17-108_k141_196041 | Not-determined |
| 17-108_k141_85929 | Not-determined |
| 17-108_k141_154887 | Not-determined |
| 17-108_k141_154890 | Not-determined |
| 17-108_k141_306810 | Not-determined |
| 17-108_k141_223884 | Not-determined |
| 17-108_k141_44540 | Not-determined |
| 17-108_k141_71774 | Low-quality |
| 17-108_k141_169014 | Not-determined |
| 17-108_k141_85941 | Low-quality |
| 17-108_k141_141029 | Not-determined |
| 17-108_k141_279250 | Not-determined |
| 17-108_k141_251288 | Not-determined |
| 17-108_k141_223902 | Not-determined |
| 17-108_k141_196059 | Low-quality |
| 17-108_k141_223904 | Not-determined |
| 17-108_k141_3301 | Not-determined |
| 17-108_k141_44558 | Low-quality |
| 17-108_k141_16953 | Not-determined |
| 17-108_k141_44563 | Not-determined |
| 17-108_k141_44565 | Not-determined |
| 17-108_k141_306836 | Not-determined |
| 17-108_k141_44574 | Not-determined |
| 17-108_k141_306839 | Not-determined |
| 17-108_k141_306844 | Low-quality |
| 17-108_k141_44581 | Not-determined |
| 17-108_k141_44582 | Not-determined |
| 17-108_k141_154913 | Not-determined |
| 17-108_k141_100052 | Low-quality |
| 17-108_k141_113085 | Not-determined |
| 17-108_k141_223914 | Not-determined |
| 17-108_k141_71796 | Not-determined |
| 17-108_k141_154916 | Not-determined |
| 17-108_k141_44584 | Not-determined |
| 17-108_k141_210263 | Not-determined |
| 17-108_k141_44589 | Not-determined |
| 17-108_k141_237973 | Not-determined |
| 17-108_k141_169040 | Not-determined |
| 17-108_k141_44595 | Not-determined |
| 17-108_k141_85980 | Not-determined |
| 17-108_k141_154925 | Not-determined |
| 17-108_k141_100062 | Not-determined |
| 17-108_k141_223919 | Not-determined |
| 17-108_k141_169042 | Not-determined |
| 17-108_k141_237985 | Low-quality |
| 17-108_k141_113100 | Not-determined |
| 17-108_k141_306868 | Not-determined |
| 17-108_k141_113105 | Not-determined |
| 17-108_k141_251323 | Not-determined |
| 17-108_k141_182326 | Not-determined |
| 17-108_k141_3309 | Not-determined |
| 17-108_k141_169056 | Not-determined |
| 17-108_k141_237992 | Not-determined |
| 17-108_k141_169061 | Not-determined |
| 17-108_k141_182339 | Not-determined |
| 17-108_k141_292363 | Low-quality |
| 17-108_k141_210298 | Not-determined |
| 17-108_k141_154937 | Not-determined |
| 17-108_k141_100080 | Not-determined |
| 17-108_k141_154938 | Not-determined |
| 17-108_k141_44616 | Not-determined |
| 17-108_k141_127280 | Not-determined |
| 17-108_k141_210307 | Not-determined |
| 17-108_k141_71828 | Not-determined |
| 17-108_k141_210311 | Not-determined |
| 17-108_k141_3327 | Not-determined |
| 17-108_k141_44625 | Not-determined |
| 17-108_k141_113114 | Not-determined |
| 17-108_k141_238004 | Not-determined |
| 17-108_k141_44628 | Low-quality |
| 17-108_k141_71835 | Not-determined |
| 17-108_k141_71839 | Not-determined |
| 17-108_k141_141063 | Not-determined |
| 17-108_k141_44632 | Not-determined |
| 17-108_k141_223945 | Not-determined |
| 17-108_k141_85996 | Not-determined |
| 17-108_k141_71842 | Not-determined |
| 17-108_k141_279306 | Not-determined |
| 17-108_k141_319917 | Not-determined |
| 17-108_k141_44638 | Not-determined |
| 17-108_k141_265151 | Not-determined |
| 17-108_k141_71846 | Not-determined |
| 17-108_k141_182371 | Not-determined |
| 17-108_k141_154960 | Not-determined |
| 17-108_k141_17009 | Not-determined |
| 17-108_k141_279313 | Not-determined |
| 17-108_k141_279316 | Not-determined |
| 17-108_k141_319935 | Not-determined |
| 17-108_k141_85999 | Low-quality |
| 17-108_k141_17024 | Not-determined |
| 17-108_k141_223958 | Not-determined |
| 17-108_k141_182386 | Not-determined |
| 17-108_k141_30886 | Not-determined |
| 17-108_k141_265167 | Not-determined |
| 17-108_k141_265168 | Not-determined |

| | |
|---|---|
| 17-108_k141_44666 | Not-determined |
| 17-108_k141_319945 | Not-determined |
| 17-108_k141_319952 | Not-determined |
| 17-108_k141_30890 | Not-determined |
| 17-108_k141_182400 | Not-determined |
| 17-108_k141_169113 | Not-determined |
| 17-108_k141_182401 | Not-determined |
| 17-108_k141_279339 | Not-determined |
| 17-108_k141_141104 | Not-determined |
| 17-108_k141_141122 | Not-determined |
| 17-108_k141_306871 | Low-quality |
| 17-108_k141_279354 | Not-determined |
| 17-108_k141_113146 | Not-determined |
| 17-108_k141_319974 | Not-determined |
| 17-108_k141_238011 | Not-determined |
| 17-108_k141_306877 | Not-determined |
| 17-108_k141_210341 | Low-quality |
| 17-108_k141_210343 | Not-determined |
| 17-108_k141_292405 | Not-determined |
| 17-108_k141_127311 | Not-determined |
| 17-108_k141_210345 | Not-determined |
| 17-108_k141_238019 | Not-determined |
| 17-108_k141_306884 | Not-determined |
| 17-108_k141_292415 | Not-determined |
| 17-108_k141_319985 | Not-determined |
| 17-108_k141_251335 | Not-determined |
| 17-108_k141_306891 | Not-determined |
| 17-108_k141_182426 | Not-determined |
| 17-108_k141_319990 | Not-determined |
| 17-108_k141_251342 | Not-determined |
| 17-108_k141_306897 | Not-determined |
| 17-108_k141_86061 | Not-determined |
| 17-108_k141_127318 | Not-determined |
| 17-108_k141_30935 | Not-determined |
| 17-108_k141_154993 | Not-determined |
| 17-108_k141_306903 | Not-determined |
| 17-108_k141_154994 | Not-determined |
| 17-108_k141_86070 | Not-determined |
| 17-108_k141_251362 | Not-determined |
| 17-108_k141_306910 | Low-quality |
| 17-108_k141_279368 | Not-determined |
| 17-108_k141_100150 | Not-determined |
| 17-108_k141_292421 | Not-determined |
| 17-108_k141_44707 | Not-determined |
| 17-108_k141_113178 | Not-determined |
| 17-108_k141_58690 | Not-determined |
| 17-108_k141_292426 | Not-determined |
| 17-108_k141_292430 | Not-determined |
| 17-108_k141_58696 | Not-determined |
| 17-108_k141_182451 | Low-quality |
| 17-108_k141_306924 | Not-determined |
| 17-108_k141_58703 | Not-determined |
| 17-108_k141_3422 | Not-determined |
| 17-108_k141_3423 | Not-determined |
| 17-108_k141_320019 | Not-determined |
| 17-108_k141_223990 | Not-determined |
| 17-108_k141_17039 | Not-determined |
| 17-108_k141_17042 | Not-determined |
| 17-108_k141_223995 | Not-determined |
| 17-108_k141_223998 | Not-determined |
| 17-108_k141_210374 | Not-determined |
| 17-108_k141_306949 | Not-determined |
| 17-108_k141_17046 | Not-determined |
| 17-108_k141_100180 | Not-determined |
| 17-108_k141_100181 | Low-quality |
| 17-108_k141_17048 | Not-determined |
| 17-108_k141_113200 | Not-determined |
| 17-108_k141_100184 | Not-determined |
| 17-108_k141_251395 | Low-quality |
| 17-108_k141_279389 | Not-determined |
| 17-108_k141_279390 | Not-determined |
| 17-108_k141_58724 | Not-determined |
| 17-108_k141_169167 | Not-determined |
| 17-108_k141_169173 | Not-determined |
| 17-108_k141_292449 | Low-quality |
| 17-108_k141_58728 | Not-determined |
| 17-108_k141_306968 | Low-quality |
| 17-108_k141_238086 | Not-determined |
| 17-108_k141_279402 | Not-determined |
| 17-108_k141_292450 | Not-determined |
| 17-108_k141_238087 | Not-determined |
| 17-108_k141_44736 | Not-determined |
| 17-108_k141_265209 | Not-determined |
| 17-108_k141_58740 | Not-determined |
| 17-108_k141_265211 | Not-determined |
| 17-108_k141_86098 | Not-determined |
| 17-108_k141_224020 | Not-determined |
| 17-108_k141_265212 | Not-determined |
| 17-108_k141_58743 | Not-determined |
| 17-108_k141_182483 | Not-determined |
| 17-108_k141_44747 | Not-determined |
| 17-108_k141_224025 | Not-determined |
| 17-108_k141_44749 | Low-quality |
| 17-108_k141_292466 | Not-determined |
| 17-108_k141_292469 | Not-determined |
| 17-108_k141_265221 | Not-determined |
| 17-108_k141_86123 | Not-determined |
| 17-108_k141_155073 | Not-determined |
| 17-108_k141_320060 | Not-determined |
| 17-108_k141_210386 | Low-quality |
| 17-108_k141_58768 | Not-determined |
| 17-108_k141_3448 | Not-determined |
| 17-108_k141_169222 | Not-determined |
| 17-108_k141_320067 | Low-quality |
| 17-108_k141_251411 | Not-determined |
| 17-108_k141_169229 | Not-determined |
| 17-108_k141_155087 | Not-determined |
| 17-108_k141_169230 | Not-determined |
| 17-108_k141_169233 | Not-determined |
| 17-108_k141_210394 | Not-determined |
| 17-108_k141_44752 | Not-determined |
| 17-108_k141_292488 | Not-determined |
| 17-108_k141_320081 | Not-determined |
| 17-108_k141_320082 | Not-determined |
| 17-108_k141_169241 | Not-determined |
| 17-108_k141_224071 | Not-determined |
| 17-108_k141_127377 | Low-quality |
| 17-108_k141_86151 | Not-determined |
| 17-108_k141_113229 | Not-determined |
| 17-108_k141_320089 | Not-determined |
| 17-108_k141_155111 | Not-determined |
| 17-108_k141_169250 | Not-determined |
| 17-108_k141_17057 | Not-determined |
| 17-108_k141_44785 | Not-determined |
| 17-108_k141_113237 | Not-determined |
| 17-108_k141_71874 | Not-determined |
| 17-108_k141_100242 | Not-determined |
| 17-108_k141_58803 | Not-determined |
| 17-108_k141_182536 | Not-determined |
| 17-108_k141_86168 | Not-determined |
| 17-108_k141_58805 | Low-quality |
| 17-108_k141_3456 | Not-determined |
| 17-108_k141_44796 | Not-determined |
| 17-108_k141_169267 | Not-determined |
| 17-108_k141_44808 | Not-determined |
| 17-108_k141_210432 | Not-determined |
| 17-108_k141_292520 | Not-determined |
| 17-108_k141_224084 | Not-determined |
| 17-108_k141_292524 | Not-determined |
| 17-108_k141_113248 | Not-determined |
| 17-108_k141_279408 | Not-determined |
| 17-108_k141_169283 | Not-determined |
| 17-108_k141_292527 | Not-determined |
| 17-108_k141_30972 | Not-determined |
| 17-108_k141_224093 | Not-determined |
| 17-108_k141_3479 | Not-determined |
| 17-108_k141_265268 | Low-quality |
| 17-108_k141_44825 | Not-determined |
| 17-108_k141_86197 | Not-determined |
| 17-108_k141_86198 | Not-determined |
| 17-108_k141_86199 | Not-determined |
| 17-108_k141_100262 | Not-determined |
| 17-108_k141_30985 | Not-determined |
| 17-108_k141_86211 | Not-determined |
| 17-108_k141_58833 | Not-determined |
| 17-108_k141_306980 | Not-determined |
| 17-108_k141_58842 | Not-determined |
| 17-108_k141_182576 | Not-determined |
| 17-108_k141_58846 | Not-determined |
| 17-108_k141_30995 | Not-determined |
| 17-108_k141_210471 | Not-determined |
| 17-108_k141_58853 | Not-determined |
| 17-108_k141_71901 | Not-determined |
| 17-108_k141_292559 | Not-determined |
| 17-108_k141_279429 | Not-determined |
| 17-108_k141_292564 | Not-determined |
| 17-108_k141_224128 | Not-determined |
| 17-108_k141_58872 | Low-quality |
| 17-108_k141_31015 | Not-determined |
| 17-108_k141_86251 | Low-quality |
| 17-108_k141_292567 | Not-determined |
| 17-108_k141_210482 | Not-determined |
| 17-108_k141_224133 | Not-determined |
| 17-108_k141_113286 | Not-determined |
| 17-108_k141_279432 | Not-determined |
| 17-108_k141_307014 | Not-determined |
| 17-108_k141_279436 | Not-determined |
| 17-108_k141_86259 | Not-determined |
| 17-108_k141_307016 | Not-determined |
| 17-108_k141_224142 | Not-determined |
| 17-108_k141_224143 | Not-determined |
| 17-108_k141_238169 | Not-determined |
| 17-108_k141_127444 | Not-determined |
| 17-108_k141_141219 | Low-quality |
| 17-108_k141_320106 | Not-determined |
| 17-108_k141_279441 | Not-determined |
| 17-108_k141_141238 | Not-determined |
| 17-108_k141_44860 | Not-determined |
| 17-108_k141_292598 | Not-determined |
| 17-108_k141_86269 | Not-determined |
| 17-108_k141_210501 | Not-determined |
| 17-108_k141_224157 | Not-determined |
| 17-108_k141_320128 | Not-determined |
| 17-108_k141_320130 | Not-determined |
| 17-108_k141_292603 | Not-determined |
| 17-108_k141_141253 | Not-determined |
| 17-108_k141_44875 | Not-determined |
| 17-108_k141_86277 | Not-determined |
| 17-108_k141_127483 | Not-determined |
| 17-108_k141_155147 | Low-quality |
| 17-108_k141_238206 | Not-determined |
| 17-108_k141_155154 | Not-determined |
| 17-108_k141_292613 | Not-determined |
| 17-108_k141_182625 | Not-determined |
| 17-108_k141_292614 | Low-quality |
| 17-108_k141_238213 | Not-determined |
| 17-108_k141_210533 | Not-determined |
| 17-108_k141_17129 | Not-determined |
| 17-108_k141_238216 | Not-determined |
| 17-108_k141_279460 | Not-determined |
| 17-108_k141_210543 | Not-determined |
| 17-108_k141_17137 | Not-determined |
| 17-108_k141_71945 | Not-determined |
| 17-108_k141_100346 | Not-determined |
| 17-108_k141_224163 | Not-determined |
| 17-108_k141_31044 | Not-determined |
| 17-108_k141_31051 | Not-determined |
| 17-108_k141_71955 | Not-determined |
| 17-108_k141_44904 | Not-determined |
| 17-108_k141_127497 | Not-determined |
| 17-108_k141_86310 | Not-determined |
| 17-108_k141_224173 | Low-quality |
| 17-108_k141_320150 | Not-determined |
| 17-108_k141_210564 | Not-determined |
| 17-108_k141_44915 | Not-determined |
| 17-108_k141_320159 | Not-determined |
| 17-108_k141_86328 | Not-determined |
| 17-108_k141_127515 | Low-quality |
| 17-108_k141_155212 | Not-determined |
| 17-108_k141_113322 | Not-determined |
| 17-108_k141_113325 | Not-determined |
| 17-108_k141_279492 | Not-determined |
| 17-108_k141_86338 | Not-determined |
| 17-108_k141_44927 | Not-determined |
| 17-108_k141_155215 | Not-determined |
| 17-108_k141_196128 | Not-determined |
| 17-108_k141_44932 | Low-quality |
| 17-108_k141_251442 | Not-determined |
| 17-108_k141_127540 | Not-determined |
| 17-108_k141_196141 | Not-determined |
| 17-108_k141_72009 | Not-determined |
| 17-108_k141_100411 | Low-quality |
| 17-108_k141_100421 | Low-quality |
| 17-108_k141_210606 | Not-determined |
| 17-108_k141_196151 | Not-determined |
| 17-108_k141_210607 | Not-determined |
| 17-108_k141_182679 | Not-determined |
| 17-108_k141_113335 | Low-quality |
| 17-108_k141_196153 | Not-determined |
| 17-108_k141_141303 | Not-determined |
| 17-108_k141_86367 | Not-determined |
| 17-108_k141_127547 | Not-determined |
| 17-108_k141_251457 | Not-determined |
| 17-108_k141_182689 | Not-determined |
| 17-108_k141_141310 | Not-determined |
| 17-108_k141_320210 | Not-determined |
| 17-108_k141_100435 | Not-determined |
| 17-108_k141_127556 | Not-determined |
| 17-108_k141_224225 | Not-determined |
| 17-108_k141_31070 | Low-quality |
| 17-108_k141_251462 | Not-determined |
| 17-108_k141_292723 | Not-determined |
| 17-108_k141_44962 | Not-determined |
| 17-108_k141_182703 | Not-determined |
| 17-108_k141_72021 | Low-quality |
| 17-108_k141_155234 | Not-determined |
| 17-108_k141_100447 | Not-determined |
| 17-108_k141_31076 | Not-determined |
| 17-108_k141_44971 | Low-quality |
| 17-108_k141_86394 | Not-determined |
| 17-108_k141_72031 | Not-determined |
| 17-108_k141_17151 | Not-determined |
| 17-108_k141_44975 | Not-determined |
| 17-108_k141_307074 | Not-determined |
| 17-108_k141_17155 | Low-quality |
| 17-108_k141_58900 | Not-determined |
| 17-108_k141_196186 | Not-determined |
| 17-108_k141_320229 | Not-determined |
| 17-108_k141_182722 | Not-determined |
| 17-108_k141_292734 | Not-determined |
| 17-108_k141_210649 | Not-determined |
| 17-108_k141_292743 | Not-determined |
| 17-108_k141_17175 | Not-determined |
| 17-108_k141_17176 | Not-determined |
| 17-108_k141_58922 | Not-determined |
| 17-108_k141_86423 | Not-determined |
| 17-108_k141_3573 | Not-determined |
| 17-108_k141_3574 | Not-determined |
| 17-108_k141_31107 | Not-determined |
| 17-108_k141_86436 | Not-determined |
| 17-108_k141_307112 | Not-determined |
| 17-108_k141_292765 | Not-determined |
| 17-108_k141_17186 | Not-determined |
| 17-108_k141_320258 | Not-determined |
| 17-108_k141_307128 | Not-determined |
| 17-108_k141_196218 | Not-determined |
| 17-108_k141_58941 | Not-determined |
| 17-108_k141_196222 | Not-determined |
| 17-108_k141_292796 | Not-determined |
| 17-108_k141_86466 | Not-determined |
| 17-108_k141_58948 | Not-determined |
| 17-108_k141_279564 | Not-determined |
| 17-108_k141_265287 | Not-determined |
| 17-108_k141_3577 | Not-determined |
| 17-108_k141_113377 | Not-determined |
| 17-108_k141_238240 | Not-determined |
| 17-108_k141_58959 | Low-quality |
| 17-108_k141_279568 | Not-determined |
| 17-108_k141_58962 | Not-determined |
| 17-108_k141_238252 | Not-determined |
| 17-108_k141_182757 | Low-quality |
| 17-108_k141_292826 | Not-determined |
| 17-108_k141_127602 | Low-quality |
| 17-108_k141_292829 | Not-determined |
| 17-108_k141_251528 | Not-determined |
| 17-108_k141_45051 | Low-quality |
| 17-108_k141_251530 | Not-determined |
| 17-108_k141_265304 | Not-determined |
| 17-108_k141_100489 | Not-determined |
| 17-108_k141_100490 | Not-determined |
| 17-108_k141_265316 | Not-determined |
| 17-108_k141_265318 | Low-quality |
| 17-108_k141_45067 | Not-determined |
| 17-108_k141_251547 | Low-quality |
| 17-108_k141_251555 | Not-determined |
| 17-108_k141_320307 | Not-determined |
| 17-108_k141_141363 | Not-determined |
| 17-108_k141_279619 | Not-determined |
| 17-108_k141_59009 | Not-determined |
| 17-108_k141_210718 | Not-determined |
| 17-108_k141_59011 | Low-quality |
| 17-108_k141_320315 | Not-determined |
| 17-108_k141_292859 | Not-determined |
| 17-108_k141_31133 | Not-determined |
| 17-108_k141_100524 | Not-determined |
| 17-108_k141_86497 | Not-determined |
| 17-108_k141_100525 | Not-determined |
| 17-108_k141_127644 | Not-determined |
| 17-108_k141_100526 | Not-determined |
| 17-108_k141_127645 | Low-quality |
| 17-108_k141_127647 | Not-determined |
| 17-108_k141_141379 | Not-determined |
| 17-108_k141_265345 | Not-determined |
| 17-108_k141_141381 | Not-determined |
| 17-108_k141_59018 | Not-determined |
| 17-108_k141_127655 | Not-determined |
| 17-108_k141_127656 | Not-determined |
| 17-108_k141_182800 | Not-determined |
| 17-108_k141_182805 | Not-determined |
| 17-108_k141_210727 | Not-determined |
| 17-108_k141_127667 | Not-determined |
| 17-108_k141_59028 | Not-determined |
| 17-108_k141_86512 | Not-determined |
| 17-108_k141_113409 | Not-determined |
| 17-108_k141_141395 | Not-determined |
| 17-108_k141_86516 | Low-quality |
| 17-108_k141_196245 | Not-determined |
| 17-108_k141_265359 | Not-determined |
| 17-108_k141_59037 | Not-determined |
| 17-108_k141_31146 | Not-determined |
| 17-108_k141_59038 | Not-determined |
| 17-108_k141_59041 | Not-determined |
| 17-108_k141_59043 | Not-determined |
| 17-108_k141_141413 | Low-quality |
| 17-108_k141_17254 | Not-determined |
| 17-108_k141_265371 | Not-determined |
| 17-108_k141_86536 | Not-determined |
| 17-108_k141_59048 | Not-determined |
| 17-108_k141_141424 | Not-determined |
| 17-108_k141_210755 | Not-determined |
| 17-108_k141_86545 | Not-determined |
| 17-108_k141_279665 | Not-determined |
| 17-108_k141_86548 | Not-determined |
| 17-108_k141_292888 | Not-determined |
| 17-108_k141_292891 | Low-quality |
| 17-108_k141_182838 | Not-determined |
| 17-108_k141_169450 | Not-determined |
| 17-108_k141_100585 | Not-determined |
| 17-108_k141_169452 | Not-determined |
| 17-108_k141_292894 | Low-quality |
| 17-108_k141_210770 | Not-determined |
| 17-108_k141_72068 | Low-quality |
| 17-108_k141_307160 | Low-quality |
| 17-108_k141_113439 | Not-determined |
| 17-108_k141_155344 | Not-determined |
| 17-108_k141_59050 | Not-determined |
| 17-108_k141_127684 | Not-determined |
| 17-108_k141_320360 | Not-determined |
| 17-108_k141_265419 | Not-determined |
| 17-108_k141_251594 | Not-determined |
| 17-108_k141_196305 | Not-determined |
| 17-108_k141_265422 | Not-determined |
| 17-108_k141_31194 | Not-determined |
| 17-108_k141_113450 | Not-determined |
| 17-108_k141_113452 | Not-determined |
| 17-108_k141_251603 | Not-determined |
| 17-108_k141_59060 | Not-determined |
| 17-108_k141_320381 | Low-quality |
| 17-108_k141_210810 | Not-determined |
| 17-108_k141_45133 | Not-determined |
| 17-108_k141_320387 | Not-determined |
| 17-108_k141_265429 | Not-determined |
| 17-108_k141_265432 | Not-determined |
| 17-108_k141_182874 | Low-quality |
| 17-108_k141_210818 | Not-determined |
| 17-108_k141_279708 | Low-quality |
| 17-108_k141_72090 | Low-quality |
| 17-108_k141_265447 | Not-determined |
| 17-108_k141_72097 | Not-determined |
| 17-108_k141_251620 | Low-quality |
| 17-108_k141_292936 | Not-determined |
| 17-108_k141_279714 | Not-determined |
| 17-108_k141_210835 | Not-determined |
| 17-108_k141_155367 | Not-determined |
| 17-108_k141_292943 | Not-determined |
| 17-108_k141_292944 | Not-determined |
| 17-108_k141_169485 | Not-determined |
| 17-108_k141_307176 | Low-quality |
| 17-108_k141_279727 | Not-determined |
| 17-108_k141_3628 | Not-determined |
| 17-108_k141_196321 | Not-determined |
| 17-108_k141_182892 | Not-determined |
| 17-108_k141_86613 | Not-determined |
| 17-108_k141_265470 | Low-quality |
| 17-108_k141_320416 | Not-determined |
| 17-108_k141_292950 | Low-quality |
| 17-108_k141_86623 | Not-determined |
| 17-108_k141_72111 | Not-determined |
| 17-108_k141_155387 | Not-determined |
| 17-108_k141_251644 | Not-determined |
| 17-108_k141_224343 | Not-determined |
| 17-108_k141_100638 | Not-determined |
| 17-108_k141_320421 | Not-determined |
| 17-108_k141_224349 | Not-determined |
| 17-108_k141_210873 | Not-determined |
| 17-108_k141_307186 | Not-determined |
| 17-108_k141_251661 | Low-quality |
| 17-108_k141_59090 | Not-determined |
| 17-108_k141_224362 | Not-determined |
| 17-108_k141_320439 | Low-quality |
| 17-108_k141_86639 | Not-determined |
| 17-108_k141_86643 | Not-determined |
| 17-108_k141_292985 | Not-determined |
| 17-108_k141_292986 | Not-determined |
| 17-108_k141_169534 | Not-determined |
| 17-108_k141_320452 | Not-determined |
| 17-108_k141_251681 | Not-determined |
| 17-108_k141_169538 | Low-quality |
| 17-108_k141_320464 | Not-determined |
| 17-108_k141_307220 | Not-determined |
| 17-108_k141_251685 | Not-determined |
| 17-108_k141_31226 | Not-determined |
| 17-108_k141_307224 | Not-determined |
| 17-108_k141_31229 | Not-determined |
| 17-108_k141_320473 | Not-determined |
| 17-108_k141_224375 | Not-determined |
| 17-108_k141_86661 | Not-determined |
| 17-108_k141_182924 | Not-determined |
| 17-108_k141_292994 | Not-determined |
| 17-108_k141_45146 | Not-determined |
| 17-108_k141_31246 | Not-determined |
| 17-108_k141_293002 | Low-quality |
| 17-108_k141_72149 | Not-determined |
| 17-108_k141_72155 | Not-determined |
| 17-108_k141_224395 | Not-determined |
| 17-108_k141_31250 | Not-determined |
| 17-108_k141_72158 | Not-determined |
| 17-108_k141_224397 | Not-determined |
| 17-108_k141_72166 | Not-determined |
| 17-108_k141_320491 | Not-determined |
| 17-108_k141_320495 | Not-determined |
| 17-108_k141_265514 | Low-quality |
| 17-108_k141_141525 | Not-determined |
| 17-108_k141_45169 | Not-determined |
| 17-108_k141_31263 | Not-determined |
| 17-108_k141_196336 | Not-determined |
| 17-108_k141_31277 | Not-determined |
| 17-108_k141_265529 | Not-determined |
| 17-108_k141_45176 | Not-determined |
| 17-108_k141_72179 | Not-determined |
| 17-108_k141_251701 | Not-determined |
| 17-108_k141_31282 | Low-quality |
| 17-108_k141_59170 | Not-determined |
| 17-108_k141_72185 | Not-determined |
| 17-108_k141_238305 | Not-determined |
| 17-108_k141_251706 | Not-determined |
| 17-108_k141_31289 | Not-determined |
| 17-108_k141_238311 | Low-quality |
| 17-108_k141_72194 | Not-determined |
| 17-108_k141_210946 | Not-determined |
| 17-108_k141_59176 | Low-quality |
| 17-108_k141_224430 | Not-determined |
| 17-108_k141_251718 | Not-determined |
| 17-108_k141_224433 | Not-determined |
| 17-108_k141_196345 | Not-determined |
| 17-108_k141_238322 | Not-determined |
| 17-108_k141_279802 | Not-determined |
| 17-108_k141_155471 | Not-determined |
| 17-108_k141_169589 | Not-determined |
| 17-108_k141_113483 | Not-determined |
| 17-108_k141_169590 | Low-quality |
| 17-108_k141_141553 | Not-determined |
| 17-108_k141_196350 | Not-determined |
| 17-108_k141_210963 | Not-determined |
| 17-108_k141_100688 | Not-determined |
| 17-108_k141_307242 | Not-determined |
| 17-108_k141_100691 | Not-determined |
| 17-108_k141_72198 | Low-quality |
| 17-108_k141_238342 | Low-quality |
| 17-108_k141_3723 | Not-determined |
| 17-108_k141_279819 | Not-determined |
| 17-108_k141_265545 | Not-determined |
| 17-108_k141_307255 | Not-determined |
| 17-108_k141_141578 | Low-quality |
| 17-108_k141_31308 | Low-quality |
| 17-108_k141_169601 | Not-determined |
| 17-108_k141_224449 | Not-determined |
| 17-108_k141_45182 | Not-determined |
| 17-108_k141_17346 | Low-quality |
| 17-108_k141_141581 | Low-quality |
| 17-108_k141_141583 | Low-quality |
| 17-108_k141_279833 | Not-determined |
| 17-108_k141_155485 | Not-determined |
| 17-108_k141_72220 | Low-quality |
| 17-108_k141_224452 | Not-determined |
| 17-108_k141_224458 | Not-determined |
| 17-108_k141_113518 | Not-determined |
| 17-108_k141_72231 | Low-quality |
| 17-108_k141_307275 | Not-determined |
| 17-108_k141_72236 | Not-determined |
| 17-108_k141_127816 | Not-determined |
| 17-108_k141_224460 | Not-determined |
| 17-108_k141_169622 | Not-determined |
| 17-108_k141_86713 | Not-determined |
| 17-108_k141_72256 | Low-quality |
| 17-108_k141_307304 | Not-determined |
| 17-108_k141_59230 | Not-determined |
| 17-108_k141_293020 | Not-determined |
| 17-108_k141_17370 | Not-determined |
| 17-108_k141_307308 | Not-determined |
| 17-108_k141_196391 | Not-determined |
| 17-108_k141_59235 | Not-determined |
| 17-108_k141_210994 | Not-determined |
| 17-108_k141_293031 | Not-determined |
| 17-108_k141_293034 | Not-determined |
| 17-108_k141_293036 | Not-determined |
| 17-108_k141_251728 | Not-determined |
| 17-108_k141_307317 | Not-determined |
| 17-108_k141_141607 | Low-quality |
| 17-108_k141_307319 | Not-determined |
| 17-108_k141_45204 | Not-determined |
| 17-108_k141_251742 | Not-determined |
| 17-108_k141_320514 | Not-determined |
| 17-108_k141_155511 | Not-determined |
| 17-108_k141_251748 | Not-determined |
| 17-108_k141_251751 | Not-determined |
| 17-108_k141_86741 | Not-determined |
| 17-108_k141_251754 | Not-determined |
| 17-108_k141_265569 | Not-determined |
| 17-108_k141_169650 | Not-determined |
| 17-108_k141_320521 | Not-determined |
| 17-108_k141_251765 | Not-determined |
| 17-108_k141_169652 | Low-quality |
| 17-108_k141_155523 | Not-determined |
| 17-108_k141_279855 | Not-determined |
| 17-108_k141_59282 | Low-quality |
| 17-108_k141_251783 | Not-determined |
| 17-108_k141_320543 | Not-determined |
| 17-108_k141_320544 | Low-quality |
| 17-108_k141_293091 | Not-determined |
| 17-108_k141_3799 | Not-determined |
| 17-108_k141_141663 | Not-determined |
| 17-108_k141_293098 | Not-determined |
| 17-108_k141_141674 | Not-determined |
| 17-108_k141_293104 | Not-determined |
| 17-108_k141_169677 | Not-determined |
| 17-108_k141_238382 | Not-determined |
| 17-108_k141_113560 | Not-determined |
| 17-108_k141_141679 | Low-quality |
| 17-108_k141_45238 | Low-quality |
| 17-108_k141_238397 | Not-determined |
| 17-108_k141_307366 | Not-determined |
| 17-108_k141_224472 | Low-quality |
| 17-108_k141_182967 | Not-determined |
| 17-108_k141_86802 | Not-determined |
| 17-108_k141_211046 | Not-determined |
| 17-108_k141_251810 | Not-determined |
| 17-108_k141_211049 | Not-determined |
| 17-108_k141_86807 | Not-determined |

| | |
|---|---|
| 17-108_k141_169696 | Not-determined |
| 17-108_k141_141692 | Not-determined |
| 17-108_k141_196468 | Not-determined |
| 17-108_k141_307375 | Not-determined |
| 17-108_k141_307376 | Not-determined |
| 17-108_k141_59305 | Not-determined |
| 17-108_k141_72304 | Not-determined |
| 17-108_k141_169705 | Not-determined |
| 17-108_k141_224485 | Not-determined |
| 17-108_k141_293129 | Low-quality |
| 17-108_k141_182987 | Not-determined |
| 17-108_k141_211064 | Not-determined |
| 17-108_k141_45257 | Not-determined |
| 17-108_k141_224489 | Not-determined |
| 17-108_k141_100727 | Not-determined |
| 17-108_k141_86817 | Not-determined |
| 18-101_k141_102342 | Medium-quality |
| 17-108_k141_320561 | Not-determined |
| 17-108_k141_238423 | Not-determined |
| 17-108_k141_265620 | Not-determined |
| 17-108_k141_320568 | Not-determined |
| 17-108_k141_59322 | Not-determined |
| 17-108_k141_251822 | Not-determined |
| 17-108_k141_17446 | Low-quality |
| 17-108_k141_307403 | Not-determined |
| 17-108_k141_100738 | Not-determined |
| 17-108_k141_251830 | Not-determined |
| 17-108_k141_265631 | Not-determined |
| 17-108_k141_86823 | Not-determined |
| 17-108_k141_3864 | Not-determined |
| 17-108_k141_169725 | Not-determined |
| 17-108_k141_320582 | Not-determined |
| 17-108_k141_17456 | Not-determined |
| 17-108_k141_31393 | Not-determined |
| 17-108_k141_265639 | Not-determined |
| 17-108_k141_31409 | Not-determined |
| 17-108_k141_211099 | Not-determined |
| 17-108_k141_211102 | Not-determined |
| 17-108_k141_17466 | Low-quality |
| 17-108_k141_45297 | Low-quality |
| 17-108_k141_183034 | Not-determined |
| 17-108_k141_183038 | Not-determined |
| 17-108_k141_265652 | Not-determined |
| 17-108_k141_155607 | Not-determined |
| 17-108_k141_141768 | Not-determined |
| 17-108_k141_31428 | Not-determined |
| 17-108_k141_86856 | Low-quality |
| 17-108_k141_183043 | Not-determined |
| 17-108_k141_238496 | Not-determined |
| 17-108_k141_45322 | Low-quality |
| 17-108_k141_127887 | Not-determined |
| 17-108_k141_17482 | Not-determined |
| 17-108_k141_127894 | Not-determined |
| 17-108_k141_211136 | Not-determined |
| 17-108_k141_211139 | Not-determined |
| 17-108_k141_155622 | Not-determined |
| 17-108_k141_127909 | Not-determined |
| 17-108_k141_113595 | Not-determined |
| 17-108_k141_196520 | Low-quality |
| 17-108_k141_45335 | Not-determined |
| 17-108_k141_224497 | Not-determined |
| 17-108_k141_169751 | Not-determined |
| 17-108_k141_45348 | Not-determined |
| 17-108_k141_45350 | Not-determined |
| 17-108_k141_86894 | Not-determined |
| 17-108_k141_86895 | Not-determined |
| 17-108_k141_196524 | Not-determined |
| 17-108_k141_3898 | Not-determined |
| 17-108_k141_127920 | Not-determined |
| 17-108_k141_3901 | Not-determined |
| 17-108_k141_45362 | Low-quality |
| 17-108_k141_169757 | Not-determined |
| 17-108_k141_196535 | Low-quality |
| 17-108_k141_45364 | Not-determined |
| 17-108_k141_31435 | Not-determined |
| 17-108_k141_196539 | Not-determined |
| 17-108_k141_3906 | Not-determined |
| 17-108_k141_196541 | Not-determined |
| 17-108_k141_127923 | Not-determined |
| 17-108_k141_224515 | Not-determined |
| 17-108_k141_169759 | Low-quality |
| 17-108_k141_141799 | Not-determined |
| 17-108_k141_320625 | Low-quality |
| 17-108_k141_113625 | Not-determined |
| 17-108_k141_279987 | Not-determined |
| 17-108_k141_31448 | Not-determined |
| 17-108_k141_72375 | Not-determined |
| 17-108_k141_279998 | Low-quality |
| 17-108_k141_59364 | Not-determined |
| 17-108_k141_127937 | Not-determined |
| 17-108_k141_141804 | Low-quality |
| 17-108_k141_169776 | Not-determined |
| 17-108_k141_59370 | Not-determined |
| 17-108_k141_86906 | Not-determined |
| 17-108_k141_224534 | Not-determined |
| 17-108_k141_251927 | Not-determined |
| 17-108_k141_251928 | Not-determined |
| 17-108_k141_238543 | Not-determined |
| 17-108_k141_127959 | Not-determined |
| 17-108_k141_169790 | Not-determined |
| 17-108_k141_211180 | Not-determined |
| 17-108_k141_238553 | Not-determined |
| 17-108_k141_45394 | Not-determined |
| 17-108_k141_196565 | Low-quality |
| 17-108_k141_59393 | Not-determined |
| 17-108_k141_45398 | Not-determined |
| 17-108_k141_113648 | Not-determined |
| 17-108_k141_280033 | Not-determined |
| 17-108_k141_265747 | Not-determined |
| 17-108_k141_155670 | Not-determined |
| 17-108_k141_113654 | Not-determined |
| 17-108_k141_307478 | Not-determined |
| 17-108_k141_72396 | Not-determined |
| 17-108_k141_113661 | Not-determined |
| 17-108_k141_238571 | Not-determined |
| 17-108_k141_17508 | Not-determined |
| 17-108_k141_45409 | Not-determined |
| 17-108_k141_113668 | Low-quality |
| 17-108_k141_265763 | Not-determined |
| 17-108_k141_224566 | Low-quality |
| 17-108_k141_169805 | Low-quality |
| 17-108_k141_224575 | Not-determined |
| 17-108_k141_86916 | Not-determined |
| 17-108_k141_211201 | Low-quality |
| 17-108_k141_113688 | Not-determined |
| 17-108_k141_211206 | Not-determined |
| 17-108_k141_307509 | Not-determined |
| 17-108_k141_183123 | Not-determined |
| 17-108_k141_307513 | Not-determined |
| 17-108_k141_3955 | Not-determined |
| 17-108_k141_86917 | Low-quality |
| 17-108_k141_86921 | Not-determined |
| 17-108_k141_3959 | Not-determined |
| 17-108_k141_86922 | Not-determined |
| 17-108_k141_86924 | Not-determined |
| 17-108_k141_17533 | Not-determined |
| 17-108_k141_307518 | Not-determined |
| 17-108_k141_251955 | Not-determined |
| 17-108_k141_251957 | Low-quality |
| 17-108_k141_17542 | Not-determined |
| 17-108_k141_86934 | Low-quality |
| 17-108_k141_17543 | Not-determined |
| 17-108_k141_265796 | Not-determined |
| 17-108_k141_169815 | Not-determined |
| 17-108_k141_72428 | Not-determined |
| 17-108_k141_17547 | Not-determined |
| 17-108_k141_238624 | Not-determined |
| 17-108_k141_224595 | Not-determined |
| 17-108_k141_155691 | Low-quality |
| 17-108_k141_17549 | Not-determined |
| 17-108_k141_17550 | Not-determined |
| 17-108_k141_141822 | Not-determined |
| 17-108_k141_265804 | Low-quality |
| 17-108_k141_238628 | Not-determined |
| 17-108_k141_265807 | Not-determined |
| 17-108_k141_155696 | Not-determined |
| 17-108_k141_141824 | Low-quality |
| 17-108_k141_113702 | Low-quality |
| 17-108_k141_3965 | Low-quality |
| 17-108_k141_183131 | Low-quality |
| 17-108_k141_211220 | Not-determined |
| 17-108_k141_265821 | Not-determined |
| 17-108_k141_141828 | Low-quality |
| 17-108_k141_113718 | Not-determined |
| 17-108_k141_141832 | Not-determined |
| 17-108_k141_280085 | Not-determined |
| 17-108_k141_113721 | Not-determined |
| 17-108_k141_293200 | Not-determined |
| 17-108_k141_280090 | Not-determined |
| 17-108_k141_211243 | Not-determined |
| 17-108_k141_211245 | Low-quality |
| 17-108_k141_45434 | Not-determined |
| 17-108_k141_169835 | Not-determined |
| 17-108_k141_211258 | Not-determined |
| 17-108_k141_141851 | Not-determined |
| 17-108_k141_100815 | Not-determined |
| 17-108_k141_141859 | Not-determined |
| 17-108_k141_211279 | Not-determined |
| 17-108_k141_141865 | Not-determined |
| 17-108_k141_251990 | Not-determined |
| 17-108_k141_100820 | Not-determined |
| 17-108_k141_86975 | Not-determined |
| 17-108_k141_127989 | Not-determined |
| 17-108_k141_280127 | Low-quality |
| 17-108_k141_31508 | Not-determined |
| 17-108_k141_127994 | Not-determined |
| 17-108_k141_251995 | Not-determined |
| 17-108_k141_31512 | Not-determined |
| 17-108_k141_280131 | Low-quality |
| 17-108_k141_141870 | Not-determined |
| 17-108_k141_238691 | Not-determined |
| 17-108_k141_211309 | Not-determined |
| 17-108_k141_252003 | Not-determined |
| 17-108_k141_238693 | Not-determined |
| 17-108_k141_238698 | Not-determined |
| 17-108_k141_169851 | Not-determined |
| 17-108_k141_3985 | Not-determined |
| 17-108_k141_238702 | Not-determined |
| 17-108_k141_169859 | Not-determined |
| 17-108_k141_211322 | Not-determined |
| 17-108_k141_87006 | Not-determined |
| 17-108_k141_238711 | Not-determined |
| 17-108_k141_87016 | Not-determined |
| 17-108_k141_238717 | Low-quality |
| 17-108_k141_169887 | Not-determined |
| 17-108_k141_141895 | Not-determined |
| 17-108_k141_265869 | Low-quality |
| 17-108_k141_252028 | Not-determined |
| 17-108_k141_238734 | Not-determined |
| 17-108_k141_265877 | Not-determined |
| 17-108_k141_252037 | Not-determined |
| 17-108_k141_211356 | Not-determined |
| 17-108_k141_224654 | Not-determined |
| 17-108_k141_238739 | Not-determined |
| 17-108_k141_169907 | Not-determined |
| 17-108_k141_265882 | Not-determined |
| 17-108_k141_211362 | Not-determined |
| 17-108_k141_211368 | Not-determined |
| 17-108_k141_141920 | Not-determined |
| 17-108_k141_45483 | Not-determined |
| 17-108_k141_211370 | Not-determined |
| 17-108_k141_169911 | Not-determined |
| 17-108_k141_113744 | Not-determined |
| 17-108_k141_224671 | Not-determined |
| 17-108_k141_307552 | Not-determined |
| 17-108_k141_211375 | Not-determined |
| 17-108_k141_87020 | Not-determined |
| 17-108_k141_59423 | Not-determined |
| 17-108_k141_31593 | Not-determined |
| 17-108_k141_141933 | Not-determined |
| 17-108_k141_169920 | Not-determined |
| 17-108_k141_307565 | Not-determined |
| 17-108_k141_211387 | Not-determined |
| 17-108_k141_196631 | Not-determined |
| 17-108_k141_211389 | Not-determined |
| 17-108_k141_100921 | Low-quality |
| 17-108_k141_252055 | Low-quality |
| 17-108_k141_59431 | Low-quality |
| 17-108_k141_72449 | Not-determined |
| 17-108_k141_45505 | Not-determined |
| 17-108_k141_72452 | Not-determined |
| 17-108_k141_252061 | Not-determined |
| 17-108_k141_59435 | Not-determined |
| 17-108_k141_320667 | Not-determined |
| 17-108_k141_265913 | Not-determined |
| 17-108_k141_224689 | Not-determined |
| 17-108_k141_87045 | Not-determined |
| 17-108_k141_238770 | Not-determined |
| 17-108_k141_87055 | Not-determined |
| 17-108_k141_211414 | Not-determined |
| 17-108_k141_183208 | Not-determined |
| 17-108_k141_169933 | Low-quality |
| 17-108_k141_238785 | Not-determined |
| 17-108_k141_252065 | Not-determined |
| 17-108_k141_265940 | Not-determined |
| 17-108_k141_183218 | Not-determined |
| 17-108_k141_183224 | Not-determined |
| 17-108_k141_196667 | Low-quality |
| 17-108_k141_224724 | Low-quality |
| 17-108_k141_45537 | Not-determined |
| 17-108_k141_128028 | Not-determined |
| 17-108_k141_72490 | Not-determined |
| 17-108_k141_169940 | Low-quality |
| 17-108_k141_320679 | Low-quality |
| 17-108_k141_169941 | Not-determined |
| 17-108_k141_280161 | Low-quality |
| 17-108_k141_280162 | Low-quality |
| 17-108_k141_141953 | Not-determined |
| 17-108_k141_320700 | Not-determined |
| 17-108_k141_252087 | Low-quality |
| 17-108_k141_72497 | Not-determined |
| 17-108_k141_183234 | Not-determined |
| 17-108_k141_72506 | Not-determined |
| 17-108_k141_72508 | Not-determined |
| 17-108_k141_141970 | Low-quality |
| 17-108_k141_293357 | Not-determined |
| 17-108_k141_4026 | Not-determined |
| 17-108_k141_169955 | Not-determined |
| 17-108_k141_141974 | Low-quality |
| 17-108_k141_45565 | Not-determined |
| 17-108_k141_72517 | Low-quality |
| 17-108_k141_100971 | Not-determined |
| 17-108_k141_45569 | Low-quality |
| 17-108_k141_183252 | Not-determined |
| 17-108_k141_87122 | Not-determined |
| 17-108_k141_196707 | Not-determined |
| 17-108_k141_211462 | Not-determined |
| 17-108_k141_196708 | Not-determined |
| 17-108_k141_211464 | Not-determined |
| 17-108_k141_155714 | Not-determined |
| 17-108_k141_72537 | Not-determined |
| 17-108_k141_183269 | Not-determined |
| 17-108_k141_280205 | Not-determined |
| 17-108_k141_196715 | Not-determined |
| 17-108_k141_45589 | Not-determined |
| 17-108_k141_72543 | Not-determined |
| 17-108_k141_45594 | Not-determined |
| 17-108_k141_266009 | Not-determined |
| 17-108_k141_100981 | Not-determined |
| 17-108_k141_31611 | Not-determined |
| 17-108_k141_72561 | Low-quality |
| 17-108_k141_252123 | Not-determined |
| 17-108_k141_87142 | Not-determined |
| 17-108_k141_155742 | Low-quality |
| 17-108_k141_142007 | Not-determined |
| 17-108_k141_293399 | Low-quality |
| 17-108_k141_4055 | Not-determined |
| 17-108_k141_72569 | Not-determined |
| 17-108_k141_183290 | Not-determined |
| 17-108_k141_155745 | Not-determined |
| 17-108_k141_280237 | Low-quality |
| 17-108_k141_72571 | Not-determined |
| 17-108_k141_169983 | Not-determined |
| 17-108_k141_142022 | Not-determined |
| 17-108_k141_307602 | Not-determined |
| 17-108_k141_169987 | Low-quality |
| 17-108_k141_169991 | Not-determined |
| 17-108_k141_113750 | Not-determined |
| 17-108_k141_293420 | Not-determined |
| 17-108_k141_169996 | Not-determined |
| 17-108_k141_4064 | Not-determined |
| 17-108_k141_169998 | Not-determined |
| 17-108_k141_252146 | Not-determined |
| 17-108_k141_183328 | Not-determined |
| 17-108_k141_252148 | Not-determined |
| 17-108_k141_142035 | Not-determined |
| 17-108_k141_183331 | Not-determined |
| 17-108_k141_142038 | Not-determined |
| 17-108_k141_31639 | Low-quality |
| 17-108_k141_307611 | Not-determined |
| 17-108_k141_280248 | Not-determined |
| 17-108_k141_31644 | Not-determined |
| 17-108_k141_266025 | Not-determined |
| 17-108_k141_280251 | Not-determined |
| 17-108_k141_196725 | Not-determined |
| 17-108_k141_320726 | Not-determined |
| 17-108_k141_183350 | Not-determined |
| 17-108_k141_59464 | Not-determined |
| 17-108_k141_142048 | Not-determined |
| 17-108_k141_238868 | Not-determined |
| 17-108_k141_280266 | Not-determined |
| 17-108_k141_320738 | Not-determined |
| 17-108_k141_45636 | Not-determined |
| 17-108_k141_320741 | Not-determined |
| 17-108_k141_280276 | Not-determined |
| 17-108_k141_252182 | Low-quality |
| 17-108_k141_224774 | Low-quality |
| 17-108_k141_17595 | Not-determined |
| 17-108_k141_17597 | Not-determined |
| 17-108_k141_252186 | Not-determined |
| 17-108_k141_155790 | Not-determined |
| 17-108_k141_87185 | Low-quality |
| 17-108_k141_31680 | Not-determined |
| 17-108_k141_211512 | Not-determined |
| 17-108_k141_211516 | Not-determined |
| 17-108_k141_307651 | Not-determined |
| 17-108_k141_196776 | Not-determined |
| 17-108_k141_307657 | Not-determined |
| 17-108_k141_155815 | Not-determined |
| 17-108_k141_238874 | Not-determined |
| 17-108_k141_45679 | Not-determined |
| 17-108_k141_196792 | Not-determined |
| 17-108_k141_17623 | Low-quality |
| 17-108_k141_45694 | Not-determined |
| 17-108_k141_238899 | Not-determined |
| 17-108_k141_183376 | Not-determined |
| 17-108_k141_113780 | Low-quality |
| 17-108_k141_238904 | Not-determined |
| 17-108_k141_280315 | Not-determined |
| 17-108_k141_113787 | Not-determined |
| 17-108_k141_266066 | Not-determined |
| 17-108_k141_266072 | Not-determined |
| 17-108_k141_266074 | Not-determined |
| 17-108_k141_155832 | Not-determined |
| 17-108_k141_113805 | Not-determined |
| 17-108_k141_183390 | Not-determined |
| 17-108_k141_280322 | Low-quality |
| 17-108_k141_280323 | Not-determined |
| 17-108_k141_252222 | Not-determined |
| 17-108_k141_183399 | Not-determined |
| 17-108_k141_59527 | Not-determined |
| 17-108_k141_266088 | Not-determined |
| 17-108_k141_170093 | Not-determined |
| 17-108_k141_307671 | Not-determined |
| 17-108_k141_142080 | Not-determined |
| 17-108_k141_155847 | Not-determined |
| 17-108_k141_320767 | Low-quality |
| 17-108_k141_45732 | Not-determined |
| 17-108_k141_170102 | Not-determined |
| 17-108_k141_252236 | Not-determined |
| 17-108_k141_170104 | Not-determined |
| 17-108_k141_252243 | Not-determined |
| 17-108_k141_211560 | Not-determined |
| 17-108_k141_252245 | Not-determined |
| 17-108_k141_155867 | Low-quality |
| 17-108_k141_183420 | Not-determined |
| 17-108_k141_252254 | Not-determined |
| 17-108_k141_4101 | Not-determined |
| 17-108_k141_266107 | Not-determined |
| 17-108_k141_266108 | Not-determined |
| 17-108_k141_87231 | Not-determined |
| 17-108_k141_238959 | Not-determined |
| 17-108_k141_238966 | Not-determined |
| 17-108_k141_183430 | Not-determined |
| 17-108_k141_238969 | Not-determined |
| 17-108_k141_320801 | Not-determined |
| 17-108_k141_252272 | Not-determined |
| 17-108_k141_211578 | Not-determined |
| 17-108_k141_128155 | Not-determined |
| 17-108_k141_238975 | Not-determined |
| 17-108_k141_224858 | Low-quality |
| 17-108_k141_320805 | Low-quality |
| 17-108_k141_307707 | Not-determined |
| 17-108_k141_113858 | Not-determined |
| 17-108_k141_87255 | Not-determined |
| 17-108_k141_87256 | Not-determined |
| 17-108_k141_101057 | Not-determined |
| 17-108_k141_266127 | Not-determined |
| 17-108_k141_101058 | Not-determined |
| 17-108_k141_87260 | Not-determined |
| 17-108_k141_113869 | Low-quality |
| 17-108_k141_87265 | Not-determined |
| 17-108_k141_307722 | Not-determined |
| 17-108_k141_307729 | Not-determined |
| 17-108_k141_307732 | Not-determined |
| 17-108_k141_113875 | Low-quality |
| 17-108_k141_101071 | Not-determined |
| 17-108_k141_113876 | Not-determined |
| 17-108_k141_4150 | Not-determined |
| 17-108_k141_31742 | Not-determined |
| 17-108_k141_101076 | Not-determined |
| 17-108_k141_155896 | Not-determined |
| 17-108_k141_142107 | Low-quality |
| 17-108_k141_239008 | Not-determined |
| 17-108_k141_266138 | Not-determined |
| 17-108_k141_196846 | Not-determined |
| 17-108_k141_196848 | Not-determined |
| 17-108_k141_266148 | Not-determined |
| 17-108_k141_293573 | Not-determined |
| 17-108_k141_307742 | Not-determined |
| 17-108_k141_128208 | Not-determined |
| 17-108_k141_4155 | Low-quality |
| 17-108_k141_72592 | Not-determined |
| 17-108_k141_155908 | Low-quality |
| 17-108_k141_155910 | Not-determined |
| 17-108_k141_155912 | Not-determined |
| 17-108_k141_239018 | Low-quality |
| 17-108_k141_113898 | Not-determined |
| 17-108_k141_142122 | Not-determined |
| 17-108_k141_142125 | Low-quality |
| 17-108_k141_128221 | Not-determined |
| 17-108_k141_239026 | Not-determined |
| 17-108_k141_101102 | Not-determined |
| 17-108_k141_59596 | Not-determined |
| 17-108_k141_101107 | Not-determined |
| 17-108_k141_252308 | Not-determined |
| 17-108_k141_307756 | Low-quality |
| 17-108_k141_155921 | Not-determined |
| 17-108_k141_307758 | Not-determined |
| 17-108_k141_17660 | Not-determined |
| 17-108_k141_101116 | Not-determined |
| 17-108_k141_266164 | Not-determined |
| 17-108_k141_113912 | Not-determined |
| 17-108_k141_266166 | Not-determined |
| 17-108_k141_45796 | Not-determined |
| 17-108_k141_45797 | Low-quality |
| 17-108_k141_211616 | Not-determined |
| 17-108_k141_72605 | Not-determined |
| 17-108_k141_252324 | Not-determined |
| 17-108_k141_142151 | Not-determined |
| 17-108_k141_183452 | Not-determined |
| 17-108_k141_4187 | Not-determined |
| 17-108_k141_280425 | Not-determined |
| 17-108_k141_239058 | Not-determined |
| 17-108_k141_72624 | Not-determined |
| 17-108_k141_4200 | Not-determined |
| 17-108_k141_59611 | Low-quality |
| 17-108_k141_280430 | Not-determined |
| 17-108_k141_45819 | Low-quality |
| 17-108_k141_31751 | Not-determined |
| 17-108_k141_266184 | Not-determined |
| 17-108_k141_128253 | Not-determined |
| 17-108_k141_59629 | Not-determined |
| 17-108_k141_320852 | Not-determined |
| 17-108_k141_45823 | Not-determined |
| 17-108_k141_45824 | Not-determined |
| 17-108_k141_320857 | Not-determined |
| 17-108_k141_211636 | Not-determined |
| 17-108_k141_239068 | Not-determined |
| 18-101-final-viral-combined_k141_102342\|\|full | Medium-quality |
| 17-108_k141_31767 | Low-quality |
| 17-108_k141_183487 | Not-determined |
| 17-108_k141_170158 | Not-determined |
| 17-108_k141_280443 | Not-determined |
| 17-108_k141_280444 | Not-determined |
| 17-108_k141_266200 | Not-determined |
| 17-108_k141_266203 | Not-determined |
| 17-108_k141_128276 | Not-determined |
| 17-108_k141_59653 | Not-determined |
| 17-108_k141_4222 | Low-quality |
| 17-108_k141_307801 | Not-determined |
| 17-108_k141_266209 | Not-determined |
| 17-108_k141_31784 | Not-determined |
| 17-108_k141_183505 | Low-quality |
| 17-108_k141_4230 | Not-determined |
| 17-108_k141_45845 | Not-determined |
| 17-108_k141_59660 | Not-determined |
| 17-108_k141_307812 | Not-determined |
| 17-108_k141_280452 | Not-determined |
| 17-108_k141_170177 | Not-determined |
| 17-108_k141_320875 | Not-determined |
| 17-108_k141_196910 | Not-determined |
| 17-108_k141_320878 | Not-determined |
| 17-108_k141_239098 | Not-determined |
| 17-108_k141_183519 | Not-determined |
| 17-108_k141_87300 | Not-determined |
| 17-108_k141_101193 | Not-determined |
| 17-108_k141_307822 | Not-determined |
| 17-108_k141_4246 | Not-determined |
| 17-108_k141_170184 | Not-determined |
| 17-108_k141_320888 | Not-determined |
| 17-108_k141_252365 | Not-determined |
| 17-108_k141_72682 | Not-determined |
| 17-108_k141_87309 | Not-determined |
| 17-108_k141_183522 | Not-determined |
| 17-108_k141_307832 | Not-determined |
| 17-108_k141_155947 | Not-determined |
| 17-108_k141_155951 | Not-determined |
| 17-108_k141_128293 | Not-determined |
| 17-108_k141_266247 | Not-determined |
| 17-108_k141_266252 | Not-determined |
| 17-108_k141_266253 | Not-determined |
| 17-108_k141_183539 | Not-determined |
| 17-108_k141_280484 | Not-determined |
| 17-108_k141_128312 | Not-determined |
| 17-108_k141_31799 | Not-determined |
| 17-108_k141_307859 | Not-determined |
| 17-108_k141_224966 | Low-quality |
| 17-108_k141_196932 | Not-determined |
| 17-108_k141_280495 | Not-determined |
| 17-108_k141_239147 | Not-determined |
| 17-108_k141_155964 | Not-determined |
| 17-108_k141_224972 | Not-determined |
| 17-108_k141_155966 | Not-determined |
| 17-108_k141_31813 | Not-determined |
| 17-108_k141_307869 | Not-determined |
| 17-108_k141_239155 | Not-determined |
| 17-108_k141_307876 | Not-determined |
| 17-108_k141_101222 | Not-determined |
| 17-108_k141_280519 | Not-determined |
| 17-108_k141_293690 | Low-quality |
| 17-108_k141_45896 | Not-determined |
| 17-108_k141_45900 | Not-determined |
| 17-108_k141_293694 | Not-determined |
| 17-108_k141_307887 | Not-determined |
| 17-108_k141_128321 | Not-determined |
| 17-108_k141_72751 | Low-quality |
| 17-108_k141_211646 | Low-quality |
| 17-108_k141_307898 | Not-determined |
| 17-108_k141_128326 | Not-determined |
| 17-108_k141_170233 | Not-determined |
| 17-108_k141_4287 | Not-determined |
| 17-108_k141_17713 | Low-quality |
| 17-108_k141_113952 | Not-determined |
| 17-108_k141_101240 | Not-determined |
| 17-108_k141_4293 | Not-determined |
| 17-108_k141_293711 | Not-determined |
| 17-108_k141_101256 | Not-determined |
| 17-108_k141_252388 | Not-determined |
| 17-108_k141_142247 | Not-determined |
| 17-108_k141_31847 | Not-determined |
| 17-108_k141_101262 | Not-determined |
| 17-108_k141_266315 | Not-determined |
| 17-108_k141_307920 | Not-determined |
| 17-108_k141_87331 | Not-determined |
| 17-108_k141_87332 | Not-determined |
| 17-108_k141_87334 | Not-determined |
| 17-108_k141_280551 | Not-determined |
| 17-108_k141_211662 | Not-determined |
| 17-108_k141_280556 | Low-quality |
| 17-108_k141_31864 | Not-determined |
| 17-108_k141_113964 | Not-determined |
| 17-108_k141_307932 | Not-determined |
| 17-108_k141_307936 | Not-determined |
| 17-108_k141_101281 | Not-determined |
| 17-108_k141_31865 | Not-determined |
| 17-108_k141_307938 | Low-quality |
| 17-108_k141_72758 | Not-determined |
| 17-108_k141_113976 | Not-determined |

| | |
|---|---|
| 17-108_k141_266343 | Not-determined |
| 17-108_k141_142257 | Low-quality |
| 17-108_k141_101285 | Not-determined |
| 17-108_k141_293733 | Not-determined |
| 17-108_k141_266348 | Not-determined |
| 17-108_k141_266349 | Not-determined |
| 17-108_k141_4328 | Not-determined |
| 17-108_k141_113991 | Low-quality |
| 17-108_k141_197007 | Not-determined |
| 17-108_k141_293746 | Not-determined |
| 17-108_k141_266368 | Not-determined |
| 17-108_k141_72766 | Not-determined |
| 17-108_k141_293756 | Not-determined |
| 17-108_k141_4344 | Not-determined |
| 17-108_k141_266375 | Not-determined |
| 17-108_k141_197022 | Not-determined |
| 17-108_k141_183569 | Not-determined |
| 17-108_k141_72773 | Not-determined |
| 17-108_k141_45955 | Not-determined |
| 17-108_k141_252441 | Not-determined |
| 17-108_k141_252444 | Not-determined |
| 17-108_k141_72781 | Not-determined |
| 17-108_k141_293765 | Low-quality |
| 17-108_k141_252449 | Not-determined |
| 17-108_k141_31931 | Not-determined |
| 17-108_k141_183588 | Not-determined |
| 17-108_k141_170250 | Not-determined |
| 17-108_k141_239186 | Not-determined |
| 17-108_k141_266396 | Not-determined |
| 17-108_k141_45976 | Not-determined |
| 17-108_k141_101331 | Not-determined |
| 17-108_k141_59717 | Not-determined |
| 17-108_k141_72805 | Not-determined |
| 17-108_k141_59730 | Not-determined |
| 17-108_k141_72809 | Not-determined |
| 17-108_k141_72812 | Not-determined |
| 17-108_k141_45986 | Not-determined |
| 17-108_k141_320964 | Low-quality |
| 17-108_k141_45988 | Not-determined |
| 17-108_k141_183598 | Not-determined |
| 17-108_k141_320969 | Not-determined |
| 17-108_k141_128391 | Not-determined |
| 17-108_k141_72821 | Not-determined |
| 17-108_k141_128392 | Not-determined |
| 17-108_k141_17780 | Not-determined |
| 17-108_k141_17782 | Not-determined |
| 17-108_k141_156025 | Not-determined |
| 17-108_k141_156026 | Not-determined |
| 17-108_k141_72830 | Not-determined |
| 17-108_k141_320979 | Not-determined |
| 17-108_k141_4375 | Not-determined |
| 17-108_k141_320982 | Not-determined |
| 17-108_k141_320984 | Not-determined |
| 17-108_k141_156038 | Low-quality |
| 17-108_k141_156039 | Not-determined |
| 17-108_k141_46002 | Low-quality |
| 17-108_k141_114048 | Not-determined |
| 17-108_k141_4378 | Not-determined |
| 17-108_k141_142319 | Not-determined |
| 18-102-final-viral-combined_k141_191613\|\|full | Medium-quality |
| 17-108_k141_156045 | Not-determined |
| 17-108_k141_156046 | Not-determined |
| 17-108_k141_142325 | Not-determined |
| 17-108_k141_170293 | Not-determined |
| 17-108_k141_72838 | Not-determined |
| 17-108_k141_31963 | Not-determined |
| 17-108_k141_170296 | Not-determined |
| 17-108_k141_183635 | Not-determined |
| 17-108_k141_211690 | Not-determined |
| 17-108_k141_170300 | Not-determined |
| 17-108_k141_4400 | Not-determined |
| 17-108_k141_156056 | Not-determined |
| 17-108_k141_114067 | Not-determined |
| 17-108_k141_72850 | Not-determined |
| 17-108_k141_183640 | Not-determined |
| 17-108_k141_225097 | Not-determined |
| 17-108_k141_225102 | Not-determined |
| 17-108_k141_72855 | Not-determined |
| 17-108_k141_72856 | Not-determined |
| 17-108_k141_46011 | Not-determined |
| 17-108_k141_225105 | Low-quality |
| 17-108_k141_17813 | Not-determined |
| 17-108_k141_183656 | Not-determined |
| 17-108_k141_211705 | Not-determined |
| 17-108_k141_211707 | Not-determined |
| 17-108_k141_225123 | Not-determined |
| 17-108_k141_170327 | Not-determined |
| 17-108_k141_321006 | Not-determined |
| 17-108_k141_170330 | Not-determined |
| 17-108_k141_252528 | Not-determined |
| 17-108_k141_170333 | Not-determined |
| 17-108_k141_114112 | Not-determined |
| 17-108_k141_114113 | Not-determined |
| 17-108_k141_239235 | Low-quality |
| 17-108_k141_197066 | Low-quality |
| 17-108_k141_225140 | Not-determined |
| 17-108_k141_170338 | Not-determined |
| 17-108_k141_252537 | Not-determined |
| 17-108_k141_128430 | Not-determined |
| 17-108_k141_280684 | Low-quality |
| 17-108_k141_72885 | Low-quality |
| 17-108_k141_4418 | Not-determined |
| 17-108_k141_59754 | Not-determined |
| 17-108_k141_142357 | Not-determined |
| 17-108_k141_114138 | Not-determined |
| 17-108_k141_156087 | Not-determined |
| 17-108_k141_46049 | Not-determined |
| 17-108_k141_183699 | Not-determined |
| 17-108_k141_59774 | Not-determined |
| 17-108_k141_87372 | Not-determined |
| 17-108_k141_87374 | Not-determined |
| 17-108_k141_46060 | Not-determined |
| 17-108_k141_101351 | Not-determined |
| 17-108_k141_101352 | Not-determined |
| 17-108_k141_59784 | Low-quality |
| 17-108_k141_17858 | Not-determined |
| 17-108_k141_211759 | Not-determined |
| 17-108_k141_46063 | Not-determined |
| 17-108_k141_31994 | Not-determined |
| 17-108_k141_307954 | Not-determined |
| 17-108_k141_32001 | Not-determined |
| 17-108_k141_211771 | Not-determined |
| 17-108_k141_183717 | Not-determined |
| 17-108_k141_225184 | Not-determined |
| 17-108_k141_225185 | Low-quality |
| 17-108_k141_46066 | Not-determined |
| 17-108_k141_225186 | Not-determined |
| 17-108_k141_307968 | Not-determined |
| 17-108_k141_183731 | Not-determined |
| 17-108_k141_156097 | Not-determined |
| 17-108_k141_17871 | Not-determined |
| 17-108_k141_266450 | Not-determined |
| 17-108_k141_142414 | Not-determined |
| 17-108_k141_321053 | Low-quality |
| 17-108_k141_225202 | Not-determined |
| 17-108_k141_87388 | Not-determined |
| 17-108_k141_307979 | Not-determined |
| 17-108_k141_114168 | Not-determined |
| 17-108_k141_266457 | Not-determined |
| 17-108_k141_266458 | Not-determined |
| 17-108_k141_156111 | Not-determined |
| 17-108_k141_183752 | Not-determined |
| 17-108_k141_156113 | Not-determined |
| 17-108_k141_101373 | Not-determined |
| 17-108_k141_197110 | Not-determined |
| 17-108_k141_211791 | Not-determined |
| 17-108_k141_72925 | Not-determined |
| 17-108_k141_183759 | Not-determined |
| 17-108_k141_252610 | Not-determined |
| 17-108_k141_308000 | Low-quality |
| 17-108_k141_46105 | Not-determined |
| 17-108_k141_87414 | Not-determined |
| 17-108_k141_183771 | Not-determined |
| 17-108_k141_142424 | Not-determined |
| 17-108_k141_46106 | Not-determined |
| 17-108_k141_142427 | Not-determined |
| 17-108_k141_225236 | Not-determined |
| 17-108_k141_46112 | Low-quality |
| 17-108_k141_4475 | Not-determined |
| 17-108_k141_114187 | Not-determined |
| 17-108_k141_72958 | Not-determined |
| 17-108_k141_128477 | Not-determined |
| 17-108_k141_183779 | Not-determined |
| 17-108_k141_183780 | Not-determined |
| 17-108_k141_197121 | Not-determined |
| 17-108_k141_72962 | Not-determined |
| 17-108_k141_225246 | Not-determined |
| 17-108_k141_59803 | Not-determined |
| 17-108_k141_280707 | Not-determined |
| 17-108_k141_211834 | Not-determined |
| 17-108_k141_114208 | Not-determined |
| 17-108_k141_128509 | Not-determined |
| 17-108_k141_211846 | Not-determined |
| 17-108_k141_128512 | Not-determined |
| 17-108_k141_266487 | Not-determined |
| 17-108_k141_87465 | Not-determined |
| 17-108_k141_266492 | Not-determined |
| 17-108_k141_321073 | Low-quality |
| 17-108_k141_128523 | Low-quality |
| 17-108_k141_87473 | Not-determined |
| 17-108_k141_4510 | Not-determined |
| 17-108_k141_46130 | Not-determined |
| 17-108_k141_266498 | Not-determined |
| 17-108_k141_72988 | Not-determined |
| 17-108_k141_170398 | Not-determined |
| 17-108_k141_87483 | Not-determined |
| 17-108_k141_308041 | Not-determined |
| 17-108_k141_293812 | Not-determined |
| 17-108_k141_308043 | Not-determined |
| 17-108_k141_59818 | Not-determined |
| 17-108_k141_321084 | Not-determined |
| 17-108_k141_32082 | Not-determined |
| 17-108_k141_239281 | Not-determined |
| 17-108_k141_87494 | Not-determined |
| 17-108_k141_32085 | Not-determined |
| 17-108_k141_321089 | Not-determined |
| 17-108_k141_4528 | Not-determined |
| 17-108_k141_101415 | Not-determined |
| 17-108_k141_321092 | Not-determined |
| 17-108_k141_197143 | Not-determined |
| 17-108_k141_280720 | Not-determined |
| 17-108_k141_280724 | Not-determined |
| 17-108_k141_197149 | Not-determined |
| 17-108_k141_87508 | Not-determined |
| 17-108_k141_4533 | Not-determined |
| 17-108_k141_156163 | Not-determined |
| 17-108_k141_225269 | Not-determined |
| 17-108_k141_197160 | Not-determined |
| 17-108_k141_308065 | Not-determined |
| 17-108_k141_239287 | Not-determined |
| 17-108_k141_225272 | Not-determined |
| 17-108_k141_4537 | Not-determined |
| 17-108_k141_197163 | Not-determined |
| 17-108_k141_128538 | Not-determined |
| 17-108_k141_170420 | Not-determined |
| 17-108_k141_4545 | Not-determined |
| 17-108_k141_59831 | Not-determined |
| 17-108_k141_32108 | Not-determined |
| 17-108_k141_46166 | Not-determined |
| 17-108_k141_17893 | Not-determined |
| 17-108_k141_73033 | Not-determined |
| 17-108_k141_128540 | Low-quality |
| 17-108_k141_32112 | Low-quality |
| 17-108_k141_239299 | Not-determined |
| 17-108_k141_17900 | Not-determined |
| 17-108_k141_87515 | Not-determined |
| 17-108_k141_59845 | Not-determined |
| 17-108_k141_197182 | Not-determined |
| 17-108_k141_4563 | Low-quality |
| 17-108_k141_17904 | Not-determined |
| 17-108_k141_101455 | Not-determined |
| 17-108_k141_128546 | Not-determined |
| 17-108_k141_128547 | Not-determined |
| 17-108_k141_170429 | Not-determined |
| 17-108_k141_239310 | Not-determined |
| 17-108_k141_73051 | Not-determined |
| 17-108_k141_156190 | Not-determined |
| 17-108_k141_46186 | Not-determined |
| 17-108_k141_183803 | Not-determined |
| 17-108_k141_101465 | Not-determined |
| 17-108_k141_17917 | Low-quality |
| 17-108_k141_239324 | Not-determined |
| 17-108_k141_280758 | Not-determined |
| 17-108_k141_32131 | Not-determined |
| 17-108_k141_17922 | Low-quality |
| 17-108_k141_225306 | Not-determined |
| 17-108_k141_280761 | Not-determined |
| 17-108_k141_87539 | Not-determined |
| 17-108_k141_170443 | Not-determined |
| 17-108_k141_142513 | Not-determined |
| 17-108_k141_211897 | Not-determined |
| 17-108_k141_142519 | Not-determined |
| 17-108_k141_32142 | Not-determined |
| 17-108_k141_197216 | Not-determined |
| 17-108_k141_280777 | Not-determined |
| 17-108_k141_101475 | Not-determined |
| 17-108_k141_293841 | Not-determined |
| 17-108_k141_293842 | Not-determined |
| 17-108_k141_170457 | Not-determined |
| 17-108_k141_197227 | Not-determined |
| 17-108_k141_197231 | Not-determined |
| 17-108_k141_266533 | Not-determined |
| 17-108_k141_293853 | Not-determined |
| 17-108_k141_197238 | Not-determined |
| 17-108_k141_308114 | Not-determined |
| 17-108_k141_225347 | Not-determined |
| 17-108_k141_266537 | Not-determined |
| 17-108_k141_225348 | Not-determined |
| 17-108_k141_17935 | Not-determined |
| 17-108_k141_142537 | Not-determined |
| 17-108_k141_4602 | Not-determined |
| 17-108_k141_59869 | Not-determined |
| 17-108_k141_17938 | Not-determined |
| 17-108_k141_266539 | Not-determined |
| 17-108_k141_156206 | Low-quality |
| 17-108_k141_197244 | Not-determined |
| 17-108_k141_114214 | Low-quality |
| 17-108_k141_293860 | Not-determined |
| 17-108_k141_252672 | Low-quality |
| 17-108_k141_59874 | Not-determined |
| 17-108_k141_321162 | Not-determined |
| 17-108_k141_266552 | Not-determined |
| 17-108_k141_59881 | Low-quality |
| 17-108_k141_321168 | Low-quality |
| 17-108_k141_170483 | Not-determined |
| 17-108_k141_46210 | Not-determined |
| 17-108_k141_321171 | Not-determined |
| 17-108_k141_156220 | Not-determined |
| 17-108_k141_321173 | Not-determined |
| 17-108_k141_225371 | Not-determined |
| 17-108_k141_183829 | Not-determined |
| 17-108_k141_183836 | Not-determined |
| 17-108_k141_101487 | Not-determined |
| 17-108_k141_239343 | Not-determined |
| 17-108_k141_46222 | Not-determined |
| 17-108_k141_114221 | Not-determined |
| 17-108_k141_280787 | Not-determined |
| 17-108_k141_225379 | Not-determined |
| 17-108_k141_197262 | Not-determined |
| 17-108_k141_128602 | Not-determined |
| 17-108_k141_4626 | Not-determined |
| 17-108_k141_101494 | Not-determined |
| 17-108_k141_225393 | Not-determined |
| 17-108_k141_183845 | Not-determined |
| 17-108_k141_239358 | Not-determined |
| 17-108_k141_101510 | Not-determined |
| 17-108_k141_308145 | Low-quality |
| 17-108_k141_114235 | Not-determined |
| 17-108_k141_239361 | Not-determined |
| 17-108_k141_114242 | Not-determined |
| 17-108_k141_321176 | Not-determined |
| 17-108_k141_114244 | Not-determined |
| 17-108_k141_170499 | Not-determined |
| 17-108_k141_239363 | Not-determined |
| 17-108_k141_183865 | Not-determined |
| 17-108_k141_183866 | Not-determined |
| 17-108_k141_321180 | Low-quality |
| 17-108_k141_239365 | Not-determined |
| 17-108_k141_101516 | Low-quality |
| 17-108_k141_170502 | Not-determined |
| 17-108_k141_156262 | Low-quality |
| 17-108_k141_183872 | Not-determined |
| 17-108_k141_211964 | Not-determined |
| 17-108_k141_225407 | Not-determined |
| 17-108_k141_293888 | Low-quality |
| 17-108_k141_128609 | Not-determined |
| 17-108_k141_170511 | Not-determined |
| 17-108_k141_128618 | Not-determined |
| 17-108_k141_156268 | Low-quality |
| 17-108_k141_156270 | Not-determined |
| 17-108_k141_101523 | Not-determined |
| 17-108_k141_321186 | Low-quality |
| 17-108_k141_156277 | Not-determined |
| 17-108_k141_114260 | Not-determined |
| 17-108_k141_225433 | Not-determined |
| 17-108_k141_59920 | Low-quality |
| 17-108_k141_308167 | Not-determined |
| 17-108_k141_321196 | Not-determined |
| 17-108_k141_293910 | Not-determined |
| 17-108_k141_293911 | Not-determined |
| 17-108_k141_308169 | Not-determined |
| 17-108_k141_321198 | Not-determined |
| 17-108_k141_114270 | Not-determined |
| 17-108_k141_114271 | Not-determined |
| 17-108_k141_280794 | Not-determined |
| 17-108_k141_114275 | Not-determined |
| 17-108_k141_183879 | Not-determined |
| 17-108_k141_114284 | Low-quality |
| 17-108_k141_101535 | Not-determined |
| 17-108_k141_280801 | Not-determined |
| 17-108_k141_293923 | Not-determined |
| 17-108_k141_4650 | Not-determined |
| 17-108_k141_46273 | Not-determined |
| 17-108_k141_252732 | Not-determined |
| 17-108_k141_280820 | Not-determined |
| 17-108_k141_170538 | Not-determined |
| 17-108_k141_170542 | Not-determined |
| 17-108_k141_87582 | Low-quality |
| 17-108_k141_101552 | Not-determined |
| 17-108_k141_17965 | Not-determined |
| 17-108_k141_17969 | Not-determined |
| 17-108_k141_128631 | Not-determined |
| 17-108_k141_321232 | Not-determined |
| 17-108_k141_17974 | Not-determined |
| 17-108_k141_114300 | Not-determined |
| 17-108_k141_252748 | Not-determined |
| 17-108_k141_101563 | Not-determined |
| 17-108_k141_114302 | Not-determined |
| 17-108_k141_101567 | Not-determined |
| 17-108_k141_101568 | Not-determined |
| 17-108_k141_280848 | Low-quality |
| 17-108_k141_59975 | Not-determined |
| 17-108_k141_321242 | Low-quality |
| 17-108_k141_59978 | Low-quality |
| 17-108_k141_212037 | Low-quality |
| 17-108_k141_183898 | Low-quality |
| 17-108_k141_225481 | Not-determined |
| 17-108_k141_142633 | Not-determined |
| 17-108_k141_266620 | Not-determined |
| 17-108_k141_266622 | Not-determined |
| 17-108_k141_46298 | Not-determined |
| 17-108_k141_321255 | Not-determined |
| 17-108_k141_142637 | Not-determined |
| 17-108_k141_239462 | Not-determined |
| 17-108_k141_183911 | Not-determined |
| 17-108_k141_183912 | Not-determined |
| 17-108_k141_321259 | Not-determined |
| 17-108_k141_321262 | Not-determined |
| 17-108_k141_183913 | Not-determined |
| 17-108_k141_46309 | Low-quality |
| 17-108_k141_225496 | Not-determined |
| 17-108_k141_142640 | Not-determined |
| 17-108_k141_46312 | Not-determined |
| 17-108_k141_17998 | Not-determined |
| 17-108_k141_128670 | Not-determined |
| 17-108_k141_170591 | Not-determined |
| 17-108_k141_101589 | Not-determined |
| 17-108_k141_128674 | Not-determined |
| 17-108_k141_197332 | Low-quality |
| 17-108_k141_197334 | Not-determined |
| 17-108_k141_225508 | Not-determined |
| 17-108_k141_142650 | Not-determined |
| 17-108_k141_197338 | Not-determined |
| 17-108_k141_4688 | Not-determined |
| 17-108_k141_252777 | Not-determined |
| 17-108_k141_321291 | Not-determined |
| 17-108_k141_18016 | Not-determined |
| 17-108_k141_142659 | Not-determined |
| 17-108_k141_142661 | Not-determined |
| 17-108_k141_114327 | Low-quality |
| 17-108_k141_170621 | Low-quality |
| 17-108_k141_170626 | Not-determined |
| 17-108_k141_142676 | Not-determined |
| 17-108_k141_142678 | Not-determined |
| 17-108_k141_170631 | Not-determined |
| 17-108_k141_142680 | Not-determined |
| 17-108_k141_156352 | Not-determined |
| 17-108_k141_114343 | Not-determined |
| 17-108_k141_212068 | Low-quality |
| 17-108_k141_197350 | Not-determined |
| 17-108_k141_280912 | Not-determined |
| 17-108_k141_73116 | Not-determined |
| 17-108_k141_4704 | Low-quality |
| 17-108_k141_156357 | Not-determined |
| 17-108_k141_142681 | Not-determined |
| 17-108_k141_142682 | Not-determined |
| 17-108_k141_18043 | Low-quality |
| 17-108_k141_114348 | Not-determined |
| 17-108_k141_170635 | Low-quality |
| 17-108_k141_114350 | Not-determined |
| 17-108_k141_128710 | Not-determined |
| 17-108_k141_294017 | Not-determined |
| 17-108_k141_46329 | Not-determined |
| 17-108_k141_114355 | Not-determined |
| 17-108_k141_294020 | Not-determined |
| 17-108_k141_197352 | Not-determined |
| 17-108_k141_225511 | Low-quality |
| 17-108_k141_239475 | Not-determined |
| 17-108_k141_156374 | Low-quality |
| 17-108_k141_46343 | Not-determined |
| 17-108_k141_212074 | Low-quality |
| 17-108_k141_18045 | Not-determined |
| 17-108_k141_4722 | Not-determined |
| 17-108_k141_46354 | Not-determined |
| 17-108_k141_239489 | Not-determined |
| 17-108_k141_32268 | Not-determined |
| 17-108_k141_142705 | Not-determined |
| 17-108_k141_170647 | Not-determined |
| 17-108_k141_280947 | Not-determined |
| 17-108_k141_170651 | Not-determined |
| 17-108_k141_183931 | Not-determined |
| 17-108_k141_4741 | Not-determined |
| 17-108_k141_156403 | Not-determined |
| 17-108_k141_32279 | Not-determined |
| 17-108_k141_170657 | Not-determined |
| 17-108_k141_225524 | Not-determined |
| 17-108_k141_183939 | Not-determined |
| 17-108_k141_128767 | Not-determined |
| 17-108_k141_101646 | Not-determined |
| 17-108_k141_197384 | Not-determined |
| 17-108_k141_239510 | Not-determined |
| 17-108_k141_239512 | Not-determined |
| 17-108_k141_197386 | Not-determined |
| 17-108_k141_170673 | Low-quality |
| 17-108_k141_156423 | Not-determined |
| 17-108_k141_128776 | Not-determined |
| 17-108_k141_60026 | Not-determined |
| 17-108_k141_212097 | Not-determined |
| 17-108_k141_212098 | Not-determined |
| 17-108_k141_101653 | Not-determined |
| 17-108_k141_252845 | Not-determined |
| 17-108_k141_252846 | Not-determined |
| 17-108_k141_321346 | Not-determined |
| 17-108_k141_87681 | Low-quality |
| 17-108_k141_321348 | Not-determined |
| 17-108_k141_266670 | Not-determined |
| 17-108_k141_321349 | Not-determined |
| 17-108_k141_225556 | Not-determined |
| 17-108_k141_101664 | Not-determined |
| 17-108_k141_73179 | Not-determined |
| 17-108_k141_32306 | Not-determined |
| 17-108_k141_46367 | Not-determined |
| 17-108_k141_225561 | Not-determined |
| 17-108_k141_239543 | Not-determined |
| 17-108_k141_46371 | Not-determined |
| 17-108_k141_156451 | Not-determined |
| 17-108_k141_156453 | Not-determined |
| 17-108_k141_156456 | Not-determined |
| 17-108_k141_142743 | Not-determined |
| 17-108_k141_128803 | Not-determined |
| 17-108_k141_294099 | Not-determined |
| 17-108_k141_321358 | Not-determined |
| 17-108_k141_239559 | Low-quality |
| 17-108_k141_252869 | Not-determined |
| 17-108_k141_170707 | Not-determined |
| 17-108_k141_128816 | Not-determined |
| 17-108_k141_239562 | Not-determined |
| 17-108_k141_128821 | Not-determined |
| 17-108_k141_239565 | Not-determined |
| 17-108_k141_73190 | Not-determined |
| 17-108_k141_239566 | Low-quality |
| 17-108_k141_170713 | Not-determined |
| 17-108_k141_156461 | Not-determined |
| 17-108_k141_114438 | Not-determined |
| 17-108_k141_183959 | Not-determined |
| 17-108_k141_239582 | Not-determined |
| 17-108_k141_225585 | Low-quality |
| 17-108_k141_197417 | Not-determined |
| 17-108_k141_183967 | Not-determined |
| 17-108_k141_252888 | Not-determined |
| 17-108_k141_239590 | Low-quality |
| 17-108_k141_252892 | Not-determined |
| 17-108_k141_252896 | Not-determined |
| 17-108_k141_32317 | Not-determined |
| 17-108_k141_225596 | Not-determined |
| 17-108_k141_321378 | Not-determined |
| 17-108_k141_18115 | Not-determined |
| 17-108_k141_252902 | Not-determined |
| 17-108_k141_212167 | Not-determined |
| 17-108_k141_183991 | Not-determined |
| 17-108_k141_142784 | Not-determined |
| 17-108_k141_114477 | Not-determined |
| 17-108_k141_266732 | Not-determined |
| 17-108_k141_101718 | Not-determined |
| 17-108_k141_197436 | Not-determined |
| 17-108_k141_281070 | Not-determined |
| 17-108_k141_321394 | Not-determined |
| 17-108_k141_142798 | Not-determined |
| 17-108_k141_142799 | Low-quality |
| 17-108_k141_239623 | Not-determined |
| 17-108_k141_266744 | Not-determined |
| 17-108_k141_101735 | Not-determined |
| 17-108_k141_32355 | Not-determined |
| 17-108_k141_294125 | Not-determined |
| 17-108_k141_114494 | Not-determined |
| 17-108_k141_239625 | Low-quality |
| 17-108_k141_308276 | Not-determined |
| 17-108_k141_114498 | Low-quality |
| 17-108_k141_170729 | Not-determined |
| 17-108_k141_4770 | Not-determined |
| 17-108_k141_308283 | Not-determined |
| 17-108_k141_156484 | Not-determined |
| 17-108_k141_114508 | Not-determined |
| 17-108_k141_101757 | Not-determined |
| 17-108_k141_156492 | Not-determined |
| 17-108_k141_87714 | Not-determined |
| 17-108_k141_87715 | Not-determined |
| 17-108_k141_281095 | Not-determined |
| 17-108_k141_101763 | Not-determined |
| 17-108_k141_225642 | Not-determined |
| 17-108_k141_101765 | Not-determined |
| 17-108_k141_101766 | Not-determined |
| 17-108_k141_156498 | Low-quality |
| 17-108_k141_128830 | Not-determined |
| 17-108_k141_252933 | Not-determined |
| 17-108_k141_252934 | Not-determined |
| 17-108_k141_183997 | Not-determined |
| 17-108_k141_156506 | Not-determined |
| 17-108_k141_142811 | Low-quality |
| 17-108_k141_114533 | Not-determined |
| 17-108_k141_281108 | Not-determined |
| 17-108_k141_156517 | Not-determined |
| 17-108_k141_142814 | Not-determined |
| 17-108_k141_239629 | Not-determined |
| 17-108_k141_4796 | Not-determined |
| 17-108_k141_197447 | Not-determined |
| 17-108_k141_18130 | Not-determined |
| 17-108_k141_184021 | Low-quality |
| 17-108_k141_128842 | Low-quality |
| 17-108_k141_114548 | Not-determined |
| 17-108_k141_46400 | Not-determined |
| 17-108_k141_46402 | Not-determined |
| 17-108_k141_4805 | Low-quality |
| 17-108_k141_60075 | Not-determined |
| 17-108_k141_212242 | Not-determined |
| 17-108_k141_252955 | Not-determined |
| 17-108_k141_266791 | Not-determined |
| 17-108_k141_142826 | Not-determined |
| 17-108_k141_197469 | Not-determined |
| 17-108_k141_46410 | Not-determined |
| 17-108_k141_142833 | Not-determined |
| 17-108_k141_184034 | Not-determined |
| 17-108_k141_114562 | Not-determined |
| 17-108_k141_73215 | Not-determined |
| 17-108_k141_197477 | Not-determined |
| 17-108_k141_281135 | Not-determined |
| 17-108_k141_18139 | Not-determined |
| 17-108_k141_281139 | Not-determined |
| 17-108_k141_184036 | Not-determined |

| | |
|---|---|
| 17-108_k141_87738 | Not-determined |
| 17-108_k141_4823 | Not-determined |
| 17-108_k141_128870 | Not-determined |
| 17-108_k141_156526 | Not-determined |
| 17-108_k141_114582 | Not-determined |
| 17-108_k141_32418 | Not-determined |
| 17-108_k141_87740 | Not-determined |
| 17-108_k141_170749 | Not-determined |
| 17-108_k141_170750 | Not-determined |
| 17-108_k141_321415 | Not-determined |
| 17-108_k141_32428 | Not-determined |
| 17-108_k141_32434 | Not-determined |
| 17-108_k141_252967 | Not-determined |
| 17-108_k141_239636 | Not-determined |
| 17-108_k141_101805 | Not-determined |
| 17-108_k141_60109 | Not-determined |
| 17-108_k141_128898 | Not-determined |
| 17-108_k141_114586 | Not-determined |
| 17-108_k141_114588 | Not-determined |
| 17-108_k141_101819 | Low-quality |
| 17-108_k141_197501 | Not-determined |
| 17-108_k141_101824 | Not-determined |
| 17-108_k141_239645 | Not-determined |
| 17-108_k141_197503 | Not-determined |
| 17-108_k141_197504 | Not-determined |
| 17-108_k141_32456 | Low-quality |
| 17-108_k141_239652 | Not-determined |
| 17-108_k141_18180 | Not-determined |
| 17-108_k141_197517 | Not-determined |
| 17-108_k141_46440 | Not-determined |
| 17-108_k141_60146 | Not-determined |
| 17-108_k141_60147 | Not-determined |
| 17-108_k141_18185 | Not-determined |
| 17-108_k141_197526 | Low-quality |
| 17-108_k141_142842 | Not-determined |
| 17-108_k141_184070 | Not-determined |
| 17-108_k141_60150 | Not-determined |
| 17-108_k141_142847 | Not-determined |
| 17-108_k141_4835 | Not-determined |
| 17-108_k141_73232 | Not-determined |
| 17-108_k141_101863 | Not-determined |
| 17-108_k141_184084 | Not-determined |
| 17-108_k141_142854 | Not-determined |
| 17-108_k141_32479 | Not-determined |
| 17-108_k141_281207 | Not-determined |
| 17-108_k141_281208 | Not-determined |
| 17-108_k141_294144 | Not-determined |
| 17-108_k141_60171 | Low-quality |
| 17-108_k141_239667 | Not-determined |
| 17-108_k141_46457 | Not-determined |
| 17-108_k141_46458 | Not-determined |
| 17-108_k141_87789 | Not-determined |
| 17-108_k141_114619 | Not-determined |
| 17-108_k141_294153 | Not-determined |
| 17-108_k141_170772 | Not-determined |
| 17-108_k141_321450 | Not-determined |
| 17-108_k141_73246 | Not-determined |
| 17-108_k141_4855 | Not-determined |
| 17-108_k141_170773 | Not-determined |
| 17-108_k141_281212 | Not-determined |
| 17-108_k141_142867 | Not-determined |
| 17-108_k141_60178 | Not-determined |
| 17-108_k141_197556 | Not-determined |
| 17-108_k141_266829 | Not-determined |
| 17-108_k141_239690 | Not-determined |
| 17-108_k141_252990 | Not-determined |
| 17-108_k141_128918 | Not-determined |
| 17-108_k141_101896 | Not-determined |
| 17-108_k141_73253 | Not-determined |
| 17-108_k141_18216 | Not-determined |
| 17-108_k141_32496 | Not-determined |
| 17-108_k141_281218 | Not-determined |
| 17-108_k141_46471 | Not-determined |
| 17-108_k141_101902 | Low-quality |
| 17-108_k141_266837 | Not-determined |
| 17-108_k141_170794 | Not-determined |
| 17-108_k141_128930 | Not-determined |
| 17-108_k141_60192 | Not-determined |
| 17-108_k141_294167 | Not-determined |
| 17-108_k141_73273 | Not-determined |
| 17-108_k141_101907 | Not-determined |
| 17-108_k141_101908 | Not-determined |
| 17-108_k141_294174 | Not-determined |
| 17-108_k141_60200 | Not-determined |
| 17-108_k141_184111 | Not-determined |
| 17-108_k141_170813 | Low-quality |
| 17-108_k141_142891 | Low-quality |
| 17-108_k141_142896 | Not-determined |
| 17-108_k141_308422 | Not-determined |
| 17-108_k141_308423 | Not-determined |
| 17-108_k141_308424 | Low-quality |
| 17-108_k141_212295 | Not-determined |
| 17-108_k141_128966 | Low-quality |
| 17-108_k141_18249 | Low-quality |
| 17-108_k141_128969 | Not-determined |
| 17-108_k141_294182 | Not-determined |
| 17-108_k141_73306 | Not-determined |
| 17-108_k141_32531 | Not-determined |
| 17-108_k141_212298 | Not-determined |
| 17-108_k141_73308 | Not-determined |
| 17-108_k141_18252 | Not-determined |
| 17-108_k141_294186 | Not-determined |
| 17-108_k141_294187 | Not-determined |
| 17-108_k141_170816 | Low-quality |
| 17-108_k141_101929 | Not-determined |
| 17-108_k141_101930 | Not-determined |
| 17-108_k141_294190 | Not-determined |
| 17-108_k141_184124 | Low-quality |
| 17-108_k141_101936 | Not-determined |
| 17-108_k141_184127 | Not-determined |
| 17-108_k141_239759 | Not-determined |
| 17-108_k141_184129 | Not-determined |
| 17-108_k141_308430 | Not-determined |
| 17-108_k141_156566 | Not-determined |
| 17-108_k141_170827 | Not-determined |
| 17-108_k141_253022 | Not-determined |
| 17-108_k141_4878 | Not-determined |
| 17-108_k141_32545 | Not-determined |
| 17-108_k141_142912 | Not-determined |
| 17-108_k141_253031 | Not-determined |
| 17-108_k141_128976 | Not-determined |
| 17-108_k141_212303 | Not-determined |
| 17-108_k141_212305 | Not-determined |
| 17-108_k141_142919 | Not-determined |
| 17-108_k141_46486 | Not-determined |
| 17-108_k141_184149 | Not-determined |
| 17-108_k141_142925 | Not-determined |
| 17-108_k141_156591 | Not-determined |
| 17-108_k141_46494 | Not-determined |
| 17-108_k141_142932 | Not-determined |
| 17-108_k141_281296 | Not-determined |
| 17-108_k141_184157 | Not-determined |
| 17-108_k141_156595 | Low-quality |
| 17-108_k141_266905 | Not-determined |
| 17-108_k141_156596 | Not-determined |
| 17-108_k141_156598 | Low-quality |
| 17-108_k141_308455 | Not-determined |
| 17-108_k141_266909 | Not-determined |
| 17-108_k141_73340 | Not-determined |
| 17-108_k141_60244 | Low-quality |
| 17-108_k141_281303 | Not-determined |
| 17-108_k141_170859 | Not-determined |
| 17-108_k141_60247 | Not-determined |
| 17-108_k141_73346 | Not-determined |
| 17-108_k141_266915 | Not-determined |
| 17-108_k141_239799 | Not-determined |
| 17-108_k141_46508 | Not-determined |
| 17-108_k141_239800 | Not-determined |
| 17-108_k141_321484 | Not-determined |
| 17-108_k141_114693 | Not-determined |
| 17-108_k141_170873 | Not-determined |
| 17-108_k141_253074 | Not-determined |
| 17-108_k141_60268 | Not-determined |
| 17-108_k141_281327 | Not-determined |
| 17-108_k141_281333 | Not-determined |
| 17-108_k141_18292 | Not-determined |
| 17-108_k141_225724 | Low-quality |
| 17-108_k141_321505 | Not-determined |
| 17-108_k141_101984 | Not-determined |
| 17-108_k141_142957 | Not-determined |
| 17-108_k141_142959 | Not-determined |
| 17-108_k141_170899 | Not-determined |
| 17-108_k141_253093 | Not-determined |
| 17-108_k141_142963 | Not-determined |
| 17-108_k141_225730 | Not-determined |
| 17-108_k141_142966 | Not-determined |
| 17-108_k141_87877 | Not-determined |
| 17-108_k141_101999 | Not-determined |
| 17-108_k141_142972 | Not-determined |
| 17-108_k141_225738 | Not-determined |
| 17-108_k141_239853 | Not-determined |
| 17-108_k141_197623 | Low-quality |
| 17-108_k141_253116 | Not-determined |
| 17-108_k141_197625 | Not-determined |
| 17-108_k141_18310 | Not-determined |
| 17-108_k141_212333 | Not-determined |
| 17-108_k141_294267 | Not-determined |
| 17-108_k141_129008 | Low-quality |
| 17-108_k141_32628 | Low-quality |
| 17-108_k141_129010 | Not-determined |
| 17-108_k141_239860 | Not-determined |
| 17-108_k141_102015 | Low-quality |
| 17-108_k141_321520 | Not-determined |
| 17-108_k141_321521 | Low-quality |
| 17-108_k141_32636 | Not-determined |
| 17-108_k141_281348 | Not-determined |
| 17-108_k141_225755 | Not-determined |
| 17-108_k141_102037 | Not-determined |
| 17-108_k141_253128 | Not-determined |
| 17-108_k141_225779 | Not-determined |
| 17-108_k141_143005 | Not-determined |
| 17-108_k141_225780 | Not-determined |
| 17-108_k141_102054 | Not-determined |
| 17-108_k141_46559 | Not-determined |
| 17-108_k141_170909 | Low-quality |
| 17-108_k141_197659 | Not-determined |
| 17-108_k141_102067 | Not-determined |
| 17-108_k141_294306 | Not-determined |
| 17-108_k141_281379 | Not-determined |
| 17-108_k141_102070 | Not-determined |
| 17-108_k141_114724 | Not-determined |
| 17-108_k141_170915 | Not-determined |
| 17-108_k141_129049 | Not-determined |
| 17-108_k141_102076 | Not-determined |
| 17-108_k141_197670 | Not-determined |
| 17-108_k141_321551 | Not-determined |
| 17-108_k141_170921 | Not-determined |
| 17-108_k141_170923 | Low-quality |
| 17-108_k141_102080 | Not-determined |
| 17-108_k141_4976 | Not-determined |
| 17-108_k141_266925 | Low-quality |
| 17-108_k141_239899 | Not-determined |
| 17-108_k141_197674 | Not-determined |
| 17-108_k141_321556 | Not-determined |
| 17-108_k141_308547 | Not-determined |
| 17-108_k141_253164 | Not-determined |
| 17-108_k141_197681 | Not-determined |
| 17-108_k141_4979 | Not-determined |
| 17-108_k141_32688 | Low-quality |
| 17-108_k141_32690 | Not-determined |
| 17-108_k141_184167 | Not-determined |
| 17-108_k141_308563 | Not-determined |
| 17-108_k141_225802 | Not-determined |
| 17-108_k141_87956 | Not-determined |
| 17-108_k141_281407 | Not-determined |
| 17-108_k141_308574 | Not-determined |
| 17-108_k141_225812 | Not-determined |
| 17-108_k141_46570 | Not-determined |
| 17-108_k141_281418 | Not-determined |
| 17-108_k141_197692 | Not-determined |
| 17-108_k141_156616 | Not-determined |
| 17-108_k141_156617 | Not-determined |
| 17-108_k141_266944 | Not-determined |
| 17-108_k141_87968 | Not-determined |
| 17-108_k141_4998 | Not-determined |
| 17-108_k141_253186 | Not-determined |
| 17-108_k141_129067 | Not-determined |
| 17-108_k141_129068 | Not-determined |
| 17-108_k141_321582 | Not-determined |
| 17-108_k141_87972 | Not-determined |
| 17-108_k141_129074 | Not-determined |
| 17-108_k141_60325 | Not-determined |
| 17-108_k141_294330 | Not-determined |
| 17-108_k141_294331 | Not-determined |
| 17-108_k141_87985 | Not-determined |
| 17-108_k141_46581 | Not-determined |
| 17-108_k141_60336 | Not-determined |
| 17-108_k141_102108 | Not-determined |
| 17-108_k141_294337 | Not-determined |
| 17-108_k141_129086 | Not-determined |
| 17-108_k141_5031 | Not-determined |
| 17-108_k141_321605 | Low-quality |
| 17-108_k141_294340 | Not-determined |
| 17-108_k141_321607 | Not-determined |
| 17-108_k141_266976 | Not-determined |
| 17-108_k141_212399 | Not-determined |
| 17-108_k141_156633 | Not-determined |
| 17-108_k141_170962 | Not-determined |
| 17-108_k141_60349 | Not-determined |
| 17-108_k141_170967 | Not-determined |
| 17-108_k141_87995 | Not-determined |
| 17-108_k141_281447 | Not-determined |
| 17-108_k141_294349 | Not-determined |
| 17-108_k141_294351 | Not-determined |
| 17-108_k141_321624 | Not-determined |
| 17-108_k141_294353 | Not-determined |
| 17-108_k141_129100 | Not-determined |
| 17-108_k141_18374 | Not-determined |
| 17-108_k141_60370 | Not-determined |
| 17-108_k141_294370 | Not-determined |
| 17-108_k141_129107 | Not-determined |
| 17-108_k141_156649 | Low-quality |
| 17-108_k141_114777 | Not-determined |
| 17-108_k141_170981 | Not-determined |
| 17-108_k141_88002 | Low-quality |
| 17-108_k141_60376 | Not-determined |
| 17-108_k141_143052 | Not-determined |
| 17-108_k141_129110 | Not-determined |
| 17-108_k141_294377 | Not-determined |
| 17-108_k141_73381 | Not-determined |
| 17-108_k141_170985 | Not-determined |
| 17-108_k141_156657 | Not-determined |
| 17-108_k141_88011 | Low-quality |
| 17-108_k141_239961 | Not-determined |
| 17-108_k141_294387 | Low-quality |
| 17-108_k141_102155 | Not-determined |
| 17-108_k141_294390 | Not-determined |
| 17-108_k141_18395 | Low-quality |
| 17-108_k141_225840 | Not-determined |
| 17-108_k141_308621 | Not-determined |
| 17-108_k141_225847 | Not-determined |
| 17-108_k141_197710 | Low-quality |
| 17-108_k141_60396 | Not-determined |
| 17-108_k141_294399 | Not-determined |
| 17-108_k141_225850 | Not-determined |
| 17-108_k141_212419 | Not-determined |
| 17-108_k141_212420 | Not-determined |
| 17-108_k141_171000 | Not-determined |
| 17-108_k141_281485 | Not-determined |
| 17-108_k141_281487 | Not-determined |
| 17-108_k141_184213 | Not-determined |
| 17-108_k141_88025 | Not-determined |
| 17-108_k141_46612 | Not-determined |
| 17-108_k141_225870 | Not-determined |
| 17-108_k141_212423 | Low-quality |
| 17-108_k141_18403 | Not-determined |
| 17-108_k141_197723 | Low-quality |
| 17-108_k141_18405 | Not-determined |
| 17-108_k141_225878 | Not-determined |
| 17-108_k141_156679 | Not-determined |
| 17-108_k141_184217 | Not-determined |
| 17-108_k141_46620 | Not-determined |
| 17-108_k141_156686 | Not-determined |
| 17-108_k141_60404 | Not-determined |
| 17-108_k141_239987 | Not-determined |
| 17-108_k141_171004 | Not-determined |
| 17-108_k141_281503 | Not-determined |
| 17-108_k141_156697 | Not-determined |
| 17-108_k141_197742 | Not-determined |
| 17-108_k141_281506 | Not-determined |
| 17-108_k141_18416 | Not-determined |
| 17-108_k141_143102 | Not-determined |
| 17-108_k141_212433 | Not-determined |
| 17-108_k141_88048 | Not-determined |
| 17-108_k141_143112 | Not-determined |
| 17-108_k141_281518 | Not-determined |
| 17-108_k141_88052 | Not-determined |
| 17-108_k141_60418 | Not-determined |
| 17-108_k141_308651 | Not-determined |
| 17-108_k141_197762 | Not-determined |
| 17-108_k141_308655 | Not-determined |
| 17-108_k141_102163 | Not-determined |
| 17-108_k141_294430 | Not-determined |
| 17-108_k141_171026 | Not-determined |
| 17-108_k141_102169 | Not-determined |
| 17-108_k141_267044 | Not-determined |
| 17-108_k141_308659 | Not-determined |
| 17-108_k141_5124 | Not-determined |
| 17-108_k141_46657 | Not-determined |
| 17-108_k141_114823 | Not-determined |
| 17-108_k141_5132 | Not-determined |
| 17-108_k141_197778 | Not-determined |
| 17-108_k141_184230 | Not-determined |
| 17-108_k141_281530 | Not-determined |
| 17-108_k141_308665 | Not-determined |
| 17-108_k141_18456 | Low-quality |
| 17-108_k141_308670 | Not-determined |
| 17-108_k141_102186 | Not-determined |
| 17-108_k141_308678 | Not-determined |
| 17-108_k141_114830 | Not-determined |
| 17-108_k141_102188 | Not-determined |
| 17-108_k141_197796 | Not-determined |
| 17-108_k141_102190 | Not-determined |
| 17-108_k141_18468 | Not-determined |
| 17-108_k141_156720 | Not-determined |
| 17-108_k141_102195 | Not-determined |
| 17-108_k141_156721 | Not-determined |
| 17-108_k141_88061 | Not-determined |
| 17-108_k141_156723 | Not-determined |
| 17-108_k141_102198 | Low-quality |
| 17-108_k141_212466 | Not-determined |
| 17-108_k141_114838 | Not-determined |
| 17-108_k141_102201 | Not-determined |
| 17-108_k141_60465 | Low-quality |
| 17-108_k141_88069 | Low-quality |
| 17-108_k141_102208 | Not-determined |
| 17-108_k141_212469 | Not-determined |
| 17-108_k141_308686 | Not-determined |
| 17-108_k141_129145 | Not-determined |
| 17-108_k141_197815 | Not-determined |
| 17-108_k141_114848 | Not-determined |
| 17-108_k141_212475 | Not-determined |
| 17-108_k141_267088 | Not-determined |
| 17-108_k141_171044 | Not-determined |
| 17-108_k141_129154 | Not-determined |
| 17-108_k141_197822 | Not-determined |
| 17-108_k141_281558 | Not-determined |
| 17-108_k141_308703 | Not-determined |
| 17-108_k141_5162 | Not-determined |
| 17-108_k141_184277 | Not-determined |
| 17-108_k141_225922 | Not-determined |
| 17-108_k141_46674 | Low-quality |
| 17-108_k141_88079 | Not-determined |
| 17-108_k141_18486 | Not-determined |
| 17-109-final-viral-combined_k141_147628\|\|full | Complete |
| 17-108_k141_321674 | Not-determined |
| 17-108_k141_143149 | Not-determined |
| 17-108_k141_114870 | Not-determined |
| 17-108_k141_73444 | Not-determined |
| 17-108_k141_88085 | Not-determined |
| 17-108_k141_60489 | Not-determined |
| 17-108_k141_308715 | Not-determined |
| 17-108_k141_114874 | Low-quality |
| 17-108_k141_308719 | Not-determined |
| 17-108_k141_73452 | Not-determined |
| 17-108_k141_197845 | Not-determined |
| 17-108_k141_102222 | Not-determined |
| 17-108_k141_184298 | Not-determined |
| 17-108_k141_18492 | Not-determined |
| 17-108_k141_102226 | Not-determined |
| 17-108_k141_102228 | Not-determined |
| 17-108_k141_102232 | Not-determined |
| 17-108_k141_294465 | Not-determined |
| 17-108_k141_267125 | Not-determined |
| 17-108_k141_184304 | Not-determined |
| 17-108_k141_281589 | Not-determined |
| 17-108_k141_156756 | Not-determined |
| 17-108_k141_197857 | Not-determined |
| 17-108_k141_308724 | Not-determined |
| 17-108_k141_156758 | Not-determined |
| 17-108_k141_267127 | Low-quality |
| 17-108_k141_5190 | Not-determined |
| 17-108_k141_294475 | Not-determined |
| 17-108_k141_114902 | Not-determined |
| 17-108_k141_114903 | Not-determined |
| 17-108_k141_240046 | Not-determined |
| 17-108_k141_294479 | Not-determined |
| 17-108_k141_197867 | Not-determined |
| 17-108_k141_114908 | Not-determined |
| 17-108_k141_225951 | Not-determined |
| 17-108_k141_294487 | Not-determined |
| 17-108_k141_73472 | Not-determined |
| 17-108_k141_197875 | Not-determined |
| 17-108_k141_32727 | Not-determined |
| 17-108_k141_281617 | Not-determined |
| 17-108_k141_281620 | Not-determined |
| 17-108_k141_267139 | Not-determined |
| 17-108_k141_197893 | Not-determined |
| 17-108_k141_114928 | Not-determined |
| 17-108_k141_281635 | Low-quality |
| 17-108_k141_129201 | Not-determined |
| 17-108_k141_143205 | Not-determined |
| 17-108_k141_114947 | Not-determined |
| 17-108_k141_308750 | Low-quality |
| 17-108_k141_253250 | Not-determined |
| 17-108_k141_32748 | Not-determined |
| 17-108_k141_114953 | Not-determined |
| 17-108_k141_308758 | Not-determined |
| 17-108_k141_281648 | Low-quality |
| 17-108_k141_129212 | Not-determined |
| 17-108_k141_197922 | Not-determined |
| 17-108_k141_240093 | Not-determined |
| 17-108_k141_308770 | Not-determined |
| 17-108_k141_308774 | Not-determined |
| 17-108_k141_46742 | Not-determined |
| 17-108_k141_197935 | Not-determined |
| 17-108_k141_253278 | Not-determined |
| 17-108_k141_212577 | Not-determined |
| 17-108_k141_18554 | Not-determined |
| 17-108_k141_88119 | Not-determined |
| 17-108_k141_102282 | Not-determined |
| 17-108_k141_240104 | Low-quality |
| 17-108_k141_102284 | Not-determined |
| 17-108_k141_240107 | Not-determined |
| 17-108_k141_114960 | Not-determined |
| 17-108_k141_212592 | Not-determined |
| 17-108_k141_253304 | Low-quality |
| 17-108_k141_5222 | Not-determined |
| 17-108_k141_5228 | Low-quality |
| 17-108_k141_32781 | Not-determined |
| 17-108_k141_114969 | Not-determined |
| 17-108_k141_114976 | Not-determined |
| 17-108_k141_184348 | Not-determined |
| 17-108_k141_281679 | Low-quality |
| 17-108_k141_32788 | Not-determined |
| 17-108_k141_281684 | Not-determined |
| 17-108_k141_184354 | Not-determined |
| 17-108_k141_88136 | Not-determined |
| 17-108_k141_18564 | Not-determined |
| 17-108_k141_60524 | Not-determined |
| 17-108_k141_184358 | Not-determined |
| 17-108_k141_156823 | Not-determined |
| 17-108_k141_184366 | Not-determined |
| 17-108_k141_18574 | Not-determined |
| 17-108_k141_32794 | Not-determined |
| 17-108_k141_73514 | Low-quality |
| 17-108_k141_225980 | Low-quality |
| 17-108_k141_5251 | Not-determined |
| 17-108_k141_240138 | Not-determined |
| 17-108_k141_184368 | Not-determined |
| 17-108_k141_225985 | Not-determined |
| 17-108_k141_184370 | Not-determined |
| 17-108_k141_115002 | Not-determined |
| 17-108_k141_225987 | Not-determined |
| 17-108_k141_18582 | Not-determined |
| 17-108_k141_184376 | Not-determined |
| 17-108_k141_129265 | Not-determined |
| 17-108_k141_88152 | Not-determined |
| 17-108_k141_88154 | Not-determined |
| 17-108_k141_60538 | Not-determined |
| 17-108_k141_197961 | Not-determined |
| 17-108_k141_253351 | Not-determined |
| 17-108_k141_212638 | Not-determined |
| 17-108_k141_197964 | Low-quality |
| 17-108_k141_156840 | Not-determined |
| 17-108_k141_321704 | Not-determined |
| 17-108_k141_308841 | Not-determined |
| 17-108_k141_240147 | Low-quality |
| 17-108_k141_143268 | Not-determined |
| 17-108_k141_240150 | Not-determined |
| 17-108_k141_184386 | Low-quality |
| 17-108_k141_156845 | Not-determined |
| 17-108_k141_88173 | Not-determined |
| 17-108_k141_102309 | Not-determined |
| 17-108_k141_143274 | Not-determined |
| 17-108_k141_129280 | Not-determined |
| 17-108_k141_129283 | Not-determined |
| 17-108_k141_73539 | Low-quality |
| 17-108_k141_88181 | Not-determined |
| 17-108_k141_321715 | Low-quality |
| 17-108_k141_129289 | Not-determined |
| 17-108_k141_308861 | Not-determined |
| 17-108_k141_129292 | Not-determined |
| 17-108_k141_294511 | Not-determined |
| 17-108_k141_115044 | Not-determined |
| 17-108_k141_240184 | Not-determined |
| 17-108_k141_321725 | Low-quality |
| 17-108_k141_143280 | Not-determined |
| 17-108_k141_102334 | Not-determined |
| 17-108_k141_212666 | Not-determined |
| 17-108_k141_5295 | Not-determined |
| 17-108_k141_294527 | Not-determined |
| 17-108_k141_156871 | Not-determined |
| 17-108_k141_156874 | Not-determined |
| 17-108_k141_115061 | Low-quality |
| 17-108_k141_73571 | Not-determined |
| 17-108_k141_46800 | Not-determined |
| 17-108_k141_184408 | Not-determined |
| 17-108_k141_226016 | Not-determined |
| 17-108_k141_212681 | Not-determined |
| 17-108_k141_156889 | Not-determined |
| 17-108_k141_129333 | Not-determined |
| 17-108_k141_156892 | Not-determined |
| 17-108_k141_60568 | Not-determined |
| 17-108_k141_73602 | Low-quality |
| 17-108_k141_88185 | Not-determined |
| 17-108_k141_102357 | Not-determined |
| 17-108_k141_60571 | Not-determined |
| 17-108_k141_226042 | Not-determined |
| 17-108_k141_197986 | Not-determined |
| 17-108_k141_102368 | Not-determined |
| 17-108_k141_197996 | Not-determined |
| 17-108_k141_88208 | Not-determined |
| 17-108_k141_267177 | Not-determined |
| 17-108_k141_267184 | Not-determined |
| 17-108_k141_184436 | Not-determined |
| 17-108_k141_267185 | Not-determined |
| 17-108_k141_212723 | Not-determined |
| 17-108_k141_102393 | Not-determined |
| 17-108_k141_198012 | Not-determined |
| 17-108_k141_115103 | Not-determined |
| 17-108_k141_321750 | Not-determined |
| 17-108_k141_102396 | Low-quality |
| 17-108_k141_308897 | Not-determined |
| 17-108_k141_102397 | Not-determined |
| 17-108_k141_32895 | Low-quality |
| 17-108_k141_226077 | Not-determined |
| 17-108_k141_156919 | Low-quality |
| 17-108_k141_308904 | Low-quality |
| 17-108_k141_198020 | Not-determined |
| 17-108_k141_294577 | Low-quality |
| 17-108_k141_73643 | Not-determined |
| 17-108_k141_212741 | Not-determined |
| 17-108_k141_281688 | Low-quality |
| 17-108_k141_73646 | Not-determined |
| 17-108_k141_308908 | Not-determined |
| 17-108_k141_253426 | Not-determined |
| 17-108_k141_115129 | Not-determined |
| 17-108_k141_156936 | Not-determined |
| 17-108_k141_240236 | Not-determined |
| 17-108_k141_156938 | Not-determined |
| 17-108_k141_102416 | Not-determined |
| 17-108_k141_240239 | Not-determined |
| 17-108_k141_115141 | Not-determined |
| 17-108_k141_46826 | Not-determined |
| 17-108_k141_321756 | Not-determined |
| 17-108_k141_240247 | Not-determined |
| 17-108_k141_46830 | Not-determined |
| 17-108_k141_102431 | Not-determined |
| 17-108_k141_321762 | Not-determined |
| 17-108_k141_321764 | Not-determined |
| 17-108_k141_88226 | Not-determined |
| 17-108_k141_60598 | Low-quality |
| 17-108_k141_253437 | Not-determined |
| 17-108_k141_46837 | Not-determined |
| 17-108_k141_184465 | Not-determined |
| 17-108_k141_46839 | Not-determined |
| 17-108_k141_32929 | Low-quality |
| 17-108_k141_5380 | Not-determined |

| | |
|---|---|
| 17-108_k141_73680 | Not-determined |
| 17-108_k141_198047 | Not-determined |
| 17-108_k141_198051 | Not-determined |
| 17-108_k141_184476 | Not-determined |
| 17-108_k141_5388 | Not-determined |
| 17-108_k141_198058 | Not-determined |
| 17-108_k141_184485 | Not-determined |
| 17-108_k141_102437 | Not-determined |
| 17-108_k141_115170 | Not-determined |
| 17-108_k141_184490 | Low-quality |
| 17-108_k141_32940 | Not-determined |
| 17-108_k141_102440 | Not-determined |
| 17-108_k141_253455 | Not-determined |
| 17-108_k141_198068 | Not-determined |
| 17-108_k141_88252 | Not-determined |
| 17-108_k141_88253 | Not-determined |
| 17-108_k141_156978 | Not-determined |
| 17-108_k141_102449 | Not-determined |
| 17-108_k141_267198 | Low-quality |
| 17-108_k141_184507 | Not-determined |
| 17-108_k141_32975 | Not-determined |
| 17-108_k141_143311 | Not-determined |
| 17-108_k141_143315 | Not-determined |
| 17-108_k141_184520 | Not-determined |
| 17-108_k141_253471 | Not-determined |
| 17-108_k141_240313 | Not-determined |
| 17-108_k141_102470 | Not-determined |
| 17-108_k141_171112 | Not-determined |
| 17-108_k141_143328 | Not-determined |
| 17-108_k141_212784 | Not-determined |
| 17-108_k141_32994 | Not-determined |
| 17-108_k141_143331 | Not-determined |
| 17-108_k141_171119 | Not-determined |
| 17-108_k141_73706 | Not-determined |
| 17-108_k141_253473 | Not-determined |
| 17-108_k141_33002 | Not-determined |
| 17-108_k141_171121 | Not-determined |
| 17-108_k141_184533 | Low-quality |
| 17-108_k141_184535 | Not-determined |
| 17-108_k141_240327 | Not-determined |
| 17-108_k141_171127 | Not-determined |
| 17-108_k141_73718 | Not-determined |
| 17-108_k141_73722 | Not-determined |
| 17-108_k141_156994 | Not-determined |
| 17-108_k141_88276 | Not-determined |
| 17-108_k141_294676 | Not-determined |
| 17-108_k141_321779 | Not-determined |
| 17-108_k141_88279 | Not-determined |
| 17-108_k141_267221 | Not-determined |
| 17-108_k141_18591 | Not-determined |
| 17-108_k141_294688 | Low-quality |
| 17-108_k141_240356 | Not-determined |
| 17-108_k141_281737 | Not-determined |
| 17-108_k141_102515 | Not-determined |
| 17-108_k141_308958 | Not-determined |
| 17-108_k141_143372 | Not-determined |
| 17-108_k141_143374 | Not-determined |
| 17-108_k141_5461 | Not-determined |
| 17-108_k141_212802 | Not-determined |
| 17-108_k141_294705 | Not-determined |
| 17-108_k141_212805 | Not-determined |
| 17-108_k141_171145 | Not-determined |
| 17-108_k141_171147 | Not-determined |
| 17-108_k141_198120 | Not-determined |
| 17-108_k141_198123 | Not-determined |
| 17-108_k141_171152 | Not-determined |
| 17-108_k141_294719 | Not-determined |
| 17-108_k141_102531 | Not-determined |
| 17-108_k141_102532 | Not-determined |
| 17-108_k141_294724 | Not-determined |
| 17-108_k141_5475 | Not-determined |
| 17-108_k141_143394 | Low-quality |
| 17-108_k141_46900 | Not-determined |
| 17-108_k141_5481 | Not-determined |
| 17-108_k141_253515 | Not-determined |
| 17-108_k141_143402 | Not-determined |
| 17-108_k141_129414 | Not-determined |
| 17-108_k141_157024 | Not-determined |
| 17-108_k141_73747 | Not-determined |
| 17-108_k141_309009 | Not-determined |
| 17-108_k141_240398 | Not-determined |
| 17-108_k141_157035 | Not-determined |
| 17-108_k141_129424 | Not-determined |
| 17-108_k141_60671 | Not-determined |
| 17-108_k141_129427 | Low-quality |
| 17-108_k141_184613 | Not-determined |
| 17-108_k141_184614 | Not-determined |
| 17-108_k141_309020 | Not-determined |
| 17-108_k141_294743 | Not-determined |
| 17-108_k141_46922 | Low-quality |
| 17-108_k141_253537 | Low-quality |
| 17-108_k141_309027 | Not-determined |
| 17-108_k141_115219 | Not-determined |
| 17-108_k141_184620 | Low-quality |
| 17-108_k141_267267 | Not-determined |
| 17-108_k141_73768 | Not-determined |
| 17-108_k141_73771 | Not-determined |
| 17-108_k141_294755 | Not-determined |
| 17-108_k141_171179 | Not-determined |
| 17-108_k141_171180 | Low-quality |
| 17-108_k141_46933 | Not-determined |
| 17-108_k141_212817 | Not-determined |
| 17-108_k141_60703 | Not-determined |
| 17-108_k141_18643 | Low-quality |
| 17-108_k141_60708 | Not-determined |
| 17-108_k141_198169 | Not-determined |
| 17-108_k141_73790 | Not-determined |
| 17-108_k141_309047 | Not-determined |
| 17-108_k141_143444 | Not-determined |
| 17-108_k141_267280 | Not-determined |
| 17-108_k141_60714 | Not-determined |
| 17-108_k141_129456 | Low-quality |
| 17-108_k141_60719 | Not-determined |
| 17-108_k141_157052 | Not-determined |
| 17-108_k141_309055 | Not-determined |
| 17-108_k141_253558 | Not-determined |
| 17-108_k141_46952 | Not-determined |
| 17-108_k141_60724 | Not-determined |
| 17-108_k141_267292 | Not-determined |
| 17-108_k141_253565 | Not-determined |
| 17-108_k141_171201 | Not-determined |
| 17-108_k141_309068 | Not-determined |
| 17-108_k141_157062 | Low-quality |
| 17-108_k141_309070 | Not-determined |
| 17-108_k141_102591 | Not-determined |
| 17-108_k141_129476 | Not-determined |
| 17-108_k141_60736 | Not-determined |
| 17-108_k141_253580 | Not-determined |
| 17-108_k141_115256 | Low-quality |
| 17-108_k141_294761 | Not-determined |
| 17-108_k141_294764 | Not-determined |
| 17-108_k141_171224 | Not-determined |
| 17-108_k141_115266 | Not-determined |
| 17-108_k141_212854 | Not-determined |
| 17-108_k141_115268 | Not-determined |
| 17-108_k141_157076 | Not-determined |
| 17-108_k141_226111 | Not-determined |
| 17-108_k141_73841 | Low-quality |
| 17-108_k141_115273 | Not-determined |
| 17-108_k141_226113 | Low-quality |
| 17-108_k141_60756 | Not-determined |
| 17-108_k141_60758 | Not-determined |
| 17-108_k141_143476 | Not-determined |
| 17-108_k141_157092 | Not-determined |
| 17-108_k141_240467 | Not-determined |
| 17-108_k141_102619 | Not-determined |
| 17-108_k141_267309 | Not-determined |
| 17-108_k141_184664 | Not-determined |
| 17-108_k141_73856 | Not-determined |
| 17-108_k141_184667 | Not-determined |
| 17-108_k141_253611 | Not-determined |
| 17-108_k141_47010 | Not-determined |
| 17-108_k141_294776 | Low-quality |
| 17-108_k141_47017 | Not-determined |
| 17-108_k141_73861 | Not-determined |
| 17-108_k141_267316 | Low-quality |
| 17-108_k141_309110 | Not-determined |
| 17-108_k141_102636 | Low-quality |
| 17-108_k141_73868 | Not-determined |
| 17-108_k141_73871 | Not-determined |
| 17-108_k141_18679 | Not-determined |
| 17-108_k141_171245 | Not-determined |
| 17-108_k141_253631 | Not-determined |
| 17-108_k141_321849 | Not-determined |
| 17-108_k141_212869 | Low-quality |
| 17-108_k141_294800 | Not-determined |
| 17-108_k141_143492 | Not-determined |
| 17-108_k141_226145 | Not-determined |
| 17-108_k141_267333 | Low-quality |
| 17-108_k141_47034 | Not-determined |
| 17-108_k141_281780 | Not-determined |
| 17-108_k141_129493 | Not-determined |
| 17-108_k141_253637 | Low-quality |
| 17-108_k141_171252 | Not-determined |
| 17-108_k141_226153 | Not-determined |
| 17-108_k141_143496 | Not-determined |
| 17-108_k141_294805 | Low-quality |
| 17-108_k141_18692 | Not-determined |
| 17-108_k141_18700 | Not-determined |
| 17-108_k141_129503 | Not-determined |
| 17-108_k141_88381 | Not-determined |
| 17-108_k141_294810 | Not-determined |
| 17-108_k141_198222 | Low-quality |
| 17-108_k141_253654 | Not-determined |
| 17-108_k141_47054 | Not-determined |
| 17-108_k141_73892 | Not-determined |
| 17-108_k141_73893 | Not-determined |
| 17-108_k141_33089 | Not-determined |
| 17-108_k141_18714 | Not-determined |
| 17-108_k141_267350 | Not-determined |
| 17-108_k141_47071 | Not-determined |
| 17-108_k141_18720 | Not-determined |
| 17-108_k141_5514 | Not-determined |
| 17-108_k141_73908 | Not-determined |
| 17-108_k141_47077 | Not-determined |
| 17-108_k141_60769 | Low-quality |
| 17-108_k141_60774 | Not-determined |
| 17-108_k141_240510 | Not-determined |
| 17-108_k141_18729 | Low-quality |
| 17-108_k141_129527 | Low-quality |
| 17-108_k141_73917 | Not-determined |
| 17-108_k141_129529 | Not-determined |
| 17-108_k141_18735 | Not-determined |
| 17-108_k141_184690 | Not-determined |
| 17-108_k141_253664 | Not-determined |
| 17-108_k141_143538 | Not-determined |
| 17-108_k141_5529 | Low-quality |
| 17-108_k141_212898 | Not-determined |
| 17-108_k141_294838 | Not-determined |
| 17-108_k141_198255 | Not-determined |
| 17-108_k141_198256 | Not-determined |
| 17-108_k141_321866 | Not-determined |
| 17-108_k141_309155 | Not-determined |
| 17-108_k141_321877 | Low-quality |
| 17-108_k141_73938 | Not-determined |
| 17-108_k141_18758 | Not-determined |
| 17-108_k141_5547 | Not-determined |
| 17-108_k141_309170 | Not-determined |
| 17-108_k141_18764 | Not-determined |
| 17-108_k141_171299 | Low-quality |
| 17-108_k141_240529 | Not-determined |
| 17-108_k141_198282 | Not-determined |
| 17-108_k141_18771 | Low-quality |
| 17-108_k141_129560 | Not-determined |
| 17-108_k141_198288 | Not-determined |
| 17-108_k141_143572 | Not-determined |
| 17-108_k141_102707 | Not-determined |
| 17-108_k141_309178 | Not-determined |
| 17-108_k141_212920 | Not-determined |
| 17-108_k141_47120 | Not-determined |
| 17-108_k141_171300 | Not-determined |
| 17-108_k141_321895 | Not-determined |
| 17-108_k141_321900 | Not-determined |
| 17-108_k141_253688 | Not-determined |
| 17-108_k141_309194 | Not-determined |
| 17-108_k141_321908 | Not-determined |
| 17-108_k141_321909 | Not-determined |
| 17-108_k141_321914 | Not-determined |
| 17-108_k141_47138 | Not-determined |
| 17-108_k141_143594 | Not-determined |
| 17-108_k141_129583 | Not-determined |
| 17-108_k141_240535 | Not-determined |
| 17-108_k141_294864 | Not-determined |
| 17-108_k141_253691 | Not-determined |
| 17-108_k141_294867 | Not-determined |
| 17-108_k141_281844 | Not-determined |
| 17-108_k141_73959 | Low-quality |
| 17-108_k141_226170 | Not-determined |
| 17-108_k141_294873 | Not-determined |
| 17-108_k141_294875 | Not-determined |
| 17-108_k141_171313 | Not-determined |
| 17-108_k141_321927 | Not-determined |
| 17-108_k141_33141 | Not-determined |
| 17-108_k141_240549 | Not-determined |
| 17-108_k141_60824 | Not-determined |
| 17-108_k141_267371 | Low-quality |
| 17-108_k141_226184 | Not-determined |
| 17-108_k141_184739 | Not-determined |
| 17-108_k141_171317 | Not-determined |
| 17-108_k141_267374 | Not-determined |
| 17-108_k141_157159 | Not-determined |
| 17-108_k141_5573 | Not-determined |
| 17-108_k141_226194 | Not-determined |
| 17-108_k141_47152 | Not-determined |
| 17-108_k141_240559 | Not-determined |
| 17-108_k141_309221 | Not-determined |
| 17-108_k141_226203 | Not-determined |
| 17-108_k141_212959 | Not-determined |
| 17-108_k141_73990 | Not-determined |
| 17-108_k141_267388 | Not-determined |
| 17-108_k141_171335 | Not-determined |
| 17-108_k141_60858 | Not-determined |
| 17-108_k141_294900 | Not-determined |
| 17-108_k141_309232 | Not-determined |
| 17-108_k141_157175 | Not-determined |
| 17-108_k141_253715 | Low-quality |
| 17-108_k141_102718 | Not-determined |
| 17-108_k141_129597 | Not-determined |
| 17-108_k141_240575 | Not-determined |
| 17-108_k141_240576 | Not-determined |
| 17-108_k141_309244 | Low-quality |
| 17-108_k141_129608 | Not-determined |
| 17-108_k141_171349 | Not-determined |
| 17-108_k141_88453 | Not-determined |
| 17-108_k141_184777 | Not-determined |
| 17-108_k141_171350 | Not-determined |
| 17-108_k141_171355 | Not-determined |
| 17-108_k141_88459 | Not-determined |
| 17-108_k141_47169 | Not-determined |
| 17-108_k141_88461 | Not-determined |
| 17-108_k141_129612 | Low-quality |
| 17-108_k141_184786 | Not-determined |
| 17-108_k141_198313 | Not-determined |
| 17-108_k141_102726 | Not-determined |
| 17-108_k141_198316 | Not-determined |
| 17-108_k141_157199 | Not-determined |
| 17-108_k141_212984 | Low-quality |
| 17-108_k141_88472 | Not-determined |
| 17-108_k141_102743 | Not-determined |
| 17-108_k141_171377 | Not-determined |
| 17-108_k141_60886 | Not-determined |
| 17-108_k141_47183 | Not-determined |
| 17-108_k141_102749 | Not-determined |
| 17-108_k141_171380 | Not-determined |
| 17-108_k141_240618 | Not-determined |
| 17-108_k141_184794 | Not-determined |
| 17-108_k141_102751 | Not-determined |
| 17-108_k141_212994 | Not-determined |
| 17-108_k141_33167 | Low-quality |
| 17-108_k141_309258 | Not-determined |
| 17-108_k141_5601 | Low-quality |
| 17-108_k141_88484 | Not-determined |
| 17-108_k141_33181 | Not-determined |
| 17-108_k141_198352 | Not-determined |
| 17-108_k141_88493 | Not-determined |
| 17-108_k141_309271 | Not-determined |
| 17-108_k141_198357 | Not-determined |
| 17-108_k141_321943 | Not-determined |
| 17-108_k141_33195 | Not-determined |
| 17-108_k141_294935 | Not-determined |
| 17-108_k141_294944 | Not-determined |
| 17-108_k141_321948 | Low-quality |
| 17-108_k141_129630 | Not-determined |
| 17-108_k141_253764 | Not-determined |
| 17-108_k141_226241 | Not-determined |
| 17-108_k141_33211 | Low-quality |
| 17-108_k141_60924 | Not-determined |
| 17-108_k141_281880 | Not-determined |
| 17-108_k141_198365 | Not-determined |
| 17-108_k141_171402 | Not-determined |
| 17-108_k141_60928 | Not-determined |
| 17-108_k141_267471 | Not-determined |
| 17-108_k141_267472 | Not-determined |
| 17-108_k141_198375 | Not-determined |
| 17-108_k141_171406 | Not-determined |
| 17-108_k141_60929 | Low-quality |
| 17-108_k141_60931 | Not-determined |
| 17-108_k141_240644 | Low-quality |
| 17-108_k141_74031 | Not-determined |
| 17-108_k141_198381 | Not-determined |
| 17-108_k141_321968 | Not-determined |
| 17-108_k141_198384 | Not-determined |
| 17-108_k141_74040 | Not-determined |
| 17-108_k141_253793 | Not-determined |
| 17-108_k141_5620 | Not-determined |
| 17-108_k141_47213 | Not-determined |
| 17-108_k141_281907 | Not-determined |
| 17-108_k141_294967 | Not-determined |
| 17-108_k141_88512 | Not-determined |
| 17-108_k141_226266 | Not-determined |
| 17-108_k141_281926 | Not-determined |
| 17-108_k141_74053 | Not-determined |
| 17-108_k141_321990 | Low-quality |
| 17-108_k141_281933 | Not-determined |
| 17-108_k141_281934 | Not-determined |
| 17-108_k141_33254 | Not-determined |
| 17-108_k141_226272 | Not-determined |
| 17-108_k141_281942 | Not-determined |
| 17-108_k141_157241 | Low-quality |
| 17-108_k141_253809 | Not-determined |
| 17-108_k141_294984 | Not-determined |
| 17-108_k141_281951 | Not-determined |
| 17-108_k141_102805 | Not-determined |
| 17-108_k141_294987 | Not-determined |
| 17-108_k141_143608 | Not-determined |
| 17-108_k141_253815 | Not-determined |
| 17-108_k141_143611 | Not-determined |
| 17-108_k141_198419 | Not-determined |
| 17-108_k141_294994 | Not-determined |
| 17-108_k141_143616 | Not-determined |
| 17-108_k141_309285 | Low-quality |
| 17-108_k141_309289 | Not-determined |
| 17-108_k141_295002 | Not-determined |
| 17-108_k141_295003 | Not-determined |
| 17-108_k141_309296 | Not-determined |
| 17-108_k141_157263 | Not-determined |
| 17-108_k141_157264 | Not-determined |
| 17-108_k141_309298 | Not-determined |
| 17-108_k141_198432 | Not-determined |
| 17-108_k141_157269 | Not-determined |
| 17-108_k141_213055 | Not-determined |
| 17-108_k141_309311 | Not-determined |
| 17-108_k141_295013 | Low-quality |
| 17-108_k141_47246 | Not-determined |
| 17-108_k141_18846 | Not-determined |
| 17-108_k141_322027 | Not-determined |
| 17-108_k141_157290 | Not-determined |
| 17-108_k141_33288 | Not-determined |
| 17-108_k141_47261 | Not-determined |
| 17-108_k141_60988 | Not-determined |
| 17-108_k141_309324 | Not-determined |
| 17-108_k141_253862 | Low-quality |
| 17-108_k141_226313 | Not-determined |
| 17-108_k141_295019 | Not-determined |
| 17-108_k141_33296 | Not-determined |
| 17-108_k141_33299 | Not-determined |
| 17-108_k141_253870 | Not-determined |
| 17-108_k141_240687 | Not-determined |
| 17-108_k141_74103 | Not-determined |
| 17-108_k141_240693 | Not-determined |
| 17-108_k141_267514 | Not-determined |
| 17-108_k141_198449 | Not-determined |
| 17-108_k141_213082 | Not-determined |
| 17-108_k141_143662 | Not-determined |
| 17-108_k141_198458 | Not-determined |
| 17-108_k141_213089 | Not-determined |
| 17-108_k141_143672 | Not-determined |
| 17-108_k141_295041 | Not-determined |
| 17-108_k141_240703 | Not-determined |
| 17-108_k141_267533 | Not-determined |
| 17-108_k141_61024 | Not-determined |
| 17-108_k141_213100 | Not-determined |
| 17-108_k141_282029 | Not-determined |
| 17-108_k141_47298 | Not-determined |
| 17-108_k141_157328 | Not-determined |
| 17-108_k141_102849 | Not-determined |
| 17-108_k141_5678 | Not-determined |
| 17-108_k141_5679 | Not-determined |
| 17-108_k141_129680 | Not-determined |
| 17-108_k141_88537 | Not-determined |
| 17-108_k141_102859 | Not-determined |
| 17-108_k141_47307 | Not-determined |
| 17-108_k141_322035 | Not-determined |
| 17-108_k141_143698 | Not-determined |
| 17-108_k141_74131 | Not-determined |
| 17-108_k141_226352 | Not-determined |
| 17-108_k141_322037 | Low-quality |
| 17-108_k141_33319 | Not-determined |
| 17-108_k141_102876 | Low-quality |
| 17-108_k141_226357 | Not-determined |
| 17-108_k141_5696 | Not-determined |
| 17-108_k141_267564 | Not-determined |
| 17-108_k141_171432 | Not-determined |
| 17-108_k141_171433 | Not-determined |
| 17-108_k141_171434 | Not-determined |
| 17-108_k141_198474 | Not-determined |
| 17-108_k141_129705 | Not-determined |
| 17-108_k141_33337 | Not-determined |
| 17-108_k141_102887 | Not-determined |
| 17-108_k141_213132 | Not-determined |
| 17-108_k141_47335 | Not-determined |
| 17-108_k141_267575 | Not-determined |
| 17-108_k141_322046 | Low-quality |
| 17-108_k141_322049 | Not-determined |
| 17-108_k141_322051 | Not-determined |
| 17-108_k141_282055 | Not-determined |
| 17-108_k141_213137 | Not-determined |
| 17-108_k141_240740 | Not-determined |
| 17-108_k141_129717 | Not-determined |
| 17-108_k141_157369 | Not-determined |
| 17-108_k141_184799 | Not-determined |
| 17-108_k141_115354 | Low-quality |
| 17-108_k141_213149 | Not-determined |
| 17-108_k141_322059 | Not-determined |
| 17-108_k141_198489 | Low-quality |
| 17-108_k141_88574 | Not-determined |
| 17-108_k141_267592 | Low-quality |
| 17-108_k141_47347 | Not-determined |
| 17-108_k141_33352 | Not-determined |
| 17-108_k141_129729 | Not-determined |
| 17-108_k141_129734 | Not-determined |
| 17-108_k141_253944 | Not-determined |
| 17-108_k141_253945 | Low-quality |
| 17-108_k141_18883 | Not-determined |
| 17-108_k141_115361 | Not-determined |
| 17-108_k141_171457 | Not-determined |
| 17-108_k141_102902 | Low-quality |
| 17-108_k141_267604 | Not-determined |
| 17-108_k141_240761 | Not-determined |
| 17-108_k141_253954 | Not-determined |
| 17-108_k141_240766 | Not-determined |
| 17-108_k141_47370 | Not-determined |
| 17-108_k141_61040 | Not-determined |
| 17-108_k141_33369 | Not-determined |
| 17-108_k141_115372 | Not-determined |
| 17-108_k141_157398 | Not-determined |
| 17-108_k141_18893 | Not-determined |
| 17-108_k141_309429 | Not-determined |
| 17-108_k141_184810 | Not-determined |
| 17-108_k141_184811 | Not-determined |
| 17-108_k141_309436 | Not-determined |
| 17-108_k141_115381 | Not-determined |
| 17-108_k141_184817 | Low-quality |
| 17-108_k141_33379 | Low-quality |
| 17-108_k141_226413 | Not-determined |
| 17-108_k141_240785 | Not-determined |
| 17-108_k141_253982 | Not-determined |
| 17-108_k141_115395 | Not-determined |
| 17-108_k141_5726 | Not-determined |
| 17-108_k141_213183 | Low-quality |
| 17-108_k141_309453 | Not-determined |
| 17-108_k141_5729 | Not-determined |
| 17-108_k141_309454 | Not-determined |
| 17-108_k141_184834 | Not-determined |
| 17-108_k141_198500 | Not-determined |
| 17-108_k141_102933 | Not-determined |
| 17-108_k141_74201 | Not-determined |
| 17-108_k141_184838 | Low-quality |
| 17-108_k141_226433 | Not-determined |
| 17-108_k141_254000 | Low-quality |
| 17-108_k141_226435 | Not-determined |
| 17-108_k141_33401 | Not-determined |
| 17-108_k141_213198 | Not-determined |
| 17-108_k141_282110 | Not-determined |
| 17-108_k141_295107 | Not-determined |
| 17-108_k141_184843 | Low-quality |
| 17-108_k141_213208 | Not-determined |
| 17-108_k141_18938 | Not-determined |
| 17-108_k141_213217 | Not-determined |
| 17-108_k141_129803 | Not-determined |
| 17-108_k141_61067 | Not-determined |
| 17-108_k141_143747 | Not-determined |
| 17-108_k141_171487 | Not-determined |
| 17-108_k141_267668 | Not-determined |
| 17-108_k141_33415 | Not-determined |
| 17-108_k141_129813 | Not-determined |
| 17-108_k141_47395 | Not-determined |
| 17-108_k141_171495 | Not-determined |
| 17-108_k141_129816 | Not-determined |
| 17-108_k141_18951 | Not-determined |
| 17-108_k141_33422 | Not-determined |
| 17-108_k141_184857 | Not-determined |
| 17-108_k141_184858 | Not-determined |
| 17-108_k141_295126 | Not-determined |
| 17-108_k141_240809 | Not-determined |
| 17-108_k141_5757 | Not-determined |
| 17-108_k141_115440 | Low-quality |
| 17-108_k141_184869 | Low-quality |
| 17-108_k141_129827 | Not-determined |
| 17-108_k141_254015 | Not-determined |
| 17-108_k141_143761 | Not-determined |
| 17-108_k141_295128 | Not-determined |
| 17-108_k141_295129 | Not-determined |
| 17-108_k141_33441 | Not-determined |
| 17-108_k141_61089 | Not-determined |
| 17-108_k141_171503 | Not-determined |
| 17-108_k141_295132 | Not-determined |
| 17-108_k141_282128 | Not-determined |
| 17-108_k141_33444 | Not-determined |
| 17-108_k141_322105 | Low-quality |
| 17-108_k141_322106 | Not-determined |
| 17-108_k141_88626 | Not-determined |
| 17-108_k141_115450 | Not-determined |
| 17-108_k141_33446 | Not-determined |
| 17-108_k141_282137 | Not-determined |
| 17-108_k141_254030 | Not-determined |
| 17-108_k141_88631 | Not-determined |
| 17-108_k141_226464 | Low-quality |
| 17-108_k141_309461 | Not-determined |
| 17-108_k141_47398 | Not-determined |
| 17-108_k141_157461 | Low-quality |
| 17-108_k141_226476 | Not-determined |
| 17-108_k141_143771 | Not-determined |
| 17-108_k141_143772 | Not-determined |
| 17-108_k141_267694 | Not-determined |
| 17-108_k141_254054 | Not-determined |
| 17-108_k141_309471 | Not-determined |
| 17-108_k141_254059 | Not-determined |
| 17-108_k141_254062 | Not-determined |
| 17-108_k141_33462 | Low-quality |
| 17-108_k141_198525 | Not-determined |
| 17-108_k141_102965 | Not-determined |
| 17-108_k141_171523 | Not-determined |
| 17-108_k141_88650 | Not-determined |
| 17-108_k141_254067 | Not-determined |
| 17-108_k141_102967 | Not-determined |
| 17-108_k141_102968 | Not-determined |
| 17-108_k141_198530 | Not-determined |
| 17-108_k141_254071 | Low-quality |
| 17-108_k141_47420 | Not-determined |
| 17-108_k141_102972 | Not-determined |
| 17-108_k141_102973 | Not-determined |
| 17-108_k141_184893 | Not-determined |
| 17-108_k141_198540 | Not-determined |
| 17-108_k141_213269 | Not-determined |
| 17-108_k141_267728 | Not-determined |
| 17-108_k141_282159 | Not-determined |
| 17-108_k141_226507 | Not-determined |
| 17-108_k141_184918 | Not-determined |
| 17-108_k141_213274 | Low-quality |
| 17-108_k141_61122 | Not-determined |
| 17-108_k141_184924 | Not-determined |
| 17-108_k141_184925 | Not-determined |
| 17-108_k141_61126 | Not-determined |
| 17-108_k141_171540 | Not-determined |
| 17-108_k141_184931 | Not-determined |
| 17-108_k141_267761 | Not-determined |
| 17-108_k141_88698 | Not-determined |
| 17-108_k141_171542 | Not-determined |

| | |
|---|---|
| 17-108_k141_184934 | Not-determined |
| 17-108_k141_61134 | Not-determined |
| 17-108_k141_33501 | Not-determined |
| 17-108_k141_184944 | Not-determined |
| 17-108_k141_5795 | Not-determined |
| 17-108_k141_267776 | Not-determined |
| 17-108_k141_5797 | Not-determined |
| 17-108_k141_74301 | Not-determined |
| 17-108_k141_309519 | Not-determined |
| 17-108_k141_295135 | Not-determined |
| 17-108_k141_322165 | Not-determined |
| 17-108_k141_115481 | Not-determined |
| 17-108_k141_267784 | Not-determined |
| 17-108_k141_295140 | Not-determined |
| 17-108_k141_240834 | Not-determined |
| 17-108_k141_74309 | Not-determined |
| 17-108_k141_309529 | Not-determined |
| 17-108_k141_5807 | Not-determined |
| 17-108_k141_240840 | Not-determined |
| 17-108_k141_226518 | Not-determined |
| 17-108_k141_295145 | Low-quality |
| 17-108_k141_213305 | Not-determined |
| 17-108_k141_171562 | Not-determined |
| 17-108_k141_226524 | Not-determined |
| 17-108_k141_226527 | Not-determined |
| 17-108_k141_267796 | Not-determined |
| 17-108_k141_171565 | Not-determined |
| 17-108_k141_74330 | Not-determined |
| 17-108_k141_309540 | Not-determined |
| 17-108_k141_267802 | Not-determined |
| 17-108_k141_267803 | Low-quality |
| 17-108_k141_198576 | Not-determined |
| 17-108_k141_171570 | Not-determined |
| 17-108_k141_115492 | Not-determined |
| 17-108_k141_88704 | Low-quality |
| 17-108_k141_143815 | Low-quality |
| 17-108_k141_267810 | Not-determined |
| 17-108_k141_184972 | Not-determined |
| 17-108_k141_184975 | Not-determined |
| 17-108_k141_254133 | Not-determined |
| 17-108_k141_254136 | Not-determined |
| 17-108_k141_143823 | Not-determined |
| 17-108_k141_322190 | Not-determined |
| 17-108_k141_88709 | Not-determined |
| 17-108_k141_47429 | Not-determined |
| 17-108_k141_5825 | Not-determined |
| 17-108_k141_143827 | Not-determined |
| 17-108_k141_5827 | Not-determined |
| 17-108_k141_143828 | Not-determined |
| 17-108_k141_5829 | Not-determined |
| 17-108_k141_103016 | Not-determined |
| 17-108_k141_19025 | Not-determined |
| 17-108_k141_33522 | Not-determined |
| 17-108_k141_129882 | Not-determined |
| 17-108_k141_115524 | Not-determined |
| 17-108_k141_282216 | Not-determined |
| 17-108_k141_198586 | Not-determined |
| 17-108_k141_171579 | Not-determined |
| 17-108_k141_19034 | Not-determined |
| 17-108_k141_47453 | Not-determined |
| 17-108_k141_143844 | Not-determined |
| 17-108_k141_226552 | Not-determined |
| 17-108_k141_198590 | Not-determined |
| 17-108_k141_103032 | Not-determined |
| 17-108_k141_129893 | Not-determined |
| 17-108_k141_226554 | Not-determined |
| 17-108_k141_103036 | Low-quality |
| 17-108_k141_5856 | Not-determined |
| 17-108_k141_143852 | Not-determined |
| 17-108_k141_322211 | Not-determined |
| 17-108_k141_103041 | Not-determined |
| 17-108_k141_61172 | Not-determined |
| 17-108_k141_322214 | Not-determined |
| 17-108_k141_103051 | Not-determined |
| 17-108_k141_254174 | Not-determined |
| 17-108_k141_129904 | Not-determined |
| 17-108_k141_115535 | Not-determined |
| 17-108_k141_322217 | Not-determined |
| 17-108_k141_157607 | Not-determined |
| 17-108_k141_33540 | Not-determined |
| 17-108_k141_295203 | Not-determined |
| 17-108_k141_115547 | Not-determined |
| 17-108_k141_213340 | Not-determined |
| 17-108_k141_295212 | Not-determined |
| 17-108_k141_322234 | Not-determined |
| 17-108_k141_19069 | Not-determined |
| 17-108_k141_33553 | Not-determined |
| 17-108_k141_198604 | Not-determined |
| 17-108_k141_19075 | Not-determined |
| 17-108_k141_61200 | Low-quality |
| 17-108_k141_157621 | Not-determined |
| 17-108_k141_309608 | Not-determined |
| 17-108_k141_143870 | Not-determined |
| 17-108_k141_5880 | Not-determined |
| 17-108_k141_115564 | Low-quality |
| 17-108_k141_213346 | Not-determined |
| 17-108_k141_240901 | Low-quality |
| 17-108_k141_115575 | Not-determined |
| 17-108_k141_240909 | Low-quality |
| 17-108_k141_157630 | Not-determined |
| 17-108_k141_103087 | Not-determined |
| 17-108_k141_240914 | Not-determined |
| 17-108_k141_115584 | Low-quality |
| 17-108_k141_240915 | Not-determined |
| 17-108_k141_88720 | Not-determined |
| 17-108_k141_240917 | Not-determined |
| 17-108_k141_171643 | Not-determined |
| 17-108_k141_19102 | Not-determined |
| 17-108_k141_282253 | Not-determined |
| 17-108_k141_88726 | Not-determined |
| 17-108_k141_240926 | Not-determined |
| 17-108_k141_129948 | Not-determined |
| 17-108_k141_240928 | Not-determined |
| 17-108_k141_282268 | Not-determined |
| 17-108_k141_61215 | Not-determined |
| 17-108_k141_115609 | Not-determined |
| 17-108_k141_129957 | Not-determined |
| 17-108_k141_74379 | Low-quality |
| 17-108_k141_240943 | Not-determined |
| 17-108_k141_198641 | Not-determined |
| 17-108_k141_282279 | Low-quality |
| 17-108_k141_282284 | Not-determined |
| 17-108_k141_240947 | Not-determined |
| 17-108_k141_47506 | Not-determined |
| 17-108_k141_226577 | Not-determined |
| 17-108_k141_5904 | Not-determined |
| 17-108_k141_115633 | Not-determined |
| 17-108_k141_115636 | Not-determined |
| 17-108_k141_47520 | Not-determined |
| 17-108_k141_47521 | Not-determined |
| 17-108_k141_226583 | Not-determined |
| 17-108_k141_74399 | Not-determined |
| 17-108_k141_322285 | Not-determined |
| 17-108_k141_47527 | Not-determined |
| 17-108_k141_143888 | Low-quality |
| 17-108_k141_198658 | Low-quality |
| 17-108_k141_5913 | Low-quality |
| 17-108_k141_254232 | Not-determined |
| 17-108_k141_267863 | Not-determined |
| 17-108_k141_267868 | Low-quality |
| 17-108_k141_267869 | Not-determined |
| 17-108_k141_309635 | Not-determined |
| 17-108_k141_47537 | Not-determined |
| 17-108_k141_74409 | Not-determined |
| 17-108_k141_295231 | Not-determined |
| 17-108_k141_267875 | Low-quality |
| 17-108_k141_322291 | Not-determined |
| 17-108_k141_322292 | Not-determined |
| 17-108_k141_198670 | Not-determined |
| 17-108_k141_143897 | Low-quality |
| 17-108_k141_115663 | Not-determined |
| 17-108_k141_171652 | Not-determined |
| 17-108_k141_61231 | Low-quality |
| 17-108_k141_267886 | Not-determined |
| 17-108_k141_33623 | Not-determined |
| 17-108_k141_267891 | Not-determined |
| 17-108_k141_61238 | Not-determined |
| 17-108_k141_157711 | Not-determined |
| 17-108_k141_226611 | Not-determined |
| 17-108_k141_295248 | Not-determined |
| 17-108_k141_143922 | Not-determined |
| 17-108_k141_47568 | Not-determined |
| 17-108_k141_226613 | Low-quality |
| 17-108_k141_282312 | Low-quality |
| 17-108_k141_309645 | Not-determined |
| 17-108_k141_240965 | Not-determined |
| 17-108_k141_295253 | Not-determined |
| 17-108_k141_322318 | Not-determined |
| 17-108_k141_322323 | Not-determined |
| 17-108_k141_47569 | Not-determined |
| 17-108_k141_198693 | Not-determined |
| 17-108_k141_5945 | Not-determined |
| 17-108_k141_254267 | Not-determined |
| 17-108_k141_61260 | Not-determined |
| 17-108_k141_157727 | Not-determined |
| 17-108_k141_157729 | Not-determined |
| 17-108_k141_157730 | Not-determined |
| 17-108_k141_47588 | Not-determined |
| 17-108_k141_295270 | Not-determined |
| 17-108_k141_143936 | Not-determined |
| 17-108_k141_103199 | Not-determined |
| 17-108_k141_103203 | Not-determined |
| 17-108_k141_103204 | Not-determined |
| 17-108_k141_33642 | Not-determined |
| 17-108_k141_88733 | Not-determined |
| 17-108_k141_295279 | Not-determined |
| 17-108_k141_74458 | Not-determined |
| 17-108_k141_157745 | Not-determined |
| 17-108_k141_88740 | Not-determined |
| 17-108_k141_198719 | Not-determined |
| 17-108_k141_295283 | Not-determined |
| 17-108_k141_322348 | Not-determined |
| 17-108_k141_171685 | Not-determined |
| 17-108_k141_171686 | Not-determined |
| 17-108_k141_157750 | Not-determined |
| 17-108_k141_61291 | Not-determined |
| 17-108_k141_295289 | Not-determined |
| 17-108_k141_295290 | Low-quality |
| 17-108_k141_61297 | Not-determined |
| 17-108_k141_130033 | Not-determined |
| 17-108_k141_213399 | Low-quality |
| 17-108_k141_322359 | Not-determined |
| 17-108_k141_322360 | Not-determined |
| 17-108_k141_322364 | Not-determined |
| 17-108_k141_130039 | Not-determined |
| 17-108_k141_185049 | Not-determined |
| 17-108_k141_19205 | Not-determined |
| 17-108_k141_19208 | Not-determined |
| 17-108_k141_157776 | Not-determined |
| 17-108_k141_309682 | Not-determined |
| 17-108_k141_309688 | Not-determined |
| 17-108_k141_115698 | Not-determined |
| 17-108_k141_282366 | Not-determined |
| 17-108_k141_88768 | Not-determined |
| 17-108_k141_267930 | Not-determined |
| 17-108_k141_295305 | Not-determined |
| 17-108_k141_33647 | Not-determined |
| 17-108_k141_254306 | Not-determined |
| 17-108_k141_143947 | Not-determined |
| 17-108_k141_267937 | Not-determined |
| 17-108_k141_322390 | Not-determined |
| 17-108_k141_115713 | Low-quality |
| 17-108_k141_88778 | Not-determined |
| 17-108_k141_33654 | Not-determined |
| 17-108_k141_88781 | Not-determined |
| 17-108_k141_226662 | Not-determined |
| 17-108_k141_115719 | Not-determined |
| 17-108_k141_171712 | Not-determined |
| 17-108_k141_6006 | Low-quality |
| 17-108_k141_185064 | Not-determined |
| 17-108_k141_171715 | Low-quality |
| 17-108_k141_61321 | Not-determined |
| 17-108_k141_6015 | Not-determined |
| 17-108_k141_171729 | Not-determined |
| 17-108_k141_6018 | Low-quality |
| 17-108_k141_157811 | Not-determined |
| 17-108_k141_241018 | Not-determined |
| 17-108_k141_226672 | Not-determined |
| 17-108_k141_6020 | Not-determined |
| 17-108_k141_6021 | Not-determined |
| 17-108_k141_33670 | Not-determined |
| 17-108_k141_115736 | Not-determined |
| 17-108_k141_309726 | Low-quality |
| 17-108_k141_130078 | Not-determined |
| 17-108_k141_157814 | Low-quality |
| 17-108_k141_241023 | Not-determined |
| 17-108_k141_143975 | Not-determined |
| 17-108_k141_282390 | Not-determined |
| 17-108_k141_130083 | Not-determined |
| 17-108_k141_282391 | Not-determined |
| 17-108_k141_282392 | Not-determined |
| 17-108_k141_19215 | Not-determined |
| 17-108_k141_254338 | Not-determined |
| 17-108_k141_282394 | Not-determined |
| 17-108_k141_103215 | Not-determined |
| 17-108_k141_322426 | Not-determined |
| 17-108_k141_33679 | Not-determined |
| 17-108_k141_198749 | Not-determined |
| 17-108_k141_130091 | Not-determined |
| 17-108_k141_213436 | Not-determined |
| 17-108_k141_74477 | Not-determined |
| 17-108_k141_309751 | Not-determined |
| 17-108_k141_226707 | Not-determined |
| 17-108_k141_267972 | Not-determined |
| 17-108_k141_185089 | Not-determined |
| 17-108_k141_309756 | Not-determined |
| 17-108_k141_226716 | Low-quality |
| 17-108_k141_267973 | Not-determined |
| 17-108_k141_267975 | Not-determined |
| 17-108_k141_295340 | Not-determined |
| 17-108_k141_143988 | Low-quality |
| 17-108_k141_115749 | Not-determined |
| 17-108_k141_185096 | Low-quality |
| 17-108_k141_33688 | Not-determined |
| 17-108_k141_6048 | Not-determined |
| 17-108_k141_115755 | Not-determined |
| 17-108_k141_115757 | Not-determined |
| 17-108_k141_88826 | Low-quality |
| 17-108_k141_322444 | Not-determined |
| 17-108_k141_157844 | Low-quality |
| 17-108_k141_254357 | Low-quality |
| 17-108_k141_267987 | Not-determined |
| 17-108_k141_254360 | Not-determined |
| 17-108_k141_295349 | Not-determined |
| 17-108_k141_103235 | Not-determined |
| 17-108_k141_295352 | Low-quality |
| 17-108_k141_157849 | Not-determined |
| 17-108_k141_171772 | Not-determined |
| 17-108_k141_157853 | Not-determined |
| 17-108_k141_322447 | Not-determined |
| 17-108_k141_33698 | Not-determined |
| 17-108_k141_144001 | Not-determined |
| 17-108_k141_309803 | Not-determined |
| 17-108_k141_309805 | Not-determined |
| 17-108_k141_322448 | Low-quality |
| 17-108_k141_254368 | Not-determined |
| 17-108_k141_144008 | Not-determined |
| 17-108_k141_19251 | Not-determined |
| 17-108_k141_157867 | Low-quality |
| 17-108_k141_88838 | Low-quality |
| 17-108_k141_61409 | Not-determined |
| 17-108_k141_19259 | Not-determined |
| 17-108_k141_267990 | Not-determined |
| 17-108_k141_282439 | Not-determined |
| 17-108_k141_282455 | Not-determined |
| 17-108_k141_19265 | Not-determined |
| 17-108_k141_88852 | Not-determined |
| 17-108_k141_61427 | Not-determined |
| 17-108_k141_74523 | Not-determined |
| 17-108_k141_198765 | Not-determined |
| 17-108_k141_88859 | Not-determined |
| 17-108_k141_254389 | Not-determined |
| 17-108_k141_157885 | Not-determined |
| 17-108_k141_322489 | Not-determined |
| 17-108_k141_254393 | Low-quality |
| 17-108_k141_213470 | Not-determined |
| 17-108_k141_88863 | Not-determined |
| 17-108_k141_144033 | Not-determined |
| 17-108_k141_115796 | Not-determined |
| 17-108_k141_19278 | Not-determined |
| 17-108_k141_115804 | Not-determined |
| 17-108_k141_185135 | Not-determined |
| 17-108_k141_144040 | Not-determined |
| 17-108_k141_213485 | Not-determined |
| 17-108_k141_198797 | Not-determined |
| 17-108_k141_88872 | Low-quality |
| 17-108_k141_61452 | Not-determined |
| 17-108_k141_213494 | Not-determined |
| 17-108_k141_213495 | Not-determined |
| 17-108_k141_309863 | Not-determined |
| 17-108_k141_115824 | Low-quality |
| 17-108_k141_309865 | Not-determined |
| 17-108_k141_144059 | Not-determined |
| 17-108_k141_6132 | Not-determined |
| 17-108_k141_171810 | Not-determined |
| 17-108_k141_157917 | Not-determined |
| 17-108_k141_6138 | Not-determined |
| 17-108_k141_6141 | Not-determined |
| 17-108_k141_88887 | Not-determined |
| 17-108_k141_157922 | Not-determined |
| 17-108_k141_282482 | Not-determined |
| 17-108_k141_47694 | Not-determined |
| 17-108_k141_213510 | Not-determined |
| 17-108_k141_171822 | Not-determined |
| 17-108_k141_198829 | Not-determined |
| 17-108_k141_171827 | Not-determined |
| 17-108_k141_295410 | Not-determined |
| 17-108_k141_47705 | Not-determined |
| 17-108_k141_185158 | Not-determined |
| 17-108_k141_74561 | Not-determined |
| 17-108_k141_115875 | Not-determined |
| 17-108_k141_295420 | Not-determined |
| 17-108_k141_115879 | Not-determined |
| 17-108_k141_213517 | Not-determined |
| 17-108_k141_254417 | Low-quality |
| 17-108_k141_61476 | Not-determined |
| 17-108_k141_226814 | Not-determined |
| 17-108_k141_254421 | Not-determined |
| 17-108_k141_61478 | Not-determined |
| 17-108_k141_213525 | Not-determined |
| 17-108_k141_19314 | Not-determined |
| 17-108_k141_254424 | Not-determined |
| 17-108_k141_198852 | Not-determined |
| 17-108_k141_157936 | Not-determined |
| 17-108_k141_322549 | Low-quality |
| 17-108_k141_115886 | Not-determined |
| 17-108_k141_33778 | Not-determined |
| 17-108_k141_171832 | Not-determined |
| 17-108_k141_213532 | Not-determined |
| 17-108_k141_226827 | Not-determined |
| 17-108_k141_115891 | Not-determined |
| 17-108_k141_322559 | Not-determined |
| 17-108_k141_88934 | Not-determined |
| 17-108_k141_282500 | Low-quality |
| 17-108_k141_88935 | Not-determined |
| 17-108_k141_226839 | Low-quality |
| 17-108_k141_282505 | Not-determined |
| 17-108_k141_115902 | Not-determined |
| 17-108_k141_115903 | Not-determined |
| 17-108_k141_282507 | Low-quality |
| 17-108_k141_267996 | Not-determined |
| 17-108_k141_157959 | Not-determined |
| 17-108_k141_282512 | Not-determined |
| 17-108_k141_144111 | Not-determined |
| 17-108_k141_282519 | Not-determined |
| 17-108_k141_213561 | Not-determined |
| 17-108_k141_6188 | Not-determined |
| 17-108_k141_6189 | Not-determined |
| 17-108_k141_88950 | Not-determined |
| 17-108_k141_61506 | Low-quality |
| 17-108_k141_6191 | Not-determined |
| 17-108_k141_322585 | Not-determined |
| 17-108_k141_322586 | Not-determined |
| 17-108_k141_322588 | Not-determined |
| 17-108_k141_171838 | Not-determined |
| 17-108_k141_268014 | Not-determined |
| 17-108_k141_115924 | Not-determined |
| 17-108_k141_241187 | Not-determined |
| 17-108_k141_61521 | Not-determined |
| 17-108_k141_241193 | Not-determined |
| 17-108_k141_198867 | Not-determined |
| 17-108_k141_33811 | Not-determined |
| 17-108_k141_115933 | Not-determined |
| 17-108_k141_157984 | Not-determined |
| 17-108_k141_61528 | Not-determined |
| 17-108_k141_115942 | Not-determined |
| 17-108_k141_61534 | Not-determined |
| 17-108_k141_226868 | Not-determined |
| 17-108_k141_226873 | Not-determined |
| 17-108_k141_295483 | Not-determined |
| 17-108_k141_115949 | Not-determined |
| 17-108_k141_268043 | Not-determined |
| 17-108_k141_282544 | Not-determined |
| 17-108_k141_115957 | Not-determined |
| 17-108_k141_254448 | Not-determined |
| 17-108_k141_144138 | Not-determined |
| 17-108_k141_254455 | Not-determined |
| 17-108_k141_74589 | Not-determined |
| 17-108_k141_226886 | Not-determined |
| 17-108_k141_144145 | Not-determined |
| 17-108_k141_226888 | Low-quality |
| 17-108_k141_185168 | Not-determined |
| 17-108_k141_74606 | Not-determined |
| 17-108_k141_33830 | Not-determined |
| 17-108_k141_115979 | Not-determined |
| 17-108_k141_309918 | Not-determined |
| 17-108_k141_282588 | Not-determined |
| 17-108_k141_254473 | Not-determined |
| 17-108_k141_144163 | Not-determined |
| 17-108_k141_61574 | Not-determined |
| 17-108_k141_185185 | Not-determined |
| 17-108_k141_322640 | Not-determined |
| 17-108_k141_282600 | Not-determined |
| 17-108_k141_88997 | Not-determined |
| 17-108_k141_322649 | Not-determined |
| 18-106-final-viral-combined_k141_39021\|\|full | Complete |
| 17-108_k141_282607 | Not-determined |
| 17-108_k141_158032 | Not-determined |
| 17-108_k141_254488 | Not-determined |
| 17-108_k141_158038 | Not-determined |
| 17-108_k141_171877 | Not-determined |
| 17-108_k141_19356 | Low-quality |
| 17-108_k141_19357 | Not-determined |
| 17-108_k141_74628 | Not-determined |
| 17-108_k141_268063 | Not-determined |
| 17-108_k141_241278 | Not-determined |
| 17-108_k141_103341 | Not-determined |
| 17-108_k141_103344 | Not-determined |
| 17-108_k141_268070 | Low-quality |
| 17-108_k141_322665 | Not-determined |
| 17-108_k141_282622 | Not-determined |
| 17-108_k141_254508 | Not-determined |
| 17-108_k141_282627 | Not-determined |
| 17-108_k141_185219 | Not-determined |
| 17-108_k141_61600 | Low-quality |
| 17-108_k141_33857 | Not-determined |
| 17-108_k141_6219 | Not-determined |
| 17-108_k141_254526 | Not-determined |
| 17-108_k141_33861 | Not-determined |
| 17-108_k141_226940 | Not-determined |
| 17-108_k141_19370 | Not-determined |
| 17-108_k141_74646 | Not-determined |
| 17-108_k141_185232 | Not-determined |
| 17-108_k141_33869 | Not-determined |
| 17-108_k141_282634 | Low-quality |
| 17-108_k141_33873 | Not-determined |
| 17-108_k141_158057 | Not-determined |
| 17-108_k141_185243 | Not-determined |
| 17-108_k141_282641 | Not-determined |
| 17-108_k141_309932 | Not-determined |
| 17-108_k141_282642 | Not-determined |
| 17-108_k141_241293 | Not-determined |
| 17-108_k141_158062 | Not-determined |
| 17-108_k141_171909 | Not-determined |
| 17-108_k141_282644 | Not-determined |
| 17-108_k141_254541 | Low-quality |
| 17-108_k141_19386 | Not-determined |
| 17-108_k141_103363 | Not-determined |
| 17-108_k141_61636 | Not-determined |
| 17-108_k141_282651 | Not-determined |
| 17-108_k141_103368 | Not-determined |
| 17-108_k141_144200 | Not-determined |
| 17-108_k141_19395 | Not-determined |
| 17-108_k141_268108 | Low-quality |
| 17-108_k141_6260 | Not-determined |
| 17-108_k141_47732 | Not-determined |
| 17-108_k141_116050 | Not-determined |
| 17-108_k141_309962 | Not-determined |
| 17-108_k141_241313 | Low-quality |
| 17-108_k141_61660 | Not-determined |
| 17-108_k141_158091 | Not-determined |
| 17-108_k141_213659 | Not-determined |
| 17-108_k141_171937 | Not-determined |
| 17-108_k141_213662 | Low-quality |
| 17-108_k141_268127 | Not-determined |
| 17-108_k141_130142 | Not-determined |
| 17-108_k141_226961 | Not-determined |
| 17-108_k141_254568 | Not-determined |
| 17-108_k141_268133 | Low-quality |
| 17-108_k141_185267 | Not-determined |
| 17-108_k141_254573 | Not-determined |
| 17-108_k141_171949 | Not-determined |
| 17-108_k141_254577 | Not-determined |
| 17-108_k141_74676 | Not-determined |
| 17-108_k141_19436 | Not-determined |
| 17-108_k141_130151 | Not-determined |
| 17-108_k141_19439 | Low-quality |
| 17-108_k141_322740 | Not-determined |
| 17-108_k141_6303 | Not-determined |
| 17-108_k141_19444 | Not-determined |
| 17-108_k141_185280 | Not-determined |
| 17-108_k141_226986 | Not-determined |
| 17-108_k141_198938 | Not-determined |
| 17-108_k141_144246 | Not-determined |
| 17-108_k141_144247 | Not-determined |
| 17-108_k141_295501 | Not-determined |
| 17-108_k141_33892 | Low-quality |
| 17-108_k141_130163 | Not-determined |
| 17-108_k141_198945 | Not-determined |
| 17-108_k141_158113 | Not-determined |
| 17-108_k141_322747 | Low-quality |
| 17-108_k141_6313 | Not-determined |
| 17-108_k141_33905 | Not-determined |
| 17-108_k141_322755 | Not-determined |
| 17-108_k141_185299 | Not-determined |
| 17-108_k141_158117 | Not-determined |
| 17-108_k141_158120 | Not-determined |
| 17-108_k141_61680 | Not-determined |
| 17-108_k141_33914 | Not-determined |
| 17-108_k141_171978 | Not-determined |
| 17-108_k141_144273 | Low-quality |
| 17-108_k141_198972 | Not-determined |
| 17-108_k141_322770 | Not-determined |
| 17-108_k141_268174 | Not-determined |
| 17-108_k141_322771 | Not-determined |
| 17-108_k141_103432 | Not-determined |
| 17-108_k141_130192 | Low-quality |
| 17-108_k141_171993 | Not-determined |
| 17-108_k141_103434 | Low-quality |
| 17-108_k141_310005 | Low-quality |
| 17-108_k141_268188 | Not-determined |
| 17-108_k141_213690 | Not-determined |
| 17-108_k141_213692 | Not-determined |
| 17-108_k141_295530 | Not-determined |
| 17-108_k141_213693 | Not-determined |
| 17-108_k141_213694 | Not-determined |
| 17-108_k141_144291 | Not-determined |
| 17-108_k141_19459 | Not-determined |
| 17-108_k141_185318 | Not-determined |
| 17-108_k141_282688 | Not-determined |
| 17-108_k141_310018 | Not-determined |
| 17-108_k141_19463 | Not-determined |
| 17-108_k141_310026 | Low-quality |
| 17-108_k141_254633 | Not-determined |
| 17-108_k141_310028 | Not-determined |
| 17-108_k141_47779 | Not-determined |
| 17-108_k141_282703 | Not-determined |
| 17-108_k141_295547 | Not-determined |
| 17-108_k141_268197 | Low-quality |
| 17-108_k141_268200 | Not-determined |
| 17-108_k141_6343 | Not-determined |
| 17-108_k141_172022 | Not-determined |
| 17-108_k141_226999 | Low-quality |
| 17-108_k141_198985 | Not-determined |
| 17-108_k141_227007 | Low-quality |
| 17-108_k141_47794 | Not-determined |
| 17-108_k141_47798 | Not-determined |
| 17-108_k141_6355 | Low-quality |
| 17-108_k141_185336 | Not-determined |
| 17-108_k141_295562 | Not-determined |
| 17-108_k141_172039 | Not-determined |
| 17-108_k141_185339 | Low-quality |
| 17-108_k141_322781 | Not-determined |
| 17-108_k141_295566 | Not-determined |
| 17-108_k141_241454 | Not-determined |
| 17-108_k141_172050 | Not-determined |
| 17-108_k141_61750 | Low-quality |
| 17-108_k141_241456 | Not-determined |
| 17-108_k141_74704 | Not-determined |
| 17-108_k141_130232 | Not-determined |
| 17-108_k141_310071 | Not-determined |
| 17-108_k141_6375 | Not-determined |
| 17-108_k141_6378 | Not-determined |
| 17-108_k141_185362 | Not-determined |
| 17-108_k141_89093 | Not-determined |
| 17-108_k141_19478 | Not-determined |
| 17-108_k141_310074 | Not-determined |
| 17-108_k141_89097 | Not-determined |
| 17-108_k141_74724 | Low-quality |

| | |
|---|---|
| 17-108_k141_241483 | Not-determined |
| 17-108_k141_268254 | Not-determined |
| 17-108_k141_185368 | Not-determined |
| 17-108_k141_282722 | Not-determined |
| 17-108_k141_19485 | Not-determined |
| 17-108_k141_268258 | Not-determined |
| 17-108_k141_322799 | Not-determined |
| 17-108_k141_213720 | Not-determined |
| 17-108_k141_268260 | Not-determined |
| 17-108_k141_19489 | Not-determined |
| 17-108_k141_268261 | Not-determined |
| 17-108_k141_6402 | Not-determined |
| 17-108_k141_6403 | Not-determined |
| 17-108_k141_172089 | Not-determined |
| 17-108_k141_172092 | Not-determined |
| 17-108_k141_19493 | Not-determined |
| 17-108_k141_172095 | Not-determined |
| 17-108_k141_241506 | Not-determined |
| 17-108_k141_6412 | Not-determined |
| 17-108_k141_213731 | Not-determined |
| 17-108_k141_19498 | Not-determined |
| 17-108_k141_310089 | Not-determined |
| 17-108_k141_322807 | Not-determined |
| 17-108_k141_130244 | Not-determined |
| 17-108_k141_158193 | Not-determined |
| 17-108_k141_310094 | Not-determined |
| 17-108_k141_130246 | Not-determined |
| 17-108_k141_227018 | Low-quality |
| 17-108_k141_19499 | Not-determined |
| 17-108_k141_172097 | Not-determined |
| 17-108_k141_227020 | Not-determined |
| 17-108_k141_310100 | Not-determined |
| 17-108_k141_116074 | Not-determined |
| 17-108_k141_227025 | Not-determined |
| 17-108_k141_172103 | Not-determined |
| 17-108_k141_130251 | Low-quality |
| 17-108_k141_130252 | Not-determined |
| 17-108_k141_185385 | Not-determined |
| 17-108_k141_158205 | Not-determined |
| 17-108_k141_89113 | Not-determined |
| 17-108_k141_6431 | Not-determined |
| 17-108_k141_310110 | Not-determined |
| 17-108_k141_74755 | Not-determined |
| 17-108_k141_116086 | Not-determined |
| 17-108_k141_310114 | Low-quality |
| 17-108_k141_130256 | Not-determined |
| 17-108_k141_116089 | Not-determined |
| 17-108_k141_227039 | Not-determined |
| 17-108_k141_254692 | Not-determined |
| 17-108_k141_61797 | Not-determined |
| 17-108_k141_61798 | Not-determined |
| 17-108_k141_310122 | Not-determined |
| 17-108_k141_185391 | Not-determined |
| 17-108_k141_227042 | Low-quality |
| 17-108_k141_227043 | Not-determined |
| 17-108_k141_74760 | Not-determined |
| 17-108_k141_322830 | Not-determined |
| 17-108_k141_74761 | Not-determined |
| 17-108_k141_172118 | Not-determined |
| 17-108_k141_116099 | Not-determined |
| 17-108_k141_103480 | Not-determined |
| 17-108_k141_61804 | Not-determined |
| 17-108_k141_213759 | Not-determined |
| 17-108_k141_61808 | Not-determined |
| 17-108_k141_61809 | Not-determined |
| 17-108_k141_199040 | Not-determined |
| 17-108_k141_199046 | Not-determined |
| 17-108_k141_61811 | Not-determined |
| 17-108_k141_310138 | Not-determined |
| 17-108_k141_61815 | Not-determined |
| 17-108_k141_61818 | Not-determined |
| 17-108_k141_6463 | Not-determined |
| 17-108_k141_61821 | Not-determined |
| 17-108_k141_74772 | Not-determined |
| 17-108_k141_268309 | Low-quality |
| 17-108_k141_74786 | Not-determined |
| 17-108_k141_33983 | Not-determined |
| 17-108_k141_89144 | Not-determined |
| 17-108_k141_103497 | Not-determined |
| 17-108_k141_103500 | Not-determined |
| 17-108_k141_47843 | Not-determined |
| 17-108_k141_89163 | Not-determined |
| 17-108_k141_213778 | Low-quality |
| 17-108_k141_74800 | Not-determined |
| 17-108_k141_74801 | Not-determined |
| 17-108_k141_172139 | Not-determined |
| 17-108_k141_227090 | Not-determined |
| 17-108_k141_158272 | Not-determined |
| 17-108_k141_34006 | Not-determined |
| 17-108_k141_227093 | Not-determined |
| 17-108_k141_172144 | Not-determined |
| 17-108_k141_34008 | Not-determined |
| 17-108_k141_241519 | Not-determined |
| 17-108_k141_19506 | Not-determined |
| 17-108_k141_172148 | Not-determined |
| 17-108_k141_227099 | Not-determined |
| 17-108_k141_227103 | Not-determined |
| 17-108_k141_103534 | Not-determined |
| 17-108_k141_227108 | Not-determined |
| 17-108_k141_227112 | Not-determined |
| 17-108_k141_185412 | Low-quality |
| 17-108_k141_199082 | Not-determined |
| 17-108_k141_295614 | Not-determined |
| 17-108_k141_89183 | Not-determined |
| 17-108_k141_19518 | Low-quality |
| 17-108_k141_213805 | Not-determined |
| 17-108_k141_89187 | Not-determined |
| 17-108_k141_19524 | Not-determined |
| 17-108_k141_310162 | Low-quality |
| 17-108_k141_295622 | Not-determined |
| 17-108_k141_74821 | Not-determined |
| 17-108_k141_61835 | Not-determined |
| 17-108_k141_227122 | Not-determined |
| 17-108_k141_227123 | Low-quality |
| 17-108_k141_227124 | Not-determined |
| 17-108_k141_74823 | Not-determined |
| 17-108_k141_241541 | Not-determined |
| 17-108_k141_213817 | Not-determined |
| 17-108_k141_254730 | Not-determined |
| 17-108_k141_61842 | Not-determined |
| 17-108_k141_19530 | Not-determined |
| 17-108_k141_241544 | Not-determined |
| 17-108_k141_295627 | Not-determined |
| 17-108_k141_295628 | Not-determined |
| 17-108_k141_61855 | Not-determined |
| 17-108_k141_268323 | Not-determined |
| 17-108_k141_254740 | Not-determined |
| 17-108_k141_322867 | Not-determined |
| 17-108_k141_61861 | Not-determined |
| 17-108_k141_19548 | Not-determined |
| 17-108_k141_322868 | Not-determined |
| 17-108_k141_254744 | Not-determined |
| 17-108_k141_74832 | Not-determined |
| 17-108_k141_213836 | Not-determined |
| 17-108_k141_103564 | Not-determined |
| 17-108_k141_295643 | Not-determined |
| 17-108_k141_185458 | Not-determined |
| 17-108_k141_185460 | Not-determined |
| 17-108_k141_295652 | Not-determined |
| 17-108_k141_47864 | Not-determined |
| 17-108_k141_282748 | Low-quality |
| 17-108_k141_254763 | Not-determined |
| 17-108_k141_158323 | Not-determined |
| 17-108_k141_310211 | Not-determined |
| 17-108_k141_172186 | Not-determined |
| 17-108_k141_227131 | Not-determined |
| 17-108_k141_158336 | Not-determined |
| 17-108_k141_268339 | Not-determined |
| 17-108_k141_116154 | Not-determined |
| 17-108_k141_268342 | Not-determined |
| 17-108_k141_227135 | Not-determined |
| 17-108_k141_158347 | Not-determined |
| 17-108_k141_47879 | Not-determined |
| 17-108_k141_282752 | Not-determined |
| 17-108_k141_6559 | Not-determined |
| 17-108_k141_172196 | Not-determined |
| 17-108_k141_254775 | Not-determined |
| 17-108_k141_213857 | Not-determined |
| 17-108_k141_282756 | Not-determined |
| 17-108_k141_185498 | Low-quality |
| 17-108_k141_241558 | Not-determined |
| 17-108_k141_268348 | Not-determined |
| 17-108_k141_74859 | Not-determined |
| 17-108_k141_322897 | Not-determined |
| 17-108_k141_322904 | Not-determined |
| 17-108_k141_295661 | Not-determined |
| 17-108_k141_103596 | Low-quality |
| 17-108_k141_172206 | Low-quality |
| 17-108_k141_116164 | Not-determined |
| 17-108_k141_310240 | Not-determined |
| 17-108_k141_185521 | Not-determined |
| 17-108_k141_144399 | Not-determined |
| 17-108_k141_310249 | Not-determined |
| 17-108_k141_144402 | Not-determined |
| 17-108_k141_227165 | Not-determined |
| 17-108_k141_116174 | Low-quality |
| 17-108_k141_282779 | Low-quality |
| 17-108_k141_254805 | Low-quality |
| 17-108_k141_241600 | Low-quality |
| 17-108_k141_227179 | Not-determined |
| 17-108_k141_158381 | Not-determined |
| 17-108_k141_310263 | Not-determined |
| 17-108_k141_34079 | Not-determined |
| 17-108_k141_47921 | Not-determined |
| 17-108_k141_310267 | Not-determined |
| 17-108_k141_172226 | Not-determined |
| 17-108_k141_310268 | Not-determined |
| 17-108_k141_241607 | Not-determined |
| 17-108_k141_74881 | Not-determined |
| 17-108_k141_172229 | Not-determined |
| 17-108_k141_199115 | Not-determined |
| 17-108_k141_19606 | Not-determined |
| 17-108_k141_34092 | Not-determined |
| 17-108_k141_254823 | Not-determined |
| 17-108_k141_241614 | Low-quality |
| 17-108_k141_172234 | Not-determined |
| 17-108_k141_172237 | Not-determined |
| 17-108_k141_116207 | Not-determined |
| 17-108_k141_172242 | Not-determined |
| 17-108_k141_172244 | Not-determined |
| 17-108_k141_310283 | Not-determined |
| 17-108_k141_74893 | Low-quality |
| 17-108_k141_158401 | Not-determined |
| 17-108_k141_158402 | Not-determined |
| 17-108_k141_74898 | Not-determined |
| 17-108_k141_172249 | Not-determined |
| 17-108_k141_268382 | Not-determined |
| 17-108_k141_19622 | Low-quality |
| 17-108_k141_310290 | Not-determined |
| 17-108_k141_158414 | Not-determined |
| 17-108_k141_61949 | Low-quality |
| 17-108_k141_19633 | Not-determined |
| 17-108_k141_295671 | Not-determined |
| 17-108_k141_213873 | Not-determined |
| 17-108_k141_89260 | Not-determined |
| 17-108_k141_6577 | Not-determined |
| 17-108_k141_213877 | Not-determined |
| 17-108_k141_34127 | Not-determined |
| 17-108_k141_34128 | Not-determined |
| 17-108_k141_74915 | Not-determined |
| 17-108_k141_61959 | Not-determined |
| 17-108_k141_19645 | Low-quality |
| 17-108_k141_144425 | Low-quality |
| 17-108_k141_158427 | Not-determined |
| 17-108_k141_241639 | Not-determined |
| 17-108_k141_172266 | Low-quality |
| 17-108_k141_158433 | Not-determined |
| 17-108_k141_172267 | Not-determined |
| 17-108_k141_158436 | Not-determined |
| 17-108_k141_310315 | Low-quality |
| 17-108_k141_282825 | Low-quality |
| 17-108_k141_158442 | Not-determined |
| 17-108_k141_282828 | Not-determined |
| 17-108_k141_172278 | Not-determined |
| 17-108_k141_34142 | Low-quality |
| 17-108_k141_172281 | Not-determined |
| 17-108_k141_282838 | Not-determined |
| 17-108_k141_89268 | Not-determined |
| 17-108_k141_268393 | Not-determined |
| 17-108_k141_227210 | Not-determined |
| 17-108_k141_74957 | Not-determined |
| 17-108_k141_213885 | Not-determined |
| 17-108_k141_116245 | Not-determined |
| 17-108_k141_322923 | Not-determined |
| 17-108_k141_241660 | Low-quality |
| 17-108_k141_322926 | Not-determined |
| 17-108_k141_34155 | Not-determined |
| 17-108_k141_199148 | Low-quality |
| 17-108_k141_322932 | Not-determined |
| 17-108_k141_74964 | Not-determined |
| 17-108_k141_322934 | Not-determined |
| 17-108_k141_227223 | Not-determined |
| 17-108_k141_158461 | Not-determined |
| 17-108_k141_130345 | Not-determined |
| 17-108_k141_103662 | Not-determined |
| 17-108_k141_158481 | Not-determined |
| 17-108_k141_295732 | Not-determined |
| 17-108_k141_144480 | Not-determined |
| 17-108_k141_47940 | Not-determined |
| 17-108_k141_144484 | Not-determined |
| 17-108_k141_34181 | Low-quality |
| 17-108_k141_103668 | Not-determined |
| 17-108_k141_241691 | Not-determined |
| 17-108_k141_103680 | Not-determined |
| 17-108_k141_158511 | Not-determined |
| 17-108_k141_227263 | Not-determined |
| 17-108_k141_172349 | Not-determined |
| 17-108_k141_144508 | Not-determined |
| 17-108_k141_47952 | Not-determined |
| 17-108_k141_6637 | Not-determined |
| 17-108_k141_47953 | Not-determined |
| 17-108_k141_130366 | Low-quality |
| 17-108_k141_116275 | Not-determined |
| 17-108_k141_130370 | Not-determined |
| 17-108_k141_185548 | Not-determined |
| 17-108_k141_213922 | Not-determined |
| 17-108_k141_144521 | Not-determined |
| 17-108_k141_158541 | Not-determined |
| 17-108_k141_199195 | Low-quality |
| 17-108_k141_116302 | Not-determined |
| 17-108_k141_116304 | Not-determined |
| 17-108_k141_295762 | Not-determined |
| 17-108_k141_116315 | Low-quality |
| 17-108_k141_282879 | Not-determined |
| 17-108_k141_19711 | Low-quality |
| 17-108_k141_241711 | Not-determined |
| 17-108_k141_241714 | Not-determined |
| 17-108_k141_130393 | Not-determined |
| 17-108_k141_268439 | Not-determined |
| 17-108_k141_62017 | Not-determined |
| 17-108_k141_62018 | Not-determined |
| 17-108_k141_130406 | Not-determined |
| 17-108_k141_268443 | Not-determined |
| 17-108_k141_62021 | Not-determined |
| 17-108_k141_144553 | Low-quality |
| 17-108_k141_268446 | Low-quality |
| 17-108_k141_227307 | Not-determined |
| 17-108_k141_268449 | Not-determined |
| 17-108_k141_268451 | Not-determined |
| 17-108_k141_62032 | Not-determined |
| 17-108_k141_62033 | Not-determined |
| 17-108_k141_103697 | Not-determined |
| 17-108_k141_172363 | Low-quality |
| 17-108_k141_295785 | Not-determined |
| 17-108_k141_62043 | Not-determined |
| 17-108_k141_241738 | Not-determined |
| 17-108_k141_6686 | Not-determined |
| 17-108_k141_241739 | Not-determined |
| 17-108_k141_254953 | Not-determined |
| 17-108_k141_227314 | Low-quality |
| 17-108_k141_254957 | Not-determined |
| 17-108_k141_74998 | Not-determined |
| 17-108_k141_172374 | Not-determined |
| 17-108_k141_62051 | Not-determined |
| 17-108_k141_241744 | Not-determined |
| 17-108_k141_172376 | Not-determined |
| 17-108_k141_213952 | Low-quality |
| 17-108_k141_199239 | Not-determined |
| 17-108_k141_227331 | Not-determined |
| 17-108_k141_185573 | Not-determined |
| 17-108_k141_19751 | Not-determined |
| 17-108_k141_75002 | Not-determined |
| 17-108_k141_6698 | Not-determined |
| 17-108_k141_213961 | Not-determined |
| 17-108_k141_75005 | Not-determined |
| 17-108_k141_103722 | Not-determined |
| 17-108_k141_116346 | Not-determined |
| 17-108_k141_227349 | Not-determined |
| 17-108_k141_75012 | Not-determined |
| 17-108_k141_282916 | Not-determined |
| 17-108_k141_282917 | Not-determined |
| 17-108_k141_322964 | Low-quality |
| 17-108_k141_47992 | Not-determined |
| 17-108_k141_199253 | Not-determined |
| 17-108_k141_47994 | Not-determined |
| 17-108_k141_34226 | Not-determined |
| 17-108_k141_268496 | Not-determined |
| 17-108_k141_116361 | Not-determined |
| 17-108_k141_227374 | Not-determined |
| 17-108_k141_116366 | Not-determined |
| 17-108_k141_48003 | Not-determined |
| 17-108_k141_268503 | Not-determined |
| 17-108_k141_75035 | Not-determined |
| 17-108_k141_116370 | Not-determined |
| 17-108_k141_6729 | Not-determined |
| 17-108_k141_62078 | Not-determined |
| 17-108_k141_62080 | Not-determined |
| 17-108_k141_199266 | Not-determined |
| 17-108_k141_185614 | Not-determined |
| 17-108_k141_213987 | Not-determined |
| 17-108_k141_19783 | Not-determined |
| 17-108_k141_89390 | Not-determined |
| 17-108_k141_227388 | Not-determined |
| 17-108_k141_116379 | Not-determined |
| 17-108_k141_75054 | Low-quality |
| 17-108_k141_282942 | Not-determined |
| 17-108_k141_116381 | Not-determined |
| 17-108_k141_199273 | Not-determined |
| 17-108_k141_310419 | Not-determined |
| 17-108_k141_282954 | Not-determined |
| 17-108_k141_282955 | Not-determined |
| 17-108_k141_214001 | Not-determined |
| 17-108_k141_34250 | Not-determined |
| 17-108_k141_268534 | Not-determined |
| 17-108_k141_295843 | Not-determined |
| 17-108_k141_268538 | Not-determined |
| 17-108_k141_103748 | Not-determined |
| 17-108_k141_62108 | Not-determined |
| 17-108_k141_19801 | Not-determined |
| 17-108_k141_255009 | Not-determined |
| 17-108_k141_130430 | Not-determined |
| 17-108_k141_62113 | Not-determined |
| 17-108_k141_130432 | Not-determined |
| 17-108_k141_34256 | Not-determined |
| 17-108_k141_130436 | Low-quality |
| 17-108_k141_172406 | Not-determined |
| 17-108_k141_322971 | Not-determined |
| 17-108_k141_103753 | Not-determined |
| 17-108_k141_34272 | Not-determined |
| 17-108_k141_214014 | Not-determined |
| 17-108_k141_295857 | Not-determined |
| 17-108_k141_19819 | Not-determined |
| 17-108_k141_19821 | Not-determined |
| 17-108_k141_268564 | Not-determined |
| 17-108_k141_19829 | Low-quality |
| 17-108_k141_144615 | Not-determined |
| 17-108_k141_130462 | Low-quality |
| 17-108_k141_172421 | Not-determined |
| 17-108_k141_48032 | Not-determined |
| 17-108_k141_295878 | Not-determined |
| 17-108_k141_295880 | Not-determined |
| 17-108_k141_295882 | Not-determined |
| 17-108_k141_255054 | Not-determined |
| 17-108_k141_255055 | Not-determined |
| 17-108_k141_6797 | Not-determined |
| 17-108_k141_144633 | Not-determined |
| 17-108_k141_62166 | Not-determined |
| 17-108_k141_268578 | Not-determined |
| 17-108_k141_185659 | Not-determined |
| 17-108_k141_89451 | Not-determined |
| 17-108_k141_62168 | Low-quality |
| 17-108_k141_185662 | Not-determined |
| 17-108_k141_89454 | Not-determined |
| 17-108_k141_103789 | Not-determined |
| 17-108_k141_185671 | Not-determined |
| 17-108_k141_282977 | Not-determined |
| 17-108_k141_214023 | Not-determined |
| 17-108_k141_75075 | Not-determined |
| 17-108_k141_103800 | Not-determined |
| 17-108_k141_295908 | Low-quality |
| 17-108_k141_89465 | Not-determined |
| 17-108_k141_34321 | Low-quality |
| 17-108_k141_295912 | Not-determined |
| 17-108_k141_89475 | Not-determined |
| 17-108_k141_19874 | Not-determined |
| 17-108_k141_227448 | Not-determined |
| 17-108_k141_19882 | Not-determined |
| 17-108_k141_322994 | Low-quality |
| 17-108_k141_322995 | Not-determined |
| 17-108_k141_34328 | Not-determined |
| 17-108_k141_158664 | Not-determined |
| 17-108_k141_214047 | Not-determined |
| 17-108_k141_227465 | Not-determined |
| 17-108_k141_227467 | Not-determined |
| 17-108_k141_310494 | Not-determined |
| 17-108_k141_158673 | Not-determined |
| 17-108_k141_241809 | Not-determined |
| 17-108_k141_227474 | Not-determined |
| 17-108_k141_62201 | Not-determined |
| 17-108_k141_75118 | Not-determined |
| 17-108_k141_75120 | Low-quality |
| 17-108_k141_34364 | Not-determined |
| 17-108_k141_75124 | Not-determined |
| 17-108_k141_62208 | Not-determined |
| 17-108_k141_227486 | Not-determined |
| 17-108_k141_6829 | Not-determined |
| 17-108_k141_144696 | Low-quality |
| 17-108_k141_144697 | Not-determined |
| 17-108_k141_144698 | Not-determined |
| 17-108_k141_241828 | Low-quality |
| 17-108_k141_89503 | Not-determined |
| 17-108_k141_158692 | Not-determined |
| 17-108_k141_214070 | Not-determined |
| 17-108_k141_103844 | Not-determined |
| 17-108_k141_62219 | Not-determined |
| 17-108_k141_130471 | Not-determined |
| 17-108_k141_130475 | Not-determined |
| 17-108_k141_310518 | Not-determined |
| 17-108_k141_227505 | Low-quality |
| 17-108_k141_89520 | Not-determined |
| 17-108_k141_144711 | Not-determined |
| 17-108_k141_323013 | Not-determined |
| 17-108_k141_158702 | Not-determined |
| 17-108_k141_158704 | Not-determined |
| 17-108_k141_227508 | Not-determined |
| 17-108_k141_48099 | Not-determined |
| 17-108_k141_103853 | Not-determined |
| 17-108_k141_227510 | Not-determined |
| 17-108_k141_62234 | Not-determined |
| 17-108_k141_130486 | Not-determined |
| 17-108_k141_62242 | Not-determined |
| 17-108_k141_75170 | Not-determined |
| 17-108_k141_62245 | Not-determined |
| 17-108_k141_75172 | Not-determined |
| 17-108_k141_144722 | Not-determined |
| 17-108_k141_116479 | Not-determined |
| 17-108_k141_116481 | Not-determined |
| 17-108_k141_144724 | Not-determined |
| 17-108_k141_214083 | Not-determined |
| 17-108_k141_214084 | Not-determined |
| 17-108_k141_214085 | Not-determined |
| 17-108_k141_34387 | Not-determined |
| 17-108_k141_103870 | Not-determined |
| 17-108_k141_199315 | Not-determined |
| 17-108_k141_6864 | Not-determined |
| 17-108_k141_199318 | Not-determined |
| 17-108_k141_310550 | Not-determined |
| 17-108_k141_199322 | Not-determined |
| 17-108_k141_75190 | Not-determined |
| 17-108_k141_34402 | Not-determined |
| 17-108_k141_199326 | Not-determined |
| 17-108_k141_34407 | Not-determined |
| 17-108_k141_34410 | Not-determined |
| 17-108_k141_214097 | Not-determined |
| 17-108_k141_268640 | Not-determined |
| 17-108_k141_199342 | Not-determined |
| 17-108_k141_48127 | Not-determined |
| 17-108_k141_310567 | Low-quality |
| 17-108_k141_241853 | Not-determined |
| 17-108_k141_48130 | Not-determined |
| 17-108_k141_310569 | Low-quality |
| 17-108_k141_116497 | Low-quality |
| 17-108_k141_255116 | Not-determined |
| 17-108_k141_214112 | Not-determined |
| 17-108_k141_172452 | Low-quality |
| 17-108_k141_214117 | Not-determined |
| 17-108_k141_144736 | Not-determined |
| 17-108_k141_172460 | Not-determined |
| 17-108_k141_323050 | Not-determined |
| 17-108_k141_19927 | Not-determined |
| 17-108_k141_255132 | Not-determined |
| 17-108_k141_310592 | Low-quality |
| 17-108_k141_62290 | Not-determined |
| 17-108_k141_130513 | Not-determined |
| 17-108_k141_323056 | Low-quality |
| 17-108_k141_62306 | Not-determined |
| 17-108_k141_103906 | Not-determined |
| 17-108_k141_199359 | Not-determined |
| 17-108_k141_310607 | Not-determined |
| 17-108_k141_172476 | Not-determined |
| 17-108_k141_116530 | Not-determined |
| 17-108_k141_214144 | Not-determined |
| 17-108_k141_6901 | Not-determined |
| 17-108_k141_34440 | Not-determined |
| 17-108_k141_185712 | Not-determined |
| 17-108_k141_89551 | Not-determined |
| 17-108_k141_282987 | Not-determined |
| 17-108_k141_158744 | Not-determined |
| 17-108_k141_89561 | Not-determined |
| 17-108_k141_323065 | Not-determined |
| 17-108_k141_103932 | Not-determined |
| 17-108_k141_241899 | Not-determined |
| 17-108_k141_199390 | Not-determined |
| 17-108_k141_19967 | Not-determined |
| 17-108_k141_130544 | Not-determined |
| 17-108_k141_6911 | Low-quality |
| 17-108_k141_268678 | Not-determined |
| 17-108_k141_227577 | Not-determined |
| 17-108_k141_116549 | Not-determined |
| 17-108_k141_89571 | Not-determined |
| 17-108_k141_214166 | Not-determined |
| 17-108_k141_172499 | Not-determined |
| 17-108_k141_214171 | Not-determined |
| 17-108_k141_130559 | Not-determined |
| 17-108_k141_19983 | Not-determined |
| 17-108_k141_48186 | Not-determined |
| 17-108_k141_130563 | Low-quality |
| 17-108_k141_62328 | Not-determined |
| 17-108_k141_310648 | Low-quality |
| 17-108_k141_19990 | Not-determined |
| 17-108_k141_255178 | Not-determined |
| 17-108_k141_172510 | Not-determined |
| 17-108_k141_172512 | Not-determined |
| 17-108_k141_310651 | Not-determined |
| 17-108_k141_214185 | Not-determined |
| 17-108_k141_144773 | Not-determined |
| 17-108_k141_323085 | Not-determined |
| 17-108_k141_295963 | Not-determined |
| 17-108_k141_48202 | Not-determined |
| 17-108_k141_48203 | Not-determined |
| 17-108_k141_89594 | Not-determined |
| 17-108_k141_268711 | Not-determined |
| 17-108_k141_6931 | Not-determined |
| 17-108_k141_48215 | Not-determined |
| 17-108_k141_6933 | Not-determined |
| 17-108_k141_227605 | Not-determined |
| 17-108_k141_89607 | Not-determined |
| 17-108_k141_130583 | Not-determined |
| 17-108_k141_130586 | Not-determined |
| 17-108_k141_75244 | Not-determined |
| 17-108_k141_241928 | Not-determined |
| 17-108_k141_255204 | Not-determined |
| 17-108_k141_144801 | Not-determined |
| 17-108_k141_295985 | Not-determined |
| 17-108_k141_172529 | Not-determined |
| 17-108_k141_144806 | Not-determined |
| 17-108_k141_255211 | Not-determined |
| 17-108_k141_172537 | Not-determined |
| 17-108_k141_62383 | Not-determined |
| 17-108_k141_283044 | Not-determined |
| 17-108_k141_283047 | Not-determined |
| 17-108_k141_34457 | Not-determined |
| 17-108_k141_241939 | Not-determined |
| 17-108_k141_323109 | Not-determined |
| 17-108_k141_199449 | Not-determined |
| 17-108_k141_310695 | Low-quality |
| 17-108_k141_62395 | Not-determined |
| 17-108_k141_268738 | Not-determined |
| 17-108_k141_34466 | Not-determined |
| 17-108_k141_75266 | Not-determined |
| 17-108_k141_214232 | Low-quality |
| 17-108_k141_144823 | Not-determined |
| 17-108_k141_199458 | Low-quality |
| 17-108_k141_283056 | Not-determined |
| 17-108_k141_185746 | Not-determined |
| 17-108_k141_172558 | Not-determined |
| 17-108_k141_20043 | Not-determined |
| 17-108_k141_75278 | Low-quality |
| 17-108_k141_323132 | Not-determined |

| | |
|---|---|
| 17-108_k141_62405 | Not-determined |
| 17-108_k141_310727 | Not-determined |
| 17-108_k141_310732 | Not-determined |
| 17-108_k141_283072 | Not-determined |
| 17-108_k141_172578 | Not-determined |
| 17-108_k141_6972 | Not-determined |
| 17-108_k141_104011 | Not-determined |
| 17-108_k141_20062 | Low-quality |
| 17-108_k141_241961 | Not-determined |
| 17-108_k141_241962 | Not-determined |
| 17-108_k141_34522 | Low-quality |
| 17-108_k141_89639 | Not-determined |
| 17-108_k141_323158 | Low-quality |
| 17-108_k141_296013 | Low-quality |
| 17-108_k141_283100 | Not-determined |
| 17-108_k141_323163 | Not-determined |
| 17-108_k141_89646 | Not-determined |
| 17-108_k141_255229 | Low-quality |
| 17-108_k141_185758 | Not-determined |
| 17-108_k141_323170 | Not-determined |
| 17-108_k141_185764 | Not-determined |
| 17-108_k141_255234 | Not-determined |
| 17-108_k141_130595 | Not-determined |
| 17-108_k141_34535 | Not-determined |
| 17-108_k141_34536 | Low-quality |
| 17-108_k141_89660 | Not-determined |
| 17-108_k141_89662 | Not-determined |
| 17-108_k141_144870 | Not-determined |
| 17-108_k141_241980 | Not-determined |
| 17-108_k141_323189 | Not-determined |
| 17-108_k141_241985 | Not-determined |
| 17-108_k141_62434 | Not-determined |
| 17-108_k141_283111 | Not-determined |
| 17-108_k141_241989 | Not-determined |
| 17-108_k141_255248 | Not-determined |
| 17-108_k141_296037 | Not-determined |
| 17-108_k141_241993 | Low-quality |
| 17-108_k141_283116 | Not-determined |
| 17-108_k141_185772 | Low-quality |
| 17-108_k141_62450 | Not-determined |
| 17-108_k141_268787 | Not-determined |
| 17-108_k141_104039 | Not-determined |
| 17-108_k141_62454 | Not-determined |
| 17-108_k141_104044 | Not-determined |
| 17-108_k141_268790 | Not-determined |
| 17-108_k141_104047 | Not-determined |
| 17-108_k141_227668 | Not-determined |
| 17-108_k141_172639 | Not-determined |
| 17-108_k141_242014 | Low-quality |
| 17-108_k141_34558 | Not-determined |
| 17-108_k141_89668 | Not-determined |
| 17-108_k141_310797 | Not-determined |
| 17-108_k141_268795 | Not-determined |
| 17-108_k141_268799 | Not-determined |
| 17-108_k141_323198 | Not-determined |
| 17-108_k141_7030 | Not-determined |
| 17-108_k141_89678 | Not-determined |
| 17-108_k141_255271 | Not-determined |
| 17-108_k141_172656 | Not-determined |
| 17-108_k141_34576 | Not-determined |
| 17-108_k141_242024 | Not-determined |
| 17-108_k141_199514 | Not-determined |
| 17-108_k141_20086 | Not-determined |
| 17-108_k141_130625 | Not-determined |
| 17-108_k141_255279 | Not-determined |
| 17-108_k141_20092 | Not-determined |
| 17-108_k141_185789 | Not-determined |
| 17-108_k141_199530 | Not-determined |
| 17-108_k141_310807 | Not-determined |
| 17-108_k141_310808 | Not-determined |
| 17-108_k141_296084 | Not-determined |
| 17-108_k141_310809 | Not-determined |
| 17-108_k141_310811 | Not-determined |
| 17-108_k141_75328 | Not-determined |
| 17-108_k141_199534 | Low-quality |
| 17-108_k141_89689 | Not-determined |
| 17-108_k141_89690 | Low-quality |
| 17-108_k141_310817 | Low-quality |
| 17-108_k141_130637 | Not-determined |
| 17-108_k141_62471 | Not-determined |
| 17-108_k141_199542 | Not-determined |
| 17-108_k141_199544 | Not-determined |
| 17-108_k141_199548 | Not-determined |
| 17-108_k141_172679 | Not-determined |
| 17-108_k141_283171 | Not-determined |
| 17-108_k141_172680 | Not-determined |
| 17-108_k141_34604 | Not-determined |
| 17-108_k141_227721 | Not-determined |
| 17-108_k141_242052 | Not-determined |
| 17-108_k141_227725 | Not-determined |
| 17-108_k141_172685 | Not-determined |
| 17-108_k141_185804 | Not-determined |
| 17-108_k141_158772 | Low-quality |
| 17-108_k141_20127 | Not-determined |
| 17-108_k141_7061 | Not-determined |
| 17-108_k141_158780 | Low-quality |
| 17-108_k141_62474 | Not-determined |
| 17-108_k141_62478 | Not-determined |
| 17-108_k141_20129 | Not-determined |
| 17-108_k141_7070 | Not-determined |
| 17-108_k141_20133 | Not-determined |
| 17-108_k141_130653 | Not-determined |
| 17-108_k141_20136 | Not-determined |
| 17-108_k141_199569 | Not-determined |
| 17-108_k141_255284 | Not-determined |
| 17-108_k141_185831 | Not-determined |
| 17-108_k141_7083 | Not-determined |
| 17-108_k141_20140 | Not-determined |
| 17-108_k141_255289 | Not-determined |
| 17-108_k141_283205 | Not-determined |
| 17-108_k141_130660 | Not-determined |
| 17-108_k141_158788 | Not-determined |
| 17-108_k141_214284 | Not-determined |
| 17-108_k141_48254 | Not-determined |
| 17-108_k141_310850 | Low-quality |
| 17-108_k141_242066 | Low-quality |
| 17-108_k141_242068 | Not-determined |
| 17-108_k141_255299 | Not-determined |
| 17-108_k141_296134 | Low-quality |
| 17-108_k141_158801 | Not-determined |
| 17-108_k141_172736 | Not-determined |
| 17-108_k141_48268 | Low-quality |
| 17-108_k141_268826 | Not-determined |
| 17-108_k141_62498 | Not-determined |
| 17-108_k141_268832 | Not-determined |
| 17-108_k141_62504 | Not-determined |
| 17-108_k141_48276 | Not-determined |
| 17-108_k141_158822 | Not-determined |
| 17-108_k141_20163 | Not-determined |
| 17-108_k141_104103 | Not-determined |
| 17-108_k141_268845 | Not-determined |
| 17-108_k141_242096 | Not-determined |
| 17-108_k141_158826 | Not-determined |
| 17-108_k141_214328 | Not-determined |
| 17-108_k141_75372 | Not-determined |
| 17-108_k141_7111 | Not-determined |
| 17-108_k141_34666 | Low-quality |
| 17-108_k141_7113 | Not-determined |
| 17-108_k141_268853 | Not-determined |
| 17-108_k141_104111 | Not-determined |
| 17-108_k141_268857 | Not-determined |
| 17-108_k141_255327 | Not-determined |
| 17-108_k141_104120 | Not-determined |
| 17-108_k141_158838 | Low-quality |
| 17-108_k141_323273 | Not-determined |
| 17-108_k141_130710 | Not-determined |
| 17-108_k141_255334 | Not-determined |
| 17-108_k141_214335 | Not-determined |
| 17-108_k141_89737 | Not-determined |
| 17-108_k141_34681 | Low-quality |
| 17-108_k141_89743 | Not-determined |
| 17-108_k141_89745 | Not-determined |
| 17-108_k141_158846 | Not-determined |
| 17-108_k141_172780 | Not-determined |
| 17-108_k141_158851 | Not-determined |
| 17-108_k141_199588 | Not-determined |
| 17-108_k141_34698 | Not-determined |
| 17-108_k141_62523 | Low-quality |
| 17-108_k141_130719 | Not-determined |
| 17-108_k141_310879 | Not-determined |
| 17-108_k141_185861 | Not-determined |
| 17-108_k141_227747 | Not-determined |
| 17-108_k141_185865 | Low-quality |
| 17-108_k141_172796 | Not-determined |
| 17-108_k141_255351 | Low-quality |
| 17-108_k141_310895 | Not-determined |
| 17-108_k141_48323 | Not-determined |
| 17-108_k141_89753 | Not-determined |
| 17-108_k141_145003 | Not-determined |
| 17-108_k141_268890 | Not-determined |
| 17-108_k141_89761 | Not-determined |
| 17-108_k141_172800 | Not-determined |
| 17-108_k141_62545 | Not-determined |
| 17-108_k141_227767 | Not-determined |
| 17-108_k141_242103 | Not-determined |
| 17-108_k141_20210 | Not-determined |
| 17-108_k141_145015 | Low-quality |
| 17-108_k141_34726 | Not-determined |
| 17-108_k141_145018 | Low-quality |
| 17-108_k141_20213 | Low-quality |
| 17-108_k141_310914 | Not-determined |
| 17-108_k141_62565 | Not-determined |
| 17-108_k141_242120 | Low-quality |
| 17-108_k141_199607 | Not-determined |
| 17-108_k141_242124 | Not-determined |
| 17-108_k141_172824 | Low-quality |
| 17-108_k141_34733 | Not-determined |
| 17-108_k141_104175 | Not-determined |
| 17-108_k141_172827 | Not-determined |
| 17-108_k141_323285 | Not-determined |
| 17-108_k141_116648 | Not-determined |
| 17-108_k141_62576 | Not-determined |
| 17-108_k141_48362 | Not-determined |
| 17-108_k141_242132 | Not-determined |
| 17-108_k141_89789 | Not-determined |
| 17-108_k141_116655 | Not-determined |
| 17-108_k141_242135 | Not-determined |
| 17-108_k141_48366 | Not-determined |
| 17-108_k141_323304 | Not-determined |
| 17-108_k141_20230 | Not-determined |
| 17-108_k141_227798 | Not-determined |
| 17-108_k141_145054 | Not-determined |
| 17-108_k141_145057 | Low-quality |
| 17-108_k141_7156 | Not-determined |
| 17-108_k141_116663 | Low-quality |
| 17-108_k141_7160 | Not-determined |
| 17-108_k141_20234 | Not-determined |
| 17-108_k141_116667 | Low-quality |
| 17-108_k141_242138 | Not-determined |
| 17-108_k141_172849 | Not-determined |
| 17-108_k141_283234 | Not-determined |
| 17-108_k141_323308 | Low-quality |
| 17-108_k141_283241 | Not-determined |
| 17-108_k141_20247 | Not-determined |
| 17-108_k141_283245 | Not-determined |
| 17-108_k141_268900 | Not-determined |
| 17-108_k141_242148 | Not-determined |
| 17-108_k141_116673 | Not-determined |
| 17-108_k141_7173 | Not-determined |
| 17-108_k141_242151 | Not-determined |
| 17-108_k141_7177 | Not-determined |
| 17-108_k141_75422 | Not-determined |
| 17-108_k141_34766 | Not-determined |
| 17-108_k141_296168 | Not-determined |
| 17-108_k141_48398 | Low-quality |
| 17-108_k141_75430 | Not-determined |
| 17-108_k141_268909 | Not-determined |
| 17-108_k141_62593 | Not-determined |
| 17-108_k141_172868 | Not-determined |
| 17-108_k141_145072 | Low-quality |
| 17-108_k141_62598 | Not-determined |
| 17-108_k141_116687 | Not-determined |
| 17-108_k141_296178 | Not-determined |
| 17-108_k141_255440 | Low-quality |
| 17-108_k141_20253 | Not-determined |
| 17-108_k141_268920 | Not-determined |
| 17-108_k141_242173 | Not-determined |
| 17-108_k141_75440 | Not-determined |
| 17-108_k141_199651 | Not-determined |
| 17-108_k141_227835 | Not-determined |
| 17-108_k141_62620 | Not-determined |
| 17-108_k141_227843 | Not-determined |
| 17-108_k141_242179 | Not-determined |
| 17-108_k141_48422 | Not-determined |
| 17-108_k141_242184 | Not-determined |
| 17-108_k141_296188 | Not-determined |
| 17-108_k141_242187 | Not-determined |
| 17-108_k141_75454 | Not-determined |
| 17-108_k141_158876 | Not-determined |
| 17-108_k141_130764 | Not-determined |
| 17-108_k141_242193 | Not-determined |
| 17-108_k141_75458 | Not-determined |
| 17-108_k141_7219 | Not-determined |
| 17-108_k141_34789 | Not-determined |
| 17-108_k141_199669 | Not-determined |
| 17-108_k141_62646 | Not-determined |
| 17-108_k141_296199 | Not-determined |
| 17-108_k141_116713 | Low-quality |
| 17-108_k141_227862 | Not-determined |
| 17-108_k141_255468 | Not-determined |
| 17-108_k141_48443 | Not-determined |
| 17-108_k141_75470 | Not-determined |
| 17-108_k141_255471 | Not-determined |
| 17-108_k141_214426 | Not-determined |
| 17-108_k141_283266 | Not-determined |
| 17-108_k141_296203 | Low-quality |
| 17-108_k141_48453 | Not-determined |
| 17-108_k141_158895 | Not-determined |
| 17-108_k141_48455 | Not-determined |
| 17-108_k141_48459 | Low-quality |
| 17-108_k141_242203 | Low-quality |
| 17-108_k141_323330 | Not-determined |
| 17-108_k141_130784 | Not-determined |
| 17-108_k141_227881 | Not-determined |
| 17-108_k141_296211 | Not-determined |
| 17-108_k141_62660 | Not-determined |
| 17-108_k141_62661 | Not-determined |
| 17-108_k141_242218 | Not-determined |
| 17-108_k141_296214 | Not-determined |
| 17-108_k141_116743 | Not-determined |
| 17-108_k141_214431 | Not-determined |
| 17-108_k141_255502 | Not-determined |
| 17-108_k141_34829 | Not-determined |
| 17-108_k141_75495 | Not-determined |
| 17-108_k141_7246 | Not-determined |
| 17-108_k141_7249 | Not-determined |
| 17-108_k141_130808 | Not-determined |
| 17-108_k141_75503 | Not-determined |
| 17-108_k141_116752 | Not-determined |
| 17-108_k141_116759 | Not-determined |
| 17-108_k141_185927 | Not-determined |
| 17-108_k141_283299 | Not-determined |
| 17-108_k141_214443 | Not-determined |
| 17-108_k141_310930 | Low-quality |
| 17-108_k141_214447 | Not-determined |
| 17-108_k141_310931 | Low-quality |
| 17-108_k141_323355 | Not-determined |
| 17-108_k141_185937 | Low-quality |
| 17-108_k141_62689 | Not-determined |
| 17-108_k141_310934 | Not-determined |
| 17-108_k141_158914 | Low-quality |
| 17-108_k141_310936 | Not-determined |
| 17-108_k141_323357 | Not-determined |
| 17-108_k141_62693 | Not-determined |
| 17-108_k141_214456 | Not-determined |
| 17-108_k141_255519 | Not-determined |
| 17-108_k141_185948 | Not-determined |
| 17-108_k141_310947 | Not-determined |
| 17-108_k141_34847 | Not-determined |
| 17-108_k141_255522 | Not-determined |
| 17-108_k141_172901 | Low-quality |
| 17-108_k141_296258 | Not-determined |
| 17-108_k141_20293 | Not-determined |
| 17-108_k141_242259 | Not-determined |
| 17-108_k141_172904 | Not-determined |
| 17-108_k141_242260 | Not-determined |
| 17-108_k141_75536 | Not-determined |
| 17-108_k141_62708 | Low-quality |
| 17-108_k141_185957 | Not-determined |
| 17-108_k141_242264 | Not-determined |
| 17-108_k141_283306 | Low-quality |
| 17-108_k141_145159 | Not-determined |
| 17-108_k141_89852 | Not-determined |
| 17-108_k141_214466 | Not-determined |
| 17-108_k141_172914 | Not-determined |
| 17-108_k141_130846 | Not-determined |
| 17-108_k141_104212 | Not-determined |
| 17-108_k141_62718 | Not-determined |
| 17-108_k141_158939 | Not-determined |
| 17-108_k141_185974 | Not-determined |
| 17-108_k141_20308 | Not-determined |
| 17-108_k141_214483 | Not-determined |
| 17-108_k141_48485 | Not-determined |
| 17-108_k141_172918 | Not-determined |
| 17-108_k141_104216 | Not-determined |
| 17-108_k141_130850 | Not-determined |
| 17-108_k141_185983 | Not-determined |
| 17-108_k141_20320 | Not-determined |
| 17-108_k141_172929 | Not-determined |
| 17-108_k141_199691 | Not-determined |
| 17-108_k141_20323 | Not-determined |
| 17-108_k141_48495 | Not-determined |
| 17-108_k141_20326 | Not-determined |
| 17-108_k141_20327 | Low-quality |
| 17-108_k141_255564 | Low-quality |
| 17-108_k141_268964 | Not-determined |
| 17-108_k141_20332 | Not-determined |
| 17-108_k141_48500 | Not-determined |
| 17-108_k141_89875 | Not-determined |
| 17-108_k141_75577 | Not-determined |
| 17-108_k141_296286 | Not-determined |
| 17-108_k141_104239 | Not-determined |
| 17-108_k141_323416 | Not-determined |
| 17-108_k141_158960 | Not-determined |
| 17-108_k141_145191 | Not-determined |
| 17-108_k141_172943 | Not-determined |
| 17-108_k141_145197 | Not-determined |
| 17-108_k141_227965 | Not-determined |
| 17-108_k141_104253 | Not-determined |
| 17-108_k141_34894 | Not-determined |
| 17-108_k141_323425 | Not-determined |
| 17-108_k141_7287 | Not-determined |
| 17-108_k141_323428 | Not-determined |
| 17-108_k141_255588 | Not-determined |
| 17-108_k141_323429 | Not-determined |
| 17-108_k141_34899 | Not-determined |
| 17-108_k141_62743 | Not-determined |
| 17-108_k141_214505 | Not-determined |
| 17-108_k141_323431 | Not-determined |
| 17-108_k141_89907 | Not-determined |
| 17-108_k141_227974 | Not-determined |
| 17-108_k141_255594 | Not-determined |
| 17-108_k141_199714 | Not-determined |
| 17-108_k141_48506 | Not-determined |
| 17-108_k141_268979 | Low-quality |
| 17-108_k141_116843 | Not-determined |
| 17-108_k141_172972 | Not-determined |
| 17-108_k141_89931 | Not-determined |
| 17-108_k141_214526 | Low-quality |
| 17-108_k141_214528 | Not-determined |
| 17-108_k141_116854 | Not-determined |
| 17-108_k141_62766 | Not-determined |
| 17-108_k141_159004 | Not-determined |
| 17-108_k141_89934 | Not-determined |
| 17-108_k141_48518 | Not-determined |
| 17-108_k141_214530 | Not-determined |
| 17-108_k141_34921 | Not-determined |
| 17-108_k141_48529 | Not-determined |
| 17-108_k141_62773 | Not-determined |
| 17-108_k141_255620 | Not-determined |
| 17-108_k141_227997 | Not-determined |
| 17-108_k141_172980 | Not-determined |
| 17-108_k141_199741 | Not-determined |
| 17-108_k141_214541 | Not-determined |
| 17-108_k141_34943 | Not-determined |
| 17-108_k141_269003 | Not-determined |
| 17-108_k141_255623 | Not-determined |
| 17-108_k141_255624 | Not-determined |
| 17-108_k141_75650 | Not-determined |
| 17-108_k141_130875 | Not-determined |
| 17-108_k141_242314 | Not-determined |
| 17-108_k141_20383 | Not-determined |
| 17-108_k141_75657 | Not-determined |
| 17-108_k141_228014 | Low-quality |
| 17-108_k141_255636 | Not-determined |
| 17-108_k141_62794 | Not-determined |
| 17-108_k141_199761 | Not-determined |
| 17-108_k141_130881 | Not-determined |
| 17-108_k141_7325 | Not-determined |
| 17-108_k141_228018 | Not-determined |
| 17-108_k141_228020 | Not-determined |
| 17-108_k141_75659 | Not-determined |
| 17-108_k141_255642 | Not-determined |
| 17-108_k141_228021 | Not-determined |
| 17-108_k141_62802 | Not-determined |
| 17-108_k141_255646 | Not-determined |
| 17-108_k141_283392 | Not-determined |
| 17-108_k141_269019 | Not-determined |
| 17-108_k141_296357 | Not-determined |
| 17-108_k141_255654 | Not-determined |
| 17-108_k141_296359 | Not-determined |
| 17-108_k141_75663 | Low-quality |
| 17-108_k141_75665 | Not-determined |
| 17-108_k141_199775 | Not-determined |
| 17-108_k141_199778 | Not-determined |
| 17-108_k141_283402 | Not-determined |
| 17-108_k141_255671 | Not-determined |
| 17-108_k141_104300 | Not-determined |
| 17-108_k141_323467 | Not-determined |
| 17-108_k141_323469 | Not-determined |
| 17-108_k141_116893 | Not-determined |
| 17-108_k141_48578 | Not-determined |
| 17-108_k141_145274 | Not-determined |
| 17-108_k141_116897 | Not-determined |
| 17-108_k141_214553 | Not-determined |
| 17-108_k141_89971 | Not-determined |
| 17-108_k141_255683 | Not-determined |
| 17-108_k141_296379 | Not-determined |
| 17-108_k141_186069 | Not-determined |
| 17-108_k141_186071 | Not-determined |
| 17-108_k141_214565 | Not-determined |
| 17-108_k141_242338 | Low-quality |
| 17-108_k141_296382 | Not-determined |
| 17-108_k141_130914 | Low-quality |
| 17-108_k141_269044 | Not-determined |
| 17-108_k141_228045 | Not-determined |
| 17-108_k141_214573 | Not-determined |
| 17-108_k141_228047 | Not-determined |
| 17-108_k141_242352 | Low-quality |
| 17-108_k141_186084 | Not-determined |
| 17-108_k141_311038 | Not-determined |
| 17-108_k141_186091 | Not-determined |
| 17-108_k141_35001 | Not-determined |
| 17-108_k141_20428 | Not-determined |
| 17-108_k141_311051 | Not-determined |
| 17-108_k141_116923 | Not-determined |
| 17-108_k141_255688 | Not-determined |
| 17-108_k141_228072 | Not-determined |
| 17-108_k141_311053 | Low-quality |
| 17-108_k141_296398 | Not-determined |
| 17-108_k141_283445 | Not-determined |
| 17-108_k141_283448 | Not-determined |
| 17-108_k141_35007 | Not-determined |
| 17-108_k141_104340 | Not-determined |
| 17-108_k141_283449 | Not-determined |
| 17-108_k141_311060 | Not-determined |
| 17-108_k141_323483 | Not-determined |
| 17-108_k141_323486 | Not-determined |
| 17-108_k141_145304 | Not-determined |
| 17-108_k141_104342 | Not-determined |
| 17-108_k141_311070 | Not-determined |
| 17-108_k141_35017 | Not-determined |
| 17-108_k141_311075 | Low-quality |
| 17-108_k141_255704 | Not-determined |
| 17-108_k141_173087 | Not-determined |
| 17-108_k141_296420 | Low-quality |
| 17-108_k141_20432 | Not-determined |
| 17-108_k141_130953 | Not-determined |
| 17-108_k141_104354 | Not-determined |
| 17-108_k141_159030 | Not-determined |
| 17-108_k141_228082 | Not-determined |
| 17-108_k141_130959 | Not-determined |
| 17-108_k141_323507 | Not-determined |
| 17-108_k141_255720 | Not-determined |
| 17-108_k141_159051 | Low-quality |
| 17-108_k141_255731 | Not-determined |
| 17-108_k141_242399 | Not-determined |
| 17-108_k141_35047 | Not-determined |
| 17-108_k141_228090 | Low-quality |
| 17-108_k141_199839 | Not-determined |
| 17-108_k141_311101 | Low-quality |
| 17-108_k141_311103 | Not-determined |
| 17-108_k141_214618 | Not-determined |
| 17-108_k141_116959 | Low-quality |
| 17-108_k141_159067 | Not-determined |
| 17-108_k141_159068 | Not-determined |
| 17-108_k141_199850 | Not-determined |
| 17-108_k141_90029 | Not-determined |
| 17-108_k141_116962 | Not-determined |
| 17-108_k141_311109 | Not-determined |
| 17-108_k141_269077 | Not-determined |
| 17-108_k141_159081 | Not-determined |
| 17-108_k141_116971 | Not-determined |
| 17-108_k141_199864 | Not-determined |
| 17-108_k141_20453 | Not-determined |
| 17-108_k141_242425 | Not-determined |
| 17-108_k141_269086 | Not-determined |
| 17-108_k141_228108 | Not-determined |
| 17-108_k141_242429 | Not-determined |
| 17-108_k141_131003 | Not-determined |
| 17-108_k141_228114 | Not-determined |
| 17-108_k141_323521 | Not-determined |
| 17-108_k141_255754 | Not-determined |
| 17-108_k141_323522 | Not-determined |
| 17-108_k141_311128 | Not-determined |
| 17-108_k141_116986 | Not-determined |
| 17-108_k141_311130 | Not-determined |
| 17-108_k141_242439 | Not-determined |
| 17-108_k141_311135 | Not-determined |
| 17-108_k141_323524 | Not-determined |
| 17-108_k141_48649 | Not-determined |
| 17-108_k141_242453 | Not-determined |
| 17-108_k141_242458 | Not-determined |
| 17-108_k141_48661 | Not-determined |
| 17-108_k141_228134 | Not-determined |
| 17-108_k141_117001 | Not-determined |
| 17-108_k141_75735 | Not-determined |
| 17-108_k141_311150 | Not-determined |
| 17-108_k141_131036 | Not-determined |
| 17-108_k141_242469 | Not-determined |
| 17-108_k141_186146 | Not-determined |
| 17-108_k141_311157 | Not-determined |
| 17-108_k141_20473 | Not-determined |
| 17-108_k141_75740 | Low-quality |
| 17-108_k141_311160 | Not-determined |
| 17-108_k141_131045 | Not-determined |
| 17-108_k141_75752 | Not-determined |
| 17-108_k141_90065 | Not-determined |
| 17-108_k141_48684 | Not-determined |
| 17-108_k141_75756 | Not-determined |
| 17-108_k141_20480 | Not-determined |
| 17-108_k141_173161 | Low-quality |
| 17-108_k141_283495 | Not-determined |
| 17-108_k141_269113 | Not-determined |
| 17-108_k141_311170 | Not-determined |
| 17-108_k141_48693 | Not-determined |
| 17-108_k141_20488 | Not-determined |
| 17-108_k141_269115 | Not-determined |
| 17-108_k141_269117 | Not-determined |
| 17-108_k141_283506 | Low-quality |
| 17-108_k141_269123 | Low-quality |
| 17-108_k141_296500 | Not-determined |
| 17-108_k141_173178 | Not-determined |
| 17-108_k141_62897 | Not-determined |
| 17-108_k141_199928 | Not-determined |
| 17-108_k141_35115 | Not-determined |
| 17-108_k141_90096 | Not-determined |
| 17-108_k141_62900 | Not-determined |
| 17-108_k141_296506 | Not-determined |
| 17-108_k141_269134 | Not-determined |
| 17-108_k141_35122 | Not-determined |
| 17-108_k141_228173 | Not-determined |
| 17-108_k141_173181 | Low-quality |
| 17-108_k141_173185 | Not-determined |
| 17-108_k141_296522 | Not-determined |
| 17-108_k141_311201 | Not-determined |
| 17-108_k141_75782 | Not-determined |
| 17-108_k141_186157 | Not-determined |
| 17-108_k141_90106 | Not-determined |
| 17-108_k141_62911 | Not-determined |
| 17-108_k141_62912 | Not-determined |
| 17-108_k141_173188 | Not-determined |
| 17-108_k141_296530 | Not-determined |
| 17-108_k141_104393 | Not-determined |
| 17-108_k141_186160 | Low-quality |
| 17-108_k141_75792 | Not-determined |
| 17-108_k141_186161 | Not-determined |
| 17-108_k141_104397 | Not-determined |
| 17-108_k141_242476 | Not-determined |
| 17-108_k141_48720 | Not-determined |
| 17-108_k141_255818 | Not-determined |
| 17-108_k141_269154 | Not-determined |
| 17-108_k141_186177 | Low-quality |
| 17-108_k141_296543 | Not-determined |
| 17-108_k141_228189 | Not-determined |
| 17-108_k141_186181 | Not-determined |
| 17-108_k141_199952 | Not-determined |
| 17-108_k141_228195 | Low-quality |
| 17-108_k141_296554 | Not-determined |

| | |
|---|---|
| 17-108_k141_242500 | Not-determined |
| 17-108_k141_242502 | Not-determined |
| 17-108_k141_186190 | Not-determined |
| 17-108_k141_186193 | Low-quality |
| 17-108_k141_296573 | Low-quality |
| 17-108_k141_48732 | Not-determined |
| 17-108_k141_283529 | Not-determined |
| 17-108_k141_48736 | Not-determined |
| 17-108_k141_48737 | Not-determined |
| 17-108_k141_323586 | Low-quality |
| 17-108_k141_323587 | Not-determined |
| 17-108_k141_159140 | Not-determined |
| 17-108_k141_323593 | Not-determined |
| 17-108_k141_214674 | Not-determined |
| 17-108_k141_323599 | Not-determined |
| 17-108_k141_117058 | Not-determined |
| 17-108_k141_48750 | Not-determined |
| 17-108_k141_159157 | Not-determined |
| 17-108_k141_7483 | Not-determined |
| 17-108_k141_48755 | Not-determined |
| 17-108_k141_242523 | Low-quality |
| 17-108_k141_35192 | Not-determined |
| 17-108_k141_62950 | Not-determined |
| 17-108_k141_296593 | Low-quality |
| 17-108_k141_48766 | Not-determined |
| 17-108_k141_48767 | Not-determined |
| 17-108_k141_242527 | Not-determined |
| 17-108_k141_145366 | Not-determined |
| 17-108_k141_269194 | Not-determined |
| 17-108_k141_20547 | Not-determined |
| 17-108_k141_199992 | Not-determined |
| 17-108_k141_255868 | Not-determined |
| 17-108_k141_173224 | Not-determined |
| 17-108_k141_117076 | Not-determined |
| 17-108_k141_269199 | Not-determined |
| 17-108_k141_117078 | Not-determined |
| 17-108_k141_35208 | Not-determined |
| 17-108_k141_323618 | Not-determined |
| 17-108_k141_159174 | Low-quality |
| 17-108_k141_200001 | Not-determined |
| 17-108_k141_200002 | Not-determined |
| 17-108_k141_242547 | Not-determined |
| 17-108_k141_62981 | Not-determined |
| 17-108_k141_242549 | Not-determined |
| 17-108_k141_35219 | Not-determined |
| 17-108_k141_269214 | Not-determined |
| 17-108_k141_186225 | Not-determined |
| 17-108_k141_311240 | Low-quality |
| 17-108_k141_255888 | Not-determined |
| 17-108_k141_48789 | Not-determined |
| 17-108_k141_90189 | Not-determined |
| 17-108_k141_35232 | Low-quality |
| 17-108_k141_75822 | Not-determined |
| 17-108_k141_228201 | Not-determined |
| 17-108_k141_75829 | Not-determined |
| 17-108_k141_228205 | Not-determined |
| 17-108_k141_214687 | Not-determined |
| 17-108_k141_269248 | Not-determined |
| 17-108_k141_35241 | Not-determined |
| 17-108_k141_214691 | Not-determined |
| 17-108_k141_90205 | Not-determined |
| 17-108_k141_35251 | Not-determined |
| 17-108_k141_48813 | Not-determined |
| 17-108_k141_173272 | Not-determined |
| 17-108_k141_283562 | Not-determined |
| 17-108_k141_311267 | Not-determined |
| 17-108_k141_200019 | Not-determined |
| 17-108_k141_159200 | Not-determined |
| 17-108_k141_35256 | Low-quality |
| 17-108_k141_48822 | Not-determined |
| 17-108_k141_242585 | Not-determined |
| 17-108_k141_186260 | Not-determined |
| 17-108_k141_90216 | Not-determined |
| 17-108_k141_186267 | Not-determined |
| 17-108_k141_200037 | Not-determined |
| 17-108_k141_269272 | Low-quality |
| 17-108_k141_145399 | Not-determined |
| 17-108_k141_200046 | Low-quality |
| 17-108_k141_186274 | Not-determined |
| 17-108_k141_117148 | Not-determined |
| 17-108_k141_90241 | Not-determined |
| 17-108_k141_117153 | Not-determined |
| 17-108_k141_283587 | Low-quality |
| 17-108_k141_145412 | Not-determined |
| 17-108_k141_145414 | Not-determined |
| 17-108_k141_214708 | Not-determined |
| 17-108_k141_35261 | Not-determined |
| 17-108_k141_90251 | Not-determined |
| 17-108_k141_145417 | Not-determined |
| 17-108_k141_228242 | Not-determined |
| 17-108_k141_200056 | Not-determined |
| 17-108_k141_255919 | Not-determined |
| 17-108_k141_242618 | Not-determined |
| 17-108_k141_228245 | Not-determined |
| 17-108_k141_255924 | Not-determined |
| 17-108_k141_20606 | Not-determined |
| 17-108_k141_75857 | Low-quality |
| 17-108_k141_20608 | Not-determined |
| 17-108_k141_145421 | Not-determined |
| 17-108_k141_75859 | Not-determined |
| 17-108_k141_75862 | Low-quality |
| 17-108_k141_75867 | Not-determined |
| 17-108_k141_200073 | Not-determined |
| 17-108_k141_35285 | Not-determined |
| 17-108_k141_173320 | Not-determined |
| 17-108_k141_20626 | Not-determined |
| 17-108_k141_323678 | Not-determined |
| 17-108_k141_228258 | Low-quality |
| 17-108_k141_90272 | Not-determined |
| 17-108_k141_173327 | Not-determined |
| 17-108_k141_20634 | Not-determined |
| 17-108_k141_228264 | Not-determined |
| 17-108_k141_131184 | Not-determined |
| 17-108_k141_48846 | Not-determined |
| 17-108_k141_173334 | Not-determined |
| 17-108_k141_90280 | Not-determined |
| 17-108_k141_255946 | Not-determined |
| 17-108_k141_186315 | Not-determined |
| 17-108_k141_48853 | Not-determined |
| 17-108_k141_186316 | Not-determined |
| 17-108_k141_255952 | Not-determined |
| 17-108_k141_35310 | Not-determined |
| 17-108_k141_35312 | Not-determined |
| 17-108_k141_255955 | Not-determined |
| 17-108_k141_228280 | Not-determined |
| 17-108_k141_104422 | Not-determined |
| 17-108_k141_228282 | Not-determined |
| 17-108_k141_255960 | Not-determined |
| 17-108_k141_228285 | Not-determined |
| 17-108_k141_228287 | Not-determined |
| 17-108_k141_186332 | Not-determined |
| 17-108_k141_173344 | Not-determined |
| 17-108_k141_311320 | Not-determined |
| 17-108_k141_269294 | Not-determined |
| 17-108_k141_255977 | Not-determined |
| 17-108_k141_228298 | Low-quality |
| 17-108_k141_159258 | Not-determined |
| 17-108_k141_228300 | Not-determined |
| 17-108_k141_104432 | Not-determined |
| 17-108_k141_104434 | Not-determined |
| 17-108_k141_145452 | Not-determined |
| 17-108_k141_117187 | Not-determined |
| 17-108_k141_75908 | Not-determined |
| 17-108_k141_117191 | Not-determined |
| 17-108_k141_159274 | Not-determined |
| 17-108_k141_7539 | Not-determined |
| 17-108_k141_20675 | Low-quality |
| 17-108_k141_159276 | Not-determined |
| 17-108_k141_200087 | Not-determined |
| 17-108_k141_228312 | Not-determined |
| 17-108_k141_117193 | Not-determined |
| 17-108_k141_228314 | Not-determined |
| 17-108_k141_104464 | Not-determined |
| 17-108_k141_20677 | Not-determined |
| 17-108_k141_228320 | Not-determined |
| 17-108_k141_200095 | Not-determined |
| 17-108_k141_131220 | Not-determined |
| 17-108_k141_48878 | Not-determined |
| 17-108_k141_323699 | Not-determined |
| 17-108_k141_48883 | Not-determined |
| 17-108_k141_214742 | Not-determined |
| 17-108_k141_75925 | Not-determined |
| 17-108_k141_200111 | Not-determined |
| 17-108_k141_200114 | Not-determined |
| 17-108_k141_269313 | Not-determined |
| 17-108_k141_296664 | Not-determined |
| 17-108_k141_131245 | Not-determined |
| 17-108_k141_7558 | Not-determined |
| 17-108_k141_228339 | Not-determined |
| 17-108_k141_214751 | Not-determined |
| 17-108_k141_296675 | Not-determined |
| 17-108_k141_48889 | Not-determined |
| 17-108_k141_7564 | Not-determined |
| 17-108_k141_75954 | Not-determined |
| 17-108_k141_323730 | Not-determined |
| 17-108_k141_75957 | Not-determined |
| 17-108_k141_269326 | Low-quality |
| 17-108_k141_311345 | Not-determined |
| 17-108_k141_200122 | Low-quality |
| 17-108_k141_159307 | Not-determined |
| 17-108_k141_159317 | Not-determined |
| 17-108_k141_159318 | Not-determined |
| 17-108_k141_117222 | Not-determined |
| 17-108_k141_311348 | Low-quality |
| 17-108_k141_311351 | Not-determined |
| 17-108_k141_296698 | Low-quality |
| 17-108_k141_283593 | Not-determined |
| 17-108_k141_283597 | Not-determined |
| 17-108_k141_104534 | Not-determined |
| 17-108_k141_35410 | Not-determined |
| 17-108_k141_173430 | Not-determined |
| 17-108_k141_242734 | Low-quality |
| 17-108_k141_186395 | Low-quality |
| 17-108_k141_214797 | Low-quality |
| 17-108_k141_90377 | Not-determined |
| 17-108_k141_63084 | Not-determined |
| 17-108_k141_186414 | Low-quality |
| 17-108_k141_63089 | Not-determined |
| 17-108_k141_159349 | Low-quality |
| 17-108_k141_269386 | Not-determined |
| 17-108_k141_269387 | Not-determined |
| 17-108_k141_63107 | Not-determined |
| 17-108_k141_159363 | Not-determined |
| 17-108_k141_104550 | Not-determined |
| 17-108_k141_104556 | Not-determined |
| 17-108_k141_256044 | Not-determined |
| 17-108_k141_269396 | Not-determined |
| 17-108_k141_75993 | Not-determined |
| 17-108_k141_296739 | Low-quality |
| 17-108_k141_214826 | Not-determined |
| 17-108_k141_145463 | Not-determined |
| 17-108_k141_269403 | Not-determined |
| 17-108_k141_296743 | Not-determined |
| 17-108_k141_145466 | Not-determined |
| 17-108_k141_186442 | Not-determined |
| 17-108_k141_145471 | Not-determined |
| 17-108_k141_200136 | Not-determined |
| 17-108_k141_296753 | Not-determined |
| 17-108_k141_145474 | Not-determined |
| 17-108_k141_145477 | Not-determined |
| 17-108_k141_269415 | Not-determined |
| 17-108_k141_228373 | Not-determined |
| 17-108_k141_35449 | Not-determined |
| 17-108_k141_323775 | Not-determined |
| 17-108_k141_311390 | Not-determined |
| 17-108_k141_104592 | Not-determined |
| 17-108_k141_256052 | Not-determined |
| 17-108_k141_269432 | Not-determined |
| 17-108_k141_104594 | Not-determined |
| 17-108_k141_242824 | Not-determined |
| 17-108_k141_311403 | Not-determined |
| 17-108_k141_104600 | Not-determined |
| 17-108_k141_186465 | Not-determined |
| 17-108_k141_200149 | Not-determined |
| 17-108_k141_104605 | Not-determined |
| 17-108_k141_214839 | Not-determined |
| 17-108_k141_296762 | Not-determined |
| 17-108_k141_104609 | Not-determined |
| 17-108_k141_90428 | Not-determined |
| 17-108_k141_35466 | Not-determined |
| 17-108_k141_242829 | Not-determined |
| 17-108_k141_186473 | Not-determined |
| 17-108_k141_159377 | Not-determined |
| 17-108_k141_104618 | Not-determined |
| 17-108_k141_159380 | Not-determined |
| 17-108_k141_214851 | Not-determined |
| 17-108_k141_214852 | Not-determined |
| 17-108_k141_90438 | Not-determined |
| 17-108_k141_256064 | Low-quality |
| 17-108_k141_242835 | Not-determined |
| 17-108_k141_35485 | Low-quality |
| 17-108_k141_104629 | Not-determined |
| 17-108_k141_90441 | Not-determined |
| 17-108_k141_256067 | Not-determined |
| 17-108_k141_7662 | Not-determined |
| 17-108_k141_214863 | Not-determined |
| 17-108_k141_256070 | Not-determined |
| 17-108_k141_311436 | Not-determined |
| 17-108_k141_200176 | Low-quality |
| 17-108_k141_159395 | Not-determined |
| 17-108_k141_228404 | Not-determined |
| 17-108_k141_228406 | Not-determined |
| 17-108_k141_214875 | Not-determined |
| 17-108_k141_145494 | Not-determined |
| 17-108_k141_256082 | Not-determined |
| 17-108_k141_90451 | Not-determined |
| 17-108_k141_173510 | Not-determined |
| 17-108_k141_214880 | Not-determined |
| 17-108_k141_269450 | Not-determined |
| 17-108_k141_63164 | Low-quality |
| 17-108_k141_296778 | Not-determined |
| 17-108_k141_269454 | Not-determined |
| 17-108_k141_159404 | Not-determined |
| 17-108_k141_20691 | Not-determined |
| 17-108_k141_269458 | Not-determined |
| 17-108_k141_269460 | Not-determined |
| 17-108_k141_145510 | Not-determined |
| 17-108_k141_63178 | Not-determined |
| 17-108_k141_63187 | Not-determined |
| 17-108_k141_117286 | Low-quality |
| 17-108_k141_7702 | Not-determined |
| 17-108_k141_296800 | Not-determined |
| 17-108_k141_296803 | Not-determined |
| 17-108_k141_186499 | Not-determined |
| 17-108_k141_7713 | Not-determined |
| 17-108_k141_117297 | Not-determined |
| 17-108_k141_49014 | Not-determined |
| 17-108_k141_7718 | Not-determined |
| 17-108_k141_214893 | Not-determined |
| 17-108_k141_283636 | Not-determined |
| 17-108_k141_296817 | Not-determined |
| 17-108_k141_283640 | Not-determined |
| 17-108_k141_200224 | Not-determined |
| 17-108_k141_90460 | Not-determined |
| 17-108_k141_159435 | Low-quality |
| 17-108_k141_90463 | Not-determined |
| 17-108_k141_200234 | Low-quality |
| 17-108_k141_200237 | Not-determined |
| 17-108_k141_269471 | Not-determined |
| 17-108_k141_214908 | Not-determined |
| 17-108_k141_20701 | Not-determined |
| 17-108_k141_200242 | Low-quality |
| 17-108_k141_7733 | Not-determined |
| 17-108_k141_117321 | Not-determined |
| 17-108_k141_104687 | Not-determined |
| 17-108_k141_200252 | Not-determined |
| 17-108_k141_90471 | Not-determined |
| 17-108_k141_76047 | Not-determined |
| 17-108_k141_228422 | Not-determined |
| 17-108_k141_214927 | Not-determined |
| 17-108_k141_200254 | Not-determined |
| 17-108_k141_90481 | Not-determined |
| 17-108_k141_283661 | Not-determined |
| 17-108_k141_7758 | Not-determined |
| 17-108_k141_186529 | Not-determined |
| 17-108_k141_173540 | Not-determined |
| 17-108_k141_283667 | Not-determined |
| 17-108_k141_7764 | Not-determined |
| 17-108_k141_242852 | Not-determined |
| 17-108_k141_173546 | Not-determined |
| 17-108_k141_76054 | Not-determined |
| 17-108_k141_76058 | Not-determined |
| 17-108_k141_159452 | Low-quality |
| 17-108_k141_173550 | Low-quality |
| 17-108_k141_117333 | Not-determined |
| 17-108_k141_7772 | Not-determined |
| 17-108_k141_159459 | Not-determined |
| 17-108_k141_7774 | Not-determined |
| 17-108_k141_145563 | Not-determined |
| 17-108_k141_256167 | Low-quality |
| 17-108_k141_35513 | Not-determined |
| 17-108_k141_269507 | Not-determined |
| 17-108_k141_269510 | Not-determined |
| 17-108_k141_117343 | Not-determined |
| 17-108_k141_283673 | Low-quality |
| 17-108_k141_35522 | Not-determined |
| 17-108_k141_173563 | Not-determined |
| 17-108_k141_269516 | Not-determined |
| 17-108_k141_173572 | Not-determined |
| 17-108_k141_7787 | Low-quality |
| 17-108_k141_323826 | Not-determined |
| 17-108_k141_90498 | Not-determined |
| 17-108_k141_63244 | Not-determined |
| 17-108_k141_145582 | Not-determined |
| 17-108_k141_283691 | Not-determined |
| 17-108_k141_269520 | Not-determined |
| 17-108_k141_186549 | Low-quality |
| 17-108_k141_145594 | Not-determined |
| 17-108_k141_269523 | Not-determined |
| 17-108_k141_63258 | Not-determined |
| 17-108_k141_269527 | Not-determined |
| 17-108_k141_117361 | Not-determined |
| 17-108_k141_228474 | Not-determined |
| 17-108_k141_35536 | Not-determined |
| 17-108_k141_117369 | Not-determined |
| 17-108_k141_90515 | Not-determined |
| 17-108_k141_104756 | Not-determined |
| 17-108_k141_159505 | Not-determined |
| 17-108_k141_145608 | Not-determined |
| 17-108_k141_311477 | Not-determined |
| 17-108_k141_214980 | Not-determined |
| 17-108_k141_256207 | Not-determined |
| 17-108_k141_117382 | Not-determined |
| 17-108_k141_145618 | Not-determined |
| 17-108_k141_242882 | Not-determined |
| 17-108_k141_35557 | Not-determined |
| 17-108_k141_159514 | Low-quality |
| 17-108_k141_159516 | Not-determined |
| 17-108_k141_35560 | Not-determined |
| 17-108_k141_186565 | Not-determined |
| 17-108_k141_186568 | Not-determined |
| 17-108_k141_49089 | Not-determined |
| 17-108_k141_323852 | Not-determined |
| 17-108_k141_173591 | Not-determined |
| 17-108_k141_283729 | Not-determined |
| 17-108_k141_159526 | Not-determined |
| 17-108_k141_200285 | Low-quality |
| 17-108_k141_228483 | Not-determined |
| 17-108_k141_323855 | Not-determined |
| 17-108_k141_200291 | Low-quality |
| 17-108_k141_117399 | Not-determined |
| 17-108_k141_117403 | Not-determined |
| 17-108_k141_242903 | Low-quality |
| 17-108_k141_186581 | Not-determined |
| 17-108_k141_186582 | Not-determined |
| 17-108_k141_7858 | Not-determined |
| 17-108_k141_49106 | Not-determined |
| 17-108_k141_7859 | Not-determined |
| 17-108_k141_186586 | Not-determined |
| 17-108_k141_323871 | Not-determined |
| 17-108_k141_145654 | Not-determined |
| 17-108_k141_35582 | Not-determined |
| 17-108_k141_76082 | Low-quality |
| 17-108_k141_283742 | Not-determined |
| 17-108_k141_323875 | Not-determined |
| 17-108_k141_104789 | Not-determined |
| 17-108_k141_63271 | Not-determined |
| 17-108_k141_200309 | Low-quality |
| 17-108_k141_159554 | Not-determined |
| 17-108_k141_186593 | Not-determined |
| 17-108_k141_269558 | Not-determined |
| 17-108_k141_131308 | Not-determined |
| 17-108_k141_159558 | Not-determined |
| 17-108_k141_311492 | Not-determined |
| 17-108_k141_269563 | Not-determined |
| 17-108_k141_159563 | Not-determined |
| 17-108_k141_186606 | Low-quality |
| 17-108_k141_159564 | Not-determined |
| 17-108_k141_7883 | Not-determined |
| 17-108_k141_214999 | Not-determined |
| 17-108_k141_269576 | Not-determined |
| 17-108_k141_215007 | Not-determined |
| 17-108_k141_7895 | Low-quality |
| 17-108_k141_104814 | Not-determined |
| 17-108_k141_63294 | Not-determined |
| 17-108_k141_7897 | Not-determined |
| 17-108_k141_90548 | Not-determined |
| 17-108_k141_90550 | Not-determined |
| 17-108_k141_200336 | Not-determined |
| 17-108_k141_35610 | Not-determined |
| 17-108_k141_49142 | Not-determined |
| 17-108_k141_7909 | Not-determined |
| 17-108_k141_145671 | Low-quality |
| 17-108_k141_311518 | Not-determined |
| 17-108_k141_242945 | Not-determined |
| 17-108_k141_228513 | Not-determined |
| 17-108_k141_35617 | Not-determined |
| 17-108_k141_242949 | Not-determined |
| 17-108_k141_228522 | Not-determined |
| 17-108_k141_215024 | Not-determined |
| 17-108_k141_228524 | Not-determined |
| 17-108_k141_269614 | Not-determined |
| 17-108_k141_323895 | Not-determined |
| 17-108_k141_104841 | Not-determined |
| 17-108_k141_104842 | Not-determined |
| 17-108_k141_228529 | Not-determined |
| 17-108_k141_296900 | Not-determined |
| 17-108_k141_117411 | Not-determined |
| 17-108_k141_296903 | Not-determined |
| 17-108_k141_323902 | Not-determined |
| 17-108_k141_269621 | Low-quality |
| 17-108_k141_323905 | Not-determined |
| 17-108_k141_131353 | Not-determined |
| 17-108_k141_311538 | Not-determined |
| 17-108_k141_311539 | Not-determined |
| 17-108_k141_7947 | Not-determined |
| 17-108_k141_296908 | Not-determined |
| 17-108_k141_186621 | Low-quality |
| 17-108_k141_90567 | Not-determined |
| 17-108_k141_117431 | Not-determined |
| 17-108_k141_269628 | Not-determined |
| 17-108_k141_296921 | Low-quality |
| 17-108_k141_323918 | Not-determined |
| 17-108_k141_228561 | Not-determined |
| 17-108_k141_296926 | Not-determined |
| 17-108_k141_76117 | Not-determined |
| 17-108_k141_311555 | Not-determined |
| 17-108_k141_7950 | Not-determined |
| 17-108_k141_7952 | Not-determined |
| 17-108_k141_228567 | Not-determined |
| 17-108_k141_76124 | Not-determined |
| 17-108_k141_283784 | Not-determined |
| 17-108_k141_256225 | Not-determined |
| 17-108_k141_76129 | Not-determined |
| 17-108_k141_200400 | Not-determined |
| 17-108_k141_76131 | Not-determined |
| 17-108_k141_90585 | Not-determined |
| 17-108_k141_311575 | Not-determined |
| 17-108_k141_117455 | Not-determined |
| 17-108_k141_269637 | Not-determined |
| 17-108_k141_269638 | Not-determined |
| 17-108_k141_296946 | Not-determined |
| 17-108_k141_35647 | Low-quality |
| 17-108_k141_296948 | Low-quality |
| 17-108_k141_200413 | Not-determined |
| 17-108_k141_228598 | Not-determined |
| 17-108_k141_76143 | Not-determined |
| 17-108_k141_7980 | Not-determined |
| 17-108_k141_63333 | Not-determined |
| 17-108_k141_35656 | Not-determined |
| 17-108_k141_200422 | Low-quality |
| 17-108_k141_7982 | Not-determined |
| 17-108_k141_63336 | Not-determined |
| 17-108_k141_296966 | Low-quality |
| 17-108_k141_200443 | Not-determined |
| 17-108_k141_296967 | Not-determined |
| 17-108_k141_20780 | Not-determined |
| 17-108_k141_173655 | Low-quality |
| 17-108_k141_269671 | Not-determined |
| 17-108_k141_311601 | Not-determined |
| 17-108_k141_269674 | Not-determined |
| 17-108_k141_8000 | Not-determined |
| 17-108_k141_200452 | Not-determined |
| 17-108_k141_35677 | Low-quality |
| 17-108_k141_117515 | Not-determined |
| 17-108_k141_8016 | Not-determined |
| 17-108_k141_8017 | Not-determined |
| 17-108_k141_256261 | Not-determined |
| 17-108_k141_49195 | Not-determined |
| 17-108_k141_228641 | Not-determined |
| 17-108_k141_228642 | Not-determined |
| 17-108_k141_90622 | Not-determined |
| 17-108_k141_145711 | Not-determined |
| 17-108_k141_256273 | Not-determined |
| 17-108_k141_323959 | Not-determined |
| 17-108_k141_35689 | Not-determined |
| 17-108_k141_63368 | Not-determined |
| 17-108_k141_145724 | Not-determined |
| 17-108_k141_228649 | Not-determined |
| 17-108_k141_131410 | Not-determined |
| 17-108_k141_145729 | Not-determined |
| 17-108_k141_242987 | Low-quality |
| 17-108_k141_49202 | Not-determined |
| 17-108_k141_242988 | Not-determined |
| 17-108_k141_311622 | Not-determined |
| 17-108_k141_35695 | Not-determined |
| 17-108_k141_323971 | Low-quality |
| 17-108_k141_283788 | Not-determined |
| 17-108_k141_131415 | Not-determined |
| 17-108_k141_311627 | Not-determined |
| 17-108_k141_200483 | Not-determined |
| 17-108_k141_242998 | Not-determined |
| 17-108_k141_49211 | Not-determined |
| 17-108_k141_200485 | Not-determined |
| 17-108_k141_117550 | Not-determined |
| 17-108_k141_35698 | Low-quality |
| 17-108_k141_243008 | Not-determined |
| 17-108_k141_243010 | Not-determined |
| 17-108_k141_228667 | Not-determined |
| 17-108_k141_90645 | Low-quality |
| 17-108_k141_35702 | Low-quality |
| 17-108_k141_159663 | Not-determined |
| 17-108_k141_104897 | Not-determined |
| 17-108_k141_186690 | Not-determined |
| 17-108_k141_131422 | Not-determined |
| 17-108_k141_243021 | Not-determined |
| 17-108_k141_104901 | Low-quality |
| 17-108_k141_90659 | Not-determined |
| 17-108_k141_20845 | Not-determined |
| 17-108_k141_76200 | Not-determined |
| 17-108_k141_35716 | Not-determined |
| 17-108_k141_131437 | Not-determined |
| 17-108_k141_8044 | Not-determined |
| 17-108_k141_228674 | Not-determined |
| 17-108_k141_228675 | Not-determined |
| 17-108_k141_35723 | Not-determined |
| 17-108_k141_323985 | Not-determined |
| 17-108_k141_228682 | Not-determined |
| 17-108_k141_283819 | Not-determined |
| 17-108_k141_323986 | Not-determined |
| 17-108_k141_323987 | Not-determined |
| 17-108_k141_117596 | Not-determined |
| 17-108_k141_117602 | Not-determined |
| 17-108_k141_228691 | Not-determined |
| 17-108_k141_228692 | Not-determined |
| 17-108_k141_131466 | Not-determined |
| 17-108_k141_243036 | Not-determined |
| 17-108_k141_131468 | Not-determined |
| 17-108_k141_117607 | Not-determined |
| 17-108_k141_200509 | Not-determined |
| 17-108_k141_117612 | Not-determined |
| 17-108_k141_90695 | Not-determined |
| 17-108_k141_131474 | Not-determined |
| 17-108_k141_20878 | Not-determined |
| 17-108_k141_269742 | Not-determined |
| 17-108_k141_8082 | Not-determined |
| 17-108_k141_90701 | Not-determined |
| 17-108_k141_63390 | Not-determined |
| 17-108_k141_256307 | Not-determined |
| 17-108_k141_20883 | Low-quality |
| 17-108_k141_269752 | Not-determined |
| 17-108_k141_269756 | Not-determined |
| 17-108_k141_256315 | Not-determined |
| 17-108_k141_49255 | Not-determined |
| 17-108_k141_311657 | Not-determined |
| 17-108_k141_228714 | Not-determined |
| 17-108_k141_269759 | Low-quality |
| 17-108_k141_63409 | Not-determined |
| 17-108_k141_131501 | Not-determined |
| 17-108_k141_283852 | Not-determined |
| 17-108_k141_283853 | Not-determined |
| 17-108_k141_324003 | Not-determined |
| 17-108_k141_311662 | Not-determined |
| 17-108_k141_145787 | Not-determined |
| 17-108_k141_63415 | Not-determined |
| 17-108_k141_8103 | Not-determined |
| 17-108_k141_324007 | Not-determined |
| 17-108_k141_63419 | Not-determined |
| 17-108_k141_200533 | Not-determined |

| | |
|---|---|
| 17-108_k141_145795 | Not-determined |
| 17-108_k141_243061 | Not-determined |
| 17-108_k141_117634 | Low-quality |
| 17-108_k141_76264 | Not-determined |
| 17-108_k141_63431 | Not-determined |
| 17-108_k141_283874 | Not-determined |
| 17-108_k141_76276 | Not-determined |
| 17-108_k141_8133 | Not-determined |
| 17-108_k141_186747 | Not-determined |
| 17-108_k141_104981 | Not-determined |
| 17-108_k141_90742 | Not-determined |
| 17-108_k141_186758 | Not-determined |
| 17-108_k141_90746 | Not-determined |
| 17-108_k141_215117 | Not-determined |
| 17-108_k141_311701 | Not-determined |
| 17-108_k141_186763 | Low-quality |
| 17-108_k141_243084 | Not-determined |
| 17-108_k141_173719 | Low-quality |
| 17-108_k141_20897 | Not-determined |
| 17-108_k141_269800 | Not-determined |
| 17-108_k141_269803 | Not-determined |
| 17-108_k141_311704 | Not-determined |
| 17-108_k141_283899 | Not-determined |
| 17-108_k141_76291 | Low-quality |
| 17-108_k141_283903 | Not-determined |
| 17-108_k141_20907 | Not-determined |
| 17-108_k141_173730 | Not-determined |
| 17-108_k141_283909 | Not-determined |
| 17-108_k141_228737 | Not-determined |
| 17-108_k141_173740 | Not-determined |
| 17-108_k141_35754 | Not-determined |
| 17-108_k141_243092 | Not-determined |
| 17-108_k141_228745 | Not-determined |
| 17-108_k141_283917 | Low-quality |
| 17-108_k141_173746 | Not-determined |
| 17-108_k141_243096 | Not-determined |
| 17-108_k141_35762 | Not-determined |
| 17-108_k141_311717 | Not-determined |
| 17-108_k141_283922 | Not-determined |
| 17-108_k141_186787 | Not-determined |
| 17-108_k141_173751 | Not-determined |
| 17-108_k141_200583 | Not-determined |
| 17-108_k141_283925 | Not-determined |
| 17-108_k141_186789 | Low-quality |
| 17-108_k141_35768 | Not-determined |
| 17-108_k141_159695 | Not-determined |
| 17-108_k141_131546 | Not-determined |
| 17-108_k141_186791 | Not-determined |
| 17-108_k141_20929 | Not-determined |
| 17-108_k141_131548 | Not-determined |
| 17-108_k141_243104 | Not-determined |
| 17-108_k141_186795 | Not-determined |
| 17-108_k141_35774 | Not-determined |
| 17-108_k141_311737 | Not-determined |
| 17-108_k141_283939 | Not-determined |
| 17-108_k141_117668 | Not-determined |
| 17-108_k141_173759 | Low-quality |
| 17-108_k141_35786 | Low-quality |
| 17-108_k141_283947 | Not-determined |
| 17-108_k141_186812 | Low-quality |
| 17-108_k141_159708 | Low-quality |
| 17-108_k141_131558 | Not-determined |
| 17-108_k141_105009 | Not-determined |
| 17-108_k141_76333 | Not-determined |
| 17-108_k141_76337 | Not-determined |
| 17-108_k141_311752 | Not-determined |
| 17-108_k141_311756 | Not-determined |
| 17-108_k141_20969 | Not-determined |
| 17-108_k141_269860 | Not-determined |
| 17-108_k141_117680 | Not-determined |
| 17-108_k141_8187 | Not-determined |
| 17-108_k141_76349 | Not-determined |
| 17-108_k141_76350 | Not-determined |
| 17-108_k141_49279 | Not-determined |
| 17-108_k141_324037 | Not-determined |
| 17-108_k141_131566 | Not-determined |
| 17-108_k141_186817 | Not-determined |
| 17-108_k141_311781 | Not-determined |
| 17-108_k141_311789 | Not-determined |
| 17-108_k141_49297 | Low-quality |
| 17-108_k141_117707 | Not-determined |
| 17-108_k141_117713 | Not-determined |
| 17-108_k141_131574 | Not-determined |
| 17-108_k141_145873 | Not-determined |
| 17-108_k141_159730 | Not-determined |
| 17-108_k141_269885 | Not-determined |
| 17-108_k141_256429 | Not-determined |
| 17-108_k141_186844 | Not-determined |
| 17-108_k141_269888 | Not-determined |
| 17-108_k141_35840 | Not-determined |
| 17-108_k141_49322 | Not-determined |
| 17-108_k141_186855 | Not-determined |
| 17-108_k141_311801 | Not-determined |
| 17-108_k141_76396 | Not-determined |
| 17-108_k141_76402 | Not-determined |
| 17-108_k141_311803 | Low-quality |
| 17-108_k141_284004 | Not-determined |
| 17-108_k141_8256 | Not-determined |
| 17-108_k141_159744 | Low-quality |
| 17-108_k141_90806 | Not-determined |
| 17-108_k141_284007 | Not-determined |
| 17-108_k141_63459 | Low-quality |
| 17-108_k141_49340 | Not-determined |
| 17-108_k141_76406 | Not-determined |
| 17-108_k141_297103 | Not-determined |
| 17-108_k141_131602 | Not-determined |
| 17-108_k141_131604 | Not-determined |
| 17-108_k141_49350 | Low-quality |
| 17-108_k141_311814 | Low-quality |
| 17-108_k141_8270 | Not-determined |
| 17-108_k141_105068 | Not-determined |
| 17-108_k141_105072 | Low-quality |
| 17-108_k141_324060 | Not-determined |
| 17-108_k141_311833 | Not-determined |
| 17-108_k141_284014 | Not-determined |
| 17-108_k141_269905 | Not-determined |
| 17-108_k141_76412 | Not-determined |
| 17-108_k141_200610 | Not-determined |
| 17-108_k141_145911 | Not-determined |
| 17-108_k141_35893 | Low-quality |
| 17-108_k141_186883 | Not-determined |
| 17-108_k141_324071 | Not-determined |
| 17-108_k141_324072 | Not-determined |
| 17-108_k141_145921 | Not-determined |
| 17-108_k141_284034 | Not-determined |
| 17-108_k141_117735 | Not-determined |
| 17-108_k141_186891 | Not-determined |
| 17-108_k141_186895 | Not-determined |
| 17-108_k141_215183 | Not-determined |
| 17-108_k141_228791 | Not-determined |
| 17-108_k141_311867 | Not-determined |
| 17-108_k141_8290 | Not-determined |
| 17-108_k141_269922 | Not-determined |
| 17-108_k141_8294 | Not-determined |
| 17-108_k141_90839 | Not-determined |
| 17-108_k141_243157 | Not-determined |
| 17-108_k141_284057 | Not-determined |
| 17-108_k141_243158 | Not-determined |
| 17-108_k141_131632 | Not-determined |
| 17-108_k141_311875 | Not-determined |
| 17-108_k141_35910 | Not-determined |
| 17-108_k141_215201 | Not-determined |
| 17-108_k141_49395 | Not-determined |
| 17-108_k141_173837 | Not-determined |
| 17-108_k141_145953 | Not-determined |
| 17-108_k141_215205 | Not-determined |
| 17-108_k141_284061 | Not-determined |
| 17-108_k141_215211 | Not-determined |
| 17-108_k141_200636 | Not-determined |
| 17-108_k141_324112 | Not-determined |
| 17-108_k141_35914 | Not-determined |
| 17-108_k141_145964 | Not-determined |
| 17-108_k141_105098 | Not-determined |
| 17-108_k141_35917 | Low-quality |
| 17-108_k141_21000 | Not-determined |
| 17-108_k141_200652 | Not-determined |
| 17-108_k141_324115 | Low-quality |
| 17-108_k141_76466 | Not-determined |
| 17-108_k141_49413 | Not-determined |
| 17-108_k141_35925 | Not-determined |
| 17-108_k141_35926 | Not-determined |
| 17-108_k141_297136 | Not-determined |
| 17-108_k141_105099 | Not-determined |
| 17-108_k141_8311 | Not-determined |
| 17-108_k141_76469 | Not-determined |
| 17-108_k141_243185 | Not-determined |
| 17-108_k141_324126 | Not-determined |
| 17-108_k141_173863 | Not-determined |
| 17-108_k141_243190 | Not-determined |
| 17-108_k141_173871 | Not-determined |
| 17-108_k141_173873 | Not-determined |
| 17-108_k141_297145 | Not-determined |
| 17-108_k141_297150 | Not-determined |
| 17-108_k141_35935 | Not-determined |
| 17-108_k141_131660 | Not-determined |
| 17-108_k141_63508 | Low-quality |
| 17-108_k141_63511 | Not-determined |
| 17-108_k141_8324 | Not-determined |
| 17-108_k141_8325 | Not-determined |
| 17-108_k141_269963 | Not-determined |
| 17-108_k141_243213 | Not-determined |
| 17-108_k141_145993 | Not-determined |
| 17-108_k141_63516 | Not-determined |
| 17-108_k141_131668 | Not-determined |
| 17-108_k141_215232 | Not-determined |
| 17-108_k141_131670 | Not-determined |
| 17-108_k141_297158 | Not-determined |
| 17-108_k141_145996 | Not-determined |
| 17-108_k141_284095 | Not-determined |
| 17-108_k141_284100 | Not-determined |
| 17-108_k141_200688 | Not-determined |
| 17-108_k141_243219 | Not-determined |
| 17-108_k141_8352 | Not-determined |
| 17-108_k141_90888 | Not-determined |
| 17-108_k141_243223 | Not-determined |
| 17-108_k141_105114 | Not-determined |
| 17-108_k141_105115 | Not-determined |
| 17-108_k141_105125 | Not-determined |
| 17-108_k141_146027 | Not-determined |
| 17-108_k141_200693 | Low-quality |
| 17-108_k141_269988 | Not-determined |
| 17-108_k141_49439 | Not-determined |
| 17-108_k141_269989 | Low-quality |
| 17-108_k141_200696 | Not-determined |
| 17-108_k141_256514 | Not-determined |
| 17-108_k141_63542 | Not-determined |
| 17-108_k141_63543 | Not-determined |
| 17-108_k141_256517 | Low-quality |
| 17-108_k141_117784 | Low-quality |
| 17-108_k141_200709 | Not-determined |
| 17-108_k141_90909 | Not-determined |
| 17-108_k141_200710 | Not-determined |
| 17-108_k141_117794 | Not-determined |
| 17-108_k141_173885 | Not-determined |
| 17-108_k141_200718 | Not-determined |
| 17-108_k141_284126 | Not-determined |
| 17-108_k141_105148 | Not-determined |
| 17-108_k141_243252 | Not-determined |
| 17-108_k141_173894 | Not-determined |
| 17-108_k141_63568 | Not-determined |
| 17-108_k141_270021 | Not-determined |
| 17-108_k141_8388 | Not-determined |
| 17-108_k141_284136 | Not-determined |
| 17-108_k141_105164 | Not-determined |
| 17-108_k141_243261 | Not-determined |
| 17-108_k141_21073 | Low-quality |
| 17-108_k141_243262 | Not-determined |
| 17-108_k141_63575 | Not-determined |
| 17-108_k141_243270 | Not-determined |
| 17-108_k141_284151 | Not-determined |
| 17-108_k141_297231 | Not-determined |
| 17-108_k141_215274 | Not-determined |
| 17-108_k141_215278 | Not-determined |
| 17-108_k141_284156 | Not-determined |
| 17-108_k141_8406 | Not-determined |
| 17-108_k141_284162 | Not-determined |
| 17-108_k141_284164 | Not-determined |
| 17-108_k141_63597 | Not-determined |
| 17-108_k141_200751 | Not-determined |
| 17-108_k141_90940 | Not-determined |
| 17-108_k141_200754 | Not-determined |
| 17-108_k141_228824 | Not-determined |
| 17-108_k141_297240 | Not-determined |
| 17-108_k141_200763 | Not-determined |
| 17-108_k141_200764 | Not-determined |
| 17-108_k141_256555 | Not-determined |
| 17-108_k141_90949 | Not-determined |
| 17-108_k141_159884 | Not-determined |
| 17-108_k141_76543 | Not-determined |
| 17-108_k141_8444 | Not-determined |
| 17-108_k141_117839 | Not-determined |
| 17-108_k141_76549 | Low-quality |
| 17-108_k141_117841 | Not-determined |
| 17-108_k141_186980 | Not-determined |
| 17-108_k141_105205 | Not-determined |
| 17-108_k141_297249 | Not-determined |
| 17-108_k141_49469 | Not-determined |
| 17-108_k141_173945 | Not-determined |
| 17-108_k141_186987 | Not-determined |
| 17-108_k141_284180 | Not-determined |
| 17-108_k141_35981 | Not-determined |
| 17-108_k141_173949 | Not-determined |
| 17-108_k141_284184 | Not-determined |
| 17-108_k141_90961 | Not-determined |
| 17-108_k141_105216 | Not-determined |
| 17-108_k141_49476 | Not-determined |
| 17-108_k141_90964 | Not-determined |
| 17-108_k141_284187 | Not-determined |
| 17-108_k141_76561 | Not-determined |
| 17-108_k141_284193 | Not-determined |
| 17-108_k141_117849 | Not-determined |
| 17-108_k141_35987 | Not-determined |
| 17-108_k141_63624 | Not-determined |
| 17-108_k141_49483 | Not-determined |
| 17-108_k141_63626 | Not-determined |
| 17-108_k141_284202 | Not-determined |
| 17-108_k141_105228 | Not-determined |
| 17-108_k141_117852 | Not-determined |
| 17-108_k141_105231 | Not-determined |
| 17-108_k141_8467 | Not-determined |
| 17-108_k141_105235 | Not-determined |
| 17-108_k141_187007 | Not-determined |
| 17-108_k141_105237 | Not-determined |
| 17-108_k141_146134 | Not-determined |
| 17-108_k141_105247 | Not-determined |
| 17-108_k141_105248 | Not-determined |
| 17-108_k141_187031 | Not-determined |
| 17-108_k141_215315 | Not-determined |
| 17-108_k141_63662 | Not-determined |
| 17-108_k141_8488 | Not-determined |
| 17-108_k141_117873 | Low-quality |
| 17-108_k141_159934 | Not-determined |
| 17-108_k141_215339 | Not-determined |
| 17-108_k141_8494 | Not-determined |
| 17-108_k141_256624 | Low-quality |
| 17-108_k141_324214 | Not-determined |
| 17-108_k141_159950 | Not-determined |
| 17-108_k141_284229 | Not-determined |
| 17-108_k141_228892 | Not-determined |
| 17-108_k141_200801 | Not-determined |
| 17-108_k141_105258 | Not-determined |
| 17-108_k141_63673 | Not-determined |
| 17-108_k141_90997 | Low-quality |
| 17-108_k141_324221 | Not-determined |
| 17-108_k141_90999 | Low-quality |
| 17-108_k141_76599 | Not-determined |
| 17-108_k141_105263 | Not-determined |
| 17-108_k141_173969 | Not-determined |
| 17-108_k141_91006 | Not-determined |
| 17-108_k141_76612 | Not-determined |
| 17-108_k141_21127 | Not-determined |
| 17-108_k141_215367 | Not-determined |
| 17-108_k141_21129 | Not-determined |
| 17-108_k141_76619 | Not-determined |
| 17-108_k141_200826 | Not-determined |
| 17-108_k141_159962 | Not-determined |
| 17-108_k141_21135 | Not-determined |
| 17-108_k141_91018 | Not-determined |
| 17-108_k141_187071 | Not-determined |
| 17-108_k141_36028 | Not-determined |
| 17-108_k141_187075 | Not-determined |
| 17-108_k141_187080 | Not-determined |
| 17-108_k141_36030 | Low-quality |
| 17-108_k141_91028 | Not-determined |
| 17-108_k141_36033 | Not-determined |
| 17-108_k141_270068 | Low-quality |
| 17-108_k141_36036 | Not-determined |
| 17-108_k141_187095 | Not-determined |
| 17-108_k141_243348 | Not-determined |
| 17-108_k141_117898 | Not-determined |
| 17-108_k141_284264 | Not-determined |
| 17-108_k141_21159 | Not-determined |
| 17-108_k141_187102 | Not-determined |
| 17-108_k141_324257 | Not-determined |
| 17-108_k141_187111 | Not-determined |
| 17-108_k141_200851 | Not-determined |
| 17-108_k141_215402 | Not-determined |
| 17-108_k141_297373 | Not-determined |
| 17-108_k141_21177 | Not-determined |
| 17-108_k141_63695 | Not-determined |
| 17-108_k141_49533 | Not-determined |
| 17-108_k141_228970 | Not-determined |
| 17-108_k141_174009 | Low-quality |
| 17-108_k141_117931 | Not-determined |
| 17-108_k141_270085 | Not-determined |
| 17-108_k141_8565 | Not-determined |
| 17-108_k141_256689 | Not-determined |
| 17-108_k141_8569 | Not-determined |
| 17-108_k141_284303 | Not-determined |
| 17-108_k141_131785 | Not-determined |
| 17-108_k141_36067 | Not-determined |
| 17-108_k141_146151 | Low-quality |
| 17-108_k141_117946 | Not-determined |
| 17-108_k141_21215 | Not-determined |
| 17-108_k141_105325 | Not-determined |
| 17-108_k141_76667 | Not-determined |
| 17-108_k141_284305 | Not-determined |
| 17-108_k141_8581 | Not-determined |
| 17-108_k141_243352 | Not-determined |
| 17-108_k141_228983 | Not-determined |
| 17-108_k141_117957 | Not-determined |
| 17-108_k141_36076 | Not-determined |
| 17-108_k141_63730 | Not-determined |
| 17-108_k141_8585 | Low-quality |
| 17-108_k141_76682 | Not-determined |
| 17-108_k141_160020 | Not-determined |
| 17-108_k141_76685 | Not-determined |
| 17-108_k141_36082 | Low-quality |
| 17-108_k141_8589 | Not-determined |
| 17-108_k141_297399 | Not-determined |
| 17-108_k141_174035 | Not-determined |
| 17-108_k141_284319 | Not-determined |
| 17-108_k141_131811 | Not-determined |
| 17-108_k141_284324 | Not-determined |
| 17-108_k141_63750 | Not-determined |
| 17-108_k141_49578 | Not-determined |
| 17-108_k141_297402 | Not-determined |
| 17-108_k141_256711 | Not-determined |
| 17-108_k141_76698 | Not-determined |
| 17-108_k141_256714 | Not-determined |
| 17-108_k141_8604 | Not-determined |
| 17-108_k141_8605 | Not-determined |
| 17-108_k141_243376 | Not-determined |
| 17-108_k141_228997 | Low-quality |
| 17-108_k141_131817 | Not-determined |
| 17-108_k141_187160 | Not-determined |
| 17-108_k141_243387 | Not-determined |
| 17-108_k141_76705 | Low-quality |
| 17-108_k141_297407 | Not-determined |
| 17-108_k141_256724 | Not-determined |
| 17-108_k141_187163 | Low-quality |
| 17-108_k141_243409 | Not-determined |
| 17-108_k141_131829 | Not-determined |
| 17-108_k141_160037 | Low-quality |
| 17-108_k141_200876 | Not-determined |
| 17-108_k141_131835 | Not-determined |
| 17-108_k141_49619 | Not-determined |
| 17-108_k141_36128 | Not-determined |
| 17-108_k141_21254 | Not-determined |
| 17-108_k141_324297 | Not-determined |
| 17-108_k141_146175 | Not-determined |
| 17-108_k141_21258 | Not-determined |
| 17-108_k141_21261 | Not-determined |
| 17-108_k141_131836 | Low-quality |
| 17-108_k141_131842 | Not-determined |
| 17-108_k141_284360 | Not-determined |
| 17-108_k141_297429 | Not-determined |
| 17-108_k141_146200 | Not-determined |
| 17-108_k141_131843 | Low-quality |
| 17-108_k141_187174 | Not-determined |
| 17-108_k141_324321 | Not-determined |
| 17-108_k141_324322 | Not-determined |
| 17-108_k141_146206 | Not-determined |
| 17-108_k141_311999 | Not-determined |
| 17-108_k141_229045 | Not-determined |
| 17-108_k141_215452 | Not-determined |
| 17-108_k141_160074 | Not-determined |
| 17-108_k141_215460 | Not-determined |
| 17-108_k141_297437 | Not-determined |
| 17-108_k141_174087 | Not-determined |
| 17-108_k141_200919 | Not-determined |
| 17-108_k141_312009 | Not-determined |
| 17-108_k141_284380 | Low-quality |
| 17-108_k141_312012 | Not-determined |
| 17-108_k141_215463 | Not-determined |
| 17-108_k141_215464 | Not-determined |
| 17-108_k141_200930 | Not-determined |
| 17-108_k141_324339 | Not-determined |
| 17-108_k141_312028 | Not-determined |
| 17-108_k141_200935 | Not-determined |
| 17-108_k141_324354 | Not-determined |
| 17-108_k141_215476 | Not-determined |
| 17-108_k141_243420 | Low-quality |
| 17-108_k141_215479 | Not-determined |
| 17-108_k141_243421 | Not-determined |
| 17-108_k141_187204 | Not-determined |
| 17-108_k141_63767 | Not-determined |
| 17-108_k141_200946 | Not-determined |
| 17-108_k141_118026 | Not-determined |
| 17-108_k141_284394 | Not-determined |
| 17-108_k141_160118 | Not-determined |
| 17-108_k141_8701 | Not-determined |
| 17-108_k141_118037 | Not-determined |
| 17-108_k141_131860 | Not-determined |
| 17-108_k141_200951 | Not-determined |
| 17-108_k141_76741 | Not-determined |
| 17-108_k141_174121 | Not-determined |
| 17-108_k141_215490 | Not-determined |
| 17-108_k141_256802 | Not-determined |
| 17-108_k141_160127 | Not-determined |
| 17-108_k141_215495 | Not-determined |
| 17-108_k141_312048 | Not-determined |
| 17-108_k141_131867 | Not-determined |
| 17-108_k141_174134 | Not-determined |
| 17-108_k141_49679 | Not-determined |
| 17-108_k141_131872 | Not-determined |
| 17-108_k141_256815 | Not-determined |
| 17-108_k141_49685 | Not-determined |
| 17-108_k141_256820 | Not-determined |
| 17-108_k141_160137 | Not-determined |
| 17-108_k141_284419 | Not-determined |
| 17-108_k141_270122 | Not-determined |
| 17-108_k141_200969 | Not-determined |
| 17-108_k141_243465 | Not-determined |
| 17-108_k141_76769 | Not-determined |
| 17-108_k141_270124 | Low-quality |
| 17-108_k141_174151 | Low-quality |
| 17-108_k141_160149 | Not-determined |
| 17-108_k141_76783 | Not-determined |
| 17-108_k141_118083 | Low-quality |
| 17-108_k141_229094 | Not-determined |
| 17-108_k141_36230 | Not-determined |
| 17-108_k141_174159 | Not-determined |
| 17-108_k141_36238 | Not-determined |
| 17-108_k141_36241 | Not-determined |
| 17-108_k141_200999 | Not-determined |
| 17-108_k141_8758 | Not-determined |
| 17-108_k141_36244 | Not-determined |
| 17-108_k141_201004 | Not-determined |
| 17-108_k141_49732 | Not-determined |
| 17-108_k141_21342 | Not-determined |
| 17-108_k141_297466 | Not-determined |
| 17-108_k141_76788 | Not-determined |
| 17-108_k141_187225 | Not-determined |
| 17-108_k141_270128 | Not-determined |
| 17-108_k141_229102 | Not-determined |
| 17-108_k141_187231 | Not-determined |
| 17-108_k141_229105 | Not-determined |
| 17-108_k141_91128 | Low-quality |
| 17-108_k141_284450 | Not-determined |
| 17-108_k141_256878 | Not-determined |
| 17-108_k141_215520 | Not-determined |
| 17-108_k141_131903 | Not-determined |
| 17-108_k141_91132 | Not-determined |
| 17-108_k141_324367 | Not-determined |
| 17-108_k141_160188 | Not-determined |
| 17-108_k141_229113 | Not-determined |
| 17-108_k141_91136 | Not-determined |
| 17-108_k141_215523 | Not-determined |
| 17-108_k141_187239 | Not-determined |
| 17-108_k141_160193 | Not-determined |
| 17-108_k141_229120 | Not-determined |
| 17-108_k141_174188 | Not-determined |
| 17-108_k141_256885 | Not-determined |
| 17-108_k141_76802 | Not-determined |
| 17-108_k141_256886 | Not-determined |
| 17-108_k141_229129 | Not-determined |
| 17-108_k141_146305 | Not-determined |
| 17-108_k141_36272 | Not-determined |
| 17-108_k141_201032 | Not-determined |
| 17-108_k141_297482 | Not-determined |
| 17-108_k141_174204 | Not-determined |
| 17-108_k141_118100 | Not-determined |
| 17-108_k141_146313 | Not-determined |
| 17-108_k141_256891 | Not-determined |
| 17-108_k141_229138 | Not-determined |
| 17-108_k141_36284 | Not-determined |
| 17-108_k141_229143 | Not-determined |
| 17-108_k141_49757 | Not-determined |
| 17-108_k141_284469 | Not-determined |
| 17-108_k141_174214 | Not-determined |
| 17-108_k141_118114 | Not-determined |
| 17-108_k141_105473 | Not-determined |
| 17-108_k141_49773 | Not-determined |
| 17-108_k141_36299 | Not-determined |
| 17-108_k141_324396 | Not-determined |
| 17-108_k141_105484 | Not-determined |
| 17-108_k141_118129 | Not-determined |
| 17-108_k141_201050 | Not-determined |
| 17-108_k141_215549 | Low-quality |
| 17-108_k141_324401 | Not-determined |
| 17-108_k141_76838 | Not-determined |
| 17-108_k141_229178 | Not-determined |
| 17-108_k141_105493 | Not-determined |
| 17-108_k141_215555 | Not-determined |
| 17-108_k141_229190 | Low-quality |
| 17-108_k141_160232 | Not-determined |
| 17-108_k141_76846 | Not-determined |
| 17-108_k141_160236 | Not-determined |
| 17-108_k141_105511 | Not-determined |
| 17-108_k141_21373 | Not-determined |
| 17-108_k141_49803 | Not-determined |
| 17-108_k141_160242 | Not-determined |
| 17-108_k141_105516 | Not-determined |
| 17-108_k141_105518 | Not-determined |
| 17-108_k141_256932 | Not-determined |
| 17-108_k141_160246 | Not-determined |
| 17-108_k141_49809 | Not-determined |
| 17-108_k141_312120 | Not-determined |
| 17-108_k141_187275 | Not-determined |
| 17-108_k141_243500 | Not-determined |
| 17-108_k141_243501 | Not-determined |
| 17-108_k141_256940 | Not-determined |
| 17-108_k141_312125 | Not-determined |
| 17-108_k141_160249 | Not-determined |
| 17-108_k141_21384 | Not-determined |
| 17-108_k141_174231 | Not-determined |
| 17-108_k141_21389 | Not-determined |
| 17-108_k141_118147 | Not-determined |
| 17-108_k141_76885 | Not-determined |
| 17-108_k141_284520 | Not-determined |
| 17-108_k141_160254 | Not-determined |
| 17-108_k141_49837 | Not-determined |
| 17-108_k141_49838 | Not-determined |
| 17-108_k141_160260 | Not-determined |
| 17-108_k141_49841 | Not-determined |
| 17-108_k141_215577 | Not-determined |
| 17-108_k141_229217 | Not-determined |
| 17-108_k141_215579 | Not-determined |
| 17-108_k141_8819 | Not-determined |
| 17-108_k141_256960 | Not-determined |
| 17-108_k141_229220 | Not-determined |
| 17-108_k141_21403 | Not-determined |
| 17-108_k141_297490 | Not-determined |
| 17-108_k141_256967 | Not-determined |
| 17-108_k141_21408 | Not-determined |
| 17-108_k141_146378 | Not-determined |
| 17-108_k141_91215 | Not-determined |
| 17-108_k141_312137 | Not-determined |
| 17-108_k141_105544 | Not-determined |
| 17-108_k141_297500 | Low-quality |
| 17-108_k141_8835 | Not-determined |
| 17-108_k141_229238 | Low-quality |
| 17-108_k141_324468 | Low-quality |
| 17-108_k141_21415 | Not-determined |
| 17-108_k141_105550 | Not-determined |
| 17-108_k141_201064 | Low-quality |
| 17-108_k141_312144 | Not-determined |
| 17-108_k141_174273 | Not-determined |

| | |
|---|---|
| 17-108_k141_146403 | Not-determined |
| 17-108_k141_8845 | Not-determined |
| 17-108_k141_243529 | Low-quality |
| 17-108_k141_160266 | Low-quality |
| 17-108_k141_160270 | Not-determined |
| 17-108_k141_324474 | Not-determined |
| 17-108_k141_160271 | Not-determined |
| 17-108_k141_49875 | Not-determined |
| 17-108_k141_243535 | Not-determined |
| 17-108_k141_8849 | Not-determined |
| 17-108_k141_297511 | Low-quality |
| 17-108_k141_270167 | Low-quality |
| 17-108_k141_8855 | Not-determined |
| 17-108_k141_297514 | Not-determined |
| 17-108_k141_270168 | Not-determined |
| 17-108_k141_215617 | Not-determined |
| 17-108_k141_215619 | Not-determined |
| 17-108_k141_174285 | Low-quality |
| 17-108_k141_270172 | Not-determined |
| 17-108_k141_187293 | Not-determined |
| 17-108_k141_215628 | Not-determined |
| 17-108_k141_91242 | Not-determined |
| 17-108_k141_146430 | Low-quality |
| 17-108_k141_256987 | Not-determined |
| 17-108_k141_146431 | Not-determined |
| 17-108_k141_36373 | Not-determined |
| 17-108_k141_63826 | Not-determined |
| 17-108_k141_201101 | Not-determined |
| 17-108_k141_243555 | Low-quality |
| 17-108_k141_243556 | Not-determined |
| 17-108_k141_63829 | Low-quality |
| 17-108_k141_76912 | Low-quality |
| 17-108_k141_256994 | Not-determined |
| 17-108_k141_187305 | Not-determined |
| 17-108_k141_201105 | Not-determined |
| 17-108_k141_215634 | Not-determined |
| 17-108_k141_105574 | Not-determined |
| 17-108_k141_270192 | Not-determined |
| 17-108_k141_49904 | Not-determined |
| 17-108_k141_76920 | Not-determined |
| 17-108_k141_36391 | Not-determined |
| 17-108_k141_160304 | Not-determined |
| 17-108_k141_63849 | Not-determined |
| 17-108_k141_63850 | Not-determined |
| 17-108_k141_243573 | Not-determined |
| 17-108_k141_36394 | Not-determined |
| 17-108_k141_63851 | Not-determined |
| 17-108_k141_49910 | Not-determined |
| 17-108_k141_297523 | Not-determined |
| 17-108_k141_36402 | Not-determined |
| 17-108_k141_118188 | Not-determined |
| 17-108_k141_8895 | Not-determined |
| 17-108_k141_229278 | Not-determined |
| 17-108_k141_243588 | Not-determined |
| 17-108_k141_118189 | Not-determined |
| 17-108_k141_243590 | Not-determined |
| 17-108_k141_118191 | Not-determined |
| 17-108_k141_174295 | Not-determined |
| 17-108_k141_270197 | Not-determined |
| 17-108_k141_63858 | Not-determined |
| 17-108_k141_8905 | Not-determined |
| 17-108_k141_270199 | Not-determined |
| 17-108_k141_160324 | Not-determined |
| 17-108_k141_243598 | Not-determined |
| 17-108_k141_270203 | Not-determined |
| 17-108_k141_105601 | Low-quality |
| 17-108_k141_201119 | Not-determined |
| 17-108_k141_131921 | Not-determined |
| 17-108_k141_91268 | Not-determined |
| 17-108_k141_49927 | Not-determined |
| 17-108_k141_229291 | Not-determined |
| 17-108_k141_270218 | Not-determined |
| 17-108_k141_174306 | Low-quality |
| 17-108_k141_201123 | Not-determined |
| 17-108_k141_215661 | Not-determined |
| 17-108_k141_174314 | Not-determined |
| 17-108_k141_229305 | Not-determined |
| 17-108_k141_243624 | Low-quality |
| 17-108_k141_243625 | Not-determined |
| 17-108_k141_160332 | Not-determined |
| 17-108_k141_105628 | Not-determined |
| 17-108_k141_243628 | Not-determined |
| 17-108_k141_312172 | Not-determined |
| 17-108_k141_215677 | Not-determined |
| 17-108_k141_105637 | Not-determined |
| 17-108_k141_160333 | Not-determined |
| 17-108_k141_243637 | Not-determined |
| 17-108_k141_174332 | Not-determined |
| 17-108_k141_8951 | Not-determined |
| 17-108_k141_63887 | Not-determined |
| 17-108_k141_118235 | Low-quality |
| 17-108_k141_63889 | Not-determined |
| 17-108_k141_243641 | Not-determined |
| 17-108_k141_146480 | Not-determined |
| 17-108_k141_105651 | Not-determined |
| 17-108_k141_215694 | Not-determined |
| 17-108_k141_215695 | Not-determined |
| 17-108_k141_243649 | Not-determined |
| 17-108_k141_105653 | Not-determined |
| 17-108_k141_8961 | Low-quality |
| 17-108_k141_131930 | Not-determined |
| 17-108_k141_146494 | Low-quality |
| 17-108_k141_146497 | Not-determined |
| 17-108_k141_229327 | Not-determined |
| 17-108_k141_160345 | Low-quality |
| 17-108_k141_201162 | Not-determined |
| 17-108_k141_63909 | Not-determined |
| 17-108_k141_187347 | Not-determined |
| 17-108_k141_201165 | Not-determined |
| 17-108_k141_105668 | Not-determined |
| 17-108_k141_76943 | Not-determined |
| 17-108_k141_201173 | Not-determined |
| 17-108_k141_243677 | Not-determined |
| 17-108_k141_257047 | Not-determined |
| 17-108_k141_297555 | Not-determined |
| 17-108_k141_118261 | Not-determined |
| 17-108_k141_243684 | Not-determined |
| 17-108_k141_243687 | Not-determined |
| 17-108_k141_257051 | Not-determined |
| 17-108_k141_312219 | Not-determined |
| 17-108_k141_21483 | Not-determined |
| 17-108_k141_257056 | Not-determined |
| 17-108_k141_131938 | Not-determined |
| 17-108_k141_49972 | Not-determined |
| 17-108_k141_243696 | Low-quality |
| 17-108_k141_105694 | Not-determined |
| 17-108_k141_187363 | Not-determined |
| 17-108_k141_257064 | Not-determined |
| 17-108_k141_49984 | Not-determined |
| 17-108_k141_312234 | Low-quality |
| 17-108_k141_284533 | Not-determined |
| 17-108_k141_76960 | Not-determined |
| 17-108_k141_76965 | Not-determined |
| 17-108_k141_270239 | Not-determined |
| 17-108_k141_174350 | Not-determined |
| 17-108_k141_297590 | Not-determined |
| 17-108_k141_187376 | Not-determined |
| 17-108_k141_270247 | Not-determined |
| 17-108_k141_49994 | Not-determined |
| 17-108_k141_297599 | Not-determined |
| 17-108_k141_187381 | Not-determined |
| 17-108_k141_297601 | Not-determined |
| 17-108_k141_63931 | Not-determined |
| 17-108_k141_284548 | Not-determined |
| 17-108_k141_257069 | Not-determined |
| 17-108_k141_324523 | Not-determined |
| 17-108_k141_174371 | Not-determined |
| 17-108_k141_63942 | Not-determined |
| 17-108_k141_9023 | Not-determined |
| 17-108_k141_9026 | Not-determined |
| 17-108_k141_76986 | Not-determined |
| 17-108_k141_284563 | Not-determined |
| 17-108_k141_312264 | Not-determined |
| 17-108_k141_229369 | Not-determined |
| 17-108_k141_201237 | Low-quality |
| 17-108_k141_243729 | Not-determined |
| 17-108_k141_243730 | Not-determined |
| 17-108_k141_160371 | Not-determined |
| 17-108_k141_160373 | Not-determined |
| 17-108_k141_160377 | Not-determined |
| 17-108_k141_76994 | Not-determined |
| 17-108_k141_297630 | Not-determined |
| 17-108_k141_324527 | Not-determined |
| 17-108_k141_324531 | Not-determined |
| 17-108_k141_324542 | Not-determined |
| 17-108_k141_312280 | Not-determined |
| 17-108_k141_77009 | Not-determined |
| 17-108_k141_229385 | Not-determined |
| 17-108_k141_201252 | Not-determined |
| 17-108_k141_324547 | Not-determined |
| 17-108_k141_312286 | Not-determined |
| 17-108_k141_77014 | Low-quality |
| 17-108_k141_270297 | Not-determined |
| 17-108_k141_270299 | Not-determined |
| 17-108_k141_312292 | Not-determined |
| 17-108_k141_105721 | Not-determined |
| 17-108_k141_36430 | Not-determined |
| 17-108_k141_312295 | Not-determined |
| 17-108_k141_324568 | Not-determined |
| 17-108_k141_257081 | Not-determined |
| 17-108_k141_201265 | Not-determined |
| 17-108_k141_215724 | Not-determined |
| 17-108_k141_9061 | Low-quality |
| 17-108_k141_270322 | Not-determined |
| 17-108_k141_50060 | Not-determined |
| 17-108_k141_187442 | Not-determined |
| 17-108_k141_215729 | Not-determined |
| 17-108_k141_118307 | Not-determined |
| 17-108_k141_201273 | Not-determined |
| 17-108_k141_201279 | Not-determined |
| 17-108_k141_160413 | Not-determined |
| 17-108_k141_9080 | Not-determined |
| 17-108_k141_91347 | Not-determined |
| 17-108_k141_9081 | Low-quality |
| 17-108_k141_64018 | Not-determined |
| 17-108_k141_64019 | Not-determined |
| 17-108_k141_312324 | Not-determined |
| 17-108_k141_9092 | Low-quality |
| 17-108_k141_64024 | Not-determined |
| 17-108_k141_215740 | Not-determined |
| 17-108_k141_105750 | Not-determined |
| 17-108_k141_91366 | Not-determined |
| 17-108_k141_312337 | Not-determined |
| 17-108_k141_146512 | Low-quality |
| 17-108_k141_312343 | Not-determined |
| 17-108_k141_284626 | Not-determined |
| 17-108_k141_146518 | Not-determined |
| 17-108_k141_105758 | Not-determined |
| 17-108_k141_174416 | Not-determined |
| 17-108_k141_36439 | Not-determined |
| 17-108_k141_105766 | Not-determined |
| 17-108_k141_324624 | Not-determined |
| 17-108_k141_297682 | Low-quality |
| 17-108_k141_229411 | Not-determined |
| 17-108_k141_174428 | Not-determined |
| 17-108_k141_257094 | Not-determined |
| 17-108_k141_146529 | Not-determined |
| 17-108_k141_91373 | Low-quality |
| 17-108_k141_36455 | Not-determined |
| 17-108_k141_174437 | Not-determined |
| 17-108_k141_91375 | Not-determined |
| 17-108_k141_64063 | Not-determined |
| 17-108_k141_91376 | Not-determined |
| 17-108_k141_187462 | Not-determined |
| 17-108_k141_9115 | Not-determined |
| 17-108_k141_270364 | Not-determined |
| 17-108_k141_21535 | Not-determined |
| 17-108_k141_21536 | Not-determined |
| 17-108_k141_270368 | Not-determined |
| 17-108_k141_64074 | Not-determined |
| 17-108_k141_312363 | Not-determined |
| 17-108_k141_257100 | Not-determined |
| 17-108_k141_215776 | Not-determined |
| 17-108_k141_21538 | Low-quality |
| 17-108_k141_146538 | Low-quality |
| 17-108_k141_297707 | Not-determined |
| 17-108_k141_118346 | Low-quality |
| 17-108_k141_91393 | Not-determined |
| 17-108_k141_50126 | Not-determined |
| 17-108_k141_50129 | Not-determined |
| 17-108_k141_229438 | Not-determined |
| 17-108_k141_187483 | Not-determined |
| 17-108_k141_270382 | Not-determined |
| 17-108_k141_257116 | Not-determined |
| 17-108_k141_284660 | Not-determined |
| 17-108_k141_21545 | Not-determined |
| 17-108_k141_187490 | Not-determined |
| 17-108_k141_243772 | Not-determined |
| 17-108_k141_215786 | Not-determined |
| 17-108_k141_201344 | Not-determined |
| 17-108_k141_118362 | Not-determined |
| 17-108_k141_229449 | Low-quality |
| 17-108_k141_118371 | Not-determined |
| 17-108_k141_201352 | Not-determined |
| 17-108_k141_77079 | Not-determined |
| 17-108_k141_50149 | Not-determined |
| 17-108_k141_160488 | Not-determined |
| 17-108_k141_9165 | Not-determined |
| 17-108_k141_146553 | Not-determined |
| 17-108_k141_9171 | Not-determined |
| 17-108_k141_118384 | Not-determined |
| 17-108_k141_297738 | Not-determined |
| 17-108_k141_118386 | Not-determined |
| 17-108_k141_257139 | Not-determined |
| 17-108_k141_36475 | Not-determined |
| 17-108_k141_21572 | Not-determined |
| 17-108_k141_21577 | Not-determined |
| 17-108_k141_146568 | Not-determined |
| 17-108_k141_160504 | Not-determined |
| 17-108_k141_160506 | Not-determined |
| 17-108_k141_229460 | Not-determined |
| 17-108_k141_215822 | Not-determined |
| 17-108_k141_229462 | Not-determined |
| 17-108_k141_174486 | Low-quality |
| 17-108_k141_201382 | Not-determined |
| 17-108_k141_50196 | Not-determined |
| 17-108_k141_312420 | Not-determined |
| 17-108_k141_229470 | Low-quality |
| 17-108_k141_174491 | Not-determined |
| 17-108_k141_243811 | Not-determined |
| 17-108_k141_174498 | Not-determined |
| 17-108_k141_243824 | Not-determined |
| 17-108_k141_160523 | Not-determined |
| 17-108_k141_284699 | Not-determined |
| 17-108_k141_257171 | Not-determined |
| 17-108_k141_174503 | Not-determined |
| 17-108_k141_284707 | Not-determined |
| 17-108_k141_21613 | Not-determined |
| 17-108_k141_105809 | Not-determined |
| 17-108_k141_9205 | Not-determined |
| 17-108_k141_160548 | Not-determined |
| 17-108_k141_21616 | Not-determined |
| 17-108_k141_50216 | Not-determined |
| 17-108_k141_201401 | Not-determined |
| 17-108_k141_284713 | Not-determined |
| 17-108_k141_215862 | Not-determined |
| 17-108_k141_174516 | Not-determined |
| 17-108_k141_324658 | Low-quality |
| 17-108_k141_229494 | Low-quality |
| 17-108_k141_270395 | Low-quality |
| 17-108_k141_9214 | Not-determined |
| 17-108_k141_270396 | Not-determined |
| 17-108_k141_174521 | Not-determined |
| 17-108_k141_50229 | Not-determined |
| 17-108_k141_21622 | Not-determined |
| 17-108_k141_297779 | Not-determined |
| 17-108_k141_50239 | Not-determined |
| 17-108_k141_257218 | Not-determined |
| 17-108_k141_105830 | Not-determined |
| 17-108_k141_187555 | Not-determined |
| 17-108_k141_77108 | Not-determined |
| 17-108_k141_297793 | Not-determined |
| 17-108_k141_187557 | Not-determined |
| 17-108_k141_297794 | Not-determined |
| 17-108_k141_91486 | Not-determined |
| 17-108_k141_215877 | Not-determined |
| 17-108_k141_215878 | Not-determined |
| 17-108_k141_297796 | Low-quality |
| 17-108_k141_201411 | Not-determined |
| 17-108_k141_187561 | Not-determined |
| 17-108_k141_229518 | Not-determined |
| 17-108_k141_187563 | Not-determined |
| 17-108_k141_50249 | Not-determined |
| 17-108_k141_187570 | Not-determined |
| 17-108_k141_50262 | Not-determined |
| 17-108_k141_201429 | Not-determined |
| 17-108_k141_9258 | Low-quality |
| 17-108_k141_297808 | Not-determined |
| 17-108_k141_91510 | Not-determined |
| 17-108_k141_215905 | Low-quality |
| 17-108_k141_270427 | Not-determined |
| 17-108_k141_77127 | Not-determined |
| 17-108_k141_91515 | Not-determined |
| 17-108_k141_312462 | Not-determined |
| 17-108_k141_257262 | Not-determined |
| 17-108_k141_160595 | Not-determined |
| 17-108_k141_270443 | Not-determined |
| 17-108_k141_9270 | Not-determined |
| 17-108_k141_187588 | Not-determined |
| 17-108_k141_9271 | Low-quality |
| 17-108_k141_118435 | Not-determined |
| 17-108_k141_50284 | Not-determined |
| 17-108_k141_146600 | Not-determined |
| 17-108_k141_50294 | Not-determined |
| 17-108_k141_160614 | Not-determined |
| 17-108_k141_201462 | Not-determined |
| 17-108_k141_174557 | Not-determined |
| 17-108_k141_201464 | Not-determined |
| 17-108_k141_21686 | Not-determined |
| 17-108_k141_118451 | Not-determined |
| 17-108_k141_105894 | Not-determined |
| 17-108_k141_36535 | Not-determined |
| 17-108_k141_297837 | Not-determined |
| 17-108_k141_118453 | Not-determined |
| 17-108_k141_105896 | Low-quality |
| 17-108_k141_36536 | Not-determined |
| 17-108_k141_118456 | Not-determined |
| 17-108_k141_201469 | Not-determined |
| 17-108_k141_146607 | Not-determined |
| 17-108_k141_312473 | Not-determined |
| 17-108_k141_36541 | Not-determined |
| 17-108_k141_284762 | Not-determined |
| 17-108_k141_64162 | Low-quality |
| 17-108_k141_270475 | Not-determined |
| 17-108_k141_324753 | Not-determined |
| 17-108_k141_118463 | Not-determined |
| 17-108_k141_118469 | Not-determined |
| 17-108_k141_50309 | Not-determined |
| 17-108_k141_146616 | Not-determined |
| 17-108_k141_297845 | Not-determined |
| 17-108_k141_324760 | Not-determined |
| 17-108_k141_146620 | Low-quality |
| 17-108_k141_50317 | Not-determined |
| 17-108_k141_312486 | Not-determined |
| 17-108_k141_64173 | Not-determined |
| 17-108_k141_324771 | Not-determined |
| 17-108_k141_324774 | Low-quality |
| 17-108_k141_50325 | Not-determined |
| 17-108_k141_324776 | Not-determined |
| 17-108_k141_64176 | Not-determined |
| 17-108_k141_50327 | Not-determined |
| 17-108_k141_91541 | Low-quality |
| 17-108_k141_229574 | Not-determined |
| 17-108_k141_91546 | Not-determined |
| 17-108_k141_91547 | Not-determined |
| 17-108_k141_270494 | Not-determined |
| 17-108_k141_201493 | Not-determined |
| 17-108_k141_118487 | Not-determined |
| 17-108_k141_50346 | Not-determined |
| 17-108_k141_187624 | Not-determined |
| 17-108_k141_118492 | Not-determined |
| 17-108_k141_105931 | Not-determined |
| 17-108_k141_50349 | Not-determined |
| 17-108_k141_229584 | Low-quality |
| 17-108_k141_270507 | Not-determined |
| 17-108_k141_9308 | Not-determined |
| 17-108_k141_324807 | Not-determined |
| 17-108_k141_174584 | Low-quality |
| 17-108_k141_91571 | Not-determined |
| 17-108_k141_77160 | Not-determined |
| 17-108_k141_64194 | Not-determined |
| 17-108_k141_284809 | Not-determined |
| 17-108_k141_324811 | Not-determined |
| 17-108_k141_174589 | Low-quality |
| 17-108_k141_284813 | Not-determined |
| 17-108_k141_243883 | Not-determined |
| 17-108_k141_243887 | Not-determined |
| 17-108_k141_243888 | Not-determined |
| 17-108_k141_243889 | Low-quality |
| 17-108_k141_257326 | Low-quality |
| 17-108_k141_324823 | Not-determined |
| 17-108_k141_284823 | Not-determined |
| 17-108_k141_118521 | Low-quality |
| 17-108_k141_105942 | Not-determined |
| 17-108_k141_64213 | Not-determined |
| 17-108_k141_324825 | Not-determined |
| 17-108_k141_270527 | Low-quality |
| 17-108_k141_118528 | Not-determined |
| 17-108_k141_270530 | Low-quality |
| 17-108_k141_9328 | Not-determined |
| 17-108_k141_146662 | Not-determined |
| 17-108_k141_284839 | Not-determined |
| 17-108_k141_9332 | Not-determined |
| 17-108_k141_284840 | Not-determined |
| 17-108_k141_9336 | Not-determined |
| 17-108_k141_64227 | Not-determined |
| 17-108_k141_297887 | Not-determined |
| 17-108_k141_229626 | Low-quality |
| 17-108_k141_118553 | Not-determined |
| 17-108_k141_201509 | Not-determined |
| 17-108_k141_131999 | Not-determined |
| 17-108_k141_91611 | Not-determined |
| 17-108_k141_50375 | Not-determined |
| 17-108_k141_118562 | Not-determined |
| 17-108_k141_146685 | Not-determined |
| 17-108_k141_21703 | Low-quality |
| 17-108_k141_270537 | Not-determined |
| 17-108_k141_118568 | Low-quality |
| 17-108_k141_132012 | Not-determined |
| 17-108_k141_229649 | Not-determined |
| 17-108_k141_229651 | Not-determined |
| 17-108_k141_146698 | Not-determined |
| 17-108_k141_201527 | Not-determined |
| 17-108_k141_118583 | Not-determined |
| 17-108_k141_201531 | Not-determined |
| 17-108_k141_243928 | Not-determined |
| 17-108_k141_229654 | Not-determined |
| 17-108_k141_36550 | Not-determined |
| 17-108_k141_9399 | Not-determined |
| 17-108_k141_284869 | Not-determined |
| 17-108_k141_91639 | Not-determined |
| 17-108_k141_174605 | Not-determined |
| 17-108_k141_187686 | Not-determined |
| 17-108_k141_187687 | Not-determined |
| 17-108_k141_174607 | Not-determined |
| 17-108_k141_146715 | Not-determined |
| 17-108_k141_106011 | Not-determined |
| 17-108_k141_243932 | Low-quality |
| 17-108_k141_243933 | Not-determined |
| 17-108_k141_91653 | Not-determined |
| 17-108_k141_215909 | Not-determined |
| 17-108_k141_64247 | Not-determined |
| 17-108_k141_146734 | Not-determined |
| 17-108_k141_270543 | Not-determined |
| 17-108_k141_324863 | Low-quality |
| 17-108_k141_106031 | Not-determined |
| 17-108_k141_64250 | Not-determined |
| 17-108_k141_146749 | Not-determined |
| 17-108_k141_118613 | Not-determined |
| 17-108_k141_270561 | Not-determined |
| 17-108_k141_146758 | Not-determined |
| 17-108_k141_257387 | Not-determined |
| 17-108_k141_118626 | Not-determined |
| 17-108_k141_160723 | Not-determined |
| 17-108_k141_324885 | Not-determined |
| 17-108_k141_77228 | Not-determined |
| 17-108_k141_77229 | Not-determined |
| 17-108_k141_270583 | Not-determined |
| 17-108_k141_324888 | Not-determined |
| 17-108_k141_9405 | Not-determined |
| 17-108_k141_132057 | Not-determined |
| 17-108_k141_257410 | Not-determined |
| 17-108_k141_257412 | Not-determined |
| 17-108_k141_257422 | Not-determined |
| 17-108_k141_9417 | Not-determined |
| 17-108_k141_146782 | Not-determined |
| 17-108_k141_91672 | Low-quality |
| 17-108_k141_132073 | Not-determined |
| 17-108_k141_146785 | Not-determined |
| 17-108_k141_106095 | Not-determined |
| 17-108_k141_215931 | Not-determined |
| 17-108_k141_146787 | Not-determined |
| 17-108_k141_146789 | Not-determined |
| 17-108_k141_106102 | Low-quality |
| 17-108_k141_146791 | Not-determined |
| 17-108_k141_91687 | Not-determined |
| 17-108_k141_215947 | Not-determined |
| 17-108_k141_257458 | Not-determined |
| 17-108_k141_187759 | Not-determined |
| 17-108_k141_187760 | Not-determined |
| 17-108_k141_284955 | Not-determined |
| 17-108_k141_201606 | Low-quality |
| 17-108_k141_174631 | Not-determined |
| 17-108_k141_132103 | Not-determined |
| 17-108_k141_50421 | Not-determined |
| 17-108_k141_21708 | Not-determined |
| 17-108_k141_174640 | Not-determined |
| 17-108_k141_132111 | Not-determined |
| 17-108_k141_201613 | Not-determined |
| 17-108_k141_201615 | Not-determined |
| 17-108_k141_201617 | Not-determined |
| 17-108_k141_284969 | Not-determined |
| 17-108_k141_215956 | Not-determined |
| 17-108_k141_284971 | Not-determined |
| 17-108_k141_201621 | Not-determined |
| 17-108_k141_146819 | Not-determined |
| 17-108_k141_297913 | Low-quality |
| 17-108_k141_36637 | Not-determined |
| 17-108_k141_174655 | Not-determined |
| 17-108_k141_36641 | Not-determined |
| 17-108_k141_146828 | Not-determined |
| 17-108_k141_229749 | Not-determined |
| 17-108_k141_77303 | Low-quality |
| 17-108_k141_160793 | Not-determined |
| 17-108_k141_50428 | Not-determined |
| 17-108_k141_174658 | Not-determined |
| 17-108_k141_132123 | Low-quality |
| 17-108_k141_77310 | Not-determined |
| 17-108_k141_36655 | Not-determined |
| 17-108_k141_297921 | Not-determined |
| 17-108_k141_77316 | Not-determined |
| 17-108_k141_215962 | Not-determined |
| 17-108_k141_243973 | Not-determined |
| 17-108_k141_324915 | Not-determined |
| 17-108_k141_36668 | Not-determined |
| 17-108_k141_229768 | Not-determined |
| 17-108_k141_187815 | Low-quality |
| 17-108_k141_132136 | Not-determined |
| 17-108_k141_9467 | Not-determined |
| 17-108_k141_64289 | Not-determined |
| 17-108_k141_106127 | Not-determined |
| 17-108_k141_187820 | Low-quality |
| 17-108_k141_187823 | Low-quality |
| 17-108_k141_257474 | Low-quality |
| 17-108_k141_77337 | Not-determined |
| 17-108_k141_297935 | Not-determined |
| 17-108_k141_243985 | Not-determined |
| 17-108_k141_297940 | Low-quality |
| 17-108_k141_257478 | Not-determined |
| 17-108_k141_201669 | Not-determined |
| 17-108_k141_243992 | Not-determined |
| 17-108_k141_324939 | Not-determined |
| 17-108_k141_9479 | Not-determined |
| 17-108_k141_118665 | Not-determined |
| 17-108_k141_118666 | Not-determined |
| 17-108_k141_229782 | Not-determined |
| 17-108_k141_257486 | Low-quality |
| 17-108_k141_91747 | Not-determined |
| 17-108_k141_187834 | Not-determined |
| 17-108_k141_50459 | Not-determined |
| 17-108_k141_285019 | Not-determined |
| 17-108_k141_50460 | Not-determined |
| 17-108_k141_229790 | Not-determined |
| 17-108_k141_9493 | Not-determined |
| 17-108_k141_257500 | Low-quality |
| 17-108_k141_9494 | Not-determined |
| 17-108_k141_50466 | Not-determined |
| 17-108_k141_9500 | Not-determined |
| 17-108_k141_257502 | Not-determined |
| 17-108_k141_229797 | Not-determined |
| 17-108_k141_324964 | Not-determined |
| 17-108_k141_201703 | Low-quality |
| 17-108_k141_132160 | Not-determined |
| 17-108_k141_36709 | Not-determined |
| 17-108_k141_50478 | Not-determined |
| 17-108_k141_106186 | Low-quality |
| 17-108_k141_50483 | Not-determined |
| 17-108_k141_36722 | Low-quality |
| 17-108_k141_187859 | Not-determined |
| 17-108_k141_257508 | Not-determined |
| 17-108_k141_146857 | Low-quality |
| 17-108_k141_201716 | Not-determined |
| 17-108_k141_146861 | Not-determined |
| 17-108_k141_257514 | Not-determined |
| 17-108_k141_257515 | Not-determined |
| 17-108_k141_257519 | Not-determined |
| 17-108_k141_9529 | Not-determined |
| 17-108_k141_297985 | Not-determined |
| 17-108_k141_324978 | Not-determined |

| | |
|---|---|
| 17-108_k141_324982 | Not-determined |
| 17-108_k141_77353 | Not-determined |
| 17-108_k141_174722 | Low-quality |
| 17-108_k141_21736 | Not-determined |
| 17-108_k141_324988 | Not-determined |
| 17-108_k141_21737 | Not-determined |
| 17-108_k141_36737 | Not-determined |
| 17-108_k141_160805 | Not-determined |
| 17-108_k141_36742 | Not-determined |
| 17-108_k141_229817 | Not-determined |
| 17-108_k141_160817 | Not-determined |
| 17-108_k141_187890 | Not-determined |
| 17-108_k141_244050 | Not-determined |
| 17-108_k141_132191 | Not-determined |
| 17-108_k141_270681 | Not-determined |
| 17-108_k141_146897 | Not-determined |
| 17-108_k141_160824 | Not-determined |
| 17-108_k141_106223 | Not-determined |
| 17-108_k141_21764 | Not-determined |
| 17-108_k141_174743 | Not-determined |
| 17-108_k141_174744 | Not-determined |
| 17-108_k141_160838 | Not-determined |
| 17-108_k141_312656 | Not-determined |
| 17-108_k141_9539 | Not-determined |
| 17-108_k141_132198 | Not-determined |
| 17-108_k141_297996 | Not-determined |
| 17-108_k141_64367 | Not-determined |
| 17-108_k141_106233 | Not-determined |
| 17-108_k141_91797 | Not-determined |
| 17-108_k141_160851 | Not-determined |
| 17-108_k141_160854 | Not-determined |
| 17-108_k141_160855 | Not-determined |
| 17-108_k141_244075 | Not-determined |
| 17-108_k141_146916 | Low-quality |
| 17-108_k141_325025 | Low-quality |
| 17-108_k141_50526 | Not-determined |
| 17-108_k141_244078 | Not-determined |
| 17-108_k141_215983 | Not-determined |
| 17-108_k141_9563 | Not-determined |
| 17-108_k141_9565 | Not-determined |
| 17-108_k141_201778 | Low-quality |
| 17-108_k141_132212 | Not-determined |
| 17-108_k141_201782 | Low-quality |
| 17-108_k141_201783 | Not-determined |
| 17-108_k141_229859 | Not-determined |
| 17-108_k141_312675 | Not-determined |
| 17-108_k141_285075 | Not-determined |
| 17-108_k141_106259 | Not-determined |
| 17-108_k141_187921 | Not-determined |
| 17-108_k141_312680 | Not-determined |
| 17-108_k141_201796 | Not-determined |
| 17-108_k141_325034 | Not-determined |
| 17-108_k141_132217 | Low-quality |
| 17-108_k141_36785 | Not-determined |
| 17-108_k141_244099 | Not-determined |
| 17-108_k141_257552 | Not-determined |
| 17-108_k141_187937 | Not-determined |
| 17-108_k141_298028 | Not-determined |
| 17-108_k141_312699 | Not-determined |
| 17-108_k141_257558 | Not-determined |
| 17-108_k141_187941 | Not-determined |
| 17-108_k141_118739 | Not-determined |
| 17-108_k141_244102 | Low-quality |
| 17-108_k141_325062 | Not-determined |
| 17-108_k141_201814 | Not-determined |
| 17-108_k141_160873 | Not-determined |
| 17-108_k141_285082 | Not-determined |
| 17-108_k141_312713 | Not-determined |
| 17-108_k141_77430 | Not-determined |
| 17-108_k141_216007 | Not-determined |
| 17-108_k141_118747 | Not-determined |
| 17-108_k141_257581 | Not-determined |
| 17-108_k141_50557 | Not-determined |
| 17-108_k141_106296 | Not-determined |
| 17-108_k141_77437 | Not-determined |
| 17-108_k141_285094 | Not-determined |
| 17-108_k141_9590 | Low-quality |
| 17-108_k141_50564 | Low-quality |
| 17-108_k141_257589 | Not-determined |
| 17-108_k141_216016 | Not-determined |
| 17-108_k141_257591 | Not-determined |
| 17-108_k141_50568 | Not-determined |
| 17-108_k141_9599 | Not-determined |
| 17-108_k141_132238 | Low-quality |
| 17-108_k141_50571 | Not-determined |
| 17-108_k141_201836 | Not-determined |
| 17-108_k141_229894 | Not-determined |
| 17-108_k141_118753 | Not-determined |
| 17-108_k141_257598 | Not-determined |
| 17-108_k141_146952 | Not-determined |
| 17-108_k141_160884 | Not-determined |
| 17-108_k141_187982 | Not-determined |
| 17-108_k141_216031 | Not-determined |
| 17-108_k141_146961 | Not-determined |
| 17-108_k141_187990 | Not-determined |
| 17-108_k141_9620 | Not-determined |
| 17-108_k141_91834 | Not-determined |
| 17-108_k141_160887 | Not-determined |
| 17-108_k141_50596 | Not-determined |
| 17-108_k141_174784 | Not-determined |
| 17-108_k141_160893 | Not-determined |
| 17-108_k141_257610 | Not-determined |
| 17-108_k141_270762 | Not-determined |
| 17-108_k141_118770 | Not-determined |
| 17-108_k141_146975 | Not-determined |
| 17-108_k141_118775 | Not-determined |
| 17-108_k141_50608 | Not-determined |
| 17-108_k141_244130 | Not-determined |
| 17-108_k141_36813 | Not-determined |
| 17-108_k141_9634 | Not-determined |
| 17-108_k141_244135 | Not-determined |
| 17-108_k141_118785 | Not-determined |
| 17-108_k141_21847 | Not-determined |
| 17-108_k141_270774 | Not-determined |
| 17-108_k141_36832 | Not-determined |
| 17-108_k141_91858 | Not-determined |
| 17-108_k141_36840 | Not-determined |
| 17-108_k141_325133 | Not-determined |
| 17-108_k141_244152 | Not-determined |
| 17-108_k141_312751 | Not-determined |
| 17-108_k141_91866 | Not-determined |
| 17-108_k141_298092 | Not-determined |
| 17-108_k141_270797 | Not-determined |
| 17-108_k141_118792 | Not-determined |
| 17-108_k141_201857 | Not-determined |
| 17-108_k141_160944 | Not-determined |
| 17-108_k141_325143 | Not-determined |
| 17-108_k141_188008 | Low-quality |
| 17-108_k141_285163 | Not-determined |
| 17-108_k141_91876 | Not-determined |
| 17-108_k141_244175 | Not-determined |
| 17-108_k141_147008 | Not-determined |
| 17-108_k141_21871 | Low-quality |
| 17-108_k141_285179 | Not-determined |
| 17-108_k141_298105 | Not-determined |
| 17-108_k141_77491 | Not-determined |
| 17-108_k141_298108 | Not-determined |
| 17-108_k141_312780 | Low-quality |
| 17-108_k141_36878 | Not-determined |
| 17-108_k141_312781 | Not-determined |
| 17-108_k141_160959 | Not-determined |
| 17-108_k141_298120 | Not-determined |
| 17-108_k141_50654 | Low-quality |
| 17-108_k141_21887 | Not-determined |
| 17-108_k141_118818 | Not-determined |
| 17-108_k141_285193 | Not-determined |
| 17-108_k141_50660 | Not-determined |
| 17-108_k141_257619 | Not-determined |
| 17-108_k141_9682 | Not-determined |
| 17-108_k141_244190 | Not-determined |
| 17-108_k141_229921 | Low-quality |
| 17-108_k141_160982 | Not-determined |
| 17-108_k141_216114 | Not-determined |
| 17-108_k141_201880 | Not-determined |
| 17-108_k141_201884 | Not-determined |
| 17-108_k141_36905 | Low-quality |
| 17-108_k141_174838 | Not-determined |
| 17-108_k141_50684 | Not-determined |
| 17-108_k141_106320 | Not-determined |
| 17-108_k141_64395 | Not-determined |
| 17-108_k141_312794 | Low-quality |
| 17-108_k141_312795 | Not-determined |
| 17-108_k141_64396 | Not-determined |
| 17-108_k141_64400 | Not-determined |
| 17-108_k141_118841 | Not-determined |
| 17-108_k141_118842 | Not-determined |
| 17-108_k141_201917 | Not-determined |
| 17-108_k141_298139 | Not-determined |
| 17-108_k141_161007 | Not-determined |
| 17-108_k141_64410 | Not-determined |
| 17-108_k141_244217 | Not-determined |
| 17-108_k141_174859 | Low-quality |
| 17-108_k141_285228 | Not-determined |
| 17-108_k141_285229 | Not-determined |
| 17-108_k141_285232 | Low-quality |
| 17-108_k141_298159 | Not-determined |
| 17-108_k141_50722 | Not-determined |
| 17-108_k141_201929 | Not-determined |
| 17-108_k141_161025 | Not-determined |
| 17-108_k141_201930 | Not-determined |
| 17-108_k141_216125 | Not-determined |
| 17-108_k141_174872 | Not-determined |
| 17-108_k141_118864 | Low-quality |
| 17-108_k141_50735 | Not-determined |
| 17-108_k141_298171 | Not-determined |
| 17-108_k141_9702 | Not-determined |
| 17-108_k141_298177 | Low-quality |
| 17-108_k141_244232 | Low-quality |
| 17-108_k141_161054 | Not-determined |
| 17-108_k141_9717 | Not-determined |
| 17-108_k141_161057 | Not-determined |
| 17-108_k141_147071 | Not-determined |
| 17-108_k141_174892 | Not-determined |
| 17-108_k141_118888 | Not-determined |
| 17-108_k141_9722 | Not-determined |
| 17-108_k141_201961 | Not-determined |
| 17-108_k141_201962 | Not-determined |
| 17-108_k141_174896 | Not-determined |
| 17-108_k141_312812 | Not-determined |
| 17-108_k141_147077 | Not-determined |
| 17-108_k141_174899 | Not-determined |
| 17-108_k141_132273 | Not-determined |
| 17-108_k141_132274 | Not-determined |
| 17-108_k141_312825 | Not-determined |
| 17-108_k141_312828 | Not-determined |
| 17-108_k141_229931 | Not-determined |
| 17-108_k141_201978 | Not-determined |
| 17-108_k141_201979 | Not-determined |
| 17-108_k141_201983 | Low-quality |
| 17-108_k141_9744 | Not-determined |
| 17-108_k141_36928 | Not-determined |
| 17-108_k141_201984 | Not-determined |
| 17-108_k141_257680 | Not-determined |
| 17-108_k141_36933 | Low-quality |
| 17-108_k141_216165 | Not-determined |
| 17-108_k141_244250 | Not-determined |
| 17-108_k141_270848 | Not-determined |
| 17-108_k141_132288 | Not-determined |
| 17-108_k141_229946 | Not-determined |
| 17-108_k141_106378 | Not-determined |
| 17-108_k141_216174 | Not-determined |
| 17-108_k141_50783 | Not-determined |
| 17-108_k141_270852 | Not-determined |
| 17-108_k141_174905 | Not-determined |
| 17-108_k141_312849 | Not-determined |
| 17-108_k141_36952 | Not-determined |
| 17-108_k141_201993 | Not-determined |
| 17-108_k141_257696 | Not-determined |
| 17-108_k141_298190 | Not-determined |
| 17-108_k141_118914 | Not-determined |
| 17-108_k141_216186 | Not-determined |
| 17-108_k141_147113 | Not-determined |
| 17-108_k141_257702 | Not-determined |
| 17-108_k141_132302 | Not-determined |
| 17-108_k141_312856 | Low-quality |
| 17-108_k141_147114 | Not-determined |
| 17-108_k141_36957 | Not-determined |
| 17-108_k141_202006 | Not-determined |
| 17-108_k141_147119 | Not-determined |
| 17-108_k141_91971 | Not-determined |
| 17-108_k141_91974 | Not-determined |
| 17-108_k141_229962 | Not-determined |
| 17-108_k141_188079 | Not-determined |
| 17-108_k141_285312 | Not-determined |
| 17-108_k141_244286 | Not-determined |
| 17-108_k141_118938 | Not-determined |
| 17-108_k141_285323 | Not-determined |
| 17-108_k141_298209 | Not-determined |
| 17-108_k141_106417 | Not-determined |
| 17-108_k141_36964 | Not-determined |
| 17-108_k141_216207 | Not-determined |
| 17-108_k141_188090 | Not-determined |
| 17-108_k141_188091 | Not-determined |
| 17-108_k141_312879 | Not-determined |
| 17-108_k141_298213 | Not-determined |
| 17-108_k141_132332 | Not-determined |
| 17-108_k141_174940 | Not-determined |
| 17-108_k141_312882 | Not-determined |
| 17-108_k141_325187 | Not-determined |
| 17-108_k141_325190 | Not-determined |
| 17-108_k141_64486 | Low-quality |
| 17-108_k141_174951 | Not-determined |
| 17-108_k141_118956 | Not-determined |
| 17-108_k141_216225 | Not-determined |
| 17-108_k141_202054 | Not-determined |
| 17-108_k141_92008 | Not-determined |
| 17-108_k141_77617 | Not-determined |
| 17-108_k141_202056 | Not-determined |
| 17-108_k141_270881 | Not-determined |
| 17-108_k141_132349 | Not-determined |
| 17-108_k141_147161 | Not-determined |
| 17-108_k141_174959 | Not-determined |
| 17-108_k141_188122 | Not-determined |
| 17-108_k141_312902 | Not-determined |
| 17-108_k141_325209 | Low-quality |
| 17-108_k141_64501 | Not-determined |
| 17-108_k141_312906 | Not-determined |
| 17-108_k141_118960 | Not-determined |
| 17-108_k141_36985 | Not-determined |
| 17-108_k141_9785 | Not-determined |
| 17-108_k141_257754 | Not-determined |
| 17-108_k141_298239 | Not-determined |
| 17-108_k141_298240 | Not-determined |
| 17-108_k141_106433 | Not-determined |
| 17-108_k141_64506 | Not-determined |
| 17-108_k141_312918 | Not-determined |
| 17-108_k141_147177 | Not-determined |
| 17-108_k141_147180 | Not-determined |
| 17-108_k141_147181 | Not-determined |
| 17-108_k141_216257 | Not-determined |
| 17-108_k141_216258 | Low-quality |
| 17-108_k141_188141 | Not-determined |
| 17-108_k141_147193 | Low-quality |
| 17-108_k141_325237 | Not-determined |
| 17-108_k141_132375 | Not-determined |
| 17-108_k141_230004 | Low-quality |
| 17-108_k141_230008 | Not-determined |
| 17-108_k141_132381 | Not-determined |
| 17-108_k141_92035 | Not-determined |
| 17-108_k141_77651 | Not-determined |
| 17-108_k141_244323 | Low-quality |
| 17-108_k141_202100 | Not-determined |
| 17-108_k141_257782 | Not-determined |
| 17-108_k141_298270 | Not-determined |
| 17-108_k141_175010 | Not-determined |
| 17-108_k141_175011 | Not-determined |
| 17-108_k141_188172 | Not-determined |
| 17-108_k141_230016 | Not-determined |
| 17-108_k141_285368 | Not-determined |
| 17-108_k141_175015 | Not-determined |
| 17-108_k141_175018 | Not-determined |
| 17-108_k141_118997 | Not-determined |
| 17-108_k141_50872 | Not-determined |
| 17-108_k141_106474 | Not-determined |
| 17-108_k141_175032 | Not-determined |
| 17-108_k141_132401 | Not-determined |
| 17-108_k141_119014 | Not-determined |
| 17-108_k141_202130 | Not-determined |
| 17-108_k141_77693 | Not-determined |
| 17-108_k141_37024 | Not-determined |
| 17-108_k141_106491 | Not-determined |
| 17-108_k141_106494 | Not-determined |
| 17-108_k141_230035 | Not-determined |
| 17-108_k141_230036 | Not-determined |
| 17-108_k141_202139 | Not-determined |
| 17-108_k141_175051 | Not-determined |
| 17-108_k141_175052 | Not-determined |
| 17-108_k141_119023 | Low-quality |
| 17-108_k141_106507 | Not-determined |
| 17-108_k141_175059 | Not-determined |
| 17-108_k141_312975 | Not-determined |
| 17-108_k141_77715 | Not-determined |
| 17-108_k141_257827 | Not-determined |
| 17-108_k141_257829 | Low-quality |
| 17-108_k141_285374 | Not-determined |
| 17-108_k141_257834 | Not-determined |
| 17-108_k141_244342 | Low-quality |
| 17-108_k141_216316 | Not-determined |
| 17-108_k141_312982 | Not-determined |
| 17-108_k141_175074 | Not-determined |
| 17-108_k141_312987 | Not-determined |
| 17-108_k141_77723 | Not-determined |
| 17-108_k141_216327 | Not-determined |
| 17-108_k141_64583 | Not-determined |
| 17-108_k141_312990 | Not-determined |
| 17-108_k141_37054 | Not-determined |
| 17-108_k141_9809 | Low-quality |
| 17-108_k141_257848 | Not-determined |
| 17-108_k141_216338 | Not-determined |
| 17-108_k141_106525 | Not-determined |
| 17-108_k141_64595 | Not-determined |
| 17-108_k141_257853 | Not-determined |
| 17-108_k141_119049 | Low-quality |
| 17-108_k141_92057 | Not-determined |
| 17-108_k141_257858 | Not-determined |
| 17-108_k141_285401 | Not-determined |
| 17-108_k141_270901 | Not-determined |
| 17-108_k141_37070 | Not-determined |
| 17-108_k141_270907 | Not-determined |
| 17-108_k141_325283 | Not-determined |
| 17-108_k141_298357 | Not-determined |
| 17-108_k141_37079 | Not-determined |
| 17-108_k141_21952 | Not-determined |
| 17-108_k141_119063 | Not-determined |
| 17-108_k141_244375 | Not-determined |
| 17-108_k141_37082 | Not-determined |
| 17-108_k141_147233 | Not-determined |
| 17-108_k141_285418 | Not-determined |
| 17-108_k141_313030 | Not-determined |
| 17-108_k141_147238 | Not-determined |
| 17-108_k141_161170 | Not-determined |
| 17-108_k141_216361 | Not-determined |
| 17-108_k141_147244 | Not-determined |
| 17-108_k141_119081 | Not-determined |
| 17-108_k141_313034 | Not-determined |
| 17-108_k141_325301 | Not-determined |
| 17-108_k141_92088 | Not-determined |
| 17-108_k141_244388 | Low-quality |
| 17-108_k141_9854 | Not-determined |
| 17-108_k141_161184 | Not-determined |
| 17-108_k141_188212 | Low-quality |
| 17-108_k141_202174 | Not-determined |
| 17-108_k141_64604 | Low-quality |
| 17-108_k141_147257 | Not-determined |
| 17-108_k141_270950 | Not-determined |
| 17-108_k141_147263 | Not-determined |
| 17-108_k141_92109 | Not-determined |
| 17-108_k141_270955 | Not-determined |
| 17-108_k141_64612 | Not-determined |
| 17-108_k141_92110 | Not-determined |
| 17-108_k141_64614 | Not-determined |
| 17-108_k141_313059 | Not-determined |
| 17-108_k141_161200 | Not-determined |
| 17-108_k141_313062 | Not-determined |
| 17-108_k141_244412 | Not-determined |
| 17-108_k141_202183 | Not-determined |
| 17-108_k141_244416 | Not-determined |
| 17-108_k141_147276 | Not-determined |
| 17-108_k141_9873 | Not-determined |
| 17-108_k141_147278 | Not-determined |
| 17-108_k141_161210 | Not-determined |
| 17-108_k141_132466 | Low-quality |
| 17-108_k141_175111 | Not-determined |
| 17-108_k141_325317 | Not-determined |
| 17-108_k141_9877 | Not-determined |
| 17-108_k141_21977 | Not-determined |
| 17-108_k141_132472 | Not-determined |
| 17-108_k141_285446 | Not-determined |
| 17-108_k141_257889 | Not-determined |
| 17-108_k141_21984 | Not-determined |
| 17-108_k141_244427 | Not-determined |
| 17-108_k141_244428 | Not-determined |
| 17-108_k141_244430 | Not-determined |
| 17-108_k141_285453 | Not-determined |
| 17-108_k141_21990 | Not-determined |
| 17-108_k141_202196 | Not-determined |
| 17-108_k141_270970 | Not-determined |
| 17-108_k141_270972 | Low-quality |
| 17-108_k141_285457 | Not-determined |
| 17-108_k141_285460 | Not-determined |
| 17-108_k141_230077 | Low-quality |
| 17-108_k141_119113 | Low-quality |
| 17-108_k141_230080 | Not-determined |
| 17-108_k141_77752 | Not-determined |
| 17-108_k141_202206 | Not-determined |
| 17-108_k141_325340 | Not-determined |
| 17-108_k141_132487 | Not-determined |
| 17-108_k141_325342 | Not-determined |
| 17-108_k141_325347 | Not-determined |
| 17-108_k141_257908 | Not-determined |
| 17-108_k141_285473 | Not-determined |
| 17-108_k141_9894 | Not-determined |
| 17-108_k141_325352 | Not-determined |
| 17-108_k141_270985 | Not-determined |
| 17-108_k141_270986 | Not-determined |
| 17-108_k141_175135 | Not-determined |
| 17-108_k141_50993 | Not-determined |
| 17-108_k141_244464 | Not-determined |
| 17-108_k141_22007 | Not-determined |
| 17-108_k141_22010 | Not-determined |
| 17-108_k141_257929 | Not-determined |
| 17-108_k141_147325 | Not-determined |
| 17-108_k141_325353 | Low-quality |
| 17-108_k141_244476 | Not-determined |
| 17-108_k141_147330 | Not-determined |
| 17-108_k141_257935 | Not-determined |
| 17-108_k141_202229 | Not-determined |
| 17-108_k141_119137 | Not-determined |
| 17-108_k141_9908 | Not-determined |
| 17-108_k141_64640 | Not-determined |
| 17-108_k141_175149 | Not-determined |
| 17-108_k141_51027 | Not-determined |
| 17-108_k141_230101 | Not-determined |
| 17-108_k141_161242 | Not-determined |
| 17-108_k141_244498 | Not-determined |
| 17-108_k141_298388 | Not-determined |
| 17-108_k141_175166 | Not-determined |
| 17-108_k141_271015 | Not-determined |
| 17-108_k141_271020 | Not-determined |
| 17-108_k141_175175 | Not-determined |
| 17-108_k141_202241 | Not-determined |
| 17-108_k141_298394 | Low-quality |
| 17-108_k141_161255 | Not-determined |
| 17-108_k141_313091 | Not-determined |
| 17-108_k141_22031 | Low-quality |
| 17-108_k141_202246 | Not-determined |
| 17-108_k141_298400 | Not-determined |
| 17-108_k141_257966 | Not-determined |
| 17-108_k141_64653 | Low-quality |
| 17-108_k141_119171 | Low-quality |
| 17-108_k141_119172 | Not-determined |
| 17-108_k141_51042 | Not-determined |
| 17-108_k141_51043 | Not-determined |
| 17-108_k141_257971 | Not-determined |
| 17-108_k141_216394 | Not-determined |
| 17-108_k141_161264 | Not-determined |
| 17-108_k141_244519 | Not-determined |
| 17-108_k141_325406 | Not-determined |
| 17-108_k141_216415 | Not-determined |
| 17-108_k141_161279 | Not-determined |
| 17-108_k141_313125 | Not-determined |
| 17-108_k141_216423 | Not-determined |
| 17-108_k141_230143 | Not-determined |
| 17-108_k141_313128 | Not-determined |
| 17-108_k141_325419 | Not-determined |
| 17-108_k141_161296 | Not-determined |
| 17-108_k141_202275 | Not-determined |
| 17-108_k141_188310 | Not-determined |
| 17-108_k141_188313 | Not-determined |
| 17-108_k141_92143 | Not-determined |
| 17-108_k141_202286 | Not-determined |
| 17-108_k141_147360 | Not-determined |
| 17-108_k141_92147 | Not-determined |
| 17-108_k141_77779 | Not-determined |
| 17-108_k141_313141 | Low-quality |
| 17-108_k141_258010 | Not-determined |
| 17-108_k141_258012 | Not-determined |
| 17-108_k141_147368 | Not-determined |
| 17-108_k141_106558 | Not-determined |
| 17-108_k141_106562 | Not-determined |
| 17-108_k141_147373 | Not-determined |
| 17-108_k141_92159 | Not-determined |
| 17-108_k141_313153 | Low-quality |
| 17-108_k141_313157 | Not-determined |
| 17-108_k141_313158 | Not-determined |
| 17-108_k141_325437 | Not-determined |
| 17-108_k141_244526 | Not-determined |
| 17-108_k141_285524 | Low-quality |
| 17-108_k141_92167 | Not-determined |
| 17-108_k141_313166 | Not-determined |
| 17-108_k141_325443 | Not-determined |
| 17-108_k141_64672 | Low-quality |
| 17-108_k141_175198 | Not-determined |
| 17-108_k141_51104 | Not-determined |
| 17-108_k141_325448 | Not-determined |
| 17-108_k141_22044 | Not-determined |
| 17-108_k141_161317 | Low-quality |
| 17-108_k141_22048 | Not-determined |
| 17-108_k141_313176 | Not-determined |
| 17-108_k141_147390 | Low-quality |
| 17-108_k141_132545 | Not-determined |
| 17-108_k141_51113 | Not-determined |
| 17-108_k141_244533 | Not-determined |
| 17-108_k141_92177 | Not-determined |
| 17-108_k141_77809 | Not-determined |
| 17-108_k141_313191 | Not-determined |
| 17-108_k141_51126 | Not-determined |
| 17-108_k141_132554 | Not-determined |
| 17-108_k141_285540 | Not-determined |
| 17-108_k141_285541 | Low-quality |
| 17-108_k141_161324 | Not-determined |
| 17-108_k141_132557 | Not-determined |
| 17-108_k141_161327 | Not-determined |
| 17-108_k141_175221 | Low-quality |
| 17-108_k141_132558 | Not-determined |
| 17-108_k141_147409 | Not-determined |
| 17-108_k141_188346 | Not-determined |
| 17-108_k141_298468 | Not-determined |
| 17-108_k141_37181 | Not-determined |
| 17-108_k141_37182 | Not-determined |
| 17-108_k141_147411 | Not-determined |
| 17-108_k141_37186 | Not-determined |
| 17-108_k141_77822 | Not-determined |
| 17-108_k141_119226 | Not-determined |
| 17-108_k141_298475 | Not-determined |
| 17-108_k141_92194 | Not-determined |
| 17-108_k141_37189 | Not-determined |
| 17-108_k141_188357 | Not-determined |
| 17-108_k141_64701 | Not-determined |
| 17-108_k141_230196 | Not-determined |
| 17-108_k141_285552 | Not-determined |
| 17-108_k141_37193 | Not-determined |
| 17-108_k141_51155 | Not-determined |
| 17-108_k141_37195 | Not-determined |
| 17-108_k141_298489 | Not-determined |
| 17-108_k141_147440 | Not-determined |
| 17-108_k141_313205 | Not-determined |
| 17-108_k141_147448 | Low-quality |
| 17-108_k141_92211 | Not-determined |
| 17-108_k141_230202 | Not-determined |
| 17-108_k141_230203 | Low-quality |
| 17-108_k141_51175 | Not-determined |
| 17-108_k141_258059 | Low-quality |
| 17-108_k141_51180 | Not-determined |
| 17-108_k141_258060 | Not-determined |
| 17-108_k141_64726 | Not-determined |
| 17-108_k141_216465 | Low-quality |
| 17-108_k141_37224 | Not-determined |
| 17-108_k141_37227 | Not-determined |
| 17-108_k141_298515 | Not-determined |
| 17-108_k141_64736 | Not-determined |
| 17-108_k141_202339 | Not-determined |
| 17-108_k141_285572 | Not-determined |
| 17-108_k141_175239 | Not-determined |
| 17-108_k141_77828 | Not-determined |
| 17-108_k141_106628 | Not-determined |
| 17-108_k141_188369 | Not-determined |
| 17-108_k141_147487 | Low-quality |
| 17-108_k141_325502 | Not-determined |
| 17-108_k141_216486 | Not-determined |
| 17-108_k141_230222 | Not-determined |
| 17-108_k141_106638 | Not-determined |
| 17-108_k141_298530 | Low-quality |
| 17-108_k141_202353 | Not-determined |
| 17-108_k141_244555 | Not-determined |
| 17-108_k141_147500 | Not-determined |
| 17-108_k141_258085 | Not-determined |
| 17-108_k141_188382 | Low-quality |
| 17-108_k141_106644 | Not-determined |

| | |
|---|---|
| 17-108_k141_51211 | Not-determined |
| 17-108_k141_244559 | Not-determined |
| 17-108_k141_64765 | Not-determined |
| 17-108_k141_147509 | Not-determined |
| 17-108_k141_285595 | Low-quality |
| 17-108_k141_258094 | Not-determined |
| 17-108_k141_106651 | Not-determined |
| 17-108_k141_119265 | Not-determined |
| 17-108_k141_51214 | Not-determined |
| 17-108_k141_132601 | Not-determined |
| 17-108_k141_132602 | Not-determined |
| 17-108_k141_161343 | Low-quality |
| 17-108_k141_175268 | Low-quality |
| 17-108_k141_244577 | Not-determined |
| 17-108_k141_161347 | Not-determined |
| 17-108_k141_313225 | Not-determined |
| 17-108_k141_325509 | Not-determined |
| 17-108_k141_37268 | Not-determined |
| 17-108_k141_51219 | Not-determined |
| 17-108_k141_202379 | Not-determined |
| 17-108_k141_132615 | Not-determined |
| 17-108_k141_132616 | Not-determined |
| 17-108_k141_132618 | Not-determined |
| 17-108_k141_216506 | Not-determined |
| 17-108_k141_244589 | Not-determined |
| 17-108_k141_298569 | Not-determined |
| 17-108_k141_132628 | Not-determined |
| 17-108_k141_244593 | Not-determined |
| 17-108_k141_244597 | Not-determined |
| 17-108_k141_64777 | Not-determined |
| 17-108_k141_64778 | Not-determined |
| 17-108_k141_106683 | Low-quality |
| 17-108_k141_298585 | Not-determined |
| 17-108_k141_188405 | Not-determined |
| 17-108_k141_188406 | Not-determined |
| 17-108_k141_64787 | Not-determined |
| 17-108_k141_285628 | Not-determined |
| 17-108_k141_244608 | Not-determined |
| 17-108_k141_51242 | Low-quality |
| 17-108_k141_202403 | Not-determined |
| 17-108_k141_244615 | Not-determined |
| 17-108_k141_132640 | Not-determined |
| 17-108_k141_298601 | Not-determined |
| 17-108_k141_285634 | Not-determined |
| 17-108_k141_216532 | Not-determined |
| 17-108_k141_106709 | Not-determined |
| 17-108_k141_9972 | Not-determined |
| 17-108_k141_77882 | Not-determined |
| 17-108_k141_9975 | Not-determined |
| 17-108_k141_175294 | Not-determined |
| 17-108_k141_92311 | Not-determined |
| 17-108_k141_64809 | Not-determined |
| 17-108_k141_271110 | Not-determined |
| 17-108_k141_37309 | Not-determined |
| 17-108_k141_202427 | Low-quality |
| 17-108_k141_119312 | Not-determined |
| 17-108_k141_9993 | Not-determined |
| 17-108_k141_244649 | Not-determined |
| 17-108_k141_313278 | Not-determined |
| 17-108_k141_106727 | Not-determined |
| 17-108_k141_230303 | Not-determined |
| 17-108_k141_147552 | Not-determined |
| 17-108_k141_271142 | Not-determined |
| 17-108_k141_202437 | Not-determined |
| 17-108_k141_188451 | Not-determined |
| 17-108_k141_202440 | Not-determined |
| 17-108_k141_244664 | Not-determined |
| 17-108_k141_244665 | Not-determined |
| 17-108_k141_298626 | Not-determined |
| 17-108_k141_202445 | Not-determined |
| 17-108_k141_132663 | Not-determined |
| 17-108_k141_175300 | Low-quality |
| 17-108_k141_119347 | Not-determined |
| 17-108_k141_64861 | Not-determined |
| 17-108_k141_216599 | Not-determined |
| 17-108_k141_216600 | Low-quality |
| 17-108_k141_106732 | Not-determined |
| 17-108_k141_64871 | Not-determined |
| 17-108_k141_230334 | Not-determined |
| 17-108_k141_298633 | Not-determined |
| 17-108_k141_106745 | Not-determined |
| 17-108_k141_230335 | Not-determined |
| 17-108_k141_298637 | Not-determined |
| 17-108_k141_285663 | Not-determined |
| 17-108_k141_285665 | Not-determined |
| 17-108_k141_147576 | Low-quality |
| 17-108_k141_161383 | Not-determined |
| 17-108_k141_10008 | Not-determined |
| 17-108_k141_37340 | Not-determined |
| 17-108_k141_51311 | Not-determined |
| 17-108_k141_285670 | Low-quality |
| 17-108_k141_147583 | Low-quality |
| 17-108_k141_37343 | Not-determined |
| 17-108_k141_216618 | Not-determined |
| 17-108_k141_64883 | Not-determined |
| 17-108_k141_298647 | Not-determined |
| 17-108_k141_298649 | Not-determined |
| 17-108_k141_285676 | Not-determined |
| 17-108_k141_230349 | Not-determined |
| 17-108_k141_22169 | Low-quality |
| 17-108_k141_313287 | Not-determined |
| 17-108_k141_92382 | Not-determined |
| 17-108_k141_64896 | Not-determined |
| 17-108_k141_92387 | Not-determined |
| 17-108_k141_10024 | Not-determined |
| 17-108_k141_92388 | Not-determined |
| 17-108_k141_64899 | Not-determined |
| 17-108_k141_175334 | Not-determined |
| 17-108_k141_132676 | Not-determined |
| 17-108_k141_258182 | Not-determined |
| 17-108_k141_298658 | Not-determined |
| 17-108_k141_22184 | Not-determined |
| 17-108_k141_119381 | Not-determined |
| 17-108_k141_147605 | Not-determined |
| 17-108_k141_230361 | Not-determined |
| 17-108_k141_313298 | Low-quality |
| 17-108_k141_188491 | Not-determined |
| 17-108_k141_325535 | Not-determined |
| 17-108_k141_285694 | Not-determined |
| 17-108_k141_216635 | Not-determined |
| 17-108_k141_77923 | Not-determined |
| 17-108_k141_325537 | Not-determined |
| 17-108_k141_188495 | Not-determined |
| 17-108_k141_298665 | Not-determined |
| 17-108_k141_147622 | Not-determined |
| 17-108_k141_77928 | Not-determined |
| 17-108_k141_147625 | Low-quality |
| 17-108_k141_161403 | Not-determined |
| 17-108_k141_77933 | Not-determined |
| 17-108_k141_244753 | Not-determined |
| 17-108_k141_271157 | Not-determined |
| 17-108_k141_77935 | Not-determined |
| 17-108_k141_77936 | Not-determined |
| 17-108_k141_132700 | Not-determined |
| 17-108_k141_132702 | Not-determined |
| 17-108_k141_298682 | Not-determined |
| 17-108_k141_77940 | Not-determined |
| 17-108_k141_244772 | Not-determined |
| 17-108_k141_325553 | Low-quality |
| 17-108_k141_106772 | Not-determined |
| 17-108_k141_325554 | Not-determined |
| 17-108_k141_132717 | Not-determined |
| 17-108_k141_325563 | Low-quality |
| 17-108_k141_271187 | Not-determined |
| 17-108_k141_298696 | Not-determined |
| 17-108_k141_244778 | Not-determined |
| 17-108_k141_37405 | Not-determined |
| 17-108_k141_37411 | Low-quality |
| 17-108_k141_175388 | Not-determined |
| 17-108_k141_77960 | Not-determined |
| 17-108_k141_51338 | Not-determined |
| 17-108_k141_37418 | Not-determined |
| 17-108_k141_22205 | Not-determined |
| 17-108_k141_147658 | Not-determined |
| 17-108_k141_147661 | Not-determined |
| 17-108_k141_37421 | Not-determined |
| 17-108_k141_161422 | Not-determined |
| 17-108_k141_258242 | Low-quality |
| 17-108_k141_132739 | Not-determined |
| 17-108_k141_22228 | Not-determined |
| 17-108_k141_216677 | Not-determined |
| 17-108_k141_216679 | Not-determined |
| 17-108_k141_298720 | Not-determined |
| 17-108_k141_22234 | Not-determined |
| 17-108_k141_161430 | Not-determined |
| 17-108_k141_161436 | Not-determined |
| 17-108_k141_22250 | Not-determined |
| 17-108_k141_106812 | Not-determined |
| 17-108_k141_175427 | Not-determined |
| 17-108_k141_51350 | Not-determined |
| 17-108_k141_106816 | Not-determined |
| 17-108_k141_64935 | Not-determined |
| 17-108_k141_92400 | Not-determined |
| 17-108_k141_22269 | Not-determined |
| 17-108_k141_175439 | Not-determined |
| 17-108_k141_132760 | Not-determined |
| 17-108_k141_78008 | Not-determined |
| 17-108_k141_161469 | Not-determined |
| 17-108_k141_285733 | Not-determined |
| 17-108_k141_202507 | Not-determined |
| 17-108_k141_230377 | Low-quality |
| 17-108_k141_202513 | Not-determined |
| 17-108_k141_161482 | Not-determined |
| 17-108_k141_106832 | Not-determined |
| 17-108_k141_22295 | Not-determined |
| 17-108_k141_51383 | Not-determined |
| 17-108_k141_258273 | Not-determined |
| 17-108_k141_92438 | Not-determined |
| 17-108_k141_298768 | Not-determined |
| 17-108_k141_258278 | Not-determined |
| 17-108_k141_106838 | Not-determined |
| 17-108_k141_106839 | Not-determined |
| 17-108_k141_285753 | Not-determined |
| 17-108_k141_313346 | Not-determined |
| 17-108_k141_188515 | Not-determined |
| 17-108_k141_22311 | Not-determined |
| 17-108_k141_64980 | Not-determined |
| 17-108_k141_51395 | Not-determined |
| 17-108_k141_325618 | Not-determined |
| 17-108_k141_202536 | Not-determined |
| 17-108_k141_244800 | Not-determined |
| 17-108_k141_258300 | Not-determined |
| 17-108_k141_22318 | Not-determined |
| 17-108_k141_64984 | Not-determined |
| 17-108_k141_230400 | Low-quality |
| 17-108_k141_244806 | Not-determined |
| 17-108_k141_51400 | Not-determined |
| 17-108_k141_64991 | Not-determined |
| 17-108_k141_230404 | Not-determined |
| 17-108_k141_147710 | Not-determined |
| 17-108_k141_92463 | Not-determined |
| 17-108_k141_216739 | Not-determined |
| 17-108_k141_51403 | Not-determined |
| 17-108_k141_230414 | Not-determined |
| 17-108_k141_175506 | Not-determined |
| 17-108_k141_106857 | Not-determined |
| 17-108_k141_202543 | Not-determined |
| 17-108_k141_22328 | Low-quality |
| 17-108_k141_37515 | Not-determined |
| 17-108_k141_161524 | Not-determined |
| 17-108_k141_313377 | Not-determined |
| 17-108_k141_161525 | Not-determined |
| 17-108_k141_37517 | Low-quality |
| 17-108_k141_202548 | Not-determined |
| 17-108_k141_313381 | Not-determined |
| 17-108_k141_313382 | Not-determined |
| 17-108_k141_92491 | Low-quality |
| 17-108_k141_244829 | Not-determined |
| 17-108_k141_78084 | Low-quality |
| 17-108_k141_175527 | Not-determined |
| 17-108_k141_175528 | Not-determined |
| 17-108_k141_244835 | Not-determined |
| 17-108_k141_132814 | Low-quality |
| 17-108_k141_92500 | Not-determined |
| 17-108_k141_188539 | Not-determined |
| 17-108_k141_202555 | Low-quality |
| 17-108_k141_202556 | Not-determined |
| 17-108_k141_298787 | Not-determined |
| 17-108_k141_244847 | Not-determined |
| 17-108_k141_188549 | Not-determined |
| 17-108_k141_22345 | Not-determined |
| 17-108_k141_119489 | Not-determined |
| 17-108_k141_161555 | Not-determined |
| 17-108_k141_51445 | Not-determined |
| 17-108_k141_230439 | Not-determined |
| 17-108_k141_285794 | Not-determined |
| 17-108_k141_147746 | Low-quality |
| 17-108_k141_147750 | Not-determined |
| 17-108_k141_51454 | Not-determined |
| 17-108_k141_119503 | Low-quality |
| 17-108_k141_119510 | Not-determined |
| 17-108_k141_313402 | Not-determined |
| 17-108_k141_106906 | Not-determined |
| 17-108_k141_216790 | Not-determined |
| 17-108_k141_65045 | Not-determined |
| 17-108_k141_22387 | Not-determined |
| 17-108_k141_10116 | Not-determined |
| 17-108_k141_10120 | Not-determined |
| 17-108_k141_202572 | Not-determined |
| 17-108_k141_313411 | Not-determined |
| 17-108_k141_106913 | Not-determined |
| 17-108_k141_230477 | Not-determined |
| 17-108_k141_106916 | Not-determined |
| 17-108_k141_313414 | Not-determined |
| 17-108_k141_106919 | Not-determined |
| 17-108_k141_22390 | Not-determined |
| 17-108_k141_202582 | Not-determined |
| 17-108_k141_10123 | Not-determined |
| 17-108_k141_51458 | Not-determined |
| 17-108_k141_51460 | Low-quality |
| 17-108_k141_132871 | Not-determined |
| 17-108_k141_230488 | Not-determined |
| 17-108_k141_230489 | Not-determined |
| 17-108_k141_271306 | Low-quality |
| 17-108_k141_10144 | Not-determined |
| 17-108_k141_258337 | Low-quality |
| 17-108_k141_51468 | Not-determined |
| 17-108_k141_271314 | Not-determined |
| 17-108_k141_298807 | Not-determined |
| 17-108_k141_37538 | Not-determined |
| 17-108_k141_230502 | Not-determined |
| 17-108_k141_216822 | Not-determined |
| 17-108_k141_37545 | Not-determined |
| 17-108_k141_230512 | Not-determined |
| 17-108_k141_92517 | Not-determined |
| 17-108_k141_271325 | Low-quality |
| 17-108_k141_119534 | Not-determined |
| 17-108_k141_37550 | Not-determined |
| 17-108_k141_51477 | Low-quality |
| 17-108_k141_132893 | Not-determined |
| 17-108_k141_119535 | Not-determined |
| 17-108_k141_325647 | Not-determined |
| 17-108_k141_119541 | Not-determined |
| 17-108_k141_51489 | Not-determined |
| 17-108_k141_10169 | Not-determined |
| 17-108_k141_202609 | Not-determined |
| 17-108_k141_132900 | Not-determined |
| 17-108_k141_258370 | Low-quality |
| 17-108_k141_119560 | Not-determined |
| 17-108_k141_147808 | Not-determined |
| 17-108_k141_161587 | Not-determined |
| 17-108_k141_313467 | Not-determined |
| 17-108_k141_132920 | Not-determined |
| 17-108_k141_271350 | Not-determined |
| 17-108_k141_161591 | Not-determined |
| 17-108_k141_92564 | Not-determined |
| 17-108_k141_51525 | Not-determined |
| 17-108_k141_119578 | Not-determined |
| 17-108_k141_258385 | Not-determined |
| 17-108_k141_51527 | Not-determined |
| 17-108_k141_92575 | Not-determined |
| 17-108_k141_51532 | Not-determined |
| 17-108_k141_258391 | Not-determined |
| 17-108_k141_119592 | Not-determined |
| 17-108_k141_230534 | Not-determined |
| 17-108_k141_271361 | Not-determined |
| 17-108_k141_161611 | Not-determined |
| 17-108_k141_161612 | Not-determined |
| 17-108_k141_51539 | Not-determined |
| 17-108_k141_78185 | Not-determined |
| 17-108_k141_119596 | Low-quality |
| 17-108_k141_271374 | Not-determined |
| 17-108_k141_244871 | Not-determined |
| 17-108_k141_119597 | Low-quality |
| 17-108_k141_230547 | Not-determined |
| 17-108_k141_51551 | Not-determined |
| 17-108_k141_230551 | Not-determined |
| 17-108_k141_132936 | Not-determined |
| 17-108_k141_65124 | Not-determined |
| 17-108_k141_92595 | Low-quality |
| 17-108_k141_78198 | Not-determined |
| 17-108_k141_313487 | Low-quality |
| 17-108_k141_119607 | Not-determined |
| 17-108_k141_22453 | Not-determined |
| 17-108_k141_22454 | Low-quality |
| 17-108_k141_271385 | Not-determined |
| 17-108_k141_202656 | Not-determined |
| 17-108_k141_216836 | Not-determined |
| 17-108_k141_22456 | Low-quality |
| 17-108_k141_202659 | Not-determined |
| 17-108_k141_325688 | Not-determined |
| 17-108_k141_51564 | Not-determined |
| 17-108_k141_22467 | Not-determined |
| 17-108_k141_132953 | Not-determined |
| 17-108_k141_51575 | Not-determined |
| 17-108_k141_271395 | Not-determined |
| 17-108_k141_271398 | Not-determined |
| 17-108_k141_22475 | Not-determined |
| 17-108_k141_51579 | Not-determined |
| 17-108_k141_161621 | Not-determined |
| 17-108_k141_161623 | Not-determined |
| 17-108_k141_147827 | Not-determined |
| 17-108_k141_230573 | Not-determined |
| 17-108_k141_298847 | Not-determined |
| 17-108_k141_230576 | Not-determined |
| 17-108_k141_175596 | Low-quality |
| 17-108_k141_202685 | Not-determined |
| 17-108_k141_132970 | Not-determined |
| 17-108_k141_22490 | Low-quality |
| 17-108_k141_10182 | Not-determined |
| 17-108_k141_51593 | Not-determined |
| 17-108_k141_161654 | Not-determined |
| 17-108_k141_175612 | Not-determined |
| 17-108_k141_298857 | Not-determined |
| 17-108_k141_271417 | Not-determined |
| 17-108_k141_285861 | Not-determined |
| 17-108_k141_325704 | Not-determined |
| 17-108_k141_22499 | Low-quality |
| 17-108_k141_10217 | Low-quality |
| 17-108_k141_37630 | Not-determined |
| 17-108_k141_10220 | Low-quality |
| 17-108_k141_258451 | Not-determined |
| 17-108_k141_10222 | Low-quality |
| 17-108_k141_51607 | Not-determined |
| 17-108_k141_298871 | Not-determined |
| 17-108_k141_147853 | Low-quality |
| 17-108_k141_37642 | Not-determined |
| 17-108_k141_271425 | Not-determined |
| 17-108_k141_161656 | Not-determined |
| 17-108_k141_202715 | Not-determined |
| 17-108_k141_271429 | Not-determined |
| 17-108_k141_92628 | Not-determined |
| 17-108_k141_51615 | Not-determined |
| 17-108_k141_161660 | Not-determined |
| 17-108_k141_258472 | Not-determined |
| 17-108_k141_325717 | Not-determined |
| 17-108_k141_216853 | Not-determined |
| 17-108_k141_216854 | Not-determined |
| 17-108_k141_202726 | Low-quality |
| 17-108_k141_10244 | Not-determined |
| 17-108_k141_51626 | Low-quality |
| 17-108_k141_51636 | Not-determined |
| 17-108_k141_271440 | Not-determined |
| 17-108_k141_22535 | Not-determined |
| 17-108_k141_313535 | Not-determined |
| 17-108_k141_78266 | Not-determined |
| 17-108_k141_65180 | Not-determined |
| 17-108_k141_147877 | Not-determined |
| 17-108_k141_65190 | Not-determined |
| 17-108_k141_92640 | Not-determined |
| 17-108_k141_107013 | Not-determined |
| 17-108_k141_78271 | Not-determined |
| 17-108_k141_119658 | Not-determined |
| 17-108_k141_271453 | Low-quality |
| 17-108_k141_325743 | Not-determined |
| 17-108_k141_51651 | Not-determined |
| 17-108_k141_188677 | Not-determined |
| 17-108_k141_188678 | Not-determined |
| 17-108_k141_271455 | Low-quality |
| 17-108_k141_188682 | Not-determined |
| 17-108_k141_271457 | Not-determined |
| 17-108_k141_285904 | Not-determined |
| 17-108_k141_271461 | Not-determined |
| 17-108_k141_51662 | Not-determined |
| 17-108_k141_107038 | Not-determined |
| 17-108_k141_271466 | Low-quality |
| 17-108_k141_298905 | Not-determined |
| 17-108_k141_216893 | Not-determined |
| 17-108_k141_119680 | Low-quality |
| 17-108_k141_298911 | Not-determined |
| 17-108_k141_298913 | Not-determined |
| 17-108_k141_119685 | Not-determined |
| 17-108_k141_188691 | Not-determined |
| 17-108_k141_202761 | Not-determined |
| 17-108_k141_161704 | Not-determined |
| 17-108_k141_313560 | Not-determined |
| 17-108_k141_188697 | Not-determined |
| 17-108_k141_325756 | Not-determined |
| 17-108_k141_202771 | Not-determined |
| 17-108_k141_161709 | Not-determined |
| 17-108_k141_161710 | Not-determined |
| 17-108_k141_244950 | Not-determined |
| 17-108_k141_271481 | Not-determined |
| 17-108_k141_258488 | Not-determined |
| 17-108_k141_216902 | Not-determined |
| 17-108_k141_175671 | Not-determined |
| 17-108_k141_313568 | Not-determined |
| 17-108_k141_244957 | Not-determined |
| 17-108_k141_285931 | Not-determined |
| 17-108_k141_37667 | Not-determined |
| 17-108_k141_325767 | Not-determined |
| 17-108_k141_188722 | Not-determined |
| 17-108_k141_244963 | Not-determined |
| 17-108_k141_188725 | Not-determined |
| 17-108_k141_244966 | Not-determined |
| 17-108_k141_51691 | Not-determined |
| 17-108_k141_51692 | Not-determined |
| 17-108_k141_161721 | Not-determined |
| 17-108_k141_271491 | Not-determined |
| 17-108_k141_271496 | Low-quality |
| 17-108_k141_161726 | Not-determined |
| 17-108_k141_65252 | Not-determined |
| 17-108_k141_161727 | Not-determined |
| 17-108_k141_298937 | Not-determined |
| 17-108_k141_107070 | Not-determined |
| 17-108_k141_22577 | Not-determined |
| 17-108_k141_244979 | Not-determined |
| 17-108_k141_216913 | Not-determined |
| 17-108_k141_51706 | Not-determined |
| 17-108_k141_92668 | Not-determined |
| 17-108_k141_325778 | Not-determined |
| 17-108_k141_313578 | Not-determined |
| 17-108_k141_258513 | Not-determined |
| 17-108_k141_65260 | Low-quality |
| 17-108_k141_37687 | Not-determined |
| 17-108_k141_51723 | Not-determined |
| 17-108_k141_51724 | Not-determined |
| 17-108_k141_78315 | Not-determined |
| 17-108_k141_271525 | Low-quality |
| 17-108_k141_133056 | Not-determined |
| 17-108_k141_298951 | Not-determined |
| 17-108_k141_51732 | Not-determined |
| 17-108_k141_313598 | Not-determined |
| 17-108_k141_51735 | Not-determined |
| 17-108_k141_147927 | Not-determined |
| 17-108_k141_78326 | Not-determined |
| 17-108_k141_51738 | Low-quality |
| 17-108_k141_271536 | Not-determined |
| 17-108_k141_244995 | Not-determined |
| 17-108_k141_202810 | Low-quality |
| 17-108_k141_188758 | Not-determined |
| 17-108_k141_119735 | Not-determined |
| 17-108_k141_216933 | Not-determined |
| 17-108_k141_107112 | Not-determined |
| 17-108_k141_216937 | Not-determined |
| 17-108_k141_202819 | Not-determined |
| 17-108_k141_188763 | Not-determined |
| 17-108_k141_22597 | Not-determined |
| 17-108_k141_51751 | Low-quality |
| 17-108_k141_37712 | Not-determined |
| 17-108_k141_245007 | Not-determined |
| 17-108_k141_271544 | Not-determined |
| 17-108_k141_188770 | Not-determined |
| 17-108_k141_92708 | Low-quality |
| 17-108_k141_313619 | Low-quality |
| 17-108_k141_216951 | Not-determined |
| 17-108_k141_10371 | Not-determined |
| 17-108_k141_161750 | Not-determined |
| 17-108_k141_175714 | Not-determined |
| 17-108_k141_271551 | Not-determined |
| 17-108_k141_119752 | Not-determined |
| 17-108_k141_258559 | Not-determined |
| 17-108_k141_51765 | Not-determined |
| 17-108_k141_65302 | Not-determined |
| 17-108_k141_175725 | Not-determined |
| 17-108_k141_271559 | Not-determined |
| 17-108_k141_271561 | Not-determined |
| 17-108_k141_216972 | Low-quality |
| 17-108_k141_271565 | Not-determined |
| 17-108_k141_175729 | Not-determined |
| 17-108_k141_216983 | Not-determined |
| 17-108_k141_216985 | Not-determined |
| 17-108_k141_258577 | Not-determined |
| 17-108_k141_298990 | Not-determined |
| 17-108_k141_230702 | Not-determined |
| 17-108_k141_175735 | Low-quality |
| 17-108_k141_119772 | Not-determined |
| 17-108_k141_37750 | Low-quality |
| 17-108_k141_92743 | Not-determined |
| 17-108_k141_22647 | Not-determined |
| 17-108_k141_133070 | Not-determined |
| 17-108_k141_133074 | Not-determined |
| 17-108_k141_175743 | Not-determined |
| 17-108_k141_147965 | Not-determined |
| 17-108_k141_285933 | Not-determined |
| 17-108_k141_161782 | Not-determined |
| 17-108_k141_285935 | Low-quality |
| 17-108_k141_299006 | Low-quality |
| 17-108_k141_10393 | Not-determined |
| 17-108_k141_37761 | Not-determined |
| 17-108_k141_258593 | Low-quality |
| 17-108_k141_230715 | Not-determined |
| 17-108_k141_202864 | Not-determined |
| 17-108_k141_285939 | Not-determined |
| 17-108_k141_216999 | Not-determined |
| 17-108_k141_37767 | Not-determined |
| 17-108_k141_133087 | Not-determined |
| 17-108_k141_119798 | Not-determined |
| 17-108_k141_133089 | Not-determined |
| 17-108_k141_258605 | Not-determined |
| 17-108_k141_65331 | Not-determined |
| 17-108_k141_78396 | Not-determined |
| 17-108_k141_271593 | Not-determined |
| 17-108_k141_299016 | Not-determined |
| 17-108_k141_133097 | Not-determined |
| 17-108_k141_217004 | Not-determined |
| 17-108_k141_107160 | Not-determined |
| 17-108_k141_299020 | Not-determined |
| 17-108_k141_37775 | Not-determined |
| 17-108_k141_147992 | Not-determined |
| 17-108_k141_78412 | Not-determined |
| 17-108_k141_147995 | Not-determined |
| 17-108_k141_175770 | Not-determined |
| 17-108_k141_92775 | Low-quality |
| 17-108_k141_245045 | Not-determined |
| 17-108_k141_78419 | Not-determined |
| 17-108_k141_133106 | Not-determined |
| 17-108_k141_245047 | Not-determined |
| 17-108_k141_313679 | Not-determined |
| 17-108_k141_245051 | Not-determined |
| 17-108_k141_92780 | Not-determined |
| 17-108_k141_230740 | Not-determined |
| 17-108_k141_22662 | Not-determined |
| 17-108_k141_92783 | Not-determined |
| 17-108_k141_133113 | Not-determined |
| 17-108_k141_37804 | Low-quality |
| 17-108_k141_230745 | Not-determined |
| 17-108_k141_78428 | Not-determined |
| 17-108_k141_133115 | Not-determined |
| 17-108_k141_107178 | Low-quality |
| 17-108_k141_148006 | Not-determined |
| 17-108_k141_299047 | Not-determined |
| 17-108_k141_245059 | Not-determined |
| 17-108_k141_299054 | Not-determined |
| 17-108_k141_245062 | Not-determined |
| 17-108_k141_133130 | Not-determined |
| 17-108_k141_299058 | Low-quality |
| 17-108_k141_202923 | Not-determined |
| 17-108_k141_107189 | Not-determined |
| 17-108_k141_325879 | Low-quality |
| 17-108_k141_325881 | Not-determined |
| 17-108_k141_325883 | Not-determined |
| 17-108_k141_175782 | Not-determined |
| 17-108_k141_245078 | Low-quality |
| 17-108_k141_188839 | Not-determined |
| 17-108_k141_175789 | Not-determined |
| 17-108_k141_78442 | Not-determined |
| 17-108_k141_10465 | Not-determined |
| 17-108_k141_175792 | Not-determined |

| | |
|---|---|
| 17-108_k141_313710 | Not-determined |
| 17-108_k141_10474 | Not-determined |
| 17-108_k141_299088 | Not-determined |
| 17-108_k141_148037 | Low-quality |
| 17-108_k141_161811 | Not-determined |
| 17-108_k141_271644 | Not-determined |
| 17-108_k141_230776 | Not-determined |
| 17-108_k141_258633 | Not-determined |
| 17-108_k141_78459 | Not-determined |
| 17-108_k141_271649 | Not-determined |
| 17-108_k141_65393 | Not-determined |
| 17-108_k141_202962 | Not-determined |
| 17-108_k141_37858 | Not-determined |
| 17-108_k141_230785 | Not-determined |
| 17-108_k141_119805 | Not-determined |
| 17-108_k141_230793 | Not-determined |
| 17-108_k141_271658 | Not-determined |
| 17-108_k141_271659 | Not-determined |
| 17-108_k141_148047 | Not-determined |
| 17-108_k141_119811 | Not-determined |
| 17-108_k141_188856 | Not-determined |
| 17-108_k141_271669 | Not-determined |
| 17-108_k141_245118 | Not-determined |
| 17-108_k141_65414 | Not-determined |
| 17-108_k141_65415 | Low-quality |
| 17-108_k141_202984 | Not-determined |
| 17-108_k141_148056 | Not-determined |
| 17-108_k141_217072 | Not-determined |
| 17-108_k141_65419 | Not-determined |
| 17-108_k141_325904 | Not-determined |
| 17-108_k141_230804 | Not-determined |
| 17-108_k141_78484 | Not-determined |
| 17-108_k141_325910 | Not-determined |
| 17-108_k141_22725 | Not-determined |
| 17-108_k141_148070 | Not-determined |
| 17-108_k141_175839 | Not-determined |
| 17-108_k141_161849 | Not-determined |
| 17-108_k141_161850 | Not-determined |
| 17-108_k141_258682 | Low-quality |
| 17-108_k141_107237 | Not-determined |
| 17-108_k141_148086 | Not-determined |
| 17-108_k141_78509 | Not-determined |
| 17-108_k141_148089 | Not-determined |
| 17-108_k141_119841 | Not-determined |
| 17-108_k141_148091 | Not-determined |
| 17-108_k141_107240 | Not-determined |
| 17-108_k141_65451 | Not-determined |
| 17-108_k141_22747 | Not-determined |
| 17-108_k141_37890 | Not-determined |
| 17-108_k141_148099 | Not-determined |
| 17-108_k141_148101 | Not-determined |
| 17-108_k141_203001 | Not-determined |
| 17-108_k141_37895 | Not-determined |
| 17-108_k141_92798 | Not-determined |
| 17-108_k141_230825 | Not-determined |
| 17-108_k141_175870 | Not-determined |
| 17-108_k141_217081 | Not-determined |
| 17-108_k141_133152 | Not-determined |
| 17-108_k141_22760 | Low-quality |
| 17-108_k141_217084 | Not-determined |
| 17-108_k141_51894 | Not-determined |
| 17-108_k141_133167 | Not-determined |
| 17-108_k141_65471 | Not-determined |
| 17-108_k141_22776 | Low-quality |
| 17-108_k141_286043 | Not-determined |
| 17-108_k141_22782 | Not-determined |
| 17-108_k141_133175 | Not-determined |
| 17-108_k141_133176 | Not-determined |
| 17-108_k141_325935 | Not-determined |
| 17-108_k141_286045 | Not-determined |
| 17-108_k141_92820 | Not-determined |
| 17-108_k141_188913 | Not-determined |
| 17-108_k141_188916 | Not-determined |
| 17-108_k141_10511 | Not-determined |
| 17-108_k141_286051 | Not-determined |
| 17-108_k141_217116 | Low-quality |
| 17-108_k141_203028 | Not-determined |
| 17-108_k141_230851 | Not-determined |
| 17-108_k141_10516 | Not-determined |
| 17-108_k141_203032 | Not-determined |
| 17-108_k141_22789 | Not-determined |
| 17-108_k141_10521 | Low-quality |
| 17-108_k141_217121 | Not-determined |
| 17-108_k141_107265 | Not-determined |
| 17-108_k141_22797 | Not-determined |
| 17-108_k141_148128 | Not-determined |
| 17-108_k141_92852 | Not-determined |
| 17-108_k141_119876 | Not-determined |
| 17-108_k141_37933 | Not-determined |
| 17-108_k141_119882 | Not-determined |
| 17-108_k141_119884 | Not-determined |
| 17-108_k141_286065 | Not-determined |
| 17-108_k141_161909 | Not-determined |
| 17-108_k141_22805 | Not-determined |
| 17-108_k141_188942 | Not-determined |
| 17-108_k141_258706 | Not-determined |
| 17-108_k141_37941 | Not-determined |
| 17-108_k141_286074 | Not-determined |
| 17-108_k141_299197 | Not-determined |
| 17-108_k141_203053 | Not-determined |
| 17-108_k141_119896 | Not-determined |
| 17-108_k141_92889 | Not-determined |
| 17-108_k141_119900 | Not-determined |
| 17-108_k141_230858 | Not-determined |
| 17-108_k141_325949 | Not-determined |
| 17-108_k141_65523 | Not-determined |
| 17-108_k141_92898 | Not-determined |
| 17-108_k141_188973 | Not-determined |
| 17-108_k141_133211 | Low-quality |
| 17-108_k141_188974 | Not-determined |
| 17-108_k141_230867 | Not-determined |
| 17-108_k141_22810 | Not-determined |
| 17-108_k141_325956 | Not-determined |
| 17-108_k141_37977 | Not-determined |
| 17-108_k141_92910 | Not-determined |
| 17-108_k141_92912 | Not-determined |
| 17-108_k141_286096 | Not-determined |
| 17-108_k141_325966 | Not-determined |
| 17-108_k141_299209 | Not-determined |
| 17-108_k141_161953 | Not-determined |
| 17-108_k141_271752 | Not-determined |
| 17-108_k141_37993 | Not-determined |
| 17-108_k141_10558 | Not-determined |
| 17-108_k141_22832 | Not-determined |
| 17-108_k141_203098 | Not-determined |
| 17-108_k141_203102 | Not-determined |
| 17-108_k141_119922 | Not-determined |
| 17-108_k141_119924 | Not-determined |
| 17-108_k141_245205 | Low-quality |
| 17-108_k141_133246 | Low-quality |
| 17-108_k141_325968 | Not-determined |
| 17-108_k141_245212 | Not-determined |
| 17-108_k141_258739 | Not-determined |
| 17-108_k141_119926 | Not-determined |
| 17-108_k141_175923 | Not-determined |
| 17-108_k141_258742 | Not-determined |
| 17-108_k141_22848 | Not-determined |
| 17-108_k141_271764 | Not-determined |
| 17-108_k141_22850 | Not-determined |
| 17-108_k141_107320 | Not-determined |
| 17-108_k141_299242 | Not-determined |
| 17-108_k141_65556 | Not-determined |
| 17-108_k141_107326 | Not-determined |
| 17-108_k141_230905 | Not-determined |
| 17-108_k141_217188 | Not-determined |
| 17-108_k141_258761 | Not-determined |
| 17-108_k141_22857 | Not-determined |
| 17-108_k141_230910 | Not-determined |
| 17-108_k141_38038 | Low-quality |
| 17-108_k141_38039 | Not-determined |
| 17-108_k141_258765 | Not-determined |
| 17-108_k141_38044 | Not-determined |
| 17-108_k141_258768 | Not-determined |
| 17-108_k141_52018 | Low-quality |
| 17-108_k141_217196 | Not-determined |
| 17-108_k141_258772 | Not-determined |
| 17-108_k141_299273 | Not-determined |
| 17-108_k141_299278 | Not-determined |
| 17-108_k141_325984 | Not-determined |
| 17-108_k141_325988 | Not-determined |
| 17-108_k141_78562 | Not-determined |
| 17-108_k141_38065 | Not-determined |
| 17-108_k141_217209 | Not-determined |
| 17-108_k141_162007 | Not-determined |
| 17-108_k141_175960 | Not-determined |
| 17-108_k141_203115 | Not-determined |
| 17-108_k141_245235 | Not-determined |
| 17-108_k141_203119 | Not-determined |
| 17-108_k141_325998 | Not-determined |
| 17-108_k141_92959 | Not-determined |
| 17-108_k141_119962 | Not-determined |
| 17-108_k141_119963 | Not-determined |
| 17-108_k141_286137 | Not-determined |
| 17-108_k141_92964 | Not-determined |
| 17-108_k141_162016 | Not-determined |
| 17-108_k141_133276 | Not-determined |
| 17-108_k141_245258 | Not-determined |
| 17-108_k141_38070 | Not-determined |
| 17-108_k141_203145 | Not-determined |
| 17-108_k141_119974 | Not-determined |
| 17-108_k141_326002 | Low-quality |
| 17-108_k141_299308 | Not-determined |
| 17-108_k141_133289 | Not-determined |
| 17-108_k141_119983 | Not-determined |
| 17-108_k141_119984 | Low-quality |
| 17-108_k141_299313 | Not-determined |
| 17-108_k141_230940 | Not-determined |
| 17-108_k141_38080 | Not-determined |
| 17-108_k141_22877 | Low-quality |
| 17-108_k141_189035 | Not-determined |
| 17-108_k141_22879 | Not-determined |
| 17-108_k141_175969 | Not-determined |
| 17-108_k141_299316 | Not-determined |
| 17-108_k141_175972 | Low-quality |
| 17-108_k141_162032 | Not-determined |
| 17-108_k141_230949 | Not-determined |
| 17-108_k141_133296 | Not-determined |
| 17-108_k141_38086 | Not-determined |
| 17-108_k141_22882 | Not-determined |
| 17-108_k141_38091 | Not-determined |
| 17-108_k141_245288 | Not-determined |
| 17-108_k141_326005 | Not-determined |
| 17-108_k141_10578 | Not-determined |
| 17-108_k141_175984 | Not-determined |
| 17-108_k141_133301 | Not-determined |
| 17-108_k141_38093 | Not-determined |
| 17-108_k141_92998 | Not-determined |
| 17-108_k141_203170 | Low-quality |
| 17-108_k141_65653 | Not-determined |
| 17-108_k141_326014 | Not-determined |
| 17-108_k141_93004 | Low-quality |
| 17-108_k141_10588 | Not-determined |
| 17-108_k141_78616 | Not-determined |
| 17-108_k141_93007 | Not-determined |
| 17-108_k141_148158 | Not-determined |
| 17-108_k141_10595 | Not-determined |
| 17-108_k141_313868 | Not-determined |
| 17-108_k141_258827 | Low-quality |
| 17-108_k141_326029 | Not-determined |
| 17-108_k141_189067 | Not-determined |
| 17-108_k141_78626 | Not-determined |
| 17-108_k141_313878 | Not-determined |
| 17-108_k141_286170 | Not-determined |
| 17-108_k141_258836 | Not-determined |
| 17-108_k141_22917 | Not-determined |
| 17-108_k141_286182 | Not-determined |
| 17-108_k141_22921 | Not-determined |
| 17-108_k141_107396 | Not-determined |
| 17-108_k141_52088 | Not-determined |
| 17-108_k141_313893 | Low-quality |
| 17-108_k141_299354 | Not-determined |
| 17-108_k141_299357 | Not-determined |
| 17-108_k141_93048 | Not-determined |
| 17-108_k141_120021 | Not-determined |
| 17-108_k141_230985 | Low-quality |
| 17-108_k141_78649 | Not-determined |
| 17-108_k141_120026 | Not-determined |
| 17-108_k141_78656 | Not-determined |
| 17-108_k141_93050 | Not-determined |
| 17-108_k141_120031 | Not-determined |
| 17-108_k141_107431 | Not-determined |
| 17-108_k141_65692 | Not-determined |
| 17-108_k141_93056 | Not-determined |
| 17-108_k141_93057 | Not-determined |
| 17-108_k141_162095 | Not-determined |
| 17-108_k141_299392 | Not-determined |
| 17-108_k141_148210 | Not-determined |
| 17-108_k141_231017 | Not-determined |
| 17-108_k141_107437 | Low-quality |
| 17-108_k141_148216 | Not-determined |
| 17-108_k141_120041 | Not-determined |
| 17-108_k141_38175 | Not-determined |
| 17-108_k141_38176 | Not-determined |
| 17-108_k141_38177 | Not-determined |
| 17-108_k141_162120 | Not-determined |
| 17-108_k141_52128 | Not-determined |
| 17-108_k141_133361 | Low-quality |
| 17-108_k141_203245 | Not-determined |
| 17-108_k141_52131 | Not-determined |
| 17-108_k141_258893 | Not-determined |
| 17-108_k141_93075 | Not-determined |
| 17-108_k141_271806 | Not-determined |
| 17-108_k141_231031 | Not-determined |
| 17-108_k141_203249 | Not-determined |
| 17-108_k141_162134 | Not-determined |
| 17-108_k141_93082 | Low-quality |
| 17-108_k141_148228 | Not-determined |
| 17-108_k141_299422 | Low-quality |
| 17-108_k141_299424 | Not-determined |
| 17-108_k141_52143 | Not-determined |
| 17-108_k141_22943 | Not-determined |
| 17-108_k141_326044 | Low-quality |
| 17-108_k141_203259 | Low-quality |
| 17-108_k141_133381 | Not-determined |
| 17-108_k141_313899 | Not-determined |
| 17-108_k141_22951 | Not-determined |
| 17-108_k141_38210 | Not-determined |
| 17-108_k141_176089 | Not-determined |
| 17-108_k141_326056 | Not-determined |
| 17-108_k141_271830 | Not-determined |
| 17-108_k141_271831 | Low-quality |
| 17-108_k141_271834 | Not-determined |
| 17-108_k141_38222 | Low-quality |
| 17-108_k141_133388 | Not-determined |
| 17-108_k141_133389 | Not-determined |
| 17-108_k141_162158 | Not-determined |
| 17-108_k141_133395 | Not-determined |
| 17-108_k141_176103 | Not-determined |
| 17-108_k141_326067 | Low-quality |
| 17-108_k141_107484 | Not-determined |
| 17-108_k141_313921 | Not-determined |
| 17-108_k141_245381 | Not-determined |
| 17-108_k141_326069 | Not-determined |
| 17-108_k141_107487 | Not-determined |
| 17-108_k141_133404 | Not-determined |
| 17-108_k141_217291 | Not-determined |
| 17-108_k141_133405 | Not-determined |
| 17-108_k141_217292 | Not-determined |
| 17-108_k141_326073 | Low-quality |
| 17-108_k141_148245 | Low-quality |
| 17-108_k141_162177 | Not-determined |
| 17-108_k141_10629 | Not-determined |
| 17-108_k141_10631 | Low-quality |
| 17-108_k141_10632 | Not-determined |
| 17-108_k141_52181 | Not-determined |
| 17-108_k141_286267 | Low-quality |
| 17-108_k141_258927 | Not-determined |
| 17-108_k141_176121 | Not-determined |
| 17-108_k141_326085 | Not-determined |
| 17-108_k141_10642 | Not-determined |
| 17-108_k141_189086 | Not-determined |
| 17-108_k141_299472 | Not-determined |
| 17-108_k141_313936 | Not-determined |
| 17-108_k141_10643 | Low-quality |
| 17-108_k141_133444 | Not-determined |
| 17-108_k141_313941 | Not-determined |
| 17-108_k141_258936 | Not-determined |
| 17-108_k141_107528 | Not-determined |
| 17-108_k141_286280 | Not-determined |
| 17-108_k141_326093 | Not-determined |
| 17-108_k141_162197 | Not-determined |
| 17-108_k141_286284 | Not-determined |
| 17-108_k141_286285 | Not-determined |
| 17-108_k141_258945 | Not-determined |
| 17-108_k141_258949 | Not-determined |
| 17-108_k141_286292 | Low-quality |
| 17-108_k141_107548 | Not-determined |
| 17-108_k141_52214 | Not-determined |
| 17-108_k141_107551 | Low-quality |
| 17-108_k141_22994 | Low-quality |
| 17-108_k141_162209 | Not-determined |
| 17-108_k141_22999 | Low-quality |
| 17-108_k141_271886 | Not-determined |
| 17-108_k141_133470 | Not-determined |
| 17-108_k141_189118 | Not-determined |
| 17-108_k141_52228 | Not-determined |
| 17-108_k141_78703 | Low-quality |
| 17-108_k141_189121 | Not-determined |
| 17-108_k141_299501 | Not-determined |
| 17-108_k141_52235 | Not-determined |
| 17-108_k141_65744 | Low-quality |
| 17-108_k141_65745 | Not-determined |
| 17-108_k141_52237 | Low-quality |
| 17-108_k141_245433 | Not-determined |
| 17-108_k141_258982 | Not-determined |
| 17-108_k141_176128 | Low-quality |
| 17-108_k141_162240 | Not-determined |
| 17-108_k141_78724 | Not-determined |
| 17-108_k141_65769 | Not-determined |
| 17-108_k141_258992 | Not-determined |
| 17-108_k141_148282 | Low-quality |
| 17-108_k141_231107 | Not-determined |
| 17-108_k141_52249 | Not-determined |
| 17-108_k141_162249 | Not-determined |
| 17-108_k141_258996 | Not-determined |
| 17-108_k141_93146 | Not-determined |
| 17-108_k141_107584 | Not-determined |
| 17-108_k141_65778 | Not-determined |
| 17-108_k141_231113 | Not-determined |
| 17-108_k141_299524 | Low-quality |
| 17-108_k141_299530 | Not-determined |
| 17-108_k141_286323 | Low-quality |
| 17-108_k141_65787 | Not-determined |
| 17-108_k141_93166 | Not-determined |
| 17-108_k141_38248 | Not-determined |
| 17-108_k141_162271 | Not-determined |
| 17-108_k141_326137 | Low-quality |
| 17-108_k141_78742 | Not-determined |
| 17-108_k141_65819 | Not-determined |
| 17-108_k141_189161 | Not-determined |
| 17-108_k141_245472 | Not-determined |
| 17-108_k141_189167 | Low-quality |
| 17-108_k141_299562 | Not-determined |
| 17-108_k141_299565 | Not-determined |
| 17-108_k141_107618 | Not-determined |
| 17-108_k141_271934 | Low-quality |
| 17-108_k141_38275 | Not-determined |
| 17-108_k141_203325 | Not-determined |
| 17-108_k141_23041 | Not-determined |
| 17-108_k141_38280 | Not-determined |
| 17-108_k141_231150 | Not-determined |
| 17-108_k141_38282 | Not-determined |
| 17-108_k141_259055 | Low-quality |
| 17-108_k141_93211 | Not-determined |
| 17-108_k141_93212 | Not-determined |
| 17-108_k141_148310 | Not-determined |
| 17-108_k141_245491 | Not-determined |
| 17-108_k141_107633 | Not-determined |
| 17-108_k141_10687 | Not-determined |
| 17-108_k141_107639 | Not-determined |
| 17-108_k141_93226 | Not-determined |
| 17-108_k141_203340 | Not-determined |
| 17-108_k141_65840 | Low-quality |
| 17-108_k141_148329 | Not-determined |
| 17-108_k141_286368 | Not-determined |
| 17-108_k141_286370 | Not-determined |
| 17-108_k141_203348 | Not-determined |
| 17-108_k141_286373 | Not-determined |
| 17-108_k141_133535 | Not-determined |
| 17-108_k141_78765 | Not-determined |
| 17-108_k141_162280 | Not-determined |
| 17-108_k141_52281 | Not-determined |
| 17-108_k141_217340 | Not-determined |
| 17-108_k141_326172 | Not-determined |
| 17-108_k141_38314 | Low-quality |
| 17-108_k141_271955 | Not-determined |
| 17-108_k141_133553 | Not-determined |
| 17-108_k141_313949 | Not-determined |
| 17-108_k141_10720 | Not-determined |
| 17-108_k141_107663 | Not-determined |
| 17-108_k141_78785 | Not-determined |
| 17-108_k141_245524 | Not-determined |
| 17-108_k141_313956 | Not-determined |
| 17-108_k141_107672 | Not-determined |
| 17-108_k141_162301 | Low-quality |
| 17-108_k141_271970 | Low-quality |
| 17-108_k141_245533 | Not-determined |
| 17-108_k141_148349 | Not-determined |
| 17-108_k141_65871 | Not-determined |
| 17-108_k141_65872 | Not-determined |
| 17-108_k141_23058 | Not-determined |
| 17-108_k141_107691 | Not-determined |
| 17-108_k141_93274 | Not-determined |
| 17-108_k141_326203 | Not-determined |
| 17-108_k141_326204 | Not-determined |
| 17-108_k141_23076 | Not-determined |
| 17-108_k141_38325 | Not-determined |
| 17-108_k141_231187 | Not-determined |
| 17-108_k141_231190 | Not-determined |
| 17-108_k141_217387 | Not-determined |
| 17-108_k141_271996 | Not-determined |
| 17-108_k141_162334 | Not-determined |
| 17-108_k141_23086 | Not-determined |
| 17-108_k141_245556 | Not-determined |
| 17-108_k141_148369 | Not-determined |
| 17-108_k141_217403 | Not-determined |
| 17-108_k141_326225 | Not-determined |
| 17-108_k141_38329 | Low-quality |
| 17-108_k141_189201 | Not-determined |
| 17-108_k141_148380 | Not-determined |
| 17-108_k141_245564 | Not-determined |
| 17-108_k141_286429 | Not-determined |
| 17-108_k141_217411 | Not-determined |
| 17-108_k141_148386 | Not-determined |
| 17-108_k141_23106 | Not-determined |
| 17-108_k141_326233 | Not-determined |
| 17-108_k141_313990 | Not-determined |
| 17-108_k141_148390 | Not-determined |
| 17-108_k141_286436 | Not-determined |
| 17-108_k141_162354 | Not-determined |
| 17-108_k141_176242 | Not-determined |
| 17-108_k141_38352 | Not-determined |
| 17-108_k141_231222 | Not-determined |
| 17-108_k141_217424 | Not-determined |
| 17-108_k141_231226 | Not-determined |
| 17-108_k141_93321 | Not-determined |
| 17-108_k141_245596 | Not-determined |
| 17-108_k141_231234 | Not-determined |
| 17-108_k141_314006 | Not-determined |
| 17-108_k141_189232 | Not-determined |
| 17-108_k141_245606 | Not-determined |
| 17-108_k141_176255 | Not-determined |
| 17-108_k141_299595 | Not-determined |
| 17-108_k141_10753 | Not-determined |
| 17-108_k141_286455 | Not-determined |
| 17-108_k141_65923 | Not-determined |
| 17-108_k141_23135 | Not-determined |
| 17-108_k141_120061 | Not-determined |
| 17-108_k141_23143 | Not-determined |
| 17-108_k141_107705 | Not-determined |
| 17-108_k141_10765 | Low-quality |
| 17-108_k141_120070 | Not-determined |
| 17-108_k141_10768 | Not-determined |
| 17-108_k141_231254 | Not-determined |
| 17-108_k141_189256 | Not-determined |
| 17-108_k141_245620 | Not-determined |
| 17-108_k141_217465 | Not-determined |
| 17-108_k141_176288 | Not-determined |
| 17-108_k141_93353 | Not-determined |
| 17-108_k141_314028 | Not-determined |
| 17-108_k141_326266 | Low-quality |
| 17-108_k141_162396 | Not-determined |
| 17-108_k141_38411 | Not-determined |
| 17-108_k141_203357 | Not-determined |
| 17-108_k141_120085 | Not-determined |
| 17-108_k141_189293 | Not-determined |
| 17-108_k141_133673 | Not-determined |
| 17-108_k141_189299 | Low-quality |
| 17-108_k141_162416 | Not-determined |
| 17-108_k141_314055 | Low-quality |
| 17-108_k141_231268 | Not-determined |
| 17-108_k141_107739 | Not-determined |
| 17-108_k141_231274 | Not-determined |
| 17-108_k141_286497 | Not-determined |
| 17-108_k141_93385 | Not-determined |
| 17-108_k141_203377 | Not-determined |
| 17-108_k141_93386 | Not-determined |
| 17-108_k141_107744 | Low-quality |
| 17-108_k141_314064 | Not-determined |
| 17-108_k141_286499 | Not-determined |
| 17-108_k141_245645 | Low-quality |
| 17-108_k141_65988 | Not-determined |
| 17-108_k141_162439 | Not-determined |
| 17-108_k141_314070 | Not-determined |
| 17-108_k141_259105 | Not-determined |
| 17-108_k141_217497 | Not-determined |
| 17-108_k141_52349 | Not-determined |
| 17-108_k141_52352 | Not-determined |
| 17-108_k141_176318 | Not-determined |
| 17-108_k141_299650 | Low-quality |
| 17-108_k141_66005 | Not-determined |
| 17-108_k141_93406 | Not-determined |
| 17-108_k141_272035 | Not-determined |
| 17-108_k141_217501 | Not-determined |
| 17-108_k141_286507 | Not-determined |
| 17-108_k141_286513 | Not-determined |
| 17-108_k141_259127 | Low-quality |
| 17-108_k141_133717 | Low-quality |
| 17-108_k141_120114 | Low-quality |
| 17-108_k141_314098 | Not-determined |
| 17-108_k141_176327 | Not-determined |
| 17-108_k141_326303 | Low-quality |
| 17-108_k141_38430 | Not-determined |
| 17-108_k141_286518 | Low-quality |
| 17-108_k141_272042 | Not-determined |
| 17-108_k141_217516 | Not-determined |
| 17-108_k141_326311 | Low-quality |
| 17-108_k141_217518 | Not-determined |
| 17-108_k141_259143 | Not-determined |
| 17-108_k141_52373 | Not-determined |
| 17-108_k141_52379 | Not-determined |
| 17-108_k141_52382 | Not-determined |
| 17-108_k141_78804 | Low-quality |
| 17-108_k141_66046 | Not-determined |
| 17-108_k141_217532 | Low-quality |
| 17-108_k141_162471 | Not-determined |
| 17-108_k141_52386 | Not-determined |
| 17-108_k141_299693 | Not-determined |
| 17-108_k141_66050 | Not-determined |
| 17-108_k141_107769 | Not-determined |
| 17-108_k141_133739 | Not-determined |
| 17-108_k141_217536 | Not-determined |
| 17-108_k141_162479 | Not-determined |
| 17-108_k141_176358 | Not-determined |
| 17-108_k141_259161 | Not-determined |
| 17-108_k141_10805 | Not-determined |
| 17-108_k141_23215 | Low-quality |
| 17-108_k141_38455 | Not-determined |
| 17-108_k141_52400 | Not-determined |
| 17-108_k141_176360 | Not-determined |
| 17-108_k141_52401 | Not-determined |
| 17-108_k141_93445 | Not-determined |
| 17-108_k141_107781 | Not-determined |
| 17-108_k141_272047 | Not-determined |
| 17-108_k141_120150 | Low-quality |
| 17-108_k141_38465 | Not-determined |
| 17-108_k141_107792 | Not-determined |
| 17-108_k141_10822 | Not-determined |
| 17-108_k141_120159 | Not-determined |
| 17-108_k141_314126 | Not-determined |
| 17-108_k141_176375 | Not-determined |
| 17-108_k141_148497 | Not-determined |
| 17-108_k141_299716 | Not-determined |
| 17-108_k141_23227 | Not-determined |
| 17-108_k141_23234 | Not-determined |
| 17-108_k141_162485 | Not-determined |
| 17-108_k141_162487 | Not-determined |
| 17-108_k141_203424 | Not-determined |
| 17-108_k141_120180 | Not-determined |
| 17-108_k141_38489 | Not-determined |
| 17-108_k141_217562 | Not-determined |
| 17-108_k141_259197 | Not-determined |
| 17-108_k141_217565 | Not-determined |
| 17-108_k141_286547 | Not-determined |
| 17-108_k141_10849 | Not-determined |
| 17-108_k141_23249 | Not-determined |
| 17-108_k141_259203 | Not-determined |
| 17-108_k141_23251 | Low-quality |
| 17-108_k141_93469 | Low-quality |
| 17-108_k141_217570 | Not-determined |
| 17-108_k141_78857 | Not-determined |
| 17-108_k141_162498 | Not-determined |
| 17-108_k141_162499 | Not-determined |
| 17-108_k141_120187 | Not-determined |
| 17-108_k141_38504 | Not-determined |
| 17-108_k141_120188 | Not-determined |
| 17-108_k141_78864 | Not-determined |
| 17-108_k141_52433 | Not-determined |
| 17-108_k141_286558 | Not-determined |

| | |
|---|---|
| 17-108_k141_162506 | Not-determined |
| 17-108_k141_120194 | Not-determined |
| 17-108_k141_133795 | Not-determined |
| 17-108_k141_231306 | Not-determined |
| 17-108_k141_52441 | Not-determined |
| 17-108_k141_189316 | Not-determined |
| 17-108_k141_66111 | Low-quality |
| 17-108_k141_66114 | Not-determined |
| 17-108_k141_245691 | Not-determined |
| 17-108_k141_78886 | Not-determined |
| 17-108_k141_203443 | Not-determined |
| 17-108_k141_245699 | Not-determined |
| 17-108_k141_245700 | Low-quality |
| 17-108_k141_93500 | Not-determined |
| 17-108_k141_107818 | Not-determined |
| 17-108_k141_23265 | Not-determined |
| 17-108_k141_259221 | Not-determined |
| 17-108_k141_259223 | Low-quality |
| 17-108_k141_10877 | Low-quality |
| 17-108_k141_23272 | Not-determined |
| 17-108_k141_259227 | Not-determined |
| 17-108_k141_93508 | Not-determined |
| 17-108_k141_245714 | Not-determined |
| 17-108_k141_148558 | Not-determined |
| 17-108_k141_286592 | Not-determined |
| 17-108_k141_286595 | Not-determined |
| 17-108_k141_162534 | Low-quality |
| 17-108_k141_272099 | Low-quality |
| 17-108_k141_217590 | Not-determined |
| 17-108_k141_203465 | Not-determined |
| 17-108_k141_203467 | Low-quality |
| 17-108_k141_148563 | Not-determined |
| 17-108_k141_52477 | Not-determined |
| 17-108_k141_299766 | Low-quality |
| 17-108_k141_286604 | Not-determined |
| 17-108_k141_52485 | Not-determined |
| 17-108_k141_259253 | Not-determined |
| 17-108_k141_148577 | Not-determined |
| 17-108_k141_217596 | Not-determined |
| 17-108_k141_326355 | Not-determined |
| 17-108_k141_314171 | Low-quality |
| 17-108_k141_189355 | Not-determined |
| 17-108_k141_52502 | Not-determined |
| 17-108_k141_286614 | Not-determined |
| 17-108_k141_314182 | Not-determined |
| 17-108_k141_162563 | Not-determined |
| 17-108_k141_314185 | Not-determined |
| 17-108_k141_272105 | Not-determined |
| 17-108_k141_93539 | Not-determined |
| 17-108_k141_10904 | Not-determined |
| 17-108_k141_10905 | Not-determined |
| 17-108_k141_231351 | Not-determined |
| 17-108_k141_148595 | Not-determined |
| 17-108_k141_10911 | Not-determined |
| 17-108_k141_93549 | Not-determined |
| 17-108_k141_148602 | Low-quality |
| 17-108_k141_314201 | Low-quality |
| 17-108_k141_314202 | Not-determined |
| 17-108_k141_133857 | Low-quality |
| 17-108_k141_326390 | Not-determined |
| 17-108_k141_107851 | Not-determined |
| 17-108_k141_162577 | Not-determined |
| 17-108_k141_133860 | Not-determined |
| 17-108_k141_314208 | Not-determined |
| 17-108_k141_259281 | Not-determined |
| 17-108_k141_78943 | Not-determined |
| 17-108_k141_107863 | Not-determined |
| 17-108_k141_107866 | Not-determined |
| 17-108_k141_203490 | Not-determined |
| 17-108_k141_189368 | Not-determined |
| 17-108_k141_272133 | Not-determined |
| 17-108_k141_259294 | Not-determined |
| 17-108_k141_133875 | Not-determined |
| 17-108_k141_231368 | Not-determined |
| 17-108_k141_272146 | Not-determined |
| 17-108_k141_93560 | Not-determined |
| 17-108_k141_107875 | Not-determined |
| 17-108_k141_189371 | Not-determined |
| 17-108_k141_10943 | Not-determined |
| 17-108_k141_326412 | Low-quality |
| 17-108_k141_133887 | Not-determined |
| 17-108_k141_38531 | Not-determined |
| 17-108_k141_38532 | Not-determined |
| 17-108_k141_52544 | Not-determined |
| 17-108_k141_259303 | Not-determined |
| 17-108_k141_162595 | Not-determined |
| 17-108_k141_93566 | Not-determined |
| 17-108_k141_299810 | Not-determined |
| 17-108_k141_133904 | Not-determined |
| 17-108_k141_38541 | Low-quality |
| 17-108_k141_10959 | Not-determined |
| 17-108_k141_299818 | Low-quality |
| 17-108_k141_245804 | Not-determined |
| 17-108_k141_245806 | Not-determined |
| 17-108_k141_148611 | Not-determined |
| 17-108_k141_176416 | Not-determined |
| 17-108_k141_176419 | Not-determined |
| 17-108_k141_38547 | Not-determined |
| 17-108_k141_217619 | Not-determined |
| 17-108_k141_133921 | Not-determined |
| 17-108_k141_38554 | Not-determined |
| 17-108_k141_272170 | Not-determined |
| 17-108_k141_231403 | Low-quality |
| 17-108_k141_93570 | Not-determined |
| 17-108_k141_259324 | Not-determined |
| 17-108_k141_217624 | Not-determined |
| 17-108_k141_217626 | Not-determined |
| 17-108_k141_259334 | Not-determined |
| 17-108_k141_203531 | Not-determined |
| 17-108_k141_259336 | Not-determined |
| 17-108_k141_203532 | Not-determined |
| 17-108_k141_93589 | Not-determined |
| 17-108_k141_78996 | Low-quality |
| 17-108_k141_326450 | Not-determined |
| 17-108_k141_272181 | Not-determined |
| 17-108_k141_259342 | Not-determined |
| 17-108_k141_272190 | Not-determined |
| 17-108_k141_11008 | Not-determined |
| 17-108_k141_107932 | Not-determined |
| 17-108_k141_107937 | Not-determined |
| 17-108_k141_23359 | Not-determined |
| 17-108_k141_189401 | Not-determined |
| 17-108_k141_189403 | Low-quality |
| 17-108_k141_176451 | Not-determined |
| 17-108_k141_286680 | Not-determined |
| 17-108_k141_326461 | Not-determined |
| 17-108_k141_107943 | Not-determined |
| 17-108_k141_272204 | Not-determined |
| 17-108_k141_107950 | Not-determined |
| 17-108_k141_79023 | Not-determined |
| 17-108_k141_52597 | Not-determined |
| 17-108_k141_189408 | Low-quality |
| 17-108_k141_176466 | Not-determined |
| 17-108_k141_162638 | Not-determined |
| 17-108_k141_11030 | Not-determined |
| 17-108_k141_189417 | Not-determined |
| 17-108_k141_162643 | Not-determined |
| 17-108_k141_66174 | Not-determined |
| 17-108_k141_107955 | Not-determined |
| 17-108_k141_79032 | Not-determined |
| 17-108_k141_286706 | Not-determined |
| 17-108_k141_299864 | Not-determined |
| 17-108_k141_79039 | Not-determined |
| 17-108_k141_107964 | Not-determined |
| 17-108_k141_23378 | Not-determined |
| 17-108_k141_162670 | Not-determined |
| 17-108_k141_231440 | Not-determined |
| 17-108_k141_299875 | Not-determined |
| 17-108_k141_299876 | Not-determined |
| 17-108_k141_107971 | Not-determined |
| 17-108_k141_11041 | Not-determined |
| 17-108_k141_231448 | Not-determined |
| 17-108_k141_259357 | Not-determined |
| 17-108_k141_326498 | Not-determined |
| 17-108_k141_259360 | Not-determined |
| 17-108_k141_203593 | Not-determined |
| 17-108_k141_259362 | Not-determined |
| 17-108_k141_217663 | Low-quality |
| 17-108_k141_259365 | Not-determined |
| 17-108_k141_52641 | Not-determined |
| 17-108_k141_259374 | Not-determined |
| 17-108_k141_176488 | Low-quality |
| 17-108_k141_231470 | Not-determined |
| 17-108_k141_217682 | Not-determined |
| 17-108_k141_176490 | Not-determined |
| 17-108_k141_217683 | Not-determined |
| 17-108_k141_299890 | Not-determined |
| 17-108_k141_217687 | Not-determined |
| 17-108_k141_217689 | Not-determined |
| 17-108_k141_66188 | Not-determined |
| 17-108_k141_120259 | Not-determined |
| 17-108_k141_162706 | Not-determined |
| 17-108_k141_79060 | Not-determined |
| 17-108_k141_176498 | Not-determined |
| 17-108_k141_79062 | Not-determined |
| 17-108_k141_162709 | Not-determined |
| 17-108_k141_133991 | Not-determined |
| 17-108_k141_52677 | Not-determined |
| 17-108_k141_245827 | Not-determined |
| 17-108_k141_133995 | Not-determined |
| 17-108_k141_162716 | Not-determined |
| 17-108_k141_259390 | Low-quality |
| 17-108_k141_11073 | Not-determined |
| 17-108_k141_231490 | Not-determined |
| 17-108_k141_314239 | Low-quality |
| 17-108_k141_203627 | Not-determined |
| 17-108_k141_259397 | Not-determined |
| 17-108_k141_38655 | Not-determined |
| 17-108_k141_134004 | Not-determined |
| 17-108_k141_79076 | Not-determined |
| 17-108_k141_203635 | Not-determined |
| 17-108_k141_107985 | Not-determined |
| 17-108_k141_52694 | Not-determined |
| 17-108_k141_107986 | Not-determined |
| 17-108_k141_79085 | Not-determined |
| 17-108_k141_52697 | Not-determined |
| 17-108_k141_231512 | Not-determined |
| 17-108_k141_52699 | Low-quality |
| 17-108_k141_314247 | Not-determined |
| 17-108_k141_134017 | Not-determined |
| 17-108_k141_93673 | Not-determined |
| 17-108_k141_189474 | Not-determined |
| 17-108_k141_286721 | Not-determined |
| 17-108_k141_299935 | Not-determined |
| 17-108_k141_148632 | Low-quality |
| 17-108_k141_299944 | Not-determined |
| 17-108_k141_299946 | Not-determined |
| 17-108_k141_259408 | Not-determined |
| 17-108_k141_203662 | Not-determined |
| 17-108_k141_203663 | Not-determined |
| 17-108_k141_203664 | Not-determined |
| 17-108_k141_245860 | Low-quality |
| 17-108_k141_162751 | Not-determined |
| 17-108_k141_299953 | Not-determined |
| 17-108_k141_134024 | Not-determined |
| 17-108_k141_259414 | Not-determined |
| 17-108_k141_107990 | Not-determined |
| 17-108_k141_259417 | Not-determined |
| 17-108_k141_259418 | Not-determined |
| 17-108_k141_107995 | Not-determined |
| 17-108_k141_38687 | Not-determined |
| 17-108_k141_107997 | Not-determined |
| 17-108_k141_286748 | Not-determined |
| 17-108_k141_107999 | Not-determined |
| 17-108_k141_231533 | Not-determined |
| 17-108_k141_162769 | Not-determined |
| 17-108_k141_134033 | Not-determined |
| 17-108_k141_108004 | Not-determined |
| 17-108_k141_203693 | Not-determined |
| 17-108_k141_23419 | Low-quality |
| 17-108_k141_38703 | Not-determined |
| 17-108_k141_259421 | Not-determined |
| 17-108_k141_120305 | Not-determined |
| 17-108_k141_23426 | Not-determined |
| 17-108_k141_148642 | Not-determined |
| 17-108_k141_162780 | Not-determined |
| 17-108_k141_314263 | Not-determined |
| 17-108_k141_38711 | Not-determined |
| 17-108_k141_134049 | Low-quality |
| 17-108_k141_52749 | Not-determined |
| 17-108_k141_134051 | Not-determined |
| 17-108_k141_38718 | Not-determined |
| 17-108_k141_299956 | Not-determined |
| 17-108_k141_148657 | Not-determined |
| 17-108_k141_217700 | Not-determined |
| 17-108_k141_11122 | Not-determined |
| 17-108_k141_11125 | Not-determined |
| 17-108_k141_176566 | Not-determined |
| 17-108_k141_299961 | Not-determined |
| 17-108_k141_299963 | Not-determined |
| 17-108_k141_52760 | Not-determined |
| 17-108_k141_134068 | Not-determined |
| 17-108_k141_120346 | Not-determined |
| 17-108_k141_299979 | Not-determined |
| 17-108_k141_162792 | Not-determined |
| 17-108_k141_120353 | Not-determined |
| 17-108_k141_203737 | Not-determined |
| 17-108_k141_162798 | Not-determined |
| 17-108_k141_134081 | Not-determined |
| 17-108_k141_79134 | Not-determined |
| 17-108_k141_23470 | Not-determined |
| 17-108_k141_245914 | Not-determined |
| 17-108_k141_120365 | Not-determined |
| 17-108_k141_120369 | Not-determined |
| 17-108_k141_148690 | Not-determined |
| 17-108_k141_286783 | Not-determined |
| 17-108_k141_11153 | Not-determined |
| 17-108_k141_176599 | Not-determined |
| 17-108_k141_272298 | Not-determined |
| 17-108_k141_259436 | Low-quality |
| 17-108_k141_134108 | Not-determined |
| 17-108_k141_272302 | Not-determined |
| 17-108_k141_299994 | Not-determined |
| 17-108_k141_52809 | Not-determined |
| 17-108_k141_217739 | Not-determined |
| 17-108_k141_38751 | Low-quality |
| 17-108_k141_176604 | Not-determined |
| 17-108_k141_11162 | Not-determined |
| 17-108_k141_23477 | Not-determined |
| 17-108_k141_23479 | Not-determined |
| 17-108_k141_52813 | Not-determined |
| 17-108_k141_259448 | Not-determined |
| 17-108_k141_38765 | Not-determined |
| 17-108_k141_66258 | Not-determined |
| 17-108_k141_38772 | Not-determined |
| 17-108_k141_176612 | Not-determined |
| 17-108_k141_231620 | Low-quality |
| 17-108_k141_176615 | Not-determined |
| 17-108_k141_272317 | Low-quality |
| 17-108_k141_38777 | Low-quality |
| 17-108_k141_259459 | Low-quality |
| 17-108_k141_79182 | Not-determined |
| 17-108_k141_314313 | Not-determined |
| 17-108_k141_120379 | Not-determined |
| 17-108_k141_38782 | Not-determined |
| 17-108_k141_93732 | Not-determined |
| 17-108_k141_162849 | Not-determined |
| 17-108_k141_300025 | Not-determined |
| 17-108_k141_52820 | Not-determined |
| 17-108_k141_148728 | Not-determined |
| 17-108_k141_66272 | Not-determined |
| 17-108_k141_66275 | Not-determined |
| 17-108_k141_272332 | Low-quality |
| 17-108_k141_326536 | Not-determined |
| 17-108_k141_176637 | Low-quality |
| 17-108_k141_259472 | Not-determined |
| 17-108_k141_108072 | Not-determined |
| 17-108_k141_52832 | Not-determined |
| 17-108_k141_176640 | Not-determined |
| 17-108_k141_231650 | Not-determined |
| 17-108_k141_108073 | Not-determined |
| 17-108_k141_272336 | Not-determined |
| 17-108_k141_231653 | Not-determined |
| 17-108_k141_38805 | Not-determined |
| 17-108_k141_286809 | Not-determined |
| 17-108_k141_176650 | Not-determined |
| 17-108_k141_176655 | Not-determined |
| 17-108_k141_93761 | Not-determined |
| 17-108_k141_134140 | Low-quality |
| 17-108_k141_245989 | Not-determined |
| 17-108_k141_300054 | Not-determined |
| 17-108_k141_148749 | Not-determined |
| 17-108_k141_286822 | Not-determined |
| 17-108_k141_286825 | Not-determined |
| 17-108_k141_108090 | Low-quality |
| 17-108_k141_23506 | Not-determined |
| 17-108_k141_108091 | Not-determined |
| 17-108_k141_314336 | Not-determined |
| 17-108_k141_108093 | Not-determined |
| 17-108_k141_11186 | Not-determined |
| 17-108_k141_148758 | Not-determined |
| 17-108_k141_314339 | Not-determined |
| 17-108_k141_93767 | Not-determined |
| 17-108_k141_11190 | Not-determined |
| 17-108_k141_326569 | Not-determined |
| 17-108_k141_231676 | Not-determined |
| 17-108_k141_66315 | Not-determined |
| 17-108_k141_11196 | Not-determined |
| 17-108_k141_108102 | Not-determined |
| 17-108_k141_23523 | Not-determined |
| 17-108_k141_23527 | Not-determined |
| 17-108_k141_259511 | Not-determined |
| 17-108_k141_300066 | Not-determined |
| 17-108_k141_11202 | Not-determined |
| 17-108_k141_231697 | Not-determined |
| 17-108_k141_203776 | Not-determined |
| 17-108_k141_120422 | Low-quality |
| 17-108_k141_11210 | Not-determined |
| 17-108_k141_11212 | Not-determined |
| 17-108_k141_203783 | Not-determined |
| 17-108_k141_134160 | Not-determined |
| 17-108_k141_189584 | Not-determined |
| 17-108_k141_272364 | Not-determined |
| 17-108_k141_286868 | Not-determined |
| 17-108_k141_246038 | Not-determined |
| 17-108_k141_231716 | Not-determined |
| 17-108_k141_286874 | Not-determined |
| 17-108_k141_326606 | Not-determined |
| 17-108_k141_52896 | Low-quality |
| 17-108_k141_326608 | Not-determined |
| 17-108_k141_259536 | Not-determined |
| 17-108_k141_148811 | Not-determined |
| 17-108_k141_23539 | Not-determined |
| 17-108_k141_162902 | Not-determined |
| 17-108_k141_162906 | Not-determined |
| 17-108_k141_326615 | Not-determined |
| 17-108_k141_23543 | Not-determined |
| 17-108_k141_176692 | Not-determined |
| 17-108_k141_108112 | Not-determined |
| 17-108_k141_176695 | Not-determined |
| 17-108_k141_108115 | Not-determined |
| 17-108_k141_23550 | Not-determined |
| 17-108_k141_286898 | Not-determined |
| 17-108_k141_314365 | Not-determined |
| 17-108_k141_134180 | Not-determined |
| 17-108_k141_314369 | Not-determined |
| 17-108_k141_120444 | Not-determined |
| 17-108_k141_286903 | Not-determined |
| 17-108_k141_300104 | Not-determined |
| 17-108_k141_38862 | Low-quality |
| 17-108_k141_148829 | Not-determined |
| 17-108_k141_120449 | Not-determined |
| 17-108_k141_79242 | Not-determined |
| 17-108_k141_326627 | Not-determined |
| 17-108_k141_203817 | Not-determined |
| 17-108_k141_326630 | Not-determined |
| 17-108_k141_231726 | Not-determined |
| 17-108_k141_300109 | Not-determined |
| 17-108_k141_231729 | Not-determined |
| 17-108_k141_326635 | Not-determined |
| 17-108_k141_134189 | Not-determined |
| 17-108_k141_79249 | Not-determined |
| 17-108_k141_148832 | Not-determined |
| 17-108_k141_203829 | Not-determined |
| 17-108_k141_148835 | Not-determined |
| 17-108_k141_79251 | Not-determined |
| 17-108_k141_148836 | Not-determined |
| 17-108_k141_66318 | Not-determined |
| 17-108_k141_79256 | Not-determined |
| 17-108_k141_66322 | Not-determined |
| 17-108_k141_259575 | Not-determined |
| 17-108_k141_203839 | Not-determined |
| 17-108_k141_326644 | Not-determined |
| 17-108_k141_108143 | Not-determined |
| 17-108_k141_148842 | Not-determined |
| 17-108_k141_108145 | Not-determined |
| 17-108_k141_120484 | Not-determined |
| 17-108_k141_259587 | Low-quality |
| 17-108_k141_314376 | Low-quality |
| 17-108_k141_93786 | Not-determined |
| 17-108_k141_314380 | Not-determined |
| 17-108_k141_231734 | Not-determined |
| 17-108_k141_314381 | Not-determined |
| 17-108_k141_93789 | Low-quality |
| 17-108_k141_272391 | Not-determined |
| 17-108_k141_189623 | Not-determined |
| 17-108_k141_108156 | Not-determined |
| 17-108_k141_66341 | Not-determined |
| 17-108_k141_93798 | Not-determined |
| 17-108_k141_108165 | Not-determined |
| 17-108_k141_326661 | Not-determined |
| 17-108_k141_148855 | Not-determined |
| 17-108_k141_326666 | Not-determined |
| 17-108_k141_286949 | Not-determined |
| 17-108_k141_148875 | Not-determined |
| 17-108_k141_148879 | Not-determined |
| 17-108_k141_300158 | Not-determined |
| 17-108_k141_286956 | Not-determined |
| 17-108_k141_120505 | Not-determined |
| 17-108_k141_66364 | Not-determined |
| 17-108_k141_286962 | Not-determined |
| 17-108_k141_66367 | Low-quality |
| 17-108_k141_217794 | Not-determined |
| 17-108_k141_217796 | Not-determined |
| 17-108_k141_326681 | Low-quality |
| 17-108_k141_231758 | Not-determined |
| 17-108_k141_38902 | Low-quality |
| 17-108_k141_231770 | Not-determined |
| 17-108_k141_326684 | Low-quality |
| 17-108_k141_66380 | Not-determined |
| 17-108_k141_79316 | Not-determined |
| 17-108_k141_272433 | Not-determined |
| 17-108_k141_314401 | Not-determined |
| 17-108_k141_79320 | Not-determined |
| 17-108_k141_300179 | Not-determined |
| 17-108_k141_79324 | Not-determined |
| 17-108_k141_286979 | Not-determined |
| 17-108_k141_246076 | Not-determined |
| 17-108_k141_79328 | Low-quality |
| 17-108_k141_246079 | Not-determined |
| 17-108_k141_79329 | Not-determined |
| 17-108_k141_286984 | Not-determined |
| 17-108_k141_300190 | Not-determined |
| 17-108_k141_231796 | Not-determined |
| 17-108_k141_272461 | Not-determined |
| 17-108_k141_11284 | Not-determined |
| 17-108_k141_11285 | Not-determined |
| 17-108_k141_287001 | Not-determined |
| 17-108_k141_176723 | Not-determined |
| 17-108_k141_231802 | Low-quality |
| 17-108_k141_162976 | Not-determined |
| 17-108_k141_203924 | Not-determined |
| 17-108_k141_272483 | Not-determined |
| 17-108_k141_176737 | Not-determined |
| 17-108_k141_176742 | Not-determined |
| 17-108_k141_300216 | Not-determined |
| 17-108_k141_272489 | Not-determined |
| 17-108_k141_176744 | Low-quality |
| 17-108_k141_176745 | Not-determined |
| 17-108_k141_66401 | Not-determined |
| 17-108_k141_287017 | Not-determined |
| 17-108_k141_66408 | Not-determined |
| 17-108_k141_259613 | Not-determined |
| 17-108_k141_231812 | Not-determined |
| 17-108_k141_120525 | Not-determined |
| 17-108_k141_38940 | Not-determined |
| 17-108_k141_66411 | Not-determined |
| 17-108_k141_66412 | Not-determined |
| 17-108_k141_79362 | Not-determined |
| 17-108_k141_52951 | Not-determined |
| 17-108_k141_148902 | Not-determined |
| 17-108_k141_11321 | Not-determined |
| 17-108_k141_11323 | Not-determined |
| 17-108_k141_108222 | Not-determined |
| 17-108_k141_287046 | Not-determined |
| 17-108_k141_231827 | Not-determined |
| 17-108_k141_203943 | Not-determined |
| 17-108_k141_108228 | Not-determined |
| 17-108_k141_176764 | Not-determined |
| 17-108_k141_11331 | Not-determined |
| 17-108_k141_108236 | Not-determined |
| 17-108_k141_176773 | Low-quality |
| 17-108_k141_23575 | Not-determined |
| 17-108_k141_176776 | Not-determined |
| 17-108_k141_23577 | Not-determined |
| 17-108_k141_176778 | Not-determined |
| 17-108_k141_11338 | Not-determined |
| 17-108_k141_23584 | Not-determined |
| 17-108_k141_246136 | Not-determined |
| 17-108_k141_66448 | Not-determined |
| 17-108_k141_52993 | Not-determined |
| 17-108_k141_326709 | Not-determined |
| 17-108_k141_300254 | Not-determined |
| 17-108_k141_272525 | Not-determined |
| 17-108_k141_23607 | Not-determined |
| 17-108_k141_52999 | Not-determined |
| 17-108_k141_203979 | Not-determined |
| 17-108_k141_53001 | Not-determined |
| 17-108_k141_246156 | Low-quality |
| 17-108_k141_272533 | Not-determined |
| 17-108_k141_120562 | Not-determined |
| 17-108_k141_326721 | Not-determined |
| 17-108_k141_326723 | Low-quality |
| 17-108_k141_246159 | Not-determined |
| 17-108_k141_66468 | Not-determined |
| 17-108_k141_79393 | Not-determined |
| 17-108_k141_79394 | Low-quality |
| 17-108_k141_120570 | Not-determined |
| 17-108_k141_79395 | Low-quality |
| 17-108_k141_203999 | Not-determined |
| 17-108_k141_326732 | Not-determined |
| 17-108_k141_53014 | Not-determined |
| 17-108_k141_66478 | Not-determined |
| 17-108_k141_148956 | Not-determined |
| 17-108_k141_93876 | Not-determined |
| 17-108_k141_93878 | Not-determined |
| 17-108_k141_231861 | Low-quality |
| 17-108_k141_79408 | Low-quality |
| 17-108_k141_108286 | Not-determined |
| 17-108_k141_120589 | Not-determined |
| 17-108_k141_231870 | Not-determined |
| 17-108_k141_189688 | Not-determined |
| 17-108_k141_108297 | Not-determined |
| 17-108_k141_23633 | Not-determined |
| 17-108_k141_189694 | Not-determined |
| 17-108_k141_259690 | Not-determined |
| 17-108_k141_79422 | Low-quality |
| 17-108_k141_108305 | Not-determined |
| 17-108_k141_134243 | Not-determined |
| 17-108_k141_53033 | Not-determined |
| 17-108_k141_66517 | Not-determined |
| 17-108_k141_326764 | Low-quality |
| 17-108_k141_204045 | Low-quality |
| 17-108_k141_134254 | Not-determined |
| 17-108_k141_204051 | Not-determined |
| 17-108_k141_189715 | Not-determined |
| 17-108_k141_246210 | Not-determined |
| 17-108_k141_326774 | Not-determined |
| 17-108_k141_163035 | Not-determined |
| 17-108_k141_204060 | Not-determined |
| 17-108_k141_79438 | Not-determined |
| 17-108_k141_23648 | Low-quality |
| 17-108_k141_66537 | Not-determined |
| 17-108_k141_259704 | Not-determined |
| 17-108_k141_134276 | Not-determined |
| 17-108_k141_134282 | Not-determined |
| 17-108_k141_326784 | Not-determined |
| 17-108_k141_163054 | Not-determined |
| 17-108_k141_259723 | Not-determined |
| 17-108_k141_287092 | Not-determined |
| 17-108_k141_134296 | Not-determined |
| 17-108_k141_134300 | Not-determined |
| 17-108_k141_134302 | Not-determined |
| 17-108_k141_287107 | Not-determined |
| 17-108_k141_134305 | Not-determined |
| 17-108_k141_300275 | Not-determined |
| 17-108_k141_272608 | Not-determined |
| 17-108_k141_259750 | Not-determined |
| 17-108_k141_272610 | Not-determined |
| 17-108_k141_272613 | Not-determined |
| 17-108_k141_259751 | Low-quality |
| 17-108_k141_108328 | Not-determined |
| 17-108_k141_148999 | Not-determined |
| 17-108_k141_287120 | Not-determined |
| 17-108_k141_176802 | Not-determined |
| 17-108_k141_189738 | Not-determined |
| 17-108_k141_79493 | Not-determined |
| 17-108_k141_66568 | Low-quality |
| 17-108_k141_246251 | Not-determined |
| 17-108_k141_38965 | Not-determined |
| 17-108_k141_287126 | Not-determined |
| 17-108_k141_38968 | Not-determined |
| 17-108_k141_300286 | Not-determined |
| 17-108_k141_287131 | Not-determined |
| 17-108_k141_246263 | Not-determined |
| 17-108_k141_149012 | Not-determined |
| 17-108_k141_287137 | Not-determined |
| 17-108_k141_149021 | Not-determined |
| 17-108_k141_66588 | Low-quality |

| | |
|---|---|
| 17-108_k141_176810 | Not-determined |
| 17-108_k141_53109 | Low-quality |
| 17-108_k141_38979 | Not-determined |
| 17-108_k141_38985 | Not-determined |
| 17-108_k141_246275 | Not-determined |
| 17-108_k141_176831 | Not-determined |
| 17-108_k141_120651 | Not-determined |
| 17-108_k141_300322 | Not-determined |
| 17-108_k141_163112 | Not-determined |
| 17-108_k141_259773 | Low-quality |
| 17-108_k141_246288 | Low-quality |
| 17-108_k141_163120 | Not-determined |
| 17-108_k141_176843 | Not-determined |
| 17-108_k141_246292 | Not-determined |
| 17-108_k141_79529 | Not-determined |
| 17-108_k141_163128 | Not-determined |
| 17-108_k141_120663 | Not-determined |
| 17-108_k141_53137 | Not-determined |
| 17-108_k141_79539 | Not-determined |
| 17-108_k141_93906 | Not-determined |
| 17-108_k141_11358 | Not-determined |
| 17-108_k141_326836 | Not-determined |
| 17-108_k141_149033 | Low-quality |
| 17-108_k141_93913 | Not-determined |
| 17-108_k141_120677 | Not-determined |
| 17-108_k141_259789 | Low-quality |
| 17-108_k141_93920 | Not-determined |
| 17-108_k141_11368 | Not-determined |
| 17-108_k141_163143 | Not-determined |
| 17-108_k141_23761 | Not-determined |
| 17-108_k141_163147 | Low-quality |
| 17-108_k141_108376 | Not-determined |
| 17-108_k141_53167 | Not-determined |
| 17-108_k141_163148 | Not-determined |
| 17-108_k141_314435 | Not-determined |
| 17-108_k141_176874 | Not-determined |
| 17-108_k141_53173 | Not-determined |
| 17-108_k141_314441 | Not-determined |
| 17-108_k141_66656 | Low-quality |
| 17-108_k141_176882 | Not-determined |
| 17-108_k141_66660 | Not-determined |
| 17-108_k141_287145 | Low-quality |
| 17-108_k141_287146 | Not-determined |
| 17-108_k141_217883 | Not-determined |
| 17-108_k141_287147 | Not-determined |
| 17-108_k141_326853 | Low-quality |
| 17-108_k141_134349 | Not-determined |
| 17-108_k141_108393 | Not-determined |
| 17-108_k141_217885 | Not-determined |
| 17-108_k141_66665 | Not-determined |
| 17-108_k141_231934 | Not-determined |
| 17-108_k141_120710 | Not-determined |
| 17-108_k141_120719 | Not-determined |
| 17-108_k141_23784 | Not-determined |
| 17-108_k141_53199 | Not-determined |
| 17-108_k141_272694 | Not-determined |
| 17-108_k141_300379 | Not-determined |
| 17-108_k141_134369 | Low-quality |
| 17-108_k141_217889 | Low-quality |
| 17-108_k141_314469 | Not-determined |
| 17-108_k141_314471 | Not-determined |
| 17-108_k141_189794 | Low-quality |
| 17-108_k141_300384 | Not-determined |
| 17-108_k141_272701 | Not-determined |
| 17-108_k141_204085 | Not-determined |
| 17-108_k141_108409 | Not-determined |
| 17-108_k141_287173 | Not-determined |
| 17-108_k141_272705 | Not-determined |
| 17-108_k141_108413 | Not-determined |
| 17-108_k141_272711 | Low-quality |
| 17-108_k141_231971 | Not-determined |
| 17-108_k141_326869 | Not-determined |
| 17-108_k141_217893 | Not-determined |
| 17-108_k141_176922 | Not-determined |
| 17-108_k141_120746 | Not-determined |
| 17-108_k141_189809 | Not-determined |
| 17-108_k141_287188 | Not-determined |
| 17-108_k141_272730 | Not-determined |
| 17-108_k141_287192 | Not-determined |
| 17-108_k141_246360 | Not-determined |
| 17-108_k141_246363 | Not-determined |
| 17-108_k141_272738 | Not-determined |
| 17-108_k141_149060 | Not-determined |
| 17-108_k141_314496 | Not-determined |
| 17-108_k141_326877 | Not-determined |
| 17-108_k141_134395 | Not-determined |
| 17-108_k141_108439 | Not-determined |
| 17-108_k141_246377 | Not-determined |
| 17-108_k141_300407 | Not-determined |
| 17-108_k141_287197 | Not-determined |
| 17-108_k141_300414 | Not-determined |
| 17-108_k141_204116 | Not-determined |
| 17-108_k141_176949 | Not-determined |
| 17-108_k141_66707 | Low-quality |
| 17-108_k141_23820 | Not-determined |
| 17-108_k141_149091 | Not-determined |
| 17-108_k141_246390 | Not-determined |
| 17-108_k141_11422 | Low-quality |
| 17-108_k141_149101 | Not-determined |
| 17-108_k141_39091 | Not-determined |
| 17-108_k141_39092 | Not-determined |
| 17-108_k141_204135 | Not-determined |
| 17-108_k141_326907 | Not-determined |
| 17-108_k141_134424 | Not-determined |
| 17-108_k141_300440 | Not-determined |
| 17-108_k141_232012 | Low-quality |
| 17-108_k141_300444 | Not-determined |
| 17-108_k141_232016 | Not-determined |
| 17-108_k141_314514 | Not-determined |
| 17-108_k141_108469 | Not-determined |
| 17-108_k141_300447 | Not-determined |
| 17-108_k141_11432 | Not-determined |
| 17-108_k141_39096 | Not-determined |
| 17-108_k141_108476 | Not-determined |
| 17-108_k141_176977 | Not-determined |
| 17-108_k141_326919 | Not-determined |
| 17-108_k141_246411 | Not-determined |
| 17-108_k141_53222 | Low-quality |
| 17-108_k141_300451 | Not-determined |
| 17-108_k141_39106 | Low-quality |
| 17-108_k141_11448 | Not-determined |
| 17-108_k141_53227 | Not-determined |
| 17-108_k141_246424 | Not-determined |
| 17-108_k141_246427 | Not-determined |
| 17-108_k141_326934 | Not-determined |
| 17-108_k141_272793 | Not-determined |
| 17-108_k141_176990 | Low-quality |
| 17-108_k141_134460 | Not-determined |
| 17-108_k141_176993 | Not-determined |
| 17-108_k141_108493 | Not-determined |
| 17-108_k141_246433 | Not-determined |
| 17-108_k141_149140 | Not-determined |
| 17-108_k141_272796 | Not-determined |
| 17-108_k141_204166 | Not-determined |
| 17-108_k141_120779 | Low-quality |
| 17-108_k141_108504 | Not-determined |
| 17-108_k141_39126 | Not-determined |
| 17-108_k141_23847 | Not-determined |
| 17-108_k141_272805 | Not-determined |
| 17-108_k141_66742 | Not-determined |
| 17-108_k141_300480 | Not-determined |
| 17-108_k141_217928 | Not-determined |
| 17-108_k141_39136 | Not-determined |
| 17-108_k141_232052 | Not-determined |
| 17-108_k141_287245 | Low-quality |
| 17-108_k141_189881 | Not-determined |
| 17-108_k141_177016 | Not-determined |
| 17-108_k141_272812 | Not-determined |
| 17-108_k141_259873 | Not-determined |
| 17-108_k141_53240 | Not-determined |
| 17-108_k141_326954 | Not-determined |
| 17-108_k141_326955 | Not-determined |
| 17-108_k141_134483 | Not-determined |
| 17-108_k141_120797 | Not-determined |
| 17-108_k141_134486 | Not-determined |
| 17-108_k141_149157 | Not-determined |
| 17-108_k141_39149 | Not-determined |
| 17-108_k141_39150 | Not-determined |
| 17-108_k141_66745 | Low-quality |
| 17-108_k141_23863 | Not-determined |
| 17-108_k141_23866 | Not-determined |
| 17-108_k141_177032 | Not-determined |
| 17-108_k141_232078 | Not-determined |
| 17-108_k141_134510 | Not-determined |
| 17-108_k141_232080 | Not-determined |
| 17-108_k141_259895 | Not-determined |
| 17-108_k141_23871 | Not-determined |
| 17-108_k141_66758 | Not-determined |
| 17-108_k141_300506 | Not-determined |
| 17-108_k141_23882 | Not-determined |
| 17-108_k141_53263 | Not-determined |
| 17-108_k141_300509 | Not-determined |
| 17-108_k141_163227 | Not-determined |
| 17-108_k141_246471 | Not-determined |
| 17-108_k141_149180 | Not-determined |
| 17-108_k141_66765 | Not-determined |
| 17-108_k141_149184 | Not-determined |
| 17-108_k141_287287 | Not-determined |
| 17-108_k141_259912 | Not-determined |
| 17-108_k141_120828 | Not-determined |
| 17-108_k141_11483 | Not-determined |
| 17-108_k141_120834 | Not-determined |
| 17-108_k141_259915 | Not-determined |
| 17-108_k141_120837 | Not-determined |
| 17-108_k141_177059 | Not-determined |
| 17-108_k141_79647 | Not-determined |
| 17-108_k141_163251 | Not-determined |
| 17-108_k141_108550 | Not-determined |
| 17-108_k141_39194 | Not-determined |
| 17-108_k141_11500 | Not-determined |
| 17-108_k141_163262 | Not-determined |
| 17-108_k141_120855 | Not-determined |
| 17-108_k141_259933 | Not-determined |
| 17-108_k141_163266 | Not-determined |
| 17-108_k141_79665 | Not-determined |
| 17-108_k141_120859 | Not-determined |
| 17-108_k141_326992 | Not-determined |
| 17-108_k141_120861 | Not-determined |
| 17-108_k141_66793 | Not-determined |
| 17-108_k141_11514 | Not-determined |
| 17-108_k141_272855 | Not-determined |
| 17-108_k141_108561 | Not-determined |
| 17-108_k141_39208 | Low-quality |
| 17-108_k141_134547 | Not-determined |
| 17-108_k141_149216 | Not-determined |
| 17-108_k141_232097 | Low-quality |
| 17-108_k141_120871 | Low-quality |
| 17-108_k141_232100 | Not-determined |
| 17-108_k141_11520 | Not-determined |
| 17-108_k141_300527 | Not-determined |
| 17-108_k141_11525 | Not-determined |
| 17-108_k141_79679 | Not-determined |
| 17-108_k141_11529 | Not-determined |
| 17-108_k141_149233 | Not-determined |
| 17-108_k141_11530 | Not-determined |
| 17-108_k141_11531 | Not-determined |
| 17-108_k141_11533 | Not-determined |
| 17-108_k141_11536 | Not-determined |
| 17-108_k141_246506 | Not-determined |
| 17-108_k141_314535 | Not-determined |
| 17-108_k141_66821 | Not-determined |
| 17-108_k141_134586 | Not-determined |
| 17-108_k141_272886 | Not-determined |
| 17-108_k141_108595 | Not-determined |
| 17-108_k141_66827 | Low-quality |
| 17-108_k141_177082 | Not-determined |
| 17-108_k141_232124 | Not-determined |
| 17-108_k141_300546 | Not-determined |
| 17-108_k141_66839 | Not-determined |
| 17-108_k141_39229 | Not-determined |
| 17-108_k141_232131 | Not-determined |
| 17-108_k141_11557 | Not-determined |
| 17-108_k141_23911 | Not-determined |
| 17-108_k141_177089 | Not-determined |
| 17-108_k141_232136 | Not-determined |
| 17-108_k141_11565 | Not-determined |
| 17-108_k141_23916 | Not-determined |
| 17-108_k141_23918 | Not-determined |
| 17-108_k141_218019 | Not-determined |
| 17-108_k141_246544 | Not-determined |
| 17-108_k141_327028 | Not-determined |
| 17-108_k141_149260 | Not-determined |
| 17-108_k141_259965 | Not-determined |
| 17-108_k141_120917 | Not-determined |
| 17-108_k141_259973 | Not-determined |
| 17-108_k141_163329 | Not-determined |
| 17-108_k141_79732 | Not-determined |
| 17-108_k141_259977 | Not-determined |
| 17-108_k141_79735 | Not-determined |
| 17-108_k141_189911 | Low-quality |
| 17-108_k141_246565 | Not-determined |
| 17-108_k141_53269 | Not-determined |
| 17-108_k141_134627 | Not-determined |
| 17-108_k141_134629 | Low-quality |
| 17-108_k141_53272 | Not-determined |
| 17-108_k141_246572 | Not-determined |
| 17-108_k141_218044 | Low-quality |
| 17-108_k141_108644 | Not-determined |
| 17-108_k141_53281 | Not-determined |
| 17-108_k141_177121 | Not-determined |
| 17-108_k141_232164 | Not-determined |
| 17-108_k141_327051 | Not-determined |
| 17-108_k141_23947 | Not-determined |
| 17-108_k141_53283 | Not-determined |
| 17-108_k141_120934 | Not-determined |
| 17-108_k141_108671 | Not-determined |
| 17-108_k141_246595 | Not-determined |
| 17-108_k141_79762 | Not-determined |
| 17-108_k141_327059 | Not-determined |
| 17-108_k141_246602 | Not-determined |
| 17-108_k141_149301 | Not-determined |
| 17-108_k141_189941 | Low-quality |
| 17-108_k141_149302 | Not-determined |
| 17-108_k141_189942 | Not-determined |
| 17-108_k141_39298 | Not-determined |
| 17-108_k141_53301 | Not-determined |
| 17-108_k141_53302 | Not-determined |
| 17-108_k141_11605 | Not-determined |
| 17-108_k141_246612 | Not-determined |
| 17-108_k141_79780 | Low-quality |
| 17-108_k141_287352 | Low-quality |
| 17-108_k141_79788 | Not-determined |
| 17-108_k141_79789 | Not-determined |
| 17-108_k141_108700 | Not-determined |
| 17-108_k141_218069 | Not-determined |
| 17-108_k141_120971 | Not-determined |
| 17-108_k141_218075 | Not-determined |
| 17-108_k141_149316 | Low-quality |
| 17-108_k141_232202 | Not-determined |
| 17-108_k141_163361 | Not-determined |
| 17-108_k141_149321 | Not-determined |
| 17-108_k141_93964 | Not-determined |
| 17-108_k141_218079 | Not-determined |
| 17-108_k141_177145 | Not-determined |
| 17-108_k141_163377 | Low-quality |
| 17-108_k141_108717 | Not-determined |
| 17-108_k141_23973 | Not-determined |
| 17-108_k141_134663 | Not-determined |
| 17-108_k141_149348 | Not-determined |
| 17-108_k141_232226 | Not-determined |
| 17-108_k141_121000 | Not-determined |
| 17-108_k141_23989 | Low-quality |
| 17-108_k141_121002 | Not-determined |
| 17-108_k141_314598 | Low-quality |
| 17-108_k141_246637 | Not-determined |
| 17-108_k141_23994 | Not-determined |
| 17-108_k141_79834 | Not-determined |
| 17-108_k141_204190 | Not-determined |
| 17-108_k141_246639 | Not-determined |
| 17-108_k141_260014 | Not-determined |
| 17-108_k141_79839 | Not-determined |
| 17-108_k141_246651 | Not-determined |
| 17-108_k141_204199 | Not-determined |
| 17-108_k141_24006 | Not-determined |
| 17-108_k141_149377 | Not-determined |
| 17-108_k141_24008 | Not-determined |
| 17-108_k141_93997 | Not-determined |
| 17-108_k141_24012 | Not-determined |
| 17-108_k141_189987 | Not-determined |
| 17-108_k141_149385 | Not-determined |
| 17-108_k141_11638 | Not-determined |
| 17-108_k141_24016 | Not-determined |
| 17-108_k141_163408 | Not-determined |
| 17-108_k141_260021 | Not-determined |
| 17-108_k141_24017 | Not-determined |
| 17-108_k141_204210 | Not-determined |
| 17-108_k141_218117 | Not-determined |
| 17-108_k141_246664 | Not-determined |
| 17-108_k141_177192 | Not-determined |
| 17-108_k141_163414 | Not-determined |
| 17-108_k141_24021 | Not-determined |
| 17-108_k141_39356 | Not-determined |
| 17-108_k141_204217 | Not-determined |
| 17-108_k141_314625 | Not-determined |
| 17-108_k141_149400 | Not-determined |
| 17-108_k141_260030 | Not-determined |
| 17-108_k141_246666 | Not-determined |
| 17-108_k141_246667 | Not-determined |
| 17-108_k141_39372 | Not-determined |
| 17-108_k141_149404 | Not-determined |
| 17-108_k141_327105 | Not-determined |
| 17-108_k141_260046 | Not-determined |
| 17-108_k141_24038 | Not-determined |
| 17-108_k141_53359 | Not-determined |
| 17-108_k141_327119 | Low-quality |
| 17-108_k141_327123 | Not-determined |
| 17-108_k141_218138 | Not-determined |
| 17-108_k141_260061 | Not-determined |
| 17-108_k141_327130 | Not-determined |
| 17-108_k141_260067 | Not-determined |
| 17-108_k141_39398 | Not-determined |
| 17-108_k141_108751 | Not-determined |
| 17-108_k141_66885 | Not-determined |
| 17-108_k141_163439 | Not-determined |
| 17-108_k141_121021 | Not-determined |
| 17-108_k141_246710 | Not-determined |
| 17-108_k141_11692 | Not-determined |
| 17-108_k141_121026 | Not-determined |
| 17-108_k141_53382 | Not-determined |
| 17-108_k141_177215 | Not-determined |
| 17-108_k141_24065 | Not-determined |
| 17-108_k141_246720 | Not-determined |
| 17-108_k141_218168 | Not-determined |
| 17-108_k141_24083 | Not-determined |
| 17-108_k141_204260 | Not-determined |
| 17-108_k141_108761 | Low-quality |
| 17-108_k141_149411 | Not-determined |
| 17-108_k141_11716 | Not-determined |
| 17-108_k141_121034 | Not-determined |
| 17-108_k141_177222 | Not-determined |
| 17-108_k141_53411 | Not-determined |
| 17-108_k141_190032 | Not-determined |
| 17-108_k141_66915 | Not-determined |
| 17-108_k141_287410 | Not-determined |
| 17-108_k141_287412 | Not-determined |
| 17-108_k141_272956 | Low-quality |
| 17-108_k141_94015 | Not-determined |
| 17-108_k141_190040 | Not-determined |
| 17-108_k141_39427 | Not-determined |
| 17-108_k141_39428 | Not-determined |
| 17-108_k141_121052 | Not-determined |
| 17-108_k141_53419 | Not-determined |
| 17-108_k141_94025 | Not-determined |
| 17-108_k141_108782 | Not-determined |
| 17-108_k141_204282 | Not-determined |
| 17-108_k141_66927 | Low-quality |
| 17-108_k141_272966 | Not-determined |
| 17-108_k141_177243 | Not-determined |
| 17-108_k141_53427 | Not-determined |
| 17-108_k141_108786 | Not-determined |
| 17-108_k141_287417 | Low-quality |
| 17-108_k141_94035 | Not-determined |
| 17-108_k141_94037 | Not-determined |
| 17-108_k141_108792 | Not-determined |
| 17-108_k141_246742 | Not-determined |
| 17-108_k141_204290 | Low-quality |
| 17-108_k141_287420 | Not-determined |
| 17-108_k141_53431 | Not-determined |
| 17-108_k141_39437 | Not-determined |
| 17-108_k141_190050 | Not-determined |
| 17-108_k141_218197 | Not-determined |
| 17-108_k141_272980 | Low-quality |
| 17-108_k141_314693 | Not-determined |
| 17-108_k141_149439 | Not-determined |
| 17-108_k141_300602 | Not-determined |
| 17-108_k141_11765 | Not-determined |
| 17-108_k141_204303 | Not-determined |
| 17-108_k141_204305 | Not-determined |
| 17-108_k141_204308 | Low-quality |
| 17-108_k141_24107 | Not-determined |
| 17-108_k141_177256 | Not-determined |
| 17-108_k141_287440 | Not-determined |
| 17-108_k141_204316 | Not-determined |
| 17-108_k141_190073 | Not-determined |
| 17-108_k141_300623 | Not-determined |
| 17-108_k141_39443 | Not-determined |
| 17-108_k141_287443 | Not-determined |
| 17-108_k141_24118 | Not-determined |
| 17-108_k141_314717 | Not-determined |
| 17-108_k141_11780 | Not-determined |
| 17-108_k141_204323 | Not-determined |
| 17-108_k141_94065 | Not-determined |
| 17-108_k141_121069 | Not-determined |
| 17-108_k141_24133 | Not-determined |
| 17-108_k141_327180 | Not-determined |
| 17-108_k141_24135 | Not-determined |
| 17-108_k141_163506 | Not-determined |
| 17-108_k141_204338 | Not-determined |
| 17-108_k141_163512 | Not-determined |
| 17-108_k141_163514 | Not-determined |
| 17-108_k141_204341 | Not-determined |
| 17-108_k141_272996 | Not-determined |
| 17-108_k141_53478 | Low-quality |
| 17-108_k141_11789 | Not-determined |
| 17-108_k141_108818 | Not-determined |
| 17-108_k141_287460 | Not-determined |
| 17-108_k141_246780 | Not-determined |
| 17-108_k141_300638 | Not-determined |
| 17-108_k141_24152 | Not-determined |
| 17-108_k141_246784 | Not-determined |
| 17-108_k141_246785 | Not-determined |
| 17-108_k141_177281 | Not-determined |
| 17-108_k141_327205 | Not-determined |
| 17-108_k141_11796 | Low-quality |
| 17-108_k141_232297 | Not-determined |
| 17-108_k141_66960 | Not-determined |
| 17-108_k141_287465 | Not-determined |
| 17-108_k141_287467 | Not-determined |
| 17-108_k141_24161 | Not-determined |
| 17-108_k141_39466 | Not-determined |
| 17-108_k141_108832 | Not-determined |
| 17-108_k141_79883 | Not-determined |
| 17-108_k141_24169 | Not-determined |
| 17-108_k141_94098 | Not-determined |
| 17-108_k141_314724 | Not-determined |
| 17-108_k141_149495 | Not-determined |
| 17-108_k141_24174 | Not-determined |
| 17-108_k141_149498 | Not-determined |
| 17-108_k141_177301 | Not-determined |
| 17-108_k141_314727 | Not-determined |
| 17-108_k141_163549 | Not-determined |
| 17-108_k141_24182 | Not-determined |
| 17-108_k141_79906 | Not-determined |
| 17-108_k141_232310 | Not-determined |
| 17-108_k141_246788 | Not-determined |
| 17-108_k141_79914 | Not-determined |
| 17-108_k141_94124 | Not-determined |
| 17-108_k141_246795 | Not-determined |
| 17-108_k141_24197 | Not-determined |
| 17-108_k141_204358 | Low-quality |
| 17-108_k141_79919 | Not-determined |
| 17-108_k141_204360 | Not-determined |
| 17-108_k141_273036 | Not-determined |
| 17-108_k141_287505 | Not-determined |
| 17-108_k141_177311 | Not-determined |
| 17-108_k141_232323 | Not-determined |
| 17-108_k141_300677 | Not-determined |
| 17-108_k141_273049 | Not-determined |
| 17-108_k141_66992 | Not-determined |
| 17-108_k141_273058 | Not-determined |
| 17-108_k141_177321 | Not-determined |
| 17-108_k141_273059 | Not-determined |
| 17-108_k141_260127 | Not-determined |
| 17-108_k141_79938 | Not-determined |
| 17-108_k141_134828 | Not-determined |
| 17-108_k141_273063 | Not-determined |
| 17-108_k141_232333 | Not-determined |
| 17-108_k141_273067 | Low-quality |
| 17-108_k141_24207 | Low-quality |
| 17-108_k141_300684 | Not-determined |
| 17-108_k141_260131 | Not-determined |
| 17-108_k141_260132 | Not-determined |
| 17-108_k141_94148 | Not-determined |
| 17-108_k141_232340 | Not-determined |
| 17-108_k141_190121 | Not-determined |
| 17-108_k141_300686 | Not-determined |
| 17-108_k141_39521 | Not-determined |
| 17-108_k141_24212 | Not-determined |
| 17-108_k141_260134 | Not-determined |
| 17-108_k141_177332 | Not-determined |
| 17-108_k141_287535 | Not-determined |
| 17-108_k141_134846 | Not-determined |
| 17-108_k141_94160 | Not-determined |
| 17-108_k141_94162 | Not-determined |
| 17-108_k141_24218 | Not-determined |
| 17-108_k141_24219 | Not-determined |
| 17-108_k141_108860 | Not-determined |
| 17-108_k141_24222 | Not-determined |
| 17-108_k141_204408 | Not-determined |
| 17-108_k141_218231 | Not-determined |
| 17-108_k141_300701 | Not-determined |
| 17-108_k141_163569 | Not-determined |
| 17-108_k141_218239 | Not-determined |
| 17-108_k141_273100 | Not-determined |
| 17-108_k141_260158 | Not-determined |
| 17-108_k141_300716 | Not-determined |
| 17-108_k141_11836 | Not-determined |
| 17-108_k141_190151 | Not-determined |
| 17-108_k141_218248 | Not-determined |
| 17-108_k141_163577 | Not-determined |
| 17-108_k141_218249 | Not-determined |
| 17-108_k141_108874 | Not-determined |
| 17-108_k141_327253 | Not-determined |
| 17-108_k141_260171 | Not-determined |
| 17-108_k141_79984 | Not-determined |
| 17-108_k141_218254 | Not-determined |
| 17-108_k141_218256 | Not-determined |
| 17-108_k141_287556 | Not-determined |
| 17-108_k141_204431 | Not-determined |
| 17-108_k141_39568 | Not-determined |
| 17-108_k141_190163 | Not-determined |
| 17-108_k141_94177 | Not-determined |
| 17-108_k141_190168 | Not-determined |
| 17-108_k141_11853 | Not-determined |
| 17-108_k141_134856 | Not-determined |
| 17-108_k141_67046 | Not-determined |
| 17-108_k141_300746 | Not-determined |
| 17-108_k141_79996 | Not-determined |
| 17-108_k141_94185 | Not-determined |
| 17-108_k141_24238 | Low-quality |
| 17-108_k141_53551 | Not-determined |
| 17-108_k141_232359 | Not-determined |
| 17-108_k141_121150 | Not-determined |
| 17-108_k141_121151 | Not-determined |
| 17-108_k141_11874 | Not-determined |
| 17-108_k141_24255 | Not-determined |
| 17-108_k141_24257 | Not-determined |
| 17-108_k141_24258 | Not-determined |
| 17-108_k141_11876 | Not-determined |
| 17-108_k141_39587 | Not-determined |
| 17-108_k141_24259 | Not-determined |
| 17-108_k141_204443 | Not-determined |
| 17-108_k141_11877 | Not-determined |
| 17-108_k141_177385 | Not-determined |
| 17-108_k141_204445 | Not-determined |
| 17-108_k141_11880 | Not-determined |
| 17-108_k141_204447 | Not-determined |
| 17-108_k141_218295 | Low-quality |
| 17-108_k141_314801 | Not-determined |
| 17-108_k141_273109 | Not-determined |
| 17-108_k141_53562 | Not-determined |
| 17-108_k141_134883 | Not-determined |
| 17-108_k141_11887 | Not-determined |
| 17-108_k141_39603 | Not-determined |
| 17-108_k141_260199 | Low-quality |
| 17-108_k141_108901 | Low-quality |
| 17-108_k141_121175 | Not-determined |
| 17-108_k141_260207 | Low-quality |
| 17-108_k141_121183 | Not-determined |
| 17-108_k141_80022 | Not-determined |
| 17-108_k141_121185 | Not-determined |
| 17-108_k141_300787 | Not-determined |
| 17-108_k141_94233 | Not-determined |
| 17-108_k141_327298 | Not-determined |
| 17-108_k141_204469 | Not-determined |
| 17-108_k141_190229 | Not-determined |
| 17-108_k141_218312 | Not-determined |
| 17-108_k141_300796 | Not-determined |
| 17-108_k141_53592 | Not-determined |
| 17-108_k141_246849 | Not-determined |
| 17-108_k141_177416 | Not-determined |
| 17-108_k141_149606 | Not-determined |
| 17-108_k141_80040 | Not-determined |
| 17-108_k141_327311 | Not-determined |
| 17-108_k141_190238 | Not-determined |
| 17-108_k141_11898 | Low-quality |
| 17-108_k141_80042 | Not-determined |
| 17-108_k141_134904 | Not-determined |
| 17-108_k141_300803 | Not-determined |

| | |
|---|---|
| 17-108_k141_232391 | Not-determined |
| 17-108_k141_134907 | Not-determined |
| 17-108_k141_67113 | Not-determined |
| 17-108_k141_177425 | Not-determined |
| 17-108_k141_260222 | Not-determined |
| 17-108_k141_177426 | Not-determined |
| 17-108_k141_177432 | Not-determined |
| 17-108_k141_24291 | Not-determined |
| 17-108_k141_24292 | Not-determined |
| 17-108_k141_273130 | Not-determined |
| 17-108_k141_260231 | Not-determined |
| 17-108_k141_121211 | Not-determined |
| 17-108_k141_246871 | Not-determined |
| 17-108_k141_80060 | Not-determined |
| 17-108_k141_177441 | Not-determined |
| 17-108_k141_287630 | Not-determined |
| 17-108_k141_190255 | Not-determined |
| 17-108_k141_163631 | Not-determined |
| 17-108_k141_11928 | Not-determined |
| 17-108_k141_163632 | Not-determined |
| 17-108_k141_163635 | Not-determined |
| 17-108_k141_260238 | Not-determined |
| 17-108_k141_53620 | Not-determined |
| 17-108_k141_246885 | Not-determined |
| 17-108_k141_177450 | Not-determined |
| 17-108_k141_121224 | Not-determined |
| 17-108_k141_163642 | Not-determined |
| 17-108_k141_39648 | Not-determined |
| 17-108_k141_121226 | Not-determined |
| 17-108_k141_94278 | Not-determined |
| 17-108_k141_134945 | Not-determined |
| 17-108_k141_80070 | Not-determined |
| 17-108_k141_314838 | Not-determined |
| 17-108_k141_80071 | Not-determined |
| 17-108_k141_11943 | Not-determined |
| 17-108_k141_67126 | Not-determined |
| 17-108_k141_94284 | Not-determined |
| 17-108_k141_80083 | Not-determined |
| 17-108_k141_94295 | Not-determined |
| 17-108_k141_108924 | Not-determined |
| 17-108_k141_163659 | Not-determined |
| 17-108_k141_24327 | Low-quality |
| 17-108_k141_24331 | Not-determined |
| 17-108_k141_11964 | Not-determined |
| 17-108_k141_80099 | Not-determined |
| 17-108_k141_80101 | Not-determined |
| 17-108_k141_327357 | Not-determined |
| 17-108_k141_80103 | Not-determined |
| 17-108_k141_177479 | Low-quality |
| 17-108_k141_177482 | Not-determined |
| 17-108_k141_314847 | Not-determined |
| 17-108_k141_163676 | Not-determined |
| 17-108_k141_80124 | Not-determined |
| 17-108_k141_246919 | Not-determined |
| 17-108_k141_314854 | Not-determined |
| 17-108_k141_149645 | Not-determined |
| 17-108_k141_80127 | Not-determined |
| 17-108_k141_11976 | Low-quality |
| 17-108_k141_53654 | Not-determined |
| 17-108_k141_149648 | Not-determined |
| 17-108_k141_80131 | Low-quality |
| 17-108_k141_67132 | Not-determined |
| 17-108_k141_134977 | Not-determined |
| 17-108_k141_39682 | Not-determined |
| 17-108_k141_39684 | Not-determined |
| 17-108_k141_314870 | Not-determined |
| 17-108_k141_11986 | Not-determined |
| 17-108_k141_53666 | Not-determined |
| 17-108_k141_163684 | Not-determined |
| 17-108_k141_53670 | Not-determined |
| 17-108_k141_232412 | Low-quality |
| 17-108_k141_134984 | Low-quality |
| 17-108_k141_287690 | Low-quality |
| 17-108_k141_53672 | Not-determined |
| 17-108_k141_232419 | Not-determined |
| 17-108_k141_53676 | Not-determined |
| 17-108_k141_246951 | Not-determined |
| 17-108_k141_134991 | Not-determined |
| 17-108_k141_260272 | Not-determined |
| 17-108_k141_121247 | Not-determined |
| 17-108_k141_12006 | Not-determined |
| 17-108_k141_218379 | Not-determined |
| 17-108_k141_287695 | Low-quality |
| 17-108_k141_273183 | Not-determined |
| 17-108_k141_12010 | Low-quality |
| 17-108_k141_314894 | Not-determined |
| 17-108_k141_135007 | Not-determined |
| 17-108_k141_218393 | Not-determined |
| 17-108_k141_12020 | Not-determined |
| 17-108_k141_260296 | Not-determined |
| 17-108_k141_135014 | Not-determined |
| 17-108_k141_12021 | Low-quality |
| 17-108_k141_12022 | Not-determined |
| 17-108_k141_327430 | Not-determined |
| 17-108_k141_135021 | Not-determined |
| 17-108_k141_287720 | Not-determined |
| 17-108_k141_12025 | Not-determined |
| 17-108_k141_232438 | Not-determined |
| 17-108_k141_67186 | Not-determined |
| 17-108_k141_287728 | Not-determined |
| 17-108_k141_108973 | Not-determined |
| 17-108_k141_327436 | Not-determined |
| 17-108_k141_67189 | Not-determined |
| 17-108_k141_67191 | Not-determined |
| 17-108_k141_135030 | Not-determined |
| 17-108_k141_24395 | Not-determined |
| 17-108_k141_108977 | Not-determined |
| 17-108_k141_327442 | Not-determined |
| 17-108_k141_135036 | Not-determined |
| 17-108_k141_314921 | Not-determined |
| 17-108_k141_121256 | Not-determined |
| 17-108_k141_163735 | Not-determined |
| 17-108_k141_163736 | Low-quality |
| 17-108_k141_24402 | Not-determined |
| 17-108_k141_260322 | Not-determined |
| 17-108_k141_260323 | Not-determined |
| 17-108_k141_260326 | Not-determined |
| 17-108_k141_149692 | Low-quality |
| 17-108_k141_121268 | Not-determined |
| 17-108_k141_273200 | Not-determined |
| 17-108_k141_273201 | Not-determined |
| 17-108_k141_260334 | Not-determined |
| 17-108_k141_232468 | Not-determined |
| 17-108_k141_121279 | Not-determined |
| 17-108_k141_327471 | Not-determined |
| 17-108_k141_163755 | Not-determined |
| 17-108_k141_67217 | Not-determined |
| 17-108_k141_94382 | Not-determined |
| 17-108_k141_24425 | Not-determined |
| 17-108_k141_314928 | Not-determined |
| 17-108_k141_109022 | Not-determined |
| 17-108_k141_163763 | Not-determined |
| 17-108_k141_94389 | Not-determined |
| 17-108_k141_94390 | Not-determined |
| 17-108_k141_232486 | Not-determined |
| 17-108_k141_149716 | Not-determined |
| 17-108_k141_300840 | Low-quality |
| 17-108_k141_232500 | Not-determined |
| 17-108_k141_135075 | Not-determined |
| 17-108_k141_300843 | Not-determined |
| 17-108_k141_53734 | Not-determined |
| 17-108_k141_149730 | Not-determined |
| 17-108_k141_327496 | Not-determined |
| 17-108_k141_314938 | Low-quality |
| 17-108_k141_327497 | Not-determined |
| 17-108_k141_260356 | Not-determined |
| 17-108_k141_24458 | Not-determined |
| 17-108_k141_121319 | Not-determined |
| 17-108_k141_135094 | Not-determined |
| 17-108_k141_232514 | Not-determined |
| 17-108_k141_39777 | Not-determined |
| 17-108_k141_135104 | Not-determined |
| 17-108_k141_94401 | Not-determined |
| 17-108_k141_121330 | Not-determined |
| 17-108_k141_177586 | Not-determined |
| 17-108_k141_94407 | Not-determined |
| 17-108_k141_149741 | Not-determined |
| 17-108_k141_300868 | Not-determined |
| 17-108_k141_287815 | Not-determined |
| 17-108_k141_190329 | Not-determined |
| 17-108_k141_94424 | Not-determined |
| 17-108_k141_94426 | Not-determined |
| 17-108_k141_135118 | Not-determined |
| 17-108_k141_190347 | Not-determined |
| 17-108_k141_39803 | Not-determined |
| 17-108_k141_24487 | Not-determined |
| 17-108_k141_24488 | Not-determined |
| 17-108_k141_300885 | Not-determined |
| 17-108_k141_177615 | Not-determined |
| 17-108_k141_287837 | Not-determined |
| 17-108_k141_109062 | Not-determined |
| 17-108_k141_314956 | Not-determined |
| 17-108_k141_149769 | Not-determined |
| 17-108_k141_314957 | Not-determined |
| 17-108_k141_314958 | Not-determined |
| 17-108_k141_218441 | Not-determined |
| 17-108_k141_80159 | Not-determined |
| 17-108_k141_314959 | Not-determined |
| 17-108_k141_163836 | Not-determined |
| 17-108_k141_163838 | Not-determined |
| 17-108_k141_149783 | Not-determined |
| 17-108_k141_273259 | Not-determined |
| 17-108_k141_53751 | Not-determined |
| 17-108_k141_204503 | Not-determined |
| 17-108_k141_80186 | Not-determined |
| 17-108_k141_204504 | Not-determined |
| 17-108_k141_273272 | Not-determined |
| 17-108_k141_287870 | Not-determined |
| 17-108_k141_204506 | Not-determined |
| 17-108_k141_163859 | Low-quality |
| 17-108_k141_163860 | Not-determined |
| 17-108_k141_287875 | Not-determined |
| 17-108_k141_287876 | Not-determined |
| 17-108_k141_247069 | Not-determined |
| 17-108_k141_53767 | Not-determined |
| 17-108_k141_260359 | Not-determined |
| 17-108_k141_39828 | Not-determined |
| 17-108_k141_300922 | Not-determined |
| 17-108_k141_247077 | Low-quality |
| 17-108_k141_273285 | Not-determined |
| 17-108_k141_109083 | Not-determined |
| 17-108_k141_327539 | Not-determined |
| 17-108_k141_232527 | Low-quality |
| 17-108_k141_260375 | Low-quality |
| 17-108_k141_190396 | Not-determined |
| 17-108_k141_190399 | Not-determined |
| 17-108_k141_300932 | Low-quality |
| 17-108_k141_80211 | Not-determined |
| 17-108_k141_177654 | Low-quality |
| 17-108_k141_327547 | Not-determined |
| 17-108_k141_232543 | Not-determined |
| 17-108_k141_327551 | Not-determined |
| 17-108_k141_300941 | Not-determined |
| 17-108_k141_80219 | Not-determined |
| 17-108_k141_273309 | Not-determined |
| 17-108_k141_121421 | Not-determined |
| 17-108_k141_80222 | Not-determined |
| 17-108_k141_39858 | Not-determined |
| 17-108_k141_177671 | Not-determined |
| 17-108_k141_39863 | Not-determined |
| 17-108_k141_314996 | Not-determined |
| 17-108_k141_314998 | Not-determined |
| 17-108_k141_149839 | Low-quality |
| 17-108_k141_163908 | Not-determined |
| 17-108_k141_327564 | Not-determined |
| 17-108_k141_177678 | Not-determined |
| 17-108_k141_315003 | Not-determined |
| 17-108_k141_247103 | Low-quality |
| 17-108_k141_109114 | Not-determined |
| 17-108_k141_273340 | Not-determined |
| 17-108_k141_190427 | Not-determined |
| 17-108_k141_121439 | Not-determined |
| 17-108_k141_109121 | Not-determined |
| 17-108_k141_232574 | Not-determined |
| 17-108_k141_232576 | Not-determined |
| 17-108_k141_232577 | Not-determined |
| 17-108_k141_53788 | Low-quality |
| 17-108_k141_247109 | Not-determined |
| 17-108_k141_260385 | Not-determined |
| 17-108_k141_121441 | Not-determined |
| 17-108_k141_327580 | Not-determined |
| 17-108_k141_260392 | Not-determined |
| 17-108_k141_327582 | Not-determined |
| 17-108_k141_39899 | Not-determined |
| 17-108_k141_327590 | Not-determined |
| 17-108_k141_53797 | Not-determined |
| 17-108_k141_39904 | Not-determined |
| 17-108_k141_39905 | Not-determined |
| 17-108_k141_109146 | Not-determined |
| 17-108_k141_190457 | Not-determined |
| 17-108_k141_24533 | Low-quality |
| 17-108_k141_218548 | Low-quality |
| 17-108_k141_149867 | Not-determined |
| 17-108_k141_149869 | Not-determined |
| 17-108_k141_315034 | Not-determined |
| 17-108_k141_163937 | Not-determined |
| 17-108_k141_190467 | Not-determined |
| 17-108_k141_12070 | Not-determined |
| 17-108_k141_80243 | Not-determined |
| 17-108_k141_24550 | Not-determined |
| 17-108_k141_315043 | Not-determined |
| 17-108_k141_121481 | Not-determined |
| 17-108_k141_121483 | Not-determined |
| 17-108_k141_53821 | Not-determined |
| 17-108_k141_109154 | Not-determined |
| 17-108_k141_163949 | Not-determined |
| 17-108_k141_232600 | Not-determined |
| 17-108_k141_39926 | Not-determined |
| 17-108_k141_260405 | Not-determined |
| 17-108_k141_109157 | Not-determined |
| 17-108_k141_247128 | Low-quality |
| 17-108_k141_53827 | Low-quality |
| 17-108_k141_24569 | Not-determined |
| 17-108_k141_327624 | Not-determined |
| 17-108_k141_109163 | Low-quality |
| 17-108_k141_12092 | Not-determined |
| 17-108_k141_190493 | Low-quality |
| 17-108_k141_232615 | Not-determined |
| 17-108_k141_121522 | Not-determined |
| 17-108_k141_149883 | Not-determined |
| 17-108_k141_232619 | Not-determined |
| 17-108_k141_177735 | Not-determined |
| 17-108_k141_232621 | Not-determined |
| 17-108_k141_39946 | Low-quality |
| 17-108_k141_300999 | Not-determined |
| 17-108_k141_204529 | Low-quality |
| 17-108_k141_12109 | Not-determined |
| 17-108_k141_177741 | Not-determined |
| 17-108_k141_53836 | Not-determined |
| 17-108_k141_135161 | Not-determined |
| 17-108_k141_67290 | Not-determined |
| 17-108_k141_232628 | Not-determined |
| 17-108_k141_301005 | Not-determined |
| 17-108_k141_94523 | Not-determined |
| 17-108_k141_177748 | Not-determined |
| 17-108_k141_80276 | Low-quality |
| 17-108_k141_260437 | Not-determined |
| 17-108_k141_53840 | Not-determined |
| 17-108_k141_80278 | Not-determined |
| 17-108_k141_53841 | Not-determined |
| 17-108_k141_109184 | Not-determined |
| 17-108_k141_39963 | Not-determined |
| 17-108_k141_260443 | Not-determined |
| 17-108_k141_39964 | Not-determined |
| 17-108_k141_204553 | Low-quality |
| 17-108_k141_121545 | Not-determined |
| 17-108_k141_287983 | Low-quality |
| 17-108_k141_260448 | Not-determined |
| 17-108_k141_67298 | Not-determined |
| 17-108_k141_24603 | Not-determined |
| 17-108_k141_67305 | Not-determined |
| 17-108_k141_67306 | Not-determined |
| 17-108_k141_121560 | Not-determined |
| 17-108_k141_315077 | Not-determined |
| 17-108_k141_327661 | Not-determined |
| 17-108_k141_135179 | Not-determined |
| 17-108_k141_135180 | Not-determined |
| 17-108_k141_204563 | Low-quality |
| 17-108_k141_24622 | Not-determined |
| 17-108_k141_94567 | Not-determined |
| 17-108_k141_94568 | Not-determined |
| 17-108_k141_94569 | Not-determined |
| 17-108_k141_12124 | Not-determined |
| 17-108_k141_315090 | Low-quality |
| 17-108_k141_164012 | Not-determined |
| 17-108_k141_273386 | Not-determined |
| 17-108_k141_67324 | Low-quality |
| 17-108_k141_164017 | Not-determined |
| 17-108_k141_135195 | Not-determined |
| 17-108_k141_288016 | Not-determined |
| 17-108_k141_67335 | Not-determined |
| 17-108_k141_315092 | Not-determined |
| 17-108_k141_204583 | Not-determined |
| 17-108_k141_247192 | Not-determined |
| 17-108_k141_288029 | Not-determined |
| 17-108_k141_232672 | Not-determined |
| 17-108_k141_53861 | Not-determined |
| 17-108_k141_121564 | Not-determined |
| 17-108_k141_53862 | Not-determined |
| 17-108_k141_204587 | Low-quality |
| 17-108_k141_204589 | Not-determined |
| 17-108_k141_149896 | Not-determined |
| 17-108_k141_94588 | Not-determined |
| 17-108_k141_135218 | Not-determined |
| 17-108_k141_232688 | Not-determined |
| 17-108_k141_53873 | Not-determined |
| 17-108_k141_12158 | Not-determined |
| 17-108_k141_204594 | Not-determined |
| 17-108_k141_12159 | Not-determined |
| 17-108_k141_288051 | Not-determined |
| 17-108_k141_121573 | Not-determined |
| 17-108_k141_164040 | Not-determined |
| 17-108_k141_121574 | Not-determined |
| 17-108_k141_53883 | Not-determined |
| 17-108_k141_190543 | Low-quality |
| 17-108_k141_94600 | Not-determined |
| 17-108_k141_121580 | Not-determined |
| 17-108_k141_164047 | Low-quality |
| 17-108_k141_315126 | Not-determined |
| 17-108_k141_315127 | Not-determined |
| 17-108_k141_24648 | Not-determined |
| 17-108_k141_12175 | Not-determined |
| 17-108_k141_218652 | Low-quality |
| 17-108_k141_80344 | Not-determined |
| 17-108_k141_53894 | Not-determined |
| 17-108_k141_135239 | Not-determined |
| 17-108_k141_288066 | Not-determined |
| 17-108_k141_135242 | Not-determined |
| 17-108_k141_121597 | Not-determined |
| 17-108_k141_273429 | Not-determined |
| 17-108_k141_80348 | Not-determined |
| 17-108_k141_53901 | Not-determined |
| 17-108_k141_149922 | Low-quality |
| 17-108_k141_67391 | Not-determined |
| 17-108_k141_327683 | Not-determined |
| 17-108_k141_190573 | Not-determined |
| 17-108_k141_121608 | Not-determined |
| 17-108_k141_40013 | Not-determined |
| 17-108_k141_273444 | Not-determined |
| 17-108_k141_260507 | Not-determined |
| 17-108_k141_260508 | Not-determined |
| 17-108_k141_40016 | Not-determined |
| 17-108_k141_288089 | Not-determined |
| 17-108_k141_190581 | Not-determined |
| 17-108_k141_190582 | Not-determined |
| 17-108_k141_149942 | Not-determined |
| 17-108_k141_12201 | Not-determined |
| 17-108_k141_12202 | Not-determined |
| 17-108_k141_177822 | Not-determined |
| 17-108_k141_315150 | Not-determined |
| 17-108_k141_260516 | Not-determined |
| 17-108_k141_288103 | Not-determined |
| 17-108_k141_204624 | Not-determined |
| 17-108_k141_177837 | Not-determined |
| 17-108_k141_80362 | Low-quality |
| 17-108_k141_260530 | Not-determined |
| 17-108_k141_135261 | Low-quality |
| 17-108_k141_135264 | Not-determined |
| 17-108_k141_177851 | Not-determined |
| 17-108_k141_67406 | Not-determined |
| 17-108_k141_177854 | Not-determined |
| 17-108_k141_232763 | Not-determined |
| 17-108_k141_247270 | Not-determined |
| 17-108_k141_288132 | Not-determined |
| 17-108_k141_135283 | Not-determined |
| 17-108_k141_288135 | Not-determined |
| 17-108_k141_327708 | Not-determined |
| 17-108_k141_24683 | Low-quality |
| 17-108_k141_40063 | Not-determined |
| 17-108_k141_121640 | Not-determined |
| 17-108_k141_24691 | Not-determined |
| 17-108_k141_121642 | Not-determined |
| 17-108_k141_12231 | Not-determined |
| 17-108_k141_12235 | Not-determined |
| 17-108_k141_247290 | Not-determined |
| 17-108_k141_247291 | Not-determined |
| 17-108_k141_121651 | Not-determined |
| 17-108_k141_109211 | Not-determined |
| 17-108_k141_260561 | Low-quality |
| 17-108_k141_109218 | Not-determined |
| 17-108_k141_12257 | Low-quality |
| 17-108_k141_109222 | Low-quality |
| 17-108_k141_80415 | Not-determined |
| 17-108_k141_12264 | Not-determined |
| 17-108_k141_12265 | Not-determined |
| 17-108_k141_12267 | Not-determined |
| 17-108_k141_232780 | Not-determined |
| 17-108_k141_150000 | Not-determined |
| 17-108_k141_121672 | Not-determined |
| 17-108_k141_327732 | Not-determined |
| 17-108_k141_315193 | Not-determined |
| 17-108_k141_315195 | Not-determined |
| 17-108_k141_121674 | Not-determined |
| 17-108_k141_177900 | Not-determined |
| 17-108_k141_12284 | Not-determined |
| 17-108_k141_232790 | Not-determined |
| 17-108_k141_40097 | Not-determined |
| 17-108_k141_150015 | Low-quality |
| 17-108_k141_121682 | Not-determined |
| 17-108_k141_177904 | Not-determined |
| 17-108_k141_150018 | Not-determined |
| 17-108_k141_273472 | Not-determined |
| 17-108_k141_260580 | Low-quality |
| 17-108_k141_94652 | Not-determined |
| 17-108_k141_40108 | Low-quality |
| 17-108_k141_40112 | Not-determined |
| 17-108_k141_204665 | Not-determined |
| 17-108_k141_260591 | Not-determined |
| 17-108_k141_204667 | Not-determined |
| 17-108_k141_260594 | Low-quality |
| 17-108_k141_121699 | Not-determined |
| 17-108_k141_150035 | Not-determined |
| 17-108_k141_94663 | Not-determined |
| 17-108_k141_273482 | Not-determined |
| 17-108_k141_40114 | Not-determined |
| 17-108_k141_273484 | Not-determined |
| 17-108_k141_260601 | Not-determined |
| 17-108_k141_177917 | Not-determined |
| 17-108_k141_327748 | Not-determined |
| 17-108_k141_12302 | Not-determined |
| 17-108_k141_273492 | Not-determined |
| 17-108_k141_260604 | Not-determined |
| 17-108_k141_260610 | Not-determined |
| 17-108_k141_80448 | Not-determined |
| 17-108_k141_190660 | Not-determined |
| 17-108_k141_121726 | Not-determined |
| 17-108_k141_190664 | Not-determined |
| 17-108_k141_12318 | Not-determined |
| 17-108_k141_177930 | Not-determined |
| 17-108_k141_24697 | Not-determined |
| 17-108_k141_40147 | Not-determined |
| 17-108_k141_247327 | Not-determined |
| 17-108_k141_150049 | Not-determined |
| 17-108_k141_232839 | Not-determined |
| 17-108_k141_327764 | Not-determined |
| 17-108_k141_80464 | Not-determined |
| 17-108_k141_12326 | Not-determined |
| 17-108_k141_67450 | Not-determined |
| 17-108_k141_327770 | Not-determined |
| 17-108_k141_109254 | Not-determined |
| 17-108_k141_327774 | Not-determined |
| 17-108_k141_109260 | Not-determined |
| 17-108_k141_24709 | Not-determined |
| 17-108_k141_218724 | Not-determined |
| 17-108_k141_109267 | Not-determined |
| 17-108_k141_260634 | Not-determined |
| 17-108_k141_315242 | Not-determined |
| 17-108_k141_177957 | Not-determined |
| 17-108_k141_190677 | Not-determined |
| 17-108_k141_24719 | Not-determined |
| 17-108_k141_177962 | Not-determined |
| 17-108_k141_40163 | Not-determined |
| 17-108_k141_177966 | Low-quality |
| 17-108_k141_273520 | Not-determined |
| 17-108_k141_204705 | Not-determined |
| 17-108_k141_24733 | Not-determined |
| 17-108_k141_135375 | Not-determined |
| 17-108_k141_315260 | Not-determined |
| 17-108_k141_260656 | Low-quality |
| 17-108_k141_218747 | Not-determined |
| 17-108_k141_67487 | Not-determined |
| 17-108_k141_260666 | Not-determined |
| 17-108_k141_40176 | Not-determined |
| 17-108_k141_218756 | Not-determined |
| 17-108_k141_12361 | Not-determined |
| 17-108_k141_135404 | Not-determined |
| 17-108_k141_327800 | Not-determined |
| 17-108_k141_260679 | Not-determined |
| 17-108_k141_273541 | Low-quality |
| 17-108_k141_273543 | Not-determined |
| 17-108_k141_260687 | Not-determined |
| 17-108_k141_150105 | Not-determined |
| 17-108_k141_24764 | Not-determined |
| 17-108_k141_204743 | Low-quality |
| 17-108_k141_204745 | Not-determined |
| 17-108_k141_177988 | Not-determined |
| 17-108_k141_260691 | Not-determined |
| 17-108_k141_24772 | Not-determined |
| 17-108_k141_177994 | Not-determined |
| 17-108_k141_315287 | Not-determined |
| 17-108_k141_204754 | Not-determined |
| 17-108_k141_67520 | Not-determined |
| 17-108_k141_67522 | Not-determined |
| 17-108_k141_218783 | Not-determined |
| 17-108_k141_288203 | Not-determined |
| 17-108_k141_301098 | Not-determined |
| 17-108_k141_94715 | Not-determined |
| 17-108_k141_178006 | Not-determined |
| 17-108_k141_232899 | Not-determined |
| 17-108_k141_260706 | Not-determined |
| 17-108_k141_109340 | Not-determined |
| 17-108_k141_135443 | Not-determined |
| 17-108_k141_301115 | Not-determined |
| 17-108_k141_24801 | Not-determined |
| 17-108_k141_135447 | Not-determined |
| 17-108_k141_109351 | Not-determined |
| 17-108_k141_190769 | Not-determined |
| 17-108_k141_40210 | Not-determined |
| 17-108_k141_12414 | Not-determined |
| 17-108_k141_94741 | Low-quality |
| 17-108_k141_273579 | Not-determined |
| 17-108_k141_150137 | Not-determined |
| 17-108_k141_178042 | Not-determined |
| 17-108_k141_94748 | Not-determined |
| 17-108_k141_315321 | Not-determined |
| 17-108_k141_109377 | Not-determined |
| 17-108_k141_190777 | Low-quality |
| 17-108_k141_53984 | Not-determined |
| 17-108_k141_135457 | Low-quality |
| 17-108_k141_260725 | Not-determined |
| 17-108_k141_53988 | Not-determined |
| 17-108_k141_135459 | Not-determined |
| 17-108_k141_67558 | Not-determined |
| 17-108_k141_12432 | Not-determined |
| 17-108_k141_109391 | Not-determined |
| 17-108_k141_53993 | Not-determined |
| 17-108_k141_150150 | Not-determined |
| 17-108_k141_109397 | Not-determined |
| 17-108_k141_218815 | Low-quality |
| 17-108_k141_315330 | Not-determined |
| 17-108_k141_218819 | Low-quality |
| 17-108_k141_109403 | Not-determined |
| 17-108_k141_40247 | Low-quality |
| 17-108_k141_273587 | Not-determined |
| 17-108_k141_315342 | Not-determined |
| 17-108_k141_273589 | Not-determined |
| 17-108_k141_135477 | Not-determined |
| 17-108_k141_135483 | Not-determined |
| 17-108_k141_260756 | Not-determined |
| 17-108_k141_121746 | Low-quality |
| 17-108_k141_94782 | Not-determined |
| 17-108_k141_260762 | Not-determined |
| 17-108_k141_12461 | Not-determined |
| 17-108_k141_204799 | Not-determined |
| 17-108_k141_260772 | Not-determined |
| 17-108_k141_204800 | Not-determined |
| 17-108_k141_273611 | Low-quality |
| 17-108_k141_150170 | Low-quality |
| 17-108_k141_204805 | Not-determined |
| 17-108_k141_135497 | Not-determined |
| 17-108_k141_54034 | Not-determined |
| 17-108_k141_40283 | Not-determined |
| 17-108_k141_135500 | Not-determined |
| 17-108_k141_54048 | Not-determined |
| 17-108_k141_315377 | Low-quality |
| 17-108_k141_315382 | Not-determined |
| 17-108_k141_315383 | Not-determined |
| 17-108_k141_218834 | Not-determined |
| 17-108_k141_40295 | Not-determined |

| | |
|---|---|
| 17-108_k141_67594 | Not-determined |
| 17-108_k141_247362 | Not-determined |
| 17-108_k141_247363 | Not-determined |
| 17-108_k141_218843 | Low-quality |
| 17-108_k141_150178 | Not-determined |
| 17-108_k141_135528 | Not-determined |
| 17-108_k141_204831 | Not-determined |
| 17-108_k141_315401 | Not-determined |
| 17-108_k141_40315 | Not-determined |
| 17-108_k141_273627 | Not-determined |
| 17-108_k141_273628 | Not-determined |
| 17-108_k141_247381 | Not-determined |
| 17-108_k141_204841 | Not-determined |
| 17-108_k141_24870 | Not-determined |
| 17-108_k141_327818 | Not-determined |
| 17-108_k141_273642 | Not-determined |
| 17-108_k141_232995 | Not-determined |
| 17-108_k141_150197 | Not-determined |
| 17-108_k141_247403 | Not-determined |
| 17-108_k141_121792 | Not-determined |
| 17-108_k141_204854 | Low-quality |
| 17-108_k141_80518 | Not-determined |
| 17-108_k141_150201 | Not-determined |
| 17-108_k141_260814 | Not-determined |
| 17-108_k141_109482 | Not-determined |
| 17-108_k141_315433 | Not-determined |
| 17-108_k141_327831 | Low-quality |
| 17-108_k141_80533 | Not-determined |
| 17-108_k141_327835 | Not-determined |
| 17-108_k141_247417 | Low-quality |
| 17-108_k141_54104 | Not-determined |
| 17-108_k141_54105 | Not-determined |
| 17-108_k141_327841 | Low-quality |
| 17-108_k141_315456 | Not-determined |
| 17-108_k141_315457 | Not-determined |
| 17-108_k141_135567 | Not-determined |
| 17-108_k141_315459 | Not-determined |
| 17-108_k141_135571 | Not-determined |
| 17-108_k141_301147 | Not-determined |
| 17-108_k141_233039 | Not-determined |
| 17-108_k141_327851 | Not-determined |
| 17-108_k141_164092 | Not-determined |
| 17-108_k141_247422 | Low-quality |
| 17-108_k141_94817 | Low-quality |
| 17-108_k141_150233 | Not-determined |
| 17-108_k141_109518 | Not-determined |
| 17-108_k141_260841 | Not-determined |
| 17-108_k141_54123 | Not-determined |
| 17-108_k141_247433 | Not-determined |
| 17-108_k141_135585 | Not-determined |
| 17-108_k141_327862 | Not-determined |
| 17-108_k141_260847 | Not-determined |
| 17-108_k141_315484 | Not-determined |
| 17-108_k141_109528 | Not-determined |
| 17-108_k141_190885 | Low-quality |
| 17-108_k141_109532 | Not-determined |
| 17-108_k141_178098 | Not-determined |
| 17-108_k141_54146 | Not-determined |
| 17-108_k141_273682 | Not-determined |
| 17-108_k141_233067 | Low-quality |
| 17-108_k141_233069 | Not-determined |
| 17-108_k141_54148 | Low-quality |
| 17-108_k141_121836 | Not-determined |
| 17-108_k141_94833 | Not-determined |
| 17-108_k141_12527 | Not-determined |
| 17-108_k141_67672 | Not-determined |
| 17-108_k141_301181 | Not-determined |
| 17-108_k141_327874 | Low-quality |
| 17-108_k141_121849 | Not-determined |
| 17-108_k141_233075 | Not-determined |
| 17-108_k141_301185 | Not-determined |
| 17-108_k141_327884 | Not-determined |
| 17-108_k141_178105 | Not-determined |
| 17-108_k141_247463 | Not-determined |
| 17-108_k141_178110 | Low-quality |
| 17-108_k141_204871 | Not-determined |
| 17-108_k141_135620 | Not-determined |
| 17-108_k141_67684 | Not-determined |
| 17-108_k141_135622 | Not-determined |
| 17-108_k141_150279 | Not-determined |
| 17-108_k141_40394 | Not-determined |
| 17-108_k141_190918 | Not-determined |
| 17-108_k141_94844 | Not-determined |
| 17-108_k141_150285 | Not-determined |
| 17-108_k141_40401 | Not-determined |
| 17-108_k141_218869 | Not-determined |
| 17-108_k141_273718 | Not-determined |
| 17-108_k141_150292 | Not-determined |
| 17-108_k141_150296 | Not-determined |
| 17-108_k141_273724 | Not-determined |
| 17-108_k141_301206 | Not-determined |
| 17-108_k141_273726 | Low-quality |
| 17-108_k141_288249 | Low-quality |
| 17-108_k141_301207 | Not-determined |
| 17-108_k141_150300 | Not-determined |
| 17-108_k141_288257 | Not-determined |
| 17-108_k141_178140 | Not-determined |
| 17-108_k141_12568 | Not-determined |
| 17-108_k141_204894 | Not-determined |
| 17-108_k141_288262 | Not-determined |
| 17-108_k141_288263 | Not-determined |
| 17-108_k141_135635 | Low-quality |
| 17-108_k141_273730 | Not-determined |
| 17-108_k141_273733 | Not-determined |
| 17-108_k141_135637 | Not-determined |
| 17-108_k141_94864 | Not-determined |
| 17-108_k141_315513 | Not-determined |
| 17-108_k141_218887 | Low-quality |
| 17-108_k141_67719 | Not-determined |
| 17-108_k141_204910 | Low-quality |
| 17-108_k141_135652 | Not-determined |
| 17-108_k141_273757 | Not-determined |
| 17-108_k141_40406 | Not-determined |
| 17-108_k141_301237 | Low-quality |
| 17-108_k141_109588 | Not-determined |
| 17-108_k141_178156 | Not-determined |
| 17-108_k141_218908 | Not-determined |
| 17-108_k141_327911 | Not-determined |
| 17-108_k141_40430 | Not-determined |
| 17-108_k141_178171 | Not-determined |
| 17-108_k141_54168 | Not-determined |
| 17-108_k141_218917 | Not-determined |
| 17-108_k141_178173 | Not-determined |
| 17-108_k141_273781 | Not-determined |
| 17-108_k141_109595 | Not-determined |
| 17-108_k141_190954 | Low-quality |
| 17-108_k141_190955 | Not-determined |
| 17-108_k141_164208 | Not-determined |
| 17-108_k141_164210 | Not-determined |
| 17-108_k141_24911 | Not-determined |
| 17-108_k141_247523 | Not-determined |
| 17-108_k141_273794 | Not-determined |
| 17-108_k141_150355 | Not-determined |
| 17-108_k141_247528 | Not-determined |
| 17-108_k141_135689 | Not-determined |
| 17-108_k141_150362 | Not-determined |
| 17-108_k141_24935 | Not-determined |
| 17-108_k141_121923 | Not-determined |
| 17-108_k141_204927 | Not-determined |
| 17-108_k141_67776 | Not-determined |
| 17-108_k141_150368 | Not-determined |
| 17-108_k141_109626 | Not-determined |
| 17-108_k141_12603 | Not-determined |
| 17-108_k141_40472 | Not-determined |
| 17-108_k141_94914 | Not-determined |
| 17-108_k141_233175 | Not-determined |
| 17-108_k141_121925 | Not-determined |
| 17-108_k141_24939 | Not-determined |
| 17-108_k141_109633 | Not-determined |
| 17-108_k141_135705 | Not-determined |
| 17-108_k141_164238 | Not-determined |
| 17-108_k141_150380 | Low-quality |
| 17-108_k141_273825 | Not-determined |
| 17-108_k141_109636 | Not-determined |
| 17-108_k141_40486 | Not-determined |
| 17-108_k141_301285 | Not-determined |
| 17-108_k141_12609 | Not-determined |
| 17-108_k141_260946 | Not-determined |
| 17-108_k141_150386 | Not-determined |
| 17-108_k141_54222 | Not-determined |
| 17-108_k141_164249 | Not-determined |
| 17-108_k141_12617 | Not-determined |
| 17-108_k141_301289 | Not-determined |
| 17-108_k141_301296 | Not-determined |
| 17-108_k141_40500 | Not-determined |
| 17-108_k141_233179 | Not-determined |
| 17-108_k141_94935 | Low-quality |
| 17-108_k141_273849 | Low-quality |
| 17-108_k141_67822 | Not-determined |
| 17-108_k141_94940 | Not-determined |
| 17-108_k141_109655 | Low-quality |
| 17-108_k141_288352 | Not-determined |
| 17-108_k141_191030 | Not-determined |
| 17-108_k141_40509 | Not-determined |
| 17-108_k141_94945 | Not-determined |
| 17-108_k141_260973 | Not-determined |
| 17-108_k141_121953 | Not-determined |
| 17-108_k141_67829 | Not-determined |
| 17-108_k141_164275 | Not-determined |
| 17-108_k141_24957 | Not-determined |
| 17-108_k141_135732 | Not-determined |
| 17-108_k141_164280 | Not-determined |
| 17-108_k141_164281 | Not-determined |
| 17-108_k141_12634 | Not-determined |
| 17-108_k141_327972 | Low-quality |
| 17-108_k141_247566 | Not-determined |
| 17-108_k141_288369 | Not-determined |
| 17-108_k141_218988 | Low-quality |
| 17-108_k141_24966 | Not-determined |
| 17-108_k141_24969 | Not-determined |
| 17-108_k141_109680 | Not-determined |
| 17-108_k141_12639 | Not-determined |
| 17-108_k141_327988 | Not-determined |
| 17-108_k141_204945 | Not-determined |
| 17-108_k141_12641 | Not-determined |
| 17-108_k141_191049 | Not-determined |
| 17-108_k141_67854 | Not-determined |
| 17-108_k141_54254 | Low-quality |
| 17-108_k141_233191 | Not-determined |
| 17-108_k141_24973 | Not-determined |
| 17-108_k141_12648 | Low-quality |
| 17-108_k141_109691 | Not-determined |
| 17-108_k141_80598 | Not-determined |
| 17-108_k141_40529 | Not-determined |
| 17-108_k141_178224 | Not-determined |
| 17-108_k141_219007 | Not-determined |
| 17-108_k141_219008 | Not-determined |
| 17-108_k141_67867 | Not-determined |
| 17-108_k141_178228 | Not-determined |
| 17-108_k141_164316 | Not-determined |
| 17-108_k141_204962 | Not-determined |
| 17-108_k141_80610 | Not-determined |
| 17-108_k141_164321 | Not-determined |
| 17-108_k141_12662 | Not-determined |
| 17-108_k141_247592 | Not-determined |
| 17-108_k141_122002 | Not-determined |
| 17-108_k141_40538 | Low-quality |
| 17-108_k141_233218 | Not-determined |
| 17-108_k141_261021 | Not-determined |
| 17-108_k141_219017 | Low-quality |
| 17-108_k141_12674 | Not-determined |
| 17-108_k141_233220 | Not-determined |
| 17-108_k141_80624 | Not-determined |
| 17-108_k141_191070 | Not-determined |
| 17-108_k141_40544 | Not-determined |
| 17-108_k141_261027 | Not-determined |
| 17-108_k141_219019 | Low-quality |
| 17-108_k141_204985 | Not-determined |
| 17-108_k141_191075 | Not-determined |
| 17-108_k141_261034 | Not-determined |
| 17-108_k141_247605 | Not-determined |
| 17-108_k141_67885 | Not-determined |
| 17-108_k141_273894 | Not-determined |
| 17-108_k141_273896 | Not-determined |
| 17-108_k141_12683 | Not-determined |
| 17-108_k141_122015 | Not-determined |
| 17-108_k141_191088 | Not-determined |
| 17-108_k141_94956 | Low-quality |
| 17-108_k141_12688 | Not-determined |
| 17-108_k141_80639 | Low-quality |
| 17-108_k141_219030 | Not-determined |
| 17-108_k141_67891 | Not-determined |
| 17-108_k141_164340 | Not-determined |
| 17-108_k141_247623 | Not-determined |
| 17-108_k141_178246 | Not-determined |
| 17-108_k141_288406 | Not-determined |
| 17-108_k141_94965 | Not-determined |
| 17-108_k141_54288 | Not-determined |
| 17-108_k141_233222 | Not-determined |
| 17-108_k141_12701 | Not-determined |
| 17-108_k141_94970 | Not-determined |
| 17-108_k141_150439 | Not-determined |
| 17-108_k141_315543 | Not-determined |
| 17-108_k141_315545 | Not-determined |
| 17-108_k141_261061 | Low-quality |
| 17-108_k141_288418 | Not-determined |
| 17-108_k141_150449 | Not-determined |
| 17-108_k141_54299 | Not-determined |
| 17-108_k141_178263 | Not-determined |
| 17-108_k141_109748 | Not-determined |
| 17-108_k141_164352 | Not-determined |
| 17-108_k141_178269 | Not-determined |
| 17-108_k141_219045 | Not-determined |
| 17-108_k141_94989 | Not-determined |
| 17-108_k141_219046 | Not-determined |
| 17-108_k141_67909 | Not-determined |
| 17-108_k141_67912 | Not-determined |
| 17-108_k141_80660 | Not-determined |
| 17-108_k141_233239 | Not-determined |
| 17-108_k141_12723 | Not-determined |
| 17-108_k141_135757 | Not-determined |
| 17-108_k141_67921 | Not-determined |
| 17-108_k141_261084 | Not-determined |
| 17-108_k141_178286 | Not-determined |
| 17-108_k141_247646 | Low-quality |
| 17-108_k141_109770 | Not-determined |
| 17-108_k141_247652 | Not-determined |
| 17-108_k141_135769 | Not-determined |
| 17-108_k141_135774 | Not-determined |
| 17-108_k141_247658 | Not-determined |
| 17-108_k141_247661 | Not-determined |
| 17-108_k141_247662 | Low-quality |
| 17-108_k141_135779 | Not-determined |
| 17-108_k141_80690 | Not-determined |
| 17-108_k141_233264 | Not-determined |
| 17-108_k141_328028 | Not-determined |
| 17-108_k141_164393 | Not-determined |
| 17-108_k141_219059 | Low-quality |
| 17-108_k141_273939 | Not-determined |
| 17-108_k141_273940 | Not-determined |
| 17-108_k141_178300 | Not-determined |
| 17-108_k141_247679 | Not-determined |
| 17-108_k141_95011 | Not-determined |
| 17-108_k141_328040 | Not-determined |
| 17-108_k141_40588 | Not-determined |
| 17-108_k141_178311 | Not-determined |
| 17-108_k141_80709 | Not-determined |
| 17-108_k141_12746 | Not-determined |
| 17-108_k141_164401 | Not-determined |
| 17-108_k141_67957 | Not-determined |
| 17-108_k141_150502 | Not-determined |
| 17-108_k141_12748 | Low-quality |
| 17-108_k141_328052 | Not-determined |
| 17-108_k141_54338 | Low-quality |
| 17-108_k141_191098 | Not-determined |
| 17-108_k141_150518 | Not-determined |
| 17-108_k141_273975 | Not-determined |
| 17-108_k141_164409 | Not-determined |
| 17-108_k141_219075 | Not-determined |
| 17-108_k141_219076 | Not-determined |
| 17-108_k141_164413 | Not-determined |
| 17-108_k141_315606 | Not-determined |
| 17-108_k141_150526 | Not-determined |
| 17-108_k141_150528 | Not-determined |
| 17-108_k141_205067 | Not-determined |
| 17-108_k141_205068 | Not-determined |
| 17-108_k141_164418 | Not-determined |
| 17-108_k141_40615 | Not-determined |
| 17-108_k141_95042 | Not-determined |
| 17-108_k141_25068 | Not-determined |
| 17-108_k141_40617 | Not-determined |
| 17-108_k141_40618 | Low-quality |
| 17-108_k141_205071 | Not-determined |
| 17-108_k141_205072 | Not-determined |
| 17-108_k141_25073 | Not-determined |
| 17-108_k141_80735 | Not-determined |
| 17-108_k141_247706 | Not-determined |
| 17-108_k141_54362 | Not-determined |
| 17-108_k141_109830 | Low-quality |
| 17-108_k141_135824 | Not-determined |
| 17-108_k141_233282 | Not-determined |
| 17-108_k141_288497 | Not-determined |
| 17-108_k141_205087 | Not-determined |
| 17-108_k141_233288 | Low-quality |
| 17-108_k141_315630 | Not-determined |
| 17-108_k141_95047 | Not-determined |
| 17-108_k141_12788 | Not-determined |
| 17-108_k141_135841 | Not-determined |
| 17-108_k141_261131 | Not-determined |
| 17-108_k141_205095 | Not-determined |
| 17-108_k141_164424 | Not-determined |
| 17-108_k141_95056 | Not-determined |
| 17-108_k141_95062 | Not-determined |
| 17-108_k141_315644 | Not-determined |
| 17-108_k141_205101 | Not-determined |
| 17-108_k141_301394 | Not-determined |
| 17-108_k141_135857 | Not-determined |
| 17-108_k141_135861 | Not-determined |
| 17-108_k141_178363 | Not-determined |
| 17-108_k141_288530 | Not-determined |
| 17-108_k141_54402 | Not-determined |
| 17-108_k141_261139 | Not-determined |
| 17-108_k141_301401 | Not-determined |
| 17-108_k141_95069 | Not-determined |
| 17-108_k141_288536 | Not-determined |
| 17-108_k141_247745 | Not-determined |
| 17-108_k141_261144 | Not-determined |
| 17-108_k141_135870 | Not-determined |
| 17-108_k141_135871 | Not-determined |
| 17-108_k141_205109 | Not-determined |
| 17-108_k141_191150 | Not-determined |
| 17-108_k141_95072 | Not-determined |
| 17-108_k141_109852 | Low-quality |
| 17-108_k141_247751 | Not-determined |
| 17-108_k141_135879 | Not-determined |
| 17-108_k141_150562 | Not-determined |
| 17-108_k141_12814 | Not-determined |
| 17-108_k141_122082 | Not-determined |
| 17-108_k141_219098 | Not-determined |
| 17-108_k141_80760 | Not-determined |
| 17-108_k141_25088 | Not-determined |
| 17-108_k141_178374 | Not-determined |
| 17-108_k141_178375 | Not-determined |
| 17-108_k141_233331 | Not-determined |
| 17-108_k141_178377 | Not-determined |
| 17-108_k141_328077 | Not-determined |
| 17-108_k141_40687 | Not-determined |
| 17-108_k141_328087 | Not-determined |
| 17-108_k141_80777 | Low-quality |
| 17-108_k141_178390 | Not-determined |
| 17-108_k141_164433 | Not-determined |
| 17-108_k141_80781 | Not-determined |
| 17-108_k141_135903 | Not-determined |
| 17-108_k141_328091 | Not-determined |
| 17-108_k141_301425 | Low-quality |
| 17-108_k141_80785 | Low-quality |
| 17-108_k141_191172 | Low-quality |
| 17-108_k141_247783 | Not-determined |
| 17-108_k141_95117 | Not-determined |
| 17-108_k141_301432 | Not-determined |
| 17-108_k141_109867 | Not-determined |
| 17-108_k141_164441 | Not-determined |
| 17-108_k141_40692 | Not-determined |
| 17-108_k141_40693 | Not-determined |
| 17-108_k141_191183 | Low-quality |
| 17-108_k141_178395 | Not-determined |
| 17-108_k141_68043 | Not-determined |
| 17-108_k141_40700 | Not-determined |
| 17-108_k141_150600 | Low-quality |
| 17-108_k141_40706 | Not-determined |
| 17-108_k141_135922 | Not-determined |
| 17-108_k141_68054 | Low-quality |
| 17-108_k141_80812 | Not-determined |
| 17-108_k141_247800 | Not-determined |
| 17-108_k141_109889 | Not-determined |
| 17-108_k141_135932 | Not-determined |
| 17-108_k141_219130 | Not-determined |
| 17-108_k141_95137 | Not-determined |
| 17-108_k141_261153 | Low-quality |
| 17-108_k141_247811 | Low-quality |
| 17-108_k141_54465 | Not-determined |
| 17-108_k141_219135 | Not-determined |
| 17-108_k141_219139 | Not-determined |
| 17-108_k141_95138 | Not-determined |
| 17-108_k141_12854 | Not-determined |
| 17-108_k141_80819 | Not-determined |
| 17-108_k141_12857 | Not-determined |
| 17-108_k141_261161 | Not-determined |
| 17-108_k141_178425 | Not-determined |
| 17-108_k141_25123 | Not-determined |
| 17-108_k141_122099 | Not-determined |
| 17-108_k141_12867 | Not-determined |
| 17-108_k141_95144 | Not-determined |
| 17-108_k141_247829 | Not-determined |
| 17-108_k141_315694 | Low-quality |
| 17-108_k141_95147 | Low-quality |
| 17-108_k141_315704 | Not-determined |
| 17-108_k141_301467 | Low-quality |
| 17-108_k141_122104 | Not-determined |
| 17-108_k141_25132 | Low-quality |
| 17-108_k141_164479 | Not-determined |
| 17-108_k141_164481 | Not-determined |
| 17-108_k141_178436 | Not-determined |
| 17-108_k141_164482 | Not-determined |
| 17-108_k141_205113 | Not-determined |
| 17-108_k141_178440 | Not-determined |
| 17-108_k141_80840 | Not-determined |
| 17-108_k141_247837 | Not-determined |
| 17-108_k141_205129 | Not-determined |
| 17-108_k141_164493 | Low-quality |
| 17-108_k141_247842 | Not-determined |
| 17-108_k141_233382 | Not-determined |
| 17-108_k141_12891 | Not-determined |
| 17-108_k141_150631 | Not-determined |
| 17-108_k141_178447 | Not-determined |
| 17-108_k141_95171 | Not-determined |
| 17-108_k141_274083 | Not-determined |
| 17-108_k141_233394 | Low-quality |
| 17-108_k141_25159 | Not-determined |
| 17-108_k141_301493 | Not-determined |
| 17-108_k141_247867 | Not-determined |
| 17-108_k141_80851 | Not-determined |
| 17-108_k141_68089 | Not-determined |
| 17-108_k141_219181 | Low-quality |
| 17-108_k141_25174 | Not-determined |
| 17-108_k141_233402 | Not-determined |
| 17-108_k141_68095 | Not-determined |
| 17-108_k141_191226 | Not-determined |
| 17-108_k141_12910 | Not-determined |
| 17-108_k141_135962 | Not-determined |
| 17-108_k141_68099 | Not-determined |
| 17-108_k141_109937 | Low-quality |
| 17-108_k141_150649 | Not-determined |
| 17-108_k141_95192 | Not-determined |
| 17-108_k141_261187 | Not-determined |
| 17-108_k141_54529 | Low-quality |
| 17-108_k141_54530 | Not-determined |
| 17-108_k141_315747 | Low-quality |
| 17-108_k141_40791 | Not-determined |
| 17-108_k141_233422 | Not-determined |
| 17-108_k141_274103 | Not-determined |
| 17-108_k141_219193 | Not-determined |
| 17-108_k141_315753 | Not-determined |
| 17-108_k141_25201 | Not-determined |
| 17-108_k141_150668 | Not-determined |
| 17-108_k141_328132 | Not-determined |
| 17-108_k141_205190 | Not-determined |
| 17-108_k141_40800 | Low-quality |
| 17-108_k141_95208 | Not-determined |
| 17-108_k141_12930 | Not-determined |
| 17-108_k141_40807 | Low-quality |
| 17-108_k141_191256 | Not-determined |
| 17-108_k141_164528 | Not-determined |
| 17-108_k141_205202 | Not-determined |
| 17-108_k141_247889 | Low-quality |
| 17-108_k141_40811 | Not-determined |
| 17-108_k141_191267 | Not-determined |
| 17-108_k141_25223 | Not-determined |
| 17-108_k141_301513 | Low-quality |
| 17-108_k141_233440 | Low-quality |
| 17-108_k141_178513 | Low-quality |
| 17-108_k141_219205 | Not-determined |
| 17-108_k141_205212 | Not-determined |
| 17-108_k141_25229 | Not-determined |
| 17-108_k141_219209 | Not-determined |
| 17-108_k141_150713 | Not-determined |
| 17-108_k141_25235 | Low-quality |
| 17-108_k141_12945 | Low-quality |
| 17-108_k141_205218 | Not-determined |
| 17-108_k141_191282 | Not-determined |
| 17-108_k141_40834 | Not-determined |
| 17-108_k141_205223 | Not-determined |
| 17-108_k141_328161 | Low-quality |
| 17-108_k141_233448 | Not-determined |
| 17-108_k141_233449 | Not-determined |
| 17-108_k141_54543 | Low-quality |
| 17-108_k141_164553 | Not-determined |
| 17-108_k141_191296 | Not-determined |
| 17-108_k141_25242 | Not-determined |
| 17-108_k141_54550 | Not-determined |
| 17-108_k141_261208 | Not-determined |
| 17-108_k141_233458 | Not-determined |
| 17-108_k141_54552 | Not-determined |
| 17-108_k141_54553 | Not-determined |
| 17-108_k141_191299 | Not-determined |
| 17-108_k141_150729 | Not-determined |
| 17-108_k141_122238 | Not-determined |
| 17-108_k141_122239 | Not-determined |
| 17-108_k141_110000 | Not-determined |
| 17-108_k141_191307 | Not-determined |
| 17-108_k141_328181 | Not-determined |
| 17-108_k141_233462 | Not-determined |
| 17-108_k141_191312 | Low-quality |
| 17-108_k141_315776 | Not-determined |
| 17-108_k141_191321 | Not-determined |
| 17-108_k141_12968 | Not-determined |
| 17-108_k141_288650 | Low-quality |
| 17-108_k141_191329 | Not-determined |
| 17-108_k141_261222 | Not-determined |
| 17-108_k141_178532 | Low-quality |
| 17-108_k141_95235 | Not-determined |
| 17-108_k141_68173 | Not-determined |
| 17-108_k141_288655 | Not-determined |
| 17-108_k141_288658 | Low-quality |
| 17-108_k141_247901 | Not-determined |
| 17-108_k141_233484 | Not-determined |
| 17-108_k141_315796 | Not-determined |
| 17-108_k141_68178 | Low-quality |
| 17-108_k141_191338 | Not-determined |
| 17-108_k141_68181 | Not-determined |
| 17-108_k141_95248 | Not-determined |
| 17-108_k141_261233 | Low-quality |
| 17-108_k141_315803 | Not-determined |
| 17-108_k141_315804 | Not-determined |
| 17-108_k141_150759 | Not-determined |
| 17-108_k141_233491 | Not-determined |
| 17-108_k141_261236 | Not-determined |
| 17-108_k141_122275 | Not-determined |
| 17-108_k141_301523 | Not-determined |
| 17-108_k141_261239 | Not-determined |
| 17-108_k141_261241 | Not-determined |
| 17-108_k141_40888 | Not-determined |
| 17-108_k141_219251 | Not-determined |
| 17-108_k141_205254 | Not-determined |
| 17-108_k141_261243 | Not-determined |
| 17-108_k141_315816 | Not-determined |
| 17-108_k141_122287 | Not-determined |
| 17-108_k141_205261 | Not-determined |
| 17-108_k141_25279 | Not-determined |
| 17-108_k141_25283 | Not-determined |
| 17-108_k141_247928 | Not-determined |
| 17-108_k141_274175 | Not-determined |
| 17-108_k141_25285 | Not-determined |
| 17-108_k141_247936 | Not-determined |
| 17-108_k141_136045 | Not-determined |
| 17-108_k141_178548 | Not-determined |
| 17-108_k141_261256 | Not-determined |
| 17-108_k141_40898 | Not-determined |
| 17-108_k141_164606 | Not-determined |
| 17-108_k141_95268 | Not-determined |
| 17-108_k141_274182 | Low-quality |
| 17-108_k141_233514 | Not-determined |
| 17-108_k141_40904 | Not-determined |
| 17-108_k141_328240 | Low-quality |
| 17-108_k141_164620 | Not-determined |
| 17-108_k141_164622 | Not-determined |
| 17-108_k141_205283 | Not-determined |
| 17-108_k141_301541 | Not-determined |
| 17-108_k141_150782 | Not-determined |
| 17-108_k141_261268 | Not-determined |
| 17-108_k141_274188 | Not-determined |
| 17-108_k141_164627 | Not-determined |
| 17-108_k141_25316 | Not-determined |
| 17-108_k141_110071 | Not-determined |
| 17-108_k141_164632 | Not-determined |
| 17-108_k141_150794 | Not-determined |
| 17-108_k141_328250 | Not-determined |
| 17-108_k141_328251 | Not-determined |
| 17-108_k141_25326 | Not-determined |

| | |
|---|---|
| 17-108_k141_25328 | Not-determined |
| 17-108_k141_54609 | Low-quality |
| 17-108_k141_150800 | Not-determined |
| 17-108_k141_122313 | Not-determined |
| 17-108_k141_25332 | Not-determined |
| 17-108_k141_54614 | Not-determined |
| 17-108_k141_315827 | Not-determined |
| 17-108_k141_150811 | Not-determined |
| 17-108_k141_288687 | Not-determined |
| 17-108_k141_301556 | Not-determined |
| 17-108_k141_150817 | Low-quality |
| 17-108_k141_274214 | Not-determined |
| 17-108_k141_110085 | Low-quality |
| 17-108_k141_233548 | Not-determined |
| 17-108_k141_110087 | Not-determined |
| 17-108_k141_164653 | Not-determined |
| 17-108_k141_122315 | Not-determined |
| 17-108_k141_150825 | Low-quality |
| 17-108_k141_13020 | Not-determined |
| 17-108_k141_191402 | Not-determined |
| 17-108_k141_247957 | Not-determined |
| 17-108_k141_122319 | Not-determined |
| 17-108_k141_233564 | Not-determined |
| 17-108_k141_122320 | Not-determined |
| 17-108_k141_247963 | Not-determined |
| 17-108_k141_80946 | Not-determined |
| 17-108_k141_261290 | Not-determined |
| 17-108_k141_288702 | Low-quality |
| 17-108_k141_233577 | Not-determined |
| 17-108_k141_150838 | Not-determined |
| 17-108_k141_328289 | Not-determined |
| 17-108_k141_219324 | Low-quality |
| 17-108_k141_164675 | Low-quality |
| 17-108_k141_13044 | Not-determined |
| 17-108_k141_40970 | Low-quality |
| 17-108_k141_80954 | Not-determined |
| 17-108_k141_328297 | Not-determined |
| 17-108_k141_247972 | Not-determined |
| 17-108_k141_122338 | Low-quality |
| 17-108_k141_233587 | Not-determined |
| 17-108_k141_274242 | Not-determined |
| 17-108_k141_150856 | Not-determined |
| 17-108_k141_25361 | Not-determined |
| 17-108_k141_301592 | Not-determined |
| 17-108_k141_178601 | Not-determined |
| 17-108_k141_164695 | Not-determined |
| 17-108_k141_261310 | Not-determined |
| 17-108_k141_13074 | Not-determined |
| 17-108_k141_136057 | Not-determined |
| 17-108_k141_122354 | Not-determined |
| 17-108_k141_301607 | Not-determined |
| 17-108_k141_136063 | Not-determined |
| 17-108_k141_95355 | Low-quality |
| 17-108_k141_95357 | Not-determined |
| 17-108_k141_13080 | Not-determined |
| 17-108_k141_13085 | Not-determined |
| 17-108_k141_178612 | Not-determined |
| 17-108_k141_178624 | Not-determined |
| 17-108_k141_247991 | Not-determined |
| 17-108_k141_136093 | Not-determined |
| 17-108_k141_164710 | Not-determined |
| 17-108_k141_54674 | Not-determined |
| 17-108_k141_219345 | Not-determined |
| 17-108_k141_80978 | Not-determined |
| 17-108_k141_136109 | Not-determined |
| 17-108_k141_68208 | Not-determined |
| 17-108_k141_178645 | Not-determined |
| 17-108_k141_68212 | Not-determined |
| 17-108_k141_178647 | Not-determined |
| 17-108_k141_54687 | Not-determined |
| 17-108_k141_54689 | Not-determined |
| 17-108_k141_261347 | Not-determined |
| 17-108_k141_80996 | Not-determined |
| 17-108_k141_68215 | Low-quality |
| 17-108_k141_164718 | Not-determined |
| 17-108_k141_150866 | Not-determined |
| 17-108_k141_315853 | Not-determined |
| 17-108_k141_315854 | Not-determined |
| 17-108_k141_261355 | Not-determined |
| 17-108_k141_68219 | Not-determined |
| 17-108_k141_164724 | Not-determined |
| 17-108_k141_178652 | Low-quality |
| 17-108_k141_248024 | Not-determined |
| 17-108_k141_219364 | Low-quality |
| 17-108_k141_122370 | Not-determined |
| 17-108_k141_54706 | Not-determined |
| 17-108_k141_54710 | Low-quality |
| 17-108_k141_205337 | Not-determined |
| 17-108_k141_150887 | Not-determined |
| 17-108_k141_80997 | Not-determined |
| 17-108_k141_54715 | Not-determined |
| 17-108_k141_178667 | Not-determined |
| 17-108_k141_233631 | Not-determined |
| 17-108_k141_205351 | Not-determined |
| 17-108_k141_233633 | Not-determined |
| 17-108_k141_288732 | Low-quality |
| 17-108_k141_150896 | Not-determined |
| 17-108_k141_301628 | Not-determined |
| 17-108_k141_110189 | Not-determined |
| 17-108_k141_25416 | Not-determined |
| 17-108_k141_219400 | Low-quality |
| 17-108_k141_219401 | Not-determined |
| 17-108_k141_54736 | Not-determined |
| 17-108_k141_54740 | Not-determined |
| 17-108_k141_54741 | Not-determined |
| 17-108_k141_328324 | Not-determined |
| 17-108_k141_205357 | Not-determined |
| 17-108_k141_150915 | Not-determined |
| 17-108_k141_288750 | Not-determined |
| 17-108_k141_191462 | Not-determined |
| 17-108_k141_164764 | Not-determined |
| 17-108_k141_288754 | Not-determined |
| 17-108_k141_233652 | Low-quality |
| 17-108_k141_328327 | Not-determined |
| 17-108_k141_54756 | Not-determined |
| 17-108_k141_248052 | Not-determined |
| 17-108_k141_25427 | Not-determined |
| 17-108_k141_68283 | Not-determined |
| 17-108_k141_122379 | Not-determined |
| 17-108_k141_178679 | Not-determined |
| 17-108_k141_136154 | Not-determined |
| 17-108_k141_95427 | Not-determined |
| 17-108_k141_288772 | Not-determined |
| 17-108_k141_68293 | Not-determined |
| 17-108_k141_288776 | Not-determined |
| 17-108_k141_68296 | Not-determined |
| 17-108_k141_68297 | Not-determined |
| 17-108_k141_54768 | Not-determined |
| 17-108_k141_40999 | Not-determined |
| 17-108_k141_122394 | Not-determined |
| 17-108_k141_233677 | Not-determined |
| 17-108_k141_122397 | Not-determined |
| 17-108_k141_219435 | Not-determined |
| 17-108_k141_136177 | Not-determined |
| 17-108_k141_164779 | Not-determined |
| 17-108_k141_205365 | Not-determined |
| 17-108_k141_110230 | Not-determined |
| 17-108_k141_13196 | Not-determined |
| 17-108_k141_54776 | Not-determined |
| 17-108_k141_136187 | Not-determined |
| 17-108_k141_178695 | Not-determined |
| 17-108_k141_315920 | Not-determined |
| 17-108_k141_122402 | Not-determined |
| 17-108_k141_122406 | Not-determined |
| 17-108_k141_25465 | Low-quality |
| 17-108_k141_54791 | Not-determined |
| 17-108_k141_54792 | Not-determined |
| 17-108_k141_301663 | Not-determined |
| 17-108_k141_191488 | Not-determined |
| 17-108_k141_233705 | Not-determined |
| 17-108_k141_301667 | Not-determined |
| 17-108_k141_178711 | Not-determined |
| 17-108_k141_301669 | Low-quality |
| 17-108_k141_261445 | Not-determined |
| 17-108_k141_178716 | Not-determined |
| 17-108_k141_288806 | Not-determined |
| 17-108_k141_25472 | Not-determined |
| 17-108_k141_233710 | Not-determined |
| 17-108_k141_205387 | Not-determined |
| 17-108_k141_25474 | Not-determined |
| 17-108_k141_95457 | Not-determined |
| 17-108_k141_205391 | Not-determined |
| 17-108_k141_261457 | Not-determined |
| 17-108_k141_110246 | Not-determined |
| 17-108_k141_315927 | Not-determined |
| 17-108_k141_95462 | Not-determined |
| 17-108_k141_41025 | Low-quality |
| 17-108_k141_219453 | Not-determined |
| 17-108_k141_25476 | Low-quality |
| 17-108_k141_191508 | Not-determined |
| 17-108_k141_219456 | Not-determined |
| 17-108_k141_13227 | Low-quality |
| 17-108_k141_328352 | Not-determined |
| 17-108_k141_13229 | Not-determined |
| 17-108_k141_25480 | Not-determined |
| 17-108_k141_219462 | Not-determined |
| 17-108_k141_54821 | Not-determined |
| 17-108_k141_233718 | Not-determined |
| 17-108_k141_274352 | Not-determined |
| 17-108_k141_205397 | Low-quality |
| 17-108_k141_150932 | Not-determined |
| 17-108_k141_95476 | Low-quality |
| 17-108_k141_191526 | Not-determined |
| 17-108_k141_205408 | Not-determined |
| 17-108_k141_81081 | Not-determined |
| 17-108_k141_261467 | Not-determined |
| 17-108_k141_41040 | Not-determined |
| 17-108_k141_13244 | Not-determined |
| 17-108_k141_164797 | Not-determined |
| 17-108_k141_233737 | Not-determined |
| 17-108_k141_95485 | Not-determined |
| 17-108_k141_95486 | Not-determined |
| 17-108_k141_205415 | Not-determined |
| 17-108_k141_315964 | Not-determined |
| 17-108_k141_219485 | Low-quality |
| 17-108_k141_25507 | Low-quality |
| 17-108_k141_95489 | Not-determined |
| 17-108_k141_13251 | Not-determined |
| 17-108_k141_122465 | Not-determined |
| 17-108_k141_164814 | Not-determined |
| 17-108_k141_164815 | Not-determined |
| 17-108_k141_288847 | Not-determined |
| 17-108_k141_13259 | Not-determined |
| 17-108_k141_13261 | Not-determined |
| 17-108_k141_95499 | Not-determined |
| 17-108_k141_164826 | Not-determined |
| 17-108_k141_233748 | Not-determined |
| 17-108_k141_41056 | Not-determined |
| 17-108_k141_219498 | Not-determined |
| 17-108_k141_68335 | Not-determined |
| 17-108_k141_68336 | Not-determined |
| 17-108_k141_25517 | Not-determined |
| 17-108_k141_25525 | Not-determined |
| 17-108_k141_301730 | Not-determined |
| 17-108_k141_95515 | Low-quality |
| 17-108_k141_81112 | Not-determined |
| 17-108_k141_233753 | Not-determined |
| 17-108_k141_95517 | Not-determined |
| 17-108_k141_41072 | Low-quality |
| 17-108_k141_150982 | Not-determined |
| 17-108_k141_233757 | Not-determined |
| 17-108_k141_288857 | Not-determined |
| 17-108_k141_315978 | Not-determined |
| 17-108_k141_95529 | Not-determined |
| 17-108_k141_164833 | Not-determined |
| 17-108_k141_288868 | Not-determined |
| 17-108_k141_315981 | Not-determined |
| 17-108_k141_150992 | Not-determined |
| 17-108_k141_25546 | Not-determined |
| 17-108_k141_288870 | Not-determined |
| 17-108_k141_191560 | Not-determined |
| 17-108_k141_68342 | Not-determined |
| 17-108_k141_315989 | Not-determined |
| 17-108_k141_54841 | Not-determined |
| 17-108_k141_191566 | Not-determined |
| 17-108_k141_191569 | Not-determined |
| 17-108_k141_68349 | Not-determined |
| 17-108_k141_261518 | Not-determined |
| 17-108_k141_151010 | Low-quality |
| 17-108_k141_233779 | Not-determined |
| 17-108_k141_164863 | Low-quality |
| 17-108_k141_68362 | Not-determined |
| 17-108_k141_95545 | Not-determined |
| 17-108_k141_95547 | Not-determined |
| 17-108_k141_41101 | Not-determined |
| 17-108_k141_68365 | Not-determined |
| 17-108_k141_54853 | Not-determined |
| 17-108_k141_233784 | Not-determined |
| 17-108_k141_191587 | Not-determined |
| 17-108_k141_68371 | Not-determined |
| 17-108_k141_233787 | Low-quality |
| 17-108_k141_136310 | Not-determined |
| 17-108_k141_81125 | Not-determined |
| 17-108_k141_301783 | Not-determined |
| 17-108_k141_164878 | Not-determined |
| 17-108_k141_178765 | Not-determined |
| 17-108_k141_136313 | Not-determined |
| 17-108_k141_274366 | Low-quality |
| 17-108_k141_288891 | Not-determined |
| 17-108_k141_136316 | Not-determined |
| 17-108_k141_95556 | Not-determined |
| 17-108_k141_81138 | Not-determined |
| 17-108_k141_81142 | Not-determined |
| 17-108_k141_288908 | Not-determined |
| 17-108_k141_13288 | Not-determined |
| 17-108_k141_178773 | Not-determined |
| 17-108_k141_81149 | Not-determined |
| 17-108_k141_122517 | Not-determined |
| 17-108_k141_122521 | Not-determined |
| 17-108_k141_13293 | Not-determined |
| 17-108_k141_122522 | Not-determined |
| 17-108_k141_81160 | Not-determined |
| 17-108_k141_151035 | Not-determined |
| 17-108_k141_288916 | Not-determined |
| 17-108_k141_178787 | Low-quality |
| 17-108_k141_301826 | Not-determined |
| 17-108_k141_151042 | Not-determined |
| 17-108_k141_274391 | Not-determined |
| 17-108_k141_301830 | Not-determined |
| 17-108_k141_151048 | Low-quality |
| 17-108_k141_13310 | Not-determined |
| 17-108_k141_122536 | Low-quality |
| 17-108_k141_328439 | Not-determined |
| 17-108_k141_13314 | Not-determined |
| 17-108_k141_205454 | Not-determined |
| 17-108_k141_191643 | Not-determined |
| 17-108_k141_25580 | Not-determined |
| 17-108_k141_13317 | Not-determined |
| 17-108_k141_191645 | Not-determined |
| 17-108_k141_191647 | Not-determined |
| 17-108_k141_261573 | Not-determined |
| 17-108_k141_205460 | Not-determined |
| 17-108_k141_191648 | Not-determined |
| 17-108_k141_261575 | Not-determined |
| 17-108_k141_219595 | Not-determined |
| 17-108_k141_205468 | Not-determined |
| 17-108_k141_301854 | Not-determined |
| 17-108_k141_68414 | Not-determined |
| 17-108_k141_25598 | Not-determined |
| 17-108_k141_68418 | Not-determined |
| 17-108_k141_191660 | Not-determined |
| 17-108_k141_122558 | Not-determined |
| 17-108_k141_54883 | Not-determined |
| 17-108_k141_233832 | Not-determined |
| 17-108_k141_233833 | Not-determined |
| 17-108_k141_233835 | Not-determined |
| 17-108_k141_205476 | Not-determined |
| 17-108_k141_191671 | Low-quality |
| 17-108_k141_205482 | Not-determined |
| 17-108_k141_301885 | Not-determined |
| 17-108_k141_68431 | Not-determined |
| 17-108_k141_205485 | Not-determined |
| 17-108_k141_233845 | Not-determined |
| 17-108_k141_301890 | Not-determined |
| 17-108_k141_301895 | Not-determined |
| 17-108_k141_274418 | Not-determined |
| 17-108_k141_122566 | Not-determined |
| 17-108_k141_205499 | Not-determined |
| 17-108_k141_219614 | Not-determined |
| 17-108_k141_68438 | Not-determined |
| 17-108_k141_164920 | Not-determined |
| 17-108_k141_301901 | Not-determined |
| 17-108_k141_301905 | Low-quality |
| 17-108_k141_205513 | Not-determined |
| 17-108_k141_274426 | Low-quality |
| 17-108_k141_13356 | Not-determined |
| 17-108_k141_122584 | Not-determined |
| 17-108_k141_288946 | Not-determined |
| 17-108_k141_136375 | Not-determined |
| 17-108_k141_191685 | Not-determined |
| 17-108_k141_95620 | Not-determined |
| 17-108_k141_316000 | Not-determined |
| 17-108_k141_122589 | Low-quality |
| 17-108_k141_219626 | Not-determined |
| 17-108_k141_261610 | Not-determined |
| 17-108_k141_136381 | Not-determined |
| 17-108_k141_301925 | Not-determined |
| 17-108_k141_288959 | Not-determined |
| 17-108_k141_191692 | Not-determined |
| 17-108_k141_164945 | Not-determined |
| 17-108_k141_54900 | Not-determined |
| 17-108_k141_288967 | Not-determined |
| 17-108_k141_81187 | Not-determined |
| 17-108_k141_219637 | Not-determined |
| 17-108_k141_54904 | Not-determined |
| 17-108_k141_301932 | Not-determined |
| 17-108_k141_178831 | Not-determined |
| 17-108_k141_274448 | Not-determined |
| 17-108_k141_95622 | Not-determined |
| 17-108_k141_316029 | Not-determined |
| 17-108_k141_178833 | Not-determined |
| 17-108_k141_54910 | Not-determined |
| 17-108_k141_54912 | Not-determined |
| 17-108_k141_54913 | Not-determined |
| 17-108_k141_191712 | Not-determined |
| 17-108_k141_25662 | Not-determined |
| 17-108_k141_136403 | Not-determined |
| 17-108_k141_164957 | Not-determined |
| 17-108_k141_191716 | Not-determined |
| 17-108_k141_261626 | Not-determined |
| 17-108_k141_274468 | Not-determined |
| 17-108_k141_274469 | Not-determined |
| 17-108_k141_151058 | Not-determined |
| 17-108_k141_191725 | Not-determined |
| 17-108_k141_233905 | Not-determined |
| 17-108_k141_191733 | Not-determined |
| 17-108_k141_41146 | Not-determined |
| 17-108_k141_13409 | Not-determined |
| 17-108_k141_13412 | Not-determined |
| 17-108_k141_122635 | Not-determined |
| 17-108_k141_219669 | Not-determined |
| 17-108_k141_288977 | Not-determined |
| 17-108_k141_164977 | Not-determined |
| 17-108_k141_164982 | Not-determined |
| 17-108_k141_81219 | Not-determined |
| 17-108_k141_164991 | Low-quality |
| 17-108_k141_136411 | Not-determined |
| 17-108_k141_41163 | Not-determined |
| 17-108_k141_81230 | Not-determined |
| 17-108_k141_13435 | Low-quality |
| 17-108_k141_151077 | Not-determined |
| 17-108_k141_205556 | Not-determined |
| 17-108_k141_54966 | Not-determined |
| 17-108_k141_122659 | Low-quality |
| 17-108_k141_95651 | Not-determined |
| 17-108_k141_288995 | Not-determined |
| 17-108_k141_136425 | Not-determined |
| 17-108_k141_41190 | Not-determined |
| 17-108_k141_81255 | Not-determined |
| 17-108_k141_95653 | Not-determined |
| 17-108_k141_95655 | Not-determined |
| 17-108_k141_95657 | Not-determined |
| 17-108_k141_95659 | Low-quality |
| 17-108_k141_81262 | Not-determined |
| 17-108_k141_81263 | Not-determined |
| 17-108_k141_289014 | Not-determined |
| 17-108_k141_233954 | Not-determined |
| 17-108_k141_178848 | Not-determined |
| 17-108_k141_316098 | Not-determined |
| 17-108_k141_41200 | Not-determined |
| 17-108_k141_41202 | Low-quality |
| 17-108_k141_261668 | Not-determined |
| 17-108_k141_122681 | Not-determined |
| 17-108_k141_233959 | Not-determined |
| 17-108_k141_54991 | Not-determined |
| 17-108_k141_81275 | Not-determined |
| 17-108_k141_328490 | Not-determined |
| 17-108_k141_316112 | Not-determined |
| 17-108_k141_151102 | Not-determined |
| 17-108_k141_316119 | Not-determined |
| 17-108_k141_233979 | Not-determined |
| 17-108_k141_316121 | Not-determined |
| 17-108_k141_191816 | Not-determined |
| 17-108_k141_302020 | Not-determined |
| 17-108_k141_191819 | Not-determined |
| 17-108_k141_289039 | Not-determined |
| 17-108_k141_136453 | Not-determined |
| 17-108_k141_316128 | Not-determined |
| 17-108_k141_191820 | Low-quality |
| 17-108_k141_316141 | Not-determined |
| 17-108_k141_274514 | Not-determined |
| 17-108_k141_25740 | Not-determined |
| 17-108_k141_13482 | Not-determined |
| 17-108_k141_165035 | Not-determined |
| 17-108_k141_81303 | Not-determined |
| 17-108_k141_151113 | Not-determined |
| 17-108_k141_55008 | Not-determined |
| 17-108_k141_25753 | Not-determined |
| 17-108_k141_122718 | Not-determined |
| 17-108_k141_302040 | Not-determined |
| 17-108_k141_274526 | Not-determined |
| 17-108_k141_81312 | Not-determined |
| 17-108_k141_178883 | Not-determined |
| 17-108_k141_165056 | Not-determined |
| 17-108_k141_151126 | Not-determined |
| 17-108_k141_234008 | Not-determined |
| 17-108_k141_122736 | Not-determined |
| 17-108_k141_81322 | Low-quality |
| 17-108_k141_151135 | Not-determined |
| 17-108_k141_41247 | Not-determined |
| 17-108_k141_274531 | Not-determined |
| 17-108_k141_165072 | Low-quality |
| 17-108_k141_274537 | Not-determined |
| 17-108_k141_328538 | Not-determined |
| 17-108_k141_328539 | Low-quality |
| 17-108_k141_234020 | Not-determined |
| 17-108_k141_122755 | Not-determined |
| 17-108_k141_219744 | Not-determined |
| 17-108_k141_289064 | Not-determined |
| 17-108_k141_178902 | Not-determined |
| 17-108_k141_302054 | Low-quality |
| 17-108_k141_219747 | Not-determined |
| 17-108_k141_302056 | Not-determined |
| 17-108_k141_234026 | Not-determined |
| 17-108_k141_205607 | Not-determined |
| 17-108_k141_95721 | Not-determined |
| 17-108_k141_219750 | Not-determined |
| 17-108_k141_95727 | Not-determined |
| 17-108_k141_95732 | Not-determined |
| 17-108_k141_274550 | Not-determined |
| 17-108_k141_122767 | Not-determined |
| 17-108_k141_13508 | Not-determined |
| 17-108_k141_178908 | Not-determined |
| 17-108_k141_302063 | Not-determined |
| 17-108_k141_95743 | Low-quality |
| 17-108_k141_55049 | Not-determined |
| 17-108_k141_136521 | Not-determined |
| 17-108_k141_95751 | Low-quality |
| 17-108_k141_41276 | Not-determined |
| 17-108_k141_68547 | Not-determined |
| 17-108_k141_261753 | Not-determined |
| 17-108_k141_219767 | Not-determined |
| 17-108_k141_219768 | Not-determined |
| 17-108_k141_25801 | Not-determined |
| 17-108_k141_302072 | Not-determined |
| 17-108_k141_205617 | Not-determined |
| 17-108_k141_95773 | Not-determined |
| 17-108_k141_95774 | Not-determined |
| 17-108_k141_178931 | Low-quality |
| 17-108_k141_55071 | Not-determined |
| 17-108_k141_68558 | Not-determined |
| 17-108_k141_41298 | Not-determined |
| 17-108_k141_41301 | Not-determined |
| 17-108_k141_234050 | Not-determined |
| 17-108_k141_289080 | Low-quality |
| 17-108_k141_55074 | Not-determined |
| 17-108_k141_41305 | Not-determined |
| 17-108_k141_302105 | Not-determined |
| 17-108_k141_81391 | Not-determined |
| 17-108_k141_68567 | Low-quality |
| 17-108_k141_328576 | Not-determined |
| 17-108_k141_205643 | Low-quality |
| 17-108_k141_205645 | Not-determined |
| 17-108_k141_151190 | Not-determined |
| 17-108_k141_81406 | Low-quality |
| 17-108_k141_122799 | Not-determined |
| 17-108_k141_205653 | Not-determined |
| 17-108_k141_274596 | Not-determined |
| 17-108_k141_234077 | Not-determined |
| 17-108_k141_25844 | Not-determined |
| 17-108_k141_151200 | Low-quality |
| 17-108_k141_316203 | Not-determined |
| 17-108_k141_205661 | Not-determined |
| 17-108_k141_151204 | Not-determined |
| 17-108_k141_274605 | Not-determined |
| 17-108_k141_302119 | Not-determined |
| 17-108_k141_289113 | Not-determined |
| 17-108_k141_41317 | Not-determined |
| 17-108_k141_178949 | Not-determined |
| 17-108_k141_95807 | Not-determined |
| 17-108_k141_151211 | Low-quality |
| 17-108_k141_289124 | Low-quality |
| 17-108_k141_191847 | Not-determined |
| 17-108_k141_95813 | Not-determined |
| 17-108_k141_234103 | Not-determined |
| 17-108_k141_95822 | Not-determined |
| 17-108_k141_219795 | Not-determined |
| 17-108_k141_289132 | Low-quality |
| 17-108_k141_178962 | Not-determined |
| 17-108_k141_316228 | Not-determined |
| 17-108_k141_68598 | Not-determined |
| 17-108_k141_205687 | Not-determined |
| 17-108_k141_165162 | Not-determined |
| 17-108_k141_136549 | Not-determined |
| 17-108_k141_81427 | Not-determined |
| 17-108_k141_122838 | Not-determined |
| 17-108_k141_81431 | Not-determined |
| 17-108_k141_328624 | Not-determined |
| 17-108_k141_13559 | Not-determined |
| 17-108_k141_219815 | Not-determined |
| 17-108_k141_302161 | Low-quality |
| 17-108_k141_151240 | Not-determined |
| 17-108_k141_41342 | Low-quality |
| 17-108_k141_302167 | Not-determined |
| 17-108_k141_328633 | Not-determined |
| 17-108_k141_81434 | Low-quality |
| 17-108_k141_178995 | Not-determined |
| 17-108_k141_13577 | Not-determined |
| 17-108_k141_95852 | Not-determined |
| 17-108_k141_95853 | Not-determined |
| 17-108_k141_289163 | Not-determined |
| 17-108_k141_122855 | Not-determined |
| 17-108_k141_219826 | Not-determined |
| 17-108_k141_205709 | Not-determined |
| 17-108_k141_95859 | Low-quality |
| 17-108_k141_316247 | Not-determined |
| 17-108_k141_234146 | Not-determined |
| 17-108_k141_13591 | Not-determined |
| 17-108_k141_205712 | Not-determined |
| 17-108_k141_165193 | Not-determined |
| 17-108_k141_205713 | Not-determined |
| 17-108_k141_302195 | Not-determined |
| 17-108_k141_191856 | Not-determined |
| 17-108_k141_81445 | Not-determined |
| 17-108_k141_165198 | Not-determined |
| 17-108_k141_179015 | Not-determined |
| 17-108_k141_25911 | Not-determined |
| 17-108_k141_316259 | Low-quality |
| 17-108_k141_13600 | Not-determined |
| 17-108_k141_302216 | Not-determined |
| 17-108_k141_234166 | Not-determined |
| 17-108_k141_136600 | Not-determined |
| 17-108_k141_234180 | Not-determined |
| 17-108_k141_13611 | Not-determined |
| 17-108_k141_179033 | Not-determined |
| 17-108_k141_205741 | Not-determined |
| 17-108_k141_234187 | Not-determined |
| 17-108_k141_179036 | Not-determined |
| 17-108_k141_13621 | Not-determined |
| 17-108_k141_316281 | Not-determined |
| 17-108_k141_316282 | Not-determined |
| 17-108_k141_165217 | Low-quality |
| 17-108_k141_234191 | Not-determined |
| 17-108_k141_205750 | Not-determined |
| 17-108_k141_165225 | Not-determined |
| 17-108_k141_55103 | Low-quality |
| 17-108_k141_179050 | Not-determined |
| 17-108_k141_302237 | Not-determined |
| 17-108_k141_234203 | Not-determined |
| 17-108_k141_234204 | Not-determined |
| 17-108_k141_316295 | Not-determined |
| 17-108_k141_81490 | Not-determined |
| 17-108_k141_13630 | Not-determined |
| 17-108_k141_179058 | Not-determined |
| 17-108_k141_151276 | Not-determined |
| 17-108_k141_328672 | Not-determined |
| 17-108_k141_179063 | Not-determined |
| 17-108_k141_179064 | Not-determined |
| 17-108_k141_219855 | Not-determined |

| | |
|---|---|
| 17-108_k141_205774 | Not-determined |
| 17-108_k141_219862 | Not-determined |
| 17-108_k141_151289 | Not-determined |
| 17-108_k141_302248 | Not-determined |
| 17-108_k141_95902 | Not-determined |
| 17-108_k141_81503 | Not-determined |
| 17-108_k141_136617 | Not-determined |
| 17-108_k141_219871 | Not-determined |
| 17-108_k141_136619 | Not-determined |
| 17-108_k141_234216 | Low-quality |
| 17-108_k141_302262 | Not-determined |
| 17-108_k141_205777 | Not-determined |
| 17-108_k141_234219 | Not-determined |
| 17-108_k141_205778 | Not-determined |
| 17-108_k141_274736 | Not-determined |
| 17-108_k141_165258 | Not-determined |
| 17-108_k141_302273 | Not-determined |
| 17-108_k141_289229 | Not-determined |
| 17-108_k141_179075 | Not-determined |
| 17-108_k141_136635 | Low-quality |
| 17-108_k141_151310 | Not-determined |
| 17-108_k141_219901 | Not-determined |
| 17-108_k141_81525 | Not-determined |
| 17-108_k141_122936 | Not-determined |
| 17-108_k141_328696 | Not-determined |
| 17-108_k141_151317 | Not-determined |
| 17-108_k141_219903 | Low-quality |
| 17-108_k141_234233 | Not-determined |
| 17-108_k141_81533 | Not-determined |
| 17-108_k141_302291 | Not-determined |
| 17-108_k141_191914 | Not-determined |
| 17-108_k141_205797 | Not-determined |
| 17-108_k141_179095 | Not-determined |
| 17-108_k141_191919 | Not-determined |
| 17-108_k141_165277 | Not-determined |
| 17-108_k141_165279 | Not-determined |
| 17-108_k141_165280 | Not-determined |
| 17-108_k141_165284 | Not-determined |
| 17-108_k141_289253 | Not-determined |
| 17-108_k141_205812 | Not-determined |
| 17-108_k141_136646 | Not-determined |
| 17-108_k141_328707 | Not-determined |
| 17-108_k141_81541 | Not-determined |
| 17-108_k141_234243 | Not-determined |
| 17-108_k141_191931 | Not-determined |
| 17-108_k141_95920 | Low-quality |
| 17-108_k141_68677 | Not-determined |
| 17-108_k141_289272 | Not-determined |
| 17-108_k141_328718 | Not-determined |
| 17-108_k141_95932 | Not-determined |
| 17-108_k141_26007 | Not-determined |
| 17-108_k141_328726 | Not-determined |
| 17-108_k141_81558 | Low-quality |
| 17-108_k141_151356 | Not-determined |
| 17-108_k141_316317 | Not-determined |
| 17-108_k141_328728 | Not-determined |
| 17-108_k141_13683 | Not-determined |
| 17-108_k141_122972 | Not-determined |
| 17-108_k141_122974 | Not-determined |
| 17-108_k141_328733 | Not-determined |
| 17-108_k141_219937 | Not-determined |
| 17-108_k141_274770 | Not-determined |
| 17-108_k141_13689 | Not-determined |
| 17-108_k141_13694 | Not-determined |
| 17-108_k141_26025 | Not-determined |
| 17-108_k141_302335 | Not-determined |
| 17-108_k141_165319 | Not-determined |
| 17-108_k141_179127 | Not-determined |
| 17-108_k141_165321 | Not-determined |
| 17-108_k141_151376 | Not-determined |
| 17-108_k141_165324 | Not-determined |
| 17-108_k141_151382 | Not-determined |
| 17-108_k141_26035 | Not-determined |
| 17-108_k141_151387 | Not-determined |
| 17-108_k141_316340 | Low-quality |
| 17-108_k141_151398 | Not-determined |
| 17-108_k141_234274 | Low-quality |
| 17-108_k141_274794 | Not-determined |
| 17-108_k141_289305 | Not-determined |
| 17-108_k141_26046 | Not-determined |
| 17-108_k141_289312 | Low-quality |
| 17-108_k141_165339 | Not-determined |
| 17-108_k141_151408 | Not-determined |
| 17-108_k141_165344 | Not-determined |
| 17-108_k141_289322 | Not-determined |
| 17-108_k141_123027 | Not-determined |
| 17-108_k141_26053 | Not-determined |
| 17-108_k141_151414 | Not-determined |
| 17-108_k141_316357 | Not-determined |
| 17-108_k141_302364 | Not-determined |
| 17-108_k141_13730 | Not-determined |
| 17-108_k141_81605 | Not-determined |
| 17-108_k141_13736 | Not-determined |
| 17-108_k141_219962 | Low-quality |
| 17-108_k141_151419 | Not-determined |
| 17-108_k141_165353 | Low-quality |
| 17-108_k141_219967 | Not-determined |
| 17-108_k141_26070 | Not-determined |
| 17-108_k141_316359 | Not-determined |
| 17-108_k141_95975 | Not-determined |
| 17-108_k141_136722 | Not-determined |
| 17-108_k141_289340 | Not-determined |
| 17-108_k141_136723 | Not-determined |
| 17-108_k141_316368 | Not-determined |
| 17-108_k141_179150 | Not-determined |
| 17-108_k141_68686 | Not-determined |
| 17-108_k141_289343 | Not-determined |
| 17-108_k141_136733 | Not-determined |
| 17-108_k141_191999 | Not-determined |
| 17-108_k141_289355 | Not-determined |
| 17-108_k141_95995 | Not-determined |
| 17-108_k141_68701 | Not-determined |
| 17-108_k141_123036 | Not-determined |
| 17-108_k141_13754 | Not-determined |
| 17-108_k141_26093 | Not-determined |
| 17-108_k141_26094 | Not-determined |
| 17-108_k141_68709 | Not-determined |
| 17-108_k141_205873 | Not-determined |
| 17-108_k141_274838 | Not-determined |
| 17-108_k141_274842 | Not-determined |
| 17-108_k141_302417 | Not-determined |
| 17-108_k141_13769 | Not-determined |
| 17-108_k141_205885 | Not-determined |
| 17-108_k141_316375 | Not-determined |
| 17-108_k141_13778 | Not-determined |
| 17-108_k141_302432 | Low-quality |
| 17-108_k141_302433 | Not-determined |
| 17-108_k141_302434 | Not-determined |
| 17-108_k141_205895 | Not-determined |
| 17-108_k141_136778 | Not-determined |
| 17-108_k141_205898 | Low-quality |
| 17-108_k141_26120 | Not-determined |
| 17-108_k141_192023 | Not-determined |
| 17-108_k141_219991 | Not-determined |
| 17-108_k141_274852 | Not-determined |
| 17-108_k141_289388 | Not-determined |
| 17-108_k141_274853 | Not-determined |
| 17-108_k141_316389 | Not-determined |
| 17-108_k141_123075 | Not-determined |
| 17-108_k141_96055 | Not-determined |
| 17-108_k141_136808 | Not-determined |
| 17-108_k141_136813 | Not-determined |
| 17-108_k141_328766 | Not-determined |
| 17-108_k141_192056 | Not-determined |
| 17-108_k141_192065 | Not-determined |
| 17-108_k141_68749 | Not-determined |
| 17-108_k141_302482 | Not-determined |
| 17-108_k141_136831 | Not-determined |
| 17-108_k141_302489 | Low-quality |
| 17-108_k141_96091 | Not-determined |
| 17-108_k141_26155 | Not-determined |
| 17-108_k141_302492 | Not-determined |
| 17-108_k141_136841 | Not-determined |
| 17-108_k141_26164 | Not-determined |
| 17-108_k141_123120 | Not-determined |
| 17-108_k141_274890 | Not-determined |
| 17-108_k141_26166 | Not-determined |
| 17-108_k141_96102 | Not-determined |
| 17-108_k141_136852 | Not-determined |
| 17-108_k141_26170 | Not-determined |
| 17-108_k141_302502 | Not-determined |
| 17-108_k141_328788 | Not-determined |
| 17-108_k141_274907 | Not-determined |
| 17-108_k141_205972 | Not-determined |
| 17-108_k141_328795 | Not-determined |
| 17-108_k141_123126 | Not-determined |
| 17-108_k141_123127 | Not-determined |
| 17-108_k141_328797 | Not-determined |
| 17-108_k141_96105 | Not-determined |
| 17-108_k141_192088 | Not-determined |
| 17-108_k141_205983 | Not-determined |
| 17-108_k141_205984 | Not-determined |
| 17-108_k141_205989 | Not-determined |
| 17-108_k141_26215 | Not-determined |
| 17-108_k141_220073 | Not-determined |
| 17-108_k141_96124 | Not-determined |
| 17-108_k141_81722 | Low-quality |
| 17-108_k141_81727 | Not-determined |
| 17-108_k141_68769 | Not-determined |
| 17-108_k141_302522 | Low-quality |
| 17-108_k141_151435 | Not-determined |
| 17-108_k141_328826 | Not-determined |
| 17-108_k141_220090 | Not-determined |
| 17-108_k141_81739 | Not-determined |
| 17-108_k141_220095 | Not-determined |
| 17-108_k141_151440 | Not-determined |
| 17-108_k141_220103 | Not-determined |
| 17-108_k141_96158 | Not-determined |
| 17-108_k141_68780 | Not-determined |
| 17-108_k141_26243 | Not-determined |
| 17-108_k141_302544 | Not-determined |
| 17-108_k141_123145 | Low-quality |
| 17-108_k141_316487 | Not-determined |
| 17-108_k141_220115 | Not-determined |
| 17-108_k141_302556 | Not-determined |
| 17-108_k141_81762 | Not-determined |
| 17-108_k141_26258 | Not-determined |
| 17-108_k141_192129 | Not-determined |
| 17-108_k141_136900 | Not-determined |
| 17-108_k141_136906 | Not-determined |
| 17-108_k141_206028 | Not-determined |
| 17-108_k141_192136 | Not-determined |
| 17-108_k141_206030 | Not-determined |
| 17-108_k141_220126 | Not-determined |
| 17-108_k141_81783 | Not-determined |
| 17-108_k141_96195 | Not-determined |
| 17-108_k141_220133 | Not-determined |
| 17-108_k141_81790 | Not-determined |
| 17-108_k141_151458 | Not-determined |
| 17-108_k141_151461 | Not-determined |
| 17-108_k141_302597 | Not-determined |
| 17-108_k141_26278 | Not-determined |
| 17-108_k141_192145 | Not-determined |
| 17-108_k141_151476 | Not-determined |
| 17-108_k141_96215 | Not-determined |
| 17-108_k141_136943 | Not-determined |
| 17-108_k141_151478 | Not-determined |
| 17-108_k141_206049 | Low-quality |
| 17-108_k141_151486 | Low-quality |
| 17-108_k141_302618 | Low-quality |
| 17-108_k141_123205 | Not-determined |
| 17-108_k141_123206 | Not-determined |
| 17-108_k141_96227 | Not-determined |
| 17-108_k141_206059 | Not-determined |
| 17-108_k141_192165 | Not-determined |
| 17-108_k141_274995 | Not-determined |
| 17-108_k141_96235 | Not-determined |
| 17-108_k141_302629 | Not-determined |
| 17-108_k141_302632 | Not-determined |
| 17-108_k141_220170 | Not-determined |
| 17-108_k141_206083 | Not-determined |
| 17-108_k141_275008 | Not-determined |
| 17-108_k141_206084 | Low-quality |
| 17-108_k141_151491 | Not-determined |
| 17-108_k141_316518 | Not-determined |
| 17-108_k141_192185 | Not-determined |
| 17-108_k141_26308 | Not-determined |
| 17-108_k141_220187 | Not-determined |
| 17-108_k141_26312 | Not-determined |
| 17-108_k141_275033 | Not-determined |
| 17-108_k141_26314 | Not-determined |
| 17-108_k141_26317 | Not-determined |
| 17-108_k141_302655 | Not-determined |
| 17-108_k141_302656 | Not-determined |
| 17-108_k141_316530 | Not-determined |
| 17-108_k141_316533 | Not-determined |
| 17-108_k141_81838 | Not-determined |
| 17-108_k141_316539 | Not-determined |
| 17-108_k141_68820 | Not-determined |
| 17-108_k141_26327 | Not-determined |
| 17-108_k141_81842 | Low-quality |
| 17-108_k141_275054 | Not-determined |
| 17-108_k141_26335 | Not-determined |
| 17-108_k141_192203 | Not-determined |
| 17-108_k141_26338 | Not-determined |
| 17-108_k141_123256 | Not-determined |
| 17-108_k141_328875 | Not-determined |
| 17-108_k141_81865 | Not-determined |
| 17-108_k141_192222 | Not-determined |
| 17-108_k141_68857 | Not-determined |
| 17-108_k141_81871 | Not-determined |
| 17-108_k141_68858 | Not-determined |
| 17-108_k141_220193 | Not-determined |
| 17-108_k141_302687 | Not-determined |
| 17-108_k141_96286 | Not-determined |
| 17-108_k141_151511 | Not-determined |
| 17-108_k141_123277 | Not-determined |
| 17-108_k141_151515 | Not-determined |
| 17-108_k141_81885 | Not-determined |
| 17-108_k141_123284 | Not-determined |
| 17-108_k141_26349 | Not-determined |
| 17-108_k141_302716 | Not-determined |
| 17-108_k141_192244 | Not-determined |
| 17-108_k141_206101 | Not-determined |
| 17-108_k141_275089 | Not-determined |
| 17-108_k141_302727 | Not-determined |
| 17-108_k141_220219 | Not-determined |
| 17-108_k141_220221 | Not-determined |
| 17-108_k141_275094 | Not-determined |
| 17-108_k141_192261 | Not-determined |
| 17-108_k141_192262 | Not-determined |
| 17-108_k141_206111 | Not-determined |
| 17-108_k141_206112 | Not-determined |
| 17-108_k141_137041 | Not-determined |
| 17-108_k141_96308 | Not-determined |
| 17-108_k141_192268 | Not-determined |
| 17-108_k141_151552 | Not-determined |
| 17-108_k141_123317 | Not-determined |
| 17-108_k141_192272 | Not-determined |
| 17-108_k141_302749 | Not-determined |
| 17-108_k141_328921 | Not-determined |
| 17-108_k141_137052 | Low-quality |
| 17-108_k141_192288 | Not-determined |
| 17-108_k141_123327 | Not-determined |
| 17-108_k141_137054 | Not-determined |
| 17-108_k141_192292 | Not-determined |
| 17-108_k141_220247 | Not-determined |
| 17-108_k141_151576 | Not-determined |
| 17-108_k141_96334 | Not-determined |
| 17-108_k141_96336 | Not-determined |
| 17-108_k141_192302 | Not-determined |
| 17-108_k141_123349 | Not-determined |
| 17-108_k141_302785 | Not-determined |
| 17-108_k141_192305 | Not-determined |
| 17-108_k141_302794 | Not-determined |
| 17-108_k141_302798 | Not-determined |
| 17-108_k141_123358 | Not-determined |
| 17-108_k141_137095 | Not-determined |
| 17-108_k141_26413 | Not-determined |
| 17-108_k141_192328 | Not-determined |
| 17-108_k141_192340 | Not-determined |
| 17-108_k141_206124 | Not-determined |
| 17-108_k141_192344 | Not-determined |
| 17-108_k141_220269 | Not-determined |
| 17-108_k141_328960 | Not-determined |
| 17-108_k141_26431 | Not-determined |
| 17-108_k141_328963 | Not-determined |
| 17-108_k141_206130 | Not-determined |
| 17-108_k141_206131 | Not-determined |
| 17-108_k141_328969 | Not-determined |
| 17-108_k141_328971 | Not-determined |
| 17-108_k141_123392 | Not-determined |
| 17-108_k141_302808 | Not-determined |
| 17-108_k141_275157 | Not-determined |
| 17-108_k141_206138 | Not-determined |
| 17-108_k141_275158 | Not-determined |
| 17-108_k141_192367 | Not-determined |
| 17-108_k141_275161 | Not-determined |
| 17-108_k141_123404 | Low-quality |
| 17-108_k141_123408 | Not-determined |
| 17-108_k141_137136 | Not-determined |
| 17-108_k141_206145 | Not-determined |
| 17-108_k141_302811 | Not-determined |
| 17-108_k141_137141 | Not-determined |
| 17-108_k141_275179 | Not-determined |
| 17-108_k141_275180 | Not-determined |
| 17-108_k141_275184 | Not-determined |
| 17-108_k141_26473 | Low-quality |
| 17-108_k141_220291 | Not-determined |
| 17-108_k141_26484 | Low-quality |
| 17-108_k141_123428 | Not-determined |
| 17-108_k141_26501 | Not-determined |
| 17-108_k141_275196 | Not-determined |
| 17-108_k141_220318 | Not-determined |
| 17-108_k141_26504 | Not-determined |
| 17-108_k141_81999 | Not-determined |
| 17-108_k141_316577 | Not-determined |
| 17-108_k141_275207 | Low-quality |
| 17-108_k141_275208 | Not-determined |
| 17-108_k141_192404 | Not-determined |
| 17-108_k141_26514 | Not-determined |
| 17-108_k141_26516 | Not-determined |
| 17-108_k141_302830 | Not-determined |
| 17-108_k141_302834 | Not-determined |
| 17-108_k141_316589 | Not-determined |
| 17-108_k141_220341 | Not-determined |
| 17-108_k141_123468 | Not-determined |
| 17-108_k141_192415 | Not-determined |
| 17-108_k141_82014 | Low-quality |
| 17-108_k141_137192 | Not-determined |
| 17-108_k141_96420 | Not-determined |
| 17-108_k141_302844 | Not-determined |
| 17-108_k141_137199 | Not-determined |
| 17-108_k141_137205 | Not-determined |
| 17-108_k141_302849 | Not-determined |
| 17-108_k141_316611 | Not-determined |
| 17-108_k141_302851 | Not-determined |
| 17-108_k141_82031 | Not-determined |
| 17-108_k141_82034 | Not-determined |
| 17-108_k141_123477 | Not-determined |
| 17-108_k141_123480 | Not-determined |
| 17-108_k141_206176 | Not-determined |
| 17-108_k141_302872 | Low-quality |
| 17-108_k141_137228 | Low-quality |
| 17-108_k141_316632 | Not-determined |
| 17-108_k141_137231 | Not-determined |
| 17-108_k141_26566 | Low-quality |
| 17-108_k141_328973 | Not-determined |
| 17-108_k141_82059 | Not-determined |
| 18-102-final-viral-combined_k141_110215\|\|full | High-quality |
| 17-108_k141_26576 | Not-determined |
| 17-108_k141_275268 | Not-determined |
| 17-108_k141_302883 | Not-determined |
| 17-108_k141_328984 | Not-determined |
| 17-108_k141_68881 | Not-determined |
| 17-108_k141_302888 | Not-determined |
| 17-108_k141_328990 | Not-determined |
| 17-108_k141_275282 | Low-quality |
| 17-108_k141_192429 | Not-determined |
| 17-108_k141_192431 | Not-determined |
| 17-108_k141_192433 | Not-determined |
| 17-108_k141_275287 | Not-determined |
| 17-108_k141_192436 | Not-determined |
| 17-108_k141_68900 | Not-determined |
| 17-108_k141_192441 | Not-determined |
| 17-108_k141_302904 | Not-determined |
| 17-108_k141_137263 | Not-determined |
| 17-108_k141_192451 | Not-determined |
| 17-108_k141_275302 | Not-determined |
| 17-108_k141_302914 | Not-determined |
| 17-108_k141_137269 | Not-determined |
| 17-108_k141_137270 | Not-determined |
| 17-108_k141_302916 | Not-determined |
| 17-108_k141_220369 | Not-determined |
| 17-108_k141_329016 | Not-determined |
| 17-108_k141_220375 | Not-determined |
| 17-108_k141_275314 | Not-determined |
| 17-108_k141_26594 | Not-determined |
| 17-108_k141_220377 | Not-determined |
| 17-108_k141_137281 | Not-determined |
| 17-108_k141_192461 | Not-determined |
| 17-108_k141_329027 | Not-determined |
| 17-108_k141_123571 | Not-determined |
| 17-108_k141_123572 | Not-determined |
| 17-108_k141_192480 | Not-determined |
| 17-108_k141_192484 | Not-determined |
| 17-108_k141_137313 | Low-quality |
| 17-108_k141_192491 | Not-determined |
| 17-108_k141_275324 | Not-determined |
| 17-108_k141_302950 | Not-determined |
| 17-108_k141_137324 | Not-determined |
| 17-108_k141_137329 | Not-determined |
| 17-108_k141_302956 | Not-determined |
| 17-108_k141_192508 | Not-determined |
| 17-108_k141_82101 | Not-determined |
| 17-108_k141_220416 | Not-determined |
| 17-108_k141_26655 | Not-determined |
| 17-108_k141_275356 | Not-determined |
| 17-108_k141_329071 | Not-determined |
| 17-108_k141_329073 | Not-determined |
| 17-108_k141_220433 | Not-determined |
| 17-108_k141_26667 | Not-determined |
| 17-108_k141_302987 | Not-determined |
| 17-108_k141_220436 | Not-determined |
| 17-108_k141_316667 | Not-determined |
| 17-108_k141_275375 | Not-determined |
| 17-108_k141_192540 | Low-quality |
| 17-108_k141_275383 | Not-determined |
| 17-108_k141_329085 | Not-determined |
| 17-108_k141_192556 | Not-determined |
| 17-108_k141_316692 | Not-determined |
| 17-108_k141_220460 | Not-determined |
| 17-108_k141_123626 | Not-determined |
| 17-108_k141_220466 | Not-determined |
| 17-108_k141_316704 | Not-determined |
| 17-108_k141_316707 | Not-determined |
| 17-108_k141_26685 | Not-determined |
| 17-108_k141_26686 | Low-quality |
| 17-108_k141_137412 | Not-determined |
| 17-108_k141_192601 | Not-determined |
| 17-108_k141_316721 | Low-quality |
| 17-108_k141_192609 | Not-determined |
| 17-108_k141_220475 | Not-determined |
| 17-108_k141_316731 | Low-quality |
| 17-108_k141_192620 | Not-determined |
| 17-108_k141_192622 | Not-determined |
| 17-108_k141_329106 | Low-quality |
| 17-108_k141_26708 | Not-determined |
| 17-108_k141_329110 | Not-determined |
| 17-108_k141_123660 | Not-determined |
| 17-108_k141_82202 | Not-determined |
| 17-108_k141_82203 | Not-determined |
| 17-108_k141_26726 | Low-quality |
| 17-108_k141_82213 | Not-determined |
| 17-108_k141_192652 | Not-determined |
| 17-108_k141_26729 | Not-determined |
| 17-108_k141_316749 | Not-determined |
| 17-108_k141_275423 | Not-determined |
| 17-108_k141_137424 | Not-determined |
| 17-108_k141_26744 | Not-determined |
| 17-108_k141_192676 | Not-determined |
| 17-108_k141_123678 | Low-quality |
| 17-108_k141_192681 | Not-determined |
| 17-108_k141_137441 | Not-determined |
| 17-108_k141_329132 | Not-determined |
| 17-108_k141_82251 | Not-determined |
| 17-108_k141_329140 | Not-determined |
| 17-108_k141_303088 | Not-determined |
| 17-108_k141_316794 | Not-determined |
| 17-108_k141_316797 | Not-determined |
| 17-108_k141_316806 | Not-determined |
| 17-108_k141_316807 | Not-determined |
| 17-108_k141_303104 | Not-determined |
| 17-108_k141_137487 | Not-determined |
| 17-108_k141_137492 | Not-determined |
| 17-108_k141_26802 | Low-quality |
| 17-108_k141_26804 | Not-determined |
| 17-108_k141_26810 | Low-quality |
| 17-108_k141_303112 | Not-determined |
| 17-108_k141_303114 | Not-determined |
| 17-108_k141_303116 | Low-quality |
| 17-108_k141_303123 | Not-determined |
| 17-108_k141_316841 | Low-quality |
| 17-108_k141_303129 | Not-determined |
| 17-108_k141_82349 | Not-determined |
| 17-108_k141_192711 | Not-determined |
| 17-108_k141_82355 | Not-determined |
| 17-108_k141_275463 | Not-determined |
| 17-108_k141_192723 | Not-determined |
| 17-108_k141_316865 | Not-determined |
| 17-108_k141_303151 | Not-determined |
| 17-108_k141_329194 | Not-determined |
| 17-108_k141_316875 | Not-determined |
| 17-108_k141_192746 | Not-determined |
| 17-108_k141_329205 | Not-determined |
| 17-108_k141_275482 | Not-determined |
| 17-108_k141_316883 | Low-quality |
| 17-108_k141_123802 | Not-determined |
| 17-108_k141_192768 | Not-determined |
| 17-108_k141_26870 | Not-determined |
| 17-108_k141_275492 | Not-determined |
| 17-108_k141_137594 | Not-determined |
| 17-108_k141_329227 | Not-determined |
| 17-108_k141_206329 | Not-determined |
| 17-108_k141_82396 | Not-determined |
| 17-108_k141_82397 | Not-determined |
| 17-108_k141_123828 | Not-determined |
| 17-108_k141_206338 | Not-determined |
| 17-108_k141_137605 | Not-determined |
| 17-108_k141_82409 | Not-determined |
| 17-108_k141_82412 | Not-determined |
| 17-108_k141_82413 | Not-determined |
| 17-108_k141_275512 | Not-determined |
| 17-108_k141_275518 | Not-determined |
| 17-108_k141_329238 | Low-quality |
| 17-108_k141_275521 | Not-determined |
| 17-108_k141_329243 | Not-determined |
| 17-108_k141_123858 | Low-quality |
| 17-108_k141_26888 | Not-determined |
| 17-108_k141_26890 | Not-determined |
| 17-108_k141_137639 | Not-determined |
| 17-108_k141_82440 | Not-determined |
| 17-108_k141_206369 | Not-determined |
| 17-108_k141_123872 | Not-determined |
| 17-108_k141_26900 | Not-determined |
| 17-108_k141_82447 | Not-determined |
| 17-108_k141_192831 | Low-quality |
| 17-108_k141_82452 | Not-determined |
| 17-108_k141_206393 | Not-determined |
| 17-108_k141_206409 | Not-determined |
| 17-108_k141_123898 | Not-determined |
| 17-108_k141_329287 | Not-determined |
| 17-108_k141_329289 | Not-determined |
| 17-108_k141_192854 | Not-determined |
| 17-108_k141_316934 | Not-determined |
| 17-108_k141_206418 | Not-determined |
| 17-108_k141_26943 | Not-determined |
| 17-108_k141_26952 | Not-determined |
| 17-108_k141_123917 | Not-determined |
| 17-108_k141_206429 | Low-quality |
| 17-108_k141_123920 | Not-determined |
| 17-108_k141_275569 | Not-determined |
| 17-108_k141_137659 | Not-determined |
| 17-108_k141_137661 | Not-determined |
| 17-108_k141_206436 | Not-determined |
| 17-108_k141_206441 | Not-determined |
| 17-108_k141_123928 | Not-determined |
| 17-108_k141_329308 | Not-determined |
| 17-108_k141_26967 | Not-determined |
| 17-108_k141_82498 | Not-determined |
| 17-108_k141_82499 | Not-determined |
| 17-108_k141_123947 | Not-determined |
| 17-108_k141_123954 | Not-determined |
| 17-108_k141_123955 | Not-determined |
| 17-108_k141_329317 | Not-determined |
| 17-108_k141_137703 | Low-quality |
| 17-108_k141_329330 | Not-determined |
| 17-108_k141_275614 | Not-determined |
| 17-108_k141_206476 | Not-determined |
| 17-108_k141_137718 | Not-determined |
| 17-108_k141_137719 | Not-determined |
| 17-108_k141_206481 | Low-quality |
| 17-108_k141_123961 | Not-determined |
| 17-108_k141_26974 | Not-determined |
| 17-108_k141_206487 | Low-quality |
| 17-108_k141_137734 | Not-determined |
| 17-108_k141_82529 | Not-determined |
| 17-108_k141_82537 | Not-determined |
| 17-108_k141_206524 | Low-quality |
| 17-108_k141_206525 | Not-determined |
| 17-108_k141_26994 | Not-determined |
| 17-108_k141_137763 | Not-determined |
| 17-108_k141_137767 | Not-determined |
| 17-108_k141_82547 | Not-determined |
| 17-108_k141_206549 | Not-determined |
| 17-108_k141_137787 | Not-determined |
| 17-108_k141_82552 | Not-determined |
| 17-108_k141_27032 | Not-determined |
| 17-108_k141_27033 | Not-determined |

| | |
|---|---|
| 17-108_k141_27036 | Not-determined |
| 17-108_k141_27039 | Not-determined |
| 17-108_k141_82559 | Not-determined |
| 17-108_k141_124028 | Not-determined |
| 17-108_k141_329429 | Not-determined |
| 17-108_k141_137795 | Not-determined |
| 17-108_k141_137801 | Not-determined |
| 17-108_k141_206618 | Not-determined |
| 17-108_k141_27090 | Not-determined |
| 17-108_k141_27091 | Not-determined |
| 17-108_k141_27094 | Not-determined |
| 17-108_k141_329460 | Not-determined |
| 17-108_k141_329462 | Not-determined |
| 17-108_k141_192906 | Not-determined |
| 17-108_k141_329478 | Not-determined |
| 17-108_k141_82601 | Not-determined |
| 17-108_k141_27112 | Low-quality |
| 17-108_k141_27113 | Not-determined |
| 17-108_k141_329484 | Low-quality |
| 17-108_k141_27117 | Not-determined |
| 17-108_k141_329491 | Not-determined |
| 17-108_k141_27139 | Not-determined |
| 17-108_k141_206689 | Not-determined |
| 17-108_k141_329504 | Not-determined |
| 17-108_k141_82679 | Not-determined |
| 17-108_k141_329513 | Not-determined |
| 17-108_k141_329533 | Not-determined |
| 17-108_k141_27207 | Not-determined |
| 17-108_k141_329563 | Not-determined |
| 17-108_k141_329576 | Not-determined |
| 17-108_k141_329581 | Low-quality |
| 17-108_k141_329587 | Not-determined |
| 17-108_k141_329605 | Not-determined |
| 17-108_k141_329623 | Not-determined |
| 17-108_k141_329633 | Not-determined |
| 17-108_k141_27287 | Not-determined |
| 17-108_k141_329650 | Not-determined |
| 17-108_k141_27289 | Not-determined |
| 17-108_k141_27300 | Not-determined |
| 17-108_k141_27318 | Low-quality |
| 17-108_k141_329689 | Not-determined |
| 17-108_k141_27345 | Not-determined |
| 17-108_k141_27348 | Not-determined |
| 17-108_k141_329721 | Not-determined |
| 17-108_k141_27362 | Not-determined |
| 17-108_k141_27367 | Not-determined |
| 17-108_k141_329789 | Not-determined |
| 17-108_k141_27440 | Not-determined |
| 17-108_k141_27441 | Not-determined |
| 17-108_k141_329839 | Not-determined |
| 17-108_k141_329841 | Not-determined |
| 17-108_k141_329855 | Not-determined |
| 17-108_k141_329856 | Not-determined |
| 17-108_k141_27467 | Not-determined |
| 17-108_k141_27471 | Not-determined |
| 17-108_k141_329894 | Not-determined |
| 17-108_k141_27488 | Not-determined |
| 17-108_k141_329938 | Not-determined |
| 17-108_k141_329939 | Not-determined |
| 17-108_k141_329944 | Not-determined |
| 17-108_k141_329979 | Not-determined |
| 17-108_k141_329984 | Low-quality |
| 17-108_k141_27532 | Not-determined |
| 17-108_k141_27542 | Not-determined |
| 17-108_k141_27543 | Not-determined |
| 17-108_k141_27546 | Not-determined |
| 17-108_k141_330048 | Not-determined |
| 17-108_k141_330074 | Not-determined |
| 17-108_k141_330079 | Not-determined |
| 17-108_k141_330081 | Not-determined |
| 17-108_k141_330082 | Not-determined |
| 17-108_k141_330096 | Low-quality |
| 17-108_k141_27560 | Not-determined |
| 17-108_k141_330115 | Not-determined |
| 17-108_k141_330164 | Not-determined |
| 17-108_k141_330178 | Not-determined |
| 17-108_k141_330202 | Low-quality |
| 17-108_k141_330217 | Not-determined |
| 17-108_k141_330232 | Not-determined |
| 17-108_k141_330244 | Not-determined |
| 17-108_k141_330245 | Not-determined |
| 17-108_k141_330278 | Not-determined |
| 17-108_k141_330339 | Not-determined |
| 17-108_k141_330340 | Not-determined |
| 17-108_k141_330348 | Not-determined |
| 17-108_k141_330403 | Not-determined |
| 17-108_k141_330449 | Not-determined |
| 17-108_k141_330477 | Not-determined |
| 17-108-final-viral-combined_k141_101337\|\|full | Medium-quality |
| 17-108_k141_330482 | Not-determined |
| 17-108_k141_330484 | Not-determined |
| 17-108_k141_330503 | Low-quality |
| 17-108_k141_330506 | Not-determined |
| 17-108_k141_330513 | Not-determined |
| 17-108_k141_330535 | Not-determined |
| 17-108_k141_330538 | Not-determined |
| 17-108_k141_330542 | Not-determined |
| 17-108_k141_330552 | Not-determined |
| 17-108_k141_330562 | Not-determined |
| 17-108_k141_330564 | Not-determined |
| 17-108_k141_330573 | Not-determined |
| 17-108_k141_330576 | Not-determined |
| 17-108_k141_330585 | Not-determined |
| 17-108_k141_330594 | Low-quality |
| 17-108_k141_330596 | Not-determined |
| 17-108_k141_330599 | Not-determined |
| 17-108_k141_330603 | Not-determined |
| 17-108_k141_330606 | Not-determined |
| 17-108_k141_330627 | Not-determined |
| 17-108_k141_330630 | Not-determined |
| 17-108_k141_330631 | Not-determined |
| 17-108_k141_330643 | Not-determined |
| 17-108_k141_330660 | Not-determined |
| 17-108_k141_330661 | Not-determined |
| 17-108_k141_330667 | Not-determined |
| 17-108_k141_330674 | Not-determined |
| 17-108_k141_330676 | Not-determined |
| 17-108_k141_330677 | Not-determined |
| 17-108_k141_330691 | Not-determined |
| 17-108_k141_330699 | Not-determined |
| 17-108_k141_330701 | Not-determined |
| 17-108_k141_330711 | Not-determined |
| 17-108_k141_330713 | Not-determined |
| 17-108_k141_330714 | Not-determined |
| 17-108_k141_330715 | Not-determined |
| 17-108_k141_330716 | Not-determined |
| 17-108_k141_330717 | Not-determined |
| 17-108_k141_330719 | Low-quality |
| 17-108_k141_330720 | Not-determined |
| 17-108_k141_330723 | Not-determined |
| 17-108_k141_330724 | Not-determined |
| 17-108_k141_330726 | Not-determined |
| 17-108_k141_330727 | Not-determined |
| 17-108_k141_330731 | Not-determined |
| 17-108_k141_330735 | Not-determined |
| 17-108_k141_330736 | Not-determined |
| 17-108_k141_330738 | Not-determined |
| 17-108_k141_330739 | Not-determined |
| 18-101-final-viral-combined_k141_263323\|\|full | Not-determined |
| 18-101-final-viral-combined_k141_533\|\|full | Low-quality |
| 18-101-final-viral-combined_k141_346502\|\|full | Not-determined |
| 18-101-final-viral-combined_k141_100830\|\|full | Low-quality |
| 18-101-final-viral-combined_k141_184415\|\|full | Low-quality |
| 18-101-final-viral-combined_k141_70298\|\|full | Not-determined |
| 18-110-final-viral-combined_k141_200168\|\|full | High-quality |
| 18-101-final-viral-combined_k141_359778\|\|full | Not-determined |
| 18-101-final-viral-combined_k141_285012\|\|full | High-quality |
| 18-101-final-viral-combined_k141_63574\|\|full | Low-quality |
| 18-101-final-viral-combined_k141_146954\|\|full | Not-determined |
| 18-102-final-viral-combined_k141_218002\|\|full | Medium-quality |
| 18-101-final-viral-combined_k141_394572\|\|full | Not-determined |
| 17-109-final-viral-combined_k141_23517\|\|full | High-quality |
| 18-101-final-viral-combined_k141_394592\|\|full | Low-quality |
| 18-101-final-viral-combined_k141_394600\|\|full | Not-determined |
| 18-102-final-viral-combined_k141_30750\|\|full | Medium-quality |
| 18-101-final-viral-combined_k141_394643\|\|full | Not-determined |
| 18-101-final-viral-combined_k141_65815\|\|full | Not-determined |
| 18-101-final-viral-combined_k141_164534\|\|full | Not-determined |
| 18-101-final-viral-combined_k141_148079\|\|full | Low-quality |
| 18-101-final-viral-combined_k141_180985\|\|full | Not-determined |
| 18-101-final-viral-combined_k141_345521\|\|full | Low-quality |
| 18-101-final-viral-combined_k141_115184\|\|full | Low-quality |
| 18-101-final-viral-combined_k141_49376\|\|full | Not-determined |
| 18-101-final-viral-combined_k141_164556\|\|full | Not-determined |
| 18-101-final-viral-combined_k141_115188\|\|full | Low-quality |
| 18-101-final-viral-combined_k141_329082\|\|full | Low-quality |
| 18-101-final-viral-combined_k141_279715\|\|full | Not-determined |
| 18-101-final-viral-combined_k141_131658\|\|full | Not-determined |
| 18-101-final-viral-combined_k141_345530\|\|full | Low-quality |
| 18-101-final-viral-combined_k141_49387\|\|full | Not-determined |
| 18-101-final-viral-combined_k141_197448\|\|full | Low-quality |
| 18-101-final-viral-combined_k141_98744\|\|full | Low-quality |
| 18-101-final-viral-combined_k141_345549\|\|full | Not-determined |
| 18-101-final-viral-combined_k141_361999\|\|full | Not-determined |
| 18-101-final-viral-combined_k141_164561\|\|full | Not-determined |
| 18-101-final-viral-combined_k141_82290\|\|full | Low-quality |
| 18-101-final-viral-combined_k141_46\|\|full | Not-determined |
| 18-101-final-viral-combined_k141_131675\|\|full | Low-quality |
| 18-101-final-viral-combined_k141_312651\|\|full | Not-determined |
| 18-101-final-viral-combined_k141_49400\|\|full | Not-determined |
| 18-101-final-viral-combined_k141_164566\|\|full | Not-determined |
| 18-101-final-viral-combined_k141_263274\|\|full | Not-determined |
| 18-101-final-viral-combined_k141_49406\|\|full | Low-quality |
| 18-101-final-viral-combined_k141_16472\|\|full | Not-determined |
| 18-101-final-viral-combined_k141_32956\|\|full | Low-quality |
| 18-101-final-viral-combined_k141_49429\|\|full | Not-determined |
| 18-101-final-viral-combined_k141_32969\|\|full | Not-determined |
| 18-101-final-viral-combined_k141_148133\|\|full | Not-determined |
| 18-101-final-viral-combined_k141_82325\|\|full | Not-determined |
| 18-101-final-viral-combined_k141_49448\|\|full | Low-quality |
| 18-101-final-viral-combined_k141_362022\|\|full | Not-determined |
| 18-101-final-viral-combined_k141_82328\|\|full | Low-quality |
| 18-101-final-viral-combined_k141_86\|\|full | Low-quality |
| 18-101-final-viral-combined_k141_312688\|\|full | Low-quality |
| 18-101-final-viral-combined_k141_98772\|\|full | Low-quality |
| 18-101-final-viral-combined_k141_329114\|\|full | Not-determined |
| 18-101-final-viral-combined_k141_16499\|\|full | Low-quality |
| 18-101-final-viral-combined_k141_16500\|\|full | Low-quality |
| 18-101-final-viral-combined_k141_115219\|\|full | Low-quality |
| 18-101-final-viral-combined_k141_345591\|\|full | Not-determined |
| 18-101-final-viral-combined_k141_100\|\|full | Not-determined |
| 18-101-final-viral-combined_k141_213924\|\|full | Not-determined |
| 18-101-final-viral-combined_k141_329133\|\|full | Not-determined |
| 18-101-final-viral-combined_k141_263299\|\|full | Not-determined |
| 18-101-final-viral-combined_k141_362029\|\|full | Low-quality |
| 18-101-final-viral-combined_k141_98785\|\|full | Low-quality |
| 18-101-final-viral-combined_k141_230402\|\|full | Low-quality |
| 18-101-final-viral-combined_k141_131740\|\|full | Not-determined |
| 18-101-final-viral-combined_k141_345605\|\|full | Low-quality |
| 18-101-final-viral-combined_k141_312720\|\|full | Low-quality |
| 18-101-final-viral-combined_k141_312722\|\|full | Low-quality |
| 18-101-final-viral-combined_k141_131748\|\|full | Low-quality |
| 18-101-final-viral-combined_k141_98791\|\|full | Not-determined |
| 18-101-final-viral-combined_k141_148159\|\|full | Low-quality |
| 18-101-final-viral-combined_k141_123\|\|full | Not-determined |
| 18-101-final-viral-combined_k141_246824\|\|full | Low-quality |
| 18-101-final-viral-combined_k141_98797\|\|full | Low-quality |
| 18-101-final-viral-combined_k141_378525\|\|full | Not-determined |
| 18-101-final-viral-combined_k141_98799\|\|full | Low-quality |
| 18-101-final-viral-combined_k141_213935\|\|full | Low-quality |
| 18-101-final-viral-combined_k141_362058\|\|full | Not-determined |
| 18-101-final-viral-combined_k141_115273\|\|full | Not-determined |
| 18-101-final-viral-combined_k141_246834\|\|full | Low-quality |
| 18-101-final-viral-combined_k141_49489\|\|full | Not-determined |
| 18-101-final-viral-combined_k141_246845\|\|full | Not-determined |
| 18-101-final-viral-combined_k141_33039\|\|full | Not-determined |
| 18-101-final-viral-combined_k141_230433\|\|full | Low-quality |
| 18-101-final-viral-combined_k141_131776\|\|full | Low-quality |
| 18-101-final-viral-combined_k141_49494\|\|full | Low-quality |
| 18-101-final-viral-combined_k141_33042\|\|full | Low-quality |
| 18-101-final-viral-combined_k141_362074\|\|full | Not-determined |
| 18-101-final-viral-combined_k141_164599\|\|full | Not-determined |
| 18-101-final-viral-combined_k141_98816\|\|full | Low-quality |
| 18-101-final-viral-combined_k141_230440\|\|full | Not-determined |
| 18-101-final-viral-combined_k141_246847\|\|full | Not-determined |
| 18-101-final-viral-combined_k141_345613\|\|full | Low-quality |
| 18-101-final-viral-combined_k141_131786\|\|full | Low-quality |
| 18-101-final-viral-combined_k141_115301\|\|full | Not-determined |
| 18-101-final-viral-combined_k141_159\|\|full | Low-quality |
| 18-101-final-viral-combined_k141_115308\|\|full | Not-determined |
| 18-101-final-viral-combined_k141_163\|\|full | Low-quality |
| 18-101-final-viral-combined_k141_213971\|\|full | Not-determined |
| 18-101-final-viral-combined_k141_164618\|\|full | Not-determined |
| 18-101-final-viral-combined_k141_362096\|\|full | Low-quality |
| 18-101-final-viral-combined_k141_16528\|\|full | Not-determined |
| 18-101-final-viral-combined_k141_16529\|\|full | Not-determined |
| 18-101-final-viral-combined_k141_246864\|\|full | Not-determined |
| 18-101-final-viral-combined_k141_362105\|\|full | Low-quality |
| 18-101-final-viral-combined_k141_148194\|\|full | Low-quality |
| 18-101-final-viral-combined_k141_312780\|\|full | Not-determined |
| 18-101-final-viral-combined_k141_345645\|\|full | Not-determined |
| 18-101-final-viral-combined_k141_214008\|\|full | Not-determined |
| 18-101-final-viral-combined_k141_193\|\|full | Low-quality |
| 18-101-final-viral-combined_k141_345650\|\|full | Not-determined |
| 18-101-final-viral-combined_k141_279822\|\|full | Low-quality |
| 18-101-final-viral-combined_k141_164641\|\|full | Not-determined |
| 18-101-final-viral-combined_k141_214013\|\|full | Low-quality |
| 18-101-final-viral-combined_k141_49525\|\|full | Not-determined |
| 18-101-final-viral-combined_k141_131829\|\|full | Low-quality |
| 18-101-final-viral-combined_k141_279828\|\|full | Low-quality |
| 18-101-final-viral-combined_k141_214018\|\|full | Low-quality |
| 18-101-final-viral-combined_k141_82437\|\|full | Low-quality |
| 18-101-final-viral-combined_k141_181133\|\|full | Low-quality |
| 18-101-final-viral-combined_k141_362117\|\|full | Low-quality |
| 18-101-final-viral-combined_k141_345662\|\|full | Not-determined |
| 18-101-final-viral-combined_k141_279838\|\|full | Not-determined |
| 18-101-final-viral-combined_k141_296311\|\|full | Not-determined |
| 18-101-final-viral-combined_k141_215\|\|full | Not-determined |
| 18-101-final-viral-combined_k141_312828\|\|full | Not-determined |
| 18-101-final-viral-combined_k141_263316\|\|full | Not-determined |
| 18-101-final-viral-combined_k141_16569\|\|full | Low-quality |
| 18-101-final-viral-combined_k141_98846\|\|full | Not-determined |
| 18-101-final-viral-combined_k141_312847\|\|full | Not-determined |
| 18-101-final-viral-combined_k141_362143\|\|full | Low-quality |
| 18-101-final-viral-combined_k141_115376\|\|full | Not-determined |
| 18-101-final-viral-combined_k141_329213\|\|full | Not-determined |
| 18-101-final-viral-combined_k141_131842\|\|full | Not-determined |
| 18-101-final-viral-combined_k141_65832\|\|full | Low-quality |
| 18-101-final-viral-combined_k141_197551\|\|full | Not-determined |
| 18-101-final-viral-combined_k141_246918\|\|full | Not-determined |
| 18-101-final-viral-combined_k141_16599\|\|full | Not-determined |
| 18-101-final-viral-combined_k141_329225\|\|full | Low-quality |
| 18-101-final-viral-combined_k141_230510\|\|full | Not-determined |
| 18-101-final-viral-combined_k141_230513\|\|full | Not-determined |
| 18-101-final-viral-combined_k141_296326\|\|full | Low-quality |
| 18-101-final-viral-combined_k141_65844\|\|full | Not-determined |
| 18-101-final-viral-combined_k141_131856\|\|full | Not-determined |
| 18-101-final-viral-combined_k141_263337\|\|full | Low-quality |
| 18-101-final-viral-combined_k141_378608\|\|full | Low-quality |
| 18-101-final-viral-combined_k141_197561\|\|full | Not-determined |
| 18-101-final-viral-combined_k141_82496\|\|full | Low-quality |
| 18-101-final-viral-combined_k141_65855\|\|full | Not-determined |
| 18-101-final-viral-combined_k141_230541\|\|full | Low-quality |
| 18-101-final-viral-combined_k141_214078\|\|full | Not-determined |
| 18-101-final-viral-combined_k141_181157\|\|full | Not-determined |
| 18-101-final-viral-combined_k141_131873\|\|full | Not-determined |
| 18-101-final-viral-combined_k141_214085\|\|full | Low-quality |
| 18-101-final-viral-combined_k141_312862\|\|full | Not-determined |
| 18-101-final-viral-combined_k141_362174\|\|full | Not-determined |
| 18-101-final-viral-combined_k141_65862\|\|full | Not-determined |
| 18-101-final-viral-combined_k141_16621\|\|full | Not-determined |
| 18-101-final-viral-combined_k141_181163\|\|full | Not-determined |
| 18-101-final-viral-combined_k141_49560\|\|full | Low-quality |
| 18-101-final-viral-combined_k141_302166\|\|full | Medium-quality |
| 18-101-final-viral-combined_k141_164693\|\|full | Not-determined |
| 18-101-final-viral-combined_k141_246925\|\|full | Low-quality |
| 18-101-final-viral-combined_k141_230553\|\|full | Not-determined |
| 18-101-final-viral-combined_k141_362181\|\|full | Not-determined |
| 18-101-final-viral-combined_k141_131889\|\|full | Not-determined |
| 18-101-final-viral-combined_k141_296349\|\|full | Not-determined |
| 18-101-final-viral-combined_k141_329259\|\|full | Not-determined |
| 18-101-final-viral-combined_k141_378628\|\|full | Low-quality |
| 18-101-final-viral-combined_k141_148299\|\|full | Low-quality |
| 18-101-final-viral-combined_k141_296353\|\|full | Low-quality |
| 18-101-final-viral-combined_k141_131904\|\|full | Not-determined |
| 18-101-final-viral-combined_k141_318\|\|full | Not-determined |
| 18-101-final-viral-combined_k141_329266\|\|full | Low-quality |
| 18-101-final-viral-combined_k141_246953\|\|full | Not-determined |
| 18-101-final-viral-combined_k141_329269\|\|full | Not-determined |
| 18-101-final-viral-combined_k141_131908\|\|full | Low-quality |
| 18-101-final-viral-combined_k141_197611\|\|full | Low-quality |
| 18-101-final-viral-combined_k141_230577\|\|full | Low-quality |
| 18-101-final-viral-combined_k141_65873\|\|full | Not-determined |
| 18-101-final-viral-combined_k141_230583\|\|full | Low-quality |
| 18-101-final-viral-combined_k141_263384\|\|full | Low-quality |
| 18-101-final-viral-combined_k141_263385\|\|full | Low-quality |
| 18-101-final-viral-combined_k141_65884\|\|full | Low-quality |
| 18-101-final-viral-combined_k141_98902\|\|full | Low-quality |
| 18-101-final-viral-combined_k141_197622\|\|full | Not-determined |
| 18-101-final-viral-combined_k141_65890\|\|full | Low-quality |
| 18-101-final-viral-combined_k141_197635\|\|full | Low-quality |
| 18-101-final-viral-combined_k141_296374\|\|full | Not-determined |
| 18-101-final-viral-combined_k141_214136\|\|full | Not-determined |
| 18-101-final-viral-combined_k141_377\|\|full | Low-quality |
| 18-101-final-viral-combined_k141_246979\|\|full | Low-quality |
| 18-101-final-viral-combined_k141_296379\|\|full | Not-determined |
| 18-101-final-viral-combined_k141_263400\|\|full | Not-determined |
| 18-101-final-viral-combined_k141_82527\|\|full | Low-quality |
| 18-101-final-viral-combined_k141_197653\|\|full | Low-quality |
| 18-101-final-viral-combined_k141_16705\|\|full | Low-quality |
| 18-101-final-viral-combined_k141_279969\|\|full | Not-determined |
| 18-101-final-viral-combined_k141_115429\|\|full | Not-determined |
| 18-101-final-viral-combined_k141_296413\|\|full | Not-determined |
| 18-101-final-viral-combined_k141_395\|\|full | Not-determined |
| 18-101-final-viral-combined_k141_345719\|\|full | Not-determined |
| 18-101-final-viral-combined_k141_399\|\|full | Low-quality |
| 18-101-final-viral-combined_k141_115431\|\|full | Not-determined |
| 18-101-final-viral-combined_k141_181242\|\|full | Not-determined |
| 18-101-final-viral-combined_k141_345728\|\|full | Not-determined |
| 18-101-final-viral-combined_k141_65913\|\|full | Low-quality |
| 18-101-final-viral-combined_k141_230642\|\|full | Low-quality |
| 18-101-final-viral-combined_k141_181248\|\|full | Not-determined |
| 18-101-final-viral-combined_k141_131964\|\|full | Not-determined |
| 18-101-final-viral-combined_k141_329293\|\|full | Low-quality |
| 18-101-final-viral-combined_k141_164775\|\|full | Not-determined |
| 18-101-final-viral-combined_k141_131979\|\|full | Not-determined |
| 18-101-final-viral-combined_k141_435\|\|full | Low-quality |
| 18-101-final-viral-combined_k141_197708\|\|full | Not-determined |
| 18-101-final-viral-combined_k141_164781\|\|full | Not-determined |
| 18-101-final-viral-combined_k141_98968\|\|full | Not-determined |
| 18-101-final-viral-combined_k141_131989\|\|full | Not-determined |
| 18-101-final-viral-combined_k141_362249\|\|full | Low-quality |
| 18-101-final-viral-combined_k141_312939\|\|full | Low-quality |
| 18-101-final-viral-combined_k141_378698\|\|full | Low-quality |
| 18-101-final-viral-combined_k141_164803\|\|full | Low-quality |
| 18-101-final-viral-combined_k141_49597\|\|full | Not-determined |
| 18-101-final-viral-combined_k141_230689\|\|full | Not-determined |
| 18-101-final-viral-combined_k141_457\|\|full | Low-quality |
| 18-101-final-viral-combined_k141_16783\|\|full | Not-determined |
| 18-101-final-viral-combined_k141_181275\|\|full | Not-determined |
| 18-101-final-viral-combined_k141_33072\|\|full | Low-quality |
| 18-101-final-viral-combined_k141_362284\|\|full | Not-determined |
| 18-101-final-viral-combined_k141_164818\|\|full | Not-determined |
| 18-101-final-viral-combined_k141_362286\|\|full | Low-quality |
| 18-101-final-viral-combined_k141_362288\|\|full | Low-quality |
| 18-101-final-viral-combined_k141_181277\|\|full | Not-determined |
| 18-101-final-viral-combined_k141_345790\|\|full | Not-determined |
| 18-101-final-viral-combined_k141_247084\|\|full | Low-quality |
| 18-101-final-viral-combined_k141_247086\|\|full | Low-quality |
| 18-101-final-viral-combined_k141_115473\|\|full | Not-determined |
| 18-101-final-viral-combined_k141_82669\|\|full | Not-determined |
| 18-101-final-viral-combined_k141_82674\|\|full | Not-determined |
| 18-101-final-viral-combined_k141_329318\|\|full | Not-determined |
| 18-101-final-viral-combined_k141_520\|\|full | Not-determined |
| 18-101-final-viral-combined_k141_296526\|\|full | Not-determined |
| 18-101-final-viral-combined_k141_49641\|\|full | Not-determined |
| 18-101-final-viral-combined_k141_329334\|\|full | Not-determined |
| 18-101-final-viral-combined_k141_148362\|\|full | Not-determined |
| 18-101-final-viral-combined_k141_214205\|\|full | Low-quality |
| 18-101-final-viral-combined_k141_528\|\|full | Low-quality |
| 18-101-final-viral-combined_k141_378728\|\|full | Not-determined |
| 18-101-final-viral-combined_k141_132065\|\|full | Not-determined |
| 18-101-final-viral-combined_k141_230768\|\|full | Not-determined |
| 18-101-final-viral-combined_k141_345795\|\|full | Not-determined |
| 18-101-final-viral-combined_k141_181306\|\|full | Not-determined |
| 18-101-final-viral-combined_k141_82699\|\|full | Low-quality |
| 18-101-final-viral-combined_k141_247126\|\|full | Low-quality |
| 18-101-final-viral-combined_k141_280109\|\|full | Not-determined |
| 18-101-final-viral-combined_k141_296563\|\|full | Not-determined |
| 18-101-final-viral-combined_k141_378749\|\|full | Not-determined |
| 18-101-final-viral-combined_k141_329369\|\|full | Low-quality |
| 18-101-final-viral-combined_k141_49659\|\|full | Not-determined |
| 18-101-final-viral-combined_k141_33113\|\|full | Low-quality |
| 18-101-final-viral-combined_k141_329379\|\|full | Low-quality |
| 18-101-final-viral-combined_k141_329381\|\|full | Low-quality |
| 18-101-final-viral-combined_k141_329386\|\|full | Low-quality |
| 18-101-final-viral-combined_k141_148420\|\|full | Low-quality |
| 18-101-final-viral-combined_k141_569\|\|full | Not-determined |
| 18-101-final-viral-combined_k141_362379\|\|full | Not-determined |
| 18-101-final-viral-combined_k141_82747\|\|full | Low-quality |
| 18-101-final-viral-combined_k141_132092\|\|full | Low-quality |
| 18-101-final-viral-combined_k141_66014\|\|full | Low-quality |
| 18-101-final-viral-combined_k141_280149\|\|full | Low-quality |
| 18-101-final-viral-combined_k141_579\|\|full | Low-quality |
| 18-101-final-viral-combined_k141_313044\|\|full | Not-determined |
| 18-101-final-viral-combined_k141_362385\|\|full | Not-determined |
| 18-101-final-viral-combined_k141_49692\|\|full | Low-quality |
| 18-101-final-viral-combined_k141_16793\|\|full | Not-determined |
| 18-101-final-viral-combined_k141_247161\|\|full | Low-quality |
| 18-101-final-viral-combined_k141_115565\|\|full | Low-quality |
| 18-101-final-viral-combined_k141_49696\|\|full | Low-quality |
| 18-101-final-viral-combined_k141_313062\|\|full | Low-quality |
| 18-105-final-viral-combined_k141_134381\|\|full | Medium-quality |
| 18-101-final-viral-combined_k141_115571\|\|full | Not-determined |
| 18-101-final-viral-combined_k141_33145\|\|full | Not-determined |
| 18-101-final-viral-combined_k141_49700\|\|full | Not-determined |
| 18-101-final-viral-combined_k141_247167\|\|full | Not-determined |
| 18-101-final-viral-combined_k141_362395\|\|full | Not-determined |
| 18-101-final-viral-combined_k141_362396\|\|full | Not-determined |
| 18-101-final-viral-combined_k141_263569\|\|full | Low-quality |
| 18-101-final-viral-combined_k141_230822\|\|full | Not-determined |
| 18-101-final-viral-combined_k141_362407\|\|full | Low-quality |
| 18-101-final-viral-combined_k141_132120\|\|full | Not-determined |
| 18-101-final-viral-combined_k141_197846\|\|full | Not-determined |
| 18-101-final-viral-combined_k141_99095\|\|full | Not-determined |
| 17-108-final-viral-combined_k141_267993\|\|full | High-quality |
| 18-101-final-viral-combined_k141_280157\|\|full | Not-determined |
| 18-101-final-viral-combined_k141_115603\|\|full | Low-quality |
| 18-101-final-viral-combined_k141_214294\|\|full | Low-quality |
| 18-101-final-viral-combined_k141_280172\|\|full | Not-determined |
| 18-101-final-viral-combined_k141_16840\|\|full | Low-quality |
| 18-101-final-viral-combined_k141_132131\|\|full | Low-quality |
| 18-101-final-viral-combined_k141_33184\|\|full | Not-determined |
| 18-101-final-viral-combined_k141_49735\|\|full | Low-quality |
| 18-101-final-viral-combined_k141_329408\|\|full | Low-quality |
| 18-101-final-viral-combined_k141_313080\|\|full | Low-quality |
| 18-101-final-viral-combined_k141_280181\|\|full | Low-quality |
| 18-101-final-viral-combined_k141_181411\|\|full | Low-quality |
| 18-101-final-viral-combined_k141_378797\|\|full | Low-quality |
| 18-101-final-viral-combined_k141_635\|\|full | Not-determined |
| 18-101-final-viral-combined_k141_638\|\|full | Not-determined |
| 18-101-final-viral-combined_k141_99120\|\|full | Low-quality |
| 18-101-final-viral-combined_k141_99124\|\|full | Not-determined |
| 18-101-final-viral-combined_k141_263600\|\|full | Not-determined |
| 18-101-final-viral-combined_k141_197887\|\|full | Not-determined |
| 18-101-final-viral-combined_k141_66039\|\|full | Not-determined |
| 18-101-final-viral-combined_k141_181420\|\|full | Low-quality |
| 18-101-final-viral-combined_k141_16874\|\|full | Not-determined |
| 18-101-final-viral-combined_k141_296577\|\|full | Not-determined |
| 18-101-final-viral-combined_k141_132134\|\|full | Not-determined |
| 18-101-final-viral-combined_k141_33199\|\|full | Not-determined |
| 18-101-final-viral-combined_k141_181436\|\|full | Not-determined |
| 18-101-final-viral-combined_k141_49766\|\|full | Not-determined |
| 18-101-final-viral-combined_k141_99148\|\|full | Low-quality |
| 18-101-final-viral-combined_k141_329440\|\|full | Low-quality |
| 18-101-final-viral-combined_k141_313100\|\|full | Low-quality |
| 18-101-final-viral-combined_k141_99154\|\|full | Not-determined |
| 18-101-final-viral-combined_k141_329445\|\|full | Not-determined |
| 18-101-final-viral-combined_k141_16890\|\|full | Not-determined |
| 18-101-final-viral-combined_k141_280210\|\|full | Not-determined |
| 18-101-final-viral-combined_k141_313110\|\|full | Low-quality |
| 18-101-final-viral-combined_k141_181457\|\|full | Not-determined |
| 18-101-final-viral-combined_k141_280217\|\|full | Not-determined |
| 18-101-final-viral-combined_k141_230849\|\|full | Not-determined |
| 18-101-final-viral-combined_k141_230856\|\|full | Not-determined |
| 18-101-final-viral-combined_k141_115656\|\|full | Not-determined |
| 18-101-final-viral-combined_k141_148484\|\|full | Low-quality |
| 18-101-final-viral-combined_k141_378865\|\|full | Low-quality |
| 18-101-final-viral-combined_k141_296624\|\|full | Not-determined |
| 18-101-final-viral-combined_k141_164947\|\|full | Low-quality |
| 18-101-final-viral-combined_k141_313129\|\|full | Not-determined |
| 18-101-final-viral-combined_k141_263644\|\|full | Not-determined |
| 18-101-final-viral-combined_k141_230860\|\|full | Not-determined |
| 18-101-final-viral-combined_k141_378876\|\|full | Not-determined |
| 18-101-final-viral-combined_k141_378882\|\|full | Not-determined |
| 18-101-final-viral-combined_k141_148488\|\|full | Not-determined |
| 18-101-final-viral-combined_k141_230872\|\|full | Not-determined |
| 18-101-final-viral-combined_k141_164968\|\|full | Low-quality |
| 18-101-final-viral-combined_k141_378891\|\|full | Not-determined |
| 18-101-final-viral-combined_k141_99204\|\|full | Not-determined |
| 18-101-final-viral-combined_k141_115697\|\|full | Not-determined |
| 18-101-final-viral-combined_k141_16922\|\|full | Not-determined |
| 18-101-final-viral-combined_k141_329484\|\|full | Low-quality |
| 18-101-final-viral-combined_k141_132190\|\|full | Not-determined |
| 18-101-final-viral-combined_k141_345966\|\|full | Low-quality |
| 18-101-final-viral-combined_k141_82829\|\|full | Low-quality |
| 18-101-final-viral-combined_k141_115702\|\|full | Not-determined |
| 18-101-final-viral-combined_k141_329493\|\|full | Not-determined |
| 18-101-final-viral-combined_k141_82836\|\|full | Not-determined |
| 18-101-final-viral-combined_k141_99231\|\|full | Not-determined |
| 18-101-final-viral-combined_k141_82837\|\|full | Low-quality |
| 18-101-final-viral-combined_k141_132226\|\|full | Not-determined |
| 18-101-final-viral-combined_k141_66077\|\|full | Low-quality |
| 18-101-final-viral-combined_k141_115723\|\|full | Not-determined |
| 18-101-final-viral-combined_k141_66078\|\|full | Not-determined |
| 18-101-final-viral-combined_k141_148551\|\|full | Low-quality |
| 18-101-final-viral-combined_k141_296678\|\|full | Not-determined |
| 18-101-final-viral-combined_k141_82882\|\|full | Low-quality |
| 18-101-final-viral-combined_k141_660\|\|full | Low-quality |
| 18-101-final-viral-combined_k141_115750\|\|full | Not-determined |
| 18-101-final-viral-combined_k141_82900\|\|full | Low-quality |
| 18-101-final-viral-combined_k141_66112\|\|full | Low-quality |
| 18-101-final-viral-combined_k141_329562\|\|full | Not-determined |
| 18-101-final-viral-combined_k141_296707\|\|full | Not-determined |
| 18-101-final-viral-combined_k141_82924\|\|full | Low-quality |
| 18-101-final-viral-combined_k141_181565\|\|full | Not-determined |
| 18-101-final-viral-combined_k141_99311\|\|full | Low-quality |
| 18-101-final-viral-combined_k141_685\|\|full | Low-quality |
| 18-101-final-viral-combined_k141_99312\|\|full | Low-quality |
| 18-101-final-viral-combined_k141_230930\|\|full | Low-quality |
| 18-101-final-viral-combined_k141_99315\|\|full | Not-determined |
| 18-101-final-viral-combined_k141_181586\|\|full | Not-determined |
| 18-101-final-viral-combined_k141_296730\|\|full | Low-quality |
| 18-101-final-viral-combined_k141_692\|\|full | Low-quality |
| 18-101-final-viral-combined_k141_66136\|\|full | Not-determined |
| 18-101-final-viral-combined_k141_230940\|\|full | Low-quality |
| 18-101-final-viral-combined_k141_165087\|\|full | Not-determined |
| 18-101-final-viral-combined_k141_181601\|\|full | Not-determined |
| 18-101-final-viral-combined_k141_197974\|\|full | Low-quality |
| 18-101-final-viral-combined_k141_181611\|\|full | Low-quality |
| 18-101-final-viral-combined_k141_214465\|\|full | Low-quality |
| 18-101-final-viral-combined_k141_296744\|\|full | Not-determined |
| 18-101-final-viral-combined_k141_197980\|\|full | Low-quality |
| 18-101-final-viral-combined_k141_329578\|\|full | Low-quality |
| 18-101-final-viral-combined_k141_66159\|\|full | Not-determined |
| 18-101-final-viral-combined_k141_263783\|\|full | Not-determined |
| 18-101-final-viral-combined_k141_181626\|\|full | Not-determined |
| 18-101-final-viral-combined_k141_346049\|\|full | Not-determined |
| 18-101-final-viral-combined_k141_115808\|\|full | Not-determined |
| 18-101-final-viral-combined_k141_132329\|\|full | Low-quality |
| 18-101-final-viral-combined_k141_115820\|\|full | Not-determined |
| 18-101-final-viral-combined_k141_66176\|\|full | Not-determined |
| 18-101-final-viral-combined_k141_16991\|\|full | Low-quality |
| 18-101-final-viral-combined_k141_362505\|\|full | Not-determined |
| 18-101-final-viral-combined_k141_296777\|\|full | Not-determined |
| 18-101-final-viral-combined_k141_148607\|\|full | Low-quality |
| 18-101-final-viral-combined_k141_132334\|\|full | Not-determined |
| 18-101-final-viral-combined_k141_17010\|\|full | Low-quality |
| 18-101-final-viral-combined_k141_148617\|\|full | Not-determined |
| 18-101-final-viral-combined_k141_214504\|\|full | Not-determined |
| 18-101-final-viral-combined_k141_214507\|\|full | Low-quality |
| 18-101-final-viral-combined_k141_115838\|\|full | Low-quality |
| 18-101-final-viral-combined_k141_198030\|\|full | Not-determined |
| 18-101-final-viral-combined_k141_198034\|\|full | Low-quality |
| 18-101-final-viral-combined_k141_362547\|\|full | Low-quality |
| 18-101-final-viral-combined_k141_724\|\|full | Low-quality |
| 18-101-final-viral-combined_k141_82976\|\|full | Low-quality |
| 18-101-final-viral-combined_k141_115877\|\|full | Not-determined |
| 18-101-final-viral-combined_k141_82984\|\|full | Not-determined |
| 18-101-final-viral-combined_k141_181634\|\|full | Low-quality |
| 18-101-final-viral-combined_k141_17057\|\|full | Not-determined |
| 18-101-final-viral-combined_k141_17061\|\|full | Low-quality |
| 18-101-final-viral-combined_k141_198055\|\|full | Not-determined |
| 18-101-final-viral-combined_k141_17065\|\|full | Not-determined |
| 18-101-final-viral-combined_k141_132379\|\|full | Low-quality |
| 18-101-final-viral-combined_k141_379035\|\|full | Low-quality |
| 18-101-final-viral-combined_k141_263835\|\|full | Not-determined |
| 18-101-final-viral-combined_k141_132385\|\|full | Not-determined |
| 18-101-final-viral-combined_k141_17088\|\|full | Low-quality |
| 18-101-final-viral-combined_k141_346185\|\|full | Not-determined |
| 18-101-final-viral-combined_k141_362573\|\|full | Not-determined |
| 18-101-final-viral-combined_k141_165143\|\|full | Not-determined |
| 18-101-final-viral-combined_k141_198056\|\|full | Low-quality |
| 18-101-final-viral-combined_k141_247413\|\|full | Not-determined |
| 18-101-final-viral-combined_k141_247416\|\|full | Not-determined |
| 18-101-final-viral-combined_k141_214619\|\|full | Low-quality |
| 18-101-final-viral-combined_k141_165148\|\|full | Low-quality |
| 18-101-final-viral-combined_k141_362577\|\|full | Low-quality |
| 18-101-final-viral-combined_k141_764\|\|full | Low-quality |
| 18-101-final-viral-combined_k141_115950\|\|full | Not-determined |
| 18-101-final-viral-combined_k141_263858\|\|full | Low-quality |
| 18-101-final-viral-combined_k141_346203\|\|full | Low-quality |
| 18-101-final-viral-combined_k141_165159\|\|full | Not-determined |
| 18-101-final-viral-combined_k141_362591\|\|full | Low-quality |
| 18-101-final-viral-combined_k141_346214\|\|full | Not-determined |
| 18-101-final-viral-combined_k141_247428\|\|full | Not-determined |
| 18-101-final-viral-combined_k141_148665\|\|full | Not-determined |
| 18-101-final-viral-combined_k141_148667\|\|full | Low-quality |
| 18-101-final-viral-combined_k141_198097\|\|full | Low-quality |

| | |
|---|---|
| 18-101-final-viral-combined_k141_811\|\|full | Low-quality |
| 18-101-final-viral-combined_k141_329671\|\|full | Low-quality |
| 18-101-final-viral-combined_k141_83004\|\|full | Low-quality |
| 18-101-final-viral-combined_k141_346228\|\|full | Low-quality |
| 18-101-final-viral-combined_k141_230950\|\|full | Low-quality |
| 18-101-final-viral-combined_k141_280401\|\|full | Low-quality |
| 18-101-final-viral-combined_k141_362600\|\|full | Low-quality |
| 18-101-final-viral-combined_k141_839\|\|full | Not-determined |
| 18-101-final-viral-combined_k141_346236\|\|full | Not-determined |
| 18-101-final-viral-combined_k141_280413\|\|full | Not-determined |
| 18-101-final-viral-combined_k141_198110\|\|full | Low-quality |
| 18-101-final-viral-combined_k141_165188\|\|full | Low-quality |
| 18-101-final-viral-combined_k141_329694\|\|full | Not-determined |
| 18-101-final-viral-combined_k141_362619\|\|full | Low-quality |
| 18-101-final-viral-combined_k141_346243\|\|full | Not-determined |
| 18-101-final-viral-combined_k141_230977\|\|full | Low-quality |
| 18-101-final-viral-combined_k141_362628\|\|full | Low-quality |
| 18-101-final-viral-combined_k141_247437\|\|full | Low-quality |
| 18-101-final-viral-combined_k141_116000\|\|full | Not-determined |
| 18-101-final-viral-combined_k141_148693\|\|full | Low-quality |
| 18-101-final-viral-combined_k141_17147\|\|full | Low-quality |
| 18-101-final-viral-combined_k141_165204\|\|full | Not-determined |
| 18-101-final-viral-combined_k141_296801\|\|full | Not-determined |
| 18-101-final-viral-combined_k141_362655\|\|full | Low-quality |
| 18-101-final-viral-combined_k141_296812\|\|full | Low-quality |
| 18-101-final-viral-combined_k141_148726\|\|full | Not-determined |
| 18-101-final-viral-combined_k141_66238\|\|full | Not-determined |
| 18-101-final-viral-combined_k141_379037\|\|full | Not-determined |
| 18-101-final-viral-combined_k141_99434\|\|full | Low-quality |
| 18-101-final-viral-combined_k141_165225\|\|full | Not-determined |
| 18-101-final-viral-combined_k141_362665\|\|full | Low-quality |
| 18-101-final-viral-combined_k141_263952\|\|full | Low-quality |
| 18-101-final-viral-combined_k141_247507\|\|full | Low-quality |
| 18-101-final-viral-combined_k141_362683\|\|full | Not-determined |
| 18-101-final-viral-combined_k141_280459\|\|full | Not-determined |
| 18-101-final-viral-combined_k141_83108\|\|full | Not-determined |
| 18-101-final-viral-combined_k141_116043\|\|full | Low-quality |
| 18-101-final-viral-combined_k141_379081\|\|full | Low-quality |
| 18-101-final-viral-combined_k141_231001\|\|full | Not-determined |
| 18-101-final-viral-combined_k141_927\|\|full | Not-determined |
| 18-101-final-viral-combined_k141_165273\|\|full | Low-quality |
| 18-101-final-viral-combined_k141_17181\|\|full | Not-determined |
| 18-101-final-viral-combined_k141_83124\|\|full | Not-determined |
| 18-101-final-viral-combined_k141_280478\|\|full | Low-quality |
| 18-101-final-viral-combined_k141_181847\|\|full | Not-determined |
| 18-101-final-viral-combined_k141_99468\|\|full | Low-quality |
| 18-101-final-viral-combined_k141_132471\|\|full | Low-quality |
| 18-101-final-viral-combined_k141_346300\|\|full | Not-determined |
| 18-101-final-viral-combined_k141_17195\|\|full | Not-determined |
| 18-101-final-viral-combined_k141_33413\|\|full | Low-quality |
| 18-101-final-viral-combined_k141_346320\|\|full | Low-quality |
| 18-101-final-viral-combined_k141_280514\|\|full | Low-quality |
| 18-101-final-viral-combined_k141_313256\|\|full | Not-determined |
| 17-109-final-viral-combined_k141_299293\|\|full | Complete |
| 18-101-final-viral-combined_k141_116087\|\|full | Low-quality |
| 18-101-final-viral-combined_k141_247550\|\|full | Low-quality |
| 18-101-final-viral-combined_k141_33421\|\|full | Low-quality |
| 18-101-final-viral-combined_k141_116093\|\|full | Not-determined |
| 18-101-final-viral-combined_k141_198227\|\|full | Not-determined |
| 18-101-final-viral-combined_k141_247557\|\|full | Not-determined |
| 18-101-final-viral-combined_k141_313279\|\|full | Not-determined |
| 18-101-final-viral-combined_k141_231062\|\|full | Low-quality |
| 18-101-final-viral-combined_k141_280547\|\|full | Not-determined |
| 18-101-final-viral-combined_k141_83183\|\|full | Low-quality |
| 18-101-final-viral-combined_k141_264022\|\|full | Not-determined |
| 18-101-final-viral-combined_k141_313301\|\|full | Low-quality |
| 18-101-final-viral-combined_k141_313304\|\|full | Low-quality |
| 18-101-final-viral-combined_k141_33440\|\|full | Low-quality |
| 18-101-final-viral-combined_k141_346363\|\|full | Not-determined |
| 18-101-final-viral-combined_k141_280562\|\|full | Low-quality |
| 18-101-final-viral-combined_k141_247577\|\|full | Not-determined |
| 18-101-final-viral-combined_k141_132529\|\|full | Low-quality |
| 18-101-final-viral-combined_k141_247578\|\|full | Not-determined |
| 18-101-final-viral-combined_k141_346382\|\|full | Not-determined |
| 18-101-final-viral-combined_k141_33448\|\|full | Low-quality |
| 18-101-final-viral-combined_k141_362812\|\|full | Not-determined |
| 18-101-final-viral-combined_k141_346392\|\|full | Not-determined |
| 18-101-final-viral-combined_k141_247593\|\|full | Not-determined |
| 18-101-final-viral-combined_k141_362822\|\|full | Low-quality |
| 18-101-final-viral-combined_k141_1014\|\|full | Not-determined |
| 18-101-final-viral-combined_k141_214883\|\|full | Not-determined |
| 18-101-final-viral-combined_k141_313366\|\|full | Not-determined |
| 18-101-final-viral-combined_k141_280581\|\|full | Low-quality |
| 18-101-final-viral-combined_k141_313369\|\|full | Not-determined |
| 18-101-final-viral-combined_k141_296921\|\|full | Not-determined |
| 18-101-final-viral-combined_k141_231080\|\|full | Low-quality |
| 18-101-final-viral-combined_k141_231082\|\|full | Low-quality |
| 18-101-final-viral-combined_k141_116180\|\|full | Low-quality |
| 18-101-final-viral-combined_k141_231084\|\|full | Not-determined |
| 18-101-final-viral-combined_k141_148824\|\|full | Not-determined |
| 18-101-final-viral-combined_k141_280590\|\|full | Not-determined |
| 18-101-final-viral-combined_k141_116199\|\|full | Not-determined |
| 18-101-final-viral-combined_k141_280592\|\|full | Low-quality |
| 18-101-final-viral-combined_k141_231087\|\|full | Low-quality |
| 18-101-final-viral-combined_k141_116216\|\|full | Low-quality |
| 18-101-final-viral-combined_k141_214898\|\|full | Not-determined |
| 18-101-final-viral-combined_k141_329783\|\|full | Not-determined |
| 18-101-final-viral-combined_k141_264062\|\|full | Low-quality |
| 18-101-final-viral-combined_k141_1068\|\|full | Low-quality |
| 18-101-final-viral-combined_k141_247635\|\|full | Low-quality |
| 18-101-final-viral-combined_k141_296962\|\|full | Low-quality |
| 18-101-final-viral-combined_k141_280621\|\|full | Low-quality |
| 18-101-final-viral-combined_k141_148844\|\|full | Low-quality |
| 18-101-final-viral-combined_k141_1080\|\|full | Not-determined |
| 18-101-final-viral-combined_k141_280623\|\|full | Not-determined |
| 18-101-final-viral-combined_k141_165343\|\|full | Low-quality |
| 18-101-final-viral-combined_k141_362850\|\|full | Low-quality |
| 18-101-final-viral-combined_k141_346473\|\|full | Low-quality |
| 18-101-final-viral-combined_k141_148858\|\|full | Low-quality |
| 18-101-final-viral-combined_k141_296974\|\|full | Low-quality |
| 18-101-final-viral-combined_k141_231131\|\|full | Not-determined |
| 18-101-final-viral-combined_k141_296985\|\|full | Not-determined |
| 18-101-final-viral-combined_k141_116263\|\|full | Low-quality |
| 18-101-final-viral-combined_k141_280648\|\|full | Low-quality |
| 18-101-final-viral-combined_k141_1114\|\|full | Not-determined |
| 18-101-final-viral-combined_k141_17256\|\|full | Not-determined |
| 18-101-final-viral-combined_k141_1118\|\|full | Low-quality |
| 18-101-final-viral-combined_k141_132644\|\|full | Low-quality |
| 18-101-final-viral-combined_k141_116283\|\|full | Low-quality |
| 18-101-final-viral-combined_k141_346507\|\|full | Not-determined |
| 18-101-final-viral-combined_k141_379264\|\|full | Not-determined |
| 18-101-final-viral-combined_k141_148884\|\|full | Low-quality |
| 18-101-final-viral-combined_k141_346514\|\|full | Low-quality |
| 18-101-final-viral-combined_k141_247704\|\|full | Low-quality |
| 18-101-final-viral-combined_k141_313409\|\|full | Low-quality |
| 18-101-final-viral-combined_k141_231184\|\|full | Low-quality |
| 18-101-final-viral-combined_k141_329814\|\|full | Not-determined |
| 18-101-final-viral-combined_k141_198359\|\|full | Not-determined |
| 18-101-final-viral-combined_k141_214966\|\|full | Low-quality |
| 18-101-final-viral-combined_k141_116322\|\|full | Not-determined |
| 18-101-final-viral-combined_k141_132662\|\|full | Not-determined |
| 18-101-final-viral-combined_k141_264107\|\|full | Low-quality |
| 18-101-final-viral-combined_k141_280695\|\|full | Low-quality |
| 18-101-final-viral-combined_k141_247727\|\|full | Not-determined |
| 18-101-final-viral-combined_k141_99541\|\|full | Low-quality |
| 18-101-final-viral-combined_k141_182015\|\|full | Low-quality |
| 18-101-final-viral-combined_k141_362926\|\|full | Low-quality |
| 18-101-final-viral-combined_k141_231210\|\|full | Not-determined |
| 18-101-final-viral-combined_k141_379289\|\|full | Not-determined |
| 18-101-final-viral-combined_k141_215020\|\|full | Low-quality |
| 18-101-final-viral-combined_k141_132719\|\|full | Low-quality |
| 18-101-final-viral-combined_k141_49799\|\|full | Low-quality |
| 18-101-final-viral-combined_k141_362955\|\|full | Not-determined |
| 18-101-final-viral-combined_k141_215036\|\|full | Low-quality |
| 18-101-final-viral-combined_k141_1173\|\|full | Not-determined |
| 18-101-final-viral-combined_k141_329842\|\|full | Not-determined |
| 18-101-final-viral-combined_k141_148939\|\|full | Low-quality |
| 18-101-final-viral-combined_k141_215037\|\|full | Not-determined |
| 18-101-final-viral-combined_k141_182040\|\|full | Low-quality |
| 18-101-final-viral-combined_k141_313457\|\|full | Low-quality |
| 18-101-final-viral-combined_k141_280718\|\|full | Not-determined |
| 18-101-final-viral-combined_k141_66406\|\|full | Not-determined |
| 18-101-final-viral-combined_k141_247755\|\|full | Low-quality |
| 18-101-final-viral-combined_k141_313462\|\|full | Low-quality |
| 18-101-final-viral-combined_k141_66424\|\|full | Low-quality |
| 18-101-final-viral-combined_k141_165393\|\|full | Low-quality |
| 18-101-final-viral-combined_k141_313471\|\|full | Low-quality |
| 18-101-final-viral-combined_k141_132770\|\|full | Not-determined |
| 18-109-final-viral-combined_k141_147158\|\|0_partial | Medium-quality |
| 18-101-final-viral-combined_k141_264146\|\|full | Low-quality |
| 18-101-final-viral-combined_k141_33581\|\|full | Not-determined |
| 18-101-final-viral-combined_k141_116384\|\|full | Low-quality |
| 18-101-final-viral-combined_k141_165397\|\|full | Not-determined |
| 18-101-final-viral-combined_k141_148963\|\|full | Low-quality |
| 18-101-final-viral-combined_k141_66458\|\|full | Not-determined |
| 18-101-final-viral-combined_k141_66459\|\|full | Not-determined |
| 18-101-final-viral-combined_k141_363002\|\|full | Not-determined |
| 18-101-final-viral-combined_k141_264174\|\|full | Not-determined |
| 18-101-final-viral-combined_k141_363003\|\|full | Low-quality |
| 18-101-final-viral-combined_k141_264182\|\|full | Low-quality |
| 18-101-final-viral-combined_k141_313516\|\|full | Low-quality |
| 18-101-final-viral-combined_k141_198425\|\|full | Not-determined |
| 18-101-final-viral-combined_k141_99599\|\|full | Not-determined |
| 18-101-final-viral-combined_k141_17327\|\|full | Low-quality |
| 18-101-final-viral-combined_k141_198434\|\|full | Not-determined |
| 18-101-final-viral-combined_k141_231276\|\|full | Low-quality |
| 18-101-final-viral-combined_k141_313526\|\|full | Low-quality |
| 18-101-final-viral-combined_k141_66473\|\|full | Low-quality |
| 18-101-final-viral-combined_k141_49896\|\|full | Not-determined |
| 18-101-final-viral-combined_k141_17345\|\|full | Low-quality |
| 18-101-final-viral-combined_k141_66476\|\|full | Not-determined |
| 18-101-final-viral-combined_k141_66480\|\|full | Not-determined |
| 18-101-final-viral-combined_k141_215089\|\|full | Low-quality |
| 18-101-final-viral-combined_k141_33611\|\|full | Not-determined |
| 18-101-final-viral-combined_k141_17366\|\|full | Not-determined |
| 18-101-final-viral-combined_k141_346613\|\|full | Not-determined |
| 18-101-final-viral-combined_k141_363082\|\|full | Not-determined |
| 18-101-final-viral-combined_k141_297180\|\|full | Not-determined |
| 18-101-final-viral-combined_k141_215114\|\|full | Low-quality |
| 18-101-final-viral-combined_k141_17380\|\|full | Not-determined |
| 18-101-final-viral-combined_k141_379362\|\|full | Low-quality |
| 18-101-final-viral-combined_k141_247813\|\|full | Not-determined |
| 18-101-final-viral-combined_k141_346635\|\|full | Low-quality |
| 18-101-final-viral-combined_k141_379367\|\|full | Low-quality |
| 18-101-final-viral-combined_k141_33638\|\|full | Low-quality |
| 18-101-final-viral-combined_k141_66520\|\|full | Low-quality |
| 18-101-final-viral-combined_k141_33639\|\|full | Not-determined |
| 18-101-final-viral-combined_k141_17397\|\|full | Not-determined |
| 18-101-final-viral-combined_k141_215135\|\|full | Low-quality |
| 18-101-final-viral-combined_k141_1271\|\|full | Not-determined |
| 18-101-final-viral-combined_k141_148973\|\|full | Not-determined |
| 18-101-final-viral-combined_k141_264254\|\|full | Low-quality |
| 18-101-final-viral-combined_k141_132823\|\|full | Low-quality |
| 18-101-final-viral-combined_k141_148975\|\|full | Low-quality |
| 18-101-final-viral-combined_k141_280797\|\|full | Low-quality |
| 18-101-final-viral-combined_k141_379381\|\|full | Not-determined |
| 18-101-final-viral-combined_k141_83362\|\|full | Not-determined |
| 18-101-final-viral-combined_k141_165469\|\|full | Not-determined |
| 18-101-final-viral-combined_k141_264264\|\|full | Not-determined |
| 18-101-final-viral-combined_k141_49936\|\|full | Not-determined |
| 18-101-final-viral-combined_k141_329891\|\|full | Not-determined |
| 18-101-final-viral-combined_k141_280807\|\|full | Not-determined |
| 18-101-final-viral-combined_k141_165479\|\|full | Low-quality |
| 18-101-final-viral-combined_k141_280811\|\|full | Not-determined |
| 18-101-final-viral-combined_k141_346665\|\|full | Not-determined |
| 18-101-final-viral-combined_k141_1311\|\|full | Low-quality |
| 18-101-final-viral-combined_k141_149014\|\|full | Not-determined |
| 18-101-final-viral-combined_k141_1318\|\|full | Not-determined |
| 18-101-final-viral-combined_k141_33687\|\|full | Low-quality |
| 18-101-final-viral-combined_k141_231354\|\|full | Not-determined |
| 18-101-final-viral-combined_k141_313630\|\|full | Low-quality |
| 18-101-final-viral-combined_k141_329924\|\|full | Low-quality |
| 18-101-final-viral-combined_k141_231366\|\|full | Not-determined |
| 18-101-final-viral-combined_k141_1349\|\|full | Not-determined |
| 18-101-final-viral-combined_k141_83405\|\|full | Not-determined |
| 18-101-final-viral-combined_k141_264291\|\|full | Not-determined |
| 18-101-final-viral-combined_k141_280817\|\|full | Low-quality |
| 18-101-final-viral-combined_k141_149018\|\|full | Low-quality |
| 18-101-final-viral-combined_k141_1359\|\|full | Not-determined |
| 18-101-final-viral-combined_k141_297288\|\|full | Not-determined |
| 18-101-final-viral-combined_k141_198541\|\|full | Not-determined |
| 18-101-final-viral-combined_k141_1364\|\|full | Not-determined |
| 18-101-final-viral-combined_k141_17449\|\|full | Not-determined |
| 18-101-final-viral-combined_k141_116432\|\|full | Low-quality |
| 18-101-final-viral-combined_k141_346687\|\|full | Low-quality |
| 18-101-final-viral-combined_k141_182089\|\|full | Low-quality |
| 18-101-final-viral-combined_k141_280825\|\|full | Not-determined |
| 18-101-final-viral-combined_k141_149040\|\|full | Low-quality |
| 18-101-final-viral-combined_k141_198558\|\|full | Low-quality |
| 18-101-final-viral-combined_k141_247919\|\|full | Low-quality |
| 18-101-final-viral-combined_k141_379425\|\|full | Low-quality |
| 18-101-final-viral-combined_k141_379427\|\|full | Low-quality |
| 18-101-final-viral-combined_k141_49977\|\|full | Low-quality |
| 18-101-final-viral-combined_k141_247921\|\|full | Not-determined |
| 18-101-final-viral-combined_k141_33725\|\|full | Low-quality |
| 18-101-final-viral-combined_k141_379433\|\|full | Not-determined |
| 18-101-final-viral-combined_k141_99716\|\|full | Low-quality |
| 18-101-final-viral-combined_k141_280856\|\|full | Low-quality |
| 18-101-final-viral-combined_k141_329981\|\|full | Not-determined |
| 18-101-final-viral-combined_k141_280858\|\|full | Low-quality |
| 18-101-final-viral-combined_k141_280863\|\|full | Not-determined |
| 18-101-final-viral-combined_k141_231404\|\|full | Not-determined |
| 18-101-final-viral-combined_k141_149079\|\|full | Low-quality |
| 18-101-final-viral-combined_k141_379439\|\|full | Low-quality |
| 18-101-final-viral-combined_k141_116478\|\|full | Not-determined |
| 18-101-final-viral-combined_k141_215255\|\|full | Low-quality |
| 18-101-final-viral-combined_k141_1415\|\|full | Not-determined |
| 18-101-final-viral-combined_k141_280879\|\|full | Not-determined |
| 18-110-final-viral-combined_k141_68780\|\|full | High-quality |
| 18-101-final-viral-combined_k141_379453\|\|full | Low-quality |
| 18-101-final-viral-combined_k141_215267\|\|full | Low-quality |
| 18-101-final-viral-combined_k141_247988\|\|full | Low-quality |
| 18-101-final-viral-combined_k141_198600\|\|full | Not-determined |
| 18-101-final-viral-combined_k141_280887\|\|full | Not-determined |
| 18-101-final-viral-combined_k141_99746\|\|full | Low-quality |
| 18-101-final-viral-combined_k141_99749\|\|full | Not-determined |
| 18-102_k141_6424 | Complete |
| 18-101-final-viral-combined_k141_99759\|\|full | Not-determined |
| 18-101-final-viral-combined_k141_280898\|\|full | Low-quality |
| 18-101-final-viral-combined_k141_66554\|\|full | Not-determined |
| 18-101-final-viral-combined_k141_198625\|\|full | Low-quality |
| 18-101-final-viral-combined_k141_198626\|\|full | Not-determined |
| 18-101-final-viral-combined_k141_99766\|\|full | Low-quality |
| 18-101-final-viral-combined_k141_198633\|\|full | Not-determined |
| 18-101-final-viral-combined_k141_198634\|\|full | Low-quality |
| 18-101-final-viral-combined_k141_248007\|\|full | Not-determined |
| 18-101-final-viral-combined_k141_248010\|\|full | Not-determined |
| 18-101-final-viral-combined_k141_248018\|\|full | Low-quality |
| 18-101-final-viral-combined_k141_33813\|\|full | Not-determined |
| 18-101-final-viral-combined_k141_198656\|\|full | Low-quality |
| 18-101-final-viral-combined_k141_379455\|\|full | Low-quality |
| 18-101-final-viral-combined_k141_132983\|\|full | Not-determined |
| 18-101-final-viral-combined_k141_313743\|\|full | Low-quality |
| 18-101-final-viral-combined_k141_215337\|\|full | Not-determined |
| 18-101-final-viral-combined_k141_33837\|\|full | Low-quality |
| 18-101-final-viral-combined_k141_182154\|\|full | Not-determined |
| 18-101-final-viral-combined_k141_50047\|\|full | Not-determined |
| 18-101-final-viral-combined_k141_50049\|\|full | Not-determined |
| 18-101-final-viral-combined_k141_363124\|\|full | Low-quality |
| 18-101-final-viral-combined_k141_215349\|\|full | Not-determined |
| 18-101-final-viral-combined_k141_363128\|\|full | Low-quality |
| 18-101-final-viral-combined_k141_346771\|\|full | Low-quality |
| 18-101-final-viral-combined_k141_33854\|\|full | Low-quality |
| 18-101-final-viral-combined_k141_313762\|\|full | Low-quality |
| 18-101-final-viral-combined_k141_313763\|\|full | Not-determined |
| 18-101-final-viral-combined_k141_313767\|\|full | Not-determined |
| 18-101-final-viral-combined_k141_17569\|\|full | Low-quality |
| 18-101-final-viral-combined_k141_297422\|\|full | Low-quality |
| 18-101-final-viral-combined_k141_346805\|\|full | Low-quality |
| 18-101-final-viral-combined_k141_248087\|\|full | Not-determined |
| 18-101-final-viral-combined_k141_1470\|\|full | Low-quality |
| 18-101-final-viral-combined_k141_346817\|\|full | Low-quality |
| 18-101-final-viral-combined_k141_182241\|\|full | Not-determined |
| 18-101-final-viral-combined_k141_33908\|\|full | Not-determined |
| 18-101-final-viral-combined_k141_133010\|\|full | Low-quality |
| 18-101-final-viral-combined_k141_313824\|\|full | Not-determined |
| 18-101-final-viral-combined_k141_231467\|\|full | Low-quality |
| 18-101-final-viral-combined_k141_33912\|\|full | Low-quality |
| 18-101-final-viral-combined_k141_281012\|\|full | Not-determined |
| 18-101-final-viral-combined_k141_346858\|\|full | Low-quality |
| 18-101-final-viral-combined_k141_165561\|\|full | Not-determined |
| 18-101-final-viral-combined_k141_281018\|\|full | Not-determined |
| 18-101-final-viral-combined_k141_346878\|\|full | Not-determined |
| 18-101-final-viral-combined_k141_346881\|\|full | Not-determined |
| 18-101-final-viral-combined_k141_346882\|\|full | Low-quality |
| 18-101-final-viral-combined_k141_281033\|\|full | Not-determined |
| 18-101-final-viral-combined_k141_99833\|\|full | Low-quality |
| 18-101-final-viral-combined_k141_165607\|\|full | Not-determined |
| 18-101-final-viral-combined_k141_165608\|\|full | Not-determined |
| 18-101-final-viral-combined_k141_116551\|\|full | Low-quality |
| 18-101-final-viral-combined_k141_297452\|\|full | Low-quality |
| 18-101-final-viral-combined_k141_116554\|\|full | Not-determined |
| 18-101-final-viral-combined_k141_116555\|\|full | Not-determined |
| 18-101-final-viral-combined_k141_116556\|\|full | Not-determined |
| 18-101-final-viral-combined_k141_116562\|\|full | Not-determined |
| 18-101-final-viral-combined_k141_83440\|\|full | Low-quality |
| 18-101-final-viral-combined_k141_281052\|\|full | Not-determined |
| 18-101-final-viral-combined_k141_50126\|\|full | Low-quality |
| 18-101-final-viral-combined_k141_149201\|\|full | Low-quality |
| 18-101-final-viral-combined_k141_133074\|\|full | Low-quality |
| 18-101-final-viral-combined_k141_231492\|\|full | Not-determined |
| 18-101-final-viral-combined_k141_297483\|\|full | Low-quality |
| 18-101-final-viral-combined_k141_281062\|\|full | Not-determined |
| 18-101-final-viral-combined_k141_83452\|\|full | Low-quality |
| 18-110-final-viral-combined_k141_26087\|\|full | High-quality |
| 18-101-final-viral-combined_k141_149211\|\|full | Low-quality |
| 18-101-final-viral-combined_k141_363233\|\|full | Not-determined |
| 18-101-final-viral-combined_k141_165641\|\|full | Not-determined |
| 18-101-final-viral-combined_k141_297485\|\|full | Not-determined |
| 18-101-final-viral-combined_k141_363234\|\|full | Low-quality |
| 18-101-final-viral-combined_k141_215400\|\|full | Not-determined |
| 18-101-final-viral-combined_k141_215401\|\|full | Not-determined |
| 18-102-final-viral-combined_k141_6424\|\|full | High-quality |
| 18-101-final-viral-combined_k141_297501\|\|full | Low-quality |
| 18-101-final-viral-combined_k141_99903\|\|full | Low-quality |
| 18-101-final-viral-combined_k141_297503\|\|full | Low-quality |
| 18-101-final-viral-combined_k141_83483\|\|full | Not-determined |
| 18-101-final-viral-combined_k141_313865\|\|full | Not-determined |
| 18-101-final-viral-combined_k141_1583\|\|full | Not-determined |
| 18-101-final-viral-combined_k141_17683\|\|full | Not-determined |
| 18-101-final-viral-combined_k141_99918\|\|full | Not-determined |
| 18-101-final-viral-combined_k141_17689\|\|full | Not-determined |
| 18-101-final-viral-combined_k141_116594\|\|full | Low-quality |
| 18-101-final-viral-combined_k141_83511\|\|full | Low-quality |
| 18-101-final-viral-combined_k141_50160\|\|full | Not-determined |
| 18-101-final-viral-combined_k141_330199\|\|full | Not-determined |
| 18-101-final-viral-combined_k141_116595\|\|full | Low-quality |
| 18-101-final-viral-combined_k141_297508\|\|full | Low-quality |
| 18-101-final-viral-combined_k141_165672\|\|full | Low-quality |
| 18-101-final-viral-combined_k141_34002\|\|full | Low-quality |
| 18-101-final-viral-combined_k141_231562\|\|full | Not-determined |
| 18-101-final-viral-combined_k141_165677\|\|full | Not-determined |
| 18-101-final-viral-combined_k141_50180\|\|full | Not-determined |
| 18-101-final-viral-combined_k141_66632\|\|full | Low-quality |
| 18-101-final-viral-combined_k141_116633\|\|full | Not-determined |
| 18-101-final-viral-combined_k141_297511\|\|full | Not-determined |
| 18-101-final-viral-combined_k141_1602\|\|full | Not-determined |
| 18-101-final-viral-combined_k141_133144\|\|full | Not-determined |
| 18-101-final-viral-combined_k141_182370\|\|full | Not-determined |
| 18-101-final-viral-combined_k141_149249\|\|full | Low-quality |
| 18-101-final-viral-combined_k141_182372\|\|full | Not-determined |
| 18-101-final-viral-combined_k141_83525\|\|full | Low-quality |
| 18-101-final-viral-combined_k141_116651\|\|full | Low-quality |
| 18-101-final-viral-combined_k141_83530\|\|full | Low-quality |
| 18-101-final-viral-combined_k141_34057\|\|full | Low-quality |
| 18-101-final-viral-combined_k141_1622\|\|full | Not-determined |
| 18-101-final-viral-combined_k141_198889\|\|full | Low-quality |
| 18-101-final-viral-combined_k141_264352\|\|full | Not-determined |
| 18-101-final-viral-combined_k141_133175\|\|full | Low-quality |
| 18-101-final-viral-combined_k141_133188\|\|full | Not-determined |
| 18-101-final-viral-combined_k141_198905\|\|full | Low-quality |
| 18-101-final-viral-combined_k141_149272\|\|full | Not-determined |
| 18-101-final-viral-combined_k141_281180\|\|full | Not-determined |
| 18-101-final-viral-combined_k141_248139\|\|full | Low-quality |
| 18-101-final-viral-combined_k141_1644\|\|full | Low-quality |
| 18-101-final-viral-combined_k141_50193\|\|full | Not-determined |
| 18-101-final-viral-combined_k141_133193\|\|full | Not-determined |
| 18-101-final-viral-combined_k141_379564\|\|full | Not-determined |
| 18-101-final-viral-combined_k141_347072\|\|full | Low-quality |
| 18-101-final-viral-combined_k141_83571\|\|full | Not-determined |
| 18-101-final-viral-combined_k141_330284\|\|full | Not-determined |
| 18-101-final-viral-combined_k141_379587\|\|full | Not-determined |
| 18-101-final-viral-combined_k141_50204\|\|full | Low-quality |
| 18-101-final-viral-combined_k141_281215\|\|full | Not-determined |
| 18-101-final-viral-combined_k141_149309\|\|full | Low-quality |
| 18-101-final-viral-combined_k141_330288\|\|full | Low-quality |
| 18-101-final-viral-combined_k141_347093\|\|full | Low-quality |
| 18-101-final-viral-combined_k141_83585\|\|full | Not-determined |
| 18-101-final-viral-combined_k141_198931\|\|full | Not-determined |
| 18-101-final-viral-combined_k141_83588\|\|full | Low-quality |
| 18-101-final-viral-combined_k141_379598\|\|full | Not-determined |
| 18-101-final-viral-combined_k141_363303\|\|full | Not-determined |
| 18-101-final-viral-combined_k141_116691\|\|full | Not-determined |
| 18-101-final-viral-combined_k141_1685\|\|full | Not-determined |
| 18-101-final-viral-combined_k141_313966\|\|full | Low-quality |
| 18-101-final-viral-combined_k141_313971\|\|full | Low-quality |
| 18-101-final-viral-combined_k141_83628\|\|full | Low-quality |
| 18-101-final-viral-combined_k141_231572\|\|full | Not-determined |
| 18-101-final-viral-combined_k141_17774\|\|full | Low-quality |
| 18-104-final-viral-combined_k141_17284\|\|full | Complete |
| 18-101-final-viral-combined_k141_264420\|\|full | Low-quality |
| 18-101-final-viral-combined_k141_198975\|\|full | Low-quality |
| 18-101-final-viral-combined_k141_314006\|\|full | Not-determined |
| 18-101-final-viral-combined_k141_17781\|\|full | Not-determined |
| 18-101-final-viral-combined_k141_149340\|\|full | Not-determined |
| 18-101-final-viral-combined_k141_17784\|\|full | Not-determined |
| 18-101-final-viral-combined_k141_281269\|\|full | Low-quality |
| 18-101-final-viral-combined_k141_99972\|\|full | Low-quality |
| 18-101-final-viral-combined_k141_231600\|\|full | Low-quality |
| 18-101-final-viral-combined_k141_34176\|\|full | Not-determined |
| 18-101-final-viral-combined_k141_231608\|\|full | Low-quality |
| 18-101-final-viral-combined_k141_314036\|\|full | Low-quality |
| 18-101-final-viral-combined_k141_231614\|\|full | Not-determined |
| 18-101-final-viral-combined_k141_347139\|\|full | Not-determined |
| 18-101-final-viral-combined_k141_297613\|\|full | Low-quality |
| 18-101-final-viral-combined_k141_50280\|\|full | Not-determined |
| 18-101-final-viral-combined_k141_314062\|\|full | Low-quality |
| 18-105-final-viral-combined_k141_94786\|\|full | Medium-quality |
| 18-101-final-viral-combined_k141_215465\|\|full | Low-quality |
| 18-101-final-viral-combined_k141_314066\|\|full | Low-quality |
| 18-101-final-viral-combined_k141_330332\|\|full | Low-quality |
| 18-101-final-viral-combined_k141_83684\|\|full | Not-determined |
| 18-101-final-viral-combined_k141_330333\|\|full | Not-determined |
| 18-101-final-viral-combined_k141_34223\|\|full | Low-quality |
| 18-101-final-viral-combined_k141_199012\|\|full | Not-determined |
| 18-101-final-viral-combined_k141_182446\|\|full | Not-determined |
| 18-101-final-viral-combined_k141_50297\|\|full | Not-determined |
| 18-101-final-viral-combined_k141_50304\|\|full | Not-determined |
| 18-101-final-viral-combined_k141_1769\|\|full | Not-determined |
| 18-101-final-viral-combined_k141_66712\|\|full | Low-quality |
| 18-101-final-viral-combined_k141_50306\|\|full | Not-determined |
| 18-101-final-viral-combined_k141_83701\|\|full | Not-determined |
| 18-101-final-viral-combined_k141_379732\|\|full | Low-quality |
| 18-101-final-viral-combined_k141_215482\|\|full | Not-determined |
| 18-101-final-viral-combined_k141_133285\|\|full | Not-determined |
| 18-101-final-viral-combined_k141_66722\|\|full | Not-determined |
| 18-101-final-viral-combined_k141_149378\|\|full | Not-determined |
| 18-101-final-viral-combined_k141_83716\|\|full | Low-quality |
| 18-101-final-viral-combined_k141_149381\|\|full | Low-quality |
| 18-101-final-viral-combined_k141_133286\|\|full | Low-quality |
| 18-101-final-viral-combined_k141_149386\|\|full | Low-quality |
| 18-101-final-viral-combined_k141_314080\|\|full | Not-determined |
| 18-101-final-viral-combined_k141_149399\|\|full | Low-quality |
| 18-101-final-viral-combined_k141_1778\|\|full | Low-quality |
| 18-101-final-viral-combined_k141_297659\|\|full | Not-determined |
| 18-101-final-viral-combined_k141_215511\|\|full | Not-determined |
| 18-101-final-viral-combined_k141_182475\|\|full | Low-quality |
| 18-101-final-viral-combined_k141_347233\|\|full | Not-determined |
| 18-101-final-viral-combined_k141_34278\|\|full | Not-determined |
| 18-101-final-viral-combined_k141_182481\|\|full | Low-quality |
| 18-101-final-viral-combined_k141_1801\|\|full | Low-quality |
| 18-101-final-viral-combined_k141_83726\|\|full | Not-determined |
| 18-101-final-viral-combined_k141_83735\|\|full | Low-quality |
| 18-101-final-viral-combined_k141_149420\|\|full | Not-determined |
| 18-101-final-viral-combined_k141_363364\|\|full | Low-quality |
| 18-101-final-viral-combined_k141_149430\|\|full | Low-quality |
| 18-101-final-viral-combined_k141_215526\|\|full | Low-quality |
| 18-101-final-viral-combined_k141_363373\|\|full | Not-determined |
| 18-101-final-viral-combined_k141_297709\|\|full | Not-determined |
| 18-101-final-viral-combined_k141_314129\|\|full | Not-determined |
| 18-101-final-viral-combined_k141_314147\|\|full | Not-determined |
| 18-101-final-viral-combined_k141_215575\|\|full | Not-determined |
| 18-101-final-viral-combined_k141_66770\|\|full | Not-determined |
| 18-101-final-viral-combined_k141_215580\|\|full | Not-determined |
| 18-101-final-viral-combined_k141_215584\|\|full | Low-quality |
| 18-101-final-viral-combined_k141_149462\|\|full | Low-quality |
| 18-101-final-viral-combined_k141_17907\|\|full | Low-quality |
| 18-101-final-viral-combined_k141_248221\|\|full | Not-determined |
| 18-101-final-viral-combined_k141_83783\|\|full | Not-determined |
| 18-101-final-viral-combined_k141_165771\|\|full | Not-determined |
| 18-101-final-viral-combined_k141_133373\|\|full | Low-quality |
| 18-101-final-viral-combined_k141_50386\|\|full | Not-determined |
| 18-101-final-viral-combined_k141_248240\|\|full | Not-determined |
| 18-101-final-viral-combined_k141_248241\|\|full | Low-quality |
| 18-101-final-viral-combined_k141_231792\|\|full | Not-determined |
| 18-101-final-viral-combined_k141_297754\|\|full | Not-determined |
| 18-101-final-viral-combined_k141_297756\|\|full | Not-determined |
| 18-101-final-viral-combined_k141_314198\|\|full | Not-determined |
| 18-101-final-viral-combined_k141_231798\|\|full | Not-determined |
| 18-101-final-viral-combined_k141_149465\|\|full | Not-determined |
| 18-101-final-viral-combined_k141_297766\|\|full | Low-quality |
| 18-101-final-viral-combined_k141_248280\|\|full | Low-quality |
| 18-101-final-viral-combined_k141_50405\|\|full | Not-determined |
| 18-101-final-viral-combined_k141_215650\|\|full | Not-determined |
| 18-101-final-viral-combined_k141_1861\|\|full | Low-quality |
| 18-101-final-viral-combined_k141_248293\|\|full | Not-determined |
| 18-101-final-viral-combined_k141_281362\|\|full | Low-quality |
| 18-101-final-viral-combined_k141_199140\|\|full | Not-determined |
| 18-101-final-viral-combined_k141_149472\|\|full | Not-determined |
| 18-101-final-viral-combined_k141_34316\|\|full | Not-determined |
| 18-101-final-viral-combined_k141_281370\|\|full | Not-determined |
| 18-101-final-viral-combined_k141_248309\|\|full | Low-quality |
| 18-101-final-viral-combined_k141_314247\|\|full | Not-determined |
| 18-109_k141_244425 | Medium-quality |
| 18-101-final-viral-combined_k141_248315\|\|full | Not-determined |
| 18-101-final-viral-combined_k141_330509\|\|full | Not-determined |
| 18-101-final-viral-combined_k141_149498\|\|full | Not-determined |
| 18-101-final-viral-combined_k141_116790\|\|full | Low-quality |
| 18-101-final-viral-combined_k141_330512\|\|full | Not-determined |
| 18-101-final-viral-combined_k141_116795\|\|full | Low-quality |
| 18-101-final-viral-combined_k141_248343\|\|full | Low-quality |
| 18-101-final-viral-combined_k141_182581\|\|full | Low-quality |
| 18-101-final-viral-combined_k141_199157\|\|full | Low-quality |
| 18-101-final-viral-combined_k141_379919\|\|full | Not-determined |
| 18-101-final-viral-combined_k141_116808\|\|full | Not-determined |
| 18-101-final-viral-combined_k141_215676\|\|full | Low-quality |
| 18-101-final-viral-combined_k141_231839\|\|full | Not-determined |
| 18-101-final-viral-combined_k141_264526\|\|full | Low-quality |
| 18-101-final-viral-combined_k141_149506\|\|full | Not-determined |
| 18-101-final-viral-combined_k141_199170\|\|full | Low-quality |
| 18-101-final-viral-combined_k141_149508\|\|full | Not-determined |
| 18-101-final-viral-combined_k141_100062\|\|full | Not-determined |
| 18-101-final-viral-combined_k141_199194\|\|full | Not-determined |
| 18-101-final-viral-combined_k141_100082\|\|full | Not-determined |
| 18-101-final-viral-combined_k141_379959\|\|full | Not-determined |
| 18-101-final-viral-combined_k141_50436\|\|full | Not-determined |
| 18-101-final-viral-combined_k141_34429\|\|full | Not-determined |
| 18-101-final-viral-combined_k141_182621\|\|full | Low-quality |
| 18-101-final-viral-combined_k141_264578\|\|full | Not-determined |
| 18-101-final-viral-combined_k141_264580\|\|full | Low-quality |
| 18-101-final-viral-combined_k141_281425\|\|full | Not-determined |
| 18-101-final-viral-combined_k141_182634\|\|full | Not-determined |
| 18-101-final-viral-combined_k141_182636\|\|full | Low-quality |
| 18-101-final-viral-combined_k141_149574\|\|full | Low-quality |
| 18-101-final-viral-combined_k141_248392\|\|full | Not-determined |
| 18-101-final-viral-combined_k141_248394\|\|full | Not-determined |
| 18-101-final-viral-combined_k141_182643\|\|full | Low-quality |
| 18-101-final-viral-combined_k141_2019\|\|full | Not-determined |
| 18-101-final-viral-combined_k141_297882\|\|full | Low-quality |
| 18-101-final-viral-combined_k141_2020\|\|full | Low-quality |
| 18-101-final-viral-combined_k141_50450\|\|full | Low-quality |
| 18-101-final-viral-combined_k141_199213\|\|full | Low-quality |
| 18-101-final-viral-combined_k141_100096\|\|full | Low-quality |
| 18-101-final-viral-combined_k141_133538\|\|full | Not-determined |
| 18-101-final-viral-combined_k141_2030\|\|full | Not-determined |
| 18-101-final-viral-combined_k141_182676\|\|full | Not-determined |
| 18-101-final-viral-combined_k141_281477\|\|full | Not-determined |
| 18-101-final-viral-combined_k141_50469\|\|full | Not-determined |
| 18-101-final-viral-combined_k141_199239\|\|full | Not-determined |
| 18-101-final-viral-combined_k141_18089\|\|full | Not-determined |
| 18-101-final-viral-combined_k141_133562\|\|full | Not-determined |
| 18-101-final-viral-combined_k141_314329\|\|full | Low-quality |
| 18-101-final-viral-combined_k141_231917\|\|full | Not-determined |
| 18-101-final-viral-combined_k141_330606\|\|full | Not-determined |
| 18-101-final-viral-combined_k141_50483\|\|full | Not-determined |
| 18-101-final-viral-combined_k141_379998\|\|full | Low-quality |
| 18-101-final-viral-combined_k141_83918\|\|full | Not-determined |
| 18-101-final-viral-combined_k141_116855\|\|full | Not-determined |
| 18-101-final-viral-combined_k141_231924\|\|full | Not-determined |
| 18-101-final-viral-combined_k141_380008\|\|full | Low-quality |
| 18-101-final-viral-combined_k141_231927\|\|full | Not-determined |
| 18-101-final-viral-combined_k141_116860\|\|full | Low-quality |
| 18-101-final-viral-combined_k141_347356\|\|full | Low-quality |
| 18-101-final-viral-combined_k141_248457\|\|full | Not-determined |
| 18-101-final-viral-combined_k141_231935\|\|full | Not-determined |
| 18-101-final-viral-combined_k141_248460\|\|full | Not-determined |
| 18-101-final-viral-combined_k141_116870\|\|full | Not-determined |
| 18-101-final-viral-combined_k141_83936\|\|full | Low-quality |
| 18-101-final-viral-combined_k141_18104\|\|full | Not-determined |
| 18-101-final-viral-combined_k141_199268\|\|full | Not-determined |
| 18-101-final-viral-combined_k141_50508\|\|full | Not-determined |
| 18-101-final-viral-combined_k141_281534\|\|full | Not-determined |
| 18-101-final-viral-combined_k141_297977\|\|full | Low-quality |
| 18-101-final-viral-combined_k141_100196\|\|full | Not-determined |
| 18-101-final-viral-combined_k141_133606\|\|full | Not-determined |
| 18-101-final-viral-combined_k141_18130\|\|full | Low-quality |
| 18-101-final-viral-combined_k141_314352\|\|full | Not-determined |
| 18-101-final-viral-combined_k141_298004\|\|full | Low-quality |
| 18-101-final-viral-combined_k141_380061\|\|full | Low-quality |
| 18-101-final-viral-combined_k141_149656\|\|full | Not-determined |
| 18-101-final-viral-combined_k141_18149\|\|full | Low-quality |
| 18-101-final-viral-combined_k141_18150\|\|full | Low-quality |
| 18-101-final-viral-combined_k141_380064\|\|full | Not-determined |
| 18-101-final-viral-combined_k141_298027\|\|full | Low-quality |
| 18-101-final-viral-combined_k141_363666\|\|full | Not-determined |
| 18-101-final-viral-combined_k141_2113\|\|full | Low-quality |
| 18-101-final-viral-combined_k141_298028\|\|full | Low-quality |
| 18-101-final-viral-combined_k141_314356\|\|full | Low-quality |
| 18-101-final-viral-combined_k141_314357\|\|full | Low-quality |

| | |
|---|---|
| 18-101-final-viral-combined_k141_116925\|\|full | Low-quality |
| 18-101-final-viral-combined_k141_165809\|\|full | Not-determined |
| 18-101-final-viral-combined_k141_116927\|\|full | Not-determined |
| 18-101-final-viral-combined_k141_149671\|\|full | Not-determined |
| 18-101-final-viral-combined_k141_50540\|\|full | Low-quality |
| 18-101-final-viral-combined_k141_281605\|\|full | Not-determined |
| 18-101-final-viral-combined_k141_66851\|\|full | Not-determined |
| 18-101-final-viral-combined_k141_215796\|\|full | Not-determined |
| 18-101-final-viral-combined_k141_83988\|\|full | Low-quality |
| 18-101-final-viral-combined_k141_347432\|\|full | Low-quality |
| 18-101-final-viral-combined_k141_363693\|\|full | Not-determined |
| 18-101-final-viral-combined_k141_330691\|\|full | Not-determined |
| 18-101-final-viral-combined_k141_100258\|\|full | Not-determined |
| 18-101-final-viral-combined_k141_314370\|\|full | Low-quality |
| 18-101-final-viral-combined_k141_231995\|\|full | Not-determined |
| 18-101-final-viral-combined_k141_347441\|\|full | Not-determined |
| 18-101-final-viral-combined_k141_314372\|\|full | Low-quality |
| 18-101-final-viral-combined_k141_18177\|\|full | Low-quality |
| 18-101-final-viral-combined_k141_215806\|\|full | Not-determined |
| 18-101-final-viral-combined_k141_330694\|\|full | Not-determined |
| 18-101-final-viral-combined_k141_50586\|\|full | Low-quality |
| 18-101-final-viral-combined_k141_66882\|\|full | Low-quality |
| 18-101-final-viral-combined_k141_165834\|\|full | Not-determined |
| 18-101-final-viral-combined_k141_18186\|\|full | Not-determined |
| 18-101-final-viral-combined_k141_182799\|\|full | Low-quality |
| 18-101-final-viral-combined_k141_100287\|\|full | Not-determined |
| 18-101-final-viral-combined_k141_215846\|\|full | Not-determined |
| 18-101-final-viral-combined_k141_330703\|\|full | Low-quality |
| 18-101-final-viral-combined_k141_133680\|\|full | Not-determined |
| 18-101-final-viral-combined_k141_281660\|\|full | Low-quality |
| 18-101-final-viral-combined_k141_100303\|\|full | Low-quality |
| 18-101-final-viral-combined_k141_248588\|\|full | Low-quality |
| 18-101-final-viral-combined_k141_314398\|\|full | Low-quality |
| 18-101-final-viral-combined_k141_347481\|\|full | Low-quality |
| 18-101-final-viral-combined_k141_165878\|\|full | Low-quality |
| 18-101-final-viral-combined_k141_380145\|\|full | Not-determined |
| 18-109-final-viral-combined_k141_244425\|\|full | Medium-quality |
| 18-101-final-viral-combined_k141_363720\|\|full | Not-determined |
| 18-101-final-viral-combined_k141_100316\|\|full | Not-determined |
| 18-101-final-viral-combined_k141_2178\|\|full | Low-quality |
| 18-101-final-viral-combined_k141_281674\|\|full | Not-determined |
| 18-101-final-viral-combined_k141_2186\|\|full | Not-determined |
| 18-101-final-viral-combined_k141_281685\|\|full | Not-determined |
| 18-101-final-viral-combined_k141_215880\|\|full | Not-determined |
| 18-101-final-viral-combined_k141_215882\|\|full | Low-quality |
| 18-101-final-viral-combined_k141_182835\|\|full | Not-determined |
| 18-105-final-viral-combined_k141_117213\|\|0_partial | High-quality |
| 18-101-final-viral-combined_k141_182838\|\|full | Low-quality |
| 18-101-final-viral-combined_k141_215896\|\|full | Not-determined |
| 18-101-final-viral-combined_k141_182839\|\|full | Not-determined |
| 18-101-final-viral-combined_k141_149751\|\|full | Low-quality |
| 18-101-final-viral-combined_k141_330737\|\|full | Not-determined |
| 18-101-final-viral-combined_k141_2222\|\|full | Not-determined |
| 18-101-final-viral-combined_k141_281705\|\|full | Not-determined |
| 18-101-final-viral-combined_k141_330745\|\|full | Not-determined |
| 18-101-final-viral-combined_k141_264703\|\|full | Low-quality |
| 18-101-final-viral-combined_k141_50621\|\|full | Low-quality |
| 18-101-final-viral-combined_k141_264709\|\|full | Not-determined |
| 18-110-final-viral-combined_k141_40666\|\|full | Medium-quality |
| 18-101-final-viral-combined_k141_100347\|\|full | Not-determined |
| 18-101-final-viral-combined_k141_199398\|\|full | Not-determined |
| 18-101-final-viral-combined_k141_18251\|\|full | Low-quality |
| 18-101-final-viral-combined_k141_66977\|\|full | Not-determined |
| 18-101-final-viral-combined_k141_232050\|\|full | Low-quality |
| 18-101-final-viral-combined_k141_116993\|\|full | Not-determined |
| 18-101-final-viral-combined_k141_232060\|\|full | Low-quality |
| 18-101-final-viral-combined_k141_84061\|\|full | Low-quality |
| 18-101-final-viral-combined_k141_298129\|\|full | Low-quality |
| 18-101-final-viral-combined_k141_182900\|\|full | Not-determined |
| 18-101-final-viral-combined_k141_182907\|\|full | Low-quality |
| 18-101-final-viral-combined_k141_18272\|\|full | Low-quality |
| 18-101-final-viral-combined_k141_264734\|\|full | Low-quality |
| 18-101-final-viral-combined_k141_165889\|\|full | Not-determined |
| 18-101-final-viral-combined_k141_298154\|\|full | Low-quality |
| 18-101-final-viral-combined_k141_50640\|\|full | Not-determined |
| 18-101-final-viral-combined_k141_347563\|\|full | Not-determined |
| 18-101-final-viral-combined_k141_314438\|\|full | Not-determined |
| 18-101-final-viral-combined_k141_133763\|\|full | Not-determined |
| 18-101-final-viral-combined_k141_67009\|\|full | Not-determined |
| 18-101-final-viral-combined_k141_34689\|\|full | Low-quality |
| 18-101-final-viral-combined_k141_165897\|\|full | Low-quality |
| 18-101-final-viral-combined_k141_84130\|\|full | Low-quality |
| 18-101-final-viral-combined_k141_281743\|\|full | Not-determined |
| 18-101-final-viral-combined_k141_380165\|\|full | Not-determined |
| 18-101-final-viral-combined_k141_18312\|\|full | Not-determined |
| 18-101-final-viral-combined_k141_84141\|\|full | Low-quality |
| 18-101-final-viral-combined_k141_281746\|\|full | Not-determined |
| 18-101-final-viral-combined_k141_67035\|\|full | Not-determined |
| 18-101-final-viral-combined_k141_133794\|\|full | Not-determined |
| 18-101-final-viral-combined_k141_133795\|\|full | Not-determined |
| 18-101-final-viral-combined_k141_133799\|\|full | Low-quality |
| 18-101-final-viral-combined_k141_18334\|\|full | Low-quality |
| 18-101-final-viral-combined_k141_380184\|\|full | Not-determined |
| 18-101-final-viral-combined_k141_67049\|\|full | Not-determined |
| 18-101-final-viral-combined_k141_50688\|\|full | Not-determined |
| 18-101-final-viral-combined_k141_133811\|\|full | Not-determined |
| 18-101-final-viral-combined_k141_380204\|\|full | Not-determined |
| 18-101-final-viral-combined_k141_215928\|\|full | Low-quality |
| 18-101-final-viral-combined_k141_232109\|\|full | Low-quality |
| 18-101-final-viral-combined_k141_232112\|\|full | Low-quality |
| 18-101-final-viral-combined_k141_347614\|\|full | Low-quality |
| 18-101-final-viral-combined_k141_67061\|\|full | Low-quality |
| 18-101-final-viral-combined_k141_50694\|\|full | Not-determined |
| 18-101-final-viral-combined_k141_363829\|\|full | Low-quality |
| 18-101-final-viral-combined_k141_117096\|\|full | Low-quality |
| 18-102-final-viral-combined_k141_244564\|\|full | High-quality |
| 18-101-final-viral-combined_k141_314503\|\|full | Not-determined |
| 18-101-final-viral-combined_k141_248682\|\|full | Not-determined |
| 18-101-final-viral-combined_k141_84201\|\|full | Not-determined |
| 18-101-final-viral-combined_k141_165981\|\|full | Not-determined |
| 18-101-final-viral-combined_k141_363837\|\|full | Low-quality |
| 18-101-final-viral-combined_k141_199470\|\|full | Low-quality |
| 18-101-final-viral-combined_k141_248696\|\|full | Not-determined |
| 18-101-final-viral-combined_k141_67087\|\|full | Not-determined |
| 18-101-final-viral-combined_k141_232122\|\|full | Low-quality |
| 18-101-final-viral-combined_k141_182958\|\|full | Not-determined |
| 18-101-final-viral-combined_k141_67089\|\|full | Not-determined |
| 18-101-final-viral-combined_k141_182963\|\|full | Low-quality |
| 18-101-final-viral-combined_k141_363851\|\|full | Not-determined |
| 18-101-final-viral-combined_k141_314547\|\|full | Low-quality |
| 18-101-final-viral-combined_k141_363853\|\|full | Not-determined |
| 18-101-final-viral-combined_k141_34814\|\|full | Low-quality |
| 18-101-final-viral-combined_k141_18367\|\|full | Not-determined |
| 18-101-final-viral-combined_k141_298237\|\|full | Not-determined |
| 18-101-final-viral-combined_k141_2290\|\|full | Low-quality |
| 18-101-final-viral-combined_k141_84238\|\|full | Not-determined |
| 18-101-final-viral-combined_k141_232159\|\|full | Not-determined |
| 18-101-final-viral-combined_k141_347641\|\|full | Not-determined |
| 18-101-final-viral-combined_k141_34823\|\|full | Low-quality |
| 18-101-final-viral-combined_k141_149844\|\|full | Not-determined |
| 18-101-final-viral-combined_k141_84250\|\|full | Not-determined |
| 18-101-final-viral-combined_k141_117161\|\|full | Low-quality |
| 18-101-final-viral-combined_k141_314581\|\|full | Low-quality |
| 18-101-final-viral-combined_k141_363895\|\|full | Not-determined |
| 18-101-final-viral-combined_k141_50804\|\|full | Not-determined |
| 18-101-final-viral-combined_k141_84278\|\|full | Not-determined |
| 18-101-final-viral-combined_k141_100469\|\|full | Low-quality |
| 18-101-final-viral-combined_k141_347664\|\|full | Low-quality |
| 18-101-final-viral-combined_k141_363910\|\|full | Low-quality |
| 18-101-final-viral-combined_k141_149876\|\|full | Low-quality |
| 18-101-final-viral-combined_k141_166012\|\|full | Not-determined |
| 18-101-final-viral-combined_k141_314615\|\|full | Not-determined |
| 18-101-final-viral-combined_k141_133890\|\|full | Low-quality |
| 18-101-final-viral-combined_k141_133892\|\|full | Low-quality |
| 18-101-final-viral-combined_k141_166022\|\|full | Low-quality |
| 18-101-final-viral-combined_k141_330799\|\|full | Not-determined |
| 18-101-final-viral-combined_k141_298315\|\|full | Low-quality |
| 18-101-final-viral-combined_k141_84309\|\|full | Low-quality |
| 18-101-final-viral-combined_k141_149890\|\|full | Low-quality |
| 18-101-final-viral-combined_k141_232213\|\|full | Low-quality |
| 18-101-final-viral-combined_k141_363915\|\|full | Not-determined |
| 18-101-final-viral-combined_k141_133906\|\|full | Low-quality |
| 18-101-final-viral-combined_k141_380237\|\|full | Low-quality |
| 18-101-final-viral-combined_k141_84323\|\|full | Not-determined |
| 18-101-final-viral-combined_k141_166047\|\|full | Not-determined |
| 18-101-final-viral-combined_k141_133924\|\|full | Not-determined |
| 18-101-final-viral-combined_k141_363924\|\|full | Low-quality |
| 18-101-final-viral-combined_k141_183059\|\|full | Not-determined |
| 18-101-final-viral-combined_k141_298341\|\|full | Low-quality |
| 18-101-final-viral-combined_k141_166054\|\|full | Low-quality |
| 18-101-final-viral-combined_k141_149900\|\|full | Low-quality |
| 18-101-final-viral-combined_k141_84331\|\|full | Not-determined |
| 18-101-final-viral-combined_k141_248802\|\|full | Not-determined |
| 18-101-final-viral-combined_k141_347705\|\|full | Not-determined |
| 18-101-final-viral-combined_k141_232267\|\|full | Low-quality |
| 18-101-final-viral-combined_k141_298364\|\|full | Not-determined |
| 18-101-final-viral-combined_k141_133954\|\|full | Low-quality |
| 18-101-final-viral-combined_k141_166078\|\|full | Low-quality |
| 18-101-final-viral-combined_k141_347718\|\|full | Low-quality |
| 18-101-final-viral-combined_k141_183066\|\|full | Low-quality |
| 18-101-final-viral-combined_k141_363959\|\|full | Not-determined |
| 18-101-final-viral-combined_k141_216125\|\|full | Low-quality |
| 18-101-final-viral-combined_k141_298369\|\|full | Not-determined |
| 18-101-final-viral-combined_k141_347725\|\|full | Low-quality |
| 18-101-final-viral-combined_k141_298371\|\|full | Not-determined |
| 18-101-final-viral-combined_k141_133968\|\|full | Low-quality |
| 18-101-final-viral-combined_k141_232273\|\|full | Low-quality |
| 18-101-final-viral-combined_k141_298377\|\|full | Not-determined |
| 18-101-final-viral-combined_k141_50899\|\|full | Not-determined |
| 18-101-final-viral-combined_k141_281849\|\|full | Not-determined |
| 18-101-final-viral-combined_k141_183091\|\|full | Low-quality |
| 18-101-final-viral-combined_k141_67194\|\|full | Not-determined |
| 18-101-final-viral-combined_k141_67199\|\|full | Low-quality |
| 18-101-final-viral-combined_k141_264932\|\|full | Not-determined |
| 18-101-final-viral-combined_k141_149946\|\|full | Not-determined |
| 18-101-final-viral-combined_k141_84385\|\|full | Low-quality |
| 18-101-final-viral-combined_k141_134002\|\|full | Not-determined |
| 18-101-final-viral-combined_k141_264947\|\|full | Low-quality |
| 18-101-final-viral-combined_k141_232312\|\|full | Not-determined |
| 18-101-final-viral-combined_k141_166110\|\|full | Low-quality |
| 18-101-final-viral-combined_k141_380300\|\|full | Not-determined |
| 18-101-final-viral-combined_k141_18432\|\|full | Low-quality |
| 18-101-final-viral-combined_k141_166112\|\|full | Low-quality |
| 18-101-final-viral-combined_k141_100575\|\|full | Not-determined |
| 18-101-final-viral-combined_k141_67226\|\|full | Low-quality |
| 18-101-final-viral-combined_k141_216142\|\|full | Not-determined |
| 18-101-final-viral-combined_k141_199567\|\|full | Low-quality |
| 18-101-final-viral-combined_k141_248879\|\|full | Low-quality |
| 18-101-final-viral-combined_k141_314718\|\|full | Not-determined |
| 18-101-final-viral-combined_k141_363977\|\|full | Low-quality |
| 18-101-final-viral-combined_k141_166126\|\|full | Low-quality |
| 18-101-final-viral-combined_k141_34881\|\|full | Not-determined |
| 18-101-final-viral-combined_k141_34885\|\|full | Low-quality |
| 18-101-final-viral-combined_k141_314739\|\|full | Not-determined |
| 18-101-final-viral-combined_k141_183128\|\|full | Not-determined |
| 18-101-final-viral-combined_k141_248922\|\|full | Not-determined |
| 18-101-final-viral-combined_k141_232342\|\|full | Not-determined |
| 18-101-final-viral-combined_k141_2418\|\|full | Low-quality |
| 18-101-final-viral-combined_k141_150023\|\|full | Not-determined |
| 18-101-final-viral-combined_k141_2423\|\|full | Low-quality |
| 18-101-final-viral-combined_k141_380353\|\|full | Not-determined |
| 18-101-final-viral-combined_k141_150026\|\|full | Not-determined |
| 18-101-final-viral-combined_k141_34894\|\|full | Low-quality |
| 18-101-final-viral-combined_k141_183133\|\|full | Low-quality |
| 18-101-final-viral-combined_k141_232363\|\|full | Not-determined |
| 18-101-final-viral-combined_k141_380365\|\|full | Low-quality |
| 18-101-final-viral-combined_k141_314776\|\|full | Not-determined |
| 18-101-final-viral-combined_k141_150039\|\|full | Low-quality |
| 18-101-final-viral-combined_k141_84451\|\|full | Low-quality |
| 18-101-final-viral-combined_k141_248961\|\|full | Not-determined |
| 18-101-final-viral-combined_k141_216202\|\|full | Not-determined |
| 18-101-final-viral-combined_k141_199638\|\|full | Low-quality |
| 18-101-final-viral-combined_k141_117307\|\|full | Low-quality |
| 18-101-final-viral-combined_k141_248980\|\|full | Not-determined |
| 18-101-final-viral-combined_k141_232418\|\|full | Low-quality |
| 18-101-final-viral-combined_k141_67298\|\|full | Low-quality |
| 18-101-final-viral-combined_k141_347832\|\|full | Low-quality |
| 18-101-final-viral-combined_k141_199643\|\|full | Low-quality |
| 18-101-final-viral-combined_k141_364030\|\|full | Not-determined |
| 18-101-final-viral-combined_k141_248992\|\|full | Not-determined |
| 18-101-final-viral-combined_k141_183180\|\|full | Not-determined |
| 18-101-final-viral-combined_k141_249007\|\|full | Not-determined |
| 18-101-final-viral-combined_k141_134118\|\|full | Low-quality |
| 18-101-final-viral-combined_k141_314820\|\|full | Not-determined |
| 18-101-final-viral-combined_k141_216260\|\|full | Not-determined |
| 18-101-final-viral-combined_k141_100645\|\|full | Low-quality |
| 18-101-final-viral-combined_k141_314836\|\|full | Low-quality |
| 18-101-final-viral-combined_k141_134149\|\|full | Not-determined |
| 18-101-final-viral-combined_k141_51109\|\|full | Not-determined |
| 18-101-final-viral-combined_k141_67346\|\|full | Low-quality |
| 18-101-final-viral-combined_k141_330923\|\|full | Not-determined |
| 18-101-final-viral-combined_k141_51117\|\|full | Not-determined |
| 18-101-final-viral-combined_k141_34950\|\|full | Low-quality |
| 18-101-final-viral-combined_k141_134168\|\|full | Low-quality |
| 18-101-final-viral-combined_k141_51135\|\|full | Low-quality |
| 18-101-final-viral-combined_k141_2529\|\|full | Not-determined |
| 18-101-final-viral-combined_k141_298421\|\|full | Not-determined |
| 18-101-final-viral-combined_k141_282012\|\|full | Not-determined |
| 18-101-final-viral-combined_k141_199708\|\|full | Low-quality |
| 18-101-final-viral-combined_k141_84504\|\|full | Not-determined |
| 18-101-final-viral-combined_k141_249068\|\|full | Not-determined |
| 18-101-final-viral-combined_k141_67392\|\|full | Low-quality |
| 18-101-final-viral-combined_k141_216301\|\|full | Not-determined |
| 18-101-final-viral-combined_k141_249070\|\|full | Not-determined |
| 18-101-final-viral-combined_k141_183239\|\|full | Not-determined |
| 18-101-final-viral-combined_k141_232496\|\|full | Not-determined |
| 18-101-final-viral-combined_k141_364086\|\|full | Not-determined |
| 18-101-final-viral-combined_k141_199714\|\|full | Not-determined |
| 18-101-final-viral-combined_k141_84514\|\|full | Low-quality |
| 18-101-final-viral-combined_k141_380467\|\|full | Not-determined |
| 18-101-final-viral-combined_k141_364091\|\|full | Not-determined |
| 18-101-final-viral-combined_k141_100686\|\|full | Not-determined |
| 18-101-final-viral-combined_k141_84515\|\|full | Low-quality |
| 18-101-final-viral-combined_k141_67413\|\|full | Not-determined |
| 18-101-final-viral-combined_k141_100695\|\|full | Low-quality |
| 18-101-final-viral-combined_k141_347841\|\|full | Low-quality |
| 18-101-final-viral-combined_k141_265058\|\|full | Not-determined |
| 18-101-final-viral-combined_k141_314856\|\|full | Low-quality |
| 18-101-final-viral-combined_k141_347842\|\|full | Low-quality |
| 18-101-final-viral-combined_k141_216341\|\|full | Not-determined |
| 18-101-final-viral-combined_k141_298484\|\|full | Not-determined |
| 18-101-final-viral-combined_k141_100701\|\|full | Low-quality |
| 18-101-final-viral-combined_k141_298499\|\|full | Not-determined |
| 18-101-final-viral-combined_k141_364131\|\|full | Not-determined |
| 18-101-final-viral-combined_k141_216372\|\|full | Low-quality |
| 18-101-final-viral-combined_k141_100703\|\|full | Not-determined |
| 18-101-final-viral-combined_k141_100709\|\|full | Not-determined |
| 18-101-final-viral-combined_k141_100712\|\|full | Low-quality |
| 18-101-final-viral-combined_k141_347869\|\|full | Low-quality |
| 18-101-final-viral-combined_k141_232513\|\|full | Low-quality |
| 18-101-final-viral-combined_k141_314889\|\|full | Low-quality |
| 18-101-final-viral-combined_k141_35017\|\|full | Low-quality |
| 18-101-final-viral-combined_k141_364145\|\|full | Low-quality |
| 18-101-final-viral-combined_k141_298524\|\|full | Not-determined |
| 18-101-final-viral-combined_k141_298528\|\|full | Low-quality |
| 18-101-final-viral-combined_k141_314906\|\|full | Not-determined |
| 18-101-final-viral-combined_k141_232526\|\|full | Low-quality |
| 18-101-final-viral-combined_k141_18536\|\|full | Not-determined |
| 18-101-final-viral-combined_k141_314909\|\|full | Not-determined |
| 18-101-final-viral-combined_k141_232545\|\|full | Not-determined |
| 18-101-final-viral-combined_k141_380531\|\|full | Low-quality |
| 18-101-final-viral-combined_k141_166301\|\|full | Not-determined |
| 18-101-final-viral-combined_k141_314917\|\|full | Low-quality |
| 18-101-final-viral-combined_k141_347925\|\|full | Not-determined |
| 18-101-final-viral-combined_k141_18560\|\|full | Low-quality |
| 18-101-final-viral-combined_k141_314933\|\|full | Low-quality |
| 18-101-final-viral-combined_k141_51246\|\|full | Not-determined |
| 18-101-final-viral-combined_k141_51259\|\|full | Not-determined |
| 18-101-final-viral-combined_k141_347934\|\|full | Low-quality |
| 18-101-final-viral-combined_k141_35066\|\|full | Low-quality |
| 18-101-final-viral-combined_k141_314950\|\|full | Not-determined |
| 18-101-final-viral-combined_k141_298562\|\|full | Not-determined |
| 18-101-final-viral-combined_k141_216435\|\|full | Low-quality |
| 18-101-final-viral-combined_k141_314958\|\|full | Not-determined |
| 18-101-final-viral-combined_k141_35082\|\|full | Low-quality |
| 18-101-final-viral-combined_k141_380551\|\|full | Not-determined |
| 18-101-final-viral-combined_k141_216456\|\|full | Low-quality |
| 18-101-final-viral-combined_k141_232610\|\|full | Not-determined |
| 18-101-final-viral-combined_k141_18586\|\|full | Not-determined |
| 18-101-final-viral-combined_k141_216457\|\|full | Not-determined |
| 18-101-final-viral-combined_k141_84663\|\|full | Low-quality |
| 18-101-final-viral-combined_k141_84664\|\|full | Low-quality |
| 18-101-final-viral-combined_k141_282083\|\|full | Not-determined |
| 18-101-final-viral-combined_k141_117457\|\|full | Not-determined |
| 18-101-final-viral-combined_k141_18591\|\|full | Not-determined |
| 18-101-final-viral-combined_k141_380562\|\|full | Not-determined |
| 18-101-final-viral-combined_k141_183359\|\|full | Not-determined |
| 18-101-final-viral-combined_k141_117464\|\|full | Not-determined |
| 18-101-final-viral-combined_k141_364235\|\|full | Not-determined |
| 18-101-final-viral-combined_k141_216462\|\|full | Low-quality |
| 18-101-final-viral-combined_k141_51301\|\|full | Not-determined |
| 18-101-final-viral-combined_k141_35144\|\|full | Low-quality |
| 18-101-final-viral-combined_k141_67538\|\|full | Low-quality |
| 18-101-final-viral-combined_k141_150182\|\|full | Low-quality |
| 18-101-final-viral-combined_k141_150185\|\|full | Not-determined |
| 18-101-final-viral-combined_k141_298619\|\|full | Not-determined |
| 18-101-final-viral-combined_k141_100721\|\|full | Low-quality |
| 18-101-final-viral-combined_k141_35165\|\|full | Not-determined |
| 18-101-final-viral-combined_k141_18631\|\|full | Low-quality |
| 18-101-final-viral-combined_k141_67547\|\|full | Not-determined |
| 18-101-final-viral-combined_k141_18632\|\|full | Low-quality |
| 18-101-final-viral-combined_k141_199908\|\|full | Not-determined |
| 18-101-final-viral-combined_k141_84686\|\|full | Low-quality |
| 18-101-final-viral-combined_k141_2619\|\|full | Low-quality |
| 18-101-final-viral-combined_k141_166368\|\|full | Low-quality |
| 18-101-final-viral-combined_k141_84694\|\|full | Low-quality |
| 18-101-final-viral-combined_k141_35170\|\|full | Low-quality |
| 18-101-final-viral-combined_k141_67568\|\|full | Low-quality |
| 18-101-final-viral-combined_k141_265210\|\|full | Not-determined |
| 18-101-final-viral-combined_k141_51345\|\|full | Not-determined |
| 18-101-final-viral-combined_k141_18646\|\|full | Not-determined |
| 18-101-final-viral-combined_k141_2636\|\|full | Low-quality |
| 18-101-final-viral-combined_k141_216511\|\|full | Not-determined |
| 18-101-final-viral-combined_k141_199922\|\|full | Not-determined |
| 18-101-final-viral-combined_k141_134341\|\|full | Not-determined |
| 18-101-final-viral-combined_k141_380613\|\|full | Not-determined |
| 18-101-final-viral-combined_k141_364260\|\|full | Not-determined |
| 18-101-final-viral-combined_k141_166375\|\|full | Not-determined |
| 18-101-final-viral-combined_k141_232661\|\|full | Not-determined |
| 18-101-final-viral-combined_k141_380623\|\|full | Low-quality |
| 18-101-final-viral-combined_k141_166385\|\|full | Low-quality |
| 18-101-final-viral-combined_k141_18667\|\|full | Not-determined |
| 18-101-final-viral-combined_k141_314986\|\|full | Low-quality |
| 18-101-final-viral-combined_k141_35203\|\|full | Not-determined |
| 18-101-final-viral-combined_k141_67577\|\|full | Low-quality |
| 18-101-final-viral-combined_k141_265224\|\|full | Not-determined |
| 18-101-final-viral-combined_k141_134368\|\|full | Low-quality |
| 18-101-final-viral-combined_k141_265233\|\|full | Low-quality |
| 18-101-final-viral-combined_k141_232687\|\|full | Not-determined |
| 18-101-final-viral-combined_k141_249288\|\|full | Not-determined |
| 18-101-final-viral-combined_k141_134375\|\|full | Low-quality |
| 18-101-final-viral-combined_k141_67591\|\|full | Not-determined |
| 18-101-final-viral-combined_k141_2685\|\|full | Not-determined |
| 18-101-final-viral-combined_k141_2688\|\|full | Low-quality |
| 18-101-final-viral-combined_k141_298661\|\|full | Not-determined |
| 18-101-final-viral-combined_k141_249299\|\|full | Not-determined |
| KM-final-viral-combined_k141_203701\|\|full | Medium-quality |
| 18-101-final-viral-combined_k141_315003\|\|full | Low-quality |
| 18-101-final-viral-combined_k141_315005\|\|full | Not-determined |
| 18-101-final-viral-combined_k141_100737\|\|full | Not-determined |
| 18-101-final-viral-combined_k141_199957\|\|full | Not-determined |
| 18-101-final-viral-combined_k141_67598\|\|full | Not-determined |
| 18-101-final-viral-combined_k141_134398\|\|full | Low-quality |
| 18-101-final-viral-combined_k141_315018\|\|full | Not-determined |
| 18-101-final-viral-combined_k141_380661\|\|full | Low-quality |
| 18-101-final-viral-combined_k141_2706\|\|full | Low-quality |
| 18-101-final-viral-combined_k141_380671\|\|full | Not-determined |
| 18-101-final-viral-combined_k141_2710\|\|full | Not-determined |
| 18-101-final-viral-combined_k141_150263\|\|full | Low-quality |
| 18-101-final-viral-combined_k141_117569\|\|full | Low-quality |
| 18-101-final-viral-combined_k141_51405\|\|full | Not-determined |
| 18-101-final-viral-combined_k141_249364\|\|full | Not-determined |
| 18-101-final-viral-combined_k141_117571\|\|full | Low-quality |
| 18-101-final-viral-combined_k141_232694\|\|full | Low-quality |
| 18-101-final-viral-combined_k141_199981\|\|full | Not-determined |
| 18-101-final-viral-combined_k141_150289\|\|full | Low-quality |
| 18-101-final-viral-combined_k141_2755\|\|full | Low-quality |
| 18-101-final-viral-combined_k141_2758\|\|full | Low-quality |
| 18-101-final-viral-combined_k141_315090\|\|full | Not-determined |
| 18-101-final-viral-combined_k141_150311\|\|full | Low-quality |
| 18-101-final-viral-combined_k141_134438\|\|full | Low-quality |
| 18-101-final-viral-combined_k141_117606\|\|full | Low-quality |
| 18-101-final-viral-combined_k141_249405\|\|full | Low-quality |
| 18-101-final-viral-combined_k141_35301\|\|full | Low-quality |
| 18-101-final-viral-combined_k141_249408\|\|full | Low-quality |
| 18-101-final-viral-combined_k141_216655\|\|full | Low-quality |
| 18-101-final-viral-combined_k141_100788\|\|full | Low-quality |
| 18-101-final-viral-combined_k141_232722\|\|full | Not-determined |
| 18-101-final-viral-combined_k141_35313\|\|full | Low-quality |
| 18-101-final-viral-combined_k141_232725\|\|full | Low-quality |
| 18-101-final-viral-combined_k141_84824\|\|full | Not-determined |
| 17-108-final-viral-combined_k141_10571\|\|full | Medium-quality |
| 18-101-final-viral-combined_k141_134484\|\|full | Low-quality |
| 18-101-final-viral-combined_k141_2792\|\|full | Not-determined |
| 18-101-final-viral-combined_k141_134486\|\|full | Low-quality |
| 18-101-final-viral-combined_k141_364383\|\|full | Low-quality |
| 18-101-final-viral-combined_k141_331161\|\|full | Not-determined |
| 18-101-final-viral-combined_k141_364387\|\|full | Not-determined |
| 18-101-final-viral-combined_k141_2800\|\|full | Low-quality |
| 18-101-final-viral-combined_k141_298765\|\|full | Not-determined |
| 18-101-final-viral-combined_k141_2801\|\|full | Not-determined |
| 18-101-final-viral-combined_k141_380734\|\|full | Not-determined |
| 18-101-final-viral-combined_k141_84831\|\|full | Low-quality |
| 18-101-final-viral-combined_k141_298784\|\|full | Not-determined |
| 18-101-final-viral-combined_k141_232747\|\|full | Not-determined |
| 18-101-final-viral-combined_k141_282251\|\|full | Low-quality |
| 18-101-final-viral-combined_k141_2812\|\|full | Low-quality |
| 18-101-final-viral-combined_k141_232767\|\|full | Low-quality |
| 18-101-final-viral-combined_k141_265302\|\|full | Not-determined |
| 18-101-final-viral-combined_k141_150348\|\|full | Not-determined |
| 18-101-final-viral-combined_k141_232771\|\|full | Low-quality |
| 18-101-final-viral-combined_k141_18767\|\|full | Low-quality |
| 18-101-final-viral-combined_k141_298822\|\|full | Not-determined |
| 18-101-final-viral-combined_k141_380763\|\|full | Low-quality |
| 18-101-final-viral-combined_k141_232781\|\|full | Low-quality |
| 18-101-final-viral-combined_k141_331200\|\|full | Low-quality |
| 18-101-final-viral-combined_k141_67725\|\|full | Low-quality |
| 18-101-final-viral-combined_k141_166494\|\|full | Not-determined |
| 18-101-final-viral-combined_k141_150363\|\|full | Low-quality |
| 18-101-final-viral-combined_k141_364446\|\|full | Not-determined |
| 18-101-final-viral-combined_k141_117669\|\|full | Not-determined |
| 18-101-final-viral-combined_k141_282286\|\|full | Not-determined |
| 18-101-final-viral-combined_k141_331225\|\|full | Not-determined |
| 18-101-final-viral-combined_k141_380771\|\|full | Low-quality |
| 18-101-final-viral-combined_k141_2838\|\|full | Low-quality |
| 18-101-final-viral-combined_k141_100856\|\|full | Not-determined |
| 18-101-final-viral-combined_k141_200032\|\|full | Low-quality |
| 18-101-final-viral-combined_k141_18785\|\|full | Low-quality |
| 18-101-final-viral-combined_k141_134515\|\|full | Low-quality |
| 18-101-final-viral-combined_k141_282300\|\|full | Not-determined |
| 18-101-final-viral-combined_k141_298853\|\|full | Low-quality |
| 18-101-final-viral-combined_k141_315164\|\|full | Low-quality |
| 18-101-final-viral-combined_k141_298862\|\|full | Not-determined |
| 18-101-final-viral-combined_k141_200067\|\|full | Low-quality |
| 18-101-final-viral-combined_k141_232823\|\|full | Low-quality |
| 18-101-final-viral-combined_k141_315180\|\|full | Low-quality |
| 18-101-final-viral-combined_k141_249419\|\|full | Low-quality |
| 18-101-final-viral-combined_k141_282312\|\|full | Not-determined |
| 18-101-final-viral-combined_k141_298881\|\|full | Low-quality |
| 18-101-final-viral-combined_k141_298887\|\|full | Low-quality |
| 18-101-final-viral-combined_k141_166512\|\|full | Not-determined |
| 18-101-final-viral-combined_k141_249439\|\|full | Low-quality |
| 18-101-final-viral-combined_k141_364484\|\|full | Low-quality |
| 18-101-final-viral-combined_k141_166516\|\|full | Low-quality |
| 18-101-final-viral-combined_k141_232865\|\|full | Not-determined |
| 18-101-final-viral-combined_k141_216761\|\|full | Not-determined |
| 18-101-final-viral-combined_k141_134565\|\|full | Not-determined |
| 18-101-final-viral-combined_k141_67786\|\|full | Low-quality |
| 18-101-final-viral-combined_k141_249453\|\|full | Not-determined |
| 18-101-final-viral-combined_k141_84956\|\|full | Low-quality |
| 18-101-final-viral-combined_k141_282339\|\|full | Low-quality |
| 18-101-final-viral-combined_k141_183559\|\|full | Low-quality |
| 18-101-final-viral-combined_k141_282352\|\|full | Not-determined |
| 18-101-final-viral-combined_k141_183566\|\|full | Low-quality |
| 18-101-final-viral-combined_k141_117733\|\|full | Low-quality |
| 18-101-final-viral-combined_k141_51436\|\|full | Low-quality |
| 18-101-final-viral-combined_k141_150431\|\|full | Low-quality |
| 18-101-final-viral-combined_k141_249462\|\|full | Low-quality |
| 18-101-final-viral-combined_k141_183576\|\|full | Not-determined |
| 18-101-final-viral-combined_k141_100911\|\|full | Low-quality |
| 18-101-final-viral-combined_k141_100914\|\|full | Low-quality |
| 18-101-final-viral-combined_k141_100923\|\|full | Not-determined |
| 18-101-final-viral-combined_k141_331326\|\|full | Not-determined |
| 18-101-final-viral-combined_k141_200140\|\|full | Low-quality |
| 18-101-final-viral-combined_k141_315214\|\|full | Not-determined |
| 18-101-final-viral-combined_k141_134610\|\|full | Not-determined |
| 18-101-final-viral-combined_k141_348150\|\|full | Low-quality |
| 18-101-final-viral-combined_k141_282370\|\|full | Not-determined |
| 18-101-final-viral-combined_k141_100929\|\|full | Not-determined |
| 18-101-final-viral-combined_k141_67833\|\|full | Not-determined |
| 18-101-final-viral-combined_k141_200146\|\|full | Not-determined |
| 18-101-final-viral-combined_k141_150484\|\|full | Not-determined |
| 18-101-final-viral-combined_k141_100934\|\|full | Not-determined |
| 18-101-final-viral-combined_k141_35429\|\|full | Not-determined |
| 18-101-final-viral-combined_k141_35438\|\|full | Not-determined |
| 18-101-final-viral-combined_k141_315245\|\|full | Not-determined |
| 18-101-final-viral-combined_k141_51519\|\|full | Low-quality |
| 18-101-final-viral-combined_k141_364547\|\|full | Low-quality |
| 18-101-final-viral-combined_k141_134637\|\|full | Low-quality |
| 18-101-final-viral-combined_k141_85001\|\|full | Not-determined |
| 18-101-final-viral-combined_k141_18900\|\|full | Not-determined |
| 18-101-final-viral-combined_k141_216840\|\|full | Not-determined |
| 18-101-final-viral-combined_k141_315265\|\|full | Low-quality |
| 18-101-final-viral-combined_k141_282383\|\|full | Low-quality |
| 18-101-final-viral-combined_k141_232916\|\|full | Low-quality |
| 18-101-final-viral-combined_k141_265367\|\|full | Low-quality |
| 18-101-final-viral-combined_k141_85004\|\|full | Not-determined |
| 18-101-final-viral-combined_k141_18918\|\|full | Low-quality |
| 18-101-final-viral-combined_k141_117799\|\|full | Not-determined |
| 18-101-final-viral-combined_k141_232927\|\|full | Not-determined |
| 18-101-final-viral-combined_k141_134649\|\|full | Low-quality |
| 18-101-final-viral-combined_k141_51543\|\|full | Low-quality |
| 18-101-final-viral-combined_k141_117811\|\|full | Not-determined |
| 18-101-final-viral-combined_k141_18934\|\|full | Not-determined |
| 18-101-final-viral-combined_k141_364587\|\|full | Low-quality |
| 18-101-final-viral-combined_k141_265385\|\|full | Not-determined |
| 18-101-final-viral-combined_k141_85015\|\|full | Low-quality |
| 18-101-final-viral-combined_k141_348182\|\|full | Not-determined |
| 18-101-final-viral-combined_k141_18941\|\|full | Low-quality |
| 18-101-final-viral-combined_k141_85020\|\|full | Not-determined |
| 18-101-final-viral-combined_k141_18942\|\|full | Low-quality |
| 18-101-final-viral-combined_k141_18943\|\|full | Not-determined |
| 18-101-final-viral-combined_k141_265395\|\|full | Low-quality |
| 18-101-final-viral-combined_k141_282406\|\|full | Not-determined |
| 18-101-final-viral-combined_k141_249560\|\|full | Low-quality |
| 18-101-final-viral-combined_k141_232944\|\|full | Low-quality |
| 18-101-final-viral-combined_k141_249563\|\|full | Low-quality |
| 18-101-final-viral-combined_k141_331403\|\|full | Not-determined |
| 18-101-final-viral-combined_k141_150531\|\|full | Low-quality |
| 18-101-final-viral-combined_k141_2964\|\|full | Not-determined |
| 18-101-final-viral-combined_k141_67893\|\|full | Not-determined |
| 18-101-final-viral-combined_k141_117861\|\|full | Low-quality |
| 18-101-final-viral-combined_k141_348199\|\|full | Low-quality |
| 18-101-final-viral-combined_k141_232966\|\|full | Low-quality |
| 18-101-final-viral-combined_k141_364641\|\|full | Not-determined |
| 18-101-final-viral-combined_k141_298963\|\|full | Not-determined |
| 18-101-final-viral-combined_k141_348217\|\|full | Low-quality |
| 18-101-final-viral-combined_k141_232989\|\|full | Low-quality |
| 18-101-final-viral-combined_k141_2986\|\|full | Not-determined |
| 18-101-final-viral-combined_k141_216905\|\|full | Not-determined |
| 18-101-final-viral-combined_k141_331457\|\|full | Low-quality |
| 18-101-final-viral-combined_k141_249603\|\|full | Not-determined |
| 18-101-final-viral-combined_k141_51620\|\|full | Low-quality |
| 18-101-final-viral-combined_k141_200206\|\|full | Low-quality |
| 18-101-final-viral-combined_k141_249607\|\|full | Not-determined |
| 18-101-final-viral-combined_k141_117876\|\|full | Not-determined |
| 18-101-final-viral-combined_k141_265462\|\|full | Low-quality |
| 18-101-final-viral-combined_k141_298996\|\|full | Low-quality |
| 18-101-final-viral-combined_k141_200210\|\|full | Low-quality |
| 18-101-final-viral-combined_k141_35541\|\|full | Low-quality |
| 18-101-final-viral-combined_k141_19017\|\|full | Not-determined |
| 18-101-final-viral-combined_k141_35544\|\|full | Not-determined |
| 18-101-final-viral-combined_k141_282465\|\|full | Not-determined |
| 18-101-final-viral-combined_k141_315376\|\|full | Low-quality |
| 18-101-final-viral-combined_k141_51665\|\|full | Not-determined |
| 18-101-final-viral-combined_k141_35571\|\|full | Low-quality |
| 18-101-final-viral-combined_k141_150617\|\|full | Low-quality |
| 18-101-final-viral-combined_k141_249666\|\|full | Low-quality |
| 18-101-final-viral-combined_k141_249674\|\|full | Not-determined |
| 18-101-final-viral-combined_k141_134759\|\|full | Low-quality |
| 18-101-final-viral-combined_k141_101108\|\|full | Not-determined |
| 18-101-final-viral-combined_k141_315406\|\|full | Low-quality |
| 18-101-final-viral-combined_k141_364710\|\|full | Not-determined |
| 18-101-final-viral-combined_k141_35582\|\|full | Low-quality |
| 17-109-final-viral-combined_k141_37250\|\|full | High-quality |
| 18-101-final-viral-combined_k141_35585\|\|full | Not-determined |
| 18-101-final-viral-combined_k141_134769\|\|full | Not-determined |
| 18-101-final-viral-combined_k141_35590\|\|full | Low-quality |
| 18-101-final-viral-combined_k141_101127\|\|full | Not-determined |
| 18-101-final-viral-combined_k141_348338\|\|full | Not-determined |
| 18-101-final-viral-combined_k141_101131\|\|full | Not-determined |
| 18-101-final-viral-combined_k141_35591\|\|full | Low-quality |
| 18-101-final-viral-combined_k141_117926\|\|full | Low-quality |
| 18-101-final-viral-combined_k141_134796\|\|full | Not-determined |
| 18-101-final-viral-combined_k141_183840\|\|full | Low-quality |
| 18-101-final-viral-combined_k141_117936\|\|full | Not-determined |
| 18-101-final-viral-combined_k141_249737\|\|full | Low-quality |
| 18-101-final-viral-combined_k141_150685\|\|full | Not-determined |
| 18-101-final-viral-combined_k141_19076\|\|full | Low-quality |
| 18-101-final-viral-combined_k141_282546\|\|full | Not-determined |
| 18-101-final-viral-combined_k141_150710\|\|full | Low-quality |
| 18-101-final-viral-combined_k141_117951\|\|full | Not-determined |
| 18-101-final-viral-combined_k141_282557\|\|full | Not-determined |
| 18-101-final-viral-combined_k141_85124\|\|full | Not-determined |
| 18-101-final-viral-combined_k141_150725\|\|full | Low-quality |
| 18-101-final-viral-combined_k141_265542\|\|full | Not-determined |
| 18-101-final-viral-combined_k141_19097\|\|full | Not-determined |
| 18-101-final-viral-combined_k141_67972\|\|full | Not-determined |
| 18-101-final-viral-combined_k141_233047\|\|full | Not-determined |
| 18-101-final-viral-combined_k141_348379\|\|full | Not-determined |
| 18-101-final-viral-combined_k141_282570\|\|full | Low-quality |
| 18-101-final-viral-combined_k141_348384\|\|full | Low-quality |
| 18-101-final-viral-combined_k141_19104\|\|full | Low-quality |
| 18-101-final-viral-combined_k141_19109\|\|full | Low-quality |
| 18-101-final-viral-combined_k141_216981\|\|full | Low-quality |
| 18-101-final-viral-combined_k141_150757\|\|full | Low-quality |
| 18-101-final-viral-combined_k141_299172\|\|full | Not-determined |
| 18-101-final-viral-combined_k141_200256\|\|full | Low-quality |
| 18-101-final-viral-combined_k141_348406\|\|full | Low-quality |

| | |
|---|---|
| 18-101-final-viral-combined_k141_51732\|\|full | Low-quality |
| 18-101-final-viral-combined_k141_348422\|\|full | Not-determined |
| 18-101-final-viral-combined_k141_166678\|\|full | Not-determined |
| 18-101-final-viral-combined_k141_183901\|\|full | Not-determined |
| 18-101-final-viral-combined_k141_51737\|\|full | Not-determined |
| 18-101-final-viral-combined_k141_249800\|\|full | Not-determined |
| 18-101-final-viral-combined_k141_282627\|\|full | Low-quality |
| 18-101-final-viral-combined_k141_381004\|\|full | Low-quality |
| 18-101-final-viral-combined_k141_35647\|\|full | Not-determined |
| 18-101-final-viral-combined_k141_348451\|\|full | Low-quality |
| 18-101-final-viral-combined_k141_249825\|\|full | Not-determined |
| 18-101-final-viral-combined_k141_233083\|\|full | Low-quality |
| 18-101-final-viral-combined_k141_364786\|\|full | Not-determined |
| 18-101-final-viral-combined_k141_348455\|\|full | Not-determined |
| 18-101-final-viral-combined_k141_150798\|\|full | Not-determined |
| 18-101-final-viral-combined_k141_35652\|\|full | Not-determined |
| 18-101-final-viral-combined_k141_200274\|\|full | Low-quality |
| 18-101-final-viral-combined_k141_315474\|\|full | Not-determined |
| 18-101-final-viral-combined_k141_265560\|\|full | Low-quality |
| 18-101-final-viral-combined_k141_265562\|\|full | Not-determined |
| 18-101-final-viral-combined_k141_282664\|\|full | Low-quality |
| 18-101-final-viral-combined_k141_200283\|\|full | Not-determined |
| 18-101-final-viral-combined_k141_166713\|\|full | Not-determined |
| 18-101-final-viral-combined_k141_183965\|\|full | Not-determined |
| 18-101-final-viral-combined_k141_200289\|\|full | Not-determined |
| 18-101-final-viral-combined_k141_19176\|\|full | Low-quality |
| 18-101-final-viral-combined_k141_315503\|\|full | Not-determined |
| 18-101-final-viral-combined_k141_282680\|\|full | Low-quality |
| 18-101-final-viral-combined_k141_315507\|\|full | Not-determined |
| 18-101-final-viral-combined_k141_183975\|\|full | Not-determined |
| 18-101-final-viral-combined_k141_364829\|\|full | Not-determined |
| 18-101-final-viral-combined_k141_35686\|\|full | Not-determined |
| 18-101-final-viral-combined_k141_315517\|\|full | Not-determined |
| 18-101-final-viral-combined_k141_35687\|\|full | Low-quality |
| 18-101-final-viral-combined_k141_282686\|\|full | Not-determined |
| 18-101-final-viral-combined_k141_200308\|\|full | Low-quality |
| 18-101-final-viral-combined_k141_315535\|\|full | Low-quality |
| 18-101-final-viral-combined_k141_134913\|\|full | Low-quality |
| 18-101-final-viral-combined_k141_35710\|\|full | Not-determined |
| 18-101-final-viral-combined_k141_134914\|\|full | Not-determined |
| 18-101-final-viral-combined_k141_348516\|\|full | Not-determined |
| 18-101-final-viral-combined_k141_381089\|\|full | Not-determined |
| 18-101-final-viral-combined_k141_348520\|\|full | Not-determined |
| 18-101-final-viral-combined_k141_381093\|\|full | Not-determined |
| 18-101-final-viral-combined_k141_348530\|\|full | Not-determined |
| 18-101-final-viral-combined_k141_134917\|\|full | Not-determined |
| 18-101-final-viral-combined_k141_265614\|\|full | Low-quality |
| 18-101-final-viral-combined_k141_299335\|\|full | Not-determined |
| 18-101-final-viral-combined_k141_265618\|\|full | Low-quality |
| 18-101-final-viral-combined_k141_101252\|\|full | Not-determined |
| 18-101-final-viral-combined_k141_68105\|\|full | Low-quality |
| 18-101-final-viral-combined_k141_85217\|\|full | Low-quality |
| 18-101-final-viral-combined_k141_150879\|\|full | Not-determined |
| 18-101-final-viral-combined_k141_118064\|\|full | Not-determined |
| 18-101-final-viral-combined_k141_315593\|\|full | Not-determined |
| 18-101-final-viral-combined_k141_166737\|\|full | Not-determined |
| 18-101-final-viral-combined_k141_265637\|\|full | Low-quality |
| 18-101-final-viral-combined_k141_200344\|\|full | Low-quality |
| 18-101-final-viral-combined_k141_299356\|\|full | Low-quality |
| 18-101-final-viral-combined_k141_282745\|\|full | Low-quality |
| 18-101-final-viral-combined_k141_315604\|\|full | Not-determined |
| 18-101-final-viral-combined_k141_200349\|\|full | Not-determined |
| 18-101-final-viral-combined_k141_233138\|\|full | Low-quality |
| 18-101-final-viral-combined_k141_299371\|\|full | Low-quality |
| 18-101-final-viral-combined_k141_233149\|\|full | Low-quality |
| 18-101-final-viral-combined_k141_233150\|\|full | Not-determined |
| 18-101-final-viral-combined_k141_118100\|\|full | Low-quality |
| 18-101-final-viral-combined_k141_249928\|\|full | Not-determined |
| 18-101-final-viral-combined_k141_51810\|\|full | Not-determined |
| 18-101-final-viral-combined_k141_166759\|\|full | Not-determined |
| 18-101-final-viral-combined_k141_233170\|\|full | Low-quality |
| 18-101-final-viral-combined_k141_299404\|\|full | Not-determined |
| 18-101-final-viral-combined_k141_233182\|\|full | Low-quality |
| 18-101-final-viral-combined_k141_299412\|\|full | Not-determined |
| 18-101-final-viral-combined_k141_364955\|\|full | Not-determined |
| 18-101-final-viral-combined_k141_265689\|\|full | Low-quality |
| 18-101-final-viral-combined_k141_150933\|\|full | Not-determined |
| 18-101-final-viral-combined_k141_184140\|\|full | Low-quality |
| 18-101-final-viral-combined_k141_331615\|\|full | Low-quality |
| 18-101-final-viral-combined_k141_249955\|\|full | Low-quality |
| 18-101-final-viral-combined_k141_217065\|\|full | Low-quality |
| 18-101-final-viral-combined_k141_35798\|\|full | Not-determined |
| 18-101-final-viral-combined_k141_217068\|\|full | Low-quality |
| 18-101-final-viral-combined_k141_249960\|\|full | Low-quality |
| 18-101-final-viral-combined_k141_184157\|\|full | Low-quality |
| 18-101-final-viral-combined_k141_35803\|\|full | Low-quality |
| 18-101-final-viral-combined_k141_51856\|\|full | Not-determined |
| 18-101-final-viral-combined_k141_217082\|\|full | Low-quality |
| 18-101-final-viral-combined_k141_381142\|\|full | Not-determined |
| 18-101-final-viral-combined_k141_134958\|\|full | Low-quality |
| 18-101-final-viral-combined_k141_166825\|\|full | Low-quality |
| 18-101-final-viral-combined_k141_101293\|\|full | Not-determined |
| 18-101-final-viral-combined_k141_381152\|\|full | Low-quality |
| 18-101-final-viral-combined_k141_217105\|\|full | Not-determined |
| 18-101-final-viral-combined_k141_315670\|\|full | Not-determined |
| 18-101-final-viral-combined_k141_3084\|\|full | Not-determined |
| 18-101-final-viral-combined_k141_51880\|\|full | Not-determined |
| 18-101-final-viral-combined_k141_101299\|\|full | Not-determined |
| 18-101-final-viral-combined_k141_364989\|\|full | Low-quality |
| 18-101-final-viral-combined_k141_184184\|\|full | Low-quality |
| 18-101-final-viral-combined_k141_217110\|\|full | Low-quality |
| 18-101-final-viral-combined_k141_381168\|\|full | Low-quality |
| 18-101-final-viral-combined_k141_134973\|\|full | Low-quality |
| 18-101-final-viral-combined_k141_19306\|\|full | Low-quality |
| 18-101-final-viral-combined_k141_265741\|\|full | Not-determined |
| 18-101-final-viral-combined_k141_299457\|\|full | Low-quality |
| 18-101-final-viral-combined_k141_68197\|\|full | Low-quality |
| 18-101-final-viral-combined_k141_101303\|\|full | Low-quality |
| 18-101-final-viral-combined_k141_19316\|\|full | Not-determined |
| 18-101-final-viral-combined_k141_51893\|\|full | Low-quality |
| 18-101-final-viral-combined_k141_134992\|\|full | Low-quality |
| 18-101-final-viral-combined_k141_365002\|\|full | Low-quality |
| 18-101-final-viral-combined_k141_348632\|\|full | Low-quality |
| 18-101-final-viral-combined_k141_101310\|\|full | Low-quality |
| 18-101-final-viral-combined_k141_348641\|\|full | Not-determined |
| 18-101-final-viral-combined_k141_282764\|\|full | Not-determined |
| 18-101-final-viral-combined_k141_51908\|\|full | Not-determined |
| 18-101-final-viral-combined_k141_118160\|\|full | Not-determined |
| 18-101-final-viral-combined_k141_135004\|\|full | Low-quality |
| 18-101-final-viral-combined_k141_118161\|\|full | Low-quality |
| 18-101-final-viral-combined_k141_184217\|\|full | Not-determined |
| 18-101-final-viral-combined_k141_348653\|\|full | Low-quality |
| 18-101-final-viral-combined_k141_51914\|\|full | Low-quality |
| 18-101-final-viral-combined_k141_184223\|\|full | Not-determined |
| 18-101-final-viral-combined_k141_184228\|\|full | Not-determined |
| 18-101-final-viral-combined_k141_68219\|\|full | Not-determined |
| 18-101-final-viral-combined_k141_51930\|\|full | Low-quality |
| 18-101-final-viral-combined_k141_51932\|\|full | Not-determined |
| 18-101-final-viral-combined_k141_299503\|\|full | Low-quality |
| 18-101-final-viral-combined_k141_184231\|\|full | Low-quality |
| 18-101-final-viral-combined_k141_51933\|\|full | Low-quality |
| 18-101-final-viral-combined_k141_348682\|\|full | Low-quality |
| 18-101-final-viral-combined_k141_184239\|\|full | Not-determined |
| 18-101-final-viral-combined_k141_101361\|\|full | Not-determined |
| 18-101-final-viral-combined_k141_135020\|\|full | Low-quality |
| 18-101-final-viral-combined_k141_51945\|\|full | Low-quality |
| 18-101-final-viral-combined_k141_315698\|\|full | Not-determined |
| 18-101-final-viral-combined_k141_184241\|\|full | Low-quality |
| 18-101-final-viral-combined_k141_299522\|\|full | Low-quality |
| 18-101-final-viral-combined_k141_282791\|\|full | Not-determined |
| 18-101-final-viral-combined_k141_3151\|\|full | Not-determined |
| 18-101-final-viral-combined_k141_381219\|\|full | Not-determined |
| 18-101-final-viral-combined_k141_217183\|\|full | Low-quality |
| 18-101-final-viral-combined_k141_3157\|\|full | Low-quality |
| 18-101-final-viral-combined_k141_217186\|\|full | Low-quality |
| 18-101-final-viral-combined_k141_35925\|\|full | Not-determined |
| 18-101-final-viral-combined_k141_51959\|\|full | Low-quality |
| 18-101-final-viral-combined_k141_151032\|\|full | Not-determined |
| 18-101-final-viral-combined_k141_381237\|\|full | Not-determined |
| 18-109-final-viral-combined_k141_7205\|\|full | High-quality |
| 18-101-final-viral-combined_k141_265816\|\|full | Low-quality |
| 18-101-final-viral-combined_k141_265833\|\|full | Not-determined |
| 18-101-final-viral-combined_k141_348744\|\|full | Not-determined |
| 18-101-final-viral-combined_k141_118179\|\|full | Not-determined |
| 18-101-final-viral-combined_k141_365071\|\|full | Not-determined |
| 18-101-final-viral-combined_k141_299558\|\|full | Not-determined |
| 18-101-final-viral-combined_k141_200489\|\|full | Not-determined |
| 18-101-final-viral-combined_k141_331757\|\|full | Low-quality |
| 18-101-final-viral-combined_k141_68271\|\|full | Low-quality |
| 18-101-final-viral-combined_k141_118185\|\|full | Low-quality |
| 18-101-final-viral-combined_k141_166838\|\|full | Not-determined |
| 18-101-final-viral-combined_k141_166839\|\|full | Low-quality |
| 18-101-final-viral-combined_k141_331775\|\|full | Not-determined |
| 18-101-final-viral-combined_k141_315722\|\|full | Not-determined |
| 18-101-final-viral-combined_k141_68288\|\|full | Not-determined |
| 18-101-final-viral-combined_k141_184280\|\|full | Not-determined |
| 18-101-final-viral-combined_k141_166846\|\|full | Not-determined |
| 18-101-final-viral-combined_k141_282867\|\|full | Low-quality |
| 18-101-final-viral-combined_k141_381254\|\|full | Not-determined |
| 18-101-final-viral-combined_k141_19376\|\|full | Not-determined |
| 18-101-final-viral-combined_k141_68293\|\|full | Low-quality |
| 18-101-final-viral-combined_k141_299574\|\|full | Not-determined |
| 18-101-final-viral-combined_k141_19378\|\|full | Low-quality |
| 18-101-final-viral-combined_k141_68294\|\|full | Low-quality |
| 18-101-final-viral-combined_k141_217253\|\|full | Low-quality |
| 18-101-final-viral-combined_k141_217254\|\|full | Low-quality |
| 18-101-final-viral-combined_k141_135058\|\|full | Not-determined |
| 18-101-final-viral-combined_k141_19406\|\|full | Not-determined |
| 18-101-final-viral-combined_k141_3187\|\|full | Not-determined |
| 18-101-final-viral-combined_k141_184295\|\|full | Low-quality |
| 18-101-final-viral-combined_k141_101441\|\|full | Not-determined |
| 18-101-final-viral-combined_k141_184317\|\|full | Not-determined |
| 18-101-final-viral-combined_k141_315775\|\|full | Not-determined |
| 18-101-final-viral-combined_k141_166892\|\|full | Not-determined |
| 18-101-final-viral-combined_k141_52059\|\|full | Not-determined |
| 18-101-final-viral-combined_k141_250078\|\|full | Low-quality |
| 18-101-final-viral-combined_k141_3202\|\|full | Low-quality |
| 18-101-final-viral-combined_k141_365161\|\|full | Not-determined |
| 18-101-final-viral-combined_k141_35989\|\|full | Low-quality |
| 18-101-final-viral-combined_k141_151062\|\|full | Not-determined |
| 18-101-final-viral-combined_k141_135108\|\|full | Not-determined |
| 18-101-final-viral-combined_k141_135117\|\|full | Not-determined |
| 18-101-final-viral-combined_k141_19422\|\|full | Low-quality |
| 18-101-final-viral-combined_k141_331801\|\|full | Not-determined |
| 18-101-final-viral-combined_k141_315802\|\|full | Low-quality |
| 18-101-final-viral-combined_k141_348833\|\|full | Not-determined |
| 18-101-final-viral-combined_k141_233346\|\|full | Low-quality |
| 18-101-final-viral-combined_k141_315808\|\|full | Not-determined |
| 18-101-final-viral-combined_k141_250101\|\|full | Low-quality |
| 18-101-final-viral-combined_k141_331826\|\|full | Not-determined |
| 18-101-final-viral-combined_k141_19446\|\|full | Low-quality |
| 18-101-final-viral-combined_k141_250104\|\|full | Not-determined |
| 18-101-final-viral-combined_k141_166940\|\|full | Not-determined |
| 18-101-final-viral-combined_k141_166944\|\|full | Low-quality |
| 18-101-final-viral-combined_k141_299598\|\|full | Low-quality |
| 18-101-final-viral-combined_k141_233371\|\|full | Low-quality |
| 18-101-final-viral-combined_k141_315820\|\|full | Not-determined |
| 18-101-final-viral-combined_k141_348843\|\|full | Not-determined |
| 18-101-final-viral-combined_k141_299600\|\|full | Not-determined |
| 18-101-final-viral-combined_k141_331848\|\|full | Not-determined |
| 18-101-final-viral-combined_k141_135162\|\|full | Low-quality |
| 18-101-final-viral-combined_k141_166959\|\|full | Not-determined |
| 18-101-final-viral-combined_k141_233389\|\|full | Low-quality |
| 18-101-final-viral-combined_k141_52097\|\|full | Low-quality |
| 18-101-final-viral-combined_k141_315851\|\|full | Low-quality |
| 18-101-final-viral-combined_k141_36015\|\|full | Low-quality |
| 18-101-final-viral-combined_k141_151102\|\|full | Low-quality |
| 18-101-final-viral-combined_k141_166975\|\|full | Low-quality |
| 18-101-final-viral-combined_k141_85244\|\|full | Not-determined |
| 18-101-final-viral-combined_k141_118306\|\|full | Not-determined |
| 18-101-final-viral-combined_k141_381366\|\|full | Not-determined |
| 18-101-final-viral-combined_k141_118308\|\|full | Not-determined |
| 18-101-final-viral-combined_k141_85248\|\|full | Low-quality |
| 18-101-final-viral-combined_k141_283014\|\|full | Not-determined |
| 18-101-final-viral-combined_k141_166985\|\|full | Low-quality |
| 18-101-final-viral-combined_k141_381368\|\|full | Not-determined |
| 18-101-final-viral-combined_k141_118314\|\|full | Low-quality |
| 18-101-final-viral-combined_k141_135193\|\|full | Not-determined |
| 18-101-final-viral-combined_k141_283024\|\|full | Not-determined |
| 18-101-final-viral-combined_k141_118317\|\|full | Not-determined |
| KM-final-viral-combined_k141_2234\|\|full | High-quality |
| 18-101-final-viral-combined_k141_118338\|\|full | Not-determined |
| 18-101-final-viral-combined_k141_36062\|\|full | Not-determined |
| 18-101-final-viral-combined_k141_381385\|\|full | Low-quality |
| 18-101-final-viral-combined_k141_381386\|\|full | Low-quality |
| 18-101-final-viral-combined_k141_381390\|\|full | Low-quality |
| 18-101-final-viral-combined_k141_299691\|\|full | Not-determined |
| 18-101-final-viral-combined_k141_184468\|\|full | Not-determined |
| 18-101-final-viral-combined_k141_135223\|\|full | Low-quality |
| 18-101-final-viral-combined_k141_52159\|\|full | Not-determined |
| 18-101-final-viral-combined_k141_151126\|\|full | Not-determined |
| 18-101-final-viral-combined_k141_19500\|\|full | Not-determined |
| 18-101-final-viral-combined_k141_381402\|\|full | Low-quality |
| 18-101-final-viral-combined_k141_85278\|\|full | Low-quality |
| 18-101-final-viral-combined_k141_151131\|\|full | Low-quality |
| 18-101-final-viral-combined_k141_19507\|\|full | Low-quality |
| 18-101-final-viral-combined_k141_365216\|\|full | Not-determined |
| 18-101-final-viral-combined_k141_381406\|\|full | Low-quality |
| 18-101-final-viral-combined_k141_381408\|\|full | Not-determined |
| 18-101-final-viral-combined_k141_118405\|\|full | Low-quality |
| 18-101-final-viral-combined_k141_250168\|\|full | Not-determined |
| 18-101-final-viral-combined_k141_381418\|\|full | Low-quality |
| 18-101-final-viral-combined_k141_184510\|\|full | Not-determined |
| 18-101-final-viral-combined_k141_315918\|\|full | Low-quality |
| 18-101-final-viral-combined_k141_299723\|\|full | Not-determined |
| 18-101-final-viral-combined_k141_217443\|\|full | Not-determined |
| 18-101-final-viral-combined_k141_167055\|\|full | Not-determined |
| 18-101-final-viral-combined_k141_250190\|\|full | Low-quality |
| 18-101-final-viral-combined_k141_331939\|\|full | Low-quality |
| 18-101-final-viral-combined_k141_135275\|\|full | Low-quality |
| 18-101-final-viral-combined_k141_331942\|\|full | Low-quality |
| 18-101-final-viral-combined_k141_118458\|\|full | Low-quality |
| 18-101-final-viral-combined_k141_3323\|\|full | Not-determined |
| 18-101-final-viral-combined_k141_217470\|\|full | Not-determined |
| 18-101-final-viral-combined_k141_200684\|\|full | Not-determined |
| 18-101-final-viral-combined_k141_85367\|\|full | Low-quality |
| 18-101-final-viral-combined_k141_118476\|\|full | Not-determined |
| 18-101-final-viral-combined_k141_381464\|\|full | Not-determined |
| 18-101-final-viral-combined_k141_200690\|\|full | Not-determined |
| 18-101-final-viral-combined_k141_118478\|\|full | Low-quality |
| 18-101-final-viral-combined_k141_19565\|\|full | Not-determined |
| 18-101-final-viral-combined_k141_36139\|\|full | Not-determined |
| 18-101-final-viral-combined_k141_85384\|\|full | Not-determined |
| 18-101-final-viral-combined_k141_283072\|\|full | Not-determined |
| 18-101-final-viral-combined_k141_167080\|\|full | Low-quality |
| 18-101-final-viral-combined_k141_19573\|\|full | Not-determined |
| 18-101-final-viral-combined_k141_331972\|\|full | Not-determined |
| 18-101-final-viral-combined_k141_167084\|\|full | Low-quality |
| 18-101-final-viral-combined_k141_135330\|\|full | Not-determined |
| 18-101-final-viral-combined_k141_52256\|\|full | Low-quality |
| 18-101-final-viral-combined_k141_217493\|\|full | Low-quality |
| 18-101-final-viral-combined_k141_348971\|\|full | Low-quality |
| 18-101-final-viral-combined_k141_217500\|\|full | Not-determined |
| 18-101-final-viral-combined_k141_331983\|\|full | Not-determined |
| 18-101-final-viral-combined_k141_184555\|\|full | Not-determined |
| 18-101-final-viral-combined_k141_36181\|\|full | Low-quality |
| 18-101-final-viral-combined_k141_331993\|\|full | Low-quality |
| 18-101-final-viral-combined_k141_184559\|\|full | Not-determined |
| 18-101-final-viral-combined_k141_151228\|\|full | Low-quality |
| 18-101-final-viral-combined_k141_167127\|\|full | Not-determined |
| 18-101-final-viral-combined_k141_250281\|\|full | Not-determined |
| 18-101-final-viral-combined_k141_331999\|\|full | Low-quality |
| 18-101-final-viral-combined_k141_200726\|\|full | Not-determined |
| 18-101-final-viral-combined_k141_85413\|\|full | Low-quality |
| 18-101-final-viral-combined_k141_52311\|\|full | Not-determined |
| 18-101-final-viral-combined_k141_3363\|\|full | Low-quality |
| 18-101-final-viral-combined_k141_184582\|\|full | Low-quality |
| 18-101-final-viral-combined_k141_85425\|\|full | Low-quality |
| 18-101-final-viral-combined_k141_3372\|\|full | Not-determined |
| 18-101-final-viral-combined_k141_36208\|\|full | Low-quality |
| 18-101-final-viral-combined_k141_332026\|\|full | Low-quality |
| 18-101-final-viral-combined_k141_184595\|\|full | Not-determined |
| 18-101-final-viral-combined_k141_349004\|\|full | Not-determined |
| 18-101-final-viral-combined_k141_3375\|\|full | Not-determined |
| 18-101-final-viral-combined_k141_365357\|\|full | Not-determined |
| 18-101-final-viral-combined_k141_266038\|\|full | Low-quality |
| 18-101-final-viral-combined_k141_250283\|\|full | Not-determined |
| 18-101-final-viral-combined_k141_68461\|\|full | Low-quality |
| 18-101-final-viral-combined_k141_381525\|\|full | Not-determined |
| 18-101-final-viral-combined_k141_266045\|\|full | Not-determined |
| 18-101-final-viral-combined_k141_36216\|\|full | Not-determined |
| 18-101-final-viral-combined_k141_167149\|\|full | Low-quality |
| 18-101-final-viral-combined_k141_151256\|\|full | Low-quality |
| 18-101-final-viral-combined_k141_299770\|\|full | Low-quality |
| 18-101-final-viral-combined_k141_101528\|\|full | Not-determined |
| 18-101-final-viral-combined_k141_184606\|\|full | Not-determined |
| 18-101-final-viral-combined_k141_250297\|\|full | Not-determined |
| 18-101-final-viral-combined_k141_332035\|\|full | Low-quality |
| 18-101-final-viral-combined_k141_332037\|\|full | Not-determined |
| 18-101-final-viral-combined_k141_316024\|\|full | Not-determined |
| 18-101-final-viral-combined_k141_36237\|\|full | Not-determined |
| 18-101-final-viral-combined_k141_85439\|\|full | Low-quality |
| 18-101-final-viral-combined_k141_167167\|\|full | Not-determined |
| 18-101-final-viral-combined_k141_233529\|\|full | Not-determined |
| 18-101-final-viral-combined_k141_283139\|\|full | Not-determined |
| 18-101-final-viral-combined_k141_184627\|\|full | Low-quality |
| 18-101-final-viral-combined_k141_135380\|\|full | Low-quality |
| 18-101-final-viral-combined_k141_332051\|\|full | Not-determined |
| 18-101-final-viral-combined_k141_184637\|\|full | Not-determined |
| 18-101-final-viral-combined_k141_233551\|\|full | Not-determined |
| 18-101-final-viral-combined_k141_233556\|\|full | Low-quality |
| 18-101-final-viral-combined_k141_381578\|\|full | Not-determined |
| 18-101-final-viral-combined_k141_36269\|\|full | Low-quality |
| 18-101-final-viral-combined_k141_200783\|\|full | Low-quality |
| 18-101-final-viral-combined_k141_167206\|\|full | Not-determined |
| 18-101-final-viral-combined_k141_151322\|\|full | Not-determined |
| 18-101-final-viral-combined_k141_36273\|\|full | Low-quality |
| 18-101-final-viral-combined_k141_283188\|\|full | Not-determined |
| 18-101-final-viral-combined_k141_316076\|\|full | Not-determined |
| 18-101-final-viral-combined_k141_135414\|\|full | Low-quality |
| 18-101-final-viral-combined_k141_101601\|\|full | Not-determined |
| 18-101-final-viral-combined_k141_250335\|\|full | Low-quality |
| 18-101-final-viral-combined_k141_135415\|\|full | Not-determined |
| 18-101-final-viral-combined_k141_365392\|\|full | Low-quality |
| 18-101-final-viral-combined_k141_184685\|\|full | Not-determined |
| 18-101-final-viral-combined_k141_68539\|\|full | Not-determined |
| 18-101-final-viral-combined_k141_184694\|\|full | Low-quality |
| 18-101-final-viral-combined_k141_266136\|\|full | Not-determined |
| 18-101-final-viral-combined_k141_365418\|\|full | Not-determined |
| 18-101-final-viral-combined_k141_184708\|\|full | Not-determined |
| 18-101-final-viral-combined_k141_68555\|\|full | Not-determined |
| 18-101-final-viral-combined_k141_316121\|\|full | Low-quality |
| 18-101-final-viral-combined_k141_135468\|\|full | Not-determined |
| 18-101-final-viral-combined_k141_118613\|\|full | Not-determined |
| 18-101-final-viral-combined_k141_381624\|\|full | Low-quality |
| 18-101-final-viral-combined_k141_135474\|\|full | Low-quality |
| 18-101-final-viral-combined_k141_36332\|\|full | Not-determined |
| 18-101-final-viral-combined_k141_381637\|\|full | Low-quality |
| 18-101-final-viral-combined_k141_135497\|\|full | Not-determined |
| 18-101-final-viral-combined_k141_151392\|\|full | Low-quality |
| 18-101-final-viral-combined_k141_266202\|\|full | Low-quality |
| 18-101-final-viral-combined_k141_299830\|\|full | Low-quality |
| 18-101-final-viral-combined_k141_266214\|\|full | Low-quality |
| 18-101-final-viral-combined_k141_283237\|\|full | Not-determined |
| 18-101-final-viral-combined_k141_365473\|\|full | Low-quality |
| 18-101-final-viral-combined_k141_3464\|\|full | Low-quality |
| 18-101-final-viral-combined_k141_299851\|\|full | Low-quality |
| 18-101-final-viral-combined_k141_316165\|\|full | Low-quality |
| 18-101-final-viral-combined_k141_85495\|\|full | Not-determined |
| 18-101-final-viral-combined_k141_233643\|\|full | Low-quality |
| 18-101-final-viral-combined_k141_316184\|\|full | Not-determined |
| 18-101-final-viral-combined_k141_266237\|\|full | Low-quality |
| 18-101-final-viral-combined_k141_151426\|\|full | Not-determined |
| 18-101-final-viral-combined_k141_217574\|\|full | Low-quality |
| 18-101-final-viral-combined_k141_85518\|\|full | Not-determined |
| 18-101-final-viral-combined_k141_36355\|\|full | Low-quality |
| 18-101-final-viral-combined_k141_52420\|\|full | Low-quality |
| 18-101-final-viral-combined_k141_118705\|\|full | Low-quality |
| 18-101-final-viral-combined_k141_135556\|\|full | Low-quality |
| 18-101-final-viral-combined_k141_332161\|\|full | Low-quality |
| 18-101-final-viral-combined_k141_365526\|\|full | Not-determined |
| 18-101-final-viral-combined_k141_233690\|\|full | Not-determined |
| 18-101-final-viral-combined_k141_217596\|\|full | Not-determined |
| 18-101-final-viral-combined_k141_151461\|\|full | Low-quality |
| 18-101-final-viral-combined_k141_3510\|\|full | Low-quality |
| 18-101-final-viral-combined_k141_381703\|\|full | Not-determined |
| 18-101-final-viral-combined_k141_349100\|\|full | Not-determined |
| 18-101-final-viral-combined_k141_299948\|\|full | Not-determined |
| 18-101-final-viral-combined_k141_52449\|\|full | Low-quality |
| 18-101-final-viral-combined_k141_184811\|\|full | Not-determined |
| 18-101-final-viral-combined_k141_299951\|\|full | Low-quality |
| 18-101-final-viral-combined_k141_200927\|\|full | Not-determined |
| 18-101-final-viral-combined_k141_3527\|\|full | Low-quality |
| 18-101-final-viral-combined_k141_266291\|\|full | Not-determined |
| 18-101-final-viral-combined_k141_217597\|\|full | Low-quality |
| 18-101-final-viral-combined_k141_299967\|\|full | Not-determined |
| 18-101-final-viral-combined_k141_135574\|\|full | Low-quality |
| 18-101-final-viral-combined_k141_36388\|\|full | Low-quality |
| 18-101-final-viral-combined_k141_68609\|\|full | Low-quality |
| 18-101-final-viral-combined_k141_85571\|\|full | Low-quality |
| 18-101-final-viral-combined_k141_151508\|\|full | Not-determined |
| 18-110-final-viral-combined_k141_108917\|\|full | High-quality |
| 18-101-final-viral-combined_k141_151522\|\|full | Low-quality |
| 18-101-final-viral-combined_k141_381758\|\|full | Not-determined |
| 18-101-final-viral-combined_k141_52457\|\|full | Low-quality |
| 18-101-final-viral-combined_k141_3549\|\|full | Low-quality |
| 18-101-final-viral-combined_k141_217618\|\|full | Low-quality |
| 18-101-final-viral-combined_k141_52463\|\|full | Not-determined |
| 18-101-final-viral-combined_k141_3559\|\|full | Low-quality |
| 18-101-final-viral-combined_k141_365587\|\|full | Low-quality |
| 18-101-final-viral-combined_k141_19809\|\|full | Not-determined |
| 18-101-final-viral-combined_k141_151536\|\|full | Not-determined |
| 18-101-final-viral-combined_k141_250503\|\|full | Not-determined |
| 18-101-final-viral-combined_k141_19821\|\|full | Not-determined |
| 18-101-final-viral-combined_k141_266340\|\|full | Low-quality |
| 18-101-final-viral-combined_k141_3583\|\|full | Not-determined |
| 18-101-final-viral-combined_k141_332232\|\|full | Low-quality |
| 18-101-final-viral-combined_k141_19828\|\|full | Low-quality |
| 18-101-final-viral-combined_k141_151559\|\|full | Low-quality |
| 18-101-final-viral-combined_k141_332233\|\|full | Not-determined |
| 18-101-final-viral-combined_k141_266361\|\|full | Not-determined |
| 18-101-final-viral-combined_k141_19846\|\|full | Not-determined |
| 18-101-final-viral-combined_k141_19850\|\|full | Not-determined |
| 18-101-final-viral-combined_k141_250537\|\|full | Not-determined |
| 18-101-final-viral-combined_k141_200958\|\|full | Low-quality |
| 18-101-final-viral-combined_k141_36449\|\|full | Low-quality |
| 18-101-final-viral-combined_k141_283370\|\|full | Low-quality |
| 18-101-final-viral-combined_k141_151589\|\|full | Not-determined |
| 18-101-final-viral-combined_k141_118763\|\|full | Not-determined |
| 18-101-final-viral-combined_k141_135632\|\|full | Low-quality |
| 18-101-final-viral-combined_k141_101775\|\|full | Low-quality |
| 18-101-final-viral-combined_k141_118766\|\|full | Not-determined |
| 18-101-final-viral-combined_k141_101777\|\|full | Low-quality |
| 18-101-final-viral-combined_k141_19877\|\|full | Low-quality |
| 18-102_k141_130687 | High-quality |
| 18-101-final-viral-combined_k141_283377\|\|full | Low-quality |
| 18-101-final-viral-combined_k141_332239\|\|full | Not-determined |
| 18-101-final-viral-combined_k141_118771\|\|full | Not-determined |
| 18-101-final-viral-combined_k141_151600\|\|full | Not-determined |
| 18-101-final-viral-combined_k141_332245\|\|full | Not-determined |
| 18-101-final-viral-combined_k141_101795\|\|full | Low-quality |
| 18-101-final-viral-combined_k141_300044\|\|full | Low-quality |
| 18-101-final-viral-combined_k141_233837\|\|full | Not-determined |
| 18-101-final-viral-combined_k141_283393\|\|full | Low-quality |
| 18-101-final-viral-combined_k141_151614\|\|full | Low-quality |
| 18-101-final-viral-combined_k141_349212\|\|full | Not-determined |
| 18-101-final-viral-combined_k141_118806\|\|full | Low-quality |
| 18-101-final-viral-combined_k141_151634\|\|full | Not-determined |
| 18-101-final-viral-combined_k141_300080\|\|full | Low-quality |
| 18-101-final-viral-combined_k141_52510\|\|full | Not-determined |
| 18-101-final-viral-combined_k141_52514\|\|full | Low-quality |
| 18-101-final-viral-combined_k141_151647\|\|full | Not-determined |
| 18-101-final-viral-combined_k141_365635\|\|full | Low-quality |
| 18-101-final-viral-combined_k141_151648\|\|full | Low-quality |
| 18-101-final-viral-combined_k141_365640\|\|full | Not-determined |
| 18-101-final-viral-combined_k141_300102\|\|full | Not-determined |
| 18-101-final-viral-combined_k141_52535\|\|full | Low-quality |
| 18-101-final-viral-combined_k141_283401\|\|full | Not-determined |
| 18-101-final-viral-combined_k141_118841\|\|full | Low-quality |
| 18-101-final-viral-combined_k141_3623\|\|full | Low-quality |
| 18-101-final-viral-combined_k141_250635\|\|full | Low-quality |
| 18-101-final-viral-combined_k141_283406\|\|full | Not-determined |
| 18-101-final-viral-combined_k141_381793\|\|full | Not-determined |
| 18-101-final-viral-combined_k141_52552\|\|full | Not-determined |
| 18-101-final-viral-combined_k141_233894\|\|full | Low-quality |
| 18-101-final-viral-combined_k141_250666\|\|full | Not-determined |
| 18-101-final-viral-combined_k141_19953\|\|full | Low-quality |
| 18-101-final-viral-combined_k141_283441\|\|full | Low-quality |
| 18-101-final-viral-combined_k141_19954\|\|full | Not-determined |
| 18-101-final-viral-combined_k141_365658\|\|full | Not-determined |
| 18-101-final-viral-combined_k141_365659\|\|full | Not-determined |
| 18-101-final-viral-combined_k141_167419\|\|full | Low-quality |
| 18-101-final-viral-combined_k141_381811\|\|full | Not-determined |
| 18-101-final-viral-combined_k141_151691\|\|full | Not-determined |
| 18-101-final-viral-combined_k141_201020\|\|full | Not-determined |
| 18-101-final-viral-combined_k141_316409\|\|full | Low-quality |
| 18-101-final-viral-combined_k141_85645\|\|full | Low-quality |
| 18-101-final-viral-combined_k141_118882\|\|full | Low-quality |
| 18-101-final-viral-combined_k141_52592\|\|full | Not-determined |
| 18-101-final-viral-combined_k141_365670\|\|full | Low-quality |
| 18-101-final-viral-combined_k141_381812\|\|full | Low-quality |
| 18-101-final-viral-combined_k141_85652\|\|full | Low-quality |
| 18-101-final-viral-combined_k141_184924\|\|full | Not-determined |
| 18-101-final-viral-combined_k141_3702\|\|full | Not-determined |
| 18-101-final-viral-combined_k141_250698\|\|full | Not-determined |
| 18-101-final-viral-combined_k141_52604\|\|full | Low-quality |
| 18-101-final-viral-combined_k141_250699\|\|full | Low-quality |
| 18-101-final-viral-combined_k141_250700\|\|full | Not-determined |
| 18-101-final-viral-combined_k141_68649\|\|full | Not-determined |
| 18-101-final-viral-combined_k141_300169\|\|full | Low-quality |
| 18-101-final-viral-combined_k141_118903\|\|full | Not-determined |
| 18-101-final-viral-combined_k141_135666\|\|full | Low-quality |
| 18-101-final-viral-combined_k141_316446\|\|full | Not-determined |
| 18-101-final-viral-combined_k141_167448\|\|full | Low-quality |
| 18-101-final-viral-combined_k141_68663\|\|full | Low-quality |
| 18-101-final-viral-combined_k141_283463\|\|full | Not-determined |
| 18-101-final-viral-combined_k141_184936\|\|full | Not-determined |
| 18-101-final-viral-combined_k141_167466\|\|full | Low-quality |
| 18-101-final-viral-combined_k141_300199\|\|full | Not-determined |
| 18-101-final-viral-combined_k141_135673\|\|full | Not-determined |
| 18-101-final-viral-combined_k141_349257\|\|full | Low-quality |
| 18-101-final-viral-combined_k141_300212\|\|full | Not-determined |
| 18-101-final-viral-combined_k141_233933\|\|full | Not-determined |
| 18-101-final-viral-combined_k141_20001\|\|full | Low-quality |
| 18-101-final-viral-combined_k141_184948\|\|full | Not-determined |
| 18-101-final-viral-combined_k141_316511\|\|full | Low-quality |
| 18-101-final-viral-combined_k141_52615\|\|full | Not-determined |
| 18-101-final-viral-combined_k141_233954\|\|full | Not-determined |
| 18-101-final-viral-combined_k141_52627\|\|full | Not-determined |
| 18-101-final-viral-combined_k141_184976\|\|full | Not-determined |
| 18-101-final-viral-combined_k141_250766\|\|full | Not-determined |
| 18-101-final-viral-combined_k141_307323\|\|0_partial | Medium-quality |
| 18-101-final-viral-combined_k141_52637\|\|full | Low-quality |
| 18-101-final-viral-combined_k141_68728\|\|full | Low-quality |
| 18-101-final-viral-combined_k141_283497\|\|full | Low-quality |
| 18-101-final-viral-combined_k141_185011\|\|full | Low-quality |
| 18-101-final-viral-combined_k141_283509\|\|full | Not-determined |
| 18-101-final-viral-combined_k141_233991\|\|full | Not-determined |
| 18-101-final-viral-combined_k141_332403\|\|full | Low-quality |
| 18-101-final-viral-combined_k141_283516\|\|full | Not-determined |
| 18-101-final-viral-combined_k141_151720\|\|full | Low-quality |
| 18-101-final-viral-combined_k141_101906\|\|full | Low-quality |
| 18-101-final-viral-combined_k141_20026\|\|full | Low-quality |
| 18-101-final-viral-combined_k141_332409\|\|full | Not-determined |
| 18-101-final-viral-combined_k141_283520\|\|full | Not-determined |
| 18-101-final-viral-combined_k141_20029\|\|full | Low-quality |
| KM-final-viral-combined_k141_259811\|\|full | Medium-quality |
| 18-101-final-viral-combined_k141_101919\|\|full | Low-quality |
| 18-101-final-viral-combined_k141_266575\|\|full | Low-quality |
| 18-101-final-viral-combined_k141_68775\|\|full | Low-quality |
| 18-101-final-viral-combined_k141_349307\|\|full | Low-quality |
| 18-101-final-viral-combined_k141_349308\|\|full | Not-determined |
| 18-101-final-viral-combined_k141_217640\|\|full | Not-determined |
| 18-101-final-viral-combined_k141_101951\|\|full | Not-determined |
| 18-101-final-viral-combined_k141_332425\|\|full | Low-quality |
| 18-101-final-viral-combined_k141_365739\|\|full | Low-quality |
| 18-101-final-viral-combined_k141_332439\|\|full | Not-determined |
| 18-101-final-viral-combined_k141_300327\|\|full | Low-quality |
| 18-101-final-viral-combined_k141_201130\|\|full | Low-quality |
| 18-101-final-viral-combined_k141_101958\|\|full | Not-determined |
| 18-101-final-viral-combined_k141_118921\|\|full | Not-determined |
| 18-101-final-viral-combined_k141_266599\|\|full | Not-determined |
| 18-101-final-viral-combined_k141_217685\|\|full | Low-quality |
| 18-101-final-viral-combined_k141_118928\|\|full | Not-determined |
| 18-101-final-viral-combined_k141_283600\|\|full | Not-determined |
| 18-101-final-viral-combined_k141_332485\|\|full | Not-determined |
| 18-101-final-viral-combined_k141_266623\|\|full | Not-determined |
| 18-101-final-viral-combined_k141_135740\|\|full | Not-determined |
| 18-101-final-viral-combined_k141_201170\|\|full | Not-determined |
| 18-105-final-viral-combined_k141_98202\|\|0_partial | Medium-quality |
| 18-101-final-viral-combined_k141_52701\|\|full | Not-determined |
| 18-101-final-viral-combined_k141_52705\|\|full | Low-quality |
| 18-101-final-viral-combined_k141_316660\|\|full | Low-quality |
| 18-101-final-viral-combined_k141_300382\|\|full | Low-quality |
| 18-101-final-viral-combined_k141_135784\|\|full | Low-quality |
| 18-101-final-viral-combined_k141_266657\|\|full | Low-quality |
| 18-101-final-viral-combined_k141_300391\|\|full | Low-quality |
| 18-101-final-viral-combined_k141_266662\|\|full | Low-quality |
| 18-101-final-viral-combined_k141_151789\|\|full | Not-determined |
| 18-101-final-viral-combined_k141_167543\|\|full | Low-quality |
| 18-101-final-viral-combined_k141_300402\|\|full | Low-quality |
| 18-101-final-viral-combined_k141_85824\|\|full | Low-quality |
| 18-101-final-viral-combined_k141_151799\|\|full | Low-quality |
| 18-101-final-viral-combined_k141_250847\|\|full | Low-quality |
| 18-101-final-viral-combined_k141_250855\|\|full | Low-quality |
| 18-101-final-viral-combined_k141_118960\|\|full | Low-quality |
| 18-101-final-viral-combined_k141_300414\|\|full | Low-quality |
| 18-101-final-viral-combined_k141_118963\|\|full | Not-determined |
| 18-101-final-viral-combined_k141_266673\|\|full | Not-determined |
| 18-101-final-viral-combined_k141_332505\|\|full | Not-determined |
| 18-101-final-viral-combined_k141_167554\|\|full | Not-determined |
| 18-101-final-viral-combined_k141_201201\|\|full | Not-determined |
| 18-101-final-viral-combined_k141_118971\|\|full | Not-determined |
| 18-101-final-viral-combined_k141_201204\|\|full | Low-quality |
| 18-101-final-viral-combined_k141_201208\|\|full | Low-quality |
| 18-101-final-viral-combined_k141_167575\|\|full | Low-quality |
| 18-106-final-viral-combined_k141_126236\|\|full | Medium-quality |
| 18-101-final-viral-combined_k141_217721\|\|full | Not-determined |
| 18-101-final-viral-combined_k141_185075\|\|full | Not-determined |
| 18-101-final-viral-combined_k141_300421\|\|full | Low-quality |
| 18-101-final-viral-combined_k141_250878\|\|full | Low-quality |
| 18-101-final-viral-combined_k141_52728\|\|full | Not-determined |
| 18-101-final-viral-combined_k141_349409\|\|full | Not-determined |
| 18-101-final-viral-combined_k141_382039\|\|full | Not-determined |
| 18-101-final-viral-combined_k141_234095\|\|full | Low-quality |
| 18-101-final-viral-combined_k141_234097\|\|full | Low-quality |
| 18-101-final-viral-combined_k141_250889\|\|full | Not-determined |
| 18-101-final-viral-combined_k141_167587\|\|full | Not-determined |
| 18-101-final-viral-combined_k141_234102\|\|full | Not-determined |

| | |
|---|---|
| 18-101-final-viral-combined_k141_20168\|\|full | Low-quality |
| 18-101-final-viral-combined_k141_201261\|\|full | Not-determined |
| 18-101-final-viral-combined_k141_266706\|\|full | Not-determined |
| 18-101-final-viral-combined_k141_382060\|\|full | Not-determined |
| 18-101-final-viral-combined_k141_349423\|\|full | Low-quality |
| 18-101-final-viral-combined_k141_234121\|\|full | Low-quality |
| 18-101-final-viral-combined_k141_266715\|\|full | Low-quality |
| 18-101-final-viral-combined_k141_151845\|\|full | Not-determined |
| 18-101-final-viral-combined_k141_135868\|\|full | Low-quality |
| 18-101-final-viral-combined_k141_102017\|\|full | Not-determined |
| 18-101-final-viral-combined_k141_266731\|\|full | Not-determined |
| 18-101-final-viral-combined_k141_185118\|\|full | Not-determined |
| 18-101-final-viral-combined_k141_300433\|\|full | Low-quality |
| 18-101-final-viral-combined_k141_382081\|\|full | Not-determined |
| 18-101-final-viral-combined_k141_332614\|\|full | Low-quality |
| 18-101-final-viral-combined_k141_300439\|\|full | Low-quality |
| 18-101-final-viral-combined_k141_300442\|\|full | Not-determined |
| 18-101-final-viral-combined_k141_266735\|\|full | Low-quality |
| 18-101-final-viral-combined_k141_119051\|\|full | Low-quality |
| 18-101-final-viral-combined_k141_217737\|\|full | Not-determined |
| 18-101-final-viral-combined_k141_3774\|\|full | Low-quality |
| 18-101-final-viral-combined_k141_234133\|\|full | Low-quality |
| 18-101-final-viral-combined_k141_151873\|\|full | Low-quality |
| 18-101-final-viral-combined_k141_382092\|\|full | Low-quality |
| 18-101-final-viral-combined_k141_382093\|\|full | Low-quality |
| 18-101-final-viral-combined_k141_332626\|\|full | Not-determined |
| 18-101-final-viral-combined_k141_266749\|\|full | Not-determined |
| 18-101-final-viral-combined_k141_349464\|\|full | Low-quality |
| 18-101-final-viral-combined_k141_300462\|\|full | Not-determined |
| 18-101-final-viral-combined_k141_52817\|\|full | Not-determined |
| 18-101-final-viral-combined_k141_332633\|\|full | Not-determined |
| 18-101-final-viral-combined_k141_185155\|\|full | Not-determined |
| 18-101-final-viral-combined_k141_119081\|\|full | Low-quality |
| 18-101-final-viral-combined_k141_119088\|\|full | Low-quality |
| 18-101-final-viral-combined_k141_36544\|\|full | Not-determined |
| 18-101-final-viral-combined_k141_52833\|\|full | Not-determined |
| 18-101-final-viral-combined_k141_250985\|\|full | Not-determined |
| 18-101-final-viral-combined_k141_266767\|\|full | Low-quality |
| 18-101-final-viral-combined_k141_3800\|\|full | Not-determined |
| 18-101-final-viral-combined_k141_382127\|\|full | Low-quality |
| 18-101-final-viral-combined_k141_68799\|\|full | Low-quality |
| 18-101-final-viral-combined_k141_135895\|\|full | Not-determined |
| 18-101-final-viral-combined_k141_36556\|\|full | Low-quality |
| 18-101-final-viral-combined_k141_316835\|\|full | Not-determined |
| 18-101-final-viral-combined_k141_316836\|\|full | Not-determined |
| [illegible] | [illegible] |

| | |
|---|---|
| 18-101-final-viral-combined_k141_284667\|\|full | Low-quality |
| 18-101-final-viral-combined_k141_86644\|\|full | Not-determined |
| 18-101-final-viral-combined_k141_152756\|\|full | Not-determined |
| 18-101-final-viral-combined_k141_120296\|\|full | Not-determined |
| 18-101-final-viral-combined_k141_366632\|\|full | Not-determined |
| 18-101-final-viral-combined_k141_120300\|\|full | Not-determined |
| 18-101-final-viral-combined_k141_366639\|\|full | Not-determined |
| 18-101-final-viral-combined_k141_267813\|\|full | Low-quality |
| 18-101-final-viral-combined_k141_102920\|\|full | Not-determined |
| 18-101-final-viral-combined_k141_120311\|\|full | Not-determined |
| 18-101-final-viral-combined_k141_21333\|\|full | Not-determined |
| 18-101-final-viral-combined_k141_366652\|\|full | Low-quality |
| 18-101-final-viral-combined_k141_267815\|\|full | Low-quality |
| 18-101-final-viral-combined_k141_317882\|\|full | Low-quality |
| 18-101-final-viral-combined_k141_120324\|\|full | Not-determined |
| 18-101-final-viral-combined_k141_120326\|\|full | Low-quality |
| 18-101-final-viral-combined_k141_53869\|\|full | Not-determined |
| 18-101-final-viral-combined_k141_317890\|\|full | Not-determined |
| 18-101-final-viral-combined_k141_21366\|\|full | Low-quality |
| 18-101-final-viral-combined_k141_53870\|\|full | Low-quality |
| 18-101-final-viral-combined_k141_235083\|\|full | Not-determined |
| 18-101-final-viral-combined_k141_120348\|\|full | Not-determined |
| 18-101-final-viral-combined_k141_168633\|\|full | Low-quality |
| 18-101-final-viral-combined_k141_267831\|\|full | Low-quality |
| 18-101-final-viral-combined_k141_383133\|\|full | Not-determined |
| 18-101-final-viral-combined_k141_152828\|\|full | Not-determined |
| 18-101-final-viral-combined_k141_120356\|\|full | Low-quality |
| 18-101-final-viral-combined_k141_267840\|\|full | Not-determined |
| 18-101-final-viral-combined_k141_350658\|\|full | Low-quality |
| 18-101-final-viral-combined_k141_301628\|\|full | Not-determined |
| 18-101-final-viral-combined_k141_21377\|\|full | Not-determined |
| 18-101-final-viral-combined_k141_317913\|\|full | Not-determined |
| 18-101-final-viral-combined_k141_168672\|\|full | Low-quality |
| 18-101-final-viral-combined_k141_202311\|\|full | Not-determined |
| 18-101-final-viral-combined_k141_317918\|\|full | Low-quality |
| 18-101-final-viral-combined_k141_86684\|\|full | Low-quality |
| 18-101-final-viral-combined_k141_284761\|\|full | Not-determined |
| 18-101-final-viral-combined_k141_168694\|\|full | Low-quality |
| 18-101-final-viral-combined_k141_350690\|\|full | Low-quality |
| 18-101-final-viral-combined_k141_350691\|\|full | Not-determined |
| 18-101-final-viral-combined_k141_383172\|\|full | Not-determined |
| 18-101-final-viral-combined_k141_86692\|\|full | Not-determined |
| 18-101-final-viral-combined_k141_136948\|\|full | Not-determined |
| 18-101-final-viral-combined_k141_136950\|\|full | Not-determined |
| 18-101-final-viral-combined_k141_168717\|\|full | Low-quality |
| 18-101-final-viral-combined_k141_186102\|\|full | Not-determined |
| 18-101-final-viral-combined_k141_366710\|\|full | Not-determined |
| 18-101-final-viral-combined_k141_4653\|\|full | Not-determined |
| 18-101-final-viral-combined_k141_218650\|\|full | Not-determined |
| 18-102-final-viral-combined_k141_208559\|\|full | Medium-quality |
| 18-101-final-viral-combined_k141_202359\|\|full | Not-determined |
| 18-101-final-viral-combined_k141_168728\|\|full | Low-quality |
| 18-101-final-viral-combined_k141_168731\|\|full | Not-determined |
| 18-101-final-viral-combined_k141_252079\|\|full | Not-determined |
| 18-101-final-viral-combined_k141_69630\|\|full | Low-quality |
| 18-101-final-viral-combined_k141_218670\|\|full | Low-quality |
| 18-101-final-viral-combined_k141_4665\|\|full | Low-quality |
| 18-101-final-viral-combined_k141_301690\|\|full | Low-quality |
| 18-101-final-viral-combined_k141_53913\|\|full | Low-quality |
| 18-101-final-viral-combined_k141_366730\|\|full | Not-determined |
| 18-101-final-viral-combined_k141_4673\|\|full | Low-quality |
| 18-101-final-viral-combined_k141_86725\|\|full | Not-determined |
| 18-101-final-viral-combined_k141_86737\|\|full | Low-quality |
| 18-101-final-viral-combined_k141_4679\|\|full | Not-determined |
| 18-101-final-viral-combined_k141_120456\|\|full | Not-determined |
| 18-101-final-viral-combined_k141_152916\|\|full | Low-quality |
| 18-101-final-viral-combined_k141_218685\|\|full | Low-quality |
| 18-101-final-viral-combined_k141_186133\|\|full | Low-quality |
| 18-101-final-viral-combined_k141_383229\|\|full | Low-quality |
| 18-101-final-viral-combined_k141_301746\|\|full | Low-quality |
| 18-101-final-viral-combined_k141_4703\|\|full | Low-quality |
| 18-101-final-viral-combined_k141_202425\|\|full | Not-determined |
| 18-101-final-viral-combined_k141_235209\|\|full | Not-determined |
| 18-101-final-viral-combined_k141_168803\|\|full | Low-quality |
| 18-101-final-viral-combined_k141_218702\|\|full | Not-determined |
| 18-101-final-viral-combined_k141_202434\|\|full | Low-quality |
| 18-101-final-viral-combined_k141_103012\|\|full | Low-quality |
| 18-101-final-viral-combined_k141_333520\|\|full | Low-quality |
| 18-101-final-viral-combined_k141_202439\|\|full | Not-determined |
| 18-101-final-viral-combined_k141_186188\|\|full | Not-determined |
| 18-101-final-viral-combined_k141_252176\|\|full | Low-quality |
| 18-101-final-viral-combined_k141_186214\|\|full | Not-determined |
| 18-101-final-viral-combined_k141_120484\|\|full | Low-quality |
| 18-101-final-viral-combined_k141_69758\|\|full | Low-quality |
| 18-101-final-viral-combined_k141_186240\|\|full | Low-quality |
| 18-101-final-viral-combined_k141_235276\|\|full | Low-quality |
| 18-101-final-viral-combined_k141_267872\|\|full | Low-quality |
| 18-101-final-viral-combined_k141_333553\|\|full | Not-determined |
| 18-101-final-viral-combined_k141_202474\|\|full | Not-determined |
| 18-101-final-viral-combined_k141_37681\|\|full | Low-quality |
| 18-101-final-viral-combined_k141_350783\|\|full | Low-quality |
| 18-101-final-viral-combined_k141_235294\|\|full | Not-determined |
| 18-101-final-viral-combined_k141_383344\|\|full | Low-quality |
| 18-101-final-viral-combined_k141_333564\|\|full | Not-determined |
| 17-109_k141_352846 | High-quality |
| 18-101-final-viral-combined_k141_152993\|\|full | Low-quality |
| 18-101-final-viral-combined_k141_120545\|\|full | Not-determined |
| 18-101-final-viral-combined_k141_267892\|\|full | Not-determined |
| 18-101-final-viral-combined_k141_318109\|\|full | Low-quality |
| 18-101-final-viral-combined_k141_86765\|\|full | Low-quality |
| 18-101-final-viral-combined_k141_120551\|\|full | Low-quality |
| 18-101-final-viral-combined_k141_318124\|\|full | Low-quality |
| 18-101-final-viral-combined_k141_53945\|\|full | Not-determined |
| 18-101-final-viral-combined_k141_333589\|\|full | Not-determined |
| 18-101-final-viral-combined_k141_53953\|\|full | Low-quality |
| 18-101-final-viral-combined_k141_86801\|\|full | Low-quality |
| 18-101-final-viral-combined_k141_333605\|\|full | Low-quality |
| 18-101-final-viral-combined_k141_318143\|\|full | Low-quality |
| 18-101-final-viral-combined_k141_333613\|\|full | Low-quality |
| 18-101-final-viral-combined_k141_4794\|\|full | Low-quality |
| 18-101-final-viral-combined_k141_69814\|\|full | Low-quality |
| 18-101-final-viral-combined_k141_202534\|\|full | Low-quality |
| 18-101-final-viral-combined_k141_186319\|\|full | Low-quality |
| 18-101-final-viral-combined_k141_202535\|\|full | Low-quality |
| 18-101-final-viral-combined_k141_383405\|\|full | Low-quality |
| 18-101-final-viral-combined_k141_186326\|\|full | Low-quality |
| 18-101-final-viral-combined_k141_4827\|\|full | Low-quality |
| 18-101-final-viral-combined_k141_366858\|\|full | Not-determined |
| 18-101-final-viral-combined_k141_202550\|\|full | Low-quality |
| 18-101-final-viral-combined_k141_267920\|\|full | Not-determined |
| 18-101-final-viral-combined_k141_350873\|\|full | Not-determined |
| 18-101-final-viral-combined_k141_120584\|\|full | Low-quality |
| 18-101-final-viral-combined_k141_202576\|\|full | Not-determined |
| 18-101-final-viral-combined_k141_37791\|\|full | Low-quality |
| 18-101-final-viral-combined_k141_366878\|\|full | Low-quality |
| 18-101-final-viral-combined_k141_137107\|\|full | Not-determined |
| 18-101-final-viral-combined_k141_186336\|\|full | Low-quality |
| 18-101-final-viral-combined_k141_4847\|\|full | Not-determined |
| 18-101-final-viral-combined_k141_350901\|\|full | Low-quality |
| 18-101-final-viral-combined_k141_137111\|\|full | Low-quality |
| 18-101-final-viral-combined_k141_366891\|\|full | Low-quality |
| 18-101-final-viral-combined_k141_69837\|\|full | Not-determined |
| 18-101-final-viral-combined_k141_267982\|\|full | Low-quality |
| 18-101-final-viral-combined_k141_137134\|\|full | Not-determined |
| 18-101-final-viral-combined_k141_86865\|\|full | Not-determined |
| 18-101-final-viral-combined_k141_4867\|\|full | Not-determined |
| 18-101-final-viral-combined_k141_86880\|\|full | Not-determined |
| 18-101-final-viral-combined_k141_37843\|\|full | Low-quality |
| 18-101-final-viral-combined_k141_218900\|\|full | Low-quality |
| 18-101-final-viral-combined_k141_54025\|\|full | Low-quality |
| 18-101-final-viral-combined_k141_318222\|\|full | Not-determined |
| 18-101-final-viral-combined_k141_37856\|\|full | Low-quality |
| 18-101-final-viral-combined_k141_218925\|\|full | Not-determined |
| 18-101-final-viral-combined_k141_120663\|\|full | Low-quality |
| 18-101-final-viral-combined_k141_169012\|\|full | Not-determined |
| 18-101-final-viral-combined_k141_120667\|\|full | Low-quality |
| 18-101-final-viral-combined_k141_103140\|\|full | Low-quality |
| 18-101-final-viral-combined_k141_235395\|\|full | Not-determined |
| 18-101-final-viral-combined_k141_302001\|\|full | Not-determined |
| 18-101-final-viral-combined_k141_318236\|\|full | Low-quality |
| 18-101-final-viral-combined_k141_218938\|\|full | Not-determined |
| 18-101-final-viral-combined_k141_103146\|\|full | Not-determined |
| 18-101-final-viral-combined_k141_302003\|\|full | Not-determined |
| 18-101-final-viral-combined_k141_120673\|\|full | Low-quality |
| 18-101-final-viral-combined_k141_366982\|\|full | Low-quality |
| 18-101-final-viral-combined_k141_137196\|\|full | Not-determined |
| 18-101-final-viral-combined_k141_120682\|\|full | Not-determined |
| 18-101-final-viral-combined_k141_137203\|\|full | Low-quality |
| 18-101-final-viral-combined_k141_284782\|\|full | Not-determined |
| 18-101-final-viral-combined_k141_103157\|\|full | Not-determined |
| 18-101-final-viral-combined_k141_350963\|\|full | Low-quality |
| 18-101-final-viral-combined_k141_383458\|\|full | Low-quality |
| 18-101-final-viral-combined_k141_202667\|\|full | Low-quality |
| 18-101-final-viral-combined_k141_268055\|\|full | Low-quality |
| 18-101-final-viral-combined_k141_366994\|\|full | Low-quality |
| 18-101-final-viral-combined_k141_318268\|\|full | Not-determined |
| 18-101-final-viral-combined_k141_350983\|\|full | Low-quality |
| 18-101-final-viral-combined_k141_202678\|\|full | Not-determined |
| 18-101-final-viral-combined_k141_318278\|\|full | Not-determined |
| 18-101-final-viral-combined_k141_4934\|\|full | Not-determined |
| 18-101-final-viral-combined_k141_54040\|\|full | Low-quality |
| 18-101-final-viral-combined_k141_284811\|\|full | Not-determined |
| 18-101-final-viral-combined_k141_284815\|\|full | Not-determined |
| 18-101-final-viral-combined_k141_69882\|\|full | Not-determined |
| 18-101-final-viral-combined_k141_235446\|\|full | Not-determined |
| 18-101-final-viral-combined_k141_4950\|\|full | Low-quality |
| 18-101-final-viral-combined_k141_37914\|\|full | Not-determined |
| 18-101-final-viral-combined_k141_103204\|\|full | Not-determined |
| 18-101-final-viral-combined_k141_54051\|\|full | Low-quality |
| 18-101-final-viral-combined_k141_137274\|\|full | Low-quality |
| 18-101-final-viral-combined_k141_4969\|\|full | Not-determined |
| 18-101-final-viral-combined_k141_302048\|\|full | Not-determined |
| 18-101-final-viral-combined_k141_367013\|\|full | Not-determined |
| 18-101-final-viral-combined_k141_302061\|\|full | Not-determined |
| 18-101-final-viral-combined_k141_54076\|\|full | Not-determined |
| 18-101-final-viral-combined_k141_302073\|\|full | Not-determined |
| 18-101-final-viral-combined_k141_202715\|\|full | Not-determined |
| 18-101-final-viral-combined_k141_218985\|\|full | Not-determined |
| 18-101-final-viral-combined_k141_302077\|\|full | Low-quality |
| 18-101-final-viral-combined_k141_153149\|\|full | Low-quality |
| 18-101-final-viral-combined_k141_4987\|\|full | Low-quality |
| 18-101-final-viral-combined_k141_252339\|\|full | Not-determined |
| 18-101-final-viral-combined_k141_202719\|\|full | Low-quality |
| 18-101-final-viral-combined_k141_103224\|\|full | Low-quality |
| 18-101-final-viral-combined_k141_4992\|\|full | Low-quality |
| 18-101-final-viral-combined_k141_169069\|\|full | Low-quality |
| 18-101-final-viral-combined_k141_4996\|\|full | Low-quality |
| 18-101-final-viral-combined_k141_318313\|\|full | Low-quality |
| 18-101-final-viral-combined_k141_5003\|\|full | Not-determined |
| 18-101-final-viral-combined_k141_351030\|\|full | Low-quality |
| 18-101-final-viral-combined_k141_169086\|\|full | Not-determined |
| 18-101-final-viral-combined_k141_318328\|\|full | Low-quality |
| 18-101-final-viral-combined_k141_284864\|\|full | Not-determined |
| 18-101-final-viral-combined_k141_383544\|\|full | Not-determined |
| 18-101-final-viral-combined_k141_252357\|\|full | Low-quality |
| 18-101-final-viral-combined_k141_284869\|\|full | Not-determined |
| 18-101-final-viral-combined_k141_383545\|\|full | Low-quality |
| 18-101-final-viral-combined_k141_284875\|\|full | Not-determined |
| 18-101-final-viral-combined_k141_86969\|\|full | Not-determined |
| 18-101-final-viral-combined_k141_169101\|\|full | Not-determined |
| 18-101-final-viral-combined_k141_318352\|\|full | Low-quality |
| 18-101-final-viral-combined_k141_120757\|\|full | Not-determined |
| 18-101-final-viral-combined_k141_103281\|\|full | Not-determined |
| 18-101-final-viral-combined_k141_367076\|\|full | Low-quality |
| 18-101-final-viral-combined_k141_268120\|\|full | Low-quality |
| 18-101-final-viral-combined_k141_252397\|\|full | Not-determined |
| 18-101-final-viral-combined_k141_383571\|\|full | Low-quality |
| 18-101-final-viral-combined_k141_37955\|\|full | Low-quality |
| 18-101-final-viral-combined_k141_383579\|\|full | Not-determined |
| 18-101-final-viral-combined_k141_120792\|\|full | Low-quality |
| 18-101-final-viral-combined_k141_383581\|\|full | Not-determined |
| 18-101-final-viral-combined_k141_252402\|\|full | Low-quality |
| 18-101-final-viral-combined_k141_235508\|\|full | Not-determined |
| 18-101-final-viral-combined_k141_284908\|\|full | Low-quality |
| 18-101-final-viral-combined_k141_235519\|\|full | Not-determined |
| 18-101-final-viral-combined_k141_21475\|\|full | Not-determined |
| 18-101-final-viral-combined_k141_235522\|\|full | Low-quality |
| 18-101-final-viral-combined_k141_284933\|\|full | Low-quality |
| 18-101-final-viral-combined_k141_37997\|\|full | Low-quality |
| 18-101-final-viral-combined_k141_333758\|\|full | Not-determined |
| 18-101-final-viral-combined_k141_383616\|\|full | Low-quality |
| 18-101-final-viral-combined_k141_137346\|\|full | Low-quality |
| 18-101-final-viral-combined_k141_333769\|\|full | Low-quality |
| 18-109-final-viral-combined_k141_107679\|\|0_partial | Medium-quality |
| 18-101-final-viral-combined_k141_186459\|\|full | Low-quality |
| 18-101-final-viral-combined_k141_268194\|\|full | Not-determined |
| 18-101-final-viral-combined_k141_333776\|\|full | Not-determined |
| 18-101-final-viral-combined_k141_54177\|\|full | Not-determined |
| 18-101-final-viral-combined_k141_202751\|\|full | Not-determined |
| 18-101-final-viral-combined_k141_351087\|\|full | Low-quality |
| 18-101-final-viral-combined_k141_202752\|\|full | Low-quality |
| 18-101-final-viral-combined_k141_202753\|\|full | Low-quality |
| 18-101-final-viral-combined_k141_54180\|\|full | Low-quality |
| 18-101-final-viral-combined_k141_318431\|\|full | Not-determined |
| 18-101-final-viral-combined_k141_302145\|\|full | Not-determined |
| 18-101-final-viral-combined_k141_5064\|\|full | Low-quality |
| 18-101-final-viral-combined_k141_318434\|\|full | Low-quality |
| 18-101-final-viral-combined_k141_351117\|\|full | Not-determined |
| 18-101-final-viral-combined_k141_137388\|\|full | Low-quality |
| 18-101-final-viral-combined_k141_383662\|\|full | Not-determined |
| 18-101-final-viral-combined_k141_153242\|\|full | Not-determined |
| 18-101-final-viral-combined_k141_268231\|\|full | Low-quality |
| 18-101-final-viral-combined_k141_219156\|\|full | Not-determined |
| 18-101-final-viral-combined_k141_69972\|\|full | Not-determined |
| 18-101-final-viral-combined_k141_120892\|\|full | Low-quality |
| 18-101-final-viral-combined_k141_383680\|\|full | Not-determined |
| 18-101-final-viral-combined_k141_21499\|\|full | Not-determined |
| 18-101-final-viral-combined_k141_137417\|\|full | Not-determined |
| 18-101-final-viral-combined_k141_318450\|\|full | Not-determined |
| 18-101-final-viral-combined_k141_351155\|\|full | Not-determined |
| 18-101-final-viral-combined_k141_186532\|\|full | Not-determined |
| 18-101-final-viral-combined_k141_137424\|\|full | Not-determined |
| 18-101-final-viral-combined_k141_186535\|\|full | Not-determined |
| 18-101-final-viral-combined_k141_54236\|\|full | Low-quality |
| 18-101-final-viral-combined_k141_219195\|\|full | Low-quality |
| 18-101-final-viral-combined_k141_186536\|\|full | Not-determined |
| 18-101-final-viral-combined_k141_87102\|\|full | Low-quality |
| 18-101-final-viral-combined_k141_153292\|\|full | Low-quality |
| 18-101-final-viral-combined_k141_120927\|\|full | Low-quality |
| 18-101-final-viral-combined_k141_21520\|\|full | Low-quality |
| 18-101-final-viral-combined_k141_284991\|\|full | Low-quality |
| 18-101-final-viral-combined_k141_5119\|\|full | Not-determined |
| 18-101-final-viral-combined_k141_120941\|\|full | Low-quality |
| 18-101-final-viral-combined_k141_5124\|\|full | Not-determined |
| 18-101-final-viral-combined_k141_284995\|\|full | Not-determined |
| 18-101-final-viral-combined_k141_252538\|\|full | Low-quality |
| 18-101-final-viral-combined_k141_54278\|\|full | Not-determined |
| 18-101-final-viral-combined_k141_318519\|\|full | Not-determined |
| 18-101-final-viral-combined_k141_153336\|\|full | Not-determined |
| 18-110-final-viral-combined_k141_190394\|\|full | High-quality |
| 18-101-final-viral-combined_k141_5134\|\|full | Low-quality |
| 18-101-final-viral-combined_k141_333919\|\|full | Low-quality |
| 18-101-final-viral-combined_k141_70016\|\|full | Low-quality |
| 18-101-final-viral-combined_k141_252551\|\|full | Low-quality |
| 18-101-final-viral-combined_k141_70019\|\|full | Not-determined |
| 18-101-final-viral-combined_k141_367256\|\|full | Not-determined |
| 18-101-final-viral-combined_k141_70033\|\|full | Low-quality |
| 18-101-final-viral-combined_k141_21536\|\|full | Low-quality |
| 18-101-final-viral-combined_k141_333937\|\|full | Not-determined |
| 18-101-final-viral-combined_k141_268308\|\|full | Low-quality |
| 18-101-final-viral-combined_k141_219300\|\|full | Low-quality |
| 18-101-final-viral-combined_k141_351236\|\|full | Not-determined |
| 18-101-final-viral-combined_k141_367267\|\|full | Low-quality |
| 18-101-final-viral-combined_k141_21552\|\|full | Low-quality |
| 18-101-final-viral-combined_k141_235621\|\|full | Not-determined |
| 18-101-final-viral-combined_k141_383769\|\|full | Not-determined |
| 18-101-final-viral-combined_k141_70049\|\|full | Not-determined |
| 18-101-final-viral-combined_k141_186627\|\|full | Low-quality |
| 18-101-final-viral-combined_k141_169159\|\|full | Low-quality |
| 18-101-final-viral-combined_k141_38101\|\|full | Not-determined |
| 18-101-final-viral-combined_k141_87156\|\|full | Not-determined |
| 18-101-final-viral-combined_k141_153375\|\|full | Not-determined |
| 18-101-final-viral-combined_k141_351275\|\|full | Low-quality |
| 18-101-final-viral-combined_k141_5171\|\|full | Not-determined |
| 18-101-final-viral-combined_k141_186651\|\|full | Low-quality |
| 18-101-final-viral-combined_k141_367301\|\|full | Low-quality |
| 18-101-final-viral-combined_k141_186660\|\|full | Not-determined |
| 18-101-final-viral-combined_k141_186661\|\|full | Low-quality |
| 18-101-final-viral-combined_k141_5176\|\|full | Not-determined |
| 18-101-final-viral-combined_k141_186662\|\|full | Low-quality |
| 18-101-final-viral-combined_k141_38118\|\|full | Low-quality |
| 18-101-final-viral-combined_k141_21578\|\|full | Low-quality |
| 18-101-final-viral-combined_k141_252575\|\|full | Low-quality |
| 18-101-final-viral-combined_k141_153389\|\|full | Not-determined |
| 18-101-final-viral-combined_k141_121029\|\|full | Low-quality |
| 18-101-final-viral-combined_k141_21596\|\|full | Not-determined |
| 18-101-final-viral-combined_k141_186679\|\|full | Low-quality |
| 18-101-final-viral-combined_k141_70078\|\|full | Not-determined |
| 18-104_k141_73990 | Medium-quality |
| 18-101-final-viral-combined_k141_219363\|\|full | Not-determined |
| 18-101-final-viral-combined_k141_202905\|\|full | Low-quality |
| 18-101-final-viral-combined_k141_318563\|\|full | Low-quality |
| 18-101-final-viral-combined_k141_21608\|\|full | Not-determined |
| 18-101-final-viral-combined_k141_54344\|\|full | Not-determined |
| 18-101-final-viral-combined_k141_5205\|\|full | Not-determined |
| 18-101-final-viral-combined_k141_70100\|\|full | Low-quality |
| 18-101-final-viral-combined_k141_137518\|\|full | Low-quality |
| 18-101-final-viral-combined_k141_302231\|\|full | Not-determined |
| 18-101-final-viral-combined_k141_186704\|\|full | Low-quality |
| 18-101-final-viral-combined_k141_38171\|\|full | Not-determined |
| 18-101-final-viral-combined_k141_268356\|\|full | Low-quality |
| 18-101-final-viral-combined_k141_153411\|\|full | Not-determined |
| 18-101-final-viral-combined_k141_367335\|\|full | Not-determined |
| 18-101-final-viral-combined_k141_186714\|\|full | Low-quality |
| 18-101-final-viral-combined_k141_121070\|\|full | Not-determined |
| 18-101-final-viral-combined_k141_351316\|\|full | Not-determined |
| 18-101-final-viral-combined_k141_137547\|\|full | Low-quality |
| 18-101-final-viral-combined_k141_5234\|\|full | Low-quality |
| 18-101-final-viral-combined_k141_70121\|\|full | Low-quality |
| 18-101-final-viral-combined_k141_333984\|\|full | Low-quality |
| 18-101-final-viral-combined_k141_5235\|\|full | Low-quality |
| 18-101-final-viral-combined_k141_87181\|\|full | Low-quality |
| 18-101-final-viral-combined_k141_54369\|\|full | Not-determined |
| 18-101-final-viral-combined_k141_302249\|\|full | Not-determined |
| 18-101-final-viral-combined_k141_21638\|\|full | Low-quality |
| 18-101-final-viral-combined_k141_169259\|\|full | Not-determined |
| 18-101-final-viral-combined_k141_318604\|\|full | Low-quality |
| 18-101-final-viral-combined_k141_70132\|\|full | Low-quality |
| 18-101-final-viral-combined_k141_202970\|\|full | Low-quality |
| 18-101-final-viral-combined_k141_169261\|\|full | Not-determined |
| 18-101-final-viral-combined_k141_186763\|\|full | Not-determined |
| 18-101-final-viral-combined_k141_202974\|\|full | Not-determined |
| 18-101-final-viral-combined_k141_21659\|\|full | Not-determined |
| 18-101-final-viral-combined_k141_121083\|\|full | Low-quality |
| 18-101-final-viral-combined_k141_285073\|\|full | Low-quality |
| 18-101-final-viral-combined_k141_5250\|\|full | Low-quality |
| 18-101-final-viral-combined_k141_202986\|\|full | Not-determined |
| 18-101-final-viral-combined_k141_137574\|\|full | Not-determined |
| 18-101-final-viral-combined_k141_87229\|\|full | Low-quality |
| 18-101-final-viral-combined_k141_252648\|\|full | Not-determined |
| 18-101-final-viral-combined_k141_285085\|\|full | Low-quality |
| 18-101-final-viral-combined_k141_235758\|\|full | Low-quality |
| 18-101-final-viral-combined_k141_186802\|\|full | Not-determined |
| 18-101-final-viral-combined_k141_235777\|\|full | Low-quality |
| 18-101-final-viral-combined_k141_383802\|\|full | Low-quality |
| 18-101-final-viral-combined_k141_87251\|\|full | Not-determined |
| 18-101-final-viral-combined_k141_302310\|\|full | Low-quality |
| 18-101-final-viral-combined_k141_334016\|\|full | Low-quality |
| 18-101-final-viral-combined_k141_203026\|\|full | Not-determined |
| 18-101-final-viral-combined_k141_137626\|\|full | Not-determined |
| 18-101-final-viral-combined_k141_285138\|\|full | Low-quality |
| 18-101-final-viral-combined_k141_252711\|\|full | Not-determined |
| 18-101-final-viral-combined_k141_153489\|\|full | Not-determined |
| 18-101-final-viral-combined_k141_137627\|\|full | Not-determined |
| 18-101-final-viral-combined_k141_285168\|\|full | Low-quality |
| 18-101-final-viral-combined_k141_302360\|\|full | Not-determined |
| 18-101-final-viral-combined_k141_351441\|\|full | Not-determined |
| 18-101-final-viral-combined_k141_70179\|\|full | Low-quality |
| 18-101-final-viral-combined_k141_351447\|\|full | Not-determined |
| 18-101-final-viral-combined_k141_21721\|\|full | Low-quality |
| 18-104-final-viral-combined_k141_73990\|\|full | Medium-quality |
| 18-101-final-viral-combined_k141_137654\|\|full | Low-quality |
| 18-101-final-viral-combined_k141_285189\|\|full | Low-quality |
| 18-101-final-viral-combined_k141_137658\|\|full | Not-determined |
| 18-101-final-viral-combined_k141_318666\|\|full | Not-determined |
| 18-101-final-viral-combined_k141_268439\|\|full | Low-quality |
| 18-101-final-viral-combined_k141_318672\|\|full | Not-determined |
| 18-101-final-viral-combined_k141_186887\|\|full | Low-quality |
| 18-101-final-viral-combined_k141_285192\|\|full | Not-determined |
| 18-101-final-viral-combined_k141_54451\|\|full | Low-quality |
| 18-101-final-viral-combined_k141_203104\|\|full | Not-determined |
| 18-101-final-viral-combined_k141_153526\|\|full | Low-quality |
| 18-101-final-viral-combined_k141_121223\|\|full | Not-determined |
| 18-101-final-viral-combined_k141_334103\|\|full | Low-quality |
| 18-101-final-viral-combined_k141_186931\|\|full | Not-determined |
| 18-101-final-viral-combined_k141_334105\|\|full | Not-determined |
| 18-101-final-viral-combined_k141_318703\|\|full | Not-determined |
| 18-101-final-viral-combined_k141_38210\|\|full | Low-quality |
| 18-101-final-viral-combined_k141_121242\|\|full | Not-determined |
| 18-101-final-viral-combined_k141_103364\|\|full | Low-quality |
| 18-101-final-viral-combined_k141_203140\|\|full | Low-quality |
| 18-101-final-viral-combined_k141_21767\|\|full | Low-quality |
| 18-101-final-viral-combined_k141_302386\|\|full | Low-quality |
| 18-101-final-viral-combined_k141_121253\|\|full | Not-determined |
| 18-101-final-viral-combined_k141_87365\|\|full | Low-quality |
| 18-101-final-viral-combined_k141_38241\|\|full | Low-quality |
| 18-101-final-viral-combined_k141_21773\|\|full | Not-determined |
| 18-101-final-viral-combined_k141_5332\|\|full | Not-determined |
| 18-101-final-viral-combined_k141_219519\|\|full | Low-quality |
| 18-101-final-viral-combined_k141_38244\|\|full | Low-quality |
| 18-101-final-viral-combined_k141_235891\|\|full | Not-determined |
| 18-101-final-viral-combined_k141_137713\|\|full | Low-quality |
| 18-101-final-viral-combined_k141_5340\|\|full | Low-quality |
| 18-101-final-viral-combined_k141_153559\|\|full | Low-quality |
| 18-101-final-viral-combined_k141_351543\|\|full | Not-determined |
| 18-101-final-viral-combined_k141_21789\|\|full | Low-quality |
| 18-101-final-viral-combined_k141_235906\|\|full | Not-determined |
| 18-101-final-viral-combined_k141_38264\|\|full | Not-determined |
| 18-101-final-viral-combined_k141_252804\|\|full | Not-determined |
| 18-101-final-viral-combined_k141_103403\|\|full | Not-determined |
| 18-101-final-viral-combined_k141_103405\|\|full | Low-quality |
| 18-101-final-viral-combined_k141_103409\|\|full | Low-quality |
| 18-101-final-viral-combined_k141_169303\|\|full | Not-determined |
| 18-101-final-viral-combined_k141_302449\|\|full | Not-determined |
| 18-101-final-viral-combined_k141_169306\|\|full | Low-quality |
| 18-101-final-viral-combined_k141_383904\|\|full | Low-quality |
| 18-101-final-viral-combined_k141_334150\|\|full | Low-quality |
| 18-101-final-viral-combined_k141_87388\|\|full | Low-quality |
| 18-101-final-viral-combined_k141_203176\|\|full | Not-determined |
| 18-101-final-viral-combined_k141_87398\|\|full | Not-determined |
| 18-101-final-viral-combined_k141_137762\|\|full | Low-quality |
| 18-101-final-viral-combined_k141_252864\|\|full | Not-determined |
| 18-101-final-viral-combined_k141_70327\|\|full | Not-determined |
| 18-101-final-viral-combined_k141_235947\|\|full | Low-quality |
| 18-101-final-viral-combined_k141_203196\|\|full | Low-quality |
| 18-101-final-viral-combined_k141_285365\|\|full | Not-determined |
| 18-101-final-viral-combined_k141_103435\|\|full | Not-determined |
| 18-101-final-viral-combined_k141_252871\|\|full | Not-determined |
| 18-101-final-viral-combined_k141_351604\|\|full | Low-quality |
| 18-101-final-viral-combined_k141_70348\|\|full | Not-determined |
| 18-101-final-viral-combined_k141_235971\|\|full | Not-determined |
| 18-101-final-viral-combined_k141_268568\|\|full | Low-quality |
| 18-101-final-viral-combined_k141_54553\|\|full | Not-determined |
| 18-101-final-viral-combined_k141_285390\|\|full | Low-quality |
| 18-101-final-viral-combined_k141_137784\|\|full | Low-quality |
| 18-101-final-viral-combined_k141_383983\|\|full | Low-quality |
| 18-101-final-viral-combined_k141_252897\|\|full | Low-quality |
| 18-101-final-viral-combined_k141_85225\|\|0_partial | High-quality |
| 18-101-final-viral-combined_k141_70365\|\|full | Not-determined |
| 18-101-final-viral-combined_k141_70374\|\|full | Not-determined |
| 18-101-final-viral-combined_k141_219596\|\|full | Low-quality |
| 18-101-final-viral-combined_k141_169393\|\|full | Low-quality |
| 18-101-final-viral-combined_k141_219601\|\|full | Low-quality |
| 18-101-final-viral-combined_k141_367649\|\|full | Not-determined |
| 18-101-final-viral-combined_k141_334198\|\|full | Not-determined |
| 18-101-final-viral-combined_k141_318775\|\|full | Low-quality |
| 18-101-final-viral-combined_k141_70392\|\|full | Not-determined |
| 18-101-final-viral-combined_k141_70394\|\|full | Not-determined |
| 18-101-final-viral-combined_k141_153635\|\|full | Not-determined |
| 18-101-final-viral-combined_k141_302539\|\|full | Low-quality |
| 18-101-final-viral-combined_k141_153636\|\|full | Not-determined |
| 18-101-final-viral-combined_k141_302547\|\|full | Low-quality |
| 18-101-final-viral-combined_k141_21867\|\|full | Low-quality |
| 18-101-final-viral-combined_k141_203252\|\|full | Not-determined |
| 18-101-final-viral-combined_k141_285434\|\|full | Not-determined |
| 18-101-final-viral-combined_k141_236024\|\|full | Not-determined |
| 18-101-final-viral-combined_k141_203255\|\|full | Not-determined |
| 18-101-final-viral-combined_k141_38346\|\|full | Not-determined |
| 18-101-final-viral-combined_k141_236027\|\|full | Low-quality |
| 18-101-final-viral-combined_k141_219614\|\|full | Low-quality |
| 18-101-final-viral-combined_k141_268607\|\|full | Low-quality |
| 18-101-final-viral-combined_k141_351619\|\|full | Not-determined |
| 18-101-final-viral-combined_k141_103479\|\|full | Low-quality |
| 18-101-final-viral-combined_k141_70432\|\|full | Low-quality |
| 18-101-final-viral-combined_k141_203268\|\|full | Low-quality |
| 18-101-final-viral-combined_k141_70437\|\|full | Not-determined |
| 18-101-final-viral-combined_k141_187136\|\|full | Low-quality |
| 18-101-final-viral-combined_k141_103482\|\|full | Low-quality |
| 18-101-final-viral-combined_k141_121284\|\|full | Low-quality |
| 18-101-final-viral-combined_k141_38380\|\|full | Not-determined |
| 18-101-final-viral-combined_k141_187155\|\|full | Low-quality |
| 18-101-final-viral-combined_k141_103497\|\|full | Not-determined |
| 18-101-final-viral-combined_k141_236067\|\|full | Not-determined |
| 18-101-final-viral-combined_k141_367727\|\|full | Low-quality |
| 18-101-final-viral-combined_k141_5453\|\|full | Not-determined |
| 18-101-final-viral-combined_k141_187168\|\|full | Low-quality |
| 18-101-final-viral-combined_k141_334238\|\|full | Not-determined |
| 18-101-final-viral-combined_k141_302618\|\|full | Not-determined |
| 18-101-final-viral-combined_k141_203294\|\|full | Low-quality |
| 18-101-final-viral-combined_k141_121313\|\|full | Not-determined |
| 18-101-final-viral-combined_k141_87489\|\|full | Low-quality |
| 18-101-final-viral-combined_k141_318859\|\|full | Low-quality |
| 18-101-final-viral-combined_k141_103512\|\|full | Low-quality |
| 17-108-final-viral-combined_k141_125176\|\|full | Medium-quality |
| 18-101-final-viral-combined_k141_334269\|\|full | Not-determined |
| 18-101-final-viral-combined_k141_70493\|\|full | Low-quality |
| 18-101-final-viral-combined_k141_203315\|\|full | Not-determined |
| 18-101-final-viral-combined_k141_21942\|\|full | Not-determined |
| 18-101-final-viral-combined_k141_103520\|\|full | Not-determined |
| 18-101-final-viral-combined_k141_70494\|\|full | Low-quality |
| 18-101-final-viral-combined_k141_121334\|\|full | Low-quality |
| 18-101-final-viral-combined_k141_268632\|\|full | Not-determined |
| 18-101-final-viral-combined_k141_219707\|\|full | Not-determined |
| 18-101-final-viral-combined_k141_285489\|\|full | Not-determined |
| 18-101-final-viral-combined_k141_70503\|\|full | Not-determined |
| 18-101-final-viral-combined_k141_203335\|\|full | Not-determined |
| 18-101-final-viral-combined_k141_203343\|\|full | Low-quality |
| 18-101-final-viral-combined_k141_268648\|\|full | Not-determined |
| 18-101-final-viral-combined_k141_203347\|\|full | Low-quality |
| 18-101-final-viral-combined_k141_187235\|\|full | Not-determined |
| 18-101-final-viral-combined_k141_384091\|\|full | Not-determined |
| 18-101-final-viral-combined_k141_252972\|\|full | Not-determined |
| 18-101-final-viral-combined_k141_351660\|\|full | Low-quality |
| 18-101-final-viral-combined_k141_38464\|\|full | Not-determined |
| 18-101-final-viral-combined_k141_121339\|\|full | Not-determined |
| 18-101-final-viral-combined_k141_5507\|\|full | Not-determined |
| 18-101-final-viral-combined_k141_351665\|\|full | Not-determined |
| 18-101-final-viral-combined_k141_187260\|\|full | Low-quality |
| 18-101-final-viral-combined_k141_169512\|\|full | Not-determined |
| 18-101-final-viral-combined_k141_252977\|\|full | Low-quality |
| 18-101-final-viral-combined_k141_5519\|\|full | Not-determined |
| 18-101-final-viral-combined_k141_203367\|\|full | Low-quality |
| 18-101-final-viral-combined_k141_384120\|\|full | Not-determined |
| 18-105-final-viral-combined_k141_206792\|\|full | Complete |
| 18-101-final-viral-combined_k141_121350\|\|full | Not-determined |
| 18-101-final-viral-combined_k141_334278\|\|full | Low-quality |
| 18-101-final-viral-combined_k141_5525\|\|full | Not-determined |
| 18-101-final-viral-combined_k141_5534\|\|full | Low-quality |
| 18-101-final-viral-combined_k141_187275\|\|full | Not-determined |
| 18-101-final-viral-combined_k141_268688\|\|full | Low-quality |
| 18-101-final-viral-combined_k141_285537\|\|full | Not-determined |
| 18-101-final-viral-combined_k141_268695\|\|full | Not-determined |
| 18-101-final-viral-combined_k141_236127\|\|full | Low-quality |
| 18-101-final-viral-combined_k141_384152\|\|full | Not-determined |
| 18-101-final-viral-combined_k141_285542\|\|full | Not-determined |
| 18-101-final-viral-combined_k141_103592\|\|full | Low-quality |
| 18-101-final-viral-combined_k141_236135\|\|full | Not-determined |
| 18-101-final-viral-combined_k141_318869\|\|full | Low-quality |
| 18-101-final-viral-combined_k141_87549\|\|full | Not-determined |
| 18-101-final-viral-combined_k141_268709\|\|full | Low-quality |
| 18-101-final-viral-combined_k141_367877\|\|full | Not-determined |
| 18-101-final-viral-combined_k141_121379\|\|full | Low-quality |
| 18-101-final-viral-combined_k141_268719\|\|full | Low-quality |
| 18-101-final-viral-combined_k141_351734\|\|full | Not-determined |
| 18-101-final-viral-combined_k141_187333\|\|full | Low-quality |
| 18-101-final-viral-combined_k141_5585\|\|full | Not-determined |
| 18-101-final-viral-combined_k141_268724\|\|full | Low-quality |
| 18-101-final-viral-combined_k141_187335\|\|full | Low-quality |
| 18-101-final-viral-combined_k141_384202\|\|full | Not-determined |
| 18-101-final-viral-combined_k141_169566\|\|full | Not-determined |
| 18-101-final-viral-combined_k141_70607\|\|full | Low-quality |
| 18-101-final-viral-combined_k141_253045\|\|full | Not-determined |
| 18-101-final-viral-combined_k141_70609\|\|full | Low-quality |
| 18-101-final-viral-combined_k141_38540\|\|full | Low-quality |
| 18-101-final-viral-combined_k141_5600\|\|full | Not-determined |
| 18-101-final-viral-combined_k141_38542\|\|full | Not-determined |
| 18-101-final-viral-combined_k141_38544\|\|full | Low-quality |
| 18-101-final-viral-combined_k141_70616\|\|full | Not-determined |
| 18-101-final-viral-combined_k141_38547\|\|full | Not-determined |
| 18-101-final-viral-combined_k141_137853\|\|full | Low-quality |
| 18-101-final-viral-combined_k141_236184\|\|full | Not-determined |
| 18-101-final-viral-combined_k141_22006\|\|full | Low-quality |
| 18-101-final-viral-combined_k141_121413\|\|full | Low-quality |
| 18-101-final-viral-combined_k141_367920\|\|full | Low-quality |
| 18-101-final-viral-combined_k141_268763\|\|full | Not-determined |
| 18-101-final-viral-combined_k141_268764\|\|full | Low-quality |
| 18-101-final-viral-combined_k141_318944\|\|full | Low-quality |
| 18-101-final-viral-combined_k141_384221\|\|full | Low-quality |
| 18-101-final-viral-combined_k141_384223\|\|full | Not-determined |
| 18-101-final-viral-combined_k141_54767\|\|full | Not-determined |
| 18-101-final-viral-combined_k141_268766\|\|full | Not-determined |
| 18-101-final-viral-combined_k141_5639\|\|full | Not-determined |
| 18-101-final-viral-combined_k141_203505\|\|full | Not-determined |
| 18-101-final-viral-combined_k141_87637\|\|full | Not-determined |
| 18-101-final-viral-combined_k141_87640\|\|full | Not-determined |
| 18-101-final-viral-combined_k141_367935\|\|full | Not-determined |
| 18-101-final-viral-combined_k141_203509\|\|full | Low-quality |
| 18-101-final-viral-combined_k141_285618\|\|full | Not-determined |
| 18-101-final-viral-combined_k141_351768\|\|full | Low-quality |
| 18-101-final-viral-combined_k141_384250\|\|full | Low-quality |
| 18-101-final-viral-combined_k141_187378\|\|full | Not-determined |
| 18-101-final-viral-combined_k141_121446\|\|full | Low-quality |
| 18-101-final-viral-combined_k141_367939\|\|full | Low-quality |
| 18-101-final-viral-combined_k141_219857\|\|full | Low-quality |
| 18-101-final-viral-combined_k141_384264\|\|full | Not-determined |
| 18-101-final-viral-combined_k141_351782\|\|full | Low-quality |
| 18-101-final-viral-combined_k141_268808\|\|full | Low-quality |
| 18-101-final-viral-combined_k141_351784\|\|full | Not-determined |
| 18-101-final-viral-combined_k141_318963\|\|full | Low-quality |

| | |
|---|---|
| 18-101-final-viral-combined_k141_5694\|\|full | Low-quality |
| 18-101-final-viral-combined_k141_187402\|\|full | Low-quality |
| 18-101-final-viral-combined_k141_5704\|\|full | Not-determined |
| 18-101-final-viral-combined_k141_268833\|\|full | Low-quality |
| 18-101-final-viral-combined_k141_54830\|\|full | Low-quality |
| 18-101-final-viral-combined_k141_285642\|\|full | Low-quality |
| 18-101-final-viral-combined_k141_203547\|\|full | Not-determined |
| 18-101-final-viral-combined_k141_318993\|\|full | Not-determined |
| 18-101-final-viral-combined_k141_38621\|\|full | Low-quality |
| 18-101-final-viral-combined_k141_187445\|\|full | Not-determined |
| 18-101-final-viral-combined_k141_351819\|\|full | Not-determined |
| 18-101-final-viral-combined_k141_54859\|\|full | Not-determined |
| 18-101-final-viral-combined_k141_38627\|\|full | Not-determined |
| 18-101-final-viral-combined_k141_187452\|\|full | Low-quality |
| 18-101-final-viral-combined_k141_285671\|\|full | Low-quality |
| 18-101-final-viral-combined_k141_368001\|\|full | Low-quality |
| 18-101-final-viral-combined_k141_169647\|\|full | Low-quality |
| 18-101-final-viral-combined_k141_70701\|\|full | Not-determined |
| 18-101-final-viral-combined_k141_368023\|\|full | Not-determined |
| 18-101-final-viral-combined_k141_334328\|\|full | Low-quality |
| 18-101-final-viral-combined_k141_22044\|\|full | Low-quality |
| 18-101-final-viral-combined_k141_121517\|\|full | Not-determined |
| 18-101-final-viral-combined_k141_103736\|\|full | Low-quality |
| 18-101-final-viral-combined_k141_334336\|\|full | Low-quality |
| 18-101-final-viral-combined_k141_203572\|\|full | Not-determined |
| 18-101-final-viral-combined_k141_219971\|\|full | Low-quality |
| 18-101-final-viral-combined_k141_22065\|\|full | Low-quality |
| 18-101-final-viral-combined_k141_285690\|\|full | Low-quality |
| 18-101-final-viral-combined_k141_219988\|\|full | Low-quality |
| 18-101-final-viral-combined_k141_351883\|\|full | Low-quality |
| 18-101-final-viral-combined_k141_219989\|\|full | Low-quality |
| 18-101-final-viral-combined_k141_253142\|\|full | Low-quality |
| 18-101-final-viral-combined_k141_187472\|\|full | Low-quality |
| 18-101-final-viral-combined_k141_87726\|\|full | Not-determined |
| 18-101-final-viral-combined_k141_153704\|\|full | Not-determined |
| 18-101-final-viral-combined_k141_253159\|\|full | Not-determined |
| 18-101-final-viral-combined_k141_22082\|\|full | Low-quality |
| 18-101-final-viral-combined_k141_319063\|\|full | Low-quality |
| 18-101-final-viral-combined_k141_153727\|\|full | Not-determined |
| 18-101-final-viral-combined_k141_103787\|\|full | Low-quality |
| KM-final-viral-combined_k141_126774\|\|full | High-quality |
| 18-101-final-viral-combined_k141_153728\|\|full | Not-determined |
| 18-110-final-viral-combined_k141_48046\|\|full | Medium-quality |
| 18-101-final-viral-combined_k141_268923\|\|full | Low-quality |
| 18-101-final-viral-combined_k141_236291\|\|full | Not-determined |
| 18-101-final-viral-combined_k141_368077\|\|full | Low-quality |
| 18-101-final-viral-combined_k141_169722\|\|full | Low-quality |
| 18-101-final-viral-combined_k141_153736\|\|full | Low-quality |
| 18-101-final-viral-combined_k141_236324\|\|full | Not-determined |
| 18-101-final-viral-combined_k141_236325\|\|full | Low-quality |
| 18-101-final-viral-combined_k141_103819\|\|full | Low-quality |
| 18-101-final-viral-combined_k141_285722\|\|full | Low-quality |
| 18-101-final-viral-combined_k141_153776\|\|full | Not-determined |
| 18-101-final-viral-combined_k141_319126\|\|full | Not-determined |
| 18-101-final-viral-combined_k141_70836\|\|full | Not-determined |
| 18-101-final-viral-combined_k141_253188\|\|full | Low-quality |
| 18-101-final-viral-combined_k141_87808\|\|full | Not-determined |
| 18-101-final-viral-combined_k141_384458\|\|full | Low-quality |
| 18-101-final-viral-combined_k141_153822\|\|full | Low-quality |
| 18-101-final-viral-combined_k141_351941\|\|full | Not-determined |
| 18-101-final-viral-combined_k141_203650\|\|full | Low-quality |
| 18-101-final-viral-combined_k141_236368\|\|full | Low-quality |
| 18-101-final-viral-combined_k141_103865\|\|full | Low-quality |
| 18-101-final-viral-combined_k141_121680\|\|full | Low-quality |
| 18-101-final-viral-combined_k141_334430\|\|full | Low-quality |
| 18-101-final-viral-combined_k141_121682\|\|full | Low-quality |
| 18-101-final-viral-combined_k141_169754\|\|full | Not-determined |
| 18-101-final-viral-combined_k141_334434\|\|full | Low-quality |
| 18-101-final-viral-combined_k141_268989\|\|full | Not-determined |
| 18-101-final-viral-combined_k141_87862\|\|full | Low-quality |
| 18-101-final-viral-combined_k141_153836\|\|full | Low-quality |
| 18-101-final-viral-combined_k141_87868\|\|full | Not-determined |
| 18-101-final-viral-combined_k141_203683\|\|full | Not-determined |
| 18-101-final-viral-combined_k141_203686\|\|full | Low-quality |
| 18-101-final-viral-combined_k141_351948\|\|full | Not-determined |
| 18-101-final-viral-combined_k141_169785\|\|full | Low-quality |
| 18-101-final-viral-combined_k141_302896\|\|full | Low-quality |
| 18-101-final-viral-combined_k141_38736\|\|full | Low-quality |
| 18-101-final-viral-combined_k141_5832\|\|full | Not-determined |
| 18-101-final-viral-combined_k141_384498\|\|full | Low-quality |
| 18-110-final-viral-combined_k141_215102\|\|full | High-quality |
| 18-101-final-viral-combined_k141_236401\|\|full | Not-determined |
| 18-101-final-viral-combined_k141_351965\|\|full | Not-determined |
| 18-101-final-viral-combined_k141_334456\|\|full | Low-quality |
| 18-101-final-viral-combined_k141_169801\|\|full | Not-determined |
| 18-101-final-viral-combined_k141_153871\|\|full | Not-determined |
| 18-101-final-viral-combined_k141_203707\|\|full | Not-determined |
| 18-101-final-viral-combined_k141_351983\|\|full | Not-determined |
| 18-101-final-viral-combined_k141_253246\|\|full | Not-determined |
| 18-101-final-viral-combined_k141_253248\|\|full | Not-determined |
| 18-101-final-viral-combined_k141_253254\|\|full | Not-determined |
| 18-101-final-viral-combined_k141_87903\|\|full | Low-quality |
| 18-101-final-viral-combined_k141_153900\|\|full | Not-determined |
| 18-101-final-viral-combined_k141_138114\|\|full | Low-quality |
| 18-101-final-viral-combined_k141_70920\|\|full | Low-quality |
| 18-101-final-viral-combined_k141_87910\|\|full | Low-quality |
| 18-101-final-viral-combined_k141_384519\|\|full | Not-determined |
| 18-101-final-viral-combined_k141_352017\|\|full | Low-quality |
| 18-101-final-viral-combined_k141_302929\|\|full | Low-quality |
| 18-101-final-viral-combined_k141_169835\|\|full | Not-determined |
| 18-101-final-viral-combined_k141_87919\|\|full | Not-determined |
| 18-101-final-viral-combined_k141_236435\|\|full | Low-quality |
| 18-101-final-viral-combined_k141_236437\|\|full | Not-determined |
| 18-101-final-viral-combined_k141_87928\|\|full | Not-determined |
| 18-101-final-viral-combined_k141_70953\|\|full | Low-quality |
| 18-101-final-viral-combined_k141_87929\|\|full | Not-determined |
| 18-101-final-viral-combined_k141_236450\|\|full | Not-determined |
| 18-101-final-viral-combined_k141_5850\|\|full | Low-quality |
| 18-101-final-viral-combined_k141_269061\|\|full | Not-determined |
| 18-101-final-viral-combined_k141_87935\|\|full | Not-determined |
| 18-101-final-viral-combined_k141_203729\|\|full | Not-determined |
| 18-102-final-viral-combined_k141_135154\|\|full | Medium-quality |
| 18-101-final-viral-combined_k141_334525\|\|full | Low-quality |
| 18-101-final-viral-combined_k141_253316\|\|full | Not-determined |
| 18-101-final-viral-combined_k141_368145\|\|full | Not-determined |
| 18-101-final-viral-combined_k141_169852\|\|full | Not-determined |
| 18-101-final-viral-combined_k141_203744\|\|full | Not-determined |
| 18-101-final-viral-combined_k141_253319\|\|full | Not-determined |
| 18-101-final-viral-combined_k141_253320\|\|full | Low-quality |
| 18-101-final-viral-combined_k141_269073\|\|full | Not-determined |
| 18-101-final-viral-combined_k141_121774\|\|full | Not-determined |
| 18-101-final-viral-combined_k141_319282\|\|full | Low-quality |
| 18-101-final-viral-combined_k141_334552\|\|full | Low-quality |
| 18-101-final-viral-combined_k141_269081\|\|full | Low-quality |
| 18-101-final-viral-combined_k141_38861\|\|full | Not-determined |
| 18-101-final-viral-combined_k141_253354\|\|full | Low-quality |
| 18-101-final-viral-combined_k141_187523\|\|full | Not-determined |
| 18-101-final-viral-combined_k141_70997\|\|full | Low-quality |
| 18-101-final-viral-combined_k141_285804\|\|full | Not-determined |
| 18-101-final-viral-combined_k141_22215\|\|full | Low-quality |
| 18-101-final-viral-combined_k141_38889\|\|full | Not-determined |
| 18-101-final-viral-combined_k141_138174\|\|full | Low-quality |
| 18-101-final-viral-combined_k141_5917\|\|full | Not-determined |
| 18-101-final-viral-combined_k141_236524\|\|full | Not-determined |
| 18-101-final-viral-combined_k141_187533\|\|full | Low-quality |
| 17-109-final-viral-combined_k141_352846\|\|full | High-quality |
| 18-101-final-viral-combined_k141_220063\|\|full | Not-determined |
| 18-101-final-viral-combined_k141_187538\|\|full | Not-determined |
| 18-101-final-viral-combined_k141_269146\|\|full | Not-determined |
| 18-101-final-viral-combined_k141_104034\|\|full | Not-determined |
| 18-101-final-viral-combined_k141_121817\|\|full | Not-determined |
| 18-101-final-viral-combined_k141_285847\|\|full | Not-determined |
| 18-101-final-viral-combined_k141_269160\|\|full | Not-determined |
| 18-101-final-viral-combined_k141_55181\|\|full | Low-quality |
| 18-101-final-viral-combined_k141_121818\|\|full | Not-determined |
| 18-101-final-viral-combined_k141_220103\|\|full | Not-determined |
| 18-101-final-viral-combined_k141_285855\|\|full | Not-determined |
| 18-101-final-viral-combined_k141_334609\|\|full | Not-determined |
| 18-101-final-viral-combined_k141_352129\|\|full | Not-determined |
| 18-101-final-viral-combined_k141_269178\|\|full | Low-quality |
| 18-101-final-viral-combined_k141_334611\|\|full | Low-quality |
| 18-101-final-viral-combined_k141_384672\|\|full | Low-quality |
| 18-101-final-viral-combined_k141_55191\|\|full | Not-determined |
| 18-101-final-viral-combined_k141_22266\|\|full | Not-determined |
| 18-101-final-viral-combined_k141_187571\|\|full | Low-quality |
| 18-101-final-viral-combined_k141_384688\|\|full | Low-quality |
| 18-101-final-viral-combined_k141_285876\|\|full | Not-determined |
| 18-101-final-viral-combined_k141_169950\|\|full | Low-quality |
| 18-101-final-viral-combined_k141_334632\|\|full | Not-determined |
| 18-101-final-viral-combined_k141_352156\|\|full | Low-quality |
| 18-101-final-viral-combined_k141_87987\|\|full | Low-quality |
| 18-101-final-viral-combined_k141_220129\|\|full | Not-determined |
| 18-101-final-viral-combined_k141_302982\|\|full | Not-determined |
| 18-101-final-viral-combined_k141_71052\|\|full | Not-determined |
| 18-101-final-viral-combined_k141_220135\|\|full | Not-determined |
| 18-101-final-viral-combined_k141_121827\|\|full | Not-determined |
| 18-101-final-viral-combined_k141_253446\|\|full | Low-quality |
| 18-101-final-viral-combined_k141_302985\|\|full | Low-quality |
| 18-101-final-viral-combined_k141_55231\|\|full | Not-determined |
| 18-101-final-viral-combined_k141_38965\|\|full | Low-quality |
| 18-101-final-viral-combined_k141_303005\|\|full | Not-determined |
| 18-101-final-viral-combined_k141_88028\|\|full | Low-quality |
| 18-101-final-viral-combined_k141_88030\|\|full | Low-quality |
| 18-101-final-viral-combined_k141_71073\|\|full | Low-quality |
| 18-101-final-viral-combined_k141_6009\|\|full | Low-quality |
| 18-101-final-viral-combined_k141_104096\|\|full | Low-quality |
| 18-101-final-viral-combined_k141_285944\|\|full | Not-determined |
| 18-101-final-viral-combined_k141_121860\|\|full | Not-determined |
| 18-101-final-viral-combined_k141_6016\|\|full | Not-determined |
| 18-101-final-viral-combined_k141_319432\|\|full | Not-determined |
| 18-101-final-viral-combined_k141_104104\|\|full | Not-determined |
| 18-101-final-viral-combined_k141_55245\|\|full | Low-quality |
| 18-101-final-viral-combined_k141_22297\|\|full | Low-quality |
| 18-101-final-viral-combined_k141_71089\|\|full | Not-determined |
| 18-101-final-viral-combined_k141_121866\|\|full | Not-determined |
| 18-101-final-viral-combined_k141_352215\|\|full | Low-quality |
| 18-101-final-viral-combined_k141_319448\|\|full | Not-determined |
| 18-101-final-viral-combined_k141_104121\|\|full | Low-quality |
| 18-101-final-viral-combined_k141_352221\|\|full | Not-determined |
| 18-101-final-viral-combined_k141_319466\|\|full | Low-quality |
| 18-101-final-viral-combined_k141_170003\|\|full | Not-determined |
| 18-101-final-viral-combined_k141_170005\|\|full | Low-quality |
| 18-101-final-viral-combined_k141_22336\|\|full | Not-determined |
| 18-101-final-viral-combined_k141_285988\|\|full | Low-quality |
| 18-101-final-viral-combined_k141_170016\|\|full | Not-determined |
| 18-101-final-viral-combined_k141_88045\|\|full | Low-quality |
| 18-101-final-viral-combined_k141_104145\|\|full | Low-quality |
| 18-101-final-viral-combined_k141_55278\|\|full | Low-quality |
| 18-101-final-viral-combined_k141_88053\|\|full | Not-determined |
| 18-101-final-viral-combined_k141_170029\|\|full | Low-quality |
| 18-101-final-viral-combined_k141_269247\|\|full | Low-quality |
| 18-101-final-viral-combined_k141_170041\|\|full | Low-quality |
| 18-101-final-viral-combined_k141_253540\|\|full | Not-determined |
| 18-101-final-viral-combined_k141_71163\|\|full | Not-determined |
| 18-101-final-viral-combined_k141_187756\|\|full | Low-quality |
| 18-101-final-viral-combined_k141_269281\|\|full | Not-determined |
| 18-101-final-viral-combined_k141_220191\|\|full | Low-quality |
| 18-101-final-viral-combined_k141_6053\|\|full | Low-quality |
| 18-101-final-viral-combined_k141_55324\|\|full | Not-determined |
| 18-101-final-viral-combined_k141_121969\|\|full | Not-determined |
| 18-101-final-viral-combined_k141_187774\|\|full | Low-quality |
| 18-101-final-viral-combined_k141_104184\|\|full | Not-determined |
| 18-101-final-viral-combined_k141_334743\|\|full | Not-determined |
| 18-101-final-viral-combined_k141_71183\|\|full | Low-quality |
| 18-101-final-viral-combined_k141_286050\|\|full | Low-quality |
| 18-101-final-viral-combined_k141_154040\|\|full | Not-determined |
| 18-101-final-viral-combined_k141_22445\|\|full | Not-determined |
| 18-101-final-viral-combined_k141_253629\|\|full | Not-determined |
| 18-101-final-viral-combined_k141_286065\|\|full | Not-determined |
| 18-101-final-viral-combined_k141_286070\|\|full | Not-determined |
| 18-101-final-viral-combined_k141_253642\|\|full | Not-determined |
| 18-101-final-viral-combined_k141_352308\|\|full | Low-quality |
| 18-101-final-viral-combined_k141_55395\|\|full | Low-quality |
| 18-101-final-viral-combined_k141_88158\|\|full | Low-quality |
| 18-101-final-viral-combined_k141_88160\|\|full | Low-quality |
| 18-101-final-viral-combined_k141_352329\|\|full | Not-determined |
| 18-101-final-viral-combined_k141_253673\|\|full | Low-quality |
| 18-101-final-viral-combined_k141_253685\|\|full | Low-quality |
| 18-101-final-viral-combined_k141_55415\|\|full | Not-determined |
| 18-101-final-viral-combined_k141_39062\|\|full | Not-determined |
| 18-101-final-viral-combined_k141_122063\|\|full | Low-quality |
| 18-101-final-viral-combined_k141_154095\|\|full | Not-determined |
| 18-101-final-viral-combined_k141_122069\|\|full | Low-quality |
| 18-101-final-viral-combined_k141_352353\|\|full | Low-quality |
| 18-101-final-viral-combined_k141_352357\|\|full | Low-quality |
| 18-101-final-viral-combined_k141_6158\|\|full | Not-determined |
| 18-101-final-viral-combined_k141_154109\|\|full | Not-determined |
| 18-101-final-viral-combined_k141_204065\|\|full | Not-determined |
| 18-101-final-viral-combined_k141_352371\|\|full | Not-determined |
| 18-101-final-viral-combined_k141_187812\|\|full | Low-quality |
| 18-101-final-viral-combined_k141_170104\|\|full | Low-quality |
| 18-101-final-viral-combined_k141_104279\|\|full | Low-quality |
| 18-101-final-viral-combined_k141_104282\|\|full | Low-quality |
| 18-101-final-viral-combined_k141_39107\|\|full | Low-quality |
| 18-101-final-viral-combined_k141_170112\|\|full | Low-quality |
| 18-101-final-viral-combined_k141_104297\|\|full | Low-quality |
| 18-101-final-viral-combined_k141_122100\|\|full | Low-quality |
| 18-101-final-viral-combined_k141_204097\|\|full | Not-determined |
| 18-101-final-viral-combined_k141_154138\|\|full | Not-determined |
| 18-101-final-viral-combined_k141_104315\|\|full | Low-quality |
| 18-101-final-viral-combined_k141_88224\|\|full | Not-determined |
| 18-101-final-viral-combined_k141_269362\|\|full | Low-quality |
| 18-101-final-viral-combined_k141_122127\|\|full | Low-quality |
| 18-101-final-viral-combined_k141_71260\|\|full | Not-determined |
| 18-101-final-viral-combined_k141_319608\|\|full | Low-quality |
| 18-101-final-viral-combined_k141_55515\|\|full | Not-determined |
| 18-101-final-viral-combined_k141_352417\|\|full | Not-determined |
| 18-101-final-viral-combined_k141_71284\|\|full | Low-quality |
| 18-102-final-viral-combined_k141_24549\|\|full | Medium-quality |
| 18-101-final-viral-combined_k141_384880\|\|full | Not-determined |
| 18-101-final-viral-combined_k141_204134\|\|full | Low-quality |
| 18-101-final-viral-combined_k141_71292\|\|full | Not-determined |
| 18-101-final-viral-combined_k141_170125\|\|full | Not-determined |
| 18-101-final-viral-combined_k141_187855\|\|full | Low-quality |
| 18-101-final-viral-combined_k141_204139\|\|full | Not-determined |
| 18-101-final-viral-combined_k141_138328\|\|full | Low-quality |
| 18-101-final-viral-combined_k141_187867\|\|full | Not-determined |
| 18-101-final-viral-combined_k141_334900\|\|full | Not-determined |
| 18-101-final-viral-combined_k141_71304\|\|full | Not-determined |
| 18-101-final-viral-combined_k141_6228\|\|full | Low-quality |
| 18-101-final-viral-combined_k141_55554\|\|full | Low-quality |
| 18-101-final-viral-combined_k141_253790\|\|full | Not-determined |
| 18-101-final-viral-combined_k141_55558\|\|full | Low-quality |
| 18-101-final-viral-combined_k141_319648\|\|full | Low-quality |
| 18-101-final-viral-combined_k141_303217\|\|full | Low-quality |
| 18-101-final-viral-combined_k141_236738\|\|full | Not-determined |
| 18-101-final-viral-combined_k141_170161\|\|full | Not-determined |
| 18-101-final-viral-combined_k141_269432\|\|full | Low-quality |
| 18-101-final-viral-combined_k141_104344\|\|full | Low-quality |
| 18-101-final-viral-combined_k141_71342\|\|full | Not-determined |
| 18-101-final-viral-combined_k141_88292\|\|full | Low-quality |
| 18-101-final-viral-combined_k141_104347\|\|full | Low-quality |
| 18-101-final-viral-combined_k141_253804\|\|full | Low-quality |
| 18-101-final-viral-combined_k141_269442\|\|full | Not-determined |
| 18-101-final-viral-combined_k141_6241\|\|full | Not-determined |
| 18-101-final-viral-combined_k141_55576\|\|full | Not-determined |
| 18-101-final-viral-combined_k141_352436\|\|full | Not-determined |
| 18-101-final-viral-combined_k141_319675\|\|full | Low-quality |
| 18-101-final-viral-combined_k141_71349\|\|full | Not-determined |
| 18-101-final-viral-combined_k141_170191\|\|full | Low-quality |
| 18-101-final-viral-combined_k141_39170\|\|full | Low-quality |
| 18-101-final-viral-combined_k141_236773\|\|full | Low-quality |
| 18-101-final-viral-combined_k141_39174\|\|full | Low-quality |
| 18-101-final-viral-combined_k141_352445\|\|full | Low-quality |
| 18-101-final-viral-combined_k141_334958\|\|full | Not-determined |
| 18-101-final-viral-combined_k141_6287\|\|full | Not-determined |
| 18-101-final-viral-combined_k141_122213\|\|full | Low-quality |
| 18-101-final-viral-combined_k141_269492\|\|full | Low-quality |
| 18-101-final-viral-combined_k141_88326\|\|full | Low-quality |
| 18-101-final-viral-combined_k141_104373\|\|full | Low-quality |
| 18-101-final-viral-combined_k141_368164\|\|full | Not-determined |
| 18-101-final-viral-combined_k141_39209\|\|full | Low-quality |
| 18-101-final-viral-combined_k141_138381\|\|full | Not-determined |
| 18-101-final-viral-combined_k141_204207\|\|full | Low-quality |
| 18-101-final-viral-combined_k141_236798\|\|full | Low-quality |
| 18-101-final-viral-combined_k141_122230\|\|full | Low-quality |
| 18-101-final-viral-combined_k141_55595\|\|full | Low-quality |
| 18-101-final-viral-combined_k141_88337\|\|full | Not-determined |
| 18-101-final-viral-combined_k141_138393\|\|full | Not-determined |
| 18-101-final-viral-combined_k141_220334\|\|full | Low-quality |
| 18-101-final-viral-combined_k141_88342\|\|full | Not-determined |
| 18-101-final-viral-combined_k141_352490\|\|full | Not-determined |
| 18-101-final-viral-combined_k141_122244\|\|full | Low-quality |
| 18-101-final-viral-combined_k141_55607\|\|full | Low-quality |
| 18-101-final-viral-combined_k141_55612\|\|full | Not-determined |
| 18-101-final-viral-combined_k141_220350\|\|full | Low-quality |
| 18-101-final-viral-combined_k141_104412\|\|full | Low-quality |
| 18-101-final-viral-combined_k141_187950\|\|full | Low-quality |
| 18-101-final-viral-combined_k141_88380\|\|full | Not-determined |
| 18-101-final-viral-combined_k141_368244\|\|full | Not-determined |
| 18-101-final-viral-combined_k141_236835\|\|full | Low-quality |
| 18-101-final-viral-combined_k141_204228\|\|full | Not-determined |
| 18-101-final-viral-combined_k141_71442\|\|full | Not-determined |
| 18-101-final-viral-combined_k141_335020\|\|full | Low-quality |
| 18-101-final-viral-combined_k141_71457\|\|full | Not-determined |
| 18-101-final-viral-combined_k141_204255\|\|full | Low-quality |
| 18-101-final-viral-combined_k141_220363\|\|full | Low-quality |
| 18-101-final-viral-combined_k141_71467\|\|full | Not-determined |
| 18-101-final-viral-combined_k141_71468\|\|full | Low-quality |
| 18-101-final-viral-combined_k141_335033\|\|full | Not-determined |
| 18-101-final-viral-combined_k141_253917\|\|full | Low-quality |
| 18-101-final-viral-combined_k141_154292\|\|full | Low-quality |
| 18-101-final-viral-combined_k141_220375\|\|full | Low-quality |
| 18-101-final-viral-combined_k141_220378\|\|full | Not-determined |
| 18-101-final-viral-combined_k141_303326\|\|full | Low-quality |
| 18-101-final-viral-combined_k141_188004\|\|full | Not-determined |
| 18-101-final-viral-combined_k141_138500\|\|full | Not-determined |
| 18-101-final-viral-combined_k141_220386\|\|full | Not-determined |
| 18-101-final-viral-combined_k141_269559\|\|full | Not-determined |
| 18-101-final-viral-combined_k141_352575\|\|full | Low-quality |
| 18-101-final-viral-combined_k141_368277\|\|full | Not-determined |
| 18-101-final-viral-combined_k141_204283\|\|full | Not-determined |
| 18-101-final-viral-combined_k141_154304\|\|full | Low-quality |
| 18-101-final-viral-combined_k141_188011\|\|full | Low-quality |
| 18-101-final-viral-combined_k141_319764\|\|full | Low-quality |
| 18-101-final-viral-combined_k141_352581\|\|full | Not-determined |
| 18-101-final-viral-combined_k141_352592\|\|full | Not-determined |
| 18-101-final-viral-combined_k141_335054\|\|full | Not-determined |
| 18-101-final-viral-combined_k141_6378\|\|full | Low-quality |
| 18-101-final-viral-combined_k141_319772\|\|full | Not-determined |
| 18-101-final-viral-combined_k141_170265\|\|full | Not-determined |
| 18-101-final-viral-combined_k141_385001\|\|full | Not-determined |
| 18-101-final-viral-combined_k141_253948\|\|full | Not-determined |
| 18-101-final-viral-combined_k141_385010\|\|full | Low-quality |
| 18-101-final-viral-combined_k141_319785\|\|full | Low-quality |
| 18-101-final-viral-combined_k141_236949\|\|full | Not-determined |
| 18-101-final-viral-combined_k141_204326\|\|full | Low-quality |
| 18-101-final-viral-combined_k141_352634\|\|full | Low-quality |
| 18-101-final-viral-combined_k141_204333\|\|full | Low-quality |
| 18-101-final-viral-combined_k141_154324\|\|full | Low-quality |
| 18-101-final-viral-combined_k141_352639\|\|full | Low-quality |
| 18-101-final-viral-combined_k141_236957\|\|full | Not-determined |
| 18-101-final-viral-combined_k141_352643\|\|full | Low-quality |
| 18-101-final-viral-combined_k141_204354\|\|full | Low-quality |
| 18-101-final-viral-combined_k141_204356\|\|full | Not-determined |
| 18-101-final-viral-combined_k141_236972\|\|full | Not-determined |
| 18-101-final-viral-combined_k141_22755\|\|full | Not-determined |
| 18-101-final-viral-combined_k141_138561\|\|full | Low-quality |
| 18-101-final-viral-combined_k141_352672\|\|full | Not-determined |
| 18-101-final-viral-combined_k141_253957\|\|full | Low-quality |
| 18-101-final-viral-combined_k141_154360\|\|full | Low-quality |
| 18-101-final-viral-combined_k141_236997\|\|full | Not-determined |
| 18-101-final-viral-combined_k141_269581\|\|full | Not-determined |
| 18-101-final-viral-combined_k141_122383\|\|full | Low-quality |
| 18-101-final-viral-combined_k141_237008\|\|full | Low-quality |
| 18-101-final-viral-combined_k141_319868\|\|full | Not-determined |
| 18-101-final-viral-combined_k141_55688\|\|full | Low-quality |
| 18-101-final-viral-combined_k141_39344\|\|full | Not-determined |
| 18-101-final-viral-combined_k141_368329\|\|full | Not-determined |
| 18-101-final-viral-combined_k141_188126\|\|full | Low-quality |
| 18-101-final-viral-combined_k141_122414\|\|full | Not-determined |
| 18-101-final-viral-combined_k141_286280\|\|full | Not-determined |
| 18-101-final-viral-combined_k141_22802\|\|full | Not-determined |
| 18-101-final-viral-combined_k141_122415\|\|full | Not-determined |
| 18-101-final-viral-combined_k141_6495\|\|full | Low-quality |
| 18-101-final-viral-combined_k141_6499\|\|full | Not-determined |
| 18-101-final-viral-combined_k141_122439\|\|full | Low-quality |
| 18-101-final-viral-combined_k141_55716\|\|full | Not-determined |
| 18-101-final-viral-combined_k141_385175\|\|full | Not-determined |
| 18-101-final-viral-combined_k141_335103\|\|full | Low-quality |
| 18-101-final-viral-combined_k141_269609\|\|full | Not-determined |
| 18-101-final-viral-combined_k141_220465\|\|full | Not-determined |
| 18-101-final-viral-combined_k141_22841\|\|full | Low-quality |
| 18-101-final-viral-combined_k141_220467\|\|full | Not-determined |
| 18-101-final-viral-combined_k141_6512\|\|full | Low-quality |
| 18-101-final-viral-combined_k141_269614\|\|full | Not-determined |
| 18-101-final-viral-combined_k141_104476\|\|full | Low-quality |
| 18-101-final-viral-combined_k141_385189\|\|full | Low-quality |
| 18-101-final-viral-combined_k141_39365\|\|full | Not-determined |
| 18-101-final-viral-combined_k141_385192\|\|full | Low-quality |
| 18-101-final-viral-combined_k141_204444\|\|full | Not-determined |
| 18-101-final-viral-combined_k141_71573\|\|full | Low-quality |
| 18-101-final-viral-combined_k141_6516\|\|full | Not-determined |
| 18-101-final-viral-combined_k141_237077\|\|full | Not-determined |
| 18-101-final-viral-combined_k141_188168\|\|full | Low-quality |
| 18-101-final-viral-combined_k141_352733\|\|full | Low-quality |
| 18-101-final-viral-combined_k141_55767\|\|full | Not-determined |
| 18-101-final-viral-combined_k141_254042\|\|full | Not-determined |
| 18-101-final-viral-combined_k141_319909\|\|full | Not-determined |
| 18-101-final-viral-combined_k141_254044\|\|full | Low-quality |
| 18-101-final-viral-combined_k141_188182\|\|full | Low-quality |
| 18-101-final-viral-combined_k141_22885\|\|full | Not-determined |
| 18-101-final-viral-combined_k141_55776\|\|full | Low-quality |
| 18-101-final-viral-combined_k141_188183\|\|full | Low-quality |
| 18-101-final-viral-combined_k141_55779\|\|full | Low-quality |
| 18-101-final-viral-combined_k141_55781\|\|full | Low-quality |
| 18-101-final-viral-combined_k141_237111\|\|full | Low-quality |
| 18-101-final-viral-combined_k141_138686\|\|full | Not-determined |
| 18-101-final-viral-combined_k141_220519\|\|full | Not-determined |
| 18-101-final-viral-combined_k141_6541\|\|full | Not-determined |
| 18-101-final-viral-combined_k141_188200\|\|full | Low-quality |
| 18-101-final-viral-combined_k141_6542\|\|full | Not-determined |
| 18-101-final-viral-combined_k141_319959\|\|full | Not-determined |
| 18-101-final-viral-combined_k141_385247\|\|full | Low-quality |
| 18-101-final-viral-combined_k141_385256\|\|full | Not-determined |
| 18-101-final-viral-combined_k141_368422\|\|full | Low-quality |
| 18-101-final-viral-combined_k141_170398\|\|full | Low-quality |
| 18-101-final-viral-combined_k141_55824\|\|full | Not-determined |
| 18-101-final-viral-combined_k141_55825\|\|full | Low-quality |
| 18-101-final-viral-combined_k141_6574\|\|full | Low-quality |
| 18-101-final-viral-combined_k141_71645\|\|full | Not-determined |
| 18-101-final-viral-combined_k141_138722\|\|full | Low-quality |
| 18-101-final-viral-combined_k141_188216\|\|full | Low-quality |
| 18-101-final-viral-combined_k141_55832\|\|full | Low-quality |
| 18-101-final-viral-combined_k141_319985\|\|full | Low-quality |
| 18-101-final-viral-combined_k141_122545\|\|full | Low-quality |
| 18-101-final-viral-combined_k141_319988\|\|full | Not-determined |
| 18-101-final-viral-combined_k141_39461\|\|full | Not-determined |
| 18-101-final-viral-combined_k141_220573\|\|full | Not-determined |
| 18-101-final-viral-combined_k141_385304\|\|full | Low-quality |
| 18-101-final-viral-combined_k141_170428\|\|full | Not-determined |
| 18-101-final-viral-combined_k141_237175\|\|full | Not-determined |
| 18-101-final-viral-combined_k141_368467\|\|full | Low-quality |
| 18-101-final-viral-combined_k141_22904\|\|full | Low-quality |
| 18-101-final-viral-combined_k141_254135\|\|full | Low-quality |
| 18-101-final-viral-combined_k141_122576\|\|full | Not-determined |
| 18-101-final-viral-combined_k141_154423\|\|full | Not-determined |
| 18-101-final-viral-combined_k141_22917\|\|full | Low-quality |
| 18-101-final-viral-combined_k141_385327\|\|full | Not-determined |
| 18-101-final-viral-combined_k141_237198\|\|full | Low-quality |
| 18-101-final-viral-combined_k141_220613\|\|full | Low-quality |
| 18-101-final-viral-combined_k141_335205\|\|full | Not-determined |
| 18-101-final-viral-combined_k141_71697\|\|full | Not-determined |
| 18-101-final-viral-combined_k141_154436\|\|full | Low-quality |
| 18-101-final-viral-combined_k141_22934\|\|full | Low-quality |
| 18-101-final-viral-combined_k141_154449\|\|full | Low-quality |
| 18-101-final-viral-combined_k141_204572\|\|full | Not-determined |
| 18-101-final-viral-combined_k141_122607\|\|full | Not-determined |
| 18-101-final-viral-combined_k141_39505\|\|full | Not-determined |
| 18-101-final-viral-combined_k141_154461\|\|full | Low-quality |
| 18-101-final-viral-combined_k141_71711\|\|full | Not-determined |
| 18-101-final-viral-combined_k141_385364\|\|full | Not-determined |
| 18-101-final-viral-combined_k141_154463\|\|full | Not-determined |
| 18-101-final-viral-combined_k141_335225\|\|full | Not-determined |
| 18-101-final-viral-combined_k141_385367\|\|full | Low-quality |
| 18-101-final-viral-combined_k141_269797\|\|full | Low-quality |
| 18-101-final-viral-combined_k141_237233\|\|full | Low-quality |
| 18-101-final-viral-combined_k141_352880\|\|full | Not-determined |
| 18-101-final-viral-combined_k141_303547\|\|full | Low-quality |
| 18-101-final-viral-combined_k141_122635\|\|full | Not-determined |
| 18-101-final-viral-combined_k141_320039\|\|full | Not-determined |
| 18-101-final-viral-combined_k141_254204\|\|full | Low-quality |
| 18-101-final-viral-combined_k141_22972\|\|full | Low-quality |
| 18-101-final-viral-combined_k141_254211\|\|full | Not-determined |
| 18-101-final-viral-combined_k141_303562\|\|full | Not-determined |
| 18-101-final-viral-combined_k141_55879\|\|full | Not-determined |
| 18-101-final-viral-combined_k141_138779\|\|full | Not-determined |
| 18-101-final-viral-combined_k141_385406\|\|full | Not-determined |
| 18-101-final-viral-combined_k141_55885\|\|full | Not-determined |
| 18-101-final-viral-combined_k141_237252\|\|full | Not-determined |
| 18-101-final-viral-combined_k141_320066\|\|full | Low-quality |
| 18-101-final-viral-combined_k141_368523\|\|full | Low-quality |
| 18-101-final-viral-combined_k141_320070\|\|full | Not-determined |
| 18-101-final-viral-combined_k141_88754\|\|full | Not-determined |
| 18-101-final-viral-combined_k141_138794\|\|full | Low-quality |
| 18-101-final-viral-combined_k141_368526\|\|full | Low-quality |
| 18-101-final-viral-combined_k141_23000\|\|full | Low-quality |
| 18-101-final-viral-combined_k141_104636\|\|full | Low-quality |
| 18-101-final-viral-combined_k141_71754\|\|full | Not-determined |
| 18-101-final-viral-combined_k141_368528\|\|full | Not-determined |
| 18-101-final-viral-combined_k141_254235\|\|full | Low-quality |
| 18-101-final-viral-combined_k141_368535\|\|full | Low-quality |
| 18-101-final-viral-combined_k141_138808\|\|full | Low-quality |
| 18-101-final-viral-combined_k141_104641\|\|full | Not-determined |
| 18-101-final-viral-combined_k141_39556\|\|full | Low-quality |
| 18-101-final-viral-combined_k141_204651\|\|full | Not-determined |
| 18-101-final-viral-combined_k141_254239\|\|full | Not-determined |
| 18-101-final-viral-combined_k141_55914\|\|full | Not-determined |
| 18-101-final-viral-combined_k141_269856\|\|full | Low-quality |
| 18-101-final-viral-combined_k141_55923\|\|full | Low-quality |
| 18-101-final-viral-combined_k141_303598\|\|full | Not-determined |
| 18-101-final-viral-combined_k141_55930\|\|full | Low-quality |
| 18-101-final-viral-combined_k141_88783\|\|full | Not-determined |
| 18-101-final-viral-combined_k141_138826\|\|full | Not-determined |
| 18-101-final-viral-combined_k141_335285\|\|full | Low-quality |
| 18-101-final-viral-combined_k141_6714\|\|full | Not-determined |
| 18-101-final-viral-combined_k141_204676\|\|full | Low-quality |
| 18-101-final-viral-combined_k141_269868\|\|full | Not-determined |
| 18-101-final-viral-combined_k141_55939\|\|full | Not-determined |
| 18-101-final-viral-combined_k141_122680\|\|full | Not-determined |
| 18-101-final-viral-combined_k141_55948\|\|full | Not-determined |
| 18-101-final-viral-combined_k141_23049\|\|full | Not-determined |
| 18-101-final-viral-combined_k141_352971\|\|full | Low-quality |
| 18-101-final-viral-combined_k141_23065\|\|full | Not-determined |
| 18-101-final-viral-combined_k141_237312\|\|full | Low-quality |
| 18-101-final-viral-combined_k141_104689\|\|full | Not-determined |
| 18-101-final-viral-combined_k141_71812\|\|full | Low-quality |
| 18-101-final-viral-combined_k141_320142\|\|full | Not-determined |
| 18-101-final-viral-combined_k141_320146\|\|full | Not-determined |
| 18-101-final-viral-combined_k141_286411\|\|full | Not-determined |
| 18-101-final-viral-combined_k141_286412\|\|full | Low-quality |
| 18-101-final-viral-combined_k141_154543\|\|full | Not-determined |
| 18-101-final-viral-combined_k141_170521\|\|full | Not-determined |
| 18-101-final-viral-combined_k141_237340\|\|full | Low-quality |
| 18-101-final-viral-combined_k141_368609\|\|full | Low-quality |
| 18-101-final-viral-combined_k141_204736\|\|full | Low-quality |
| 18-101-final-viral-combined_k141_154567\|\|full | Not-determined |
| 18-101-final-viral-combined_k141_170529\|\|full | Not-determined |
| 18-101-final-viral-combined_k141_220777\|\|full | Not-determined |
| 18-101-final-viral-combined_k141_170537\|\|full | Not-determined |
| 18-101-final-viral-combined_k141_104716\|\|full | Not-determined |
| 18-101-final-viral-combined_k141_104718\|\|full | Low-quality |
| 18-101-final-viral-combined_k141_6742\|\|full | Low-quality |
| 18-101-final-viral-combined_k141_385429\|\|full | Not-determined |
| 18-101-final-viral-combined_k141_170552\|\|full | Not-determined |
| 18-101-final-viral-combined_k141_303697\|\|full | Not-determined |
| 18-101-final-viral-combined_k141_286466\|\|full | Low-quality |
| 18-101-final-viral-combined_k141_303703\|\|full | Not-determined |
| 18-101-final-viral-combined_k141_320208\|\|full | Low-quality |
| 18-101-final-viral-combined_k141_368653\|\|full | Not-determined |
| 18-101-final-viral-combined_k141_385431\|\|full | Low-quality |
| 18-101-final-viral-combined_k141_368654\|\|full | Low-quality |
| 18-101-final-viral-combined_k141_303709\|\|full | Not-determined |
| 18-101-final-viral-combined_k141_6755\|\|full | Not-determined |
| 18-101-final-viral-combined_k141_353054\|\|full | Low-quality |
| 18-101-final-viral-combined_k141_353056\|\|full | Low-quality |
| 18-101-final-viral-combined_k141_56006\|\|full | Not-determined |
| 18-101-final-viral-combined_k141_286482\|\|full | Not-determined |
| 18-101-final-viral-combined_k141_56014\|\|full | Not-determined |
| 18-101-final-viral-combined_k141_170590\|\|full | Low-quality |
| 18-101-final-viral-combined_k141_170592\|\|full | Low-quality |
| 18-101-final-viral-combined_k141_56023\|\|full | Not-determined |
| 18-101-final-viral-combined_k141_6781\|\|full | Low-quality |
| 18-101-final-viral-combined_k141_122761\|\|full | Not-determined |
| 18-101-final-viral-combined_k141_254315\|\|full | Not-determined |
| 18-101-final-viral-combined_k141_368679\|\|full | Low-quality |
| 18-101-final-viral-combined_k141_254319\|\|full | Low-quality |
| 18-101-final-viral-combined_k141_170610\|\|full | Low-quality |
| 18-101-final-viral-combined_k141_122801\|\|full | Low-quality |
| 18-101-final-viral-combined_k141_56069\|\|full | Not-determined |
| 18-101-final-viral-combined_k141_170623\|\|full | Low-quality |
| 18-101-final-viral-combined_k141_204830\|\|full | Not-determined |
| 18-101-final-viral-combined_k141_385471\|\|full | Low-quality |
| 18-101-final-viral-combined_k141_335374\|\|full | Low-quality |
| 18-101-final-viral-combined_k141_254361\|\|full | Not-determined |
| 18-101-final-viral-combined_k141_88935\|\|full | Not-determined |
| 18-101-final-viral-combined_k141_303765\|\|full | Low-quality |
| 18-101-final-viral-combined_k141_335375\|\|full | Low-quality |
| 18-101-final-viral-combined_k141_56099\|\|full | Not-determined |
| 18-101-final-viral-combined_k141_303783\|\|full | Low-quality |
| 18-101-final-viral-combined_k141_320311\|\|full | Low-quality |
| 18-101-final-viral-combined_k141_220828\|\|full | Not-determined |
| 18-101-final-viral-combined_k141_188384\|\|full | Low-quality |
| 18-101-final-viral-combined_k141_188388\|\|full | Low-quality |
| 18-101-final-viral-combined_k141_39616\|\|full | Not-determined |
| 18-101-final-viral-combined_k141_104873\|\|full | Low-quality |
| 18-101-final-viral-combined_k141_204849\|\|full | Low-quality |
| 18-101-final-viral-combined_k141_320324\|\|full | Not-determined |
| 18-101-final-viral-combined_k141_188400\|\|full | Low-quality |
| 18-101-final-viral-combined_k141_154618\|\|full | Not-determined |
| 18-101-final-viral-combined_k141_320328\|\|full | Low-quality |
| 18-101-final-viral-combined_k141_353113\|\|full | Not-determined |
| 18-101-final-viral-combined_k141_23211\|\|full | Low-quality |
| 18-101-final-viral-combined_k141_320339\|\|full | Not-determined |
| 18-101-final-viral-combined_k141_39641\|\|full | Not-determined |
| 18-101-final-viral-combined_k141_269987\|\|full | Low-quality |
| 18-101-final-viral-combined_k141_220842\|\|full | Low-quality |
| 18-101-final-viral-combined_k141_237424\|\|full | Not-determined |
| 18-101-final-viral-combined_k141_220843\|\|full | Not-determined |

| | |
|---|---|
| 18-101-final-viral-combined_k141_122896\|\|full | Not-determined |
| 18-101-final-viral-combined_k141_254386\|\|full | Low-quality |
| 18-101-final-viral-combined_k141_303839\|\|full | Low-quality |
| 18-101-final-viral-combined_k141_353133\|\|full | Low-quality |
| 18-101-final-viral-combined_k141_56158\|\|full | Not-determined |
| 18-101-final-viral-combined_k141_138938\|\|full | Not-determined |
| 18-101-final-viral-combined_k141_6880\|\|full | Not-determined |
| 18-101-final-viral-combined_k141_122910\|\|full | Not-determined |
| 18-101-final-viral-combined_k141_368751\|\|full | Not-determined |
| 18-101-final-viral-combined_k141_23246\|\|full | Low-quality |
| 18-101-final-viral-combined_k141_353139\|\|full | Low-quality |
| 18-101-final-viral-combined_k141_237447\|\|full | Low-quality |
| 18-101-final-viral-combined_k141_335455\|\|full | Not-determined |
| 18-101-final-viral-combined_k141_270009\|\|full | Low-quality |
| 18-101-final-viral-combined_k141_71977\|\|full | Low-quality |
| 18-101-final-viral-combined_k141_122914\|\|full | Low-quality |
| 18-101-final-viral-combined_k141_270013\|\|full | Low-quality |
| 18-101-final-viral-combined_k141_353148\|\|full | Not-determined |
| 18-101-final-viral-combined_k141_320362\|\|full | Low-quality |
| 18-101-final-viral-combined_k141_188429\|\|full | Not-determined |
| 18-101-final-viral-combined_k141_303854\|\|full | Low-quality |
| 18-101-final-viral-combined_k141_188439\|\|full | Not-determined |
| 18-101-final-viral-combined_k141_204891\|\|full | Not-determined |
| 18-101-final-viral-combined_k141_303870\|\|full | Low-quality |
| 18-101-final-viral-combined_k141_154694\|\|full | Low-quality |
| 18-101-final-viral-combined_k141_303884\|\|full | Not-determined |
| 18-101-final-viral-combined_k141_303886\|\|full | Not-determined |
| 18-101-final-viral-combined_k141_368784\|\|full | Low-quality |
| 18-101-final-viral-combined_k141_154710\|\|full | Not-determined |
| 18-101-final-viral-combined_k141_88984\|\|full | Not-determined |
| 18-101-final-viral-combined_k141_237518\|\|full | Low-quality |
| 18-101-final-viral-combined_k141_353179\|\|full | Low-quality |
| 18-101-final-viral-combined_k141_368811\|\|full | Not-determined |
| 18-101-final-viral-combined_k141_188475\|\|full | Not-determined |
| 18-101-final-viral-combined_k141_286614\|\|full | Not-determined |
| 18-101-final-viral-combined_k141_6934\|\|full | Not-determined |
| 18-101-final-viral-combined_k141_170761\|\|full | Low-quality |
| 18-101-final-viral-combined_k141_237542\|\|full | Low-quality |
| 18-101-final-viral-combined_k141_254472\|\|full | Low-quality |
| 18-101-final-viral-combined_k141_270026\|\|full | Low-quality |
| 18-101-final-viral-combined_k141_154731\|\|full | Low-quality |
| 18-101-final-viral-combined_k141_89012\|\|full | Low-quality |
| 18-101-final-viral-combined_k141_237555\|\|full | Low-quality |
| 18-101-final-viral-combined_k141_56221\|\|full | Low-quality |
| 18-101-final-viral-combined_k141_154742\|\|full | Low-quality |
| 18-101-final-viral-combined_k141_72054\|\|full | Not-determined |
| 18-106_k141_140743 | Complete |
| 18-101-final-viral-combined_k141_353198\|\|full | Not-determined |
| 18-101-final-viral-combined_k141_204929\|\|full | Not-determined |
| 18-101-final-viral-combined_k141_286642\|\|full | Low-quality |
| 18-101-final-viral-combined_k141_139011\|\|full | Not-determined |
| 18-101-final-viral-combined_k141_254511\|\|full | Low-quality |
| 18-101-final-viral-combined_k141_220911\|\|full | Low-quality |
| 18-101-final-viral-combined_k141_204943\|\|full | Not-determined |
| 18-101-final-viral-combined_k141_56246\|\|full | Low-quality |
| 18-101-final-viral-combined_k141_39709\|\|full | Low-quality |
| 18-101-final-viral-combined_k141_188509\|\|full | Not-determined |
| 18-101-final-viral-combined_k141_6985\|\|full | Not-determined |
| 18-101-final-viral-combined_k141_170816\|\|full | Not-determined |
| 18-101-final-viral-combined_k141_320496\|\|full | Low-quality |
| 18-101-final-viral-combined_k141_123011\|\|full | Low-quality |
| 18-101-final-viral-combined_k141_320506\|\|full | Not-determined |
| 18-101-final-viral-combined_k141_303925\|\|full | Low-quality |
| 18-101-final-viral-combined_k141_89055\|\|full | Low-quality |
| 18-101-final-viral-combined_k141_335515\|\|full | Not-determined |
| 18-101-final-viral-combined_k141_303928\|\|full | Not-determined |
| 18-101-final-viral-combined_k141_188532\|\|full | Low-quality |
| 18-101-final-viral-combined_k141_7027\|\|full | Low-quality |
| 18-101-final-viral-combined_k141_286676\|\|full | Not-determined |
| 18-101-final-viral-combined_k141_154779\|\|full | Not-determined |
| 18-101-final-viral-combined_k141_237598\|\|full | Low-quality |
| 18-101-final-viral-combined_k141_7045\|\|full | Low-quality |
| 18-101-final-viral-combined_k141_303952\|\|full | Low-quality |
| 18-101-final-viral-combined_k141_237609\|\|full | Not-determined |
| 18-101-final-viral-combined_k141_188545\|\|full | Not-determined |
| 18-101-final-viral-combined_k141_353246\|\|full | Not-determined |
| 18-101-final-viral-combined_k141_303981\|\|full | Not-determined |
| 18-101-final-viral-combined_k141_7081\|\|full | Not-determined |
| 18-101-final-viral-combined_k141_286747\|\|full | Not-determined |
| 18-101-final-viral-combined_k141_205003\|\|full | Not-determined |
| 18-101-final-viral-combined_k141_286781\|\|full | Low-quality |
| 18-101-final-viral-combined_k141_237642\|\|full | Not-determined |
| 18-101-final-viral-combined_k141_139149\|\|full | Not-determined |
| 18-101-final-viral-combined_k141_335579\|\|full | Not-determined |
| 18-101-final-viral-combined_k141_188615\|\|full | Not-determined |
| 18-101-final-viral-combined_k141_188622\|\|full | Low-quality |
| 18-101-final-viral-combined_k141_286810\|\|full | Not-determined |
| 18-101-final-viral-combined_k141_353292\|\|full | Not-determined |
| 18-101-final-viral-combined_k141_205018\|\|full | Low-quality |
| 18-101-final-viral-combined_k141_72097\|\|full | Not-determined |
| 18-101-final-viral-combined_k141_270130\|\|full | Not-determined |
| 18-101-final-viral-combined_k141_237668\|\|full | Not-determined |
| 18-101-final-viral-combined_k141_23324\|\|full | Low-quality |
| 18-101-final-viral-combined_k141_270156\|\|full | Low-quality |
| 18-101-final-viral-combined_k141_385639\|\|full | Not-determined |
| 18-101-final-viral-combined_k141_139183\|\|full | Low-quality |
| 18-101-final-viral-combined_k141_89144\|\|full | Low-quality |
| 18-101-final-viral-combined_k141_72127\|\|full | Low-quality |
| 18-101-final-viral-combined_k141_72128\|\|full | Not-determined |
| 18-101-final-viral-combined_k141_254645\|\|full | Not-determined |
| 18-101-final-viral-combined_k141_139193\|\|full | Low-quality |
| 18-101-final-viral-combined_k141_237695\|\|full | Not-determined |
| 18-101-final-viral-combined_k141_254654\|\|full | Not-determined |
| 18-101-final-viral-combined_k141_23362\|\|full | Not-determined |
| 18-101-final-viral-combined_k141_270183\|\|full | Not-determined |
| 18-101-final-viral-combined_k141_188651\|\|full | Not-determined |
| 18-101-final-viral-combined_k141_254665\|\|full | Not-determined |
| 18-101-final-viral-combined_k141_286854\|\|full | Not-determined |
| 18-101-final-viral-combined_k141_72143\|\|full | Not-determined |
| 18-101-final-viral-combined_k141_72148\|\|full | Not-determined |
| 18-101-final-viral-combined_k141_188668\|\|full | Low-quality |
| 18-101-final-viral-combined_k141_254690\|\|full | Not-determined |
| 18-101-final-viral-combined_k141_89159\|\|full | Not-determined |
| 18-101-final-viral-combined_k141_385689\|\|full | Low-quality |
| 18-101-final-viral-combined_k141_89161\|\|full | Low-quality |
| 18-101-final-viral-combined_k141_237750\|\|full | Low-quality |
| 18-101-final-viral-combined_k141_385707\|\|full | Low-quality |
| 18-101-final-viral-combined_k141_205067\|\|full | Not-determined |
| 18-101-final-viral-combined_k141_353336\|\|full | Not-determined |
| 18-101-final-viral-combined_k141_205068\|\|full | Not-determined |
| 18-101-final-viral-combined_k141_39761\|\|full | Low-quality |
| 18-101-final-viral-combined_k141_154896\|\|full | Not-determined |
| 18-101-final-viral-combined_k141_123174\|\|full | Low-quality |
| 18-101-final-viral-combined_k141_188697\|\|full | Not-determined |
| 18-106-final-viral-combined_k141_140743\|\|full | High-quality |
| 18-101-final-viral-combined_k141_89203\|\|full | Not-determined |
| 18-101-final-viral-combined_k141_286923\|\|full | Not-determined |
| 18-101-final-viral-combined_k141_56444\|\|full | Low-quality |
| 18-101-final-viral-combined_k141_335664\|\|full | Low-quality |
| 18-101-final-viral-combined_k141_220987\|\|full | Low-quality |
| 18-101-final-viral-combined_k141_104942\|\|full | Not-determined |
| 18-101-final-viral-combined_k141_270302\|\|full | Low-quality |
| 18-101-final-viral-combined_k141_39805\|\|full | Not-determined |
| 18-101-final-viral-combined_k141_56461\|\|full | Low-quality |
| 18-101-final-viral-combined_k141_7160\|\|full | Not-determined |
| 18-101-final-viral-combined_k141_170988\|\|full | Low-quality |
| 18-101-final-viral-combined_k141_123208\|\|full | Low-quality |
| 18-101-final-viral-combined_k141_56488\|\|full | Low-quality |
| 18-101-final-viral-combined_k141_56490\|\|full | Not-determined |
| 18-101-final-viral-combined_k141_385740\|\|full | Not-determined |
| 18-101-final-viral-combined_k141_335678\|\|full | Low-quality |
| 18-101-final-viral-combined_k141_286946\|\|full | Not-determined |
| 18-101-final-viral-combined_k141_56493\|\|full | Low-quality |
| 18-101-final-viral-combined_k141_23477\|\|full | Not-determined |
| 18-101-final-viral-combined_k141_23489\|\|full | Not-determined |
| 18-101-final-viral-combined_k141_89250\|\|full | Low-quality |
| 18-101-final-viral-combined_k141_237828\|\|full | Low-quality |
| 18-101-final-viral-combined_k141_320815\|\|full | Low-quality |
| 18-101-final-viral-combined_k141_72223\|\|full | Not-determined |
| 18-101-final-viral-combined_k141_320823\|\|full | Not-determined |
| 18-101-final-viral-combined_k141_188776\|\|full | Low-quality |
| 18-101-final-viral-combined_k141_139369\|\|full | Low-quality |
| 18-101-final-viral-combined_k141_286996\|\|full | Not-determined |
| 18-101-final-viral-combined_k141_171035\|\|full | Low-quality |
| 18-101-final-viral-combined_k141_39814\|\|full | Not-determined |
| 18-101-final-viral-combined_k141_286998\|\|full | Low-quality |
| 18-101-final-viral-combined_k141_23497\|\|full | Not-determined |
| 18-101-final-viral-combined_k141_287014\|\|full | Low-quality |
| 18-101-final-viral-combined_k141_139377\|\|full | Not-determined |
| 18-101-final-viral-combined_k141_368962\|\|full | Not-determined |
| 18-101-final-viral-combined_k141_154996\|\|full | Not-determined |
| 18-101-final-viral-combined_k141_205178\|\|full | Low-quality |
| 18-101-final-viral-combined_k141_368990\|\|full | Low-quality |
| 18-101-final-viral-combined_k141_105032\|\|full | Low-quality |
| 18-101-final-viral-combined_k141_7169\|\|full | Low-quality |
| 18-101-final-viral-combined_k141_171074\|\|full | Low-quality |
| 18-101-final-viral-combined_k141_39843\|\|full | Low-quality |
| 18-101-final-viral-combined_k141_7191\|\|full | Low-quality |
| 18-101-final-viral-combined_k141_39849\|\|full | Not-determined |
| 18-101-final-viral-combined_k141_39855\|\|full | Not-determined |
| 18-101-final-viral-combined_k141_205225\|\|full | Not-determined |
| 18-101-final-viral-combined_k141_72333\|\|full | Low-quality |
| 18-101-final-viral-combined_k141_89361\|\|full | Low-quality |
| 18-101-final-viral-combined_k141_205232\|\|full | Not-determined |
| 18-101-final-viral-combined_k141_72339\|\|full | Not-determined |
| 18-101-final-viral-combined_k141_89367\|\|full | Low-quality |
| 18-101-final-viral-combined_k141_205235\|\|full | Low-quality |
| 18-101-final-viral-combined_k141_188852\|\|full | Not-determined |
| 18-101-final-viral-combined_k141_155036\|\|full | Not-determined |
| 18-101-final-viral-combined_k141_56520\|\|full | Not-determined |
| 18-101-final-viral-combined_k141_155044\|\|full | Not-determined |
| 18-101-final-viral-combined_k141_23553\|\|full | Not-determined |
| 18-101-final-viral-combined_k141_155048\|\|full | Low-quality |
| 18-101-final-viral-combined_k141_171103\|\|full | Not-determined |
| 18-101-final-viral-combined_k141_221154\|\|full | Low-quality |
| 18-101-final-viral-combined_k141_335825\|\|full | Not-determined |
| 18-101-final-viral-combined_k141_105078\|\|full | Low-quality |
| 18-101-final-viral-combined_k141_105082\|\|full | Low-quality |
| 18-101-final-viral-combined_k141_56565\|\|full | Low-quality |
| 18-101-final-viral-combined_k141_270381\|\|full | Low-quality |
| 18-101-final-viral-combined_k141_221181\|\|full | Low-quality |
| 18-101-final-viral-combined_k141_320968\|\|full | Not-determined |
| 18-101-final-viral-combined_k141_72382\|\|full | Low-quality |
| 18-101-final-viral-combined_k141_304182\|\|full | Low-quality |
| 18-101-final-viral-combined_k141_39945\|\|full | Not-determined |
| 18-101-final-viral-combined_k141_353480\|\|full | Low-quality |
| 18-101-final-viral-combined_k141_238007\|\|full | Low-quality |
| 18-101-final-viral-combined_k141_335855\|\|full | Not-determined |
| 18-101-final-viral-combined_k141_304191\|\|full | Not-determined |
| 18-101-final-viral-combined_k141_123338\|\|full | Low-quality |
| 18-101-final-viral-combined_k141_320991\|\|full | Not-determined |
| 18-101-final-viral-combined_k141_72395\|\|full | Not-determined |
| 18-101-final-viral-combined_k141_155051\|\|full | Low-quality |
| 18-101-final-viral-combined_k141_205299\|\|full | Low-quality |
| 18-101-final-viral-combined_k141_123352\|\|full | Low-quality |
| 18-101-final-viral-combined_k141_23558\|\|full | Low-quality |
| 18-101-final-viral-combined_k141_254826\|\|full | Low-quality |
| 18-101-final-viral-combined_k141_72409\|\|full | Low-quality |
| 18-101-final-viral-combined_k141_123357\|\|full | Not-determined |
| 18-101-final-viral-combined_k141_105116\|\|full | Low-quality |
| 18-101-final-viral-combined_k141_171111\|\|full | Low-quality |
| 18-101-final-viral-combined_k141_72421\|\|full | Low-quality |
| 18-101-final-viral-combined_k141_254840\|\|full | Not-determined |
| 18-101-final-viral-combined_k141_139442\|\|full | Low-quality |
| 18-101-final-viral-combined_k141_353510\|\|full | Not-determined |
| 18-101-final-viral-combined_k141_7307\|\|full | Low-quality |
| 18-101-final-viral-combined_k141_369073\|\|full | Low-quality |
| 18-101-final-viral-combined_k141_23590\|\|full | Not-determined |
| 18-101-final-viral-combined_k141_304236\|\|full | Not-determined |
| 18-101-final-viral-combined_k141_123387\|\|full | Not-determined |
| 18-101-final-viral-combined_k141_254858\|\|full | Not-determined |
| 18-101-final-viral-combined_k141_188911\|\|full | Low-quality |
| 18-101-final-viral-combined_k141_221262\|\|full | Not-determined |
| 18-101-final-viral-combined_k141_56592\|\|full | Low-quality |
| 18-101-final-viral-combined_k141_254881\|\|full | Low-quality |
| 18-101-final-viral-combined_k141_123406\|\|full | Not-determined |
| 18-101-final-viral-combined_k141_205326\|\|full | Low-quality |
| 18-101-final-viral-combined_k141_155113\|\|full | Not-determined |
| 18-101-final-viral-combined_k141_23609\|\|full | Not-determined |
| 18-101-final-viral-combined_k141_254887\|\|full | Low-quality |
| 18-101-final-viral-combined_k141_56609\|\|full | Not-determined |
| 18-101-final-viral-combined_k141_89473\|\|full | Low-quality |
| 18-101-final-viral-combined_k141_89478\|\|full | Not-determined |
| 18-101-final-viral-combined_k141_123434\|\|full | Low-quality |
| 18-101-final-viral-combined_k141_353558\|\|full | Not-determined |
| 18-101-final-viral-combined_k141_385968\|\|full | Not-determined |
| 18-101-final-viral-combined_k141_287033\|\|full | Low-quality |
| 18-101-final-viral-combined_k141_321061\|\|full | Not-determined |
| 18-101-final-viral-combined_k141_238083\|\|full | Not-determined |
| 18-101-final-viral-combined_k141_287038\|\|full | Low-quality |
| 18-101-final-viral-combined_k141_105166\|\|full | Not-determined |
| 18-101-final-viral-combined_k141_40023\|\|full | Not-determined |
| 18-101-final-viral-combined_k141_40025\|\|full | Not-determined |
| 18-101-final-viral-combined_k141_369143\|\|full | Not-determined |
| 18-101-final-viral-combined_k141_254921\|\|full | Not-determined |
| 18-101-final-viral-combined_k141_89500\|\|full | Low-quality |
| 18-101-final-viral-combined_k141_56627\|\|full | Not-determined |
| 18-101-final-viral-combined_k141_7397\|\|full | Not-determined |
| 18-101-final-viral-combined_k141_369146\|\|full | Low-quality |
| 18-101-final-viral-combined_k141_254935\|\|full | Not-determined |
| 18-101-final-viral-combined_k141_238091\|\|full | Not-determined |
| 18-101-final-viral-combined_k141_7400\|\|full | Not-determined |
| 18-101-final-viral-combined_k141_353589\|\|full | Low-quality |
| 18-101-final-viral-combined_k141_155154\|\|full | Not-determined |
| 18-101-final-viral-combined_k141_155170\|\|full | Low-quality |
| 18-101-final-viral-combined_k141_238111\|\|full | Low-quality |
| 18-101-final-viral-combined_k141_386016\|\|full | Not-determined |
| 18-101-final-viral-combined_k141_171227\|\|full | Not-determined |
| 18-101-final-viral-combined_k141_335887\|\|full | Low-quality |
| 18-101-final-viral-combined_k141_238122\|\|full | Not-determined |
| 18-101-final-viral-combined_k141_123512\|\|full | Low-quality |
| 18-101-final-viral-combined_k141_270567\|\|full | Low-quality |
| 18-101-final-viral-combined_k141_238125\|\|full | Not-determined |
| 18-101-final-viral-combined_k141_171240\|\|full | Low-quality |
| 18-101-final-viral-combined_k141_386046\|\|full | Not-determined |
| 18-101-final-viral-combined_k141_23655\|\|full | Low-quality |
| 18-101-final-viral-combined_k141_321178\|\|full | Low-quality |
| 18-101-final-viral-combined_k141_335918\|\|full | Not-determined |
| 18-101-final-viral-combined_k141_56710\|\|full | Not-determined |
| 18-101-final-viral-combined_k141_255003\|\|full | Not-determined |
| 18-101-final-viral-combined_k141_386079\|\|full | Not-determined |
| 18-101-final-viral-combined_k141_56729\|\|full | Not-determined |
| 18-101-final-viral-combined_k141_56737\|\|full | Low-quality |
| 18-101-final-viral-combined_k141_105294\|\|full | Low-quality |
| 18-101-final-viral-combined_k141_56745\|\|full | Low-quality |
| 18-101-final-viral-combined_k141_155188\|\|full | Low-quality |
| 18-101-final-viral-combined_k141_205474\|\|full | Not-determined |
| 17-108-final-viral-combined_k141_215723\|\|full | High-quality |
| 18-101-final-viral-combined_k141_221393\|\|full | Low-quality |
| 18-101-final-viral-combined_k141_335950\|\|full | Low-quality |
| 18-101-final-viral-combined_k141_238188\|\|full | Not-determined |
| 18-101-final-viral-combined_k141_221399\|\|full | Low-quality |
| 18-101-final-viral-combined_k141_221401\|\|full | Not-determined |
| 18-101-final-viral-combined_k141_105311\|\|full | Not-determined |
| 18-101-final-viral-combined_k141_255037\|\|full | Low-quality |
| 18-101-final-viral-combined_k141_304343\|\|full | Not-determined |
| 18-101-final-viral-combined_k141_105325\|\|full | Not-determined |
| 18-101-final-viral-combined_k141_287147\|\|full | Not-determined |
| 18-101-final-viral-combined_k141_321236\|\|full | Not-determined |
| 18-101-final-viral-combined_k141_139608\|\|full | Not-determined |
| 18-101-final-viral-combined_k141_123535\|\|full | Low-quality |
| 18-101-final-viral-combined_k141_304369\|\|full | Not-determined |
| 18-101-final-viral-combined_k141_155203\|\|full | Low-quality |
| 18-101-final-viral-combined_k141_123536\|\|full | Low-quality |
| 18-101-final-viral-combined_k141_139619\|\|full | Low-quality |
| 18-101-final-viral-combined_k141_56794\|\|full | Low-quality |
| 18-101-final-viral-combined_k141_123547\|\|full | Low-quality |
| 18-101-final-viral-combined_k141_287155\|\|full | Low-quality |
| 18-101-final-viral-combined_k141_270668\|\|full | Not-determined |
| 18-101-final-viral-combined_k141_72563\|\|full | Low-quality |
| 18-101-final-viral-combined_k141_188991\|\|full | Low-quality |
| 18-101-final-viral-combined_k141_123553\|\|full | Low-quality |
| 18-101-final-viral-combined_k141_238221\|\|full | Not-determined |
| 18-101-final-viral-combined_k141_369276\|\|full | Not-determined |
| 18-101-final-viral-combined_k141_72565\|\|full | Not-determined |
| 18-101-final-viral-combined_k141_221449\|\|full | Low-quality |
| 18-101-final-viral-combined_k141_321268\|\|full | Low-quality |
| 18-101-final-viral-combined_k141_369279\|\|full | Low-quality |
| 18-101-final-viral-combined_k141_270684\|\|full | Not-determined |
| 18-101-final-viral-combined_k141_72579\|\|full | Not-determined |
| 18-101-final-viral-combined_k141_205530\|\|full | Not-determined |
| 18-101-final-viral-combined_k141_238241\|\|full | Not-determined |
| 18-101-final-viral-combined_k141_139670\|\|full | Not-determined |
| 18-101-final-viral-combined_k141_105385\|\|full | Not-determined |
| 18-101-final-viral-combined_k141_369291\|\|full | Low-quality |
| 18-101-final-viral-combined_k141_89607\|\|full | Not-determined |
| 18-101-final-viral-combined_k141_270711\|\|full | Not-determined |
| 18-101-final-viral-combined_k141_287212\|\|full | Low-quality |
| 18-101-final-viral-combined_k141_335965\|\|full | Low-quality |
| 18-101-final-viral-combined_k141_40166\|\|full | Not-determined |
| 18-101-final-viral-combined_k141_171343\|\|full | Low-quality |
| 18-101-final-viral-combined_k141_221488\|\|full | Low-quality |
| 18-101-final-viral-combined_k141_72605\|\|full | Not-determined |
| 18-101-final-viral-combined_k141_205552\|\|full | Low-quality |
| 18-101-final-viral-combined_k141_23694\|\|full | Not-determined |
| 18-101-final-viral-combined_k141_40170\|\|full | Low-quality |
| 18-101-final-viral-combined_k141_23695\|\|full | Not-determined |
| 18-101-final-viral-combined_k141_56856\|\|full | Low-quality |
| 18-101-final-viral-combined_k141_205553\|\|full | Low-quality |
| 18-101-final-viral-combined_k141_155280\|\|full | Not-determined |
| 18-101-final-viral-combined_k141_23698\|\|full | Low-quality |
| 18-101-final-viral-combined_k141_369324\|\|full | Not-determined |
| 18-101-final-viral-combined_k141_221507\|\|full | Low-quality |
| 18-101-final-viral-combined_k141_105412\|\|full | Low-quality |
| 18-101-final-viral-combined_k141_40195\|\|full | Low-quality |
| 18-101-final-viral-combined_k141_336004\|\|full | Low-quality |
| 18-101-final-viral-combined_k141_155331\|\|full | Not-determined |
| 18-101-final-viral-combined_k141_221529\|\|full | Not-determined |
| 18-101-final-viral-combined_k141_7602\|\|full | Not-determined |
| 18-101-final-viral-combined_k141_40201\|\|full | Not-determined |
| 17-109-final-viral-combined_k141_359190\|\|full | Medium-quality |
| 18-101-final-viral-combined_k141_89651\|\|full | Not-determined |
| 18-101-final-viral-combined_k141_386146\|\|full | Not-determined |
| 18-101-final-viral-combined_k141_105434\|\|full | Low-quality |
| 18-101-final-viral-combined_k141_321302\|\|full | Not-determined |
| 18-101-final-viral-combined_k141_155374\|\|full | Not-determined |
| 18-101-final-viral-combined_k141_304404\|\|full | Not-determined |
| 18-101-final-viral-combined_k141_189101\|\|full | Not-determined |
| 18-109-final-viral-combined_k141_14267\|\|full | Medium-quality |
| 18-101-final-viral-combined_k141_7635\|\|full | Low-quality |
| 18-101-final-viral-combined_k141_105444\|\|full | Low-quality |
| 18-101-final-viral-combined_k141_171395\|\|full | Not-determined |
| 18-101-final-viral-combined_k141_321328\|\|full | Low-quality |
| 18-101-final-viral-combined_k141_221553\|\|full | Not-determined |
| 18-101-final-viral-combined_k141_123653\|\|full | Low-quality |
| 18-101-final-viral-combined_k141_56913\|\|full | Not-determined |
| 18-101-final-viral-combined_k141_155398\|\|full | Low-quality |
| 18-101-final-viral-combined_k141_189145\|\|full | Low-quality |
| 18-101-final-viral-combined_k141_72675\|\|full | Not-determined |
| 18-101-final-viral-combined_k141_23765\|\|full | Low-quality |
| 18-101-final-viral-combined_k141_105471\|\|full | Low-quality |
| 18-101-final-viral-combined_k141_205654\|\|full | Not-determined |
| 18-101-final-viral-combined_k141_7669\|\|full | Not-determined |
| 18-101-final-viral-combined_k141_386207\|\|full | Not-determined |
| 18-101-final-viral-combined_k141_321354\|\|full | Low-quality |
| 18-101-final-viral-combined_k141_321361\|\|full | Not-determined |
| 18-101-final-viral-combined_k141_56934\|\|full | Low-quality |
| 18-101-final-viral-combined_k141_189176\|\|full | Low-quality |
| 18-101-final-viral-combined_k141_353801\|\|full | Not-determined |
| 18-101-final-viral-combined_k141_189195\|\|full | Not-determined |
| 18-101-final-viral-combined_k141_353806\|\|full | Low-quality |
| 18-101-final-viral-combined_k141_23795\|\|full | Not-determined |
| 18-101-final-viral-combined_k141_56955\|\|full | Low-quality |
| 18-101-final-viral-combined_k141_304481\|\|full | Low-quality |
| 18-101-final-viral-combined_k141_56964\|\|full | Not-determined |
| 18-101-final-viral-combined_k141_369407\|\|full | Not-determined |
| 18-101-final-viral-combined_k141_287343\|\|full | Not-determined |
| 18-101-final-viral-combined_k141_386248\|\|full | Low-quality |
| 18-101-final-viral-combined_k141_304491\|\|full | Not-determined |
| 18-101-final-viral-combined_k141_105476\|\|full | Low-quality |
| 18-101-final-viral-combined_k141_221630\|\|full | Low-quality |
| 18-101-final-viral-combined_k141_171472\|\|full | Not-determined |
| 18-101-final-viral-combined_k141_304508\|\|full | Low-quality |
| 18-101-final-viral-combined_k141_23829\|\|full | Not-determined |
| 18-101-final-viral-combined_k141_89728\|\|full | Not-determined |
| 18-101-final-viral-combined_k141_386270\|\|full | Not-determined |
| 18-101-final-viral-combined_k141_287376\|\|full | Not-determined |
| 18-101-final-viral-combined_k141_221644\|\|full | Low-quality |
| 18-101-final-viral-combined_k141_105504\|\|full | Low-quality |
| 18-101-final-viral-combined_k141_287386\|\|full | Not-determined |
| 18-101-final-viral-combined_k141_139765\|\|full | Low-quality |
| 18-101-final-viral-combined_k141_171493\|\|full | Low-quality |
| 18-101-final-viral-combined_k141_336087\|\|full | Not-determined |
| 18-101-final-viral-combined_k141_221653\|\|full | Not-determined |
| 18-101-final-viral-combined_k141_221654\|\|full | Not-determined |
| 18-101-final-viral-combined_k141_123711\|\|full | Low-quality |
| 18-101-final-viral-combined_k141_171494\|\|full | Low-quality |
| 18-101-final-viral-combined_k141_139771\|\|full | Low-quality |
| 18-101-final-viral-combined_k141_123715\|\|full | Not-determined |
| 18-101-final-viral-combined_k141_139774\|\|full | Not-determined |
| 18-101-final-viral-combined_k141_221664\|\|full | Not-determined |
| 18-101-final-viral-combined_k141_369483\|\|full | Not-determined |
| 18-101-final-viral-combined_k141_139779\|\|full | Low-quality |
| 18-101-final-viral-combined_k141_139783\|\|full | Not-determined |
| 18-101-final-viral-combined_k141_155451\|\|full | Not-determined |
| 18-101-final-viral-combined_k141_139785\|\|full | Not-determined |
| 18-101-final-viral-combined_k141_7718\|\|full | Low-quality |
| 18-101-final-viral-combined_k141_205769\|\|full | Low-quality |
| 18-101-final-viral-combined_k141_287402\|\|full | Not-determined |
| 18-101-final-viral-combined_k141_139789\|\|full | Low-quality |
| 18-101-final-viral-combined_k141_353876\|\|full | Low-quality |
| 18-101-final-viral-combined_k141_205779\|\|full | Low-quality |
| 18-101-final-viral-combined_k141_304570\|\|full | Low-quality |
| 18-101-final-viral-combined_k141_386329\|\|full | Not-determined |
| 18-101-final-viral-combined_k141_205783\|\|full | Low-quality |
| 18-101-final-viral-combined_k141_105545\|\|full | Not-determined |
| 18-101-final-viral-combined_k141_304575\|\|full | Low-quality |
| 18-101-final-viral-combined_k141_336112\|\|full | Low-quality |
| 18-101-final-viral-combined_k141_23870\|\|full | Not-determined |
| 18-101-final-viral-combined_k141_369505\|\|full | Low-quality |
| 18-101-final-viral-combined_k141_304590\|\|full | Not-determined |
| 18-101-final-viral-combined_k141_105556\|\|full | Not-determined |
| 18-101-final-viral-combined_k141_189269\|\|full | Low-quality |
| 18-101-final-viral-combined_k141_205793\|\|full | Low-quality |
| 18-101-final-viral-combined_k141_336130\|\|full | Low-quality |
| 18-101-final-viral-combined_k141_89778\|\|full | Not-determined |
| 18-101-final-viral-combined_k141_369517\|\|full | Low-quality |
| 18-101-final-viral-combined_k141_221700\|\|full | Low-quality |
| 18-101-final-viral-combined_k141_369520\|\|full | Not-determined |
| 18-101-final-viral-combined_k141_139808\|\|full | Not-determined |
| 18-101-final-viral-combined_k141_23896\|\|full | Low-quality |
| 18-101-final-viral-combined_k141_139812\|\|full | Not-determined |
| 18-101-final-viral-combined_k141_369537\|\|full | Not-determined |
| 18-101-final-viral-combined_k141_7772\|\|full | Low-quality |
| 18-101-final-viral-combined_k141_123774\|\|full | Low-quality |
| 18-101-final-viral-combined_k141_336145\|\|full | Not-determined |
| 18-101-final-viral-combined_k141_23926\|\|full | Not-determined |
| 18-101-final-viral-combined_k141_369564\|\|full | Not-determined |
| 18-101-final-viral-combined_k141_221728\|\|full | Low-quality |
| 18-101-final-viral-combined_k141_221729\|\|full | Not-determined |
| 18-101-final-viral-combined_k141_105608\|\|full | Low-quality |
| 18-101-final-viral-combined_k141_386392\|\|full | Low-quality |
| 18-101-final-viral-combined_k141_23944\|\|full | Not-determined |
| 18-101-final-viral-combined_k141_336180\|\|full | Not-determined |
| 18-101-final-viral-combined_k141_7821\|\|full | Not-determined |
| 18-101-final-viral-combined_k141_72763\|\|full | Low-quality |
| 18-109-final-viral-combined_k141_134452\|\|full | Medium-quality |
| 18-101-final-viral-combined_k141_89845\|\|full | Low-quality |
| 18-101-final-viral-combined_k141_221752\|\|full | Low-quality |
| 18-101-final-viral-combined_k141_353978\|\|full | Not-determined |
| 18-101-final-viral-combined_k141_23957\|\|full | Low-quality |
| 18-101-final-viral-combined_k141_221757\|\|full | Not-determined |
| 18-101-final-viral-combined_k141_89851\|\|full | Low-quality |
| 18-101-final-viral-combined_k141_57130\|\|full | Low-quality |
| 18-101-final-viral-combined_k141_72771\|\|full | Low-quality |
| 18-101-final-viral-combined_k141_40359\|\|full | Not-determined |
| 18-101-final-viral-combined_k141_72777\|\|full | Low-quality |
| 18-101-final-viral-combined_k141_336187\|\|full | Not-determined |
| 18-101-final-viral-combined_k141_155495\|\|full | Low-quality |
| 18-101-final-viral-combined_k141_205886\|\|full | Low-quality |
| 18-101-final-viral-combined_k141_105650\|\|full | Low-quality |
| 18-101-final-viral-combined_k141_89876\|\|full | Low-quality |
| 18-101-final-viral-combined_k141_72814\|\|full | Not-determined |
| 18-101-final-viral-combined_k141_270983\|\|full | Not-determined |
| 18-101-final-viral-combined_k141_72817\|\|full | Not-determined |
| 18-101-final-viral-combined_k141_171534\|\|full | Not-determined |
| 18-101-final-viral-combined_k141_238300\|\|full | Not-determined |
| 18-101-final-viral-combined_k141_221792\|\|full | Not-determined |
| 18-101-final-viral-combined_k141_105698\|\|full | Low-quality |
| 18-101-final-viral-combined_k141_386415\|\|full | Low-quality |
| 18-101-final-viral-combined_k141_369607\|\|full | Low-quality |
| 18-101-final-viral-combined_k141_270991\|\|full | Low-quality |
| 18-101-final-viral-combined_k141_155544\|\|full | Not-determined |
| 18-101-final-viral-combined_k141_72854\|\|full | Low-quality |
| 18-101-final-viral-combined_k141_386428\|\|full | Not-determined |
| 18-101-final-viral-combined_k141_155567\|\|full | Not-determined |
| 18-101-final-viral-combined_k141_271001\|\|full | Low-quality |
| 18-101-final-viral-combined_k141_189438\|\|full | Low-quality |
| 18-101-final-viral-combined_k141_23998\|\|full | Low-quality |
| 18-101-final-viral-combined_k141_238335\|\|full | Low-quality |
| 18-101-final-viral-combined_k141_155586\|\|full | Not-determined |
| 18-101-final-viral-combined_k141_221834\|\|full | Not-determined |
| 18-101-final-viral-combined_k141_354024\|\|full | Not-determined |
| 18-101-final-viral-combined_k141_123886\|\|full | Low-quality |
| 18-101-final-viral-combined_k141_271026\|\|full | Low-quality |
| 18-101-final-viral-combined_k141_155602\|\|full | Not-determined |
| 18-101-final-viral-combined_k141_238357\|\|full | Not-determined |
| 18-101-final-viral-combined_k141_155604\|\|full | Not-determined |
| 18-101-final-viral-combined_k141_105759\|\|full | Not-determined |
| 18-101-final-viral-combined_k141_155606\|\|full | Low-quality |
| 18-101-final-viral-combined_k141_354048\|\|full | Not-determined |
| 18-101-final-viral-combined_k141_189459\|\|full | Not-determined |
| 18-101-final-viral-combined_k141_386492\|\|full | Low-quality |
| 18-101-final-viral-combined_k141_57147\|\|full | Not-determined |
| 18-101-final-viral-combined_k141_221863\|\|full | Not-determined |
| 18-101-final-viral-combined_k141_336285\|\|full | Low-quality |
| 18-101-final-viral-combined_k141_105773\|\|full | Low-quality |
| 18-101-final-viral-combined_k141_238376\|\|full | Low-quality |
| 18-101-final-viral-combined_k141_57149\|\|full | Low-quality |
| 18-101-final-viral-combined_k141_155639\|\|full | Not-determined |
| 18-101-final-viral-combined_k141_139972\|\|full | Not-determined |
| 18-101-final-viral-combined_k141_321604\|\|full | Low-quality |
| 18-101-final-viral-combined_k141_287548\|\|full | Low-quality |
| 18-101-final-viral-combined_k141_105783\|\|full | Low-quality |
| 18-101-final-viral-combined_k141_89958\|\|full | Not-determined |
| 18-101-final-viral-combined_k141_255309\|\|full | Low-quality |
| 18-101-final-viral-combined_k141_40501\|\|full | Low-quality |
| 18-101-final-viral-combined_k141_238388\|\|full | Not-determined |
| 18-101-final-viral-combined_k141_255314\|\|full | Not-determined |
| 18-101-final-viral-combined_k141_7963\|\|full | Not-determined |
| 18-101-final-viral-combined_k141_369647\|\|full | Not-determined |
| 18-101-final-viral-combined_k141_271072\|\|full | Not-determined |
| 18-101-final-viral-combined_k141_369649\|\|full | Not-determined |
| 18-101-final-viral-combined_k141_369652\|\|full | Not-determined |
| 18-101-final-viral-combined_k141_386520\|\|full | Not-determined |
| 18-101-final-viral-combined_k141_287559\|\|full | Low-quality |
| 18-101-final-viral-combined_k141_105792\|\|full | Not-determined |
| 18-101-final-viral-combined_k141_205908\|\|full | Low-quality |
| 18-101-final-viral-combined_k141_7973\|\|full | Low-quality |
| 18-101-final-viral-combined_k141_189507\|\|full | Not-determined |
| 18-101-final-viral-combined_k141_89986\|\|full | Not-determined |
| 18-101-final-viral-combined_k141_205918\|\|full | Low-quality |
| 18-101-final-viral-combined_k141_105804\|\|full | Low-quality |
| 18-101-final-viral-combined_k141_171610\|\|full | Low-quality |
| 18-101-final-viral-combined_k141_255336\|\|full | Low-quality |
| 18-101-final-viral-combined_k141_238435\|\|full | Not-determined |
| 18-101-final-viral-combined_k141_189530\|\|full | Not-determined |
| 18-101-final-viral-combined_k141_155694\|\|full | Not-determined |
| 18-101-final-viral-combined_k141_386569\|\|full | Low-quality |
| 18-101-final-viral-combined_k141_304687\|\|full | Low-quality |
| 18-101-final-viral-combined_k141_105813\|\|full | Low-quality |
| 18-101-final-viral-combined_k141_238443\|\|full | Not-determined |
| 18-101-final-viral-combined_k141_123964\|\|full | Not-determined |
| 18-101-final-viral-combined_k141_105818\|\|full | Low-quality |
| 18-101-final-viral-combined_k141_238452\|\|full | Not-determined |
| 18-101-final-viral-combined_k141_386580\|\|full | Not-determined |
| 18-101-final-viral-combined_k141_321653\|\|full | Low-quality |
| 18-101-final-viral-combined_k141_8013\|\|full | Low-quality |
| 18-101-final-viral-combined_k141_123973\|\|full | Not-determined |
| 18-101-final-viral-combined_k141_221894\|\|full | Low-quality |
| 18-101-final-viral-combined_k141_255368\|\|full | Low-quality |
| 18-101-final-viral-combined_k141_287607\|\|full | Not-determined |
| 18-101-final-viral-combined_k141_221900\|\|full | Not-determined |
| 18-101-final-viral-combined_k141_255378\|\|full | Not-determined |
| 18-101-final-viral-combined_k141_155736\|\|full | Not-determined |
| 18-101-final-viral-combined_k141_336359\|\|full | Not-determined |
| 18-101-final-viral-combined_k141_369702\|\|full | Not-determined |
| 18-101-final-viral-combined_k141_189547\|\|full | Low-quality |
| 18-101-final-viral-combined_k141_72980\|\|full | Low-quality |
| 18-101-final-viral-combined_k141_238493\|\|full | Not-determined |
| 18-101-final-viral-combined_k141_238496\|\|full | Low-quality |
| 18-101-final-viral-combined_k141_321697\|\|full | Not-determined |
| 18-101-final-viral-combined_k141_171669\|\|full | Low-quality |
| 18-101-final-viral-combined_k141_40606\|\|full | Low-quality |
| 18-101-final-viral-combined_k141_304753\|\|full | Low-quality |
| 18-101-final-viral-combined_k141_287643\|\|full | Not-determined |
| 18-102-final-viral-combined_k141_197577\|\|full | Medium-quality |
| 18-101-final-viral-combined_k141_189589\|\|full | Not-determined |
| 18-101-final-viral-combined_k141_8058\|\|full | Not-determined |
| 18-101-final-viral-combined_k141_57201\|\|full | Low-quality |
| 18-101-final-viral-combined_k141_24146\|\|full | Low-quality |
| 18-101-final-viral-combined_k141_73034\|\|full | Low-quality |
| 18-101-final-viral-combined_k141_336393\|\|full | Low-quality |
| 18-101-final-viral-combined_k141_189610\|\|full | Low-quality |
| 18-101-final-viral-combined_k141_304778\|\|full | Low-quality |
| 18-101-final-viral-combined_k141_336399\|\|full | Low-quality |
| 18-101-final-viral-combined_k141_321748\|\|full | Low-quality |
| 18-101-final-viral-combined_k141_206015\|\|full | Not-determined |
| 18-101-final-viral-combined_k141_287671\|\|full | Low-quality |
| 18-101-final-viral-combined_k141_386635\|\|full | Low-quality |
| 18-101-final-viral-combined_k141_189619\|\|full | Not-determined |
| 18-101-final-viral-combined_k141_73069\|\|full | Not-determined |
| 18-101-final-viral-combined_k141_271227\|\|full | Low-quality |
| 18-101-final-viral-combined_k141_57221\|\|full | Low-quality |
| 18-101-final-viral-combined_k141_336423\|\|full | Low-quality |
| 18-101-final-viral-combined_k141_8069\|\|full | Not-determined |
| 18-101-final-viral-combined_k141_8074\|\|full | Low-quality |
| 18-101-final-viral-combined_k141_304828\|\|full | Low-quality |
| 18-101-final-viral-combined_k141_321781\|\|full | Not-determined |
| 18-101-final-viral-combined_k141_369761\|\|full | Not-determined |
| 18-101-final-viral-combined_k141_24193\|\|full | Low-quality |
| 18-101-final-viral-combined_k141_304839\|\|full | Not-determined |
| 18-101-final-viral-combined_k141_24198\|\|full | Not-determined |
| 18-101-final-viral-combined_k141_336443\|\|full | Low-quality |
| 18-101-final-viral-combined_k141_73082\|\|full | Not-determined |
| 18-101-final-viral-combined_k141_189663\|\|full | Low-quality |
| 18-101-final-viral-combined_k141_206044\|\|full | Not-determined |
| 18-101-final-viral-combined_k141_8107\|\|full | Not-determined |
| 18-101-final-viral-combined_k141_321793\|\|full | Low-quality |

| | |
|---|---|
| 18-101-final-viral-combined_k141_238579\|\|full | Low-quality |
| 18-101-final-viral-combined_k141_386667\|\|full | Not-determined |
| 18-101-final-viral-combined_k141_336466\|\|full | Not-determined |
| 18-101-final-viral-combined_k141_321800\|\|full | Low-quality |
| 18-101-final-viral-combined_k141_206060\|\|full | Not-determined |
| 18-101-final-viral-combined_k141_238583\|\|full | Low-quality |
| 18-101-final-viral-combined_k141_369787\|\|full | Not-determined |
| 18-101-final-viral-combined_k141_206064\|\|full | Not-determined |
| 18-101-final-viral-combined_k141_386694\|\|full | Low-quality |
| 18-101-final-viral-combined_k141_321833\|\|full | Low-quality |
| 18-101-final-viral-combined_k141_189692\|\|full | Not-determined |
| 18-101-final-viral-combined_k141_57306\|\|full | Low-quality |
| 18-101-final-viral-combined_k141_304890\|\|full | Low-quality |
| 18-101-final-viral-combined_k141_206076\|\|full | Low-quality |
| 18-101-final-viral-combined_k141_336515\|\|full | Low-quality |
| 18-101-final-viral-combined_k141_304895\|\|full | Not-determined |
| 18-101-final-viral-combined_k141_57314\|\|full | Not-determined |
| 18-101-final-viral-combined_k141_369808\|\|full | Low-quality |
| 18-101-final-viral-combined_k141_321851\|\|full | Not-determined |
| 18-101-final-viral-combined_k141_73113\|\|full | Low-quality |
| 18-101-final-viral-combined_k141_57326\|\|full | Low-quality |
| 18-101-final-viral-combined_k141_336541\|\|full | Not-determined |
| 18-101-final-viral-combined_k141_90125\|\|full | Low-quality |
| 18-101-final-viral-combined_k141_238647\|\|full | Low-quality |
| 18-101-final-viral-combined_k141_8174\|\|full | Low-quality |
| 18-101-final-viral-combined_k141_336555\|\|full | Not-determined |
| 18-101-final-viral-combined_k141_287800\|\|full | Low-quality |
| 18-101-final-viral-combined_k141_73122\|\|full | Low-quality |
| 18-101-final-viral-combined_k141_57336\|\|full | Low-quality |
| 18-101-final-viral-combined_k141_238667\|\|full | Low-quality |
| 18-101-final-viral-combined_k141_73131\|\|full | Not-determined |
| 18-101-final-viral-combined_k141_8205\|\|full | Not-determined |
| 18-101-final-viral-combined_k141_105989\|\|full | Low-quality |
| 18-101-final-viral-combined_k141_238680\|\|full | Not-determined |
| 18-101-final-viral-combined_k141_386742\|\|full | Low-quality |
| 18-101-final-viral-combined_k141_336588\|\|full | Low-quality |
| 18-101-final-viral-combined_k141_206144\|\|full | Low-quality |
| 18-101-final-viral-combined_k141_221981\|\|full | Low-quality |
| 18-101-final-viral-combined_k141_8242\|\|full | Low-quality |
| 18-101-final-viral-combined_k141_354135\|\|full | Not-determined |
| 18-101-final-viral-combined_k141_255501\|\|full | Not-determined |
| 18-101-final-viral-combined_k141_386781\|\|full | Low-quality |
| 18-101-final-viral-combined_k141_124144\|\|full | Low-quality |
| 18-101-final-viral-combined_k141_354136\|\|full | Low-quality |
| 18-101-final-viral-combined_k141_140027\|\|full | Not-determined |
| 18-101-final-viral-combined_k141_171795\|\|full | Low-quality |
| 18-101-final-viral-combined_k141_255513\|\|full | Not-determined |
| 18-101-final-viral-combined_k141_386790\|\|full | Low-quality |
| 18-101-final-viral-combined_k141_222008\|\|full | Not-determined |
| 18-101-final-viral-combined_k141_386793\|\|full | Low-quality |
| 18-101-final-viral-combined_k141_8264\|\|full | Not-determined |
| 18-101-final-viral-combined_k141_336640\|\|full | Not-determined |
| 18-101-final-viral-combined_k141_124153\|\|full | Low-quality |
| 18-101-final-viral-combined_k141_238726\|\|full | Low-quality |
| 18-101-final-viral-combined_k141_222027\|\|full | Low-quality |
| 18-101-final-viral-combined_k141_238728\|\|full | Low-quality |
| 18-101-final-viral-combined_k141_354165\|\|full | Low-quality |
| 18-101-final-viral-combined_k141_189784\|\|full | Not-determined |
| 18-101-final-viral-combined_k141_255537\|\|full | Not-determined |
| 18-101-final-viral-combined_k141_271270\|\|full | Not-determined |
| 18-101-final-viral-combined_k141_305016\|\|full | Not-determined |
| 18-101-final-viral-combined_k141_189791\|\|full | Not-determined |
| 18-101-final-viral-combined_k141_106017\|\|full | Low-quality |
| 18-101-final-viral-combined_k141_189796\|\|full | Not-determined |
| 18-101-final-viral-combined_k141_287883\|\|full | Not-determined |
| 18-101-final-viral-combined_k141_305028\|\|full | Not-determined |
| 18-101-final-viral-combined_k141_206213\|\|full | Not-determined |
| 18-101-final-viral-combined_k141_124184\|\|full | Not-determined |
| 18-101-final-viral-combined_k141_386841\|\|full | Not-determined |
| 18-101-final-viral-combined_k141_271282\|\|full | Not-determined |
| 18-101-final-viral-combined_k141_255565\|\|full | Low-quality |
| 18-101-final-viral-combined_k141_189815\|\|full | Not-determined |
| 18-101-final-viral-combined_k141_238768\|\|full | Low-quality |
| 18-101-final-viral-combined_k141_287907\|\|full | Not-determined |
| 18-101-final-viral-combined_k141_8317\|\|full | Not-determined |
| 18-101-final-viral-combined_k141_189829\|\|full | Not-determined |
| 18-101-final-viral-combined_k141_140052\|\|full | Not-determined |
| 18-101-final-viral-combined_k141_73202\|\|full | Low-quality |
| 18-101-final-viral-combined_k141_124209\|\|full | Low-quality |
| 18-101-final-viral-combined_k141_369976\|\|full | Not-determined |
| 18-101-final-viral-combined_k141_271316\|\|full | Not-determined |
| 18-101-final-viral-combined_k141_171841\|\|full | Not-determined |
| 18-101-final-viral-combined_k141_106034\|\|full | Low-quality |
| 18-101-final-viral-combined_k141_255611\|\|full | Not-determined |
| 18-101-final-viral-combined_k141_222078\|\|full | Not-determined |
| 18-101-final-viral-combined_k141_189869\|\|full | Not-determined |
| 18-101-final-viral-combined_k141_106049\|\|full | Low-quality |
| 18-101-final-viral-combined_k141_271332\|\|full | Low-quality |
| 18-101-final-viral-combined_k141_238845\|\|full | Not-determined |
| 18-101-final-viral-combined_k141_354276\|\|full | Not-determined |
| 18-101-final-viral-combined_k141_206266\|\|full | Low-quality |
| 18-101-final-viral-combined_k141_238850\|\|full | Not-determined |
| 18-101-final-viral-combined_k141_106069\|\|full | Low-quality |
| 18-101-final-viral-combined_k141_171862\|\|full | Not-determined |
| 18-101-final-viral-combined_k141_321985\|\|full | Low-quality |
| 18-101-final-viral-combined_k141_222105\|\|full | Not-determined |
| 18-101-final-viral-combined_k141_73245\|\|full | Not-determined |
| 18-101-final-viral-combined_k141_369991\|\|full | Low-quality |
| 18-101-final-viral-combined_k141_386903\|\|full | Not-determined |
| 18-101-final-viral-combined_k141_73271\|\|full | Not-determined |
| 18-101-final-viral-combined_k141_90233\|\|full | Low-quality |
| 18-101-final-viral-combined_k141_386913\|\|full | Low-quality |
| 18-101-final-viral-combined_k141_24380\|\|full | Low-quality |
| 18-101-final-viral-combined_k141_73277\|\|full | Low-quality |
| 18-102_k141_248315 | Complete |
| 18-101-final-viral-combined_k141_222156\|\|full | Low-quality |
| 18-101-final-viral-combined_k141_40724\|\|full | Low-quality |
| 18-101-final-viral-combined_k141_171909\|\|full | Not-determined |
| 18-101-final-viral-combined_k141_287984\|\|full | Low-quality |
| 18-101-final-viral-combined_k141_124331\|\|full | Low-quality |
| 18-101-final-viral-combined_k141_238876\|\|full | Low-quality |
| 18-101-final-viral-combined_k141_8417\|\|full | Low-quality |
| 18-101-final-viral-combined_k141_90249\|\|full | Low-quality |
| 18-101-final-viral-combined_k141_370021\|\|full | Not-determined |
| 18-101-final-viral-combined_k141_271402\|\|full | Not-determined |
| 18-101-final-viral-combined_k141_370027\|\|full | Low-quality |
| 18-101-final-viral-combined_k141_24416\|\|full | Not-determined |
| 18-101-final-viral-combined_k141_354380\|\|full | Not-determined |
| 18-101-final-viral-combined_k141_8432\|\|full | Low-quality |
| 18-101-final-viral-combined_k141_386941\|\|full | Low-quality |
| 18-101-final-viral-combined_k141_238891\|\|full | Low-quality |
| 18-101-final-viral-combined_k141_190010\|\|full | Low-quality |
| 18-101-final-viral-combined_k141_354384\|\|full | Low-quality |
| 18-101-final-viral-combined_k141_354386\|\|full | Not-determined |
| 18-101-final-viral-combined_k141_255737\|\|full | Not-determined |
| 18-102-final-viral-combined_k141_248315\|\|full | High-quality |
| 18-101-final-viral-combined_k141_288013\|\|full | Low-quality |
| 18-101-final-viral-combined_k141_386975\|\|full | Not-determined |
| 18-101-final-viral-combined_k141_206381\|\|full | Not-determined |
| 18-101-final-viral-combined_k141_57433\|\|full | Low-quality |
| 18-101-final-viral-combined_k141_106131\|\|full | Low-quality |
| 18-101-final-viral-combined_k141_288022\|\|full | Low-quality |
| 18-101-final-viral-combined_k141_40752\|\|full | Not-determined |
| 18-101-final-viral-combined_k141_90307\|\|full | Not-determined |
| 18-101-final-viral-combined_k141_90311\|\|full | Not-determined |
| 18-101-final-viral-combined_k141_24464\|\|full | Low-quality |
| 18-101-final-viral-combined_k141_305143\|\|full | Low-quality |
| 18-101-final-viral-combined_k141_271463\|\|full | Low-quality |
| 18-101-final-viral-combined_k141_387021\|\|full | Not-determined |
| 18-101-final-viral-combined_k141_387026\|\|full | Not-determined |
| 18-105-final-viral-combined_k141_29272\|\|full | High-quality |
| 18-101-final-viral-combined_k141_90332\|\|full | Not-determined |
| 18-101-final-viral-combined_k141_222287\|\|full | Low-quality |
| 18-101-final-viral-combined_k141_238901\|\|full | Low-quality |
| 18-101-final-viral-combined_k141_90333\|\|full | Not-determined |
| 18-101-final-viral-combined_k141_24491\|\|full | Not-determined |
| 18-101-final-viral-combined_k141_288034\|\|full | Not-determined |
| 18-101-final-viral-combined_k141_73305\|\|full | Not-determined |
| 18-101-final-viral-combined_k141_24492\|\|full | Low-quality |
| 18-101-final-viral-combined_k141_73309\|\|full | Low-quality |
| 18-101-final-viral-combined_k141_387052\|\|full | Low-quality |
| 18-101-final-viral-combined_k141_24494\|\|full | Not-determined |
| 18-101-final-viral-combined_k141_354463\|\|full | Not-determined |
| 18-101-final-viral-combined_k141_124443\|\|full | Low-quality |
| 18-101-final-viral-combined_k141_305185\|\|full | Low-quality |
| 18-101-final-viral-combined_k141_238907\|\|full | Low-quality |
| 18-101-final-viral-combined_k141_73310\|\|full | Low-quality |
| 18-101-final-viral-combined_k141_255823\|\|full | Not-determined |
| 18-101-final-viral-combined_k141_238910\|\|full | Not-determined |
| 18-101-final-viral-combined_k141_322161\|\|full | Not-determined |
| 18-101-final-viral-combined_k141_124452\|\|full | Not-determined |
| 18-101-final-viral-combined_k141_90346\|\|full | Low-quality |
| 18-101-final-viral-combined_k141_288044\|\|full | Low-quality |
| 18-101-final-viral-combined_k141_271512\|\|full | Not-determined |
| 18-101-final-viral-combined_k141_370128\|\|full | Not-determined |
| 18-101-final-viral-combined_k141_387082\|\|full | Low-quality |
| 18-101-final-viral-combined_k141_206425\|\|full | Low-quality |
| 18-101-final-viral-combined_k141_106194\|\|full | Low-quality |
| 18-101-final-viral-combined_k141_255845\|\|full | Low-quality |
| 18-101-final-viral-combined_k141_24515\|\|full | Not-determined |
| 18-101-final-viral-combined_k141_255852\|\|full | Low-quality |
| 18-101-final-viral-combined_k141_8450\|\|full | Low-quality |
| 18-101-final-viral-combined_k141_255854\|\|full | Not-determined |
| 18-101-final-viral-combined_k141_288059\|\|full | Not-determined |
| 18-101-final-viral-combined_k141_124482\|\|full | Not-determined |
| 18-101-final-viral-combined_k141_271545\|\|full | Low-quality |
| 18-101-final-viral-combined_k141_106220\|\|full | Low-quality |
| 18-101-final-viral-combined_k141_171968\|\|full | Not-determined |
| 18-101-final-viral-combined_k141_238931\|\|full | Not-determined |
| 18-101-final-viral-combined_k141_190157\|\|full | Not-determined |
| 18-101-final-viral-combined_k141_124486\|\|full | Low-quality |
| 18-101-final-viral-combined_k141_238933\|\|full | Low-quality |
| 18-101-final-viral-combined_k141_305244\|\|full | Low-quality |
| 18-101-final-viral-combined_k141_40822\|\|full | Not-determined |
| 18-101-final-viral-combined_k141_354511\|\|full | Low-quality |
| 18-101-final-viral-combined_k141_124496\|\|full | Not-determined |
| 18-101-final-viral-combined_k141_73338\|\|full | Not-determined |
| 18-101-final-viral-combined_k141_370168\|\|full | Low-quality |
| 18-101-final-viral-combined_k141_222360\|\|full | Low-quality |
| 18-101-final-viral-combined_k141_73347\|\|full | Low-quality |
| 18-101-final-viral-combined_k141_255900\|\|full | Low-quality |
| 18-101-final-viral-combined_k141_106251\|\|full | Low-quality |
| 18-101-final-viral-combined_k141_387116\|\|full | Low-quality |
| 18-101-final-viral-combined_k141_24555\|\|full | Low-quality |
| 18-101-final-viral-combined_k141_40863\|\|full | Not-determined |
| 18-101-final-viral-combined_k141_354537\|\|full | Low-quality |
| 18-101-final-viral-combined_k141_255935\|\|full | Not-determined |
| 18-101-final-viral-combined_k141_90409\|\|full | Low-quality |
| 18-101-final-viral-combined_k141_8498\|\|full | Low-quality |
| 18-101-final-viral-combined_k141_24572\|\|full | Not-determined |
| 18-101-final-viral-combined_k141_8502\|\|full | Not-determined |
| 18-101-final-viral-combined_k141_172018\|\|full | Low-quality |
| 18-101-final-viral-combined_k141_238973\|\|full | Low-quality |
| 18-101-final-viral-combined_k141_238977\|\|full | Low-quality |
| 18-101-final-viral-combined_k141_124528\|\|full | Not-determined |
| 18-101-final-viral-combined_k141_40895\|\|full | Low-quality |
| 18-101-final-viral-combined_k141_305278\|\|full | Low-quality |
| 18-101-final-viral-combined_k141_255946\|\|full | Low-quality |
| 18-101-final-viral-combined_k141_206478\|\|full | Low-quality |
| 18-101-final-viral-combined_k141_90416\|\|full | Not-determined |
| 18-101-final-viral-combined_k141_73393\|\|full | Low-quality |
| 18-101-final-viral-combined_k141_40900\|\|full | Low-quality |
| 18-101-final-viral-combined_k141_271625\|\|full | Not-determined |
| 18-101-final-viral-combined_k141_155883\|\|full | Not-determined |
| 18-101-final-viral-combined_k141_40905\|\|full | Low-quality |
| 18-101-final-viral-combined_k141_190228\|\|full | Low-quality |
| 18-101-final-viral-combined_k141_172038\|\|full | Low-quality |
| 18-101-final-viral-combined_k141_271638\|\|full | Low-quality |
| 18-101-final-viral-combined_k141_305287\|\|full | Not-determined |
| 18-101-final-viral-combined_k141_238991\|\|full | Not-determined |
| 18-101-final-viral-combined_k141_206505\|\|full | Not-determined |
| 18-101-final-viral-combined_k141_370206\|\|full | Low-quality |
| 18-101-final-viral-combined_k141_40930\|\|full | Not-determined |
| 18-101-final-viral-combined_k141_206527\|\|full | Low-quality |
| 18-101-final-viral-combined_k141_190258\|\|full | Not-determined |
| 18-101-final-viral-combined_k141_387141\|\|full | Low-quality |
| 18-101-final-viral-combined_k141_73444\|\|full | Not-determined |
| 18-101-final-viral-combined_k141_336781\|\|full | Low-quality |
| 18-101-final-viral-combined_k141_8560\|\|full | Low-quality |
| 18-101-final-viral-combined_k141_155915\|\|full | Not-determined |
| 18-101-final-viral-combined_k141_255975\|\|full | Not-determined |
| 18-101-final-viral-combined_k141_106283\|\|full | Low-quality |
| 18-101-final-viral-combined_k141_322246\|\|full | Not-determined |
| 18-101-final-viral-combined_k141_387162\|\|full | Low-quality |
| 18-101-final-viral-combined_k141_271693\|\|full | Not-determined |
| 18-101-final-viral-combined_k141_8575\|\|full | Low-quality |
| 18-101-final-viral-combined_k141_336818\|\|full | Not-determined |
| 18-101-final-viral-combined_k141_8576\|\|full | Not-determined |
| 18-101-final-viral-combined_k141_40941\|\|full | Low-quality |
| 18-101-final-viral-combined_k141_8578\|\|full | Low-quality |
| 18-101-final-viral-combined_k141_124564\|\|full | Low-quality |
| 18-101-final-viral-combined_k141_370270\|\|full | Low-quality |
| 18-101-final-viral-combined_k141_239054\|\|full | Low-quality |
| 18-101-final-viral-combined_k141_354637\|\|full | Low-quality |
| 18-101-final-viral-combined_k141_336847\|\|full | Not-determined |
| 18-101-final-viral-combined_k141_370277\|\|full | Low-quality |
| 18-101-final-viral-combined_k141_370285\|\|full | Low-quality |
| 18-101-final-viral-combined_k141_90511\|\|full | Not-determined |
| 18-101-final-viral-combined_k141_190317\|\|full | Low-quality |
| 18-101-final-viral-combined_k141_40968\|\|full | Not-determined |
| 18-101-final-viral-combined_k141_288156\|\|full | Not-determined |
| 18-101-final-viral-combined_k141_124601\|\|full | Not-determined |
| 18-101-final-viral-combined_k141_387206\|\|full | Low-quality |
| 18-101-final-viral-combined_k141_124608\|\|full | Not-determined |
| 18-101-final-viral-combined_k141_370308\|\|full | Low-quality |
| 18-101-final-viral-combined_k141_106319\|\|full | Low-quality |
| 18-101-final-viral-combined_k141_322319\|\|full | Low-quality |
| 18-101-final-viral-combined_k141_106320\|\|full | Low-quality |
| 18-101-final-viral-combined_k141_90532\|\|full | Low-quality |
| 18-101-final-viral-combined_k141_106339\|\|full | Low-quality |
| 18-101-final-viral-combined_k141_90538\|\|full | Low-quality |
| 18-101-final-viral-combined_k141_8648\|\|full | Low-quality |
| 18-101-final-viral-combined_k141_73519\|\|full | Not-determined |
| 18-101-final-viral-combined_k141_305382\|\|full | Low-quality |
| 18-101-final-viral-combined_k141_322336\|\|full | Not-determined |
| 18-101-final-viral-combined_k141_57597\|\|full | Not-determined |
| 18-101-final-viral-combined_k141_288174\|\|full | Low-quality |
| 18-101-final-viral-combined_k141_57600\|\|full | Low-quality |
| 18-101-final-viral-combined_k141_239123\|\|full | Not-determined |
| 18-101-final-viral-combined_k141_90559\|\|full | Low-quality |
| 18-101-final-viral-combined_k141_288178\|\|full | Not-determined |
| 18-101-final-viral-combined_k141_305400\|\|full | Not-determined |
| 18-101-final-viral-combined_k141_239133\|\|full | Not-determined |
| 18-101-final-viral-combined_k141_57613\|\|full | Not-determined |
| 18-101-final-viral-combined_k141_206600\|\|full | Low-quality |
| 18-101-final-viral-combined_k141_370332\|\|full | Not-determined |
| 18-101-final-viral-combined_k141_40977\|\|full | Not-determined |
| 18-101-final-viral-combined_k141_370335\|\|full | Low-quality |
| 18-101-final-viral-combined_k141_190381\|\|full | Low-quality |
| 18-101-final-viral-combined_k141_155936\|\|full | Low-quality |
| 18-101-final-viral-combined_k141_40979\|\|full | Not-determined |
| 18-101-final-viral-combined_k141_387282\|\|full | Low-quality |
| 18-101-final-viral-combined_k141_124678\|\|full | Low-quality |
| 18-101-final-viral-combined_k141_387285\|\|full | Not-determined |
| 18-101-final-viral-combined_k141_8717\|\|full | Not-determined |
| 18-101-final-viral-combined_k141_155950\|\|full | Not-determined |
| 18-101-final-viral-combined_k141_24776\|\|full | Not-determined |
| 18-101-final-viral-combined_k141_206632\|\|full | Not-determined |
| 18-101-final-viral-combined_k141_124697\|\|full | Low-quality |
| 18-101-final-viral-combined_k141_73543\|\|full | Low-quality |
| 18-101-final-viral-combined_k141_155968\|\|full | Not-determined |
| 18-101-final-viral-combined_k141_305434\|\|full | Low-quality |
| 18-101-final-viral-combined_k141_370367\|\|full | Low-quality |
| 18-101-final-viral-combined_k141_8748\|\|full | Not-determined |
| 18-101-final-viral-combined_k141_190423\|\|full | Low-quality |
| 18-101-final-viral-combined_k141_90635\|\|full | Low-quality |
| 18-101-final-viral-combined_k141_370377\|\|full | Low-quality |
| 18-101-final-viral-combined_k141_222507\|\|full | Not-determined |
| 18-101-final-viral-combined_k141_24809\|\|full | Not-determined |
| 18-101-final-viral-combined_k141_336998\|\|full | Not-determined |
| 18-101-final-viral-combined_k141_387310\|\|full | Not-determined |
| 18-101-final-viral-combined_k141_190441\|\|full | Not-determined |
| 18-101-final-viral-combined_k141_354651\|\|full | Not-determined |
| 18-101-final-viral-combined_k141_370387\|\|full | Low-quality |
| 18-101-final-viral-combined_k141_387324\|\|full | Not-determined |
| 18-101-final-viral-combined_k141_337013\|\|full | Not-determined |
| 18-101-final-viral-combined_k141_222530\|\|full | Low-quality |
| 18-101-final-viral-combined_k141_337018\|\|full | Low-quality |
| 18-101-final-viral-combined_k141_73553\|\|full | Low-quality |
| 18-101-final-viral-combined_k141_305458\|\|full | Low-quality |
| 18-101-final-viral-combined_k141_190451\|\|full | Not-determined |
| 18-101-final-viral-combined_k141_41039\|\|full | Low-quality |
| 18-101-final-viral-combined_k141_354676\|\|full | Low-quality |
| 18-101-final-viral-combined_k141_288255\|\|full | Not-determined |
| 18-101-final-viral-combined_k141_255996\|\|full | Not-determined |
| 18-101-final-viral-combined_k141_140273\|\|full | Low-quality |
| 18-101-final-viral-combined_k141_73562\|\|full | Not-determined |
| 18-101-final-viral-combined_k141_370410\|\|full | Not-determined |
| 18-101-final-viral-combined_k141_387362\|\|full | Low-quality |
| 18-101-final-viral-combined_k141_8796\|\|full | Low-quality |
| 18-101-final-viral-combined_k141_322413\|\|full | Not-determined |
| 18-101-final-viral-combined_k141_354705\|\|full | Low-quality |
| 18-101-final-viral-combined_k141_206672\|\|full | Not-determined |
| 18-101-final-viral-combined_k141_322416\|\|full | Not-determined |
| 18-101-final-viral-combined_k141_256029\|\|full | Not-determined |
| 18-101-final-viral-combined_k141_305477\|\|full | Not-determined |
| 18-101-final-viral-combined_k141_140299\|\|full | Low-quality |
| 18-101-final-viral-combined_k141_370443\|\|full | Not-determined |
| 18-101-final-viral-combined_k141_124731\|\|full | Low-quality |
| 18-101-final-viral-combined_k141_206687\|\|full | Low-quality |
| 18-101-final-viral-combined_k141_90665\|\|full | Low-quality |
| 18-101-final-viral-combined_k141_73611\|\|full | Low-quality |
| 18-101-final-viral-combined_k141_73614\|\|full | Low-quality |
| 18-101-final-viral-combined_k141_41079\|\|full | Not-determined |
| 18-101-final-viral-combined_k141_24864\|\|full | Not-determined |
| 18-101-final-viral-combined_k141_8806\|\|full | Not-determined |
| 18-101-final-viral-combined_k141_172156\|\|full | Low-quality |
| 18-101-final-viral-combined_k141_222622\|\|full | Not-determined |
| 18-101-final-viral-combined_k141_271801\|\|full | Low-quality |
| 18-101-final-viral-combined_k141_41088\|\|full | Low-quality |
| 18-101-final-viral-combined_k141_106396\|\|full | Not-determined |
| 18-101-final-viral-combined_k141_239248\|\|full | Low-quality |
| 18-101-final-viral-combined_k141_222635\|\|full | Not-determined |
| 18-101-final-viral-combined_k141_190531\|\|full | Not-determined |
| 18-101-final-viral-combined_k141_271817\|\|full | Not-determined |
| 18-101-final-viral-combined_k141_222638\|\|full | Low-quality |
| 18-101-final-viral-combined_k141_172169\|\|full | Low-quality |
| 18-101-final-viral-combined_k141_387422\|\|full | Low-quality |
| 18-101-final-viral-combined_k141_288322\|\|full | Not-determined |
| 18-101-final-viral-combined_k141_90688\|\|full | Not-determined |
| 18-101-final-viral-combined_k141_239263\|\|full | Low-quality |
| 18-101-final-viral-combined_k141_288324\|\|full | Low-quality |
| 18-101-final-viral-combined_k141_239271\|\|full | Low-quality |
| 18-101-final-viral-combined_k141_41123\|\|full | Low-quality |
| 18-101-final-viral-combined_k141_90701\|\|full | Not-determined |
| 18-101-final-viral-combined_k141_140351\|\|full | Not-determined |
| 18-101-final-viral-combined_k141_172188\|\|full | Low-quality |
| 18-101-final-viral-combined_k141_256122\|\|full | Low-quality |
| 18-101-final-viral-combined_k141_354724\|\|full | Not-determined |
| 18-101-final-viral-combined_k141_288357\|\|full | Not-determined |
| 18-101-final-viral-combined_k141_140356\|\|full | Not-determined |
| 18-101-final-viral-combined_k141_305584\|\|full | Low-quality |
| 18-101-final-viral-combined_k141_256130\|\|full | Not-determined |
| 18-101-final-viral-combined_k141_370543\|\|full | Not-determined |
| 18-101-final-viral-combined_k141_41148\|\|full | Low-quality |
| 18-101-final-viral-combined_k141_370545\|\|full | Low-quality |
| 18-101-final-viral-combined_k141_271851\|\|full | Low-quality |
| 18-101-final-viral-combined_k141_222676\|\|full | Low-quality |
| 18-101-final-viral-combined_k141_370551\|\|full | Not-determined |
| 18-101-final-viral-combined_k141_140372\|\|full | Not-determined |
| 18-101-final-viral-combined_k141_370556\|\|full | Low-quality |
| 18-101-final-viral-combined_k141_24921\|\|full | Low-quality |
| 18-101-final-viral-combined_k141_222683\|\|full | Not-determined |
| 18-101-final-viral-combined_k141_156075\|\|full | Low-quality |
| 18-101-final-viral-combined_k141_106447\|\|full | Low-quality |
| 18-101-final-viral-combined_k141_288400\|\|full | Low-quality |
| 18-101-final-viral-combined_k141_337175\|\|full | Low-quality |
| 18-101-final-viral-combined_k141_271862\|\|full | Low-quality |
| 18-101-final-viral-combined_k141_354739\|\|full | Not-determined |
| 18-101-final-viral-combined_k141_305629\|\|full | Low-quality |
| 18-101-final-viral-combined_k141_337184\|\|full | Low-quality |
| 18-101-final-viral-combined_k141_222691\|\|full | Low-quality |
| 18-101-final-viral-combined_k141_370587\|\|full | Low-quality |
| 18-101-final-viral-combined_k141_90760\|\|full | Low-quality |
| 18-101-final-viral-combined_k141_337191\|\|full | Low-quality |
| 18-101-final-viral-combined_k141_106474\|\|full | Low-quality |
| 18-101-final-viral-combined_k141_140412\|\|full | Not-determined |
| 18-101-final-viral-combined_k141_288436\|\|full | Not-determined |
| 18-101-final-viral-combined_k141_156117\|\|full | Low-quality |
| 18-101-final-viral-combined_k141_24987\|\|full | Not-determined |
| 18-101-final-viral-combined_k141_190626\|\|full | Not-determined |
| 18-101-final-viral-combined_k141_271870\|\|full | Low-quality |
| 18-101-final-viral-combined_k141_288448\|\|full | Low-quality |
| 18-101-final-viral-combined_k141_190636\|\|full | Not-determined |
| 18-101-final-viral-combined_k141_239362\|\|full | Not-determined |
| 18-101-final-viral-combined_k141_354782\|\|full | Not-determined |
| 18-101-final-viral-combined_k141_222708\|\|full | Not-determined |
| 18-101-final-viral-combined_k141_124868\|\|full | Low-quality |
| 18-101-final-viral-combined_k141_190661\|\|full | Not-determined |
| 18-101-final-viral-combined_k141_222721\|\|full | Low-quality |
| 18-101-final-viral-combined_k141_57756\|\|full | Low-quality |
| 18-101-final-viral-combined_k141_206852\|\|full | Low-quality |
| 18-101-final-viral-combined_k141_172284\|\|full | Not-determined |
| 18-101-final-viral-combined_k141_124896\|\|full | Low-quality |
| 18-101-final-viral-combined_k141_57760\|\|full | Not-determined |
| 18-101-final-viral-combined_k141_90822\|\|full | Low-quality |
| 18-101-final-viral-combined_k141_41288\|\|full | Not-determined |
| 18-101-final-viral-combined_k141_370684\|\|full | Not-determined |
| 18-101-final-viral-combined_k141_271958\|\|full | Not-determined |
| 18-101-final-viral-combined_k141_124908\|\|full | Low-quality |
| 18-101-final-viral-combined_k141_25031\|\|full | Low-quality |
| 18-101-final-viral-combined_k141_124910\|\|full | Low-quality |
| 18-101-final-viral-combined_k141_172314\|\|full | Not-determined |
| 18-101-final-viral-combined_k141_271969\|\|full | Low-quality |
| 18-101-final-viral-combined_k141_206881\|\|full | Low-quality |
| 18-101-final-viral-combined_k141_25046\|\|full | Low-quality |
| 18-101-final-viral-combined_k141_271975\|\|full | Low-quality |
| 18-101-final-viral-combined_k141_140505\|\|full | Low-quality |
| 18-101-final-viral-combined_k141_256250\|\|full | Low-quality |
| 18-101-final-viral-combined_k141_239427\|\|full | Low-quality |
| 18-101-final-viral-combined_k141_354876\|\|full | Low-quality |
| 18-101-final-viral-combined_k141_222745\|\|full | Low-quality |
| 18-101-final-viral-combined_k141_57827\|\|full | Not-determined |
| 18-101-final-viral-combined_k141_387585\|\|full | Low-quality |
| 18-101-final-viral-combined_k141_354887\|\|full | Low-quality |
| 18-101-final-viral-combined_k141_288519\|\|full | Low-quality |
| 18-101-final-viral-combined_k141_57839\|\|full | Not-determined |
| 18-101-final-viral-combined_k141_222761\|\|full | Low-quality |
| 18-101-final-viral-combined_k141_190697\|\|full | Low-quality |
| 18-101-final-viral-combined_k141_140577\|\|full | Not-determined |
| 17-106-final-viral-combined_k141_26410\|\|full | High-quality |
| 18-101-final-viral-combined_k141_322501\|\|full | Not-determined |
| 18-101-final-viral-combined_k141_239461\|\|full | Not-determined |
| 18-101-final-viral-combined_k141_190702\|\|full | Not-determined |
| 18-101-final-viral-combined_k141_222786\|\|full | Not-determined |
| 18-101-final-viral-combined_k141_25092\|\|full | Not-determined |
| 18-101-final-viral-combined_k141_370744\|\|full | Low-quality |
| 18-101-final-viral-combined_k141_370755\|\|full | Not-determined |
| 18-101-final-viral-combined_k141_140615\|\|full | Low-quality |
| 18-101-final-viral-combined_k141_190727\|\|full | Not-determined |
| 18-101-final-viral-combined_k141_140618\|\|full | Low-quality |
| 18-101-final-viral-combined_k141_172388\|\|full | Not-determined |
| 18-101-final-viral-combined_k141_156205\|\|full | Not-determined |
| 18-101-final-viral-combined_k141_354936\|\|full | Not-determined |
| 18-101-final-viral-combined_k141_272083\|\|full | Not-determined |
| 18-101-final-viral-combined_k141_288611\|\|full | Low-quality |
| 18-101-final-viral-combined_k141_288614\|\|full | Not-determined |
| 18-101-final-viral-combined_k141_73770\|\|full | Not-determined |
| 18-101-final-viral-combined_k141_106563\|\|full | Not-determined |
| 18-101-final-viral-combined_k141_156234\|\|full | Not-determined |
| 18-102-final-viral-combined_k141_7756\|\|0_partial | Medium-quality |
| 18-101-final-viral-combined_k141_354987\|\|full | Low-quality |
| 18-101-final-viral-combined_k141_337256\|\|full | Low-quality |
| 18-101-final-viral-combined_k141_322577\|\|full | Not-determined |
| 18-101-final-viral-combined_k141_322579\|\|full | Low-quality |
| 18-101-final-viral-combined_k141_73801\|\|full | Low-quality |
| 18-101-final-viral-combined_k141_239540\|\|full | Low-quality |
| 18-101-final-viral-combined_k141_106579\|\|full | Not-determined |
| 18-101-final-viral-combined_k141_25136\|\|full | Low-quality |
| 18-101-final-viral-combined_k141_370831\|\|full | Not-determined |
| 18-101-final-viral-combined_k141_239545\|\|full | Not-determined |
| 18-101-final-viral-combined_k141_106584\|\|full | Low-quality |
| 18-101-final-viral-combined_k141_256361\|\|full | Not-determined |
| 18-101-final-viral-combined_k141_125057\|\|full | Not-determined |
| 18-101-final-viral-combined_k141_256371\|\|full | Not-determined |
| 18-101-final-viral-combined_k141_106601\|\|full | Low-quality |
| 18-101-final-viral-combined_k141_91001\|\|full | Not-determined |
| 18-101-final-viral-combined_k141_57929\|\|full | Not-determined |
| 18-101-final-viral-combined_k141_140700\|\|full | Not-determined |
| 18-101-final-viral-combined_k141_370867\|\|full | Low-quality |
| 18-101-final-viral-combined_k141_337276\|\|full | Low-quality |
| 18-101-final-viral-combined_k141_9033\|\|full | Low-quality |
| 18-101-final-viral-combined_k141_125075\|\|full | Not-determined |
| 18-101-final-viral-combined_k141_140706\|\|full | Low-quality |
| 18-101-final-viral-combined_k141_322606\|\|full | Not-determined |
| 18-101-final-viral-combined_k141_387667\|\|full | Low-quality |
| 18-101-final-viral-combined_k141_41379\|\|full | Not-determined |
| 18-101-final-viral-combined_k141_288677\|\|full | Not-determined |
| 18-101-final-viral-combined_k141_41387\|\|full | Not-determined |
| 18-101-final-viral-combined_k141_156272\|\|full | Not-determined |
| 18-101-final-viral-combined_k141_370906\|\|full | Low-quality |
| 18-101-final-viral-combined_k141_322615\|\|full | Not-determined |
| 18-101-final-viral-combined_k141_73852\|\|full | Low-quality |
| 18-101-final-viral-combined_k141_355035\|\|full | Low-quality |
| 18-101-final-viral-combined_k141_9055\|\|full | Not-determined |
| 18-101-final-viral-combined_k141_256411\|\|full | Low-quality |
| 18-101-final-viral-combined_k141_125111\|\|full | Not-determined |
| 18-101-final-viral-combined_k141_57965\|\|full | Low-quality |
| 18-101-final-viral-combined_k141_370912\|\|full | Low-quality |
| 18-101-final-viral-combined_k141_387706\|\|full | Not-determined |
| 18-101-final-viral-combined_k141_73863\|\|full | Not-determined |
| 18-101-final-viral-combined_k141_207052\|\|full | Not-determined |
| 18-101-final-viral-combined_k141_73865\|\|full | Not-determined |
| 18-101-final-viral-combined_k141_125132\|\|full | Low-quality |
| 18-101-final-viral-combined_k141_337325\|\|full | Not-determined |
| 18-101-final-viral-combined_k141_57992\|\|full | Low-quality |
| 18-101-final-viral-combined_k141_239603\|\|full | Not-determined |
| 18-101-final-viral-combined_k141_256430\|\|full | Not-determined |
| 18-101-final-viral-combined_k141_9085\|\|full | Not-determined |
| 18-101-final-viral-combined_k141_125146\|\|full | Not-determined |
| 18-101-final-viral-combined_k141_156333\|\|full | Low-quality |
| 18-101-final-viral-combined_k141_272193\|\|full | Not-determined |
| 18-101-final-viral-combined_k141_387749\|\|full | Low-quality |
| 18-101-final-viral-combined_k141_387752\|\|full | Not-determined |
| 18-101-final-viral-combined_k141_305953\|\|full | Not-determined |
| 18-101-final-viral-combined_k141_305954\|\|full | Low-quality |
| 18-101-final-viral-combined_k141_156356\|\|full | Not-determined |
| 18-101-final-viral-combined_k141_305958\|\|full | Low-quality |
| 18-101-final-viral-combined_k141_355095\|\|full | Low-quality |
| 18-101-final-viral-combined_k141_239630\|\|full | Low-quality |
| 18-101-final-viral-combined_k141_25253\|\|full | Low-quality |
| 18-101-final-viral-combined_k141_370973\|\|full | Not-determined |
| 18-101-final-viral-combined_k141_140803\|\|full | Low-quality |
| 18-101-final-viral-combined_k141_25263\|\|full | Not-determined |
| 18-101-final-viral-combined_k141_25270\|\|full | Not-determined |
| 18-101-final-viral-combined_k141_106706\|\|full | Not-determined |
| 18-101-final-viral-combined_k141_58067\|\|full | Low-quality |
| 18-101-final-viral-combined_k141_322691\|\|full | Not-determined |
| 18-101-final-viral-combined_k141_337370\|\|full | Not-determined |
| 18-101-final-viral-combined_k141_322699\|\|full | Not-determined |
| 18-101-final-viral-combined_k141_172478\|\|full | Not-determined |
| 18-101-final-viral-combined_k141_256539\|\|full | Low-quality |
| 18-101-final-viral-combined_k141_207116\|\|full | Not-determined |
| 18-101-final-viral-combined_k141_322706\|\|full | Not-determined |
| 18-101-final-viral-combined_k141_106730\|\|full | Not-determined |
| 18-101-final-viral-combined_k141_9182\|\|full | Not-determined |
| 18-101-final-viral-combined_k141_9188\|\|full | Low-quality |
| 18-101-final-viral-combined_k141_207121\|\|full | Low-quality |
| 18-101-final-viral-combined_k141_322714\|\|full | Low-quality |
| 18-101-final-viral-combined_k141_106732\|\|full | Not-determined |
| 18-101-final-viral-combined_k141_371001\|\|full | Not-determined |
| 18-101-final-viral-combined_k141_91107\|\|full | Low-quality |
| 18-101-final-viral-combined_k141_106745\|\|full | Low-quality |
| 18-101-final-viral-combined_k141_156401\|\|full | Low-quality |
| 18-101-final-viral-combined_k141_125205\|\|full | Not-determined |
| 18-101-final-viral-combined_k141_305987\|\|full | Not-determined |
| 18-101-final-viral-combined_k141_156429\|\|full | Low-quality |
| 18-101-final-viral-combined_k141_288750\|\|full | Low-quality |
| 18-101-final-viral-combined_k141_222956\|\|full | Low-quality |
| 18-101-final-viral-combined_k141_58132\|\|full | Low-quality |
| 18-101-final-viral-combined_k141_25336\|\|full | Low-quality |
| 18-109-final-viral-combined_k141_190281\|\|full | High-quality |
| 18-101-final-viral-combined_k141_41503\|\|full | Not-determined |
| 18-101-final-viral-combined_k141_172548\|\|full | Low-quality |
| 18-101-final-viral-combined_k141_25342\|\|full | Low-quality |
| 18-101-final-viral-combined_k141_9218\|\|full | Low-quality |
| 18-101-final-viral-combined_k141_272307\|\|full | Not-determined |
| 18-101-final-viral-combined_k141_322768\|\|full | Low-quality |
| 18-101-final-viral-combined_k141_140868\|\|full | Not-determined |
| 18-101-final-viral-combined_k141_222988\|\|full | Not-determined |
| 18-101-final-viral-combined_k141_125241\|\|full | Low-quality |
| 18-101-final-viral-combined_k141_337405\|\|full | Low-quality |
| 18-101-final-viral-combined_k141_25376\|\|full | Not-determined |
| 18-101-final-viral-combined_k141_140875\|\|full | Low-quality |
| 18-101-final-viral-combined_k141_222997\|\|full | Not-determined |
| 18-101-final-viral-combined_k141_74001\|\|full | Not-determined |
| 18-101-final-viral-combined_k141_25386\|\|full | Low-quality |
| 18-101-final-viral-combined_k141_156458\|\|full | Low-quality |
| 18-101-final-viral-combined_k141_140894\|\|full | Not-determined |
| 18-101-final-viral-combined_k141_322808\|\|full | Not-determined |
| 18-101-final-viral-combined_k141_337423\|\|full | Low-quality |
| 18-101-final-viral-combined_k141_106759\|\|full | Low-quality |
| 18-101-final-viral-combined_k141_190879\|\|full | Not-determined |
| 18-101-final-viral-combined_k141_223024\|\|full | Not-determined |
| 18-101-final-viral-combined_k141_288806\|\|full | Not-determined |
| 18-101-final-viral-combined_k141_91224\|\|full | Low-quality |
| 18-101-final-viral-combined_k141_387896\|\|full | Not-determined |
| 18-101-final-viral-combined_k141_25435\|\|full | Low-quality |
| 18-101-final-viral-combined_k141_256651\|\|full | Low-quality |
| 18-101-final-viral-combined_k141_41550\|\|full | Low-quality |
| 18-101-final-viral-combined_k141_223052\|\|full | Not-determined |
| 18-101-final-viral-combined_k141_91252\|\|full | Low-quality |
| 18-101-final-viral-combined_k141_41561\|\|full | Low-quality |
| 18-101-final-viral-combined_k141_172615\|\|full | Low-quality |
| 18-101-final-viral-combined_k141_288835\|\|full | Not-determined |
| 18-101-final-viral-combined_k141_156495\|\|full | Low-quality |
| 18-101-final-viral-combined_k141_125336\|\|full | Low-quality |
| 18-101-final-viral-combined_k141_58197\|\|full | Not-determined |
| 18-101-final-viral-combined_k141_337458\|\|full | Not-determined |
| 18-101-final-viral-combined_k141_106797\|\|full | Not-determined |
| 18-101-final-viral-combined_k141_156506\|\|full | Not-determined |
| 18-101-final-viral-combined_k141_371108\|\|full | Low-quality |
| 18-101-final-viral-combined_k141_207263\|\|full | Not-determined |
| 18-101-final-viral-combined_k141_190922\|\|full | Low-quality |
| 18-101-final-viral-combined_k141_106810\|\|full | Not-determined |
| 18-101-final-viral-combined_k141_156514\|\|full | Not-determined |
| 18-101-final-viral-combined_k141_256676\|\|full | Not-determined |
| 18-101-final-viral-combined_k141_91291\|\|full | Low-quality |
| 18-101-final-viral-combined_k141_25454\|\|full | Low-quality |

| | |
|---|---|
| 18-101-final-viral-combined_k141_156523\|\|full | Not-determined |
| 18-101-final-viral-combined_k141_322854\|\|full | Low-quality |
| 18-101-final-viral-combined_k141_91293\|\|full | Low-quality |
| 18-101-final-viral-combined_k141_172627\|\|full | Low-quality |
| 18-101-final-viral-combined_k141_172632\|\|full | Not-determined |
| 18-101-final-viral-combined_k141_239790\|\|full | Not-determined |
| 18-101-final-viral-combined_k141_387931\|\|full | Low-quality |
| 18-101-final-viral-combined_k141_371137\|\|full | Low-quality |
| 18-101-final-viral-combined_k141_172636\|\|full | Not-determined |
| 18-101-final-viral-combined_k141_322861\|\|full | Not-determined |
| 18-101-final-viral-combined_k141_337514\|\|full | Low-quality |
| 18-101-final-viral-combined_k141_172644\|\|full | Not-determined |
| 18-101-final-viral-combined_k141_322872\|\|full | Low-quality |
| 18-101-final-viral-combined_k141_306142\|\|full | Not-determined |
| 18-101-final-viral-combined_k141_106867\|\|full | Not-determined |
| 18-101-final-viral-combined_k141_58268\|\|full | Not-determined |
| 18-101-final-viral-combined_k141_322878\|\|full | Low-quality |
| 18-101-final-viral-combined_k141_9350\|\|full | Not-determined |
| 18-101-final-viral-combined_k141_172663\|\|full | Not-determined |
| 18-101-final-viral-combined_k141_322880\|\|full | Low-quality |
| 18-101-final-viral-combined_k141_223081\|\|full | Not-determined |
| 18-101-final-viral-combined_k141_207327\|\|full | Not-determined |
| 18-101-final-viral-combined_k141_355195\|\|full | Low-quality |
| 18-101-final-viral-combined_k141_91342\|\|full | Not-determined |
| 18-101-final-viral-combined_k141_322906\|\|full | Low-quality |
| 18-101-final-viral-combined_k141_337549\|\|full | Not-determined |
| 18-101-final-viral-combined_k141_337559\|\|full | Not-determined |
| 18-101-final-viral-combined_k141_172708\|\|full | Not-determined |
| 18-101-final-viral-combined_k141_239826\|\|full | Low-quality |
| 18-101-final-viral-combined_k141_355216\|\|full | Low-quality |
| 18-101-final-viral-combined_k141_156571\|\|full | Not-determined |
| 18-101-final-viral-combined_k141_9387\|\|full | Low-quality |
| 18-101-final-viral-combined_k141_337568\|\|full | Low-quality |
| 18-101-final-viral-combined_k141_141031\|\|full | Low-quality |
| 18-101-final-viral-combined_k141_207362\|\|full | Low-quality |
| 18-101-final-viral-combined_k141_41630\|\|full | Low-quality |
| 18-101-final-viral-combined_k141_125352\|\|full | Not-determined |
| 18-101-final-viral-combined_k141_322919\|\|full | Not-determined |
| 18-101-final-viral-combined_k141_106935\|\|full | Low-quality |
| 18-101-final-viral-combined_k141_371166\|\|full | Not-determined |
| 18-101-final-viral-combined_k141_355243\|\|full | Not-determined |
| 18-101-final-viral-combined_k141_256746\|\|full | Low-quality |
| 18-101-final-viral-combined_k141_41648\|\|full | Not-determined |
| 18-101-final-viral-combined_k141_172749\|\|full | Low-quality |
| 18-101-final-viral-combined_k141_207417\|\|full | Low-quality |
| 18-101-final-viral-combined_k141_371190\|\|full | Not-determined |
| 18-101-final-viral-combined_k141_41664\|\|full | Not-determined |
| 18-101-final-viral-combined_k141_191030\|\|full | Not-determined |
| 18-101-final-viral-combined_k141_207439\|\|full | Low-quality |
| 18-101-final-viral-combined_k141_355283\|\|full | Low-quality |
| 18-101-final-viral-combined_k141_41690\|\|full | Not-determined |
| 18-101-final-viral-combined_k141_191034\|\|full | Low-quality |
| 18-101-final-viral-combined_k141_371214\|\|full | Low-quality |
| 18-101-final-viral-combined_k141_306247\|\|full | Not-determined |
| 18-101-final-viral-combined_k141_141108\|\|full | Low-quality |
| 18-101-final-viral-combined_k141_272479\|\|full | Not-determined |
| 18-101-final-viral-combined_k141_74182\|\|full | Not-determined |
| 18-101-final-viral-combined_k141_288954\|\|full | Not-determined |
| 18-101-final-viral-combined_k141_371221\|\|full | Not-determined |
| 18-101-final-viral-combined_k141_207452\|\|full | Not-determined |
| 18-101-final-viral-combined_k141_91430\|\|full | Low-quality |
| 18-101-final-viral-combined_k141_172801\|\|full | Low-quality |
| 18-101-final-viral-combined_k141_288957\|\|full | Not-determined |
| 18-109-final-viral-combined_k141_14529\|\|full | Medium-quality |
| 18-101-final-viral-combined_k141_256825\|\|full | Not-determined |
| 18-101-final-viral-combined_k141_74208\|\|full | Not-determined |
| 18-101-final-viral-combined_k141_223210\|\|full | Not-determined |
| 18-101-final-viral-combined_k141_74220\|\|full | Not-determined |
| KM-final-viral-combined_k141_106940\|\|full | Medium-quality |
| 18-101-final-viral-combined_k141_9480\|\|full | Not-determined |
| 18-101-final-viral-combined_k141_172831\|\|full | Low-quality |
| 18-101-final-viral-combined_k141_74233\|\|full | Not-determined |
| 18-101-final-viral-combined_k141_41739\|\|full | Not-determined |
| 18-101-final-viral-combined_k141_141152\|\|full | Not-determined |
| 18-101-final-viral-combined_k141_207495\|\|full | Not-determined |
| 18-101-final-viral-combined_k141_223243\|\|full | Low-quality |
| 18-101-final-viral-combined_k141_156653\|\|full | Low-quality |
| 18-101-final-viral-combined_k141_223253\|\|full | Not-determined |
| 18-101-final-viral-combined_k141_272540\|\|full | Low-quality |
| 18-101-final-viral-combined_k141_91492\|\|full | Low-quality |
| 18-101-final-viral-combined_k141_106984\|\|full | Low-quality |
| 18-101-final-viral-combined_k141_25500\|\|full | Low-quality |
| 18-101-final-viral-combined_k141_371277\|\|full | Low-quality |
| 18-101-final-viral-combined_k141_387998\|\|full | Not-determined |
| 18-101-final-viral-combined_k141_106996\|\|full | Low-quality |
| 18-101-final-viral-combined_k141_25503\|\|full | Low-quality |
| 18-101-final-viral-combined_k141_74265\|\|full | Not-determined |
| 18-101-final-viral-combined_k141_107001\|\|full | Low-quality |
| 18-101-final-viral-combined_k141_256865\|\|full | Low-quality |
| 18-101-final-viral-combined_k141_355408\|\|full | Not-determined |
| 18-101-final-viral-combined_k141_322982\|\|full | Not-determined |
| 18-101-final-viral-combined_k141_207518\|\|full | Low-quality |
| 18-101-final-viral-combined_k141_172876\|\|full | Low-quality |
| 18-101-final-viral-combined_k141_223296\|\|full | Not-determined |
| 18-101-final-viral-combined_k141_388049\|\|full | Not-determined |
| 18-101-final-viral-combined_k141_125392\|\|full | Not-determined |
| 18-101-final-viral-combined_k141_289029\|\|full | Low-quality |
| 18-101-final-viral-combined_k141_74298\|\|full | Low-quality |
| 18-101-final-viral-combined_k141_207548\|\|full | Low-quality |
| 18-101-final-viral-combined_k141_141196\|\|full | Not-determined |
| 18-101-final-viral-combined_k141_58377\|\|full | Not-determined |
| 18-101-final-viral-combined_k141_107040\|\|full | Not-determined |
| 18-101-final-viral-combined_k141_172897\|\|full | Not-determined |
| 18-101-final-viral-combined_k141_125416\|\|full | Low-quality |
| 18-101-final-viral-combined_k141_172923\|\|full | Low-quality |
| 18-101-final-viral-combined_k141_371321\|\|full | Low-quality |
| 18-101-final-viral-combined_k141_207568\|\|full | Low-quality |
| 18-101-final-viral-combined_k141_9511\|\|full | Not-determined |
| 18-101-final-viral-combined_k141_156733\|\|full | Not-determined |
| 18-101-final-viral-combined_k141_207569\|\|full | Not-determined |
| 18-101-final-viral-combined_k141_272598\|\|full | Low-quality |
| 18-101-final-viral-combined_k141_156747\|\|full | Low-quality |
| 18-101-final-viral-combined_k141_9524\|\|full | Low-quality |
| 18-101-final-viral-combined_k141_156749\|\|full | Low-quality |
| 18-101-final-viral-combined_k141_223319\|\|full | Low-quality |
| 18-101-final-viral-combined_k141_58411\|\|full | Not-determined |
| 18-101-final-viral-combined_k141_9534\|\|full | Not-determined |
| 18-101-final-viral-combined_k141_239998\|\|full | Not-determined |
| 18-101-final-viral-combined_k141_223324\|\|full | Not-determined |
| 18-101-final-viral-combined_k141_74342\|\|full | Low-quality |
| 18-101-final-viral-combined_k141_272620\|\|full | Low-quality |
| 18-101-final-viral-combined_k141_289113\|\|full | Low-quality |
| 18-101-final-viral-combined_k141_240006\|\|full | Not-determined |
| 18-101-final-viral-combined_k141_371355\|\|full | Low-quality |
| 18-102-final-viral-combined_k141_101826\|\|full | Complete |
| 18-101-final-viral-combined_k141_125463\|\|full | Low-quality |
| 18-101-final-viral-combined_k141_337676\|\|full | Low-quality |
| 18-101-final-viral-combined_k141_388114\|\|full | Not-determined |
| 18-101-final-viral-combined_k141_289120\|\|full | Not-determined |
| 18-101-final-viral-combined_k141_191141\|\|full | Low-quality |
| 18-101-final-viral-combined_k141_141232\|\|full | Not-determined |
| 18-101-final-viral-combined_k141_107102\|\|full | Not-determined |
| 18-101-final-viral-combined_k141_9568\|\|full | Not-determined |
| 18-101-final-viral-combined_k141_306287\|\|full | Low-quality |
| 18-101-final-viral-combined_k141_125471\|\|full | Not-determined |
| 18-101-final-viral-combined_k141_74361\|\|full | Not-determined |
| 18-101-final-viral-combined_k141_388132\|\|full | Not-determined |
| 18-101-final-viral-combined_k141_173016\|\|full | Low-quality |
| 18-101-final-viral-combined_k141_191183\|\|full | Not-determined |
| 18-101-final-viral-combined_k141_41851\|\|full | Not-determined |
| 18-101-final-viral-combined_k141_156785\|\|full | Low-quality |
| 18-101-final-viral-combined_k141_337706\|\|full | Low-quality |
| 18-101-final-viral-combined_k141_371391\|\|full | Low-quality |
| 18-101-final-viral-combined_k141_388155\|\|full | Low-quality |
| 18-101-final-viral-combined_k141_371397\|\|full | Low-quality |
| 18-101-final-viral-combined_k141_74412\|\|full | Not-determined |
| 18-101-final-viral-combined_k141_74413\|\|full | Low-quality |
| 18-101-final-viral-combined_k141_337711\|\|full | Not-determined |
| 18-101-final-viral-combined_k141_156796\|\|full | Not-determined |
| 18-101-final-viral-combined_k141_388173\|\|full | Low-quality |
| 18-101-final-viral-combined_k141_74424\|\|full | Not-determined |
| 18-101-final-viral-combined_k141_337725\|\|full | Low-quality |
| 18-101-final-viral-combined_k141_240048\|\|full | Low-quality |
| 18-101-final-viral-combined_k141_25725\|\|full | Low-quality |
| 18-101-final-viral-combined_k141_141285\|\|full | Not-determined |
| 18-101-final-viral-combined_k141_306336\|\|full | Not-determined |
| 18-101-final-viral-combined_k141_91687\|\|full | Low-quality |
| 18-101-final-viral-combined_k141_289179\|\|full | Not-determined |
| 18-101-final-viral-combined_k141_74441\|\|full | Low-quality |
| 18-101-final-viral-combined_k141_125544\|\|full | Not-determined |
| 18-101-final-viral-combined_k141_107140\|\|full | Low-quality |
| 18-101-final-viral-combined_k141_223356\|\|full | Not-determined |
| 18-101-final-viral-combined_k141_371448\|\|full | Not-determined |
| 18-101-final-viral-combined_k141_191206\|\|full | Not-determined |
| 18-101-final-viral-combined_k141_9677\|\|full | Not-determined |
| 18-101-final-viral-combined_k141_240090\|\|full | Low-quality |
| 18-101-final-viral-combined_k141_355665\|\|full | Not-determined |
| 18-101-final-viral-combined_k141_388241\|\|full | Not-determined |
| 18-101-final-viral-combined_k141_141318\|\|full | Low-quality |
| 18-101-final-viral-combined_k141_156842\|\|full | Low-quality |
| 18-101-final-viral-combined_k141_337775\|\|full | Low-quality |
| 18-101-final-viral-combined_k141_191223\|\|full | Low-quality |
| 18-101-final-viral-combined_k141_371493\|\|full | Not-determined |
| 18-101-final-viral-combined_k141_388267\|\|full | Not-determined |
| 18-101-final-viral-combined_k141_9727\|\|full | Low-quality |
| 18-101-final-viral-combined_k141_388278\|\|full | Not-determined |
| 18-101-final-viral-combined_k141_41971\|\|full | Not-determined |
| 18-101-final-viral-combined_k141_41986\|\|full | Low-quality |
| 18-101-final-viral-combined_k141_323197\|\|full | Not-determined |
| 18-101-final-viral-combined_k141_173095\|\|full | Low-quality |
| 18-101-final-viral-combined_k141_74528\|\|full | Not-determined |
| 18-101-final-viral-combined_k141_58467\|\|full | Not-determined |
| 18-101-final-viral-combined_k141_9760\|\|full | Not-determined |
| 18-101-final-viral-combined_k141_91800\|\|full | Not-determined |
| 18-101-final-viral-combined_k141_173115\|\|full | Not-determined |
| 18-101-final-viral-combined_k141_91809\|\|full | Not-determined |
| 18-101-final-viral-combined_k141_289221\|\|full | Low-quality |
| 18-101-final-viral-combined_k141_289223\|\|full | Not-determined |
| 18-101-final-viral-combined_k141_74545\|\|full | Not-determined |
| 18-101-final-viral-combined_k141_25812\|\|full | Low-quality |
| 18-101-final-viral-combined_k141_323240\|\|full | Not-determined |
| 18-101-final-viral-combined_k141_323242\|\|full | Low-quality |
| 18-101-final-viral-combined_k141_323250\|\|full | Low-quality |
| 18-101-final-viral-combined_k141_74561\|\|full | Low-quality |
| 18-101-final-viral-combined_k141_289236\|\|full | Not-determined |
| 18-101-final-viral-combined_k141_42013\|\|full | Not-determined |
| 18-101-final-viral-combined_k141_191296\|\|full | Not-determined |
| 18-101-final-viral-combined_k141_156897\|\|full | Not-determined |
| 18-101-final-viral-combined_k141_42023\|\|full | Not-determined |
| 18-101-final-viral-combined_k141_289252\|\|full | Not-determined |
| 18-101-final-viral-combined_k141_173137\|\|full | Not-determined |
| 18-101-final-viral-combined_k141_156909\|\|full | Low-quality |
| 18-101-final-viral-combined_k141_9830\|\|full | Low-quality |
| 18-101-final-viral-combined_k141_207771\|\|full | Low-quality |
| 18-101-final-viral-combined_k141_207776\|\|full | Not-determined |
| 18-101-final-viral-combined_k141_257156\|\|full | Not-determined |
| 18-101-final-viral-combined_k141_323295\|\|full | Not-determined |
| 18-101-final-viral-combined_k141_141431\|\|full | Not-determined |
| 18-101-final-viral-combined_k141_289291\|\|full | Not-determined |
| 18-101-final-viral-combined_k141_173169\|\|full | Low-quality |
| 18-101-final-viral-combined_k141_306481\|\|full | Not-determined |
| 18-101-final-viral-combined_k141_25859\|\|full | Not-determined |
| 18-101-final-viral-combined_k141_289301\|\|full | Not-determined |
| 18-101-final-viral-combined_k141_91841\|\|full | Low-quality |
| 18-101-final-viral-combined_k141_125627\|\|full | Low-quality |
| 18-101-final-viral-combined_k141_91843\|\|full | Low-quality |
| 18-101-final-viral-combined_k141_355791\|\|full | Low-quality |
| 18-101-final-viral-combined_k141_257165\|\|full | Not-determined |
| 18-101-final-viral-combined_k141_141449\|\|full | Not-determined |
| 18-101-final-viral-combined_k141_58534\|\|full | Low-quality |
| 18-101-final-viral-combined_k141_156935\|\|full | Low-quality |
| 18-101-final-viral-combined_k141_306499\|\|full | Not-determined |
| 18-101-final-viral-combined_k141_58543\|\|full | Low-quality |
| 18-101-final-viral-combined_k141_191364\|\|full | Low-quality |
| 18-101-final-viral-combined_k141_141459\|\|full | Low-quality |
| 18-101-final-viral-combined_k141_257176\|\|full | Not-determined |
| 18-101-final-viral-combined_k141_240223\|\|full | Low-quality |
| 18-101-final-viral-combined_k141_191383\|\|full | Low-quality |
| 18-101-final-viral-combined_k141_323361\|\|full | Not-determined |
| 18-101-final-viral-combined_k141_107367\|\|full | Low-quality |
| 18-101-final-viral-combined_k141_355800\|\|full | Low-quality |
| 18-101-final-viral-combined_k141_289369\|\|full | Not-determined |
| 18-101-final-viral-combined_k141_74632\|\|full | Low-quality |
| 18-101-final-viral-combined_k141_207842\|\|full | Not-determined |
| 18-101-final-viral-combined_k141_156991\|\|full | Not-determined |
| 18-101-final-viral-combined_k141_207856\|\|full | Low-quality |
| 18-101-final-viral-combined_k141_306555\|\|full | Not-determined |
| 18-101-final-viral-combined_k141_74656\|\|full | Not-determined |
| 18-101-final-viral-combined_k141_257254\|\|full | Low-quality |
| 18-101-final-viral-combined_k141_240282\|\|full | Not-determined |
| 18-101-final-viral-combined_k141_355830\|\|full | Low-quality |
| 18-101-final-viral-combined_k141_337961\|\|full | Not-determined |
| 18-101-final-viral-combined_k141_223398\|\|full | Low-quality |
| 18-101-final-viral-combined_k141_141510\|\|full | Not-determined |
| 18-101-final-viral-combined_k141_91860\|\|full | Low-quality |
| 18-101-final-viral-combined_k141_337972\|\|full | Not-determined |
| 18-101-final-viral-combined_k141_289428\|\|full | Not-determined |
| 18-101-final-viral-combined_k141_173207\|\|full | Not-determined |
| 18-101-final-viral-combined_k141_223414\|\|full | Not-determined |
| 18-101-final-viral-combined_k141_191447\|\|full | Not-determined |
| 18-101-final-viral-combined_k141_125676\|\|full | Not-determined |
| 18-101-final-viral-combined_k141_240307\|\|full | Low-quality |
| 18-101-final-viral-combined_k141_207894\|\|full | Low-quality |
| 18-101-final-viral-combined_k141_355849\|\|full | Low-quality |
| 18-101-final-viral-combined_k141_91884\|\|full | Low-quality |
| 18-101-final-viral-combined_k141_323418\|\|full | Low-quality |
| 18-101-final-viral-combined_k141_223426\|\|full | Not-determined |
| 18-101-final-viral-combined_k141_107426\|\|full | Low-quality |
| 18-101-final-viral-combined_k141_191473\|\|full | Not-determined |
| 18-101-final-viral-combined_k141_42129\|\|full | Not-determined |
| 18-101-final-viral-combined_k141_191475\|\|full | Low-quality |
| 18-101-final-viral-combined_k141_223439\|\|full | Not-determined |
| 18-101-final-viral-combined_k141_207919\|\|full | Low-quality |
| 18-101-final-viral-combined_k141_289473\|\|full | Low-quality |
| 18-101-final-viral-combined_k141_289478\|\|full | Not-determined |
| 18-101-final-viral-combined_k141_125700\|\|full | Not-determined |
| 18-101-final-viral-combined_k141_42142\|\|full | Low-quality |
| 18-101-final-viral-combined_k141_9952\|\|full | Low-quality |
| 18-101-final-viral-combined_k141_191497\|\|full | Not-determined |
| 18-101-final-viral-combined_k141_355917\|\|full | Not-determined |
| 18-101-final-viral-combined_k141_125729\|\|full | Low-quality |
| 18-101-final-viral-combined_k141_306652\|\|full | Low-quality |
| 18-101-final-viral-combined_k141_272921\|\|full | Low-quality |
| 18-101-final-viral-combined_k141_223487\|\|full | Not-determined |
| 18-101-final-viral-combined_k141_289508\|\|full | Low-quality |
| 18-101-final-viral-combined_k141_371624\|\|full | Low-quality |
| 18-101-final-viral-combined_k141_125749\|\|full | Low-quality |
| 18-101-final-viral-combined_k141_208007\|\|full | Not-determined |
| 18-101-final-viral-combined_k141_125754\|\|full | Low-quality |
| 18-101-final-viral-combined_k141_208011\|\|full | Low-quality |
| 18-101-final-viral-combined_k141_58668\|\|full | Not-determined |
| 18-101-final-viral-combined_k141_107506\|\|full | Not-determined |
| 18-101-final-viral-combined_k141_371633\|\|full | Low-quality |
| 18-101-final-viral-combined_k141_223512\|\|full | Low-quality |
| 18-101-final-viral-combined_k141_371637\|\|full | Not-determined |
| 18-101-final-viral-combined_k141_371642\|\|full | Not-determined |
| 18-101-final-viral-combined_k141_338044\|\|full | Low-quality |
| 18-101-final-viral-combined_k141_289530\|\|full | Low-quality |
| 18-101-final-viral-combined_k141_371653\|\|full | Not-determined |
| 18-101-final-viral-combined_k141_257326\|\|full | Not-determined |
| 18-101-final-viral-combined_k141_289545\|\|full | Not-determined |
| 18-101-final-viral-combined_k141_323459\|\|full | Low-quality |
| 18-101-final-viral-combined_k141_26072\|\|full | Low-quality |
| 18-101-final-viral-combined_k141_157083\|\|full | Not-determined |
| 18-101-final-viral-combined_k141_42182\|\|full | Low-quality |
| 18-101-final-viral-combined_k141_191590\|\|full | Not-determined |
| 18-101-final-viral-combined_k141_272928\|\|full | Low-quality |
| 18-101-final-viral-combined_k141_240407\|\|full | Low-quality |
| 18-101-final-viral-combined_k141_323508\|\|full | Low-quality |
| 18-101-final-viral-combined_k141_371685\|\|full | Not-determined |
| 18-101-final-viral-combined_k141_257356\|\|full | Low-quality |
| 18-101-final-viral-combined_k141_74714\|\|full | Not-determined |
| 18-101-final-viral-combined_k141_157140\|\|full | Low-quality |
| 18-101-final-viral-combined_k141_92003\|\|full | Not-determined |
| 18-101-final-viral-combined_k141_10069\|\|full | Not-determined |
| 18-101-final-viral-combined_k141_306743\|\|full | Not-determined |
| 18-101-final-viral-combined_k141_42225\|\|full | Not-determined |
| 18-101-final-viral-combined_k141_289629\|\|full | Low-quality |
| 18-101-final-viral-combined_k141_92013\|\|full | Not-determined |
| 18-101-final-viral-combined_k141_338133\|\|full | Low-quality |
| 18-101-final-viral-combined_k141_289631\|\|full | Low-quality |
| 18-101-final-viral-combined_k141_26125\|\|full | Not-determined |
| 18-101-final-viral-combined_k141_323547\|\|full | Low-quality |
| 18-101-final-viral-combined_k141_223601\|\|full | Low-quality |
| 18-101-final-viral-combined_k141_92035\|\|full | Not-determined |
| 18-101-final-viral-combined_k141_10093\|\|full | Not-determined |
| 18-101-final-viral-combined_k141_338175\|\|full | Not-determined |
| 18-101-final-viral-combined_k141_157175\|\|full | Low-quality |
| 18-101-final-viral-combined_k141_257376\|\|full | Low-quality |
| 18-101-final-viral-combined_k141_289640\|\|full | Low-quality |
| 18-101-final-viral-combined_k141_107632\|\|full | Not-determined |
| 18-101-final-viral-combined_k141_240481\|\|full | Not-determined |
| 18-101-final-viral-combined_k141_306778\|\|full | Low-quality |
| 18-101-final-viral-combined_k141_10105\|\|full | Low-quality |
| 18-101-final-viral-combined_k141_323584\|\|full | Not-determined |
| 18-101-final-viral-combined_k141_173326\|\|full | Not-determined |
| 17-106-final-viral-combined_k141_53991\|\|full | High-quality |
| 18-101-final-viral-combined_k141_173328\|\|full | Not-determined |
| 18-101-final-viral-combined_k141_10116\|\|full | Not-determined |
| 18-101-final-viral-combined_k141_107647\|\|full | Low-quality |
| 18-101-final-viral-combined_k141_157201\|\|full | Low-quality |
| 18-101-final-viral-combined_k141_289664\|\|full | Not-determined |
| 18-101-final-viral-combined_k141_371708\|\|full | Low-quality |
| 18-101-final-viral-combined_k141_257408\|\|full | Low-quality |
| 18-101-final-viral-combined_k141_125837\|\|full | Not-determined |
| 18-101-final-viral-combined_k141_10131\|\|full | Low-quality |
| 18-101-final-viral-combined_k141_173350\|\|full | Not-determined |
| 18-101-final-viral-combined_k141_125841\|\|full | Not-determined |
| 18-101-final-viral-combined_k141_323591\|\|full | Low-quality |
| 18-101-final-viral-combined_k141_355979\|\|full | Not-determined |
| 18-101-final-viral-combined_k141_371734\|\|full | Low-quality |
| 18-109-final-viral-combined_k141_84688\|\|full | Complete |
| 18-101-final-viral-combined_k141_338224\|\|full | Not-determined |
| 18-101-final-viral-combined_k141_371753\|\|full | Not-determined |
| 18-101-final-viral-combined_k141_173376\|\|full | Not-determined |
| 18-101-final-viral-combined_k141_191640\|\|full | Not-determined |
| 18-101-final-viral-combined_k141_223643\|\|full | Not-determined |
| 18-101-final-viral-combined_k141_306814\|\|full | Low-quality |
| 18-101-final-viral-combined_k141_141550\|\|full | Low-quality |
| 18-101-final-viral-combined_k141_356030\|\|full | Not-determined |
| 18-101-final-viral-combined_k141_338245\|\|full | Low-quality |
| 18-101-final-viral-combined_k141_273001\|\|full | Low-quality |
| 18-101-final-viral-combined_k141_157257\|\|full | Not-determined |
| 18-101-final-viral-combined_k141_141568\|\|full | Not-determined |
| 18-101-final-viral-combined_k141_125857\|\|full | Low-quality |
| 18-101-final-viral-combined_k141_157270\|\|full | Not-determined |
| 18-101-final-viral-combined_k141_289702\|\|full | Not-determined |
| 18-101-final-viral-combined_k141_191650\|\|full | Not-determined |
| 18-101-final-viral-combined_k141_356053\|\|full | Not-determined |
| 18-101-final-viral-combined_k141_289704\|\|full | Not-determined |
| 18-101-final-viral-combined_k141_273006\|\|full | Low-quality |
| 18-101-final-viral-combined_k141_223663\|\|full | Low-quality |
| 18-101-final-viral-combined_k141_173400\|\|full | Low-quality |
| 18-101-final-viral-combined_k141_107666\|\|full | Not-determined |
| 18-101-final-viral-combined_k141_173406\|\|full | Low-quality |
| 18-101-final-viral-combined_k141_371760\|\|full | Low-quality |
| 18-101-final-viral-combined_k141_125888\|\|full | Low-quality |
| 18-101-final-viral-combined_k141_58861\|\|full | Low-quality |
| 18-101-final-viral-combined_k141_338282\|\|full | Low-quality |
| 18-101-final-viral-combined_k141_141578\|\|full | Not-determined |
| 18-101-final-viral-combined_k141_323644\|\|full | Low-quality |
| 18-101-final-viral-combined_k141_257496\|\|full | Not-determined |
| 18-101-final-viral-combined_k141_306863\|\|full | Not-determined |
| 18-101-final-viral-combined_k141_240533\|\|full | Low-quality |
| 18-101-final-viral-combined_k141_141588\|\|full | Not-determined |
| 18-101-final-viral-combined_k141_289714\|\|full | Not-determined |
| 18-101-final-viral-combined_k141_208248\|\|full | Low-quality |
| 18-101-final-viral-combined_k141_356106\|\|full | Not-determined |
| 18-101-final-viral-combined_k141_240544\|\|full | Not-determined |
| 18-101-final-viral-combined_k141_306879\|\|full | Not-determined |
| 18-101-final-viral-combined_k141_371791\|\|full | Not-determined |
| 18-101-final-viral-combined_k141_306898\|\|full | Low-quality |
| 18-101-final-viral-combined_k141_257516\|\|full | Not-determined |
| 18-101-final-viral-combined_k141_92191\|\|full | Low-quality |
| 18-101-final-viral-combined_k141_356114\|\|full | Low-quality |
| 18-101-final-viral-combined_k141_208276\|\|full | Not-determined |
| 18-101-final-viral-combined_k141_223700\|\|full | Low-quality |
| 18-101-final-viral-combined_k141_356123\|\|full | Low-quality |
| 18-101-final-viral-combined_k141_58921\|\|full | Not-determined |
| 18-101-final-viral-combined_k141_306919\|\|full | Low-quality |
| 18-101-final-viral-combined_k141_356133\|\|full | Low-quality |
| 18-101-final-viral-combined_k141_338346\|\|full | Low-quality |
| 18-101-final-viral-combined_k141_92192\|\|full | Low-quality |
| 18-101-final-viral-combined_k141_26337\|\|full | Low-quality |
| 18-101-final-viral-combined_k141_42397\|\|full | Not-determined |
| 18-101-final-viral-combined_k141_74781\|\|full | Not-determined |
| 18-101-final-viral-combined_k141_74782\|\|full | Not-determined |
| 18-101-final-viral-combined_k141_26353\|\|full | Low-quality |
| 18-101-final-viral-combined_k141_42403\|\|full | Not-determined |
| 18-101-final-viral-combined_k141_208307\|\|full | Not-determined |
| 18-101-final-viral-combined_k141_289752\|\|full | Low-quality |
| 18-101-final-viral-combined_k141_306933\|\|full | Low-quality |
| 18-101-final-viral-combined_k141_107751\|\|full | Low-quality |
| 18-101-final-viral-combined_k141_141626\|\|full | Low-quality |
| 18-101-final-viral-combined_k141_240594\|\|full | Low-quality |
| 18-101-final-viral-combined_k141_141627\|\|full | Not-determined |
| 18-101-final-viral-combined_k141_107757\|\|full | Not-determined |
| 18-101-final-viral-combined_k141_191743\|\|full | Not-determined |
| 18-101-final-viral-combined_k141_257584\|\|full | Not-determined |
| 18-101-final-viral-combined_k141_141646\|\|full | Low-quality |
| 18-101-final-viral-combined_k141_191754\|\|full | Low-quality |
| 18-101-final-viral-combined_k141_289787\|\|full | Not-determined |
| 18-101-final-viral-combined_k141_42445\|\|full | Not-determined |
| 18-101-final-viral-combined_k141_10203\|\|full | Low-quality |
| 18-101-final-viral-combined_k141_273054\|\|full | Not-determined |
| 18-101-final-viral-combined_k141_289796\|\|full | Low-quality |
| 18-101-final-viral-combined_k141_125983\|\|full | Low-quality |
| 18-101-final-viral-combined_k141_58968\|\|full | Low-quality |
| 18-101-final-viral-combined_k141_74851\|\|full | Not-determined |
| 18-101-final-viral-combined_k141_26367\|\|full | Low-quality |
| 18-101-final-viral-combined_k141_356210\|\|full | Not-determined |
| 18-101-final-viral-combined_k141_257617\|\|full | Not-determined |
| 18-101-final-viral-combined_k141_273088\|\|full | Not-determined |
| 18-101-final-viral-combined_k141_257634\|\|full | Not-determined |
| 18-101-final-viral-combined_k141_257635\|\|full | Low-quality |
| 18-101-final-viral-combined_k141_42460\|\|full | Low-quality |
| 18-101-final-viral-combined_k141_173467\|\|full | Not-determined |
| 18-101-final-viral-combined_k141_338393\|\|full | Not-determined |
| 18-101-final-viral-combined_k141_289836\|\|full | Not-determined |
| 18-101-final-viral-combined_k141_58986\|\|full | Low-quality |
| 18-101-final-viral-combined_k141_141716\|\|full | Low-quality |
| 18-101-final-viral-combined_k141_273106\|\|full | Low-quality |
| 18-101-final-viral-combined_k141_208347\|\|full | Low-quality |
| 18-106-final-viral-combined_k141_119138\|\|full | Medium-quality |
| 18-101-final-viral-combined_k141_240655\|\|full | Not-determined |
| 18-101-final-viral-combined_k141_356255\|\|full | Not-determined |
| 18-101-final-viral-combined_k141_92329\|\|full | Not-determined |
| 18-101-final-viral-combined_k141_306989\|\|full | Low-quality |
| 18-101-final-viral-combined_k141_157453\|\|full | Low-quality |
| 18-101-final-viral-combined_k141_371924\|\|full | Low-quality |
| 18-101-final-viral-combined_k141_92333\|\|full | Low-quality |
| 18-101-final-viral-combined_k141_42482\|\|full | Not-determined |
| 18-101-final-viral-combined_k141_240679\|\|full | Not-determined |
| 18-101-final-viral-combined_k141_141757\|\|full | Not-determined |
| 18-101-final-viral-combined_k141_289873\|\|full | Low-quality |
| 18-101-final-viral-combined_k141_257680\|\|full | Low-quality |
| 18-101-final-viral-combined_k141_42505\|\|full | Low-quality |
| 18-101-final-viral-combined_k141_257686\|\|full | Low-quality |
| 18-101-final-viral-combined_k141_273155\|\|full | Not-determined |
| 18-101-final-viral-combined_k141_257700\|\|full | Not-determined |
| 18-101-final-viral-combined_k141_26394\|\|full | Not-determined |
| 18-101-final-viral-combined_k141_191829\|\|full | Low-quality |
| 18-101-final-viral-combined_k141_273159\|\|full | Low-quality |
| 18-101-final-viral-combined_k141_223907\|\|full | Low-quality |
| 18-101-final-viral-combined_k141_223909\|\|full | Low-quality |
| 18-101-final-viral-combined_k141_191839\|\|full | Not-determined |
| 18-101-final-viral-combined_k141_92368\|\|full | Not-determined |
| 18-101-final-viral-combined_k141_388394\|\|full | Low-quality |
| 18-101-final-viral-combined_k141_323830\|\|full | Low-quality |
| 18-101-final-viral-combined_k141_289920\|\|full | Low-quality |
| 18-101-final-viral-combined_k141_59018\|\|full | Not-determined |
| 18-101-final-viral-combined_k141_173515\|\|full | Low-quality |
| 18-101-final-viral-combined_k141_59021\|\|full | Not-determined |
| 18-101-final-viral-combined_k141_92377\|\|full | Low-quality |
| 18-101-final-viral-combined_k141_10264\|\|full | Not-determined |
| 18-101-final-viral-combined_k141_173523\|\|full | Not-determined |
| 18-101-final-viral-combined_k141_388411\|\|full | Low-quality |
| 18-101-final-viral-combined_k141_240740\|\|full | Not-determined |
| 18-101-final-viral-combined_k141_173531\|\|full | Low-quality |
| 18-101-final-viral-combined_k141_356345\|\|full | Not-determined |
| 18-101-final-viral-combined_k141_173534\|\|full | Low-quality |
| 18-101-final-viral-combined_k141_191846\|\|full | Not-determined |
| 18-101-final-viral-combined_k141_173536\|\|full | Not-determined |
| 18-101-final-viral-combined_k141_323856\|\|full | Not-determined |
| 18-101-final-viral-combined_k141_240744\|\|full | Low-quality |
| 18-101-final-viral-combined_k141_338441\|\|full | Low-quality |
| 18-101-final-viral-combined_k141_59047\|\|full | Low-quality |
| 18-101-final-viral-combined_k141_208446\|\|full | Low-quality |
| 18-101-final-viral-combined_k141_173544\|\|full | Low-quality |
| 18-101-final-viral-combined_k141_372006\|\|full | Low-quality |
| 18-102-final-viral-combined_k141_174241\|\|full | Medium-quality |
| 18-101-final-viral-combined_k141_356352\|\|full | Not-determined |
| 18-101-final-viral-combined_k141_289954\|\|full | Not-determined |
| 18-101-final-viral-combined_k141_157541\|\|full | Not-determined |
| 18-101-final-viral-combined_k141_240763\|\|full | Low-quality |
| 18-101-final-viral-combined_k141_338458\|\|full | Low-quality |
| 18-101-final-viral-combined_k141_240766\|\|full | Not-determined |
| 18-101-final-viral-combined_k141_191869\|\|full | Not-determined |
| 18-101-final-viral-combined_k141_157544\|\|full | Low-quality |
| 18-101-final-viral-combined_k141_74953\|\|full | Low-quality |
| 18-101-final-viral-combined_k141_257785\|\|full | Low-quality |
| 18-101-final-viral-combined_k141_141832\|\|full | Not-determined |
| 18-101-final-viral-combined_k141_289965\|\|full | Not-determined |
| 18-101-final-viral-combined_k141_223988\|\|full | Not-determined |
| 18-101-final-viral-combined_k141_273226\|\|full | Low-quality |
| 18-101-final-viral-combined_k141_223989\|\|full | Not-determined |
| 18-101-final-viral-combined_k141_173568\|\|full | Not-determined |
| 18-101-final-viral-combined_k141_26417\|\|full | Not-determined |
| 18-101-final-viral-combined_k141_126047\|\|full | Not-determined |
| 18-101-final-viral-combined_k141_273233\|\|full | Low-quality |
| 18-101-final-viral-combined_k141_126049\|\|full | Not-determined |
| 18-101-final-viral-combined_k141_257788\|\|full | Low-quality |
| 18-101-final-viral-combined_k141_59084\|\|full | Not-determined |
| 18-101-final-viral-combined_k141_42601\|\|full | Not-determined |
| 18-101-final-viral-combined_k141_289979\|\|full | Not-determined |
| 18-101-final-viral-combined_k141_42605\|\|full | Low-quality |
| 18-101-final-viral-combined_k141_59085\|\|full | Not-determined |
| 18-101-final-viral-combined_k141_273236\|\|full | Not-determined |
| 18-101-final-viral-combined_k141_224033\|\|full | Low-quality |
| 18-101-final-viral-combined_k141_307146\|\|full | Low-quality |
| 18-101-final-viral-combined_k141_26447\|\|full | Not-determined |
| 18-101-final-viral-combined_k141_323918\|\|full | Low-quality |
| 18-101-final-viral-combined_k141_372053\|\|full | Low-quality |
| 18-101-final-viral-combined_k141_273238\|\|full | Low-quality |
| 18-101-final-viral-combined_k141_10286\|\|full | Not-determined |
| 18-101-final-viral-combined_k141_356368\|\|full | Not-determined |
| 18-101-final-viral-combined_k141_323922\|\|full | Low-quality |
| 18-101-final-viral-combined_k141_141867\|\|full | Low-quality |
| 18-101-final-viral-combined_k141_372074\|\|full | Not-determined |
| 18-101-final-viral-combined_k141_257827\|\|full | Low-quality |
| 18-101-final-viral-combined_k141_126086\|\|full | Low-quality |
| 18-101-final-viral-combined_k141_372078\|\|full | Not-determined |
| 18-101-final-viral-combined_k141_75002\|\|full | Low-quality |
| 18-101-final-viral-combined_k141_338499\|\|full | Low-quality |
| 18-101-final-viral-combined_k141_224057\|\|full | Low-quality |
| 18-101-final-viral-combined_k141_59103\|\|full | Low-quality |
| 18-101-final-viral-combined_k141_338503\|\|full | Low-quality |
| 18-101-final-viral-combined_k141_356369\|\|full | Low-quality |
| 18-101-final-viral-combined_k141_307184\|\|full | Not-determined |
| 18-101-final-viral-combined_k141_126103\|\|full | Not-determined |
| 18-101-final-viral-combined_k141_75019\|\|full | Not-determined |
| 18-101-final-viral-combined_k141_42645\|\|full | Not-determined |
| 18-101-final-viral-combined_k141_26465\|\|full | Not-determined |
| 18-101-final-viral-combined_k141_323942\|\|full | Low-quality |
| 18-101-final-viral-combined_k141_257851\|\|full | Not-determined |
| 18-101-final-viral-combined_k141_356387\|\|full | Not-determined |
| 18-101-final-viral-combined_k141_141883\|\|full | Low-quality |
| 18-101-final-viral-combined_k141_126124\|\|full | Low-quality |
| 18-101-final-viral-combined_k141_323952\|\|full | Low-quality |
| 18-101-final-viral-combined_k141_257856\|\|full | Not-determined |
| 18-101-final-viral-combined_k141_290074\|\|full | Low-quality |
| 18-101-final-viral-combined_k141_157632\|\|full | Not-determined |
| 18-101-final-viral-combined_k141_356401\|\|full | Not-determined |
| 18-101-final-viral-combined_k141_372134\|\|full | Not-determined |
| 18-101-final-viral-combined_k141_224072\|\|full | Low-quality |
| 18-101-final-viral-combined_k141_92497\|\|full | Low-quality |
| 18-101-final-viral-combined_k141_59122\|\|full | Not-determined |
| 18-101-final-viral-combined_k141_356411\|\|full | Low-quality |
| 18-101-final-viral-combined_k141_338549\|\|full | Not-determined |
| 18-101-final-viral-combined_k141_126170\|\|full | Not-determined |
| 18-101-final-viral-combined_k141_257885\|\|full | Not-determined |
| 18-101-final-viral-combined_k141_307207\|\|full | Not-determined |
| 18-101-final-viral-combined_k141_10362\|\|full | Low-quality |
| 18-101-final-viral-combined_k141_356422\|\|full | Not-determined |
| 18-101-final-viral-combined_k141_26524\|\|full | Low-quality |
| 18-101-final-viral-combined_k141_273282\|\|full | Low-quality |
| 18-101-final-viral-combined_k141_75087\|\|full | Not-determined |
| 18-101-final-viral-combined_k141_388490\|\|full | Low-quality |
| 18-101-final-viral-combined_k141_338569\|\|full | Not-determined |
| 18-101-final-viral-combined_k141_157669\|\|full | Low-quality |
| 18-101-final-viral-combined_k141_173712\|\|full | Low-quality |
| 18-101-final-viral-combined_k141_257896\|\|full | Low-quality |
| 18-101-final-viral-combined_k141_372179\|\|full | Low-quality |
| 18-101-final-viral-combined_k141_173733\|\|full | Not-determined |
| 18-101-final-viral-combined_k141_173737\|\|full | Not-determined |
| 18-101-final-viral-combined_k141_26544\|\|full | Not-determined |
| 18-101-final-viral-combined_k141_388509\|\|full | Low-quality |
| 18-101-final-viral-combined_k141_338582\|\|full | Not-determined |
| 18-101-final-viral-combined_k141_107990\|\|full | Not-determined |
| 18-101-final-viral-combined_k141_42694\|\|full | Not-determined |
| 18-101-final-viral-combined_k141_240940\|\|full | Not-determined |
| 18-101-final-viral-combined_k141_257934\|\|full | Low-quality |
| 18-101-final-viral-combined_k141_208550\|\|full | Not-determined |
| 18-101-final-viral-combined_k141_141993\|\|full | Not-determined |
| 17-109-final-viral-combined_k141_313953\|\|full | High-quality |
| 18-101-final-viral-combined_k141_224136\|\|full | Not-determined |
| 18-101-final-viral-combined_k141_173761\|\|full | Low-quality |
| 18-101-final-viral-combined_k141_26559\|\|full | Low-quality |
| 18-101-final-viral-combined_k141_273362\|\|full | Not-determined |
| 18-101-final-viral-combined_k141_307242\|\|full | Low-quality |
| 18-101-final-viral-combined_k141_75145\|\|full | Not-determined |
| 18-101-final-viral-combined_k141_356498\|\|full | Not-determined |
| 18-101-final-viral-combined_k141_257962\|\|full | Not-determined |
| 18-101-final-viral-combined_k141_307250\|\|full | Not-determined |
| 18-101-final-viral-combined_k141_224160\|\|full | Not-determined |
| 18-101-final-viral-combined_k141_208588\|\|full | Not-determined |
| 18-101-final-viral-combined_k141_173782\|\|full | Not-determined |
| 18-101-final-viral-combined_k141_126302\|\|full | Not-determined |
| 18-101-final-viral-combined_k141_324069\|\|full | Low-quality |

18-101-final-viral-combined_k141_173800||full Low-quality
18-101-final-viral-combined_k141_257972||full Not-determined
18-101-final-viral-combined_k141_372268||full Not-determined
18-101-final-viral-combined_k141_59189||full Not-determined
18-101-final-viral-combined_k141_338624||full Low-quality
18-101-final-viral-combined_k141_388554||full Low-quality
18-101-final-viral-combined_k141_108051||full Low-quality
18-101-final-viral-combined_k141_191963||full Low-quality
18-101-final-viral-combined_k141_108068||full Low-quality
18-101-final-viral-combined_k141_42771||full Low-quality
18-101-final-viral-combined_k141_240967||full Low-quality
18-101-final-viral-combined_k141_59204||full Not-determined
18-101-final-viral-combined_k141_108071||full Not-determined
18-101-final-viral-combined_k141_372283||full Low-quality
18-101-final-viral-combined_k141_324115||full Not-determined
18-101-final-viral-combined_k141_388583||full Not-determined
18-101-final-viral-combined_k141_307299||full Not-determined
18-101-final-viral-combined_k141_26606||full Not-determined
18-101-final-viral-combined_k141_142045||full Not-determined
18-101-final-viral-combined_k141_388584||full Not-determined
18-101-final-viral-combined_k141_290203||full Not-determined
18-101-final-viral-combined_k141_59226||full Not-determined
18-101-final-viral-combined_k141_388587||full Low-quality
18-101-final-viral-combined_k141_224189||full Not-determined
18-101-final-viral-combined_k141_26615||full Not-determined
18-101-final-viral-combined_k141_324141||full Not-determined
18-101-final-viral-combined_k141_208661||full Not-determined
18-101-final-viral-combined_k141_258020||full Low-quality
18-101-final-viral-combined_k141_92636||full Not-determined
18-101-final-viral-combined_k141_142067||full Not-determined
18-101-final-viral-combined_k141_388606||full Not-determined
18-101-final-viral-combined_k141_324147||full Low-quality
18-101-final-viral-combined_k141_356572||full Not-determined
18-101-final-viral-combined_k141_290214||full Low-quality
18-101-final-viral-combined_k141_240987||full Not-determined
18-101-final-viral-combined_k141_324150||full Low-quality
18-101-final-viral-combined_k141_240990||full Not-determined
18-101-final-viral-combined_k141_241002||full Not-determined
18-101-final-viral-combined_k141_192004||full Not-determined
18-101-final-viral-combined_k141_290233||full Not-determined
18-101-final-viral-combined_k141_59261||full Not-determined
18-101-final-viral-combined_k141_372347||full Low-quality
18-101-final-viral-combined_k141_108099||full Not-determined
18-101-final-viral-combined_k141_208687||full Not-determined
18-101-final-viral-combined_k141_92671||full Low-quality
18-101-final-viral-combined_k141_208691||full Not-determined
18-101-final-viral-combined_k141_224229||full Not-determined
18-101-final-viral-combined_k141_192021||full Low-quality
18-101-final-viral-combined_k141_92681||full Low-quality
18-101-final-viral-combined_k141_338697||full Not-determined
18-101-final-viral-combined_k141_208701||full Not-determined
18-101-final-viral-combined_k141_372362||full Low-quality
18-101-final-viral-combined_k141_258081||full Low-quality
18-101-final-viral-combined_k141_108121||full Not-determined
18-101-final-viral-combined_k141_372368||full Low-quality
18-101-final-viral-combined_k141_208706||full Not-determined
18-101-final-viral-combined_k141_42834||full Low-quality
18-101-final-viral-combined_k141_59281||full Low-quality
18-101-final-viral-combined_k141_324199||full Low-quality
18-101-final-viral-combined_k141_258096||full Not-determined
18-101-final-viral-combined_k141_157791||full Low-quality
18-101-final-viral-combined_k141_173897||full Low-quality
18-101-final-viral-combined_k141_192068||full Low-quality
18-101-final-viral-combined_k141_208724||full Not-determined
18-101-final-viral-combined_k141_192089||full Not-determined
18-101-final-viral-combined_k141_92728||full Low-quality
18-101-final-viral-combined_k141_108157||full Not-determined
18-101-final-viral-combined_k141_59310||full Low-quality
18-101-final-viral-combined_k141_241034||full Low-quality
18-101-final-viral-combined_k141_59315||full Not-determined
18-101-final-viral-combined_k141_258121||full Low-quality
18-101-final-viral-combined_k141_92752||full Low-quality
18-101-final-viral-combined_k141_59321||full Low-quality
18-101-final-viral-combined_k141_338748||full Not-determined
18-101-final-viral-combined_k141_372429||full Not-determined
18-101-final-viral-combined_k141_157822||full Not-determined
18-101-final-viral-combined_k141_372440||full Low-quality
18-101-final-viral-combined_k141_157834||full Not-determined
18-101-final-viral-combined_k141_208752||full Low-quality
18-101-final-viral-combined_k141_126470||full Not-determined
18-101-final-viral-combined_k141_356662||full Not-determined
18-101-final-viral-combined_k141_372461||full Low-quality
18-101-final-viral-combined_k141_208764||full Not-determined
18-101-final-viral-combined_k141_157861||full Low-quality
18-101-final-viral-combined_k141_356683||full Low-quality
18-101-final-viral-combined_k141_142187||full Low-quality
18-101-final-viral-combined_k141_208775||full Not-determined
18-101-final-viral-combined_k141_241111||full Not-determined
18-101-final-viral-combined_k141_126510||full Low-quality
18-101-final-viral-combined_k141_338785||full Not-determined
18-101-final-viral-combined_k141_173982||full Low-quality
18-101-final-viral-combined_k141_59383||full Low-quality
18-101-final-viral-combined_k141_42968||full Low-quality
18-101-final-viral-combined_k141_126519||full Low-quality
18-101-final-viral-combined_k141_59399||full Low-quality
18-101-final-viral-combined_k141_173997||full Low-quality
18-101-final-viral-combined_k141_388731||full Not-determined
18-101-final-viral-combined_k141_258197||full Not-determined
18-101-final-viral-combined_k141_388737||full Low-quality
18-101-final-viral-combined_k141_258199||full Not-determined
18-101-final-viral-combined_k141_258203||full Not-determined
18-101-final-viral-combined_k141_388742||full Not-determined
18-101-final-viral-combined_k141_157923||full Low-quality
18-101-final-viral-combined_k141_290285||full Low-quality
18-101-final-viral-combined_k141_338816||full Not-determined
18-101-final-viral-combined_k141_388769||full Low-quality
18-101-final-viral-combined_k141_338818||full Low-quality
18-101-final-viral-combined_k141_290298||full Not-determined
18-101-final-viral-combined_k141_356732||full Low-quality
18-101-final-viral-combined_k141_388771||full Not-determined
18-101-final-viral-combined_k141_43000||full Low-quality
18-101-final-viral-combined_k141_356733||full Not-determined
18-101-final-viral-combined_k141_192205||full Low-quality
18-101-final-viral-combined_k141_224402||full Not-determined
18-101-final-viral-combined_k141_273527||full Not-determined
18-101-final-viral-combined_k141_241188||full Not-determined
18-101-final-viral-combined_k141_338837||full Not-determined
18-101-final-viral-combined_k141_192209||full Low-quality
18-101-final-viral-combined_k141_59467||full Low-quality
18-101-final-viral-combined_k141_142293||full Low-quality
18-101-final-viral-combined_k141_208837||full Not-determined
18-101-final-viral-combined_k141_290347||full Not-determined
18-101-final-viral-combined_k141_157954||full Not-determined
18-101-final-viral-combined_k141_290350||full Not-determined
18-101-final-viral-combined_k141_372498||full Low-quality
18-101-final-viral-combined_k141_192250||full Low-quality
18-101-final-viral-combined_k141_324293||full Not-determined
18-101-final-viral-combined_k141_241218||full Low-quality
18-101-final-viral-combined_k141_241222||full Low-quality
18-101-final-viral-combined_k141_307324||full Not-determined
18-101-final-viral-combined_k141_258249||full Not-determined
18-101-final-viral-combined_k141_10426||full Not-determined
18-101-final-viral-combined_k141_258255||full Low-quality
18-101-final-viral-combined_k141_241242||full Low-quality
18-101-final-viral-combined_k141_241248||full Not-determined
18-101-final-viral-combined_k141_126617||full Not-determined
18-101-final-viral-combined_k141_126618||full Not-determined
18-101-final-viral-combined_k141_258282||full Not-determined
18-101-final-viral-combined_k141_258290||full Low-quality
18-105_k141_85650 High-quality
18-101-final-viral-combined_k141_157998||full Not-determined
18-101-final-viral-combined_k141_224446||full Low-quality
18-101-final-viral-combined_k141_43075||full Low-quality
18-101-final-viral-combined_k141_324331||full Low-quality
18-101-final-viral-combined_k141_388854||full Not-determined
18-101-final-viral-combined_k141_126672||full Low-quality
18-101-final-viral-combined_k141_388862||full Not-determined
18-101-final-viral-combined_k141_142368||full Not-determined
18-101-final-viral-combined_k141_142374||full Not-determined
18-101-final-viral-combined_k141_241318||full Low-quality
18-101-final-viral-combined_k141_43109||full Low-quality
18-101-final-viral-combined_k141_59551||full Not-determined
18-101-final-viral-combined_k141_108364||full Not-determined
18-101-final-viral-combined_k141_10502||full Not-determined
18-101-final-viral-combined_k141_208925||full Low-quality
18-101-final-viral-combined_k141_192321||full Not-determined
18-101-final-viral-combined_k141_142395||full Not-determined
18-101-final-viral-combined_k141_290423||full Not-determined
18-101-final-viral-combined_k141_372596||full Not-determined
18-101-final-viral-combined_k141_307388||full Not-determined
18-101-final-viral-combined_k141_356816||full Not-determined
18-101-final-viral-combined_k141_258313||full Not-determined
18-101-final-viral-combined_k141_93004||full Not-determined
18-101-final-viral-combined_k141_174133||full Low-quality
18-101-final-viral-combined_k141_258316||full Low-quality
18-101-final-viral-combined_k141_142408||full Low-quality
18-101-final-viral-combined_k141_59579||full Low-quality
18-101-final-viral-combined_k141_273637||full Not-determined
18-101-final-viral-combined_k141_356823||full Not-determined
18-101-final-viral-combined_k141_10521||full Low-quality
18-101-final-viral-combined_k141_158066||full Not-determined
18-101-final-viral-combined_k141_126717||full Low-quality
18-101-final-viral-combined_k141_158073||full Low-quality
18-101-final-viral-combined_k141_258317||full Not-determined
18-101-final-viral-combined_k141_108394||full Low-quality
18-101-final-viral-combined_k141_75464||full Not-determined
18-101-final-viral-combined_k141_258321||full Low-quality
18-101-final-viral-combined_k141_356848||full Low-quality
18-101-final-viral-combined_k141_26768||full Not-determined
18-101-final-viral-combined_k141_273667||full Low-quality
18-101-final-viral-combined_k141_93038||full Not-determined
18-101-final-viral-combined_k141_43153||full Low-quality
18-101-final-viral-combined_k141_356859||full Not-determined
18-101-final-viral-combined_k141_258341||full Low-quality
18-101-final-viral-combined_k141_43167||full Not-determined
18-101-final-viral-combined_k141_356867||full Not-determined
18-101-final-viral-combined_k141_142451||full Low-quality
18-101-final-viral-combined_k141_26797||full Low-quality
18-101-final-viral-combined_k141_388889||full Not-determined
18-101-final-viral-combined_k141_93047||full Not-determined
18-101-final-viral-combined_k141_307405||full Not-determined
18-101-final-viral-combined_k141_43177||full Low-quality
18-101-final-viral-combined_k141_142454||full Not-determined
18-101-final-viral-combined_k141_43178||full Not-determined
18-101-final-viral-combined_k141_10571||full Not-determined
18-101-final-viral-combined_k141_108430||full Low-quality
18-101-final-viral-combined_k141_273678||full Low-quality
18-101-final-viral-combined_k141_174173||full Not-determined
18-101-final-viral-combined_k141_192398||full Not-determined
18-101-final-viral-combined_k141_93065||full Not-determined
18-101-final-viral-combined_k141_290494||full Not-determined
18-101-final-viral-combined_k141_108452||full Not-determined
18-101-final-viral-combined_k141_26821||full Low-quality
18-101-final-viral-combined_k141_324368||full Not-determined
18-101-final-viral-combined_k141_93073||full Not-determined
18-101-final-viral-combined_k141_59653||full Not-determined
18-101-final-viral-combined_k141_158110||full Low-quality
18-101-final-viral-combined_k141_174209||full Not-determined
18-101-final-viral-combined_k141_273689||full Low-quality
18-101-final-viral-combined_k141_26840||full Not-determined
18-101-final-viral-combined_k141_142461||full Not-determined
18-101-final-viral-combined_k141_174221||full Low-quality
18-101-final-viral-combined_k141_126778||full Not-determined
18-101-final-viral-combined_k141_10609||full Not-determined
18-101-final-viral-combined_k141_388920||full Not-determined
18-101-final-viral-combined_k141_388921||full Low-quality
18-101-final-viral-combined_k141_209016||full Low-quality
18-101-final-viral-combined_k141_372636||full Not-determined
18-101-final-viral-combined_k141_388929||full Not-determined
18-101-final-viral-combined_k141_93106||full Low-quality
18-101-final-viral-combined_k141_142467||full Low-quality
18-101-final-viral-combined_k141_10641||full Low-quality
18-101-final-viral-combined_k141_126796||full Not-determined
18-101-final-viral-combined_k141_142473||full Low-quality
18-101-final-viral-combined_k141_290537||full Low-quality
18-101-final-viral-combined_k141_356908||full Not-determined
18-101-final-viral-combined_k141_273732||full Not-determined
18-101-final-viral-combined_k141_10650||full Not-determined
18-101-final-viral-combined_k141_290543||full Low-quality
18-101-final-viral-combined_k141_142486||full Not-determined
18-101-final-viral-combined_k141_209064||full Low-quality
18-101-final-viral-combined_k141_93121||full Low-quality
18-101-final-viral-combined_k141_273761||full Low-quality
18-102-final-viral-combined_k141_62377||full High-quality
18-101-final-viral-combined_k141_338951||full Not-determined
18-101-final-viral-combined_k141_108511||full Low-quality
18-101-final-viral-combined_k141_26890||full Low-quality
18-101-final-viral-combined_k141_307508||full Not-determined
18-101-final-viral-combined_k141_192472||full Not-determined
18-101-final-viral-combined_k141_158185||full Low-quality
18-101-final-viral-combined_k141_338965||full Low-quality
18-101-final-viral-combined_k141_192487||full Not-determined
18-101-final-viral-combined_k141_158200||full Not-determined
18-101-final-viral-combined_k141_126852||full Low-quality
18-101-final-viral-combined_k141_108538||full Low-quality
18-101-final-viral-combined_k141_258450||full Not-determined
18-101-final-viral-combined_k141_126855||full Not-determined
18-101-final-viral-combined_k141_93144||full Not-determined
18-101-final-viral-combined_k141_338985||full Low-quality
18-101-final-viral-combined_k141_142532||full Low-quality
18-101-final-viral-combined_k141_75603||full Not-determined
18-101-final-viral-combined_k141_10701||full Not-determined
18-101-final-viral-combined_k141_356923||full Not-determined
18-101-final-viral-combined_k141_10704||full Low-quality
18-101-final-viral-combined_k141_224588||full Low-quality
18-101-final-viral-combined_k141_10724||full Not-determined
18-101-final-viral-combined_k141_241380||full Not-determined
18-101-final-viral-combined_k141_126891||full Low-quality
18-101-final-viral-combined_k141_93184||full Low-quality
18-101-final-viral-combined_k141_307592||full Low-quality
18-101-final-viral-combined_k141_209142||full Low-quality
18-101-final-viral-combined_k141_307604||full Not-determined
18-101-final-viral-combined_k141_372742||full Not-determined
18-101-final-viral-combined_k141_126927||full Not-determined
18-101-final-viral-combined_k141_10764||full Low-quality
18-101-final-viral-combined_k141_192530||full Low-quality
18-101-final-viral-combined_k141_142569||full Not-determined
18-101-final-viral-combined_k141_307608||full Not-determined
18-101-final-viral-combined_k141_372747||full Not-determined
18-101-final-viral-combined_k141_59776||full Low-quality
18-101-final-viral-combined_k141_10770||full Not-determined
18-101-final-viral-combined_k141_356970||full Low-quality
18-101-final-viral-combined_k141_356971||full Low-quality
18-101-final-viral-combined_k141_75632||full Not-determined
18-101-final-viral-combined_k141_290607||full Low-quality
18-101-final-viral-combined_k141_59781||full Low-quality
18-101-final-viral-combined_k141_192534||full Not-determined
18-101-final-viral-combined_k141_241449||full Low-quality
18-101-final-viral-combined_k141_26996||full Low-quality
18-101-final-viral-combined_k141_126938||full Low-quality
18-101-final-viral-combined_k141_290621||full Not-determined
18-101-final-viral-combined_k141_339105||full Not-determined
18-101-final-viral-combined_k141_389058||full Low-quality
18-101-final-viral-combined_k141_93226||full Not-determined
18-101-final-viral-combined_k141_192563||full Low-quality
18-101-final-viral-combined_k141_10801||full Not-determined
18-101-final-viral-combined_k141_126948||full Not-determined
18-101-final-viral-combined_k141_10819||full Low-quality
18-101-final-viral-combined_k141_241482||full Low-quality
18-101-final-viral-combined_k141_142592||full Not-determined
18-101-final-viral-combined_k141_209205||full Not-determined
18-101-final-viral-combined_k141_93258||full Not-determined
18-101-final-viral-combined_k141_290643||full Not-determined
18-101-final-viral-combined_k141_142613||full Low-quality
18-101-final-viral-combined_k141_209216||full Low-quality
18-101-final-viral-combined_k141_224718||full Not-determined
18-101-final-viral-combined_k141_158309||full Not-determined
18-101-final-viral-combined_k141_174326||full Low-quality
18-101-final-viral-combined_k141_108606||full Low-quality
18-101-final-viral-combined_k141_241508||full Not-determined
18-101-final-viral-combined_k141_258587||full Not-determined
18-101-final-viral-combined_k141_307681||full Not-determined
18-101-final-viral-combined_k141_43378||full Not-determined
18-101-final-viral-combined_k141_126965||full Not-determined
18-101-final-viral-combined_k141_59807||full Low-quality
18-101-final-viral-combined_k141_290681||full Low-quality
18-101-final-viral-combined_k141_339182||full Low-quality
18-101-final-viral-combined_k141_27085||full Low-quality
18-101-final-viral-combined_k141_372867||full Not-determined
18-101-final-viral-combined_k141_27092||full Not-determined
18-101-final-viral-combined_k141_10887||full Low-quality
18-101-final-viral-combined_k141_224752||full Low-quality
18-101-final-viral-combined_k141_324625||full Low-quality
18-101-final-viral-combined_k141_59847||full Not-determined
18-101-final-viral-combined_k141_10888||full Not-determined
18-101-final-viral-combined_k141_389133||full Low-quality
18-101-final-viral-combined_k141_27106||full Low-quality
18-101-final-viral-combined_k141_43398||full Low-quality
18-101-final-viral-combined_k141_324647||full Not-determined
18-101-final-viral-combined_k141_357034||full Not-determined
18-101-final-viral-combined_k141_258647||full Low-quality
18-101-final-viral-combined_k141_10920||full Low-quality
18-101-final-viral-combined_k141_93331||full Not-determined
18-101-final-viral-combined_k141_59870||full Not-determined
18-101-final-viral-combined_k141_158350||full Not-determined
18-101-final-viral-combined_k141_307742||full Not-determined
18-101-final-viral-combined_k141_389161||full Not-determined
18-101-final-viral-combined_k141_75659||full Low-quality
18-101-final-viral-combined_k141_339228||full Low-quality
18-101-final-viral-combined_k141_290738||full Low-quality
18-101-final-viral-combined_k141_372880||full Not-determined
18-101-final-viral-combined_k141_241575||full Not-determined
18-101-final-viral-combined_k141_258671||full Not-determined
18-101-final-viral-combined_k141_209319||full Low-quality
18-101-final-viral-combined_k141_258677||full Low-quality
18-101-final-viral-combined_k141_43440||full Low-quality
18-101-final-viral-combined_k141_357066||full Not-determined
18-101-final-viral-combined_k141_339243||full Not-determined
18-101-final-viral-combined_k141_75696||full Low-quality
18-101-final-viral-combined_k141_142699||full Not-determined
18-101-final-viral-combined_k141_174381||full Low-quality
18-101-final-viral-combined_k141_59904||full Not-determined
18-101-final-viral-combined_k141_273920||full Not-determined
18-101-final-viral-combined_k141_174390||full Not-determined
18-101-final-viral-combined_k141_258713||full Not-determined
18-101-final-viral-combined_k141_158392||full Low-quality
18-101-final-viral-combined_k141_75714||full Low-quality
18-101-final-viral-combined_k141_339278||full Low-quality
18-101-final-viral-combined_k141_224840||full Low-quality
18-101-final-viral-combined_k141_192675||full Low-quality
18-101-final-viral-combined_k141_127056||full Not-determined
18-101-final-viral-combined_k141_389210||full Not-determined
18-101-final-viral-combined_k141_10996||full Low-quality
18-101-final-viral-combined_k141_241618||full Not-determined
18-101-final-viral-combined_k141_174420||full Low-quality
18-101-final-viral-combined_k141_290804||full Not-determined
18-101-final-viral-combined_k141_158428||full Not-determined
18-101-final-viral-combined_k141_258750||full Not-determined
18-101-final-viral-combined_k141_324685||full Low-quality
18-101-final-viral-combined_k141_11011||full Not-determined
18-101-final-viral-combined_k141_142721||full Not-determined
18-101-final-viral-combined_k141_339289||full Not-determined
18-101-final-viral-combined_k141_389228||full Not-determined
18-101-final-viral-combined_k141_324691||full Low-quality
18-101-final-viral-combined_k141_258764||full Not-determined
18-101-final-viral-combined_k141_224859||full Low-quality
18-101-final-viral-combined_k141_75767||full Low-quality
18-101-final-viral-combined_k141_324699||full Not-determined
18-101-final-viral-combined_k141_241653||full Low-quality
18-101-final-viral-combined_k141_158451||full Not-determined
18-101-final-viral-combined_k141_75779||full Not-determined
18-101-final-viral-combined_k141_108755||full Low-quality
18-101-final-viral-combined_k141_224870||full Not-determined
18-101-final-viral-combined_k141_357126||full Low-quality
18-101-final-viral-combined_k141_290848||full Not-determined
18-101-final-viral-combined_k141_43468||full Low-quality
18-101-final-viral-combined_k141_59925||full Not-determined
18-101-final-viral-combined_k141_209398||full Low-quality
18-101-final-viral-combined_k141_324717||full Not-determined
18-101-final-viral-combined_k141_108769||full Low-quality
18-101-final-viral-combined_k141_43485||full Low-quality
18-101-final-viral-combined_k141_209411||full Not-determined
18-101-final-viral-combined_k141_27169||full Low-quality
18-101-final-viral-combined_k141_241675||full Low-quality
18-101-final-viral-combined_k141_224901||full Not-determined
18-101-final-viral-combined_k141_108775||full Low-quality
18-101-final-viral-combined_k141_59940||full Low-quality
18-101-final-viral-combined_k141_75831||full Not-determined
18-101-final-viral-combined_k141_108788||full Not-determined
18-101-final-viral-combined_k141_389300||full Low-quality
18-101-final-viral-combined_k141_75847||full Low-quality
18-101-final-viral-combined_k141_11080||full Low-quality
18-101-final-viral-combined_k141_273985||full Not-determined
18-101-final-viral-combined_k141_241722||full Low-quality
18-101-final-viral-combined_k141_75865||full Not-determined
18-101-final-viral-combined_k141_93481||full Not-determined
18-101-final-viral-combined_k141_389319||full Not-determined
18-101-final-viral-combined_k141_241731||full Low-quality
18-101-final-viral-combined_k141_27198||full Low-quality
18-101-final-viral-combined_k141_373051||full Not-determined
18-101-final-viral-combined_k141_27202||full Low-quality
18-101-final-viral-combined_k141_224943||full Not-determined
18-101-final-viral-combined_k141_43538||full Not-determined
18-101-final-viral-combined_k141_93512||full Not-determined
18-101-final-viral-combined_k141_192703||full Low-quality
18-101-final-viral-combined_k141_127181||full Low-quality
18-101-final-viral-combined_k141_241763||full Not-determined
18-101-final-viral-combined_k141_59973||full Low-quality
18-101-final-viral-combined_k141_290959||full Low-quality
18-101-final-viral-combined_k141_224958||full Not-determined
18-101-final-viral-combined_k141_11115||full Low-quality
18-101-final-viral-combined_k141_174539||full Low-quality
18-101-final-viral-combined_k141_27227||full Not-determined
18-101-final-viral-combined_k141_108867||full Low-quality
18-101-final-viral-combined_k141_290967||full Not-determined
18-101-final-viral-combined_k141_174545||full Not-determined
18-101-final-viral-combined_k141_357148||full Not-determined
18-101-final-viral-combined_k141_174550||full Low-quality
18-101-final-viral-combined_k141_59989||full Not-determined
18-101-final-viral-combined_k141_307987||full Low-quality
18-101-final-viral-combined_k141_290976||full Low-quality
18-101-final-viral-combined_k141_389355||full Low-quality
18-101-final-viral-combined_k141_357156||full Low-quality
18-101-final-viral-combined_k141_93536||full Low-quality
18-101-final-viral-combined_k141_60006||full Low-quality
18-101-final-viral-combined_k141_224984||full Not-determined
18-101-final-viral-combined_k141_324830||full Low-quality
18-101-final-viral-combined_k141_127209||full Low-quality
18-101-final-viral-combined_k141_43587||full Not-determined
18-101-final-viral-combined_k141_357178||full Not-determined
18-101-final-viral-combined_k141_339423||full Not-determined
18-101-final-viral-combined_k141_258917||full Low-quality
18-101-final-viral-combined_k141_209523||full Low-quality
18-101-final-viral-combined_k141_209524||full Not-determined
18-101-final-viral-combined_k141_93574||full Not-determined
18-101-final-viral-combined_k141_209525||full Low-quality
18-101-final-viral-combined_k141_308030||full Low-quality
18-101-final-viral-combined_k141_308032||full Not-determined
18-101-final-viral-combined_k141_43603||full Low-quality
18-101-final-viral-combined_k141_291020||full Low-quality
18-101-final-viral-combined_k141_43606||full Low-quality
18-101-final-viral-combined_k141_43610||full Not-determined
18-101-final-viral-combined_k141_291028||full Low-quality
18-101-final-viral-combined_k141_308049||full Low-quality
18-101-final-viral-combined_k141_60060||full Low-quality
18-101-final-viral-combined_k141_339446||full Low-quality
18-101-final-viral-combined_k141_158528||full Not-determined
18-101-final-viral-combined_k141_308053||full Low-quality
18-101-final-viral-combined_k141_43614||full Not-determined
18-101-final-viral-combined_k141_258963||full Low-quality
18-101-final-viral-combined_k141_241850||full Low-quality
18-101-final-viral-combined_k141_225049||full Not-determined
18-101-final-viral-combined_k141_76010||full Not-determined
18-101-final-viral-combined_k141_174621||full Low-quality
18-101-final-viral-combined_k141_357211||full Low-quality
18-101-final-viral-combined_k141_357229||full Low-quality
18-101-final-viral-combined_k141_174622||full Low-quality
18-101-final-viral-combined_k141_225074||full Low-quality
18-101-final-viral-combined_k141_174636||full Low-quality
18-101-final-viral-combined_k141_274095||full Not-determined
18-101-final-viral-combined_k141_192780||full Low-quality
18-101-final-viral-combined_k141_60093||full Not-determined
18-101-final-viral-combined_k141_43654||full Not-determined
18-101-final-viral-combined_k141_274118||full Low-quality
18-101-final-viral-combined_k141_11153||full Low-quality
18-101-final-viral-combined_k141_339468||full Not-determined
18-101-final-viral-combined_k141_11155||full Not-determined
18-101-final-viral-combined_k141_241905||full Low-quality
18-101-final-viral-combined_k141_339477||full Low-quality
18-101-final-viral-combined_k141_259053||full Not-determined
18-101-final-viral-combined_k141_43666||full Low-quality
18-101-final-viral-combined_k141_308127||full Low-quality
18-101-final-viral-combined_k141_76038||full Not-determined
18-101-final-viral-combined_k141_291106||full Not-determined
18-101-final-viral-combined_k141_339483||full Not-determined
18-101-final-viral-combined_k141_142819||full Low-quality
18-101-final-viral-combined_k141_324904||full Low-quality
18-101-final-viral-combined_k141_60131||full Low-quality
18-101-final-viral-combined_k141_274143||full Not-determined
18-101-final-viral-combined_k141_158589||full Low-quality
18-101-final-viral-combined_k141_192842||full Not-determined
18-101-final-viral-combined_k141_142832||full Low-quality
18-101-final-viral-combined_k141_291127||full Not-determined
18-101-final-viral-combined_k141_241935||full Not-determined
18-101-final-viral-combined_k141_357319||full Low-quality
18-101-final-viral-combined_k141_142857||full Not-determined
18-101-final-viral-combined_k141_291147||full Low-quality
18-101-final-viral-combined_k141_142869||full Not-determined
18-101-final-viral-combined_k141_291153||full Not-determined
18-101-final-viral-combined_k141_389486||full Low-quality
18-101-final-viral-combined_k141_43736||full Not-determined
18-101-final-viral-combined_k141_93706||full Low-quality
18-101-final-viral-combined_k141_108980||full Low-quality
18-101-final-viral-combined_k141_142878||full Low-quality
18-101-final-viral-combined_k141_108984||full Low-quality
18-101-final-viral-combined_k141_158637||full Low-quality
18-101-final-viral-combined_k141_11214||full Low-quality
18-101-final-viral-combined_k141_308135||full Not-determined
18-101-final-viral-combined_k141_76101||full Not-determined
18-101-final-viral-combined_k141_174732||full Low-quality
18-101-final-viral-combined_k141_60211||full Low-quality
18-101-final-viral-combined_k141_109001||full Low-quality
18-101-final-viral-combined_k141_324961||full Not-determined
18-101-final-viral-combined_k141_11240||full Low-quality
18-101-final-viral-combined_k141_27321||full Low-quality
18-101-final-viral-combined_k141_291193||full Low-quality
18-101-final-viral-combined_k141_158651||full Not-determined
18-101-final-viral-combined_k141_209536||full Not-determined
18-101-final-viral-combined_k141_27327||full Not-determined
18-101-final-viral-combined_k141_225210||full Not-determined
18-101-final-viral-combined_k141_76129||full Low-quality
18-101-final-viral-combined_k141_174758||full Not-determined
18-101-final-viral-combined_k141_174759||full Low-quality
18-101-final-viral-combined_k141_11249||full Low-quality
18-101-final-viral-combined_k141_76130||full Not-determined
18-101-final-viral-combined_k141_225225||full Low-quality
18-101-final-viral-combined_k141_308160||full Not-determined
18-101-final-viral-combined_k141_389538||full Low-quality
18-101-final-viral-combined_k141_389541||full Not-determined
18-101-final-viral-combined_k141_43787||full Not-determined
18-101-final-viral-combined_k141_209557||full Not-determined
18-101-final-viral-combined_k141_291219||full Low-quality
18-101-final-viral-combined_k141_158676||full Not-determined
18-101-final-viral-combined_k141_155480||full High-quality
18-101-final-viral-combined_k141_291225||full Not-determined
18-101-final-viral-combined_k141_60253||full Low-quality
18-101-final-viral-combined_k141_158679||full Low-quality
18-101-final-viral-combined_k141_209580||full Low-quality
18-101-final-viral-combined_k141_357408||full Not-determined
18-101-final-viral-combined_k141_142958||full Low-quality
18-101-final-viral-combined_k141_339560||full Not-determined
18-101-final-viral-combined_k141_225273||full Not-determined
18-101-final-viral-combined_k141_373226||full Not-determined
18-101-final-viral-combined_k141_209600||full Low-quality
18-101-final-viral-combined_k141_158702||full Not-determined
18-101-final-viral-combined_k141_357424||full Not-determined
18-101-final-viral-combined_k141_389588||full Low-quality
18-101-final-viral-combined_k141_109044||full Low-quality
18-101-final-viral-combined_k141_76201||full Low-quality
18-101-final-viral-combined_k141_339577||full Low-quality
18-101-final-viral-combined_k141_60282||full Low-quality
18-101-final-viral-combined_k141_192956||full Low-quality
18-101-final-viral-combined_k141_109055||full Not-determined
18-101-final-viral-combined_k141_373236||full Not-determined
18-101-final-viral-combined_k141_389605||full Low-quality
18-101-final-viral-combined_k141_291295||full Low-quality
18-101-final-viral-combined_k141_109061||full Low-quality
18-101-final-viral-combined_k141_339583||full Not-determined
18-101-final-viral-combined_k141_242076||full Not-determined
18-101-final-viral-combined_k141_274296||full Low-quality
18-101-final-viral-combined_k141_225298||full Not-determined
18-101-final-viral-combined_k141_27404||full Low-quality
18-101-final-viral-combined_k141_291299||full Low-quality
18-101-final-viral-combined_k141_11293||full Not-determined
18-101-final-viral-combined_k141_142988||full Not-determined
18-101-final-viral-combined_k141_142995||full Not-determined
18-101-final-viral-combined_k141_158714||full Low-quality
18-101-final-viral-combined_k141_11319||full Low-quality
18-101-final-viral-combined_k141_209665||full Low-quality
18-101-final-viral-combined_k141_27433||full Not-determined
18-101-final-viral-combined_k141_242105||full Low-quality
18-101-final-viral-combined_k141_60368||full Not-determined
18-101-final-viral-combined_k141_76249||full Low-quality
18-101-final-viral-combined_k141_291346||full Low-quality
18-101-final-viral-combined_k141_109127||full Low-quality
18-101-final-viral-combined_k141_225360||full Low-quality
18-101-final-viral-combined_k141_193020||full Not-determined
18-101-final-viral-combined_k141_158770||full Low-quality
18-101-final-viral-combined_k141_93891||full Not-determined
18-101-final-viral-combined_k141_291361||full Low-quality
18-101-final-viral-combined_k141_127526||full Low-quality
18-101-final-viral-combined_k141_373292||full Low-quality
18-101-final-viral-combined_k141_127534||full Not-determined
18-101-final-viral-combined_k141_389654||full Not-determined
18-101-final-viral-combined_k141_373295||full Low-quality
18-101-final-viral-combined_k141_93906||full Low-quality
18-101-final-viral-combined_k141_193064||full Low-quality
18-101-final-viral-combined_k141_60403||full Not-determined
18-101-final-viral-combined_k141_291380||full Low-quality
18-101-final-viral-combined_k141_193068||full Not-determined
18-101-final-viral-combined_k141_174918||full Not-determined
18-101-final-viral-combined_k141_109153||full Low-quality
18-101-final-viral-combined_k141_76289||full Not-determined
18-101-final-viral-combined_k141_389674||full Not-determined
18-101-final-viral-combined_k141_291405||full Not-determined
18-101-final-viral-combined_k141_193101||full Low-quality
18-101-final-viral-combined_k141_389679||full Low-quality
18-101-final-viral-combined_k141_259234||full Not-determined
18-109-final-viral-combined_k141_174935||full Medium-quality

| | |
|---|---|
| 18-101-final-viral-combined_k141_76303\|\|full | Low-quality |
| 18-101-final-viral-combined_k141_93949\|\|full | Not-determined |
| 18-101-final-viral-combined_k141_93952\|\|full | Not-determined |
| 18-101-final-viral-combined_k141_325093\|\|full | Not-determined |
| 18-101-final-viral-combined_k141_158852\|\|full | Low-quality |
| 18-101-final-viral-combined_k141_373316\|\|full | Not-determined |
| 18-101-final-viral-combined_k141_76307\|\|full | Low-quality |
| 18-101-final-viral-combined_k141_225454\|\|full | Low-quality |
| 18-101-final-viral-combined_k141_225467\|\|full | Not-determined |
| 18-101-final-viral-combined_k141_357579\|\|full | Low-quality |
| 18-101-final-viral-combined_k141_373338\|\|full | Low-quality |
| 18-101-final-viral-combined_k141_357597\|\|full | Not-determined |
| 18-101-final-viral-combined_k141_357601\|\|full | Low-quality |
| 18-101-final-viral-combined_k141_109189\|\|full | Not-determined |
| 18-101-final-viral-combined_k141_93978\|\|full | Not-determined |
| 18-101-final-viral-combined_k141_308339\|\|full | Not-determined |
| 18-101-final-viral-combined_k141_357608\|\|full | Not-determined |
| 18-101-final-viral-combined_k141_325139\|\|full | Not-determined |
| 18-101-final-viral-combined_k141_308365\|\|full | Low-quality |
| 18-101-final-viral-combined_k141_389726\|\|full | Low-quality |
| 18-101-final-viral-combined_k141_242169\|\|full | Low-quality |
| 18-101-final-viral-combined_k141_209777\|\|full | Low-quality |
| 18-101-final-viral-combined_k141_109209\|\|full | Not-determined |
| 18-101-final-viral-combined_k141_242176\|\|full | Not-determined |
| 18-101-final-viral-combined_k141_27551\|\|full | Low-quality |
| 18-101-final-viral-combined_k141_242177\|\|full | Not-determined |
| 18-101-final-viral-combined_k141_242184\|\|full | Not-determined |
| 18-101-final-viral-combined_k141_143150\|\|full | Low-quality |
| 18-101-final-viral-combined_k141_259291\|\|full | Low-quality |
| 18-101-final-viral-combined_k141_209794\|\|full | Low-quality |
| 18-101-final-viral-combined_k141_43970\|\|full | Not-determined |
| 18-101-final-viral-combined_k141_143154\|\|full | Low-quality |
| 18-101-final-viral-combined_k141_259295\|\|full | Not-determined |
| 18-101-final-viral-combined_k141_209795\|\|full | Not-determined |
| 18-101-final-viral-combined_k141_325187\|\|full | Not-determined |
| 18-105-final-viral-combined_k141_85650\|\|full | High-quality |
| 18-101-final-viral-combined_k141_373395\|\|full | Low-quality |
| 18-101-final-viral-combined_k141_389753\|\|full | Not-determined |
| 18-101-final-viral-combined_k141_27606\|\|full | Low-quality |
| 18-101-final-viral-combined_k141_143184\|\|full | Not-determined |
| 18-101-final-viral-combined_k141_225513\|\|full | Not-determined |
| 18-101-final-viral-combined_k141_193228\|\|full | Not-determined |
| 18-101-final-viral-combined_k141_94055\|\|full | Not-determined |
| 18-101-final-viral-combined_k141_389755\|\|full | Low-quality |
| 18-101-final-viral-combined_k141_274450\|\|full | Low-quality |
| 18-101-final-viral-combined_k141_109260\|\|full | Low-quality |
| 18-101-final-viral-combined_k141_175059\|\|full | Not-determined |
| 18-101-final-viral-combined_k141_373408\|\|full | Not-determined |
| 18-101-final-viral-combined_k141_373412\|\|full | Low-quality |
| 18-101-final-viral-combined_k141_94073\|\|full | Not-determined |
| 18-101-final-viral-combined_k141_291566\|\|full | Not-determined |
| 18-101-final-viral-combined_k141_11429\|\|full | Low-quality |
| 18-101-final-viral-combined_k141_274459\|\|full | Low-quality |
| 18-101-final-viral-combined_k141_43992\|\|full | Not-determined |
| 18-101-final-viral-combined_k141_109272\|\|full | Low-quality |
| 18-101-final-viral-combined_k141_76368\|\|full | Low-quality |
| 18-101-final-viral-combined_k141_76369\|\|full | Low-quality |
| 18-101-final-viral-combined_k141_373439\|\|full | Low-quality |
| 18-101-final-viral-combined_k141_259366\|\|full | Low-quality |
| 18-101-final-viral-combined_k141_158942\|\|full | Not-determined |
| 18-101-final-viral-combined_k141_357705\|\|full | Low-quality |
| 18-101-final-viral-combined_k141_94103\|\|full | Not-determined |
| 18-101-final-viral-combined_k141_158950\|\|full | Not-determined |
| 18-101-final-viral-combined_k141_44029\|\|full | Not-determined |
| 18-101-final-viral-combined_k141_193282\|\|full | Low-quality |
| 18-101-final-viral-combined_k141_225538\|\|full | Low-quality |
| 18-101-final-viral-combined_k141_225546\|\|full | Low-quality |
| 18-101-final-viral-combined_k141_325276\|\|full | Not-determined |
| 18-101-final-viral-combined_k141_193296\|\|full | Not-determined |
| 18-101-final-viral-combined_k141_357734\|\|full | Low-quality |
| 18-101-final-viral-combined_k141_109305\|\|full | Low-quality |
| 18-101-final-viral-combined_k141_209886\|\|full | Not-determined |
| 18-101-final-viral-combined_k141_175133\|\|full | Not-determined |
| 18-101-final-viral-combined_k141_193300\|\|full | Not-determined |
| 18-101-final-viral-combined_k141_44065\|\|full | Low-quality |
| 18-101-final-viral-combined_k141_158978\|\|full | Not-determined |
| 18-101-final-viral-combined_k141_109320\|\|full | Not-determined |
| 18-101-final-viral-combined_k141_225570\|\|full | Low-quality |
| 18-101-final-viral-combined_k141_291651\|\|full | Not-determined |
| 18-101-final-viral-combined_k141_274508\|\|full | Not-determined |
| 18-101-final-viral-combined_k141_76412\|\|full | Not-determined |
| 18-101-final-viral-combined_k141_109335\|\|full | Not-determined |
| 18-101-final-viral-combined_k141_76413\|\|full | Low-quality |
| 18-101-final-viral-combined_k141_143289\|\|full | Low-quality |
| 18-101-final-viral-combined_k141_94161\|\|full | Low-quality |
| 18-101-final-viral-combined_k141_259424\|\|full | Not-determined |
| 18-101-final-viral-combined_k141_44101\|\|full | Low-quality |
| 18-101-final-viral-combined_k141_274526\|\|full | Not-determined |
| 18-101-final-viral-combined_k141_373463\|\|full | Low-quality |
| 18-101-final-viral-combined_k141_60615\|\|full | Not-determined |
| 18-101-final-viral-combined_k141_44107\|\|full | Not-determined |
| 18-101-final-viral-combined_k141_291681\|\|full | Low-quality |
| 18-101-final-viral-combined_k141_193328\|\|full | Not-determined |
| 18-101-final-viral-combined_k141_94176\|\|full | Low-quality |
| 18-101-final-viral-combined_k141_175144\|\|full | Low-quality |
| 18-101-final-viral-combined_k141_357755\|\|full | Low-quality |
| 18-101-final-viral-combined_k141_143322\|\|full | Not-determined |
| 18-101-final-viral-combined_k141_373481\|\|full | Not-determined |
| 18-101-final-viral-combined_k141_357758\|\|full | Not-determined |
| 18-101-final-viral-combined_k141_357765\|\|full | Not-determined |
| 18-101-final-viral-combined_k141_274533\|\|full | Not-determined |
| 18-101-final-viral-combined_k141_127718\|\|full | Low-quality |
| 18-101-final-viral-combined_k141_94197\|\|full | Low-quality |
| 18-101-final-viral-combined_k141_242230\|\|full | Not-determined |
| 18-101-final-viral-combined_k141_76465\|\|full | Low-quality |
| 18-101-final-viral-combined_k141_109362\|\|full | Low-quality |
| 18-101-final-viral-combined_k141_159030\|\|full | Not-determined |
| 18-101-final-viral-combined_k141_109365\|\|full | Low-quality |
| 18-101-final-viral-combined_k141_259474\|\|full | Low-quality |
| 18-101-final-viral-combined_k141_291710\|\|full | Not-determined |
| 18-101-final-viral-combined_k141_339819\|\|full | Not-determined |
| 18-101-final-viral-combined_k141_259475\|\|full | Not-determined |
| 18-101-final-viral-combined_k141_109381\|\|full | Low-quality |
| 18-101-final-viral-combined_k141_357790\|\|full | Not-determined |
| 18-101-final-viral-combined_k141_175193\|\|full | Not-determined |
| 18-101-final-viral-combined_k141_308515\|\|full | Low-quality |
| 18-101-final-viral-combined_k141_225657\|\|full | Low-quality |
| 18-101-final-viral-combined_k141_209949\|\|full | Low-quality |
| 18-101-final-viral-combined_k141_325353\|\|full | Not-determined |
| 18-101-final-viral-combined_k141_94227\|\|full | Low-quality |
| 18-101-final-viral-combined_k141_109393\|\|full | Not-determined |
| 18-101-final-viral-combined_k141_193412\|\|full | Low-quality |
| 18-101-final-viral-combined_k141_27715\|\|full | Low-quality |
| 18-101-final-viral-combined_k141_143351\|\|full | Low-quality |
| 18-101-final-viral-combined_k141_193427\|\|full | Not-determined |
| 18-101-final-viral-combined_k141_259508\|\|full | Not-determined |
| 18-101-final-viral-combined_k141_159083\|\|full | Low-quality |
| 18-101-final-viral-combined_k141_389912\|\|full | Low-quality |
| 18-101-final-viral-combined_k141_94257\|\|full | Not-determined |
| 18-101-final-viral-combined_k141_209982\|\|full | Low-quality |
| 18-101-final-viral-combined_k141_109429\|\|full | Not-determined |
| 18-101-final-viral-combined_k141_11562\|\|full | Not-determined |
| 18-101-final-viral-combined_k141_225670\|\|full | Low-quality |
| 18-101-final-viral-combined_k141_143382\|\|full | Not-determined |
| 18-101-final-viral-combined_k141_175223\|\|full | Low-quality |
| 18-101-final-viral-combined_k141_94270\|\|full | Not-determined |
| 18-101-final-viral-combined_k141_11576\|\|full | Low-quality |
| 18-101-final-viral-combined_k141_308595\|\|full | Not-determined |
| 18-101-final-viral-combined_k141_291803\|\|full | Not-determined |
| 18-101-final-viral-combined_k141_143404\|\|full | Not-determined |
| 18-101-final-viral-combined_k141_259563\|\|full | Low-quality |
| 18-101-final-viral-combined_k141_210015\|\|full | Low-quality |
| 18-101-final-viral-combined_k141_127736\|\|full | Low-quality |
| 18-101-final-viral-combined_k141_308609\|\|full | Low-quality |
| 18-101-final-viral-combined_k141_109457\|\|full | Low-quality |
| 18-101-final-viral-combined_k141_193460\|\|full | Low-quality |
| 18-101-final-viral-combined_k141_11616\|\|full | Not-determined |
| 18-101-final-viral-combined_k141_193472\|\|full | Low-quality |
| 18-101-final-viral-combined_k141_242378\|\|full | Not-determined |
| 18-101-final-viral-combined_k141_259602\|\|full | Not-determined |
| 18-101-final-viral-combined_k141_389969\|\|full | Low-quality |
| 18-101-final-viral-combined_k141_210059\|\|full | Low-quality |
| 18-101-final-viral-combined_k141_143453\|\|full | Low-quality |
| 18-101-final-viral-combined_k141_373640\|\|full | Low-quality |
| 18-101-final-viral-combined_k141_76546\|\|full | Not-determined |
| 18-101-final-viral-combined_k141_225734\|\|full | Low-quality |
| 18-101-final-viral-combined_k141_242386\|\|full | Not-determined |
| 18-101-final-viral-combined_k141_291866\|\|full | Not-determined |
| 18-101-final-viral-combined_k141_76548\|\|full | Not-determined |
| 18-101-final-viral-combined_k141_193485\|\|full | Not-determined |
| 18-101-final-viral-combined_k141_175281\|\|full | Low-quality |
| 18-101-final-viral-combined_k141_60782\|\|full | Not-determined |
| 18-101-final-viral-combined_k141_175299\|\|full | Low-quality |
| 18-101-final-viral-combined_k141_389999\|\|full | Low-quality |
| 18-101-final-viral-combined_k141_357943\|\|full | Not-determined |
| 18-101-final-viral-combined_k141_390005\|\|full | Low-quality |
| 18-101-final-viral-combined_k141_373692\|\|full | Low-quality |
| 18-101-final-viral-combined_k141_44300\|\|full | Not-determined |
| 18-101-final-viral-combined_k141_109527\|\|full | Low-quality |
| 18-110-final-viral-combined_k141_72911\|\|full | Medium-quality |
| 18-101-final-viral-combined_k141_159141\|\|full | Low-quality |
| 18-101-final-viral-combined_k141_259639\|\|full | Low-quality |
| 18-101-final-viral-combined_k141_127794\|\|full | Low-quality |
| 18-101-final-viral-combined_k141_259641\|\|full | Low-quality |
| 18-101-final-viral-combined_k141_193534\|\|full | Not-determined |
| 18-101-final-viral-combined_k141_390031\|\|full | Not-determined |
| 18-101-final-viral-combined_k141_109563\|\|full | Low-quality |
| 18-101-final-viral-combined_k141_357992\|\|full | Low-quality |
| 18-101-final-viral-combined_k141_259646\|\|full | Low-quality |
| 18-101-final-viral-combined_k141_11702\|\|full | Low-quality |
| 18-101-final-viral-combined_k141_175346\|\|full | Not-determined |
| 18-101-final-viral-combined_k141_11704\|\|full | Not-determined |
| 18-101-final-viral-combined_k141_325510\|\|full | Not-determined |
| 18-101-final-viral-combined_k141_143485\|\|full | Not-determined |
| 18-101-final-viral-combined_k141_109586\|\|full | Not-determined |
| 18-101-final-viral-combined_k141_259664\|\|full | Not-determined |
| 18-101-final-viral-combined_k141_339877\|\|full | Not-determined |
| 18-101-final-viral-combined_k141_325516\|\|full | Not-determined |
| 18-101-final-viral-combined_k141_259667\|\|full | Not-determined |
| 18-101-final-viral-combined_k141_225832\|\|full | Low-quality |
| 18-101-final-viral-combined_k141_44358\|\|full | Low-quality |
| 18-101-final-viral-combined_k141_274732\|\|full | Not-determined |
| 18-101-final-viral-combined_k141_259675\|\|full | Low-quality |
| 18-101-final-viral-combined_k141_358016\|\|full | Not-determined |
| 18-101-final-viral-combined_k141_94446\|\|full | Not-determined |
| 18-101-final-viral-combined_k141_339904\|\|full | Not-determined |
| 18-101-final-viral-combined_k141_339909\|\|full | Not-determined |
| 18-101-final-viral-combined_k141_358032\|\|full | Low-quality |
| 18-101-final-viral-combined_k141_259695\|\|full | Not-determined |
| 18-101-final-viral-combined_k141_11736\|\|full | Low-quality |
| 18-101-final-viral-combined_k141_225862\|\|full | Low-quality |
| 18-101-final-viral-combined_k141_27965\|\|full | Low-quality |
| 18-101-final-viral-combined_k141_76600\|\|full | Low-quality |
| 18-101-final-viral-combined_k141_76601\|\|full | Not-determined |
| 18-101-final-viral-combined_k141_109605\|\|full | Low-quality |
| 18-101-final-viral-combined_k141_210155\|\|full | Not-determined |
| 18-101-final-viral-combined_k141_76605\|\|full | Low-quality |
| 18-101-final-viral-combined_k141_325559\|\|full | Low-quality |
| 18-101-final-viral-combined_k141_210162\|\|full | Low-quality |
| 18-101-final-viral-combined_k141_308752\|\|full | Not-determined |
| 18-101-final-viral-combined_k141_11754\|\|full | Not-determined |
| 18-101-final-viral-combined_k141_94487\|\|full | Not-determined |
| 18-101-final-viral-combined_k141_60912\|\|full | Low-quality |
| 18-101-final-viral-combined_k141_339949\|\|full | Not-determined |
| 18-101-final-viral-combined_k141_210164\|\|full | Low-quality |
| 18-101-final-viral-combined_k141_76632\|\|full | Not-determined |
| 18-101-final-viral-combined_k141_11768\|\|full | Not-determined |
| 18-101-final-viral-combined_k141_325581\|\|full | Not-determined |
| 18-101-final-viral-combined_k141_259730\|\|full | Not-determined |
| 18-101-final-viral-combined_k141_259731\|\|full | Not-determined |
| 18-101-final-viral-combined_k141_325590\|\|full | Not-determined |
| 18-101-final-viral-combined_k141_27999\|\|full | Low-quality |
| 18-101-final-viral-combined_k141_76650\|\|full | Low-quality |
| 18-101-final-viral-combined_k141_76654\|\|full | Not-determined |
| 18-101-final-viral-combined_k141_292019\|\|full | Not-determined |
| 18-101-final-viral-combined_k141_339976\|\|full | Not-determined |
| 18-101-final-viral-combined_k141_28013\|\|full | Low-quality |
| 18-101-final-viral-combined_k141_76665\|\|full | Not-determined |
| 18-101-final-viral-combined_k141_159242\|\|full | Not-determined |
| 18-101-final-viral-combined_k141_225915\|\|full | Not-determined |
| 18-101-final-viral-combined_k141_325624\|\|full | Not-determined |
| 18-101-final-viral-combined_k141_143607\|\|full | Not-determined |
| 18-101-final-viral-combined_k141_390109\|\|full | Not-determined |
| 18-101-final-viral-combined_k141_242525\|\|full | Not-determined |
| 18-101-final-viral-combined_k141_127973\|\|full | Not-determined |
| 18-101-final-viral-combined_k141_340006\|\|full | Not-determined |
| 18-101-final-viral-combined_k141_109670\|\|full | Not-determined |
| 18-101-final-viral-combined_k141_175395\|\|full | Not-determined |
| 18-101-final-viral-combined_k141_44463\|\|full | Not-determined |
| 18-101-final-viral-combined_k141_175411\|\|full | Not-determined |
| 18-101-final-viral-combined_k141_210227\|\|full | Low-quality |
| 18-101-final-viral-combined_k141_28078\|\|full | Not-determined |
| 18-101-final-viral-combined_k141_193687\|\|full | Not-determined |
| 18-101-final-viral-combined_k141_242553\|\|full | Low-quality |
| 18-101-final-viral-combined_k141_159306\|\|full | Low-quality |
| 18-101-final-viral-combined_k141_94540\|\|full | Not-determined |
| 18-101-final-viral-combined_k141_390159\|\|full | Not-determined |
| 18-101-final-viral-combined_k141_390161\|\|full | Low-quality |
| 18-101-final-viral-combined_k141_373803\|\|full | Low-quality |
| 18-101-final-viral-combined_k141_373807\|\|full | Low-quality |
| 18-101-final-viral-combined_k141_159314\|\|full | Not-determined |
| 18-101-final-viral-combined_k141_175425\|\|full | Not-determined |
| 18-101-final-viral-combined_k141_94560\|\|full | Low-quality |
| 18-102-final-viral-combined_k141_74387\|\|full | Complete |
| 18-110_k141_127191 | High-quality |
| 18-101-final-viral-combined_k141_175439\|\|full | Not-determined |
| 18-101-final-viral-combined_k141_159320\|\|full | Low-quality |
| 18-101-final-viral-combined_k141_325658\|\|full | Not-determined |
| 18-101-final-viral-combined_k141_11881\|\|full | Low-quality |
| 18-101-final-viral-combined_k141_373837\|\|full | Not-determined |
| 18-101-final-viral-combined_k141_193767\|\|full | Not-determined |
| 18-101-final-viral-combined_k141_143680\|\|full | Low-quality |
| 18-101-final-viral-combined_k141_225988\|\|full | Low-quality |
| 18-101-final-viral-combined_k141_94598\|\|full | Not-determined |
| 18-101-final-viral-combined_k141_61064\|\|full | Low-quality |
| 18-101-final-viral-combined_k141_340095\|\|full | Low-quality |
| 18-101-final-viral-combined_k141_28120\|\|full | Low-quality |
| 18-101-final-viral-combined_k141_109733\|\|full | Low-quality |
| 18-101-final-viral-combined_k141_11911\|\|full | Low-quality |
| 18-101-final-viral-combined_k141_308897\|\|full | Not-determined |
| 18-101-final-viral-combined_k141_94600\|\|full | Not-determined |
| 18-101-final-viral-combined_k141_175484\|\|full | Not-determined |
| 18-101-final-viral-combined_k141_373878\|\|full | Low-quality |
| 18-101-final-viral-combined_k141_274876\|\|full | Low-quality |
| 18-101-final-viral-combined_k141_128014\|\|full | Low-quality |
| 18-101-final-viral-combined_k141_44540\|\|full | Low-quality |
| 18-101-final-viral-combined_k141_28134\|\|full | Low-quality |
| 18-101-final-viral-combined_k141_292137\|\|full | Not-determined |
| 18-101-final-viral-combined_k141_259876\|\|full | Not-determined |
| 18-101-final-viral-combined_k141_109753\|\|full | Not-determined |
| 18-101-final-viral-combined_k141_143736\|\|full | Not-determined |
| 18-101-final-viral-combined_k141_159363\|\|full | Low-quality |
| 18-101-final-viral-combined_k141_128029\|\|full | Not-determined |
| 18-101-final-viral-combined_k141_325734\|\|full | Not-determined |
| 18-101-final-viral-combined_k141_76819\|\|full | Not-determined |
| 18-101-final-viral-combined_k141_143749\|\|full | Not-determined |
| 18-101-final-viral-combined_k141_128038\|\|full | Not-determined |
| 18-101-final-viral-combined_k141_61104\|\|full | Low-quality |
| 18-101-final-viral-combined_k141_340119\|\|full | Not-determined |
| 18-101-final-viral-combined_k141_210322\|\|full | Not-determined |
| 18-101-final-viral-combined_k141_358290\|\|full | Not-determined |
| 18-101-final-viral-combined_k141_109767\|\|full | Low-quality |
| 18-101-final-viral-combined_k141_325744\|\|full | Not-determined |
| 18-101-final-viral-combined_k141_109769\|\|full | Low-quality |
| 18-101-final-viral-combined_k141_292172\|\|full | Not-determined |
| 18-101-final-viral-combined_k141_109782\|\|full | Not-determined |
| 18-105-final-viral-combined_k141_94482\|\|full | High-quality |
| 18-101-final-viral-combined_k141_226001\|\|full | Low-quality |
| 18-101-final-viral-combined_k141_373923\|\|full | Low-quality |
| 18-101-final-viral-combined_k141_226005\|\|full | Low-quality |
| 18-101-final-viral-combined_k141_325758\|\|full | Not-determined |
| 18-101-final-viral-combined_k141_28194\|\|full | Low-quality |
| 18-101-final-viral-combined_k141_358306\|\|full | Low-quality |
| 18-101-final-viral-combined_k141_61147\|\|full | Low-quality |
| 18-101-final-viral-combined_k141_390210\|\|full | Not-determined |
| 18-101-final-viral-combined_k141_143794\|\|full | Low-quality |
| 18-101-final-viral-combined_k141_94640\|\|full | Not-determined |
| 18-101-final-viral-combined_k141_325770\|\|full | Not-determined |
| 18-101-final-viral-combined_k141_11974\|\|full | Not-determined |
| 18-101-final-viral-combined_k141_109826\|\|full | Low-quality |
| 18-101-final-viral-combined_k141_76881\|\|full | Low-quality |
| 18-101-final-viral-combined_k141_44628\|\|full | Low-quality |
| 18-101-final-viral-combined_k141_109850\|\|full | Low-quality |
| 18-101-final-viral-combined_k141_210400\|\|full | Low-quality |
| 18-101-final-viral-combined_k141_226052\|\|full | Not-determined |
| 18-101-final-viral-combined_k141_292229\|\|full | Low-quality |
| 18-101-final-viral-combined_k141_226060\|\|full | Not-determined |
| 18-101-final-viral-combined_k141_94683\|\|full | Low-quality |
| 18-101-final-viral-combined_k141_128072\|\|full | Low-quality |
| 18-101-final-viral-combined_k141_175523\|\|full | Low-quality |
| 18-101-final-viral-combined_k141_274988\|\|full | Not-determined |
| 18-101-final-viral-combined_k141_76919\|\|full | Not-determined |
| 18-101-final-viral-combined_k141_340191\|\|full | Low-quality |
| 18-101-final-viral-combined_k141_308994\|\|full | Not-determined |
| 18-101-final-viral-combined_k141_275003\|\|full | Not-determined |
| 18-101-final-viral-combined_k141_340195\|\|full | Not-determined |
| 18-101-final-viral-combined_k141_292254\|\|full | Not-determined |
| 18-101-final-viral-combined_k141_109895\|\|full | Not-determined |
| 18-101-final-viral-combined_k141_390276\|\|full | Low-quality |
| 18-101-final-viral-combined_k141_128107\|\|full | Not-determined |
| 18-101-final-viral-combined_k141_340220\|\|full | Not-determined |
| 18-101-final-viral-combined_k141_109915\|\|full | Low-quality |
| 18-101-final-viral-combined_k141_143857\|\|full | Low-quality |
| 18-101-final-viral-combined_k141_128137\|\|full | Low-quality |
| 18-101-final-viral-combined_k141_226126\|\|full | Low-quality |
| 18-101-final-viral-combined_k141_128147\|\|full | Low-quality |
| 18-101-final-viral-combined_k141_325836\|\|full | Not-determined |
| 18-101-final-viral-combined_k141_374057\|\|full | Not-determined |
| 18-101-final-viral-combined_k141_259967\|\|full | Not-determined |
| 18-101-final-viral-combined_k141_76963\|\|full | Not-determined |
| 18-101-final-viral-combined_k141_12090\|\|full | Not-determined |
| 18-101-final-viral-combined_k141_210508\|\|full | Not-determined |
| 18-101-final-viral-combined_k141_76974\|\|full | Low-quality |
| 18-101-final-viral-combined_k141_358456\|\|full | Not-determined |
| 18-101-final-viral-combined_k141_61308\|\|full | Not-determined |
| 18-101-final-viral-combined_k141_143889\|\|full | Not-determined |
| 18-101-final-viral-combined_k141_175599\|\|full | Not-determined |
| 18-101-final-viral-combined_k141_275097\|\|full | Low-quality |
| 18-101-final-viral-combined_k141_358474\|\|full | Low-quality |
| 18-101-final-viral-combined_k141_175609\|\|full | Low-quality |
| 18-101-final-viral-combined_k141_159466\|\|full | Not-determined |
| 18-101-final-viral-combined_k141_44761\|\|full | Not-determined |
| 18-101-final-viral-combined_k141_12154\|\|full | Not-determined |
| 18-101-final-viral-combined_k141_390370\|\|full | Not-determined |
| 18-101-final-viral-combined_k141_292292\|\|full | Low-quality |
| 18-101-final-viral-combined_k141_143905\|\|full | Low-quality |
| 18-101-final-viral-combined_k141_159490\|\|full | Not-determined |
| 18-101-final-viral-combined_k141_12158\|\|full | Low-quality |
| 18-101-final-viral-combined_k141_12169\|\|full | Not-determined |
| 18-101-final-viral-combined_k141_260038\|\|full | Low-quality |
| 18-101-final-viral-combined_k141_226179\|\|full | Not-determined |
| 18-101-final-viral-combined_k141_340270\|\|full | Low-quality |
| 18-101-final-viral-combined_k141_340271\|\|full | Not-determined |
| 18-101-final-viral-combined_k141_242761\|\|full | Low-quality |
| 18-101-final-viral-combined_k141_374112\|\|full | Low-quality |
| 18-101-final-viral-combined_k141_309110\|\|full | Low-quality |
| 18-101-final-viral-combined_k141_94699\|\|full | Not-determined |
| 18-101-final-viral-combined_k141_143925\|\|full | Not-determined |
| 18-101-final-viral-combined_k141_374118\|\|full | Low-quality |
| 18-101-final-viral-combined_k141_374121\|\|full | Not-determined |
| 18-101-final-viral-combined_k141_309123\|\|full | Low-quality |
| 18-101-final-viral-combined_k141_242795\|\|full | Not-determined |
| 18-101-final-viral-combined_k141_340301\|\|full | Low-quality |
| 18-101-final-viral-combined_k141_210573\|\|full | Not-determined |
| 18-101-final-viral-combined_k141_242800\|\|full | Not-determined |
| 18-101-final-viral-combined_k141_94731\|\|full | Low-quality |
| 18-101-final-viral-combined_k141_292365\|\|full | Low-quality |
| 18-101-final-viral-combined_k141_12222\|\|full | Not-determined |
| 18-101-final-viral-combined_k141_358535\|\|full | Low-quality |
| 18-101-final-viral-combined_k141_340316\|\|full | Not-determined |
| 18-101-final-viral-combined_k141_226252\|\|full | Not-determined |
| 18-101-final-viral-combined_k141_77019\|\|full | Low-quality |
| 18-101-final-viral-combined_k141_77020\|\|full | Not-determined |
| 18-101-final-viral-combined_k141_275179\|\|full | Low-quality |
| 18-101-final-viral-combined_k141_159520\|\|full | Not-determined |
| 18-101-final-viral-combined_k141_390446\|\|full | Not-determined |
| 18-101-final-viral-combined_k141_159538\|\|full | Low-quality |
| 18-101-final-viral-combined_k141_292398\|\|full | Low-quality |
| 18-101-final-viral-combined_k141_128221\|\|full | Low-quality |
| 18-101-final-viral-combined_k141_260128\|\|full | Low-quality |
| 18-101-final-viral-combined_k141_275217\|\|full | Low-quality |
| 18-101-final-viral-combined_k141_28390\|\|full | Low-quality |
| 18-101-final-viral-combined_k141_61430\|\|full | Not-determined |
| 18-101-final-viral-combined_k141_77053\|\|full | Not-determined |
| 18-101-final-viral-combined_k141_358592\|\|full | Low-quality |
| 18-110-final-viral-combined_k141_127191\|\|full | High-quality |
| 18-101-final-viral-combined_k141_242858\|\|full | Not-determined |
| 18-101-final-viral-combined_k141_260149\|\|full | Low-quality |
| 18-101-final-viral-combined_k141_260154\|\|full | Low-quality |
| 18-101-final-viral-combined_k141_28427\|\|full | Low-quality |
| 18-101-final-viral-combined_k141_292442\|\|full | Low-quality |
| 18-101-final-viral-combined_k141_390523\|\|full | Low-quality |
| 18-101-final-viral-combined_k141_128247\|\|full | Low-quality |
| 18-101-final-viral-combined_k141_12293\|\|full | Low-quality |
| 18-101-final-viral-combined_k141_159588\|\|full | Low-quality |
| 18-101-final-viral-combined_k141_44852\|\|full | Low-quality |
| 18-101-final-viral-combined_k141_94756\|\|full | Not-determined |
| 18-101-final-viral-combined_k141_309197\|\|full | Not-determined |
| 18-101-final-viral-combined_k141_61490\|\|full | Not-determined |
| 18-101-final-viral-combined_k141_260188\|\|full | Not-determined |
| 18-101-final-viral-combined_k141_358623\|\|full | Low-quality |
| 18-101-final-viral-combined_k141_309199\|\|full | Low-quality |
| 18-101-final-viral-combined_k141_193815\|\|full | Low-quality |
| 18-101-final-viral-combined_k141_242902\|\|full | Low-quality |
| 18-101-final-viral-combined_k141_144008\|\|full | Low-quality |
| 18-101-final-viral-combined_k141_226314\|\|full | Not-determined |
| 18-101-final-viral-combined_k141_226316\|\|full | Low-quality |
| 18-101-final-viral-combined_k141_28458\|\|full | Not-determined |
| 18-101-final-viral-combined_k141_77099\|\|full | Not-determined |
| 18-101-final-viral-combined_k141_309209\|\|full | Not-determined |
| 18-101-final-viral-combined_k141_309211\|\|full | Not-determined |
| 18-101-final-viral-combined_k141_226324\|\|full | Not-determined |
| 18-101-final-viral-combined_k141_77105\|\|full | Not-determined |
| 18-101-final-viral-combined_k141_12334\|\|full | Low-quality |
| 18-101-final-viral-combined_k141_326008\|\|full | Not-determined |
| 18-101-final-viral-combined_k141_226338\|\|full | Low-quality |
| 18-101-final-viral-combined_k141_144043\|\|full | Low-quality |
| 18-101-final-viral-combined_k141_390548\|\|full | Low-quality |
| 18-101_k141_212940 | High-quality |
| 18-101-final-viral-combined_k141_226358\|\|full | Not-determined |
| 18-101-final-viral-combined_k141_193839\|\|full | Low-quality |
| 18-101-final-viral-combined_k141_374230\|\|full | Low-quality |
| 18-101-final-viral-combined_k141_12369\|\|full | Not-determined |
| 18-101-final-viral-combined_k141_374231\|\|full | Low-quality |
| 18-101-final-viral-combined_k141_292516\|\|full | Not-determined |
| 18-101-final-viral-combined_k141_358683\|\|full | Not-determined |
| 18-101-final-viral-combined_k141_12377\|\|full | Not-determined |
| 18-101-final-viral-combined_k141_61590\|\|full | Low-quality |
| 18-101-final-viral-combined_k141_242923\|\|full | Low-quality |
| 18-101-final-viral-combined_k141_144089\|\|full | Not-determined |
| 18-101-final-viral-combined_k141_61598\|\|full | Low-quality |
| 18-101-final-viral-combined_k141_242932\|\|full | Not-determined |
| 18-101-final-viral-combined_k141_144092\|\|full | Not-determined |
| 18-101-final-viral-combined_k141_390558\|\|full | Not-determined |
| 18-101-final-viral-combined_k141_358696\|\|full | Not-determined |
| 18-101-final-viral-combined_k141_390566\|\|full | Not-determined |
| 18-101-final-viral-combined_k141_374267\|\|full | Low-quality |
| 18-101-final-viral-combined_k141_28498\|\|full | Not-determined |
| 18-101-final-viral-combined_k141_358716\|\|full | Low-quality |
| 18-101-final-viral-combined_k141_193884\|\|full | Not-determined |
| 18-101-final-viral-combined_k141_340420\|\|full | Not-determined |
| 18-101-final-viral-combined_k141_390580\|\|full | Low-quality |
| 18-101-final-viral-combined_k141_210713\|\|full | Low-quality |
| 18-101-final-viral-combined_k141_144101\|\|full | Low-quality |
| 18-101-final-viral-combined_k141_12434\|\|full | Not-determined |
| 18-101-final-viral-combined_k141_340436\|\|full | Not-determined |
| 18-101-final-viral-combined_k141_77188\|\|full | Not-determined |
| 18-101-final-viral-combined_k141_77191\|\|full | Low-quality |
| 18-101-final-viral-combined_k141_175759\|\|full | Low-quality |
| 18-101-final-viral-combined_k141_340440\|\|full | Low-quality |
| 18-101-final-viral-combined_k141_292576\|\|full | Low-quality |
| 18-101-final-viral-combined_k141_61626\|\|full | Not-determined |
| 18-101-final-viral-combined_k141_159641\|\|full | Low-quality |
| 18-101-final-viral-combined_k141_326038\|\|full | Not-determined |
| 18-101-final-viral-combined_k141_374301\|\|full | Low-quality |
| 18-101-final-viral-combined_k141_12453\|\|full | Not-determined |
| 18-101-final-viral-combined_k141_28536\|\|full | Not-determined |
| 18-101-final-viral-combined_k141_128366\|\|full | Not-determined |
| 18-101-final-viral-combined_k141_28557\|\|full | Low-quality |
| 18-101-final-viral-combined_k141_159670\|\|full | Not-determined |
| 18-101-final-viral-combined_k141_109947\|\|full | Low-quality |
| 18-101-final-viral-combined_k141_61643\|\|full | Low-quality |
| 18-101-final-viral-combined_k141_128375\|\|full | Low-quality |
| 18-101-final-viral-combined_k141_260366\|\|full | Not-determined |
| 18-101-final-viral-combined_k141_260367\|\|full | Not-determined |
| 18-101-final-viral-combined_k141_340458\|\|full | Low-quality |
| 18-101-final-viral-combined_k141_358777\|\|full | Not-determined |
| 18-101-final-viral-combined_k141_144157\|\|full | Low-quality |
| 18-101-final-viral-combined_k141_374360\|\|full | Low-quality |
| 18-101-final-viral-combined_k141_28568\|\|full | Low-quality |
| 18-101-final-viral-combined_k141_390670\|\|full | Low-quality |
| 18-101-final-viral-combined_k141_28572\|\|full | Low-quality |
| 18-101-final-viral-combined_k141_175820\|\|full | Low-quality |
| 18-101-final-viral-combined_k141_61657\|\|full | Not-determined |
| 18-101-final-viral-combined_k141_109981\|\|full | Not-determined |
| 18-101-final-viral-combined_k141_390677\|\|full | Not-determined |
| 18-101-final-viral-combined_k141_144171\|\|full | Not-determined |
| 18-101-final-viral-combined_k141_94851\|\|full | Not-determined |
| 18-101-final-viral-combined_k141_292658\|\|full | Low-quality |
| 18-101-final-viral-combined_k141_275322\|\|full | Low-quality |
| 18-101-final-viral-combined_k141_159695\|\|full | Low-quality |
| 18-101-final-viral-combined_k141_326103\|\|full | Not-determined |
| 18-101-final-viral-combined_k141_28609\|\|full | Low-quality |
| 18-101-final-viral-combined_k141_358813\|\|full | Not-determined |
| 18-101-final-viral-combined_k141_275332\|\|full | Low-quality |
| 18-101-final-viral-combined_k141_28615\|\|full | Low-quality |
| 18-101-final-viral-combined_k141_159711\|\|full | Not-determined |
| 18-101-final-viral-combined_k141_45039\|\|full | Not-determined |
| 18-101-final-viral-combined_k141_144188\|\|full | Low-quality |
| 18-101-final-viral-combined_k141_194042\|\|full | Low-quality |
| 18-101-final-viral-combined_k141_226517\|\|full | Not-determined |
| 18-101-final-viral-combined_k141_390740\|\|full | Low-quality |
| 18-101-final-viral-combined_k141_110004\|\|full | Not-determined |
| 18-101-final-viral-combined_k141_309347\|\|full | Low-quality |
| 18-101-final-viral-combined_k141_226521\|\|full | Low-quality |
| 18-101-final-viral-combined_k141_326139\|\|full | Low-quality |
| 18-101-final-viral-combined_k141_374392\|\|full | Not-determined |
| 18-101-final-viral-combined_k141_28645\|\|full | Low-quality |
| 18-101-final-viral-combined_k141_159733\|\|full | Not-determined |
| 18-101-final-viral-combined_k141_159734\|\|full | Not-determined |
| 18-101-final-viral-combined_k141_243092\|\|full | Low-quality |
| 18-101-final-viral-combined_k141_309353\|\|full | Low-quality |
| 18-101-final-viral-combined_k141_374410\|\|full | Not-determined |
| 18-101-final-viral-combined_k141_243095\|\|full | Not-determined |
| 18-101-final-viral-combined_k141_110023\|\|full | Not-determined |
| 18-101-final-viral-combined_k141_110024\|\|full | Low-quality |
| 18-101-final-viral-combined_k141_159744\|\|full | Low-quality |
| 18-101-final-viral-combined_k141_12549\|\|full | Low-quality |
| 18-101-final-viral-combined_k141_159746\|\|full | Not-determined |
| 18-101-final-viral-combined_k141_45089\|\|full | Not-determined |
| 18-101-final-viral-combined_k141_28652\|\|full | Low-quality |
| 18-101-final-viral-combined_k141_374442\|\|full | Not-determined |
| 18-101-final-viral-combined_k141_275350\|\|full | Not-determined |
| 18-101-final-viral-combined_k141_45094\|\|full | Not-determined |
| 18-101-final-viral-combined_k141_110042\|\|full | Not-determined |
| 18-101-final-viral-combined_k141_144214\|\|full | Low-quality |
| 18-101-final-viral-combined_k141_12566\|\|full | Low-quality |
| 18-101-final-viral-combined_k141_144225\|\|full | Low-quality |
| 18-101-final-viral-combined_k141_94880\|\|full | Not-determined |
| 18-101-final-viral-combined_k141_226570\|\|full | Not-determined |
| 18-101-final-viral-combined_k141_159765\|\|full | Not-determined |
| 18-101-final-viral-combined_k141_61767\|\|full | Low-quality |
| 18-101-final-viral-combined_k141_340546\|\|full | Not-determined |
| 18-101-final-viral-combined_k141_275374\|\|full | Not-determined |
| 18-101-final-viral-combined_k141_210736\|\|full | Not-determined |
| 18-101-final-viral-combined_k141_210746\|\|full | Low-quality |
| 18-101-final-viral-combined_k141_326225\|\|full | Not-determined |
| 18-101-final-viral-combined_k141_243144\|\|full | Not-determined |
| 18-101-final-viral-combined_k141_144244\|\|full | Low-quality |
| 18-101-final-viral-combined_k141_94919\|\|full | Low-quality |
| 18-101-final-viral-combined_k141_94922\|\|full | Low-quality |
| 18-101-final-viral-combined_k141_77320\|\|full | Not-determined |
| 18-101-final-viral-combined_k141_12590\|\|full | Low-quality |
| 18-101-final-viral-combined_k141_292778\|\|full | Low-quality |
| 18-101-final-viral-combined_k141_128553\|\|full | Not-determined |
| 18-101-final-viral-combined_k141_326250\|\|full | Not-determined |
| 18-101-final-viral-combined_k141_212940\|\|full | High-quality |
| 18-101-final-viral-combined_k141_194124\|\|full | Not-determined |
| 18-101-final-viral-combined_k141_210791\|\|full | Not-determined |
| 18-101-final-viral-combined_k141_77327\|\|full | Not-determined |
| 18-101-final-viral-combined_k141_28676\|\|full | Not-determined |
| 18-101-final-viral-combined_k141_358934\|\|full | Not-determined |
| 18-101-final-viral-combined_k141_275444\|\|full | Not-determined |
| 18-101-final-viral-combined_k141_77329\|\|full | Not-determined |
| 18-101-final-viral-combined_k141_243185\|\|full | Low-quality |
| 18-101-final-viral-combined_k141_292811\|\|full | Not-determined |
| 18-101-final-viral-combined_k141_12640\|\|full | Not-determined |
| 18-101-final-viral-combined_k141_275457\|\|full | Not-determined |
| 18-101-final-viral-combined_k141_292822\|\|full | Low-quality |
| 18-101-final-viral-combined_k141_358970\|\|full | Low-quality |
| 18-101-final-viral-combined_k141_275459\|\|full | Low-quality |
| 18-101-final-viral-combined_k141_260479\|\|full | Low-quality |
| 18-101-final-viral-combined_k141_340600\|\|full | Not-determined |
| 18-101-final-viral-combined_k141_128611\|\|full | Not-determined |
| 18-101-final-viral-combined_k141_226672\|\|full | Low-quality |
| 18-101-final-viral-combined_k141_292836\|\|full | Low-quality |
| 18-101-final-viral-combined_k141_358989\|\|full | Low-quality |
| 18-101-final-viral-combined_k141_374570\|\|full | Low-quality |
| 18-101-final-viral-combined_k141_194177\|\|full | Low-quality |
| 18-101-final-viral-combined_k141_194180\|\|full | Not-determined |
| 18-101-final-viral-combined_k141_77380\|\|full | Not-determined |
| 18-101-final-viral-combined_k141_210843\|\|full | Low-quality |
| 18-101-final-viral-combined_k141_12679\|\|full | Low-quality |
| 18-101-final-viral-combined_k141_340616\|\|full | Not-determined |
| 18-101-final-viral-combined_k141_77389\|\|full | Not-determined |
| 18-101-final-viral-combined_k141_77391\|\|full | Low-quality |
| 18-101-final-viral-combined_k141_226687\|\|full | Low-quality |
| 18-101-final-viral-combined_k141_61896\|\|full | Low-quality |
| 18-101-final-viral-combined_k141_12688\|\|full | Not-determined |
| 18-101-final-viral-combined_k141_210866\|\|full | Low-quality |
| 18-101-final-viral-combined_k141_340631\|\|full | Not-determined |
| 18-101-final-viral-combined_k141_159835\|\|full | Low-quality |
| 18-101-final-viral-combined_k141_359010\|\|full | Not-determined |
| 18-101-final-viral-combined_k141_194239\|\|full | Not-determined |
| 18-101-final-viral-combined_k141_159851\|\|full | Low-quality |
| 18-101-final-viral-combined_k141_374609\|\|full | Low-quality |
| 18-101-final-viral-combined_k141_194245\|\|full | Low-quality |
| 18-101-final-viral-combined_k141_340648\|\|full | Low-quality |
| 18-101-final-viral-combined_k141_340649\|\|full | Low-quality |

18-101-final-viral-combined_k141_243249||full Low-quality
18-101-final-viral-combined_k141_77444||full Low-quality
18-101-final-viral-combined_k141_45195||full Low-quality
18-101-final-viral-combined_k141_374616||full Not-determined
18-101-final-viral-combined_k141_340654||full Not-determined
18-101-final-viral-combined_k141_61946||full Low-quality
18-101-final-viral-combined_k141_374624||full Not-determined
18-101-final-viral-combined_k141_390870||full Low-quality
18-101-final-viral-combined_k141_390871||full Not-determined
18-101-final-viral-combined_k141_326323||full Low-quality
18-101-final-viral-combined_k141_326324||full Not-determined
18-101-final-viral-combined_k141_326325||full Low-quality
18-101-final-viral-combined_k141_77471||full Low-quality
18-101-final-viral-combined_k141_28759||full Low-quality
18-101-final-viral-combined_k141_359068||full Not-determined
18-101-final-viral-combined_k141_194280||full Low-quality
18-101-final-viral-combined_k141_159859||full Low-quality
18-101-final-viral-combined_k141_45228||full Not-determined
18-101-final-viral-combined_k141_210914||full Low-quality
18-101-final-viral-combined_k141_194288||full Not-determined
18-101-final-viral-combined_k141_340696||full Low-quality
18-101-final-viral-combined_k141_194304||full Low-quality
18-101-final-viral-combined_k141_45249||full Low-quality
18-101-final-viral-combined_k141_28784||full Low-quality
18-101-final-viral-combined_k141_61974||full Not-determined
18-101-final-viral-combined_k141_77537||full Low-quality
18-101-final-viral-combined_k141_77544||full Not-determined
18-101-final-viral-combined_k141_28797||full Not-determined
18-101-final-viral-combined_k141_390923||full Low-quality
18-101-final-viral-combined_k141_128750||full Not-determined
18-101-final-viral-combined_k141_61997||full Not-determined
18-101-final-viral-combined_k141_77560||full Not-determined
18-101-final-viral-combined_k141_194335||full Not-determined
18-101-final-viral-combined_k141_292934||full Not-determined
18-101-final-viral-combined_k141_340750||full Not-determined
18-101-final-viral-combined_k141_62026||full Not-determined
18-101-final-viral-combined_k141_243343||full Not-determined
18-101-final-viral-combined_k141_28829||full Not-determined
18-101-final-viral-combined_k141_275634||full Not-determined
18-101-final-viral-combined_k141_359134||full Low-quality
18-101-final-viral-combined_k141_77591||full Not-determined
18-101-final-viral-combined_k141_226824||full Not-determined
18-101-final-viral-combined_k141_77592||full Not-determined
18-101-final-viral-combined_k141_359141||full Low-quality
18-101-final-viral-combined_k141_77598||full Not-determined
18-101-final-viral-combined_k141_28839||full Not-determined
18-101-final-viral-combined_k141_309607||full Low-quality
18-101-final-viral-combined_k141_309608||full Not-determined
18-101-final-viral-combined_k141_243362||full Low-quality
18-101-final-viral-combined_k141_309613||full Low-quality
18-101-final-viral-combined_k141_260691||full Low-quality
18-101-final-viral-combined_k141_292971||full Not-determined
18-101-final-viral-combined_k141_243377||full Low-quality
18-101-final-viral-combined_k141_226848||full Low-quality
18-101-final-viral-combined_k141_144320||full Low-quality
18-101-final-viral-combined_k141_275674||full Not-determined
18-101-final-viral-combined_k141_326451||full Low-quality
18-101-final-viral-combined_k141_275678||full Low-quality
18-101-final-viral-combined_k141_326463||full Not-determined
18-101-final-viral-combined_k141_359174||full Not-determined
18-101-final-viral-combined_k141_110052||full Not-determined
18-101-final-viral-combined_k141_326470||full Low-quality
18-101-final-viral-combined_k141_62080||full Low-quality
18-101-final-viral-combined_k141_293004||full Not-determined
18-101-final-viral-combined_k141_391007||full Not-determined
18-101-final-viral-combined_k141_144330||full Low-quality
18-101-final-viral-combined_k141_260716||full Not-determined
18-101-final-viral-combined_k141_260730||full Low-quality
18-101-final-viral-combined_k141_391029||full Not-determined
18-101-final-viral-combined_k141_293030||full Not-determined
18-101-final-viral-combined_k141_128806||full Low-quality
18-101-final-viral-combined_k141_326497||full Low-quality
18-101-final-viral-combined_k141_293050||full Not-determined
18-101-final-viral-combined_k141_226891||full Not-determined
18-101-final-viral-combined_k141_95099||full Not-determined
18-101-final-viral-combined_k141_293054||full Low-quality
18-101-final-viral-combined_k141_194461||full Not-determined
18-101-final-viral-combined_k141_243443||full Not-determined
18-101-final-viral-combined_k141_62124||full Not-determined
18-101-final-viral-combined_k141_159970||full Not-determined
18-101-final-viral-combined_k141_293075||full Low-quality
18-101-final-viral-combined_k141_359229||full Low-quality
18-101-final-viral-combined_k141_128847||full Not-determined
18-101-final-viral-combined_k141_144409||full Low-quality
18-101-final-viral-combined_k141_62134||full Not-determined
18-101-final-viral-combined_k141_309685||full Low-quality
18-101-final-viral-combined_k141_95140||full Not-determined
18-101-final-viral-combined_k141_12792||full Low-quality
18-101-final-viral-combined_k141_110134||full Low-quality
18-101-final-viral-combined_k141_211062||full Not-determined
18-101-final-viral-combined_k141_326505||full Not-determined
18-101-final-viral-combined_k141_211068||full Low-quality
18-101-final-viral-combined_k141_128895||full Not-determined
18-101-final-viral-combined_k141_374730||full Not-determined
18-101-final-viral-combined_k141_340882||full Not-determined
18-101-final-viral-combined_k141_194503||full Not-determined
18-101-final-viral-combined_k141_211079||full Not-determined
18-101-final-viral-combined_k141_144429||full Not-determined
18-101-final-viral-combined_k141_110147||full Low-quality
18-101-final-viral-combined_k141_144432||full Not-determined
18-101-final-viral-combined_k141_28904||full Not-determined
18-101-final-viral-combined_k141_293113||full Not-determined
18-101-final-viral-combined_k141_77710||full Not-determined
18-101-final-viral-combined_k141_95169||full Low-quality
18-101-final-viral-combined_k141_243488||full Low-quality
18-101-final-viral-combined_k141_309717||full Low-quality
18-101-final-viral-combined_k141_243499||full Low-quality
18-101-final-viral-combined_k141_62183||full Not-determined
18-101-final-viral-combined_k141_275741||full Low-quality
18-101-final-viral-combined_k141_28941||full Not-determined
18-101-final-viral-combined_k141_340922||full Low-quality
18-101-final-viral-combined_k141_62207||full Not-determined
18-101-final-viral-combined_k141_144465||full Not-determined
18-101-final-viral-combined_k141_391062||full Low-quality
18-101-final-viral-combined_k141_359287||full Low-quality
18-101-final-viral-combined_k141_95202||full Low-quality
18-101-final-viral-combined_k141_160060||full Low-quality
18-101-final-viral-combined_k141_28946||full Low-quality
18-101-final-viral-combined_k141_293171||full Not-determined
18-101-final-viral-combined_k141_275762||full Low-quality
18-101-final-viral-combined_k141_391075||full Not-determined
18-101-final-viral-combined_k141_359295||full Low-quality
18-101-final-viral-combined_k141_226939||full Not-determined
18-101-final-viral-combined_k141_12872||full Not-determined
18-101-final-viral-combined_k141_211161||full Low-quality
18-101-final-viral-combined_k141_194589||full Not-determined
18-101-final-viral-combined_k141_12877||full Low-quality
18-101-final-viral-combined_k141_95226||full Not-determined
18-101-final-viral-combined_k141_260827||full Not-determined
18-101-final-viral-combined_k141_194599||full Not-determined
18-101-final-viral-combined_k141_275787||full Not-determined
18-101-final-viral-combined_k141_176034||full Not-determined
18-101-final-viral-combined_k141_275794||full Not-determined
18-101-final-viral-combined_k141_77779||full Not-determined
18-101-final-viral-combined_k141_211188||full Not-determined
18-101-final-viral-combined_k141_391112||full Low-quality
18-101-final-viral-combined_k141_243589||full Not-determined
18-101-final-viral-combined_k141_144502||full Not-determined
18-101-final-viral-combined_k141_29011||full Not-determined
18-101-final-viral-combined_k141_128966||full Low-quality
18-101-final-viral-combined_k141_29013||full Low-quality
18-101-final-viral-combined_k141_77803||full Not-determined
18-101-final-viral-combined_k141_391125||full Low-quality
18-101-final-viral-combined_k141_144535||full Not-determined
18-101-final-viral-combined_k141_160108||full Not-determined
18-101-final-viral-combined_k141_211218||full Not-determined
18-101-final-viral-combined_k141_110220||full Not-determined
17-106-final-viral-combined_k141_28805||full Medium-quality
18-101-final-viral-combined_k141_341003||full Not-determined
18-101-final-viral-combined_k141_374867||full Not-determined
18-101-final-viral-combined_k141_243633||full Not-determined
18-101-final-viral-combined_k141_45388||full Not-determined
18-101-final-viral-combined_k141_243642||full Low-quality
18-101-final-viral-combined_k141_260913||full Not-determined
18-101-final-viral-combined_k141_243643||full Low-quality
18-101-final-viral-combined_k141_260914||full Not-determined
18-101-final-viral-combined_k141_129017||full Not-determined
18-101-final-viral-combined_k141_359375||full Low-quality
18-101-final-viral-combined_k141_260922||full Low-quality
18-101-final-viral-combined_k141_62326||full Low-quality
18-101-final-viral-combined_k141_194669||full Low-quality
18-101-final-viral-combined_k141_374892||full Not-determined
18-101-final-viral-combined_k141_243666||full Not-determined
18-101-final-viral-combined_k141_45395||full Low-quality
18-101-final-viral-combined_k141_275871||full Not-determined
18-101-final-viral-combined_k141_160176||full Not-determined
18-101-final-viral-combined_k141_211260||full Not-determined
18-101-final-viral-combined_k141_211270||full Not-determined
18-101-final-viral-combined_k141_12952||full Not-determined
18-101-final-viral-combined_k141_326687||full Not-determined
18-101-final-viral-combined_k141_176149||full Low-quality
18-101-final-viral-combined_k141_160204||full Not-determined
18-101-final-viral-combined_k141_12958||full Not-determined
18-101-final-viral-combined_k141_211281||full Low-quality
18-101-final-viral-combined_k141_45410||full Not-determined
18-101-final-viral-combined_k141_129093||full Low-quality
18-101-final-viral-combined_k141_194724||full Low-quality
18-101-final-viral-combined_k141_341080||full Low-quality
18-101-final-viral-combined_k141_176160||full Not-determined
18-101-final-viral-combined_k141_77819||full Low-quality
18-101-final-viral-combined_k141_293288||full Low-quality
18-101-final-viral-combined_k141_309869||full Not-determined
18-101-final-viral-combined_k141_45442||full Low-quality
18-101-final-viral-combined_k141_160248||full Not-determined
18-101-final-viral-combined_k141_211326||full Low-quality
18-101-final-viral-combined_k141_359429||full Not-determined
18-101-final-viral-combined_k141_12987||full Low-quality
18-101-final-viral-combined_k141_374958||full Low-quality
18-101-final-viral-combined_k141_261044||full Not-determined
18-101-final-viral-combined_k141_129149||full Not-determined
18-101-final-viral-combined_k141_359450||full Low-quality
18-101-final-viral-combined_k141_227156||full Low-quality
18-101-final-viral-combined_k141_29100||full Not-determined
18-101-final-viral-combined_k141_77868||full Low-quality
18-101-final-viral-combined_k141_211351||full Low-quality
18-101-final-viral-combined_k141_160269||full Not-determined
18-101-final-viral-combined_k141_77878||full Low-quality
18-101-final-viral-combined_k141_374963||full Low-quality
18-101-final-viral-combined_k141_293308||full Low-quality
18-101-final-viral-combined_k141_77881||full Not-determined
18-101-final-viral-combined_k141_359476||full Low-quality
18-101-final-viral-combined_k141_293333||full Not-determined
18-101-final-viral-combined_k141_261089||full Low-quality
18-101-final-viral-combined_k141_309919||full Low-quality
18-101-final-viral-combined_k141_160304||full Low-quality
18-101-final-viral-combined_k141_95403||full Not-determined
18-101-final-viral-combined_k141_243826||full Low-quality
18-101-final-viral-combined_k141_45489||full Not-determined
18-101-final-viral-combined_k141_326800||full Not-determined
18-101-final-viral-combined_k141_243837||full Low-quality
18-101-final-viral-combined_k141_211403||full Not-determined
18-101-final-viral-combined_k141_144713||full Not-determined
18-101-final-viral-combined_k141_29160||full Not-determined
18-101-final-viral-combined_k141_391222||full Not-determined
18-101-final-viral-combined_k141_211406||full Not-determined
18-101-final-viral-combined_k141_13078||full Low-quality
18-101-final-viral-combined_k141_309943||full Not-determined
18-101-final-viral-combined_k141_211412||full Low-quality
18-101-final-viral-combined_k141_13090||full Not-determined
18-101-final-viral-combined_k141_194818||full Low-quality
18-101-final-viral-combined_k141_341146||full Low-quality
18-101-final-viral-combined_k141_326824||full Low-quality
18-101-final-viral-combined_k141_77943||full Low-quality
18-101-final-viral-combined_k141_309963||full Low-quality
18-101-final-viral-combined_k141_261144||full Not-determined
18-101-final-viral-combined_k141_110420||full Low-quality
18-101-final-viral-combined_k141_194827||full Low-quality
18-101-final-viral-combined_k141_95460||full Low-quality
18-101-final-viral-combined_k141_13091||full Low-quality
18-101-final-viral-combined_k141_227203||full Not-determined
18-101-final-viral-combined_k141_275961||full Not-determined
18-101-final-viral-combined_k141_227206||full Not-determined
18-101-final-viral-combined_k141_227209||full Low-quality
18-101-final-viral-combined_k141_275978||full Not-determined
18-101-final-viral-combined_k141_194848||full Not-determined
18-101-final-viral-combined_k141_110447||full Low-quality
18-101-final-viral-combined_k141_261163||full Low-quality
18-101-final-viral-combined_k141_194852||full Low-quality
18-101-final-viral-combined_k141_95486||full Not-determined
18-101-final-viral-combined_k141_211467||full Not-determined
18-101-final-viral-combined_k141_275989||full Low-quality
18-101-final-viral-combined_k141_326880||full Low-quality
18-101-final-viral-combined_k141_176320||full Not-determined
18-101-final-viral-combined_k141_275995||full Not-determined
18-101-final-viral-combined_k141_243919||full Not-determined
18-101-final-viral-combined_k141_144772||full Not-determined
18-101-final-viral-combined_k141_129288||full Not-determined
18-101-final-viral-combined_k141_77993||full Not-determined
18-101-final-viral-combined_k141_194879||full Not-determined
18-101-final-viral-combined_k141_391296||full Not-determined
18-101-final-viral-combined_k141_194880||full Not-determined
18-101-final-viral-combined_k141_77995||full Not-determined
18-101-final-viral-combined_k141_176340||full Not-determined
18-101-final-viral-combined_k141_62410||full Low-quality
18-101-final-viral-combined_k141_276014||full Low-quality
18-101-final-viral-combined_k141_211506||full Not-determined
18-101-final-viral-combined_k141_310044||full Not-determined
18-101-final-viral-combined_k141_227265||full Not-determined
18-101-final-viral-combined_k141_211508||full Low-quality
18-101-final-viral-combined_k141_194926||full Not-determined
18-101-final-viral-combined_k141_45590||full Low-quality
18-101-final-viral-combined_k141_129338||full Not-determined
18-101-final-viral-combined_k141_176375||full Low-quality
18-101-final-viral-combined_k141_341227||full Not-determined
18-101-final-viral-combined_k141_45594||full Not-determined
18-101-final-viral-combined_k141_144834||full Low-quality
18-101-final-viral-combined_k141_13156||full Low-quality
18-101-final-viral-combined_k141_194928||full Low-quality
18-101-final-viral-combined_k141_293451||full Low-quality
18-101-final-viral-combined_k141_211536||full Not-determined
18-101-final-viral-combined_k141_293456||full Low-quality
18-101-final-viral-combined_k141_310088||full Not-determined
18-101-final-viral-combined_k141_276092||full Low-quality
18-101-final-viral-combined_k141_293477||full Low-quality
18-101-final-viral-combined_k141_13183||full Low-quality
18-101-final-viral-combined_k141_129384||full Not-determined
18-101-final-viral-combined_k141_261273||full Low-quality
18-101-final-viral-combined_k141_375197||full Low-quality
18-101-final-viral-combined_k141_129401||full Low-quality
18-101-final-viral-combined_k141_326953||full Not-determined
18-101-final-viral-combined_k141_211581||full Not-determined
18-101-final-viral-combined_k141_13200||full Not-determined
18-101-final-viral-combined_k141_176412||full Low-quality
18-101-final-viral-combined_k141_261287||full Not-determined
18-101-final-viral-combined_k141_129411||full Low-quality
18-101-final-viral-combined_k141_13210||full Not-determined
18-101-final-viral-combined_k141_326978||full Not-determined
18-101-final-viral-combined_k141_45629||full Low-quality
18-101-final-viral-combined_k141_160546||full Low-quality
18-101-final-viral-combined_k141_375225||full Not-determined
18-101-final-viral-combined_k141_95553||full Not-determined
18-101-final-viral-combined_k141_375233||full Low-quality
18-101-final-viral-combined_k141_45647||full Not-determined
18-101-final-viral-combined_k141_375241||full Not-determined
18-101-final-viral-combined_k141_176441||full Low-quality
18-101-final-viral-combined_k141_129450||full Low-quality
18-101-final-viral-combined_k141_78024||full Not-determined
18-101-final-viral-combined_k141_244041||full Not-determined
18-101-final-viral-combined_k141_29183||full Low-quality
18-101-final-viral-combined_k141_276164||full Not-determined
18-101-final-viral-combined_k141_375253||full Low-quality
18-101-final-viral-combined_k141_293554||full Not-determined
18-101-final-viral-combined_k141_13242||full Not-determined
18-101-final-viral-combined_k141_310152||full Not-determined
18-101-final-viral-combined_k141_375254||full Low-quality
18-101-final-viral-combined_k141_227359||full Not-determined
18-101-final-viral-combined_k141_276180||full Low-quality
18-101-final-viral-combined_k141_211682||full Low-quality
18-101-final-viral-combined_k141_95582||full Not-determined
18-101-final-viral-combined_k141_144933||full Not-determined
18-101-final-viral-combined_k141_62481||full Low-quality
18-101-final-viral-combined_k141_244081||full Low-quality
18-101-final-viral-combined_k141_195010||full Not-determined
18-101-final-viral-combined_k141_176469||full Not-determined
18-101-final-viral-combined_k141_13289||full Not-determined
18-101-final-viral-combined_k141_211726||full Not-determined
18-101-final-viral-combined_k141_95609||full Not-determined
18-101-final-viral-combined_k141_29198||full Low-quality
18-101-final-viral-combined_k141_176479||full Low-quality
18-101-final-viral-combined_k141_176480||full Not-determined
18-101-final-viral-combined_k141_29201||full Not-determined
18-101-final-viral-combined_k141_129528||full Not-determined
18-101-final-viral-combined_k141_195044||full Low-quality
18-101-final-viral-combined_k141_211738||full Not-determined
18-101-final-viral-combined_k141_391453||full Low-quality
18-101-final-viral-combined_k141_375313||full Low-quality
18-101-final-viral-combined_k141_160635||full Low-quality
18-101-final-viral-combined_k141_211746||full Low-quality
18-101-final-viral-combined_k141_62540||full Low-quality
18-101-final-viral-combined_k141_276250||full Not-determined
18-101-final-viral-combined_k141_160638||full Not-determined
18-101-final-viral-combined_k141_145002||full Not-determined
18-101-final-viral-combined_k141_391471||full Low-quality
18-101-final-viral-combined_k141_195085||full Low-quality
18-101-final-viral-combined_k141_359658||full Not-determined
18-101-final-viral-combined_k141_310218||full Low-quality
18-101-final-viral-combined_k141_62576||full Low-quality
18-101-final-viral-combined_k141_375331||full Not-determined
18-101-final-viral-combined_k141_227437||full Low-quality
18-101-final-viral-combined_k141_160675||full Low-quality
18-101-final-viral-combined_k141_145022||full Low-quality
18-101-final-viral-combined_k141_244188||full Not-determined
18-101-final-viral-combined_k141_341429||full Not-determined
18-101-final-viral-combined_k141_244190||full Low-quality
18-101-final-viral-combined_k141_176535||full Not-determined
18-101-final-viral-combined_k141_62595||full Low-quality
18-101-final-viral-combined_k141_129613||full Low-quality
18-101-final-viral-combined_k141_95686||full Low-quality
18-101-final-viral-combined_k141_176552||full Not-determined
18-101-final-viral-combined_k141_160702||full Not-determined
18-101-final-viral-combined_k141_227480||full Not-determined
18-101-final-viral-combined_k141_276303||full Low-quality
18-101-final-viral-combined_k141_160720||full Not-determined
18-101-final-viral-combined_k141_261411||full Low-quality
18-101-final-viral-combined_k141_341466||full Low-quality
18-101-final-viral-combined_k141_227494||full Low-quality
18-101-final-viral-combined_k141_110720||full Not-determined
18-101-final-viral-combined_k141_244238||full Not-determined
18-101-final-viral-combined_k141_62626||full Low-quality
18-101-final-viral-combined_k141_244241||full Not-determined
18-101-final-viral-combined_k141_160733||full Low-quality
18-101-final-viral-combined_k141_341489||full Low-quality
18-101-final-viral-combined_k141_45762||full Not-determined
18-101-final-viral-combined_k141_45768||full Not-determined
18-101-final-viral-combined_k141_391538||full Not-determined
18-101-final-viral-combined_k141_359690||full Not-determined
18-101-final-viral-combined_k141_261473||full Not-determined
18-101-final-viral-combined_k141_78110||full Not-determined
18-101-final-viral-combined_k141_276375||full Not-determined
18-101-final-viral-combined_k141_293701||full Low-quality
18-101-final-viral-combined_k141_62633||full Low-quality
18-101-final-viral-combined_k141_95787||full Low-quality
18-101-final-viral-combined_k141_195174||full Not-determined
18-101-final-viral-combined_k141_341508||full Not-determined
18-101-final-viral-combined_k141_391551||full Low-quality
18-101-final-viral-combined_k141_211897||full Not-determined
18-101-final-viral-combined_k141_261501||full Low-quality
18-101-final-viral-combined_k141_62638||full Low-quality
18-101-final-viral-combined_k141_227515||full Low-quality
18-101-final-viral-combined_k141_276405||full Not-determined
18-101-final-viral-combined_k141_276406||full Low-quality
18-101-final-viral-combined_k141_391561||full Not-determined
18-101-final-viral-combined_k141_359734||full Low-quality
18-101-final-viral-combined_k141_195191||full Low-quality
18-101-final-viral-combined_k141_129690||full Not-determined
18-101-final-viral-combined_k141_195193||full Low-quality
18-101-final-viral-combined_k141_78167||full Low-quality
18-101-final-viral-combined_k141_145154||full Not-determined
18-101-final-viral-combined_k141_110752||full Not-determined
18-101-final-viral-combined_k141_276419||full Not-determined
18-101-final-viral-combined_k141_110753||full Low-quality
18-101-final-viral-combined_k141_145157||full Not-determined
18-101-final-viral-combined_k141_129700||full Low-quality
18-101-final-viral-combined_k141_375368||full Low-quality
18-101-final-viral-combined_k141_327085||full Not-determined
18-101-final-viral-combined_k141_160798||full Low-quality
18-101-final-viral-combined_k141_45806||full Low-quality
18-101-final-viral-combined_k141_341554||full Low-quality
18-101-final-viral-combined_k141_176656||full Not-determined
18-101-final-viral-combined_k141_375389||full Not-determined
18-101-final-viral-combined_k141_276447||full Low-quality
18-101-final-viral-combined_k141_13470||full Not-determined
18-101-final-viral-combined_k141_176664||full Not-determined
18-101-final-viral-combined_k141_293800||full Not-determined
18-101-final-viral-combined_k141_45826||full Not-determined
18-101-final-viral-combined_k141_375397||full Not-determined
18-101-final-viral-combined_k141_341574||full Low-quality
18-101-final-viral-combined_k141_29271||full Low-quality
18-101-final-viral-combined_k141_62680||full Not-determined
18-101-final-viral-combined_k141_110786||full Not-determined
18-101-final-viral-combined_k141_227542||full Low-quality
18-101-final-viral-combined_k141_276483||full Not-determined
18-101-final-viral-combined_k141_62703||full Not-determined
18-101-final-viral-combined_k141_244380||full Not-determined
18-101-final-viral-combined_k141_195247||full Not-determined
18-101-final-viral-combined_k141_375441||full Low-quality
18-101-final-viral-combined_k141_310295||full Not-determined
18-101-final-viral-combined_k141_293843||full Not-determined
18-101-final-viral-combined_k141_276507||full Low-quality
18-101-final-viral-combined_k141_195260||full Low-quality
18-101-final-viral-combined_k141_62714||full Not-determined
18-101-final-viral-combined_k141_45881||full Low-quality
18-101-final-viral-combined_k141_129753||full Low-quality
18-101-final-viral-combined_k141_293849||full Not-determined
18-101-final-viral-combined_k141_227591||full Not-determined
18-101-final-viral-combined_k141_95908||full Not-determined
18-101-final-viral-combined_k141_341634||full Not-determined
18-101-final-viral-combined_k141_62739||full Low-quality
18-101-final-viral-combined_k141_145290||full Low-quality
18-101-final-viral-combined_k141_391684||full Not-determined
18-101-final-viral-combined_k141_145292||full Low-quality
KM-final-viral-combined_k141_199746||full High-quality
18-101-final-viral-combined_k141_293865||full Not-determined
18-101-final-viral-combined_k141_212057||full Not-determined
18-101-final-viral-combined_k141_145295||full Low-quality
18-101-final-viral-combined_k141_129799||full Low-quality
18-101-final-viral-combined_k141_13568||full Not-determined
18-101-final-viral-combined_k141_62759||full Not-determined
18-101-final-viral-combined_k141_95944||full Low-quality
18-101-final-viral-combined_k141_244453||full Not-determined
18-101-final-viral-combined_k141_129804||full Not-determined
18-101-final-viral-combined_k141_176753||full Not-determined
18-101-final-viral-combined_k141_145309||full Low-quality
18-101-final-viral-combined_k141_95955||full Low-quality
18-101-final-viral-combined_k141_391725||full Not-determined
18-101-final-viral-combined_k141_359862||full Low-quality
18-101-final-viral-combined_k141_95965||full Not-determined
18-101-final-viral-combined_k141_176776||full Low-quality
18-101-final-viral-combined_k141_29345||full Not-determined
18-101-final-viral-combined_k141_227663||full Not-determined
18-101-final-viral-combined_k141_45945||full Low-quality
18-101-final-viral-combined_k141_341686||full Not-determined
18-101-final-viral-combined_k141_212089||full Low-quality
18-101-final-viral-combined_k141_29364||full Not-determined
18-101-final-viral-combined_k141_13617||full Not-determined
18-101-final-viral-combined_k141_261551||full Low-quality
18-101-final-viral-combined_k141_244499||full Not-determined
18-101-final-viral-combined_k141_78357||full Not-determined
18-101-final-viral-combined_k141_110909||full Not-determined
18-101-final-viral-combined_k141_391732||full Not-determined
18-101-final-viral-combined_k141_78362||full Not-determined
18-101-final-viral-combined_k141_96007||full Not-determined
18-101-final-viral-combined_k141_244507||full Not-determined
18-101-final-viral-combined_k141_13635||full Low-quality
18-101-final-viral-combined_k141_227680||full Not-determined
18-101-final-viral-combined_k141_310323||full Not-determined
18-101-final-viral-combined_k141_227689||full Not-determined
18-101-final-viral-combined_k141_13643||full Not-determined
18-101-final-viral-combined_k141_212110||full Not-determined
18-101-final-viral-combined_k141_160964||full Not-determined
18-101-final-viral-combined_k141_212113||full Low-quality
18-101-final-viral-combined_k141_160972||full Low-quality
18-101-final-viral-combined_k141_359901||full Not-determined
18-101-final-viral-combined_k141_13662||full Not-determined
18-101-final-viral-combined_k141_46023||full Not-determined
18-101-final-viral-combined_k141_96028||full Not-determined
18-101-final-viral-combined_k141_78379||full Not-determined
18-101-final-viral-combined_k141_244530||full Low-quality
18-101-final-viral-combined_k141_78382||full Not-determined
18-101-final-viral-combined_k141_341726||full Low-quality
18-101-final-viral-combined_k141_129871||full Not-determined
18-101-final-viral-combined_k141_375527||full Not-determined
18-101-final-viral-combined_k141_341729||full Low-quality
18-101-final-viral-combined_k141_145378||full Low-quality
18-101-final-viral-combined_k141_46048||full Not-determined
18-101-final-viral-combined_k141_341739||full Not-determined
18-101-final-viral-combined_k141_161005||full Low-quality
18-101-final-viral-combined_k141_46059||full Not-determined
18-101-final-viral-combined_k141_227745||full Not-determined
18-101-final-viral-combined_k141_145419||full Not-determined
18-101-final-viral-combined_k141_341767||full Low-quality
18-101-final-viral-combined_k141_375567||full Low-quality
18-101-final-viral-combined_k141_129920||full Low-quality
18-101-final-viral-combined_k141_261620||full Not-determined
18-101-final-viral-combined_k141_244617||full Low-quality
18-101-final-viral-combined_k141_195464||full Not-determined
18-101-final-viral-combined_k141_359954||full Low-quality
18-101-final-viral-combined_k141_110985||full Not-determined
18-101-final-viral-combined_k141_195473||full Low-quality
18-101-final-viral-combined_k141_145442||full Low-quality
18-101-final-viral-combined_k141_227795||full Not-determined
18-101-final-viral-combined_k141_78426||full Low-quality
18-101-final-viral-combined_k141_293970||full Not-determined
18-101-final-viral-combined_k141_212187||full Low-quality
18-101-final-viral-combined_k141_129962||full Low-quality
18-101-final-viral-combined_k141_244629||full Not-determined
18-101-final-viral-combined_k141_13736||full Low-quality
18-101-final-viral-combined_k141_145467||full Low-quality
18-101-final-viral-combined_k141_195487||full Not-determined
18-101-final-viral-combined_k141_310386||full Not-determined
18-101-final-viral-combined_k141_261653||full Low-quality
18-101-final-viral-combined_k141_78447||full Low-quality
18-101-final-viral-combined_k141_341815||full Not-determined
18-101-final-viral-combined_k141_244654||full Not-determined
18-101-final-viral-combined_k141_195492||full Low-quality
18-101-final-viral-combined_k141_212200||full Low-quality
18-101-final-viral-combined_k141_293998||full Not-determined
18-101-final-viral-combined_k141_359993||full Low-quality
18-101-final-viral-combined_k141_212204||full Low-quality
18-101-final-viral-combined_k141_129997||full Low-quality
18-101-final-viral-combined_k141_261681||full Not-determined
18-101-final-viral-combined_k141_195504||full Low-quality
18-101-final-viral-combined_k141_310419||full Low-quality
18-101-final-viral-combined_k141_62943||full Not-determined
18-101-final-viral-combined_k141_111051||full Low-quality
18-101-final-viral-combined_k141_13773||full Low-quality
18-101-final-viral-combined_k141_29388||full Low-quality
18-101-final-viral-combined_k141_111063||full Not-determined
18-101-final-viral-combined_k141_244662||full Low-quality
18-101-final-viral-combined_k141_195529||full Low-quality
18-101-final-viral-combined_k141_276653||full Low-quality
18-101-final-viral-combined_k141_62976||full Not-determined
18-101-final-viral-combined_k141_195557||full Not-determined
18-101-final-viral-combined_k141_391816||full Not-determined
18-101-final-viral-combined_k141_130051||full Low-quality
18-101-final-viral-combined_k141_227824||full Low-quality
18-101-final-viral-combined_k141_391822||full Not-determined
18-101-final-viral-combined_k141_13812||full Not-determined
18-101-final-viral-combined_k141_195570||full Not-determined
18-101-final-viral-combined_k141_310467||full Low-quality
18-101-final-viral-combined_k141_29451||full Not-determined
18-101-final-viral-combined_k141_63027||full Not-determined
18-101-final-viral-combined_k141_13823||full Low-quality
18-101-final-viral-combined_k141_360061||full Low-quality
18-101-final-viral-combined_k141_145577||full Not-determined
18-101-final-viral-combined_k141_310488||full Low-quality
18-101-final-viral-combined_k141_130071||full Not-determined
18-101-final-viral-combined_k141_78510||full Low-quality
18-101-final-viral-combined_k141_276702||full Low-quality
18-101-final-viral-combined_k141_360069||full Low-quality
18-101-final-viral-combined_k141_244754||full Not-determined
18-101-final-viral-combined_k141_111128||full Low-quality
18-101-final-viral-combined_k141_244758||full Low-quality
18-101-final-viral-combined_k141_212307||full Low-quality
18-101-final-viral-combined_k141_111135||full Low-quality
18-101-final-viral-combined_k141_13847||full Not-determined
18-101-final-viral-combined_k141_177023||full Low-quality
18-101-final-viral-combined_k141_46228||full Not-determined
18-101-final-viral-combined_k141_177026||full Not-determined
18-101-final-viral-combined_k141_276728||full Low-quality
18-101-final-viral-combined_k141_195608||full Not-determined
18-101-final-viral-combined_k141_375745||full Not-determined
18-101-final-viral-combined_k141_161251||full Low-quality
18-101-final-viral-combined_k141_212338||full Not-determined
18-101-final-viral-combined_k141_227879||full Low-quality
18-101-final-viral-combined_k141_111158||full Low-quality
18-101-final-viral-combined_k141_13874||full Low-quality
18-101-final-viral-combined_k141_261839||full Not-determined
18-101-final-viral-combined_k141_375782||full Not-determined
18-101-final-viral-combined_k141_111169||full Not-determined
18-101-final-viral-combined_k141_111179||full Low-quality
18-101-final-viral-combined_k141_161293||full Low-quality
18-101-final-viral-combined_k141_276750||full Low-quality
18-101-final-viral-combined_k141_130164||full Low-quality
18-101-final-viral-combined_k141_111200||full Not-determined
18-101-final-viral-combined_k141_130175||full Not-determined
18-101-final-viral-combined_k141_341944||full Not-determined
18-101-final-viral-combined_k141_391908||full Not-determined
18-101-final-viral-combined_k141_276778||full Low-quality
18-101-final-viral-combined_k141_111214||full Not-determined
18-101-final-viral-combined_k141_161315||full Low-quality
18-101-final-viral-combined_k141_212344||full Low-quality
18-101-final-viral-combined_k141_13927||full Low-quality
18-101-final-viral-combined_k141_29523||full Low-quality
18-101-final-viral-combined_k141_391910||full Not-determined
18-101-final-viral-combined_k141_195697||full Low-quality
18-101-final-viral-combined_k141_63151||full Not-determined
18-101-final-viral-combined_k141_177145||full Low-quality
18-101-final-viral-combined_k141_391913||full Not-determined

| | |
|---|---|
| 18-101-final-viral-combined_k141_276808||full | Low-quality |
| 18-101-final-viral-combined_k141_212378||full | Not-determined |
| 18-101-final-viral-combined_k141_276816||full | Not-determined |
| 17-109-final-viral-combined_k141_263377||full | High-quality |
| 18-101-final-viral-combined_k141_391923||full | Low-quality |
| 18-101-final-viral-combined_k141_276817||full | Not-determined |
| 18-101-final-viral-combined_k141_177159||full | Low-quality |
| 18-101-final-viral-combined_k141_96143||full | Not-determined |
| 18-110_k141_48935 | High-quality |
| 18-101-final-viral-combined_k141_111236||full | Low-quality |
| 18-101-final-viral-combined_k141_310533||full | Not-determined |
| 18-101-final-viral-combined_k141_212407||full | Not-determined |
| 18-101-final-viral-combined_k141_96145||full | Low-quality |
| 18-101-final-viral-combined_k141_227929||full | Not-determined |
| 18-101-final-viral-combined_k141_244857||full | Not-determined |
| 18-101-final-viral-combined_k141_261970||full | Not-determined |
| 18-101-final-viral-combined_k141_227934||full | Low-quality |
| 18-101-final-viral-combined_k141_244858||full | Not-determined |
| 18-101-final-viral-combined_k141_111255||full | Low-quality |
| 18-101-final-viral-combined_k141_96163||full | Low-quality |
| 18-101-final-viral-combined_k141_360189||full | Not-determined |
| 18-101-final-viral-combined_k141_244871||full | Not-determined |
| 18-101-final-viral-combined_k141_13979||full | Low-quality |
| 18-101-final-viral-combined_k141_63190||full | Low-quality |
| 18-101-final-viral-combined_k141_391955||full | Not-determined |
| 18-101-final-viral-combined_k141_78575||full | Low-quality |
| 18-101-final-viral-combined_k141_342020||full | Not-determined |
| 18-101-final-viral-combined_k141_391956||full | Low-quality |
| 18-101-final-viral-combined_k141_161386||full | Not-determined |
| 18-101-final-viral-combined_k141_161394||full | Low-quality |
| 18-101-final-viral-combined_k141_262004||full | Low-quality |
| 18-101-final-viral-combined_k141_29625||full | Not-determined |
| 18-101-final-viral-combined_k141_391970||full | Not-determined |
| 18-101-final-viral-combined_k141_96202||full | Not-determined |
| 18-101-final-viral-combined_k141_342044||full | Not-determined |
| 18-101-final-viral-combined_k141_111297||full | Not-determined |
| 18-101-final-viral-combined_k141_391971||full | Low-quality |
| 18-101-final-viral-combined_k141_342046||full | Low-quality |
| 18-101-final-viral-combined_k141_244903||full | Low-quality |
| 18-101-final-viral-combined_k141_46265||full | Low-quality |
| 18-101-final-viral-combined_k141_391974||full | Not-determined |
| 18-101-final-viral-combined_k141_195799||full | Not-determined |
| 18-101-final-viral-combined_k141_310594||full | Not-determined |
| 18-101-final-viral-combined_k141_212493||full | Not-determined |
| 18-101-final-viral-combined_k141_14004||full | Not-determined |
| 18-101-final-viral-combined_k141_130259||full | Low-quality |
| 18-101-final-viral-combined_k141_360269||full | Not-determined |
| 18-101-final-viral-combined_k141_310617||full | Not-determined |
| 18-101-final-viral-combined_k141_375923||full | Low-quality |
| 18-101-final-viral-combined_k141_262040||full | Not-determined |
| 18-101-final-viral-combined_k141_276911||full | Not-determined |
| 18-101-final-viral-combined_k141_212516||full | Not-determined |
| 18-101-final-viral-combined_k141_294200||full | Low-quality |
| 18-101-final-viral-combined_k141_111341||full | Low-quality |
| 18-101-final-viral-combined_k141_63216||full | Not-determined |
| 18-101-final-viral-combined_k141_14015||full | Not-determined |
| 18-101-final-viral-combined_k141_342095||full | Not-determined |
| 18-101-final-viral-combined_k141_29667||full | Low-quality |
| 18-101-final-viral-combined_k141_212536||full | Low-quality |
| 18-101-final-viral-combined_k141_262061||full | Low-quality |
| 18-101-final-viral-combined_k141_195867||full | Not-determined |
| 18-101-final-viral-combined_k141_228010||full | Low-quality |
| 18-101-final-viral-combined_k141_375959||full | Low-quality |
| 18-101-final-viral-combined_k141_294212||full | Low-quality |
| 18-101-final-viral-combined_k141_342101||full | Low-quality |
| 18-101-final-viral-combined_k141_111366||full | Not-determined |
| 18-101-final-viral-combined_k141_161468||full | Low-quality |
| 18-101-final-viral-combined_k141_161470||full | Not-determined |
| 18-101-final-viral-combined_k141_375981||full | Not-determined |
| 18-101-final-viral-combined_k141_161472||full | Not-determined |
| 18-101-final-viral-combined_k141_244986||full | Not-determined |
| 18-101-final-viral-combined_k141_161477||full | Low-quality |
| 18-101-final-viral-combined_k141_195909||full | Not-determined |
| 18-101-final-viral-combined_k141_310670||full | Low-quality |
| 18-101-final-viral-combined_k141_327240||full | Not-determined |
| 18-102_k141_124416 | Medium-quality |
| 18-101-final-viral-combined_k141_111382||full | Not-determined |
| 18-101-final-viral-combined_k141_262091||full | Not-determined |
| 18-101-final-viral-combined_k141_78618||full | Low-quality |
| 18-101-final-viral-combined_k141_130297||full | Not-determined |
| 18-101-final-viral-combined_k141_245020||full | Low-quality |
| 18-101-final-viral-combined_k141_29703||full | Not-determined |
| 18-101-final-viral-combined_k141_78621||full | Not-determined |
| 18-101-final-viral-combined_k141_342141||full | Low-quality |
| 18-101-final-viral-combined_k141_130302||full | Low-quality |
| 18-101-final-viral-combined_k141_161509||full | Low-quality |
| 18-101-final-viral-combined_k141_294232||full | Low-quality |
| 18-101-final-viral-combined_k141_111391||full | Low-quality |
| 18-101-final-viral-combined_k141_262114||full | Low-quality |
| 18-101-final-viral-combined_k141_46294||full | Low-quality |
| 18-101-final-viral-combined_k141_78639||full | Low-quality |
| 18-101-final-viral-combined_k141_46296||full | Low-quality |
| 18-101-final-viral-combined_k141_145813||full | Not-determined |
| 18-101-final-viral-combined_k141_376022||full | Low-quality |
| 18-101-final-viral-combined_k141_14061||full | Not-determined |
| 18-101-final-viral-combined_k141_376024||full | Not-determined |
| 18-101-final-viral-combined_k141_46305||full | Low-quality |
| 18-101-final-viral-combined_k141_130331||full | Low-quality |
| 18-101-final-viral-combined_k141_360351||full | Low-quality |
| 18-101-final-viral-combined_k141_310716||full | Not-determined |
| 18-101-final-viral-combined_k141_161563||full | Low-quality |
| 18-101-final-viral-combined_k141_294277||full | Not-determined |
| 18-101-final-viral-combined_k141_29754||full | Not-determined |
| 18-101-final-viral-combined_k141_195983||full | Not-determined |
| 18-101-final-viral-combined_k141_294289||full | Not-determined |
| 18-101-final-viral-combined_k141_212638||full | Not-determined |
| 18-101-final-viral-combined_k141_310739||full | Not-determined |
| 18-101-final-viral-combined_k141_327301||full | Low-quality |
| 18-101-final-viral-combined_k141_177415||full | Low-quality |
| 18-101-final-viral-combined_k141_195996||full | Not-determined |
| 18-101-final-viral-combined_k141_145848||full | Not-determined |
| 18-101-final-viral-combined_k141_78689||full | Low-quality |
| 18-101-final-viral-combined_k141_196001||full | Not-determined |
| 18-101-final-viral-combined_k141_14110||full | Not-determined |
| 18-101-final-viral-combined_k141_310758||full | Low-quality |
| 18-101-final-viral-combined_k141_262173||full | Not-determined |
| 18-101-final-viral-combined_k141_177440||full | Not-determined |
| 18-101-final-viral-combined_k141_294318||full | Not-determined |
| 18-101-final-viral-combined_k141_196022||full | Not-determined |
| 18-101-final-viral-combined_k141_177445||full | Low-quality |
| 18-101-final-viral-combined_k141_310784||full | Not-determined |
| 18-101-final-viral-combined_k141_277046||full | Low-quality |
| 18-101-final-viral-combined_k141_96311||full | Not-determined |
| 18-101-final-viral-combined_k141_14138||full | Low-quality |
| 18-101-final-viral-combined_k141_327343||full | Not-determined |
| 18-101-final-viral-combined_k141_46361||full | Low-quality |
| 18-101-final-viral-combined_k141_228215||full | Not-determined |
| 18-101-final-viral-combined_k141_14151||full | Low-quality |
| 18-101-final-viral-combined_k141_96322||full | Not-determined |
| 18-101-final-viral-combined_k141_161607||full | Low-quality |
| 18-101-final-viral-combined_k141_14154||full | Not-determined |
| 18-101-final-viral-combined_k141_14162||full | Low-quality |
| 18-101-final-viral-combined_k141_14163||full | Not-determined |
| 18-101-final-viral-combined_k141_111446||full | Low-quality |
| 18-101-final-viral-combined_k141_130382||full | Not-determined |
| 18-101-final-viral-combined_k141_14169||full | Not-determined |
| 18-101-final-viral-combined_k141_46389||full | Not-determined |
| 18-101-final-viral-combined_k141_294370||full | Not-determined |
| 18-101-final-viral-combined_k141_14173||full | Not-determined |
| 18-101-final-viral-combined_k141_29833||full | Low-quality |
| 18-101-final-viral-combined_k141_78784||full | Low-quality |
| 18-101-final-viral-combined_k141_111476||full | Not-determined |
| 18-101-final-viral-combined_k141_78792||full | Low-quality |
| 18-101-final-viral-combined_k141_294390||full | Not-determined |
| 18-101-final-viral-combined_k141_212742||full | Low-quality |
| 18-101-final-viral-combined_k141_145949||full | Not-determined |
| 18-101-final-viral-combined_k141_111483||full | Not-determined |
| 18-101-final-viral-combined_k141_196097||full | Low-quality |
| 18-101-final-viral-combined_k141_196101||full | Not-determined |
| 18-101-final-viral-combined_k141_310860||full | Low-quality |
| 18-101-final-viral-combined_k141_96351||full | Low-quality |
| 18-101-final-viral-combined_k141_310870||full | Not-determined |
| 18-101-final-viral-combined_k141_376155||full | Low-quality |
| 18-101-final-viral-combined_k141_376156||full | Low-quality |
| 18-101-final-viral-combined_k141_310886||full | Low-quality |
| 18-101-final-viral-combined_k141_63465||full | Low-quality |
| 18-101-final-viral-combined_k141_212786||full | Low-quality |
| 18-101-final-viral-combined_k141_360449||full | Low-quality |
| 18-101-final-viral-combined_k141_78848||full | Not-determined |
| 18-101-final-viral-combined_k141_212796||full | Low-quality |
| 18-101-final-viral-combined_k141_327445||full | Low-quality |
| 18-101-final-viral-combined_k141_342300||full | Low-quality |
| 18-101-final-viral-combined_k141_146001||full | Not-determined |
| 18-101-final-viral-combined_k141_327465||full | Not-determined |
| 18-101-final-viral-combined_k141_63497||full | Low-quality |
| 18-101-final-viral-combined_k141_342314||full | Not-determined |
| 18-101-final-viral-combined_k141_78885||full | Not-determined |
| 18-101-final-viral-combined_k141_196149||full | Not-determined |
| 18-101-final-viral-combined_k141_310902||full | Low-quality |
| 18-101-final-viral-combined_k141_360485||full | Not-determined |
| 18-101-final-viral-combined_k141_262289||full | Low-quality |
| 18-101-final-viral-combined_k141_327482||full | Not-determined |
| 18-101-final-viral-combined_k141_63515||full | Not-determined |
| 18-101-final-viral-combined_k141_277170||full | Low-quality |
| 18-101-final-viral-combined_k141_177585||full | Low-quality |
| 18-101-final-viral-combined_k141_29927||full | Low-quality |
| 18-101-final-viral-combined_k141_360488||full | Not-determined |
| 18-101-final-viral-combined_k141_245166||full | Not-determined |
| 18-101-final-viral-combined_k141_161705||full | Low-quality |
| 18-101-final-viral-combined_k141_63533||full | Not-determined |
| 18-101-final-viral-combined_k141_96416||full | Low-quality |
| 18-101-final-viral-combined_k141_310904||full | Low-quality |
| 18-101-final-viral-combined_k141_376253||full | Low-quality |
| 18-101-final-viral-combined_k141_46507||full | Low-quality |
| 18-101-final-viral-combined_k141_360494||full | Not-determined |
| 18-101-final-viral-combined_k141_46509||full | Not-determined |
| 18-101-final-viral-combined_k141_29950||full | Low-quality |
| 18-101-final-viral-combined_k141_294534||full | Not-determined |
| 18-101-final-viral-combined_k141_14284||full | Not-determined |
| 18-101-final-viral-combined_k141_63549||full | Not-determined |
| 18-101-final-viral-combined_k141_177606||full | Low-quality |
| 18-101-final-viral-combined_k141_146020||full | Low-quality |
| 18-101-final-viral-combined_k141_177618||full | Low-quality |
| 18-101-final-viral-combined_k141_146027||full | Low-quality |
| 18-101-final-viral-combined_k141_294545||full | Not-determined |
| 18-101-final-viral-combined_k141_177630||full | Low-quality |
| 18-101-final-viral-combined_k141_245194||full | Not-determined |
| 18-101-final-viral-combined_k141_111587||full | Low-quality |
| 18-101-final-viral-combined_k141_177643||full | Low-quality |
| 18-101-final-viral-combined_k141_130456||full | Low-quality |
| 18-101-final-viral-combined_k141_111595||full | Not-determined |
| 18-101-final-viral-combined_k141_177646||full | Low-quality |
| 18-101-final-viral-combined_k141_342349||full | Not-determined |
| 18-101-final-viral-combined_k141_392228||full | Not-determined |
| 18-101-final-viral-combined_k141_161742||full | Low-quality |
| 18-101-final-viral-combined_k141_294563||full | Low-quality |
| 18-101-final-viral-combined_k141_262356||full | Low-quality |
| 18-101-final-viral-combined_k141_146048||full | Not-determined |
| 18-101-final-viral-combined_k141_262365||full | Not-determined |
| 18-101-final-viral-combined_k141_376280||full | Low-quality |
| 18-101-final-viral-combined_k141_392237||full | Not-determined |
| 18-101-final-viral-combined_k141_30006||full | Low-quality |
| 18-101-final-viral-combined_k141_360530||full | Not-determined |
| 18-101-final-viral-combined_k141_78967||full | Low-quality |
| 18-101-final-viral-combined_k141_360544||full | Not-determined |
| 18-101-final-viral-combined_k141_196245||full | Not-determined |
| 18-101-final-viral-combined_k141_342385||full | Not-determined |
| 18-101-final-viral-combined_k141_294595||full | Low-quality |
| 18-101-final-viral-combined_k141_177679||full | Not-determined |
| 18-101-final-viral-combined_k141_161777||full | Not-determined |
| 18-101-final-viral-combined_k141_130517||full | Not-determined |
| 18-101-final-viral-combined_k141_78972||full | Low-quality |
| 18-101-final-viral-combined_k141_130518||full | Low-quality |
| 18-101-final-viral-combined_k141_161787||full | Not-determined |
| 18-101-final-viral-combined_k141_294609||full | Not-determined |
| 18-101-final-viral-combined_k141_342407||full | Not-determined |
| 18-101-final-viral-combined_k141_327575||full | Low-quality |
| 18-101-final-viral-combined_k141_161810||full | Not-determined |
| 18-101-final-viral-combined_k141_360571||full | Not-determined |
| 18-101-final-viral-combined_k141_245264||full | Low-quality |
| 18-101-final-viral-combined_k141_327587||full | Low-quality |
| 18-101-final-viral-combined_k141_294618||full | Low-quality |
| 18-101-final-viral-combined_k141_311018||full | Low-quality |
| 18-101-final-viral-combined_k141_277294||full | Not-determined |
| 18-101-final-viral-combined_k141_360582||full | Low-quality |
| 18-101-final-viral-combined_k141_130541||full | Not-determined |
| 18-101-final-viral-combined_k141_262449||full | Low-quality |
| 18-101-final-viral-combined_k141_342429||full | Not-determined |
| 18-101-final-viral-combined_k141_63621||full | Not-determined |
| 18-101-final-viral-combined_k141_96569||full | Not-determined |
| 18-101-final-viral-combined_k141_96590||full | Low-quality |
| 18-101-final-viral-combined_k141_342454||full | Low-quality |
| 18-101-final-viral-combined_k141_392304||full | Not-determined |
| 18-101-final-viral-combined_k141_30095||full | Not-determined |
| 18-101-final-viral-combined_k141_245334||full | Low-quality |
| 18-101-final-viral-combined_k141_294659||full | Not-determined |
| 18-101-final-viral-combined_k141_228415||full | Not-determined |
| 18-101-final-viral-combined_k141_63673||full | Low-quality |
| 18-102-final-viral-combined_k141_124416||full | Medium-quality |
| 18-101-final-viral-combined_k141_146086||full | Low-quality |
| 18-101-final-viral-combined_k141_245362||full | Low-quality |
| 18-101-final-viral-combined_k141_342497||full | Not-determined |
| 18-101-final-viral-combined_k141_294693||full | Low-quality |
| 18-101-final-viral-combined_k141_161854||full | Low-quality |
| 18-101-final-viral-combined_k141_146096||full | Low-quality |
| 18-101-final-viral-combined_k141_294694||full | Not-determined |
| 18-101-final-viral-combined_k141_212913||full | Low-quality |
| 18-101-final-viral-combined_k141_311131||full | Low-quality |
| 18-101-final-viral-combined_k141_146113||full | Low-quality |
| 18-101-final-viral-combined_k141_196382||full | Not-determined |
| 18-101-final-viral-combined_k141_392331||full | Low-quality |
| 18-101-final-viral-combined_k141_177784||full | Low-quality |
| 18-101-final-viral-combined_k141_245411||full | Not-determined |
| 18-101-final-viral-combined_k141_79035||full | Not-determined |
| 18-101-final-viral-combined_k141_342553||full | Not-determined |
| 18-101-final-viral-combined_k141_294729||full | Not-determined |
| 18-101-final-viral-combined_k141_30137||full | Not-determined |
| 18-101-final-viral-combined_k141_262512||full | Not-determined |
| 18-101-final-viral-combined_k141_96704||full | Not-determined |
| 18-101-final-viral-combined_k141_14374||full | Not-determined |
| 18-101-final-viral-combined_k141_262514||full | Low-quality |
| 18-101-final-viral-combined_k141_96708||full | Not-determined |
| 18-101-final-viral-combined_k141_14386||full | Low-quality |
| 18-101-final-viral-combined_k141_392369||full | Low-quality |
| 18-101-final-viral-combined_k141_14389||full | Low-quality |
| 18-101-final-viral-combined_k141_245453||full | Not-determined |
| 18-101-final-viral-combined_k141_327722||full | Not-determined |
| 18-101-final-viral-combined_k141_177849||full | Low-quality |
| 18-101-final-viral-combined_k141_96718||full | Low-quality |
| 18-101-final-viral-combined_k141_327726||full | Low-quality |
| 18-101-final-viral-combined_k141_79058||full | Not-determined |
| 18-101-final-viral-combined_k141_376395||full | Not-determined |
| 18-101-final-viral-combined_k141_161928||full | Not-determined |
| 18-101-final-viral-combined_k141_294751||full | Low-quality |
| 18-101-final-viral-combined_k141_360646||full | Not-determined |
| 18-101-final-viral-combined_k141_30190||full | Not-determined |
| 18-101-final-viral-combined_k141_14432||full | Low-quality |
| 18-101-final-viral-combined_k141_96744||full | Not-determined |
| 18-101-final-viral-combined_k141_79081||full | Not-determined |
| 18-101-final-viral-combined_k141_14442||full | Not-determined |
| 18-101-final-viral-combined_k141_277382||full | Low-quality |
| 18-101-final-viral-combined_k141_111694||full | Not-determined |
| 18-101-final-viral-combined_k141_130661||full | Not-determined |
| 18-101-final-viral-combined_k141_30231||full | Low-quality |
| 18-101-final-viral-combined_k141_177882||full | Not-determined |
| 18-101-final-viral-combined_k141_177887||full | Not-determined |
| 18-101-final-viral-combined_k141_311261||full | Low-quality |
| 18-101-final-viral-combined_k141_130673||full | Not-determined |
| 18-101-final-viral-combined_k141_360693||full | Low-quality |
| 18-101-final-viral-combined_k141_392442||full | Not-determined |
| 18-101-final-viral-combined_k141_327763||full | Not-determined |
| 18-110-final-viral-combined_k141_48935||full | High-quality |
| 18-101-final-viral-combined_k141_262573||full | Low-quality |
| 18-101-final-viral-combined_k141_392456||full | Low-quality |
| 18-101-final-viral-combined_k141_30268||full | Not-determined |
| 18-101-final-viral-combined_k141_376463||full | Low-quality |
| 18-101-final-viral-combined_k141_130707||full | Low-quality |
| 18-101-final-viral-combined_k141_96832||full | Low-quality |
| 18-101-final-viral-combined_k141_130710||full | Low-quality |
| 18-101-final-viral-combined_k141_245583||full | Low-quality |
| 18-101-final-viral-combined_k141_96848||full | Not-determined |
| 18-101-final-viral-combined_k141_162035||full | Not-determined |
| 18-101-final-viral-combined_k141_30292||full | Low-quality |
| 18-101-final-viral-combined_k141_228423||full | Low-quality |
| 18-101-final-viral-combined_k141_392473||full | Low-quality |
| 18-101-final-viral-combined_k141_14519||full | Not-determined |
| 18-101-final-viral-combined_k141_63789||full | Not-determined |
| 18-101-final-viral-combined_k141_14531||full | Low-quality |
| 18-101-final-viral-combined_k141_277430||full | Not-determined |
| 18-101-final-viral-combined_k141_228450||full | Low-quality |
| 18-101-final-viral-combined_k141_63799||full | Low-quality |
| 18-101-final-viral-combined_k141_177981||full | Not-determined |
| 18-101-final-viral-combined_k141_63802||full | Low-quality |
| 18-101-final-viral-combined_k141_96868||full | Low-quality |
| 18-101-final-viral-combined_k141_14545||full | Not-determined |
| 18-101-final-viral-combined_k141_30315||full | Low-quality |
| 18-101-final-viral-combined_k141_162071||full | Not-determined |
| 18-101-final-viral-combined_k141_96885||full | Low-quality |
| 18-101-final-viral-combined_k141_245656||full | Not-determined |
| 18-101-final-viral-combined_k141_111757||full | Not-determined |
| 18-101-final-viral-combined_k141_245665||full | Not-determined |
| 18-101-final-viral-combined_k141_392511||full | Not-determined |
| 18-101-final-viral-combined_k141_294887||full | Low-quality |
| 18-101-final-viral-combined_k141_14568||full | Not-determined |
| 18-101-final-viral-combined_k141_63831||full | Low-quality |
| 18-101-final-viral-combined_k141_327830||full | Low-quality |
| 18-101-final-viral-combined_k141_392515||full | Not-determined |
| 18-101-final-viral-combined_k141_360771||full | Not-determined |
| 18-101-final-viral-combined_k141_262668||full | Not-determined |
| 18-101-final-viral-combined_k141_30353||full | Low-quality |
| 18-101-final-viral-combined_k141_376546||full | Not-determined |
| 18-101-final-viral-combined_k141_360787||full | Not-determined |
| 18-101-final-viral-combined_k141_392539||full | Low-quality |
| 18-101-final-viral-combined_k141_342773||full | Not-determined |
| 18-101-final-viral-combined_k141_327855||full | Not-determined |
| 18-101-final-viral-combined_k141_360796||full | Not-determined |
| 18-101-final-viral-combined_k141_277494||full | Not-determined |
| 18-101-final-viral-combined_k141_376559||full | Low-quality |
| 18-101-final-viral-combined_k141_178034||full | Low-quality |
| 18-101-final-viral-combined_k141_327862||full | Low-quality |
| 18-101-final-viral-combined_k141_162158||full | Not-determined |
| 18-101-final-viral-combined_k141_311423||full | Not-determined |
| 18-101-final-viral-combined_k141_111802||full | Low-quality |
| 18-101-final-viral-combined_k141_178042||full | Not-determined |
| 18-101-final-viral-combined_k141_294923||full | Not-determined |
| 18-101-final-viral-combined_k141_146235||full | Low-quality |
| 18-101-final-viral-combined_k141_162165||full | Low-quality |
| 18-101-final-viral-combined_k141_14609||full | Not-determined |
| 18-101-final-viral-combined_k141_376589||full | Not-determined |
| 18-101-final-viral-combined_k141_392563||full | Not-determined |
| 18-101-final-viral-combined_k141_360846||full | Low-quality |
| 18-101-final-viral-combined_k141_96933||full | Not-determined |
| 18-101-final-viral-combined_k141_277548||full | Low-quality |
| 18-101-final-viral-combined_k141_14620||full | Not-determined |
| 18-101-final-viral-combined_k141_262756||full | Not-determined |
| 18-101-final-viral-combined_k141_277572||full | Low-quality |
| 18-101-final-viral-combined_k141_277574||full | Not-determined |
| 18-101-final-viral-combined_k141_342839||full | Not-determined |
| 18-101-final-viral-combined_k141_63925||full | Low-quality |
| 18-101-final-viral-combined_k141_196505||full | Low-quality |
| 18-101-final-viral-combined_k141_111833||full | Low-quality |
| 18-101-final-viral-combined_k141_130851||full | Low-quality |
| 18-101-final-viral-combined_k141_294977||full | Low-quality |
| 18-101-final-viral-combined_k141_342862||full | Not-determined |
| 18-101-final-viral-combined_k141_311501||full | Low-quality |
| 18-101-final-viral-combined_k141_63940||full | Low-quality |
| 18-101-final-viral-combined_k141_96959||full | Low-quality |
| 18-101-final-viral-combined_k141_111858||full | Low-quality |
| 18-101-final-viral-combined_k141_30471||full | Not-determined |
| 18-101-final-viral-combined_k141_96973||full | Not-determined |
| 18-101-final-viral-combined_k141_360908||full | Not-determined |
| 18-101-final-viral-combined_k141_245808||full | Low-quality |
| 18-101-final-viral-combined_k141_96976||full | Not-determined |
| 18-101-final-viral-combined_k141_130899||full | Not-determined |
| 18-101-final-viral-combined_k141_79241||full | Low-quality |
| 18-101-final-viral-combined_k141_96982||full | Not-determined |
| 18-101-final-viral-combined_k141_262781||full | Not-determined |
| 18-101-final-viral-combined_k141_327960||full | Low-quality |
| 18-101-final-viral-combined_k141_342920||full | Low-quality |
| 18-101-final-viral-combined_k141_79258||full | Low-quality |
| 18-101-final-viral-combined_k141_111903||full | Low-quality |
| 18-101-final-viral-combined_k141_295022||full | Low-quality |
| 18-101-final-viral-combined_k141_360919||full | Not-determined |
| 18-101-final-viral-combined_k141_342935||full | Low-quality |
| 18-101-final-viral-combined_k141_111905||full | Not-determined |
| 18-101-final-viral-combined_k141_162285||full | Low-quality |
| 18-101-final-viral-combined_k141_327975||full | Low-quality |
| 18-101-final-viral-combined_k141_342937||full | Not-determined |
| 18-101-final-viral-combined_k141_245874||full | Low-quality |
| 18-101-final-viral-combined_k141_79297||full | Low-quality |
| 18-101-final-viral-combined_k141_376631||full | Low-quality |
| 18-101-final-viral-combined_k141_30525||full | Low-quality |
| 18-101-final-viral-combined_k141_392674||full | Not-determined |
| 18-101-final-viral-combined_k141_178182||full | Not-determined |
| 18-101-final-viral-combined_k141_228648||full | Low-quality |
| 18-101-final-viral-combined_k141_178184||full | Low-quality |
| 18-101-final-viral-combined_k141_14762||full | Not-determined |
| 18-101-final-viral-combined_k141_14767||full | Low-quality |
| 18-101-final-viral-combined_k141_178192||full | Not-determined |
| 18-101-final-viral-combined_k141_130951||full | Not-determined |
| 18-101-final-viral-combined_k141_213008||full | Not-determined |
| 18-101-final-viral-combined_k141_111943||full | Not-determined |
| 18-101-final-viral-combined_k141_213012||full | Low-quality |
| 18-101-final-viral-combined_k141_376664||full | Low-quality |
| 18-101-final-viral-combined_k141_97053||full | Low-quality |
| 18-101-final-viral-combined_k141_162337||full | Not-determined |
| 18-101-final-viral-combined_k141_342949||full | Low-quality |
| 18-101-final-viral-combined_k141_392693||full | Low-quality |
| 18-101-final-viral-combined_k141_245894||full | Low-quality |
| 18-101-final-viral-combined_k141_178208||full | Low-quality |
| 18-101-final-viral-combined_k141_30562||full | Not-determined |
| 18-101-final-viral-combined_k141_311613||full | Low-quality |
| 18-101-final-viral-combined_k141_196594||full | Not-determined |
| 18-101-final-viral-combined_k141_14799||full | Not-determined |
| 18-101-final-viral-combined_k141_79320||full | Low-quality |
| 18-101-final-viral-combined_k141_392702||full | Low-quality |
| 18-101-final-viral-combined_k141_14803||full | Low-quality |
| 18-101-final-viral-combined_k141_196600||full | Not-determined |
| 18-101-final-viral-combined_k141_392709||full | Low-quality |
| 18-101-final-viral-combined_k141_79338||full | Low-quality |
| 18-101-final-viral-combined_k141_213047||full | Not-determined |
| 18-101-final-viral-combined_k141_97086||full | Low-quality |
| 18-101-final-viral-combined_k141_130989||full | Low-quality |
| 18-101-final-viral-combined_k141_178240||full | Low-quality |
| 18-101-final-viral-combined_k141_245934||full | Low-quality |
| 18-101-final-viral-combined_k141_245935||full | Not-determined |
| 18-101-final-viral-combined_k141_228683||full | Not-determined |
| 18-101-final-viral-combined_k141_295109||full | Low-quality |
| 18-101-final-viral-combined_k141_376712||full | Not-determined |
| 18-101-final-viral-combined_k141_146421||full | Not-determined |
| 18-101-final-viral-combined_k141_111992||full | Low-quality |
| 18-101-final-viral-combined_k141_178244||full | Not-determined |
| 18-101-final-viral-combined_k141_228698||full | Low-quality |
| 18-101-final-viral-combined_k141_245937||full | Low-quality |
| 18-101-final-viral-combined_k141_392733||full | Low-quality |
| 18-101-final-viral-combined_k141_162364||full | Not-determined |
| 18-101-final-viral-combined_k141_14849||full | Low-quality |
| 18-101-final-viral-combined_k141_328078||full | Not-determined |
| 18-101-final-viral-combined_k141_228716||full | Low-quality |
| 18-101-final-viral-combined_k141_311686||full | Low-quality |
| 18-101-final-viral-combined_k141_245946||full | Not-determined |
| 18-101-final-viral-combined_k141_262801||full | Low-quality |
| 18-101-final-viral-combined_k141_311699||full | Not-determined |
| 18-101-final-viral-combined_k141_245953||full | Low-quality |
| 18-101-final-viral-combined_k141_343034||full | Low-quality |
| 18-101-final-viral-combined_k141_376779||full | Low-quality |
| 18-101-final-viral-combined_k141_376781||full | Not-determined |
| 18-101-final-viral-combined_k141_228729||full | Not-determined |
| 18-101-final-viral-combined_k141_97135||full | Low-quality |
| 18-101-final-viral-combined_k141_97138||full | Not-determined |
| 18-101-final-viral-combined_k141_361004||full | Not-determined |
| 18-101-final-viral-combined_k141_162420||full | Not-determined |
| 18-101-final-viral-combined_k141_328125||full | Not-determined |
| 18-101-final-viral-combined_k141_311738||full | Not-determined |
| 18-101-final-viral-combined_k141_277765||full | Low-quality |
| 18-101-final-viral-combined_k141_392782||full | Not-determined |
| 18-101-final-viral-combined_k141_178332||full | Low-quality |
| 18-101-final-viral-combined_k141_196677||full | Not-determined |
| 18-101-final-viral-combined_k141_343076||full | Low-quality |
| 18-101-final-viral-combined_k141_196682||full | Low-quality |
| 18-101-final-viral-combined_k141_311756||full | Not-determined |
| 18-101-final-viral-combined_k141_131047||full | Low-quality |
| 18-101-final-viral-combined_k141_79479||full | Not-determined |
| 18-101-final-viral-combined_k141_311761||full | Not-determined |
| 18-101-final-viral-combined_k141_262826||full | Not-determined |
| 18-101-final-viral-combined_k141_277787||full | Low-quality |
| 18-101-final-viral-combined_k141_343111||full | Low-quality |
| 18-101-final-viral-combined_k141_262833||full | Not-determined |
| 18-101-final-viral-combined_k141_311781||full | Not-determined |
| 18-101-final-viral-combined_k141_262837||full | Not-determined |
| 18-101-final-viral-combined_k141_361045||full | Not-determined |
| 18-101-final-viral-combined_k141_376849||full | Low-quality |
| 18-101-final-viral-combined_k141_97188||full | Low-quality |
| 18-101-final-viral-combined_k141_162482||full | Low-quality |
| 18-101-final-viral-combined_k141_162483||full | Low-quality |
| 18-101-final-viral-combined_k141_361049||full | Low-quality |
| 18-101-final-viral-combined_k141_262853||full | Low-quality |
| 18-101-final-viral-combined_k141_14885||full | Not-determined |
| 18-101-final-viral-combined_k141_328153||full | Low-quality |
| 18-101-final-viral-combined_k141_131088||full | Not-determined |
| 18-101-final-viral-combined_k141_46585||full | Low-quality |
| 18-101-final-viral-combined_k141_277817||full | Not-determined |
| 18-101-final-viral-combined_k141_262877||full | Low-quality |
| 18-101-final-viral-combined_k141_213093||full | Not-determined |
| 18-101-final-viral-combined_k141_228825||full | Low-quality |
| 18-101-final-viral-combined_k141_213096||full | Low-quality |
| 18-101-final-viral-combined_k141_46595||full | Low-quality |
| 18-101-final-viral-combined_k141_14924||full | Not-determined |
| 18-101-final-viral-combined_k141_146519||full | Low-quality |
| 18-101-final-viral-combined_k141_112027||full | Low-quality |
| 18-101-final-viral-combined_k141_311831||full | Low-quality |
| 18-101-final-viral-combined_k141_311832||full | Not-determined |
| 18-101-final-viral-combined_k141_97251||full | Low-quality |
| 18-101-final-viral-combined_k141_112036||full | Low-quality |
| 18-101-final-viral-combined_k141_131134||full | Not-determined |
| 18-101-final-viral-combined_k141_311837||full | Not-determined |
| 18-101-final-viral-combined_k141_112041||full | Not-determined |
| 18-101-final-viral-combined_k141_361083||full | Low-quality |
| 18-101-final-viral-combined_k141_213128||full | Low-quality |
| 18-101-final-viral-combined_k141_146542||full | Not-determined |
| 18-101-final-viral-combined_k141_376912||full | Not-determined |
| 18-101-final-viral-combined_k141_97264||full | Low-quality |
| 18-101-final-viral-combined_k141_277857||full | Low-quality |
| 18-101-final-viral-combined_k141_97272||full | Low-quality |
| 18-101-final-viral-combined_k141_277866||full | Low-quality |
| 18-101-final-viral-combined_k141_328223||full | Low-quality |
| 18-101-final-viral-combined_k141_376929||full | Low-quality |
| 18-101-final-viral-combined_k141_277867||full | Not-determined |
| 18-101-final-viral-combined_k141_277869||full | Not-determined |
| 18-101-final-viral-combined_k141_343170||full | Not-determined |
| 18-101-final-viral-combined_k141_328239||full | Low-quality |
| 18-101-final-viral-combined_k141_112081||full | Low-quality |
| 18-101-final-viral-combined_k141_112091||full | Low-quality |
| 18-101-final-viral-combined_k141_178504||full | Not-determined |
| 18-101-final-viral-combined_k141_361097||full | Low-quality |
| 18-101-final-viral-combined_k141_14986||full | Low-quality |
| 18-101-final-viral-combined_k141_178516||full | Low-quality |
| 18-101-final-viral-combined_k141_46643||full | Low-quality |
| 18-101-final-viral-combined_k141_146604||full | Not-determined |
| 18-101-final-viral-combined_k141_14991||full | Not-determined |
| 18-101-final-viral-combined_k141_213196||full | Low-quality |
| 18-101-final-viral-combined_k141_146617||full | Low-quality |
| 18-101-final-viral-combined_k141_311915||full | Not-determined |
| 18-101-final-viral-combined_k141_15006||full | Not-determined |
| 18-101-final-viral-combined_k141_196775||full | Low-quality |
| 18-101-final-viral-combined_k141_311924||full | Not-determined |
| 18-101-final-viral-combined_k141_131180||full | Not-determined |
| 18-101-final-viral-combined_k141_277956||full | Not-determined |
| 18-101-final-viral-combined_k141_392822||full | Not-determined |
| 18-101-final-viral-combined_k141_30797||full | Not-determined |
| 18-101-final-viral-combined_k141_131200||full | Low-quality |
| 18-101-final-viral-combined_k141_196806||full | Low-quality |
| 18-101-final-viral-combined_k141_277969||full | Low-quality |
| 18-101-final-viral-combined_k141_246071||full | Not-determined |
| 18-101-final-viral-combined_k141_377005||full | Not-determined |
| 18-101-final-viral-combined_k141_162599||full | Not-determined |
| 18-101-final-viral-combined_k141_79706||full | Low-quality |
| 18-101-final-viral-combined_k141_262984||full | Low-quality |
| 18-101-final-viral-combined_k141_178551||full | Low-quality |
| 18-101-final-viral-combined_k141_178553||full | Not-determined |
| 18-101-final-viral-combined_k141_30840||full | Low-quality |
| 18-101-final-viral-combined_k141_343240||full | Low-quality |
| 18-101-final-viral-combined_k141_196823||full | Not-determined |
| 18-101-final-viral-combined_k141_246087||full | Not-determined |
| 18-101-final-viral-combined_k141_30856||full | Low-quality |
| 18-101-final-viral-combined_k141_196828||full | Not-determined |
| 18-101-final-viral-combined_k141_343254||full | Not-determined |
| 18-101-final-viral-combined_k141_263032||full | Not-determined |
| 18-101-final-viral-combined_k141_64242||full | Low-quality |
| 18-101-final-viral-combined_k141_162643||full | Low-quality |
| 18-101-final-viral-combined_k141_64244||full | Not-determined |
| 18-101-final-viral-combined_k141_162645||full | Low-quality |
| 18-101-final-viral-combined_k141_278006||full | Low-quality |
| 18-101-final-viral-combined_k141_46780||full | Not-determined |
| 18-101-final-viral-combined_k141_377044||full | Not-determined |
| 18-101-final-viral-combined_k141_343282||full | Low-quality |
| 18-101-final-viral-combined_k141_97326||full | Not-determined |
| 18-101-final-viral-combined_k141_343285||full | Not-determined |
| 18-101-final-viral-combined_k141_79745||full | Not-determined |
| 18-101-final-viral-combined_k141_178615||full | Low-quality |
| 18-101-final-viral-combined_k141_146705||full | Low-quality |
| 18-101-final-viral-combined_k141_131267||full | Low-quality |
| 18-101-final-viral-combined_k141_196834||full | Low-quality |
| 18-101-final-viral-combined_k141_246138||full | Low-quality |
| 18-101-final-viral-combined_k141_15076||full | Low-quality |
| 18-101-final-viral-combined_k141_295384||full | Not-determined |
| 18-101-final-viral-combined_k141_295400||full | Low-quality |
| 18-101-final-viral-combined_k141_392866||full | Not-determined |
| 18-101-final-viral-combined_k141_46820||full | Low-quality |
| 18-101-final-viral-combined_k141_392873||full | Low-quality |
| 18-101-final-viral-combined_k141_97358||full | Not-determined |
| 18-101-final-viral-combined_k141_97361||full | Low-quality |
| 18-101-final-viral-combined_k141_15107||full | Low-quality |
| 18-101-final-viral-combined_k141_343320||full | Not-determined |
| 18-101-final-viral-combined_k141_295438||full | Low-quality |
| 17-109-final-viral-combined_k141_117628||full | Medium-quality |
| 18-101-final-viral-combined_k141_278071||full | Not-determined |
| 18-101-final-viral-combined_k141_15111||full | Not-determined |
| 18-101-final-viral-combined_k141_178650||full | Low-quality |

18-101-final-viral-combined_k141_162741||full Low-quality
18-101-final-viral-combined_k141_30931||full Low-quality
18-101-final-viral-combined_k141_229029||full Low-quality
18-101-final-viral-combined_k141_361274||full Low-quality
18-101-final-viral-combined_k141_97394||full Not-determined
18-101-final-viral-combined_k141_328479||full Not-determined
18-101-final-viral-combined_k141_30954||full Not-determined
18-101-final-viral-combined_k141_392909||full Low-quality
18-101-final-viral-combined_k141_263102||full Not-determined
18-101-final-viral-combined_k141_30961||full Not-determined
18-101-final-viral-combined_k141_97406||full Not-determined
18-101-final-viral-combined_k141_97408||full Not-determined
18-101-final-viral-combined_k141_263106||full Low-quality
18-101-final-viral-combined_k141_64309||full Low-quality
18-101-final-viral-combined_k141_361307||full Not-determined
18-101-final-viral-combined_k141_229081||full Low-quality
18-101-final-viral-combined_k141_162810||full Low-quality
18-101-final-viral-combined_k141_328503||full Not-determined
18-101-final-viral-combined_k141_213332||full Low-quality
18-101-final-viral-combined_k141_97446||full Not-determined
18-101-final-viral-combined_k141_162832||full Not-determined
18-101-final-viral-combined_k141_343383||full Not-determined
18-101-final-viral-combined_k141_278162||full Low-quality
18-101-final-viral-combined_k141_178754||full Low-quality
18-101-final-viral-combined_k141_30994||full Low-quality
18-101-final-viral-combined_k141_295527||full Not-determined
18-101-final-viral-combined_k141_178757||full Low-quality
18-101-final-viral-combined_k141_79885||full Not-determined
18-101-final-viral-combined_k141_295531||full Not-determined
18-101-final-viral-combined_k141_343398||full Low-quality
18-101-final-viral-combined_k141_361347||full Low-quality
18-101-final-viral-combined_k141_196983||full Not-determined
18-101-final-viral-combined_k141_162864||full Low-quality
18-101-final-viral-combined_k141_97486||full Low-quality
18-101-final-viral-combined_k141_278178||full Not-determined
18-101-final-viral-combined_k141_213333||full Not-determined
18-101-final-viral-combined_k141_97495||full Low-quality
18-101-final-viral-combined_k141_146820||full Not-determined
18-101-final-viral-combined_k141_377215||full Not-determined
18-101-final-viral-combined_k141_377222||full Low-quality
18-101-final-viral-combined_k141_343422||full Low-quality
18-101-final-viral-combined_k141_46968||full Low-quality
18-101-final-viral-combined_k141_392978||full Not-determined
18-101-final-viral-combined_k141_213346||full Low-quality
18-101-final-viral-combined_k141_97527||full Low-quality
18-101-final-viral-combined_k141_377233||full Not-determined
18-101-final-viral-combined_k141_343426||full Not-determined
18-101-final-viral-combined_k141_377236||full Not-determined
18-101-final-viral-combined_k141_295578||full Not-determined
18-101-final-viral-combined_k141_197010||full Low-quality
18-101-final-viral-combined_k141_229196||full Low-quality
18-101-final-viral-combined_k141_343445||full Low-quality
18-101-final-viral-combined_k141_263130||full Not-determined
18-101-final-viral-combined_k141_312157||full Not-determined
18-101-final-viral-combined_k141_263143||full Not-determined
18-101-final-viral-combined_k141_64377||full Low-quality
18-101-final-viral-combined_k141_213385||full Low-quality
18-101-final-viral-combined_k141_15174||full Low-quality
18-101-final-viral-combined_k141_178868||full Not-determined
18-101-final-viral-combined_k141_146866||full Low-quality
18-101-final-viral-combined_k141_229217||full Low-quality
18-101-final-viral-combined_k141_197012||full Low-quality
18-101-final-viral-combined_k141_131364||full Not-determined
18-101-final-viral-combined_k141_393034||full Low-quality
18-101-final-viral-combined_k141_178900||full Not-determined
18-101-final-viral-combined_k141_146894||full Low-quality
18-101-final-viral-combined_k141_229228||full Not-determined
18-101-final-viral-combined_k141_328545||full Not-determined
18-101-final-viral-combined_k141_377299||full Not-determined
18-101-final-viral-combined_k141_64440||full Not-determined
18-101-final-viral-combined_k141_377311||full Not-determined
18-101-final-viral-combined_k141_295611||full Not-determined
18-101-final-viral-combined_k141_178932||full Not-determined
18-101-final-viral-combined_k141_15241||full Not-determined
18-101-final-viral-combined_k141_146934||full Low-quality
18-101-final-viral-combined_k141_295613||full Low-quality
18-101-final-viral-combined_k141_213445||full Not-determined
18-101-final-viral-combined_k141_295614||full Not-determined
18-101-final-viral-combined_k141_393055||full Low-quality
18-101-final-viral-combined_k141_377326||full Not-determined
18-101-final-viral-combined_k141_213447||full Not-determined
18-101-final-viral-combined_k141_197064||full Low-quality
18-101-final-viral-combined_k141_15256||full Not-determined
18-101-final-viral-combined_k141_377333||full Not-determined
18-101-final-viral-combined_k141_213459||full Low-quality
18-101-final-viral-combined_k141_47030||full Not-determined
18-101-final-viral-combined_k141_97642||full Low-quality
18-101-final-viral-combined_k141_112235||full Not-determined
18-101-final-viral-combined_k141_64484||full Not-determined
18-101-final-viral-combined_k141_97652||full Low-quality
18-101-final-viral-combined_k141_361490||full Low-quality
18-101-final-viral-combined_k141_361494||full Not-determined
18-101-final-viral-combined_k141_47055||full Low-quality
18-101-final-viral-combined_k141_393092||full Low-quality
18-101-final-viral-combined_k141_246261||full Low-quality
18-101-final-viral-combined_k141_64499||full Low-quality
18-101-final-viral-combined_k141_229306||full Not-determined
18-101-final-viral-combined_k141_328594||full Not-determined
18-101-final-viral-combined_k141_246266||full Not-determined
18-101-final-viral-combined_k141_178979||full Not-determined
18-101-final-viral-combined_k141_263231||full Not-determined
18-101-final-viral-combined_k141_97684||full Not-determined
18-101-final-viral-combined_k141_246276||full Low-quality
18-101-final-viral-combined_k141_377359||full Low-quality
18-101-final-viral-combined_k141_229323||full Not-determined
18-101-final-viral-combined_k141_343541||full Not-determined
18-101-final-viral-combined_k141_393095||full Not-determined
18-101-final-viral-combined_k141_213472||full Not-determined
18-101-final-viral-combined_k141_31166||full Not-determined
18-101-final-viral-combined_k141_263243||full Low-quality
18-101-final-viral-combined_k141_80066||full Low-quality
18-101-final-viral-combined_k141_112272||full Low-quality
18-101-final-viral-combined_k141_312226||full Low-quality
18-101-final-viral-combined_k141_213475||full Low-quality
18-101-final-viral-combined_k141_31177||full Not-determined
18-101-final-viral-combined_k141_64542||full Not-determined
18-101-final-viral-combined_k141_64548||full Low-quality
17-109_k141_12658 High-quality
18-101-final-viral-combined_k141_229344||full Not-determined
18-101-final-viral-combined_k141_80086||full Low-quality
18-101-final-viral-combined_k141_377395||full Low-quality
18-101-final-viral-combined_k141_328652||full Not-determined
18-101-final-viral-combined_k141_131383||full Low-quality
18-101-final-viral-combined_k141_15332||full Low-quality
18-101-final-viral-combined_k141_213493||full Low-quality
18-101-final-viral-combined_k141_197139||full Low-quality
18-101-final-viral-combined_k141_393121||full Low-quality
18-101-final-viral-combined_k141_343569||full Low-quality
18-101-final-viral-combined_k141_343573||full Low-quality
18-101-final-viral-combined_k141_97737||full Not-determined
18-101-final-viral-combined_k141_97742||full Low-quality
18-101-final-viral-combined_k141_163070||full Not-determined
18-101-final-viral-combined_k141_131390||full Not-determined
18-101-final-viral-combined_k141_229381||full Low-quality
18-101-final-viral-combined_k141_343586||full Not-determined
18-101-final-viral-combined_k141_15368||full Low-quality
18-101-final-viral-combined_k141_47131||full Not-determined
18-101-final-viral-combined_k141_146974||full Low-quality
18-101-final-viral-combined_k141_213525||full Low-quality
18-101-final-viral-combined_k141_112310||full Not-determined
18-101-final-viral-combined_k141_246318||full Low-quality
18-101-final-viral-combined_k141_64598||full Not-determined
18-101-final-viral-combined_k141_97767||full Low-quality
18-101-final-viral-combined_k141_393162||full Low-quality
18-101-final-viral-combined_k141_146985||full Not-determined
18-101-final-viral-combined_k141_47175||full Not-determined
18-101-final-viral-combined_k141_31272||full Low-quality
18-101-final-viral-combined_k141_31274||full Low-quality
18-101-final-viral-combined_k141_147001||full Low-quality
18-101-final-viral-combined_k141_64631||full Low-quality
18-101-final-viral-combined_k141_64632||full Not-determined
18-101-final-viral-combined_k141_112374||full Low-quality
18-101-final-viral-combined_k141_213590||full Low-quality
18-101-final-viral-combined_k141_15423||full Low-quality
18-101-final-viral-combined_k141_64646||full Not-determined
18-101-final-viral-combined_k141_112388||full Low-quality
18-101-final-viral-combined_k141_328754||full Low-quality
18-101-final-viral-combined_k141_312273||full Not-determined
18-101-final-viral-combined_k141_147025||full Low-quality
18-101-final-viral-combined_k141_147030||full Not-determined
18-101-final-viral-combined_k141_47220||full Not-determined
18-101-final-viral-combined_k141_179105||full Not-determined
18-101-final-viral-combined_k141_377516||full Not-determined
18-101-final-viral-combined_k141_163114||full Low-quality
18-101-final-viral-combined_k141_328772||full Low-quality
18-101-final-viral-combined_k141_64679||full Low-quality
18-101-final-viral-combined_k141_393177||full Low-quality
18-101-final-viral-combined_k141_328780||full Not-determined
18-101-final-viral-combined_k141_377526||full Low-quality
18-101-final-viral-combined_k141_97843||full Not-determined
18-101-final-viral-combined_k141_147042||full Low-quality
18-101-final-viral-combined_k141_312305||full Low-quality
18-101-final-viral-combined_k141_377535||full Low-quality
18-101-final-viral-combined_k141_393186||full Not-determined
18-101-final-viral-combined_k141_197214||full Low-quality
18-101-final-viral-combined_k141_80234||full Not-determined
18-101-final-viral-combined_k141_278285||full Low-quality
18-101-final-viral-combined_k141_47266||full Not-determined
18-101-final-viral-combined_k141_97861||full Not-determined
18-101-final-viral-combined_k141_246371||full Low-quality
18-101-final-viral-combined_k141_328816||full Low-quality
18-101-final-viral-combined_k141_97870||full Low-quality
18-101-final-viral-combined_k141_64731||full Low-quality
18-101-final-viral-combined_k141_213652||full Not-determined
18-101-final-viral-combined_k141_295817||full Low-quality
18-101-final-viral-combined_k141_197265||full Not-determined
18-101-final-viral-combined_k141_97901||full Not-determined
18-101-final-viral-combined_k141_131500||full Low-quality
18-101-final-viral-combined_k141_328849||full Not-determined
18-101-final-viral-combined_k141_80297||full Low-quality
18-101-final-viral-combined_k141_97910||full Low-quality
18-101-final-viral-combined_k141_15542||full Not-determined
18-101-final-viral-combined_k141_163172||full Not-determined
18-101-final-viral-combined_k141_343680||full Low-quality
18-101-final-viral-combined_k141_393219||full Low-quality
18-101-final-viral-combined_k141_213693||full Low-quality
18-101-final-viral-combined_k141_393220||full Low-quality
18-101-final-viral-combined_k141_361639||full Low-quality
18-101-final-viral-combined_k141_295836||full Not-determined
18-101-final-viral-combined_k141_393222||full Low-quality
18-101-final-viral-combined_k141_80302||full Low-quality
18-101-final-viral-combined_k141_64782||full Low-quality
18-101-final-viral-combined_k141_147071||full Low-quality
18-101-final-viral-combined_k141_15568||full Not-determined
18-101-final-viral-combined_k141_97966||full Low-quality
18-101-final-viral-combined_k141_112425||full Low-quality
18-101-final-viral-combined_k141_213721||full Low-quality
18-101-final-viral-combined_k141_147093||full Not-determined
18-101-final-viral-combined_k141_312413||full Low-quality
18-101-final-viral-combined_k141_343723||full Not-determined
18-101-final-viral-combined_k141_179302||full Not-determined
18-101-final-viral-combined_k141_131545||full Not-determined
18-101-final-viral-combined_k141_15606||full Not-determined
18-101-final-viral-combined_k141_147130||full Low-quality
18-101-final-viral-combined_k141_377661||full Low-quality
18-101-final-viral-combined_k141_278337||full Not-determined
18-101-final-viral-combined_k141_377664||full Low-quality
18-101-final-viral-combined_k141_361674||full Not-determined
18-101-final-viral-combined_k141_197296||full Low-quality
18-101-final-viral-combined_k141_295904||full Not-determined
18-101-final-viral-combined_k141_64831||full Low-quality
18-101-final-viral-combined_k141_163265||full Not-determined
18-101-final-viral-combined_k141_98058||full Not-determined
18-101-final-viral-combined_k141_131576||full Low-quality
18-101-final-viral-combined_k141_64844||full Not-determined
18-101-final-viral-combined_k141_112477||full Low-quality
18-101-final-viral-combined_k141_147146||full Low-quality
18-101-final-viral-combined_k141_64868||full Low-quality
18-101-final-viral-combined_k141_329009||full Low-quality
18-101-final-viral-combined_k141_47351||full Low-quality
18-101-final-viral-combined_k141_131597||full Not-determined
18-101-final-viral-combined_k141_147176||full Low-quality
18-101-final-viral-combined_k141_147180||full Low-quality
18-101-final-viral-combined_k141_131600||full Low-quality
18-101-final-viral-combined_k141_197339||full Low-quality
18-101-final-viral-combined_k141_112514||full Low-quality
18-101-final-viral-combined_k141_80424||full Not-determined
18-101-final-viral-combined_k141_278405||full Not-determined
18-101-final-viral-combined_k141_47381||full Low-quality
18-101-final-viral-combined_k141_312504||full Not-determined
18-101-final-viral-combined_k141_278415||full Low-quality
18-101-final-viral-combined_k141_361731||full Low-quality
18-101-final-viral-combined_k141_197349||full Low-quality
18-101-final-viral-combined_k141_213767||full Low-quality
18-101-final-viral-combined_k141_312515||full Not-determined
18-101-final-viral-combined_k141_197351||full Not-determined
18-101-final-viral-combined_k141_361735||full Not-determined
18-101-final-viral-combined_k141_131608||full Low-quality
18-101-final-viral-combined_k141_47390||full Low-quality
18-101-final-viral-combined_k141_197355||full Not-determined
18-101-final-viral-combined_k141_80476||full Low-quality
18-101-final-viral-combined_k141_64894||full Low-quality
18-101-final-viral-combined_k141_361745||full Low-quality
18-101-final-viral-combined_k141_213791||full Not-determined
18-101-final-viral-combined_k141_179316||full Not-determined
18-101-final-viral-combined_k141_246456||full Low-quality
18-101-final-viral-combined_k141_213804||full Low-quality
18-101-final-viral-combined_k141_31355||full Not-determined
18-101-final-viral-combined_k141_361752||full Not-determined
18-101-final-viral-combined_k141_147237||full Not-determined
18-101-final-viral-combined_k141_377767||full Low-quality
18-101-final-viral-combined_k141_64919||full Low-quality
18-101-final-viral-combined_k141_213822||full Low-quality
18-101-final-viral-combined_k141_229525||full Low-quality
18-101-final-viral-combined_k141_98122||full Low-quality
18-101-final-viral-combined_k141_64942||full Not-determined
18-101-final-viral-combined_k141_229531||full Not-determined
18-101-final-viral-combined_k141_343811||full Low-quality
18-101-final-viral-combined_k141_197382||full Not-determined
18-101-final-viral-combined_k141_31407||full Not-determined
18-101-final-viral-combined_k141_197391||full Low-quality
18-101-final-viral-combined_k141_15714||full Not-determined
18-101-final-viral-combined_k141_98143||full Low-quality
18-101-final-viral-combined_k141_15726||full Low-quality
18-101-final-viral-combined_k141_377773||full Low-quality
18-101-final-viral-combined_k141_80574||full Not-determined
18-101-final-viral-combined_k141_64969||full Not-determined
18-101-final-viral-combined_k141_80577||full Low-quality
18-101-final-viral-combined_k141_343831||full Not-determined
18-101-final-viral-combined_k141_31434||full Low-quality
18-101-final-viral-combined_k141_361785||full Not-determined
18-101-final-viral-combined_k141_47461||full Low-quality
18-101-final-viral-combined_k141_377814||full Not-determined
18-101-final-viral-combined_k141_147288||full Low-quality
18-101-final-viral-combined_k141_15786||full Low-quality
18-101-final-viral-combined_k141_163363||full Not-determined
18-101-final-viral-combined_k141_179384||full Low-quality
18-101-final-viral-combined_k141_47493||full Low-quality
17-108_k141_55610 Medium-quality
18-101-final-viral-combined_k141_163377||full Low-quality
18-101-final-viral-combined_k141_393408||full Low-quality
18-101-final-viral-combined_k141_296047||full Not-determined
18-101-final-viral-combined_k141_163378||full Low-quality
18-101-final-viral-combined_k141_179395||full Not-determined
18-101-final-viral-combined_k141_296060||full Not-determined
18-101-final-viral-combined_k141_377851||full Low-quality
18-101-final-viral-combined_k141_163388||full Not-determined
18-101-final-viral-combined_k141_213881||full Low-quality
18-101-final-viral-combined_k141_65053||full Not-determined
18-101-final-viral-combined_k141_229564||full Not-determined
18-101-final-viral-combined_k141_361829||full Not-determined
18-101-final-viral-combined_k141_343898||full Low-quality
18-101-final-viral-combined_k141_296076||full Low-quality
18-101-final-viral-combined_k141_393428||full Not-determined
18-101-final-viral-combined_k141_179428||full Low-quality
18-101-final-viral-combined_k141_80688||full Low-quality
18-101-final-viral-combined_k141_80691||full Low-quality
18-101-final-viral-combined_k141_98229||full Low-quality
18-101-final-viral-combined_k141_80695||full Low-quality
18-101-final-viral-combined_k141_393451||full Low-quality
18-101-final-viral-combined_k141_296087||full Low-quality
18-101-final-viral-combined_k141_163434||full Not-determined
18-101-final-viral-combined_k141_147332||full Not-determined
18-101-final-viral-combined_k141_229585||full Low-quality
18-101-final-viral-combined_k141_179474||full Low-quality
18-101-final-viral-combined_k141_147340||full Low-quality
18-101-final-viral-combined_k141_296126||full Not-determined
18-101-final-viral-combined_k141_80722||full Not-determined
18-101-final-viral-combined_k141_15827||full Not-determined
18-101-final-viral-combined_k141_377904||full Not-determined
18-101-final-viral-combined_k141_47549||full Not-determined
18-101-final-viral-combined_k141_296151||full Low-quality
18-101-final-viral-combined_k141_147380||full Low-quality
18-101-final-viral-combined_k141_98268||full Not-determined
18-101-final-viral-combined_k141_15850||full Low-quality
18-101-final-viral-combined_k141_98280||full Low-quality
18-101-final-viral-combined_k141_278676||full Not-determined
18-101-final-viral-combined_k141_98282||full Low-quality
18-101-final-viral-combined_k141_80772||full Not-determined
18-101-final-viral-combined_k141_361884||full Not-determined
18-101-final-viral-combined_k141_278694||full Low-quality
18-101-final-viral-combined_k141_47603||full Low-quality
18-101-final-viral-combined_k141_278701||full Not-determined
18-101-final-viral-combined_k141_31461||full Not-determined
18-101-final-viral-combined_k141_377976||full Low-quality
18-101-final-viral-combined_k141_163498||full Low-quality
18-101-final-viral-combined_k141_15886||full Low-quality
18-101-final-viral-combined_k141_112658||full Not-determined
18-101-final-viral-combined_k141_65149||full Low-quality
18-101-final-viral-combined_k141_278715||full Low-quality
18-101-final-viral-combined_k141_112660||full Not-determined
18-101-final-viral-combined_k141_393569||full Low-quality
18-101-final-viral-combined_k141_393570||full Not-determined
18-101-final-viral-combined_k141_163526||full Not-determined
18-101-final-viral-combined_k141_246628||full Not-determined
18-101-final-viral-combined_k141_47634||full Low-quality
18-101-final-viral-combined_k141_15910||full Low-quality
18-101-final-viral-combined_k141_278753||full Low-quality
18-101-final-viral-combined_k141_31487||full Not-determined
18-101-final-viral-combined_k141_229640||full Low-quality
18-101-final-viral-combined_k141_80801||full Not-determined
18-101-final-viral-combined_k141_229646||full Low-quality
18-101-final-viral-combined_k141_47677||full Low-quality
18-101-final-viral-combined_k141_246665||full Low-quality
18-101-final-viral-combined_k141_393603||full Not-determined
18-101-final-viral-combined_k141_278799||full Low-quality
18-101-final-viral-combined_k141_179639||full Low-quality
18-101-final-viral-combined_k141_15947||full Not-determined
18-101-final-viral-combined_k141_31527||full Low-quality
18-101-final-viral-combined_k141_246677||full Low-quality
18-101-final-viral-combined_k141_31535||full Not-determined
18-101-final-viral-combined_k141_278812||full Not-determined
18-101-final-viral-combined_k141_47702||full Low-quality
18-101-final-viral-combined_k141_378018||full Not-determined
18-101-final-viral-combined_k141_246683||full Not-determined
18-101-final-viral-combined_k141_15988||full Low-quality
18-101-final-viral-combined_k141_278831||full Not-determined
18-101-final-viral-combined_k141_163600||full Low-quality
18-101-final-viral-combined_k141_16003||full Not-determined
18-101-final-viral-combined_k141_246694||full Not-determined
18-101-final-viral-combined_k141_163625||full Not-determined
18-101-final-viral-combined_k141_393651||full Not-determined
18-101-final-viral-combined_k141_47745||full Not-determined
18-101-final-viral-combined_k141_112803||full Not-determined
18-101-final-viral-combined_k141_147547||full Low-quality
18-101-final-viral-combined_k141_163646||full Low-quality
18-101-final-viral-combined_k141_147553||full Not-determined
18-101-final-viral-combined_k141_278866||full Low-quality
18-101-final-viral-combined_k141_246721||full Low-quality
18-101-final-viral-combined_k141_147557||full Not-determined
18-101-final-viral-combined_k141_344130||full Not-determined
18-101-final-viral-combined_k141_65270||full Low-quality
18-101-final-viral-combined_k141_112833||full Not-determined
18-101-final-viral-combined_k141_278877||full Not-determined
18-101-final-viral-combined_k141_80950||full Not-determined
18-101-final-viral-combined_k141_163677||full Not-determined
18-101-final-viral-combined_k141_278883||full Low-quality
18-101-final-viral-combined_k141_229765||full Not-determined
18-101-final-viral-combined_k141_179759||full Low-quality
18-101-final-viral-combined_k141_229783||full Low-quality
18-101-final-viral-combined_k141_393680||full Not-determined
18-101-final-viral-combined_k141_393681||full Low-quality
18-101-final-viral-combined_k141_47786||full Low-quality
18-101-final-viral-combined_k141_98448||full Low-quality
18-101-final-viral-combined_k141_98452||full Not-determined
18-101-final-viral-combined_k141_179779||full Low-quality
18-101-final-viral-combined_k141_98454||full Low-quality
18-101-final-viral-combined_k141_98472||full Low-quality
18-101-final-viral-combined_k141_344220||full Low-quality
18-101-final-viral-combined_k141_16043||full Not-determined
18-101-final-viral-combined_k141_163749||full Low-quality
18-101-final-viral-combined_k141_47830||full Low-quality
18-101-final-viral-combined_k141_393733||full Not-determined
18-101-final-viral-combined_k141_31634||full Low-quality
18-101-final-viral-combined_k141_246788||full Low-quality
18-101-final-viral-combined_k141_65310||full Low-quality
18-101-final-viral-combined_k141_16071||full Low-quality
18-101-final-viral-combined_k141_65315||full Low-quality
18-101-final-viral-combined_k141_47855||full Low-quality
18-101-final-viral-combined_k141_112909||full Not-determined
18-101-final-viral-combined_k141_47860||full Low-quality
18-101-final-viral-combined_k141_163768||full Low-quality
18-101-final-viral-combined_k141_47862||full Low-quality
18-101-final-viral-combined_k141_344259||full Low-quality
18-101-final-viral-combined_k141_65340||full Not-determined
18-101-final-viral-combined_k141_81068||full Not-determined
18-101-final-viral-combined_k141_393758||full Not-determined
18-101-final-viral-combined_k141_112929||full Not-determined
18-101-final-viral-combined_k141_229859||full Not-determined
18-101-final-viral-combined_k141_163782||full Low-quality
18-101-final-viral-combined_k141_65362||full Low-quality
18-101-final-viral-combined_k141_278989||full Not-determined
18-101-final-viral-combined_k141_65391||full Low-quality
18-101-final-viral-combined_k141_278991||full Low-quality
18-101-final-viral-combined_k141_31701||full Low-quality
18-101-final-viral-combined_k141_16114||full Not-determined
18-101-final-viral-combined_k141_31718||full Low-quality
18-101-final-viral-combined_k141_393808||full Not-determined
18-101-final-viral-combined_k141_344339||full Not-determined
18-101-final-viral-combined_k141_179910||full Low-quality
18-101-final-viral-combined_k141_279021||full Not-determined
18-101-final-viral-combined_k141_112967||full Not-determined
18-101-final-viral-combined_k141_65407||full Not-determined
18-101-final-viral-combined_k141_81149||full Not-determined
18-101-final-viral-combined_k141_16151||full Low-quality
18-101-final-viral-combined_k141_16164||full Not-determined
18-101-final-viral-combined_k141_47967||full Not-determined
18-101-final-viral-combined_k141_31745||full Low-quality
18-101-final-viral-combined_k141_16175||full Not-determined
18-101-final-viral-combined_k141_279043||full Low-quality
18-101-final-viral-combined_k141_98598||full Low-quality
18-101-final-viral-combined_k141_81197||full Low-quality
18-101-final-viral-combined_k141_47982||full Not-determined
18-101-final-viral-combined_k141_378237||full Low-quality
18-101-final-viral-combined_k141_47986||full Not-determined
18-101-final-viral-combined_k141_378244||full Low-quality
18-101-final-viral-combined_k141_179995||full Low-quality
18-101-final-viral-combined_k141_31760||full Not-determined
18-101-final-viral-combined_k141_65480||full Low-quality
18-101-final-viral-combined_k141_393855||full Not-determined
18-101-final-viral-combined_k141_65482||full Low-quality
18-101-final-viral-combined_k141_180007||full Not-determined
18-101-final-viral-combined_k141_48016||full Low-quality
18-101-final-viral-combined_k141_81262||full Not-determined
18-101-final-viral-combined_k141_81266||full Not-determined
18-101-final-viral-combined_k141_393889||full Not-determined
18-101-final-viral-combined_k141_229995||full Low-quality
18-101-final-viral-combined_k141_163901||full Low-quality
18-101-final-viral-combined_k141_230006||full Not-determined
18-101-final-viral-combined_k141_180037||full Low-quality
18-101-final-viral-combined_k141_378291||full Not-determined
18-101-final-viral-combined_k141_393911||full Not-determined
18-101-final-viral-combined_k141_147674||full Low-quality
18-101-final-viral-combined_k141_279137||full Low-quality
18-101-final-viral-combined_k141_31849||full Low-quality
18-101-final-viral-combined_k141_98643||full Not-determined
18-101-final-viral-combined_k141_98647||full Low-quality
18-101-final-viral-combined_k141_113138||full Low-quality
18-101-final-viral-combined_k141_230061||full Low-quality
18-101-final-viral-combined_k141_163945||full Low-quality
18-101-final-viral-combined_k141_344487||full Not-determined
18-101-final-viral-combined_k141_98669||full Not-determined
18-101-final-viral-combined_k141_180082||full Not-determined
18-101-final-viral-combined_k141_163975||full Low-quality
18-101-final-viral-combined_k141_230082||full Low-quality
18-101-final-viral-combined_k141_48098||full Not-determined
18-101-final-viral-combined_k141_163982||full Low-quality
18-101-final-viral-combined_k141_344538||full Not-determined
18-101-final-viral-combined_k141_163991||full Low-quality
18-101-final-viral-combined_k141_65577||full Not-determined
18-101-final-viral-combined_k141_230112||full Not-determined
18-101-final-viral-combined_k141_147787||full Low-quality
18-101-final-viral-combined_k141_98682||full Not-determined
18-101-final-viral-combined_k141_344559||full Not-determined
18-101-final-viral-combined_k141_344565||full Not-determined
18-101-final-viral-combined_k141_394044||full Not-determined
18-101-final-viral-combined_k141_164063||full Low-quality
18-101-final-viral-combined_k141_147807||full Not-determined
18-101-final-viral-combined_k141_164071||full Not-determined
18-101-final-viral-combined_k141_16338||full Not-determined
18-101-final-viral-combined_k141_16340||full Not-determined
18-101-final-viral-combined_k141_65587||full Not-determined
18-101-final-viral-combined_k141_65589||full Low-quality
18-101-final-viral-combined_k141_164086||full Low-quality
18-101-final-viral-combined_k141_31984||full Not-determined
18-101-final-viral-combined_k141_16347||full Not-determined
18-101-final-viral-combined_k141_147832||full Low-quality
18-101-final-viral-combined_k141_394078||full Low-quality
18-101-final-viral-combined_k141_230157||full Low-quality
18-101-final-viral-combined_k141_147844||full Not-determined
18-101-final-viral-combined_k141_48152||full Low-quality
18-101-final-viral-combined_k141_344607||full Low-quality
18-101-final-viral-combined_k141_230199||full Not-determined
18-101-final-viral-combined_k141_279260||full Not-determined
18-101-final-viral-combined_k141_16388||full Not-determined
18-101-final-viral-combined_k141_180198||full Not-determined
18-101-final-viral-combined_k141_279287||full Not-determined
18-101-final-viral-combined_k141_344624||full Low-quality
18-101-final-viral-combined_k141_164133||full Not-determined
18-101-final-viral-combined_k141_164141||full Not-determined
18-101-final-viral-combined_k141_113368||full Not-determined
18-101-final-viral-combined_k141_81490||full Low-quality
18-101-final-viral-combined_k141_81501||full Not-determined
18-101-final-viral-combined_k141_230265||full Low-quality
18-101-final-viral-combined_k141_230272||full Not-determined
18-101-final-viral-combined_k141_180265||full Not-determined
18-101-final-viral-combined_k141_16448||full Not-determined
18-101-final-viral-combined_k141_81544||full Not-determined
18-101-final-viral-combined_k141_164204||full Low-quality
18-101-final-viral-combined_k141_113420||full Low-quality
18-101-final-viral-combined_k141_180273||full Low-quality
18-101-final-viral-combined_k141_32084||full Not-determined
18-101-final-viral-combined_k141_113423||full Low-quality
18-101-final-viral-combined_k141_113427||full Low-quality
18-101-final-viral-combined_k141_81584||full Low-quality
18-101-final-viral-combined_k141_164218||full Not-determined
18-101-final-viral-combined_k141_230331||full Not-determined
18-101-final-viral-combined_k141_180292||full Low-quality
18-101-final-viral-combined_k141_344770||full Low-quality
18-101-final-viral-combined_k141_48284||full Low-quality
18-101-final-viral-combined_k141_32158||full Not-determined
18-101-final-viral-combined_k141_164281||full Low-quality
18-101-final-viral-combined_k141_48302||full Low-quality
18-101-final-viral-combined_k141_148071||full Low-quality
18-101-final-viral-combined_k141_378389||full Not-determined
18-101-final-viral-combined_k141_81669||full Low-quality
18-101-final-viral-combined_k141_32191||full Not-determined
18-101-final-viral-combined_k141_180381||full Low-quality
18-101-final-viral-combined_k141_65702||full Not-determined
18-101-final-viral-combined_k141_113540||full Low-quality
18-101-final-viral-combined_k141_394259||full Low-quality
18-101-final-viral-combined_k141_180399||full Not-determined
18-101-final-viral-combined_k141_344835||full Low-quality
18-101-final-viral-combined_k141_164325||full Low-quality
18-101-final-viral-combined_k141_180418||full Not-determined
18-101-final-viral-combined_k141_344848||full Low-quality
18-101-final-viral-combined_k141_344850||full Low-quality
18-101-final-viral-combined_k141_81730||full Not-determined
18-101-final-viral-combined_k141_344859||full Low-quality
18-101-final-viral-combined_k141_344860||full Low-quality
18-101-final-viral-combined_k141_32272||full Not-determined
18-101-final-viral-combined_k141_344861||full Low-quality
18-101-final-viral-combined_k141_394326||full Not-determined
18-101-final-viral-combined_k141_394327||full Not-determined
18-101-final-viral-combined_k141_48326||full Low-quality
18-101-final-viral-combined_k141_344889||full Not-determined
18-101-final-viral-combined_k141_394332||full Not-determined
18-101-final-viral-combined_k141_164380||full Low-quality
18-101-final-viral-combined_k141_81739||full Not-determined
18-101-final-viral-combined_k141_344897||full Low-quality
18-101-final-viral-combined_k141_279418||full Not-determined
18-101-final-viral-combined_k141_180459||full Low-quality
18-101-final-viral-combined_k141_164399||full Low-quality
18-101-final-viral-combined_k141_164401||full Not-determined
18-101-final-viral-combined_k141_180468||full Not-determined
18-101-final-viral-combined_k141_48374||full Low-quality
18-101-final-viral-combined_k141_113633||full Not-determined
18-101-final-viral-combined_k141_394372||full Low-quality
18-101-final-viral-combined_k141_48389||full Not-determined
18-101-final-viral-combined_k141_279480||full Not-determined
18-101-final-viral-combined_k141_180490||full Low-quality
18-101-final-viral-combined_k141_48411||full Low-quality
18-101-final-viral-combined_k141_394384||full Not-determined
18-101-final-viral-combined_k141_48419||full Low-quality
18-101-final-viral-combined_k141_344967||full Low-quality
18-101-final-viral-combined_k141_32382||full Not-determined
18-101-final-viral-combined_k141_344970||full Low-quality
18-101-final-viral-combined_k141_394391||full Not-determined
18-101-final-viral-combined_k141_394396||full Not-determined
18-101-final-viral-combined_k141_344987||full Low-quality
17-108-final-viral-combined_k141_55610||full Medium-quality
18-101-final-viral-combined_k141_48451||full Low-quality
18-101-final-viral-combined_k141_81854||full Not-determined
18-101-final-viral-combined_k141_32406||full Low-quality
18-101-final-viral-combined_k141_48480||full Low-quality
18-101-final-viral-combined_k141_32438||full Low-quality
18-101-final-viral-combined_k141_180558||full Not-determined
18-101-final-viral-combined_k141_345059||full Not-determined
18-101-final-viral-combined_k141_180603||full Not-determined

| | |
|---|---|
| 18-101-final-viral-combined_k141_394456\|\|full | Low-quality |
| 18-101-final-viral-combined_k141_32497\|\|full | Not-determined |
| 18-101-final-viral-combined_k141_48530\|\|full | Not-determined |
| 18-101-final-viral-combined_k141_279528\|\|full | Low-quality |
| 18-101-final-viral-combined_k141_32504\|\|full | Low-quality |
| 18-101-final-viral-combined_k141_180622\|\|full | Low-quality |
| 18-101-final-viral-combined_k141_113793\|\|full | Low-quality |
| 18-101-final-viral-combined_k141_48545\|\|full | Low-quality |
| 18-101-final-viral-combined_k141_345092\|\|full | Low-quality |
| 18-101-final-viral-combined_k141_32547\|\|full | Not-determined |
| 18-101-final-viral-combined_k141_113823\|\|full | Low-quality |
| 18-101-final-viral-combined_k141_65777\|\|full | Low-quality |
| 18-101-final-viral-combined_k141_345112\|\|full | Not-determined |
| 18-101-final-viral-combined_k141_345129\|\|full | Not-determined |
| 18-101-final-viral-combined_k141_113846\|\|full | Not-determined |
| 18-101-final-viral-combined_k141_164436\|\|full | Not-determined |
| 18-101-final-viral-combined_k141_164439\|\|full | Not-determined |
| 18-101-final-viral-combined_k141_81942\|\|full | Low-quality |
| 18-101-final-viral-combined_k141_81947\|\|full | Not-determined |
| 18-101-final-viral-combined_k141_164467\|\|full | Low-quality |
| 18-101-final-viral-combined_k141_394532\|\|full | Not-determined |
| 18-105_k141_127218 | High-quality |
| 18-101-final-viral-combined_k141_164486\|\|full | Low-quality |
| 18-101-final-viral-combined_k141_48676\|\|full | Not-determined |
| 18-101-final-viral-combined_k141_32651\|\|full | Not-determined |
| 18-101-final-viral-combined_k141_180742\|\|full | Low-quality |
| 18-101-final-viral-combined_k141_32669\|\|full | Low-quality |
| 18-101-final-viral-combined_k141_113935\|\|full | Low-quality |
| 18-101-final-viral-combined_k141_164517\|\|full | Low-quality |
| 18-101-final-viral-combined_k141_279611\|\|full | Not-determined |
| 18-101-final-viral-combined_k141_180779\|\|full | Not-determined |
| 18-101-final-viral-combined_k141_180781\|\|full | Not-determined |
| 18-101-final-viral-combined_k141_279629\|\|full | Low-quality |
| 18-101-final-viral-combined_k141_345168\|\|full | Not-determined |
| 18-101-final-viral-combined_k141_82011\|\|full | Low-quality |
| 18-101-final-viral-combined_k141_32738\|\|full | Not-determined |
| 18-101-final-viral-combined_k141_180806\|\|full | Not-determined |
| 18-101-final-viral-combined_k141_180807\|\|full | Not-determined |
| 18-101-final-viral-combined_k141_32789\|\|full | Low-quality |
| 18-101-final-viral-combined_k141_345238\|\|full | Not-determined |
| 18-101-final-viral-combined_k141_114071\|\|full | Low-quality |
| 18-101-final-viral-combined_k141_345249\|\|full | Not-determined |
| 18-101-final-viral-combined_k141_114080\|\|full | Not-determined |
| 18-101-final-viral-combined_k141_82151\|\|full | Not-determined |
| 18-101-final-viral-combined_k141_345300\|\|full | Low-quality |
| 18-101-final-viral-combined_k141_48809\|\|full | Not-determined |
| 18-101-final-viral-combined_k141_48811\|\|full | Low-quality |
| 18-101-final-viral-combined_k141_48813\|\|full | Low-quality |
| 18-101-final-viral-combined_k141_114145\|\|full | Not-determined |
| 18-101-final-viral-combined_k141_82187\|\|full | Not-determined |
| 18-101-final-viral-combined_k141_114158\|\|full | Low-quality |
| 18-101-final-viral-combined_k141_114162\|\|full | Low-quality |
| 18-101-final-viral-combined_k141_345325\|\|full | Not-determined |
| 18-101-final-viral-combined_k141_82205\|\|full | Not-determined |
| 18-101-final-viral-combined_k141_48883\|\|full | Not-determined |
| 18-101-final-viral-combined_k141_345351\|\|full | Not-determined |
| 18-101-final-viral-combined_k141_48905\|\|full | Low-quality |
| 18-101-final-viral-combined_k141_345383\|\|full | Not-determined |
| 18-101-final-viral-combined_k141_48930\|\|full | Low-quality |
| 18-101-final-viral-combined_k141_114239\|\|full | Low-quality |
| 17-109-final-viral-combined_k141_12658\|\|full | High-quality |
| 18-101-final-viral-combined_k141_48966\|\|full | Not-determined |
| 18-101-final-viral-combined_k141_32868\|\|full | Not-determined |
| 18-101-final-viral-combined_k141_114263\|\|full | Low-quality |
| 18-101-final-viral-combined_k141_114282\|\|full | Not-determined |
| 18-101-final-viral-combined_k141_345404\|\|full | Not-determined |
| 18-101-final-viral-combined_k141_114291\|\|full | Low-quality |
| 18-101-final-viral-combined_k141_345429\|\|full | Not-determined |
| 18-102-final-viral-combined_k141_25727\|\|full | Medium-quality |
| 18-101-final-viral-combined_k141_114352\|\|full | Low-quality |
| 18-101-final-viral-combined_k141_345468\|\|full | Low-quality |
| 18-101-final-viral-combined_k141_32883\|\|full | Not-determined |
| 18-101-final-viral-combined_k141_114382\|\|full | Not-determined |
| 18-101-final-viral-combined_k141_114460\|\|full | Low-quality |
| 18-101-final-viral-combined_k141_114474\|\|full | Not-determined |
| 18-101-final-viral-combined_k141_49100\|\|full | Low-quality |
| 18-101-final-viral-combined_k141_114478\|\|full | Low-quality |
| 18-101-final-viral-combined_k141_49114\|\|full | Low-quality |
| 18-101-final-viral-combined_k141_114506\|\|full | Low-quality |
| 18-101-final-viral-combined_k141_114540\|\|full | Low-quality |
| 18-101-final-viral-combined_k141_49157\|\|full | Low-quality |
| 18-101-final-viral-combined_k141_49236\|\|full | Not-determined |
| 18-101-final-viral-combined_k141_49249\|\|full | Not-determined |
| 18-101-final-viral-combined_k141_114627\|\|full | Low-quality |
| 18-101-final-viral-combined_k141_49284\|\|full | Not-determined |
| 18-101-final-viral-combined_k141_114663\|\|full | Low-quality |
| 18-101-final-viral-combined_k141_114673\|\|full | Low-quality |
| 18-101-final-viral-combined_k141_114686\|\|full | Low-quality |
| 18-101-final-viral-combined_k141_114690\|\|full | Low-quality |
| 18-101-final-viral-combined_k141_114773\|\|full | Low-quality |
| 18-101-final-viral-combined_k141_114784\|\|full | Not-determined |
| 18-101-final-viral-combined_k141_114796\|\|full | Low-quality |
| 18-101-final-viral-combined_k141_114798\|\|full | Low-quality |
| 18-101-final-viral-combined_k141_114803\|\|full | Not-determined |
| 18-101-final-viral-combined_k141_114832\|\|full | Low-quality |
| 18-101-final-viral-combined_k141_114978\|\|full | Not-determined |
| 18-101-final-viral-combined_k141_115088\|\|full | Not-determined |
| 18-101-final-viral-combined_k141_115104\|\|full | Not-determined |
| 18-101-final-viral-combined_k141_313149\|\|0_partial | Low-quality |
| 18-101-final-viral-combined_k141_49772\|\|0_partial | Low-quality |
| 18-101-final-viral-combined_k141_264065\|\|0_partial | Low-quality |
| 18-101-final-viral-combined_k141_116429\|\|0_partial | Low-quality |
| 18-101-final-viral-combined_k141_49960\|\|0_partial | Low-quality |
| 18-101-final-viral-combined_k141_18465\|\|0_partial | Low-quality |
| 18-101-final-viral-combined_k141_2551\|\|0_partial | Not-determined |
| 18-101-final-viral-combined_k141_2595\|\|0_partial | Low-quality |
| 18-101-final-viral-combined_k141_282151\|\|0_partial | Low-quality |
| 18-101-final-viral-combined_k141_216719\|\|0_partial | Low-quality |
| 18-110-final-viral-combined_k141_49369\|\|full | High-quality |
| 18-101-final-viral-combined_k141_216985\|\|0_partial | Low-quality |
| 18-101-final-viral-combined_k141_68332\|\|0_partial | Not-determined |
| 18-101-final-viral-combined_k141_265860\|\|0_partial | Not-determined |
| 18-105-final-viral-combined_k141_207291\|\|full | Medium-quality |
| 18-101-final-viral-combined_k141_85258\|\|0_partial | Low-quality |
| 18-101-final-viral-combined_k141_36114\|\|0_partial | Not-determined |
| 18-101-final-viral-combined_k141_250445\|\|0_partial | Low-quality |
| 18-101-final-viral-combined_k141_52605\|\|0_partial | Low-quality |
| 18-101-final-viral-combined_k141_118937\|\|0_partial | Low-quality |
| 18-101-final-viral-combined_k141_332829\|\|0_partial | Low-quality |
| 18-109-final-viral-combined_k141_134458\|\|full | Medium-quality |
| 18-101-final-viral-combined_k141_267657\|\|0_partial | Not-determined |
| 18-101-final-viral-combined_k141_168463\|\|0_partial | Not-determined |
| 18-101-final-viral-combined_k141_284778\|\|0_partial | Not-determined |
| 18-101-final-viral-combined_k141_37915\|\|0_partial | Low-quality |
| 18-101-final-viral-combined_k141_302144\|\|0_partial | Not-determined |
| 18-101-final-viral-combined_k141_169130\|\|0_partial | Low-quality |
| 18-101-final-viral-combined_k141_38039\|\|0_partial | Low-quality |
| 18-101-final-viral-combined_k141_169272\|\|0_partial | Not-determined |
| 18-101-final-viral-combined_k141_219533\|\|0_partial | Not-determined |
| 18-101-final-viral-combined_k141_121278\|\|0_partial | Not-determined |
| 18-101-final-viral-combined_k141_54654\|\|0_partial | Low-quality |
| 18-101-final-viral-combined_k141_219763\|\|0_partial | Not-determined |
| 18-101-final-viral-combined_k141_5759\|\|0_partial | Low-quality |
| 18-101-final-viral-combined_k141_38714\|\|0_partial | Low-quality |
| 18-101-final-viral-combined_k141_187505\|\|0_partial | Low-quality |
| 18-101-final-viral-combined_k141_154009\|\|0_partial | Low-quality |
| 18-101-final-viral-combined_k141_23096\|\|0_partial | Low-quality |
| 18-101-final-viral-combined_k141_138958\|\|0_partial | Not-determined |
| 18-101-final-viral-combined_k141_255289\|\|0_partial | Low-quality |
| 18-101-final-viral-combined_k141_255289\|\|1_partial | Not-determined |
| 18-101-final-viral-combined_k141_221925\|\|0_partial | Low-quality |
| 18-101-final-viral-combined_k141_155743\|\|0_partial | Not-determined |
| 18-101-final-viral-combined_k141_172194\|\|0_partial | Low-quality |
| 18-101-final-viral-combined_k141_273012\|\|0_partial | Not-determined |
| 18-101-final-viral-combined_k141_126022\|\|0_partial | Low-quality |
| 18-105-final-viral-combined_k141_206050\|\|full | Medium-quality |
| 18-101-final-viral-combined_k141_224684\|\|0_partial | Low-quality |
| 18-101-final-viral-combined_k141_127310\|\|0_partial | Low-quality |
| 18-101-final-viral-combined_k141_175512\|\|0_partial | Low-quality |
| 18-101-final-viral-combined_k141_193811\|\|0_partial | Low-quality |
| 18-101-final-viral-combined_k141_326034\|\|0_partial | Not-determined |
| 18-105-final-viral-combined_k141_127218\|\|full | High-quality |
| KM-final-viral-combined_k141_106262\|\|full | High-quality |
| 18-101-final-viral-combined_k141_95394\|\|0_partial | Low-quality |
| 18-101-final-viral-combined_k141_375477\|\|0_partial | Not-determined |
| 18-101-final-viral-combined_k141_46244\|\|0_partial | Low-quality |
| 18-101-final-viral-combined_k141_294159\|\|0_partial | Not-determined |
| 18-101-final-viral-combined_k141_294159\|\|1_partial | Not-determined |
| 18-101-final-viral-combined_k141_343763\|\|0_partial | Not-determined |
| 18-101-final-viral-combined_k141_31949\|\|0_partial | Low-quality |
| 18-101-final-viral-combined_k141_180675\|\|0_partial | Low-quality |
| 18-101-final-viral-combined_k141_49364\|\|lt2gene | Low-quality |
| 18-101-final-viral-combined_k141_82293\|\|lt2gene | Low-quality |
| 18-101-final-viral-combined_k141_16474\|\|lt2gene | Not-determined |
| 18-101-final-viral-combined_k141_148124\|\|lt2gene | Low-quality |
| 18-101-final-viral-combined_k141_148131\|\|lt2gene | Not-determined |
| 18-101-final-viral-combined_k141_329136\|\|lt2gene | Low-quality |
| 18-101-final-viral-combined_k141_378509\|\|lt2gene | Not-determined |
| 18-101-final-viral-combined_k141_132\|\|lt2gene | Low-quality |
| 18-101-final-viral-combined_k141_312757\|\|lt2gene | Not-determined |
| 18-101-final-viral-combined_k141_115303\|\|lt2gene | Not-determined |
| 18-101-final-viral-combined_k141_162\|\|lt2gene | Low-quality |
| 18-101-final-viral-combined_k141_279781\|\|lt2gene | Not-determined |
| 18-101-final-viral-combined_k141_82418\|\|lt2gene | Low-quality |
| 18-101-final-viral-combined_k141_115318\|\|lt2gene | Low-quality |
| 18-101-final-viral-combined_k141_214015\|\|lt2gene | Low-quality |
| 18-101-final-viral-combined_k141_148232\|\|lt2gene | Not-determined |
| 18-101-final-viral-combined_k141_197536\|\|lt2gene | Low-quality |
| 18-101-final-viral-combined_k141_234\|\|lt2gene | Not-determined |
| 18-101-final-viral-combined_k141_362148\|\|lt2gene | Low-quality |
| 18-101-final-viral-combined_k141_261\|\|lt2gene | Low-quality |
| 18-101-final-viral-combined_k141_65846\|\|lt2gene | Not-determined |
| 18-101-final-viral-combined_k141_65859\|\|lt2gene | Not-determined |
| 18-101-final-viral-combined_k141_292\|\|lt2gene | Not-determined |
| 18-101-final-viral-combined_k141_279898\|\|lt2gene | Low-quality |
| 18-101-final-viral-combined_k141_82518\|\|lt2gene | Low-quality |
| 18-101-final-viral-combined_k141_279928\|\|lt2gene | Low-quality |
| 18-101-final-viral-combined_k141_279937\|\|lt2gene | Low-quality |
| 18-101-final-viral-combined_k141_164720\|\|lt2gene | Not-determined |
| 18-101-final-viral-combined_k141_296380\|\|lt2gene | Not-determined |
| 18-101-final-viral-combined_k141_230607\|\|lt2gene | Not-determined |
| 18-101-final-viral-combined_k141_82533\|\|lt2gene | Low-quality |
| 18-101-final-viral-combined_k141_378680\|\|lt2gene | Low-quality |
| 18-101-final-viral-combined_k141_230638\|\|lt2gene | Low-quality |
| 18-101-final-viral-combined_k141_16711\|\|lt2gene | Low-quality |
| 18-101-final-viral-combined_k141_131980\|\|lt2gene | Low-quality |
| 18-101-final-viral-combined_k141_296447\|\|lt2gene | Low-quality |
| 18-101-final-viral-combined_k141_296448\|\|lt2gene | Low-quality |
| 18-101-final-viral-combined_k141_65921\|\|lt2gene | Not-determined |
| 18-101-final-viral-combined_k141_148351\|\|lt2gene | Not-determined |
| 18-101-final-viral-combined_k141_345753\|\|lt2gene | Low-quality |
| 18-101-final-viral-combined_k141_247060\|\|lt2gene | Low-quality |
| 18-101-final-viral-combined_k141_49602\|\|lt2gene | Low-quality |
| 18-101-final-viral-combined_k141_82626\|\|lt2gene | Low-quality |
| 18-101-final-viral-combined_k141_49607\|\|lt2gene | Low-quality |
| 18-101-final-viral-combined_k141_132027\|\|lt2gene | Low-quality |
| 18-101-final-viral-combined_k141_214199\|\|lt2gene | Not-determined |
| 18-101-final-viral-combined_k141_263527\|\|lt2gene | Low-quality |
| 18-101-final-viral-combined_k141_296535\|\|lt2gene | Not-determined |
| 18-101-final-viral-combined_k141_345803\|\|lt2gene | Low-quality |
| 18-101-final-viral-combined_k141_181309\|\|lt2gene | Low-quality |
| 18-101-final-viral-combined_k141_99025\|\|lt2gene | Low-quality |
| 18-101-final-viral-combined_k141_247144\|\|lt2gene | Low-quality |
| 18-101-final-viral-combined_k141_263564\|\|lt2gene | Low-quality |
| 18-101-final-viral-combined_k141_598\|\|lt2gene | Low-quality |
| 18-101-final-viral-combined_k141_115581\|\|lt2gene | Low-quality |
| 18-101-final-viral-combined_k141_345878\|\|lt2gene | Low-quality |
| 18-101-final-viral-combined_k141_247193\|\|lt2gene | Low-quality |
| 18-101-final-viral-combined_k141_197856\|\|lt2gene | Low-quality |
| 18-101-final-viral-combined_k141_214290\|\|lt2gene | Low-quality |
| 18-101-final-viral-combined_k141_362435\|\|lt2gene | Low-quality |
| 18-101-final-viral-combined_k141_82797\|\|lt2gene | Not-determined |
| 18-101-final-viral-combined_k141_181421\|\|lt2gene | Not-determined |
| 18-101-final-viral-combined_k141_280227\|\|lt2gene | Low-quality |
| 18-101-final-viral-combined_k141_214336\|\|lt2gene | Low-quality |
| 18-101-final-viral-combined_k141_115680\|\|lt2gene | Low-quality |
| 18-101-final-viral-combined_k141_313140\|\|lt2gene | Low-quality |
| 18-101-final-viral-combined_k141_16926\|\|lt2gene | Low-quality |
| 18-101-final-viral-combined_k141_362485\|\|lt2gene | Low-quality |
| 18-101-final-viral-combined_k141_115732\|\|lt2gene | Low-quality |
| 18-101-final-viral-combined_k141_296677\|\|lt2gene | Low-quality |
| 18-101-final-viral-combined_k141_132243\|\|lt2gene | Low-quality |
| 18-101-final-viral-combined_k141_66144\|\|lt2gene | Low-quality |
| 18-101-final-viral-combined_k141_214463\|\|lt2gene | Not-determined |
| 18-101-final-viral-combined_k141_197989\|\|lt2gene | Low-quality |
| 18-101-final-viral-combined_k141_296762\|\|lt2gene | Low-quality |
| 18-101-final-viral-combined_k141_132323\|\|lt2gene | Low-quality |
| 18-101-final-viral-combined_k141_198005\|\|lt2gene | Not-determined |
| 18-101-final-viral-combined_k141_198022\|\|lt2gene | Low-quality |
| 18-101-final-viral-combined_k141_346125\|\|lt2gene | Low-quality |
| 18-101-final-viral-combined_k141_82969\|\|lt2gene | Low-quality |
| 18-101-final-viral-combined_k141_214550\|\|lt2gene | Not-determined |
| 18-101-final-viral-combined_k141_115879\|\|lt2gene | Low-quality |
| 18-101-final-viral-combined_k141_746\|\|lt2gene | Not-determined |
| 18-101-final-viral-combined_k141_280339\|\|lt2gene | Low-quality |
| 18-101-final-viral-combined_k141_148631\|\|lt2gene | Low-quality |
| 18-101-final-viral-combined_k141_148633\|\|lt2gene | Not-determined |
| 18-101-final-viral-combined_k141_214642\|\|lt2gene | Low-quality |
| 18-101-final-viral-combined_k141_115975\|\|lt2gene | Low-quality |
| 18-101-final-viral-combined_k141_181747\|\|lt2gene | Not-determined |
| 18-101-final-viral-combined_k141_802\|\|lt2gene | Low-quality |
| 18-101-final-viral-combined_k141_99384\|\|lt2gene | Low-quality |
| 18-101-final-viral-combined_k141_313182\|\|lt2gene | Low-quality |
| 18-101-final-viral-combined_k141_181785\|\|lt2gene | Low-quality |
| 18-101-final-viral-combined_k141_214707\|\|lt2gene | Not-determined |
| 18-101-final-viral-combined_k141_198156\|\|lt2gene | Low-quality |
| 18-101-final-viral-combined_k141_895\|\|lt2gene | Low-quality |
| 18-101-final-viral-combined_k141_362676\|\|lt2gene | Low-quality |
| 18-101-final-viral-combined_k141_329745\|\|lt2gene | Not-determined |
| 18-101-final-viral-combined_k141_362699\|\|lt2gene | Low-quality |
| 18-101-final-viral-combined_k141_116052\|\|lt2gene | Not-determined |
| 18-101-final-viral-combined_k141_116054\|\|lt2gene | Not-determined |
| 18-101-final-viral-combined_k141_181839\|\|lt2gene | Low-quality |
| 18-101-final-viral-combined_k141_83126\|\|lt2gene | Not-determined |
| 18-101-final-viral-combined_k141_181844\|\|lt2gene | Low-quality |
| 18-101-final-viral-combined_k141_247531\|\|lt2gene | Not-determined |
| 18-101-final-viral-combined_k141_362745\|\|lt2gene | Low-quality |
| 18-101-final-viral-combined_k141_33415\|\|lt2gene | Low-quality |
| 18-101-final-viral-combined_k141_132481\|\|lt2gene | Low-quality |
| 18-101-final-viral-combined_k141_132485\|\|lt2gene | Low-quality |
| 18-101-final-viral-combined_k141_379104\|\|lt2gene | Low-quality |
| 18-101-final-viral-combined_k141_346340\|\|lt2gene | Low-quality |
| 18-101-final-viral-combined_k141_264009\|\|lt2gene | Low-quality |
| 18-101-final-viral-combined_k141_296875\|\|lt2gene | Low-quality |
| 18-101-final-viral-combined_k141_116120\|\|lt2gene | Low-quality |
| 18-101-final-viral-combined_k141_379131\|\|lt2gene | Low-quality |
| 18-101-final-viral-combined_k141_346383\|\|lt2gene | Low-quality |
| 18-101-final-viral-combined_k141_379162\|\|lt2gene | Low-quality |
| 18-101-final-viral-combined_k141_296926\|\|lt2gene | Not-determined |
| 18-101-final-viral-combined_k141_296992\|\|lt2gene | Low-quality |
| 18-101-final-viral-combined_k141_346504\|\|lt2gene | Low-quality |
| 18-101-final-viral-combined_k141_313411\|\|lt2gene | Low-quality |
| 18-101-final-viral-combined_k141_165371\|\|lt2gene | Low-quality |
| 18-101-final-viral-combined_k141_247722\|\|lt2gene | Low-quality |
| 18-101-final-viral-combined_k141_231222\|\|lt2gene | Low-quality |
| 18-101-final-viral-combined_k141_66402\|\|lt2gene | Low-quality |
| 18-101-final-viral-combined_k141_49804\|\|lt2gene | Not-determined |
| 18-101-final-viral-combined_k141_99558\|\|lt2gene | Low-quality |
| 18-101-final-viral-combined_k141_66448\|\|lt2gene | Low-quality |
| 18-101-final-viral-combined_k141_231264\|\|lt2gene | Low-quality |
| 18-101-final-viral-combined_k141_99600\|\|lt2gene | Low-quality |
| 18-101-final-viral-combined_k141_264200\|\|lt2gene | Not-determined |
| 18-101-final-viral-combined_k141_313536\|\|lt2gene | Low-quality |
| 18-101-final-viral-combined_k141_99619\|\|lt2gene | Not-determined |
| 18-101-final-viral-combined_k141_66518\|\|lt2gene | Low-quality |
| 18-101-final-viral-combined_k141_297223\|\|lt2gene | Low-quality |
| 18-101-final-viral-combined_k141_329884\|\|lt2gene | Not-determined |
| 18-101-final-viral-combined_k141_329904\|\|lt2gene | Low-quality |
| 18-101-final-viral-combined_k141_83384\|\|lt2gene | Low-quality |
| 18-101-final-viral-combined_k141_132873\|\|lt2gene | Low-quality |
| 18-101-final-viral-combined_k141_116447\|\|lt2gene | Not-determined |
| 18-101-final-viral-combined_k141_1397\|\|lt2gene | Low-quality |
| 18-101-final-viral-combined_k141_297322\|\|lt2gene | Low-quality |
| 18-101-final-viral-combined_k141_330026\|\|lt2gene | Not-determined |
| 18-101-final-viral-combined_k141_346736\|\|lt2gene | Low-quality |
| 18-101-final-viral-combined_k141_248015\|\|lt2gene | Low-quality |
| 18-101-final-viral-combined_k141_66565\|\|lt2gene | Not-determined |
| 18-101-final-viral-combined_k141_182131\|\|lt2gene | Not-determined |
| 18-101-final-viral-combined_k141_313736\|\|lt2gene | Low-quality |
| 18-101-final-viral-combined_k141_50040\|\|lt2gene | Low-quality |
| 18-101-final-viral-combined_k141_132989\|\|lt2gene | Not-determined |
| 18-101-final-viral-combined_k141_33850\|\|lt2gene | Low-quality |
| 18-101-final-viral-combined_k141_33864\|\|lt2gene | Low-quality |
| 18-101-final-viral-combined_k141_116549\|\|lt2gene | Not-determined |
| 18-101-final-viral-combined_k141_99874\|\|lt2gene | Low-quality |
| 18-101-final-viral-combined_k141_330154\|\|lt2gene | Low-quality |
| 18-101-final-viral-combined_k141_297488\|\|lt2gene | Low-quality |
| 18-101-final-viral-combined_k141_198784\|\|lt2gene | Low-quality |
| 18-101-final-viral-combined_k141_182328\|\|lt2gene | Low-quality |
| 18-101-final-viral-combined_k141_313877\|\|lt2gene | Not-determined |
| 18-101-final-viral-combined_k141_83486\|\|lt2gene | Not-determined |
| 18-101-final-viral-combined_k141_133127\|\|lt2gene | Low-quality |
| 18-101-final-viral-combined_k141_182349\|\|lt2gene | Low-quality |
| 18-101-final-viral-combined_k141_231532\|\|lt2gene | Low-quality |
| 18-101-final-viral-combined_k141_281129\|\|lt2gene | Low-quality |
| 18-101-final-viral-combined_k141_17734\|\|lt2gene | Not-determined |
| 18-101-final-viral-combined_k141_363288\|\|lt2gene | Not-determined |
| 18-101-final-viral-combined_k141_116644\|\|lt2gene | Low-quality |
| 18-101-final-viral-combined_k141_198878\|\|lt2gene | Not-determined |
| 18-101-final-viral-combined_k141_347046\|\|lt2gene | Low-quality |
| 18-101-final-viral-combined_k141_50200\|\|lt2gene | Not-determined |
| 18-101-final-viral-combined_k141_313959\|\|lt2gene | Not-determined |
| 18-101-final-viral-combined_k141_149315\|\|lt2gene | Low-quality |
| 18-101-final-viral-combined_k141_347122\|\|lt2gene | Not-determined |
| 18-101-final-viral-combined_k141_379621\|\|lt2gene | Low-quality |
| 18-101-final-viral-combined_k141_34120\|\|lt2gene | Low-quality |
| 18-101-final-viral-combined_k141_50237\|\|lt2gene | Not-determined |
| 18-101-final-viral-combined_k141_50292\|\|lt2gene | Low-quality |
| 18-101-final-viral-combined_k141_347190\|\|lt2gene | Low-quality |
| 18-101-final-viral-combined_k141_330357\|\|lt2gene | Low-quality |
| 18-101-final-viral-combined_k141_199023\|\|lt2gene | Not-determined |
| 18-101-final-viral-combined_k141_330383\|\|lt2gene | Not-determined |
| 18-101-final-viral-combined_k141_297696\|\|lt2gene | Low-quality |
| 18-101-final-viral-combined_k141_231707\|\|lt2gene | Not-determined |
| 18-101-final-viral-combined_k141_1805\|\|lt2gene | Low-quality |
| 18-101-final-viral-combined_k141_215565\|\|lt2gene | Low-quality |
| 18-101-final-viral-combined_k141_165741\|\|lt2gene | Not-determined |
| 18-101-final-viral-combined_k141_363420\|\|lt2gene | Low-quality |
| 18-101-final-viral-combined_k141_66796\|\|lt2gene | Not-determined |
| 18-101-final-viral-combined_k141_379877\|\|lt2gene | Low-quality |
| 18-101-final-viral-combined_k141_17936\|\|lt2gene | Not-determined |
| 18-101-final-viral-combined_k141_215652\|\|lt2gene | Low-quality |
| 18-101-final-viral-combined_k141_1888\|\|lt2gene | Low-quality |
| 18-101-final-viral-combined_k141_133439\|\|lt2gene | Low-quality |
| 18-101-final-viral-combined_k141_116779\|\|lt2gene | Low-quality |
| 18-101-final-viral-combined_k141_34356\|\|lt2gene | Low-quality |
| 18-101-final-viral-combined_k141_347276\|\|lt2gene | Low-quality |
| 18-101-final-viral-combined_k141_281409\|\|lt2gene | Low-quality |
| 18-101-final-viral-combined_k141_199166\|\|lt2gene | Low-quality |
| 18-101-final-viral-combined_k141_83825\|\|lt2gene | Low-quality |
| 18-101-final-viral-combined_k141_363552\|\|lt2gene | Low-quality |
| 18-101-final-viral-combined_k141_264563\|\|lt2gene | Not-determined |
| 18-101-final-viral-combined_k141_1965\|\|lt2gene | Low-quality |
| 18-101-final-viral-combined_k141_34439\|\|lt2gene | Not-determined |
| 18-101-final-viral-combined_k141_314281\|\|lt2gene | Not-determined |
| 18-101-final-viral-combined_k141_330555\|\|lt2gene | Not-determined |
| 18-101-final-viral-combined_k141_149575\|\|lt2gene | Low-quality |
| 18-101-final-viral-combined_k141_330595\|\|lt2gene | Low-quality |
| 18-101-final-viral-combined_k141_215726\|\|lt2gene | Low-quality |
| 18-101-final-viral-combined_k141_2061\|\|lt2gene | Low-quality |
| 18-101-final-viral-combined_k141_281530\|\|lt2gene | Low-quality |
| 18-101-final-viral-combined_k141_297975\|\|lt2gene | Low-quality |
| 18-101-final-viral-combined_k141_100187\|\|lt2gene | Not-determined |
| 18-101-final-viral-combined_k141_330668\|\|lt2gene | Low-quality |
| 18-101-final-viral-combined_k141_66812\|\|lt2gene | Not-determined |
| 18-101-final-viral-combined_k141_380086\|\|lt2gene | Low-quality |
| 18-101-final-viral-combined_k141_50587\|\|lt2gene | Low-quality |
| 18-101-final-viral-combined_k141_347468\|\|lt2gene | Low-quality |
| 18-101-final-viral-combined_k141_66887\|\|lt2gene | Low-quality |
| 18-101-final-viral-combined_k141_281690\|\|lt2gene | Low-quality |
| 18-101-final-viral-combined_k141_50604\|\|lt2gene | Low-quality |
| 18-101-final-viral-combined_k141_264731\|\|lt2gene | Not-determined |
| 18-101-final-viral-combined_k141_149780\|\|lt2gene | Not-determined |
| 18-101-final-viral-combined_k141_347583\|\|lt2gene | Low-quality |
| 18-101-final-viral-combined_k141_232089\|\|lt2gene | Low-quality |
| 18-101-final-viral-combined_k141_149819\|\|lt2gene | Not-determined |
| 18-101-final-viral-combined_k141_117076\|\|lt2gene | Not-determined |
| 18-101-final-viral-combined_k141_264801\|\|lt2gene | Low-quality |
| 18-101-final-viral-combined_k141_117106\|\|lt2gene | Low-quality |
| 18-101-final-viral-combined_k141_314524\|\|lt2gene | Low-quality |
| 18-101-final-viral-combined_k141_215956\|\|lt2gene | Not-determined |
| 18-101-final-viral-combined_k141_117136\|\|lt2gene | Low-quality |
| 18-101-final-viral-combined_k141_84237\|\|lt2gene | Low-quality |
| 18-101-final-viral-combined_k141_248746\|\|lt2gene | Not-determined |
| 18-101-final-viral-combined_k141_2310\|\|lt2gene | Not-determined |
| 18-101-final-viral-combined_k141_117170\|\|lt2gene | Low-quality |
| 18-101-final-viral-combined_k141_298296\|\|lt2gene | Low-quality |
| 18-101-final-viral-combined_k141_183015\|\|lt2gene | Low-quality |
| 18-101-final-viral-combined_k141_232201\|\|lt2gene | Low-quality |
| 18-101-final-viral-combined_k141_84314\|\|lt2gene | Low-quality |
| 18-101-final-viral-combined_k141_314627\|\|lt2gene | Low-quality |
| 18-101-final-viral-combined_k141_380245\|\|lt2gene | Not-determined |
| 18-101-final-viral-combined_k141_100499\|\|lt2gene | Low-quality |
| 18-101-final-viral-combined_k141_2348\|\|lt2gene | Not-determined |
| 18-101-final-viral-combined_k141_298376\|\|lt2gene | Low-quality |
| 18-101-final-viral-combined_k141_232277\|\|lt2gene | Low-quality |
| 18-101-final-viral-combined_k141_216131\|\|lt2gene | Low-quality |
| 18-101-final-viral-combined_k141_264929\|\|lt2gene | Low-quality |
| 18-101-final-viral-combined_k141_166109\|\|lt2gene | Not-determined |
| 18-101-final-viral-combined_k141_380306\|\|lt2gene | Low-quality |
| 18-101-final-viral-combined_k141_216150\|\|lt2gene | Low-quality |
| 18-101-final-viral-combined_k141_50989\|\|lt2gene | Low-quality |
| 18-101-final-viral-combined_k141_84429\|\|lt2gene | Low-quality |
| 18-101-final-viral-combined_k141_51000\|\|lt2gene | Low-quality |
| 18-101-final-viral-combined_k141_248933\|\|lt2gene | Low-quality |
| 18-101-final-viral-combined_k141_248940\|\|lt2gene | Not-determined |
| 18-101-final-viral-combined_k141_199607\|\|lt2gene | Not-determined |
| 18-101-final-viral-combined_k141_67267\|\|lt2gene | Not-determined |
| 18-101-final-viral-combined_k141_166184\|\|lt2gene | Low-quality |
| 18-101-final-viral-combined_k141_2463\|\|lt2gene | Not-determined |
| 18-101-final-viral-combined_k141_183202\|\|lt2gene | Low-quality |
| 18-101-final-viral-combined_k141_282030\|\|lt2gene | Low-quality |
| 18-101-final-viral-combined_k141_51154\|\|lt2gene | Not-determined |
| 18-101-final-viral-combined_k141_183260\|\|lt2gene | Low-quality |
| 18-101-final-viral-combined_k141_199733\|\|lt2gene | Low-quality |
| 18-101-final-viral-combined_k141_216359\|\|lt2gene | Low-quality |
| 18-101-final-viral-combined_k141_347882\|\|lt2gene | Low-quality |
| 18-101-final-viral-combined_k141_18526\|\|lt2gene | Low-quality |
| 18-101-final-viral-combined_k141_117396\|\|lt2gene | Low-quality |
| 18-101-final-viral-combined_k141_199811\|\|lt2gene | Low-quality |
| 18-101-final-viral-combined_k141_84624\|\|lt2gene | Low-quality |
| 18-101-final-viral-combined_k141_249148\|\|lt2gene | Low-quality |
| 18-101-final-viral-combined_k141_166333\|\|lt2gene | Low-quality |
| 18-101-final-viral-combined_k141_199871\|\|lt2gene | Low-quality |
| 18-101-final-viral-combined_k141_67484\|\|lt2gene | Not-determined |
| 18-101-final-viral-combined_k141_67516\|\|lt2gene | Not-determined |
| 18-101-final-viral-combined_k141_2632\|\|lt2gene | Low-quality |
| 18-101-final-viral-combined_k141_249256\|\|lt2gene | Low-quality |
| 18-101-final-viral-combined_k141_166378\|\|lt2gene | Not-determined |
| 18-101-final-viral-combined_k141_216538\|\|lt2gene | Low-quality |
| 18-101-final-viral-combined_k141_216556\|\|lt2gene | Low-quality |
| 18-101-final-viral-combined_k141_249287\|\|lt2gene | Low-quality |
| 18-101-final-viral-combined_k141_84745\|\|lt2gene | Low-quality |
| 18-101-final-viral-combined_k141_380674\|\|lt2gene | Not-determined |
| 18-101-final-viral-combined_k141_315080\|\|lt2gene | Low-quality |
| 18-101-final-viral-combined_k141_232700\|\|lt2gene | Low-quality |
| 18-101-final-viral-combined_k141_298722\|\|lt2gene | Low-quality |
| 18-101-final-viral-combined_k141_232732\|\|lt2gene | Low-quality |
| 18-101-final-viral-combined_k141_166461\|\|lt2gene | Low-quality |
| 18-101-final-viral-combined_k141_298778\|\|lt2gene | Low-quality |
| 18-101-final-viral-combined_k141_265294\|\|lt2gene | Low-quality |
| 18-101-final-viral-combined_k141_232769\|\|lt2gene | Low-quality |
| 18-101-final-viral-combined_k141_380778\|\|lt2gene | Low-quality |
| 18-101-final-viral-combined_k141_2857\|\|lt2gene | Low-quality |
| 18-101-final-viral-combined_k141_200063\|\|lt2gene | Not-determined |
| 18-101-final-viral-combined_k141_200073\|\|lt2gene | Low-quality |
| 18-101-final-viral-combined_k141_315185\|\|lt2gene | Not-determined |
| 18-101-final-viral-combined_k141_200089\|\|lt2gene | Low-quality |
| 18-101-final-viral-combined_k141_364494\|\|lt2gene | Low-quality |
| 18-101-final-viral-combined_k141_134589\|\|lt2gene | Low-quality |
| 18-101-final-viral-combined_k141_51446\|\|lt2gene | Not-determined |
| 18-101-final-viral-combined_k141_150444\|\|lt2gene | Not-determined |
| 18-101-final-viral-combined_k141_331325\|\|lt2gene | Low-quality |
| 18-101-final-viral-combined_k141_183595\|\|lt2gene | Low-quality |
| 18-101-final-viral-combined_k141_183601\|\|lt2gene | Low-quality |
| 18-101-final-viral-combined_k141_249485\|\|lt2gene | Not-determined |
| 18-101-final-viral-combined_k141_134626\|\|lt2gene | Low-quality |
| 18-101-final-viral-combined_k141_51544\|\|lt2gene | Low-quality |
| 18-101-final-viral-combined_k141_364578\|\|lt2gene | Not-determined |
| 18-101-final-viral-combined_k141_364598\|\|lt2gene | Low-quality |
| 18-101-final-viral-combined_k141_249534\|\|lt2gene | Low-quality |
| 18-101-final-viral-combined_k141_35489\|\|lt2gene | Low-quality |
| 18-101-final-viral-combined_k141_67883\|\|lt2gene | Low-quality |
| 18-101-final-viral-combined_k141_35493\|\|lt2gene | Low-quality |
| 18-101-final-viral-combined_k141_249564\|\|lt2gene | Low-quality |
| 18-101-final-viral-combined_k141_216876\|\|lt2gene | Low-quality |
| 18-101-final-viral-combined_k141_331427\|\|lt2gene | Not-determined |
| 18-101-final-viral-combined_k141_348211\|\|lt2gene | Not-determined |
| 18-101-final-viral-combined_k141_18985\|\|lt2gene | Low-quality |
| 18-101-final-viral-combined_k141_265434\|\|lt2gene | Low-quality |
| 18-101-final-viral-combined_k141_265445\|\|lt2gene | Low-quality |
| 18-101-final-viral-combined_k141_348221\|\|lt2gene | Not-determined |
| 18-101-final-viral-combined_k141_3005\|\|lt2gene | Low-quality |
| 18-101-final-viral-combined_k141_150632\|\|lt2gene | Low-quality |
| 18-101-final-viral-combined_k141_331533\|\|lt2gene | Low-quality |
| 18-101-final-viral-combined_k141_183843\|\|lt2gene | Low-quality |
| 18-101-final-viral-combined_k141_249752\|\|lt2gene | Low-quality |
| 18-101-final-viral-combined_k141_19089\|\|lt2gene | Low-quality |
| 18-101-final-viral-combined_k141_249787\|\|lt2gene | Low-quality |
| 18-101-final-viral-combined_k141_299189\|\|lt2gene | Low-quality |
| 18-101-final-viral-combined_k141_150779\|\|lt2gene | Low-quality |
| 18-101-final-viral-combined_k141_150781\|\|lt2gene | Low-quality |
| 18-101-final-viral-combined_k141_364774\|\|lt2gene | Not-determined |
| 18-101-final-viral-combined_k141_150810\|\|lt2gene | Low-quality |
| 18-101-final-viral-combined_k141_299231\|\|lt2gene | Low-quality |
| 18-101-final-viral-combined_k141_265559\|\|lt2gene | Low-quality |
| 18-101-final-viral-combined_k141_183947\|\|lt2gene | Low-quality |
| 18-101-final-viral-combined_k141_183950\|\|lt2gene | Not-determined |
| 18-101-final-viral-combined_k141_233096\|\|lt2gene | Low-quality |
| 18-101-final-viral-combined_k141_85179\|\|lt2gene | Low-quality |
| 18-101-final-viral-combined_k141_265579\|\|lt2gene | Low-quality |
| 18-101-final-viral-combined_k141_315527\|\|lt2gene | Low-quality |
| 18-101-final-viral-combined_k141_134905\|\|lt2gene | Low-quality |
| 18-101-final-viral-combined_k141_299305\|\|lt2gene | Low-quality |
| 18-101-final-viral-combined_k141_364896\|\|lt2gene | Low-quality |
| 18-101-final-viral-combined_k141_249910\|\|lt2gene | Not-determined |
| 18-101-final-viral-combined_k141_166760\|\|lt2gene | Low-quality |
| 18-101-final-viral-combined_k141_150912\|\|lt2gene | Low-quality |
| 18-101-final-viral-combined_k141_200380\|\|lt2gene | Low-quality |
| 18-101-final-viral-combined_k141_364949\|\|lt2gene | Low-quality |
| 18-101-final-viral-combined_k141_364952\|\|lt2gene | Low-quality |
| 18-101-final-viral-combined_k141_68153\|\|lt2gene | Low-quality |
| 18-101-final-viral-combined_k141_150924\|\|lt2gene | Low-quality |
| 18-101-final-viral-combined_k141_331605\|\|lt2gene | Low-quality |
| 18-101-final-viral-combined_k141_150948\|\|lt2gene | Low-quality |
| 18-101-final-viral-combined_k141_249968\|\|lt2gene | Low-quality |
| 18-101-final-viral-combined_k141_184180\|\|lt2gene | Not-determined |
| 18-101-final-viral-combined_k141_381167\|\|lt2gene | Low-quality |
| 18-101-final-viral-combined_k141_19314\|\|lt2gene | Low-quality |
| 18-101-final-viral-combined_k141_282757\|\|lt2gene | Low-quality |
| 18-101-final-viral-combined_k141_51903\|\|lt2gene | Not-determined |
| 18-101-final-viral-combined_k141_265762\|\|lt2gene | Low-quality |
| 18-101-final-viral-combined_k141_35885\|\|lt2gene | Not-determined |
| 18-101-final-viral-combined_k141_299521\|\|lt2gene | Low-quality |
| 18-101-final-viral-combined_k141_217187\|\|lt2gene | Low-quality |
| 18-101-final-viral-combined_k141_35946\|\|lt2gene | Not-determined |
| 18-101-final-viral-combined_k141_365097\|\|lt2gene | Not-determined |
| 18-101-final-viral-combined_k141_315724\|\|lt2gene | Low-quality |
| 18-101-final-viral-combined_k141_299575\|\|lt2gene | Not-determined |
| 18-101-final-viral-combined_k141_315753\|\|lt2gene | Low-quality |
| 18-101-final-viral-combined_k141_200516\|\|lt2gene | Not-determined |
| 18-101-final-viral-combined_k141_217284\|\|lt2gene | Low-quality |
| 18-101-final-viral-combined_k141_35978\|\|lt2gene | Low-quality |
| 18-101-final-viral-combined_k141_217296\|\|lt2gene | Low-quality |
| 18-101-final-viral-combined_k141_217300\|\|lt2gene | Not-determined |
| 18-101-final-viral-combined_k141_3222\|\|lt2gene | Not-determined |
| 18-101-final-viral-combined_k141_184342\|\|lt2gene | Low-quality |
| 18-101-final-viral-combined_k141_3234\|\|lt2gene | Low-quality |
| 18-101-final-viral-combined_k141_166936\|\|lt2gene | Low-quality |
| 18-101-final-viral-combined_k141_282938\|\|lt2gene | Low-quality |
| 18-101-final-viral-combined_k141_282945\|\|lt2gene | Low-quality |
| 18-101-final-viral-combined_k141_250117\|\|lt2gene | Low-quality |
| 18-101-final-viral-combined_k141_217341\|\|lt2gene | Low-quality |
| 18-101-final-viral-combined_k141_331859\|\|lt2gene | Low-quality |
| 18-101-final-viral-combined_k141_282976\|\|lt2gene | Low-quality |
| 18-101-final-viral-combined_k141_299620\|\|lt2gene | Not-determined |
| 18-101-final-viral-combined_k141_250125\|\|lt2gene | Low-quality |
| 18-101-final-viral-combined_k141_166977\|\|lt2gene | Not-determined |
| 18-101-final-viral-combined_k141_135188\|\|lt2gene | Low-quality |
| 18-101-final-viral-combined_k141_184448\|\|lt2gene | Not-determined |
| 18-101-final-viral-combined_k141_283039\|\|lt2gene | Low-quality |
| 18-101-final-viral-combined_k141_135232\|\|lt2gene | Low-quality |
| 18-101-final-viral-combined_k141_85314\|\|lt2gene | Low-quality |
| 18-101-final-viral-combined_k141_283041\|\|lt2gene | Not-determined |
| 18-101-final-viral-combined_k141_118455\|\|lt2gene | Low-quality |
| 18-101-final-viral-combined_k141_265943\|\|lt2gene | Low-quality |
| 18-101-final-viral-combined_k141_265954\|\|lt2gene | Low-quality |
| 18-101-final-viral-combined_k141_52264\|\|lt2gene | Low-quality |
| 18-101-final-viral-combined_k141_118516\|\|lt2gene | Not-determined |
| 18-101-final-viral-combined_k141_381514\|\|lt2gene | Not-determined |
| 18-101-final-viral-combined_k141_217523\|\|lt2gene | Not-determined |
| 18-101-final-viral-combined_k141_167169\|\|lt2gene | Low-quality |
| 18-101-final-viral-combined_k141_151290\|\|lt2gene | Low-quality |
| 18-101-final-viral-combined_k141_85444\|\|lt2gene | Low-quality |
| 18-101-final-viral-combined_k141_283146\|\|lt2gene | Low-quality |
| 18-101-final-viral-combined_k141_299799\|\|lt2gene | Not-determined |
| 18-101-final-viral-combined_k141_68492\|\|lt2gene | Low-quality |
| 18-101-final-viral-combined_k141_332078\|\|lt2gene | Low-quality |
| 18-101-final-viral-combined_k141_167205\|\|lt2gene | Low-quality |
| 18-101-final-viral-combined_k141_266086\|\|lt2gene | Low-quality |
| 18-101-final-viral-combined_k141_283192\|\|lt2gene | Low-quality |
| 18-101-final-viral-combined_k141_151337\|\|lt2gene | Low-quality |
| 18-101-final-viral-combined_k141_332107\|\|lt2gene | Low-quality |
| 18-101-final-viral-combined_k141_283217\|\|lt2gene | Low-quality |
| 18-101-final-viral-combined_k141_19687\|\|lt2gene | Not-determined |
| 18-101-final-viral-combined_k141_184688\|\|lt2gene | Not-determined |
| 18-101-final-viral-combined_k141_3419\|\|lt2gene | Low-quality |
| 18-101-final-viral-combined_k141_365426\|\|lt2gene | Low-quality |
| 18-101-final-viral-combined_k141_167241\|\|lt2gene | Not-determined |
| 18-101-final-viral-combined_k141_167247\|\|lt2gene | Not-determined |
| 18-101-final-viral-combined_k141_365441\|\|lt2gene | Low-quality |
| 18-101-final-viral-combined_k141_381630\|\|lt2gene | Not-determined |
| 18-101-final-viral-combined_k141_266196\|\|lt2gene | Low-quality |
| 18-101-final-viral-combined_k141_250422\|\|lt2gene | Not-determined |
| 18-101-final-viral-combined_k141_135522\|\|lt2gene | Low-quality |
| 18-101-final-viral-combined_k141_85511\|\|lt2gene | Low-quality |
| 18-101-final-viral-combined_k141_381669\|\|lt2gene | Low-quality |
| 18-101-final-viral-combined_k141_85524\|\|lt2gene | Low-quality |
| 18-101-final-viral-combined_k141_217583\|\|lt2gene | Low-quality |

18-101-final-viral-combined_k141_299910||lt2gene Not-determined
18-101-final-viral-combined_k141_118709||lt2gene Low-quality
18-101-final-viral-combined_k141_316222||lt2gene Low-quality
18-101-final-viral-combined_k141_332183||lt2gene Not-determined
18-101-final-viral-combined_k141_167259||lt2gene Not-determined
18-101-final-viral-combined_k141_200911||lt2gene Low-quality
18-101-final-viral-combined_k141_299959||lt2gene Low-quality
18-101-final-viral-combined_k141_381740||lt2gene Not-determined
18-101-final-viral-combined_k141_167288||lt2gene Low-quality
18-101-final-viral-combined_k141_299985||lt2gene Not-determined
18-101-final-viral-combined_k141_283351||lt2gene Low-quality
18-101-final-viral-combined_k141_151566||lt2gene Low-quality
18-101-final-viral-combined_k141_300012||lt2gene Low-quality
18-101-final-viral-combined_k141_365625||lt2gene Low-quality
18-101-final-viral-combined_k141_184879||lt2gene Low-quality
18-101-final-viral-combined_k141_3598||lt2gene Low-quality
18-101-final-viral-combined_k141_349201||lt2gene Not-determined
18-101-final-viral-combined_k141_332262||lt2gene Not-determined
18-101-final-viral-combined_k141_349214||lt2gene Low-quality
18-101-final-viral-combined_k141_19902||lt2gene Low-quality
18-101-final-viral-combined_k141_250614||lt2gene Low-quality
18-101-final-viral-combined_k141_300082||lt2gene Low-quality
18-101-final-viral-combined_k141_52524||lt2gene Low-quality
18-101-final-viral-combined_k141_283402||lt2gene Low-quality
18-101-final-viral-combined_k141_3641||lt2gene Low-quality
18-101-final-viral-combined_k141_85629||lt2gene Low-quality
18-101-final-viral-combined_k141_167413||lt2gene Not-determined
18-101-final-viral-combined_k141_250674||lt2gene Low-quality
18-101-final-viral-combined_k141_201024||lt2gene Not-determined
18-101-final-viral-combined_k141_118887||lt2gene Not-determined
18-101-final-viral-combined_k141_85667||lt2gene Low-quality
18-101-final-viral-combined_k141_101859||lt2gene Not-determined
18-101-final-viral-combined_k141_266522||lt2gene Not-determined
18-101-final-viral-combined_k141_300216||lt2gene Low-quality
18-101-final-viral-combined_k141_332377||lt2gene Not-determined
18-101-final-viral-combined_k141_349273||lt2gene Low-quality
18-101-final-viral-combined_k141_266560||lt2gene Low-quality
18-101-final-viral-combined_k141_300325||lt2gene Low-quality
18-101-final-viral-combined_k141_381978||lt2gene Low-quality
18-101-final-viral-combined_k141_300347||lt2gene Not-determined
18-101-final-viral-combined_k141_20085||lt2gene Low-quality
18-101-final-viral-combined_k141_201190||lt2gene Low-quality
18-101-final-viral-combined_k141_167541||lt2gene Low-quality
18-101-final-viral-combined_k141_349369||lt2gene Low-quality
18-101-final-viral-combined_k141_20116||lt2gene Low-quality
18-101-final-viral-combined_k141_151797||lt2gene Low-quality
18-101-final-viral-combined_k141_316708||lt2gene Low-quality
18-101-final-viral-combined_k141_250859||lt2gene Low-quality
18-101-final-viral-combined_k141_185079||lt2gene Not-determined
18-101-final-viral-combined_k141_185086||lt2gene Low-quality
18-101-final-viral-combined_k141_20169||lt2gene Not-determined
18-101-final-viral-combined_k141_382056||lt2gene Not-determined
18-101-final-viral-combined_k141_332584||lt2gene Not-determined
18-101-final-viral-combined_k141_151839||lt2gene Low-quality
18-101-final-viral-combined_k141_167609||lt2gene Low-quality
18-101-final-viral-combined_k141_316814||lt2gene Low-quality
18-101-final-viral-combined_k141_119111||lt2gene Low-quality
18-101-final-viral-combined_k141_85873||lt2gene Not-determined
18-101-final-viral-combined_k141_201392||lt2gene Low-quality
18-101-final-viral-combined_k141_185206||lt2gene Low-quality
18-101-final-viral-combined_k141_201411||lt2gene Low-quality
18-101-final-viral-combined_k141_20227||lt2gene Not-determined
18-101-final-viral-combined_k141_167659||lt2gene Low-quality
18-101-final-viral-combined_k141_151984||lt2gene Low-quality
18-101-final-viral-combined_k141_20271||lt2gene Low-quality
18-101-final-viral-combined_k141_201439||lt2gene Low-quality
18-101-final-viral-combined_k141_266863||lt2gene Low-quality
18-101-final-viral-combined_k141_283752||lt2gene Low-quality
18-101-final-viral-combined_k141_68833||lt2gene Not-determined
18-101-final-viral-combined_k141_234242||lt2gene Low-quality
18-101-final-viral-combined_k141_234246||lt2gene Low-quality
18-101-final-viral-combined_k141_3891||lt2gene Low-quality
18-101-final-viral-combined_k141_36591||lt2gene Low-quality
18-101-final-viral-combined_k141_36592||lt2gene Low-quality
18-101-final-viral-combined_k141_283828||lt2gene Low-quality
18-101-final-viral-combined_k141_266937||lt2gene Not-determined
18-101-final-viral-combined_k141_3923||lt2gene Not-determined
18-101-final-viral-combined_k141_53013||lt2gene Not-determined
18-101-final-viral-combined_k141_234319||lt2gene Low-quality
18-101-final-viral-combined_k141_332818||lt2gene Low-quality
18-101-final-viral-combined_k141_349623||lt2gene Low-quality
18-101-final-viral-combined_k141_36671||lt2gene Not-determined
18-101-final-viral-combined_k141_68946||lt2gene Not-determined
18-101-final-viral-combined_k141_217915||lt2gene Not-determined
18-101-final-viral-combined_k141_317046||lt2gene Low-quality
18-101-final-viral-combined_k141_317070||lt2gene Low-quality
18-101-final-viral-combined_k141_234399||lt2gene Low-quality
18-101-final-viral-combined_k141_234454||lt2gene Not-determined
18-101-final-viral-combined_k141_283894||lt2gene Low-quality
18-101-final-viral-combined_k141_332894||lt2gene Low-quality
18-101-final-viral-combined_k141_283909||lt2gene Low-quality
18-101-final-viral-combined_k141_185511||lt2gene Low-quality
18-101-final-viral-combined_k141_234523||lt2gene Low-quality
18-101-final-viral-combined_k141_167867||lt2gene Low-quality
18-101-final-viral-combined_k141_251404||lt2gene Not-determined
18-101-final-viral-combined_k141_332966||lt2gene Low-quality
18-101-final-viral-combined_k141_251484||lt2gene Not-determined
18-101-final-viral-combined_k141_167952||lt2gene Not-determined
18-101-final-viral-combined_k141_332994||lt2gene Low-quality
18-101-final-viral-combined_k141_218125||lt2gene Low-quality
18-101-final-viral-combined_k141_86186||lt2gene Low-quality
18-101-final-viral-combined_k141_267260||lt2gene Low-quality
18-101-final-viral-combined_k141_350001||lt2gene Low-quality
18-101-final-viral-combined_k141_218172||lt2gene Not-determined
18-101-final-viral-combined_k141_168024||lt2gene Low-quality
18-101-final-viral-combined_k141_350007||lt2gene Low-quality
18-101-final-viral-combined_k141_333059||lt2gene Low-quality
18-101-final-viral-combined_k141_136174||lt2gene Low-quality
18-101-final-viral-combined_k141_136187||lt2gene Low-quality
18-101-final-viral-combined_k141_119637||lt2gene Not-determined
18-101-final-viral-combined_k141_168077||lt2gene Not-determined
18-101-final-viral-combined_k141_185675||lt2gene Not-determined
18-101-final-viral-combined_k141_152305||lt2gene Not-determined
18-101-final-viral-combined_k141_86241||lt2gene Low-quality
18-101-final-viral-combined_k141_366169||lt2gene Low-quality
18-101-final-viral-combined_k141_53240||lt2gene Low-quality
18-101-final-viral-combined_k141_317310||lt2gene Low-quality
18-101-final-viral-combined_k141_185733||lt2gene Low-quality
18-101-final-viral-combined_k141_267450||lt2gene Low-quality
18-101-final-viral-combined_k141_152335||lt2gene Low-quality
18-101-final-viral-combined_k141_185804||lt2gene Not-determined
18-101-final-viral-combined_k141_251700||lt2gene Not-determined
18-101-final-viral-combined_k141_284203||lt2gene Not-determined
18-101-final-viral-combined_k141_20811||lt2gene Low-quality
18-101-final-viral-combined_k141_284226||lt2gene Low-quality
18-101-final-viral-combined_k141_53379||lt2gene Low-quality
18-101-final-viral-combined_k141_168164||lt2gene Low-quality
18-101-final-viral-combined_k141_37139||lt2gene Low-quality
18-101-final-viral-combined_k141_37146||lt2gene Not-determined
18-101-final-viral-combined_k141_37171||lt2gene Low-quality
18-101-final-viral-combined_k141_69250||lt2gene Low-quality
18-101-final-viral-combined_k141_119907||lt2gene Low-quality
18-101-final-viral-combined_k141_382663||lt2gene Low-quality
18-101-final-viral-combined_k141_234768||lt2gene Low-quality
18-101-final-viral-combined_k141_218209||lt2gene Low-quality
18-101-final-viral-combined_k141_86434||lt2gene Low-quality
18-101-final-viral-combined_k141_152458||lt2gene Low-quality
18-101-final-viral-combined_k141_102592||lt2gene Low-quality
18-101-final-viral-combined_k141_301293||lt2gene Not-determined
18-101-final-viral-combined_k141_53566||lt2gene Not-determined
18-101-final-viral-combined_k141_333376||lt2gene Low-quality
18-101-final-viral-combined_k141_350376||lt2gene Not-determined
18-101-final-viral-combined_k141_251782||lt2gene Not-determined
18-101-final-viral-combined_k141_234793||lt2gene Not-determined
18-101-final-viral-combined_k141_301390||lt2gene Low-quality
18-101-final-viral-combined_k141_382792||lt2gene Not-determined
18-101-final-viral-combined_k141_284422||lt2gene Low-quality
18-101-final-viral-combined_k141_317633||lt2gene Low-quality
18-101-final-viral-combined_k141_284440||lt2gene Not-determined
18-101-final-viral-combined_k141_317641||lt2gene Low-quality
18-101-final-viral-combined_k141_234853||lt2gene Not-determined
18-101-final-viral-combined_k141_382834||lt2gene Low-quality
18-101-final-viral-combined_k141_53671||lt2gene Not-determined
18-101-final-viral-combined_k141_350428||lt2gene Low-quality
18-101-final-viral-combined_k141_86550||lt2gene Low-quality
18-101-final-viral-combined_k141_366524||lt2gene Low-quality
18-101-final-viral-combined_k141_21153||lt2gene Low-quality
18-101-final-viral-combined_k141_202169||lt2gene Not-determined
18-101-final-viral-combined_k141_301485||lt2gene Not-determined
18-101-final-viral-combined_k141_235005||lt2gene Not-determined
18-101-final-viral-combined_k141_21210||lt2gene Low-quality
18-101-final-viral-combined_k141_21221||lt2gene Low-quality
18-101-final-viral-combined_k141_86594||lt2gene Low-quality
18-101-final-viral-combined_k141_350641||lt2gene Not-determined
18-101-final-viral-combined_k141_301569||lt2gene Low-quality
18-101-final-viral-combined_k141_152783||lt2gene Low-quality
18-101-final-viral-combined_k141_252045||lt2gene Low-quality
18-101-final-viral-combined_k141_120347||lt2gene Not-determined
18-101-final-viral-combined_k141_168652||lt2gene Low-quality
18-101-final-viral-combined_k141_4630||lt2gene Not-determined
18-101-final-viral-combined_k141_284764||lt2gene Low-quality
18-101-final-viral-combined_k141_136945||lt2gene Low-quality
18-101-final-viral-combined_k141_152893||lt2gene Low-quality
18-101-final-viral-combined_k141_120422||lt2gene Low-quality
18-101-final-viral-combined_k141_37587||lt2gene Low-quality
18-101-final-viral-combined_k141_37588||lt2gene Low-quality
18-101-final-viral-combined_k141_317973||lt2gene Low-quality
18-101-final-viral-combined_k141_37603||lt2gene Low-quality
18-101-final-viral-combined_k141_168797||lt2gene Not-determined
18-101-final-viral-combined_k141_252152||lt2gene Not-determined
18-101-final-viral-combined_k141_103015||lt2gene Not-determined
18-101-final-viral-combined_k141_186209||lt2gene Low-quality
18-101-final-viral-combined_k141_301786||lt2gene Not-determined
18-101-final-viral-combined_k141_366824||lt2gene Not-determined
18-101-final-viral-combined_k141_350822||lt2gene Low-quality
18-101-final-viral-combined_k141_383391||lt2gene Not-determined
18-101-final-viral-combined_k141_252305||lt2gene Not-determined
18-101-final-viral-combined_k141_252314||lt2gene Low-quality
18-101-final-viral-combined_k141_53980||lt2gene Low-quality
18-101-final-viral-combined_k141_202582||lt2gene Low-quality
18-101-final-viral-combined_k141_53992||lt2gene Low-quality
18-101-final-viral-combined_k141_53997||lt2gene Low-quality
18-101-final-viral-combined_k141_4853||lt2gene Not-determined
18-101-final-viral-combined_k141_69846||lt2gene Not-determined
18-101-final-viral-combined_k141_54001||lt2gene Low-quality
18-101-final-viral-combined_k141_137142||lt2gene Low-quality
18-101-final-viral-combined_k141_202627||lt2gene Low-quality
18-101-final-viral-combined_k141_218929||lt2gene Not-determined
18-101-final-viral-combined_k141_350947||lt2gene Low-quality
18-101-final-viral-combined_k141_202639||lt2gene Low-quality
18-101-final-viral-combined_k141_235399||lt2gene Not-determined
18-101-final-viral-combined_k141_366985||lt2gene Low-quality
18-101-final-viral-combined_k141_383483||lt2gene Not-determined
18-101-final-viral-combined_k141_235432||lt2gene Not-determined
18-101-final-viral-combined_k141_284833||lt2gene Low-quality
18-101-final-viral-combined_k141_268095||lt2gene Low-quality
18-101-final-viral-combined_k141_5016||lt2gene Not-determined
18-101-final-viral-combined_k141_351058||lt2gene Low-quality
18-101-final-viral-combined_k141_21457||lt2gene Low-quality
18-101-final-viral-combined_k141_219068||lt2gene Low-quality
18-101-final-viral-combined_k141_367148||lt2gene Low-quality
18-101-final-viral-combined_k141_383607||lt2gene Low-quality
18-101-final-viral-combined_k141_69933||lt2gene Low-quality
18-101-final-viral-combined_k141_120872||lt2gene Low-quality
18-101-final-viral-combined_k141_219127||lt2gene Low-quality
18-101-final-viral-combined_k141_5083||lt2gene Low-quality
18-101-final-viral-combined_k141_69986||lt2gene Low-quality
18-101-final-viral-combined_k141_383702||lt2gene Not-determined
18-101-final-viral-combined_k141_268269||lt2gene Not-determined
18-101-final-viral-combined_k141_333869||lt2gene Low-quality
18-101-final-viral-combined_k141_120943||lt2gene Low-quality
18-101-final-viral-combined_k141_351213||lt2gene Low-quality
18-101-final-viral-combined_k141_70010||lt2gene Low-quality
18-101-final-viral-combined_k141_202841||lt2gene Not-determined
18-101-final-viral-combined_k141_202847||lt2gene Low-quality
18-101-final-viral-combined_k141_120988||lt2gene Low-quality
18-101-final-viral-combined_k141_38072||lt2gene Not-determined
18-101-final-viral-combined_k141_186603||lt2gene Low-quality
18-101-final-viral-combined_k141_38086||lt2gene Low-quality
18-101-final-viral-combined_k141_169156||lt2gene Low-quality
18-101-final-viral-combined_k141_70085||lt2gene Low-quality
18-101-final-viral-combined_k141_54343||lt2gene Not-determined
18-101-final-viral-combined_k141_169227||lt2gene Low-quality
18-101-final-viral-combined_k141_153409||lt2gene Low-quality
18-101-final-viral-combined_k141_137537||lt2gene Low-quality
18-101-final-viral-combined_k141_54356||lt2gene Low-quality
18-101-final-viral-combined_k141_54360||lt2gene Low-quality
18-101-final-viral-combined_k141_186734||lt2gene Not-determined
18-101-final-viral-combined_k141_5239||lt2gene Low-quality
18-101-final-viral-combined_k141_318597||lt2gene Low-quality
18-101-final-viral-combined_k141_70128||lt2gene Low-quality
18-101-final-viral-combined_k141_202967||lt2gene Low-quality
18-101-final-viral-combined_k141_186766||lt2gene Low-quality
18-101-final-viral-combined_k141_302286||lt2gene Low-quality
18-101-final-viral-combined_k141_87242||lt2gene Low-quality
18-101-final-viral-combined_k141_235762||lt2gene Low-quality
18-101-final-viral-combined_k141_186808||lt2gene Low-quality
18-101-final-viral-combined_k141_302302||lt2gene Not-determined
18-101-final-viral-combined_k141_87256||lt2gene Low-quality
18-101-final-viral-combined_k141_318627||lt2gene Not-determined
18-101-final-viral-combined_k141_351405||lt2gene Low-quality
18-101-final-viral-combined_k141_383829||lt2gene Not-determined
18-101-final-viral-combined_k141_318657||lt2gene Low-quality
18-101-final-viral-combined_k141_87278||lt2gene Low-quality
18-101-final-viral-combined_k141_302329||lt2gene Low-quality
18-101-final-viral-combined_k141_70198||lt2gene Not-determined
18-101-final-viral-combined_k141_87343||lt2gene Low-quality
18-101-final-viral-combined_k141_219509||lt2gene Low-quality
18-101-final-viral-combined_k141_70236||lt2gene Low-quality
18-101-final-viral-combined_k141_252829||lt2gene Not-determined
18-101-final-viral-combined_k141_268556||lt2gene Low-quality
18-101-final-viral-combined_k141_285371||lt2gene Not-determined
18-101-final-viral-combined_k141_252875||lt2gene Low-quality
18-101-final-viral-combined_k141_219564||lt2gene Low-quality
18-101-final-viral-combined_k141_268576||lt2gene Low-quality
18-101-final-viral-combined_k141_169388||lt2gene Not-determined
18-101-final-viral-combined_k141_169392||lt2gene Not-determined
18-101-final-viral-combined_k141_334183||lt2gene Low-quality
18-101-final-viral-combined_k141_54572||lt2gene Low-quality
18-101-final-viral-combined_k141_70395||lt2gene Low-quality
18-101-final-viral-combined_k141_187084||lt2gene Low-quality
18-101-final-viral-combined_k141_285421||lt2gene Low-quality
18-101-final-viral-combined_k141_367681||lt2gene Low-quality
18-101-final-viral-combined_k141_169434||lt2gene Low-quality
18-101-final-viral-combined_k141_285452||lt2gene Low-quality
18-101-final-viral-combined_k141_5434||lt2gene Low-quality
18-101-final-viral-combined_k141_169446||lt2gene Low-quality
18-101-final-viral-combined_k141_367715||lt2gene Low-quality
18-101-final-viral-combined_k141_367718||lt2gene Low-quality
18-101-final-viral-combined_k141_367720||lt2gene Low-quality
18-101-final-viral-combined_k141_367721||lt2gene Low-quality
18-101-final-viral-combined_k141_169453||lt2gene Low-quality
18-101-final-viral-combined_k141_103506||lt2gene Low-quality
18-101-final-viral-combined_k141_302614||lt2gene Not-determined
18-101-final-viral-combined_k141_103508||lt2gene Not-determined
18-101-final-viral-combined_k141_38410||lt2gene Low-quality
18-101-final-viral-combined_k141_302641||lt2gene Low-quality
18-101-final-viral-combined_k141_367832||lt2gene Low-quality
18-101-final-viral-combined_k141_203385||lt2gene Low-quality
18-101-final-viral-combined_k141_5538||lt2gene Low-quality
18-101-final-viral-combined_k141_302675||lt2gene Low-quality
18-101-final-viral-combined_k141_285548||lt2gene Not-determined
18-101-final-viral-combined_k141_384171||lt2gene Low-quality
18-101-final-viral-combined_k141_38515||lt2gene Low-quality
18-101-final-viral-combined_k141_169564||lt2gene Not-determined
18-101-final-viral-combined_k141_121400||lt2gene Low-quality
18-101-final-viral-combined_k141_219801||lt2gene Low-quality
18-101-final-viral-combined_k141_219842||lt2gene Not-determined
18-101-final-viral-combined_k141_384286||lt2gene Low-quality
18-101-final-viral-combined_k141_318978||lt2gene Low-quality
18-101-final-viral-combined_k141_302821||lt2gene Low-quality
18-101-final-viral-combined_k141_137939||lt2gene Not-determined
18-101-final-viral-combined_k141_334333||lt2gene Low-quality
18-101-final-viral-combined_k141_121519||lt2gene Low-quality
18-101-final-viral-combined_k141_384357||lt2gene Low-quality
18-101-final-viral-combined_k141_368050||lt2gene Not-determined
18-101-final-viral-combined_k141_236275||lt2gene Low-quality
18-101-final-viral-combined_k141_121579||lt2gene Low-quality
18-101-final-viral-combined_k141_319129||lt2gene Not-determined
18-101-final-viral-combined_k141_334403||lt2gene Not-determined
18-101-final-viral-combined_k141_253216||lt2gene Low-quality
18-101-final-viral-combined_k141_236393||lt2gene Low-quality
18-101-final-viral-combined_k141_153857||lt2gene Low-quality
18-101-final-viral-combined_k141_302921||lt2gene Not-determined
18-101-final-viral-combined_k141_253258||lt2gene Low-quality
18-101-final-viral-combined_k141_121748||lt2gene Not-determined
18-101-final-viral-combined_k141_38823||lt2gene Not-determined
18-101-final-viral-combined_k141_169845||lt2gene Low-quality
18-101-final-viral-combined_k141_203738||lt2gene Not-determined
18-101-final-viral-combined_k141_368150||lt2gene Low-quality
18-101-final-viral-combined_k141_352056||lt2gene Not-determined
18-101-final-viral-combined_k141_38863||lt2gene Low-quality
18-101-final-viral-combined_k141_269096||lt2gene Low-quality
18-101-final-viral-combined_k141_103977||lt2gene Not-determined
18-101-final-viral-combined_k141_55113||lt2gene Not-determined
18-101-final-viral-combined_k141_138184||lt2gene Not-determined
18-101-final-viral-combined_k141_138199||lt2gene Low-quality
18-101-final-viral-combined_k141_220113||lt2gene Not-determined
18-101-final-viral-combined_k141_38939||lt2gene Not-determined
18-101-final-viral-combined_k141_334629||lt2gene Low-quality
18-101-final-viral-combined_k141_334633||lt2gene Low-quality
18-101-final-viral-combined_k141_169953||lt2gene Low-quality
18-101-final-viral-combined_k141_384701||lt2gene Low-quality
18-101-final-viral-combined_k141_319417||lt2gene Low-quality
18-101-final-viral-combined_k141_71061||lt2gene Low-quality
18-101-final-viral-combined_k141_384707||lt2gene Not-determined
18-101-final-viral-combined_k141_384710||lt2gene Low-quality
18-101-final-viral-combined_k141_38982||lt2gene Not-determined
18-101-final-viral-combined_k141_203884||lt2gene Low-quality
18-101-final-viral-combined_k141_154027||lt2gene Low-quality
18-101-final-viral-combined_k141_88041||lt2gene Not-determined
18-101-final-viral-combined_k141_22342||lt2gene Low-quality
18-101-final-viral-combined_k141_121928||lt2gene Not-determined
18-101-final-viral-combined_k141_88060||lt2gene Low-quality
18-101-final-viral-combined_k141_88074||lt2gene Low-quality
18-101-final-viral-combined_k141_187745||lt2gene Low-quality
18-101-final-viral-combined_k141_6031||lt2gene Low-quality
18-101-final-viral-combined_k141_220195||lt2gene Low-quality
18-101-final-viral-combined_k141_6065||lt2gene Low-quality
18-101-final-viral-combined_k141_6068||lt2gene Low-quality
18-101-final-viral-combined_k141_6100||lt2gene Low-quality
18-101-final-viral-combined_k141_253637||lt2gene Low-quality
18-101-final-viral-combined_k141_204033||lt2gene Low-quality
18-101-final-viral-combined_k141_286083||lt2gene Not-determined
18-101-final-viral-combined_k141_104277||lt2gene Low-quality
18-101-final-viral-combined_k141_88222||lt2gene Low-quality
18-101-final-viral-combined_k141_334843||lt2gene Low-quality
18-101-final-viral-combined_k141_253756||lt2gene Low-quality
18-101-final-viral-combined_k141_22543||lt2gene Low-quality
18-101-final-viral-combined_k141_352411||lt2gene Low-quality
18-101-final-viral-combined_k141_253769||lt2gene Not-determined
18-101-final-viral-combined_k141_286120||lt2gene Not-determined
18-101-final-viral-combined_k141_55536||lt2gene Low-quality
18-101-final-viral-combined_k141_22572||lt2gene Low-quality
18-101-final-viral-combined_k141_55540||lt2gene Low-quality
18-101-final-viral-combined_k141_204140||lt2gene Low-quality
18-101-final-viral-combined_k141_88274||lt2gene Low-quality
18-101-final-viral-combined_k141_88276||lt2gene Low-quality
18-101-final-viral-combined_k141_122176||lt2gene Low-quality
18-101-final-viral-combined_k141_269467||lt2gene Not-determined
18-101-final-viral-combined_k141_170203||lt2gene Low-quality
18-101-final-viral-combined_k141_55594||lt2gene Low-quality
18-101-final-viral-combined_k141_55597||lt2gene Low-quality
18-101-final-viral-combined_k141_236805||lt2gene Low-quality
18-101-final-viral-combined_k141_384978||lt2gene Not-determined
18-101-final-viral-combined_k141_39230||lt2gene Low-quality
18-101-final-viral-combined_k141_122299||lt2gene Low-quality
18-101-final-viral-combined_k141_236843||lt2gene Low-quality
18-101-final-viral-combined_k141_6331||lt2gene Low-quality
18-101-final-viral-combined_k141_368260||lt2gene Low-quality
18-101-final-viral-combined_k141_122322||lt2gene Low-quality
18-101-final-viral-combined_k141_22699||lt2gene Low-quality
18-101-final-viral-combined_k141_122348||lt2gene Low-quality
18-101-final-viral-combined_k141_22740||lt2gene Low-quality
18-101-final-viral-combined_k141_39268||lt2gene Low-quality
18-101-final-viral-combined_k141_170295||lt2gene Not-determined
18-101-final-viral-combined_k141_237010||lt2gene Low-quality
18-101-final-viral-combined_k141_368328||lt2gene Low-quality
18-101-final-viral-combined_k141_368334||lt2gene Low-quality
18-101-final-viral-combined_k141_253984||lt2gene Low-quality
18-101-final-viral-combined_k141_22800||lt2gene Not-determined
18-101-final-viral-combined_k141_55732||lt2gene Low-quality
18-101-final-viral-combined_k141_269629||lt2gene Low-quality
18-101-final-viral-combined_k141_303382||lt2gene Not-determined
18-101-final-viral-combined_k141_220497||lt2gene Low-quality
18-101-final-viral-combined_k141_335152||lt2gene Low-quality
18-101-final-viral-combined_k141_39412||lt2gene Low-quality
18-101-final-viral-combined_k141_319942||lt2gene Low-quality
18-101-final-viral-combined_k141_170397||lt2gene Low-quality
18-101-final-viral-combined_k141_385249||lt2gene Not-determined
18-101-final-viral-combined_k141_88613||lt2gene Not-determined
18-101-final-viral-combined_k141_319963||lt2gene Low-quality
18-101-final-viral-combined_k141_6575||lt2gene Not-determined
18-101-final-viral-combined_k141_104589||lt2gene Low-quality
18-101-final-viral-combined_k141_104590||lt2gene Not-determined
18-101-final-viral-combined_k141_104591||lt2gene Low-quality
18-101-final-viral-combined_k141_71684||lt2gene Not-determined
18-101-final-viral-combined_k141_122597||lt2gene Not-determined
18-101-final-viral-combined_k141_254160||lt2gene Low-quality
18-101-final-viral-combined_k141_71703||lt2gene Low-quality
18-101-final-viral-combined_k141_22938||lt2gene Low-quality
18-101-final-viral-combined_k141_138755||lt2gene Low-quality
18-101-final-viral-combined_k141_352894||lt2gene Low-quality
18-101-final-viral-combined_k141_6658||lt2gene Low-quality
18-101-final-viral-combined_k141_71752||lt2gene Low-quality
18-101-final-viral-combined_k141_154513||lt2gene Low-quality
18-101-final-viral-combined_k141_39539||lt2gene Low-quality
18-101-final-viral-combined_k141_269846||lt2gene Low-quality
18-101-final-viral-combined_k141_138804||lt2gene Low-quality
18-101-final-viral-combined_k141_188243||lt2gene Low-quality
18-101-final-viral-combined_k141_204683||lt2gene Low-quality
18-101-final-viral-combined_k141_204686||lt2gene Not-determined
18-101-final-viral-combined_k141_368584||lt2gene Not-determined
18-101-final-viral-combined_k141_188301||lt2gene Low-quality
18-101-final-viral-combined_k141_170507||lt2gene Low-quality
18-101-final-viral-combined_k141_254308||lt2gene Not-determined
18-101-final-viral-combined_k141_303665||lt2gene Low-quality
18-101-final-viral-combined_k141_104723||lt2gene Low-quality
18-101-final-viral-combined_k141_204753||lt2gene Not-determined
18-101-final-viral-combined_k141_237361||lt2gene Not-determined
18-101-final-viral-combined_k141_320197||lt2gene Not-determined
18-101-final-viral-combined_k141_204775||lt2gene Low-quality
18-101-final-viral-combined_k141_170567||lt2gene Low-quality
18-101-final-viral-combined_k141_286489||lt2gene Not-determined
18-101-final-viral-combined_k141_286513||lt2gene Low-quality
18-101-final-viral-combined_k141_23147||lt2gene Not-determined
18-101-final-viral-combined_k141_368708||lt2gene Low-quality
18-101-final-viral-combined_k141_71894||lt2gene Low-quality
18-101-final-viral-combined_k141_254354||lt2gene Not-determined
18-101-final-viral-combined_k141_254358||lt2gene Low-quality
18-101-final-viral-combined_k141_254359||lt2gene Not-determined
18-101-final-viral-combined_k141_303773||lt2gene Low-quality
18-101-final-viral-combined_k141_71935||lt2gene Not-determined
18-101-final-viral-combined_k141_335404||lt2gene Low-quality
18-101-final-viral-combined_k141_71943||lt2gene Low-quality
18-101-final-viral-combined_k141_138906||lt2gene Low-quality
18-101-final-viral-combined_k141_303814||lt2gene Not-determined
18-101-final-viral-combined_k141_303815||lt2gene Not-determined
18-101-final-viral-combined_k141_39637||lt2gene Not-determined
18-101-final-viral-combined_k141_320338||lt2gene Low-quality
18-101-final-viral-combined_k141_72003||lt2gene Low-quality
18-101-final-viral-combined_k141_385522||lt2gene Low-quality
18-101-final-viral-combined_k141_385554||lt2gene Low-quality
18-101-final-viral-combined_k141_270021||lt2gene Low-quality
18-101-final-viral-combined_k141_353200||lt2gene Not-determined
18-101-final-viral-combined_k141_23256||lt2gene Low-quality
18-101-final-viral-combined_k141_39685||lt2gene Low-quality
18-101-final-viral-combined_k141_6968||lt2gene Low-quality
18-101-final-viral-combined_k141_139031||lt2gene Low-quality
18-101-final-viral-combined_k141_335499||lt2gene Low-quality
18-101-final-viral-combined_k141_368841||lt2gene Low-quality
18-101-final-viral-combined_k141_139048||lt2gene Not-determined
18-101-final-viral-combined_k141_123028||lt2gene Low-quality
18-101-final-viral-combined_k141_89087||lt2gene Low-quality
18-101-final-viral-combined_k141_237636||lt2gene Low-quality
18-101-final-viral-combined_k141_139138||lt2gene Low-quality
18-101-final-viral-combined_k141_286795||lt2gene Low-quality
18-101-final-viral-combined_k141_72078||lt2gene Low-quality
18-101-final-viral-combined_k141_385595||lt2gene Low-quality
18-101-final-viral-combined_k141_23305||lt2gene Low-quality
18-101-final-viral-combined_k141_286817||lt2gene Low-quality
18-101-final-viral-combined_k141_237677||lt2gene Low-quality
18-101-final-viral-combined_k141_286836||lt2gene Not-determined
18-101-final-viral-combined_k141_220958||lt2gene Low-quality
18-101-final-viral-combined_k141_139260||lt2gene Not-determined
18-101-final-viral-combined_k141_270289||lt2gene Low-quality
18-101-final-viral-combined_k141_23454||lt2gene Not-determined
18-101-final-viral-combined_k141_72204||lt2gene Low-quality
18-101-final-viral-combined_k141_286967||lt2gene Low-quality
18-101-final-viral-combined_k141_89267||lt2gene Not-determined
18-101-final-viral-combined_k141_89276||lt2gene Low-quality
18-101-final-viral-combined_k141_139393||lt2gene Low-quality
18-101-final-viral-combined_k141_254751||lt2gene Low-quality
18-101-final-viral-combined_k141_123273||lt2gene Low-quality
18-101-final-viral-combined_k141_205186||lt2gene Not-determined
18-101-final-viral-combined_k141_89318||lt2gene Low-quality
18-101-final-viral-combined_k141_335778||lt2gene Low-quality
18-101-final-viral-combined_k141_188817||lt2gene Not-determined
18-101-final-viral-combined_k141_105045||lt2gene Low-quality
18-101-final-viral-combined_k141_188822||lt2gene Not-determined
18-101-final-viral-combined_k141_237917||lt2gene Low-quality
18-101-final-viral-combined_k141_56504||lt2gene Low-quality
18-101-final-viral-combined_k141_335809||lt2gene Not-determined
18-101-final-viral-combined_k141_39876||lt2gene Low-quality
18-101-final-viral-combined_k141_56527||lt2gene Low-quality
18-101-final-viral-combined_k141_254814||lt2gene Low-quality
18-101-final-viral-combined_k141_205260||lt2gene Low-quality
18-101-final-viral-combined_k141_72350||lt2gene Not-determined
18-101-final-viral-combined_k141_105086||lt2gene Low-quality
18-101-final-viral-combined_k141_237992||lt2gene Not-determined
18-101-final-viral-combined_k141_385861||lt2gene Low-quality
18-101-final-viral-combined_k141_89415||lt2gene Low-quality
18-101-final-viral-combined_k141_72393||lt2gene Low-quality
18-101-final-viral-combined_k141_221209||lt2gene Not-determined
18-101-final-viral-combined_k141_304214||lt2gene Low-quality
18-101-final-viral-combined_k141_23583||lt2gene Not-determined
18-101-final-viral-combined_k141_254854||lt2gene Low-quality
18-101-final-viral-combined_k141_188898||lt2gene Not-determined
18-101-final-viral-combined_k141_7316||lt2gene Low-quality
18-101-final-viral-combined_k141_353536||lt2gene Low-quality
18-101-final-viral-combined_k141_270448||lt2gene Not-determined
18-101-final-viral-combined_k141_270451||lt2gene Low-quality
18-101-final-viral-combined_k141_56604||lt2gene Low-quality
18-101-final-viral-combined_k141_155124||lt2gene Low-quality
18-101-final-viral-combined_k141_123440||lt2gene Not-determined
18-101-final-viral-combined_k141_221319||lt2gene Low-quality
18-101-final-viral-combined_k141_7409||lt2gene Low-quality
18-101-final-viral-combined_k141_89529||lt2gene Low-quality
18-101-final-viral-combined_k141_89533||lt2gene Not-determined
18-101-final-viral-combined_k141_353591||lt2gene Low-quality
18-101-final-viral-combined_k141_56651||lt2gene Low-quality
18-101-final-viral-combined_k141_72490||lt2gene Low-quality
18-101-final-viral-combined_k141_171238||lt2gene Low-quality
18-101-final-viral-combined_k141_386073||lt2gene Low-quality
18-101-final-viral-combined_k141_321202||lt2gene Low-quality
18-101-final-viral-combined_k141_171274||lt2gene Low-quality
18-101-final-viral-combined_k141_72524||lt2gene Low-quality
18-101-final-viral-combined_k141_221388||lt2gene Low-quality
18-101-final-viral-combined_k141_255030||lt2gene Not-determined
18-101-final-viral-combined_k141_238205||lt2gene Low-quality
18-101-final-viral-combined_k141_139618||lt2gene Low-quality
18-101-final-viral-combined_k141_155221||lt2gene Low-quality
18-101-final-viral-combined_k141_353733||lt2gene Low-quality
18-101-final-viral-combined_k141_221472||lt2gene Low-quality
18-101-final-viral-combined_k141_221476||lt2gene Not-determined
18-101-final-viral-combined_k141_7565||lt2gene Not-determined
18-101-final-viral-combined_k141_287217||lt2gene Not-determined
18-101-final-viral-combined_k141_287237||lt2gene Low-quality
18-101-final-viral-combined_k141_23717||lt2gene Low-quality
18-101-final-viral-combined_k141_7598||lt2gene Low-quality
18-101-final-viral-combined_k141_40214||lt2gene Low-quality
18-101-final-viral-combined_k141_189102||lt2gene Low-quality
18-101-final-viral-combined_k141_205607||lt2gene Low-quality
18-101-final-viral-combined_k141_270786||lt2gene Not-determined
18-101-final-viral-combined_k141_336052||lt2gene Low-quality
18-101-final-viral-combined_k141_205633||lt2gene Low-quality
18-101-final-viral-combined_k141_255271||lt2gene Low-quality
18-101-final-viral-combined_k141_89706||lt2gene Low-quality
18-101-final-viral-combined_k141_270857||lt2gene Low-quality
18-101-final-viral-combined_k141_56983||lt2gene Low-quality
18-101-final-viral-combined_k141_353831||lt2gene Low-quality
18-101-final-viral-combined_k141_205712||lt2gene Low-quality
18-101-final-viral-combined_k141_40257||lt2gene Low-quality
18-101-final-viral-combined_k141_270885||lt2gene Low-quality
18-101-final-viral-combined_k141_171502||lt2gene Low-quality
18-101-final-viral-combined_k141_369485||lt2gene Low-quality
18-101-final-viral-combined_k141_353875||lt2gene Low-quality
18-101-final-viral-combined_k141_353916||lt2gene Low-quality
18-101-final-viral-combined_k141_369513||lt2gene Low-quality
18-101-final-viral-combined_k141_89788||lt2gene Low-quality
18-101-final-viral-combined_k141_336196||lt2gene Low-quality
18-101-final-viral-combined_k141_336197||lt2gene Low-quality
18-101-final-viral-combined_k141_123849||lt2gene Low-quality
18-101-final-viral-combined_k141_40385||lt2gene Low-quality
18-101-final-viral-combined_k141_72822||lt2gene Not-determined
18-101-final-viral-combined_k141_321559||lt2gene Low-quality
18-101-final-viral-combined_k141_40394||lt2gene Low-quality
18-101-final-viral-combined_k141_221802||lt2gene Low-quality
18-101-final-viral-combined_k141_189417||lt2gene Not-determined
18-101-final-viral-combined_k141_72876||lt2gene Not-determined
18-101-final-viral-combined_k141_354022||lt2gene Low-quality
18-101-final-viral-combined_k141_238337||lt2gene Not-determined
18-101-final-viral-combined_k141_7908||lt2gene Low-quality
18-101-final-viral-combined_k141_40448||lt2gene Low-quality
18-101-final-viral-combined_k141_287528||lt2gene Low-quality
18-101-final-viral-combined_k141_72901||lt2gene Not-determined
18-101-final-viral-combined_k141_238374||lt2gene Low-quality
18-101-final-viral-combined_k141_271051||lt2gene Low-quality
18-101-final-viral-combined_k141_386506||lt2gene Low-quality
18-101-final-viral-combined_k141_7957||lt2gene Not-determined
18-101-final-viral-combined_k141_40500||lt2gene Low-quality
18-101-final-viral-combined_k141_321617||lt2gene Low-quality
18-101-final-viral-combined_k141_336311||lt2gene Low-quality
18-101-final-viral-combined_k141_369669||lt2gene Not-determined
18-101-final-viral-combined_k141_369676||lt2gene Not-determined
18-101-final-viral-combined_k141_238467||lt2gene Low-quality
18-101-final-viral-combined_k141_287617||lt2gene Low-quality
18-101-final-viral-combined_k141_171653||lt2gene Low-quality
18-101-final-viral-combined_k141_205978||lt2gene Low-quality

| | |
|---|---|
| 18-101-final-viral-combined_k141_189580|||lt2gene | Low-quality |
| 18-101-final-viral-combined_k141_238519|||lt2gene | Low-quality |
| 18-101-final-viral-combined_k141_321725|||lt2gene | Low-quality |
| 18-101-final-viral-combined_k141_8063||lt2gene | Low-quality |
| 18-101-final-viral-combined_k141_206000|||lt2gene | Low-quality |
| 18-101-final-viral-combined_k141_287667|||lt2gene | Not-determined |
| 18-101-final-viral-combined_k141_73058|||lt2gene | Low-quality |
| 18-101-final-viral-combined_k141_386634|||lt2gene | Not-determined |
| 18-101-final-viral-combined_k141_24179|||lt2gene | Not-determined |
| 18-101-final-viral-combined_k141_90029|||lt2gene | Low-quality |
| 18-101-final-viral-combined_k141_24217|||lt2gene | Not-determined |
| 18-101-final-viral-combined_k141_24220|||lt2gene | Not-determined |
| 18-101-final-viral-combined_k141_105909|||lt2gene | Not-determined |
| 18-101-final-viral-combined_k141_336489|||lt2gene | Not-determined |
| 18-101-final-viral-combined_k141_304883|||lt2gene | Low-quality |
| 18-101-final-viral-combined_k141_287752|||lt2gene | Low-quality |
| 18-101-final-viral-combined_k141_321848|||lt2gene | Low-quality |
| 18-101-final-viral-combined_k141_386715|||lt2gene | Not-determined |
| 18-101-final-viral-combined_k141_8158||lt2gene | Low-quality |
| 18-101-final-viral-combined_k141_287767|||lt2gene | Not-determined |
| 18-101-final-viral-combined_k141_105954|||lt2gene | Low-quality |
| 18-101-final-viral-combined_k141_336554|||lt2gene | Low-quality |
| 18-101-final-viral-combined_k141_57334|||lt2gene | Low-quality |
| 18-101-final-viral-combined_k141_287822|||lt2gene | Low-quality |
| 18-101-final-viral-combined_k141_321885|||lt2gene | Low-quality |
| 18-101-final-viral-combined_k141_171771|||lt2gene | Low-quality |
| 18-101-final-viral-combined_k141_386755|||lt2gene | Low-quality |
| 18-101-final-viral-combined_k141_189736|||lt2gene | Low-quality |
| 18-101-final-viral-combined_k141_221985|||lt2gene | Low-quality |
| 18-101-final-viral-combined_k141_206167|||lt2gene | Low-quality |
| 18-101-final-viral-combined_k141_206169|||lt2gene | Low-quality |
| 18-101-final-viral-combined_k141_8253||lt2gene | Low-quality |
| 18-101-final-viral-combined_k141_8259||lt2gene | Low-quality |
| 18-101-final-viral-combined_k141_305024|||lt2gene | Not-determined |
| 18-101-final-viral-combined_k141_238750|||lt2gene | Low-quality |
| 18-101-final-viral-combined_k141_238800|||lt2gene | Low-quality |
| 18-101-final-viral-combined_k141_238827|||lt2gene | Not-determined |
| 18-101-final-viral-combined_k141_386884|||lt2gene | Not-determined |
| 18-101-final-viral-combined_k141_386898|||lt2gene | Low-quality |
| 18-101-final-viral-combined_k141_171901|||lt2gene | Not-determined |
| 18-101-final-viral-combined_k141_255669|||lt2gene | Not-determined |
| 18-101-final-viral-combined_k141_354333|||lt2gene | Low-quality |
| 18-101-final-viral-combined_k141_90242|||lt2gene | Low-quality |
| 18-101-final-viral-combined_k141_386926|||lt2gene | Low-quality |
| 18-101-final-viral-combined_k141_386935|||lt2gene | Low-quality |
| 18-101-final-viral-combined_k141_57392|||lt2gene | Low-quality |
| 18-101-final-viral-combined_k141_190006|||lt2gene | Low-quality |
| 18-101-final-viral-combined_k141_124396|||lt2gene | Low-quality |
| 18-101-final-viral-combined_k141_171954|||lt2gene | Not-determined |
| 18-101-final-viral-combined_k141_73314|||lt2gene | Low-quality |
| 18-101-final-viral-combined_k141_206424|||lt2gene | Low-quality |
| 18-101-final-viral-combined_k141_155803|||lt2gene | Low-quality |
| 18-101-final-viral-combined_k141_271522|||lt2gene | Low-quality |
| 18-101-final-viral-combined_k141_155807|||lt2gene | Low-quality |
| 18-101-final-viral-combined_k141_255847|||lt2gene | Low-quality |
| 18-101-final-viral-combined_k141_40802|||lt2gene | Low-quality |
| 18-101-final-viral-combined_k141_106208|||lt2gene | Low-quality |
| 18-101-final-viral-combined_k141_8452||lt2gene | Low-quality |
| 18-101-final-viral-combined_k141_90396|||lt2gene | Low-quality |
| 18-101-final-viral-combined_k141_40862|||lt2gene | Low-quality |
| 18-101-final-viral-combined_k141_255934|||lt2gene | Low-quality |
| 18-101-final-viral-combined_k141_238969|||lt2gene | Low-quality |
| 18-101-final-viral-combined_k141_238970|||lt2gene | Low-quality |
| 18-101-final-viral-combined_k141_8519||lt2gene | Low-quality |
| 18-101-final-viral-combined_k141_90424|||lt2gene | Not-determined |
| 18-101-final-viral-combined_k141_90432|||lt2gene | Low-quality |
| 18-101-final-viral-combined_k141_8545||lt2gene | Low-quality |
| 18-101-final-viral-combined_k141_190265|||lt2gene | Low-quality |
| 18-101-final-viral-combined_k141_24618|||lt2gene | Low-quality |
| 18-101-final-viral-combined_k141_206538|||lt2gene | Low-quality |
| 18-101-final-viral-combined_k141_354598|||lt2gene | Not-determined |
| 18-101-final-viral-combined_k141_354599|||lt2gene | Not-determined |
| 18-101-final-viral-combined_k141_140113|||lt2gene | Low-quality |
| 18-101-final-viral-combined_k141_57537|||lt2gene | Low-quality |
| 18-101-final-viral-combined_k141_90475|||lt2gene | Low-quality |
| 18-101-final-viral-combined_k141_336830|||lt2gene | Low-quality |
| 18-101-final-viral-combined_k141_57558|||lt2gene | Low-quality |
| 18-101-final-viral-combined_k141_140152|||lt2gene | Not-determined |
| 18-101-final-viral-combined_k141_190295|||lt2gene | Low-quality |
| 18-101-final-viral-combined_k141_40962|||lt2gene | Low-quality |
| 18-101-final-viral-combined_k141_172072|||lt2gene | Low-quality |
| 18-101-final-viral-combined_k141_336862|||lt2gene | Not-determined |
| 18-101-final-viral-combined_k141_336896|||lt2gene | Low-quality |
| 18-101-final-viral-combined_k141_8667||lt2gene | Not-determined |
| 18-101-final-viral-combined_k141_57614|||lt2gene | Low-quality |
| 18-101-final-viral-combined_k141_90575|||lt2gene | Not-determined |
| 18-101-final-viral-combined_k141_90615|||lt2gene | Low-quality |
| 18-101-final-viral-combined_k141_336965|||lt2gene | Low-quality |
| 18-101-final-viral-combined_k141_222492|||lt2gene | Low-quality |
| 18-101-final-viral-combined_k141_336982|||lt2gene | Low-quality |
| 18-101-final-viral-combined_k141_190426|||lt2gene | Low-quality |
| 18-101-final-viral-combined_k141_336992|||lt2gene | Low-quality |
| 18-101-final-viral-combined_k141_90650|||lt2gene | Low-quality |
| 18-101-final-viral-combined_k141_370402|||lt2gene | Not-determined |
| 18-101-final-viral-combined_k141_156015|||lt2gene | Low-quality |
| 18-101-final-viral-combined_k141_156024|||lt2gene | Low-quality |
| 18-101-final-viral-combined_k141_370414|||lt2gene | Low-quality |
| 18-101-final-viral-combined_k141_222595|||lt2gene | Not-determined |
| 18-101-final-viral-combined_k141_140336|||lt2gene | Low-quality |
| 18-101-final-viral-combined_k141_124764|||lt2gene | Low-quality |
| 18-101-final-viral-combined_k141_387434|||lt2gene | Low-quality |
| 18-101-final-viral-combined_k141_370515|||lt2gene | Low-quality |
| 18-101-final-viral-combined_k141_206781|||lt2gene | Low-quality |
| 18-101-final-viral-combined_k141_370552|||lt2gene | Low-quality |
| 18-101-final-viral-combined_k141_24943|||lt2gene | Not-determined |
| 18-101-final-viral-combined_k141_156083|||lt2gene | Not-determined |
| 18-101-final-viral-combined_k141_24948|||lt2gene | Not-determined |
| 18-101-final-viral-combined_k141_57723|||lt2gene | Low-quality |
| 18-101-final-viral-combined_k141_172206|||lt2gene | Not-determined |
| 18-101-final-viral-combined_k141_24973|||lt2gene | Low-quality |
| 18-101-final-viral-combined_k141_256177|||lt2gene | Low-quality |
| 18-101-final-viral-combined_k141_156120|||lt2gene | Low-quality |
| 18-101-final-viral-combined_k141_387513|||lt2gene | Low-quality |
| 18-101-final-viral-combined_k141_271910|||lt2gene | Not-determined |
| 18-101-final-viral-combined_k141_239392|||lt2gene | Low-quality |
| 18-101-final-viral-combined_k141_305699|||lt2gene | Low-quality |
| 18-101-final-viral-combined_k141_206897|||lt2gene | Not-determined |
| 18-101-final-viral-combined_k141_354871|||lt2gene | Low-quality |
| 18-101-final-viral-combined_k141_354872|||lt2gene | Not-determined |
| 18-101-final-viral-combined_k141_206905|||lt2gene | Low-quality |
| 18-101-final-viral-combined_k141_8924||lt2gene | Low-quality |
| 18-101-final-viral-combined_k141_354878|||lt2gene | Low-quality |
| 18-101-final-viral-combined_k141_190663|||lt2gene | Not-determined |
| 18-101-final-viral-combined_k141_256279|||lt2gene | Low-quality |
| 18-101-final-viral-combined_k141_387610|||lt2gene | Low-quality |
| 18-101-final-viral-combined_k141_288526|||lt2gene | Not-determined |
| 18-101-final-viral-combined_k141_57846|||lt2gene | Low-quality |
| 18-101-final-viral-combined_k141_190691|||lt2gene | Low-quality |
| 18-101-final-viral-combined_k141_387631|||lt2gene | Not-determined |
| 18-101-final-viral-combined_k141_305795|||lt2gene | Low-quality |
| 18-101-final-viral-combined_k141_90937|||lt2gene | Not-determined |
| 18-101-final-viral-combined_k141_140645|||lt2gene | Not-determined |
| 18-101-final-viral-combined_k141_190772|||lt2gene | Low-quality |
| 18-101-final-viral-combined_k141_256360|||lt2gene | Not-determined |
| 18-101-final-viral-combined_k141_272127|||lt2gene | Low-quality |
| 18-101-final-viral-combined_k141_25150|||lt2gene | Low-quality |
| 18-101-final-viral-combined_k141_125074|||lt2gene | Low-quality |
| 18-101-final-viral-combined_k141_355019|||lt2gene | Low-quality |
| 18-101-final-viral-combined_k141_322608|||lt2gene | Low-quality |
| 18-101-final-viral-combined_k141_140713|||lt2gene | Low-quality |
| 18-101-final-viral-combined_k141_73843|||lt2gene | Low-quality |
| 18-101-final-viral-combined_k141_387688|||lt2gene | Low-quality |
| 18-101-final-viral-combined_k141_305905|||lt2gene | Low-quality |
| 18-101-final-viral-combined_k141_387698|||lt2gene | Low-quality |
| 18-101-final-viral-combined_k141_337301|||lt2gene | Not-determined |
| 18-101-final-viral-combined_k141_387701|||lt2gene | Low-quality |
| 18-101-final-viral-combined_k141_239601|||lt2gene | Low-quality |
| 18-101-final-viral-combined_k141_172440|||lt2gene | Not-determined |
| 18-101-final-viral-combined_k141_256454|||lt2gene | Not-determined |
| 18-101-final-viral-combined_k141_322652|||lt2gene | Not-determined |
| 18-101-final-viral-combined_k141_41462|||lt2gene | Low-quality |
| 18-101-final-viral-combined_k141_207080|||lt2gene | Not-determined |
| 18-101-final-viral-combined_k141_355096|||lt2gene | Low-quality |
| 18-101-final-viral-combined_k141_256488|||lt2gene | Low-quality |
| 18-101-final-viral-combined_k141_355110|||lt2gene | Not-determined |
| 18-101-final-viral-combined_k141_272207|||lt2gene | Low-quality |
| 18-101-final-viral-combined_k141_272229|||lt2gene | Low-quality |
| 18-101-final-viral-combined_k141_156423|||lt2gene | Low-quality |
| 18-101-final-viral-combined_k141_9215||lt2gene | Low-quality |
| 18-101-final-viral-combined_k141_41508|||lt2gene | Low-quality |
| 18-101-final-viral-combined_k141_306016|||lt2gene | Not-determined |
| 18-101-final-viral-combined_k141_306018|||lt2gene | Not-determined |
| 18-101-final-viral-combined_k141_337449|||lt2gene | Low-quality |
| 18-101-final-viral-combined_k141_306079|||lt2gene | Low-quality |
| 18-101-final-viral-combined_k141_58188|||lt2gene | Low-quality |
| 18-101-final-viral-combined_k141_272372|||lt2gene | Low-quality |
| 18-101-final-viral-combined_k141_58203|||lt2gene | Low-quality |
| 18-101-final-viral-combined_k141_91290|||lt2gene | Low-quality |
| 18-101-final-viral-combined_k141_256682|||lt2gene | Low-quality |
| 18-101-final-viral-combined_k141_156540|||lt2gene | Not-determined |
| 18-101-final-viral-combined_k141_91313|||lt2gene | Not-determined |
| 18-101-final-viral-combined_k141_306148|||lt2gene | Low-quality |
| 18-101-final-viral-combined_k141_223079|||lt2gene | Low-quality |
| 18-101-final-viral-combined_k141_91349|||lt2gene | Low-quality |
| 18-101-final-viral-combined_k141_239819|||lt2gene | Low-quality |
| 18-101-final-viral-combined_k141_91357|||lt2gene | Low-quality |
| 18-101-final-viral-combined_k141_239835|||lt2gene | Low-quality |
| 18-101-final-viral-combined_k141_256718|||lt2gene | Low-quality |
| 18-101-final-viral-combined_k141_322916|||lt2gene | Not-determined |
| 18-101-final-viral-combined_k141_106941|||lt2gene | Low-quality |
| 18-101-final-viral-combined_k141_272451|||lt2gene | Low-quality |
| 18-101-final-viral-combined_k141_337601|||lt2gene | Low-quality |
| 18-101-final-viral-combined_k141_41644|||lt2gene | Low-quality |
| 18-101-final-viral-combined_k141_91380|||lt2gene | Low-quality |
| 18-101-final-viral-combined_k141_355296|||lt2gene | Low-quality |
| 18-101-final-viral-combined_k141_41703|||lt2gene | Low-quality |
| 18-101-final-viral-combined_k141_272517|||lt2gene | Low-quality |
| 18-101-final-viral-combined_k141_387981|||lt2gene | Low-quality |
| 18-101-final-viral-combined_k141_355381|||lt2gene | Low-quality |
| 18-101-final-viral-combined_k141_172870|||lt2gene | Not-determined |
| 18-101-final-viral-combined_k141_172873|||lt2gene | Not-determined |
| 18-101-final-viral-combined_k141_239924|||lt2gene | Low-quality |
| 18-101-final-viral-combined_k141_25533|||lt2gene | Low-quality |
| 18-101-final-viral-combined_k141_223292|||lt2gene | Low-quality |
| 18-101-final-viral-combined_k141_74291|||lt2gene | Low-quality |
| 18-101-final-viral-combined_k141_272579|||lt2gene | Not-determined |
| 18-101-final-viral-combined_k141_156701|||lt2gene | Not-determined |
| 18-101-final-viral-combined_k141_107046|||lt2gene | Low-quality |
| 18-101-final-viral-combined_k141_172902|||lt2gene | Not-determined |
| 18-101-final-viral-combined_k141_289072|||lt2gene | Low-quality |
| 18-101-final-viral-combined_k141_289073|||lt2gene | Low-quality |
| 18-101-final-viral-combined_k141_337640|||lt2gene | Not-determined |
| 18-101-final-viral-combined_k141_289087|||lt2gene | Not-determined |
| 18-101-final-viral-combined_k141_25596|||lt2gene | Low-quality |
| 18-101-final-viral-combined_k141_25604|||lt2gene | Low-quality |
| 18-101-final-viral-combined_k141_9532||lt2gene | Low-quality |
| 18-101-final-viral-combined_k141_25609|||lt2gene | Low-quality |
| 18-101-final-viral-combined_k141_272610|||lt2gene | Low-quality |
| 18-101-final-viral-combined_k141_207593|||lt2gene | Low-quality |
| 18-101-final-viral-combined_k141_191165|||lt2gene | Low-quality |
| 18-101-final-viral-combined_k141_306304|||lt2gene | Low-quality |
| 18-101-final-viral-combined_k141_191190|||lt2gene | Low-quality |
| 18-101-final-viral-combined_k141_41859|||lt2gene | Low-quality |
| 18-101-final-viral-combined_k141_41867|||lt2gene | Not-determined |
| 18-101-final-viral-combined_k141_9656||lt2gene | Low-quality |
| 18-101-final-viral-combined_k141_337745|||lt2gene | Low-quality |
| 18-101-final-viral-combined_k141_74436|||lt2gene | Low-quality |
| 18-101-final-viral-combined_k141_41908|||lt2gene | Low-quality |
| 18-101-final-viral-combined_k141_240074|||lt2gene | Low-quality |
| 18-101-final-viral-combined_k141_207628|||lt2gene | Low-quality |
| 18-101-final-viral-combined_k141_74459|||lt2gene | Not-determined |
| 18-101-final-viral-combined_k141_191214|||lt2gene | Low-quality |
| 18-101-final-viral-combined_k141_272714|||lt2gene | Low-quality |
| 18-101-final-viral-combined_k141_74485|||lt2gene | Not-determined |
| 18-101-final-viral-combined_k141_58452|||lt2gene | Low-quality |
| 18-101-final-viral-combined_k141_107234|||lt2gene | Low-quality |
| 18-101-final-viral-combined_k141_141374|||lt2gene | Low-quality |
| 18-101-final-viral-combined_k141_9799||lt2gene | Low-quality |
| 18-101-final-viral-combined_k141_306417|||lt2gene | Low-quality |
| 18-101-final-viral-combined_k141_107264|||lt2gene | Low-quality |
| 18-101-final-viral-combined_k141_107275|||lt2gene | Low-quality |
| 18-101-final-viral-combined_k141_207753|||lt2gene | Not-determined |
| 18-101-final-viral-combined_k141_58513|||lt2gene | Low-quality |
| 18-101-final-viral-combined_k141_125622|||lt2gene | Not-determined |
| 18-101-final-viral-combined_k141_355786|||lt2gene | Low-quality |
| 18-101-final-viral-combined_k141_371581|||lt2gene | Low-quality |
| 18-101-final-viral-combined_k141_207827|||lt2gene | Low-quality |
| 18-101-final-viral-combined_k141_156946|||lt2gene | Low-quality |
| 18-101-final-viral-combined_k141_306522|||lt2gene | Low-quality |
| 18-101-final-viral-combined_k141_156970|||lt2gene | Low-quality |
| 18-101-final-viral-combined_k141_156989|||lt2gene | Not-determined |
| 18-101-final-viral-combined_k141_289387|||lt2gene | Low-quality |
| 18-101-final-viral-combined_k141_257238|||lt2gene | Low-quality |
| 18-101-final-viral-combined_k141_355822|||lt2gene | Low-quality |
| 18-101-final-viral-combined_k141_58588|||lt2gene | Low-quality |
| 18-101-final-viral-combined_k141_191437|||lt2gene | Not-determined |
| 18-101-final-viral-combined_k141_272821|||lt2gene | Low-quality |
| 18-101-final-viral-combined_k141_141520|||lt2gene | Low-quality |
| 18-101-final-viral-combined_k141_323405|||lt2gene | Low-quality |
| 18-101-final-viral-combined_k141_240309|||lt2gene | Low-quality |
| 18-101-final-viral-combined_k141_355851|||lt2gene | Low-quality |
| 18-101-final-viral-combined_k141_355852|||lt2gene | Not-determined |
| 18-101-final-viral-combined_k141_173221|||lt2gene | Not-determined |
| 18-101-final-viral-combined_k141_289467|||lt2gene | Not-determined |
| 18-101-final-viral-combined_k141_107452|||lt2gene | Low-quality |
| 18-101-final-viral-combined_k141_207963|||lt2gene | Not-determined |
| 18-101-final-viral-combined_k141_207967|||lt2gene | Low-quality |
| 18-101-final-viral-combined_k141_272887|||lt2gene | Low-quality |
| 18-101-final-viral-combined_k141_306627|||lt2gene | Low-quality |
| 18-101-final-viral-combined_k141_157018|||lt2gene | Not-determined |
| 18-101-final-viral-combined_k141_306665|||lt2gene | Low-quality |
| 18-101-final-viral-combined_k141_306679|||lt2gene | Low-quality |
| 18-101-final-viral-combined_k141_240396|||lt2gene | Low-quality |
| 18-101-final-viral-combined_k141_306707|||lt2gene | Not-determined |
| 18-101-final-viral-combined_k141_240414|||lt2gene | Not-determined |
| 18-101-final-viral-combined_k141_157115|||lt2gene | Not-determined |
| 18-101-final-viral-combined_k141_223596|||lt2gene | Low-quality |
| 18-101-final-viral-combined_k141_107617|||lt2gene | Not-determined |
| 18-101-final-viral-combined_k141_323588|||lt2gene | Not-determined |
| 18-101-final-viral-combined_k141_355968|||lt2gene | Low-quality |
| 18-101-final-viral-combined_k141_125838|||lt2gene | Not-determined |
| 18-101-final-viral-combined_k141_26208|||lt2gene | Low-quality |
| 18-101-final-viral-combined_k141_355981|||lt2gene | Low-quality |
| 18-101-final-viral-combined_k141_26229|||lt2gene | Not-determined |
| 18-101-final-viral-combined_k141_157236|||lt2gene | Low-quality |
| 18-101-final-viral-combined_k141_208182|||lt2gene | Not-determined |
| 18-101-final-viral-combined_k141_58819|||lt2gene | Low-quality |
| 18-101-final-viral-combined_k141_42351|||lt2gene | Low-quality |
| 18-101-final-viral-combined_k141_338285|||lt2gene | Low-quality |
| 18-101-final-viral-combined_k141_58869|||lt2gene | Not-determined |
| 18-101-final-viral-combined_k141_356103|||lt2gene | Low-quality |
| 18-101-final-viral-combined_k141_223679|||lt2gene | Low-quality |
| 18-101-final-viral-combined_k141_289721|||lt2gene | Low-quality |
| 18-101-final-viral-combined_k141_26325|||lt2gene | Low-quality |
| 18-101-final-viral-combined_k141_42365|||lt2gene | Not-determined |
| 18-101-final-viral-combined_k141_26331|||lt2gene | Low-quality |
| 18-101-final-viral-combined_k141_42394|||lt2gene | Not-determined |
| 18-101-final-viral-combined_k141_58956|||lt2gene | Low-quality |
| 18-101-final-viral-combined_k141_92237|||lt2gene | Not-determined |
| 18-101-final-viral-combined_k141_157389|||lt2gene | Not-determined |
| 18-101-final-viral-combined_k141_208319|||lt2gene | Low-quality |
| 18-101-final-viral-combined_k141_92271|||lt2gene | Low-quality |
| 18-101-final-viral-combined_k141_289821|||lt2gene | Low-quality |
| 18-101-final-viral-combined_k141_323733|||lt2gene | Low-quality |
| 18-101-final-viral-combined_k141_323747|||lt2gene | Low-quality |
| 18-101-final-viral-combined_k141_338399|||lt2gene | Not-determined |
| 18-101-final-viral-combined_k141_323788|||lt2gene | Not-determined |
| 18-101-final-viral-combined_k141_42518|||lt2gene | Low-quality |
| 18-101-final-viral-combined_k141_173509|||lt2gene | Not-determined |
| 18-101-final-viral-combined_k141_338418|||lt2gene | Low-quality |
| 18-101-final-viral-combined_k141_388424|||lt2gene | Low-quality |
| 18-101-final-viral-combined_k141_323862|||lt2gene | Low-quality |
| 18-101-final-viral-combined_k141_273211|||lt2gene | Not-determined |
| 18-101-final-viral-combined_k141_126029|||lt2gene | Low-quality |
| 18-101-final-viral-combined_k141_223982|||lt2gene | Not-determined |
| 18-101-final-viral-combined_k141_157548|||lt2gene | Low-quality |
| 18-101-final-viral-combined_k141_307142|||lt2gene | Not-determined |
| 18-101-final-viral-combined_k141_323914|||lt2gene | Low-quality |
| 18-101-final-viral-combined_k141_289994|||lt2gene | Low-quality |
| 18-101-final-viral-combined_k141_307147|||lt2gene | Low-quality |
| 18-101-final-viral-combined_k141_307151|||lt2gene | Low-quality |
| 18-101-final-viral-combined_k141_107923|||lt2gene | Low-quality |
| 18-101-final-viral-combined_k141_240835|||lt2gene | Low-quality |
| 18-101-final-viral-combined_k141_224064|||lt2gene | Low-quality |
| 18-101-final-viral-combined_k141_173626|||lt2gene | Low-quality |
| 18-101-final-viral-combined_k141_26462|||lt2gene | Low-quality |
| 18-101-final-viral-combined_k141_75017|||lt2gene | Low-quality |
| 18-101-final-viral-combined_k141_224077|||lt2gene | Not-determined |
| 18-101-final-viral-combined_k141_224093|||lt2gene | Low-quality |
| 18-101-final-viral-combined_k141_372157|||lt2gene | Low-quality |
| 18-101-final-viral-combined_k141_92506|||lt2gene | Low-quality |
| 18-101-final-viral-combined_k141_240907|||lt2gene | Low-quality |
| 18-101-final-viral-combined_k141_290102|||lt2gene | Low-quality |
| 18-101-final-viral-combined_k141_372220|||lt2gene | Low-quality |
| 18-101-final-viral-combined_k141_290112|||lt2gene | Not-determined |
| 18-101-final-viral-combined_k141_208571|||lt2gene | Low-quality |
| 18-101-final-viral-combined_k141_240949|||lt2gene | Low-quality |
| 18-101-final-viral-combined_k141_290134|||lt2gene | Not-determined |
| 18-101-final-viral-combined_k141_290156|||lt2gene | Low-quality |
| 18-101-final-viral-combined_k141_257973|||lt2gene | Low-quality |
| 18-101-final-viral-combined_k141_26597|||lt2gene | Not-determined |
| 18-101-final-viral-combined_k141_191955|||lt2gene | Not-determined |
| 18-101-final-viral-combined_k141_126361|||lt2gene | Low-quality |
| 18-101-final-viral-combined_k141_307303|||lt2gene | Low-quality |
| 18-101-final-viral-combined_k141_258011|||lt2gene | Low-quality |
| 18-101-final-viral-combined_k141_307306|||lt2gene | Not-determined |
| 18-101-final-viral-combined_k141_388601|||lt2gene | Low-quality |
| 18-101-final-viral-combined_k141_258018|||lt2gene | Not-determined |
| 18-101-final-viral-combined_k141_92633|||lt2gene | Not-determined |
| 18-101-final-viral-combined_k141_192013|||lt2gene | Not-determined |
| 18-101-final-viral-combined_k141_126419|||lt2gene | Not-determined |
| 18-101-final-viral-combined_k141_372386|||lt2gene | Low-quality |
| 18-101-final-viral-combined_k141_173913|||lt2gene | Not-determined |
| 18-101-final-viral-combined_k141_241023|||lt2gene | Low-quality |
| 18-101-final-viral-combined_k141_241024|||lt2gene | Not-determined |
| 18-101-final-viral-combined_k141_208736|||lt2gene | Low-quality |
| 18-101-final-viral-combined_k141_241052|||lt2gene | Not-determined |
| 18-101-final-viral-combined_k141_324247|||lt2gene | Low-quality |
| 18-101-final-viral-combined_k141_224305|||lt2gene | Not-determined |
| 18-101-final-viral-combined_k141_108219|||lt2gene | Low-quality |
| 18-101-final-viral-combined_k141_157888|||lt2gene | Low-quality |
| 18-101-final-viral-combined_k141_142217|||lt2gene | Not-determined |
| 18-101-final-viral-combined_k141_224355|||lt2gene | Not-determined |
| 18-101-final-viral-combined_k141_258195|||lt2gene | Low-quality |
| 18-101-final-viral-combined_k141_356725|||lt2gene | Low-quality |
| 18-101-final-viral-combined_k141_157920|||lt2gene | Low-quality |
| 18-101-final-viral-combined_k141_290291|||lt2gene | Low-quality |
| 18-101-final-viral-combined_k141_126580|||lt2gene | Low-quality |
| 18-101-final-viral-combined_k141_338826|||lt2gene | Not-determined |
| 18-101-final-viral-combined_k141_290302|||lt2gene | Not-determined |
| 18-101-final-viral-combined_k141_241206|||lt2gene | Low-quality |
| 18-101-final-viral-combined_k141_75335|||lt2gene | Not-determined |
| 18-101-final-viral-combined_k141_157981|||lt2gene | Low-quality |
| 18-101-final-viral-combined_k141_290402|||lt2gene | Low-quality |
| 18-101-final-viral-combined_k141_258296|||lt2gene | Low-quality |
| 18-101-final-viral-combined_k141_92950|||lt2gene | Low-quality |
| 18-101-final-viral-combined_k141_224466|||lt2gene | Low-quality |
| 18-101-final-viral-combined_k141_356788|||lt2gene | Low-quality |
| 18-101-final-viral-combined_k141_75434|||lt2gene | Low-quality |
| 18-101-final-viral-combined_k141_356800|||lt2gene | Low-quality |
| 18-101-final-viral-combined_k141_92989|||lt2gene | Not-determined |
| 18-101-final-viral-combined_k141_307399|||lt2gene | Low-quality |
| 18-101-final-viral-combined_k141_273656|||lt2gene | Low-quality |
| 18-101-final-viral-combined_k141_126742|||lt2gene | Low-quality |
| 18-101-final-viral-combined_k141_174169|||lt2gene | Low-quality |
| 18-101-final-viral-combined_k141_93063|||lt2gene | Not-determined |
| 18-101-final-viral-combined_k141_26828|||lt2gene | Low-quality |
| 18-101-final-viral-combined_k141_93093|||lt2gene | Not-determined |
| 18-101-final-viral-combined_k141_174246|||lt2gene | Low-quality |
| 18-101-final-viral-combined_k141_75552|||lt2gene | Low-quality |
| 18-101-final-viral-combined_k141_388948|||lt2gene | Not-determined |
| 18-101-final-viral-combined_k141_324438|||lt2gene | Not-determined |
| 18-101-final-viral-combined_k141_93126|||lt2gene | Low-quality |
| 18-101-final-viral-combined_k141_158177|||lt2gene | Low-quality |
| 18-101-final-viral-combined_k141_324458|||lt2gene | Low-quality |
| 18-101-final-viral-combined_k141_75580|||lt2gene | Low-quality |
| 18-101-final-viral-combined_k141_75582|||lt2gene | Low-quality |
| 18-101-final-viral-combined_k141_75597|||lt2gene | Low-quality |
| 18-101-final-viral-combined_k141_338995|||lt2gene | Low-quality |
| 18-101-final-viral-combined_k141_307535|||lt2gene | Low-quality |
| 18-101-final-viral-combined_k141_93173|||lt2gene | Low-quality |
| 18-101-final-viral-combined_k141_192514|||lt2gene | Low-quality |
| 18-101-final-viral-combined_k141_389008|||lt2gene | Low-quality |
| 18-101-final-viral-combined_k141_10739|||lt2gene | Low-quality |
| 18-101-final-viral-combined_k141_158277|||lt2gene | Not-determined |
| 18-101-final-viral-combined_k141_209181|||lt2gene | Low-quality |
| 18-101-final-viral-combined_k141_93247|||lt2gene | Low-quality |
| 18-101-final-viral-combined_k141_389112|||lt2gene | Low-quality |
| 18-101-final-viral-combined_k141_142612|||lt2gene | Low-quality |
| 18-101-final-viral-combined_k141_339158|||lt2gene | Low-quality |
| 18-101-final-viral-combined_k141_59805|||lt2gene | Low-quality |
| 18-101-final-viral-combined_k141_10860|||lt2gene | Low-quality |
| 18-101-final-viral-combined_k141_43386|||lt2gene | Low-quality |
| 18-101-final-viral-combined_k141_59840|||lt2gene | Low-quality |
| 18-101-final-viral-combined_k141_59866|||lt2gene | Not-determined |
| 18-101-final-viral-combined_k141_339209|||lt2gene | Low-quality |
| 18-101-final-viral-combined_k141_10904|||lt2gene | Low-quality |
| 18-101-final-viral-combined_k141_389156|||lt2gene | Not-determined |
| 18-101-final-viral-combined_k141_59871|||lt2gene | Low-quality |
| 18-101-final-viral-combined_k141_93355|||lt2gene | Not-determined |
| 18-101-final-viral-combined_k141_273919|||lt2gene | Low-quality |
| 18-101-final-viral-combined_k141_290774|||lt2gene | Low-quality |
| 18-101-final-viral-combined_k141_273926|||lt2gene | Low-quality |
| 18-101-final-viral-combined_k141_273937|||lt2gene | Not-determined |
| 18-101-final-viral-combined_k141_372949|||lt2gene | Low-quality |
| 18-101-final-viral-combined_k141_192678|||lt2gene | Not-determined |
| 18-101-final-viral-combined_k141_389217|||lt2gene | Low-quality |
| 18-101-final-viral-combined_k141_75763|||lt2gene | Low-quality |
| 18-101-final-viral-combined_k141_389229|||lt2gene | Low-quality |
| 18-101-final-viral-combined_k141_43461|||lt2gene | Not-determined |
| 18-101-final-viral-combined_k141_357103|||lt2gene | Low-quality |
| 18-101-final-viral-combined_k141_339306|||lt2gene | Low-quality |
| 18-101-final-viral-combined_k141_127094|||lt2gene | Low-quality |
| 18-101-final-viral-combined_k141_290834|||lt2gene | Not-determined |
| 18-101-final-viral-combined_k141_158447|||lt2gene | Not-determined |
| 18-101-final-viral-combined_k141_209380|||lt2gene | Low-quality |
| 18-101-final-viral-combined_k141_241671|||lt2gene | Low-quality |
| 18-101-final-viral-combined_k141_142767|||lt2gene | Low-quality |
| 18-101-final-viral-combined_k141_127117|||lt2gene | Low-quality |
| 18-101-final-viral-combined_k141_127124|||lt2gene | Low-quality |
| 18-101-final-viral-combined_k141_241686|||lt2gene | Not-determined |
| 18-101-final-viral-combined_k141_93443|||lt2gene | Low-quality |
| 18-101-final-viral-combined_k141_339364|||lt2gene | Low-quality |
| 18-101-final-viral-combined_k141_389302|||lt2gene | Low-quality |
| 18-101-final-viral-combined_k141_127161|||lt2gene | Low-quality |
| 18-101-final-viral-combined_k141_241764|||lt2gene | Low-quality |
| 18-101-final-viral-combined_k141_59996|||lt2gene | Low-quality |
| 18-101-final-viral-combined_k141_192733|||lt2gene | Not-determined |
| 18-101-final-viral-combined_k141_158488|||lt2gene | Low-quality |
| 18-101-final-viral-combined_k141_389393|||lt2gene | Low-quality |
| 18-101-final-viral-combined_k141_192742|||lt2gene | Not-determined |
| 18-101-final-viral-combined_k141_357194|||lt2gene | Low-quality |
| 18-101-final-viral-combined_k141_75964|||lt2gene | Low-quality |
| 18-101-final-viral-combined_k141_158522|||lt2gene | Low-quality |
| 18-101-final-viral-combined_k141_43615|||lt2gene | Not-determined |
| 18-101-final-viral-combined_k141_158537|||lt2gene | Low-quality |
| 18-101-final-viral-combined_k141_389416|||lt2gene | Low-quality |
| 18-101-final-viral-combined_k141_192764|||lt2gene | Low-quality |
| 18-101-final-viral-combined_k141_357253|||lt2gene | Low-quality |
| 18-101-final-viral-combined_k141_11143|||lt2gene | Low-quality |
| 18-101-final-viral-combined_k141_225140|||lt2gene | Low-quality |
| 18-101-final-viral-combined_k141_357297|||lt2gene | Low-quality |
| 18-101-final-viral-combined_k141_76056|||lt2gene | Not-determined |
| 18-101-final-viral-combined_k141_93673|||lt2gene | Not-determined |
| 18-101-final-viral-combined_k141_324941|||lt2gene | Low-quality |
| 18-101-final-viral-combined_k141_76077|||lt2gene | Low-quality |
| 18-101-final-viral-combined_k141_142872|||lt2gene | Not-determined |
| 18-101-final-viral-combined_k141_225181|||lt2gene | Low-quality |
| 18-101-final-viral-combined_k141_241994|||lt2gene | Not-determined |
| 18-101-final-viral-combined_k141_324969|||lt2gene | Not-determined |
| 18-101-final-viral-combined_k141_27326|||lt2gene | Not-determined |
| 18-101-final-viral-combined_k141_209545|||lt2gene | Not-determined |
| 18-101-final-viral-combined_k141_242032|||lt2gene | Low-quality |
| 18-101-final-viral-combined_k141_43782|||lt2gene | Low-quality |
| 18-101-final-viral-combined_k141_76160|||lt2gene | Not-determined |
| 18-101-final-viral-combined_k141_109018|||lt2gene | Low-quality |
| 18-101-final-viral-combined_k141_43814|||lt2gene | Low-quality |
| 18-101-final-viral-combined_k141_274243|||lt2gene | Not-determined |
| 18-101-final-viral-combined_k141_127420|||lt2gene | Low-quality |
| 18-101-final-viral-combined_k141_389585|||lt2gene | Low-quality |
| 18-101-final-viral-combined_k141_43849|||lt2gene | Low-quality |
| 18-101-final-viral-combined_k141_76217|||lt2gene | Not-determined |
| 18-101-final-viral-combined_k141_158710|||lt2gene | Low-quality |
| 18-101-final-viral-combined_k141_27418|||lt2gene | Low-quality |
| 18-101-final-viral-combined_k141_174866|||lt2gene | Not-determined |
| 18-101-final-viral-combined_k141_225309|||lt2gene | Low-quality |
| 18-101-final-viral-combined_k141_308223|||lt2gene | Low-quality |
| 18-101-final-viral-combined_k141_291349|||lt2gene | Low-quality |
| 18-101-final-viral-combined_k141_325030|||lt2gene | Low-quality |
| 18-101-final-viral-combined_k141_225375|||lt2gene | Low-quality |
| 18-101-final-viral-combined_k141_225380|||lt2gene | Not-determined |
| 18-101-final-viral-combined_k141_325050|||lt2gene | Low-quality |
| 18-101-final-viral-combined_k141_291386|||lt2gene | Not-determined |
| 18-101-final-viral-combined_k141_143090|||lt2gene | Low-quality |
| 18-101-final-viral-combined_k141_143092|||lt2gene | Not-determined |
| 18-101-final-viral-combined_k141_158843|||lt2gene | Not-determined |
| 18-101-final-viral-combined_k141_259254|||lt2gene | Low-quality |
| 18-101-final-viral-combined_k141_158861|||lt2gene | Low-quality |
| 18-101-final-viral-combined_k141_274376|||lt2gene | Low-quality |
| 18-101-final-viral-combined_k141_27533|||lt2gene | Low-quality |
| 18-101-final-viral-combined_k141_193191|||lt2gene | Low-quality |
| 18-101-final-viral-combined_k141_325182|||lt2gene | Not-determined |
| 18-101-final-viral-combined_k141_60467|||lt2gene | Low-quality |
| 18-101-final-viral-combined_k141_158913|||lt2gene | Low-quality |
| 18-101-final-viral-combined_k141_193215|||lt2gene | Not-determined |
| 18-101-final-viral-combined_k141_259305|||lt2gene | Low-quality |
| 18-101-final-viral-combined_k141_225517|||lt2gene | Low-quality |
| 18-101-final-viral-combined_k141_11423|||lt2gene | Low-quality |
| 18-101-final-viral-combined_k141_389765|||lt2gene | Low-quality |
| 18-101-final-viral-combined_k141_325216|||lt2gene | Not-determined |
| 18-101-final-viral-combined_k141_373418|||lt2gene | Not-determined |
| 18-101-final-viral-combined_k141_357691|||lt2gene | Not-determined |
| 18-101-final-viral-combined_k141_175077|||lt2gene | Low-quality |
| 18-101-final-viral-combined_k141_94092|||lt2gene | Not-determined |
| 18-101-final-viral-combined_k141_259374|||lt2gene | Low-quality |
| 18-101-final-viral-combined_k141_44035|||lt2gene | Low-quality |
| 18-101-final-viral-combined_k141_259386|||lt2gene | Low-quality |
| 18-101-final-viral-combined_k141_209881|||lt2gene | Not-determined |
| 18-101-final-viral-combined_k141_44071|||lt2gene | Not-determined |
| 18-101-final-viral-combined_k141_339783|||lt2gene | Low-quality |
| 18-101-final-viral-combined_k141_274522|||lt2gene | Not-determined |
| 18-101-final-viral-combined_k141_11501|||lt2gene | Low-quality |
| 18-101-final-viral-combined_k141_27643|||lt2gene | Low-quality |
| 18-101-final-viral-combined_k141_175150|||lt2gene | Low-quality |
| 18-101-final-viral-combined_k141_308493|||lt2gene | Low-quality |
| 18-101-final-viral-combined_k141_325338|||lt2gene | Not-determined |
| 18-101-final-viral-combined_k141_44146|||lt2gene | Low-quality |
| 18-101-final-viral-combined_k141_159049|||lt2gene | Not-determined |
| 18-101-final-viral-combined_k141_193425|||lt2gene | Low-quality |
| 18-101-final-viral-combined_k141_274600|||lt2gene | Low-quality |
| 18-101-final-viral-combined_k141_373559|||lt2gene | Low-quality |
| 18-101-final-viral-combined_k141_389921|||lt2gene | Low-quality |
| 18-101-final-viral-combined_k141_209993|||lt2gene | Not-determined |
| 18-101-final-viral-combined_k141_225706|||lt2gene | Not-determined |
| 18-101-final-viral-combined_k141_210029|||lt2gene | Not-determined |
| 18-101-final-viral-combined_k141_175252|||lt2gene | Not-determined |
| 18-101-final-viral-combined_k141_389957|||lt2gene | Not-determined |
| 18-101-final-viral-combined_k141_389971|||lt2gene | Low-quality |
| 18-101-final-viral-combined_k141_259606|||lt2gene | Low-quality |
| 18-101-final-viral-combined_k141_373664|||lt2gene | Low-quality |
| 18-101-final-viral-combined_k141_27828|||lt2gene | Low-quality |
| 18-101-final-viral-combined_k141_210077|||lt2gene | Low-quality |
| 18-101-final-viral-combined_k141_27838|||lt2gene | Low-quality |
| 18-101-final-viral-combined_k141_325498|||lt2gene | Low-quality |
| 18-101-final-viral-combined_k141_225814|||lt2gene | Low-quality |
| 18-101-final-viral-combined_k141_225815|||lt2gene | Low-quality |
| 18-101-final-viral-combined_k141_27901|||lt2gene | Not-determined |
| 18-101-final-viral-combined_k141_242481|||lt2gene | Low-quality |
| 18-101-final-viral-combined_k141_225848|||lt2gene | Low-quality |
| 18-101-final-viral-combined_k141_143534|||lt2gene | Not-determined |
| 18-101-final-viral-combined_k141_373758|||lt2gene | Not-determined |
| 18-101-final-viral-combined_k141_94499|||lt2gene | Low-quality |
| 18-101-final-viral-combined_k141_210172|||lt2gene | Low-quality |
| 18-101-final-viral-combined_k141_127942|||lt2gene | Low-quality |
| 18-101-final-viral-combined_k141_193647|||lt2gene | Low-quality |
| 18-101-final-viral-combined_k141_193649|||lt2gene | Low-quality |
| 18-101-final-viral-combined_k141_60974|||lt2gene | Low-quality |
| 18-101-final-viral-combined_k141_109682|||lt2gene | Not-determined |
| 18-101-final-viral-combined_k141_109685|||lt2gene | Not-determined |
| 18-101-final-viral-combined_k141_373786|||lt2gene | Low-quality |
| 18-101-final-viral-combined_k141_274831|||lt2gene | Low-quality |
| 18-101-final-viral-combined_k141_340043|||lt2gene | Not-determined |
| 18-101-final-viral-combined_k141_242554|||lt2gene | Low-quality |
| 18-101-final-viral-combined_k141_292097|||lt2gene | Not-determined |
| 18-101-final-viral-combined_k141_210263|||lt2gene | Low-quality |
| 18-101-final-viral-combined_k141_259843|||lt2gene | Low-quality |
| 18-101-final-viral-combined_k141_143693|||lt2gene | Low-quality |
| 18-101-final-viral-combined_k141_109750|||lt2gene | Low-quality |
| 18-101-final-viral-combined_k141_11916|||lt2gene | Not-determined |
| 18-101-final-viral-combined_k141_210313|||lt2gene | Not-determined |
| 18-101-final-viral-combined_k141_128022|||lt2gene | Not-determined |
| 18-101-final-viral-combined_k141_11922|||lt2gene | Low-quality |
| 18-101-final-viral-combined_k141_274914|||lt2gene | Not-determined |
| 18-101-final-viral-combined_k141_259891|||lt2gene | Not-determined |
| 18-101-final-viral-combined_k141_259900|||lt2gene | Not-determined |
| 18-101-final-viral-combined_k141_210327|||lt2gene | Low-quality |
| 18-101-final-viral-combined_k141_340121|||lt2gene | Low-quality |
| 18-101-final-viral-combined_k141_76832|||lt2gene | Low-quality |
| 18-101-final-viral-combined_k141_358309|||lt2gene | Not-determined |
| 18-101-final-viral-combined_k141_325763|||lt2gene | Low-quality |
| 18-101-final-viral-combined_k141_159402|||lt2gene | Low-quality |
| 18-101-final-viral-combined_k141_292208|||lt2gene | Not-determined |
| 18-101-final-viral-combined_k141_109862|||lt2gene | Not-determined |
| 18-101-final-viral-combined_k141_175521|||lt2gene | Low-quality |
| 18-101-final-viral-combined_k141_109876|||lt2gene | Low-quality |
| 18-101-final-viral-combined_k141_308985|||lt2gene | Low-quality |
| 18-101-final-viral-combined_k141_358378|||lt2gene | Low-quality |
| 18-101-final-viral-combined_k141_358386|||lt2gene | Not-determined |
| 18-101-final-viral-combined_k141_374012|||lt2gene | Low-quality |
| 18-101-final-viral-combined_k141_76932|||lt2gene | Low-quality |
| 18-101-final-viral-combined_k141_109906|||lt2gene | Low-quality |
| 18-101-final-viral-combined_k141_175539|||lt2gene | Low-quality |
| 18-101-final-viral-combined_k141_44703|||lt2gene | Low-quality |
| 18-101-final-viral-combined_k141_44716|||lt2gene | Low-quality |
| 18-101-final-viral-combined_k141_242705|||lt2gene | Low-quality |
| 18-101-final-viral-combined_k141_193776|||lt2gene | Not-determined |
| 18-101-final-viral-combined_k141_374081|||lt2gene | Low-quality |
| 18-101-final-viral-combined_k141_175619|||lt2gene | Not-determined |
| 18-101-final-viral-combined_k141_61339|||lt2gene | Low-quality |

18-101-final-viral-combined_k141_44771||lt2gene Low-quality
18-101-final-viral-combined_k141_325898||lt2gene Low-quality
18-101-final-viral-combined_k141_44778||lt2gene Low-quality
18-101-final-viral-combined_k141_390385||lt2gene Not-determined
18-101-final-viral-combined_k141_242758||lt2gene Low-quality
18-101-final-viral-combined_k141_175645||lt2gene Low-quality
18-101-final-viral-combined_k141_143932||lt2gene Not-determined
18-101-final-viral-combined_k141_374133||lt2gene Low-quality
18-101-final-viral-combined_k141_292363||lt2gene Low-quality
18-101-final-viral-combined_k141_390418||lt2gene Low-quality
18-101-final-viral-combined_k141_309144||lt2gene Low-quality
18-101-final-viral-combined_k141_374161||lt2gene Low-quality
18-101-final-viral-combined_k141_374162||lt2gene Low-quality
18-101-final-viral-combined_k141_159536||lt2gene Low-quality
18-101-final-viral-combined_k141_390483||lt2gene Low-quality
18-101-final-viral-combined_k141_159548||lt2gene Low-quality
18-101-final-viral-combined_k141_159552||lt2gene Not-determined
18-101-final-viral-combined_k141_94744||lt2gene Low-quality
18-101-final-viral-combined_k141_159562||lt2gene Low-quality
18-101-final-viral-combined_k141_94751||lt2gene Not-determined
18-101-final-viral-combined_k141_28439||lt2gene Not-determined
18-101-final-viral-combined_k141_61500||lt2gene Not-determined
18-101-final-viral-combined_k141_292482||lt2gene Low-quality
18-101-final-viral-combined_k141_358687||lt2gene Not-determined
18-101-final-viral-combined_k141_12395||lt2gene Low-quality
18-101-final-viral-combined_k141_340428||lt2gene Low-quality
18-101-final-viral-combined_k141_159627||lt2gene Not-determined
18-101-final-viral-combined_k141_12443||lt2gene Low-quality
18-101-final-viral-combined_k141_292586||lt2gene Low-quality
18-101-final-viral-combined_k141_44950||lt2gene Not-determined
18-101-final-viral-combined_k141_374322||lt2gene Not-determined
18-101-final-viral-combined_k141_226461||lt2gene Low-quality
18-101-final-viral-combined_k141_12491||lt2gene Low-quality
18-101-final-viral-combined_k141_193981||lt2gene Not-determined
18-101-final-viral-combined_k141_159706||lt2gene Low-quality
18-101-final-viral-combined_k141_326114||lt2gene Low-quality
18-101-final-viral-combined_k141_326136||lt2gene Not-determined
18-101-final-viral-combined_k141_374401||lt2gene Low-quality
18-101-final-viral-combined_k141_128457||lt2gene Not-determined
18-101-final-viral-combined_k141_144211||lt2gene Low-quality
18-101-final-viral-combined_k141_45086||lt2gene Low-quality
18-101-final-viral-combined_k141_28659||lt2gene Low-quality
18-101-final-viral-combined_k141_292726||lt2gene Low-quality
18-101-final-viral-combined_k141_358905||lt2gene Low-quality
18-101-final-viral-combined_k141_292743||lt2gene Low-quality
18-101-final-viral-combined_k141_210748||lt2gene Low-quality
18-101-final-viral-combined_k141_226599||lt2gene Not-determined
18-101-final-viral-combined_k141_374489||lt2gene Not-determined
18-101-final-viral-combined_k141_309462||lt2gene Low-quality
18-101-final-viral-combined_k141_128565||lt2gene Not-determined
18-101-final-viral-combined_k141_340565||lt2gene Low-quality
18-101-final-viral-combined_k141_128574||lt2gene Not-determined
18-101-final-viral-combined_k141_260461||lt2gene Not-determined
18-101-final-viral-combined_k141_159788||lt2gene Not-determined
18-101-final-viral-combined_k141_210819||lt2gene Not-determined
18-101-final-viral-combined_k141_243219||lt2gene Low-quality
18-101-final-viral-combined_k141_61865||lt2gene Low-quality
18-101-final-viral-combined_k141_128614||lt2gene Low-quality
18-101-final-viral-combined_k141_28705||lt2gene Low-quality
18-101-final-viral-combined_k141_210850||lt2gene Low-quality
18-101-final-viral-combined_k141_128640||lt2gene Not-determined
18-101-final-viral-combined_k141_340637||lt2gene Low-quality
18-101-final-viral-combined_k141_28719||lt2gene Not-determined
18-101-final-viral-combined_k141_128692||lt2gene Low-quality
18-101-final-viral-combined_k141_28755||lt2gene Not-determined
18-101-final-viral-combined_k141_326344||lt2gene Low-quality
18-101-final-viral-combined_k141_28763||lt2gene Low-quality
18-101-final-viral-combined_k141_226739||lt2gene Low-quality
18-101-final-viral-combined_k141_340678||lt2gene Low-quality
18-101-final-viral-combined_k141_340679||lt2gene Low-quality
18-101-final-viral-combined_k141_326352||lt2gene Low-quality
18-101-final-viral-combined_k141_260598||lt2gene Low-quality
18-101-final-viral-combined_k141_128738||lt2gene Low-quality
18-101-final-viral-combined_k141_309564||lt2gene Low-quality
18-101-final-viral-combined_k141_292915||lt2gene Low-quality
18-101-final-viral-combined_k141_77551||lt2gene Not-determined
18-101-final-viral-combined_k141_45257||lt2gene Low-quality
18-101-final-viral-combined_k141_210975||lt2gene Not-determined
18-101-final-viral-combined_k141_77574||lt2gene Low-quality
18-101-final-viral-combined_k141_175952||lt2gene Low-quality
18-101-final-viral-combined_k141_275697||lt2gene Not-determined
18-101-final-viral-combined_k141_128826||lt2gene Low-quality
18-101-final-viral-combined_k141_144369||lt2gene Low-quality
18-101-final-viral-combined_k141_62116||lt2gene Low-quality
18-101-final-viral-combined_k141_176003||lt2gene Low-quality
18-101-final-viral-combined_k141_293085||lt2gene Low-quality
18-101-final-viral-combined_k141_12814||lt2gene Low-quality
18-101-final-viral-combined_k141_77735||lt2gene Low-quality
18-101-final-viral-combined_k141_194550||lt2gene Low-quality
18-101-final-viral-combined_k141_309742||lt2gene Low-quality
18-101-final-viral-combined_k141_12849||lt2gene Not-determined
18-101-final-viral-combined_k141_62217||lt2gene Low-quality
18-101-final-viral-combined_k141_359291||lt2gene Low-quality
18-101-final-viral-combined_k141_211141||lt2gene Low-quality
18-101-final-viral-combined_k141_309758||lt2gene Not-determined
18-101-final-viral-combined_k141_28965||lt2gene Low-quality
18-101-final-viral-combined_k141_275774||lt2gene Not-determined
18-101-final-viral-combined_k141_260839||lt2gene Low-quality
18-101-final-viral-combined_k141_28990||lt2gene Low-quality
18-101-final-viral-combined_k141_374827||lt2gene Not-determined
18-101-final-viral-combined_k141_374835||lt2gene Low-quality
18-101-final-viral-combined_k141_95286||lt2gene Low-quality
18-101-final-viral-combined_k141_62270||lt2gene Low-quality
18-101-final-viral-combined_k141_62271||lt2gene Not-determined
18-101-final-viral-combined_k141_29037||lt2gene Low-quality
18-101-final-viral-combined_k141_62284||lt2gene Low-quality
18-101-final-viral-combined_k141_275840||lt2gene Low-quality
18-101-final-viral-combined_k141_374875||lt2gene Low-quality
18-101-final-viral-combined_k141_359367||lt2gene Low-quality
18-101-final-viral-combined_k141_243648||lt2gene Not-determined
18-101-final-viral-combined_k141_243650||lt2gene Low-quality
18-101-final-viral-combined_k141_211238||lt2gene Not-determined
18-101-final-viral-combined_k141_144545||lt2gene Low-quality
18-101-final-viral-combined_k141_129040||lt2gene Low-quality
18-101-final-viral-combined_k141_227055||lt2gene Low-quality
18-101-final-viral-combined_k141_95384||lt2gene Low-quality
18-101-final-viral-combined_k141_227065||lt2gene Not-determined
18-101-final-viral-combined_k141_12936||lt2gene Not-determined
18-101-final-viral-combined_k141_341071||lt2gene Low-quality
18-101-final-viral-combined_k141_260983||lt2gene Low-quality
18-101-final-viral-combined_k141_211306||lt2gene Low-quality
18-101-final-viral-combined_k141_110338||lt2gene Not-determined
18-101-final-viral-combined_k141_144636||lt2gene Low-quality
18-101-final-viral-combined_k141_129144||lt2gene Not-determined
18-101-final-viral-combined_k141_144643||lt2gene Low-quality
18-101-final-viral-combined_k141_293303||lt2gene Low-quality
18-101-final-viral-combined_k141_261103||lt2gene Low-quality
18-101-final-viral-combined_k141_211404||lt2gene Low-quality
18-101-final-viral-combined_k141_359542||lt2gene Low-quality
18-101-final-viral-combined_k141_194831||lt2gene Low-quality
18-101-final-viral-combined_k141_341163||lt2gene Low-quality
18-101-final-viral-combined_k141_129252||lt2gene Low-quality
18-101-final-viral-combined_k141_129256||lt2gene Low-quality
18-101-final-viral-combined_k141_326874||lt2gene Low-quality
18-101-final-viral-combined_k141_227224||lt2gene Not-determined
18-101-final-viral-combined_k141_144755||lt2gene Low-quality
18-101-final-viral-combined_k141_326890||lt2gene Low-quality
18-101-final-viral-combined_k141_326891||lt2gene Low-quality
18-101-final-viral-combined_k141_227239||lt2gene Low-quality
18-101-final-viral-combined_k141_45582||lt2gene Low-quality
18-101-final-viral-combined_k141_194886||lt2gene Not-determined
18-101-final-viral-combined_k141_261191||lt2gene Low-quality
18-101-final-viral-combined_k141_160413||lt2gene Low-quality
18-101-final-viral-combined_k141_160414||lt2gene Low-quality
18-101-final-viral-combined_k141_160422||lt2gene Low-quality
18-101-final-viral-combined_k141_261231||lt2gene Not-determined
18-101-final-viral-combined_k141_227313||lt2gene Not-determined
18-101-final-viral-combined_k141_391318||lt2gene Low-quality
18-101-final-viral-combined_k141_160504||lt2gene Not-determined
18-101-final-viral-combined_k141_276111||lt2gene Low-quality
18-101-final-viral-combined_k141_326950||lt2gene Low-quality
18-101-final-viral-combined_k141_341295||lt2gene Low-quality
18-101-final-viral-combined_k141_326964||lt2gene Low-quality
18-101-final-viral-combined_k141_244012||lt2gene Low-quality
18-101-final-viral-combined_k141_176419||lt2gene Low-quality
18-101-final-viral-combined_k141_110631||lt2gene Low-quality
18-101-final-viral-combined_k141_276178||lt2gene Not-determined
18-101-final-viral-combined_k141_110667||lt2gene Low-quality
18-101-final-viral-combined_k141_211687||lt2gene Low-quality
18-101-final-viral-combined_k141_391407||lt2gene Low-quality
18-101-final-viral-combined_k141_144937||lt2gene Low-quality
18-101-final-viral-combined_k141_261344||lt2gene Not-determined
18-101-final-viral-combined_k141_227384||lt2gene Not-determined
18-101-final-viral-combined_k141_359596||lt2gene Low-quality
18-101-final-viral-combined_k141_95607||lt2gene Not-determined
18-101-final-viral-combined_k141_95613||lt2gene Low-quality
18-101-final-viral-combined_k141_227407||lt2gene Low-quality
18-101-final-viral-combined_k141_244112||lt2gene Low-quality
18-101-final-viral-combined_k141_261361||lt2gene Low-quality
18-101-final-viral-combined_k141_261365||lt2gene Low-quality
18-101-final-viral-combined_k141_244130||lt2gene Low-quality
18-101-final-viral-combined_k141_160650||lt2gene Low-quality
18-101-final-viral-combined_k141_13321||lt2gene Low-quality
18-101-final-viral-combined_k141_276267||lt2gene Not-determined
18-101-final-viral-combined_k141_244174||lt2gene Low-quality
18-101-final-viral-combined_k141_244187||lt2gene Low-quality
18-101-final-viral-combined_k141_341461||lt2gene Low-quality
18-101-final-viral-combined_k141_227499||lt2gene Low-quality
18-101-final-viral-combined_k141_62608||lt2gene Not-determined
18-101-final-viral-combined_k141_227508||lt2gene Low-quality
18-101-final-viral-combined_k141_293677||lt2gene Low-quality
18-101-final-viral-combined_k141_129680||lt2gene Not-determined
18-101-final-viral-combined_k141_244281||lt2gene Low-quality
18-101-final-viral-combined_k141_391558||lt2gene Low-quality
18-101-final-viral-combined_k141_78143||lt2gene Not-determined
18-101-final-viral-combined_k141_211909||lt2gene Low-quality
18-101-final-viral-combined_k141_110742||lt2gene Low-quality
18-101-final-viral-combined_k141_293787||lt2gene Low-quality
18-101-final-viral-combined_k141_129727||lt2gene Not-determined
18-101-final-viral-combined_k141_78205||lt2gene Low-quality
18-101-final-viral-combined_k141_211971||lt2gene Low-quality
18-101-final-viral-combined_k141_391657||lt2gene Low-quality
18-101-final-viral-combined_k141_359804||lt2gene Low-quality
18-101-final-viral-combined_k141_261505||lt2gene Low-quality
18-101-final-viral-combined_k141_29324||lt2gene Low-quality
18-101-final-viral-combined_k141_261541||lt2gene Low-quality
18-101-final-viral-combined_k141_227660||lt2gene Low-quality
18-101-final-viral-combined_k141_45949||lt2gene Low-quality
18-101-final-viral-combined_k141_212087||lt2gene Low-quality
18-101-final-viral-combined_k141_45994||lt2gene Not-determined
18-101-final-viral-combined_k141_195391||lt2gene Low-quality
18-101-final-viral-combined_k141_145366||lt2gene Low-quality
18-101-final-viral-combined_k141_13666||lt2gene Low-quality
18-101-final-viral-combined_k141_391777||lt2gene Low-quality
18-101-final-viral-combined_k141_13698||lt2gene Not-determined
18-101-final-viral-combined_k141_276589||lt2gene Low-quality
18-101-final-viral-combined_k141_227756||lt2gene Low-quality
18-101-final-viral-combined_k141_341755||lt2gene Not-determined
18-101-final-viral-combined_k141_276613||lt2gene Low-quality
18-101-final-viral-combined_k141_13727||lt2gene Low-quality
18-101-final-viral-combined_k141_176906||lt2gene Low-quality
18-101-final-viral-combined_k141_78442||lt2gene Low-quality
18-101-final-viral-combined_k141_13762||lt2gene Low-quality
18-101-final-viral-combined_k141_375626||lt2gene Low-quality
18-101-final-viral-combined_k141_244666||lt2gene Not-determined
18-101-final-viral-combined_k141_130021||lt2gene Low-quality
18-101-final-viral-combined_k141_276661||lt2gene Low-quality
18-101-final-viral-combined_k141_46195||lt2gene Low-quality
18-101-final-viral-combined_k141_13809||lt2gene Low-quality
18-101-final-viral-combined_k141_227823||lt2gene Not-determined
18-101-final-viral-combined_k141_78475||lt2gene Low-quality
18-101-final-viral-combined_k141_244738||lt2gene Low-quality
18-101-final-viral-combined_k141_111112||lt2gene Low-quality
18-101-final-viral-combined_k141_375734||lt2gene Low-quality
18-101-final-viral-combined_k141_261808||lt2gene Low-quality
18-101-final-viral-combined_k141_13863||lt2gene Low-quality
18-101-final-viral-combined_k141_111151||lt2gene Low-quality
18-101-final-viral-combined_k141_261828||lt2gene Not-determined
18-101-final-viral-combined_k141_244799||lt2gene Not-determined
18-101-final-viral-combined_k141_375803||lt2gene Not-determined
18-101-final-viral-combined_k141_375819||lt2gene Low-quality
18-101-final-viral-combined_k141_294136||lt2gene Low-quality
18-101-final-viral-combined_k141_63123||lt2gene Low-quality
18-101-final-viral-combined_k141_341941||lt2gene Low-quality
18-101-final-viral-combined_k141_261903||lt2gene Not-determined
18-101-final-viral-combined_k141_29525||lt2gene Low-quality
18-101-final-viral-combined_k141_161345||lt2gene Low-quality
18-101-final-viral-combined_k141_360139||lt2gene Not-determined
18-101-final-viral-combined_k141_130209||lt2gene Not-determined
18-101-final-viral-combined_k141_96130||lt2gene Low-quality
18-101-final-viral-combined_k141_244840||lt2gene Not-determined
18-101-final-viral-combined_k141_96141||lt2gene Not-determined
18-101-final-viral-combined_k141_212395||lt2gene Low-quality
18-101-final-viral-combined_k141_310526||lt2gene Low-quality
18-101-final-viral-combined_k141_145644||lt2gene Low-quality
18-101-final-viral-combined_k141_375845||lt2gene Low-quality
18-101-final-viral-combined_k141_96161||lt2gene Low-quality
18-101-final-viral-combined_k141_177200||lt2gene Low-quality
18-101-final-viral-combined_k141_96175||lt2gene Low-quality
18-101-final-viral-combined_k141_276888||lt2gene Not-determined
18-101-final-viral-combined_k141_96228||lt2gene Low-quality
18-101-final-viral-combined_k141_360238||lt2gene Low-quality
18-101-final-viral-combined_k141_29657||lt2gene Low-quality
18-101-final-viral-combined_k141_342100||lt2gene Low-quality
18-101-final-viral-combined_k141_276946||lt2gene Not-determined
18-101-final-viral-combined_k141_327232||lt2gene Low-quality
18-101-final-viral-combined_k141_212592||lt2gene Low-quality
18-101-final-viral-combined_k141_294241||lt2gene Not-determined
18-101-final-viral-combined_k141_360331||lt2gene Low-quality
18-101-final-viral-combined_k141_63314||lt2gene Low-quality
18-101-final-viral-combined_k141_78730||lt2gene Low-quality
18-101-final-viral-combined_k141_327341||lt2gene Low-quality
18-101-final-viral-combined_k141_262209||lt2gene Low-quality
18-101-final-viral-combined_k141_327363||lt2gene Low-quality
18-101-final-viral-combined_k141_327370||lt2gene Low-quality
18-101-final-viral-combined_k141_145921||lt2gene Not-determined
18-101-final-viral-combined_k141_228236||lt2gene Low-quality
18-101-final-viral-combined_k141_228243||lt2gene Low-quality
18-101-final-viral-combined_k141_145934||lt2gene Low-quality
18-101-final-viral-combined_k141_342222||lt2gene Not-determined
18-101-final-viral-combined_k141_294403||lt2gene Low-quality
18-101-final-viral-combined_k141_262259||lt2gene Low-quality
18-101-final-viral-combined_k141_46442||lt2gene Not-determined
18-101-final-viral-combined_k141_228276||lt2gene Low-quality
18-101-final-viral-combined_k141_177542||lt2gene Low-quality
18-101-final-viral-combined_k141_96369||lt2gene Low-quality
18-101-final-viral-combined_k141_376180||lt2gene Low-quality
18-101-final-viral-combined_k141_29909||lt2gene Low-quality
18-101-final-viral-combined_k141_29912||lt2gene Not-determined
18-101-final-viral-combined_k141_360489||lt2gene Not-determined
18-101-final-viral-combined_k141_310954||lt2gene Low-quality
18-101-final-viral-combined_k141_212863||lt2gene Low-quality
18-101-final-viral-combined_k141_130509||lt2gene Low-quality
18-101-final-viral-combined_k141_196247||lt2gene Low-quality
18-101-final-viral-combined_k141_177670||lt2gene Low-quality
18-101-final-viral-combined_k141_376304||lt2gene Low-quality
18-101-final-viral-combined_k141_63616||lt2gene Low-quality
18-101-final-viral-combined_k141_327571||lt2gene Low-quality
18-101-final-viral-combined_k141_161820||lt2gene Low-quality
18-101-final-viral-combined_k141_360578||lt2gene Low-quality
18-101-final-viral-combined_k141_294622||lt2gene Low-quality
18-101-final-viral-combined_k141_161840||lt2gene Not-determined
18-101-final-viral-combined_k141_196338||lt2gene Low-quality
18-101-final-viral-combined_k141_212895||lt2gene Not-determined
18-101-final-viral-combined_k141_96612||lt2gene Low-quality
18-101-final-viral-combined_k141_245365||lt2gene Not-determined
18-101-final-viral-combined_k141_245395||lt2gene Low-quality
18-101-final-viral-combined_k141_146117||lt2gene Not-determined
18-101-final-viral-combined_k141_130624||lt2gene Low-quality
18-101-final-viral-combined_k141_177812||lt2gene Not-determined
18-101-final-viral-combined_k141_161916||lt2gene Low-quality
18-101-final-viral-combined_k141_14406||lt2gene Low-quality
18-101-final-viral-combined_k141_14409||lt2gene Low-quality
18-101-final-viral-combined_k141_30178||lt2gene Low-quality
18-101-final-viral-combined_k141_30181||lt2gene Low-quality
18-101-final-viral-combined_k141_311230||lt2gene Low-quality
18-101-final-viral-combined_k141_311245||lt2gene Not-determined
18-101-final-viral-combined_k141_161966||lt2gene Low-quality
18-101-final-viral-combined_k141_30235||lt2gene Low-quality
18-101-final-viral-combined_k141_96843||lt2gene Not-determined
18-101-final-viral-combined_k141_245604||lt2gene Low-quality
18-101-final-viral-combined_k141_63804||lt2gene Not-determined
18-101-final-viral-combined_k141_311373||lt2gene Low-quality
18-101-final-viral-combined_k141_30340||lt2gene Low-quality
18-101-final-viral-combined_k141_162094||lt2gene Low-quality
18-101-final-viral-combined_k141_30341||lt2gene Not-determined
18-101-final-viral-combined_k141_376556||lt2gene Low-quality
18-101-final-viral-combined_k141_111787||lt2gene Low-quality
18-101-final-viral-combined_k141_30410||lt2gene Low-quality
18-101-final-viral-combined_k141_146275||lt2gene Low-quality
18-101-final-viral-combined_k141_130887||lt2gene Not-determined
18-101-final-viral-combined_k141_392645||lt2gene Low-quality
18-101-final-viral-combined_k141_79248||lt2gene Low-quality
18-101-final-viral-combined_k141_295028||lt2gene Not-determined
18-101-final-viral-combined_k141_327980||lt2gene Low-quality
18-101-final-viral-combined_k141_130937||lt2gene Low-quality
18-101-final-viral-combined_k141_14798||lt2gene Low-quality
18-101-final-viral-combined_k141_14802||lt2gene Low-quality
18-101-final-viral-combined_k141_111962||lt2gene Low-quality
18-101-final-viral-combined_k141_277714||lt2gene Low-quality
18-101-final-viral-combined_k141_79354||lt2gene Low-quality
18-101-final-viral-combined_k141_328064||lt2gene Not-determined
18-101-final-viral-combined_k141_277742||lt2gene Not-determined
18-101-final-viral-combined_k141_277753||lt2gene Low-quality
18-101-final-viral-combined_k141_311677||lt2gene Low-quality
18-101-final-viral-combined_k141_213066||lt2gene Not-determined
18-101-final-viral-combined_k141_343038||lt2gene Not-determined
18-101-final-viral-combined_k141_64080||lt2gene Low-quality
18-101-final-viral-combined_k141_295150||lt2gene Low-quality
18-101-final-viral-combined_k141_131025||lt2gene Not-determined
18-101-final-viral-combined_k141_178350||lt2gene Not-determined
18-101-final-viral-combined_k141_97179||lt2gene Low-quality
18-101-final-viral-combined_k141_228767||lt2gene Low-quality
18-101-final-viral-combined_k141_311794||lt2gene Low-quality
18-101-final-viral-combined_k141_64162||lt2gene Low-quality
18-101-final-viral-combined_k141_262857||lt2gene Not-determined
18-101-final-viral-combined_k141_30650||lt2gene Not-determined
18-101-final-viral-combined_k141_46590||lt2gene Low-quality
18-101-final-viral-combined_k141_30676||lt2gene Not-determined
18-101-final-viral-combined_k141_328181||lt2gene Low-quality
18-101-final-viral-combined_k141_131127||lt2gene Low-quality
18-101-final-viral-combined_k141_328207||lt2gene Low-quality
18-101-final-viral-combined_k141_277864||lt2gene Not-determined
18-101-final-viral-combined_k141_262888||lt2gene Low-quality
18-101-final-viral-combined_k141_178506||lt2gene Not-determined
18-101-final-viral-combined_k141_328256||lt2gene Not-determined
18-101-final-viral-combined_k141_146603||lt2gene Low-quality
18-101-final-viral-combined_k141_146608||lt2gene Not-determined
18-101-final-viral-combined_k141_196762||lt2gene Low-quality
18-101-final-viral-combined_k141_295310||lt2gene Low-quality
18-101-final-viral-combined_k141_311932||lt2gene Low-quality
18-101-final-viral-combined_k141_376988||lt2gene Low-quality
18-101-final-viral-combined_k141_79689||lt2gene Not-determined
18-101-final-viral-combined_k141_328342||lt2gene Low-quality
18-101-final-viral-combined_k141_228955||lt2gene Low-quality
18-101-final-viral-combined_k141_311955||lt2gene Low-quality
18-101-final-viral-combined_k141_131229||lt2gene Low-quality
18-101-final-viral-combined_k141_178609||lt2gene Not-determined
18-101-final-viral-combined_k141_178617||lt2gene Not-determined
18-101-final-viral-combined_k141_278030||lt2gene Not-determined
18-101-final-viral-combined_k141_79760||lt2gene Not-determined
18-101-final-viral-combined_k141_79764||lt2gene Not-determined
18-101-final-viral-combined_k141_64278||lt2gene Low-quality
18-101-final-viral-combined_k141_46808||lt2gene Not-determined
18-101-final-viral-combined_k141_162701||lt2gene Not-determined
18-101-final-viral-combined_k141_46818||lt2gene Low-quality
18-101-final-viral-combined_k141_196869||lt2gene Low-quality
18-101-final-viral-combined_k141_79816||lt2gene Low-quality
18-101-final-viral-combined_k141_229024||lt2gene Low-quality
18-101-final-viral-combined_k141_263089||lt2gene Not-determined
18-101-final-viral-combined_k141_97402||lt2gene Low-quality
18-101-final-viral-combined_k141_295475||lt2gene Not-determined
18-101-final-viral-combined_k141_295485||lt2gene Low-quality
18-101-final-viral-combined_k141_312047||lt2gene Not-determined
18-101-final-viral-combined_k141_15146||lt2gene Low-quality
18-101-final-viral-combined_k141_263116||lt2gene Low-quality
18-101-final-viral-combined_k141_278170||lt2gene Low-quality
18-101-final-viral-combined_k141_79894||lt2gene Low-quality
18-101-final-viral-combined_k141_229132||lt2gene Low-quality
18-101-final-viral-combined_k141_213344||lt2gene Low-quality
18-101-final-viral-combined_k141_361376||lt2gene Low-quality
18-101-final-viral-combined_k141_79918||lt2gene Low-quality
18-101-final-viral-combined_k141_229186||lt2gene Not-determined
18-101-final-viral-combined_k141_97546||lt2gene Not-determined
18-101-final-viral-combined_k141_361413||lt2gene Low-quality
18-101-final-viral-combined_k141_131341||lt2gene Not-determined
18-101-final-viral-combined_k141_213417||lt2gene Not-determined
18-101-final-viral-combined_k141_197027||lt2gene Low-quality
18-101-final-viral-combined_k141_79996||lt2gene Not-determined
18-101-final-viral-combined_k141_64477||lt2gene Low-quality
18-101-final-viral-combined_k141_162989||lt2gene Low-quality
18-101-final-viral-combined_k141_97649||lt2gene Low-quality
18-101-final-viral-combined_k141_163015||lt2gene Low-quality
18-101-final-viral-combined_k141_64550||lt2gene Not-determined
18-101-final-viral-combined_k141_197132||lt2gene Not-determined
18-101-final-viral-combined_k141_163062||lt2gene Low-quality
18-101-final-viral-combined_k141_295713||lt2gene Not-determined
18-101-final-viral-combined_k141_146975||lt2gene Low-quality
18-101-final-viral-combined_k141_15371||lt2gene Low-quality
18-101-final-viral-combined_k141_295727||lt2gene Low-quality
18-101-final-viral-combined_k141_97776||lt2gene Not-determined
18-101-final-viral-combined_k141_229414||lt2gene Low-quality
18-101-final-viral-combined_k141_163101||lt2gene Low-quality
18-101-final-viral-combined_k141_147009||lt2gene Low-quality
18-101-final-viral-combined_k141_64638||lt2gene Not-determined
18-101-final-viral-combined_k141_229438||lt2gene Low-quality
18-101-final-viral-combined_k141_112406||lt2gene Low-quality
18-101-final-viral-combined_k141_112418||lt2gene Not-determined
18-101-final-viral-combined_k141_179140||lt2gene Not-determined
18-101-final-viral-combined_k141_179178||lt2gene Low-quality
18-101-final-viral-combined_k141_377608||lt2gene Low-quality
18-101-final-viral-combined_k141_179254||lt2gene Not-determined
18-101-final-viral-combined_k141_229473||lt2gene Low-quality
18-101-final-viral-combined_k141_312385||lt2gene Not-determined
18-101-final-viral-combined_k141_343699||lt2gene Not-determined
18-101-final-viral-combined_k141_328919||lt2gene Low-quality
18-101-final-viral-combined_k141_328966||lt2gene Low-quality
18-101-final-viral-combined_k141_393284||lt2gene Low-quality
18-101-final-viral-combined_k141_377665||lt2gene Low-quality
18-101-final-viral-combined_k141_278346||lt2gene Not-determined
18-101-final-viral-combined_k141_377683||lt2gene Low-quality
18-101-final-viral-combined_k141_312485||lt2gene Not-determined
18-101-final-viral-combined_k141_131596||lt2gene Not-determined
18-101-final-viral-combined_k141_80413||lt2gene Low-quality
18-101-final-viral-combined_k141_147187||lt2gene Low-quality
18-101-final-viral-combined_k141_377731||lt2gene Low-quality
18-101-final-viral-combined_k141_329046||lt2gene Not-determined
18-101-final-viral-combined_k141_179324||lt2gene Low-quality
18-101-final-viral-combined_k141_246497||lt2gene Low-quality
18-101-final-viral-combined_k141_15689||lt2gene Not-determined
18-101-final-viral-combined_k141_278470||lt2gene Not-determined
18-101-final-viral-combined_k141_393338||lt2gene Low-quality
18-101-final-viral-combined_k141_98130||lt2gene Low-quality
18-101-final-viral-combined_k141_377777||lt2gene Low-quality
18-101-final-viral-combined_k141_213847||lt2gene Low-quality
18-101-final-viral-combined_k141_377835||lt2gene Low-quality
18-101-final-viral-combined_k141_65032||lt2gene Low-quality
18-101-final-viral-combined_k141_65079||lt2gene Low-quality
18-101-final-viral-combined_k141_179466||lt2gene Not-determined
18-101-final-viral-combined_k141_343923||lt2gene Low-quality
18-101-final-viral-combined_k141_65085||lt2gene Low-quality
18-101-final-viral-combined_k141_278643||lt2gene Low-quality
18-101-final-viral-combined_k141_278660||lt2gene Low-quality
18-101-final-viral-combined_k141_278665||lt2gene Low-quality
18-101-final-viral-combined_k141_229623||lt2gene Low-quality
18-101-final-viral-combined_k141_47591||lt2gene Low-quality
18-101-final-viral-combined_k141_47597||lt2gene Not-determined
18-101-final-viral-combined_k141_15883||lt2gene Low-quality
18-101-final-viral-combined_k141_179576||lt2gene Low-quality
18-101-final-viral-combined_k141_278791||lt2gene Low-quality
18-101-final-viral-combined_k141_98348||lt2gene Low-quality
18-101-final-viral-combined_k141_65220||lt2gene Not-determined
18-101-final-viral-combined_k141_393620||lt2gene Low-quality
18-101-final-viral-combined_k141_31554||lt2gene Not-determined
18-101-final-viral-combined_k141_179707||lt2gene Low-quality
18-101-final-viral-combined_k141_278879||lt2gene Not-determined
18-101-final-viral-combined_k141_163670||lt2gene Low-quality
18-101-final-viral-combined_k141_278888||lt2gene Low-quality
18-101-final-viral-combined_k141_80967||lt2gene Low-quality
18-101-final-viral-combined_k141_98420||lt2gene Not-determined
18-101-final-viral-combined_k141_112857||lt2gene Low-quality
18-101-final-viral-combined_k141_179775||lt2gene Low-quality
18-101-final-viral-combined_k141_112879||lt2gene Low-quality
18-101-final-viral-combined_k141_179794||lt2gene Not-determined
18-101-final-viral-combined_k141_163744||lt2gene Low-quality
18-101-final-viral-combined_k141_163753||lt2gene Low-quality
18-101-final-viral-combined_k141_65317||lt2gene Low-quality
18-101-final-viral-combined_k141_98513||lt2gene Not-determined
18-101-final-viral-combined_k141_112916||lt2gene Low-quality
18-101-final-viral-combined_k141_65368||lt2gene Low-quality
18-101-final-viral-combined_k141_81134||lt2gene Low-quality
18-101-final-viral-combined_k141_47930||lt2gene Low-quality
18-101-final-viral-combined_k141_112982||lt2gene Not-determined
18-101-final-viral-combined_k141_16170||lt2gene Low-quality
18-101-final-viral-combined_k141_279078||lt2gene Low-quality
18-101-final-viral-combined_k141_81241||lt2gene Low-quality
18-101-final-viral-combined_k141_393871||lt2gene Low-quality
18-101-final-viral-combined_k141_279102||lt2gene Low-quality
18-101-final-viral-combined_k141_16219||lt2gene Low-quality
18-101-final-viral-combined_k141_147681||lt2gene Low-quality
18-101-final-viral-combined_k141_31879||lt2gene Not-determined
18-101-final-viral-combined_k141_378332||lt2gene Low-quality
18-101-final-viral-combined_k141_344521||lt2gene Low-quality
18-101-final-viral-combined_k141_147777||lt2gene Low-quality
18-101-final-viral-combined_k141_48119||lt2gene Not-determined
18-101-final-viral-combined_k141_180146||lt2gene Low-quality
18-101-final-viral-combined_k141_279200||lt2gene Low-quality
18-101-final-viral-combined_k141_16337||lt2gene Low-quality
18-101-final-viral-combined_k141_48207||lt2gene Not-determined
18-101-final-viral-combined_k141_48213||lt2gene Low-quality
18-101-final-viral-combined_k141_344732||lt2gene Low-quality
18-101-final-viral-combined_k141_180283||lt2gene Low-quality
18-101-final-viral-combined_k141_48258||lt2gene Low-quality
18-101-final-viral-combined_k141_164216||lt2gene Low-quality
18-101-final-viral-combined_k141_32094||lt2gene Low-quality
18-101-final-viral-combined_k141_113438||lt2gene Not-determined
18-101-final-viral-combined_k141_164241||lt2gene Low-quality
18-101-final-viral-combined_k141_32172||lt2gene Not-determined
18-101-final-viral-combined_k141_180357||lt2gene Low-quality
18-101-final-viral-combined_k141_48310||lt2gene Not-determined
18-101-final-viral-combined_k141_394238||lt2gene Low-quality
18-101-final-viral-combined_k141_81688||lt2gene Low-quality
18-101-final-viral-combined_k141_180403||lt2gene Low-quality
18-101-final-viral-combined_k141_344884||lt2gene Low-quality
18-101-final-viral-combined_k141_48375||lt2gene Not-determined
18-101-final-viral-combined_k141_279482||lt2gene Low-quality
18-101-final-viral-combined_k141_48403||lt2gene Not-determined
18-101-final-viral-combined_k141_32380||lt2gene Low-quality
18-101-final-viral-combined_k141_394403||lt2gene Low-quality
18-101-final-viral-combined_k141_345007||lt2gene Low-quality
18-101-final-viral-combined_k141_279518||lt2gene Low-quality
18-101-final-viral-combined_k141_81891||lt2gene Low-quality
18-101-final-viral-combined_k141_81920||lt2gene Not-determined
18-101-final-viral-combined_k141_48620||lt2gene Low-quality
18-101-final-viral-combined_k141_48722||lt2gene Low-quality
18-101-final-viral-combined_k141_164527||lt2gene Low-quality
18-101-final-viral-combined_k141_180791||lt2gene Low-quality
18-101-final-viral-combined_k141_32717||lt2gene Low-quality
18-101-final-viral-combined_k141_180802||lt2gene Low-quality
18-101-final-viral-combined_k141_82036||lt2gene Low-quality
18-101-final-viral-combined_k141_180812||lt2gene Low-quality
18-101-final-viral-combined_k141_180857||lt2gene Low-quality
18-101-final-viral-combined_k141_345247||lt2gene Low-quality
18-101-final-viral-combined_k141_82127||lt2gene Low-quality
18-101-final-viral-combined_k141_345267||lt2gene Low-quality
18-101-final-viral-combined_k141_32823||lt2gene Low-quality
18-101-final-viral-combined_k141_114101||lt2gene Low-quality
18-101-final-viral-combined_k141_180949||lt2gene Low-quality
18-101-final-viral-combined_k141_82220||lt2gene Low-quality
18-101-final-viral-combined_k141_114323||lt2gene Not-determined
18-101-final-viral-combined_k141_114351||lt2gene Low-quality
18-101-final-viral-combined_k141_49107||lt2gene Not-determined
18-101-final-viral-combined_k141_114584||lt2gene Not-determined
18-101-final-viral-combined_k141_114941||lt2gene Low-quality
18-101-final-viral-combined_k141_114999||lt2gene Not-determined
18-101_k141_180983 Not-determined
18-101_k141_329059 Not-determined
18-101_k141_246796 Not-determined
18-101_k141_131626 Not-determined
18-101_k141_263251 Not-determined
18-101_k141_3 Not-determined
18-101_k141_279704 Not-determined
18-101_k141_279705 Not-determined
18-101_k141_361966 Not-determined
18-101_k141_16462 Not-determined
18-101_k141_296158 Not-determined
18-101_k141_49367 Not-determined
18-101_k141_213891 Not-determined
18-101_k141_115179 Not-determined
18-101_k141_329070 Not-determined
18-101_k141_65824 Not-determined
18-101_k141_361978 Low-quality
18-101_k141_148088 Not-determined
18-101_k141_329079 Not-determined
18-101_k141_49376 Not-determined
18-101_k141_230358 Not-determined
18-101_k141_98728 Not-determined
18-101_k141_378447 Not-determined
18-101_k141_230360 Not-determined
18-101_k141_98731 Not-determined
18-101_k141_230363 Not-determined
18-101_k141_230365 Not-determined
18-101_k141_329087 Not-determined
18-101_k141_49386 Not-determined
18-101_k141_312637 Not-determined
18-101_k141_296166 Not-determined
18-101_k141_345535 Not-determined
18-101_k141_312638 Not-determined
18-101_k141_279723 Not-determined
18-101_k141_16464 Not-determined
18-101_k141_49388 Not-determined
18-101_k141_38 Low-quality
18-101_k141_312644 Not-determined
18-101_k141_197447 Not-determined
18-101_k141_180997 Not-determined
18-101_k141_82286 Not-determined
18-101_k141_329103 Not-determined
18-101_k141_82289 Not-determined
18-101_k141_32929 Not-determined

| | |
|---|---|
| 18-101_k141_82293 | Low-quality |
| 18-101_k141_362001 | Not-determined |
| 18-101_k141_32935 | Not-determined |
| 18-101_k141_362002 | Not-determined |
| 18-101_k141_32938 | Not-determined |
| 18-101_k141_181010 | Not-determined |
| 18-101_k141_148104 | Not-determined |
| 18-101_k141_345557 | Low-quality |
| 18-101_k141_230376 | Not-determined |
| 18-101_k141_164567 | Not-determined |
| 18-101_k141_312656 | Not-determined |
| 18-101_k141_51 | Not-determined |
| 18-101_k141_49410 | Not-determined |
| 18-101_k141_362009 | Not-determined |
| 18-101_k141_16475 | Not-determined |
| 18-101_k141_32948 | Not-determined |
| 18-101_k141_16481 | Not-determined |
| 18-101_k141_345573 | Not-determined |
| 18-101_k141_16485 | Not-determined |
| 18-101_k141_16486 | Not-determined |
| 18-101_k141_378477 | Not-determined |
| 18-101_k141_312671 | Not-determined |
| 18-101_k141_49427 | Not-determined |
| 18-101_k141_197462 | Not-determined |
| 18-101_k141_16492 | Not-determined |
| 18-101_k141_164576 | Not-determined |
| 18-101_k141_148127 | Not-determined |
| 18-101_k141_197470 | Not-determined |
| 18-101_k141_164583 | Not-determined |
| 18-101_k141_312676 | Not-determined |
| 18-101_k141_345584 | Not-determined |
| 18-101_k141_131693 | Not-determined |
| 18-101_k141_131695 | Not-determined |
| 18-101_k141_296204 | Not-determined |
| 18-101_k141_197480 | Low-quality |
| 18-101_k141_197481 | Not-determined |
| 18-101_k141_131700 | Not-determined |
| 18-101_k141_82317 | Not-determined |
| 18-101_k141_197486 | Not-determined |
| 18-101_k141_49438 | Not-determined |
| 18-101_k141_75 | Not-determined |
| 18-101_k141_197488 | Not-determined |
| 18-101_k141_362016 | Not-determined |
| 18-101_k141_362020 | Not-determined |
| 18-101_k141_378484 | Not-determined |
| 18-101_k141_32983 | Not-determined |
| 18-101_k141_362023 | Not-determined |
| 18-101_k141_329107 | Low-quality |
| 18-101_k141_197495 | Not-determined |
| 18-101_k141_98772 | Low-quality |
| 18-101_k141_92 | Not-determined |
| 18-101_k141_246805 | Not-determined |
| 18-101_k141_378494 | Not-determined |
| 18-101_k141_246806 | Not-determined |
| 18-101_k141_329114 | Not-determined |
| 18-101_k141_246807 | Not-determined |
| 18-101_k141_115213 | Not-determined |
| 18-101_k141_131714 | Not-determined |
| 18-101_k141_33005 | Not-determined |
| 18-101_k141_164589 | Low-quality |
| 18-101_k141_230384 | Not-determined |
| 18-101_k141_197509 | Low-quality |
| 18-101_k141_246814 | Not-determined |
| 18-101_k141_115230 | Not-determined |
| 18-101_k141_263297 | Not-determined |
| 18-101_k141_106 | Not-determined |
| 18-101_k141_329136 | Low-quality |
| 18-101_k141_312707 | Not-determined |
| 18-101_k141_107 | Not-determined |
| 18-101_k141_329140 | Not-determined |
| 18-101_k141_263305 | Not-determined |
| 18-101_k141_296228 | Not-determined |
| 18-101_k141_378496 | Not-determined |
| 18-101_k141_115246 | Not-determined |
| 18-101_k141_230399 | Not-determined |
| 18-101_k141_312716 | Not-determined |
| 18-101_k141_148157 | Not-determined |
| 18-101_k141_345605 | Low-quality |
| 18-101_k141_82353 | Not-determined |
| 18-101_k141_181065 | Not-determined |
| 18-101_k141_181066 | Not-determined |
| 18-101_k141_312726 | Not-determined |
| 18-101_k141_312727 | Not-determined |
| 18-101_k141_131753 | Not-determined |
| 18-101_k141_378514 | Not-determined |
| 18-101_k141_164590 | Not-determined |
| 18-101_k141_131763 | Not-determined |
| 18-101_k141_148170 | Not-determined |
| 18-101_k141_279746 | Not-determined |
| 18-101_k141_131766 | Not-determined |
| 18-101_k141_98804 | Not-determined |
| 18-101_k141_329152 | Not-determined |
| 18-101_k141_33033 | Not-determined |
| 18-101_k141_82382 | Not-determined |
| 18-101_k141_129 | Not-determined |
| 18-101_k141_132 | Low-quality |
| 18-101_k141_246843 | Not-determined |
| 18-101_k141_135 | Not-determined |
| 18-101_k141_296251 | Not-determined |
| 18-101_k141_246844 | Not-determined |
| 18-101_k141_98812 | Not-determined |
| 18-101_k141_329161 | Not-determined |
| 18-101_k141_131776 | Low-quality |
| 18-101_k141_312753 | Not-determined |
| 18-101_k141_152 | Not-determined |
| 18-101_k141_49499 | Not-determined |
| 18-101_k141_115292 | Not-determined |
| 18-101_k141_362074 | Not-determined |
| 18-101_k141_362075 | Not-determined |
| 18-101_k141_279763 | Not-determined |
| 18-101_k141_115296 | Not-determined |
| 18-101_k141_181092 | Not-determined |
| 18-101_k141_181094 | Not-determined |
| 18-101_k141_82389 | Not-determined |
| 18-101_k141_312771 | Not-determined |
| 18-101_k141_164604 | Not-determined |
| 18-101_k141_230450 | Not-determined |
| 18-101_k141_230452 | Not-determined |
| 18-101_k141_296279 | Not-determined |
| 18-101_k141_213957 | Not-determined |
| 18-101_k141_230459 | Not-determined |
| 18-101_k141_164612 | Not-determined |
| 18-101_k141_181105 | Not-determined |
| 18-101_k141_362087 | Not-determined |
| 18-101_k141_213962 | Not-determined |
| 18-101_k141_213963 | Not-determined |
| 18-101_k141_213964 | Not-determined |
| 18-101_k141_148182 | Not-determined |
| 18-101_k141_296285 | Not-determined |
| 18-101_k141_362092 | Not-determined |
| 18-101_k141_345622 | Not-determined |
| 18-101_k141_164621 | Not-determined |
| 18-101_k141_279797 | Not-determined |
| 18-101_k141_213981 | Not-determined |
| 18-101_k141_213982 | Not-determined |
| 18-101_k141_246864 | Not-determined |
| 18-101_k141_173 | Not-determined |
| 18-101_k141_329173 | Not-determined |
| 18-101_k141_131809 | Not-determined |
| 18-101_k141_82417 | Low-quality |
| 18-101_k141_329174 | Low-quality |
| 18-101_k141_296300 | Not-determined |
| 18-101_k141_279810 | Not-determined |
| 18-101_k141_181116 | Not-determined |
| 18-101_k141_115317 | Not-determined |
| 18-101_k141_16540 | Not-determined |
| 18-101_k141_312780 | Not-determined |
| 18-101_k141_180 | Not-determined |
| 18-101_k141_181 | Not-determined |
| 18-101_k141_312788 | Not-determined |
| 18-101_k141_131815 | Not-determined |
| 18-101_k141_312793 | Not-determined |
| 18-101_k141_115323 | Not-determined |
| 18-101_k141_378558 | Not-determined |
| 18-101_k141_345654 | Not-determined |
| 18-101_k141_345655 | Not-determined |
| 18-101_k141_196 | Not-determined |
| 18-101_k141_148223 | Not-determined |
| 18-101_k141_164646 | Not-determined |
| 18-101_k141_82436 | Not-determined |
| 18-101_k141_378559 | Low-quality |
| 18-101_k141_279834 | Not-determined |
| 18-101_k141_98823 | Not-determined |
| 18-101_k141_214026 | Not-determined |
| 18-101_k141_181139 | Not-determined |
| 18-101_k141_164655 | Not-determined |
| 18-101_k141_115343 | Not-determined |
| 18-101_k141_164656 | Not-determined |
| 18-101_k141_214037 | Not-determined |
| 18-101_k141_197516 | Low-quality |
| 18-101_k141_197518 | Not-determined |
| 18-101_k141_378565 | Not-determined |
| 18-101_k141_214048 | Not-determined |
| 18-101_k141_197524 | Not-determined |
| 18-101_k141_115353 | Not-determined |
| 18-101_k141_228 | Not-determined |
| 18-101_k141_197529 | Not-determined |
| 18-101_k141_362130 | Low-quality |
| 18-101_k141_197533 | Not-determined |
| 18-101_k141_16570 | Not-determined |
| 18-101_k141_312837 | Not-determined |
| 18-101_k141_232 | Not-determined |
| 18-101_k141_82468 | Low-quality |
| 18-101_k141_148250 | Not-determined |
| 18-101_k141_16574 | Not-determined |
| 18-101_k141_246894 | Not-determined |
| 18-101_k141_263323 | Not-determined |
| 18-101_k141_115370 | Not-determined |
| 18-101_k141_246903 | Not-determined |
| 18-101_k141_148259 | Not-determined |
| 18-101_k141_148260 | Not-determined |
| 18-101_k141_115374 | Not-determined |
| 18-101_k141_246908 | Not-determined |
| 18-101_k141_16588 | Not-determined |
| 18-101_k141_329207 | Not-determined |
| 18-101_k141_378589 | Low-quality |
| 18-101_k141_378591 | Not-determined |
| 18-101_k141_378594 | Not-determined |
| 18-101_k141_230504 | Not-determined |
| 18-101_k141_279865 | Not-determined |
| 18-101_k141_197556 | Low-quality |
| 18-101_k141_279868 | Not-determined |
| 18-101_k141_345676 | Not-determined |
| 18-101_k141_197558 | Not-determined |
| 18-101_k141_329225 | Low-quality |
| 18-101_k141_255 | Not-determined |
| 18-101_k141_378597 | Not-determined |
| 18-101_k141_65838 | Low-quality |
| 18-101_k141_263329 | Low-quality |
| 18-101_k141_263 | Not-determined |
| 18-101_k141_263333 | Not-determined |
| 18-101_k141_264 | Not-determined |
| 18-101_k141_345682 | Not-determined |
| 18-101_k141_296327 | Not-determined |
| 18-101_k141_263334 | Not-determined |
| 18-101_k141_263335 | Not-determined |
| 18-101_k141_345686 | Not-determined |
| 18-101_k141_65845 | Not-determined |
| 18-101_k141_131857 | Not-determined |
| 18-101_k141_148268 | Not-determined |
| 18-101_k141_279883 | Not-determined |
| 18-101_k141_230531 | Not-determined |
| 18-101_k141_16606 | Not-determined |
| 18-101_k141_197567 | Not-determined |
| 18-101_k141_82493 | Not-determined |
| 18-101_k141_131869 | Not-determined |
| 18-101_k141_362171 | Not-determined |
| 18-101_k141_131870 | Not-determined |
| 18-101_k141_65859 | Not-determined |
| 18-101_k141_98874 | Not-determined |
| 18-101_k141_181157 | Not-determined |
| 18-101_k141_291 | Not-determined |
| 18-101_k141_16618 | Not-determined |
| 18-101_k141_16620 | Not-determined |
| 18-101_k141_295 | Not-determined |
| 18-101_k141_181161 | Not-determined |
| 18-101_k141_214091 | Not-determined |
| 18-101_k141_148277 | Not-determined |
| 18-101_k141_230548 | Not-determined |
| 18-101_k141_131885 | Not-determined |
| 18-101_k141_16626 | Not-determined |
| 18-101_k141_49560 | Low-quality |
| 18-101_k141_230552 | Not-determined |
| 18-101_k141_246926 | Not-determined |
| 18-101_k141_148285 | Not-determined |
| 18-101_k141_131887 | Not-determined |
| 18-101_k141_279892 | Not-determined |
| 18-101_k141_246930 | Not-determined |
| 18-101_k141_246932 | Not-determined |
| 18-101_k141_303 | Not-determined |
| 18-101_k141_246937 | Not-determined |
| 18-101_k141_148294 | Not-determined |
| 18-101_k141_312872 | Not-determined |
| 18-101_k141_263360 | Not-determined |
| 18-101_k141_197596 | Not-determined |
| 18-101_k141_131897 | Not-determined |
| 18-101_k141_307 | Not-determined |
| 18-101_k141_148296 | Not-determined |
| 18-101_k141_279900 | Not-determined |
| 18-101_k141_181176 | Not-determined |
| 18-101_k141_16636 | Not-determined |
| 18-101_k141_317 | Not-determined |
| 18-101_k141_230568 | Not-determined |
| 18-101_k141_329268 | Not-determined |
| 18-101_k141_181180 | Not-determined |
| 18-101_k141_378631 | Not-determined |
| 18-101_k141_296360 | Not-determined |
| 18-101_k141_378635 | Not-determined |
| 18-101_k141_214103 | Not-determined |
| 18-101_k141_312888 | Not-determined |
| 18-101_k141_197611 | Low-quality |
| 18-101_k141_230576 | Not-determined |
| 18-101_k141_279914 | Not-determined |
| 18-101_k141_332 | Not-determined |
| 18-101_k141_296368 | Low-quality |
| 18-101_k141_197619 | Not-determined |
| 18-101_k141_181193 | Not-determined |
| 18-101_k141_263383 | Not-determined |
| 18-101_k141_312899 | Not-determined |
| 18-101_k141_230587 | Not-determined |
| 18-101_k141_16657 | Not-determined |
| 18-101_k141_16661 | Not-determined |
| 18-101_k141_296370 | Low-quality |
| 18-101_k141_214115 | Low-quality |
| 18-101_k141_378649 | Low-quality |
| 18-101_k141_263389 | Low-quality |
| 18-101_k141_181198 | Not-determined |
| 18-101_k141_16666 | Not-determined |
| 18-101_k141_329277 | Not-determined |
| 18-101_k141_353 | Not-determined |
| 18-101_k141_329279 | Not-determined |
| 18-101_k141_115407 | Low-quality |
| 18-101_k141_197626 | Not-determined |
| 18-101_k141_214118 | Not-determined |
| 18-101_k141_279937 | Low-quality |
| 18-101_k141_181207 | Not-determined |
| 18-101_k141_279939 | Not-determined |
| 18-101_k141_214122 | Not-determined |
| 18-101_k141_164716 | Not-determined |
| 18-101_k141_164718 | Not-determined |
| 18-101_k141_164724 | Not-determined |
| 18-101_k141_148314 | Not-determined |
| 18-101_k141_98917 | Not-determined |
| 18-101_k141_230595 | Not-determined |
| 18-101_k141_362202 | Not-determined |
| 18-101_k141_181222 | Not-determined |
| 18-101_k141_148324 | Not-determined |
| 18-101_k141_148325 | Not-determined |
| 18-101_k141_16697 | Not-determined |
| 18-101_k141_230602 | Not-determined |
| 18-101_k141_279947 | Not-determined |
| 18-101_k141_279948 | Not-determined |
| 18-101_k141_164736 | Low-quality |
| 18-101_k141_164737 | Not-determined |
| 18-101_k141_214139 | Not-determined |
| 18-101_k141_378677 | Not-determined |
| 18-101_k141_296392 | Not-determined |
| 18-101_k141_230612 | Not-determined |
| 18-101_k141_230614 | Low-quality |
| 18-101_k141_230617 | Not-determined |
| 18-101_k141_197654 | Not-determined |
| 18-101_k141_16701 | Not-determined |
| 18-101_k141_279961 | Low-quality |
| 18-101_k141_378685 | Not-determined |
| 18-101_k141_362205 | Not-determined |
| 18-101_k141_82545 | Not-determined |
| 18-101_k141_33049 | Not-determined |
| 18-101_k141_197658 | Not-determined |
| 18-101_k141_98943 | Not-determined |
| 18-101_k141_279967 | Low-quality |
| 18-101_k141_98944 | Not-determined |
| 18-101_k141_49585 | Low-quality |
| 18-101_k141_98945 | Not-determined |
| 18-101_k141_16705 | Low-quality |
| 18-101_k141_362207 | Not-determined |
| 18-101_k141_345716 | Not-determined |
| 18-101_k141_181239 | Not-determined |
| 18-101_k141_98951 | Not-determined |
| 18-101_k141_164745 | Not-determined |
| 18-101_k141_65908 | Not-determined |
| 18-101_k141_362208 | Low-quality |
| 18-101_k141_247009 | Not-determined |
| 18-101_k141_214145 | Not-determined |
| 18-101_k141_98957 | Not-determined |
| 18-101_k141_131945 | Not-determined |
| 18-101_k141_164752 | Not-determined |
| 18-101_k141_279974 | Not-determined |
| 18-101_k141_279977 | Not-determined |
| 18-101_k141_230630 | Not-determined |
| 18-101_k141_131950 | Not-determined |
| 18-101_k141_279979 | Not-determined |
| 18-101_k141_378695 | Not-determined |
| 18-101_k141_115435 | Not-determined |
| 18-101_k141_115436 | Not-determined |
| 18-101_k141_197681 | Not-determined |
| 18-101_k141_345725 | Not-determined |
| 18-101_k141_214148 | Not-determined |
| 18-101_k141_410 | Not-determined |
| 18-101_k141_296429 | Not-determined |
| 18-101_k141_230643 | Not-determined |
| 18-101_k141_65917 | Not-determined |
| 18-101_k141_247027 | Not-determined |
| 18-101_k141_312925 | Not-determined |
| 18-101_k141_164759 | Not-determined |
| 18-101_k141_181248 | Not-determined |
| 18-101_k141_16719 | Not-determined |
| 18-101_k141_131964 | Not-determined |
| 18-101_k141_197694 | Not-determined |
| 18-101_k141_312929 | Not-determined |
| 18-101_k141_181253 | Not-determined |
| 18-101_k141_362226 | Not-determined |
| 18-101_k141_164769 | Not-determined |
| 18-101_k141_362234 | Not-determined |
| 18-101_k141_148335 | Not-determined |
| 18-101_k141_16733 | Not-determined |
| 18-101_k141_148338 | Not-determined |
| 18-101_k141_164777 | Not-determined |
| 18-101_k141_432 | Not-determined |
| 18-101_k141_362243 | Not-determined |
| 18-101_k141_263449 | Not-determined |
| 18-101_k141_247042 | Not-determined |
| 18-101_k141_16742 | Not-determined |
| 18-101_k141_98969 | Not-determined |
| 18-101_k141_16748 | Low-quality |
| 18-101_k141_296453 | Not-determined |
| 18-101_k141_16755 | Not-determined |
| 18-101_k141_296457 | Not-determined |
| 18-101_k141_65927 | Low-quality |
| 18-101_k141_131994 | Not-determined |
| 18-101_k141_148344 | Not-determined |
| 18-101_k141_296461 | Not-determined |
| 18-101_k141_65930 | Not-determined |
| 18-101_k141_280017 | Not-determined |
| 18-101_k141_362250 | Low-quality |
| 18-101_k141_280019 | Not-determined |
| 18-101_k141_65931 | Low-quality |
| 18-101_k141_230666 | Not-determined |
| 18-101_k141_312939 | Low-quality |
| 18-101_k141_362259 | Not-determined |
| 18-101_k141_131997 | Not-determined |
| 18-101_k141_263476 | Not-determined |
| 18-101_k141_65939 | Not-determined |
| 18-101_k141_65940 | Not-determined |
| 18-101_k141_362269 | Not-determined |
| 18-101_k141_230681 | Not-determined |
| 18-101_k141_280032 | Low-quality |
| 18-101_k141_280039 | Not-determined |
| 18-101_k141_230693 | Low-quality |
| 18-101_k141_115448 | Not-determined |
| 18-101_k141_362278 | Not-determined |
| 18-101_k141_312969 | Not-determined |
| 18-101_k141_460 | Not-determined |
| 18-101_k141_312972 | Not-determined |
| 18-101_k141_214152 | Low-quality |
| 18-101_k141_82629 | Not-determined |
| 18-101_k141_280049 | Not-determined |
| 18-101_k141_471 | Not-determined |
| 18-101_k141_164817 | Not-determined |
| 18-101_k141_132016 | Not-determined |
| 18-101_k141_230702 | Not-determined |
| 18-101_k141_312980 | Not-determined |
| 18-101_k141_476 | Not-determined |
| 18-101_k141_132021 | Low-quality |
| 18-101_k141_164825 | Not-determined |
| 18-101_k141_247075 | Low-quality |
| 18-101_k141_296483 | Not-determined |
| 18-101_k141_296486 | Not-determined |
| 18-101_k141_132028 | Low-quality |
| 18-101_k141_345789 | Not-determined |
| 18-101_k141_378711 | Low-quality |
| 18-101_k141_164835 | Not-determined |
| 18-101_k141_115465 | Not-determined |
| 18-101_k141_82644 | Not-determined |
| 18-101_k141_498 | Not-determined |
| 18-101_k141_33088 | Not-determined |
| 18-101_k141_247084 | Low-quality |
| 18-101_k141_115468 | Not-determined |
| 18-101_k141_115469 | Not-determined |
| 18-101_k141_132034 | Low-quality |
| 18-101_k141_98985 | Low-quality |
| 18-101_k141_362299 | Not-determined |
| 18-101_k141_49622 | Not-determined |
| 18-101_k141_115475 | Not-determined |
| 18-101_k141_82668 | Not-determined |
| 18-101_k141_115477 | Not-determined |
| 18-101_k141_296508 | Not-determined |
| 18-101_k141_98991 | Not-determined |
| 18-101_k141_247095 | Not-determined |
| 18-101_k141_164843 | Not-determined |
| 18-101_k141_329315 | Not-determined |
| 18-101_k141_98998 | Not-determined |
| 18-101_k141_98999 | Not-determined |
| 18-101_k141_115484 | Not-determined |
| 18-101_k141_513 | Not-determined |
| 18-101_k141_362303 | Not-determined |
| 18-101_k141_313008 | Not-determined |
| 18-101_k141_115492 | Not-determined |
| 18-101_k141_329321 | Not-determined |
| 18-101_k141_82679 | Not-determined |
| 18-101_k141_99007 | Not-determined |
| 18-101_k141_214201 | Not-determined |
| 18-101_k141_524 | Not-determined |
| 18-101_k141_280077 | Not-determined |
| 18-101_k141_263527 | Low-quality |
| 18-101_k141_527 | Not-determined |
| 18-101_k141_230767 | Not-determined |
| 18-101_k141_296535 | Not-determined |
| 18-101_k141_214210 | Not-determined |
| 18-101_k141_362315 | Low-quality |
| 18-101_k141_197760 | Not-determined |
| 18-101_k141_132060 | Not-determined |
| 18-101_k141_65962 | Not-determined |
| 18-101_k141_528 | Low-quality |
| 18-101_k141_82689 | Not-determined |
| 18-101_k141_148367 | Not-determined |
| 18-101_k141_82691 | Not-determined |
| 18-101_k141_378730 | Not-determined |
| 18-101_k141_296543 | Not-determined |
| 18-101_k141_378732 | Not-determined |
| 18-101_k141_362326 | Not-determined |
| 18-101_k141_296545 | Not-determined |
| 18-101_k141_362332 | Not-determined |
| 18-101_k141_115499 | Not-determined |
| 18-101_k141_533 | Low-quality |
| 18-101_k141_230774 | Low-quality |
| 18-101_k141_82697 | Not-determined |
| 18-101_k141_99018 | Not-determined |
| 18-101_k141_230777 | Not-determined |
| 18-101_k141_197772 | Not-determined |
| 18-101_k141_230779 | Low-quality |
| 18-101_k141_263531 | Not-determined |
| 18-101_k141_65988 | Not-determined |
| 18-101_k141_82703 | Not-determined |
| 18-101_k141_329358 | Not-determined |
| 18-101_k141_65992 | Not-determined |
| 18-101_k141_263541 | Not-determined |
| 18-101_k141_181316 | Not-determined |
| 18-101_k141_362352 | Not-determined |
| 18-101_k141_296567 | Not-determined |
| 18-101_k141_66007 | Not-determined |
| 18-101_k141_66009 | Not-determined |
| 18-101_k141_181330 | Not-determined |
| 18-101_k141_560 | Not-determined |
| 18-101_k141_99029 | Not-determined |
| 18-101_k141_181334 | Not-determined |
| 18-101_k141_33107 | Not-determined |
| 18-101_k141_49659 | Not-determined |
| 18-101_k141_148400 | Not-determined |
| 18-101_k141_49660 | Not-determined |
| 18-101_k141_181338 | Not-determined |
| 18-101_k141_280125 | Not-determined |
| 18-101_k141_230801 | Not-determined |
| 18-101_k141_181339 | Not-determined |
| 18-101_k141_33111 | Not-determined |
| 18-101_k141_49664 | Not-determined |
| 18-101_k141_132084 | Not-determined |
| 18-101_k141_132085 | Not-determined |
| 18-101_k141_99039 | Not-determined |
| 18-101_k141_345826 | Not-determined |
| 18-101_k141_345829 | Low-quality |
| 18-101_k141_329386 | Low-quality |
| 18-101_k141_115527 | Not-determined |
| 18-101_k141_148425 | Not-determined |
| 18-101_k141_378763 | Not-determined |
| 18-101_k141_164877 | Not-determined |
| 18-101_k141_378769 | Not-determined |
| 18-101_k141_132103 | Not-determined |
| 18-101_k141_49684 | Low-quality |
| 18-101_k141_82755 | Not-determined |
| 18-101_k141_33133 | Low-quality |
| 18-101_k141_33134 | Not-determined |
| 18-101_k141_345846 | Not-determined |
| 18-101_k141_214244 | Not-determined |
| 18-101_k141_115552 | Not-determined |
| 18-101_k141_579 | Low-quality |
| 18-101_k141_181362 | Not-determined |
| 18-101_k141_280152 | Not-determined |
| 18-101_k141_164887 | Not-determined |
| 18-101_k141_247158 | Not-determined |
| 18-101_k141_49689 | Not-determined |
| 18-101_k141_115556 | Not-determined |
| 18-101_k141_115559 | Not-determined |
| 18-101_k141_33138 | Not-determined |
| 18-101_k141_313050 | Not-determined |
| 18-101_k141_214258 | Not-determined |
| 18-101_k141_345862 | Not-determined |
| 18-101_k141_345864 | Not-determined |
| 18-101_k141_362387 | Low-quality |
| 18-101_k141_345865 | Not-determined |
| 18-101_k141_362388 | Not-determined |
| 18-101_k141_33140 | Not-determined |
| 18-101_k141_33142 | Not-determined |
| 18-101_k141_148450 | Not-determined |
| 18-101_k141_16794 | Not-determined |
| 18-101_k141_313060 | Not-determined |
| 18-101_k141_82765 | Low-quality |
| 18-101_k141_115567 | Not-determined |
| 18-101_k141_181384 | Not-determined |
| 18-101_k141_263563 | Not-determined |
| 18-101_k141_181385 | Not-determined |
| 18-101_k141_263564 | Low-quality |
| 18-101_k141_313066 | Low-quality |
| 18-101_k141_592 | Not-determined |
| 18-101_k141_596 | Not-determined |
| 18-101_k141_99075 | Not-determined |
| 18-101_k141_597 | Not-determined |
| 18-101_k141_247167 | Not-determined |
| 18-101_k141_345875 | Not-determined |
| 18-101_k141_132114 | Low-quality |
| 18-101_k141_263569 | Not-determined |
| 18-101_k141_247183 | Not-determined |
| 18-101_k141_263570 | Not-determined |
| 18-101_k141_313072 | Low-quality |
| 18-101_k141_214271 | Not-determined |
| 18-101_k141_345879 | Not-determined |
| 18-101_k141_230826 | Not-determined |
| 18-101_k141_115597 | Not-determined |
| 18-101_k141_33150 | Not-determined |
| 18-101_k141_115600 | Not-determined |
| 18-101_k141_362417 | Not-determined |
| 18-101_k141_49713 | Not-determined |
| 18-101_k141_345889 | Not-determined |
| 18-101_k141_263576 | Low-quality |
| 18-101_k141_197851 | Not-determined |
| 18-101_k141_99097 | Not-determined |
| 18-101_k141_115603 | Low-quality |
| 18-101_k141_49722 | Not-determined |
| 18-101_k141_247218 | Not-determined |
| 18-101_k141_49727 | Not-determined |

| | |
|---|---|
| 18-101_k141_115609 | Not-determined |
| 18-101_k141_33174 | Not-determined |
| 18-101_k141_99110 | Not-determined |
| 18-101_k141_148462 | Not-determined |
| 18-101_k141_280173 | Not-determined |
| 18-101_k141_362431 | Not-determined |
| 18-101_k141_280176 | Not-determined |
| 18-101_k141_247230 | Not-determined |
| 18-101_k141_49730 | Not-determined |
| 18-101_k141_620 | Not-determined |
| 18-101_k141_313078 | Not-determined |
| 18-101_k141_148474 | Not-determined |
| 18-101_k141_313080 | Low-quality |
| 18-101_k141_378796 | Not-determined |
| 18-101_k141_148478 | Not-determined |
| 18-101_k141_247237 | Not-determined |
| 18-101_k141_164896 | Not-determined |
| 18-101_k141_329414 | Not-determined |
| 18-101_k141_16854 | Not-determined |
| 18-101_k141_99116 | Not-determined |
| 18-101_k141_313086 | Not-determined |
| 18-101_k141_634 | Not-determined |
| 18-101_k141_164899 | Not-determined |
| 18-101_k141_636 | Low-quality |
| 18-101_k141_313091 | Not-determined |
| 18-101_k141_197883 | Not-determined |
| 18-101_k141_99126 | Not-determined |
| 18-101_k141_247254 | Not-determined |
| 18-101_k141_362457 | Not-determined |
| 18-101_k141_164907 | Not-determined |
| 18-101_k141_99134 | Not-determined |
| 18-101_k141_33197 | Not-determined |
| 18-101_k141_296578 | Not-determined |
| 18-101_k141_296582 | Not-determined |
| 18-101_k141_132138 | Not-determined |
| 18-101_k141_329440 | Low-quality |
| 18-101_k141_313098 | Not-determined |
| 18-101_k141_99149 | Not-determined |
| 18-101_k141_115621 | Not-determined |
| 18-101_k141_329441 | Not-determined |
| 18-101_k141_16881 | Not-determined |
| 18-101_k141_115624 | Not-determined |
| 18-101_k141_115625 | Not-determined |
| 18-101_k141_16883 | Low-quality |
| 18-101_k141_378825 | Not-determined |
| 18-101_k141_378826 | Not-determined |
| 18-101_k141_164923 | Not-determined |
| 18-101_k141_263612 | Not-determined |
| 18-101_k141_230843 | Not-determined |
| 18-101_k141_378837 | Not-determined |
| 18-101_k141_181452 | Low-quality |
| 18-101_k141_82811 | Not-determined |
| 18-101_k141_263618 | Not-determined |
| 18-101_k141_181455 | Not-determined |
| 18-101_k141_115638 | Not-determined |
| 18-101_k141_214325 | Not-determined |
| 18-101_k141_214327 | Not-determined |
| 18-101_k141_345932 | Not-determined |
| 18-101_k141_214331 | Not-determined |
| 18-101_k141_313113 | Not-determined |
| 18-101_k141_115645 | Low-quality |
| 18-101_k141_296603 | Not-determined |
| 18-101_k141_230852 | Not-determined |
| 18-101_k141_230854 | Not-determined |
| 18-101_k141_66067 | Not-determined |
| 18-101_k141_280227 | Low-quality |
| 18-101_k141_16917 | Not-determined |
| 18-101_k141_132154 | Low-quality |
| 18-101_k141_247272 | Not-determined |
| 18-101_k141_197904 | Not-determined |
| 18-101_k141_280229 | Not-determined |
| 18-101_k141_197905 | Not-determined |
| 18-101_k141_164938 | Not-determined |
| 18-101_k141_197906 | Not-determined |
| 18-101_k141_164941 | Not-determined |
| 18-101_k141_329453 | Not-determined |
| 18-101_k141_164944 | Not-determined |
| 18-101_k141_197912 | Not-determined |
| 18-101_k141_197914 | Not-determined |
| 18-101_k141_247288 | Not-determined |
| 18-101_k141_247289 | Not-determined |
| 18-101_k141_197916 | Not-determined |
| 18-101_k141_247291 | Not-determined |
| 18-101_k141_164949 | Not-determined |
| 18-101_k141_197930 | Not-determined |
| 18-101_k141_115674 | Not-determined |
| 18-101_k141_181481 | Not-determined |
| 18-101_k141_164956 | Not-determined |
| 18-101_k141_313132 | Low-quality |
| 18-101_k141_378877 | Not-determined |
| 18-101_k141_378881 | Not-determined |
| 18-101_k141_115684 | Not-determined |
| 18-101_k141_230864 | Not-determined |
| 18-101_k141_164963 | Not-determined |
| 18-101_k141_313140 | Low-quality |
| 18-101_k141_378891 | Not-determined |
| 18-101_k141_345958 | Not-determined |
| 18-101_k141_247298 | Not-determined |
| 18-101_k141_263659 | Not-determined |
| 18-101_k141_378901 | Not-determined |
| 18-101_k141_378902 | Not-determined |
| 18-101_k141_197951 | Not-determined |
| 18-101_k141_33288 | Not-determined |
| 18-101_k141_345962 | Not-determined |
| 18-101_k141_247307 | Not-determined |
| 18-101_k141_329482 | Not-determined |
| 18-101_k141_16927 | Not-determined |
| 18-101_k141_16928 | Not-determined |
| 18-101_k141_33300 | Not-determined |
| 18-101_k141_33301 | Not-determined |
| 18-101_k141_230883 | Not-determined |
| 18-101_k141_378912 | Not-determined |
| 18-101_k141_263675 | Not-determined |
| 18-101_k141_345973 | Not-determined |
| 18-101_k141_33314 | Not-determined |
| 18-101_k141_247315 | Not-determined |
| 18-101_k141_280270 | Not-determined |
| [illegible] | [illegible] |

| | |
|---|---|
| 18-101_k141_214936 | Not-determined |
| 18-101_k141_346490 | Not-determined |
| 18-101_k141_296978 | Not-determined |
| 18-101_k141_116262 | Not-determined |
| 18-101_k141_198315 | Not-determined |
| 18-101_k141_66297 | Not-determined |
| 18-101_k141_1103 | Not-determined |
| 18-101_k141_198318 | Not-determined |
| 18-101_k141_264066 | Not-determined |
| 18-101_k141_132628 | Not-determined |
| 18-101_k141_165361 | Not-determined |
| 18-101_k141_280649 | Not-determined |
| 18-101_k141_165362 | Not-determined |
| 18-101_k141_296997 | Not-determined |
| 18-101_k141_264080 | Not-determined |
| 18-101_k141_17257 | Not-determined |
| 18-101_k141_280653 | Not-determined |
| 18-101_k141_346500 | Not-determined |
| 18-101_k141_66324 | Low-quality |
| 18-101_k141_264086 | Not-determined |
| 18-101_k141_148874 | Not-determined |
| 18-101_k141_132644 | Low-quality |
| 18-101_k141_181973 | Not-determined |
| 18-101_k141_346510 | Not-determined |
| 18-101_k141_247683 | Not-determined |
| 18-101_k141_346512 | Not-determined |
| 18-101_k141_231161 | Not-determined |
| 18-101_k141_280671 | Not-determined |
| 18-101_k141_116297 | Not-determined |
| 18-101_k141_116298 | Low-quality |
| 18-101_k141_83304 | Not-determined |
| 18-101_k141_116299 | Not-determined |
| 18-101_k141_148884 | Low-quality |
| 18-101_k141_148888 | Not-determined |
| 18-101_k141_346514 | Low-quality |
| 18-101_k141_346519 | Low-quality |
| 18-101_k141_231171 | Not-determined |
| 18-101_k141_379272 | Not-determined |
| 18-101_k141_148896 | Not-determined |
| 18-101_k141_362899 | Low-quality |
| 18-101_k141_379275 | Not-determined |
| 18-101_k141_297010 | Not-determined |
| 18-101_k141_247702 | Not-determined |
| 18-101_k141_116307 | Low-quality |
| 18-101_k141_33577 | Not-determined |
| 18-101_k141_83324 | Not-determined |
| 18-101_k141_297017 | Not-determined |
| 18-101_k141_198354 | Low-quality |
| 18-101_k141_116312 | Not-determined |
| 18-101_k141_247707 | Not-determined |
| 18-101_k141_313411 | Low-quality |
| 18-101_k141_346529 | Not-determined |
| 18-101_k141_297025 | Not-determined |
| 18-101_k141_181981 | Not-determined |
| 18-101_k141_231189 | Not-determined |
| 18-101_k141_247719 | Not-determined |
| 18-101_k141_116323 | Not-determined |
| 18-101_k141_329820 | Not-determined |
| 18-101_k141_247721 | Low-quality |
| 18-101_k141_346536 | Not-determined |
| 18-101_k141_313415 | Not-determined |
| 18-101_k141_214974 | Not-determined |
| 18-101_k141_198368 | Low-quality |
| 18-101_k141_329822 | Not-determined |
| 18-101_k141_99526 | Not-determined |
| 18-101_k141_214978 | Not-determined |
| 18-101_k141_362916 | Not-determined |
| 18-101_k141_181997 | Not-determined |
| 18-101_k141_132673 | Not-determined |
| 18-101_k141_280703 | Not-determined |
| 18-101_k141_99533 | Low-quality |
| 18-101_k141_214985 | Not-determined |
| 18-101_k141_99536 | Not-determined |
| 18-101_k141_99538 | Not-determined |
| 18-101_k141_297040 | Not-determined |
| 18-101_k141_99542 | Not-determined |
| 18-101_k141_1160 | Not-determined |
| 18-101_k141_215000 | Not-determined |
| 18-101_k141_132691 | Not-determined |
| 18-101_k141_313424 | Not-determined |
| 18-101_k141_182019 | Not-determined |
| 18-101_k141_379288 | Not-determined |
| 18-101_k141_116337 | Not-determined |
| 18-101_k141_116339 | Not-determined |
| 18-101_k141_313429 | Not-determined |
| 18-101_k141_313430 | Not-determined |
| 18-101_k141_215011 | Not-determined |
| 18-101_k141_66390 | Not-determined |
| 18-101_k141_247737 | Not-determined |
| 18-101_k141_17283 | Not-determined |
| 18-101_k141_346549 | Not-determined |
| 18-101_k141_132713 | Not-determined |
| 18-101_k141_132715 | Not-determined |
| 18-101_k141_362947 | Not-determined |
| 18-101_k141_116357 | Low-quality |
| 18-101_k141_231223 | Low-quality |
| 18-101_k141_116358 | Not-determined |
| 18-101_k141_49793 | Not-determined |
| 18-101_k141_165383 | Not-determined |
| 18-101_k141_165384 | Not-determined |
| 18-101_k141_379296 | Not-determined |
| 18-101_k141_165386 | Not-determined |
| 18-101_k141_379299 | Not-determined |
| 18-101_k141_231232 | Not-determined |
| 18-101_k141_198386 | Not-determined |
| 18-101_k141_215036 | Low-quality |
| 18-101_k141_198387 | Not-determined |
| 18-101_k141_1174 | Not-determined |
| 18-101_k141_148939 | Low-quality |
| 18-101_k141_99549 | Not-determined |
| 18-101_k141_215040 | Not-determined |
| 18-101_k141_215044 | Not-determined |
| 18-101_k141_182045 | Not-determined |
| 18-101_k141_280723 | Not-determined |
| 18-101_k141_264129 | Not-determined |
| 18-101_k141_1192 | Not-determined |
| 18-101_k141_66414 | Not-determined |
| 18-101_k141_99554 | Not-determined |
| 18-101_k141_313462 | Low-quality |
| 18-101_k141_280736 | Not-determined |
| 18-101_k141_247763 | Not-determined |
| 18-101_k141_297073 | Not-determined |
| 18-101_k141_148951 | Not-determined |
| 18-101_k141_362977 | Not-determined |
| 18-101_k141_49835 | Not-determined |
| 18-101_k141_1205 | Not-determined |
| 18-101_k141_264144 | Not-determined |
| 18-101_k141_17288 | Not-determined |
| 18-101_k141_313479 | Not-determined |
| 18-101_k141_329852 | Not-determined |
| 18-101_k141_346564 | Not-determined |
| 18-101_k141_313482 | Not-determined |
| 18-101_k141_362982 | Low-quality |
| 18-101_k141_264146 | Low-quality |
| 18-101_k141_231242 | Not-determined |
| 18-101_k141_297078 | Not-determined |
| 18-101_k141_264149 | Not-determined |
| 18-101_k141_116384 | Low-quality |
| 18-101_k141_362985 | Not-determined |
| 18-101_k141_99572 | Not-determined |
| 18-101_k141_17290 | Low-quality |
| 18-101_k141_297087 | Not-determined |
| 18-101_k141_148962 | Not-determined |
| 18-101_k141_231248 | Not-determined |
| 18-101_k141_346582 | Not-determined |
| 18-101_k141_297094 | Not-determined |
| 18-101_k141_346587 | Low-quality |
| 18-101_k141_297098 | Not-determined |
| 18-101_k141_17302 | Not-determined |
| 18-101_k141_264169 | Not-determined |
| 18-101_k141_231266 | Not-determined |
| 18-101_k141_66460 | Not-determined |
| 18-101_k141_49852 | Not-determined |
| 18-101_k141_132796 | Not-determined |
| 18-101_k141_17307 | Low-quality |
| 18-101_k141_297130 | Not-determined |
| 18-101_k141_297134 | Not-determined |
| 18-101_k141_297136 | Not-determined |
| 18-101_k141_17314 | Low-quality |
| 18-101_k141_297137 | Not-determined |
| 18-101_k141_363013 | Not-determined |
| 18-101_k141_17319 | Not-determined |
| 18-101_k141_49866 | Not-determined |
| 18-101_k141_297144 | Not-determined |
| 18-101_k141_198431 | Not-determined |
| 18-101_k141_99601 | Not-determined |
| 18-101_k141_198433 | Not-determined |
| 18-101_k141_17327 | Low-quality |
| 18-101_k141_17329 | Not-determined |
| 18-101_k141_17331 | Not-determined |
| 18-101_k141_198435 | Not-determined |
| 18-101_k141_198438 | Not-determined |
| 18-101_k141_297157 | Not-determined |
| 18-101_k141_1223 | Not-determined |
| 18-101_k141_1224 | Not-determined |
| 18-101_k141_17336 | Not-determined |
| 18-101_k141_17337 | Not-determined |
| 18-101_k141_49885 | Not-determined |
| 18-101_k141_215058 | Not-determined |
| 18-101_k141_99614 | Not-determined |
| 18-101_k141_49888 | Not-determined |
| 18-101_k141_198447 | Not-determined |
| 18-101_k141_49891 | Not-determined |
| 18-101_k141_247808 | Not-determined |
| 18-101_k141_264193 | Not-determined |
| 18-101_k141_215074 | Not-determined |
| 18-101_k141_379343 | Not-determined |
| 18-101_k141_379344 | Not-determined |
| 18-101_k141_215078 | Not-determined |
| 18-101_k141_215079 | Low-quality |
| 18-101_k141_49902 | Not-determined |
| 18-101_k141_264204 | Not-determined |
| 18-101_k141_297174 | Not-determined |
| 18-101_k141_379346 | Not-determined |
| 18-101_k141_264205 | Not-determined |
| 18-101_k141_313528 | Not-determined |
| 18-101_k141_17355 | Not-determined |
| 18-101_k141_280758 | Not-determined |
| 18-101_k141_99621 | Not-determined |
| 18-101_k141_264214 | Not-determined |
| 18-101_k141_1237 | Not-determined |
| 18-101_k141_363067 | Not-determined |
| 18-101_k141_33612 | Not-determined |
| 18-101_k141_1240 | Not-determined |
| 18-101_k141_66493 | Not-determined |
| 18-101_k141_1244 | Not-determined |
| 18-101_k141_363071 | Not-determined |
| 18-101_k141_363072 | Not-determined |
| 18-101_k141_33617 | Not-determined |
| 18-101_k141_215101 | Not-determined |
| 18-101_k141_33619 | Not-determined |
| 18-101_k141_215103 | Not-determined |
| 18-101_k141_363075 | Not-determined |
| 18-101_k141_280770 | Not-determined |
| 18-101_k141_346618 | Not-determined |
| 18-101_k141_313550 | Not-determined |
| 18-101_k141_132810 | Not-determined |
| 18-101_k141_329865 | Not-determined |
| 18-101_k141_264239 | Not-determined |
| 18-101_k141_66505 | Not-determined |
| 18-101_k141_1260 | Not-determined |
| 18-101_k141_83336 | Not-determined |
| 18-101_k141_132820 | Not-determined |
| 18-101_k141_379361 | Not-determined |
| 18-101_k141_165445 | Low-quality |
| 18-101_k141_83344 | Not-determined |
| 18-101_k141_280780 | Not-determined |
| 18-101_k141_379366 | Not-determined |
| 18-101_k141_313558 | Not-determined |
| 18-101_k141_33633 | Not-determined |
| 18-101_k141_49927 | Not-determined |
| 18-101_k141_280787 | Not-determined |
| 18-101_k141_280793 | Not-determined |
| 18-101_k141_297197 | Not-determined |
| 18-101_k141_215135 | Low-quality |
| 18-101_k141_379369 | Not-determined |
| 18-101_k141_215137 | Not-determined |
| 18-101_k141_297200 | Not-determined |
| 18-101_k141_33646 | Not-determined |
| 18-101_k141_297204 | Not-determined |
| 18-101_k141_182060 | Not-determined |
| 18-101_k141_198486 | Not-determined |
| 18-101_k141_1282 | Not-determined |
| 18-101_k141_198488 | Low-quality |
| 18-101_k141_313581 | Not-determined |
| 18-101_k141_313582 | Not-determined |
| 18-101_k141_297213 | Not-determined |
| 18-101_k141_379373 | Not-determined |
| 18-101_k141_33658 | Not-determined |
| 18-101_k141_329876 | Not-determined |
| 18-101_k141_148975 | Low-quality |
| 18-101_k141_132827 | Not-determined |
| 18-101_k141_17414 | Not-determined |
| 18-101_k141_148978 | Not-determined |
| 18-101_k141_99645 | Not-determined |
| 18-101_k141_99649 | Not-determined |
| 18-101_k141_231309 | Not-determined |
| 18-101_k141_182070 | Low-quality |
| 18-101_k141_165468 | Not-determined |
| 18-101_k141_247851 | Not-determined |
| 18-101_k141_215147 | Not-determined |
| 18-101_k141_231317 | Low-quality |
| 18-101_k141_99653 | Low-quality |
| 18-101_k141_99655 | Not-determined |
| 18-101_k141_313606 | Not-determined |
| 18-101_k141_379389 | Not-determined |
| 18-101_k141_99661 | Not-determined |
| 18-101_k141_49937 | Not-determined |
| 18-101_k141_379397 | Not-determined |
| 18-101_k141_17423 | Not-determined |
| 18-101_k141_297227 | Not-determined |
| 18-101_k141_329897 | Not-determined |
| 18-101_k141_215155 | Not-determined |
| 18-101_k141_231335 | Not-determined |
| 18-101_k141_264282 | Not-determined |
| 18-101_k141_215157 | Not-determined |
| 18-101_k141_1309 | Not-determined |
| 18-101_k141_215159 | Not-determined |
| 18-101_k141_346669 | Not-determined |
| 18-101_k141_231344 | Not-determined |
| 18-101_k141_215168 | Not-determined |
| 18-101_k141_231346 | Not-determined |
| 18-101_k141_149015 | Not-determined |
| 18-101_k141_1316 | Not-determined |
| 18-101_k141_66536 | Not-determined |
| 18-101_k141_132848 | Not-determined |
| 18-101_k141_99689 | Not-determined |
| 18-101_k141_247871 | Not-determined |
| 18-101_k141_215176 | Not-determined |
| 18-101_k141_379405 | Not-determined |
| 18-101_k141_1322 | Not-determined |
| 18-101_k141_99692 | Not-determined |
| 18-101_k141_198517 | Not-determined |
| 18-101_k141_198520 | Not-determined |
| 18-101_k141_33689 | Not-determined |
| 18-101_k141_313629 | Low-quality |
| 18-101_k141_247890 | Not-determined |
| 18-101_k141_66540 | Not-determined |
| 18-101_k141_215192 | Not-determined |
| 18-101_k141_132863 | Not-determined |
| 18-101_k141_33699 | Not-determined |
| 18-101_k141_231371 | Not-determined |
| 18-101_k141_313647 | Not-determined |
| 18-101_k141_132873 | Low-quality |
| 18-101_k141_33707 | Not-determined |
| 18-101_k141_17445 | Not-determined |
| 18-101_k141_99715 | Not-determined |
| 18-101_k141_1352 | Not-determined |
| 18-101_k141_329931 | Not-determined |
| 18-101_k141_1353 | Not-determined |
| 18-101_k141_149020 | Not-determined |
| 18-101_k141_264294 | Not-determined |
| 18-101_k141_132888 | Not-determined |
| 18-101_k141_149024 | Not-determined |
| 18-101_k141_346686 | Not-determined |
| 18-101_k141_297294 | Not-determined |
| 18-101_k141_116432 | Low-quality |
| 18-101_k141_132897 | Not-determined |
| 18-101_k141_116437 | Not-determined |
| 18-101_k141_33716 | Not-determined |
| 18-101_k141_313668 | Not-determined |
| 18-101_k141_313670 | Not-determined |
| 18-101_k141_165491 | Not-determined |
| 18-101_k141_132904 | Not-determined |
| 18-101_k141_182098 | Not-determined |
| 18-101_k141_198563 | Not-determined |
| 18-101_k141_329951 | Not-determined |
| 18-101_k141_116457 | Not-determined |
| 18-101_k141_116460 | Not-determined |
| 18-101_k141_379430 | Low-quality |
| 18-101_k141_329961 | Not-determined |
| 18-101_k141_313688 | Not-determined |
| 18-101_k141_198571 | Not-determined |
| 18-101_k141_17452 | Not-determined |
| 18-101_k141_49980 | Not-determined |
| 18-101_k141_198574 | Not-determined |
| 18-101_k141_1377 | Not-determined |
| 18-101_k141_49984 | Not-determined |
| 18-101_k141_231392 | Not-determined |
| 18-101_k141_165507 | Not-determined |
| 18-101_k141_346706 | Not-determined |
| 18-101_k141_231394 | Not-determined |
| 18-101_k141_165509 | Not-determined |
| 18-101_k141_329973 | Not-determined |
| 18-101_k141_1384 | Not-determined |
| 18-101_k141_329978 | Not-determined |
| 18-101_k141_215234 | Not-determined |
| 18-101_k141_33737 | Low-quality |
| 18-101_k141_99716 | Low-quality |
| 18-101_k141_149058 | Not-determined |
| 18-101_k141_280867 | Not-determined |
| 18-101_k141_346726 | Not-determined |
| 18-101_k141_329993 | Not-determined |
| 18-101_k141_329996 | Not-determined |
| 18-101_k141_280871 | Not-determined |
| 18-101_k141_346732 | Low-quality |
| 18-101_k141_1407 | Not-determined |
| 18-101_k141_247964 | Not-determined |
| 18-101_k141_215255 | Low-quality |
| 18-101_k141_99733 | Not-determined |
| 18-101_k141_247976 | Not-determined |
| 18-101_k141_363087 | Not-determined |
| 18-101_k141_99744 | Not-determined |
| 18-101_k141_215260 | Not-determined |
| 18-101_k141_330017 | Low-quality |
| 18-101_k141_149098 | Not-determined |
| 18-101_k141_149099 | Not-determined |
| 18-101_k141_116499 | Low-quality |
| 18-101_k141_99747 | Not-determined |
| 18-101_k141_99748 | Not-determined |
| 18-101_k141_297322 | Low-quality |
| 18-101_k141_17500 | Not-determined |
| 18-101_k141_280898 | Low-quality |
| 18-101_k141_33797 | Not-determined |
| 18-101_k141_363100 | Not-determined |
| 18-101_k141_280902 | Not-determined |
| 18-101_k141_33802 | Not-determined |
| 18-101_k141_198623 | Not-determined |
| 18-101_k141_215291 | Not-determined |
| 18-101_k141_50003 | Not-determined |
| 18-101_k141_17516 | Low-quality |
| 18-101_k141_50004 | Not-determined |
| 18-101_k141_33806 | Not-determined |
| 18-101_k141_50005 | Low-quality |
| 18-101_k141_363106 | Not-determined |
| 18-101_k141_165528 | Low-quality |
| 18-101_k141_297337 | Not-determined |
| 18-101_k141_99771 | Not-determined |
| 18-101_k141_297340 | Not-determined |
| 18-101_k141_50012 | Not-determined |
| 18-101_k141_280915 | Not-determined |
| 18-101_k141_346736 | Low-quality |
| 18-101_k141_297351 | Not-determined |
| 18-101_k141_346741 | Not-determined |
| 18-101_k141_198648 | Not-determined |
| 18-101_k141_248018 | Low-quality |
| 18-101_k141_215315 | Not-determined |
| 18-101_k141_182130 | Low-quality |
| 18-101_k141_215320 | Not-determined |
| 18-101_k141_313736 | Low-quality |
| 18-101_k141_215328 | Low-quality |
| 18-101_k141_198659 | Low-quality |
| 18-101_k141_297366 | Not-determined |
| 18-101_k141_346751 | Not-determined |
| 18-101_k141_297371 | Not-determined |
| 18-101_k141_346757 | Not-determined |
| 18-101_k141_379460 | Not-determined |
| 18-101_k141_149104 | Not-determined |
| 18-101_k141_33828 | Not-determined |
| 18-101_k141_297379 | Not-determined |
| 18-101_k141_379465 | Not-determined |
| 18-101_k141_66568 | Not-determined |
| 18-101_k141_132984 | Not-determined |
| 18-101_k141_33833 | Not-determined |
| 18-101_k141_149109 | Not-determined |
| 18-101_k141_379469 | Not-determined |
| 18-101_k141_149112 | Not-determined |
| 18-101_k141_313746 | Low-quality |
| 18-101_k141_17520 | Low-quality |
| 18-101_k141_313748 | Not-determined |
| 18-101_k141_313750 | Low-quality |
| 18-101_k141_313751 | Not-determined |
| 18-101_k141_1441 | Not-determined |
| 18-101_k141_1442 | Not-determined |
| 18-101_k141_149123 | Not-determined |
| 18-101_k141_198678 | Not-determined |
| 18-101_k141_33847 | Not-determined |
| 18-101_k141_313761 | Not-determined |
| 18-101_k141_99786 | Not-determined |
| 18-101_k141_99789 | Not-determined |
| 18-101_k141_379491 | Not-determined |
| 18-101_k141_17545 | Not-determined |
| 18-101_k141_330097 | Not-determined |
| 18-101_k141_182181 | Not-determined |
| 18-101_k141_33853 | Not-determined |
| 18-101_k141_182185 | Not-determined |
| 18-101_k141_17553 | Not-determined |
| 18-101_k141_66572 | Not-determined |
| 18-101_k141_17556 | Not-determined |
| 18-101_k141_248058 | Not-determined |
| 18-101_k141_297399 | Not-determined |
| 18-101_k141_182191 | Not-determined |
| 18-101_k141_1449 | Not-determined |
| 18-101_k141_182195 | Not-determined |
| 18-101_k141_248067 | Not-determined |
| 18-101_k141_149138 | Not-determined |
| 18-101_k141_149140 | Not-determined |
| 18-101_k141_231455 | Not-determined |
| 18-101_k141_248073 | Not-determined |
| 18-101_k141_313770 | Not-determined |
| 18-101_k141_198720 | Not-determined |
| 18-101_k141_66593 | Not-determined |
| 18-101_k141_132997 | Not-determined |
| 18-101_k141_346782 | Not-determined |
| 18-101_k141_346788 | Not-determined |
| 18-101_k141_50068 | Low-quality |
| 18-101_k141_379498 | Not-determined |
| 18-101_k141_379506 | Low-quality |
| 18-101_k141_346805 | Low-quality |
| 18-101_k141_182218 | Not-determined |
| 18-101_k141_313804 | Not-determined |
| 18-101_k141_379512 | Not-determined |
| 18-101_k141_33891 | Not-determined |
| 18-101_k141_50077 | Not-determined |
| 18-101_k141_313809 | Not-determined |
| 18-101_k141_313815 | Not-determined |
| 18-101_k141_280976 | Not-determined |
| 18-101_k141_231464 | Not-determined |
| 18-101_k141_198749 | Not-determined |
| 18-101_k141_133010 | Low-quality |
| 18-101_k141_116518 | Not-determined |
| 18-101_k141_33917 | Not-determined |
| 18-101_k141_182256 | Not-determined |
| 18-101_k141_280996 | Not-determined |
| 18-101_k141_346837 | Not-determined |
| 18-101_k141_346838 | Not-determined |
| 18-101_k141_280999 | Not-determined |
| 18-101_k141_363171 | Not-determined |
| 18-101_k141_379527 | Not-determined |
| 18-101_k141_346852 | Not-determined |
| 18-101_k141_17612 | Not-determined |
| 18-101_k141_17614 | Not-determined |
| 18-101_k141_346861 | Not-determined |
| 18-101_k141_281017 | Not-determined |
| 18-101_k141_99810 | Not-determined |
| 18-101_k141_363180 | Not-determined |
| 18-101_k141_346871 | Not-determined |
| 18-101_k141_281020 | Not-determined |
| 18-101_k141_165576 | Not-determined |
| 18-101_k141_346873 | Not-determined |
| 18-101_k141_165583 | Not-determined |
| 18-101_k141_363193 | Not-determined |
| 18-101_k141_165591 | Not-determined |
| 18-101_k141_363196 | Not-determined |
| 18-101_k141_297440 | Not-determined |
| 18-101_k141_33923 | Low-quality |
| 18-101_k141_99826 | Not-determined |
| 18-101_k141_17622 | Not-determined |
| 18-101_k141_149188 | Not-determined |
| 18-101_k141_33935 | Not-determined |
| 18-101_k141_231485 | Not-determined |
| 18-101_k141_149193 | Not-determined |
| 18-101_k141_116546 | Not-determined |
| 18-101_k141_33941 | Not-determined |
| 18-101_k141_33942 | Not-determined |
| 18-101_k141_149199 | Not-determined |
| 18-101_k141_83431 | Low-quality |
| 18-101_k141_330138 | Low-quality |
| 18-101_k141_346901 | Not-determined |
| 18-101_k141_1508 | Not-determined |
| 18-101_k141_17628 | Not-determined |
| 18-101_k141_66599 | Not-determined |
| 18-101_k141_116555 | Not-determined |
| 18-101_k141_1513 | Not-determined |
| 18-101_k141_50115 | Not-determined |
| 18-101_k141_17634 | Not-determined |
| 18-101_k141_116562 | Not-determined |
| 18-101_k141_346912 | Not-determined |
| 18-101_k141_363228 | Not-determined |
| 18-101_k141_297464 | Not-determined |
| 18-101_k141_346918 | Not-determined |
| 18-101_k141_215378 | Not-determined |
| 18-101_k141_50126 | Low-quality |
| 18-101_k141_281055 | Not-determined |
| 18-101_k141_248108 | Not-determined |
| 18-101_k141_165619 | Not-determined |
| 18-101_k141_297478 | Low-quality |
| 18-101_k141_133073 | Not-determined |
| 18-101_k141_215385 | Not-determined |
| 18-101_k141_33979 | Not-determined |
| 18-101_k141_50131 | Not-determined |
| 18-101_k141_99885 | Not-determined |
| 18-101_k141_231498 | Not-determined |
| 18-101_k141_83454 | Low-quality |
| 18-101_k141_149215 | Not-determined |
| 18-101_k141_215387 | Not-determined |
| 18-101_k141_33991 | Not-determined |
| 18-101_k141_149221 | Not-determined |
| 18-101_k141_215391 | Not-determined |
| 18-101_k141_83460 | Not-determined |
| 18-101_k141_297490 | Not-determined |
| 18-101_k141_83461 | Not-determined |
| 18-101_k141_83465 | Not-determined |
| 18-101_k141_215402 | Not-determined |
| 18-101_k141_1567 | Not-determined |
| 18-101_k141_17669 | Not-determined |
| 18-101_k141_133102 | Not-determined |
| 18-101_k141_1576 | Not-determined |
| 18-101_k141_182318 | Not-determined |
| 18-101_k141_198791 | Not-determined |
| 18-101_k141_198795 | Not-determined |
| 18-101_k141_346944 | Not-determined |
| 18-101_k141_346945 | Not-determined |
| 18-101_k141_17678 | Not-determined |
| 18-101_k141_363244 | Not-determined |
| 18-101_k141_182326 | Not-determined |
| 18-101_k141_99912 | Not-determined |
| 18-101_k141_182334 | Not-determined |
| 18-101_k141_363254 | Not-determined |
| 18-101_k141_182336 | Not-determined |
| 18-101_k141_198808 | Not-determined |
| 18-101_k141_313883 | Not-determined |
| 18-101_k141_50140 | Not-determined |
| 18-101_k141_182345 | Not-determined |
| 18-101_k141_83500 | Not-determined |
| 18-101_k141_264309 | Not-determined |
| 18-101_k141_182349 | Low-quality |
| 18-101_k141_182353 | Not-determined |
| 18-101_k141_99932 | Not-determined |
| 18-101_k141_231523 | Not-determined |
| 18-101_k141_99933 | Not-determined |
| 18-101_k141_165654 | Not-determined |
| 18-101_k141_99938 | Not-determined |
| 18-101_k141_99939 | Not-determined |
| 18-101_k141_198822 | Not-determined |
| 18-101_k141_346977 | Not-determined |
| 18-101_k141_281104 | Not-determined |
| 18-101_k141_330192 | Not-determined |
| 18-101_k141_83518 | Not-determined |
| 18-101_k141_330201 | Not-determined |
| 18-101_k141_363266 | Not-determined |
| 18-101_k141_133132 | Not-determined |
| 18-101_k141_346986 | Not-determined |
| 18-101_k141_330207 | Not-determined |
| 18-101_k141_17715 | Not-determined |
| 18-101_k141_346990 | Not-determined |
| 18-101_k141_330209 | Not-determined |
| 18-101_k141_66617 | Not-determined |
| 18-101_k141_165673 | Not-determined |

| | |
|---|---|
| 18-101_k141_313902 | Not-determined |
| 18-101_k141_24002 | Low-quality |
| 18-101_k141_231561 | Not-determined |
| 18-101_k141_17719 | Not-determined |
| 18-101_k141_116607 | Not-determined |
| 18-101_k141_133138 | Not-determined |
| 18-101_k141_116610 | Not-determined |
| 18-101_k141_281128 | Not-determined |
| 18-101_k141_24018 | Not-determined |
| 18-101_k141_313911 | Not-determined |
| 18-101_k141_330223 | Not-determined |
| 18-101_k141_24021 | Not-determined |
| 18-101_k141_17732 | Not-determined |
| 18-101_k141_1596 | Not-determined |
| 18-101_k141_1599 | Not-determined |
| 18-101_k141_17738 | Not-determined |
| 18-101_k141_24029 | Not-determined |
| 18-101_k141_281138 | Not-determined |
| 18-101_k141_17741 | Not-determined |
| 18-101_k141_17746 | Not-determined |
| 18-101_k141_1606 | Not-determined |
| 18-101_k141_198860 | Not-determined |
| 18-101_k141_297516 | Not-determined |
| 18-101_k141_363285 | Not-determined |
| 18-101_k141_133160 | Not-determined |
| 18-101_k141_363290 | Not-determined |
| 18-101_k141_116646 | Not-determined |
| 18-101_k141_24052 | Not-determined |
| 18-101_k141_363294 | Low-quality |
| 18-101_k141_83525 | Low-quality |
| 18-101_k141_198877 | Not-determined |
| 18-101_k141_264339 | Low-quality |
| 18-101_k141_281165 | Low-quality |
| 18-101_k141_17751 | Not-determined |
| 18-101_k141_281168 | Not-determined |
| 18-101_k141_264344 | Not-determined |
| 18-101_k141_133170 | Not-determined |
| 18-101_k141_24060 | Not-determined |
| 18-101_k141_182382 | Not-determined |
| 18-101_k141_264353 | Not-determined |
| 18-101_k141_133175 | Low-quality |
| 18-101_k141_50187 | Low-quality |
| 18-101_k141_133177 | Not-determined |
| 18-101_k141_24069 | Low-quality |
| 18-101_k141_198891 | Low-quality |
| [illegible] | [illegible] |

| | |
|---|---|
| 18-101_k141_133662 | Not-determined |
| 18-101_k141_363680 | Not-determined |
| 18-101_k141_50538 | Not-determined |
| 18-101_k141_133665 | Not-determined |
| 18-101_k141_215792 | Not-determined |
| 18-101_k141_149674 | Not-determined |
| 18-101_k141_34594 | Not-determined |
| 18-101_k141_363683 | Low-quality |
| 18-101_k141_199309 | Not-determined |
| 18-101_k141_231977 | Low-quality |
| 18-101_k141_347422 | Not-determined |
| 18-101_k141_83985 | Not-determined |
| 18-101_k141_281610 | Not-determined |
| 18-101_k141_34603 | Not-determined |
| 18-101_k141_149683 | Not-determined |
| 18-101_k141_314365 | Not-determined |
| 18-101_k141_149686 | Not-determined |
| 18-101_k141_380099 | Not-determined |
| 18-101_k141_347432 | Low-quality |
| 18-101_k141_330690 | Not-determined |
| 18-101_k141_34608 | Not-determined |
| 18-101_k141_199324 | Not-determined |
| 18-101_k141_50553 | Not-determined |
| 18-101_k141_182769 | Not-determined |
| 18-101_k141_66867 | Not-determined |
| 18-101_k141_231990 | Not-determined |
| 18-101_k141_199333 | Not-determined |
| 18-101_k141_18172 | Not-determined |
| 18-101_k141_380109 | Not-determined |
| 18-101_k141_231997 | Not-determined |
| 18-101_k141_330693 | Not-determined |
| 18-101_k141_264623 | Not-determined |
| 18-101_k141_84021 | Not-determined |
| 18-101_k141_66869 | Not-determined |
| 18-101_k141_264627 | Not-determined |
| 18-101_k141_2140 | Not-determined |
| 18-101_k141_215814 | Not-determined |
| 18-101_k141_298041 | Not-determined |
| 18-101_k141_347453 | Low-quality |
| 18-101_k141_215822 | Not-determined |
| 18-101_k141_215823 | Not-determined |
| 18-101_k141_215824 | Not-determined |
| 18-101_k141_199350 | Not-determined |
| 18-101_k141_182788 | Not-determined |
| 18-101_k141_330699 | Low-quality |
| 18-101_k141_347463 | Not-determined |
| 18-101_k141_314379 | Not-determined |
| 18-101_k141_182796 | Not-determined |
| 18-101_k141_116936 | Low-quality |
| 18-101_k141_66886 | Low-quality |
| 18-101_k141_66889 | Not-determined |
| 18-101_k141_298049 | Not-determined |
| 18-101_k141_100286 | Low-quality |
| 18-101_k141_165848 | Not-determined |
| 18-101_k141_2171 | Not-determined |
| 18-101_k141_182804 | Not-determined |
| 18-101_k141_165858 | Not-determined |
| 18-101_k141_380136 | Not-determined |
| 18-101_k141_133673 | Not-determined |
| 18-101_k141_298063 | Not-determined |
| 18-101_k141_66900 | Not-determined |
| 18-101_k141_133681 | Not-determined |
| 18-101_k141_330713 | Not-determined |
| 18-101_k141_182809 | Not-determined |
| 18-101_k141_100305 | Not-determined |
| 18-101_k141_133686 | Not-determined |
| 18-101_k141_165873 | Not-determined |
| 18-101_k141_314398 | Low-quality |
| 18-101_k141_264655 | Not-determined |
| 18-101_k141_264656 | Not-determined |
| 18-101_k141_298076 | Not-determined |
| 18-101_k141_248598 | Not-determined |
| 18-101_k141_380146 | Not-determined |
| 18-101_k141_363720 | Not-determined |
| 18-101_k141_2175 | Low-quality |
| 18-101_k141_215866 | Not-determined |
| 18-101_k141_66904 | Not-determined |
| 18-101_k141_330729 | Not-determined |
| 18-101_k141_100319 | Not-determined |
| 18-101_k141_2185 | Not-determined |
| 18-101_k141_84050 | Not-determined |
| 18-101_k141_347496 | Not-determined |
| 18-101_k141_2193 | Not-determined |
| 18-101_k141_182813 | Not-determined |
| 18-101_k141_363741 | Not-determined |
| 18-101_k141_2194 | Not-determined |
| 18-101_k141_133704 | Not-determined |
| 18-101_k141_281683 | Not-determined |
| 18-101_k141_281684 | Not-determined |
| 18-101_k141_182828 | Not-determined |
| 18-101_k141_34663 | Low-quality |
| 18-101_k141_34664 | Not-determined |
| 18-101_k141_182830 | Not-determined |
| 18-101_k141_66919 | Low-quality |
| 18-101_k141_149738 | Not-determined |
| 18-101_k141_149739 | Not-determined |
| 18-101_k141_116955 | Not-determined |
| 18-101_k141_116962 | Not-determined |
| 18-101_k141_363771 | Not-determined |
| 18-101_k141_182838 | Low-quality |
| 18-101_k141_149751 | Low-quality |
| 18-101_k141_182843 | Not-determined |
| 18-101_k141_330739 | Not-determined |
| 18-101_k141_66939 | Not-determined |
| 18-101_k141_66942 | Not-determined |
| 18-101_k141_66944 | Not-determined |
| 18-101_k141_66945 | Not-determined |
| 18-101_k141_2224 | Not-determined |
| 18-101_k141_100342 | Not-determined |
| 18-101_k141_34669 | Not-determined |
| 18-101_k141_2227 | Not-determined |
| 18-101_k141_182851 | Not-determined |
| 18-101_k141_347524 | Not-determined |
| 18-101_k141_66953 | Not-determined |
| 18-101_k141_2231 | Low-quality |
| 18-101_k141_182855 | Not-determined |
| 18-101_k141_116978 | Not-determined |
| 18-101_k141_314415 | Not-determined |
| 18-101_k141_66962 | Not-determined |
| 18-101_k141_347543 | Not-determined |
| 18-101_k141_347546 | Not-determined |
| 18-101_k141_116985 | Not-determined |
| 18-101_k141_116986 | Not-determined |
| 18-101_k141_199404 | Not-determined |
| 18-101_k141_116987 | Not-determined |
| 18-101_k141_182880 | Not-determined |
| 18-101_k141_264726 | Not-determined |
| 18-101_k141_314422 | Not-determined |
| 18-101_k141_149755 | Not-determined |
| 18-101_k141_100363 | Not-determined |
| 18-101_k141_18261 | Not-determined |
| 18-101_k141_298129 | Low-quality |
| 18-101_k141_18268 | Low-quality |
| 18-101_k141_182906 | Not-determined |
| 18-101_k141_18272 | Low-quality |
| 18-101_k141_149761 | Not-determined |
| 18-101_k141_298136 | Not-determined |
| 18-101_k141_281720 | Low-quality |
| 18-101_k141_281721 | Not-determined |
| 18-101_k141_149766 | Not-determined |
| 18-101_k141_281724 | Not-determined |
| 18-101_k141_149768 | Not-determined |
| 18-101_k141_314433 | Not-determined |
| 18-101_k141_18294 | Not-determined |
| 18-101_k141_133753 | Not-determined |
| 18-101_k141_84093 | Not-determined |
| 18-101_k141_84098 | Not-determined |
| 18-101_k141_50645 | Not-determined |
| 18-101_k141_149782 | Not-determined |
| 18-101_k141_50655 | Not-determined |
| 18-101_k141_232070 | Not-determined |
| 18-101_k141_298158 | Not-determined |
| 18-101_k141_100391 | Not-determined |
| 18-101_k141_117016 | Not-determined |
| 18-101_k141_100399 | Not-determined |
| 18-101_k141_117036 | Not-determined |
| 18-101_k141_100405 | Not-determined |
| 18-101_k141_84120 | Not-determined |
| 18-101_k141_182938 | Not-determined |
| 18-101_k141_314455 | Not-determined |
| 18-101_k141_182939 | Not-determined |
| 18-101_k141_165897 | Low-quality |
| 18-101_k141_165899 | Not-determined |
| 18-101_k141_165901 | Not-determined |
| 18-101_k141_117042 | Not-determined |
| 18-101_k141_34699 | Not-determined |
| 18-101_k141_100412 | Not-determined |
| 18-101_k141_165915 | Not-determined |
| 18-101_k141_117054 | Not-determined |
| 18-101_k141_117055 | Not-determined |
| 18-101_k141_67027 | Not-determined |
| 18-101_k141_165916 | Not-determined |
| 18-101_k141_314467 | Not-determined |
| 18-101_k141_149802 | Not-determined |
| 18-101_k141_380172 | Not-determined |
| 18-101_k141_281746 | Not-determined |
| 18-101_k141_363792 | Not-determined |
| 18-101_k141_281747 | Not-determined |
| 18-101_k141_363793 | Not-determined |
| 18-101_k141_84146 | Not-determined |
| 18-101_k141_281749 | Not-determined |
| 18-101_k141_232080 | Low-quality |
| 18-101_k141_264767 | Not-determined |
| 18-101_k141_363797 | Not-determined |
| 18-101_k141_232083 | Not-determined |
| 18-101_k141_165932 | Not-determined |
| 18-101_k141_232089 | Low-quality |
| 18-101_k141_149817 | Low-quality |
| 18-101_k141_117072 | Not-determined |
| 18-101_k141_363799 | Not-determined |
| 18-101_k141_67048 | Not-determined |
| 18-101_k141_330767 | Not-determined |
| 18-101_k141_380190 | Not-determined |
| 18-101_k141_84161 | Not-determined |
| 18-101_k141_18342 | Not-determined |
| 18-101_k141_215917 | Not-determined |
| 18-101_k141_165946 | Not-determined |
| 18-101_k141_117083 | Not-determined |
| 18-101_k141_347604 | Not-determined |
| 18-101_k141_117087 | Not-determined |
| 18-101_k141_165953 | Not-determined |
| 18-101_k141_215927 | Low-quality |
| 18-101_k141_67059 | Not-determined |
| 18-101_k141_2245 | Not-determined |
| 18-101_k141_199433 | Not-determined |
| 18-101_k141_264795 | Low-quality |
| 18-101_k141_149824 | Not-determined |
| 18-101_k141_215932 | Not-determined |
| 18-101_k141_199434 | Not-determined |
| 18-101_k141_314498 | Not-determined |
| 18-101_k141_232114 | Not-determined |
| 18-101_k141_264801 | Low-quality |
| 18-101_k141_50697 | Not-determined |
| 18-101_k141_84189 | Not-determined |
| 18-101_k141_50698 | Not-determined |
| 18-101_k141_363828 | Not-determined |
| 18-101_k141_50702 | Not-determined |
| 18-101_k141_199450 | Not-determined |
| 18-101_k141_248672 | Not-determined |
| 18-101_k141_34760 | Not-determined |
| 18-101_k141_363831 | Low-quality |
| 18-101_k141_67069 | Not-determined |
| 18-101_k141_84200 | Not-determined |
| 18-101_k141_298202 | Not-determined |
| 18-101_k141_165981 | Not-determined |
| 18-101_k141_117106 | Low-quality |
| 18-101_k141_314513 | Not-determined |
| 18-101_k141_182948 | Not-determined |
| 18-101_k141_84204 | Not-determined |
| 18-101_k141_117109 | Not-determined |
| 18-101_k141_133830 | Not-determined |
| 18-101_k141_2265 | Low-quality |
| 18-101_k141_50723 | Not-determined |
| 18-101_k141_232121 | Not-determined |
| 18-101_k141_117116 | Not-determined |
| 18-101_k141_100451 | Not-determined |
| 18-101_k141_215944 | Not-determined |
| 18-101_k141_84212 | Not-determined |
| 18-101_k141_215948 | Low-quality |
| 18-101_k141_314528 | Low-quality |
| 18-101_k141_18357 | Low-quality |
| 18-101_k141_50731 | Not-determined |
| 18-101_k141_18358 | Not-determined |
| 18-101_k141_199478 | Low-quality |
| 18-101_k141_248700 | Not-determined |
| 18-101_k141_84213 | Not-determined |
| 18-101_k141_34791 | Not-determined |
| 18-101_k141_215952 | Not-determined |
| 18-101_k141_67089 | Not-determined |
| 18-101_k141_133838 | Not-determined |
| 18-101_k141_100455 | Not-determined |
| 18-101_k141_2277 | Not-determined |
| 18-101_k141_314545 | Not-determined |
| 18-101_k141_34804 | Not-determined |
| 18-101_k141_18362 | Not-determined |
| 18-101_k141_232139 | Not-determined |
| 18-101_k141_182969 | Not-determined |
| 18-101_k141_50747 | Not-determined |
| 18-101_k141_281771 | Not-determined |
| 18-101_k141_298231 | Not-determined |
| 18-101_k141_182975 | Not-determined |
| 18-101_k141_182976 | Not-determined |
| 18-101_k141_363858 | Not-determined |
| 18-101_k141_50758 | Not-determined |
| 18-101_k141_363859 | Not-determined |
| 18-101_k141_84224 | Not-determined |
| 18-101_k141_363863 | Not-determined |
| 18-101_k141_363866 | Low-quality |
| 18-101_k141_84231 | Not-determined |
| 18-101_k141_84236 | Not-determined |
| 18-101_k141_248734 | Not-determined |
| 18-101_k141_2290 | Low-quality |
| 18-101_k141_232158 | Not-determined |
| 18-101_k141_298243 | Not-determined |
| 18-101_k141_84241 | Not-determined |
| 18-101_k141_2298 | Not-determined |
| 18-101_k141_248742 | Not-determined |
| 18-101_k141_215981 | Not-determined |
| 18-101_k141_264840 | Not-determined |
| 18-101_k141_117148 | Not-determined |
| 18-101_k141_133861 | Not-determined |
| 18-101_k141_264843 | Not-determined |
| 18-101_k141_347642 | Not-determined |
| 18-101_k141_50784 | Not-determined |
| 18-101_k141_264847 | Not-determined |
| 18-101_k141_182981 | Not-determined |
| 18-101_k141_117156 | Not-determined |
| 18-101_k141_347649 | Not-determined |
| 18-101_k141_314571 | Not-determined |
| 18-101_k141_182984 | Not-determined |
| 18-101_k141_117160 | Low-quality |
| 18-101_k141_182990 | Not-determined |
| 18-101_k141_298269 | Not-determined |
| 18-101_k141_84250 | Not-determined |
| 18-101_k141_216003 | Not-determined |
| 18-101_k141_363877 | Not-determined |
| 18-101_k141_363880 | Not-determined |
| 18-101_k141_298280 | Not-determined |
| 18-101_k141_363888 | Not-determined |
| 18-101_k141_314578 | Not-determined |
| 18-101_k141_314579 | Not-determined |
| 18-101_k141_117168 | Not-determined |
| 18-101_k141_216015 | Not-determined |
| 18-101_k141_182998 | Not-determined |
| 18-101_k141_281789 | Not-determined |
| 18-101_k141_133875 | Not-determined |
| 18-101_k141_2319 | Not-determined |
| 18-101_k141_183003 | Not-determined |
| 18-101_k141_183006 | Low-quality |
| 18-101_k141_100468 | Not-determined |
| 18-101_k141_34849 | Not-determined |
| 18-101_k141_133882 | Not-determined |
| 18-101_k141_34851 | Not-determined |
| 18-101_k141_232190 | Not-determined |
| 18-101_k141_314596 | Not-determined |
| 18-101_k141_314599 | Not-determined |
| 18-101_k141_50810 | Not-determined |
| 18-101_k141_183009 | Not-determined |
| 18-101_k141_84292 | Not-determined |
| 18-101_k141_117186 | Not-determined |
| 18-101_k141_298301 | Not-determined |
| 18-101_k141_216053 | Not-determined |
| 18-101_k141_298304 | Not-determined |
| 18-101_k141_149882 | Low-quality |
| 18-101_k141_314617 | Not-determined |
| 18-101_k141_100483 | Not-determined |
| 18-101_k141_133893 | Not-determined |
| 18-101_k141_232195 | Not-determined |
| 18-101_k141_166021 | Not-determined |
| 18-101_k141_380226 | Not-determined |
| 18-101_k141_216057 | Not-determined |
| 18-101_k141_84310 | Not-determined |
| 18-101_k141_298319 | Not-determined |
| 18-101_k141_314625 | Not-determined |
| 18-101_k141_100493 | Not-determined |
| 18-101_k141_347669 | Not-determined |
| 18-101_k141_166031 | Not-determined |
| 18-101_k141_380233 | Not-determined |
| 18-101_k141_347671 | Not-determined |
| 18-101_k141_363915 | Not-determined |
| 18-101_k141_133906 | Low-quality |
| 18-101_k141_298328 | Not-determined |
| 18-101_k141_363917 | Not-determined |
| 18-101_k141_18387 | Not-determined |
| 18-101_k141_298333 | Not-determined |
| 18-101_k141_183044 | Not-determined |
| 18-101_k141_363922 | Not-determined |
| 18-101_k141_183045 | Not-determined |
| 18-101_k141_216077 | Not-determined |
| 18-101_k141_117205 | Not-determined |
| 18-101_k141_100497 | Not-determined |
| 18-101_k141_232231 | Not-determined |
| 18-101_k141_183055 | Not-determined |
| 18-101_k141_380253 | Not-determined |
| 18-101_k141_248792 | Not-determined |
| 18-101_k141_248793 | Not-determined |
| 18-101_k141_133926 | Not-determined |
| 18-101_k141_347694 | Not-determined |
| 18-101_k141_298346 | Not-determined |
| 18-101_k141_100511 | Low-quality |
| 18-101_k141_149899 | Not-determined |
| 18-101_k141_298348 | Not-determined |
| 18-101_k141_232245 | Not-determined |
| 18-101_k141_298350 | Not-determined |
| 18-101_k141_149903 | Not-determined |
| 18-101_k141_84335 | Not-determined |
| 18-101_k141_281837 | Not-determined |
| 18-101_k141_2334 | Not-determined |
| 18-101_k141_84337 | Not-determined |
| 18-101_k141_2335 | Not-determined |
| 18-101_k141_2337 | Not-determined |
| 18-101_k141_100521 | Not-determined |
| 18-101_k141_50867 | Not-determined |
| 18-101_k141_232260 | Not-determined |
| 18-101_k141_363933 | Not-determined |
| 18-101_k141_133936 | Low-quality |
| 18-101_k141_133937 | Not-determined |
| 18-101_k141_314660 | Not-determined |
| 18-101_k141_314662 | Not-determined |
| 18-101_k141_100532 | Not-determined |
| 18-101_k141_149923 | Not-determined |
| 18-101_k141_149924 | Not-determined |
| 18-101_k141_347713 | Low-quality |
| 18-101_k141_216109 | Not-determined |
| 18-101_k141_363948 | Not-determined |
| 18-101_k141_100536 | Not-determined |
| 18-101_k141_84356 | Not-determined |
| 18-101_k141_264893 | Low-quality |
| 18-101_k141_363951 | Low-quality |
| 18-101_k141_67145 | Not-determined |
| 18-101_k141_248817 | Low-quality |
| 18-101_k141_2348 | Not-determined |
| 18-101_k141_166082 | Not-determined |
| 18-101_k141_347723 | Not-determined |
| 18-101_k141_166089 | Low-quality |
| 18-101_k141_100554 | Not-determined |
| 18-101_k141_166091 | Not-determined |
| 18-101_k141_133969 | Not-determined |
| 18-101_k141_264900 | Not-determined |
| 18-101_k141_232275 | Not-determined |
| 18-101_k141_166102 | Not-determined |
| 18-101_k141_298377 | Not-determined |
| 18-101_k141_67171 | Not-determined |
| 18-101_k141_84363 | Not-determined |
| 18-101_k141_183083 | Not-determined |
| 18-101_k141_232280 | Not-determined |
| 18-101_k141_84372 | Not-determined |
| 18-101_k141_347736 | Not-determined |
| 18-101_k141_133984 | Not-determined |
| 18-101_k141_232287 | Not-determined |
| 18-101_k141_67182 | Not-determined |
| 18-101_k141_248844 | Not-determined |
| 18-101_k141_248845 | Not-determined |
| 18-101_k141_281846 | Not-determined |
| 18-101_k141_50911 | Not-determined |
| 18-101_k141_183092 | Not-determined |
| 18-101_k141_183093 | Not-determined |
| 18-101_k141_281853 | Not-determined |
| 18-101_k141_2363 | Not-determined |
| 18-101_k141_232297 | Not-determined |
| 18-101_k141_18402 | Not-determined |
| 18-101_k141_264936 | Not-determined |
| 18-101_k141_50932 | Not-determined |
| 18-101_k141_50933 | Not-determined |
| 18-101_k141_2369 | Not-determined |
| 18-101_k141_50936 | Not-determined |
| 18-101_k141_67212 | Not-determined |
| 18-101_k141_232309 | Not-determined |
| 18-101_k141_199547 | Low-quality |
| 18-101_k141_264952 | Not-determined |
| 18-101_k141_248856 | Not-determined |
| 18-101_k141_166109 | Not-determined |
| 18-101_k141_2373 | Not-determined |
| 18-101_k141_248858 | Not-determined |
| 18-101_k141_50946 | Not-determined |
| 18-101_k141_100571 | Not-determined |
| 18-101_k141_149956 | Not-determined |
| 18-101_k141_199558 | Not-determined |
| 18-101_k141_18432 | Low-quality |
| 18-101_k141_18433 | Not-determined |
| 18-101_k141_232317 | Low-quality |
| 18-101_k141_281865 | Low-quality |
| 18-101_k141_18435 | Not-determined |
| 18-101_k141_84391 | Not-determined |
| 18-101_k141_314705 | Not-determined |
| 18-101_k141_67226 | Low-quality |
| 18-101_k141_50957 | Not-determined |
| 18-101_k141_67233 | Not-determined |
| 18-101_k141_50959 | Not-determined |
| 18-101_k141_67235 | Not-determined |
| 18-101_k141_199567 | Low-quality |
| 18-101_k141_84398 | Not-determined |
| 18-101_k141_50968 | Not-determined |
| 18-101_k141_347765 | Low-quality |
| 18-101_k141_183104 | Low-quality |
| 18-101_k141_18455 | Not-determined |
| 18-101_k141_380329 | Low-quality |
| 18-101_k141_232320 | Not-determined |
| 18-101_k141_149975 | Not-determined |
| 18-101_k141_216163 | Not-determined |
| 18-101_k141_149979 | Not-determined |
| 18-101_k141_18460 | Not-determined |
| 18-101_k141_314720 | Not-determined |
| 18-101_k141_166122 | Not-determined |
| 18-101_k141_314722 | Not-determined |
| 18-101_k141_2385 | Not-determined |
| 18-101_k141_166127 | Not-determined |
| 18-101_k141_34881 | Not-determined |
| 18-101_k141_248895 | Not-determined |
| 18-101_k141_281880 | Not-determined |
| 18-101_k141_2393 | Not-determined |
| 18-101_k141_248904 | Not-determined |
| 18-101_k141_199584 | Not-determined |
| 18-101_k141_347776 | Not-determined |
| 18-101_k141_363986 | Not-determined |
| 18-101_k141_149997 | Not-determined |
| 18-101_k141_2407 | Not-determined |
| 18-101_k141_150001 | Not-determined |
| 18-101_k141_380339 | Not-determined |
| 18-101_k141_2409 | Not-determined |
| 18-101_k141_2410 | Not-determined |
| 18-101_k141_134044 | Not-determined |
| 18-101_k141_2413 | Not-determined |
| 18-101_k141_2415 | Not-determined |
| 18-101_k141_84423 | Not-determined |
| 18-101_k141_347788 | Low-quality |
| 18-101_k141_117235 | Not-determined |
| 18-101_k141_150009 | Low-quality |
| 18-101_k141_347792 | Not-determined |
| 18-101_k141_150013 | Not-determined |
| 18-101_k141_264964 | Not-determined |
| 18-101_k141_50999 | Not-determined |
| 18-101_k141_84429 | Low-quality |
| 18-101_k141_117244 | Not-determined |
| 18-101_k141_150021 | Not-determined |
| 18-101_k141_2418 | Low-quality |
| 18-101_k141_380350 | Not-determined |
| 18-101_k141_347800 | Not-determined |
| 18-101_k141_232347 | Low-quality |
| 18-101_k141_264974 | Not-determined |
| 18-101_k141_51011 | Not-determined |
| 18-101_k141_2429 | Not-determined |
| 18-101_k141_199587 | Not-determined |
| 18-101_k141_199588 | Not-determined |
| 18-101_k141_2432 | Low-quality |
| 18-101_k141_347810 | Not-determined |
| 18-101_k141_2434 | Not-determined |
| 18-101_k141_281899 | Low-quality |
| 18-101_k141_232355 | Not-determined |
| 18-101_k141_264980 | Low-quality |
| 18-101_k141_51024 | Not-determined |
| 18-101_k141_248933 | Low-quality |
| 18-101_k141_34897 | Not-determined |
| 18-101_k141_2441 | Not-determined |
| 18-101_k141_281904 | Not-determined |
| 18-101_k141_2447 | Not-determined |
| 18-101_k141_51031 | Low-quality |
| 18-101_k141_117270 | Not-determined |
| 18-101_k141_232365 | Not-determined |
| 18-101_k141_232366 | Not-determined |
| 18-101_k141_117282 | Not-determined |
| 18-101_k141_84438 | Not-determined |
| 18-101_k141_330851 | Not-determined |
| 18-101_k141_364012 | Not-determined |
| 18-101_k141_232385 | Not-determined |
| 18-101_k141_166175 | Not-determined |
| 18-101_k141_166176 | Not-determined |
| 18-101_k141_347820 | Not-determined |
| 18-101_k141_134063 | Not-determined |
| 18-101_k141_2451 | Not-determined |
| 18-101_k141_51053 | Not-determined |
| 18-101_k141_117293 | Not-determined |
| 18-101_k141_232396 | Not-determined |
| 18-101_k141_183152 | Not-determined |
| 18-101_k141_134068 | Not-determined |
| 18-101_k141_51057 | Not-determined |
| 18-101_k141_134072 | Not-determined |
| 18-101_k141_117301 | Not-determined |
| 18-101_k141_380378 | Not-determined |
| 18-101_k141_347823 | Not-determined |
| 18-101_k141_248969 | Not-determined |
| 18-101_k141_199635 | Not-determined |
| 18-101_k141_380379 | Not-determined |
| 18-101_k141_2459 | Not-determined |
| 18-101_k141_216207 | Low-quality |
| 18-101_k141_67296 | Not-determined |
| 18-101_k141_84469 | Low-quality |
| 18-101_k141_84470 | Not-determined |
| 18-101_k141_216223 | Not-determined |
| 18-101_k141_100642 | Not-determined |
| 18-101_k141_216226 | Not-determined |
| 18-101_k141_51076 | Not-determined |
| 18-101_k141_166202 | Not-determined |
| 18-101_k141_330893 | Not-determined |
| 18-101_k141_330894 | Not-determined |
| 18-101_k141_67299 | Low-quality |
| 18-101_k141_364026 | Not-determined |
| 18-101_k141_347832 | Low-quality |
| 18-101_k141_216235 | Not-determined |
| 18-101_k141_216237 | Not-determined |
| 18-101_k141_216239 | Not-determined |
| 18-101_k141_67304 | Not-determined |
| 18-101_k141_314803 | Not-determined |
| 18-101_k141_67306 | Not-determined |
| 18-101_k141_183171 | Not-determined |
| 18-101_k141_117326 | Not-determined |
| 18-101_k141_117330 | Not-determined |
| 18-101_k141_216254 | Not-determined |
| 18-101_k141_134102 | Not-determined |
| 18-101_k141_51089 | Not-determined |
| 18-101_k141_281927 | Not-determined |
| 18-101_k141_67317 | Not-determined |
| 18-101_k141_51099 | Not-determined |
| 18-101_k141_150048 | Not-determined |
| 18-101_k141_67322 | Not-determined |
| 18-101_k141_150051 | Not-determined |
| 18-101_k141_2484 | Low-quality |
| 18-101_k141_150054 | Not-determined |
| 18-101_k141_183189 | Not-determined |
| 18-101_k141_183191 | Not-determined |
| 18-101_k141_150057 | Low-quality |
| 18-101_k141_314828 | Not-determined |
| 18-101_k141_84474 | Not-determined |
| 18-101_k141_281946 | Not-determined |
| 18-101_k141_84476 | Not-determined |
| 18-101_k141_2491 | Not-determined |
| 18-101_k141_216261 | Not-determined |
| 18-101_k141_216262 | Not-determined |
| 18-101_k141_100646 | Not-determined |

| | |
|---|---|
| 18-101_k141_281951 | Not-determined |
| 18-101_k141_2499 | Low-quality |
| 18-101_k141_216264 | Low-quality |
| 18-101_k141_216265 | Not-determined |
| 18-101_k141_183198 | Not-determined |
| 18-101_k141_232464 | Not-determined |
| 18-101_k141_2504 | Low-quality |
| 18-101_k141_232467 | Not-determined |
| 18-101_k141_150071 | Not-determined |
| 18-101_k141_34928 | Not-determined |
| 18-101_k141_232471 | Not-determined |
| 18-101_k141_34935 | Not-determined |
| 18-101_k141_232474 | Not-determined |
| 18-101_k141_67346 | Low-quality |
| 18-101_k141_2511 | Not-determined |
| 18-101_k141_380418 | Not-determined |
| 18-101_k141_232479 | Not-determined |
| 18-101_k141_380422 | Low-quality |
| 18-101_k141_281979 | Not-determined |
| 18-101_k141_330923 | Not-determined |
| 18-101_k141_330925 | Not-determined |
| 18-101_k141_51124 | Not-determined |
| 18-101_k141_51129 | Not-determined |
| 18-101_k141_67354 | Not-determined |
| 18-101_k141_34957 | Not-determined |
| 18-101_k141_134169 | Not-determined |
| 18-101_k141_199673 | Not-determined |
| 18-101_k141_84495 | Not-determined |
| 18-101_k141_281997 | Not-determined |
| 18-101_k141_34964 | Low-quality |
| 18-101_k141_199692 | Not-determined |
| 18-101_k141_282003 | Not-determined |
| 18-101_k141_34969 | Not-determined |
| 18-101_k141_216289 | Not-determined |
| 18-101_k141_2530 | Not-determined |
| 18-101_k141_298418 | Not-determined |
| 18-101_k141_364064 | Not-determined |
| 18-101_k141_166236 | Not-determined |
| 18-101_k141_249058 | Not-determined |
| 18-101_k141_166238 | Not-determined |
| 18-101_k141_134198 | Not-determined |
| 18-101_k141_298426 | Low-quality |
| 18-101_k141_199708 | Low-quality |
| 18-101_k141_265009 | Not-determined |
| 18-101_k141_67383 | Not-determined |
| 18-101_k141_34980 | Not-determined |
| 18-101_k141_34981 | Not-determined |
| 18-101_k141_380444 | Not-determined |
| 18-101_k141_380445 | Not-determined |
| 18-101_k141_265012 | Not-determined |
| 18-101_k141_134221 | Not-determined |
| 18-101_k141_330957 | Not-determined |
| 18-101_k141_298446 | Not-determined |
| 18-101_k141_380454 | Not-determined |
| 18-101_k141_216313 | Not-determined |
| 18-101_k141_265019 | Not-determined |
| 18-101_k141_249071 | Not-determined |
| 18-101_k141_249074 | Not-determined |
| 18-101_k141_249077 | Not-determined |
| 18-101_k141_282041 | Not-determined |
| 18-101_k141_298457 | Not-determined |
| 18-101_k141_282043 | Not-determined |
| 18-101_k141_183252 | Not-determined |
| 18-101_k141_298461 | Not-determined |
| 18-101_k141_34986 | Not-determined |
| 18-101_k141_199720 | Not-determined |
| 18-101_k141_34987 | Not-determined |
| 18-101_k141_364095 | Not-determined |
| 18-101_k141_199721 | Not-determined |
| 18-101_k141_67405 | Not-determined |
| 18-101_k141_18477 | Not-determined |
| 18-101_k141_84515 | Low-quality |
| 18-101_k141_380470 | Low-quality |
| 18-101_k141_330984 | Not-determined |
| 18-101_k141_216328 | Not-determined |
| 18-101_k141_265053 | Not-determined |
| 18-101_k141_249091 | Not-determined |
| 18-101_k141_216335 | Not-determined |
| 18-101_k141_84524 | Not-determined |
| 18-101_k141_84532 | Not-determined |
| 18-101_k141_150113 | Not-determined |
| 18-101_k141_265070 | Not-determined |
| 18-101_k141_67426 | Not-determined |
| 18-101_k141_67427 | Not-determined |
| 18-101_k141_331007 | Low-quality |
| 18-101_k141_84536 | Not-determined |
| 18-101_k141_216351 | Not-determined |
| 18-101_k141_331010 | Not-determined |
| 18-101_k141_380490 | Not-determined |
| 18-101_k141_100700 | Low-quality |
| 18-101_k141_282073 | Not-determined |
| 18-101_k141_134265 | Not-determined |
| 18-101_k141_364123 | Low-quality |
| 18-101_k141_216360 | Not-determined |
| 18-101_k141_298494 | Not-determined |
| 18-101_k141_35000 | Not-determined |
| 18-101_k141_18502 | Not-determined |
| 18-101_k141_331016 | Not-determined |
| 18-101_k141_298501 | Not-determined |
| 18-101_k141_18506 | Not-determined |
| 18-101_k141_18507 | Not-determined |
| 18-101_k141_35006 | Not-determined |
| 18-101_k141_100705 | Not-determined |
| 18-101_k141_100707 | Not-determined |
| 18-101_k141_18511 | Not-determined |
| 18-101_k141_199748 | Not-determined |
| 18-101_k141_199750 | Not-determined |
| 18-101_k141_35011 | Not-determined |
| 18-101_k141_216384 | Not-determined |
| 18-101_k141_150127 | Not-determined |
| 18-101_k141_199758 | Not-determined |
| 18-101_k141_380510 | Not-determined |
| 18-101_k141_51186 | Not-determined |
| 18-101_k141_347872 | Low-quality |
| 18-101_k141_199763 | Not-determined |
| 18-101_k141_166273 | Not-determined |
| 18-101_k141_199765 | Not-determined |
| 18-101_k141_347874 | Not-determined |
| 18-101_k141_35021 | Not-determined |
| 18-101_k141_347875 | Not-determined |
| 18-101_k141_347877 | Not-determined |
| 18-101_k141_314896 | Not-determined |
| 18-101_k141_347879 | Not-determined |
| 18-101_k141_298525 | Not-determined |
| 18-101_k141_216409 | Not-determined |
| 18-101_k141_216412 | Not-determined |
| 18-101_k141_117374 | Not-determined |
| 18-101_k141_117379 | Not-determined |
| 18-101_k141_18532 | Low-quality |
| 18-101_k141_84556 | Not-determined |
| 18-101_k141_150144 | Not-determined |
| 18-101_k141_298535 | Not-determined |
| 18-101_k141_150154 | Not-determined |
| 18-101_k141_117387 | Not-determined |
| 18-101_k141_166290 | Not-determined |
| 18-101_k141_150157 | Not-determined |
| 18-101_k141_18538 | Not-determined |
| 18-101_k141_18539 | Low-quality |
| 18-101_k141_347901 | Not-determined |
| 18-101_k141_347905 | Not-determined |
| 18-101_k141_347908 | Not-determined |
| 18-101_k141_347909 | Not-determined |
| 18-101_k141_314912 | Not-determined |
| 18-101_k141_84575 | Not-determined |
| 18-101_k141_134301 | Low-quality |
| 18-101_k141_314915 | Not-determined |
| 18-101_k141_347910 | Not-determined |
| 18-101_k141_314916 | Not-determined |
| 18-101_k141_134302 | Not-determined |
| 18-101_k141_380529 | Not-determined |
| 18-101_k141_84579 | Not-determined |
| 18-101_k141_232549 | Not-determined |
| 18-101_k141_84581 | Not-determined |
| 18-101_k141_347914 | Not-determined |
| 18-101_k141_84582 | Not-determined |
| 18-101_k141_134305 | Not-determined |
| 18-101_k141_18555 | Low-quality |
| 18-101_k141_347917 | Not-determined |
| 18-101_k141_298544 | Not-determined |
| 18-101_k141_298547 | Not-determined |
| 18-101_k141_298549 | Not-determined |
| 18-101_k141_347921 | Not-determined |
| 18-101_k141_2571 | Not-determined |
| 18-101_k141_51237 | Not-determined |
| 18-101_k141_2576 | Not-determined |
| 18-101_k141_84597 | Not-determined |
| 18-101_k141_199815 | Not-determined |
| 18-101_k141_84599 | Not-determined |
| 18-101_k141_35031 | Not-determined |
| 18-101_k141_35033 | Not-determined |
| 18-101_k141_347931 | Not-determined |
| 18-101_k141_265110 | Not-determined |
| 18-101_k141_84608 | Not-determined |
| 18-101_k141_232567 | Not-determined |
| 18-101_k141_199822 | Not-determined |
| 18-101_k141_314940 | Not-determined |
| 18-101_k141_199826 | Low-quality |
| 18-101_k141_199830 | Low-quality |
| 18-101_k141_84617 | Not-determined |
| 18-101_k141_84618 | Not-determined |
| 18-101_k141_35045 | Low-quality |
| 18-101_k141_84624 | Low-quality |
| 18-101_k141_199840 | Not-determined |
| 18-101_k141_216424 | Not-determined |
| 18-101_k141_314943 | Not-determined |
| 18-101_k141_183306 | Not-determined |
| 18-101_k141_84630 | Not-determined |
| 18-101_k141_134310 | Not-determined |
| 18-101_k141_84632 | Not-determined |
| 18-101_k141_364192 | Not-determined |
| 18-101_k141_134314 | Not-determined |
| 18-101_k141_265136 | Not-determined |
| 18-101_k141_265137 | Not-determined |
| 18-101_k141_199849 | Not-determined |
| 18-101_k141_265142 | Not-determined |
| 18-101_k141_347948 | Not-determined |
| 18-101_k141_265144 | Not-determined |
| 18-101_k141_67462 | Not-determined |
| 18-101_k141_249136 | Not-determined |
| 18-101_k141_380547 | Low-quality |
| 18-101_k141_298563 | Not-determined |
| 18-101_k141_199857 | Not-determined |
| 18-101_k141_364198 | Not-determined |
| 18-101_k141_364199 | Not-determined |
| 18-101_k141_298566 | Not-determined |
| 18-101_k141_347953 | Not-determined |
| 18-101_k141_265148 | Not-determined |
| 18-101_k141_347954 | Not-determined |
| 18-101_k141_232594 | Not-determined |
| 18-101_k141_67470 | Not-determined |
| 18-101_k141_35079 | Not-determined |
| 18-101_k141_35080 | Not-determined |
| 18-101_k141_216437 | Not-determined |
| 18-101_k141_216442 | Not-determined |
| 18-101_k141_216444 | Not-determined |
| 18-101_k141_166329 | Not-determined |
| 18-101_k141_249148 | Low-quality |
| 18-101_k141_216452 | Not-determined |
| 18-101_k141_265160 | Not-determined |
| 18-101_k141_117432 | Not-determined |
| 18-101_k141_35093 | Not-determined |
| 18-101_k141_298568 | Not-determined |
| 18-101_k141_364203 | Not-determined |
| 18-101_k141_117447 | Not-determined |
| 18-101_k141_84655 | Not-determined |
| 18-101_k141_232610 | Not-determined |
| 18-101_k141_298580 | Low-quality |
| 18-101_k141_117449 | Not-determined |
| 18-101_k141_51274 | Not-determined |
| 18-101_k141_134336 | Low-quality |
| 18-101_k141_18587 | Not-determined |
| 18-101_k141_51279 | Not-determined |
| 18-101_k141_364225 | Not-determined |
| 18-101_k141_380554 | Low-quality |
| 18-101_k141_183343 | Not-determined |
| 18-101_k141_183345 | Not-determined |
| 18-101_k141_183348 | Not-determined |
| 18-101_k141_331074 | Not-determined |
| 18-101_k141_347971 | Not-determined |
| 18-101_k141_282083 | Not-determined |
| 18-101_k141_347974 | Not-determined |
| 18-101_k141_347975 | Not-determined |
| 18-101_k141_35119 | Not-determined |
| 18-101_k141_232626 | Not-determined |
| 18-101_k141_183355 | Not-determined |
| 18-101_k141_35125 | Not-determined |
| 18-101_k141_232631 | Not-determined |
| 18-101_k141_35127 | Not-determined |
| 18-101_k141_18593 | Not-determined |
| 18-101_k141_18594 | Not-determined |
| 18-101_k141_35130 | Not-determined |
| 18-101_k141_282098 | Not-determined |
| 18-101_k141_216462 | Low-quality |
| 18-101_k141_364236 | Not-determined |
| 18-101_k141_347983 | Not-determined |
| 18-101_k141_35141 | Not-determined |
| 18-101_k141_35142 | Not-determined |
| 18-101_k141_249209 | Not-determined |
| 18-101_k141_35143 | Not-determined |
| 18-101_k141_51302 | Not-determined |
| 18-101_k141_347988 | Not-determined |
| 18-101_k141_183372 | Not-determined |
| 18-101_k141_347989 | Not-determined |
| 18-101_k141_18603 | Not-determined |
| 18-101_k141_380578 | Not-determined |
| 18-101_k141_35148 | Not-determined |
| 18-101_k141_117474 | Not-determined |
| 18-101_k141_249224 | Not-determined |
| 18-101_k141_18613 | Not-determined |
| 18-101_k141_183381 | Not-determined |
| 18-101_k141_183382 | Not-determined |
| 18-101_k141_166350 | Low-quality |
| 18-101_k141_166351 | Not-determined |
| 18-101_k141_35159 | Not-determined |
| 18-101_k141_2613 | Not-determined |
| 18-101_k141_331086 | Not-determined |
| 18-101_k141_265185 | Not-determined |
| 18-101_k141_117478 | Not-determined |
| 18-101_k141_18621 | Not-determined |
| 18-101_k141_51318 | Not-determined |
| 18-101_k141_298619 | Not-determined |
| 18-101_k141_265193 | Not-determined |
| 18-101_k141_183386 | Not-determined |
| 18-101_k141_183388 | Not-determined |
| 18-101_k141_51322 | Not-determined |
| 18-101_k141_51323 | Not-determined |
| 18-101_k141_282133 | Not-determined |
| 18-101_k141_364238 | Not-determined |
| 18-101_k141_67549 | Not-determined |
| 18-101_k141_199904 | Not-determined |
| 18-101_k141_249241 | Not-determined |
| 18-101_k141_216487 | Not-determined |
| 18-101_k141_232639 | Low-quality |
| 18-101_k141_35167 | Low-quality |
| 18-101_k141_183395 | Not-determined |
| 18-101_k141_2622 | Not-determined |
| 18-101_k141_183400 | Not-determined |
| 18-101_k141_166370 | Not-determined |
| 18-101_k141_67565 | Not-determined |
| 18-101_k141_380601 | Not-determined |
| 18-101_k141_84696 | Not-determined |
| 18-101_k141_380603 | Not-determined |
| 18-101_k141_35170 | Low-quality |
| 18-101_k141_249250 | Not-determined |
| 18-101_k141_380609 | Not-determined |
| 18-101_k141_216509 | Not-determined |
| 18-101_k141_51350 | Not-determined |
| 18-101_k141_249258 | Not-determined |
| 18-101_k141_265215 | Not-determined |
| 18-101_k141_216514 | Not-determined |
| 18-101_k141_282142 | Not-determined |
| 18-101_k141_150207 | Not-determined |
| 18-101_k141_51356 | Not-determined |
| 18-101_k141_282143 | Not-determined |
| 18-101_k141_134343 | Not-determined |
| 18-101_k141_199930 | Not-determined |
| 18-101_k141_282148 | Not-determined |
| 18-101_k141_216529 | Not-determined |
| 18-101_k141_380621 | Not-determined |
| 18-101_k141_364270 | Not-determined |
| 18-101_k141_2652 | Not-determined |
| 18-101_k141_364272 | Not-determined |
| 18-101_k141_84708 | Not-determined |
| 18-101_k141_232669 | Not-determined |
| 18-101_k141_2656 | Not-determined |
| 18-101_k141_134351 | Not-determined |
| 18-101_k141_183435 | Low-quality |
| 18-101_k141_380633 | Not-determined |
| 18-101_k141_232676 | Not-determined |
| 18-101_k141_35204 | Low-quality |
| 18-101_k141_232678 | Low-quality |
| 18-101_k141_67578 | Not-determined |
| 18-101_k141_298647 | Not-determined |
| 18-101_k141_166391 | Not-determined |
| 18-101_k141_2668 | Not-determined |
| 18-101_k141_150239 | Not-determined |
| 18-101_k141_265229 | Not-determined |
| 18-101_k141_134368 | Low-quality |
| 18-101_k141_2669 | Not-determined |
| 18-101_k141_35214 | Low-quality |
| 18-101_k141_117499 | Not-determined |
| 18-101_k141_183450 | Not-determined |
| 18-101_k141_117505 | Not-determined |
| 18-101_k141_364284 | Not-determined |
| 18-101_k141_315000 | Not-determined |
| 18-101_k141_117510 | Low-quality |
| 18-101_k141_134385 | Not-determined |
| 18-101_k141_134386 | Not-determined |
| 18-101_k141_134388 | Not-determined |
| 18-101_k141_249299 | Not-determined |
| 18-101_k141_364293 | Not-determined |
| 18-101_k141_249304 | Not-determined |
| 18-101_k141_18690 | Not-determined |
| 18-101_k141_315007 | Not-determined |
| 18-101_k141_134397 | Not-determined |
| 18-101_k141_265251 | Not-determined |
| 18-101_k141_18704 | Not-determined |
| 18-101_k141_117527 | Not-determined |
| 18-101_k141_364305 | Low-quality |
| 18-101_k141_166405 | Not-determined |
| 18-101_k141_100751 | Not-determined |
| 18-101_k141_2699 | Not-determined |
| 18-101_k141_2700 | Not-determined |
| 18-101_k141_199960 | Not-determined |
| 18-101_k141_364307 | Not-determined |
| 18-101_k141_150251 | Not-determined |
| 18-101_k141_2703 | Not-determined |
| 18-101_k141_249314 | Not-determined |
| 18-101_k141_249315 | Not-determined |
| 18-101_k141_315023 | Not-determined |
| 18-101_k141_51386 | Not-determined |
| 18-101_k141_315025 | Not-determined |
| 18-101_k141_67610 | Not-determined |
| 18-101_k141_117535 | Not-determined |
| 18-101_k141_117536 | Not-determined |
| 18-101_k141_67613 | Not-determined |
| 18-101_k141_249324 | Not-determined |
| 18-101_k141_100763 | Not-determined |
| 18-101_k141_364312 | Not-determined |
| 18-101_k141_51395 | Not-determined |
| 18-101_k141_232689 | Not-determined |
| 18-101_k141_51398 | Not-determined |
| 18-101_k141_249329 | Not-determined |
| 18-101_k141_67624 | Not-determined |
| 18-101_k141_35251 | Not-determined |
| 18-101_k141_84760 | Low-quality |
| 18-101_k141_117549 | Low-quality |
| 18-101_k141_2734 | Not-determined |
| 18-101_k141_2735 | Not-determined |
| 18-101_k141_150266 | Not-determined |
| 18-101_k141_199971 | Not-determined |
| 18-101_k141_298680 | Not-determined |
| 18-101_k141_18714 | Not-determined |
| 18-101_k141_249355 | Not-determined |
| 18-101_k141_298692 | Not-determined |
| 18-101_k141_18720 | Not-determined |
| 18-101_k141_51404 | Not-determined |
| 18-101_k141_265274 | Not-determined |
| 18-101_k141_84776 | Not-determined |
| 18-101_k141_380683 | Not-determined |
| 18-101_k141_67640 | Not-determined |
| 18-101_k141_117569 | Low-quality |
| 18-101_k141_348007 | Not-determined |
| 18-101_k141_150282 | Not-determined |
| 18-101_k141_348009 | Not-determined |
| 18-101_k141_100767 | Not-determined |
| 18-101_k141_265277 | Low-quality |
| 18-101_k141_298705 | Not-determined |
| 18-101_k141_298707 | Not-determined |
| 18-101_k141_315077 | Not-determined |
| 18-101_k141_298708 | Not-determined |
| 18-101_k141_315079 | Not-determined |
| 18-101_k141_150293 | Not-determined |
| 18-101_k141_150294 | Not-determined |
| 18-101_k141_84795 | Not-determined |
| 18-101_k141_84798 | Not-determined |
| 18-101_k141_84799 | Not-determined |
| 18-101_k141_117581 | Not-determined |
| 18-101_k141_199989 | Not-determined |
| 18-101_k141_84802 | Not-determined |
| 18-101_k141_331125 | Not-determined |
| 18-101_k141_348024 | Not-determined |
| 18-101_k141_166428 | Low-quality |
| 18-101_k141_18739 | Low-quality |
| 18-101_k141_315091 | Not-determined |
| 18-101_k141_331131 | Not-determined |
| 18-101_k141_315094 | Not-determined |
| 18-101_k141_84808 | Not-determined |
| 18-101_k141_364361 | Not-determined |
| 18-101_k141_166431 | Not-determined |
| 18-101_k141_100773 | Not-determined |
| 18-101_k141_117588 | Not-determined |
| 18-101_k141_216642 | Not-determined |
| 18-101_k141_117595 | Not-determined |
| 18-101_k141_35292 | Not-determined |
| 18-101_k141_232707 | Not-determined |
| 18-101_k141_18750 | Not-determined |
| 18-101_k141_18751 | Low-quality |
| 18-101_k141_331144 | Not-determined |
| 18-101_k141_216648 | Not-determined |
| 18-101_k141_232712 | Not-determined |
| 18-101_k141_166440 | Not-determined |
| 18-101_k141_18754 | Not-determined |
| 18-101_k141_35301 | Low-quality |
| 18-101_k141_331147 | Not-determined |
| 18-101_k141_117610 | Not-determined |
| 18-101_k141_134452 | Not-determined |
| 18-101_k141_249413 | Not-determined |
| 18-101_k141_200002 | Not-determined |
| 18-101_k141_232723 | Not-determined |
| 18-101_k141_35316 | Low-quality |
| 18-101_k141_134460 | Not-determined |
| 18-101_k141_117619 | Not-determined |
| 18-101_k141_232726 | Not-determined |
| 18-101_k141_380707 | Not-determined |
| 18-101_k141_380708 | Not-determined |
| 17-109-final-viral-combined_k141_358010\|\|full | High-quality |
| 18-101_k141_380711 | Not-determined |
| 18-101_k141_18764 | Not-determined |
| 18-101_k141_348045 | Not-determined |
| 18-101_k141_298756 | Not-determined |
| 18-101_k141_315124 | Not-determined |
| 18-101_k141_84826 | Not-determined |
| 18-101_k141_100821 | Not-determined |
| 18-101_k141_364383 | Low-quality |
| 18-101_k141_298760 | Low-quality |
| 18-101_k141_2796 | Not-determined |
| 18-101_k141_67708 | Not-determined |
| 18-101_k141_331162 | Not-determined |
| 18-101_k141_380723 | Not-determined |
| 18-101_k141_282238 | Not-determined |
| 18-101_k141_298765 | Not-determined |
| 18-101_k141_348066 | Not-determined |
| 18-101_k141_282247 | Not-determined |
| 18-101_k141_298772 | Not-determined |
| 18-101_k141_315132 | Not-determined |
| 18-101_k141_232737 | Not-determined |
| 18-101_k141_364399 | Not-determined |
| 18-101_k141_315134 | Not-determined |
| 18-101_k141_84831 | Low-quality |
| 18-101_k141_35361 | Not-determined |
| 18-101_k141_35363 | Not-determined |
| 18-101_k141_166470 | Not-determined |
| 18-101_k141_232757 | Not-determined |
| 18-101_k141_166473 | Not-determined |
| 18-101_k141_183482 | Not-determined |
| 18-101_k141_282251 | Low-quality |
| 18-101_k141_364417 | Not-determined |
| 18-101_k141_298799 | Not-determined |
| 18-101_k141_331173 | Not-determined |
| 18-101_k141_298808 | Not-determined |
| 18-101_k141_315154 | Not-determined |
| 18-101_k141_166480 | Not-determined |
| 18-101_k141_84857 | Not-determined |
| 18-101_k141_282260 | Not-determined |
| 18-101_k141_265308 | Not-determined |
| 18-101_k141_100841 | Not-determined |
| 18-101_k141_84860 | Not-determined |
| 18-101_k141_2828 | Not-determined |
| 18-101_k141_84873 | Not-determined |
| 18-101_k141_298824 | Not-determined |
| 18-101_k141_348084 | Not-determined |
| 18-101_k141_200014 | Low-quality |
| 18-101_k141_166490 | Not-determined |
| 18-101_k141_200019 | Low-quality |
| 18-101_k141_150361 | Not-determined |
| 18-101_k141_298831 | Not-determined |
| 18-101_k141_331218 | Low-quality |
| 18-101_k141_166497 | Low-quality |
| 18-101_k141_84890 | Not-determined |
| 18-101_k141_183497 | Not-determined |
| 18-101_k141_117671 | Not-determined |
| 18-101_k141_183500 | Not-determined |
| 18-101_k141_183501 | Not-determined |
| 18-101_k141_200025 | Not-determined |
| 18-101_k141_117680 | Low-quality |
| 18-101_k141_84897 | Not-determined |
| 18-101_k141_331232 | Not-determined |
| 18-101_k141_18786 | Not-determined |
| 18-101_k141_2845 | Not-determined |
| 18-101_k141_380776 | Not-determined |
| 18-101_k141_298849 | Not-determined |
| 18-101_k141_67728 | Low-quality |
| 18-101_k141_200039 | Not-determined |
| 18-101_k141_84913 | Not-determined |
| 18-101_k141_35390 | Not-determined |
| 18-101_k141_200044 | Not-determined |
| 18-101_k141_315160 | Not-determined |
| 18-101_k141_331249 | Not-determined |
| 18-101_k141_200047 | Not-determined |
| 18-101_k141_200049 | Not-determined |
| 18-101_k141_117701 | Low-quality |
| 18-101_k141_67745 | Low-quality |
| 18-101_k141_2857 | Low-quality |
| 18-101_k141_100860 | Not-determined |
| 18-101_k141_84931 | Not-determined |
| 18-101_k141_315169 | Not-determined |
| 18-101_k141_150374 | Not-determined |
| 18-101_k141_35396 | Not-determined |
| 18-101_k141_84938 | Not-determined |
| 18-101_k141_315178 | Low-quality |
| 18-101_k141_200070 | Not-determined |
| 18-101_k141_166510 | Not-determined |
| 18-101_k141_150384 | Not-determined |
| 18-101_k141_200076 | Not-determined |
| 18-101_k141_150390 | Not-determined |
| 18-101_k141_249423 | Not-determined |
| 18-101_k141_200083 | Not-determined |
| 18-101_k141_67750 | Low-quality |
| 18-101_k141_117714 | Not-determined |
| 18-101_k141_232839 | Not-determined |
| 18-101_k141_67752 | Not-determined |
| 18-101_k141_249426 | Not-determined |
| 18-101_k141_298881 | Low-quality |
| 18-101_k141_150397 | Not-determined |
| 18-101_k141_117723 | Not-determined |
| 18-101_k141_150400 | Not-determined |
| 18-101_k141_298886 | Not-determined |
| 18-101_k141_216735 | Not-determined |
| 18-101_k141_200090 | Not-determined |
| 18-101_k141_249439 | Low-quality |
| 18-101_k141_298891 | Not-determined |
| 18-101_k141_67768 | Not-determined |
| 18-101_k141_18822 | Low-quality |
| 18-101_k141_134555 | Not-determined |
| 18-101_k141_216747 | Not-determined |
| 18-101_k141_232853 | Not-determined |
| 18-101_k141_249446 | Not-determined |
| 18-101_k141_216750 | Not-determined |
| 18-101_k141_183540 | Not-determined |
| 18-101_k141_18826 | Not-determined |
| 18-101_k141_364484 | Low-quality |
| 18-101_k141_315191 | Not-determined |
| 18-101_k141_364488 | Not-determined |
| 18-101_k141_331270 | Not-determined |
| 18-101_k141_183547 | Not-determined |
| 18-101_k141_183548 | Not-determined |
| 18-101_k141_150418 | Not-determined |
| 18-101_k141_364496 | Not-determined |
| 18-101_k141_364498 | Not-determined |
| 18-101_k141_364499 | Low-quality |
| 18-101_k141_232873 | Not-determined |
| 18-101_k141_183559 | Low-quality |
| 18-101_k141_166529 | Not-determined |
| 18-101_k141_331287 | Not-determined |
| 18-101_k141_216774 | Not-determined |
| 18-101_k141_216775 | Not-determined |
| 18-101_k141_282343 | Not-determined |
| 18-101_k141_380816 | Not-determined |

| | |
|---|---|
| 18-101_k141_282349 | Not-determined |
| 18-101_k141_380817 | Not-determined |
| 18-101_k141_364506 | Not-determined |
| 18-101_k141_216786 | Not-determined |
| 18-101_k141_84964 | Not-determined |
| 18-101_k141_84965 | Not-determined |
| 18-101_k141_117734 | Not-determined |
| 18-101_k141_150429 | Low-quality |
| 18-101_k141_216798 | Not-determined |
| 18-101_k141_150430 | Not-determined |
| 18-101_k141_216800 | Not-determined |
| 18-101_k141_380822 | Not-determined |
| 18-101_k141_150431 | Low-quality |
| 18-101_k141_364517 | Not-determined |
| 18-101_k141_331308 | Not-determined |
| 18-101_k141_67806 | Not-determined |
| 18-101_k141_315195 | Not-determined |
| 18-101_k141_100911 | Low-quality |
| 18-101_k141_380840 | Not-determined |
| 18-101_k141_67813 | Not-determined |
| 18-101_k141_249468 | Not-determined |
| 18-101_k141_100916 | Not-determined |
| 18-101_k141_100919 | Not-determined |
| 18-101_k141_100920 | Not-determined |
| 18-101_k141_100921 | Not-determined |
| 18-101_k141_315201 | Not-determined |
| 18-101_k141_216810 | Low-quality |
| 18-101_k141_84980 | Not-determined |
| 18-101_k141_117741 | Low-quality |
| 18-101_k141_117742 | Not-determined |
| 18-101_k141_67822 | Not-determined |
| 18-101_k141_331329 | Not-determined |
| 18-101_k141_67825 | Not-determined |
| 18-101_k141_331332 | Not-determined |
| 18-101_k141_117748 | Not-determined |
| 18-101_k141_216817 | Not-determined |
| 18-101_k141_331335 | Not-determined |
| 18-101_k141_249483 | Not-determined |
| 18-101_k141_51467 | Not-determined |
| 18-101_k141_249488 | Not-determined |
| 18-101_k141_249489 | Not-determined |
| 18-101_k141_51469 | Not-determined |
| 18-101_k141_249492 | Not-determined |
| 18-101_k141_51471 | Not-determined |
| 18-101_k141_331342 | Low-quality |
| 18-101_k141_150465 | Not-determined |
| 18-101_k141_134613 | Not-determined |
| 18-101_k141_150468 | Not-determined |
| 18-101_k141_150470 | Not-determined |
| 18-101_k141_150471 | Not-determined |
| 18-101_k141_331344 | Not-determined |
| 18-101_k141_150477 | Not-determined |
| 18-101_k141_315220 | Not-determined |
| 18-101_k141_150481 | Not-determined |
| 18-101_k141_150484 | Not-determined |
| 18-101_k141_315228 | Not-determined |
| 18-101_k141_364542 | Not-determined |
| 18-101_k141_315230 | Not-determined |
| 18-101_k141_364543 | Not-determined |
| 18-101_k141_315234 | Not-determined |
| 18-101_k141_117766 | Not-determined |
| 18-101_k141_315237 | Not-determined |
| 18-101_k141_166537 | Not-determined |
| 18-101_k141_117776 | Not-determined |
| 18-101_k141_18884 | Not-determined |
| 18-101_k141_134619 | Not-determined |
| 18-101_k141_282379 | Not-determined |
| 18-101_k141_150499 | Not-determined |
| 18-101_k141_315242 | Low-quality |
| 18-101_k141_51506 | Not-determined |
| 18-101_k141_183626 | Not-determined |
| 18-101_k141_315244 | Not-determined |
| 18-101_k141_216827 | Not-determined |
| 18-101_k141_35442 | Low-quality |
| 18-101_k141_331359 | Not-determined |
| 18-101_k141_51519 | Low-quality |
| 18-101_k141_183630 | Not-determined |
| 18-101_k141_85000 | Not-determined |
| 18-101_k141_2892 | Not-determined |
| 18-101_k141_183634 | Not-determined |
| 18-101_k141_134637 | Low-quality |
| 18-101_k141_100975 | Not-determined |
| 18-101_k141_2895 | Not-determined |
| 18-101_k141_315263 | Not-determined |
| 18-101_k141_364560 | Not-determined |
| 18-101_k141_51531 | Not-determined |
| 18-101_k141_51532 | Not-determined |
| 18-101_k141_100980 | Not-determined |
| 18-101_k141_117789 | Not-determined |
| 18-101_k141_265362 | Not-determined |
| 18-101_k141_117798 | Low-quality |
| 18-101_k141_134643 | Not-determined |
| 18-101_k141_265370 | Not-determined |
| 18-101_k141_232922 | Not-determined |
| 18-101_k141_117806 | Not-determined |
| 18-101_k141_18922 | Not-determined |
| 18-101_k141_85012 | Not-determined |
| 18-101_k141_18927 | Not-determined |
| 18-101_k141_134654 | Not-determined |
| 18-101_k141_331370 | Not-determined |
| 18-101_k141_101003 | Not-determined |
| 18-101_k141_216851 | Not-determined |
| 18-101_k141_183674 | Not-determined |
| 18-101_k141_298908 | Not-determined |
| 18-101_k141_85015 | Low-quality |
| 18-101_k141_364599 | Not-determined |
| 18-101_k141_85017 | Not-determined |
| 18-101_k141_67872 | Not-determined |
| 18-101_k141_18941 | Low-quality |
| 18-101_k141_2918 | Not-determined |
| 18-101_k141_85021 | Not-determined |
| 18-101_k141_85022 | Not-determined |
| 18-101_k141_67874 | Not-determined |
| 18-101_k141_51565 | Not-determined |
| 18-101_k141_249542 | Not-determined |
| 18-101_k141_35481 | Not-determined |
| 18-101_k141_35484 | Not-determined |
| 18-101_k141_331389 | Not-determined |
| 18-101_k141_282404 | Not-determined |
| 18-101_k141_134681 | Not-determined |
| 18-101_k141_380859 | Not-determined |
| 18-101_k141_348196 | Not-determined |
| 18-101_k141_134684 | Not-determined |
| 18-101_k141_183691 | Not-determined |
| 18-101_k141_298921 | Not-determined |
| 18-101_k141_150516 | Not-determined |
| 18-101_k141_150518 | Not-determined |
| 18-101_k141_150519 | Not-determined |
| 18-101_k141_282411 | Not-determined |
| 18-101_k141_249560 | Low-quality |
| 18-101_k141_265402 | Not-determined |
| 18-101_k141_232944 | Low-quality |
| 18-101_k141_249564 | Low-quality |
| 18-101_k141_249566 | Not-determined |
| 18-101_k141_51576 | Not-determined |
| 18-101_k141_282419 | Not-determined |
| 18-101_k141_232954 | Not-determined |
| 18-101_k141_35504 | Not-determined |
| 18-101_k141_183694 | Not-determined |
| 18-101_k141_249576 | Not-determined |
| 18-101_k141_265412 | Not-determined |
| 18-101_k141_331413 | Not-determined |
| 18-101_k141_67887 | Not-determined |
| 18-101_k141_183700 | Not-determined |
| 18-101_k141_315288 | Not-determined |
| 18-101_k141_117862 | Not-determined |
| 18-101_k141_216880 | Not-determined |
| 18-101_k141_35509 | Not-determined |
| 18-101_k141_150541 | Not-determined |
| 18-101_k141_183705 | Low-quality |
| 18-101_k141_348202 | Not-determined |
| 18-101_k141_18972 | Not-determined |
| 18-101_k141_380881 | Not-determined |
| 18-101_k141_117867 | Not-determined |
| 18-101_k141_331426 | Not-determined |
| 18-101_k141_117869 | Not-determined |
| 18-101_k141_216884 | Not-determined |
| 18-101_k141_117870 | Not-determined |
| 18-101_k141_200157 | Not-determined |
| 18-101_k141_315297 | Not-determined |
| 18-101_k141_51604 | Not-determined |
| 18-101_k141_265425 | Not-determined |
| 18-101_k141_183713 | Low-quality |
| 18-101_k141_150551 | Not-determined |
| 18-101_k141_18986 | Not-determined |
| 18-101_k141_183722 | Not-determined |
| 18-101_k141_200178 | Not-determined |
| 18-101_k141_232989 | Low-quality |
| 18-101_k141_331445 | Not-determined |
| 18-101_k141_67912 | Not-determined |
| 18-101_k141_348236 | Not-determined |
| 18-101_k141_232991 | Not-determined |
| 18-101_k141_216901 | Not-determined |
| 18-101_k141_315340 | Not-determined |
| 18-101_k141_183746 | Not-determined |
| 18-101_k141_150566 | Not-determined |
| 18-101_k141_134702 | Not-determined |
| 18-101_k141_67924 | Not-determined |
| 18-101_k141_67925 | Not-determined |
| 18-101_k141_183751 | Not-determined |
| 18-101_k141_67930 | Not-determined |
| 18-101_k141_315348 | Not-determined |
| 18-101_k141_249605 | Not-determined |
| 18-101_k141_2995 | Not-determined |
| 18-101_k141_19007 | Not-determined |
| 18-101_k141_331461 | Not-determined |
| 18-101_k141_2996 | Low-quality |
| 18-101_k141_51621 | Not-determined |
| 18-101_k141_282456 | Not-determined |
| 18-101_k141_249612 | Not-determined |
| 18-101_k141_117880 | Not-determined |
| 18-101_k141_19012 | Not-determined |
| 18-101_k141_35538 | Not-determined |
| 18-101_k141_315367 | Not-determined |
| 18-101_k141_348253 | Not-determined |
| 18-101_k141_19016 | Not-determined |
| 18-101_k141_200218 | Not-determined |
| 18-101_k141_364661 | Not-determined |
| 18-101_k141_51642 | Low-quality |
| 18-101_k141_134705 | Not-determined |
| 18-101_k141_348257 | Not-determined |
| 18-101_k141_364663 | Not-determined |
| 18-101_k141_85046 | Not-determined |
| 18-101_k141_51647 | Not-determined |
| 18-101_k141_134707 | Not-determined |
| 18-101_k141_200227 | Not-determined |
| 18-101_k141_134708 | Not-determined |
| 18-101_k141_364669 | Not-determined |
| 18-101_k141_134716 | Not-determined |
| 18-101_k141_364676 | Not-determined |
| 18-101_k141_364678 | Not-determined |
| 18-101_k141_85061 | Not-determined |
| 18-101_k141_85062 | Not-determined |
| 18-101_k141_85065 | Low-quality |
| 18-101_k141_315372 | Not-determined |
| 18-101_k141_249637 | Not-determined |
| 18-101_k141_331478 | Not-determined |
| 18-101_k141_35552 | Not-determined |
| 18-101_k141_265482 | Not-determined |
| 18-101_k141_134724 | Not-determined |
| 18-101_k141_249638 | Not-determined |
| 18-101_k141_117894 | Not-determined |
| 18-101_k141_166569 | Not-determined |
| 18-101_k141_364690 | Not-determined |
| 18-101_k141_101047 | Not-determined |
| 18-101_k141_299020 | Not-determined |
| 18-101_k141_134730 | Not-determined |
| 18-101_k141_315390 | Low-quality |
| 18-101_k141_150594 | Not-determined |
| 18-101_k141_265497 | Low-quality |
| 18-101_k141_315394 | Not-determined |
| 18-101_k141_348278 | Not-determined |
| 18-101_k141_183769 | Not-determined |
| 18-101_k141_348282 | Not-determined |
| 18-101_k141_331483 | Not-determined |
| 18-101_k141_380912 | Not-determined |
| 18-101_k141_331485 | Not-determined |
| 18-101_k141_249651 | Not-determined |
| 18-101_k141_348289 | Not-determined |
| 18-101_k141_101059 | Not-determined |
| 18-101_k141_249657 | Not-determined |
| 18-101_k141_249663 | Not-determined |
| 18-101_k141_101074 | Not-determined |
| 18-101_k141_282480 | Not-determined |
| 18-101_k141_150618 | Not-determined |
| 18-101_k141_265501 | Not-determined |
| 18-101_k141_249668 | Not-determined |
| 18-101_k141_380935 | Low-quality |
| 18-101_k141_249673 | Not-determined |
| 18-101_k141_380941 | Not-determined |
| 18-101_k141_166591 | Not-determined |
| 18-101_k141_134757 | Not-determined |
| 18-101_k141_134761 | Not-determined |
| 18-101_k141_101108 | Not-determined |
| 18-101_k141_166604 | Not-determined |
| 18-101_k141_200234 | Not-determined |
| 18-101_k141_265509 | Not-determined |
| 18-101_k141_200239 | Not-determined |
| 18-101_k141_282488 | Not-determined |
| 18-101_k141_67960 | Not-determined |
| 18-101_k141_150640 | Not-determined |
| 18-101_k141_35582 | Low-quality |
| 18-101_k141_35583 | Not-determined |
| 18-101_k141_19033 | Not-determined |
| 18-101_k141_249698 | Not-determined |
| 18-101_k141_134772 | Not-determined |
| 18-101_k141_364714 | Low-quality |
| 18-101_k141_101123 | Not-determined |
| 18-101_k141_117912 | Not-determined |
| 18-101_k141_331519 | Not-determined |
| 18-101_k141_117915 | Not-determined |
| 18-101_k141_348341 | Not-determined |
| 18-101_k141_364729 | Not-determined |
| 18-101_k141_19046 | Not-determined |
| 18-101_k141_183822 | Not-determined |
| 18-101_k141_265525 | Not-determined |
| 18-101_k141_331527 | Not-determined |
| 18-101_k141_364736 | Not-determined |
| 18-101_k141_183828 | Not-determined |
| 18-101_k141_19057 | Not-determined |
| 18-101_k141_265533 | Not-determined |
| 18-101_k141_183839 | Not-determined |
| 18-101_k141_150674 | Not-determined |
| 18-101_k141_249732 | Not-determined |
| 18-101_k141_35600 | Not-determined |
| 18-101_k141_315435 | Not-determined |
| 18-101_k141_150680 | Not-determined |
| 18-101_k141_282522 | Not-determined |
| 18-101_k141_364740 | Not-determined |
| 18-101_k141_282523 | Low-quality |
| 18-101_k141_3016 | Not-determined |
| 18-101_k141_101141 | Not-determined |
| 18-101_k141_282530 | Not-determined |
| 18-101_k141_85108 | Not-determined |
| 18-101_k141_183852 | Not-determined |
| 18-101_k141_315445 | Not-determined |
| 18-101_k141_150692 | Not-determined |
| 18-101_k141_299106 | Not-determined |
| 18-101_k141_150694 | Not-determined |
| 18-101_k141_150697 | Not-determined |
| 18-101_k141_150700 | Not-determined |
| 18-101_k141_150705 | Not-determined |
| 18-101_k141_216941 | Not-determined |
| 18-101_k141_216944 | Low-quality |
| 18-101_k141_150712 | Low-quality |
| 18-101_k141_51706 | Not-determined |
| 18-101_k141_299119 | Not-determined |
| 18-101_k141_51707 | Not-determined |
| 18-101_k141_101160 | Not-determined |
| 18-101_k141_249745 | Not-determined |
| 18-101_k141_282555 | Not-determined |
| 18-101_k141_85120 | Not-determined |
| 18-101_k141_51714 | Not-determined |
| 18-101_k141_380945 | Not-determined |
| 18-101_k141_150721 | Not-determined |
| 18-101_k141_150722 | Not-determined |
| 18-101_k141_348359 | Not-determined |
| 18-101_k141_101164 | Not-determined |
| 18-101_k141_35621 | Low-quality |
| 18-101_k141_249752 | Low-quality |
| 18-101_k141_150723 | Not-determined |
| 18-101_k141_3030 | Not-determined |
| 18-101_k141_3034 | Not-determined |
| 18-101_k141_85132 | Not-determined |
| 18-101_k141_85136 | Not-determined |
| 18-101_k141_101175 | Low-quality |
| 18-101_k141_265552 | Not-determined |
| 18-101_k141_166635 | Not-determined |
| 18-101_k141_348374 | Low-quality |
| 18-101_k141_67976 | Not-determined |
| 18-101_k141_85140 | Not-determined |
| 18-101_k141_166642 | Low-quality |
| 18-101_k141_166644 | Not-determined |
| 18-101_k141_249760 | Not-determined |
| 18-101_k141_282579 | Not-determined |
| 18-101_k141_19107 | Not-determined |
| 18-101_k141_282582 | Not-determined |
| 18-101_k141_183873 | Not-determined |
| 18-101_k141_348391 | Not-determined |
| 18-101_k141_299167 | Not-determined |
| 18-101_k141_134859 | Not-determined |
| 18-101_k141_19112 | Not-determined |
| 18-101_k141_299173 | Not-determined |
| 18-101_k141_249771 | Not-determined |
| 18-101_k141_282588 | Not-determined |
| 18-101_k141_380981 | Not-determined |
| 18-101_k141_19121 | Not-determined |
| 18-101_k141_380982 | Not-determined |
| 18-101_k141_348399 | Not-determined |
| 18-101_k141_183880 | Not-determined |
| 18-101_k141_249776 | Not-determined |
| 18-101_k141_348406 | Low-quality |
| 18-101_k141_249787 | Low-quality |
| 18-101_k141_117975 | Not-determined |
| 18-101_k141_101208 | Not-determined |
| 18-101_k141_282606 | Not-determined |
| 18-101_k141_233062 | Not-determined |
| 18-101_k141_348417 | Not-determined |
| 18-101_k141_150770 | Not-determined |
| 18-101_k141_348422 | Not-determined |
| 18-101_k141_101216 | Not-determined |
| 18-101_k141_101220 | Not-determined |
| 18-101_k141_150772 | Not-determined |
| 18-101_k141_166681 | Not-determined |
| 18-101_k141_348429 | Not-determined |
| 18-101_k141_381003 | Not-determined |
| 18-101_k141_51738 | Not-determined |
| 18-101_k141_348431 | Not-determined |
| 18-101_k141_249803 | Not-determined |
| 18-101_k141_348432 | Low-quality |
| 18-101_k141_19144 | Not-determined |
| 18-101_k141_166688 | Not-determined |
| 18-101_k141_183918 | Not-determined |
| 18-101_k141_315462 | Not-determined |
| 18-101_k141_183922 | Not-determined |
| 18-101_k141_51757 | Not-determined |
| 18-101_k141_249824 | Not-determined |
| 18-101_k141_35649 | Not-determined |
| 18-101_k141_348451 | Low-quality |
| 18-101_k141_249825 | Not-determined |
| 18-101_k141_150790 | Not-determined |
| 18-101_k141_381019 | Low-quality |
| 18-101_k141_117990 | Not-determined |
| 18-101_k141_68024 | Not-determined |
| 18-101_k141_150792 | Not-determined |
| 18-101_k141_68027 | Not-determined |
| 18-101_k141_150796 | Not-determined |
| 18-101_k141_51773 | Not-determined |
| 18-101_k141_150798 | Not-determined |
| 18-101_k141_381023 | Not-determined |
| 18-101_k141_35650 | Low-quality |
| 18-101_k141_315471 | Not-determined |
| 18-101_k141_364795 | Not-determined |
| 18-101_k141_364797 | Not-determined |
| 18-101_k141_166710 | Not-determined |
| 18-101_k141_150807 | Not-determined |
| 18-101_k141_150809 | Not-determined |
| 18-101_k141_19155 | Not-determined |
| 18-101_k141_117998 | Not-determined |
| 18-101_k141_150812 | Not-determined |
| 18-101_k141_35659 | Not-determined |
| 18-101_k141_299230 | Not-determined |
| 18-101_k141_265560 | Low-quality |
| 18-101_k141_249844 | Not-determined |
| 18-101_k141_348470 | Not-determined |
| 18-101_k141_200280 | Not-determined |
| 18-101_k141_315482 | Not-determined |
| 18-101_k141_348471 | Low-quality |
| 18-101_k141_68047 | Not-determined |
| 18-101_k141_315486 | Not-determined |
| 18-101_k141_19163 | Low-quality |
| 18-101_k141_249852 | Not-determined |
| 18-101_k141_315493 | Not-determined |
| 18-101_k141_299239 | Not-determined |
| 18-101_k141_282674 | Not-determined |
| 18-101_k141_35670 | Not-determined |
| 18-101_k141_348482 | Not-determined |
| 18-101_k141_134869 | Not-determined |
| 18-101_k141_249863 | Not-determined |
| 18-101_k141_233100 | Not-determined |
| 18-101_k141_85189 | Not-determined |
| 18-101_k141_85190 | Not-determined |
| 18-101_k141_134875 | Not-determined |
| 18-101_k141_299249 | Not-determined |
| 18-101_k141_118002 | Not-determined |
| 18-101_k141_364823 | Not-determined |
| 18-101_k141_282681 | Not-determined |
| 18-101_k141_299252 | Not-determined |
| 18-101_k141_348487 | Not-determined |
| 18-101_k141_68068 | Not-determined |
| 18-101_k141_249880 | Not-determined |
| 18-101_k141_249883 | Not-determined |
| 18-101_k141_118005 | Not-determined |
| 18-101_k141_299265 | Not-determined |
| 18-101_k141_299275 | Not-determined |
| 18-101_k141_200306 | Not-determined |
| 18-101_k141_35688 | Not-determined |
| 18-101_k141_200307 | Not-determined |
| 18-101_k141_85206 | Not-determined |
| 18-101_k141_85207 | Not-determined |
| 18-101_k141_85208 | Low-quality |
| 18-101_k141_134898 | Not-determined |
| 18-101_k141_364842 | Not-determined |
| 18-101_k141_200308 | Low-quality |
| 18-101_k141_183984 | Not-determined |
| 18-101_k141_265582 | Not-determined |
| 18-101_k141_19202 | Not-determined |
| 18-101_k141_315527 | Low-quality |
| 18-101_k141_200310 | Not-determined |
| 18-101_k141_183992 | Not-determined |
| 18-101_k141_381063 | Not-determined |
| 18-101_k141_315533 | Not-determined |
| 18-101_k141_348500 | Not-determined |
| 18-101_k141_200316 | Not-determined |
| 18-101_k141_200317 | Not-determined |
| 18-101_k141_348503 | Not-determined |
| 18-101_k141_315538 | Not-determined |
| 18-101_k141_265588 | Not-determined |
| 18-101_k141_200324 | Not-determined |
| 18-101_k141_233106 | Not-determined |
| 18-101_k141_364858 | Not-determined |
| 18-101_k141_166724 | Low-quality |
| 18-101_k141_19210 | Not-determined |
| 18-101_k141_315544 | Not-determined |
| 18-101_k141_265603 | Not-determined |
| 18-101_k141_299305 | Low-quality |
| 18-101_k141_348523 | Not-determined |
| 18-101_k141_299309 | Not-determined |
| 18-101_k141_35724 | Not-determined |
| 18-101_k141_315551 | Not-determined |
| 18-101_k141_68101 | Not-determined |
| 18-101_k141_315555 | Not-determined |
| 18-101_k141_265609 | Not-determined |
| 18-101_k141_19229 | Not-determined |
| 18-101_k141_299326 | Not-determined |
| 18-101_k141_315564 | Not-determined |
| 18-101_k141_282719 | Not-determined |
| 18-101_k141_299333 | Not-determined |
| 18-101_k141_51791 | Not-determined |
| 18-101_k141_51792 | Not-determined |
| 18-101_k141_3051 | Not-determined |
| 18-101_k141_184039 | Low-quality |
| 18-101_k141_118058 | Not-determined |
| 18-101_k141_184044 | Not-determined |
| 18-101_k141_315574 | Not-determined |
| 18-101_k141_51803 | Not-determined |
| 18-101_k141_315576 | Not-determined |
| 18-101_k141_315578 | Not-determined |
| 18-101_k141_315579 | Not-determined |
| 18-101_k141_381114 | Not-determined |
| 18-101_k141_68105 | Low-quality |
| 18-101_k141_101257 | Not-determined |
| 18-101_k141_101258 | Not-determined |
| 18-101_k141_68110 | Not-determined |
| 18-101_k141_68111 | Not-determined |
| 18-101_k141_184057 | Not-determined |
| 18-101_k141_184059 | Not-determined |
| 18-101_k141_348557 | Not-determined |
| 18-101_k141_150880 | Not-determined |
| 18-101_k141_348560 | Not-determined |
| 18-101_k141_265631 | Not-determined |
| 18-101_k141_200341 | Not-determined |
| 18-101_k141_184072 | Not-determined |
| 18-101_k141_150890 | Not-determined |
| 18-101_k141_282743 | Not-determined |
| 18-101_k141_233129 | Not-determined |
| 18-101_k141_233130 | Not-determined |
| 18-101_k141_364896 | Low-quality |
| 18-101_k141_200351 | Not-determined |
| 18-101_k141_200353 | Not-determined |
| 18-101_k141_315609 | Not-determined |
| 18-101_k141_249910 | Not-determined |
| 18-101_k141_348577 | Not-determined |
| 18-101_k141_364905 | Low-quality |
| 18-101_k141_184085 | Not-determined |
| 18-101_k141_150903 | Not-determined |
| 18-101_k141_299379 | Not-determined |
| 18-101_k141_348584 | Not-determined |
| 18-101_k141_68134 | Not-determined |
| 18-101_k141_364912 | Not-determined |
| 18-101_k141_364913 | Not-determined |
| 18-101_k141_364914 | Not-determined |
| 18-101_k141_233150 | Low-quality |
| 18-101_k141_200360 | Not-determined |
| 18-101_k141_233154 | Not-determined |
| 18-101_k141_35744 | Not-determined |
| 18-101_k141_19254 | Not-determined |
| 18-101_k141_35747 | Not-determined |
| 18-101_k141_19258 | Not-determined |
| 18-101_k141_51811 | Not-determined |
| 18-101_k141_331580 | Not-determined |
| 18-101_k141_19260 | Not-determined |
| 18-101_k141_35752 | Not-determined |
| 18-101_k141_166762 | Not-determined |
| 18-101_k141_348607 | Not-determined |
| 18-101_k141_217014 | Not-determined |
| 18-101_k141_299402 | Not-determined |
| 18-101_k141_299403 | Not-determined |
| 18-101_k141_217016 | Not-determined |
| 18-101_k141_200380 | Low-quality |
| 18-101_k141_184120 | Low-quality |
| 18-101_k141_184125 | Not-determined |
| 18-101_k141_35757 | Not-determined |
| 18-101_k141_51839 | Not-determined |
| 18-101_k141_299406 | Not-determined |
| 18-101_k141_51846 | Not-determined |
| 18-101_k141_217027 | Not-determined |
| 18-101_k141_331606 | Not-determined |
| 18-101_k141_217043 | Not-determined |
| 18-101_k141_150928 | Not-determined |
| 18-101_k141_265689 | Low-quality |
| 18-101_k141_68161 | Not-determined |
| 18-101_k141_364962 | Not-determined |
| 18-101_k141_150930 | Low-quality |
| 18-101_k141_68165 | Not-determined |
| 18-101_k141_233195 | Low-quality |
| 18-101_k141_184140 | Low-quality |
| 18-101_k141_19272 | Not-determined |
| 18-101_k141_35788 | Low-quality |
| 18-101_k141_150939 | Not-determined |
| 18-101_k141_118124 | Not-determined |
| 18-101_k141_35791 | Not-determined |
| 18-101_k141_299416 | Not-determined |
| 18-101_k141_166796 | Not-determined |
| 18-101_k141_134934 | Not-determined |
| 18-101_k141_265706 | Not-determined |
| 18-101_k141_331633 | Not-determined |
| 18-101_k141_299419 | Not-determined |
| 18-101_k141_265710 | Not-determined |
| 18-101_k141_331639 | Not-determined |
| 18-101_k141_51850 | Not-determined |
| 18-101_k141_331641 | Not-determined |
| 18-101_k141_265716 | Not-determined |
| 18-101_k141_265717 | Not-determined |
| 18-101_k141_299425 | Not-determined |
| 18-101_k141_233204 | Not-determined |
| 18-101_k141_134943 | Not-determined |
| 18-101_k141_200403 | Not-determined |
| 18-101_k141_315658 | Not-determined |
| 18-101_k141_249975 | Not-determined |
| 18-101_k141_101280 | Not-determined |
| 18-101_k141_299434 | Not-determined |
| 18-101_k141_315662 | Not-determined |
| 18-101_k141_381142 | Not-determined |
| 18-101_k141_217093 | Not-determined |
| 18-101_k141_19280 | Not-determined |
| 18-101_k141_217098 | Not-determined |
| 18-101_k141_134954 | Not-determined |
| 18-101_k141_166821 | Not-determined |

| | |
|---|---|
| 18-101_k141_51868 | Not-determined |
| 18-101_k141_166823 | Not-determined |
| 18-101_k141_51870 | Not-determined |
| 18-101_k141_19289 | Not-determined |
| 18-101_k141_249980 | Not-determined |
| 18-101_k141_134963 | Not-determined |
| 18-101_k141_166829 | Not-determined |
| 18-101_k141_184173 | Not-determined |
| 18-101_k141_200414 | Not-determined |
| 18-101_k141_150974 | Not-determined |
| 18-101_k141_217107 | Not-determined |
| 18-101_k141_364982 | Not-determined |
| 18-101_k141_3085 | Not-determined |
| 18-101_k141_315674 | Not-determined |
| 18-101_k141_19294 | Not-determined |
| 18-101_k141_331676 | Not-determined |
| 18-101_k141_184184 | Low-quality |
| 18-101_k141_118143 | Not-determined |
| 18-101_k141_331677 | Low-quality |
| 18-101_k141_381166 | Not-determined |
| 18-101_k141_134973 | Low-quality |
| 18-101_k141_381169 | Not-determined |
| 18-101_k141_19307 | Not-determined |
| 18-101_k141_249996 | Not-determined |
| 18-101_k141_299455 | Not-determined |
| 18-101_k141_299458 | Not-determined |
| 18-101_k141_101303 | Low-quality |
| 18-101_k141_51889 | Not-determined |
| 18-101_k141_51892 | Not-determined |
| 18-101_k141_217118 | Not-determined |
| 18-101_k141_51893 | Low-quality |
| 18-101_k141_35821 | Not-determined |
| 18-101_k141_348631 | Not-determined |
| 18-101_k141_184207 | Not-determined |
| 18-101_k141_250011 | Not-determined |
| 18-101_k141_265757 | Not-determined |
| 18-101_k141_233209 | Not-determined |
| 18-101_k141_35831 | Not-determined |
| 18-101_k141_51905 | Not-determined |
| 18-101_k141_299479 | Not-determined |
| 18-101_k141_35833 | Not-determined |
| 18-101_k141_51906 | Not-determined |
| 18-101_k141_35836 | Not-determined |
| 18-101_k141_184212 | Not-determined |
| 18-101_k141_365029 | Not-determined |
| 18-101_k141_217127 | Not-determined |
| 18-101_k141_3120 | Not-determined |
| 18-101_k141_118162 | Not-determined |
| 18-101_k141_217133 | Not-determined |
| 18-101_k141_3122 | Not-determined |
| 18-101_k141_348651 | Not-determined |
| 18-101_k141_299482 | Not-determined |
| 18-101_k141_135016 | Not-determined |
| 18-101_k141_365034 | Not-determined |
| 18-101_k141_101322 | Not-determined |
| 18-101_k141_184221 | Not-determined |
| 18-101_k141_348654 | Not-determined |
| 18-101_k141_101324 | Not-determined |
| 18-101_k141_35852 | Not-determined |
| 18-101_k141_348656 | Not-determined |
| 18-101_k141_35855 | Not-determined |
| 18-101_k141_200438 | Low-quality |
| 18-101_k141_348658 | Not-determined |
| 18-101_k141_348660 | Not-determined |
| 18-101_k141_35861 | Not-determined |
| 18-101_k141_150994 | Not-determined |
| 18-101_k141_35862 | Not-determined |
| 18-101_k141_68218 | Not-determined |
| 18-101_k141_3125 | Not-determined |
| 18-101_k141_35865 | Not-determined |
| 18-101_k141_265779 | Not-determined |
| 18-101_k141_250029 | Not-determined |
| 18-101_k141_348668 | Low-quality |
| 18-101_k141_265783 | Not-determined |
| 18-101_k141_35874 | Not-determined |
| 18-101_k141_19338 | Not-determined |
| 18-101_k141_233229 | Not-determined |
| 18-101_k141_101343 | Not-determined |
| 18-101_k141_19344 | Not-determined |
| 18-101_k141_331717 | Not-determined |
| 18-101_k141_51930 | Low-quality |
| 18-101_k141_19347 | Not-determined |
| 18-101_k141_51933 | Low-quality |
| 18-101_k141_282771 | Not-determined |
| 18-101_k141_200448 | Not-determined |
| 18-101_k141_282772 | Not-determined |
| 18-101_k141_184235 | Not-determined |
| 18-101_k141_184236 | Not-determined |
| 18-101_k141_381199 | Low-quality |
| 18-101_k141_365063 | Not-determined |
| 18-101_k141_51941 | Not-determined |
| 18-101_k141_101361 | Not-determined |
| 18-101_k141_35885 | Not-determined |
| 18-101_k141_51945 | Low-quality |
| 18-101_k141_282786 | Not-determined |
| 18-101_k141_348689 | Not-determined |
| 18-101_k141_282787 | Not-determined |
| 18-101_k141_151010 | Not-determined |
| 18-101_k141_348691 | Not-determined |
| 18-101_k141_101368 | Not-determined |
| 18-101_k141_381208 | Not-determined |
| 18-101_k141_282788 | Not-determined |
| 18-101_k141_151011 | Not-determined |
| 18-101_k141_315699 | Not-determined |
| 18-101_k141_315700 | Not-determined |
| 18-101_k141_35891 | Not-determined |
| 18-101_k141_365068 | Not-determined |
| 18-101_k141_35894 | Not-determined |
| 18-101_k141_348699 | Not-determined |
| 18-101_k141_3150 | Low-quality |
| 18-101_k141_348701 | Low-quality |
| 18-101_k141_35902 | Not-determined |
| 18-101_k141_265794 | Not-determined |
| 18-101_k141_331741 | Not-determined |
| 18-101_k141_200453 | Not-determined |
| 18-101_k141_233264 | Not-determined |
| 18-101_k141_233266 | Not-determined |
| 18-101_k141_200459 | Not-determined |
| 18-101_k141_101391 | Not-determined |
| 18-101_k141_282812 | Not-determined |
| 18-101_k141_282816 | Not-determined |
| 18-101_k141_51959 | Low-quality |
| 18-101_k141_217200 | Not-determined |
| 18-101_k141_184249 | Not-determined |
| 18-101_k141_265816 | Low-quality |
| 18-101_k141_299543 | Not-determined |
| 18-101_k141_299544 | Not-determined |
| 18-101_k141_282827 | Low-quality |
| 18-101_k141_217210 | Not-determined |
| 18-101_k141_331751 | Not-determined |
| 18-101_k141_299554 | Not-determined |
| 18-101_k141_265834 | Not-determined |
| 18-101_k141_365070 | Not-determined |
| 18-101_k141_118177 | Not-determined |
| 18-101_k141_184259 | Not-determined |
| 18-101_k141_282839 | Not-determined |
| 18-101_k141_200489 | Not-determined |
| 18-101_k141_135025 | Not-determined |
| 18-101_k141_331759 | Not-determined |
| 18-101_k141_3169 | Not-determined |
| 18-101_k141_381245 | Not-determined |
| 18-101_k141_381246 | Not-determined |
| 18-101_k141_381249 | Not-determined |
| 18-101_k141_118185 | Low-quality |
| 18-101_k141_35948 | Not-determined |
| 18-101_k141_265845 | Low-quality |
| 18-101_k141_365090 | Not-determined |
| 18-101_k141_365091 | Not-determined |
| 18-101_k141_101421 | Not-determined |
| 18-101_k141_331770 | Not-determined |
| 18-101_k141_331772 | Not-determined |
| 18-101_k141_217231 | Low-quality |
| 18-101_k141_348758 | Not-determined |
| 18-101_k141_299572 | Not-determined |
| 18-101_k141_348761 | Not-determined |
| 18-101_k141_381252 | Low-quality |
| 18-101_k141_331779 | Not-determined |
| 18-101_k141_166848 | Not-determined |
| 18-101_k141_184286 | Not-determined |
| 18-101_k141_282869 | Not-determined |
| 18-101_k141_348764 | Not-determined |
| 18-101_k141_184288 | Not-determined |
| 18-101_k141_19378 | Low-quality |
| 18-101_k141_365106 | Not-determined |
| 18-101_k141_19381 | Not-determined |
| 18-101_k141_348768 | Not-determined |
| 18-101_k141_52022 | Not-determined |
| 18-101_k141_381265 | Not-determined |
| 18-101_k141_68299 | Not-determined |
| 18-101_k141_68301 | Not-determined |
| 18-101_k141_381270 | Not-determined |
| 18-101_k141_315743 | Low-quality |
| 18-101_k141_101428 | Not-determined |
| 18-101_k141_348779 | Not-determined |
| 18-101_k141_348781 | Not-determined |
| 18-101_k141_166857 | Not-determined |
| 18-101_k141_68311 | Not-determined |
| 18-101_k141_3182 | Not-determined |
| 18-101_k141_68314 | Not-determined |
| 18-101_k141_381273 | Not-determined |
| 18-101_k141_19399 | Not-determined |
| 18-101_k141_348784 | Not-determined |
| 18-101_k141_68320 | Not-determined |
| 18-101_k141_200520 | Not-determined |
| 18-101_k141_348789 | Not-determined |
| 18-101_k141_348794 | Not-determined |
| 18-101_k141_381286 | Not-determined |
| 18-101_k141_217265 | Not-determined |
| 18-101_k141_200531 | Not-determined |
| 18-101_k141_348801 | Low-quality |
| 18-101_k141_348802 | Not-determined |
| 18-101_k141_315763 | Not-determined |
| 18-101_k141_381301 | Low-quality |
| 18-101_k141_233297 | Not-determined |
| 18-101_k141_184297 | Not-determined |
| 18-101_k141_118196 | Not-determined |
| 18-101_k141_184301 | Not-determined |
| 18-101_k141_35964 | Not-determined |
| 18-101_k141_233307 | Not-determined |
| 18-101_k141_315768 | Not-determined |
| 18-101_k141_101447 | Not-determined |
| 18-101_k141_365146 | Not-determined |
| 18-101_k141_365148 | Not-determined |
| 18-101_k141_217279 | Not-determined |
| 18-101_k141_135086 | Not-determined |
| 18-101_k141_166886 | Not-determined |
| 18-101_k141_166887 | Not-determined |
| 18-101_k141_118215 | Not-determined |
| 18-101_k141_151042 | Not-determined |
| 18-101_k141_151045 | Not-determined |
| 18-101_k141_52058 | Not-determined |
| 18-101_k141_233315 | Not-determined |
| 18-101_k141_282898 | Low-quality |
| 18-101_k141_118222 | Not-determined |
| 18-101_k141_118224 | Not-determined |
| 18-101_k141_282905 | Not-determined |
| 18-101_k141_52061 | Not-determined |
| 18-101_k141_118234 | Low-quality |
| 18-101_k141_118235 | Not-determined |
| 18-101_k141_282910 | Not-determined |
| 18-101_k141_233327 | Not-determined |
| 18-101_k141_365165 | Not-determined |
| 18-101_k141_184333 | Not-determined |
| 18-101_k141_101459 | Not-determined |
| 18-101_k141_118241 | Not-determined |
| 18-101_k141_365171 | Not-determined |
| 18-101_k141_217299 | Not-determined |
| 18-101_k141_265864 | Not-determined |
| 18-101_k141_19414 | Low-quality |
| 18-101_k141_200540 | Not-determined |
| 18-101_k141_3223 | Not-determined |
| 18-101_k141_135106 | Not-determined |
| 18-101_k141_315782 | Not-determined |
| 18-101_k141_282919 | Low-quality |
| 18-101_k141_184342 | Low-quality |
| 18-101_k141_35993 | Not-determined |
| 18-101_k141_381324 | Not-determined |
| 18-101_k141_348823 | Not-determined |
| 18-101_k141_135111 | Not-determined |
| 18-101_k141_282929 | Not-determined |
| 18-101_k141_265870 | Not-determined |
| 18-101_k141_331793 | Not-determined |
| 18-101_k141_233340 | Not-determined |
| 18-101_k141_265877 | Not-determined |
| 18-101_k141_348827 | Not-determined |
| 18-101_k141_348831 | Not-determined |
| 18-101_k141_381340 | Not-determined |
| 18-101_k141_381342 | Not-determined |
| 18-101_k141_166929 | Not-determined |
| 18-101_k141_217314 | Not-determined |
| 18-101_k141_331823 | Low-quality |
| 18-101_k141_381349 | Not-determined |
| 18-101_k141_3239 | Not-determined |
| 18-101_k141_233353 | Not-determined |
| 18-101_k141_217315 | Not-determined |
| 18-101_k141_135140 | Low-quality |
| 18-101_k141_381363 | Not-determined |
| 18-101_k141_52074 | Not-determined |
| 18-101_k141_331828 | Not-determined |
| 18-101_k141_233363 | Not-determined |
| 18-101_k141_233369 | Not-determined |
| 18-101_k141_135149 | Not-determined |
| 18-101_k141_299590 | Not-determined |
| 18-101_k141_315816 | Not-determined |
| 18-101_k141_135152 | Not-determined |
| 18-101_k141_166942 | Not-determined |
| 18-101_k141_282943 | Not-determined |
| 18-101_k141_135160 | Not-determined |
| 18-101_k141_233371 | Low-quality |
| 18-101_k141_184395 | Not-determined |
| 18-101_k141_233378 | Low-quality |
| 18-101_k141_282953 | Not-determined |
| 18-101_k141_250116 | Not-determined |
| 18-101_k141_217335 | Not-determined |
| 18-101_k141_299600 | Not-determined |
| 18-101_k141_299601 | Not-determined |
| 18-101_k141_233382 | Low-quality |
| 18-101_k141_331850 | Not-determined |
| 18-101_k141_233385 | Not-determined |
| 18-101_k141_233386 | Not-determined |
| 18-101_k141_315839 | Not-determined |
| 18-101_k141_166963 | Not-determined |
| 18-101_k141_52095 | Low-quality |
| 18-101_k141_331859 | Low-quality |
| 18-101_k141_233394 | Not-determined |
| 18-101_k141_331863 | Not-determined |
| 18-101_k141_233397 | Not-determined |
| 18-101_k141_282972 | Not-determined |
| 18-101_k141_331866 | Not-determined |
| 18-101_k141_118294 | Not-determined |
| 18-101_k141_365187 | Not-determined |
| 18-101_k141_233401 | Not-determined |
| 18-101_k141_233402 | Not-determined |
| 18-101_k141_282976 | Low-quality |
| 18-101_k141_118297 | Not-determined |
| 18-101_k141_233409 | Not-determined |
| 18-101_k141_135170 | Not-determined |
| 18-101_k141_135172 | Not-determined |
| 18-101_k141_118300 | Not-determined |
| 18-101_k141_315860 | Not-determined |
| 18-101_k141_282988 | Not-determined |
| 18-101_k141_282991 | Not-determined |
| 18-101_k141_331873 | Not-determined |
| 18-101_k141_151102 | Low-quality |
| 18-101_k141_299640 | Not-determined |
| 18-101_k141_3279 | Low-quality |
| 18-101_k141_265902 | Not-determined |
| 18-101_k141_265905 | Not-determined |
| 18-101_k141_331883 | Not-determined |
| 18-101_k141_184418 | Not-determined |
| 18-101_k141_331887 | Not-determined |
| 18-101_k141_52113 | Not-determined |
| 18-101_k141_200564 | Not-determined |
| 18-101_k141_36040 | Not-determined |
| 18-101_k141_283018 | Not-determined |
| 18-101_k141_299652 | Not-determined |
| 18-101_k141_283022 | Not-determined |
| 18-101_k141_118316 | Low-quality |
| 18-101_k141_250159 | Not-determined |
| 18-101_k141_299657 | Not-determined |
| 18-101_k141_52126 | Low-quality |
| 18-101_k141_52127 | Not-determined |
| 18-101_k141_184438 | Not-determined |
| 18-101_k141_135202 | Not-determined |
| 18-101_k141_348869 | Not-determined |
| 18-101_k141_200576 | Not-determined |
| 18-101_k141_299660 | Not-determined |
| 18-101_k141_166997 | Not-determined |
| 18-101_k141_52141 | Not-determined |
| 18-101_k141_348872 | Not-determined |
| 18-101_k141_52147 | Not-determined |
| 18-101_k141_217371 | Not-determined |
| 18-101_k141_217373 | Not-determined |
| 18-101_k141_381384 | Not-determined |
| 18-101_k141_118341 | Not-determined |
| 18-101_k141_151115 | Not-determined |
| 18-101_k141_381386 | Low-quality |
| 18-101_k141_299678 | Not-determined |
| 18-101_k141_151117 | Not-determined |
| 18-101_k141_118344 | Not-determined |
| 18-101_k141_151120 | Not-determined |
| 18-101_k141_299684 | Not-determined |
| 18-101_k141_52151 | Not-determined |
| 18-101_k141_365205 | Not-determined |
| 18-101_k141_85259 | Not-determined |
| 18-101_k141_167022 | Not-determined |
| 18-101_k141_151125 | Not-determined |
| 18-101_k141_52154 | Not-determined |
| 18-101_k141_118369 | Not-determined |
| 18-101_k141_36067 | Not-determined |
| 18-101_k141_365210 | Not-determined |
| 18-101_k141_135242 | Not-determined |
| 18-101_k141_217384 | Not-determined |
| 18-101_k141_118377 | Not-determined |
| 18-101_k141_118380 | Not-determined |
| 18-101_k141_217386 | Not-determined |
| 18-101_k141_233420 | Not-determined |
| 18-101_k141_299709 | Not-determined |
| 18-101_k141_200596 | Not-determined |
| 18-101_k141_233422 | Not-determined |
| 18-101_k141_315901 | Not-determined |
| 18-101_k141_233423 | Not-determined |
| 18-101_k141_85283 | Not-determined |
| 18-101_k141_85285 | Not-determined |
| 18-101_k141_151136 | Not-determined |
| 18-101_k141_36079 | Not-determined |
| 18-101_k141_315902 | Low-quality |
| 18-101_k141_184491 | Not-determined |
| 18-101_k141_118391 | Not-determined |
| 18-101_k141_19516 | Not-determined |
| 18-101_k141_315907 | Not-determined |
| 18-101_k141_19518 | Not-determined |
| 18-101_k141_315908 | Not-determined |
| 18-101_k141_184497 | Not-determined |
| 18-101_k141_36101 | Not-determined |
| 18-101_k141_85300 | Not-determined |
| 18-101_k141_381413 | Low-quality |
| 18-101_k141_85302 | Not-determined |
| 18-101_k141_151146 | Not-determined |
| 18-101_k141_118402 | Not-determined |
| 18-101_k141_200620 | Not-determined |
| 18-101_k141_200621 | Not-determined |
| 18-101_k141_68381 | Not-determined |
| 18-101_k141_217418 | Not-determined |
| 18-101_k141_118406 | Not-determined |
| 18-101_k141_85314 | Low-quality |
| 18-101_k141_85317 | Low-quality |
| 18-101_k141_250173 | Not-determined |
| 18-101_k141_250174 | Not-determined |
| 18-101_k141_85322 | Not-determined |
| 18-101_k141_365242 | Not-determined |
| 18-101_k141_217429 | Not-determined |
| 18-101_k141_118418 | Not-determined |
| 18-101_k141_184518 | Not-determined |
| 18-101_k141_167049 | Not-determined |
| 18-101_k141_167050 | Not-determined |
| 18-101_k141_68406 | Not-determined |
| 18-101_k141_381436 | Not-determined |
| 18-101_k141_68408 | Not-determined |
| 18-101_k141_265920 | Not-determined |
| 18-101_k141_68409 | Not-determined |
| 18-101_k141_265921 | Not-determined |
| 18-101_k141_265924 | Not-determined |
| 18-101_k141_200643 | Not-determined |
| 18-101_k141_348916 | Not-determined |
| 18-101_k141_381441 | Not-determined |
| 18-101_k141_19536 | Not-determined |
| 18-101_k141_200645 | Not-determined |
| 18-101_k141_36109 | Not-determined |
| 18-101_k141_331935 | Not-determined |
| 18-101_k141_200649 | Not-determined |
| 18-101_k141_52187 | Not-determined |
| 18-101_k141_315928 | Not-determined |
| 18-101_k141_118440 | Not-determined |
| 18-101_k141_135262 | Not-determined |
| 18-101_k141_283058 | Not-determined |
| 18-101_k141_348927 | Not-determined |
| 18-101_k141_118443 | Not-determined |
| 18-101_k141_200662 | Not-determined |
| 18-101_k141_52203 | Not-determined |
| 18-101_k141_331939 | Low-quality |
| 18-101_k141_348936 | Not-determined |
| 18-101_k141_118451 | Not-determined |
| 18-101_k141_315937 | Not-determined |
| 18-101_k141_85339 | Not-determined |
| 18-101_k141_200667 | Not-determined |
| 18-101_k141_265938 | Not-determined |
| 18-101_k141_265939 | Low-quality |
| 18-101_k141_265941 | Not-determined |
| 18-101_k141_19544 | Not-determined |
| 18-101_k141_85352 | Not-determined |
| 18-101_k141_250211 | Not-determined |
| 18-101_k141_365262 | Not-determined |
| 18-101_k141_265945 | Not-determined |
| 18-101_k141_365265 | Not-determined |
| 18-101_k141_85355 | Not-determined |
| 18-101_k141_217467 | Not-determined |
| 18-101_k141_118462 | Not-determined |
| 18-101_k141_217468 | Not-determined |
| 18-101_k141_118463 | Not-determined |
| 18-101_k141_315947 | Not-determined |
| 18-101_k141_118466 | Not-determined |
| 18-101_k141_118467 | Not-determined |
| 18-101_k141_265953 | Not-determined |
| 18-101_k141_348946 | Not-determined |
| 18-101_k141_265954 | Low-quality |
| 18-101_k141_381452 | Not-determined |
| 18-101_k141_348949 | Not-determined |
| 18-101_k141_151165 | Not-determined |
| 18-101_k141_331958 | Not-determined |
| 18-101_k141_200682 | Not-determined |
| 18-101_k141_118474 | Not-determined |
| 18-101_k141_250235 | Not-determined |
| 18-101_k141_135297 | Not-determined |
| 18-101_k141_19555 | Low-quality |
| 18-101_k141_217470 | Not-determined |
| 18-101_k141_265963 | Not-determined |
| 18-101_k141_200689 | Not-determined |
| 18-101_k141_250246 | Not-determined |
| 18-101_k141_151173 | Not-determined |
| 18-101_k141_36135 | Not-determined |
| 18-101_k141_365288 | Not-determined |
| 18-101_k141_52231 | Not-determined |
| 18-101_k141_85381 | Not-determined |
| 18-101_k141_52234 | Not-determined |
| 18-101_k141_52235 | Not-determined |
| 18-101_k141_135317 | Not-determined |
| 18-101_k141_68427 | Not-determined |
| 18-101_k141_68429 | Low-quality |
| 18-101_k141_283070 | Not-determined |
| 18-101_k141_151190 | Not-determined |
| 18-101_k141_36143 | Not-determined |
| 18-101_k141_315957 | Not-determined |
| 18-101_k141_151197 | Not-determined |
| 18-101_k141_217483 | Not-determined |
| 18-101_k141_118485 | Not-determined |
| 18-101_k141_233460 | Not-determined |
| 18-101_k141_365306 | Not-determined |
| 18-101_k141_118487 | Not-determined |
| 18-101_k141_331974 | Not-determined |
| 18-101_k141_250266 | Not-determined |
| 18-101_k141_315965 | Not-determined |
| 18-101_k141_3343 | Not-determined |
| 18-101_k141_315967 | Not-determined |
| 18-101_k141_36157 | Not-determined |
| 18-101_k141_283077 | Not-determined |
| 18-101_k141_365308 | Not-determined |
| 18-101_k141_68445 | Not-determined |
| 18-101_k141_36168 | Not-determined |
| 18-101_k141_167097 | Low-quality |
| 18-101_k141_135334 | Not-determined |
| 18-101_k141_135335 | Not-determined |
| 18-101_k141_365313 | Not-determined |
| 18-101_k141_52265 | Not-determined |
| 18-101_k141_52266 | Not-determined |
| 18-101_k141_167103 | Not-determined |
| 18-101_k141_52271 | Not-determined |
| 18-101_k141_217501 | Not-determined |
| 18-101_k141_151213 | Not-determined |
| 18-101_k141_19596 | Not-determined |
| 18-101_k141_200709 | Not-determined |
| 18-101_k141_283093 | Not-determined |
| 18-101_k141_200717 | Not-determined |
| 18-101_k141_283100 | Not-determined |
| 18-101_k141_101493 | Not-determined |
| 18-101_k141_348986 | Not-determined |
| 18-101_k141_85403 | Not-determined |
| 18-101_k141_85404 | Not-determined |
| 18-101_k141_85405 | Not-determined |
| 18-101_k141_101501 | Not-determined |
| 18-101_k141_266011 | Low-quality |
| 18-101_k141_3355 | Not-determined |
| 18-101_k141_3356 | Not-determined |
| 18-101_k141_167123 | Not-determined |
| 18-101_k141_52304 | Low-quality |
| 18-101_k141_52307 | Not-determined |
| 18-101_k141_19604 | Not-determined |
| 18-101_k141_101509 | Not-determined |
| 18-101_k141_299751 | Low-quality |
| 18-101_k141_167129 | Not-determined |
| 18-101_k141_233485 | Low-quality |
| 18-101_k141_266017 | Not-determined |
| 18-101_k141_332004 | Not-determined |
| 18-101_k141_332006 | Not-determined |
| 18-101_k141_200730 | Not-determined |
| 18-101_k141_52311 | Not-determined |
| 18-101_k141_349003 | Not-determined |
| 18-101_k141_233494 | Not-determined |
| 18-101_k141_332012 | Not-determined |
| 18-101_k141_36204 | Not-determined |
| 18-101_k141_365340 | Not-determined |
| 18-101_k141_151246 | Not-determined |
| 18-101_k141_167134 | Not-determined |
| 18-101_k141_52314 | Not-determined |
| 18-101_k141_200735 | Low-quality |
| 18-101_k141_184586 | Not-determined |
| 18-101_k141_200736 | Not-determined |
| 18-101_k141_118532 | Not-determined |
| 18-101_k141_217510 | Not-determined |
| 18-101_k141_36208 | Low-quality |
| 18-101_k141_381520 | Not-determined |
| 18-101_k141_135344 | Not-determined |
| 18-101_k141_266028 | Not-determined |
| 18-101_k141_233506 | Not-determined |
| 18-101_k141_68456 | Not-determined |
| 18-101_k141_19623 | Not-determined |
| 18-101_k141_299766 | Not-determined |
| 18-101_k141_381523 | Not-determined |
| 18-101_k141_19626 | Not-determined |
| 18-101_k141_36216 | Not-determined |
| 18-101_k141_381528 | Not-determined |
| 18-101_k141_381529 | Not-determined |
| 18-101_k141_36220 | Low-quality |
| 18-101_k141_68468 | Not-determined |
| 18-101_k141_299774 | Not-determined |
| 18-101_k141_118552 | Not-determined |
| 18-101_k141_36226 | Not-determined |
| 18-101_k141_101531 | Not-determined |
| 18-101_k141_184607 | Not-determined |
| 18-101_k141_250289 | Not-determined |
| 18-101_k141_184611 | Not-determined |
| 18-101_k141_250293 | Not-determined |
| 18-101_k141_36231 | Not-determined |
| 18-101_k141_19637 | Not-determined |
| 18-101_k141_283128 | Not-determined |
| 18-101_k141_381543 | Not-determined |
| 18-101_k141_316028 | Not-determined |
| 18-101_k141_233526 | Low-quality |
| 18-101_k141_381553 | Not-determined |
| 18-101_k141_316031 | Not-determined |
| 18-101_k141_167168 | Not-determined |
| 18-101_k141_68478 | Not-determined |
| 18-101_k141_283139 | Not-determined |
| 18-101_k141_118569 | Not-determined |
| 18-101_k141_151295 | Not-determined |
| 18-101_k141_381561 | Not-determined |
| 18-101_k141_299787 | Not-determined |
| 18-101_k141_184636 | Not-determined |
| 18-101_k141_135388 | Not-determined |
| 18-101_k141_151305 | Not-determined |
| 18-101_k141_68483 | Low-quality |
| 18-101_k141_167186 | Not-determined |
| 18-101_k141_316042 | Not-determined |
| 18-101_k141_36257 | Not-determined |
| 18-101_k141_167188 | Not-determined |
| 18-101_k141_316045 | Low-quality |
| 18-101_k141_316046 | Not-determined |
| 18-101_k141_233550 | Not-determined |
| 18-101_k141_266051 | Not-determined |

| | |
|---|---|
| 18-101_k141_365373 | Not-determined |
| 18-101_k141_36258 | Not-determined |
| 18-101_k141_85465 | Low-quality |
| 18-101_k141_233553 | Not-determined |
| 18-101_k141_283160 | Not-determined |
| 18-101_k141_250315 | Not-determined |
| 18-101_k141_266063 | Not-determined |
| 18-101_k141_36266 | Not-determined |
| 18-101_k141_283164 | Low-quality |
| 18-101_k141_266066 | Not-determined |
| 18-101_k141_68497 | Not-determined |
| 18-101_k141_250318 | Not-determined |
| 18-101_k141_151316 | Not-determined |
| 18-101_k141_316060 | Low-quality |
| 18-101_k141_167205 | Low-quality |
| 18-101_k141_283179 | Not-determined |
| 18-101_k141_101581 | Not-determined |
| 18-101_k141_283184 | Not-determined |
| 18-101_k141_36273 | Low-quality |
| 18-101_k141_36275 | Not-determined |
| 18-101_k141_250329 | Not-determined |
| 18-101_k141_151329 | Not-determined |
| 18-101_k141_68505 | Not-determined |
| 18-101_k141_332088 | Not-determined |
| 18-101_k141_101594 | Not-determined |
| 18-101_k141_266096 | Not-determined |
| 18-101_k141_332095 | Not-determined |
| 18-101_k141_151337 | Low-quality |
| 18-101_k141_332104 | Not-determined |
| 18-101_k141_52354 | Not-determined |
| 18-101_k141_184666 | Low-quality |
| 18-101_k141_365388 | Not-determined |
| 18-101_k141_36286 | Not-determined |
| 18-101_k141_332110 | Not-determined |
| 18-101_k141_101606 | Not-determined |
| 18-101_k141_365392 | Low-quality |
| 18-101_k141_283207 | Low-quality |
| 18-101_k141_68526 | Not-determined |
| 18-101_k141_118576 | Not-determined |
| 18-101_k141_68530 | Not-determined |
| 18-101_k141_19681 | Not-determined |
| 18-101_k141_250346 | Low-quality |
| 18-101_k141_266122 | Not-determined |
| 18-101_k141_250349 | Not-determined |
| 18-101_k141_52367 | Not-determined |
| 18-101_k141_101621 | Not-determined |
| 18-101_k141_52372 | Not-determined |
| 18-101_k141_19686 | Low-quality |
| 18-101_k141_52374 | Not-determined |
| 18-101_k141_316094 | Low-quality |
| 18-101_k141_184686 | Not-determined |
| 18-101_k141_68538 | Not-determined |
| 18-101_k141_167216 | Not-determined |
| 18-101_k141_316098 | Not-determined |
| 18-101_k141_316099 | Not-determined |
| 18-101_k141_316104 | Not-determined |
| 18-101_k141_250359 | Low-quality |
| 18-101_k141_266131 | Not-determined |
| 18-101_k141_68541 | Not-determined |
| 18-101_k141_250360 | Not-determined |
| 18-101_k141_266132 | Not-determined |
| 18-101_k141_250362 | Not-determined |
| 18-101_k141_184694 | Low-quality |
| 18-101_k141_381599 | Not-determined |
| 18-101_k141_52390 | Not-determined |
| 18-101_k141_101632 | Not-determined |
| 18-101_k141_68546 | Low-quality |
| 18-101_k141_101635 | Not-determined |
| 18-101_k141_283225 | Not-determined |
| 18-101_k141_266143 | Not-determined |
| 18-101_k141_101642 | Not-determined |
| 18-101_k141_118597 | Not-determined |
| 18-101_k141_101644 | Not-determined |
| 18-101_k141_250382 | Not-determined |
| 18-101_k141_3418 | Not-determined |
| 18-101_k141_3421 | Not-determined |
| 18-101_k141_266153 | Low-quality |
| 18-101_k141_3422 | Low-quality |
| 18-101_k141_299818 | Not-determined |
| 18-101_k141_266156 | Not-determined |
| 18-101_k141_316119 | Not-determined |
| 18-101_k141_266161 | Not-determined |
| 18-101_k141_266162 | Not-determined |
| 18-101_k141_365430 | Not-determined |
| 18-101_k141_266165 | Not-determined |
| 18-101_k141_36318 | Not-determined |
| 18-101_k141_167251 | Not-determined |
| 18-101_k141_118616 | Not-determined |
| 18-101_k141_151389 | Not-determined |
| 18-101_k141_135474 | Low-quality |
| 18-101_k141_266183 | Low-quality |
| 18-101_k141_101658 | Not-determined |
| 18-101_k141_135490 | Not-determined |
| 18-101_k141_3439 | Not-determined |
| 18-101_k141_316145 | Not-determined |
| 18-101_k141_19716 | Not-determined |
| 18-101_k141_266200 | Not-determined |
| 18-101_k141_250409 | Not-determined |
| 18-101_k141_299833 | Not-determined |
| 18-101_k141_316154 | Not-determined |
| 18-101_k141_184730 | Not-determined |
| 18-101_k141_217548 | Not-determined |
| 18-101_k141_135502 | Low-quality |
| 18-101_k141_118643 | Not-determined |
| 18-101_k141_365467 | Not-determined |
| 18-101_k141_266221 | Not-determined |
| 18-101_k141_118646 | Not-determined |
| 18-101_k141_184747 | Not-determined |
| 18-101_k141_349057 | Not-determined |
| 18-101_k141_184749 | Not-determined |
| 18-101_k141_118648 | Not-determined |
| 18-101_k141_233634 | Not-determined |
| 18-101_k141_200821 | Not-determined |
| 18-101_k141_101693 | Not-determined |
| 18-101_k141_299850 | Not-determined |
| 18-101_k141_118650 | Not-determined |
| 18-101_k141_3464 | Low-quality |
| 18-101_k141_266226 | Not-determined |
| 18-101_k141_200825 | Not-determined |
| 18-101_k141_3466 | Not-determined |
| 18-101_k141_68566 | Not-determined |
| 18-101_k141_250427 | Not-determined |
| 18-101_k141_349064 | Low-quality |
| 18-101_k141_151410 | Not-determined |
| 18-101_k141_233639 | Not-determined |
| 18-101_k141_68576 | Not-determined |
| 18-101_k141_200835 | Not-determined |
| 18-101_k141_250435 | Not-determined |
| 18-101_k141_217551 | Not-determined |
| 18-101_k141_299870 | Not-determined |
| 18-101_k141_250442 | Not-determined |
| 18-101_k141_381652 | Not-determined |
| 18-101_k141_217559 | Not-determined |
| 18-101_k141_266232 | Low-quality |
| 18-101_k141_85504 | Not-determined |
| 18-101_k141_283268 | Not-determined |
| 18-101_k141_200857 | Not-determined |
| 18-101_k141_118671 | Not-determined |
| 18-101_k141_233653 | Not-determined |
| 18-101_k141_200862 | Not-determined |
| 18-101_k141_381660 | Low-quality |
| 18-101_k141_200863 | Not-determined |
| 18-101_k141_381663 | Not-determined |
| 18-101_k141_233663 | Not-determined |
| 18-101_k141_283276 | Not-determined |
| 18-101_k141_19745 | Not-determined |
| 18-101_k141_52407 | Not-determined |
| 18-101_k141_118684 | Not-determined |
| 18-101_k141_135537 | Not-determined |
| 18-101_k141_365501 | Not-determined |
| 18-101_k141_316195 | Not-determined |
| 18-101_k141_316196 | Not-determined |
| 18-101_k141_3473 | Not-determined |
| 18-101_k141_349090 | Not-determined |
| 18-101_k141_118692 | Low-quality |
| 18-101_k141_299903 | Not-determined |
| 18-101_k141_316205 | Not-determined |
| 18-101_k141_85524 | Low-quality |
| 18-101_k141_365510 | Low-quality |
| 18-101_k141_200874 | Not-determined |
| 18-101_k141_151433 | Not-determined |
| 18-101_k141_52420 | Low-quality |
| 18-101_k141_200880 | Not-determined |
| 18-101_k141_200883 | Not-determined |
| 18-101_k141_365520 | Not-determined |
| 18-101_k141_266255 | Not-determined |
| 18-101_k141_332157 | Low-quality |
| 18-101_k141_381674 | Not-determined |
| 18-101_k141_200890 | Not-determined |
| 18-101_k141_381675 | Not-determined |
| 18-101_k141_299915 | Not-determined |
| 18-101_k141_332164 | Not-determined |
| 18-101_k141_381681 | Not-determined |
| 18-101_k141_3503 | Not-determined |
| 18-101_k141_332183 | Not-determined |
| 18-101_k141_85533 | Low-quality |
| 18-101_k141_135562 | Not-determined |
| 18-101_k141_85539 | Not-determined |
| 18-101_k141_151462 | Not-determined |
| 18-101_k141_332191 | Low-quality |
| 18-101_k141_3512 | Low-quality |
| 18-101_k141_381708 | Not-determined |
| 18-101_k141_200906 | Low-quality |
| 18-101_k141_118731 | Not-determined |
| 18-101_k141_151473 | Not-determined |
| 18-101_k141_85568 | Not-determined |
| 18-101_k141_200911 | Low-quality |
| 18-101_k141_151478 | Not-determined |
| 18-101_k141_151480 | Not-determined |
| 18-101_k141_151481 | Not-determined |
| 18-101_k141_316227 | Not-determined |
| 18-101_k141_266268 | Not-determined |
| 18-101_k141_200913 | Not-determined |
| 18-101_k141_316233 | Not-determined |
| 18-101_k141_316235 | Not-determined |
| 18-101_k141_266278 | Not-determined |
| 18-101_k141_200923 | Not-determined |
| 18-101_k141_349115 | Not-determined |
| 18-101_k141_349120 | Not-determined |
| 18-101_k141_250457 | Not-determined |
| 18-101_k141_266286 | Not-determined |
| 18-101_k141_266287 | Not-determined |
| 18-101_k141_316243 | Not-determined |
| 18-101_k141_266289 | Not-determined |
| 18-101_k141_299958 | Not-determined |
| 18-101_k141_233722 | Not-determined |
| 18-101_k141_299959 | Low-quality |
| 18-101_k141_19783 | Not-determined |
| 18-101_k141_365560 | Not-determined |
| 18-101_k141_299965 | Not-determined |
| 18-101_k141_381734 | Not-determined |
| 18-101_k141_299970 | Not-determined |
| 18-101_k141_68607 | Not-determined |
| 18-101_k141_381737 | Not-determined |
| 18-101_k141_68609 | Low-quality |
| 18-101_k141_381739 | Not-determined |
| 18-101_k141_381740 | Not-determined |
| 18-101_k141_36392 | Not-determined |
| 18-101_k141_349141 | Not-determined |
| 18-101_k141_151507 | Low-quality |
| 18-101_k141_85577 | Not-determined |
| 18-101_k141_3548 | Not-determined |
| 18-101_k141_233737 | Not-determined |
| 18-101_k141_36401 | Not-determined |
| 18-101_k141_184830 | Not-determined |
| 18-101_k141_151519 | Not-determined |
| 18-101_k141_381756 | Not-determined |
| 18-101_k141_349156 | Low-quality |
| 18-101_k141_365581 | Low-quality |
| 18-101_k141_365582 | Low-quality |
| 18-101_k141_283334 | Not-determined |
| 18-101_k141_332202 | Not-determined |
| 18-101_k141_349160 | Not-determined |
| 18-101_k141_266319 | Not-determined |
| 18-101_k141_349163 | Not-determined |
| 18-101_k141_3556 | Not-determined |
| 18-101_k141_217619 | Not-determined |
| 18-101_k141_365587 | Low-quality |
| 18-101_k141_36411 | Not-determined |
| 18-101_k141_332221 | Not-determined |
| 18-101_k141_184841 | Not-determined |
| 18-101_k141_266336 | Not-determined |
| 18-101_k141_381783 | Not-determined |
| 18-101_k141_167288 | Low-quality |
| 18-101_k141_332231 | Not-determined |
| 18-101_k141_266340 | Low-quality |
| 18-101_k141_250508 | Not-determined |
| 18-101_k141_283347 | Not-determined |
| 18-101_k141_299985 | Not-determined |
| 18-101_k141_283349 | Not-determined |
| 18-101_k141_151555 | Not-determined |
| 18-101_k141_299990 | Not-determined |
| 18-101_k141_184845 | Not-determined |
| 18-101_k141_184847 | Low-quality |
| 18-101_k141_250526 | Not-determined |
| 18-101_k141_151559 | Low-quality |
| 18-101_k141_250528 | Not-determined |
| 18-101_k141_266352 | Not-determined |
| 18-101_k141_266354 | Not-determined |
| 18-101_k141_184851 | Not-determined |
| 18-101_k141_365605 | Not-determined |
| 18-101_k141_332233 | Not-determined |
| 18-101_k141_118737 | Low-quality |
| 18-101_k141_151564 | Not-determined |
| 18-101_k141_151568 | Not-determined |
| 18-101_k141_36439 | Not-determined |
| 18-101_k141_19839 | Not-determined |
| 18-101_k141_184859 | Not-determined |
| 18-101_k141_19843 | Not-determined |
| 18-101_k141_300005 | Not-determined |
| 18-101_k141_118741 | Low-quality |
| 18-101_k141_365620 | Not-determined |
| 18-101_k141_167311 | Not-determined |
| 18-101_k141_300010 | Not-determined |
| 18-101_k141_19849 | Not-determined |
| 18-101_k141_365621 | Not-determined |
| 18-101_k141_365623 | Not-determined |
| 18-101_k141_200956 | Not-determined |
| 18-101_k141_184868 | Not-determined |
| 18-101_k141_300019 | Not-determined |
| 18-101_k141_266371 | Not-determined |
| 18-101_k141_36451 | Not-determined |
| 18-101_k141_250541 | Not-determined |
| 18-101_k141_118756 | Not-determined |
| 18-101_k141_19860 | Not-determined |
| 18-101_k141_3596 | Not-determined |
| 18-101_k141_3598 | Low-quality |
| 18-101_k141_3599 | Not-determined |
| 18-101_k141_316286 | Not-determined |
| 18-101_k141_316287 | Not-determined |
| 18-101_k141_167317 | Not-determined |
| 18-101_k141_200975 | Not-determined |
| 18-101_k141_3612 | Not-determined |
| 18-101_k141_200979 | Not-determined |
| 18-101_k141_200982 | Not-determined |
| 18-101_k141_19871 | Not-determined |
| 18-101_k141_101777 | Low-quality |
| 18-101_k141_349184 | Not-determined |
| 18-101_k141_316313 | Not-determined |
| 18-101_k141_19880 | Not-determined |
| 18-101_k141_283384 | Not-determined |
| 18-101_k141_167342 | Not-determined |
| 18-101_k141_250573 | Not-determined |
| 18-101_k141_101800 | Not-determined |
| 18-101_k141_300044 | Low-quality |
| 18-101_k141_300045 | Not-determined |
| 18-101_k141_19883 | Low-quality |
| 18-101_k141_332249 | Not-determined |
| 18-101_k141_332252 | Not-determined |
| 18-101_k141_36468 | Not-determined |
| 18-101_k141_332253 | Not-determined |
| 18-101_k141_266398 | Not-determined |
| 18-101_k141_332256 | Not-determined |
| 18-101_k141_135645 | Not-determined |
| 18-101_k141_151619 | Not-determined |
| 18-101_k141_135648 | Not-determined |
| 18-101_k141_250587 | Not-determined |
| 18-101_k141_250588 | Not-determined |
| 18-101_k141_118797 | Not-determined |
| 18-101_k141_349213 | Not-determined |
| 18-101_k141_349216 | Not-determined |
| 18-101_k141_250599 | Not-determined |
| 18-101_k141_135652 | Not-determined |
| 18-101_k141_349220 | Not-determined |
| 18-101_k141_349221 | Not-determined |
| 18-101_k141_250600 | Not-determined |
| 18-101_k141_19900 | Low-quality |
| 18-101_k141_151630 | Not-determined |
| 18-101_k141_300071 | Not-determined |
| 18-101_k141_151633 | Not-determined |
| 18-101_k141_52506 | Not-determined |
| 18-101_k141_19903 | Not-determined |
| 18-101_k141_118810 | Not-determined |
| 18-101_k141_250607 | Not-determined |
| 18-101_k141_300076 | Not-determined |
| 18-101_k141_19909 | Not-determined |
| 18-101_k141_19911 | Not-determined |
| 18-101_k141_300084 | Not-determined |
| 18-101_k141_300089 | Not-determined |
| 18-101_k141_300090 | Not-determined |
| 18-101_k141_52520 | Not-determined |
| 18-101_k141_365635 | Low-quality |
| 18-101_k141_52521 | Not-determined |
| 18-101_k141_52524 | Low-quality |
| 18-101_k141_300100 | Not-determined |
| 18-101_k141_167371 | Not-determined |
| 18-101_k141_365640 | Not-determined |
| 18-101_k141_201004 | Not-determined |
| 18-101_k141_250631 | Not-determined |
| 18-101_k141_52533 | Not-determined |
| 18-101_k141_300107 | Not-determined |
| 18-101_k141_300108 | Not-determined |
| 18-101_k141_250633 | Not-determined |
| 18-101_k141_85595 | Not-determined |
| 18-101_k141_52538 | Not-determined |
| 18-101_k141_118841 | Low-quality |
| 18-101_k141_381787 | Not-determined |
| 18-101_k141_3628 | Not-determined |
| 18-101_k141_283403 | Not-determined |
| 18-101_k141_85598 | Low-quality |
| 18-101_k141_233876 | Not-determined |
| 18-101_k141_233878 | Not-determined |
| 18-101_k141_250636 | Not-determined |
| 18-101_k141_85605 | Not-determined |
| 18-101_k141_167388 | Not-determined |
| 18-101_k141_85606 | Not-determined |
| 18-101_k141_19934 | Not-determined |
| 18-101_k141_19936 | Not-determined |
| 18-101_k141_19937 | Not-determined |
| 18-101_k141_151656 | Not-determined |
| 18-101_k141_201009 | Not-determined |
| 18-101_k141_201010 | Not-determined |
| 18-101_k141_19944 | Not-determined |
| 18-101_k141_266468 | Not-determined |
| 18-101_k141_3650 | Not-determined |
| 18-101_k141_19946 | Not-determined |
| 18-101_k141_19947 | Not-determined |
| 18-101_k141_266471 | Not-determined |
| 18-101_k141_381796 | Not-determined |
| 18-101_k141_3656 | Not-determined |
| 18-101_k141_381798 | Low-quality |
| 18-101_k141_151669 | Low-quality |
| 18-101_k141_151672 | Low-quality |
| 18-101_k141_85620 | Not-determined |
| 18-101_k141_365646 | Not-determined |
| 18-101_k141_52572 | Low-quality |
| 18-101_k141_3667 | Not-determined |
| 18-101_k141_52574 | Not-determined |
| 18-101_k141_283441 | Low-quality |
| 18-101_k141_300124 | Not-determined |
| 18-101_k141_316401 | Low-quality |
| 18-101_k141_85638 | Low-quality |
| 18-101_k141_381807 | Not-determined |
| 18-101_k141_52587 | Not-determined |
| 18-101_k141_332304 | Not-determined |
| 18-101_k141_184905 | Not-determined |
| 18-101_k141_184907 | Not-determined |
| 18-101_k141_184911 | Not-determined |
| 18-101_k141_201027 | Not-determined |
| 18-101_k141_118878 | Not-determined |
| 18-101_k141_300142 | Not-determined |
| 18-101_k141_316417 | Not-determined |
| 18-101_k141_68637 | Not-determined |
| 18-101_k141_381812 | Low-quality |
| 18-101_k141_381813 | Not-determined |
| 18-101_k141_300147 | Not-determined |
| 18-101_k141_381815 | Not-determined |
| 18-101_k141_36478 | Not-determined |
| 18-101_k141_36483 | Not-determined |
| 18-101_k141_118890 | Not-determined |
| 18-101_k141_36486 | Not-determined |
| 18-101_k141_36490 | Not-determined |
| 18-101_k141_250695 | Not-determined |
| 18-101_k141_300158 | Not-determined |
| 18-101_k141_52604 | Low-quality |
| 18-101_k141_85670 | Not-determined |
| 18-101_k141_316442 | Low-quality |
| 18-101_k141_85675 | Not-determined |
| 18-101_k141_300168 | Not-determined |
| 18-101_k141_85676 | Not-determined |
| 18-101_k141_85680 | Not-determined |
| 18-101_k141_85682 | Not-determined |
| 18-101_k141_68654 | Not-determined |
| 18-101_k141_201056 | Not-determined |
| 18-101_k141_101854 | Not-determined |
| 18-101_k141_101856 | Not-determined |
| 18-101_k141_266500 | Not-determined |
| 18-101_k141_381842 | Not-determined |
| 18-101_k141_266507 | Not-determined |
| 18-101_k141_266510 | Not-determined |
| 18-101_k141_101870 | Not-determined |
| 18-101_k141_3720 | Not-determined |
| 18-101_k141_201065 | Not-determined |
| 18-101_k141_300187 | Not-determined |
| 18-101_k141_349253 | Low-quality |
| 18-101_k141_349256 | Not-determined |
| 18-101_k141_349257 | Low-quality |
| 18-101_k141_184943 | Not-determined |
| 18-101_k141_68684 | Not-determined |
| 18-101_k141_85695 | Not-determined |
| 18-101_k141_233918 | Not-determined |
| 18-101_k141_332345 | Not-determined |
| 18-101_k141_300209 | Not-determined |
| 18-101_k141_19993 | Not-determined |
| 18-101_k141_68688 | Not-determined |
| 18-101_k141_85698 | Not-determined |
| 18-101_k141_233929 | Not-determined |
| 18-101_k141_365681 | Not-determined |
| 18-101_k141_3735 | Not-determined |
| 18-101_k141_266538 | Not-determined |
| 18-101_k141_68696 | Not-determined |
| 18-101_k141_135680 | Not-determined |
| 18-101_k141_283468 | Not-determined |
| 18-101_k141_3752 | Not-determined |
| 18-101_k141_167499 | Not-determined |
| 18-101_k141_381864 | Not-determined |
| 18-101_k141_300235 | Not-determined |
| 18-101_k141_250743 | Not-determined |
| 18-101_k141_52608 | Not-determined |
| 18-101_k141_3758 | Not-determined |
| 18-101_k141_381870 | Not-determined |
| 18-101_k141_233944 | Not-determined |
| 18-101_k141_68703 | Not-determined |
| 18-101_k141_250745 | Not-determined |
| 18-101_k141_85717 | Not-determined |
| 18-101_k141_68704 | Not-determined |
| 18-101_k141_3761 | Not-determined |
| 18-101_k141_135692 | Not-determined |
| 18-101_k141_332377 | Not-determined |
| 18-101_k141_52620 | Not-determined |
| 18-101_k141_85727 | Not-determined |
| 18-101_k141_184964 | Not-determined |
| 18-101_k141_250751 | Not-determined |
| 18-101_k141_184968 | Not-determined |
| 18-101_k141_167515 | Not-determined |
| 18-101_k141_85738 | Not-determined |
| 18-101_k141_85744 | Not-determined |
| 18-101_k141_381886 | Not-determined |
| 18-101_k141_365690 | Not-determined |
| 18-101_k141_20022 | Not-determined |
| 18-101_k141_184987 | Not-determined |
| 18-101_k141_68728 | Low-quality |
| 18-101_k141_349274 | Not-determined |
| 18-101_k141_52640 | Not-determined |
| 18-101_k141_381907 | Not-determined |
| 18-101_k141_349276 | Not-determined |
| 18-101_k141_349277 | Not-determined |
| 18-101_k141_250782 | Not-determined |
| 18-101_k141_101888 | Not-determined |
| 18-101_k141_365703 | Not-determined |
| 18-101_k141_101892 | Not-determined |
| 18-101_k141_381915 | Not-determined |
| 18-101_k141_185014 | Not-determined |
| 18-101_k141_101906 | Low-quality |
| 18-101_k141_20026 | Low-quality |
| 18-101_k141_201083 | Not-determined |
| 18-101_k141_381931 | Not-determined |
| 18-101_k141_201084 | Not-determined |
| 18-101_k141_381932 | Not-determined |
| 18-101_k141_233994 | Not-determined |
| 18-101_k141_332409 | Not-determined |
| 18-101_k141_381935 | Not-determined |
| 18-101_k141_316556 | Not-determined |
| 18-101_k141_332414 | Not-determined |
| 18-101_k141_20030 | Low-quality |
| 18-101_k141_349294 | Not-determined |
| 18-101_k141_381938 | Not-determined |
| 18-101_k141_381940 | Not-determined |
| 18-101_k141_300288 | Not-determined |
| 18-101_k141_68771 | Not-determined |
| 18-101_k141_300291 | Not-determined |
| 18-101_k141_234005 | Not-determined |
| 18-101_k141_151732 | Not-determined |
| 18-101_k141_101929 | Not-determined |
| 18-101_k141_234010 | Not-determined |
| 18-101_k141_316578 | Not-determined |
| 18-101_k141_68783 | Not-determined |
| 18-101_k141_349307 | Low-quality |
| 18-101_k141_101935 | Not-determined |
| 18-101_k141_349312 | Not-determined |
| 18-101_k141_101938 | Not-determined |
| 18-101_k141_316596 | Not-determined |
| 18-101_k141_101941 | Low-quality |
| 18-101_k141_201111 | Not-determined |
| 18-101_k141_316600 | Not-determined |
| 18-101_k141_85760 | Not-determined |
| 18-101_k141_381955 | Not-determined |
| 18-101_k141_283557 | Not-determined |
| 18-101_k141_85766 | Not-determined |
| 18-101_k141_332422 | Not-determined |
| 18-101_k141_381963 | Not-determined |
| 18-101_k141_381966 | Not-determined |
| 18-101_k141_381967 | Low-quality |
| 18-101_k141_201122 | Not-determined |
| 18-101_k141_234032 | Not-determined |
| 18-101_k141_151739 | Not-determined |
| 18-101_k141_234033 | Not-determined |
| 18-101_k141_52661 | Not-determined |
| 18-101_k141_52664 | Not-determined |
| 18-101_k141_185025 | Not-determined |
| 18-101_k141_151740 | Not-determined |
| 18-101_k141_300320 | Not-determined |
| 18-101_k141_185031 | Not-determined |
| 18-101_k141_283586 | Not-determined |
| 18-101_k141_332440 | Not-determined |
| 18-101_k141_300327 | Low-quality |
| 18-101_k141_266588 | Not-determined |
| 18-101_k141_316605 | Not-determined |
| 18-101_k141_365745 | Not-determined |
| 18-101_k141_381988 | Not-determined |
| 18-101_k141_332451 | Not-determined |
| 18-101_k141_135714 | Not-determined |
| 18-101_k141_316607 | Not-determined |
| 18-101_k141_201149 | Not-determined |
| 18-101_k141_381991 | Not-determined |
| 18-101_k141_85804 | Not-determined |
| 18-101_k141_316612 | Not-determined |
| 18-101_k141_20058 | Not-determined |
| 18-101_k141_381997 | Not-determined |
| 18-101_k141_266599 | Low-quality |
| 18-101_k141_52683 | Not-determined |
| 18-101_k141_20067 | Low-quality |
| 18-101_k141_217691 | Not-determined |
| 18-101_k141_316632 | Not-determined |
| 18-101_k141_266614 | Not-determined |
| 18-101_k141_332482 | Not-determined |
| 18-101_k141_349327 | Not-determined |
| 18-101_k141_135730 | Not-determined |
| 18-101_k141_201163 | Not-determined |
| 18-101_k141_135733 | Not-determined |
| 18-101_k141_266619 | Not-determined |
| 18-101_k141_135736 | Not-determined |
| 18-101_k141_283612 | Not-determined |
| 18-101_k141_250812 | Not-determined |
| 18-101_k141_283620 | Not-determined |
| 18-101-final-viral-combined_k141_68998\|\|full | High-quality |
| 18-101_k141_266628 | Low-quality |
| 18-101_k141_266630 | Not-determined |
| 18-101_k141_20086 | Not-determined |
| 18-101_k141_266634 | Not-determined |
| 18-101_k141_52702 | Low-quality |
| 18-101_k141_20091 | Not-determined |
| 18-101_k141_135767 | Not-determined |
| 18-101_k141_135768 | Not-determined |
| 18-101_k141_167530 | Not-determined |
| 18-101_k141_316670 | Not-determined |
| 18-101_k141_201199 | Not-determined |
| 18-101_k141_266656 | Not-determined |
| 18-101_k141_300386 | Not-determined |

| | |
|---|---|
| 18-101_k141_266658 | Not-determined |
| 18-101_k141_300389 | Low-quality |
| 18-101_k141_349361 | Not-determined |
| 18-101_k141_250834 | Low-quality |
| 18-101_k141_135797 | Not-determined |
| 18-101_k141_349369 | Low-quality |
| 18-101_k141_167543 | Low-quality |
| 18-101_k141_300402 | Low-quality |
| 18-101_k141_36499 | Not-determined |
| 18-101_k141_300411 | Not-determined |
| 18-101_k141_167552 | Not-determined |
| 18-101_k141_316709 | Not-determined |
| 18-101_k141_36512 | Not-determined |
| 18-101_k141_332502 | Not-determined |
| 18-101_k141_250859 | Low-quality |
| 18-101_k141_20122 | Not-determined |
| 18-101_k141_332505 | Not-determined |
| 18-101_k141_135808 | Not-determined |
| 18-101_k141_135810 | Not-determined |
| 18-101_k141_167558 | Not-determined |
| 18-101_k141_332514 | Not-determined |
| 18-101_k141_217704 | Not-determined |
| 18-101_k141_135821 | Not-determined |
| 18-101_k141_332519 | Not-determined |
| 18-101_k141_266683 | Not-determined |
| 18-101_k141_20138 | Not-determined |
| 18-101_k141_118979 | Not-determined |
| 18-101_k141_201208 | Low-quality |
| 18-101_k141_20146 | Not-determined |
| 18-101_k141_201215 | Not-determined |
| 18-101_k141_135837 | Not-determined |
| 18-101_k141_20150 | Not-determined |
| 18-101_k141_135838 | Not-determined |
| 18-101_k141_185075 | Not-determined |
| 18-101_k141_332541 | Not-determined |
| 18-101_k141_135850 | Not-determined |
| 18-101_k141_52715 | Not-determined |
| 18-101_k141_118987 | Not-determined |
| 18-101_k141_52721 | Not-determined |
| 18-101_k141_316728 | Not-determined |
| 18-101_k141_201229 | Low-quality |
| 18-101_k141_316732 | Not-determined |
| 18-101_k141_316734 | Not-determined |
| 18-101_k141_52727 | Not-determined |
| 18-101_k141_135854 | Not-determined |
| 18-101_k141_234085 | Not-determined |
| 18-101_k141_52731 | Not-determined |
| 18-101_k141_382038 | Not-determined |
| 18-101_k141_52733 | Not-determined |
| 18-101_k141_234088 | Not-determined |
| 18-101_k141_52734 | Not-determined |
| 18-101_k141_151813 | Low-quality |
| 18-101_k141_266693 | Not-determined |
| 18-101_k141_119013 | Not-determined |
| 18-101_k141_349415 | Not-determined |
| 18-101_k141_382045 | Not-determined |
| 18-101_k141_332567 | Not-determined |
| 18-101_k141_349417 | Not-determined |
| 18-101_k141_201251 | Not-determined |
| 18-101_k141_201255 | Not-determined |
| 18-101_k141_167595 | Not-determined |
| 18-101_k141_234106 | Not-determined |
| 18-101_k141_332579 | Not-determined |
| 18-101_k141_266703 | Not-determined |
| 18-101_k141_167603 | Not-determined |
| 18-101_k141_52751 | Not-determined |
| 18-101_k141_266708 | Not-determined |
| 18-101_k141_185095 | Not-determined |
| 18-101_k141_201272 | Not-determined |
| 18-101_k141_349424 | Not-determined |
| 18-101_k141_151840 | Low-quality |
| 18-101_k141_234126 | Not-determined |
| 18-101_k141_332591 | Not-determined |
| 18-101_k141_119026 | Not-determined |
| 18-101_k141_201293 | Not-determined |
| 18-101_k141_167611 | Not-determined |
| 18-101_k141_151847 | Not-determined |
| 18-101_k141_167612 | Not-determined |
| 18-101_k141_349434 | Not-determined |
| 18-101_k141_332603 | Not-determined |
| 18-101_k141_300426 | Not-determined |
| 18-101_k141_20184 | Not-determined |
| 18-101_k141_316758 | Not-determined |
| 18-101_k141_300439 | Low-quality |
| 18-101_k141_119047 | Not-determined |
| 18-101_k141_52789 | Not-determined |
| 18-101_k141_151865 | Not-determined |
| 18-101_k141_300450 | Not-determined |
| 18-101_k141_250942 | Not-determined |
| 18-101_k141_234134 | Not-determined |
| 18-101_k141_300454 | Low-quality |
| 18-101_k141_332624 | Not-determined |
| 18-101_k141_217741 | Not-determined |
| 18-101_k141_316769 | Not-determined |
| 18-101_k141_316770 | Not-determined |
| 18-101_k141_119064 | Not-determined |
| 18-101_k141_201316 | Not-determined |
| 18-101_k141_316780 | Not-determined |
| 18-101_k141_316782 | Not-determined |
| 18-101_k141_119068 | Not-determined |
| 18-101_k141_250951 | Not-determined |
| 18-101_k141_316785 | Not-determined |
| 18-101_k141_316786 | Not-determined |
| 18-101_k141_283661 | Not-determined |
| 18-101_k141_316789 | Not-determined |
| 18-101_k141_250954 | Not-determined |
| 18-101_k141_185136 | Not-determined |
| 18-101_k141_234144 | Not-determined |
| 18-101_k141_185140 | Not-determined |
| 18-101_k141_300468 | Not-determined |
| 18-101_k141_201323 | Not-determined |
| 18-101_k141_316796 | Not-determined |
| 18-101_k141_185145 | Not-determined |
| 18-101_k141_52823 | Not-determined |
| 18-101_k141_382111 | Not-determined |
| 18-101_k141_316800 | Not-determined |
| 18-101_k141_201333 | Low-quality |
| 18-101_k141_201334 | Not-determined |
| 18-101_k141_36532 | Not-determined |
| 18-101_k141_201337 | Not-determined |
| 18-101_k141_201339 | Not-determined |
| 18-101_k141_382115 | Not-determined |
| 18-101_k141_382116 | Not-determined |
| 18-101_k141_151895 | Not-determined |
| 18-101_k141_349473 | Not-determined |
| 18-101_k141_151897 | Not-determined |
| 18-101_k141_85842 | Not-determined |
| 18-101_k141_234158 | Not-determined |
| 18-101_k141_119086 | Not-determined |
| 18-101_k141_300481 | Not-determined |
| 18-101_k141_3790 | Not-determined |
| 18-101_k141_250981 | Not-determined |
| 18-101_k141_250983 | Not-determined |
| 18-101_k141_36548 | Not-determined |
| 18-101_k141_151909 | Not-determined |
| 18-101_k141_85855 | Not-determined |
| 18-101_k141_52837 | Not-determined |
| 18-101_k141_332649 | Not-determined |
| 18-101_k141_332651 | Not-determined |
| 18-101_k141_102042 | Not-determined |
| 18-101_k141_102045 | Not-determined |
| 18-101_k141_119107 | Not-determined |
| 18-101_k141_349492 | Not-determined |
| 18-101_k141_316833 | Not-determined |
| 18-101_k141_36554 | Not-determined |
| 18-101_k141_52845 | Not-determined |
| 18-101_k141_316834 | Not-determined |
| 18-101_k141_251004 | Not-determined |
| 18-101_k141_266779 | Not-determined |
| 18-101_k141_266780 | Not-determined |
| 18-101_k141_382134 | Not-determined |
| 18-101_k141_300485 | Not-determined |
| 18-101_k141_266787 | Not-determined |
| 18-101_k141_85866 | Not-determined |
| 18-101_k141_85867 | Not-determined |
| 18-101_k141_266789 | Not-determined |
| 18-101_k141_382141 | Not-determined |
| 18-101_k141_85886 | Not-determined |
| 18-101_k141_36560 | Not-determined |
| 18-101_k141_201365 | Not-determined |
| 18-101_k141_135911 | Not-determined |
| 18-101_k141_316840 | Not-determined |
| 18-101_k141_85890 | Not-determined |
| 18-101_k141_300504 | Not-determined |
| 18-101_k141_85898 | Not-determined |
| 18-101_k141_151916 | Not-determined |
| 18-101_k141_382153 | Not-determined |
| 18-101_k141_102068 | Not-determined |
| 18-101_k141_349503 | Not-determined |
| 18-101_k141_201384 | Not-determined |
| 18-101_k141_300511 | Not-determined |
| 18-101_k141_119120 | Not-determined |
| 18-101_k141_167623 | Not-determined |
| 18-101_k141_85907 | Low-quality |
| 18-101_k141_167624 | Not-determined |
| 18-101_k141_102077 | Not-determined |
| 18-101_k141_151927 | Not-determined |
| 18-101_k141_251031 | Not-determined |
| 18-101_k141_167628 | Not-determined |
| 18-101_k141_283705 | Not-determined |
| 18-101_k141_201396 | Not-determined |
| 18-101_k141_119135 | Low-quality |
| 18-101_k141_185212 | Not-determined |
| 18-101_k141_266809 | Not-determined |
| 18-101_k141_185214 | Not-determined |
| 18-101_k141_300518 | Not-determined |
| 18-101_k141_135936 | Not-determined |
| 18-101_k141_135939 | Not-determined |
| 18-101_k141_85911 | Low-quality |
| 18-101_k141_135942 | Not-determined |
| 18-101_k141_52884 | Not-determined |
| 18-101_k141_151955 | Not-determined |
| 18-101_k141_52889 | Not-determined |
| 18-101_k141_316858 | Not-determined |
| 18-101_k141_151962 | Not-determined |
| 18-101_k141_167644 | Not-determined |
| 18-101_k141_85919 | Not-determined |
| 18-101_k141_332685 | Low-quality |
| 18-101_k141_20214 | Not-determined |
| 18-101_k141_266816 | Not-determined |
| 18-101_k141_20217 | Not-determined |
| 18-101_k141_3805 | Not-determined |
| 18-101_k141_283711 | Not-determined |
| 18-101_k141_201434 | Not-determined |
| 18-101_k141_201435 | Not-determined |
| 18-101_k141_119138 | Low-quality |
| 18-101_k141_119139 | Not-determined |
| 18-101_k141_382175 | Not-determined |
| 18-101_k141_119142 | Not-determined |
| 18-101_k141_300546 | Not-determined |
| 18-101_k141_3811 | Low-quality |
| 18-101_k141_52900 | Not-determined |
| 18-101_k141_20244 | Not-determined |
| 18-101_k141_251057 | Not-determined |
| 18-101_k141_316890 | Low-quality |
| 18-101_k141_217772 | Not-determined |
| 18-101_k141_151984 | Low-quality |
| 18-101_k141_20255 | Not-determined |
| 18-101_k141_332706 | Not-determined |
| 18-101_k141_20256 | Not-determined |
| 18-101_k141_3825 | Not-determined |
| 18-101_k141_20259 | Not-determined |
| 18-101_k141_217774 | Not-determined |
| 18-101_k141_332716 | Not-determined |
| 18-101_k141_316902 | Not-determined |
| 18-101_k141_316903 | Not-determined |
| 18-101_k141_20268 | Not-determined |
| 18-101_k141_85943 | Not-determined |
| 18-101_k141_52922 | Not-determined |
| 18-101_k141_52923 | Not-determined |
| 18-101_k141_382190 | Not-determined |
| 18-101_k141_135974 | Not-determined |
| 18-101_k141_316910 | Not-determined |
| 18-101_k141_167670 | Not-determined |
| 18-101_k141_332733 | Not-determined |
| 18-101_k141_151988 | Not-determined |
| 18-101_k141_119173 | Not-determined |
| 18-101_k141_266862 | Not-determined |
| 18-101_k141_151995 | Not-determined |
| 18-101_k141_201447 | Not-determined |
| 18-101_k141_151997 | Not-determined |
| 18-101_k141_119184 | Not-determined |
| 18-101_k141_102101 | Not-determined |
| 18-101_k141_300559 | Not-determined |
| 18-101_k141_382202 | Not-determined |
| 18-101_k141_119187 | Not-determined |
| 18-101_k141_300565 | Not-determined |
| 18-101_k141_234195 | Not-determined |
| 18-101_k141_283759 | Low-quality |
| 18-101_k141_382206 | Not-determined |
| 18-101_k141_167674 | Not-determined |
| 18-101_k141_217814 | Not-determined |
| 18-101_k141_20293 | Low-quality |
| 18-101_k141_68837 | Not-determined |
| 18-101_k141_135995 | Not-determined |
| 18-101_k141_20294 | Low-quality |
| 18-101_k141_283767 | Not-determined |
| 18-101_k141_135997 | Not-determined |
| 18-101_k141_135999 | Not-determined |
| 18-101_k141_266880 | Not-determined |
| 18-101_k141_349523 | Not-determined |
| 18-101_k141_201471 | Not-determined |
| 18-101_k141_217819 | Not-determined |
| 18-101_k141_152013 | Not-determined |
| 18-101_k141_152014 | Not-determined |
| 18-101_k141_332754 | Not-determined |
| 18-101_k141_332755 | Not-determined |
| 18-101_k141_185265 | Not-determined |
| 18-101_k141_136011 | Not-determined |
| 18-101_k141_234210 | Not-determined |
| 18-101_k141_201473 | Not-determined |
| 18-101_k141_365768 | Not-determined |
| 18-101_k141_349527 | Not-determined |
| 18-101_k141_201478 | Not-determined |
| 18-101_k141_217827 | Not-determined |
| 18-101_k141_185271 | Not-determined |
| 18-101_k141_85990 | Not-determined |
| 18-101_k141_52929 | Not-determined |
| 18-101_k141_217831 | Not-determined |
| 18-101_k141_332764 | Not-determined |
| 18-101_k141_234224 | Not-determined |
| 18-101_k141_234225 | Not-determined |
| 18-101_k141_251119 | Not-determined |
| 18-101_k141_3868 | Not-determined |
| 18-101_k141_102124 | Not-determined |
| 18-101_k141_3871 | Not-determined |
| 18-101_k141_52943 | Not-determined |
| 18-101_k141_102127 | Not-determined |
| 18-101_k141_102129 | Not-determined |
| 18-101_k141_102132 | Not-determined |
| 18-101_k141_85998 | Not-determined |
| 18-101_k141_102138 | Not-determined |
| 18-101_k141_119216 | Not-determined |
| 18-101_k141_3879 | Not-determined |
| 18-101_k141_349540 | Not-determined |
| 18-101_k141_152030 | Not-determined |
| 18-101_k141_152032 | Low-quality |
| 18-101_k141_266890 | Not-determined |
| 18-101_k141_86006 | Not-determined |
| 18-101_k141_86007 | Not-determined |
| 18-101_k141_68868 | Low-quality |
| 18-101_k141_349544 | Not-determined |
| 18-101_k141_102154 | Not-determined |
| 18-101_k141_152036 | Not-determined |
| 18-101_k141_68871 | Not-determined |
| 18-101_k141_365799 | Not-determined |
| 18-101_k141_3884 | Not-determined |
| 18-101_k141_68877 | Not-determined |
| 18-101_k141_167688 | Not-determined |
| 18-101_k141_234250 | Low-quality |
| 18-101_k141_167689 | Not-determined |
| 18-101_k141_316974 | Low-quality |
| 18-101_k141_102157 | Not-determined |
| 18-101_k141_167691 | Not-determined |
| 18-101_k141_283799 | Not-determined |
| 18-101_k141_234256 | Low-quality |
| 18-101_k141_36571 | Not-determined |
| 18-101_k141_300612 | Not-determined |
| 18-101_k141_152052 | Not-determined |
| 18-101_k141_217845 | Not-determined |
| 18-101_k141_217846 | Not-determined |
| 18-101_k141_300618 | Not-determined |
| 18-101_k141_68887 | Not-determined |
| 18-101_k141_217850 | Not-determined |
| 18-101_k141_68890 | Not-determined |
| 18-101_k141_234263 | Not-determined |
| 18-101_k141_36582 | Not-determined |
| 18-101_k141_283811 | Not-determined |
| 18-101_k141_283812 | Not-determined |
| 18-101_k141_52977 | Low-quality |
| 18-101_k141_52978 | Not-determined |
| 18-101_k141_300628 | Not-determined |
| 18-101_k141_185294 | Not-determined |
| 18-101_k141_300632 | Not-determined |
| 18-101_k141_365806 | Not-determined |
| 18-101_k141_234271 | Not-determined |
| 18-101_k141_266928 | Not-determined |
| 18-101_k141_349581 | Not-determined |
| 18-101_k141_201486 | Not-determined |
| 18-101_k141_300638 | Not-determined |
| 18-101_k141_251166 | Low-quality |
| 18-101_k141_185304 | Not-determined |
| 18-101_k141_185305 | Low-quality |
| 18-101_k141_36602 | Not-determined |
| 18-101_k141_36604 | Not-determined |
| 18-101_k141_52990 | Not-determined |
| 18-101_k141_52994 | Not-determined |
| 18-101_k141_266931 | Low-quality |
| 18-101_k141_136058 | Not-determined |
| 18-101_k141_332781 | Not-determined |
| 18-101_k141_349596 | Not-determined |
| 18-101_k141_36613 | Not-determined |
| 18-101_k141_217863 | Low-quality |
| 18-101_k141_234307 | Not-determined |
| 18-101_k141_234308 | Not-determined |
| 18-101_k141_266941 | Not-determined |
| 18-101_k141_234309 | Not-determined |
| 18-101_k141_185323 | Not-determined |
| 18-101_k141_266943 | Not-determined |
| 18-101_k141_332791 | Not-determined |
| 18-101_k141_349600 | Not-determined |
| 18-101_k141_53006 | Not-determined |
| 18-101_k141_185325 | Not-determined |
| 18-101_k141_136068 | Not-determined |
| 18-101_k141_36621 | Not-determined |
| 18-101_k141_349613 | Not-determined |
| 18-101_k141_266950 | Low-quality |
| 18-101_k141_217876 | Not-determined |
| 18-101_k141_365830 | Low-quality |
| 18-101_k141_185334 | Not-determined |
| 18-101_k141_53011 | Low-quality |
| 18-101_k141_217885 | Not-determined |
| 18-101_k141_251188 | Not-determined |
| 18-101_k141_251190 | Not-determined |
| 18-101_k141_234319 | Low-quality |
| 18-101_k141_36628 | Not-determined |
| 18-101_k141_234321 | Not-determined |
| 18-101_k141_68908 | Low-quality |
| 18-101_k141_36632 | Not-determined |
| 18-101_k141_119252 | Not-determined |
| 18-101_k141_68913 | Not-determined |
| 18-101_k141_382215 | Not-determined |
| 18-101_k141_234327 | Not-determined |
| 18-101_k141_201522 | Low-quality |
| 18-101_k141_185353 | Not-determined |
| 18-101_k141_185356 | Not-determined |
| 18-101_k141_251196 | Not-determined |
| 18-101_k141_102202 | Not-determined |
| 18-101_k141_102204 | Low-quality |
| 18-101_k141_251202 | Not-determined |
| 18-101_k141_68926 | Not-determined |
| 18-101_k141_68927 | Not-determined |
| 18-101_k141_185374 | Not-determined |
| 18-101_k141_382245 | Not-determined |
| 18-101_k141_102214 | Not-determined |
| 18-101_k141_332822 | Not-determined |
| 18-101_k141_349627 | Not-determined |
| 18-101_k141_152073 | Not-determined |
| 18-101_k141_102218 | Not-determined |
| 18-101_k141_36650 | Not-determined |
| 18-101_k141_349631 | Not-determined |
| 18-101_k141_167733 | Not-determined |
| 18-101_k141_119261 | Not-determined |
| 18-101_k141_20409 | Low-quality |
| 18-101_k141_20410 | Not-determined |
| 18-101_k141_167736 | Not-determined |
| 18-101_k141_251217 | Not-determined |
| 18-101_k141_201542 | Not-determined |
| 18-101_k141_201543 | Not-determined |
| 18-101_k141_3936 | Not-determined |
| 18-101_k141_201544 | Not-determined |
| 18-101_k141_185395 | Not-determined |
| 18-101_k141_300690 | Not-determined |
| 18-101_k141_201552 | Not-determined |
| 18-101_k141_119272 | Not-determined |
| 18-101_k141_36670 | Not-determined |
| 18-101_k141_266972 | Not-determined |
| 18-101_k141_234358 | Not-determined |
| 18-101_k141_201562 | Not-determined |
| 18-101_k141_68945 | Not-determined |
| 18-101_k141_152101 | Not-determined |
| 18-101_k141_68946 | Not-determined |
| 18-101_k141_201570 | Not-determined |
| 18-101_k141_266977 | Not-determined |
| 18-101_k141_201572 | Low-quality |
| 18-101_k141_382255 | Not-determined |
| 18-101_k141_234378 | Not-determined |
| 18-101_k141_119293 | Not-determined |
| 18-101_k141_266986 | Not-determined |
| 18-101_k141_217915 | Not-determined |
| 18-101_k141_251246 | Not-determined |
| 18-101_k141_68949 | Not-determined |
| 18-101_k141_68950 | Not-determined |
| 18-101_k141_152117 | Not-determined |
| 18-101_k141_68951 | Not-determined |
| 18-101_k141_68952 | Not-determined |
| 18-101_k141_68954 | Not-determined |
| 18-101_k141_119301 | Not-determined |
| 18-101_k141_86082 | Not-determined |
| 18-101_k141_3964 | Not-determined |
| 18-101_k141_317044 | Low-quality |
| 18-101_k141_102261 | Not-determined |
| 18-101_k141_251264 | Not-determined |
| 18-101_k141_217935 | Not-determined |
| 18-101_k141_251268 | Not-determined |
| 18-101_k141_36692 | Not-determined |
| 18-101_k141_36696 | Not-determined |
| 18-101_k141_349657 | Not-determined |
| 18-101_k141_3979 | Not-determined |
| 18-101_k141_382280 | Not-determined |
| 18-101_k141_20424 | Not-determined |
| 18-101_k141_68980 | Not-determined |
| 18-101_k141_382283 | Low-quality |
| 18-101_k141_349668 | Not-determined |
| 18-101_k141_36703 | Not-determined |
| 18-101_k141_36706 | Not-determined |
| 18-101_k141_365867 | Not-determined |
| 18-101_k141_382285 | Not-determined |
| 18-101_k141_217940 | Low-quality |
| 18-101_k141_365871 | Low-quality |
| 18-101_k141_167749 | Low-quality |
| 18-101_k141_382288 | Not-determined |
| 18-101_k141_119323 | Not-determined |
| 18-101_k141_317070 | Low-quality |
| 18-101_k141_234399 | Low-quality |
| 18-101_k141_201588 | Not-determined |
| 18-101_k141_317073 | Not-determined |
| 18-101_k141_234403 | Not-determined |
| 18-101_k141_317074 | Not-determined |
| 18-101_k141_317075 | Not-determined |
| 18-101_k141_201591 | Not-determined |
| 18-101_k141_201593 | Not-determined |
| 18-101_k141_332834 | Not-determined |
| 18-101_k141_20434 | Low-quality |
| 18-101_k141_234412 | Not-determined |
| 18-101_k141_20443 | Not-determined |
| 18-101_k141_267056 | Not-determined |
| 18-101_k141_36716 | Not-determined |
| 18-101_k141_119330 | Not-determined |
| 18-101_k141_317088 | Not-determined |
| 18-101_k141_167762 | Not-determined |
| 18-101_k141_317090 | Not-determined |
| 18-101_k141_201597 | Low-quality |
| 18-101_k141_283852 | Not-determined |
| 18-101_k141_365879 | Low-quality |
| 18-101_k141_53059 | Low-quality |
| 18-101_k141_201600 | Low-quality |
| 18-101_k141_167766 | Not-determined |
| 18-101_k141_201603 | Not-determined |
| 18-101_k141_365886 | Not-determined |
| 18-101_k141_365888 | Not-determined |
| 18-101_k141_201606 | Not-determined |
| 18-101_k141_283861 | Not-determined |
| 18-101_k141_317102 | Not-determined |
| 18-101_k141_317103 | Not-determined |
| 18-101_k141_53081 | Not-determined |
| 18-101_k141_119343 | Not-determined |
| 18-101_k141_349725 | Not-determined |
| 18-101_k141_119348 | Not-determined |
| 18-101_k141_251302 | Not-determined |
| 18-101_k141_365901 | Low-quality |
| 18-101_k141_332860 | Not-determined |
| 18-101_k141_349728 | Not-determined |
| 18-101_k141_332862 | Not-determined |
| 18-101_k141_20467 | Not-determined |
| 18-101_k141_267071 | Not-determined |
| 18-101_k141_167787 | Not-determined |
| 18-101_k141_283882 | Not-determined |
| 18-101_k141_267076 | Not-determined |
| 18-101_k141_283884 | Not-determined |
| 18-101_k141_234434 | Not-determined |
| 18-101_k141_267079 | Low-quality |
| 18-101_k141_185475 | Not-determined |
| 18-101_k141_365913 | Not-determined |
| 18-101_k141_317122 | Not-determined |
| 18-101_k141_382301 | Not-determined |
| 18-101_k141_251311 | Not-determined |
| 18-101_k141_365919 | Not-determined |
| 18-101_k141_234452 | Not-determined |
| 18-101_k141_349752 | Not-determined |
| 18-101_k141_201637 | Not-determined |
| 18-101_k141_4027 | Not-determined |
| 18-101_k141_382305 | Not-determined |
| 18-101_k141_382306 | Not-determined |
| 18-101_k141_234457 | Not-determined |
| 18-101_k141_267099 | Not-determined |
| 18-101_k141_251321 | Low-quality |
| 18-101_k141_185489 | Not-determined |
| 18-101_k141_332888 | Not-determined |
| 18-101_k141_201641 | Not-determined |
| 18-101_k141_201642 | Low-quality |
| 18-101_k141_251330 | Not-determined |
| 18-101_k141_283900 | Not-determined |
| 18-101_k141_317129 | Not-determined |
| 18-101_k141_234465 | Not-determined |
| 18-101_k141_185502 | Not-determined |
| 18-101_k141_283904 | Not-determined |
| 18-101_k141_201649 | Low-quality |
| 18-101_k141_119398 | Not-determined |
| 18-101_k141_251343 | Not-determined |
| 18-101_k141_234471 | Not-determined |
| 18-101_k141_69012 | Not-determined |
| 18-101_k141_185507 | Not-determined |
| 18-101_k141_349763 | Low-quality |
| 18-101_k141_218000 | Not-determined |
| 18-101_k141_283921 | Not-determined |
| 18-101_k141_86127 | Not-determined |
| 18-101_k141_69022 | Not-determined |
| 18-101_k141_365949 | Not-determined |
| 18-101_k141_317134 | Not-determined |
| 18-101_k141_69029 | Not-determined |
| 18-101_k141_283925 | Not-determined |
| 18-101_k141_267124 | Low-quality |
| 18-101_k141_300711 | Low-quality |
| 18-101_k141_300716 | Not-determined |
| 18-101_k141_69040 | Not-determined |
| 18-101_k141_86142 | Not-determined |
| 18-101_k141_300718 | Not-determined |
| 18-101_k141_36755 | Not-determined |
| 18-101_k141_267138 | Not-determined |
| 18-101_k141_36759 | Not-determined |
| 18-101_k141_20490 | Not-determined |
| 18-101_k141_167835 | Not-determined |
| 18-101_k141_201687 | Low-quality |
| 18-101_k141_119431 | Not-determined |
| 18-101_k141_218025 | Not-determined |
| 18-101_k141_167841 | Not-determined |
| 18-101_k141_218035 | Not-determined |
| 18-101_k141_167844 | Not-determined |
| 18-101_k141_218040 | Not-determined |
| 18-101_k141_349800 | Low-quality |
| 18-101_k141_69059 | Low-quality |
| 18-101_k141_234500 | Not-determined |
| 18-101_k141_36773 | Not-determined |
| 18-101_k141_36774 | Not-determined |
| 18-101_k141_234511 | Not-determined |
| 18-101_k141_185535 | Not-determined |
| 18-101_k141_234513 | Not-determined |
| 18-101_k141_20504 | Not-determined |
| 18-101_k141_36778 | Not-determined |
| 18-101_k141_185541 | Not-determined |
| 18-101_k141_349817 | Low-quality |
| 18-101_k141_300724 | Not-determined |
| 18-101_k141_365974 | Not-determined |
| 18-101_k141_36792 | Not-determined |
| 18-101_k141_317149 | Not-determined |
| 18-101_k141_267162 | Not-determined |
| 18-101_k141_185548 | Not-determined |
| 18-101_k141_20516 | Not-determined |

| | |
|---|---|
| 18-101_k141_382325 | Not-determined |
| 18-101_k141_382330 | Not-determined |
| 18-101_k141_349845 | Not-determined |
| 18-101_k141_349846 | Not-determined |
| 18-101_k141_317168 | Not-determined |
| 18-101_k141_218064 | Not-determined |
| 18-101_k141_20535 | Not-determined |
| 18-101_k141_53096 | Not-determined |
| 18-101_k141_20539 | Not-determined |
| 18-101_k141_20541 | Low-quality |
| 18-101_k141_234551 | Not-determined |
| 18-101_k141_234552 | Not-determined |
| 18-101_k141_218074 | Not-determined |
| 18-101_k141_300764 | Not-determined |
| 18-101_k141_119483 | Not-determined |
| 18-101_k141_234558 | Not-determined |
| 18-101_k141_267188 | Not-determined |
| 18-101_k141_53105 | Not-determined |
| 18-101_k141_234559 | Not-determined |
| 18-101_k141_267191 | Not-determined |
| 18-101_k141_251406 | Not-determined |
| 18-101_k141_102320 | Low-quality |
| 18-101_k141_201722 | Not-determined |
| 18-101_k141_53123 | Not-determined |
| 18-101_k141_251416 | Not-determined |
| 18-101_k141_201727 | Not-determined |
| 18-101_k141_251418 | Not-determined |
| 18-101_k141_332917 | Not-determined |
| 18-101_k141_382351 | Not-determined |
| 18-101_k141_267201 | Not-determined |
| 18-109-final-viral-combined_k141_235537\|\|full | High-quality |
| 18-101_k141_234568 | Low-quality |
| 18-101_k141_332919 | Not-determined |
| 18-101_k141_366043 | Not-determined |
| 18-101_k141_332922 | Not-determined |
| 18-101_k141_332923 | Not-determined |
| 18-101_k141_20574 | Not-determined |
| 18-101_k141_185553 | Not-determined |
| 18-101_k141_382364 | Not-determined |
| 18-101_k141_69119 | Not-determined |
| 18-101_k141_332936 | Low-quality |
| 18-101_k141_267216 | Not-determined |
| 18-101_k141_185562 | Not-determined |
| 18-101_k141_251439 | Not-determined |
| 18-101_k141_102338 | Not-determined |
| 18-101_k141_185582 | Not-determined |
| 18-101_k141_152143 | Not-determined |
| 18-101_k141_349894 | Not-determined |
| 18-101_k141_349896 | Not-determined |
| 18-101_k141_152152 | Not-determined |
| 18-101_k141_152153 | Not-determined |
| 18-101_k141_332951 | Not-determined |
| 18-101_k141_332952 | Not-determined |
| 18-101_k141_152166 | Low-quality |
| 18-101_k141_152167 | Not-determined |
| 18-101_k141_36834 | Not-determined |
| 18-101_k141_349910 | Not-determined |
| 18-101_k141_152170 | Not-determined |
| 18-101_k141_201762 | Not-determined |
| 18-101_k141_332959 | Not-determined |
| 18-101_k141_332960 | Not-determined |
| 18-101_k141_36843 | Not-determined |
| 18-101_k141_136110 | Not-determined |
| 18-101_k141_136111 | Not-determined |
| 18-101_k141_152171 | Low-quality |
| 18-101_k141_152173 | Not-determined |
| 18-101_k141_136113 | Low-quality |
| 18-101_k141_332967 | Not-determined |
| 18-101_k141_152179 | Not-determined |
| 18-101_k141_267220 | Low-quality |
| 18-101_k141_300845 | Not-determined |
| 18-101_k141_300846 | Low-quality |
| 18-101_k141_136124 | Not-determined |
| 18-101_k141_349935 | Not-determined |
| 18-101_k141_218094 | Low-quality |
| 18-101_k141_251477 | Not-determined |
| 18-101_k141_167933 | Not-determined |
| 18-101_k141_36860 | Low-quality |
| 18-101_k141_36861 | Not-determined |
| 18-101_k141_167937 | Not-determined |
| 18-101_k141_332973 | Not-determined |
| 18-101_k141_36865 | Not-determined |
| 18-101_k141_53185 | Low-quality |
| 18-101_k141_119522 | Not-determined |
| 18-101_k141_201773 | Not-determined |
| 18-101_k141_366086 | Not-determined |
| 18-101_k141_201778 | Not-determined |
| 18-101_k141_36873 | Not-determined |
| 18-101_k141_251497 | Not-determined |
| 18-101_k141_332982 | Not-determined |
| 18-101_k141_332983 | Not-determined |
| 18-101_k141_366093 | Not-determined |
| 18-101_k141_382393 | Not-determined |
| 18-101_k141_53199 | Not-determined |
| 18-101_k141_267228 | Not-determined |
| 18-101_k141_4074 | Not-determined |
| 18-101_k141_267229 | Not-determined |
| 18-101_k141_284018 | Not-determined |
| 18-101_k141_167954 | Not-determined |
| 18-101_k141_36881 | Not-determined |
| 18-101_k141_36883 | Not-determined |
| 18-101_k141_20611 | Not-determined |
| 18-101_k141_332987 | Not-determined |
| 18-101_k141_53201 | Not-determined |
| 18-101_k141_332992 | Not-determined |
| 18-101_k141_382404 | Not-determined |
| 18-101_k141_349955 | Not-determined |
| 18-101_k141_284024 | Not-determined |
| 18-101_k141_53205 | Not-determined |
| 18-101_k141_382409 | Not-determined |
| 18-101_k141_349956 | Not-determined |
| 18-101_k141_218112 | Not-determined |
| 18-101_k141_119545 | Not-determined |
| 18-101_k141_382424 | Not-determined |
| 18-101_k141_136150 | Not-determined |
| 18-101_k141_86183 | Not-determined |
| 18-101_k141_284038 | Not-determined |
| 18-101_k141_284039 | Not-determined |
| 18-101_k141_136155 | Not-determined |
| 18-101_k141_136156 | Not-determined |
| 18-101_k141_366103 | Not-determined |
| 18-101_k141_167983 | Not-determined |
| 18-101_k141_267270 | Not-determined |
| 18-101_k141_284052 | Not-determined |
| 18-101_k141_366107 | Not-determined |
| 18-101_k141_36904 | Not-determined |
| 18-101_k141_86203 | Not-determined |
| 18-101_k141_36905 | Low-quality |
| 18-101_k141_167993 | Not-determined |
| 18-101_k141_152225 | Not-determined |
| 18-101_k141_20623 | Not-determined |
| 18-101_k141_251544 | Not-determined |
| 18-101_k141_284059 | Not-determined |
| 18-101_k141_36911 | Not-determined |
| 18-102-final-viral-combined_k141_93845\|\|full | Medium-quality |
| 18-101_k141_102343 | Not-determined |
| 18-101_k141_218148 | Not-determined |
| 18-101_k141_267273 | Low-quality |
| 18-101_k141_349987 | Not-determined |
| 18-101_k141_168002 | Not-determined |
| 18-101_k141_267279 | Not-determined |
| 18-101_k141_53218 | Not-determined |
| 18-101_k141_168006 | Not-determined |
| 18-101_k141_251552 | Not-determined |
| 18-101_k141_152240 | Not-determined |
| 18-101_k141_317199 | Not-determined |
| 18-101_k141_267287 | Not-determined |
| 18-101_k141_267288 | Not-determined |
| 18-101_k141_36928 | Not-determined |
| 18-101_k141_267290 | Not-determined |
| 18-101_k141_366116 | Not-determined |
| 18-101_k141_218170 | Low-quality |
| 18-101_k141_350001 | Low-quality |
| 18-101_k141_36932 | Not-determined |
| 18-101_k141_300911 | Not-determined |
| 18-101_k141_218172 | Not-determined |
| 18-101_k141_300917 | Not-determined |
| 18-101_k141_300919 | Not-determined |
| 18-101_k141_333037 | Not-determined |
| 18-101_k141_333038 | Not-determined |
| 18-101_k141_267301 | Not-determined |
| 18-101_k141_333041 | Not-determined |
| 18-101_k141_86210 | Not-determined |
| 18-101_k141_86211 | Not-determined |
| 18-101_k141_168031 | Not-determined |
| 18-101_k141_350011 | Not-determined |
| 18-101_k141_366129 | Not-determined |
| 18-101_k141_102346 | Low-quality |
| 18-101_k141_366134 | Not-determined |
| 18-101_k141_152272 | Not-determined |
| 18-101_k141_300945 | Not-determined |
| 18-101_k141_317224 | Low-quality |
| 18-101_k141_350018 | Low-quality |
| 18-101_k141_300951 | Not-determined |
| 18-101_k141_102358 | Not-determined |
| 18-101_k141_136178 | Not-determined |
| 18-101_k141_152278 | Not-determined |
| 18-101_k141_201843 | Not-determined |
| 18-101_k141_350020 | Not-determined |
| 18-101_k141_251586 | Not-determined |
| 18-101_k141_119607 | Not-determined |
| 18-101_k141_102361 | Low-quality |
| 18-101_k141_300956 | Not-determined |
| 18-101_k141_119612 | Not-determined |
| 18-101_k141_234585 | Low-quality |
| 18-101_k141_152292 | Not-determined |
| 18-101_k141_234592 | Not-determined |
| 18-101_k141_185652 | Not-determined |
| 18-101_k141_168049 | Low-quality |
| 18-101_k141_267325 | Not-determined |
| 18-101_k141_234597 | Not-determined |
| 18-101_k141_102368 | Not-determined |
| 18-101_k141_267330 | Not-determined |
| 18-101_k141_234598 | Low-quality |
| 18-101_k141_300971 | Not-determined |
| 18-101_k141_201845 | Not-determined |
| 18-101_k141_136198 | Low-quality |
| 18-101_k141_366150 | Not-determined |
| 18-101_k141_102383 | Low-quality |
| 18-101_k141_36959 | Not-determined |
| 18-101_k141_350047 | Not-determined |
| 18-101_k141_350049 | Not-determined |
| 18-101_k141_251596 | Not-determined |
| 18-101_k141_366156 | Not-determined |
| 18-101_k141_366158 | Not-determined |
| 18-101_k141_350054 | Not-determined |
| 18-101_k141_317266 | Not-determined |
| 18-101_k141_267351 | Not-determined |
| 18-101_k141_301000 | Not-determined |
| 18-101_k141_152303 | Not-determined |
| 18-101_k141_201867 | Not-determined |
| 18-101_k141_201868 | Low-quality |
| 18-101_k141_201869 | Not-determined |
| 18-101_k141_136234 | Not-determined |
| 18-101_k141_251602 | Not-determined |
| 18-101_k141_201871 | Not-determined |
| 18-101_k141_301010 | Low-quality |
| 18-101_k141_185686 | Not-determined |
| 18-101_k141_301013 | Not-determined |
| 18-101_k141_201875 | Not-determined |
| 18-101_k141_136243 | Not-determined |
| 18-101_k141_301017 | Not-determined |
| 18-101_k141_136245 | Not-determined |
| 18-101_k141_366166 | Not-determined |
| 18-101_k141_152317 | Not-determined |
| 18-101_k141_102415 | Not-determined |
| 18-101_k141_333094 | Low-quality |
| 18-101_k141_152318 | Low-quality |
| 18-101_k141_333104 | Not-determined |
| 18-101_k141_136255 | Not-determined |
| 18-101_k141_267379 | Not-determined |
| 18-101_k141_366170 | Not-determined |
| 18-101_k141_267383 | Not-determined |
| 18-101_k141_168102 | Not-determined |
| 18-101_k141_333110 | Low-quality |
| 18-101_k141_119658 | Not-determined |
| 18-101_k141_20724 | Not-determined |
| 18-101_k141_267391 | Not-determined |
| 18-101_k141_350088 | Not-determined |
| 18-101_k141_168106 | Not-determined |
| 18-101_k141_350092 | Not-determined |
| 18-101_k141_136259 | Not-determined |
| 18-101_k141_382429 | Not-determined |
| 18-101_k141_102432 | Not-determined |
| 18-101_k141_86267 | Not-determined |
| 18-101_k141_251619 | Not-determined |
| 18-101_k141_301045 | Not-determined |
| 18-101_k141_382437 | Not-determined |
| 18-101_k141_119672 | Not-determined |
| 18-101_k141_267413 | Not-determined |
| 18-101_k141_317277 | Not-determined |
| 18-101_k141_350100 | Low-quality |
| 18-101_k141_366183 | Not-determined |
| 18-101_k141_350103 | Not-determined |
| 18-101_k141_317282 | Not-determined |
| 18-101_k141_382443 | Not-determined |
| 18-101_k141_136283 | Not-determined |
| 18-101_k141_53228 | Low-quality |
| 18-101_k141_20750 | Not-determined |
| 18-101_k141_366194 | Not-determined |
| 18-101_k141_333130 | Low-quality |
| 18-101_k141_382457 | Not-determined |
| 18-101_k141_366198 | Not-determined |
| 18-101_k141_333133 | Not-determined |
| 18-101_k141_317295 | Not-determined |
| 18-101_k141_301068 | Not-determined |
| 18-101_k141_301069 | Not-determined |
| 18-101_k141_382462 | Not-determined |
| 18-101_k141_36995 | Not-determined |
| 18-101_k141_350113 | Not-determined |
| 18-101_k141_234618 | Not-determined |
| 18-101_k141_36998 | Not-determined |
| 18-101_k141_366212 | Not-determined |
| 18-101_k141_201912 | Not-determined |
| 18-101_k141_185721 | Low-quality |
| 18-101_k141_366216 | Not-determined |
| 18-101_k141_382478 | Not-determined |
| 18-101_k141_185726 | Not-determined |
| 18-101_k141_185727 | Not-determined |
| 18-101_k141_301086 | Not-determined |
| 18-101_k141_20757 | Not-determined |
| 18-101_k141_136326 | Not-determined |
| 18-101_k141_317309 | Not-determined |
| 18-101_k141_53261 | Low-quality |
| 18-101_k141_317310 | Low-quality |
| 18-101_k141_333150 | Not-determined |
| 18-101_k141_53263 | Not-determined |
| 18-101_k141_53264 | Not-determined |
| 18-101_k141_136330 | Not-determined |
| 18-101_k141_20762 | Not-determined |
| 18-101_k141_53267 | Not-determined |
| 18-101_k141_301093 | Not-determined |
| 18-101_k141_234634 | Not-determined |
| 18-101_k141_333160 | Not-determined |
| 18-101_k141_53271 | Not-determined |
| 18-101_k141_350132 | Not-determined |
| 18-101_k141_234637 | Not-determined |
| 18-101_k141_185731 | Not-determined |
| 18-101_k141_102465 | Not-determined |
| 18-101_k141_333166 | Not-determined |
| 18-101_k141_251651 | Not-determined |
| 18-101_k141_350134 | Not-determined |
| 18-101_k141_366238 | Not-determined |
| 18-101_k141_350135 | Not-determined |
| 18-101_k141_37035 | Not-determined |
| 18-101_k141_267433 | Low-quality |
| 18-101_k141_382490 | Not-determined |
| 18-101_k141_382491 | Not-determined |
| 18-101_k141_382494 | Not-determined |
| 18-101_k141_201920 | Not-determined |
| 18-101_k141_301109 | Not-determined |
| 18-101_k141_185748 | Not-determined |
| 18-101_k141_102472 | Not-determined |
| 18-101_k141_251674 | Not-determined |
| 18-101_k141_317325 | Not-determined |
| 18-101_k141_86306 | Not-determined |
| 18-101_k141_86310 | Not-determined |
| 18-101_k141_37047 | Not-determined |
| 18-101_k141_267450 | Low-quality |
| 18-101_k141_37050 | Not-determined |
| 18-101_k141_333192 | Not-determined |
| 18-101_k141_234661 | Not-determined |
| 18-101_k141_267457 | Not-determined |
| 18-101_k141_102477 | Not-determined |
| 18-101_k141_317333 | Not-determined |
| 18-101_k141_37054 | Not-determined |
| 18-101_k141_366258 | Low-quality |
| 18-101_k141_37057 | Not-determined |
| 18-101_k141_119737 | Not-determined |
| 18-101_k141_185768 | Not-determined |
| 18-101_k141_136355 | Not-determined |
| 18-101_k141_152336 | Not-determined |
| 18-101_k141_69169 | Low-quality |
| 18-101_k141_317341 | Not-determined |
| 18-101_k141_152339 | Not-determined |
| 18-101_k141_152343 | Not-determined |
| 18-101_k141_37067 | Not-determined |
| 18-101_k141_53312 | Not-determined |
| 18-101_k141_152347 | Not-determined |
| 18-101_k141_152348 | Not-determined |
| 18-101_k141_102483 | Not-determined |
| 18-101_k141_185778 | Low-quality |
| 18-101_k141_185779 | Not-determined |
| 18-101_k141_136375 | Not-determined |
| 18-101_k141_284179 | Not-determined |
| 18-101_k141_152360 | Not-determined |
| 18-101_k141_20782 | Not-determined |
| 18-101_k141_382539 | Not-determined |
| 18-101_k141_152367 | Not-determined |
| 18-101_k141_317366 | Not-determined |
| 18-101_k141_317367 | Not-determined |
| 18-101_k141_284185 | Not-determined |
| 18-101_k141_37082 | Not-determined |
| 18-101_k141_69195 | Not-determined |
| 18-101_k141_185804 | Not-determined |
| 18-101_k141_251697 | Not-determined |
| 18-101_k141_69200 | Low-quality |
| 18-101_k141_102491 | Not-determined |
| 18-101_k141_251700 | Not-determined |
| 18-101_k141_119775 | Not-determined |
| 18-101_k141_382563 | Not-determined |
| 18-101_k141_234698 | Not-determined |
| 18-101_k141_201932 | Not-determined |
| 18-101_k141_168136 | Not-determined |
| 18-101_k141_317392 | Not-determined |
| 18-101_k141_53357 | Low-quality |
| 18-101_k141_69203 | Not-determined |
| 18-101_k141_350191 | Not-determined |
| 18-101_k141_4127 | Low-quality |
| 18-101_k141_4131 | Not-determined |
| 18-101_k141_333222 | Not-determined |
| 18-101_k141_136383 | Not-determined |
| 18-101_k141_284210 | Not-determined |
| 18-101_k141_251715 | Not-determined |
| 18-101_k141_136389 | Not-determined |
| 18-101_k141_382582 | Not-determined |
| 18-101_k141_185815 | Not-determined |
| 18-101_k141_136394 | Not-determined |
| 18-101_k141_20826 | Not-determined |
| 18-101_k141_37110 | Low-quality |
| 18-101_k141_37112 | Not-determined |
| 18-101_k141_284224 | Not-determined |
| 18-101_k141_284226 | Low-quality |
| 18-101_k141_301148 | Not-determined |
| 18-101_k141_284233 | Not-determined |
| 18-101_k141_382591 | Low-quality |
| 18-101_k141_152391 | Low-quality |
| 18-101_k141_317403 | Not-determined |
| 18-101_k141_333235 | Not-determined |
| 18-101_k141_317407 | Not-determined |
| 18-101_k141_37125 | Not-determined |
| 18-101_k141_119812 | Not-determined |
| 18-101_k141_317409 | Not-determined |
| 18-101_k141_317413 | Not-determined |
| 18-101_k141_301165 | Not-determined |
| 18-101_k141_119818 | Not-determined |
| 18-101_k141_69213 | Low-quality |
| 18-101_k141_119820 | Not-determined |
| 18-101_k141_136418 | Not-determined |
| 18-101_k141_350229 | Not-determined |
| 18-101_k141_136419 | Not-determined |
| 18-101_k141_119826 | Not-determined |
| 18-101_k141_350231 | Not-determined |
| 18-101_k141_37139 | Low-quality |
| 18-101_k141_37141 | Not-determined |
| 18-101_k141_317422 | Not-determined |
| 18-101_k141_317423 | Not-determined |
| 18-101_k141_102517 | Not-determined |
| 18-101_k141_382605 | Not-determined |
| 18-101_k141_201987 | Not-determined |
| 18-101_k141_333250 | Not-determined |
| 18-101_k141_136437 | Not-determined |
| 18-101_k141_119828 | Not-determined |
| 18-101_k141_185831 | Not-determined |
| 18-101_k141_168183 | Not-determined |
| 18-101_k141_86341 | Low-quality |
| 18-101_k141_4155 | Not-determined |
| 18-101_k141_102525 | Not-determined |
| 18-101_k141_201990 | Not-determined |
| 18-101_k141_317434 | Not-determined |
| 18-101_k141_4160 | Not-determined |
| 18-101_k141_136443 | Not-determined |
| 18-101_k141_119840 | Not-determined |
| 18-101_k141_382610 | Not-determined |
| 18-101_k141_119843 | Not-determined |
| 18-101_k141_53433 | Not-determined |
| 18-101_k141_69224 | Low-quality |
| 18-101_k141_102535 | Not-determined |
| 18-101_k141_102537 | Not-determined |
| 18-101_k141_102539 | Not-determined |
| 18-101_k141_152419 | Not-determined |
| 18-101_k141_301193 | Low-quality |
| 18-101_k141_251737 | Not-determined |
| 18-101_k141_136459 | Not-determined |
| 18-101_k141_102548 | Not-determined |
| 18-101_k141_53441 | Not-determined |
| 18-101_k141_267507 | Not-determined |
| 18-101_k141_202023 | Not-determined |
| 18-101_k141_4181 | Low-quality |
| 18-101_k141_317471 | Not-determined |
| 18-101_k141_382624 | Not-determined |
| 18-101_k141_136469 | Not-determined |
| 18-101_k141_234725 | Not-determined |
| 18-101_k141_267511 | Low-quality |
| 18-101_k141_37169 | Not-determined |
| 18-101_k141_234733 | Not-determined |
| 18-101_k141_267512 | Not-determined |
| 18-101_k141_366279 | Not-determined |
| 18-101_k141_20880 | Not-determined |
| 18-101_k141_37172 | Not-determined |
| 18-101_k141_86359 | Not-determined |
| 18-101_k141_251749 | Not-determined |
| 18-101_k141_317475 | Not-determined |
| 18-101_k141_37176 | Not-determined |
| 18-101_k141_202041 | Low-quality |
| 18-101_k141_168233 | Not-determined |
| 18-101_k141_37184 | Not-determined |
| 18-101_k141_102571 | Not-determined |
| 18-101_k141_382637 | Not-determined |
| 18-101_k141_350252 | Not-determined |
| 18-101_k141_317484 | Not-determined |
| 18-101_k141_69250 | Low-quality |
| 18-101_k141_284278 | Not-determined |
| 18-101_k141_317490 | Low-quality |
| 18-101_k141_366309 | Low-quality |
| 18-101_k141_53472 | Not-determined |
| 18-101_k141_136494 | Not-determined |
| 18-101_k141_382639 | Not-determined |
| 18-101_k141_53474 | Not-determined |
| 18-101_k141_267540 | Not-determined |
| 18-101_k141_119887 | Not-determined |
| 18-101_k141_69261 | Not-determined |
| 18-101_k141_284281 | Low-quality |
| 18-101_k141_267549 | Not-determined |
| 18-101_k141_136504 | Not-determined |
| 18-101_k141_333287 | Not-determined |
| 18-101_k141_69266 | Not-determined |
| 18-101_k141_267559 | Not-determined |
| 18-101_k141_267561 | Not-determined |
| 18-101_k141_234750 | Not-determined |
| 18-101_k141_284287 | Low-quality |
| 18-101_k141_267573 | Not-determined |
| 18-101_k141_218179 | Not-determined |
| 18-101_k141_284289 | Not-determined |
| 18-101_k141_20886 | Not-determined |
| 18-101_k141_168270 | Not-determined |
| 18-101_k141_202074 | Not-determined |
| 18-101_k141_301228 | Not-determined |
| 18-101_k141_202079 | Not-determined |
| 18-101_k141_301231 | Not-determined |
| 18-101_k141_284300 | Not-determined |
| 18-101_k141_53489 | Not-determined |
| 18-101_k141_382662 | Not-determined |
| 18-101_k141_301251 | Not-determined |
| 18-101_k141_86421 | Not-determined |
| 18-101_k141_284314 | Not-determined |
| 18-101_k141_53499 | Not-determined |
| 18-101_k141_218203 | Low-quality |
| 18-101_k141_202091 | Not-determined |
| 18-101_k141_69274 | Not-determined |
| 18-101_k141_382672 | Not-determined |
| 18-101_k141_218208 | Not-determined |
| 18-101_k141_301268 | Not-determined |
| 18-101_k141_218209 | Low-quality |
| 18-101_k141_168282 | Not-determined |
| 18-101_k141_218212 | Not-determined |
| 18-101_k141_333314 | Not-determined |
| 18-101_k141_301276 | Not-determined |
| 18-101_k141_4210 | Not-determined |
| 18-101_k141_317495 | Not-determined |
| 18-101_k141_333318 | Not-determined |
| 18-101_k141_333319 | Not-determined |
| 18-101_k141_53515 | Not-determined |
| 18-101_k141_267593 | Not-determined |
| 18-101_k141_152460 | Not-determined |
| 18-101_k141_185845 | Not-determined |
| 18-101_k141_102584 | Not-determined |
| 18-101_k141_317496 | Low-quality |
| 18-101_k141_4225 | Not-determined |
| 18-101_k141_267596 | Not-determined |
| 18-101_k141_317499 | Not-determined |
| 18-101_k141_53518 | Not-determined |
| 18-101_k141_4228 | Not-determined |
| 18-101_k141_317502 | Not-determined |
| 18-101_k141_168297 | Not-determined |
| 18-101_k141_152470 | Not-determined |
| 18-101_k141_301289 | Not-determined |
| 18-101_k141_185849 | Not-determined |
| 18-101_k141_4231 | Not-determined |
| 18-101_k141_168301 | Not-determined |
| 18-101_k141_152473 | Low-quality |
| 18-101_k141_69282 | Low-quality |
| 18-101_k141_185857 | Not-determined |
| 18-101_k141_350293 | Not-determined |
| 18-101_k141_69283 | Not-determined |
| 18-101_k141_333337 | Not-determined |
| 18-101_k141_350295 | Not-determined |
| 18-101_k141_366348 | Not-determined |
| 18-101_k141_382711 | Low-quality |
| 18-101_k141_333340 | Not-determined |
| 18-101_k141_4245 | Not-determined |
| 18-101_k141_218243 | Not-determined |
| 18-101_k141_350301 | Not-determined |
| 18-101_k141_53534 | Not-determined |
| 18-101_k141_218249 | Not-determined |
| 18-101_k141_53537 | Not-determined |
| 18-101_k141_168312 | Not-determined |
| 18-101_k141_136565 | Not-determined |
| 18-101_k141_102617 | Low-quality |
| 18-101_k141_301302 | Low-quality |
| 18-101_k141_333348 | Not-determined |
| 18-101_k141_218255 | Not-determined |
| 18-101_k141_4249 | Not-determined |
| 18-101_k141_350310 | Not-determined |
| 18-101_k141_136572 | Not-determined |
| 18-101_k141_333353 | Not-determined |
| 18-101_k141_350319 | Not-determined |
| 18-101_k141_350321 | Not-determined |
| 18-101_k141_53560 | Not-determined |
| 18-101_k141_350327 | Not-determined |
| 18-101_k141_152479 | Not-determined |
| 18-101_k141_86454 | Not-determined |
| 18-101_k141_333365 | Not-determined |
| 18-101_k141_152481 | Low-quality |
| 18-101_k141_119967 | Not-determined |
| 18-101_k141_86458 | Not-determined |
| 18-101_k141_102625 | Not-determined |
| 18-101_k141_185875 | Not-determined |
| 18-101_k141_218289 | Not-determined |
| 18-101_k141_333368 | Not-determined |
| 18-101_k141_136587 | Not-determined |
| 18-101_k141_317542 | Not-determined |
| 18-101_k141_4253 | Not-determined |
| 18-101_k141_4259 | Not-determined |
| 18-101_k141_301326 | Not-determined |
| 18-101_k141_168356 | Not-determined |
| 18-101_k141_317550 | Not-determined |
| 18-101_k141_366374 | Not-determined |
| 18-101_k141_37240 | Not-determined |
| 18-101_k141_284348 | Not-determined |
| 18-101_k141_284349 | Not-determined |
| 18-101_k141_152501 | Not-determined |
| 18-101_k141_333386 | Not-determined |
| 18-101_k141_152503 | Not-determined |
| 18-101_k141_4266 | Not-determined |
| 18-101_k141_37251 | Not-determined |
| 18-101_k141_284360 | Not-determined |
| 18-101_k141_37258 | Not-determined |
| 18-101_k141_333397 | Not-determined |

| | |
|---|---|
| 18-101_k141_284362 | Not-determined |
| 18-101_k141_168384 | Not-determined |
| 18-101_k141_317567 | Not-determined |
| 18-101_k141_168387 | Not-determined |
| 18-101_k141_53581 | Not-determined |
| 18-101_k141_53582 | Not-determined |
| 18-101_k141_333405 | Not-determined |
| 18-101_k141_86475 | Not-determined |
| 18-101_k141_218309 | Not-determined |
| 18-101_k141_382745 | Not-determined |
| 18-101_k141_350342 | Low-quality |
| 18-101_k141_102637 | Not-determined |
| 18-101_k141_382751 | Not-determined |
| 18-101_k141_317580 | Not-determined |
| 18-101_k141_20984 | Not-determined |
| 18-101_k141_267614 | Not-determined |
| 18-101_k141_301355 | Not-determined |
| 18-101_k141_102650 | Not-determined |
| 18-101_k141_382762 | Not-determined |
| 18-101_k141_152540 | Not-determined |
| 18-101_k141_86498 | Not-determined |
| 18-101_k141_53602 | Not-determined |
| 18-101_k141_37285 | Not-determined |
| 18-101_k141_350361 | Not-determined |
| 18-101_k141_350364 | Not-determined |
| 18-101_k141_152546 | Not-determined |
| 18-101_k141_284384 | Not-determined |
| 18-101_k141_284385 | Not-determined |
| 18-101_k141_350367 | Not-determined |
| 18-101_k141_152549 | Not-determined |
| 18-101_k141_218328 | Not-determined |
| 18-101_k141_284389 | Low-quality |
| 18-101_k141_37291 | Not-determined |
| 18-101_k141_317590 | Not-determined |
| 18-101_k141_37292 | Not-determined |
| 18-101_k141_136616 | Not-determined |
| 18-101_k141_152554 | Not-determined |
| 18-101_k141_86503 | Not-determined |
| 18-101_k141_218336 | Not-determined |
| 18-101_k141_284398 | Not-determined |
| 18-101_k141_152559 | Not-determined |
| 18-101_k141_102669 | Not-determined |
| 18-101_k141_382771 | Not-determined |
| 18-101_k141_301383 | Not-determined |
| 18-101_k141_382775 | Not-determined |
| 18-101_k141_350380 | Not-determined |
| 18-101_k141_168426 | Not-determined |
| 18-101_k141_284409 | Not-determined |
| 18-101_k141_366420 | Not-determined |
| 18-101_k141_119979 | Not-determined |
| 18-101_k141_168432 | Not-determined |
| 18-101_k141_168433 | Not-determined |
| 18-101_k141_234796 | Low-quality |
| 18-101_k141_37300 | Low-quality |
| 18-101_k141_168434 | Not-determined |
| 18-102-final-viral-combined_k141_122457\|\|full | Medium-quality |
| 18-101_k141_234798 | Not-determined |
| 18-101_k141_218348 | Not-determined |
| 18-101_k141_218351 | Not-determined |
| 18-101_k141_317611 | Not-determined |
| 18-101_k141_267631 | Not-determined |
| 18-101_k141_69331 | Not-determined |
| 18-101_k141_136628 | Not-determined |
| 18-101_k141_267635 | Not-determined |
| 18-101_k141_382792 | Not-determined |
| 18-101_k141_284414 | Not-determined |
| 18-101_k141_53644 | Not-determined |
| 18-101_k141_366427 | Not-determined |
| 18-101_k141_234823 | Not-determined |
| 18-101_k141_382801 | Not-determined |
| 18-101_k141_284427 | Not-determined |
| 18-101_k141_168449 | Not-determined |
| 18-101_k141_218377 | Not-determined |
| 18-101_k141_234830 | Not-determined |
| 18-101_k141_284432 | Not-determined |
| 18-101_k141_382807 | Not-determined |
| 18-101_k141_317632 | Not-determined |
| 18-101_k141_366438 | Not-determined |
| 18-101_k141_120003 | Not-determined |
| 18-101_k141_152580 | Not-determined |
| 18-101_k141_317633 | Low-quality |
| 18-101_k141_234836 | Not-determined |
| 18-101_k141_366444 | Not-determined |
| 18-101_k141_234839 | Not-determined |
| 18-101_k141_168455 | Not-determined |
| 18-101_k141_317638 | Not-determined |
| 18-101_k141_21040 | Not-determined |
| 18-101_k141_284452 | Not-determined |
| 18-101_k141_102701 | Not-determined |
| 18-101_k141_382823 | Not-determined |
| 18-101_k141_333447 | Not-determined |
| 18-101_k141_120017 | Not-determined |
| 18-101_k141_4305 | Low-quality |
| 18-101_k141_4306 | Not-determined |
| 18-101_k141_317663 | Not-determined |
| 18-101_k141_120027 | Low-quality |
| 18-101_k141_120032 | Not-determined |
| 18-101_k141_120034 | Not-determined |
| 18-101_k141_382836 | Not-determined |
| 18-101_k141_102709 | Not-determined |
| 18-101_k141_120042 | Not-determined |
| 18-101_k141_251802 | Low-quality |
| 18-101_k141_4318 | Not-determined |
| 18-101_k141_350405 | Not-determined |
| 18-101_k141_382847 | Not-determined |
| 18-101_k141_382852 | Not-determined |
| 18-101_k141_120048 | Not-determined |
| 18-101_k141_136675 | Not-determined |
| 18-101_k141_53670 | Not-determined |
| 18-101_k141_102719 | Not-determined |
| 18-101_k141_366480 | Not-determined |
| 18-101_k141_152606 | Not-determined |
| 18-101_k141_37344 | Not-determined |
| 18-101_k141_53673 | Not-determined |
| 18-101_k141_21064 | Not-determined |
| 18-101_k141_120057 | Not-determined |
| 18-101_k141_267653 | Not-determined |
| 18-101_k141_267654 | Not-determined |
| 18-101_k141_284479 | Not-determined |
| 18-101_k141_382865 | Not-determined |
| 18-101_k141_382866 | Not-determined |
| 18-101_k141_120062 | Not-determined |
| 18-101_k141_251827 | Not-determined |
| 18-101_k141_102729 | Not-determined |
| 18-101_k141_382875 | Not-determined |
| 18-101_k141_234869 | Not-determined |
| 18-101_k141_69368 | Not-determined |
| 18-101_k141_234874 | Not-determined |
| 18-101_k141_185953 | Not-determined |
| 18-101_k141_4336 | Not-determined |
| 18-101_k141_120084 | Not-determined |
| 18-101_k141_301427 | Not-determined |
| 18-101_k141_284506 | Not-determined |
| 18-101_k141_234880 | Not-determined |
| 18-101_k141_382897 | Not-determined |
| 18-101_k141_366513 | Not-determined |
| 18-101_k141_37362 | Not-determined |
| 18-101_k141_4361 | Low-quality |
| 18-101_k141_382901 | Not-determined |
| 18-101_k141_69382 | Not-determined |
| 18-101_k141_102759 | Not-determined |
| 18-101_k141_284522 | Not-determined |
| 18-101_k141_234894 | Not-determined |
| 18-101_k141_317690 | Not-determined |
| 18-101_k141_234896 | Not-determined |
| 18-101_k141_69391 | Not-determined |
| 18-101_k141_185976 | Low-quality |
| 18-101_k141_136740 | Not-determined |
| 18-101_k141_86549 | Not-determined |
| 18-101_k141_86554 | Not-determined |
| 18-101_k141_4373 | Not-determined |
| 18-101_k141_317697 | Not-determined |
| 18-101_k141_4376 | Not-determined |
| 18-101_k141_251876 | Not-determined |
| 18-101_k141_251882 | Not-determined |
| 18-101_k141_21122 | Not-determined |
| 18-101_k141_102774 | Not-determined |
| 18-101_k141_251887 | Low-quality |
| 18-101_k141_234909 | Not-determined |
| 18-101_k141_185990 | Not-determined |
| 18-101_k141_251889 | Not-determined |
| 18-101_k141_86561 | Not-determined |
| 18-101_k141_185992 | Low-quality |
| 18-101_k141_251893 | Not-determined |
| 18-101_k141_21143 | Low-quality |
| 18-101_k141_350466 | Not-determined |
| 18-101_k141_21144 | Not-determined |
| 18-105-final-viral-combined_k141_83370\|\|full | Medium-quality |
| 18-101_k141_136775 | Not-determined |
| 18-101_k141_152619 | Not-determined |
| 18-101_k141_120112 | Not-determined |
| 18-101_k141_136788 | Not-determined |
| 18-101_k141_152636 | Not-determined |
| 18-101_k141_53707 | Not-determined |
| 18-101_k141_152643 | Not-determined |
| 18-101_k141_350485 | Not-determined |
| 18-101_k141_267664 | Low-quality |
| 18-101_k141_234917 | Not-determined |
| 18-101_k141_21151 | Not-determined |
| 18-101_k141_234927 | Not-determined |
| 18-101_k141_21152 | Not-determined |
| 18-101_k141_234930 | Not-determined |
| 18-101_k141_234933 | Not-determined |
| 18-101_k141_382923 | Low-quality |
| 18-101_k141_152653 | Not-determined |
| 18-101_k141_267675 | Not-determined |
| 18-101_k141_218443 | Not-determined |
| 18-101_k141_202157 | Not-determined |
| 18-101_k141_202159 | Not-determined |
| 18-101_k141_37382 | Not-determined |
| 18-101_k141_202160 | Not-determined |
| 18-101_k141_350504 | Low-quality |
| 18-101_k141_37385 | Not-determined |
| 18-101_k141_382942 | Not-determined |
| 18-101_k141_202168 | Not-determined |
| 18-101_k141_301456 | Not-determined |
| 18-101_k141_251922 | Not-determined |
| 18-101_k141_382962 | Not-determined |
| 18-101_k141_267687 | Not-determined |
| 18-101_k141_69455 | Not-determined |
| 18-101_k141_120134 | Not-determined |
| 18-101_k141_234979 | Not-determined |
| 18-101_k141_301468 | Not-determined |
| 18-101_k141_218469 | Low-quality |
| 18-101_k141_218471 | Not-determined |
| 18-101_k141_53735 | Not-determined |
| 18-101_k141_366569 | Not-determined |
| 18-101_k141_251923 | Not-determined |
| 18-101_k141_218476 | Low-quality |
| 18-101_k141_4434 | Not-determined |
| 18-101_k141_234983 | Not-determined |
| 18-101_k141_382979 | Not-determined |
| 18-101_k141_53739 | Low-quality |
| 18-101_k141_284556 | Not-determined |
| 18-101_k141_350520 | Not-determined |
| 18-101_k141_69477 | Not-determined |
| 18-101_k141_21192 | Not-determined |
| 18-101_k141_53747 | Not-determined |
| 18-101_k141_382983 | Not-determined |
| 18-101_k141_301498 | Low-quality |
| 18-101_k141_317784 | Not-determined |
| 18-101_k141_350529 | Not-determined |
| 18-101_k141_37400 | Not-determined |
| 18-101_k141_102815 | Not-determined |
| 18-101_k141_120164 | Low-quality |
| 18-101_k141_366588 | Low-quality |
| 18-101_k141_53760 | Not-determined |
| 18-101_k141_152684 | Not-determined |
| 18-101_k141_53773 | Not-determined |
| 18-101_k141_136803 | Low-quality |
| 18-101_k141_350538 | Low-quality |
| 18-101_k141_284588 | Not-determined |
| 18-101_k141_251960 | Not-determined |
| 18-101_k141_317798 | Not-determined |
| 18-101_k141_235014 | Low-quality |
| 18-101_k141_301512 | Not-determined |
| 18-101_k141_69505 | Not-determined |
| 18-101_k141_152700 | Not-determined |
| 18-101_k141_69508 | Not-determined |
| 18-101_k141_4458 | Not-determined |
| 18-101_k141_136818 | Not-determined |
| 18-101_k141_4459 | Not-determined |
| 18-101_k141_37419 | Not-determined |
| 18-101_k141_69514 | Not-determined |
| 18-101_k141_37421 | Not-determined |
| 18-101_k141_218510 | Low-quality |
| 18-101_k141_218511 | Not-determined |
| 18-101_k141_366607 | Not-determined |
| 18-101_k141_53778 | Not-determined |
| 18-101_k141_186014 | Not-determined |
| 18-101_k141_120211 | Not-determined |
| 18-101_k141_21215 | Not-determined |
| 18-101_k141_202194 | Not-determined |
| 18-101_k141_186020 | Not-determined |
| 18-101_k141_37428 | Low-quality |
| 18-101_k141_4471 | Not-determined |
| 18-101_k141_168491 | Not-determined |
| 18-101_k141_235033 | Not-determined |
| 18-101_k141_120222 | Not-determined |
| 18-101_k141_301521 | Low-quality |
| 18-101_k141_69538 | Not-determined |
| 18-101_k141_69539 | Not-determined |
| 18-101_k141_37442 | Not-determined |
| 18-101_k141_21227 | Not-determined |
| 18-101_k141_53796 | Not-determined |
| 18-101_k141_284621 | Not-determined |
| 18-101_k141_4487 | Not-determined |
| 18-101_k141_21237 | Not-determined |
| 18-101_k141_202210 | Not-determined |
| 18-101_k141_235037 | Low-quality |
| 18-101_k141_53809 | Not-determined |
| 18-101_k141_102850 | Not-determined |
| 18-101_k141_382998 | Not-determined |
| 18-101_k141_120248 | Low-quality |
| 18-101_k141_218543 | Not-determined |
| 18-101_k141_235046 | Not-determined |
| 18-101_k141_136857 | Not-determined |
| 18-101_k141_284625 | Not-determined |
| 18-101_k141_301539 | Not-determined |
| 18-101_k141_120251 | Not-determined |
| 18-101_k141_120252 | Not-determined |
| 18-101_k141_102857 | Not-determined |
| 18-101_k141_251982 | Not-determined |
| 18-101_k141_102858 | Low-quality |
| 18-101_k141_168527 | Low-quality |
| 18-101_k141_267730 | Not-determined |
| 18-101_k141_53829 | Low-quality |
| 18-101_k141_383005 | Not-determined |
| 18-101_k141_136866 | Not-determined |
| 18-101_k141_69555 | Not-determined |
| 18-101_k141_21258 | Not-determined |
| 18-101_k141_102859 | Not-determined |
| 18-101_k141_69558 | Not-determined |
| 18-101_k141_267736 | Not-determined |
| 18-101_k141_251987 | Not-determined |
| 18-101_k141_86597 | Low-quality |
| 18-101_k141_69561 | Low-quality |
| 18-101_k141_267742 | Not-determined |
| 18-101_k141_4510 | Not-determined |
| 18-101_k141_4514 | Not-determined |
| 18-101_k141_202240 | Not-determined |
| 18-101_k141_251989 | Not-determined |
| 18-101_k141_267744 | Low-quality |
| 18-101_k141_69566 | Not-determined |
| 18-101_k141_267747 | Not-determined |
| 18-101_k141_102867 | Not-determined |
| 18-101_k141_301547 | Low-quality |
| 18-101_k141_4519 | Not-determined |
| 18-101_k141_383011 | Not-determined |
| 18-101_k141_301549 | Not-determined |
| 18-101_k141_21275 | Not-determined |
| 18-101_k141_202247 | Not-determined |
| 18-101_k141_235060 | Not-determined |
| 18-101_k141_202249 | Not-determined |
| 18-101_k141_53838 | Not-determined |
| 18-101_k141_218553 | Not-determined |
| 18-101_k141_21282 | Low-quality |
| 18-101_k141_53840 | Low-quality |
| 18-101_k141_37478 | Not-determined |
| 18-101_k141_4533 | Not-determined |
| 18-101_k141_284645 | Not-determined |
| 18-101_k141_37483 | Not-determined |
| 18-101_k141_383028 | Not-determined |
| 18-101_k141_251999 | Not-determined |
| 18-101_k141_383029 | Not-determined |
| 18-101_k141_53843 | Not-determined |
| 18-101_k141_235068 | Not-determined |
| 18-101_k141_252002 | Not-determined |
| 18-101_k141_252003 | Not-determined |
| 18-101_k141_152755 | Not-determined |
| 18-101_k141_350618 | Not-determined |
| 18-101_k141_69578 | Not-determined |
| 18-101_k141_4542 | Not-determined |
| 18-101_k141_366619 | Not-determined |
| 18-101_k141_168573 | Not-determined |
| 18-101_k141_86635 | Not-determined |
| 18-101_k141_4547 | Low-quality |
| 18-101_k141_21292 | Not-determined |
| 18-101_k141_267777 | Low-quality |
| 18-101_k141_218573 | Not-determined |
| 18-101_k141_4549 | Not-determined |
| 18-101_k141_21296 | Not-determined |
| 18-101_k141_218577 | Not-determined |
| 18-101_k141_53866 | Not-determined |
| 18-101_k141_120283 | Not-determined |
| 18-101_k141_284650 | Not-determined |
| 18-101_k141_366623 | Low-quality |
| 18-101_k141_317836 | Not-determined |
| 18-101_k141_284655 | Not-determined |
| 18-101_k141_21305 | Not-determined |
| 18-101_k141_37509 | Not-determined |
| 18-101_k141_333504 | Not-determined |
| 18-101_k141_317843 | Not-determined |
| 18-101_k141_102898 | Not-determined |
| 18-101_k141_21310 | Not-determined |
| 18-101_k141_86643 | Not-determined |
| 18-101_k141_168598 | Not-determined |
| 18-101_k141_168604 | Not-determined |
| 18-101_k141_102902 | Low-quality |
| 18-101_k141_69600 | Not-determined |
| 18-101_k141_267799 | Not-determined |
| 18-101_k141_102906 | Not-determined |
| 18-101_k141_267802 | Not-determined |
| 18-101_k141_4577 | Not-determined |
| 18-101_k141_21315 | Not-determined |
| 18-101_k141_284669 | Not-determined |
| 18-101_k141_383066 | Not-determined |
| 18-101_k141_120299 | Not-determined |
| 18-101_k141_102911 | Not-determined |
| 18-101_k141_366635 | Not-determined |
| 18-101_k141_218605 | Not-determined |
| 18-101_k141_21325 | Not-determined |
| 18-101_k141_284677 | Not-determined |
| 18-101_k141_301569 | Low-quality |
| 18-101_k141_120308 | Not-determined |
| 18-101_k141_21329 | Not-determined |
| 18-101_k141_366645 | Not-determined |
| 18-101_k141_120315 | Not-determined |
| 18-101_k141_235078 | Not-determined |
| 18-101_k141_317874 | Not-determined |
| 18-101_k141_284691 | Not-determined |
| 18-101_k141_152782 | Not-determined |
| 18-101_k141_350651 | Not-determined |
| 18-101_k141_21347 | Not-determined |
| 18-101_k141_301584 | Not-determined |
| 18-101_k141_152790 | Not-determined |
| 18-101_k141_186067 | Not-determined |
| 18-101_k141_202290 | Not-determined |
| 18-101_k141_301594 | Not-determined |
| 18-101_k141_301595 | Not-determined |
| 18-101_k141_186072 | Low-quality |
| 18-101_k141_301598 | Not-determined |
| 18-101_k141_267829 | Not-determined |
| 18-101_k141_168620 | Not-determined |
| 18-101_k141_86652 | Not-determined |
| 18-101_k141_21364 | Low-quality |
| 18-101_k141_86656 | Not-determined |
| 18-101_k141_202297 | Not-determined |
| 18-101_k141_21365 | Not-determined |
| 18-101_k141_235082 | Not-determined |
| 18-101_k141_186078 | Not-determined |
| 18-101_k141_284730 | Not-determined |
| 18-101_k141_168631 | Not-determined |
| 18-101_k141_301613 | Low-quality |
| 18-101_k141_366682 | Not-determined |
| 18-101_k141_102965 | Not-determined |
| 18-101_k141_53881 | Not-determined |
| 18-101_k141_383129 | Not-determined |
| 18-101_k141_136896 | Not-determined |
| 18-101_k141_366694 | Not-determined |
| 18-101_k141_267835 | Not-determined |
| 18-101_k141_267837 | Not-determined |
| 18-101_k141_284748 | Not-determined |
| 18-101_k141_218616 | Not-determined |
| 18-101_k141_21377 | Not-determined |
| 18-101_k141_152845 | Not-determined |
| 18-101_k141_120370 | Not-determined |
| 18-101_k141_235109 | Not-determined |
| 18-101_k141_267845 | Not-determined |
| 18-101_k141_152848 | Not-determined |
| 18-101_k141_301636 | Not-determined |
| 18-101_k141_152851 | Low-quality |
| 18-101_k141_86675 | Low-quality |
| 18-101_k141_202310 | Not-determined |
| 18-101_k141_284757 | Not-determined |
| 18-101_k141_152853 | Not-determined |
| 18-101_k141_120382 | Not-determined |
| 18-101_k141_136913 | Not-determined |
| 18-101_k141_136914 | Not-determined |
| 18-101_k141_136922 | Not-determined |
| 18-101_k141_53903 | Not-determined |
| 18-101_k141_4631 | Not-determined |
| 18-101_k141_168685 | Low-quality |
| 18-101_k141_136927 | Not-determined |
| 18-101_k141_53910 | Not-determined |
| 18-101_k141_152871 | Not-determined |
| 18-101_k141_86684 | Low-quality |
| 18-101_k141_21393 | Not-determined |
| 18-101_k141_284764 | Low-quality |
| 18-101_k141_252057 | Not-determined |
| 18-101_k141_301654 | Not-determined |
| 18-101_k141_86690 | Not-determined |
| 18-101_k141_284773 | Not-determined |
| 18-101_k141_350693 | Not-determined |
| 18-101_k141_202339 | Not-determined |
| 18-101_k141_4639 | Not-determined |
| 18-101_k141_301658 | Not-determined |
| 18-101_k141_301660 | Not-determined |
| 18-101_k141_86692 | Not-determined |
| 18-101_k141_350699 | Not-determined |
| 18-101_k141_350700 | Not-determined |
| 18-101_k141_383181 | Not-determined |
| 18-101_k141_120409 | Not-determined |
| 18-101_k141_168714 | Not-determined |
| 18-101_k141_120412 | Not-determined |
| 18-101_k141_120414 | Not-determined |
| 18-101_k141_168715 | Low-quality |
| 18-101_k141_4650 | Not-determined |
| 18-101_k141_86709 | Not-determined |
| 18-101_k141_69624 | Not-determined |
| 18-101_k141_136953 | Not-determined |
| 18-101_k141_152893 | Low-quality |
| 18-101_k141_267858 | Not-determined |
| 18-101_k141_186103 | Not-determined |
| 18-101_k141_136959 | Not-determined |
| 18-101_k141_366714 | Not-determined |
| 18-101_k141_37563 | Not-determined |
| 18-101_k141_37564 | Not-determined |
| 18-101_k141_301673 | Not-determined |
| 18-101_k141_37568 | Not-determined |
| 18-101_k141_218660 | Not-determined |
| 18-101_k141_37570 | Not-determined |
| 18-101_k141_218666 | Not-determined |
| 18-101_k141_252081 | Not-determined |
| 18-101_k141_120437 | Not-determined |
| 18-101_k141_366723 | Not-determined |
| 18-101_k141_168735 | Not-determined |
| 18-101_k141_301689 | Not-determined |
| 18-101_k141_383197 | Not-determined |
| 18-101_k141_317951 | Not-determined |
| 18-101_k141_4673 | Low-quality |
| 18-101_k141_168745 | Not-determined |
| 18-101_k141_136963 | Not-determined |
| 18-101_k141_37584 | Low-quality |
| 18-101_k141_37587 | Low-quality |
| 18-101_k141_136966 | Not-determined |
| 18-101_k141_383207 | Not-determined |
| 18-101_k141_235172 | Not-determined |
| 18-101_k141_317969 | Not-determined |
| 18-101_k141_383211 | Not-determined |
| 18-101_k141_383212 | Not-determined |
| 18-101_k141_37592 | Not-determined |
| 18-101_k141_4683 | Not-determined |
| 18-101_k141_86743 | Not-determined |
| 18-101_k141_186122 | Not-determined |
| 18-101_k141_235188 | Not-determined |
| 18-101_k141_4686 | Low-quality |
| 18-101_k141_383220 | Not-determined |
| 18-101_k141_383227 | Not-determined |
| 18-101_k141_383228 | Not-determined |
| 18-101_k141_301712 | Not-determined |
| 18-101_k141_69659 | Not-determined |
| 18-101_k141_69660 | Not-determined |
| 18-101_k141_186133 | Low-quality |
| 18-101_k141_366754 | Not-determined |
| 18-101_k141_53932 | Not-determined |
| 18-101_k141_168785 | Low-quality |
| 18-101_k141_301735 | Not-determined |
| 18-101_k141_366761 | Low-quality |
| 18-101_k141_252113 | Not-determined |
| 18-101_k141_186144 | Low-quality |
| 18-101_k141_202405 | Not-determined |
| 18-101_k141_318009 | Not-determined |
| 18-101_k141_202410 | Low-quality |
| 18-101_k141_318010 | Not-determined |
| 18-101_k141_202415 | Not-determined |
| 18-101_k141_202421 | Not-determined |
| 18-101_k141_4698 | Not-determined |
| 18-101_k141_4700 | Low-quality |
| 18-101_k141_383236 | Low-quality |
| 18-101_k141_383238 | Not-determined |
| 18-101_k141_168801 | Not-determined |
| 18-101_k141_37614 | Not-determined |
| 18-101_k141_37615 | Not-determined |
| 18-101_k141_152936 | Not-determined |
| 18-101_k141_235200 | Not-determined |
| 18-101_k141_202424 | Not-determined |
| 18-101_k141_301771 | Not-determined |
| 18-101_k141_318017 | Not-determined |
| 18-101_k141_252151 | Not-determined |
| 18-101_k141_186168 | Not-determined |
| 18-101_k141_235212 | Not-determined |
| 18-101_k141_4714 | Not-determined |
| 18-101_k141_186171 | Not-determined |
| 18-101_k141_152953 | Not-determined |
| 18-101_k141_383264 | Not-determined |
| 18-101_k141_202434 | Low-quality |
| 18-101_k141_218704 | Not-determined |
| 18-101_k141_202436 | Not-determined |
| 18-101_k141_218705 | Not-determined |
| 18-101_k141_168812 | Not-determined |
| 18-101_k141_383266 | Not-determined |
| 18-101_k141_168815 | Not-determined |
| 18-101_k141_136989 | Not-determined |
| 18-101_k141_136995 | Not-determined |
| 18-101_k141_4731 | Not-determined |
| 18-101_k141_120463 | Not-determined |
| 18-101_k141_4733 | Not-determined |
| 18-101_k141_120465 | Not-determined |
| 18-101_k141_69727 | Not-determined |
| 18-101_k141_152960 | Not-determined |
| 18-101_k141_186198 | Not-determined |
| 18-101_k141_235233 | Not-determined |
| 18-101_k141_186207 | Not-determined |
| 18-101_k141_333537 | Not-determined |
| 18-101_k141_37647 | Not-determined |
| 18-101_k141_69734 | Not-determined |
| 18-101_k141_137014 | Not-determined |
| 18-101_k141_69738 | Not-determined |
| 18-101_k141_252190 | Not-determined |
| 18-101_k141_350735 | Not-determined |
| 18-101_k141_137020 | Not-determined |
| 18-101_k141_218736 | Not-determined |
| 18-101_k141_37651 | Not-determined |
| 18-101_k141_186227 | Not-determined |
| 18-101_k141_218740 | Not-determined |
| 18-101_k141_235258 | Not-determined |
| 18-101_k141_301789 | Not-determined |
| 18-101_k141_301795 | Not-determined |
| 18-101_k141_69766 | Not-determined |
| 18-101_k141_69767 | Not-determined |
| 18-101_k141_218752 | Low-quality |
| 18-101_k141_318057 | Not-determined |
| 18-101_k141_137026 | Not-determined |
| 18-101_k141_137028 | Not-determined |
| 18-101_k141_318061 | Not-determined |
| 18-101_k141_37663 | Not-determined |
| 18-101_k141_37664 | Not-determined |
| 18-101_k141_301811 | Not-determined |
| 18-101_k141_301818 | Not-determined |
| 18-101_k141_350765 | Not-determined |
| 18-101_k141_69772 | Not-determined |
| 18-101_k141_318064 | Not-determined |
| 18-101_k141_168837 | Not-determined |
| 18-101_k141_186248 | Not-determined |
| 18-101_k141_69775 | Not-determined |
| 18-101_k141_318069 | Not-determined |
| 18-101_k141_202470 | Not-determined |
| 18-101_k141_218766 | Not-determined |

| | |
|---|---|
| 18-101_k141_252229 | Not-determined |
| 18-101_k141_383326 | Not-determined |
| 18-101_k141_267887 | Not-determined |
| 18-101_k141_37679 | Not-determined |
| 18-101_k141_350776 | Not-determined |
| 18-101_k141_120527 | Not-determined |
| 18-101_k141_318078 | Not-determined |
| 18-101_k141_202480 | Low-quality |
| 18-101_k141_318085 | Not-determined |
| 18-101_k141_103086 | Not-determined |
| 18-101_k141_301828 | Not-determined |
| 18-101_k141_152979 | Not-determined |
| 18-101_k141_235294 | Not-determined |
| 18-101_k141_152982 | Not-determined |
| 18-101_k141_37691 | Not-determined |
| 18-101_k141_333574 | Not-determined |
| 18-101_k141_366807 | Not-determined |
| 18-101_k141_318096 | Not-determined |
| 18-101_k141_69779 | Not-determined |
| 18-101_k141_318098 | Not-determined |
| 18-101_k141_252248 | Not-determined |
| 18-101_k141_235315 | Not-determined |
| 18-101_k141_252251 | Not-determined |
| 18-101_k141_333577 | Not-determined |
| 18-101_k141_152995 | Not-determined |
| 18-101_k141_252254 | Not-determined |
| 18-101_k141_86762 | Not-determined |
| 18-101_k141_153002 | Not-determined |
| 18-101_k141_383369 | Not-determined |
| 18-101_k141_202489 | Not-determined |
| 18-101_k141_383379 | Not-determined |
| 18-101_k141_218816 | Not-determined |
| 18-101_k141_86773 | Not-determined |
| 18-101_k141_252279 | Low-quality |
| 18-101_k141_333588 | Not-determined |
| 18-101_k141_301879 | Not-determined |
| 18-101_k141_318125 | Not-determined |
| 18-101_k141_333592 | Not-determined |
| 18-101_k141_218827 | Not-determined |
| 18-101_k141_69789 | Not-determined |
| 18-101_k141_366824 | Not-determined |
| 18-101_k141_218830 | Not-determined |
| 18-101_k141_318132 | Not-determined |
| 18-101_k141_86792 | Not-determined |
| 18-101_k141_366827 | Not-determined |
| 18-101_k141_218837 | Not-determined |
| 18-101_k141_301888 | Not-determined |
| 18-101_k141_218839 | Not-determined |
| 18-101_k141_218842 | Not-determined |
| 18-101_k141_218843 | Not-determined |
| 18-101_k141_69795 | Not-determined |
| 18-101_k141_186298 | Not-determined |
| 18-101_k141_202510 | Not-determined |
| 18-101_k141_4790 | Not-determined |
| 18-101_k141_350824 | Not-determined |
| 18-101_k141_350825 | Not-determined |
| 18-101_k141_69810 | Not-determined |
| 18-101_k141_69811 | Low-quality |
| 18-101_k141_153059 | Not-determined |
| 18-101_k141_168867 | Not-determined |
| 18-101_k141_383395 | Not-determined |
| 18-101_k141_333614 | Not-determined |
| 18-101_k141_350837 | Not-determined |
| 18-101_k141_186314 | Not-determined |
| 18-101_k141_366841 | Not-determined |
| 18-101_k141_168882 | Not-determined |
| 18-101_k141_4805 | Not-determined |
| 18-101_k141_267912 | Not-determined |
| 18-101_k141_4809 | Not-determined |
| 18-101_k141_168890 | Not-determined |
| 18-101_k141_4811 | Not-determined |
| 18-101_k141_252304 | Not-determined |
| 18-101_k141_318155 | Not-determined |
| 18-101_k141_318160 | Not-determined |
| 18-101_k141_350843 | Not-determined |
| 18-101_k141_186319 | Low-quality |
| 18-101_k141_350845 | Not-determined |
| 18-101_k141_120569 | Not-determined |
| 18-101_k141_4822 | Not-determined |
| 18-101_k141_37746 | Not-determined |
| 18-101_k141_186326 | Low-quality |
| 18-101_k141_318169 | Not-determined |
| 18-101_k141_252310 | Not-determined |
| 18-101_k141_37751 | Not-determined |
| 18-101_k141_252311 | Low-quality |
| 18-101_k141_37754 | Not-determined |
| 18-101_k141_252315 | Not-determined |
| 18-101_k141_252317 | Not-determined |
| 18-101_k141_37760 | Not-determined |
| 18-101_k141_37761 | Not-determined |
| 18-101_k141_301925 | Not-determined |
| 18-101_k141_137065 | Low-quality |
| 18-101_k141_267929 | Not-determined |
| 18-101_k141_168914 | Not-determined |
| 18-101_k141_202556 | Not-determined |
| 18-101_k141_252324 | Not-determined |
| 18-101_k141_318180 | Not-determined |
| 18-101_k141_350856 | Not-determined |
| 18-101_k141_318181 | Not-determined |
| 18-101_k141_318183 | Not-determined |
| 18-101_k141_350860 | Low-quality |
| 18-101_k141_168921 | Not-determined |
| 18-101_k141_350862 | Not-determined |
| 18-101_k141_350863 | Low-quality |
| 18-101_k141_301928 | Not-determined |
| 18-101_k141_53965 | Not-determined |
| 18-101_k141_53966 | Not-determined |
| 18-101_k141_37770 | Not-determined |
| 18-101_k141_137078 | Not-determined |
| 18-101_k141_4829 | Not-determined |
| 18-101_k141_120575 | Not-determined |
| 18-101_k141_350873 | Not-determined |
| 18-101_k141_301944 | Not-determined |
| 18-101_k141_4838 | Not-determined |
| 18-101_k141_301946 | Not-determined |
| 18-101_k141_4841 | Not-determined |
| 18-101_k141_120579 | Not-determined |
| 18-101_k141_202569 | Not-determined |
| 18-101_k141_168932 | Not-determined |
| 18-101_k141_350886 | Not-determined |
| 18-101_k141_120586 | Not-determined |
| 18-101_k141_53990 | Not-determined |
| 18-101_k141_86836 | Not-determined |
| 18-101_k141_168942 | Not-determined |
| 18-101_k141_350893 | Not-determined |
| 18-101_k141_103094 | Not-determined |
| 18-101_k141_168957 | Not-determined |
| 18-101_k141_137111 | Low-quality |
| 18-101_k141_186342 | Not-determined |
| 18-101_k141_120600 | Not-determined |
| 18-101_k141_137115 | Not-determined |
| 18-101_k141_53992 | Low-quality |
| 18-101_k141_120604 | Not-determined |
| 18-101_k141_53995 | Not-determined |
| 18-101_k141_202598 | Not-determined |
| 18-101_k141_137116 | Not-determined |
| 18-101_k141_218874 | Not-determined |
| 18-101_k141_366900 | Not-determined |
| 18-101_k141_137121 | Not-determined |
| 18-101_k141_103102 | Not-determined |
| 18-101_k141_318210 | Not-determined |
| 18-101_k141_168966 | Not-determined |
| 18-101_k141_366908 | Low-quality |
| 18-101_k141_37827 | Not-determined |
| 18-101_k141_301958 | Not-determined |
| 18-101_k141_350915 | Not-determined |
| 18-101_k141_120620 | Not-determined |
| 18-101_k141_168972 | Not-determined |
| 18-101_k141_69856 | Not-determined |
| 18-101_k141_350928 | Not-determined |
| 18-101_k141_218890 | Not-determined |
| 18-101_k141_350930 | Not-determined |
| 18-101_k141_235363 | Not-determined |
| 18-101_k141_218894 | Low-quality |
| 18-101_k141_218895 | Not-determined |
| 18-101_k141_366925 | Not-determined |
| 18-101_k141_37837 | Not-determined |
| 18-101_k141_86872 | Not-determined |
| 18-101_k141_137152 | Not-determined |
| 18-101_k141_69873 | Not-determined |
| 18-101_k141_120631 | Not-determined |
| 18-101_k141_4874 | Not-determined |
| 18-101_k141_120635 | Not-determined |
| 18-101_k141_202611 | Not-determined |
| 18-101_k141_383437 | Not-determined |
| 18-101_k141_137160 | Not-determined |
| 18-101_k141_54025 | Low-quality |
| 18-101_k141_120646 | Not-determined |
| 18-101_k141_69877 | Not-determined |
| 18-101_k141_37844 | Not-determined |
| 18-101_k141_54028 | Not-determined |
| 18-101_k141_86894 | Not-determined |
| 18-101_k141_218909 | Not-determined |
| 18-101_k141_268003 | Not-determined |
| 18-101_k141_37854 | Low-quality |
| 18-101_k141_218913 | Not-determined |
| 18-101_k141_153095 | Not-determined |
| 18-101_k141_186368 | Not-determined |
| 18-101_k141_318225 | Not-determined |
| 18-101_k141_218921 | Not-determined |
| 18-101_k141_268009 | Not-determined |
| 18-101_k141_137170 | Not-determined |
| 18-101_k141_268013 | Not-determined |
| 18-101_k141_366965 | Not-determined |
| 18-101_k141_186380 | Not-determined |
| 18-101_k141_186381 | Not-determined |
| 18-101_k141_37864 | Not-determined |
| 18-101_k141_103141 | Not-determined |
| 18-101_k141_103145 | Not-determined |
| 18-101_k141_37873 | Not-determined |
| 18-101_k141_268025 | Not-determined |
| 18-101_k141_37874 | Not-determined |
| 18-101_k141_137187 | Not-determined |
| 18-101_k141_202650 | Not-determined |
| 18-101_k141_202653 | Low-quality |
| 18-101_k141_235402 | Not-determined |
| 18-101_k141_37880 | Not-determined |
| 18-101_k141_268034 | Not-determined |
| 18-101_k141_103153 | Not-determined |
| 18-101_k141_366984 | Not-determined |
| 18-101_k141_202655 | Not-determined |
| 18-101_k141_4919 | Not-determined |
| 18-101_k141_137199 | Not-determined |
| 18-101_k141_350961 | Low-quality |
| 18-101_k141_137200 | Not-determined |
| 18-101_k141_4924 | Not-determined |
| 18-101_k141_366987 | Low-quality |
| 18-101_k141_202660 | Not-determined |
| 18-101_k141_284783 | Not-determined |
| 18-101_k141_4932 | Not-determined |
| 18-101_k141_218947 | Not-determined |
| 18-101_k141_169029 | Not-determined |
| 18-101_k141_137208 | Not-determined |
| 18-101_k141_318264 | Not-determined |
| 18-101_k141_284792 | Not-determined |
| 18-101_k141_120698 | Not-determined |
| 18-101_k141_383468 | Not-determined |
| 18-101_k141_383469 | Not-determined |
| 18-101_k141_153107 | Not-determined |
| 18-101_k141_302022 | Not-determined |
| 18-101_k141_383471 | Not-determined |
| 18-101_k141_153110 | Not-determined |
| 18-101_k141_137221 | Not-determined |
| 18-101_k141_202674 | Not-determined |
| 18-101_k141_235413 | Not-determined |
| 18-101_k141_383476 | Not-determined |
| 18-101_k141_350979 | Low-quality |
| 18-101_k141_284807 | Not-determined |
| 18-101_k141_302032 | Not-determined |
| 18-101_k141_318281 | Not-determined |
| 18-101_k141_54040 | Low-quality |
| 18-101_k141_235432 | Not-determined |
| 18-101_k141_4937 | Not-determined |
| 18-101_k141_235434 | Not-determined |
| 18-101_k141_137247 | Not-determined |
| 18-101_k141_137250 | Not-determined |
| 18-101_k141_37902 | Not-determined |
| 18-101_k141_4943 | Not-determined |
| 18-101_k141_284822 | Not-determined |
| 18-101_k141_37905 | Not-determined |
| 18-101_k141_4944 | Low-quality |
| 18-101_k141_86929 | Not-determined |
| 18-101_k141_169059 | Not-determined |
| 18-101_k141_54049 | Not-determined |
| 18-101_k141_54059 | Not-determined |
| 18-101_k141_137270 | Not-determined |
| 18-101_k141_383508 | Not-determined |
| 18-101_k141_268065 | Not-determined |
| 18-101_k141_383511 | Not-determined |
| 18-101_k141_350988 | Not-determined |
| 18-101_k141_137277 | Low-quality |
| 18-101_k141_4971 | Not-determined |
| 18-101_k141_153116 | Not-determined |
| 18-101_k141_153117 | Not-determined |
| 18-101_k141_367016 | Not-determined |
| 18-101_k141_54069 | Not-determined |
| 18-101_k141_4981 | Not-determined |
| 18-101_k141_302058 | Not-determined |
| 18-101_k141_4982 | Not-determined |
| 18-101_k141_202699 | Not-determined |
| 18-101_k141_4985 | Not-determined |
| 18-101_k141_302062 | Low-quality |
| 18-101_k141_153129 | Not-determined |
| 18-101_k141_137282 | Not-determined |
| 18-101_k141_268079 | Not-determined |
| 18-101_k141_153135 | Not-determined |
| 18-101_k141_318309 | Not-determined |
| 18-101_k141_54078 | Not-determined |
| 18-101_k141_153146 | Not-determined |
| 18-101_k141_169065 | Not-determined |
| 18-101_k141_153153 | Not-determined |
| 18-101_k141_351013 | Not-determined |
| 18-101_k141_137293 | Not-determined |
| 18-101_k141_218994 | Not-determined |
| 18-101_k141_333652 | Not-determined |
| 18-101_k141_153162 | Not-determined |
| 18-101_k141_202094 | Not-determined |
| 18-101_k141_103233 | Not-determined |
| 18-101_k141_4997 | Not-determined |
| 18-101_k141_235466 | Not-determined |
| 18-101_k141_333665 | Not-determined |
| 18-101_k141_103245 | Not-determined |
| 18-101_k141_169085 | Not-determined |
| 18-101_k141_5002 | Not-determined |
| 18-101_k141_367048 | Not-determined |
| 18-101_k141_103255 | Not-determined |
| 18-101_k141_351037 | Not-determined |
| 18-101_k141_383539 | Not-determined |
| 18-101_k141_120729 | Not-determined |
| 18-101_k141_252359 | Not-determined |
| 18-101_k141_103260 | Not-determined |
| 18-101_k141_333687 | Not-determined |
| 18-101_k141_169096 | Not-determined |
| 18-101_k141_219015 | Not-determined |
| 18-101_k141_5030 | Not-determined |
| 18-101_k141_153202 | Not-determined |
| 18-101_k141_268097 | Not-determined |
| 18-101_k141_367061 | Not-determined |
| 18-101_k141_302105 | Not-determined |
| 18-101_k141_120753 | Not-determined |
| 18-101_k141_5039 | Not-determined |
| 18-101_k141_103270 | Not-determined |
| 18-101_k141_235488 | Low-quality |
| 18-101_k141_284894 | Not-determined |
| 18-101_k141_302118 | Not-determined |
| 18-101_k141_219030 | Not-determined |
| 18-101_k141_367076 | Low-quality |
| 18-101_k141_54103 | Not-determined |
| 18-101_k141_103294 | Not-determined |
| 18-101_k141_37928 | Not-determined |
| 18-101_k141_268114 | Not-determined |
| 18-101_k141_268116 | Not-determined |
| 18-101_k141_351070 | Not-determined |
| 18-101_k141_103301 | Not-determined |
| 18-101_k141_367104 | Low-quality |
| 18-101_k141_103317 | Not-determined |
| 18-101_k141_268137 | Not-determined |
| 18-101_k141_235506 | Not-determined |
| 18-101_k141_383572 | Not-determined |
| 18-101_k141_318371 | Not-determined |
| 18-101_k141_37949 | Not-determined |
| 18-101_k141_383575 | Not-determined |
| 18-101_k141_219050 | Not-determined |
| 18-101_k141_54111 | Not-determined |
| 18-101_k141_219055 | Not-determined |
| 18-101_k141_367114 | Not-determined |
| 18-101_k141_54115 | Low-quality |
| 18-101_k141_120791 | Not-determined |
| 18-101_k141_87019 | Not-determined |
| 18-101_k141_383583 | Not-determined |
| 18-101_k141_383586 | Not-determined |
| 18-101_k141_54124 | Not-determined |
| 18-101_k141_383587 | Not-determined |
| 18-101_k141_186411 | Not-determined |
| 18-101_k141_383592 | Not-determined |
| 18-101_k141_37960 | Not-determined |
| 18-101_k141_54127 | Not-determined |
| 18-101_k141_318377 | Low-quality |
| 18-101_k141_21456 | Not-determined |
| 18-101_k141_318381 | Not-determined |
| 18-101_k141_318382 | Not-determined |
| 18-101_k141_21460 | Not-determined |
| 18-101_k141_235512 | Not-determined |
| 18-101_k141_186421 | Not-determined |
| 18-101_k141_37968 | Not-determined |
| 18-101_k141_284917 | Not-determined |
| 18-101_k141_235517 | Not-determined |
| 18-101_k141_21468 | Not-determined |
| 18-101_k141_219071 | Not-determined |
| 18-101_k141_120814 | Not-determined |
| 18-101_k141_333746 | Not-determined |
| 18-101_k141_268149 | Not-determined |
| 18-101_k141_252425 | Not-determined |
| 18-101_k141_268151 | Not-determined |
| 18-101_k141_54149 | Not-determined |
| 18-101_k141_235524 | Not-determined |
| 18-101_k141_367143 | Not-determined |
| 18-101_k141_252429 | Not-determined |
| 18-101_k141_137343 | Not-determined |
| 18-101_k141_268158 | Not-determined |
| 18-101_k141_367150 | Not-determined |
| 18-101_k141_235525 | Not-determined |
| 18-101_k141_69908 | Not-determined |
| 18-101_k141_69909 | Not-determined |
| 18-101_k141_69912 | Not-determined |
| 18-101_k141_5042 | Not-determined |
| 18-101_k141_318410 | Not-determined |
| 18-101_k141_5043 | Not-determined |
| 18-101_k141_120821 | Not-determined |
| 18-101_k141_318413 | Not-determined |
| 18-101_k141_38004 | Not-determined |
| 18-101_k141_268171 | Not-determined |
| 18-101_k141_367164 | Not-determined |
| 18-101_k141_219095 | Not-determined |
| 18-101_k141_186448 | Not-determined |
| 18-101_k141_202727 | Not-determined |
| 18-101_k141_367167 | Low-quality |
| 18-101_k141_268174 | Not-determined |
| 18-101_k141_87035 | Not-determined |
| 18-101_k141_268179 | Not-determined |
| 18-101_k141_87042 | Not-determined |
| 18-101_k141_268181 | Not-determined |
| 18-101_k141_268183 | Not-determined |
| 18-101_k141_383616 | Low-quality |
| 18-101_k141_333767 | Not-determined |
| 18-101_k141_284937 | Not-determined |
| 18-101_k141_219109 | Not-determined |
| 18-101_k141_284942 | Not-determined |
| 18-101_k141_219111 | Not-determined |
| 18-101_k141_235541 | Not-determined |
| 18-101_k141_219113 | Not-determined |
| 18-101_k141_235543 | Not-determined |
| 18-101_k141_383628 | Not-determined |
| 18-101_k141_137359 | Not-determined |
| 18-101_k141_284952 | Not-determined |
| 18-101_k141_186463 | Not-determined |
| 18-101_k141_120850 | Not-determined |
| 18-101_k141_87065 | Not-determined |
| 18-101_k141_120856 | Not-determined |
| 18-101_k141_120858 | Not-determined |
| 18-101_k141_268204 | Not-determined |
| 18-101_k141_87068 | Not-determined |
| 18-101_k141_186470 | Not-determined |
| 18-101_k141_318427 | Not-determined |
| 18-101_k141_202753 | Low-quality |
| 18-101_k141_120860 | Not-determined |
| 18-101_k141_367172 | Not-determined |
| 18-101_k141_252451 | Not-determined |
| 18-101_k141_186481 | Not-determined |
| 18-101_k141_383642 | Not-determined |
| 18-101_k141_186486 | Low-quality |
| 18-101_k141_21481 | Not-determined |
| 18-101_k141_302146 | Not-determined |
| 18-101_k141_252459 | Not-determined |
| 18-101_k141_21488 | Not-determined |
| 18-101_k141_21489 | Not-determined |
| 18-101_k141_120881 | Not-determined |
| 18-101_k141_219131 | Not-determined |
| 18-101_k141_302150 | Not-determined |
| 18-101_k141_5064 | Low-quality |
| 18-101_k141_318434 | Low-quality |
| 18-101_k141_54188 | Not-determined |
| 18-101_k141_5070 | Not-determined |
| 18-101_k141_153229 | Not-determined |
| 18-101_k141_302164 | Not-determined |
| 18-101_k141_383656 | Not-determined |
| 18-101_k141_268222 | Not-determined |
| 18-101_k141_54194 | Not-determined |
| 18-101_k141_333819 | Not-determined |
| 18-101_k141_137376 | Not-determined |
| 18-101_k141_137380 | Not-determined |
| 18-101_k141_383659 | Not-determined |
| 18-101_k141_137383 | Not-determined |
| 18-101_k141_137386 | Not-determined |
| 18-101_k141_383660 | Low-quality |
| 18-101_k141_268227 | Not-determined |
| 18-101_k141_333821 | Not-determined |
| 18-101_k141_153244 | Not-determined |
| 18-101_k141_153245 | Not-determined |
| 18-101_k141_219153 | Not-determined |
| 18-101_k141_268237 | Not-determined |
| 18-101_k141_351127 | Not-determined |
| 18-101_k141_333824 | Not-determined |
| 18-101_k141_351133 | Not-determined |
| 18-101_k141_120888 | Not-determined |
| 18-101_k141_252479 | Not-determined |
| 18-101_k141_54214 | Not-determined |
| 18-101_k141_333831 | Not-determined |
| 18-101_k141_219159 | Not-determined |
| 18-101_k141_54222 | Not-determined |
| 18-101_k141_137402 | Not-determined |
| 18-101_k141_219162 | Low-quality |
| 18-101_k141_333839 | Not-determined |
| 18-101_k141_219163 | Not-determined |
| 18-101_k141_219167 | Not-determined |
| 18-101_k141_219169 | Not-determined |
| 18-101_k141_137409 | Not-determined |
| 18-101_k141_252487 | Not-determined |
| 18-101_k141_21501 | Not-determined |
| 18-101_k141_219179 | Not-determined |
| 18-101_k141_137421 | Not-determined |
| 18-101_k141_186530 | Not-determined |
| 18-101_k141_69984 | Not-determined |
| 18-101_k141_367218 | Low-quality |
| 18-101_k141_69986 | Low-quality |
| 18-101_k141_87089 | Not-determined |
| 18-101_k141_5092 | Not-determined |
| 18-101_k141_202798 | Not-determined |
| 18-101_k141_5094 | Not-determined |
| 18-101_k141_5095 | Not-determined |
| 18-101_k141_202800 | Not-determined |
| 18-101_k141_351163 | Low-quality |
| 18-101_k141_153288 | Not-determined |
| 18-101_k141_219195 | Low-quality |
| 18-101_k141_137425 | Not-determined |
| 18-101_k141_137426 | Not-determined |
| 18-101_k141_351170 | Not-determined |
| 18-101_k141_351171 | Not-determined |
| 18-101_k141_186541 | Not-determined |
| 18-101_k141_54243 | Not-determined |
| 18-101_k141_5102 | Not-determined |
| 18-101_k141_219204 | Not-determined |
| 18-101_k141_219210 | Not-determined |
| 18-101_k141_318465 | Not-determined |
| 18-101_k141_268267 | Not-determined |
| 18-101_k141_219214 | Not-determined |
| 18-101_k141_268270 | Not-determined |
| 18-101_k141_153302 | Not-determined |
| 18-101_k141_186550 | Low-quality |
| 18-101_k141_186553 | Not-determined |
| 18-101_k141_21516 | Not-determined |
| 18-101_k141_252519 | Not-determined |
| 18-101_k141_21518 | Not-determined |
| 18-101_k141_186559 | Not-determined |
| 18-101_k141_383715 | Not-determined |
| 18-101_k141_318474 | Not-determined |
| 18-101_k141_333869 | Low-quality |
| 18-101_k141_268279 | Not-determined |
| 18-101_k141_252526 | Not-determined |
| 18-101_k141_219229 | Not-determined |
| 18-101_k141_120934 | Not-determined |
| 18-101_k141_284994 | Not-determined |
| 18-101_k141_120937 | Low-quality |
| 18-101_k141_351187 | Not-determined |
| 18-101_k141_268283 | Not-determined |
| 18-101_k141_318485 | Not-determined |
| 18-101_k141_252532 | Not-determined |
| 18-101_k141_219240 | Not-determined |
| 18-101_k141_252534 | Not-determined |
| 18-101_k141_284995 | Not-determined |
| 18-101_k141_351203 | Not-determined |
| 18-101_k141_219247 | Not-determined |
| 18-101_k141_367225 | Not-determined |
| 18-101_k141_5131 | Not-determined |
| 18-101_k141_284997 | Not-determined |
| 18-101_k141_219259 | Not-determined |
| 18-101_k141_219260 | Not-determined |
| 18-101_k141_333899 | Not-determined |
| 18-101_k141_219261 | Not-determined |
| 18-101_k141_202828 | Not-determined |
| 18-101_k141_54271 | Not-determined |
| 18-101_k141_252540 | Not-determined |
| 18-101_k141_318511 | Not-determined |
| 18-101_k141_333907 | Low-quality |
| 18-101_k141_219269 | Not-determined |
| 18-101_k141_70001 | Not-determined |
| 18-101_k141_318513 | Not-determined |
| 18-101_k141_70007 | Not-determined |
| 18-101_k141_153326 | Not-determined |
| 18-101_k141_219277 | Not-determined |
| 18-101_k141_202838 | Not-determined |
| 18-101_k141_268293 | Not-determined |
| 18-101_k141_219279 | Not-determined |
| 18-101_k141_153329 | Not-determined |
| 18-101_k141_70010 | Low-quality |
| 18-101_k141_367237 | Low-quality |
| 18-101_k141_219286 | Not-determined |
| 18-101_k141_153341 | Not-determined |
| 18-101_k141_333919 | Low-quality |
| 18-101_k141_367243 | Not-determined |
| 18-101_k141_5137 | Not-determined |
| 18-101_k141_54291 | Not-determined |
| 18-101_k141_87113 | Not-determined |
| 18-101_k141_333926 | Not-determined |
| 18-101_k141_70018 | Not-determined |
| 18-101_k141_302183 | Not-determined |
| 18-101_k141_202841 | Not-determined |
| 18-101_k141_333928 | Not-determined |
| 18-101_k141_302186 | Not-determined |
| 18-101_k141_302187 | Not-determined |
| 18-101_k141_38049 | Not-determined |
| 18-101_k141_87117 | Not-determined |
| 18-101_k141_120961 | Not-determined |
| 18-101_k141_87118 | Not-determined |
| 18-101_k141_153351 | Not-determined |
| 18-101_k141_38057 | Not-determined |
| 18-101_k141_302190 | Not-determined |
| 18-101_k141_54303 | Not-determined |
| 18-101_k141_202856 | Not-determined |
| 18-101_k141_302194 | Not-determined |
| 18-101_k141_235591 | Not-determined |
| 18-101_k141_70033 | Low-quality |
| 18-101_k141_383748 | Not-determined |
| 18-101_k141_21536 | Low-quality |
| 18-101_k141_70036 | Not-determined |
| 18-101_k141_351228 | Not-determined |
| 18-101_k141_120980 | Not-determined |
| 18-101_k141_268309 | Not-determined |
| 18-101_k141_87133 | Not-determined |
| 18-101_k141_235596 | Not-determined |
| 18-101_k141_219304 | Not-determined |
| 18-101_k141_235603 | Not-determined |
| 18-101_k141_21539 | Not-determined |
| 18-101_k141_268320 | Not-determined |
| 18-101_k141_70047 | Not-determined |
| 18-101_k141_351239 | Not-determined |
| 18-101_k141_21552 | Low-quality |
| 18-101_k141_186603 | Low-quality |
| 18-101_k141_38083 | Not-determined |
| 18-101_k141_38084 | Not-determined |
| 18-101_k141_351249 | Not-determined |
| 18-101_k141_252559 | Not-determined |
| 18-101_k141_21561 | Not-determined |
| 18-101_k141_186613 | Not-determined |
| 18-101_k141_38092 | Not-determined |
| 18-101_k141_235625 | Not-determined |
| 18-101_k141_169155 | Not-determined |
| 18-101_k141_137452 | Not-determined |
| 18-101_k141_137453 | Not-determined |

| | |
|---|---|
| 18-101_k141_153366 | Not-determined |
| 18-101_k141_219320 | Not-determined |
| 18-101_k141_318549 | Not-determined |
| 18-101_k141_219323 | Not-determined |
| 18-101_k141_351271 | Not-determined |
| 18-101_k141_351273 | Not-determined |
| 18-101_k141_219328 | Not-determined |
| 18-101_k141_137462 | Not-determined |
| 18-101_k141_202862 | Not-determined |
| 18-101_k141_137470 | Not-determined |
| 18-101_k141_21566 | Not-determined |
| 18-101_k141_186653 | Not-determined |
| 18-101_k141_169175 | Not-determined |
| 18-101_k141_38116 | Not-determined |
| 18-101_k141_169182 | Not-determined |
| 18-101_k141_38118 | Low-quality |
| 18-101_k141_38121 | Not-determined |
| 18-101_k141_121011 | Not-determined |
| 18-101_k141_202884 | Not-determined |
| 18-101_k141_38126 | Not-determined |
| 18-101_k141_21582 | Not-determined |
| 18-101_k141_169195 | Not-determined |
| 18-101_k141_121015 | Not-determined |
| 18-101_k141_38128 | Not-determined |
| 18-101_k141_252577 | Not-determined |
| 18-101_k141_121017 | Not-determined |
| 18-101_k141_5187 | Not-determined |
| 18-101_k141_169201 | Not-determined |
| 18-101_k141_5190 | Not-determined |
| 18-101_k141_186673 | Not-determined |
| 18-101_k141_70075 | Not-determined |
| 18-101_k141_202895 | Not-determined |
| 18-101_k141_302214 | Not-determined |
| 18-101_k141_121029 | Low-quality |
| 18-101_k141_318555 | Not-determined |
| 18-101_k141_186679 | Low-quality |
| 18-101_k141_70079 | Not-determined |
| 18-101_k141_318557 | Not-determined |
| 18-101_k141_137495 | Not-determined |
| 18-101_k141_235651 | Not-determined |
| 18-101_k141_202907 | Not-determined |
| 18-101_k141_318558 | Not-determined |
| 18-101_k141_5197 | Not-determined |
| 18-101_k141_186688 | Not-determined |
| 18-101_k141_137501 | Not-determined |
| 18-101_k141_70093 | Not-determined |
| 18-101_k141_318563 | Low-quality |
| 18-101_k141_5202 | Not-determined |
| 18-101_k141_5206 | Not-determined |
| 18-101_k141_137510 | Not-determined |
| 18-101_k141_137514 | Not-determined |
| 18-101_k141_219383 | Not-determined |
| 18-101_k141_318583 | Not-determined |
| 18-101_k141_351286 | Not-determined |
| 18-101_k141_318589 | Not-determined |
| 18-101_k141_38162 | Low-quality |
| 18-101_k141_186697 | Not-determined |
| 18-101_k141_302235 | Not-determined |
| 18-101_k141_333951 | Not-determined |
| 18-101_k141_351292 | Not-determined |
| 18-101_k141_153407 | Not-determined |
| 18-101_k141_186705 | Not-determined |
| 18-101_k141_21613 | Not-determined |
| 18-101_k141_186708 | Not-determined |
| 18-101_k141_268356 | Low-quality |
| 18-101_k141_137531 | Low-quality |
| 18-101_k141_153419 | Not-determined |
| 18-101_k141_252614 | Low-quality |
| 18-101_k141_121061 | Not-determined |
| 18-101_k141_121062 | Not-determined |
| 18-101_k141_202946 | Not-determined |
| 18-101_k141_285035 | Low-quality |
| 18-101_k141_137540 | Not-determined |
| 18-101_k141_21621 | Not-determined |
| 18-101_k141_54352 | Not-determined |
| 18-101_k141_202949 | Not-determined |
| 18-101_k141_186721 | Not-determined |
| 18-101_k141_121067 | Low-quality |
| 18-101_k141_186724 | Not-determined |
| 18-101_k141_202951 | Not-determined |
| 18-101_k141_137544 | Not-determined |
| 18-101_k141_38175 | Not-determined |
| 18-101_k141_252615 | Not-determined |
| 18-101_k141_235697 | Not-determined |
| 18-101_k141_38180 | Not-determined |
| 18-101_k141_235704 | Not-determined |
| 18-101_k141_54364 | Not-determined |
| 18-101_k141_219420 | Not-determined |
| 18-101_k141_87179 | Not-determined |
| 18-101_k141_285043 | Not-determined |
| 18-101_k141_186738 | Not-determined |
| 18-101_k141_351330 | Not-determined |
| 18-101_k141_186739 | Not-determined |
| 18-101_k141_186740 | Not-determined |
| 18-101_k141_169257 | Not-determined |
| 18-101_k141_169258 | Not-determined |
| 18-101_k141_367348 | Not-determined |
| 18-101_k141_87184 | Low-quality |
| 18-101_k141_21643 | Low-quality |
| 18-101_k141_21648 | Not-determined |
| 18-101_k141_351336 | Not-determined |
| 18-101_k141_70132 | Low-quality |
| 18-101_k141_333990 | Not-determined |
| 18-101_k141_137556 | Not-determined |
| 18-101_k141_70146 | Not-determined |
| 18-101_k141_333993 | Low-quality |
| 18-101_k141_285063 | Not-determined |
| 18-101_k141_351350 | Not-determined |
| 18-101_k141_121077 | Not-determined |
| 18-101_k141_268393 | Not-determined |
| 18-101_k141_21659 | Not-determined |
| 18-101_k141_21662 | Not-determined |
| 18-101_k141_87207 | Not-determined |
| 18-101_k141_351360 | Not-determined |
| 18-101_k141_87213 | Not-determined |
| 18-101_k141_285073 | Low-quality |
| 18-101_k141_351364 | Not-determined |
| 18-101_k141_121091 | Not-determined |
| 18-101_k141_367376 | Low-quality |
| 18-101_k141_252633 | Not-determined |
| 18-101_k141_202985 | Not-determined |
| 18-101_k141_121097 | Not-determined |
| 18-101_k141_186778 | Not-determined |
| 18-101_k141_252641 | Not-determined |
| 18-101_k141_285081 | Not-determined |
| 18-101_k141_21666 | Not-determined |
| 18-101_k141_202989 | Not-determined |
| 18-101_k141_202992 | Not-determined |
| 18-101_k141_87231 | Not-determined |
| 18-101_k141_121111 | Not-determined |
| 18-101_k141_268402 | Not-determined |
| 18-101_k141_219462 | Not-determined |
| 18-101_k141_121113 | Not-determined |
| 18-101_k141_268405 | Low-quality |
| 18-101_k141_383786 | Not-determined |
| 18-101_k141_54404 | Not-determined |
| 18-101_k141_121117 | Not-determined |
| 18-101_k141_334000 | Not-determined |
| 18-101_k141_87234 | Not-determined |
| 18-101_k141_21681 | Not-determined |
| 18-101_k141_87238 | Not-determined |
| 18-101_k141_137588 | Not-determined |
| 18-101_k141_302299 | Not-determined |
| 18-101_k141_351378 | Not-determined |
| 18-101_k141_186813 | Not-determined |
| 18-101_k141_334004 | Not-determined |
| 18-101_k141_121131 | Not-determined |
| 18-101_k141_235774 | Not-determined |
| 18-101_k141_351382 | Not-determined |
| 18-101_k141_235776 | Not-determined |
| 18-101_k141_235778 | Not-determined |
| 18-101_k141_186827 | Not-determined |
| 18-101_k141_235781 | Not-determined |
| 18-101_k141_186828 | Not-determined |
| 18-101_k141_186835 | Not-determined |
| 18-101_k141_383802 | Low-quality |
| 18-101_k141_252688 | Not-determined |
| 18-101_k141_252689 | Not-determined |
| 18-101_k141_318623 | Not-determined |
| 18-101_k141_252693 | Not-determined |
| 18-101_k141_235793 | Not-determined |
| 18-101_k141_87263 | Not-determined |
| 18-101_k141_153447 | Not-determined |
| 18-101_k141_153448 | Not-determined |
| 18-101_k141_351404 | Not-determined |
| 18-101_k141_203021 | Not-determined |
| 18-101_k141_252699 | Not-determined |
| 18-101_k141_252700 | Not-determined |
| 18-101_k141_87272 | Not-determined |
| 18-101_k141_334020 | Not-determined |
| 18-101_k141_302320 | Not-determined |
| 18-101_k141_153457 | Not-determined |
| 18-101_k141_351410 | Not-determined |
| 18-101_k141_302322 | Not-determined |
| 18-101_k141_153462 | Not-determined |
| 18-101_k141_186870 | Not-determined |
| 18-101_k141_235809 | Not-determined |
| 18-101_k141_367420 | Low-quality |
| 18-101_k141_302326 | Not-determined |
| 18-101_k141_121157 | Not-determined |
| 18-101_k141_302333 | Not-determined |
| 18-101_k141_302335 | Not-determined |
| 18-101_k141_351420 | Not-determined |
| 18-101_k141_383846 | Not-determined |
| 18-101_k141_153487 | Not-determined |
| 18-101_k141_351424 | Not-determined |
| 18-101_k141_203041 | Not-determined |
| 18-101_k141_351428 | Not-determined |
| 18-101_k141_121177 | Not-determined |
| 18-101_k141_21705 | Not-determined |
| 18-101_k141_367438 | Low-quality |
| 18-101_k141_285154 | Not-determined |
| 18-101_k141_203055 | Not-determined |
| 18-101_k141_252736 | Not-determined |
| 18-101_k141_367441 | Not-determined |
| 18-101_k141_203058 | Not-determined |
| 18-101_k141_351437 | Not-determined |
| 18-101_k141_302351 | Not-determined |
| 18-101_k141_203063 | Not-determined |
| 18-101_k141_383861 | Not-determined |
| 18-101_k141_383864 | Not-determined |
| 18-101_k141_383868 | Not-determined |
| 18-101_k141_285163 | Not-determined |
| 18-101_k141_383870 | Not-determined |
| 18-101_k141_383872 | Not-determined |
| 18-101_k141_285169 | Not-determined |
| 18-101_k141_87300 | Not-determined |
| 18-101_k141_87302 | Not-determined |
| 18-101_k141_252757 | Not-determined |
| 18-101_k141_153504 | Low-quality |
| 18-101_k141_70195 | Not-determined |
| 18-101_k141_153508 | Not-determined |
| 18-101_k141_87305 | Not-determined |
| 18-101_k141_186880 | Not-determined |
| 18-101_k141_87307 | Not-determined |
| 18-101_k141_87311 | Not-determined |
| 18-101_k141_203077 | Not-determined |
| 18-101_k141_54430 | Not-determined |
| 18-101_k141_54432 | Not-determined |
| 18-101_k141_268429 | Not-determined |
| 18-101_k141_235848 | Not-determined |
| 18-101_k141_121193 | Not-determined |
| 18-101_k141_21736 | Not-determined |
| 18-101_k141_54443 | Not-determined |
| 18-101_k141_367456 | Not-determined |
| 18-101_k141_203092 | Not-determined |
| 18-101_k141_5290 | Not-determined |
| 18-101_k141_351475 | Not-determined |
| 18-101_k141_54447 | Not-determined |
| 18-101_k141_285195 | Not-determined |
| 18-101_k141_367461 | Not-determined |
| 18-101_k141_87332 | Not-determined |
| 18-101_k141_235866 | Low-quality |
| 18-101_k141_285203 | Not-determined |
| 18-101_k141_268452 | Not-determined |
| 18-101_k141_235867 | Not-determined |
| 18-101_k141_21752 | Not-determined |
| 18-101_k141_235868 | Not-determined |
| 18-101_k141_121207 | Not-determined |
| 18-101_k141_70215 | Not-determined |
| 18-101_k141_235871 | Not-determined |
| 18-101_k141_203104 | Not-determined |
| 18-101_k141_203106 | Not-determined |
| 18-101_k141_203107 | Not-determined |
| 18-101_k141_121213 | Not-determined |
| 18-101_k141_121215 | Not-determined |
| 18-101_k141_235877 | Not-determined |
| 18-101_k141_87343 | Low-quality |
| 18-101_k141_54468 | Not-determined |
| 18-101_k141_203120 | Not-determined |
| 18-101_k141_87346 | Not-determined |
| 18-101_k141_285228 | Low-quality |
| 18-101_k141_153526 | Low-quality |
| 18-101_k141_351498 | Not-determined |
| 18-101_k141_334095 | Not-determined |
| 18-101_k141_5310 | Not-determined |
| 18-101_k141_186921 | Not-determined |
| 18-101_k141_54486 | Not-determined |
| 18-101_k141_285235 | Not-determined |
| 18-101_k141_186927 | Not-determined |
| 18-101_k141_186929 | Not-determined |
| 18-101_k141_169279 | Not-determined |
| 18-101_k141_351510 | Not-determined |
| 18-101_k141_252772 | Not-determined |
| 18-101_k141_186932 | Not-determined |
| 18-101_k141_219515 | Not-determined |
| 18-101_k141_219516 | Not-determined |
| 18-101_k141_169285 | Not-determined |
| 18-101_k141_186934 | Low-quality |
| 18-101_k141_54495 | Not-determined |
| 18-101_k141_54502 | Not-determined |
| 18-101_k141_54503 | Not-determined |
| 18-101_k141_103352 | Low-quality |
| 18-101_k141_302375 | Not-determined |
| 18-101_k141_351527 | Not-determined |
| 18-101_k141_318713 | Low-quality |
| 18-101_k141_268484 | Not-determined |
| 18-101_k141_285258 | Not-determined |
| 18-101_k141_367501 | Not-determined |
| 18-101_k141_103364 | Low-quality |
| 18-101_k141_203140 | Low-quality |
| 18-101_k141_186942 | Low-quality |
| 18-101_k141_367503 | Not-determined |
| 18-101_k141_38225 | Not-determined |
| 18-101_k141_38230 | Low-quality |
| 18-101_k141_137680 | Low-quality |
| 18-101_k141_38231 | Not-determined |
| 18-101_k141_137682 | Not-determined |
| 18-101_k141_351536 | Not-determined |
| 18-101_k141_351537 | Not-determined |
| 18-101_k141_103378 | Not-determined |
| 18-101_k141_21771 | Not-determined |
| 18-101_k141_367514 | Not-determined |
| 18-101_k141_186952 | Not-determined |
| 18-101_k141_5325 | Not-determined |
| 18-101_k141_70248 | Not-determined |
| 18-101_k141_70252 | Not-determined |
| 18-101_k141_87368 | Not-determined |
| 18-101_k141_186961 | Not-determined |
| 18-101_k141_137701 | Not-determined |
| 18-101_k141_137703 | Not-determined |
| 18-101_k141_38243 | Not-determined |
| 18-101_k141_103383 | Not-determined |
| 18-101_k141_219519 | Low-quality |
| 18-101_k141_5334 | Not-determined |
| 18-101_k141_121267 | Not-determined |
| 18-101_k141_153548 | Low-quality |
| 18-101_k141_38247 | Low-quality |
| 18-101_k141_153550 | Not-determined |
| 18-101_k141_137713 | Low-quality |
| 18-101_k141_153554 | Not-determined |
| 18-101_k141_38248 | Not-determined |
| 18-101_k141_153557 | Not-determined |
| 18-101_k141_137718 | Not-determined |
| 18-101_k141_38258 | Not-determined |
| 18-101_k141_351546 | Not-determined |
| 18-101_k141_103402 | Not-determined |
| 18-101_k141_203173 | Not-determined |
| 18-101_k141_367538 | Not-determined |
| 18-101_k141_235909 | Not-determined |
| 18-101_k141_367543 | Not-determined |
| 18-101_k141_285297 | Not-determined |
| 18-101_k141_367547 | Not-determined |
| 18-101_k141_38283 | Not-determined |
| 18-101_k141_367549 | Not-determined |
| 18-101_k141_169296 | Not-determined |
| 18-101_k141_103405 | Low-quality |
| 18-101_k141_285305 | Not-determined |
| 18-101_k141_21811 | Not-determined |
| 18-101_k141_21816 | Not-determined |
| 18-101_k141_252820 | Not-determined |
| 18-101_k141_367556 | Not-determined |
| 18-101_k141_367559 | Not-determined |
| 18-101_k141_21824 | Not-determined |
| 18-101_k141_285320 | Not-determined |
| 18-101_k141_137745 | Not-determined |
| 18-101_k141_302451 | Not-determined |
| 18-101_k141_70297 | Not-determined |
| 18-101_k141_38295 | Not-determined |
| 18-101_k141_383905 | Not-determined |
| 18-101_k141_334148 | Not-determined |
| 18-101_k141_285325 | Not-determined |
| 18-101_k141_285327 | Not-determined |
| 18-101_k141_302458 | Not-determined |
| 18-101_k141_153583 | Not-determined |
| 18-101_k141_87384 | Not-determined |
| 18-101_k141_252840 | Not-determined |
| 18-101_k141_367572 | Not-determined |
| 18-101_k141_383921 | Not-determined |
| 18-101_k141_268515 | Not-determined |
| 18-101_k141_235932 | Not-determined |
| 18-101_k141_87387 | Not-determined |
| 18-101_k141_187003 | Not-determined |
| 18-101_k141_235936 | Not-determined |
| 18-101_k141_5367 | Not-determined |
| 18-101_k141_235938 | Not-determined |
| 18-101_k141_383932 | Not-determined |
| 18-101_k141_87395 | Not-determined |
| 18-101_k141_252848 | Not-determined |
| 18-101_k141_252854 | Not-determined |
| 18-101_k141_268526 | Not-determined |
| 18-101_k141_137751 | Not-determined |
| 18-101_k141_137755 | Not-determined |
| 18-101_k141_54523 | Not-determined |
| 18-101_k141_187011 | Not-determined |
| 18-101_k141_137758 | Not-determined |
| 18-101_k141_252863 | Not-determined |
| 18-101_k141_54531 | Not-determined |
| 18-101_k141_285353 | Not-determined |
| 18-101_k141_5383 | Not-determined |
| 18-101_k141_54533 | Not-determined |
| 18-101_k141_70324 | Not-determined |
| 18-101_k141_21850 | Low-quality |
| 18-101_k141_54536 | Not-determined |
| 18-101_k141_302483 | Not-determined |
| 18-101_k141_21852 | Not-determined |
| 18-101_k141_252867 | Not-determined |
| 18-101_k141_103432 | Not-determined |
| 18-101_k141_103433 | Not-determined |
| 18-101_k141_334158 | Not-determined |
| 18-101_k141_70331 | Not-determined |
| 18-101_k141_203200 | Not-determined |
| 18-101_k141_187031 | Not-determined |
| 18-101_k141_235957 | Not-determined |
| 18-101_k141_169355 | Not-determined |
| 18-101_k141_285369 | Not-determined |
| 18-101_k141_219552 | Not-determined |
| 18-101_k141_334172 | Not-determined |
| 18-101_k141_219555 | Not-determined |
| 18-101_k141_187041 | Not-determined |
| 18-101_k141_235966 | Not-determined |
| 18-101_k141_70343 | Not-determined |
| 18-101_k141_219561 | Not-determined |
| 18-101_k141_268563 | Not-determined |
| 18-101_k141_203215 | Not-determined |
| 18-101_k141_252880 | Not-determined |
| 18-101_k141_285379 | Not-determined |
| 18-101_k141_383973 | Not-determined |
| 18-101_k141_302494 | Not-determined |
| 18-101_k141_383978 | Not-determined |
| 18-101_k141_203240 | Not-determined |
| 18-101_k141_137782 | Not-determined |
| 18-101_k141_203242 | Not-determined |
| 18-101_k141_153604 | Not-determined |
| 18-101_k141_153610 | Not-determined |
| 18-101_k141_367611 | Not-determined |
| 18-101_k141_137786 | Not-determined |
| 18-101_k141_153613 | Not-determined |
| 18-101_k141_318746 | Not-determined |
| 18-101_k141_302506 | Not-determined |
| 18-101_k141_302509 | Not-determined |
| 17-109-final-viral-combined_k141_368462\|\|full | High-quality |
| 18-101_k141_302511 | Not-determined |
| 18-101_k141_318752 | Low-quality |
| 18-101_k141_187066 | Not-determined |
| 18-101_k141_318753 | Not-determined |
| 18-101_k141_318754 | Not-determined |
| 18-101_k141_383991 | Not-determined |
| 18-101_k141_153617 | Low-quality |
| 18-101_k141_5388 | Not-determined |
| 18-101_k141_70370 | Not-determined |
| 18-101_k141_70371 | Not-determined |
| 18-101_k141_367624 | Not-determined |
| 18-101_k141_302518 | Not-determined |
| 18-101_k141_318759 | Not-determined |
| 18-101_k141_137796 | Not-determined |
| 18-101_k141_5393 | Not-determined |
| 18-101_k141_5394 | Not-determined |
| 18-101_k141_219592 | Not-determined |
| 18-101_k141_318764 | Not-determined |
| 18-101_k141_219594 | Not-determined |
| 18-101_k141_318766 | Not-determined |
| 18-101_k141_103463 | Not-determined |
| 18-101_k141_268589 | Not-determined |
| 18-101_k141_87437 | Not-determined |
| 18-101_k141_87438 | Not-determined |
| 18-101_k141_268591 | Not-determined |
| 18-101_k141_187070 | Not-determined |
| 18-101_k141_334181 | Not-determined |
| 18-101_k141_169394 | Not-determined |
| 18-101_k141_219598 | Not-determined |
| 18-101_k141_5399 | Not-determined |
| 18-101_k141_5400 | Not-determined |
| 18-101_k141_5403 | Not-determined |
| 18-101_k141_285408 | Not-determined |
| 18-101_k141_87447 | Low-quality |
| 18-101_k141_302538 | Not-determined |
| 18-101_k141_38309 | Not-determined |
| 18-101_k141_367651 | Not-determined |
| 18-101_k141_70392 | Not-determined |
| 18-101_k141_54574 | Not-determined |
| 18-101_k141_54575 | Not-determined |
| 18-101_k141_87450 | Not-determined |
| 18-101_k141_285412 | Not-determined |
| 18-101_k141_70399 | Low-quality |
| 18-101_k141_70400 | Not-determined |
| 18-101_k141_318791 | Not-determined |
| 18-101_k141_367669 | Not-determined |
| 18-101_k141_153644 | Not-determined |
| 18-101_k141_153647 | Not-determined |
| 18-101_k141_153648 | Not-determined |
| 18-101_k141_384002 | Not-determined |
| 18-101_k141_21869 | Not-determined |
| 18-101_k141_302555 | Not-determined |
| 18-101_k141_187097 | Low-quality |
| 18-101_k141_54591 | Low-quality |
| 18-101_k141_302560 | Not-determined |
| 18-101_k141_70415 | Not-determined |
| 18-101_k141_203254 | Not-determined |
| 18-101_k141_21876 | Not-determined |
| 18-101_k141_334226 | Not-determined |
| 18-101_k141_70421 | Not-determined |
| 18-101_k141_236030 | Not-determined |
| 18-101_k141_384022 | Not-determined |
| 18-101_k141_38351 | Not-determined |
| 18-101_k141_5426 | Not-determined |
| 18-101_k141_54605 | Not-determined |
| 18-101_k141_21887 | Not-determined |
| 18-101_k141_367690 | Low-quality |
| 18-101_k141_384029 | Not-determined |
| 18-101_k141_21890 | Not-determined |
| 18-101_k141_38359 | Not-determined |
| 18-101_k141_169435 | Not-determined |
| 18-101_k141_302571 | Not-determined |
| 18-101_k141_203263 | Low-quality |
| 18-101_k141_285451 | Not-determined |
| 18-101_k141_21903 | Not-determined |
| 18-101_k141_38364 | Not-determined |
| 18-101_k141_70432 | Low-quality |
| 18-101_k141_203268 | Low-quality |
| 18-101_k141_285457 | Low-quality |
| 18-101_k141_219629 | Not-determined |
| 18-101_k141_70436 | Not-determined |
| 18-101_k141_302585 | Not-determined |
| 18-101_k141_302586 | Not-determined |
| 18-101_k141_5439 | Not-determined |
| 18-101_k141_38375 | Not-determined |
| 18-101_k141_268609 | Not-determined |
| 18-101_k141_103483 | Not-determined |
| 18-101_k141_70442 | Not-determined |
| 18-101_k141_5448 | Not-determined |
| 18-101_k141_187144 | Not-determined |
| 18-101_k141_384039 | Not-determined |
| 18-101_k141_367717 | Not-determined |
| 18-101_k141_70455 | Not-determined |
| 18-101_k141_21921 | Not-determined |
| 18-101_k141_121291 | Not-determined |
| 18-101_k141_21923 | Low-quality |
| 18-101_k141_5451 | Not-determined |
| 18-101_k141_187155 | Low-quality |
| 18-101_k141_367725 | Not-determined |
| 18-101_k141_70459 | Not-determined |
| 18-101_k141_219656 | Not-determined |
| 18-101_k141_21936 | Not-determined |
| 18-101_k141_252917 | Not-determined |
| 18-101_k141_252918 | Not-determined |
| 18-101_k141_5455 | Not-determined |
| 18-101_k141_121300 | Not-determined |
| 18-101_k141_302605 | Not-determined |
| 18-101_k141_302606 | Not-determined |
| 18-101_k141_70467 | Not-determined |
| 18-101_k141_302607 | Not-determined |
| 18-101_k141_137804 | Not-determined |
| 18-101_k141_318837 | Not-determined |
| 18-101_k141_219670 | Not-determined |
| 18-101_k141_103504 | Not-determined |
| 18-101_k141_121308 | Not-determined |
| 18-101_k141_285486 | Not-determined |
| 18-101_k141_5465 | Not-determined |
| 18-101_k141_334246 | Not-determined |
| 18-101_k141_334248 | Not-determined |
| 18-101_k141_38410 | Low-quality |
| 18-101_k141_38414 | Not-determined |
| 18-101_k141_187188 | Not-determined |
| 18-101_k141_268624 | Not-determined |
| 18-101_k141_87492 | Not-determined |
| 18-101_k141_38419 | Not-determined |
| 18-101_k141_318856 | Not-determined |
| 18-101_k141_252937 | Not-determined |
| 18-101_k141_54651 | Not-determined |
| 18-101_k141_70493 | Low-quality |
| 18-101_k141_334270 | Not-determined |
| 18-101_k141_219688 | Not-determined |
| 18-101_k141_121321 | Not-determined |
| 18-101_k141_203320 | Not-determined |
| 18-101_k141_334277 | Not-determined |
| 18-101_k141_367781 | Not-determined |
| 18-101_k141_121333 | Not-determined |
| 18-101_k141_236096 | Not-determined |
| 18-101_k141_5474 | Not-determined |
| 18-101_k141_302636 | Not-determined |
| 18-101_k141_153684 | Not-determined |
| 18-101_k141_187223 | Not-determined |
| 18-101_k141_103524 | Not-determined |
| 18-101_k141_219712 | Not-determined |
| 18-101_k141_367789 | Not-determined |
| 18-101_k141_367790 | Not-determined |
| 18-101_k141_87497 | Not-determined |
| 18-101_k141_219716 | Not-determined |
| 18-101_k141_285493 | Not-determined |
| 18-101_k141_268647 | Not-determined |
| 18-101_k141_38435 | Not-determined |
| 18-101_k141_351646 | Not-determined |
| 18-101_k141_351647 | Not-determined |
| 18-101_k141_236104 | Not-determined |
| 18-101_k141_5497 | Not-determined |
| 18-101_k141_187236 | Not-determined |
| 18-101_k141_187237 | Not-determined |
| 18-101_k141_367809 | Low-quality |
| 18-101_k141_285502 | Not-determined |
| 18-101_k141_285503 | Not-determined |
| 18-101_k141_384095 | Not-determined |
| 18-101_k141_285506 | Low-quality |
| 18-101_k141_21965 | Not-determined |
| 18-101_k141_203355 | Not-determined |
| 18-101_k141_203358 | Not-determined |
| 18-101_k141_38450 | Not-determined |
| 18-101_k141_38451 | Low-quality |
| 18-101_k141_268663 | Not-determined |
| 18-101_k141_121337 | Not-determined |
| 18-101_k141_103540 | Not-determined |
| 18-101_k141_285521 | Not-determined |
| 18-101_k141_252976 | Not-determined |
| 18-101_k141_5509 | Not-determined |
| 18-101_k141_169510 | Not-determined |
| 18-101_k141_21979 | Not-determined |
| 18-101_k141_351666 | Not-determined |
| 18-101_k141_121346 | Not-determined |
| 18-101_k141_5511 | Not-determined |

| | |
|---|---|
| 18-101_k141_285525 | Not-determined |
| 18-101_k141_5516 | Not-determined |
| 18-101_k141_187268 | Not-determined |
| 18-101_k141_219756 | Not-determined |
| 18-101_k141_103557 | Not-determined |
| 18-101_k141_5521 | Not-determined |
| 18-101_k141_169516 | Not-determined |
| 18-109-final-viral-combined_k141_72070\|\|0_partial | High-quality |
| 18-101_k141_367824 | Not-determined |
| 18-101_k141_351677 | Not-determined |
| 18-101_k141_351679 | Low-quality |
| 18-101_k141_367831 | Not-determined |
| 18-101_k141_285529 | Not-determined |
| 18-101_k141_203384 | Not-determined |
| 18-101_k141_367836 | Not-determined |
| 18-101_k141_268682 | Not-determined |
| 18-101_k141_54676 | Not-determined |
| 18-101_k141_203386 | Not-determined |
| 18-101_k141_268684 | Low-quality |
| 18-101_k141_5536 | Low-quality |
| 18-101_k141_285530 | Low-quality |
| 18-101_k141_367840 | Not-determined |
| 18-101_k141_103568 | Not-determined |
| 18-101_k141_285535 | Low-quality |
| 18-101_k141_103569 | Not-determined |
| 18-101_k141_169540 | Not-determined |
| 18-101_k141_103570 | Not-determined |
| 18-101_k141_268697 | Low-quality |
| 18-101_k141_187282 | Not-determined |
| 18-101_k141_384152 | Not-determined |
| 18-101_k141_302676 | Not-determined |
| 18-101_k141_187284 | Not-determined |
| 18-101_k141_367857 | Not-determined |
| 18-101_k141_268701 | Not-determined |
| 18-101_k141_5552 | Not-determined |
| 18-101_k141_103582 | Low-quality |
| 18-101_k141_54690 | Low-quality |
| 18-101_k141_203405 | Not-determined |
| 18-101_k141_285543 | Not-determined |
| 18-101_k141_38485 | Not-determined |
| 18-101_k141_38486 | Not-determined |
| 18-101_k141_187295 | Not-determined |
| 18-101_k141_70540 | Not-determined |
| 18-101_k141_384161 | Not-determined |
| 18-101_k141_87534 | Not-determined |
| 18-101_k141_203419 | Not-determined |
| 18-101_k141_38494 | Low-quality |
| 18-101_k141_187304 | Not-determined |
| 18-101_k141_38495 | Not-determined |
| 18-101_k141_121370 | Not-determined |
| 18-101_k141_384171 | Low-quality |
| 18-101_k141_187308 | Not-determined |
| 18-101_k141_302709 | Not-determined |
| 18-101_k141_5570 | Not-determined |
| 18-101_k141_384176 | Not-determined |
| 18-101_k141_103605 | Not-determined |
| 18-101_k141_384177 | Not-determined |
| 18-101_k141_384178 | Not-determined |
| 18-101_k141_70561 | Not-determined |
| 18-101_k141_38498 | Not-determined |
| 18-101_k141_203430 | Not-determined |
| 18-101_k141_38502 | Low-quality |
| 18-101_k141_137808 | Not-determined |
| 18-101_k141_351706 | Not-determined |
| 18-101_k141_87550 | Not-determined |
| 18-101_k141_38504 | Not-determined |
| 18-101_k141_367878 | Not-determined |
| 18-101_k141_103608 | Not-determined |
| 18-101_k141_318872 | Not-determined |
| 18-101_k141_87553 | Not-determined |
| 18-101_k141_187319 | Not-determined |
| 18-101_k141_38506 | Not-determined |
| 18-101_k141_87556 | Low-quality |
| 18-101_k141_137819 | Not-determined |
| 18-101_k141_87559 | Not-determined |
| 18-101_k141_187326 | Not-determined |
| 18-101_k141_203434 | Low-quality |
| 18-101_k141_38515 | Low-quality |
| 18-101_k141_203435 | Not-determined |
| 18-101_k141_38516 | Not-determined |
| 18-101_k141_302721 | Not-determined |
| 18-101_k141_203436 | Low-quality |
| 18-101_k141_38517 | Not-determined |
| 18-101_k141_351729 | Not-determined |
| 18-101_k141_253021 | Not-determined |
| 18-101_k141_351731 | Not-determined |
| 18-101_k141_121381 | Not-determined |
| 18-101_k141_70579 | Not-determined |
| 18-101_k141_318881 | Not-determined |
| 18-101_k141_70580 | Not-determined |
| 18-101_k141_318882 | Not-determined |
| 18-101_k141_351738 | Not-determined |
| 18-101_k141_318883 | Not-determined |
| 18-101_k141_384192 | Not-determined |
| 18-101_k141_54719 | Not-determined |
| 18-101_k141_103622 | Not-determined |
| 18-101_k141_351743 | Not-determined |
| 18-101_k141_121397 | Not-determined |
| 18-101_k141_70594 | Not-determined |
| 18-101_k141_70595 | Not-determined |
| 18-101_k141_187335 | Low-quality |
| 18-101_k141_367889 | Not-determined |
| 18-101_k141_318896 | Not-determined |
| 18-101_k141_38525 | Not-determined |
| 18-101_k141_384200 | Not-determined |
| 18-101_k141_70602 | Not-determined |
| 18-101_k141_187339 | Not-determined |
| 18-101_k141_87602 | Not-determined |
| 18-101_k141_367894 | Not-determined |
| 18-101_k141_187343 | Not-determined |
| 18-101_k141_70609 | Low-quality |
| 18-101_k141_253047 | Not-determined |
| 18-101_k141_302752 | Not-determined |
| 18-101_k141_219781 | Not-determined |
| 18-101_k141_219784 | Not-determined |
| 18-101_k141_236152 | Low-quality |
| 18-101_k141_203470 | Not-determined |
| 18-101_k141_5608 | Not-determined |
| 18-101_k141_38547 | Not-determined |
| 18-101_k141_318925 | Not-determined |
| 18-101_k141_54737 | Not-determined |
| 18-101_k141_38549 | Not-determined |
| 18-101_k141_268745 | Not-determined |
| 18-101_k141_318929 | Not-determined |
| 18-101_k141_21997 | Low-quality |
| 18-101_k141_21998 | Not-determined |
| 18-101_k141_203488 | Not-determined |
| 18-101_k141_302776 | Not-determined |
| 18-101_k141_203490 | Not-determined |
| 18-101_k141_54746 | Not-determined |
| 18-101_k141_367916 | Not-determined |
| 18-101_k141_236182 | Not-determined |
| 18-101_k141_268758 | Not-determined |
| 18-101_k141_121411 | Not-determined |
| 18-101_k141_285600 | Not-determined |
| 18-101_k141_219826 | Not-determined |
| 18-101_k141_203503 | Low-quality |
| 18-101_k141_384219 | Not-determined |
| 18-101_k141_121421 | Not-determined |
| 18-101_k141_384225 | Not-determined |
| 18-101_k141_54772 | Not-determined |
| 18-101_k141_384229 | Not-determined |
| 18-101_k141_121427 | Not-determined |
| 18-101_k141_5640 | Not-determined |
| 18-101_k141_268772 | Not-determined |
| 18-101_k141_54777 | Not-determined |
| 18-101_k141_103649 | Not-determined |
| 18-101_k141_103664 | Not-determined |
| 18-101_k141_318951 | Not-determined |
| 18-101_k141_87643 | Not-determined |
| 18-101_k141_384253 | Not-determined |
| 18-101_k141_54782 | Not-determined |
| 18-101_k141_5664 | Not-determined |
| 18-101_k141_367939 | Low-quality |
| 18-101_k141_384257 | Low-quality |
| 18-101_k141_384258 | Not-determined |
| 18-101_k141_268792 | Not-determined |
| 18-101_k141_268794 | Not-determined |
| 18-101_k141_351774 | Not-determined |
| 18-101_k141_54793 | Not-determined |
| 18-101_k141_54794 | Not-determined |
| 18-101_k141_236216 | Not-determined |
| 18-101_k141_5682 | Not-determined |
| 18-101_k141_351781 | Not-determined |
| 18-101_k141_318957 | Not-determined |
| 18-101_k141_87670 | Not-determined |
| 18-101_k141_70655 | Not-determined |
| 18-101_k141_285629 | Low-quality |
| 18-101_k141_236219 | Not-determined |
| 18-101_k141_285630 | Not-determined |
| 18-101_k141_367952 | Not-determined |
| 18-101_k141_219875 | Not-determined |
| 18-101_k141_384281 | Not-determined |
| 18-101_k141_54804 | Not-determined |
| 18-101_k141_219880 | Not-determined |
| 18-101_k141_367956 | Not-determined |
| 18-101_k141_103691 | Not-determined |
| 18-101_k141_137899 | Not-determined |
| 18-101_k141_351790 | Not-determined |
| 18-101_k141_302802 | Not-determined |
| 18-101_k141_219887 | Not-determined |
| 18-101_k141_253091 | Not-determined |
| 18-101_k141_137905 | Not-determined |
| 18-101_k141_268821 | Not-determined |
| 18-101_k141_253094 | Low-quality |
| 18-101_k141_70674 | Not-determined |
| 18-101_k141_137910 | Not-determined |
| 18-101_k141_302812 | Not-determined |
| 18-101_k141_384290 | Not-determined |
| 18-101_k141_54820 | Not-determined |
| 18-101_k141_384298 | Not-determined |
| 18-101_k141_219904 | Not-determined |
| 18-101_k141_384299 | Not-determined |
| 18-101_k141_384300 | Not-determined |
| 18-101_k141_5710 | Not-determined |
| 18-101_k141_367978 | Not-determined |
| 18-101_k141_5714 | Not-determined |
| 18-101_k141_5715 | Not-determined |
| 18-101_k141_367982 | Not-determined |
| 18-101_k141_253105 | Not-determined |
| 18-101_k141_219918 | Not-determined |
| 18-101_k141_219919 | Not-determined |
| 18-101_k141_318985 | Low-quality |
| 18-101_k141_54834 | Not-determined |
| 18-101_k141_253112 | Low-quality |
| 18-101_k141_38608 | Not-determined |
| 18-101_k141_285647 | Not-determined |
| 18-101_k141_121468 | Low-quality |
| 18-101_k141_54839 | Not-determined |
| 18-101_k141_187430 | Not-determined |
| 18-101_k141_54842 | Not-determined |
| 18-101_k141_187436 | Not-determined |
| 18-101_k141_137930 | Not-determined |
| 18-101_k141_121472 | Not-determined |
| 18-101_k141_302821 | Low-quality |
| 18-101_k141_54846 | Not-determined |
| 18-101_k141_187440 | Not-determined |
| 18-101_k141_38616 | Not-determined |
| 18-101_k141_54850 | Not-determined |
| 18-101_k141_5740 | Not-determined |
| 18-101_k141_38622 | Not-determined |
| 18-101_k141_54856 | Not-determined |
| 18-101_k141_54859 | Not-determined |
| 18-101_k141_54860 | Low-quality |
| 18-101_k141_22026 | Not-determined |
| 18-101_k141_103732 | Not-determined |
| 18-101_k141_334293 | Not-determined |
| 18-101_k141_38630 | Not-determined |
| 18-101_k141_384321 | Not-determined |
| 18-101_k141_70686 | Not-determined |
| 18-101_k141_70687 | Not-determined |
| 18-101_k141_22032 | Not-determined |
| 18-101_k141_268854 | Not-determined |
| 18-101_k141_268856 | Not-determined |
| 18-101_k141_219927 | Not-determined |
| 18-101_k141_70695 | Not-determined |
| 18-101_k141_368004 | Not-determined |
| 18-101_k141_38644 | Not-determined |
| 18-101_k141_169648 | Not-determined |
| 18-101_k141_351829 | Not-determined |
| 18-101_k141_351830 | Not-determined |
| 18-101_k141_268868 | Not-determined |
| 18-101_k141_54870 | Not-determined |
| 18-101_k141_121498 | Not-determined |
| 18-101_k141_54874 | Not-determined |
| 18-101_k141_319009 | Not-determined |
| 18-101_k141_334311 | Not-determined |
| 18-101_k141_54880 | Not-determined |
| 18-101_k141_236261 | Not-determined |
| 18-101_k141_268882 | Not-determined |
| 18-101_k141_70710 | Low-quality |
| 18-101_k141_236263 | Not-determined |
| 18-101_k141_285687 | Not-determined |
| 18-101_k141_351853 | Not-determined |
| 18-101_k141_285689 | Not-determined |
| 18-101_k141_203554 | Not-determined |
| 18-101_k141_334331 | Not-determined |
| 18-101_k141_203556 | Not-determined |
| 18-101_k141_70716 | Not-determined |
| 18-101_k141_54893 | Not-determined |
| 18-101_k141_137976 | Not-determined |
| 18-101_k141_319018 | Low-quality |
| 18-101_k141_70723 | Not-determined |
| 18-101_k141_236266 | Not-determined |
| 18-101_k141_219950 | Not-determined |
| 18-101_k141_121528 | Low-quality |
| 18-101_k141_302825 | Not-determined |
| 18-101_k141_384354 | Not-determined |
| 18-101_k141_253124 | Not-determined |
| 18-101_k141_334344 | Not-determined |
| 18-101_k141_253127 | Not-determined |
| 18-101_k141_319020 | Not-determined |
| 18-101_k141_302829 | Not-determined |
| 18-101_k141_351866 | Low-quality |
| 18-101_k141_319025 | Not-determined |
| 18-101_k141_334353 | Not-determined |
| 18-101_k141_54898 | Not-determined |
| 18-101_k141_54901 | Not-determined |
| 18-101_k141_70746 | Not-determined |
| 18-101_k141_368038 | Not-determined |
| 18-101_k141_319035 | Not-determined |
| 18-101_k141_70752 | Not-determined |
| 18-101_k141_302832 | Not-determined |
| 18-101_k141_219973 | Not-determined |
| 18-101_k141_302833 | Not-determined |
| 18-101_k141_70758 | Not-determined |
| 18-101_k141_219980 | Not-determined |
| 18-101_k141_54910 | Not-determined |
| 18-101_k141_219986 | Not-determined |
| 18-101_k141_384383 | Not-determined |
| 18-101_k141_351881 | Not-determined |
| 18-101_k141_22068 | Not-determined |
| 18-101_k141_187464 | Not-determined |
| 18-101_k141_384388 | Not-determined |
| 18-101_k141_334365 | Not-determined |
| 18-101_k141_54923 | Low-quality |
| 18-101_k141_87721 | Low-quality |
| 18-101_k141_285699 | Not-determined |
| 18-101_k141_368049 | Not-determined |
| 18-101_k141_285706 | Not-determined |
| 18-101_k141_253140 | Not-determined |
| 18-101_k141_384403 | Not-determined |
| 18-101_k141_121556 | Not-determined |
| 18-101_k141_121560 | Not-determined |
| 18-101_k141_153692 | Not-determined |
| 18-101_k141_121563 | Not-determined |
| 18-101_k141_169680 | Not-determined |
| 18-101_k141_187481 | Not-determined |
| 18-101_k141_236276 | Not-determined |
| 18-101_k141_220006 | Not-determined |
| 18-101_k141_187499 | Not-determined |
| 18-101_k141_236280 | Not-determined |
| 18-101_k141_169697 | Not-determined |
| 18-101_k141_137996 | Not-determined |
| 18-101_k141_169701 | Not-determined |
| 18-101_k141_153725 | Not-determined |
| 18-101_k141_54968 | Not-determined |
| 18-101_k141_220010 | Not-determined |
| 18-101_k141_87759 | Not-determined |
| 18-101_k141_70783 | Not-determined |
| 18-101_k141_220016 | Not-determined |
| 18-101_k141_236287 | Not-determined |
| 18-101_k141_220017 | Not-determined |
| 18-101_k141_70784 | Not-determined |
| 18-101_k141_302840 | Not-determined |
| 18-101_k141_302841 | Not-determined |
| 18-101_k141_70789 | Low-quality |
| 18-101_k141_87767 | Not-determined |
| 18-101_k141_319084 | Not-determined |
| 18-101_k141_236292 | Not-determined |
| 18-101_k141_253171 | Not-determined |
| 18-101_k141_103793 | Not-determined |
| 18-101_k141_87772 | Not-determined |
| 18-101_k141_253172 | Not-determined |
| 18-101_k141_103800 | Not-determined |
| 18-101_k141_121603 | Low-quality |
| 18-101_k141_236298 | Not-determined |
| 18-101_k141_368077 | Low-quality |
| 18-101_k141_169725 | Not-determined |
| 18-101_k141_121609 | Not-determined |
| 18-101_k141_121612 | Not-determined |
| 18-101_k141_268932 | Not-determined |
| 18-101_k141_268933 | Not-determined |
| 18-101_k141_70803 | Not-determined |
| 18-101_k141_351901 | Not-determined |
| 18-101_k141_334388 | Not-determined |
| 18-101_k141_368086 | Not-determined |
| 18-101_k141_268942 | Not-determined |
| 18-101_k141_153746 | Not-determined |
| 18-101_k141_70814 | Not-determined |
| 18-101_k141_351912 | Not-determined |
| 18-101_k141_368091 | Not-determined |
| 18-101_k141_351916 | Not-determined |
| 18-101_k141_153757 | Not-determined |
| 18-101_k141_121626 | Not-determined |
| 18-101_k141_268946 | Not-determined |
| 18-101_k141_70825 | Not-determined |
| 18-101_k141_253181 | Not-determined |
| 18-101_k141_384442 | Not-determined |
| 18-101_k141_103822 | Not-determined |
| 18-101_k141_153774 | Not-determined |
| 18-101_k141_70828 | Not-determined |
| 18-101_k141_70832 | Low-quality |
| 18-101_k141_38669 | Not-determined |
| 18-101_k141_70837 | Not-determined |
| 18-101_k141_302847 | Not-determined |
| 18-101_k141_169740 | Low-quality |
| 18-101_k141_302851 | Not-determined |
| 18-101_k141_138037 | Not-determined |
| 18-101_k141_268961 | Not-determined |
| 18-101_k141_103838 | Not-determined |
| 18-101_k141_22097 | Low-quality |
| 18-101_k141_302857 | Not-determined |
| 18-101_k141_54992 | Not-determined |
| 18-101_k141_253188 | Low-quality |
| 18-101_k141_236354 | Not-determined |
| 18-101_k141_253190 | Not-determined |
| 18-101_k141_103848 | Not-determined |
| 18-101_k141_285737 | Low-quality |
| 18-101_k141_121652 | Not-determined |
| 18-101_k141_368104 | Low-quality |
| 18-101_k141_121653 | Not-determined |
| 18-101_k141_121654 | Low-quality |
| 18-101_k141_253200 | Not-determined |
| 18-101_k141_22111 | Not-determined |
| 18-101_k141_5766 | Not-determined |
| 18-101_k141_334403 | Not-determined |
| 18-101_k141_334405 | Not-determined |
| 18-101_k141_22119 | Not-determined |
| 18-101_k141_334416 | Not-determined |
| 18-101_k141_203645 | Not-determined |
| 18-101_k141_334417 | Not-determined |
| 18-101_k141_334420 | Low-quality |
| 18-101_k141_153819 | Not-determined |
| 18-101_k141_55014 | Not-determined |
| 18-101_k141_384457 | Not-determined |
| 18-101_k141_5782 | Not-determined |
| 18-101_k141_22131 | Not-determined |
| 18-101_k141_70869 | Not-determined |
| 18-101_k141_38708 | Not-determined |
| 18-101_k141_268978 | Not-determined |
| 18-101_k141_203650 | Low-quality |
| 18-101_k141_236365 | Not-determined |
| 18-101_k141_334426 | Not-determined |
| 18-101_k141_22142 | Not-determined |
| 18-101_k141_203652 | Not-determined |
| 18-101_k141_121680 | Low-quality |
| 18-101_k141_5801 | Not-determined |
| 18-101_k141_334431 | Not-determined |
| 18-101_k141_334432 | Not-determined |
| 18-101_k141_138073 | Not-determined |
| 18-101_k141_334433 | Not-determined |
| 18-101_k141_55037 | Not-determined |
| 18-101_k141_334436 | Not-determined |
| 18-101_k141_334437 | Not-determined |
| 18-101_k141_268995 | Not-determined |
| 18-101_k141_138090 | Not-determined |
| 18-101_k141_302862 | Not-determined |
| 18-101_k141_103876 | Not-determined |
| 18-101_k141_236385 | Not-determined |
| 18-101_k141_236386 | Not-determined |
| 18-101_k141_169766 | Not-determined |
| 18-101_k141_103881 | Not-determined |
| 18-101_k141_203671 | Not-determined |
| 18-101_k141_121690 | Not-determined |
| 18-101_k141_302869 | Not-determined |
| 18-101_k141_103884 | Not-determined |
| 18-101_k141_5824 | Low-quality |
| 18-101_k141_302874 | Not-determined |
| 18-101_k141_87859 | Low-quality |
| 18-101_k141_121698 | Not-determined |
| 18-101_k141_87864 | Not-determined |
| 18-101_k141_169771 | Not-determined |
| 18-101_k141_169775 | Not-determined |
| 18-101_k141_103897 | Not-determined |
| 18-101_k141_236389 | Not-determined |
| 18-101_k141_87869 | Not-determined |
| 18-101_k141_55058 | Not-determined |
| 18-101_k141_55059 | Not-determined |
| 18-101_k141_169781 | Low-quality |
| 18-101_k141_169782 | Not-determined |
| 18-101_k141_169783 | Not-determined |
| 18-101_k141_302888 | Not-determined |
| 18-101_k141_55064 | Not-determined |
| 18-101_k141_236392 | Low-quality |
| 18-101_k141_236393 | Low-quality |
| 18-101_k141_38737 | Low-quality |
| 18-101_k141_319151 | Not-determined |
| 18-101_k141_22167 | Not-determined |
| 18-101_k141_203694 | Not-determined |
| 18-101_k141_319159 | Not-determined |
| 18-101_k141_319164 | Not-determined |
| 18-101_k141_38745 | Low-quality |
| 18-101_k141_351966 | Not-determined |
| 18-101_k141_203697 | Not-determined |
| 18-101_k141_334457 | Not-determined |
| 18-101_k141_70905 | Not-determined |
| 18-101_k141_153866 | Not-determined |
| 18-101_k141_351978 | Low-quality |
| 18-101_k141_203702 | Not-determined |
| 18-101_k141_55098 | Not-determined |
| 18-101_k141_121713 | Not-determined |
| 18-101_k141_334470 | Not-determined |
| 18-101_k141_103921 | Not-determined |
| 18-101_k141_103922 | Not-determined |
| 18-101_k141_203722 | Not-determined |
| 18-101_k141_103925 | Not-determined |
| 18-101_k141_121725 | Not-determined |
| 18-101_k141_121727 | Not-determined |
| 18-101_k141_334482 | Not-determined |
| 18-101_k141_253258 | Low-quality |
| 18-101_k141_121739 | Not-determined |
| 18-101_k141_169822 | Not-determined |
| 18-101_k141_269032 | Not-determined |
| 18-101_k141_253272 | Not-determined |
| 18-101_k141_121746 | Not-determined |
| 18-101_k141_138115 | Not-determined |
| 18-101_k141_38766 | Not-determined |
| 18-101_k141_253279 | Not-determined |
| 18-101_k141_319208 | Not-determined |
| 18-101_k141_269039 | Not-determined |
| 18-101_k141_319213 | Not-determined |
| 18-101_k141_334494 | Not-determined |
| 18-101_k141_70934 | Not-determined |
| 18-101_k141_285748 | Not-determined |
| 18-101_k141_302932 | Not-determined |
| 18-101_k141_70937 | Not-determined |
| 18-101_k141_285750 | Not-determined |
| 18-101_k141_352026 | Not-determined |
| 18-101_k141_352029 | Not-determined |
| 18-101_k141_352030 | Not-determined |
| 18-101_k141_285754 | Not-determined |
| 18-101_k141_302942 | Low-quality |
| 18-101_k141_302943 | Not-determined |
| 18-101_k141_285757 | Not-determined |
| 18-101_k141_319230 | Not-determined |
| 18-101_k141_285758 | Not-determined |
| 18-101_k141_236431 | Not-determined |
| 18-101_k141_302949 | Not-determined |
| 18-101_k141_153920 | Low-quality |
| 18-101_k141_153924 | Not-determined |
| 18-101_k141_334498 | Not-determined |
| 18-101_k141_5846 | Not-determined |
| 18-101_k141_334500 | Not-determined |
| 18-101_k141_334502 | Not-determined |
| 18-101_k141_334503 | Not-determined |
| 18-101_k141_70957 | Low-quality |
| 18-101_k141_38809 | Not-determined |
| 18-101_k141_253299 | Not-determined |
| 18-101_k141_38810 | Not-determined |
| 18-101_k141_138129 | Not-determined |
| 18-101_k141_38814 | Not-determined |
| 18-101_k141_285777 | Not-determined |
| 18-101_k141_269062 | Not-determined |
| 18-101_k141_253303 | Not-determined |
| 18-101_k141_220029 | Not-determined |
| 18-101_k141_169841 | Not-determined |
| 18-101_k141_121753 | Not-determined |
| 18-101_k141_253309 | Not-determined |
| 18-101_k141_169843 | Not-determined |
| 18-101_k141_38837 | Not-determined |
| 18-101_k141_352054 | Not-determined |
| 18-101_k141_203738 | Not-determined |
| 18-101_k141_121766 | Not-determined |
| 18-101_k141_319264 | Not-determined |
| 18-101_k141_203746 | Not-determined |
| 18-101_k141_384545 | Not-determined |
| 18-101_k141_203748 | Not-determined |
| 18-101_k141_38850 | Low-quality |
| 18-101_k141_22196 | Not-determined |
| 18-101_k141_352056 | Not-determined |
| 18-101_k141_253328 | Not-determined |
| 18-101_k141_319277 | Not-determined |
| 18-101_k141_253329 | Not-determined |
| 18-101_k141_253331 | Not-determined |
| 18-101_k141_70967 | Not-determined |
| 18-101_k141_138151 | Not-determined |
| 18-101_k141_153927 | Not-determined |
| 18-101_k141_169865 | Not-determined |
| 18-101_k141_138153 | Not-determined |
| 18-101_k141_169866 | Not-determined |
| 18-101_k141_203760 | Not-determined |
| 18-101_k141_319289 | Not-determined |
| 18-101_k141_153933 | Not-determined |
| 18-101_k141_169870 | Not-determined |
| 18-101_k141_334555 | Not-determined |
| 18-101_k141_220032 | Not-determined |
| 18-101_k141_384565 | Not-determined |
| 18-101_k141_138161 | Not-determined |
| 18-101_k141_70988 | Not-determined |
| 18-101_k141_5886 | Not-determined |
| 18-101_k141_103970 | Not-determined |
| 18-101_k141_187510 | Not-determined |
| 18-101_k141_269097 | Not-determined |
| 18-101_k141_38875 | Not-determined |
| 18-101_k141_352072 | Not-determined |
| 18-101_k141_38876 | Low-quality |
| 18-101_k141_236500 | Not-determined |
| 18-101_k141_285801 | Not-determined |
| 18-101_k141_38885 | Not-determined |
| 18-101_k141_269119 | Not-determined |
| 18-101_k141_334578 | Not-determined |
| 18-101_k141_22208 | Not-determined |
| 18-101_k141_70997 | Low-quality |
| 18-101_k141_103992 | Not-determined |
| 18-101_k141_153949 | Not-determined |
| 18-101_k141_103994 | Not-determined |
| 18-101_k141_103996 | Not-determined |
| 18-101_k141_103997 | Not-determined |
| 18-101_k141_285813 | Not-determined |
| 18-101_k141_103998 | Not-determined |
| 18-101_k141_5908 | Not-determined |
| 18-101_k141_220058 | Not-determined |
| 18-101_k141_384618 | Not-determined |
| 18-101_k141_71008 | Not-determined |
| 18-101_k141_5914 | Not-determined |
| 18-101_k141_55136 | Not-determined |
| 18-101_k141_138174 | Low-quality |
| 18-101_k141_236523 | Not-determined |
| 18-101_k141_285822 | Not-determined |
| 18-101_k141_285823 | Not-determined |
| 18-101_k141_153960 | Not-determined |
| 18-101_k141_352083 | Not-determined |
| 18-101_k141_187537 | Not-determined |
| 18-101_k141_104020 | Not-determined |
| 18-101_k141_253383 | Not-determined |
| 18-101_k141_121799 | Not-determined |
| 18-101_k141_22222 | Not-determined |
| 18-101_k141_55155 | Not-determined |

| | |
|---|---|
| 18-101_k141_138181 | Not-determined |
| 18-101_k141_104027 | Not-determined |
| 18-101_k141_352095 | Not-determined |
| 18-101_k141_352099 | Not-determined |
| 18-101_k141_104028 | Not-determined |
| 18-101_k141_22233 | Not-determined |
| 18-101_k141_285839 | Not-determined |
| 18-101_k141_285840 | Not-determined |
| 18-101_k141_5941 | Not-determined |
| 18-101_k141_269147 | Not-determined |
| 18-101_k141_38913 | Not-determined |
| 18-101_k141_71015 | Not-determined |
| 18-101_k141_203795 | Not-determined |
| 18-101_k141_187541 | Not-determined |
| 18-101_k141_334594 | Not-determined |
| 18-101_k141_220093 | Not-determined |
| 18-101_k141_253392 | Not-determined |
| 18-101_k141_384637 | Not-determined |
| 18-101_k141_169898 | Not-determined |
| 18-101_k141_334597 | Not-determined |
| 18-101_k141_38917 | Not-determined |
| 18-101_k141_121814 | Not-determined |
| 18-101_k141_104038 | Not-determined |
| 18-101_k141_55173 | Not-determined |
| 18-101_k141_285846 | Not-determined |
| 18-101_k141_187551 | Not-determined |
| 18-101_k141_203806 | Not-determined |
| 18-101_k141_187555 | Not-determined |
| 18-101_k141_319346 | Not-determined |
| 18-101_k141_187556 | Not-determined |
| 18-101_k141_104042 | Not-determined |
| 18-101_k141_384647 | Not-determined |
| 18-101_k141_121818 | Not-determined |
| 18-101_k141_319350 | Not-determined |
| 18-101_k141_104044 | Not-determined |
| 18-101_k141_203816 | Not-determined |
| 18-101_k141_169912 | Not-determined |
| 18-101_k141_220101 | Low-quality |
| 18-101_k141_187564 | Not-determined |
| 18-101_k141_187565 | Not-determined |
| 18-101_k141_253405 | Not-determined |
| 18-101_k141_22246 | Not-determined |
| 18-101_k141_334604 | Not-determined |
| 18-101_k141_253406 | Not-determined |
| 18-101_k141_384659 | Not-determined |
| 18-101_k141_104047 | Not-determined |
| 18-101_k141_253407 | Not-determined |
| 18-101_k141_352133 | Not-determined |
| 18-101_k141_55189 | Not-determined |
| 18-101_k141_169923 | Low-quality |
| 18-101_k141_285864 | Not-determined |
| 18-101_k141_203831 | Low-quality |
| 18-101_k141_384675 | Low-quality |
| 18-101_k141_55193 | Not-determined |
| 18-101_k141_55194 | Not-determined |
| 18-101_k141_169931 | Not-determined |
| 18-101_k141_55196 | Not-determined |
| 18-101_k141_236542 | Not-determined |
| 18-101_k141_22265 | Not-determined |
| 18-101_k141_169937 | Not-determined |
| 18-101_k141_236547 | Not-determined |
| 18-101_k141_38936 | Not-determined |
| 18-101_k141_187571 | Low-quality |
| 18-101_k141_253418 | Not-determined |
| 18-101_k141_153980 | Low-quality |
| 18-101_k141_87975 | Not-determined |
| 18-101_k141_253424 | Not-determined |
| 18-101_k141_22274 | Not-determined |
| 18-101_k141_384688 | Low-quality |
| 18-101_k141_153985 | Not-determined |
| 18-101_k141_384689 | Not-determined |
| 18-101_k141_22278 | Low-quality |
| 18-101_k141_153987 | Not-determined |
| 18-101_k141_285877 | Not-determined |
| 18-101_k141_38944 | Not-determined |
| 18-101_k141_253429 | Not-determined |
| 18-101_k141_236560 | Not-determined |
| 18-101_k141_38946 | Not-determined |
| 18-101_k141_153991 | Not-determined |
| 18-101_k141_38949 | Not-determined |
| 18-101_k141_5961 | Not-determined |
| 18-101_k141_236565 | Low-quality |
| 18-101_k141_236566 | Not-determined |
| 18-101_k141_22285 | Not-determined |
| 18-101_k141_87984 | Not-determined |
| 18-101_k141_285885 | Not-determined |
| 18-101_k141_104063 | Not-determined |
| 18-101_k141_285890 | Not-determined |
| 18-101_k141_203856 | Not-determined |
| 18-101_k141_153994 | Not-determined |
| 18-101_k141_236568 | Not-determined |
| 18-101_k141_55210 | Not-determined |
| 18-101_k141_153996 | Not-determined |
| 18-101_k141_236570 | Low-quality |
| 18-101_k141_22289 | Not-determined |
| 18-101_k141_104072 | Not-determined |
| 18-101_k141_352158 | Not-determined |
| 18-101_k141_302979 | Not-determined |
| 18-101_k141_22293 | Not-determined |
| 18-101_k141_319410 | Not-determined |
| 18-101_k141_319411 | Not-determined |
| 18-101_k141_187603 | Not-determined |
| 18-101_k141_187604 | Not-determined |
| 18-101_k141_55225 | Low-quality |
| 18-101_k141_352175 | Not-determined |
| 18-101_k141_71055 | Not-determined |
| 18-101_k141_121831 | Not-determined |
| 18-101_k141_121832 | Not-determined |
| 18-101_k141_253450 | Not-determined |
| 18-101_k141_121833 | Low-quality |
| 18-101_k141_121834 | Not-determined |
| 18-101_k141_236594 | Not-determined |
| 18-101_k141_220138 | Low-quality |
| 18-101_k141_220139 | Not-determined |
| 18-101_k141_352178 | Not-determined |
| 18-101_k141_384702 | Not-determined |
| 18-101_k141_5984 | Not-determined |
| 18-101_k141_5985 | Not-determined |
| 18-101_k141_88001 | Not-determined |
| 18-101_k141_5986 | Not-determined |
| 18-101_k141_236598 | Not-determined |
| 18-101_k141_285897 | Low-quality |
| 18-101_k141_285898 | Not-determined |
| 18-101_k141_319420 | Not-determined |
| 18-101_k141_203866 | Not-determined |
| 18-101_k141_203870 | Not-determined |
| 18-101_k141_203872 | Not-determined |
| 18-101_k141_88015 | Not-determined |
| 18-101_k141_38968 | Not-determined |
| 18-101_k141_38969 | Not-determined |
| 18-101_k141_38972 | Not-determined |
| 18-101_k141_352182 | Not-determined |
| 18-101_k141_319423 | Not-determined |
| 18-101_k141_303007 | Not-determined |
| 18-101_k141_55236 | Not-determined |
| 18-101_k141_71061 | Low-quality |
| 18-101_k141_88023 | Not-determined |
| 18-101_k141_6002 | Not-determined |
| 18-101_k141_352192 | Not-determined |
| 18-101_k141_6005 | Not-determined |
| 18-101_k141_319428 | Not-determined |
| 18-101_k141_121849 | Not-determined |
| 18-101_k141_38982 | Not-determined |
| 18-101_k141_334654 | Not-determined |
| 18-101_k141_203881 | Not-determined |
| 18-101_k141_334655 | Low-quality |
| 18-101_k141_384720 | Not-determined |
| 18-101_k141_303015 | Not-determined |
| 18-101_k141_154017 | Not-determined |
| 18-101_k141_236604 | Not-determined |
| 18-101_k141_6015 | Not-determined |
| 18-101_k141_88037 | Not-determined |
| 18-101_k141_38983 | Not-determined |
| 18-101_k141_285949 | Not-determined |
| 18-101_k141_169970 | Not-determined |
| 18-101_k141_121862 | Not-determined |
| 18-101_k141_71083 | Not-determined |
| 18-101_k141_303018 | Not-determined |
| 18-101_k141_187656 | Not-determined |
| 18-101_k141_55245 | Low-quality |
| 18-101_k141_71087 | Not-determined |
| 18-101_k141_55246 | Not-determined |
| 18-101_k141_154027 | Low-quality |
| 18-101_k141_104109 | Not-determined |
| 18-101_k141_169980 | Not-determined |
| 18-101_k141_203893 | Not-determined |
| 18-101_k141_303024 | Not-determined |
| 18-101_k141_121869 | Not-determined |
| 18-101_k141_71100 | Not-determined |
| 18-101_k141_187669 | Not-determined |
| 18-101_k141_169988 | Not-determined |
| 18-101_k141_334674 | Not-determined |
| 18-101_k141_334675 | Not-determined |
| 18-101_k141_22305 | Not-determined |
| 18-101_k141_203903 | Not-determined |
| 18-101_k141_187677 | Not-determined |
| 18-101_k141_352211 | Not-determined |
| 18-101_k141_303040 | Not-determined |
| 18-101_k141_334684 | Not-determined |
| 18-101_k141_22313 | Not-determined |
| 18-101_k141_71105 | Not-determined |
| 18-101_k141_352215 | Low-quality |
| 18-101_k141_55267 | Not-determined |
| 18-101_k141_55268 | Low-quality |
| 18-101_k141_285962 | Not-determined |
| 18-101_k141_71108 | Not-determined |
| 18-101_k141_285964 | Not-determined |
| 18-101_k141_203917 | Not-determined |
| 18-101_k141_303046 | Low-quality |
| 18-101_k141_352220 | Not-determined |
| 18-101_k141_285967 | Low-quality |
| 18-101_k141_187687 | Not-determined |
| 18-101_k141_334698 | Not-determined |
| 18-101_k141_285972 | Not-determined |
| 18-101_k141_104127 | Not-determined |
| 18-101_k141_253506 | Not-determined |
| 18-101_k141_104128 | Not-determined |
| 18-101_k141_203925 | Not-determined |
| 18-101_k141_187694 | Not-determined |
| 18-101_k141_187695 | Not-determined |
| 18-101_k141_203931 | Not-determined |
| 18-101_k141_303057 | Not-determined |
| 18-101_k141_170007 | Not-determined |
| 18-101_k141_384764 | Not-determined |
| 18-101_k141_384768 | Not-determined |
| 18-101_k141_384769 | Not-determined |
| 18-101_k141_187701 | Not-determined |
| 18-101_k141_121909 | Not-determined |
| 18-101_k141_303064 | Not-determined |
| 18-101_k141_104143 | Low-quality |
| 18-101_k141_88048 | Not-determined |
| 18-101_k141_319484 | Not-determined |
| 18-101_k141_187710 | Not-determined |
| 18-101_k141_384778 | Not-determined |
| 18-101_k141_203943 | Not-determined |
| 18-101_k141_203944 | Not-determined |
| 18-101_k141_121921 | Not-determined |
| 18-101_k141_187717 | Not-determined |
| 18-101_k141_352259 | Not-determined |
| 18-101_k141_104151 | Not-determined |
| 18-101_k141_121928 | Not-determined |
| 18-101_k141_22363 | Not-determined |
| 18-101_k141_187725 | Not-determined |
| 18-101_k141_187726 | Low-quality |
| 18-101_k141_121931 | Not-determined |
| 18-101_k141_203955 | Not-determined |
| 18-101_k141_88058 | Not-determined |
| 18-101_k141_220171 | Not-determined |
| 18-101_k141_170040 | Not-determined |
| 18-101_k141_121933 | Low-quality |
| 18-101_k141_303081 | Not-determined |
| 18-101_k141_303082 | Not-determined |
| 18-101_k141_39014 | Low-quality |
| 18-101_k141_121935 | Not-determined |
| 18-101_k141_303084 | Low-quality |
| 18-101_k141_319502 | Not-determined |
| 18-101_k141_71141 | Not-determined |
| 18-101_k141_334719 | Low-quality |
| 18-101_k141_104168 | Not-determined |
| 18-101_k141_203961 | Not-determined |
| 18-101_k141_22373 | Not-determined |
| 18-101_k141_187737 | Not-determined |
| 18-101_k141_253540 | Not-determined |
| 18-101_k141_253548 | Not-determined |
| 18-101_k141_170045 | Not-determined |
| 18-101_k141_303096 | Not-determined |
| 18-101_k141_269268 | Not-determined |
| 18-101_k141_253560 | Not-determined |
| 18-101_k141_253563 | Not-determined |
| 18-101_k141_269273 | Not-determined |
| 18-101_k141_71155 | Not-determined |
| 18-101_k141_203984 | Not-determined |
| 18-101_k141_253575 | Not-determined |
| 18-101_k141_55308 | Not-determined |
| 18-101_k141_121953 | Not-determined |
| 18-101_k141_55314 | Not-determined |
| 18-101_k141_55318 | Not-determined |
| 18-101_k141_22400 | Not-determined |
| 18-101_k141_55322 | Not-determined |
| 18-101_k141_88108 | Not-determined |
| 18-101_k141_121962 | Not-determined |
| 18-101_k141_269285 | Not-determined |
| 18-101_k141_253586 | Not-determined |
| 18-101_k141_22408 | Not-determined |
| 18-101_k141_138266 | Not-determined |
| 18-101_k141_22413 | Low-quality |
| 18-101_k141_187773 | Not-determined |
| 18-101_k141_55336 | Not-determined |
| 18-101_k141_6061 | Not-determined |
| 18-101_k141_121976 | Not-determined |
| 18-101_k141_220212 | Not-determined |
| 18-101_k141_71175 | Not-determined |
| 18-101_k141_104185 | Not-determined |
| 18-101_k141_104186 | Not-determined |
| 18-101_k141_138279 | Not-determined |
| 18-101_k141_170064 | Not-determined |
| 18-101_k141_22428 | Not-determined |
| 18-101_k141_104194 | Not-determined |
| 18-101_k141_71183 | Low-quality |
| 18-101_k141_138282 | Not-determined |
| 18-101_k141_55360 | Low-quality |
| 18-101_k141_220226 | Not-determined |
| 18-101_k141_122007 | Not-determined |
| 18-101_k141_352293 | Not-determined |
| 18-101_k141_154035 | Not-determined |
| 18-101_k141_303112 | Not-determined |
| 18-101_k141_253624 | Not-determined |
| 18-101_k141_6092 | Not-determined |
| 18-101_k141_303115 | Not-determined |
| 18-101_k141_352297 | Not-determined |
| 18-101_k141_6096 | Not-determined |
| 18-101_k141_104213 | Not-determined |
| 18-101_k141_187794 | Not-determined |
| 18-101_k141_154041 | Not-determined |
| 18-101_k141_154047 | Not-determined |
| 18-101_k141_286063 | Not-determined |
| 18-101_k141_6100 | Low-quality |
| 18-101_k141_6102 | Not-determined |
| 18-101_k141_253638 | Not-determined |
| 18-101_k141_122027 | Not-determined |
| 18-101_k141_154052 | Not-determined |
| 18-101_k141_22452 | Not-determined |
| 18-101_k141_352309 | Not-determined |
| 18-101_k141_71200 | Not-determined |
| 18-101_k141_39030 | Not-determined |
| 18-101_k141_88148 | Not-determined |
| 18-101_k141_154061 | Not-determined |
| 18-101_k141_6116 | Not-determined |
| 18-101_k141_122042 | Not-determined |
| 18-101_k141_122043 | Not-determined |
| 18-101_k141_88155 | Not-determined |
| 18-101_k141_384789 | Not-determined |
| 18-101_k141_88157 | Not-determined |
| 18-101_k141_352324 | Not-determined |
| 18-101_k141_384791 | Not-determined |
| 18-101_k141_88158 | Low-quality |
| 18-101_k141_6125 | Not-determined |
| 18-101_k141_236638 | Not-determined |
| 18-101_k141_6131 | Not-determined |
| 18-101_k141_88163 | Not-determined |
| 18-101_k141_236641 | Not-determined |
| 18-101_k141_6133 | Not-determined |
| 18-101_k141_253667 | Not-determined |
| 18-101_k141_39053 | Not-determined |
| 18-101_k141_154080 | Not-determined |
| 18-101_k141_253673 | Low-quality |
| 18-101_k141_253674 | Not-determined |
| 18-101_k141_187804 | Not-determined |
| 18-101_k141_187809 | Not-determined |
| 18-101_k141_88171 | Not-determined |
| 18-101_k141_204018 | Not-determined |
| 18-101_k141_253682 | Not-determined |
| 18-101_k141_88174 | Not-determined |
| 18-101_k141_204021 | Not-determined |
| 18-101_k141_384811 | Not-determined |
| 18-101_k141_71220 | Not-determined |
| 18-101_k141_55412 | Low-quality |
| 18-101_k141_352344 | Not-determined |
| 18-101_k141_154097 | Not-determined |
| 18-101_k141_303137 | Not-determined |
| 18-101_k141_334754 | Not-determined |
| 18-101_k141_204041 | Low-quality |
| 18-101_k141_204043 | Not-determined |
| 18-101_k141_122073 | Not-determined |
| 18-101_k141_334757 | Not-determined |
| 18-101_k141_303141 | Not-determined |
| 18-101_k141_303145 | Not-determined |
| 18-101_k141_88209 | Not-determined |
| 18-101_k141_104268 | Not-determined |
| 18-101_k141_384827 | Not-determined |
| 18-101_k141_334761 | Not-determined |
| 18-101_k141_154107 | Low-quality |
| 18-101_k141_319526 | Not-determined |
| 18-101_k141_319527 | Not-determined |
| 18-101_k141_319528 | Low-quality |
| 18-101_k141_204065 | Not-determined |
| 18-101_k141_39094 | Low-quality |
| 18-101_k141_204066 | Low-quality |
| 18-101_k141_236646 | Not-determined |
| 18-101_k141_187812 | Low-quality |
| 18-101_k141_55436 | Not-determined |
| 18-101_k141_334770 | Not-determined |
| 18-101_k141_55441 | Not-determined |
| 18-101_k141_204075 | Not-determined |
| 18-101_k141_104277 | Low-quality |
| 18-101_k141_104278 | Not-determined |
| 18-101_k141_236658 | Not-determined |
| 18-101_k141_170109 | Low-quality |
| 18-101_k141_170111 | Not-determined |
| 18-101_k141_39105 | Low-quality |
| 18-101_k141_303150 | Not-determined |
| 18-101_k141_334775 | Not-determined |
| 18-101_k141_104289 | Not-determined |
| 18-101_k141_39107 | Low-quality |
| 18-101_k141_334779 | Not-determined |
| 18-101_k141_334780 | Low-quality |
| 18-101_k141_269335 | Not-determined |
| 18-101_k141_236667 | Not-determined |
| 18-101_k141_170112 | Low-quality |
| 18-101_k141_39114 | Not-determined |
| 18-101_k141_384842 | Not-determined |
| 18-101_k141_253708 | Not-determined |
| 18-101_k141_71223 | Not-determined |
| 18-101_k141_236671 | Not-determined |
| 18-101_k141_204099 | Low-quality |
| 18-101_k141_334791 | Not-determined |
| 18-101_k141_71232 | Not-determined |
| 18-101_k141_22510 | Not-determined |
| 18-101_k141_154137 | Not-determined |
| 18-101_k141_334796 | Low-quality |
| 18-101_k141_122110 | Not-determined |
| 18-101_k141_6181 | Low-quality |
| 18-101_k141_39123 | Not-determined |
| 18-101_k141_22516 | Not-determined |
| 18-101_k141_39127 | Not-determined |
| 18-101_k141_319564 | Not-determined |
| 18-101_k141_384861 | Not-determined |
| 18-101_k141_187829 | Not-determined |
| 18-101_k141_384864 | Not-determined |
| 18-101_k141_384865 | Not-determined |
| 18-101_k141_319567 | Not-determined |
| 18-101_k141_104319 | Not-determined |
| 18-101_k141_319571 | Not-determined |
| 18-101_k141_6193 | Low-quality |
| 18-101_k141_71245 | Not-determined |
| 18-101_k141_319574 | Not-determined |
| 18-101_k141_319576 | Not-determined |
| 18-101_k141_319577 | Not-determined |
| 18-101_k141_154140 | Not-determined |
| 18-101_k141_303170 | Not-determined |
| 18-101_k141_303172 | Not-determined |
| 18-101_k141_253733 | Not-determined |
| 18-101_k141_187839 | Not-determined |
| 18-101_k141_253736 | Not-determined |
| 18-101_k141_253742 | Not-determined |
| 18-101_k141_55498 | Not-determined |
| 18-101_k141_269362 | Low-quality |
| 18-101_k141_204111 | Not-determined |
| 18-101_k141_269364 | Not-determined |
| 18-101_k141_22541 | Not-determined |
| 18-101_k141_269367 | Not-determined |
| 18-101_k141_269368 | Not-determined |
| 18-101_k141_55503 | Not-determined |
| 18-101_k141_204118 | Not-determined |
| 18-101_k141_269372 | Not-determined |
| 18-101_k141_154163 | Not-determined |
| 18-101_k141_286107 | Not-determined |
| 18-101_k141_384876 | Not-determined |
| 18-101_k141_319600 | Not-determined |
| 18-101_k141_269375 | Not-determined |
| 18-101_k141_303193 | Not-determined |
| 18-101_k141_319604 | Not-determined |
| 18-101_k141_303197 | Low-quality |
| 18-101_k141_352408 | Not-determined |
| 18-101_k141_352411 | Low-quality |
| 18-101_k141_122146 | Not-determined |
| 18-101_k141_122147 | Not-determined |
| 18-101_k141_236685 | Not-determined |
| 18-101_k141_22548 | Not-determined |
| 18-101_k141_22550 | Not-determined |
| 18-101_k141_55522 | Not-determined |
| 18-101_k141_253771 | Not-determined |
| 18-101_k141_253772 | Not-determined |
| 17-109-final-viral-combined_k141_323943\|\|full | High-quality |
| 18-101_k141_22561 | Not-determined |
| 18-101_k141_334877 | Not-determined |
| 18-101_k141_22564 | Not-determined |
| 18-101_k141_71289 | Not-determined |
| 18-101_k141_154186 | Not-determined |
| 18-101_k141_187852 | Low-quality |
| 18-101_k141_286132 | Not-determined |
| 18-101_k141_286133 | Not-determined |
| 18-101_k141_6203 | Not-determined |
| 18-101_k141_6206 | Not-determined |
| 18-101_k141_334889 | Not-determined |
| 18-101_k141_204136 | Not-determined |
| 18-101_k141_55541 | Not-determined |
| 18-101_k141_71292 | Not-determined |
| 18-101_k141_88258 | Not-determined |
| 18-101_k141_6210 | Not-determined |
| 18-101_k141_269396 | Not-determined |
| 18-101_k141_71294 | Not-determined |
| 18-101_k141_303203 | Not-determined |
| 18-101_k141_187858 | Not-determined |
| 18-101_k141_220247 | Not-determined |
| 18-101_k141_187861 | Not-determined |
| 18-101_k141_104336 | Not-determined |
| 18-101_k141_204142 | Not-determined |
| 18-101_k141_22591 | Not-determined |
| 18-101_k141_236720 | Not-determined |
| 18-101_k141_269415 | Not-determined |
| 18-101_k141_88269 | Not-determined |
| 18-101_k141_187870 | Not-determined |
| 18-101_k141_220263 | Not-determined |
| 18-101_k141_319632 | Not-determined |
| 18-101_k141_220274 | Not-determined |
| 18-101_k141_122165 | Not-determined |
| 18-101_k141_55554 | Low-quality |
| 18-101_k141_6230 | Not-determined |
| 18-101_k141_286157 | Not-determined |
| 18-101_k141_253793 | Not-determined |
| 18-101_k141_319646 | Not-determined |
| 18-101_k141_122171 | Not-determined |
| 18-101_k141_88280 | Not-determined |
| 18-101_k141_6233 | Not-determined |
| 18-101_k141_269426 | Not-determined |
| 18-101_k141_319649 | Not-determined |
| 18-101_k141_170161 | Not-determined |
| 18-101_k141_334914 | Low-quality |
| 18-101_k141_154201 | Not-determined |
| 18-101_k141_170162 | Not-determined |
| 18-101_k141_71338 | Not-determined |
| 18-101_k141_204159 | Not-determined |
| 18-101_k141_334919 | Not-determined |
| 18-101_k141_6236 | Not-determined |
| 18-101_k141_384921 | Not-determined |
| 18-101_k141_220294 | Not-determined |
| 18-101_k141_122181 | Not-determined |
| 18-101_k141_220297 | Not-determined |
| 18-101_k141_220300 | Not-determined |
| 18-101_k141_6242 | Not-determined |
| 18-101_k141_303230 | Not-determined |
| 18-101_k141_253808 | Low-quality |
| 18-101_k141_104357 | Not-determined |
| 18-101_k141_303233 | Not-determined |
| 18-101_k141_303236 | Not-determined |
| 18-101_k141_334935 | Not-determined |
| 18-101_k141_170164 | Not-determined |
| 18-101_k141_286189 | Not-determined |
| 18-101_k141_88300 | Not-determined |
| 18-101_k141_122190 | Not-determined |
| 18-101_k141_104359 | Low-quality |
| 18-101_k141_170172 | Not-determined |
| 18-101_k141_104362 | Not-determined |
| 18-101_k141_303246 | Not-determined |
| 18-101_k141_384937 | Not-determined |
| 18-101_k141_187900 | Not-determined |
| 18-101_k141_170177 | Not-determined |
| 18-101_k141_352432 | Not-determined |
| 18-101_k141_384939 | Not-determined |
| 18-101_k141_334945 | Not-determined |
| 18-101_k141_170182 | Not-determined |
| 18-101_k141_170187 | Not-determined |
| 18-101_k141_384944 | Not-determined |
| 18-101_k141_352438 | Not-determined |
| 18-101_k141_319675 | Low-quality |
| 18-101_k141_55582 | Not-determined |
| 18-101_k141_88306 | Not-determined |
| 18-101_k141_352440 | Not-determined |
| 18-101_k141_104371 | Not-determined |
| 18-101_k141_88314 | Not-determined |
| 18-101_k141_39176 | Low-quality |
| 18-101_k141_303276 | Not-determined |
| 18-101_k141_319682 | Low-quality |
| 18-101_k141_352445 | Low-quality |
| 18-101_k141_253832 | Not-determined |
| 18-101_k141_352449 | Not-determined |
| 18-101_k141_204192 | Not-determined |
| 18-101_k141_138359 | Not-determined |
| 18-101_k141_170204 | Not-determined |
| 18-101_k141_122211 | Not-determined |
| 18-101_k141_303288 | Not-determined |
| 18-101_k141_71360 | Low-quality |
| 18-101_k141_334962 | Not-determined |
| 18-101_k141_253844 | Not-determined |
| 18-101_k141_253845 | Not-determined |
| 18-101_k141_71362 | Not-determined |
| 18-101_k141_269490 | Not-determined |
| 18-101_k141_6291 | Not-determined |
| 18-101_k141_71373 | Not-determined |
| 18-101_k141_352479 | Not-determined |
| 18-101_k141_104373 | Low-quality |
| 18-101_k141_352483 | Not-determined |
| 18-101_k141_170211 | Not-determined |
| 18-101_k141_204198 | Not-determined |
| 18-101_k141_334971 | Not-determined |
| 18-101_k141_384963 | Low-quality |
| 18-101_k141_170214 | Not-determined |
| 18-101_k141_104376 | Not-determined |
| 18-101_k141_334974 | Not-determined |
| 18-101_k141_334975 | Not-determined |
| 18-101_k141_236792 | Not-determined |
| 18-101_k141_334976 | Not-determined |
| 18-101_k141_204205 | Not-determined |
| 18-101_k141_269498 | Not-determined |
| 18-101_k141_236795 | Not-determined |
| 18-101_k141_39214 | Not-determined |
| 18-101_k141_384966 | Not-determined |
| 18-101_k141_368173 | Not-determined |
| 18-101_k141_39217 | Not-determined |
| 18-101_k141_236798 | Low-quality |
| 18-101_k141_334981 | Not-determined |
| 18-101_k141_204210 | Not-determined |
| 18-101_k141_6297 | Not-determined |
| 18-101_k141_55594 | Low-quality |
| 18-101_k141_220334 | Low-quality |
| 18-101_k141_334993 | Not-determined |
| 18-101_k141_204212 | Not-determined |
| 18-101_k141_204214 | Not-determined |
| 18-101_k141_88346 | Not-determined |
| 18-101_k141_335003 | Not-determined |
| 18-101_k141_303302 | Not-determined |
| 18-101_k141_138406 | Not-determined |
| 18-101_k141_71398 | Not-determined |
| 18-101_k141_104388 | Not-determined |
| 18-101_k141_368193 | Not-determined |
| 18-101_k141_154235 | Not-determined |
| 18-101_k141_39227 | Not-determined |
| 18-101_k141_170217 | Not-determined |

| | |
|---|---|
| 18-101_k141_104397 | Not-determined |
| 18-101_k141_303312 | Not-determined |
| 18-101_k141_319707 | Not-determined |
| 18-101_k141_88355 | Not-determined |
| 18-101_k141_122259 | Not-determined |
| 18-101_k141_6312 | Not-determined |
| 18-101_k141_88359 | Not-determined |
| 18-101_k141_122264 | Low-quality |
| 18-101_k141_187939 | Not-determined |
| 18-101_k141_71416 | Not-determined |
| 18-101_k141_220350 | Low-quality |
| 18-101_k141_368211 | Not-determined |
| 18-101_k141_39235 | Not-determined |
| 18-101_k141_368212 | Low-quality |
| 18-101_k141_71423 | Not-determined |
| 18-101_k141_187944 | Not-determined |
| 18-101_k141_187950 | Low-quality |
| 18-101_k141_253864 | Not-determined |
| 18-101_k141_236815 | Not-determined |
| 18-101_k141_187958 | Not-determined |
| 18-101_k141_71427 | Not-determined |
| 18-101_k141_352508 | Not-determined |
| 18-101_k141_71432 | Not-determined |
| 18-101_k141_138430 | Not-determined |
| 18-101_k141_319717 | Not-determined |
| 18-101_k141_368245 | Not-determined |
| 18-101_k141_55640 | Not-determined |
| 18-101_k141_122297 | Not-determined |
| 18-101_k141_55643 | Not-determined |
| 18-101_k141_352525 | Not-determined |
| 18-101_k141_39244 | Not-determined |
| 18-101_k141_122304 | Not-determined |
| 18-101_k141_71443 | Not-determined |
| 18-101_k141_204236 | Not-determined |
| 18-101_k141_319726 | Not-determined |
| 18-101_k141_236851 | Not-determined |
| 18-101_k141_253887 | Not-determined |
| 18-101_k141_335020 | Low-quality |
| 18-101_k141_154261 | Not-determined |
| 18-101_k141_368257 | Not-determined |
| 18-101_k141_352532 | Not-determined |
| 18-101_k141_71456 | Not-determined |
| 18-101_k141_236859 | Not-determined |
| 18-101_k141_6339 | Not-determined |
| 18-101_k141_368262 | Not-determined |
| 18-101_k141_253902 | Not-determined |
| 18-101_k141_88413 | Low-quality |
| 18-101_k141_22645 | Not-determined |
| 18-101_k141_319743 | Not-determined |
| 18-101_k141_384985 | Not-determined |
| 18-101_k141_138476 | Not-determined |
| 18-101_k141_71468 | Low-quality |
| 18-101_k141_352552 | Low-quality |
| 18-101_k141_253915 | Not-determined |
| 18-101_k141_220371 | Not-determined |
| 18-101_k141_236879 | Not-determined |
| 18-101_k141_204266 | Low-quality |
| 18-101_k141_88428 | Not-determined |
| 18-101_k141_71472 | Low-quality |
| 18-101_k141_236884 | Not-determined |
| 18-101_k141_138487 | Not-determined |
| 18-101_k141_88433 | Not-determined |
| 18-101_k141_236885 | Not-determined |
| 18-101_k141_154294 | Not-determined |
| 18-101_k141_269552 | Not-determined |
| 18-101_k141_88441 | Not-determined |
| 18-101_k141_236890 | Not-determined |
| 18-101_k141_22663 | Not-determined |
| 18-101_k141_22665 | Not-determined |
| 18-101_k141_188009 | Not-determined |
| 18-101_k141_55652 | Not-determined |
| 18-101_k141_253932 | Not-determined |
| 18-101_k141_220388 | Not-determined |
| 18-101_k141_138506 | Not-determined |
| 18-101_k141_368276 | Not-determined |
| 18-101_k141_204280 | Not-determined |
| 18-101_k141_236903 | Not-determined |
| 18-101_k141_352575 | Low-quality |
| 18-101_k141_88453 | Not-determined |
| 18-101_k141_352576 | Not-determined |
| 18-101_k141_204283 | Not-determined |
| 18-101_k141_154304 | Low-quality |
| 18-101_k141_220391 | Not-determined |
| 18-101_k141_269561 | Not-determined |
| 18-101_k141_236913 | Not-determined |
| 18-101_k141_154305 | Low-quality |
| 18-101_k141_170246 | Not-determined |
| 18-101_k141_220396 | Not-determined |
| 18-101_k141_55663 | Not-determined |
| 18-101_k141_188016 | Not-determined |
| 18-101_k141_88467 | Not-determined |
| 18-101_k141_138520 | Not-determined |
| 18-101_k141_88471 | Not-determined |
| 18-101_k141_236927 | Not-determined |
| 18-101_k141_335055 | Not-determined |
| 18-101_k141_22680 | Not-determined |
| 18-101_k141_22681 | Not-determined |
| 18-101_k141_22684 | Not-determined |
| 18-101_k141_319770 | Not-determined |
| 18-101_k141_335063 | Not-determined |
| 18-101_k141_170264 | Not-determined |
| 18-101_k141_236941 | Not-determined |
| 18-101_k141_253943 | Not-determined |
| 18-101_k141_319773 | Not-determined |
| 18-101_k141_204312 | Not-determined |
| 18-101_k141_204314 | Not-determined |
| 18-101_k141_71492 | Not-determined |
| 18-101_k141_22698 | Low-quality |
| 18-101_k141_22699 | Low-quality |
| 18-101_k141_71498 | Not-determined |
| 18-101_k141_204318 | Not-determined |
| 18-101_k141_188038 | Not-determined |
| 18-101_k141_352618 | Not-determined |
| 18-101_k141_253955 | Not-determined |
| 18-101_k141_204322 | Not-determined |
| 18-101_k141_188042 | Not-determined |
| 18-101_k141_188043 | Low-quality |
| 18-101_k141_352626 | Not-determined |
| 18-101_k141_236950 | Not-determined |
| 18-101_k141_335074 | Not-determined |
| 18-101_k141_6409 | Not-determined |
| 18-101_k141_352633 | Not-determined |
| 18-101_k141_385020 | Not-determined |
| 18-101_k141_204333 | Low-quality |
| 18-101_k141_319808 | Not-determined |
| 18-101_k141_104438 | Not-determined |
| 18-101_k141_385027 | Not-determined |
| 18-101_k141_154328 | Low-quality |
| 18-101_k141_170275 | Not-determined |
| 18-101_k141_6422 | Not-determined |
| 18-101_k141_335094 | Not-determined |
| 18-101_k141_6423 | Not-determined |
| 18-101_k141_6425 | Not-determined |
| 18-101_k141_154337 | Not-determined |
| 18-101_k141_319813 | Not-determined |
| 18-101_k141_6429 | Not-determined |
| 18-101_k141_22740 | Low-quality |
| 18-101_k141_39267 | Not-determined |
| 18-101_k141_39268 | Low-quality |
| 18-101_k141_170289 | Not-determined |
| 18-101_k141_220406 | Not-determined |
| 18-101_k141_39270 | Not-determined |
| 18-101_k141_269577 | Not-determined |
| 18-101_k141_319825 | Not-determined |
| 18-101_k141_88507 | Not-determined |
| 18-101_k141_88508 | Not-determined |
| 18-101_k141_236970 | Not-determined |
| 18-101_k141_220420 | Not-determined |
| 18-101_k141_39284 | Not-determined |
| 18-101_k141_39285 | Not-determined |
| 18-101_k141_22748 | Not-determined |
| 18-101_k141_22751 | Not-determined |
| 18-101_k141_319833 | Not-determined |
| 18-101_k141_88516 | Not-determined |
| 18-101_k141_39291 | Not-determined |
| 18-101_k141_352666 | Not-determined |
| 18-101_k141_104465 | Not-determined |
| 18-101_k141_385075 | Not-determined |
| 18-101_k141_104470 | Low-quality |
| 18-101_k141_170314 | Not-determined |
| 18-101_k141_22760 | Not-determined |
| 18-101_k141_170317 | Not-determined |
| 18-101_k141_22762 | Not-determined |
| 18-101_k141_188086 | Not-determined |
| 18-101_k141_368314 | Not-determined |
| 18-101_k141_138568 | Not-determined |
| 18-101_k141_352677 | Not-determined |
| 18-101_k141_237003 | Not-determined |
| 18-101_k141_6452 | Low-quality |
| 18-101_k141_6453 | Not-determined |
| 18-101_k141_6457 | Not-determined |
| 18-101_k141_237007 | Not-determined |
| 18-101_k141_71546 | Not-determined |
| 18-101_k141_286259 | Not-determined |
| 18-101_k141_39323 | Not-determined |
| 18-101_k141_286262 | Not-determined |
| 18-101_k141_319864 | Not-determined |
| 18-101_k141_237013 | Not-determined |
| 18-101_k141_319869 | Not-determined |
| 18-101_k141_319870 | Not-determined |
| 18-101_k141_39334 | Not-determined |
| 18-101_k141_385114 | Not-determined |
| 18-101_k141_39336 | Not-determined |
| 18-101_k141_39338 | Not-determined |
| 18-101_k141_39343 | Not-determined |
| 18-101_k141_352682 | Not-determined |
| 18-101_k141_352684 | Not-determined |
| 18-101_k141_237025 | Not-determined |
| 18-101_k141_138594 | Not-determined |
| 18-101_k141_385123 | Not-determined |
| 18-101_k141_88546 | Not-determined |
| 18-101_k141_303335 | Not-determined |
| 18-101_k141_138601 | Low-quality |
| 18-101_k141_286268 | Not-determined |
| 18-101_k141_319891 | Not-determined |
| 18-101_k141_368334 | Low-quality |
| 18-101_k141_368335 | Not-determined |
| 18-101_k141_253973 | Not-determined |
| 18-101_k141_22788 | Not-determined |
| 18-101_k141_188124 | Not-determined |
| 18-101_k141_303347 | Not-determined |
| 18-101_k141_286279 | Not-determined |
| 18-101_k141_204399 | Not-determined |
| 18-101_k141_352704 | Not-determined |
| 18-101_k141_237037 | Not-determined |
| 18-101_k141_286283 | Not-determined |
| 18-101_k141_237040 | Not-determined |
| 18-101_k141_352708 | Not-determined |
| 18-101_k141_220447 | Not-determined |
| 18-101_k141_385158 | Low-quality |
| 18-101_k141_385161 | Not-determined |
| 18-101_k141_269592 | Not-determined |
| 18-101_k141_385163 | Not-determined |
| 18-101_k141_170339 | Not-determined |
| 18-101_k141_385167 | Not-determined |
| 18-101_k141_253994 | Not-determined |
| 18-101_k141_22828 | Low-quality |
| 18-101_k141_204421 | Not-determined |
| 18-101_k141_269601 | Not-determined |
| 18-101_k141_237052 | Not-determined |
| 18-101_k141_6504 | Not-determined |
| 18-101_k141_204427 | Not-determined |
| 18-101_k141_220457 | Not-determined |
| 18-101_k141_204428 | Low-quality |
| 18-101_k141_204429 | Not-determined |
| 18-101_k141_269605 | Low-quality |
| 18-101_k141_154403 | Not-determined |
| 18-101_k141_55720 | Not-determined |
| 18-101_k141_88588 | Not-determined |
| 18-101_k141_269613 | Not-determined |
| 18-101_k141_204435 | Not-determined |
| 18-101_k141_6508 | Not-determined |
| 18-101_k141_237068 | Not-determined |
| 18-101_k141_237071 | Not-determined |
| 18-101_k141_6512 | Not-determined |
| 18-101_k141_204442 | Not-determined |
| 18-101_k141_88596 | Not-determined |
| 18-101_k141_55729 | Not-determined |
| 18-101_k141_254006 | Not-determined |
| 18-101_k141_88599 | Not-determined |
| 18-101_k141_335113 | Not-determined |
| 18-101_k141_104477 | Not-determined |
| 18-101_k141_352715 | Not-determined |
| 18-101_k141_39369 | Not-determined |
| 18-101_k141_286315 | Not-determined |
| 18-101_k141_188158 | Not-determined |
| 18-101_k141_385192 | Low-quality |
| 18-101_k141_170356 | Not-determined |
| 18-101_k141_352718 | Not-determined |
| 18-101_k141_303378 | Not-determined |
| 18-101_k141_204447 | Not-determined |
| 18-101_k141_71575 | Not-determined |
| 18-101_k141_220475 | Not-determined |
| 18-101_k141_237080 | Not-determined |
| 18-101_k141_385200 | Not-determined |
| 18-101_k141_269629 | Low-quality |
| 18-101_k141_269632 | Not-determined |
| 18-101_k141_335125 | Not-determined |
| 18-101_k141_237087 | Not-determined |
| 18-101_k141_303384 | Not-determined |
| 18-101_k141_269637 | Not-determined |
| 18-101_k141_154411 | Low-quality |
| 18-101_k141_269641 | Not-determined |
| 18-101_k141_39384 | Not-determined |
| 18-101_k141_138643 | Not-determined |
| 18-101_k141_368364 | Not-determined |
| 18-101_k141_254036 | Not-determined |
| 18-101_k141_352732 | Not-determined |
| 18-101_k141_254037 | Not-determined |
| 18-101_k141_55764 | Not-determined |
| 18-101_k141_303392 | Not-determined |
| 18-101_k141_39392 | Not-determined |
| 18-101_k141_303394 | Not-determined |
| 18-101_k141_39396 | Not-determined |
| 18-101_k141_22875 | Not-determined |
| 18-101_k141_286336 | Not-determined |
| 18-101_k141_286338 | Not-determined |
| 18-101_k141_188178 | Not-determined |
| 18-101_k141_39406 | Not-determined |
| 18-101_k141_385205 | Not-determined |
| 18-101_k141_220502 | Not-determined |
| 18-101_k141_368389 | Not-determined |
| 18-101_k141_385207 | Not-determined |
| 18-101_k141_385208 | Not-determined |
| 18-101_k141_71605 | Not-determined |
| 18-101_k141_254049 | Not-determined |
| 18-101_k141_55776 | Low-quality |
| 18-101_k141_368393 | Not-determined |
| 18-101_k141_237102 | Not-determined |
| 18-101_k141_286346 | Not-determined |
| 18-101_k141_303413 | Not-determined |
| 18-101_k141_204482 | Not-determined |
| 18-101_k141_237105 | Not-determined |
| 18-101_k141_170368 | Not-determined |
| 18-101_k141_6535 | Not-determined |
| 18-101_k141_71614 | Not-determined |
| 18-101_k141_204488 | Not-determined |
| 18-101_k141_335165 | Not-determined |
| 18-101_k141_303414 | Not-determined |
| 18-101_k141_237112 | Not-determined |
| 18-101_k141_335170 | Not-determined |
| 18-101_k141_303419 | Not-determined |
| 18-101_k141_335171 | Not-determined |
| 18-101_k141_39413 | Not-determined |
| 18-101_k141_138697 | Not-determined |
| 18-101_k141_188200 | Low-quality |
| 18-101_k141_55791 | Not-determined |
| 18-101_k141_319934 | Not-determined |
| 18-101_k141_385221 | Low-quality |
| 18-101_k141_319938 | Not-determined |
| 18-101_k141_22894 | Not-determined |
| 18-101_k141_39419 | Not-determined |
| 18-101_k141_220529 | Low-quality |
| 18-101_k141_303428 | Not-determined |
| 18-101_k141_55799 | Not-determined |
| 18-101_k141_71635 | Not-determined |
| 18-101_k141_39426 | Low-quality |
| 18-101_k141_385232 | Low-quality |
| 18-101_k141_319949 | Not-determined |
| 18-101_k141_122506 | Not-determined |
| 18-101_k141_170386 | Not-determined |
| 18-101_k141_385234 | Not-determined |
| 18-101_k141_71640 | Not-determined |
| 18-101_k141_254074 | Not-determined |
| 18-101_k141_71642 | Not-determined |
| 18-101_k141_55808 | Not-determined |
| 18-101_k141_220544 | Not-determined |
| 18-101_k141_170397 | Low-quality |
| 18-101_k141_104511 | Low-quality |
| 18-101_k141_368410 | Not-determined |
| 18-101_k141_6563 | Not-determined |
| 18-101_k141_385243 | Not-determined |
| 18-101_k141_303456 | Not-determined |
| 18-101_k141_368414 | Not-determined |
| 18-101_k141_104516 | Not-determined |
| 18-101_k141_55819 | Not-determined |
| 18-101_k141_385257 | Not-determined |
| 18-101_k141_188204 | Not-determined |
| 18-101_k141_55821 | Low-quality |
| 18-101_k141_188206 | Not-determined |
| 18-101_k141_254088 | Low-quality |
| 18-101_k141_254089 | Not-determined |
| 18-101_k141_204504 | Not-determined |
| 18-101_k141_88621 | Not-determined |
| 18-101_k141_88622 | Not-determined |
| 18-101_k141_55828 | Not-determined |
| 18-101_k141_55830 | Not-determined |
| 18-101_k141_204519 | Not-determined |
| 18-101_k141_254098 | Low-quality |
| 18-101_k141_71651 | Not-determined |
| 18-101_k141_39447 | Not-determined |
| 18-101_k141_220553 | Not-determined |
| 18-101_k141_122532 | Not-determined |
| 18-101_k141_104532 | Not-determined |
| 18-101_k141_170410 | Low-quality |
| 18-101_k141_368430 | Not-determined |
| 18-101_k141_204529 | Not-determined |
| 18-101_k141_122535 | Not-determined |
| 18-101_k141_335180 | Not-determined |
| 18-101_k141_88631 | Not-determined |
| 18-101_k141_352761 | Not-determined |
| 18-101_k141_335184 | Low-quality |
| 18-101_k141_352764 | Not-determined |
| 18-101_k141_220562 | Not-determined |
| 18-101_k141_138736 | Not-determined |
| 18-101_k141_39454 | Not-determined |
| 18-101_k141_138737 | Not-determined |
| 18-101_k141_385288 | Not-determined |
| 18-101_k141_138740 | Not-determined |
| 18-101_k141_138742 | Not-determined |
| 18-101_k141_6605 | Not-determined |
| 18-101_k141_254106 | Not-determined |
| 18-101_k141_335195 | Not-determined |
| 18-101_k141_104550 | Not-determined |
| 18-101_k141_352772 | Not-determined |
| 18-101_k141_170421 | Not-determined |
| 18-101_k141_88645 | Not-determined |
| 18-101_k141_71663 | Not-determined |
| 18-101_k141_6607 | Not-determined |
| 18-101_k141_170425 | Not-determined |
| 18-101_k141_88649 | Not-determined |
| 18-101_k141_385304 | Low-quality |
| 18-101_k141_385305 | Low-quality |
| 18-101_k141_254119 | Not-determined |
| 18-101_k141_88651 | Not-determined |
| 18-101_k141_368456 | Not-determined |
| 18-101_k141_39471 | Not-determined |
| 18-101_k141_122560 | Not-determined |
| 18-101_k141_104568 | Not-determined |
| 18-101_k141_319993 | Not-determined |
| 18-101_k141_104577 | Not-determined |
| 18-101_k141_220585 | Low-quality |
| 18-101_k141_204543 | Not-determined |
| 18-101_k141_55871 | Not-determined |
| 18-101_k141_220587 | Not-determined |
| 18-101_k141_104586 | Not-determined |
| 18-101_k141_237181 | Not-determined |
| 18-101_k141_104589 | Low-quality |
| 18-101_k141_269756 | Not-determined |
| 18-101_k141_71673 | Not-determined |
| 18-101_k141_352806 | Not-determined |
| 18-101_k141_254138 | Not-determined |
| 18-101_k141_204553 | Not-determined |
| 18-101_k141_220590 | Not-determined |
| 18-101_k141_254141 | Not-determined |
| 18-101_k141_6631 | Not-determined |
| 18-101_k141_269760 | Not-determined |
| 18-101_k141_220594 | Not-determined |
| 18-101_k141_385328 | Not-determined |
| 18-101_k141_385330 | Not-determined |
| 18-101_k141_385331 | Not-determined |
| 18-101_k141_220600 | Not-determined |
| 18-101_k141_104605 | Not-determined |
| 18-101_k141_104606 | Not-determined |
| 18-101_k141_352817 | Not-determined |
| 18-101_k141_22920 | Not-determined |
| 18-101_k141_220605 | Not-determined |
| 18-101_k141_254161 | Not-determined |
| 18-101_k141_122600 | Not-determined |
| 18-101_k141_352827 | Low-quality |
| 18-101_k141_220612 | Not-determined |
| 18-101_k141_237202 | Not-determined |
| 18-101_k141_88699 | Low-quality |
| 18-101_k141_352836 | Not-determined |
| 18-101_k141_352837 | Not-determined |
| 18-101_k141_385352 | Not-determined |
| 18-101_k141_154442 | Not-determined |
| 18-101_k141_220618 | Not-determined |
| 18-101_k141_88709 | Not-determined |
| 18-101_k141_88710 | Not-determined |
| 18-101_k141_71700 | Low-quality |
| 18-101_k141_154446 | Not-determined |
| 18-101_k141_352845 | Not-determined |
| 18-101_k141_320024 | Not-determined |
| 18-101_k141_22937 | Not-determined |
| 18-101_k141_22940 | Not-determined |
| 18-101_k141_352850 | Not-determined |
| 18-101_k141_352851 | Not-determined |
| 18-101_k141_204570 | Not-determined |
| 18-101_k141_352853 | Low-quality |
| 18-101_k141_71708 | Not-determined |
| 18-101_k141_368506 | Not-determined |
| 18-101_k141_39501 | Not-determined |
| 18-101_k141_39507 | Not-determined |
| 18-101_k141_352863 | Not-determined |
| 18-101_k141_39508 | Not-determined |
| 18-101_k141_154461 | Low-quality |
| 18-101_k141_220637 | Not-determined |
| 18-101_k141_368512 | Not-determined |
| 18-101_k141_352865 | Not-determined |
| 18-101_k141_352867 | Not-determined |
| 18-101_k141_303535 | Not-determined |
| 18-101_k141_254176 | Low-quality |
| 18-101_k141_220641 | Not-determined |
| 18-101_k141_385366 | Not-determined |
| 18-101_k141_154467 | Not-determined |
| 18-101_k141_237230 | Not-determined |
| 18-101_k141_22951 | Not-determined |
| 18-101_k141_254182 | Not-determined |
| 18-101_k141_352879 | Not-determined |
| 18-101_k141_254183 | Not-determined |
| 18-101_k141_138755 | Low-quality |
| 18-101_k141_122621 | Not-determined |
| 18-101_k141_335234 | Not-determined |
| 18-101_k141_154472 | Not-determined |
| 18-101_k141_22954 | Not-determined |
| 18-101_k141_71719 | Not-determined |
| 18-101_k141_122628 | Not-determined |
| 18-101_k141_254194 | Not-determined |
| 18-101_k141_220655 | Not-determined |
| 18-101_k141_303546 | Not-determined |
| 18-101_k141_254196 | Not-determined |
| 18-101_k141_71722 | Not-determined |
| 18-101_k141_385384 | Not-determined |
| 18-101_k141_154481 | Not-determined |
| 18-101_k141_368515 | Low-quality |
| 18-101_k141_22965 | Not-determined |
| 18-101_k141_385390 | Not-determined |
| 18-101_k141_154487 | Not-determined |
| 18-101_k141_88730 | Not-determined |
| 18-101_k141_104617 | Not-determined |
| 18-101_k141_204609 | Low-quality |
| 18-101_k141_352904 | Not-determined |
| 18-101_k141_22973 | Not-determined |
| 18-101_k141_335261 | Not-determined |
| 18-101_k141_269823 | Not-determined |
| 18-101_k141_352914 | Not-determined |
| 18-101_k141_122647 | Not-determined |
| 18-101_k141_104624 | Not-determined |
| 18-101_k141_22980 | Not-determined |
| 18-101_k141_122650 | Not-determined |
| 18-101_k141_320059 | Not-determined |
| 18-101_k141_154506 | Not-determined |
| 18-101_k141_71752 | Low-quality |
| 18-101_k141_88745 | Not-determined |
| 18-101_k141_154509 | Not-determined |
| 18-101_k141_237253 | Not-determined |
| 18-101_k141_303576 | Not-determined |
| 18-101_k141_385414 | Not-determined |
| 18-101_k141_204628 | Not-determined |
| 18-101_k141_154513 | Low-quality |
| 18-101_k141_154514 | Not-determined |
| 18-101_k141_88748 | Not-determined |
| 18-101_k141_39532 | Not-determined |
| 18-101_k141_22995 | Not-determined |
| 18-101_k141_88753 | Not-determined |
| 18-101_k141_104632 | Not-determined |
| 18-101_k141_88755 | Not-determined |
| 18-101_k141_39538 | Not-determined |
| 18-101_k141_320073 | Not-determined |
| 18-101_k141_122658 | Not-determined |
| 18-101_k141_237260 | Not-determined |
| 18-101_k141_39540 | Not-determined |
| 18-101_k141_55893 | Not-determined |
| 18-101_k141_204639 | Not-determined |
| 18-101_k141_39544 | Not-determined |
| 18-101_k141_269846 | Low-quality |
| 18-101_k141_71759 | Not-determined |
| 18-101_k141_368530 | Not-determined |
| 18-101_k141_55902 | Not-determined |
| 18-101_k141_6684 | Not-determined |
| 18-101_k141_320080 | Not-determined |
| 18-101_k141_204650 | Not-determined |
| 18-101_k141_188241 | Low-quality |
| 18-101_k141_6693 | Not-determined |
| 18-101_k141_6695 | Not-determined |
| 18-101_k141_6696 | Not-determined |
| 18-101_k141_138821 | Not-determined |
| 18-101_k141_55920 | Not-determined |
| 18-101_k141_88771 | Not-determined |
| 18-101_k141_320094 | Low-quality |
| 18-101_k141_170452 | Not-determined |
| 18-101_k141_55923 | Low-quality |
| 18-101_k141_23031 | Not-determined |
| 18-101_k141_368544 | Not-determined |
| 18-101_k141_254254 | Not-determined |
| 18-101_k141_71768 | Not-determined |
| 18-101_k141_88782 | Not-determined |
| 18-101_k141_188257 | Not-determined |
| 18-101_k141_320103 | Low-quality |
| 18-101_k141_335284 | Not-determined |
| 18-101_k141_188266 | Not-determined |
| 18-101_k141_71776 | Not-determined |
| 18-101_k141_71779 | Not-determined |
| 18-101_k141_88785 | Not-determined |
| 18-101_k141_320109 | Low-quality |
| 18-101_k141_170469 | Not-determined |
| 18-101_k141_204671 | Not-determined |
| 18-101_k141_335285 | Low-quality |
| 18-101_k141_55933 | Not-determined |
| 18-101_k141_220717 | Not-determined |
| 18-101_k141_352948 | Not-determined |
| 18-101_k141_6715 | Low-quality |
| 18-101_k141_204676 | Low-quality |
| 18-101_k141_170478 | Not-determined |
| 18-101_k141_104665 | Not-determined |
| 18-101_k141_204678 | Not-determined |
| 18-101_k141_55940 | Not-determined |
| 18-101_k141_220726 | Not-determined |
| 18-101_k141_368561 | Not-determined |
| 18-101_k141_352954 | Not-determined |
| 18-101_k141_170484 | Not-determined |
| 18-101_k141_368566 | Not-determined |
| 18-101_k141_6725 | Not-determined |
| 18-101_k141_6729 | Not-determined |
| 18-101_k141_368574 | Not-determined |
| 18-101_k141_204688 | Not-determined |
| 18-101_k141_188281 | Not-determined |
| 18-101_k141_122684 | Low-quality |
| 18-101_k141_88814 | Not-determined |
| 18-101_k141_286385 | Not-determined |
| 18-101_k141_88817 | Not-determined |
| 18-101_k141_104676 | Not-determined |
| 18-101_k141_23054 | Not-determined |
| 18-101_k141_55966 | Not-determined |
| 18-101_k141_220737 | Not-determined |
| 18-101_k141_88823 | Not-determined |
| 18-101_k141_122699 | Not-determined |
| 18-101_k141_204701 | Not-determined |
| 18-101_k141_23067 | Not-determined |
| 18-101_k141_170498 | Not-determined |
| 18-101_k141_286399 | Not-determined |
| 18-101_k141_303614 | Not-determined |
| 18-101_k141_368586 | Not-determined |
| 18-101_k141_303615 | Not-determined |
| 18-101_k141_368587 | Not-determined |
| 18-101_k141_88826 | Not-determined |
| 18-101_k141_188299 | Not-determined |
| 18-101_k141_237317 | Not-determined |

| | |
|---|---|
| 18-101_k141_220748 | Not-determined |
| 18-101_k141_170507 | Low-quality |
| 18-101_k141_188309 | Not-determined |
| 18-101_k141_254300 | Not-determined |
| 18-101_k141_352989 | Not-determined |
| 18-101_k141_23072 | Not-determined |
| 18-101_k141_220756 | Low-quality |
| 18-101_k141_188313 | Not-determined |
| 18-101_k141_220758 | Not-determined |
| 18-101_k141_220760 | Not-determined |
| 18-101_k141_220761 | Not-determined |
| 18-101_k141_154529 | Not-determined |
| 18-101_k141_254303 | Not-determined |
| 18-101_k141_269887 | Not-determined |
| 18-101_k141_220770 | Not-determined |
| 18-101_k141_320152 | Low-quality |
| 18-101_k141_269888 | Not-determined |
| 18-101_k141_220772 | Not-determined |
| 18-101_k141_286413 | Not-determined |
| 18-101_k141_23074 | Not-determined |
| 18-101_k141_353000 | Not-determined |
| 18-101_k141_303637 | Not-determined |
| 18-101_k141_286417 | Not-determined |
| 18-101_k141_71821 | Not-determined |
| 18-101_k141_104710 | Not-determined |
| 18-101_k141_71824 | Not-determined |
| 18-101_k141_220774 | Not-determined |
| 18-101_k141_353003 | Not-determined |
| 18-101_k141_368601 | Not-determined |
| 18-101_k141_154544 | Not-determined |
| 18-101_k141_368607 | Not-determined |
| 18-101_k141_286424 | Not-determined |
| 18-101_k141_353009 | Not-determined |
| 18-101_k141_286426 | Not-determined |
| 18-101_k141_335307 | Not-determined |
| 18-101_k141_303648 | Not-determined |
| 18-101_k141_335312 | Not-determined |
| 18-101_k141_353013 | Not-determined |
| 18-101_k141_335316 | Not-determined |
| 18-101_k141_71834 | Not-determined |
| 18-101_k141_286433 | Not-determined |
| 18-101_k141_154561 | Not-determined |
| 18-101_k141_154564 | Not-determined |
| 18-101_k141_303661 | Not-determined |
| 18-101_k141_286440 | Not-determined |
| 18-101_k141_335317 | Not-determined |
| 18-101_k141_368616 | Not-determined |
| 18-101_k141_71844 | Not-determined |
| 18-101_k141_170533 | Low-quality |
| 18-101_k141_170539 | Not-determined |
| 18-101_k141_286450 | Not-determined |
| 18-101_k141_303676 | Not-determined |
| 18-101_k141_71855 | Not-determined |
| 18-101_k141_353025 | Not-determined |
| 18-101_k141_6735 | Not-determined |
| 18-101_k141_286456 | Not-determined |
| 18-101_k141_303680 | Not-determined |
| 18-101_k141_269919 | Not-determined |
| 18-101_k141_320173 | Not-determined |
| 18-101_k141_320174 | Not-determined |
| 18-101_k141_320175 | Not-determined |
| 18-101_k141_6742 | Not-determined |
| 18-101_k141_303685 | Not-determined |
| 18-101_k141_320183 | Not-determined |
| 18-101_k141_154588 | Not-determined |
| 18-101_k141_353026 | Not-determined |
| 18-101_k141_122708 | Not-determined |
| 18-101_k141_385430 | Not-determined |
| 18-101_k141_204759 | Not-determined |
| 18-101_k141_237361 | Not-determined |
| 18-101_k141_104733 | Not-determined |
| 18-101_k141_237366 | Low-quality |
| 18-101_k141_303694 | Not-determined |
| 18-101_k141_320192 | Not-determined |
| 18-101_k141_353034 | Not-determined |
| 18-101_k141_286465 | Not-determined |
| 18-101_k141_204772 | Not-determined |
| 18-101_k141_303700 | Not-determined |
| 18-101_k141_170559 | Not-determined |
| 18-101_k141_122728 | Not-determined |
| 18-101_k141_353040 | Low-quality |
| 18-101_k141_320205 | Not-determined |
| 18-101_k141_138837 | Not-determined |
| 18-101_k141_6754 | Not-determined |
| 18-101_k141_104761 | Not-determined |
| 18-101_k141_88858 | Not-determined |
| 18-101_k141_320216 | Not-determined |
| 18-101_k141_353056 | Low-quality |
| 18-101_k141_237377 | Not-determined |
| 18-101_k141_138853 | Not-determined |
| 18-101_k141_88869 | Not-determined |
| 18-101_k141_237382 | Not-determined |
| 18-101_k141_122750 | Not-determined |
| 18-101_k141_122754 | Not-determined |
| 18-101_k141_368673 | Not-determined |
| 18-101_k141_237388 | Not-determined |
| 18-101_k141_353071 | Not-determined |
| 18-101_k141_237393 | Not-determined |
| 18-101_k141_353080 | Not-determined |
| 18-101_k141_6782 | Not-determined |
| 18-101_k141_6784 | Not-determined |
| 18-101_k141_6785 | Not-determined |
| 18-101_k141_23107 | Not-determined |
| 18-101_k141_56031 | Not-determined |
| 18-101_k141_269939 | Not-determined |
| 18-101_k141_269940 | Not-determined |
| 18-101_k141_204796 | Not-determined |
| 18-101_k141_254310 | Not-determined |
| 18-101_k141_204799 | Not-determined |
| 18-101_k141_122762 | Not-determined |
| 18-101_k141_6792 | Not-determined |
| 18-101_k141_269942 | Low-quality |
| 18-101_k141_88895 | Not-determined |
| 18-101_k141_122771 | Not-determined |
| 18-101_k141_56044 | Not-determined |
| 18-101_k141_122774 | Not-determined |
| 18-101_k141_23124 | Not-determined |
| 18-101_k141_23126 | Not-determined |
| 18-101_k141_122779 | Not-determined |
| 18-101_k141_220794 | Not-determined |
| 18-101_k141_220795 | Not-determined |
| 18-101_k141_204801 | Not-determined |
| 18-101_k141_204802 | Not-determined |
| 18-101_k141_204804 | Not-determined |
| 18-101_k141_385442 | Not-determined |
| 18-101_k141_104810 | Low-quality |
| 18-101_k141_254320 | Low-quality |
| 18-101_k141_286496 | Not-determined |
| 18-101_k141_122789 | Not-determined |
| 18-101_k141_170612 | Not-determined |
| 18-101_k141_286501 | Not-determined |
| 18-101_k141_204811 | Not-determined |
| 18-101_k141_6802 | Not-determined |
| 18-101_k141_23140 | Low-quality |
| 18-101_k141_220809 | Not-determined |
| 18-101_k141_6805 | Not-determined |
| 18-101_k141_104822 | Not-determined |
| 18-101_k141_56068 | Not-determined |
| 18-101_k141_188353 | Not-determined |
| 18-101_k141_170614 | Not-determined |
| 18-101_k141_88917 | Not-determined |
| 18-101_k141_220813 | Not-determined |
| 18-101_k141_122805 | Not-determined |
| 18-101_k141_303744 | Not-determined |
| 18-101_k141_122810 | Not-determined |
| 18-101_k141_385460 | Not-determined |
| 18-101_k141_23148 | Not-determined |
| 18-101_k141_6818 | Not-determined |
| 18-101_k141_303749 | Not-determined |
| 18-101_k141_303750 | Low-quality |
| 18-101_k141_104839 | Not-determined |
| 18-101_k141_104840 | Not-determined |
| 18-101_k141_6823 | Low-quality |
| 18-101_k141_23153 | Not-determined |
| 18-101_k141_6826 | Not-determined |
| 18-101_k141_368707 | Not-determined |
| 18-101_k141_254352 | Not-determined |
| 18-101_k141_39583 | Not-determined |
| 18-101_k141_204827 | Not-determined |
| 18-101_k141_254354 | Not-determined |
| 18-101_k141_254358 | Low-quality |
| 18-101_k141_335373 | Not-determined |
| 18-101_k141_23164 | Not-determined |
| 18-101_k141_320251 | Not-determined |
| 18-101_k141_254359 | Not-determined |
| 18-101_k141_385473 | Not-determined |
| 18-101_k141_204835 | Not-determined |
| 18-101_k141_254361 | Not-determined |
| 18-101_k141_254362 | Not-determined |
| 18-101_k141_254369 | Not-determined |
| 18-101_k141_104847 | Not-determined |
| 18-101_k141_220818 | Not-determined |
| 18-101_k141_385482 | Not-determined |
| 18-101_k141_385483 | Not-determined |
| 18-101_k141_71906 | Low-quality |
| 18-101_k141_122838 | Not-determined |
| 18-101_k141_71915 | Not-determined |
| 18-101_k141_170654 | Not-determined |
| 18-101_k141_303771 | Low-quality |
| 18-101_k141_320283 | Not-determined |
| 18-101_k141_56099 | Not-determined |
| 18-101_k141_71921 | Not-determined |
| 18-101_k141_138880 | Not-determined |
| 18-101_k141_71925 | Not-determined |
| 18-101_k141_303777 | Not-determined |
| 18-101_k141_154615 | Not-determined |
| 18-101_k141_170665 | Not-determined |
| 18-101_k141_138888 | Not-determined |
| 18-101_k141_23185 | Not-determined |
| 18-101_k141_303786 | Not-determined |
| 18-101_k141_56115 | Not-determined |
| 18-101_k141_303788 | Not-determined |
| 18-101_k141_303790 | Not-determined |
| 18-101_k141_320312 | Not-determined |
| 18-101_k141_122862 | Low-quality |
| 18-101_k141_353103 | Not-determined |
| 18-101_k141_303797 | Not-determined |
| 18-101_k141_269966 | Not-determined |
| 18-101_k141_237413 | Not-determined |
| 18-101_k141_56129 | Not-determined |
| 18-101_k141_303814 | Not-determined |
| 18-101_k141_303815 | Not-determined |
| 18-101_k141_269975 | Not-determined |
| 18-101_k141_204853 | Not-determined |
| 18-101_k141_122869 | Low-quality |
| 18-101_k141_303820 | Not-determined |
| 18-101_k141_335417 | Low-quality |
| 18-101_k141_269978 | Not-determined |
| 18-101_k141_154622 | Not-determined |
| 18-101_k141_104895 | Not-determined |
| 18-101_k141_303825 | Not-determined |
| 18-101_k141_368739 | Not-determined |
| 18-101_k141_104900 | Not-determined |
| 18-101_k141_320344 | Not-determined |
| 18-101_k141_335424 | Low-quality |
| 18-101_k141_335425 | Not-determined |
| 18-101_k141_170700 | Not-determined |
| 18-101_k141_71954 | Not-determined |
| 18-101_k141_23229 | Not-determined |
| 18-101_k141_39650 | Not-determined |
| 18-101_k141_122896 | Not-determined |
| 18-101_k141_154632 | Not-determined |
| 18-101_k141_254391 | Not-determined |
| 18-101_k141_204865 | Not-determined |
| 18-101_k141_188426 | Not-determined |
| 18-101_k141_254395 | Not-determined |
| 18-101_k141_138940 | Not-determined |
| 18-101_k141_237440 | Not-determined |
| 18-101_k141_204866 | Not-determined |
| 18-101_k141_237445 | Not-determined |
| 18-101_k141_71966 | Not-determined |
| 18-101_k141_237453 | Low-quality |
| 18-101_k141_220852 | Not-determined |
| 18-101_k141_237458 | Not-determined |
| 18-101_k141_335462 | Not-determined |
| 18-101_k141_270011 | Not-determined |
| 18-101_k141_220859 | Not-determined |
| 18-101_k141_122916 | Not-determined |
| 18-101_k141_71984 | Not-determined |
| 18-101_k141_254421 | Not-determined |
| 18-101_k141_6900 | Low-quality |
| 18-101_k141_368770 | Not-determined |
| 18-101_k141_237477 | Not-determined |
| 18-101_k141_335476 | Not-determined |
| 18-101_k141_303856 | Not-determined |
| 18-101_k141_353156 | Not-determined |
| 18-101_k141_188431 | Low-quality |
| 18-101_k141_154675 | Not-determined |
| 18-101_k141_170712 | Not-determined |
| 18-101_k141_320365 | Not-determined |
| 18-101_k141_122930 | Not-determined |
| 18-101_k141_237481 | Not-determined |
| 18-101_k141_368775 | Not-determined |
| 18-101_k141_237485 | Not-determined |
| 18-101_k141_353167 | Low-quality |
| 18-101_k141_368780 | Not-determined |
| 18-101_k141_368783 | Not-determined |
| 18-101_k141_122934 | Not-determined |
| 18-101_k141_154687 | Not-determined |
| 18-101_k141_72015 | Not-determined |
| 18-101_k141_188447 | Not-determined |
| 18-101_k141_303870 | Low-quality |
| 18-101_k141_303871 | Not-determined |
| 18-101_k141_385519 | Not-determined |
| 18-101_k141_237495 | Not-determined |
| 18-101_k141_72027 | Not-determined |
| 18-101_k141_385522 | Low-quality |
| 18-101_k141_303880 | Low-quality |
| 18-101_k141_154699 | Low-quality |
| 18-101_k141_237501 | Not-determined |
| 18-101_k141_154705 | Not-determined |
| 18-101_k141_368784 | Low-quality |
| 18-101_k141_368785 | Not-determined |
| 18-101_k141_254453 | Not-determined |
| 18-101_k141_170731 | Not-determined |
| 18-101_k141_320401 | Not-determined |
| 18-101_k141_170734 | Not-determined |
| 18-101_k141_368791 | Not-determined |
| 18-101_k141_154719 | Not-determined |
| 18-101_k141_320407 | Not-determined |
| 18-101_k141_237513 | Not-determined |
| 18-101_k141_320410 | Not-determined |
| 18-101_k141_138961 | Not-determined |
| 18-101_k141_220874 | Not-determined |
| 18-101_k141_237517 | Not-determined |
| 18-101_k141_56193 | Not-determined |
| 18-101_k141_6903 | Not-determined |
| 18-101_k141_385550 | Not-determined |
| 18-101_k141_56202 | Not-determined |
| 18-101_k141_188474 | Not-determined |
| 18-101_k141_56205 | Not-determined |
| 18-101_k141_368811 | Not-determined |
| 18-101_k141_220889 | Not-determined |
| 18-101_k141_88990 | Not-determined |
| 18-101_k141_368817 | Not-determined |
| 18-101_k141_286609 | Not-determined |
| 18-101_k141_188484 | Not-determined |
| 18-101_k141_56216 | Not-determined |
| 18-101_k141_368822 | Not-determined |
| 18-101_k141_89001 | Not-determined |
| 18-101_k141_286614 | Not-determined |
| 18-101_k141_6936 | Not-determined |
| 18-101_k141_270019 | Not-determined |
| 18-101_k141_204898 | Not-determined |
| 18-101_k141_385554 | Low-quality |
| 18-101_k141_6939 | Not-determined |
| 18-101_k141_303897 | Not-determined |
| 18-101_k141_237543 | Not-determined |
| 18-101_k141_237546 | Low-quality |
| 18-101_k141_56219 | Not-determined |
| 18-101_k141_39659 | Not-determined |
| 18-101_k141_254474 | Not-determined |
| 18-101_k141_204906 | Not-determined |
| 18-101_k141_39667 | Not-determined |
| 18-101_k141_89017 | Not-determined |
| 18-101_k141_89018 | Not-determined |
| 18-101_k141_353187 | Not-determined |
| 18-101_k141_254484 | Not-determined |
| 18-101_k141_237567 | Not-determined |
| 18-101_k141_188489 | Not-determined |
| 18-101_k141_89029 | Not-determined |
| 18-101_k141_320461 | Not-determined |
| 18-101_k141_154750 | Not-determined |
| 18-101_k141_39682 | Not-determined |
| 18-101_k141_139001 | Not-determined |
| 18-101_k141_23256 | Low-quality |
| 18-101_k141_139004 | Not-determined |
| 18-101_k141_237580 | Not-determined |
| 18-101_k141_204930 | Low-quality |
| 18-101_k141_122980 | Not-determined |
| 18-101_k141_23261 | Not-determined |
| 18-101_k141_139017 | Not-determined |
| 18-101_k141_170789 | Not-determined |
| 18-101_k141_204942 | Not-determined |
| 18-101_k141_89034 | Not-determined |
| 18-101_k141_170794 | Low-quality |
| 18-101_k141_188505 | Not-determined |
| 18-101_k141_170796 | Not-determined |
| 18-101_k141_270042 | Not-determined |
| 18-101_k141_254515 | Not-determined |
| 18-101_k141_335499 | Low-quality |
| 18-101_k141_39703 | Not-determined |
| 18-101_k141_39707 | Not-determined |
| 18-101_k141_56255 | Not-determined |
| 18-101_k141_254521 | Not-determined |
| 18-101_k141_237594 | Not-determined |
| 18-101_k141_188517 | Low-quality |
| 18-101_k141_368836 | Not-determined |
| 18-101_k141_254535 | Not-determined |
| 18-101_k141_335507 | Not-determined |
| 18-101_k141_170828 | Not-determined |
| 18-101_k141_123013 | Low-quality |
| 18-101_k141_335510 | Not-determined |
| 18-101_k141_139038 | Not-determined |
| 18-101_k141_7012 | Not-determined |
| 18-101_k141_368840 | Not-determined |
| 18-101_k141_89055 | Low-quality |
| 18-101_k141_7013 | Not-determined |
| 18-101_k141_368841 | Low-quality |
| 18-101_k141_254544 | Not-determined |
| 18-101_k141_139043 | Low-quality |
| 18-101_k141_368849 | Not-determined |
| 18-101_k141_368851 | Not-determined |
| 18-101_k141_139044 | Not-determined |
| 18-101_k141_335520 | Not-determined |
| 18-101_k141_154763 | Not-determined |
| 18-101_k141_335522 | Not-determined |
| 18-101_k141_385582 | Not-determined |
| 18-101_k141_39719 | Not-determined |
| 18-101_k141_89071 | Not-determined |
| 18-101_k141_154770 | Not-determined |
| 18-101_k141_154772 | Not-determined |
| 18-101_k141_154773 | Not-determined |
| 18-101_k141_154775 | Not-determined |
| 18-101_k141_154777 | Not-determined |
| 18-101_k141_254567 | Not-determined |
| 18-101_k141_320517 | Not-determined |
| 18-101_k141_254573 | Not-determined |
| 18-101_k141_7037 | Not-determined |
| 18-101_k141_303940 | Not-determined |
| 18-101_k141_154788 | Not-determined |
| 18-101_k141_286687 | Not-determined |
| 18-101_k141_368877 | Not-determined |
| 18-101_k141_123055 | Not-determined |
| 18-101_k141_7042 | Not-determined |
| 18-101_k141_7043 | Not-determined |
| 18-101_k141_56266 | Not-determined |
| 18-101_k141_139064 | Not-determined |
| 18-101_k141_270060 | Not-determined |
| 18-101_k141_7044 | Not-determined |
| 18-101_k141_368890 | Low-quality |
| 18-101_k141_237602 | Not-determined |
| 18-101_k141_204973 | Not-determined |
| 18-101_k141_7048 | Not-determined |
| 18-101_k141_7052 | Not-determined |
| 18-101_k141_303957 | Not-determined |
| 18-101_k141_270068 | Not-determined |
| 18-101_k141_237616 | Not-determined |
| 18-101_k141_170851 | Not-determined |
| 18-101_k141_139090 | Not-determined |
| 18-101_k141_188557 | Not-determined |
| 18-101_k141_320543 | Not-determined |
| 18-101_k141_320547 | Low-quality |
| 18-101_k141_320548 | Not-determined |
| 18-101_k141_139098 | Not-determined |
| 18-101_k141_353230 | Not-determined |
| 18-101_k141_204976 | Not-determined |
| 18-101_k141_204977 | Not-determined |
| 18-101_k141_286722 | Not-determined |
| 18-101_k141_139102 | Not-determined |
| 18-101_k141_320556 | Not-determined |
| 18-101_k141_188570 | Low-quality |
| 18-101_k141_286730 | Not-determined |
| 18-101_k141_188573 | Not-determined |
| 18-101_k141_188575 | Not-determined |
| 18-101_k141_270080 | Low-quality |
| 18-101_k141_320565 | Not-determined |
| 18-101_k141_7079 | Not-determined |
| 18-101_k141_303982 | Not-determined |
| 18-101_k141_353255 | Not-determined |
| 18-101_k141_188585 | Not-determined |
| 18-101_k141_320577 | Not-determined |
| 18-101_k141_286750 | Not-determined |
| 18-101_k141_56288 | Not-determined |
| 18-101_k141_39739 | Not-determined |
| 18-101_k141_286754 | Not-determined |
| 18-101_k141_286755 | Not-determined |
| 18-101_k141_123060 | Not-determined |
| 18-101_k141_286760 | Not-determined |
| 18-101_k141_237638 | Not-determined |
| 18-101_k141_286765 | Not-determined |
| 18-101_k141_303994 | Not-determined |
| 18-101_k141_303996 | Not-determined |
| 18-101_k141_56308 | Not-determined |
| 18-101_k141_368907 | Not-determined |
| 18-101_k141_335564 | Not-determined |
| 18-101_k141_286770 | Not-determined |
| 18-101_k141_89104 | Not-determined |
| 18-101_k141_56315 | Not-determined |
| 18-101_k141_270101 | Not-determined |
| 18-101_k141_89110 | Not-determined |
| 18-101_k141_56320 | Not-determined |
| 18-101_k141_89114 | Not-determined |
| 18-101_k141_286790 | Not-determined |
| 18-101_k141_368925 | Not-determined |
| 18-101_k141_154821 | Not-determined |
| 18-101_k141_56325 | Not-determined |
| 18-101_k141_353277 | Not-determined |
| 18-101_k141_123088 | Not-determined |
| 18-101_k141_320590 | Not-determined |
| 18-101_k141_72078 | Low-quality |
| 18-101_k141_385591 | Low-quality |
| 18-101_k141_286800 | Not-determined |
| 18-101_k141_320596 | Not-determined |
| 18-101_k141_320600 | Not-determined |
| 18-101_k141_23294 | Not-determined |
| 18-101_k141_320603 | Not-determined |
| 18-101_k141_304015 | Not-determined |
| 18-101_k141_188613 | Not-determined |
| 18-101_k141_353282 | Not-determined |
| 18-101_k141_353284 | Not-determined |
| 18-101_k141_170908 | Not-determined |
| 18-101_k141_72087 | Not-determined |
| 18-101_k141_254619 | Not-determined |
| 18-101_k141_385618 | Not-determined |
| 18-101_k141_304024 | Not-determined |
| 18-101_k141_72094 | Low-quality |
| 18-101_k141_170913 | Not-determined |
| 18-101_k141_304028 | Not-determined |
| 18-101_k141_72099 | Not-determined |
| 18-101_k141_237664 | Not-determined |
| 18-101_k141_237667 | Not-determined |
| 18-101_k141_254628 | Not-determined |
| 18-101_k141_72114 | Not-determined |
| 18-101_k141_237672 | Low-quality |
| 18-101_k141_188633 | Not-determined |
| 18-101_k141_188635 | Not-determined |
| 18-101_k141_385636 | Not-determined |
| 18-101_k141_139173 | Not-determined |
| 18-101_k141_23324 | Low-quality |
| 18-101_k141_23325 | Not-determined |
| 18-101_k141_23330 | Not-determined |
| 18-101_k141_237679 | Not-determined |
| 18-101_k141_254633 | Not-determined |
| 18-101_k141_23332 | Not-determined |
| 18-101_k141_139180 | Not-determined |
| 18-101_k141_237680 | Not-determined |
| 18-101_k141_254638 | Not-determined |
| 18-101_k141_286823 | Not-determined |
| 18-101_k141_286825 | Low-quality |
| 18-101_k141_23342 | Not-determined |
| 18-101_k141_320615 | Not-determined |
| 18-101_k141_56347 | Not-determined |
| 18-101_k141_56348 | Not-determined |
| 18-101_k141_320617 | Not-determined |
| 18-101_k141_56350 | Not-determined |
| 18-101_k141_56356 | Not-determined |
| 18-101_k141_254648 | Low-quality |
| 18-101_k141_56358 | Not-determined |
| 18-101_k141_188648 | Not-determined |
| 18-101_k141_254653 | Not-determined |
| 18-101_k141_170958 | Not-determined |
| 18-101_k141_237700 | Not-determined |
| 18-101_k141_254657 | Not-determined |
| 18-101_k141_23369 | Not-determined |
| 18-101_k141_270196 | Not-determined |
| 18-101_k141_139195 | Not-determined |
| 18-101_k141_188656 | Not-determined |
| 18-101_k141_188657 | Not-determined |
| 18-101_k141_188660 | Not-determined |
| 18-101_k141_188661 | Not-determined |
| 18-101_k141_139201 | Not-determined |
| 18-101_k141_188663 | Not-determined |
| 18-101_k141_154860 | Not-determined |
| 18-101_k141_320645 | Not-determined |
| 18-101_k141_335636 | Not-determined |
| 18-101_k141_385665 | Not-determined |
| 18-101_k141_385667 | Not-determined |
| 18-101_k141_385670 | Not-determined |
| 18-101_k141_320653 | Low-quality |
| 18-101_k141_385675 | Not-determined |
| 18-101_k141_139208 | Not-determined |
| 18-101_k141_320658 | Not-determined |
| 18-101_k141_139210 | Not-determined |
| 18-101_k141_353332 | Not-determined |
| 18-101_k141_139213 | Not-determined |
| 18-101_k141_56389 | Not-determined |
| 18-101_k141_139220 | Not-determined |
| 18-101_k141_270216 | Not-determined |
| 18-101_k141_286879 | Not-determined |
| 18-101_k141_270220 | Not-determined |
| 18-101_k141_56394 | Not-determined |
| 18-101_k141_286883 | Not-determined |
| 18-101_k141_220925 | Not-determined |
| 18-101_k141_237739 | Not-determined |
| 18-101_k141_304073 | Not-determined |
| 18-101_k141_254682 | Not-determined |
| 18-101_k141_123132 | Not-determined |
| 18-101_k141_123134 | Not-determined |
| 18-101_k141_320679 | Not-determined |
| 18-101_k141_304077 | Low-quality |
| 18-101_k141_72153 | Not-determined |
| 18-101_k141_320684 | Not-determined |
| 18-101_k141_320685 | Low-quality |
| 18-101_k141_254694 | Not-determined |
| 18-101_k141_89159 | Not-determined |
| 18-101_k141_72166 | Not-determined |
| 18-101_k141_385689 | Low-quality |
| 18-101_k141_188687 | Not-determined |
| 18-101_k141_237750 | Low-quality |
| 18-101_k141_237751 | Not-determined |
| 18-101_k141_205063 | Not-determined |
| 18-101_k141_270240 | Not-determined |
| 18-101_k141_123147 | Not-determined |
| 18-101_k141_270245 | Not-determined |
| 18-101_k141_56399 | Not-determined |
| 18-101_k141_220949 | Not-determined |
| 18-101_k141_123150 | Not-determined |
| 18-101_k141_270262 | Not-determined |
| 18-101_k141_123162 | Not-determined |
| 18-101_k141_270264 | Not-determined |
| 18-101_k141_353345 | Not-determined |
| 18-101_k141_205078 | Low-quality |
| 18-101_k141_56428 | Not-determined |
| 18-101_k141_89188 | Not-determined |
| 18-101_k141_23418 | Not-determined |
| 18-101_k141_154904 | Not-determined |
| 18-101_k141_286915 | Low-quality |
| 18-101_k141_237782 | Not-determined |
| 18-101_k141_39783 | Not-determined |
| 18-101_k141_23432 | Not-determined |
| 18-101_k141_89201 | Low-quality |
| 18-101_k141_205094 | Not-determined |
| 18-101_k141_286924 | Not-determined |
| 18-101_k141_89208 | Not-determined |
| 18-101_k141_270281 | Not-determined |
| 18-101_k141_154916 | Not-determined |
| 18-101_k141_154920 | Not-determined |
| 18-101_k141_154922 | Not-determined |
| 18-101_k141_270293 | Not-determined |
| 18-101_k141_89218 | Not-determined |
| 18-101_k141_320740 | Low-quality |
| 18-101_k141_39797 | Not-determined |
| 18-101_k141_72176 | Not-determined |
| 18-101_k141_205108 | Not-determined |
| 18-101_k141_139281 | Not-determined |
| 18-101_k141_320748 | Not-determined |
| 18-101_k141_139285 | Not-determined |
| 18-101_k141_335670 | Not-determined |

| | |
|---|---|
| 18-101_k141_320749 | Not-determined |
| 18-101_k141_270302 | Low-quality |
| 18-101_k141_170976 | Not-determined |
| 18-101_k141_56459 | Not-determined |
| 18-101_k141_139293 | Low-quality |
| 18-101_k141_139294 | Not-determined |
| 18-101_k141_188736 | Not-determined |
| 18-101_k141_320761 | Not-determined |
| 18-101_k141_205125 | Not-determined |
| 18-101_k141_221006 | Not-determined |
| 18-101_k141_56462 | Not-determined |
| 18-101_k141_221008 | Not-determined |
| 18-101_k141_353371 | Not-determined |
| 18-101_k141_7163 | Not-determined |
| 18-101_k141_170985 | Not-determined |
| 18-101_k141_7164 | Not-determined |
| 18-101_k141_385728 | Not-determined |
| 18-101_k141_89220 | Not-determined |
| 18-101_k141_56471 | Low-quality |
| 18-101_k141_237786 | Not-determined |
| 18-101_k141_104969 | Not-determined |
| 18-101_k141_139312 | Not-determined |
| 18-101_k141_89227 | Not-determined |
| 18-101_k141_104974 | Not-determined |
| 18-101_k141_221021 | Not-determined |
| 18-101_k141_286932 | Not-determined |
| 18-101_k141_139317 | Not-determined |
| 18-101_k141_72179 | Not-determined |
| 18-101_k141_353380 | Low-quality |
| 18-101_k141_154945 | Not-determined |
| 18-101_k141_353383 | Not-determined |
| 18-101_k141_221025 | Low-quality |
| 18-101_k141_221026 | Not-determined |
| 18-101_k141_237796 | Not-determined |
| 18-101_k141_335676 | Not-determined |
| 18-101_k141_385738 | Not-determined |
| 18-101_k141_237801 | Not-determined |
| 18-101_k141_335678 | Low-quality |
| 18-101_k141_335679 | Not-determined |
| 18-101_k141_270309 | Not-determined |
| 18-101_k141_237814 | Not-determined |
| 18-101_k141_56496 | Not-determined |
| 18-101_k141_56497 | Not-determined |
| 18-101_k141_72193 | Low-quality |
| 18-101_k141_171000 | Not-determined |
| 18-101_k141_320790 | Not-determined |
| 18-101_k141_335683 | Not-determined |
| 18-101_k141_286953 | Not-determined |
| 18-101_k141_320797 | Not-determined |
| 18-101_k141_270324 | Not-determined |
| 18-101_k141_335689 | Not-determined |
| 18-101_k141_23480 | Not-determined |
| 18-101_k141_23481 | Not-determined |
| 18-101_k141_72204 | Low-quality |
| 18-101_k141_72206 | Not-determined |
| 18-101_k141_72207 | Not-determined |
| 18-101_k141_353413 | Not-determined |
| 18-101_k141_320813 | Not-determined |
| 18-101_k141_353416 | Not-determined |
| 18-101_k141_171005 | Not-determined |
| 18-101_k141_270331 | Not-determined |
| 18-101_k141_237832 | Not-determined |
| 18-101_k141_270336 | Not-determined |
| 18-101_k141_139345 | Not-determined |
| 18-101_k141_286967 | Low-quality |
| 18-101_k141_139347 | Not-determined |
| 18-101_k141_353425 | Not-determined |
| 18-101_k141_353426 | Not-determined |
| 18-101_k141_72233 | Not-determined |
| 18-101_k141_270350 | Not-determined |
| 18-101_k141_139358 | Not-determined |
| 18-101_k141_154959 | Not-determined |
| 18-101_k141_72239 | Not-determined |
| 18-101_k141_270357 | Not-determined |
| 18-101_k141_72244 | Not-determined |
| 18-101_k141_171031 | Not-determined |
| 18-101_k141_123240 | Not-determined |
| 18-101_k141_335715 | Not-determined |
| 18-101_k141_154968 | Not-determined |
| 18-101_k141_320840 | Not-determined |
| 18-101_k141_154975 | Not-determined |
| 18-101_k141_72259 | Not-determined |
| 18-101_k141_89280 | Not-determined |
| 18-101_k141_72262 | Not-determined |
| 18-101_k141_39815 | Not-determined |
| 18-101_k141_320843 | Not-determined |
| 18-101_k141_286999 | Not-determined |
| 18-101_k141_171039 | Not-determined |
| 18-101_k141_89283 | Not-determined |
| 18-101_k141_287004 | Not-determined |
| 18-101_k141_287006 | Not-determined |
| 18-101_k141_320854 | Not-determined |
| 18-101_k141_171044 | Not-determined |
| 18-101_k141_72276 | Not-determined |
| 18-101_k141_237868 | Not-determined |
| 18-101_k141_237873 | Not-determined |
| 18-101_k141_335742 | Not-determined |
| 18-101_k141_154994 | Not-determined |
| 18-101_k141_320856 | Not-determined |
| 18-101_k141_39830 | Not-determined |
| 18-101_k141_237876 | Not-determined |
| 18-101_k141_368971 | Not-determined |
| 18-101_k141_39837 | Not-determined |
| 18-101_k141_23511 | Not-determined |
| 18-101_k141_154999 | Not-determined |
| 18-101_k141_254747 | Not-determined |
| 18-101_k141_139398 | Not-determined |
| 18-101_k141_368978 | Not-determined |
| 18-101_k141_254751 | Low-quality |
| 18-101_k141_368983 | Not-determined |
| 18-101_k141_254764 | Not-determined |
| 18-101_k141_304109 | Not-determined |
| 18-101_k141_72304 | Not-determined |
| 18-101_k141_385782 | Not-determined |
| 18-101_k141_72310 | Not-determined |
| 18-101_k141_72316 | Not-determined |
| 18-101_k141_335769 | Not-determined |
| 18-101_k141_368991 | Not-determined |
| 18-101_k141_205195 | Not-determined |
| 18-101_k141_171072 | Not-determined |
| 18-101_k141_335772 | Not-determined |
| 18-101_k141_155003 | Not-determined |
| 18-101_k141_171074 | Low-quality |
| 18-101_k141_205198 | Not-determined |
| 18-101_k141_205201 | Not-determined |
| 18-101_k141_72322 | Not-determined |
| 18-101_k141_39843 | Low-quality |
| 18-101_k141_123291 | Not-determined |
| 18-101_k141_155010 | Not-determined |
| 18-101_k141_385802 | Low-quality |
| 18-101_k141_139414 | Not-determined |
| 18-101_k141_221110 | Low-quality |
| 18-101_k141_89329 | Not-determined |
| 18-101_k141_23522 | Not-determined |
| 18-101_k141_7187 | Not-determined |
| 18-101_k141_254786 | Not-determined |
| 18-101_k141_139416 | Not-determined |
| 18-101_k141_205217 | Not-determined |
| 18-101_k141_89334 | Not-determined |
| 18-101_k141_320873 | Not-determined |
| 18-101_k141_205221 | Low-quality |
| 18-101_k141_89349 | Not-determined |
| 18-101_k141_105043 | Not-determined |
| 18-101_k141_320887 | Not-determined |
| 18-101_k141_105045 | Low-quality |
| 18-101_k141_320892 | Not-determined |
| 18-101_k141_287027 | Not-determined |
| 18-101_k141_89358 | Not-determined |
| 18-101_k141_320896 | Not-determined |
| 18-101_k141_205228 | Not-determined |
| [illegible] | [illegible] |

| | |
|---|---|
| 18-101_k141_270712 | Not-determined |
| 18-101_k141_155250 | Not-determined |
| 18-101_k141_221484 | Not-determined |
| 18-101_k141_270713 | Not-determined |
| 18-101_k141_238268 | Not-determined |
| 18-101_k141_56847 | Not-determined |
| 18-101_k141_386112 | Not-determined |
| 18-101_k141_155258 | Not-determined |
| 18-101_k141_155264 | Not-determined |
| 18-101_k141_40167 | Not-determined |
| 18-101_k141_123567 | Not-determined |
| 18-101_k141_287217 | Not-determined |
| 18-101_k141_40169 | Not-determined |
| 18-101_k141_238281 | Not-determined |
| 18-101_k141_40170 | Low-quality |
| 18-101_k141_189033 | Not-determined |
| 18-101_k141_72610 | Not-determined |
| 18-101_k141_287225 | Not-determined |
| 18-101_k141_221494 | Not-determined |
| 18-101_k141_123576 | Not-determined |
| 18-101_k141_205558 | Not-determined |
| 18-101_k141_56864 | Not-determined |
| 18-101_k141_155283 | Not-determined |
| 18-101_k141_171364 | Not-determined |
| 18-101_k141_40178 | Not-determined |
| 18-101_k141_287237 | Low-quality |
| 18-101_k141_105397 | Not-determined |
| 18-101_k141_171366 | Not-determined |
| 18-101_k141_287239 | Not-determined |
| 18-101_k141_171373 | Not-determined |
| 18-101_k141_40187 | Not-determined |
| 18-101_k141_105404 | Not-determined |
| 18-101_k141_7582 | Not-determined |
| 18-101_k141_123587 | Not-determined |
| 18-101_k141_123588 | Not-determined |
| 18-101_k141_221507 | Low-quality |
| 18-101_k141_335986 | Low-quality |
| 18-101_k141_369331 | Not-determined |
| 18-101_k141_335988 | Not-determined |
| 18-101_k141_369332 | Not-determined |
| 18-101_k141_287245 | Not-determined |
| 18-101_k141_40193 | Not-determined |
| 18-101_k141_23712 | Not-determined |
| 18-101_k141_336001 | Not-determined |
| 18-101_k141_105418 | Not-determined |
| 18-101_k141_155317 | Not-determined |
| 18-101_k141_23717 | Low-quality |
| 18-101_k141_105422 | Not-determined |
| 18-101_k141_56886 | Not-determined |
| 18-101_k141_123596 | Not-determined |
| 18-101_k141_123600 | Not-determined |
| 18-101_k141_287252 | Not-determined |
| 18-101_k141_287253 | Not-determined |
| 18-101_k141_221519 | Not-determined |
| 18-101_k141_205572 | Not-determined |
| 18-101_k141_255156 | Not-determined |
| 18-101_k141_23732 | Not-determined |
| 18-101_k141_336013 | Not-determined |
| 18-101_k141_155329 | Not-determined |
| 18-101_k141_336018 | Not-determined |
| 18-101_k141_255165 | Not-determined |
| 18-101_k141_386132 | Not-determined |
| 18-101_k141_386133 | Not-determined |
| 18-101_k141_40197 | Not-determined |
| 18-101_k141_287275 | Not-determined |
| 18-101_k141_155335 | Not-determined |
| 18-101_k141_72621 | Not-determined |
| 18-101_k141_336021 | Not-determined |
| 18-101_k141_270748 | Not-determined |
| 18-101_k141_336023 | Not-determined |
| 18-101_k141_336024 | Not-determined |
| 18-101_k141_221532 | Not-determined |
| 18-101_k141_336028 | Not-determined |
| 18-101_k141_336029 | Low-quality |
| 18-101_k141_72624 | Low-quality |
| 18-101_k141_270756 | Not-determined |
| 18-101_k141_23744 | Not-determined |
| 18-101_k141_304379 | Not-determined |
| 18-101_k141_72628 | Low-quality |
| 18-101_k141_7612 | Not-determined |
| 18-101_k141_56902 | Not-determined |
| 18-101_k141_255193 | Not-determined |
| 18-101_k141_72634 | Not-determined |
| 18-101_k141_255195 | Not-determined |
| 18-101_k141_189082 | Not-determined |
| 18-101_k141_72638 | Not-determined |
| 18-101_k141_155361 | Not-determined |
| 18-101_k141_89661 | Not-determined |
| 18-101_k141_72642 | Not-determined |
| 18-101_k141_369358 | Not-determined |
| 18-101_k141_72644 | Low-quality |
| 18-101_k141_386148 | Not-determined |
| 18-101_k141_72648 | Not-determined |
| 18-101_k141_7616 | Not-determined |
| 18-101_k141_72653 | Not-determined |
| 18-101_k141_386155 | Not-determined |
| 18-101_k141_40224 | Not-determined |
| 18-101_k141_304403 | Not-determined |
| 18-101_k141_321309 | Not-determined |
| 18-101_k141_255218 | Not-determined |
| 18-101_k141_304408 | Low-quality |
| 18-101_k141_255221 | Not-determined |
| 18-101_k141_7633 | Not-determined |
| 18-101_k141_123616 | Not-determined |
| 18-101_k141_189107 | Not-determined |
| 18-101_k141_321317 | Not-determined |
| 18-101_k141_155378 | Not-determined |
| 18-101_k141_321319 | Not-determined |
| 18-101_k141_205607 | Low-quality |
| 18-101_k141_205611 | Not-determined |
| 18-101_k141_171394 | Not-determined |
| 18-101_k141_369366 | Not-determined |
| 18-101_k141_205612 | Low-quality |
| 18-101_k141_205614 | Not-determined |
| 18-101_k141_255242 | Not-determined |
| 18-101_k141_89665 | Not-determined |
| 18-101_k141_189137 | Not-determined |
| 18-101_k141_89666 | Not-determined |
| 18-101_k141_189138 | Not-determined |
| 18-101_k141_7644 | Not-determined |
| 18-101_k141_89668 | Not-determined |
| 18-101_k141_353772 | Not-determined |
| 18-101_k141_72663 | Not-determined |
| 18-101_k141_353773 | Not-determined |
| 18-101_k141_336055 | Not-determined |
| 18-101_k141_7648 | Not-determined |
| 18-101_k141_89671 | Not-determined |
| 18-101_k141_89672 | Not-determined |
| 18-101_k141_205621 | Not-determined |
| 18-101_k141_123648 | Not-determined |
| 18-101_k141_255258 | Not-determined |
| 18-101_k141_205628 | Not-determined |
| 18-101_k141_7653 | Not-determined |
| 18-101_k141_105455 | Not-determined |
| 18-101_k141_123655 | Not-determined |
| 18-101_k141_155397 | Not-determined |
| 18-101_k141_255268 | Not-determined |
| 18-101_k141_89686 | Not-determined |
| 18-101_k141_205642 | Not-determined |
| 18-101_k141_171416 | Not-determined |
| 18-101_k141_155405 | Not-determined |
| 18-101_k141_189145 | Low-quality |
| 18-101_k141_304442 | Low-quality |
| 18-101_k141_105468 | Not-determined |
| 18-101_k141_386198 | Low-quality |
| 18-101_k141_321350 | Not-determined |
| 18-101_k141_89694 | Not-determined |
| 18-101_k141_221576 | Not-determined |
| 18-101_k141_205649 | Not-determined |
| 18-101_k141_386201 | Not-determined |
| 18-101_k141_221580 | Not-determined |
| 18-101_k141_72684 | Not-determined |
| 18-101_k141_56921 | Not-determined |
| 18-101_k141_123664 | Not-determined |
| 18-101_k141_255288 | Not-determined |
| 18-101_k141_189158 | Not-determined |
| 18-101_k141_304445 | Not-determined |
| 18-101_k141_205657 | Not-determined |
| 18-101_k141_171431 | Not-determined |
| 18-101_k141_205660 | Not-determined |
| 18-101_k141_171440 | Not-determined |
| 18-101_k141_56932 | Not-determined |
| 18-101_k141_321362 | Not-determined |
| 18-101_k141_221595 | Not-determined |
| 18-101_k141_386222 | Not-determined |
| 18-101_k141_304463 | Not-determined |
| 18-101_k141_189176 | Low-quality |
| 18-101_k141_353799 | Not-determined |
| 18-101_k141_304472 | Not-determined |
| 18-101_k141_221598 | Not-determined |
| 18-101_k141_89715 | Not-determined |
| 18-101_k141_56941 | Not-determined |
| 18-101_k141_72707 | Not-determined |
| 18-101_k141_321381 | Not-determined |
| 18-101_k141_270825 | Not-determined |
| 18-101_k141_189199 | Not-determined |
| 18-101_k141_23794 | Not-determined |
| 18-101_k141_155416 | Not-determined |
| 18-101_k141_321387 | Not-determined |
| 18-101_k141_189207 | Not-determined |
| 18-101_k141_287329 | Not-determined |
| 18-101_k141_139700 | Not-determined |
| 18-101_k141_386234 | Not-determined |
| 18-101_k141_189209 | Not-determined |
| 18-101_k141_7695 | Not-determined |
| 18-101_k141_205696 | Low-quality |
| 18-101_k141_304480 | Not-determined |
| 18-101_k141_56959 | Not-determined |
| 18-101_k141_205697 | Not-determined |
| 18-101_k141_321392 | Not-determined |
| 18-101_k141_56962 | Not-determined |
| 18-101_k141_386242 | Not-determined |
| 18-101_k141_287340 | Not-determined |
| 18-101_k141_56965 | Not-determined |
| 18-101_k141_386246 | Not-determined |
| 18-101_k141_23813 | Not-determined |
| 18-101_k141_23815 | Not-determined |
| 18-101_k141_221619 | Not-determined |
| 18-101_k141_221620 | Not-determined |
| 18-101_k141_304489 | Not-determined |
| 18-101_k141_353817 | Not-determined |
| 18-101_k141_171462 | Not-determined |
| 18-101_k141_270858 | Not-determined |
| 18-101_k141_270865 | Not-determined |
| 18-101_k141_369418 | Not-determined |
| 18-101_k141_386255 | Not-determined |
| 18-101_k141_386258 | Not-determined |
| 18-101_k141_369421 | Not-determined |
| 18-101_k141_270869 | Not-determined |
| 18-101_k141_221626 | Not-determined |
| 18-101_k141_72730 | Not-determined |
| 18-101_k141_369440 | Not-determined |
| 18-101_k141_304509 | Not-determined |
| 18-101_k141_270879 | Not-determined |
| 18-101_k141_304512 | Not-determined |
| 18-101_k141_40254 | Not-determined |
| 18-101_k141_40257 | Low-quality |
| 18-101_k141_40261 | Not-determined |
| 18-101_k141_304521 | Not-determined |
| 18-101_k141_56998 | Not-determined |
| 18-101_k141_270894 | Not-determined |
| 18-101_k141_7700 | Not-determined |
| 18-101_k141_57001 | Not-determined |
| 18-101_k141_386274 | Not-determined |
| 18-101_k141_189234 | Not-determined |
| 18-101_k141_139758 | Not-determined |
| 18-101_k141_336073 | Low-quality |
| 18-101_k141_171482 | Not-determined |
| 18-101_k141_57007 | Not-determined |
| 18-101_k141_221644 | Low-quality |
| 18-101_k141_336081 | Not-determined |
| 18-101_k141_287386 | Not-determined |
| 18-101_k141_287388 | Not-determined |
| 18-101_k141_336085 | Not-determined |
| 18-101_k141_336086 | Low-quality |
| 18-101_k141_205761 | Not-determined |
| 18-101_k141_171493 | Low-quality |
| 18-101_k141_386291 | Not-determined |
| 18-101_k141_23841 | Not-determined |
| 18-101_k141_270914 | Not-determined |
| 18-101_k141_40277 | Not-determined |
| 18-101_k141_105512 | Not-determined |
| 18-101_k141_105513 | Not-determined |
| 18-101_k141_155438 | Not-determined |
| 18-101_k141_221657 | Not-determined |
| 18-101_k141_221658 | Not-determined |
| 18-101_k141_57022 | Not-determined |
| 18-101_k141_105518 | Not-determined |
| 18-101_k141_171496 | Not-determined |
| 18-101_k141_221662 | Not-determined |
| 18-101_k141_353862 | Not-determined |
| 18-101_k141_205764 | Not-determined |
| 18-101_k141_40285 | Not-determined |
| 18-101_k141_205766 | Not-determined |
| 18-101_k141_304542 | Not-determined |
| 18-101_k141_287393 | Low-quality |
| 18-101_k141_57032 | Not-determined |
| 18-101_k141_23847 | Not-determined |
| 18-101_k141_386301 | Not-determined |
| 18-101_k141_386314 | Not-determined |
| 18-101_k141_304552 | Not-determined |
| 18-101_k141_386316 | Not-determined |
| 18-101_k141_139786 | Not-determined |
| 18-101_k141_7718 | Low-quality |
| 18-101_k141_369485 | Low-quality |
| 18-101_k141_205769 | Low-quality |
| 18-101_k141_7727 | Not-determined |
| 18-101_k141_386323 | Not-determined |
| 18-101_k141_57055 | Not-determined |
| 18-101_k141_57056 | Not-determined |
| 18-101_k141_287408 | Not-determined |
| 18-101_k141_205780 | Not-determined |
| 18-101_k141_7731 | Not-determined |
| 18-101_k141_386328 | Not-determined |
| 18-101_k141_369498 | Low-quality |
| 18-101_k141_7737 | Not-determined |
| 18-101_k141_336111 | Not-determined |
| 18-101_k141_270920 | Not-determined |
| 18-101_k141_155467 | Not-determined |
| 18-101_k141_57068 | Not-determined |
| 18-101_k141_353896 | Low-quality |
| 18-101_k141_57069 | Not-determined |
| 18-101_k141_270926 | Not-determined |
| 18-101_k141_304582 | Not-determined |
| 18-101_k141_189258 | Not-determined |
| 18-101_k141_304583 | Not-determined |
| 18-101_k141_189260 | Not-determined |
| 18-101_k141_7744 | Not-determined |
| 18-101_k141_369505 | Low-quality |
| 18-101_k141_336121 | Not-determined |
| 18-101_k141_369509 | Not-determined |
| 18-101_k141_57084 | Not-determined |
| 18-101_k141_336128 | Not-determined |
| 18-101_k141_221684 | Low-quality |
| 18-101_k141_205794 | Not-determined |
| 18-101_k141_221686 | Not-determined |
| 18-101_k141_89765 | Not-determined |
| 18-101_k141_89768 | Not-determined |
| 18-101_k141_386357 | Not-determined |
| 18-101_k141_353916 | Low-quality |
| 18-101_k141_89770 | Not-determined |
| 18-101_k141_139800 | Not-determined |
| 18-101_k141_205802 | Not-determined |
| 18-101_k141_386360 | Not-determined |
| 18-101_k141_105572 | Not-determined |
| 18-101_k141_205814 | Not-determined |
| 18-101_k141_40330 | Not-determined |
| 18-101_k141_205817 | Not-determined |
| 18-101_k141_57096 | Not-determined |
| 18-101_k141_386372 | Not-determined |
| 18-101_k141_89787 | Not-determined |
| 18-101_k141_23896 | Low-quality |
| 18-101_k141_89789 | Not-determined |
| 18-101_k141_205826 | Not-determined |
| 18-101_k141_270954 | Not-determined |
| 18-101_k141_205829 | Not-determined |
| 18-101_k141_270955 | Not-determined |
| 18-101_k141_369532 | Not-determined |
| 18-101_k141_7766 | Not-determined |
| 18-101_k141_221706 | Not-determined |
| 18-101_k141_336137 | Not-determined |
| 18-101_k141_369543 | Not-determined |
| 18-101_k141_7772 | Low-quality |
| 18-101_k141_336141 | Not-determined |
| 18-101_k141_336142 | Not-determined |
| 18-101_k141_336146 | Not-determined |
| 18-101_k141_353950 | Not-determined |
| 18-101_k141_304624 | Not-determined |
| 18-101_k141_7779 | Not-determined |
| 18-101_k141_336151 | Not-determined |
| 18-101_k141_139819 | Not-determined |
| 18-101_k141_189299 | Not-determined |
| 18-101_k141_287459 | Not-determined |
| 18-101_k141_139823 | Not-determined |
| 18-101_k141_205853 | Not-determined |
| 18-101_k141_189305 | Not-determined |
| 18-101_k141_189309 | Not-determined |
| 18-101_k141_7793 | Not-determined |
| 18-101_k141_105605 | Low-quality |
| 18-101_k141_89816 | Not-determined |
| 18-101_k141_336165 | Not-determined |
| 18-101_k141_221728 | Low-quality |
| 18-101_k141_189324 | Low-quality |
| 18-101_k141_40341 | Not-determined |
| 18-101_k141_189328 | Not-determined |
| 18-101_k141_23937 | Low-quality |
| 18-101_k141_221731 | Not-determined |
| 18-101_k141_336171 | Not-determined |
| 18-101_k141_23939 | Not-determined |
| 18-101_k141_89827 | Not-determined |
| 18-101_k141_221735 | Not-determined |
| 18-101_k141_336175 | Not-determined |
| 18-101_k141_386385 | Not-determined |
| 18-101_k141_123788 | Not-determined |
| 18-101_k141_321498 | Not-determined |
| 18-101_k141_353974 | Not-determined |
| 18-101_k141_123791 | Not-determined |
| 18-101_k141_336177 | Not-determined |
| 18-101_k141_7818 | Not-determined |
| 18-101_k141_321504 | Not-determined |
| 18-101_k141_72760 | Not-determined |
| 18-101_k141_105622 | Not-determined |
| 18-101_k141_123804 | Not-determined |
| 18-101_k141_57123 | Not-determined |
| 18-101_k141_7821 | Not-determined |
| 18-101_k141_72766 | Low-quality |
| 18-101_k141_221757 | Not-determined |
| 18-101_k141_321518 | Not-determined |
| 18-101_k141_353989 | Not-determined |
| 18-101_k141_57132 | Not-determined |
| 18-101_k141_57133 | Not-determined |
| 18-101_k141_270973 | Not-determined |
| 18-101_k141_123827 | Not-determined |
| 18-101_k141_7839 | Not-determined |
| 18-101_k141_89863 | Not-determined |
| 18-101_k141_72779 | Not-determined |
| 18-101_k141_123829 | Not-determined |
| 18-101_k141_7844 | Not-determined |
| 18-101_k141_189376 | Not-determined |
| 18-101_k141_7845 | Not-determined |
| 18-101_k141_72782 | Not-determined |
| 18-101_k141_139871 | Not-determined |
| 18-101_k141_189383 | Not-determined |
| 18-101_k141_72794 | Not-determined |
| 18-101_k141_89874 | Not-determined |
| 18-101_k141_72797 | Not-determined |
| 18-101_k141_72799 | Not-determined |
| 18-101_k141_23963 | Not-determined |
| 18-101_k141_336205 | Not-determined |
| 18-101_k141_40374 | Not-determined |
| 18-101_k141_155513 | Not-determined |
| 18-101_k141_189392 | Not-determined |
| 18-101_k141_205894 | Not-determined |
| 18-101_k141_89884 | Not-determined |
| 18-101_k141_238291 | Not-determined |
| 18-101_k141_105675 | Not-determined |
| 18-101_k141_171525 | Not-determined |
| 18-101_k141_369590 | Not-determined |
| 18-101_k141_369591 | Not-determined |
| 18-101_k141_155523 | Not-determined |
| 18-101_k141_139894 | Not-determined |
| 18-101_k141_139895 | Not-determined |
| 18-101_k141_89890 | Not-determined |
| 18-101_k141_89893 | Not-determined |
| 18-101_k141_238292 | Not-determined |
| 18-101_k141_238293 | Not-determined |
| 18-101_k141_238297 | Not-determined |
| 18-101_k141_270986 | Not-determined |
| 18-101_k141_72822 | Not-determined |
| 18-101_k141_89905 | Not-determined |
| 18-101_k141_386408 | Not-determined |
| 18-101_k141_369601 | Not-determined |
| 18-101_k141_89910 | Not-determined |
| 18-101_k141_89915 | Not-determined |
| 18-101_k141_7870 | Not-determined |
| 18-101_k141_40388 | Not-determined |
| 18-101_k141_40390 | Not-determined |
| 18-101_k141_7874 | Not-determined |
| 18-101_k141_321564 | Not-determined |
| 18-101_k141_89920 | Not-determined |
| 18-101_k141_105702 | Not-determined |
| 18-101_k141_238310 | Not-determined |
| 18-101_k141_336219 | Not-determined |
| 18-101_k141_7884 | Not-determined |
| 18-101_k141_171548 | Low-quality |
| 18-101_k141_72851 | Not-determined |
| 18-101_k141_386425 | Not-determined |
| 18-101_k141_7888 | Not-determined |
| 18-101_k141_321574 | Not-determined |
| 18-101_k141_321576 | Not-determined |
| 18-101_k141_336233 | Not-determined |
| 18-101_k141_23995 | Not-determined |
| 18-101_k141_221812 | Not-determined |
| 18-101_k141_189417 | Not-determined |
| 18-101_k141_354014 | Not-determined |
| 18-101_k141_189419 | Not-determined |
| 18-101_k141_40412 | Not-determined |
| 18-101_k141_139920 | Not-determined |
| 18-101_k141_139924 | Not-determined |
| 18-101_k141_271001 | Low-quality |
| 18-101_k141_271002 | Not-determined |
| 18-101_k141_189427 | Not-determined |
| 18-101_k141_139931 | Not-determined |
| 18-101_k141_321593 | Not-determined |
| 18-101_k141_139933 | Low-quality |
| 18-101_k141_89928 | Not-determined |
| 18-101_k141_40429 | Not-determined |
| 18-101_k141_386466 | Not-determined |
| 18-101_k141_354022 | Low-quality |
| 18-101_k141_155577 | Not-determined |
| 18-101_k141_189442 | Not-determined |
| 18-101_k141_238340 | Not-determined |
| 18-101_k141_24009 | Not-determined |
| 18-101_k141_221830 | Not-determined |
| 18-101_k141_238344 | Not-determined |
| 18-101_k141_221834 | Not-determined |
| 18-101_k141_7909 | Low-quality |
| 18-101_k141_271026 | Low-quality |
| 18-101_k141_89942 | Not-determined |
| 18-101_k141_369614 | Not-determined |
| 18-101_k141_7913 | Not-determined |
| 18-101_k141_89949 | Not-determined |
| 18-101_k141_287507 | Not-determined |
| 18-101_k141_221848 | Not-determined |
| 18-101_k141_7917 | Not-determined |
| 18-101_k141_40456 | Not-determined |
| 18-101_k141_369617 | Not-determined |
| 18-101_k141_7918 | Not-determined |
| 18-101_k141_72881 | Not-determined |
| 18-101_k141_123893 | Not-determined |
| 18-101_k141_354035 | Not-determined |
| 18-101_k141_40461 | Not-determined |
| 18-101_k141_304636 | Not-determined |
| 18-101_k141_155604 | Not-determined |
| 18-101_k141_40465 | Not-determined |
| 18-101_k141_304640 | Not-determined |
| 18-101_k141_336261 | Not-determined |
| 18-101_k141_139968 | Not-determined |
| 18-101_k141_255295 | Not-determined |
| 18-101_k141_24034 | Not-determined |
| 18-101_k141_189452 | Not-determined |
| 18-101_k141_336267 | Not-determined |
| 18-101_k141_304647 | Not-determined |
| 18-101_k141_255303 | Not-determined |
| 18-101_k141_24041 | Not-determined |
| 18-101_k141_221860 | Not-determined |
| 18-101_k141_24046 | Not-determined |
| 18-101_k141_155619 | Low-quality |
| 18-101_k141_24050 | Not-determined |
| 18-101_k141_155623 | Not-determined |
| 18-101_k141_72908 | Not-determined |
| 18-101_k141_238373 | Not-determined |
| 18-101_k141_24052 | Not-determined |
| 18-101_k141_72913 | Not-determined |
| 18-101_k141_89951 | Not-determined |
| 18-101_k141_221864 | Not-determined |
| 18-101_k141_105774 | Not-determined |
| 18-101_k141_271055 | Not-determined |
| 18-101_k141_155642 | Not-determined |
| 18-101_k141_155644 | Not-determined |
| 18-101_k141_287546 | Not-determined |
| 18-101_k141_369637 | Not-determined |
| 18-101_k141_336292 | Not-determined |
| 18-101_k141_155653 | Not-determined |
| 18-101_k141_321607 | Not-determined |
| 18-101_k141_155655 | Not-determined |
| 18-101_k141_189489 | Not-determined |
| 18-101_k141_304662 | Not-determined |
| 18-101_k141_321611 | Not-determined |
| 18-101_k141_7959 | Not-determined |
| 18-101_k141_369639 | Not-determined |
| 18-101_k141_304664 | Not-determined |
| 18-101_k141_57156 | Not-determined |
| 18-101_k141_40501 | Low-quality |
| 18-101_k141_336305 | Not-determined |
| 18-101_k141_123917 | Not-determined |
| 18-101_k141_287554 | Low-quality |
| 18-101_k141_7963 | Not-determined |
| 18-101_k141_24080 | Not-determined |
| 18-101_k141_7964 | Low-quality |
| 18-101_k141_304670 | Not-determined |
| 18-101_k141_57162 | Not-determined |
| 18-101_k141_336312 | Not-determined |
| 18-101_k141_369649 | Not-determined |
| 18-101_k141_386519 | Not-determined |
| 18-101_k141_336315 | Low-quality |
| 18-101_k141_189500 | Not-determined |
| 18-101_k141_189502 | Not-determined |
| 18-101_k141_7968 | Not-determined |
| 18-101_k141_24087 | Not-determined |
| 18-101_k141_287561 | Not-determined |
| 18-101_k141_24089 | Not-determined |
| 18-101_k141_123933 | Not-determined |
| 18-101_k141_238402 | Not-determined |
| 18-101_k141_24096 | Not-determined |
| 18-101_k141_205911 | Not-determined |
| 18-101_k141_336327 | Not-determined |
| 18-101_k141_155672 | Not-determined |
| 18-101_k141_40522 | Not-determined |
| 18-101_k141_189508 | Not-determined |
| 18-101_k141_105804 | Low-quality |
| 18-101_k141_105807 | Not-determined |
| 18-101_k141_7979 | Not-determined |
| 18-101_k141_238419 | Not-determined |
| 18-101_k141_40545 | Not-determined |
| 18-101_k141_271093 | Not-determined |
| 18-101_k141_171611 | Not-determined |
| 18-101_k141_238431 | Low-quality |
| 18-101_k141_238432 | Not-determined |
| 18-101_k141_171614 | Not-determined |
| 18-101_k141_40562 | Not-determined |
| 18-101_k141_40563 | Not-determined |
| 18-101_k141_386565 | Not-determined |
| 18-101_k141_40567 | Not-determined |
| 18-101_k141_24114 | Not-determined |
| 18-101_k141_7999 | Not-determined |
| 18-101_k141_321646 | Not-determined |
| 18-101_k141_238450 | Not-determined |
| 18-101_k141_8003 | Not-determined |
| 18-101_k141_221879 | Not-determined |
| 18-101_k141_123965 | Not-determined |
| 18-101_k141_105818 | Low-quality |
| 18-101_k141_205946 | Not-determined |
| 18-101_k141_105819 | Not-determined |
| 18-101_k141_123969 | Not-determined |
| 18-101_k141_369678 | Not-determined |
| 18-101_k141_271114 | Not-determined |
| 18-101_k141_336346 | Not-determined |
| 18-101_k141_238458 | Low-quality |
| 18-101_k141_369683 | Not-determined |
| 18-101_k141_321654 | Low-quality |
| 18-101_k141_271119 | Low-quality |
| 18-101_k141_155712 | Not-determined |
| 18-101_k141_369684 | Not-determined |
| 18-101_k141_8020 | Not-determined |
| 18-101_k141_8022 | Not-determined |
| 18-101_k141_238468 | Not-determined |
| 18-101_k141_123981 | Not-determined |
| 18-101_k141_271133 | Not-determined |
| 18-101_k141_24125 | Not-determined |
| 18-101_k141_72958 | Not-determined |
| 18-101_k141_155723 | Not-determined |
| 18-101_k141_105829 | Not-determined |
| 18-101_k141_72962 | Not-determined |
| 18-101_k141_369693 | Not-determined |
| 18-101_k141_321664 | Not-determined |
| 18-101_k141_287612 | Not-determined |
| 18-101_k141_155729 | Not-determined |
| 18-101_k141_123992 | Not-determined |

| | |
|---|---|
| 18-101_k141_123993 | Not-determined |
| 18-101_k141_238473 | Not-determined |
| 18-101_k141_238475 | Not-determined |
| 18-101_k141_105841 | Not-determined |
| 18-101_k141_354094 | Not-determined |
| 18-101_k141_155734 | Not-determined |
| 18-101_k141_221912 | Low-quality |
| 18-101_k141_171650 | Not-determined |
| 18-101_k141_271152 | Not-determined |
| 18-101_k141_105844 | Not-determined |
| 18-101_k141_386608 | Not-determined |
| 18-101_k141_238482 | Not-determined |
| 18-101_k141_336360 | Not-determined |
| 18-101_k141_271153 | Not-determined |
| 18-101_k141_271154 | Not-determined |
| 18-101_k141_171655 | Not-determined |
| 18-101_k141_321680 | Not-determined |
| 18-101_k141_386611 | Not-determined |
| 18-101_k141_321682 | Not-determined |
| 18-101_k141_155741 | Not-determined |
| 18-101_k141_255383 | Not-determined |
| 18-101_k141_255393 | Not-determined |
| 18-101_k141_255394 | Not-determined |
| 18-101_k141_321687 | Low-quality |
| 18-101_k141_304728 | Not-determined |
| 18-101_k141_72990 | Not-determined |
| 18-101_k141_189556 | Not-determined |
| 18-101_k141_321691 | Not-determined |
| 18-101_k141_24134 | Not-determined |
| 18-101_k141_40597 | Not-determined |
| 18-101_k141_8044 | Not-determined |
| 18-101_k141_255399 | Not-determined |
| 18-101_k141_255402 | Not-determined |
| 18-101_k141_124025 | Not-determined |
| 18-101_k141_255403 | Low-quality |
| 18-101_k141_171666 | Not-determined |
| 18-101_k141_304746 | Not-determined |
| 18-101_k141_57173 | Not-determined |
| 18-101_k141_321702 | Not-determined |
| 18-101_k141_73011 | Not-determined |
| 18-101_k141_73014 | Not-determined |
| 18-101_k141_57178 | Not-determined |
| 18-101_k141_238512 | Not-determined |
| 18-101_k141_57182 | Low-quality |
| 18-101_k141_105869 | Not-determined |
| 18-101_k141_57185 | Not-determined |
| 18-101_k141_238518 | Not-determined |
| 18-101_k141_304762 | Not-determined |
| 18-101_k141_321713 | Low-quality |
| 18-101_k141_271170 | Low-quality |
| 18-101_k141_124038 | Not-determined |
| 18-101_k141_271172 | Not-determined |
| 18-101_k141_40618 | Low-quality |
| 18-101_k141_336387 | Not-determined |
| 18-101_k141_287648 | Not-determined |
| 18-101_k141_8056 | Not-determined |
| 18-101_k141_189594 | Not-determined |
| 18-101_k141_124041 | Not-determined |
| 18-101_k141_124042 | Not-determined |
| 18-101_k141_304770 | Not-determined |
| 18-101_k141_57201 | Low-quality |
| 18-101_k141_205998 | Not-determined |
| 18-101_k141_206004 | Low-quality |
| 18-101_k141_271193 | Not-determined |
| 18-101_k141_189606 | Not-determined |
| 18-101_k141_304776 | Not-determined |
| 18-101_k141_189610 | Low-quality |
| 18-101_k141_189611 | Not-determined |
| 18-101_k141_24166 | Not-determined |
| 18-101_k141_304782 | Low-quality |
| 18-101_k141_321749 | Low-quality |
| 18-101_k141_336401 | Not-determined |
| 18-101_k141_287669 | Not-determined |
| 18-101_k141_304791 | Not-determined |
| 18-101_k141_369740 | Not-determined |
| 18-101_k141_73066 | Not-determined |
| 18-101_k141_105877 | Not-determined |
| 18-101_k141_189620 | Not-determined |
| 18-101_k141_287679 | Not-determined |
| 18-101_k141_57214 | Not-determined |
| 18-101_k141_73071 | Not-determined |
| 18-101_k141_189622 | Not-determined |
| 18-101_k141_321763 | Not-determined |
| 18-101_k141_386640 | Not-determined |
| 18-101_k141_271227 | Low-quality |
| 18-101_k141_189623 | Not-determined |
| 18-101_k141_238549 | Not-determined |
| 18-101_k141_271231 | Not-determined |
| 18-101_k141_304816 | Not-determined |
| 18-101_k141_105889 | Not-determined |
| 18-101_k141_271238 | Not-determined |
| 18-101_k141_57230 | Not-determined |
| 18-101_k141_57235 | Not-determined |
| 18-101_k141_57236 | Not-determined |
| 18-101_k141_57238 | Not-determined |
| 18-101_k141_8073 | Not-determined |
| 18-101_k141_8074 | Low-quality |
| 18-101_k141_321779 | Low-quality |
| 18-101_k141_24187 | Not-determined |
| 18-101_k141_255433 | Not-determined |
| 18-101_k141_255434 | Not-determined |
| 18-101_k141_304831 | Not-determined |
| 18-101_k141_8081 | Not-determined |
| 18-101_k141_8082 | Not-determined |
| 18-101_k141_369764 | Not-determined |
| 18-101_k141_171695 | Not-determined |
| 18-101_k141_57248 | Not-determined |
| 18-101_k141_57249 | Not-determined |
| 18-101_k141_171700 | Not-determined |
| 18-101_k141_57251 | Not-determined |
| 18-101_k141_369767 | Not-determined |
| 18-101_k141_124080 | Not-determined |
| 18-101_k141_57256 | Not-determined |
| 18-101_k141_336443 | Low-quality |
| 18-101_k141_321789 | Not-determined |
| 18-101_k141_73084 | Not-determined |
| 18-101_k141_73086 | Not-determined |
| 18-101_k141_90064 | Not-determined |
| 18-101_k141_90066 | Not-determined |
| 18-101_k141_73090 | Not-determined |
| 18-101_k141_57267 | Not-determined |
| 18-101_k141_73099 | Not-determined |
| 18-101_k141_8111 | Not-determined |
| 18-101_k141_287700 | Not-determined |
| 18-101_k141_287701 | Not-determined |
| 18-101_k141_206052 | Not-determined |
| 18-101_k141_189676 | Not-determined |
| 18-101_k141_321793 | Low-quality |
| 18-101_k141_206053 | Not-determined |
| 18-101_k141_304853 | Not-determined |
| 18-101_k141_238579 | Low-quality |
| 18-101_k141_321801 | Not-determined |
| 18-101_k141_206061 | Not-determined |
| 18-101_k141_206062 | Not-determined |
| 18-101_k141_336469 | Not-determined |
| 18-101_k141_369784 | Not-determined |
| 18-101_k141_24217 | Not-determined |
| 18-101_k141_336479 | Not-determined |
| 18-101_k141_24227 | Not-determined |
| 18-101_k141_24229 | Not-determined |
| 18-101_k141_40674 | Not-determined |
| 18-101_k141_90097 | Not-determined |
| 18-101_k141_304861 | Not-determined |
| 18-101_k141_40677 | Not-determined |
| 18-101_k141_40679 | Not-determined |
| 18-101_k141_171726 | Not-determined |
| 18-101_k141_57300 | Not-determined |
| 18-101_k141_369797 | Not-determined |
| 18-101_k141_304870 | Not-determined |
| 18-101_k141_90110 | Not-determined |
| 18-101_k141_304874 | Not-determined |
| 18-101_k141_40687 | Not-determined |
| 18-101_k141_304882 | Not-determined |
| 18-101_k141_238611 | Not-determined |
| 18-101_k141_40690 | Not-determined |
| 18-101_k141_238614 | Not-determined |
| 18-101_k141_321835 | Not-determined |
| 18-101_k141_238617 | Not-determined |
| 18-101_k141_57311 | Not-determined |
| 18-101_k141_8149 | Not-determined |
| 18-101_k141_238624 | Not-determined |
| 18-101_k141_321843 | Not-determined |
| 18-101_k141_321847 | Not-determined |
| 18-101_k141_336515 | Low-quality |
| 18-101_k141_321850 | Not-determined |
| 18-101_k141_287755 | Not-determined |
| 18-101_k141_73102 | Not-determined |
| 18-101_k141_386714 | Not-determined |
| 18-101_k141_105940 | Not-determined |
| 18-101_k141_386716 | Not-determined |
| 18-101_k141_287762 | Not-determined |
| 18-101_k141_336528 | Not-determined |
| 18-101_k141_238640 | Not-determined |
| 18-101_k141_105948 | Not-determined |
| 18-101_k141_105949 | Not-determined |
| 18-101_k141_189714 | Not-determined |
| 18-101_k141_189716 | Not-determined |
| 18-101_k141_171753 | Not-determined |
| 18-101_k141_304917 | Not-determined |
| 18-101_k141_105953 | Not-determined |
| 18-101_k141_321864 | Low-quality |
| 18-101_k141_8164 | Not-determined |
| 18-101_k141_321865 | Not-determined |
| 18-101_k141_369819 | Not-determined |
| 18-101_k141_105954 | Low-quality |
| 18-101_k141_386724 | Not-determined |
| 18-101_k141_255480 | Not-determined |
| 18-101_k141_90124 | Not-determined |
| 18-101_k141_8169 | Not-determined |
| 18-101_k141_336544 | Not-determined |
| 18-101_k141_336546 | Not-determined |
| 18-101_k141_206094 | Not-determined |
| 18-101_k141_221946 | Not-determined |
| 18-101_k141_238645 | Not-determined |
| 18-101_k141_304925 | Not-determined |
| 18-101_k141_90126 | Not-determined |
| 18-101_k141_336553 | Not-determined |
| 18-101_k141_336554 | Low-quality |
| 18-101_k141_304928 | Not-determined |
| 18-101_k141_255491 | Not-determined |
| 18-101_k141_90132 | Not-determined |
| 18-101_k141_8184 | Not-determined |
| 18-101_k141_24264 | Not-determined |
| 18-101_k141_105961 | Low-quality |
| 18-101_k141_24267 | Not-determined |
| 18-101_k141_24268 | Not-determined |
| 18-101_k141_287803 | Not-determined |
| 18-101_k141_73124 | Not-determined |
| 18-101_k141_369824 | Not-determined |
| 18-101_k141_287809 | Not-determined |
| 18-101_k141_206126 | Not-determined |
| 18-101_k141_386737 | Not-determined |
| 18-101_k141_369839 | Not-determined |
| 18-101_k141_238660 | Not-determined |
| 18-101_k141_105980 | Not-determined |
| 18-101_k141_238662 | Not-determined |
| 18-101_k141_304949 | Not-determined |
| 18-101_k141_24290 | Not-determined |
| 18-101_k141_90161 | Not-determined |
| 18-101_k141_171764 | Not-determined |
| 18-101_k141_90168 | Not-determined |
| 18-101_k141_8203 | Not-determined |
| 18-101_k141_304956 | Not-determined |
| 18-101_k141_90182 | Not-determined |
| 18-101_k141_336574 | Not-determined |
| 18-101_k141_304960 | Not-determined |
| 18-101_k141_124107 | Not-determined |
| 18-101_k141_57354 | Not-determined |
| 18-101_k141_124109 | Not-determined |
| 18-101_k141_386742 | Low-quality |
| 18-101_k141_386745 | Not-determined |
| 18-101_k141_90188 | Not-determined |
| 18-101_k141_124111 | Not-determined |
| 18-101_k141_287829 | Not-determined |
| 18-101_k141_287830 | Not-determined |
| 18-101_k141_171785 | Not-determined |
| 18-101_k141_287836 | Low-quality |
| 18-101_k141_106011 | Not-determined |
| 18-101_k141_73152 | Low-quality |
| 18-101_k141_369856 | Not-determined |
| 18-101_k141_73154 | Low-quality |
| 18-101_k141_369857 | Not-determined |
| 18-101_k141_140009 | Not-determined |
| 18-101_k141_287842 | Not-determined |
| 18-101_k141_73156 | Not-determined |
| 18-101_k141_369861 | Not-determined |
| 18-101_k141_287845 | Not-determined |
| 18-101_k141_140014 | Not-determined |
| 18-101_k141_238688 | Not-determined |
| 18-101_k141_369862 | Not-determined |
| 18-101_k141_189733 | Not-determined |
| 18-101_k141_287848 | Not-determined |
| 18-101_k141_73160 | Low-quality |
| 18-101_k141_189738 | Not-determined |
| 18-101_k141_73164 | Not-determined |
| 18-101_k141_221981 | Low-quality |
| 18-101_k141_124132 | Not-determined |
| 18-101_k141_189742 | Not-determined |
| 18-101_k141_124138 | Low-quality |
| 18-101_k141_321919 | Not-determined |
| 18-101_k141_336616 | Not-determined |
| 18-101_k141_287855 | Not-determined |
| 18-101_k141_221996 | Not-determined |
| 18-101_k141_255503 | Not-determined |
| 18-101_k141_73166 | Not-determined |
| 18-101_k141_206168 | Not-determined |
| 18-101_k141_189757 | Not-determined |
| 18-101_k141_369879 | Not-determined |
| 18-101_k141_8250 | Not-determined |
| 18-101_k141_8253 | Low-quality |
| 18-101_k141_336630 | Not-determined |
| 18-101_k141_24299 | Not-determined |
| 18-101_k141_321936 | Not-determined |
| 18-101_k141_369892 | Not-determined |
| 18-101_k141_124151 | Not-determined |
| 18-101_k141_369895 | Not-determined |
| 18-101_k141_238716 | Not-determined |
| 18-101_k141_222008 | Not-determined |
| 18-101_k141_189770 | Not-determined |
| 18-101_k141_238720 | Not-determined |
| 18-101_k141_222015 | Not-determined |
| 18-101_k141_336638 | Not-determined |
| 18-101_k141_304988 | Not-determined |
| 18-101_k141_255523 | Not-determined |
| 18-101_k141_189775 | Not-determined |
| 18-101_k141_189776 | Not-determined |
| 18-101_k141_255529 | Not-determined |
| 18-101_k141_354165 | Low-quality |
| 18-101_k141_189783 | Not-determined |
| 18-101_k141_369924 | Not-determined |
| 18-101_k141_73176 | Not-determined |
| 18-101_k141_206196 | Not-determined |
| 18-101_k141_369929 | Not-determined |
| 18-101_k141_238732 | Not-determined |
| 18-101_k141_369933 | Low-quality |
| 18-101_k141_369935 | Not-determined |
| 18-101_k141_255547 | Not-determined |
| 18-101_k141_386817 | Not-determined |
| 18-101_k141_369936 | Not-determined |
| 18-101_k141_305016 | Not-determined |
| 18-101_k141_222037 | Not-determined |
| 18-101_k141_287875 | Not-determined |
| 18-101_k141_124173 | Not-determined |
| 18-101_k141_354176 | Not-determined |
| 18-101_k141_8289 | Not-determined |
| 18-101_k141_305026 | Not-determined |
| 18-101_k141_369944 | Not-determined |
| 18-101_k141_124176 | Not-determined |
| 18-101_k141_124177 | Not-determined |
| 18-101_k141_73190 | Not-determined |
| 18-101_k141_238750 | Low-quality |
| 18-101_k141_354193 | Not-determined |
| 18-101_k141_354198 | Not-determined |
| 18-101_k141_90194 | Not-determined |
| 18-101_k141_287896 | Not-determined |
| 18-101_k141_305049 | Not-determined |
| 18-101_k141_238775 | Not-determined |
| 18-101_k141_255578 | Low-quality |
| 18-101_k141_369959 | Not-determined |
| 18-101_k141_287920 | Not-determined |
| 18-101_k141_386856 | Low-quality |
| 18-101_k141_354218 | Not-determined |
| 18-101_k141_369965 | Not-determined |
| 18-101_k141_189831 | Not-determined |
| 18-101_k141_124207 | Not-determined |
| 18-101_k141_171809 | Not-determined |
| 18-101_k141_287933 | Not-determined |
| 18-101_k141_140057 | Not-determined |
| 18-101_k141_73204 | Not-determined |
| 18-101_k141_369972 | Not-determined |
| 18-101_k141_321947 | Not-determined |
| 18-101_k141_354230 | Not-determined |
| 18-101_k141_189838 | Not-determined |
| 18-101_k141_386871 | Not-determined |
| 18-101_k141_140068 | Low-quality |
| 18-101_k141_321954 | Not-determined |
| 18-101_k141_171824 | Not-determined |
| 18-101_k141_106026 | Not-determined |
| 18-101_k141_189847 | Not-determined |
| 18-101_k141_189853 | Not-determined |
| 18-101_k141_140080 | Not-determined |
| 18-101_k141_140083 | Not-determined |
| 18-101_k141_140084 | Low-quality |
| 18-101_k141_171835 | Not-determined |
| 18-101_k141_222057 | Not-determined |
| 18-101_k141_369982 | Not-determined |
| 18-101_k141_171840 | Not-determined |
| 18-101_k141_189860 | Not-determined |
| 18-101_k141_238811 | Not-determined |
| 18-101_k141_238812 | Low-quality |
| 18-101_k141_24308 | Not-determined |
| 18-101_k141_255604 | Not-determined |
| 18-101_k141_271326 | Low-quality |
| 18-101_k141_271328 | Low-quality |
| 18-101_k141_222068 | Low-quality |
| 18-101_k141_271329 | Not-determined |
| 18-101_k141_321969 | Not-determined |
| 18-101_k141_222072 | Not-determined |
| 18-101_k141_24318 | Not-determined |
| 18-101_k141_106039 | Not-determined |
| 18-101_k141_238823 | Not-determined |
| 18-101_k141_354254 | Not-determined |
| 18-101_k141_124244 | Not-determined |
| 18-101_k141_189871 | Not-determined |
| 18-101_k141_222080 | Low-quality |
| 18-101_k141_336691 | Not-determined |
| 18-101_k141_171859 | Not-determined |
| 18-101_k141_124253 | Not-determined |
| 18-101_k141_206262 | Not-determined |
| 18-101_k141_255628 | Not-determined |
| 18-101_k141_124257 | Not-determined |
| 18-101_k141_354275 | Not-determined |
| 18-101_k141_106068 | Low-quality |
| 18-101_k141_189891 | Not-determined |
| 18-101_k141_354279 | Not-determined |
| 18-101_k141_305078 | Not-determined |
| 18-101_k141_206273 | Not-determined |
| 18-101_k141_287944 | Not-determined |
| 18-101_k141_255635 | Not-determined |
| 18-101_k141_321984 | Not-determined |
| 18-101_k141_73226 | Not-determined |
| 18-101_k141_73228 | Not-determined |
| 18-101_k141_386884 | Not-determined |
| 18-101_k141_206281 | Not-determined |
| 18-101_k141_287951 | Low-quality |
| 18-101_k141_73231 | Not-determined |
| 18-101_k141_222104 | Not-determined |
| 18-101_k141_90201 | Not-determined |
| 18-101_k141_90203 | Not-determined |
| 18-101_k141_171872 | Not-determined |
| 18-101_k141_321990 | Not-determined |
| 18-101_k141_206286 | Not-determined |
| 18-101_k141_271359 | Not-determined |
| 18-101_k141_305085 | Not-determined |
| 18-101_k141_24340 | Low-quality |
| 18-101_k141_171877 | Not-determined |
| 18-101_k141_222112 | Not-determined |
| 18-101_k141_40712 | Not-determined |
| 18-101_k141_321999 | Not-determined |
| 18-101_k141_124278 | Not-determined |
| 18-101_k141_189913 | Not-determined |
| 18-101_k141_106088 | Not-determined |
| 18-101_k141_189914 | Low-quality |
| 18-101_k141_90216 | Not-determined |
| 18-101_k141_124287 | Not-determined |
| 18-101_k141_255659 | Not-determined |
| 18-101_k141_90222 | Not-determined |
| 18-101_k141_8374 | Not-determined |
| 18-101_k141_73257 | Not-determined |
| 18-101_k141_171894 | Low-quality |
| 18-101_k141_271376 | Not-determined |
| 18-101_k141_124296 | Not-determined |
| 18-101_k141_206303 | Not-determined |
| 18-101_k141_222135 | Not-determined |
| 18-101_k141_24369 | Not-determined |
| 18-101_k141_24371 | Not-determined |
| 18-101_k141_386907 | Not-determined |
| 18-101_k141_189939 | Not-determined |
| 18-101_k141_124303 | Not-determined |
| 18-101_k141_271381 | Not-determined |
| 18-101_k141_354323 | Not-determined |
| 18-101_k141_386908 | Not-determined |
| 18-101_k141_73271 | Not-determined |
| 18-101_k141_386913 | Low-quality |
| 18-101_k141_40717 | Not-determined |
| 18-101_k141_222154 | Not-determined |
| 18-101_k141_40720 | Not-determined |
| 18-101_k141_40722 | Not-determined |
| 18-101_k141_305091 | Not-determined |
| 18-101_k141_238863 | Low-quality |
| 18-101_k141_8402 | Not-determined |
| 18-101_k141_305100 | Not-determined |
| 18-101_k141_40724 | Low-quality |
| 18-101_k141_206307 | Not-determined |
| 18-101_k141_189968 | Not-determined |
| 18-101_k141_206310 | Not-determined |
| 18-101_k141_8410 | Not-determined |
| 18-101_k141_386931 | Not-determined |
| 18-101_k141_189973 | Not-determined |
| 18-101_k141_8417 | Low-quality |
| 18-101_k141_124335 | Not-determined |
| 18-101_k141_322030 | Not-determined |
| 18-101_k141_354370 | Low-quality |
| 18-101_k141_24408 | Not-determined |
| 18-101_k141_106111 | Not-determined |
| 18-101_k141_24409 | Not-determined |
| 18-101_k141_189993 | Not-determined |
| 18-101_k141_222172 | Not-determined |
| 18-101_k141_255722 | Not-determined |
| 18-101_k141_238887 | Not-determined |
| 18-101_k141_8434 | Not-determined |
| 18-101_k141_57409 | Not-determined |
| 18-101_k141_8435 | Not-determined |
| 18-101_k141_287986 | Not-determined |
| 18-101_k141_386947 | Not-determined |
| 18-101_k141_90264 | Not-determined |
| 18-101_k141_90267 | Not-determined |
| 18-101_k141_124340 | Not-determined |
| 18-101_k141_287998 | Not-determined |
| 18-101_k141_322039 | Not-determined |
| 18-101_k141_57424 | Low-quality |
| 18-101_k141_57426 | Not-determined |
| 18-101_k141_354385 | Not-determined |
| 18-101_k141_90276 | Low-quality |
| 18-101_k141_255742 | Not-determined |
| 18-101_k141_322054 | Low-quality |
| 18-101_k141_24438 | Not-determined |
| 18-101_k141_322057 | Not-determined |
| 18-101_k141_73288 | Not-determined |
| 18-101_k141_73290 | Not-determined |
| 18-101_k141_90282 | Not-determined |
| 18-101_k141_206367 | Not-determined |
| 18-101_k141_271428 | Not-determined |
| 18-101_k141_255752 | Not-determined |
| 18-101_k141_222218 | Not-determined |
| 18-101_k141_322070 | Not-determined |
| 18-101_k141_155757 | Not-determined |
| 17-106_k141_19025 | High-quality |
| 18-101_k141_288013 | Low-quality |
| 18-101_k141_255757 | Not-determined |
| 18-101_k141_106116 | Not-determined |
| 18-101_k141_305104 | Not-determined |
| 18-101_k141_322077 | Not-determined |
| 18-101_k141_322082 | Not-determined |
| 18-101_k141_124362 | Not-determined |
| 18-101_k141_370040 | Not-determined |
| 18-101_k141_206380 | Not-determined |
| 18-101_k141_57430 | Not-determined |
| 18-101_k141_106125 | Not-determined |
| 18-101_k141_190028 | Not-determined |
| 18-101_k141_106127 | Not-determined |
| 18-101_k141_206386 | Not-determined |
| 18-101_k141_370051 | Not-determined |
| 18-101_k141_305117 | Not-determined |
| 18-101_k141_305119 | Not-determined |
| 18-101_k141_305120 | Not-determined |
| 18-101_k141_305123 | Not-determined |
| 18-101_k141_386984 | Not-determined |
| 18-101_k141_305125 | Low-quality |
| 18-101_k141_305126 | Not-determined |
| 18-101_k141_222238 | Not-determined |
| 18-101_k141_222239 | Not-determined |
| 18-101_k141_305129 | Not-determined |
| 18-101_k141_386989 | Low-quality |
| 18-101_k141_222240 | Not-determined |
| 18-101_k141_206400 | Not-determined |
| 18-101_k141_171938 | Not-determined |
| 18-101_k141_322098 | Not-determined |
| 18-101_k141_222247 | Not-determined |
| 18-101_k141_24453 | Not-determined |
| 18-101_k141_106138 | Low-quality |
| 18-101_k141_370063 | Not-determined |
| 18-101_k141_222260 | Low-quality |
| 18-101_k141_370068 | Not-determined |
| 18-101_k141_40761 | Not-determined |
| 18-101_k141_305143 | Low-quality |
| 18-101_k141_271463 | Low-quality |
| 18-101_k141_370073 | Not-determined |
| 18-101_k141_238896 | Not-determined |
| 18-101_k141_305152 | Not-determined |
| 18-101_k141_124396 | Low-quality |
| 18-101_k141_124399 | Not-determined |
| 18-101_k141_370082 | Not-determined |
| 18-101_k141_40770 | Not-determined |
| 18-101_k141_57450 | Not-determined |
| 18-101_k141_57451 | Not-determined |
| 18-101_k141_73297 | Not-determined |
| 18-101_k141_40775 | Not-determined |
| 18-101_k141_73300 | Not-determined |
| 18-101_k141_354454 | Low-quality |
| 18-101_k141_190079 | Not-determined |
| 18-101_k141_238905 | Not-determined |
| 18-101_k141_73305 | Not-determined |
| 18-101_k141_106167 | Not-determined |
| 18-101_k141_124430 | Not-determined |
| 18-101_k141_40784 | Not-determined |
| 18-101_k141_354461 | Not-determined |
| 18-101_k141_73309 | Low-quality |
| 18-101_k141_387052 | Low-quality |
| 18-101_k141_106171 | Not-determined |
| 18-101_k141_387054 | Not-determined |
| 18-101_k141_124437 | Not-determined |
| 18-101_k141_106174 | Not-determined |
| 18-101_k141_271500 | Not-determined |
| 18-101_k141_106177 | Not-determined |
| 18-101_k141_387059 | Not-determined |
| 18-101_k141_40789 | Not-determined |
| 18-101_k141_322153 | Not-determined |
| 18-101_k141_124444 | Not-determined |
| 18-101_k141_238907 | Low-quality |
| 18-101_k141_106182 | Not-determined |
| 18-101_k141_387063 | Not-determined |
| 18-101_k141_255827 | Not-determined |
| 18-101_k141_238909 | Not-determined |
| 18-101_k141_387064 | Not-determined |
| 18-101_k141_387067 | Not-determined |
| 18-101_k141_90345 | Not-determined |
| 18-101_k141_387069 | Not-determined |
| 18-101_k141_271509 | Not-determined |
| 18-101_k141_155799 | Not-determined |
| 18-101_k141_387073 | Not-determined |
| 18-101_k141_370125 | Not-determined |
| 18-101_k141_190107 | Not-determined |
| 18-101_k141_206422 | Not-determined |
| 18-101_k141_190108 | Not-determined |
| 18-101_k141_106191 | Not-determined |
| 18-101_k141_354483 | Low-quality |
| 18-101_k141_238919 | Not-determined |
| 18-101_k141_305202 | Not-determined |
| 18-101_k141_106193 | Not-determined |
| 18-101_k141_238923 | Not-determined |
| 18-101_k141_40796 | Not-determined |
| 18-101_k141_171961 | Not-determined |
| 18-101_k141_190118 | Not-determined |
| 18-101_k141_106198 | Not-determined |
| 18-101_k141_255844 | Not-determined |
| 18-101_k141_305209 | Not-determined |
| 18-101_k141_106201 | Not-determined |
| 18-101_k141_354489 | Not-determined |
| 18-101_k141_40802 | Low-quality |
| 18-101_k141_24517 | Not-determined |
| 18-101_k141_40803 | Not-determined |
| 18-101_k141_206432 | Not-determined |
| 18-101_k141_106204 | Low-quality |
| 18-101_k141_305219 | Not-determined |
| 18-101_k141_288051 | Not-determined |
| 18-101_k141_24524 | Not-determined |
| 18-101_k141_288055 | Not-determined |
| 18-101_k141_155813 | Not-determined |

| | |
|---|---|
| 18-101_k141_8452 | Low-quality |
| 18-101_k141_222321 | Not-determined |
| 18-101_k141_255855 | Not-determined |
| 18-101_k141_354498 | Not-determined |
| 18-101_k141_370145 | Not-determined |
| 18-101_k141_322178 | Not-determined |
| 18-101_k141_190153 | Not-determined |
| 18-101_k141_322180 | Not-determined |
| 18-101_k141_40817 | Low-quality |
| 18-101_k141_190156 | Not-determined |
| 18-101_k141_206441 | Low-quality |
| 18-101_k141_24530 | Not-determined |
| 18-101_k141_255875 | Not-determined |
| 18-101_k141_255878 | Low-quality |
| 18-101_k141_124488 | Not-determined |
| 18-101_k141_190163 | Not-determined |
| 18-101_k141_222338 | Not-determined |
| 18-101_k141_57495 | Not-determined |
| 18-101_k141_8460 | Not-determined |
| 18-101_k141_206444 | Not-determined |
| 18-101_k141_190174 | Not-determined |
| 18-101_k141_106233 | Not-determined |
| 18-101_k141_24543 | Not-determined |
| 18-101_k141_90382 | Not-determined |
| 18-101_k141_206448 | Not-determined |
| 18-101_k141_222347 | Not-determined |
| 18-101_k141_124502 | Not-determined |
| 18-101_k141_370165 | Not-determined |
| 18-101_k141_354516 | Not-determined |
| 18-101_k141_171989 | Not-determined |
| 18-101_k141_155839 | Not-determined |
| 18-101_k141_255888 | Not-determined |
| 18-101_k141_24547 | Not-determined |
| 18-101_k141_40835 | Not-determined |
| 18-101_k141_222357 | Not-determined |
| 18-101_k141_387099 | Not-determined |
| 18-101_k141_73349 | Not-determined |
| 18-101_k141_40844 | Not-determined |
| 18-101_k141_73354 | Not-determined |
| 18-101_k141_255907 | Not-determined |
| 18-101_k141_288091 | Not-determined |
| 18-101_k141_40850 | Not-determined |
| 18-101_k141_155841 | Not-determined |
| 18-101_k141_190198 | Not-determined |
| 18-101_k141_90396 | Low-quality |
| 18-101_k141_8474 | Not-determined |
| 18-101_k141_387116 | Low-quality |
| 18-101_k141_8478 | Not-determined |
| 18-101_k141_8485 | Not-determined |
| 18-101_k141_57521 | Low-quality |
| 18-101_k141_190209 | Low-quality |
| 18-101_k141_40865 | Not-determined |
| 18-101_k141_336721 | Not-determined |
| 18-101_k141_155854 | Low-quality |
| 18-101_k141_322205 | Not-determined |
| 18-101_k141_73377 | Not-determined |
| 18-101_k141_271597 | Not-determined |
| 18-101_k141_238956 | Not-determined |
| 18-101_k141_322209 | Not-determined |
| 18-101_k141_172005 | Not-determined |
| 18-101_k141_40873 | Not-determined |
| 18-101_k141_238961 | Not-determined |
| 18-101_k141_172009 | Not-determined |
| 18-101_k141_155861 | Not-determined |
| 18-101_k141_206460 | Not-determined |
| 18-101_k141_288100 | Not-determined |
| 18-101_k141_370190 | Not-determined |
| 18-101_k141_288103 | Not-determined |
| 18-101_k141_238970 | Low-quality |
| 18-101_k141_288105 | Not-determined |
| 18-101_k141_271607 | Not-determined |
| 18-101_k141_8505 | Low-quality |
| 18-101_k141_238976 | Not-determined |
| 18-101_k141_370196 | Not-determined |
| 18-101_k141_40886 | Not-determined |
| 18-101_k141_40888 | Not-determined |
| 18-101_k141_172028 | Not-determined |
| 18-101_k141_40896 | Not-determined |
| 18-101_k141_305279 | Not-determined |
| 18-101_k141_73391 | Not-determined |
| 18-101_k141_124538 | Not-determined |
| 18-101_k141_222386 | Not-determined |
| 18-101_k141_222387 | Not-determined |
| 18-101_k141_336742 | Not-determined |
| 18-101_k141_73395 | Not-determined |
| 18-101_k141_8530 | Not-determined |
| 18-101_k141_40901 | Not-determined |
| 18-101_k141_73399 | Not-determined |
| 18-101_k141_40902 | Not-determined |
| 18-101_k141_24580 | Not-determined |
| 18-101_k141_8533 | Not-determined |
| 18-101_k141_155884 | Not-determined |
| 18-101_k141_106266 | Not-determined |
| 18-101_k141_222392 | Low-quality |
| 18-101_k141_140091 | Low-quality |
| 18-101_k141_73403 | Not-determined |
| 18-101_k141_222393 | Not-determined |
| 18-101_k141_8536 | Not-determined |
| 18-101_k141_106271 | Not-determined |
| 18-101_k141_40905 | Low-quality |
| 18-101_k141_322215 | Not-determined |
| 18-101_k141_90429 | Not-determined |
| 18-101_k141_40912 | Not-determined |
| 18-101_k141_370208 | Not-determined |
| 18-101_k141_172043 | Not-determined |
| 18-101_k141_370209 | Not-determined |
| 18-101_k141_73414 | Not-determined |
| 18-101_k141_305296 | Not-determined |
| 18-101_k141_370220 | Not-determined |
| 18-101_k141_322236 | Not-determined |
| 18-101_k141_73426 | Not-determined |
| 18-101_k141_190247 | Not-determined |
| 18-101_k141_354550 | Not-determined |
| 18-101_k141_354553 | Not-determined |
| 18-101_k141_155900 | Not-determined |
| 18-101_k141_336764 | Not-determined |
| 18-101_k141_73433 | Not-determined |
| 18-101_k141_354555 | Not-determined |
| 18-101_k141_90443 | Not-determined |
| 18-101_k141_271657 | Not-determined |
| 18-101_k141_354563 | Not-determined |
| 18-101_k141_255961 | Not-determined |
| 18-101_k141_222418 | Not-determined |
| 18-101_k141_190259 | Not-determined |
| 18-101_k141_90453 | Not-determined |
| 18-101_k141_271664 | Not-determined |
| 18-101_k141_255969 | Low-quality |
| 18-101_k141_206533 | Not-determined |
| 18-101_k141_387142 | Not-determined |
| 18-101_k141_155909 | Not-determined |
| 18-101_k141_155911 | Not-determined |
| 18-101_k141_222441 | Not-determined |
| 18-101_k141_370246 | Not-determined |
| 18-101_k141_155914 | Not-determined |
| 18-101_k141_336786 | Not-determined |
| 18-101_k141_271680 | Not-determined |
| 18-101_k141_239016 | Not-determined |
| 18-101_k141_354594 | Not-determined |
| 18-101_k141_8569 | Not-determined |
| 18-101_k141_239020 | Not-determined |
| 18-101_k141_239023 | Not-determined |
| 18-101_k141_73466 | Not-determined |
| 18-101_k141_24641 | Not-determined |
| 18-101_k141_354599 | Not-determined |
| 18-101_k141_387153 | Not-determined |
| 18-101_k141_305313 | Not-determined |
| 18-101_k141_354603 | Not-determined |
| 18-101_k141_222450 | Not-determined |
| 18-101_k141_57537 | Low-quality |
| 18-101_k141_354605 | Not-determined |
| 18-101_k141_90478 | Not-determined |
| 18-101_k141_322248 | Not-determined |
| 18-101_k141_305321 | Not-determined |
| 18-101_k141_305324 | Not-determined |
| 18-101_k141_239043 | Not-determined |
| 18-101_k141_106287 | Not-determined |
| 18-101_k141_305329 | Not-determined |
| 18-101_k141_73477 | Not-determined |
| 18-101_k141_322253 | Not-determined |
| 18-101_k141_206565 | Low-quality |
| 18-101_k141_73481 | Not-determined |
| 18-101_k141_57550 | Not-determined |
| 18-101_k141_336824 | Not-determined |
| 18-101_k141_354625 | Not-determined |
| 18-101_k141_206571 | Not-determined |
| 18-101_k141_172050 | Not-determined |
| 18-101_k141_140139 | Not-determined |
| 18-101_k141_90482 | Not-determined |
| 18-101_k141_8581 | Not-determined |
| 18-101_k141_8582 | Not-determined |
| 18-101_k141_288132 | Not-determined |
| 18-101_k141_222471 | Not-determined |
| 18-101_k141_288134 | Not-determined |
| 18-101_k141_40943 | Not-determined |
| 18-101_k141_222475 | Not-determined |
| 18-101_k141_288138 | Not-determined |
| 18-101_k141_40952 | Not-determined |
| 18-101_k141_8591 | Not-determined |
| 18-101_k141_90492 | Not-determined |
| 18-101_k141_124570 | Not-determined |
| 18-101_k141_40954 | Not-determined |
| 18-101_k141_8606 | Not-determined |
| 18-101_k141_90500 | Not-determined |
| 18-101_k141_140156 | Not-determined |
| 18-101_k141_40955 | Not-determined |
| 18-101_k141_40956 | Not-determined |
| 18-101_k141_190295 | Low-quality |
| 18-101_k141_370280 | Not-determined |
| 18-101_k141_40959 | Not-determined |
| 18-101_k141_140158 | Not-determined |
| 18-101_k141_40961 | Not-determined |
| 18-101_k141_322288 | Not-determined |
| 18-101_k141_172064 | Not-determined |
| 18-101_k141_387172 | Not-determined |
| 18-101_k141_106292 | Not-determined |
| 18-101_k141_387173 | Not-determined |
| 18-101_k141_239088 | Not-determined |
| 18-101_k141_387178 | Not-determined |
| 18-101_k141_370285 | Low-quality |
| 18-101_k141_106297 | Not-determined |
| 18-101_k141_172069 | Not-determined |
| 18-101_k141_172072 | Low-quality |
| 18-101_k141_106300 | Not-determined |
| 18-101_k141_336858 | Not-determined |
| 18-101_k141_370291 | Not-determined |
| 18-101_k141_387188 | Not-determined |
| 18-101_k141_24707 | Not-determined |
| 18-101_k141_322310 | Not-determined |
| 18-101_k141_140166 | Not-determined |
| 18-101_k141_336873 | Not-determined |
| 18-101_k141_24712 | Not-determined |
| 18-101_k141_24717 | Not-determined |
| 18-101_k141_271712 | Not-determined |
| 18-101_k141_387211 | Not-determined |
| 18-101_k141_140183 | Not-determined |
| 18-101_k141_305374 | Not-determined |
| 18-101_k141_206587 | Not-determined |
| 18-101_k141_140185 | Not-determined |
| 18-101_k141_239108 | Not-determined |
| 18-101_k141_271715 | Not-determined |
| 18-101_k141_271716 | Not-determined |
| 18-101_k141_73506 | Not-determined |
| 18-101_k141_206593 | Not-determined |
| 18-101_k141_106320 | Low-quality |
| 18-101_k141_322320 | Not-determined |
| 18-101_k141_73510 | Low-quality |
| 18-101_k141_190331 | Not-determined |
| 18-101_k141_305376 | Not-determined |
| 18-101_k141_90532 | Low-quality |
| 18-101_k141_387227 | Not-determined |
| 18-101_k141_322322 | Low-quality |
| 18-101_k141_387231 | Not-determined |
| 18-101_k141_90538 | Low-quality |
| 18-101_k141_387236 | Not-determined |
| 18-101_k141_90541 | Not-determined |
| 18-101_k141_140202 | Not-determined |
| 18-101_k141_8648 | Low-quality |
| 18-101_k141_8649 | Not-determined |
| 18-101_k141_73521 | Not-determined |
| 18-101_k141_336902 | Low-quality |
| 18-101_k141_24722 | Not-determined |
| 18-101_k141_190347 | Not-determined |
| 18-101_k141_155927 | Not-determined |
| 18-101_k141_190349 | Not-determined |
| 18-101_k141_24727 | Not-determined |
| 18-101_k141_73536 | Not-determined |
| 18-101_k141_124633 | Not-determined |
| 18-101_k141_336922 | Not-determined |
| 18-101_k141_387252 | Low-quality |
| 18-101_k141_387253 | Not-determined |
| 18-101_k141_387256 | Not-determined |
| 18-101_k141_24745 | Not-determined |
| 18-101_k141_370321 | Not-determined |
| 18-101_k141_8668 | Not-determined |
| 18-101_k141_387262 | Not-determined |
| 18-101_k141_387266 | Not-determined |
| 18-101_k141_190364 | Not-determined |
| 18-101_k141_90570 | Not-determined |
| 18-101_k141_8683 | Not-determined |
| 18-101_k141_336935 | Not-determined |
| 18-101_k141_24755 | Low-quality |
| 18-101_k141_57621 | Not-determined |
| 18-101_k141_239138 | Not-determined |
| 18-101_k141_336940 | Not-determined |
| 18-101_k141_40975 | Not-determined |
| 18-101_k141_206601 | Not-determined |
| 18-101_k141_8696 | Not-determined |
| 18-101_k141_336944 | Not-determined |
| 18-101_k141_288203 | Not-determined |
| 18-101_k141_206607 | Not-determined |
| 18-101_k141_206610 | Not-determined |
| 18-101_k141_8705 | Low-quality |
| 18-101_k141_124665 | Not-determined |
| 18-101_k141_90591 | Not-determined |
| 18-101_k141_305415 | Not-determined |
| 18-101_k141_90597 | Not-determined |
| 18-101_k141_155949 | Not-determined |
| 18-101_k141_239151 | Not-determined |
| 18-101_k141_124679 | Not-determined |
| 18-101_k141_387290 | Not-determined |
| 18-101_k141_8725 | Not-determined |
| 18-101_k141_288229 | Not-determined |
| 18-101_k141_172086 | Not-determined |
| 18-101_k141_190413 | Not-determined |
| 18-101_k141_206626 | Not-determined |
| 18-101_k141_90616 | Not-determined |
| 18-101_k141_40999 | Not-determined |
| 18-101_k141_24781 | Not-determined |
| 18-101_k141_305429 | Low-quality |
| 18-101_k141_24788 | Not-determined |
| 18-101_k141_41007 | Not-determined |
| 18-101_k141_124698 | Not-determined |
| 18-101_k141_322364 | Not-determined |
| 18-101_k141_305433 | Not-determined |
| 18-101_k141_90626 | Not-determined |
| 18-101_k141_336977 | Not-determined |
| 18-101_k141_90627 | Not-determined |
| 18-101_k141_172097 | Not-determined |
| 18-101_k141_222498 | Not-determined |
| 18-101_k141_24795 | Not-determined |
| 18-101_k141_190426 | Low-quality |
| 18-101_k141_24801 | Not-determined |
| 18-101_k141_90634 | Not-determined |
| 18-101_k141_172106 | Not-determined |
| 18-101_k141_140246 | Not-determined |
| 18-101_k141_387305 | Not-determined |
| 18-101_k141_90637 | Not-determined |
| 18-101_k141_24805 | Not-determined |
| 18-101_k141_190438 | Low-quality |
| 18-101_k141_8760 | Not-determined |
| 18-101_k141_140257 | Not-determined |
| 18-101_k141_322392 | Not-determined |
| 18-101_k141_24813 | Not-determined |
| 18-101_k141_8763 | Not-determined |
| 18-101_k141_222515 | Not-determined |
| 18-101_k141_387319 | Low-quality |
| 18-101_k141_24816 | Not-determined |
| 18-101_k141_239174 | Not-determined |
| 18-101_k141_41035 | Not-determined |
| 18-101_k141_387333 | Not-determined |
| 18-101_k141_370395 | Not-determined |
| 18-101_k141_90654 | Not-determined |
| 18-101_k141_156008 | Not-determined |
| 18-101_k141_90655 | Low-quality |
| 18-101_k141_255988 | Not-determined |
| 18-101_k141_190458 | Not-determined |
| 18-101_k141_73561 | Not-determined |
| 18-101_k141_337029 | Not-determined |
| 18-101_k141_222537 | Low-quality |
| 18-101_k141_222539 | Not-determined |
| 18-101_k141_190461 | Not-determined |
| 18-101_k141_222541 | Not-determined |
| 18-101_k141_41041 | Not-determined |
| 18-101_k141_140273 | Low-quality |
| 18-101_k141_24845 | Not-determined |
| 18-101_k141_206646 | Not-determined |
| 18-101_k141_354694 | Not-determined |
| 18-101_k141_222550 | Not-determined |
| 18-101_k141_41050 | Not-determined |
| 18-101_k141_24850 | Not-determined |
| 18-101_k141_387360 | Not-determined |
| 18-101_k141_41052 | Not-determined |
| 18-101_k141_8791 | Not-determined |
| 18-101_k141_239187 | Not-determined |
| 18-101_k141_156036 | Not-determined |
| 18-101_k141_370421 | Not-determined |
| 18-101_k141_337047 | Not-determined |
| 18-101_k141_239192 | Not-determined |
| 18-101_k141_172116 | Not-determined |
| 18-101_k141_239194 | Not-determined |
| 18-101_k141_256017 | Not-determined |
| 18-101_k141_140291 | Not-determined |
| 18-101_k141_239200 | Not-determined |
| 18-101_k141_140293 | Not-determined |
| 18-101_k141_124720 | Not-determined |
| 18-101_k141_256024 | Not-determined |
| 18-101_k141_124722 | Not-determined |
| 18-101_k141_172124 | Not-determined |
| 18-101_k141_172126 | Low-quality |
| 18-101_k141_288275 | Low-quality |
| 18-101_k141_190486 | Not-determined |
| 18-101_k141_305476 | Not-determined |
| 18-101_k141_106356 | Not-determined |
| 18-101_k141_370440 | Not-determined |
| 18-101_k141_370441 | Not-determined |
| 18-101_k141_322424 | Not-determined |
| 18-101_k141_337072 | Not-determined |
| 18-101_k141_322425 | Not-determined |
| 18-101_k141_239218 | Not-determined |
| 18-101_k141_24860 | Not-determined |
| 18-101_k141_222582 | Not-determined |
| 18-101_k141_256041 | Not-determined |
| 18-101_k141_190498 | Not-determined |
| 18-101_k141_90663 | Not-determined |
| 18-101_k141_140311 | Not-determined |
| 18-101_k141_256047 | Not-determined |
| 18-101_k141_337085 | Not-determined |
| 18-101_k141_256048 | Not-determined |
| 18-101_k141_256055 | Not-determined |
| 18-101_k141_337089 | Low-quality |
| 18-101_k141_239228 | Not-determined |
| 18-101_k141_271788 | Not-determined |
| 18-101_k141_337097 | Not-determined |
| 18-101_k141_206693 | Not-determined |
| 18-101_k141_387390 | Not-determined |
| 18-101_k141_106373 | Not-determined |
| 18-101_k141_222604 | Not-determined |
| 18-101_k141_337099 | Low-quality |
| 18-101_k141_271796 | Low-quality |
| 18-101_k141_337105 | Not-determined |
| 18-101_k141_239236 | Not-determined |
| 18-101_k141_271797 | Not-determined |
| 18-101_k141_8805 | Not-determined |
| 18-101_k141_222613 | Not-determined |
| 18-101_k141_73618 | Not-determined |
| 18-101_k141_106386 | Not-determined |
| 18-101_k141_305498 | Not-determined |
| 18-101_k141_73629 | Not-determined |
| 18-101_k141_305499 | Not-determined |
| 18-101_k141_271802 | Not-determined |
| 18-101_k141_41092 | Not-determined |
| 18-101_k141_140331 | Not-determined |
| 18-101_k141_190529 | Low-quality |
| 18-101_k141_41101 | Not-determined |
| 18-101_k141_305511 | Not-determined |
| 18-101_k141_124752 | Not-determined |
| 18-101_k141_305517 | Low-quality |
| 18-101_k141_41106 | Not-determined |
| 18-101_k141_206720 | Not-determined |
| 18-101_k141_206721 | Not-determined |
| 18-101_k141_57669 | Not-determined |
| 18-101_k141_172168 | Not-determined |
| 18-101_k141_73645 | Not-determined |
| 18-101_k141_41108 | Low-quality |
| 18-101_k141_370479 | Not-determined |
| 18-101_k141_222642 | Not-determined |
| 18-101_k141_8824 | Not-determined |
| 18-101_k141_370484 | Not-determined |
| 18-101_k141_305525 | Not-determined |
| 18-101_k141_305526 | Not-determined |
| 18-101_k141_73659 | Not-determined |
| 18-101_k141_172178 | Not-determined |
| 18-101_k141_172179 | Not-determined |
| 18-101_k141_172180 | Not-determined |
| 18-101_k141_41117 | Not-determined |
| 18-101_k141_305532 | Not-determined |
| 18-101_k141_305533 | Not-determined |
| 18-101_k141_370496 | Not-determined |
| 18-101_k141_305535 | Not-determined |
| 18-101_k141_90695 | Not-determined |
| 18-101_k141_90696 | Not-determined |
| 18-101_k141_288324 | Low-quality |
| 18-101_k141_8844 | Not-determined |
| 18-101_k141_24885 | Not-determined |
| 18-101_k141_140344 | Low-quality |
| 18-101_k141_24890 | Low-quality |
| 18-101_k141_370505 | Not-determined |
| 18-101_k141_239271 | Low-quality |
| 18-101_k141_90701 | Not-determined |
| 18-101_k141_90702 | Not-determined |
| 18-101_k141_156050 | Not-determined |
| 18-101_k141_156052 | Not-determined |
| 18-101_k141_41125 | Not-determined |
| 18-101_k141_305551 | Not-determined |
| 18-101_k141_354719 | Not-determined |
| 18-101_k141_206752 | Not-determined |
| 18-101_k141_90710 | Not-determined |
| 18-101_k141_190535 | Not-determined |
| 18-101_k141_305553 | Not-determined |
| 18-101_k141_305555 | Not-determined |
| 18-101_k141_41134 | Not-determined |
| 18-101_k141_305557 | Not-determined |
| 18-101_k141_354720 | Not-determined |
| 18-101_k141_387446 | Not-determined |
| 18-101_k141_256127 | Not-determined |
| 18-101_k141_124790 | Not-determined |
| 18-101_k141_337145 | Not-determined |
| 18-101_k141_271822 | Not-determined |
| 18-101_k141_190544 | Not-determined |
| 18-101_k141_41136 | Not-determined |
| 18-101_k141_124794 | Not-determined |
| 18-101_k141_271825 | Not-determined |
| 18-101_k141_305564 | Not-determined |
| 18-101_k141_305566 | Not-determined |
| 18-101_k141_288356 | Not-determined |
| 18-101_k141_370521 | Not-determined |
| 18-101_k141_73678 | Not-determined |
| 18-101_k141_190558 | Not-determined |
| 18-101_k141_370527 | Not-determined |
| 18-101_k141_288362 | Not-determined |
| 18-101_k141_370530 | Not-determined |
| 18-101_k141_305580 | Not-determined |
| 18-101_k141_288374 | Low-quality |
| 18-101_k141_370539 | Not-determined |
| 18-101_k141_41138 | Not-determined |
| 18-101_k141_24912 | Not-determined |
| 18-101_k141_24913 | Not-determined |
| 18-101_k141_222667 | Not-determined |
| 18-101_k141_206789 | Low-quality |
| 18-101_k141_24915 | Not-determined |
| 18-101_k141_305592 | Not-determined |
| 18-101_k141_24919 | Not-determined |
| 18-101_k141_305595 | Not-determined |
| 18-101_k141_370550 | Not-determined |
| 18-101_k141_140364 | Not-determined |
| 18-101_k141_140369 | Not-determined |
| 18-101_k141_322452 | Low-quality |
| 18-101_k141_370552 | Low-quality |
| 18-101_k141_106429 | Not-determined |
| 18-101_k141_206798 | Not-determined |
| 18-101_k141_239317 | Not-determined |
| 18-101_k141_124820 | Low-quality |
| 18-101_k141_305608 | Not-determined |
| 18-101_k141_305610 | Not-determined |
| 18-101_k141_124821 | Not-determined |
| 18-101_k141_41171 | Not-determined |
| 18-101_k141_206805 | Not-determined |
| 18-101_k141_156075 | Low-quality |
| 18-101_k141_222686 | Not-determined |
| 18-101_k141_90720 | Not-determined |
| 18-101_k141_24934 | Not-determined |
| 18-101_k141_24938 | Low-quality |
| 18-101_k141_288392 | Not-determined |
| 18-101_k141_305617 | Low-quality |
| 18-101_k141_354729 | Not-determined |
| 18-101_k141_106443 | Not-determined |
| 18-101_k141_288399 | Not-determined |
| 18-101_k141_271859 | Not-determined |
| 18-101_k141_106448 | Not-determined |
| 18-101_k141_190598 | Not-determined |
| 18-101_k141_337167 | Not-determined |
| 18-101_k141_239344 | Not-determined |
| 18-101_k141_256163 | Not-determined |
| 18-101_k141_57719 | Not-determined |
| 18-101_k141_57720 | Not-determined |
| 18-101_k141_156094 | Not-determined |
| 18-101_k141_387472 | Not-determined |
| 18-101_k141_387473 | Not-determined |
| 18-101_k141_90748 | Not-determined |
| 18-101_k141_106456 | Not-determined |
| 18-101_k141_24957 | Not-determined |
| 18-101_k141_206811 | Not-determined |
| 18-101_k141_140401 | Not-determined |
| 18-101_k141_106462 | Not-determined |
| 18-101_k141_337187 | Not-determined |
| 18-101_k141_90757 | Not-determined |
| 18-101_k141_222697 | Not-determined |
| 18-101_k141_222698 | Not-determined |
| 18-101_k141_370591 | Not-determined |
| 18-101_k141_305639 | Not-determined |
| 18-101_k141_24968 | Not-determined |
| 18-101_k141_172220 | Not-determined |
| 18-101_k141_24973 | Low-quality |
| 18-101_k141_190615 | Not-determined |
| 18-101_k141_256176 | Not-determined |
| 18-101_k141_172225 | Not-determined |
| 18-101_k141_90767 | Not-determined |
| 18-101_k141_57738 | Not-determined |
| 18-101_k141_354768 | Not-determined |
| 18-101_k141_90771 | Not-determined |
| 18-101_k141_172232 | Not-determined |
| 18-101_k141_90776 | Not-determined |
| 18-101_k141_140413 | Not-determined |
| 18-101_k141_140418 | Not-determined |
| 18-101_k141_305643 | Low-quality |
| 18-101_k141_370617 | Not-determined |
| 18-101_k141_387501 | Not-determined |
| 18-101_k141_172245 | Not-determined |
| 18-101_k141_271870 | Low-quality |
| 18-101_k141_305650 | Low-quality |
| 18-101_k141_370627 | Not-determined |
| 18-101_k141_172253 | Not-determined |
| 18-101_k141_370628 | Not-determined |
| 18-101_k141_387511 | Not-determined |
| 18-101_k141_305655 | Not-determined |
| 18-101_k141_354777 | Not-determined |
| 18-101_k141_25003 | Not-determined |
| 18-101_k141_124860 | Not-determined |
| 18-101_k141_305657 | Not-determined |
| 18-101_k141_305658 | Not-determined |
| 18-101_k141_172255 | Low-quality |
| 18-101_k141_140437 | Not-determined |
| 18-101_k141_124864 | Not-determined |
| 18-101_k141_271887 | Not-determined |
| 18-101_k141_288463 | Not-determined |
| 18-101_k141_206827 | Not-determined |
| 18-101_k141_305662 | Low-quality |
| 18-101_k141_222712 | Not-determined |
| 18-101_k141_271896 | Not-determined |
| 18-101_k141_25006 | Not-determined |
| 18-101_k141_387524 | Low-quality |
| 18-101_k141_8881 | Not-determined |
| 18-101_k141_206831 | Not-determined |
| 18-101_k141_256209 | Not-determined |
| 18-101_k141_256211 | Not-determined |
| 18-101_k141_387529 | Not-determined |
| 18-101_k141_271903 | Not-determined |
| 18-101_k141_305667 | Not-determined |
| 18-101_k141_370654 | Not-determined |
| 18-101_k141_124874 | Not-determined |
| 18-101_k141_124877 | Not-determined |
| 18-101_k141_271913 | Not-determined |
| 18-101_k141_370658 | Not-determined |
| 18-101_k141_239387 | Not-determined |
| 18-101_k141_190656 | Not-determined |
| 18-101_k141_73716 | Not-determined |
| 18-101_k141_172276 | Not-determined |
| 18-101_k141_106494 | Not-determined |
| 18-101_k141_124888 | Not-determined |

| | |
|---|---|
| 18-101_k141_124889 | Not-determined |
| 18-101_k141_239399 | Not-determined |
| 18-101_k141_57752 | Not-determined |
| 18-101_k141_106498 | Not-determined |
| 18-101_k141_90810 | Not-determined |
| 18-101_k141_57754 | Not-determined |
| 18-101_k141_57755 | Not-determined |
| 18-101_k141_222720 | Not-determined |
| 18-101_k141_25013 | Not-determined |
| 18-101_k141_124898 | Not-determined |
| 18-101_k141_140466 | Not-determined |
| 18-101_k141_206859 | Not-determined |
| 18-101_k141_222728 | Not-determined |
| 18-101_k141_140475 | Not-determined |
| 18-101_k141_354828 | Not-determined |
| 18-101_k141_387541 | Not-determined |
| 18-101_k141_172302 | Not-determined |
| 18-101_k141_271944 | Low-quality |
| 18-101_k141_206864 | Not-determined |
| 18-101_k141_387543 | Not-determined |
| 18-101_k141_271948 | Not-determined |
| 18-101_k141_41289 | Not-determined |
| 18-101_k141_354837 | Not-determined |
| 18-101_k141_57781 | Not-determined |
| 18-101_k141_387545 | Low-quality |
| 18-101_k141_57784 | Not-determined |
| 18-101_k141_41293 | Not-determined |
| 18-101_k141_206874 | Not-determined |
| 18-101_k141_370686 | Not-determined |
| 18-101_k141_90836 | Not-determined |
| 18-101_k141_106515 | Not-determined |
| 18-101_k141_8902 | Low-quality |
| 18-101_k141_239426 | Not-determined |
| 18-101_k141_25032 | Not-determined |
| 18-101_k141_370695 | Not-determined |
| 18-101_k141_25035 | Not-determined |
| 18-101_k141_57797 | Not-determined |
| 18-101_k141_370696 | Not-determined |
| 18-101_k141_354853 | Not-determined |
| 18-101_k141_90842 | Not-determined |
| 18-101_k141_57800 | Not-determined |
| 18-101_k141_305711 | Not-determined |
| 18-101_k141_124914 | Not-determined |
| 18-101_k141_41304 | Not-determined |
| 18-101_k141_206881 | Low-quality |
| 18-101_k141_172319 | Not-determined |
| 18-101_k141_73727 | Not-determined |
| 18-101_k141_271973 | Not-determined |
| 18-101_k141_206887 | Not-determined |
| 18-101_k141_90850 | Not-determined |
| 18-101_k141_106536 | Not-determined |
| 18-101_k141_106537 | Not-determined |
| 18-101_k141_305723 | Not-determined |
| 18-101_k141_57807 | Not-determined |
| 18-101_k141_8916 | Not-determined |
| 18-101_k141_256250 | Low-quality |
| 18-101_k141_222733 | Not-determined |
| 18-101_k141_106541 | Not-determined |
| 18-101_k141_288505 | Not-determined |
| 18-101_k141_271985 | Not-determined |
| 18-101_k141_387563 | Not-determined |
| 18-101_k141_172340 | Not-determined |
| 18-101_k141_387567 | Not-determined |
| 18-101_k141_172344 | Not-determined |
| 18-101_k141_172345 | Not-determined |
| 18-101_k141_106553 | Not-determined |
| 18-101_k141_222737 | Low-quality |
| 18-101_k141_387572 | Not-determined |
| 18-101_k141_90853 | Not-determined |
| 18-101_k141_354872 | Not-determined |
| 18-101_k141_106559 | Not-determined |
| 18-101_k141_140518 | Not-determined |
| 18-101_k141_256267 | Not-determined |
| 18-101_k141_239438 | Not-determined |
| 18-101_k141_90859 | Not-determined |
| 18-101_k141_25057 | Not-determined |
| 18-101_k141_222745 | Low-quality |
| 18-101_k141_272000 | Not-determined |
| 18-101_k141_288518 | Not-determined |
| 18-101_k141_140523 | Low-quality |
| 18-101_k141_256281 | Not-determined |
| 18-101_k141_387587 | Not-determined |
| 18-101_k141_305740 | Not-determined |
| 18-101_k141_322477 | Not-determined |
| 18-101_k141_387589 | Not-determined |
| 18-101_k141_387590 | Not-determined |
| 18-101_k141_140531 | Not-determined |
| 18-101_k141_354889 | Not-determined |
| 18-101_k141_140546 | Not-determined |
| 18-101_k141_206919 | Not-determined |
| 18-101_k141_354894 | Not-determined |
| 18-101_k141_206921 | Low-quality |
| 18-101_k141_90882 | Low-quality |
| 18-101_k141_387604 | Not-determined |
| 18-101_k141_272015 | Not-determined |
| 18-101_k141_73743 | Not-determined |
| 18-101_k141_387610 | Low-quality |
| 18-101_k141_57841 | Low-quality |
| 18-101_k141_288527 | Not-determined |
| 18-101_k141_272022 | Not-determined |
| 18-101_k141_288529 | Not-determined |
| 18-101_k141_156146 | Not-determined |
| 18-101_k141_140562 | Not-determined |
| 18-101_k141_156152 | Not-determined |
| 18-101_k141_239451 | Not-determined |
| 18-101_k141_156157 | Not-determined |
| 18-101_k141_370739 | Not-determined |
| 18-101_k141_140569 | Not-determined |
| 18-101_k141_124957 | Not-determined |
| 18-101_k141_272031 | Not-determined |
| 18-101_k141_140575 | Not-determined |
| 18-101_k141_124961 | Not-determined |
| 18-101_k141_156162 | Not-determined |
| 18-101_k141_190701 | Not-determined |
| 18-101_k141_57862 | Not-determined |
| 18-101_k141_222782 | Not-determined |
| 18-101_k141_288545 | Not-determined |
| 18-101_k141_124968 | Not-determined |
| 18-101_k141_8955 | Not-determined |
| 18-101_k141_90898 | Not-determined |
| 18-101_k141_140591 | Low-quality |
| 18-101_k141_124973 | Not-determined |
| 18-101_k141_272038 | Not-determined |
| 18-101_k141_156174 | Not-determined |
| 18-101_k141_206954 | Not-determined |
| 18-101_k141_272040 | Not-determined |
| 18-101_k141_354904 | Not-determined |
| 18-101_k141_370747 | Not-determined |
| 18-101_k141_25095 | Not-determined |
| 18-101_k141_190715 | Not-determined |
| 18-101_k141_25099 | Not-determined |
| 18-101_k141_370752 | Not-determined |
| 18-101_k141_272047 | Not-determined |
| 18-101_k141_140601 | Not-determined |
| 18-101_k141_354912 | Not-determined |
| 18-101_k141_288564 | Not-determined |
| 18-101_k141_90920 | Not-determined |
| 18-101_k141_156190 | Not-determined |
| 18-101_k141_354918 | Not-determined |
| 18-101_k141_354919 | Not-determined |
| 18-101_k141_41362 | Not-determined |
| 18-101_k141_305787 | Not-determined |
| 18-101_k141_354928 | Not-determined |
| 18-101_k141_140618 | Low-quality |
| 18-101_k141_190732 | Not-determined |
| 18-101_k141_305796 | Not-determined |
| 18-101_k141_90927 | Not-determined |
| 18-101_k141_305797 | Not-determined |
| 18-101_k141_322530 | Low-quality |
| 18-101_k141_387647 | Not-determined |
| 18-101_k141_256296 | Not-determined |
| 18-101_k141_387648 | Not-determined |
| 18-101_k141_387649 | Not-determined |
| 18-101_k141_206968 | Not-determined |
| 18-101_k141_140621 | Not-determined |
| 18-101_k141_90933 | Not-determined |
| 18-101_k141_256312 | Not-determined |
| 18-101_k141_8987 | Not-determined |
| 18-101_k141_288588 | Not-determined |
| 18-101_k141_140628 | Not-determined |
| 18-101_k141_140629 | Low-quality |
| 18-101_k141_370771 | Not-determined |
| 18-101_k141_140631 | Not-determined |
| 18-101_k141_354944 | Not-determined |
| 18-101_k141_156219 | Not-determined |
| 18-101_k141_322548 | Not-determined |
| 18-101_k141_57893 | Not-determined |
| 18-101_k141_305817 | Not-determined |
| 18-101_k141_156222 | Not-determined |
| 18-101_k141_370780 | Not-determined |
| 18-101_k141_322552 | Not-determined |
| 18-101_k141_156224 | Low-quality |
| 18-101_k141_156226 | Not-determined |
| 18-101_k141_73747 | Not-determined |
| 18-101_k141_140644 | Not-determined |
| 18-101_k141_8998 | Not-determined |
| 18-101_k141_125020 | Not-determined |
| 18-101_k141_337231 | Not-determined |
| 18-101_k141_305822 | Not-determined |
| 18-101_k141_354967 | Low-quality |
| 18-101_k141_337236 | Low-quality |
| 18-101_k141_125031 | Not-determined |
| 18-101_k141_9012 | Not-determined |
| 18-101_k141_125033 | Not-determined |
| 18-101_k141_305829 | Not-determined |
| 18-101_k141_337251 | Not-determined |
| 18-101_k141_140652 | Not-determined |
| 18-101_k141_90961 | Not-determined |
| 18-101_k141_73773 | Not-determined |
| 18-101_k141_288629 | Not-determined |
| 18-101_k141_206998 | Not-determined |
| 18-101_k141_156236 | Not-determined |
| 18-101_k141_73781 | Not-determined |
| 18-101_k141_57924 | Not-determined |
| 18-101_k141_370814 | Not-determined |
| 18-101_k141_73789 | Not-determined |
| 18-101_k141_90974 | Not-determined |
| 18-101_k141_73801 | Low-quality |
| 18-101_k141_256349 | Not-determined |
| 18-101_k141_256352 | Not-determined |
| 18-101_k141_106579 | Not-determined |
| 18-101_k141_288636 | Not-determined |
| 18-101_k141_305842 | Not-determined |
| 18-101_k141_322586 | Not-determined |
| 18-101_k141_305843 | Not-determined |
| 18-101_k141_222797 | Not-determined |
| 18-101_k141_140677 | Not-determined |
| 18-101_k141_256360 | Not-determined |
| 18-101_k141_90987 | Not-determined |
| 18-101_k141_222799 | Not-determined |
| 18-101_k141_125040 | Not-determined |
| 18-101_k141_239545 | Not-determined |
| 18-101_k141_190785 | Not-determined |
| 18-101_k141_125043 | Not-determined |
| 18-101_k141_370839 | Not-determined |
| 18-101_k141_370840 | Not-determined |
| 18-101_k141_73814 | Not-determined |
| 18-101_k141_25145 | Not-determined |
| 18-101_k141_25146 | Not-determined |
| 18-101_k141_106590 | Not-determined |
| 18-101_k141_272127 | Low-quality |
| 18-101_k141_25152 | Not-determined |
| 18-101_k141_272137 | Not-determined |
| 18-101_k141_222814 | Not-determined |
| 18-101_k141_288652 | Not-determined |
| 18-101_k141_125054 | Not-determined |
| 18-101_k141_305861 | Not-determined |
| 18-101_k141_25159 | Not-determined |
| 18-101_k141_106596 | Low-quality |
| 18-101_k141_41373 | Low-quality |
| 18-101_k141_305867 | Not-determined |
| 18-101_k141_239571 | Not-determined |
| 18-101_k141_73825 | Not-determined |
| 18-101_k141_41377 | Not-determined |
| 18-101_k141_9030 | Not-determined |
| 18-101_k141_256377 | Not-determined |
| 18-101_k141_355010 | Not-determined |
| 18-101_k141_222822 | Not-determined |
| 18-101_k141_239576 | Not-determined |
| 18-101_k141_355019 | Low-quality |
| 18-101_k141_370868 | Not-determined |
| 18-101_k141_256380 | Not-determined |
| 18-101_k141_322598 | Not-determined |
| 18-101_k141_9033 | Low-quality |
| 18-101_k141_57938 | Not-determined |
| 18-101_k141_73834 | Not-determined |
| 18-101_k141_9034 | Not-determined |
| 18-101_k141_57940 | Not-determined |
| 18-101_k141_57942 | Not-determined |
| 18-101_k141_322603 | Not-determined |
| 18-101_k141_239582 | Not-determined |
| 18-101_k141_125076 | Not-determined |
| 18-101_k141_387667 | Low-quality |
| 18-101_k141_9046 | Not-determined |
| 18-101_k141_25184 | Not-determined |
| 18-101_k141_125086 | Not-determined |
| 18-101_k141_337284 | Not-determined |
| 18-101_k141_337287 | Not-determined |
| 18-101_k141_337289 | Not-determined |
| 18-101_k141_370893 | Not-determined |
| 18-101_k141_222845 | Not-determined |
| 18-101_k141_9050 | Not-determined |
| 18-101_k141_370896 | Not-determined |
| 18-101_k141_125095 | Not-determined |
| 18-101_k141_106629 | Low-quality |
| 18-101_k141_222853 | Not-determined |
| 18-101_k141_41395 | Not-determined |
| 18-101_k141_222854 | Not-determined |
| 18-101_k141_125103 | Not-determined |
| 18-101_k141_322616 | Not-determined |
| 18-101_k141_305897 | Not-determined |
| 18-101_k141_140733 | Not-determined |
| 18-101_k141_222864 | Not-determined |
| 18-101_k141_387694 | Not-determined |
| 18-101_k141_57963 | Not-determined |
| 18-101_k141_256411 | Low-quality |
| 18-101_k141_9059 | Not-determined |
| 18-101_k141_207037 | Not-determined |
| 18-101_k141_207038 | Not-determined |
| 18-101_k141_256417 | Not-determined |
| 18-101_k141_387698 | Low-quality |
| 18-101_k141_305911 | Not-determined |
| 18-101_k141_57974 | Not-determined |
| 18-101_k141_140738 | Not-determined |
| 18-101_k141_387701 | Low-quality |
| 18-101_k141_288699 | Not-determined |
| 18-101_k141_207046 | Not-determined |
| 18-101_k141_9065 | Not-determined |
| 18-101_k141_73859 | Not-determined |
| 18-101_k141_140740 | Not-determined |
| 18-101_k141_140741 | Not-determined |
| 18-101_k141_370915 | Not-determined |
| 18-101_k141_222890 | Not-determined |
| 18-101_k141_9074 | Not-determined |
| 18-101_k141_125125 | Not-determined |
| 18-101_k141_41408 | Not-determined |
| 18-101_k141_337320 | Not-determined |
| 18-101_k141_125128 | Not-determined |
| 18-101_k141_156314 | Not-determined |
| 18-101_k141_57988 | Not-determined |
| 18-101_k141_305918 | Not-determined |
| 18-101_k141_239598 | Not-determined |
| 18-101_k141_272161 | Not-determined |
| 18-101_k141_41412 | Not-determined |
| 18-101_k141_41414 | Not-determined |
| 18-101_k141_222897 | Not-determined |
| 18-101_k141_172428 | Not-determined |
| 18-101_k141_222898 | Not-determined |
| 18-101_k141_57997 | Not-determined |
| 18-101_k141_140749 | Low-quality |
| 18-101_k141_140750 | Not-determined |
| 18-101_k141_387725 | Not-determined |
| 18-101_k141_41431 | Low-quality |
| 18-101_k141_156318 | Not-determined |
| 18-101_k141_140753 | Not-determined |
| 18-101_k141_41433 | Not-determined |
| 18-101_k141_156323 | Not-determined |
| 18-101_k141_272176 | Not-determined |
| 18-101_k141_272178 | Not-determined |
| 18-101_k141_73882 | Not-determined |
| 18-101_k141_58013 | Not-determined |
| 18-101_k141_41440 | Not-determined |
| 18-101_k141_272188 | Not-determined |
| 18-101_k141_25208 | Not-determined |
| 18-101_k141_337342 | Not-determined |
| 18-101_k141_272190 | Not-determined |
| 18-101_k141_355078 | Not-determined |
| 18-101_k141_387736 | Not-determined |
| 18-101_k141_9104 | Not-determined |
| 18-101_k141_387739 | Not-determined |
| 18-101_k141_140773 | Not-determined |
| 18-101_k141_9106 | Not-determined |
| 18-101_k141_387742 | Not-determined |
| 18-101_k141_25215 | Not-determined |
| 18-101_k141_387744 | Not-determined |
| 18-101_k141_73890 | Not-determined |
| 18-101_k141_387745 | Not-determined |
| 18-101_k141_125166 | Not-determined |
| 18-101_k141_25217 | Not-determined |
| 18-101_k141_172448 | Not-determined |
| 18-101_k141_73893 | Not-determined |
| 18-101_k141_272194 | Not-determined |
| 18-101_k141_256451 | Not-determined |
| 18-101_k141_305949 | Not-determined |
| 18-101_k141_73895 | Not-determined |
| 18-101_k141_305952 | Not-determined |
| 18-101_k141_322645 | Not-determined |
| 18-101_k141_25233 | Not-determined |
| 18-101_k141_305954 | Low-quality |
| 18-101_k141_9124 | Not-determined |
| 18-101_k141_256461 | Not-determined |
| 18-101_k141_9125 | Not-determined |
| 18-101_k141_256462 | Not-determined |
| 18-101_k141_322651 | Not-determined |
| 18-101_k141_73901 | Not-determined |
| 18-101_k141_156358 | Not-determined |
| 18-101_k141_91040 | Not-determined |
| 18-101_k141_322656 | Not-determined |
| 18-101_k141_91042 | Low-quality |
| 18-101_k141_91043 | Not-determined |
| 18-101_k141_370963 | Low-quality |
| 18-101_k141_256472 | Not-determined |
| 18-101_k141_91045 | Not-determined |
| 18-101_k141_370964 | Not-determined |
| 18-101_k141_73912 | Not-determined |
| 18-101_k141_73916 | Not-determined |
| 18-101_k141_156364 | Low-quality |
| 18-101_k141_41470 | Not-determined |
| 18-101_k141_9144 | Not-determined |
| 18-101_k141_355094 | Low-quality |
| 18-101_k141_156369 | Not-determined |
| 18-101_k141_156370 | Not-determined |
| 18-101_k141_370970 | Not-determined |
| 18-101_k141_256485 | Not-determined |
| 18-101_k141_125184 | Not-determined |
| 18-101_k141_25259 | Not-determined |
| 18-101_k141_207086 | Not-determined |
| 18-101_k141_140803 | Low-quality |
| 18-101_k141_58054 | Not-determined |
| 18-101_k141_106695 | Not-determined |
| 18-101_k141_140806 | Not-determined |
| 18-101_k141_125195 | Not-determined |
| 18-101_k141_58058 | Not-determined |
| 18-101_k141_91057 | Not-determined |
| 18-101_k141_322675 | Not-determined |
| 18-101_k141_355102 | Not-determined |
| 18-101_k141_172465 | Not-determined |
| 18-101_k141_322680 | Not-determined |
| 18-101_k141_106702 | Not-determined |
| 18-101_k141_239665 | Not-determined |
| 18-101_k141_91070 | Not-determined |
| 18-101_k141_172470 | Not-determined |
| 18-101_k141_25274 | Not-determined |
| 18-101_k141_387779 | Not-determined |
| 18-101_k141_140819 | Not-determined |
| 18-101_k141_91076 | Not-determined |
| 18-101_k141_140824 | Not-determined |
| 18-101_k141_140825 | Not-determined |
| 18-101_k141_140829 | Not-determined |
| 18-101_k141_272219 | Not-determined |
| 18-101_k141_322696 | Not-determined |
| 18-101_k141_190801 | Not-determined |
| 18-101_k141_256525 | Not-determined |
| 18-101_k141_25289 | Not-determined |
| 18-101_k141_387796 | Not-determined |
| 18-101_k141_239669 | Not-determined |
| 18-101_k141_239670 | Not-determined |
| 18-101_k141_337368 | Not-determined |
| 18-101_k141_140836 | Not-determined |
| 18-101_k141_106726 | Not-determined |
| 18-101_k141_156385 | Not-determined |
| 18-101_k141_73938 | Not-determined |
| 18-101_k141_387799 | Not-determined |
| 18-101_k141_239674 | Not-determined |
| 18-101_k141_207110 | Not-determined |
| 18-101_k141_156392 | Not-determined |
| 18-101_k141_190810 | Not-determined |
| 18-101_k141_256539 | Low-quality |
| 18-101_k141_239679 | Not-determined |
| 18-101_k141_91092 | Not-determined |
| 18-101_k141_190812 | Not-determined |
| 18-101_k141_172484 | Not-determined |
| 18-101_k141_387806 | Low-quality |
| 18-101_k141_288723 | Not-determined |
| 18-101_k141_25304 | Not-determined |
| 18-101_k141_322708 | Not-determined |
| 18-101_k141_322714 | Not-determined |
| 18-101_k141_207123 | Not-determined |
| 18-101_k141_106734 | Not-determined |
| 18-101_k141_387822 | Not-determined |
| 18-101_k141_371005 | Not-determined |
| 18-101_k141_256554 | Not-determined |
| 18-101_k141_288732 | Low-quality |
| 18-101_k141_371008 | Not-determined |
| 18-101_k141_106743 | Not-determined |
| 18-101_k141_25311 | Not-determined |
| 18-101_k141_256559 | Not-determined |
| 18-101_k141_73956 | Not-determined |
| 18-101_k141_322728 | Not-determined |
| 18-101_k141_322729 | Not-determined |
| 18-101_k141_322730 | Not-determined |
| 18-101_k141_256567 | Not-determined |
| 18-101_k141_207138 | Not-determined |
| 18-101_k141_58101 | Not-determined |
| 18-101_k141_288741 | Not-determined |
| 18-101_k141_73961 | Not-determined |
| 18-101_k141_172518 | Not-determined |
| 18-101_k141_207148 | Not-determined |
| 18-101_k141_222937 | Not-determined |
| 18-101_k141_91135 | Not-determined |
| 18-101_k141_58109 | Low-quality |
| 18-101_k141_305986 | Not-determined |
| 18-101_k141_272262 | Not-determined |
| 18-101_k141_172528 | Not-determined |
| 18-101_k141_207162 | Not-determined |
| 18-101_k141_25319 | Not-determined |
| 18-101_k141_222953 | Not-determined |
| 18-101_k141_272270 | Not-determined |
| 18-101_k141_272272 | Not-determined |
| 18-101_k141_322744 | Not-determined |
| 18-101_k141_73986 | Not-determined |
| 18-101_k141_305996 | Not-determined |
| 18-101_k141_207178 | Not-determined |
| 18-101_k141_272281 | Not-determined |
| 18-101_k141_222958 | Not-determined |
| 18-101_k141_272283 | Not-determined |
| 18-101_k141_256582 | Not-determined |
| 18-101_k141_172546 | Low-quality |
| 18-101_k141_306006 | Not-determined |
| 18-101_k141_272291 | Not-determined |
| 18-101_k141_25336 | Low-quality |
| 18-101_k141_322751 | Not-determined |
| 18-101_k141_322753 | Not-determined |
| 18-101_k141_272293 | Not-determined |
| 18-101_k141_41507 | Not-determined |
| 18-101_k141_172548 | Low-quality |
| 18-101_k141_190859 | Not-determined |
| 18-101_k141_306015 | Not-determined |
| 18-101_k141_288772 | Low-quality |
| 18-101_k141_222976 | Not-determined |
| 18-101_k141_322759 | Not-determined |
| 18-101_k141_106752 | Not-determined |
| 18-101_k141_156446 | Not-determined |
| 18-101_k141_222983 | Not-determined |
| 18-101_k141_387850 | Not-determined |
| 18-101_k141_172563 | Not-determined |
| 18-101_k141_125231 | Not-determined |
| 18-101_k141_41519 | Not-determined |
| 18-101_k141_371035 | Not-determined |
| 18-101_k141_222987 | Not-determined |
| 18-101_k141_9223 | Not-determined |
| 18-101_k141_25365 | Not-determined |
| 18-101_k141_387857 | Not-determined |
| 18-101_k141_25366 | Not-determined |
| 18-101_k141_371037 | Not-determined |
| 18-101_k141_91179 | Not-determined |
| 18-101_k141_41526 | Not-determined |
| 18-101_k141_172565 | Not-determined |
| 18-101_k141_172566 | Not-determined |
| 18-101_k141_125247 | Not-determined |
| 18-101_k141_91188 | Not-determined |
| 18-101_k141_371046 | Not-determined |
| 18-101_k141_125249 | Not-determined |
| 18-101_k141_207192 | Not-determined |
| 18-101_k141_9231 | Not-determined |
| 18-101_k141_306041 | Not-determined |
| 18-101_k141_25374 | Low-quality |
| 18-101_k141_207196 | Low-quality |
| 18-101_k141_125256 | Not-determined |
| 18-101_k141_306044 | Not-determined |
| 18-101_k141_306049 | Not-determined |
| 18-101_k141_41534 | Not-determined |
| 18-101_k141_58166 | Not-determined |
| 18-101_k141_9241 | Not-determined |
| 18-101_k141_306051 | Not-determined |
| 18-101_k141_322786 | Not-determined |
| 18-101_k141_207205 | Not-determined |
| 18-101_k141_41536 | Not-determined |
| 18-101_k141_9249 | Not-determined |
| 18-101_k141_272325 | Not-determined |
| 18-101_k141_387868 | Low-quality |
| 18-101_k141_272326 | Not-determined |
| 18-101_k141_25384 | Not-determined |
| 18-101_k141_207211 | Not-determined |
| 18-101_k141_207213 | Not-determined |
| 18-101_k141_322791 | Not-determined |
| 18-101_k141_25390 | Not-determined |
| 18-101_k141_58177 | Not-determined |
| 18-101_k141_25394 | Not-determined |
| 18-101_k141_125271 | Not-determined |
| 18-101_k141_337411 | Low-quality |
| 18-101_k141_337412 | Not-determined |
| 18-101_k141_140882 | Not-determined |
| 18-101_k141_322798 | Not-determined |
| 18-101_k141_256626 | Not-determined |
| 18-101_k141_306056 | Not-determined |
| 18-101_k141_25406 | Not-determined |
| 18-101_k141_190871 | Low-quality |
| 18-101_k141_207222 | Not-determined |
| 18-101_k141_58185 | Not-determined |
| 18-101_k141_172589 | Low-quality |
| 18-101_k141_9260 | Not-determined |
| 18-101_k141_322805 | Low-quality |
| 18-101_k141_337421 | Not-determined |
| 18-101_k141_172591 | Not-determined |
| 18-101_k141_172595 | Not-determined |
| 18-101_k141_337422 | Not-determined |
| 18-101_k141_125286 | Not-determined |
| 18-101_k141_256632 | Not-determined |
| 18-101_k141_256633 | Not-determined |
| 18-101_k141_125292 | Not-determined |
| 18-101_k141_9273 | Not-determined |
| 18-101_k141_9279 | Not-determined |
| 18-101_k141_156470 | Not-determined |
| 18-101_k141_337434 | Not-determined |
| 18-101_k141_156473 | Not-determined |
| 18-101_k141_371057 | Not-determined |
| 18-101_k141_387886 | Not-determined |
| 18-101_k141_9288 | Not-determined |
| 18-101_k141_9290 | Not-determined |
| 18-101_k141_9291 | Not-determined |
| 18-101_k141_74026 | Not-determined |
| 18-101_k141_91232 | Not-determined |
| 18-101_k141_125312 | Not-determined |
| 18-101_k141_25433 | Not-determined |
| 18-101_k141_9296 | Not-determined |
| 18-101_k141_91234 | Not-determined |
| 18-101_k141_106777 | Not-determined |
| 18-101_k141_223034 | Not-determined |
| 18-101_k141_91241 | Not-determined |
| 18-101_k141_58188 | Low-quality |
| 18-101_k141_256651 | Low-quality |
| 18-101_k141_106782 | Not-determined |
| 18-101_k141_41551 | Not-determined |
| 18-101_k141_74036 | Not-determined |
| 18-101_k141_306084 | Not-determined |
| 18-101_k141_371083 | Low-quality |
| 18-101_k141_106786 | Not-determined |
| 18-101_k141_256654 | Not-determined |
| 18-101_k141_355114 | Not-determined |
| 18-101_k141_190900 | Not-determined |
| 18-101_k141_272376 | Not-determined |
| 18-101_k141_371090 | Not-determined |
| 18-101_k141_387902 | Not-determined |
| 18-101_k141_106791 | Not-determined |
| 18-101_k141_140929 | Not-determined |
| 18-101_k141_272381 | Not-determined |
| 18-101_k141_9304 | Not-determined |
| 18-101_k141_239749 | Low-quality |

| | |
|---|---|
| 18-101_k141_190904 | Not-determined |
| 18-101_k141_355122 | Not-determined |
| 18-101_k141_125336 | Low-quality |
| 18-101_k141_223061 | Not-determined |
| 18-101_k141_207244 | Not-determined |
| 18-101_k141_190909 | Not-determined |
| 18-101_k141_387906 | Not-determined |
| 18-101_k141_207245 | Not-determined |
| 18-101_k141_355123 | Not-determined |
| 18-101_k141_58201 | Not-determined |
| 18-101_k141_256667 | Not-determined |
| 18-101_k141_58202 | Not-determined |
| 18-101_k141_9313 | Not-determined |
| 18-101_k141_337460 | Low-quality |
| 18-101_k141_140941 | Not-determined |
| 18-101_k141_322834 | Not-determined |
| 18-101_k141_207259 | Not-determined |
| 18-101_k141_337463 | Not-determined |
| 18-101_k141_355136 | Not-determined |
| 18-101_k141_207260 | Not-determined |
| 18-101_k141_387910 | Not-determined |
| 18-101_k141_25448 | Not-determined |
| 18-101_k141_355139 | Not-determined |
| 18-101_k141_156508 | Not-determined |
| 18-101_k141_337468 | Not-determined |
| 18-101_k141_9320 | Low-quality |
| 18-101_k141_387915 | Not-determined |
| 18-101_k141_140950 | Low-quality |
| 18-101_k141_322840 | Not-determined |
| 18-101_k141_190926 | Not-determined |
| 18-101_k141_9325 | Not-determined |
| 18-101_k141_190927 | Not-determined |
| 18-101_k141_156517 | Not-determined |
| 18-101_k141_190932 | Not-determined |
| 18-101_k141_9327 | Not-determined |
| 18-101_k141_172618 | Not-determined |
| 18-101_k141_207273 | Not-determined |
| 18-101_k141_239778 | Not-determined |
| 18-101_k141_355150 | Not-determined |
| 18-101_k141_288877 | Not-determined |
| 18-101_k141_239781 | Low-quality |
| 18-101_k141_337489 | Not-determined |
| 18-101_k141_207277 | Not-determined |
| 18-101_k141_172621 | Low-quality |
| 18-101_k141_371121 | Not-determined |
| 18-101_k141_25455 | Not-determined |
| 18-101_k141_239786 | Not-determined |
| 18-101_k141_25457 | Not-determined |
| 18-101_k141_106826 | Not-determined |
| 18-101_k141_256694 | Not-determined |
| 18-101_k141_156526 | Not-determined |
| 18-101_k141_106831 | Not-determined |
| 18-101_k141_91293 | Low-quality |
| 18-101_k141_371129 | Not-determined |
| 18-101_k141_172631 | Not-determined |
| 18-101_k141_9341 | Not-determined |
| 18-101_k141_207281 | Not-determined |
| 18-101_k141_91296 | Not-determined |
| 18-101_k141_272400 | Not-determined |
| 18-101_k141_272401 | Not-determined |
| 18-101_k141_288887 | Not-determined |
| 18-101_k141_74046 | Not-determined |
| 18-101_k141_106837 | Not-determined |
| 18-101_k141_337502 | Not-determined |
| 18-101_k141_387931 | Low-quality |
| 18-101_k141_106848 | Not-determined |
| 18-101_k141_337504 | Not-determined |
| 18-101_k141_207299 | Not-determined |
| 18-101_k141_41602 | Not-determined |
| 18-101_k141_91309 | Not-determined |
| 18-101_k141_337509 | Not-determined |
| 18-101_k141_25472 | Not-determined |
| 18-101_k141_9349 | Not-determined |
| 18-101_k141_58243 | Not-determined |
| 18-101_k141_41606 | Not-determined |
| 18-101_k141_91313 | Not-determined |
| 18-101_k141_41608 | Not-determined |
| 18-101_k141_140973 | Not-determined |
| 18-101_k141_207307 | Not-determined |
| 18-101_k141_190948 | Not-determined |
| 18-101_k141_58252 | Not-determined |
| 18-101_k141_106853 | Not-determined |
| 18-101_k141_306136 | Not-determined |
| 18-101_k141_322866 | Low-quality |
| 18-101_k141_58255 | Not-determined |
| 18-101_k141_41614 | Not-determined |
| 18-101_k141_172648 | Not-determined |
| 18-101_k141_306141 | Not-determined |
| 18-101_k141_322869 | Not-determined |
| 18-101_k141_106857 | Not-determined |
| 18-101_k141_106858 | Not-determined |
| 18-101_k141_322871 | Low-quality |
| 18-101_k141_288894 | Not-determined |
| 18-101_k141_106864 | Not-determined |
| 18-101_k141_172652 | Low-quality |
| 18-101_k141_25488 | Not-determined |
| 18-101_k141_74080 | Not-determined |
| 18-101_k141_387941 | Not-determined |
| 18-101_k141_140975 | Not-determined |
| 18-101_k141_140976 | Not-determined |
| 18-101_k141_172658 | Not-determined |
| 18-101_k141_172660 | Not-determined |
| 18-101_k141_322878 | Low-quality |
| 18-101_k141_91341 | Not-determined |
| 18-101_k141_172665 | Not-determined |
| 18-101_k141_272409 | Not-determined |
| 18-101_k141_172666 | Not-determined |
| 18-101_k141_140986 | Not-determined |
| 18-101_k141_106874 | Not-determined |
| 18-101_k141_106878 | Not-determined |
| 18-101_k141_272415 | Not-determined |
| 18-101_k141_322890 | Not-determined |
| 18-101_k141_74092 | Not-determined |
| 18-101_k141_41624 | Not-determined |
| 18-101_k141_355195 | Low-quality |
| 18-101_k141_140996 | Not-determined |
| 18-101_k141_172678 | Not-determined |
| 18-101_k141_106892 | Not-determined |
| 18-101_k141_9363 | Not-determined |
| 18-101_k141_74103 | Not-determined |
| 18-101_k141_106900 | Not-determined |
| 18-101_k141_172686 | Not-determined |
| 18-101_k141_106901 | Low-quality |
| 18-101_k141_172687 | Not-determined |
| 18-101_k141_306173 | Not-determined |
| 18-101_k141_172691 | Not-determined |
| 18-101_k141_172695 | Not-determined |
| 18-101_k141_288913 | Not-determined |
| 18-101_k141_337552 | Low-quality |
| 18-101_k141_172698 | Not-determined |
| 18-101_k141_239821 | Not-determined |
| 18-101_k141_207348 | Not-determined |
| 18-101_k141_207356 | Not-determined |
| 18-101_k141_9382 | Not-determined |
| 18-101_k141_9384 | Not-determined |
| 18-101_k141_223117 | Not-determined |
| 18-101_k141_223120 | Not-determined |
| 18-101_k141_239834 | Not-determined |
| 18-101_k141_207358 | Not-determined |
| 18-101_k141_106924 | Not-determined |
| 18-101_k141_74124 | Not-determined |
| 18-101_k141_91366 | Not-determined |
| 18-101_k141_172716 | Not-determined |
| 18-101_k141_256718 | Low-quality |
| 18-101_k141_288921 | Not-determined |
| 18-101_k141_106929 | Not-determined |
| 18-101_k141_41632 | Not-determined |
| 18-101_k141_337576 | Low-quality |
| 18-101_k141_106931 | Not-determined |
| 18-101_k141_125353 | Low-quality |
| 18-101_k141_106933 | Not-determined |
| 18-101_k141_322917 | Low-quality |
| 18-101_k141_125355 | Not-determined |
| 18-101_k141_272433 | Not-determined |
| 18-101_k141_371154 | Not-determined |
| 18-101_k141_272438 | Not-determined |
| 18-101_k141_190987 | Not-determined |
| 18-101_k141_141045 | Not-determined |
| 18-101_k141_306195 | Not-determined |
| 18-101_k141_256730 | Not-determined |
| 18-101_k141_106938 | Not-determined |
| 18-101_k141_141053 | Not-determined |
| 18-101_k141_106939 | Not-determined |
| 18-101_k141_371169 | Not-determined |
| 18-101_k141_256739 | Low-quality |
| 18-101_k141_288924 | Not-determined |
| 18-101_k141_256742 | Not-determined |
| 18-101_k141_74136 | Not-determined |
| 18-101_k141_223166 | Low-quality |
| 18-101_k141_41647 | Not-determined |
| 18-101_k141_91386 | Not-determined |
| 18-101_k141_256748 | Not-determined |
| 18-101_k141_172749 | Low-quality |
| 18-101_k141_156593 | Not-determined |
| 18-101_k141_207414 | Not-determined |
| 18-101_k141_156595 | Not-determined |
| 18-101_k141_191011 | Not-determined |
| 18-101_k141_355253 | Not-determined |
| 18-101_k141_58276 | Not-determined |
| 18-101_k141_207415 | Not-determined |
| 18-101_k141_355256 | Not-determined |
| 18-101_k141_141084 | Low-quality |
| 18-101_k141_191014 | Not-determined |
| 18-101_k141_355261 | Not-determined |
| 18-101_k141_337616 | Not-determined |
| 18-101_k141_355271 | Not-determined |
| 18-101_k141_74160 | Not-determined |
| 18-101_k141_272470 | Not-determined |
| 18-101_k141_74167 | Not-determined |
| 18-101_k141_74169 | Low-quality |
| 18-101_k141_41675 | Not-determined |
| 18-101_k141_91409 | Not-determined |
| 18-101_k141_41681 | Not-determined |
| 18-101_k141_387962 | Not-determined |
| 18-101_k141_58288 | Not-determined |
| 18-101_k141_58289 | Not-determined |
| 18-101_k141_355283 | Low-quality |
| 18-101_k141_371208 | Not-determined |
| 18-101_k141_371209 | Not-determined |
| 18-101_k141_91415 | Not-determined |
| 18-101_k141_355286 | Not-determined |
| 18-101_k141_239842 | Not-determined |
| 18-101_k141_256778 | Not-determined |
| 18-101_k141_239845 | Not-determined |
| 18-101_k141_288943 | Not-determined |
| 18-101_k141_58295 | Not-determined |
| 18-101_k141_172792 | Not-determined |
| 18-101_k141_207444 | Not-determined |
| 18-101_k141_355301 | Not-determined |
| 18-101_k141_156609 | Not-determined |
| 18-101_k141_239856 | Not-determined |
| 18-101_k141_272481 | Not-determined |
| 18-101_k141_288954 | Not-determined |
| 18-101_k141_355306 | Not-determined |
| 18-101_k141_172800 | Not-determined |
| 18-101_k141_239862 | Not-determined |
| 18-101_k141_239865 | Not-determined |
| 18-101_k141_74193 | Not-determined |
| 18-101_k141_256798 | Not-determined |
| 18-101_k141_355314 | Low-quality |
| 18-101_k141_191053 | Not-determined |
| 18-101_k141_256800 | Not-determined |
| 18-101_k141_239869 | Not-determined |
| 18-101_k141_239871 | Not-determined |
| 18-101_k141_256806 | Not-determined |
| 18-101_k141_172805 | Not-determined |
| 18-101_k141_239875 | Not-determined |
| 18-101_k141_239879 | Not-determined |
| 18-101_k141_355326 | Not-determined |
| 18-101_k141_371225 | Not-determined |
| 18-101_k141_207467 | Not-determined |
| 18-101_k141_355331 | Not-determined |
| 18-101_k141_371228 | Not-determined |
| 18-101_k141_41700 | Not-determined |
| 18-101_k141_306254 | Not-determined |
| 18-101_k141_191085 | Not-determined |
| 18-101_k141_355342 | Not-determined |
| 18-101_k141_355343 | Not-determined |
| 18-101_k141_74212 | Not-determined |
| 18-101_k141_9477 | Not-determined |
| 18-101_k141_41712 | Not-determined |
| 18-101_k141_9478 | Not-determined |
| 18-101_k141_41714 | Not-determined |
| 18-101_k141_355349 | Not-determined |
| 18-101_k141_355350 | Not-determined |
| 18-101_k141_239901 | Not-determined |
| 18-101_k141_239902 | Not-determined |
| 18-101_k141_141119 | Not-determined |
| 18-101_k141_207480 | Not-determined |
| 18-101_k141_74223 | Low-quality |
| 18-101_k141_239905 | Low-quality |
| 18-101_k141_172827 | Not-determined |
| 18-101_k141_141122 | Low-quality |
| 18-101_k141_172828 | Not-determined |
| 18-101_k141_272507 | Not-determined |
| 18-101_k141_272510 | Not-determined |
| 18-101_k141_141129 | Not-determined |
| 18-101_k141_156642 | Not-determined |
| 18-101_k141_172831 | Low-quality |
| 18-101_k141_172836 | Not-determined |
| 18-101_k141_141140 | Not-determined |
| 18-101_k141_74231 | Not-determined |
| 18-101_k141_74232 | Not-determined |
| 18-101_k141_74239 | Not-determined |
| 18-101_k141_371256 | Not-determined |
| 18-101_k141_74242 | Not-determined |
| 18-101_k141_91477 | Not-determined |
| 18-101_k141_207495 | Not-determined |
| 18-101_k141_272532 | Not-determined |
| 18-101_k141_355382 | Not-determined |
| 18-101_k141_355383 | Not-determined |
| 18-101_k141_141160 | Not-determined |
| 18-101_k141_322949 | Low-quality |
| 18-101_k141_141162 | Not-determined |
| 18-101_k141_207500 | Not-determined |
| 18-101_k141_272539 | Not-determined |
| 18-101_k141_272540 | Low-quality |
| 18-101_k141_172854 | Not-determined |
| 18-101_k141_41748 | Not-determined |
| 18-101_k141_74253 | Not-determined |
| 18-101_k141_322957 | Not-determined |
| 18-101_k141_371268 | Not-determined |
| 18-101_k141_41758 | Not-determined |
| 18-101_k141_239920 | Low-quality |
| 18-101_k141_91492 | Low-quality |
| 18-101_k141_106984 | Low-quality |
| 18-101_k141_172862 | Not-determined |
| 18-101_k141_371273 | Not-determined |
| 18-101_k141_256845 | Not-determined |
| 18-101_k141_41764 | Low-quality |
| 18-101_k141_25500 | Low-quality |
| 18-101_k141_58334 | Not-determined |
| 18-101_k141_106994 | Not-determined |
| 18-101_k141_172871 | Not-determined |
| 18-101_k141_256855 | Not-determined |
| 18-101_k141_25506 | Not-determined |
| 18-101_k141_272552 | Not-determined |
| 18-101_k141_58342 | Not-determined |
| 18-101_k141_388004 | Not-determined |
| 18-101_k141_388007 | Not-determined |
| 18-101_k141_41776 | Not-determined |
| 18-101_k141_207507 | Not-determined |
| 18-101_k141_239925 | Not-determined |
| 18-101_k141_322978 | Not-determined |
| 18-101_k141_256869 | Not-determined |
| 18-101_k141_156678 | Not-determined |
| 18-101_k141_91531 | Not-determined |
| 18-101_k141_207513 | Not-determined |
| 18-101_k141_355413 | Not-determined |
| 18-101_k141_25524 | Not-determined |
| 18-101_k141_207519 | Not-determined |
| 18-101_k141_74275 | Not-determined |
| 18-101_k141_388026 | Not-determined |
| 18-101_k141_207522 | Not-determined |
| 18-101_k141_107013 | Not-determined |
| 18-101_k141_74281 | Not-determined |
| 18-101_k141_91544 | Not-determined |
| 18-101_k141_207531 | Not-determined |
| 18-101_k141_207533 | Not-determined |
| 18-101_k141_239941 | Not-determined |
| 18-101_k141_322996 | Not-determined |
| 18-101_k141_25541 | Not-determined |
| 18-101_k141_25543 | Not-determined |
| 18-101_k141_289021 | Not-determined |
| 18-101_k141_156695 | Not-determined |
| 18-101_k141_355438 | Not-determined |
| 18-101_k141_355439 | Not-determined |
| 18-101_k141_125389 | Not-determined |
| 18-101_k141_107027 | Not-determined |
| 18-101_k141_371309 | Not-determined |
| 18-101_k141_355444 | Not-determined |
| 18-101_k141_289027 | Not-determined |
| 18-101_k141_91564 | Not-determined |
| 18-101_k141_125394 | Low-quality |
| 18-101_k141_371312 | Not-determined |
| 18-101_k141_223310 | Not-determined |
| 18-101_k141_223312 | Not-determined |
| 18-101_k141_289033 | Low-quality |
| 18-101_k141_323023 | Not-determined |
| 18-101_k141_355456 | Not-determined |
| 18-101_k141_355457 | Not-determined |
| 18-101_k141_207553 | Not-determined |
| 18-101_k141_25566 | Not-determined |
| 18-101_k141_289042 | Not-determined |
| 18-101_k141_289043 | Not-determined |
| 18-101_k141_355467 | Not-determined |
| 18-101_k141_107046 | Low-quality |
| 18-101_k141_272593 | Not-determined |
| 18-101_k141_107047 | Not-determined |
| 18-101_k141_91576 | Not-determined |
| 18-101_k141_107048 | Not-determined |
| 18-101_k141_156714 | Not-determined |
| 18-101_k141_107051 | Not-determined |
| 18-101_k141_58390 | Low-quality |
| 18-101_k141_172900 | Not-determined |
| 18-101_k141_355473 | Not-determined |
| 18-101_k141_156721 | Not-determined |
| 18-101_k141_91583 | Not-determined |
| 18-101_k141_207558 | Not-determined |
| 18-101_k141_289059 | Not-determined |
| 18-101_k141_172909 | Not-determined |
| 18-101_k141_107062 | Not-determined |
| 18-101_k141_172910 | Not-determined |
| 18-101_k141_172911 | Not-determined |
| 18-101_k141_388075 | Not-determined |
| 18-101_k141_191134 | Not-determined |
| 18-101_k141_74314 | Not-determined |
| 18-101_k141_156725 | Not-determined |
| 18-101_k141_156731 | Not-determined |
| 18-101_k141_91595 | Not-determined |
| 18-101_k141_125421 | Not-determined |
| 18-101_k141_107069 | Low-quality |
| 18-101_k141_323041 | Not-determined |
| 18-101_k141_371324 | Not-determined |
| 18-101_k141_323042 | Not-determined |
| 18-101_k141_25587 | Not-determined |
| 18-101_k141_74319 | Not-determined |
| 18-101_k141_272596 | Not-determined |
| 18-101_k141_74320 | Not-determined |
| 18-101_k141_289084 | Not-determined |
| 18-101_k141_74324 | Not-determined |
| 18-101_k141_371333 | Not-determined |
| 18-101_k141_25593 | Not-determined |
| 18-101_k141_289090 | Not-determined |
| 18-101_k141_272605 | Not-determined |
| 18-101_k141_256910 | Low-quality |
| 18-101_k141_74329 | Not-determined |
| 18-101_k141_388084 | Not-determined |
| 18-101_k141_41813 | Not-determined |
| 18-101_k141_388089 | Not-determined |
| 18-101_k141_191139 | Not-determined |
| 18-101_k141_207582 | Not-determined |
| 18-101_k141_58407 | Not-determined |
| 18-101_k141_9530 | Not-determined |
| 18-101_k141_355494 | Not-determined |
| 18-101_k141_125430 | Not-determined |
| 18-101_k141_125432 | Not-determined |
| 18-101_k141_125433 | Not-determined |
| 18-101_k141_172935 | Not-determined |
| 18-101_k141_337647 | Low-quality |
| 18-101_k141_125438 | Not-determined |
| 18-101_k141_337651 | Not-determined |
| 18-101_k141_25615 | Low-quality |
| 18-101_k141_256921 | Not-determined |
| 18-101_k141_388100 | Low-quality |
| 18-101_k141_355509 | Not-determined |
| 18-101_k141_172942 | Not-determined |
| 18-101_k141_337663 | Not-determined |
| 18-101_k141_9538 | Not-determined |
| 18-101_k141_107083 | Not-determined |
| 18-101_k141_107084 | Not-determined |
| 18-101_k141_125462 | Not-determined |
| 18-101_k141_74345 | Not-determined |
| 18-101_k141_9547 | Not-determined |
| 18-101_k141_272637 | Not-determined |
| 18-101_k141_388117 | Not-determined |
| 18-101_k141_306270 | Not-determined |
| 18-101_k141_9556 | Low-quality |
| 18-101_k141_107096 | Not-determined |
| 18-101_k141_107097 | Not-determined |
| 18-101_k141_289121 | Not-determined |
| 18-101_k141_172986 | Not-determined |
| 18-101_k141_355536 | Not-determined |
| 18-101_k141_256962 | Not-determined |
| 18-101_k141_25634 | Not-determined |
| 18-101_k141_172991 | Not-determined |
| 18-101_k141_306285 | Not-determined |
| 18-101_k141_323059 | Not-determined |
| 18-101_k141_272656 | Not-determined |
| 18-101_k141_9575 | Not-determined |
| 18-101_k141_337692 | Not-determined |
| 18-101_k141_156772 | Not-determined |
| 18-101_k141_91609 | Not-determined |
| 18-101_k141_256977 | Not-determined |
| 18-101_k141_355551 | Not-determined |
| 18-101_k141_191165 | Low-quality |
| 18-101_k141_207606 | Not-determined |
| 18-101_k141_173001 | Not-determined |
| 18-101_k141_191171 | Not-determined |
| 18-101_k141_306301 | Not-determined |
| 18-101_k141_107118 | Not-determined |
| 18-101_k141_91620 | Not-determined |
| 18-101_k141_337698 | Not-determined |
| 18-101_k141_256991 | Not-determined |
| 18-101_k141_125482 | Low-quality |
| 18-101_k141_91622 | Not-determined |
| 18-101_k141_191176 | Not-determined |
| 18-101_k141_25649 | Not-determined |
| 18-101_k141_371383 | Not-determined |
| 18-101_k141_25656 | Not-determined |
| 18-101_k141_173015 | Not-determined |
| 18-101_k141_388134 | Not-determined |
| 18-101_k141_25659 | Not-determined |
| 18-101_k141_25661 | Not-determined |
| 18-101_k141_9594 | Not-determined |
| 18-101_k141_91632 | Low-quality |
| 18-101_k141_74381 | Not-determined |
| 18-101_k141_388137 | Not-determined |
| 18-101_k141_41848 | Not-determined |
| 18-101_k141_74384 | Not-determined |
| 18-101_k141_191187 | Not-determined |
| 18-101_k141_58432 | Not-determined |
| 18-101_k141_257012 | Not-determined |
| 18-101_k141_9608 | Not-determined |
| 18-101_k141_74395 | Not-determined |
| 18-101_k141_9610 | Not-determined |
| 18-101_k141_91637 | Not-determined |
| 18-101_k141_41857 | Not-determined |
| 18-101_k141_323078 | Not-determined |
| 18-101_k141_125508 | Not-determined |
| 18-101_k141_91647 | Not-determined |
| 18-101_k141_107129 | Not-determined |
| 18-101_k141_91649 | Not-determined |
| 18-101_k141_371398 | Low-quality |
| 18-101_k141_41872 | Not-determined |
| 18-101_k141_323082 | Not-determined |
| 18-101_k141_240019 | Not-determined |
| 18-101_k141_25694 | Not-determined |
| 18-101_k141_9633 | Not-determined |
| 18-101_k141_306314 | Low-quality |
| 18-101_k141_9635 | Not-determined |
| 18-101_k141_388167 | Not-determined |
| 18-101_k141_371407 | Low-quality |
| 18-101_k141_306316 | Not-determined |
| 18-101_k141_223335 | Not-determined |
| 18-101_k141_355591 | Not-determined |
| 18-101_k141_25702 | Not-determined |
| 18-101_k141_257035 | Not-determined |
| 18-101_k141_173041 | Not-determined |
| 18-101_k141_41882 | Not-determined |
| 18-101_k141_371412 | Not-determined |
| 18-101_k141_9648 | Not-determined |
| 18-101_k141_323096 | Not-determined |
| 18-101_k141_306325 | Not-determined |
| 18-101_k141_289169 | Not-determined |
| 18-101_k141_240039 | Not-determined |
| 18-101_k141_272671 | Not-determined |
| 18-101_k141_240040 | Not-determined |
| 18-101_k141_240041 | Not-determined |
| 18-101_k141_125519 | Not-determined |
| 18-101_k141_257044 | Not-determined |
| 18-101_k141_141277 | Not-determined |
| 18-101_k141_25712 | Not-determined |
| 18-101_k141_156814 | Not-determined |
| 18-101_k141_25714 | Not-determined |
| 18-101_k141_25720 | Not-determined |
| 18-101_k141_337731 | Not-determined |
| 18-101_k141_156820 | Not-determined |
| 18-101_k141_141281 | Not-determined |
| 18-101_k141_257055 | Not-determined |
| 18-101_k141_337738 | Not-determined |
| 18-101_k141_25725 | Low-quality |
| 18-101_k141_371434 | Not-determined |
| 18-101_k141_41898 | Not-determined |
| 18-101_k141_25727 | Not-determined |
| 18-101_k141_9656 | Low-quality |
| 18-101_k141_371442 | Not-determined |
| 18-101_k141_74425 | Not-determined |
| 18-101_k141_323121 | Not-determined |
| 18-101_k141_355621 | Not-determined |
| 18-101_k141_337749 | Low-quality |
| 18-101_k141_25740 | Not-determined |
| 18-101_k141_323123 | Not-determined |
| 18-101_k141_337753 | Not-determined |
| 18-101_k141_240059 | Not-determined |
| 18-101_k141_355631 | Low-quality |
| 18-101_k141_74434 | Not-determined |
| 18-101_k141_91689 | Low-quality |
| 18-101_k141_371445 | Not-determined |
| 18-101_k141_388199 | Not-determined |
| 18-101_k141_74436 | Low-quality |
| 18-101_k141_74442 | Not-determined |
| 18-101_k141_74443 | Not-determined |
| 18-101_k141_257064 | Not-determined |
| 18-101_k141_355645 | Not-determined |
| 18-101_k141_9663 | Not-determined |
| 18-101_k141_388205 | Not-determined |
| 18-101_k141_355650 | Not-determined |
| 18-101_k141_355651 | Not-determined |
| 18-101_k141_388212 | Not-determined |
| 18-101_k141_289191 | Not-determined |
| 18-101_k141_107142 | Not-determined |
| 18-101_k141_240083 | Low-quality |
| 18-101_k141_272690 | Not-determined |
| 18-101_k141_41916 | Not-determined |
| 18-101_k141_207639 | Not-determined |
| 18-101_k141_388220 | Not-determined |
| 18-101_k141_289195 | Not-determined |
| 18-101_k141_289196 | Not-determined |
| 18-101_k141_355658 | Not-determined |
| 18-101_k141_41918 | Not-determined |
| 18-101_k141_74471 | Not-determined |
| 18-101_k141_306362 | Not-determined |
| 18-101_k141_272701 | Not-determined |
| 18-101_k141_240090 | Low-quality |
| 18-101_k141_388238 | Not-determined |
| 18-101_k141_91709 | Not-determined |
| 18-101_k141_9684 | Not-determined |
| 18-101_k141_25766 | Not-determined |
| 18-101_k141_91718 | Not-determined |
| 18-101_k141_41933 | Not-determined |
| 18-101_k141_156842 | Low-quality |
| 18-101_k141_141321 | Not-determined |
| 18-101_k141_306368 | Not-determined |
| 18-101_k141_355668 | Not-determined |
| 18-101_k141_306370 | Not-determined |
| 18-101_k141_388246 | Not-determined |
| 18-101_k141_9698 | Not-determined |
| 18-101_k141_41943 | Not-determined |
| 18-101_k141_388250 | Low-quality |
| 18-101_k141_355675 | Not-determined |
| 18-101_k141_323166 | Not-determined |
| 18-101_k141_337780 | Not-determined |
| 18-101_k141_9708 | Not-determined |
| 18-101_k141_388252 | Not-determined |
| 18-101_k141_41947 | Not-determined |
| 18-101_k141_141330 | Low-quality |
| 18-101_k141_91732 | Not-determined |
| 18-101_k141_9712 | Not-determined |
| 18-101_k141_240105 | Low-quality |
| 18-101_k141_240115 | Not-determined |
| 18-101_k141_240118 | Not-determined |
| 18-101_k141_74504 | Not-determined |
| 18-101_k141_107169 | Not-determined |
| 18-101_k141_306380 | Not-determined |
| 18-101_k141_107171 | Not-determined |
| 18-101_k141_355705 | Not-determined |
| 18-101_k141_355706 | Not-determined |
| 18-101_k141_323173 | Not-determined |

| | |
|---|---|
| 18-101_k141_191225 | Low-quality |
| 18-101_k141_323184 | Not-determined |
| 18-101_k141_156857 | Not-determined |
| 18-101_k141_337807 | Not-determined |
| 18-101_k141_41968 | Not-determined |
| 18-101_k141_25780 | Not-determined |
| 18-101_k141_289203 | Not-determined |
| 18-101_k141_41970 | Not-determined |
| 18-101_k141_323190 | Not-determined |
| 18-101_k141_355712 | Not-determined |
| 18-101_k141_156870 | Not-determined |
| 18-101_k141_207672 | Not-determined |
| 18-101_k141_388286 | Not-determined |
| 18-101_k141_74510 | Not-determined |
| 18-101_k141_355717 | Not-determined |
| 18-101_k141_355718 | Not-determined |
| 18-101_k141_74513 | Not-determined |
| 18-101_k141_371501 | Not-determined |
| 18-101_k141_272741 | Not-determined |
| 18-101_k141_272742 | Not-determined |
| 18-101_k141_207680 | Not-determined |
| 18-101_k141_74518 | Not-determined |
| 18-101_k141_355727 | Not-determined |
| KM-final-viral-combined_k141_47860\|\|full | Medium-quality |
| 18-101_k141_388294 | Not-determined |
| 18-101_k141_91778 | Not-determined |
| 18-101_k141_91779 | Not-determined |
| 18-101_k141_141345 | Low-quality |
| 18-101_k141_141346 | Not-determined |
| 18-101_k141_306385 | Not-determined |
| 18-101_k141_173098 | Not-determined |
| 18-101_k141_207686 | Not-determined |
| 18-101_k141_91785 | Not-determined |
| 18-101_k141_74528 | Not-determined |
| 18-101_k141_58468 | Not-determined |
| 18-101_k141_355737 | Not-determined |
| 18-101_k141_74530 | Not-determined |
| 18-101_k141_141356 | Not-determined |
| 18-101_k141_289208 | Not-determined |
| 18-101_k141_107221 | Not-determined |
| 18-101_k141_9761 | Not-determined |
| 18-101_k141_91793 | Not-determined |
| 18-101_k141_107222 | Not-determined |
| 18-101_k141_388315 | Not-determined |
| 18-101_k141_141368 | Not-determined |
| 18-101_k141_74540 | Not-determined |
| 18-101_k141_191262 | Low-quality |
| 18-101_k141_306397 | Not-determined |
| 18-101_k141_191270 | Not-determined |
| 18-101_k141_323230 | Not-determined |
| 18-101_k141_257095 | Not-determined |
| 18-101_k141_306402 | Not-determined |
| 18-101_k141_58476 | Not-determined |
| 18-101_k141_306403 | Not-determined |
| 18-101_k141_337862 | Not-determined |
| 18-101_k141_257111 | Not-determined |
| 18-101_k141_257112 | Low-quality |
| 18-101_k141_371537 | Not-determined |
| 18-101_k141_323241 | Not-determined |
| 18-101_k141_9797 | Not-determined |
| 18-101_k141_41996 | Low-quality |
| 18-101_k141_41997 | Not-determined |
| 18-101_k141_323249 | Not-determined |
| 18-101_k141_74558 | Not-determined |
| 18-101_k141_388325 | Not-determined |
| 18-101_k141_58488 | Not-determined |
| 18-101_k141_107250 | Not-determined |
| 18-101_k141_337873 | Low-quality |
| 18-101_k141_191287 | Not-determined |
| 18-101_k141_74566 | Not-determined |
| 18-101_k141_306428 | Not-determined |
| 18-101_k141_272757 | Not-determined |
| 18-101_k141_257122 | Not-determined |
| 18-101_k141_141394 | Not-determined |
| 18-101_k141_272760 | Not-determined |
| 18-101_k141_289247 | Not-determined |
| 18-101_k141_355744 | Not-determined |
| 18-101_k141_125557 | Not-determined |
| 18-101_k141_42016 | Not-determined |
| 18-101_k141_207733 | Not-determined |
| 18-101_k141_42020 | Not-determined |
| 18-101_k141_42021 | Low-quality |
| 18-101_k141_289249 | Not-determined |
| 18-101_k141_337893 | Not-determined |
| 18-101_k141_323260 | Not-determined |
| 18-101_k141_337896 | Low-quality |
| 18-101_k141_289254 | Not-determined |
| 18-101_k141_355756 | Not-determined |
| 18-101_k141_141409 | Not-determined |
| 18-101_k141_323271 | Not-determined |
| 18-101_k141_371544 | Not-determined |
| 18-101_k141_323274 | Not-determined |
| 18-101_k141_257134 | Not-determined |
| 18-101_k141_289260 | Not-determined |
| 18-101_k141_306453 | Not-determined |
| 18-101_k141_371554 | Not-determined |
| 18-101_k141_355765 | Not-determined |
| 18-101_k141_58501 | Not-determined |
| 18-101_k141_107280 | Not-determined |
| 18-101_k141_156914 | Not-determined |
| 18-101_k141_207762 | Low-quality |
| 18-101_k141_257141 | Not-determined |
| 18-101_k141_257142 | Not-determined |
| 18-101_k141_257143 | Not-determined |
| 18-101_k141_257145 | Not-determined |
| 18-101_k141_240179 | Not-determined |
| 18-101_k141_25844 | Not-determined |
| 18-101_k141_25845 | Not-determined |
| 18-101_k141_25847 | Not-determined |
| 18-101_k141_9836 | Not-determined |
| 18-101_k141_141426 | Not-determined |
| 18-101_k141_173156 | Not-determined |
| 18-101_k141_388337 | Not-determined |
| 18-101_k141_173160 | Not-determined |
| 18-101_k141_25853 | Not-determined |
| 18-101_k141_25854 | Not-determined |
| 18-101_k141_25856 | Not-determined |
| 18-101_k141_25858 | Not-determined |
| 18-101_k141_191328 | Not-determined |
| 18-101_k141_272802 | Not-determined |
| 18-101_k141_58516 | Not-determined |
| 18-101_k141_91824 | Not-determined |
| 18-101_k141_91825 | Not-determined |
| 18-101_k141_191333 | Not-determined |
| 18-101_k141_191336 | Not-determined |
| 18-101_k141_91830 | Not-determined |
| 18-101_k141_125613 | Not-determined |
| 18-101_k141_191342 | Not-determined |
| 18-101_k141_91835 | Not-determined |
| 18-101_k141_289302 | Not-determined |
| 18-101_k141_107322 | Not-determined |
| 18-101_k141_207803 | Not-determined |
| 18-101_k141_125622 | Not-determined |
| 18-101_k141_125627 | Low-quality |
| 18-101_k141_91845 | Not-determined |
| 18-101_k141_9862 | Not-determined |
| 18-101_k141_388361 | Low-quality |
| 18-101_k141_207813 | Not-determined |
| 18-101_k141_388365 | Not-determined |
| 18-101_k141_9871 | Not-determined |
| 18-101_k141_9872 | Not-determined |
| 18-101_k141_141453 | Not-determined |
| 18-101_k141_141454 | Not-determined |
| 18-101_k141_306501 | Not-determined |
| 18-101_k141_191363 | Not-determined |
| 18-101_k141_323330 | Not-determined |
| 18-101_k141_371585 | Not-determined |
| 18-101_k141_156941 | Not-determined |
| 18-101_k141_156943 | Not-determined |
| 18-101_k141_141458 | Not-determined |
| 18-101_k141_207826 | Not-determined |
| 18-101_k141_191366 | Not-determined |
| 18-101_k141_191367 | Not-determined |
| 18-101_k141_289334 | Not-determined |
| 18-101_k141_191369 | Low-quality |
| 18-101_k141_125634 | Not-determined |
| 18-101_k141_156950 | Not-determined |
| 18-101_k141_25895 | Not-determined |
| 18-101_k141_371595 | Not-determined |
| 18-101_k141_289342 | Not-determined |
| 18-101_k141_289343 | Not-determined |
| 18-101_k141_125648 | Not-determined |
| 18-101_k141_25902 | Not-determined |
| 18-101_k141_337942 | Not-determined |
| 18-101_k141_9884 | Not-determined |
| 18-101_k141_9887 | Not-determined |
| 18-101_k141_58567 | Not-determined |
| 18-101_k141_240235 | Not-determined |
| 18-101_k141_289352 | Not-determined |
| 18-101_k141_107346 | Not-determined |
| 18-101_k141_107351 | Not-determined |
| 18-101_k141_42037 | Not-determined |
| 18-101_k141_306526 | Not-determined |
| 18-101_k141_9905 | Not-determined |
| 18-101_k141_191398 | Not-determined |
| 18-101_k141_223374 | Not-determined |
| 18-101_k141_25921 | Not-determined |
| 18-101_k141_323368 | Not-determined |
| 18-101_k141_42050 | Not-determined |
| 18-101_k141_306534 | Not-determined |
| 18-101_k141_306542 | Not-determined |
| 18-101_k141_107369 | Not-determined |
| 18-101_k141_207838 | Not-determined |
| 18-101_k141_289376 | Low-quality |
| 18-101_k141_9923 | Not-determined |
| 18-101_k141_74637 | Not-determined |
| 18-101_k141_355812 | Low-quality |
| 18-101_k141_240273 | Not-determined |
| 18-101_k141_240278 | Not-determined |
| 18-101_k141_25953 | Not-determined |
| 18-101_k141_191419 | Low-quality |
| 18-101_k141_141492 | Not-determined |
| 18-101_k141_191426 | Not-determined |
| 18-101_k141_74659 | Not-determined |
| 18-101_k141_306560 | Not-determined |
| 18-101_k141_9933 | Not-determined |
| 18-101_k141_337955 | Not-determined |
| 18-101_k141_240283 | Not-determined |
| 18-101_k141_9935 | Not-determined |
| 18-101_k141_306561 | Not-determined |
| 18-101_k141_25968 | Not-determined |
| 18-101_k141_25969 | Not-determined |
| 18-101_k141_306563 | Not-determined |
| 18-101_k141_141507 | Not-determined |
| 18-101_k141_74672 | Not-determined |
| 18-101_k141_289413 | Not-determined |
| 18-101_k141_191433 | Not-determined |
| 18-101_k141_207875 | Not-determined |
| 18-101_k141_191435 | Not-determined |
| 18-101_k141_306571 | Not-determined |
| 18-101_k141_107382 | Not-determined |
| 18-101_k141_355836 | Not-determined |
| 18-101_k141_240294 | Not-determined |
| 18-101_k141_191439 | Not-determined |
| 18-101_k141_25976 | Not-determined |
| 18-101_k141_257271 | Not-determined |
| 18-101_k141_107386 | Not-determined |
| 18-101_k141_107388 | Not-determined |
| 18-101_k141_173204 | Not-determined |
| 18-101_k141_58596 | Not-determined |
| 18-101_k141_223403 | Not-determined |
| 18-101_k141_223414 | Not-determined |
| 18-101_k141_141520 | Low-quality |
| 18-101_k141_223420 | Not-determined |
| 18-101_k141_207894 | Low-quality |
| 18-101_k141_91871 | Not-determined |
| 18-101_k141_207897 | Not-determined |
| 18-101_k141_42112 | Not-determined |
| 18-101_k141_107416 | Low-quality |
| 18-101_k141_107417 | Not-determined |
| 18-101_k141_58608 | Not-determined |
| 18-101_k141_107418 | Low-quality |
| 18-101_k141_58609 | Not-determined |
| 18-101_k141_125683 | Not-determined |
| 18-101_k141_306582 | Low-quality |
| 18-101_k141_272838 | Not-determined |
| 18-101_k141_125688 | Not-determined |
| 18-101_k141_42118 | Not-determined |
| 18-101_k141_355856 | Low-quality |
| 18-101_k141_173222 | Not-determined |
| 18-101_k141_125689 | Not-determined |
| 18-101_k141_173224 | Not-determined |
| 18-101_k141_42126 | Low-quality |
| 18-101_k141_107426 | Low-quality |
| 18-101_k141_272854 | Not-determined |
| 18-101_k141_306588 | Not-determined |
| 18-101_k141_272855 | Not-determined |
| 18-101_k141_107430 | Not-determined |
| 18-101_k141_337995 | Not-determined |
| 18-101_k141_306589 | Not-determined |
| 18-101_k141_107431 | Not-determined |
| 18-101_k141_207914 | Not-determined |
| 18-101_k141_207915 | Not-determined |
| 18-101_k141_74690 | Not-determined |
| 18-101_k141_74692 | Not-determined |
| 18-101_k141_355878 | Not-determined |
| 18-101_k141_125697 | Not-determined |
| 18-101_k141_74694 | Low-quality |
| 18-101_k141_173237 | Not-determined |
| 18-101_k141_191477 | Not-determined |
| 18-101_k141_289475 | Not-determined |
| 18-101_k141_306606 | Not-determined |
| 18-101_k141_9946 | Not-determined |
| 18-101_k141_191488 | Not-determined |
| 18-101_k141_355886 | Not-determined |
| 18-101_k141_125704 | Not-determined |
| 18-101_k141_25997 | Not-determined |
| 18-101_k141_338006 | Not-determined |
| 18-101_k141_9953 | Not-determined |
| 18-101_k141_323427 | Low-quality |
| 18-101_k141_91904 | Not-determined |
| 18-101_k141_338012 | Not-determined |
| 18-101_k141_207953 | Not-determined |
| 18-101_k141_223466 | Not-determined |
| 18-101_k141_125713 | Not-determined |
| 18-101_k141_91911 | Not-determined |
| 18-101_k141_125714 | Not-determined |
| 18-101_k141_355900 | Not-determined |
| 18-101_k141_355906 | Not-determined |
| 18-101_k141_355908 | Not-determined |
| 18-101_k141_355909 | Not-determined |
| 18-101_k141_207968 | Not-determined |
| 18-101_k141_355912 | Not-determined |
| 18-101_k141_272887 | Low-quality |
| 18-101_k141_125725 | Not-determined |
| 18-101_k141_125726 | Not-determined |
| 18-101_k141_355922 | Not-determined |
| 18-101_k141_371607 | Not-determined |
| 18-101_k141_371609 | Not-determined |
| 18-101_k141_207979 | Not-determined |
| 18-101_k141_289491 | Not-determined |
| 18-101_k141_107476 | Not-determined |
| 18-101_k141_371611 | Not-determined |
| 18-101_k141_26007 | Low-quality |
| 18-101_k141_223473 | Not-determined |
| 18-101_k141_223474 | Not-determined |
| 18-101_k141_306639 | Not-determined |
| 18-101_k141_240333 | Not-determined |
| 18-101_k141_223476 | Not-determined |
| 18-101_k141_207990 | Not-determined |
| 18-101_k141_223478 | Not-determined |
| 18-101_k141_306643 | Not-determined |
| 18-101_k141_58649 | Not-determined |
| 18-101_k141_272911 | Not-determined |
| 18-101_k141_191522 | Not-determined |
| 18-101_k141_9977 | Not-determined |
| 18-101_k141_58652 | Not-determined |
| 18-101_k141_58654 | Not-determined |
| 18-101_k141_107485 | Not-determined |
| 18-101_k141_371613 | Not-determined |
| 18-101_k141_191526 | Low-quality |
| 18-101_k141_107492 | Low-quality |
| 18-101_k141_223487 | Not-determined |
| 18-101_k141_371622 | Not-determined |
| 18-101_k141_157016 | Not-determined |
| 18-101_k141_306658 | Not-determined |
| 18-101_k141_157017 | Low-quality |
| 18-101_k141_125747 | Not-determined |
| 18-101_k141_355942 | Not-determined |
| 18-101_k141_289510 | Not-determined |
| 18-101_k141_223493 | Not-determined |
| 18-101_k141_157028 | Not-determined |
| 18-101_k141_191537 | Not-determined |
| 18-101_k141_208012 | Not-determined |
| 18-101_k141_208013 | Not-determined |
| 18-101_k141_208014 | Not-determined |
| 18-101_k141_107506 | Not-determined |
| 18-101_k141_371633 | Low-quality |
| 18-101_k141_371636 | Not-determined |
| 18-101_k141_91949 | Not-determined |
| 18-101_k141_10006 | Not-determined |
| 18-101_k141_208026 | Not-determined |
| 18-101_k141_157048 | Not-determined |
| 18-101_k141_157049 | Not-determined |
| 18-101_k141_91957 | Not-determined |
| 18-101_k141_26045 | Not-determined |
| 18-101_k141_208038 | Not-determined |
| 18-101_k141_223516 | Not-determined |
| 18-101_k141_26053 | Not-determined |
| 18-101_k141_240357 | Not-determined |
| 18-101_k141_208046 | Not-determined |
| 18-101_k141_338050 | Not-determined |
| 18-101_k141_240361 | Not-determined |
| 18-101_k141_26058 | Not-determined |
| 18-101_k141_91962 | Not-determined |
| 18-101_k141_289530 | Low-quality |
| 18-101_k141_191563 | Not-determined |
| 18-101_k141_10012 | Not-determined |
| 18-101_k141_91966 | Not-determined |
| 18-101_k141_289538 | Not-determined |
| 18-101_k141_208056 | Not-determined |
| 18-101_k141_223527 | Not-determined |
| 18-101_k141_191571 | Not-determined |
| 18-101_k141_338065 | Not-determined |
| 18-101_k141_157078 | Not-determined |
| 18-101_k141_323463 | Not-determined |
| 18-101_k141_157081 | Not-determined |
| 18-101_k141_26076 | Not-determined |
| 18-101_k141_240383 | Not-determined |
| 18-101_k141_74706 | Low-quality |
| 18-101_k141_323473 | Not-determined |
| 18-101_k141_107534 | Not-determined |
| 18-101_k141_191587 | Not-determined |
| 18-101_k141_289561 | Not-determined |
| 18-101_k141_208063 | Not-determined |
| 18-101_k141_26082 | Not-determined |
| 18-101_k141_323485 | Not-determined |
| 18-101_k141_91995 | Low-quality |
| 18-101_k141_240403 | Not-determined |
| 18-101_k141_26086 | Not-determined |
| 18-101_k141_173294 | Not-determined |
| 18-101_k141_26089 | Not-determined |
| 18-101_k141_323487 | Not-determined |
| 18-101_k141_58700 | Not-determined |
| 18-101_k141_306705 | Not-determined |
| 18-101_k141_323490 | Not-determined |
| 18-101_k141_208064 | Not-determined |
| 18-101_k141_240407 | Low-quality |
| 18-101_k141_208067 | Not-determined |
| 18-101_k141_371670 | Not-determined |
| 18-101_k141_10038 | Not-determined |
| 18-101_k141_157098 | Not-determined |
| 18-101_k141_240411 | Not-determined |
| 18-101_k141_107550 | Not-determined |
| 18-101_k141_323495 | Not-determined |
| 18-101_k141_323499 | Not-determined |
| 18-101_k141_223537 | Not-determined |
| 18-101_k141_157105 | Not-determined |
| 18-101_k141_191599 | Not-determined |
| 18-101_k141_10047 | Not-determined |
| 18-101_k141_42218 | Not-determined |
| 18-101_k141_157114 | Low-quality |
| 18-101_k141_289593 | Not-determined |
| 18-101_k141_323508 | Low-quality |
| 18-101_k141_157125 | Low-quality |
| 18-101_k141_338109 | Not-determined |
| 18-101_k141_323523 | Not-determined |
| 18-101_k141_323524 | Not-determined |
| 18-101_k141_257357 | Not-determined |
| 18-101_k141_272945 | Low-quality |
| 18-101_k141_191613 | Not-determined |
| 18-101_k141_257365 | Not-determined |
| 18-101_k141_289607 | Low-quality |
| 18-101_k141_10061 | Not-determined |
| 18-101_k141_107577 | Not-determined |
| 18-101_k141_208108 | Not-determined |
| 18-101_k141_257373 | Not-determined |
| 18-101_k141_74717 | Not-determined |
| 18-101_k141_107588 | Not-determined |
| 18-101_k141_74718 | Not-determined |
| 18-101_k141_107590 | Not-determined |
| 18-101_k141_223584 | Not-determined |
| 18-101_k141_157153 | Not-determined |
| 18-101_k141_157156 | Not-determined |
| 18-101_k141_10083 | Low-quality |
| 18-101_k141_125775 | Not-determined |
| 18-101_k141_157158 | Not-determined |
| 18-101_k141_157159 | Not-determined |
| 18-101_k141_240436 | Not-determined |
| 18-101_k141_338137 | Not-determined |
| 18-101_k141_92023 | Low-quality |
| 18-101_k141_223592 | Not-determined |
| 18-101_k141_306751 | Not-determined |
| 18-101_k141_338151 | Not-determined |
| 18-101_k141_223601 | Low-quality |
| 18-101_k141_58766 | Not-determined |
| 18-101_k141_125784 | Not-determined |
| 18-101_k141_223605 | Not-determined |
| 18-101_k141_208129 | Not-determined |
| 18-101_k141_157173 | Not-determined |
| 18-101_k141_208131 | Not-determined |
| 18-101_k141_240460 | Not-determined |
| 18-101_k141_208132 | Not-determined |
| 18-101_k141_107624 | Low-quality |
| 18-101_k141_26148 | Not-determined |
| 18-101_k141_125802 | Not-determined |
| 18-101_k141_272954 | Not-determined |
| 18-101_k141_125805 | Not-determined |
| 18-101_k141_208136 | Not-determined |
| 18-101_k141_58778 | Low-quality |
| 18-101_k141_257377 | Not-determined |
| 18-101_k141_157178 | Not-determined |
| 18-101_k141_42242 | Low-quality |
| 18-101_k141_125813 | Not-determined |
| 18-101_k141_323581 | Not-determined |
| 18-101_k141_240481 | Not-determined |
| 18-101_k141_157181 | Not-determined |
| 18-101_k141_42248 | Not-determined |
| 18-101_k141_107636 | Not-determined |
| 18-101_k141_92059 | Not-determined |
| 18-101_k141_323587 | Not-determined |
| 18-101_k141_323588 | Not-determined |
| 18-101_k141_26171 | Not-determined |
| 18-101_k141_257394 | Not-determined |
| 18-101_k141_173329 | Not-determined |
| 18-101_k141_92062 | Not-determined |
| 18-101_k141_338191 | Not-determined |
| 18-101_k141_92063 | Not-determined |
| 18-101_k141_26184 | Not-determined |
| 18-101_k141_92069 | Not-determined |
| 18-101_k141_42269 | Not-determined |
| 18-101_k141_371709 | Not-determined |
| 18-101_k141_173335 | Not-determined |
| 18-101_k141_173339 | Not-determined |
| 18-101_k141_371724 | Low-quality |
| 18-101_k141_355968 | Low-quality |
| 18-101_k141_173348 | Not-determined |
| 18-101_k141_157228 | Not-determined |
| 18-101_k141_92076 | Not-determined |
| 18-101_k141_10131 | Low-quality |
| 18-101_k141_355974 | Not-determined |
| 18-101_k141_92083 | Low-quality |
| 18-101_k141_323593 | Low-quality |
| 18-101_k141_371733 | Not-determined |
| 18-101_k141_173357 | Not-determined |
| 18-101_k141_323595 | Not-determined |
| 18-101_k141_42290 | Not-determined |
| 17-106-final-viral-combined_k141_19025\|\|full | High-quality |
| 18-101_k141_355990 | Not-determined |
| 18-101_k141_355992 | Not-determined |
| 18-101_k141_58790 | Not-determined |
| 18-101_k141_208171 | Not-determined |
| 18-101_k141_272976 | Low-quality |
| 18-101_k141_223639 | Not-determined |
| 18-101_k141_223641 | Not-determined |
| 18-101_k141_338225 | Not-determined |
| 18-101_k141_92108 | Not-determined |
| 18-101_k141_289685 | Not-determined |
| 18-101_k141_92111 | Not-determined |
| 18-101_k141_356002 | Not-determined |
| 18-101_k141_92112 | Not-determined |
| 18-101_k141_26239 | Not-determined |
| 18-101_k141_92115 | Not-determined |
| 18-101_k141_338233 | Not-determined |
| 18-101_k141_157238 | Not-determined |
| 18-101_k141_356011 | Not-determined |
| 18-101_k141_257430 | Not-determined |
| 18-101_k141_223643 | Not-determined |
| 18-101_k141_323622 | Not-determined |
| 18-101_k141_58811 | Not-determined |
| 18-101_k141_58813 | Not-determined |
| 18-101_k141_58814 | Not-determined |
| 18-101_k141_92117 | Not-determined |
| 18-101_k141_257433 | Not-determined |
| 18-101_k141_323628 | Not-determined |
| 18-101_k141_356021 | Not-determined |
| 18-101_k141_257438 | Not-determined |
| 18-101_k141_257440 | Not-determined |
| 18-101_k141_58824 | Not-determined |
| 18-101_k141_173385 | Not-determined |
| 18-101_k141_356037 | Not-determined |
| 18-101_k141_208193 | Not-determined |
| 18-101_k141_306832 | Not-determined |
| 18-101_k141_257447 | Not-determined |
| 18-101_k141_356048 | Not-determined |
| 18-101_k141_191646 | Not-determined |
| 18-101_k141_107659 | Not-determined |
| 18-101_k141_58830 | Not-determined |
| 18-101_k141_257450 | Low-quality |
| 18-101_k141_157271 | Not-determined |
| 18-101_k141_356056 | Not-determined |
| 18-101_k141_157274 | Not-determined |
| 18-101_k141_306841 | Not-determined |
| 18-101_k141_107662 | Not-determined |
| 18-101_k141_257456 | Not-determined |
| 18-101_k141_208205 | Not-determined |
| 18-101_k141_26266 | Not-determined |
| 18-101_k141_157284 | Not-determined |
| 18-101_k141_306849 | Not-determined |
| 18-101_k141_356059 | Not-determined |
| 18-101_k141_125889 | Low-quality |
| 18-101_k141_58856 | Not-determined |
| 18-101_k141_240512 | Not-determined |
| 18-101_k141_323641 | Not-determined |
| 18-101_k141_58858 | Not-determined |
| 18-101_k141_257483 | Not-determined |
| 18-101_k141_371774 | Not-determined |
| 18-101_k141_26284 | Not-determined |
| 18-101_k141_371778 | Not-determined |
| 18-101_k141_208228 | Not-determined |
| 18-101_k141_356080 | Not-determined |
| 18-101_k141_240529 | Not-determined |
| 18-101_k141_157299 | Not-determined |
| 18-101_k141_107682 | Not-determined |
| 18-101_k141_191679 | Not-determined |
| 18-101_k141_58871 | Not-determined |
| 18-101_k141_107686 | Not-determined |
| 18-101_k141_356091 | Not-determined |
| 18-101_k141_208229 | Low-quality |
| 18-101_k141_26302 | Not-determined |
| 18-101_k141_191688 | Not-determined |
| 18-101_k141_208234 | Not-determined |
| 18-101_k141_356094 | Not-determined |
| 18-101_k141_257502 | Not-determined |
| 18-101_k141_125910 | Not-determined |
| 18-101_k141_208241 | Not-determined |
| 18-101_k141_208242 | Not-determined |
| 18-101_k141_356100 | Not-determined |
| 18-101_k141_107693 | Not-determined |
| 18-101_k141_157309 | Not-determined |
| 18-101_k141_157312 | Not-determined |
| 18-101_k141_74743 | Not-determined |
| 18-101_k141_323655 | Not-determined |
| 18-101_k141_208253 | Not-determined |
| 18-101_k141_306879 | Not-determined |
| 18-101_k141_306884 | Not-determined |
| 18-101_k141_240548 | Not-determined |
| 18-101_k141_208261 | Low-quality |
| 18-101_k141_92188 | Not-determined |
| 18-101_k141_306894 | Not-determined |
| 18-101_k141_26322 | Not-determined |
| 18-101_k141_338319 | Not-determined |
| 18-101_k141_306901 | Not-determined |
| 18-101_k141_26325 | Low-quality |
| 18-101_k141_157322 | Not-determined |
| 18-101_k141_125936 | Not-determined |
| 18-101_k141_306903 | Not-determined |
| 18-101_k141_92191 | Low-quality |
| 18-101_k141_223682 | Not-determined |
| 18-101_k141_289726 | Not-determined |
| 18-101_k141_157329 | Not-determined |
| 18-101_k141_208275 | Low-quality |
| 18-101_k141_125940 | Not-determined |
| 18-101_k141_42368 | Not-determined |
| 18-101_k141_26331 | Low-quality |
| 18-101_k141_289730 | Not-determined |
| 18-101_k141_289732 | Not-determined |
| 18-101_k141_125944 | Not-determined |
| 18-101_k141_223694 | Not-determined |

| | |
|---|---|
| 18-101_k141_356114 | Low-quality |
| 18-101_k141_223698 | Not-determined |
| 18-101_k141_356121 | Not-determined |
| 18-101_k141_157343 | Not-determined |
| 18-101_k141_338337 | Not-determined |
| 18-101_k141_356124 | Not-determined |
| 18-101_k141_306919 | Low-quality |
| 18-101_k141_257533 | Not-determined |
| 18-101_k141_74763 | Not-determined |
| 18-101_k141_125955 | Not-determined |
| 18-101_k141_157356 | Not-determined |
| 18-101_k141_338347 | Not-determined |
| 18-101_k141_191710 | Not-determined |
| 18-101_k141_338348 | Not-determined |
| 18-101_k141_107726 | Not-determined |
| 18-101_k141_42385 | Not-determined |
| 18-101_k141_191714 | Not-determined |
| 18-101_k141_306925 | Not-determined |
| 18-101_k141_338349 | Low-quality |
| 18-101_k141_26335 | Not-determined |
| 18-101_k141_125963 | Not-determined |
| 18-101_k141_173438 | Not-determined |
| 18-101_k141_257541 | Not-determined |
| 18-101_k141_208289 | Low-quality |
| 18-101_k141_58932 | Not-determined |
| 18-101_k141_208291 | Not-determined |
| 18-101_k141_257544 | Not-determined |
| 18-101_k141_157360 | Not-determined |
| 18-101_k141_42394 | Not-determined |
| 18-101_k141_208297 | Not-determined |
| 18-101_k141_371807 | Not-determined |
| 18-101_k141_356150 | Not-determined |
| 18-101_k141_306931 | Not-determined |
| 18-101_k141_92203 | Not-determined |
| 18-101_k141_223715 | Not-determined |
| 18-101_k141_208306 | Not-determined |
| 18-101_k141_26350 | Not-determined |
| 18-101_k141_223717 | Not-determined |
| 18-101_k141_157376 | Not-determined |
| 18-101_k141_141610 | Not-determined |
| 18-101_k141_10175 | Not-determined |
| 18-101_k141_208307 | Not-determined |
| 18-101_k141_92212 | Not-determined |
| 18-101_k141_289756 | Not-determined |
| 18-101_k141_191734 | Not-determined |
| 18-101_k141_74789 | Not-determined |
| 18-101_k141_289757 | Not-determined |
| 18-101_k141_306933 | Low-quality |
| 18-101_k141_107751 | Low-quality |
| 18-101_k141_223727 | Not-determined |
| 18-101_k141_223735 | Not-determined |
| 18-101_k141_257568 | Not-determined |
| 18-101_k141_223739 | Not-determined |
| 18-101_k141_42422 | Not-determined |
| 18-101_k141_42425 | Not-determined |
| 18-101_k141_141629 | Not-determined |
| 18-101_k141_42428 | Not-determined |
| 18-101_k141_240605 | Low-quality |
| 18-101_k141_141633 | Not-determined |
| 18-101_k141_371832 | Not-determined |
| 18-101_k141_191740 | Not-determined |
| 18-101_k141_240608 | Low-quality |
| 18-101_k141_371835 | Not-determined |
| 18-101_k141_371837 | Not-determined |
| 18-101_k141_191746 | Not-determined |
| 18-101_k141_92225 | Not-determined |
| 18-101_k141_323685 | Low-quality |
| 18-101_k141_273034 | Low-quality |
| 18-101_k141_10195 | Not-determined |
| 18-101_k141_356184 | Not-determined |
| 18-101_k141_141657 | Not-determined |
| 18-101_k141_92235 | Not-determined |
| 18-101_k141_371862 | Not-determined |
| 18-101_k141_191761 | Not-determined |
| 18-101_k141_356193 | Not-determined |
| 18-101_k141_92241 | Not-determined |
| 18-101_k141_92243 | Not-determined |
| 18-101_k141_356197 | Not-determined |
| 18-101_k141_10206 | Not-determined |
| 18-101_k141_306955 | Not-determined |
| 18-101_k141_356200 | Not-determined |
| 18-101_k141_208312 | Not-determined |
| 18-101_k141_92247 | Not-determined |
| 18-101_k141_289802 | Not-determined |
| 18-101_k141_223778 | Not-determined |
| 18-101_k141_125984 | Not-determined |
| 18-101_k141_125985 | Not-determined |
| 18-101_k141_223781 | Not-determined |
| 18-101_k141_240622 | Not-determined |
| 18-101_k141_141680 | Not-determined |
| 18-101_k141_273069 | Not-determined |
| 18-101_k141_371878 | Not-determined |
| 18-101_k141_289808 | Not-determined |
| 18-101_k141_323716 | Low-quality |
| 18-101_k141_338362 | Not-determined |
| 18-101_k141_323718 | Not-determined |
| 18-101_k141_125994 | Not-determined |
| 18-101_k141_240627 | Not-determined |
| 18-101_k141_371882 | Not-determined |
| 18-101_k141_58975 | Not-determined |
| 18-101_k141_74860 | Not-determined |
| 18-101_k141_26370 | Not-determined |
| 18-101_k141_323724 | Not-determined |
| 18-101_k141_240639 | Not-determined |
| 18-101_k141_273080 | Not-determined |
| 18-101_k141_289817 | Not-determined |
| 18-101_k141_191789 | Not-determined |
| 18-101_k141_191791 | Not-determined |
| 18-101_k141_191792 | Not-determined |
| 18-101_k141_191795 | Not-determined |
| 18-101_k141_26377 | Not-determined |
| 18-101_k141_356224 | Not-determined |
| 18-101_k141_273086 | Not-determined |
| 18-101_k141_126000 | Not-determined |
| 18-101_k141_371893 | Not-determined |
| 18-101_k141_191800 | Not-determined |
| 18-101_k141_191801 | Not-determined |
| 18-101_k141_257630 | Not-determined |
| 18-101_k141_240642 | Not-determined |
| 18-101_k141_240643 | Low-quality |
| 18-101_k141_240645 | Not-determined |
| 18-101_k141_92285 | Not-determined |
| 18-101_k141_338379 | Not-determined |
| 18-101_k141_323733 | Low-quality |
| 18-101_k141_356228 | Not-determined |
| 18-101_k141_173460 | Low-quality |
| 18-101_k141_92289 | Not-determined |
| 18-101_k141_157412 | Not-determined |
| 18-101_k141_126014 | Not-determined |
| 18-101_k141_257635 | Low-quality |
| 18-101_k141_223792 | Not-determined |
| 18-101_k141_257638 | Not-determined |
| 18-101_k141_208338 | Not-determined |
| 18-101_k141_157417 | Not-determined |
| 18-101_k141_42460 | Low-quality |
| 18-101_k141_306973 | Low-quality |
| 18-101_k141_306974 | Not-determined |
| 18-101_k141_306975 | Not-determined |
| 18-101_k141_289832 | Not-determined |
| 18-101_k141_208343 | Not-determined |
| 18-101_k141_323751 | Not-determined |
| 18-101_k141_306977 | Not-determined |
| 18-101_k141_26380 | Not-determined |
| 18-101_k141_223809 | Not-determined |
| 18-101_k141_26388 | Not-determined |
| 18-101_k141_157430 | Not-determined |
| 18-101_k141_58990 | Not-determined |
| 18-101_k141_173481 | Not-determined |
| 18-101_k141_141724 | Not-determined |
| 18-101_k141_92326 | Not-determined |
| 18-101_k141_58992 | Not-determined |
| 18-101_k141_223820 | Not-determined |
| 18-101_k141_208349 | Not-determined |
| 18-101_k141_208352 | Not-determined |
| 18-101_k141_356253 | Not-determined |
| 18-101_k141_42475 | Not-determined |
| 18-101_k141_257651 | Not-determined |
| 18-101_k141_240656 | Not-determined |
| 18-101_k141_289843 | Not-determined |
| 18-101_k141_240659 | Not-determined |
| 18-101_k141_173489 | Not-determined |
| 18-101_k141_338405 | Not-determined |
| 18-101_k141_223834 | Not-determined |
| 18-101_k141_338406 | Not-determined |
| 18-101_k141_223835 | Not-determined |
| 18-101_k141_323771 | Not-determined |
| 18-101_k141_289850 | Not-determined |
| 18-101_k141_289853 | Low-quality |
| 18-101_k141_306982 | Not-determined |
| 18-101_k141_273123 | Not-determined |
| 18-101_k141_58995 | Not-determined |
| 18-101_k141_107782 | Not-determined |
| 18-101_k141_58997 | Not-determined |
| 18-101_k141_157453 | Low-quality |
| 18-101_k141_42482 | Not-determined |
| 18-101_k141_42483 | Not-determined |
| 18-101_k141_273125 | Low-quality |
| 18-101_k141_141746 | Not-determined |
| 18-101_k141_240674 | Not-determined |
| 18-101_k141_208365 | Not-determined |
| 18-101_k141_157462 | Not-determined |
| 18-101_k141_157465 | Not-determined |
| 18-101_k141_141757 | Not-determined |
| 18-101_k141_208376 | Not-determined |
| 18-101_k141_42499 | Not-determined |
| 18-101_k141_307010 | Not-determined |
| 18-101_k141_240691 | Not-determined |
| 18-101_k141_257678 | Not-determined |
| 18-101_k141_107806 | Not-determined |
| 18-101_k141_223868 | Not-determined |
| 18-101_k141_289880 | Not-determined |
| 18-101_k141_356285 | Not-determined |
| 18-101_k141_323811 | Not-determined |
| 18-101_k141_257686 | Low-quality |
| 18-101_k141_10231 | Not-determined |
| 18-101_k141_307022 | Not-determined |
| 18-101_k141_141775 | Not-determined |
| 18-101_k141_141776 | Not-determined |
| 18-101_k141_371955 | Not-determined |
| 18-101_k141_157477 | Not-determined |
| 18-101_k141_107824 | Not-determined |
| 18-101_k141_42518 | Low-quality |
| 18-101_k141_107827 | Not-determined |
| 18-101_k141_157481 | Not-determined |
| 18-101_k141_307031 | Not-determined |
| 18-101_k141_356303 | Not-determined |
| 18-101_k141_356304 | Not-determined |
| 18-101_k141_208405 | Not-determined |
| 18-101_k141_371966 | Not-determined |
| 18-101_k141_26391 | Not-determined |
| 18-101_k141_42523 | Not-determined |
| 18-101_k141_371970 | Not-determined |
| 18-101_k141_223895 | Not-determined |
| 18-101_k141_10253 | Not-determined |
| 18-101_k141_26400 | Not-determined |
| 18-101_k141_141783 | Not-determined |
| 18-101_k141_273159 | Low-quality |
| 18-101_k141_289911 | Not-determined |
| 18-101_k141_323821 | Low-quality |
| 18-101_k141_257713 | Not-determined |
| 18-101_k141_356327 | Not-determined |
| 18-101_k141_240722 | Not-determined |
| 18-101_k141_356329 | Not-determined |
| 18-101_k141_107848 | Not-determined |
| 18-101_k141_157496 | Not-determined |
| 18-101_k141_92371 | Low-quality |
| 18-101_k141_307062 | Not-determined |
| 18-101_k141_338418 | Low-quality |
| 18-101_k141_289922 | Not-determined |
| 18-101_k141_257726 | Not-determined |
| 18-101_k141_173519 | Not-determined |
| 18-101_k141_191844 | Not-determined |
| 18-101_k141_257735 | Not-determined |
| 18-101_k141_371977 | Low-quality |
| 18-101_k141_323839 | Not-determined |
| 18-101_k141_157508 | Not-determined |
| 18-101_k141_208424 | Not-determined |
| 18-101_k141_157509 | Not-determined |
| 18-101_k141_173524 | Not-determined |
| 18-101_k141_173525 | Not-determined |
| 18-101_k141_10268 | Not-determined |
| 18-101_k141_92383 | Not-determined |
| 18-101_k141_157514 | Not-determined |
| 18-101_k141_338427 | Not-determined |
| 18-101_k141_42534 | Low-quality |
| 18-101_k141_307075 | Not-determined |
| 18-101_k141_338429 | Not-determined |
| 18-101_k141_323844 | Not-determined |
| 18-101_k141_307077 | Not-determined |
| 18-101_k141_388417 | Not-determined |
| 18-101_k141_338432 | Not-determined |
| 18-101_k141_323846 | Not-determined |
| 18-101_k141_157516 | Not-determined |
| 18-101_k141_173531 | Low-quality |
| 18-101_k141_42546 | Not-determined |
| 18-101_k141_356343 | Not-determined |
| 18-101_k141_141806 | Not-determined |
| 18-101_k141_141809 | Not-determined |
| 18-101_k141_323853 | Not-determined |
| 18-101_k141_141810 | Not-determined |
| 18-101_k141_191849 | Not-determined |
| 18-101_k141_273197 | Not-determined |
| 18-101_k141_323856 | Not-determined |
| 18-101_k141_323858 | Not-determined |
| 18-101_k141_59044 | Not-determined |
| 18-101_k141_371983 | Not-determined |
| 18-101_k141_273201 | Not-determined |
| 18-101_k141_223947 | Not-determined |
| 18-101_k141_223950 | Not-determined |
| 18-101_k141_173542 | Not-determined |
| 18-101_k141_371988 | Not-determined |
| 18-101_k141_191855 | Not-determined |
| 18-101_k141_59048 | Not-determined |
| 18-101_k141_240755 | Not-determined |
| 18-101_k141_371998 | Not-determined |
| 18-101_k141_59053 | Not-determined |
| 18-101_k141_157540 | Not-determined |
| 18-101_k141_240758 | Not-determined |
| 18-101_k141_372003 | Not-determined |
| 18-101_k141_92404 | Not-determined |
| 18-101_k141_223966 | Not-determined |
| 18-101_k141_59059 | Not-determined |
| 18-101_k141_240762 | Not-determined |
| 18-101_k141_10285 | Not-determined |
| 18-101_k141_307103 | Not-determined |
| 18-101_k141_173551 | Not-determined |
| 18-101_k141_372012 | Not-determined |
| 18-101_k141_372013 | Not-determined |
| 18-101_k141_307106 | Not-determined |
| 18-101_k141_141826 | Not-determined |
| 18-101_k141_257772 | Not-determined |
| 18-101_k141_372014 | Not-determined |
| 18-101_k141_59066 | Not-determined |
| 18-101_k141_141830 | Not-determined |
| 18-101_k141_74956 | Not-determined |
| 18-101_k141_173555 | Not-determined |
| 18-101_k141_223987 | Not-determined |
| 18-101_k141_307121 | Not-determined |
| 18-101_k141_338469 | Not-determined |
| 18-101_k141_273225 | Not-determined |
| 18-101_k141_92412 | Not-determined |
| 18-101_k141_141836 | Not-determined |
| 18-101_k141_338472 | Not-determined |
| 18-101_k141_338473 | Low-quality |
| 18-101_k141_126044 | Not-determined |
| 18-101_k141_273227 | Not-determined |
| 18-101_k141_126045 | Not-determined |
| 18-101_k141_42590 | Not-determined |
| 18-101_k141_240789 | Not-determined |
| 18-101_k141_191879 | Not-determined |
| 18-101_k141_74966 | Not-determined |
| 18-101_k141_289967 | Not-determined |
| 18-101_k141_240790 | Not-determined |
| 18-101_k141_191883 | Not-determined |
| 18-101_k141_307127 | Not-determined |
| 18-101_k141_307131 | Not-determined |
| 18-101_k141_372033 | Not-determined |
| 18-101_k141_74979 | Not-determined |
| 18-101_k141_126055 | Not-determined |
| 18-101_k141_42607 | Not-determined |
| 18-101_k141_240811 | Low-quality |
| 18-101_k141_42612 | Not-determined |
| 18-101_k141_191889 | Not-determined |
| 18-101_k141_289990 | Not-determined |
| 18-101_k141_92435 | Not-determined |
| 18-101_k141_323914 | Low-quality |
| 18-101_k141_224025 | Not-determined |
| 18-101_k141_126070 | Not-determined |
| 18-101_k141_157576 | Not-determined |
| 18-101_k141_307145 | Not-determined |
| 18-101_k141_191902 | Not-determined |
| 18-101_k141_126078 | Not-determined |
| 18-101_k141_240828 | Not-determined |
| 18-101_k141_92449 | Not-determined |
| 18-101_k141_92451 | Not-determined |
| 18-101_k141_290008 | Low-quality |
| 18-101_k141_10294 | Low-quality |
| 18-101_k141_372066 | Not-determined |
| 18-101_k141_208475 | Not-determined |
| 18-101_k141_372068 | Not-determined |
| 18-101_k141_273249 | Low-quality |
| 18-101_k141_141861 | Not-determined |
| 18-101_k141_240830 | Not-determined |
| 18-101_k141_338489 | Not-determined |
| 18-101_k141_356368 | Not-determined |
| 18-101_k141_388467 | Not-determined |
| 18-101_k141_307160 | Not-determined |
| 18-101_k141_126084 | Not-determined |
| 18-101_k141_157592 | Not-determined |
| 18-101_k141_126086 | Low-quality |
| 18-101_k141_10310 | Not-determined |
| 18-101_k141_126089 | Not-determined |
| 18-101_k141_372080 | Not-determined |
| 18-101_k141_338495 | Not-determined |
| 18-101_k141_224055 | Not-determined |
| 18-101_k141_224057 | Low-quality |
| 18-101_k141_92459 | Not-determined |
| 18-101_k141_338500 | Not-determined |
| 18-101_k141_224058 | Not-determined |
| 18-101_k141_59103 | Low-quality |
| 18-101_k141_257835 | Low-quality |
| 18-101_k141_191925 | Not-determined |
| 18-101_k141_126102 | Not-determined |
| 18-101_k141_191927 | Not-determined |
| 18-101_k141_224064 | Low-quality |
| 18-101_k141_356369 | Low-quality |
| 18-101_k141_290032 | Not-determined |
| 18-101_k141_290033 | Not-determined |
| 18-101_k141_240841 | Not-determined |
| 18-101_k141_26456 | Not-determined |
| 18-101_k141_338511 | Not-determined |
| 18-101_k141_290037 | Not-determined |
| 18-101_k141_141875 | Not-determined |
| 18-101_k141_257841 | Not-determined |
| 18-101_k141_372098 | Not-determined |
| 18-101_k141_257842 | Not-determined |
| 18-101_k141_75016 | Not-determined |
| 18-101_k141_26464 | Not-determined |
| 18-101_k141_75019 | Not-determined |
| 18-101_k141_240846 | Not-determined |
| 18-101_k141_173633 | Not-determined |
| 18-101_k141_107928 | Not-determined |
| 18-101_k141_290052 | Not-determined |
| 18-101_k141_173638 | Not-determined |
| 18-101_k141_323949 | Not-determined |
| 18-101_k141_26472 | Not-determined |
| 18-101_k141_126124 | Low-quality |
| 18-101_k141_356394 | Not-determined |
| 18-101_k141_173645 | Not-determined |
| 18-101_k141_141889 | Low-quality |
| 18-101_k141_240856 | Not-determined |
| 18-101_k141_26481 | Not-determined |
| 18-101_k141_240858 | Not-determined |
| 18-101_k141_126136 | Not-determined |
| 18-101_k141_257857 | Not-determined |
| 18-101_k141_141894 | Not-determined |
| 18-101_k141_75051 | Not-determined |
| 18-101_k141_126139 | Not-determined |
| 18-101_k141_290068 | Not-determined |
| 18-101_k141_290072 | Not-determined |
| 18-101_k141_173660 | Not-determined |
| 18-101_k141_290074 | Low-quality |
| 18-101_k141_141901 | Not-determined |
| 18-101_k141_257869 | Not-determined |
| 18-101_k141_257870 | Not-determined |
| 18-101_k141_59109 | Not-determined |
| 18-101_k141_290083 | Not-determined |
| 18-101_k141_208513 | Not-determined |
| 18-101_k141_323980 | Not-determined |
| 18-101_k141_208516 | Not-determined |
| 18-101_k141_26505 | Not-determined |
| 18-101_k141_141917 | Low-quality |
| 18-101_k141_224074 | Not-determined |
| 18-101_k141_141920 | Not-determined |
| 18-101_k141_42658 | Low-quality |
| 18-101_k141_75071 | Low-quality |
| 18-101_k141_224078 | Not-determined |
| 18-101_k141_42663 | Not-determined |
| 18-101_k141_372145 | Not-determined |
| 18-101_k141_126167 | Not-determined |
| 18-101_k141_356412 | Not-determined |
| 18-101_k141_290088 | Not-determined |
| 18-101_k141_107970 | Not-determined |
| 18-101_k141_26515 | Not-determined |
| 18-101_k141_10353 | Not-determined |
| 18-101_k141_356419 | Not-determined |
| 18-101_k141_126174 | Not-determined |
| 18-101_k141_240873 | Not-determined |
| 18-101_k141_141932 | Not-determined |
| 18-101_k141_10364 | Not-determined |
| 18-101_k141_141935 | Not-determined |
| 18-101_k141_224093 | Low-quality |
| 18-101_k141_338554 | Not-determined |
| 18-101_k141_372158 | Not-determined |
| 18-101_k141_307211 | Not-determined |
| 18-101_k141_224096 | Not-determined |
| 18-101_k141_42666 | Not-determined |
| 18-101_k141_75084 | Not-determined |
| 18-101_k141_173695 | Not-determined |
| 18-101_k141_107981 | Not-determined |
| 18-101_k141_323999 | Not-determined |
| 18-101_k141_224098 | Not-determined |
| 18-101_k141_307219 | Not-determined |
| 18-101_k141_173699 | Not-determined |
| 18-101_k141_324003 | Not-determined |
| 18-101_k141_126185 | Not-determined |
| 18-101_k141_126186 | Not-determined |
| 18-101_k141_126187 | Not-determined |
| 18-101_k141_388489 | Not-determined |
| 18-101_k141_356427 | Not-determined |
| 18-101_k141_126189 | Not-determined |
| 18-101_k141_338566 | Not-determined |
| 18-101_k141_126191 | Not-determined |
| 18-101_k141_240894 | Not-determined |
| 18-101_k141_26531 | Not-determined |
| 18-101_k141_273284 | Not-determined |
| 18-101_k141_173711 | Not-determined |
| 18-101_k141_42675 | Not-determined |
| 18-101_k141_372172 | Low-quality |
| 18-101_k141_324013 | Not-determined |
| 18-101_k141_324014 | Not-determined |
| 18-101_k141_324022 | Not-determined |
| 18-101_k141_324025 | Not-determined |
| 18-101_k141_157678 | Not-determined |
| 18-101_k141_240906 | Not-determined |
| 18-101_k141_240907 | Low-quality |
| 18-101_k141_141947 | Not-determined |
| 18-101_k141_141948 | Not-determined |
| 18-101_k141_92520 | Not-determined |
| 18-101_k141_224118 | Not-determined |
| 18-101_k141_141957 | Not-determined |
| 18-101_k141_157691 | Not-determined |
| 18-101_k141_173736 | Not-determined |
| 18-101_k141_273308 | Not-determined |
| 18-101_k141_372187 | Not-determined |
| 18-101_k141_273316 | Not-determined |
| 18-101_k141_173738 | Not-determined |
| 18-101_k141_126220 | Not-determined |
| 18-101_k141_42688 | Not-determined |
| 18-101_k141_173743 | Not-determined |
| 18-101_k141_257930 | Not-determined |
| 18-101_k141_240931 | Not-determined |
| 18-101_k141_372208 | Not-determined |
| 18-101_k141_388514 | Not-determined |
| 18-101_k141_208542 | Not-determined |
| 18-101_k141_75122 | Not-determined |
| 18-101_k141_126229 | Not-determined |
| 18-101_k141_208545 | Not-determined |
| 18-101_k141_75124 | Not-determined |
| 18-101_k141_42705 | Not-determined |
| 18-101_k141_126233 | Not-determined |
| 18-101_k141_290107 | Not-determined |
| 18-101_k141_108002 | Not-determined |
| 18-101_k141_290110 | Not-determined |
| 18-101_k141_59130 | Not-determined |
| 18-101_k141_126244 | Not-determined |
| 18-101_k141_141986 | Not-determined |
| 18-101_k141_356460 | Not-determined |
| 18-101_k141_141988 | Not-determined |
| 18-101_k141_108008 | Not-determined |
| 18-101_k141_208554 | Low-quality |
| 18-101_k141_108011 | Not-determined |
| 18-101_k141_141995 | Not-determined |
| 18-101_k141_273343 | Not-determined |
| 18-101_k141_224133 | Not-determined |
| 18-101_k141_173761 | Low-quality |
| 18-101_k141_273351 | Not-determined |
| 18-101_k141_338598 | Not-determined |
| 18-101_k141_257959 | Not-determined |
| 18-101_k141_240953 | Not-determined |
| 18-101_k141_108026 | Not-determined |
| 18-101_k141_307242 | Low-quality |
| 18-101_k141_224144 | Low-quality |
| 18-101_k141_75146 | Not-determined |
| 18-101_k141_75147 | Not-determined |
| 18-101_k141_92542 | Not-determined |
| 18-101_k141_173769 | Not-determined |
| 18-101_k141_224149 | Not-determined |
| 18-101_k141_372238 | Not-determined |
| 18-101_k141_75152 | Not-determined |
| 18-101_k141_372251 | Not-determined |
| 18-101_k141_257964 | Not-determined |
| 18-101_k141_92550 | Not-determined |
| 18-101_k141_338617 | Not-determined |
| 18-101_k141_257966 | Not-determined |
| 18-101_k141_388532 | Not-determined |
| 18-101_k141_92556 | Not-determined |
| 18-101_k141_356509 | Not-determined |
| 18-101_k141_290142 | Not-determined |
| 18-101_k141_157723 | Not-determined |
| 18-101_k141_59165 | Not-determined |
| 18-101_k141_126304 | Low-quality |
| 18-101_k141_324064 | Not-determined |
| 18-101_k141_307261 | Not-determined |
| 18-101_k141_59172 | Not-determined |
| 18-101_k141_307262 | Not-determined |
| 18-101_k141_173798 | Not-determined |
| 18-101_k141_290158 | Not-determined |
| 18-101_k141_257972 | Not-determined |
| 18-101_k141_126308 | Not-determined |
| 18-101_k141_126309 | Not-determined |
| 18-101_k141_126310 | Not-determined |
| 18-101_k141_307269 | Not-determined |
| 18-101_k141_126312 | Not-determined |
| 18-101_k141_75175 | Not-determined |
| 18-101_k141_388555 | Not-determined |
| 18-101_k141_388557 | Not-determined |
| 18-101_k141_307275 | Not-determined |
| 18-101_k141_388560 | Not-determined |
| 18-101_k141_324088 | Not-determined |
| 18-101_k141_307277 | Not-determined |
| 18-101_k141_126331 | Not-determined |
| 18-101_k141_388563 | Not-determined |
| 18-101_k141_173816 | Not-determined |
| 18-101_k141_126335 | Not-determined |
| 18-101_k141_108051 | Low-quality |
| 18-101_k141_208625 | Low-quality |
| 18-101_k141_208627 | Not-determined |
| 18-101_k141_257989 | Not-determined |
| 18-101_k141_307287 | Not-determined |
| 18-101_k141_324100 | Not-determined |
| 18-101_k141_324101 | Not-determined |
| 18-101_k141_75198 | Not-determined |
| 18-101_k141_42769 | Not-determined |
| 18-101_k141_307295 | Not-determined |
| 18-101_k141_290188 | Not-determined |
| 18-101_k141_142022 | Not-determined |
| 18-101_k141_126353 | Not-determined |
| 18-101_k141_191968 | Not-determined |
| 18-101_k141_240967 | Low-quality |
| 18-101_k141_126354 | Low-quality |
| 18-101_k141_142025 | Not-determined |
| 18-101_k141_338639 | Low-quality |
| 18-101_k141_142028 | Not-determined |
| 18-101_k141_191974 | Not-determined |
| 18-101_k141_59208 | Not-determined |
| 18-101_k141_59209 | Low-quality |
| 18-101_k141_324121 | Not-determined |
| 18-101_k141_258005 | Low-quality |
| 18-101_k141_173845 | Not-determined |
| 18-101_k141_307301 | Not-determined |
| 18-101_k141_307304 | Not-determined |
| 18-101_k141_372301 | Not-determined |
| 18-101_k141_372302 | Not-determined |
| 18-101_k141_92619 | Not-determined |
| 18-101_k141_307309 | Not-determined |
| 18-101_k141_356557 | Not-determined |
| 18-101_k141_324125 | Not-determined |
| 18-101_k141_75214 | Not-determined |

| | |
|---|---|
| 18-101_k141_92624 | Not-determined |
| 18-101_k141_59226 | Not-determined |
| 18-101_k141_42780 | Not-determined |
| 18-101_k141_388587 | Low-quality |
| 18-101_k141_92629 | Not-determined |
| 18-101_k141_338668 | Not-determined |
| 18-101_k141_224193 | Not-determined |
| 18-101_k141_324139 | Not-determined |
| 18-101_k141_26613 | Not-determined |
| 18-101_k141_92631 | Not-determined |
| 18-101_k141_59237 | Not-determined |
| 18-101_k141_173857 | Not-determined |
| 18-101_k141_173858 | Not-determined |
| 18-101_k141_59251 | Not-determined |
| 18-101_k141_324148 | Not-determined |
| 18-101_k141_388607 | Low-quality |
| 18-101_k141_290215 | Not-determined |
| 18-101_k141_142071 | Not-determined |
| 18-101_k141_92645 | Not-determined |
| 18-101_k141_92646 | Not-determined |
| 18-101_k141_372332 | Not-determined |
| 18-101_k141_372333 | Not-determined |
| 18-101_k141_388613 | Not-determined |
| 18-101_k141_324158 | Not-determined |
| 18-101_k141_92656 | Not-determined |
| 18-101_k141_372336 | Not-determined |
| 18-101_k141_173866 | Not-determined |
| 18-101_k141_191993 | Not-determined |
| 18-101_k141_224204 | Not-determined |
| 18-101_k141_191999 | Not-determined |
| 18-101_k141_92661 | Not-determined |
| 18-101_k141_142093 | Not-determined |
| 18-101_k141_258046 | Not-determined |
| 18-101_k141_173868 | Not-determined |
| 18-101_k141_241003 | Not-determined |
| 18-101_k141_338684 | Not-determined |
| 18-101_k141_290235 | Not-determined |
| 18-101_k141_108097 | Not-determined |
| 18-101_k141_142102 | Not-determined |
| 18-101_k141_108098 | Not-determined |
| 18-101_k141_142105 | Not-determined |
| 18-101_k141_142106 | Low-quality |
| 18-101_k141_173874 | Not-determined |
| 18-101_k141_324173 | Not-determined |
| 18-101_k141_192013 | Not-determined |
| 18-101_k141_258061 | Not-determined |
| 18-101_k141_108111 | Not-determined |
| 18-101_k141_192021 | Low-quality |
| 18-101_k141_224234 | Not-determined |
| 18-101_k141_75230 | Not-determined |
| 18-101_k141_192027 | Low-quality |
| 18-101_k141_192033 | Not-determined |
| 18-101_k141_142115 | Not-determined |
| 18-101_k141_208703 | Not-determined |
| 18-101_k141_42819 | Low-quality |
| 18-101_k141_42820 | Low-quality |
| 18-101_k141_208704 | Not-determined |
| 18-101_k141_192040 | Not-determined |
| 18-101_k141_324182 | Not-determined |
| 18-101_k141_92686 | Not-determined |
| 18-101_k141_338703 | Not-determined |
| 18-101_k141_338709 | Low-quality |
| 18-101_k141_324190 | Not-determined |
| 18-101_k141_157782 | Not-determined |
| 18-101_k141_173888 | Low-quality |
| 18-101_k141_192051 | Not-determined |
| 18-101_k141_273400 | Not-determined |
| 18-101_k141_273403 | Not-determined |
| 18-101_k141_258097 | Not-determined |
| 18-101_k141_192053 | Not-determined |
| 18-101_k141_142129 | Low-quality |
| 18-101_k141_92706 | Not-determined |
| 18-101_k141_157794 | Not-determined |
| 18-101_k141_59284 | Not-determined |
| 18-101_k141_324203 | Not-determined |
| 18-101_k141_208715 | Not-determined |
| 18-101_k141_372374 | Not-determined |
| 18-101_k141_59286 | Not-determined |
| 18-101_k141_75247 | Not-determined |
| 18-101_k141_224258 | Not-determined |
| 18-101_k141_42837 | Not-determined |
| 18-101_k141_42839 | Not-determined |
| 18-101_k141_258099 | Not-determined |
| 18-101_k141_108142 | Not-determined |
| 18-101_k141_192067 | Low-quality |
| 18-101_k141_42843 | Not-determined |
| 18-101_k141_59293 | Not-determined |
| 18-101_k141_75256 | Not-determined |
| 18-101_k141_273413 | Not-determined |
| 18-101_k141_372381 | Not-determined |
| 18-101_k141_142134 | Not-determined |
| 18-101_k141_42847 | Not-determined |
| 18-101_k141_273423 | Not-determined |
| 18-101_k141_356583 | Not-determined |
| 18-101_k141_108146 | Low-quality |
| 18-101_k141_173906 | Not-determined |
| 18-101_k141_59299 | Not-determined |
| 18-101_k141_92721 | Not-determined |
| 18-101_k141_26647 | Not-determined |
| 18-101_k141_192081 | Not-determined |
| 18-101_k141_126418 | Not-determined |
| 18-101_k141_372386 | Low-quality |
| 18-101_k141_92723 | Low-quality |
| 18-101_k141_126421 | Not-determined |
| 18-101_k141_42859 | Not-determined |
| 18-101_k141_338723 | Not-determined |
| 18-101_k141_273428 | Not-determined |
| 18-101_k141_157803 | Not-determined |
| 18-101_k141_173921 | Not-determined |
| 18-101_k141_126434 | Not-determined |
| 18-101_k141_356608 | Not-determined |
| 18-101_k141_108168 | Not-determined |
| 18-101_k141_241036 | Not-determined |
| 18-101_k141_290266 | Not-determined |
| 18-101_k141_273434 | Not-determined |
| 18-101_k141_338737 | Not-determined |
| 18-101_k141_290269 | Not-determined |
| 18-101_k141_356613 | Not-determined |
| 18-101_k141_356615 | Not-determined |
| 18-101_k141_208745 | Not-determined |
| 18-101_k141_273440 | Low-quality |
| 18-101_k141_356619 | Not-determined |
| 18-101_k141_192092 | Not-determined |
| 18-101_k141_142140 | Not-determined |
| 18-101_k141_338745 | Not-determined |
| 18-101_k141_192097 | Low-quality |
| 18-101_k141_75279 | Not-determined |
| 18-101_k141_157818 | Low-quality |
| 18-101_k141_192100 | Low-quality |
| 18-101_k141_92748 | Not-determined |
| 18-101_k141_192101 | Low-quality |
| 18-101_k141_241052 | Not-determined |
| 18-101_k141_372427 | Not-determined |
| 18-101_k141_192103 | Low-quality |
| 18-101_k141_75289 | Not-determined |
| 18-101_k141_388666 | Not-determined |
| 18-101_k141_126464 | Not-determined |
| 18-101_k141_241057 | Not-determined |
| 18-101_k141_59328 | Not-determined |
| 18-101_k141_372439 | Not-determined |
| 18-101_k141_59329 | Low-quality |
| 18-101_k141_388675 | Not-determined |
| 18-101_k141_224274 | Not-determined |
| 18-101_k141_42903 | Not-determined |
| 18-101_k141_224276 | Not-determined |
| 18-101_k141_42905 | Not-determined |
| 18-101_k141_42906 | Low-quality |
| 18-101_k141_59331 | Low-quality |
| 18-101_k141_324248 | Not-determined |
| 18-101_k141_224284 | Not-determined |
| 18-101_k141_192117 | Not-determined |
| 18-101_k141_142159 | Not-determined |
| 18-101_k141_126468 | Not-determined |
| 18-101_k141_42911 | Not-determined |
| 18-101_k141_208752 | Low-quality |
| 18-101_k141_192120 | Not-determined |
| 18-101_k141_372451 | Not-determined |
| 18-101_k141_92776 | Not-determined |
| 18-101_k141_92777 | Not-determined |
| 18-101_k141_224299 | Low-quality |
| 18-101_k141_241077 | Low-quality |
| 18-101_k141_224300 | Not-determined |
| 18-101_k141_157845 | Not-determined |
| 18-101_k141_157846 | Not-determined |
| 18-101_k141_356665 | Not-determined |
| 18-101_k141_224304 | Not-determined |
| 18-101_k141_258139 | Not-determined |
| 18-101_k141_126480 | Not-determined |
| 18-101_k141_224305 | Not-determined |
| 18-101_k141_241091 | Not-determined |
| 18-101_k141_388693 | Not-determined |
| 18-101_k141_338767 | Not-determined |
| 18-101_k141_192145 | Not-determined |
| 18-101_k141_273497 | Not-determined |
| 18-101_k141_108189 | Not-determined |
| 18-101_k141_157861 | Low-quality |
| 18-101_k141_59352 | Not-determined |
| 18-101_k141_241098 | Not-determined |
| 18-101_k141_192158 | Not-determined |
| 18-101_k141_142187 | Low-quality |
| 18-101_k141_224327 | Not-determined |
| 18-101_k141_192164 | Not-determined |
| 18-101_k141_356689 | Low-quality |
| 18-101_k141_108201 | Not-determined |
| 18-101_k141_92801 | Not-determined |
| 18-101_k141_42948 | Not-determined |
| 18-101_k141_192170 | Not-determined |
| 18-101_k141_356693 | Not-determined |
| 18-101_k141_173970 | Not-determined |
| 18-101_k141_356694 | Low-quality |
| 18-101_k141_273512 | Not-determined |
| 18-101_k141_241113 | Not-determined |
| 18-101_k141_108208 | Low-quality |
| 18-101_k141_92803 | Not-determined |
| 18-101_k141_26680 | Not-determined |
| 18-101_k141_75316 | Not-determined |
| 18-101_k141_388707 | Not-determined |
| 18-101_k141_26685 | Not-determined |
| 18-101_k141_241127 | Not-determined |
| 18-101_k141_59373 | Not-determined |
| 18-101_k141_157887 | Not-determined |
| 18-101_k141_59375 | Low-quality |
| 18-101_k141_258172 | Not-determined |
| 18-101_k141_258173 | Not-determined |
| 18-101_k141_42962 | Not-determined |
| 18-101_k141_173977 | Not-determined |
| 18-101_k141_241131 | Not-determined |
| 18-101_k141_157892 | Not-determined |
| 18-101_k141_42965 | Not-determined |
| 18-101_k141_388713 | Not-determined |
| 18-101_k141_173982 | Low-quality |
| 18-101_k141_92809 | Not-determined |
| 18-101_k141_142225 | Not-determined |
| 18-101_k141_108230 | Low-quality |
| 18-101_k141_126518 | Not-determined |
| 18-101_k141_356714 | Not-determined |
| 18-101_k141_108237 | Not-determined |
| 18-101_k141_92815 | Not-determined |
| 18-101_k141_92816 | Not-determined |
| 18-101_k141_142229 | Low-quality |
| 18-101_k141_42972 | Not-determined |
| 18-101_k141_356718 | Not-determined |
| 18-101_k141_356720 | Not-determined |
| 18-101_k141_59399 | Low-quality |
| 18-101_k141_42980 | Not-determined |
| 18-101_k141_224365 | Not-determined |
| 18-101_k141_108249 | Not-determined |
| 18-101_k141_59404 | Not-determined |
| 18-101_k141_108255 | Not-determined |
| 18-101_k141_92825 | Not-determined |
| 18-101_k141_142238 | Not-determined |
| 18-101_k141_142239 | Not-determined |
| 18-101_k141_126539 | Not-determined |
| 18-101_k141_92829 | Not-determined |
| 18-101_k141_59412 | Not-determined |
| 18-101_k141_108266 | Not-determined |
| 18-101_k141_388746 | Low-quality |
| 18-101_k141_174002 | Not-determined |
| 18-101_k141_108272 | Not-determined |
| 18-101_k141_126552 | Not-determined |
| 18-101_k141_388748 | Not-determined |
| 18-101_k141_388749 | Not-determined |
| 18-101_k141_10401 | Not-determined |
| 18-101_k141_388751 | Not-determined |
| 18-101_k141_92843 | Not-determined |
| 18-101_k141_142250 | Low-quality |
| 18-101_k141_388756 | Not-determined |
| 18-101_k141_92852 | Not-determined |
| 18-101_k141_290288 | Low-quality |
| 18-101_k141_174013 | Not-determined |
| 18-101_k141_42989 | Not-determined |
| 18-101_k141_388763 | Not-determined |
| 18-101_k141_388765 | Not-determined |
| 18-101_k141_192188 | Low-quality |
| 18-101_k141_338816 | Not-determined |
| 18-101_k141_59433 | Not-determined |
| 18-101_k141_208789 | Not-determined |
| 18-101_k141_59435 | Not-determined |
| 18-101_k141_338818 | Low-quality |
| 18-101_k141_42997 | Not-determined |
| 18-101_k141_142268 | Not-determined |
| 18-101_k141_59448 | Not-determined |
| 18-101_k141_142272 | Not-determined |
| 18-101_k141_388771 | Not-determined |
| 18-101_k141_142273 | Not-determined |
| 18-101_k141_208798 | Not-determined |
| 18-101_k141_290300 | Not-determined |
| 18-101_k141_92873 | Not-determined |
| 18-101_k141_126584 | Not-determined |
| 18-101_k141_208802 | Not-determined |
| 18-101_k141_388774 | Not-determined |
| 18-101_k141_174024 | Not-determined |
| 18-101_k141_59453 | Not-determined |
| 18-101_k141_92876 | Not-determined |
| 18-101_k141_241169 | Not-determined |
| 18-101_k141_290310 | Not-determined |
| 18-101_k141_356733 | Not-determined |
| 18-101_k141_192202 | Not-determined |
| 18-101_k141_388779 | Not-determined |
| 18-101_k141_273518 | Not-determined |
| 18-101_k141_241177 | Not-determined |
| 18-101_k141_273520 | Not-determined |
| 18-101_k141_208811 | Not-determined |
| 18-101_k141_174038 | Not-determined |
| 18-101_k141_208814 | Not-determined |
| 18-101_k141_356742 | Not-determined |
| 18-101_k141_273527 | Not-determined |
| 18-101_k141_388789 | Not-determined |
| 18-101_k141_59463 | Not-determined |
| 18-101_k141_356748 | Not-determined |
| 18-101_k141_108285 | Not-determined |
| 18-101_k141_59469 | Not-determined |
| 18-101_k141_356757 | Not-determined |
| 18-101_k141_241198 | Not-determined |
| 18-101_k141_92887 | Not-determined |
| 18-101_k141_59474 | Not-determined |
| 18-101_k141_372483 | Not-determined |
| 18-101_k141_59477 | Not-determined |
| 18-101_k141_157948 | Not-determined |
| 18-101_k141_241206 | Low-quality |
| 18-101_k141_372489 | Not-determined |
| 18-101_k141_142296 | Not-determined |
| 18-101_k141_157950 | Not-determined |
| 18-101_k141_174061 | Not-determined |
| 18-101_k141_59485 | Not-determined |
| 18-101_k141_59486 | Not-determined |
| 18-101_k141_92901 | Not-determined |
| 18-101_k141_92902 | Not-determined |
| 18-101_k141_108289 | Not-determined |
| 18-101_k141_174066 | Not-determined |
| 18-101_k141_157958 | Not-determined |
| 18-101_k141_273552 | Not-determined |
| 18-101_k141_192251 | Not-determined |
| 18-101_k141_208847 | Not-determined |
| 18-101_k141_75328 | Not-determined |
| 18-101_k141_388809 | Not-determined |
| 18-101_k141_142301 | Not-determined |
| 18-101_k141_157971 | Not-determined |
| 18-101_k141_192260 | Not-determined |
| 18-101_k141_324298 | Not-determined |
| 18-101_k141_338866 | Not-determined |
| 18-101_k141_258247 | Not-determined |
| 18-101_k141_307326 | Not-determined |
| 18-101_k141_307328 | Not-determined |
| 18-101_k141_10426 | Not-determined |
| 18-101_k141_208863 | Not-determined |
| 18-101_k141_10427 | Not-determined |
| 18-101_k141_174091 | Low-quality |
| 18-101_k141_126601 | Not-determined |
| 18-101_k141_43016 | Not-determined |
| 18-101_k141_174095 | Not-determined |
| 18-101_k141_324306 | Not-determined |
| 18-101_k141_157984 | Not-determined |
| 18-101_k141_192274 | Low-quality |
| 18-101_k141_241238 | Not-determined |
| 18-101_k141_372520 | Not-determined |
| 18-101_k141_59504 | Low-quality |
| 18-101_k141_208872 | Not-determined |
| 18-101_k141_59506 | Not-determined |
| 18-101_k141_290381 | Low-quality |
| 18-101_k141_356763 | Not-determined |
| 18-101_k141_241245 | Not-determined |
| 18-101_k141_174106 | Low-quality |
| 18-101_k141_388822 | Not-determined |
| 18-101_k141_174108 | Not-determined |
| 18-101_k141_372528 | Not-determined |
| 18-101_k141_208874 | Not-determined |
| 18-101_k141_75350 | Not-determined |
| 18-101_k141_241248 | Not-determined |
| 18-101_k141_75362 | Not-determined |
| 18-101_k141_108299 | Not-determined |
| 18-101_k141_224420 | Not-determined |
| 18-101_k141_75363 | Not-determined |
| 18-101_k141_356778 | Not-determined |
| 18-101_k141_241256 | Not-determined |
| 18-101_k141_388836 | Not-determined |
| 18-101_k141_324312 | Low-quality |
| 18-101_k141_241262 | Not-determined |
| 18-101_k141_324313 | Low-quality |
| 18-101_k141_324314 | Not-determined |
| 18-101_k141_241263 | Not-determined |
| 18-101_k141_372531 | Not-determined |
| 18-101_k141_59523 | Not-determined |
| 18-101_k141_307338 | Not-determined |
| 18-101_k141_126620 | Not-determined |
| 18-101_k141_126621 | Not-determined |
| 18-101_k141_43045 | Not-determined |
| 18-101_k141_241267 | Not-determined |
| 18-101_k141_108313 | Not-determined |
| 18-101_k141_388844 | Not-determined |
| 18-101_k141_92943 | Low-quality |
| 18-101_k141_372536 | Not-determined |
| 18-101_k141_75381 | Not-determined |
| 18-101_k141_157994 | Not-determined |
| 18-101_k141_142333 | Not-determined |
| 18-101_k141_372543 | Not-determined |
| 18-101_k141_157997 | Not-determined |
| 17-108-final-viral-combined_k141_140009\|\|full | High-quality |
| 18-101_k141_307363 | Not-determined |
| 18-101_k141_126637 | Not-determined |
| 18-101_k141_142343 | Not-determined |
| 18-101_k141_290405 | Not-determined |
| 18-101_k141_290406 | Not-determined |
| 18-101_k141_273570 | Not-determined |
| 18-101_k141_208878 | Not-determined |
| 18-101_k141_108323 | Not-determined |
| 18-101_k141_43071 | Not-determined |
| 18-101_k141_92947 | Not-determined |
| 18-101_k141_10473 | Not-determined |
| 18-101_k141_192296 | Not-determined |
| 18-101_k141_192300 | Not-determined |
| 18-101_k141_43078 | Low-quality |
| 18-101_k141_108332 | Not-determined |
| 18-101_k141_338917 | Not-determined |
| 18-101_k141_372559 | Not-determined |
| 18-101_k141_43079 | Not-determined |
| 18-101_k141_338923 | Not-determined |
| 18-101_k141_158015 | Not-determined |
| 18-101_k141_372562 | Low-quality |
| 18-101_k141_75401 | Not-determined |
| 18-101_k141_75403 | Not-determined |
| 18-101_k141_388859 | Not-determined |
| 18-101_k141_307378 | Low-quality |
| 18-101_k141_307382 | Not-determined |
| 18-101_k141_142363 | Low-quality |
| 18-101_k141_126672 | Low-quality |
| 18-101_k141_224464 | Not-determined |
| 18-101_k141_75416 | Not-determined |
| 18-101_k141_75418 | Not-determined |
| 18-101_k141_10496 | Not-determined |
| 18-101_k141_108360 | Not-determined |
| 18-101_k141_388864 | Not-determined |
| 18-101_k141_273594 | Not-determined |
| 18-101_k141_158020 | Not-determined |
| 18-101_k141_142373 | Not-determined |
| 18-101_k141_92969 | Not-determined |
| 18-101_k141_10501 | Not-determined |
| 18-101_k141_208918 | Not-determined |
| 18-101_k141_192315 | Not-determined |
| 18-101_k141_75428 | Not-determined |
| 18-101_k141_241317 | Not-determined |
| 18-101_k141_43110 | Not-determined |
| 18-101_k141_59553 | Not-determined |
| 18-101_k141_108364 | Not-determined |
| 18-101_k141_59557 | Not-determined |
| 18-101_k141_372583 | Not-determined |
| 18-101_k141_241323 | Not-determined |
| 18-101_k141_158041 | Not-determined |
| 18-101_k141_208927 | Not-determined |
| 18-101_k141_43121 | Not-determined |
| 18-101_k141_192323 | Not-determined |
| 18-101_k141_126707 | Not-determined |
| 18-101_k141_10508 | Not-determined |
| 18-101_k141_10510 | Not-determined |
| 18-101_k141_273621 | Not-determined |
| 18-101_k141_241333 | Not-determined |
| 18-101_k141_75441 | Not-determined |
| 18-101_k141_192331 | Not-determined |
| 18-101_k141_158048 | Not-determined |
| 18-101_k141_59568 | Not-determined |
| 18-101_k141_158051 | Not-determined |
| 18-101_k141_208939 | Not-determined |
| 18-101_k141_43132 | Not-determined |
| 18-101_k141_59575 | Not-determined |
| 18-101_k141_356815 | Not-determined |
| 18-101_k141_174128 | Not-determined |
| 18-101_k141_192340 | Not-determined |
| 18-101_k141_93004 | Not-determined |
| 18-101_k141_93005 | Not-determined |
| 18-101_k141_356819 | Not-determined |
| 18-101_k141_26760 | Not-determined |
| 18-101_k141_290434 | Not-determined |
| 18-101_k141_174137 | Not-determined |
| 18-101_k141_59579 | Low-quality |
| 18-101_k141_93015 | Not-determined |
| 18-101_k141_356826 | Not-determined |
| 18-101_k141_356827 | Not-determined |
| 18-101_k141_356830 | Not-determined |
| 18-101_k141_290442 | Not-determined |
| 18-101_k141_10532 | Not-determined |
| 18-101_k141_10533 | Not-determined |
| 18-101_k141_93029 | Not-determined |
| 18-101_k141_356840 | Low-quality |
| 18-101_k141_142428 | Not-determined |
| 18-101_k141_43145 | Not-determined |
| 18-101_k141_258331 | Not-determined |
| 18-101_k141_258333 | Not-determined |
| 18-101_k141_174152 | Not-determined |
| 18-101_k141_290464 | Not-determined |
| 18-101_k141_174154 | Not-determined |
| 18-101_k141_10550 | Not-determined |
| 18-101_k141_273668 | Not-determined |
| 18-101_k141_142438 | Not-determined |
| 18-101_k141_224487 | Not-determined |
| 18-101_k141_93039 | Low-quality |
| 18-101_k141_43160 | Not-determined |
| 18-101_k141_192383 | Not-determined |
| 18-101_k141_26779 | Not-determined |
| 18-101_k141_356859 | Not-determined |
| 18-101_k141_208977 | Low-quality |
| 18-101_k141_192387 | Not-determined |
| 18-101_k141_324347 | Not-determined |
| 18-101_k141_158087 | Not-determined |
| 18-101_k141_388884 | Not-determined |
| 18-101_k141_26791 | Not-determined |
| 18-101_k141_142449 | Not-determined |
| 18-101_k141_258347 | Not-determined |
| 18-101_k141_43170 | Not-determined |
| 18-101_k141_142451 | Low-quality |
| 18-101_k141_43171 | Not-determined |
| 18-101_k141_290480 | Not-determined |
| 18-101_k141_126735 | Not-determined |
| 18-101_k141_258350 | Not-determined |
| 18-101_k141_324350 | Not-determined |
| 18-101_k141_356876 | Not-determined |
| 18-101_k141_26800 | Not-determined |
| 18-101_k141_26802 | Not-determined |
| 18-101_k141_258359 | Not-determined |
| 18-101_k141_324353 | Not-determined |
| 18-101_k141_93054 | Not-determined |
| 18-101_k141_59628 | Not-determined |
| 18-101_k141_388897 | Not-determined |
| 18-101_k141_273678 | Low-quality |
| 18-101_k141_324358 | Not-determined |
| 18-101_k141_59630 | Not-determined |
| 18-101_k141_290490 | Not-determined |
| 18-101_k141_26812 | Not-determined |
| 18-101_k141_324363 | Not-determined |
| 18-101_k141_258367 | Not-determined |
| 18-101_k141_26818 | Low-quality |
| 18-101_k141_108441 | Not-determined |
| 18-101_k141_108445 | Not-determined |
| 18-101_k141_192401 | Not-determined |
| 18-101_k141_43186 | Not-determined |
| 18-101_k141_10574 | Not-determined |
| 18-101_k141_59637 | Not-determined |
| 18-101_k141_356880 | Not-determined |
| 18-101_k141_10581 | Not-determined |
| 18-101_k141_93071 | Not-determined |
| 18-101_k141_59643 | Not-determined |
| 18-101_k141_192408 | Not-determined |
| 18-101_k141_241363 | Not-determined |
| 18-101_k141_59646 | Not-determined |
| 18-101_k141_75498 | Not-determined |
| 18-101_k141_10585 | Not-determined |
| 18-101_k141_192409 | Not-determined |
| 18-101_k141_108464 | Not-determined |
| 18-101_k141_59649 | Not-determined |
| 18-101_k141_93073 | Not-determined |
| 18-101_k141_93076 | Not-determined |
| 18-101_k141_158105 | Not-determined |
| 18-101_k141_59652 | Not-determined |
| 18-101_k141_108469 | Not-determined |
| 18-101_k141_26835 | Not-determined |
| 18-101_k141_174204 | Not-determined |
| 18-101_k141_174205 | Not-determined |
| 18-101_k141_43187 | Not-determined |
| 18-101_k141_208998 | Not-determined |
| 18-101_k141_174210 | Not-determined |
| 18-101_k141_158122 | Not-determined |
| 18-101_k141_10601 | Not-determined |
| 18-101_k141_158126 | Low-quality |
| 18-101_k141_388904 | Not-determined |
| 18-101_k141_174214 | Not-determined |
| 18-101_k141_158128 | Low-quality |
| 18-101_k141_93093 | Not-determined |
| 18-101_k141_75521 | Not-determined |
| 18-101_k141_158137 | Not-determined |
| 18-101_k141_258383 | Not-determined |
| 18-101_k141_209013 | Low-quality |
| 18-101_k141_126780 | Not-determined |
| 18-101_k141_258388 | Not-determined |
| 18-101_k141_75527 | Low-quality |
| 18-101_k141_158142 | Low-quality |
| 18-101_k141_388917 | Not-determined |
| 18-101_k141_192426 | Not-determined |
| 18-101_k141_388919 | Not-determined |
| 18-101_k141_10616 | Not-determined |
| 18-101_k141_43205 | Not-determined |
| 18-101_k141_224550 | Not-determined |
| 18-101_k141_209017 | Not-determined |
| 18-101_k141_224553 | Not-determined |
| 18-101_k141_75536 | Not-determined |
| 18-101_k141_75537 | Not-determined |
| 18-101_k141_43211 | Not-determined |
| 18-101_k141_26850 | Not-determined |
| 18-101_k141_224555 | Not-determined |
| 18-101_k141_258393 | Not-determined |
| 18-101_k141_307448 | Not-determined |
| 18-101_k141_388926 | Not-determined |
| 18-101_k141_372641 | Low-quality |
| 18-101_k141_158157 | Not-determined |
| 18-101_k141_224566 | Not-determined |
| 18-101_k141_126792 | Not-determined |
| 18-101_k141_174244 | Not-determined |
| 18-101_k141_273717 | Not-determined |
| 18-101_k141_174246 | Low-quality |
| 18-101_k141_158162 | Not-determined |
| 18-101_k141_10635 | Not-determined |
| 18-101_k141_307462 | Not-determined |
| 18-101_k141_388936 | Not-determined |
| 18-101_k141_108487 | Low-quality |
| 18-101_k141_356896 | Not-determined |
| 18-101_k141_388938 | Not-determined |
| 18-101_k141_126798 | Not-determined |
| 18-101_k141_75554 | Not-determined |
| 18-101_k141_372652 | Not-determined |
| 18-101_k141_59661 | Not-determined |
| 18-101_k141_43222 | Low-quality |

| | |
|---|---|
| 18-101_k141_258413 | Not-determined |
| 18-101_k141_258414 | Not-determined |
| 18-101_k141_209058 | Not-determined |
| 18-101_k141_338936 | Not-determined |
| 18-101_k141_43226 | Not-determined |
| 18-101_k141_43227 | Not-determined |
| 18-101_k141_307479 | Not-determined |
| 18-101_k141_126824 | Not-determined |
| 18-101_k141_192447 | Not-determined |
| 18-101_k141_324449 | Not-determined |
| 18-101_k141_388959 | Not-determined |
| 18-101_k141_10666 | Not-determined |
| 18-101_k141_59673 | Not-determined |
| 18-101_k141_192452 | Not-determined |
| 18-101_k141_290564 | Not-determined |
| 18-101_k141_388963 | Not-determined |
| 18-101_k141_43235 | Not-determined |
| 18-101_k141_338944 | Not-determined |
| 18-101_k141_338945 | Not-determined |
| 18-101_k141_26869 | Not-determined |
| 18-101_k141_26871 | Low-quality |
| 18-101_k141_126831 | Not-determined |
| 18-101_k141_26874 | Low-quality |
| 18-101_k141_93129 | Not-determined |
| 18-101_k141_26881 | Not-determined |
| 18-101_k141_258439 | Not-determined |
| 18-101_k141_142498 | Low-quality |
| 18-101_k141_258443 | Not-determined |
| 18-101_k141_75582 | Low-quality |
| 18-101_k141_338953 | Not-determined |
| 18-101_k141_142505 | Not-determined |
| 18-101_k141_324465 | Not-determined |
| 18-101_k141_372669 | Not-determined |
| 18-101_k141_108519 | Not-determined |
| 18-101_k141_142509 | Not-determined |
| 18-101_k141_290582 | Not-determined |
| 18-101_k141_108522 | Not-determined |
| 18-101_k141_108528 | Not-determined |
| 18-101_k141_224579 | Not-determined |
| 18-101_k141_338965 | Low-quality |
| 18-101_k141_93131 | Not-determined |
| 18-101_k141_372681 | Not-determined |
| 18-101_k141_158192 | Not-determined |
| 18-101_k141_43248 | Not-determined |
| 18-101_k141_26903 | Not-determined |
| 18-101_k141_93136 | Not-determined |
| 18-101_k141_338976 | Not-determined |
| 18-101_k141_388974 | Not-determined |
| 18-101_k141_372688 | Not-determined |
| 18-101_k141_26909 | Not-determined |
| 18-101_k141_158200 | Not-determined |
| 18-101_k141_10678 | Not-determined |
| 18-101_k141_324483 | Not-determined |
| 18-101_k141_209081 | Not-determined |
| 18-101_k141_142529 | Not-determined |
| 18-101_k141_142530 | Not-determined |
| 18-101_k141_10685 | Not-determined |
| 18-101_k141_108538 | Low-quality |
| 18-101_k141_59698 | Not-determined |
| 18-101_k141_324493 | Not-determined |
| 18-101_k141_26915 | Not-determined |
| 18-101_k141_209084 | Low-quality |
| 18-101_k141_26918 | Not-determined |
| 18-101_k141_108543 | Not-determined |
| 18-101_k141_209086 | Not-determined |
| 18-101_k141_10691 | Not-determined |
| 18-101_k141_59702 | Not-determined |
| 18-101_k141_356918 | Not-determined |
| 18-101_k141_59705 | Not-determined |
| 18-101_k141_59709 | Not-determined |
| 18-101_k141_307532 | Not-determined |
| 18-101_k141_75598 | Low-quality |
| 18-101_k141_75600 | Not-determined |
| 18-101_k141_209102 | Not-determined |
| 18-101_k141_158217 | Not-determined |
| 18-101_k141_339001 | Not-determined |
| 18-101_k141_356924 | Not-determined |
| 18-101_k141_258471 | Low-quality |
| 18-101_k141_59724 | Not-determined |
| 18-101_k141_258479 | Not-determined |
| 18-101_k141_192508 | Not-determined |
| 18-101_k141_224588 | Low-quality |
| 18-101_k141_324499 | Not-determined |
| 18-101_k141_339014 | Not-determined |
| 18-101_k141_93169 | Not-determined |
| 18-101_k141_192509 | Not-determined |
| 18-101_k141_192510 | Not-determined |
| 18-101_k141_192512 | Not-determined |
| 18-101_k141_209123 | Not-determined |
| 18-101_k141_307556 | Not-determined |
| 18-101_k141_26947 | Not-determined |
| 18-101_k141_43283 | Not-determined |
| 18-101_k141_324507 | Not-determined |
| 18-101_k141_324510 | Not-determined |
| 18-101_k141_26954 | Not-determined |
| 18-101_k141_26956 | Not-determined |
| 18-101_k141_324512 | Not-determined |
| 18-101_k141_241380 | Not-determined |
| 18-101_k141_126893 | Not-determined |
| 18-101_k141_307570 | Not-determined |
| 18-101_k141_26962 | Not-determined |
| 18-101_k141_59740 | Low-quality |
| 18-101_k141_93184 | Low-quality |
| 18-101_k141_241384 | Not-determined |
| 18-101_k141_93188 | Not-determined |
| 18-101_k141_209129 | Not-determined |
| 18-101_k141_389008 | Low-quality |
| 18-101_k141_339046 | Not-determined |
| 18-101_k141_26965 | Not-determined |
| 18-101_k141_307583 | Not-determined |
| 18-101_k141_126905 | Not-determined |
| 18-101_k141_307585 | Low-quality |
| 18-101_k141_339048 | Not-determined |
| 18-101_k141_158232 | Not-determined |
| 18-101_k141_126908 | Not-determined |
| 18-101_k141_158234 | Not-determined |
| 18-101_k141_258485 | Not-determined |
| 18-101_k141_258488 | Not-determined |
| 18-101_k141_258491 | Not-determined |
| 18-101_k141_59747 | Not-determined |
| 18-101_k141_324534 | Not-determined |
| 18-101_k141_10739 | Low-quality |
| 18-101_k141_59756 | Not-determined |
| 18-101_k141_59758 | Not-determined |
| 18-101_k141_307592 | Low-quality |
| 18-101_k141_258500 | Not-determined |
| 18-101_k141_224640 | Not-determined |
| 18-101_k141_43294 | Not-determined |
| 18-101_k141_224641 | Not-determined |
| 18-101_k141_192519 | Not-determined |
| 18-101_k141_43299 | Not-determined |
| 18-101_k141_209142 | Low-quality |
| 18-101_k141_59770 | Not-determined |
| 18-101_k141_126925 | Not-determined |
| 18-101_k141_224663 | Not-determined |
| 18-101_k141_356961 | Not-determined |
| 18-101_k141_142569 | Not-determined |
| 18-101_k141_209158 | Not-determined |
| 18-101_k141_224666 | Not-determined |
| 18-101_k141_43321 | Not-determined |
| 18-101_k141_258516 | Not-determined |
| 18-101_k141_307614 | Not-determined |
| 18-101_k141_372756 | Not-determined |
| 18-101_k141_241430 | Not-determined |
| 18-101_k141_75631 | Not-determined |
| 18-101_k141_10772 | Not-determined |
| 18-101_k141_307617 | Not-determined |
| 18-101_k141_26990 | Not-determined |
| 18-101_k141_290607 | Low-quality |
| 18-101_k141_324560 | Not-determined |
| 18-101_k141_389054 | Not-determined |
| 18-101_k141_372768 | Not-determined |
| 18-101_k141_324562 | Not-determined |
| 18-101_k141_339096 | Not-determined |
| 18-101_k141_307623 | Not-determined |
| 18-101_k141_258524 | Not-determined |
| 18-101_k141_126934 | Not-determined |
| 18-101_k141_192536 | Not-determined |
| 18-101_k141_158280 | Low-quality |
| 18-101_k141_224670 | Not-determined |
| 18-101_k141_158281 | Not-determined |
| 18-101_k141_372775 | Not-determined |
| 18-101_k141_75639 | Not-determined |
| 18-101_k141_192538 | Not-determined |
| 18-101_k141_174266 | Not-determined |
| 18-101_k141_43333 | Not-determined |
| 18-101_k141_126941 | Not-determined |
| 18-101_k141_27002 | Not-determined |
| 18-101_k141_372781 | Not-determined |
| 18-101_k141_192547 | Not-determined |
| 18-101_k141_258538 | Not-determined |
| 18-101_k141_258539 | Not-determined |
| 18-101_k141_339105 | Not-determined |
| 18-101_k141_324578 | Not-determined |
| 18-101_k141_372786 | Not-determined |
| 18-101_k141_290625 | Not-determined |
| 18-101_k141_290626 | Not-determined |
| 18-101_k141_142580 | Not-determined |
| 18-101_k141_192564 | Not-determined |
| 18-101_k141_174293 | Not-determined |
| 18-101_k141_126943 | Not-determined |
| 18-101_k141_192567 | Not-determined |
| 18-101_k141_27013 | Not-determined |
| 18-101_k141_224690 | Not-determined |
| 18-101_k141_307628 | Not-determined |
| 18-101_k141_372808 | Not-determined |
| 18-101_k141_389074 | Not-determined |
| 18-101_k141_10812 | Not-determined |
| 18-101_k141_27019 | Not-determined |
| 18-101_k141_142584 | Not-determined |
| 18-101_k141_10816 | Not-determined |
| 18-101_k141_10817 | Not-determined |
| 18-101_k141_174302 | Not-determined |
| 18-101_k141_324587 | Not-determined |
| 18-101_k141_307637 | Not-determined |
| 18-101_k141_324588 | Not-determined |
| 18-101_k141_324591 | Not-determined |
| 18-101_k141_389087 | Not-determined |
| 18-101_k141_339133 | Not-determined |
| 18-101_k141_158290 | Not-determined |
| 18-101_k141_241485 | Not-determined |
| 18-101_k141_27034 | Not-determined |
| 18-101_k141_372816 | Not-determined |
| 18-101_k141_258556 | Not-determined |
| 18-101_k141_339145 | Not-determined |
| 18-101_k141_324601 | Not-determined |
| 18-101_k141_93257 | Not-determined |
| 18-101_k141_158301 | Not-determined |
| 18-101_k141_339148 | Not-determined |
| 18-101_k141_258560 | Not-determined |
| 18-101_k141_174312 | Not-determined |
| 18-101_k141_389112 | Low-quality |
| 18-101_k141_339156 | Not-determined |
| 18-101_k141_93273 | Not-determined |
| 18-101_k141_59790 | Not-determined |
| 18-101_k141_59794 | Not-determined |
| 18-101_k141_209216 | Low-quality |
| 18-101_k141_10841 | Low-quality |
| 18-101_k141_241501 | Not-determined |
| 18-101_k141_209223 | Not-determined |
| 18-101_k141_290658 | Not-determined |
| 18-101_k141_59803 | Not-determined |
| 18-101_k141_372830 | Not-determined |
| 18-101_k141_290661 | Not-determined |
| 18-101_k141_142622 | Not-determined |
| 18-101_k141_10848 | Not-determined |
| 18-101_k141_241507 | Not-determined |
| 18-101_k141_27067 | Not-determined |
| 18-101_k141_290666 | Not-determined |
| 18-101_k141_108606 | Low-quality |
| 18-101_k141_389114 | Low-quality |
| 18-101_k141_389117 | Not-determined |
| 18-101_k141_307677 | Not-determined |
| 18-101_k141_241509 | Not-determined |
| 18-101_k141_290673 | Not-determined |
| 18-101_k141_389124 | Not-determined |
| 18-101_k141_241513 | Not-determined |
| 18-101_k141_224722 | Not-determined |
| 18-101_k141_142630 | Not-determined |
| 18-101_k141_273839 | Not-determined |
| 18-101_k141_43379 | Not-determined |
| 18-101_k141_142632 | Not-determined |
| 18-101_k141_290680 | Not-determined |
| 18-101_k141_209256 | Not-determined |
| 18-101_k141_27081 | Low-quality |
| 18-101_k141_59810 | Not-determined |
| 18-101_k141_389130 | Not-determined |
| 18-101_k141_224738 | Low-quality |
| 18-101_k141_324618 | Not-determined |
| 18-101_k141_290689 | Not-determined |
| 18-101_k141_192584 | Not-determined |
| 18-101_k141_126973 | Not-determined |
| 18-101_k141_59821 | Not-determined |
| 18-101_k141_389132 | Not-determined |
| 18-101_k141_59822 | Not-determined |
| 18-101_k141_290690 | Not-determined |
| 18-101_k141_324620 | Not-determined |
| 18-101_k141_307688 | Not-determined |
| 18-101_k141_258606 | Not-determined |
| 18-101_k141_126977 | Not-determined |
| 18-101_k141_43387 | Not-determined |
| 18-101_k141_43388 | Not-determined |
| 18-101_k141_241544 | Not-determined |
| 18-101_k141_59838 | Not-determined |
| 18-101_k141_339191 | Not-determined |
| 18-101_k141_174330 | Not-determined |
| 18-101_k141_324624 | Not-determined |
| 18-101_k141_307704 | Not-determined |
| 18-101_k141_27101 | Not-determined |
| 18-101_k141_290704 | Not-determined |
| 18-101_k141_126987 | Not-determined |
| 18-101_k141_339200 | Low-quality |
| 18-101_k141_357010 | Not-determined |
| 18-101_k141_258623 | Not-determined |
| 18-101_k141_59856 | Not-determined |
| 18-101_k141_307713 | Not-determined |
| 18-101_k141_273858 | Not-determined |
| 18-101_k141_389136 | Not-determined |
| 18-101_k141_241556 | Not-determined |
| 18-101_k141_241557 | Not-determined |
| 18-101_k141_59860 | Not-determined |
| 18-101_k141_258634 | Not-determined |
| 18-101_k141_273865 | Not-determined |
| 18-101_k141_241559 | Not-determined |
| 18-101_k141_389141 | Not-determined |
| 18-101_k141_192602 | Not-determined |
| 18-101_k141_389143 | Not-determined |
| 18-101_k141_209295 | Not-determined |
| 18-101_k141_43401 | Not-determined |
| 18-101_k141_158338 | Not-determined |
| 18-101_k141_258648 | Not-determined |
| 18-101_k141_10916 | Not-determined |
| 18-101_k141_158344 | Not-determined |
| 18-101_k141_290728 | Not-determined |
| 18-101_k141_209301 | Low-quality |
| 18-101_k141_273880 | Not-determined |
| 18-101_k141_10927 | Not-determined |
| 18-101_k141_10928 | Not-determined |
| 18-101_k141_192620 | Not-determined |
| 18-101_k141_273884 | Not-determined |
| 18-101_k141_192626 | Not-determined |
| 18-101_k141_357042 | Not-determined |
| 18-101_k141_43418 | Not-determined |
| 18-101_k141_158360 | Not-determined |
| 18-101_k141_290738 | Low-quality |
| 18-101_k141_59873 | Low-quality |
| 18-101_k141_209311 | Not-determined |
| 18-101_k141_93345 | Not-determined |
| 18-101_k141_10943 | Not-determined |
| 18-101_k141_75667 | Not-determined |
| 18-101_k141_224776 | Not-determined |
| 18-101_k141_174353 | Not-determined |
| 18-101_k141_290748 | Not-determined |
| 18-101_k141_10948 | Not-determined |
| 18-101_k141_192631 | Not-determined |
| 18-101_k141_192632 | Not-determined |
| 18-101_k141_339240 | Not-determined |
| 18-101_k141_142685 | Not-determined |
| 18-101_k141_224781 | Not-determined |
| 18-101_k141_93354 | Not-determined |
| 18-101_k141_43437 | Not-determined |
| 18-101_k141_209319 | Low-quality |
| 18-101_k141_258674 | Not-determined |
| 18-101_k141_75682 | Not-determined |
| 18-101_k141_224795 | Not-determined |
| 18-101_k141_209321 | Not-determined |
| 18-101_k141_209327 | Not-determined |
| 18-101_k141_258686 | Not-determined |
| 18-101_k141_59887 | Not-determined |
| 18-101_k141_59889 | Not-determined |
| 18-101_k141_357069 | Not-determined |
| 18-101_k141_108684 | Not-determined |
| 18-101_k141_142698 | Low-quality |
| 18-101_k141_241581 | Low-quality |
| 18-101_k141_59892 | Not-determined |
| 18-101_k141_43456 | Not-determined |
| 18-101_k141_108689 | Not-determined |
| 18-101_k141_59894 | Not-determined |
| 18-101_k141_43457 | Not-determined |
| 18-101_k141_192642 | Not-determined |
| 18-101_k141_307770 | Not-determined |
| 18-101_k141_192643 | Low-quality |
| 18-101_k141_10971 | Not-determined |
| 18-101_k141_174377 | Not-determined |
| 18-101_k141_174381 | Low-quality |
| 18-101_k141_224818 | Not-determined |
| 18-101_k141_59903 | Not-determined |
| 18-101_k141_324661 | Not-determined |
| 18-101_k141_339250 | Not-determined |
| 18-101_k141_258697 | Not-determined |
| 18-101_k141_372924 | Not-determined |
| 18-101_k141_389196 | Not-determined |
| 18-101_k141_174389 | Not-determined |
| 18-101_k141_10975 | Not-determined |
| 18-101_k141_307790 | Not-determined |
| 18-101_k141_307791 | Not-determined |
| 18-101_k141_258714 | Not-determined |
| 18-101_k141_209341 | Not-determined |
| 18-101_k141_158392 | Low-quality |
| 18-101_k141_59915 | Not-determined |
| 18-101_k141_241590 | Not-determined |
| 18-101_k141_224821 | Not-determined |
| 18-101_k141_174403 | Not-determined |
| 18-101_k141_59920 | Not-determined |
| 18-101_k141_174405 | Not-determined |
| 18-101_k141_357082 | Not-determined |
| 18-101_k141_27145 | Not-determined |
| 18-101_k141_372937 | Not-determined |
| 18-101_k141_258722 | Not-determined |
| 18-101_k141_290787 | Not-determined |
| 18-101_k141_158401 | Not-determined |
| 18-101_k141_241603 | Not-determined |
| 18-101_k141_339277 | Not-determined |
| 18-101_k141_174410 | Not-determined |
| 18-101_k141_192655 | Not-determined |
| 18-101_k141_192656 | Not-determined |
| 18-101_k141_108712 | Not-determined |
| 18-101_k141_158406 | Not-determined |
| 18-101_k141_75724 | Not-determined |
| 18-101_k141_27155 | Not-determined |
| 18-101_k141_258729 | Low-quality |
| 18-101_k141_174413 | Not-determined |
| 18-101_k141_75727 | Not-determined |
| 18-101_k141_307798 | Not-determined |
| 18-101_k141_389205 | Not-determined |
| 18-101_k141_93382 | Not-determined |
| 18-101_k141_158416 | Not-determined |
| 18-101_k141_158418 | Not-determined |
| 18-101_k141_209360 | Not-determined |
| 18-101_k141_174418 | Low-quality |
| 18-101_k141_209362 | Not-determined |
| 18-101_k141_142720 | Not-determined |
| 18-101_k141_158425 | Not-determined |
| 18-101_k141_127059 | Not-determined |
| 18-101_k141_258739 | Not-determined |
| 18-101_k141_224849 | Not-determined |
| 18-101_k141_258741 | Not-determined |
| 18-101_k141_93398 | Not-determined |
| 18-101_k141_93399 | Not-determined |
| 18-101_k141_108720 | Not-determined |
| 18-101_k141_10999 | Not-determined |
| 18-101_k141_11004 | Not-determined |
| 18-101_k141_11005 | Not-determined |
| 18-101_k141_11007 | Not-determined |
| 18-101_k141_192681 | Not-determined |
| 18-101_k141_174434 | Not-determined |
| 18-101_k141_241627 | Not-determined |
| 18-101_k141_307823 | Not-determined |
| 18-101_k141_389217 | Low-quality |
| 18-101_k141_324685 | Low-quality |
| 18-101_k141_93410 | Not-determined |
| 18-101_k141_241635 | Not-determined |
| 18-101_k141_307828 | Not-determined |
| 18-101_k141_174439 | Not-determined |
| 18-101_k141_93413 | Not-determined |
| 18-101_k141_241637 | Not-determined |
| 18-101_k141_75761 | Not-determined |
| 18-101_k141_258758 | Not-determined |
| 18-101_k141_241640 | Not-determined |
| 18-101_k141_258759 | Not-determined |
| 18-101_k141_241641 | Not-determined |
| 18-101_k141_224854 | Not-determined |
| 18-101_k141_127081 | Not-determined |
| 18-101_k141_127083 | Not-determined |
| 18-101_k141_224858 | Not-determined |
| 18-101_k141_11018 | Not-determined |
| 18-101_k141_142731 | Not-determined |
| 18-101_k141_290812 | Not-determined |
| 18-101_k141_339296 | Not-determined |
| 18-101_k141_174447 | Low-quality |
| 18-101_k141_389237 | Not-determined |
| 18-101_k141_389239 | Not-determined |
| 18-101_k141_127088 | Low-quality |
| 18-101_k141_127089 | Not-determined |
| 18-101_k141_224859 | Low-quality |
| 18-101_k141_241647 | Not-determined |
| 18-101_k141_174452 | Not-determined |
| 18-101_k141_307839 | Not-determined |
| 18-101_k141_339306 | Low-quality |
| 18-101_k141_290823 | Not-determined |
| 18-101_k141_357111 | Not-determined |
| 18-101_k141_307846 | Not-determined |
| 18-101_k141_108743 | Not-determined |
| 18-101_k141_372975 | Not-determined |
| 18-101_k141_290838 | Not-determined |
| 18-101_k141_357122 | Not-determined |
| 18-101_k141_307850 | Low-quality |
| 18-101_k141_357125 | Not-determined |
| 18-101_k141_75779 | Not-determined |
| 18-101_k141_290842 | Not-determined |
| 18-101_k141_339320 | Not-determined |
| 18-101_k141_158459 | Not-determined |
| 18-101_k141_209389 | Not-determined |
| 18-101_k141_258794 | Not-determined |
| 18-101_k141_339326 | Not-determined |
| 18-101_k141_209394 | Not-determined |
| 18-101_k141_11048 | Not-determined |
| 18-101_k141_158469 | Not-determined |
| 18-101_k141_373001 | Not-determined |
| 18-101_k141_241662 | Not-determined |
| 18-101_k141_258805 | Not-determined |
| 18-101_k141_290850 | Not-determined |
| 18-101_k141_75798 | Not-determined |
| 18-101_k141_43470 | Not-determined |
| 18-101_k141_373008 | Not-determined |
| 18-101_k141_307855 | Not-determined |
| 18-101_k141_75805 | Not-determined |
| 18-101_k141_142757 | Not-determined |
| 18-101_k141_373012 | Not-determined |
| 18-101_k141_389265 | Not-determined |
| 18-101_k141_258809 | Not-determined |
| 18-101_k141_241668 | Not-determined |
| 18-101_k141_43481 | Not-determined |
| 18-101_k141_324724 | Not-determined |
| 18-101_k141_389270 | Not-determined |
| 18-101_k141_127115 | Not-determined |
| 18-101_k141_142767 | Low-quality |
| 18-101_k141_127117 | Low-quality |
| 18-101_k141_324728 | Not-determined |
| 18-101_k141_224896 | Not-determined |
| 18-101_k141_192686 | Not-determined |
| 18-101_k141_324733 | Not-determined |
| 18-101_k141_290862 | Not-determined |
| 18-101_k141_389278 | Not-determined |
| 18-101_k141_209420 | Not-determined |
| 18-101_k141_241687 | Not-determined |
| 18-101_k141_127126 | Not-determined |
| 18-101_k141_209421 | Not-determined |
| 18-101_k141_339357 | Not-determined |
| 18-101_k141_11065 | Not-determined |
| 18-101_k141_373022 | Not-determined |
| 18-101_k141_59947 | Not-determined |
| 18-101_k141_93446 | Not-determined |
| 18-101_k141_241697 | Not-determined |
| 18-101_k141_59948 | Not-determined |
| 18-101_k141_43491 | Not-determined |
| 18-101_k141_93450 | Not-determined |
| 18-101_k141_174477 | Not-determined |
| 18-101_k141_174478 | Not-determined |
| 18-101_k141_307894 | Not-determined |
| 18-101_k141_324749 | Not-determined |
| 18-101_k141_75837 | Low-quality |
| 18-101_k141_290884 | Not-determined |
| 18-101_k141_389298 | Not-determined |
| 18-101_k141_373034 | Not-determined |
| 18-101_k141_290886 | Not-determined |
| 18-101_k141_174486 | Not-determined |
| 18-101_k141_209429 | Not-determined |
| 18-101_k141_373037 | Low-quality |
| 18-101_k141_290894 | Not-determined |
| 18-101_k141_174488 | Not-determined |
| 18-101_k141_258836 | Not-determined |
| 18-101_k141_339382 | Not-determined |
| 18-101_k141_108799 | Not-determined |
| 18-101_k141_142781 | Not-determined |
| 18-101_k141_27195 | Not-determined |
| 18-101_k141_108805 | Not-determined |
| 18-101_k141_93463 | Not-determined |
| 18-101_k141_43507 | Not-determined |
| 18-101_k141_290903 | Not-determined |
| 18-101_k141_11075 | Not-determined |
| 18-101_k141_389310 | Not-determined |
| 18-101_k141_224910 | Not-determined |
| 18-101_k141_43510 | Not-determined |
| 18-101_k141_224914 | Not-determined |
| 18-101_k141_307924 | Low-quality |
| 18-101_k141_108815 | Not-determined |
| 18-101_k141_273981 | Not-determined |
| 18-101_k141_108820 | Not-determined |
| 18-101_k141_127143 | Not-determined |
| 18-101_k141_273984 | Not-determined |
| 18-101_k141_75859 | Not-determined |
| 18-101_k141_174516 | Not-determined |
| 18-101_k141_324770 | Not-determined |
| 18-101_k141_75864 | Low-quality |
| 18-101_k141_307935 | Not-determined |
| 18-101_k141_324779 | Not-determined |
| 18-101_k141_258862 | Not-determined |
| 18-101_k141_258864 | Not-determined |
| 18-101_k141_108832 | Not-determined |
| 18-101_k141_258872 | Not-determined |
| 18-101_k141_290928 | Not-determined |
| 18-101_k141_224928 | Not-determined |
| 18-101_k141_224929 | Not-determined |
| 18-101_k141_108838 | Not-determined |
| 18-101_k141_209461 | Not-determined |
| 18-101_k141_274001 | Not-determined |
| 18-101_k141_43531 | Not-determined |
| 18-101_k141_108843 | Not-determined |
| 18-101_k141_43534 | Low-quality |
| 18-101_k141_290934 | Not-determined |
| 18-101_k141_127153 | Not-determined |
| 18-101_k141_43535 | Not-determined |
| 18-101_k141_389328 | Not-determined |
| 18-101_k141_224933 | Not-determined |
| 18-101_k141_324787 | Not-determined |
| 18-101_k141_75874 | Not-determined |
| 18-101_k141_307958 | Not-determined |
| 18-101_k141_241739 | Not-determined |
| 18-101_k141_93496 | Not-determined |
| 18-101_k141_290944 | Not-determined |
| 18-101_k141_11095 | Not-determined |
| 18-101_k141_389342 | Low-quality |
| 18-101_k141_59972 | Not-determined |
| 18-101_k141_192699 | Low-quality |
| 18-101_k141_11103 | Low-quality |
| 18-101_k141_241753 | Not-determined |
| 18-101_k141_11104 | Not-determined |
| 18-101_k141_43545 | Not-determined |
| 18-101_k141_192702 | Not-determined |
| 18-101_k141_11106 | Not-determined |
| 18-101_k141_373070 | Not-determined |
| 18-101_k141_290951 | Not-determined |
| 18-101_k141_224954 | Not-determined |
| 18-101_k141_241761 | Not-determined |
| 18-101_k141_75905 | Not-determined |
| 18-101_k141_93519 | Not-determined |
| 18-101_k141_258880 | Not-determined |
| 18-101_k141_43552 | Not-determined |
| 18-101_k141_290959 | Low-quality |
| 18-101_k141_174534 | Not-determined |
| 18-101_k141_209467 | Not-determined |
| 18-101_k141_241769 | Not-determined |
| 18-101_k141_11118 | Not-determined |
| 18-101_k141_75911 | Not-determined |
| 18-101_k141_108868 | Not-determined |
| 18-101_k141_59984 | Not-determined |
| 18-101_k141_307980 | Not-determined |
| 18-101_k141_373083 | Not-determined |

| | |
|---|---|
| 18-101_k141_241781 | Not-determined |
| 18-101_k141_59988 | Not-determined |
| 18-101_k141_43569 | Not-determined |
| 18-101_k141_373087 | Not-determined |
| 18-101_k141_357148 | Not-determined |
| 18-101_k141_389344 | Low-quality |
| 18-101_k141_127195 | Not-determined |
| 18-101_k141_75913 | Low-quality |
| 18-101_k141_192725 | Low-quality |
| 18-101_k141_389346 | Not-determined |
| 18-101_k141_307987 | Low-quality |
| 18-101_k141_93528 | Low-quality |
| 18-101_k141_389351 | Not-determined |
| 18-101_k141_307990 | Low-quality |
| 18-101_k141_324814 | Not-determined |
| 18-101_k141_75921 | Not-determined |
| 18-101_k141_174556 | Not-determined |
| 18-101_k141_307994 | Not-determined |
| 18-101_k141_127201 | Not-determined |
| 18-101_k141_324816 | Not-determined |
| 18-101_k141_224979 | Not-determined |
| 18-101_k141_27251 | Not-determined |
| 18-101_k141_75933 | Not-determined |
| 18-101_k141_258896 | Low-quality |
| 18-101_k141_127205 | Not-determined |
| 18-101_k141_174563 | Not-determined |
| 18-101_k141_357163 | Not-determined |
| 18-101_k141_224983 | Not-determined |
| 18-101_k141_357165 | Not-determined |
| 18-101_k141_43579 | Not-determined |
| 18-101_k141_27257 | Not-determined |
| 18-101_k141_290990 | Not-determined |
| 18-101_k141_324829 | Not-determined |
| 18-101_k141_43584 | Not-determined |
| 18-101_k141_373101 | Not-determined |
| 18-101_k141_27260 | Not-determined |
| 18-101_k141_224990 | Not-determined |
| 18-101_k141_373107 | Not-determined |
| 18-101_k141_127210 | Not-determined |
| 18-101_k141_308019 | Not-determined |
| 18-101_k141_357177 | Low-quality |
| 18-101_k141_241805 | Not-determined |
| 18-101_k141_324843 | Not-determined |
| 18-101_k141_290997 | Not-determined |
| 18-101_k141_60032 | Not-determined |
| 18-101_k141_241813 | Not-determined |
| 18-101_k141_373120 | Not-determined |
| 18-101_k141_373123 | Not-determined |
| 18-101_k141_174587 | Not-determined |
| 18-101_k141_225015 | Not-determined |
| 18-101_k141_291005 | Not-determined |
| 18-101_k141_389394 | Not-determined |
| 18-101_k141_75950 | Not-determined |
| 18-101_k141_357185 | Not-determined |
| 18-101_k141_225018 | Not-determined |
| 18-101_k141_291017 | Not-determined |
| 18-101_k141_357190 | Not-determined |
| 18-101_k141_93580 | Not-determined |
| 18-101_k141_158503 | Not-determined |
| 18-101_k141_291019 | Not-determined |
| 18-101_k141_43603 | Low-quality |
| 18-101_k141_174599 | Low-quality |
| 18-101_k141_158507 | Not-determined |
| 18-101_k141_75968 | Not-determined |
| 18-101_k141_241826 | Not-determined |
| 18-101_k141_158515 | Not-determined |
| 18-101_k141_274044 | Not-determined |
| 18-101_k141_158518 | Not-determined |
| 18-101_k141_291028 | Low-quality |
| 18-101_k141_339440 | Not-determined |
| 18-101_k141_308051 | Not-determined |
| 18-101_k141_274045 | Not-determined |
| 18-101_k141_127218 | Not-determined |
| 18-101_k141_274050 | Not-determined |
| 18-101_k141_127225 | Not-determined |
| 18-101_k141_357199 | Not-determined |
| 18-101_k141_225038 | Not-determined |
| 18-101_k141_174605 | Not-determined |
| 18-101_k141_127229 | Not-determined |
| 18-101_k141_274057 | Not-determined |
| 18-101_k141_308054 | Not-determined |
| 18-101_k141_389410 | Low-quality |
| 18-101_k141_291047 | Low-quality |
| 18-101_k141_174610 | Not-determined |
| 18-101_k141_308059 | Not-determined |
| 18-101_k141_308060 | Not-determined |
| 18-101_k141_308061 | Not-determined |
| 18-101_k141_93600 | Not-determined |
| 18-101_k141_225048 | Not-determined |
| 18-101_k141_174613 | Not-determined |
| 18-101_k141_308063 | Not-determined |
| 18-101_k141_274062 | Not-determined |
| 18-101_k141_192759 | Not-determined |
| 18-101_k141_174617 | Not-determined |
| 18-101_k141_174618 | Not-determined |
| 18-101_k141_291056 | Not-determined |
| 18-101_k141_225054 | Not-determined |
| 18-101_k141_258977 | Not-determined |
| 18-101_k141_291063 | Not-determined |
| 18-101_k141_192768 | Not-determined |
| 18-101_k141_158541 | Low-quality |
| 18-101_k141_241853 | Not-determined |
| 18-101_k141_192769 | Not-determined |
| 18-101_k141_308072 | Not-determined |
| 18-101_k141_127256 | Not-determined |
| 18-101_k141_192772 | Not-determined |
| 18-101_k141_339457 | Not-determined |
| 18-101_k141_108901 | Not-determined |
| 18-101_k141_258996 | Not-determined |
| 18-101_k141_60074 | Low-quality |
| 18-101_k141_225073 | Not-determined |
| 18-101_k141_158545 | Not-determined |
| 18-101_k141_241860 | Low-quality |
| 18-101_k141_241865 | Not-determined |
| 18-101_k141_174631 | Not-determined |
| 18-101_k141_274084 | Not-determined |
| 18-101_k141_93630 | Not-determined |
| 18-101_k141_324862 | Not-determined |
| 18-101_k141_127262 | Not-determined |
| 18-101_k141_291077 | Not-determined |
| 18-101_k141_259007 | Not-determined |
| 18-101_k141_274092 | Not-determined |
| 18-101_k141_174636 | Low-quality |
| 18-101_k141_373166 | Not-determined |
| 18-101_k141_241873 | Low-quality |
| 18-101_k141_76019 | Not-determined |
| 18-101_k141_11137 | Not-determined |
| 18-101_k141_11139 | Not-determined |
| 18-101_k141_225081 | Not-determined |
| 18-101_k141_308092 | Not-determined |
| 18-101_k141_60093 | Not-determined |
| 18-101_k141_60094 | Not-determined |
| 18-101_k141_389453 | Not-determined |
| 18-101_k141_60098 | Not-determined |
| 18-101_k141_225093 | Not-determined |
| 18-101_k141_108933 | Not-determined |
| 18-101_k141_192787 | Not-determined |
| 18-101_k141_192792 | Not-determined |
| 18-101_k141_60105 | Not-determined |
| 18-101_k141_259033 | Not-determined |
| 18-101_k141_324875 | Not-determined |
| 18-101_k141_225117 | Not-determined |
| 18-101_k141_274118 | Low-quality |
| 18-101_k141_43659 | Not-determined |
| 18-101_k141_308120 | Not-determined |
| 18-101_k141_339470 | Not-determined |
| 18-101_k141_241905 | Low-quality |
| 18-101_k141_192811 | Not-determined |
| 18-101_k141_76034 | Not-determined |
| 18-101_k141_324892 | Not-determined |
| 18-101_k141_324893 | Not-determined |
| 18-101_k141_11171 | Not-determined |
| 18-101_k141_339477 | Low-quality |
| 18-101_k141_241915 | Not-determined |
| 18-101_k141_225133 | Low-quality |
| 18-101_k141_174651 | Not-determined |
| 18-101_k141_357284 | Not-determined |
| 18-101_k141_225138 | Not-determined |
| 18-101_k141_324900 | Not-determined |
| 18-101_k141_127297 | Not-determined |
| 18-101_k141_291107 | Not-determined |
| 18-101_k141_291108 | Not-determined |
| 18-101_k141_259056 | Not-determined |
| 18-101_k141_259057 | Not-determined |
| 18-101_k141_274140 | Not-determined |
| 18-101_k141_324907 | Not-determined |
| 18-101_k141_108947 | Low-quality |
| 18-101_k141_324909 | Low-quality |
| 18-101_k141_291114 | Not-determined |
| 18-101_k141_339491 | Not-determined |
| 18-101_k141_274144 | Not-determined |
| 18-101_k141_43679 | Not-determined |
| 18-101_k141_241921 | Not-determined |
| 18-101_k141_43680 | Not-determined |
| 18-101_k141_60135 | Not-determined |
| 18-101_k141_60137 | Not-determined |
| 18-101_k141_158589 | Low-quality |
| 18-101_k141_291124 | Not-determined |
| 18-101_k141_108956 | Not-determined |
| 18-101_k141_389483 | Not-determined |
| 18-101_k141_357303 | Not-determined |
| 18-101_k141_93661 | Not-determined |
| 18-101_k141_60143 | Not-determined |
| 18-101_k141_93663 | Not-determined |
| 18-101_k141_158598 | Not-determined |
| 18-101_k141_43695 | Not-determined |
| 18-101_k141_108967 | Not-determined |
| 18-101_k141_291132 | Not-determined |
| 18-101_k141_174680 | Not-determined |
| 18-101_k141_174681 | Not-determined |
| 18-101_k141_43699 | Low-quality |
| 18-101_k141_291136 | Not-determined |
| 18-101_k141_259085 | Not-determined |
| 18-101_k141_43703 | Not-determined |
| 18-101_k141_259086 | Not-determined |
| 18-101_k141_225170 | Not-determined |
| 18-101_k141_43705 | Not-determined |
| 18-101_k141_76056 | Not-determined |
| 18-101_k141_241942 | Not-determined |
| 18-101_k141_60155 | Low-quality |
| 18-101_k141_357315 | Low-quality |
| 18-101_k141_93668 | Not-determined |
| 18-101_k141_93669 | Not-determined |
| 18-101_k141_93670 | Not-determined |
| 18-101_k141_339502 | Not-determined |
| 18-101_k141_241947 | Not-determined |
| 18-101_k141_158611 | Not-determined |
| 18-101_k141_158620 | Not-determined |
| 18-101_k141_339512 | Not-determined |
| 18-101_k141_373170 | Not-determined |
| 18-101_k141_43719 | Not-determined |
| 18-101_k141_142859 | Low-quality |
| 18-101_k141_76079 | Not-determined |
| 18-101_k141_373174 | Not-determined |
| 18-101_k141_11198 | Not-determined |
| 18-101_k141_373176 | Not-determined |
| 18-101_k141_76084 | Not-determined |
| 18-101_k141_127314 | Not-determined |
| 18-101_k141_158627 | Not-determined |
| 18-101_k141_27293 | Not-determined |
| 18-101_k141_274158 | Not-determined |
| 18-101_k141_389486 | Low-quality |
| 18-101_k141_274165 | Not-determined |
| 18-101_k141_60193 | Not-determined |
| 18-101_k141_127325 | Not-determined |
| 18-101_k141_259131 | Not-determined |
| 18-101_k141_274174 | Not-determined |
| 18-101_k141_60198 | Not-determined |
| 18-101_k141_174719 | Low-quality |
| 18-101_k141_108987 | Not-determined |
| 18-101_k141_291170 | Not-determined |
| 18-101_k141_158637 | Low-quality |
| 18-101_k141_192878 | Not-determined |
| 18-101_k141_192879 | Not-determined |
| 18-101_k141_108994 | Not-determined |
| 18-101_k141_43743 | Not-determined |
| 18-101_k141_225183 | Not-determined |
| 18-101_k141_108998 | Not-determined |
| 18-101_k141_241994 | Not-determined |
| 18-101_k141_60208 | Not-determined |
| 18-101_k141_225190 | Not-determined |
| 18-101_k141_174732 | Low-quality |
| 18-101_k141_27310 | Low-quality |
| 18-101_k141_43749 | Not-determined |
| 18-101_k141_389499 | Not-determined |
| 18-101_k141_142885 | Not-determined |
| 18-101_k141_274191 | Not-determined |
| 18-101_k141_142888 | Not-determined |
| 18-101_k141_27312 | Not-determined |
| 18-101_k141_225195 | Not-determined |
| 18-101_k141_174738 | Not-determined |
| 18-101_k141_225196 | Not-determined |
| 18-101_k141_142891 | Not-determined |
| 18-101_k141_11228 | Not-determined |
| 18-101_k141_27314 | Not-determined |
| 18-101_k141_11229 | Not-determined |
| 18-101_k141_192883 | Not-determined |
| 18-101_k141_93714 | Low-quality |
| 18-101_k141_127347 | Not-determined |
| 18-101_k141_76118 | Not-determined |
| 18-101_k141_291190 | Not-determined |
| 18-101_k141_274203 | Not-determined |
| 18-101_k141_209530 | Low-quality |
| 18-101_k141_324970 | Not-determined |
| 18-101_k141_158650 | Not-determined |
| 18-101_k141_209533 | Low-quality |
| 18-101_k141_274209 | Not-determined |
| 18-101_k141_192898 | Not-determined |
| 18-101_k141_142907 | Not-determined |
| 18-101_k141_389518 | Not-determined |
| 18-101_k141_389519 | Not-determined |
| 18-101_k141_324981 | Not-determined |
| 18-101_k141_357341 | Low-quality |
| 18-101_k141_389521 | Not-determined |
| 18-101_k141_158658 | Not-determined |
| 18-101_k141_389522 | Not-determined |
| 18-101_k141_389523 | Not-determined |
| 18-101_k141_60223 | Not-determined |
| 18-101_k141_11246 | Not-determined |
| 18-101_k141_174759 | Low-quality |
| 18-101_k141_389527 | Not-determined |
| 18-101_k141_174760 | Low-quality |
| 18-101_k141_389528 | Not-determined |
| 18-101_k141_357346 | Not-determined |
| 18-101_k141_209545 | Not-determined |
| 18-101_k141_93733 | Not-determined |
| 18-101_k141_127370 | Not-determined |
| 18-101_k141_27329 | Not-determined |
| 18-101_k141_93738 | Not-determined |
| 18-101_k141_142924 | Not-determined |
| 18-101_k141_357358 | Not-determined |
| 18-101_k141_158667 | Not-determined |
| 18-101_k141_43776 | Not-determined |
| 18-101_k141_142930 | Not-determined |
| 18-101_k141_43781 | Not-determined |
| 18-101_k141_259144 | Not-determined |
| 18-101_k141_158671 | Not-determined |
| 18-101_k141_76149 | Not-determined |
| 18-101_k141_43784 | Not-determined |
| 18-101_k141_357365 | Not-determined |
| 18-101_k141_158674 | Not-determined |
| 18-101_k141_192927 | Not-determined |
| 18-101_k141_43786 | Not-determined |
| 18-101_k141_389548 | Not-determined |
| 18-101_k141_357372 | Not-determined |
| 18-101_k141_43791 | Not-determined |
| 18-101_k141_27338 | Not-determined |
| 18-101_k141_291223 | Not-determined |
| 18-101_k141_158677 | Not-determined |
| 18-101_k141_339539 | Not-determined |
| 18-101_k141_60253 | Low-quality |
| 18-101_k141_339540 | Not-determined |
| 18-101_k141_209565 | Low-quality |
| 18-101_k141_339545 | Low-quality |
| 18-101_k141_339548 | Not-determined |
| 18-101_k141_225243 | Not-determined |
| 18-101_k141_158679 | Low-quality |
| 18-101_k141_339552 | Not-determined |
| 18-101_k141_27345 | Not-determined |
| 18-101_k141_339556 | Not-determined |
| 18-101_k141_357388 | Not-determined |
| 18-101_k141_27348 | Not-determined |
| 18-101_k141_142948 | Low-quality |
| 18-101_k141_259163 | Not-determined |
| 18-101_k141_43816 | Not-determined |
| 18-101_k141_357397 | Not-determined |
| 18-101_k141_357403 | Not-determined |
| 18-101_k141_127402 | Low-quality |
| 18-101_k141_225256 | Not-determined |
| 18-101_k141_259173 | Not-determined |
| 18-101_k141_274245 | Not-determined |
| 18-101_k141_339558 | Not-determined |
| 18-101_k141_174823 | Low-quality |
| 18-101_k141_389576 | Not-determined |
| 18-101_k141_373217 | Not-determined |
| 18-101_k141_109038 | Low-quality |
| 18-101_k141_308192 | Not-determined |
| 18-101_k141_209596 | Not-determined |
| 18-101_k141_60261 | Not-determined |
| 18-101_k141_60262 | Not-determined |
| 18-101_k141_291265 | Not-determined |
| 18-101_k141_27371 | Not-determined |
| 18-101_k141_389584 | Not-determined |
| 18-101_k141_142968 | Not-determined |
| 18-101_k141_291267 | Not-determined |
| 18-101_k141_127420 | Low-quality |
| 18-101_k141_174839 | Low-quality |
| 18-101_k141_76186 | Low-quality |
| 18-101_k141_274252 | Not-determined |
| 18-101_k141_209606 | Not-determined |
| 18-101_k141_43844 | Not-determined |
| 18-101_k141_109044 | Low-quality |
| 18-101_k141_93780 | Low-quality |
| 18-101_k141_93783 | Not-determined |
| 18-101_k141_174844 | Not-determined |
| 18-101_k141_109050 | Not-determined |
| 18-101_k141_357428 | Not-determined |
| 18-101_k141_43849 | Low-quality |
| 18-101_k141_291284 | Not-determined |
| 18-101_k141_357432 | Not-determined |
| 18-101_k141_142973 | Not-determined |
| 18-101_k141_43850 | Not-determined |
| 18-101_k141_242052 | Not-determined |
| 18-101_k141_93796 | Not-determined |
| 18-101_k141_109053 | Not-determined |
| 18-101_k141_291290 | Not-determined |
| 18-101_k141_43857 | Not-determined |
| 18-101_k141_209624 | Not-determined |
| 18-101_k141_43858 | Not-determined |
| 18-101_k141_209625 | Not-determined |
| 18-101_k141_127430 | Not-determined |
| 18-101_k141_76206 | Low-quality |
| 18-101_k141_76210 | Not-determined |
| 18-101_k141_93802 | Not-determined |
| 18-101_k141_274274 | Not-determined |
| 18-101_k141_274276 | Not-determined |
| 18-101_k141_357447 | Not-determined |
| 18-101_k141_60302 | Not-determined |
| 18-101_k141_209633 | Not-determined |
| 18-101_k141_76213 | Not-determined |
| 18-101_k141_209634 | Not-determined |
| 18-101_k141_76217 | Not-determined |
| 18-101_k141_76218 | Not-determined |
| 18-101_k141_192968 | Not-determined |
| 18-101_k141_11277 | Not-determined |
| 18-101_k141_274291 | Not-determined |
| 18-101_k141_11281 | Not-determined |
| 18-101_k141_225291 | Not-determined |
| 18-101_k141_93814 | Not-determined |
| 18-101_k141_60312 | Not-determined |
| 18-101_k141_60314 | Not-determined |
| 18-101_k141_357464 | Not-determined |
| 18-101_k141_209643 | Not-determined |
| 18-101_k141_225299 | Not-determined |
| 18-101_k141_127456 | Not-determined |
| 18-101_k141_225300 | Not-determined |
| 18-101_k141_225301 | Not-determined |
| 18-101_k141_11292 | Not-determined |
| 18-101_k141_242082 | Not-determined |
| 18-101_k141_93820 | Not-determined |
| 18-101_k141_291300 | Not-determined |
| 18-101_k141_109075 | Not-determined |
| 18-101_k141_209648 | Not-determined |
| 18-101_k141_11296 | Not-determined |
| 18-101_k141_389625 | Not-determined |
| 18-101_k141_225303 | Not-determined |
| 18-101_k141_291303 | Not-determined |
| 18-101_k141_174854 | Not-determined |
| 18-101_k141_109081 | Not-determined |
| 18-101_k141_158711 | Not-determined |
| 18-101_k141_27415 | Not-determined |
| 18-101_k141_76234 | Not-determined |
| 18-101_k141_274302 | Not-determined |
| 18-101_k141_209658 | Not-determined |
| 18-101_k141_389638 | Not-determined |
| 18-101_k141_109089 | Not-determined |
| 18-101_k141_174866 | Not-determined |
| 18-101_k141_127466 | Not-determined |
| 18-101_k141_109090 | Not-determined |
| 18-101_k141_109093 | Not-determined |
| 18-101_k141_127470 | Not-determined |
| 18-101_k141_225309 | Low-quality |
| 18-101_k141_193001 | Not-determined |
| 18-101_k141_308199 | Not-determined |
| 18-101_k141_109095 | Not-determined |
| 18-101_k141_308203 | Not-determined |
| 18-101_k141_143003 | Not-determined |
| 18-101_k141_274314 | Not-determined |
| 18-101_k141_259199 | Not-determined |
| 18-101_k141_225317 | Not-determined |
| 18-101_k141_60337 | Not-determined |
| 18-101_k141_308212 | Not-determined |
| 18-101_k141_158728 | Not-determined |
| 18-101_k141_274317 | Not-determined |
| 18-101_k141_158730 | Not-determined |
| 18-101_k141_127484 | Not-determined |
| 18-101_k141_27432 | Not-determined |
| 18-101_k141_242088 | Not-determined |
| 18-101_k141_43893 | Low-quality |
| 18-101_k141_242089 | Not-determined |
| 18-101_k141_325010 | Low-quality |
| 18-101_k141_325011 | Low-quality |
| 18-101_k141_259204 | Not-determined |
| 18-101_k141_93848 | Not-determined |
| 18-101_k141_60353 | Not-determined |
| 18-101_k141_308217 | Not-determined |
| 18-101_k141_308223 | Low-quality |
| 18-101_k141_27445 | Low-quality |
| 18-101_k141_209682 | Not-determined |
| 18-101_k141_109123 | Not-determined |
| 18-101_k141_274333 | Not-determined |
| 18-101_k141_225340 | Not-determined |
| 18-101_k141_339597 | Not-determined |
| 18-101_k141_242106 | Not-determined |
| 18-101_k141_209688 | Low-quality |
| 18-101_k141_242112 | Low-quality |
| 18-101_k141_76245 | Not-determined |
| 18-101_k141_76249 | Low-quality |
| 18-101_k141_308241 | Not-determined |
| 18-101_k141_259217 | Not-determined |
| 18-101_k141_76253 | Not-determined |
| 18-101_k141_291349 | Low-quality |
| 18-101_k141_339606 | Not-determined |
| 18-101_k141_158756 | Not-determined |
| 18-101_k141_158757 | Not-determined |
| 18-101_k141_339613 | Not-determined |
| 18-101_k141_76266 | Not-determined |
| 18-101_k141_93862 | Not-determined |
| 18-101_k141_11327 | Not-determined |
| 18-101_k141_291354 | Not-determined |
| 18-101_k141_325027 | Not-determined |
| 18-101_k141_339618 | Not-determined |
| 18-101_k141_11331 | Not-determined |
| 18-101_k141_225375 | Low-quality |
| 18-101_k141_93881 | Not-determined |
| 18-101_k141_193029 | Not-determined |
| 18-101_k141_193030 | Not-determined |
| 18-101_k141_373268 | Not-determined |
| 18-101_k141_339630 | Not-determined |
| 18-101_k141_193036 | Not-determined |
| 18-101_k141_76274 | Not-determined |
| 18-101_k141_143034 | Not-determined |
| 18-101_k141_193041 | Not-determined |
| 18-101_k141_291363 | Not-determined |
| 18-101_k141_209699 | Not-determined |
| 18-101_k141_60388 | Not-determined |
| 18-101_k141_325044 | Not-determined |
| 18-101_k141_373285 | Not-determined |
| 18-101_k141_325045 | Not-determined |
| 18-101_k141_109140 | Not-determined |
| 18-101_k141_93899 | Not-determined |
| 18-101_k141_373286 | Not-determined |
| 18-101_k141_308248 | Not-determined |
| 18-101_k141_143046 | Not-determined |
| 18-101_k141_143052 | Not-determined |
| 18-101_k141_225418 | Not-determined |
| 18-101_k141_357537 | Not-determined |
| 18-101_k141_143055 | Not-determined |
| 18-101_k141_60405 | Not-determined |
| 18-101_k141_373298 | Not-determined |
| 18-101_k141_127552 | Not-determined |
| 18-101_k141_43932 | Not-determined |
| 18-101_k141_109149 | Not-determined |
| 18-101_k141_193079 | Not-determined |
| 18-101_k141_109151 | Not-determined |
| 18-101_k141_325055 | Not-determined |
| 18-101_k141_43934 | Not-determined |
| 18-101_k141_174923 | Not-determined |
| 18-101_k141_225430 | Not-determined |
| 18-101_k141_158813 | Low-quality |
| 18-101_k141_27489 | Not-determined |
| 18-101_k141_373307 | Not-determined |
| 18-101_k141_291391 | Not-determined |
| 18-101_k141_174930 | Not-determined |
| 18-101_k141_76291 | Not-determined |
| 18-101_k141_109164 | Low-quality |
| 18-101_k141_158824 | Not-determined |
| 18-101_k141_242135 | Not-determined |
| 18-101_k141_109170 | Not-determined |
| 18-101_k141_109171 | Not-determined |
| 18-101_k141_109173 | Not-determined |
| 18-101_k141_93935 | Not-determined |
| 18-101_k141_225449 | Low-quality |
| 18-101_k141_174939 | Not-determined |
| 18-101_k141_339637 | Not-determined |
| 18-101_k141_143079 | Not-determined |
| 18-101_k141_11367 | Not-determined |
| 18-101_k141_389677 | Low-quality |
| 18-101_k141_193102 | Not-determined |
| 18-101_k141_308284 | Not-determined |
| 18-101_k141_193103 | Not-determined |
| 18-101_k141_325079 | Not-determined |
| 18-101_k141_339644 | Not-determined |
| 18-101_k141_291413 | Not-determined |
| 18-101_k141_308287 | Not-determined |
| 18-101_k141_76302 | Not-determined |
| 18-101_k141_357544 | Not-determined |
| 18-101_k141_357545 | Not-determined |
| 18-101_k141_308290 | Not-determined |
| 18-101_k141_193111 | Not-determined |
| 18-101_k141_11373 | Not-determined |
| 18-101_k141_308292 | Not-determined |
| 18-101_k141_259237 | Not-determined |
| 18-101_k141_259239 | Not-determined |
| 18-101_k141_308295 | Not-determined |
| 18-101_k141_11378 | Not-determined |
| 18-101_k141_357553 | Not-determined |
| 18-101_k141_259251 | Not-determined |
| 18-101_k141_291415 | Not-determined |
| 18-101_k141_158848 | Not-determined |
| 18-101_k141_357557 | Not-determined |
| 18-101_k141_76305 | Not-determined |
| 18-101_k141_291421 | Not-determined |
| 18-101_k141_209726 | Not-determined |
| 18-101_k141_27497 | Not-determined |
| 18-101_k141_291423 | Not-determined |
| 18-101_k141_373322 | Not-determined |
| 18-101_k141_357563 | Not-determined |
| 18-101_k141_174958 | Not-determined |
| 18-101_k141_158856 | Not-determined |
| 18-101_k141_76308 | Not-determined |
| 18-101_k141_339659 | Not-determined |
| 18-101_k141_291429 | Not-determined |
| 18-101_k141_76311 | Not-determined |
| 18-101_k141_274374 | Not-determined |
| 18-101_k141_357570 | Not-determined |
| 18-101_k141_291438 | Not-determined |
| 18-101_k141_242144 | Not-determined |
| 18-101_k141_373330 | Not-determined |
| 18-101_k141_339670 | Low-quality |
| 18-101_k141_357577 | Not-determined |
| 18-101_k141_76321 | Not-determined |
| 18-101_k141_325103 | Low-quality |
| 18-101_k141_143107 | Not-determined |
| 18-101_k141_339677 | Not-determined |
| 18-101_k141_259270 | Not-determined |
| 18-101_k141_325109 | Not-determined |
| 18-101_k141_242149 | Not-determined |
| 18-101_k141_357580 | Not-determined |
| 18-101_k141_174969 | Not-determined |
| 18-101_k141_291453 | Not-determined |
| 18-101_k141_389690 | Not-determined |
| 18-101_k141_158885 | Not-determined |
| 18-101_k141_291460 | Not-determined |
| 18-101_k141_193136 | Not-determined |
| 18-101_k141_274383 | Not-determined |
| 18-101_k141_193140 | Not-determined |
| 18-101_k141_60431 | Not-determined |
| 18-101_k141_274384 | Not-determined |

| | |
|---|---|
| 18-101_k141_373338 | Low-quality |
| 18-101_k141_27517 | Not-determined |
| 18-101_k141_158891 | Not-determined |
| 18-101_k141_308324 | Not-determined |
| 18-101_k141_109179 | Not-determined |
| 18-101_k141_109182 | Not-determined |
| 18-101_k141_291467 | Not-determined |
| 18-101_k141_158895 | Low-quality |
| 18-101_k141_308331 | Not-determined |
| 18-101_k141_109187 | Not-determined |
| 18-101_k141_291471 | Not-determined |
| 18-101_k141_274400 | Not-determined |
| 18-101_k141_389708 | Not-determined |
| 18-101_k141_225487 | Not-determined |
| 18-101_k141_27524 | Not-determined |
| 18-101_k141_11389 | Not-determined |
| 18-101_k141_274402 | Not-determined |
| 18-101_k141_339705 | Not-determined |
| 18-101_k141_308340 | Not-determined |
| 18-101_k141_127574 | Not-determined |
| 18-101_k141_11394 | Not-determined |
| 18-101_k141_389711 | Not-determined |
| 18-101_k141_174998 | Low-quality |
| 18-101_k141_242164 | Not-determined |
| 18-101_k141_175000 | Not-determined |
| 18-101_k141_27534 | Not-determined |
| 18-101_k141_60455 | Not-determined |
| 18-101_k141_291492 | Not-determined |
| 18-101_k141_325141 | Not-determined |
| 18-101_k141_308363 | Low-quality |
| 18-101_k141_127581 | Not-determined |
| 18-101_k141_11400 | Not-determined |
| 18-101_k141_373356 | Not-determined |
| 18-101_k141_143130 | Not-determined |
| 18-101_k141_274413 | Not-determined |
| 18-101_k141_308365 | Low-quality |
| 18-101_k141_76350 | Not-determined |
| 18-101_k141_339715 | Not-determined |
| 18-101_k141_291494 | Not-determined |
| 18-101_k141_291496 | Not-determined |
| 18-101_k141_308369 | Not-determined |
| 18-101_k141_27544 | Not-determined |
| 18-101_k141_27546 | Not-determined |
| 18-101_k141_325158 | Not-determined |
| 18-101_k141_325161 | Not-determined |
| 18-101_k141_175006 | Not-determined |
| 18-101_k141_339721 | Not-determined |
| 18-101_k141_209785 | Not-determined |
| 18-101_k141_339725 | Not-determined |
| 18-101_k141_193181 | Not-determined |
| 18-101_k141_193182 | Not-determined |
| 18-101_k141_308384 | Not-determined |
| 18-101_k141_193186 | Not-determined |
| 18-101_k141_193187 | Not-determined |
| 18-101_k141_389739 | Not-determined |
| 18-101_k141_175021 | Not-determined |
| 18-101_k141_259291 | Low-quality |
| 18-101_k141_274429 | Not-determined |
| 18-101_k141_27570 | Not-determined |
| 18-101_k141_193195 | Not-determined |
| 18-101_k141_27571 | Not-determined |
| 18-101_k141_389751 | Not-determined |
| 18-101_k141_60465 | Not-determined |
| 18-101_k141_94008 | Not-determined |
| 18-101_k141_109224 | Not-determined |
| 18-101_k141_27577 | Low-quality |
| 18-101_k141_94023 | Not-determined |
| 18-101_k141_225500 | Not-determined |
| 18-101_k141_175040 | Not-determined |
| 18-101_k141_242204 | Not-determined |
| 18-101_k141_27589 | Not-determined |
| 18-101_k141_373390 | Not-determined |
| 18-101_k141_259302 | Not-determined |
| 18-101_k141_339735 | Not-determined |
| 18-101_k141_94036 | Not-determined |
| 18-101_k141_242209 | Not-determined |
| 18-101_k141_308409 | Not-determined |
| 18-101_k141_357658 | Not-determined |
| 18-101_k141_291537 | Not-determined |
| 18-101_k141_193212 | Low-quality |
| 18-101_k141_158917 | Not-determined |
| 18-101_k141_291539 | Not-determined |
| 18-101_k141_291543 | Not-determined |
| 18-101_k141_109253 | Not-determined |
| 18-101_k141_109254 | Not-determined |
| 18-101_k141_193228 | Not-determined |
| 18-101_k141_274445 | Not-determined |
| 18-101_k141_325202 | Not-determined |
| 18-101_k141_259319 | Not-determined |
| 18-101_k141_259321 | Not-determined |
| 18-101_k141_193236 | Low-quality |
| 18-101_k141_127643 | Not-determined |
| 18-101_k141_274458 | Not-determined |
| 18-101_k141_127645 | Not-determined |
| 18-101_k141_127646 | Not-determined |
| 18-101_k141_242223 | Not-determined |
| 18-101_k141_242224 | Not-determined |
| 18-101_k141_94071 | Not-determined |
| 18-101_k141_94072 | Not-determined |
| 18-101_k141_357669 | Not-determined |
| 18-101_k141_209827 | Not-determined |
| 18-101_k141_357673 | Not-determined |
| 18-101_k141_357674 | Not-determined |
| 18-101_k141_193252 | Not-determined |
| 18-101_k141_357679 | Not-determined |
| 18-101_k141_357682 | Not-determined |
| 18-101_k141_109269 | Not-determined |
| 18-101_k141_339764 | Not-determined |
| 18-101_k141_60507 | Low-quality |
| 18-101_k141_357689 | Not-determined |
| 18-101_k141_60510 | Not-determined |
| 18-101_k141_209845 | Not-determined |
| 18-101_k141_357691 | Not-determined |
| 18-101_k141_94081 | Not-determined |
| 18-101_k141_143225 | Not-determined |
| 18-101_k141_94082 | Not-determined |
| 18-101_k141_60517 | Not-determined |
| 18-101_k141_143226 | Not-determined |
| 18-101_k141_291580 | Not-determined |
| 18-101_k141_94087 | Not-determined |
| 18-101_k141_175081 | Not-determined |
| 18-101_k141_175083 | Not-determined |
| 18-101_k141_109280 | Not-determined |
| 18-101_k141_60523 | Not-determined |
| 18-101_k141_259351 | Not-determined |
| 18-101_k141_175085 | Not-determined |
| 18-101_k141_325228 | Not-determined |
| 18-101_k141_11442 | Not-determined |
| 18-101_k141_11445 | Not-determined |
| 18-101_k141_389778 | Not-determined |
| 18-101_k141_291586 | Not-determined |
| 18-101_k141_44010 | Not-determined |
| 18-101_k141_325238 | Not-determined |
| 18-101_k141_60536 | Not-determined |
| 18-101_k141_76369 | Low-quality |
| 18-101_k141_259366 | Low-quality |
| 18-101_k141_143238 | Not-determined |
| 18-101_k141_60541 | Not-determined |
| 18-101_k141_94102 | Not-determined |
| 18-101_k141_193271 | Not-determined |
| 18-101_k141_127674 | Not-determined |
| 18-101_k141_259370 | Not-determined |
| 18-101_k141_143242 | Not-determined |
| 18-101_k141_94106 | Not-determined |
| 18-101_k141_44021 | Not-determined |
| 18-101_k141_291600 | Not-determined |
| 18-101_k141_209865 | Not-determined |
| 18-101_k141_11461 | Low-quality |
| 18-101_k141_209866 | Not-determined |
| 18-101_k141_109287 | Not-determined |
| 18-101_k141_274485 | Not-determined |
| 18-101_k141_109289 | Not-determined |
| 18-101_k141_325253 | Not-determined |
| 18-101_k141_175104 | Not-determined |
| 18-101_k141_109291 | Not-determined |
| 18-101_k141_373448 | Not-determined |
| 18-101_k141_389787 | Not-determined |
| 18-101_k141_259382 | Not-determined |
| 18-101_k141_44029 | Not-determined |
| 18-101_k141_325257 | Not-determined |
| 18-101_k141_175109 | Not-determined |
| 18-101_k141_175112 | Not-determined |
| 18-101_k141_94123 | Not-determined |
| 18-101_k141_76382 | Low-quality |
| 18-101_k141_325264 | Not-determined |
| 18-101_k141_158951 | Not-determined |
| 18-101_k141_274492 | Not-determined |
| 18-101_k141_274496 | Not-determined |
| 18-101_k141_109303 | Not-determined |
| 18-101_k141_291616 | Not-determined |
| 18-101_k141_60555 | Not-determined |
| 18-101_k141_175122 | Not-determined |
| 18-101_k141_209883 | Not-determined |
| 18-101_k141_325278 | Not-determined |
| 18-101_k141_325283 | Low-quality |
| 18-101_k141_339773 | Not-determined |
| 18-101_k141_109307 | Not-determined |
| 18-101_k141_291631 | Not-determined |
| 18-101_k141_60572 | Not-determined |
| 18-101_k141_259396 | Not-determined |
| 18-101_k141_143258 | Not-determined |
| 18-101_k141_158975 | Not-determined |
| 18-101_k141_127688 | Not-determined |
| 18-101_k141_109312 | Not-determined |
| 18-101_k141_109313 | Not-determined |
| 18-101_k141_143262 | Low-quality |
| 18-101_k141_193304 | Not-determined |
| 18-101_k141_308451 | Not-determined |
| 18-101_k141_225562 | Not-determined |
| 18-101_k141_158977 | Not-determined |
| 18-101_k141_44066 | Low-quality |
| 18-101_k141_225568 | Not-determined |
| 18-101_k141_109321 | Not-determined |
| 18-101_k141_274499 | Not-determined |
| 18-101_k141_158981 | Not-determined |
| 18-101_k141_389790 | Not-determined |
| 18-101_k141_11491 | Not-determined |
| 18-101_k141_60578 | Not-determined |
| 18-101_k141_76394 | Not-determined |
| 18-101_k141_339788 | Not-determined |
| 18-101_k141_76399 | Not-determined |
| 18-101_k141_357738 | Not-determined |
| 18-101_k141_44081 | Not-determined |
| 18-101_k141_60593 | Not-determined |
| 18-101_k141_94146 | Not-determined |
| 18-101_k141_76412 | Not-determined |
| 18-101_k141_274520 | Not-determined |
| 18-101_k141_44089 | Not-determined |
| 18-101_k141_109335 | Not-determined |
| 18-101_k141_44091 | Low-quality |
| 18-101_k141_76416 | Not-determined |
| 18-101_k141_44097 | Not-determined |
| 18-101_k141_259419 | Not-determined |
| 18-101_k141_259420 | Not-determined |
| 18-101_k141_76421 | Not-determined |
| 18-101_k141_94161 | Low-quality |
| 18-101_k141_76424 | Not-determined |
| 18-101_k141_76426 | Not-determined |
| 18-101_k141_76427 | Not-determined |
| 18-101_k141_357745 | Not-determined |
| 18-101_k141_143303 | Not-determined |
| 18-101_k141_44102 | Not-determined |
| 18-101_k141_158989 | Not-determined |
| 18-101_k141_373463 | Low-quality |
| 18-101_k141_76438 | Not-determined |
| 18-101_k141_76441 | Not-determined |
| 18-101_k141_143312 | Not-determined |
| 18-101_k141_291681 | Low-quality |
| 18-101_k141_259436 | Not-determined |
| 18-101_k141_373473 | Not-determined |
| 18-101_k141_127690 | Low-quality |
| 18-101_k141_76447 | Not-determined |
| 18-101_k141_291690 | Not-determined |
| 18-101_k141_60621 | Not-determined |
| 18-101_k141_225614 | Not-determined |
| 18-101_k141_27642 | Not-determined |
| 18-101_k141_357751 | Not-determined |
| 18-101_k141_308461 | Not-determined |
| 18-101_k141_76453 | Not-determined |
| 18-101_k141_308467 | Not-determined |
| 18-101_k141_159003 | Not-determined |
| 18-101_k141_373480 | Not-determined |
| 18-101_k141_209893 | Not-determined |
| 18-101_k141_143324 | Not-determined |
| 18-101_k141_209895 | Not-determined |
| 18-101_k141_209896 | Not-determined |
| 18-101_k141_259454 | Not-determined |
| 18-101_k141_159006 | Not-determined |
| 18-101_k141_127702 | Not-determined |
| 18-101_k141_389819 | Not-determined |
| 18-101_k141_94184 | Not-determined |
| 18-101_k141_373485 | Not-determined |
| 18-101_k141_159011 | Not-determined |
| 18-101_k141_373487 | Not-determined |
| 18-101_k141_94186 | Not-determined |
| 18-101_k141_127709 | Not-determined |
| 18-101_k141_373494 | Not-determined |
| 18-101_k141_274533 | Not-determined |
| 18-101_k141_339803 | Not-determined |
| 18-101_k141_76464 | Not-determined |
| 18-101_k141_339804 | Not-determined |
| 18-101_k141_339805 | Not-determined |
| 18-101_k141_357770 | Not-determined |
| 18-101_k141_339807 | Not-determined |
| 18-101_k141_27659 | Not-determined |
| 18-101_k141_44125 | Not-determined |
| 18-101_k141_127712 | Not-determined |
| 18-101_k141_357771 | Not-determined |
| 18-101_k141_373501 | Not-determined |
| 18-101_k141_209910 | Not-determined |
| 18-101_k141_127718 | Low-quality |
| 18-101_k141_291706 | Low-quality |
| 18-101_k141_94197 | Low-quality |
| 18-101_k141_209913 | Not-determined |
| 18-101_k141_27673 | Not-determined |
| 18-101_k141_373509 | Not-determined |
| 18-101_k141_339808 | Not-determined |
| 18-101_k141_175165 | Not-determined |
| 18-101_k141_44135 | Not-determined |
| 18-101_k141_373513 | Not-determined |
| 18-101_k141_94207 | Not-determined |
| 18-101_k141_94208 | Not-determined |
| 18-101_k141_44139 | Not-determined |
| 18-101_k141_339813 | Not-determined |
| 18-101_k141_339814 | Not-determined |
| 18-101_k141_339816 | Not-determined |
| 18-101_k141_159033 | Not-determined |
| 18-101_k141_76471 | Low-quality |
| 18-101_k141_389829 | Not-determined |
| 18-101_k141_193370 | Low-quality |
| 18-101_k141_175176 | Not-determined |
| 18-101_k141_109375 | Not-determined |
| 18-101_k141_373526 | Not-determined |
| 18-101_k141_109376 | Not-determined |
| 18-101_k141_259480 | Not-determined |
| 18-101_k141_259481 | Not-determined |
| 18-101_k141_259484 | Not-determined |
| 18-101_k141_291715 | Not-determined |
| 18-101_k141_44142 | Not-determined |
| 18-101_k141_109383 | Not-determined |
| 18-101_k141_389841 | Not-determined |
| 18-101_k141_291721 | Not-determined |
| 18-101_k141_44145 | Not-determined |
| 18-101_k141_325338 | Not-determined |
| 18-101_k141_389847 | Not-determined |
| 18-101_k141_44148 | Not-determined |
| 18-101_k141_308509 | Not-determined |
| 18-101_k141_175192 | Not-determined |
| 18-101_k141_357794 | Not-determined |
| 18-101_k141_193391 | Not-determined |
| 18-101_k141_291728 | Not-determined |
| 18-101_k141_60658 | Not-determined |
| 18-101_k141_159049 | Not-determined |
| 18-101_k141_291735 | Not-determined |
| 18-101_k141_94223 | Low-quality |
| 18-101_k141_389859 | Not-determined |
| 18-101_k141_274578 | Not-determined |
| 18-101_k141_94232 | Not-determined |
| 18-101_k141_259489 | Not-determined |
| 18-101_k141_159057 | Not-determined |
| 18-101_k141_357807 | Not-determined |
| 18-101_k141_159061 | Low-quality |
| 18-101_k141_259501 | Not-determined |
| 18-101_k141_193412 | Low-quality |
| 18-101_k141_373548 | Not-determined |
| 18-101_k141_27708 | Not-determined |
| 18-101_k141_373557 | Not-determined |
| 18-101_k141_193419 | Low-quality |
| 18-101_k141_109404 | Not-determined |
| 18-101_k141_325374 | Not-determined |
| 18-101_k141_325375 | Not-determined |
| 18-101_k141_389901 | Not-determined |
| 18-101_k141_159079 | Not-determined |
| 18-101_k141_109408 | Not-determined |
| 18-101_k141_193425 | Low-quality |
| 18-101_k141_389902 | Not-determined |
| 18-101_k141_11544 | Not-determined |
| 18-101_k141_389903 | Not-determined |
| 18-101_k141_143362 | Not-determined |
| 18-101_k141_143363 | Not-determined |
| 18-101_k141_60685 | Not-determined |
| 18-101_k141_209963 | Not-determined |
| 18-101_k141_325378 | Low-quality |
| 18-101_k141_389908 | Not-determined |
| 18-101_k141_109416 | Not-determined |
| 18-101_k141_291776 | Not-determined |
| 18-101_k141_308565 | Not-determined |
| 18-101_k141_357842 | Not-determined |
| 18-101_k141_259516 | Not-determined |
| 18-101_k141_175205 | Not-determined |
| 18-101_k141_259518 | Not-determined |
| 18-101_k141_308575 | Not-determined |
| 18-101_k141_389910 | Not-determined |
| 18-101_k141_357849 | Not-determined |
| 18-101_k141_193437 | Not-determined |
| 18-101_k141_209977 | Not-determined |
| 18-101_k141_242274 | Not-determined |
| 18-101_k141_291782 | Not-determined |
| 18-101_k141_143380 | Not-determined |
| 18-101_k141_209980 | Low-quality |
| 18-101_k141_242277 | Not-determined |
| 18-101_k141_94259 | Not-determined |
| 18-101_k141_143381 | Not-determined |
| 18-101_k141_76491 | Not-determined |
| 18-101_k141_325393 | Low-quality |
| 18-101_k141_357850 | Not-determined |
| 18-101_k141_373559 | Low-quality |
| 18-101_k141_11561 | Not-determined |
| 18-101_k141_242286 | Not-determined |
| 18-101_k141_209989 | Not-determined |
| 18-101_k141_11562 | Not-determined |
| 18-101_k141_159100 | Not-determined |
| 18-101_k141_11563 | Not-determined |
| 18-101_k141_159102 | Not-determined |
| 18-101_k141_60702 | Not-determined |
| 18-101_k141_209992 | Not-determined |
| 18-101_k141_60704 | Not-determined |
| 18-101_k141_357857 | Not-determined |
| 18-101_k141_76499 | Not-determined |
| 18-101_k141_159105 | Low-quality |
| 18-101_k141_143386 | Low-quality |
| 18-101_k141_225675 | Not-determined |
| 18-101_k141_357862 | Not-determined |
| 18-101_k141_11572 | Not-determined |
| 18-101_k141_357864 | Not-determined |
| 18-101_k141_357865 | Not-determined |
| 18-101_k141_242304 | Low-quality |
| 18-101_k141_11575 | Not-determined |
| 18-101_k141_308592 | Low-quality |
| 18-101_k141_109438 | Not-determined |
| 18-101_k141_11576 | Low-quality |
| 18-101_k141_225691 | Not-determined |
| 18-101_k141_373573 | Not-determined |
| 18-101_k141_259547 | Not-determined |
| 18-101_k141_11585 | Not-determined |
| 18-101_k141_127727 | Not-determined |
| 18-101_k141_76518 | Not-determined |
| 18-101_k141_76521 | Not-determined |
| 18-101_k141_143407 | Not-determined |
| 18-101_k141_325419 | Not-determined |
| 18-101_k141_11592 | Not-determined |
| 18-101_k141_308603 | Not-determined |
| 18-101_k141_225697 | Low-quality |
| 18-101_k141_291815 | Not-determined |
| 18-101_k141_291816 | Not-determined |
| 18-101_k141_325426 | Not-determined |
| 18-101_k141_210015 | Low-quality |
| 18-101_k141_60726 | Not-determined |
| 18-101_k141_27789 | Not-determined |
| 18-101_k141_193455 | Not-determined |
| 18-101_k141_193456 | Not-determined |
| 18-101_k141_44222 | Not-determined |
| 18-101_k141_27794 | Not-determined |
| 18-101_k141_193459 | Not-determined |
| 18-101_k141_225720 | Not-determined |
| 18-101_k141_357882 | Not-determined |
| 18-101_k141_76537 | Not-determined |
| 18-101_k141_76538 | Not-determined |
| 18-101_k141_94286 | Not-determined |
| 18-101_k141_94288 | Not-determined |
| 18-101_k141_60733 | Low-quality |
| 18-101_k141_76543 | Not-determined |
| 18-101_k141_242351 | Not-determined |
| 18-101_k141_291847 | Not-determined |
| 18-101_k141_143437 | Not-determined |
| 18-101_k141_325442 | Not-determined |
| 18-101_k141_94301 | Not-determined |
| 18-101_k141_308625 | Not-determined |
| 18-101_k141_44241 | Not-determined |
| 18-101_k141_193466 | Not-determined |
| 18-101_k141_175259 | Not-determined |
| 18-101_k141_373617 | Not-determined |
| 18-101_k141_373620 | Not-determined |
| 18-101_k141_389960 | Not-determined |
| 18-101_k141_357891 | Not-determined |
| 18-101_k141_193469 | Not-determined |
| 18-101_k141_210046 | Not-determined |
| 18-101_k141_143448 | Not-determined |
| 18-101_k141_325457 | Not-determined |
| 18-101_k141_210048 | Not-determined |
| 18-101_k141_94307 | Not-determined |
| 18-101_k141_373625 | Not-determined |
| 18-101_k141_193475 | Not-determined |
| 18-101_k141_210052 | Not-determined |
| 18-101_k141_210054 | Not-determined |
| 18-101_k141_175270 | Low-quality |
| 18-101_k141_11632 | Not-determined |
| 18-101_k141_11634 | Not-determined |
| 18-101_k141_60746 | Not-determined |
| 18-101_k141_109464 | Not-determined |
| 18-101_k141_127772 | Not-determined |
| 18-101_k141_389971 | Low-quality |
| 18-101_k141_143457 | Not-determined |
| 18-101_k141_76549 | Not-determined |
| 18-101_k141_274632 | Not-determined |
| 18-101_k141_193487 | Not-determined |
| 18-101_k141_175280 | Not-determined |
| 18-101_k141_389976 | Not-determined |
| 18-101_k141_44260 | Not-determined |
| 18-101_k141_44261 | Not-determined |
| 18-101_k141_109473 | Not-determined |
| 18-101_k141_373645 | Not-determined |
| 18-101_k141_308637 | Not-determined |
| 18-101_k141_193495 | Not-determined |
| 18-101_k141_259605 | Not-determined |
| 18-101_k141_76561 | Not-determined |
| 18-101_k141_60767 | Not-determined |
| 18-101_k141_175292 | Not-determined |
| 18-101_k141_27819 | Not-determined |
| 18-101_k141_259611 | Not-determined |
| 18-101_k141_357926 | Not-determined |
| 18-101_k141_308653 | Not-determined |
| 18-101_k141_242399 | Not-determined |
| 18-101_k141_109488 | Not-determined |
| 18-101_k141_94341 | Not-determined |
| 18-101_k141_389988 | Not-determined |
| 18-101_k141_225750 | Not-determined |
| 18-101_k141_60780 | Not-determined |
| 18-101_k141_109497 | Not-determined |
| 18-101_k141_193502 | Not-determined |
| 18-101_k141_109499 | Not-determined |
| 18-101_k141_325471 | Not-determined |
| 18-101_k141_193504 | Not-determined |
| 18-101_k141_44271 | Not-determined |
| 18-101_k141_76579 | Not-determined |
| 18-101_k141_325476 | Not-determined |
| 18-101_k141_325478 | Not-determined |
| 18-101_k141_127775 | Not-determined |
| 18-101_k141_44278 | Not-determined |
| 18-101_k141_225766 | Not-determined |
| 18-101_k141_274652 | Low-quality |
| 18-101_k141_94350 | Not-determined |
| 18-101_k141_225773 | Not-determined |
| 18-101_k141_274654 | Not-determined |
| 18-101_k141_11660 | Not-determined |
| 18-101_k141_27835 | Not-determined |
| 18-101_k141_27836 | Not-determined |
| 18-101_k141_210077 | Low-quality |
| 18-101_k141_76594 | Not-determined |
| 18-101_k141_76595 | Not-determined |
| 18-101_k141_109515 | Not-determined |
| 18-101_k141_373683 | Not-determined |
| 18-101_k141_242417 | Not-determined |
| 18-101_k141_11670 | Not-determined |
| 18-101_k141_210083 | Not-determined |
| 18-101_k141_27842 | Not-determined |
| 18-101_k141_44295 | Not-determined |
| 18-101_k141_357955 | Not-determined |
| 18-101_k141_225778 | Not-determined |
| 18-101_k141_357957 | Not-determined |
| 18-101_k141_44298 | Not-determined |
| 18-101_k141_11678 | Not-determined |
| 18-101_k141_44299 | Low-quality |
| 18-101_k141_259621 | Not-determined |
| 18-101_k141_357958 | Not-determined |
| 18-101_k141_259622 | Not-determined |
| 18-101_k141_127790 | Not-determined |
| 18-101_k141_109527 | Low-quality |
| 18-101_k141_210097 | Not-determined |
| 18-101_k141_357962 | Not-determined |
| 18-101_k141_339829 | Not-determined |
| 18-101_k141_242431 | Not-determined |
| 18-101_k141_109532 | Not-determined |
| 18-101_k141_109533 | Not-determined |
| 18-101_k141_143460 | Not-determined |
| 18-101_k141_274681 | Not-determined |
| 18-101_k141_242436 | Not-determined |
| 18-101_k141_210103 | Not-determined |
| 18-101_k141_225789 | Not-determined |
| 18-101_k141_109542 | Not-determined |
| 18-101_k141_60824 | Not-determined |
| 18-101_k141_159141 | Low-quality |
| 18-101_k141_27855 | Not-determined |
| 18-101_k141_109545 | Not-determined |
| 18-101_k141_193526 | Not-determined |
| 18-101_k141_27861 | Not-determined |
| 18-101_k141_109551 | Not-determined |
| 18-101_k141_259639 | Low-quality |
| 18-101_k141_390025 | Not-determined |
| 18-101_k141_357979 | Not-determined |
| 18-101_k141_27871 | Not-determined |
| 18-101_k141_127795 | Not-determined |
| 18-101_k141_60841 | Not-determined |
| 18-101_k141_225805 | Not-determined |
| 18-101_k141_210120 | Not-determined |
| 18-101_k141_159154 | Not-determined |
| 18-101_k141_27878 | Not-determined |
| 18-101_k141_60849 | Not-determined |
| 18-101_k141_60850 | Not-determined |
| 18-101_k141_159158 | Not-determined |
| 18-101_k141_27881 | Not-determined |
| 18-101_k141_44320 | Not-determined |
| 18-101_k141_159162 | Not-determined |
| 18-101_k141_291907 | Not-determined |
| 18-101_k141_308662 | Not-determined |
| 18-101_k141_27886 | Not-determined |
| 18-101_k141_159164 | Not-determined |
| 18-101_k141_193542 | Not-determined |
| 18-101_k141_390038 | Not-determined |
| 18-101_k141_308666 | Not-determined |
| 18-101_k141_27891 | Not-determined |
| 18-101_k141_225814 | Low-quality |
| 18-101_k141_308671 | Not-determined |
| 18-101_k141_27898 | Not-determined |
| 18-101_k141_193555 | Not-determined |
| 18-101_k141_175341 | Not-determined |
| 18-101_k141_390043 | Low-quality |
| 18-101_k141_44334 | Not-determined |
| 18-101_k141_390046 | Not-determined |
| 18-101_k141_357998 | Not-determined |
| 18-101_k141_259657 | Not-determined |
| 18-101_k141_225820 | Not-determined |
| 18-101_k141_159169 | Not-determined |
| 18-101_k141_291930 | Not-determined |
| 18-101_k141_291931 | Not-determined |
| 18-101_k141_94390 | Not-determined |
| 18-101_k141_308692 | Not-determined |
| 18-101_k141_308693 | Not-determined |
| 18-101_k141_291936 | Not-determined |
| 18-101_k141_339867 | Not-determined |
| 18-101_k141_308695 | Not-determined |
| 18-101_k141_193567 | Not-determined |
| 18-101_k141_325513 | Not-determined |
| 18-101_k141_274723 | Not-determined |
| 18-101_k141_259665 | Not-determined |
| 18-101_k141_127832 | Not-determined |
| 18-101_k141_94401 | Not-determined |
| 18-101_k141_339884 | Not-determined |
| 18-101_k141_274731 | Not-determined |

| | |
|---|---|
| 18-101_k141_242450 | Not-determined |
| 18-101_k141_44355 | Not-determined |
| 18-101_k141_325524 | Not-determined |
| 18-101_k141_94407 | Not-determined |
| 18-101_k141_143493 | Not-determined |
| 18-101_k141_308706 | Not-determined |
| 18-101_k141_143495 | Not-determined |
| 18-101_k141_210136 | Not-determined |
| 18-101_k141_210138 | Not-determined |
| 18-101_k141_44362 | Not-determined |
| 18-101_k141_143503 | Not-determined |
| 18-101_k141_143505 | Not-determined |
| 18-101_k141_44371 | Not-determined |
| 18-101_k141_11726 | Low-quality |
| 18-101_k141_94420 | Not-determined |
| 18-101_k141_27931 | Not-determined |
| 18-101_k141_11733 | Not-determined |
| 18-101_k141_225836 | Not-determined |
| 18-101_k141_325529 | Not-determined |
| 18-101_k141_242477 | Not-determined |
| 18-101_k141_94435 | Low-quality |
| 18-101_k141_274749 | Not-determined |
| 18-101_k141_325538 | Not-determined |
| 18-101_k141_242483 | Not-determined |
| 18-101_k141_94447 | Not-determined |
| 18-101_k141_94449 | Not-determined |
| 18-101_k141_358028 | Not-determined |
| 18-101_k141_159211 | Not-determined |
| 18-101_k141_339905 | Not-determined |
| 18-101_k141_159212 | Not-determined |
| 18-101_k141_193597 | Not-determined |
| 18-101_k141_373728 | Not-determined |
| 18-101_k141_308730 | Not-determined |
| 18-101_k141_259693 | Not-determined |
| 18-101_k141_210145 | Not-determined |
| 18-101_k141_60866 | Not-determined |
| 18-101_k141_94455 | Not-determined |
| 18-101_k141_60868 | Not-determined |
| 18-101_k141_143533 | Low-quality |
| 18-101_k141_210150 | Low-quality |
| 18-101_k141_210151 | Not-determined |
| 18-101_k141_225862 | Low-quality |
| 18-101_k141_193604 | Not-determined |
| 18-101_k141_11743 | Not-determined |
| 18-101_k141_325556 | Not-determined |
| 18-101_k141_325557 | Not-determined |
| 18-101_k141_60882 | Not-determined |
| 18-101_k141_373748 | Not-determined |
| 18-101_k141_225873 | Not-determined |
| 18-101_k141_373750 | Not-determined |
| 18-101_k141_373752 | Not-determined |
| 18-101_k141_109601 | Not-determined |
| 18-101_k141_373753 | Not-determined |
| 18-101_k141_225875 | Not-determined |
| 18-101_k141_373755 | Not-determined |
| 18-101_k141_339934 | Not-determined |
| 18-101_k141_358048 | Not-determined |
| 18-101_k141_60895 | Not-determined |
| 18-101_k141_143554 | Not-determined |
| 18-101_k141_225883 | Not-determined |
| 18-101_k141_210162 | Low-quality |
| 18-101_k141_358054 | Not-determined |
| 18-101_k141_76610 | Not-determined |
| 18-101_k141_94482 | Low-quality |
| 18-101_k141_109613 | Not-determined |
| 18-101_k141_291981 | Not-determined |
| 18-101_k141_127894 | Not-determined |
| 18-101_k141_27982 | Not-determined |
| 18-101_k141_291987 | Not-determined |
| 18-101_k141_373761 | Not-determined |
| 18-101_k141_325569 | Not-determined |
| 18-101_k141_76616 | Not-determined |
| 18-101_k141_358056 | Not-determined |
| 18-101_k141_127897 | Not-determined |
| 18-101_k141_60909 | Not-determined |
| 18-101_k141_259711 | Not-determined |
| 18-101_k141_60912 | Low-quality |
| 18-101_k141_291996 | Not-determined |
| 18-101_k141_291998 | Not-determined |
| 18-101_k141_11758 | Not-determined |
| 18-101_k141_11763 | Not-determined |
| 18-101_k141_225901 | Not-determined |
| 18-101_k141_109628 | Not-determined |
| 18-101_k141_60916 | Not-determined |
| 18-101_k141_308765 | Not-determined |
| 18-101_k141_76633 | Not-determined |
| 18-101_k141_339955 | Not-determined |
| 18-101_k141_76639 | Not-determined |
| 18-101_k141_76642 | Not-determined |
| 18-101_k141_127921 | Not-determined |
| 18-101_k141_44414 | Not-determined |
| 18-101_k141_127924 | Low-quality |
| 18-101_k141_143576 | Not-determined |
| 18-101_k141_390075 | Not-determined |
| 18-101_k141_76651 | Not-determined |
| 18-101_k141_339969 | Not-determined |
| 18-101_k141_339971 | Not-determined |
| 18-101_k141_44429 | Not-determined |
| 18-101_k141_127930 | Not-determined |
| 18-101_k141_28012 | Not-determined |
| 18-101_k141_274779 | Low-quality |
| 18-101_k141_292020 | Not-determined |
| 18-101_k141_358108 | Not-determined |
| 18-101_k141_308782 | Not-determined |
| 18-101_k141_390088 | Low-quality |
| 18-101_k141_11797 | Not-determined |
| 18-101_k141_358111 | Not-determined |
| 18-101_k141_292028 | Not-determined |
| 18-101_k141_358112 | Not-determined |
| 18-101_k141_339981 | Not-determined |
| 18-101_k141_308795 | Not-determined |
| 18-101_k141_159243 | Not-determined |
| 18-101_k141_60942 | Not-determined |
| 18-101_k141_292034 | Not-determined |
| 18-101_k141_159247 | Not-determined |
| 18-101_k141_60946 | Not-determined |
| 18-101_k141_60949 | Not-determined |
| 18-101_k141_60950 | Not-determined |
| 18-101_k141_94504 | Not-determined |
| 18-101_k141_292046 | Not-determined |
| 18-101_k141_76680 | Not-determined |
| 18-101_k141_193626 | Not-determined |
| 18-101_k141_308802 | Not-determined |
| 18-101_k141_175376 | Not-determined |
| 18-101_k141_308804 | Not-determined |
| 18-101_k141_159264 | Not-determined |
| 18-101_k141_11814 | Not-determined |
| 18-101_k141_159266 | Not-determined |
| 18-101_k141_193642 | Not-determined |
| 18-101_k141_242524 | Not-determined |
| 18-101_k141_274796 | Not-determined |
| 18-101_k141_274799 | Not-determined |
| 18-101_k141_11820 | Not-determined |
| 18-101_k141_11821 | Not-determined |
| 18-101_k141_242526 | Not-determined |
| 18-101_k141_390116 | Not-determined |
| 18-101_k141_358133 | Not-determined |
| 18-101_k141_127971 | Not-determined |
| 18-101_k141_175387 | Not-determined |
| 18-101_k141_225923 | Not-determined |
| 18-101_k141_193649 | Low-quality |
| 18-101_k141_259771 | Not-determined |
| 18-101_k141_325634 | Not-determined |
| 18-101_k141_109671 | Not-determined |
| 18-101_k141_292076 | Not-determined |
| 18-101_k141_143622 | Low-quality |
| 18-101_k141_193658 | Not-determined |
| 18-101_k141_109673 | Not-determined |
| 18-101_k141_127982 | Not-determined |
| 18-101_k141_109674 | Not-determined |
| 18-101_k141_127984 | Not-determined |
| 18-101_k141_210204 | Low-quality |
| 18-101_k141_109677 | Not-determined |
| 18-101_k141_159279 | Not-determined |
| 18-101_k141_28043 | Not-determined |
| 18-101_k141_358139 | Not-determined |
| 18-101_k141_193669 | Not-determined |
| 18-101_k141_28050 | Not-determined |
| 18-101_k141_127991 | Not-determined |
| 18-101_k141_308823 | Not-determined |
| 18-101_k141_127993 | Not-determined |
| 18-101_k141_44459 | Not-determined |
| 18-101_k141_373785 | Low-quality |
| 18-101_k141_109689 | Not-determined |
| 18-101_k141_193675 | Low-quality |
| 18-101_k141_60986 | Low-quality |
| 18-101_k141_274831 | Low-quality |
| 18-101_k141_210214 | Not-determined |
| 18-101_k141_193681 | Not-determined |
| 18-101_k141_390141 | Not-determined |
| 18-101_k141_308829 | Not-determined |
| 18-101_k141_44465 | Not-determined |
| 18-101_k141_28067 | Not-determined |
| 18-101_k141_340034 | Not-determined |
| 18-101_k141_128007 | Not-determined |
| 18-101_k141_358167 | Not-determined |
| 18-101_k141_340038 | Not-determined |
| 18-101_k141_358169 | Not-determined |
| 18-101_k141_358170 | Not-determined |
| 18-101_k141_340042 | Not-determined |
| 18-101_k141_28079 | Not-determined |
| 18-101_k141_159309 | Not-determined |
| 18-101_k141_44487 | Not-determined |
| 18-101_k141_60993 | Not-determined |
| 18-101_k141_60996 | Not-determined |
| 18-101_k141_143660 | Not-determined |
| 18-101_k141_94549 | Not-determined |
| 18-101_k141_94552 | Not-determined |
| 18-101_k141_193707 | Not-determined |
| 18-101_k141_193713 | Not-determined |
| 18-101_k141_61000 | Not-determined |
| 18-101_k141_373808 | Not-determined |
| 18-101_k141_28086 | Not-determined |
| 18-101_k141_259788 | Not-determined |
| 18-101_k141_373813 | Not-determined |
| 18-101_k141_11858 | Not-determined |
| 18-101_k141_259791 | Not-determined |
| 18-101_k141_340068 | Not-determined |
| 18-101_k141_159315 | Low-quality |
| 18-101_k141_76742 | Not-determined |
| 18-101_k141_11861 | Not-determined |
| 18-101_k141_175426 | Not-determined |
| 18-101_k141_210235 | Not-determined |
| 18-101_k141_340073 | Not-determined |
| 18-101_k141_225958 | Not-determined |
| 18-101_k141_340075 | Not-determined |
| 18-101_k141_340077 | Not-determined |
| 18-101_k141_44500 | Not-determined |
| 18-101_k141_109699 | Not-determined |
| 18-101_k141_358208 | Not-determined |
| 18-101_k141_390170 | Low-quality |
| 18-101_k141_259805 | Not-determined |
| 18-101_k141_193742 | Not-determined |
| 18-101_k141_390173 | Not-determined |
| 18-101_k141_292090 | Not-determined |
| 18-101_k141_44514 | Not-determined |
| 18-101_k141_61029 | Not-determined |
| 18-101_k141_193750 | Not-determined |
| 18-101_k141_292096 | Not-determined |
| 18-101_k141_390181 | Not-determined |
| 18-101_k141_373829 | Not-determined |
| 18-101_k141_242567 | Not-determined |
| 18-101_k141_259823 | Not-determined |
| 18-101_k141_143667 | Not-determined |
| 18-101_k141_175444 | Not-determined |
| 18-101_k141_175445 | Not-determined |
| 18-101_k141_143671 | Not-determined |
| 18-101_k141_109720 | Not-determined |
| 18-101_k141_242574 | Not-determined |
| 18-101_k141_259835 | Low-quality |
| 18-101_k141_109721 | Not-determined |
| 18-101_k141_242576 | Not-determined |
| 18-101_k141_76778 | Not-determined |
| 18-101_k141_292100 | Not-determined |
| 18-101_k141_210262 | Not-determined |
| 18-101_k141_210263 | Low-quality |
| 18-101_k141_76781 | Not-determined |
| 18-101_k141_292103 | Not-determined |
| 18-101_k141_210264 | Low-quality |
| 18-101_k141_11890 | Not-determined |
| 18-101_k141_274869 | Not-determined |
| 18-101_k141_143679 | Not-determined |
| 18-101_k141_94582 | Not-determined |
| 18-101_k141_373849 | Not-determined |
| 18-101_k141_175455 | Low-quality |
| 18-101_k141_94586 | Not-determined |
| 18-101_k141_358238 | Low-quality |
| 18-101_k141_11903 | Not-determined |
| 18-101_k141_94594 | Low-quality |
| 18-101_k141_308880 | Not-determined |
| 18-101_k141_308882 | Not-determined |
| 18-101_k141_373861 | Not-determined |
| 18-101_k141_308885 | Not-determined |
| 18-101_k141_225991 | Not-determined |
| 18-101_k141_358244 | Not-determined |
| 18-101_k141_373866 | Not-determined |
| 18-101_k141_225993 | Not-determined |
| 18-101_k141_175469 | Not-determined |
| 18-101_k141_143696 | Not-determined |
| 18-101_k141_175472 | Not-determined |
| 18-101_k141_61070 | Not-determined |
| 18-101_k141_143700 | Not-determined |
| 18-101_k141_143706 | Not-determined |
| 18-101_k141_143707 | Not-determined |
| 18-101_k141_109734 | Not-determined |
| 18-101_k141_143714 | Not-determined |
| 18-101_k141_373880 | Not-determined |
| 18-101_k141_210296 | Not-determined |
| 18-101_k141_242605 | Not-determined |
| 18-101_k141_308905 | Not-determined |
| 18-101_k141_109739 | Not-determined |
| 18-101_k141_292124 | Not-determined |
| 18-101_k141_28129 | Not-determined |
| 18-101_k141_109742 | Not-determined |
| 18-101_k141_325710 | Not-determined |
| 18-101_k141_44536 | Not-determined |
| 18-101_k141_358260 | Not-determined |
| 18-101_k141_292133 | Not-determined |
| 18-101_k141_128018 | Not-determined |
| 18-101_k141_210306 | Not-determined |
| 18-101_k141_210307 | Not-determined |
| 18-101_k141_94612 | Not-determined |
| 18-101_k141_159358 | Not-determined |
| 18-101_k141_373893 | Not-determined |
| 18-101_k141_210313 | Not-determined |
| 18-101_k141_373894 | Not-determined |
| 18-101_k141_390199 | Not-determined |
| 18-101_k141_61091 | Not-determined |
| 18-101_k141_159361 | Not-determined |
| 18-101_k141_128025 | Not-determined |
| 18-101_k141_325726 | Low-quality |
| 18-101_k141_373897 | Not-determined |
| 18-101_k141_308910 | Not-determined |
| 18-101_k141_373899 | Not-determined |
| 18-101_k141_128028 | Not-determined |
| 18-101_k141_274895 | Not-determined |
| 18-101_k141_61099 | Not-determined |
| 18-101_k141_159371 | Low-quality |
| 18-101_k141_358284 | Not-determined |
| 18-101_k141_28161 | Not-determined |
| 18-101_k141_274896 | Not-determined |
| 18-101_k141_128034 | Not-determined |
| 18-101_k141_340108 | Not-determined |
| 18-101_k141_44563 | Low-quality |
| 18-101_k141_340109 | Not-determined |
| 18-101_k141_274901 | Not-determined |
| 18-101_k141_274906 | Not-determined |
| 18-101_k141_44570 | Not-determined |
| 18-101_k141_292156 | Not-determined |
| 18-101_k141_242627 | Not-determined |
| 18-101_k141_44575 | Not-determined |
| 18-101_k141_292161 | Not-determined |
| 18-101_k141_94636 | Not-determined |
| 18-101_k141_76827 | Not-determined |
| 18-101_k141_143762 | Not-determined |
| 18-101_k141_259898 | Not-determined |
| 18-101_k141_109771 | Not-determined |
| 18-101_k141_325747 | Not-determined |
| 18-101_k141_274923 | Not-determined |
| 18-101_k141_259906 | Not-determined |
| 18-101_k141_143778 | Not-determined |
| 18-101_k141_109779 | Not-determined |
| 18-101_k141_358305 | Not-determined |
| 18-101_k141_44596 | Not-determined |
| 17-109-final-viral-combined_k141_17617\|\|full | Medium-quality |
| 18-101_k141_274929 | Not-determined |
| 18-101_k141_373919 | Not-determined |
| 18-101_k141_292184 | Not-determined |
| 18-101_k141_61132 | Not-determined |
| 18-101_k141_76844 | Not-determined |
| 18-101_k141_292185 | Not-determined |
| 18-101_k141_373924 | Not-determined |
| 18-101_k141_76846 | Not-determined |
| 18-101_k141_373926 | Not-determined |
| 18-101_k141_390202 | Low-quality |
| 18-101_k141_61136 | Not-determined |
| 18-101_k141_61137 | Not-determined |
| 18-101_k141_390208 | Not-determined |
| 18-101_k141_308948 | Not-determined |
| 18-101_k141_28198 | Not-determined |
| 18-101_k141_325764 | Not-determined |
| 18-101_k141_109803 | Not-determined |
| 18-101_k141_159380 | Low-quality |
| 18-101_k141_28203 | Not-determined |
| 18-101_k141_28204 | Not-determined |
| 18-101_k141_143794 | Low-quality |
| 18-101_k141_11964 | Not-determined |
| 18-101_k141_76856 | Not-determined |
| 18-101_k141_373947 | Not-determined |
| 18-101_k141_358319 | Not-determined |
| 18-101_k141_390214 | Not-determined |
| 18-101_k141_390215 | Not-determined |
| 18-101_k141_274955 | Not-determined |
| 18-101_k141_292190 | Not-determined |
| 18-101_k141_226017 | Not-determined |
| 18-101_k141_292192 | Not-determined |
| 18-101_k141_390216 | Not-determined |
| 18-101_k141_259930 | Not-determined |
| 18-101_k141_325776 | Not-determined |
| 18-101_k141_259931 | Not-determined |
| 18-101_k141_373960 | Not-determined |
| 18-101_k141_61167 | Not-determined |
| 18-101_k141_226021 | Not-determined |
| 18-101_k141_128046 | Not-determined |
| 18-101_k141_28216 | Not-determined |
| 18-101_k141_226025 | Not-determined |
| 18-101_k141_28219 | Low-quality |
| 18-101_k141_11979 | Not-determined |
| 18-101_k141_76870 | Not-determined |
| 18-101_k141_210375 | Not-determined |
| 18-101_k141_76871 | Not-determined |
| 18-101_k141_226027 | Not-determined |
| 18-101_k141_109830 | Not-determined |
| 18-101_k141_109831 | Not-determined |
| 18-101_k141_358336 | Not-determined |
| 18-101_k141_390235 | Not-determined |
| 18-101_k141_94661 | Not-determined |
| 18-101_k141_373968 | Not-determined |
| 18-101_k141_11987 | Not-determined |
| 18-101_k141_109838 | Not-determined |
| 18-101_k141_109842 | Not-determined |
| 18-101_k141_226041 | Low-quality |
| 18-101_k141_11993 | Not-determined |
| 18-101_k141_340157 | Not-determined |
| 18-101_k141_210389 | Not-determined |
| 18-101_k141_128067 | Not-determined |
| 18-101_k141_390247 | Not-determined |
| 18-101_k141_226043 | Not-determined |
| 18-101_k141_28224 | Not-determined |
| 18-101_k141_340164 | Not-determined |
| 18-101_k141_390255 | Not-determined |
| 18-101_k141_159412 | Not-determined |
| 18-101_k141_390257 | Not-determined |
| 18-101_k141_210399 | Not-determined |
| 18-101_k141_373984 | Not-determined |
| 18-101_k141_76895 | Not-determined |
| 18-101_k141_292231 | Not-determined |
| 18-101_k141_76896 | Not-determined |
| 18-101_k141_226055 | Not-determined |
| 18-101_k141_44638 | Not-determined |
| 18-101_k141_274981 | Not-determined |
| 18-101_k141_242653 | Not-determined |
| 18-101_k141_94678 | Not-determined |
| 18-101_k141_109866 | Not-determined |
| 18-101_k141_28242 | Not-determined |
| 18-101_k141_159417 | Not-determined |
| 18-101_k141_44643 | Not-determined |
| 18-101_k141_210411 | Not-determined |
| 18-101_k141_242656 | Not-determined |
| 18-101_k141_274987 | Not-determined |
| 18-101_k141_358365 | Not-determined |
| 18-101_k141_210414 | Low-quality |
| 18-101_k141_44646 | Not-determined |
| 18-101_k141_358369 | Not-determined |
| 18-101_k141_292244 | Not-determined |
| 18-101_k141_358370 | Not-determined |
| 18-101_k141_292247 | Not-determined |
| 18-101_k141_308984 | Not-determined |
| 18-101_k141_226068 | Not-determined |
| 18-101_k141_175528 | Not-determined |
| 18-101_k141_340180 | Not-determined |
| 18-101_k141_143823 | Not-determined |
| 18-101_k141_128080 | Not-determined |
| 18-101_k141_374008 | Not-determined |
| 18-101_k141_61224 | Not-determined |
| 18-101_k141_340185 | Not-determined |
| 18-101_k141_340186 | Low-quality |
| 18-101_k141_109883 | Not-determined |
| 18-101_k141_109884 | Not-determined |
| 18-101_k141_12019 | Not-determined |
| 18-101_k141_94689 | Not-determined |
| 18-101_k141_44659 | Low-quality |
| 18-101_k141_308997 | Not-determined |
| 18-101_k141_226095 | Not-determined |
| 18-101_k141_374022 | Not-determined |
| 18-101_k141_275007 | Not-determined |
| 18-101_k141_61242 | Not-determined |
| 18-101_k141_44664 | Not-determined |
| 18-101_k141_325810 | Not-determined |
| 18-101_k141_275013 | Not-determined |
| 18-101_k141_390271 | Not-determined |
| 18-101_k141_12033 | Not-determined |
| 18-101_k141_275015 | Not-determined |
| 18-101_k141_292265 | Not-determined |
| 18-101_k141_44674 | Not-determined |
| 18-101_k141_143848 | Not-determined |
| 18-101_k141_143850 | Not-determined |
| 18-101_k141_390277 | Not-determined |
| 18-101_k141_340210 | Not-determined |
| 18-101_k141_242676 | Not-determined |
| 18-101_k141_242678 | Not-determined |
| 18-101_k141_143856 | Not-determined |
| 18-101_k141_340219 | Not-determined |
| 18-101_k141_210450 | Not-determined |
| 18-101_k141_128115 | Not-determined |
| 18-101_k141_390292 | Not-determined |
| 18-101_k141_128123 | Not-determined |
| 18-101_k141_210460 | Not-determined |
| 18-101_k141_44699 | Not-determined |
| 18-101_k141_390296 | Not-determined |
| 18-101_k141_390297 | Not-determined |
| 18-101_k141_175541 | Not-determined |
| 18-101_k141_358418 | Not-determined |
| 18-101_k141_28277 | Not-determined |
| 18-101_k141_61279 | Not-determined |
| 18-101_k141_210471 | Not-determined |
| 18-101_k141_390307 | Not-determined |
| 18-101_k141_61282 | Not-determined |
| 18-101_k141_143858 | Not-determined |
| 18-101_k141_76955 | Not-determined |
| 18-101_k141_210476 | Not-determined |
| 18-101_k141_76960 | Not-determined |
| 18-101_k141_61287 | Not-determined |
| 18-101_k141_28287 | Low-quality |
| 18-101_k141_175556 | Not-determined |
| 18-101_k141_275045 | Not-determined |
| 18-101_k141_12079 | Not-determined |
| 18-101_k141_44719 | Not-determined |
| 18-101_k141_28296 | Not-determined |
| 18-101_k141_12085 | Not-determined |
| 18-101_k141_44730 | Not-determined |
| 18-101_k141_374065 | Not-determined |
| 18-101_k141_193773 | Not-determined |
| 18-101_k141_193776 | Not-determined |
| 18-101_k141_128157 | Not-determined |
| 18-101_k141_12093 | Not-determined |
| 18-101_k141_390332 | Not-determined |
| 18-101_k141_275059 | Not-determined |
| 18-101_k141_259976 | Not-determined |
| 18-101_k141_358450 | Not-determined |
| 18-101_k141_275061 | Not-determined |
| 18-101_k141_12095 | Not-determined |
| 18-101_k141_210508 | Not-determined |
| 18-101_k141_175582 | Not-determined |
| 18-101_k141_175584 | Not-determined |
| 18-101_k141_61296 | Not-determined |
| 18-101_k141_143871 | Not-determined |
| 18-101_k141_259985 | Not-determined |
| 18-101_k141_390344 | Not-determined |
| 18-101_k141_61302 | Not-determined |
| 18-101_k141_175588 | Not-determined |
| 18-101_k141_193786 | Not-determined |
| 18-101_k141_143881 | Not-determined |
| 18-101_k141_128168 | Not-determined |
| 18-101_k141_193792 | Not-determined |
| 18-101_k141_193794 | Not-determined |
| 18-101_k141_12123 | Not-determined |
| 18-101_k141_159452 | Not-determined |
| 18-101_k141_193806 | Not-determined |
| 18-101_k141_128182 | Not-determined |
| 18-101_k141_275097 | Low-quality |
| 18-101_k141_12130 | Not-determined |
| 18-101_k141_358474 | Low-quality |
| 18-101_k141_76992 | Not-determined |
| 18-101_k141_12131 | Not-determined |
| 18-101_k141_358476 | Not-determined |
| 18-101_k141_28310 | Not-determined |
| 18-101_k141_175607 | Not-determined |
| 18-101_k141_159464 | Not-determined |
| 18-101_k141_325875 | Not-determined |
| 18-101_k141_159469 | Not-determined |
| 18-101_k141_340242 | Not-determined |
| 18-101_k141_390364 | Not-determined |
| 18-101_k141_159471 | Not-determined |
| 18-101_k141_340245 | Not-determined |
| 18-101_k141_175616 | Not-determined |
| 18-101_k141_175620 | Not-determined |
| 18-101_k141_309064 | Not-determined |
| 18-101_k141_159481 | Not-determined |
| 18-101_k141_292285 | Not-determined |
| 18-101_k141_159486 | Not-determined |
| 18-101_k141_128194 | Not-determined |
| 18-101_k141_44762 | Not-determined |
| 18-101_k141_260019 | Not-determined |
| 18-101_k141_143905 | Low-quality |
| 18-101_k141_260024 | Not-determined |
| 18-101_k141_275119 | Not-determined |
| 18-101_k141_28320 | Not-determined |
| 18-101_k141_275120 | Not-determined |
| 18-101_k141_358487 | Not-determined |
| 18-101_k141_61343 | Not-determined |
| 18-101_k141_44771 | Low-quality |
| 18-101_k141_61345 | Not-determined |
| 18-101_k141_292303 | Not-determined |
| 18-101_k141_61347 | Not-determined |
| 18-101_k141_292306 | Not-determined |
| 18-101_k141_175628 | Low-quality |
| 18-101_k141_325898 | Low-quality |
| 18-101_k141_175630 | Not-determined |
| 18-101_k141_44775 | Not-determined |
| 18-101_k141_242737 | Not-determined |
| 18-101_k141_242742 | Not-determined |
| 18-101_k141_44780 | Not-determined |
| 18-101_k141_44781 | Not-determined |
| 18-101_k141_275136 | Not-determined |
| 18-101_k141_12172 | Not-determined |
| 18-101_k141_128205 | Not-determined |
| 18-101_k141_292312 | Not-determined |
| 18-101_k141_44791 | Not-determined |
| 18-101_k141_292313 | Not-determined |
| 18-101_k141_226182 | Not-determined |
| 18-101_k141_175641 | Not-determined |
| 18-101_k141_390390 | Not-determined |
| 18-101_k141_226184 | Low-quality |
| 18-101_k141_309108 | Not-determined |
| 18-101_k141_226188 | Not-determined |
| 18-101_k141_390399 | Not-determined |
| 18-101_k141_242759 | Not-determined |
| 18-101_k141_175645 | Low-quality |
| 18-101_k141_260059 | Not-determined |
| 18-101_k141_374112 | Low-quality |
| 18-101_k141_325905 | Not-determined |
| 18-101_k141_340280 | Low-quality |
| 18-101_k141_374113 | Not-determined |
| 18-101_k141_275144 | Not-determined |
| 18-101_k141_390408 | Not-determined |
| 18-101_k141_309112 | Not-determined |
| 18-101_k141_358503 | Not-determined |
| 18-101_k141_143925 | Not-determined |
| 18-101_k141_242770 | Not-determined |
| 18-101_k141_325909 | Not-determined |
| 18-101_k141_175650 | Not-determined |
| 18-101_k141_94705 | Not-determined |
| 18-101_k141_143931 | Not-determined |
| 18-101_k141_374119 | Not-determined |
| 18-101_k141_44817 | Not-determined |
| 18-101_k141_94719 | Low-quality |
| 18-101_k141_374123 | Not-determined |
| 18-101_k141_28346 | Not-determined |

| | |
|---|---|
| 18-101_k141_260082 | Not-determined |
| 18-101_k141_28351 | Not-determined |
| 18-101_k141_340302 | Not-determined |
| 18-101_k141_260087 | Not-determined |
| 18-101_k141_325927 | Not-determined |
| 18-101_k141_325929 | Not-determined |
| 18-101_k141_292358 | Not-determined |
| 18-101_k141_61362 | Not-determined |
| 18-101_k141_275173 | Low-quality |
| 18-101_k141_210577 | Not-determined |
| 18-101_k141_260099 | Not-determined |
| 18-101_k141_340307 | Not-determined |
| 18-101_k141_340311 | Not-determined |
| 18-101_k141_28369 | Not-determined |
| 18-101_k141_210586 | Not-determined |
| 18-101_k141_12216 | Not-determined |
| 18-101_k141_374138 | Not-determined |
| 18-101_k141_61376 | Not-determined |
| 18-101_k141_143950 | Not-determined |
| 18-101_k141_61377 | Not-determined |
| 18-101_k141_143952 | Not-determined |
| 18-101_k141_226242 | Not-determined |
| 18-101_k141_143953 | Not-determined |
| 18-101_k141_226245 | Not-determined |
| 18-101_k141_12223 | Not-determined |
| 18-101_k141_143959 | Not-determined |
| 18-101_k141_210594 | Not-determined |
| 18-101_k141_358538 | Not-determined |
| 18-101_k141_143961 | Not-determined |
| 18-101_k141_390425 | Not-determined |
| 18-101_k141_374146 | Not-determined |
| 18-101_k141_390430 | Not-determined |
| 18-101_k141_77019 | Low-quality |
| 18-101_k141_309143 | Not-determined |
| 18-101_k141_292382 | Not-determined |
| 18-101_k141_275179 | Low-quality |
| 18-101_k141_159522 | Not-determined |
| 18-101_k141_12233 | Not-determined |
| 18-101_k141_374161 | Low-quality |
| 18-101_k141_390444 | Not-determined |
| 18-101_k141_210602 | Not-determined |
| 18-101_k141_275193 | Not-determined |
| 18-101_k141_325959 | Not-determined |
| 18-101_k141_242821 | Not-determined |
| 18-101_k141_159538 | Low-quality |
| 18-101_k141_358563 | Not-determined |
| 18-101_k141_390460 | Not-determined |
| 18-101_k141_358568 | Not-determined |
| 18-101_k141_390470 | Not-determined |
| 18-101_k141_61413 | Low-quality |
| 18-101_k141_61416 | Not-determined |
| 18-101_k141_260136 | Not-determined |
| 18-101_k141_242835 | Not-determined |
| 18-101_k141_390483 | Low-quality |
| 18-101_k141_77052 | Not-determined |
| 18-101_k141_242843 | Not-determined |
| 18-101_k141_390493 | Not-determined |
| 18-101_k141_61433 | Not-determined |
| 18-101_k141_28405 | Not-determined |
| 18-101_k141_61438 | Not-determined |
| 18-101_k141_61444 | Not-determined |
| 18-101_k141_77061 | Low-quality |
| 18-101_k141_159552 | Not-determined |
| 18-101_k141_358604 | Not-determined |
| 18-101_k141_390507 | Not-determined |
| 18-101_k141_159566 | Not-determined |
| 18-101_k141_77070 | Not-determined |
| 18-101_k141_159572 | Not-determined |
| 18-101_k141_292439 | Not-determined |
| 18-101_k141_128246 | Not-determined |
| 18-101_k141_28429 | Not-determined |
| 18-101_k141_94751 | Not-determined |
| 18-101_k141_28434 | Low-quality |
| 18-101_k141_143975 | Not-determined |
| 18-101_k141_374185 | Low-quality |
| 18-101_k141_28439 | Not-determined |
| 18-101_k141_143980 | Not-determined |
| 18-101_k141_210639 | Not-determined |
| 18-101_k141_159585 | Not-determined |
| 18-101_k141_292447 | Not-determined |
| 18-101_k141_374190 | Not-determined |
| 18-101_k141_128247 | Low-quality |
| 18-101_k141_94753 | Not-determined |
| 18-101_k141_44849 | Not-determined |
| 18-101_k141_210643 | Not-determined |
| 18-101_k141_143985 | Not-determined |
| 18-101_k141_358613 | Not-determined |
| 18-101_k141_77081 | Not-determined |
| 18-101_k141_309189 | Not-determined |
| 18-101_k141_94761 | Not-determined |
| 18-101_k141_210651 | Not-determined |
| 18-101_k141_61502 | Not-determined |
| 18-101_k141_159593 | Not-determined |
| 18-101_k141_242894 | Not-determined |
| 18-101_k141_358630 | Not-determined |
| 18-101_k141_374209 | Not-determined |
| 18-101_k141_242902 | Low-quality |
| 18-101_k141_309204 | Not-determined |
| 18-101_k141_242903 | Not-determined |
| 18-101_k141_193818 | Not-determined |
| 18-101_k141_28450 | Not-determined |
| 18-101_k141_260194 | Not-determined |
| 18-101_k141_94782 | Not-determined |
| 18-101_k141_144012 | Not-determined |
| 18-101_k141_193824 | Not-determined |
| 18-101_k141_77104 | Not-determined |
| 18-101_k141_358652 | Low-quality |
| 18-101_k141_358653 | Not-determined |
| 18-101_k141_326006 | Not-determined |
| 18-101_k141_94798 | Low-quality |
| 18-101_k141_226329 | Not-determined |
| 18-101_k141_44871 | Not-determined |
| 18-101_k141_260207 | Low-quality |
| 18-101_k141_128267 | Not-determined |
| 18-101_k141_94801 | Low-quality |
| 18-101_k141_12342 | Not-determined |
| 18-101_k141_358659 | Not-determined |
| 18-101_k141_94803 | Not-determined |
| 18-101_k141_340340 | Not-determined |
| 18-101_k141_340341 | Not-determined |
| 18-101_k141_44878 | Not-determined |
| 18-101_k141_94805 | Not-determined |
| 18-101_k141_292493 | Not-determined |
| 18-101_k141_77118 | Not-determined |
| 18-101_k141_77120 | Not-determined |
| 18-101_k141_390541 | Not-determined |
| 18-101_k141_340345 | Not-determined |
| 18-101_k141_28480 | Not-determined |
| 18-101_k141_144051 | Not-determined |
| 18-101_k141_128272 | Not-determined |
| 18-101_k141_128273 | Not-determined |
| 18-101_k141_226350 | Not-determined |
| 18-101_k141_340349 | Not-determined |
| 18-101_k141_340350 | Not-determined |
| 18-101_k141_390549 | Not-determined |
| 18-101_k141_61558 | Not-determined |
| 18-101_k141_12353 | Not-determined |
| 18-101_k141_193836 | Low-quality |
| 18-101_k141_61560 | Not-determined |
| 18-101_k141_193838 | Not-determined |
| 18-101_k141_326028 | Not-determined |
| 18-101_k141_292501 | Not-determined |
| 18-101_k141_309236 | Not-determined |
| 18-101_k141_260234 | Not-determined |
| 18-101_k141_374228 | Not-determined |
| 18-101_k141_12360 | Low-quality |
| 18-101_k141_210658 | Not-determined |
| 18-101_k141_175711 | Not-determined |
| 18-101_k141_144069 | Not-determined |
| 18-101_k141_260244 | Not-determined |
| 18-101_k141_358676 | Not-determined |
| 18-101_k141_77132 | Not-determined |
| 18-101_k141_210668 | Not-determined |
| 18-101_k141_260253 | Not-determined |
| 18-101_k141_309255 | Not-determined |
| 18-101_k141_340378 | Not-determined |
| 18-101_k141_193848 | Not-determined |
| 18-101_k141_340384 | Low-quality |
| 18-101_k141_358686 | Not-determined |
| 18-101_k141_292521 | Not-determined |
| 18-101_k141_144085 | Not-determined |
| 18-101_k141_374241 | Not-determined |
| 18-101_k141_61597 | Not-determined |
| 18-101_k141_175725 | Low-quality |
| 18-101_k141_242929 | Not-determined |
| 18-101_k141_242930 | Not-determined |
| 18-101_k141_210672 | Not-determined |
| 18-101_k141_226380 | Not-determined |
| 18-101_k141_242934 | Not-determined |
| 18-101_k141_12388 | Not-determined |
| 18-101_k141_12389 | Not-determined |
| 18-101_k141_210678 | Not-determined |
| 18-101_k141_340404 | Not-determined |
| 18-101_k141_210679 | Not-determined |
| 18-101_k141_242940 | Not-determined |
| 18-101_k141_175736 | Not-determined |
| 18-101_k141_12393 | Not-determined |
| 18-101_k141_210682 | Not-determined |
| 18-101_k141_374250 | Not-determined |
| 18-101_k141_12395 | Low-quality |
| 18-101_k141_374255 | Not-determined |
| 18-101_k141_260269 | Not-determined |
| 18-101_k141_340410 | Low-quality |
| 18-101_k141_390565 | Not-determined |
| 18-101_k141_128316 | Not-determined |
| 18-101_k141_390569 | Not-determined |
| 18-101_k141_374264 | Not-determined |
| 18-101_k141_128317 | Low-quality |
| 18-101_k141_260277 | Not-determined |
| 18-101_k141_390572 | Not-determined |
| 18-101_k141_12410 | Not-determined |
| 18-101_k141_144100 | Not-determined |
| 18-101_k141_210700 | Not-determined |
| 18-101_k141_358713 | Not-determined |
| 18-101_k141_260283 | Not-determined |
| 18-101_k141_358714 | Not-determined |
| 18-101_k141_210702 | Not-determined |
| 18-101_k141_12419 | Not-determined |
| 18-101_k141_309268 | Not-determined |
| 18-101_k141_28501 | Not-determined |
| 18-101_k141_12422 | Not-determined |
| 18-101_k141_193886 | Low-quality |
| 18-101_k141_340422 | Low-quality |
| 18-101_k141_390582 | Not-determined |
| 18-101_k141_292544 | Not-determined |
| 18-101_k141_193896 | Not-determined |
| 18-101_k141_44918 | Not-determined |
| 18-101_k141_358721 | Low-quality |
| 18-101_k141_159626 | Not-determined |
| 18-101_k141_144101 | Low-quality |
| 18-101_k141_44920 | Not-determined |
| 18-101_k141_260305 | Not-determined |
| 18-101_k141_242969 | Not-determined |
| 18-101_k141_358728 | Low-quality |
| 18-101_k141_358729 | Not-determined |
| 18-101_k141_226427 | Not-determined |
| 18-101_k141_128334 | Not-determined |
| 18-101_k141_226429 | Not-determined |
| 18-101_k141_309284 | Not-determined |
| 18-101_k141_390607 | Not-determined |
| 18-101_k141_242983 | Not-determined |
| 18-101_k141_77196 | Not-determined |
| 18-101_k141_144118 | Not-determined |
| 18-101_k141_242984 | Not-determined |
| 18-101_k141_159639 | Not-determined |
| 18-101_k141_12447 | Not-determined |
| 18-101_k141_144123 | Not-determined |
| 18-101_k141_193906 | Not-determined |
| 18-101_k141_159641 | Low-quality |
| 18-101_k141_374297 | Low-quality |
| 18-101_k141_292582 | Not-determined |
| 18-101_k141_242991 | Not-determined |
| 18-101_k141_77207 | Not-determined |
| 18-101_k141_128352 | Low-quality |
| 18-101_k141_159652 | Not-determined |
| 18-101_k141_374302 | Not-determined |
| 18-101_k141_358752 | Not-determined |
| 18-101_k141_292594 | Not-determined |
| 18-101_k141_193929 | Not-determined |
| 18-101_k141_374305 | Not-determined |
| 18-101_k141_292596 | Not-determined |
| 18-101_k141_326043 | Not-determined |
| 18-101_k141_260311 | Not-determined |
| 18-101_k141_193931 | Not-determined |
| 18-101_k141_326047 | Not-determined |
| 18-101_k141_340447 | Not-determined |
| 18-101_k141_128358 | Not-determined |
| 18-101_k141_12467 | Not-determined |
| 18-101_k141_374313 | Not-determined |
| 18-101_k141_175793 | Not-determined |
| 18-101_k141_374316 | Not-determined |
| 18-101_k141_175798 | Not-determined |
| 18-101_k141_326053 | Not-determined |
| 18-101_k141_326058 | Not-determined |
| 18-101_k141_326060 | Not-determined |
| 18-101_k141_275256 | Not-determined |
| 18-101_k141_275257 | Not-determined |
| 18-101_k141_243018 | Not-determined |
| 18-101_k141_159667 | Not-determined |
| 18-101_k141_193957 | Not-determined |
| 18-101_k141_309293 | Not-determined |
| 18-101_k141_28557 | Low-quality |
| 18-101_k141_326070 | Not-determined |
| 18-101_k141_275260 | Not-determined |
| 18-101_k141_275261 | Not-determined |
| 18-101_k141_243027 | Not-determined |
| 18-101_k141_326077 | Not-determined |
| 18-101_k141_358762 | Not-determined |
| 18-101_k141_390659 | Not-determined |
| 18-101_k141_159671 | Not-determined |
| 18-101_k141_109949 | Not-determined |
| 18-101_k141_275272 | Not-determined |
| 18-101_k141_193971 | Not-determined |
| 18-101_k141_260361 | Not-determined |
| 18-101_k141_292622 | Not-determined |
| 18-101_k141_193978 | Not-determined |
| 18-101_k141_109955 | Not-determined |
| 18-101_k141_243047 | Not-determined |
| 18-101_k141_275285 | Not-determined |
| 18-101_k141_358773 | Not-determined |
| 18-101_k141_292629 | Not-determined |
| 18-101_k141_77221 | Not-determined |
| 18-101_k141_374352 | Not-determined |
| 18-101_k141_340458 | Low-quality |
| 18-101_k141_175819 | Not-determined |
| 18-101_k141_358778 | Not-determined |
| 18-101_k141_193986 | Not-determined |
| 18-101_k141_193987 | Not-determined |
| 18-101_k141_374360 | Low-quality |
| 18-101_k141_193988 | Not-determined |
| 18-101_k141_292638 | Not-determined |
| 18-101_k141_260377 | Not-determined |
| 18-101_k141_275297 | Not-determined |
| 18-101_k141_159686 | Not-determined |
| 18-101_k141_28576 | Not-determined |
| 18-101_k141_358790 | Not-determined |
| 18-101_k141_326100 | Not-determined |
| 18-101_k141_175826 | Low-quality |
| 18-101_k141_275306 | Not-determined |
| 18-101_k141_94847 | Not-determined |
| 18-101_k141_45011 | Not-determined |
| 18-101_k141_28590 | Not-determined |
| 18-101_k141_390679 | Not-determined |
| 18-101_k141_128402 | Not-determined |
| 18-101_k141_292655 | Not-determined |
| 18-101_k141_340479 | Not-determined |
| 18-101_k141_226485 | Not-determined |
| 18-101_k141_390687 | Not-determined |
| 18-101_k141_292661 | Not-determined |
| 18-101_k141_175832 | Not-determined |
| 18-101_k141_292662 | Not-determined |
| 18-101_k141_175835 | Not-determined |
| 18-101_k141_358794 | Not-determined |
| 18-101_k141_374371 | Not-determined |
| 18-101_k141_374373 | Not-determined |
| 18-101_k141_159696 | Not-determined |
| 18-101_k141_175842 | Not-determined |
| 18-101_k141_309311 | Not-determined |
| 18-101_k141_309312 | Not-determined |
| 18-101_k141_374377 | Not-determined |
| 18-101_k141_28609 | Low-quality |
| 18-101_k141_28610 | Not-determined |
| 18-101_k141_226490 | Not-determined |
| 18-101_k141_292674 | Not-determined |
| 18-101_k141_159705 | Not-determined |
| 18-101_k141_390703 | Not-determined |
| 18-101_k141_28616 | Not-determined |
| 18-101_k141_77255 | Not-determined |
| 18-101_k141_358818 | Not-determined |
| 18-101_k141_45035 | Not-determined |
| 18-101_k141_390709 | Not-determined |
| 18-101_k141_260402 | Low-quality |
| 18-101_k141_292688 | Not-determined |
| 18-101_k141_194025 | Not-determined |
| 18-101_k141_309320 | Not-determined |
| 18-101_k141_309322 | Not-determined |
| 18-101_k141_226501 | Not-determined |
| 18-101_k141_144188 | Low-quality |
| 18-101_k141_109987 | Not-determined |
| 18-101_k141_12510 | Not-determined |
| 18-101_k141_12513 | Not-determined |
| 18-101_k141_194037 | Not-determined |
| 18-101_k141_309334 | Not-determined |
| 18-101_k141_292693 | Not-determined |
| 18-101_k141_159715 | Not-determined |
| 18-101_k141_159718 | Not-determined |
| 18-101_k141_309338 | Not-determined |
| 18-101_k141_77261 | Not-determined |
| 18-101_k141_159720 | Not-determined |
| 18-101_k141_77265 | Low-quality |
| 18-101_k141_194046 | Not-determined |
| 18-101_k141_226516 | Not-determined |
| 18-101_k141_28634 | Not-determined |
| 18-101_k141_326138 | Not-determined |
| 18-101_k141_309345 | Not-determined |
| 18-101_k141_390737 | Not-determined |
| 18-101_k141_260424 | Not-determined |
| 18-101_k141_340525 | Not-determined |
| 18-101_k141_340529 | Not-determined |
| 18-101_k141_390745 | Not-determined |
| 18-101_k141_340533 | Not-determined |
| 18-101_k141_326139 | Low-quality |
| 18-101_k141_226525 | Not-determined |
| 18-101_k141_374393 | Not-determined |
| 18-101_k141_309349 | Not-determined |
| 18-101_k141_194056 | Not-determined |
| 18-101_k141_28645 | Low-quality |
| 18-101_k141_309352 | Not-determined |
| 18-101_k141_28646 | Not-determined |
| 18-101_k141_374406 | Not-determined |
| 18-101_k141_144201 | Not-determined |
| 18-101_k141_45060 | Not-determined |
| 18-101_k141_12527 | Not-determined |
| 18-101_k141_144207 | Not-determined |
| 18-101_k141_374413 | Not-determined |
| 18-101_k141_28651 | Not-determined |
| 18-101_k141_358862 | Not-determined |
| 18-101_k141_45064 | Low-quality |
| 18-101_k141_374426 | Not-determined |
| 18-101_k141_45069 | Not-determined |
| 18-101_k141_45072 | Not-determined |
| 18-101_k141_260442 | Not-determined |
| 18-101_k141_358875 | Low-quality |
| 18-101_k141_358878 | Not-determined |
| 18-101_k141_110031 | Not-determined |
| 18-101_k141_358879 | Not-determined |
| 18-101_k141_390771 | Not-determined |
| 18-101_k141_275338 | Not-determined |
| 18-101_k141_77274 | Not-determined |
| 18-101_k141_194073 | Not-determined |
| 18-101_k141_275340 | Not-determined |
| 18-101_k141_77278 | Not-determined |
| 18-101_k141_159749 | Not-determined |
| 18-101_k141_226539 | Not-determined |
| 18-101_k141_326177 | Not-determined |
| 18-101_k141_194079 | Not-determined |
| 18-101_k141_45090 | Not-determined |
| 18-101_k141_94870 | Not-determined |
| 18-101_k141_28657 | Not-determined |
| 18-101_k141_226550 | Not-determined |
| 18-101_k141_128472 | Not-determined |
| 18-101_k141_340539 | Not-determined |
| 18-101_k141_358890 | Not-determined |
| 18-101_k141_45103 | Not-determined |
| 18-101_k141_128474 | Not-determined |
| 18-101_k141_340543 | Not-determined |
| 18-101_k141_309387 | Not-determined |
| 18-101_k141_358893 | Not-determined |
| 18-101_k141_243107 | Not-determined |
| 18-101_k141_144215 | Not-determined |
| 18-101_k141_309388 | Low-quality |
| 18-101_k141_309392 | Not-determined |
| 18-101_k141_358898 | Not-determined |
| 18-101_k141_159763 | Not-determined |
| 18-101_k141_194088 | Not-determined |
| 18-101_k141_128491 | Not-determined |
| 18-101_k141_28673 | Not-determined |
| 18-101_k141_292741 | Not-determined |
| 18-101_k141_61771 | Not-determined |
| 18-101_k141_128501 | Not-determined |
| 18-101_k141_210734 | Not-determined |
| 18-101_k141_194105 | Not-determined |
| 18-101_k141_292745 | Not-determined |
| 18-101_k141_61776 | Not-determined |
| 18-101_k141_226586 | Not-determined |
| 18-101_k141_210738 | Not-determined |
| 18-101_k141_210741 | Not-determined |
| 18-101_k141_144231 | Not-determined |
| 18-101_k141_374469 | Low-quality |
| 18-101_k141_292754 | Not-determined |
| 18-101_k141_128513 | Not-determined |
| 18-101_k141_94904 | Not-determined |
| 18-101_k141_210746 | Low-quality |
| 18-101_k141_326224 | Not-determined |
| 18-101_k141_77303 | Not-determined |
| 18-101_k141_128517 | Not-determined |
| 18-101_k141_94907 | Not-determined |
| 18-101_k141_326229 | Not-determined |
| 18-101_k141_374480 | Not-determined |
| 18-101_k141_226602 | Not-determined |
| 18-101_k141_77314 | Not-determined |
| 18-101_k141_77319 | Not-determined |
| 18-101_k141_159784 | Not-determined |
| 18-101_k141_144247 | Not-determined |
| 18-101_k141_243147 | Not-determined |
| 18-101_k141_275407 | Not-determined |
| 18-101_k141_61794 | Not-determined |
| 18-101_k141_243149 | Not-determined |
| 18-101_k141_309450 | Not-determined |
| 18-101_k141_243153 | Not-determined |
| 18-101_k141_128551 | Not-determined |
| 18-101_k141_292778 | Low-quality |
| 18-101_k141_12597 | Not-determined |
| 18-101_k141_309460 | Not-determined |
| 18-101_k141_12598 | Not-determined |
| 18-101_k141_243165 | Not-determined |
| 18-101_k141_309465 | Low-quality |
| 18-101_k141_12605 | Not-determined |
| 18-101_k141_61812 | Not-determined |
| 18-101_k141_309468 | Not-determined |
| 18-101_k141_374510 | Not-determined |
| 18-101_k141_128566 | Not-determined |
| 18-101_k141_340563 | Not-determined |
| 18-101_k141_61817 | Not-determined |
| 18-101_k141_326256 | Not-determined |
| 18-101_k141_326259 | Not-determined |
| 18-101_k141_326262 | Not-determined |
| 18-101_k141_358923 | Not-determined |
| 18-101_k141_12613 | Not-determined |
| 18-101_k141_144261 | Not-determined |
| 18-101_k141_292799 | Low-quality |
| 18-101_k141_243183 | Not-determined |
| 18-101_k141_12618 | Not-determined |
| 18-101_k141_226652 | Not-determined |
| 18-101_k141_326269 | Not-determined |
| 18-101_k141_77330 | Low-quality |
| 18-101_k141_326271 | Not-determined |
| 18-101_k141_77332 | Not-determined |
| 18-101_k141_358941 | Not-determined |
| 18-101_k141_358943 | Not-determined |
| 18-101_k141_61829 | Not-determined |
| 18-101_k141_128585 | Not-determined |
| 18-101_k141_61832 | Not-determined |
| 18-101_k141_61833 | Not-determined |
| 18-101_k141_243196 | Not-determined |
| 18-101_k141_358955 | Low-quality |
| 18-101_k141_374529 | Not-determined |
| 18-101_k141_292817 | Not-determined |
| 18-101_k141_358956 | Not-determined |
| 18-101_k141_260461 | Not-determined |
| 18-101_k141_159788 | Not-determined |
| 18-101_k141_374541 | Not-determined |
| 18-101_k141_292821 | Not-determined |
| 18-101_k141_94971 | Not-determined |
| 18-101_k141_194163 | Not-determined |
| 18-101_k141_94974 | Not-determined |
| 18-101_k141_275457 | Not-determined |
| 18-101_k141_260467 | Not-determined |
| 18-101_k141_390818 | Not-determined |
| 18-101_k141_194171 | Not-determined |
| 18-101_k141_61850 | Not-determined |
| 18-101_k141_194173 | Not-determined |
| 18-101_k141_12654 | Not-determined |
| 18-101_k141_159804 | Not-determined |
| 18-101_k141_292829 | Not-determined |
| 18-101_k141_243222 | Not-determined |
| 18-101_k141_61860 | Not-determined |
| 18-101_k141_374555 | Not-determined |
| 18-101_k141_390827 | Not-determined |
| 18-101_k141_374557 | Not-determined |
| 18-101_k141_390828 | Not-determined |
| 18-101_k141_292834 | Not-determined |
| 18-101_k141_144277 | Not-determined |
| 18-101_k141_374561 | Not-determined |
| 18-101_k141_144279 | Not-determined |
| 18-101_k141_144280 | Low-quality |
| 18-101_k141_374562 | Not-determined |
| 18-101_k141_226672 | Low-quality |
| 18-101_k141_226675 | Not-determined |
| 18-101_k141_374567 | Not-determined |
| 18-101_k141_260486 | Not-determined |
| 18-101_k141_358990 | Not-determined |
| 18-101_k141_12666 | Not-determined |
| 18-101_k141_374570 | Low-quality |
| 18-101_k141_61877 | Not-determined |
| 18-101_k141_194177 | Low-quality |
| 18-101_k141_292842 | Low-quality |
| 18-101_k141_12670 | Not-determined |
| 18-101_k141_61881 | Not-determined |
| 18-101_k141_340604 | Not-determined |
| 18-101_k141_260494 | Not-determined |
| 18-101_k141_309486 | Not-determined |
| 18-101_k141_275485 | Not-determined |
| 18-101_k141_128624 | Not-determined |
| 18-101_k141_260506 | Not-determined |
| 18-101_k141_194196 | Not-determined |
| 18-101_k141_45166 | Not-determined |
| 18-101_k141_210850 | Low-quality |
| 18-101_k141_159827 | Not-determined |
| 18-101_k141_45168 | Low-quality |
| 18-101_k141_61892 | Not-determined |
| 18-101_k141_292865 | Not-determined |
| 18-101_k141_326274 | Not-determined |
| 18-101_k141_210862 | Not-determined |
| 18-101_k141_210864 | Not-determined |
| 18-101_k141_210865 | Not-determined |
| 18-101_k141_292873 | Not-determined |
| 18-101_k141_309489 | Not-determined |
| 18-101_k141_194219 | Not-determined |
| 18-101_k141_194221 | Not-determined |
| 18-101_k141_309493 | Not-determined |
| 18-101_k141_159837 | Not-determined |
| 18-101_k141_309494 | Not-determined |
| 18-101_k141_45177 | Low-quality |
| 18-101_k141_340637 | Low-quality |
| 18-101_k141_210877 | Low-quality |
| 18-101_k141_28708 | Low-quality |
| 18-101_k141_194231 | Not-determined |
| 18-101_k141_61924 | Not-determined |
| 18-101_k141_275526 | Not-determined |
| 18-101_k141_275527 | Not-determined |
| 18-101_k141_28711 | Not-determined |
| 18-101_k141_326281 | Not-determined |
| 18-101_k141_275528 | Not-determined |
| 18-101_k141_275529 | Not-determined |
| 18-101_k141_210885 | Not-determined |
| 18-101_k141_340642 | Low-quality |
| 18-101_k141_260519 | Not-determined |
| 18-101_k141_326292 | Not-determined |
| 18-101_k141_359024 | Not-determined |
| 18-101_k141_359026 | Not-determined |
| 18-101_k141_210904 | Not-determined |
| 18-101_k141_210906 | Not-determined |
| 18-101_k141_390863 | Low-quality |
| 18-101_k141_390864 | Not-determined |
| 18-101_k141_292899 | Not-determined |
| 18-101_k141_61931 | Not-determined |
| 18-101_k141_326309 | Not-determined |
| 18-101_k141_77443 | Not-determined |
| 18-101_k141_275542 | Not-determined |
| 18-101_k141_260552 | Not-determined |
| 18-101_k141_61936 | Not-determined |
| 18-101_k141_359036 | Low-quality |
| 18-101_k141_28730 | Not-determined |
| 18-101_k141_194254 | Not-determined |
| 18-101_k141_275544 | Not-determined |
| 18-101_k141_260562 | Not-determined |
| 18-101_k141_309519 | Not-determined |
| 18-101_k141_309523 | Not-determined |

| | |
|---|---|
| 18-101_k141_61943 | Low-quality |
| 18-101_k141_359046 | Low-quality |
| 18-101_k141_61946 | Low-quality |
| 18-101_k141_12714 | Low-quality |
| 18-101_k141_61953 | Not-determined |
| 18-101_k141_28746 | Not-determined |
| 18-101_k141_390872 | Not-determined |
| 18-101_k141_275565 | Not-determined |
| 18-101_k141_128701 | Not-determined |
| 18-101_k141_359050 | Not-determined |
| 18-101_k141_128703 | Not-determined |
| 18-101_k141_61956 | Not-determined |
| 18-101_k141_243274 | Not-determined |
| 18-101_k141_390875 | Low-quality |
| 18-101_k141_243275 | Low-quality |
| 18-101_k141_226720 | Not-determined |
| 18-101_k141_77464 | Not-determined |
| 18-101_k141_275573 | Not-determined |
| 18-101_k141_374634 | Not-determined |
| 18-101_k141_128717 | Not-determined |
| 18-101_k141_175857 | Not-determined |
| 18-101_k141_275578 | Not-determined |
| 18-101_k141_77475 | Not-determined |
| 18-101_k141_128720 | Not-determined |
| 18-101_k141_374645 | Not-determined |
| 18-101_k141_128722 | Not-determined |
| 18-101_k141_128723 | Not-determined |
| 18-101_k141_128724 | Not-determined |
| 18-101_k141_175863 | Not-determined |
| 18-101_k141_226733 | Not-determined |
| 18-101_k141_28764 | Not-determined |
| 18-101_k141_326346 | Not-determined |
| 18-101_k141_390889 | Not-determined |
| 18-101_k141_243297 | Not-determined |
| 18-101_k141_359079 | Not-determined |
| 18-101_k141_226740 | Not-determined |
| 18-101_k141_374657 | Not-determined |
| 18-101_k141_340678 | Low-quality |
| 18-101_k141_12729 | Not-determined |
| 18-101_k141_77501 | Not-determined |
| 18-101_k141_309537 | Not-determined |
| 18-101_k141_45228 | Not-determined |
| 18-101_k141_77506 | Not-determined |
| 18-101_k141_243304 | Not-determined |
| 18-101_k141_326361 | Not-determined |
| 18-101_k141_226751 | Not-determined |
| 18-101_k141_326363 | Not-determined |
| 18-101_k141_226753 | Not-determined |
| 18-101_k141_243307 | Not-determined |
| 18-101_k141_128731 | Not-determined |
| 18-101_k141_390901 | Not-determined |
| 18-101_k141_226757 | Not-determined |
| 18-101_k141_194297 | Not-determined |
| 18-101_k141_45241 | Not-determined |
| 18-101_k141_194303 | Not-determined |
| 18-101_k141_77523 | Not-determined |
| 18-101_k141_210922 | Not-determined |
| 18-101_k141_340697 | Not-determined |
| 18-101_k141_175894 | Not-determined |
| 18-101_k141_45250 | Not-determined |
| 18-101_k141_309560 | Not-determined |
| 18-101_k141_309561 | Not-determined |
| 18-101_k141_128738 | Low-quality |
| 18-101_k141_194311 | Not-determined |
| 18-101_k141_260609 | Not-determined |
| 18-101_k141_275594 | Not-determined |
| 18-101_k141_326365 | Not-determined |
| 18-101_k141_340711 | Not-determined |
| 18-101_k141_359104 | Not-determined |
| 18-101_k141_28791 | Not-determined |
| 18-101_k141_326368 | Not-determined |
| 18-101_k141_194313 | Not-determined |
| 18-101_k141_175908 | Not-determined |
| 18-101_k141_340720 | Not-determined |
| 18-101_k141_175911 | Not-determined |
| 18-101_k141_210949 | Not-determined |
| 18-101_k141_340723 | Not-determined |
| 18-101_k141_12749 | Not-determined |
| 18-101_k141_340724 | Not-determined |
| 18-101_k141_359114 | Not-determined |
| 18-101_k141_340726 | Not-determined |
| 18-101_k141_260629 | Not-determined |
| 18-101_k141_12752 | Not-determined |
| 18-101_k141_390926 | Not-determined |
| 18-101_k141_194321 | Not-determined |
| 18-101_k141_210957 | Not-determined |
| 18-101_k141_210959 | Not-determined |
| 18-101_k141_62006 | Not-determined |
| 18-101_k141_194328 | Not-determined |
| 18-101_k141_226800 | Not-determined |
| 18-101_k141_194336 | Not-determined |
| 18-101_k141_159898 | Not-determined |
| 18-101_k141_340754 | Not-determined |
| 18-101_k141_175928 | Not-determined |
| 18-101_k141_390943 | Not-determined |
| 18-101_k141_340763 | Not-determined |
| 18-101_k141_210966 | Not-determined |
| 18-101_k141_275617 | Not-determined |
| 18-101_k141_95001 | Not-determined |
| 18-101_k141_210971 | Not-determined |
| 18-101_k141_260660 | Not-determined |
| 18-101_k141_326389 | Not-determined |
| 18-101_k141_243339 | Not-determined |
| 18-101_k141_210973 | Not-determined |
| 18-101_k141_62034 | Low-quality |
| 18-101_k141_340777 | Not-determined |
| 18-101_k141_390946 | Not-determined |
| 18-101_k141_243346 | Not-determined |
| 18-101_k141_210987 | Not-determined |
| 18-101_k141_194350 | Not-determined |
| 18-101_k141_159917 | Not-determined |
| 18-101_k141_45261 | Not-determined |
| 18-101_k141_12772 | Not-determined |
| 18-101_k141_275630 | Not-determined |
| 18-101_k141_243350 | Not-determined |
| 18-101_k141_12774 | Not-determined |
| 18-101_k141_326402 | Not-determined |
| 18-101_k141_390957 | Not-determined |
| 18-101_k141_95018 | Not-determined |
| 18-101_k141_292952 | Not-determined |
| 18-101_k141_194360 | Not-determined |
| 18-101_k141_275636 | Not-determined |
| 18-101_k141_77590 | Not-determined |
| 18-101_k141_326411 | Not-determined |
| 18-101_k141_359139 | Low-quality |
| 18-101_k141_326413 | Not-determined |
| 18-101_k141_194368 | Not-determined |
| 18-101_k141_292964 | Not-determined |
| 18-101_k141_226828 | Not-determined |
| 18-101_k141_95037 | Not-determined |
| 18-101_k141_326416 | Not-determined |
| 18-101_k141_359149 | Not-determined |
| 18-101_k141_175960 | Not-determined |
| 18-101_k141_390965 | Not-determined |
| 18-101_k141_95041 | Not-determined |
| 18-101_k141_309603 | Not-determined |
| 18-101_k141_77607 | Low-quality |
| 18-101_k141_28838 | Not-determined |
| 18-101_k141_144300 | Not-determined |
| 18-101_k141_144302 | Not-determined |
| 18-101_k141_340782 | Not-determined |
| 18-101_k141_292970 | Not-determined |
| 18-101_k141_211001 | Not-determined |
| 18-101_k141_77616 | Not-determined |
| 18-101_k141_326431 | Not-determined |
| 18-101_k141_226841 | Not-determined |
| 18-101_k141_243371 | Not-determined |
| 18-101_k141_309614 | Not-determined |
| 18-101_k141_260692 | Not-determined |
| 18-101_k141_175977 | Not-determined |
| 18-101_k141_175978 | Not-determined |
| 18-101_k141_309619 | Not-determined |
| 18-101_k141_211014 | Not-determined |
| 18-101_k141_28856 | Not-determined |
| 18-101_k141_292974 | Not-determined |
| 18-101_k141_275672 | Not-determined |
| 18-101_k141_95063 | Not-determined |
| 18-101_k141_194409 | Not-determined |
| 18-101_k141_194411 | Not-determined |
| 18-101_k141_275680 | Not-determined |
| 18-101_k141_110046 | Not-determined |
| 18-101_k141_275686 | Not-determined |
| 18-101_k141_275687 | Not-determined |
| 18-101_k141_326469 | Not-determined |
| 18-101_k141_62081 | Not-determined |
| 18-101_k141_293004 | Not-determined |
| 18-101_k141_391011 | Not-determined |
| 18-101_k141_260713 | Not-determined |
| 18-101_k141_62086 | Not-determined |
| 18-101_k141_194417 | Not-determined |
| 18-101_k141_77626 | Low-quality |
| 18-101_k141_144331 | Not-determined |
| 18-101_k141_340810 | Not-determined |
| 18-101_k141_293014 | Not-determined |
| 18-101_k141_326479 | Not-determined |
| 18-101_k141_340816 | Not-determined |
| 18-101_k141_293017 | Not-determined |
| 18-101_k141_110068 | Not-determined |
| 18-101_k141_243405 | Not-determined |
| 18-101_k141_391022 | Not-determined |
| 18-101_k141_359196 | Low-quality |
| 18-101_k141_326483 | Not-determined |
| 18-101_k141_77638 | Low-quality |
| 18-101_k141_110070 | Not-determined |
| 18-101_k141_194439 | Not-determined |
| 18-101_k141_194441 | Not-determined |
| 18-101_k141_359209 | Not-determined |
| 18-101_k141_144342 | Not-determined |
| 18-101_k141_391037 | Not-determined |
| 18-101_k141_194446 | Not-determined |
| 18-101_k141_144345 | Low-quality |
| 18-101_k141_391039 | Not-determined |
| 18-101_k141_260741 | Not-determined |
| 18-101_k141_309635 | Not-determined |
| 18-101_k141_62103 | Not-determined |
| 18-101_k141_175994 | Not-determined |
| 18-101_k141_211030 | Not-determined |
| 18-101_k141_243416 | Not-determined |
| 18-101_k141_211031 | Not-determined |
| 18-101_k141_243417 | Not-determined |
| 18-101_k141_62105 | Not-determined |
| 18-101_k141_110084 | Not-determined |
| 18-101_k141_194450 | Not-determined |
| 18-101_k141_110088 | Not-determined |
| 18-101_k141_128813 | Not-determined |
| 18-101_k141_211035 | Low-quality |
| 18-101_k141_45291 | Not-determined |
| 18-101_k141_340842 | Low-quality |
| 18-101_k141_95104 | Not-determined |
| 18-101_k141_110102 | Not-determined |
| 18-101_k141_309655 | Not-determined |
| 18-101_k141_110103 | Low-quality |
| 18-101_k141_275705 | Not-determined |
| 18-101_k141_62116 | Low-quality |
| 18-101_k141_159958 | Not-determined |
| 18-101_k141_128832 | Not-determined |
| 18-101_k141_260767 | Not-determined |
| 18-101_k141_260768 | Not-determined |
| 18-101_k141_144376 | Not-determined |
| 18-101_k141_128837 | Not-determined |
| 18-101_k141_243446 | Not-determined |
| 18-101_k141_128839 | Not-determined |
| 18-101_k141_144381 | Not-determined |
| 18-101_k141_293071 | Not-determined |
| 18-101_k141_62125 | Low-quality |
| 18-101_k141_159969 | Not-determined |
| 18-101_k141_62126 | Not-determined |
| 18-101_k141_309667 | Not-determined |
| 18-101_k141_260779 | Low-quality |
| 18-101_k141_45296 | Not-determined |
| 18-101_k141_128843 | Not-determined |
| 18-101_k141_62127 | Not-determined |
| 18-101_k141_176002 | Not-determined |
| 18-101_k141_45302 | Not-determined |
| 18-101_k141_309673 | Not-determined |
| 18-101_k141_309679 | Not-determined |
| 18-101_k141_95133 | Not-determined |
| 18-101_k141_275722 | Not-determined |
| 18-101_k141_159983 | Low-quality |
| 18-101_k141_293083 | Not-determined |
| 18-101_k141_176015 | Not-determined |
| 18-101_k141_359240 | Not-determined |
| 18-101_k141_359242 | Not-determined |
| 18-101_k141_340871 | Not-determined |
| 18-101_k141_110130 | Not-determined |
| 18-101_k141_309685 | Low-quality |
| 18-101_k141_77659 | Not-determined |
| 18-101_k141_243457 | Not-determined |
| 18-101_k141_275732 | Not-determined |
| 18-101_k141_128867 | Not-determined |
| 18-101_k141_12792 | Low-quality |
| 18-101_k141_95142 | Not-determined |
| 18-101_k141_95146 | Not-determined |
| 18-101_k141_128873 | Not-determined |
| 18-101_k141_243464 | Not-determined |
| 18-101_k141_293088 | Not-determined |
| 18-101_k141_293089 | Not-determined |
| 18-101_k141_77668 | Not-determined |
| 18-101_k141_211058 | Not-determined |
| 18-101_k141_159994 | Not-determined |
| 18-101_k141_144417 | Not-determined |
| 18-101_k141_77671 | Not-determined |
| 18-101_k141_128882 | Not-determined |
| 18-101_k141_128885 | Not-determined |
| 18-101_k141_374722 | Low-quality |
| 18-101_k141_62145 | Not-determined |
| 18-101_k141_374724 | Not-determined |
| 18-101_k141_374725 | Not-determined |
| 18-101_k141_326507 | Not-determined |
| 18-101_k141_194496 | Not-determined |
| 18-101_k141_326510 | Not-determined |
| 18-101_k141_326511 | Not-determined |
| 18-101_k141_309706 | Not-determined |
| 18-101_k141_326513 | Not-determined |
| 18-101_k141_110140 | Not-determined |
| 18-101_k141_211075 | Not-determined |
| 18-101_k141_28888 | Not-determined |
| 18-101_k141_62155 | Not-determined |
| 18-101_k141_293102 | Low-quality |
| 18-101_k141_160006 | Not-determined |
| 18-101_k141_326521 | Not-determined |
| 18-101_k141_374738 | Not-determined |
| 18-101_k141_28891 | Not-determined |
| 18-101_k141_28892 | Not-determined |
| 18-101_k141_110146 | Not-determined |
| 18-101_k141_128906 | Not-determined |
| 18-101_k141_28896 | Not-determined |
| 18-101_k141_340891 | Not-determined |
| 18-101_k141_128911 | Not-determined |
| 18-101_k141_194518 | Not-determined |
| 18-101_k141_12813 | Not-determined |
| 18-101_k141_374753 | Not-determined |
| 18-101_k141_211091 | Not-determined |
| 18-101_k141_374757 | Not-determined |
| 18-101_k141_77716 | Not-determined |
| 18-101_k141_194522 | Not-determined |
| 18-101_k141_95174 | Not-determined |
| 18-101_k141_211098 | Not-determined |
| 18-101_k141_243497 | Not-determined |
| 18-101_k141_110164 | Not-determined |
| 18-101_k141_194534 | Not-determined |
| 18-101_k141_309719 | Not-determined |
| 18-101_k141_374765 | Not-determined |
| 18-101_k141_77736 | Not-determined |
| 18-101_k141_28916 | Not-determined |
| 18-101_k141_77740 | Not-determined |
| 18-101_k141_62183 | Not-determined |
| 18-101_k141_340907 | Not-determined |
| 18-101_k141_160035 | Not-determined |
| 18-101_k141_62191 | Not-determined |
| 18-101_k141_160045 | Not-determined |
| 18-101_k141_326554 | Not-determined |
| 18-101_k141_128933 | Not-determined |
| 18-101_k141_144453 | Not-determined |
| 18-101_k141_275744 | Not-determined |
| 18-101_k141_243512 | Not-determined |
| 18-101_k141_62201 | Not-determined |
| 18-101_k141_194553 | Not-determined |
| 18-101_k141_340922 | Low-quality |
| 18-101_k141_340924 | Not-determined |
| 18-101_k141_226917 | Not-determined |
| 18-101_k141_226919 | Not-determined |
| 18-101_k141_62208 | Not-determined |
| 18-101_k141_194560 | Not-determined |
| 18-101_k141_309737 | Not-determined |
| 18-101_k141_12846 | Not-determined |
| 18-101_k141_391059 | Not-determined |
| 18-101_k141_359285 | Not-determined |
| 18-101_k141_309742 | Low-quality |
| 18-101_k141_243520 | Not-determined |
| 18-101_k141_211121 | Not-determined |
| 18-101_k141_95196 | Not-determined |
| 18-101_k141_95197 | Not-determined |
| 18-101_k141_293153 | Not-determined |
| 18-101_k141_326572 | Not-determined |
| 18-101_k141_359287 | Low-quality |
| 18-101_k141_243522 | Not-determined |
| 18-101_k141_95207 | Not-determined |
| 18-101_k141_326581 | Not-determined |
| 18-101_k141_391073 | Not-determined |
| 18-101_k141_243526 | Not-determined |
| 18-101_k141_243530 | Not-determined |
| 18-101_k141_293172 | Not-determined |
| 18-101_k141_309751 | Not-determined |
| 18-101_k141_243532 | Low-quality |
| 18-101_k141_391077 | Not-determined |
| 18-101_k141_211149 | Not-determined |
| 18-101_k141_260821 | Low-quality |
| 18-101_k141_211153 | Not-determined |
| 18-101_k141_326597 | Not-determined |
| 18-101_k141_211158 | Not-determined |
| 18-101_k141_211160 | Not-determined |
| 18-101_k141_275765 | Low-quality |
| 18-101_k141_243544 | Not-determined |
| 18-101_k141_160079 | Not-determined |
| 18-101_k141_243548 | Not-determined |
| 18-101_k141_28965 | Low-quality |
| 18-101_k141_374809 | Not-determined |
| 18-101_k141_144496 | Not-determined |
| 18-101_k141_326604 | Not-determined |
| 18-101_k141_243550 | Not-determined |
| 18-101_k141_243551 | Not-determined |
| 18-101_k141_359304 | Not-determined |
| 18-101_k141_28971 | Not-determined |
| 18-101_k141_275774 | Not-determined |
| 18-101_k141_340935 | Not-determined |
| 18-101_k141_260827 | Not-determined |
| 18-101_k141_28977 | Not-determined |
| 18-101_k141_326612 | Not-determined |
| 18-101_k141_243563 | Not-determined |
| 18-101_k141_128951 | Not-determined |
| 18-101_k141_243564 | Not-determined |
| 18-101_k141_95237 | Not-determined |
| 18-101_k141_293187 | Not-determined |
| 18-101_k141_243566 | Not-determined |
| 18-101_k141_293188 | Not-determined |
| 18-101_k141_226951 | Not-determined |
| 18-101_k141_326617 | Not-determined |
| 18-101_k141_95242 | Low-quality |
| 18-101_k141_326624 | Not-determined |
| 18-101_k141_226960 | Not-determined |
| 18-101_k141_194618 | Not-determined |
| 18-101_k141_95258 | Not-determined |
| 18-101_k141_110170 | Not-determined |
| 18-101_k141_45347 | Not-determined |
| 18-101_k141_28990 | Low-quality |
| 18-101_k141_194630 | Not-determined |
| 18-101_k141_226972 | Not-determined |
| 18-101_k141_144501 | Not-determined |
| 18-101_k141_211189 | Not-determined |
| 18-101_k141_243585 | Not-determined |
| 18-101_k141_128959 | Not-determined |
| 18-101_k141_211190 | Not-determined |
| 18-101_k141_28994 | Not-determined |
| 18-101_k141_243588 | Not-determined |
| 18-101_k141_110183 | Not-determined |
| 18-101_k141_340946 | Not-determined |
| 18-101_k141_243591 | Not-determined |
| 18-101_k141_340954 | Not-determined |
| 18-101_k141_176047 | Not-determined |
| 18-101_k141_194639 | Not-determined |
| 18-101_k141_128965 | Not-determined |
| 18-101_k141_226990 | Not-determined |
| 18-101_k141_110199 | Low-quality |
| 18-101_k141_243593 | Not-determined |
| 18-101_k141_374832 | Not-determined |
| 18-101_k141_110202 | Not-determined |
| 18-101_k141_77801 | Low-quality |
| 18-101_k141_275811 | Not-determined |
| 18-101_k141_309803 | Not-determined |
| 18-101_k141_95285 | Not-determined |
| 18-101_k141_62267 | Not-determined |
| 18-101_k141_95288 | Not-determined |
| 18-101_k141_29036 | Not-determined |
| 18-101_k141_62271 | Not-determined |
| 18-101_k141_243609 | Not-determined |
| 18-101_k141_340981 | Not-determined |
| 18-101_k141_144533 | Not-determined |
| 18-101_k141_391125 | Low-quality |
| 18-101_k141_29042 | Not-determined |
| 18-101_k141_260869 | Not-determined |
| 18-101_k141_243612 | Not-determined |
| 18-101_k141_45375 | Not-determined |
| 18-101_k141_62276 | Not-determined |
| 18-101_k141_95296 | Not-determined |
| 18-101_k141_359342 | Low-quality |
| 18-101_k141_340983 | Not-determined |
| 18-101_k141_374848 | Not-determined |
| 18-101_k141_160110 | Low-quality |
| 18-101_k141_62286 | Not-determined |
| 18-101_k141_260878 | Not-determined |
| 18-101_k141_45377 | Not-determined |
| 18-101_k141_176064 | Not-determined |
| 18-101_k141_374856 | Not-determined |
| 18-101_k141_243617 | Not-determined |
| 18-101_k141_176068 | Not-determined |
| 18-101_k141_260883 | Not-determined |
| 18-101_k141_176069 | Not-determined |
| 18-101_k141_160112 | Not-determined |
| 18-101_k141_359357 | Not-determined |
| 18-101_k141_359360 | Not-determined |
| 18-101_k141_176073 | Not-determined |
| 18-101_k141_95310 | Not-determined |
| 18-101_k141_194648 | Not-determined |
| 18-101_k141_160120 | Not-determined |
| 18-101_k141_293213 | Not-determined |
| 18-101_k141_128992 | Low-quality |
| 18-101_k141_12907 | Not-determined |
| 18-101_k141_293216 | Not-determined |
| 18-101_k141_326640 | Not-determined |
| 18-101_k141_160128 | Not-determined |
| 18-101_k141_160129 | Not-determined |
| 18-101_k141_129001 | Not-determined |
| 18-101_k141_293223 | Not-determined |
| 18-101_k141_326641 | Not-determined |
| 18-101_k141_293225 | Not-determined |
| 18-101_k141_326642 | Not-determined |
| 18-101_k141_341005 | Not-determined |
| 18-101_k141_260906 | Not-determined |
| 18-101_k141_326647 | Not-determined |
| 18-101_k141_374869 | Not-determined |
| 18-101_k141_95327 | Not-determined |
| 18-101_k141_227005 | Not-determined |
| 18-101_k141_260908 | Not-determined |
| 18-101_k141_95329 | Not-determined |
| 18-101_k141_374873 | Not-determined |
| 18-101_k141_391141 | Not-determined |
| 18-101_k141_326654 | Not-determined |
| 18-101_k141_260912 | Not-determined |
| 18-101_k141_326657 | Not-determined |
| 18-101_k141_326659 | Not-determined |
| 18-101_k141_293234 | Not-determined |
| 18-101_k141_341018 | Not-determined |
| 18-101_k141_326662 | Not-determined |
| 18-101_k141_62317 | Not-determined |
| 18-101_k141_243645 | Not-determined |
| 18-101_k141_160136 | Low-quality |
| 18-101_k141_243646 | Low-quality |
| 18-101_k141_160137 | Not-determined |
| 18-101_k141_62320 | Not-determined |
| 18-101_k141_293242 | Not-determined |
| 18-101_k141_243653 | Not-determined |
| 18-101_k141_176091 | Not-determined |
| 18-101_k141_260919 | Not-determined |
| 18-101_k141_341030 | Not-determined |
| 18-101_k141_326669 | Not-determined |
| 18-101_k141_144544 | Low-quality |
| 18-101_k141_160154 | Not-determined |
| 18-101_k141_275854 | Not-determined |
| 18-101_k141_194685 | Low-quality |
| 18-101_k141_144559 | Not-determined |
| 18-101_k141_326672 | Not-determined |
| 18-101_k141_374905 | Low-quality |
| 18-101_k141_391159 | Not-determined |
| 18-101_k141_391160 | Not-determined |
| 18-101_k141_391161 | Not-determined |
| 18-101_k141_160167 | Not-determined |
| 18-101_k141_95367 | Not-determined |
| 18-101_k141_326677 | Not-determined |
| 18-101_k141_391163 | Not-determined |
| 18-101_k141_160171 | Not-determined |
| 18-101_k141_227054 | Not-determined |
| 18-101_k141_341036 | Not-determined |
| 18-101_k141_95374 | Not-determined |
| 18-101_k141_176117 | Low-quality |
| 18-101_k141_260952 | Not-determined |
| 18-101_k141_359390 | Not-determined |
| 18-101_k141_129050 | Not-determined |
| 18-101_k141_129054 | Not-determined |
| 18-101_k141_95384 | Low-quality |
| 18-101_k141_95385 | Low-quality |
| 18-101_k141_260958 | Not-determined |
| 18-101_k141_359395 | Not-determined |
| 18-101_k141_275874 | Not-determined |
| 18-101_k141_227069 | Not-determined |
| 18-101_k141_211257 | Not-determined |
| 18-101_k141_129062 | Not-determined |
| 18-101_k141_144578 | Not-determined |
| 18-101_k141_12935 | Not-determined |
| 18-101_k141_341039 | Not-determined |
| 18-101_k141_275878 | Not-determined |
| 18-101_k141_160179 | Not-determined |
| 18-101_k141_341040 | Not-determined |
| 18-101_k141_341041 | Not-determined |
| 18-101_k141_12943 | Not-determined |
| 18-101_k141_160186 | Low-quality |
| 18-101_k141_211265 | Not-determined |
| 18-101_k141_211266 | Not-determined |
| 18-101_k141_374920 | Low-quality |
| 18-101_k141_227080 | Not-determined |
| 18-101_k141_243702 | Not-determined |
| 18-101_k141_129074 | Not-determined |
| 18-101_k141_374926 | Not-determined |
| 18-101_k141_12948 | Not-determined |
| 18-101_k141_341047 | Not-determined |
| 18-101_k141_341048 | Not-determined |
| 18-101_k141_374930 | Not-determined |
| 18-101_k141_227085 | Not-determined |
| 18-101_k141_309830 | Not-determined |
| 18-101_k141_309831 | Not-determined |
| 18-101_k141_359405 | Not-determined |
| 18-101_k141_243709 | Not-determined |
| 18-101_k141_160195 | Not-determined |
| 18-101_k141_341057 | Low-quality |
| 18-101_k141_110298 | Not-determined |
| 18-101_k141_227093 | Not-determined |
| 18-101_k141_243717 | Not-determined |
| 18-101_k141_45399 | Not-determined |
| 18-101_k141_45403 | Low-quality |
| 18-101_k141_12953 | Not-determined |
| 18-101_k141_62358 | Not-determined |
| 18-101_k141_374939 | Not-determined |
| 18-101_k141_12957 | Low-quality |
| 18-101_k141_194706 | Not-determined |
| 18-101_k141_110307 | Not-determined |
| 18-101_k141_326690 | Not-determined |
| 18-101_k141_374942 | Not-determined |
| 18-101_k141_110309 | Not-determined |
| 18-101_k141_275891 | Not-determined |
| 18-101_k141_176153 | Not-determined |
| 18-101_k141_326693 | Not-determined |
| 18-101_k141_110311 | Not-determined |
| 18-101_k141_194709 | Not-determined |
| 18-101_k141_160212 | Not-determined |
| 18-101_k141_194715 | Not-determined |
| 18-101_k141_260983 | Low-quality |
| 18-101_k141_176158 | Not-determined |
| 18-101_k141_309849 | Low-quality |
| 18-101_k141_160224 | Not-determined |
| 18-101_k141_341083 | Not-determined |
| 18-101_k141_341085 | Not-determined |
| 18-101_k141_275904 | Not-determined |
| 18-101_k141_29050 | Not-determined |
| 18-101_k141_129101 | Not-determined |
| 18-101_k141_326712 | Not-determined |
| 18-101_k141_129105 | Not-determined |
| 18-101_k141_211308 | Not-determined |
| 18-101_k141_194740 | Not-determined |
| 18-101_k141_77822 | Not-determined |
| 18-101_k141_194742 | Not-determined |
| 18-101_k141_160239 | Low-quality |
| 18-101_k141_309862 | Not-determined |
| 18-101_k141_309864 | Not-determined |
| 18-101_k141_326727 | Not-determined |
| 18-101_k141_77828 | Not-determined |
| 18-101_k141_77829 | Not-determined |
| 18-101_k141_326729 | Not-determined |
| 18-101_k141_160242 | Low-quality |
| 18-101_k141_309867 | Not-determined |

| | |
|---|---|
| 18-101_k141_293294 | Not-determined |
| 18-101_k141_29063 | Not-determined |
| 18-101_k141_144628 | Not-determined |
| 18-101_k141_326733 | Not-determined |
| 18-101_k141_275920 | Not-determined |
| 18-101_k141_110334 | Not-determined |
| 18-101_k141_176185 | Not-determined |
| 18-101_k141_110335 | Not-determined |
| 18-101_k141_211320 | Not-determined |
| 18-101_k141_129126 | Not-determined |
| 18-101_k141_62366 | Not-determined |
| 18-101_k141_309870 | Low-quality |
| 18-101_k141_309871 | Not-determined |
| 18-101_k141_129129 | Not-determined |
| 18-101_k141_129131 | Not-determined |
| 18-101_k141_275930 | Not-determined |
| 18-101_k141_391166 | Not-determined |
| 18-101_k141_261029 | Not-determined |
| 18-101_k141_62367 | Not-determined |
| 18-101_k141_29073 | Low-quality |
| 18-101_k141_129136 | Not-determined |
| 18-101_k141_359432 | Not-determined |
| 18-101_k141_243761 | Not-determined |
| 18-101_k141_211331 | Not-determined |
| 18-101_k141_211335 | Not-determined |
| 18-101_k141_29086 | Not-determined |
| 18-101_k141_110356 | Not-determined |
| 18-101_k141_391175 | Low-quality |
| 18-101_k141_144635 | Not-determined |
| 18-101_k141_144636 | Low-quality |
| 18-101_k141_309888 | Not-determined |
| 18-101_k141_243767 | Not-determined |
| 18-101_k141_110359 | Not-determined |
| 18-101_k141_374957 | Not-determined |
| 18-101_k141_391178 | Not-determined |
| 18-101_k141_144643 | Low-quality |
| 18-101_k141_160256 | Not-determined |
| 18-101_k141_160258 | Not-determined |
| 18-101_k141_243773 | Not-determined |
| 18-101_k141_45449 | Not-determined |
| 18-101_k141_309895 | Not-determined |
| 18-101_k141_62371 | Not-determined |
| 18-101_k141_359450 | Low-quality |
| 18-101_k141_77863 | Not-determined |
| 18-101_k141_77865 | Not-determined |
| 18-101_k141_309905 | Not-determined |
| 18-101_k141_77867 | Not-determined |
| 18-101_k141_176207 | Not-determined |
| 18-101_k141_194785 | Not-determined |
| 18-101_k141_29107 | Not-determined |
| 18-101_k141_144660 | Not-determined |
| 18-101_k141_194788 | Low-quality |
| 18-101_k141_326762 | Not-determined |
| 18-101_k141_293303 | Low-quality |
| 18-101_k141_129161 | Not-determined |
| 18-101_k141_227165 | Not-determined |
| 18-101_k141_309914 | Not-determined |
| 18-101_k141_359459 | Not-determined |
| 18-101_k141_144671 | Not-determined |
| 18-101_k141_227169 | Low-quality |
| 18-101_k141_45470 | Not-determined |
| 18-101_k141_45471 | Not-determined |
| 18-101_k141_261062 | Not-determined |
| 18-101_k141_374967 | Not-determined |
| 18-101_k141_160288 | Not-determined |
| 18-101_k141_211366 | Not-determined |
| 18-101_k141_359469 | Not-determined |
| 18-101_k141_293313 | Not-determined |
| 18-101_k141_374974 | Not-determined |
| 18-101_k141_326769 | Not-determined |
| 18-101_k141_261074 | Not-determined |
| 18-101_k141_129168 | Low-quality |
| 18-101_k141_359475 | Not-determined |
| 18-101_k141_129170 | Low-quality |
| 18-101_k141_77888 | Not-determined |
| 18-101_k141_176227 | Not-determined |
| 18-101_k141_359482 | Not-determined |
| 18-101_k141_211379 | Not-determined |
| 18-101_k141_261083 | Not-determined |
| 18-101_k141_129179 | Not-determined |
| 18-101_k141_29140 | Not-determined |
| 18-101_k141_77901 | Low-quality |
| 18-101_k141_129182 | Not-determined |
| 18-101_k141_77905 | Low-quality |
| 18-101_k141_129191 | Not-determined |
| 18-101_k141_29149 | Not-determined |
| 18-101_k141_129193 | Not-determined |
| 18-101_k141_261092 | Not-determined |
| 18-101_k141_129197 | Not-determined |
| 18-101_k141_211393 | Not-determined |
| 18-101_k141_160305 | Not-determined |
| 18-101_k141_309922 | Not-determined |
| 18-101_k141_176251 | Not-determined |
| 18-101_k141_293345 | Not-determined |
| 18-101_k141_326796 | Not-determined |
| 18-101_k141_13049 | Not-determined |
| 18-101_k141_243828 | Not-determined |
| 18-101_k141_341120 | Low-quality |
| 18-101_k141_309927 | Not-determined |
| 18-101_k141_341122 | Not-determined |
| 18-101_k141_160311 | Not-determined |
| 18-101_k141_293349 | Not-determined |
| 18-101_k141_13051 | Not-determined |
| 18-101_k141_160313 | Not-determined |
| 18-101_k141_176259 | Not-determined |
| 18-101_k141_129218 | Not-determined |
| 18-101_k141_95415 | Not-determined |
| 18-101_k141_176268 | Not-determined |
| 18-101_k141_391221 | Not-determined |
| 18-101_k141_243849 | Not-determined |
| 18-101_k141_341137 | Low-quality |
| 18-101_k141_391222 | Not-determined |
| 18-101_k141_243853 | Not-determined |
| 18-101_k141_359496 | Not-determined |
| 18-101_k141_359497 | Not-determined |
| 18-101_k141_359498 | Not-determined |
| 18-101_k141_326819 | Not-determined |
| 18-101_k141_375025 | Not-determined |
| 18-101_k141_45502 | Not-determined |
| 18-101_k141_375027 | Not-determined |
| 18-101_k141_391227 | Not-determined |
| 18-101_k141_293362 | Not-determined |
| 18-101_k141_176274 | Not-determined |
| 18-101_k141_13085 | Low-quality |
| 18-101_k141_13086 | Not-determined |
| 18-101_k141_309946 | Not-determined |
| 18-101_k141_160341 | Not-determined |
| 18-101_k141_243865 | Not-determined |
| 18-101_k141_293372 | Not-determined |
| 18-101_k141_391243 | Not-determined |
| 18-101_k141_77938 | Not-determined |
| 18-101_k141_359516 | Not-determined |
| 18-101_k141_95434 | Not-determined |
| 18-101_k141_391247 | Not-determined |
| 18-101_k141_309950 | Not-determined |
| 18-101_k141_160345 | Not-determined |
| 18-101_k141_375042 | Not-determined |
| 18-101_k141_309954 | Not-determined |
| 18-101_k141_391250 | Not-determined |
| 18-101_k141_243872 | Not-determined |
| 18-101_k141_144719 | Not-determined |
| 18-101_k141_309960 | Not-determined |
| 18-101_k141_95449 | Not-determined |
| 18-101_k141_227193 | Not-determined |
| 18-101_k141_243885 | Not-determined |
| 18-101_k141_359530 | Not-determined |
| 18-101_k141_176293 | Not-determined |
| 18-101_k141_359531 | Not-determined |
| 18-101_k141_243890 | Not-determined |
| 18-101_k141_309963 | Low-quality |
| 18-101_k141_326835 | Not-determined |
| 18-101_k141_326840 | Not-determined |
| 18-101_k141_326841 | Not-determined |
| 18-101_k141_359534 | Not-determined |
| 18-101_k141_77952 | Not-determined |
| 18-101_k141_144730 | Not-determined |
| 18-101_k141_375056 | Not-determined |
| 18-101_k141_110424 | Not-determined |
| 18-101_k141_275954 | Not-determined |
| 18-101_k141_243899 | Not-determined |
| 18-101_k141_275956 | Low-quality |
| 18-101_k141_176306 | Not-determined |
| 18-101_k141_391259 | Low-quality |
| 18-101_k141_95461 | Not-determined |
| 18-101_k141_144740 | Not-determined |
| 18-101_k141_375061 | Not-determined |
| 18-101_k141_211441 | Not-determined |
| 18-101_k141_194833 | Not-determined |
| 18-101_k141_144744 | Not-determined |
| 18-101_k141_194834 | Not-determined |
| 18-101_k141_129248 | Not-determined |
| 18-101_k141_160374 | Not-determined |
| 18-101_k141_129249 | Not-determined |
| 18-101_k141_243905 | Not-determined |
| 18-101_k141_194844 | Not-determined |
| 18-101_k141_194846 | Low-quality |
| 18-101_k141_391275 | Not-determined |
| 18-101_k141_243907 | Not-determined |
| 18-101_k141_391276 | Not-determined |
| 18-101_k141_375072 | Not-determined |
| 18-101_k141_95476 | Low-quality |
| 18-101_k141_227214 | Low-quality |
| 18-101_k141_227218 | Not-determined |
| 18-101_k141_375081 | Not-determined |
| 18-101_k141_13101 | Not-determined |
| 18-101_k141_160388 | Not-determined |
| 18-101_k141_275984 | Not-determined |
| 18-101_k141_176309 | Not-determined |
| 18-101_k141_95485 | Not-determined |
| 18-101_k141_13104 | Not-determined |
| 18-101_k141_391283 | Low-quality |
| 18-101_k141_211466 | Not-determined |
| 18-101_k141_309989 | Not-determined |
| 18-101_k141_326877 | Not-determined |
| 18-101_k141_391284 | Not-determined |
| 18-101_k141_211467 | Not-determined |
| 18-101_k141_275989 | Low-quality |
| 18-101_k141_176319 | Not-determined |
| 18-101_k141_211469 | Not-determined |
| 18-101_k141_194856 | Not-determined |
| 18-101_k141_194857 | Not-determined |
| 18-101_k141_45552 | Not-determined |
| 18-101_k141_375093 | Not-determined |
| 18-101_k141_243920 | Not-determined |
| 18-101_k141_375105 | Not-determined |
| 18-101_k141_144774 | Not-determined |
| 18-101_k141_45566 | Not-determined |
| 18-101_k141_194871 | Not-determined |
| 18-101_k141_45570 | Not-determined |
| 18-101_k141_326895 | Not-determined |
| 18-101_k141_45573 | Not-determined |
| 18-101_k141_293391 | Not-determined |
| 18-101_k141_391292 | Not-determined |
| 18-101_k141_326902 | Not-determined |
| 18-101_k141_391294 | Not-determined |
| 18-101_k141_129294 | Not-determined |
| 18-101_k141_293400 | Not-determined |
| 18-101_k141_310026 | Not-determined |
| 18-101_k141_310027 | Not-determined |
| 18-101_k141_129301 | Not-determined |
| 18-101_k141_293406 | Not-determined |
| 18-101_k141_160400 | Not-determined |
| 18-101_k141_194890 | Not-determined |
| 18-101_k141_341190 | Not-determined |
| 18-101_k141_110502 | Low-quality |
| 18-101_k141_144798 | Not-determined |
| 18-101_k141_110510 | Not-determined |
| 18-101_k141_227257 | Not-determined |
| 18-101_k141_276017 | Not-determined |
| 18-101_k141_326920 | Not-determined |
| 18-101_k141_227260 | Not-determined |
| 18-101_k141_211507 | Not-determined |
| 18-101_k141_129314 | Not-determined |
| 18-101_k141_227264 | Not-determined |
| 18-101_k141_13142 | Not-determined |
| 18-101_k141_160417 | Not-determined |
| 18-101_k141_160423 | Not-determined |
| 18-101_k141_194915 | Not-determined |
| 18-101_k141_276040 | Not-determined |
| 18-101_k141_227274 | Not-determined |
| 18-101_k141_243950 | Not-determined |
| 18-101_k141_29170 | Not-determined |
| 18-101_k141_160434 | Not-determined |
| 18-101_k141_293423 | Low-quality |
| 18-101_k141_29173 | Not-determined |
| 18-101_k141_227280 | Low-quality |
| 18-101_k141_110519 | Not-determined |
| 18-101_k141_110523 | Not-determined |
| 18-101_k141_276051 | Low-quality |
| 18-101_k141_276052 | Not-determined |
| 18-101_k141_341226 | Not-determined |
| 18-101_k141_45592 | Not-determined |
| 18-101_k141_293431 | Not-determined |
| 18-101_k141_45593 | Not-determined |
| 18-101_k141_375142 | Not-determined |
| 18-101_k141_13155 | Not-determined |
| 18-101_k141_95513 | Not-determined |
| 18-101_k141_45595 | Not-determined |
| 18-101_k141_176380 | Not-determined |
| 18-101_k141_144828 | Low-quality |
| 18-101_k141_310064 | Not-determined |
| 18-101_k141_144830 | Not-determined |
| 18-101_k141_176386 | Not-determined |
| 18-101_k141_276063 | Not-determined |
| 18-101_k141_227293 | Not-determined |
| 18-101_k141_110540 | Not-determined |
| 18-101_k141_13160 | Not-determined |
| 18-101_k141_243960 | Not-determined |
| 18-101_k141_110545 | Not-determined |
| 18-101_k141_227298 | Not-determined |
| 18-101_k141_375151 | Not-determined |
| 18-101_k141_341240 | Not-determined |
| 18-101_k141_375157 | Not-determined |
| 18-101_k141_176396 | Not-determined |
| 18-101_k141_293451 | Low-quality |
| 18-101_k141_129361 | Not-determined |
| 18-101_k141_194940 | Not-determined |
| 18-101_k141_227312 | Not-determined |
| 18-101_k141_391318 | Low-quality |
| 18-101_k141_326929 | Not-determined |
| 18-101_k141_129364 | Not-determined |
| 18-101_k141_227319 | Not-determined |
| 18-101_k141_243965 | Not-determined |
| 18-101_k141_326930 | Not-determined |
| 18-101_k141_160488 | Not-determined |
| 18-101_k141_261259 | Not-determined |
| 18-101_k141_391325 | Not-determined |
| 18-101_k141_144861 | Not-determined |
| 18-101_k141_129374 | Low-quality |
| 18-101_k141_144862 | Not-determined |
| 18-101_k141_176397 | Not-determined |
| 18-101_k141_13180 | Not-determined |
| 18-101_k141_375185 | Not-determined |
| 18-101_k141_144868 | Not-determined |
| 18-101_k141_176404 | Not-determined |
| 18-101_k141_243978 | Not-determined |
| 18-101_k141_326941 | Not-determined |
| 18-101_k141_276104 | Not-determined |
| 18-101_k141_211560 | Not-determined |
| 18-101_k141_129387 | Not-determined |
| 18-101_k141_243987 | Not-determined |
| 18-101_k141_110566 | Not-determined |
| 18-101_k141_341274 | Not-determined |
| 18-101_k141_276111 | Low-quality |
| 18-101_k141_129393 | Not-determined |
| 18-101_k141_194973 | Not-determined |
| 18-101_k141_194975 | Not-determined |
| 18-101_k141_129397 | Not-determined |
| 18-101_k141_310108 | Not-determined |
| 18-101_k141_211572 | Not-determined |
| 18-101_k141_310109 | Not-determined |
| 18-101_k141_341286 | Low-quality |
| 18-101_k141_95526 | Not-determined |
| 18-101_k141_129402 | Not-determined |
| 18-101_k141_341289 | Not-determined |
| 18-101_k141_310113 | Not-determined |
| 18-101_k141_276117 | Not-determined |
| 18-101_k141_95529 | Not-determined |
| 18-101_k141_276120 | Not-determined |
| 18-101_k141_341291 | Not-determined |
| 18-101_k141_194988 | Not-determined |
| 18-101_k141_211583 | Not-determined |
| 18-101_k141_341297 | Low-quality |
| 18-101_k141_211588 | Not-determined |
| 18-101_k141_310117 | Not-determined |
| 18-101_k141_95538 | Not-determined |
| 18-101_k141_95540 | Not-determined |
| 18-101_k141_45620 | Not-determined |
| 18-101_k141_95542 | Not-determined |
| 18-101_k141_129411 | Low-quality |
| 18-101_k141_326967 | Not-determined |
| 18-101_k141_45623 | Not-determined |
| 18-101_k141_45624 | Not-determined |
| 18-101_k141_13211 | Not-determined |
| 18-101_k141_13212 | Not-determined |
| 18-101_k141_195000 | Not-determined |
| 18-101_k141_326971 | Not-determined |
| 18-101_k141_211600 | Not-determined |
| 18-101_k141_129421 | Low-quality |
| 18-101_k141_211605 | Not-determined |
| 18-101_k141_78008 | Not-determined |
| 18-101_k141_326974 | Low-quality |
| 18-101_k141_244006 | Not-determined |
| 18-101_k141_13224 | Low-quality |
| 18-101_k141_326977 | Low-quality |
| 18-101_k141_13227 | Not-determined |
| 18-101_k141_326979 | Not-determined |
| 18-101_k141_341318 | Not-determined |
| 18-101_k141_326980 | Not-determined |
| 18-101_k141_341319 | Not-determined |
| 18-101_k141_95552 | Not-determined |
| 18-101_k141_160547 | Not-determined |
| 18-101_k141_261302 | Not-determined |
| 18-101_k141_261303 | Not-determined |
| 18-101_k141_276134 | Not-determined |
| 18-101_k141_244021 | Low-quality |
| 18-101_k141_391340 | Not-determined |
| 18-101_k141_244022 | Not-determined |
| 18-101_k141_244025 | Not-determined |
| 18-101_k141_244026 | Low-quality |
| 18-101_k141_129442 | Low-quality |
| 18-101_k141_244028 | Not-determined |
| 18-101_k141_375238 | Not-determined |
| 18-101_k141_45648 | Not-determined |
| 18-101_k141_391346 | Not-determined |
| 18-101_k141_176436 | Not-determined |
| 18-101_k141_227323 | Not-determined |
| 18-101_k141_293532 | Not-determined |
| 18-101_k141_176441 | Low-quality |
| 18-101_k141_276151 | Not-determined |
| 18-101_k141_176444 | Not-determined |
| 18-101_k141_327001 | Not-determined |
| 18-101_k141_176445 | Not-determined |
| 18-101_k141_176447 | Not-determined |
| 18-101_k141_110637 | Low-quality |
| 18-101_k141_129459 | Not-determined |
| 18-101_k141_13238 | Low-quality |
| 18-101_k141_211646 | Not-determined |
| 18-101_k141_293547 | Not-determined |
| 18-101_k141_227344 | Not-determined |
| 18-101_k141_129464 | Not-determined |
| 18-101_k141_176457 | Not-determined |
| 18-101_k141_176458 | Not-determined |
| 18-101_k141_29189 | Low-quality |
| 18-101_k141_227348 | Not-determined |
| 18-101_k141_391379 | Not-determined |
| 18-101_k141_144908 | Not-determined |
| 18-101_k141_211662 | Not-determined |
| 18-101_k141_211664 | Not-determined |
| 18-101_k141_144913 | Not-determined |
| 18-101_k141_310155 | Not-determined |
| 18-101_k141_45679 | Low-quality |
| 18-101_k141_391384 | Not-determined |
| 18-101_k141_110657 | Not-determined |
| 18-101_k141_375254 | Low-quality |
| 18-101_k141_211670 | Not-determined |
| 18-101_k141_327017 | Not-determined |
| 18-101_k141_276175 | Low-quality |
| 18-101_k141_341347 | Not-determined |
| 18-101_k141_359568 | Not-determined |
| 18-101_k141_261322 | Not-determined |
| 18-101_k141_62465 | Not-determined |
| 18-101_k141_129479 | Not-determined |
| 18-101_k141_391391 | Low-quality |
| 18-101_k141_293568 | Not-determined |
| 18-101_k141_144924 | Not-determined |
| 18-101_k141_359575 | Not-determined |
| 18-101_k141_160591 | Not-determined |
| 18-101_k141_110672 | Not-determined |
| 18-101_k141_359580 | Not-determined |
| 18-101_k141_276180 | Low-quality |
| 18-101_k141_293577 | Not-determined |
| 18-101_k141_211683 | Not-determined |
| 18-101_k141_391399 | Not-determined |
| 18-101_k141_227371 | Not-determined |
| 18-101_k141_211689 | Not-determined |
| 18-101_k141_129486 | Low-quality |
| 18-101_k141_391403 | Low-quality |
| 18-101_k141_62478 | Not-determined |
| 18-101_k141_195003 | Not-determined |
| 18-101_k141_62481 | Low-quality |
| 18-101_k141_95586 | Not-determined |
| 18-101_k141_211693 | Low-quality |
| 18-101_k141_62485 | Not-determined |
| 18-101_k141_359582 | Not-determined |
| 18-101_k141_293585 | Not-determined |
| 18-101_k141_261333 | Not-determined |
| 18-101_k141_391412 | Not-determined |
| 18-101_k141_359584 | Not-determined |
| 18-101_k141_110686 | Not-determined |
| 18-101_k141_144939 | Not-determined |
| 18-101_k141_359587 | Low-quality |
| 18-101_k141_13270 | Low-quality |
| 18-101_k141_129494 | Not-determined |
| 18-101_k141_341372 | Not-determined |
| 18-101_k141_211702 | Not-determined |
| 18-101_k141_227380 | Not-determined |
| 18-101_k141_276193 | Not-determined |
| 18-101_k141_160595 | Not-determined |
| 18-101_k141_244087 | Not-determined |
| 18-101_k141_244091 | Not-determined |
| 18-101_k141_341381 | Not-determined |
| 18-101_k141_341383 | Not-determined |
| 18-101_k141_359596 | Low-quality |
| 18-101_k141_160599 | Not-determined |
| 18-101_k141_211715 | Not-determined |
| 18-101_k141_391427 | Not-determined |
| 18-101_k141_391429 | Not-determined |
| 18-101_k141_160603 | Not-determined |
| 18-101_k141_45686 | Not-determined |
| 18-101_k141_129506 | Not-determined |
| 18-101_k141_95596 | Not-determined |
| 18-101_k141_160607 | Not-determined |
| 18-101_k141_359606 | Not-determined |
| 18-101_k141_95598 | Low-quality |
| 18-101_k141_375293 | Not-determined |
| 18-101_k141_359614 | Not-determined |
| 18-101_k141_129513 | Not-determined |
| 18-101_k141_375295 | Low-quality |
| 18-101_k141_375298 | Not-determined |
| 18-101_k141_359620 | Not-determined |
| 18-101_k141_160613 | Not-determined |
| 18-101_k141_29194 | Not-determined |
| 18-101_k141_375301 | Not-determined |
| 18-101_k141_110703 | Not-determined |
| 18-101_k141_29198 | Low-quality |
| 18-101_k141_359628 | Not-determined |
| 18-101_k141_276226 | Not-determined |
| 18-101_k141_195041 | Not-determined |
| 18-101_k141_359629 | Not-determined |
| 18-101_k141_45699 | Not-determined |
| 18-101_k141_359634 | Not-determined |
| 18-101_k141_276231 | Not-determined |
| 18-101_k141_391452 | Not-determined |
| 18-101_k141_227400 | Not-determined |
| 18-101_k141_62519 | Not-determined |
| 18-101_k141_211740 | Not-determined |
| 18-101_k141_62527 | Not-determined |
| 18-101_k141_129546 | Not-determined |
| 18-101_k141_129547 | Not-determined |
| 18-101_k141_211744 | Low-quality |
| 18-101_k141_129549 | Low-quality |
| 18-101_k141_45716 | Not-determined |
| 18-101_k141_160636 | Not-determined |
| 18-101_k141_62539 | Not-determined |
| 18-101_k141_195069 | Not-determined |
| 18-101_k141_144995 | Not-determined |
| 18-101_k141_160639 | Not-determined |
| 18-101_k141_160640 | Low-quality |
| 18-101_k141_276255 | Not-determined |
| 18-101_k141_227424 | Not-determined |
| 18-101_k141_129563 | Not-determined |
| 18-101_k141_95633 | Not-determined |
| 18-101_k141_261379 | Not-determined |
| 18-101_k141_62557 | Not-determined |
| 18-101_k141_227429 | Not-determined |
| 18-101_k141_310210 | Not-determined |
| 18-101_k141_310212 | Not-determined |
| 18-101_k141_391476 | Not-determined |
| 18-101_k141_211753 | Not-determined |
| 18-101_k141_195088 | Not-determined |
| 18-101_k141_95639 | Not-determined |
| 18-101_k141_176502 | Not-determined |
| 18-101_k141_211757 | Not-determined |
| 18-101_k141_276263 | Not-determined |
| 18-101_k141_78042 | Not-determined |
| 18-101_k141_129587 | Not-determined |
| 18-101_k141_129589 | Not-determined |
| 18-101_k141_129593 | Not-determined |
| 18-101_k141_176510 | Low-quality |
| 18-101_k141_211769 | Low-quality |
| 18-101_k141_95656 | Not-determined |
| 18-101_k141_211770 | Not-determined |
| 18-101_k141_375336 | Not-determined |
| 18-101_k141_13334 | Not-determined |
| 18-101_k141_293625 | Not-determined |
| 18-101_k141_293629 | Not-determined |
| 18-101_k141_244189 | Not-determined |
| 18-101_k141_145025 | Not-determined |
| 18-101_k141_176515 | Not-determined |
| 18-101_k141_293631 | Low-quality |
| 18-101_k141_211791 | Not-determined |
| 18-101_k141_176518 | Not-determined |
| 18-101_k141_341420 | Not-determined |
| 18-101_k141_391485 | Not-determined |
| 18-101_k141_359674 | Not-determined |
| 18-101_k141_375347 | Low-quality |
| 18-101_k141_391489 | Low-quality |
| 18-101_k141_341426 | Not-determined |
| 18-101_k141_375353 | Not-determined |
| 18-101_k141_62589 | Low-quality |
| 18-101_k141_145042 | Not-determined |
| 18-101_k141_276282 | Not-determined |
| 18-101_k141_244191 | Not-determined |
| 18-101_k141_95676 | Not-determined |
| 18-101_k141_95677 | Not-determined |
| 18-101_k141_341437 | Not-determined |
| 18-101_k141_62594 | Not-determined |
| 18-101_k141_244196 | Not-determined |
| 18-101_k141_341440 | Not-determined |
| 18-101_k141_276289 | Not-determined |
| 18-101_k141_244200 | Not-determined |
| 18-101_k141_244203 | Not-determined |
| 18-101_k141_160691 | Not-determined |
| 18-101_k141_13341 | Not-determined |
| 18-101_k141_13343 | Not-determined |
| 18-101_k141_160695 | Not-determined |
| 18-101_k141_211808 | Not-determined |
| 18-101_k141_13345 | Not-determined |
| 18-101_k141_145064 | Not-determined |
| 18-101_k141_95687 | Not-determined |
| 18-101_k141_195134 | Not-determined |
| 18-101_k141_227475 | Not-determined |
| 18-101_k141_195140 | Not-determined |
| 18-101_k141_95692 | Not-determined |
| 18-101_k141_261393 | Not-determined |
| 18-101_k141_211819 | Not-determined |
| 18-101_k141_391502 | Not-determined |
| 18-101_k141_391503 | Not-determined |
| 18-101_k141_176554 | Not-determined |
| 18-101_k141_211824 | Not-determined |
| 18-101_k141_211825 | Not-determined |
| 18-101_k141_129624 | Not-determined |
| 18-101_k141_261401 | Not-determined |
| 18-101_k141_145077 | Not-determined |
| 18-101_k141_78066 | Not-determined |
| 18-101_k141_276317 | Not-determined |
| 18-101_k141_244221 | Not-determined |
| 18-101_k141_176572 | Not-determined |
| 18-101_k141_341471 | Low-quality |
| 18-101_k141_95726 | Low-quality |
| 18-101_k141_227508 | Low-quality |
| 18-101_k141_95734 | Not-determined |
| 18-101_k141_95735 | Not-determined |
| 18-101_k141_327027 | Not-determined |
| 18-101_k141_129637 | Not-determined |
| 18-101_k141_391515 | Not-determined |
| 18-101_k141_341483 | Not-determined |
| 18-101_k141_244239 | Low-quality |
| 18-101_k141_244240 | Not-determined |
| 18-101_k141_211830 | Not-determined |
| 18-101_k141_62623 | Not-determined |
| 18-101_k141_13386 | Not-determined |
| 18-101_k141_45749 | Not-determined |
| 18-101_k141_176593 | Not-determined |
| 18-101_k141_95750 | Not-determined |
| 18-101_k141_13388 | Not-determined |
| 18-101_k141_195158 | Not-determined |
| 18-101_k141_261441 | Not-determined |

| | |
|---|---|
| 18-101_k141_195160 | Not-determined |
| 18-101_k141_261450 | Not-determined |
| 18-101_k141_176603 | Not-determined |
| 18-101_k141_95757 | Low-quality |
| 18-101_k141_211846 | Low-quality |
| 18-101_k141_160733 | Low-quality |
| 18-101_k141_160739 | Not-determined |
| 18-101_k141_160741 | Not-determined |
| 18-101_k141_195166 | Not-determined |
| 18-101_k141_293684 | Not-determined |
| 18-101_k141_195168 | Not-determined |
| 18-101_k141_244255 | Not-determined |
| 18-101_k141_160749 | Not-determined |
| 18-101_k141_145105 | Not-determined |
| 18-101_k141_211859 | Not-determined |
| 18-101_k141_160753 | Low-quality |
| 18-101_k141_293696 | Not-determined |
| 18-101_k141_95764 | Not-determined |
| 18-101_k141_391538 | Not-determined |
| 18-101_k141_327042 | Not-determined |
| 18-101_k141_327043 | Not-determined |
| 18-101_k141_244264 | Not-determined |
| 18-101_k141_261477 | Not-determined |
| 18-101_k141_359698 | Not-determined |
| 18-101_k141_327052 | Not-determined |
| 18-101_k141_13398 | Not-determined |
| 18-101_k141_13400 | Not-determined |
| 18-101_k141_129676 | Not-determined |
| 18-101_k141_327059 | Not-determined |
| 18-101_k141_95781 | Not-determined |
| 18-101_k141_95785 | Not-determined |
| 18-101_k141_261492 | Not-determined |
| 18-101_k141_211888 | Not-determined |
| 18-101_k141_13408 | Not-determined |
| 18-101_k141_391549 | Low-quality |
| 18-101_k141_211894 | Not-determined |
| 18-101_k141_145114 | Not-determined |
| 18-101_k141_29208 | Not-determined |
| 18-101_k141_95793 | Not-determined |
| 18-101_k141_391552 | Not-determined |
| 18-101_k141_95795 | Not-determined |
| 18-101_k141_261501 | Low-quality |
| 18-101_k141_13414 | Not-determined |
| 18-101_k141_78133 | Not-determined |
| 18-101_k141_359726 | Not-determined |
| 18-101_k141_62638 | Low-quality |
| 18-101_k141_391556 | Not-determined |
| 18-101_k141_391559 | Not-determined |
| 18-101_k141_327075 | Not-determined |
| 18-101_k141_391560 | Not-determined |
| 18-101_k141_327077 | Not-determined |
| 18-101_k141_13418 | Low-quality |
| 18-101_k141_391561 | Not-determined |
| 18-101_k141_244302 | Not-determined |
| 18-101_k141_195191 | Low-quality |
| 18-101_k141_391565 | Not-determined |
| 18-101_k141_78153 | Not-determined |
| 18-101_k141_244305 | Not-determined |
| 18-101_k141_145139 | Low-quality |
| 18-101_k141_293732 | Not-determined |
| 18-101_k141_78160 | Not-determined |
| 18-101_k141_45790 | Not-determined |
| 18-101_k141_176627 | Not-determined |
| 18-101_k141_145149 | Not-determined |
| 18-101_k141_13428 | Not-determined |
| 18-101_k141_129695 | Not-determined |
| 18-101_k141_195193 | Low-quality |
| 18-101_k141_211937 | Not-determined |
| 18-101_k141_211938 | Not-determined |
| 18-101_k141_341530 | Not-determined |
| 18-101_k141_244321 | Not-determined |
| 18-101_k141_62648 | Not-determined |
| 18-101_k141_244325 | Not-determined |
| 18-101_k141_110753 | Low-quality |
| 18-101_k141_129700 | Low-quality |
| 18-101_k141_45797 | Not-determined |
| 18-101_k141_391569 | Low-quality |
| 18-101_k141_244329 | Not-determined |
| 18-101_k141_95834 | Not-determined |
| 18-101_k141_391571 | Not-determined |
| 18-101_k141_341544 | Not-determined |
| 18-101_k141_293772 | Not-determined |
| 18-101_k141_45805 | Not-determined |
| 18-101_k141_391576 | Not-determined |
| 18-101_k141_310220 | Not-determined |
| 18-101_k141_276430 | Not-determined |
| 18-101_k141_391578 | Not-determined |
| 18-101_k141_95842 | Not-determined |
| 18-101_k141_276432 | Not-determined |
| 18-101_k141_293779 | Not-determined |
| 18-101_k141_13445 | Not-determined |
| 18-101_k141_160798 | Low-quality |
| 18-101_k141_391581 | Not-determined |
| 18-101_k141_145166 | Not-determined |
| 18-101_k141_293781 | Not-determined |
| 18-101_k141_78182 | Low-quality |
| 18-101_k141_176650 | Not-determined |
| 18-101_k141_391583 | Not-determined |
| 18-101_k141_310227 | Not-determined |
| 18-101_k141_13452 | Low-quality |
| 18-101_k141_145172 | Not-determined |
| 18-101_k141_160804 | Not-determined |
| 18-101_k141_359773 | Not-determined |
| 18-101_k141_293785 | Not-determined |
| 18-101_k141_375388 | Not-determined |
| 18-101_k141_13463 | Not-determined |
| 18-101_k141_310235 | Not-determined |
| 18-101_k141_95852 | Low-quality |
| 18-101_k141_327107 | Not-determined |
| 18-101_k141_293794 | Not-determined |
| 18-101_k141_45816 | Not-determined |
| 18-101_k141_145187 | Not-determined |
| 18-101_k141_276456 | Not-determined |
| 18-101_k141_110762 | Not-determined |
| 18-101_k141_391599 | Not-determined |
| 18-101_k141_375395 | Not-determined |
| 18-101_k141_29263 | Low-quality |
| 18-101_k141_45823 | Not-determined |
| 18-101_k141_160819 | Not-determined |
| 18-101_k141_13483 | Low-quality |
| 18-101_k141_78203 | Not-determined |
| 18-101_k141_160822 | Not-determined |
| 18-101_k141_145192 | Not-determined |
| 18-101_k141_78208 | Not-determined |
| 18-101_k141_391603 | Not-determined |
| 18-101_k141_110769 | Not-determined |
| 18-101_k141_276461 | Not-determined |
| 18-101_k141_110771 | Not-determined |
| 18-101_k141_375398 | Not-determined |
| 18-101_k141_13490 | Not-determined |
| 18-101_k141_45830 | Not-determined |
| 18-101_k141_45831 | Low-quality |
| 18-101_k141_391609 | Not-determined |
| 18-101_k141_110779 | Not-determined |
| 18-101_k141_391613 | Not-determined |
| 18-101_k141_211955 | Not-determined |
| 18-101_k141_211956 | Not-determined |
| 18-101_k141_375404 | Not-determined |
| 18-101_k141_375405 | Not-determined |
| 18-101_k141_375406 | Not-determined |
| 18-101_k141_29277 | Not-determined |
| 18-101_k141_293807 | Not-determined |
| 18-101_k141_375411 | Not-determined |
| 18-101_k141_176677 | Not-determined |
| 18-101_k141_310265 | Not-determined |
| 18-101_k141_244354 | Not-determined |
| 18-101_k141_29282 | Not-determined |
| 18-101_k141_276475 | Low-quality |
| 18-101_k141_45842 | Not-determined |
| 18-101_k141_244357 | Not-determined |
| 18-101_k141_13502 | Not-determined |
| 18-101_k141_375418 | Not-determined |
| 18-101_k141_227544 | Not-determined |
| 18-101_k141_375422 | Not-determined |
| 18-101_k141_78232 | Not-determined |
| 18-101_k141_13506 | Not-determined |
| 18-101_k141_244363 | Not-determined |
| 18-101_k141_341599 | Not-determined |
| 18-101_k141_227547 | Low-quality |
| 18-101_k141_13512 | Not-determined |
| 18-101_k141_62702 | Not-determined |
| 18-101_k141_244370 | Not-determined |
| 18-101_k141_110811 | Not-determined |
| 18-101_k141_176690 | Not-determined |
| 18-101_k141_176692 | Not-determined |
| 18-101_k141_310288 | Not-determined |
| 18-101_k141_110815 | Not-determined |
| 18-101_k141_276495 | Not-determined |
| 18-101_k141_211978 | Not-determined |
| 18-101_k141_160847 | Low-quality |
| 18-101_k141_45860 | Not-determined |
| 18-101_k141_341607 | Not-determined |
| 18-101_k141_45861 | Not-determined |
| 18-101_k141_176703 | Not-determined |
| 18-101_k141_310295 | Not-determined |
| 18-101_k141_327161 | Not-determined |
| 18-101_k141_341616 | Not-determined |
| 18-101_k141_176706 | Not-determined |
| 18-101_k141_211989 | Not-determined |
| 18-101_k141_375449 | Not-determined |
| 18-101_k141_391659 | Not-determined |
| 18-101_k141_13535 | Not-determined |
| 18-101_k141_211991 | Not-determined |
| 18-101_k141_211993 | Not-determined |
| 18-101_k141_293847 | Not-determined |
| 18-101_k141_95887 | Not-determined |
| 18-101_k141_375450 | Not-determined |
| 18-101_k141_95889 | Not-determined |
| 18-101_k141_195265 | Not-determined |
| 18-101_k141_359786 | Not-determined |
| 18-101_k141_13545 | Not-determined |
| 18-101_k141_145247 | Not-determined |
| 18-101_k141_212002 | Not-determined |
| 18-101_k141_110831 | Not-determined |
| 18-101_k141_45885 | Not-determined |
| 18-101_k141_129754 | Not-determined |
| 18-101_k141_110834 | Not-determined |
| 18-101_k141_45887 | Not-determined |
| 18-101_k141_13552 | Not-determined |
| 18-101_k141_129758 | Not-determined |
| 18-101_k141_293858 | Not-determined |
| 18-101_k141_276525 | Not-determined |
| 18-101_k141_359803 | Not-determined |
| 18-101_k141_341624 | Not-determined |
| 18-101_k141_160879 | Not-determined |
| 18-101_k141_341625 | Not-determined |
| 18-101_k141_62722 | Not-determined |
| 18-101_k141_375475 | Not-determined |
| 18-101_k141_62730 | Not-determined |
| 18-101_k141_195289 | Not-determined |
| 18-101_k141_95923 | Not-determined |
| 18-101_k141_62734 | Not-determined |
| 18-101_k141_341640 | Not-determined |
| 18-101_k141_62739 | Low-quality |
| 18-101_k141_212038 | Not-determined |
| 18-101_k141_62740 | Not-determined |
| 18-101_k141_195300 | Not-determined |
| 18-101_k141_212042 | Not-determined |
| 18-101_k141_391688 | Not-determined |
| 18-101_k141_359823 | Not-determined |
| 18-101_k141_110868 | Not-determined |
| 18-101_k141_359824 | Not-determined |
| 18-101_k141_160909 | Not-determined |
| 17-109-final-viral-combined_k141_270839\|\|0_partial | Complete |
| 18-101_k141_160911 | Not-determined |
| 18-101_k141_391693 | Low-quality |
| 18-101_k141_45921 | Not-determined |
| 18-101_k141_359832 | Not-determined |
| 18-101_k141_29296 | Not-determined |
| 18-101_k141_359838 | Not-determined |
| 18-101_k141_212057 | Not-determined |
| 18-101_k141_391702 | Not-determined |
| 18-101_k141_29302 | Low-quality |
| 18-101_k141_13559 | Not-determined |
| 18-101_k141_341663 | Low-quality |
| 18-101_k141_261505 | Low-quality |
| 18-101_k141_341664 | Not-determined |
| 18-101_k141_45927 | Not-determined |
| 18-101_k141_78310 | Not-determined |
| 18-101_k141_29306 | Not-determined |
| 18-101_k141_78314 | Not-determined |
| 18-101_k141_391710 | Not-determined |
| 18-101_k141_62756 | Low-quality |
| 18-101_k141_145304 | Not-determined |
| 18-101_k141_244443 | Not-determined |
| 18-101_k141_13567 | Not-determined |
| 18-101_k141_276539 | Not-determined |
| 18-101_k141_45937 | Not-determined |
| 18-101_k141_244450 | Not-determined |
| 18-101_k141_78325 | Not-determined |
| 18-101_k141_78329 | Not-determined |
| 18-101_k141_391717 | Not-determined |
| 18-101_k141_227637 | Low-quality |
| 18-101_k141_45942 | Not-determined |
| 18-101_k141_95950 | Not-determined |
| 18-101_k141_95951 | Not-determined |
| 18-101_k141_78334 | Not-determined |
| 18-101_k141_261529 | Not-determined |
| 18-101_k141_176757 | Not-determined |
| 18-101_k141_391722 | Not-determined |
| 18-101_k141_244460 | Not-determined |
| 18-101_k141_29324 | Low-quality |
| 18-101_k141_276545 | Not-determined |
| 18-101_k141_95959 | Not-determined |
| 18-101_k141_13595 | Not-determined |
| 18-101_k141_29332 | Low-quality |
| 18-101_k141_359864 | Not-determined |
| 18-101_k141_391726 | Low-quality |
| 18-101_k141_195333 | Not-determined |
| 18-101_k141_341681 | Not-determined |
| 18-101_k141_129827 | Not-determined |
| 18-101_k141_45946 | Not-determined |
| 18-101_k141_95971 | Not-determined |
| 18-101_k141_45950 | Not-determined |
| 18-101_k141_29347 | Low-quality |
| 18-101_k141_176785 | Not-determined |
| 18-101_k141_244486 | Not-determined |
| 18-101_k141_212081 | Not-determined |
| 18-101_k141_13604 | Not-determined |
| 18-101_k141_145331 | Not-determined |
| 18-101_k141_95978 | Not-determined |
| 18-101_k141_29355 | Not-determined |
| 18-101_k141_110892 | Not-determined |
| 18-101_k141_212089 | Low-quality |
| 18-101_k141_13613 | Not-determined |
| 18-101_k141_244493 | Not-determined |
| 18-101_k141_45965 | Not-determined |
| 18-101_k141_110900 | Low-quality |
| 18-101_k141_95986 | Not-determined |
| 18-101_k141_261551 | Low-quality |
| 18-101_k141_110905 | Not-determined |
| 18-101_k141_310308 | Not-determined |
| 18-101_k141_110908 | Not-determined |
| 18-101_k141_391729 | Not-determined |
| 18-101_k141_95997 | Not-determined |
| 18-101_k141_96001 | Not-determined |
| 18-101_k141_227669 | Not-determined |
| 18-101_k141_261559 | Not-determined |
| 18-101_k141_310314 | Not-determined |
| 18-101_k141_45980 | Not-determined |
| 18-101_k141_227674 | Not-determined |
| 18-101_k141_176806 | Not-determined |
| 18-101_k141_227680 | Not-determined |
| 18-101_k141_310324 | Not-determined |
| 18-101_k141_45991 | Not-determined |
| 18-101_k141_13638 | Not-determined |
| 18-101_k141_375485 | Not-determined |
| 18-101_k141_227687 | Not-determined |
| 18-101_k141_62823 | Not-determined |
| 18-101_k141_195362 | Not-determined |
| 18-101_k141_375489 | Not-determined |
| 18-101_k141_145344 | Not-determined |
| 18-101_k141_310330 | Not-determined |
| 18-101_k141_110933 | Not-determined |
| 18-101_k141_341692 | Not-determined |
| 18-101_k141_13650 | Not-determined |
| 18-101_k141_13652 | Not-determined |
| 18-101_k141_46004 | Not-determined |
| 18-101_k141_341696 | Not-determined |
| 18-101_k141_78367 | Not-determined |
| 18-101_k141_96012 | Not-determined |
| 18-101_k141_62827 | Not-determined |
| 18-101_k141_96017 | Not-determined |
| 18-101_k141_310340 | Low-quality |
| 18-101_k141_145358 | Not-determined |
| 18-101_k141_160968 | Not-determined |
| 18-101_k141_375505 | Not-determined |
| 18-101_k141_96021 | Not-determined |
| 18-101_k141_375509 | Not-determined |
| 18-101_k141_391745 | Not-determined |
| 18-101_k141_195391 | Low-quality |
| 18-101_k141_227713 | Not-determined |
| 18-101_k141_13664 | Not-determined |
| 18-101_k141_310356 | Not-determined |
| 18-101_k141_375514 | Not-determined |
| 18-101_k141_96031 | Not-determined |
| 18-101_k141_391755 | Not-determined |
| 18-101_k141_96032 | Not-determined |
| 18-101_k141_391759 | Low-quality |
| 18-101_k141_244533 | Not-determined |
| 18-101_k141_160985 | Not-determined |
| 18-101_k141_62831 | Not-determined |
| 18-101_k141_13679 | Not-determined |
| 18-101_k141_46033 | Not-determined |
| 18-101_k141_78383 | Not-determined |
| 18-101_k141_78385 | Not-determined |
| 18-101_k141_341726 | Low-quality |
| 18-101_k141_78388 | Not-determined |
| 18-101_k141_78390 | Not-determined |
| 18-101_k141_46037 | Not-determined |
| 18-101_k141_391770 | Not-determined |
| 18-101_k141_110939 | Not-determined |
| 18-101_k141_160989 | Not-determined |
| 18-101_k141_46042 | Not-determined |
| 18-101_k141_244547 | Not-determined |
| 18-101_k141_129878 | Not-determined |
| 18-101_k141_375529 | Not-determined |
| 18-101_k141_96044 | Not-determined |
| 18-101_k141_276565 | Not-determined |
| 18-101_k141_212123 | Not-determined |
| 18-101_k141_276568 | Not-determined |
| 18-101_k141_212126 | Not-determined |
| 18-101_k141_110946 | Low-quality |
| 18-101_k141_391775 | Not-determined |
| 18-101_k141_375536 | Not-determined |
| 18-101_k141_391777 | Low-quality |
| 18-101_k141_96052 | Not-determined |
| 18-101_k141_227736 | Not-determined |
| 18-101_k141_244562 | Not-determined |
| 18-101_k141_310375 | Not-determined |
| 18-101_k141_341737 | Not-determined |
| 18-101_k141_129887 | Not-determined |
| 18-101_k141_261592 | Not-determined |
| 18-101_k141_341742 | Low-quality |
| 18-101_k141_145393 | Not-determined |
| 18-101_k141_145395 | Not-determined |
| 18-101_k141_176870 | Not-determined |
| 18-101_k141_244587 | Not-determined |
| 18-101_k141_13719 | Not-determined |
| 18-101_k141_261599 | Not-determined |
| 18-101_k141_161016 | Not-determined |
| 18-101_k141_359932 | Not-determined |
| 18-101_k141_341755 | Not-determined |
| 18-101_k141_341759 | Not-determined |
| 18-101_k141_195430 | Not-determined |
| 18-101_k141_129907 | Not-determined |
| 18-101_k141_145415 | Not-determined |
| 18-101_k141_227764 | Not-determined |
| 18-101_k141_375566 | Not-determined |
| 18-101_k141_129914 | Not-determined |
| 18-101_k141_145419 | Not-determined |
| 18-101_k141_261617 | Not-determined |
| 18-101_k141_341774 | Not-determined |
| 18-101_k141_244599 | Not-determined |
| 18-101_k141_62863 | Not-determined |
| 18-101_k141_161038 | Not-determined |
| 18-101_k141_195455 | Low-quality |
| 18-101_k141_244607 | Not-determined |
| 18-101_k141_195463 | Not-determined |
| 18-101_k141_227782 | Not-determined |
| 18-101_k141_293946 | Not-determined |
| 18-101_k141_129937 | Not-determined |
| 18-101_k141_129939 | Not-determined |
| 18-101_k141_145434 | Not-determined |
| 18-101_k141_293960 | Not-determined |
| 18-101_k141_227794 | Not-determined |
| 18-101_k141_359957 | Not-determined |
| 18-101_k141_293965 | Low-quality |
| 18-101_k141_129952 | Not-determined |
| 18-101_k141_359962 | Not-determined |
| 18-101_k141_129954 | Not-determined |
| 18-101_k141_176898 | Not-determined |
| 18-101_k141_145440 | Not-determined |
| 18-101_k141_195473 | Low-quality |
| 18-101_k141_145443 | Not-determined |
| 18-101_k141_212185 | Not-determined |
| 18-101_k141_145446 | Not-determined |
| 18-101_k141_176904 | Not-determined |
| 18-101_k141_261635 | Low-quality |
| 18-101_k141_359970 | Not-determined |
| 18-101_k141_145451 | Not-determined |
| 18-101_k141_29372 | Low-quality |
| 18-101_k141_145459 | Not-determined |
| 18-101_k141_62887 | Not-determined |
| 18-101_k141_359974 | Not-determined |
| 18-101_k141_212188 | Low-quality |
| 18-101_k141_195477 | Low-quality |
| 18-101_k141_129967 | Not-determined |
| 18-101_k141_13736 | Low-quality |
| 18-101_k141_244641 | Not-determined |
| 18-101_k141_13741 | Not-determined |
| 18-101_k141_176907 | Low-quality |
| 18-101_k141_111004 | Not-determined |
| 18-101_k141_244646 | Not-determined |
| 18-101_k141_13747 | Not-determined |
| 18-101_k141_111009 | Not-determined |
| 18-101_k141_161085 | Not-determined |
| 18-101_k141_375593 | Not-determined |
| 18-101_k141_293985 | Not-determined |
| 18-101_k141_62909 | Low-quality |
| 18-101_k141_212198 | Not-determined |
| 18-101_k141_129976 | Not-determined |
| 18-101_k141_161092 | Not-determined |
| 18-101_k141_62912 | Not-determined |
| 18-101_k141_129979 | Not-determined |
| 18-101_k141_310401 | Not-determined |
| 18-101_k141_341824 | Not-determined |
| 18-101_k141_111018 | Not-determined |
| 18-101_k141_46113 | Not-determined |
| 18-101_k141_310402 | Not-determined |
| 18-101_k141_46114 | Not-determined |
| 18-101_k141_129986 | Not-determined |
| 18-101_k141_62924 | Not-determined |
| 18-101_k141_341832 | Not-determined |
| 18-101_k141_129991 | Not-determined |
| 18-101_k141_111025 | Not-determined |
| 18-101_k141_46124 | Not-determined |
| 18-101_k141_176936 | Not-determined |
| 18-101_k141_261676 | Not-determined |
| 18-101_k141_62929 | Not-determined |
| 18-101_k141_195503 | Not-determined |
| 18-101_k141_310407 | Not-determined |
| 18-101_k141_341844 | Not-determined |
| 18-101_k141_129998 | Not-determined |
| 18-101_k141_310410 | Not-determined |
| 18-101_k141_341849 | Not-determined |
| 18-101_k141_375629 | Not-determined |
| 18-101_k141_176946 | Not-determined |
| 18-101_k141_310413 | Not-determined |
| 18-101_k141_261689 | Not-determined |
| 18-101_k141_195504 | Low-quality |
| 18-101_k141_294019 | Not-determined |
| 18-101_k141_310420 | Not-determined |
| 18-101_k141_360012 | Not-determined |
| 18-101_k141_145507 | Not-determined |
| 18-101_k141_111051 | Low-quality |
| 18-101_k141_375639 | Not-determined |
| 18-101_k141_294027 | Not-determined |
| 18-101_k141_13773 | Low-quality |
| 18-101_k141_161120 | Not-determined |
| 18-101_k141_13774 | Not-determined |
| 18-101_k141_161121 | Not-determined |
| 18-101_k141_145513 | Not-determined |
| 18-101_k141_261706 | Not-determined |
| 18-101_k141_29398 | Not-determined |
| 18-101_k141_360033 | Not-determined |
| 18-101_k141_161133 | Not-determined |
| 18-101_k141_195527 | Not-determined |
| 18-101_k141_360037 | Low-quality |
| 18-101_k141_261711 | Not-determined |
| 18-101_k141_111070 | Not-determined |
| 18-101_k141_276642 | Not-determined |
| 18-101_k141_391799 | Not-determined |
| 18-101_k141_29407 | Low-quality |
| 18-101_k141_195531 | Not-determined |
| 18-101_k141_176964 | Not-determined |
| 18-101_k141_391806 | Not-determined |
| 18-101_k141_244669 | Not-determined |
| 18-101_k141_176969 | Not-determined |
| 18-101_k141_276648 | Not-determined |
| 18-101_k141_310449 | Not-determined |
| 18-101_k141_176974 | Not-determined |
| 18-101_k141_130033 | Not-determined |
| 18-101_k141_212229 | Not-determined |
| 18-101_k141_46178 | Not-determined |
| 18-101_k141_176978 | Not-determined |
| 18-101_k141_13787 | Not-determined |
| 18-101_k141_261735 | Not-determined |
| 18-101_k141_13788 | Not-determined |
| 18-101_k141_13790 | Not-determined |
| 18-101_k141_145547 | Low-quality |
| 18-101_k141_244687 | Not-determined |
| 18-101_k141_145548 | Not-determined |
| 18-101_k141_111082 | Not-determined |
| 18-101_k141_161155 | Not-determined |
| 18-101_k141_244692 | Not-determined |
| 18-101_k141_244693 | Not-determined |
| 18-101_k141_29433 | Low-quality |
| 18-101_k141_176981 | Not-determined |
| 18-101_k141_62984 | Not-determined |
| 18-101_k141_13801 | Not-determined |
| 18-101_k141_227812 | Low-quality |
| 18-101_k141_212244 | Not-determined |
| 18-101_k141_310460 | Low-quality |
| 18-101_k141_13809 | Low-quality |
| 18-101_k141_130051 | Low-quality |
| 18-101_k141_294057 | Not-determined |
| 18-101_k141_276671 | Not-determined |
| 18-101_k141_261746 | Not-determined |
| 18-101_k141_195565 | Not-determined |
| 18-101_k141_227824 | Low-quality |
| 18-101_k141_62996 | Not-determined |
| 18-101_k141_375674 | Not-determined |
| 18-101_k141_244713 | Not-determined |
| 18-101_k141_13812 | Low-quality |
| 18-101_k141_78472 | Not-determined |
| 18-101_k141_276677 | Not-determined |
| 18-101_k141_212272 | Not-determined |
| 18-101_k141_212273 | Low-quality |
| 18-101_k141_310466 | Not-determined |
| 18-101_k141_227834 | Not-determined |
| 18-101_k141_375684 | Not-determined |
| 18-101_k141_310467 | Low-quality |
| 18-101_k141_341876 | Not-determined |
| 18-101_k141_294068 | Not-determined |
| 18-101_k141_161183 | Not-determined |
| 18-101_k141_244730 | Not-determined |
| 18-101_k141_375691 | Not-determined |
| 18-101_k141_130069 | Not-determined |
| 18-101_k141_29462 | Not-determined |
| 18-101_k141_78484 | Not-determined |
| 18-101_k141_360056 | Not-determined |
| 18-101_k141_63022 | Not-determined |
| 18-101_k141_78489 | Not-determined |
| 18-101_k141_261759 | Not-determined |
| 18-101_k141_227847 | Not-determined |
| 18-101_k141_360060 | Not-determined |
| 18-101_k141_13823 | Low-quality |
| 18-101_k141_261766 | Not-determined |
| 18-101_k141_310483 | Not-determined |
| 18-101_k141_176997 | Not-determined |
| 18-101_k141_341893 | Not-determined |
| 18-101_k141_360065 | Not-determined |
| 18-101_k141_195575 | Not-determined |
| 18-101_k141_276693 | Not-determined |
| 18-101_k141_195576 | Not-determined |
| 18-101_k141_78499 | Not-determined |
| 18-101_k141_161201 | Not-determined |
| 18-101_k141_261777 | Not-determined |
| 18-101_k141_111120 | Not-determined |
| 18-101_k141_63041 | Not-determined |
| 18-101_k141_276696 | Not-determined |
| 18-101_k141_212297 | Low-quality |
| 18-101_k141_78512 | Not-determined |
| 18-101_k141_212301 | Not-determined |
| 18-101_k141_212302 | Not-determined |
| 18-101_k141_130084 | Not-determined |
| 18-101_k141_130085 | Not-determined |
| 18-101_k141_78518 | Not-determined |
| 18-101_k141_78519 | Not-determined |
| 18-101_k141_261792 | Not-determined |
| 18-101_k141_360071 | Not-determined |
| 18-101_k141_13842 | Not-determined |
| 18-101_k141_360073 | Not-determined |
| 18-101_k141_145596 | Not-determined |
| 18-101_k141_212307 | Low-quality |
| 18-101_k141_391847 | Not-determined |
| 18-101_k141_360076 | Not-determined |
| 18-101_k141_195588 | Not-determined |

| | |
|---|---|
| 18-101_k141_78524 | Low-quality |
| 18-101_k141_161224 | Not-determined |
| 18-101_k141_212309 | Not-determined |
| 18-101_k141_195590 | Not-determined |
| 18-101_k141_375725 | Not-determined |
| 18-101_k141_375726 | Not-determined |
| 18-101_k141_177016 | Low-quality |
| 18-101_k141_177018 | Not-determined |
| 18-101_k141_227849 | Not-determined |
| 18-101_k141_130102 | Not-determined |
| 18-101_k141_276718 | Not-determined |
| 18-101_k141_375736 | Not-determined |
| 18-101_k141_161232 | Low-quality |
| 18-101_k141_375737 | Not-determined |
| 18-101_k141_294099 | Not-determined |
| 18-101_k141_276722 | Not-determined |
| 18-101_k141_13857 | Not-determined |
| 18-101_k141_276725 | Not-determined |
| 18-101_k141_294103 | Not-determined |
| 18-101_k141_145605 | Not-determined |
| 18-101_k141_46230 | Not-determined |
| 18-101_k141_294109 | Not-determined |
| 18-101_k141_130110 | Low-quality |
| 18-101_k141_375748 | Not-determined |
| 18-101_k141_29503 | Not-determined |
| 18-101_k141_161247 | Not-determined |
| 18-101_k141_145610 | Not-determined |
| 18-101_k141_212335 | Not-determined |
| 18-101_k141_63083 | Not-determined |
| 18-101_k141_276733 | Not-determined |
| 18-101_k141_111152 | Not-determined |
| 18-101_k141_161252 | Not-determined |
| 18-101_k141_391851 | Not-determined |
| 18-101_k141_227872 | Not-determined |
| 18-101_k141_244791 | Low-quality |
| 18-101_k141_195618 | Not-determined |
| 18-101_k141_177061 | Not-determined |
| 18-101_k141_96070 | Not-determined |
| 18-101_k141_63105 | Not-determined |
| 18-101_k141_96079 | Not-determined |
| 18-101_k141_161274 | Not-determined |
| 18-101_k141_177074 | Not-determined |
| 18-101_k141_111171 | Not-determined |
| 18-101_k141_177077 | Not-determined |
| 18-101_k141_111174 | Not-determined |
| 18-101_k141_244805 | Not-determined |
| 18-101_k141_391871 | Not-determined |
| 18-101_k141_195642 | Not-determined |
| 18-101_k141_111181 | Not-determined |
| 18-101_k141_244814 | Not-determined |
| 18-101_k141_195647 | Not-determined |
| 18-101_k141_375804 | Not-determined |
| 18-101_k141_360082 | Not-determined |
| 18-101_k141_195652 | Not-determined |
| 18-101_k141_96092 | Not-determined |
| 18-101_k141_130155 | Not-determined |
| 18-101_k141_96093 | Not-determined |
| 18-101_k141_244822 | Not-determined |
| 18-101_k141_161293 | Low-quality |
| 18-101_k141_195659 | Not-determined |
| 18-101_k141_177093 | Not-determined |
| 18-101_k141_63112 | Not-determined |
| 18-101_k141_244826 | Not-determined |
| 18-101_k141_276753 | Not-determined |
| 18-101_k141_341923 | Not-determined |
| 18-101_k141_294133 | Not-determined |
| 18-101_k141_360094 | Not-determined |
| 18-101_k141_294139 | Not-determined |
| 18-101_k141_261867 | Not-determined |
| 18-101_k141_276759 | Not-determined |
| 18-101_k141_177108 | Not-determined |
| 18-101_k141_276766 | Not-determined |
| 18-101_k141_375828 | Not-determined |
| 18-101_k141_261879 | Not-determined |
| 18-101_k141_261881 | Not-determined |
| 18-101_k141_244833 | Not-determined |
| 18-101_k141_360105 | Not-determined |
| 18-101_k141_341941 | Low-quality |
| 18-101_k141_227897 | Not-determined |
| 18-101_k141_261886 | Not-determined |
| 18-101_k141_63128 | Not-determined |
| 18-101_k141_375835 | Not-determined |
| 18-101_k141_130175 | Not-determined |
| 18-101_k141_78535 | Not-determined |
| 18-101_k141_360114 | Not-determined |
| 18-101_k141_360116 | Not-determined |
| 18-101_k141_195682 | Not-determined |
| 18-101_k141_130181 | Not-determined |
| 18-101_k141_161316 | Not-determined |
| 18-101_k141_161317 | Not-determined |
| 18-101_k141_29520 | Not-determined |
| 18-101_k141_212345 | Not-determined |
| 18-101_k141_212346 | Not-determined |
| 18-101_k141_13927 | Low-quality |
| 18-101_k141_294148 | Not-determined |
| 18-101_k141_161328 | Not-determined |
| 18-101_k141_261910 | Not-determined |
| 18-101_k141_360130 | Not-determined |
| 18-101_k141_341950 | Not-determined |
| 18-101_k141_63145 | Not-determined |
| 18-101_k141_261915 | Not-determined |
| 18-101_k141_29531 | Not-determined |
| 18-101_k141_212354 | Not-determined |
| 18-101_k141_177143 | Not-determined |
| 18-101_k141_195699 | Not-determined |
| 18-101_k141_177146 | Not-determined |
| 18-101_k141_195703 | Not-determined |
| 18-101_k141_261924 | Not-determined |
| 18-101_k141_63154 | Not-determined |
| 18-101_k141_212364 | Not-determined |
| 18-101_k141_261926 | Not-determined |
| 18-101_k141_63157 | Not-determined |
| 18-101_k141_360145 | Not-determined |
| 18-101_k141_261931 | Not-determined |
| 18-101_k141_276805 | Not-determined |
| 18-101_k141_161366 | Not-determined |
| 18-101_k141_391917 | Not-determined |
| 18-101_k141_212377 | Not-determined |
| 18-101_k141_96130 | Low-quality |
| 18-101_k141_261940 | Not-determined |
| 18-109-final-viral-combined_k141_71573\|\|full | High-quality |
| 18-101_k141_391923 | Low-quality |
| 18-101_k141_195733 | Not-determined |
| 18-101_k141_29548 | Not-determined |
| 18-101_k141_244839 | Low-quality |
| 18-101_k141_212395 | Low-quality |
| 18-101_k141_145633 | Not-determined |
| 18-101_k141_177159 | Low-quality |
| 18-101_k141_391933 | Not-determined |
| 18-101_k141_341984 | Not-determined |
| 18-101_k141_13934 | Not-determined |
| 18-101_k141_96144 | Not-determined |
| 18-101_k141_261960 | Not-determined |
| 18-101_k141_227920 | Low-quality |
| 18-101_k141_177163 | Low-quality |
| 18-101_k141_161372 | Not-determined |
| 18-101_k141_161373 | Not-determined |
| 18-101_k141_341991 | Not-determined |
| 18-101_k141_261963 | Not-determined |
| 18-101_k141_161375 | Not-determined |
| 18-101_k141_78556 | Low-quality |
| 18-101_k141_145644 | Low-quality |
| 18-101_k141_145645 | Not-determined |
| 18-101_k141_212405 | Not-determined |
| 18-101_k141_227925 | Not-determined |
| 18-101_k141_227928 | Not-determined |
| 18-101_k141_96145 | Low-quality |
| 18-101_k141_391943 | Not-determined |
| 18-101_k141_212415 | Not-determined |
| 18-101_k141_310548 | Not-determined |
| 18-101_k141_276842 | Not-determined |
| 18-101_k141_29586 | Not-determined |
| 18-101_k141_276846 | Not-determined |
| 18-101_k141_360177 | Not-determined |
| 18-101_k141_244859 | Not-determined |
| 18-101_k141_111257 | Not-determined |
| 18-101_k141_96156 | Low-quality |
| 18-101_k141_96157 | Low-quality |
| 18-101_k141_360180 | Not-determined |
| 18-101_k141_29594 | Not-determined |
| 18-101_k141_342012 | Not-determined |
| 18-101_k141_375851 | Low-quality |
| 18-101_k141_276851 | Not-determined |
| 18-101_k141_111263 | Not-determined |
| 18-101_k141_227944 | Not-determined |
| 18-101_k141_195748 | Not-determined |
| 18-101_k141_96161 | Low-quality |
| 18-101_k141_177194 | Not-determined |
| 18-101_k141_227950 | Not-determined |
| 18-101_k141_276855 | Not-determined |
| 18-101_k141_244868 | Not-determined |
| 18-101_k141_111273 | Not-determined |
| 18-101_k141_276859 | Low-quality |
| 18-101_k141_177200 | Low-quality |
| 18-101_k141_212430 | Not-determined |
| 18-101_k141_391948 | Not-determined |
| 18-101_k141_29597 | Not-determined |
| 18-101_k141_375858 | Low-quality |
| 18-101_k141_375859 | Not-determined |
| 18-101_k141_29603 | Not-determined |
| 18-101_k141_261991 | Not-determined |
| 18-101_k141_161382 | Not-determined |
| 18-101_k141_391955 | Not-determined |
| 18-101_k141_29611 | Not-determined |
| 18-101_k141_177206 | Not-determined |
| 18-101_k141_276870 | Not-determined |
| 18-101_k141_177209 | Not-determined |
| 18-101_k141_310562 | Not-determined |
| 18-101_k141_96173 | Low-quality |
| 17-108-final-viral-combined_k141_217790\|\|full | Medium-quality |
| 18-101_k141_29616 | Not-determined |
| 18-101_k141_244878 | Not-determined |
| 18-101_k141_261998 | Not-determined |
| 18-101_k141_96190 | Not-determined |
| 18-101_k141_391964 | Not-determined |
| 18-101_k141_29624 | Not-determined |
| 18-101_k141_195770 | Not-determined |
| 18-101_k141_130222 | Not-determined |
| 18-101_k141_227976 | Not-determined |
| 18-101_k141_294162 | Not-determined |
| 18-101_k141_262007 | Not-determined |
| 18-101_k141_111288 | Not-determined |
| 18-101_k141_262010 | Not-determined |
| 18-101_k141_96198 | Not-determined |
| 18-101_k141_29626 | Not-determined |
| 18-101_k141_195787 | Not-determined |
| 18-101_k141_212451 | Not-determined |
| 18-101_k141_212455 | Low-quality |
| 18-101_k141_276884 | Not-determined |
| 18-101_k141_262022 | Low-quality |
| 18-101_k141_294179 | Not-determined |
| 18-101_k141_360218 | Not-determined |
| 18-101_k141_227982 | Not-determined |
| 18-101_k141_96214 | Not-determined |
| 18-101_k141_46267 | Low-quality |
| 18-101_k141_391983 | Not-determined |
| 18-101_k141_375899 | Not-determined |
| 18-101_k141_46269 | Not-determined |
| 18-101_k141_29629 | Not-determined |
| 18-101_k141_130247 | Not-determined |
| 18-101_k141_342058 | Not-determined |
| 18-101_k141_244918 | Not-determined |
| 18-101_k141_111304 | Not-determined |
| 18-101_k141_177232 | Not-determined |
| 18-101_k141_145688 | Not-determined |
| 18-101_k141_145690 | Not-determined |
| 18-101_k141_145694 | Not-determined |
| 18-101_k141_29645 | Not-determined |
| 18-101_k141_310593 | Not-determined |
| 18-101_k141_342066 | Not-determined |
| 18-101_k141_360250 | Not-determined |
| 18-101_k141_29649 | Not-determined |
| 18-101_k141_342070 | Not-determined |
| 18-101_k141_342073 | Not-determined |
| 18-101_k141_130256 | Not-determined |
| 18-101_k141_111310 | Not-determined |
| 18-101_k141_375905 | Not-determined |
| 18-101_k141_392001 | Not-determined |
| 18-101_k141_111311 | Not-determined |
| 18-101_k141_145708 | Not-determined |
| 18-101_k141_111319 | Not-determined |
| 18-101_k141_276908 | Not-determined |
| 18-101_k141_227996 | Not-determined |
| 18-101_k141_276909 | Not-determined |
| 18-101_k141_111322 | Not-determined |
| 18-101_k141_392008 | Not-determined |
| 18-101_k141_161418 | Not-determined |
| 18-101_k141_262033 | Not-determined |
| 18-101_k141_375922 | Not-determined |
| 18-101_k141_96258 | Not-determined |
| 18-101_k141_262034 | Not-determined |
| 18-101_k141_360274 | Not-determined |
| 18-101_k141_262036 | Not-determined |
| 18-101_k141_294194 | Not-determined |
| 18-101_k141_310618 | Not-determined |
| 18-101_k141_262037 | Not-determined |
| 18-101_k141_14007 | Low-quality |
| 18-101_k141_294198 | Not-determined |
| 18-101_k141_63209 | Not-determined |
| 18-101_k141_145716 | Not-determined |
| 18-101_k141_29657 | Low-quality |
| 18-101_k141_177251 | Not-determined |
| 18-101_k141_212515 | Not-determined |
| 18-101_k141_177256 | Not-determined |
| 18-101_k141_161425 | Not-determined |
| 18-101_k141_29658 | Not-determined |
| 18-101_k141_375936 | Not-determined |
| 18-101_k141_375942 | Not-determined |
| 18-101_k141_111350 | Not-determined |
| 18-101_k141_161437 | Not-determined |
| 18-101_k141_276923 | Not-determined |
| 18-101_k141_195858 | Not-determined |
| 18-101_k141_212536 | Low-quality |
| 18-101_k141_212537 | Not-determined |
| 18-101_k141_145733 | Not-determined |
| 18-101_k141_327192 | Not-determined |
| 18-101_k141_177275 | Not-determined |
| 18-101_k141_212540 | Not-determined |
| 18-101_k141_342100 | Low-quality |
| 18-101_k141_276929 | Not-determined |
| 18-101_k141_161440 | Not-determined |
| 18-101_k141_161443 | Not-determined |
| 18-101_k141_195868 | Not-determined |
| 18-101_k141_375958 | Not-determined |
| 18-101_k141_177279 | Not-determined |
| 18-101_k141_14020 | Not-determined |
| 18-101_k141_244966 | Not-determined |
| 18-101_k141_375964 | Not-determined |
| 18-101_k141_177290 | Not-determined |
| 18-101_k141_294212 | Low-quality |
| 18-101_k141_327213 | Not-determined |
| 18-101_k141_310658 | Not-determined |
| 18-101_k141_212561 | Not-determined |
| 18-101_k141_310660 | Not-determined |
| 18-101_k141_46274 | Not-determined |
| 18-101_k141_78580 | Not-determined |
| 18-101_k141_212566 | Not-determined |
| 18-101_k141_63224 | Not-determined |
| 18-101_k141_161469 | Not-determined |
| 18-101_k141_244984 | Not-determined |
| 18-101_k141_111373 | Not-determined |
| 18-101_k141_161474 | Not-determined |
| 18-101_k141_177301 | Not-determined |
| 18-101_k141_244989 | Not-determined |
| 18-101_k141_244991 | Not-determined |
| 18-101_k141_327226 | Not-determined |
| 18-101_k141_327227 | Not-determined |
| 18-101_k141_262086 | Low-quality |
| 18-101_k141_161481 | Not-determined |
| 18-101_k141_161482 | Not-determined |
| 18-101_k141_212580 | Not-determined |
| 18-101_k141_262089 | Not-determined |
| 18-101_k141_327232 | Low-quality |
| 18-101_k141_228047 | Not-determined |
| 18-101_k141_195908 | Not-determined |
| 18-101_k141_29682 | Not-determined |
| 18-101_k141_327236 | Not-determined |
| 18-101_k141_245003 | Low-quality |
| 18-101_k141_327238 | Not-determined |
| 18-101_k141_195912 | Not-determined |
| 18-101_k141_212600 | Not-determined |
| 18-101_k141_145766 | Not-determined |
| 18-101_k141_29685 | Not-determined |
| 18-101_k141_212603 | Not-determined |
| 18-101_k141_177328 | Not-determined |
| 18-101_k141_327246 | Low-quality |
| 18-101_k141_212614 | Not-determined |
| 18-101_k141_376000 | Not-determined |
| 18-101_k141_145773 | Not-determined |
| 18-101_k141_360297 | Not-determined |
| 18-101_k141_212620 | Not-determined |
| 18-101_k141_29695 | Not-determined |
| 18-101_k141_177339 | Not-determined |
| 18-101_k141_360301 | Not-determined |
| 18-101_k141_145784 | Not-determined |
| 18-101_k141_212627 | Not-determined |
| 18-101_k141_29702 | Not-determined |
| 18-101_k141_145786 | Not-determined |
| 18-101_k141_145787 | Not-determined |
| 18-101_k141_342137 | Not-determined |
| 18-101_k141_245023 | Not-determined |
| 18-101_k141_276965 | Not-determined |
| 18-101_k141_228078 | Not-determined |
| 18-101_k141_360312 | Not-determined |
| 18-101_k141_161506 | Not-determined |
| 18-101_k141_276969 | Not-determined |
| 18-101_k141_130302 | Low-quality |
| 18-101_k141_276973 | Not-determined |
| 18-101_k141_276976 | Not-determined |
| 18-101_k141_376009 | Not-determined |
| 18-101_k141_276978 | Not-determined |
| 18-101_k141_276980 | Not-determined |
| 18-101_k141_392018 | Not-determined |
| 18-101_k141_327266 | Not-determined |
| 18-101_k141_262118 | Not-determined |
| 18-101_k141_327270 | Not-determined |
| 18-101_k141_228087 | Not-determined |
| 18-101_k141_360327 | Not-determined |
| 18-101_k141_276987 | Low-quality |
| 18-101_k141_63278 | Not-determined |
| 18-101_k141_376019 | Low-quality |
| 18-101_k141_376020 | Low-quality |
| 18-101_k141_327278 | Not-determined |
| 18-101_k141_327279 | Not-determined |
| 18-101_k141_130313 | Not-determined |
| 18-101_k141_63288 | Not-determined |
| 18-101_k141_276992 | Low-quality |
| 18-101_k141_111399 | Not-determined |
| 18-101_k141_276994 | Not-determined |
| 18-101_k141_145816 | Not-determined |
| 18-101_k141_228099 | Not-determined |
| 18-101_k141_177367 | Not-determined |
| 18-101_k141_342164 | Not-determined |
| 18-101_k141_294251 | Not-determined |
| 18-101_k141_145823 | Not-determined |
| 18-101_k141_392034 | Not-determined |
| 18-101_k141_161538 | Not-determined |
| 18-101_k141_228102 | Low-quality |
| 18-101_k141_14063 | Not-determined |
| 18-101_k141_392038 | Not-determined |
| 18-101_k141_277003 | Not-determined |
| 18-101_k141_327288 | Not-determined |
| 18-101_k141_145828 | Not-determined |
| 18-101_k141_46305 | Low-quality |
| 18-101_k141_327291 | Not-determined |
| 18-101_k141_29737 | Low-quality |
| 18-101_k141_360345 | Not-determined |
| 18-101_k141_63303 | Not-determined |
| 18-101_k141_130336 | Not-determined |
| 18-101_k141_63304 | Not-determined |
| 18-101_k141_376028 | Not-determined |
| 18-101_k141_195969 | Not-determined |
| 18-101_k141_360351 | Low-quality |
| 18-101_k141_310713 | Not-determined |
| 18-101_k141_360352 | Not-determined |
| 18-101_k141_294262 | Not-determined |
| 18-101_k141_63310 | Not-determined |
| 18-101_k141_392044 | Not-determined |
| 18-101_k141_29744 | Not-determined |
| 18-101_k141_195976 | Not-determined |
| 18-101_k141_277010 | Not-determined |
| 18-101_k141_161558 | Not-determined |
| 18-101_k141_29748 | Not-determined |
| 18-101_k141_228123 | Not-determined |
| 18-101_k141_63314 | Low-quality |
| 18-101_k141_177395 | Not-determined |
| 18-101_k141_161565 | Not-determined |
| 18-101_k141_78675 | Not-determined |
| 18-101_k141_262147 | Not-determined |
| 18-101_k141_294286 | Not-determined |
| 18-101_k141_46324 | Not-determined |
| 18-101_k141_63320 | Not-determined |
| 18-101_k141_310727 | Not-determined |
| 18-101_k141_177403 | Not-determined |
| 18-101_k141_195983 | Not-determined |
| 18-101_k141_63322 | Not-determined |
| 18-101_k141_177404 | Not-determined |
| 18-101_k141_46327 | Not-determined |
| 18-101_k141_177407 | Not-determined |
| 18-101_k141_392057 | Not-determined |
| 18-101_k141_392061 | Not-determined |
| 18-101_k141_360353 | Not-determined |
| 18-101_k141_294292 | Not-determined |
| 18-101_k141_14094 | Not-determined |
| 18-101_k141_78677 | Not-determined |
| 18-101_k141_14095 | Not-determined |
| 18-101_k141_228144 | Low-quality |
| 18-101_k141_392071 | Not-determined |
| 18-101_k141_161570 | Not-determined |
| 18-101_k141_294295 | Low-quality |
| 18-101_k141_161573 | Not-determined |
| 18-101_k141_177413 | Not-determined |
| 18-101_k141_78685 | Not-determined |
| 18-101_k141_277035 | Not-determined |
| 18-101_k141_294303 | Not-determined |
| 18-101_k141_63334 | Not-determined |
| 18-101_k141_29772 | Not-determined |
| 18-101_k141_78690 | Not-determined |
| 18-101_k141_111408 | Not-determined |
| 18-101_k141_310755 | Not-determined |
| 18-101_k141_29775 | Not-determined |
| 18-101_k141_29777 | Low-quality |
| 18-101_k141_111412 | Not-determined |
| 18-101_k141_294306 | Not-determined |
| 18-101_k141_111413 | Low-quality |
| 18-101_k141_327316 | Not-determined |
| 18-101_k141_96285 | Not-determined |
| 18-101_k141_130351 | Not-determined |
| 18-101_k141_29785 | Not-determined |
| 18-101_k141_78703 | Not-determined |
| 18-101_k141_111418 | Not-determined |
| 18-101_k141_245044 | Not-determined |
| 18-101_k141_327324 | Not-determined |
| 18-101_k141_212664 | Not-determined |
| 18-101_k141_29795 | Not-determined |
| 18-101_k141_196019 | Not-determined |
| 18-101_k141_29798 | Not-determined |
| 18-101_k141_29799 | Not-determined |
| 18-101_k141_29800 | Not-determined |
| 18-101_k141_29802 | Not-determined |
| 18-101_k141_63344 | Not-determined |
| 18-101_k141_145867 | Not-determined |
| 18-101_k141_29803 | Not-determined |
| 18-101_k141_96295 | Not-determined |
| 18-101_k141_14127 | Not-determined |
| 18-101_k141_262179 | Not-determined |
| 18-101_k141_161586 | Not-determined |
| 18-101_k141_342188 | Low-quality |
| 18-101_k141_96302 | Not-determined |
| 18-101_k141_228188 | Not-determined |
| 18-101_k141_212682 | Not-determined |
| 18-101_k141_212683 | Not-determined |
| 18-101_k141_96306 | Not-determined |
| 18-101_k141_212686 | Not-determined |
| 18-101_k141_196030 | Not-determined |
| 18-101_k141_212688 | Not-determined |
| 18-101_k141_96308 | Low-quality |
| 18-101_k141_262190 | Not-determined |
| 18-101_k141_96309 | Not-determined |
| 18-101_k141_196031 | Not-determined |
| 18-101_k141_376037 | Not-determined |
| 18-101_k141_63355 | Not-determined |
| 18-101_k141_196034 | Not-determined |
| 18-101_k141_14136 | Not-determined |
| 18-101_k141_196035 | Not-determined |
| 18-101_k141_161599 | Not-determined |
| 18-101_k141_262193 | Not-determined |
| 18-101_k141_29818 | Not-determined |
| 18-101_k141_212701 | Not-determined |
| 18-101_k141_78736 | Low-quality |
| 18-101_k141_245059 | Not-determined |
| 18-101_k141_262200 | Not-determined |
| 18-101_k141_376049 | Not-determined |
| 18-101_k141_277057 | Not-determined |
| 18-101_k141_327344 | Not-determined |
| 18-101_k141_14149 | Not-determined |
| 18-101_k141_294343 | Not-determined |
| 18-101_k141_277060 | Not-determined |
| 18-101_k141_376062 | Not-determined |
| 18-101_k141_78752 | Not-determined |
| 18-101_k141_63376 | Not-determined |
| 18-101_k141_46367 | Not-determined |
| 18-101_k141_78762 | Not-determined |
| 18-101_k141_360391 | Low-quality |
| 18-101_k141_78764 | Not-determined |
| 18-101_k141_96333 | Not-determined |
| 18-101_k141_376071 | Not-determined |
| 18-101_k141_376074 | Not-determined |
| 18-101_k141_294358 | Not-determined |
| 18-101_k141_376076 | Not-determined |
| 18-101_k141_46375 | Not-determined |
| 18-101_k141_262224 | Not-determined |
| 18-101_k141_262225 | Not-determined |
| 18-101_k141_130366 | Low-quality |
| 18-101_k141_262227 | Not-determined |
| 18-101_k141_130370 | Not-determined |
| 18-101_k141_161620 | Not-determined |
| 18-101_k141_327378 | Not-determined |
| 18-101_k141_161624 | Not-determined |
| 18-101_k141_63385 | Not-determined |
| 18-101_k141_310817 | Not-determined |
| 18-101_k141_196080 | Not-determined |
| 18-101_k141_63393 | Not-determined |
| 18-101_k141_310821 | Not-determined |
| 18-101_k141_294370 | Not-determined |
| 18-101_k141_177490 | Low-quality |
| 18-101_k141_376106 | Not-determined |
| 18-101_k141_161646 | Not-determined |
| 18-101_k141_145930 | Low-quality |
| 18-101_k141_277084 | Low-quality |
| 18-101_k141_177495 | Not-determined |
| 18-101_k141_310835 | Not-determined |
| 18-101_k141_111460 | Low-quality |
| 18-101_k141_327395 | Not-determined |
| 18-101_k141_145933 | Not-determined |
| 18-101_k141_294379 | Not-determined |
| 18-101_k141_212735 | Not-determined |
| 18-101_k141_228247 | Not-determined |
| 18-101_k141_177510 | Not-determined |
| 18-101_k141_327402 | Not-determined |
| 18-101_k141_342220 | Not-determined |
| 18-101_k141_310845 | Not-determined |
| 18-101_k141_342222 | Not-determined |
| 18-101_k141_327407 | Not-determined |
| 18-101_k141_161658 | Low-quality |
| 18-101_k141_14189 | Not-determined |
| 18-101_k141_161661 | Not-determined |
| 18-101_k141_376125 | Not-determined |
| 18-101_k141_63416 | Not-determined |
| 18-101_k141_78792 | Low-quality |
| 18-101_k141_161664 | Low-quality |
| 18-101_k141_310852 | Not-determined |
| 18-101_k141_78797 | Not-determined |
| 18-101_k141_392096 | Not-determined |
| 18-101_k141_96342 | Not-determined |
| 18-101_k141_262248 | Not-determined |
| 18-101_k141_262249 | Not-determined |
| 18-101_k141_212746 | Not-determined |
| 18-101_k141_294396 | Not-determined |
| 18-101_k141_277094 | Not-determined |
| 18-101_k141_294397 | Not-determined |
| 18-101_k141_294398 | Not-determined |
| 18-101_k141_78805 | Not-determined |
| 18-101_k141_46428 | Not-determined |
| 18-101_k141_228264 | Not-determined |
| 18-101_k141_78808 | Not-determined |
| 18-101_k141_392103 | Not-determined |
| 18-101_k141_212753 | Not-determined |
| 18-101_k141_294406 | Low-quality |
| 18-101_k141_63438 | Not-determined |
| 18-101_k141_327423 | Not-determined |
| 18-101_k141_277106 | Not-determined |
| 18-101_k141_111490 | Not-determined |
| 18-101_k141_63444 | Not-determined |
| 18-101_k141_161684 | Not-determined |
| 18-101_k141_161685 | Not-determined |
| 18-101_k141_310867 | Not-determined |
| 18-101_k141_46437 | Not-determined |
| 18-101_k141_130387 | Not-determined |
| 18-101_k141_262255 | Not-determined |
| 18-101_k141_63452 | Not-determined |
| 18-101_k141_161689 | Not-determined |
| 18-101_k141_145976 | Not-determined |
| 18-101_k141_342252 | Not-determined |
| 18-101_k141_228280 | Low-quality |
| 18-101_k141_78825 | Not-determined |
| 18-101_k141_245080 | Not-determined |
| 18-101_k141_29857 | Not-determined |
| 18-101_k141_376153 | Not-determined |

| | |
|---|---|
| 18-101_k141_310878 | Not-determined |
| 18-101_k141_392105 | Not-determined |
| 18-101_k141_392106 | Not-determined |
| 18-101_k141_342265 | Not-determined |
| 18-101_k141_96372 | Not-determined |
| 18-101_k141_376155 | Low-quality |
| 18-101_k141_392108 | Low-quality |
| 18-101_k141_392109 | Not-determined |
| 18-101_k141_96375 | Not-determined |
| 18-101_k141_294436 | Not-determined |
| 18-101_k141_29877 | Not-determined |
| 18-101_k141_46470 | Not-determined |
| 18-101_k141_196118 | Not-determined |
| 18-101_k141_46471 | Not-determined |
| 18-101_k141_228300 | Not-determined |
| 18-101_k141_294439 | Not-determined |
| 18-101_k141_228303 | Not-determined |
| 18-101_k141_392116 | Not-determined |
| 18-101_k141_46476 | Not-determined |
| 18-101_k141_294447 | Not-determined |
| 18-101_k141_294448 | Low-quality |
| 18-101_k141_78847 | Low-quality |
| 18-101_k141_392123 | Not-determined |
| 18-101_k141_376170 | Not-determined |
| 18-101_k141_342290 | Not-determined |
| 18-101_k141_392126 | Not-determined |
| 18-101_k141_360450 | Not-determined |
| 18-101_k141_46482 | Not-determined |
| 18-101_k141_360456 | Not-determined |
| 18-101_k141_327444 | Not-determined |
| 18-101_k141_46483 | Not-determined |
| 18-101_k141_212796 | Low-quality |
| 18-101_k141_145992 | Not-determined |
| 18-101_k141_212797 | Not-determined |
| 18-101_k141_245120 | Not-determined |
| 18-101_k141_14234 | Not-determined |
| 18-101_k141_46487 | Not-determined |
| 18-101_k141_212800 | Not-determined |
| 18-101_k141_327453 | Not-determined |
| 18-101_k141_196129 | Not-determined |
| 18-101_k141_277146 | Not-determined |
| 18-101_k141_78865 | Not-determined |
| 18-101_k141_245129 | Not-determined |
| 18-101_k141_177565 | Not-determined |
| 18-101_k141_310898 | Not-determined |
| 18-101_k141_177568 | Not-determined |
| 18-101_k141_376188 | Not-determined |
| 18-101_k141_392139 | Not-determined |
| 18-101_k141_342306 | Not-determined |
| 18-101_k141_212808 | Not-determined |
| 18-101_k141_392141 | Not-determined |
| 18-101_k141_294466 | Not-determined |
| 18-101_k141_245138 | Not-determined |
| 18-101_k141_177577 | Not-determined |
| 18-101_k141_342311 | Not-determined |
| 18-101_k141_29898 | Not-determined |
| 18-101_k141_130406 | Not-determined |
| 18-101_k141_245140 | Not-determined |
| 18-101_k141_327470 | Not-determined |
| 18-101_k141_262279 | Not-determined |
| 18-101_k141_14248 | Not-determined |
| 18-101_k141_14249 | Not-determined |
| 18-101_k141_245143 | Not-determined |
| 18-101_k141_327475 | Not-determined |
| 18-101_k141_96394 | Not-determined |
| 18-101_k141_392151 | Not-determined |
| 18-101_k141_294482 | Not-determined |
| 18-101_k141_360477 | Not-determined |
| 18-101_k141_14252 | Not-determined |
| 18-101_k141_376211 | Not-determined |
| 18-101_k141_360479 | Not-determined |
| 18-101_k141_96398 | Not-determined |
| 18-101_k141_29906 | Not-determined |
| 18-101_k141_96401 | Not-determined |
| 18-101_k141_161697 | Not-determined |
| 18-101_k141_29912 | Not-determined |
| 18-101_k141_130419 | Low-quality |
| 18-101_k141_130420 | Not-determined |
| 18-101_k141_376221 | Not-determined |
| 18-101_k141_360484 | Not-determined |
| 18-101_k141_262286 | Low-quality |
| 18-101_k141_196156 | Not-determined |
| 18-101_k141_376223 | Not-determined |
| 18-101_k141_376225 | Not-determined |
| 18-101_k141_161698 | Not-determined |
| 18-101_k141_294503 | Not-determined |
| 18-101_k141_392158 | Not-determined |
| 18-101_k141_196163 | Not-determined |
| 18-101_k141_96406 | Not-determined |
| 18-101_k141_342325 | Not-determined |
| 18-101_k141_245157 | Not-determined |
| 18-101_k141_392160 | Not-determined |
| 18-101_k141_96409 | Not-determined |
| 18-101_k141_177585 | Low-quality |
| 18-101_k141_342328 | Not-determined |
| 18-101_k141_392162 | Not-determined |
| 18-101_k141_177586 | Not-determined |
| 18-101_k141_29923 | Not-determined |
| 18-101_k141_277173 | Low-quality |
| 18-101_k141_327486 | Not-determined |
| 18-101_k141_245159 | Not-determined |
| 18-101_k141_342331 | Not-determined |
| 18-101_k141_245161 | Not-determined |
| 18-101_k141_262302 | Not-determined |
| 18-101_k141_294512 | Not-determined |
| 18-101_k141_245162 | Not-determined |
| 18-101_k141_245164 | Not-determined |
| 18-101_k141_78903 | Not-determined |
| 18-101_k141_262307 | Low-quality |
| 18-101_k141_327495 | Not-determined |
| 18-101_k141_177596 | Not-determined |
| 18-101_k141_161705 | Low-quality |
| 18-101_k141_392176 | Not-determined |
| 18-101_k141_29936 | Not-determined |
| 18-101_k141_277190 | Not-determined |
| 18-101_k141_392184 | Not-determined |
| 18-101_k141_310905 | Low-quality |
| 18-101_k141_29940 | Low-quality |
| 18-101_k141_212813 | Not-determined |
| 18-101_k141_29943 | Not-determined |
| 18-101_k141_327500 | Not-determined |
| 18-101_k141_212816 | Not-determined |
| 18-101_k141_262323 | Not-determined |
| 18-101_k141_277200 | Not-determined |
| 18-101_k141_96426 | Not-determined |
| 18-101_k141_245183 | Not-determined |
| 18-101_k141_277204 | Not-determined |
| 18-101_k141_392198 | Not-determined |
| 18-101_k141_161721 | Not-determined |
| 18-101_k141_212823 | Not-determined |
| 18-101_k141_277209 | Not-determined |
| 18-101_k141_14279 | Not-determined |
| 18-101_k141_392200 | Not-determined |
| 18-101_k141_392201 | Not-determined |
| 18-101_k141_360499 | Not-determined |
| 18-101_k141_29956 | Not-determined |
| 18-101_k141_78928 | Not-determined |
| 18-101_k141_78929 | Not-determined |
| 18-101_k141_46513 | Not-determined |
| 18-101_k141_130440 | Low-quality |
| 18-101_k141_262327 | Not-determined |
| 18-101_k141_310932 | Not-determined |
| 18-101_k141_130445 | Not-determined |
| 18-101_k141_146020 | Low-quality |
| 18-101_k141_146021 | Not-determined |
| 18-101_k141_130447 | Not-determined |
| 18-101_k141_161740 | Not-determined |
| 18-101_k141_14296 | Not-determined |
| 18-101_k141_262333 | Not-determined |
| 18-101_k141_262336 | Not-determined |
| 18-101_k141_196193 | Not-determined |
| 18-101_k141_46530 | Low-quality |
| 18-101_k141_360516 | Not-determined |
| 18-101_k141_327529 | Low-quality |
| 18-101_k141_196197 | Not-determined |
| 18-101_k141_360519 | Not-determined |
| 18-101_k141_196198 | Not-determined |
| 18-101_k141_310945 | Low-quality |
| 18-101_k141_327532 | Not-determined |
| 18-101_k141_46540 | Not-determined |
| 18-101_k141_376256 | Not-determined |
| 18-101_k141_327536 | Not-determined |
| 18-101_k141_177636 | Low-quality |
| 18-101_k141_310948 | Not-determined |
| 18-101_k141_196210 | Not-determined |
| 18-101_k141_376261 | Low-quality |
| 18-101_k141_196215 | Not-determined |
| 18-101_k141_96465 | Not-determined |
| 18-101_k141_96467 | Low-quality |
| 18-101_k141_392219 | Not-determined |
| 18-101_k141_327540 | Not-determined |
| 18-101_k141_111591 | Not-determined |
| 18-101_k141_277239 | Not-determined |
| 18-101_k141_196225 | Not-determined |
| 18-101_k141_111602 | Not-determined |
| 18-101_k141_111603 | Low-quality |
| 18-101_k141_262349 | Not-determined |
| 18-101_k141_146041 | Not-determined |
| 18-101_k141_46557 | Not-determined |
| 18-101_k141_96478 | Not-determined |
| 18-101_k141_63569 | Not-determined |
| 18-101_k141_29987 | Not-determined |
| 18-101_k141_228326 | Not-determined |
| 18-101_k141_262362 | Not-determined |
| 18-101_k141_29993 | Not-determined |
| 18-101_k141_96487 | Not-determined |
| 18-101_k141_262369 | Not-determined |
| 18-101_k141_111613 | Not-determined |
| 18-101_k141_78957 | Not-determined |
| 18-101_k141_228341 | Not-determined |
| 18-101_k141_262373 | Not-determined |
| 18-101_k141_96494 | Low-quality |
| 18-101_k141_342367 | Not-determined |
| 18-101_k141_177647 | Not-determined |
| 18-101_k141_63583 | Not-determined |
| 18-101_k141_392245 | Not-determined |
| 18-101_k141_63585 | Not-determined |
| 18-101_k141_392248 | Not-determined |
| 18-101_k141_130495 | Not-determined |
| 18-101_k141_14324 | Not-determined |
| 18-101_k141_111618 | Not-determined |
| 18-101_k141_130498 | Not-determined |
| 18-101_k141_360535 | Not-determined |
| 18-101_k141_342370 | Low-quality |
| 18-101_k141_262390 | Not-determined |
| 18-101_k141_196242 | Low-quality |
| 18-101_k141_196243 | Not-determined |
| 18-101_k141_342382 | Not-determined |
| 18-101_k141_376291 | Not-determined |
| 18-101_k141_376292 | Not-determined |
| 18-101_k141_262403 | Not-determined |
| 18-101_k141_262404 | Not-determined |
| 18-101_k141_177665 | Not-determined |
| 18-101_k141_161759 | Not-determined |
| 18-101_k141_196247 | Low-quality |
| 18-101_k141_360550 | Not-determined |
| 18-101_k141_360551 | Not-determined |
| 18-101_k141_294592 | Not-determined |
| 18-101_k141_360552 | Not-determined |
| 18-101_k141_196253 | Not-determined |
| 18-101_k141_327557 | Not-determined |
| 18-101_k141_262411 | Not-determined |
| 18-101_k141_177678 | Not-determined |
| 18-101_k141_294596 | Not-determined |
| 18-101_k141_63608 | Not-determined |
| 18-101_k141_262414 | Not-determined |
| 18-101_k141_196262 | Not-determined |
| 18-101_k141_161774 | Not-determined |
| 18-101_k141_161775 | Not-determined |
| 18-101_k141_196267 | Not-determined |
| 18-101_k141_245236 | Not-determined |
| 18-101_k141_245237 | Not-determined |
| 18-101_k141_294603 | Not-determined |
| 18-101_k141_245243 | Not-determined |
| 18-101_k141_177692 | Not-determined |
| 18-101_k141_327569 | Not-determined |
| 18-101_k141_196277 | Not-determined |
| 18-101_k141_228369 | Not-determined |
| 18-101_k141_196279 | Not-determined |
| 18-101_k141_228372 | Not-determined |
| 18-101_k141_327574 | Not-determined |
| 18-101_k141_311010 | Not-determined |
| 18-101_k141_161803 | Not-determined |
| 18-101_k141_277280 | Not-determined |
| 18-101_k141_311017 | Low-quality |
| 18-101_k141_78987 | Not-determined |
| 18-101_k141_196285 | Not-determined |
| 18-101_k141_360572 | Not-determined |
| 18-101_k141_245265 | Not-determined |
| 18-101_k141_177714 | Not-determined |
| 18-101_k141_30050 | Not-determined |
| 18-101_k141_146069 | Not-determined |
| 18-101_k141_161817 | Not-determined |
| 18-101_k141_262436 | Not-determined |
| 18-101_k141_196292 | Not-determined |
| 18-101_k141_262437 | Not-determined |
| 18-101_k141_177729 | Not-determined |
| 18-101_k141_262438 | Not-determined |
| 18-101_k141_392268 | Not-determined |
| 18-101_k141_277290 | Not-determined |
| 18-101_k141_196295 | Not-determined |
| 18-101_k141_161823 | Not-determined |
| 18-101_k141_376315 | Low-quality |
| 18-101_k141_311018 | Low-quality |
| 18-101_k141_360577 | Not-determined |
| 18-101_k141_327591 | Not-determined |
| 18-101_k141_311020 | Not-determined |
| 18-101_k141_327599 | Not-determined |
| 18-101_k141_30067 | Not-determined |
| 18-101_k141_327603 | Not-determined |
| 18-101_k141_327609 | Not-determined |
| 18-101_k141_342432 | Not-determined |
| 18-101_k141_245286 | Not-determined |
| 18-101_k141_228411 | Not-determined |
| 18-101_k141_360589 | Not-determined |
| 18-101_k141_96562 | Not-determined |
| 18-101_k141_360590 | Not-determined |
| 18-101_k141_392285 | Not-determined |
| 18-101_k141_196300 | Not-determined |
| 18-101_k141_177741 | Not-determined |
| 18-101_k141_392289 | Not-determined |
| 18-101_k141_262465 | Not-determined |
| 18-101_k141_311028 | Not-determined |
| 18-101_k141_294631 | Not-determined |
| 18-101_k141_63631 | Low-quality |
| 18-101_k141_96570 | Not-determined |
| 18-101_k141_311033 | Low-quality |
| 18-101_k141_30080 | Not-determined |
| 18-101_k141_196316 | Not-determined |
| 18-101_k141_262477 | Not-determined |
| 18-101_k141_63641 | Not-determined |
| 18-101_k141_196320 | Not-determined |
| 18-101_k141_196326 | Not-determined |
| 18-101_k141_63650 | Not-determined |
| 18-101_k141_96591 | Not-determined |
| 18-101_k141_177750 | Not-determined |
| 18-101_k141_196340 | Not-determined |
| 18-101_k141_30093 | Not-determined |
| 18-101_k141_392305 | Not-determined |
| 18-101_k141_342457 | Not-determined |
| 18-101_k141_196344 | Not-determined |
| 18-101_k141_14347 | Not-determined |
| 18-101_k141_311061 | Not-determined |
| 18-101_k141_63663 | Not-determined |
| 18-101_k141_342466 | Not-determined |
| 18-101_k141_14348 | Not-determined |
| 18-101_k141_96600 | Not-determined |
| 18-101_k141_245333 | Not-determined |
| 18-101_k141_96601 | Not-determined |
| 18-101_k141_327622 | Not-determined |
| 18-101_k141_342475 | Not-determined |
| 18-101_k141_277322 | Not-determined |
| 18-101_k141_245344 | Not-determined |
| 18-101_k141_311076 | Not-determined |
| 18-101_k141_342480 | Not-determined |
| 18-101_k141_327630 | Not-determined |
| 18-101_k141_277326 | Not-determined |
| 18-101_k141_30105 | Not-determined |
| 18-101_k141_161841 | Not-determined |
| 18-101_k141_212895 | Not-determined |
| 18-101_k141_196366 | Not-determined |
| 18-101_k141_196369 | Not-determined |
| 18-101_k141_130561 | Not-determined |
| 18-101_k141_161844 | Not-determined |
| 18-101_k141_63682 | Not-determined |
| 18-101_k141_96615 | Not-determined |
| 18-101_k141_212905 | Not-determined |
| 18-101_k141_146089 | Not-determined |
| 18-101_k141_96625 | Not-determined |
| 18-101_k141_212907 | Not-determined |
| 18-101_k141_392318 | Not-determined |
| 18-101_k141_161854 | Low-quality |
| 18-101_k141_277344 | Not-determined |
| 18-101_k141_327640 | Not-determined |
| 18-101_k141_146099 | Not-determined |
| 18-101_k141_146101 | Not-determined |
| 18-101_k141_392321 | Not-determined |
| 18-101_k141_245375 | Not-determined |
| 18-101_k141_294700 | Not-determined |
| 18-101_k141_342507 | Not-determined |
| 18-101_k141_177759 | Not-determined |
| 18-101_k141_96642 | Not-determined |
| 18-101_k141_245391 | Not-determined |
| 18-101_k141_245395 | Low-quality |
| 18-101_k141_177770 | Not-determined |
| 18-101_k141_392330 | Not-determined |
| 18-101_k141_161867 | Not-determined |
| 18-101_k141_161868 | Not-determined |
| 18-101_k141_96652 | Not-determined |
| 18-101_k141_376342 | Not-determined |
| 18-101_k141_130589 | Not-determined |
| 18-101_k141_262489 | Not-determined |
| 18-101_k141_245404 | Not-determined |
| 18-101_k141_130592 | Not-determined |
| 18-101_k141_342537 | Low-quality |
| 18-101_k141_294712 | Not-determined |
| 18-101_k141_360603 | Not-determined |
| 18-101_k141_294714 | Not-determined |
| 18-101_k141_130594 | Not-determined |
| 18-101_k141_63710 | Not-determined |
| 18-101_k141_96668 | Low-quality |
| 18-101_k141_376349 | Not-determined |
| 18-101_k141_311148 | Not-determined |
| 18-101_k141_96676 | Not-determined |
| 18-101_k141_130600 | Low-quality |
| 18-101_k141_161878 | Not-determined |
| 18-101_k141_96680 | Not-determined |
| 18-101_k141_262497 | Not-determined |
| 18-101_k141_177792 | Not-determined |
| 18-101_k141_311162 | Not-determined |
| 18-101_k141_96685 | Not-determined |
| 18-101_k141_96686 | Not-determined |
| 18-101_k141_342547 | Not-determined |
| 18-101_k141_196385 | Not-determined |
| 18-101_k141_262507 | Not-determined |
| 18-101_k141_79029 | Not-determined |
| 18-101_k141_130615 | Not-determined |
| 18-101_k141_63727 | Not-determined |
| 18-101_k141_196392 | Not-determined |
| 18-101_k141_327680 | Not-determined |
| 18-101_k141_130621 | Not-determined |
| 18-101_k141_327682 | Not-determined |
| 18-101_k141_130623 | Not-determined |
| 18-101_k141_196395 | Not-determined |
| 18-101_k141_130626 | Not-determined |
| 18-101_k141_311179 | Not-determined |
| 18-101_k141_79040 | Not-determined |
| 18-101_k141_212924 | Not-determined |
| 18-101_k141_161884 | Not-determined |
| 18-101_k141_392354 | Not-determined |
| 18-101_k141_392358 | Not-determined |
| 18-101_k141_14374 | Not-determined |
| 18-101_k141_14377 | Not-determined |
| 18-101_k141_161888 | Not-determined |
| 18-101_k141_294736 | Not-determined |
| 18-101_k141_262515 | Low-quality |
| 18-101_k141_177812 | Not-determined |
| 18-101_k141_14382 | Not-determined |
| 18-101_k141_262516 | Not-determined |
| 18-101_k141_79053 | Not-determined |
| 18-101_k141_196400 | Not-determined |
| 18-101_k141_311193 | Not-determined |
| 18-101_k141_360609 | Not-determined |
| 18-101_k141_311198 | Low-quality |
| 18-101_k141_342568 | Not-determined |
| 18-101_k141_14392 | Not-determined |
| 18-101_k141_177826 | Not-determined |
| 18-101_k141_360619 | Not-determined |
| 18-101_k141_196406 | Not-determined |
| 18-101_k141_245454 | Not-determined |
| 18-101_k141_177831 | Not-determined |
| 18-101_k141_327716 | Not-determined |
| 18-101_k141_161913 | Not-determined |
| 18-101_k141_360624 | Not-determined |
| 18-101_k141_392380 | Not-determined |
| 18-101_k141_161916 | Low-quality |
| 18-101_k141_245464 | Not-determined |
| 18-101_k141_161921 | Not-determined |
| 18-101_k141_245468 | Not-determined |
| 18-101_k141_14409 | Low-quality |
| 18-101_k141_111634 | Low-quality |
| 18-101_k141_161924 | Not-determined |
| 18-101_k141_14414 | Not-determined |
| 18-101_k141_360632 | Not-determined |
| 18-101_k141_245472 | Not-determined |
| 18-101_k141_30170 | Not-determined |
| 18-101_k141_245478 | Not-determined |
| 18-101_k141_96720 | Not-determined |
| 18-101_k141_177856 | Not-determined |
| 18-101_k141_262531 | Not-determined |
| 18-101_k141_196412 | Not-determined |
| 18-101_k141_30180 | Low-quality |
| 18-101_k141_196418 | Not-determined |
| 18-101_k141_245494 | Not-determined |
| 18-101_k141_96725 | Not-determined |
| 18-101_k141_294751 | Not-determined |
| 18-101_k141_294756 | Not-determined |
| 18-101_k141_311230 | Low-quality |
| 18-101_k141_327733 | Not-determined |
| 18-101_k141_14422 | Not-determined |
| 18-101_k141_245501 | Not-determined |
| 18-101_k141_360647 | Not-determined |
| 18-101_k141_245503 | Not-determined |
| 18-101_k141_14425 | Not-determined |
| 18-101_k141_63732 | Not-determined |
| 18-101_k141_130628 | Not-determined |
| 18-101_k141_262543 | Low-quality |
| 18-101_k141_161943 | Not-determined |
| 18-101_k141_360654 | Not-determined |
| 18-101_k141_30196 | Not-determined |
| 18-101_k141_327739 | Not-determined |
| 18-101_k141_161947 | Not-determined |
| 18-101_k141_96743 | Not-determined |
| 18-101_k141_376418 | Not-determined |
| 18-101_k141_196436 | Not-determined |
| 18-101_k141_63743 | Not-determined |
| 18-101_k141_63744 | Not-determined |
| 18-101_k141_311249 | Not-determined |
| 18-101_k141_311251 | Not-determined |
| 18-101_k141_376420 | Not-determined |
| 18-101_k141_196439 | Not-determined |
| 18-101_k141_30215 | Not-determined |
| 18-101_k141_79081 | Not-determined |
| 18-101_k141_30218 | Not-determined |
| 18-101_k141_79086 | Not-determined |
| 18-101_k141_14441 | Not-determined |
| 18-101_k141_196445 | Not-determined |
| 18-101_k141_196446 | Not-determined |
| 18-101_k141_294785 | Not-determined |
| 18-101_k141_360672 | Not-determined |
| 18-101_k141_360674 | Not-determined |
| 18-101_k141_177873 | Low-quality |
| 18-101_k141_294794 | Not-determined |
| 18-101_k141_177877 | Not-determined |
| 18-101_k141_130661 | Low-quality |
| 18-101_k141_161983 | Not-determined |
| 18-101_k141_196458 | Not-determined |
| 18-101_k141_392422 | Not-determined |
| 18-101_k141_30231 | Low-quality |
| 18-101_k141_277397 | Not-determined |
| 18-101_k141_196462 | Not-determined |
| 18-101_k141_30235 | Low-quality |
| 18-101_k141_245541 | Not-determined |
| 18-101_k141_392432 | Not-determined |
| 18-101_k141_130672 | Not-determined |
| 18-101_k141_245550 | Not-determined |
| 18-101_k141_342616 | Not-determined |
| 18-101_k141_342618 | Not-determined |
| 18-101_k141_30247 | Not-determined |
| 18-101_k141_177895 | Not-determined |
| 18-101_k141_177897 | Not-determined |
| 18-101_k141_245563 | Not-determined |
| 18-101_k141_14472 | Not-determined |
| 18-101_k141_245565 | Not-determined |
| 18-101_k141_311278 | Not-determined |
| 18-101_k141_327751 | Low-quality |
| 18-101_k141_130686 | Not-determined |
| 18-101_k141_245570 | Low-quality |
| 18-101_k141_111718 | Not-determined |
| 18-101_k141_162009 | Not-determined |
| 18-101_k141_245572 | Not-determined |
| 18-101_k141_392447 | Not-determined |
| 18-101_k141_342628 | Not-determined |
| 18-101_k141_96815 | Not-determined |
| 18-101_k141_130697 | Not-determined |
| 18-110_k141_111056 | Medium-quality |
| 18-101_k141_342631 | Not-determined |
| 18-101_k141_360711 | Not-determined |
| 18-101_k141_212945 | Not-determined |
| 18-101_k141_360714 | Not-determined |
| 18-101_k141_130703 | Not-determined |
| 18-101_k141_63756 | Not-determined |
| 18-101_k141_212949 | Not-determined |
| 18-101_k141_130706 | Not-determined |
| 18-101_k141_162026 | Not-determined |
| 18-101_k141_262578 | Not-determined |
| 18-101_k141_111730 | Not-determined |
| 18-101_k141_342648 | Not-determined |
| 18-101_k141_96829 | Not-determined |
| 18-101_k141_130707 | Low-quality |
| 18-101_k141_311303 | Not-determined |
| 18-101_k141_294822 | Not-determined |
| 18-101_k141_130712 | Not-determined |
| 18-101_k141_130713 | Not-determined |
| 18-101_k141_294825 | Not-determined |
| 18-101_k141_14497 | Not-determined |
| 18-101_k141_277402 | Low-quality |
| 18-101_k141_245583 | Low-quality |
| 18-101_k141_96839 | Not-determined |
| 18-101_k141_14500 | Not-determined |
| 18-101_k141_212967 | Not-determined |
| 18-101_k141_262596 | Not-determined |
| 18-101_k141_177935 | Not-determined |
| 18-101_k141_162028 | Not-determined |
| 18-101_k141_277410 | Not-determined |
| 18-101_k141_30280 | Not-determined |
| 18-101_k141_294832 | Not-determined |
| 18-101_k141_360724 | Not-determined |
| 18-101_k141_360725 | Not-determined |
| 18-101_k141_311317 | Not-determined |
| 18-101_k141_376478 | Not-determined |
| 18-101_k141_63780 | Not-determined |
| 18-101_k141_245599 | Not-determined |
| 18-101_k141_376485 | Not-determined |
| 18-101_k141_79124 | Not-determined |
| 18-101_k141_63786 | Not-determined |
| 18-101_k141_96852 | Not-determined |
| 18-101_k141_96854 | Not-determined |
| 18-101_k141_277421 | Not-determined |
| 18-101_k141_294839 | Not-determined |
| 18-101_k141_96857 | Not-determined |
| 18-101_k141_262606 | Not-determined |
| 18-101_k141_96860 | Low-quality |
| 18-101_k141_228423 | Low-quality |
| 18-101_k141_392472 | Not-determined |
| 18-101_k141_294843 | Not-determined |
| 18-101_k141_30301 | Not-determined |
| 18-101_k141_342689 | Not-determined |
| 18-101_k141_177965 | Not-determined |
| 18-101_k141_228439 | Not-determined |
| 18-101_k141_392479 | Not-determined |
| 18-101_k141_79140 | Not-determined |
| 18-101_k141_360750 | Not-determined |
| 18-101_k141_360751 | Not-determined |
| 18-101_k141_130739 | Not-determined |
| 18-101_k141_245630 | Low-quality |
| 18-101_k141_177981 | Not-determined |
| 18-101_k141_376500 | Not-determined |
| 18-101_k141_360755 | Not-determined |
| 18-101_k141_63803 | Not-determined |
| 18-101_k141_327785 | Not-determined |
| 18-101_k141_376508 | Not-determined |
| 18-101_k141_245639 | Not-determined |
| 18-101_k141_130748 | Not-determined |
| 18-101_k141_327791 | Not-determined |
| 18-101_k141_96876 | Low-quality |
| 18-101_k141_63813 | Not-determined |
| 18-101_k141_277443 | Not-determined |
| 18-101_k141_376519 | Not-determined |
| 18-101_k141_392490 | Not-determined |
| 18-101_k141_294869 | Not-determined |
| 18-101_k141_360762 | Not-determined |
| 18-101_k141_262643 | Not-determined |
| 18-101_k141_360763 | Not-determined |
| 18-101_k141_130767 | Not-determined |
| 18-101_k141_311371 | Not-determined |
| 18-101_k141_130769 | Not-determined |
| 18-101_k141_111753 | Not-determined |
| 18-101_k141_376527 | Not-determined |

| | |
|---|---|
| 18-101_k141_294879 | Not-determined |
| 18-101_k141_311377 | Not-determined |
| 18-101_k141_392505 | Not-determined |
| 18-101_k141_294880 | Not-determined |
| 18-101_k141_262659 | Not-determined |
| 18-101_k141_14561 | Not-determined |
| 18-101_k141_146154 | Not-determined |
| 18-101_k141_262662 | Not-determined |
| 18-101_k141_14563 | Not-determined |
| 18-101_k141_311385 | Not-determined |
| 18-101_k141_162094 | Low-quality |
| 18-101_k141_245668 | Not-determined |
| 18-101_k141_79157 | Not-determined |
| 18-101_k141_178015 | Not-determined |
| 18-101_k141_146167 | Not-determined |
| 18-101_k141_376532 | Not-determined |
| 18-101_k141_130782 | Not-determined |
| 18-101_k141_146172 | Not-determined |
| 18-101_k141_311393 | Not-determined |
| 18-101_k141_111770 | Not-determined |
| 18-101_k141_63840 | Not-determined |
| 18-101_k141_63842 | Not-determined |
| 18-101_k141_277468 | Not-determined |
| 18-101_k141_327844 | Not-determined |
| 18-101_k141_360774 | Not-determined |
| 18-101_k141_277472 | Not-determined |
| 18-101_k141_130788 | Not-determined |
| 18-101_k141_162114 | Low-quality |
| 18-101_k141_392529 | Not-determined |
| 18-101_k141_277479 | Not-determined |
| 18-101_k141_277483 | Not-determined |
| 18-101_k141_130794 | Not-determined |
| 18-101_k141_146197 | Not-determined |
| 18-101_k141_311408 | Not-determined |
| 18-101_k141_294896 | Not-determined |
| 18-101_k141_14585 | Not-determined |
| 18-101_k141_342758 | Not-determined |
| 18-101_k141_294898 | Not-determined |
| 18-101_k141_14587 | Not-determined |
| 18-101_k141_245674 | Not-determined |
| 18-101_k141_327852 | Not-determined |
| 18-101_k141_342765 | Low-quality |
| 18-101_k141_146210 | Not-determined |
| 18-101_k141_63851 | Not-determined |
| 18-101_k141_376553 | Not-determined |
| 18-101_k141_79169 | Not-determined |
| 18-101_k141_342769 | Not-determined |
| 18-101_k141_162134 | Not-determined |
| 18-101_k141_277486 | Low-quality |
| 18-101_k141_146214 | Not-determined |
| 18-101_k141_392548 | Not-determined |
| 18-101_k141_342775 | Not-determined |
| 18-101_k141_245681 | Not-determined |
| 18-101_k141_111786 | Not-determined |
| 18-101_k141_130813 | Not-determined |
| 18-101_k141_130817 | Not-determined |
| 18-101_k141_63871 | Not-determined |
| 18-101_k141_63873 | Not-determined |
| 18-101_k141_30382 | Not-determined |
| 18-101_k141_277497 | Not-determined |
| 18-101_k141_228502 | Not-determined |
| 18-101_k141_376562 | Not-determined |
| 18-101_k141_130826 | Not-determined |
| 18-101_k141_228505 | Not-determined |
| 18-101_k141_327861 | Not-determined |
| 18-101_k141_360809 | Not-determined |
| 18-101_k141_14596 | Not-determined |
| 18-101_k141_162157 | Not-determined |
| 18-101_k141_376568 | Not-determined |
| 18-101_k141_376572 | Not-determined |
| 18-101_k141_130833 | Not-determined |
| 18-101_k141_277512 | Not-determined |
| 18-101_k141_130836 | Not-determined |
| 18-101_k141_262711 | Not-determined |
| 18-101_k141_327866 | Not-determined |
| 18-101_k141_327867 | Not-determined |
| 18-101_k141_162161 | Not-determined |
| 18-101_k141_327872 | Not-determined |
| 18-101_k141_311432 | Not-determined |
| 18-101_k141_130847 | Not-determined |
| 18-101_k141_14602 | Not-determined |
| 18-101_k141_311433 | Not-determined |
| 18-101_k141_96914 | Not-determined |
| 18-101_k141_277523 | Not-determined |
| 18-101_k141_360835 | Not-determined |
| 18-101_k141_360836 | Not-determined |
| 18-101_k141_262720 | Not-determined |
| 18-101_k141_96922 | Not-determined |
| 18-101_k141_262723 | Not-determined |
| 18-101_k141_360842 | Not-determined |
| 18-101_k141_311444 | Not-determined |
| 18-101_k141_96930 | Not-determined |
| 18-101_k141_327883 | Not-determined |
| 18-101_k141_277534 | Not-determined |
| 18-101_k141_178057 | Not-determined |
| 18-101_k141_277536 | Not-determined |
| 18-101_k141_277537 | Not-determined |
| 18-101_k141_294928 | Not-determined |
| 18-101_k141_146243 | Not-determined |
| 18-101_k141_63914 | Not-determined |
| 18-101_k141_392573 | Not-determined |
| 18-101_k141_392574 | Not-determined |
| 18-101_k141_30407 | Not-determined |
| 18-101_k141_392578 | Not-determined |
| 18-101_k141_327894 | Not-determined |
| 18-101_k141_342819 | Not-determined |
| 18-101_k141_360851 | Not-determined |
| 18-101_k141_262744 | Not-determined |
| 18-101_k141_63923 | Not-determined |
| 18-101_k141_178064 | Not-determined |
| 18-101_k141_392586 | Low-quality |
| 18-101_k141_178065 | Not-determined |
| 18-101_k141_277554 | Not-determined |
| 18-101_k141_327900 | Not-determined |
| 18-101_k141_327901 | Not-determined |
| 18-101_k141_262754 | Not-determined |
| 18-101_k141_342829 | Low-quality |
| 18-101_k141_228513 | Not-determined |
| 18-101_k141_392589 | Not-determined |
| 18-101_k141_14633 | Not-determined |
| 18-101_k141_277562 | Not-determined |
| 18-101_k141_178079 | Not-determined |
| 18-101_k141_360859 | Not-determined |
| 18-101_k141_14641 | Not-determined |
| 18-101_k141_311458 | Not-determined |
| 18-101_k141_392595 | Not-determined |
| 18-101_k141_311460 | Not-determined |
| 18-101_k141_294952 | Not-determined |
| 18-101_k141_360865 | Not-determined |
| 18-101_k141_392600 | Not-determined |
| 18-101_k141_162184 | Not-determined |
| 18-101_k141_178089 | Not-determined |
| 18-101_k141_392604 | Not-determined |
| 18-101_k141_360866 | Not-determined |
| 18-101_k141_228528 | Not-determined |
| 18-101_k141_360867 | Not-determined |
| 18-101_k141_311473 | Not-determined |
| 18-101_k141_162196 | Not-determined |
| 18-101_k141_96941 | Not-determined |
| 18-101_k141_360872 | Not-determined |
| 18-101_k141_14654 | Not-determined |
| 18-101_k141_14656 | Not-determined |
| 18-101_k141_277579 | Low-quality |
| 18-101_k141_162208 | Not-determined |
| 18-101_k141_196508 | Not-determined |
| 18-101_k141_342845 | Not-determined |
| 18-101_k141_14661 | Not-determined |
| 18-101_k141_311489 | Not-determined |
| 18-101_k141_311491 | Not-determined |
| 18-101_k141_311492 | Not-determined |
| 18-101_k141_327915 | Not-determined |
| 18-101_k141_111836 | Not-determined |
| 18-101_k141_228536 | Not-determined |
| 18-101_k141_196512 | Not-determined |
| 18-101_k141_111840 | Not-determined |
| 18-101_k141_327920 | Not-determined |
| 18-101_k141_162216 | Not-determined |
| 18-101_k141_130854 | Not-determined |
| 18-101_k141_262767 | Not-determined |
| 18-101_k141_277592 | Not-determined |
| 18-101_k141_130855 | Not-determined |
| 18-101_k141_130856 | Not-determined |
| 18-101_k141_294983 | Not-determined |
| 18-101_k141_196519 | Not-determined |
| 18-101_k141_162228 | Not-determined |
| 18-101_k141_162229 | Not-determined |
| 18-101_k141_178112 | Not-determined |
| 18-101_k141_178114 | Not-determined |
| 18-101_k141_79213 | Not-determined |
| 18-101_k141_96958 | Not-determined |
| 18-101_k141_360891 | Not-determined |
| 18-101_k141_162235 | Not-determined |
| 18-101_k141_360893 | Not-determined |
| 18-101_k141_327924 | Not-determined |
| 18-101_k141_212976 | Not-determined |
| 18-101_k141_178118 | Not-determined |
| 18-101_k141_294998 | Low-quality |
| 18-101_k141_212978 | Not-determined |
| 18-101_k141_342878 | Not-determined |
| 18-101_k141_327934 | Low-quality |
| 18-101_k141_63946 | Not-determined |
| 18-101_k141_63947 | Not-determined |
| 18-101_k141_130871 | Not-determined |
| 18-101_k141_63948 | Not-determined |
| 18-101_k141_96966 | Not-determined |
| 18-101_k141_311510 | Not-determined |
| 18-101_k141_311511 | Not-determined |
| 18-101_k141_79223 | Not-determined |
| 18-101_k141_130875 | Low-quality |
| 18-101_k141_63960 | Not-determined |
| 18-101_k141_96972 | Not-determined |
| 18-101_k141_228560 | Not-determined |
| 18-101_k141_111859 | Not-determined |
| 18-101_k141_360904 | Not-determined |
| 18-101_k141_228562 | Not-determined |
| 18-101_k141_262774 | Not-determined |
| 18-101_k141_392639 | Not-determined |
| 18-101_k141_262776 | Not-determined |
| 18-101_k141_360909 | Not-determined |
| 18-101_k141_392641 | Not-determined |
| 18-101_k141_245811 | Not-determined |
| 18-101_k141_162244 | Not-determined |
| 18-101_k141_162245 | Not-determined |
| 18-101_k141_79239 | Not-determined |
| 18-101_k141_392645 | Low-quality |
| 18-101_k141_311524 | Not-determined |
| 18-101_k141_146321 | Not-determined |
| 18-101_k141_311527 | Not-determined |
| 18-101_k141_146322 | Not-determined |
| 18-101_k141_327945 | Not-determined |
| 18-101_k141_228575 | Not-determined |
| 18-101_k141_262779 | Not-determined |
| 18-101_k141_295021 | Not-determined |
| 18-101_k141_178123 | Low-quality |
| 18-101_k141_342907 | Not-determined |
| 18-101_k141_96986 | Not-determined |
| 18-101_k141_30474 | Not-determined |
| 18-101_k141_327953 | Not-determined |
| 18-101_k141_327955 | Not-determined |
| 18-101_k141_30476 | Not-determined |
| 18-101_k141_79249 | Not-determined |
| 18-101_k141_111889 | Not-determined |
| 18-101_k141_311542 | Not-determined |
| 18-101_k141_376598 | Not-determined |
| 18-101_k141_342916 | Not-determined |
| 18-101_k141_228591 | Not-determined |
| 18-101_k141_111893 | Not-determined |
| 18-101_k141_228596 | Not-determined |
| 18-101_k141_311553 | Not-determined |
| 18-101_k141_342923 | Not-determined |
| 18-101_k141_30490 | Not-determined |
| 18-101_k141_196543 | Not-determined |
| 18-101_k141_111903 | Low-quality |
| 18-101_k141_79270 | Not-determined |
| 18-101_k141_196551 | Not-determined |
| 18-101_k141_295023 | Not-determined |
| 18-101_k141_311569 | Not-determined |
| 18-101_k141_130929 | Not-determined |
| 18-101_k141_228621 | Not-determined |
| 18-101_k141_228626 | Not-determined |
| 18-101_k141_97007 | Not-determined |
| 18-101_k141_30508 | Not-determined |
| 18-101_k141_295035 | Not-determined |
| 18-101_k141_295037 | Not-determined |
| 18-101_k141_97009 | Not-determined |
| 18-101_k141_14739 | Not-determined |
| 18-101_k141_146343 | Not-determined |
| 18-101_k141_376625 | Not-determined |
| 18-101_k141_360937 | Not-determined |
| 18-101_k141_376628 | Not-determined |
| 18-101_k141_79300 | Not-determined |
| 18-101_k141_228643 | Not-determined |
| 18-101_k141_228645 | Not-determined |
| 18-101_k141_196576 | Not-determined |
| 18-101_k141_63989 | Not-determined |
| 18-101_k141_14753 | Not-determined |
| 18-101_k141_311577 | Not-determined |
| 18-101_k141_14754 | Not-determined |
| 18-101_k141_178182 | Not-determined |
| 18-101_k141_376641 | Low-quality |
| 18-101_k141_196584 | Not-determined |
| 18-101_k141_212993 | Not-determined |
| 18-101_k141_97023 | Not-determined |
| 18-101_k141_212994 | Not-determined |
| 18-101_k141_196587 | Low-quality |
| 18-101_k141_311587 | Low-quality |
| 18-101_k141_64004 | Not-determined |
| 18-101_k141_213004 | Not-determined |
| 18-101_k141_14768 | Low-quality |
| 18-101_k141_213007 | Not-determined |
| 18-101_k141_130951 | Not-determined |
| 18-101_k141_311595 | Not-determined |
| 18-101_k141_392682 | Not-determined |
| 18-101_k141_392683 | Not-determined |
| 18-101_k141_130956 | Not-determined |
| 18-101_k141_392687 | Not-determined |
| 18-101_k141_328013 | Not-determined |
| 18-101_k141_245880 | Not-determined |
| 18-101_k141_277687 | Not-determined |
| 18-101_k141_228677 | Not-determined |
| 18-101_k141_376657 | Not-determined |
| 18-101_k141_111945 | Low-quality |
| 18-101_k141_328014 | Not-determined |
| 18-101_k141_162333 | Not-determined |
| 18-101_k141_376664 | Low-quality |
| 18-101_k141_277697 | Not-determined |
| 18-101_k141_97053 | Low-quality |
| 18-101_k141_97054 | Not-determined |
| 18-101_k141_213019 | Not-determined |
| 18-101_k141_178204 | Not-determined |
| 18-101_k141_295077 | Low-quality |
| 18-101_k141_178206 | Not-determined |
| 18-101_k141_342952 | Not-determined |
| 18-101_k141_30555 | Not-determined |
| 18-101_k141_162338 | Not-determined |
| 18-101_k141_146381 | Not-determined |
| 18-101_k141_111951 | Not-determined |
| 18-101_k141_277705 | Not-determined |
| 18-101_k141_311614 | Not-determined |
| 18-101_k141_213029 | Not-determined |
| 18-101_k141_14800 | Not-determined |
| 18-101_k141_311617 | Not-determined |
| 18-101_k141_79323 | Not-determined |
| 18-101_k141_130967 | Not-determined |
| 18-101_k141_277712 | Not-determined |
| 18-101_k141_213031 | Not-determined |
| 18-101_k141_392701 | Not-determined |
| 18-101_k141_111961 | Low-quality |
| 18-101_k141_111962 | Low-quality |
| 18-101_k141_311620 | Not-determined |
| 18-101_k141_392702 | Low-quality |
| 18-101_k141_178216 | Not-determined |
| 18-101_k141_146386 | Not-determined |
| 18-101_k141_277714 | Low-quality |
| 18-101_k141_146389 | Not-determined |
| 18-101_k141_14808 | Not-determined |
| 18-101_k141_64041 | Not-determined |
| 18-101_k141_392707 | Not-determined |
| 18-101_k141_97073 | Not-determined |
| 18-101_k141_162343 | Not-determined |
| 18-101_k141_79326 | Not-determined |
| 18-101_k141_64046 | Not-determined |
| 18-101_k141_79328 | Not-determined |
| 18-101_k141_64047 | Not-determined |
| 18-101_k141_14815 | Not-determined |
| 18-101_k141_245916 | Not-determined |
| 18-101_k141_79330 | Low-quality |
| 18-101_k141_162347 | Not-determined |
| 18-101_k141_342964 | Not-determined |
| 18-101_k141_14818 | Not-determined |
| 18-101_k141_392713 | Not-determined |
| 18-101_k141_146397 | Not-determined |
| 18-101_k141_14822 | Low-quality |
| 18-101_k141_196618 | Not-determined |
| 18-101_k141_146398 | Not-determined |
| 18-101_k141_146400 | Low-quality |
| 18-101_k141_111968 | Not-determined |
| 18-101_k141_14824 | Not-determined |
| 18-101_k141_178236 | Not-determined |
| 18-101_k141_328047 | Not-determined |
| 18-101_k141_277732 | Not-determined |
| 18-101_k141_146405 | Not-determined |
| 18-101_k141_79347 | Not-determined |
| 18-101_k141_79348 | Not-determined |
| 18-101_k141_146412 | Not-determined |
| 18-101_k141_111977 | Not-determined |
| 18-101_k141_245933 | Not-determined |
| 18-101_k141_295098 | Not-determined |
| 18-101_k141_342971 | Not-determined |
| 18-101_k141_328051 | Not-determined |
| 18-101_k141_392727 | Not-determined |
| 18-101_k141_178242 | Not-determined |
| 18-101_k141_146415 | Not-determined |
| 18-101_k141_228683 | Not-determined |
| 18-101_k141_79364 | Not-determined |
| 18-101_k141_328055 | Not-determined |
| 18-101_k141_311639 | Not-determined |
| 18-101_k141_97092 | Not-determined |
| 18-101_k141_376709 | Not-determined |
| 18-101_k141_111989 | Not-determined |
| 18-101_k141_311648 | Not-determined |
| 18-101_k141_213049 | Not-determined |
| 18-101_k141_376713 | Not-determined |
| 18-101_k141_342989 | Low-quality |
| 18-101_k141_277736 | Not-determined |
| 18-101_k141_311652 | Not-determined |
| 18-101_k141_146424 | Not-determined |
| 18-101_k141_342993 | Low-quality |
| 18-101_k141_79371 | Not-determined |
| 18-101_k141_228694 | Not-determined |
| 18-101_k141_311657 | Not-determined |
| 18-101_k141_392732 | Not-determined |
| 18-101_k141_342997 | Not-determined |
| 18-101_k141_97106 | Not-determined |
| 18-101_k141_178245 | Not-determined |
| 18-101_k141_376725 | Not-determined |
| 18-101_k141_311669 | Not-determined |
| 18-101_k141_131012 | Low-quality |
| 18-101_k141_162357 | Not-determined |
| 18-101_k141_131013 | Not-determined |
| 18-101_k141_376728 | Not-determined |
| 18-101_k141_131017 | Not-determined |
| 18-101_k141_146428 | Not-determined |
| 18-101_k141_196632 | Not-determined |
| 18-101_k141_228700 | Not-determined |
| 18-101_k141_213059 | Not-determined |
| 18-101_k141_79386 | Not-determined |
| 18-101_k141_228701 | Not-determined |
| 18-101_k141_178249 | Not-determined |
| 18-101_k141_343015 | Not-determined |
| 18-101_k141_343016 | Not-determined |
| 18-101_k141_343017 | Not-determined |
| 18-101_k141_228709 | Not-determined |
| 18-101_k141_295128 | Not-determined |
| 18-101_k141_328073 | Not-determined |
| 18-101_k141_311676 | Not-determined |
| 18-101_k141_162365 | Not-determined |
| 18-101_k141_343020 | Not-determined |
| 18-101_k141_311677 | Low-quality |
| 18-101_k141_343032 | Not-determined |
| 18-101_k141_360984 | Not-determined |
| 18-101_k141_196641 | Not-determined |
| 18-101_k141_97118 | Not-determined |
| 18-101_k141_213066 | Not-determined |
| 18-101_k141_311692 | Not-determined |
| 18-101_k141_79411 | Not-determined |
| 18-101_k141_162374 | Not-determined |
| 18-101_k141_162376 | Not-determined |
| 18-101_k141_245951 | Not-determined |
| 18-101_k141_245952 | Not-determined |
| 18-101_k141_376771 | Not-determined |
| 18-101_k141_64073 | Not-determined |
| 18-101_k141_162381 | Not-determined |
| 18-101_k141_343034 | Low-quality |
| 18-101_k141_79418 | Not-determined |
| 18-101_k141_178281 | Not-determined |
| 18-101_k141_328094 | Not-determined |
| 18-101_k141_392753 | Not-determined |
| 18-101_k141_97124 | Not-determined |
| 18-101_k141_376779 | Low-quality |
| 18-101_k141_328099 | Not-determined |
| 18-101_k141_360997 | Not-determined |
| 18-101_k141_311718 | Not-determined |
| 18-101_k141_178286 | Not-determined |
| 18-101_k141_178293 | Not-determined |
| 18-101_k141_376791 | Not-determined |
| 18-101_k141_295154 | Not-determined |
| 18-101_k141_30575 | Not-determined |
| 18-101_k141_178301 | Not-determined |
| 18-101_k141_131025 | Not-determined |
| 18-101_k141_30578 | Not-determined |
| 18-101_k141_178303 | Not-determined |
| 18-101_k141_178304 | Not-determined |
| 18-101_k141_97148 | Not-determined |
| 18-101_k141_97150 | Not-determined |
| 18-101_k141_196648 | Not-determined |
| 18-101_k141_196649 | Not-determined |
| 18-101_k141_97152 | Not-determined |
| 18-101_k141_162422 | Low-quality |
| 18-101_k141_30587 | Not-determined |
| 18-101_k141_30588 | Not-determined |
| 18-101_k141_228745 | Not-determined |
| 18-101_k141_97155 | Not-determined |
| 18-101_k141_178314 | Not-determined |
| 18-101_k141_328122 | Not-determined |
| 18-101_k141_79433 | Not-determined |
| 18-101_k141_178317 | Not-determined |
| 18-101_k141_392779 | Not-determined |
| 18-101_k141_262816 | Not-determined |
| 18-101_k141_146470 | Not-determined |
| 18-101_k141_361016 | Not-determined |
| 18-101_k141_196668 | Not-determined |
| 18-101_k141_196671 | Not-determined |
| 18-101_k141_228758 | Not-determined |
| 18-101_k141_228760 | Not-determined |
| 18-101_k141_64128 | Not-determined |
| 18-101_k141_79456 | Not-determined |
| 18-101_k141_178337 | Not-determined |
| 18-101_k141_30606 | Not-determined |
| 18-101_k141_64130 | Not-determined |
| 18-101_k141_343076 | Low-quality |
| 18-101_k141_295166 | Not-determined |
| 18-101_k141_392793 | Not-determined |
| 18-101_k141_311756 | Not-determined |
| 18-101_k141_311758 | Not-determined |
| 18-101_k141_295172 | Low-quality |
| 18-101_k141_97180 | Not-determined |
| 18-101_k141_79479 | Not-determined |
| 18-101_k141_295173 | Low-quality |
| 18-101_k141_295175 | Not-determined |
| 18-101_k141_178353 | Not-determined |
| 18-101_k141_196697 | Not-determined |
| 18-101_k141_131060 | Not-determined |
| 18-101_k141_228777 | Not-determined |
| 18-101_k141_343105 | Low-quality |
| 18-101_k141_311773 | Not-determined |
| 18-101_k141_343106 | Not-determined |
| 18-101_k141_343108 | Not-determined |
| 18-101_k141_343109 | Not-determined |
| 18-101_k141_64141 | Not-determined |
| 18-101_k141_178365 | Not-determined |
| 18-101_k141_262835 | Not-determined |
| 18-101_k141_131072 | Not-determined |
| 18-101_k141_196703 | Low-quality |
| 18-101_k141_228795 | Not-determined |
| 18-101_k141_162477 | Not-determined |
| 18-101_k141_97188 | Low-quality |
| 18-101_k141_361048 | Not-determined |
| 18-101_k141_146481 | Not-determined |
| 18-101_k141_162479 | Not-determined |
| 18-101_k141_376854 | Not-determined |
| 18-101_k141_178375 | Not-determined |
| 18-101_k141_262846 | Not-determined |
| 18-101_k141_245962 | Low-quality |
| 18-101_k141_64155 | Not-determined |
| 18-101_k141_162483 | Low-quality |
| 18-101_k141_14883 | Not-determined |
| 18-101_k141_361051 | Not-determined |
| 18-101_k141_361052 | Not-determined |
| 18-101_k141_295210 | Not-determined |
| 18-101_k141_64167 | Not-determined |
| 18-101_k141_262855 | Not-determined |
| 18-101_k141_228802 | Not-determined |
| 18-101_k141_146495 | Not-determined |
| 18-101_k141_262865 | Not-determined |
| 18-101_k141_295212 | Not-determined |
| 18-101_k141_146497 | Not-determined |
| 18-101_k141_14898 | Not-determined |
| 18-101_k141_46583 | Not-determined |
| 18-101_k141_30650 | Not-determined |
| 18-101_k141_213084 | Not-determined |
| 18-101_k141_295216 | Not-determined |
| 18-101_k141_64177 | Not-determined |
| 18-101_k141_79522 | Not-determined |
| 18-101_k141_213086 | Not-determined |
| 18-101_k141_262873 | Not-determined |
| 18-101_k141_30656 | Not-determined |
| 18-101_k141_131100 | Not-determined |
| 18-101_k141_213091 | Not-determined |
| 18-101_k141_178403 | Not-determined |
| 18-101_k141_97218 | Not-determined |
| 18-101_k141_311815 | Not-determined |
| 18-101_k141_311817 | Not-determined |
| 18-101_k141_162512 | Not-determined |
| 18-101_k141_178416 | Not-determined |
| 18-101_k141_213095 | Not-determined |
| 18-101_k141_131112 | Not-determined |
| 18-101_k141_311823 | Not-determined |
| 18-101_k141_131115 | Low-quality |
| 18-101_k141_146509 | Low-quality |
| 18-101_k141_228829 | Not-determined |
| 18-101_k141_14919 | Not-determined |
| 18-101_k141_79550 | Not-determined |
| 18-101_k141_14922 | Not-determined |
| 18-101_k141_97235 | Not-determined |
| 18-101_k141_146517 | Not-determined |
| 18-101_k141_97237 | Low-quality |
| 18-101_k141_79556 | Not-determined |
| 18-101_k141_178437 | Not-determined |
| 18-101_k141_295218 | Not-determined |
| 18-101_k141_361066 | Not-determined |
| 18-101_k141_79559 | Not-determined |
| 18-101_k141_146528 | Low-quality |
| 18-101_k141_246008 | Not-determined |
| 18-101_k141_361075 | Low-quality |
| 18-101_k141_246009 | Not-determined |
| 18-101_k141_295226 | Not-determined |
| 18-101_k141_112034 | Not-determined |
| 18-101_k141_213118 | Not-determined |
| 18-101_k141_295232 | Not-determined |
| 18-101_k141_328200 | Not-determined |
| 18-101_k141_131137 | Not-determined |
| 18-101_k141_112042 | Not-determined |
| 18-101_k141_112043 | Not-determined |
| 18-101_k141_14938 | Not-determined |
| 18-101_k141_46612 | Not-determined |
| 18-101_k141_295234 | Not-determined |
| 18-101_k141_213124 | Not-determined |
| 18-101_k141_328205 | Not-determined |
| 18-101_k141_79574 | Not-determined |
| 18-101_k141_295237 | Low-quality |
| 18-101_k141_311840 | Low-quality |
| 18-101_k141_361083 | Low-quality |
| 18-101_k141_295242 | Not-determined |
| 18-101_k141_311843 | Not-determined |
| 18-101_k141_376912 | Not-determined |
| 18-101_k141_311849 | Not-determined |
| 18-101_k141_30719 | Not-determined |
| 18-101_k141_79585 | Not-determined |
| 18-101_k141_311858 | Not-determined |
| 18-101_k141_328221 | Not-determined |
| 18-101_k141_246018 | Not-determined |
| 18-101_k141_376924 | Not-determined |
| 18-101_k141_131145 | Not-determined |
| 18-101_k141_97275 | Not-determined |
| 18-101_k141_131148 | Not-determined |
| 18-101_k141_376931 | Not-determined |
| 18-101_k141_112069 | Not-determined |
| 18-101_k141_112070 | Not-determined |
| 18-101_k141_178479 | Low-quality |
| 18-101_k141_213152 | Not-determined |
| 18-101_k141_112075 | Not-determined |
| 18-101_k141_295259 | Not-determined |
| 18-101_k141_328234 | Not-determined |
| 18-101_k141_14958 | Not-determined |
| 18-101_k141_79605 | Not-determined |
| 18-101_k141_277883 | Not-determined |
| 18-101_k141_64203 | Not-determined |
| 18-101_k141_79608 | Not-determined |

| | |
|---|---|
| 18-101_k141_64211 | Not-determined |
| 18-101_k141_178488 | Low-quality |
| 18-101_k141_79615 | Not-determined |
| 18-101_k141_178493 | Not-determined |
| 18-101_k141_97302 | Not-determined |
| 18-101_k141_97303 | Not-determined |
| 18-101_k141_376957 | Not-determined |
| 18-101_k141_262883 | Low-quality |
| 18-101_k141_376959 | Not-determined |
| 18-101_k141_328249 | Not-determined |
| 18-101_k141_376961 | Not-determined |
| 18-101_k141_343190 | Low-quality |
| 18-101_k141_131156 | Not-determined |
| 18-101_k141_112099 | Not-determined |
| 18-101_k141_178501 | Not-determined |
| 18-101_k141_262886 | Low-quality |
| 18-101_k141_146575 | Low-quality |
| 18-101_k141_277900 | Not-determined |
| 18-101_k141_328256 | Not-determined |
| 18-101_k141_328258 | Low-quality |
| 18-101_k141_146585 | Not-determined |
| 18-101_k141_376978 | Not-determined |
| 18-101_k141_131167 | Not-determined |
| 18-101_k141_228908 | Not-determined |
| 18-101_k141_196739 | Not-determined |
| 18-101_k141_112119 | Not-determined |
| 18-101_k141_196742 | Low-quality |
| 18-101_k141_328268 | Not-determined |
| 18-101_k141_46638 | Not-determined |
| 18-101_k141_328272 | Not-determined |
| 18-101_k141_277916 | Not-determined |
| 18-101_k141_112125 | Not-determined |
| 18-101_k141_361107 | Not-determined |
| 18-101_k141_146593 | Not-determined |
| 18-101_k141_146595 | Not-determined |
| 18-101_k141_146601 | Not-determined |
| 18-101_k141_295282 | Not-determined |
| 18-101_k141_178517 | Not-determined |
| 18-101_k141_328277 | Not-determined |
| 18-101_k141_361118 | Not-determined |
| 18-101_k141_46650 | Not-determined |
| 18-101_k141_14995 | Not-determined |
| 18-101_k141_196754 | Not-determined |
| 18-101_k141_328286 | Not-determined |
| 18-101_k141_196758 | Not-determined |
| 18-101_k141_328292 | Not-determined |
| 18-101_k141_361125 | Not-determined |
| 18-101_k141_328294 | Not-determined |
| 18-101_k141_112138 | Not-determined |
| 18-101_k141_328296 | Low-quality |
| 18-101_k141_196762 | Low-quality |
| 18-101_k141_30776 | Not-determined |
| 18-101_k141_146613 | Not-determined |
| 18-101_k141_146614 | Not-determined |
| 18-101_k141_311905 | Low-quality |
| 18-101_k141_311906 | Not-determined |
| 18-101_k141_262927 | Low-quality |
| 18-101_k141_79655 | Low-quality |
| 18-101_k141_79658 | Not-determined |
| 18-101_k141_262933 | Low-quality |
| 18-101_k141_46667 | Not-determined |
| 18-101_k141_311918 | Not-determined |
| 18-101_k141_295314 | Not-determined |
| 18-101_k141_328302 | Not-determined |
| 18-101_k141_262947 | Not-determined |
| 18-101_k141_228920 | Not-determined |
| 18-101_k141_228921 | Not-determined |
| 18-101_k141_30788 | Not-determined |
| 18-101_k141_30789 | Not-determined |
| 18-101_k141_392814 | Not-determined |
| 18-101_k141_30791 | Not-determined |
| 18-101_k141_361157 | Not-determined |
| 18-101_k141_178529 | Not-determined |
| 18-101_k141_178531 | Not-determined |
| 18-101_k141_15020 | Not-determined |
| 18-101_k141_162574 | Not-determined |
| 18-101_k141_15024 | Not-determined |
| 18-101_k141_262957 | Not-determined |
| 18-101_k141_131183 | Not-determined |
| 18-101_k141_277960 | Not-determined |
| 18-101_k141_277963 | Not-determined |
| 18-101_k141_131189 | Not-determined |
| 18-101_k141_46696 | Not-determined |
| 18-101_k141_392828 | Not-determined |
| 18-101_k141_376986 | Not-determined |
| 18-101_k141_376987 | Not-determined |
| 18-101_k141_178542 | Not-determined |
| 18-101_k141_15027 | Not-determined |
| 18-101_k141_376989 | Not-determined |
| 18-101_k141_196796 | Not-determined |
| 18-101_k141_146646 | Not-determined |
| 18-101_k141_328336 | Not-determined |
| 18-101_k141_311941 | Not-determined |
| 18-101_k141_311942 | Not-determined |
| 18-101_k141_15037 | Not-determined |
| 18-101_k141_328339 | Low-quality |
| 18-101_k141_64223 | Not-determined |
| 18-101_k141_246062 | Not-determined |
| 18-101_k141_228951 | Not-determined |
| 18-101_k141_146653 | Not-determined |
| 18-101_k141_46709 | Not-determined |
| 18-101_k141_377000 | Not-determined |
| 18-101_k141_46711 | Not-determined |
| 18-101_k141_112172 | Not-determined |
| 18-101_k141_377004 | Not-determined |
| 18-101_k141_162594 | Not-determined |
| 18-101_k141_30820 | Not-determined |
| 18-101_k141_262977 | Not-determined |
| 18-101_k141_46726 | Not-determined |
| 18-101_k141_146664 | Not-determined |
| 18-101_k141_228963 | Not-determined |
| 18-101_k141_196815 | Not-determined |
| 18-101_k141_131213 | Not-determined |
| 18-101_k141_131215 | Not-determined |
| 18-101_k141_46739 | Not-determined |
| 18-101_k141_343237 | Not-determined |
| 18-101_k141_377010 | Not-determined |
| 18-101_k141_377012 | Low-quality |
| 18-101_k141_30842 | Not-determined |
| 18-101_k141_30843 | Not-determined |
| 18-101_k141_377016 | Not-determined |
| 18-101_k141_343240 | Low-quality |
| 18-101_k141_262998 | Not-determined |
| 18-101_k141_361160 | Not-determined |
| 18-101_k141_361162 | Low-quality |
| 18-101_k141_263006 | Not-determined |
| 18-101_k141_131229 | Low-quality |
| 18-101_k141_361169 | Not-determined |
| 18-101_k141_343246 | Not-determined |
| 18-101_k141_46754 | Not-determined |
| 18-101_k141_178566 | Not-determined |
| 18-101_k141_46760 | Not-determined |
| 18-101_k141_328362 | Not-determined |
| 18-101_k141_162619 | Not-determined |
| 18-101_k141_246085 | Not-determined |
| 18-101_k141_295329 | Not-determined |
| 18-101_k141_46767 | Not-determined |
| 18-101_k141_131238 | Low-quality |
| 18-101_k141_178582 | Not-determined |
| 18-101_k141_263022 | Not-determined |
| 18-101_k141_131240 | Not-determined |
| 18-101_k141_46771 | Not-determined |
| 18-101_k141_196829 | Not-determined |
| 18-101_k141_178585 | Not-determined |
| 18-101_k141_178586 | Not-determined |
| 18-101_k141_178589 | Not-determined |
| 18-101_k141_246097 | Not-determined |
| 18-101_k141_178595 | Not-determined |
| 18-101_k141_295336 | Not-determined |
| 18-101_k141_328375 | Not-determined |
| 18-101_k141_328377 | Not-determined |
| 18-101_k141_343262 | Not-determined |
| 18-101_k141_131250 | Not-determined |
| 18-101_k141_178603 | Not-determined |
| 18-101_k141_246115 | Not-determined |
| 18-101_k141_343265 | Not-determined |
| 18-101_k141_343266 | Low-quality |
| 18-101_k141_328384 | Not-determined |
| 18-101_k141_328385 | Not-determined |
| 18-101_k141_79736 | Not-determined |
| 18-101_k141_146690 | Not-determined |
| 18-101_k141_64245 | Not-determined |
| 18-101_k141_343278 | Not-determined |
| 18-101_k141_328396 | Not-determined |
| 18-101_k141_278006 | Low-quality |
| 18-101_k141_328400 | Not-determined |
| 18-101_k141_97327 | Not-determined |
| 18-101_k141_131263 | Not-determined |
| 18-101_k141_213226 | Not-determined |
| 18-101_k141_263051 | Not-determined |
| 18-101_k141_278015 | Not-determined |
| 18-101_k141_328406 | Not-determined |
| 18-101_k141_46788 | Not-determined |
| 18-101_k141_97333 | Not-determined |
| 18-101_k141_295357 | Not-determined |
| 18-101_k141_246134 | Not-determined |
| 18-101_k141_162656 | Not-determined |
| 18-101_k141_196834 | Low-quality |
| 18-101_k141_246136 | Not-determined |
| 18-101_k141_295361 | Not-determined |
| 18-101_k141_361208 | Not-determined |
| 18-101_k141_295366 | Not-determined |
| 18-101_k141_361211 | Not-determined |
| 18-101_k141_162660 | Not-determined |
| 18-101_k141_246140 | Not-determined |
| 18-101_k141_178623 | Not-determined |
| 18-101_k141_213242 | Not-determined |
| 18-101_k141_196839 | Not-determined |
| 18-101_k141_328412 | Not-determined |
| 18-101_k141_30886 | Not-determined |
| 18-101_k141_295372 | Not-determined |
| 18-101_k141_228971 | Not-determined |
| 18-101_k141_162670 | Not-determined |
| 18-101_k141_146717 | Not-determined |
| 18-101_k141_278030 | Not-determined |
| 18-101_k141_79759 | Not-determined |
| 18-101_k141_79761 | Not-determined |
| 18-101_k141_213254 | Not-determined |
| 18-101_k141_64272 | Not-determined |
| 18-101_k141_392849 | Not-determined |
| 18-101_k141_246151 | Not-determined |
| 18-101_k141_64275 | Not-determined |
| 18-101_k141_79767 | Not-determined |
| 18-101_k141_246153 | Not-determined |
| 18-101_k141_97346 | Not-determined |
| 18-101_k141_361223 | Low-quality |
| 18-101_k141_295382 | Not-determined |
| 18-101_k141_46808 | Not-determined |
| 18-101_k141_15079 | Not-determined |
| 18-101_k141_278039 | Low-quality |
| 18-101_k141_15083 | Not-determined |
| 18-101_k141_278042 | Not-determined |
| 18-101_k141_15084 | Not-determined |
| 18-101_k141_377080 | Not-determined |
| 18-101_k141_146736 | Low-quality |
| 18-101_k141_328431 | Low-quality |
| 18-101_k141_213263 | Not-determined |
| 18-101_k141_15092 | Not-determined |
| 18-101_k141_196862 | Low-quality |
| 18-101_k141_79792 | Not-determined |
| 18-101_k141_79793 | Not-determined |
| 18-101_k141_162701 | Not-determined |
| 18-101_k141_229001 | Not-determined |
| 18-101_k141_162706 | Not-determined |
| 18-101_k141_15101 | Not-determined |
| 18-101_k141_343306 | Low-quality |
| 18-101_k141_392872 | Low-quality |
| 18-101_k141_46824 | Low-quality |
| 18-101_k141_295413 | Not-determined |
| 18-101_k141_162714 | Not-determined |
| 18-101_k141_146739 | Not-determined |
| 18-101_k141_97359 | Not-determined |
| 18-101_k141_196871 | Not-determined |
| 18-101_k141_162723 | Not-determined |
| 18-101_k141_196874 | Not-determined |
| 18-101_k141_79814 | Not-determined |
| 18-101_k141_377111 | Not-determined |
| 18-101_k141_79817 | Not-determined |
| 18-101_k141_30915 | Low-quality |
| 18-101_k141_146752 | Not-determined |
| 18-101_k141_196886 | Not-determined |
| 18-101_k141_343324 | Not-determined |
| 18-101_k141_328459 | Not-determined |
| 18-101_k141_328461 | Not-determined |
| 18-101_k141_229016 | Not-determined |
| 18-101_k141_278070 | Not-determined |
| 18-101_k141_311974 | Not-determined |
| 18-101_k141_311980 | Not-determined |
| 18-101_k141_64293 | Not-determined |
| 18-101_k141_343339 | Not-determined |
| 18-101_k141_162743 | Not-determined |
| 18-101_k141_15120 | Not-determined |
| 18-101_k141_46845 | Not-determined |
| 18-101_k141_263089 | Not-determined |
| 18-101_k141_361266 | Low-quality |
| 18-101_k141_229035 | Not-determined |
| 18-101_k141_343354 | Low-quality |
| 18-101_k141_295457 | Not-determined |
| 18-101_k141_162755 | Not-determined |
| 18-101_k141_295459 | Not-determined |
| 18-101_k141_178669 | Not-determined |
| 18-101_k141_97389 | Not-determined |
| 18-101_k141_46863 | Not-determined |
| 18-101_k141_229045 | Not-determined |
| 18-101_k141_97394 | Not-determined |
| 18-101_k141_30944 | Not-determined |
| 18-101_k141_229047 | Not-determined |
| 18-101_k141_30945 | Not-determined |
| 18-101_k141_30947 | Not-determined |
| 18-101_k141_229048 | Not-determined |
| 18-101_k141_131275 | Not-determined |
| 18-101_k141_312008 | Not-determined |
| 18-101_k141_46869 | Not-determined |
| 18-101_k141_278112 | Low-quality |
| 18-101_k141_178691 | Not-determined |
| 18-101_k141_30954 | Not-determined |
| 18-101_k141_278115 | Not-determined |
| 18-101_k141_377144 | Low-quality |
| 18-101_k141_30960 | Not-determined |
| 18-101_k141_295464 | Not-determined |
| 18-101_k141_229057 | Not-determined |
| 18-101_k141_295465 | Not-determined |
| 18-101_k141_46883 | Not-determined |
| 18-101_k141_162774 | Not-determined |
| 18-101_k141_15128 | Not-determined |
| 18-101_k141_263096 | Not-determined |
| 18-101_k141_15129 | Not-determined |
| 18-101_k141_97405 | Low-quality |
| 18-101_k141_46888 | Not-determined |
| 18-101_k141_162781 | Not-determined |
| 18-101_k141_361292 | Not-determined |
| 18-101_k141_263104 | Not-determined |
| 18-101_k141_328489 | Not-determined |
| 18-101_k141_361299 | Not-determined |
| 18-101_k141_178710 | Not-determined |
| 18-101_k141_162789 | Not-determined |
| 18-101_k141_312030 | Not-determined |
| 18-101_k141_178713 | Not-determined |
| 18-101_k141_162793 | Not-determined |
| 18-101_k141_15140 | Not-determined |
| 18-101_k141_162801 | Not-determined |
| 18-101_k141_377162 | Not-determined |
| 18-101_k141_213320 | Not-determined |
| 18-101_k141_213321 | Not-determined |
| 18-101_k141_178721 | Not-determined |
| 18-101_k141_343356 | Not-determined |
| 18-101_k141_312045 | Not-determined |
| 18-101_k141_312047 | Not-determined |
| 18-101_k141_263111 | Low-quality |
| 18-101_k141_361312 | Not-determined |
| 18-101_k141_229081 | Low-quality |
| 18-101_k141_97428 | Not-determined |
| 18-101_k141_46913 | Not-determined |
| 18-101_k141_162812 | Not-determined |
| 18-101_k141_263113 | Not-determined |
| 18-101_k141_178736 | Not-determined |
| 18-101_k141_162825 | Not-determined |
| 18-101_k141_97439 | Not-determined |
| 18-101_k141_328507 | Low-quality |
| 18-101_k141_15156 | Not-determined |
| 18-101_k141_343380 | Not-determined |
| 18-101_k141_196963 | Not-determined |
| 18-101_k141_196965 | Not-determined |
| 18-101_k141_178745 | Not-determined |
| 18-101_k141_178746 | Not-determined |
| 18-101_k141_196968 | Not-determined |
| 18-101_k141_162830 | Not-determined |
| 18-101_k141_295520 | Not-determined |
| 18-101_k141_278158 | Not-determined |
| 18-101_k141_178752 | Not-determined |
| 18-101_k141_328518 | Low-quality |
| 18-101_k141_343391 | Not-determined |
| 18-101_k141_162838 | Not-determined |
| 18-101_k141_162839 | Not-determined |
| 18-101_k141_229110 | Not-determined |
| 18-101_k141_178757 | Low-quality |
| 18-101_k141_312098 | Not-determined |
| 18-101_k141_278171 | Not-determined |
| 18-101_k141_343399 | Not-determined |
| 18-101_k141_97472 | Not-determined |
| 18-101_k141_79893 | Low-quality |
| 18-101_k141_79894 | Low-quality |
| 18-101_k141_162866 | Not-determined |
| 18-101_k141_392940 | Not-determined |
| 18-101_k141_79903 | Not-determined |
| 18-101_k141_392946 | Not-determined |
| 18-101_k141_392948 | Not-determined |
| 18-101_k141_79907 | Not-determined |
| 18-101_k141_178778 | Not-determined |
| 18-101_k141_46936 | Not-determined |
| 18-101_k141_79912 | Not-determined |
| 18-101_k141_146815 | Not-determined |
| 18-101_k141_146818 | Not-determined |
| 18-101_k141_178790 | Not-determined |
| 18-101_k141_178794 | Not-determined |
| 18-101_k141_361367 | Not-determined |
| 18-101_k141_178798 | Not-determined |
| 18-101_k141_229143 | Not-determined |
| 18-101_k141_146825 | Not-determined |
| 18-101_k141_392965 | Not-determined |
| 18-101_k141_31025 | Not-determined |
| 18-101_k141_343420 | Not-determined |
| 18-101_k141_178819 | Not-determined |
| 18-101_k141_196994 | Not-determined |
| 18-101_k141_343422 | Low-quality |
| 18-101_k141_312127 | Not-determined |
| 18-101_k141_392977 | Not-determined |
| 18-101_k141_361380 | Not-determined |
| 18-101_k141_295557 | Low-quality |
| 18-101_k141_377231 | Not-determined |
| 18-101_k141_392979 | Not-determined |
| 18-101_k141_79917 | Not-determined |
| 18-101_k141_79920 | Not-determined |
| 18-101_k141_46975 | Not-determined |
| 18-101_k141_213352 | Low-quality |
| 18-101_k141_131307 | Not-determined |
| 18-101_k141_377234 | Not-determined |
| 18-101_k141_361396 | Not-determined |
| 18-101_k141_361398 | Not-determined |
| 18-101_k141_112177 | Not-determined |
| 18-101_k141_229182 | Not-determined |
| 18-101_k141_79928 | Not-determined |
| 18-101_k141_162877 | Not-determined |
| 18-101_k141_263127 | Not-determined |
| 18-101_k141_178835 | Not-determined |
| 18-101_k141_213359 | Low-quality |
| 18-101_k141_213362 | Not-determined |
| 18-101_k141_178844 | Not-determined |
| 18-101_k141_361406 | Not-determined |
| 18-101_k141_343439 | Not-determined |
| 18-101_k141_229192 | Not-determined |
| 18-101_k141_178846 | Not-determined |
| 18-101_k141_162890 | Not-determined |
| 18-101_k141_295583 | Not-determined |
| 18-101_k141_328527 | Not-determined |
| 18-101_k141_229195 | Not-determined |
| 18-101_k141_131331 | Not-determined |
| 18-101_k141_31065 | Not-determined |
| 18-101_k141_328528 | Not-determined |
| 18-101_k141_295591 | Not-determined |
| 18-101_k141_343445 | Low-quality |
| 18-101_k141_295596 | Not-determined |
| 18-101_k141_312152 | Not-determined |
| 18-101_k141_328533 | Low-quality |
| 18-101_k141_312160 | Not-determined |
| 18-101_k141_343456 | Not-determined |
| 18-101_k141_263138 | Not-determined |
| 18-101_k141_131355 | Not-determined |
| 18-101_k141_162909 | Not-determined |
| 18-101_k141_263146 | Not-determined |
| 18-101_k141_328540 | Not-determined |
| 18-101_k141_178866 | Not-determined |
| 18-101_k141_162914 | Not-determined |
| 18-101_k141_64384 | Not-determined |
| 18-101_k141_64387 | Not-determined |
| 18-101_k141_393014 | Not-determined |
| 18-101_k141_162915 | Not-determined |
| 18-101_k141_97563 | Not-determined |
| 18-101_k141_393015 | Not-determined |
| 18-101_k141_146862 | Not-determined |
| 18-101_k141_178868 | Not-determined |
| 18-101_k141_47001 | Not-determined |
| 18-101_k141_178872 | Not-determined |
| 18-101_k141_97569 | Not-determined |
| 18-101_k141_31092 | Not-determined |
| 18-101_k141_146864 | Low-quality |
| 18-101_k141_64401 | Not-determined |
| 18-101_k141_146867 | Not-determined |
| 18-101_k141_47010 | Not-determined |
| 18-101_k141_15189 | Not-determined |
| 18-101_k141_47012 | Not-determined |
| 18-101_k141_229218 | Not-determined |
| 18-101_k141_146870 | Not-determined |
| 18-101_k141_146871 | Not-determined |
| 18-101_k141_47014 | Not-determined |
| 18-101_k141_393028 | Not-determined |
| 18-101_k141_31105 | Not-determined |
| 18-101_k141_97576 | Not-determined |
| 18-101_k141_278201 | Not-determined |
| 18-101_k141_229224 | Not-determined |
| 18-101_k141_31109 | Not-determined |
| 18-101_k141_393031 | Not-determined |
| 18-101_k141_178891 | Not-determined |
| 18-101_k141_146887 | Not-determined |
| 18-101_k141_213410 | Not-determined |
| 18-101_k141_377287 | Not-determined |
| 18-101_k141_79939 | Not-determined |
| 18-101_k141_377293 | Not-determined |
| 18-101_k141_79947 | Not-determined |
| 18-101_k141_64426 | Not-determined |
| 18-101_k141_64429 | Not-determined |
| 18-101_k141_393040 | Not-determined |
| 18-101_k141_377299 | Not-determined |
| 18-101_k141_162946 | Not-determined |
| 18-101_k141_162947 | Not-determined |
| 18-101_k141_377303 | Not-determined |
| 18-101_k141_377306 | Not-determined |
| 18-101_k141_213417 | Not-determined |
| 18-101_k141_197027 | Low-quality |
| 18-101_k141_162951 | Low-quality |
| 18-101_k141_146908 | Not-determined |
| 18-101_k141_146909 | Not-determined |
| 18-101_k141_246182 | Not-determined |
| 18-101_k141_97601 | Not-determined |
| 18-101_k141_213422 | Not-determined |
| 18-101_k141_197040 | Not-determined |
| 18-101_k141_246185 | Not-determined |
| 18-101_k141_278241 | Not-determined |
| 18-101_k141_146917 | Not-determined |
| 18-101_k141_146918 | Not-determined |
| 18-101_k141_97611 | Not-determined |
| 18-101_k141_146920 | Not-determined |
| 18-101_k141_361447 | Not-determined |
| 18-101_k141_377313 | Not-determined |
| 18-101_k141_197050 | Not-determined |
| 18-101_k141_263192 | Low-quality |
| 18-101_k141_393048 | Not-determined |
| 18-101_k141_79962 | Low-quality |
| 18-101_k141_377318 | Not-determined |
| 18-101_k141_112199 | Not-determined |
| 18-101_k141_146930 | Not-determined |
| 18-101_k141_178937 | Not-determined |
| 18-101_k141_112201 | Not-determined |
| 18-101_k141_15236 | Not-determined |
| 18-101_k141_15237 | Not-determined |
| 18-101_k141_112203 | Not-determined |
| 18-101_k141_112204 | Not-determined |
| 18-101_k141_343474 | Not-determined |
| 18-101_k141_343477 | Not-determined |
| 18-101_k141_64461 | Not-determined |
| 18-101_k141_328576 | Not-determined |
| 18-101_k141_162973 | Not-determined |
| 18-101_k141_64462 | Not-determined |
| 18-101_k141_328577 | Not-determined |
| 18-101_k141_178943 | Not-determined |
| 18-101_k141_79978 | Not-determined |
| 18-101_k141_393055 | Low-quality |
| 18-101_k141_295615 | Not-determined |
| 18-101_k141_312174 | Not-determined |
| 18-101_k141_343481 | Not-determined |
| 18-101_k141_213447 | Not-determined |
| 18-101_k141_97634 | Not-determined |
| 18-101_k141_328581 | Not-determined |
| 18-101_k141_47021 | Not-determined |
| 18-101_k141_79983 | Not-determined |
| 18-101_k141_47024 | Low-quality |
| 18-101_k141_328583 | Low-quality |
| 18-101_k141_246216 | Not-determined |
| 18-101_k141_246217 | Not-determined |
| 18-101_k141_229275 | Not-determined |
| 18-101_k141_377341 | Not-determined |
| 18-101_k141_197078 | Not-determined |
| 18-101_k141_393063 | Not-determined |
| 18-101_k141_112224 | Not-determined |
| 18-101_k141_278275 | Not-determined |
| 18-101_k141_31127 | Not-determined |
| 18-101_k141_31128 | Not-determined |
| 18-101_k141_229282 | Not-determined |
| 18-101_k141_146947 | Not-determined |
| 18-101_k141_146948 | Not-determined |
| 18-101_k141_47030 | Not-determined |
| 18-101_k141_64476 | Not-determined |
| 18-101_k141_361472 | Not-determined |
| 18-101_k141_197079 | Low-quality |
| 18-101_k141_47033 | Not-determined |
| 18-101_k141_343496 | Not-determined |
| 18-101_k141_64479 | Not-determined |
| 18-101_k141_197081 | Not-determined |
| 18-101_k141_295628 | Not-determined |
| 18-101_k141_80006 | Low-quality |
| 18-101_k141_229292 | Not-determined |
| 18-101_k141_229295 | Not-determined |
| 18-101_k141_31136 | Not-determined |
| 18-101_k141_263215 | Not-determined |
| 18-101_k141_246239 | Not-determined |
| 18-101_k141_163003 | Not-determined |
| 18-101_k141_97658 | Not-determined |
| 18-101_k141_178959 | Low-quality |
| 18-101_k141_15268 | Not-determined |
| 18-101_k141_163006 | Not-determined |
| 18-101_k141_312193 | Not-determined |
| 18-101_k141_97664 | Not-determined |
| 18-101_k141_15274 | Not-determined |
| 18-101_k141_361493 | Not-determined |
| 18-101_k141_312198 | Not-determined |
| 18-101_k141_97670 | Not-determined |
| 18-101_k141_15280 | Not-determined |
| 18-101_k141_163017 | Not-determined |
| 18-101_k141_64493 | Not-determined |
| 18-101_k141_97677 | Not-determined |
| 18-101_k141_97678 | Low-quality |
| 18-101_k141_328593 | Not-determined |
| 18-101_k141_178978 | Not-determined |
| 18-101_k141_377351 | Not-determined |
| 18-101_k141_361507 | Not-determined |
| 18-101_k141_229316 | Not-determined |
| 18-101_k141_377355 | Not-determined |
| 18-101_k141_31155 | Not-determined |
| 18-101_k141_64522 | Not-determined |
| 18-101_k141_213467 | Not-determined |
| 18-101_k141_31159 | Not-determined |
| 18-101_k141_80052 | Not-determined |
| 18-101_k141_361519 | Not-determined |
| 18-101_k141_263235 | Not-determined |
| 18-101_k141_295646 | Not-determined |
| 18-101_k141_377368 | Not-determined |
| 18-101_k141_64532 | Not-determined |
| 18-101_k141_15313 | Not-determined |
| 18-101_k141_361523 | Not-determined |
| 18-101_k141_229328 | Low-quality |
| 18-101_k141_163025 | Not-determined |
| 18-101_k141_312222 | Not-determined |
| 18-101_k141_328617 | Not-determined |
| 18-101_k141_80063 | Not-determined |
| 18-101_k141_31170 | Not-determined |
| 18-101_k141_47084 | Not-determined |
| 18-101_k141_361537 | Not-determined |
| 18-101_k141_112275 | Low-quality |
| 18-101_k141_97696 | Not-determined |
| 18-101_k141_246291 | Not-determined |
| 18-101_k141_80076 | Low-quality |
| 18-101_k141_328632 | Not-determined |
| 18-101_k141_178983 | Not-determined |
| 18-101_k141_328635 | Not-determined |
| 18-101_k141_178985 | Low-quality |
| 18-101_k141_213478 | Low-quality |
| 18-101_k141_213480 | Not-determined |
| 18-101_k141_31184 | Not-determined |
| 18-101_k141_343560 | Not-determined |

| | |
|---|---|
| 18-101_k141_31188 | Not-determined |
| 18-101_k141_31189 | Not-determined |
| 18-101_k141_295681 | Not-determined |
| 18-101_k141_246294 | Low-quality |
| 18-101_k141_31191 | Not-determined |
| 18-101_k141_246297 | Not-determined |
| 18-101_k141_131374 | Not-determined |
| 18-101_k141_47102 | Not-determined |
| 18-101_k141_328651 | Not-determined |
| 18-101_k141_295684 | Not-determined |
| 18-101_k141_197124 | Not-determined |
| 18-101_k141_80089 | Not-determined |
| 18-101_k141_377395 | Low-quality |
| 18-101_k141_31201 | Not-determined |
| 18-101_k141_15332 | Low-quality |
| 18-101_k141_328658 | Not-determined |
| 18-101_k141_213495 | Not-determined |
| 18-101_k141_31212 | Not-determined |
| 18-101_k141_163057 | Not-determined |
| 18-101_k141_328663 | Not-determined |
| 18-101_k141_393125 | Not-determined |
| 18-101_k141_15336 | Not-determined |
| 18-101_k141_80099 | Not-determined |
| 18-101_k141_393128 | Not-determined |
| 18-101_k141_393129 | Not-determined |
| 18-101_k141_163062 | Low-quality |
| 18-101_k141_97739 | Not-determined |
| 18-101_k141_328668 | Not-determined |
| 18-101_k141_197148 | Not-determined |
| 18-101_k141_80104 | Not-determined |
| 18-101_k141_229370 | Not-determined |
| 18-101_k141_15352 | Not-determined |
| 18-101_k141_377414 | Not-determined |
| 18-101_k141_393136 | Low-quality |
| 18-101_k141_295712 | Not-determined |
| 18-101_k141_377421 | Not-determined |
| 18-101_k141_131389 | Not-determined |
| 18-101_k141_15359 | Not-determined |
| 18-101_k141_179015 | Not-determined |
| 18-101_k141_163071 | Low-quality |
| 18-101_k141_179017 | Not-determined |
| 18-101_k141_47121 | Not-determined |
| 18-101_k141_328685 | Not-determined |
| 18-101_k141_229380 | Not-determined |
| 18-101_k141_328688 | Not-determined |
| 18-101_k141_97753 | Not-determined |
| 18-101_k141_246302 | Low-quality |
| 18-101_k141_112300 | Not-determined |
| 18-101_k141_229388 | Not-determined |
| 18-101_k141_97757 | Not-determined |
| 18-101_k141_80120 | Not-determined |
| 18-101_k141_31242 | Not-determined |
| 18-101_k141_31247 | Not-determined |
| 18-101_k141_343594 | Not-determined |
| 18-101_k141_97766 | Not-determined |
| 18-101_k141_31250 | Not-determined |
| 18-101_k141_393152 | Not-determined |
| 18-101_k141_47141 | Not-determined |
| 18-101_k141_31252 | Not-determined |
| 18-101_k141_295723 | Not-determined |
| 18-101_k141_295724 | Not-determined |
| 18-101_k141_393158 | Not-determined |
| 18-101_k141_163084 | Not-determined |
| 18-101_k141_163085 | Not-determined |
| 18-101_k141_15374 | Low-quality |
| 18-101_k141_377452 | Not-determined |
| 18-101_k141_163087 | Not-determined |
| 18-101_k141_47147 | Not-determined |
| 18-101_k141_64599 | Not-determined |
| 18-101_k141_328720 | Not-determined |
| 18-101_k141_64602 | Not-determined |
| 18-101_k141_146985 | Not-determined |
| 18-101_k141_80141 | Not-determined |
| 18-101_k141_15388 | Low-quality |
| 18-101_k141_31256 | Not-determined |
| 18-101_k141_393165 | Not-determined |
| 18-101_k141_295733 | Not-determined |
| 18-101_k141_47163 | Not-determined |
| 18-101_k141_393166 | Not-determined |
| 18-101_k141_31261 | Not-determined |
| 18-101_k141_246331 | Not-determined |
| 18-101_k141_343615 | Not-determined |
| 18-101_k141_97775 | Not-determined |
| 18-101_k141_213539 | Not-determined |
| 18-101_k141_97779 | Not-determined |
| 18-101_k141_213545 | Not-determined |
| 18-101_k141_197172 | Not-determined |
| 18-101_k141_377467 | Low-quality |
| 18-101_k141_213547 | Not-determined |
| 18-101_k141_229414 | Low-quality |
| 18-101_k141_97788 | Low-quality |
| 18-101_k141_197176 | Not-determined |
| 18-101_k141_328736 | Not-determined |
| 18-101_k141_163103 | Not-determined |
| 18-101_k141_15416 | Low-quality |
| 18-101_k141_47185 | Not-determined |
| 18-101_k141_64624 | Not-determined |
| 18-101_k141_229423 | Low-quality |
| 18-101_k141_213570 | Not-determined |
| 18-101_k141_295750 | Not-determined |
| 18-101_k141_179076 | Not-determined |
| 18-101_k141_147002 | Not-determined |
| 18-101_k141_64631 | Low-quality |
| 18-101_k141_295759 | Not-determined |
| 18-101_k141_47203 | Not-determined |
| 18-101_k141_147009 | Low-quality |
| 18-101_k141_47209 | Not-determined |
| 18-101_k141_229440 | Not-determined |
| 18-101_k141_312265 | Not-determined |
| 18-101_k141_47211 | Not-determined |
| 18-101_k141_229441 | Not-determined |
| 18-101_k141_361549 | Not-determined |
| 18-101_k141_229443 | Not-determined |
| 18-101_k141_295768 | Not-determined |
| 18-101_k141_295770 | Not-determined |
| 18-101_k141_312272 | Not-determined |
| 18-101_k141_179092 | Not-determined |
| 18-101_k141_377493 | Not-determined |
| 18-101_k141_97793 | Not-determined |
| 18-101_k141_295780 | Not-determined |
| 18-101_k141_295782 | Not-determined |
| 18-101_k141_97804 | Not-determined |
| 18-101_k141_295785 | Not-determined |
| 18-101_k141_47219 | Not-determined |
| 18-101_k141_147026 | Not-determined |
| 18-101_k141_179102 | Not-determined |
| 18-101_k141_147028 | Not-determined |
| 18-101_k141_377505 | Not-determined |
| 18-101_k141_15438 | Not-determined |
| 18-101_k141_328761 | Not-determined |
| 18-101_k141_64664 | Not-determined |
| 18-101_k141_15446 | Not-determined |
| 18-101_k141_64666 | Not-determined |
| 18-101_k141_47229 | Not-determined |
| 18-101_k141_80184 | Low-quality |
| 18-101_k141_31293 | Not-determined |
| 18-101_k141_361570 | Not-determined |
| 18-101_k141_15453 | Not-determined |
| 18-101_k141_31298 | Not-determined |
| 18-101_k141_312283 | Not-determined |
| 18-101_k141_295798 | Not-determined |
| 18-101_k141_15454 | Not-determined |
| 18-101_k141_312289 | Not-determined |
| 18-101_k141_15460 | Not-determined |
| 18-101_k141_47240 | Not-determined |
| 18-101_k141_295804 | Not-determined |
| 18-101_k141_97821 | Not-determined |
| 18-101_k141_343621 | Low-quality |
| 18-101_k141_393172 | Not-determined |
| 18-101_k141_15466 | Not-determined |
| 18-101_k141_179117 | Not-determined |
| 18-101_k141_97826 | Not-determined |
| 18-101_k141_163116 | Not-determined |
| 18-101_k141_328777 | Not-determined |
| 18-101_k141_213606 | Not-determined |
| 18-101_k141_64684 | Not-determined |
| 18-101_k141_213609 | Not-determined |
| 18-101_k141_112413 | Not-determined |
| 18-101_k141_361574 | Not-determined |
| 18-101_k141_213612 | Low-quality |
| 18-101_k141_15478 | Not-determined |
| 18-101_k141_213616 | Not-determined |
| 18-101_k141_80218 | Not-determined |
| 18-101_k141_147043 | Not-determined |
| 18-101_k141_343633 | Not-determined |
| 18-101_k141_343635 | Not-determined |
| 18-101_k141_147045 | Not-determined |
| 18-101_k141_15488 | Not-determined |
| 18-101_k141_377534 | Not-determined |
| 18-101_k141_15490 | Low-quality |
| 18-101_k141_328785 | Low-quality |
| 18-101_k141_328792 | Not-determined |
| 18-101_k141_179143 | Not-determined |
| 18-101_k141_393191 | Not-determined |
| 18-101_k141_312313 | Not-determined |
| 18-101_k141_312314 | Not-determined |
| 18-101_k141_80233 | Low-quality |
| 18-101_k141_64706 | Not-determined |
| 18-101_k141_312316 | Not-determined |
| 18-101_k141_312318 | Not-determined |
| 18-101_k141_64711 | Not-determined |
| 18-101_k141_361586 | Not-determined |
| 18-101_k141_47268 | Low-quality |
| 18-101_k141_328804 | Low-quality |
| 18-101_k141_47271 | Not-determined |
| 18-101_k141_361590 | Not-determined |
| 18-101_k141_47273 | Not-determined |
| 18-101_k141_361592 | Low-quality |
| 18-101_k141_97863 | Not-determined |
| 18-101_k141_343649 | Not-determined |
| 18-101_k141_80252 | Not-determined |
| 18-101_k141_131473 | Not-determined |
| 18-101_k141_213636 | Not-determined |
| 18-101_k141_213638 | Not-determined |
| 18-101_k141_64734 | Not-determined |
| 18-101_k141_213642 | Not-determined |
| 18-101_k141_213646 | Not-determined |
| 18-101_k141_343654 | Not-determined |
| 18-101_k141_131483 | Not-determined |
| 18-101_k141_47289 | Not-determined |
| 18-101_k141_47292 | Not-determined |
| 18-101_k141_361610 | Not-determined |
| 18-101_k141_197257 | Not-determined |
| 18-101_k141_97898 | Not-determined |
| 18-101_k141_80274 | Not-determined |
| 18-101_k141_393208 | Not-determined |
| 18-101_k141_393210 | Not-determined |
| 18-101_k141_131493 | Low-quality |
| 18-101_k141_47302 | Not-determined |
| 18-101_k141_179204 | Not-determined |
| 18-101_k141_343661 | Not-determined |
| 18-101_k141_213657 | Not-determined |
| 18-101_k141_295818 | Not-determined |
| 18-101_k141_343663 | Not-determined |
| 18-101_k141_15519 | Not-determined |
| 18-101_k141_246389 | Not-determined |
| 18-101_k141_47308 | Low-quality |
| 18-101_k141_197268 | Not-determined |
| 18-101_k141_246391 | Not-determined |
| 18-101_k141_361624 | Not-determined |
| 18-101_k141_246396 | Not-determined |
| 18-101_k141_312346 | Not-determined |
| 18-101_k141_80287 | Not-determined |
| 18-101_k141_131505 | Not-determined |
| 18-101_k141_15524 | Not-determined |
| 18-101_k141_361628 | Not-determined |
| 18-101_k141_80296 | Not-determined |
| 18-101_k141_131509 | Not-determined |
| 18-101_k141_15535 | Not-determined |
| 18-101_k141_64753 | Not-determined |
| 18-101_k141_179224 | Not-determined |
| 18-101_k141_15538 | Not-determined |
| 18-101_k141_131512 | Not-determined |
| 18-101_k141_64759 | Not-determined |
| 18-101_k141_343677 | Not-determined |
| 18-101_k141_343678 | Not-determined |
| 18-101_k141_213685 | Not-determined |
| 18-101_k141_64762 | Not-determined |
| 18-101_k141_97920 | Not-determined |
| 18-101_k141_97923 | Not-determined |
| 18-101_k141_393219 | Low-quality |
| 18-101_k141_328867 | Not-determined |
| 18-101_k141_361639 | Low-quality |
| 18-101_k141_246403 | Not-determined |
| 18-101_k141_179241 | Not-determined |
| 18-101_k141_80302 | Low-quality |
| 18-101_k141_64782 | Low-quality |
| 18-101_k141_328884 | Not-determined |
| 18-101_k141_312369 | Not-determined |
| 18-101_k141_64788 | Not-determined |
| 18-101_k141_80307 | Not-determined |
| 18-101_k141_197282 | Not-determined |
| 18-101_k141_213707 | Not-determined |
| 18-101_k141_197284 | Not-determined |
| 18-101_k141_179246 | Not-determined |
| 18-101_k141_179248 | Not-determined |
| 18-101_k141_97952 | Low-quality |
| 18-101_k141_295840 | Not-determined |
| 18-101_k141_97954 | Not-determined |
| 18-101_k141_179252 | Not-determined |
| 18-101_k141_393240 | Not-determined |
| 18-101_k141_163202 | Not-determined |
| 18-101_k141_64800 | Not-determined |
| 18-101_k141_295842 | Not-determined |
| 18-101_k141_312378 | Not-determined |
| 18-101_k141_64802 | Not-determined |
| 18-101_k141_246415 | Not-determined |
| 18-101_k141_163216 | Not-determined |
| 18-101_k141_15582 | Not-determined |
| 18-101_k141_64812 | Not-determined |
| 18-101_k141_377626 | Not-determined |
| 18-101_k141_64815 | Not-determined |
| 18-101_k141_377627 | Low-quality |
| 18-101_k141_97979 | Not-determined |
| 18-101_k141_312396 | Not-determined |
| 18-101_k141_343702 | Not-determined |
| 18-101_k141_229493 | Not-determined |
| 18-101_k141_97988 | Not-determined |
| 18-101_k141_213723 | Not-determined |
| 18-101_k141_312399 | Not-determined |
| 18-101_k141_295859 | Low-quality |
| 18-101_k141_131530 | Not-determined |
| 18-101_k141_393246 | Not-determined |
| 18-101_k141_328926 | Not-determined |
| 18-101_k141_131534 | Not-determined |
| 18-101_k141_393254 | Not-determined |
| 18-101_k141_131538 | Not-determined |
| 18-101_k141_312404 | Not-determined |
| 18-101_k141_393255 | Not-determined |
| 18-101_k141_15596 | Not-determined |
| 18-101_k141_312408 | Not-determined |
| 18-101_k141_112443 | Not-determined |
| 18-101_k141_328936 | Not-determined |
| 18-101_k141_295872 | Not-determined |
| 18-101_k141_179292 | Not-determined |
| 18-101_k141_312414 | Not-determined |
| 18-101_k141_278317 | Not-determined |
| 18-101_k141_312416 | Not-determined |
| 18-101_k141_147105 | Not-determined |
| 18-101_k141_80334 | Not-determined |
| 18-101_k141_147110 | Low-quality |
| 18-101_k141_377644 | Not-determined |
| 18-101_k141_179297 | Not-determined |
| 18-101_k141_278319 | Not-determined |
| 18-101_k141_147115 | Not-determined |
| 18-101_k141_393269 | Not-determined |
| 18-101_k141_393271 | Not-determined |
| 18-101_k141_112452 | Not-determined |
| 18-101_k141_98026 | Not-determined |
| 18-101_k141_393273 | Not-determined |
| 18-101_k141_112457 | Not-determined |
| 18-101_k141_131546 | Not-determined |
| 18-101_k141_377653 | Not-determined |
| 18-101_k141_278326 | Not-determined |
| 18-101_k141_278332 | Not-determined |
| 18-101_k141_393275 | Not-determined |
| 18-101_k141_295898 | Not-determined |
| 18-101_k141_295899 | Not-determined |
| 18-101_k141_361669 | Not-determined |
| 18-101_k141_295900 | Not-determined |
| 18-101_k141_393283 | Not-determined |
| 18-101_k141_377661 | Low-quality |
| 18-101_k141_163239 | Not-determined |
| 18-101_k141_328971 | Not-determined |
| 18-101_k141_377662 | Not-determined |
| 18-101_k141_131563 | Not-determined |
| 18-101_k141_64827 | Not-determined |
| 18-101_k141_98036 | Not-determined |
| 18-101_k141_197296 | Low-quality |
| 18-101_k141_98039 | Not-determined |
| 18-101_k141_343732 | Not-determined |
| 18-101_k141_343736 | Not-determined |
| 18-101_k141_64834 | Not-determined |
| 18-101_k141_377676 | Not-determined |
| 18-101_k141_312435 | Not-determined |
| 18-101_k141_80367 | Not-determined |
| 18-101_k141_295908 | Not-determined |
| 18-101_k141_47344 | Not-determined |
| 18-101_k141_47345 | Low-quality |
| 18-101_k141_377685 | Low-quality |
| 18-101_k141_343740 | Not-determined |
| 18-101_k141_328983 | Not-determined |
| 18-101_k141_343745 | Not-determined |
| 18-101_k141_197312 | Not-determined |
| 18-101_k141_343749 | Not-determined |
| 18-101_k141_278357 | Not-determined |
| 18-101_k141_343758 | Not-determined |
| 18-101_k141_343759 | Low-quality |
| 18-101_k141_295928 | Low-quality |
| 18-101_k141_312450 | Not-determined |
| 18-101_k141_112474 | Low-quality |
| 18-101_k141_278370 | Not-determined |
| 18-101_k141_312454 | Not-determined |
| 18-101_k141_197322 | Not-determined |
| 18-101_k141_361694 | Not-determined |
| 18-101_k141_64847 | Not-determined |
| 18-101_k141_377712 | Not-determined |
| 18-101_k141_377714 | Not-determined |
| 18-101_k141_278376 | Not-determined |
| 18-101_k141_197326 | Not-determined |
| 18-101_k141_15610 | Not-determined |
| 18-101_k141_197328 | Not-determined |
| 18-101_k141_64855 | Not-determined |
| 18-101_k141_64857 | Not-determined |
| 18-101_k141_98077 | Not-determined |
| 18-101_k141_163280 | Not-determined |
| 18-101_k141_147149 | Not-determined |
| 18-101_k141_15621 | Not-determined |
| 18-101_k141_98082 | Not-determined |
| 18-101_k141_361698 | Not-determined |
| 18-101_k141_278393 | Not-determined |
| 18-101_k141_131590 | Not-determined |
| 18-101_k141_278394 | Not-determined |
| 18-101_k141_361709 | Not-determined |
| 18-101_k141_112504 | Not-determined |
| 18-101_k141_361711 | Low-quality |
| 18-101_k141_278404 | Not-determined |
| 18-101_k141_47351 | Low-quality |
| 18-101_k141_312486 | Not-determined |
| 18-101_k141_47352 | Not-determined |
| 18-101_k141_98094 | Not-determined |
| 18-101_k141_47355 | Not-determined |
| 18-101_k141_47358 | Not-determined |
| 18-101_k141_47360 | Not-determined |
| 18-101_k141_147175 | Not-determined |
| 18-101_k141_80415 | Not-determined |
| 18-101_k141_197340 | Not-determined |
| 18-101_k141_197341 | Not-determined |
| 18-101_k141_112514 | Low-quality |
| 18-101_k141_377726 | Not-determined |
| 18-101_k141_112518 | Not-determined |
| 18-101_k141_361726 | Not-determined |
| 18-101_k141_377729 | Low-quality |
| 18-101_k141_278407 | Not-determined |
| 18-101_k141_47376 | Low-quality |
| 18-101_k141_47379 | Not-determined |
| 18-101_k141_64882 | Not-determined |
| 18-101_k141_329038 | Not-determined |
| 18-101_k141_329040 | Not-determined |
| 18-101_k141_47387 | Not-determined |
| 18-101_k141_197349 | Low-quality |
| 18-101_k141_80457 | Not-determined |
| 18-101_k141_80461 | Not-determined |
| 18-101_k141_131609 | Not-determined |
| 18-101_k141_64890 | Not-determined |
| 18-101_k141_80472 | Not-determined |
| 18-101_k141_343765 | Not-determined |
| 18-101_k141_329055 | Not-determined |
| 18-101_k141_295962 | Not-determined |
| 18-101_k141_343770 | Not-determined |
| 18-101_k141_15673 | Not-determined |
| 18-101_k141_15675 | Not-determined |
| 18-101_k141_278419 | Not-determined |
| 18-101_k141_147222 | Not-determined |
| 18-101_k141_295971 | Not-determined |
| 18-101_k141_295973 | Not-determined |
| 18-101_k141_197368 | Not-determined |
| 18-101_k141_213784 | Low-quality |
| 18-101_k141_343779 | Not-determined |
| 18-101_k141_229504 | Not-determined |
| 18-101_k141_229505 | Not-determined |
| 18-101_k141_343788 | Not-determined |
| 18-101_k141_278433 | Not-determined |
| 18-101_k141_229510 | Not-determined |
| 18-101_k141_393295 | Not-determined |
| 18-101_k141_213791 | Not-determined |
| 18-101_k141_80498 | Not-determined |
| 18-101_k141_179314 | Not-determined |
| 18-101_k141_393302 | Not-determined |
| 18-101_k141_47424 | Not-determined |
| 18-101_k141_229513 | Not-determined |
| 18-101_k141_246459 | Not-determined |
| 18-101_k141_295984 | Not-determined |
| 18-101_k141_278443 | Not-determined |
| 18-101_k141_147231 | Not-determined |
| 18-101_k141_393312 | Not-determined |
| 18-101_k141_343803 | Not-determined |
| 18-101_k141_312526 | Not-determined |
| 18-101_k141_213805 | Not-determined |
| 18-101_k141_31356 | Not-determined |
| 18-101_k141_343809 | Not-determined |
| 18-101_k141_31359 | Not-determined |
| 18-101_k141_31360 | Not-determined |
| 18-101_k141_246477 | Not-determined |
| 18-101_k141_312531 | Not-determined |
| 18-101_k141_361762 | Not-determined |
| 18-101_k141_246490 | Not-determined |
| 18-101_k141_147247 | Not-determined |
| 18-101_k141_278454 | Not-determined |
| 18-101_k141_361775 | Not-determined |
| 18-101_k141_98114 | Not-determined |
| 18-101_k141_393320 | Not-determined |
| 18-101_k141_179362 | Not-determined |
| 18-101_k141_80529 | Not-determined |
| 18-101_k141_147251 | Not-determined |
| 18-101_k141_15692 | Not-determined |
| 18-101_k141_278467 | Not-determined |
| 18-101_k141_80534 | Not-determined |
| 18-101_k141_47439 | Low-quality |
| 18-101_k141_80540 | Not-determined |
| 18-101_k141_31400 | Not-determined |
| 18-101_k141_197384 | Not-determined |
| 18-101_k141_80552 | Not-determined |
| 18-101_k141_64953 | Not-determined |
| 18-101_k141_343815 | Not-determined |
| 18-101_k141_163290 | Not-determined |
| 18-101_k141_393336 | Not-determined |
| 18-101_k141_47453 | Not-determined |
| 18-101_k141_47454 | Not-determined |
| 18-101_k141_393338 | Low-quality |
| 18-101_k141_64967 | Not-determined |
| 18-101_k141_393343 | Not-determined |
| 18-101_k141_98136 | Not-determined |
| 18-101_k141_15721 | Not-determined |
| 18-101_k141_163304 | Not-determined |
| 18-101_k141_15726 | Low-quality |
| 18-101_k141_377775 | Not-determined |
| 18-101_k141_377777 | Low-quality |
| 18-101_k141_163314 | Low-quality |
| 18-101_k141_163316 | Low-quality |
| 18-101_k141_393353 | Not-determined |
| 18-101_k141_15736 | Not-determined |
| 18-101_k141_64973 | Not-determined |
| 18-101_k141_64974 | Not-determined |
| 18-101_k141_64977 | Not-determined |
| 18-101_k141_147265 | Not-determined |
| 18-101_k141_147267 | Not-determined |
| 18-101_k141_98153 | Not-determined |
| 18-101_k141_147268 | Not-determined |
| 18-101_k141_377794 | Not-determined |
| 18-101_k141_15747 | Low-quality |
| 18-101_k141_98155 | Not-determined |
| 18-101_k141_343839 | Not-determined |
| 18-101_k141_98156 | Not-determined |
| 18-101_k141_163328 | Not-determined |
| 18-101_k141_80587 | Not-determined |
| 18-101_k141_80590 | Not-determined |
| 18-101_k141_147273 | Not-determined |
| 18-101_k141_213840 | Not-determined |
| 18-101_k141_80593 | Not-determined |
| 18-101_k141_15761 | Not-determined |
| 18-101_k141_163340 | Not-determined |
| 18-101_k141_213847 | Low-quality |
| 18-101_k141_47479 | Not-determined |
| 18-101_k141_377810 | Not-determined |
| 18-101_k141_296011 | Not-determined |
| 18-101_k141_15773 | Not-determined |
| 18-101_k141_98179 | Not-determined |
| 18-101_k141_80614 | Not-determined |
| 18-101_k141_64982 | Not-determined |
| 18-101_k141_15782 | Not-determined |
| 18-101_k141_147291 | Not-determined |
| 18-101_k141_98187 | Not-determined |
| 18-101_k141_64997 | Not-determined |
| 18-101_k141_312570 | Not-determined |
| 18-101_k141_163370 | Not-determined |
| 18-101_k141_163371 | Not-determined |
| 18-101_k141_296035 | Not-determined |
| 18-101_k141_163375 | Not-determined |
| 18-101_k141_65009 | Not-determined |
| 18-101_k141_163376 | Low-quality |
| 18-101_k141_147305 | Not-determined |
| 18-110-final-viral-combined_k141_111056\|\|full | Medium-quality |
| 18-101_k141_343868 | Low-quality |
| 18-101_k141_98205 | Not-determined |
| 18-101_k141_278561 | Not-determined |
| 18-101_k141_65019 | Not-determined |
| 18-101_k141_65021 | Not-determined |
| 18-101_k141_343875 | Not-determined |
| 18-101_k141_65036 | Not-determined |
| 18-101_k141_80668 | Low-quality |
| 18-101_k141_246526 | Not-determined |
| 18-101_k141_163385 | Not-determined |
| 18-101_k141_246528 | Not-determined |
| 18-101_k141_213878 | Not-determined |
| 18-101_k141_163391 | Not-determined |
| 18-101_k141_377857 | Not-determined |
| 18-101_k141_47524 | Not-determined |
| 18-101_k141_278564 | Not-determined |
| 18-101_k141_47526 | Low-quality |
| 18-101_k141_361831 | Not-determined |
| 18-101_k141_361835 | Not-determined |
| 18-101_k141_197420 | Low-quality |
| 18-101_k141_278569 | Low-quality |
| 18-101_k141_343901 | Not-determined |
| 18-101_k141_163410 | Not-determined |
| 18-101_k141_229573 | Not-determined |
| 18-101_k141_65065 | Not-determined |
| 18-101_k141_229574 | Not-determined |
| 18-101_k141_112525 | Not-determined |
| 18-101_k141_278574 | Not-determined |
| 18-101_k141_393427 | Not-determined |
| 18-101_k141_98221 | Not-determined |
| 18-101_k141_393429 | Not-determined |
| 18-101_k141_80693 | Not-determined |
| 18-101_k141_377864 | Not-determined |
| 18-101_k141_361845 | Not-determined |
| 18-101_k141_361847 | Not-determined |
| 18-101_k141_98247 | Not-determined |
| 18-101_k141_377879 | Not-determined |
| 18-101_k141_278601 | Not-determined |
| 18-101_k141_112553 | Low-quality |
| 18-101_k141_246562 | Not-determined |
| 18-101_k141_343922 | Not-determined |
| 18-101_k141_179475 | Not-determined |
| 18-101_k141_65088 | Not-determined |
| 18-101_k141_98256 | Not-determined |
| 18-101_k141_98259 | Not-determined |
| 18-101_k141_98262 | Not-determined |
| 18-101_k141_65094 | Not-determined |
| 18-101_k141_393467 | Not-determined |
| 18-101_k141_65096 | Not-determined |
| 18-101_k141_296109 | Not-determined |
| 18-101_k141_278632 | Low-quality |
| 18-101_k141_147342 | Not-determined |
| 18-101_k141_246580 | Not-determined |
| 18-101_k141_179488 | Not-determined |
| 18-101_k141_377899 | Low-quality |
| 18-101_k141_246590 | Not-determined |
| 18-101_k141_147353 | Not-determined |
| 18-101_k141_343944 | Not-determined |
| 18-101_k141_15821 | Not-determined |
| 18-101_k141_147359 | Not-determined |
| 18-101_k141_296126 | Not-determined |
| 18-101_k141_147362 | Not-determined |
| 18-101_k141_343952 | Not-determined |
| 18-101_k141_278655 | Not-determined |
| 18-101_k141_343953 | Not-determined |
| 18-101_k141_80727 | Not-determined |

| | |
|---|---|
| 18-101_k141_179514 | Not-determined |
| 18-101_k141_246602 | Not-determined |
| 18-101_k141_278663 | Not-determined |
| 18-101_k141_15829 | Not-determined |
| 18-101_k141_15830 | Not-determined |
| 18-101_k141_15831 | Not-determined |
| 18-101_k141_393477 | Not-determined |
| 18-101_k141_278672 | Not-determined |
| 18-101_k141_229615 | Not-determined |
| 18-101_k141_229617 | Not-determined |
| 18-101_k141_147370 | Not-determined |
| 18-101_k141_163457 | Low-quality |
| 18-101_k141_296146 | Not-determined |
| 18-101_k141_343978 | Not-determined |
| 18-101_k141_15840 | Low-quality |
| 18-101_k141_393485 | Not-determined |
| 18-101_k141_15842 | Not-determined |
| 18-101_k141_112586 | Not-determined |
| 18-101_k141_229623 | Low-quality |
| 18-101_k141_112589 | Low-quality |
| 18-101_k141_361863 | Not-determined |
| 18-101_k141_112594 | Not-determined |
| 18-101_k141_377916 | Not-determined |
| 18-101_k141_98269 | Not-determined |
| 18-101_k141_343990 | Not-determined |
| 18-101_k141_343992 | Not-determined |
| 18-101_k141_393503 | Not-determined |
| 18-101_k141_47568 | Not-determined |
| 18-101_k141_147392 | Low-quality |
| 18-101_k141_361875 | Not-determined |
| 18-101_k141_80765 | Low-quality |
| 18-101_k141_15866 | Not-determined |
| 18-101_k141_344014 | Not-determined |
| 18-101_k141_80769 | Not-determined |
| 18-101_k141_278687 | Not-determined |
| 18-101_k141_377944 | Not-determined |
| 18-101_k141_393525 | Not-determined |
| 18-101_k141_393532 | Not-determined |
| 18-101_k141_147412 | Not-determined |
| 18-101_k141_147414 | Not-determined |
| 18-101_k141_278693 | Not-determined |
| 18-101_k141_179556 | Not-determined |
| 18-101_k141_179559 | Not-determined |
| 18-101_k141_163490 | Not-determined |
| 18-101_k141_15877 | Low-quality |
| 18-101_k141_163494 | Low-quality |
| 18-101_k141_278697 | Not-determined |
| 18-101_k141_98298 | Not-determined |
| 18-101_k141_278700 | Not-determined |
| 18-101_k141_47605 | Low-quality |
| 18-101_k141_278707 | Not-determined |
| 18-101_k141_393550 | Not-determined |
| 18-101_k141_278709 | Not-determined |
| 18-101_k141_179582 | Not-determined |
| 18-101_k141_147431 | Not-determined |
| 18-101_k141_163502 | Not-determined |
| 18-101_k141_393564 | Not-determined |
| 18-101_k141_112664 | Not-determined |
| 18-101_k141_65155 | Not-determined |
| 18-101_k141_31474 | Not-determined |
| 18-101_k141_393567 | Not-determined |
| 18-101_k141_278720 | Not-determined |
| 18-101_k141_65156 | Low-quality |
| 18-101_k141_112667 | Not-determined |
| 18-101_k141_393569 | Low-quality |
| 18-101_k141_147442 | Not-determined |
| 18-101_k141_112671 | Not-determined |
| 18-101_k141_47619 | Not-determined |
| 18-101_k141_361896 | Not-determined |
| 18-101_k141_393571 | Not-determined |
| 18-101_k141_163513 | Not-determined |
| 18-101_k141_163515 | Not-determined |
| 18-101_k141_163518 | Not-determined |
| 18-101_k141_278731 | Not-determined |
| 18-101_k141_361903 | Not-determined |
| 18-101_k141_163521 | Low-quality |
| 18-101_k141_344024 | Not-determined |
| 18-101_k141_147455 | Not-determined |
| 18-101_k141_47627 | Not-determined |
| 18-101_k141_278743 | Not-determined |
| 18-101_k141_15899 | Not-determined |
| 18-101_k141_163538 | Not-determined |
| 18-101_k141_163539 | Not-determined |
| 18-101_k141_31482 | Not-determined |
| 18-101_k141_47640 | Not-determined |
| 18-101_k141_246633 | Not-determined |
| 18-101_k141_179611 | Not-determined |
| 18-101_k141_344029 | Not-determined |
| 18-101_k141_246641 | Low-quality |
| 18-101_k141_31485 | Not-determined |
| 18-101_k141_163544 | Not-determined |
| 18-101_k141_246642 | Low-quality |
| 18-101_k141_344038 | Not-determined |
| 18-101_k141_147459 | Not-determined |
| 18-101_k141_246644 | Not-determined |
| 18-101_k141_278753 | Low-quality |
| 18-101_k141_112695 | Not-determined |
| 18-101_k141_393575 | Not-determined |
| 18-101_k141_179617 | Not-determined |
| 18-101_k141_147460 | Low-quality |
| 18-101_k141_47653 | Not-determined |
| 18-101_k141_31498 | Low-quality |
| 18-101_k141_361926 | Not-determined |
| 18-101_k141_179624 | Not-determined |
| 18-101_k141_147468 | Not-determined |
| 18-101_k141_98324 | Not-determined |
| 18-101_k141_112705 | Not-determined |
| 18-101_k141_163564 | Not-determined |
| 18-101_k141_98329 | Not-determined |
| 18-101_k141_31505 | Not-determined |
| 18-101_k141_229664 | Not-determined |
| 18-101_k141_344060 | Not-determined |
| 18-101_k141_344061 | Not-determined |
| 18-101_k141_98338 | Not-determined |
| 18-101_k141_15941 | Not-determined |
| 18-101_k141_393602 | Not-determined |
| 18-101_k141_31516 | Low-quality |
| 18-101_k141_31520 | Not-determined |
| 18-101_k141_65210 | Not-determined |
| 18-101_k141_393605 | Low-quality |
| 18-101_k141_163575 | Not-determined |
| 18-101_k141_278805 | Not-determined |
| 18-101_k141_344074 | Not-determined |
| 18-101_k141_393613 | Not-determined |
| 18-101_k141_15954 | Not-determined |
| 18-101_k141_278807 | Not-determined |
| 18-101_k141_15955 | Not-determined |
| 18-101_k141_393617 | Not-determined |
| 18-101_k141_98348 | Low-quality |
| 18-101_k141_65218 | Not-determined |
| 18-101_k141_147492 | Not-determined |
| 18-101_k141_344088 | Not-determined |
| 18-101_k141_112733 | Not-determined |
| 18-101_k141_112736 | Not-determined |
| 18-101_k141_229690 | Not-determined |
| 18-101_k141_98357 | Not-determined |
| 18-101_k141_47703 | Not-determined |
| 18-101_k141_98358 | Not-determined |
| 18-101_k141_393620 | Low-quality |
| 18-101_k141_80863 | Not-determined |
| 18-101_k141_163587 | Not-determined |
| 18-101_k141_47711 | Not-determined |
| 18-101_k141_278826 | Not-determined |
| 18-101_k141_147507 | Not-determined |
| 18-101_k141_393625 | Not-determined |
| 18-101_k141_163598 | Not-determined |
| 18-101_k141_163601 | Not-determined |
| 18-101_k141_378050 | Not-determined |
| 18-101_k141_80890 | Not-determined |
| 18-101_k141_31551 | Not-determined |
| 18-101_k141_31555 | Not-determined |
| 18-101_k141_163608 | Not-determined |
| 18-101_k141_163611 | Not-determined |
| 18-101_k141_278846 | Not-determined |
| 18-101_k141_112778 | Not-determined |
| 18-101_k141_147522 | Not-determined |
| 18-101_k141_16003 | Not-determined |
| 18-101_k141_393640 | Not-determined |
| 18-101_k141_229714 | Not-determined |
| 18-101_k141_31570 | Not-determined |
| 18-101_k141_229717 | Not-determined |
| 18-101_k141_163619 | Not-determined |
| 18-101_k141_393643 | Not-determined |
| 18-101_k141_147529 | Not-determined |
| 18-101_k141_31576 | Not-determined |
| 18-101_k141_47733 | Low-quality |
| 18-101_k141_47734 | Not-determined |
| 18-101_k141_47741 | Not-determined |
| 18-101_k141_179691 | Not-determined |
| 18-101_k141_112798 | Not-determined |
| 18-101_k141_393654 | Not-determined |
| 18-101_k141_147533 | Not-determined |
| 18-101_k141_147537 | Not-determined |
| 18-101_k141_31585 | Not-determined |
| 18-101_k141_147539 | Low-quality |
| 18-101_k141_31590 | Not-determined |
| 18-101_k141_163634 | Not-determined |
| 18-101_k141_393667 | Not-determined |
| 18-101_k141_229740 | Not-determined |
| 18-101_k141_80929 | Not-determined |
| 18-101_k141_112823 | Not-determined |
| 18-101_k141_278865 | Not-determined |
| 18-101_k141_229743 | Not-determined |
| 18-101_k141_80943 | Not-determined |
| 18-101_k141_163660 | Not-determined |
| 18-101_k141_98405 | Not-determined |
| 18-101_k141_344136 | Not-determined |
| 18-101_k141_112839 | Not-determined |
| 18-101_k141_80957 | Not-determined |
| 18-101_k141_163675 | Not-determined |
| 18-101_k141_246735 | Not-determined |
| 18-101_k141_98416 | Not-determined |
| 18-101_k141_278884 | Not-determined |
| 18-101_k141_112845 | Not-determined |
| 18-101_k141_378089 | Not-determined |
| 18-101_k141_47757 | Not-determined |
| 18-101_k141_278892 | Not-determined |
| 18-101_k141_179745 | Not-determined |
| 18-101_k141_278898 | Not-determined |
| 18-101_k141_98427 | Not-determined |
| 18-101_k141_112848 | Not-determined |
| 18-101_k141_31608 | Not-determined |
| 18-101_k141_65306 | Not-determined |
| 18-101_k141_179760 | Not-determined |
| 18-101_k141_47775 | Not-determined |
| 18-101_k141_344167 | Not-determined |
| 18-101_k141_344169 | Not-determined |
| 18-101_k141_112858 | Not-determined |
| 18-101_k141_278916 | Not-determined |
| 18-101_k141_47781 | Not-determined |
| 18-101_k141_378103 | Low-quality |
| 18-101_k141_378104 | Low-quality |
| 18-101_k141_229783 | Low-quality |
| 18-101_k141_47784 | Not-determined |
| 18-101_k141_112867 | Not-determined |
| 18-101_k141_344180 | Not-determined |
| 18-101_k141_361946 | Low-quality |
| 18-101_k141_344182 | Not-determined |
| 18-101_k141_278922 | Not-determined |
| 18-101_k141_393692 | Not-determined |
| 18-101_k141_344186 | Not-determined |
| 18-101_k141_98448 | Low-quality |
| 18-101_k141_98449 | Not-determined |
| 18-101_k141_47797 | Not-determined |
| 18-101_k141_344189 | Not-determined |
| 18-101_k141_246736 | Not-determined |
| 18-101_k141_393696 | Not-determined |
| 18-101_k141_80991 | Not-determined |
| 18-101_k141_246741 | Not-determined |
| 18-101_k141_344197 | Not-determined |
| 18-101_k141_80994 | Not-determined |
| 18-101_k141_393700 | Low-quality |
| 18-101_k141_344198 | Not-determined |
| 18-101_k141_98463 | Not-determined |
| 18-101_k141_47814 | Not-determined |
| 18-101_k141_179796 | Not-determined |
| 18-101_k141_98466 | Not-determined |
| 18-101_k141_344206 | Not-determined |
| 18-101_k141_393704 | Low-quality |
| 18-101_k141_344207 | Not-determined |
| 18-101_k141_344208 | Not-determined |
| 18-101_k141_246747 | Not-determined |
| 18-101_k141_344214 | Low-quality |
| 18-101_k141_98471 | Not-determined |
| 18-101_k141_344215 | Low-quality |
| 18-101_k141_31621 | Not-determined |
| 18-101_k141_246751 | Not-determined |
| 18-101_k141_344220 | Low-quality |
| 18-101_k141_393716 | Not-determined |
| 18-101_k141_163742 | Not-determined |
| 18-101_k141_246760 | Not-determined |
| 18-101_k141_81015 | Not-determined |
| 18-101_k141_163745 | Not-determined |
| 18-101_k141_393719 | Low-quality |
| 18-101_k141_393721 | Not-determined |
| 18-101_k141_378141 | Low-quality |
| 18-101_k141_81022 | Not-determined |
| 18-101_k141_378144 | Not-determined |
| 18-101_k141_246767 | Not-determined |
| 18-101_k141_378151 | Not-determined |
| 18-101_k141_378152 | Not-determined |
| 18-101_k141_246774 | Not-determined |
| 18-101_k141_147585 | Not-determined |
| 18-101_k141_393728 | Not-determined |
| 18-101_k141_163749 | Low-quality |
| 18-101_k141_31627 | Not-determined |
| 18-101_k141_16059 | Not-determined |
| 18-101_k141_47828 | Not-determined |
| 18-101_k141_246781 | Not-determined |
| 18-101_k141_246783 | Not-determined |
| 18-101_k141_16064 | Not-determined |
| 18-101_k141_229793 | Not-determined |
| 18-101_k141_81044 | Not-determined |
| 18-101_k141_163753 | Low-quality |
| 18-101_k141_229798 | Not-determined |
| 18-101_k141_31643 | Not-determined |
| 18-101_k141_65318 | Not-determined |
| 18-101_k141_31646 | Not-determined |
| 18-101_k141_179813 | Not-determined |
| 18-101_k141_229812 | Not-determined |
| 18-101_k141_98499 | Not-determined |
| 18-101_k141_179825 | Not-determined |
| 18-101_k141_393743 | Not-determined |
| 18-101_k141_229820 | Not-determined |
| 18-101_k141_163771 | Not-determined |
| 18-101_k141_393747 | Not-determined |
| 18-101_k141_179841 | Not-determined |
| 18-101_k141_112913 | Not-determined |
| 18-101_k141_278945 | Low-quality |
| 18-101_k141_179848 | Not-determined |
| 18-101_k141_229838 | Not-determined |
| 18-101_k141_393757 | Not-determined |
| 18-101_k141_81071 | Not-determined |
| 18-101_k141_393758 | Not-determined |
| 18-101_k141_393759 | Not-determined |
| 18-101_k141_278956 | Not-determined |
| 18-101_k141_81080 | Not-determined |
| 18-101_k141_98521 | Not-determined |
| 18-101_k141_31675 | Not-determined |
| 18-101_k141_378196 | Not-determined |
| 18-101_k141_31676 | Not-determined |
| 18-101_k141_278967 | Not-determined |
| 18-101_k141_278968 | Low-quality |
| 18-101_k141_278969 | Not-determined |
| 18-101_k141_65357 | Not-determined |
| 18-101_k141_65360 | Not-determined |
| 18-101_k141_344269 | Not-determined |
| 18-101_k141_47879 | Not-determined |
| 18-101_k141_112934 | Not-determined |
| 18-101_k141_229861 | Not-determined |
| 18-101_k141_344279 | Not-determined |
| 18-101_k141_163792 | Low-quality |
| 18-101_k141_393769 | Not-determined |
| 18-101_k141_47887 | Not-determined |
| 18-101_k141_65381 | Not-determined |
| 18-101_k141_393775 | Not-determined |
| 18-101_k141_81102 | Not-determined |
| 18-101_k141_393778 | Not-determined |
| 18-101_k141_31688 | Not-determined |
| 18-101_k141_278989 | Not-determined |
| 18-101_k141_81106 | Not-determined |
| 18-101_k141_47892 | Not-determined |
| 18-101_k141_112948 | Not-determined |
| 18-101_k141_112950 | Low-quality |
| 18-101_k141_179882 | Not-determined |
| 18-101_k141_278991 | Low-quality |
| 18-101_k141_65402 | Low-quality |
| 18-101_k141_31704 | Not-determined |
| 18-101_k141_47905 | Not-determined |
| 18-101_k141_47906 | Not-determined |
| 18-101_k141_279004 | Low-quality |
| 18-101_k141_279005 | Not-determined |
| 18-101_k141_179897 | Not-determined |
| 18-101_k141_344329 | Not-determined |
| 18-101_k141_344331 | Not-determined |
| 18-101_k141_16121 | Not-determined |
| 18-101_k141_393805 | Not-determined |
| 18-101_k141_393807 | Not-determined |
| 18-101_k141_179908 | Not-determined |
| 18-101_k141_31721 | Not-determined |
| 18-101_k141_229901 | Not-determined |
| 18-101_k141_16132 | Not-determined |
| 18-101_k141_179912 | Not-determined |
| 18-101_k141_279021 | Not-determined |
| 18-101_k141_179917 | Not-determined |
| 18-101_k141_16138 | Not-determined |
| 18-101_k141_147602 | Not-determined |
| 18-101_k141_31724 | Not-determined |
| 18-101_k141_229910 | Not-determined |
| 18-101_k141_16142 | Not-determined |
| 18-101_k141_147610 | Low-quality |
| 18-101_k141_179948 | Not-determined |
| 18-101_k141_65418 | Not-determined |
| 18-101_k141_378216 | Not-determined |
| 18-101_k141_229926 | Low-quality |
| 18-101_k141_229929 | Not-determined |
| 18-101_k141_47949 | Not-determined |
| 18-101_k141_47952 | Not-determined |
| 18-101_k141_163813 | Not-determined |
| 18-101_k141_81180 | Not-determined |
| 18-101_k141_98583 | Not-determined |
| 18-101_k141_229942 | Not-determined |
| 18-101_k141_65440 | Not-determined |
| 18-101_k141_279040 | Not-determined |
| 18-101_k141_112990 | Not-determined |
| 18-101_k141_393818 | Low-quality |
| 18-101_k141_393821 | Not-determined |
| 18-101_k141_16174 | Low-quality |
| 18-101_k141_65449 | Not-determined |
| 18-101_k141_31751 | Not-determined |
| 18-101_k141_163829 | Not-determined |
| 18-101_k141_81198 | Not-determined |
| 18-101_k141_65454 | Not-determined |
| 18-101_k141_16183 | Not-determined |
| 18-101_k141_47981 | Not-determined |
| 18-101_k141_279046 | Not-determined |
| 18-101_k141_81201 | Not-determined |
| 18-101_k141_81206 | Not-determined |
| 18-101_k141_179968 | Not-determined |
| 18-101_k141_378240 | Low-quality |
| 18-101_k141_65463 | Not-determined |
| 18-101_k141_98609 | Not-determined |
| 18-101_k141_393842 | Low-quality |
| 18-101_k141_147622 | Not-determined |
| 18-101_k141_147625 | Not-determined |
| 18-101_k141_179990 | Not-determined |
| 18-101_k141_229960 | Not-determined |
| 18-101_k141_378248 | Not-determined |
| 18-101_k141_147630 | Not-determined |
| 18-101_k141_147631 | Low-quality |
| 18-101_k141_98619 | Low-quality |
| 18-101_k141_393846 | Not-determined |
| 18-101_k141_393848 | Low-quality |
| 18-101_k141_147639 | Not-determined |
| 18-101_k141_47994 | Not-determined |
| 18-101_k141_163849 | Not-determined |
| 18-101_k141_31760 | Not-determined |
| 18-101_k141_179998 | Not-determined |
| 18-101_k141_229969 | Not-determined |
| 18-101_k141_48003 | Not-determined |
| 18-101_k141_81231 | Not-determined |
| 18-101_k141_81232 | Not-determined |
| 18-101_k141_344377 | Not-determined |
| 18-101_k141_344379 | Not-determined |
| 18-101_k141_147647 | Not-determined |
| 18-101_k141_344383 | Not-determined |
| 18-101_k141_378265 | Not-determined |
| 18-101_k141_393871 | Low-quality |
| 18-101_k141_48017 | Not-determined |
| 18-101_k141_48018 | Not-determined |
| 18-101_k141_163876 | Low-quality |
| 18-101_k141_65489 | Not-determined |
| 18-101_k141_229988 | Not-determined |
| 18-101_k141_65498 | Not-determined |
| 18-101_k141_65500 | Not-determined |
| 18-101_k141_279091 | Low-quality |
| 18-101_k141_163886 | Not-determined |
| 18-101_k141_113077 | Not-determined |
| 18-101_k141_113079 | Not-determined |
| 18-101_k141_180027 | Not-determined |
| 18-101_k141_113084 | Not-determined |
| 18-101_k141_98628 | Not-determined |
| 18-101_k141_180032 | Not-determined |
| 18-101_k141_31780 | Not-determined |
| 18-101_k141_98633 | Not-determined |
| 18-101_k141_31789 | Not-determined |
| 18-101_k141_163894 | Not-determined |
| 18-101_k141_344419 | Not-determined |
| 18-101_k141_31791 | Not-determined |
| 18-101_k141_344421 | Not-determined |
| 18-101_k141_180035 | Low-quality |
| 18-101_k141_344424 | Not-determined |
| 18-101_k141_163899 | Not-determined |
| 18-101_k141_393904 | Not-determined |
| 18-101_k141_31817 | Not-determined |
| 18-101_k141_31818 | Not-determined |
| 18-101_k141_31821 | Not-determined |
| 18-101_k141_16198 | Not-determined |
| 18-101_k141_31823 | Low-quality |
| 18-101_k141_81293 | Not-determined |
| 18-101_k141_31826 | Not-determined |
| 18-101_k141_81297 | Not-determined |
| 18-101_k141_113093 | Not-determined |
| 18-101_k141_113095 | Not-determined |
| 18-101_k141_16210 | Not-determined |
| 18-101_k141_31834 | Low-quality |
| 18-101_k141_230016 | Not-determined |
| 18-101_k141_31839 | Not-determined |
| 18-101_k141_81311 | Not-determined |
| 18-101_k141_81314 | Not-determined |
| 18-101_k141_344448 | Not-determined |
| 18-101_k141_344449 | Not-determined |
| 18-101_k141_344451 | Not-determined |
| 18-101_k141_16225 | Not-determined |
| 18-101_k141_147681 | Low-quality |
| 18-101_k141_393921 | Not-determined |
| 18-101_k141_147684 | Not-determined |
| 18-101_k141_113109 | Not-determined |
| 18-101_k141_344456 | Not-determined |
| 18-101_k141_113112 | Not-determined |
| 18-101_k141_147686 | Not-determined |
| 18-101_k141_230050 | Not-determined |
| 18-101_k141_378300 | Not-determined |
| 18-101_k141_16237 | Not-determined |
| 18-101_k141_31857 | Not-determined |
| 18-101_k141_344466 | Not-determined |
| 18-101_k141_147699 | Not-determined |
| 18-101_k141_378305 | Not-determined |
| 18-101_k141_378307 | Not-determined |
| 18-101_k141_65504 | Not-determined |
| 18-101_k141_65505 | Not-determined |
| 18-101_k141_98651 | Not-determined |
| 18-101_k141_378314 | Not-determined |
| 18-101_k141_113137 | Not-determined |
| 18-101_k141_147715 | Not-determined |
| 18-101_k141_147722 | Not-determined |
| 18-101_k141_98656 | Not-determined |
| 18-101_k141_65524 | Not-determined |
| 18-101_k141_98659 | Not-determined |
| 18-101_k141_113150 | Not-determined |
| 18-101_k141_344486 | Not-determined |
| 18-101_k141_147733 | Not-determined |
| 18-101_k141_113160 | Not-determined |
| 18-101_k141_393970 | Not-determined |
| 18-101_k141_31887 | Not-determined |
| 18-101_k141_344492 | Not-determined |
| 18-101_k141_279148 | Low-quality |
| 18-101_k141_393974 | Not-determined |
| 18-101_k141_48070 | Not-determined |
| 18-101_k141_180077 | Not-determined |
| 18-101_k141_378340 | Not-determined |
| 18-101_k141_180090 | Not-determined |
| 18-101_k141_16253 | Not-determined |
| 18-101_k141_48076 | Not-determined |
| 18-101_k141_393992 | Not-determined |
| 18-101_k141_393993 | Not-determined |
| 18-101_k141_163964 | Not-determined |
| 18-101_k141_48081 | Not-determined |
| 18-101_k141_48084 | Low-quality |
| 18-101_k141_16262 | Not-determined |
| 18-101_k141_31916 | Not-determined |
| 18-101_k141_48092 | Not-determined |
| 18-101_k141_163975 | Low-quality |
| 18-101_k141_279171 | Not-determined |
| 18-101_k141_344526 | Not-determined |
| 18-101_k141_48099 | Not-determined |
| 18-101_k141_65558 | Not-determined |
| 18-101_k141_48101 | Low-quality |
| 18-101_k141_48103 | Not-determined |
| 18-101_k141_147761 | Not-determined |
| 18-101_k141_65562 | Not-determined |
| 18-101_k141_16264 | Not-determined |
| 18-101_k141_180117 | Not-determined |
| 18-101_k141_230098 | Not-determined |
| 18-101_k141_65565 | Not-determined |
| 18-101_k141_113206 | Not-determined |
| 18-101_k141_65568 | Not-determined |
| 18-101_k141_394003 | Not-determined |
| 18-101_k141_16281 | Not-determined |
| 18-101_k141_344539 | Not-determined |
| 18-101_k141_16282 | Not-determined |
| 18-101_k141_163991 | Low-quality |
| 18-101_k141_394008 | Low-quality |
| 18-101_k141_394013 | Not-determined |
| 18-101_k141_163997 | Not-determined |
| 18-101_k141_164000 | Not-determined |
| 18-101_k141_394020 | Not-determined |
| 18-101_k141_164004 | Not-determined |
| 18-101_k141_164005 | Not-determined |
| 18-101_k141_65578 | Not-determined |
| 18-101_k141_230113 | Not-determined |
| 18-101_k141_344555 | Not-determined |
| 18-101_k141_394027 | Not-determined |
| 18-101_k141_81379 | Not-determined |
| 18-101_k141_378357 | Not-determined |
| 18-101_k141_279188 | Not-determined |
| 18-101_k141_16292 | Not-determined |
| 18-101_k141_279192 | Not-determined |
| 18-101_k141_164020 | Not-determined |
| 18-101_k141_16295 | Not-determined |
| 18-101_k141_113226 | Not-determined |
| 18-101_k141_98676 | Not-determined |
| 18-101_k141_180146 | Low-quality |
| 18-101_k141_16305 | Not-determined |
| 18-101_k141_180149 | Not-determined |
| 18-101_k141_180150 | Not-determined |
| 18-101_k141_98691 | Not-determined |
| 18-101_k141_98692 | Not-determined |
| 18-101_k141_113247 | Not-determined |
| 18-101_k141_16313 | Not-determined |
| 18-101_k141_164061 | Not-determined |
| 18-101_k141_147805 | Not-determined |
| 18-101_k141_344574 | Not-determined |
| 18-101_k141_113254 | Not-determined |
| 18-101_k141_16324 | Not-determined |
| 18-101_k141_98697 | Low-quality |
| 18-101_k141_16329 | Not-determined |
| 18-101_k141_113260 | Not-determined |
| 18-101_k141_16335 | Low-quality |
| 18-101_k141_31955 | Not-determined |
| 18-101_k141_65580 | Not-determined |
| 18-101_k141_147812 | Not-determined |
| 18-101_k141_164076 | Not-determined |
| 18-101_k141_344586 | Not-determined |
| 18-101_k141_98709 | Not-determined |
| 18-101_k141_394064 | Not-determined |
| 18-101_k141_48127 | Not-determined |
| 18-101_k141_16341 | Low-quality |
| 18-101_k141_164080 | Low-quality |
| 18-101_k141_31969 | Not-determined |
| 18-101_k141_31971 | Not-determined |
| 18-101_k141_164086 | Low-quality |
| 18-101_k141_147831 | Not-determined |
| 18-101_k141_180187 | Not-determined |
| 18-101_k141_230157 | Low-quality |
| 18-101_k141_230160 | Not-determined |
| 18-101_k141_230161 | Not-determined |
| 18-101_k141_16357 | Not-determined |
| 18-101_k141_16359 | Not-determined |
| 18-101_k141_31994 | Not-determined |
| 18-101_k141_48144 | Not-determined |
| 18-101_k141_164098 | Not-determined |
| 18-101_k141_16369 | Not-determined |
| 18-101_k141_48153 | Not-determined |
| 18-101_k141_48154 | Not-determined |
| 18-101_k141_164105 | Not-determined |
| 18-101_k141_48155 | Not-determined |
| 18-101_k141_113290 | Not-determined |
| 18-101_k141_48159 | Not-determined |

| | |
|---|---|
| 18-101_k141_113304 | Not-determined |
| 18-101_k141_16377 | Not-determined |
| 18-101_k141_113305 | Not-determined |
| 18-101_k141_113306 | Not-determined |
| 18-101_k141_81413 | Not-determined |
| 18-101_k141_147866 | Not-determined |
| 18-101_k141_113313 | Not-determined |
| 18-101_k141_147876 | Not-determined |
| 18-101_k141_48170 | Low-quality |
| 18-101_k141_230204 | Not-determined |
| 18-101_k141_147877 | Not-determined |
| 18-101_k141_147878 | Not-determined |
| 18-101_k141_147879 | Not-determined |
| 18-101_k141_344613 | Not-determined |
| 18-101_k141_81426 | Not-determined |
| 18-101_k141_279283 | Not-determined |
| 18-101_k141_16393 | Not-determined |
| 18-101_k141_279285 | Not-determined |
| 18-101_k141_147886 | Low-quality |
| 18-101_k141_16402 | Not-determined |
| 18-101_k141_180213 | Not-determined |
| 18-101_k141_16407 | Not-determined |
| 18-101_k141_180220 | Not-determined |
| 18-101_k141_147896 | Not-determined |
| 18-101_k141_180222 | Not-determined |
| 18-101_k141_344628 | Not-determined |
| 18-101_k141_180227 | Not-determined |
| 18-101_k141_81461 | Not-determined |
| 18-101_k141_230225 | Not-determined |
| 18-101_k141_16418 | Not-determined |
| 18-101_k141_16422 | Not-determined |
| 18-101_k141_180244 | Not-determined |
| 18-101_k141_81481 | Not-determined |
| 18-101_k141_230238 | Low-quality |
| 18-101_k141_230241 | Not-determined |
| 18-101_k141_65617 | Not-determined |
| 18-101_k141_180250 | Not-determined |
| 18-101_k141_279327 | Not-determined |
| 18-101_k141_164143 | Not-determined |
| 18-101_k141_279332 | Not-determined |
| 18-101_k141_16438 | Not-determined |
| 18-101_k141_344652 | Not-determined |
| 18-101_k141_81491 | Not-determined |
| 18-101_k141_81493 | Low-quality |
| 18-101_k141_344653 | Not-determined |
| 18-101_k141_279340 | Not-determined |
| 18-101_k141_394098 | Not-determined |
| 18-101_k141_344659 | Not-determined |
| 18-101_k141_230253 | Not-determined |
| 18-101_k141_48197 | Not-determined |
| 18-101_k141_230259 | Not-determined |
| 18-101_k141_164171 | Not-determined |
| 18-101_k141_147942 | Not-determined |
| 18-101_k141_81510 | Not-determined |
| 18-101_k141_344688 | Not-determined |
| 18-101_k141_65626 | Low-quality |
| 18-101_k141_65627 | Not-determined |
| 18-101_k141_344699 | Low-quality |
| 18-101_k141_394117 | Not-determined |
| 18-101_k141_344701 | Not-determined |
| 18-101_k141_344706 | Not-determined |
| 18-101_k141_230272 | Low-quality |
| 18-101_k141_164187 | Not-determined |
| 18-101_k141_230274 | Not-determined |
| 18-101_k141_180264 | Not-determined |
| 18-101_k141_230275 | Not-determined |
| 18-101_k141_48223 | Not-determined |
| 18-101_k141_48227 | Not-determined |
| 18-101_k141_32053 | Not-determined |
| 18-101_k141_147968 | Not-determined |
| 18-101_k141_113398 | Not-determined |
| 18-101_k141_394126 | Not-determined |
| 18-101_k141_48238 | Not-determined |
| 18-101_k141_81554 | Not-determined |
| 18-101_k141_81556 | Not-determined |
| 18-101_k141_81558 | Not-determined |
| 18-101_k141_394129 | Not-determined |
| 18-101_k141_81560 | Not-determined |
| 18-101_k141_344732 | Low-quality |
| 18-101_k141_81563 | Not-determined |
| 18-101_k141_394131 | Not-determined |
| 18-101_k141_180271 | Not-determined |
| 18-101_k141_81569 | Not-determined |
| 18-101_k141_180275 | Not-determined |
| 18-101_k141_148001 | Low-quality |
| 18-101_k141_48259 | Not-determined |
| 18-101_k141_113425 | Not-determined |
| 18-101_k141_164216 | Low-quality |
| 18-101_k141_230306 | Not-determined |
| 18-101_k141_113434 | Not-determined |
| 18-101_k141_164220 | Not-determined |
| 18-101_k141_148017 | Not-determined |
| 18-101_k141_113443 | Not-determined |
| 18-101_k141_164222 | Not-determined |
| 18-101_k141_32098 | Not-determined |
| 18-101_k141_230311 | Not-determined |
| 18-101_k141_164234 | Not-determined |
| 18-101_k141_113462 | Not-determined |
| 18-101_k141_81619 | Not-determined |
| 18-101_k141_344751 | Not-determined |
| 18-101_k141_81623 | Not-determined |
| 18-101_k141_180294 | Not-determined |
| 18-101_k141_148039 | Not-determined |
| 18-101_k141_164261 | Not-determined |
| 18-101_k141_148044 | Not-determined |
| 18-101_k141_32114 | Not-determined |
| 18-101_k141_32115 | Not-determined |
| 18-101_k141_180302 | Not-determined |
| 18-101_k141_394168 | Low-quality |
| 18-101_k141_32124 | Not-determined |
| 18-101_k141_32130 | Not-determined |
| 18-101_k141_32133 | Not-determined |
| 18-101_k141_180313 | Not-determined |
| 18-101_k141_32135 | Low-quality |
| 18-101_k141_32140 | Not-determined |
| 18-101_k141_65652 | Not-determined |
| 18-101_k141_65653 | Not-determined |
| 18-101_k141_344783 | Not-determined |
| 18-101_k141_344784 | Low-quality |
| 18-101_k141_344787 | Not-determined |
| 18-101_k141_113492 | Not-determined |
| 18-101_k141_65660 | Not-determined |
| 18-101_k141_344794 | Not-determined |
| 18-101_k141_344795 | Not-determined |
| 18-101_k141_180330 | Not-determined |
| 18-101_k141_180336 | Not-determined |
| 18-101_k141_378381 | Low-quality |
| 18-101_k141_394191 | Not-determined |
| 18-101_k141_81647 | Not-determined |
| 18-101_k141_180347 | Not-determined |
| 18-101_k141_48302 | Low-quality |
| 18-101_k141_113510 | Not-determined |
| 18-101_k141_180357 | Low-quality |
| 18-101_k141_113514 | Not-determined |
| 18-101_k141_378385 | Not-determined |
| 18-101_k141_65667 | Not-determined |
| 18-101_k141_113517 | Not-determined |
| 18-101_k141_344807 | Not-determined |
| 18-101_k141_148074 | Not-determined |
| 18-101_k141_81665 | Not-determined |
| 18-101_k141_394223 | Not-determined |
| 18-101_k141_164290 | Low-quality |
| 18-101_k141_378396 | Low-quality |
| 18-101_k141_81675 | Not-determined |
| 18-101_k141_378397 | Not-determined |
| 18-101_k141_65684 | Not-determined |
| 18-101_k141_394237 | Not-determined |
| 18-101_k141_81679 | Not-determined |
| 18-101_k141_65697 | Not-determined |
| 18-101_k141_65698 | Not-determined |
| 18-101_k141_344816 | Not-determined |
| 18-101_k141_32216 | Not-determined |
| 18-101_k141_32218 | Low-quality |
| 18-101_k141_32220 | Not-determined |
| 18-101_k141_164317 | Not-determined |
| 18-101_k141_32225 | Not-determined |
| 18-101_k141_32226 | Not-determined |
| 18-101_k141_344822 | Not-determined |
| 18-101_k141_32229 | Not-determined |
| 18-101_k141_344825 | Not-determined |
| 18-101_k141_180401 | Not-determined |
| 18-101_k141_180403 | Low-quality |
| 18-101_k141_180406 | Not-determined |
| 18-101_k141_344837 | Not-determined |
| 18-101_k141_164327 | Not-determined |
| 18-101_k141_32248 | Not-determined |
| 18-101_k141_65705 | Not-determined |
| 18-101_k141_164340 | Not-determined |
| 18-101_k141_394283 | Not-determined |
| 18-101_k141_344846 | Not-determined |
| 18-101_k141_32255 | Not-determined |
| 18-101_k141_344850 | Low-quality |
| 18-101_k141_180432 | Not-determined |
| 18-101_k141_394307 | Not-determined |
| 18-101_k141_113553 | Not-determined |
| 18-101_k141_180434 | Not-determined |
| 18-101_k141_180435 | Not-determined |
| 18-101_k141_164351 | Not-determined |
| 18-101_k141_180442 | Not-determined |
| 18-101_k141_32267 | Not-determined |
| 18-101_k141_164357 | Not-determined |
| 18-101_k141_344861 | Low-quality |
| 18-101_k141_344864 | Not-determined |
| 18-101_k141_32277 | Not-determined |
| 18-101_k141_32281 | Not-determined |
| 18-101_k141_164358 | Not-determined |
| 18-101_k141_164359 | Not-determined |
| 18-101_k141_344873 | Not-determined |
| 18-101_k141_164366 | Not-determined |
| 18-101_k141_344881 | Not-determined |
| 18-101_k141_32304 | Not-determined |
| 18-101_k141_279401 | Not-determined |
| 18-101_k141_344897 | Low-quality |
| 18-101_k141_113573 | Not-determined |
| 18-101_k141_113578 | Not-determined |
| 18-101_k141_180449 | Not-determined |
| 18-101_k141_113579 | Not-determined |
| 18-101_k141_344899 | Not-determined |
| 18-101_k141_48340 | Not-determined |
| 18-101_k141_279418 | Not-determined |
| 18-101_k141_344906 | Not-determined |
| 18-101_k141_180459 | Low-quality |
| 18-101_k141_344914 | Not-determined |
| 18-101_k141_81776 | Not-determined |
| 18-101_k141_344917 | Not-determined |
| 18-101_k141_81778 | Not-determined |
| 18-101_k141_48349 | Not-determined |
| 18-101_k141_113601 | Not-determined |
| 18-101_k141_344921 | Not-determined |
| 18-101_k141_48358 | Not-determined |
| 18-101_k141_279430 | Not-determined |
| 18-101_k141_113604 | Low-quality |
| 18-101_k141_164399 | Low-quality |
| 18-101_k141_48361 | Not-determined |
| 18-101_k141_32336 | Not-determined |
| 18-101_k141_394351 | Not-determined |
| 18-101_k141_279440 | Not-determined |
| 18-101_k141_32340 | Not-determined |
| 18-101_k141_48367 | Low-quality |
| 18-101_k141_164404 | Not-determined |
| 18-101_k141_32348 | Not-determined |
| 18-101_k141_164415 | Not-determined |
| 18-101_k141_81792 | Not-determined |
| 18-101_k141_279455 | Not-determined |
| 18-101_k141_164426 | Not-determined |
| 18-101_k141_279479 | Not-determined |
| 18-101_k141_32362 | Not-determined |
| 18-101_k141_48400 | Not-determined |
| 18-101_k141_32371 | Not-determined |
| 18-101_k141_180492 | Low-quality |
| 18-101_k141_344954 | Not-determined |
| 18-101_k141_279488 | Not-determined |
| 18-101_k141_81796 | Not-determined |
| 18-101_k141_48413 | Low-quality |
| 18-101_k141_344964 | Not-determined |
| 18-101_k141_344965 | Not-determined |
| 18-101_k141_32379 | Not-determined |
| 18-101_k141_48434 | Not-determined |
| 18-101_k141_344978 | Not-determined |
| 18-101_k141_344982 | Not-determined |
| 18-101_k141_81834 | Not-determined |
| 18-101_k141_113682 | Not-determined |
| 18-101_k141_81838 | Not-determined |
| 18-101_k141_344987 | Low-quality |
| 18-101_k141_48448 | Not-determined |
| 18-101_k141_48449 | Not-determined |
| 18-101_k141_394403 | Low-quality |
| 18-101_k141_344994 | Not-determined |
| 18-101_k141_344996 | Not-determined |
| 18-101_k141_32403 | Not-determined |
| 18-101_k141_180528 | Low-quality |
| 18-101_k141_48455 | Not-determined |
| 18-101_k141_113697 | Not-determined |
| 18-101_k141_180540 | Not-determined |
| 18-101_k141_345006 | Not-determined |
| 18-101_k141_345010 | Not-determined |
| 18-101_k141_32416 | Not-determined |
| 18-101_k141_48470 | Not-determined |
| 18-101_k141_394414 | Low-quality |
| 18-101_k141_32424 | Not-determined |
| 18-101_k141_48484 | Not-determined |
| 18-101_k141_48487 | Not-determined |
| 18-101_k141_48488 | Not-determined |
| 18-101_k141_32453 | Not-determined |
| 18-101_k141_32459 | Not-determined |
| 18-101_k141_279508 | Not-determined |
| 18-101_k141_48491 | Not-determined |
| 18-101_k141_345043 | Not-determined |
| 18-101_k141_394436 | Not-determined |
| 18-101_k141_394437 | Not-determined |
| 18-101_k141_32464 | Not-determined |
| 18-101_k141_345053 | Not-determined |
| 18-101_k141_345057 | Not-determined |
| 18-101_k141_180583 | Not-determined |
| 18-101_k141_394446 | Not-determined |
| 18-101_k141_394451 | Not-determined |
| 18-101_k141_48508 | Not-determined |
| 18-101_k141_32478 | Not-determined |
| 18-101_k141_48513 | Not-determined |
| 18-101_k141_48517 | Not-determined |
| 18-101_k141_180601 | Not-determined |
| 18-101_k141_48522 | Not-determined |
| 18-101_k141_81900 | Low-quality |
| 18-101_k141_180604 | Not-determined |
| 18-101_k141_81901 | Not-determined |
| 18-101_k141_32493 | Not-determined |
| 18-101_k141_180611 | Not-determined |
| 18-101_k141_48532 | Not-determined |
| 18-101_k141_180617 | Not-determined |
| 18-101_k141_48538 | Not-determined |
| 18-101_k141_394475 | Not-determined |
| 18-101_k141_394478 | Not-determined |
| 18-101_k141_345079 | Not-determined |
| 18-101_k141_394480 | Low-quality |
| 18-101_k141_32523 | Not-determined |
| 18-101_k141_113797 | Not-determined |
| 18-101_k141_65753 | Not-determined |
| 18-101_k141_65767 | Not-determined |
| 18-101_k141_180652 | Not-determined |
| 18-101_k141_394491 | Not-determined |
| 18-101_k141_345105 | Not-determined |
| 18-101_k141_32549 | Not-determined |
| 18-101_k141_394503 | Not-determined |
| 18-101_k141_180663 | Not-determined |
| 18-101_k141_180664 | Low-quality |
| 18-101_k141_345118 | Not-determined |
| 18-101_k141_345119 | Not-determined |
| 18-101_k141_113835 | Not-determined |
| 18-101_k141_164430 | Low-quality |
| 18-101_k141_394513 | Not-determined |
| 18-101_k141_113845 | Not-determined |
| 18-101_k141_345131 | Not-determined |
| 18-101_k141_394520 | Not-determined |
| 18-101_k141_81939 | Not-determined |
| 18-101_k141_164438 | Not-determined |
| 18-101_k141_65790 | Not-determined |
| 18-101_k141_32568 | Not-determined |
| 18-101_k141_81947 | Not-determined |
| 18-101_k141_32571 | Not-determined |
| 18-101_k141_113863 | Not-determined |
| 18-101_k141_81960 | Not-determined |
| 18-101_k141_180678 | Not-determined |
| 18-101_k141_32584 | Not-determined |
| 18-101_k141_345148 | Not-determined |
| 18-101_k141_164454 | Not-determined |
| 18-101_k141_164455 | Not-determined |
| 18-101_k141_164466 | Not-determined |
| 18-101_k141_32608 | Low-quality |
| 18-101_k141_164470 | Not-determined |
| 18-101_k141_113906 | Not-determined |
| 18-101_k141_394530 | Not-determined |
| 18-101_k141_164482 | Not-determined |
| 18-101_k141_394535 | Not-determined |
| 18-101_k141_394537 | Not-determined |
| 18-101_k141_180699 | Not-determined |
| 18-101_k141_279548 | Not-determined |
| 18-101_k141_279550 | Not-determined |
| 18-101_k141_394542 | Not-determined |
| 18-101_k141_279554 | Not-determined |
| 18-101_k141_48656 | Not-determined |
| 18-101_k141_394551 | Not-determined |
| 18-101_k141_113924 | Not-determined |
| 18-101_k141_32641 | Low-quality |
| 18-101_k141_32645 | Not-determined |
| 18-101_k141_394563 | Low-quality |
| 18-101_k141_48681 | Not-determined |
| 18-101_k141_164492 | Not-determined |
| 18-101_k141_180738 | Not-determined |
| 18-101_k141_48685 | Not-determined |
| 18-101_k141_279587 | Not-determined |
| 18-101_k141_180744 | Not-determined |
| 18-101_k141_279593 | Not-determined |
| 18-101_k141_164500 | Not-determined |
| 18-101_k141_48700 | Not-determined |
| 18-101_k141_164502 | Not-determined |
| 18-101_k141_164503 | Not-determined |
| 18-101_k141_48708 | Not-determined |
| 18-101_k141_180756 | Not-determined |
| 18-101_k141_180758 | Not-determined |
| 18-101_k141_113935 | Low-quality |
| 18-101_k141_279603 | Not-determined |
| 18-101_k141_48724 | Not-determined |
| 18-101_k141_81983 | Not-determined |
| 18-101_k141_113958 | Low-quality |
| 18-101_k141_164529 | Not-determined |
| 18-101_k141_345165 | Not-determined |
| 18-101_k141_345166 | Not-determined |
| 18-101_k141_81990 | Not-determined |
| 18-101_k141_32714 | Low-quality |
| 18-101_k141_180791 | Low-quality |
| 18-101_k141_32725 | Not-determined |
| 18-101_k141_180795 | Not-determined |
| 18-101_k141_279636 | Not-determined |
| 18-101_k141_82005 | Not-determined |
| 18-101_k141_345184 | Not-determined |
| 18-101_k141_345186 | Not-determined |
| 18-101_k141_180798 | Not-determined |
| 18-101_k141_345191 | Not-determined |
| 18-101_k141_32750 | Not-determined |
| 18-101_k141_113980 | Not-determined |
| 18-101_k141_82034 | Not-determined |
| 18-101_k141_180812 | Low-quality |
| 18-101_k141_180813 | Low-quality |
| 18-101_k141_279652 | Not-determined |
| 18-101_k141_32758 | Not-determined |
| 18-101_k141_32760 | Not-determined |
| 18-101_k141_82038 | Not-determined |
| 18-101_k141_279659 | Low-quality |
| 18-101_k141_113984 | Not-determined |
| 18-101_k141_82044 | Not-determined |
| 18-101_k141_345214 | Not-determined |
| 18-101_k141_82050 | Not-determined |
| 18-101_k141_279675 | Not-determined |
| 18-101_k141_279676 | Not-determined |
| 18-101_k141_279679 | Not-determined |
| 18-101_k141_279684 | Not-determined |
| 18-101_k141_279685 | Not-determined |
| 18-101_k141_82072 | Low-quality |
| 18-101_k141_114025 | Not-determined |
| 18-101_k141_180861 | Not-determined |
| 18-105-final-viral-combined_k141_147531\|\|full | High-quality |
| 18-101_k141_32789 | Low-quality |
| 18-101_k141_114035 | Not-determined |
| 18-101_k141_48757 | Low-quality |
| 18-101_k141_48762 | Not-determined |
| 18-101_k141_32800 | Not-determined |
| 18-101_k141_114047 | Not-determined |
| 18-101_k141_180887 | Not-determined |
| 18-101_k141_180889 | Not-determined |
| 18-101_k141_114060 | Not-determined |
| 18-101_k141_48783 | Not-determined |
| 18-101_k141_345241 | Low-quality |
| 18-101_k141_82107 | Not-determined |
| 18-101_k141_82118 | Not-determined |
| 18-101_k141_345252 | Not-determined |
| 18-101_k141_345265 | Not-determined |
| 18-101_k141_345267 | Low-quality |
| 18-101_k141_114079 | Not-determined |
| 18-101_k141_82139 | Not-determined |
| 18-101_k141_345271 | Not-determined |
| 18-101_k141_114082 | Not-determined |
| 18-101_k141_114084 | Not-determined |
| 18-101_k141_114086 | Not-determined |
| 18-101_k141_32823 | Low-quality |
| 18-101_k141_394577 | Not-determined |
| 18-101_k141_394585 | Not-determined |
| 18-101_k141_345283 | Not-determined |
| 18-101_k141_345285 | Not-determined |
| 18-101_k141_114112 | Low-quality |
| 18-101_k141_345293 | Not-determined |
| 18-101_k141_82154 | Not-determined |
| 18-101_k141_180922 | Not-determined |
| 18-101_k141_114120 | Not-determined |
| 18-101_k141_114122 | Not-determined |
| 18-101_k141_394605 | Not-determined |
| 18-101_k141_82162 | Not-determined |
| 18-101_k141_394607 | Not-determined |
| 18-101_k141_32840 | Low-quality |
| 18-101_k141_394619 | Not-determined |
| 18-101_k141_82184 | Not-determined |
| 18-101_k141_394624 | Not-determined |
| 18-101_k141_394625 | Not-determined |
| 18-101_k141_394626 | Not-determined |
| 18-101_k141_114149 | Not-determined |
| 18-101_k141_345311 | Not-determined |
| 18-101_k141_48837 | Not-determined |
| 18-101_k141_114161 | Not-determined |
| 18-101_k141_48851 | Not-determined |
| 18-101_k141_82198 | Not-determined |
| 18-101_k141_394641 | Not-determined |
| 18-101_k141_345326 | Not-determined |
| 18-101_k141_394648 | Not-determined |
| 18-101_k141_394654 | Not-determined |
| 18-101_k141_394664 | Not-determined |
| 18-101_k141_114189 | Not-determined |
| 18-101_k141_394667 | Not-determined |
| 18-101_k141_394668 | Not-determined |
| 18-101_k141_394670 | Not-determined |
| 18-101_k141_394674 | Not-determined |
| 18-101_k141_82238 | Not-determined |
| 18-101_k141_48886 | Not-determined |
| 18-101_k141_345354 | Not-determined |
| 18-101_k141_82249 | Not-determined |
| 18-101_k141_48903 | Not-determined |
| 18-101_k141_180956 | Low-quality |
| 18-101_k141_394702 | Not-determined |
| 18-101_k141_180958 | Not-determined |
| 18-101_k141_394713 | Not-determined |
| 18-101_k141_394717 | Not-determined |
| 18-101_k141_394724 | Not-determined |
| 18-101_k141_180965 | Not-determined |
| 18-101_k141_114230 | Not-determined |
| 18-101_k141_394731 | Not-determined |
| 18-101_k141_394732 | Not-determined |
| 18-101_k141_394736 | Not-determined |
| 18-101_k141_394738 | Not-determined |
| 18-101_k141_394741 | Not-determined |
| 18-101_k141_394742 | Not-determined |
| 18-101_k141_394743 | Not-determined |
| 18-101_k141_114231 | Low-quality |
| 18-101_k141_114233 | Not-determined |
| 18-101_k141_394749 | Not-determined |
| 18-101_k141_394750 | Not-determined |
| 18-101_k141_394755 | Not-determined |
| 18-101_k141_394762 | Not-determined |
| 18-101_k141_114246 | Not-determined |
| 18-101_k141_114247 | Not-determined |
| 18-101_k141_394780 | Not-determined |
| 18-101_k141_394784 | Not-determined |
| 18-101_k141_114254 | Not-determined |
| 18-101_k141_394792 | Not-determined |
| 18-101_k141_394798 | Not-determined |
| 18-101_k141_48958 | Not-determined |
| 18-101_k141_48960 | Not-determined |
| 18-101_k141_180980 | Not-determined |
| 18-101_k141_48963 | Low-quality |
| 18-101_k141_394806 | Not-determined |
| 18-101_k141_394815 | Not-determined |
| 18-101_k141_394816 | Not-determined |
| 18-101_k141_394822 | Not-determined |
| 18-101_k141_394833 | Not-determined |
| 18-101_k141_394837 | Not-determined |
| 18-101_k141_394846 | Not-determined |
| 18-101_k141_394847 | Not-determined |
| 18-101_k141_394849 | Not-determined |
| 18-101_k141_394853 | Not-determined |
| 18-101_k141_48971 | Not-determined |
| 18-101_k141_114278 | Not-determined |
| 18-101_k141_394863 | Not-determined |
| 18-101_k141_394866 | Not-determined |
| 18-101_k141_48974 | Not-determined |
| 18-101_k141_48975 | Not-determined |
| 18-101_k141_48991 | Not-determined |
| 18-101_k141_48997 | Not-determined |
| 18-101_k141_114291 | Low-quality |
| 18-101_k141_114292 | Not-determined |
| 18-101_k141_49001 | Low-quality |
| 18-101_k141_49003 | Not-determined |
| 18-101_k141_49024 | Not-determined |
| 18-101_k141_345434 | Not-determined |
| 18-101_k141_345445 | Not-determined |
| 18-101_k141_114324 | Not-determined |
| 18-101_k141_114326 | Not-determined |
| 18-101_k141_345464 | Not-determined |
| 18-102_k141_246296 | Medium-quality |
| 18-101_k141_114357 | Not-determined |
| 18-101_k141_114365 | Not-determined |
| 18-101_k141_32885 | Not-determined |
| 18-101_k141_114413 | Not-determined |
| 18-101_k141_114417 | Not-determined |
| 18-101_k141_345501 | Not-determined |
| 18-101_k141_114431 | Not-determined |
| 18-101_k141_114433 | Not-determined |
| 18-101_k141_114439 | Not-determined |
| 18-101_k141_114452 | Not-determined |
| 18-101_k141_114459 | Not-determined |
| 18-101_k141_114467 | Low-quality |
| 18-101_k141_49107 | Not-determined |
| 18-101_k141_49111 | Not-determined |
| 18-101_k141_114486 | Not-determined |
| 18-101_k141_114498 | Not-determined |
| 18-101_k141_114500 | Not-determined |
| 18-101_k141_114506 | Low-quality |
| 18-101_k141_49134 | Not-determined |
| 18-101_k141_114510 | Low-quality |
| 18-101_k141_114533 | Not-determined |
| 18-101_k141_49147 | Not-determined |
| 18-101_k141_49149 | Not-determined |
| 18-101_k141_49155 | Not-determined |
| 18-101_k141_114574 | Not-determined |
| 18-101_k141_114583 | Not-determined |
| 18-101_k141_49202 | Not-determined |
| 18-101_k141_49203 | Not-determined |
| 18-101_k141_49204 | Not-determined |
| 18-101_k141_49205 | Not-determined |
| 18-101_k141_49231 | Low-quality |
| 18-101_k141_49236 | Not-determined |
| 18-101_k141_114633 | Not-determined |
| 18-101_k141_114637 | Not-determined |
| 18-101_k141_49268 | Not-determined |
| 18-101_k141_114654 | Low-quality |
| 18-101_k141_49290 | Not-determined |
| 18-101_k141_49331 | Not-determined |
| 18-101_k141_49353 | Not-determined |
| 18-101_k141_114681 | Not-determined |
| 18-101_k141_114687 | Not-determined |
| 18-101_k141_114688 | Not-determined |
| 18-101_k141_114694 | Not-determined |
| 18-101_k141_114721 | Not-determined |
| 18-101_k141_114753 | Not-determined |
| 18-101_k141_114756 | Not-determined |
| 18-101_k141_114759 | Not-determined |
| 18-101_k141_114774 | Not-determined |
| 18-101_k141_114795 | Not-determined |
| 18-101_k141_114802 | Not-determined |
| 18-101_k141_114841 | Not-determined |
| 18-101_k141_114843 | Not-determined |
| 18-101_k141_114844 | Not-determined |
| 18-101_k141_114851 | Not-determined |
| 18-101_k141_114860 | Not-determined |
| 18-101_k141_114867 | Low-quality |
| 18-101_k141_114886 | Not-determined |
| 18-101_k141_114904 | Not-determined |
| 18-101_k141_114906 | Not-determined |
| 18-101_k141_114918 | Not-determined |
| 18-101_k141_114952 | Not-determined |
| 18-101_k141_114986 | Not-determined |
| 18-101_k141_115010 | Not-determined |
| 18-101_k141_115031 | Not-determined |

18-101_k141_115035 Not-determined
18-101_k141_115036 Not-determined
18-101_k141_115040 Not-determined
18-101_k141_115061 Low-quality
18-101_k141_115062 Not-determined
18-101_k141_115128 Low-quality
18-101_k141_115140 Not-determined
18-101_k141_115145 Not-determined
18-101_k141_115152 Not-determined
18-101_k141_115165 Not-determined
18-102-final-viral-combined_k141_114278||0_partial Medium-quality
18-103-final-viral-combined_k141_46839||full Not-determined
18-103-final-viral-combined_k141_23880||full Low-quality
18-103-final-viral-combined_k141_128066||full Not-determined
18-103-final-viral-combined_k141_48788||full Not-determined
18-103-final-viral-combined_k141_95279||full Not-determined
18-103-final-viral-combined_k141_137874||full Not-determined
18-103-final-viral-combined_k141_137877||full Not-determined
18-103-final-viral-combined_k141_86258||full Low-quality
18-103-final-viral-combined_k141_34507||full Low-quality
18-103-final-viral-combined_k141_132259||full Not-determined
18-103-final-viral-combined_k141_5760||full Not-determined
18-103-final-viral-combined_k141_120766||full Low-quality
18-103-final-viral-combined_k141_34515||full Not-determined
18-103-final-viral-combined_k141_97763||full Not-determined
18-103-final-viral-combined_k141_109264||full Low-quality
18-103-final-viral-combined_k141_51767||full Not-determined
18-103-final-viral-combined_k141_40271||full Low-quality
18-103-final-viral-combined_k141_69021||full Low-quality
18-103-final-viral-combined_k141_57519||full Low-quality
18-103-final-viral-combined_k141_126525||full Low-quality
18-103-final-viral-combined_k141_69028||full Not-determined
18-103-final-viral-combined_k141_120780||full Not-determined
18-103-final-viral-combined_k141_115037||full Not-determined
18-103-final-viral-combined_k141_11526||full Not-determined
18-103-final-viral-combined_k141_97780||full Low-quality
18-103-final-viral-combined_k141_109290||full Not-determined
18-103-final-viral-combined_k141_23051||full Not-determined
18-103-final-viral-combined_k141_11545||full Low-quality
18-103-final-viral-combined_k141_51798||full Not-determined
18-103-final-viral-combined_k141_5798||full Not-determined
18-103-final-viral-combined_k141_74776||full Low-quality
18-103-final-viral-combined_k141_11559||full Not-determined
18-103-final-viral-combined_k141_126583||full Low-quality
18-103-final-viral-combined_k141_120835||full Low-quality
18-103-final-viral-combined_k141_86333||full Low-quality
18-103-final-viral-combined_k141_69082||full Not-determined
18-103-final-viral-combined_k141_97816||full Low-quality
18-103-final-viral-combined_k141_126601||full Not-determined
18-103-final-viral-combined_k141_63347||full Not-determined
18-103-final-viral-combined_k141_40315||full Low-quality
18-103-final-viral-combined_k141_46103||full Not-determined
18-103-final-viral-combined_k141_115104||full Not-determined
18-103-final-viral-combined_k141_34596||full Not-determined
18-103-final-viral-combined_k141_57565||full Not-determined
18-103-final-viral-combined_k141_92055||full Low-quality
18-103-final-viral-combined_k141_5829||full Low-quality
18-103-final-viral-combined_k141_17344||full Low-quality
18-103-final-viral-combined_k141_103557||full Not-determined
18-103-final-viral-combined_k141_92071||full Low-quality
18-103-final-viral-combined_k141_120890||full Not-determined
18-103-final-viral-combined_k141_103569||full Low-quality
18-103-final-viral-combined_k141_23115||full Low-quality
18-103-final-viral-combined_k141_132374||full Low-quality
18-103-final-viral-combined_k141_74835||full Low-quality
18-103-final-viral-combined_k141_115138||full Not-determined
18-103-final-viral-combined_k141_86369||full Low-quality
18-103-final-viral-combined_k141_172||full Not-determined
18-103-final-viral-combined_k141_34625||full Not-determined
18-103-final-viral-combined_k141_74837||full Low-quality
18-103-final-viral-combined_k141_132383||full Low-quality
18-103-final-viral-combined_k141_120902||full Low-quality
18-103-final-viral-combined_k141_86379||full Not-determined
18-103-final-viral-combined_k141_51849||full Not-determined
18-103-final-viral-combined_k141_5852||full Not-determined
18-103-final-viral-combined_k141_51850||full Low-quality
18-103-final-viral-combined_k141_97873||full Not-determined
18-103-final-viral-combined_k141_74848||full Not-determined
18-103-final-viral-combined_k141_103586||full Low-quality
18-103-final-viral-combined_k141_86396||full Low-quality
18-103-final-viral-combined_k141_40344||full Low-quality
18-103-final-viral-combined_k141_80585||full Not-determined
18-103-final-viral-combined_k141_17359||full Low-quality
18-103-final-viral-combined_k141_74862||full Low-quality
18-103-final-viral-combined_k141_103622||full Not-determined
18-103-final-viral-combined_k141_34665||full Not-determined
18-103-final-viral-combined_k141_126694||full Not-determined
18-103-final-viral-combined_k141_40367||full Low-quality
18-103-final-viral-combined_k141_132435||full Low-quality
18-103-final-viral-combined_k141_40371||full Low-quality
18-103-final-viral-combined_k141_23137||full Low-quality
18-103-final-viral-combined_k141_34678||full Low-quality
18-103-final-viral-combined_k141_69156||full Low-quality
18-103-final-viral-combined_k141_120957||full Low-quality
18-103-final-viral-combined_k141_51919||full Not-determined
18-103-final-viral-combined_k141_40378||full Not-determined
18-103-final-viral-combined_k141_5887||full Not-determined
18-103-final-viral-combined_k141_221||full Not-determined
18-103-final-viral-combined_k141_17387||full Low-quality
18-103-final-viral-combined_k141_46189||full Not-determined
18-103-final-viral-combined_k141_103641||full Not-determined
18-103-final-viral-combined_k141_51927||full Low-quality
18-103-final-viral-combined_k141_132450||full Not-determined
18-103-final-viral-combined_k141_40380||full Not-determined
18-103-final-viral-combined_k141_5898||full Low-quality
18-103-final-viral-combined_k141_74901||full Low-quality
18-103-final-viral-combined_k141_46198||full Low-quality
18-103-final-viral-combined_k141_103651||full Not-determined
18-103-final-viral-combined_k141_11637||full Low-quality
18-103-final-viral-combined_k141_115220||full Not-determined
18-103-final-viral-combined_k141_69190||full Not-determined
18-103-final-viral-combined_k141_97928||full Low-quality
18-103-final-viral-combined_k141_86505||full Low-quality
18-103-final-viral-combined_k141_115230||full Not-determined
18-103-final-viral-combined_k141_23202||full Low-quality
18-103-final-viral-combined_k141_92164||full Low-quality
18-103-final-viral-combined_k141_34703||full Low-quality
18-103-final-viral-combined_k141_97941||full Low-quality
18-103-final-viral-combined_k141_109486||full Low-quality
18-103-final-viral-combined_k141_92180||full Low-quality
18-103-final-viral-combined_k141_86514||full Low-quality
18-103-final-viral-combined_k141_86515||full Not-determined
18-103-final-viral-combined_k141_28899||full Low-quality
18-103-final-viral-combined_k141_120998||full Not-determined
18-103-final-viral-combined_k141_40410||full Not-determined
18-103-final-viral-combined_k141_74960||full Not-determined
18-103-final-viral-combined_k141_74961||full Low-quality
18-103-final-viral-combined_k141_51948||full Low-quality
18-103-final-viral-combined_k141_63473||full Not-determined
18-103-final-viral-combined_k141_5938||full Not-determined
18-103-final-viral-combined_k141_23249||full Not-determined
18-103-final-viral-combined_k141_126782||full Low-quality
18-103-final-viral-combined_k141_92203||full Low-quality
18-103-final-viral-combined_k141_80654||full Low-quality
18-103-final-viral-combined_k141_46256||full Not-determined
18-103-final-viral-combined_k141_80656||full Low-quality
18-103-final-viral-combined_k141_92204||full Not-determined
18-103-final-viral-combined_k141_11649||full Not-determined
18-103-final-viral-combined_k141_74975||full Not-determined
18-103-final-viral-combined_k141_132528||full Low-quality
18-103-final-viral-combined_k141_115283||full Low-quality
18-103-final-viral-combined_k141_28937||full Not-determined
18-103-final-viral-combined_k141_316||full Not-determined
18-103-final-viral-combined_k141_103726||full Not-determined
18-103-final-viral-combined_k141_57652||full Not-determined
18-103-final-viral-combined_k141_28939||full Not-determined
18-103-final-viral-combined_k141_80681||full Not-determined
18-103-final-viral-combined_k141_63491||full Low-quality
18-103-final-viral-combined_k141_98003||full Low-quality
18-103-final-viral-combined_k141_74992||full Not-determined
18-103-final-viral-combined_k141_121034||full Low-quality
18-103-final-viral-combined_k141_69215||full Low-quality
18-103-final-viral-combined_k141_103732||full Low-quality
18-103-final-viral-combined_k141_115299||full Low-quality
18-103-final-viral-combined_k141_57670||full Low-quality
18-103-final-viral-combined_k141_109567||full Low-quality
18-103-final-viral-combined_k141_121038||full Not-determined
18-103-final-viral-combined_k141_5996||full Not-determined
18-103-final-viral-combined_k141_63526||full Low-quality
18-103-final-viral-combined_k141_358||full Not-determined
18-103-final-viral-combined_k141_126808||full Low-quality
18-103-final-viral-combined_k141_126809||full Not-determined
18-103-final-viral-combined_k141_80693||full Low-quality
18-103-final-viral-combined_k141_126812||full Not-determined
18-103-final-viral-combined_k141_52014||full Low-quality
18-103-final-viral-combined_k141_103749||full Low-quality
18-103-final-viral-combined_k141_34760||full Low-quality
18-103-final-viral-combined_k141_132555||full Not-determined
18-103-final-viral-combined_k141_368||full Not-determined
18-103-final-viral-combined_k141_34784||full Not-determined
18-103-final-viral-combined_k141_17459||full Not-determined
18-103-final-viral-combined_k141_28998||full Low-quality
18-103-final-viral-combined_k141_80714||full Not-determined
18-103-final-viral-combined_k141_52045||full Low-quality
18-103-final-viral-combined_k141_40528||full Low-quality
18-103-final-viral-combined_k141_394||full Low-quality
18-103-final-viral-combined_k141_109641||full Not-determined
18-103-final-viral-combined_k141_11716||full Not-determined
18-103-final-viral-combined_k141_132594||full Not-determined
18-103-final-viral-combined_k141_121079||full Not-determined
18-103-final-viral-combined_k141_57708||full Low-quality
18-103-final-viral-combined_k141_69292||full Low-quality
18-103-final-viral-combined_k141_406||full Not-determined
18-103-final-viral-combined_k141_29013||full Low-quality
18-103-final-viral-combined_k141_17478||full Not-determined
18-103-final-viral-combined_k141_69298||full Low-quality
18-103-final-viral-combined_k141_46355||full Not-determined
18-103-final-viral-combined_k141_69299||full Low-quality
18-103-final-viral-combined_k141_52081||full Low-quality
18-103-final-viral-combined_k141_98065||full Not-determined
18-103-final-viral-combined_k141_57738||full Not-determined
18-103-final-viral-combined_k141_98069||full Low-quality
18-103-final-viral-combined_k141_52088||full Not-determined
18-103-final-viral-combined_k141_40553||full Not-determined
18-103-final-viral-combined_k141_23380||full Low-quality
18-103-final-viral-combined_k141_126865||full Not-determined
18-103-final-viral-combined_k141_63590||full Not-determined
18-103-final-viral-combined_k141_80766||full Not-determined
18-103-final-viral-combined_k141_57744||full Low-quality
18-103-final-viral-combined_k141_69319||full Low-quality
18-103-final-viral-combined_k141_40561||full Not-determined
18-103-final-viral-combined_k141_23381||full Low-quality
18-103-final-viral-combined_k141_75035||full Not-determined
18-103-final-viral-combined_k141_52108||full Not-determined
18-103-final-viral-combined_k141_86723||full Not-determined
18-103-final-viral-combined_k141_34863||full Low-quality
18-103-final-viral-combined_k141_52110||full Not-determined
18-103-final-viral-combined_k141_80789||full Low-quality
18-103-final-viral-combined_k141_132658||full Low-quality
18-103-final-viral-combined_k141_80797||full Not-determined
18-103-final-viral-combined_k141_29052||full Not-determined
18-103-final-viral-combined_k141_103826||full Low-quality
18-103-final-viral-combined_k141_92330||full Low-quality
18-103-final-viral-combined_k141_80807||full Not-determined
18-103-final-viral-combined_k141_29057||full Low-quality
18-103-final-viral-combined_k141_103845||full Not-determined
18-103-final-viral-combined_k141_126929||full Not-determined
18-103-final-viral-combined_k141_23437||full Not-determined
18-103-final-viral-combined_k141_46401||full Low-quality
18-103-final-viral-combined_k141_40590||full Low-quality
18-103-final-viral-combined_k141_40592||full Low-quality
18-103-final-viral-combined_k141_69383||full Not-determined
18-103-final-viral-combined_k141_98087||full Low-quality
18-103-final-viral-combined_k141_17572||full Not-determined
18-103-final-viral-combined_k141_23456||full Low-quality
18-103-final-viral-combined_k141_98093||full Not-determined
18-103-final-viral-combined_k141_132708||full Not-determined
18-103-final-viral-combined_k141_23463||full Not-determined
18-103-final-viral-combined_k141_17584||full Low-quality
18-103-final-viral-combined_k141_481||full Not-determined
18-103-final-viral-combined_k141_75114||full Not-determined
18-103-final-viral-combined_k141_34926||full Low-quality
18-103-final-viral-combined_k141_86780||full Not-determined
18-103-final-viral-combined_k141_490||full Not-determined
18-103-final-viral-combined_k141_17603||full Low-quality
18-103-final-viral-combined_k141_80862||full Not-determined
18-103-final-viral-combined_k141_92389||full Low-quality
18-103-final-viral-combined_k141_29087||full Low-quality
18-103-final-viral-combined_k141_75127||full Not-determined
18-103-final-viral-combined_k141_34956||full Low-quality
18-103-final-viral-combined_k141_80865||full Not-determined
18-103-final-viral-combined_k141_34957||full Low-quality
18-103-final-viral-combined_k141_80874||full Low-quality
18-103-final-viral-combined_k141_103897||full Low-quality
18-103-final-viral-combined_k141_127001||full Low-quality
18-103-final-viral-combined_k141_6190||full Not-determined
18-103-final-viral-combined_k141_98112||full Not-determined
18-103-final-viral-combined_k141_34970||full Not-determined
18-103-final-viral-combined_k141_115446||full Low-quality
18-103-final-viral-combined_k141_98117||full Not-determined
18-103-final-viral-combined_k141_132729||full Low-quality
18-103-final-viral-combined_k141_46465||full Low-quality
18-103-final-viral-combined_k141_115455||full Low-quality
18-103-final-viral-combined_k141_11848||full Low-quality
18-103-final-viral-combined_k141_11849||full Low-quality
18-103-final-viral-combined_k141_80896||full Not-determined
18-103-final-viral-combined_k141_40657||full Not-determined
18-103-final-viral-combined_k141_57849||full Not-determined
18-103-final-viral-combined_k141_6207||full Not-determined
18-103-final-viral-combined_k141_34998||full Not-determined
18-103-final-viral-combined_k141_40663||full Low-quality
18-103-final-viral-combined_k141_86807||full Not-determined
18-103-final-viral-combined_k141_63671||full Low-quality
18-103-final-viral-combined_k141_69470||full Not-determined
18-103-final-viral-combined_k141_103937||full Low-quality
18-103-final-viral-combined_k141_127047||full Low-quality
18-103-final-viral-combined_k141_23524||full Low-quality
18-103-final-viral-combined_k141_29145||full Low-quality
18-103-final-viral-combined_k141_57879||full Low-quality
18-103-final-viral-combined_k141_23530||full Low-quality
18-103-final-viral-combined_k141_98142||full Not-determined
18-103-final-viral-combined_k141_6233||full Low-quality
18-103-final-viral-combined_k141_109789||full Not-determined
18-103-final-viral-combined_k141_132783||full Not-determined
18-103-final-viral-combined_k141_6239||full Not-determined
18-103-final-viral-combined_k141_23544||full Low-quality
18-103-final-viral-combined_k141_132790||full Not-determined
18-103-final-viral-combined_k141_6243||full Low-quality
18-103-final-viral-combined_k141_6246||full Not-determined
18-103-final-viral-combined_k141_92473||full Not-determined
18-103-final-viral-combined_k141_11890||full Not-determined
18-103-final-viral-combined_k141_63715||full Low-quality
18-103-final-viral-combined_k141_11897||full Not-determined
18-103-final-viral-combined_k141_57924||full Not-determined
18-103-final-viral-combined_k141_40698||full Low-quality
18-103-final-viral-combined_k141_40702||full Low-quality
18-103-final-viral-combined_k141_52245||full Low-quality
18-103-final-viral-combined_k141_69529||full Not-determined
18-103-final-viral-combined_k141_11904||full Not-determined
18-103-final-viral-combined_k141_104032||full Not-determined
18-103-final-viral-combined_k141_6271||full Not-determined
18-103-final-viral-combined_k141_35055||full Low-quality
18-103-final-viral-combined_k141_52265||full Low-quality
18-103-final-viral-combined_k141_75205||full Not-determined
18-103-final-viral-combined_k141_127141||full Low-quality
18-103-final-viral-combined_k141_6276||full Not-determined
18-103-final-viral-combined_k141_6278||full Not-determined
18-103-final-viral-combined_k141_46601||full Low-quality
18-103-final-viral-combined_k141_599||full Low-quality
18-103-final-viral-combined_k141_35059||full Not-determined
18-103-final-viral-combined_k141_86910||full Not-determined
18-103-final-viral-combined_k141_23580||full Low-quality
18-103-final-viral-combined_k141_86913||full Not-determined
18-103-final-viral-combined_k141_29214||full Low-quality
18-103-final-viral-combined_k141_23593||full Not-determined
18-103-final-viral-combined_k141_23594||full Not-determined
18-103-final-viral-combined_k141_58000||full Not-determined
18-103-final-viral-combined_k141_127172||full Not-determined
18-103-final-viral-combined_k141_75231||full Not-determined
18-103-final-viral-combined_k141_98216||full Not-determined
18-103-final-viral-combined_k141_46638||full Not-determined
18-103-final-viral-combined_k141_40784||full Not-determined
18-103-final-viral-combined_k141_127184||full Low-quality
18-103-final-viral-combined_k141_115597||full Not-determined
18-103-final-viral-combined_k141_23598||full Not-determined
18-103-final-viral-combined_k141_52333||full Not-determined
18-103-final-viral-combined_k141_86953||full Not-determined
18-103-final-viral-combined_k141_92541||full Not-determined
18-103-final-viral-combined_k141_104129||full Low-quality
18-103-final-viral-combined_k141_6381||full Not-determined
18-103-final-viral-combined_k141_75264||full Not-determined
18-103-final-viral-combined_k141_647||full Not-determined
18-103-final-viral-combined_k141_6389||full Low-quality
18-103-final-viral-combined_k141_121309||full Low-quality
18-103-final-viral-combined_k141_46681||full Low-quality
18-103-final-viral-combined_k141_11975||full Not-determined
18-103-final-viral-combined_k141_23642||full Not-determined
18-103-final-viral-combined_k141_132903||full Not-determined
18-103-final-viral-combined_k141_40838||full Not-determined
18-103-final-viral-combined_k141_35115||full Not-determined
18-103-final-viral-combined_k141_52355||full Low-quality
18-103-final-viral-combined_k141_23651||full Not-determined
18-103-final-viral-combined_k141_52358||full Low-quality
18-103-final-viral-combined_k141_81080||full Not-determined
18-103-final-viral-combined_k141_92575||full Low-quality
18-103-final-viral-combined_k141_40857||full Not-determined
18-103-final-viral-combined_k141_132922||full Not-determined
18-103-final-viral-combined_k141_63821||full Not-determined
18-103-final-viral-combined_k141_11999||full Not-determined
18-102-final-viral-combined_k141_246296||full Medium-quality
18-103-final-viral-combined_k141_92586||full Not-determined
18-103-final-viral-combined_k141_98248||full Not-determined
18-103-final-viral-combined_k141_35146||full Not-determined
18-103-final-viral-combined_k141_132926||full Not-determined
18-103-final-viral-combined_k141_127278||full Low-quality
18-103-final-viral-combined_k141_104145||full Not-determined
18-103-final-viral-combined_k141_23677||full Not-determined
18-103-final-viral-combined_k141_132929||full Low-quality
18-103-final-viral-combined_k141_127282||full Low-quality
18-103-final-viral-combined_k141_109956||full Not-determined
18-103-final-viral-combined_k141_6447||full Low-quality
18-103-final-viral-combined_k141_46700||full Not-determined
18-103-final-viral-combined_k141_40889||full Not-determined
18-103-final-viral-combined_k141_127289||full Low-quality
18-103-final-viral-combined_k141_98257||full Low-quality
18-103-final-viral-combined_k141_127290||full Not-determined
18-103-final-viral-combined_k141_75309||full Not-determined
18-103-final-viral-combined_k141_23690||full Not-determined
18-103-final-viral-combined_k141_87036||full Low-quality
18-103-final-viral-combined_k141_69650||full Not-determined
18-103-final-viral-combined_k141_81115||full Low-quality
18-103-final-viral-combined_k141_127304||full Not-determined
18-103-final-viral-combined_k141_87050||full Low-quality
18-103-final-viral-combined_k141_46722||full Not-determined
18-103-final-viral-combined_k141_98273||full Low-quality
18-103-final-viral-combined_k141_40907||full Low-quality
18-103-final-viral-combined_k141_17845||full Low-quality
18-103-final-viral-combined_k141_69661||full Low-quality
18-103-final-viral-combined_k141_6479||full Low-quality
18-103-final-viral-combined_k141_121393||full Low-quality
18-103-final-viral-combined_k141_23714||full Not-determined
18-103-final-viral-combined_k141_6484||full Low-quality
18-103-final-viral-combined_k141_92628||full Low-quality
18-103-final-viral-combined_k141_12051||full Low-quality
18-103-final-viral-combined_k141_69679||full Not-determined
18-103-final-viral-combined_k141_92635||full Low-quality
18-103-final-viral-combined_k141_127339||full Low-quality
18-103-final-viral-combined_k141_12063||full Not-determined
18-103-final-viral-combined_k141_52442||full Low-quality
18-103-final-viral-combined_k141_121415||full Low-quality
18-103-final-viral-combined_k141_98301||full Not-determined
18-103-final-viral-combined_k141_63889||full Low-quality
18-103-final-viral-combined_k141_63890||full Low-quality
18-103-final-viral-combined_k141_92639||full Low-quality
18-103-final-viral-combined_k141_6510||full Not-determined
18-103-final-viral-combined_k141_63895||full Low-quality
18-103-final-viral-combined_k141_6521||full Not-determined
18-103-final-viral-combined_k141_63908||full Not-determined
18-103-final-viral-combined_k141_29338||full Not-determined
18-103-final-viral-combined_k141_6527||full Low-quality
18-103-final-viral-combined_k141_23727||full Not-determined
18-103-final-viral-combined_k141_87109||full Not-determined
18-103-final-viral-combined_k141_75408||full Low-quality
18-103-final-viral-combined_k141_121438||full Not-determined
18-103-final-viral-combined_k141_98328||full Low-quality
18-103-final-viral-combined_k141_52497||full Not-determined
18-103-final-viral-combined_k141_23746||full Low-quality
18-103-final-viral-combined_k141_81197||full Low-quality
18-103-final-viral-combined_k141_29368||full Not-determined
18-103-final-viral-combined_k141_81226||full Low-quality
18-103-final-viral-combined_k141_104223||full Not-determined
18-103-final-viral-combined_k141_17957||full Not-determined
18-103-final-viral-combined_k141_35260||full Low-quality
18-103-final-viral-combined_k141_75436||full Not-determined
18-103-final-viral-combined_k141_40985||full Low-quality
18-103-final-viral-combined_k141_58212||full Low-quality
18-103-final-viral-combined_k141_6582||full Not-determined
18-103-final-viral-combined_k141_17959||full Not-determined
18-103-final-viral-combined_k141_52538||full Not-determined
18-103-final-viral-combined_k141_87147||full Low-quality
18-103-final-viral-combined_k141_133056||full Low-quality
18-103-final-viral-combined_k141_75451||full Low-quality
18-103-final-viral-combined_k141_115752||full Low-quality
18-103-final-viral-combined_k141_29421||full Not-determined
18-103-final-viral-combined_k141_92733||full Low-quality
18-103-final-viral-combined_k141_104247||full Not-determined
18-103-final-viral-combined_k141_63969||full Low-quality
18-103-final-viral-combined_k141_110035||full Low-quality
18-103-final-viral-combined_k141_81275||full Not-determined
18-103-final-viral-combined_k141_17987||full Low-quality
18-103-final-viral-combined_k141_829||full Not-determined
18-103-final-viral-combined_k141_46836||full Low-quality
18-103-final-viral-combined_k141_110052||full Not-determined
18-103-final-viral-combined_k141_115762||full Low-quality
18-103-final-viral-combined_k141_29438||full Not-determined
18-103-final-viral-combined_k141_23825||full Low-quality
18-103-final-viral-combined_k141_52574||full Not-determined
18-103-final-viral-combined_k141_18003||full Not-determined
18-103-final-viral-combined_k141_12098||full Low-quality
18-103-final-viral-combined_k141_98407||full Low-quality
18-103-final-viral-combined_k141_127462||full Not-determined
18-103-final-viral-combined_k141_29468||full Low-quality
18-103-final-viral-combined_k141_98413||full Low-quality
18-103-final-viral-combined_k141_127464||full Not-determined
18-103-final-viral-combined_k141_41050||full Not-determined
18-103-final-viral-combined_k141_46886||full Not-determined
18-103-final-viral-combined_k141_75514||full Low-quality
18-103-final-viral-combined_k141_98421||full Not-determined
18-103-final-viral-combined_k141_64036||full Not-determined
18-103-final-viral-combined_k141_110095||full Not-determined
18-103-final-viral-combined_k141_58328||full Not-determined
18-103-final-viral-combined_k141_41075||full Low-quality
18-103-final-viral-combined_k141_81361||full Low-quality
18-103-final-viral-combined_k141_81366||full Not-determined
18-103-final-viral-combined_k141_898||full Not-determined
18-103-final-viral-combined_k141_900||full Not-determined
18-103-final-viral-combined_k141_81379||full Not-determined
18-103-final-viral-combined_k141_69815||full Low-quality
18-103-final-viral-combined_k141_29483||full Low-quality
18-103-final-viral-combined_k141_903||full Low-quality
18-103-final-viral-combined_k141_18042||full Not-determined
18-103-final-viral-combined_k141_41089||full Low-quality
18-103-final-viral-combined_k141_58356||full Low-quality
18-103-final-viral-combined_k141_75560||full Low-quality
18-103-final-viral-combined_k141_64076||full Not-determined
18-103-final-viral-combined_k141_133126||full Low-quality
18-103-final-viral-combined_k141_23875||full Low-quality
18-103-final-viral-combined_k141_69823||full Low-quality
18-103-final-viral-combined_k141_133131||full Not-determined
18-103-final-viral-combined_k141_127507||full Not-determined
18-103-final-viral-combined_k141_18053||full Not-determined
18-103-final-viral-combined_k141_104349||full Not-determined
18-103-final-viral-combined_k141_64096||full Not-determined
18-103-final-viral-combined_k141_75588||full Low-quality
18-103-final-viral-combined_k141_81435||full Not-determined
18-103-final-viral-combined_k141_104368||full Not-determined
18-103-final-viral-combined_k141_6689||full Not-determined
18-103-final-viral-combined_k141_121555||full Not-determined
18-103-final-viral-combined_k141_64120||full Not-determined
18-103-final-viral-combined_k141_52677||full Low-quality
18-103-final-viral-combined_k141_6704||full Low-quality
18-103-final-viral-combined_k141_35375||full Low-quality
18-103-final-viral-combined_k141_115846||full Low-quality
18-103-final-viral-combined_k141_41136||full Not-determined
18-103-final-viral-combined_k141_81464||full Not-determined
18-103-final-viral-combined_k141_41139||full Low-quality
18-103-final-viral-combined_k141_104402||full Low-quality
18-103-final-viral-combined_k141_6711||full Low-quality
18-103-final-viral-combined_k141_98471||full Low-quality
18-103-final-viral-combined_k141_35405||full Not-determined
18-103-final-viral-combined_k141_35406||full Low-quality
18-103-final-viral-combined_k141_58437||full Not-determined
18-103-final-viral-combined_k141_6730||full Low-quality
18-103-final-viral-combined_k141_75632||full Not-determined
18-103-final-viral-combined_k141_75640||full Not-determined
18-103-final-viral-combined_k141_6736||full Not-determined
18-103-final-viral-combined_k141_69881||full Not-determined
18-103-final-viral-combined_k141_121601||full Low-quality
18-103-final-viral-combined_k141_58440||full Low-quality
18-103-final-viral-combined_k141_87399||full Not-determined
18-103-final-viral-combined_k141_69896||full Low-quality
18-103-final-viral-combined_k141_121610||full Low-quality
18-103-final-viral-combined_k141_52734||full Low-quality
18-103-final-viral-combined_k141_127610||full Not-determined
18-103-final-viral-combined_k141_69906||full Not-determined
18-102-final-viral-combined_k141_196783||full Complete
18-103-final-viral-combined_k141_92947||full Not-determined
18-103-final-viral-combined_k141_58458||full Not-determined
18-103-final-viral-combined_k141_115914||full Not-determined
18-103-final-viral-combined_k141_18066||full Not-determined
18-103-final-viral-combined_k141_121631||full Low-quality
18-103-final-viral-combined_k141_81525||full Not-determined
18-103-final-viral-combined_k141_121642||full Low-quality
18-103-final-viral-combined_k141_92960||full Not-determined
18-103-final-viral-combined_k141_18073||full Not-determined
18-103-final-viral-combined_k141_75687||full Not-determined
18-103-final-viral-combined_k141_6776||full Not-determined
18-103-final-viral-combined_k141_58480||full Low-quality
18-103-final-viral-combined_k141_52765||full Not-determined
18-103-final-viral-combined_k141_133260||full Low-quality
18-103-final-viral-combined_k141_64226||full Low-quality
18-103-final-viral-combined_k141_87436||full Low-quality
18-103-final-viral-combined_k141_133261||full Not-determined
18-103-final-viral-combined_k141_127636||full Not-determined
18-103-final-viral-combined_k141_35483||full Not-determined
18-103-final-viral-combined_k141_81552||full Not-determined
18-103-final-viral-combined_k141_58488||full Low-quality
18-103-final-viral-combined_k141_98544||full Not-determined
18-103-final-viral-combined_k141_1094||full Not-determined
18-103-final-viral-combined_k141_92975||full Not-determined
18-103-final-viral-combined_k141_6823||full Low-quality
18-103-final-viral-combined_k141_98564||full Low-quality
18-103-final-viral-combined_k141_75741||full Not-determined
18-103-final-viral-combined_k141_92992||full Low-quality
18-103-final-viral-combined_k141_87486||full Low-quality
18-103-final-viral-combined_k141_35518||full Low-quality
18-103-final-viral-combined_k141_52820||full Not-determined
18-103-final-viral-combined_k141_64283||full Not-determined
18-103-final-viral-combined_k141_29661||full Low-quality
18-103-final-viral-combined_k141_1135||full Low-quality
18-103-final-viral-combined_k141_75749||full Not-determined
18-103-final-viral-combined_k141_81591||full Low-quality
18-103-final-viral-combined_k141_58556||full Not-determined
18-103-final-viral-combined_k141_69990||full Low-quality
18-103-final-viral-combined_k141_87519||full Not-determined
18-103-final-viral-combined_k141_69993||full Low-quality
18-103-final-viral-combined_k141_98621||full Not-determined
18-103-final-viral-combined_k141_81601||full Not-determined
18-103-final-viral-combined_k141_133317||full Not-determined
18-103-final-viral-combined_k141_75774||full Low-quality
18-103-final-viral-combined_k141_35554||full Low-quality
18-103-final-viral-combined_k141_75777||full Low-quality
18-103-final-viral-combined_k141_121721||full Not-determined
18-103-final-viral-combined_k141_58570||full Not-determined
18-103-final-viral-combined_k141_104557||full Not-determined
18-103-final-viral-combined_k141_64331||full Not-determined
18-103-final-viral-combined_k141_6871||full Not-determined
18-103-final-viral-combined_k141_127741||full Low-quality
18-103-final-viral-combined_k141_81623||full Low-quality
18-103-final-viral-combined_k141_24056||full Low-quality
18-103-final-viral-combined_k141_64335||full Low-quality
18-103-final-viral-combined_k141_35580||full Low-quality
18-103-final-viral-combined_k141_81631||full Low-quality
18-103-final-viral-combined_k141_70027||full Not-determined
18-103-final-viral-combined_k141_52877||full Not-determined
18-103-final-viral-combined_k141_116043||full Not-determined
18-103-final-viral-combined_k141_18197||full Not-determined
18-103-final-viral-combined_k141_58583||full Not-determined
18-103-final-viral-combined_k141_35588||full Low-quality
18-103-final-viral-combined_k141_41292||full Low-quality
18-103-final-viral-combined_k141_70034||full Low-quality
18-103-final-viral-combined_k141_127756||full Not-determined
18-103-final-viral-combined_k141_121752||full Low-quality
18-103-final-viral-combined_k141_1214||full Low-quality
18-103-final-viral-combined_k141_87562||full Low-quality
18-103-final-viral-combined_k141_24087||full Not-determined
18-103-final-viral-combined_k141_75824||full Low-quality
18-103-final-viral-combined_k141_52905||full Not-determined
18-103-final-viral-combined_k141_127779||full Not-determined
18-103-final-viral-combined_k141_127782||full Not-determined
18-103-final-viral-combined_k141_35604||full Low-quality
18-103-final-viral-combined_k141_70058||full Not-determined
18-103-final-viral-combined_k141_121778||full Not-determined
18-103-final-viral-combined_k141_12401||full Not-determined
18-103-final-viral-combined_k141_6925||full Not-determined
18-103-final-viral-combined_k141_6930||full Low-quality
18-103-final-viral-combined_k141_47214||full Low-quality
18-103-final-viral-combined_k141_64361||full Low-quality
18-103-final-viral-combined_k141_35631||full Low-quality
18-103-final-viral-combined_k141_12421||full Not-determined
18-103-final-viral-combined_k141_18246||full Low-quality
18-103-final-viral-combined_k141_18248||full Low-quality
18-103-final-viral-combined_k141_133422||full Not-determined
18-103-final-viral-combined_k141_81678||full Low-quality
18-103-final-viral-combined_k141_121801||full Low-quality
18-103-final-viral-combined_k141_133428||full Not-determined
18-103-final-viral-combined_k141_70091||full Not-determined
18-103-final-viral-combined_k141_98652||full Low-quality
18-103-final-viral-combined_k141_81704||full Low-quality

18-103-final-viral-combined_k141_47256||full Not-determined
18-103-final-viral-combined_k141_24150||full Low-quality
18-103-final-viral-combined_k141_121831||full Low-quality
18-103-final-viral-combined_k141_133455||full Not-determined
18-103-final-viral-combined_k141_116154||full Low-quality
18-103-final-viral-combined_k141_93102||full Low-quality
18-103-final-viral-combined_k141_41391||full Low-quality
18-103-final-viral-combined_k141_70116||full Low-quality
18-103-final-viral-combined_k141_12479||full Not-determined
18-103-final-viral-combined_k141_70125||full Not-determined
18-103-final-viral-combined_k141_110452||full Not-determined
18-103-final-viral-combined_k141_70129||full Not-determined
18-103-final-viral-combined_k141_52999||full Not-determined
18-103-final-viral-combined_k141_18305||full Low-quality
18-103-final-viral-combined_k141_24191||full Not-determined
18-103-final-viral-combined_k141_58669||full Not-determined
18-103-final-viral-combined_k141_93115||full Low-quality
18-103-final-viral-combined_k141_110457||full Low-quality
18-103-final-viral-combined_k141_75905||full Low-quality
18-103-final-viral-combined_k141_127875||full Low-quality
18-103-final-viral-combined_k141_70145||full Low-quality
18-103-final-viral-combined_k141_29826||full Low-quality
18-103-final-viral-combined_k141_116201||full Low-quality
18-103-final-viral-combined_k141_133482||full Not-determined
18-103-final-viral-combined_k141_24216||full Low-quality
18-103-final-viral-combined_k141_47308||full Not-determined
18-103-final-viral-combined_k141_70151||full Low-quality
18-103-final-viral-combined_k141_18341||full Low-quality
18-103-final-viral-combined_k141_70155||full Low-quality
18-103-final-viral-combined_k141_93136||full Low-quality
18-103-final-viral-combined_k141_121873||full Low-quality
18-103-final-viral-combined_k141_29844||full Not-determined
18-103-final-viral-combined_k141_127894||full Not-determined
18-103-final-viral-combined_k141_53048||full Low-quality
18-103-final-viral-combined_k141_93142||full Low-quality
18-103-final-viral-combined_k141_41460||full Not-determined
18-103-final-viral-combined_k141_81788||full Not-determined
18-103-final-viral-combined_k141_93143||full Not-determined
18-103-final-viral-combined_k141_133504||full Low-quality
18-103-final-viral-combined_k141_127910||full Low-quality
18-103-final-viral-combined_k141_110478||full Not-determined
18-103-final-viral-combined_k141_127914||full Not-determined
18-103-final-viral-combined_k141_127917||full Not-determined
18-103-final-viral-combined_k141_93171||full Not-determined
18-103-final-viral-combined_k141_18394||full Low-quality
18-103-final-viral-combined_k141_70204||full Low-quality
18-103-final-viral-combined_k141_64496||full Low-quality
18-103-final-viral-combined_k141_24287||full Low-quality
18-103-final-viral-combined_k141_81836||full Low-quality
18-103-final-viral-combined_k141_64510||full Low-quality
18-103-final-viral-combined_k141_110530||full Low-quality
18-103-final-viral-combined_k141_75989||full Low-quality
18-103-final-viral-combined_k141_53089||full Low-quality
18-103-final-viral-combined_k141_93190||full Not-determined
18-103-final-viral-combined_k141_7083||full Low-quality
18-103-final-viral-combined_k141_41491||full Low-quality
18-103-final-viral-combined_k141_58759||full Low-quality
18-103-final-viral-combined_k141_87695||full Not-determined
18-103-final-viral-combined_k141_70230||full Low-quality
18-103-final-viral-combined_k141_87700||full Not-determined
18-103-final-viral-combined_k141_35740||full Low-quality
18-103-final-viral-combined_k141_110538||full Low-quality
18-103-final-viral-combined_k141_1322||full Not-determined
18-103-final-viral-combined_k141_64528||full Not-determined
18-103-final-viral-combined_k141_47344||full Low-quality
18-103-final-viral-combined_k141_81864||full Low-quality
18-103-final-viral-combined_k141_41509||full Not-determined
18-103-final-viral-combined_k141_41510||full Not-determined
18-103-final-viral-combined_k141_133564||full Not-determined
18-103-final-viral-combined_k141_7109||full Low-quality
18-103-final-viral-combined_k141_24306||full Low-quality
18-103-final-viral-combined_k141_1339||full Low-quality
18-103-final-viral-combined_k141_116328||full Low-quality
18-103-final-viral-combined_k141_1345||full Not-determined
18-103-final-viral-combined_k141_76025||full Not-determined
18-103-final-viral-combined_k141_76027||full Low-quality
18-103-final-viral-combined_k141_104696||full Not-determined
18-103-final-viral-combined_k141_47371||full Not-determined
18-103-final-viral-combined_k141_81884||full Not-determined
18-103-final-viral-combined_k141_7115||full Low-quality
18-103-final-viral-combined_k141_93232||full Not-determined
18-103-final-viral-combined_k141_128007||full Not-determined
18-103-final-viral-combined_k141_41523||full Low-quality
18-103-final-viral-combined_k141_133599||full Low-quality
18-103-final-viral-combined_k141_58813||full Low-quality
18-103-final-viral-combined_k141_121970||full Low-quality
18-103-final-viral-combined_k141_12624||full Low-quality
18-103-final-viral-combined_k141_110589||full Low-quality
18-103-final-viral-combined_k141_58815||full Not-determined
18-103-final-viral-combined_k141_93252||full Low-quality
18-103-final-viral-combined_k141_121976||full Not-determined
18-103-final-viral-combined_k141_29947||full Low-quality
18-103-final-viral-combined_k141_87764||full Not-determined
18-103-final-viral-combined_k141_64590||full Low-quality
18-103-final-viral-combined_k141_133622||full Low-quality
18-103-final-viral-combined_k141_29954||full Low-quality
18-103-final-viral-combined_k141_76067||full Not-determined
18-103-final-viral-combined_k141_35774||full Not-determined
18-103-final-viral-combined_k141_81933||full Low-quality
18-103-final-viral-combined_k141_47421||full Low-quality
18-103-final-viral-combined_k141_24387||full Low-quality
18-103-final-viral-combined_k141_133630||full Not-determined
18-103-final-viral-combined_k141_18461||full Not-determined
18-103-final-viral-combined_k141_35783||full Not-determined
18-103-final-viral-combined_k141_18465||full Low-quality
18-103-final-viral-combined_k141_98872||full Not-determined
18-103-final-viral-combined_k141_133639||full Not-determined
18-103-final-viral-combined_k141_58871||full Not-determined
18-103-final-viral-combined_k141_98882||full Not-determined
18-103-final-viral-combined_k141_133645||full Not-determined
18-103-final-viral-combined_k141_18468||full Not-determined
18-103-final-viral-combined_k141_1423||full Low-quality
18-103-final-viral-combined_k141_58879||full Not-determined
18-103-final-viral-combined_k141_128107||full Not-determined
18-103-final-viral-combined_k141_30002||full Low-quality
18-103-final-viral-combined_k141_47440||full Low-quality
18-103-final-viral-combined_k141_41575||full Low-quality
18-103-final-viral-combined_k141_122037||full Not-determined
18-103-final-viral-combined_k141_93339||full Low-quality
18-103-final-viral-combined_k141_64621||full Not-determined
18-103-final-viral-combined_k141_7196||full Not-determined
18-103-final-viral-combined_k141_70302||full Not-determined
18-103-final-viral-combined_k141_76124||full Low-quality
18-103-final-viral-combined_k141_87807||full Not-determined
18-103-final-viral-combined_k141_7202||full Not-determined
18-103-final-viral-combined_k141_12710||full Not-determined
18-103-final-viral-combined_k141_64639||full Low-quality
18-103-final-viral-combined_k141_35845||full Low-quality
18-103-final-viral-combined_k141_70317||full Low-quality
18-103-final-viral-combined_k141_104825||full Low-quality
18-103-final-viral-combined_k141_18544||full Low-quality
18-103-final-viral-combined_k141_35868||full Low-quality
18-103-final-viral-combined_k141_41635||full Not-determined
18-103-final-viral-combined_k141_18546||full Not-determined
18-103-final-viral-combined_k141_87822||full Low-quality
18-103-final-viral-combined_k141_41638||full Low-quality
18-103-final-viral-combined_k141_30083||full Low-quality
18-103-final-viral-combined_k141_128158||full Low-quality
18-103-final-viral-combined_k141_41644||full Low-quality
18-103-final-viral-combined_k141_82027||full Low-quality
18-103-final-viral-combined_k141_87838||full Not-determined
18-103-final-viral-combined_k141_122102||full Low-quality
18-103-final-viral-combined_k141_12758||full Low-quality
18-103-final-viral-combined_k141_128165||full Low-quality
18-103-final-viral-combined_k141_110679||full Not-determined
18-103-final-viral-combined_k141_122123||full Not-determined
18-103-final-viral-combined_k141_93405||full Not-determined
18-103-final-viral-combined_k141_1507||full Not-determined
18-103-final-viral-combined_k141_12782||full Low-quality
18-103-final-viral-combined_k141_122145||full Not-determined
18-103-final-viral-combined_k141_122151||full Not-determined
18-103-final-viral-combined_k141_133763||full Low-quality
18-103-final-viral-combined_k141_12793||full Not-determined
18-103-final-viral-combined_k141_133766||full Low-quality
18-103-final-viral-combined_k141_1526||full Low-quality
18-103-final-viral-combined_k141_76230||full Not-determined
18-103-final-viral-combined_k141_41689||full Low-quality
18-103-final-viral-combined_k141_70401||full Low-quality
18-103-final-viral-combined_k141_41694||full Not-determined
18-103-final-viral-combined_k141_70408||full Low-quality
18-103-final-viral-combined_k141_18564||full Not-determined
18-103-final-viral-combined_k141_1540||full Low-quality
18-103-final-viral-combined_k141_53277||full Low-quality
18-103-final-viral-combined_k141_76242||full Low-quality
18-103-final-viral-combined_k141_47567||full Low-quality
18-103-final-viral-combined_k141_99010||full Low-quality
18-103-final-viral-combined_k141_53282||full Low-quality
18-103-final-viral-combined_k141_99014||full Not-determined
18-103-final-viral-combined_k141_99016||full Not-determined
18-103-final-viral-combined_k141_18588||full Low-quality
18-103-final-viral-combined_k141_12837||full Low-quality
18-103-final-viral-combined_k141_133811||full Low-quality
18-103-final-viral-combined_k141_12843||full Not-determined
18-103-final-viral-combined_k141_76260||full Low-quality
18-103-final-viral-combined_k141_122171||full Not-determined
18-103-final-viral-combined_k141_18597||full Low-quality
18-103-final-viral-combined_k141_1567||full Low-quality
18-103-final-viral-combined_k141_122179||full Low-quality
18-103-final-viral-combined_k141_82107||full Low-quality
18-103-final-viral-combined_k141_64791||full Low-quality
18-103-final-viral-combined_k141_12883||full Low-quality
18-103-final-viral-combined_k141_133856||full Not-determined
18-103-final-viral-combined_k141_7277||full Low-quality
18-103-final-viral-combined_k141_99054||full Low-quality
18-103-final-viral-combined_k141_64805||full Low-quality
18-103-final-viral-combined_k141_70490||full Low-quality
18-103-final-viral-combined_k141_1594||full Not-determined
18-103-final-viral-combined_k141_30119||full Not-determined
18-103-final-viral-combined_k141_110777||full Not-determined
18-103-final-viral-combined_k141_99058||full Not-determined
18-103-final-viral-combined_k141_30123||full Not-determined
18-103-final-viral-combined_k141_110778||full Not-determined
18-103-final-viral-combined_k141_99062||full Low-quality
18-103-final-viral-combined_k141_24555||full Not-determined
18-103-final-viral-combined_k141_24560||full Not-determined
18-103-final-viral-combined_k141_35965||full Low-quality
18-103-final-viral-combined_k141_64841||full Low-quality
18-103-final-viral-combined_k141_87952||full Low-quality
18-103-final-viral-combined_k141_133862||full Not-determined
18-103-final-viral-combined_k141_93541||full Not-determined
18-103-final-viral-combined_k141_47642||full Low-quality
18-103-final-viral-combined_k141_64848||full Low-quality
18-103-final-viral-combined_k141_133879||full Low-quality
18-103-final-viral-combined_k141_1635||full Low-quality
18-103-final-viral-combined_k141_128305||full Not-determined
18-103-final-viral-combined_k141_24572||full Low-quality
18-103-final-viral-combined_k141_116672||full Low-quality
18-103-final-viral-combined_k141_82165||full Not-determined
18-103-final-viral-combined_k141_110836||full Not-determined
18-103-final-viral-combined_k141_82176||full Low-quality
18-103-final-viral-combined_k141_76322||full Low-quality
18-103-final-viral-combined_k141_7341||full Low-quality
18-103-final-viral-combined_k141_116689||full Not-determined
18-103-final-viral-combined_k141_59119||full Low-quality
18-103-final-viral-combined_k141_64876||full Low-quality
18-103-final-viral-combined_k141_53432||full Not-determined
18-103-final-viral-combined_k141_88011||full Not-determined
18-103-final-viral-combined_k141_128342||full Low-quality
18-103-final-viral-combined_k141_110871||full Not-determined
18-103-final-viral-combined_k141_116719||full Low-quality
18-103-final-viral-combined_k141_70592||full Low-quality
18-103-final-viral-combined_k141_47715||full Not-determined
18-103-final-viral-combined_k141_99105||full Not-determined
18-103-final-viral-combined_k141_70598||full Not-determined
18-103-final-viral-combined_k141_99109||full Not-determined
18-103-final-viral-combined_k141_18761||full Not-determined
18-103-final-viral-combined_k141_128347||full Low-quality
18-103-final-viral-combined_k141_70604||full Not-determined
18-103-final-viral-combined_k141_105107||full Low-quality
18-103-final-viral-combined_k141_41827||full Not-determined
18-103-final-viral-combined_k141_122301||full Not-determined
18-103-final-viral-combined_k141_18781||full Not-determined
18-103-final-viral-combined_k141_116728||full Low-quality
18-103-final-viral-combined_k141_64919||full Low-quality
18-103-final-viral-combined_k141_47739||full Low-quality
18-103-final-viral-combined_k141_1718||full Low-quality
18-103-final-viral-combined_k141_59175||full Not-determined
18-103-final-viral-combined_k141_53462||full Low-quality
18-103-final-viral-combined_k141_18793||full Not-determined
18-103-final-viral-combined_k141_133958||full Low-quality
18-103-final-viral-combined_k141_7358||full Not-determined
18-103-final-viral-combined_k141_93589||full Low-quality
18-103-final-viral-combined_k141_82266||full Low-quality
18-103-final-viral-combined_k141_30245||full Low-quality
18-103-final-viral-combined_k141_36083||full Not-determined
18-103-final-viral-combined_k141_122351||full Low-quality
18-103-final-viral-combined_k141_128406||full Not-determined
18-103-final-viral-combined_k141_41882||full Not-determined
18-103-final-viral-combined_k141_1753||full Not-determined
18-103-final-viral-combined_k141_93625||full Low-quality
18-103-final-viral-combined_k141_30267||full Low-quality
18-103-final-viral-combined_k141_99144||full Low-quality
18-103-final-viral-combined_k141_122386||full Not-determined
18-103-final-viral-combined_k141_47807||full Not-determined
18-103-final-viral-combined_k141_110969||full Low-quality
18-103-final-viral-combined_k141_105176||full Not-determined
18-103-final-viral-combined_k141_76405||full Not-determined
18-103-final-viral-combined_k141_30285||full Not-determined
18-103-final-viral-combined_k141_110979||full Not-determined
18-103-final-viral-combined_k141_53497||full Not-determined
18-103-final-viral-combined_k141_88116||full Low-quality
18-103-final-viral-combined_k141_110982||full Low-quality
18-103-final-viral-combined_k141_88123||full Low-quality
18-103-final-viral-combined_k141_134035||full Not-determined
18-103-final-viral-combined_k141_128462||full Low-quality
18-103-final-viral-combined_k141_47826||full Low-quality
18-103-final-viral-combined_k141_18862||full Not-determined
18-103-final-viral-combined_k141_24705||full Not-determined
18-103-final-viral-combined_k141_93678||full Low-quality
18-103-final-viral-combined_k141_59235||full Low-quality
18-103-final-viral-combined_k141_122433||full Low-quality
18-103-final-viral-combined_k141_59238||full Not-determined
18-103-final-viral-combined_k141_122448||full Not-determined
18-103-final-viral-combined_k141_134054||full Low-quality
18-103-final-viral-combined_k141_47840||full Not-determined
18-103-final-viral-combined_k141_82369||full Low-quality
18-103-final-viral-combined_k141_128487||full Low-quality
18-103-final-viral-combined_k141_18883||full Not-determined
18-103-final-viral-combined_k141_111019||full Low-quality
18-103-final-viral-combined_k141_82376||full Low-quality
18-103-final-viral-combined_k141_116852||full Low-quality
18-103-final-viral-combined_k141_53540||full Low-quality
18-103-final-viral-combined_k141_76455||full Low-quality
18-103-final-viral-combined_k141_65067||full Low-quality
18-103-final-viral-combined_k141_134060||full Low-quality
18-103-final-viral-combined_k141_111048||full Not-determined
18-103-final-viral-combined_k141_116859||full Low-quality
18-103-final-viral-combined_k141_134077||full Low-quality
18-103-final-viral-combined_k141_111058||full Low-quality
18-103-final-viral-combined_k141_99258||full Not-determined
18-103-final-viral-combined_k141_30362||full Not-determined
18-103-final-viral-combined_k141_7521||full Low-quality
18-103-final-viral-combined_k141_116866||full Low-quality
18-103-final-viral-combined_k141_122516||full Low-quality
18-103-final-viral-combined_k141_116871||full Low-quality
18-103-final-viral-combined_k141_134099||full Low-quality
18-103-final-viral-combined_k141_70769||full Low-quality
18-103-final-viral-combined_k141_88197||full Low-quality
18-103-final-viral-combined_k141_82410||full Not-determined
18-103-final-viral-combined_k141_18936||full Not-determined
18-103-final-viral-combined_k141_134100||full Low-quality
18-103-final-viral-combined_k141_70786||full Not-determined
18-103-final-viral-combined_k141_13150||full Not-determined
18-103-final-viral-combined_k141_36216||full Low-quality
18-103-final-viral-combined_k141_59301||full Low-quality
18-103-final-viral-combined_k141_18954||full Low-quality
18-103-final-viral-combined_k141_65118||full Not-determined
18-103-final-viral-combined_k141_70794||full Not-determined
18-103-final-viral-combined_k141_1865||full Low-quality
18-103-final-viral-combined_k141_30405||full Not-determined
18-103-final-viral-combined_k141_18966||full Not-determined
18-103-final-viral-combined_k141_1872||full Not-determined
18-103-final-viral-combined_k141_70798||full Low-quality
18-103-final-viral-combined_k141_116908||full Not-determined
18-103-final-viral-combined_k141_76526||full Low-quality
18-103-final-viral-combined_k141_122557||full Low-quality
18-103-final-viral-combined_k141_1880||full Low-quality
18-103-final-viral-combined_k141_116910||full Low-quality
18-103-final-viral-combined_k141_122558||full Low-quality
18-103-final-viral-combined_k141_13172||full Not-determined
18-103-final-viral-combined_k141_128569||full Low-quality
18-103-final-viral-combined_k141_93781||full Low-quality
18-103-final-viral-combined_k141_116919||full Low-quality
18-103-final-viral-combined_k141_7550||full Low-quality
18-103-final-viral-combined_k141_128581||full Low-quality
18-103-final-viral-combined_k141_42067||full Not-determined
18-103-final-viral-combined_k141_65152||full Low-quality
18-103-final-viral-combined_k141_1896||full Low-quality
18-103-final-viral-combined_k141_42068||full Not-determined
18-103-final-viral-combined_k141_36268||full Low-quality
18-103-final-viral-combined_k141_93798||full Low-quality
18-103-final-viral-combined_k141_99318||full Low-quality
18-103-final-viral-combined_k141_13209||full Low-quality
18-103-final-viral-combined_k141_53581||full Not-determined
18-103-final-viral-combined_k141_1908||full Low-quality
18-103-final-viral-combined_k141_134168||full Low-quality
18-103-final-viral-combined_k141_13225||full Low-quality
18-103-final-viral-combined_k141_59376||full Low-quality
18-103-final-viral-combined_k141_76577||full Low-quality
18-103-final-viral-combined_k141_30468||full Not-determined
18-103-final-viral-combined_k141_47922||full Not-determined
18-103-final-viral-combined_k141_24900||full Low-quality
18-103-final-viral-combined_k141_65175||full Not-determined
18-103-final-viral-combined_k141_105395||full Not-determined
18-103-final-viral-combined_k141_30471||full Not-determined
18-103-final-viral-combined_k141_99349||full Low-quality
18-103-final-viral-combined_k141_59390||full Not-determined
18-103-final-viral-combined_k141_93810||full Not-determined
18-103-final-viral-combined_k141_88258||full Not-determined
18-103-final-viral-combined_k141_1943||full Not-determined
18-103-final-viral-combined_k141_128601||full Low-quality
18-103-final-viral-combined_k141_82509||full Not-determined
18-103-final-viral-combined_k141_105423||full Low-quality
18-103-final-viral-combined_k141_76608||full Low-quality
18-103-final-viral-combined_k141_70897||full Not-determined
18-103-final-viral-combined_k141_53639||full Not-determined
18-103-final-viral-combined_k141_42123||full Low-quality
18-103-final-viral-combined_k141_36297||full Low-quality
18-103-final-viral-combined_k141_70898||full Not-determined
18-103-final-viral-combined_k141_70903||full Low-quality
18-103-final-viral-combined_k141_82523||full Low-quality
18-103-final-viral-combined_k141_70907||full Low-quality
18-103-final-viral-combined_k141_59422||full Not-determined
18-103-final-viral-combined_k141_88270||full Low-quality
18-103-final-viral-combined_k141_65213||full Low-quality
18-103-final-viral-combined_k141_53654||full Not-determined
18-103-final-viral-combined_k141_122660||full Not-determined
18-103-final-viral-combined_k141_13278||full Not-determined
18-103-final-viral-combined_k141_53660||full Not-determined
18-103-final-viral-combined_k141_30527||full Low-quality
18-103-final-viral-combined_k141_24948||full Low-quality
18-103-final-viral-combined_k141_105446||full Not-determined
18-103-final-viral-combined_k141_99377||full Low-quality
18-103-final-viral-combined_k141_65231||full Low-quality
18-103-final-viral-combined_k141_111146||full Low-quality
18-103-final-viral-combined_k141_36317||full Low-quality
18-103-final-viral-combined_k141_42168||full Not-determined
18-103-final-viral-combined_k141_65239||full Low-quality
18-103-final-viral-combined_k141_122693||full Not-determined
18-103-final-viral-combined_k141_42178||full Low-quality
18-103-final-viral-combined_k141_82575||full Not-determined
18-103-final-viral-combined_k141_70931||full Not-determined
18-103-final-viral-combined_k141_59461||full Low-quality
18-103-final-viral-combined_k141_70935||full Low-quality
18-103-final-viral-combined_k141_70938||full Low-quality
18-103-final-viral-combined_k141_30561||full Low-quality
18-103-final-viral-combined_k141_70955||full Not-determined
18-103-final-viral-combined_k141_13264||full Not-determined
18-103-final-viral-combined_k141_2079||full Low-quality
18-103-final-viral-combined_k141_105523||full Not-determined
18-103-final-viral-combined_k141_105525||full Low-quality
18-103-final-viral-combined_k141_128705||full Low-quality
18-103-final-viral-combined_k141_7632||full Low-quality
18-103-final-viral-combined_k141_128710||full Low-quality
18-103-final-viral-combined_k141_19125||full Low-quality
18-103-final-viral-combined_k141_128717||full Low-quality
18-103-final-viral-combined_k141_2094||full Not-determined
18-103-final-viral-combined_k141_117076||full Not-determined
18-103-final-viral-combined_k141_82633||full Low-quality
18-103-final-viral-combined_k141_42244||full Low-quality
18-103-final-viral-combined_k141_36405||full Not-determined
18-103-final-viral-combined_k141_19147||full Low-quality
18-103-final-viral-combined_k141_53725||full Not-determined
18-103-final-viral-combined_k141_111204||full Not-determined
18-103-final-viral-combined_k141_105571||full Not-determined
18-103-final-viral-combined_k141_65337||full Not-determined
18-103-final-viral-combined_k141_111215||full Low-quality
18-103-final-viral-combined_k141_7659||full Not-determined
18-103-final-viral-combined_k141_19169||full Low-quality
18-103-final-viral-combined_k141_7662||full Low-quality
18-103-final-viral-combined_k141_111219||full Low-quality
18-103-final-viral-combined_k141_30643||full Low-quality
18-103-final-viral-combined_k141_2141||full Not-determined
18-103-final-viral-combined_k141_128731||full Low-quality
18-103-final-viral-combined_k141_134329||full Not-determined
18-103-final-viral-combined_k141_134333||full Not-determined
18-103-final-viral-combined_k141_7700||full Not-determined
18-103-final-viral-combined_k141_117118||full Low-quality
18-103-final-viral-combined_k141_2170||full Low-quality
18-103-final-viral-combined_k141_134349||full Not-determined
18-103-final-viral-combined_k141_122876||full Low-quality
18-103-final-viral-combined_k141_128748||full Low-quality
18-103-final-viral-combined_k141_76754||full Low-quality
18-103-final-viral-combined_k141_53780||full Low-quality
18-103-final-viral-combined_k141_30672||full Not-determined
18-103-final-viral-combined_k141_94067||full Not-determined
18-103-final-viral-combined_k141_25069||full Not-determined
18-103-final-viral-combined_k141_59557||full Low-quality
18-103-final-viral-combined_k141_76765||full Low-quality
18-103-final-viral-combined_k141_134366||full Not-determined
18-103-final-viral-combined_k141_2199||full Low-quality
18-103-final-viral-combined_k141_65354||full Low-quality
18-103-final-viral-combined_k141_53786||full Low-quality
18-103-final-viral-combined_k141_134380||full Not-determined
18-103-final-viral-combined_k141_53790||full Not-determined
18-103-final-viral-combined_k141_13473||full Not-determined
18-103-final-viral-combined_k141_88441||full Low-quality
18-103-final-viral-combined_k141_30694||full Low-quality
18-103-final-viral-combined_k141_53800||full Not-determined
18-103-final-viral-combined_k141_48185||full Not-determined
18-103-final-viral-combined_k141_117148||full Not-determined
18-103-final-viral-combined_k141_36493||full Not-determined
18-103-final-viral-combined_k141_122919||full Low-quality
18-103-final-viral-combined_k141_71111||full Not-determined
18-103-final-viral-combined_k141_59589||full Low-quality
18-103-final-viral-combined_k141_128792||full Low-quality
18-103-final-viral-combined_k141_88465||full Low-quality
18-103-final-viral-combined_k141_2234||full Low-quality
18-103-final-viral-combined_k141_19221||full Low-quality
18-103-final-viral-combined_k141_13506||full Not-determined
18-103-final-viral-combined_k141_7765||full Not-determined
18-103-final-viral-combined_k141_82698||full Not-determined
18-103-final-viral-combined_k141_30714||full Not-determined
18-103-final-viral-combined_k141_105633||full Low-quality
18-103-final-viral-combined_k141_88474||full Low-quality
18-103-final-viral-combined_k141_42347||full Not-determined
18-103-final-viral-combined_k141_88484||full Low-quality
18-103-final-viral-combined_k141_94100||full Not-determined
18-103-final-viral-combined_k141_122955||full Low-quality
18-103-final-viral-combined_k141_105648||full Low-quality
18-103-final-viral-combined_k141_82742||full Not-determined
18-103-final-viral-combined_k141_13546||full Not-determined
18-103-final-viral-combined_k141_88514||full Low-quality
18-103-final-viral-combined_k141_122971||full Low-quality
18-103-final-viral-combined_k141_25179||full Not-determined
18-103-final-viral-combined_k141_117226||full Not-determined
18-103-final-viral-combined_k141_134440||full Low-quality
18-103-final-viral-combined_k141_25185||full Not-determined
18-103-final-viral-combined_k141_99571||full Low-quality
18-103-final-viral-combined_k141_13557||full Low-quality
18-103-final-viral-combined_k141_25189||full Low-quality
18-103-final-viral-combined_k141_76821||full Not-determined
18-103-final-viral-combined_k141_13573||full Not-determined
18-103-final-viral-combined_k141_122998||full Not-determined
18-103-final-viral-combined_k141_36589||full Low-quality
18-103-final-viral-combined_k141_36595||full Not-determined
18-103-final-viral-combined_k141_99600||full Not-determined
18-103-final-viral-combined_k141_25208||full Not-determined
18-103-final-viral-combined_k141_99603||full Low-quality
18-103-final-viral-combined_k141_88556||full Low-quality
18-103-final-viral-combined_k141_59652||full Low-quality
18-103-final-viral-combined_k141_2322||full Low-quality
18-103-final-viral-combined_k141_7829||full Not-determined
18-103-final-viral-combined_k141_13594||full Not-determined
18-103-final-viral-combined_k141_42409||full Low-quality
18-103-final-viral-combined_k141_48259||full Not-determined
18-103-final-viral-combined_k141_42417||full Low-quality
18-103-final-viral-combined_k141_94206||full Low-quality
18-103-final-viral-combined_k141_99642||full Low-quality
18-103-final-viral-combined_k141_25214||full Low-quality
18-103-final-viral-combined_k141_59662||full Not-determined
18-103-final-viral-combined_k141_65468||full Not-determined
18-103-final-viral-combined_k141_94219||full Not-determined
18-103-final-viral-combined_k141_123041||full Low-quality
18-103-final-viral-combined_k141_134547||full Low-quality
18-103-final-viral-combined_k141_13629||full Not-determined
18-103-final-viral-combined_k141_7862||full Low-quality
18-103-final-viral-combined_k141_48285||full Low-quality
18-103-final-viral-combined_k141_25254||full Low-quality
18-103-final-viral-combined_k141_2372||full Low-quality
18-103-final-viral-combined_k141_36675||full Not-determined
18-103-final-viral-combined_k141_19385||full Not-determined
18-103-final-viral-combined_k141_105718||full Not-determined
18-103-final-viral-combined_k141_128936||full Low-quality
18-103-final-viral-combined_k141_30820||full Low-quality
18-103-final-viral-combined_k141_53944||full Low-quality
18-103-final-viral-combined_k141_42473||full Not-determined
18-103-final-viral-combined_k141_94245||full Not-determined
18-103-final-viral-combined_k141_2387||full Low-quality
18-103-final-viral-combined_k141_117313||full Low-quality
18-103-final-viral-combined_k141_59703||full Not-determined
18-103-final-viral-combined_k141_105734||full Not-determined
18-103-final-viral-combined_k141_30840||full Low-quality
18-103-final-viral-combined_k141_105738||full Not-determined
18-103-final-viral-combined_k141_134600||full Low-quality
18-103-final-viral-combined_k141_53965||full Not-determined
18-103-final-viral-combined_k141_76929||full Low-quality
18-103-final-viral-combined_k141_59730||full Low-quality
18-103-final-viral-combined_k141_105766||full Not-determined
18-103-final-viral-combined_k141_65538||full Not-determined
18-103-final-viral-combined_k141_82905||full Low-quality
18-103-final-viral-combined_k141_13675||full Not-determined
18-103-final-viral-combined_k141_99737||full Low-quality
18-103-final-viral-combined_k141_105773||full Not-determined
18-103-final-viral-combined_k141_99740||full Not-determined
18-103-final-viral-combined_k141_48336||full Not-determined
18-103-final-viral-combined_k141_71285||full Not-determined
18-103-final-viral-combined_k141_117359||full Low-quality
18-103-final-viral-combined_k141_42515||full Low-quality
18-103-final-viral-combined_k141_123103||full Low-quality
18-103-final-viral-combined_k141_128997||full Not-determined
18-103-final-viral-combined_k141_134646||full Not-determined
18-103-final-viral-combined_k141_71291||full Not-determined
18-103-final-viral-combined_k141_71292||full Low-quality
18-103-final-viral-combined_k141_36758||full Low-quality
18-103-final-viral-combined_k141_129003||full Low-quality
18-103-final-viral-combined_k141_42521||full Low-quality
18-103-final-viral-combined_k141_2430||full Not-determined
18-103-final-viral-combined_k141_76974||full Not-determined
18-103-final-viral-combined_k141_54011||full Low-quality
18-103-final-viral-combined_k141_123109||full Low-quality
18-103-final-viral-combined_k141_71304||full Low-quality
18-103-final-viral-combined_k141_117370||full Not-determined
18-103-final-viral-combined_k141_54012||full Not-determined
18-103-final-viral-combined_k141_48369||full Low-quality
18-103-final-viral-combined_k141_71308||full Low-quality
18-103-final-viral-combined_k141_36769||full Not-determined
18-103-final-viral-combined_k141_2446||full Low-quality
18-103-final-viral-combined_k141_42533||full Not-determined
18-103-final-viral-combined_k141_123119||full Low-quality
18-103-final-viral-combined_k141_99778||full Low-quality
18-103-final-viral-combined_k141_65566||full Low-quality
18-103-final-viral-combined_k141_59773||full Not-determined
18-103-final-viral-combined_k141_19473||full Not-determined
18-103-final-viral-combined_k141_36799||full Not-determined
18-103-final-viral-combined_k141_65576||full Low-quality
18-103-final-viral-combined_k141_48397||full Not-determined
18-103-final-viral-combined_k141_36805||full Low-quality
18-103-final-viral-combined_k141_134713||full Low-quality
18-103-final-viral-combined_k141_88758||full Not-determined
18-103-final-viral-combined_k141_117410||full Not-determined
18-103-final-viral-combined_k141_88763||full Low-quality
18-103-final-viral-combined_k141_134727||full Low-quality
18-103-final-viral-combined_k141_36827||full Not-determined
18-103-final-viral-combined_k141_82975||full Not-determined
18-103-final-viral-combined_k141_2481||full Low-quality
18-103-final-viral-combined_k141_2492||full Not-determined
18-103-final-viral-combined_k141_8012||full Low-quality
18-103-final-viral-combined_k141_111556||full Not-determined
18-103-final-viral-combined_k141_117447||full Not-determined
18-103-final-viral-combined_k141_134750||full Low-quality
18-103-final-viral-combined_k141_2508||full Low-quality
18-103-final-viral-combined_k141_111563||full Not-determined
18-103-final-viral-combined_k141_117457||full Low-quality
18-103-final-viral-combined_k141_59828||full Not-determined
18-103-final-viral-combined_k141_19532||full Not-determined
18-103-final-viral-combined_k141_30991||full Not-determined
18-103-final-viral-combined_k141_2514||full Low-quality
18-103-final-viral-combined_k141_65649||full Low-quality
18-103-final-viral-combined_k141_77078||full Not-determined
18-103-final-viral-combined_k141_19548||full Low-quality
18-103-final-viral-combined_k141_42559||full Low-quality
18-103-final-viral-combined_k141_99863||full Not-determined
18-103-final-viral-combined_k141_65651||full Not-determined
18-103-final-viral-combined_k141_105854||full Low-quality
18-103-final-viral-combined_k141_99864||full Not-determined
18-103-final-viral-combined_k141_25428||full Low-quality
18-103-final-viral-combined_k141_134778||full Not-determined
18-103-final-viral-combined_k141_2539||full Low-quality

18-103-final-viral-combined_k141_83034||full Low-quality
18-103-final-viral-combined_k141_31016||full Not-determined
18-103-final-viral-combined_k141_77106||full Low-quality
18-103-final-viral-combined_k141_129127||full Low-quality
18-103-final-viral-combined_k141_13748||full Not-determined
18-103-final-viral-combined_k141_36918||full Not-determined
18-103-final-viral-combined_k141_48466||full Not-determined
18-103-final-viral-combined_k141_77123||full Not-determined
18-103-final-viral-combined_k141_36930||full Low-quality
18-103-final-viral-combined_k141_111641||full Low-quality
18-103-final-viral-combined_k141_8062||full Low-quality
18-103-final-viral-combined_k141_31046||full Not-determined
18-103-final-viral-combined_k141_42616||full Not-determined
18-103-final-viral-combined_k141_77150||full Not-determined
18-103-final-viral-combined_k141_42619||full Not-determined
18-103-final-viral-combined_k141_99924||full Low-quality
18-103-final-viral-combined_k141_13784||full Low-quality
18-103-final-viral-combined_k141_31053||full Not-determined
18-103-final-viral-combined_k141_54117||full Low-quality
18-103-final-viral-combined_k141_71425||full Low-quality
18-103-final-viral-combined_k141_8085||full Not-determined
18-103-final-viral-combined_k141_19610||full Not-determined
18-103-final-viral-combined_k141_129181||full Low-quality
18-103-final-viral-combined_k141_105919||full Not-determined
18-103-final-viral-combined_k141_134829||full Not-determined
18-103-final-viral-combined_k141_88873||full Low-quality
18-103-final-viral-combined_k141_19614||full Not-determined
18-103-final-viral-combined_k141_8096||full Not-determined
18-103-final-viral-combined_k141_59956||full Low-quality
18-103-final-viral-combined_k141_88893||full Low-quality
18-103-final-viral-combined_k141_37004||full Low-quality
18-103-final-viral-combined_k141_48559||full Not-determined
18-103-final-viral-combined_k141_48566||full Low-quality
18-103-final-viral-combined_k141_83113||full Not-determined
18-103-final-viral-combined_k141_83119||full Low-quality
18-103-final-viral-combined_k141_65686||full Low-quality
18-103-final-viral-combined_k141_134871||full Not-determined
18-103-final-viral-combined_k141_123241||full Low-quality
18-103-final-viral-combined_k141_99961||full Not-determined
18-103-final-viral-combined_k141_134880||full Low-quality
18-103-final-viral-combined_k141_117576||full Not-determined
18-103-final-viral-combined_k141_37045||full Not-determined
18-103-final-viral-combined_k141_88939||full Low-quality
18-103-final-viral-combined_k141_105956||full Not-determined
18-103-final-viral-combined_k141_99984||full Low-quality
18-103-final-viral-combined_k141_94494||full Low-quality
18-103-final-viral-combined_k141_117590||full Not-determined
18-103-final-viral-combined_k141_25527||full Not-determined
18-103-final-viral-combined_k141_48588||full Low-quality
18-103-final-viral-combined_k141_117599||full Low-quality
18-103-final-viral-combined_k141_100007||full Low-quality
18-103-final-viral-combined_k141_31165||full Low-quality
18-103-final-viral-combined_k141_123287||full Not-determined
18-103-final-viral-combined_k141_8158||full Low-quality
18-103-final-viral-combined_k141_19732||full Not-determined
18-103-final-viral-combined_k141_129245||full Not-determined
18-103-final-viral-combined_k141_25550||full Low-quality
18-103-final-viral-combined_k141_65729||full Low-quality
18-103-final-viral-combined_k141_60057||full Low-quality
18-103-final-viral-combined_k141_111785||full Not-determined
18-103-final-viral-combined_k141_65739||full Low-quality
18-103-final-viral-combined_k141_2687||full Low-quality
18-103-final-viral-combined_k141_111788||full Not-determined
18-103-final-viral-combined_k141_2689||full Not-determined
18-103-final-viral-combined_k141_94552||full Low-quality
18-103-final-viral-combined_k141_117633||full Low-quality
18-103-final-viral-combined_k141_117636||full Not-determined
18-103-final-viral-combined_k141_2692||full Not-determined
18-103-final-viral-combined_k141_31204||full Not-determined
18-103-final-viral-combined_k141_88985||full Low-quality
18-103-final-viral-combined_k141_123315||full Low-quality
18-103-final-viral-combined_k141_88988||full Not-determined
18-103-final-viral-combined_k141_111799||full Not-determined
18-103-final-viral-combined_k141_117638||full Low-quality
18-103-final-viral-combined_k141_83200||full Low-quality
18-103-final-viral-combined_k141_89000||full Low-quality
18-103-final-viral-combined_k141_13859||full Low-quality
18-103-final-viral-combined_k141_94575||full Low-quality
18-103-final-viral-combined_k141_129295||full Not-determined
18-103-final-viral-combined_k141_129299||full Not-determined
18-103-final-viral-combined_k141_25593||full Not-determined
18-103-final-viral-combined_k141_71547||full Low-quality
18-103-final-viral-combined_k141_54251||full Low-quality
18-103-final-viral-combined_k141_65772||full Low-quality
18-103-final-viral-combined_k141_65782||full Low-quality
18-103-final-viral-combined_k141_123364||full Not-determined
18-103-final-viral-combined_k141_65785||full Low-quality
18-103-final-viral-combined_k141_8238||full Not-determined
18-103-final-viral-combined_k141_54286||full Not-determined
18-103-final-viral-combined_k141_42715||full Low-quality
18-103-final-viral-combined_k141_37117||full Low-quality
18-103-final-viral-combined_k141_25636||full Low-quality
18-103-final-viral-combined_k141_37123||full Low-quality
18-103-final-viral-combined_k141_13895||full Not-determined
18-103-final-viral-combined_k141_89054||full Not-determined
18-103-final-viral-combined_k141_83275||full Low-quality
18-103-final-viral-combined_k141_89060||full Low-quality
18-103-final-viral-combined_k141_19834||full Not-determined
18-103-final-viral-combined_k141_71587||full Low-quality
18-103-final-viral-combined_k141_106058||full Not-determined
18-103-final-viral-combined_k141_89075||full Not-determined
18-103-final-viral-combined_k141_77321||full Low-quality
18-103-final-viral-combined_k141_106065||full Not-determined
18-103-final-viral-combined_k141_42747||full Low-quality
18-103-final-viral-combined_k141_123415||full Low-quality
18-103-final-viral-combined_k141_89091||full Low-quality
18-103-final-viral-combined_k141_123426||full Low-quality
18-103-final-viral-combined_k141_71610||full Not-determined
18-103-final-viral-combined_k141_134975||full Not-determined
18-103-final-viral-combined_k141_77345||full Not-determined
18-103-final-viral-combined_k141_19861||full Low-quality
18-103-final-viral-combined_k141_19862||full Low-quality
18-103-final-viral-combined_k141_13929||full Not-determined
18-103-final-viral-combined_k141_106096||full Not-determined
18-103-final-viral-combined_k141_8295||full Low-quality
18-103-final-viral-combined_k141_94675||full Low-quality
18-103-final-viral-combined_k141_83330||full Low-quality
18-103-final-viral-combined_k141_89121||full Not-determined
18-103-final-viral-combined_k141_31349||full Not-determined
18-103-final-viral-combined_k141_60163||full Not-determined
18-103-final-viral-combined_k141_37179||full Not-determined
18-103-final-viral-combined_k141_94695||full Not-determined
18-103-final-viral-combined_k141_42789||full Not-determined
18-103-final-viral-combined_k141_100166||full Low-quality
18-103-final-viral-combined_k141_129420||full Low-quality
18-103-final-viral-combined_k141_106135||full Low-quality
18-103-final-viral-combined_k141_117738||full Not-determined
18-103-final-viral-combined_k141_129421||full Not-determined
18-103-final-viral-combined_k141_25730||full Not-determined
18-103-final-viral-combined_k141_83356||full Low-quality
18-103-final-viral-combined_k141_106139||full Low-quality
18-103-final-viral-combined_k141_8317||full Not-determined
18-103-final-viral-combined_k141_31397||full Not-determined
18-103-final-viral-combined_k141_54395||full Not-determined
18-103-final-viral-combined_k141_65873||full Not-determined
18-103-final-viral-combined_k141_8324||full Not-determined
18-103-final-viral-combined_k141_48712||full Low-quality
18-103-final-viral-combined_k141_94725||full Low-quality
18-103-final-viral-combined_k141_42812||full Not-determined
18-103-final-viral-combined_k141_60208||full Low-quality
18-103-final-viral-combined_k141_8331||full Not-determined
18-103-final-viral-combined_k141_100208||full Low-quality
18-103-final-viral-combined_k141_42825||full Not-determined
18-103-final-viral-combined_k141_54424||full Not-determined
18-103-final-viral-combined_k141_2876||full Low-quality
18-103-final-viral-combined_k141_19896||full Low-quality
18-103-final-viral-combined_k141_129466||full Low-quality
18-103-final-viral-combined_k141_19914||full Low-quality
18-103-final-viral-combined_k141_14024||full Not-determined
18-103-final-viral-combined_k141_54440||full Low-quality
18-103-final-viral-combined_k141_129470||full Not-determined
18-103-final-viral-combined_k141_2901||full Not-determined
18-103-final-viral-combined_k141_8342||full Not-determined
18-103-final-viral-combined_k141_123518||full Low-quality
18-103-final-viral-combined_k141_117805||full Not-determined
18-103-final-viral-combined_k141_100237||full Low-quality
18-103-final-viral-combined_k141_135056||full Not-determined
18-103-final-viral-combined_k141_14037||full Low-quality
18-103-final-viral-combined_k141_2921||full Not-determined
18-103-final-viral-combined_k141_77455||full Low-quality
18-103-final-viral-combined_k141_123533||full Low-quality
18-103-final-viral-combined_k141_42853||full Not-determined
18-103-final-viral-combined_k141_129488||full Not-determined
18-103-final-viral-combined_k141_31452||full Low-quality
18-103-final-viral-combined_k141_25804||full Low-quality
18-103-final-viral-combined_k141_8350||full Low-quality
18-103-final-viral-combined_k141_135079||full Low-quality
18-103-final-viral-combined_k141_129494||full Not-determined
18-103-final-viral-combined_k141_77463||full Not-determined
18-103-final-viral-combined_k141_71687||full Low-quality
18-103-final-viral-combined_k141_2933||full Low-quality
18-103-final-viral-combined_k141_77464||full Low-quality
18-103-final-viral-combined_k141_65928||full Low-quality
18-103-final-viral-combined_k141_111996||full Not-determined
18-103-final-viral-combined_k141_37267||full Low-quality
18-103-final-viral-combined_k141_14046||full Low-quality
18-103-final-viral-combined_k141_8375||full Low-quality
18-103-final-viral-combined_k141_65942||full Low-quality
18-103-final-viral-combined_k141_19978||full Not-determined
18-103-final-viral-combined_k141_89194||full Low-quality
18-103-final-viral-combined_k141_42891||full Not-determined
18-103-final-viral-combined_k141_65948||full Low-quality
18-103-final-viral-combined_k141_129528||full Not-determined
18-103-final-viral-combined_k141_89195||full Not-determined
18-103-final-viral-combined_k141_100294||full Low-quality
18-103-final-viral-combined_k141_129531||full Low-quality
18-103-final-viral-combined_k141_77483||full Low-quality
18-103-final-viral-combined_k141_83426||full Not-determined
18-103-final-viral-combined_k141_54495||full Low-quality
18-103-final-viral-combined_k141_77484||full Not-determined
18-103-final-viral-combined_k141_48813||full Low-quality
18-103-final-viral-combined_k141_123571||full Not-determined
18-103-final-viral-combined_k141_94818||full Low-quality
18-103-final-viral-combined_k141_54496||full Low-quality
18-103-final-viral-combined_k141_31489||full Low-quality
18-103-final-viral-combined_k141_123576||full Not-determined
18-103-final-viral-combined_k141_89201||full Not-determined
18-103-final-viral-combined_k141_31491||full Not-determined
18-103-final-viral-combined_k141_65972||full Not-determined
18-103-final-viral-combined_k141_48831||full Not-determined
18-103-final-viral-combined_k141_89211||full Not-determined
18-103-final-viral-combined_k141_37326||full Low-quality
18-103-final-viral-combined_k141_54504||full Low-quality
18-103-final-viral-combined_k141_123587||full Not-determined
18-103-final-viral-combined_k141_89216||full Not-determined
18-103-final-viral-combined_k141_20030||full Low-quality
18-103-final-viral-combined_k141_31497||full Low-quality
18-103-final-viral-combined_k141_48851||full Low-quality
18-103-final-viral-combined_k141_94839||full Low-quality
18-103-final-viral-combined_k141_129540||full Low-quality
18-103-final-viral-combined_k141_94841||full Low-quality
18-103-final-viral-combined_k141_112052||full Low-quality
18-103-final-viral-combined_k141_48855||full Low-quality
18-103-final-viral-combined_k141_54512||full Low-quality
18-103-final-viral-combined_k141_129549||full Low-quality
18-103-final-viral-combined_k141_135137||full Low-quality
18-103-final-viral-combined_k141_48858||full Not-determined
18-103-final-viral-combined_k141_54516||full Not-determined
18-103-final-viral-combined_k141_14115||full Not-determined
18-103-final-viral-combined_k141_3009||full Low-quality
18-103-final-viral-combined_k141_25854||full Low-quality
18-103-final-viral-combined_k141_54522||full Low-quality
18-103-final-viral-combined_k141_37355||full Not-determined
18-103-final-viral-combined_k141_31510||full Low-quality
18-103-final-viral-combined_k141_129572||full Not-determined
18-103-final-viral-combined_k141_71709||full Low-quality
18-103-final-viral-combined_k141_117908||full Low-quality
18-103-final-viral-combined_k141_60335||full Not-determined
18-103-final-viral-combined_k141_112081||full Low-quality
18-103-final-viral-combined_k141_112086||full Low-quality
18-103-final-viral-combined_k141_54557||full Low-quality
18-103-final-viral-combined_k141_60347||full Low-quality
18-103-final-viral-combined_k141_135181||full Low-quality
18-103-final-viral-combined_k141_100345||full Low-quality
18-103-final-viral-combined_k141_14132||full Not-determined
18-103-final-viral-combined_k141_94901||full Low-quality
18-103-final-viral-combined_k141_106306||full Not-determined
18-103-final-viral-combined_k141_129583||full Low-quality
18-103-final-viral-combined_k141_112111||full Low-quality
18-103-final-viral-combined_k141_60353||full Not-determined
18-103-final-viral-combined_k141_25902||full Low-quality
18-103-final-viral-combined_k141_14155||full Low-quality
18-103-final-viral-combined_k141_66055||full Low-quality
18-103-final-viral-combined_k141_20114||full Low-quality
18-103-final-viral-combined_k141_100371||full Not-determined
18-103-final-viral-combined_k141_106334||full Not-determined
18-103-final-viral-combined_k141_123665||full Low-quality
18-103-final-viral-combined_k141_60378||full Not-determined
18-103-final-viral-combined_k141_66061||full Not-determined
18-103-final-viral-combined_k141_117963||full Low-quality
18-103-final-viral-combined_k141_71749||full Low-quality
18-103-final-viral-combined_k141_89305||full Not-determined
18-103-final-viral-combined_k141_123672||full Not-determined
18-103-final-viral-combined_k141_94948||full Not-determined
18-103-final-viral-combined_k141_31585||full Low-quality
18-103-final-viral-combined_k141_123691||full Not-determined
18-103-final-viral-combined_k141_83568||full Low-quality
18-103-final-viral-combined_k141_83570||full Low-quality
18-103-final-viral-combined_k141_106366||full Low-quality
18-103-final-viral-combined_k141_8524||full Not-determined
18-103-final-viral-combined_k141_20137||full Low-quality
18-103-final-viral-combined_k141_123701||full Not-determined
18-103-final-viral-combined_k141_89333||full Not-determined
18-103-final-viral-combined_k141_83582||full Low-quality
18-103-final-viral-combined_k141_123706||full Low-quality
18-103-final-viral-combined_k141_14184||full Not-determined
18-103-final-viral-combined_k141_20150||full Not-determined
18-103-final-viral-combined_k141_43064||full Not-determined
18-103-final-viral-combined_k141_123714||full Low-quality
18-103-final-viral-combined_k141_112168||full Low-quality
18-103-final-viral-combined_k141_135247||full Not-determined
18-103-final-viral-combined_k141_60429||full Not-determined
18-103-final-viral-combined_k141_100425||full Low-quality
18-103-final-viral-combined_k141_48906||full Not-determined
18-103-final-viral-combined_k141_31597||full Not-determined
18-103-final-viral-combined_k141_106407||full Low-quality
18-103-final-viral-combined_k141_48907||full Not-determined
18-103-final-viral-combined_k141_129668||full Not-determined
18-103-final-viral-combined_k141_123729||full Low-quality
18-103-final-viral-combined_k141_118054||full Low-quality
18-103-final-viral-combined_k141_8571||full Not-determined
18-103-final-viral-combined_k141_20164||full Not-determined
18-103-final-viral-combined_k141_37481||full Low-quality
18-103-final-viral-combined_k141_112179||full Low-quality
18-103-final-viral-combined_k141_37487||full Low-quality
18-103-final-viral-combined_k141_37488||full Low-quality
18-103-final-viral-combined_k141_60444||full Low-quality
18-103-final-viral-combined_k141_77600||full Not-determined
18-103-final-viral-combined_k141_20179||full Not-determined
18-103-final-viral-combined_k141_129693||full Not-determined
18-103-final-viral-combined_k141_135292||full Low-quality
18-103-final-viral-combined_k141_123765||full Not-determined
18-103-final-viral-combined_k141_60447||full Low-quality
18-103-final-viral-combined_k141_66152||full Low-quality
18-103-final-viral-combined_k141_129704||full Not-determined
18-103-final-viral-combined_k141_60461||full Low-quality
18-103-final-viral-combined_k141_106453||full Not-determined
18-103-final-viral-combined_k141_100490||full Not-determined
18-103-final-viral-combined_k141_106458||full Low-quality
18-103-final-viral-combined_k141_129711||full Not-determined
18-103-final-viral-combined_k141_83623||full Low-quality
18-103-final-viral-combined_k141_123790||full Low-quality
18-103-final-viral-combined_k141_20213||full Not-determined
18-103-final-viral-combined_k141_8625||full Low-quality
18-103-final-viral-combined_k141_89434||full Not-determined
18-103-final-viral-combined_k141_106497||full Low-quality
18-103-final-viral-combined_k141_112237||full Not-determined
18-103-final-viral-combined_k141_89436||full Not-determined
18-103-final-viral-combined_k141_43122||full Low-quality
18-103-final-viral-combined_k141_20231||full Not-determined
18-103-final-viral-combined_k141_37545||full Low-quality
18-103-final-viral-combined_k141_71869||full Low-quality
18-103-final-viral-combined_k141_83650||full Not-determined
18-103-final-viral-combined_k141_77650||full Low-quality
18-103-final-viral-combined_k141_37553||full Low-quality
18-103-final-viral-combined_k141_14242||full Not-determined
18-103-final-viral-combined_k141_54716||full Low-quality
18-103-final-viral-combined_k141_3273||full Not-determined
18-103-final-viral-combined_k141_54727||full Not-determined
18-103-final-viral-combined_k141_71896||full Not-determined
18-103-final-viral-combined_k141_77671||full Low-quality
18-103-final-viral-combined_k141_77674||full Low-quality
18-103-final-viral-combined_k141_54737||full Not-determined
18-103-final-viral-combined_k141_89477||full Low-quality
18-103-final-viral-combined_k141_77684||full Not-determined
18-103-final-viral-combined_k141_37588||full Low-quality
18-103-final-viral-combined_k141_37591||full Not-determined
18-103-final-viral-combined_k141_25991||full Not-determined
18-103-final-viral-combined_k141_100550||full Not-determined
18-103-final-viral-combined_k141_83694||full Not-determined
18-103-final-viral-combined_k141_89499||full Not-determined
18-103-final-viral-combined_k141_118220||full Not-determined
18-103-final-viral-combined_k141_77707||full Not-determined
18-103-final-viral-combined_k141_112327||full Low-quality
18-103-final-viral-combined_k141_83708||full Low-quality
18-103-final-viral-combined_k141_26014||full Low-quality
18-103-final-viral-combined_k141_31684||full Not-determined
18-103-final-viral-combined_k141_71948||full Low-quality
18-103-final-viral-combined_k141_43195||full Not-determined
18-103-final-viral-combined_k141_100578||full Low-quality
18-103-final-viral-combined_k141_14302||full Not-determined
18-103-final-viral-combined_k141_112351||full Low-quality
18-103-final-viral-combined_k141_48963||full Low-quality
18-103-final-viral-combined_k141_95106||full Not-determined
18-103-final-viral-combined_k141_14319||full Low-quality
18-103-final-viral-combined_k141_95119||full Low-quality
18-103-final-viral-combined_k141_48986||full Not-determined
18-103-final-viral-combined_k141_48987||full Low-quality
18-103-final-viral-combined_k141_8699||full Low-quality
18-103-final-viral-combined_k141_123874||full Not-determined
18-103-final-viral-combined_k141_14323||full Not-determined
18-103-final-viral-combined_k141_71979||full Not-determined
18-103-final-viral-combined_k141_95134||full Not-determined
18-103-final-viral-combined_k141_118280||full Low-quality
18-103-final-viral-combined_k141_89549||full Low-quality
18-103-final-viral-combined_k141_83735||full Not-determined
18-103-final-viral-combined_k141_31726||full Not-determined
18-103-final-viral-combined_k141_66341||full Low-quality
18-103-final-viral-combined_k141_135415||full Not-determined
18-103-final-viral-combined_k141_54798||full Low-quality
18-103-final-viral-combined_k141_129834||full Not-determined
18-103-final-viral-combined_k141_54814||full Not-determined
18-103-final-viral-combined_k141_49029||full Not-determined
18-103-final-viral-combined_k141_43286||full Not-determined
18-103-final-viral-combined_k141_100652||full Low-quality
18-103-final-viral-combined_k141_54849||full Low-quality
18-103-final-viral-combined_k141_123906||full Low-quality
18-103-final-viral-combined_k141_43296||full Low-quality
18-103-final-viral-combined_k141_129860||full Not-determined
18-103-final-viral-combined_k141_123911||full Not-determined
18-103-final-viral-combined_k141_123912||full Low-quality
18-103-final-viral-combined_k141_8777||full Low-quality
18-103-final-viral-combined_k141_66399||full Not-determined
18-103-final-viral-combined_k141_3389||full Low-quality
18-103-final-viral-combined_k141_83786||full Low-quality
18-110-final-viral-combined_k141_186054||full High-quality
18-103-final-viral-combined_k141_123924||full Low-quality
18-103-final-viral-combined_k141_37698||full Not-determined
18-103-final-viral-combined_k141_118338||full Not-determined
18-103-final-viral-combined_k141_49064||full Low-quality
18-103-final-viral-combined_k141_14401||full Not-determined
18-103-final-viral-combined_k141_14402||full Not-determined
18-102-final-viral-combined_k141_58563||full Medium-quality
18-103-final-viral-combined_k141_49076||full Not-determined
18-103-final-viral-combined_k141_95221||full Low-quality
18-103-final-viral-combined_k141_89637||full Low-quality
18-103-final-viral-combined_k141_118367||full Low-quality
18-103-final-viral-combined_k141_89641||full Not-determined
18-103-final-viral-combined_k141_43328||full Low-quality
18-103-final-viral-combined_k141_49086||full Not-determined
18-103-final-viral-combined_k141_3421||full Not-determined
18-103-final-viral-combined_k141_20340||full Not-determined
18-103-final-viral-combined_k141_26151||full Low-quality
18-103-final-viral-combined_k141_112485||full Not-determined
18-103-final-viral-combined_k141_26170||full Not-determined
18-103-final-viral-combined_k141_95241||full Not-determined
18-103-final-viral-combined_k141_118399||full Low-quality
18-103-final-viral-combined_k141_112493||full Low-quality
18-103-final-viral-combined_k141_95249||full Low-quality
18-103-final-viral-combined_k141_123984||full Not-determined
18-103-final-viral-combined_k141_83826||full Not-determined
18-103-final-viral-combined_k141_72065||full Low-quality
18-103-final-viral-combined_k141_37755||full Low-quality
18-103-final-viral-combined_k141_123994||full Not-determined
18-103-final-viral-combined_k141_72070||full Not-determined
18-103-final-viral-combined_k141_3448||full Low-quality
18-103-final-viral-combined_k141_124006||full Low-quality
18-103-final-viral-combined_k141_135484||full Low-quality
18-103-final-viral-combined_k141_49147||full Low-quality
18-103-final-viral-combined_k141_124009||full Low-quality
18-103-final-viral-combined_k141_31813||full Not-determined
18-103-final-viral-combined_k141_60628||full Low-quality
18-103-final-viral-combined_k141_54959||full Not-determined
18-103-final-viral-combined_k141_26190||full Low-quality
18-103-final-viral-combined_k141_106671||full Not-determined
18-103-final-viral-combined_k141_118445||full Low-quality
18-103-final-viral-combined_k141_54965||full Not-determined
18-103-final-viral-combined_k141_66515||full Not-determined
18-103-final-viral-combined_k141_43394||full Low-quality
18-103-final-viral-combined_k141_135527||full Not-determined
18-103-final-viral-combined_k141_72116||full Not-determined
18-103-final-viral-combined_k141_129968||full Not-determined
18-103-final-viral-combined_k141_135534||full Not-determined
18-103-final-viral-combined_k141_66527||full Low-quality
18-103-final-viral-combined_k141_37833||full Low-quality
18-103-final-viral-combined_k141_129973||full Low-quality
18-103-final-viral-combined_k141_49197||full Not-determined
18-103-final-viral-combined_k141_14541||full Low-quality
18-103-final-viral-combined_k141_118477||full Low-quality
18-103-final-viral-combined_k141_129975||full Not-determined
18-103-final-viral-combined_k141_26249||full Not-determined
18-103-final-viral-combined_k141_3474||full Low-quality
18-103-final-viral-combined_k141_124069||full Not-determined
18-103-final-viral-combined_k141_95336||full Not-determined
18-103-final-viral-combined_k141_43443||full Low-quality
18-103-final-viral-combined_k141_3483||full Not-determined
18-103-final-viral-combined_k141_77992||full Low-quality
18-103-final-viral-combined_k141_55005||full Not-determined
18-103-final-viral-combined_k141_14568||full Not-determined
18-103-final-viral-combined_k141_130005||full Low-quality
18-103-final-viral-combined_k141_31879||full Low-quality
18-103-final-viral-combined_k141_78005||full Low-quality
18-103-final-viral-combined_k141_124100||full Low-quality
18-103-final-viral-combined_k141_124101||full Not-determined
18-103-final-viral-combined_k141_72185||full Low-quality
18-103-final-viral-combined_k141_20470||full Not-determined
18-103-final-viral-combined_k141_130018||full Not-determined
18-103-final-viral-combined_k141_72186||full Low-quality
18-103-final-viral-combined_k141_14600||full Not-determined
18-103-final-viral-combined_k141_14601||full Not-determined
18-103-final-viral-combined_k141_83988||full Not-determined
18-103-final-viral-combined_k141_112613||full Not-determined
18-103-final-viral-combined_k141_26301||full Not-determined
18-103-final-viral-combined_k141_3523||full Low-quality
18-103-final-viral-combined_k141_3525||full Not-determined
18-103-final-viral-combined_k141_3528||full Low-quality
18-103-final-viral-combined_k141_37881||full Low-quality
18-103-final-viral-combined_k141_100849||full Low-quality
18-103-final-viral-combined_k141_26328||full Low-quality
18-103-final-viral-combined_k141_112645||full Low-quality
18-103-final-viral-combined_k141_37902||full Low-quality
18-103-final-viral-combined_k141_49225||full Not-determined
18-103-final-viral-combined_k141_66594||full Not-determined
18-103-final-viral-combined_k141_72240||full Low-quality
18-103-final-viral-combined_k141_26345||full Low-quality
18-103-final-viral-combined_k141_72261||full Low-quality
18-103-final-viral-combined_k141_37940||full Not-determined
18-103-final-viral-combined_k141_66609||full Low-quality
18-103-final-viral-combined_k141_43489||full Not-determined
18-103-final-viral-combined_k141_124220||full Low-quality
18-103-final-viral-combined_k141_72275||full Not-determined
18-103-final-viral-combined_k141_60786||full Low-quality
18-103-final-viral-combined_k141_106847||full Low-quality
18-103-final-viral-combined_k141_14647||full Not-determined
18-103-final-viral-combined_k141_60797||full Not-determined
18-103-final-viral-combined_k141_135643||full Low-quality
18-103-final-viral-combined_k141_37981||full Low-quality
18-103-final-viral-combined_k141_60798||full Low-quality
18-103-final-viral-combined_k141_118619||full Low-quality
18-103-final-viral-combined_k141_95465||full Not-determined
18-103-final-viral-combined_k141_49303||full Not-determined
18-103-final-viral-combined_k141_37987||full Low-quality
18-103-final-viral-combined_k141_95467||full Not-determined
18-103-final-viral-combined_k141_130097||full Not-determined
18-103-final-viral-combined_k141_37990||full Low-quality
18-103-final-viral-combined_k141_43522||full Low-quality
18-103-final-viral-combined_k141_72295||full Low-quality
18-103-final-viral-combined_k141_43523||full Low-quality
18-103-final-viral-combined_k141_72307||full Low-quality
18-103-final-viral-combined_k141_118641||full Not-determined
18-103-final-viral-combined_k141_112701||full Low-quality
18-103-final-viral-combined_k141_112707||full Low-quality
18-103-final-viral-combined_k141_20592||full Not-determined
18-103-final-viral-combined_k141_89918||full Not-determined
18-103-final-viral-combined_k141_49341||full Not-determined
18-103-final-viral-combined_k141_66690||full Not-determined
18-103-final-viral-combined_k141_100904||full Not-determined
18-103-final-viral-combined_k141_89933||full Low-quality
18-103-final-viral-combined_k141_118662||full Not-determined
18-103-final-viral-combined_k141_89943||full Low-quality
18-103-final-viral-combined_k141_130155||full Low-quality
18-103-final-viral-combined_k141_3669||full Low-quality
18-103-final-viral-combined_k141_78113||full Not-determined
18-103-final-viral-combined_k141_135675||full Not-determined
18-103-final-viral-combined_k141_72342||full Low-quality
18-103-final-viral-combined_k141_130163||full Not-determined
18-103-final-viral-combined_k141_78115||full Not-determined
18-103-final-viral-combined_k141_84052||full Not-determined
18-103-final-viral-combined_k141_72345||full Not-determined
18-103-final-viral-combined_k141_43548||full Low-quality
18-103-final-viral-combined_k141_43552||full Low-quality
18-103-final-viral-combined_k141_100932||full Not-determined
18-103-final-viral-combined_k141_135697||full Not-determined
18-103-final-viral-combined_k141_95522||full Not-determined
18-103-final-viral-combined_k141_43567||full Low-quality
18-103-final-viral-combined_k141_130188||full Not-determined
18-103-final-viral-combined_k141_100965||full Not-determined
18-103-final-viral-combined_k141_55258||full Not-determined
18-103-final-viral-combined_k141_60874||full Not-determined
18-103-final-viral-combined_k141_43586||full Not-determined
18-103-final-viral-combined_k141_3699||full Low-quality
18-103-final-viral-combined_k141_100976||full Low-quality
18-103-final-viral-combined_k141_100979||full Not-determined
18-103-final-viral-combined_k141_112743||full Low-quality
18-103-final-viral-combined_k141_60892||full Not-determined
18-103-final-viral-combined_k141_100982||full Not-determined
18-103-final-viral-combined_k141_9102||full Not-determined
18-103-final-viral-combined_k141_124348||full Low-quality
18-103-final-viral-combined_k141_38106||full Low-quality
18-103-final-viral-combined_k141_26525||full Not-determined
18-103-final-viral-combined_k141_20708||full Not-determined
18-103-final-viral-combined_k141_32013||full Not-determined
18-103-final-viral-combined_k141_26529||full Not-determined
18-103-final-viral-combined_k141_43601||full Not-determined
18-103-final-viral-combined_k141_95555||full Not-determined
18-103-final-viral-combined_k141_32018||full Not-determined
18-103-final-viral-combined_k141_78195||full Low-quality
18-103-final-viral-combined_k141_14753||full Not-determined
18-103-final-viral-combined_k141_60924||full Low-quality
18-103-final-viral-combined_k141_112774||full Not-determined
18-103-final-viral-combined_k141_14771||full Not-determined
18-103-final-viral-combined_k141_3724||full Low-quality
18-103-final-viral-combined_k141_118746||full Not-determined
18-103-final-viral-combined_k141_20755||full Low-quality
18-103-final-viral-combined_k141_135752||full Low-quality
18-103-final-viral-combined_k141_38165||full Not-determined
18-103-final-viral-combined_k141_43638||full Not-determined
18-103-final-viral-combined_k141_9149||full Low-quality
18-103-final-viral-combined_k141_112800||full Not-determined
18-103-final-viral-combined_k141_130303||full Low-quality
18-103-final-viral-combined_k141_38170||full Low-quality
18-103-final-viral-combined_k141_130305||full Low-quality
18-103-final-viral-combined_k141_112805||full Low-quality
18-103-final-viral-combined_k141_90055||full Not-determined
18-103-final-viral-combined_k141_78227||full Not-determined
18-103-final-viral-combined_k141_55328||full Low-quality
18-103-final-viral-combined_k141_112807||full Low-quality
18-103-final-viral-combined_k141_32075||full Low-quality
18-103-final-viral-combined_k141_124408||full Not-determined
18-103-final-viral-combined_k141_78230||full Low-quality
18-103-final-viral-combined_k141_14814||full Low-quality
18-103-final-viral-combined_k141_49480||full Not-determined
18-103-final-viral-combined_k141_49481||full Low-quality
18-103-final-viral-combined_k141_20792||full Low-quality
18-103-final-viral-combined_k141_9173||full Not-determined
18-103-final-viral-combined_k141_72479||full Low-quality
18-103-final-viral-combined_k141_60968||full Low-quality
18-103-final-viral-combined_k141_66816||full Low-quality
18-103-final-viral-combined_k141_14824||full Not-determined
18-103-final-viral-combined_k141_20806||full Low-quality
18-103-final-viral-combined_k141_43686||full Low-quality
18-103-final-viral-combined_k141_66826||full Not-determined
18-103-final-viral-combined_k141_130350||full Not-determined
18-103-final-viral-combined_k141_124422||full Not-determined
18-103-final-viral-combined_k141_55367||full Not-determined
18-103-final-viral-combined_k141_3776||full Not-determined
18-103-final-viral-combined_k141_9218||full Low-quality
18-103-final-viral-combined_k141_55385||full Not-determined
18-103-final-viral-combined_k141_66850||full Low-quality
18-103-final-viral-combined_k141_90136||full Low-quality
18-103-final-viral-combined_k141_112849||full Not-determined
18-103-final-viral-combined_k141_55393||full Not-determined
17-109_k141_193565 Medium-quality
18-103-final-viral-combined_k141_90140||full Low-quality
18-103-final-viral-combined_k141_107034||full Not-determined
18-103-final-viral-combined_k141_14850||full Not-determined
18-103-final-viral-combined_k141_135808||full Not-determined
18-103-final-viral-combined_k141_49543||full Low-quality
18-103-final-viral-combined_k141_32143||full Low-quality

| | |
|---|---|
| 18-103-final-viral-combined_k141_26659\|\|full | Not-determined |
| 18-103-final-viral-combined_k141_55397\|\|full | Low-quality |
| 18-103-final-viral-combined_k141_101121\|\|full | Not-determined |
| 18-103-final-viral-combined_k141_84237\|\|full | Low-quality |
| 18-103-final-viral-combined_k141_20826\|\|full | Not-determined |
| 18-103-final-viral-combined_k141_61043\|\|full | Not-determined |
| 18-103-final-viral-combined_k141_38285\|\|full | Low-quality |
| 18-103-final-viral-combined_k141_72541\|\|full | Not-determined |
| 18-103-final-viral-combined_k141_78330\|\|full | Low-quality |
| 18-103-final-viral-combined_k141_101129\|\|full | Low-quality |
| 18-103-final-viral-combined_k141_66894\|\|full | Low-quality |
| 18-103-final-viral-combined_k141_26678\|\|full | Low-quality |
| 18-103-final-viral-combined_k141_95706\|\|full | Not-determined |
| 18-103-final-viral-combined_k141_130421\|\|full | Low-quality |
| 18-103-final-viral-combined_k141_9263\|\|full | Low-quality |
| 18-103-final-viral-combined_k141_9264\|\|full | Low-quality |
| 18-103-final-viral-combined_k141_49551\|\|full | Low-quality |
| 18-103-final-viral-combined_k141_20854\|\|full | Low-quality |
| 18-103-final-viral-combined_k141_124501\|\|full | Low-quality |
| 18-103-final-viral-combined_k141_95724\|\|full | Not-determined |
| 18-103-final-viral-combined_k141_3825\|\|full | Not-determined |
| 18-103-final-viral-combined_k141_130427\|\|full | Low-quality |
| 18-103-final-viral-combined_k141_118907\|\|full | Low-quality |
| 18-103-final-viral-combined_k141_43716\|\|full | Low-quality |
| 18-103-final-viral-combined_k141_38307\|\|full | Not-determined |
| 18-103-final-viral-combined_k141_32194\|\|full | Not-determined |
| 18-103-final-viral-combined_k141_9277\|\|full | Low-quality |
| 18-103-final-viral-combined_k141_49563\|\|full | Low-quality |
| 18-103-final-viral-combined_k141_78369\|\|full | Low-quality |
| 18-103-final-viral-combined_k141_43729\|\|full | Low-quality |
| 18-103-final-viral-combined_k141_84282\|\|full | Not-determined |
| 18-103-final-viral-combined_k141_135868\|\|full | Not-determined |
| 18-103-final-viral-combined_k141_130449\|\|full | Not-determined |
| 18-103-final-viral-combined_k141_3855\|\|full | Low-quality |
| 18-103-final-viral-combined_k141_9314\|\|full | Not-determined |
| 18-103-final-viral-combined_k141_135885\|\|full | Not-determined |
| 18-103-final-viral-combined_k141_14940\|\|full | Low-quality |
| 18-103-final-viral-combined_k141_3872\|\|full | Not-determined |
| 18-103-final-viral-combined_k141_55455\|\|full | Not-determined |
| 18-103-final-viral-combined_k141_135905\|\|full | Not-determined |
| 18-103-final-viral-combined_k141_20915\|\|full | Low-quality |
| 18-103-final-viral-combined_k141_112937\|\|full | Low-quality |
| 18-103-final-viral-combined_k141_32237\|\|full | Low-quality |
| 18-103-final-viral-combined_k141_84344\|\|full | Not-determined |
| 18-103-final-viral-combined_k141_43784\|\|full | Low-quality |
| 18-103-final-viral-combined_k141_118972\|\|full | Not-determined |
| 18-103-final-viral-combined_k141_72588\|\|full | Low-quality |
| 18-103-final-viral-combined_k141_84355\|\|full | Not-determined |
| 18-103-final-viral-combined_k141_61142\|\|full | Low-quality |
| 18-103-final-viral-combined_k141_66998\|\|full | Low-quality |
| 18-103-final-viral-combined_k141_20964\|\|full | Not-determined |
| 18-103-final-viral-combined_k141_124622\|\|full | Not-determined |
| 18-103-final-viral-combined_k141_107189\|\|full | Low-quality |
| 18-103-final-viral-combined_k141_67002\|\|full | Not-determined |
| 18-103-final-viral-combined_k141_95802\|\|full | Low-quality |
| 18-103-final-viral-combined_k141_107200\|\|full | Low-quality |
| 18-103-final-viral-combined_k141_61151\|\|full | Low-quality |
| 18-103-final-viral-combined_k141_130517\|\|full | Not-determined |
| 18-103-final-viral-combined_k141_107202\|\|full | Low-quality |
| 18-103-final-viral-combined_k141_90268\|\|full | Not-determined |
| 18-103-final-viral-combined_k141_84381\|\|full | Low-quality |
| 18-103-final-viral-combined_k141_84387\|\|full | Not-determined |
| 18-103-final-viral-combined_k141_20990\|\|full | Not-determined |
| 18-103-final-viral-combined_k141_130529\|\|full | Low-quality |
| 18-103-final-viral-combined_k141_78495\|\|full | Not-determined |
| 18-103-final-viral-combined_k141_26768\|\|full | Not-determined |
| 18-103-final-viral-combined_k141_113016\|\|full | Not-determined |
| 18-103-final-viral-combined_k141_78502\|\|full | Low-quality |
| 18-103-final-viral-combined_k141_14986\|\|full | Not-determined |
| 18-103-final-viral-combined_k141_113024\|\|full | Not-determined |
| 18-103-final-viral-combined_k141_95827\|\|full | Not-determined |
| 18-103-final-viral-combined_k141_61205\|\|full | Not-determined |
| 18-103-final-viral-combined_k141_130549\|\|full | Low-quality |
| 18-103-final-viral-combined_k141_21007\|\|full | Low-quality |
| 18-103-final-viral-combined_k141_43877\|\|full | Not-determined |
| 18-103-final-viral-combined_k141_130553\|\|full | Low-quality |
| 18-103-final-viral-combined_k141_26800\|\|full | Not-determined |
| 18-103-final-viral-combined_k141_130554\|\|full | Not-determined |
| 18-103-final-viral-combined_k141_32313\|\|full | Not-determined |
| 18-103-final-viral-combined_k141_135990\|\|full | Low-quality |
| 18-103-final-viral-combined_k141_124685\|\|full | Not-determined |
| 18-103-final-viral-combined_k141_95839\|\|full | Not-determined |
| 18-103-final-viral-combined_k141_84433\|\|full | Low-quality |
| 18-103-final-viral-combined_k141_26806\|\|full | Not-determined |
| 18-103-final-viral-combined_k141_107254\|\|full | Low-quality |
| 18-103-final-viral-combined_k141_84439\|\|full | Low-quality |
| 18-103-final-viral-combined_k141_135992\|\|full | Not-determined |
| 18-103-final-viral-combined_k141_113053\|\|full | Low-quality |
| 18-103-final-viral-combined_k141_113061\|\|full | Low-quality |
| 18-103-final-viral-combined_k141_130578\|\|full | Low-quality |
| 18-103-final-viral-combined_k141_95846\|\|full | Low-quality |
| 18-103-final-viral-combined_k141_21058\|\|full | Not-determined |
| 18-103-final-viral-combined_k141_67101\|\|full | Low-quality |
| 18-103-final-viral-combined_k141_4012\|\|full | Not-determined |
| 18-103-final-viral-combined_k141_4019\|\|full | Not-determined |
| 18-103-final-viral-combined_k141_55604\|\|full | Not-determined |
| 18-103-final-viral-combined_k141_9428\|\|full | Not-determined |
| 18-103-final-viral-combined_k141_4026\|\|full | Not-determined |
| 18-103-final-viral-combined_k141_107322\|\|full | Low-quality |
| 18-103-final-viral-combined_k141_21086\|\|full | Low-quality |
| 18-103-final-viral-combined_k141_107326\|\|full | Low-quality |
| 18-103-final-viral-combined_k141_49729\|\|full | Low-quality |
| 18-103-final-viral-combined_k141_43945\|\|full | Low-quality |
| 18-103-final-viral-combined_k141_72655\|\|full | Low-quality |
| 18-103-final-viral-combined_k141_90344\|\|full | Not-determined |
| 18-103-final-viral-combined_k141_21093\|\|full | Low-quality |
| 18-103-final-viral-combined_k141_9442\|\|full | Not-determined |
| 18-103-final-viral-combined_k141_49732\|\|full | Low-quality |
| 18-103-final-viral-combined_k141_15061\|\|full | Not-determined |
| 18-103-final-viral-combined_k141_124751\|\|full | Not-determined |
| 18-103-final-viral-combined_k141_84485\|\|full | Low-quality |
| 18-103-final-viral-combined_k141_124757\|\|full | Not-determined |
| 18-103-final-viral-combined_k141_119150\|\|full | Low-quality |
| 18-103-final-viral-combined_k141_124758\|\|full | Not-determined |
| 18-103-final-viral-combined_k141_113116\|\|full | Low-quality |
| 18-103-final-viral-combined_k141_101268\|\|full | Low-quality |
| 18-103-final-viral-combined_k141_130641\|\|full | Low-quality |
| 18-103-final-viral-combined_k141_113126\|\|full | Not-determined |
| 18-103-final-viral-combined_k141_15082\|\|full | Not-determined |
| 18-103-final-viral-combined_k141_72673\|\|full | Low-quality |
| 18-103-final-viral-combined_k141_4080\|\|full | Low-quality |
| 18-103-final-viral-combined_k141_55655\|\|full | Not-determined |
| 18-103-final-viral-combined_k141_15093\|\|full | Not-determined |
| 18-103-final-viral-combined_k141_72686\|\|full | Low-quality |
| 18-103-final-viral-combined_k141_136079\|\|full | Low-quality |
| 18-103-final-viral-combined_k141_130652\|\|full | Not-determined |
| 18-103-final-viral-combined_k141_119166\|\|full | Low-quality |
| 18-103-final-viral-combined_k141_67163\|\|full | Low-quality |
| 18-103-final-viral-combined_k141_101277\|\|full | Low-quality |
| 18-103-final-viral-combined_k141_38488\|\|full | Not-determined |
| 18-103-final-viral-combined_k141_78616\|\|full | Low-quality |
| 18-103-final-viral-combined_k141_4090\|\|full | Not-determined |
| 18-103-final-viral-combined_k141_49777\|\|full | Not-determined |
| 18-103-final-viral-combined_k141_38501\|\|full | Low-quality |
| 18-103-final-viral-combined_k141_90398\|\|full | Not-determined |
| 18-103-final-viral-combined_k141_49780\|\|full | Low-quality |
| 18-103-final-viral-combined_k141_38508\|\|full | Not-determined |
| 18-103-final-viral-combined_k141_101290\|\|full | Low-quality |
| 18-103-final-viral-combined_k141_101293\|\|full | Not-determined |
| 18-103-final-viral-combined_k141_84543\|\|full | Low-quality |
| 18-103-final-viral-combined_k141_90419\|\|full | Low-quality |
| 18-103-final-viral-combined_k141_124794\|\|full | Low-quality |
| 18-103-final-viral-combined_k141_113184\|\|full | Low-quality |
| 18-103-final-viral-combined_k141_67186\|\|full | Not-determined |
| 18-103-final-viral-combined_k141_21163\|\|full | Not-determined |
| 18-103-final-viral-combined_k141_90425\|\|full | Not-determined |
| 18-103-final-viral-combined_k141_113190\|\|full | Not-determined |
| 18-103-final-viral-combined_k141_78640\|\|full | Low-quality |
| 18-103-final-viral-combined_k141_130681\|\|full | Not-determined |
| 18-103-final-viral-combined_k141_136120\|\|full | Low-quality |
| 18-103-final-viral-combined_k141_124812\|\|full | Not-determined |
| 18-103-final-viral-combined_k141_107381\|\|full | Low-quality |
| 18-103-final-viral-combined_k141_84560\|\|full | Low-quality |
| 18-103-final-viral-combined_k141_9561\|\|full | Not-determined |
| 18-103-final-viral-combined_k141_67220\|\|full | Low-quality |
| 18-103-final-viral-combined_k141_21194\|\|full | Not-determined |
| 18-103-final-viral-combined_k141_84571\|\|full | Low-quality |
| 18-103-final-viral-combined_k141_49815\|\|full | Not-determined |
| 18-103-final-viral-combined_k141_136168\|\|full | Not-determined |
| 18-103-final-viral-combined_k141_130728\|\|full | Low-quality |
| 18-103-final-viral-combined_k141_15138\|\|full | Not-determined |
| 18-103-final-viral-combined_k141_67242\|\|full | Not-determined |
| 18-103-final-viral-combined_k141_136185\|\|full | Low-quality |
| 18-103-final-viral-combined_k141_101341\|\|full | Low-quality |
| 18-103-final-viral-combined_k141_4166\|\|full | Low-quality |
| 18-103-final-viral-combined_k141_72795\|\|full | Low-quality |
| 18-103-final-viral-combined_k141_4167\|\|full | Low-quality |
| 18-103-final-viral-combined_k141_15149\|\|full | Not-determined |
| 18-103-final-viral-combined_k141_113265\|\|full | Low-quality |
| 18-103-final-viral-combined_k141_9611\|\|full | Low-quality |
| 18-103-final-viral-combined_k141_124878\|\|full | Not-determined |
| 18-103-final-viral-combined_k141_4173\|\|full | Not-determined |
| 18-103-final-viral-combined_k141_61352\|\|full | Low-quality |
| 18-103-final-viral-combined_k141_32500\|\|full | Not-determined |
| 18-103-final-viral-combined_k141_72809\|\|full | Low-quality |
| 18-103-final-viral-combined_k141_38599\|\|full | Not-determined |
| 18-103-final-viral-combined_k141_67273\|\|full | Low-quality |
| 18-103-final-viral-combined_k141_32503\|\|full | Not-determined |
| 18-103-final-viral-combined_k141_44068\|\|full | Not-determined |
| 18-103-final-viral-combined_k141_9623\|\|full | Not-determined |
| 18-103-final-viral-combined_k141_124894\|\|full | Low-quality |
| 18-103-final-viral-combined_k141_55761\|\|full | Not-determined |
| 18-103-final-viral-combined_k141_124907\|\|full | Not-determined |
| 18-103-final-viral-combined_k141_21284\|\|full | Low-quality |
| 18-103-final-viral-combined_k141_124910\|\|full | Low-quality |
| 18-103-final-viral-combined_k141_84652\|\|full | Not-determined |
| 18-103-final-viral-combined_k141_136228\|\|full | Not-determined |
| 18-103-final-viral-combined_k141_67315\|\|full | Not-determined |
| 18-103-final-viral-combined_k141_107471\|\|full | Not-determined |
| 18-103-final-viral-combined_k141_55772\|\|full | Low-quality |
| 18-103-final-viral-combined_k141_119330\|\|full | Low-quality |
| 18-103-final-viral-combined_k141_32553\|\|full | Low-quality |
| 18-103-final-viral-combined_k141_119333\|\|full | Not-determined |
| 18-103-final-viral-combined_k141_78752\|\|full | Not-determined |
| 18-103-final-viral-combined_k141_38659\|\|full | Low-quality |
| 18-103-final-viral-combined_k141_27092\|\|full | Not-determined |
| 18-103-final-viral-combined_k141_61421\|\|full | Not-determined |
| 18-103-final-viral-combined_k141_44115\|\|full | Not-determined |
| 18-103-final-viral-combined_k141_113333\|\|full | Low-quality |
| 18-103-final-viral-combined_k141_55800\|\|full | Low-quality |
| 18-103-final-viral-combined_k141_72841\|\|full | Low-quality |
| 18-103-final-viral-combined_k141_84695\|\|full | Low-quality |
| 18-103-final-viral-combined_k141_78767\|\|full | Low-quality |
| 18-103-final-viral-combined_k141_32582\|\|full | Not-determined |
| 18-103-final-viral-combined_k141_136256\|\|full | Low-quality |
| 18-103-final-viral-combined_k141_101434\|\|full | Not-determined |
| 18-103-final-viral-combined_k141_9687\|\|full | Low-quality |
| 18-103-final-viral-combined_k141_61436\|\|full | Not-determined |
| 18-103-final-viral-combined_k141_67356\|\|full | Not-determined |
| 18-103-final-viral-combined_k141_32592\|\|full | Not-determined |
| 18-103-final-viral-combined_k141_21324\|\|full | Not-determined |
| 18-103-final-viral-combined_k141_90520\|\|full | Not-determined |
| 18-103-final-viral-combined_k141_96114\|\|full | Not-determined |
| 18-103-final-viral-combined_k141_136281\|\|full | Not-determined |
| 18-103-final-viral-combined_k141_61466\|\|full | Low-quality |
| 18-103-final-viral-combined_k141_136284\|\|full | Low-quality |
| 18-103-final-viral-combined_k141_113363\|\|full | Not-determined |
| 18-103-final-viral-combined_k141_90545\|\|full | Not-determined |
| 18-103-final-viral-combined_k141_113366\|\|full | Not-determined |
| 18-103-final-viral-combined_k141_72897\|\|full | Not-determined |
| 18-103-final-viral-combined_k141_119394\|\|full | Low-quality |
| 18-103-final-viral-combined_k141_90546\|\|full | Not-determined |
| 18-103-final-viral-combined_k141_27154\|\|full | Low-quality |
| 18-103-final-viral-combined_k141_125006\|\|full | Low-quality |
| 18-103-final-viral-combined_k141_96130\|\|full | Low-quality |
| 18-103-final-viral-combined_k141_136311\|\|full | Low-quality |
| 18-103-final-viral-combined_k141_61499\|\|full | Not-determined |
| 18-103-final-viral-combined_k141_55832\|\|full | Low-quality |
| 18-103-final-viral-combined_k141_67400\|\|full | Low-quality |
| 18-103-final-viral-combined_k141_84750\|\|full | Not-determined |
| 18-103-final-viral-combined_k141_72923\|\|full | Not-determined |
| 18-103-final-viral-combined_k141_136329\|\|full | Low-quality |
| 18-103-final-viral-combined_k141_96145\|\|full | Not-determined |
| 18-103-final-viral-combined_k141_15254\|\|full | Not-determined |
| 18-103-final-viral-combined_k141_107536\|\|full | Not-determined |
| 18-103-final-viral-combined_k141_96156\|\|full | Not-determined |
| 18-103-final-viral-combined_k141_113415\|\|full | Not-determined |
| 18-103-final-viral-combined_k141_130881\|\|full | Low-quality |
| 18-103-final-viral-combined_k141_67419\|\|full | Low-quality |
| 18-103-final-viral-combined_k141_96174\|\|full | Not-determined |
| 18-103-final-viral-combined_k141_67421\|\|full | Low-quality |
| 18-103-final-viral-combined_k141_21374\|\|full | Low-quality |
| 18-103-final-viral-combined_k141_78848\|\|full | Low-quality |
| 18-103-final-viral-combined_k141_136348\|\|full | Not-determined |
| 18-103-final-viral-combined_k141_21377\|\|full | Not-determined |
| 18-103-final-viral-combined_k141_107561\|\|full | Not-determined |
| 18-103-final-viral-combined_k141_67429\|\|full | Not-determined |
| 18-103-final-viral-combined_k141_61537\|\|full | Low-quality |
| 18-103-final-viral-combined_k141_72973\|\|full | Not-determined |
| 18-103-final-viral-combined_k141_119480\|\|full | Low-quality |
| 18-103-final-viral-combined_k141_101559\|\|full | Low-quality |
| 18-103-final-viral-combined_k141_130907\|\|full | Low-quality |
| 18-103-final-viral-combined_k141_107572\|\|full | Low-quality |
| 18-103-final-viral-combined_k141_15318\|\|full | Low-quality |
| 18-103-final-viral-combined_k141_21400\|\|full | Not-determined |
| 18-103-final-viral-combined_k141_61544\|\|full | Not-determined |
| 18-103-final-viral-combined_k141_136383\|\|full | Not-determined |
| 18-103-final-viral-combined_k141_119493\|\|full | Low-quality |
| 18-103-final-viral-combined_k141_27189\|\|full | Not-determined |
| 18-103-final-viral-combined_k141_130932\|\|full | Not-determined |
| 18-103-final-viral-combined_k141_113463\|\|full | Low-quality |
| 18-103-final-viral-combined_k141_125099\|\|full | Low-quality |
| 18-103-final-viral-combined_k141_61552\|\|full | Low-quality |
| 18-103-final-viral-combined_k141_38829\|\|full | Low-quality |
| 18-103-final-viral-combined_k141_113464\|\|full | Not-determined |
| 18-103-final-viral-combined_k141_130937\|\|full | Not-determined |
| 18-103-final-viral-combined_k141_32689\|\|full | Not-determined |
| 18-103-final-viral-combined_k141_9753\|\|full | Low-quality |
| 18-103-final-viral-combined_k141_55875\|\|full | Not-determined |
| 18-103-final-viral-combined_k141_32690\|\|full | Low-quality |
| 18-103-final-viral-combined_k141_4368\|\|full | Low-quality |
| 18-103-final-viral-combined_k141_21421\|\|full | Not-determined |
| 18-103-final-viral-combined_k141_38847\|\|full | Not-determined |
| 18-103-final-viral-combined_k141_61573\|\|full | Low-quality |
| 18-103-final-viral-combined_k141_96257\|\|full | Not-determined |
| 18-103-final-viral-combined_k141_136413\|\|full | Not-determined |
| 18-103-final-viral-combined_k141_38849\|\|full | Low-quality |
| 18-103-final-viral-combined_k141_67475\|\|full | Not-determined |
| 18-103-final-viral-combined_k141_96261\|\|full | Not-determined |
| 18-103-final-viral-combined_k141_32712\|\|full | Not-determined |
| 18-103-final-viral-combined_k141_96267\|\|full | Low-quality |
| 18-103-final-viral-combined_k141_125143\|\|full | Low-quality |
| 18-103-final-viral-combined_k141_15367\|\|full | Not-determined |
| 18-103-final-viral-combined_k141_67501\|\|full | Not-determined |
| 18-103-final-viral-combined_k141_61607\|\|full | Low-quality |
| 18-103-final-viral-combined_k141_107626\|\|full | Not-determined |
| 18-103-final-viral-combined_k141_27283\|\|full | Low-quality |
| 18-103-final-viral-combined_k141_38872\|\|full | Low-quality |
| 18-103-final-viral-combined_k141_131011\|\|full | Low-quality |
| 18-103-final-viral-combined_k141_21487\|\|full | Not-determined |
| 18-103-final-viral-combined_k141_38873\|\|full | Low-quality |
| 18-103-final-viral-combined_k141_27304\|\|full | Not-determined |
| 18-103-final-viral-combined_k141_38893\|\|full | Low-quality |
| 18-103-final-viral-combined_k141_55968\|\|full | Low-quality |
| 18-103-final-viral-combined_k141_38894\|\|full | Low-quality |
| 18-103-final-viral-combined_k141_101644\|\|full | Low-quality |
| 18-103-final-viral-combined_k141_125194\|\|full | Not-determined |
| 18-103-final-viral-combined_k141_27319\|\|full | Not-determined |
| 18-103-final-viral-combined_k141_107661\|\|full | Low-quality |
| 18-103-final-viral-combined_k141_61672\|\|full | Low-quality |
| 18-103-final-viral-combined_k141_136459\|\|full | Low-quality |
| 18-103-final-viral-combined_k141_113573\|\|full | Not-determined |
| 18-103-final-viral-combined_k141_101656\|\|full | Low-quality |
| 18-103-final-viral-combined_k141_9844\|\|full | Not-determined |
| 18-103-final-viral-combined_k141_55990\|\|full | Not-determined |
| 18-103-final-viral-combined_k141_131056\|\|full | Low-quality |
| 18-103-final-viral-combined_k141_84898\|\|full | Low-quality |
| 18-103-final-viral-combined_k141_125226\|\|full | Not-determined |
| 18-103-final-viral-combined_k141_119623\|\|full | Not-determined |
| 18-103-final-viral-combined_k141_15419\|\|full | Low-quality |
| 18-103-final-viral-combined_k141_56008\|\|full | Low-quality |
| 18-103-final-viral-combined_k141_84910\|\|full | Low-quality |
| 18-103-final-viral-combined_k141_84915\|\|full | Low-quality |
| 18-103-final-viral-combined_k141_119649\|\|full | Low-quality |
| 18-103-final-viral-combined_k141_131090\|\|full | Not-determined |
| 18-103-final-viral-combined_k141_27327\|\|full | Low-quality |
| 18-103-final-viral-combined_k141_32799\|\|full | Low-quality |
| 18-103-final-viral-combined_k141_84953\|\|full | Low-quality |
| 18-103-final-viral-combined_k141_119664\|\|full | Not-determined |
| 18-103-final-viral-combined_k141_96371\|\|full | Not-determined |
| 18-103-final-viral-combined_k141_56033\|\|full | Not-determined |
| 18-103-final-viral-combined_k141_38975\|\|full | Low-quality |
| 18-103-final-viral-combined_k141_61762\|\|full | Low-quality |
| 18-103-final-viral-combined_k141_125278\|\|full | Low-quality |
| 18-103-final-viral-combined_k141_125279\|\|full | Not-determined |
| 18-103-final-viral-combined_k141_44333\|\|full | Low-quality |
| 18-103-final-viral-combined_k141_4548\|\|full | Not-determined |
| 18-103-final-viral-combined_k141_125286\|\|full | Not-determined |
| 18-103-final-viral-combined_k141_84993\|\|full | Low-quality |
| 18-103-final-viral-combined_k141_101755\|\|full | Not-determined |
| 18-103-final-viral-combined_k141_9947\|\|full | Not-determined |
| 18-103-final-viral-combined_k141_21607\|\|full | Low-quality |
| 18-103-final-viral-combined_k141_4556\|\|full | Low-quality |
| 18-103-final-viral-combined_k141_131140\|\|full | Not-determined |
| 18-103-final-viral-combined_k141_67615\|\|full | Not-determined |
| 18-103-final-viral-combined_k141_44357\|\|full | Low-quality |
| 18-103-final-viral-combined_k141_9970\|\|full | Low-quality |
| 18-103-final-viral-combined_k141_15467\|\|full | Low-quality |
| 18-103-final-viral-combined_k141_125322\|\|full | Not-determined |
| 18-103-final-viral-combined_k141_125331\|\|full | Not-determined |
| 18-103-final-viral-combined_k141_44392\|\|full | Low-quality |
| 18-103-final-viral-combined_k141_79128\|\|full | Not-determined |
| 18-103-final-viral-combined_k141_113694\|\|full | Low-quality |
| 18-103-final-viral-combined_k141_85034\|\|full | Not-determined |
| 18-103-final-viral-combined_k141_136571\|\|full | Not-determined |
| 18-103-final-viral-combined_k141_15501\|\|full | Low-quality |
| 18-103-final-viral-combined_k141_107780\|\|full | Not-determined |
| 18-103-final-viral-combined_k141_136575\|\|full | Low-quality |
| 18-103-final-viral-combined_k141_39049\|\|full | Low-quality |
| 18-103-final-viral-combined_k141_85050\|\|full | Not-determined |
| 18-103-final-viral-combined_k141_90855\|\|full | Not-determined |
| 18-103-final-viral-combined_k141_10007\|\|full | Low-quality |
| 18-103-final-viral-combined_k141_90860\|\|full | Low-quality |
| 18-103-final-viral-combined_k141_85062\|\|full | Low-quality |
| 18-103-final-viral-combined_k141_125360\|\|full | Low-quality |
| 18-103-final-viral-combined_k141_61814\|\|full | Low-quality |
| 18-103-final-viral-combined_k141_107796\|\|full | Not-determined |
| 18-103-final-viral-combined_k141_119777\|\|full | Low-quality |
| 18-103-final-viral-combined_k141_61819\|\|full | Not-determined |
| 18-103-final-viral-combined_k141_96475\|\|full | Not-determined |
| 18-103-final-viral-combined_k141_32900\|\|full | Low-quality |
| 18-103-final-viral-combined_k141_21676\|\|full | Not-determined |
| 18-103-final-viral-combined_k141_4595\|\|full | Low-quality |
| 18-103-final-viral-combined_k141_131222\|\|full | Not-determined |
| 18-103-final-viral-combined_k141_10033\|\|full | Not-determined |
| 18-103-final-viral-combined_k141_101830\|\|full | Low-quality |
| 18-103-final-viral-combined_k141_44458\|\|full | Low-quality |
| 18-103-final-viral-combined_k141_4608\|\|full | Not-determined |
| 18-103-final-viral-combined_k141_119792\|\|full | Not-determined |
| 18-103-final-viral-combined_k141_101853\|\|full | Not-determined |
| 18-103-final-viral-combined_k141_85100\|\|full | Low-quality |
| 18-103-final-viral-combined_k141_61852\|\|full | Low-quality |
| 18-103-final-viral-combined_k141_79200\|\|full | Low-quality |
| 18-103-final-viral-combined_k141_136640\|\|full | Not-determined |
| 18-103-final-viral-combined_k141_79206\|\|full | Not-determined |
| 18-103-final-viral-combined_k141_96518\|\|full | Not-determined |
| 18-103-final-viral-combined_k141_131247\|\|full | Not-determined |
| 18-103-final-viral-combined_k141_15573\|\|full | Low-quality |
| 18-103-final-viral-combined_k141_96524\|\|full | Not-determined |
| 18-103-final-viral-combined_k141_21712\|\|full | Not-determined |
| 18-103-final-viral-combined_k141_61861\|\|full | Low-quality |
| 18-103-final-viral-combined_k141_44501\|\|full | Not-determined |
| 18-103-final-viral-combined_k141_21716\|\|full | Low-quality |
| 18-103-final-viral-combined_k141_27495\|\|full | Not-determined |
| 18-103-final-viral-combined_k141_50215\|\|full | Not-determined |
| 18-103-final-viral-combined_k141_44502\|\|full | Low-quality |
| 18-103-final-viral-combined_k141_61869\|\|full | Low-quality |
| 18-103-final-viral-combined_k141_44506\|\|full | Not-determined |
| 18-103-final-viral-combined_k141_61872\|\|full | Not-determined |
| 18-103-final-viral-combined_k141_107849\|\|full | Not-determined |
| 18-103-final-viral-combined_k141_44508\|\|full | Not-determined |
| 18-103-final-viral-combined_k141_119819\|\|full | Not-determined |
| 18-103-final-viral-combined_k141_15598\|\|full | Not-determined |
| 17-109-final-viral-combined_k141_193565\|\|full | Medium-quality |
| 18-103-final-viral-combined_k141_113803\|\|full | Low-quality |
| 18-103-final-viral-combined_k141_21739\|\|full | Not-determined |
| 18-103-final-viral-combined_k141_4667\|\|full | Low-quality |
| 18-103-final-viral-combined_k141_73265\|\|full | Low-quality |
| 18-103-final-viral-combined_k141_50255\|\|full | Not-determined |
| 18-103-final-viral-combined_k141_73270\|\|full | Not-determined |
| 18-103-final-viral-combined_k141_136676\|\|full | Low-quality |
| 18-103-final-viral-combined_k141_67705\|\|full | Low-quality |
| 18-103-final-viral-combined_k141_107890\|\|full | Low-quality |
| 18-103-final-viral-combined_k141_15619\|\|full | Low-quality |
| 18-103-final-viral-combined_k141_32972\|\|full | Not-determined |
| 18-103-final-viral-combined_k141_39144\|\|full | Not-determined |
| 18-103-final-viral-combined_k141_10133\|\|full | Low-quality |
| 18-103-final-viral-combined_k141_21786\|\|full | Low-quality |
| 18-103-final-viral-combined_k141_27572\|\|full | Not-determined |
| 18-103-final-viral-combined_k141_101919\|\|full | Low-quality |
| 18-103-final-viral-combined_k141_56257\|\|full | Low-quality |
| 18-103-final-viral-combined_k141_136701\|\|full | Low-quality |
| 18-103-final-viral-combined_k141_73308\|\|full | Not-determined |
| 18-103-final-viral-combined_k141_90965\|\|full | Not-determined |
| 18-103-final-viral-combined_k141_79300\|\|full | Low-quality |
| 18-103-final-viral-combined_k141_67750\|\|full | Low-quality |
| 18-103-final-viral-combined_k141_39172\|\|full | Not-determined |
| 18-103-final-viral-combined_k141_39180\|\|full | Low-quality |
| 18-103-final-viral-combined_k141_50306\|\|full | Low-quality |
| 18-103-final-viral-combined_k141_101946\|\|full | Not-determined |
| 18-103-final-viral-combined_k141_125518\|\|full | Not-determined |
| 18-103-final-viral-combined_k141_67773\|\|full | Low-quality |
| 18-103-final-viral-combined_k141_107958\|\|full | Not-determined |
| 18-103-final-viral-combined_k141_136742\|\|full | Not-determined |
| 18-103-final-viral-combined_k141_107960\|\|full | Not-determined |
| 18-103-final-viral-combined_k141_119894\|\|full | Low-quality |
| 18-103-final-viral-combined_k141_79348\|\|full | Not-determined |
| 18-103-final-viral-combined_k141_113881\|\|full | Low-quality |
| 18-103-final-viral-combined_k141_33034\|\|full | Low-quality |
| 18-103-final-viral-combined_k141_50344\|\|full | Not-determined |
| 18-103-final-viral-combined_k141_91042\|\|full | Low-quality |
| 18-103-final-viral-combined_k141_113913\|\|full | Low-quality |
| 18-103-final-viral-combined_k141_119931\|\|full | Not-determined |
| 18-103-final-viral-combined_k141_61992\|\|full | Low-quality |
| 18-103-final-viral-combined_k141_27677\|\|full | Not-determined |
| 18-103-final-viral-combined_k141_113927\|\|full | Low-quality |
| 18-103-final-viral-combined_k141_27682\|\|full | Low-quality |
| 18-103-final-viral-combined_k141_33060\|\|full | Not-determined |
| 18-103-final-viral-combined_k141_119947\|\|full | Low-quality |
| 18-103-final-viral-combined_k141_44636\|\|full | Low-quality |
| 18-103-final-viral-combined_k141_56360\|\|full | Low-quality |
| 18-103-final-viral-combined_k141_125569\|\|full | Low-quality |
| 18-103-final-viral-combined_k141_50393\|\|full | Not-determined |
| 18-103-final-viral-combined_k141_131372\|\|full | Low-quality |
| 18-103-final-viral-combined_k141_44648\|\|full | Not-determined |
| 18-103-final-viral-combined_k141_96673\|\|full | Not-determined |
| 18-103-final-viral-combined_k141_136808\|\|full | Not-determined |
| 18-103-final-viral-combined_k141_67844\|\|full | Not-determined |
| 18-103-final-viral-combined_k141_96681\|\|full | Low-quality |
| 18-103-final-viral-combined_k141_27710\|\|full | Not-determined |
| 18-103-final-viral-combined_k141_4808\|\|full | Not-determined |
| 18-103-final-viral-combined_k141_56387\|\|full | Not-determined |
| 18-103-final-viral-combined_k141_96686\|\|full | Low-quality |
| 18-103-final-viral-combined_k141_113967\|\|full | Not-determined |
| 18-103-final-viral-combined_k141_15737\|\|full | Not-determined |
| 18-103-final-viral-combined_k141_33108\|\|full | Not-determined |
| 18-103-final-viral-combined_k141_15739\|\|full | Not-determined |
| 18-103-final-viral-combined_k141_21937\|\|full | Not-determined |
| 18-103-final-viral-combined_k141_96699\|\|full | Low-quality |
| 18-103-final-viral-combined_k141_136828\|\|full | Low-quality |
| 18-103-final-viral-combined_k141_15745\|\|full | Low-quality |
| 18-103-final-viral-combined_k141_62057\|\|full | Not-determined |
| 18-103-final-viral-combined_k141_85250\|\|full | Low-quality |
| 18-103-final-viral-combined_k141_120024\|\|full | Low-quality |
| 18-103-final-viral-combined_k141_125621\|\|full | Low-quality |
| 18-103-final-viral-combined_k141_10303\|\|full | Not-determined |
| 18-103-final-viral-combined_k141_125628\|\|full | Low-quality |
| 18-103-final-viral-combined_k141_120035\|\|full | Not-determined |
| 18-103-final-viral-combined_k141_96716\|\|full | Low-quality |
| 18-103-final-viral-combined_k141_67912\|\|full | Low-quality |
| 18-103-final-viral-combined_k141_120053\|\|full | Low-quality |
| 18-103-final-viral-combined_k141_108076\|\|full | Low-quality |
| 18-103-final-viral-combined_k141_102038\|\|full | Low-quality |
| 18-103-final-viral-combined_k141_120060\|\|full | Not-determined |
| 18-103-final-viral-combined_k141_125658\|\|full | Low-quality |
| 18-103-final-viral-combined_k141_91129\|\|full | Not-determined |
| 18-103-final-viral-combined_k141_131461\|\|full | Low-quality |
| 18-103-final-viral-combined_k141_120071\|\|full | Not-determined |
| 18-103-final-viral-combined_k141_73467\|\|full | Low-quality |
| 18-103-final-viral-combined_k141_10332\|\|full | Low-quality |
| 18-103-final-viral-combined_k141_85297\|\|full | Not-determined |
| 18-103-final-viral-combined_k141_108101\|\|full | Not-determined |
| 18-103-final-viral-combined_k141_131466\|\|full | Not-determined |
| 18-103-final-viral-combined_k141_114039\|\|full | Not-determined |
| 18-103-final-viral-combined_k141_33191\|\|full | Low-quality |
| 18-103-final-viral-combined_k141_56462\|\|full | Low-quality |
| 18-103-final-viral-combined_k141_27793\|\|full | Not-determined |
| 18-103-final-viral-combined_k141_125695\|\|full | Not-determined |
| 18-103-final-viral-combined_k141_120093\|\|full | Not-determined |
| 18-103-final-viral-combined_k141_33201\|\|full | Low-quality |
| 18-103-final-viral-combined_k141_131496\|\|full | Low-quality |
| 18-103-final-viral-combined_k141_125707\|\|full | Not-determined |
| 18-103-final-viral-combined_k141_85325\|\|full | Not-determined |
| 18-103-final-viral-combined_k141_79510\|\|full | Not-determined |
| 18-103-final-viral-combined_k141_91160\|\|full | Low-quality |
| 18-103-final-viral-combined_k141_56484\|\|full | Not-determined |
| 18-103-final-viral-combined_k141_4899\|\|full | Not-determined |
| 18-103-final-viral-combined_k141_33234\|\|full | Not-determined |
| 18-103-final-viral-combined_k141_120133\|\|full | Low-quality |
| 18-103-final-viral-combined_k141_4908\|\|full | Not-determined |
| 18-103-final-viral-combined_k141_50536\|\|full | Not-determined |
| 18-103-final-viral-combined_k141_56510\|\|full | Low-quality |
| 18-103-final-viral-combined_k141_108148\|\|full | Low-quality |
| 18-103-final-viral-combined_k141_125733\|\|full | Low-quality |
| 18-103-final-viral-combined_k141_22066\|\|full | Not-determined |
| 18-103-final-viral-combined_k141_39399\|\|full | Low-quality |
| 18-103-final-viral-combined_k141_27857\|\|full | Low-quality |
| 18-103-final-viral-combined_k141_62178\|\|full | Low-quality |
| 18-103-final-viral-combined_k141_102096\|\|full | Low-quality |
| 18-103-final-viral-combined_k141_108166\|\|full | Not-determined |
| 18-103-final-viral-combined_k141_62179\|\|full | Not-determined |
| 18-103-final-viral-combined_k141_44793\|\|full | Not-determined |
| 18-103-final-viral-combined_k141_102103\|\|full | Low-quality |
| 18-103-final-viral-combined_k141_22094\|\|full | Not-determined |
| 18-103-final-viral-combined_k141_96831\|\|full | Not-determined |
| 18-103-final-viral-combined_k141_62189\|\|full | Low-quality |
| 18-103-final-viral-combined_k141_4961\|\|full | Low-quality |
| 18-103-final-viral-combined_k141_108175\|\|full | Low-quality |
| 18-103-final-viral-combined_k141_22108\|\|full | Not-determined |
| 18-103-final-viral-combined_k141_27891\|\|full | Not-determined |
| 18-103-final-viral-combined_k141_102124\|\|full | Not-determined |
| 18-103-final-viral-combined_k141_50590\|\|full | Low-quality |
| 18-103-final-viral-combined_k141_22117\|\|full | Low-quality |
| 18-103-final-viral-combined_k141_62207\|\|full | Low-quality |
| 18-103-final-viral-combined_k141_114126\|\|full | Low-quality |
| 18-103-final-viral-combined_k141_4969\|\|full | Low-quality |
| 18-103-final-viral-combined_k141_91243\|\|full | Low-quality |
| 18-103-final-viral-combined_k141_131595\|\|full | Not-determined |
| 18-103-final-viral-combined_k141_39450\|\|full | Not-determined |
| 18-103-final-viral-combined_k141_50602\|\|full | Not-determined |
| 18-103-final-viral-combined_k141_62219\|\|full | Low-quality |
| 18-103-final-viral-combined_k141_27911\|\|full | Not-determined |
| 18-103-final-viral-combined_k141_102138\|\|full | Low-quality |
| 18-103-final-viral-combined_k141_114132\|\|full | Not-determined |
| 18-103-final-viral-combined_k141_131601\|\|full | Low-quality |
| 18-103-final-viral-combined_k141_108215\|\|full | Low-quality |
| 18-103-final-viral-combined_k141_22133\|\|full | Not-determined |
| 18-103-final-viral-combined_k141_120208\|\|full | Low-quality |
| 18-103-final-viral-combined_k141_4994\|\|full | Low-quality |
| 18-103-final-viral-combined_k141_68101\|\|full | Not-determined |
| 18-103-final-viral-combined_k141_27928\|\|full | Not-determined |
| 18-103-final-viral-combined_k141_27930\|\|full | Low-quality |
| 18-103-final-viral-combined_k141_33338\|\|full | Not-determined |
| 18-103-final-viral-combined_k141_50624\|\|full | Low-quality |
| 18-103-final-viral-combined_k141_73613\|\|full | Not-determined |
| 18-103-final-viral-combined_k141_73614\|\|full | Low-quality |
| 18-103-final-viral-combined_k141_44876\|\|full | Low-quality |
| 18-103-final-viral-combined_k141_120242\|\|full | Not-determined |
| 18-103-final-viral-combined_k141_15987\|\|full | Low-quality |
| 18-103-final-viral-combined_k141_79582\|\|full | Low-quality |
| 18-103-final-viral-combined_k141_96922\|\|full | Not-determined |
| 18-103-final-viral-combined_k141_85477\|\|full | Low-quality |
| 18-103-final-viral-combined_k141_33355\|\|full | Low-quality |
| 18-103-final-viral-combined_k141_50649\|\|full | Low-quality |
| 18-103-final-viral-combined_k141_85491\|\|full | Low-quality |
| 18-103-final-viral-combined_k141_96935\|\|full | Low-quality |
| 18-103-final-viral-combined_k141_68151\|\|full | Low-quality |
| 18-103-final-viral-combined_k141_102181\|\|full | Not-determined |
| 18-103-final-viral-combined_k141_102182\|\|full | Low-quality |
| 18-103-final-viral-combined_k141_16010\|\|full | Not-determined |
| 18-103-final-viral-combined_k141_62294\|\|full | Low-quality |
| 18-103-final-viral-combined_k141_50660\|\|full | Not-determined |
| 18-103-final-viral-combined_k141_33377\|\|full | Not-determined |
| 18-103-final-viral-combined_k141_27975\|\|full | Low-quality |
| 18-103-final-viral-combined_k141_114187\|\|full | Low-quality |
| 18-103-final-viral-combined_k141_108272\|\|full | Not-determined |
| 18-103-final-viral-combined_k141_96945\|\|full | Not-determined |
| 18-103-final-viral-combined_k141_73649\|\|full | Not-determined |
| 18-103-final-viral-combined_k141_96950\|\|full | Not-determined |
| 18-103-final-viral-combined_k141_50673\|\|full | Low-quality |
| 18-103-final-viral-combined_k141_79630\|\|full | Not-determined |
| 18-103-final-viral-combined_k141_96952\|\|full | Not-determined |
| 18-103-final-viral-combined_k141_16045\|\|full | Not-determined |
| 18-103-final-viral-combined_k141_68189\|\|full | Low-quality |
| 18-103-final-viral-combined_k141_125874\|\|full | Not-determined |
| 18-103-final-viral-combined_k141_96961\|\|full | Low-quality |
| 18-103-final-viral-combined_k141_96962\|\|full | Not-determined |
| 18-103-final-viral-combined_k141_16052\|\|full | Not-determined |
| 18-103-final-viral-combined_k141_28006\|\|full | Low-quality |
| 18-103-final-viral-combined_k141_16053\|\|full | Not-determined |
| 18-103-final-viral-combined_k141_5095\|\|full | Low-quality |
| 18-103-final-viral-combined_k141_10567\|\|full | Low-quality |
| 18-103-final-viral-combined_k141_85543\|\|full | Low-quality |
| 18-103-final-viral-combined_k141_44915\|\|full | Low-quality |
| 18-103-final-viral-combined_k141_125893\|\|full | Low-quality |
| 18-103-final-viral-combined_k141_10580\|\|full | Low-quality |
| 18-103-final-viral-combined_k141_33438\|\|full | Not-determined |
| 18-103-final-viral-combined_k141_62337\|\|full | Low-quality |
| 18-103-final-viral-combined_k141_62341\|\|full | Not-determined |

| | |
|---|---|
| 18-103-final-viral-combined_k141_16089\|\|full | Not-determined |
| 18-103-final-viral-combined_k141_62345\|\|full | Low-quality |
| 18-103-final-viral-combined_k141_68228\|\|full | Not-determined |
| 18-103-final-viral-combined_k141_79695\|\|full | Not-determined |
| 18-103-final-viral-combined_k141_39586\|\|full | Not-determined |
| 18-103-final-viral-combined_k141_33449\|\|full | Low-quality |
| 18-103-final-viral-combined_k141_73715\|\|full | Not-determined |
| 18-103-final-viral-combined_k141_39588\|\|full | Low-quality |
| 18-103-final-viral-combined_k141_97012\|\|full | Not-determined |
| 18-103-final-viral-combined_k141_91336\|\|full | Not-determined |
| 18-103-final-viral-combined_k141_131742\|\|full | Low-quality |
| 18-103-final-viral-combined_k141_28048\|\|full | Low-quality |
| 18-103-final-viral-combined_k141_137042\|\|full | Not-determined |
| 18-103-final-viral-combined_k141_5137\|\|full | Not-determined |
| 18-103-final-viral-combined_k141_137051\|\|full | Low-quality |
| 18-103-final-viral-combined_k141_16120\|\|full | Low-quality |
| 18-103-final-viral-combined_k141_114288\|\|full | Not-determined |
| 18-103-final-viral-combined_k141_97044\|\|full | Not-determined |
| 18-103-final-viral-combined_k141_5163\|\|full | Low-quality |
| 18-103-final-viral-combined_k141_97049\|\|full | Low-quality |
| 18-103-final-viral-combined_k141_16126\|\|full | Low-quality |
| 18-103-final-viral-combined_k141_91361\|\|full | Not-determined |
| 18-103-final-viral-combined_k141_22301\|\|full | Not-determined |
| 18-103-final-viral-combined_k141_137067\|\|full | Low-quality |
| 18-103-final-viral-combined_k141_102318\|\|full | Low-quality |
| 18-103-final-viral-combined_k141_114322\|\|full | Not-determined |
| 18-103-final-viral-combined_k141_120424\|\|full | Not-determined |
| 18-103-final-viral-combined_k141_33478\|\|full | Not-determined |
| 18-103-final-viral-combined_k141_73768\|\|full | Not-determined |
| 18-103-final-viral-combined_k141_114332\|\|full | Low-quality |
| 18-103-final-viral-combined_k141_22315\|\|full | Not-determined |
| 18-103-final-viral-combined_k141_62404\|\|full | Low-quality |
| 18-103-final-viral-combined_k141_120442\|\|full | Not-determined |
| 18-103-final-viral-combined_k141_125985\|\|full | Not-determined |
| 18-103-final-viral-combined_k141_137074\|\|full | Low-quality |
| 18-103-final-viral-combined_k141_44996\|\|full | Not-determined |
| 18-103-final-viral-combined_k141_79746\|\|full | Low-quality |
| 18-103-final-viral-combined_k141_114351\|\|full | Not-determined |
| 18-103-final-viral-combined_k141_62421\|\|full | Low-quality |
| 18-103-final-viral-combined_k141_16162\|\|full | Low-quality |
| 18-103-final-viral-combined_k141_45003\|\|full | Not-determined |
| 18-103-final-viral-combined_k141_68296\|\|full | Low-quality |
| 18-103-final-viral-combined_k141_85621\|\|full | Low-quality |
| 18-103-final-viral-combined_k141_73796\|\|full | Not-determined |
| 18-103-final-viral-combined_k141_16176\|\|full | Not-determined |
| 18-103-final-viral-combined_k141_45019\|\|full | Not-determined |
| 18-103-final-viral-combined_k141_102351\|\|full | Not-determined |
| 18-103-final-viral-combined_k141_108353\|\|full | Not-determined |
| 18-103-final-viral-combined_k141_16190\|\|full | Not-determined |
| 18-103-final-viral-combined_k141_131833\|\|full | Not-determined |
| 18-103-final-viral-combined_k141_85640\|\|full | Low-quality |
| 18-103-final-viral-combined_k141_45041\|\|full | Not-determined |
| 18-103-final-viral-combined_k141_56726\|\|full | Not-determined |
| 18-103-final-viral-combined_k141_85641\|\|full | Low-quality |
| 18-103-final-viral-combined_k141_91410\|\|full | Not-determined |
| 18-103-final-viral-combined_k141_85649\|\|full | Not-determined |
| 18-103-final-viral-combined_k141_28139\|\|full | Not-determined |
| 18-103-final-viral-combined_k141_79787\|\|full | Not-determined |
| 18-103-final-viral-combined_k141_91417\|\|full | Not-determined |
| 18-103-final-viral-combined_k141_108372\|\|full | Not-determined |
| 18-103-final-viral-combined_k141_39670\|\|full | Not-determined |
| 18-103-final-viral-combined_k141_56745\|\|full | Low-quality |
| 18-103-final-viral-combined_k141_120507\|\|full | Low-quality |
| 18-103-final-viral-combined_k141_56749\|\|full | Low-quality |
| 18-103-final-viral-combined_k141_10725\|\|full | Not-determined |
| 18-103-final-viral-combined_k141_33557\|\|full | Low-quality |
| 18-103-final-viral-combined_k141_102402\|\|full | Low-quality |
| 18-103-final-viral-combined_k141_126114\|\|full | Low-quality |
| 18-103-final-viral-combined_k141_33571\|\|full | Not-determined |
| 18-103-final-viral-combined_k141_33573\|\|full | Low-quality |
| 18-103-final-viral-combined_k141_5293\|\|full | Low-quality |
| 18-103-final-viral-combined_k141_126118\|\|full | Not-determined |
| 18-103-final-viral-combined_k141_102407\|\|full | Not-determined |
| 18-103-final-viral-combined_k141_33577\|\|full | Not-determined |
| 18-103-final-viral-combined_k141_5301\|\|full | Low-quality |
| 18-103-final-viral-combined_k141_126128\|\|full | Low-quality |
| 18-103-final-viral-combined_k141_120543\|\|full | Not-determined |
| 18-103-final-viral-combined_k141_91464\|\|full | Low-quality |
| 18-103-final-viral-combined_k141_22449\|\|full | Low-quality |
| 18-103-final-viral-combined_k141_5318\|\|full | Low-quality |
| 18-103-final-viral-combined_k141_33599\|\|full | Low-quality |
| 18-103-final-viral-combined_k141_62498\|\|full | Not-determined |
| 18-103-final-viral-combined_k141_126142\|\|full | Low-quality |
| 18-103-final-viral-combined_k141_91478\|\|full | Low-quality |
| 18-103-final-viral-combined_k141_73907\|\|full | Low-quality |
| 18-103-final-viral-combined_k141_10797\|\|full | Not-determined |
| 18-103-final-viral-combined_k141_97194\|\|full | Low-quality |
| 18-103-final-viral-combined_k141_91497\|\|full | Low-quality |
| 18-103-final-viral-combined_k141_137146\|\|full | Not-determined |
| 18-103-final-viral-combined_k141_68415\|\|full | Not-determined |
| 18-103-final-viral-combined_k141_22490\|\|full | Not-determined |
| 18-103-final-viral-combined_k141_97210\|\|full | Not-determined |
| 18-103-final-viral-combined_k141_5362\|\|full | Not-determined |
| 18-103-final-viral-combined_k141_62519\|\|full | Low-quality |
| 18-103-final-viral-combined_k141_28261\|\|full | Low-quality |
| 18-103-final-viral-combined_k141_16283\|\|full | Low-quality |
| 18-103-final-viral-combined_k141_97213\|\|full | Not-determined |
| 18-103-final-viral-combined_k141_108433\|\|full | Not-determined |
| 18-103-final-viral-combined_k141_5379\|\|full | Low-quality |
| 18-103-final-viral-combined_k141_5380\|\|full | Low-quality |
| 18-103-final-viral-combined_k141_79904\|\|full | Low-quality |
| 18-103-final-viral-combined_k141_5382\|\|full | Not-determined |
| 18-103-final-viral-combined_k141_137170\|\|full | Not-determined |
| 18-103-final-viral-combined_k141_10826\|\|full | Not-determined |
| 18-103-final-viral-combined_k141_102464\|\|full | Not-determined |
| 18-103-final-viral-combined_k141_91534\|\|full | Not-determined |
| 18-103-final-viral-combined_k141_102471\|\|full | Low-quality |
| 18-103-final-viral-combined_k141_68456\|\|full | Low-quality |
| 18-103-final-viral-combined_k141_33680\|\|full | Not-determined |
| 18-103-final-viral-combined_k141_50941\|\|full | Low-quality |
| 18-103-final-viral-combined_k141_131904\|\|full | Not-determined |
| 18-103-final-viral-combined_k141_50946\|\|full | Not-determined |
| 18-103-final-viral-combined_k141_62581\|\|full | Low-quality |
| 18-103-final-viral-combined_k141_28280\|\|full | Not-determined |
| 18-103-final-viral-combined_k141_16291\|\|full | Low-quality |
| 18-103-final-viral-combined_k141_102496\|\|full | Not-determined |
| 18-103-final-viral-combined_k141_62594\|\|full | Not-determined |
| 18-103-final-viral-combined_k141_45189\|\|full | Not-determined |
| 18-103-final-viral-combined_k141_131927\|\|full | Not-determined |
| 18-103-final-viral-combined_k141_126245\|\|full | Low-quality |
| 18-103-final-viral-combined_k141_68493\|\|full | Low-quality |
| 18-103-final-viral-combined_k141_85836\|\|full | Not-determined |
| 18-103-final-viral-combined_k141_114533\|\|full | Not-determined |
| 18-103-final-viral-combined_k141_16312\|\|full | Low-quality |
| 18-103-final-viral-combined_k141_33719\|\|full | Low-quality |
| 18-103-final-viral-combined_k141_102513\|\|full | Low-quality |
| 18-103-final-viral-combined_k141_10860\|\|full | Low-quality |
| 18-103-final-viral-combined_k141_97271\|\|full | Not-determined |
| 18-103-final-viral-combined_k141_39794\|\|full | Low-quality |
| 18-103-final-viral-combined_k141_126268\|\|full | Low-quality |
| 18-103-final-viral-combined_k141_68518\|\|full | Not-determined |
| 18-103-final-viral-combined_k141_45218\|\|full | Not-determined |
| 18-103-final-viral-combined_k141_45221\|\|full | Not-determined |
| 18-103-final-viral-combined_k141_10864\|\|full | Low-quality |
| 18-103-final-viral-combined_k141_126281\|\|full | Low-quality |
| 18-103-final-viral-combined_k141_102525\|\|full | Not-determined |
| 18-103-final-viral-combined_k141_91593\|\|full | Not-determined |
| 18-103-final-viral-combined_k141_10881\|\|full | Low-quality |
| 18-103-final-viral-combined_k141_62630\|\|full | Low-quality |
| 18-103-final-viral-combined_k141_131983\|\|full | Low-quality |
| 18-103-final-viral-combined_k141_45242\|\|full | Not-determined |
| 18-103-final-viral-combined_k141_28341\|\|full | Low-quality |
| 18-103-final-viral-combined_k141_16369\|\|full | Low-quality |
| 18-103-final-viral-combined_k141_97303\|\|full | Low-quality |
| 18-103-final-viral-combined_k141_102562\|\|full | Low-quality |
| 18-103-final-viral-combined_k141_120662\|\|full | Not-determined |
| 18-103-final-viral-combined_k141_137218\|\|full | Not-determined |
| 18-103-final-viral-combined_k141_28345\|\|full | Low-quality |
| 18-103-final-viral-combined_k141_45275\|\|full | Low-quality |
| 18-103-final-viral-combined_k141_91647\|\|full | Not-determined |
| 18-103-final-viral-combined_k141_137232\|\|full | Not-determined |
| 18-103-final-viral-combined_k141_68566\|\|full | Low-quality |
| 18-103-final-viral-combined_k141_97338\|\|full | Not-determined |
| 18-103-final-viral-combined_k141_22626\|\|full | Low-quality |
| 18-103-final-viral-combined_k141_62656\|\|full | Not-determined |
| 18-103-final-viral-combined_k141_16415\|\|full | Not-determined |
| 18-103-final-viral-combined_k141_5511\|\|full | Not-determined |
| 18-103-final-viral-combined_k141_28363\|\|full | Low-quality |
| 18-103-final-viral-combined_k141_45292\|\|full | Not-determined |
| 18-103-final-viral-combined_k141_120696\|\|full | Low-quality |
| 18-103-final-viral-combined_k141_45297\|\|full | Low-quality |
| 18-103-final-viral-combined_k141_114605\|\|full | Not-determined |
| 18-103-final-viral-combined_k141_102604\|\|full | Not-determined |
| 18-103-final-viral-combined_k141_114616\|\|full | Low-quality |
| 18-103-final-viral-combined_k141_28386\|\|full | Not-determined |
| 18-103-final-viral-combined_k141_108588\|\|full | Not-determined |
| 18-103-final-viral-combined_k141_68605\|\|full | Low-quality |
| 18-103-final-viral-combined_k141_33850\|\|full | Not-determined |
| 18-103-final-viral-combined_k141_16440\|\|full | Low-quality |
| 18-103-final-viral-combined_k141_137265\|\|full | Low-quality |
| 18-103-final-viral-combined_k141_102640\|\|full | Not-determined |
| 18-103-final-viral-combined_k141_51108\|\|full | Low-quality |
| 18-103-final-viral-combined_k141_62696\|\|full | Not-determined |
| 18-103-final-viral-combined_k141_97364\|\|full | Low-quality |
| 18-103-final-viral-combined_k141_68629\|\|full | Not-determined |
| 18-103-final-viral-combined_k141_97369\|\|full | Low-quality |
| 18-103-final-viral-combined_k141_97370\|\|full | Not-determined |
| 18-103-final-viral-combined_k141_114652\|\|full | Not-determined |
| 18-101-final-viral-combined_k141_197401\|\|full | Medium-quality |
| 18-103-final-viral-combined_k141_132071\|\|full | Low-quality |
| 18-103-final-viral-combined_k141_45307\|\|full | Not-determined |
| 18-103-final-viral-combined_k141_97392\|\|full | Not-determined |
| 18-103-final-viral-combined_k141_97394\|\|full | Low-quality |
| 18-103-final-viral-combined_k141_85975\|\|full | Not-determined |
| 18-103-final-viral-combined_k141_28398\|\|full | Not-determined |
| 18-103-final-viral-combined_k141_51133\|\|full | Not-determined |
| 18-103-final-viral-combined_k141_39919\|\|full | Low-quality |
| 18-103-final-viral-combined_k141_132087\|\|full | Not-determined |
| 18-103-final-viral-combined_k141_91742\|\|full | Not-determined |
| 18-103-final-viral-combined_k141_39927\|\|full | Low-quality |
| 18-103-final-viral-combined_k141_51148\|\|full | Low-quality |
| 18-103-final-viral-combined_k141_33903\|\|full | Not-determined |
| 18-103-final-viral-combined_k141_132111\|\|full | Low-quality |
| 18-103-final-viral-combined_k141_91761\|\|full | Low-quality |
| 18-103-final-viral-combined_k141_51157\|\|full | Not-determined |
| 18-103-final-viral-combined_k141_74205\|\|full | Not-determined |
| 18-103-final-viral-combined_k141_5572\|\|full | Not-determined |
| 18-103-final-viral-combined_k141_62743\|\|full | Low-quality |
| 18-103-final-viral-combined_k141_80126\|\|full | Low-quality |
| 18-103-final-viral-combined_k141_86021\|\|full | Low-quality |
| 18-103-final-viral-combined_k141_51165\|\|full | Low-quality |
| 18-103-final-viral-combined_k141_39969\|\|full | Low-quality |
| 18-103-final-viral-combined_k141_11004\|\|full | Not-determined |
| 18-103-final-viral-combined_k141_51167\|\|full | Low-quality |
| 18-103-final-viral-combined_k141_86028\|\|full | Low-quality |
| 18-103-final-viral-combined_k141_68706\|\|full | Low-quality |
| 18-103-final-viral-combined_k141_91796\|\|full | Not-determined |
| 18-103-final-viral-combined_k141_114687\|\|full | Low-quality |
| 18-103-final-viral-combined_k141_68714\|\|full | Low-quality |
| 18-103-final-viral-combined_k141_45373\|\|full | Not-determined |
| 18-103-final-viral-combined_k141_28422\|\|full | Low-quality |
| 18-103-final-viral-combined_k141_114696\|\|full | Low-quality |
| 18-103-final-viral-combined_k141_40014\|\|full | Low-quality |
| 18-103-final-viral-combined_k141_137379\|\|full | Low-quality |
| 18-103-final-viral-combined_k141_114705\|\|full | Not-determined |
| 18-103-final-viral-combined_k141_137380\|\|full | Low-quality |
| 18-103-final-viral-combined_k141_114706\|\|full | Low-quality |
| 18-103-final-viral-combined_k141_5626\|\|full | Low-quality |
| 18-103-final-viral-combined_k141_114709\|\|full | Not-determined |
| 18-103-final-viral-combined_k141_137388\|\|full | Low-quality |
| 18-103-final-viral-combined_k141_91838\|\|full | Low-quality |
| 18-103-final-viral-combined_k141_40021\|\|full | Not-determined |
| 18-103-final-viral-combined_k141_97487\|\|full | Low-quality |
| 18-101_k141_197401 | Medium-quality |
| 18-103-final-viral-combined_k141_97490\|\|full | Not-determined |
| 18-103-final-viral-combined_k141_22751\|\|full | Not-determined |
| 18-103-final-viral-combined_k141_5647\|\|full | Low-quality |
| 18-103-final-viral-combined_k141_114732\|\|full | Low-quality |
| 18-103-final-viral-combined_k141_97496\|\|full | Low-quality |
| 18-103-final-viral-combined_k141_80182\|\|full | Not-determined |
| 18-103-final-viral-combined_k141_51230\|\|full | Not-determined |
| 18-103-final-viral-combined_k141_28454\|\|full | Not-determined |
| 18-103-final-viral-combined_k141_86076\|\|full | Not-determined |
| 18-103-final-viral-combined_k141_40040\|\|full | Not-determined |
| 18-103-final-viral-combined_k141_80194\|\|full | Not-determined |
| 18-103-final-viral-combined_k141_74275\|\|full | Not-determined |
| 18-103-final-viral-combined_k141_91870\|\|full | Low-quality |
| 18-103-final-viral-combined_k141_40045\|\|full | Not-determined |
| 18-103-final-viral-combined_k141_114752\|\|full | Low-quality |
| 18-103-final-viral-combined_k141_51247\|\|full | Low-quality |
| 18-103-final-viral-combined_k141_28477\|\|full | Not-determined |
| 18-103-final-viral-combined_k141_5666\|\|full | Low-quality |
| 18-103-final-viral-combined_k141_114759\|\|full | Low-quality |
| 18-103-final-viral-combined_k141_137426\|\|full | Not-determined |
| 18-103-final-viral-combined_k141_91896\|\|full | Low-quality |
| 18-103-final-viral-combined_k141_5683\|\|full | Low-quality |
| 18-103-final-viral-combined_k141_51275\|\|full | Not-determined |
| 18-103-final-viral-combined_k141_68777\|\|full | Low-quality |
| 18-103-final-viral-combined_k141_132228\|\|full | Low-quality |
| 18-103-final-viral-combined_k141_102800\|\|full | Not-determined |
| 18-103-final-viral-combined_k141_16610\|\|full | Low-quality |
| 18-103-final-viral-combined_k141_51299\|\|full | Low-quality |
| 18-103-final-viral-combined_k141_57173\|\|full | Not-determined |
| 18-103-final-viral-combined_k141_51302\|\|full | Not-determined |
| 18-103-final-viral-combined_k141_68798\|\|full | Low-quality |
| 18-103-final-viral-combined_k141_80234\|\|full | Low-quality |
| 18-103-final-viral-combined_k141_108786\|\|full | Not-determined |
| 18-103-final-viral-combined_k141_28568\|\|full | Low-quality |
| 18-103-final-viral-combined_k141_34124\|\|full | Low-quality |
| 18-103-final-viral-combined_k141_34127\|\|full | Not-determined |
| 18-103-final-viral-combined_k141_51346\|\|full | Not-determined |
| 18-103-final-viral-combined_k141_80271\|\|full | Low-quality |
| 18-103-final-viral-combined_k141_108812\|\|full | Not-determined |
| 18-103-final-viral-combined_k141_40112\|\|full | Low-quality |
| 18-103-final-viral-combined_k141_28597\|\|full | Not-determined |
| 18-103-final-viral-combined_k141_91989\|\|full | Low-quality |
| 18-103-final-viral-combined_k141_97620\|\|full | Not-determined |
| 18-103-final-viral-combined_k141_45477\|\|full | Low-quality |
| 18-103-final-viral-combined_k141_86156\|\|full | Low-quality |
| 18-103-final-viral-combined_k141_137528\|\|full | Low-quality |
| 18-103-final-viral-combined_k141_28618\|\|full | Not-determined |
| 18-103-final-viral-combined_k141_45481\|\|full | Low-quality |
| 18-103-final-viral-combined_k141_22893\|\|full | Low-quality |
| 18-103-final-viral-combined_k141_137541\|\|full | Not-determined |
| 18-103-final-viral-combined_k141_28641\|\|full | Low-quality |
| 18-103-final-viral-combined_k141_16731\|\|full | Not-determined |
| 18-103-final-viral-combined_k141_74466\|\|full | Not-determined |
| 18-103-final-viral-combined_k141_51393\|\|full | Not-determined |
| 18-103-final-viral-combined_k141_80293\|\|full | Low-quality |
| 18-103-final-viral-combined_k141_80302\|\|full | Not-determined |
| 18-103-final-viral-combined_k141_102922\|\|full | Not-determined |
| 18-103-final-viral-combined_k141_62906\|\|full | Low-quality |
| 18-103-final-viral-combined_k141_97680\|\|full | Not-determined |
| 18-103-final-viral-combined_k141_34182\|\|full | Low-quality |
| 18-103-final-viral-combined_k141_137571\|\|full | Not-determined |
| 18-103-final-viral-combined_k141_108852\|\|full | Not-determined |
| 18-103-final-viral-combined_k141_16751\|\|full | Not-determined |
| 18-103-final-viral-combined_k141_74494\|\|full | Low-quality |
| 18-103-final-viral-combined_k141_86217\|\|full | Not-determined |
| 18-103-final-viral-combined_k141_51419\|\|full | Low-quality |
| 18-103-final-viral-combined_k141_62929\|\|full | Not-determined |
| 18-103-final-viral-combined_k141_62945\|\|full | Low-quality |
| 18-103-final-viral-combined_k141_102973\|\|full | Not-determined |
| 18-103-final-viral-combined_k141_86236\|\|full | Not-determined |
| 18-103-final-viral-combined_k141_80345\|\|full | Low-quality |
| 18-103-final-viral-combined_k141_102977\|\|full | Not-determined |
| 18-103-final-viral-combined_k141_108891\|\|full | Not-determined |
| 18-103-final-viral-combined_k141_114968\|\|full | Low-quality |
| 18-103-final-viral-combined_k141_22993\|\|full | Low-quality |
| 18-103-final-viral-combined_k141_62971\|\|full | Low-quality |
| 18-103-final-viral-combined_k141_68912\|\|full | Not-determined |
| 18-103-final-viral-combined_k141_114975\|\|full | Low-quality |
| 18-103-final-viral-combined_k141_28707\|\|full | Low-quality |
| 18-103-final-viral-combined_k141_51441\|\|full | Not-determined |
| 18-103-final-viral-combined_k141_45560\|\|full | Not-determined |
| 18-103-final-viral-combined_k141_62989\|\|full | Low-quality |
| 18-103-final-viral-combined_k141_57364\|\|full | Not-determined |
| 18-103-final-viral-combined_k141_51465\|\|full | Not-determined |
| 18-103-final-viral-combined_k141_68950\|\|full | Low-quality |
| 18-103-final-viral-combined_k141_51471\|\|full | Not-determined |
| 18-103-final-viral-combined_k141_68953\|\|full | Not-determined |
| 18-103-final-viral-combined_k141_11354\|\|full | Low-quality |
| 18-103-final-viral-combined_k141_11357\|\|full | Not-determined |
| 18-103-final-viral-combined_k141_51488\|\|full | Low-quality |
| 18-103-final-viral-combined_k141_34309\|\|full | Not-determined |
| 18-103-final-viral-combined_k141_51493\|\|full | Low-quality |
| 18-103-final-viral-combined_k141_68969\|\|full | Low-quality |
| 18-103-final-viral-combined_k141_103080\|\|full | Low-quality |
| 18-103-final-viral-combined_k141_80464\|\|full | Not-determined |
| 18-103-final-viral-combined_k141_80477\|\|full | Not-determined |
| 18-103-final-viral-combined_k141_68979\|\|full | Low-quality |
| 18-103-final-viral-combined_k141_11380\|\|full | Not-determined |
| 18-103-final-viral-combined_k141_103088\|\|full | Low-quality |
| 18-103-final-viral-combined_k141_45648\|\|full | Low-quality |
| 18-103-final-viral-combined_k141_51515\|\|full | Not-determined |
| 18-103-final-viral-combined_k141_45658\|\|full | Not-determined |
| 18-103-final-viral-combined_k141_80493\|\|full | Not-determined |
| 18-103-final-viral-combined_k141_57455\|\|full | Not-determined |
| 18-103-final-viral-combined_k141_63078\|\|full | Not-determined |
| 18-103-final-viral-combined_k141_109013\|\|full | Not-determined |
| 18-103-final-viral-combined_k141_137686\|\|full | Low-quality |
| 18-103-final-viral-combined_k141_57461\|\|full | Low-quality |
| 18-103-final-viral-combined_k141_11444\|\|full | Low-quality |
| 18-103-final-viral-combined_k141_45698\|\|full | Low-quality |
| 18-103-final-viral-combined_k141_103118\|\|full | Not-determined |
| 18-103-final-viral-combined_k141_74599\|\|full | Not-determined |
| 18-103-final-viral-combined_k141_137733\|\|full | Not-determined |
| 18-103-final-viral-combined_k141_51566\|\|full | Low-quality |
| 18-103-final-viral-combined_k141_137744\|\|full | Not-determined |
| 18-103-final-viral-combined_k141_51575\|\|full | Not-determined |
| 18-103-final-viral-combined_k141_137764\|\|full | Not-determined |
| 18-103-final-viral-combined_k141_63157\|\|full | Not-determined |
| 18-103-final-viral-combined_k141_16999\|\|full | Not-determined |
| 18-103-final-viral-combined_k141_34429\|\|full | Not-determined |
| 18-103-final-viral-combined_k141_109117\|\|full | Not-determined |
| 18-103-final-viral-combined_k141_63192\|\|full | Low-quality |
| 18-103-final-viral-combined_k141_17037\|\|full | Not-determined |
| 18-103-final-viral-combined_k141_45763\|\|full | Low-quality |
| 18-103-final-viral-combined_k141_109160\|\|full | Not-determined |
| 18-103-final-viral-combined_k141_137799\|\|full | Low-quality |
| 18-103-final-viral-combined_k141_137800\|\|full | Low-quality |
| 18-103-final-viral-combined_k141_51632\|\|full | Not-determined |
| 18-103-final-viral-combined_k141_74719\|\|full | Low-quality |
| 18-103-final-viral-combined_k141_34475\|\|full | Low-quality |
| 18-103-final-viral-combined_k141_103184\|\|full | Not-determined |
| 18-103-final-viral-combined_k141_74753\|\|full | Not-determined |
| 18-103-final-viral-combined_k141_51664\|\|full | Low-quality |
| 18-103-final-viral-combined_k141_109199\|\|full | Not-determined |
| 18-103-final-viral-combined_k141_63238\|\|full | Not-determined |
| 18-103-final-viral-combined_k141_51680\|\|full | Low-quality |
| 18-103-final-viral-combined_k141_17103\|\|full | Low-quality |
| 18-103-final-viral-combined_k141_17105\|\|full | Low-quality |
| 18-103-final-viral-combined_k141_109218\|\|full | Not-determined |
| 18-103-final-viral-combined_k141_63256\|\|full | Not-determined |
| 18-103-final-viral-combined_k141_45814\|\|full | Not-determined |
| 18-103-final-viral-combined_k141_51708\|\|full | Low-quality |
| 18-103-final-viral-combined_k141_45823\|\|full | Not-determined |
| 18-103-final-viral-combined_k141_51736\|\|full | Not-determined |
| 18-103-final-viral-combined_k141_103291\|\|full | Not-determined |
| 18-103-final-viral-combined_k141_103293\|\|full | Not-determined |
| 18-103-final-viral-combined_k141_137869\|\|full | Low-quality |
| 18-103-final-viral-combined_k141_17186\|\|full | Not-determined |
| 18-103-final-viral-combined_k141_103322\|\|full | Not-determined |
| 18-103-final-viral-combined_k141_103334\|\|full | Low-quality |
| 18-103-final-viral-combined_k141_45917\|\|full | Low-quality |
| 18-103-final-viral-combined_k141_45986\|\|full | Low-quality |
| 18-103-final-viral-combined_k141_45987\|\|full | Not-determined |
| 18-103-final-viral-combined_k141_45996\|\|full | Low-quality |
| 18-103-final-viral-combined_k141_103482\|\|full | Not-determined |
| 18-103-final-viral-combined_k141_46694\|\|0_partial | Not-determined |
| 18-103-final-viral-combined_k141_115845\|\|0_partial | Not-determined |
| 18-103-final-viral-combined_k141_53130\|\|0_partial | Not-determined |
| 18-103-final-viral-combined_k141_7588\|\|0_partial | Low-quality |
| 18-103-final-viral-combined_k141_83057\|\|0_partial | Not-determined |
| 18-103-final-viral-combined_k141_134912\|\|0_partial | Low-quality |
| 18-103-final-viral-combined_k141_136422\|\|0_partial | Not-determined |
| 18-103-final-viral-combined_k141_120607\|\|0_partial | Low-quality |
| 18-103-final-viral-combined_k141_63265\|\|lt2gene | Not-determined |
| 18-103-final-viral-combined_k141_86268\|\|lt2gene | Low-quality |
| 18-103-final-viral-combined_k141_40300\|\|lt2gene | Not-determined |
| 18-103-final-viral-combined_k141_74775\|\|lt2gene | Low-quality |
| 18-103-final-viral-combined_k141_120826\|\|lt2gene | Not-determined |
| 18-103-final-viral-combined_k141_132329\|\|lt2gene | Low-quality |
| 18-103-final-viral-combined_k141_23092\|\|lt2gene | Low-quality |
| 18-103-final-viral-combined_k141_28819\|\|lt2gene | Low-quality |
| 18-103-final-viral-combined_k141_129\|\|lt2gene | Low-quality |
| 18-103-final-viral-combined_k141_126620\|\|lt2gene | Low-quality |
| 18-103-final-viral-combined_k141_126639\|\|lt2gene | Not-determined |
| 18-103-final-viral-combined_k141_63384\|\|lt2gene | Low-quality |
| 18-103-final-viral-combined_k141_63406\|\|lt2gene | Low-quality |
| 18-103-final-viral-combined_k141_191\|\|lt2gene | Not-determined |
| 18-103-final-viral-combined_k141_103611\|\|lt2gene | Low-quality |
| 18-103-final-viral-combined_k141_86449\|\|lt2gene | Low-quality |
| 18-103-final-viral-combined_k141_126725\|\|lt2gene | Low-quality |
| 18-103-final-viral-combined_k141_109440\|\|lt2gene | Low-quality |
| 18-103-final-viral-combined_k141_103677\|\|lt2gene | Low-quality |
| 18-103-final-viral-combined_k141_46217\|\|lt2gene | Not-determined |
| 18-103-final-viral-combined_k141_86536\|\|lt2gene | Not-determined |
| 18-103-final-viral-combined_k141_80639\|\|lt2gene | Low-quality |
| 18-103-final-viral-combined_k141_46241\|\|lt2gene | Low-quality |
| 18-103-final-viral-combined_k141_17449\|\|lt2gene | Low-quality |
| 18-103-final-viral-combined_k141_126775\|\|lt2gene | Low-quality |
| 18-103-final-viral-combined_k141_23254\|\|lt2gene | Not-determined |
| 18-103-final-viral-combined_k141_74979\|\|lt2gene | Low-quality |
| 18-103-final-viral-combined_k141_63493\|\|lt2gene | Not-determined |
| 18-103-final-viral-combined_k141_109581\|\|lt2gene | Low-quality |
| 18-103-final-viral-combined_k141_132544\|\|lt2gene | Low-quality |
| 18-103-final-viral-combined_k141_86616\|\|lt2gene | Low-quality |
| 18-103-final-viral-combined_k141_34762\|\|lt2gene | Low-quality |
| 18-103-final-viral-combined_k141_109603\|\|lt2gene | Low-quality |
| 18-103-final-viral-combined_k141_34780\|\|lt2gene | Low-quality |
| 18-103-final-viral-combined_k141_34798\|\|lt2gene | Low-quality |
| 18-103-final-viral-combined_k141_132592\|\|lt2gene | Low-quality |
| 18-103-final-viral-combined_k141_69283\|\|lt2gene | Low-quality |
| 18-103-final-viral-combined_k141_52074\|\|lt2gene | Low-quality |
| 18-103-final-viral-combined_k141_75008\|\|lt2gene | Low-quality |
| 18-103-final-viral-combined_k141_23275\|\|lt2gene | Low-quality |
| 18-103-final-viral-combined_k141_29022\|\|lt2gene | Not-determined |
| 18-103-final-viral-combined_k141_109676\|\|lt2gene | Low-quality |
| 18-103-final-viral-combined_k141_109677\|\|lt2gene | Not-determined |
| 18-103-final-viral-combined_k141_86722\|\|lt2gene | Low-quality |
| 18-103-final-viral-combined_k141_86726\|\|lt2gene | Low-quality |
| 18-103-final-viral-combined_k141_109695\|\|lt2gene | Not-determined |
| 18-103-final-viral-combined_k141_69342\|\|lt2gene | Low-quality |
| 18-103-final-viral-combined_k141_103822\|\|lt2gene | Low-quality |
| 18-103-final-viral-combined_k141_86747\|\|lt2gene | Not-determined |
| 18-103-final-viral-combined_k141_126900\|\|lt2gene | Low-quality |
| 18-103-final-viral-combined_k141_75059\|\|lt2gene | Low-quality |
| 18-103-final-viral-combined_k141_69351\|\|lt2gene | Low-quality |
| 18-103-final-viral-combined_k141_92333\|\|lt2gene | Low-quality |
| 18-103-final-viral-combined_k141_467\|\|lt2gene | Not-determined |
| 18-103-final-viral-combined_k141_80849\|\|lt2gene | Low-quality |
| 18-103-final-viral-combined_k141_103871\|\|lt2gene | Not-determined |
| 18-103-final-viral-combined_k141_6150\|\|lt2gene | Low-quality |
| 18-103-final-viral-combined_k141_498\|\|lt2gene | Not-determined |
| 18-103-final-viral-combined_k141_121161\|\|lt2gene | Low-quality |
| 18-103-final-viral-combined_k141_80875\|\|lt2gene | Low-quality |
| 18-103-final-viral-combined_k141_92404\|\|lt2gene | Low-quality |
| 18-103-final-viral-combined_k141_121188\|\|lt2gene | Low-quality |
| 18-103-final-viral-combined_k141_6202\|\|lt2gene | Not-determined |
| 18-103-final-viral-combined_k141_17652\|\|lt2gene | Low-quality |
| 18-103-final-viral-combined_k141_132778\|\|lt2gene | Not-determined |
| 18-103-final-viral-combined_k141_63701\|\|lt2gene | Low-quality |
| 18-103-final-viral-combined_k141_17667\|\|lt2gene | Low-quality |
| 18-103-final-viral-combined_k141_86857\|\|lt2gene | Low-quality |
| 18-103-final-viral-combined_k141_92468\|\|lt2gene | Not-determined |
| 18-103-final-viral-combined_k141_40691\|\|lt2gene | Not-determined |
| 18-103-final-viral-combined_k141_57910\|\|lt2gene | Low-quality |
| 18-103-final-viral-combined_k141_69498\|\|lt2gene | Not-determined |
| 18-103-final-viral-combined_k141_29177\|\|lt2gene | Low-quality |
| 18-103-final-viral-combined_k141_115511\|\|lt2gene | Low-quality |
| 18-103-final-viral-combined_k141_52238\|\|lt2gene | Low-quality |
| 18-103-final-viral-combined_k141_109841\|\|lt2gene | Not-determined |
| 18-103-final-viral-combined_k141_121233\|\|lt2gene | Low-quality |
| 18-103-final-viral-combined_k141_577\|\|lt2gene | Low-quality |
| 18-103-final-viral-combined_k141_6274\|\|lt2gene | Low-quality |
| 18-103-final-viral-combined_k141_6289\|\|lt2gene | Not-determined |
| 18-103-final-viral-combined_k141_11926\|\|lt2gene | Low-quality |
| 18-103-final-viral-combined_k141_6308\|\|lt2gene | Low-quality |
| 18-103-final-viral-combined_k141_132827\|\|lt2gene | Low-quality |
| 18-103-final-viral-combined_k141_46628\|\|lt2gene | Low-quality |
| 18-103-final-viral-combined_k141_40795\|\|lt2gene | Low-quality |
| 18-103-final-viral-combined_k141_40802\|\|lt2gene | Low-quality |
| 18-103-final-viral-combined_k141_6358\|\|lt2gene | Low-quality |
| 18-103-final-viral-combined_k141_46675\|\|lt2gene | Low-quality |
| 18-103-final-viral-combined_k141_121301\|\|lt2gene | Low-quality |
| 18-103-final-viral-combined_k141_127238\|\|lt2gene | Low-quality |
| 18-103-final-viral-combined_k141_40847\|\|lt2gene | Not-determined |
| 18-103-final-viral-combined_k141_40849\|\|lt2gene | Low-quality |
| 18-103-final-viral-combined_k141_35138\|\|lt2gene | Low-quality |
| 18-103-final-viral-combined_k141_92587\|\|lt2gene | Not-determined |
| 18-103-final-viral-combined_k141_104150\|\|lt2gene | Low-quality |
| 18-103-final-viral-combined_k141_58117\|\|lt2gene | Not-determined |
| 18-103-final-viral-combined_k141_127288\|\|lt2gene | Low-quality |
| 18-103-final-viral-combined_k141_58118\|\|lt2gene | Low-quality |
| 18-103-final-viral-combined_k141_121366\|\|lt2gene | Low-quality |
| 18-103-final-viral-combined_k141_52414\|\|lt2gene | Not-determined |
| 18-103-final-viral-combined_k141_726\|\|lt2gene | Not-determined |
| 18-103-final-viral-combined_k141_87066\|\|lt2gene | Low-quality |
| 18-103-final-viral-combined_k141_23716\|\|lt2gene | Low-quality |
| 18-103-final-viral-combined_k141_35205\|\|lt2gene | Low-quality |
| 18-103-final-viral-combined_k141_17879\|\|lt2gene | Low-quality |
| 18-103-final-viral-combined_k141_92652\|\|lt2gene | Low-quality |
| 18-103-final-viral-combined_k141_81161\|\|lt2gene | Low-quality |
| 18-103-final-viral-combined_k141_133047\|\|lt2gene | Low-quality |
| 18-103-final-viral-combined_k141_29386\|\|lt2gene | Not-determined |
| 18-103-final-viral-combined_k141_110002\|\|lt2gene | Low-quality |
| 18-103-final-viral-combined_k141_23768\|\|lt2gene | Not-determined |
| 18-103-final-viral-combined_k141_104231\|\|lt2gene | Low-quality |
| 18-103-final-viral-combined_k141_81246\|\|lt2gene | Not-determined |
| 18-103-final-viral-combined_k141_69735\|\|lt2gene | Low-quality |
| 18-103-final-viral-combined_k141_133058\|\|lt2gene | Not-determined |
| 18-103-final-viral-combined_k141_58232\|\|lt2gene | Not-determined |
| 18-103-final-viral-combined_k141_46821\|\|lt2gene | Not-determined |
| 18-103-final-viral-combined_k141_87164\|\|lt2gene | Low-quality |
| 18-103-final-viral-combined_k141_87198\|\|lt2gene | Low-quality |
| 18-103-final-viral-combined_k141_127460\|\|lt2gene | Low-quality |
| 18-103-final-viral-combined_k141_35299\|\|lt2gene | Low-quality |
| 18-103-final-viral-combined_k141_110078\|\|lt2gene | Low-quality |
| 18-103-final-viral-combined_k141_87214\|\|lt2gene | Low-quality |
| 18-103-final-viral-combined_k141_87215\|\|lt2gene | Low-quality |
| 18-103-final-viral-combined_k141_58318\|\|lt2gene | Not-determined |
| 18-103-final-viral-combined_k141_12121\|\|lt2gene | Low-quality |
| 18-103-final-viral-combined_k141_12134\|\|lt2gene | Low-quality |
| 18-103-final-viral-combined_k141_35311\|\|lt2gene | Not-determined |
| 18-103-final-viral-combined_k141_127498\|\|lt2gene | Low-quality |
| 18-103-final-viral-combined_k141_81386\|\|lt2gene | Not-determined |
| 18-103-final-viral-combined_k141_35322\|\|lt2gene | Low-quality |
| 18-103-final-viral-combined_k141_52646\|\|lt2gene | Not-determined |
| 18-103-final-viral-combined_k141_58371\|\|lt2gene | Low-quality |
| 18-103-final-viral-combined_k141_133154\|\|lt2gene | Not-determined |
| 18-103-final-viral-combined_k141_87312\|\|lt2gene | Not-determined |
| 18-103-final-viral-combined_k141_23902\|\|lt2gene | Low-quality |
| 18-103-final-viral-combined_k141_104377\|\|lt2gene | Not-determined |
| 18-103-final-viral-combined_k141_87344\|\|lt2gene | Low-quality |
| 18-103-final-viral-combined_k141_29508\|\|lt2gene | Low-quality |
| 18-103-final-viral-combined_k141_29516\|\|lt2gene | Low-quality |
| 18-103-final-viral-combined_k141_29522\|\|lt2gene | Low-quality |
| 18-103-final-viral-combined_k141_104415\|\|lt2gene | Low-quality |
| 18-103-final-viral-combined_k141_46982\|\|lt2gene | Low-quality |
| 18-103-final-viral-combined_k141_69873\|\|lt2gene | Low-quality |
| 18-103-final-viral-combined_k141_75641\|\|lt2gene | Low-quality |
| 18-103-final-viral-combined_k141_110249\|\|lt2gene | Not-determined |
| 18-103-final-viral-combined_k141_110260\|\|lt2gene | Low-quality |
| 18-103-final-viral-combined_k141_12242\|\|lt2gene | Not-determined |
| 18-103-final-viral-combined_k141_1043\|\|lt2gene | Not-determined |
| 18-103-final-viral-combined_k141_58452\|\|lt2gene | Not-determined |
| 18-103-final-viral-combined_k141_1044\|\|lt2gene | Low-quality |
| 18-103-final-viral-combined_k141_12248\|\|lt2gene | Not-determined |
| 18-103-final-viral-combined_k141_6756\|\|lt2gene | Low-quality |
| 18-103-final-viral-combined_k141_41182\|\|lt2gene | Low-quality |
| 18-103-final-viral-combined_k141_23979\|\|lt2gene | Low-quality |
| 18-103-final-viral-combined_k141_41207\|\|lt2gene | Not-determined |
| 18-103-final-viral-combined_k141_104495\|\|lt2gene | Low-quality |
| 18-103-final-viral-combined_k141_35501\|\|lt2gene | Not-determined |
| 18-103-final-viral-combined_k141_133267\|\|lt2gene | Low-quality |
| 18-103-final-viral-combined_k141_110307\|\|lt2gene | Not-determined |
| 18-103-final-viral-combined_k141_58503\|\|lt2gene | Low-quality |
| 18-103-final-viral-combined_k141_52808\|\|lt2gene | Low-quality |
| 18-103-final-viral-combined_k141_47093\|\|lt2gene | Low-quality |
| 18-103-final-viral-combined_k141_6839\|\|lt2gene | Low-quality |
| 18-103-final-viral-combined_k141_12299\|\|lt2gene | Low-quality |
| 18-103-final-viral-combined_k141_98592\|\|lt2gene | Low-quality |
| 18-103-final-viral-combined_k141_18142\|\|lt2gene | Low-quality |
| 18-103-final-viral-combined_k141_110352\|\|lt2gene | Low-quality |
| 18-103-final-viral-combined_k141_98605\|\|lt2gene | Low-quality |
| 18-103-final-viral-combined_k141_133309\|\|lt2gene | Low-quality |
| 18-103-final-viral-combined_k141_127710\|\|lt2gene | Low-quality |
| 18-103-final-viral-combined_k141_29692\|\|lt2gene | Low-quality |
| 18-103-final-viral-combined_k141_58568\|\|lt2gene | Low-quality |
| 18-103-final-viral-combined_k141_41250\|\|lt2gene | Low-quality |
| 18-103-final-viral-combined_k141_70011\|\|lt2gene | Not-determined |
| 18-103-final-viral-combined_k141_47165\|\|lt2gene | Not-determined |
| 18-103-final-viral-combined_k141_116063\|\|lt2gene | Low-quality |
| 18-103-final-viral-combined_k141_64351\|\|lt2gene | Low-quality |
| 18-103-final-viral-combined_k141_12408\|\|lt2gene | Not-determined |
| 18-103-final-viral-combined_k141_12418\|\|lt2gene | Low-quality |
| 18-103-final-viral-combined_k141_29758\|\|lt2gene | Low-quality |
| 18-103-final-viral-combined_k141_52945\|\|lt2gene | Not-determined |
| 18-103-final-viral-combined_k141_6944\|\|lt2gene | Low-quality |
| 18-103-final-viral-combined_k141_121812\|\|lt2gene | Low-quality |
| 18-103-final-viral-combined_k141_104631\|\|lt2gene | Low-quality |
| 18-103-final-viral-combined_k141_121828\|\|lt2gene | Low-quality |
| 18-103-final-viral-combined_k141_81711\|\|lt2gene | Low-quality |
| 18-103-final-viral-combined_k141_41377\|\|lt2gene | Not-determined |
| 18-103-final-viral-combined_k141_47264\|\|lt2gene | Low-quality |
| 18-103-final-viral-combined_k141_6966\|\|lt2gene | Low-quality |
| 18-103-final-viral-combined_k141_127857\|\|lt2gene | Low-quality |
| 18-103-final-viral-combined_k141_1260\|\|lt2gene | Not-determined |
| 18-103-final-viral-combined_k141_70134\|\|lt2gene | Not-determined |
| 18-103-final-viral-combined_k141_127871\|\|lt2gene | Low-quality |
| 18-103-final-viral-combined_k141_93120\|\|lt2gene | Low-quality |
| 18-103-final-viral-combined_k141_53054\|\|lt2gene | Not-determined |
| 18-103-final-viral-combined_k141_110477\|\|lt2gene | Low-quality |
| 18-103-final-viral-combined_k141_24262\|\|lt2gene | Low-quality |
| 18-103-final-viral-combined_k141_47339\|\|lt2gene | Low-quality |
| 18-103-final-viral-combined_k141_75962\|\|lt2gene | Not-determined |
| 18-103-final-viral-combined_k141_7059\|\|lt2gene | Low-quality |
| 18-103-final-viral-combined_k141_35724\|\|lt2gene | Not-determined |

18-103-final-viral-combined_k141_53077||lt2gene Low-quality
18-103-final-viral-combined_k141_24280||lt2gene Low-quality
18-103-final-viral-combined_k141_93178||lt2gene Low-quality
18-103-final-viral-combined_k141_70221||lt2gene Low-quality
18-103-final-viral-combined_k141_133541||lt2gene Low-quality
18-103-final-viral-combined_k141_116295||lt2gene Not-determined
18-103-final-viral-combined_k141_29882||lt2gene Low-quality
18-103-final-viral-combined_k141_70239||lt2gene Not-determined
18-103-final-viral-combined_k141_87703||lt2gene Low-quality
18-103-final-viral-combined_k141_12649||lt2gene Low-quality
18-103-final-viral-combined_k141_12657||lt2gene Not-determined
18-103-final-viral-combined_k141_58862||lt2gene Low-quality
18-103-final-viral-combined_k141_53176||lt2gene Low-quality
18-103-final-viral-combined_k141_122010||lt2gene Not-determined
18-103-final-viral-combined_k141_29973||lt2gene Not-determined
18-103-final-viral-combined_k141_81957||lt2gene Low-quality
18-103-final-viral-combined_k141_81961||lt2gene Low-quality
18-103-final-viral-combined_k141_87797||lt2gene Low-quality
18-103-final-viral-combined_k141_53200||lt2gene Not-determined
18-103-final-viral-combined_k141_7173||lt2gene Low-quality
18-103-final-viral-combined_k141_98911||lt2gene Low-quality
18-103-final-viral-combined_k141_7199||lt2gene Low-quality
18-103-final-viral-combined_k141_41607||lt2gene Low-quality
18-103-final-viral-combined_k141_133696||lt2gene Low-quality
18-103-final-viral-combined_k141_104818||lt2gene Low-quality
18-103-final-viral-combined_k141_76149||lt2gene Not-determined
18-103-final-viral-combined_k141_12733||lt2gene Low-quality
18-103-final-viral-combined_k141_41628||lt2gene Not-determined
18-103-final-viral-combined_k141_41631||lt2gene Low-quality
18-103-final-viral-combined_k141_122086||lt2gene Low-quality
18-103-final-viral-combined_k141_104828||lt2gene Low-quality
18-103-final-viral-combined_k141_30072||lt2gene Low-quality
18-103-final-viral-combined_k141_47515||lt2gene Low-quality
18-103-final-viral-combined_k141_53219||lt2gene Not-determined
18-103-final-viral-combined_k141_133723||lt2gene Not-determined
18-103-final-viral-combined_k141_53233||lt2gene Not-determined
18-103-final-viral-combined_k141_93410||lt2gene Low-quality
18-103-final-viral-combined_k141_104857||lt2gene Not-determined
18-103-final-viral-combined_k141_76194||lt2gene Low-quality
18-103-final-viral-combined_k141_53243||lt2gene Low-quality
18-103-final-viral-combined_k141_18550||lt2gene Low-quality
18-103-final-viral-combined_k141_18559||lt2gene Low-quality
18-103-final-viral-combined_k141_70412||lt2gene Low-quality
18-103-final-viral-combined_k141_18589||lt2gene Low-quality
18-103-final-viral-combined_k141_12849||lt2gene Low-quality
18-103-final-viral-combined_k141_12856||lt2gene Low-quality
18-103-final-viral-combined_k141_104947||lt2gene Not-determined
18-103-final-viral-combined_k141_12878||lt2gene Low-quality
18-103-final-viral-combined_k141_64794||lt2gene Low-quality
18-103-final-viral-combined_k141_133848||lt2gene Not-determined
18-103-final-viral-combined_k141_82112||lt2gene Not-determined
18-103-final-viral-combined_k141_93504||lt2gene Not-determined
18-103-final-viral-combined_k141_12892||lt2gene Low-quality
18-103-final-viral-combined_k141_116625||lt2gene Low-quality
18-103-final-viral-combined_k141_87949||lt2gene Low-quality
18-103-final-viral-combined_k141_47639||lt2gene Low-quality
18-103-final-viral-combined_k141_7329||lt2gene Low-quality
18-103-final-viral-combined_k141_128307||lt2gene Low-quality
18-103-final-viral-combined_k141_18694||lt2gene Low-quality
18-103-final-viral-combined_k141_93567||lt2gene Low-quality
18-103-final-viral-combined_k141_12969||lt2gene Low-quality
18-103-final-viral-combined_k141_18738||lt2gene Low-quality
18-103-final-viral-combined_k141_1686||lt2gene Low-quality
18-103-final-viral-combined_k141_122284||lt2gene Low-quality
18-103-final-viral-combined_k141_59158||lt2gene Low-quality
18-103-final-viral-combined_k141_99110||lt2gene Not-determined
18-103-final-viral-combined_k141_1704||lt2gene Low-quality
18-103-final-viral-combined_k141_30211||lt2gene Low-quality
18-103-final-viral-combined_k141_1717||lt2gene Low-quality
18-103-final-viral-combined_k141_93581||lt2gene Low-quality
18-103-final-viral-combined_k141_30231||lt2gene Low-quality
18-103-final-viral-combined_k141_24597||lt2gene Not-determined
18-103-final-viral-combined_k141_30247||lt2gene Low-quality
18-103-final-viral-combined_k141_105142||lt2gene Low-quality
18-103-final-viral-combined_k141_70639||lt2gene Low-quality
18-103-final-viral-combined_k141_70644||lt2gene Low-quality
18-103-final-viral-combined_k141_105157||lt2gene Low-quality
18-103-final-viral-combined_k141_24641||lt2gene Low-quality
18-103-final-viral-combined_k141_36094||lt2gene Low-quality
18-103-final-viral-combined_k141_134010||lt2gene Low-quality
18-103-final-viral-combined_k141_122375||lt2gene Low-quality
18-103-final-viral-combined_k141_24674||lt2gene Low-quality
18-103-final-viral-combined_k141_1776||lt2gene Low-quality
18-103-final-viral-combined_k141_99159||lt2gene Low-quality
18-103-final-viral-combined_k141_76409||lt2gene Low-quality
18-103-final-viral-combined_k141_65015||lt2gene Low-quality
18-103-final-viral-combined_k141_70691||lt2gene Low-quality
18-103-final-viral-combined_k141_13059||lt2gene Low-quality
18-103-final-viral-combined_k141_36134||lt2gene Low-quality
18-103-final-viral-combined_k141_24727||lt2gene Low-quality
18-103-final-viral-combined_k141_1787||lt2gene Low-quality
18-103-final-viral-combined_k141_13089||lt2gene Low-quality
18-103-final-viral-combined_k141_41992||lt2gene Low-quality
18-103-final-viral-combined_k141_105273||lt2gene Low-quality
18-103-final-viral-combined_k141_7503||lt2gene Low-quality
18-103-final-viral-combined_k141_122499||lt2gene Low-quality
18-103-final-viral-combined_k141_65094||lt2gene Low-quality
18-103-final-viral-combined_k141_7518||lt2gene Low-quality
18-103-final-viral-combined_k141_59265||lt2gene Low-quality
18-103-final-viral-combined_k141_24798||lt2gene Low-quality
18-103-final-viral-combined_k141_59271||lt2gene Low-quality
18-103-final-viral-combined_k141_59272||lt2gene Not-determined
18-103-final-viral-combined_k141_93735||lt2gene Not-determined
18-103-final-viral-combined_k141_24805||lt2gene Low-quality
18-103-final-viral-combined_k141_82447||lt2gene Low-quality
18-103-final-viral-combined_k141_18988||lt2gene Low-quality
18-103-final-viral-combined_k141_42052||lt2gene Low-quality
18-103-final-viral-combined_k141_93782||lt2gene Low-quality
18-103-final-viral-combined_k141_30425||lt2gene Low-quality
18-103-final-viral-combined_k141_42059||lt2gene Low-quality
18-103-final-viral-combined_k141_42091||lt2gene Low-quality
18-103-final-viral-combined_k141_99333||lt2gene Low-quality
18-103-final-viral-combined_k141_19029||lt2gene Low-quality
18-103-final-viral-combined_k141_105396||lt2gene Low-quality
18-103-final-viral-combined_k141_70880||lt2gene Not-determined
18-103-final-viral-combined_k141_105417||lt2gene Low-quality
18-103-final-viral-combined_k141_59406||lt2gene Low-quality
18-103-final-viral-combined_k141_24930||lt2gene Low-quality
18-103-final-viral-combined_k141_1957||lt2gene Low-quality
18-103-final-viral-combined_k141_93865||lt2gene Not-determined
18-103-final-viral-combined_k141_1976||lt2gene Low-quality
18-103-final-viral-combined_k141_99378||lt2gene Low-quality
18-103-final-viral-combined_k141_36356||lt2gene Not-determined
18-103-final-viral-combined_k141_65290||lt2gene Low-quality
18-103-final-viral-combined_k141_7622||lt2gene Low-quality
18-103-final-viral-combined_k141_65300||lt2gene Not-determined
18-103-final-viral-combined_k141_122774||lt2gene Low-quality
18-103-final-viral-combined_k141_122781||lt2gene Low-quality
18-103-final-viral-combined_k141_93952||lt2gene Low-quality
18-103-final-viral-combined_k141_19126||lt2gene Low-quality
18-103-final-viral-combined_k141_13394||lt2gene Not-determined
18-103-final-viral-combined_k141_65331||lt2gene Not-determined
18-103-final-viral-combined_k141_88380||lt2gene Not-determined
18-103-final-viral-combined_k141_7655||lt2gene Low-quality
18-103-final-viral-combined_k141_7668||lt2gene Not-determined
18-103-final-viral-combined_k141_134314||lt2gene Not-determined
18-103-final-viral-combined_k141_30650||lt2gene Not-determined
18-103-final-viral-combined_k141_42288||lt2gene Low-quality
18-103-final-viral-combined_k141_134331||lt2gene Not-determined
18-103-final-viral-combined_k141_48144||lt2gene Low-quality
18-103-final-viral-combined_k141_2211||lt2gene Not-determined
18-103-final-viral-combined_k141_105595||lt2gene Low-quality
18-103-final-viral-combined_k141_53805||lt2gene Low-quality
18-103-final-viral-combined_k141_82695||lt2gene Low-quality
18-103-final-viral-combined_k141_88476||lt2gene Low-quality
18-103-final-viral-combined_k141_76796||lt2gene Low-quality
18-103-final-viral-combined_k141_134419||lt2gene Low-quality
18-103-final-viral-combined_k141_82737||lt2gene Low-quality
18-103-final-viral-combined_k141_36537||lt2gene Low-quality
18-103-final-viral-combined_k141_53863||lt2gene Low-quality
18-103-final-viral-combined_k141_30734||lt2gene Not-determined
18-103-final-viral-combined_k141_122989||lt2gene Low-quality
18-103-final-viral-combined_k141_134464||lt2gene Not-determined
18-103-final-viral-combined_k141_94165||lt2gene Not-determined
18-103-final-viral-combined_k141_53882||lt2gene Low-quality
18-103-final-viral-combined_k141_71185||lt2gene Not-determined
18-103-final-viral-combined_k141_53887||lt2gene Low-quality
18-103-final-viral-combined_k141_111341||lt2gene Low-quality
18-103-final-viral-combined_k141_111342||lt2gene Low-quality
18-103-final-viral-combined_k141_123025||lt2gene Low-quality
18-103-final-viral-combined_k141_48270||lt2gene Not-determined
18-103-final-viral-combined_k141_105686||lt2gene Not-determined
18-103-final-viral-combined_k141_36652||lt2gene Low-quality
18-103-final-viral-combined_k141_134560||lt2gene Low-quality
18-103-final-viral-combined_k141_30803||lt2gene Not-determined
18-103-final-viral-combined_k141_42443||lt2gene Not-determined
18-103-final-viral-combined_k141_117297||lt2gene Not-determined
18-103-final-viral-combined_k141_94228||lt2gene Low-quality
18-103-final-viral-combined_k141_117309||lt2gene Low-quality
18-103-final-viral-combined_k141_48308||lt2gene Not-determined
18-103-final-viral-combined_k141_59708||lt2gene Low-quality
18-103-final-viral-combined_k141_42491||lt2gene Low-quality
18-103-final-viral-combined_k141_36730||lt2gene Low-quality
18-103-final-viral-combined_k141_134633||lt2gene Low-quality
18-103-final-viral-combined_k141_7959||lt2gene Not-determined
18-103-final-viral-combined_k141_7974||lt2gene Not-determined
18-103-final-viral-combined_k141_59791||lt2gene Not-determined
18-103-final-viral-combined_k141_71359||lt2gene Low-quality
18-103-final-viral-combined_k141_30952||lt2gene Low-quality
18-103-final-viral-combined_k141_129052||lt2gene Not-determined
18-103-final-viral-combined_k141_54054||lt2gene Low-quality
18-103-final-viral-combined_k141_134738||lt2gene Not-determined
18-103-final-viral-combined_k141_30962||lt2gene Low-quality
18-103-final-viral-combined_k141_111550||lt2gene Low-quality
18-103-final-viral-combined_k141_82989||lt2gene Not-determined
18-103-final-viral-combined_k141_71387||lt2gene Low-quality
18-103-final-viral-combined_k141_30993||lt2gene Low-quality
18-103-final-viral-combined_k141_94386||lt2gene Low-quality
18-103-final-viral-combined_k141_134762||lt2gene Low-quality
18-103-final-viral-combined_k141_83035||lt2gene Low-quality
18-103-final-viral-combined_k141_117512||lt2gene Low-quality
18-103-final-viral-combined_k141_8053||lt2gene Low-quality
18-103-final-viral-combined_k141_111645||lt2gene Low-quality
18-103-final-viral-combined_k141_123201||lt2gene Low-quality
18-103-final-viral-combined_k141_13790||lt2gene Low-quality
18-103-final-viral-combined_k141_123220||lt2gene Not-determined
18-103-final-viral-combined_k141_48531||lt2gene Not-determined
18-103-final-viral-combined_k141_71478||lt2gene Low-quality
18-103-final-viral-combined_k141_2629||lt2gene Not-determined
18-103-final-viral-combined_k141_54189||lt2gene Low-quality
18-103-final-viral-combined_k141_25515||lt2gene Not-determined
18-103-final-viral-combined_k141_19702||lt2gene Not-determined
18-103-final-viral-combined_k141_88963||lt2gene Low-quality
18-103-final-viral-combined_k141_117610||lt2gene Low-quality
18-103-final-viral-combined_k141_77221||lt2gene Low-quality
18-103-final-viral-combined_k141_117624||lt2gene Low-quality
18-103-final-viral-combined_k141_94545||lt2gene Low-quality
18-103-final-viral-combined_k141_71525||lt2gene Low-quality
18-103-final-viral-combined_k141_8194||lt2gene Low-quality
18-103-final-viral-combined_k141_123327||lt2gene Low-quality
18-103-final-viral-combined_k141_13862||lt2gene Low-quality
18-103-final-viral-combined_k141_13864||lt2gene Low-quality
18-103-final-viral-combined_k141_54244||lt2gene Low-quality
18-103-final-viral-combined_k141_65770||lt2gene Not-determined
18-103-final-viral-combined_k141_31247||lt2gene Low-quality
18-103-final-viral-combined_k141_77299||lt2gene Low-quality
18-103-final-viral-combined_k141_2747||lt2gene Low-quality
18-103-final-viral-combined_k141_71569||lt2gene Low-quality
18-103-final-viral-combined_k141_31269||lt2gene Not-determined
18-103-final-viral-combined_k141_65799||lt2gene Low-quality
18-103-final-viral-combined_k141_111863||lt2gene Low-quality
18-103-final-viral-combined_k141_19831||lt2gene Low-quality
18-103-final-viral-combined_k141_100094||lt2gene Low-quality
18-103-final-viral-combined_k141_94624||lt2gene Low-quality
18-103-final-viral-combined_k141_37131||lt2gene Low-quality
18-103-final-viral-combined_k141_19843||lt2gene Low-quality
18-103-final-viral-combined_k141_111885||lt2gene Low-quality
18-103-final-viral-combined_k141_100120||lt2gene Not-determined
18-103-final-viral-combined_k141_65825||lt2gene Not-determined
18-103-final-viral-combined_k141_134968||lt2gene Low-quality
18-103-final-viral-combined_k141_77333||lt2gene Not-determined
18-103-final-viral-combined_k141_60140||lt2gene Low-quality
18-103-final-viral-combined_k141_77355||lt2gene Not-determined
18-103-final-viral-combined_k141_71638||lt2gene Low-quality
18-103-final-viral-combined_k141_83338||lt2gene Low-quality
18-103-final-viral-combined_k141_48687||lt2gene Low-quality
18-103-final-viral-combined_k141_100167||lt2gene Not-determined
18-103-final-viral-combined_k141_117747||lt2gene Low-quality
18-103-final-viral-combined_k141_65870||lt2gene Not-determined
18-103-final-viral-combined_k141_111936||lt2gene Not-determined
18-103-final-viral-combined_k141_8336||lt2gene Low-quality
18-103-final-viral-combined_k141_54450||lt2gene Low-quality
18-103-final-viral-combined_k141_71676||lt2gene Low-quality
18-103-final-viral-combined_k141_25805||lt2gene Low-quality
18-103-final-viral-combined_k141_123551||lt2gene Low-quality
18-103-final-viral-combined_k141_60281||lt2gene Low-quality
18-103-final-viral-combined_k141_94792||lt2gene Low-quality
18-103-final-viral-combined_k141_129522||lt2gene Low-quality
18-103-final-viral-combined_k141_65945||lt2gene Not-determined
18-103-final-viral-combined_k141_19988||lt2gene Not-determined
18-103-final-viral-combined_k141_37301||lt2gene Low-quality
18-103-final-viral-combined_k141_37307||lt2gene Low-quality
18-103-final-viral-combined_k141_25823||lt2gene Not-determined
18-103-final-viral-combined_k141_37325||lt2gene Not-determined
18-103-final-viral-combined_k141_8422||lt2gene Low-quality
18-103-final-viral-combined_k141_129552||lt2gene Low-quality
18-103-final-viral-combined_k141_3005||lt2gene Low-quality
18-103-final-viral-combined_k141_129571||lt2gene Low-quality
18-103-final-viral-combined_k141_54543||lt2gene Not-determined
18-103-final-viral-combined_k141_25880||lt2gene Low-quality
18-103-final-viral-combined_k141_135174||lt2gene Low-quality
18-103-final-viral-combined_k141_77520||lt2gene Not-determined
18-103-final-viral-combined_k141_77521||lt2gene Not-determined
18-103-final-viral-combined_k141_89256||lt2gene Low-quality
18-103-final-viral-combined_k141_112085||lt2gene Low-quality
18-103-final-viral-combined_k141_31533||lt2gene Low-quality
18-103-final-viral-combined_k141_83552||lt2gene Low-quality
18-103-final-viral-combined_k141_83558||lt2gene Not-determined
18-103-final-viral-combined_k141_37411||lt2gene Not-determined
18-103-final-viral-combined_k141_89294||lt2gene Low-quality
18-103-final-viral-combined_k141_25929||lt2gene Not-determined
18-103-final-viral-combined_k141_77568||lt2gene Not-determined
18-103-final-viral-combined_k141_112146||lt2gene Not-determined
18-103-final-viral-combined_k141_83576||lt2gene Low-quality
18-103-final-viral-combined_k141_117992||lt2gene Not-determined
18-103-final-viral-combined_k141_129642||lt2gene Low-quality
18-103-final-viral-combined_k141_25955||lt2gene Not-determined
18-103-final-viral-combined_k141_135243||lt2gene Low-quality
18-103-final-viral-combined_k141_112156||lt2gene Low-quality
18-103-final-viral-combined_k141_14188||lt2gene Low-quality
18-103-final-viral-combined_k141_100413||lt2gene Low-quality
18-103-final-viral-combined_k141_94976||lt2gene Low-quality
18-103-final-viral-combined_k141_112184||lt2gene Low-quality
18-103-final-viral-combined_k141_14197||lt2gene Not-determined
18-103-final-viral-combined_k141_118131||lt2gene Not-determined
18-103-final-viral-combined_k141_89433||lt2gene Low-quality
18-103-final-viral-combined_k141_8628||lt2gene Low-quality
18-103-final-viral-combined_k141_71862||lt2gene Low-quality
18-103-final-viral-combined_k141_71864||lt2gene Not-determined
18-103-final-viral-combined_k141_89444||lt2gene Low-quality
18-103-final-viral-combined_k141_77657||lt2gene Low-quality
18-103-final-viral-combined_k141_71891||lt2gene Not-determined
18-103-final-viral-combined_k141_118167||lt2gene Low-quality
18-103-final-viral-combined_k141_129759||lt2gene Low-quality
18-103-final-viral-combined_k141_89486||lt2gene Not-determined
18-103-final-viral-combined_k141_83693||lt2gene Low-quality
18-103-final-viral-combined_k141_135354||lt2gene Low-quality
18-103-final-viral-combined_k141_89505||lt2gene Low-quality
18-103-final-viral-combined_k141_77729||lt2gene Not-determined
18-103-final-viral-combined_k141_54758||lt2gene Low-quality
18-103-final-viral-combined_k141_118260||lt2gene Low-quality
18-103-final-viral-combined_k141_89542||lt2gene Low-quality
18-103-final-viral-combined_k141_8705||lt2gene Low-quality
18-103-final-viral-combined_k141_49002||lt2gene Not-determined
18-103-final-viral-combined_k141_106577||lt2gene Low-quality
18-103-final-viral-combined_k141_8758||lt2gene Low-quality
18-103-final-viral-combined_k141_54848||lt2gene Low-quality
18-103-final-viral-combined_k141_49038||lt2gene Low-quality
18-103-final-viral-combined_k141_100660||lt2gene Not-determined
18-103-final-viral-combined_k141_100662||lt2gene Low-quality
18-103-final-viral-combined_k141_129870||lt2gene Not-determined
18-103-final-viral-combined_k141_95192||lt2gene Low-quality
18-103-final-viral-combined_k141_26116||lt2gene Low-quality
18-103-final-viral-combined_k141_89610||lt2gene Low-quality
18-103-final-viral-combined_k141_129874||lt2gene Low-quality
18-103-final-viral-combined_k141_129880||lt2gene Low-quality
18-103-final-viral-combined_k141_14394||lt2gene Not-determined
18-103-final-viral-combined_k141_14398||lt2gene Low-quality
18-103-final-viral-combined_k141_83799||lt2gene Not-determined
18-103-final-viral-combined_k141_66423||lt2gene Low-quality
18-103-final-viral-combined_k141_49070||lt2gene Low-quality
18-103-final-viral-combined_k141_3412||lt2gene Not-determined
18-103-final-viral-combined_k141_89632||lt2gene Not-determined
18-103-final-viral-combined_k141_106614||lt2gene Low-quality
18-103-final-viral-combined_k141_43345||lt2gene Not-determined
18-103-final-viral-combined_k141_89662||lt2gene Low-quality
18-103-final-viral-combined_k141_135450||lt2gene Low-quality
18-103-final-viral-combined_k141_60606||lt2gene Not-determined
18-103-final-viral-combined_k141_135481||lt2gene Low-quality
18-103-final-viral-combined_k141_77901||lt2gene Low-quality
18-103-final-viral-combined_k141_112528||lt2gene Not-determined
18-103-final-viral-combined_k141_66503||lt2gene Not-determined
18-103-final-viral-combined_k141_8866||lt2gene Low-quality
18-103-final-viral-combined_k141_95293||lt2gene Not-determined
18-103-final-viral-combined_k141_106687||lt2gene Low-quality
18-103-final-viral-combined_k141_43398||lt2gene Low-quality
18-103-final-viral-combined_k141_77945||lt2gene Low-quality
18-103-final-viral-combined_k141_89747||lt2gene Low-quality
18-103-final-viral-combined_k141_20424||lt2gene Low-quality
18-103-final-viral-combined_k141_124072||lt2gene Low-quality
18-103-final-viral-combined_k141_72157||lt2gene Not-determined
18-103-final-viral-combined_k141_100829||lt2gene Low-quality
18-103-final-viral-combined_k141_112598||lt2gene Low-quality
18-103-final-viral-combined_k141_95377||lt2gene Low-quality
18-103-final-viral-combined_k141_60717||lt2gene Not-determined
18-103-final-viral-combined_k141_124128||lt2gene Low-quality
18-103-final-viral-combined_k141_31891||lt2gene Low-quality
18-103-final-viral-combined_k141_95405||lt2gene Not-determined
18-103-final-viral-combined_k141_89821||lt2gene Low-quality
18-103-final-viral-combined_k141_66586||lt2gene Low-quality
18-103-final-viral-combined_k141_66603||lt2gene Not-determined
18-103-final-viral-combined_k141_26352||lt2gene Low-quality
18-103-final-viral-combined_k141_49241||lt2gene Low-quality
18-103-final-viral-combined_k141_43481||lt2gene Not-determined
18-103-final-viral-combined_k141_118586||lt2gene Not-determined
18-103-final-viral-combined_k141_118590||lt2gene Low-quality
18-103-final-viral-combined_k141_20551||lt2gene Low-quality
18-103-final-viral-combined_k141_89903||lt2gene Low-quality
18-103-final-viral-combined_k141_95471||lt2gene Low-quality
18-103-final-viral-combined_k141_89924||lt2gene Low-quality
18-103-final-viral-combined_k141_3657||lt2gene Low-quality
18-103-final-viral-combined_k141_60846||lt2gene Low-quality
18-103-final-viral-combined_k141_66718||lt2gene Low-quality
18-103-final-viral-combined_k141_100933||lt2gene Low-quality
18-103-final-viral-combined_k141_3692||lt2gene Low-quality
18-103-final-viral-combined_k141_124319||lt2gene Not-determined
18-103-final-viral-combined_k141_66734||lt2gene Low-quality
18-103-final-viral-combined_k141_118689||lt2gene Not-determined
18-103-final-viral-combined_k141_84085||lt2gene Low-quality
18-103-final-viral-combined_k141_124334||lt2gene Not-determined
18-103-final-viral-combined_k141_124343||lt2gene Not-determined
18-103-final-viral-combined_k141_84102||lt2gene Low-quality
18-103-final-viral-combined_k141_26519||lt2gene Not-determined
18-103-final-viral-combined_k141_55278||lt2gene Not-determined
18-103-final-viral-combined_k141_38125||lt2gene Not-determined
18-103-final-viral-combined_k141_49439||lt2gene Low-quality
18-103-final-viral-combined_k141_90027||lt2gene Low-quality
18-103-final-viral-combined_k141_101021||lt2gene Not-determined
18-103-final-viral-combined_k141_90042||lt2gene Low-quality
18-103-final-viral-combined_k141_124397||lt2gene Low-quality
18-103-final-viral-combined_k141_95598||lt2gene Low-quality
18-103-final-viral-combined_k141_3766||lt2gene Not-determined
18-103-final-viral-combined_k141_124415||lt2gene Low-quality
18-103-final-viral-combined_k141_135797||lt2gene Low-quality
18-103-final-viral-combined_k141_38212||lt2gene Low-quality
18-103-final-viral-combined_k141_26633||lt2gene Low-quality
18-103-final-viral-combined_k141_3786||lt2gene Low-quality
18-103-final-viral-combined_k141_90152||lt2gene Not-determined
18-103-final-viral-combined_k141_61035||lt2gene Not-determined
18-103-final-viral-combined_k141_112863||lt2gene Not-determined
18-103-final-viral-combined_k141_135822||lt2gene Low-quality
18-103-final-viral-combined_k141_66888||lt2gene Low-quality
18-103-final-viral-combined_k141_95704||lt2gene Not-determined
18-103-final-viral-combined_k141_78341||lt2gene Low-quality
18-103-final-viral-combined_k141_118905||lt2gene Not-determined
18-103-final-viral-combined_k141_95725||lt2gene Not-determined
18-103-final-viral-combined_k141_107078||lt2gene Not-determined
18-103-final-viral-combined_k141_49562||lt2gene Not-determined
18-103-final-viral-combined_k141_112893||lt2gene Low-quality
18-103-final-viral-combined_k141_107093||lt2gene Low-quality
18-103-final-viral-combined_k141_9311||lt2gene Not-determined
18-103-final-viral-combined_k141_135891||lt2gene Low-quality
18-103-final-viral-combined_k141_118950||lt2gene Not-determined
18-103-final-viral-combined_k141_95782||lt2gene Not-determined
18-103-final-viral-combined_k141_112982||lt2gene Not-determined
18-103-final-viral-combined_k141_78449||lt2gene Not-determined
18-103-final-viral-combined_k141_78462||lt2gene Low-quality
18-103-final-viral-combined_k141_72624||lt2gene Not-determined
18-103-final-viral-combined_k141_49636||lt2gene Low-quality
18-103-final-viral-combined_k141_9394||lt2gene Not-determined
18-103-final-viral-combined_k141_107249||lt2gene Low-quality
18-103-final-viral-combined_k141_119075||lt2gene Low-quality
18-103-final-viral-combined_k141_61228||lt2gene Low-quality
18-103-final-viral-combined_k141_21052||lt2gene Low-quality
18-103-final-viral-combined_k141_67105||lt2gene Low-quality
18-103-final-viral-combined_k141_61234||lt2gene Low-quality
18-103-final-viral-combined_k141_4041||lt2gene Low-quality
18-103-final-viral-combined_k141_55635||lt2gene Not-determined
18-103-final-viral-combined_k141_9447||lt2gene Low-quality
18-103-final-viral-combined_k141_4057||lt2gene Low-quality
18-103-final-viral-combined_k141_90363||lt2gene Not-determined
18-103-final-viral-combined_k141_32400||lt2gene Not-determined
18-103-final-viral-combined_k141_15094||lt2gene Low-quality
18-103-final-viral-combined_k141_113145||lt2gene Not-determined
18-103-final-viral-combined_k141_95920||lt2gene Not-determined
18-103-final-viral-combined_k141_113168||lt2gene Low-quality
18-103-final-viral-combined_k141_95948||lt2gene Low-quality
18-103-final-viral-combined_k141_44005||lt2gene Not-determined
18-103-final-viral-combined_k141_38522||lt2gene Low-quality
18-103-final-viral-combined_k141_44024||lt2gene Low-quality
18-103-final-viral-combined_k141_72729||lt2gene Low-quality
18-103-final-viral-combined_k141_130695||lt2gene Low-quality
18-103-final-viral-combined_k141_67222||lt2gene Low-quality
18-103-final-viral-combined_k141_4151||lt2gene Not-determined
18-103-final-viral-combined_k141_84591||lt2gene Not-determined
18-103-final-viral-combined_k141_67241||lt2gene Low-quality
18-103-final-viral-combined_k141_95995||lt2gene Low-quality
18-103-final-viral-combined_k141_32486||lt2gene Not-determined
18-103-final-viral-combined_k141_55750||lt2gene Low-quality
18-103-final-viral-combined_k141_84614||lt2gene Low-quality
18-103-final-viral-combined_k141_27033||lt2gene Low-quality
18-103-final-viral-combined_k141_119292||lt2gene Not-determined
18-103-final-viral-combined_k141_84623||lt2gene Not-determined
18-103-final-viral-combined_k141_72822||lt2gene Low-quality
18-103-final-viral-combined_k141_21275||lt2gene Not-determined
18-103-final-viral-combined_k141_136219||lt2gene Low-quality
18-103-final-viral-combined_k141_55768||lt2gene Low-quality
18-103-final-viral-combined_k141_4194||lt2gene Low-quality
18-103-final-viral-combined_k141_44100||lt2gene Low-quality
18-103-final-viral-combined_k141_27067||lt2gene Not-determined
18-103-final-viral-combined_k141_101410||lt2gene Not-determined
18-103-final-viral-combined_k141_15186||lt2gene Low-quality
18-103-final-viral-combined_k141_124952||lt2gene Not-determined
18-103-final-viral-combined_k141_107479||lt2gene Low-quality
18-103-final-viral-combined_k141_49892||lt2gene Not-determined
18-103-final-viral-combined_k141_130843||lt2gene Low-quality
18-103-final-viral-combined_k141_101448||lt2gene Low-quality
18-103-final-viral-combined_k141_61465||lt2gene Not-determined
18-103-final-viral-combined_k141_21330||lt2gene Low-quality
18-103-final-viral-combined_k141_49917||lt2gene Low-quality
18-103-final-viral-combined_k141_38717||lt2gene Low-quality
18-103-final-viral-combined_k141_101493||lt2gene Low-quality
18-103-final-viral-combined_k141_78830||lt2gene Not-determined
18-103-final-viral-combined_k141_72922||lt2gene Low-quality
18-103-final-viral-combined_k141_49945||lt2gene Low-quality
18-103-final-viral-combined_k141_15259||lt2gene Not-determined
18-103-final-viral-combined_k141_78845||lt2gene Not-determined
18-103-final-viral-combined_k141_61519||lt2gene Low-quality
18-103-final-viral-combined_k141_119452||lt2gene Low-quality
18-103-final-viral-combined_k141_101540||lt2gene Low-quality
18-103-final-viral-combined_k141_113429||lt2gene Low-quality
18-103-final-viral-combined_k141_49962||lt2gene Low-quality
18-103-final-viral-combined_k141_125054||lt2gene Low-quality
18-103-final-viral-combined_k141_90601||lt2gene Not-determined
18-103-final-viral-combined_k141_101562||lt2gene Low-quality
18-103-final-viral-combined_k141_130931||lt2gene Low-quality
18-103-final-viral-combined_k141_119497||lt2gene Low-quality
18-103-final-viral-combined_k141_136400||lt2gene Not-determined
18-103-final-viral-combined_k141_38843||lt2gene Low-quality
18-103-final-viral-combined_k141_67472||lt2gene Not-determined
18-103-final-viral-combined_k141_15325||lt2gene Low-quality
18-103-final-viral-combined_k141_38851||lt2gene Low-quality
18-103-final-viral-combined_k141_96265||lt2gene Low-quality
18-103-final-viral-combined_k141_130978||lt2gene Low-quality
18-103-final-viral-combined_k141_44256||lt2gene Not-determined
18-103-final-viral-combined_k141_101624||lt2gene Not-determined
18-103-final-viral-combined_k141_32741||lt2gene Not-determined
18-103-final-viral-combined_k141_32752||lt2gene Low-quality
18-103-final-viral-combined_k141_32756||lt2gene Low-quality
18-103-final-viral-combined_k141_55992||lt2gene Low-quality
18-103-final-viral-combined_k141_125232||lt2gene Low-quality
18-103-final-viral-combined_k141_61731||lt2gene Low-quality
18-103-final-viral-combined_k141_15431||lt2gene Low-quality
18-103-final-viral-combined_k141_61737||lt2gene Not-determined
18-103-final-viral-combined_k141_113636||lt2gene Not-determined
18-103-final-viral-combined_k141_125260||lt2gene Low-quality
18-103-final-viral-combined_k141_50138||lt2gene Low-quality
18-103-final-viral-combined_k141_131114||lt2gene Low-quality
18-103-final-viral-combined_k141_9935||lt2gene Low-quality
18-103-final-viral-combined_k141_79091||lt2gene Low-quality
18-103-final-viral-combined_k141_21622||lt2gene Not-determined
18-103-final-viral-combined_k141_96411||lt2gene Not-determined
18-103-final-viral-combined_k141_85002||lt2gene Low-quality
18-103-final-viral-combined_k141_15474||lt2gene Low-quality
18-103-final-viral-combined_k141_4563||lt2gene Low-quality
18-103-final-viral-combined_k141_113697||lt2gene Low-quality
18-103-final-viral-combined_k141_119759||lt2gene Low-quality
18-103-final-viral-combined_k141_27418||lt2gene Low-quality
18-103-final-viral-combined_k141_90866||lt2gene Not-determined
18-103-final-viral-combined_k141_4589||lt2gene Low-quality
18-103-final-viral-combined_k141_79178||lt2gene Not-determined
18-103-final-viral-combined_k141_61851||lt2gene Not-determined
18-103-final-viral-combined_k141_113765||lt2gene Not-determined
18-103-final-viral-combined_k141_32939||lt2gene Low-quality
18-103-final-viral-combined_k141_15553||lt2gene Low-quality
18-103-final-viral-combined_k141_90909||lt2gene Low-quality
18-103-final-viral-combined_k141_131250||lt2gene Not-determined
18-103-final-viral-combined_k141_85139||lt2gene Low-quality
18-103-final-viral-combined_k141_4654||lt2gene Low-quality
18-103-final-viral-combined_k141_27512||lt2gene Low-quality
18-103-final-viral-combined_k141_4657||lt2gene Low-quality
18-103-final-viral-combined_k141_96532||lt2gene Not-determined
18-103-final-viral-combined_k141_79248||lt2gene Low-quality
18-103-final-viral-combined_k141_39119||lt2gene Low-quality
18-103-final-viral-combined_k141_79259||lt2gene Not-determined
18-103-final-viral-combined_k141_79260||lt2gene Low-quality
18-103-final-viral-combined_k141_21749||lt2gene Low-quality
18-103-final-viral-combined_k141_101892||lt2gene Not-determined
18-103-final-viral-combined_k141_79270||lt2gene Not-determined
18-103-final-viral-combined_k141_101901||lt2gene Not-determined
18-103-final-viral-combined_k141_79272||lt2gene Low-quality
18-103-final-viral-combined_k141_136684||lt2gene Not-determined
18-103-final-viral-combined_k141_125454||lt2gene Not-determined
18-103-final-viral-combined_k141_4704||lt2gene Low-quality
18-103-final-viral-combined_k141_15634||lt2gene Low-quality
18-103-final-viral-combined_k141_96566||lt2gene Low-quality
18-103-final-viral-combined_k141_21790||lt2gene Not-determined
18-103-final-viral-combined_k141_131325||lt2gene Low-quality
18-103-final-viral-combined_k141_73311||lt2gene Low-quality
18-103-final-viral-combined_k141_125509||lt2gene Low-quality
18-103-final-viral-combined_k141_56283||lt2gene Low-quality
18-103-final-viral-combined_k141_79327||lt2gene Not-determined
18-103-final-viral-combined_k141_4750||lt2gene Not-determined
18-103-final-viral-combined_k141_10205||lt2gene Low-quality
18-103-final-viral-combined_k141_119921||lt2gene Low-quality
18-103-final-viral-combined_k141_73349||lt2gene Low-quality
18-103-final-viral-combined_k141_27680||lt2gene Not-determined
18-103-final-viral-combined_k141_39223||lt2gene Low-quality
18-103-final-viral-combined_k141_39226||lt2gene Low-quality
18-103-final-viral-combined_k141_62017||lt2gene Not-determined
18-103-final-viral-combined_k141_56362||lt2gene Low-quality
18-103-final-viral-combined_k141_113950||lt2gene Low-quality
18-103-final-viral-combined_k141_4796||lt2gene Not-determined
18-103-final-viral-combined_k141_79417||lt2gene Not-determined
18-103-final-viral-combined_k141_15728||lt2gene Low-quality
18-103-final-viral-combined_k141_27708||lt2gene Low-quality
18-103-final-viral-combined_k141_125590||lt2gene Not-determined
18-103-final-viral-combined_k141_125594||lt2gene Low-quality
18-103-final-viral-combined_k141_119994||lt2gene Low-quality
18-103-final-viral-combined_k141_119996||lt2gene Low-quality
18-103-final-viral-combined_k141_21943||lt2gene Not-determined
18-103-final-viral-combined_k141_56417||lt2gene Not-determined
18-103-final-viral-combined_k141_62068||lt2gene Low-quality
18-103-final-viral-combined_k141_114013||lt2gene Low-quality
18-103-final-viral-combined_k141_73440||lt2gene Low-quality
18-103-final-viral-combined_k141_67911||lt2gene Low-quality
18-103-final-viral-combined_k141_125651||lt2gene Low-quality
18-103-final-viral-combined_k141_67923||lt2gene Low-quality
18-103-final-viral-combined_k141_96750||lt2gene Low-quality
18-103-final-viral-combined_k141_15789||lt2gene Not-determined
18-103-final-viral-combined_k141_33179||lt2gene Not-determined
18-103-final-viral-combined_k141_21999||lt2gene Not-determined
18-103-final-viral-combined_k141_136862||lt2gene Low-quality
18-103-final-viral-combined_k141_136880||lt2gene Low-quality
18-103-final-viral-combined_k141_73475||lt2gene Low-quality
18-103-final-viral-combined_k141_96782||lt2gene Low-quality
18-103-final-viral-combined_k141_96786||lt2gene Low-quality
18-103-final-viral-combined_k141_131494||lt2gene Low-quality
18-103-final-viral-combined_k141_27815||lt2gene Not-determined
18-103-final-viral-combined_k141_91161||lt2gene Not-determined
18-103-final-viral-combined_k141_131515||lt2gene Not-determined
18-103-final-viral-combined_k141_62143||lt2gene Not-determined
18-103-final-viral-combined_k141_62150||lt2gene Low-quality
18-103-final-viral-combined_k141_15867||lt2gene Not-determined
18-103-final-viral-combined_k141_73528||lt2gene Low-quality
18-103-final-viral-combined_k141_4932||lt2gene Not-determined
18-103-final-viral-combined_k141_39400||lt2gene Low-quality
18-103-final-viral-combined_k141_108164||lt2gene Not-determined
18-103-final-viral-combined_k141_68038||lt2gene Not-determined

18-103-final-viral-combined_k141_27868||lt2gene Low-quality
18-103-final-viral-combined_k141_44799||lt2gene Not-determined
18-103-final-viral-combined_k141_125751||lt2gene Not-determined
18-103-final-viral-combined_k141_39415||lt2gene Low-quality
18-103-final-viral-combined_k141_50583||lt2gene Low-quality
18-103-final-viral-combined_k141_91230||lt2gene Not-determined
18-103-final-viral-combined_k141_79537||lt2gene Low-quality
18-103-final-viral-combined_k141_91233||lt2gene Low-quality
18-103-final-viral-combined_k141_56546||lt2gene Low-quality
18-103-final-viral-combined_k141_10443||lt2gene Low-quality
18-103-final-viral-combined_k141_15943||lt2gene Low-quality
18-103-final-viral-combined_k141_62232||lt2gene Low-quality
18-103-final-viral-combined_k141_15954||lt2gene Low-quality
18-103-final-viral-combined_k141_10459||lt2gene Low-quality
18-103-final-viral-combined_k141_131620||lt2gene Low-quality
18-103-final-viral-combined_k141_96902||lt2gene Low-quality
18-103-final-viral-combined_k141_10474||lt2gene Low-quality
18-103-final-viral-combined_k141_22163||lt2gene Not-determined
18-103-final-viral-combined_k141_136988||lt2gene Low-quality
18-103-final-viral-combined_k141_33354||lt2gene Not-determined
18-103-final-viral-combined_k141_56608||lt2gene Not-determined
18-103-final-viral-combined_k141_56615||lt2gene Not-determined
18-103-final-viral-combined_k141_10521||lt2gene Not-determined
18-103-final-viral-combined_k141_114209||lt2gene Not-determined
18-103-final-viral-combined_k141_85512||lt2gene Low-quality
18-103-final-viral-combined_k141_68195||lt2gene Low-quality
18-103-final-viral-combined_k141_73661||lt2gene Low-quality
18-103-final-viral-combined_k141_102220||lt2gene Low-quality
18-103-final-viral-combined_k141_22234||lt2gene Low-quality
18-103-final-viral-combined_k141_114241||lt2gene Low-quality
18-103-final-viral-combined_k141_91311||lt2gene Low-quality
18-103-final-viral-combined_k141_33435||lt2gene Low-quality
18-103-final-viral-combined_k141_50726||lt2gene Low-quality
18-103-final-viral-combined_k141_16084||lt2gene Low-quality
18-103-final-viral-combined_k141_114261||lt2gene Low-quality
18-103-final-viral-combined_k141_39578||lt2gene Low-quality
18-103-final-viral-combined_k141_91334||lt2gene Low-quality
18-103-final-viral-combined_k141_79694||lt2gene Low-quality
18-103-final-viral-combined_k141_131735||lt2gene Low-quality
18-103-final-viral-combined_k141_97009||lt2gene Not-determined
18-103-final-viral-combined_k141_97021||lt2gene Low-quality
18-103-final-viral-combined_k141_62370||lt2gene Low-quality
18-103-final-viral-combined_k141_85595||lt2gene Not-determined
18-103-final-viral-combined_k141_62410||lt2gene Low-quality
18-103-final-viral-combined_k141_10661||lt2gene Low-quality
18-103-final-viral-combined_k141_131805||lt2gene Not-determined
18-103-final-viral-combined_k141_73792||lt2gene Low-quality
18-103-final-viral-combined_k141_114355||lt2gene Not-determined
18-103-final-viral-combined_k141_137084||lt2gene Low-quality
18-103-final-viral-combined_k141_62432||lt2gene Low-quality
18-103-final-viral-combined_k141_33509||lt2gene Low-quality
18-103-final-viral-combined_k141_85632||lt2gene Not-determined
18-103-final-viral-combined_k141_102374||lt2gene Not-determined
18-103-final-viral-combined_k141_28150||lt2gene Not-determined
18-103-final-viral-combined_k141_68342||lt2gene Low-quality
18-103-final-viral-combined_k141_137125||lt2gene Low-quality
18-103-final-viral-combined_k141_5262||lt2gene Low-quality
18-103-final-viral-combined_k141_10720||lt2gene Low-quality
18-103-final-viral-combined_k141_108391||lt2gene Low-quality
18-103-final-viral-combined_k141_85701||lt2gene Low-quality
18-103-final-viral-combined_k141_126099||lt2gene Low-quality
18-103-final-viral-combined_k141_50888||lt2gene Low-quality
18-103-final-viral-combined_k141_39721||lt2gene Low-quality
18-103-final-viral-combined_k141_120561||lt2gene Low-quality
18-103-final-viral-combined_k141_79868||lt2gene Not-determined
18-103-final-viral-combined_k141_45129||lt2gene Low-quality
18-103-final-viral-combined_k141_126156||lt2gene Not-determined
18-103-final-viral-combined_k141_33619||lt2gene Low-quality
18-103-final-viral-combined_k141_16251||lt2gene Low-quality
18-103-final-viral-combined_k141_120595||lt2gene Low-quality
18-103-final-viral-combined_k141_10806||lt2gene Low-quality
18-103-final-viral-combined_k141_10807||lt2gene Low-quality
18-103-final-viral-combined_k141_62540||lt2gene Low-quality
18-103-final-viral-combined_k141_91528||lt2gene Low-quality
18-103-final-viral-combined_k141_85784||lt2gene Not-determined
18-103-final-viral-combined_k141_79935||lt2gene Low-quality
18-103-final-viral-combined_k141_73972||lt2gene Not-determined
18-103-final-viral-combined_k141_131907||lt2gene Low-quality
18-103-final-viral-combined_k141_73982||lt2gene Low-quality
18-103-final-viral-combined_k141_50955||lt2gene Low-quality
18-103-final-viral-combined_k141_126242||lt2gene Low-quality
18-103-final-viral-combined_k141_85835||lt2gene Low-quality
18-103-final-viral-combined_k141_102508||lt2gene Low-quality
18-103-final-viral-combined_k141_120608||lt2gene Low-quality
18-103-final-viral-combined_k141_74033||lt2gene Low-quality
18-103-final-viral-combined_k141_74034||lt2gene Not-determined
18-103-final-viral-combined_k141_74043||lt2gene Low-quality
18-103-final-viral-combined_k141_56918||lt2gene Not-determined
18-103-final-viral-combined_k141_126277||lt2gene Low-quality
18-103-final-viral-combined_k141_28334||lt2gene Not-determined
18-103-final-viral-combined_k141_102537||lt2gene Not-determined
18-103-final-viral-combined_k141_120643||lt2gene Low-quality
18-103-final-viral-combined_k141_5478||lt2gene Low-quality
18-103-final-viral-combined_k141_10901||lt2gene Low-quality
18-103-final-viral-combined_k141_126324||lt2gene Not-determined
18-103-final-viral-combined_k141_33789||lt2gene Low-quality
18-103-final-viral-combined_k141_137223||lt2gene Not-determined
18-103-final-viral-combined_k141_33793||lt2gene Low-quality
18-103-final-viral-combined_k141_108572||lt2gene Low-quality
18-103-final-viral-combined_k141_28359||lt2gene Not-determined
18-103-final-viral-combined_k141_80026||lt2gene Low-quality
18-103-final-viral-combined_k141_114601||lt2gene Low-quality
18-103-final-viral-combined_k141_137252||lt2gene Not-determined
18-103-final-viral-combined_k141_85922||lt2gene Low-quality
18-103-final-viral-combined_k141_80043||lt2gene Low-quality
18-103-final-viral-combined_k141_97348||lt2gene Not-determined
18-103-final-viral-combined_k141_74117||lt2gene Not-determined
18-103-final-viral-combined_k141_120716||lt2gene Low-quality
18-103-final-viral-combined_k141_33852||lt2gene Not-determined
18-103-final-viral-combined_k141_68609||lt2gene Low-quality
18-103-final-viral-combined_k141_10969||lt2gene Low-quality
18-103-final-viral-combined_k141_51109||lt2gene Low-quality
18-103-final-viral-combined_k141_97376||lt2gene Low-quality
18-103-final-viral-combined_k141_97398||lt2gene Low-quality
18-103-final-viral-combined_k141_91743||lt2gene Not-determined
18-103-final-viral-combined_k141_51138||lt2gene Low-quality
18-103-final-viral-combined_k141_86000||lt2gene Low-quality
18-103-final-viral-combined_k141_97416||lt2gene Low-quality
18-103-final-viral-combined_k141_39945||lt2gene Low-quality
18-103-final-viral-combined_k141_102697||lt2gene Low-quality
18-103-final-viral-combined_k141_137348||lt2gene Low-quality
18-103-final-viral-combined_k141_114665||lt2gene Low-quality
18-103-final-viral-combined_k141_5577||lt2gene Low-quality
18-103-final-viral-combined_k141_108668||lt2gene Not-determined
18-103-final-viral-combined_k141_68693||lt2gene Low-quality
18-103-final-viral-combined_k141_28432||lt2gene Not-determined
18-103-final-viral-combined_k141_102733||lt2gene Low-quality
18-103-final-viral-combined_k141_11059||lt2gene Low-quality
18-103-final-viral-combined_k141_28455||lt2gene Low-quality
18-103-final-viral-combined_k141_114743||lt2gene Low-quality
18-103-final-viral-combined_k141_97517||lt2gene Low-quality
18-103-final-viral-combined_k141_34035||lt2gene Low-quality
18-103-final-viral-combined_k141_51255||lt2gene Low-quality
18-103-final-viral-combined_k141_57132||lt2gene Not-determined
18-103-final-viral-combined_k141_108709||lt2gene Low-quality
18-103-final-viral-combined_k141_11114||lt2gene Low-quality
18-103-final-viral-combined_k141_40066||lt2gene Not-determined
18-103-final-viral-combined_k141_68767||lt2gene Low-quality
18-103-final-viral-combined_k141_22812||lt2gene Low-quality
18-103-final-viral-combined_k141_11145||lt2gene Not-determined
18-103-final-viral-combined_k141_62840||lt2gene Low-quality
18-103-final-viral-combined_k141_74361||lt2gene Low-quality
18-103-final-viral-combined_k141_91947||lt2gene Low-quality
18-103-final-viral-combined_k141_5742||lt2gene Low-quality
18-103-final-viral-combined_k141_34113||lt2gene Low-quality
18-103-final-viral-combined_k141_80250||lt2gene Low-quality
18-103-final-viral-combined_k141_86117||lt2gene Low-quality
18-103-final-viral-combined_k141_28585||lt2gene Low-quality
18-103-final-viral-combined_k141_102868||lt2gene Low-quality
18-103-final-viral-combined_k141_40115||lt2gene Low-quality
18-103-final-viral-combined_k141_22878||lt2gene Not-determined
18-103-final-viral-combined_k141_16701||lt2gene Low-quality
18-103-final-viral-combined_k141_97670||lt2gene Low-quality
18-103-final-viral-combined_k141_86204||lt2gene Low-quality
18-103-final-viral-combined_k141_86212||lt2gene Low-quality
18-103-final-viral-combined_k141_97700||lt2gene Not-determined
18-103-final-viral-combined_k141_16784||lt2gene Low-quality
18-103-final-viral-combined_k141_108885||lt2gene Not-determined
18-103-final-viral-combined_k141_108888||lt2gene Not-determined
18-103-final-viral-combined_k141_28696||lt2gene Not-determined
18-103-final-viral-combined_k141_108895||lt2gene Not-determined
18-103-final-viral-combined_k141_114972||lt2gene Low-quality
18-103-final-viral-combined_k141_22992||lt2gene Low-quality
18-103-final-viral-combined_k141_68907||lt2gene Low-quality
18-103-final-viral-combined_k141_16813||lt2gene Not-determined
18-103-final-viral-combined_k141_11289||lt2gene Low-quality
18-103-final-viral-combined_k141_16825||lt2gene Low-quality
18-103-final-viral-combined_k141_108927||lt2gene Low-quality
18-103-final-viral-combined_k141_103028||lt2gene Not-determined
18-103-final-viral-combined_k141_103049||lt2gene Low-quality
18-103-final-viral-combined_k141_57393||lt2gene Not-determined
18-103-final-viral-combined_k141_68962||lt2gene Low-quality
18-103-final-viral-combined_k141_68964||lt2gene Low-quality
18-103-final-viral-combined_k141_80475||lt2gene Low-quality
18-103-final-viral-combined_k141_57436||lt2gene Not-determined
18-103-final-viral-combined_k141_11401||lt2gene Not-determined
18-103-final-viral-combined_k141_45677||lt2gene Low-quality
18-103-final-viral-combined_k141_103107||lt2gene Not-determined
18-103-final-viral-combined_k141_34459||lt2gene Low-quality
18-103-final-viral-combined_k141_109163||lt2gene Low-quality
18-103-final-viral-combined_k141_51666||lt2gene Low-quality
18-103-final-viral-combined_k141_103281||lt2gene Not-determined
18-103-final-viral-combined_k141_45962||lt2gene Not-determined
18-103_k141_115007 Not-determined
18-103_k141_23004 Not-determined
18-103_k141_69009 Not-determined
18-103_k141_3 Not-determined
18-103_k141_17255 Not-determined
18-103_k141_23006 Not-determined
18-103_k141_5754 Not-determined
18-103_k141_6 Not-determined
18-103_k141_5757 Not-determined
18-103_k141_92012 Not-determined
18-103_k141_86259 Not-determined
18-103_k141_69014 Not-determined
18-103_k141_12 Low-quality
18-103_k141_80510 Not-determined
18-103_k141_126512 Not-determined
18-103_k141_109264 Low-quality
18-103_k141_46017 Not-determined
18-103_k141_46020 Low-quality
18-103_k141_51769 Not-determined
18-103_k141_126525 Low-quality
18-103_k141_69028 Not-determined
18-103_k141_86277 Not-determined
18-103_k141_126527 Not-determined
18-103_k141_126528 Not-determined
18-103_k141_40 Not-determined
18-103_k141_23026 Not-determined
18-103_k141_28787 Not-determined
18-103_k141_69040 Not-determined
18-103_k141_34533 Not-determined
18-103_k141_46038 Not-determined
18-103_k141_44 Not-determined
18-103_k141_69045 Not-determined
18-103_k141_109276 Not-determined
18-103_k141_40283 Not-determined
18-103_k141_63296 Not-determined
18-103_k141_57527 Not-determined
18-103_k141_23038 Not-determined
18-103_k141_126535 Not-determined
18-103_k141_69049 Not-determined
18-103_k141_23040 Not-determined
18-103_k141_126537 Not-determined
18-103_k141_46047 Low-quality
18-103_k141_34543 Not-determined
18-103_k141_69054 Not-determined
18-103_k141_109287 Not-determined
18-103_k141_97780 Low-quality
18-103_k141_40290 Not-determined
18-103_k141_11534 Not-determined
18-103_k141_120796 Not-determined
18-103_k141_86294 Not-determined
18-103_k141_126550 Not-determined
18-103_k141_69064 Low-quality
18-103_k141_126555 Not-determined
18-103_k141_40301 Not-determined
18-103_k141_120806 Not-determined
18-103_k141_97793 Not-determined
18-103_k141_120810 Not-determined
18-103_k141_34550 Low-quality
18-103_k141_5794 Not-determined
18-103_k141_92043 Not-determined
18-103_k141_97797 Not-determined
18-103_k141_34551 Not-determined
18-103_k141_87 Not-determined
18-103_k141_46074 Not-determined
18-103_k141_57551 Not-determined
18-103_k141_132303 Low-quality
18-103_k141_86319 Not-determined
18-103_k141_109316 Not-determined
18-103_k141_126575 Not-determined
18-103_k141_120826 Not-determined
18-103_k141_74776 Low-quality
18-103_k141_97811 Not-determined
18-103_k141_120831 Not-determined
18-103_k141_63331 Not-determined
18-103_k141_11560 Low-quality
18-103_k141_57558 Not-determined
18-103_k141_120834 Not-determined
18-103_k141_28800 Not-determined
18-103_k141_34569 Not-determined
18-103_k141_11561 Not-determined
18-103_k141_28802 Not-determined
18-103_k141_46090 Not-determined
18-103_k141_120835 Low-quality
18-103_k141_5811 Not-determined
18-103_k141_5814 Not-determined
18-103_k141_57559 Not-determined
18-103_k141_11570 Not-determined
18-103_k141_11571 Low-quality
18-103_k141_80527 Not-determined
18-103_k141_17307 Not-determined
18-103_k141_132319 Not-determined
18-103_k141_28810 Not-determined
18-103_k141_119 Not-determined
18-103_k141_28811 Not-determined
18-103_k141_132324 Not-determined
18-103_k141_46095 Not-determined
18-103_k141_97820 Not-determined
18-103_k141_11579 Low-quality
18-103_k141_28816 Not-determined
18-103_k141_23093 Low-quality
18-103_k141_63349 Not-determined
18-103_k141_5822 Not-determined
18-103_k141_129 Low-quality
18-103_k141_74815 Not-determined
18-103_k141_115106 Not-determined
18-103_k141_74819 Not-determined
18-103_k141_23098 Not-determined
18-103_k141_74822 Not-determined
18-103_k141_132352 Not-determined
18-103_k141_11585 Not-determined
18-103_k141_139 Not-determined
18-103_k141_17339 Not-determined
18-103_k141_109352 Low-quality
18-103_k141_115120 Not-determined
18-103_k141_126620 Low-quality
18-103_k141_145 Not-determined
18-103_k141_28837 Not-determined
18-103_k141_80540 Not-determined
18-103_k141_148 Not-determined
18-103_k141_92057 Not-determined
18-103_k141_120881 Not-determined
18-103_k141_57573 Not-determined
18-103_k141_5831 Low-quality
18-103_k141_97851 Not-determined
18-103_k141_28845 Not-determined
18-103_k141_126627 Not-determined
18-103_k141_97855 Not-determined
18-103_k141_23109 Not-determined
18-103_k141_74833 Not-determined
18-103_k141_103561 Not-determined
18-103_k141_69104 Low-quality
18-103_k141_103565 Not-determined
18-103_k141_92072 Not-determined
18-103_k141_109360 Not-determined
18-103_k141_51839 Not-determined
18-103_k141_11593 Not-determined
18-103_k141_5835 Not-determined
18-103_k141_74835 Low-quality
18-103_k141_5836 Not-determined
18-103_k141_132377 Not-determined
18-103_k141_86369 Low-quality
18-103_k141_63388 Not-determined
18-103_k141_5841 Not-determined
18-103_k141_86371 Low-quality
18-103_k141_5842 Low-quality
18-103_k141_74837 Low-quality
18-103_k141_46128 Not-determined
18-103_k141_5846 Not-determined
18-103_k141_103577 Not-determined
18-103_k141_103580 Not-determined
18-103_k141_97870 Not-determined
18-103_k141_109375 Not-determined
18-103_k141_11603 Not-determined
18-103_k141_80568 Not-determined
18-103_k141_5855 Not-determined
18-103_k141_109377 Not-determined
18-103_k141_80570 Not-determined
18-103_k141_80574 Not-determined
18-103_k141_51858 Not-determined
18-103_k141_69119 Not-determined
18-103_k141_103587 Not-determined
18-103_k141_97879 Not-determined
18-103_k141_103588 Not-determined
18-103_k141_51860 Not-determined
18-103_k141_120909 Not-determined
18-103_k141_120910 Not-determined
18-103_k141_46139 Not-determined
18-103_k141_126658 Not-determined
18-103_k141_120912 Not-determined
18-103_k141_97882 Not-determined
18-103_k141_74860 Not-determined
18-103_k141_97883 Not-determined
18-103_k141_186 Not-determined
18-103_k141_103597 Not-determined
18-103_k141_103598 Not-determined
18-103_k141_120917 Not-determined
18-103_k141_46150 Not-determined
18-103_k141_109388 Not-determined
18-103_k141_86398 Not-determined
18-103_k141_17355 Not-determined
18-103_k141_97888 Not-determined
18-103_k141_132410 Not-determined
18-103_k141_34648 Not-determined
18-103_k141_51878 Not-determined
18-103_k141_17359 Low-quality
18-103_k141_34650 Low-quality
18-103_k141_51882 Not-determined
18-103_k141_115166 Not-determined
18-103_k141_34652 Not-determined
18-103_k141_115168 Not-determined
18-103_k141_126681 Not-determined
18-103_k141_103614 Low-quality
18-103_k141_74862 Low-quality
18-103_k141_199 Not-determined
18-103_k141_17367 Not-determined
18-103_k141_17373 Not-determined
18-103_k141_40359 Not-determined
18-103_k141_92117 Not-determined
18-103_k141_63423 Not-determined
18-103_k141_69134 Not-determined
18-103_k141_5867 Not-determined
18-103_k141_63428 Not-determined
18-103_k141_103623 Low-quality
18-103_k141_103624 Not-determined
18-103_k141_69141 Not-determined
18-103_k141_69143 Not-determined
18-103_k141_17381 Not-determined
18-103_k141_74881 Not-determined
18-103_k141_40367 Low-quality
18-103_k141_40369 Not-determined
18-103_k141_126697 Not-determined
18-103_k141_63434 Not-determined
18-103_k141_69146 Low-quality
18-103_k141_120945 Low-quality
18-103_k141_63435 Not-determined
18-103_k141_97908 Not-determined
18-103_k141_132435 Low-quality
18-103_k141_115184 Not-determined
18-103_k141_34673 Not-determined
18-103_k141_132438 Not-determined
18-103_k141_86437 Not-determined
18-103_k141_23142 Not-determined
18-103_k141_212 Not-determined
18-103_k141_97918 Not-determined
18-103_k141_120954 Not-determined
18-103_k141_5884 Low-quality
18-103_k141_97922 Not-determined
18-103_k141_109413 Not-determined
18-103_k141_126709 Low-quality
18-103_k141_23153 Not-determined
18-103_k141_115198 Not-determined
18-103_k141_115199 Not-determined
18-103_k141_5887 Not-determined
18-103_k141_69164 Not-determined
18-103_k141_115204 Not-determined
18-103_k141_109422 Low-quality
18-103_k141_115210 Not-determined
18-103_k141_11633 Not-determined
18-103_k141_109430 Not-determined
18-103_k141_40382 Not-determined
18-103_k141_92145 Not-determined
18-103_k141_97925 Not-determined
18-103_k141_63446 Low-quality
18-103_k141_115219 Not-determined
18-103_k141_126725 Low-quality
18-103_k141_57588 Not-determined
18-103_k141_23173 Not-determined
18-103_k141_86478 Not-determined
18-103_k141_69179 Not-determined
18-103_k141_69181 Low-quality
18-103_k141_57597 Not-determined
18-103_k141_74911 Not-determined
18-103_k141_69185 Not-determined
18-103_k141_23183 Not-determined
18-103_k141_63459 Low-quality
18-103_k141_126737 Not-determined
18-103_k141_86490 Low-quality
18-103_k141_126740 Not-determined
18-103_k141_51944 Not-determined
18-103_k141_74923 Not-determined
18-103_k141_17402 Not-determined
18-103_k141_57611 Low-quality
18-103_k141_23193 Not-determined
18-103_k141_103670 Not-determined
18-103_k141_80633 Not-determined
18-103_k141_103674 Not-determined
18-103_k141_86502 Not-determined
18-103_k141_126751 Not-determined
18-103_k141_69202 Not-determined
18-103_k141_17410 Not-determined
18-103_k141_34696 Not-determined
18-103_k141_115236 Not-determined
18-103_k141_69208 Not-determined
18-103_k141_69209 Not-determined
18-103_k141_259 Not-determined
18-103_k141_120980 Not-determined
18-103_k141_92164 Low-quality
18-103_k141_261 Low-quality
18-103_k141_132474 Not-determined
18-103_k141_115243 Not-determined
18-103_k141_109469 Not-determined
18-103_k141_132476 Not-determined
18-103_k141_265 Not-determined
18-103_k141_92166 Not-determined
18-103_k141_74934 Not-determined
18-103_k141_92169 Not-determined
18-103_k141_23211 Not-determined
18-103_k141_132482 Not-determined
18-103_k141_97941 Low-quality
18-103_k141_28894 Not-determined
18-103_k141_92171 Not-determined
18-103_k141_57628 Not-determined
18-103_k141_120986 Not-determined
18-103_k141_109483 Not-determined
18-103_k141_132490 Not-determined
18-103_k141_126765 Not-determined
18-103_k141_126767 Not-determined
18-103_k141_86518 Not-determined
18-103_k141_86521 Not-determined
18-103_k141_17429 Low-quality
18-103_k141_5930 Not-determined
18-103_k141_103687 Not-determined
18-103_k141_285 Not-determined
18-103_k141_103688 Not-determined
18-103_k141_86527 Not-determined
18-103_k141_28911 Not-determined
18-103_k141_109496 Not-determined
18-103_k141_86529 Not-determined
18-103_k141_121002 Not-determined
18-103_k141_17440 Low-quality
18-103_k141_46234 Not-determined
18-103_k141_23228 Not-determined
18-103_k141_34711 Not-determined
18-103_k141_86535 Not-determined
18-103_k141_40419 Not-determined
18-103_k141_86540 Not-determined
18-103_k141_86541 Not-determined
18-103_k141_34718 Not-determined
18-103_k141_97972 Not-determined
18-103_k141_23235 Not-determined
18-103_k141_23237 Not-determined
18-103_k141_23239 Not-determined
18-103_k141_17455 Not-determined
18-103_k141_109517 Not-determined
18-103_k141_74969 Low-quality
18-103_k141_86549 Not-determined
18-103_k141_132512 Not-determined
18-103_k141_80651 Not-determined
18-103_k141_11641 Not-determined
18-103_k141_310 Not-determined
18-103_k141_126785 Not-determined
18-103_k141_312 Not-determined
18-103_k141_11645 Not-determined
18-103_k141_40423 Not-determined
18-103_k141_109530 Not-determined
18-103_k141_86557 Not-determined
18-103_k141_103717 Not-determined
18-103_k141_132523 Not-determined
18-103_k141_57651 Low-quality
18-103_k141_103719 Not-determined
18-103_k141_5945 Not-determined
18-103_k141_97991 Not-determined
18-103_k141_97992 Not-determined
18-103_k141_80665 Not-determined
18-103_k141_97997 Not-determined
18-103_k141_23267 Not-determined
18-103_k141_86563 Not-determined
18-103_k141_86565 Not-determined
18-103_k141_86572 Not-determined
18-103_k141_46276 Not-determined
18-103_k141_327 Not-determined
18-103_k141_46280 Not-determined
18-103_k141_46283 Not-determined
18-103_k141_28949 Not-determined
18-103_k141_109548 Not-determined
18-103_k141_109549 Not-determined
18-103_k141_63492 Not-determined
18-103_k141_109553 Not-determined
18-103_k141_115295 Low-quality
18-103_k141_109554 Not-determined
18-103_k141_5971 Not-determined
18-103_k141_5973 Not-determined
18-103_k141_23284 Low-quality
18-103_k141_63501 Not-determined
18-103_k141_126793 Not-determined
18-103_k141_69212 Low-quality
18-103_k141_63507 Not-determined
18-103_k141_126796 Not-determined
18-103_k141_46288 Not-determined
18-103_k141_51988 Not-determined
18-103_k141_5983 Not-determined
18-103_k141_126805 Not-determined
18-103_k141_40445 Low-quality
18-103_k141_23293 Low-quality
18-103_k141_5987 Not-determined
18-103_k141_40447 Not-determined
18-103_k141_5988 Not-determined
18-103_k141_115301 Not-determined
18-103_k141_51995 Not-determined
18-103_k141_86597 Not-determined
18-103_k141_57673 Not-determined
18-103_k141_46301 Not-determined
18-103_k141_121038 Not-determined
18-103_k141_115308 Not-determined
18-103_k141_52000 Not-determined
18-103_k141_69223 Not-determined
18-103_k141_69225 Not-determined
18-103_k141_40462 Not-determined
18-103_k141_92222 Not-determined
18-103_k141_348 Not-determined
18-103_k141_92225 Not-determined
18-103_k141_46312 Not-determined
18-103_k141_23306 Not-determined
18-103_k141_23308 Not-determined
18-103_k141_103742 Not-determined
18-103_k141_132546 Not-determined
18-103_k141_98015 Not-determined
18-103_k141_109589 Not-determined
18-103_k141_80693 Low-quality
18-103_k141_6015 Not-determined
18-103_k141_92246 Low-quality
18-103_k141_11705 Not-determined
18-103_k141_103749 Low-quality
18-103_k141_57679 Not-determined
18-103_k141_126817 Not-determined
18-103_k141_6022 Not-determined

| | |
|---|---|
| 18-103_k141_34765 | Not-determined |
| 18-103_k141_63552 | Not-determined |
| 18-103_k141_40491 | Not-determined |
| 18-103_k141_126825 | Not-determined |
| 18-103_k141_121052 | Not-determined |
| 18-103_k141_23330 | Not-determined |
| 18-103_k141_28990 | Low-quality |
| 18-103_k141_34774 | Not-determined |
| 18-103_k141_92259 | Not-determined |
| 18-103_k141_52021 | Not-determined |
| 18-103_k141_6030 | Not-determined |
| 18-103_k141_121058 | Not-determined |
| 18-103_k141_63558 | Not-determined |
| 18-103_k141_52022 | Not-determined |
| 18-103_k141_46333 | Not-determined |
| 18-103_k141_92261 | Not-determined |
| 18-103_k141_34779 | Not-determined |
| 18-103_k141_46336 | Not-determined |
| 18-103_k141_6033 | Not-determined |
| 18-103_k141_98041 | Not-determined |
| 18-103_k141_98042 | Not-determined |
| 18-103_k141_121063 | Not-determined |
| 18-103_k141_40506 | Not-determined |
| 18-103_k141_69251 | Low-quality |
| 18-103_k141_34785 | Not-determined |
| 18-103_k141_23339 | Not-determined |
| 18-103_k141_52030 | Not-determined |
| 18-103_k141_86636 | Not-determined |
| 18-103_k141_92270 | Not-determined |
| 18-103_k141_115349 | Not-determined |
| 18-103_k141_52033 | Not-determined |
| 18-103_k141_40512 | Not-determined |
| 18-103_k141_57697 | Not-determined |
| 18-103_k141_132583 | Not-determined |
| 18-103_k141_92274 | Not-determined |
| 18-103_k141_80712 | Not-determined |
| 18-103_k141_63579 | Not-determined |
| 18-103_k141_86653 | Not-determined |
| 18-103_k141_121074 | Low-quality |
| 18-103_k141_52044 | Not-determined |
| 18-103_k141_390 | Not-determined |
| 18-103_k141_34804 | Not-determined |
| 18-103_k141_63584 | Not-determined |
| 18-103_k141_40528 | Low-quality |
| 18-103_k141_69270 | Not-determined |
| 18-103_k141_121078 | Not-determined |
| 18-103_k141_86660 | Low-quality |
| 18-103_k141_69272 | Not-determined |
| 18-103_k141_6049 | Not-determined |
| 18-103_k141_6050 | Not-determined |
| 18-103_k141_57705 | Not-determined |
| 18-103_k141_103772 | Not-determined |
| 18-103_k141_29010 | Not-determined |
| 18-103_k141_132595 | Not-determined |
| 18-103_k141_402 | Not-determined |
| 18-103_k141_126850 | Not-determined |
| 18-103_k141_103776 | Not-determined |
| 18-103_k141_126852 | Not-determined |
| 18-103_k141_69290 | Not-determined |
| 18-103_k141_132601 | Not-determined |
| 18-103_k141_23359 | Not-determined |
| 18-103_k141_34822 | Low-quality |
| 18-103_k141_29013 | Low-quality |
| 18-103_k141_46351 | Not-determined |
| 18-103_k141_57722 | Not-determined |
| 18-103_k141_57725 | Not-determined |
| 18-103_k141_69298 | Low-quality |
| 18-103_k141_414 | Not-determined |
| 18-103_k141_80742 | Not-determined |
| 18-103_k141_115356 | Not-determined |
| 18-103_k141_11740 | Not-determined |
| 18-103_k141_103786 | Not-determined |
| 18-103_k141_103787 | Not-determined |
| 18-103_k141_40547 | Not-determined |
| 18-103_k141_132609 | Not-determined |
| 18-103_k141_115361 | Not-determined |
| 18-103_k141_132611 | Not-determined |
| 18-103_k141_418 | Not-determined |
| 18-103_k141_75009 | Not-determined |
| 18-103_k141_132614 | Not-determined |
| 18-103_k141_52081 | Low-quality |
| 18-103_k141_34836 | Not-determined |
| 18-103_k141_132617 | Not-determined |
| 18-103_k141_11752 | Not-determined |
| 18-103_k141_103792 | Not-determined |
| 18-103_k141_132619 | Low-quality |
| 18-103_k141_57740 | Not-determined |
| 18-103_k141_17494 | Not-determined |
| 18-103_k141_69305 | Not-determined |
| 18-103_k141_80754 | Not-determined |
| 18-103_k141_103795 | Not-determined |
| 18-103_k141_80757 | Not-determined |
| 18-103_k141_132630 | Not-determined |
| 18-103_k141_23378 | Not-determined |
| 18-103_k141_75023 | Not-determined |
| 18-103_k141_46371 | Not-determined |
| 18-103_k141_75025 | Not-determined |
| 18-103_k141_46373 | Not-determined |
| 18-103_k141_103810 | Not-determined |
| 18-103_k141_52091 | Not-determined |
| 18-103_k141_75030 | Not-determined |
| 18-103_k141_115382 | Not-determined |
| 18-103_k141_126869 | Not-determined |
| 18-103_k141_451 | Low-quality |
| 18-103_k141_86704 | Not-determined |
| 18-103_k141_80774 | Not-determined |
| 18-103_k141_29022 | Not-determined |
| 18-103_k141_23384 | Not-determined |
| 18-103_k141_52104 | Not-determined |
| 18-103_k141_52106 | Not-determined |
| 18-103_k141_109677 | Not-determined |
| 18-103_k141_29027 | Not-determined |
| 18-103_k141_109679 | Not-determined |
| 18-103_k141_92311 | Not-determined |
| 18-103_k141_86714 | Not-determined |
| 18-103_k141_75038 | Low-quality |
| 18-103_k141_29030 | Not-determined |
| 18-103_k141_46386 | Not-determined |
| 18-103_k141_86721 | Not-determined |
| 18-103_k141_57763 | Not-determined |
| 18-103_k141_52108 | Not-determined |
| 18-103_k141_80787 | Not-determined |
| 18-103_k141_86724 | Not-determined |
| 18-103_k141_57767 | Not-determined |
| 18-103_k141_109693 | Not-determined |
| 18-103_k141_29038 | Not-determined |
| 18-103_k141_126888 | Not-determined |
| 18-103_k141_86730 | Not-determined |
| 18-103_k141_75046 | Not-determined |
| 18-103_k141_86731 | Not-determined |
| 18-103_k141_126896 | Not-determined |
| 18-103_k141_75048 | Low-quality |
| 18-103_k141_63611 | Not-determined |
| 18-103_k141_75051 | Not-determined |
| 18-103_k141_69342 | Low-quality |
| 18-103_k141_23409 | Not-determined |
| 18-103_k141_98082 | Not-determined |
| 18-103_k141_57772 | Not-determined |
| 18-103_k141_126900 | Low-quality |
| 18-103_k141_80802 | Not-determined |
| 18-103_k141_6106 | Not-determined |
| 18-103_k141_132667 | Not-determined |
| 18-103_k141_121098 | Not-determined |
| 18-103_k141_115411 | Not-determined |
| 18-103_k141_6111 | Not-determined |
| 18-103_k141_69351 | Low-quality |
| 18-103_k141_461 | Not-determined |
| 18-103_k141_115414 | Not-determined |
| 18-103_k141_80807 | Not-determined |
| 18-103_k141_29057 | Low-quality |
| 18-103_k141_92333 | Low-quality |
| 18-103_k141_34891 | Not-determined |
| 18-103_k141_11778 | Not-determined |
| 18-103_k141_86756 | Not-determined |
| 18-103_k141_92338 | Low-quality |
| 18-103_k141_103842 | Not-determined |
| 18-103_k141_126920 | Not-determined |
| 18-103_k141_80817 | Not-determined |
| 18-103_k141_126923 | Not-determined |
| 18-103_k141_57780 | Not-determined |
| 18-103_k141_86760 | Low-quality |
| 18-103_k141_126927 | Low-quality |
| 18-103_k141_80823 | Not-determined |
| 18-103_k141_92352 | Low-quality |
| 18-103_k141_109708 | Not-determined |
| 18-103_k141_69369 | Not-determined |
| 18-103_k141_29070 | Not-determined |
| 18-103_k141_92355 | Not-determined |
| 18-103_k141_29071 | Not-determined |
| 18-103_k141_46401 | Low-quality |
| 18-103_k141_121126 | Not-determined |
| 18-103_k141_29075 | Not-determined |
| 18-103_k141_11800 | Not-determined |
| 18-103_k141_57792 | Not-determined |
| 18-103_k141_40590 | Low-quality |
| 18-103_k141_23451 | Not-determined |
| 18-103_k141_103859 | Not-determined |
| 18-103_k141_40591 | Not-determined |
| 18-103_k141_75087 | Not-determined |
| 18-103_k141_6134 | Not-determined |
| 18-103_k141_126943 | Not-determined |
| 18-103_k141_40596 | Not-determined |
| 18-103_k141_98089 | Not-determined |
| 18-103_k141_80847 | Not-determined |
| 18-103_k141_23456 | Low-quality |
| 18-103_k141_80848 | Not-determined |
| 18-103_k141_11818 | Not-determined |
| 18-103_k141_75100 | Not-determined |
| 18-103_k141_29081 | Not-determined |
| 18-103_k141_11822 | Not-determined |
| 18-103_k141_6140 | Not-determined |
| 18-103_k141_29085 | Not-determined |
| 18-103_k141_63638 | Not-determined |
| 18-103_k141_132712 | Not-determined |
| 18-103_k141_40607 | Not-determined |
| 18-103_k141_132714 | Not-determined |
| 18-103_k141_75110 | Not-determined |
| 18-103_k141_46425 | Not-determined |
| 18-103_k141_86774 | Not-determined |
| 18-103_k141_46428 | Not-determined |
| 18-103_k141_69398 | Not-determined |
| 18-103_k141_34924 | Not-determined |
| 18-103_k141_40612 | Not-determined |
| 18-103_k141_17590 | Not-determined |
| 18-103_k141_6152 | Not-determined |
| 18-103_k141_46439 | Not-determined |
| 18-103_k141_52130 | Not-determined |
| 18-103_k141_63651 | Not-determined |
| 18-103_k141_40621 | Not-determined |
| 18-103_k141_126963 | Not-determined |
| 18-103_k141_6159 | Not-determined |
| 18-103_k141_46445 | Not-determined |
| 18-103_k141_126971 | Not-determined |
| 18-103_k141_17608 | Not-determined |
| 18-103_k141_46451 | Low-quality |
| 18-103_k141_6171 | Low-quality |
| 18-103_k141_6172 | Not-determined |
| 18-103_k141_121157 | Not-determined |
| 18-103_k141_121160 | Not-determined |
| 18-103_k141_40633 | Not-determined |
| 18-103_k141_57823 | Not-determined |
| 18-103_k141_121162 | Not-determined |
| 18-103_k141_46462 | Not-determined |
| 18-103_k141_126981 | Not-determined |
| 18-103_k141_57827 | Not-determined |
| 18-103_k141_103889 | Not-determined |
| 18-103_k141_69422 | Low-quality |
| 18-103_k141_92395 | Not-determined |
| 18-103_k141_126987 | Not-determined |
| 18-103_k141_23484 | Not-determined |
| 18-103_k141_75134 | Not-determined |
| 18-103_k141_29095 | Not-determined |
| 18-103_k141_80874 | Low-quality |
| 18-103_k141_121173 | Not-determined |
| 18-103_k141_80875 | Low-quality |
| 18-103_k141_11838 | Not-determined |
| 18-103_k141_512 | Low-quality |
| 18-103_k141_80878 | Not-determined |
| 18-103_k141_103897 | Low-quality |
| 18-103_k141_69434 | Not-determined |
| 18-103_k141_29104 | Not-determined |
| 18-103_k141_127011 | Not-determined |
| 18-103_k141_75145 | Not-determined |
| 18-103_k141_69438 | Not-determined |
| 18-103_k141_80888 | Not-determined |
| 18-103_k141_75147 | Not-determined |
| 18-103_k141_115446 | Low-quality |
| 18-103_k141_69442 | Not-determined |
| 18-103_k141_132729 | Low-quality |
| 18-103_k141_80893 | Not-determined |
| 18-103_k141_132734 | Not-determined |
| 18-103_k141_132736 | Not-determined |
| 18-103_k141_6204 | Not-determined |
| 18-103_k141_11849 | Low-quality |
| 18-103_k141_115462 | Not-determined |
| 18-103_k141_103921 | Low-quality |
| 18-103_k141_23503 | Not-determined |
| 18-103_k141_121207 | Not-determined |
| 18-103_k141_80904 | Not-determined |
| 18-103_k141_23506 | Not-determined |
| 18-103_k141_57853 | Not-determined |
| 18-103_k141_121212 | Not-determined |
| 18-103_k141_35000 | Not-determined |
| 18-103_k141_35001 | Not-determined |
| 18-103_k141_35003 | Not-determined |
| 18-103_k141_29128 | Not-determined |
| 18-103_k141_63662 | Not-determined |
| 18-103_k141_92426 | Not-determined |
| 18-103_k141_132762 | Not-determined |
| 18-103_k141_132764 | Not-determined |
| 18-103_k141_17639 | Not-determined |
| 18-103_k141_115474 | Not-determined |
| 18-103_k141_23512 | Not-determined |
| 18-103_k141_103937 | Low-quality |
| 18-103_k141_80913 | Low-quality |
| 18-103_k141_127042 | Not-determined |
| 18-103_k141_29138 | Not-determined |
| 18-103_k141_103941 | Not-determined |
| 18-103_k141_52184 | Not-determined |
| 18-103_k141_75181 | Not-determined |
| 18-103_k141_533 | Not-determined |
| 18-103_k141_127047 | Low-quality |
| 18-103_k141_103942 | Not-determined |
| 18-103_k141_535 | Not-determined |
| 18-103_k141_132770 | Not-determined |
| 18-103_k141_86819 | Not-determined |
| 18-103_k141_132772 | Not-determined |
| 18-103_k141_57871 | Not-determined |
| 18-103_k141_121216 | Not-determined |
| 18-103_k141_92437 | Not-determined |
| 18-103_k141_92438 | Not-determined |
| 18-103_k141_103947 | Low-quality |
| 18-103_k141_57875 | Not-determined |
| 18-103_k141_127053 | Not-determined |
| 18-103_k141_17652 | Low-quality |
| 18-103_k141_121224 | Not-determined |
| 18-103_k141_57880 | Low-quality |
| 18-103_k141_52196 | Not-determined |
| 18-103_k141_11877 | Not-determined |
| 18-103_k141_63688 | Not-determined |
| 18-103_k141_80934 | Not-determined |
| 18-103_k141_75193 | Not-determined |
| 18-103_k141_75194 | Not-determined |
| 18-103_k141_127063 | Not-determined |
| 18-103_k141_46493 | Not-determined |
| 18-103_k141_63691 | Low-quality |
| 18-103_k141_98140 | Not-determined |
| 18-103_k141_63693 | Not-determined |
| 18-103_k141_6231 | Not-determined |
| 18-103_k141_103962 | Not-determined |
| 18-103_k141_40683 | Not-determined |
| 18-103_k141_98143 | Not-determined |
| 18-103_k141_46499 | Low-quality |
| 18-103_k141_46500 | Not-determined |
| 18-103_k141_80946 | Not-determined |
| 18-103_k141_127073 | Not-determined |
| 18-103_k141_132778 | Not-determined |
| 18-103_k141_46507 | Not-determined |
| 18-103_k141_92458 | Not-determined |
| 18-103_k141_92459 | Not-determined |
| 18-103_k141_115482 | Not-determined |
| 18-103_k141_35006 | Low-quality |
| 18-103_k141_6236 | Not-determined |
| 18-103_k141_11882 | Not-determined |
| 18-103_k141_63701 | Low-quality |
| 18-103_k141_57895 | Low-quality |
| 18-103_k141_127083 | Not-determined |
| 18-103_k141_46510 | Not-determined |
| 18-103_k141_23540 | Not-determined |
| 18-103_k141_103981 | Not-determined |
| 18-103_k141_63704 | Not-determined |
| 18-103_k141_35020 | Not-determined |
| 18-103_k141_127090 | Not-determined |
| 18-103_k141_23545 | Not-determined |
| 18-103_k141_92465 | Not-determined |
| 18-103_k141_46517 | Not-determined |
| 18-103_k141_98158 | Not-determined |
| 18-103_k141_127094 | Not-determined |
| 18-103_k141_46520 | Not-determined |
| 18-103_k141_40691 | Not-determined |
| 18-103_k141_63710 | Not-determined |
| 18-103_k141_92471 | Low-quality |
| 18-103_k141_127097 | Low-quality |
| 18-103_k141_98164 | Not-determined |
| 18-103_k141_98165 | Not-determined |
| 18-103_k141_98166 | Not-determined |
| 18-103_k141_17674 | Not-determined |
| 18-103_k141_52212 | Not-determined |
| 18-103_k141_57910 | Low-quality |
| 18-103_k141_69498 | Not-determined |
| 18-103_k141_52215 | Not-determined |
| 18-103_k141_92476 | Not-determined |
| 18-103_k141_52219 | Not-determined |
| 18-103_k141_69509 | Not-determined |
| 18-103_k141_52225 | Not-determined |
| 18-103_k141_109823 | Not-determined |
| 18-103_k141_109824 | Not-determined |
| 18-103_k141_92484 | Not-determined |
| 18-103_k141_109826 | Not-determined |
| 18-103_k141_92486 | Not-determined |
| 18-103_k141_115512 | Not-determined |
| 18-103_k141_46552 | Not-determined |
| 18-103_k141_46553 | Not-determined |
| 18-103_k141_6256 | Not-determined |
| 18-103_k141_104008 | Not-determined |
| 18-103_k141_17686 | Not-determined |
| 18-103_k141_80973 | Not-determined |
| 18-103_k141_127106 | Not-determined |
| 18-103_k141_17688 | Not-determined |
| 18-103_k141_98178 | Not-determined |
| 18-103_k141_46559 | Not-determined |
| 18-103_k141_104012 | Low-quality |
| 18-103_k141_109836 | Not-determined |
| 18-103_k141_69521 | Not-determined |
| 18-103_k141_92495 | Low-quality |
| 18-103_k141_17698 | Not-determined |
| 18-103_k141_115525 | Not-determined |
| 18-103_k141_46566 | Not-determined |
| 18-103_k141_35035 | Not-determined |
| 18-103_k141_46570 | Not-determined |
| 18-103_k141_63732 | Not-determined |
| 18-103_k141_80986 | Not-determined |
| 18-103_k141_17707 | Not-determined |
| 18-103_k141_40702 | Low-quality |
| 18-103_k141_86880 | Not-determined |
| 18-103_k141_63735 | Not-determined |
| 18-103_k141_40713 | Low-quality |
| 18-103_k141_92507 | Not-determined |
| 18-103_k141_127121 | Not-determined |
| 18-103_k141_104025 | Not-determined |
| 18-103_k141_92509 | Not-determined |
| 18-103_k141_69529 | Not-determined |
| 18-103_k141_40720 | Not-determined |
| 18-103_k141_569 | Not-determined |
| 18-103_k141_92515 | Not-determined |
| 18-103_k141_40721 | Not-determined |
| 18-103_k141_17718 | Not-determined |
| 18-103_k141_57944 | Not-determined |
| 18-103_k141_46584 | Not-determined |
| 18-103_k141_92519 | Not-determined |
| 18-103_k141_57948 | Not-determined |
| 18-103_k141_75201 | Not-determined |
| 18-103_k141_40724 | Not-determined |
| 18-103_k141_115546 | Not-determined |
| 18-103_k141_121235 | Not-determined |
| 18-103_k141_132803 | Not-determined |
| 18-103_k141_92528 | Not-determined |
| 18-103_k141_132806 | Not-determined |
| 18-103_k141_104042 | Not-determined |
| 18-103_k141_104047 | Not-determined |
| 18-103_k141_11918 | Not-determined |
| 18-103_k141_86892 | Not-determined |
| 18-103_k141_98194 | Low-quality |
| 18-103_k141_6278 | Not-determined |
| 18-103_k141_40729 | Not-determined |
| 18-103_k141_6283 | Not-determined |
| 18-103_k141_121255 | Not-determined |
| 18-103_k141_75215 | Not-determined |
| 18-103_k141_92530 | Not-determined |
| 18-103_k141_81009 | Not-determined |
| 18-103_k141_57976 | Not-determined |
| 18-103_k141_23573 | Not-determined |
| 18-103_k141_57978 | Not-determined |
| 18-103_k141_121262 | Not-determined |
| 18-103_k141_86906 | Not-determined |
| 18-103_k141_115559 | Not-determined |
| 18-103_k141_46608 | Not-determined |
| 18-103_k141_6297 | Not-determined |
| 18-103_k141_75223 | Not-determined |
| 18-103_k141_75224 | Not-determined |
| 18-103_k141_610 | Not-determined |
| 18-103_k141_69560 | Low-quality |
| 18-103_k141_40743 | Not-determined |
| 18-103_k141_35064 | Not-determined |
| 18-103_k141_29206 | Not-determined |
| 18-103_k141_86911 | Not-determined |
| 18-103_k141_29207 | Not-determined |
| 18-103_k141_40746 | Not-determined |
| 18-103_k141_115564 | Not-determined |
| 18-103_k141_132819 | Not-determined |
| 18-103_k141_17746 | Not-determined |
| 18-103_k141_98206 | Not-determined |
| 18-103_k141_6308 | Low-quality |
| 18-103_k141_132827 | Low-quality |
| 18-103_k141_52302 | Low-quality |
| 18-103_k141_40757 | Not-determined |
| 18-103_k141_40758 | Not-determined |
| 18-103_k141_11944 | Not-determined |
| 18-103_k141_115578 | Low-quality |
| 18-103_k141_132832 | Not-determined |
| 18-103_k141_11945 | Not-determined |
| 18-103_k141_104076 | Not-determined |
| 18-103_k141_46626 | Low-quality |
| 18-103_k141_58007 | Not-determined |
| 18-103_k141_75231 | Not-determined |
| 18-103_k141_132848 | Not-determined |
| 18-103_k141_127178 | Not-determined |
| 18-103_k141_40775 | Not-determined |
| 18-103_k141_63760 | Not-determined |
| 18-103_k141_6328 | Not-determined |
| 18-103_k141_63767 | Not-determined |
| 18-103_k141_58021 | Low-quality |
| 18-103_k141_69565 | Not-determined |
| 18-103_k141_115591 | Low-quality |
| 18-103_k141_58023 | Not-determined |
| 18-103_k141_46647 | Not-determined |
| 18-103_k141_40790 | Not-determined |
| 18-103_k141_35082 | Not-determined |
| 18-103_k141_104106 | Not-determined |
| 18-103_k141_98237 | Not-determined |
| 18-103_k141_127199 | Not-determined |
| 18-103_k141_52329 | Not-determined |
| 18-103_k141_115599 | Not-determined |
| 18-103_k141_104119 | Not-determined |
| 18-103_k141_127206 | Not-determined |
| 18-103_k141_104121 | Not-determined |
| 18-103_k141_109885 | Not-determined |
| 18-103_k141_81030 | Not-determined |
| 18-103_k141_40802 | Low-quality |
| 18-103_k141_23602 | Not-determined |
| 18-103_k141_115603 | Not-determined |
| 18-103_k141_115604 | Not-determined |
| 18-103_k141_58040 | Not-determined |
| 18-103_k141_627 | Not-determined |
| 18-103_k141_629 | Not-determined |
| 18-103_k141_109892 | Not-determined |
| 18-103_k141_115607 | Not-determined |
| 18-103_k141_630 | Low-quality |
| 18-103_k141_631 | Not-determined |
| 18-103_k141_92545 | Not-determined |
| 18-103_k141_104129 | Low-quality |
| 18-103_k141_81041 | Not-determined |
| 18-103_k141_35090 | Not-determined |
| 18-103_k141_109899 | Not-determined |
| 18-103_k141_52347 | Not-determined |
| 18-103_k141_23618 | Not-determined |
| 18-103_k141_23625 | Not-determined |
| 18-103_k141_115621 | Low-quality |
| 18-103_k141_86969 | Not-determined |
| 18-103_k141_6384 | Not-determined |
| 18-103_k141_40815 | Not-determined |
| 18-103_k141_81053 | Not-determined |
| 18-103_k141_23636 | Not-determined |
| 18-103_k141_127231 | Not-determined |
| 18-103_k141_75270 | Not-determined |
| 18-103_k141_35105 | Not-determined |
| 18-103_k141_115627 | Not-determined |
| 18-103_k141_127238 | Low-quality |
| 18-103_k141_29237 | Not-determined |
| 18-103_k141_127240 | Not-determined |
| 18-103_k141_81057 | Not-determined |
| 18-103_k141_132903 | Not-determined |
| 18-103_k141_86985 | Not-determined |
| 18-103_k141_81060 | Not-determined |
| 18-103_k141_63783 | Not-determined |
| 18-103_k141_35113 | Low-quality |
| 18-103_k141_121318 | Not-determined |
| 18-103_k141_58058 | Low-quality |
| 18-103_k141_6401 | Low-quality |
| 18-103_k141_63790 | Not-determined |
| 18-103_k141_23650 | Not-determined |
| 18-103_k141_121322 | Not-determined |
| 18-103_k141_58061 | Not-determined |
| 18-103_k141_121323 | Not-determined |
| 18-103_k141_52355 | Low-quality |
| 18-103_k141_35117 | Not-determined |
| 18-103_k141_63796 | Not-determined |
| 18-103_k141_127252 | Not-determined |
| 18-103_k141_6405 | Not-determined |
| 18-103_k141_17786 | Not-determined |
| 18-103_k141_17787 | Not-determined |
| 18-103_k141_52360 | Not-determined |
| 18-103_k141_81070 | Not-determined |
| 18-103_k141_29252 | Not-determined |
| 18-103_k141_40847 | Not-determined |
| 18-103_k141_127263 | Not-determined |
| 18-103_k141_75284 | Low-quality |
| 18-103_k141_127264 | Not-determined |
| 18-103_k141_11996 | Not-determined |
| 18-103_k141_63811 | Not-determined |
| 18-103_k141_75287 | Not-determined |
| 18-103_k141_672 | Not-determined |
| 18-103_k141_6415 | Not-determined |
| 18-103_k141_35129 | Not-determined |
| 18-103_k141_92575 | Low-quality |
| 18-103_k141_58081 | Not-determined |
| 18-103_k141_92577 | Not-determined |
| 18-103_k141_63816 | Not-determined |
| 18-103_k141_121332 | Not-determined |
| 18-103_k141_40862 | Not-determined |
| 18-103_k141_58084 | Not-determined |
| 18-103_k141_63819 | Not-determined |
| 18-103_k141_69628 | Not-determined |
| 18-103_k141_17807 | Not-determined |
| 18-103_k141_98247 | Low-quality |
| 18-103_k141_6426 | Not-determined |
| 18-103_k141_6429 | Low-quality |
| 18-103_k141_40871 | Not-determined |
| 17-109-final-viral-combined_k141_256299\|\|full | Medium-quality |
| 18-103_k141_104138 | Not-determined |
| 18-103_k141_104139 | Not-determined |
| 18-103_k141_35145 | Not-determined |
| 18-103_k141_40875 | Not-determined |
| 18-103_k141_92587 | Not-determined |
| 18-103_k141_69638 | Not-determined |
| 18-103_k141_17809 | Not-determined |
| 18-103_k141_63826 | Not-determined |
| 18-103_k141_52388 | Low-quality |
| 18-103_k141_121349 | Not-determined |
| 18-103_k141_63829 | Not-determined |
| 18-103_k141_6439 | Not-determined |
| 18-103_k141_6441 | Not-determined |
| 18-103_k141_40886 | Not-determined |
| 18-103_k141_58113 | Not-determined |
| 18-103_k141_87024 | Not-determined |
| 18-103_k141_23684 | Low-quality |
| 18-103_k141_6449 | Not-determined |
| 18-103_k141_52395 | Not-determined |
| 18-103_k141_127289 | Low-quality |
| 18-103_k141_29272 | Not-determined |
| 18-103_k141_132940 | Not-determined |
| 18-103_k141_52403 | Not-determined |
| 18-103_k141_63846 | Not-determined |
| 18-103_k141_35169 | Not-determined |
| 18-103_k141_98262 | Not-determined |
| 18-103_k141_52408 | Not-determined |
| 18-103_k141_121366 | Low-quality |
| 18-103_k141_75314 | Not-determined |
| 18-103_k141_87039 | Not-determined |

| | |
|---|---|
| 18-103_k141_12021 | Not-determined |
| 18-103_k141_132949 | Low-quality |
| 18-103_k141_132950 | Not-determined |
| 18-103_k141_46713 | Not-determined |
| 18-103_k141_75324 | Not-determined |
| 18-103_k141_46714 | Not-determined |
| 18-103_k141_46716 | Not-determined |
| 18-103_k141_87043 | Low-quality |
| 18-103_k141_81112 | Not-determined |
| 18-103_k141_81113 | Not-determined |
| 18-103_k141_40900 | Not-determined |
| 18-103_k141_46718 | Not-determined |
| 18-103_k141_132957 | Not-determined |
| 18-103_k141_40903 | Not-determined |
| 18-103_k141_104163 | Not-determined |
| 18-103_k141_109964 | Not-determined |
| 18-103_k141_87054 | Not-determined |
| 18-103_k141_6470 | Not-determined |
| 18-103_k141_127308 | Not-determined |
| 18-103_k141_46722 | Not-determined |
| 18-103_k141_121374 | Not-determined |
| 18-103_k141_17844 | Not-determined |
| 18-103_k141_121375 | Not-determined |
| 18-103_k141_121376 | Not-determined |
| 18-103_k141_40907 | Low-quality |
| 18-103_k141_121380 | Not-determined |
| 18-103_k141_109966 | Not-determined |
| 18-103_k141_92620 | Not-determined |
| 18-103_k141_63861 | Not-determined |
| 18-103_k141_17849 | Not-determined |
| 18-103_k141_98277 | Not-determined |
| 18-103_k141_40911 | Not-determined |
| 18-103_k141_46726 | Not-determined |
| 18-103_k141_127311 | Not-determined |
| 18-103_k141_87063 | Not-determined |
| 18-103_k141_732 | Not-determined |
| 18-103_k141_46732 | Not-determined |
| 18-103_k141_58138 | Not-determined |
| 18-103_k141_23702 | Not-determined |
| 18-103_k141_121389 | Not-determined |
| 18-103_k141_735 | Not-determined |
| 18-103_k141_23703 | Not-determined |
| 18-103_k141_40916 | Not-determined |
| 18-103_k141_23705 | Not-determined |
| 18-103_k141_23706 | Not-determined |
| 18-103_k141_29285 | Not-determined |
| 18-103_k141_104178 | Not-determined |
| 18-103_k141_87068 | Not-determined |
| 18-103_k141_6480 | Not-determined |
| 18-103_k141_12045 | Not-determined |
| 18-103_k141_104180 | Not-determined |
| 18-103_k141_121396 | Not-determined |
| 18-103_k141_12046 | Not-determined |
| 18-103_k141_98282 | Not-determined |
| 18-103_k141_69666 | Not-determined |
| 18-103_k141_81140 | Not-determined |
| 18-103_k141_115645 | Not-determined |
| 18-103_k141_17864 | Not-determined |
| 18-103_k141_127330 | Not-determined |
| 18-103_k141_87075 | Not-determined |
| 18-103_k141_69671 | Low-quality |
| 18-103_k141_87079 | Not-determined |
| 18-103_k141_52434 | Not-determined |
| 18-103_k141_17870 | Not-determined |
| 18-103_k141_132983 | Not-determined |
| 18-103_k141_52435 | Not-determined |
| 18-103_k141_46750 | Not-determined |
| 18-103_k141_17875 | Not-determined |
| 18-103_k141_35207 | Not-determined |
| 18-103_k141_92633 | Not-determined |
| 18-103_k141_40929 | Not-determined |
| 18-103_k141_17884 | Low-quality |
| 18-103_k141_92635 | Low-quality |
| 18-103_k141_763 | Not-determined |
| 18-103_k141_121410 | Not-determined |
| 18-103_k141_104207 | Not-determined |
| 18-103_k141_87093 | Not-determined |
| 18-103_k141_29316 | Not-determined |
| 18-103_k141_768 | Not-determined |
| 18-103_k141_75362 | Low-quality |
| 18-103_k141_81152 | Not-determined |
| 18-103_k141_29321 | Not-determined |
| 18-103_k141_58160 | Not-determined |
| 18-103_k141_98302 | Not-determined |
| 18-103_k141_58161 | Low-quality |
| 18-103_k141_772 | Not-determined |
| 18-103_k141_6506 | Not-determined |
| 18-103_k141_6507 | Not-determined |
| 18-103_k141_63890 | Low-quality |
| 18-103_k141_121422 | Not-determined |
| 18-103_k141_92642 | Not-determined |
| 18-103_k141_75374 | Not-determined |
| 18-103_k141_779 | Not-determined |
| 18-103_k141_75377 | Not-determined |
| 18-103_k141_781 | Not-determined |
| 18-103_k141_87099 | Not-determined |
| 18-103_k141_63900 | Low-quality |
| 18-103_k141_75384 | Not-determined |
| 18-103_k141_133000 | Not-determined |
| 18-103_k141_69698 | Not-determined |
| 18-103_k141_52461 | Low-quality |
| 18-103_k141_69699 | Low-quality |
| 18-103_k141_40955 | Not-determined |
| 18-103_k141_92648 | Low-quality |
| 18-103_k141_127365 | Not-determined |
| 18-103_k141_75396 | Not-determined |
| 18-103_k141_133004 | Not-determined |
| 18-103_k141_52467 | Not-determined |
| 18-103_k141_52468 | Not-determined |
| 18-103_k141_133005 | Not-determined |
| 18-103_k141_92652 | Low-quality |
| 18-103_k141_75401 | Not-determined |
| 18-103_k141_69706 | Not-determined |
| 18-103_k141_75405 | Not-determined |
| 18-103_k141_69709 | Not-determined |
| 18-103_k141_58182 | Not-determined |
| 18-103_k141_98317 | Not-determined |
| 18-103_k141_109981 | Not-determined |
| 18-103_k141_109982 | Not-determined |
| 18-103_k141_40964 | Not-determined |
| 18-103_k141_790 | Not-determined |
| 18-103_k141_52484 | Not-determined |
| 18-103_k141_40967 | Not-determined |
| 18-103_k141_121435 | Low-quality |
| 18-103_k141_29348 | Not-determined |
| 18-103_k141_58188 | Not-determined |
| 18-103_k141_58189 | Not-determined |
| 18-103_k141_69715 | Not-determined |
| 18-103_k141_796 | Not-determined |
| 18-103_k141_52490 | Not-determined |
| 18-103_k141_98324 | Not-determined |
| 18-103_k141_52492 | Low-quality |
| 18-103_k141_115690 | Not-determined |
| 18-103_k141_63922 | Not-determined |
| 18-103_k141_115693 | Not-determined |
| 18-103_k141_6538 | Not-determined |
| 18-103_k141_87115 | Not-determined |
| 18-103_k141_69724 | Not-determined |
| 18-103_k141_121451 | Low-quality |
| 18-103_k141_6545 | Not-determined |
| 18-103_k141_17933 | Not-determined |
| 18-103_k141_98341 | Not-determined |
| 18-103_k141_133036 | Not-determined |
| 18-103_k141_92678 | Not-determined |
| 18-103_k141_81198 | Not-determined |
| 18-103_k141_92680 | Not-determined |
| 18-103_k141_81200 | Not-determined |
| 18-103_k141_127381 | Not-determined |
| 18-103_k141_115718 | Not-determined |
| 18-103_k141_121461 | Low-quality |
| 18-103_k141_17948 | Not-determined |
| 18-103_k141_133046 | Not-determined |
| 18-103_k141_63943 | Not-determined |
| 18-103_k141_29387 | Not-determined |
| 18-103_k141_133048 | Not-determined |
| 18-103_k141_81219 | Not-determined |
| 18-103_k141_115729 | Not-determined |
| 18-103_k141_17952 | Not-determined |
| 18-103_k141_29389 | Not-determined |
| 18-103_k141_58203 | Not-determined |
| 18-103_k141_52519 | Low-quality |
| 18-103_k141_29391 | Not-determined |
| 18-103_k141_92700 | Not-determined |
| 18-103_k141_23754 | Not-determined |
| 18-103_k141_81226 | Low-quality |
| 18-103_k141_40981 | Not-determined |
| 18-103_k141_23760 | Not-determined |
| 18-103_k141_17955 | Not-determined |
| 18-103_k141_75432 | Not-determined |
| 18-103_k141_87136 | Not-determined |
| 18-103_k141_46801 | Not-determined |
| 18-103_k141_81236 | Not-determined |
| 18-103_k141_35260 | Low-quality |
| 18-103_k141_104227 | Not-determined |
| 18-103_k141_92710 | Not-determined |
| 18-103_k141_115738 | Not-determined |
| 18-103_k141_58215 | Not-determined |
| 18-103_k141_87141 | Low-quality |
| 18-103_k141_115740 | Not-determined |
| 18-103_k141_75443 | Not-determined |
| 18-103_k141_104231 | Low-quality |
| 18-103_k141_23773 | Low-quality |
| 18-103_k141_52532 | Not-determined |
| 18-103_k141_40988 | Not-determined |
| 18-103_k141_35266 | Low-quality |
| 18-103_k141_52535 | Not-determined |
| 18-103_k141_40989 | Low-quality |
| 18-103_k141_6590 | Not-determined |
| 18-103_k141_40991 | Not-determined |
| 18-103_k141_58223 | Not-determined |
| 18-103_k141_40995 | Not-determined |
| 18-103_k141_81252 | Not-determined |
| 18-103_k141_92725 | Not-determined |
| 18-103_k141_17970 | Not-determined |
| 18-103_k141_87147 | Low-quality |
| 18-103_k141_69733 | Not-determined |
| 18-103_k141_127405 | Not-determined |
| 18-103_k141_87149 | Not-determined |
| 18-103_k141_81257 | Not-determined |
| 18-103_k141_69738 | Not-determined |
| 18-103_k141_58232 | Not-determined |
| 18-103_k141_81262 | Low-quality |
| 18-103_k141_58234 | Not-determined |
| 18-103_k141_17977 | Not-determined |
| 18-103_k141_35276 | Low-quality |
| 18-103_k141_23790 | Not-determined |
| 18-103_k141_814 | Not-determined |
| 18-103_k141_127416 | Not-determined |
| 18-103_k141_75452 | Not-determined |
| 18-103_k141_115753 | Not-determined |
| 18-103_k141_75453 | Not-determined |
| 18-103_k141_92733 | Low-quality |
| 18-103_k141_98351 | Not-determined |
| 18-103_k141_92734 | Low-quality |
| 18-103_k141_98353 | Not-determined |
| 18-103_k141_110034 | Not-determined |
| 18-103_k141_828 | Low-quality |
| 18-103_k141_46826 | Not-determined |
| 18-103_k141_98358 | Not-determined |
| 18-103_k141_127431 | Not-determined |
| 18-103_k141_29425 | Not-determined |
| 18-103_k141_110039 | Not-determined |
| 18-103_k141_92740 | Not-determined |
| 18-103_k141_46834 | Not-determined |
| 18-103_k141_110043 | Not-determined |
| 18-103_k141_17990 | Not-determined |
| 18-103_k141_81282 | Not-determined |
| 18-103_k141_63974 | Not-determined |
| 18-103_k141_98372 | Not-determined |
| 18-103_k141_63979 | Not-determined |
| 18-103_k141_58268 | Not-determined |
| 18-103_k141_63981 | Not-determined |
| 18-103_k141_23815 | Not-determined |
| 18-103_k141_75479 | Not-determined |
| 18-103_k141_81293 | Not-determined |
| 18-103_k141_115762 | Low-quality |
| 18-103_k141_75480 | Not-determined |
| 18-103_k141_92755 | Not-determined |
| 18-103_k141_63987 | Not-determined |
| 18-103_k141_41025 | Not-determined |
| 18-103_k141_46844 | Low-quality |
| 18-103_k141_29445 | Low-quality |
| 18-103_k141_98392 | Not-determined |
| 18-103_k141_92766 | Not-determined |
| 18-103_k141_46849 | Low-quality |
| 18-103_k141_110068 | Not-determined |
| 18-103_k141_41032 | Not-determined |
| 18-103_k141_127450 | Not-determined |
| 18-103_k141_46852 | Not-determined |
| 18-103_k141_12092 | Not-determined |
| 18-103_k141_23838 | Not-determined |
| 18-103_k141_46856 | Not-determined |
| 18-103_k141_58286 | Low-quality |
| 18-103_k141_58287 | Not-determined |
| 18-103_k141_64011 | Not-determined |
| 18-103_k141_104274 | Not-determined |
| 18-103_k141_110076 | Low-quality |
| 18-103_k141_41042 | Not-determined |
| 18-103_k141_115775 | Low-quality |
| 18-103_k141_41043 | Not-determined |
| 18-103_k141_35299 | Low-quality |
| 18-103_k141_69771 | Not-determined |
| 18-103_k141_98407 | Low-quality |
| 18-103_k141_64014 | Not-determined |
| 18-103_k141_98411 | Not-determined |
| 18-103_k141_98412 | Not-determined |
| 18-103_k141_12103 | Not-determined |
| 18-103_k141_87204 | Not-determined |
| 18-103_k141_98413 | Low-quality |
| 18-103_k141_6626 | Not-determined |
| 18-103_k141_81325 | Not-determined |
| 18-103_k141_115780 | Not-determined |
| 18-103_k141_46877 | Not-determined |
| 18-103_k141_12107 | Not-determined |
| 18-103_k141_81330 | Low-quality |
| 18-103_k141_6631 | Not-determined |
| 18-103_k141_104288 | Not-determined |
| 18-103_k141_104292 | Not-determined |
| 18-103_k141_23858 | Not-determined |
| 18-103_k141_6635 | Low-quality |
| 18-103_k141_69785 | Not-determined |
| 18-103_k141_104297 | Not-determined |
| 18-103_k141_92774 | Not-determined |
| 18-103_k141_69788 | Not-determined |
| 18-103_k141_98422 | Not-determined |
| 18-103_k141_69793 | Not-determined |
| 18-103_k141_69795 | Not-determined |
| 18-103_k141_58313 | Not-determined |
| 18-103_k141_104310 | Not-determined |
| 18-103_k141_58320 | Not-determined |
| 18-103_k141_92782 | Not-determined |
| 18-103_k141_133114 | Not-determined |
| 18-103_k141_41063 | Not-determined |
| 18-103_k141_12121 | Low-quality |
| 18-103_k141_64047 | Not-determined |
| 18-103_k141_58328 | Not-determined |
| 18-103_k141_115807 | Not-determined |
| 18-103_k141_890 | Not-determined |
| 18-103_k141_81359 | Not-determined |
| 18-103_k141_52616 | Not-determined |
| 18-103_k141_52618 | Not-determined |
| 18-103_k141_115810 | Not-determined |
| 18-103_k141_121516 | Low-quality |
| 18-103_k141_64053 | Not-determined |
| 18-103_k141_41082 | Not-determined |
| 18-103_k141_69801 | Not-determined |
| 18-103_k141_92797 | Not-determined |
| 18-103_k141_58336 | Not-determined |
| 18-103_k141_58337 | Not-determined |
| 18-103_k141_58339 | Not-determined |
| 18-103_k141_92804 | Not-determined |
| 18-103_k141_127486 | Not-determined |
| 18-103_k141_12134 | Low-quality |
| 18-103_k141_127489 | Not-determined |
| 18-103_k141_110109 | Not-determined |
| 18-103_k141_23861 | Not-determined |
| 18-103_k141_35311 | Not-determined |
| 18-103_k141_29478 | Not-determined |
| 18-103_k141_35313 | Not-determined |
| 18-103_k141_64066 | Not-determined |
| 18-103_k141_52635 | Not-determined |
| 18-103_k141_75548 | Not-determined |
| 18-103_k141_46899 | Not-determined |
| 18-103_k141_81378 | Not-determined |
| 18-103_k141_115827 | Not-determined |
| 18-103_k141_12143 | Not-determined |
| 18-103_k141_115831 | Not-determined |
| 18-103_k141_75555 | Not-determined |
| 18-103_k141_46904 | Not-determined |
| 18-103_k141_29487 | Not-determined |
| 18-103_k141_87255 | Not-determined |
| 18-103_k141_104331 | Not-determined |
| 18-103_k141_12146 | Not-determined |
| 18-103_k141_12148 | Not-determined |
| 18-103_k141_12149 | Not-determined |
| 18-103_k141_58355 | Not-determined |
| 18-103_k141_46906 | Not-determined |
| 18-103_k141_18046 | Not-determined |
| 18-103_k141_64076 | Not-determined |
| 18-103_k141_6661 | Not-determined |
| 18-103_k141_87265 | Not-determined |
| 18-103_k141_914 | Not-determined |
| 18-103_k141_29498 | Low-quality |
| 18-103_k141_81397 | Not-determined |
| 18-103_k141_110125 | Not-determined |
| 18-103_k141_87270 | Not-determined |
| 18-103_k141_98434 | Not-determined |
| 18-103_k141_23881 | Not-determined |
| 18-103_k141_98436 | Not-determined |
| 18-103_k141_127508 | Not-determined |
| 18-103_k141_41098 | Low-quality |
| 18-103_k141_23884 | Not-determined |
| 18-103_k141_18055 | Not-determined |
| 18-103_k141_23885 | Not-determined |
| 18-103_k141_75572 | Not-determined |
| 18-103_k141_75573 | Not-determined |
| 18-103_k141_127515 | Not-determined |
| 18-103_k141_920 | Not-determined |
| 18-103_k141_133137 | Not-determined |
| 18-103_k141_81415 | Low-quality |
| 18-103_k141_69832 | Not-determined |
| 18-103_k141_87285 | Not-determined |
| 18-103_k141_87288 | Not-determined |
| 18-103_k141_121532 | Not-determined |
| 18-103_k141_41113 | Not-determined |
| 18-103_k141_64091 | Not-determined |
| 18-103_k141_87293 | Not-determined |
| 18-103_k141_924 | Not-determined |
| 18-103_k141_58380 | Not-determined |
| 18-103_k141_87295 | Not-determined |
| 18-103_k141_35353 | Not-determined |
| 18-103_k141_929 | Not-determined |
| 18-103_k141_52665 | Not-determined |
| 18-103_k141_64097 | Not-determined |
| 18-103_k141_92839 | Not-determined |
| 18-103_k141_933 | Not-determined |
| 18-103_k141_92843 | Not-determined |
| 18-103_k141_69846 | Not-determined |
| 18-103_k141_87311 | Not-determined |
| 18-103_k141_69847 | Not-determined |
| 18-103_k141_133160 | Not-determined |
| 18-103_k141_133161 | Not-determined |
| 18-103_k141_104370 | Not-determined |
| 18-103_k141_121552 | Not-determined |
| 18-103_k141_69851 | Low-quality |
| 18-103_k141_104372 | Not-determined |
| 18-103_k141_23903 | Not-determined |
| 18-103_k141_46939 | Not-determined |
| 18-103_k141_23904 | Not-determined |
| 18-103_k141_110172 | Not-determined |
| 18-103_k141_12173 | Not-determined |
| 18-103_k141_41122 | Not-determined |
| 18-103_k141_35365 | Not-determined |
| 18-103_k141_110173 | Not-determined |
| 18-103_k141_81447 | Not-determined |
| 18-103_k141_75598 | Not-determined |
| 18-103_k141_110176 | Not-determined |
| 18-103_k141_133168 | Not-determined |
| 18-103_k141_121558 | Not-determined |
| 18-103_k141_69859 | Low-quality |
| 18-103_k141_6700 | Not-determined |
| 18-103_k141_104383 | Not-determined |
| 18-103_k141_121562 | Not-determined |
| 18-103_k141_41134 | Not-determined |
| 18-103_k141_81458 | Not-determined |
| 18-103_k141_6704 | Low-quality |
| 18-103_k141_18059 | Not-determined |
| 18-103_k141_948 | Not-determined |
| 18-103_k141_81461 | Low-quality |
| 18-103_k141_92849 | Low-quality |
| 18-103_k141_127553 | Not-determined |
| 18-103_k141_115849 | Not-determined |
| 18-103_k141_133177 | Not-determined |
| 18-103_k141_64131 | Not-determined |
| 18-103_k141_87341 | Not-determined |
| 18-103_k141_87344 | Low-quality |
| 18-103_k141_104398 | Not-determined |
| 18-103_k141_957 | Not-determined |
| 18-103_k141_41140 | Not-determined |
| 18-103_k141_121579 | Not-determined |
| 18-103_k141_110192 | Not-determined |
| 18-103_k141_87350 | Not-determined |
| 18-103_k141_64140 | Not-determined |
| 18-103_k141_12182 | Not-determined |
| 18-103_k141_46956 | Low-quality |
| 18-103_k141_6711 | Low-quality |
| 18-103_k141_92862 | Not-determined |
| 18-103_k141_58414 | Not-determined |
| 18-103_k141_58415 | Not-determined |
| 18-103_k141_23936 | Not-determined |
| 18-103_k141_110201 | Not-determined |
| 18-103_k141_35391 | Not-determined |
| 18-103_k141_110202 | Not-determined |
| 18-103_k141_121584 | Not-determined |
| 18-103_k141_104413 | Not-determined |
| 18-103_k141_115862 | Not-determined |
| 18-103_k141_12191 | Not-determined |
| 18-103_k141_121587 | Low-quality |
| 18-103_k141_64149 | Not-determined |
| 18-103_k141_980 | Not-determined |
| 18-103_k141_58424 | Not-determined |
| 18-103_k141_12194 | Not-determined |
| 18-103_k141_35396 | Not-determined |
| 18-103_k141_92877 | Not-determined |
| 18-103_k141_988 | Not-determined |
| 18-103_k141_6719 | Not-determined |
| 18-103_k141_29530 | Not-determined |
| 18-103_k141_127562 | Not-determined |
| 18-103_k141_46969 | Not-determined |
| 18-103_k141_6722 | Not-determined |
| 18-103_k141_115873 | Low-quality |
| 18-103_k141_52680 | Not-determined |
| 18-103_k141_46971 | Not-determined |
| 18-103_k141_104418 | Not-determined |
| 18-103_k141_98484 | Not-determined |
| 18-103_k141_110216 | Not-determined |
| 18-103_k141_98485 | Not-determined |
| 18-103_k141_64161 | Not-determined |
| 18-103_k141_98488 | Not-determined |
| 18-103_k141_35409 | Not-determined |
| 18-103_k141_64164 | Not-determined |
| 18-103_k141_115879 | Low-quality |
| 18-103_k141_115885 | Not-determined |
| 18-103_k141_29545 | Not-determined |
| 18-103_k141_29547 | Not-determined |
| 18-103_k141_69866 | Not-determined |
| 18-103_k141_104430 | Not-determined |
| 18-103_k141_46985 | Not-determined |
| 18-103_k141_29552 | Not-determined |
| 18-103_k141_87382 | Not-determined |
| 18-103_k141_104436 | Not-determined |
| 18-103_k141_29557 | Not-determined |
| 18-103_k141_29558 | Low-quality |
| 18-103_k141_6735 | Not-determined |
| 18-103_k141_92903 | Not-determined |
| 18-103_k141_12215 | Not-determined |
| 18-103_k141_98503 | Low-quality |
| 18-103_k141_64184 | Not-determined |
| 18-103_k141_52696 | Low-quality |
| 18-103_k141_92909 | Not-determined |
| 18-103_k141_75641 | Low-quality |
| 18-103_k141_110240 | Not-determined |
| 18-103_k141_110241 | Not-determined |
| 18-103_k141_121592 | Not-determined |
| 18-103_k141_64187 | Not-determined |
| 18-103_k141_52703 | Not-determined |
| 18-103_k141_64190 | Not-determined |
| 18-103_k141_29570 | Not-determined |
| 18-103_k141_121596 | Not-determined |
| 18-103_k141_81484 | Low-quality |
| 18-103_k141_64192 | Not-determined |
| 18-103_k141_110249 | Not-determined |
| 18-103_k141_52708 | Not-determined |
| 18-103_k141_133211 | Not-determined |
| 18-103_k141_92921 | Not-determined |
| 18-103_k141_87398 | Not-determined |
| 18-103_k141_46997 | Not-determined |
| 18-103_k141_29575 | Not-determined |
| 18-103_k141_69898 | Low-quality |
| 18-103_k141_127600 | Not-determined |
| 18-103_k141_69902 | Low-quality |
| 18-103_k141_104455 | Not-determined |
| 18-103_k141_52726 | Not-determined |
| 18-103_k141_29583 | Not-determined |
| 18-103_k141_81495 | Not-determined |
| 18-103_k141_121612 | Not-determined |
| 18-103_k141_12240 | Not-determined |
| 18-103_k141_121613 | Not-determined |
| 18-103_k141_35450 | Not-determined |
| 18-103_k141_127607 | Not-determined |
| 18-103_k141_81505 | Not-determined |
| 18-103_k141_87415 | Not-determined |
| 18-103_k141_133228 | Not-determined |
| 18-103_k141_52736 | Low-quality |
| 18-103_k141_35456 | Not-determined |
| 18-103_k141_1044 | Low-quality |
| 18-103_k141_58453 | Not-determined |
| 18-103_k141_87417 | Not-determined |
| 18-103_k141_12247 | Not-determined |
| 18-103_k141_81510 | Not-determined |
| 18-101-final-viral-combined_k141_107766\|\|full | Medium-quality |
| 18-103_k141_87422 | Not-determined |
| 18-103_k141_1050 | Not-determined |
| 18-103_k141_81513 | Not-determined |
| 18-103_k141_1051 | Low-quality |
| 18-103_k141_133236 | Not-determined |
| 18-103_k141_52744 | Not-determined |
| 18-103_k141_35462 | Low-quality |
| 18-103_k141_92947 | Not-determined |
| 18-103_k141_64206 | Not-determined |
| 18-103_k141_47019 | Not-determined |
| 18-103_k141_58460 | Not-determined |
| 18-103_k141_1055 | Not-determined |
| 18-103_k141_69915 | Not-determined |
| 18-103_k141_29603 | Not-determined |
| 18-103_k141_23968 | Not-determined |
| 18-103_k141_104462 | Not-determined |
| 18-103_k141_6766 | Not-determined |
| 18-103_k141_58464 | Not-determined |
| 18-103_k141_121632 | Not-determined |
| 18-103_k141_81523 | Not-determined |
| 18-103_k141_92953 | Not-determined |
| 18-103_k141_87431 | Not-determined |
| 18-103_k141_1062 | Not-determined |
| 18-103_k141_87433 | Not-determined |
| 18-103_k141_64212 | Not-determined |
| 18-103_k141_29610 | Not-determined |
| 18-103_k141_69927 | Low-quality |
| 18-103_k141_127630 | Not-determined |
| 18-103_k141_58475 | Not-determined |
| 18-103_k141_58476 | Not-determined |
| 18-103_k141_121646 | Not-determined |
| 18-103_k141_29612 | Not-determined |
| 18-103_k141_127631 | Not-determined |
| 18-103_k141_58479 | Not-determined |
| 18-103_k141_110290 | Not-determined |
| 18-103_k141_18080 | Not-determined |
| 18-103_k141_121652 | Low-quality |
| 18-103_k141_41197 | Low-quality |
| 18-103_k141_6777 | Not-determined |
| 18-103_k141_64225 | Not-determined |
| 18-103_k141_98529 | Not-determined |
| 18-103_k141_64226 | Low-quality |
| 18-103_k141_104479 | Not-determined |
| 18-103_k141_110298 | Not-determined |
| 18-103_k141_69934 | Not-determined |
| 18-103_k141_104481 | Low-quality |
| 18-103_k141_12270 | Low-quality |
| 18-103_k141_87436 | Low-quality |
| 18-103_k141_18088 | Not-determined |
| 18-103_k141_64232 | Not-determined |
| 18-103_k141_121659 | Not-determined |
| 18-103_k141_98534 | Not-determined |
| 18-103_k141_115923 | Not-determined |
| 18-103_k141_115928 | Not-determined |
| 18-103_k141_92974 | Low-quality |
| 18-103_k141_87454 | Not-determined |
| 18-103_k141_81555 | Not-determined |
| 18-103_k141_115935 | Not-determined |
| 18-103_k141_41205 | Not-determined |
| 18-103_k141_12277 | Low-quality |
| 18-103_k141_104494 | Low-quality |
| 18-103_k141_58485 | Not-determined |
| 18-103_k141_87460 | Not-determined |
| 18-103_k141_121668 | Not-determined |
| 18-103_k141_115940 | Not-determined |
| 18-103_k141_12279 | Not-determined |
| 18-103_k141_12280 | Not-determined |
| 18-103_k141_1091 | Not-determined |

| | |
|---|---|
| 18-103_k141_18104 | Not-determined |
| 18-103_k141_12285 | Not-determined |
| 18-103_k141_18110 | Not-determined |
| 18-103_k141_104498 | Not-determined |
| 18-103_k141_24003 | Not-determined |
| 18-103_k141_127654 | Not-determined |
| 18-103_k141_6804 | Low-quality |
| 18-103_k141_121678 | Not-determined |
| 18-103_k141_133267 | Low-quality |
| 18-103_k141_1094 | Not-determined |
| 18-103_k141_41221 | Not-determined |
| 18-103_k141_127658 | Not-determined |
| 18-103_k141_1096 | Not-determined |
| 18-103_k141_75722 | Not-determined |
| 18-103_k141_115955 | Not-determined |
| 18-103_k141_1100 | Not-determined |
| 18-103_k141_47071 | Not-determined |
| 18-103_k141_127662 | Not-determined |
| 18-103_k141_47072 | Not-determined |
| 18-103_k141_29639 | Not-determined |
| 18-103_k141_115957 | Not-determined |
| 18-103_k141_121686 | Not-determined |
| 18-103_k141_47074 | Not-determined |
| 18-103_k141_81577 | Not-determined |
| 18-103_k141_47078 | Not-determined |
| 18-103_k141_64262 | Not-determined |
| 18-103_k141_115967 | Not-determined |
| 18-103_k141_6826 | Not-determined |
| 18-103_k141_75735 | Not-determined |
| 18-103_k141_92981 | Not-determined |
| 18-103_k141_41236 | Not-determined |
| 18-103_k141_64269 | Not-determined |
| 18-103_k141_127674 | Not-determined |
| 18-103_k141_1121 | Not-determined |
| 18-103_k141_52805 | Not-determined |
| 18-103_k141_52807 | Not-determined |
| 18-103_k141_1125 | Not-determined |
| 18-103_k141_115973 | Not-determined |
| 18-103_k141_18120 | Low-quality |
| 18-103_k141_29652 | Not-determined |
| 18-103_k141_64276 | Not-determined |
| 18-103_k141_29653 | Not-determined |
| 18-103_k141_18123 | Not-determined |
| 18-103_k141_87486 | Low-quality |
| 18-103_k141_133281 | Low-quality |
| 18-103_k141_133282 | Not-determined |
| 18-103_k141_87487 | Not-determined |
| 18-103_k141_133285 | Not-determined |
| 18-103_k141_92995 | Not-determined |
| 18-103_k141_133287 | Low-quality |
| 18-103_k141_81581 | Not-determined |
| 18-103_k141_58522 | Not-determined |
| 18-103_k141_29660 | Not-determined |
| 18-103_k141_52820 | Not-determined |
| 18-103_k141_110333 | Not-determined |
| 18-103_k141_12303 | Not-determined |
| 18-103_k141_58528 | Not-determined |
| 18-103_k141_110338 | Not-determined |
| 18-103_k141_93001 | Not-determined |
| 18-103_k141_110342 | Not-determined |
| 18-103_k141_58536 | Not-determined |
| 18-103_k141_81583 | Not-determined |
| 18-103_k141_93007 | Not-determined |
| 18-103_k141_35544 | Not-determined |
| 18-103_k141_81585 | Not-determined |
| 18-103_k141_1146 | Not-determined |
| 18-103_k141_127698 | Not-determined |
| 18-103_k141_29674 | Not-determined |
| 18-103_k141_115994 | Not-determined |
| 18-103_k141_18144 | Not-determined |
| 18-103_k141_58541 | Not-determined |
| 18-103_k141_87502 | Not-determined |
| 18-103_k141_47123 | Low-quality |
| 18-103_k141_116001 | Not-determined |
| 18-103_k141_127707 | Not-determined |
| 18-103_k141_29685 | Not-determined |
| 18-103_k141_81594 | Not-determined |
| 18-103_k141_87516 | Not-determined |
| 18-103_k141_18155 | Not-determined |
| 18-103_k141_127712 | Not-determined |
| 18-103_k141_69990 | Low-quality |
| 18-103_k141_127717 | Not-determined |
| 18-103_k141_69993 | Low-quality |
| 18-103_k141_127720 | Not-determined |
| 18-103_k141_87525 | Not-determined |
| 18-103_k141_64314 | Low-quality |
| 18-103_k141_58559 | Not-determined |
| 18-103_k141_104539 | Not-determined |
| 18-103_k141_127723 | Not-determined |
| 18-103_k141_121709 | Not-determined |
| 18-103_k141_110360 | Not-determined |
| 18-103_k141_69999 | Not-determined |
| 18-103_k141_1167 | Not-determined |
| 18-103_k141_87532 | Not-determined |
| 18-103_k141_75773 | Not-determined |
| 18-103_k141_47134 | Not-determined |
| 18-103_k141_104552 | Not-determined |
| 18-103_k141_64322 | Not-determined |
| 18-103_k141_81605 | Not-determined |
| 18-103_k141_87546 | Not-determined |
| 18-103_k141_127736 | Not-determined |
| 18-103_k141_47154 | Low-quality |
| 18-103_k141_121727 | Not-determined |
| 18-103_k141_133337 | Not-determined |
| 18-103_k141_12335 | Not-determined |
| 18-103_k141_81616 | Not-determined |
| 18-103_k141_12338 | Not-determined |
| 18-103_k141_18175 | Not-determined |
| 18-103_k141_35567 | Low-quality |
| 18-103_k141_35568 | Not-determined |
| 18-103_k141_116021 | Not-determined |
| 18-103_k141_24054 | Not-determined |
| 18-103_k141_127741 | Low-quality |
| 18-103_k141_6875 | Not-determined |
| 18-103_k141_121734 | Not-determined |
| 18-103_k141_116024 | Not-determined |
| 18-103_k141_116025 | Not-determined |
| 18-103_k141_81624 | Not-determined |
| 18-103_k141_24056 | Low-quality |
| 18-103_k141_18183 | Not-determined |
| 18-103_k141_47165 | Not-determined |
| 18-103_k141_35577 | Not-determined |
| 18-103_k141_35579 | Not-determined |
| 18-103_k141_18185 | Not-determined |
| 18-103_k141_12351 | Not-determined |
| 18-103_k141_1189 | Not-determined |
| 18-103_k141_52873 | Not-determined |
| 18-103_k141_70022 | Not-determined |
| 18-103_k141_133354 | Not-determined |
| 18-103_k141_24060 | Not-determined |
| 18-103_k141_58581 | Not-determined |
| 18-103_k141_1193 | Not-determined |
| 18-103_k141_133356 | Not-determined |
| 18-103_k141_41274 | Not-determined |
| 18-103_k141_18195 | Not-determined |
| 18-103_k141_18196 | Not-determined |
| 18-103_k141_121750 | Not-determined |
| 18-103_k141_12359 | Not-determined |
| 18-103_k141_12360 | Not-determined |
| 18-103_k141_1203 | Not-determined |
| 18-103_k141_104574 | Not-determined |
| 18-103_k141_81635 | Not-determined |
| 18-103_k141_87555 | Low-quality |
| 18-103_k141_133371 | Not-determined |
| 18-103_k141_1207 | Low-quality |
| 18-103_k141_12370 | Not-determined |
| 18-103_k141_127758 | Not-determined |
| 18-103_k141_75822 | Not-determined |
| 18-103_k141_47183 | Not-determined |
| 18-103_k141_64348 | Not-determined |
| 18-103_k141_64350 | Not-determined |
| 18-103_k141_127764 | Low-quality |
| 18-103_k141_58596 | Not-determined |
| 18-103_k141_6905 | Not-determined |
| 18-103_k141_87561 | Not-determined |
| 18-103_k141_64351 | Low-quality |
| 18-103_k141_133380 | Not-determined |
| 18-103_k141_29731 | Not-determined |
| 18-103_k141_81652 | Not-determined |
| 18-103_k141_133381 | Not-determined |
| 18-103_k141_1214 | Low-quality |
| 18-103_k141_104599 | Not-determined |
| 18-103_k141_133382 | Not-determined |
| 18-103_k141_127768 | Not-determined |
| 18-103_k141_81656 | Not-determined |
| 18-103_k141_93066 | Not-determined |
| 18-103_k141_41295 | Low-quality |
| 18-103_k141_127769 | Low-quality |
| 18-103_k141_41297 | Not-determined |
| 18-103_k141_116075 | Not-determined |
| 18-103_k141_87573 | Not-determined |
| 18-103_k141_47202 | Not-determined |
| 18-103_k141_6911 | Not-determined |
| 18-103_k141_58608 | Not-determined |
| 18-103_k141_127782 | Not-determined |
| 18-103_k141_133394 | Low-quality |
| 18-103_k141_29740 | Not-determined |
| 18-103_k141_121775 | Not-determined |
| 18-103_k141_127785 | Not-determined |
| 18-103_k141_93076 | Not-determined |
| 18-103_k141_6918 | Not-determined |
| 18-103_k141_1225 | Not-determined |
| 18-103_k141_24096 | Not-determined |
| 18-103_k141_29747 | Not-determined |
| 18-103_k141_116088 | Not-determined |
| 18-103_k141_6925 | Not-determined |
| 18-103_k141_121780 | Not-determined |
| 18-103_k141_87591 | Not-determined |
| 18-103_k141_133405 | Low-quality |
| 18-103_k141_41322 | Not-determined |
| 18-103_k141_6931 | Not-determined |
| 18-103_k141_121788 | Not-determined |
| 18-103_k141_58615 | Not-determined |
| 18-103_k141_47214 | Low-quality |
| 18-103_k141_58616 | Not-determined |
| 18-103_k141_58618 | Not-determined |
| 18-103_k141_93087 | Low-quality |
| 18-103_k141_116098 | Not-determined |
| 18-103_k141_12416 | Not-determined |
| 18-103_k141_58620 | Not-determined |
| 18-103_k141_29757 | Not-determined |
| 18-103_k141_12418 | Low-quality |
| 18-103_k141_29758 | Low-quality |
| 18-103_k141_1235 | Not-determined |
| 18-103_k141_64365 | Not-determined |
| 18-103_k141_110392 | Not-determined |
| 18-103_k141_18246 | Low-quality |
| 18-103_k141_24112 | Not-determined |
| 18-103_k141_24113 | Not-determined |
| 18-103_k141_58630 | Not-determined |
| 18-103_k141_75854 | Low-quality |
| 18-103_k141_47222 | Not-determined |
| 18-103_k141_18248 | Low-quality |
| 18-103_k141_12429 | Not-determined |
| 18-103_k141_104608 | Not-determined |
| 18-103_k141_12431 | Not-determined |
| 18-103_k141_6943 | Not-determined |
| 18-103_k141_81678 | Low-quality |
| 18-103_k141_35640 | Not-determined |
| 18-103_k141_6945 | Not-determined |
| 18-103_k141_58635 | Not-determined |
| 18-103_k141_133425 | Not-determined |
| 18-103_k141_6948 | Low-quality |
| 18-103_k141_35645 | Not-determined |
| 18-103_k141_104612 | Not-determined |
| 18-103_k141_116109 | Not-determined |
| 18-103_k141_52954 | Not-determined |
| 18-103_k141_41339 | Not-determined |
| 18-103_k141_35649 | Not-determined |
| 18-103_k141_18260 | Low-quality |
| 18-103_k141_75868 | Not-determined |
| 18-103_k141_75871 | Not-determined |
| 18-103_k141_75872 | Not-determined |
| 18-103_k141_81689 | Not-determined |
| 18-103_k141_35655 | Low-quality |
| 18-103_k141_64381 | Not-determined |
| 18-103_k141_6958 | Not-determined |
| 18-103_k141_24138 | Not-determined |
| 18-103_k141_47240 | Not-determined |
| 18-103_k141_18268 | Not-determined |
| 18-103_k141_47241 | Not-determined |
| 18-103_k141_24139 | Low-quality |
| 18-103_k141_116130 | Not-determined |
| 18-103_k141_29776 | Not-determined |
| 18-103_k141_104631 | Low-quality |
| 18-103_k141_87613 | Not-determined |
| 18-103_k141_133442 | Not-determined |
| 18-103_k141_87614 | Not-determined |
| 18-103_k141_127815 | Not-determined |
| 18-103_k141_127816 | Not-determined |
| 18-103_k141_121821 | Not-determined |
| 18-103_k141_24144 | Not-determined |
| 18-103_k141_127821 | Not-determined |
| 18-103_k141_64400 | Low-quality |
| 18-103_k141_70099 | Not-determined |
| 18-103_k141_41367 | Not-determined |
| 18-103_k141_121824 | Not-determined |
| 18-103_k141_87621 | Not-determined |
| 18-103_k141_110418 | Low-quality |
| 18-103_k141_87622 | Not-determined |
| 18-103_k141_127828 | Not-determined |
| 18-103_k141_98656 | Low-quality |
| 18-103_k141_121828 | Low-quality |
| 18-103_k141_116135 | Not-determined |
| 18-103_k141_81707 | Not-determined |
| 18-103_k141_41372 | Not-determined |
| 18-103_k141_127831 | Not-determined |
| 18-103_k141_64405 | Not-determined |
| 18-103_k141_81712 | Not-determined |
| 18-103_k141_29787 | Not-determined |
| 18-103_k141_121832 | Not-determined |
| 18-103_k141_18284 | Not-determined |
| 18-103_k141_18285 | Not-determined |
| 18-103_k141_12472 | Not-determined |
| 18-103_k141_24157 | Not-determined |
| 18-103_k141_93095 | Not-determined |
| 18-103_k141_116155 | Not-determined |
| 18-103_k141_6966 | Low-quality |
| 18-103_k141_110435 | Not-determined |
| 18-103_k141_64415 | Low-quality |
| 18-103_k141_64416 | Not-determined |
| 18-103_k141_18294 | Not-determined |
| 18-103_k141_121843 | Not-determined |
| 18-103_k141_116161 | Not-determined |
| 18-103_k141_70116 | Low-quality |
| 18-103_k141_127853 | Not-determined |
| 18-103_k141_58661 | Not-determined |
| 18-103_k141_110445 | Not-determined |
| 18-103_k141_98673 | Not-determined |
| 18-103_k141_75890 | Not-determined |
| 18-103_k141_116165 | Not-determined |
| 18-103_k141_47277 | Not-determined |
| 18-103_k141_98683 | Not-determined |
| 18-103_k141_41405 | Not-determined |
| 18-103_k141_75903 | Not-determined |
| 18-103_k141_1262 | Not-determined |
| 18-103_k141_24192 | Not-determined |
| 18-103_k141_93110 | Not-determined |
| 18-103_k141_29819 | Not-determined |
| 18-103_k141_64442 | Not-determined |
| 18-103_k141_6986 | Not-determined |
| 18-103_k141_47280 | Not-determined |
| 18-103_k141_41412 | Not-determined |
| 18-103_k141_47283 | Not-determined |
| 18-103_k141_81745 | Not-determined |
| 18-103_k141_12492 | Not-determined |
| 18-103_k141_116187 | Not-determined |
| 18-103_k141_127873 | Low-quality |
| 18-103_k141_116190 | Not-determined |
| 18-103_k141_81748 | Not-determined |
| 18-103_k141_18313 | Not-determined |
| 18-103_k141_116194 | Not-determined |
| 18-103_k141_98701 | Not-determined |
| 18-103_k141_98703 | Not-determined |
| 18-103_k141_75911 | Not-determined |
| 18-103_k141_1267 | Not-determined |
| 18-103_k141_6989 | Not-determined |
| 18-103_k141_6991 | Not-determined |
| 18-103_k141_98707 | Not-determined |
| 18-103_k141_24202 | Low-quality |
| 18-103_k141_87650 | Not-determined |
| 18-103_k141_127878 | Not-determined |
| 18-103_k141_58682 | Not-determined |
| 18-103_k141_81759 | Low-quality |
| 18-103_k141_70147 | Not-determined |
| 18-103_k141_75917 | Not-determined |
| 18-103_k141_81762 | Not-determined |
| 18-103_k141_41433 | Not-determined |
| 18-103_k141_75922 | Not-determined |
| 18-103_k141_18329 | Not-determined |
| 18-103_k141_7001 | Not-determined |
| 18-103_k141_53026 | Not-determined |
| 18-103_k141_47303 | Not-determined |
| 18-103_k141_58690 | Not-determined |
| 18-103_k141_116208 | Not-determined |
| 18-103_k141_12515 | Not-determined |
| 18-103_k141_24217 | Not-determined |
| 18-103_k141_24221 | Low-quality |
| 18-103_k141_133489 | Not-determined |
| 18-103_k141_53036 | Not-determined |
| 18-103_k141_35661 | Not-determined |
| 18-103_k141_7014 | Not-determined |
| 18-103_k141_81776 | Not-determined |
| 18-103_k141_18340 | Not-determined |
| 18-103_k141_7017 | Not-determined |
| 18-103_k141_127892 | Not-determined |
| 18-103_k141_35678 | Low-quality |
| 18-103_k141_133502 | Not-determined |
| 18-103_k141_24239 | Not-determined |
| 18-103_k141_70165 | Not-determined |
| 18-103_k141_93141 | Not-determined |
| 18-103_k141_18350 | Not-determined |
| 18-103_k141_64478 | Not-determined |
| 18-103_k141_70173 | Not-determined |
| 18-103_k141_64479 | Not-determined |
| 18-103_k141_18356 | Not-determined |
| 18-103_k141_116236 | Not-determined |
| 18-103_k141_127897 | Low-quality |
| 18-103_k141_116238 | Not-determined |
| 18-103_k141_116240 | Not-determined |
| 18-103_k141_81790 | Not-determined |
| 18-103_k141_29864 | Not-determined |
| 18-103_k141_75935 | Not-determined |
| 18-103_k141_116243 | Not-determined |
| 18-103_k141_121882 | Not-determined |
| 18-103_k141_18371 | Not-determined |
| 18-103_k141_41472 | Not-determined |
| 18-103_k141_127911 | Not-determined |
| 18-103_k141_29877 | Not-determined |
| 18-103_k141_121890 | Not-determined |
| 18-103_k141_64492 | Not-determined |
| 18-103_k141_133516 | Not-determined |
| 18-103_k141_7042 | Not-determined |
| 18-103_k141_1275 | Not-determined |
| 18-103_k141_127920 | Not-determined |
| 18-103_k141_24268 | Not-determined |
| 18-103_k141_58721 | Not-determined |
| 18-103_k141_116263 | Not-determined |
| 18-103_k141_81805 | Not-determined |
| 18-103_k141_70202 | Not-determined |
| 18-103_k141_75962 | Not-determined |
| 18-103_k141_133519 | Not-determined |
| 18-103_k141_116267 | Not-determined |
| 18-103_k141_70204 | Low-quality |
| 18-103_k141_18399 | Not-determined |
| 18-103_k141_70209 | Not-determined |
| 18-103_k141_64499 | Not-determined |
| 18-103_k141_1287 | Not-determined |
| 18-103_k141_110505 | Not-determined |
| 18-103_k141_81819 | Not-determined |
| 18-103_k141_110506 | Not-determined |
| 18-103_k141_1290 | Low-quality |
| 18-103_k141_53076 | Not-determined |
| 18-103_k141_18408 | Not-determined |
| 18-103_k141_35726 | Not-determined |
| 18-103_k141_35727 | Not-determined |
| 18-103_k141_1297 | Low-quality |
| 18-103_k141_93174 | Not-determined |
| 18-103_k141_110525 | Not-determined |
| 18-103_k141_12548 | Low-quality |
| 18-103_k141_53083 | Low-quality |
| 18-103_k141_133538 | Not-determined |
| 18-103_k141_81837 | Not-determined |
| 18-103_k141_7074 | Not-determined |
| 18-103_k141_64511 | Not-determined |
| 18-103_k141_116297 | Low-quality |
| 18-103_k141_93185 | Not-determined |
| 18-103_k141_110530 | Low-quality |
| 18-103_k141_12555 | Not-determined |
| 18-103_k141_70224 | Not-determined |
| 18-103_k141_98765 | Not-determined |
| 18-103_k141_98769 | Not-determined |
| 18-103_k141_75998 | Not-determined |
| 18-103_k141_41491 | Low-quality |
| 18-103_k141_133549 | Not-determined |
| 18-103_k141_127960 | Not-determined |
| 18-103_k141_75999 | Low-quality |
| 18-103_k141_133551 | Not-determined |
| 18-103_k141_133553 | Not-determined |
| 18-103_k141_98773 | Not-determined |
| 18-103_k141_41496 | Not-determined |
| 18-103_k141_41498 | Not-determined |
| 18-103_k141_116313 | Not-determined |
| 18-103_k141_127970 | Not-determined |
| 18-103_k141_64524 | Not-determined |
| 18-103_k141_12567 | Not-determined |
| 18-103_k141_1323 | Not-determined |
| 18-103_k141_24298 | Not-determined |
| 18-103_k141_12571 | Not-determined |
| 18-103_k141_81863 | Not-determined |
| 18-103_k141_110540 | Not-determined |
| 18-103_k141_47349 | Not-determined |
| 18-103_k141_87709 | Not-determined |
| 18-103_k141_64533 | Not-determined |
| 18-103_k141_24306 | Low-quality |
| 18-103_k141_133569 | Not-determined |
| 18-103_k141_58779 | Not-determined |
| 18-103_k141_12593 | Not-determined |
| 18-103_k141_116329 | Not-determined |
| 18-103_k141_93220 | Not-determined |
| 18-103_k141_116330 | Not-determined |
| 18-103_k141_121937 | Not-determined |
| 18-103_k141_7114 | Not-determined |
| 18-103_k141_64551 | Not-determined |
| 18-103_k141_53117 | Low-quality |
| 18-103_k141_76027 | Low-quality |
| 18-103_k141_41520 | Not-determined |
| 18-103_k141_121941 | Not-determined |
| 18-103_k141_1350 | Not-determined |
| 18-103_k141_76030 | Not-determined |
| 18-103_k141_12599 | Not-determined |
| 18-103_k141_76032 | Not-determined |
| 18-103_k141_47371 | Not-determined |
| 18-103_k141_76034 | Not-determined |
| 18-103_k141_133591 | Not-determined |
| 18-103_k141_128007 | Not-determined |
| 18-103_k141_116350 | Not-determined |
| 18-103_k141_110575 | Not-determined |
| 18-103_k141_58798 | Not-determined |
| 18-103_k141_24331 | Not-determined |
| 18-103_k141_58800 | Not-determined |
| 18-103_k141_87731 | Not-determined |
| 18-103_k141_47382 | Not-determined |
| 18-103_k141_81894 | Not-determined |
| 18-103_k141_47385 | Not-determined |
| 18-103_k141_24333 | Not-determined |
| 18-103_k141_87733 | Not-determined |
| 18-103_k141_64566 | Low-quality |
| 18-103_k141_104703 | Not-determined |
| 18-103_k141_64568 | Not-determined |
| 18-103_k141_58807 | Not-determined |
| 18-103_k141_121962 | Not-determined |
| 18-103_k141_98812 | Not-determined |
| 18-103_k141_121964 | Not-determined |
| 18-103_k141_133599 | Low-quality |
| 18-103_k141_128017 | Not-determined |
| 18-103_k141_18443 | Not-determined |
| 18-103_k141_116357 | Not-determined |
| 18-103_k141_64569 | Not-determined |
| 18-103_k141_41529 | Not-determined |
| 18-103_k141_41530 | Not-determined |
| 18-103_k141_81902 | Not-determined |
| 18-103_k141_18449 | Not-determined |
| 18-103_k141_18450 | Not-determined |
| 18-103_k141_104711 | Not-determined |
| 18-103_k141_81905 | Not-determined |
| 18-103_k141_18451 | Not-determined |
| 18-103_k141_64576 | Not-determined |
| 18-103_k141_87745 | Not-determined |
| 18-103_k141_116369 | Not-determined |
| 18-103_k141_87749 | Not-determined |
| 18-103_k141_24361 | Not-determined |
| 18-103_k141_12627 | Not-determined |
| 18-103_k141_47395 | Low-quality |
| 18-103_k141_110593 | Not-determined |
| 18-103_k141_121976 | Not-determined |
| 18-103_k141_53137 | Not-determined |
| 18-103_k141_12633 | Not-determined |
| 18-103_k141_76055 | Not-determined |
| 18-103_k141_121981 | Not-determined |
| 18-103_k141_76056 | Not-determined |
| 18-103_k141_110603 | Not-determined |
| 18-103_k141_24377 | Not-determined |
| 18-103_k141_81916 | Not-determined |
| 18-103_k141_87756 | Not-determined |
| 18-103_k141_29947 | Low-quality |
| 18-103_k141_81918 | Not-determined |
| 18-103_k141_128047 | Not-determined |
| 18-103_k141_58843 | Not-determined |
| 18-103_k141_93267 | Not-determined |
| 18-103_k141_116389 | Not-determined |
| 18-103_k141_47405 | Not-determined |
| 18-103_k141_87766 | Not-determined |
| 18-103_k141_133622 | Low-quality |
| 18-103_k141_87767 | Not-determined |
| 18-103_k141_104738 | Not-determined |
| 18-103_k141_128058 | Not-determined |
| 18-103_k141_81925 | Not-determined |
| 18-103_k141_121994 | Not-determined |
| 18-103_k141_29963 | Not-determined |
| 18-103_k141_12654 | Not-determined |
| 18-103_k141_87775 | Not-determined |
| 18-103_k141_81937 | Not-determined |
| 18-103_k141_41554 | Not-determined |
| 18-103_k141_35781 | Not-determined |
| 18-103_k141_12664 | Not-determined |
| 18-103_k141_133632 | Not-determined |
| 18-103_k141_35782 | Not-determined |
| 18-103_k141_128074 | Not-determined |
| 18-103_k141_87786 | Not-determined |
| 18-103_k141_122010 | Not-determined |
| 18-103_k141_53180 | Not-determined |
| 18-103_k141_81948 | Not-determined |
| 18-103_k141_29973 | Not-determined |
| 18-103_k141_133638 | Not-determined |
| 18-103_k141_70257 | Not-determined |
| 18-103_k141_70258 | Not-determined |
| 18-103_k141_122020 | Not-determined |
| 18-103_k141_29974 | Low-quality |
| 18-103_k141_18467 | Not-determined |
| 18-103_k141_81957 | Low-quality |
| 18-103_k141_93304 | Not-determined |
| 18-103_k141_41563 | Not-determined |
| 18-103_k141_70264 | Not-determined |
| 18-103_k141_70265 | Low-quality |
| 18-103_k141_81963 | Not-determined |
| 18-103_k141_87797 | Low-quality |
| 18-103_k141_41571 | Not-determined |
| 18-103_k141_29991 | Not-determined |
| 18-103_k141_122029 | Not-determined |
| 18-103_k141_128098 | Not-determined |
| 18-103_k141_104779 | Not-determined |
| 18-103_k141_1414 | Not-determined |
| 18-103_k141_76090 | Not-determined |
| 18-103_k141_47431 | Not-determined |
| 18-103_k141_1422 | Not-determined |
| 18-103_k141_93322 | Not-determined |
| 18-103_k141_35807 | Not-determined |
| 18-103_k141_58884 | Not-determined |
| 18-103_k141_24400 | Not-determined |
| 18-103_k141_104785 | Not-determined |
| 18-103_k141_35815 | Not-determined |
| 18-103_k141_98906 | Low-quality |
| 18-103_k141_93333 | Not-determined |
| 18-103_k141_41576 | Not-determined |
| 18-103_k141_93335 | Low-quality |
| 18-103_k141_58893 | Not-determined |
| 18-103_k141_122037 | Not-determined |
| 18-103_k141_70293 | Low-quality |
| 18-103_k141_128117 | Not-determined |
| 18-103_k141_18494 | Not-determined |
| 18-103_k141_116430 | Not-determined |
| 18-103_k141_76119 | Not-determined |
| 18-103_k141_122042 | Not-determined |
| 18-103_k141_1431 | Not-determined |
| 18-103_k141_58898 | Not-determined |
| 18-103_k141_93341 | Not-determined |
| 18-103_k141_70300 | Not-determined |
| 18-103_k141_110626 | Not-determined |
| 18-103_k141_7188 | Not-determined |
| 18-103_k141_7193 | Not-determined |
| 18-103_k141_110631 | Not-determined |
| 18-103_k141_110632 | Not-determined |
| 18-103_k141_35834 | Not-determined |
| 18-103_k141_41585 | Low-quality |
| 18-103_k141_35835 | Not-determined |
| 18-103_k141_110634 | Not-determined |
| 18-103_k141_1433 | Low-quality |
| 18-103_k141_87805 | Not-determined |

| | |
|---|---|
| 18-103_k141_35840 | Low-quality |
| 18-103_k141_30025 | Not-determined |
| 18-103_k141_41591 | Low-quality |
| 18-103_k141_41594 | Not-determined |
| 18-103_k141_41595 | Not-determined |
| 18-103_k141_41596 | Not-determined |
| 18-103_k141_104807 | Not-determined |
| 18-103_k141_12716 | Not-determined |
| 18-103_k141_41599 | Not-determined |
| 18-103_k141_30041 | Not-determined |
| 18-103_k141_64644 | Not-determined |
| 18-103_k141_18523 | Not-determined |
| 18-103_k141_122074 | Not-determined |
| 18-103_k141_30045 | Not-determined |
| 18-103_k141_116450 | Not-determined |
| 18-103_k141_128132 | Not-determined |
| 18-103_k141_76138 | Not-determined |
| 18-103_k141_110652 | Low-quality |
| 18-103_k141_41611 | Not-determined |
| 18-103_k141_64653 | Not-determined |
| 18-103_k141_128136 | Not-determined |
| 18-103_k141_110661 | Not-determined |
| 18-103_k141_104816 | Not-determined |
| 18-103_k141_30055 | Not-determined |
| 18-103_k141_110665 | Not-determined |
| 18-103_k141_133702 | Not-determined |
| 18-103_k141_110666 | Not-determined |
| 18-103_k141_128141 | Not-determined |
| 18-103_k141_116465 | Not-determined |
| 18-103_k141_41627 | Not-determined |
| 18-103_k141_12733 | Low-quality |
| 18-103_k141_41628 | Not-determined |
| 18-103_k141_128142 | Not-determined |
| 18-103_k141_47501 | Not-determined |
| 18-103_k141_133706 | Not-determined |
| 18-103_k141_47506 | Not-determined |
| 18-103_k141_116471 | Not-determined |
| 18-103_k141_64674 | Not-determined |
| 18-103_k141_12742 | Not-determined |
| 18-103_k141_87822 | Low-quality |
| 18-103_k141_53215 | Not-determined |
| 18-103_k141_70336 | Not-determined |
| 18-103_k141_87825 | Not-determined |
| 18-103_k141_98946 | Not-determined |
| 18-103_k141_41639 | Not-determined |
| 18-103_k141_53222 | Low-quality |
| 18-103_k141_128154 | Not-determined |
| 18-103_k141_70342 | Not-determined |
| 18-103_k141_47525 | Not-determined |
| 18-103_k141_128158 | Low-quality |
| 18-103_k141_30089 | Not-determined |
| 18-103_k141_30093 | Not-determined |
| 18-103_k141_47533 | Not-determined |
| 18-103_k141_87839 | Not-determined |
| 18-103_k141_41652 | Not-determined |
| 18-103_k141_12758 | Low-quality |
| 18-103_k141_35884 | Not-determined |
| 18-103_k141_58945 | Not-determined |
| 18-103_k141_87848 | Not-determined |
| 18-103_k141_64683 | Not-determined |
| 18-103_k141_128172 | Not-determined |
| 18-103_k141_53229 | Not-determined |
| 18-103_k141_116507 | Not-determined |
| 18-103_k141_110682 | Not-determined |
| 18-103_k141_58955 | Not-determined |
| 18-103_k141_116512 | Not-determined |
| 18-103_k141_76185 | Not-determined |
| 18-103_k141_110685 | Not-determined |
| 18-103_k141_110687 | Not-determined |
| 18-103_k141_24445 | Not-determined |
| 18-103_k141_7210 | Not-determined |
| 18-103_k141_128187 | Not-determined |
| 18-103_k141_7214 | Not-determined |
| 18-103_k141_116518 | Not-determined |
| 18-103_k141_128189 | Not-determined |
| 18-103_k141_104852 | Not-determined |
| 18-103_k141_87856 | Not-determined |
| 18-103_k141_87857 | Not-determined |
| 18-103_k141_104854 | Not-determined |
| 18-103_k141_116523 | Not-determined |
| 18-103_k141_110698 | Low-quality |
| 18-103_k141_7220 | Not-determined |
| 18-103_k141_122136 | Not-determined |
| 18-103_k141_12774 | Not-determined |
| 18-103_k141_87865 | Not-determined |
| 18-103_k141_128201 | Not-determined |
| 18-103_k141_87868 | Not-determined |
| 18-103_k141_116536 | Not-determined |
| 18-103_k141_110707 | Not-determined |
| 18-103_k141_12781 | Not-determined |
| 18-103_k141_41672 | Low-quality |
| 18-103_k141_53243 | Low-quality |
| 18-103_k141_12784 | Not-determined |
| 18-103_k141_128208 | Low-quality |
| 18-103_k141_47552 | Not-determined |
| 18-103_k141_24461 | Not-determined |
| 18-103_k141_128215 | Not-determined |
| 18-103_k141_53252 | Not-determined |
| 18-103_k141_1523 | Not-determined |
| 18-103_k141_47555 | Not-determined |
| 18-103_k141_128219 | Not-determined |
| 18-103_k141_64711 | Not-determined |
| 18-103_k141_128222 | Not-determined |
| 18-103_k141_24467 | Not-determined |
| 18-103_k141_133766 | Low-quality |
| 18-103_k141_18550 | Low-quality |
| 18-103_k141_24470 | Not-determined |
| 18-103_k141_133769 | Low-quality |
| 18-103_k141_58962 | Not-determined |
| 18-103_k141_87894 | Not-determined |
| 18-103_k141_128231 | Not-determined |
| 18-103_k141_98993 | Not-determined |
| 18-103_k141_128234 | Low-quality |
| 18-103_k141_128236 | Not-determined |
| 18-103_k141_1535 | Not-determined |
| 18-103_k141_98998 | Not-determined |
| 18-103_k141_99000 | Not-determined |
| 18-103_k141_70407 | Not-determined |
| 18-103_k141_18558 | Not-determined |
| 18-103_k141_87903 | Not-determined |
| 18-103_k141_87908 | Not-determined |
| 18-103_k141_82080 | Not-determined |
| 18-103_k141_133789 | Not-determined |
| 18-103_k141_1540 | Low-quality |
| 18-103_k141_53277 | Low-quality |
| 18-103_k141_70419 | Not-determined |
| 18-103_k141_24498 | Not-determined |
| 18-103_k141_70421 | Not-determined |
| 18-103_k141_64732 | Not-determined |
| 18-103_k141_116564 | Not-determined |
| 18-103_k141_76244 | Not-determined |
| 18-103_k141_70426 | Not-determined |
| 18-103_k141_64738 | Not-determined |
| 18-103_k141_82091 | Not-determined |
| 18-103_k141_12824 | Not-determined |
| 18-103_k141_41703 | Not-determined |
| 18-103_k141_41704 | Not-determined |
| 18-103_k141_70429 | Not-determined |
| 18-103_k141_24505 | Not-determined |
| 18-103_k141_104916 | Not-determined |
| 18-103_k141_41709 | Not-determined |
| 18-103_k141_104919 | Not-determined |
| 18-103_k141_87924 | Not-determined |
| 18-103_k141_64753 | Not-determined |
| 18-103_k141_53282 | Low-quality |
| 18-103_k141_70436 | Not-determined |
| 18-103_k141_41719 | Not-determined |
| 18-103_k141_1552 | Not-determined |
| 18-103_k141_104922 | Not-determined |
| 18-103_k141_87926 | Not-determined |
| 18-103_k141_58986 | Not-determined |
| 18-103_k141_1556 | Not-determined |
| 18-103_k141_64762 | Not-determined |
| 18-103_k141_76252 | Not-determined |
| 18-103_k141_18589 | Low-quality |
| 18-103_k141_110746 | Not-determined |
| 18-103_k141_12842 | Not-determined |
| 18-103_k141_7234 | Not-determined |
| 18-103_k141_104931 | Not-determined |
| 18-103_k141_122165 | Not-determined |
| 18-103_k141_58994 | Not-determined |
| 18-103_k141_133820 | Not-determined |
| 18-103_k141_18597 | Low-quality |
| 18-103_k141_59003 | Not-determined |
| 18-103_k141_70451 | Not-determined |
| 18-103_k141_70452 | Not-determined |
| 18-103_k141_1570 | Not-determined |
| 18-103_k141_122177 | Not-determined |
| 18-103_k141_104936 | Not-determined |
| 18-103_k141_59007 | Not-determined |
| 18-103_k141_116579 | Not-determined |
| 18-103_k141_18608 | Not-determined |
| 18-103_k141_104947 | Not-determined |
| 18-103_k141_87938 | Not-determined |
| 18-103_k141_59018 | Low-quality |
| 18-103_k141_47601 | Not-determined |
| 18-103_k141_18617 | Not-determined |
| 18-103_k141_116594 | Not-determined |
| 18-103_k141_1576 | Not-determined |
| 18-103_k141_116595 | Not-determined |
| 18-103_k141_116600 | Not-determined |
| 18-103_k141_128248 | Not-determined |
| 18-103_k141_116601 | Not-determined |
| 18-103_k141_93490 | Not-determined |
| 18-103_k141_133845 | Not-determined |
| 18-103_k141_116606 | Not-determined |
| 18-103_k141_82111 | Not-determined |
| 18-103_k141_133848 | Not-determined |
| 18-103_k141_82112 | Not-determined |
| 18-103_k141_70478 | Not-determined |
| 18-103_k141_104963 | Not-determined |
| 18-103_k141_47619 | Not-determined |
| 18-103_k141_59033 | Not-determined |
| 18-103_k141_59035 | Low-quality |
| 18-103_k141_104969 | Not-determined |
| 18-103_k141_116618 | Not-determined |
| 18-103_k141_7280 | Not-determined |
| 18-103_k141_64802 | Not-determined |
| 18-103_k141_53340 | Not-determined |
| 18-103_k141_64807 | Not-determined |
| 18-103_k141_53342 | Not-determined |
| 18-103_k141_93512 | Low-quality |
| 18-103_k141_82121 | Not-determined |
| 18-103_k141_12902 | Not-determined |
| 18-103_k141_18635 | Not-determined |
| 18-103_k141_18636 | Low-quality |
| 18-103_k141_64812 | Not-determined |
| 18-103_k141_18638 | Not-determined |
| 18-103_k141_70498 | Not-determined |
| 18-103_k141_116623 | Not-determined |
| 18-103_k141_122208 | Not-determined |
| 18-103_k141_64818 | Not-determined |
| 18-103_k141_82130 | Not-determined |
| 18-103_k141_116627 | Not-determined |
| 18-103_k141_99060 | Not-determined |
| 18-103_k141_53355 | Not-determined |
| 18-103_k141_93527 | Not-determined |
| 18-103_k141_1603 | Not-determined |
| 18-103_k141_76270 | Not-determined |
| 18-103_k141_18652 | Not-determined |
| 18-103_k141_110779 | Not-determined |
| 18-103_k141_1608 | Not-determined |
| 18-103_k141_64829 | Not-determined |
| 18-103_k141_82133 | Not-determined |
| 18-103_k141_18657 | Not-determined |
| 18-103_k141_99062 | Low-quality |
| 18-103_k141_122220 | Not-determined |
| 18-103_k141_93532 | Not-determined |
| 18-103_k141_53364 | Not-determined |
| 18-103_k141_116631 | Not-determined |
| 18-103_k141_30130 | Not-determined |
| 18-103_k141_116632 | Not-determined |
| 18-103_k141_87946 | Not-determined |
| 18-103_k141_116633 | Low-quality |
| 18-103_k141_30132 | Not-determined |
| 18-103_k141_1614 | Not-determined |
| 18-103_k141_116634 | Not-determined |
| 18-103_k141_18665 | Not-determined |
| 18-103_k141_53369 | Not-determined |
| 18-103_k141_110783 | Not-determined |
| 18-103_k141_59065 | Not-determined |
| 18-103_k141_116639 | Not-determined |
| 18-103_k141_87951 | Not-determined |
| 18-103_k141_35965 | Low-quality |
| 18-103_k141_110789 | Not-determined |
| 18-103_k141_110790 | Not-determined |
| 18-103_k141_1621 | Not-determined |
| 18-103_k141_87952 | Low-quality |
| 18-103_k141_30141 | Low-quality |
| 18-103_k141_110794 | Not-determined |
| 18-103_k141_59072 | Not-determined |
| 18-103_k141_30144 | Not-determined |
| 18-103_k141_47640 | Not-determined |
| 18-103_k141_76285 | Not-determined |
| 18-103_k141_35977 | Not-determined |
| 18-103_k141_87959 | Not-determined |
| 18-103_k141_47642 | Low-quality |
| 18-103_k141_35979 | Not-determined |
| 18-103_k141_93549 | Low-quality |
| 18-103_k141_18682 | Not-determined |
| 18-103_k141_18683 | Low-quality |
| 18-103_k141_47646 | Low-quality |
| 18-103_k141_76291 | Not-determined |
| 18-103_k141_53384 | Not-determined |
| 18-103_k141_110802 | Not-determined |
| 18-103_k141_53385 | Not-determined |
| 18-103_k141_99073 | Not-determined |
| 18-103_k141_7331 | Not-determined |
| 18-103_k141_110815 | Not-determined |
| 18-103_k141_128307 | Low-quality |
| 18-103_k141_116671 | Not-determined |
| 18-103_k141_99085 | Not-determined |
| 18-103_k141_76298 | Not-determined |
| 18-103_k141_87988 | Not-determined |
| 18-103_k141_105043 | Not-determined |
| 18-103_k141_122254 | Not-determined |
| 18-103_k141_1650 | Not-determined |
| 18-103_k141_70541 | Low-quality |
| 18-103_k141_18694 | Low-quality |
| 18-103_k141_105051 | Not-determined |
| 18-103_k141_59099 | Not-determined |
| 18-103_k141_76313 | Not-determined |
| 18-103_k141_59101 | Not-determined |
| 18-103_k141_82175 | Not-determined |
| 18-103_k141_110837 | Not-determined |
| 18-103_k141_110840 | Not-determined |
| 18-103_k141_133897 | Not-determined |
| 18-103_k141_64860 | Not-determined |
| 18-103_k141_128329 | Not-determined |
| 18-103_k141_133898 | Low-quality |
| 18-103_k141_110842 | Not-determined |
| 18-103_k141_87994 | Not-determined |
| 18-103_k141_18709 | Not-determined |
| 18-103_k141_53414 | Not-determined |
| 18-103_k141_7339 | Not-determined |
| 18-103_k141_18710 | Not-determined |
| 18-103_k141_41764 | Not-determined |
| 18-103_k141_18712 | Not-determined |
| 18-103_k141_82184 | Not-determined |
| 18-103_k141_59112 | Not-determined |
| 18-103_k141_18713 | Not-determined |
| 18-103_k141_122264 | Not-determined |
| 18-103_k141_35988 | Not-determined |
| 18-103_k141_30184 | Not-determined |
| 18-103_k141_76324 | Not-determined |
| 18-103_k141_7345 | Not-determined |
| 18-103_k141_116693 | Low-quality |
| 18-103_k141_110845 | Not-determined |
| 18-103_k141_18720 | Not-determined |
| 18-103_k141_133904 | Not-determined |
| 18-103_k141_64866 | Not-determined |
| 18-103_k141_12950 | Not-determined |
| 18-103_k141_41771 | Not-determined |
| 18-103_k141_70563 | Not-determined |
| 18-103_k141_128335 | Not-determined |
| 18-103_k141_41774 | Not-determined |
| 18-103_k141_1680 | Not-determined |
| 18-103_k141_59123 | Not-determined |
| 18-103_k141_30193 | Not-determined |
| 18-103_k141_12958 | Not-determined |
| 18-103_k141_64874 | Not-determined |
| 18-103_k141_133911 | Low-quality |
| 18-103_k141_110855 | Not-determined |
| 18-103_k141_59135 | Not-determined |
| 18-103_k141_36010 | Not-determined |
| 18-103_k141_36011 | Not-determined |
| 18-103_k141_18743 | Low-quality |
| 18-103_k141_36013 | Low-quality |
| 18-103_k141_116712 | Not-determined |
| 18-103_k141_70585 | Not-determined |
| 18-103_k141_110865 | Not-determined |
| 18-103_k141_82214 | Not-determined |
| 18-103_k141_18750 | Not-determined |
| 18-103_k141_128344 | Not-determined |
| 18-103_k141_82217 | Not-determined |
| 18-103_k141_105093 | Not-determined |
| 18-103_k141_76345 | Low-quality |
| 18-103_k141_76346 | Not-determined |
| 18-103_k141_1692 | Not-determined |
| 18-103_k141_53447 | Not-determined |
| 18-103_k141_110873 | Not-determined |
| 18-103_k141_59156 | Not-determined |
| 18-103_k141_12983 | Not-determined |
| 18-103_k141_128347 | Low-quality |
| 18-103_k141_99111 | Not-determined |
| 18-103_k141_70600 | Not-determined |
| 18-103_k141_128349 | Not-determined |
| 18-103_k141_122294 | Not-determined |
| 18-103_k141_128352 | Not-determined |
| 18-103_k141_133922 | Not-determined |
| 18-103_k141_47723 | Low-quality |
| 18-103_k141_18774 | Not-determined |
| 18-103_k141_133929 | Not-determined |
| 18-103_k141_133930 | Not-determined |
| 18-103_k141_133932 | Not-determined |
| 18-103_k141_110881 | Not-determined |
| 18-103_k141_110883 | Not-determined |
| 18-103_k141_47729 | Not-determined |
| 18-103_k141_82228 | Not-determined |
| 18-103_k141_64908 | Not-determined |
| 18-103_k141_30208 | Not-determined |
| 18-103_k141_110889 | Not-determined |
| 18-103_k141_30210 | Low-quality |
| 18-103_k141_122303 | Not-determined |
| 18-103_k141_110895 | Not-determined |
| 18-103_k141_24583 | Not-determined |
| 18-103_k141_122306 | Not-determined |
| 18-103_k141_1717 | Low-quality |
| 18-103_k141_88046 | Not-determined |
| 18-103_k141_116730 | Not-determined |
| 18-103_k141_47739 | Low-quality |
| 18-103_k141_82236 | Low-quality |
| 18-103_k141_70625 | Not-determined |
| 18-103_k141_18792 | Not-determined |
| 18-103_k141_76357 | Not-determined |
| 18-103_k141_82243 | Not-determined |
| 18-103_k141_122315 | Not-determined |
| 18-103_k141_110911 | Not-determined |
| 18-103_k141_47748 | Not-determined |
| 18-103_k141_7355 | Not-determined |
| 18-103_k141_122318 | Low-quality |
| 18-103_k141_93581 | Low-quality |
| 18-103_k141_122320 | Not-determined |
| 18-103_k141_30231 | Low-quality |
| 18-103_k141_116740 | Not-determined |
| 18-103_k141_1725 | Not-determined |
| 18-103_k141_18799 | Not-determined |
| 18-103_k141_116744 | Not-determined |
| 18-103_k141_30235 | Not-determined |
| 18-103_k141_88056 | Not-determined |
| 18-103_k141_128375 | Not-determined |
| 18-103_k141_76364 | Not-determined |
| 18-103_k141_64933 | Not-determined |
| 18-103_k141_47761 | Not-determined |
| 18-103_k141_82255 | Not-determined |
| 18-103_k141_110925 | Not-determined |
| 18-103_k141_36058 | Not-determined |
| 18-103_k141_36059 | Not-determined |
| 18-103_k141_82259 | Not-determined |
| 18-103_k141_24613 | Not-determined |
| 18-103_k141_122339 | Not-determined |
| 18-103_k141_76371 | Not-determined |
| 18-103_k141_59193 | Not-determined |
| 18-103_k141_105134 | Not-determined |
| 18-103_k141_47766 | Low-quality |
| 18-103_k141_53465 | Not-determined |
| 18-103_k141_12995 | Not-determined |
| 18-103_k141_7371 | Not-determined |
| 18-103_k141_53470 | Not-determined |
| 18-103_k141_53471 | Not-determined |
| 18-103_k141_116751 | Not-determined |
| 18-103_k141_116752 | Not-determined |
| 18-103_k141_105146 | Not-determined |
| 18-103_k141_70639 | Low-quality |
| 18-103_k141_13006 | Not-determined |
| 18-103_k141_64954 | Low-quality |
| 18-103_k141_110944 | Not-determined |
| 18-103_k141_133992 | Not-determined |
| 18-103_k141_116763 | Not-determined |
| 18-103_k141_7382 | Not-determined |
| 18-103_k141_105152 | Not-determined |
| 18-103_k141_116765 | Not-determined |
| 18-103_k141_133994 | Not-determined |
| 18-103_k141_18810 | Not-determined |
| 18-103_k141_93609 | Not-determined |
| 18-103_k141_70649 | Not-determined |
| 18-103_k141_133996 | Not-determined |
| 18-103_k141_41875 | Not-determined |
| 18-103_k141_70651 | Not-determined |
| 18-103_k141_128404 | Not-determined |
| 18-103_k141_36082 | Not-determined |
| 18-103_k141_59200 | Not-determined |
| 18-103_k141_88076 | Not-determined |
| 18-103_k141_59202 | Not-determined |
| 18-103_k141_93616 | Not-determined |
| 18-103_k141_36084 | Not-determined |
| 18-103_k141_70658 | Not-determined |
| 18-103_k141_99118 | Not-determined |
| 18-103_k141_1744 | Not-determined |
| 18-103_k141_99119 | Not-determined |
| 18-103_k141_122354 | Not-determined |
| 18-103_k141_41879 | Not-determined |
| 18-103_k141_64967 | Not-determined |
| 18-103_k141_24652 | Not-determined |
| 18-103_k141_88083 | Not-determined |
| 18-103_k141_1749 | Not-determined |
| 18-103_k141_30263 | Not-determined |
| 18-103_k141_47790 | Not-determined |
| 18-103_k141_105161 | Not-determined |
| 18-103_k141_128417 | Not-determined |
| 18-103_k141_93621 | Not-determined |
| 18-103_k141_134003 | Not-determined |
| 18-103_k141_41890 | Not-determined |
| 18-103_k141_110955 | Not-determined |
| 18-103_k141_36097 | Not-determined |
| 18-103_k141_105163 | Not-determined |
| 18-103_k141_53481 | Low-quality |
| 18-103_k141_64981 | Not-determined |
| 18-103_k141_53482 | Not-determined |
| 18-103_k141_24663 | Not-determined |
| 18-103_k141_1765 | Not-determined |
| 18-103_k141_122373 | Not-determined |
| 18-103_k141_93630 | Low-quality |
| 18-103_k141_18820 | Not-determined |
| 18-103_k141_41902 | Not-determined |
| 18-103_k141_122379 | Not-determined |
| 18-103_k141_7395 | Not-determined |
| 18-103_k141_88096 | Not-determined |
| 18-103_k141_24674 | Low-quality |
| 18-103_k141_24676 | Not-determined |
| 18-103_k141_76389 | Not-determined |
| 18-103_k141_76391 | Low-quality |
| 18-103_k141_7400 | Low-quality |
| 18-103_k141_110967 | Not-determined |
| 18-103_k141_30275 | Not-determined |
| 18-103_k141_99144 | Low-quality |
| 18-103_k141_1776 | Low-quality |
| 18-103_k141_47808 | Not-determined |
| 18-103_k141_30282 | Not-determined |
| 18-103_k141_128437 | Not-determined |
| 18-103_k141_70671 | Not-determined |
| 18-103_k141_76406 | Not-determined |
| 18-103_k141_76407 | Not-determined |
| 18-103_k141_93646 | Not-determined |
| 18-103_k141_99159 | Low-quality |
| 18-103_k141_36106 | Not-determined |
| 18-103_k141_47813 | Not-determined |
| 18-103_k141_30289 | Low-quality |
| 18-103_k141_53498 | Not-determined |
| 18-103_k141_53499 | Not-determined |
| 18-103_k141_7413 | Not-determined |
| 18-103_k141_99163 | Not-determined |
| 18-103_k141_70684 | Low-quality |
| 18-103_k141_105192 | Not-determined |
| 18-103_k141_65015 | Low-quality |
| 18-103_k141_99166 | Not-determined |
| 18-103_k141_105196 | Not-determined |
| 18-103_k141_122400 | Not-determined |
| 18-103_k141_99167 | Low-quality |
| 18-103_k141_82333 | Not-determined |
| 18-103_k141_70687 | Not-determined |
| 18-103_k141_128444 | Not-determined |
| 18-103_k141_105208 | Not-determined |
| 18-103_k141_13043 | Not-determined |
| 18-103_k141_13045 | Not-determined |
| 18-103_k141_70697 | Not-determined |
| 18-103_k141_30308 | Not-determined |
| 18-103_k141_7428 | Not-determined |
| 18-103_k141_111000 | Not-determined |
| 18-103_k141_111002 | Not-determined |
| 18-103_k141_24699 | Not-determined |
| 18-103_k141_70701 | Not-determined |
| 18-103_k141_105214 | Not-determined |
| 18-103_k141_116810 | Not-determined |
| 18-103_k141_128460 | Not-determined |
| 18-103_k141_65020 | Not-determined |
| 18-103_k141_116812 | Low-quality |
| 18-103_k141_47826 | Low-quality |
| 18-103_k141_24704 | Not-determined |
| 18-103_k141_128465 | Not-determined |
| 18-103_k141_36125 | Low-quality |
| 18-103_k141_105224 | Not-determined |
| 18-103_k141_18871 | Not-determined |
| 18-103_k141_105226 | Not-determined |
| 18-103_k141_18874 | Not-determined |
| 18-103_k141_70721 | Not-determined |
| 18-103_k141_59235 | Low-quality |
| 18-103_k141_7438 | Low-quality |
| 18-103_k141_122432 | Not-determined |
| 18-103_k141_116827 | Not-determined |
| 18-103_k141_82350 | Low-quality |
| 18-103_k141_82355 | Not-determined |
| 18-103_k141_65041 | Not-determined |
| 18-103_k141_24727 | Low-quality |
| 18-103_k141_41951 | Not-determined |
| 18-103_k141_70727 | Not-determined |
| 18-103_k141_88138 | Not-determined |
| 18-103_k141_65049 | Not-determined |
| 18-103_k141_116830 | Not-determined |
| 18-103_k141_105251 | Not-determined |
| 18-103_k141_41960 | Not-determined |
| 18-103_k141_7456 | Not-determined |
| 18-103_k141_99207 | Not-determined |
| 18-103_k141_116840 | Not-determined |
| 18-103_k141_128487 | Low-quality |
| 18-103_k141_70741 | Not-determined |
| 18-103_k141_53534 | Not-determined |
| 18-103_k141_59247 | Not-determined |
| 18-103_k141_99216 | Not-determined |
| 18-103_k141_111021 | Not-determined |
| 18-103_k141_47847 | Not-determined |
| 18-103_k141_7471 | Low-quality |
| 18-103_k141_122456 | Not-determined |
| 18-103_k141_116852 | Low-quality |
| 18-103_k141_99225 | Not-determined |
| 18-103_k141_76457 | Not-determined |
| 18-103_k141_116856 | Not-determined |
| 18-103_k141_1798 | Not-determined |
| 18-103_k141_47856 | Not-determined |
| 18-103_k141_47857 | Not-determined |
| 18-103_k141_82383 | Not-determined |
| 18-103_k141_99232 | Not-determined |
| 18-103_k141_18908 | Not-determined |
| 18-103_k141_122462 | Not-determined |
| 18-103_k141_53555 | Not-determined |
| 18-103_k141_122463 | Not-determined |
| 18-103_k141_111045 | Not-determined |
| 18-103_k141_128509 | Not-determined |
| 18-103_k141_82388 | Not-determined |
| 18-103_k141_41992 | Low-quality |
| 18-103_k141_47863 | Not-determined |
| 18-103_k141_82390 | Not-determined |
| 18-103_k141_128512 | Not-determined |
| 18-103_k141_7492 | Not-determined |
| 18-103_k141_47865 | Not-determined |
| 18-103_k141_82394 | Not-determined |
| 18-103_k141_82398 | Not-determined |
| 18-103_k141_122476 | Not-determined |
| 18-103_k141_93699 | Not-determined |
| 18-103_k141_42002 | Not-determined |
| 18-103_k141_105284 | Not-determined |
| 18-103_k141_122484 | Not-determined |
| 18-103_k141_122485 | Low-quality |
| 18-103_k141_93707 | Not-determined |
| 18-103_k141_7501 | Not-determined |
| 18-103_k141_30353 | Not-determined |
| 18-103_k141_93713 | Not-determined |
| 18-103_k141_7502 | Not-determined |
| 18-103_k141_24783 | Not-determined |

| | |
|---|---|
| 18-103_k141_116862 | Not-determined |
| 18-103_k141_88167 | Not-determined |
| 18-103_k141_70754 | Not-determined |
| 18-103_k141_88169 | Not-determined |
| 18-103_k141_128524 | Not-determined |
| 18-103_k141_76477 | Not-determined |
| 18-103_k141_65090 | Not-determined |
| 18-103_k141_7508 | Not-determined |
| 18-103_k141_134080 | Not-determined |
| 18-103_k141_13116 | Not-determined |
| 18-103_k141_88173 | Not-determined |
| 18-103_k141_122500 | Not-determined |
| 18-103_k141_76481 | Not-determined |
| 18-103_k141_18928 | Not-determined |
| 18-103_k141_24787 | Not-determined |
| 18-103_k141_134088 | Not-determined |
| 18-103_k141_7519 | Not-determined |
| 18-103_k141_134095 | Not-determined |
| 18-103_k141_99265 | Not-determined |
| 18-103_k141_122513 | Not-determined |
| 18-103_k141_59267 | Not-determined |
| 18-103_k141_1849 | Not-determined |
| 18-103_k141_88184 | Not-determined |
| 18-103_k141_24799 | Low-quality |
| 18-103_k141_13131 | Not-determined |
| 18-103_k141_82405 | Not-determined |
| 18-103_k141_82407 | Not-determined |
| 18-103_k141_134099 | Low-quality |
| 18-103_k141_36193 | Not-determined |
| 18-103_k141_88188 | Not-determined |
| 18-103_k141_42023 | Not-determined |
| 18-103_k141_24802 | Low-quality |
| 18-103_k141_42024 | Not-determined |
| 18-103_k141_88193 | Not-determined |
| 18-103_k141_122525 | Not-determined |
| 18-103_k141_99284 | Not-determined |
| 18-103_k141_30381 | Not-determined |
| 18-103_k141_88198 | Low-quality |
| 18-103_k141_122531 | Low-quality |
| 18-103_k141_82414 | Not-determined |
| 18-103_k141_93738 | Not-determined |
| 18-103_k141_82418 | Not-determined |
| 18-103_k141_82421 | Not-determined |
| 18-103_k141_116895 | Not-determined |
| 18-103_k141_116898 | Not-determined |
| 18-103_k141_76503 | Not-determined |
| 18-103_k141_18949 | Not-determined |
| 18-103_k141_70792 | Not-determined |
| 18-103_k141_30403 | Not-determined |
| 18-103_k141_65117 | Not-determined |
| 18-103_k141_82432 | Not-determined |
| 18-103_k141_1863 | Not-determined |
| 18-103_k141_70794 | Not-determined |
| 18-103_k141_30405 | Not-determined |
| 18-103_k141_76518 | Not-determined |
| 18-103_k141_42027 | Not-determined |
| 18-103_k141_76519 | Not-determined |
| 18-103_k141_82443 | Not-determined |
| 18-103_k141_36234 | Low-quality |
| 18-103_k141_36235 | Not-determined |
| 18-103_k141_18970 | Not-determined |
| 18-103_k141_88203 | Not-determined |
| 18-103_k141_18976 | Not-determined |
| 18-103_k141_59314 | Low-quality |
| 18-103_k141_13165 | Not-determined |
| 18-103_k141_105342 | Not-determined |
| 18-103_k141_76526 | Low-quality |
| 18-103_k141_76527 | Not-determined |
| 18-103_k141_134133 | Not-determined |
| 18-103_k141_105345 | Not-determined |
| 18-103_k141_1880 | Low-quality |
| 18-103_k141_42035 | Not-determined |
| 18-103_k141_116910 | Low-quality |
| 18-103_k141_88212 | Not-determined |
| 18-103_k141_93769 | Not-determined |
| 18-103_k141_93770 | Not-determined |
| 18-103_k141_93771 | Not-determined |
| 18-103_k141_13173 | Not-determined |
| 18-103_k141_30415 | Not-determined |
| 18-103_k141_42043 | Not-determined |
| 18-103_k141_122564 | Not-determined |
| 18-103_k141_93774 | Not-determined |
| 18-103_k141_93778 | Not-determined |
| 18-103_k141_42051 | Not-determined |
| 18-103_k141_128569 | Low-quality |
| 18-103_k141_42053 | Not-determined |
| 18-103_k141_128571 | Not-determined |
| 18-103_k141_1889 | Not-determined |
| 18-103_k141_70820 | Low-quality |
| 18-103_k141_59335 | Not-determined |
| 18-103_k141_70826 | Not-determined |
| 18-103_k141_93782 | Low-quality |
| 18-103_k141_122579 | Not-determined |
| 18-103_k141_111079 | Not-determined |
| 18-103_k141_42059 | Low-quality |
| 18-103_k141_65151 | Not-determined |
| 18-103_k141_36259 | Not-determined |
| 18-103_k141_13189 | Not-determined |
| 18-103_k141_122585 | Not-determined |
| 18-103_k141_36266 | Not-determined |
| 18-103_k141_82457 | Not-determined |
| 18-103_k141_13199 | Not-determined |
| 18-103_k141_105358 | Not-determined |
| 18-103_k141_47895 | Not-determined |
| 18-103_k141_47896 | Not-determined |
| 18-103_k141_105360 | Not-determined |
| 18-103_k141_93792 | Not-determined |
| 18-103_k141_105361 | Not-determined |
| 18-103_k141_111084 | Not-determined |
| 18-103_k141_59347 | Not-determined |
| 18-103_k141_24867 | Not-determined |
| 18-103_k141_105363 | Not-determined |
| 18-103_k141_105366 | Not-determined |
| 18-103_k141_93797 | Low-quality |
| 18-103_k141_36272 | Not-determined |
| 18-103_k141_76558 | Not-determined |
| 18-103_k141_76559 | Not-determined |
| 18-103_k141_93798 | Low-quality |
| 18-103_k141_99318 | Low-quality |
| 18-103_k141_70844 | Not-determined |
| 18-103_k141_36280 | Not-determined |
| 18-103_k141_76569 | Not-determined |
| 18-103_k141_1908 | Low-quality |
| 18-103_k141_19016 | Not-determined |
| 18-103_k141_105377 | Not-determined |
| 18-103_k141_30455 | Low-quality |
| 18-103_k141_105379 | Not-determined |
| 18-103_k141_19020 | Not-determined |
| 18-103_k141_59365 | Not-determined |
| 18-103_k141_13220 | Not-determined |
| 18-103_k141_13221 | Not-determined |
| 18-103_k141_111092 | Not-determined |
| 18-103_k141_70849 | Not-determined |
| 18-103_k141_13222 | Low-quality |
| 18-103_k141_88249 | Low-quality |
| 18-103_k141_70853 | Not-determined |
| 18-103_k141_1917 | Not-determined |
| 18-103_k141_7570 | Not-determined |
| 18-103_k141_122610 | Not-determined |
| 18-103_k141_65164 | Not-determined |
| 18-103_k141_82480 | Not-determined |
| 18-103_k141_42097 | Not-determined |
| 18-103_k141_99338 | Not-determined |
| 18-103_k141_70862 | Not-determined |
| 18-103_k141_99341 | Not-determined |
| 18-103_k141_7581 | Not-determined |
| 18-103_k141_105393 | Not-determined |
| 18-103_k141_93804 | Not-determined |
| 18-103_k141_53603 | Not-determined |
| 18-103_k141_76577 | Low-quality |
| 18-103_k141_76578 | Not-determined |
| 18-103_k141_88254 | Low-quality |
| 18-103_k141_47921 | Not-determined |
| 18-103_k141_65177 | Not-determined |
| 18-103_k141_116960 | Low-quality |
| 18-103_k141_99350 | Not-determined |
| 18-103_k141_93811 | Not-determined |
| 18-103_k141_116965 | Not-determined |
| 18-103_k141_105402 | Not-determined |
| 18-103_k141_24910 | Not-determined |
| 18-103_k141_134193 | Not-determined |
| 18-103_k141_42117 | Low-quality |
| 18-103_k141_53621 | Low-quality |
| 18-103_k141_105407 | Not-determined |
| 18-103_k141_47939 | Not-determined |
| 18-103_k141_1941 | Not-determined |
| 18-103_k141_128597 | Not-determined |
| 18-103_k141_13247 | Not-determined |
| 18-103_k141_30478 | Not-determined |
| 18-103_k141_93827 | Not-determined |
| 18-103_k141_134202 | Low-quality |
| 18-103_k141_24914 | Not-determined |
| 18-103_k141_76590 | Not-determined |
| 18-103_k141_59400 | Not-determined |
| 18-103_k141_19048 | Not-determined |
| 18-103_k141_122636 | Not-determined |
| 18-103_k141_70888 | Low-quality |
| 18-103_k141_122638 | Not-determined |
| 18-103_k141_53630 | Not-determined |
| 18-103_k141_82513 | Not-determined |
| 18-103_k141_19054 | Not-determined |
| 18-103_k141_105425 | Not-determined |
| 18-103_k141_59407 | Not-determined |
| 18-103_k141_30496 | Not-determined |
| 18-103_k141_128608 | Not-determined |
| 18-103_k141_53638 | Not-determined |
| 18-103_k141_122648 | Not-determined |
| 18-103_k141_36297 | Low-quality |
| 18-103_k141_128611 | Not-determined |
| 18-103_k141_76611 | Not-determined |
| 18-103_k141_13253 | Not-determined |
| 18-103_k141_93842 | Not-determined |
| 18-103_k141_93843 | Not-determined |
| 18-103_k141_19069 | Not-determined |
| 18-103_k141_36303 | Not-determined |
| 18-103_k141_116997 | Not-determined |
| 18-103_k141_82523 | Low-quality |
| 18-103_k141_53648 | Not-determined |
| 18-103_k141_70909 | Not-determined |
| 18-103_k141_19078 | Not-determined |
| 18-103_k141_99366 | Not-determined |
| 18-103_k141_42137 | Not-determined |
| 18-103_k141_88278 | Not-determined |
| 18-103_k141_19081 | Not-determined |
| 18-103_k141_82530 | Not-determined |
| 18-103_k141_47970 | Not-determined |
| 18-103_k141_88286 | Not-determined |
| 18-103_k141_111128 | Not-determined |
| 18-103_k141_42145 | Low-quality |
| 18-103_k141_134226 | Not-determined |
| 18-103_k141_76632 | Not-determined |
| 18-103_k141_82539 | Not-determined |
| 18-103_k141_111133 | Not-determined |
| 18-103_k141_36307 | Not-determined |
| 18-103_k141_105444 | Low-quality |
| 18-103_k141_47973 | Not-determined |
| 18-103_k141_111136 | Not-determined |
| 18-103_k141_76634 | Not-determined |
| 18-103_k141_47977 | Not-determined |
| 18-103_k141_128629 | Not-determined |
| 18-103_k141_76637 | Not-determined |
| 18-103_k141_47981 | Not-determined |
| 18-103_k141_93875 | Not-determined |
| 18-103_k141_36315 | Not-determined |
| 18-103_k141_128633 | Not-determined |
| 18-103_k141_76642 | Not-determined |
| 18-103_k141_93878 | Low-quality |
| 18-103_k141_42153 | Not-determined |
| 18-103_k141_53668 | Not-determined |
| 18-103_k141_42156 | Not-determined |
| 18-103_k141_47982 | Low-quality |
| 18-103_k141_134240 | Not-determined |
| 18-103_k141_1998 | Not-determined |
| 18-103_k141_111145 | Not-determined |
| 18-103_k141_82551 | Not-determined |
| 18-103_k141_111147 | Not-determined |
| 18-103_k141_53671 | Not-determined |
| 18-103_k141_36317 | Low-quality |
| 18-103_k141_76648 | Not-determined |
| 18-103_k141_128646 | Not-determined |
| 18-103_k141_128647 | Not-determined |
| 18-103_k141_93890 | Not-determined |
| 18-103_k141_13292 | Not-determined |
| 18-103_k141_47994 | Not-determined |
| 18-103_k141_82561 | Low-quality |
| 18-103_k141_13294 | Not-determined |
| 18-103_k141_36325 | Not-determined |
| 18-103_k141_47997 | Not-determined |
| 18-103_k141_122691 | Not-determined |
| 18-103_k141_117032 | Not-determined |
| 18-103_k141_93898 | Not-determined |
| 18-103_k141_53683 | Not-determined |
| 18-103_k141_134249 | Low-quality |
| 18-103_k141_93906 | Not-determined |
| 18-103_k141_82573 | Not-determined |
| 18-103_k141_42178 | Low-quality |
| 18-103_k141_65241 | Low-quality |
| 18-103_k141_42180 | Not-determined |
| 18-103_k141_105463 | Not-determined |
| 18-103_k141_111169 | Not-determined |
| 18-103_k141_59465 | Not-determined |
| 18-103_k141_65252 | Not-determined |
| 18-103_k141_24981 | Not-determined |
| 18-103_k141_13344 | Not-determined |
| 18-103_k141_111184 | Not-determined |
| 18-103_k141_2038 | Not-determined |
| 18-103_k141_13347 | Not-determined |
| 18-103_k141_53687 | Not-determined |
| 18-103_k141_48034 | Not-determined |
| 18-103_k141_42209 | Not-determined |
| 18-103_k141_53689 | Not-determined |
| 18-103_k141_59485 | Low-quality |
| 18-103_k141_88335 | Not-determined |
| 18-103_k141_128686 | Not-determined |
| 18-103_k141_53691 | Not-determined |
| 18-103_k141_53694 | Not-determined |
| 18-103_k141_2059 | Not-determined |
| 18-103_k141_122750 | Not-determined |
| 18-103_k141_65284 | Not-determined |
| 18-103_k141_30579 | Not-determined |
| 18-103_k141_30581 | Not-determined |
| 18-103_k141_105507 | Not-determined |
| 18-103_k141_70963 | Not-determined |
| 18-103_k141_122756 | Not-determined |
| 18-103_k141_105509 | Not-determined |
| 18-103_k141_2065 | Not-determined |
| 18-103_k141_105511 | Not-determined |
| 18-103_k141_42216 | Not-determined |
| 18-103_k141_19105 | Low-quality |
| 18-103_k141_42221 | Not-determined |
| 18-103_k141_25007 | Not-determined |
| 18-103_k141_25009 | Not-determined |
| 18-103_k141_128697 | Not-determined |
| 18-103_k141_7622 | Low-quality |
| 18-103_k141_88350 | Not-determined |
| 18-103_k141_59503 | Not-determined |
| 18-103_k141_19114 | Not-determined |
| 18-103_k141_59504 | Not-determined |
| 18-103_k141_65302 | Not-determined |
| 18-103_k141_82613 | Not-determined |
| 18-103_k141_42234 | Not-determined |
| 18-103_k141_53704 | Low-quality |
| 18-103_k141_30600 | Not-determined |
| 18-103_k141_48071 | Not-determined |
| 18-103_k141_2081 | Not-determined |
| 18-103_k141_36379 | Not-determined |
| 18-103_k141_36381 | Not-determined |
| 18-103_k141_128705 | Low-quality |
| 18-103_k141_122786 | Low-quality |
| 18-103_k141_117067 | Not-determined |
| 18-103_k141_93952 | Low-quality |
| 18-103_k141_36385 | Low-quality |
| 18-103_k141_2087 | Not-determined |
| 18-103_k141_53709 | Not-determined |
| 18-103_k141_19126 | Low-quality |
| 18-103_k141_105535 | Not-determined |
| 18-103_k141_7637 | Not-determined |
| 18-103_k141_59519 | Not-determined |
| 18-103_k141_122793 | Not-determined |
| 18-103_k141_70980 | Not-determined |
| 18-103_k141_13378 | Not-determined |
| 18-103_k141_70984 | Not-determined |
| 18-103_k141_65313 | Not-determined |
| 18-103_k141_36389 | Not-determined |
| 18-103_k141_93962 | Not-determined |
| 18-103_k141_36390 | Not-determined |
| 18-103_k141_105546 | Not-determined |
| 18-103_k141_105549 | Not-determined |
| 18-103_k141_105551 | Not-determined |
| 18-103_k141_128723 | Not-determined |
| 18-103_k141_53720 | Not-determined |
| 18-103_k141_42240 | Not-determined |
| 18-103_k141_48097 | Not-determined |
| 18-103_k141_70994 | Not-determined |
| 18-103_k141_111195 | Not-determined |
| 18-103_k141_48101 | Not-determined |
| 18-103_k141_70999 | Not-determined |
| 18-103_k141_2109 | Not-determined |
| 18-103_k141_105559 | Not-determined |
| 18-103_k141_99419 | Not-determined |
| 18-103_k141_111202 | Not-determined |
| 18-103_k141_36410 | Not-determined |
| 18-103_k141_36411 | Not-determined |
| 18-103_k141_42249 | Not-determined |
| 18-103_k141_53726 | Not-determined |
| 18-103_k141_71002 | Low-quality |
| 18-103_k141_30623 | Not-determined |
| 18-103_k141_88386 | Not-determined |
| 18-103_k141_13402 | Not-determined |
| 18-103_k141_7655 | Low-quality |
| 18-103_k141_71006 | Not-determined |
| 18-103_k141_30630 | Not-determined |
| 18-103_k141_71010 | Not-determined |
| 18-103_k141_36414 | Not-determined |
| 18-103_k141_99433 | Not-determined |
| 18-103_k141_65342 | Low-quality |
| 18-103_k141_93990 | Not-determined |
| 18-103_k141_82654 | Not-determined |
| 18-103_k141_111217 | Not-determined |
| 18-103_k141_42255 | Not-determined |
| 18-103_k141_99442 | Not-determined |
| 18-103_k141_71012 | Low-quality |
| 18-103_k141_71014 | Not-determined |
| 18-103_k141_82666 | Not-determined |
| 18-103_k141_13416 | Not-determined |
| 18-103_k141_122836 | Not-determined |
| 18-103_k141_117108 | Not-determined |
| 18-103_k141_71023 | Not-determined |
| 18-103_k141_105581 | Not-determined |
| 18-103_k141_117112 | Not-determined |
| 18-103_k141_19178 | Low-quality |
| 18-103_k141_19179 | Not-determined |
| 18-103_k141_76705 | Not-determined |
| 18-103_k141_94000 | Not-determined |
| 18-103_k141_36428 | Not-determined |
| 18-103_k141_76712 | Low-quality |
| 18-103_k141_53747 | Not-determined |
| 18-103_k141_53748 | Not-determined |
| 18-103_k141_88408 | Not-determined |
| 18-103_k141_111235 | Not-determined |
| 18-103_k141_76721 | Not-determined |
| 18-103_k141_99466 | Not-determined |
| 18-103_k141_13431 | Not-determined |
| 18-103_k141_36435 | Not-determined |
| 18-103_k141_88417 | Not-determined |
| 18-103_k141_122852 | Not-determined |
| 18-103_k141_71049 | Not-determined |
| 18-103_k141_2151 | Not-determined |
| 18-103_k141_134336 | Not-determined |
| 18-103_k141_94029 | Not-determined |
| 18-103_k141_42293 | Not-determined |
| 18-103_k141_94035 | Not-determined |
| 18-103_k141_111258 | Not-determined |
| 18-103_k141_53757 | Not-determined |
| 18-103_k141_111262 | Not-determined |
| 18-103_k141_48143 | Not-determined |
| 18-103_k141_99482 | Not-determined |
| 18-103_k141_111265 | Not-determined |
| 18-103_k141_76745 | Low-quality |
| 18-103_k141_117121 | Not-determined |
| 18-103_k141_48149 | Not-determined |
| 18-103_k141_71068 | Not-determined |
| 18-103_k141_71071 | Not-determined |
| 18-103_k141_99491 | Low-quality |
| 18-103_k141_19198 | Not-determined |
| 18-103_k141_59545 | Not-determined |
| 18-103_k141_42314 | Not-determined |
| 18-103_k141_13453 | Not-determined |
| 18-103_k141_71076 | Low-quality |
| 18-103_k141_99496 | Not-determined |
| 18-103_k141_7724 | Not-determined |
| 18-103_k141_111276 | Not-determined |
| 18-103_k141_48159 | Not-determined |
| 18-103_k141_99501 | Not-determined |
| 18-103_k141_111281 | Not-determined |
| 18-103_k141_36465 | Not-determined |
| 18-103_k141_42332 | Not-determined |
| 18-103_k141_94073 | Not-determined |
| 18-103_k141_128758 | Not-determined |
| 18-103_k141_13465 | Not-determined |
| 18-103_k141_71088 | Low-quality |
| 18-103_k141_122897 | Not-determined |
| 18-103_k141_48168 | Not-determined |
| 18-103_k141_7745 | Not-determined |
| 18-103_k141_65352 | Not-determined |
| 18-103_k141_65353 | Not-determined |
| 18-103_k141_134371 | Not-determined |
| 18-103_k141_53783 | Not-determined |
| 18-103_k141_53784 | Not-determined |
| 18-103_k141_36480 | Not-determined |
| 18-103_k141_53786 | Low-quality |
| 18-103_k141_105589 | Not-determined |
| 18-103_k141_71099 | Low-quality |
| 18-103_k141_122899 | Not-determined |
| 18-103_k141_134384 | Not-determined |
| 18-103_k141_122905 | Not-determined |
| 18-103_k141_117145 | Not-determined |
| 18-103_k141_134388 | Not-determined |
| 18-103_k141_99527 | Not-determined |
| 18-103_k141_13476 | Not-determined |
| 18-103_k141_13477 | Not-determined |
| 18-103_k141_53803 | Not-determined |
| 18-103_k141_53805 | Low-quality |
| 18-103_k141_76782 | Not-determined |
| 18-103_k141_36499 | Not-determined |
| 18-103_k141_88460 | Not-determined |
| 18-103_k141_13483 | Not-determined |
| 18-103_k141_65362 | Low-quality |
| 18-103_k141_88464 | Not-determined |
| 18-103_k141_122921 | Not-determined |
| 18-103_k141_13487 | Low-quality |
| 18-103_k141_71110 | Not-determined |
| 18-103_k141_65365 | Not-determined |
| 18-103_k141_13492 | Not-determined |
| 18-103_k141_117165 | Not-determined |
| 18-103_k141_25111 | Not-determined |
| 18-103_k141_36508 | Not-determined |
| 18-103_k141_128792 | Low-quality |
| 18-103_k141_117170 | Not-determined |
| 18-103_k141_82697 | Not-determined |
| 18-103_k141_111289 | Not-determined |
| 18-103_k141_7764 | Not-determined |
| 18-103_k141_13509 | Not-determined |
| 18-103_k141_42339 | Not-determined |
| 18-103_k141_82699 | Not-determined |
| 18-103_k141_25127 | Not-determined |
| 18-103_k141_117186 | Not-determined |
| 18-103_k141_111300 | Not-determined |
| 18-103_k141_117187 | Not-determined |
| 18-103_k141_82709 | Not-determined |
| 18-103_k141_105638 | Not-determined |
| 18-103_k141_19241 | Not-determined |
| 18-103_k141_94098 | Not-determined |
| 18-103_k141_88482 | Not-determined |
| 18-103_k141_99537 | Not-determined |
| 18-103_k141_134418 | Not-determined |
| 18-103_k141_134420 | Low-quality |
| 18-103_k141_117201 | Not-determined |
| 18-103_k141_128817 | Not-determined |
| 18-103_k141_71149 | Not-determined |
| 18-103_k141_105646 | Not-determined |
| 18-103_k141_25142 | Not-determined |
| 18-103_k141_117208 | Not-determined |
| 18-103_k141_48208 | Not-determined |
| 18-103_k141_53844 | Low-quality |
| 18-103_k141_88502 | Not-determined |
| 18-103_k141_13540 | Not-determined |
| 18-103_k141_82735 | Not-determined |
| 18-103_k141_117220 | Not-determined |
| 18-103_k141_59614 | Not-determined |
| 18-103_k141_99555 | Not-determined |
| 18-103_k141_122961 | Low-quality |
| 18-103_k141_88506 | Not-determined |
| 18-103_k141_94123 | Not-determined |
| 18-103_k141_99557 | Not-determined |
| 18-103_k141_128840 | Not-determined |
| 18-103_k141_19262 | Low-quality |
| 18-103_k141_42361 | Not-determined |
| 18-103_k141_122969 | Not-determined |
| 18-103_k141_42362 | Not-determined |
| 18-103_k141_65417 | Not-determined |
| 18-103_k141_105673 | Not-determined |
| 18-103_k141_76815 | Not-determined |
| 18-103_k141_128853 | Not-determined |
| 18-103_k141_99564 | Not-determined |
| 18-103_k141_117232 | Not-determined |
| 18-103_k141_65420 | Not-determined |
| 18-103_k141_30738 | Low-quality |
| 18-103_k141_105682 | Low-quality |
| 18-103_k141_13555 | Not-determined |
| 18-103_k141_134454 | Not-determined |
| 18-103_k141_19265 | Not-determined |
| 18-103_k141_65425 | Not-determined |
| 18-103_k141_7801 | Not-determined |
| 18-103_k141_25189 | Low-quality |
| 18-103_k141_134460 | Not-determined |
| 18-103_k141_19271 | Not-determined |
| 18-103_k141_59641 | Not-determined |
| 18-103_k141_94156 | Not-determined |
| 18-103_k141_48220 | Not-determined |
| 18-103_k141_82757 | Not-determined |
| 18-103_k141_117242 | Not-determined |
| 18-103_k141_94165 | Not-determined |
| 18-103_k141_88547 | Not-determined |
| 18-103_k141_7808 | Not-determined |
| 18-103_k141_99590 | Not-determined |
| 18-103_k141_117246 | Not-determined |
| 18-103_k141_117248 | Not-determined |
| 18-103_k141_42388 | Not-determined |
| 18-103_k141_48234 | Not-determined |
| 18-103_k141_134485 | Not-determined |
| 18-103_k141_94179 | Not-determined |
| 18-103_k141_94181 | Not-determined |
| 18-103_k141_53896 | Not-determined |
| 18-103_k141_7820 | Not-determined |
| 18-103_k141_99607 | Not-determined |
| 18-103_k141_94186 | Not-determined |
| 18-103_k141_48249 | Not-determined |
| 18-103_k141_82791 | Not-determined |
| 18-103_k141_99616 | Not-determined |
| 18-103_k141_99617 | Not-determined |
| 18-103_k141_99618 | Not-determined |
| 18-103_k141_88573 | Low-quality |
| 18-103_k141_48256 | Not-determined |
| 18-103_k141_13594 | Not-determined |
| 18-103_k141_36619 | Not-determined |
| 18-103_k141_53906 | Not-determined |
| 18-103_k141_111341 | Low-quality |
| 18-103_k141_2330 | Not-determined |
| 18-103_k141_111342 | Low-quality |
| 18-103_k141_99632 | Not-determined |
| 18-103_k141_19315 | Not-determined |
| 18-103_k141_117271 | Not-determined |
| 18-103_k141_134517 | Not-determined |
| 18-103_k141_13601 | Low-quality |
| 18-103_k141_128866 | Not-determined |
| 18-103_k141_59661 | Not-determined |
| 18-103_k141_36632 | Not-determined |
| 18-103_k141_88591 | Not-determined |
| 18-103_k141_7839 | Not-determined |
| 18-103_k141_19324 | Not-determined |
| 18-103_k141_123025 | Low-quality |
| 18-103_k141_25214 | Low-quality |
| 18-103_k141_134531 | Not-determined |
| 18-103_k141_48270 | Not-determined |
| 18-103_k141_123034 | Not-determined |
| 18-103_k141_25220 | Not-determined |
| 18-103_k141_19331 | Not-determined |
| 18-103_k141_25221 | Not-determined |
| 18-103_k141_76835 | Not-determined |
| 18-103_k141_13614 | Not-determined |
| 18-103_k141_76837 | Not-determined |
| 18-103_k141_111359 | Not-determined |
| 18-103_k141_76838 | Not-determined |
| 18-103_k141_134539 | Not-determined |
| 18-103_k141_65471 | Not-determined |
| 18-103_k141_82805 | Not-determined |
| 18-103_k141_128886 | Not-determined |
| 18-103_k141_99649 | Not-determined |
| 18-103_k141_36645 | Not-determined |
| 18-103_k141_36646 | Not-determined |
| 18-103_k141_111371 | Not-determined |
| 18-103_k141_76846 | Not-determined |
| 18-103_k141_19343 | Not-determined |
| 18-103_k141_105689 | Not-determined |
| 18-103_k141_42435 | Not-determined |
| 18-103_k141_111374 | Not-determined |
| 18-103_k141_123044 | Not-determined |
| 18-103_k141_2356 | Not-determined |

| | |
|---|---|
| 18-103_k141_111375 | Not-determined |
| 18-103_k141_19355 | Not-determined |
| 18-103_k141_82810 | Not-determined |
| 18-103_k141_105695 | Not-determined |
| 18-103_k141_36659 | Not-determined |
| 18-103_k141_36660 | Not-determined |
| 18-103_k141_82815 | Not-determined |
| 18-103_k141_25238 | Not-determined |
| 18-103_k141_19367 | Not-determined |
| 18-103_k141_82822 | Not-determined |
| 18-103_k141_111383 | Not-determined |
| 18-103_k141_111386 | Low-quality |
| 18-103_k141_82832 | Not-determined |
| 18-103_k141_134574 | Not-determined |
| 18-103_k141_2366 | Not-determined |
| 18-103_k141_7873 | Not-determined |
| 18-103_k141_42449 | Not-determined |
| 18-103_k141_105713 | Not-determined |
| 18-103_k141_88618 | Not-determined |
| 18-103_k141_7879 | Not-determined |
| 18-103_k141_94227 | Not-determined |
| 18-103_k141_128921 | Not-determined |
| 18-103_k141_59684 | Not-determined |
| 18-103_k141_36676 | Not-determined |
| 18-103_k141_7886 | Not-determined |
| 18-103_k141_53929 | Low-quality |
| 18-103_k141_94238 | Not-determined |
| 18-103_k141_42462 | Not-determined |
| 18-103_k141_82851 | Low-quality |
| 18-103_k141_88634 | Not-determined |
| 18-103_k141_134584 | Low-quality |
| 18-103_k141_42467 | Not-determined |
| 18-103_k141_76878 | Low-quality |
| 18-103_k141_123065 | Not-determined |
| 18-103_k141_42469 | Low-quality |
| 18-103_k141_42472 | Not-determined |
| 18-103_k141_134588 | Not-determined |
| 18-103_k141_128946 | Not-determined |
| 18-103_k141_88642 | Not-determined |
| 18-103_k141_105726 | Not-determined |
| 18-103_k141_134591 | Not-determined |
| 18-103_k141_53944 | Low-quality |
| 18-103_k141_99688 | Not-determined |
| 18-103_k141_59695 | Not-determined |
| 18-103_k141_59696 | Not-determined |
| 18-103_k141_48295 | Not-determined |
| 18-103_k141_94248 | Not-determined |
| 18-103_k141_65501 | Not-determined |
| 18-103_k141_111413 | Not-determined |
| 18-103_k141_25277 | Not-determined |
| 18-103_k141_82864 | Not-determined |
| 18-103_k141_71249 | Not-determined |
| 18-103_k141_25279 | Not-determined |
| 18-103_k141_82866 | Not-determined |
| 18-103_k141_48309 | Not-determined |
| 18-103_k141_76902 | Not-determined |
| 18-103_k141_30839 | Low-quality |
| 18-103_k141_59707 | Not-determined |
| 18-103_k141_2393 | Low-quality |
| 18-103_k141_94257 | Not-determined |
| 18-103_k141_94259 | Not-determined |
| 18-103_k141_111420 | Not-determined |
| 18-103_k141_134600 | Low-quality |
| 18-103_k141_19400 | Not-determined |
| 18-103_k141_82872 | Not-determined |
| 18-103_k141_36708 | Not-determined |
| 18-103_k141_65513 | Not-determined |
| 18-103_k141_65514 | Not-determined |
| 18-103_k141_48311 | Low-quality |
| 18-103_k141_99725 | Not-determined |
| 18-103_k141_65517 | Not-determined |
| 18-103_k141_99726 | Not-determined |
| 18-103_k141_30852 | Not-determined |
| 18-103_k141_53970 | Low-quality |
| 18-103_k141_13661 | Not-determined |
| 18-103_k141_105751 | Not-determined |
| 18-103_k141_88653 | Not-determined |
| 18-103_k141_88656 | Not-determined |
| 18-103_k141_48322 | Not-determined |
| 18-103_k141_30862 | Not-determined |
| 18-103_k141_48324 | Not-determined |
| 18-103_k141_13666 | Not-determined |
| 18-103_k141_2411 | Not-determined |
| 18-103_k141_88659 | Not-determined |
| 18-103_k141_123093 | Low-quality |
| 18-103_k141_111426 | Low-quality |
| 18-103_k141_88660 | Not-determined |
| 18-103_k141_76932 | Not-determined |
| 18-103_k141_123096 | Not-determined |
| 18-103_k141_76934 | Not-determined |
| 18-103_k141_123098 | Not-determined |
| 18-103_k141_105762 | Low-quality |
| 18-103_k141_42496 | Not-determined |
| 18-103_k141_105765 | Not-determined |
| 18-103_k141_134619 | Not-determined |
| 18-103_k141_30866 | Not-determined |
| 18-103_k141_42498 | Not-determined |
| 18-103_k141_111436 | Not-determined |
| 18-103_k141_134625 | Not-determined |
| 18-103_k141_71276 | Not-determined |
| 18-103_k141_71277 | Not-determined |
| 18-103_k141_53987 | Not-determined |
| 18-103_k141_88675 | Not-determined |
| 18-103_k141_65542 | Not-determined |
| 18-103_k141_105772 | Low-quality |
| 18-103_k141_134626 | Not-determined |
| 18-103_k141_59739 | Low-quality |
| 18-103_k141_65546 | Not-determined |
| 18-103_k141_65547 | Not-determined |
| 18-103_k141_25317 | Not-determined |
| 18-103_k141_36736 | Not-determined |
| 18-103_k141_36737 | Not-determined |
| 18-103_k141_53993 | Not-determined |
| 18-103_k141_25321 | Not-determined |
| 18-103_k141_82909 | Not-determined |
| 18-103_k141_94267 | Not-determined |
| 18-103_k141_134633 | Low-quality |
| 18-103_k141_134634 | Not-determined |
| 18-103_k141_88689 | Not-determined |
| 18-103_k141_82912 | Not-determined |
| 18-103_k141_76946 | Not-determined |
| 18-103_k141_36744 | Not-determined |
| 18-103_k141_2425 | Low-quality |
| 18-103_k141_71286 | Not-determined |
| 18-103_k141_111450 | Not-determined |
| 18-103_k141_30891 | Not-determined |
| 18-103_k141_105784 | Not-determined |
| 18-103_k141_123103 | Low-quality |
| 18-103_k141_123105 | Not-determined |
| 18-103_k141_13685 | Not-determined |
| 18-103_k141_123108 | Not-determined |
| 18-103_k141_94287 | Not-determined |
| 18-103_k141_88697 | Not-determined |
| 18-103_k141_71289 | Not-determined |
| 18-103_k141_105790 | Not-determined |
| 18-103_k141_129001 | Not-determined |
| 18-103_k141_71292 | Low-quality |
| 18-103_k141_48351 | Not-determined |
| 18-103_k141_30903 | Not-determined |
| 18-103_k141_117364 | Low-quality |
| 18-103_k141_2430 | Not-determined |
| 18-103_k141_48358 | Not-determined |
| 18-103_k141_2434 | Not-determined |
| 18-103_k141_30911 | Not-determined |
| 18-103_k141_123109 | Low-quality |
| 18-103_k141_129009 | Not-determined |
| 18-103_k141_129011 | Not-determined |
| 18-103_k141_111476 | Not-determined |
| 18-103_k141_54016 | Not-determined |
| 18-103_k141_88711 | Not-determined |
| 18-103_k141_54017 | Not-determined |
| 18-103_k141_117374 | Not-determined |
| 18-103_k141_76988 | Not-determined |
| 18-103_k141_36766 | Not-determined |
| 18-103_k141_129013 | Not-determined |
| 18-103_k141_65555 | Not-determined |
| 18-103_k141_19455 | Not-determined |
| 18-103_k141_48371 | Not-determined |
| 18-103_k141_71309 | Not-determined |
| 18-103_k141_94300 | Not-determined |
| 18-103_k141_76994 | Not-determined |
| 18-103_k141_88716 | Not-determined |
| 18-103_k141_2449 | Not-determined |
| 18-103_k141_48377 | Not-determined |
| 18-103_k141_76997 | Not-determined |
| 18-103_k141_129020 | Not-determined |
| 18-103_k141_94305 | Not-determined |
| 18-103_k141_105809 | Not-determined |
| 18-103_k141_117383 | Not-determined |
| 18-103_k141_36777 | Low-quality |
| 18-103_k141_54027 | Not-determined |
| 18-103_k141_59767 | Not-determined |
| 18-103_k141_19464 | Not-determined |
| 18-103_k141_65565 | Not-determined |
| 18-103_k141_99778 | Low-quality |
| 18-103_k141_99781 | Not-determined |
| 18-103_k141_111499 | Low-quality |
| 18-103_k141_99782 | Not-determined |
| 18-103_k141_2461 | Not-determined |
| 18-103_k141_65569 | Not-determined |
| 18-103_k141_82931 | Not-determined |
| 18-103_k141_25363 | Low-quality |
| 18-103_k141_99787 | Not-determined |
| 18-103_k141_82935 | Not-determined |
| 18-103_k141_30923 | Not-determined |
| 18-103_k141_25365 | Not-determined |
| 18-103_k141_99789 | Not-determined |
| 18-103_k141_2464 | Not-determined |
| 18-103_k141_30927 | Not-determined |
| 18-103_k141_123128 | Not-determined |
| 18-103_k141_25370 | Not-determined |
| 18-103_k141_30929 | Not-determined |
| 18-103_k141_7962 | Not-determined |
| 18-103_k141_25372 | Not-determined |
| 18-103_k141_30933 | Not-determined |
| 18-103_k141_65573 | Not-determined |
| 18-103_k141_25374 | Not-determined |
| 18-103_k141_30936 | Not-determined |
| 18-103_k141_19478 | Not-determined |
| 18-103_k141_94327 | Not-determined |
| 18-103_k141_2469 | Not-determined |
| 18-103_k141_59783 | Not-determined |
| 18-103_k141_77025 | Not-determined |
| 18-103_k141_65581 | Not-determined |
| 18-103_k141_88746 | Not-determined |
| 18-103_k141_111518 | Not-determined |
| 18-103_k141_65582 | Not-determined |
| 18-103_k141_2472 | Low-quality |
| 18-103_k141_71341 | Not-determined |
| 18-103_k141_65583 | Low-quality |
| 18-103_k141_123141 | Not-determined |
| 18-103_k141_25387 | Not-determined |
| 18-103_k141_105831 | Not-determined |
| 18-103_k141_36805 | Low-quality |
| 18-103_k141_129029 | Not-determined |
| 18-103_k141_82952 | Not-determined |
| 18-103_k141_59790 | Not-determined |
| 18-103_k141_7982 | Not-determined |
| 18-103_k141_65587 | Not-determined |
| 18-103_k141_123149 | Not-determined |
| 18-103_k141_48406 | Not-determined |
| 18-103_k141_59793 | Not-determined |
| 18-103_k141_54043 | Not-determined |
| 18-103_k141_65591 | Not-determined |
| 18-103_k141_25391 | Not-determined |
| 18-103_k141_134715 | Not-determined |
| 18-103_k141_2480 | Not-determined |
| 18-103_k141_99810 | Not-determined |
| 18-103_k141_36813 | Not-determined |
| 18-103_k141_77030 | Not-determined |
| 18-103_k141_30949 | Not-determined |
| 18-103_k141_30950 | Not-determined |
| 18-103_k141_48416 | Not-determined |
| 18-103_k141_19500 | Not-determined |
| 18-103_k141_30952 | Low-quality |
| 18-103_k141_82967 | Not-determined |
| 18-103_k141_88763 | Low-quality |
| 18-103_k141_117415 | Not-determined |
| 18-103_k141_111533 | Not-determined |
| 18-103_k141_117417 | Not-determined |
| 18-103_k141_59803 | Not-determined |
| 18-103_k141_82974 | Not-determined |
| 18-103_k141_88769 | Not-determined |
| 18-103_k141_71365 | Not-determined |
| 18-103_k141_88772 | Not-determined |
| 18-103_k141_111539 | Not-determined |
| 18-103_k141_54054 | Low-quality |
| 18-103_k141_82977 | Not-determined |
| 18-103_k141_99830 | Not-determined |
| 18-103_k141_117422 | Not-determined |
| 18-103_k141_129059 | Not-determined |
| 18-103_k141_134736 | Not-determined |
| 18-103_k141_65619 | Not-determined |
| 18-103_k141_36837 | Not-determined |
| 18-103_k141_77050 | Not-determined |
| 18-103_k141_88782 | Not-determined |
| 18-103_k141_88783 | Not-determined |
| 18-103_k141_36845 | Not-determined |
| 18-103_k141_19509 | Not-determined |
| 18-103_k141_8003 | Not-determined |
| 18-103_k141_77056 | Not-determined |
| 18-103_k141_134741 | Not-determined |
| 18-103_k141_65623 | Not-determined |
| 18-103_k141_117436 | Not-determined |
| 18-103_k141_71372 | Not-determined |
| 18-103_k141_36849 | Not-determined |
| 18-103_k141_65627 | Low-quality |
| 18-103_k141_54064 | Not-determined |
| 18-103_k141_94369 | Not-determined |
| 18-103_k141_8009 | Not-determined |
| 18-103_k141_129072 | Not-determined |
| 18-103_k141_65630 | Not-determined |
| 18-103_k141_36857 | Not-determined |
| 18-103_k141_30973 | Not-determined |
| 18-103_k141_129075 | Not-determined |
| 18-103_k141_19516 | Not-determined |
| 18-103_k141_99841 | Not-determined |
| 18-103_k141_129076 | Not-determined |
| 18-103_k141_129083 | Not-determined |
| 18-103_k141_2501 | Low-quality |
| 18-103_k141_88800 | Not-determined |
| 18-103_k141_117451 | Not-determined |
| 18-103_k141_71390 | Not-determined |
| 18-103_k141_77072 | Not-determined |
| 18-103_k141_129086 | Not-determined |
| 18-103_k141_77073 | Not-determined |
| 18-103_k141_8022 | Not-determined |
| 18-103_k141_65646 | Not-determined |
| 18-103_k141_8027 | Not-determined |
| 18-103_k141_48436 | Low-quality |
| 18-103_k141_88816 | Not-determined |
| 18-103_k141_2514 | Low-quality |
| 18-103_k141_65649 | Low-quality |
| 18-103_k141_111577 | Not-determined |
| 18-103_k141_2516 | Low-quality |
| 18-103_k141_134751 | Not-determined |
| 18-103_k141_117475 | Not-determined |
| 18-103_k141_36882 | Not-determined |
| 18-103_k141_42545 | Not-determined |
| 18-103_k141_36884 | Not-determined |
| 18-103_k141_59847 | Not-determined |
| 18-103_k141_36885 | Not-determined |
| 18-103_k141_19540 | Low-quality |
| 18-103_k141_42551 | Low-quality |
| 18-103_k141_117482 | Not-determined |
| 18-103_k141_134762 | Low-quality |
| 18-103_k141_54088 | Not-determined |
| 18-103_k141_83024 | Not-determined |
| 18-103_k141_42559 | Low-quality |
| 18-103_k141_77087 | Not-determined |
| 18-103_k141_19551 | Low-quality |
| 18-103_k141_77089 | Low-quality |
| 18-103_k141_65656 | Not-determined |
| 18-103_k141_19553 | Not-determined |
| 18-103_k141_54092 | Not-determined |
| 18-103_k141_94397 | Low-quality |
| 18-103_k141_77093 | Not-determined |
| 18-103_k141_94403 | Not-determined |
| 18-103_k141_13740 | Not-determined |
| 18-103_k141_36901 | Not-determined |
| 18-103_k141_25425 | Not-determined |
| 18-103_k141_129116 | Not-determined |
| 18-103_k141_117498 | Not-determined |
| 18-103_k141_117501 | Not-determined |
| 18-103_k141_134780 | Not-determined |
| 18-103_k141_111600 | Not-determined |
| 18-103_k141_77101 | Not-determined |
| 18-103_k141_83034 | Low-quality |
| 18-103_k141_88818 | Not-determined |
| 18-103_k141_83035 | Low-quality |
| 18-103_k141_31017 | Low-quality |
| 18-103_k141_134792 | Not-determined |
| 18-103_k141_31020 | Not-determined |
| 18-103_k141_48459 | Not-determined |
| 18-103_k141_77109 | Not-determined |
| 18-103_k141_111617 | Not-determined |
| 18-103_k141_129132 | Not-determined |
| 18-103_k141_2549 | Not-determined |
| 18-103_k141_88830 | Not-determined |
| 18-103_k141_19569 | Not-determined |
| 18-103_k141_13750 | Not-determined |
| 18-103_k141_48466 | Not-determined |
| 18-103_k141_129135 | Not-determined |
| 18-103_k141_88837 | Not-determined |
| 18-103_k141_48468 | Not-determined |
| 18-103_k141_105878 | Not-determined |
| 18-103_k141_59886 | Not-determined |
| 18-103_k141_19585 | Not-determined |
| 18-103_k141_48473 | Not-determined |
| 18-103_k141_129141 | Low-quality |
| 18-103_k141_88847 | Low-quality |
| 18-103_k141_71399 | Low-quality |
| 18-103_k141_31033 | Not-determined |
| 18-103_k141_105888 | Not-determined |
| 18-103_k141_111643 | Not-determined |
| 18-103_k141_88852 | Low-quality |
| 18-103_k141_99896 | Not-determined |
| 18-103_k141_48481 | Not-determined |
| 18-103_k141_99899 | Not-determined |
| 18-103_k141_59898 | Not-determined |
| 18-103_k141_123178 | Not-determined |
| 18-103_k141_123182 | Not-determined |
| 18-103_k141_129157 | Not-determined |
| 18-103_k141_2583 | Not-determined |
| 18-103_k141_13774 | Not-determined |
| 18-103_k141_111649 | Not-determined |
| 18-103_k141_123189 | Not-determined |
| 18-103_k141_8065 | Not-determined |
| 18-103_k141_71410 | Not-determined |
| 18-103_k141_88860 | Not-determined |
| 18-103_k141_19590 | Not-determined |
| 18-103_k141_71413 | Not-determined |
| 18-103_k141_71414 | Not-determined |
| 18-103_k141_123196 | Not-determined |
| 18-103_k141_117532 | Not-determined |
| 18-103_k141_77151 | Not-determined |
| 18-103_k141_88864 | Low-quality |
| 18-103_k141_99920 | Not-determined |
| 18-103_k141_123198 | Not-determined |
| 18-103_k141_129163 | Not-determined |
| 18-103_k141_2596 | Not-determined |
| 18-103_k141_99925 | Not-determined |
| 18-103_k141_42622 | Not-determined |
| 18-103_k141_19601 | Not-determined |
| 18-103_k141_36949 | Not-determined |
| 18-103_k141_65673 | Low-quality |
| 18-103_k141_65675 | Not-determined |
| 18-103_k141_94446 | Not-determined |
| 18-103_k141_13795 | Not-determined |
| 18-103_k141_54121 | Not-determined |
| 18-103_k141_83060 | Not-determined |
| 18-103_k141_59936 | Not-determined |
| 18-103_k141_54124 | Not-determined |
| 18-103_k141_59937 | Not-determined |
| 18-103_k141_94455 | Not-determined |
| 18-103_k141_54132 | Not-determined |
| 18-103_k141_111683 | Not-determined |
| 18-103_k141_48525 | Not-determined |
| 18-103_k141_88873 | Low-quality |
| 18-103_k141_117551 | Not-determined |
| 18-103_k141_94458 | Not-determined |
| 18-103_k141_36980 | Not-determined |
| 18-103_k141_129188 | Not-determined |
| 18-103_k141_88875 | Not-determined |
| 18-103_k141_36984 | Not-determined |
| 18-103_k141_48534 | Not-determined |
| 18-103_k141_83084 | Not-determined |
| 18-103_k141_48535 | Not-determined |
| 18-103_k141_54145 | Not-determined |
| 18-103_k141_83085 | Not-determined |
| 18-103_k141_59957 | Not-determined |
| 18-103_k141_59958 | Not-determined |
| 18-103_k141_134835 | Not-determined |
| 18-103_k141_19629 | Not-determined |
| 18-103_k141_83091 | Not-determined |
| 18-103_k141_42631 | Low-quality |
| 18-103_k141_99946 | Not-determined |
| 18-103_k141_111699 | Not-determined |
| 18-103_k141_8109 | Not-determined |
| 18-103_k141_129195 | Not-determined |
| 18-103_k141_99950 | Low-quality |
| 18-103_k141_111706 | Not-determined |
| 18-103_k141_54155 | Not-determined |
| 18-103_k141_77161 | Not-determined |
| 18-103_k141_19639 | Not-determined |
| 18-103_k141_83097 | Not-determined |
| 18-103_k141_77164 | Not-determined |
| 18-103_k141_48554 | Not-determined |
| 18-103_k141_8117 | Not-determined |
| 18-103_k141_134851 | Not-determined |
| 18-103_k141_19645 | Not-determined |
| 18-103_k141_31100 | Not-determined |
| 18-103_k141_83109 | Low-quality |
| 18-103_k141_83110 | Not-determined |
| 18-103_k141_111721 | Low-quality |
| 18-103_k141_94472 | Not-determined |
| 18-103_k141_54164 | Not-determined |
| 18-103_k141_25468 | Low-quality |
| 18-103_k141_19651 | Not-determined |
| 18-103_k141_94475 | Not-determined |
| 18-103_k141_94477 | Not-determined |
| 18-103_k141_111724 | Not-determined |
| 18-103_k141_19655 | Not-determined |
| 18-103_k141_37015 | Not-determined |
| 18-103_k141_19656 | Not-determined |
| 18-103_k141_111727 | Not-determined |
| 18-103_k141_77185 | Not-determined |
| 18-103_k141_2613 | Not-determined |
| 18-103_k141_65684 | Low-quality |
| 18-103_k141_2615 | Not-determined |
| 18-103_k141_13804 | Not-determined |
| 18-103_k141_48569 | Not-determined |
| 18-103_k141_8136 | Low-quality |
| 18-103_k141_13810 | Not-determined |
| 18-103_k141_123238 | Low-quality |
| 18-103_k141_42655 | Not-determined |
| 18-103_k141_134875 | Not-determined |
| 18-103_k141_59986 | Not-determined |
| 18-103_k141_123241 | Low-quality |
| 18-103_k141_94489 | Not-determined |
| 18-103_k141_123242 | Not-determined |
| 18-103_k141_134880 | Low-quality |
| 18-103_k141_59989 | Not-determined |
| 18-103_k141_99965 | Not-determined |
| 18-103_k141_13816 | Not-determined |
| 18-103_k141_54181 | Not-determined |
| 18-103_k141_8141 | Low-quality |
| 18-103_k141_13818 | Not-determined |
| 18-103_k141_88930 | Not-determined |
| 18-103_k141_99966 | Low-quality |
| 18-103_k141_99968 | Not-determined |
| 18-103_k141_54186 | Not-determined |
| 18-103_k141_111741 | Not-determined |
| 18-103_k141_31117 | Not-determined |
| 18-103_k141_37036 | Low-quality |
| 18-103_k141_105950 | Not-determined |
| 18-103_k141_25494 | Not-determined |
| 18-103_k141_83136 | Not-determined |
| 18-103_k141_13829 | Not-determined |
| 18-103_k141_54194 | Low-quality |
| 18-103_k141_54196 | Not-determined |
| 18-103_k141_71495 | Not-determined |
| 18-103_k141_54198 | Not-determined |
| 18-103_k141_25502 | Not-determined |
| 18-103_k141_99983 | Not-determined |
| 18-103_k141_111749 | Low-quality |
| 18-103_k141_60017 | Not-determined |
| 18-102-final-viral-combined_k141_71652\|\|full | Medium-quality |
| 18-103_k141_60022 | Not-determined |
| 18-103_k141_19691 | Not-determined |
| 18-103_k141_117587 | Not-determined |
| 18-103_k141_99991 | Not-determined |
| 18-103_k141_99993 | Not-determined |
| 18-103_k141_99995 | Not-determined |
| 18-103_k141_19696 | Not-determined |
| 18-103_k141_83174 | Not-determined |
| 18-103_k141_100001 | Not-determined |
| 18-103_k141_31157 | Not-determined |
| 18-103_k141_25526 | Not-determined |
| 18-103_k141_19710 | Not-determined |
| 18-103_k141_48589 | Not-determined |
| 18-103_k141_65721 | Not-determined |
| 18-103_k141_19716 | Not-determined |
| 18-103_k141_123286 | Not-determined |
| 18-103_k141_100007 | Low-quality |
| 18-103_k141_31164 | Not-determined |
| 18-103_k141_65726 | Low-quality |
| 18-103_k141_65727 | Not-determined |
| 18-103_k141_134907 | Not-determined |
| 18-103_k141_60044 | Not-determined |
| 18-103_k141_129243 | Not-determined |
| 18-103_k141_83183 | Not-determined |
| 18-103_k141_8158 | Low-quality |
| 18-103_k141_94530 | Not-determined |
| 18-103_k141_77213 | Not-determined |
| 18-103_k141_19737 | Not-determined |
| 18-103_k141_94532 | Not-determined |
| 18-103_k141_77218 | Not-determined |
| 18-103_k141_105988 | Low-quality |
| 18-103_k141_129251 | Not-determined |
| 18-103_k141_65729 | Low-quality |
| 18-103_k141_88976 | Not-determined |
| 18-103_k141_31184 | Not-determined |
| 18-103_k141_94542 | Not-determined |
| 18-103_k141_31189 | Not-determined |
| 18-103_k141_65736 | Not-determined |
| 18-103_k141_123301 | Not-determined |
| 18-103_k141_31192 | Not-determined |
| 18-103_k141_19743 | Not-determined |
| 18-103_k141_129258 | Not-determined |
| 18-103_k141_111788 | Not-determined |
| 18-103_k141_129260 | Not-determined |
| 18-103_k141_129261 | Not-determined |
| 18-103_k141_117633 | Low-quality |
| 18-103_k141_54212 | Low-quality |
| 18-103_k141_8183 | Low-quality |
| 18-103_k141_37093 | Not-determined |
| 18-103_k141_19756 | Not-determined |
| 18-103_k141_31202 | Not-determined |
| 18-103_k141_77239 | Not-determined |
| 18-103_k141_31204 | Not-determined |
| 18-103_k141_31209 | Not-determined |
| 18-103_k141_94560 | Not-determined |
| 18-103_k141_77243 | Not-determined |
| 18-103_k141_123313 | Not-determined |
| 18-103_k141_123315 | Low-quality |
| 18-103_k141_88989 | Not-determined |
| 18-103_k141_94569 | Not-determined |
| 18-103_k141_8194 | Low-quality |
| 18-103_k141_129275 | Not-determined |
| 18-103_k141_105998 | Not-determined |
| 18-103_k141_13846 | Not-determined |
| 18-103_k141_8197 | Not-determined |
| 18-103_k141_54224 | Low-quality |
| 18-103_k141_111801 | Not-determined |
| 18-103_k141_117638 | Low-quality |
| 18-103_k141_31218 | Not-determined |
| 18-103_k141_77256 | Not-determined |
| 18-103_k141_106000 | Not-determined |
| 18-103_k141_100038 | Not-determined |
| 18-103_k141_77260 | Not-determined |
| 18-103_k141_111805 | Low-quality |
| 18-103_k141_111806 | Not-determined |
| 18-103_k141_106005 | Not-determined |
| 18-103_k141_60085 | Not-determined |
| 18-103_k141_77265 | Not-determined |
| 18-103_k141_129289 | Low-quality |
| 18-103_k141_129290 | Not-determined |
| 18-103_k141_54234 | Not-determined |
| 18-103_k141_123334 | Not-determined |
| 18-103_k141_54235 | Not-determined |
| 18-103_k141_13859 | Low-quality |
| 18-103_k141_123337 | Not-determined |
| 18-103_k141_2717 | Not-determined |
| 18-103_k141_89011 | Not-determined |
| 18-103_k141_2720 | Not-determined |
| 18-103_k141_54239 | Not-determined |
| 18-103_k141_65763 | Not-determined |
| 18-103_k141_54244 | Low-quality |
| 18-103_k141_71542 | Low-quality |
| 18-103_k141_106009 | Not-determined |
| 18-103_k141_65769 | Not-determined |
| 18-103_k141_111819 | Not-determined |
| 18-103_k141_19777 | Not-determined |
| 18-103_k141_100043 | Not-determined |
| 18-103_k141_25589 | Not-determined |
| 18-103_k141_129308 | Not-determined |
| 18-103_k141_13884 | Not-determined |
| 18-103_k141_31241 | Not-determined |

| | |
|---|---|
| 18-103_k141_19782 | Not-determined |
| 18-103_k141_100052 | Not-determined |
| 18-103_k141_42693 | Low-quality |
| 18-103_k141_111831 | Not-determined |
| 18-103_k141_2727 | Not-determined |
| 18-103_k141_129312 | Not-determined |
| 18-103_k141_71552 | Not-determined |
| 18-103_k141_117664 | Not-determined |
| 18-103_k141_71554 | Not-determined |
| 18-103_k141_71555 | Not-determined |
| 18-103_k141_25608 | Not-determined |
| 18-103_k141_65779 | Not-determined |
| 18-103_k141_54262 | Not-determined |
| 18-103_k141_19793 | Not-determined |
| 18-103_k141_31256 | Not-determined |
| 18-103_k141_60099 | Not-determined |
| 18-103_k141_123360 | Not-determined |
| 18-103_k141_111841 | Not-determined |
| 18-103_k141_25617 | Not-determined |
| 18-103_k141_106017 | Low-quality |
| 18-103_k141_100065 | Low-quality |
| 18-103_k141_2740 | Low-quality |
| 18-103_k141_25622 | Not-determined |
| 18-103_k141_19800 | Not-determined |
| 18-103_k141_2742 | Not-determined |
| 18-103_k141_65785 | Low-quality |
| 18-103_k141_2744 | Not-determined |
| 18-103_k141_117683 | Not-determined |
| 18-103_k141_71565 | Not-determined |
| 18-103_k141_83252 | Low-quality |
| 18-103_k141_71569 | Low-quality |
| 18-103_k141_8242 | Low-quality |
| 18-103_k141_19814 | Not-determined |
| 18-103_k141_65804 | Not-determined |
| 18-103_k141_83264 | Not-determined |
| 18-103_k141_65805 | Not-determined |
| 18-103_k141_37117 | Low-quality |
| 18-103_k141_42721 | Not-determined |
| 18-103_k141_60109 | Not-determined |
| 18-103_k141_123384 | Not-determined |
| 18-103_k141_31277 | Not-determined |
| 18-103_k141_106037 | Not-determined |
| 18-103_k141_48622 | Not-determined |
| 18-103_k141_60117 | Low-quality |
| 18-103_k141_19827 | Not-determined |
| 18-103_k141_48632 | Not-determined |
| 18-103_k141_19831 | Low-quality |
| 18-103_k141_89062 | Not-determined |
| 18-103_k141_31290 | Not-determined |
| 18-103_k141_129348 | Not-determined |
| 18-103_k141_2773 | Low-quality |
| 18-103_k141_106058 | Not-determined |
| 18-103_k141_94630 | Not-determined |
| 18-103_k141_31295 | Low-quality |
| 18-103_k141_37139 | Not-determined |
| 18-103_k141_54310 | Not-determined |
| 18-103_k141_123407 | Not-determined |
| 18-103_k141_48648 | Not-determined |
| 18-103_k141_77321 | Low-quality |
| 18-103_k141_42740 | Not-determined |
| 18-103_k141_94636 | Not-determined |
| 18-103_k141_129366 | Low-quality |
| 18-103_k141_54322 | Not-determined |
| 18-103_k141_111885 | Low-quality |
| 18-103_k141_83300 | Not-determined |
| 18-103_k141_89083 | Low-quality |
| 18-103_k141_71603 | Not-determined |
| 18-103_k141_19851 | Not-determined |
| 18-103_k141_77330 | Not-determined |
| 18-103_k141_71608 | Not-determined |
| 18-103_k141_89091 | Low-quality |
| 18-103_k141_77337 | Not-determined |
| 18-103_k141_54335 | Not-determined |
| 18-103_k141_8283 | Not-determined |
| 18-103_k141_8284 | Not-determined |
| 18-103_k141_2789 | Not-determined |
| 18-103_k141_25684 | Not-determined |
| 18-103_k141_8289 | Not-determined |
| 18-103_k141_117698 | Not-determined |
| 18-103_k141_94654 | Not-determined |
| 18-103_k141_19861 | Low-quality |
| 18-103_k141_77346 | Not-determined |
| 18-103_k141_106089 | Not-determined |
| 18-103_k141_111902 | Not-determined |
| 18-103_k141_100125 | Not-determined |
| 18-103_k141_83321 | Low-quality |
| 18-103_k141_54345 | Not-determined |
| 18-103_k141_48667 | Not-determined |
| 18-103_k141_2806 | Not-determined |
| 18-103_k141_37159 | Not-determined |
| 18-103_k141_94665 | Not-determined |
| 18-103_k141_42758 | Not-determined |
| 18-103_k141_134983 | Not-determined |
| 18-103_k141_77351 | Low-quality |
| 18-103_k141_83324 | Not-determined |
| 18-103_k141_106099 | Low-quality |
| 18-103_k141_94667 | Not-determined |
| 18-103_k141_2813 | Not-determined |
| 18-103_k141_111913 | Low-quality |
| 18-103_k141_89115 | Not-determined |
| 18-103_k141_13938 | Low-quality |
| 18-103_k141_94671 | Not-determined |
| 18-103_k141_31340 | Not-determined |
| 18-103_k141_89118 | Not-determined |
| 18-103_k141_48679 | Not-determined |
| 18-103_k141_111915 | Low-quality |
| 18-103_k141_2817 | Low-quality |
| 18-103_k141_54350 | Low-quality |
| 18-103_k141_111918 | Not-determined |
| 18-103_k141_25697 | Not-determined |
| 18-103_k141_60161 | Not-determined |
| 18-103_k141_83330 | Low-quality |
| 18-103_k141_54354 | Not-determined |
| 18-103_k141_123444 | Not-determined |
| 18-103_k141_31349 | Not-determined |
| 18-103_k141_48682 | Not-determined |
| 18-103_k141_2824 | Not-determined |
| 18-103_k141_65849 | Not-determined |
| 18-103_k141_94681 | Not-determined |
| 18-103_k141_100144 | Not-determined |
| 18-103_k141_25706 | Not-determined |
| 18-103_k141_77364 | Not-determined |
| 18-103_k141_123452 | Not-determined |
| 18-103_k141_31354 | Not-determined |
| 18-103_k141_65854 | Low-quality |
| 18-103_k141_129400 | Not-determined |
| 18-103_k141_111919 | Not-determined |
| 18-103_k141_100146 | Low-quality |
| 18-103_k141_60164 | Not-determined |
| 18-103_k141_42775 | Not-determined |
| 18-103_k141_94685 | Not-determined |
| 18-103_k141_83348 | Not-determined |
| 18-103_k141_129404 | Not-determined |
| 18-103_k141_31368 | Not-determined |
| 18-103_k141_25709 | Not-determined |
| 18-103_k141_100157 | Not-determined |
| 18-103_k141_13964 | Not-determined |
| 18-103_k141_77379 | Not-determined |
| 18-103_k141_129410 | Low-quality |
| 18-103_k141_100164 | Not-determined |
| 18-103_k141_77384 | Not-determined |
| 18-103_k141_25717 | Not-determined |
| 18-103_k141_100166 | Low-quality |
| 18-103_k141_106131 | Not-determined |
| 18-103_k141_94702 | Not-determined |
| 18-103_k141_135004 | Not-determined |
| 18-103_k141_77390 | Not-determined |
| 18-103_k141_8307 | Not-determined |
| 18-103_k141_94704 | Low-quality |
| 18-103_k141_117740 | Not-determined |
| 18-103_k141_31388 | Not-determined |
| 18-103_k141_42800 | Not-determined |
| 18-103_k141_25730 | Not-determined |
| 18-103_k141_31389 | Low-quality |
| 18-103_k141_129428 | Low-quality |
| 18-103_k141_117746 | Not-determined |
| 18-103_k141_117748 | Not-determined |
| 18-103_k141_106140 | Not-determined |
| 18-103_k141_77394 | Not-determined |
| 18-103_k141_100179 | Low-quality |
| 18-103_k141_111935 | Low-quality |
| 18-103_k141_123480 | Not-determined |
| 18-103_k141_111936 | Not-determined |
| 18-103_k141_94721 | Not-determined |
| 18-103_k141_25741 | Not-determined |
| 18-103_k141_48714 | Low-quality |
| 18-103_k141_123491 | Not-determined |
| 18-103_k141_8329 | Not-determined |
| 18-103_k141_25744 | Not-determined |
| 18-103_k141_54402 | Not-determined |
| 18-103_k141_94732 | Not-determined |
| 18-103_k141_54404 | Low-quality |
| 18-103_k141_25746 | Not-determined |
| 18-103_k141_129438 | Low-quality |
| 18-103_k141_83373 | Not-determined |
| 18-103_k141_111947 | Not-determined |
| 18-103_k141_83375 | Not-determined |
| 18-103_k141_37204 | Not-determined |
| 18-103_k141_83377 | Not-determined |
| 18-103_k141_83378 | Low-quality |
| 18-103_k141_117766 | Not-determined |
| 18-103_k141_106169 | Low-quality |
| 18-103_k141_135034 | Not-determined |
| 18-103_k141_65887 | Not-determined |
| 18-103_k141_25759 | Low-quality |
| 18-103_k141_77417 | Not-determined |
| 18-103_k141_100203 | Not-determined |
| 18-103_k141_106173 | Not-determined |
| 18-103_k141_42816 | Not-determined |
| 18-103_k141_106176 | Not-determined |
| 18-103_k141_65894 | Not-determined |
| 18-103_k141_129456 | Not-determined |
| 18-103_k141_19885 | Not-determined |
| 18-103_k141_48736 | Not-determined |
| 18-103_k141_100214 | Not-determined |
| 18-103_k141_19891 | Not-determined |
| 18-103_k141_19893 | Not-determined |
| 18-103_k141_2875 | Not-determined |
| 18-103_k141_117785 | Not-determined |
| 18-103_k141_37225 | Not-determined |
| 18-103_k141_48746 | Not-determined |
| 18-103_k141_111963 | Not-determined |
| 18-103_k141_106190 | Not-determined |
| 18-103_k141_31425 | Not-determined |
| 18-103_k141_94759 | Not-determined |
| 18-103_k141_135047 | Not-determined |
| 18-103_k141_77438 | Not-determined |
| 18-103_k141_19903 | Not-determined |
| 18-103_k141_123515 | Not-determined |
| 18-103_k141_65910 | Not-determined |
| 18-103_k141_31426 | Not-determined |
| 18-103_k141_83395 | Low-quality |
| 18-103_k141_2886 | Not-determined |
| 18-103_k141_48754 | Not-determined |
| 18-103_k141_54439 | Not-determined |
| 18-103_k141_14021 | Low-quality |
| 18-103_k141_2891 | Not-determined |
| 18-103_k141_42838 | Not-determined |
| 18-103_k141_31434 | Not-determined |
| 18-103_k141_117804 | Not-determined |
| 18-103_k141_60225 | Low-quality |
| 18-103_k141_25780 | Not-determined |
| 18-103_k141_129472 | Not-determined |
| 18-103_k141_123518 | Low-quality |
| 18-103_k141_117807 | Not-determined |
| 18-103_k141_42848 | Not-determined |
| 18-103_k141_2906 | Not-determined |
| 18-103_k141_100237 | Low-quality |
| 18-103_k141_60234 | Not-determined |
| 18-103_k141_89147 | Not-determined |
| 18-103_k141_94779 | Not-determined |
| 18-103_k141_48765 | Not-determined |
| 18-103_k141_37255 | Not-determined |
| 18-103_k141_37256 | Not-determined |
| 18-103_k141_2919 | Not-determined |
| 18-103_k141_89156 | Not-determined |
| 18-103_k141_71680 | Not-determined |
| 18-103_k141_60248 | Not-determined |
| 18-103_k141_2925 | Not-determined |
| 18-103_k141_25795 | Not-determined |
| 18-103_k141_89161 | Not-determined |
| 18-103_k141_25798 | Not-determined |
| 18-103_k141_100254 | Low-quality |
| 18-103_k141_2928 | Not-determined |
| 18-103_k141_8346 | Not-determined |
| 18-103_k141_100259 | Not-determined |
| 18-103_k141_31450 | Not-determined |
| 18-103_k141_129489 | Not-determined |
| 18-103_k141_8349 | Not-determined |
| 18-103_k141_42857 | Not-determined |
| 18-103_k141_31452 | Low-quality |
| 18-103_k141_19950 | Not-determined |
| 18-103_k141_65923 | Not-determined |
| 18-103_k141_135077 | Not-determined |
| 18-103_k141_135080 | Not-determined |
| 18-103_k141_54464 | Not-determined |
| 18-103_k141_25815 | Not-determined |
| 18-103_k141_65929 | Not-determined |
| 18-103_k141_19962 | Not-determined |
| 18-103_k141_8363 | Not-determined |
| 18-103_k141_54471 | Not-determined |
| 18-103_k141_83405 | Not-determined |
| 18-103_k141_135091 | Not-determined |
| 18-103_k141_2946 | Low-quality |
| 18-103_k141_48796 | Not-determined |
| 18-103_k141_60274 | Not-determined |
| 18-103_k141_48797 | Not-determined |
| 18-103_k141_65935 | Not-determined |
| 18-103_k141_89186 | Not-determined |
| 18-103_k141_14054 | Not-determined |
| 18-103_k141_112009 | Not-determined |
| 18-103_k141_37277 | Not-determined |
| 18-103_k141_112010 | Not-determined |
| 18-103_k141_31478 | Not-determined |
| 18-103_k141_129516 | Low-quality |
| 18-103_k141_129519 | Not-determined |
| 18-103_k141_65942 | Low-quality |
| 18-103_k141_135104 | Not-determined |
| 18-103_k141_129522 | Low-quality |
| 18-103_k141_117830 | Not-determined |
| 18-103_k141_83422 | Not-determined |
| 18-103_k141_37287 | Not-determined |
| 18-103_k141_135111 | Not-determined |
| 18-103_k141_48806 | Not-determined |
| 18-103_k141_37290 | Not-determined |
| 18-103_k141_100294 | Low-quality |
| 18-103_k141_54495 | Low-quality |
| 18-103_k141_94808 | Not-determined |
| 18-103_k141_65955 | Not-determined |
| 18-103_k141_94810 | Not-determined |
| 18-103_k141_14075 | Not-determined |
| 18-103_k141_37300 | Not-determined |
| 18-103_k141_14076 | Not-determined |
| 18-103_k141_83429 | Not-determined |
| 18-103_k141_37306 | Not-determined |
| 18-103_k141_42903 | Low-quality |
| 18-103_k141_94816 | Not-determined |
| 18-103_k141_2969 | Not-determined |
| 18-103_k141_65965 | Low-quality |
| 18-103_k141_42911 | Not-determined |
| 18-103_k141_48822 | Not-determined |
| 18-103_k141_60293 | Not-determined |
| 18-103_k141_94821 | Not-determined |
| 18-103_k141_20005 | Not-determined |
| 18-103_k141_100303 | Not-determined |
| 18-103_k141_65966 | Low-quality |
| 18-103_k141_117839 | Not-determined |
| 18-103_k141_54501 | Not-determined |
| 18-103_k141_60302 | Not-determined |
| 18-103_k141_20016 | Not-determined |
| 18-103_k141_42921 | Not-determined |
| 18-103_k141_25823 | Not-determined |
| 18-103_k141_106219 | Not-determined |
| 18-103_k141_20025 | Not-determined |
| 18-103_k141_94828 | Low-quality |
| 18-103_k141_106223 | Not-determined |
| 18-103_k141_37320 | Not-determined |
| 18-103_k141_117856 | Not-determined |
| 18-103_k141_83451 | Not-determined |
| 18-103_k141_2992 | Not-determined |
| 18-103_k141_83452 | Not-determined |
| 18-103_k141_54504 | Low-quality |
| 18-103_k141_112041 | Low-quality |
| 18-103_k141_8416 | Not-determined |
| 18-103_k141_42941 | Not-determined |
| 18-103_k141_42944 | Not-determined |
| 18-103_k141_25833 | Not-determined |
| 18-103_k141_71696 | Not-determined |
| 18-103_k141_106239 | Not-determined |
| 18-103_k141_100333 | Not-determined |
| 18-103_k141_42946 | Not-determined |
| 18-103_k141_37344 | Not-determined |
| 18-103_k141_83466 | Not-determined |
| 18-103_k141_135134 | Not-determined |
| 18-103_k141_83468 | Not-determined |
| 18-103_k141_25842 | Low-quality |
| 18-103_k141_89222 | Low-quality |
| 18-103_k141_83470 | Not-determined |
| 18-103_k141_54512 | Low-quality |
| 18-103_k141_123600 | Not-determined |
| 18-103_k141_123603 | Not-determined |
| 18-103_k141_117876 | Not-determined |
| 18-103_k141_8433 | Low-quality |
| 18-103_k141_123606 | Not-determined |
| 18-103_k141_94856 | Not-determined |
| 18-103_k141_129560 | Not-determined |
| 18-103_k141_83485 | Not-determined |
| 18-103_k141_135148 | Not-determined |
| 18-103_k141_83487 | Not-determined |
| 18-103_k141_83489 | Not-determined |
| 18-103_k141_83491 | Not-determined |
| 18-103_k141_3010 | Not-determined |
| 18-103_k141_25854 | Low-quality |
| 18-103_k141_89231 | Not-determined |
| 18-103_k141_37355 | Not-determined |
| 18-103_k141_135154 | Not-determined |
| 18-103_k141_54527 | Not-determined |
| 18-103_k141_89238 | Not-determined |
| 18-103_k141_135156 | Not-determined |
| 18-103_k141_106280 | Not-determined |
| 18-103_k141_135159 | Not-determined |
| 18-103_k141_106282 | Not-determined |
| 18-103_k141_3022 | Not-determined |
| 18-103_k141_8453 | Not-determined |
| 18-103_k141_31516 | Not-determined |
| 18-103_k141_8457 | Not-determined |
| 18-103_k141_117903 | Not-determined |
| 18-103_k141_3029 | Not-determined |
| 18-103_k141_71709 | Low-quality |
| 18-103_k141_8461 | Not-determined |
| 18-103_k141_135168 | Not-determined |
| 18-103_k141_54547 | Not-determined |
| 18-103_k141_31523 | Low-quality |
| 18-103_k141_66010 | Low-quality |
| 18-103_k141_25879 | Not-determined |
| 18-103_k141_135173 | Not-determined |
| 18-103_k141_54553 | Not-determined |
| 18-103_k141_89253 | Not-determined |
| 18-103_k141_42965 | Not-determined |
| 18-103_k141_42966 | Not-determined |
| 18-103_k141_89256 | Low-quality |
| 18-103_k141_3042 | Not-determined |
| 18-103_k141_60340 | Not-determined |
| 18-103_k141_117924 | Not-determined |
| 18-103_k141_71721 | Not-determined |
| 18-103_k141_25883 | Not-determined |
| 18-103_k141_112083 | Not-determined |
| 18-103_k141_25884 | Not-determined |
| 18-103_k141_60341 | Not-determined |
| 18-103_k141_77525 | Not-determined |
| 18-103_k141_60342 | Not-determined |
| 18-103_k141_20084 | Not-determined |
| 18-103_k141_94889 | Not-determined |
| 18-103_k141_100341 | Not-determined |
| 18-103_k141_112089 | Not-determined |
| 18-103_k141_54557 | Low-quality |
| 18-103_k141_94894 | Not-determined |
| 18-103_k141_60348 | Not-determined |
| 18-103_k141_66024 | Not-determined |
| 18-103_k141_66025 | Not-determined |
| 18-103_k141_20092 | Not-determined |
| 18-103_k141_20093 | Not-determined |
| 18-103_k141_100345 | Low-quality |
| 18-103_k141_112102 | Not-determined |
| 18-103_k141_123623 | Not-determined |
| 18-103_k141_83522 | Not-determined |
| 18-103_k141_94901 | Low-quality |
| 18-103_k141_77528 | Not-determined |
| 18-103_k141_83525 | Low-quality |
| 18-103_k141_8465 | Not-determined |
| 18-103_k141_106310 | Not-determined |
| 18-103_k141_14146 | Not-determined |
| 18-103_k141_8467 | Not-determined |
| 18-103_k141_123629 | Not-determined |
| 18-103_k141_129587 | Not-determined |
| 18-103_k141_89275 | Not-determined |
| 18-103_k141_3075 | Not-determined |
| 18-103_k141_112116 | Not-determined |
| 18-103_k141_83534 | Not-determined |
| 18-103_k141_77534 | Not-determined |
| 18-103_k141_60353 | Not-determined |
| 18-103_k141_37384 | Not-determined |
| 18-103_k141_37385 | Not-determined |
| 18-103_k141_20102 | Not-determined |
| 18-103_k141_8477 | Not-determined |
| 18-103_k141_129591 | Not-determined |
| 18-103_k141_3089 | Not-determined |
| 18-103_k141_100359 | Not-determined |
| 18-103_k141_42988 | Not-determined |
| 18-103_k141_77545 | Not-determined |
| 18-103_k141_8483 | Not-determined |
| 18-103_k141_14156 | Not-determined |
| 18-103_k141_3092 | Not-determined |
| 18-103_k141_3093 | Not-determined |
| 18-103_k141_106321 | Not-determined |
| 18-103_k141_20111 | Not-determined |
| 18-103_k141_77551 | Not-determined |
| 18-103_k141_60366 | Not-determined |
| 18-103_k141_77552 | Not-determined |
| 18-103_k141_117943 | Not-determined |
| 18-103_k141_8489 | Not-determined |
| 18-103_k141_37404 | Not-determined |
| 18-103_k141_135221 | Not-determined |
| 18-103_k141_135222 | Not-determined |
| 18-103_k141_25922 | Not-determined |
| 18-103_k141_31569 | Not-determined |
| 18-103_k141_123663 | Not-determined |
| 18-103_k141_123664 | Not-determined |
| 18-103_k141_3108 | Not-determined |
| 18-103_k141_100373 | Not-determined |
| 18-103_k141_117958 | Not-determined |
| 18-103_k141_117960 | Not-determined |
| 18-103_k141_117962 | Not-determined |
| 18-103_k141_112142 | Not-determined |
| 18-103_k141_66065 | Not-determined |
| 18-103_k141_43009 | Not-determined |
| 18-103_k141_112145 | Not-determined |
| 18-103_k141_37418 | Not-determined |
| 18-103_k141_71753 | Not-determined |
| 18-103_k141_94944 | Not-determined |
| 18-103_k141_37433 | Not-determined |
| 18-103_k141_71767 | Not-determined |
| 18-103_k141_117983 | Not-determined |
| 18-103_k141_100390 | Not-determined |
| 18-103_k141_71773 | Not-determined |
| 18-103_k141_3124 | Not-determined |
| 18-103_k141_66083 | Not-determined |
| 18-103_k141_89317 | Low-quality |
| 18-103_k141_31585 | Low-quality |
| 18-103_k141_43033 | Not-determined |
| 18-103_k141_123692 | Not-determined |
| 18-103_k141_71778 | Not-determined |
| 18-103_k141_8525 | Low-quality |
| 18-103_k141_94964 | Not-determined |
| 18-103_k141_94965 | Low-quality |
| 18-103_k141_25944 | Not-determined |
| 18-103_k141_135227 | Not-determined |
| 18-103_k141_89328 | Low-quality |
| 18-103_k141_43044 | Not-determined |
| 18-103_k141_20139 | Not-determined |
| 18-103_k141_135232 | Not-determined |
| 18-103_k141_3137 | Not-determined |
| 18-103_k141_66096 | Low-quality |
| 18-103_k141_71784 | Not-determined |
| 18-103_k141_89335 | Not-determined |
| 18-103_k141_3140 | Low-quality |
| 18-103_k141_25955 | Not-determined |
| 18-103_k141_43052 | Not-determined |
| 18-103_k141_48892 | Not-determined |
| 18-103_k141_118011 | Low-quality |
| 18-103_k141_94968 | Not-determined |
| 18-103_k141_60410 | Not-determined |
| 18-103_k141_94970 | Not-determined |
| 18-103_k141_48895 | Not-determined |
| 18-103_k141_94971 | Not-determined |
| 18-103_k141_8537 | Not-determined |
| 18-103_k141_3144 | Not-determined |
| 18-103_k141_54592 | Not-determined |
| 18-103_k141_48897 | Not-determined |
| 18-103_k141_129649 | Not-determined |
| 18-103_k141_118019 | Not-determined |
| 18-103_k141_100405 | Not-determined |
| 18-103_k141_54596 | Not-determined |
| 18-103_k141_14184 | Not-determined |
| 18-103_k141_14186 | Not-determined |
| 18-103_k141_54598 | Not-determined |
| 18-103_k141_14188 | Low-quality |
| 18-103_k141_14189 | Not-determined |
| 18-103_k141_71791 | Not-determined |
| 18-103_k141_118024 | Not-determined |
| 18-103_k141_54601 | Not-determined |
| 18-103_k141_8542 | Not-determined |
| 18-103_k141_112165 | Not-determined |
| 18-103_k141_106392 | Not-determined |
| 18-103_k141_54604 | Not-determined |
| 18-103_k141_100414 | Not-determined |
| 18-103_k141_106393 | Not-determined |
| 18-103_k141_123715 | Not-determined |
| 18-103_k141_71799 | Not-determined |
| 18-103_k141_8544 | Not-determined |
| 18-103_k141_60423 | Not-determined |
| 18-103_k141_54607 | Not-determined |
| 18-103_k141_129658 | Not-determined |
| 18-103_k141_66108 | Not-determined |
| 18-103_k141_118032 | Not-determined |
| 18-103_k141_100422 | Not-determined |
| 18-103_k141_118034 | Not-determined |
| 18-103_k141_118035 | Not-determined |
| 18-103_k141_60427 | Not-determined |
| 18-103_k141_60429 | Low-quality |
| 18-103_k141_66113 | Not-determined |
| 18-103_k141_43076 | Not-determined |
| 18-103_k141_54617 | Low-quality |
| 18-103_k141_31600 | Not-determined |
| 18-103_k141_43080 | Not-determined |
| 18-103_k141_123725 | Not-determined |
| 18-103_k141_37469 | Low-quality |
| 18-103_k141_106410 | Low-quality |
| 18-103_k141_43084 | Not-determined |
| 18-103_k141_43085 | Not-determined |
| 18-103_k141_135262 | Not-determined |
| 18-103_k141_71813 | Low-quality |
| 18-103_k141_123734 | Not-determined |
| 18-103_k141_89363 | Not-determined |
| 18-103_k141_100436 | Not-determined |
| 18-103_k141_89367 | Not-determined |
| 18-103_k141_100439 | Low-quality |
| 18-103_k141_118058 | Not-determined |
| 18-103_k141_83606 | Not-determined |
| 18-103_k141_100444 | Not-determined |
| 18-103_k141_3163 | Not-determined |
| 18-103_k141_54635 | Not-determined |
| 18-103_k141_48923 | Low-quality |
| 18-103_k141_48924 | Not-determined |
| 18-103_k141_129690 | Not-determined |
| 18-103_k141_3169 | Not-determined |
| 18-103_k141_37487 | Low-quality |
| 18-103_k141_100452 | Not-determined |
| 18-103_k141_77593 | Not-determined |
| 18-103_k141_89376 | Not-determined |
| 18-103_k141_106427 | Not-determined |
| 18-103_k141_3174 | Not-determined |
| 18-103_k141_100461 | Not-determined |
| 18-103_k141_112191 | Not-determined |
| 18-103_k141_77604 | Not-determined |
| 18-103_k141_77605 | Not-determined |
| 18-103_k141_95008 | Not-determined |
| 18-103_k141_8589 | Not-determined |
| 18-103_k141_31612 | Not-determined |
| 18-103_k141_37497 | Not-determined |
| 18-103_k141_37500 | Low-quality |
| 18-103_k141_66140 | Not-determined |
| 18-103_k141_3193 | Not-determined |
| 18-103_k141_89398 | Not-determined |
| 18-103_k141_135299 | Not-determined |
| 18-103_k141_135302 | Not-determined |
| 18-103_k141_118101 | Low-quality |
| 18-103_k141_89400 | Not-determined |
| 18-103_k141_71830 | Not-determined |
| 18-103_k141_66147 | Not-determined |
| 18-103_k141_3202 | Not-determined |
| 18-103_k141_60455 | Not-determined |
| 18-103_k141_54656 | Not-determined |
| 18-103_k141_60457 | Not-determined |
| 18-103_k141_89408 | Not-determined |
| 18-103_k141_95024 | Not-determined |
| 18-103_k141_100485 | Low-quality |
| 18-103_k141_60463 | Not-determined |
| 18-103_k141_112201 | Not-determined |

| | |
|---|---|
| 18-103_k141_66159 | Not-determined |
| 18-103_k141_66160 | Not-determined |
| 18-103_k141_129709 | Not-determined |
| 18-103_k141_20203 | Low-quality |
| 18-103_k141_118114 | Not-determined |
| 18-103_k141_100491 | Not-determined |
| 18-103_k141_31620 | Not-determined |
| 18-103_k141_95029 | Not-determined |
| 18-103_k141_37521 | Not-determined |
| 18-103_k141_106462 | Low-quality |
| 18-103_k141_60470 | Not-determined |
| 18-103_k141_54668 | Not-determined |
| 18-103_k141_106466 | Not-determined |
| 18-103_k141_83623 | Low-quality |
| 18-103_k141_106469 | Low-quality |
| 18-103_k141_118128 | Not-determined |
| 18-103_k141_89424 | Not-determined |
| 18-103_k141_95036 | Not-determined |
| 18-103_k141_100498 | Not-determined |
| 18-103_k141_3228 | Not-determined |
| 18-103_k141_106478 | Not-determined |
| 18-103_k141_95042 | Not-determined |
| 18-103_k141_71846 | Not-determined |
| 18-103_k141_112224 | Not-determined |
| 18-103_k141_95043 | Not-determined |
| 18-103_k141_112227 | Not-determined |
| 18-103_k141_8621 | Not-determined |
| 18-103_k141_20217 | Not-determined |
| 18-103_k141_100501 | Not-determined |
| 18-103_k141_106487 | Not-determined |
| 18-103_k141_77627 | Low-quality |
| 18-103_k141_118145 | Not-determined |
| 18-103_k141_8626 | Not-determined |
| 18-103_k141_54690 | Not-determined |
| 18-103_k141_100506 | Not-determined |
| 18-103_k141_106502 | Not-determined |
| 18-103_k141_43123 | Low-quality |
| 18-103_k141_14235 | Not-determined |
| 18-103_k141_66189 | Not-determined |
| 18-103_k141_66190 | Not-determined |
| 18-103_k141_66193 | Not-determined |
| 18-103_k141_71866 | Not-determined |
| 18-103_k141_8635 | Not-determined |
| 18-103_k141_95055 | Low-quality |
| 18-103_k141_135314 | Not-determined |
| 18-103_k141_43130 | Not-determined |
| 18-103_k141_135317 | Not-determined |
| 18-103_k141_66200 | Not-determined |
| 18-103_k141_3252 | Not-determined |
| 18-103_k141_100512 | Not-determined |
| 18-103_k141_71868 | Not-determined |
| 18-103_k141_89450 | Not-determined |
| 18-103_k141_129740 | Not-determined |
| 18-103_k141_77643 | Not-determined |
| 18-103_k141_89452 | Low-quality |
| 18-103_k141_77645 | Not-determined |
| 18-103_k141_95066 | Not-determined |
| 18-103_k141_100517 | Not-determined |
| 18-103_k141_106519 | Not-determined |
| 18-103_k141_43140 | Not-determined |
| 18-103_k141_83651 | Not-determined |
| 18-103_k141_77651 | Not-determined |
| 18-103_k141_14245 | Not-determined |
| 18-103_k141_54715 | Not-determined |
| 18-103_k141_66214 | Not-determined |
| 18-103_k141_71879 | Not-determined |
| 18-103_k141_37562 | Not-determined |
| 18-103_k141_54716 | Low-quality |
| 18-103_k141_71880 | Not-determined |
| 18-103_k141_118164 | Not-determined |
| 18-103_k141_83654 | Not-determined |
| 18-103_k141_3270 | Not-determined |
| 18-103_k141_89460 | Low-quality |
| 18-103_k141_77663 | Not-determined |
| 18-103_k141_77666 | Not-determined |
| 18-103_k141_100530 | Not-determined |
| 18-103_k141_89463 | Not-determined |
| 18-103_k141_106530 | Not-determined |
| 18-103_k141_71891 | Not-determined |
| 18-103_k141_66227 | Not-determined |
| 18-103_k141_71893 | Not-determined |
| 18-103_k141_106533 | Not-determined |
| 18-103_k141_66231 | Not-determined |
| 18-103_k141_3275 | Not-determined |
| 18-103_k141_129757 | Not-determined |
| 18-103_k141_54728 | Not-determined |
| 18-103_k141_123826 | Not-determined |
| 18-103_k141_3280 | Not-determined |
| 18-103_k141_66237 | Not-determined |
| 18-103_k141_123827 | Not-determined |
| 18-103_k141_118178 | Not-determined |
| 18-103_k141_112286 | Not-determined |
| 18-103_k141_95079 | Not-determined |
| 18-103_k141_135330 | Not-determined |
| 18-103_k141_95080 | Not-determined |
| 18-103_k141_37573 | Not-determined |
| 18-103_k141_100536 | Not-determined |
| 18-103_k141_37575 | Not-determined |
| 18-103_k141_43154 | Low-quality |
| 18-103_k141_54737 | Not-determined |
| 18-103_k141_54739 | Not-determined |
| 18-103_k141_31652 | Not-determined |
| 18-103_k141_83679 | Not-determined |
| 18-103_k141_71911 | Not-determined |
| 18-103_k141_37588 | Low-quality |
| 18-103_k141_77692 | Not-determined |
| 18-103_k141_100545 | Low-quality |
| 18-103_k141_83686 | Not-determined |
| 18-103_k141_31663 | Not-determined |
| 18-103_k141_112308 | Not-determined |
| 18-103_k141_66253 | Not-determined |
| 18-103_k141_83690 | Not-determined |
| 18-103_k141_37593 | Not-determined |
| 18-103_k141_89496 | Not-determined |
| 18-103_k141_129775 | Not-determined |
| 18-103_k141_129776 | Not-determined |
| 18-103_k141_135347 | Not-determined |
| 18-103_k141_135348 | Not-determined |
| 18-103_k141_3293 | Not-determined |
| 18-103_k141_123857 | Not-determined |
| 18-103_k141_89511 | Not-determined |
| 18-103_k141_26009 | Low-quality |
| 18-103_k141_89513 | Not-determined |
| 18-103_k141_43173 | Not-determined |
| 18-103_k141_83702 | Low-quality |
| 18-103_k141_26012 | Not-determined |
| 18-103_k141_43176 | Not-determined |
| 18-103_k141_8667 | Not-determined |
| 18-103_k141_100561 | Low-quality |
| 18-103_k141_71929 | Not-determined |
| 18-103_k141_26014 | Low-quality |
| 18-103_k141_20251 | Not-determined |
| 18-103_k141_83714 | Not-determined |
| 18-103_k141_43184 | Not-determined |
| 18-103_k141_129794 | Not-determined |
| 18-103_k141_118233 | Not-determined |
| 18-103_k141_66276 | Not-determined |
| 18-103_k141_20258 | Not-determined |
| 18-103_k141_14292 | Not-determined |
| 18-103_k141_3316 | Not-determined |
| 18-103_k141_31686 | Low-quality |
| 18-103_k141_60501 | Not-determined |
| 18-103_k141_8682 | Not-determined |
| 18-103_k141_77732 | Not-determined |
| 18-103_k141_31694 | Not-determined |
| 18-103_k141_118254 | Not-determined |
| 18-103_k141_89516 | Not-determined |
| 18-103_k141_112357 | Not-determined |
| 18-103_k141_100587 | Not-determined |
| 18-103_k141_118255 | Not-determined |
| 18-103_k141_14312 | Not-determined |
| 18-103_k141_54758 | Low-quality |
| 18-103_k141_112364 | Not-determined |
| 18-103_k141_100591 | Not-determined |
| 18-103_k141_106549 | Not-determined |
| 18-103_k141_3322 | Not-determined |
| 18-103_k141_31703 | Not-determined |
| 18-103_k141_89523 | Not-determined |
| 18-103_k141_118260 | Low-quality |
| 18-103_k141_112371 | Not-determined |
| 18-103_k141_37635 | Not-determined |
| 18-103_k141_54765 | Not-determined |
| 18-103_k141_77749 | Not-determined |
| 18-103_k141_95118 | Not-determined |
| 18-103_k141_118264 | Not-determined |
| 18-103_k141_118266 | Not-determined |
| 18-103_k141_14322 | Not-determined |
| 18-103_k141_129809 | Not-determined |
| 18-103_k141_123870 | Not-determined |
| 18-103_k141_129810 | Not-determined |
| 18-103_k141_95122 | Not-determined |
| 18-103_k141_48987 | Low-quality |
| 18-103_k141_66317 | Not-determined |
| 18-103_k141_118271 | Not-determined |
| 18-103_k141_54774 | Not-determined |
| 18-103_k141_77758 | Low-quality |
| 18-103_k141_66319 | Not-determined |
| 18-103_k141_37651 | Not-determined |
| 18-103_k141_14325 | Not-determined |
| 18-103_k141_89542 | Low-quality |
| 18-103_k141_135409 | Not-determined |
| 18-103_k141_112379 | Not-determined |
| 18-103_k141_129813 | Not-determined |
| 18-103_k141_60533 | Not-determined |
| 18-103_k141_77763 | Not-determined |
| 18-103_k141_95128 | Not-determined |
| 18-103_k141_95130 | Not-determined |
| 18-103_k141_83726 | Not-determined |
| 18-103_k141_83727 | Not-determined |
| 18-103_k141_3335 | Not-determined |
| 18-103_k141_95133 | Not-determined |
| 18-103_k141_8709 | Not-determined |
| 18-103_k141_3336 | Not-determined |
| 18-103_k141_135411 | Low-quality |
| 18-103_k141_77774 | Low-quality |
| 18-103_k141_118284 | Not-determined |
| 18-103_k141_95141 | Low-quality |
| 18-103_k141_26046 | Low-quality |
| 18-103_k141_8718 | Not-determined |
| 18-103_k141_71985 | Not-determined |
| 18-103_k141_54792 | Not-determined |
| 18-103_k141_118288 | Not-determined |
| 18-103_k141_89555 | Not-determined |
| 18-103_k141_71989 | Not-determined |
| 18-103_k141_71990 | Low-quality |
| 18-103_k141_129828 | Not-determined |
| 18-103_k141_89561 | Not-determined |
| 18-103_k141_54801 | Not-determined |
| 18-103_k141_66349 | Not-determined |
| 18-103_k141_37662 | Low-quality |
| 18-103_k141_3351 | Not-determined |
| 18-103_k141_118297 | Not-determined |
| 18-103_k141_43261 | Not-determined |
| 18-103_k141_26063 | Not-determined |
| 18-103_k141_26065 | Not-determined |
| 18-103_k141_20301 | Not-determined |
| 18-103_k141_118302 | Not-determined |
| 18-103_k141_89571 | Not-determined |
| 18-103_k141_66356 | Not-determined |
| 18-103_k141_43263 | Not-determined |
| 18-103_k141_106579 | Not-determined |
| 18-103_k141_54816 | Not-determined |
| 18-103_k141_135421 | Not-determined |
| 18-103_k141_8732 | Not-determined |
| 18-103_k141_118305 | Not-determined |
| 18-103_k141_26077 | Low-quality |
| 18-103_k141_112407 | Not-determined |
| 18-103_k141_135426 | Not-determined |
| 18-103_k141_43273 | Not-determined |
| 18-103_k141_66370 | Not-determined |
| 18-103_k141_106588 | Not-determined |
| 18-103_k141_43276 | Not-determined |
| 18-103_k141_77797 | Not-determined |
| 18-103_k141_14365 | Not-determined |
| 18-103_k141_54829 | Not-determined |
| 18-103_k141_83754 | Not-determined |
| 18-103_k141_77802 | Not-determined |
| 18-103_k141_77808 | Not-determined |
| 18-103_k141_37668 | Not-determined |
| 18-103_k141_83760 | Not-determined |
| 18-103_k141_54838 | Not-determined |
| 18-103_k141_37669 | Not-determined |
| 18-103_k141_37671 | Not-determined |
| 18-103_k141_37672 | Not-determined |
| 18-103_k141_89589 | Not-determined |
| 18-103_k141_3372 | Not-determined |
| 18-103_k141_83764 | Not-determined |
| 18-103_k141_26090 | Not-determined |
| 18-103_k141_112423 | Not-determined |
| 18-103_k141_8758 | Low-quality |
| 18-103_k141_8759 | Not-determined |
| 18-103_k141_129849 | Not-determined |
| 18-103_k141_43288 | Not-determined |
| 18-103_k141_83768 | Not-determined |
| 18-103_k141_26096 | Low-quality |
| 18-103_k141_14379 | Not-determined |
| 18-103_k141_26098 | Not-determined |
| 18-103_k141_123902 | Not-determined |
| 18-103_k141_72006 | Not-determined |
| 18-103_k141_37683 | Low-quality |
| 18-103_k141_77822 | Not-determined |
| 18-103_k141_129857 | Not-determined |
| 18-103_k141_100656 | Not-determined |
| 18-103_k141_89596 | Not-determined |
| 18-103_k141_83772 | Not-determined |
| 18-103_k141_112435 | Not-determined |
| 18-103_k141_129861 | Not-determined |
| 18-103_k141_54854 | Not-determined |
| 18-103_k141_129865 | Not-determined |
| 18-103_k141_8777 | Low-quality |
| 18-103_k141_100661 | Not-determined |
| 18-103_k141_95191 | Not-determined |
| 18-103_k141_129867 | Not-determined |
| 18-103_k141_8779 | Not-determined |
| 18-103_k141_8780 | Not-determined |
| 18-103_k141_123916 | Not-determined |
| 18-103_k141_77827 | Not-determined |
| 18-103_k141_77828 | Not-determined |
| 18-103_k141_37693 | Not-determined |
| 18-103_k141_49055 | Not-determined |
| 18-103_k141_89611 | Not-determined |
| 18-103_k141_60555 | Not-determined |
| 18-103_k141_129875 | Not-determined |
| 18-103_k141_37694 | Not-determined |
| 18-103_k141_89615 | Not-determined |
| 18-103_k141_129877 | Not-determined |
| 18-103_k141_118336 | Not-determined |
| 18-103_k141_14392 | Low-quality |
| 18-103_k141_129880 | Low-quality |
| 18-103_k141_83786 | Low-quality |
| 18-103_k141_31740 | Not-determined |
| 18-103_k141_26130 | Not-determined |
| 18-103_k141_77837 | Low-quality |
| 18-103_k141_54872 | Low-quality |
| 18-103_k141_31744 | Not-determined |
| 18-103_k141_112452 | Low-quality |
| 18-103_k141_54877 | Not-determined |
| 18-103_k141_54878 | Not-determined |
| 18-103_k141_100677 | Not-determined |
| 18-103_k141_123931 | Not-determined |
| 18-103_k141_77843 | Not-determined |
| 18-103_k141_8785 | Not-determined |
| 18-103_k141_49064 | Low-quality |
| 18-103_k141_118348 | Not-determined |
| 18-103_k141_83799 | Not-determined |
| 18-103_k141_112461 | Low-quality |
| 18-103_k141_118351 | Not-determined |
| 18-103_k141_123934 | Not-determined |
| 18-103_k141_112464 | Not-determined |
| 18-103_k141_60572 | Low-quality |
| 18-103_k141_77848 | Not-determined |
| 18-103_k141_77852 | Not-determined |
| 18-103_k141_100686 | Not-determined |
| 18-103_k141_95211 | Not-determined |
| 18-103_k141_89632 | Not-determined |
| 18-103_k141_95213 | Not-determined |
| 18-103_k141_49074 | Not-determined |
| 18-103_k141_31761 | Not-determined |
| 18-103_k141_129904 | Not-determined |
| 18-103_k141_26146 | Not-determined |
| 18-103_k141_54901 | Not-determined |
| 18-103_k141_95219 | Low-quality |
| 18-103_k141_123939 | Not-determined |
| 18-103_k141_31772 | Not-determined |
| 18-103_k141_95222 | Not-determined |
| 18-103_k141_49084 | Not-determined |
| 18-103_k141_129913 | Not-determined |
| 18-103_k141_31777 | Not-determined |
| 18-103_k141_123949 | Low-quality |
| 18-103_k141_60591 | Not-determined |
| 18-103_k141_89645 | Not-determined |
| 18-103_k141_100701 | Not-determined |
| 18-103_k141_37736 | Not-determined |
| 18-103_k141_26151 | Low-quality |
| 18-103_k141_20341 | Not-determined |
| 18-103_k141_100705 | Not-determined |
| 18-103_k141_129918 | Not-determined |
| 18-103_k141_135438 | Not-determined |
| 18-103_k141_112482 | Not-determined |
| 18-103_k141_72047 | Not-determined |
| 18-103_k141_89653 | Not-determined |
| 18-103_k141_123955 | Not-determined |
| 18-103_k141_43339 | Not-determined |
| 18-103_k141_135439 | Not-determined |
| 18-103_k141_20349 | Not-determined |
| 18-103_k141_3426 | Not-determined |
| 18-103_k141_3427 | Not-determined |
| 18-103_k141_26157 | Not-determined |
| 18-103_k141_95237 | Not-determined |
| 18-103_k141_26159 | Not-determined |
| 18-103_k141_89655 | Low-quality |
| 18-103_k141_123963 | Not-determined |
| 18-103_k141_43350 | Low-quality |
| 18-103_k141_112485 | Not-determined |
| 18-103_k141_37750 | Not-determined |
| 18-103_k141_129932 | Not-determined |
| 18-103_k141_43353 | Not-determined |
| 18-103_k141_8796 | Not-determined |
| 18-103_k141_43356 | Not-determined |
| 18-103_k141_8797 | Not-determined |
| 18-103_k141_20357 | Not-determined |
| 18-103_k141_118399 | Low-quality |
| 18-103_k141_14437 | Not-determined |
| 18-103_k141_14438 | Not-determined |
| 18-103_k141_20359 | Not-determined |
| 18-103_k141_129941 | Not-determined |
| 18-103_k141_54919 | Not-determined |
| 18-103_k141_123974 | Not-determined |
| 18-103_k141_95248 | Not-determined |
| 18-103_k141_77886 | Not-determined |
| 18-103_k141_106630 | Not-determined |
| 18-103_k141_72052 | Not-determined |
| 18-103_k141_8805 | Not-determined |
| 18-103_k141_54922 | Not-determined |
| 18-103_k141_123979 | Not-determined |
| 18-103_k141_118406 | Not-determined |
| 18-103_k141_77891 | Not-determined |
| 18-103_k141_106635 | Not-determined |
| 18-103_k141_135456 | Not-determined |
| 18-103_k141_95253 | Not-determined |
| 18-103_k141_106649 | Not-determined |
| 18-103_k141_49116 | Not-determined |
| 18-103_k141_3440 | Low-quality |
| 18-103_k141_8814 | Low-quality |
| 18-103_k141_100734 | Not-determined |
| 18-103_k141_54934 | Not-determined |
| 18-103_k141_20373 | Not-determined |
| 18-103_k141_66462 | Not-determined |
| 18-103_k141_14471 | Low-quality |
| 18-103_k141_31796 | Not-determined |
| 18-103_k141_20377 | Not-determined |
| 18-103_k141_83845 | Not-determined |
| 18-103_k141_129957 | Not-determined |
| 18-103_k141_135473 | Not-determined |
| 18-103_k141_31801 | Not-determined |
| 18-103_k141_31805 | Not-determined |
| 18-103_k141_83850 | Not-determined |
| 18-103_k141_31807 | Not-determined |
| 18-103_k141_106662 | Not-determined |
| 18-103_k141_89691 | Not-determined |
| 18-103_k141_112515 | Not-determined |
| 18-103_k141_31812 | Not-determined |
| 18-103_k141_14493 | Not-determined |
| 18-103_k141_14496 | Not-determined |
| 18-103_k141_83864 | Not-determined |
| 18-103_k141_66489 | Not-determined |
| 18-103_k141_135489 | Not-determined |
| 18-103_k141_83867 | Not-determined |
| 18-103_k141_135491 | Not-determined |
| 18-103_k141_118444 | Not-determined |
| 18-103_k141_135492 | Not-determined |
| 18-103_k141_37779 | Not-determined |
| 18-103_k141_37783 | Not-determined |
| 18-103_k141_72096 | Not-determined |
| 18-103_k141_135500 | Not-determined |
| 18-103_k141_135501 | Not-determined |
| 18-103_k141_66503 | Not-determined |
| 18-103_k141_37788 | Not-determined |
| 18-103_k141_66504 | Not-determined |
| 18-103_k141_66506 | Not-determined |
| 18-103_k141_66508 | Not-determined |
| 18-103_k141_60644 | Not-determined |
| 18-103_k141_129966 | Not-determined |
| 18-103_k141_37796 | Not-determined |
| 18-103_k141_60648 | Not-determined |
| 18-103_k141_60651 | Not-determined |
| 18-103_k141_8860 | Not-determined |
| 18-103_k141_54974 | Not-determined |
| 18-103_k141_60652 | Not-determined |
| 18-103_k141_31840 | Not-determined |
| 18-103_k141_14509 | Not-determined |
| 18-103_k141_118460 | Not-determined |
| 18-103_k141_49169 | Not-determined |
| 18-103_k141_37798 | Not-determined |
| 18-103_k141_124026 | Not-determined |
| 18-103_k141_54978 | Not-determined |
| 18-103_k141_83888 | Not-determined |
| 18-103_k141_83890 | Not-determined |
| 18-103_k141_49177 | Not-determined |
| 18-103_k141_26223 | Low-quality |
| 18-103_k141_14519 | Not-determined |
| 18-103_k141_83896 | Not-determined |
| 18-103_k141_8866 | Low-quality |
| 18-103_k141_112539 | Not-determined |
| 18-103_k141_43394 | Low-quality |
| 18-103_k141_106685 | Not-determined |
| 18-103_k141_49182 | Not-determined |
| 18-103_k141_135525 | Not-determined |
| 18-103_k141_124038 | Not-determined |
| 18-103_k141_49184 | Not-determined |
| 18-103_k141_20397 | Not-determined |
| 18-103_k141_135527 | Not-determined |
| 18-103_k141_83903 | Not-determined |
| 18-103_k141_66525 | Not-determined |
| 18-103_k141_95302 | Not-determined |
| 18-103_k141_100786 | Not-determined |
| 18-103_k141_83909 | Not-determined |
| 18-103_k141_8875 | Not-determined |
| 18-103_k141_66527 | Low-quality |
| 18-103_k141_77947 | Not-determined |
| 18-103_k141_8882 | Not-determined |
| 18-103_k141_83919 | Not-determined |
| 18-103_k141_83922 | Not-determined |
| 18-103_k141_43415 | Not-determined |
| 18-103_k141_43421 | Not-determined |
| 18-103_k141_43422 | Not-determined |
| 18-103_k141_14538 | Not-determined |
| 18-103_k141_37844 | Not-determined |
| 18-103_k141_95320 | Not-determined |
| 18-103_k141_77967 | Not-determined |
| 18-103_k141_20424 | Low-quality |
| 18-103_k141_37849 | Not-determined |
| 18-103_k141_77970 | Not-determined |
| 18-103_k141_60687 | Not-determined |
| 18-103_k141_26259 | Low-quality |
| 18-103_k141_83935 | Not-determined |
| 18-103_k141_95333 | Not-determined |
| 18-103_k141_95334 | Not-determined |
| 18-103_k141_83942 | Not-determined |
| 18-103_k141_124072 | Low-quality |
| 18-103_k141_8906 | Not-determined |
| 18-103_k141_77987 | Not-determined |
| 18-103_k141_20437 | Not-determined |
| 18-103_k141_95339 | Not-determined |
| 18-103_k141_8908 | Not-determined |
| 18-103_k141_95340 | Not-determined |
| 18-103_k141_55004 | Not-determined |
| 18-103_k141_26269 | Not-determined |
| 18-103_k141_129993 | Not-determined |
| 18-103_k141_95346 | Low-quality |
| 18-103_k141_20450 | Not-determined |
| 18-103_k141_95348 | Not-determined |
| 18-103_k141_135556 | Not-determined |
| 18-103_k141_135558 | Not-determined |
| 18-103_k141_20452 | Not-determined |
| 18-103_k141_83956 | Not-determined |
| 18-103_k141_106733 | Not-determined |
| 18-103_k141_77994 | Not-determined |
| 18-103_k141_112590 | Not-determined |
| 18-103_k141_77997 | Not-determined |
| 18-103_k141_26281 | Not-determined |
| 18-103_k141_26282 | Not-determined |
| 18-103_k141_124092 | Not-determined |
| 18-103_k141_8933 | Not-determined |
| 18-103_k141_112593 | Not-determined |
| 18-103_k141_20458 | Not-determined |
| 18-103_k141_55015 | Not-determined |
| 18-103_k141_8936 | Not-determined |
| 18-103_k141_26285 | Not-determined |
| 18-103_k141_14577 | Not-determined |
| 18-103_k141_118497 | Not-determined |
| 18-103_k141_106745 | Not-determined |
| 18-103_k141_31879 | Low-quality |
| 18-103_k141_106746 | Not-determined |
| 18-103_k141_55018 | Not-determined |
| 18-103_k141_49208 | Not-determined |
| 18-103_k141_20467 | Not-determined |
| 18-103_k141_26287 | Not-determined |
| 18-103_k141_118501 | Not-determined |
| 18-103_k141_118502 | Not-determined |
| 18-103_k141_20473 | Not-determined |
| 18-103_k141_3497 | Not-determined |
| 18-103_k141_37861 | Not-determined |
| 18-103_k141_78014 | Not-determined |
| 18-103_k141_8949 | Not-determined |
| 18-103_k141_72186 | Low-quality |
| 18-103_k141_26290 | Not-determined |
| 18-103_k141_83972 | Not-determined |
| 18-103_k141_55025 | Low-quality |
| 18-103_k141_37865 | Not-determined |
| 18-103_k141_124111 | Not-determined |
| 18-103_k141_37866 | Not-determined |
| 18-103_k141_72188 | Not-determined |
| 18-103_k141_8952 | Not-determined |
| 18-103_k141_55027 | Not-determined |
| 18-103_k141_106761 | Not-determined |
| 18-103_k141_95365 | Not-determined |
| 18-103_k141_89797 | Not-determined |
| 18-103_k141_83978 | Not-determined |
| 18-103_k141_112609 | Not-determined |
| 18-103_k141_95372 | Low-quality |
| 18-103_k141_8960 | Not-determined |
| 18-103_k141_60712 | Not-determined |
| 18-103_k141_124124 | Not-determined |
| 18-103_k141_66558 | Not-determined |
| 18-103_k141_89806 | Not-determined |
| 18-103_k141_78029 | Low-quality |
| 18-103_k141_66564 | Not-determined |
| 18-103_k141_66565 | Not-determined |
| 18-103_k141_55040 | Not-determined |
| 18-103_k141_20494 | Not-determined |
| 18-103_k141_118523 | Not-determined |
| 18-103_k141_106764 | Not-determined |
| 18-103_k141_14609 | Not-determined |
| 18-103_k141_78035 | Not-determined |
| 18-103_k141_14611 | Not-determined |
| 18-103_k141_78036 | Not-determined |
| 18-103_k141_83996 | Not-determined |
| 18-103_k141_66577 | Not-determined |
| 18-103_k141_135602 | Not-determined |
| 18-103_k141_37880 | Not-determined |
| 18-103_k141_95404 | Low-quality |
| 18-103_k141_26322 | Not-determined |
| 18-103_k141_124159 | Not-determined |
| 18-103_k141_84003 | Not-determined |
| 18-103_k141_84005 | Low-quality |
| 18-103_k141_124163 | Not-determined |
| 18-103_k141_26327 | Not-determined |
| 18-103_k141_37893 | Not-determined |
| 18-103_k141_55062 | Not-determined |
| 18-103_k141_37895 | Not-determined |
| 18-103_k141_60739 | Not-determined |
| 18-103_k141_106787 | Not-determined |
| 18-103_k141_26329 | Not-determined |
| 18-103_k141_72231 | Not-determined |
| 18-103_k141_72232 | Not-determined |
| 18-103_k141_8964 | Not-determined |
| 18-103_k141_112651 | Not-determined |
| 18-103_k141_49225 | Not-determined |
| 18-103_k141_72238 | Not-determined |
| 18-103_k141_89836 | Not-determined |
| 18-103_k141_55074 | Not-determined |
| 18-103_k141_26344 | Not-determined |
| 18-103_k141_130049 | Not-determined |
| 18-103_k141_89840 | Low-quality |
| 18-103_k141_118544 | Not-determined |
| 18-103_k141_49228 | Not-determined |
| 18-103_k141_95424 | Not-determined |
| 18-103_k141_118546 | Not-determined |

| | |
|---|---|
| 18-103_k141_26345 | Low-quality |
| 18-103_k141_60755 | Not-determined |
| 18-103_k141_112662 | Not-determined |
| 18-103_k141_135624 | Not-determined |
| 18-103_k141_3550 | Not-determined |
| 18-103_k141_66598 | Not-determined |
| 18-103_k141_66600 | Not-determined |
| 18-103_k141_49232 | Not-determined |
| 18-103_k141_37921 | Not-determined |
| 18-103_k141_43471 | Not-determined |
| 18-103_k141_26352 | Low-quality |
| 18-103_k141_49237 | Not-determined |
| 18-103_k141_124192 | Not-determined |
| 18-103_k141_72257 | Not-determined |
| 18-103_k141_49241 | Low-quality |
| 18-103_k141_124195 | Not-determined |
| 18-103_k141_43479 | Not-determined |
| 18-103_k141_124198 | Not-determined |
| 18-103_k141_49246 | Not-determined |
| 18-103_k141_106805 | Low-quality |
| 18-103_k141_106806 | Not-determined |
| 18-103_k141_14628 | Not-determined |
| 18-103_k141_95434 | Not-determined |
| 18-103_k141_37939 | Not-determined |
| 18-103_k141_55090 | Not-determined |
| 18-103_k141_20525 | Not-determined |
| 18-103_k141_20526 | Not-determined |
| 18-103_k141_8989 | Not-determined |
| 18-103_k141_135637 | Not-determined |
| 18-103_k141_135638 | Not-determined |
| 18-103_k141_72266 | Not-determined |
| 18-103_k141_95438 | Not-determined |
| 18-103_k141_49255 | Not-determined |
| 18-103_k141_118586 | Not-determined |
| 18-103_k141_37949 | Not-determined |
| 18-103_k141_55107 | Low-quality |
| 18-103_k141_3594 | Not-determined |
| 18-103_k141_60780 | Not-determined |
| 18-103_k141_124221 | Not-determined |
| 18-103_k141_55115 | Not-determined |
| 18-103_k141_89882 | Not-determined |
| 18-103_k141_118596 | Not-determined |
| 18-103_k141_8998 | Not-determined |
| 18-103_k141_8999 | Not-determined |
| 18-103_k141_66624 | Not-determined |
| 18-103_k141_66626 | Not-determined |
| 18-103_k141_43495 | Not-determined |
| 18-103_k141_124233 | Not-determined |
| 18-103_k141_26389 | Not-determined |
| 18-103_k141_37967 | Not-determined |
| 18-103_k141_112681 | Not-determined |
| 18-103_k141_60792 | Not-determined |
| 18-103_k141_20540 | Not-determined |
| 18-103_k141_106847 | Low-quality |
| 18-103_k141_118604 | Not-determined |
| 18-103_k141_78058 | Not-determined |
| 18-103_k141_55138 | Not-determined |
| 18-103_k141_37976 | Not-determined |
| 18-103_k141_78060 | Not-determined |
| 18-103_k141_130082 | Low-quality |
| 18-103_k141_60796 | Not-determined |
| 18-103_k141_14649 | Not-determined |
| 18-103_k141_95458 | Not-determined |
| 18-103_k141_43506 | Not-determined |
| 18-103_k141_100889 | Not-determined |
| 18-103_k141_3607 | Not-determined |
| 18-103_k141_100892 | Low-quality |
| 18-103_k141_135645 | Not-determined |
| 18-103_k141_72283 | Not-determined |
| 18-103_k141_78068 | Not-determined |
| 18-103_k141_14654 | Not-determined |
| 18-103_k141_66636 | Not-determined |
| 18-103_k141_20558 | Not-determined |
| 18-103_k141_118621 | Low-quality |
| 18-103_k141_3617 | Not-determined |
| 18-103_k141_20563 | Not-determined |
| 18-103_k141_135652 | Not-determined |
| 18-103_k141_55151 | Low-quality |
| 18-103_k141_60810 | Not-determined |
| 18-103_k141_55152 | Not-determined |
| 18-103_k141_118625 | Not-determined |
| 18-103_k141_72292 | Not-determined |
| 18-103_k141_20566 | Not-determined |
| 18-103_k141_49307 | Not-determined |
| 18-103_k141_124259 | Not-determined |
| 18-103_k141_60812 | Not-determined |
| 18-103_k141_78074 | Not-determined |
| 18-103_k141_89904 | Not-determined |
| 18-103_k141_78075 | Not-determined |
| 18-103_k141_78077 | Not-determined |
| 18-103_k141_60816 | Not-determined |
| 18-103_k141_130102 | Not-determined |
| 18-103_k141_55167 | Not-determined |
| 18-103_k141_3629 | Not-determined |
| 18-103_k141_95472 | Not-determined |
| 18-103_k141_89911 | Not-determined |
| 18-103_k141_26423 | Low-quality |
| 18-103_k141_124270 | Not-determined |
| 18-103_k141_95475 | Not-determined |
| 18-103_k141_20585 | Not-determined |
| 18-103_k141_112701 | Low-quality |
| 18-103_k141_49324 | Not-determined |
| 18-103_k141_112704 | Not-determined |
| 18-103_k141_14674 | Not-determined |
| 18-103_k141_72312 | Low-quality |
| 18-103_k141_112707 | Low-quality |
| 18-103_k141_124276 | Not-determined |
| 18-103_k141_66676 | Not-determined |
| 18-103_k141_60833 | Not-determined |
| 18-103_k141_66681 | Not-determined |
| 18-103_k141_26428 | Not-determined |
| 18-103_k141_66686 | Not-determined |
| 18-103_k141_60840 | Not-determined |
| 18-103_k141_106852 | Not-determined |
| 18-103_k141_49344 | Not-determined |
| 18-103_k141_66691 | Not-determined |
| 18-103_k141_118661 | Not-determined |
| 18-103_k141_66696 | Not-determined |
| 18-103_k141_20604 | Not-determined |
| 18-103_k141_130138 | Not-determined |
| 18-103_k141_31962 | Not-determined |
| 18-103_k141_130140 | Not-determined |
| 18-103_k141_49359 | Not-determined |
| 18-103_k141_31967 | Not-determined |
| 18-103_k141_130145 | Not-determined |
| 18-103_k141_78106 | Not-determined |
| 18-103_k141_135669 | Not-determined |
| 18-103_k141_66706 | Not-determined |
| 18-103_k141_9043 | Not-determined |
| 18-103_k141_66709 | Not-determined |
| 18-103_k141_135674 | Not-determined |
| 18-103_k141_38017 | Not-determined |
| 18-103_k141_14688 | Not-determined |
| 18-103_k141_124302 | Not-determined |
| 18-103_k141_31977 | Not-determined |
| 18-103_k141_55209 | Not-determined |
| 18-103_k141_55210 | Not-determined |
| 18-103_k141_130155 | Low-quality |
| 18-103_k141_112723 | Not-determined |
| 18-103_k141_66712 | Not-determined |
| 18-103_k141_31981 | Not-determined |
| 18-103_k141_31982 | Not-determined |
| 18-103_k141_26457 | Not-determined |
| 18-103_k141_14696 | Not-determined |
| 18-103_k141_118666 | Not-determined |
| 18-103_k141_106880 | Not-determined |
| 18-103_k141_112728 | Not-determined |
| 18-103_k141_100918 | Not-determined |
| 18-103_k141_49372 | Not-determined |
| 18-103_k141_130166 | Not-determined |
| 18-103_k141_78117 | Not-determined |
| 18-103_k141_130168 | Not-determined |
| 18-103_k141_118676 | Not-determined |
| 18-103_k141_26460 | Not-determined |
| 18-103_k141_43546 | Not-determined |
| 18-103_k141_3675 | Not-determined |
| 18-103_k141_26462 | Not-determined |
| 18-103_k141_135683 | Not-determined |
| 18-103_k141_100923 | Low-quality |
| 18-103_k141_3683 | Not-determined |
| 18-103_k141_49375 | Not-determined |
| 18-103_k141_135684 | Not-determined |
| 18-103_k141_26472 | Not-determined |
| 18-103_k141_38038 | Low-quality |
| 18-103_k141_20644 | Not-determined |
| 18-103_k141_124317 | Low-quality |
| 18-103_k141_106894 | Not-determined |
| 18-103_k141_38043 | Not-determined |
| 18-103_k141_20645 | Not-determined |
| 18-103_k141_26476 | Not-determined |
| 18-103_k141_124319 | Not-determined |
| 18-103_k141_72354 | Not-determined |
| 18-103_k141_130179 | Not-determined |
| 18-103_k141_106897 | Not-determined |
| 18-103_k141_135688 | Not-determined |
| 18-103_k141_135689 | Not-determined |
| 18-103_k141_66733 | Low-quality |
| 18-103_k141_89963 | Not-determined |
| 18-103_k141_89964 | Not-determined |
| 18-103_k141_66734 | Low-quality |
| 18-103_k141_135695 | Not-determined |
| 18-103_k141_89968 | Not-determined |
| 18-103_k141_100944 | Not-determined |
| 18-103_k141_20659 | Not-determined |
| 18-103_k141_38053 | Not-determined |
| 18-103_k141_100947 | Not-determined |
| 18-103_k141_78142 | Not-determined |
| 18-103_k141_60853 | Not-determined |
| 18-103_k141_89973 | Not-determined |
| 18-103_k141_78145 | Not-determined |
| 18-103_k141_100949 | Not-determined |
| 18-103_k141_43567 | Low-quality |
| 18-103_k141_66742 | Not-determined |
| 18-103_k141_78147 | Not-determined |
| 18-103_k141_78149 | Not-determined |
| 18-103_k141_20672 | Not-determined |
| 18-103_k141_20673 | Not-determined |
| 18-103_k141_84081 | Not-determined |
| 18-103_k141_14724 | Not-determined |
| 18-103_k141_72368 | Not-determined |
| 18-103_k141_43572 | Not-determined |
| 18-103_k141_130195 | Not-determined |
| 18-103_k141_72371 | Low-quality |
| 18-103_k141_78155 | Low-quality |
| 18-103_k141_49400 | Low-quality |
| 18-103_k141_100959 | Not-determined |
| 18-103_k141_38066 | Not-determined |
| 18-103_k141_72376 | Not-determined |
| 18-103_k141_38067 | Not-determined |
| 18-103_k141_72380 | Not-determined |
| 18-103_k141_38070 | Not-determined |
| 18-103_k141_72384 | Not-determined |
| 18-103_k141_9084 | Not-determined |
| 18-103_k141_3694 | Not-determined |
| 18-103_k141_38075 | Not-determined |
| 18-103_k141_3697 | Not-determined |
| 18-103_k141_89994 | Not-determined |
| 18-103_k141_60876 | Not-determined |
| 18-103_k141_14738 | Not-determined |
| 18-103_k141_66748 | Not-determined |
| 18-103_k141_89997 | Not-determined |
| 18-103_k141_20690 | Low-quality |
| 18-103_k141_3702 | Not-determined |
| 18-103_k141_26509 | Not-determined |
| 18-103_k141_100976 | Low-quality |
| 18-103_k141_20698 | Not-determined |
| 18-103_k141_124338 | Not-determined |
| 18-103_k141_55268 | Not-determined |
| 18-103_k141_100983 | Not-determined |
| 18-103_k141_9102 | Not-determined |
| 18-103_k141_124346 | Not-determined |
| 18-103_k141_100987 | Not-determined |
| 18-103_k141_20705 | Not-determined |
| 18-103_k141_78180 | Not-determined |
| 18-103_k141_72404 | Not-determined |
| 18-103_k141_78183 | Not-determined |
| 18-103_k141_38104 | Not-determined |
| 18-103_k141_100993 | Not-determined |
| 18-103_k141_135723 | Not-determined |
| 18-103_k141_106918 | Not-determined |
| 18-103_k141_135724 | Low-quality |
| 18-103_k141_100999 | Not-determined |
| 18-103_k141_130229 | Not-determined |
| 18-103_k141_101000 | Not-determined |
| 18-103_k141_106921 | Not-determined |
| 18-103_k141_55275 | Not-determined |
| 18-103_k141_118717 | Not-determined |
| 18-103_k141_32012 | Low-quality |
| 18-103_k141_38109 | Not-determined |
| 18-103_k141_14751 | Low-quality |
| 18-103_k141_90019 | Not-determined |
| 18-103_k141_118728 | Not-determined |
| 18-103_k141_60910 | Not-determined |
| 18-103_k141_55285 | Not-determined |
| 18-103_k141_60911 | Not-determined |
| 18-103_k141_55286 | Not-determined |
| 18-103_k141_90022 | Not-determined |
| 18-103_k141_124370 | Not-determined |
| 18-103_k141_55287 | Not-determined |
| 18-103_k141_49427 | Not-determined |
| 18-103_k141_55288 | Not-determined |
| 18-103_k141_43607 | Not-determined |
| 18-103_k141_43608 | Not-determined |
| 18-103_k141_118734 | Not-determined |
| 18-103_k141_14754 | Not-determined |
| 18-103_k141_84119 | Not-determined |
| 18-103_k141_26538 | Low-quality |
| 18-103_k141_84120 | Not-determined |
| 18-103_k141_20726 | Not-determined |
| 18-103_k141_20727 | Not-determined |
| 18-103_k141_32028 | Not-determined |
| 18-103_k141_49438 | Not-determined |
| 18-103_k141_49439 | Low-quality |
| 18-103_k141_14757 | Not-determined |
| 18-103_k141_49440 | Not-determined |
| 18-103_k141_55294 | Not-determined |
| 18-103_k141_14759 | Not-determined |
| 18-103_k141_49441 | Low-quality |
| 18-103_k141_72428 | Not-determined |
| 18-103_k141_20739 | Not-determined |
| 18-103_k141_60937 | Not-determined |
| 18-103_k141_106952 | Not-determined |
| 18-103_k141_84129 | Not-determined |
| 18-103_k141_118742 | Not-determined |
| 18-103_k141_84132 | Not-determined |
| 18-103_k141_90029 | Not-determined |
| 18-103_k141_78208 | Not-determined |
| 18-103_k141_95578 | Not-determined |
| 18-103_k141_101022 | Not-determined |
| 18-103_k141_66784 | Not-determined |
| 18-103_k141_38142 | Not-determined |
| 18-103_k141_26563 | Not-determined |
| 18-103_k141_130269 | Not-determined |
| 18-103_k141_38144 | Not-determined |
| 18-103_k141_124380 | Not-determined |
| 18-103_k141_9124 | Not-determined |
| 18-103_k141_20750 | Not-determined |
| 18-103_k141_38151 | Low-quality |
| 18-103_k141_14776 | Not-determined |
| 18-103_k141_32044 | Not-determined |
| 18-103_k141_118750 | Not-determined |
| 18-103_k141_130281 | Not-determined |
| 18-103_k141_130282 | Not-determined |
| 18-103_k141_20758 | Not-determined |
| 18-103_k141_130284 | Not-determined |
| 18-103_k141_124394 | Not-determined |
| 18-103_k141_14783 | Not-determined |
| 18-103_k141_9139 | Not-determined |
| 18-103_k141_135749 | Low-quality |
| 18-103_k141_32055 | Not-determined |
| 18-103_k141_130288 | Not-determined |
| 18-103_k141_14787 | Low-quality |
| 18-103_k141_49461 | Not-determined |
| 18-103_k141_130291 | Not-determined |
| 18-103_k141_135754 | Not-determined |
| 18-103_k141_130300 | Not-determined |
| 18-103_k141_95602 | Not-determined |
| 18-103_k141_135762 | Not-determined |
| 18-103_k141_43645 | Not-determined |
| 18-103_k141_43647 | Not-determined |
| 18-103_k141_43648 | Not-determined |
| 18-103_k141_130305 | Low-quality |
| 18-103_k141_95606 | Not-determined |
| 18-103_k141_43650 | Not-determined |
| 18-103_k141_43652 | Not-determined |
| 18-103_k141_49471 | Not-determined |
| 18-103_k141_55326 | Not-determined |
| 18-103_k141_112806 | Low-quality |
| 18-103_k141_90056 | Not-determined |
| 18-103_k141_20788 | Not-determined |
| 18-103_k141_124408 | Not-determined |
| 18-103_k141_32077 | Not-determined |
| 18-103_k141_130309 | Not-determined |
| 18-103_k141_72469 | Not-determined |
| 18-103_k141_90062 | Not-determined |
| 18-103_k141_32080 | Not-determined |
| 18-103_k141_95614 | Not-determined |
| 18-103_k141_95615 | Not-determined |
| 18-103_k141_130315 | Not-determined |
| 18-103_k141_14814 | Low-quality |
| 18-103_k141_112814 | Not-determined |
| 18-103_k141_135776 | Low-quality |
| 18-103_k141_130319 | Not-determined |
| 18-103_k141_112816 | Not-determined |
| 18-103_k141_84165 | Not-determined |
| 18-103_k141_78237 | Not-determined |
| 18-103_k141_72477 | Not-determined |
| 18-103_k141_84169 | Not-determined |
| 18-103_k141_106989 | Not-determined |
| 18-103_k141_90078 | Not-determined |
| 18-103_k141_66815 | Not-determined |
| 18-103_k141_106991 | Not-determined |
| 18-103_k141_66816 | Low-quality |
| 18-103_k141_43674 | Not-determined |
| 18-103_k141_43675 | Not-determined |
| 18-103_k141_130334 | Low-quality |
| 18-103_k141_9183 | Not-determined |
| 18-103_k141_95623 | Not-determined |
| 18-103_k141_95624 | Not-determined |
| 18-103_k141_135787 | Low-quality |
| 18-103_k141_135788 | Not-determined |
| 18-103_k141_60977 | Not-determined |
| 18-103_k141_66821 | Not-determined |
| 18-103_k141_38205 | Not-determined |
| 18-103_k141_14831 | Not-determined |
| 18-103_k141_95633 | Not-determined |
| 18-103_k141_66823 | Not-determined |
| 18-103_k141_60985 | Not-determined |
| 18-103_k141_135797 | Low-quality |
| 18-103_k141_95637 | Not-determined |
| 18-103_k141_3774 | Not-determined |
| 18-103_k141_95638 | Not-determined |
| 18-103_k141_124419 | Low-quality |
| 18-103_k141_14836 | Not-determined |
| 18-103_k141_26618 | Not-determined |
| 18-103_k141_72497 | Not-determined |
| 18-103_k141_14842 | Not-determined |
| 18-103_k141_14844 | Not-determined |
| 18-103_k141_55366 | Not-determined |
| 18-103_k141_32111 | Not-determined |
| 18-103_k141_130358 | Not-determined |
| 18-103_k141_9193 | Not-determined |
| 18-103_k141_112830 | Not-determined |
| 18-103_k141_112831 | Not-determined |
| 18-103_k141_124426 | Not-determined |
| 18-103_k141_38224 | Not-determined |
| 18-103_k141_101095 | Not-determined |
| 18-103_k141_107015 | Not-determined |
| 18-103_k141_107016 | Not-determined |
| 18-103_k141_130365 | Not-determined |
| 18-103_k141_90111 | Not-determined |
| 18-103_k141_118808 | Not-determined |
| 18-103_k141_32118 | Not-determined |
| 18-103_k141_118814 | Not-determined |
| 18-103_k141_118817 | Not-determined |
| 18-103_k141_66835 | Not-determined |
| 18-103_k141_66836 | Not-determined |
| 18-103_k141_84204 | Not-determined |
| 18-103_k141_61015 | Not-determined |
| 18-103_k141_32125 | Not-determined |
| 18-103_k141_3785 | Not-determined |
| 18-103_k141_61020 | Not-determined |
| 18-103_k141_66843 | Not-determined |
| 18-103_k141_55382 | Not-determined |
| 18-103_k141_9220 | Not-determined |
| 18-103_k141_118830 | Not-determined |
| 18-103_k141_84214 | Not-determined |
| 18-103_k141_124445 | Low-quality |
| 18-103_k141_55391 | Not-determined |
| 18-103_k141_3794 | Not-determined |
| 18-103_k141_55392 | Not-determined |
| 18-103_k141_38248 | Not-determined |
| 18-103_k141_26644 | Not-determined |
| 18-103_k141_90144 | Not-determined |
| 18-103_k141_118844 | Not-determined |
| 18-103_k141_66864 | Not-determined |
| 18-103_k141_32137 | Low-quality |
| 18-103_k141_90147 | Not-determined |
| 18-103_k141_9230 | Not-determined |
| 18-103_k141_66869 | Not-determined |
| 18-103_k141_14857 | Not-determined |
| 18-103_k141_66870 | Not-determined |
| 18-103_k141_84221 | Not-determined |
| 18-103_k141_32148 | Not-determined |
| 18-103_k141_72524 | Not-determined |
| 18-103_k141_38271 | Not-determined |
| 18-103_k141_61032 | Not-determined |
| 18-103_k141_38274 | Not-determined |
| 18-103_k141_84226 | Not-determined |
| 18-103_k141_61035 | Not-determined |
| 18-103_k141_84227 | Not-determined |
| 18-103_k141_3803 | Not-determined |
| 18-103_k141_107047 | Not-determined |
| 18-103_k141_3804 | Not-determined |
| 18-103_k141_26661 | Not-determined |
| 18-103_k141_84231 | Low-quality |
| 18-103_k141_61039 | Not-determined |
| 18-103_k141_3809 | Not-determined |
| 18-103_k141_26671 | Not-determined |
| 18-103_k141_32161 | Not-determined |
| 18-103_k141_78317 | Not-determined |
| 18-103_k141_130405 | Not-determined |
| 18-103_k141_14876 | Not-determined |
| 18-103_k141_55403 | Not-determined |
| 18-103_k141_130411 | Not-determined |
| 18-103_k141_107062 | Not-determined |
| 18-103_k141_72539 | Not-determined |
| 18-103_k141_130413 | Not-determined |
| 18-103_k141_95691 | Low-quality |
| 18-103_k141_90173 | Not-determined |
| 18-103_k141_112874 | Not-determined |
| 18-103_k141_95694 | Not-determined |
| 18-103_k141_38287 | Not-determined |
| 18-103_k141_135832 | Not-determined |
| 18-103_k141_101129 | Low-quality |
| 18-103_k141_14882 | Not-determined |
| 18-103_k141_9259 | Not-determined |
| 18-103_k141_61046 | Not-determined |
| 18-103_k141_90180 | Not-determined |
| 18-103_k141_112881 | Not-determined |
| 18-103_k141_95705 | Not-determined |
| 18-103_k141_135837 | Not-determined |
| 18-103_k141_135840 | Not-determined |
| 18-103_k141_101135 | Not-determined |
| 18-103_k141_118889 | Not-determined |
| 18-103_k141_14886 | Not-determined |
| 18-103_k141_49550 | Not-determined |
| 18-103_k141_14891 | Low-quality |
| 18-103_k141_84267 | Not-determined |
| 18-103_k141_130423 | Not-determined |
| 18-103_k141_3816 | Not-determined |
| 18-103_k141_49551 | Low-quality |
| 18-103_k141_90188 | Not-determined |
| 18-103_k141_95717 | Not-determined |
| 18-103_k141_9268 | Not-determined |
| 18-103_k141_9269 | Not-determined |
| 18-103_k141_38298 | Not-determined |
| 18-103_k141_20857 | Not-determined |
| 18-103_k141_38300 | Not-determined |
| 18-103_k141_66913 | Not-determined |
| 18-103_k141_20865 | Low-quality |
| 18-103_k141_95725 | Not-determined |
| 18-103_k141_118907 | Low-quality |
| 18-103_k141_49561 | Low-quality |
| 18-103_k141_66916 | Not-determined |
| 18-103_k141_124509 | Not-determined |
| 18-103_k141_112893 | Low-quality |
| 18-103_k141_112894 | Not-determined |
| 18-103_k141_95732 | Not-determined |
| 18-103_k141_107084 | Not-determined |
| 18-103_k141_124511 | Not-determined |
| 18-103_k141_124512 | Not-determined |
| 18-103_k141_84278 | Not-determined |
| 18-103_k141_43721 | Not-determined |
| 18-103_k141_78364 | Low-quality |
| 18-103_k141_49563 | Low-quality |
| 18-103_k141_107091 | Not-determined |
| 18-103_k141_3844 | Not-determined |
| 18-103_k141_49566 | Not-determined |
| 18-103_k141_61059 | Not-determined |
| 18-103_k141_61060 | Not-determined |
| 18-103_k141_107097 | Not-determined |
| 18-103_k141_107099 | Not-determined |
| 18-103_k141_3853 | Low-quality |
| 18-103_k141_61062 | Not-determined |
| 18-103_k141_84284 | Not-determined |
| 18-103_k141_84285 | Not-determined |
| 18-103_k141_118925 | Not-determined |
| 18-103_k141_61068 | Low-quality |
| 18-103_k141_84288 | Not-determined |
| 18-103_k141_130448 | Not-determined |
| 18-103_k141_135869 | Not-determined |
| 18-103_k141_66928 | Low-quality |
| 18-103_k141_112912 | Not-determined |
| 18-103_k141_9295 | Not-determined |
| 18-103_k141_20892 | Not-determined |
| 18-103_k141_101165 | Not-determined |
| 18-103_k141_9299 | Low-quality |
| 18-103_k141_49578 | Not-determined |
| 18-103_k141_3855 | Low-quality |
| 18-103_k141_43735 | Not-determined |
| 18-103_k141_78378 | Not-determined |
| 18-103_k141_9304 | Not-determined |
| 18-103_k141_78381 | Not-determined |
| 18-103_k141_14930 | Not-determined |
| 18-103_k141_84292 | Not-determined |
| 18-103_k141_124538 | Not-determined |
| 18-103_k141_112927 | Not-determined |
| 18-103_k141_124541 | Not-determined |
| 18-103_k141_112930 | Not-determined |
| 18-103_k141_118942 | Not-determined |
| 18-103_k141_101179 | Not-determined |
| 18-103_k141_135885 | Not-determined |
| 18-103_k141_9317 | Not-determined |
| 18-103_k141_112931 | Not-determined |
| 18-103_k141_95757 | Not-determined |
| 18-103_k141_49587 | Not-determined |
| 18-103_k141_118945 | Low-quality |
| 18-103_k141_14941 | Not-determined |
| 18-103_k141_107118 | Not-determined |
| 18-103_k141_9320 | Low-quality |
| 18-103_k141_14947 | Not-determined |
| 18-103_k141_38336 | Not-determined |
| 18-103_k141_130467 | Not-determined |
| 18-103_k141_101187 | Not-determined |
| 18-103_k141_124555 | Not-determined |
| 18-103_k141_14954 | Not-determined |
| 18-103_k141_20911 | Not-determined |
| 18-103_k141_84317 | Low-quality |
| 18-103_k141_107128 | Not-determined |
| 18-103_k141_78392 | Not-determined |
| 18-103_k141_78393 | Not-determined |
| 18-103_k141_130470 | Not-determined |
| 18-103_k141_55459 | Not-determined |
| 18-103_k141_32231 | Low-quality |
| 18-103_k141_90205 | Not-determined |
| 18-103_k141_101194 | Low-quality |
| 18-103_k141_124561 | Not-determined |
| 18-103_k141_3887 | Not-determined |
| 18-103_k141_32233 | Not-determined |
| 18-103_k141_32234 | Low-quality |
| 18-103_k141_124563 | Not-determined |
| 18-103_k141_107139 | Not-determined |
| 18-103_k141_118951 | Not-determined |
| 18-103_k141_61089 | Not-determined |
| 18-103_k141_55468 | Not-determined |
| 18-103_k141_20915 | Low-quality |
| 18-103_k141_9341 | Not-determined |
| 18-103_k141_84337 | Not-determined |
| 18-103_k141_78404 | Not-determined |
| 18-103_k141_43769 | Not-determined |
| 18-103_k141_84339 | Not-determined |
| 18-103_k141_3903 | Not-determined |
| 18-103_k141_20919 | Not-determined |
| 18-103_k141_32237 | Low-quality |
| 18-103_k141_61095 | Not-determined |
| 18-103_k141_20923 | Low-quality |
| 18-103_k141_72577 | Low-quality |
| 18-103_k141_107143 | Not-determined |
| 18-103_k141_26700 | Not-determined |
| 18-103_k141_107146 | Not-determined |
| 18-103_k141_26705 | Not-determined |
| 18-103_k141_130486 | Low-quality |
| 18-103_k141_90220 | Not-determined |
| 18-103_k141_61105 | Not-determined |
| 18-103_k141_49605 | Not-determined |
| 18-103_k141_124575 | Not-determined |
| 18-103_k141_49611 | Not-determined |
| 18-103_k141_135920 | Not-determined |

| | |
|---|---|
| 18-103_k141_55491 | Not-determined |
| 18-103_k141_66968 | Not-determined |
| 18-103_k141_43782 | Not-determined |
| 18-103_k141_135926 | Not-determined |
| 18-103_k141_135928 | Not-determined |
| 18-103_k141_49617 | Not-determined |
| 18-103_k141_107159 | Not-determined |
| 18-103_k141_95791 | Not-determined |
| 18-103_k141_112960 | Not-determined |
| 18-103_k141_55498 | Not-determined |
| 18-103_k141_112962 | Not-determined |
| 18-103_k141_43791 | Not-determined |
| 18-103_k141_3926 | Not-determined |
| 18-103_k141_124597 | Not-determined |
| 18-103_k141_3930 | Not-determined |
| 18-103_k141_43798 | Not-determined |
| 18-103_k141_112969 | Not-determined |
| 18-103_k141_124600 | Not-determined |
| 18-103_k141_20937 | Not-determined |
| 18-103_k141_84352 | Not-determined |
| 18-103_k141_61131 | Not-determined |
| 18-103_k141_84354 | Not-determined |
| 18-103_k141_72599 | Not-determined |
| 18-103_k141_72600 | Not-determined |
| 18-103_k141_112974 | Not-determined |
| 18-103_k141_3933 | Not-determined |
| 18-103_k141_3936 | Not-determined |
| 18-103_k141_118983 | Not-determined |
| 18-103_k141_55502 | Low-quality |
| 18-103_k141_118989 | Not-determined |
| 18-103_k141_55507 | Not-determined |
| 18-103_k141_72611 | Not-determined |
| 18-103_k141_66996 | Low-quality |
| 18-103_k141_20961 | Not-determined |
| 18-103_k141_78449 | Not-determined |
| 18-103_k141_66998 | Low-quality |
| 18-103_k141_90258 | Low-quality |
| 18-103_k141_84370 | Not-determined |
| 18-103_k141_78456 | Not-determined |
| 18-103_k141_130512 | Not-determined |
| 18-103_k141_26728 | Not-determined |
| 18-103_k141_119006 | Not-determined |
| 18-103_k141_135952 | Not-determined |
| 18-103_k141_49630 | Not-determined |
| 18-103_k141_130516 | Not-determined |
| 18-103_k141_38385 | Not-determined |
| 18-103_k141_84374 | Not-determined |
| 18-103_k141_20975 | Not-determined |
| 18-103_k141_9372 | Low-quality |
| 18-103_k141_49636 | Low-quality |
| 18-103_k141_9377 | Not-determined |
| 18-103_k141_107208 | Not-determined |
| 18-103_k141_26754 | Not-determined |
| 18-103_k141_20987 | Not-determined |
| 18-103_k141_135966 | Not-determined |
| 18-103_k141_78482 | Not-determined |
| 18-103_k141_112999 | Not-determined |
| 18-103_k141_55537 | Not-determined |
| 18-103_k141_84391 | Not-determined |
| 18-103_k141_55538 | Low-quality |
| 18-103_k141_26764 | Not-determined |
| 18-103_k141_61174 | Low-quality |
| 18-103_k141_61175 | Not-determined |
| 18-103_k141_61180 | Not-determined |
| 18-103_k141_61182 | Not-determined |
| 18-103_k141_43853 | Not-determined |
| 18-103_k141_84400 | Not-determined |
| 18-103_k141_119041 | Not-determined |
| 18-103_k141_55543 | Not-determined |
| 18-103_k141_84403 | Not-determined |
| 18-103_k141_113013 | Not-determined |
| 18-103_k141_38408 | Not-determined |
| 18-103_k141_84412 | Not-determined |
| 18-103_k141_119058 | Not-determined |
| 18-103_k141_38414 | Not-determined |
| 18-103_k141_32294 | Not-determined |
| 18-103_k141_78502 | Low-quality |
| 18-103_k141_9399 | Not-determined |
| 18-103_k141_43863 | Not-determined |
| 18-103_k141_135972 | Not-determined |
| 18-103_k141_9402 | Not-determined |
| 18-103_k141_26783 | Not-determined |
| 18-103_k141_32303 | Not-determined |
| 18-103_k141_119063 | Not-determined |
| 18-103_k141_49663 | Not-determined |
| 18-103_k141_135978 | Not-determined |
| 18-103_k141_43872 | Not-determined |
| 18-103_k141_124675 | Not-determined |
| 18-103_k141_124676 | Not-determined |
| 18-103_k141_43875 | Not-determined |
| 18-103_k141_130549 | Low-quality |
| 18-103_k141_130550 | Not-determined |
| 18-103_k141_107252 | Not-determined |
| 18-103_k141_135985 | Not-determined |
| 18-103_k141_38428 | Not-determined |
| 18-103_k141_67058 | Not-determined |
| 18-103_k141_95835 | Not-determined |
| 18-103_k141_67059 | Not-determined |
| 18-103_k141_38433 | Not-determined |
| 18-103_k141_43883 | Not-determined |
| 18-103_k141_21014 | Not-determined |
| 18-103_k141_21015 | Not-determined |
| 18-103_k141_67068 | Not-determined |
| 18-103_k141_95841 | Not-determined |
| 18-103_k141_113033 | Not-determined |
| 18-103_k141_124688 | Not-determined |
| 18-103_k141_84433 | Low-quality |
| 18-103_k141_21018 | Not-determined |
| 18-103_k141_15001 | Not-determined |
| 18-103_k141_67072 | Not-determined |
| 18-103_k141_113043 | Not-determined |
| 18-103_k141_113047 | Not-determined |
| 18-103_k141_107260 | Not-determined |
| 18-103_k141_107264 | Not-determined |
| 18-103_k141_43894 | Low-quality |
| 18-103_k141_43898 | Not-determined |
| 18-103_k141_119098 | Not-determined |
| 18-103_k141_61211 | Not-determined |
| 18-103_k141_67086 | Not-determined |
| 18-103_k141_49682 | Not-determined |
| 18-103_k141_26831 | Not-determined |
| 18-103_k141_55584 | Not-determined |
| 18-103_k141_61220 | Not-determined |
| 18-103_k141_61221 | Not-determined |
| 18-103_k141_55587 | Not-determined |
| 18-103_k141_26837 | Not-determined |
| 18-103_k141_15013 | Not-determined |
| 18-103_k141_107282 | Not-determined |
| 18-103_k141_55596 | Not-determined |
| 18-103_k141_95849 | Not-determined |
| 18-103_k141_124721 | Low-quality |
| 18-103_k141_9417 | Not-determined |
| 18-103_k141_49694 | Not-determined |
| 18-103_k141_15029 | Not-determined |
| 18-103_k141_130586 | Not-determined |
| 18-103_k141_43921 | Not-determined |
| 18-103_k141_90326 | Not-determined |
| 18-103_k141_49698 | Not-determined |
| 18-103_k141_119115 | Not-determined |
| 18-103_k141_119117 | Not-determined |
| 18-103_k141_67105 | Low-quality |
| 18-103_k141_78558 | Not-determined |
| 18-103_k141_49703 | Not-determined |
| 18-103_k141_21068 | Not-determined |
| 18-103_k141_107304 | Not-determined |
| 18-103_k141_95857 | Not-determined |
| 18-103_k141_124735 | Not-determined |
| 18-103_k141_49711 | Not-determined |
| 18-103_k141_32360 | Not-determined |
| 18-103_k141_43927 | Not-determined |
| 18-103_k141_9429 | Not-determined |
| 18-103_k141_43929 | Not-determined |
| 18-103_k141_9430 | Not-determined |
| 18-103_k141_67115 | Not-determined |
| 18-103_k141_95864 | Not-determined |
| 18-103_k141_21077 | Not-determined |
| 18-103_k141_78566 | Not-determined |
| 18-103_k141_72636 | Not-determined |
| 18-103_k141_95867 | Not-determined |
| 18-103_k141_21083 | Not-determined |
| 18-103_k141_84463 | Not-determined |
| 18-103_k141_15048 | Not-determined |
| 18-103_k141_55615 | Not-determined |
| 18-103_k141_78571 | Not-determined |
| 18-103_k141_107322 | Low-quality |
| 18-103_k141_90335 | Not-determined |
| 18-103_k141_55619 | Not-determined |
| 18-103_k141_136042 | Not-determined |
| 18-103_k141_107328 | Not-determined |
| 18-103_k141_78574 | Not-determined |
| 18-103_k141_130612 | Not-determined |
| 18-103_k141_119134 | Not-determined |
| 18-103_k141_107332 | Not-determined |
| 18-103_k141_72650 | Not-determined |
| 18-103_k141_78578 | Not-determined |
| 18-103_k141_119137 | Not-determined |
| 18-103_k141_113097 | Not-determined |
| 18-103_k141_90341 | Not-determined |
| 18-103_k141_15049 | Not-determined |
| 18-103_k141_4041 | Low-quality |
| 18-103_k141_67122 | Low-quality |
| 18-103_k141_4043 | Not-determined |
| 18-103_k141_113098 | Not-determined |
| 18-103_k141_55625 | Not-determined |
| 18-103_k141_72655 | Low-quality |
| 18-103_k141_90344 | Not-determined |
| 18-103_k141_101238 | Not-determined |
| 18-103_k141_95876 | Not-determined |
| 18-103_k141_130622 | Not-determined |
| 18-103_k141_72658 | Not-determined |
| 18-103_k141_119142 | Not-determined |
| 18-103_k141_55633 | Not-determined |
| 18-103_k141_21097 | Low-quality |
| 18-103_k141_90347 | Not-determined |
| 18-103_k141_72663 | Not-determined |
| 18-103_k141_49732 | Low-quality |
| 18-103_k141_21099 | Not-determined |
| 18-103_k141_43953 | Not-determined |
| 18-103_k141_107337 | Not-determined |
| 18-103_k141_43957 | Not-determined |
| 18-103_k141_15063 | Not-determined |
| 18-103_k141_101255 | Not-determined |
| 18-103_k141_84487 | Not-determined |
| 18-103_k141_4051 | Not-determined |
| 18-103_k141_95894 | Not-determined |
| 18-103_k141_101260 | Not-determined |
| 18-103_k141_55647 | Not-determined |
| 18-103_k141_95896 | Not-determined |
| 18-103_k141_4055 | Not-determined |
| 18-103_k141_84496 | Low-quality |
| 18-103_k141_26881 | Not-determined |
| 18-103_k141_4062 | Not-determined |
| 18-103_k141_84501 | Not-determined |
| 18-103_k141_130637 | Not-determined |
| 18-103_k141_32397 | Not-determined |
| 18-103_k141_90364 | Low-quality |
| 18-103_k141_26891 | Not-determined |
| 18-103_k141_43978 | Not-determined |
| 18-103_k141_32404 | Not-determined |
| 18-103_k141_101267 | Not-determined |
| 18-103_k141_130641 | Low-quality |
| 18-103_k141_61238 | Not-determined |
| 18-103_k141_72672 | Not-determined |
| 18-103_k141_136072 | Not-determined |
| 18-103_k141_130647 | Low-quality |
| 18-103_k141_136073 | Not-determined |
| 18-103_k141_9474 | Not-determined |
| 18-103_k141_67151 | Not-determined |
| 18-103_k141_21122 | Not-determined |
| 18-103_k141_84519 | Low-quality |
| 18-103_k141_113136 | Not-determined |
| 18-103_k141_113137 | Not-determined |
| 18-103_k141_15095 | Not-determined |
| 18-103_k141_78606 | Not-determined |
| 18-103_k141_90386 | Not-determined |
| 18-103_k141_136079 | Low-quality |
| 18-103_k141_113142 | Not-determined |
| 18-103_k141_119166 | Low-quality |
| 18-103_k141_67164 | Not-determined |
| 18-103_k141_119168 | Not-determined |
| 18-103_k141_4089 | Not-determined |
| 18-103_k141_78616 | Low-quality |
| 18-103_k141_38489 | Not-determined |
| 18-103_k141_55672 | Not-determined |
| 18-103_k141_49777 | Not-determined |
| 18-103_k141_84526 | Not-determined |
| 18-103_k141_124784 | Not-determined |
| 18-103_k141_26914 | Not-determined |
| 18-103_k141_26916 | Not-determined |
| 18-103_k141_55683 | Not-determined |
| 18-103_k141_9502 | Not-determined |
| 18-103_k141_101288 | Not-determined |
| 18-103_k141_95941 | Not-determined |
| 18-103_k141_49780 | Low-quality |
| 18-103_k141_15119 | Not-determined |
| 18-103_k141_61260 | Not-determined |
| 18-103_k141_113168 | Low-quality |
| 18-103_k141_44001 | Not-determined |
| 18-103_k141_113170 | Not-determined |
| 18-103_k141_136101 | Low-quality |
| 18-103_k141_90405 | Not-determined |
| 18-103_k141_26930 | Low-quality |
| 18-103_k141_101295 | Not-determined |
| 18-103_k141_44005 | Not-determined |
| 18-103_k141_55694 | Not-determined |
| 18-103_k141_113178 | Not-determined |
| 18-103_k141_61274 | Low-quality |
| 18-103_k141_113180 | Not-determined |
| 18-103_k141_90413 | Low-quality |
| 18-103_k141_67182 | Not-determined |
| 18-103_k141_84543 | Low-quality |
| 18-103_k141_90419 | Low-quality |
| 18-103_k141_101302 | Not-determined |
| 18-103_k141_113185 | Not-determined |
| 18-103_k141_38522 | Low-quality |
| 18-103_k141_32424 | Not-determined |
| 18-103_k141_9521 | Not-determined |
| 18-103_k141_90423 | Not-determined |
| 18-103_k141_95959 | Not-determined |
| 18-103_k141_9531 | Not-determined |
| 18-103_k141_84547 | Not-determined |
| 18-103_k141_84549 | Not-determined |
| 18-103_k141_55705 | Not-determined |
| 18-103_k141_26950 | Not-determined |
| 18-103_k141_90430 | Not-determined |
| 18-103_k141_90431 | Not-determined |
| 18-103_k141_61286 | Not-determined |
| 18-103_k141_61287 | Not-determined |
| 18-103_k141_4110 | Not-determined |
| 18-103_k141_67195 | Not-determined |
| 18-103_k141_113193 | Not-determined |
| 18-103_k141_9536 | Not-determined |
| 18-103_k141_72726 | Not-determined |
| 18-103_k141_130686 | Not-determined |
| 18-103_k141_119212 | Low-quality |
| 18-103_k141_136124 | Not-determined |
| 18-103_k141_26961 | Not-determined |
| 18-103_k141_61289 | Low-quality |
| 18-103_k141_84560 | Low-quality |
| 18-103_k141_21177 | Not-determined |
| 18-103_k141_61294 | Not-determined |
| 18-103_k141_61298 | Not-determined |
| 18-103_k141_107387 | Not-determined |
| 18-103_k141_72739 | Not-determined |
| 18-103_k141_72742 | Not-determined |
| 18-103_k141_136138 | Not-determined |
| 18-103_k141_113213 | Not-determined |
| 18-103_k141_61305 | Not-determined |
| 18-103_k141_4130 | Not-determined |
| 18-103_k141_38545 | Not-determined |
| 18-103_k141_26978 | Not-determined |
| 18-103_k141_4135 | Not-determined |
| 18-103_k141_38548 | Low-quality |
| 18-103_k141_72756 | Not-determined |
| 18-103_k141_38550 | Not-determined |
| 18-103_k141_107406 | Not-determined |
| 18-103_k141_61314 | Not-determined |
| 18-103_k141_32460 | Not-determined |
| 18-103_k141_130716 | Not-determined |
| 18-103_k141_67229 | Not-determined |
| 18-103_k141_32463 | Not-determined |
| 18-103_k141_113224 | Not-determined |
| 18-103_k141_38557 | Not-determined |
| 18-103_k141_9569 | Not-determined |
| 18-103_k141_119237 | Not-determined |
| 18-103_k141_136159 | Not-determined |
| 18-103_k141_136162 | Not-determined |
| 18-103_k141_78657 | Low-quality |
| 18-103_k141_44039 | Low-quality |
| 18-103_k141_124843 | Not-determined |
| 18-103_k141_78659 | Not-determined |
| 18-103_k141_44041 | Low-quality |
| 18-103_k141_95977 | Not-determined |
| 18-103_k141_113234 | Not-determined |
| 18-103_k141_90441 | Not-determined |
| 18-103_k141_107408 | Not-determined |
| 18-103_k141_84591 | Not-determined |
| 18-103_k141_107411 | Not-determined |
| 18-103_k141_9579 | Not-determined |
| 18-103_k141_32475 | Not-determined |
| 18-103_k141_90444 | Not-determined |
| 18-103_k141_107412 | Not-determined |
| 18-103_k141_55727 | Not-determined |
| 18-103_k141_21210 | Not-determined |
| 18-103_k141_32476 | Not-determined |
| 18-103_k141_124846 | Not-determined |
| 18-103_k141_124848 | Low-quality |
| 18-103_k141_78667 | Not-determined |
| 18-103_k141_9583 | Not-determined |
| 18-103_k141_136171 | Not-determined |
| 18-103_k141_113246 | Not-determined |
| 18-103_k141_9585 | Not-determined |
| 18-103_k141_124851 | Not-determined |
| 18-103_k141_21216 | Not-determined |
| 18-103_k141_44055 | Not-determined |
| 18-103_k141_78672 | Low-quality |
| 18-103_k141_84595 | Not-determined |
| 18-103_k141_67241 | Low-quality |
| 18-103_k141_124857 | Not-determined |
| 18-103_k141_21224 | Not-determined |
| 18-103_k141_101335 | Low-quality |
| 18-103_k141_130736 | Not-determined |
| 18-103_k141_61326 | Not-determined |
| 18-103_k141_136184 | Not-determined |
| 18-103_k141_90455 | Not-determined |
| 18-103_k141_119255 | Not-determined |
| 18-103_k141_107429 | Not-determined |
| 18-103_k141_49820 | Not-determined |
| 18-103_k141_61333 | Not-determined |
| 18-103_k141_55739 | Not-determined |
| 18-103_k141_9600 | Not-determined |
| 18-103_k141_55741 | Not-determined |
| 18-103_k141_67251 | Low-quality |
| 18-103_k141_61337 | Not-determined |
| 18-103_k141_9602 | Not-determined |
| 18-103_k141_9604 | Not-determined |
| 18-103_k141_61340 | Not-determined |
| 18-103_k141_44061 | Not-determined |
| 18-103_k141_90465 | Not-determined |
| 18-103_k141_130744 | Not-determined |
| 18-103_k141_130745 | Not-determined |
| 18-103_k141_90469 | Not-determined |
| 18-103_k141_15148 | Not-determined |
| 18-103_k141_96015 | Not-determined |
| 18-103_k141_78697 | Not-determined |
| 18-103_k141_9611 | Low-quality |
| 18-103_k141_136190 | Low-quality |
| 18-103_k141_49829 | Not-determined |
| 18-103_k141_32498 | Not-determined |
| 18-103_k141_38595 | Not-determined |
| 18-103_k141_119282 | Not-determined |
| 18-103_k141_130764 | Not-determined |
| 18-103_k141_136195 | Not-determined |
| 18-103_k141_21260 | Not-determined |
| 18-103_k141_44066 | Not-determined |
| 18-103_k141_136200 | Not-determined |
| 18-103_k141_9616 | Not-determined |
| 18-103_k141_38599 | Not-determined |
| 18-103_k141_72811 | Not-determined |
| 18-103_k141_9617 | Not-determined |
| 18-103_k141_136201 | Not-determined |
| 18-103_k141_44068 | Not-determined |
| 18-103_k141_78710 | Not-determined |
| 18-103_k141_9620 | Not-determined |
| 18-103_k141_61360 | Not-determined |
| 18-103_k141_84620 | Not-determined |
| 18-103_k141_4180 | Not-determined |
| 18-103_k141_27042 | Not-determined |
| 18-103_k141_61369 | Low-quality |
| 18-103_k141_21264 | Not-determined |
| 18-103_k141_55757 | Not-determined |
| 18-103_k141_32515 | Not-determined |
| 18-103_k141_21266 | Not-determined |
| 18-103_k141_84628 | Not-determined |
| 18-103_k141_44072 | Not-determined |
| 18-103_k141_21271 | Not-determined |
| 18-103_k141_136214 | Not-determined |
| 18-103_k141_21274 | Not-determined |
| 18-103_k141_119300 | Low-quality |
| 18-103_k141_9631 | Not-determined |
| 18-103_k141_49841 | Not-determined |
| 18-103_k141_49845 | Not-determined |
| 18-103_k141_49848 | Not-determined |
| 18-103_k141_21283 | Not-determined |
| 18-103_k141_27059 | Not-determined |
| 18-103_k141_44082 | Not-determined |
| 18-103_k141_44083 | Not-determined |
| 18-103_k141_136218 | Not-determined |
| 18-103_k141_61376 | Not-determined |
| 18-103_k141_84648 | Not-determined |
| 18-103_k141_136220 | Not-determined |
| 18-103_k141_124910 | Low-quality |
| 18-103_k141_32529 | Not-determined |
| 18-103_k141_38623 | Not-determined |
| 18-103_k141_61382 | Not-determined |
| 18-103_k141_61383 | Not-determined |
| 18-103_k141_84653 | Not-determined |
| 18-103_k141_101381 | Not-determined |
| 18-103_k141_38626 | Not-determined |
| 18-103_k141_96056 | Not-determined |
| 18-103_k141_101384 | Not-determined |
| 18-103_k141_49857 | Not-determined |
| 18-103_k141_15170 | Not-determined |
| 18-103_k141_101388 | Not-determined |
| 18-103_k141_44101 | Not-determined |
| 18-103_k141_61393 | Not-determined |
| 18-103_k141_101395 | Not-determined |
| 18-103_k141_107471 | Not-determined |
| 18-103_k141_78742 | Not-determined |
| 18-103_k141_136230 | Not-determined |
| 18-103_k141_27070 | Not-determined |
| 18-103_k141_55772 | Low-quality |
| 18-103_k141_78746 | Not-determined |
| 18-103_k141_119332 | Not-determined |
| 18-103_k141_49871 | Not-determined |
| 18-103_k141_67322 | Not-determined |
| 18-103_k141_136232 | Not-determined |
| 18-103_k141_124937 | Not-determined |
| 18-103_k141_38645 | Not-determined |
| 18-103_k141_61409 | Low-quality |
| 18-103_k141_67325 | Not-determined |
| 18-103_k141_90488 | Not-determined |
| 18-103_k141_67328 | Not-determined |
| 18-103_k141_15184 | Not-determined |
| 18-103_k141_15186 | Low-quality |
| 18-103_k141_21304 | Not-determined |
| 18-103_k141_119351 | Not-determined |
| 18-103_k141_4210 | Not-determined |
| 18-103_k141_124951 | Not-determined |
| 18-103_k141_61418 | Not-determined |
| 18-103_k141_67331 | Not-determined |
| 18-103_k141_44112 | Not-determined |
| 18-103_k141_67334 | Not-determined |
| 18-103_k141_61420 | Low-quality |
| 18-103_k141_96079 | Not-determined |
| 18-103_k141_32567 | Not-determined |
| 18-103_k141_55790 | Not-determined |
| 18-103_k141_130815 | Not-determined |
| 18-103_k141_119354 | Not-determined |
| 18-103_k141_78760 | Not-determined |
| 18-103_k141_15201 | Not-determined |
| 18-103_k141_4221 | Not-determined |
| 18-103_k141_49891 | Not-determined |
| 18-103_k141_15205 | Low-quality |
| 18-103_k141_61429 | Not-determined |
| 18-103_k141_27099 | Not-determined |
| 18-103_k141_21315 | Not-determined |
| 18-103_k141_130826 | Not-determined |
| 18-103_k141_124964 | Not-determined |
| 18-103_k141_61433 | Not-determined |
| 18-103_k141_72843 | Not-determined |
| 18-103_k141_136254 | Low-quality |
| 18-103_k141_4233 | Not-determined |
| 18-103_k141_78767 | Low-quality |
| 18-103_k141_55805 | Not-determined |
| 18-103_k141_78769 | Not-determined |
| 18-103_k141_27109 | Not-determined |
| 18-103_k141_130835 | Not-determined |
| 18-103_k141_32585 | Not-determined |
| 18-103_k141_4239 | Not-determined |
| 18-103_k141_113348 | Not-determined |
| 18-103_k141_90511 | Not-determined |
| 18-103_k141_4243 | Not-determined |
| 18-103_k141_90513 | Not-determined |
| 18-103_k141_136266 | Not-determined |
| 18-103_k141_55817 | Not-determined |
| 18-103_k141_113356 | Not-determined |
| 18-103_k141_27122 | Not-determined |
| 18-103_k141_27123 | Not-determined |
| 18-103_k141_38692 | Not-determined |
| 18-103_k141_136275 | Not-determined |
| 18-103_k141_67366 | Not-determined |
| 18-103_k141_72868 | Not-determined |
| 18-103_k141_4259 | Low-quality |
| 18-103_k141_84704 | Not-determined |
| 18-103_k141_78789 | Not-determined |
| 18-103_k141_4265 | Not-determined |
| 18-103_k141_67369 | Not-determined |
| 18-103_k141_84707 | Not-determined |
| 18-103_k141_72873 | Not-determined |
| 18-103_k141_9708 | Not-determined |
| 18-103_k141_101454 | Not-determined |
| 18-103_k141_130858 | Not-determined |
| 18-103_k141_72878 | Not-determined |
| 18-103_k141_136282 | Not-determined |
| 18-103_k141_90534 | Not-determined |
| 18-103_k141_72882 | Not-determined |
| 18-103_k141_61465 | Not-determined |
| 18-103_k141_32606 | Not-determined |
| 18-103_k141_119382 | Not-determined |
| 18-103_k141_78799 | Not-determined |
| 18-103_k141_61470 | Not-determined |
| 18-103_k141_32610 | Not-determined |
| 18-103_k141_49902 | Not-determined |
| 18-103_k141_78806 | Not-determined |
| 18-103_k141_21334 | Not-determined |
| 18-103_k141_101469 | Not-determined |
| 18-103_k141_32616 | Not-determined |
| 18-103_k141_38711 | Not-determined |
| 18-103_k141_44157 | Not-determined |
| 18-103_k141_32618 | Not-determined |
| 18-103_k141_136293 | Not-determined |
| 18-103_k141_78812 | Not-determined |
| 18-103_k141_44162 | Not-determined |
| 18-103_k141_124997 | Not-determined |
| 18-103_k141_61479 | Not-determined |
| 18-103_k141_21342 | Not-determined |
| 18-103_k141_67374 | Not-determined |
| 18-103_k141_136299 | Not-determined |
| 18-103_k141_67380 | Not-determined |
| 18-103_k141_90550 | Not-determined |
| 18-103_k141_72905 | Not-determined |
| 18-103_k141_15230 | Not-determined |
| 18-103_k141_72907 | Not-determined |
| 18-103_k141_90552 | Not-determined |
| 18-103_k141_9717 | Not-determined |
| 18-103_k141_38720 | Not-determined |
| 18-103_k141_101496 | Not-determined |
| 18-103_k141_125014 | Not-determined |
| 18-103_k141_38722 | Not-determined |
| 18-103_k141_49930 | Not-determined |
| 18-103_k141_67392 | Not-determined |
| 18-103_k141_84749 | Not-determined |
| 18-103_k141_67397 | Not-determined |
| 18-103_k141_119418 | Not-determined |
| 18-103_k141_61502 | Not-determined |
| 18-103_k141_4283 | Not-determined |
| 18-103_k141_15244 | Not-determined |
| 18-103_k141_125037 | Not-determined |
| 18-103_k141_125038 | Not-determined |
| 18-103_k141_113392 | Not-determined |
| 18-103_k141_49939 | Not-determined |
| 18-103_k141_15247 | Not-determined |
| 18-103_k141_136327 | Not-determined |
| 18-103_k141_49940 | Not-determined |
| 18-103_k141_119434 | Not-determined |
| 18-103_k141_15249 | Not-determined |
| 18-103_k141_130875 | Not-determined |
| 18-103_k141_84754 | Not-determined |
| 18-103_k141_38751 | Not-determined |
| 18-103_k141_9735 | Not-determined |
| 18-103_k141_44185 | Not-determined |
| 18-103_k141_96151 | Not-determined |
| 18-103_k141_61510 | Not-determined |
| 18-103_k141_72932 | Not-determined |
| 18-103_k141_107537 | Not-determined |
| 18-103_k141_61515 | Not-determined |
| 18-103_k141_32637 | Not-determined |
| 18-103_k141_101527 | Not-determined |

| | |
|---|---|
| 18-103_k141_96157 | Not-determined |
| 18-103_k141_107540 | Not-determined |
| 18-103_k141_32644 | Not-determined |
| 18-103_k141_61520 | Not-determined |
| 18-103_k141_72943 | Not-determined |
| 18-103_k141_67410 | Not-determined |
| 18-103_k141_15271 | Not-determined |
| 18-103_k141_96166 | Not-determined |
| 18-103_k141_4292 | Not-determined |
| 18-103_k141_101537 | Not-determined |
| 18-103_k141_84779 | Not-determined |
| 18-103_k141_90575 | Not-determined |
| 18-103_k141_55845 | Low-quality |
| 18-103_k141_84781 | Not-determined |
| 18-103_k141_44201 | Not-determined |
| 18-103_k141_119452 | Low-quality |
| 18-103_k141_113422 | Not-determined |
| 18-103_k141_72949 | Not-determined |
| 18-103_k141_119453 | Not-determined |
| 18-103_k141_55847 | Not-determined |
| 18-103_k141_61530 | Not-determined |
| 18-103_k141_4297 | Not-determined |
| 18-103_k141_113424 | Low-quality |
| 18-103_k141_55848 | Low-quality |
| 18-103_k141_72954 | Not-determined |
| 18-103_k141_113428 | Not-determined |
| 18-103_k141_96176 | Not-determined |
| 18-103_k141_49956 | Not-determined |
| 18-103_k141_4303 | Not-determined |
| 18-103_k141_96183 | Not-determined |
| 18-103_k141_107558 | Not-determined |
| 18-103_k141_125050 | Low-quality |
| 18-103_k141_119463 | Not-determined |
| 18-103_k141_130894 | Not-determined |
| 18-103_k141_101546 | Not-determined |
| 18-103_k141_136348 | Not-determined |
| 18-103_k141_119466 | Not-determined |
| 18-103_k141_67426 | Not-determined |
| 18-103_k141_136352 | Not-determined |
| 18-103_k141_119469 | Not-determined |
| 18-103_k141_49971 | Not-determined |
| 18-103_k141_72959 | Not-determined |
| 18-103_k141_55866 | Not-determined |
| 18-103_k141_44231 | Not-determined |
| 18-103_k141_125071 | Not-determined |
| 18-103_k141_21386 | Not-determined |
| 18-103_k141_15297 | Not-determined |
| 18-103_k141_21388 | Not-determined |
| 18-103_k141_21389 | Not-determined |
| 18-103_k141_136359 | Not-determined |
| 18-103_k141_15303 | Not-determined |
| 18-103_k141_4328 | Not-determined |
| 18-103_k141_119481 | Not-determined |
| 18-103_k141_78864 | Not-determined |
| 18-103_k141_107572 | Low-quality |
| 18-103_k141_21395 | Not-determined |
| 18-103_k141_49995 | Not-determined |
| 18-103_k141_49998 | Low-quality |
| 18-103_k141_50001 | Low-quality |
| 18-103_k141_27182 | Not-determined |
| 18-103_k141_113460 | Not-determined |
| 18-103_k141_119486 | Not-determined |
| 18-103_k141_50003 | Not-determined |
| 18-103_k141_21399 | Not-determined |
| 18-103_k141_107577 | Not-determined |
| 18-103_k141_107579 | Not-determined |
| 18-103_k141_96207 | Low-quality |
| 18-103_k141_78872 | Not-determined |
| 18-103_k141_72981 | Not-determined |
| 18-103_k141_32673 | Not-determined |
| 18-103_k141_15318 | Low-quality |
| 18-103_k141_136378 | Not-determined |
| 18-103_k141_50005 | Not-determined |
| 18-103_k141_125094 | Not-determined |
| 18-103_k141_61547 | Not-determined |
| 18-103_k141_136383 | Low-quality |
| 18-103_k141_119493 | Low-quality |
| 18-103_k141_101572 | Not-determined |
| 18-103_k141_38822 | Not-determined |
| 18-103_k141_119495 | Not-determined |
| 18-103_k141_96225 | Not-determined |
| 18-103_k141_38826 | Not-determined |
| 18-103_k141_84800 | Not-determined |
| 18-103_k141_130937 | Not-determined |
| 18-103_k141_38832 | Not-determined |
| 18-103_k141_113468 | Not-determined |
| 18-103_k141_72997 | Not-determined |
| 18-103_k141_125107 | Not-determined |
| 18-103_k141_9753 | Low-quality |
| 18-103_k141_61563 | Not-determined |
| 18-103_k141_4355 | Not-determined |
| 18-103_k141_130945 | Low-quality |
| 18-103_k141_61567 | Not-determined |
| 18-103_k141_4363 | Not-determined |
| 18-103_k141_9761 | Not-determined |
| 18-103_k141_90629 | Not-determined |
| 18-103_k141_38843 | Low-quality |
| 18-103_k141_9766 | Not-determined |
| 18-103_k141_119514 | Low-quality |
| 18-103_k141_136402 | Not-determined |
| 18-103_k141_32694 | Not-determined |
| 18-103_k141_136404 | Not-determined |
| 18-103_k141_27219 | Not-determined |
| 18-103_k141_9771 | Not-determined |
| 18-103_k141_32698 | Not-determined |
| 18-103_k141_113484 | Not-determined |
| 18-103_k141_136408 | Not-determined |
| 18-103_k141_15323 | Low-quality |
| 18-103_k141_61573 | Low-quality |
| 18-103_k141_32704 | Low-quality |
| 18-103_k141_84822 | Not-determined |
| 18-103_k141_125123 | Not-determined |
| 18-103_k141_21425 | Low-quality |
| 18-103_k141_55889 | Low-quality |
| 18-103_k141_78921 | Not-determined |
| 18-103_k141_61578 | Low-quality |
| 18-103_k141_55891 | Not-determined |
| 18-103_k141_32711 | Not-determined |
| 18-103_k141_61584 | Not-determined |
| 18-103_k141_32714 | Not-determined |
| 18-103_k141_119538 | Not-determined |
| 18-103_k141_125135 | Not-determined |
| 18-103_k141_90654 | Not-determined |
| 18-103_k141_125138 | Not-determined |
| 18-103_k141_119544 | Not-determined |
| 18-103_k141_32719 | Not-determined |
| 18-103_k141_27243 | Not-determined |
| 18-103_k141_4393 | Not-determined |
| 18-103_k141_55906 | Not-determined |
| 18-103_k141_73026 | Not-determined |
| 18-103_k141_125143 | Low-quality |
| 18-103_k141_96271 | Not-determined |
| 18-103_k141_21447 | Not-determined |
| 18-103_k141_125147 | Not-determined |
| 18-103_k141_90669 | Not-determined |
| 18-103_k141_107610 | Not-determined |
| 18-103_k141_50047 | Not-determined |
| 18-103_k141_119558 | Not-determined |
| 18-103_k141_44250 | Not-determined |
| 18-103_k141_55925 | Not-determined |
| 18-103_k141_27266 | Low-quality |
| 18-103_k141_44253 | Low-quality |
| 18-103_k141_119565 | Not-determined |
| 18-103_k141_101619 | Not-determined |
| 18-103_k141_101624 | Not-determined |
| 18-103_k141_125161 | Not-determined |
| 18-103_k141_90681 | Not-determined |
| 18-103_k141_61618 | Not-determined |
| 18-103_k141_61620 | Not-determined |
| 18-103_k141_55935 | Not-determined |
| 18-103_k141_4418 | Not-determined |
| 18-103_k141_113536 | Not-determined |
| 18-103_k141_44265 | Not-determined |
| 18-103_k141_15378 | Low-quality |
| 18-103_k141_131002 | Not-determined |
| 18-103_k141_131009 | Not-determined |
| 18-103_k141_107632 | Not-determined |
| 18-103_k141_73064 | Not-determined |
| 18-103_k141_27293 | Low-quality |
| 18-103_k141_55947 | Not-determined |
| 18-103_k141_4428 | Not-determined |
| 18-103_k141_125177 | Low-quality |
| 18-103_k141_119577 | Not-determined |
| 18-103_k141_38876 | Not-determined |
| 18-103_k141_78951 | Not-determined |
| 18-103_k141_96310 | Not-determined |
| 18-103_k141_38878 | Low-quality |
| 18-103_k141_4431 | Not-determined |
| 18-103_k141_61641 | Not-determined |
| 18-103_k141_61642 | Not-determined |
| 18-103_k141_55956 | Not-determined |
| 18-103_k141_131016 | Not-determined |
| 18-103_k141_50069 | Not-determined |
| 18-103_k141_4439 | Not-determined |
| 18-103_k141_131019 | Not-determined |
| 18-103_k141_27309 | Not-determined |
| 18-103_k141_73070 | Not-determined |
| 18-103_k141_9819 | Not-determined |
| 18-103_k141_125191 | Not-determined |
| 18-103_k141_9821 | Not-determined |
| 18-103_k141_38893 | Low-quality |
| 18-103_k141_78968 | Not-determined |
| 18-103_k141_84851 | Not-determined |
| 18-103_k141_113559 | Not-determined |
| 18-103_k141_55969 | Not-determined |
| 18-103_k141_32746 | Not-determined |
| 18-103_k141_96323 | Not-determined |
| 18-103_k141_73083 | Low-quality |
| 18-103_k141_55974 | Not-determined |
| 18-103_k141_107661 | Low-quality |
| 18-103_k141_4455 | Not-determined |
| 18-103_k141_119592 | Not-determined |
| 18-103_k141_125201 | Low-quality |
| 18-103_k141_101649 | Not-determined |
| 18-103_k141_73087 | Not-determined |
| 18-103_k141_125204 | Not-determined |
| 18-103_k141_96329 | Not-determined |
| 18-103_k141_9839 | Not-determined |
| 18-103_k141_131036 | Not-determined |
| 18-103_k141_55983 | Not-determined |
| 18-103_k141_96330 | Not-determined |
| 18-103_k141_73093 | Not-determined |
| 18-103_k141_125217 | Not-determined |
| 18-103_k141_96334 | Low-quality |
| 18-103_k141_21509 | Not-determined |
| 18-103_k141_50100 | Not-determined |
| 18-103_k141_38912 | Low-quality |
| 18-103_k141_32763 | Low-quality |
| 18-103_k141_84878 | Not-determined |
| 18-103_k141_4469 | Not-determined |
| 18-103_k141_9849 | Not-determined |
| 18-103_k141_67552 | Not-determined |
| 18-103_k141_61694 | Not-determined |
| 18-103_k141_9853 | Not-determined |
| 18-103_k141_44273 | Not-determined |
| 18-103_k141_21519 | Not-determined |
| 18-103_k141_15400 | Not-determined |
| 18-103_k141_61697 | Low-quality |
| 18-103_k141_131055 | Not-determined |
| 18-103_k141_84883 | Not-determined |
| 18-103_k141_61701 | Not-determined |
| 18-103_k141_4477 | Not-determined |
| 18-103_k141_73100 | Not-determined |
| 18-103_k141_38923 | Not-determined |
| 18-103_k141_50102 | Not-determined |
| 18-103_k141_67563 | Not-determined |
| 18-103_k141_101673 | Low-quality |
| 18-103_k141_131059 | Not-determined |
| 18-103_k141_15401 | Not-determined |
| 18-103_k141_90701 | Not-determined |
| 18-103_k141_131063 | Not-determined |
| 18-103_k141_61709 | Not-determined |
| 18-103_k141_131068 | Low-quality |
| 18-103_k141_21540 | Not-determined |
| 18-103_k141_15411 | Not-determined |
| 18-103_k141_4490 | Not-determined |
| 18-103_k141_131072 | Not-determined |
| 18-103_k141_67568 | Not-determined |
| 18-103_k141_101690 | Not-determined |
| 18-103_k141_113608 | Not-determined |
| 18-103_k141_56007 | Not-determined |
| 18-103_k141_131075 | Not-determined |
| 18-103_k141_119630 | Low-quality |
| 18-103_k141_44280 | Not-determined |
| 18-103_k141_101694 | Low-quality |
| 18-103_k141_79022 | Not-determined |
| 18-103_k141_56010 | Not-determined |
| 18-103_k141_38936 | Not-determined |
| 18-103_k141_90717 | Not-determined |
| 18-103_k141_119643 | Not-determined |
| 18-103_k141_90721 | Not-determined |
| 18-103_k141_15425 | Not-determined |
| 18-103_k141_131082 | Not-determined |
| 18-103_k141_44293 | Not-determined |
| 18-103_k141_84919 | Not-determined |
| 18-103_k141_50112 | Not-determined |
| 18-103_k141_4512 | Not-determined |
| 18-103_k141_9896 | Not-determined |
| 18-103_k141_107709 | Not-determined |
| 18-103_k141_79028 | Not-determined |
| 18-103_k141_38948 | Low-quality |
| 18-103_k141_15438 | Not-determined |
| 18-103_k141_84929 | Not-determined |
| 18-103_k141_107711 | Not-determined |
| 18-103_k141_38951 | Not-determined |
| 18-103_k141_101714 | Not-determined |
| 18-103_k141_131089 | Not-determined |
| 18-103_k141_15444 | Not-determined |
| 18-103_k141_79033 | Not-determined |
| 18-103_k141_84939 | Not-determined |
| 18-103_k141_90742 | Not-determined |
| 18-103_k141_79037 | Not-determined |
| 18-103_k141_79038 | Not-determined |
| 18-103_k141_21557 | Not-determined |
| 18-103_k141_50122 | Low-quality |
| 18-103_k141_61745 | Not-determined |
| 18-103_k141_21559 | Not-determined |
| 18-103_k141_50123 | Not-determined |
| 18-103_k141_32795 | Not-determined |
| 18-103_k141_113633 | Not-determined |
| 18-103_k141_27327 | Low-quality |
| 18-103_k141_21565 | Low-quality |
| 18-103_k141_15459 | Not-determined |
| 18-103_k141_90750 | Not-determined |
| 18-103_k141_32799 | Low-quality |
| 18-103_k141_84951 | Not-determined |
| 18-103_k141_113636 | Not-determined |
| 18-103_k141_107723 | Not-determined |
| 18-103_k141_9909 | Not-determined |
| 18-103_k141_61752 | Not-determined |
| 18-103_k141_119661 | Not-determined |
| 18-103_k141_107728 | Not-determined |
| 18-103_k141_107731 | Not-determined |
| 18-103_k141_125258 | Not-determined |
| 18-103_k141_96371 | Not-determined |
| 18-103_k141_125262 | Not-determined |
| 18-103_k141_9920 | Not-determined |
| 18-103_k141_9921 | Not-determined |
| 18-103_k141_9922 | Low-quality |
| 18-103_k141_67576 | Not-determined |
| 18-103_k141_44320 | Not-determined |
| 18-103_k141_44321 | Not-determined |
| 18-103_k141_9928 | Not-determined |
| 18-103_k141_73127 | Not-determined |
| 18-103_k141_61766 | Not-determined |
| 18-103_k141_79065 | Low-quality |
| 18-103_k141_131114 | Low-quality |
| 18-103_k141_27347 | Not-determined |
| 18-103_k141_27348 | Low-quality |
| 18-103_k141_119671 | Not-determined |
| 18-103_k141_32802 | Low-quality |
| 18-103_k141_50150 | Not-determined |
| 18-103_k141_101743 | Not-determined |
| 18-103_k141_101747 | Not-determined |
| 18-103_k141_96388 | Not-determined |
| 18-103_k141_84989 | Not-determined |
| 18-103_k141_61789 | Not-determined |
| 18-103_k141_79085 | Not-determined |
| 18-103_k141_32821 | Not-determined |
| 18-103_k141_50155 | Not-determined |
| 18-103_k141_136530 | Not-determined |
| 18-103_k141_90778 | Not-determined |
| 18-103_k141_21602 | Not-determined |
| 18-103_k141_90780 | Not-determined |
| 18-103_k141_9945 | Not-determined |
| 18-103_k141_9946 | Not-determined |
| 18-103_k141_44335 | Not-determined |
| 18-103_k141_56069 | Not-determined |
| 18-103_k141_67605 | Low-quality |
| 18-103_k141_44344 | Not-determined |
| 18-103_k141_21616 | Not-determined |
| 18-103_k141_27375 | Not-determined |
| 18-103_k141_136537 | Not-determined |
| 18-103_k141_56086 | Not-determined |
| 18-103_k141_73148 | Not-determined |
| 18-103_k141_21622 | Not-determined |
| 18-103_k141_136541 | Not-determined |
| 18-103_k141_125315 | Not-determined |
| 18-103_k141_44345 | Low-quality |
| 18-103_k141_119685 | Not-determined |
| 18-103_k141_119688 | Not-determined |
| 18-103_k141_90799 | Not-determined |
| 18-103_k141_73158 | Not-determined |
| 18-103_k141_39008 | Not-determined |
| 18-103_k141_79097 | Not-determined |
| 18-103_k141_67619 | Not-determined |
| 18-103_k141_79098 | Not-determined |
| 18-103_k141_79099 | Not-determined |
| 18-103_k141_27392 | Not-determined |
| 18-103_k141_44359 | Low-quality |
| 18-103_k141_101776 | Low-quality |
| 18-103_k141_96417 | Not-determined |
| 18-103_k141_136549 | Low-quality |
| 18-103_k141_79105 | Not-determined |
| 18-103_k141_131160 | Low-quality |
| 18-103_k141_136551 | Not-determined |
| 18-103_k141_44368 | Not-determined |
| 18-103_k141_21629 | Low-quality |
| 18-103_k141_85001 | Not-determined |
| 18-103_k141_79110 | Not-determined |
| 18-103_k141_85004 | Not-determined |
| 18-103_k141_119710 | Not-determined |
| 18-103_k141_90812 | Not-determined |
| 18-103_k141_79115 | Not-determined |
| 18-103_k141_39023 | Not-determined |
| 18-103_k141_125323 | Not-determined |
| 18-103_k141_107769 | Not-determined |
| 18-103_k141_119721 | Not-determined |
| 18-103_k141_90825 | Not-determined |
| 18-103_k141_56108 | Not-determined |
| 18-103_k141_67639 | Not-determined |
| 18-103_k141_96447 | Not-determined |
| 18-103_k141_44392 | Low-quality |
| 18-103_k141_90833 | Not-determined |
| 18-103_k141_101787 | Not-determined |
| 18-103_k141_15491 | Not-determined |
| 18-103_k141_44398 | Not-determined |
| 18-103_k141_101789 | Not-determined |
| 18-103_k141_21655 | Not-determined |
| 18-103_k141_21656 | Not-determined |
| 18-103_k141_85034 | Not-determined |
| 18-103_k141_4560 | Not-determined |
| 18-103_k141_21658 | Not-determined |
| 18-103_k141_79141 | Not-determined |
| 18-103_k141_113696 | Not-determined |
| 18-103_k141_101795 | Not-determined |
| 18-103_k141_39041 | Not-determined |
| 18-103_k141_113699 | Not-determined |
| 18-103_k141_101800 | Not-determined |
| 18-103_k141_90850 | Not-determined |
| 18-103_k141_85039 | Not-determined |
| 18-103_k141_15505 | Not-determined |
| 18-103_k141_131200 | Not-determined |
| 18-103_k141_101804 | Low-quality |
| 18-103_k141_101805 | Not-determined |
| 18-103_k141_27412 | Not-determined |
| 18-103_k141_85046 | Not-determined |
| 18-103_k141_44419 | Not-determined |
| 18-103_k141_131203 | Low-quality |
| 18-103_k141_79158 | Not-determined |
| 18-103_k141_131204 | Not-determined |
| 18-103_k141_119755 | Not-determined |
| 18-103_k141_73207 | Not-determined |
| 18-103_k141_73208 | Not-determined |
| 18-103_k141_79164 | Not-determined |
| 18-103_k141_125352 | Not-determined |
| 18-103_k141_21669 | Not-determined |
| 18-103_k141_56131 | Low-quality |
| 18-103_k141_32875 | Low-quality |
| 18-103_k141_21670 | Not-determined |
| 18-103_k141_113724 | Not-determined |
| 18-103_k141_4572 | Not-determined |
| 18-103_k141_107792 | Not-determined |
| 18-103_k141_44434 | Not-determined |
| 18-103_k141_113729 | Not-determined |
| 18-103_k141_113730 | Not-determined |
| 18-103_k141_56136 | Not-determined |
| 18-103_k141_90866 | Not-determined |
| 18-103_k141_32886 | Not-determined |
| 18-103_k141_15509 | Not-determined |
| 18-103_k141_85064 | Not-determined |
| 18-103_k141_119775 | Not-determined |
| 18-103_k141_125360 | Low-quality |
| 18-103_k141_61814 | Low-quality |
| 18-103_k141_85071 | Low-quality |
| 18-103_k141_44443 | Not-determined |
| 18-103_k141_27427 | Not-determined |
| 18-103_k141_136585 | Not-determined |
| 18-103_k141_15517 | Not-determined |
| 18-103_k141_125367 | Not-determined |
| 18-103_k141_56146 | Not-determined |
| 18-103_k141_21674 | Not-determined |
| 18-103_k141_113736 | Not-determined |
| 18-103_k141_56147 | Not-determined |
| 18-103_k141_79170 | Not-determined |
| 18-103_k141_79173 | Low-quality |
| 18-103_k141_125373 | Low-quality |
| 18-103_k141_136596 | Not-determined |
| 18-103_k141_136597 | Low-quality |
| 18-103_k141_61826 | Not-determined |
| 18-103_k141_90874 | Not-determined |
| 18-103_k141_56154 | Not-determined |
| 18-103_k141_131225 | Not-determined |
| 18-103_k141_56159 | Not-determined |
| 18-103_k141_96482 | Not-determined |
| 18-103_k141_39085 | Not-determined |
| 18-103_k141_96489 | Not-determined |
| 18-103_k141_4605 | Low-quality |
| 18-103_k141_39089 | Not-determined |
| 18-103_k141_44458 | Low-quality |
| 18-103_k141_125387 | Not-determined |
| 18-103_k141_73248 | Not-determined |
| 18-103_k141_85088 | Not-determined |
| 18-103_k141_113752 | Not-determined |
| 18-103_k141_119790 | Not-determined |
| 18-103_k141_125393 | Not-determined |
| 18-103_k141_56165 | Not-determined |
| 18-103_k141_56168 | Not-determined |
| 18-103_k141_136615 | Not-determined |
| 18-103_k141_44464 | Low-quality |
| 18-103_k141_113756 | Not-determined |
| 18-103_k141_96494 | Not-determined |
| 18-103_k141_101848 | Not-determined |
| 18-103_k141_101849 | Not-determined |
| 18-103_k141_61843 | Not-determined |
| 18-103_k141_96498 | Not-determined |
| 18-103_k141_125400 | Not-determined |
| 18-103_k141_85098 | Not-determined |
| 18-103_k141_15547 | Not-determined |
| 18-103_k141_44471 | Not-determined |
| 18-103_k141_85100 | Low-quality |
| 18-103_k141_136623 | Not-determined |
| 18-103_k141_79194 | Not-determined |
| 18-103_k141_96504 | Low-quality |
| 18-103_k141_101857 | Not-determined |
| 18-103_k141_67699 | Not-determined |
| 18-103_k141_136631 | Not-determined |
| 18-103_k141_56179 | Not-determined |
| 18-103_k141_4619 | Not-determined |
| 18-103_k141_79198 | Not-determined |
| 18-103_k141_44476 | Not-determined |
| 18-103_k141_32940 | Not-determined |
| 18-103_k141_90898 | Not-determined |
| 18-103_k141_44480 | Not-determined |
| 18-103_k141_44482 | Not-determined |
| 18-103_k141_136638 | Not-determined |
| 18-103_k141_107831 | Not-determined |
| 18-103_k141_85116 | Not-determined |
| 18-103_k141_131244 | Not-determined |
| 18-103_k141_125418 | Not-determined |
| 18-103_k141_90909 | Low-quality |
| 18-103_k141_56194 | Low-quality |
| 18-103_k141_21710 | Not-determined |
| 18-103_k141_125426 | Not-determined |
| 18-103_k141_21715 | Not-determined |
| 18-103_k141_39100 | Not-determined |
| 18-103_k141_136659 | Not-determined |
| 18-103_k141_39101 | Not-determined |
| 18-103_k141_79225 | Not-determined |
| 18-103_k141_61868 | Not-determined |
| 18-103_k141_21718 | Not-determined |
| 18-103_k141_136662 | Not-determined |
| 18-103_k141_90923 | Not-determined |
| 18-103_k141_90924 | Not-determined |
| 18-103_k141_50218 | Not-determined |
| 18-103_k141_4640 | Not-determined |
| 18-103_k141_21726 | Not-determined |
| 18-103_k141_79232 | Not-determined |
| 18-103_k141_4644 | Not-determined |
| 18-103_k141_56209 | Not-determined |
| 18-103_k141_107845 | Low-quality |
| 18-103_k141_79235 | Not-determined |
| 18-103_k141_101872 | Low-quality |
| 18-103_k141_113789 | Not-determined |
| 18-103_k141_90932 | Not-determined |
| 18-103_k141_44507 | Not-determined |
| 18-103_k141_15585 | Not-determined |
| 18-103_k141_119817 | Not-determined |
| 18-103_k141_27508 | Not-determined |
| 18-103_k141_113795 | Low-quality |
| 18-103_k141_131272 | Not-determined |
| 18-103_k141_61885 | Not-determined |
| 18-103_k141_96531 | Not-determined |
| 18-103_k141_107867 | Not-determined |
| 18-103_k141_96534 | Not-determined |
| 18-103_k141_15602 | Not-determined |
| 18-103_k141_50247 | Not-determined |
| 18-103_k141_107868 | Not-determined |
| 18-103_k141_15603 | Not-determined |
| 18-103_k141_131282 | Not-determined |
| 18-103_k141_61893 | Not-determined |
| 18-103_k141_119829 | Not-determined |
| 18-103_k141_107872 | Not-determined |
| 18-103_k141_32952 | Not-determined |
| 18-103_k141_107874 | Not-determined |
| 18-103_k141_107875 | Not-determined |
| 18-103_k141_56225 | Not-determined |
| 18-103_k141_10087 | Not-determined |
| 18-103_k141_107877 | Not-determined |
| 18-103_k141_15613 | Not-determined |
| 18-103_k141_27532 | Not-determined |
| 18-103_k141_4681 | Not-determined |
| 18-103_k141_101892 | Not-determined |
| 18-103_k141_131296 | Low-quality |
| 18-103_k141_73277 | Not-determined |
| 18-103_k141_131299 | Not-determined |
| 18-103_k141_90950 | Not-determined |
| 18-103_k141_79263 | Not-determined |
| 18-103_k141_136676 | Low-quality |
| 18-103_k141_131300 | Not-determined |
| 18-103_k141_113819 | Not-determined |
| 18-103_k141_113820 | Not-determined |
| 18-103_k141_131301 | Low-quality |
| 18-103_k141_50269 | Not-determined |
| 18-103_k141_32962 | Not-determined |
| 18-103_k141_136678 | Low-quality |
| 18-103_k141_27541 | Not-determined |
| 18-103_k141_107887 | Not-determined |
| 18-103_k141_107888 | Not-determined |
| 18-103_k141_101899 | Not-determined |
| 18-103_k141_50272 | Low-quality |
| 18-103_k141_113827 | Not-determined |
| 18-103_k141_79270 | Not-determined |
| 18-103_k141_67710 | Not-determined |
| 18-103_k141_119835 | Not-determined |
| 18-103_k141_27549 | Not-determined |
| 18-103_k141_131304 | Low-quality |
| 18-103_k141_119836 | Not-determined |
| 18-103_k141_56245 | Not-determined |
| 18-103_k141_15619 | Low-quality |
| 18-103_k141_10106 | Not-determined |
| 18-103_k141_67714 | Not-determined |
| 18-103_k141_44547 | Not-determined |
| 18-103_k141_21774 | Not-determined |
| 18-103_k141_44549 | Not-determined |
| 18-103_k141_136688 | Not-determined |
| 18-103_k141_101905 | Not-determined |
| 18-103_k141_67721 | Not-determined |
| 18-103_k141_131311 | Not-determined |
| 18-103_k141_21780 | Not-determined |
| 18-103_k141_67723 | Not-determined |
| 18-103_k141_21781 | Not-determined |
| 18-103_k141_10117 | Not-determined |
| 18-103_k141_32974 | Not-determined |
| 18-103_k141_96558 | Not-determined |
| 18-103_k141_119845 | Not-determined |
| 18-103_k141_10122 | Not-determined |
| 18-103_k141_39146 | Not-determined |
| 18-103_k141_107914 | Not-determined |

| | |
|---|---|
| 18-103_k141_39147 | Not-determined |
| 18-103_k141_10126 | Not-determined |
| 18-103_k141_4707 | Not-determined |
| 18-103_k141_10127 | Not-determined |
| 18-103_k141_61928 | Not-determined |
| 18-103_k141_10133 | Low-quality |
| 18-103_k141_4712 | Not-determined |
| 18-103_k141_27574 | Not-determined |
| 18-103_k141_113843 | Not-determined |
| 18-103_k141_61936 | Not-determined |
| 18-103_k141_73307 | Not-determined |
| 18-103_k141_27575 | Not-determined |
| 18-103_k141_39158 | Not-determined |
| 18-103_k141_32989 | Not-determined |
| 18-103_k141_136705 | Not-determined |
| 18-103_k141_79294 | Low-quality |
| 18-103_k141_67739 | Not-determined |
| 18-103_k141_107932 | Not-determined |
| 18-103_k141_39170 | Not-determined |
| 18-103_k141_79298 | Not-determined |
| 18-103_k141_67745 | Not-determined |
| 18-103_k141_44573 | Not-determined |
| 18-103_k141_136709 | Not-determined |
| 18-103_k141_136710 | Not-determined |
| 18-103_k141_79300 | Low-quality |
| 18-103_k141_67749 | Not-determined |
| 18-103_k141_10153 | Not-determined |
| 18-103_k141_131336 | Not-determined |
| 18-103_k141_79307 | Not-determined |
| 18-103_k141_119882 | Not-determined |
| 18-103_k141_27602 | Not-determined |
| 18-103_k141_39180 | Low-quality |
| 18-103_k141_136735 | Not-determined |
| 18-103_k141_119884 | Not-determined |
| 18-103_k141_67762 | Not-determined |
| 18-103_k141_125512 | Not-determined |
| 18-103_k141_27610 | Not-determined |
| 18-103_k141_44589 | Not-determined |
| 18-103_k141_56283 | Low-quality |
| 18-103_k141_67765 | Not-determined |
| 18-103_k141_79328 | Not-determined |
| 18-103_k141_10171 | Low-quality |
| 18-103_k141_56289 | Not-determined |
| 18-103_k141_73319 | Not-determined |
| 18-103_k141_10176 | Not-determined |
| 18-103_k141_90995 | Not-determined |
| 18-103_k141_67771 | Not-determined |
| 18-103_k141_79337 | Not-determined |
| 18-103_k141_4748 | Low-quality |
| 18-103_k141_67773 | Low-quality |
| 18-103_k141_33012 | Not-determined |
| 18-103_k141_79342 | Not-determined |
| 18-103_k141_136743 | Not-determined |
| 18-103_k141_56298 | Not-determined |
| 18-103_k141_39187 | Low-quality |
| 18-103_k141_91006 | Not-determined |
| 18-103_k141_73326 | Not-determined |
| 18-103_k141_113877 | Not-determined |
| 18-103_k141_91007 | Not-determined |
| 18-103_k141_113878 | Not-determined |
| 18-103_k141_96590 | Not-determined |
| 18-103_k141_107963 | Not-determined |
| 18-103_k141_15676 | Not-determined |
| 18-103_k141_39197 | Not-determined |
| 18-103_k141_27631 | Not-determined |
| 18-103_k141_113880 | Not-determined |
| 18-103_k141_67785 | Low-quality |
| 18-103_k141_107966 | Not-determined |
| 18-103_k141_136751 | Not-determined |
| 18-103_k141_50324 | Not-determined |
| 18-103_k141_10193 | Not-determined |
| 18-103_k141_107972 | Not-determined |
| 18-103_k141_96600 | Low-quality |
| 18-103_k141_136756 | Low-quality |
| 18-103_k141_21846 | Not-determined |
| 18-103_k141_56323 | Not-determined |
| 18-103_k141_21847 | Not-determined |
| 18-103_k141_67790 | Not-determined |
| 18-103_k141_73337 | Low-quality |
| 18-103_k141_4761 | Not-determined |
| 18-103_k141_27643 | Not-determined |
| 18-103_k141_4763 | Not-determined |
| 18-103_k141_50333 | Not-determined |
| 18-103_k141_91030 | Not-determined |
| 18-103_k141_67800 | Low-quality |
| 18-103_k141_101970 | Not-determined |
| 18-103_k141_91032 | Not-determined |
| 18-103_k141_10205 | Low-quality |
| 18-103_k141_136769 | Not-determined |
| 18-103_k141_10207 | Not-determined |
| 18-103_k141_44615 | Not-determined |
| 18-103_k141_119915 | Not-determined |
| 18-103_k141_67804 | Not-determined |
| 18-103_k141_91037 | Not-determined |
| 18-103_k141_33043 | Not-determined |
| 18-103_k141_61983 | Not-determined |
| 18-103_k141_61984 | Not-determined |
| 18-103_k141_91042 | Low-quality |
| 18-103_k141_79364 | Not-determined |
| 18-103_k141_136779 | Not-determined |
| 18-103_k141_79365 | Not-determined |
| 18-103_k141_10221 | Not-determined |
| 18-103_k141_119927 | Not-determined |
| 18-103_k141_79367 | Not-determined |
| 18-103_k141_15691 | Not-determined |
| 18-103_k141_50351 | Not-determined |
| 18-103_k141_56335 | Low-quality |
| 18-103_k141_107982 | Not-determined |
| 18-103_k141_56338 | Not-determined |
| 18-103_k141_15696 | Not-determined |
| 18-103_k141_101980 | Not-determined |
| 18-103_k141_91053 | Not-determined |
| 18-103_k141_91055 | Not-determined |
| 18-103_k141_113922 | Not-determined |
| 18-103_k141_4789 | Not-determined |
| 18-103_k141_79377 | Not-determined |
| 18-103_k141_56345 | Not-determined |
| 18-103_k141_113925 | Not-determined |
| 18-103_k141_56347 | Not-determined |
| 18-103_k141_56348 | Not-determined |
| 18-103_k141_113928 | Not-determined |
| 18-103_k141_91059 | Not-determined |
| 18-103_k141_10226 | Not-determined |
| 18-103_k141_27680 | Not-determined |
| 18-103_k141_21891 | Not-determined |
| 18-103_k141_27684 | Not-determined |
| 18-103_k141_107997 | Not-determined |
| 18-103_k141_73370 | Not-determined |
| 18-103_k141_136784 | Low-quality |
| 18-103_k141_119947 | Low-quality |
| 18-103_k141_27687 | Not-determined |
| 18-103_k141_136786 | Not-determined |
| 18-103_k141_67827 | Not-determined |
| 18-103_k141_108003 | Not-determined |
| 18-103_k141_136791 | Not-determined |
| 18-103_k141_33069 | Not-determined |
| 18-103_k141_91071 | Not-determined |
| 18-103_k141_108008 | Not-determined |
| 18-103_k141_62019 | Not-determined |
| 18-103_k141_33075 | Not-determined |
| 18-103_k141_119962 | Not-determined |
| 18-103_k141_113950 | Low-quality |
| 18-103_k141_33077 | Not-determined |
| 18-103_k141_21907 | Low-quality |
| 18-103_k141_56367 | Not-determined |
| 18-103_k141_125569 | Low-quality |
| 18-103_k141_119968 | Not-determined |
| 18-103_k141_79401 | Not-determined |
| 18-103_k141_119969 | Not-determined |
| 18-103_k141_136802 | Low-quality |
| 18-103_k141_50396 | Not-determined |
| 18-103_k141_33083 | Not-determined |
| 18-103_k141_27697 | Not-determined |
| 18-103_k141_50397 | Not-determined |
| 18-103_k141_62031 | Not-determined |
| 18-103_k141_4801 | Not-determined |
| 18-103_k141_108017 | Not-determined |
| 18-103_k141_125578 | Not-determined |
| 18-103_k141_33086 | Not-determined |
| 18-103_k141_73389 | Not-determined |
| 18-103_k141_21911 | Low-quality |
| 18-103_k141_44650 | Not-determined |
| 18-103_k141_108028 | Not-determined |
| 18-103_k141_136808 | Not-determined |
| 18-103_k141_56381 | Not-determined |
| 18-103_k141_15727 | Not-determined |
| 18-103_k141_27707 | Not-determined |
| 18-103_k141_15728 | Low-quality |
| 18-103_k141_27708 | Low-quality |
| 18-103_k141_113959 | Not-determined |
| 18-103_k141_67845 | Not-determined |
| 18-103_k141_136811 | Not-determined |
| 18-103_k141_39270 | Not-determined |
| 18-103_k141_108031 | Not-determined |
| 18-103_k141_73392 | Low-quality |
| 18-103_k141_67847 | Not-determined |
| 18-103_k141_108034 | Not-determined |
| 18-103_k141_50403 | Not-determined |
| 18-103_k141_108036 | Not-determined |
| 18-103_k141_67854 | Not-determined |
| 18-103_k141_39278 | Not-determined |
| 18-103_k141_44657 | Not-determined |
| 18-103_k141_62049 | Low-quality |
| 18-103_k141_10268 | Not-determined |
| 18-103_k141_50407 | Not-determined |
| 18-103_k141_131387 | Not-determined |
| 18-103_k141_21929 | Not-determined |
| 18-103_k141_113967 | Not-determined |
| 18-103_k141_56393 | Not-determined |
| 18-103_k141_39288 | Not-determined |
| 18-103_k141_39290 | Not-determined |
| 18-103_k141_73409 | Not-determined |
| 18-103_k141_33114 | Not-determined |
| 18-103_k141_125606 | Not-determined |
| 18-103_k141_21938 | Not-determined |
| 18-103_k141_50415 | Not-determined |
| 18-103_k141_73415 | Not-determined |
| 18-103_k141_96701 | Not-determined |
| 18-103_k141_136828 | Low-quality |
| 18-103_k141_73422 | Not-determined |
| 18-103_k141_102008 | Not-determined |
| 18-103_k141_108048 | Not-determined |
| 18-103_k141_120010 | Not-determined |
| 18-103_k141_79435 | Not-determined |
| 18-103_k141_131401 | Not-determined |
| 18-103_k141_21941 | Not-determined |
| 18-103_k141_131403 | Not-determined |
| 18-103_k141_136833 | Not-determined |
| 18-103_k141_10296 | Not-determined |
| 18-103_k141_125615 | Not-determined |
| 18-103_k141_62058 | Low-quality |
| 18-103_k141_15748 | Not-determined |
| 18-103_k141_125618 | Not-determined |
| 18-103_k141_91098 | Not-determined |
| 18-103_k141_85254 | Not-determined |
| 18-103_k141_125620 | Not-determined |
| 18-103_k141_73427 | Not-determined |
| 18-103_k141_91101 | Not-determined |
| 18-103_k141_108059 | Not-determined |
| 18-103_k141_85260 | Not-determined |
| 18-103_k141_27743 | Not-determined |
| 18-103_k141_113997 | Not-determined |
| 18-103_k141_15757 | Not-determined |
| 18-103_k141_125629 | Low-quality |
| 18-103_k141_21956 | Not-determined |
| 18-103_k141_125633 | Not-determined |
| 18-103_k141_21958 | Not-determined |
| 18-103_k141_91108 | Not-determined |
| 18-103_k141_114010 | Not-determined |
| 18-103_k141_120043 | Not-determined |
| 18-103_k141_125637 | Not-determined |
| 18-103_k141_79453 | Not-determined |
| 18-103_k141_27755 | Not-determined |
| 18-103_k141_4826 | Not-determined |
| 18-103_k141_21968 | Not-determined |
| 18-103_k141_125645 | Not-determined |
| 18-103_k141_21971 | Low-quality |
| 18-103_k141_125649 | Not-determined |
| 18-103_k141_85279 | Not-determined |
| 18-103_k141_67914 | Not-determined |
| 18-103_k141_44702 | Not-determined |
| 18-103_k141_33155 | Not-determined |
| 18-103_k141_102038 | Low-quality |
| 18-103_k141_10323 | Not-determined |
| 18-103_k141_33158 | Not-determined |
| 18-103_k141_4829 | Not-determined |
| 18-103_k141_108083 | Not-determined |
| 18-103_k141_136844 | Not-determined |
| 18-103_k141_96741 | Not-determined |
| 18-103_k141_73458 | Low-quality |
| 18-103_k141_39323 | Not-determined |
| 18-103_k141_96748 | Not-determined |
| 18-103_k141_85285 | Low-quality |
| 18-103_k141_33167 | Not-determined |
| 18-103_k141_21989 | Not-determined |
| 18-103_k141_67934 | Not-determined |
| 18-103_k141_125665 | Not-determined |
| 18-103_k141_136850 | Not-determined |
| 18-103_k141_15791 | Not-determined |
| 18-103_k141_56447 | Not-determined |
| 18-103_k141_50475 | Not-determined |
| 18-103_k141_131461 | Low-quality |
| 18-103_k141_79492 | Not-determined |
| 18-103_k141_131465 | Not-determined |
| 18-103_k141_136861 | Not-determined |
| 18-103_k141_21999 | Not-determined |
| 18-103_k141_114030 | Not-determined |
| 18-103_k141_96758 | Low-quality |
| 18-103_k141_91140 | Not-determined |
| 18-103_k141_125680 | Not-determined |
| 18-103_k141_125681 | Not-determined |
| 18-103_k141_96764 | Not-determined |
| 18-103_k141_50486 | Not-determined |
| 18-103_k141_136862 | Low-quality |
| 18-103_k141_125683 | Not-determined |
| 18-103_k141_62103 | Not-determined |
| 18-103_k141_114038 | Not-determined |
| 18-103_k141_62105 | Not-determined |
| 18-103_k141_67946 | Not-determined |
| 18-103_k141_44727 | Not-determined |
| 18-103_k141_44731 | Not-determined |
| 18-103_k141_44732 | Not-determined |
| 18-103_k141_79499 | Not-determined |
| 18-103_k141_50489 | Not-determined |
| 18-103_k141_67951 | Not-determined |
| 18-103_k141_67952 | Not-determined |
| 18-103_k141_62112 | Not-determined |
| 18-103_k141_10352 | Not-determined |
| 18-103_k141_96774 | Not-determined |
| 18-103_k141_85306 | Not-determined |
| 18-103_k141_96775 | Not-determined |
| 18-103_k141_96776 | Not-determined |
| 18-103_k141_136874 | Not-determined |
| 18-103_k141_136875 | Not-determined |
| 18-103_k141_10355 | Not-determined |
| 18-103_k141_56466 | Not-determined |
| 18-103_k141_67963 | Low-quality |
| 18-103_k141_67966 | Not-determined |
| 18-103_k141_73474 | Not-determined |
| 18-103_k141_15824 | Not-determined |
| 18-103_k141_96780 | Low-quality |
| 18-103_k141_120095 | Low-quality |
| 18-103_k141_27806 | Not-determined |
| 18-103_k141_27809 | Not-determined |
| 18-103_k141_131494 | Low-quality |
| 18-103_k141_22019 | Not-determined |
| 18-103_k141_96789 | Not-determined |
| 18-103_k141_15833 | Not-determined |
| 18-103_k141_125703 | Not-determined |
| 18-103_k141_131496 | Low-quality |
| 18-103_k141_96793 | Not-determined |
| 18-103_k141_85320 | Not-determined |
| 18-103_k141_4878 | Not-determined |
| 18-103_k141_44755 | Not-determined |
| 18-103_k141_50511 | Not-determined |
| 18-103_k141_91154 | Not-determined |
| 18-103_k141_27814 | Not-determined |
| 18-103_k141_15839 | Not-determined |
| 18-103_k141_120110 | Not-determined |
| 18-103_k141_131501 | Not-determined |
| 18-103_k141_15840 | Not-determined |
| 18-103_k141_131502 | Not-determined |
| 18-103_k141_67990 | Not-determined |
| 18-103_k141_67992 | Not-determined |
| 18-103_k141_39360 | Not-determined |
| 18-103_k141_67994 | Not-determined |
| 18-103_k141_56480 | Not-determined |
| 18-103_k141_62124 | Not-determined |
| 18-103_k141_67996 | Low-quality |
| 18-103_k141_15843 | Not-determined |
| 18-103_k141_56483 | Not-determined |
| 18-103_k141_125715 | Not-determined |
| 18-103_k141_39367 | Not-determined |
| 18-103_k141_33224 | Low-quality |
| 18-103_k141_22047 | Not-determined |
| 18-103_k141_33226 | Not-determined |
| 18-103_k141_125718 | Not-determined |
| 18-103_k141_120127 | Not-determined |
| 18-103_k141_33230 | Not-determined |
| 18-103_k141_125726 | Not-determined |
| 18-103_k141_68006 | Not-determined |
| 18-103_k141_125729 | Not-determined |
| 18-103_k141_56496 | Not-determined |
| 18-103_k141_50533 | Not-determined |
| 18-103_k141_68008 | Not-determined |
| 18-103_k141_62145 | Not-determined |
| 18-103_k141_85340 | Not-determined |
| 18-103_k141_50537 | Not-determined |
| 18-103_k141_73509 | Not-determined |
| 18-103_k141_136898 | Not-determined |
| 18-103_k141_85342 | Not-determined |
| 18-103_k141_22053 | Not-determined |
| 18-103_k141_91177 | Not-determined |
| 18-103_k141_22058 | Not-determined |
| 18-103_k141_22059 | Not-determined |
| 18-103_k141_120141 | Not-determined |
| 18-103_k141_85347 | Not-determined |
| 18-103_k141_15867 | Not-determined |
| 18-103_k141_73524 | Not-determined |
| 18-103_k141_39389 | Not-determined |
| 18-103_k141_44774 | Not-determined |
| 18-103_k141_131526 | Not-determined |
| 18-103_k141_56513 | Not-determined |
| 18-103_k141_136903 | Not-determined |
| 18-103_k141_44779 | Not-determined |
| 18-103_k141_114080 | Not-determined |
| 18-103_k141_4926 | Not-determined |
| 18-103_k141_108154 | Not-determined |
| 18-103_k141_4928 | Not-determined |
| 18-103_k141_73533 | Not-determined |
| 18-103_k141_136906 | Not-determined |
| 18-103_k141_68027 | Not-determined |
| 18-103_k141_120154 | Not-determined |
| 18-103_k141_27855 | Not-determined |
| 18-103_k141_136909 | Not-determined |
| 18-103_k141_15887 | Not-determined |
| 18-103_k141_136911 | Not-determined |
| 18-103_k141_39399 | Low-quality |
| 18-103_k141_62178 | Low-quality |
| 18-103_k141_73542 | Not-determined |
| 18-103_k141_4946 | Not-determined |
| 18-103_k141_4947 | Not-determined |
| 18-103_k141_62179 | Not-determined |
| 18-103_k141_15895 | Not-determined |
| 18-103_k141_131546 | Low-quality |
| 18-103_k141_44800 | Not-determined |
| 18-103_k141_91209 | Not-determined |
| 18-103_k141_85362 | Not-determined |
| 18-103_k141_96824 | Not-determined |
| 18-103_k141_125747 | Not-determined |
| 18-103_k141_114103 | Not-determined |
| 18-103_k141_96826 | Not-determined |
| 18-103_k141_22096 | Not-determined |
| 18-103_k141_15898 | Not-determined |
| 18-103_k141_120163 | Not-determined |
| 18-103_k141_96831 | Not-determined |
| 18-103_k141_125751 | Not-determined |
| 18-103_k141_96833 | Not-determined |
| 18-103_k141_125752 | Not-determined |
| 18-103_k141_91221 | Not-determined |
| 18-103_k141_62191 | Not-determined |
| 18-103_k141_120170 | Not-determined |
| 18-103_k141_131565 | Not-determined |
| 18-103_k141_108175 | Low-quality |
| 18-103_k141_27884 | Not-determined |
| 18-103_k141_50578 | Not-determined |
| 18-103_k141_136928 | Not-determined |
| 18-103_k141_27887 | Not-determined |
| 18-103_k141_136929 | Not-determined |
| 18-103_k141_73570 | Low-quality |
| 18-103_k141_125759 | Not-determined |
| 18-103_k141_85380 | Not-determined |
| 18-103_k141_62196 | Not-determined |
| 18-103_k141_114114 | Not-determined |
| 18-103_k141_114116 | Not-determined |
| 18-103_k141_91230 | Not-determined |
| 18-103_k141_56522 | Not-determined |
| 18-103_k141_131577 | Not-determined |
| 18-103_k141_39426 | Not-determined |
| 18-103_k141_44821 | Not-determined |
| 18-103_k141_33291 | Not-determined |
| 18-103_k141_91233 | Low-quality |
| 18-103_k141_15923 | Not-determined |
| 18-103_k141_39435 | Not-determined |
| 18-103_k141_91234 | Not-determined |
| 18-103_k141_91237 | Not-determined |
| 18-103_k141_73585 | Not-determined |
| 18-103_k141_85400 | Not-determined |
| 18-103_k141_44836 | Not-determined |
| 18-103_k141_15932 | Low-quality |
| 18-103_k141_120193 | Not-determined |
| 18-103_k141_79557 | Not-determined |
| 18-103_k141_50596 | Not-determined |
| 18-103_k141_10445 | Not-determined |
| 18-103_k141_27904 | Not-determined |
| 18-103_k141_68071 | Low-quality |
| 18-103_k141_15935 | Not-determined |
| 18-103_k141_131596 | Not-determined |
| 18-103_k141_136957 | Not-determined |
| 18-103_k141_62217 | Not-determined |
| 18-103_k141_62218 | Not-determined |
| 18-103_k141_96872 | Not-determined |
| 18-103_k141_62219 | Low-quality |
| 18-103_k141_120199 | Low-quality |
| 18-103_k141_15939 | Not-determined |
| 18-103_k141_39455 | Not-determined |
| 18-103_k141_136963 | Not-determined |
| 18-103_k141_39458 | Not-determined |
| 18-103_k141_131599 | Not-determined |
| 18-103_k141_62227 | Not-determined |
| 18-103_k141_114133 | Not-determined |
| 18-103_k141_33313 | Not-determined |
| 18-103_k141_10451 | Not-determined |
| 18-103_k141_56568 | Not-determined |
| 18-103_k141_22138 | Not-determined |
| 18-103_k141_85427 | Not-determined |
| 18-103_k141_131607 | Not-determined |
| 18-103_k141_96890 | Not-determined |
| 18-103_k141_85430 | Not-determined |
| 18-103_k141_85431 | Not-determined |
| 18-103_k141_96891 | Not-determined |
| 18-103_k141_91264 | Not-determined |
| 18-103_k141_44860 | Not-determined |
| 18-103_k141_114143 | Not-determined |
| 18-103_k141_85435 | Not-determined |
| 18-103_k141_120215 | Not-determined |
| 18-103_k141_102148 | Not-determined |
| 18-103_k141_85438 | Not-determined |
| 18-103_k141_56578 | Not-determined |
| 18-103_k141_73611 | Not-determined |
| 18-103_k141_108219 | Not-determined |
| 18-103_k141_125797 | Not-determined |
| 18-103_k141_120226 | Not-determined |
| 18-103_k141_56583 | Not-determined |
| 18-103_k141_44871 | Not-determined |
| 18-103_k141_22149 | Not-determined |
| 18-103_k141_5000 | Low-quality |
| 18-103_k141_62255 | Not-determined |
| 18-103_k141_62256 | Not-determined |
| 18-103_k141_5006 | Not-determined |
| 18-103_k141_102159 | Not-determined |
| 18-103_k141_131633 | Not-determined |
| 18-103_k141_108232 | Not-determined |
| 18-103_k141_50633 | Not-determined |
| 18-103_k141_62262 | Not-determined |
| 18-103_k141_62263 | Low-quality |
| 18-103_k141_56591 | Low-quality |
| 18-103_k141_50636 | Not-determined |
| 18-103_k141_85459 | Not-determined |
| 18-103_k141_73620 | Not-determined |
| 18-103_k141_85465 | Not-determined |
| 18-103_k141_120245 | Not-determined |
| 18-103_k141_120246 | Not-determined |
| 18-103_k141_15990 | Not-determined |
| 18-103_k141_114167 | Not-determined |
| 18-103_k141_62274 | Not-determined |
| 18-103_k141_68126 | Not-determined |
| 18-103_k141_79585 | Not-determined |
| 18-103_k141_136998 | Not-determined |
| 18-103_k141_131642 | Not-determined |
| 18-103_k141_27939 | Not-determined |
| 18-103_k141_102168 | Not-determined |
| 18-103_k141_137000 | Not-determined |
| 18-103_k141_79589 | Not-determined |
| 18-103_k141_102169 | Not-determined |
| 18-103_k141_62276 | Low-quality |
| 18-103_k141_56599 | Not-determined |
| 18-103_k141_68131 | Not-determined |
| 18-103_k141_22177 | Not-determined |
| 18-103_k141_73634 | Not-determined |
| 18-103_k141_22178 | Not-determined |
| 18-103_k141_102171 | Not-determined |
| 18-103_k141_131646 | Low-quality |
| 18-103_k141_39502 | Not-determined |
| 18-103_k141_91283 | Not-determined |
| 18-103_k141_73637 | Not-determined |
| 18-103_k141_62284 | Not-determined |
| 18-103_k141_22184 | Not-determined |
| 18-103_k141_79598 | Not-determined |
| 18-103_k141_62286 | Not-determined |
| 18-103_k141_131657 | Not-determined |
| 18-103_k141_96927 | Not-determined |
| 18-103_k141_16003 | Not-determined |
| 18-103_k141_85485 | Not-determined |
| 18-103_k141_10499 | Not-determined |
| 18-103_k141_120265 | Not-determined |
| 18-103_k141_22187 | Not-determined |
| 18-103_k141_102178 | Not-determined |
| 18-103_k141_33364 | Not-determined |
| 18-103_k141_85492 | Not-determined |
| 18-103_k141_125827 | Not-determined |
| 18-103_k141_5048 | Not-determined |
| 18-103_k141_10505 | Not-determined |
| 18-103_k141_79608 | Low-quality |
| 18-103_k141_10509 | Not-determined |
| 18-103_k141_62289 | Not-determined |
| 18-103_k141_50654 | Not-determined |
| 18-103_k141_22194 | Not-determined |
| 18-103_k141_96938 | Not-determined |
| 18-103_k141_68156 | Not-determined |
| 18-103_k141_131669 | Not-determined |
| 18-103_k141_79612 | Low-quality |
| 18-103_k141_125838 | Not-determined |
| 18-103_k141_22202 | Not-determined |
| 18-103_k141_137009 | Not-determined |
| 18-103_k141_33377 | Not-determined |
| 18-103_k141_50665 | Not-determined |
| 18-103_k141_85494 | Not-determined |
| 18-103_k141_10519 | Not-determined |
| 18-103_k141_56619 | Not-determined |
| 18-103_k141_131677 | Not-determined |
| 18-103_k141_27981 | Not-determined |
| 18-103_k141_120280 | Not-determined |
| 18-103_k141_137019 | Not-determined |
| 18-103_k141_33388 | Not-determined |
| 18-103_k141_108279 | Not-determined |
| 18-103_k141_131682 | Not-determined |
| 18-103_k141_10527 | Not-determined |
| 18-103_k141_73649 | Not-determined |
| 18-103_k141_39522 | Not-determined |
| 18-103_k141_10535 | Not-determined |
| 18-103_k141_33398 | Not-determined |
| 18-103_k141_33402 | Not-determined |
| 18-103_k141_10540 | Not-determined |
| 18-103_k141_114209 | Not-determined |
| 18-103_k141_50678 | Not-determined |
| 18-103_k141_131690 | Low-quality |
| 18-103_k141_85511 | Not-determined |
| 18-103_k141_131691 | Not-determined |
| 18-103_k141_85512 | Low-quality |
| 18-103_k141_114211 | Not-determined |
| 18-103_k141_96956 | Not-determined |
| 18-103_k141_85516 | Not-determined |
| 18-103_k141_39534 | Not-determined |
| 18-103_k141_16046 | Not-determined |
| 18-103_k141_39536 | Not-determined |
| 18-103_k141_85520 | Low-quality |
| 18-103_k141_125868 | Not-determined |
| 18-103_k141_5082 | Not-determined |
| 18-103_k141_68192 | Low-quality |
| 18-103_k141_5092 | Not-determined |
| 18-103_k141_102217 | Low-quality |
| 18-103_k141_102218 | Not-determined |
| 18-103_k141_5095 | Low-quality |
| 18-103_k141_102222 | Not-determined |
| 18-103_k141_28015 | Not-determined |
| 18-103_k141_33424 | Not-determined |
| 18-103_k141_22234 | Low-quality |
| 18-103_k141_10567 | Low-quality |

| | |
|---|---|
| 18-103_k141_5100 | Not-determined |
| 18-103_k141_102230 | Not-determined |
| 18-103_k141_50705 | Not-determined |
| 18-103_k141_102232 | Not-determined |
| 18-103_k141_22237 | Not-determined |
| 18-103_k141_16065 | Not-determined |
| 18-103_k141_102236 | Not-determined |
| 18-103_k141_39560 | Not-determined |
| 18-103_k141_5108 | Not-determined |
| 18-103_k141_44912 | Not-determined |
| 18-103_k141_85546 | Not-determined |
| 18-103_k141_73683 | Not-determined |
| 18-103_k141_39565 | Not-determined |
| 18-103_k141_114245 | Not-determined |
| 18-103_k141_62330 | Not-determined |
| 18-103_k141_120333 | Not-determined |
| 18-103_k141_68219 | Not-determined |
| 18-103_k141_91314 | Not-determined |
| 18-103_k141_33439 | Not-determined |
| 18-103_k141_114254 | Not-determined |
| 18-103_k141_137022 | Not-determined |
| 18-103_k141_56628 | Low-quality |
| 18-103_k141_137023 | Not-determined |
| 18-103_k141_16081 | Not-determined |
| 18-103_k141_44921 | Not-determined |
| 18-103_k141_16083 | Not-determined |
| 18-103_k141_96992 | Not-determined |
| 18-103_k141_62343 | Not-determined |
| 18-103_k141_91325 | Low-quality |
| 18-103_k141_91326 | Not-determined |
| 18-103_k141_39574 | Not-determined |
| 18-103_k141_91329 | Not-determined |
| 18-103_k141_62345 | Low-quality |
| 18-103_k141_73695 | Not-determined |
| 18-103_k141_56643 | Not-determined |
| 18-103_k141_102254 | Not-determined |
| 18-103_k141_125909 | Not-determined |
| 18-103_k141_56644 | Not-determined |
| 18-103_k141_125910 | Not-determined |
| 18-103_k141_62349 | Not-determined |
| 18-103_k141_44935 | Low-quality |
| 18-103_k141_114266 | Not-determined |
| 18-103_k141_102258 | Not-determined |
| 18-103_k141_125914 | Not-determined |
| 18-103_k141_28039 | Not-determined |
| 18-103_k141_73707 | Not-determined |
| 18-103_k141_114270 | Not-determined |
| 18-103_k141_137036 | Not-determined |
| 18-103_k141_85566 | Not-determined |
| 18-103_k141_22270 | Not-determined |
| 18-103_k141_97007 | Low-quality |
| 18-103_k141_5135 | Not-determined |
| 18-103_k141_22274 | Not-determined |
| 18-103_k141_62360 | Not-determined |
| 18-103_k141_120379 | Not-determined |
| 18-103_k141_10610 | Not-determined |
| 18-103_k141_102274 | Not-determined |
| 18-103_k141_33451 | Not-determined |
| 18-103_k141_28047 | Not-determined |
| 18-103_k141_10612 | Not-determined |
| 18-103_k141_131744 | Not-determined |
| 18-103_k141_97016 | Not-determined |
| 18-103_k141_131745 | Not-determined |
| 18-103_k141_33457 | Not-determined |
| 18-103_k141_68242 | Not-determined |
| 18-103_k141_120385 | Not-determined |
| 18-103_k141_114278 | Not-determined |
| 18-103_k141_120386 | Not-determined |
| 18-103_k141_97021 | Low-quality |
| 18-103_k141_97022 | Not-determined |
| 18-103_k141_39593 | Not-determined |
| 18-103_k141_10621 | Not-determined |
| 18-103_k141_120390 | Not-determined |
| 18-103_k141_97026 | Not-determined |
| 18-103_k141_5143 | Low-quality |
| 18-103_k141_10625 | Low-quality |
| 18-103_k141_125925 | Not-determined |
| 18-103_k141_62380 | Not-determined |
| 18-103_k141_125933 | Not-determined |
| 18-103_k141_5154 | Not-determined |
| 18-103_k141_97039 | Not-determined |
| 18-103_k141_39606 | Not-determined |
| 18-103_k141_16120 | Low-quality |
| 18-103_k141_97041 | Not-determined |
| 18-103_k141_28070 | Not-determined |
| 18-103_k141_44949 | Not-determined |
| 18-103_k141_97043 | Not-determined |
| 18-103_k141_56679 | Not-determined |
| 18-103_k141_39611 | Not-determined |
| 18-103_k141_33466 | Not-determined |
| 18-103_k141_85586 | Not-determined |
| 18-103_k141_39614 | Not-determined |
| 18-103_k141_16124 | Not-determined |
| 18-103_k141_5163 | Low-quality |
| 18-103_k141_91356 | Not-determined |
| 18-103_k141_73739 | Not-determined |
| 18-103_k141_39617 | Not-determined |
| 18-103_k141_50758 | Not-determined |
| 18-103_k141_97050 | Not-determined |
| 18-103_k141_125947 | Not-determined |
| 18-103_k141_68252 | Not-determined |
| 18-103_k141_28076 | Not-determined |
| 18-103_k141_16128 | Not-determined |
| 18-103_k141_125949 | Not-determined |
| 18-103_k141_125951 | Not-determined |
| 18-103_k141_50759 | Not-determined |
| 18-103_k141_73747 | Not-determined |
| 18-103_k141_120410 | Not-determined |
| 18-103_k141_114303 | Not-determined |
| 18-103_k141_131772 | Not-determined |
| 18-103_k141_33476 | Low-quality |
| 18-103_k141_137067 | Low-quality |
| 18-103_k141_91369 | Not-determined |
| 18-103_k141_28082 | Not-determined |
| 18-103_k141_114320 | Not-determined |
| 18-103_k141_44971 | Not-determined |
| 18-103_k141_102322 | Not-determined |
| 18-103_k141_91372 | Not-determined |
| 18-103_k141_85601 | Not-determined |
| 18-103_k141_68267 | Not-determined |
| 18-103_k141_56692 | Not-determined |
| 18-103_k141_56694 | Not-determined |
| 18-103_k141_10647 | Not-determined |
| 18-103_k141_108308 | Not-determined |
| 18-103_k141_16149 | Not-determined |
| 18-103_k141_68274 | Not-determined |
| 18-103_k141_5186 | Not-determined |
| 18-103_k141_120432 | Not-determined |
| 18-103_k141_125975 | Not-determined |
| 18-103_k141_114333 | Not-determined |
| 18-103_k141_108310 | Not-determined |
| 18-103_k141_5190 | Not-determined |
| 18-103_k141_33486 | Not-determined |
| 18-103_k141_91381 | Not-determined |
| 18-103_k141_79733 | Not-determined |
| 18-103_k141_108314 | Not-determined |
| 18-103_k141_91383 | Not-determined |
| 18-103_k141_79735 | Low-quality |
| 18-103_k141_16156 | Not-determined |
| 18-103_k141_44982 | Not-determined |
| 18-103_k141_79736 | Not-determined |
| 18-103_k141_44984 | Not-determined |
| 18-103_k141_114341 | Not-determined |
| 18-103_k141_79739 | Not-determined |
| 18-103_k141_62410 | Low-quality |
| 18-103_k141_68287 | Not-determined |
| 18-103_k141_5198 | Not-determined |
| 18-103_k141_5199 | Not-determined |
| 18-103_k141_73793 | Not-determined |
| 18-103_k141_56708 | Low-quality |
| 18-103_k141_131811 | Not-determined |
| 18-103_k141_62427 | Not-determined |
| 18-103_k141_114360 | Not-determined |
| 18-103_k141_50789 | Not-determined |
| 18-103_k141_10673 | Not-determined |
| 18-103_k141_62432 | Low-quality |
| 18-103_k141_56712 | Low-quality |
| 18-103_k141_33510 | Not-determined |
| 18-103_k141_120456 | Not-determined |
| 18-103_k141_22338 | Not-determined |
| 18-103_k141_108333 | Not-determined |
| 18-103_k141_108334 | Not-determined |
| 18-103_k141_39649 | Not-determined |
| 18-103_k141_28114 | Not-determined |
| 18-103_k141_5221 | Not-determined |
| 18-103_k141_126020 | Not-determined |
| 18-103_k141_120461 | Not-determined |
| 18-103_k141_39652 | Not-determined |
| 18-103_k141_28116 | Not-determined |
| 18-103_k141_114369 | Not-determined |
| 18-103_k141_79758 | Not-determined |
| 18-103_k141_79759 | Not-determined |
| 18-103_k141_73798 | Low-quality |
| 18-103_k141_97072 | Not-determined |
| 18-103_k141_39658 | Not-determined |
| 18-103_k141_97073 | Not-determined |
| 18-103_k141_45022 | Low-quality |
| 18-103_k141_91391 | Not-determined |
| 18-103_k141_45025 | Not-determined |
| 18-103_k141_85630 | Not-determined |
| 18-103_k141_68313 | Not-determined |
| 18-103_k141_73803 | Not-determined |
| 18-103_k141_79768 | Not-determined |
| 18-103_k141_108349 | Not-determined |
| 18-103_k141_45032 | Not-determined |
| 18-103_k141_108356 | Not-determined |
| 18-103_k141_33534 | Not-determined |
| 18-103_k141_131831 | Not-determined |
| 18-103_k141_131832 | Not-determined |
| 18-103_k141_102361 | Not-determined |
| 18-103_k141_102362 | Low-quality |
| 18-103_k141_137093 | Not-determined |
| 18-103_k141_68328 | Not-determined |
| 18-103_k141_97079 | Not-determined |
| 18-103_k141_126045 | Not-determined |
| 18-103_k141_102367 | Not-determined |
| 18-103_k141_91410 | Not-determined |
| 18-103_k141_39666 | Not-determined |
| 18-103_k141_97086 | Not-determined |
| 18-103_k141_16197 | Not-determined |
| 18-103_k141_85650 | Not-determined |
| 18-103_k141_28140 | Not-determined |
| 18-103_k141_73816 | Not-determined |
| 18-103_k141_97088 | Not-determined |
| 18-103_k141_97090 | Not-determined |
| 18-103_k141_5247 | Low-quality |
| 18-103_k141_131847 | Not-determined |
| 18-103_k141_50828 | Low-quality |
| 18-103_k141_137105 | Not-determined |
| 18-103_k141_114413 | Not-determined |
| 18-103_k141_126061 | Not-determined |
| 18-103_k141_126062 | Not-determined |
| 18-103_k141_137110 | Not-determined |
| 18-103_k141_120493 | Low-quality |
| 18-103_k141_50835 | Not-determined |
| 18-103_k141_45065 | Not-determined |
| 18-103_k141_126069 | Not-determined |
| 18-103_k141_137119 | Not-determined |
| 18-103_k141_79797 | Not-determined |
| 18-103_k141_114421 | Not-determined |
| 18-103_k141_22388 | Not-determined |
| 18-103_k141_137121 | Not-determined |
| 18-103_k141_126072 | Not-determined |
| 18-103_k141_126073 | Not-determined |
| 18-103_k141_62467 | Not-determined |
| 18-103_k141_102386 | Not-determined |
| 18-103_k141_137125 | Low-quality |
| 18-103_k141_68343 | Not-determined |
| 18-103_k141_91429 | Not-determined |
| 18-103_k141_56744 | Not-determined |
| 18-103_k141_56745 | Low-quality |
| 18-103_k141_56746 | Not-determined |
| 18-103_k141_45077 | Not-determined |
| 18-103_k141_45079 | Not-determined |
| 18-103_k141_120508 | Low-quality |
| 18-103_k141_79809 | Not-determined |
| 18-103_k141_97127 | Not-determined |
| 18-103_k141_120511 | Not-determined |
| 18-103_k141_22401 | Not-determined |
| 18-103_k141_97132 | Not-determined |
| 18-103_k141_45084 | Not-determined |
| 18-103_k141_28172 | Not-determined |
| 18-103_k141_56754 | Not-determined |
| 18-103_k141_102390 | Not-determined |
| 18-103_k141_73850 | Not-determined |
| 18-103_k141_97138 | Not-determined |
| 18-103_k141_79814 | Not-determined |
| 18-103_k141_73857 | Not-determined |
| 18-103_k141_56762 | Not-determined |
| 18-103_k141_68357 | Not-determined |
| 18-103_k141_108399 | Not-determined |
| 18-103_k141_108403 | Not-determined |
| 18-103_k141_22414 | Not-determined |
| 18-103_k141_62487 | Not-determined |
| 18-103_k141_39700 | Not-determined |
| 18-103_k141_62490 | Not-determined |
| 18-103_k141_120524 | Not-determined |
| 18-103_k141_5286 | Not-determined |
| 18-103_k141_50872 | Not-determined |
| 18-103_k141_126105 | Not-determined |
| 18-103_k141_73865 | Not-determined |
| 18-103_k141_73867 | Not-determined |
| 18-103_k141_73869 | Not-determined |
| 18-103_k141_22430 | Not-determined |
| 18-103_k141_79832 | Not-determined |
| 18-103_k141_50880 | Not-determined |
| 18-103_k141_33573 | Low-quality |
| 18-103_k141_10746 | Not-determined |
| 18-103_k141_45101 | Not-determined |
| 18-103_k141_73874 | Low-quality |
| 18-103_k141_10752 | Not-determined |
| 18-103_k141_120542 | Not-determined |
| 18-103_k141_85719 | Not-determined |
| 18-103_k141_22439 | Not-determined |
| 18-103_k141_28198 | Not-determined |
| 18-103_k141_22441 | Low-quality |
| 18-103_k141_91461 | Not-determined |
| 18-103_k141_22446 | Not-determined |
| 18-103_k141_68375 | Low-quality |
| 18-103_k141_108414 | Not-determined |
| 18-103_k141_85726 | Not-determined |
| 18-103_k141_97164 | Not-determined |
| 18-103_k141_114460 | Not-determined |
| 18-103_k141_97166 | Not-determined |
| 18-103_k141_50897 | Not-determined |
| 18-103_k141_50898 | Not-determined |
| 18-103_k141_102417 | Not-determined |
| 18-103_k141_45114 | Not-determined |
| 18-103_k141_56784 | Not-determined |
| 18-103_k141_91464 | Low-quality |
| 18-103_k141_33596 | Low-quality |
| 18-103_k141_79854 | Not-determined |
| 18-103_k141_73889 | Not-determined |
| 18-103_k141_79857 | Not-determined |
| 18-103_k141_102423 | Not-determined |
| 18-103_k141_97176 | Not-determined |
| 18-103_k141_114468 | Not-determined |
| 18-103_k141_102429 | Not-determined |
| 18-103_k141_102430 | Not-determined |
| 18-103_k141_73891 | Not-determined |
| 18-103_k141_131869 | Not-determined |
| 18-103_k141_45128 | Low-quality |
| 18-103_k141_79870 | Not-determined |
| 18-103_k141_62500 | Not-determined |
| 18-103_k141_10780 | Not-determined |
| 18-103_k141_79875 | Not-determined |
| 18-103_k141_126145 | Not-determined |
| 18-103_k141_28230 | Not-determined |
| 18-103_k141_10786 | Low-quality |
| 18-103_k141_126147 | Not-determined |
| 18-103_k141_73898 | Not-determined |
| 18-103_k141_16234 | Not-determined |
| 18-103_k141_85744 | Not-determined |
| 18-103_k141_22467 | Not-determined |
| 18-103_k141_114484 | Low-quality |
| 18-103_k141_16241 | Not-determined |
| 18-103_k141_137141 | Not-determined |
| 18-103_k141_73908 | Not-determined |
| 18-103_k141_97182 | Not-determined |
| 18-103_k141_120584 | Not-determined |
| 18-103_k141_16249 | Not-determined |
| 18-103_k141_16252 | Not-determined |
| 18-103_k141_28239 | Not-determined |
| 18-103_k141_114491 | Not-determined |
| 18-103_k141_114492 | Not-determined |
| 18-103_k141_5338 | Not-determined |
| 18-103_k141_114494 | Not-determined |
| 18-103_k141_120591 | Not-determined |
| 18-103_k141_126165 | Not-determined |
| 18-103_k141_16258 | Not-determined |
| 18-103_k141_28249 | Low-quality |
| 18-103_k141_97197 | Not-determined |
| 18-103_k141_5348 | Not-determined |
| 18-103_k141_16261 | Low-quality |
| 18-103_k141_22480 | Not-determined |
| 18-103_k141_126171 | Not-determined |
| 18-103_k141_120595 | Low-quality |
| 18-103_k141_16271 | Low-quality |
| 18-103_k141_28257 | Not-determined |
| 18-103_k141_5357 | Not-determined |
| 18-103_k141_114506 | Not-determined |
| 18-103_k141_39757 | Not-determined |
| 18-103_k141_33638 | Not-determined |
| 18-103_k141_79887 | Not-determined |
| 18-103_k141_39762 | Not-determined |
| 18-103_k141_62523 | Not-determined |
| 18-103_k141_16284 | Not-determined |
| 18-103_k141_102444 | Not-determined |
| 18-103_k141_102445 | Not-determined |
| 18-103_k141_137157 | Low-quality |
| 18-103_k141_45139 | Not-determined |
| 18-103_k141_91517 | Not-determined |
| 18-103_k141_22494 | Not-determined |
| 18-103_k141_62530 | Not-determined |
| 18-103_k141_131889 | Not-determined |
| 18-103_k141_85777 | Not-determined |
| 18-103_k141_62533 | Not-determined |
| 18-103_k141_79895 | Not-determined |
| 18-103_k141_16290 | Not-determined |
| 18-103_k141_33650 | Not-determined |
| 18-103_k141_10812 | Not-determined |
| 18-103_k141_56853 | Not-determined |
| 18-103_k141_62540 | Low-quality |
| 18-103_k141_39774 | Not-determined |
| 18-103_k141_33654 | Low-quality |
| 18-103_k141_45153 | Not-determined |
| 18-103_k141_126197 | Low-quality |
| 18-103_k141_50922 | Not-determined |
| 18-103_k141_68449 | Not-determined |
| 18-103_k141_126201 | Not-determined |
| 18-103_k141_79913 | Not-determined |
| 18-103_k141_91533 | Not-determined |
| 18-103_k141_126206 | Not-determined |
| 18-103_k141_79920 | Not-determined |
| 18-103_k141_10830 | Not-determined |
| 18-103_k141_62558 | Not-determined |
| 18-103_k141_102465 | Not-determined |
| 18-103_k141_108457 | Low-quality |
| 18-103_k141_45164 | Not-determined |
| 18-103_k141_22517 | Not-determined |
| 18-103_k141_137176 | Not-determined |
| 18-103_k141_10834 | Not-determined |
| 18-103_k141_85802 | Not-determined |
| 18-103_k141_50935 | Not-determined |
| 18-103_k141_73964 | Not-determined |
| 18-103_k141_45170 | Not-determined |
| 18-103_k141_108463 | Not-determined |
| 18-103_k141_22527 | Not-determined |
| 18-103_k141_62567 | Not-determined |
| 18-103_k141_91546 | Not-determined |
| 18-103_k141_102477 | Not-determined |
| 18-103_k141_22529 | Not-determined |
| 18-103_k141_68459 | Not-determined |
| 18-103_k141_73972 | Not-determined |
| 18-103_k141_73975 | Not-determined |
| 18-103_k141_10842 | Not-determined |
| 18-103_k141_62574 | Not-determined |
| 18-103_k141_45173 | Not-determined |
| 18-103_k141_131906 | Not-determined |
| 18-103_k141_131907 | Low-quality |
| 18-103_k141_22537 | Low-quality |
| 18-103_k141_68467 | Not-determined |
| 18-103_k141_108473 | Not-determined |
| 18-103_k141_79945 | Not-determined |
| 18-103_k141_22540 | Low-quality |
| 18-103_k141_85818 | Not-determined |
| 18-103_k141_68478 | Not-determined |
| 18-103_k141_97244 | Not-determined |
| 18-103_k141_73993 | Low-quality |
| 18-103_k141_33699 | Not-determined |
| 18-103_k141_28284 | Not-determined |
| 18-103_k141_16297 | Not-determined |
| 18-103_k141_5415 | Not-determined |
| 18-103_k141_97253 | Low-quality |
| 18-103_k141_126242 | Low-quality |
| 18-103_k141_91553 | Not-determined |
| 18-103_k141_74005 | Not-determined |
| 18-103_k141_131928 | Not-determined |
| 18-103_k141_39780 | Not-determined |
| 18-103_k141_91562 | Not-determined |
| 18-103_k141_33714 | Not-determined |
| 18-103_k141_102508 | Low-quality |
| 18-103_k141_45199 | Not-determined |
| 18-103_k141_68501 | Not-determined |
| 18-103_k141_50978 | Not-determined |
| 18-103_k141_114535 | Not-determined |
| 18-103_k141_50980 | Not-determined |
| 18-103_k141_85840 | Low-quality |
| 18-103_k141_68509 | Not-determined |
| 18-103_k141_68510 | Not-determined |
| 18-103_k141_5436 | Not-determined |
| 18-103_k141_50988 | Not-determined |
| 18-103_k141_91575 | Not-determined |
| 18-103_k141_108486 | Not-determined |
| 18-103_k141_45213 | Not-determined |
| 18-103_k141_126267 | Not-determined |
| 18-103_k141_74035 | Not-determined |
| 18-103_k141_79984 | Not-determined |
| 18-103_k141_114548 | Not-determined |
| 18-103_k141_91581 | Not-determined |
| 18-103_k141_68518 | Low-quality |
| 18-103_k141_114551 | Not-determined |
| 18-103_k141_68522 | Not-determined |
| 18-103_k141_51003 | Not-determined |
| 18-103_k141_45220 | Not-determined |
| 18-103_k141_56915 | Not-determined |
| 18-103_k141_108499 | Not-determined |
| 18-103_k141_16337 | Not-determined |
| 18-103_k141_79995 | Not-determined |
| 18-103_k141_10864 | Low-quality |
| 18-103_k141_10866 | Not-determined |
| 18-103_k141_56923 | Not-determined |
| 18-103_k141_10870 | Not-determined |
| 18-103_k141_108503 | Not-determined |
| 18-103_k141_33733 | Not-determined |
| 18-103_k141_10873 | Not-determined |
| 18-103_k141_33737 | Low-quality |
| 18-103_k141_126280 | Not-determined |
| 18-103_k141_108513 | Not-determined |
| 18-103_k141_102521 | Not-determined |
| 18-103_k141_51009 | Not-determined |
| 18-103_k141_91592 | Not-determined |
| 18-103_k141_10877 | Not-determined |
| 18-103_k141_126290 | Not-determined |
| 18-103_k141_16356 | Not-determined |
| 18-103_k141_108527 | Not-determined |
| 18-103_k141_102533 | Not-determined |
| 18-103_k141_56930 | Not-determined |
| 18-103_k141_33749 | Not-determined |
| 18-103_k141_91597 | Not-determined |
| 18-103_k141_28337 | Not-determined |
| 18-103_k141_102541 | Not-determined |
| 18-103_k141_16364 | Not-determined |
| 18-103_k141_126296 | Not-determined |
| 18-103_k141_102544 | Not-determined |
| 18-103_k141_45246 | Not-determined |
| 18-103_k141_5476 | Not-determined |
| 18-103_k141_126301 | Not-determined |
| 18-103_k141_126303 | Not-determined |
| 18-103_k141_45250 | Not-determined |
| 18-103_k141_5478 | Low-quality |
| 18-103_k141_126307 | Not-determined |
| 18-103_k141_51030 | Not-determined |
| 18-103_k141_10897 | Low-quality |
| 18-103_k141_108537 | Not-determined |
| 18-103_k141_16370 | Not-determined |
| 18-103_k141_45253 | Not-determined |
| 18-103_k141_51035 | Not-determined |
| 18-103_k141_51038 | Not-determined |
| 18-103_k141_68550 | Not-determined |
| 18-103_k141_131999 | Not-determined |
| 18-103_k141_126321 | Not-determined |
| 18-103_k141_16378 | Not-determined |
| 18-103_k141_91618 | Not-determined |
| 18-103_k141_132006 | Not-determined |
| 18-103_k141_45263 | Not-determined |
| 18-103_k141_97310 | Not-determined |
| 18-103_k141_22601 | Not-determined |
| 18-103_k141_137216 | Not-determined |
| 18-103_k141_91628 | Not-determined |
| 18-103_k141_33770 | Not-determined |
| 18-103_k141_33772 | Not-determined |
| 18-103_k141_33775 | Not-determined |
| 18-103_k141_102569 | Not-determined |
| 18-103_k141_102570 | Low-quality |
| 18-103_k141_74070 | Not-determined |
| 18-103_k141_97321 | Not-determined |
| 18-103_k141_16391 | Not-determined |
| 18-103_k141_33781 | Not-determined |
| 18-103_k141_126335 | Not-determined |
| 18-103_k141_33783 | Not-determined |
| 18-103_k141_132012 | Not-determined |
| 18-103_k141_39840 | Not-determined |
| 18-103_k141_33787 | Not-determined |
| 18-103_k141_137223 | Not-determined |
| 18-103_k141_22616 | Not-determined |
| 18-103_k141_120676 | Not-determined |
| 18-103_k141_126341 | Not-determined |
| 18-103_k141_28345 | Low-quality |
| 18-103_k141_68560 | Not-determined |
| 18-103_k141_74078 | Not-determined |
| 18-103_k141_16400 | Not-determined |
| 18-103_k141_97329 | Not-determined |
| 18-103_k141_45274 | Not-determined |
| 18-103_k141_39851 | Not-determined |
| 18-103_k141_85877 | Low-quality |
| 18-103_k141_132023 | Not-determined |
| 18-103_k141_91645 | Not-determined |
| 18-103_k141_85880 | Not-determined |
| 18-103_k141_22618 | Not-determined |
| 18-103_k141_132028 | Not-determined |
| 18-103_k141_62646 | Not-determined |
| 18-103_k141_102585 | Not-determined |
| 18-103_k141_22624 | Not-determined |
| 18-103_k141_85887 | Not-determined |
| 18-103_k141_114578 | Not-determined |
| 18-103_k141_62650 | Not-determined |
| 18-103_k141_62652 | Not-determined |
| 18-103_k141_120684 | Not-determined |
| 18-103_k141_114582 | Not-determined |
| 18-103_k141_39866 | Not-determined |
| 18-103_k141_97345 | Not-determined |
| 18-103_k141_56977 | Not-determined |
| 18-103_k141_80022 | Not-determined |
| 18-103_k141_39869 | Not-determined |
| 18-103_k141_56982 | Not-determined |
| 18-103_k141_16415 | Not-determined |
| 18-103_k141_51062 | Not-determined |
| 18-103_k141_91660 | Not-determined |
| 18-103_k141_5511 | Not-determined |
| 18-103_k141_33813 | Not-determined |
| 18-103_k141_16417 | Not-determined |
| 18-103_k141_16418 | Low-quality |
| 18-103_k141_91663 | Not-determined |
| 18-103_k141_22632 | Not-determined |
| 18-103_k141_74096 | Not-determined |
| 18-103_k141_51065 | Low-quality |
| 18-103_k141_80033 | Not-determined |
| 18-103_k141_74098 | Not-determined |
| 18-103_k141_45297 | Low-quality |
| 18-103_k141_126369 | Not-determined |
| 18-103_k141_68580 | Not-determined |
| 18-103_k141_114601 | Low-quality |
| 18-103_k141_137252 | Not-determined |
| 18-103_k141_114603 | Not-determined |
| 18-103_k141_137254 | Not-determined |
| 18-103_k141_22643 | Not-determined |
| 18-103_k141_114605 | Not-determined |
| 18-103_k141_62668 | Not-determined |
| 18-103_k141_85909 | Not-determined |
| 18-103_k141_51076 | Not-determined |
| 18-103_k141_33835 | Not-determined |
| 18-103_k141_16427 | Not-determined |
| 18-103_k141_16429 | Not-determined |
| 18-103_k141_102605 | Not-determined |
| 18-103_k141_5522 | Not-determined |
| 18-103_k141_68596 | Not-determined |
| 18-103_k141_80043 | Low-quality |
| 18-103_k141_28380 | Not-determined |
| 18-103_k141_22661 | Not-determined |
| 18-103_k141_85930 | Not-determined |
| 18-103_k141_22662 | Low-quality |
| 18-103_k141_68603 | Low-quality |
| 18-103_k141_120718 | Not-determined |
| 18-103_k141_97352 | Not-determined |
| 18-103_k141_102618 | Not-determined |
| 18-103_k141_97355 | Not-determined |
| 18-103_k141_102623 | Low-quality |
| 18-103_k141_91696 | Low-quality |

| | |
|---|---|
| 18-103_k141_120726 | Not-determined |
| 18-103_k141_132036 | Not-determined |
| 18-103_k141_85940 | Not-determined |
| 18-103_k141_97359 | Not-determined |
| 18-103_k141_91702 | Not-determined |
| 18-103_k141_114628 | Not-determined |
| 18-103_k141_80065 | Not-determined |
| 18-103_k141_5545 | Not-determined |
| 18-103_k141_85951 | Not-determined |
| 18-103_k141_62695 | Not-determined |
| 18-103_k141_120735 | Not-determined |
| 18-103_k141_33864 | Not-determined |
| 18-103_k141_51102 | Not-determined |
| 18-103_k141_39874 | Low-quality |
| 18-103_k141_39876 | Not-determined |
| 18-103_k141_85954 | Not-determined |
| 18-103_k141_108603 | Not-determined |
| 18-103_k141_39880 | Not-determined |
| 18-103_k141_68624 | Not-determined |
| 18-103_k141_39882 | Not-determined |
| 18-103_k141_57032 | Not-determined |
| 18-103_k141_51108 | Low-quality |
| 18-103_k141_132052 | Low-quality |
| 18-103_k141_137275 | Low-quality |
| 18-103_k141_39891 | Not-determined |
| 18-103_k141_97369 | Low-quality |
| 18-103_k141_74149 | Not-determined |
| 18-103_k141_74150 | Low-quality |
| 18-103_k141_132057 | Not-determined |
| 18-103_k141_114641 | Not-determined |
| 18-103_k141_68633 | Not-determined |
| 18-103_k141_114644 | Not-determined |
| 18-103_k141_22696 | Not-determined |
| 18-103_k141_51116 | Not-determined |
| 18-103_k141_51122 | Not-determined |
| 18-103_k141_137287 | Not-determined |
| 18-103_k141_57045 | Not-determined |
| 18-103_k141_57046 | Not-determined |
| 18-103_k141_85969 | Not-determined |
| 18-103_k141_68638 | Not-determined |
| 18-103_k141_137290 | Not-determined |
| 18-103_k141_137291 | Not-determined |
| 18-103_k141_126390 | Not-determined |
| 18-103_k141_68642 | Low-quality |
| 18-103_k141_57050 | Not-determined |
| 18-103_k141_91730 | Not-determined |
| 18-103_k141_74160 | Not-determined |
| 18-103_k141_74162 | Not-determined |
| 18-103_k141_5561 | Not-determined |
| 18-103_k141_85975 | Not-determined |
| 18-103_k141_45312 | Not-determined |
| 18-103_k141_45313 | Low-quality |
| 18-103_k141_137305 | Not-determined |
| 18-103_k141_126404 | Not-determined |
| 18-103_k141_45317 | Not-determined |
| 18-103_k141_68649 | Not-determined |
| 18-103_k141_97401 | Not-determined |
| 18-103_k141_68651 | Not-determined |
| 18-103_k141_102662 | Not-determined |
| 18-103_k141_68653 | Not-determined |
| 18-103_k141_85984 | Not-determined |
| 18-103_k141_33888 | Not-determined |
| 18-103_k141_22714 | Not-determined |
| 18-103_k141_137313 | Not-determined |
| 18-103_k141_33895 | Not-determined |
| 18-103_k141_97411 | Not-determined |
| 18-103_k141_132096 | Not-determined |
| 18-103_k141_39927 | Low-quality |
| 18-103_k141_91750 | Not-determined |
| 18-103_k141_108625 | Not-determined |
| 18-103_k141_126423 | Not-determined |
| 18-103_k141_80105 | Not-determined |
| 18-103_k141_22723 | Not-determined |
| 18-103_k141_45327 | Low-quality |
| 18-103_k141_74182 | Not-determined |
| 18-103_k141_68665 | Not-determined |
| 18-103_k141_132109 | Not-determined |
| 18-103_k141_57073 | Not-determined |
| 18-103_k141_137332 | Not-determined |
| 18-103_k141_16485 | Low-quality |
| 18-103_k141_137335 | Not-determined |
| 18-103_k141_108646 | Not-determined |
| 18-103_k141_74195 | Not-determined |
| 18-103_k141_137340 | Not-determined |
| 18-103_k141_97424 | Not-determined |
| 18-103_k141_16493 | Not-determined |
| 18-103_k141_74198 | Not-determined |
| 18-103_k141_22739 | Not-determined |
| 18-103_k141_108655 | Not-determined |
| 18-103_k141_5573 | Not-determined |
| 18-103_k141_80125 | Not-determined |
| 18-103_k141_80126 | Low-quality |
| 18-103_k141_74210 | Not-determined |
| 18-103_k141_62746 | Not-determined |
| 18-103_k141_108664 | Not-determined |
| 18-103_k141_80128 | Not-determined |
| 18-103_k141_108668 | Not-determined |
| 18-103_k141_68687 | Not-determined |
| 18-103_k141_51165 | Low-quality |
| 18-103_k141_126445 | Not-determined |
| 18-103_k141_39969 | Low-quality |
| 18-103_k141_5586 | Not-determined |
| 18-103_k141_137363 | Not-determined |
| 18-103_k141_5587 | Not-determined |
| 18-103_k141_51167 | Low-quality |
| 18-103_k141_126459 | Not-determined |
| 18-103_k141_132142 | Not-determined |
| 18-103_k141_51171 | Not-determined |
| 18-103_k141_62757 | Not-determined |
| 18-103_k141_114676 | Not-determined |
| 18-103_k141_126462 | Not-determined |
| 18-103_k141_51177 | Not-determined |
| 18-103_k141_126467 | Low-quality |
| 18-103_k141_114685 | Not-determined |
| 18-103_k141_68711 | Not-determined |
| 18-103_k141_74217 | Not-determined |
| 18-103_k141_126472 | Not-determined |
| 18-103_k141_33950 | Not-determined |
| 18-103_k141_68713 | Not-determined |
| 18-103_k141_16513 | Low-quality |
| 18-103_k141_97461 | Not-determined |
| 18-103_k141_11024 | Not-determined |
| 18-103_k141_51188 | Not-determined |
| 18-103_k141_86041 | Not-determined |
| 18-103_k141_97468 | Not-determined |
| 18-103_k141_51192 | Not-determined |
| 18-103_k141_51193 | Not-determined |
| 18-103_k141_62774 | Not-determined |
| 18-103_k141_114694 | Not-determined |
| 18-103_k141_80145 | Not-determined |
| 18-103_k141_68727 | Not-determined |
| 18-103_k141_80149 | Not-determined |
| 18-103_k141_126490 | Not-determined |
| 18-103_k141_45376 | Low-quality |
| 18-103_k141_114696 | Low-quality |
| 18-103_k141_40006 | Not-determined |
| 18-103_k141_33966 | Not-determined |
| 18-103_k141_91818 | Not-determined |
| 18-103_k141_108675 | Not-determined |
| 18-103_k141_108677 | Not-determined |
| 18-103_k141_40017 | Not-determined |
| 18-103_k141_97481 | Not-determined |
| 18-103_k141_102728 | Not-determined |
| 18-103_k141_126501 | Not-determined |
| 18-103_k141_62791 | Not-determined |
| 18-103_k141_91825 | Not-determined |
| 18-103_k141_74235 | Not-determined |
| 18-103_k141_80163 | Not-determined |
| 18-103_k141_57093 | Not-determined |
| 18-103_k141_5628 | Not-determined |
| 18-103_k141_137387 | Not-determined |
| 18-103_k141_5629 | Not-determined |
| 18-103_k141_62799 | Low-quality |
| 18-103_k141_57100 | Not-determined |
| 18-103_k141_132184 | Not-determined |
| 18-103_k141_80174 | Low-quality |
| 18-103_k141_91842 | Not-determined |
| 18-103_k141_51214 | Not-determined |
| 18-103_k141_132186 | Not-determined |
| 18-103_k141_57105 | Low-quality |
| 18-103_k141_5643 | Not-determined |
| 17-106-final-viral-combined_k141_10990\|\|full | Medium-quality |
| 18-103_k141_80178 | Not-determined |
| 18-103_k141_28444 | Not-determined |
| 18-103_k141_33993 | Not-determined |
| 18-103_k141_102743 | Not-determined |
| 18-103_k141_11061 | Not-determined |
| 18-103_k141_114732 | Low-quality |
| 18-103_k141_86065 | Not-determined |
| 18-103_k141_86068 | Not-determined |
| 18-103_k141_114735 | Not-determined |
| 18-103_k141_22758 | Not-determined |
| 18-103_k141_80187 | Not-determined |
| 18-103_k141_114738 | Not-determined |
| 18-103_k141_16535 | Not-determined |
| 18-103_k141_45399 | Not-determined |
| 18-103_k141_40030 | Not-determined |
| 18-103_k141_11068 | Not-determined |
| 18-103_k141_51236 | Not-determined |
| 18-103_k141_86077 | Not-determined |
| 18-103_k141_45402 | Not-determined |
| 18-103_k141_86080 | Not-determined |
| 18-103_k141_34017 | Not-determined |
| 18-103_k141_11078 | Not-determined |
| 18-103_k141_86084 | Not-determined |
| 18-103_k141_102762 | Not-determined |
| 18-103_k141_45409 | Not-determined |
| 18-103_k141_11080 | Not-determined |
| 18-103_k141_102766 | Not-determined |
| 18-103_k141_22776 | Not-determined |
| 18-103_k141_22780 | Not-determined |
| 18-103_k141_11087 | Not-determined |
| 18-103_k141_74282 | Not-determined |
| 18-103_k141_40045 | Not-determined |
| 18-103_k141_97524 | Not-determined |
| 18-103_k141_51247 | Low-quality |
| 18-103_k141_102778 | Not-determined |
| 18-103_k141_102781 | Not-determined |
| 18-103_k141_34033 | Not-determined |
| 18-103_k141_57121 | Not-determined |
| 18-103_k141_137408 | Not-determined |
| 18-103_k141_16565 | Not-determined |
| 18-103_k141_28480 | Not-determined |
| 18-103_k141_51255 | Low-quality |
| 18-103_k141_132194 | Not-determined |
| 18-103_k141_137415 | Not-determined |
| 18-103_k141_97533 | Not-determined |
| 18-103_k141_5671 | Not-determined |
| 18-103_k141_80208 | Not-determined |
| 18-103_k141_80209 | Low-quality |
| 18-103_k141_5676 | Not-determined |
| 18-103_k141_114762 | Not-determined |
| 18-103_k141_16571 | Not-determined |
| 18-103_k141_114764 | Not-determined |
| 18-103_k141_62811 | Not-determined |
| 18-103_k141_97542 | Not-determined |
| 18-103_k141_108707 | Not-determined |
| 18-103_k141_91894 | Not-determined |
| 18-103_k141_11114 | Low-quality |
| 18-103_k141_97547 | Not-determined |
| 18-103_k141_80218 | Not-determined |
| 18-103_k141_51267 | Not-determined |
| 18-103_k141_22790 | Not-determined |
| 18-103_k141_74318 | Not-determined |
| 18-103_k141_74319 | Not-determined |
| 18-103_k141_68762 | Not-determined |
| 18-103_k141_132216 | Not-determined |
| 18-103_k141_5683 | Low-quality |
| 18-103_k141_62816 | Not-determined |
| 18-103_k141_102791 | Not-determined |
| 18-103_k141_57142 | Not-determined |
| 18-103_k141_137429 | Low-quality |
| 18-103_k141_137430 | Not-determined |
| 18-103_k141_62820 | Not-determined |
| 18-103_k141_68769 | Not-determined |
| 18-103_k141_51276 | Not-determined |
| 18-103_k141_22802 | Not-determined |
| 18-103_k141_62827 | Not-determined |
| 18-103_k141_62829 | Low-quality |
| 18-103_k141_22804 | Not-determined |
| 18-103_k141_51278 | Low-quality |
| 18-103_k141_108733 | Low-quality |
| 18-103_k141_28508 | Not-determined |
| 18-103_k141_16598 | Low-quality |
| 18-103_k141_132228 | Low-quality |
| 18-103_k141_137441 | Not-determined |
| 18-103_k141_132231 | Not-determined |
| 18-103_k141_5702 | Not-determined |
| 18-103_k141_132232 | Not-determined |
| 18-103_k141_45427 | Not-determined |
| 18-103_k141_16608 | Not-determined |
| 18-103_k141_11141 | Not-determined |
| 18-103_k141_108748 | Low-quality |
| 18-103_k141_51292 | Not-determined |
| 18-103_k141_28528 | Low-quality |
| 18-103_k141_108752 | Not-determined |
| 18-103_k141_22816 | Not-determined |
| 18-103_k141_137459 | Not-determined |
| 18-103_k141_28533 | Not-determined |
| 18-103_k141_132246 | Not-determined |
| 18-103_k141_91924 | Low-quality |
| 18-103_k141_28537 | Not-determined |
| 18-103_k141_114800 | Not-determined |
| 18-103_k141_34078 | Not-determined |
| 18-103_k141_11157 | Not-determined |
| 18-103_k141_68794 | Not-determined |
| 18-103_k141_62843 | Not-determined |
| 18-103_k141_57172 | Not-determined |
| 18-103_k141_22827 | Not-determined |
| 18-103_k141_22829 | Not-determined |
| 18-103_k141_22834 | Low-quality |
| 18-103_k141_22835 | Low-quality |
| 18-103_k141_68802 | Not-determined |
| 18-103_k141_22839 | Not-determined |
| 18-103_k141_80234 | Low-quality |
| 18-103_k141_28550 | Not-determined |
| 18-103_k141_108784 | Not-determined |
| 18-103_k141_91948 | Not-determined |
| 18-103_k141_68808 | Not-determined |
| 18-103_k141_86100 | Not-determined |
| 18-103_k141_34098 | Low-quality |
| 18-103_k141_114824 | Not-determined |
| 18-103_k141_97572 | Not-determined |
| 18-103_k141_5742 | Low-quality |
| 18-103_k141_91950 | Not-determined |
| 18-103_k141_102823 | Not-determined |
| 18-103_k141_86102 | Not-determined |
| 18-103_k141_16641 | Not-determined |
| 18-103_k141_137487 | Not-determined |
| 18-103_k141_68817 | Not-determined |
| 18-103_k141_86106 | Not-determined |
| 18-103_k141_5748 | Not-determined |
| 18-103_k141_34106 | Not-determined |
| 18-103_k141_51323 | Not-determined |
| 18-103_k141_68827 | Not-determined |
| 18-103_k141_57200 | Not-determined |
| 18-103_k141_28573 | Not-determined |
| 18-103_k141_51327 | Not-determined |
| 18-103_k141_102842 | Low-quality |
| 18-103_k141_68833 | Not-determined |
| 18-103_k141_51342 | Not-determined |
| 18-103_k141_57211 | Low-quality |
| 18-103_k141_45448 | Not-determined |
| 18-103_k141_57214 | Not-determined |
| 18-103_k141_40088 | Not-determined |
| 18-103_k141_40089 | Not-determined |
| 18-103_k141_86131 | Not-determined |
| 18-103_k141_68841 | Not-determined |
| 18-103_k141_11179 | Not-determined |
| 18-103_k141_86137 | Low-quality |
| 18-103_k141_68847 | Not-determined |
| 18-103_k141_45455 | Not-determined |
| 18-103_k141_68848 | Not-determined |
| 18-103_k141_45460 | Not-determined |
| 18-103_k141_45462 | Not-determined |
| 18-103_k141_114850 | Not-determined |
| 18-103_k141_137515 | Not-determined |
| 18-103_k141_62851 | Not-determined |
| 18-103_k141_108814 | Not-determined |
| 18-103_k141_108816 | Low-quality |
| 18-103_k141_57228 | Low-quality |
| 18-103_k141_11184 | Not-determined |
| 18-103_k141_68850 | Not-determined |
| 18-103_k141_45471 | Not-determined |
| 18-103_k141_102884 | Not-determined |
| 18-103_k141_45477 | Low-quality |
| 18-103_k141_74425 | Not-determined |
| 18-103_k141_86156 | Low-quality |
| 18-103_k141_97631 | Not-determined |
| 18-103_k141_74432 | Not-determined |
| 18-103_k141_68863 | Low-quality |
| 18-103_k141_137526 | Not-determined |
| 18-103_k141_57243 | Not-determined |
| 18-103_k141_16710 | Not-determined |
| 18-103_k141_40131 | Not-determined |
| 18-103_k141_40133 | Not-determined |
| 18-103_k141_11187 | Not-determined |
| 18-103_k141_91998 | Not-determined |
| 18-103_k141_91999 | Low-quality |
| 18-103_k141_45481 | Low-quality |
| 18-103_k141_86173 | Not-determined |
| 18-103_k141_28622 | Not-determined |
| 18-103_k141_28623 | Not-determined |
| 18-103_k141_137538 | Not-determined |
| 18-103_k141_22897 | Not-determined |
| 18-103_k141_16726 | Not-determined |
| 18-103_k141_80280 | Not-determined |
| 18-103_k141_62879 | Not-determined |
| 18-103_k141_97651 | Not-determined |
| 18-103_k141_62880 | Not-determined |
| 18-103_k141_28632 | Not-determined |
| 18-103_k141_97653 | Not-determined |
| 18-103_k141_28635 | Low-quality |
| 18-103_k141_86184 | Low-quality |
| 18-103_k141_22902 | Not-determined |
| 18-103_k141_57253 | Low-quality |
| 18-103_k141_57256 | Low-quality |
| 18-103_k141_102909 | Not-determined |
| 18-103_k141_51389 | Not-determined |
| 18-103_k141_51391 | Not-determined |
| 18-103_k141_102911 | Not-determined |
| 18-103_k141_28647 | Not-determined |
| 18-103_k141_74469 | Not-determined |
| 18-103_k141_97672 | Not-determined |
| 18-103_k141_114891 | Not-determined |
| 18-103_k141_22919 | Not-determined |
| 18-103_k141_34168 | Not-determined |
| 18-103_k141_11215 | Not-determined |
| 18-103_k141_97676 | Not-determined |
| 18-103_k141_51401 | Low-quality |
| 18-103_k141_51403 | Not-determined |
| 18-103_k141_28652 | Not-determined |
| 18-103_k141_22930 | Not-determined |
| 18-103_k141_62908 | Not-determined |
| 18-103_k141_102927 | Not-determined |
| 18-103_k141_114903 | Not-determined |
| 18-103_k141_45514 | Not-determined |
| 18-103_k141_40157 | Not-determined |
| 18-103_k141_16744 | Not-determined |
| 18-103_k141_86203 | Not-determined |
| 18-103_k141_11221 | Not-determined |
| 18-103_k141_45520 | Not-determined |
| 18-103_k141_68872 | Not-determined |
| 18-103_k141_68873 | Not-determined |
| 18-103_k141_114910 | Not-determined |
| 18-103_k141_114911 | Not-determined |
| 18-103_k141_80312 | Not-determined |
| 18-103_k141_86211 | Not-determined |
| 18-103_k141_80313 | Not-determined |
| 18-103_k141_80314 | Low-quality |
| 18-103_k141_114915 | Not-determined |
| 18-103_k141_68879 | Not-determined |
| 18-103_k141_57274 | Not-determined |
| 18-103_k141_11234 | Not-determined |
| 18-103_k141_22949 | Not-determined |
| 18-103_k141_80321 | Not-determined |
| 18-103_k141_34192 | Low-quality |
| 18-103_k141_137579 | Low-quality |
| 18-103_k141_114931 | Not-determined |
| 18-103_k141_11237 | Not-determined |
| 18-103_k141_34196 | Low-quality |
| 18-103_k141_97705 | Not-determined |
| 18-103_k141_16759 | Not-determined |
| 18-103_k141_16760 | Not-determined |
| 18-103_k141_74503 | Not-determined |
| 18-103_k141_40170 | Not-determined |
| 18-103_k141_34202 | Low-quality |
| 18-103_k141_68884 | Not-determined |
| 18-103_k141_16765 | Not-determined |
| 18-103_k141_86226 | Not-determined |
| 18-103_k141_86227 | Not-determined |
| 18-103_k141_22958 | Not-determined |
| 18-103_k141_114943 | Not-determined |
| 18-103_k141_80334 | Not-determined |
| 18-103_k141_22960 | Not-determined |
| 18-103_k141_16772 | Not-determined |
| 18-103_k141_57299 | Not-determined |
| 18-103_k141_114952 | Not-determined |
| 18-103_k141_22969 | Not-determined |
| 18-103_k141_86232 | Not-determined |
| 18-103_k141_11249 | Not-determined |
| 18-103_k141_86233 | Not-determined |
| 18-103_k141_97720 | Low-quality |
| 18-103_k141_28681 | Not-determined |
| 18-103_k141_22975 | Not-determined |
| 18-103_k141_62950 | Not-determined |
| 18-103_k141_62951 | Not-determined |
| 18-103_k141_114964 | Low-quality |
| 18-103_k141_97722 | Not-determined |
| 18-103_k141_62952 | Not-determined |
| 18-103_k141_74529 | Not-determined |
| 18-103_k141_80345 | Low-quality |
| 18-103_k141_62955 | Not-determined |
| 18-103_k141_16791 | Not-determined |
| 18-103_k141_114965 | Low-quality |
| 18-103_k141_102978 | Not-determined |
| 18-103_k141_97734 | Low-quality |
| 18-103_k141_97735 | Not-determined |
| 18-103_k141_97737 | Not-determined |
| 18-103_k141_28692 | Not-determined |
| 18-103_k141_51433 | Not-determined |
| 18-103_k141_102980 | Not-determined |
| 18-103_k141_40196 | Not-determined |
| 18-103_k141_57333 | Not-determined |
| 18-103_k141_16796 | Low-quality |
| 18-103_k141_45549 | Not-determined |
| 18-103_k141_57334 | Not-determined |
| 18-103_k141_34221 | Not-determined |
| 18-103_k141_11268 | Not-determined |
| 18-103_k141_57336 | Not-determined |
| 18-103_k141_11272 | Not-determined |
| 18-103_k141_62971 | Low-quality |
| 18-103_k141_11282 | Not-determined |
| 18-103_k141_45555 | Not-determined |
| 18-103_k141_62977 | Not-determined |
| 18-103_k141_102999 | Not-determined |
| 18-103_k141_80377 | Not-determined |
| 18-103_k141_40214 | Not-determined |
| 18-103_k141_62979 | Not-determined |
| 18-103_k141_11288 | Not-determined |
| 18-103_k141_103001 | Not-determined |
| 18-103_k141_97755 | Not-determined |
| 18-103_k141_74544 | Not-determined |
| 18-103_k141_103008 | Not-determined |
| 18-103_k141_80383 | Not-determined |
| 18-103_k141_80389 | Not-determined |
| 18-103_k141_45567 | Not-determined |
| 18-103_k141_74549 | Not-determined |
| 18-103_k141_80393 | Not-determined |
| 18-103_k141_80394 | Not-determined |
| 18-103_k141_45570 | Low-quality |
| 18-103_k141_74554 | Not-determined |
| 18-103_k141_80402 | Not-determined |
| 18-103_k141_51447 | Not-determined |
| 18-103_k141_28730 | Not-determined |
| 18-103_k141_11308 | Not-determined |
| 18-103_k141_74558 | Not-determined |
| 18-103_k141_108923 | Low-quality |
| 18-103_k141_103027 | Not-determined |
| 18-103_k141_80412 | Low-quality |
| 18-103_k141_80413 | Not-determined |
| 18-103_k141_45576 | Not-determined |
| 18-103_k141_40233 | Not-determined |
| 18-103_k141_45577 | Not-determined |
| 18-103_k141_28745 | Not-determined |
| 18-103_k141_11317 | Not-determined |
| 18-103_k141_68939 | Not-determined |
| 18-103_k141_57361 | Not-determined |
| 18-103_k141_40236 | Not-determined |
| 18-103_k141_40239 | Not-determined |
| 18-103_k141_103039 | Not-determined |
| 18-103_k141_11323 | Low-quality |
| 18-103_k141_45585 | Not-determined |
| 18-103_k141_45587 | Not-determined |
| 18-103_k141_108943 | Not-determined |
| 18-103_k141_137631 | Low-quality |
| 18-103_k141_45592 | Not-determined |
| 18-103_k141_16838 | Not-determined |
| 18-103_k141_16841 | Not-determined |
| 18-103_k141_57376 | Not-determined |
| 18-103_k141_57377 | Not-determined |
| 18-103_k141_103046 | Not-determined |
| 18-103_k141_51469 | Not-determined |
| 18-103_k141_11338 | Not-determined |
| 18-103_k141_51480 | Not-determined |
| 18-103_k141_63024 | Not-determined |
| 18-103_k141_40253 | Not-determined |
| 18-103_k141_57385 | Not-determined |
| 18-103_k141_16850 | Not-determined |
| 18-103_k141_16851 | Not-determined |
| 18-103_k141_45612 | Not-determined |
| 18-103_k141_34299 | Not-determined |
| 18-103_k141_16854 | Not-determined |
| 18-103_k141_16857 | Not-determined |
| 18-103_k141_103064 | Not-determined |
| 18-103_k141_68957 | Not-determined |
| 18-103_k141_80440 | Not-determined |
| 18-103_k141_45616 | Not-determined |
| 18-103_k141_45618 | Not-determined |
| 18-103_k141_108973 | Not-determined |
| 18-103_k141_68966 | Low-quality |
| 18-103_k141_16866 | Not-determined |
| 18-103_k141_51491 | Not-determined |
| 18-103_k141_80453 | Low-quality |
| 18-103_k141_51492 | Low-quality |
| 18-103_k141_80454 | Not-determined |
| 18-103_k141_51494 | Not-determined |
| 18-103_k141_57414 | Low-quality |
| 18-103_k141_137646 | Not-determined |
| 18-103_k141_80459 | Not-determined |
| 18-103_k141_16872 | Low-quality |
| 18-103_k141_45632 | Not-determined |
| 18-103_k141_57417 | Not-determined |
| 18-103_k141_137648 | Not-determined |
| 18-103_k141_68975 | Not-determined |
| 18-103_k141_63042 | Low-quality |
| 18-103_k141_16881 | Not-determined |
| 18-103_k141_80470 | Not-determined |
| 18-103_k141_57429 | Not-determined |
| 18-103_k141_16885 | Not-determined |
| 18-103_k141_63049 | Not-determined |
| 18-103_k141_80480 | Not-determined |
| 18-103_k141_51503 | Low-quality |
| 18-103_k141_63053 | Not-determined |
| 18-103_k141_11378 | Not-determined |
| 18-103_k141_57440 | Not-determined |
| 18-103_k141_63058 | Not-determined |
| 18-103_k141_103088 | Low-quality |
| 18-103_k141_63064 | Not-determined |
| 18-103_k141_68984 | Low-quality |
| 18-103_k141_16892 | Low-quality |
| 18-103_k141_80489 | Not-determined |
| 18-103_k141_16898 | Not-determined |
| 18-103_k141_68988 | Low-quality |
| 18-103_k141_63075 | Not-determined |
| 18-103_k141_51523 | Not-determined |
| 18-103_k141_11401 | Not-determined |
| 18-103_k141_68996 | Not-determined |
| 18-103_k141_45666 | Not-determined |
| 18-103_k141_11409 | Not-determined |
| 18-103_k141_109008 | Not-determined |
| 18-103_k141_137680 | Not-determined |
| 18-103_k141_80500 | Not-determined |
| 18-103_k141_45673 | Not-determined |
| 18-103_k141_63085 | Low-quality |
| 18-103_k141_137686 | Low-quality |
| 18-103_k141_34358 | Not-determined |
| 18-103_k141_63089 | Not-determined |
| 18-103_k141_63093 | Not-determined |
| 18-103_k141_109026 | Not-determined |
| 18-103_k141_57470 | Not-determined |
| 18-103_k141_11430 | Not-determined |
| 18-103_k141_57473 | Not-determined |
| 18-103_k141_57475 | Not-determined |
| 18-103_k141_63097 | Not-determined |
| 18-103_k141_137701 | Not-determined |
| 18-103_k141_45682 | Not-determined |
| 18-103_k141_63099 | Low-quality |
| 18-103_k141_16911 | Not-determined |
| 18-103_k141_109033 | Not-determined |
| 18-103_k141_137706 | Not-determined |
| 18-103_k141_16916 | Not-determined |
| 18-103_k141_74567 | Not-determined |
| 18-103_k141_63115 | Not-determined |
| 18-103_k141_137709 | Not-determined |
| 18-103_k141_11453 | Not-determined |
| 18-103_k141_45697 | Not-determined |
| 18-103_k141_16929 | Not-determined |
| 18-103_k141_109060 | Not-determined |

| | |
|---|---|
| 18-103_k141_109063 | Not-determined |
| 18-103_k141_74581 | Not-determined |
| 18-103_k141_45706 | Not-determined |
| 18-103_k141_11470 | Not-determined |
| 18-103_k141_34362 | Low-quality |
| 18-103_k141_45716 | Not-determined |
| 18-103_k141_63127 | Not-determined |
| 18-103_k141_63128 | Not-determined |
| 18-103_k141_51538 | Low-quality |
| 18-103_k141_57501 | Not-determined |
| 18-103_k141_51540 | Not-determined |
| 18-103_k141_137731 | Not-determined |
| 18-103_k141_16962 | Not-determined |
| 18-103_k141_103148 | Not-determined |
| 18-103_k141_137739 | Not-determined |
| 18-103_k141_137742 | Not-determined |
| 18-103_k141_51565 | Not-determined |
| 18-103_k141_137745 | Not-determined |
| 18-103_k141_74632 | Not-determined |
| 18-103_k141_74640 | Not-determined |
| 18-103_k141_74643 | Not-determined |
| 18-103_k141_34403 | Not-determined |
| 18-103_k141_74645 | Not-determined |
| 18-103_k141_137758 | Not-determined |
| 18-103_k141_137759 | Not-determined |
| 18-103_k141_51589 | Not-determined |
| 18-103_k141_34414 | Not-determined |
| 18-103_k141_74663 | Not-determined |
| 18-103_k141_137770 | Not-determined |
| 18-103_k141_74666 | Not-determined |
| 18-103_k141_109097 | Not-determined |
| 18-103_k141_74669 | Not-determined |
| 18-103_k141_74670 | Not-determined |
| 18-103_k141_74674 | Not-determined |
| 18-103_k141_74677 | Not-determined |
| 18-103_k141_63164 | Not-determined |
| 18-103_k141_63165 | Not-determined |
| 18-103_k141_109109 | Not-determined |
| 18-103_k141_63167 | Not-determined |
| 18-103_k141_45738 | Not-determined |
| 18-103_k141_34458 | Not-determined |
| 18-103_k141_137782 | Not-determined |
| 18-103_k141_109133 | Not-determined |
| 18-103_k141_34463 | Not-determined |
| 18-103_k141_74700 | Low-quality |
| 18-103_k141_45754 | Not-determined |
| 18-103_k141_63198 | Not-determined |
| 18-103_k141_109142 | Not-determined |
| 18-103_k141_17041 | Not-determined |
| 18-103_k141_109149 | Not-determined |
| 18-103_k141_45762 | Not-determined |
| 18-103_k141_51617 | Not-determined |
| 18-103_k141_74707 | Not-determined |
| 18-103_k141_45773 | Not-determined |
| 18-103_k141_109158 | Not-determined |
| 18-103_k141_63214 | Not-determined |
| 18-103_k141_17049 | Not-determined |
| 18-103_k141_109162 | Not-determined |
| 18-103_k141_51632 | Not-determined |
| 18-103_k141_45778 | Not-determined |
| 18-103_k141_45783 | Low-quality |
| 18-103_k141_17058 | Not-determined |
| 18-103_k141_109169 | Not-determined |
| 18-103_k141_17061 | Not-determined |
| 18-103_k141_34474 | Not-determined |
| 18-103_k141_109172 | Not-determined |
| 18-103_k141_17070 | Not-determined |
| 18-103_k141_109186 | Not-determined |
| 18-103_k141_34485 | Not-determined |
| 18-103_k141_51642 | Not-determined |
| 18-103_k141_74740 | Not-determined |
| 18-103_k141_63230 | Not-determined |
| 18-103_k141_109191 | Low-quality |
| 18-103_k141_63234 | Not-determined |
| 18-103_k141_51652 | Not-determined |
| 18-103_k141_17089 | Not-determined |
| 18-103_k141_74751 | Not-determined |
| 18-103_k141_51658 | Not-determined |
| 18-103_k141_51661 | Not-determined |
| 18-103_k141_103204 | Not-determined |
| 18-103_k141_51664 | Low-quality |
| 18-103_k141_103216 | Not-determined |
| 18-103_k141_109201 | Low-quality |
| 18-103_k141_103224 | Not-determined |
| 18-103_k141_103225 | Not-determined |
| 18-103_k141_109203 | Not-determined |
| 18-103_k141_103227 | Not-determined |
| 18-103_k141_103230 | Not-determined |
| 18-103_k141_63246 | Not-determined |
| 18-103_k141_63247 | Not-determined |
| 18-103_k141_63249 | Not-determined |
| 18-103_k141_17104 | Low-quality |
| 18-103_k141_17105 | Low-quality |
| 18-103_k141_63254 | Not-determined |
| 18-103_k141_51688 | Not-determined |
| 18-103_k141_137846 | Not-determined |
| 18-103_k141_51698 | Not-determined |
| 18-103_k141_45802 | Not-determined |
| 18-103_k141_17139 | Not-determined |
| 18-103_k141_103257 | Not-determined |
| 18-103_k141_51711 | Not-determined |
| 18-103_k141_51713 | Not-determined |
| 18-103_k141_109237 | Low-quality |
| 18-103_k141_45828 | Not-determined |
| 18-103_k141_51732 | Not-determined |
| 18-103_k141_51738 | Not-determined |
| 18-103_k141_45842 | Low-quality |
| 18-103_k141_45844 | Not-determined |
| 18-103_k141_45847 | Not-determined |
| 18-103_k141_51754 | Not-determined |
| 18-103_k141_45863 | Not-determined |
| 18-103_k141_103299 | Not-determined |
| 18-103_k141_45866 | Not-determined |
| 18-103_k141_45885 | Low-quality |
| 18-103_k141_103311 | Not-determined |
| 18-103_k141_45888 | Not-determined |
| 18-103_k141_17196 | Not-determined |
| 18-103_k141_137877 | Not-determined |
| 18-103_k141_137878 | Low-quality |
| 18-103_k141_45902 | Not-determined |
| 18-103_k141_45908 | Not-determined |
| 18-103_k141_137880 | Not-determined |
| 18-103_k141_137882 | Not-determined |
| 18-103_k141_137884 | Not-determined |
| 18-103_k141_137885 | Not-determined |
| 18-103_k141_137886 | Not-determined |
| 18-103_k141_137887 | Not-determined |
| 18-103_k141_137895 | Not-determined |
| 18-103_k141_137901 | Not-determined |
| 18-103_k141_17205 | Not-determined |
| 18-103_k141_137906 | Not-determined |
| 18-103_k141_137910 | Not-determined |
| 18-103_k141_45917 | Low-quality |
| 18-103_k141_17211 | Not-determined |
| 18-103_k141_137924 | Low-quality |
| 18-103_k141_137925 | Not-determined |
| 18-103_k141_137927 | Not-determined |
| 18-103_k141_17218 | Not-determined |
| 18-103_k141_137937 | Not-determined |
| 18-103_k141_137939 | Not-determined |
| 18-103_k141_137940 | Not-determined |
| 18-103_k141_137947 | Not-determined |
| 18-103_k141_137952 | Not-determined |
| 18-103_k141_137953 | Not-determined |
| 18-103_k141_137954 | Not-determined |
| 18-103_k141_137955 | Not-determined |
| 18-103_k141_137956 | Not-determined |
| 18-103_k141_137964 | Not-determined |
| 18-103_k141_137970 | Not-determined |
| 18-103_k141_137972 | Not-determined |
| 18-103_k141_137973 | Not-determined |
| 18-103_k141_137974 | Not-determined |
| 18-103_k141_137984 | Not-determined |
| 18-103_k141_137987 | Not-determined |
| 18-103_k141_137991 | Not-determined |
| 18-103_k141_137992 | Not-determined |
| 18-103_k141_137996 | Not-determined |
| 18-103_k141_138001 | Not-determined |
| 18-103_k141_138002 | Not-determined |
| 18-103_k141_138003 | Not-determined |
| 18-103_k141_138006 | Not-determined |
| 18-103_k141_45921 | Not-determined |
| 18-103_k141_103384 | Not-determined |
| 18-103_k141_45925 | Not-determined |
| 18-103_k141_45936 | Not-determined |
| 18-103_k141_45939 | Not-determined |
| 18-103_k141_103433 | Low-quality |
| 18-103_k141_45971 | Low-quality |
| 18-103_k141_45974 | Not-determined |
| 18-103_k141_103453 | Not-determined |
| 18-103_k141_103459 | Not-determined |
| 18-103_k141_103473 | Not-determined |
| 18-103_k141_45999 | Not-determined |
| 18-103_k141_103482 | Not-determined |
| 18-103_k141_103483 | Not-determined |
| 18-103_k141_46005 | Not-determined |
| 18-103_k141_103495 | Not-determined |
| 18-103_k141_103499 | Not-determined |
| 18-105-final-viral-combined_k141_53805\|\|full | Not-determined |
| 18-105-final-viral-combined_k141_18175\|\|full | Not-determined |
| 18-105-final-viral-combined_k141_36163\|\|full | Not-determined |
| 18-105-final-viral-combined_k141_90077\|\|full | Not-determined |
| 18-105-final-viral-combined_k141_63959\|\|full | Not-determined |
| 18-105-final-viral-combined_k141_162029\|\|full | Not-determined |
| 18-105-final-viral-combined_k141_28935\|\|full | Not-determined |
| 18-105-final-viral-combined_k141_38184\|\|full | Not-determined |
| 18-105-final-viral-combined_k141_3752\|\|full | Not-determined |
| 18-105-final-viral-combined_k141_181624\|\|full | Not-determined |
| 18-105-final-viral-combined_k141_85957\|\|full | Not-determined |
| 18-105-final-viral-combined_k141_23116\|\|full | Not-determined |
| 18-105-final-viral-combined_k141_124632\|\|full | Not-determined |
| 17-109-final-viral-combined_k141_375343\|\|full | Medium-quality |
| 18-105-final-viral-combined_k141_213583\|\|full | Not-determined |
| 18-105-final-viral-combined_k141_213600\|\|full | Not-determined |
| 18-105-final-viral-combined_k141_44546\|\|full | Not-determined |
| 18-105-final-viral-combined_k141_53463\|\|full | Low-quality |
| 18-105-final-viral-combined_k141_71287\|\|full | Low-quality |
| 18-105-final-viral-combined_k141_115846\|\|full | Not-determined |
| 18-105-final-viral-combined_k141_142545\|\|full | Low-quality |
| 18-105-final-viral-combined_k141_124753\|\|full | Not-determined |
| 18-105-final-viral-combined_k141_169278\|\|full | Not-determined |
| 18-105-final-viral-combined_k141_151474\|\|full | Not-determined |
| 18-105-final-viral-combined_k141_142555\|\|full | Not-determined |
| 18-105-final-viral-combined_k141_71291\|\|full | Low-quality |
| 18-105-final-viral-combined_k141_47\|\|full | Low-quality |
| 18-105-final-viral-combined_k141_35659\|\|full | Low-quality |
| 18-105-final-viral-combined_k141_115870\|\|full | Low-quality |
| 18-105-final-viral-combined_k141_26779\|\|full | Low-quality |
| 18-105-final-viral-combined_k141_89126\|\|full | Not-determined |
| 18-105-final-viral-combined_k141_8986\|\|full | Low-quality |
| 18-105-final-viral-combined_k141_151492\|\|full | Low-quality |
| 18-105-final-viral-combined_k141_80238\|\|full | Low-quality |
| 18-105-final-viral-combined_k141_62414\|\|full | Not-determined |
| 18-105-final-viral-combined_k141_35672\|\|full | Low-quality |
| 18-105-final-viral-combined_k141_69\|\|full | Not-determined |
| 18-105-final-viral-combined_k141_80244\|\|full | Low-quality |
| 18-105-final-viral-combined_k141_151518\|\|full | Low-quality |
| 18-105-final-viral-combined_k141_169295\|\|full | Low-quality |
| 18-105-final-viral-combined_k141_151530\|\|full | Low-quality |
| 18-105-final-viral-combined_k141_53484\|\|full | Low-quality |
| 18-105-final-viral-combined_k141_196018\|\|full | Low-quality |
| 18-105-final-viral-combined_k141_17869\|\|full | Low-quality |
| 18-105-final-viral-combined_k141_133716\|\|full | Low-quality |
| 18-105-final-viral-combined_k141_160416\|\|full | Low-quality |
| 18-105-final-viral-combined_k141_151552\|\|full | Low-quality |
| 18-105-final-viral-combined_k141_196058\|\|full | Not-determined |
| 18-105-final-viral-combined_k141_178183\|\|full | Low-quality |
| 18-105-final-viral-combined_k141_71384\|\|full | Not-determined |
| 18-105-final-viral-combined_k141_124834\|\|full | Not-determined |
| 18-105-final-viral-combined_k141_71388\|\|full | Not-determined |
| 18-105-final-viral-combined_k141_160464\|\|full | Low-quality |
| 18-105-final-viral-combined_k141_115916\|\|full | Not-determined |
| 18-105-final-viral-combined_k141_137\|\|full | Not-determined |
| 18-105-final-viral-combined_k141_196092\|\|full | Low-quality |
| 18-105-final-viral-combined_k141_142601\|\|full | Low-quality |
| 18-105-final-viral-combined_k141_115927\|\|full | Not-determined |
| 18-105-final-viral-combined_k141_106988\|\|full | Not-determined |
| 18-105-final-viral-combined_k141_9082\|\|full | Not-determined |
| 18-105-final-viral-combined_k141_107002\|\|full | Not-determined |
| 18-105-final-viral-combined_k141_151607\|\|full | Not-determined |
| 18-105-final-viral-combined_k141_9091\|\|full | Not-determined |
| 18-105-final-viral-combined_k141_107012\|\|full | Low-quality |
| 18-105-final-viral-combined_k141_178193\|\|full | Low-quality |
| 18-105-final-viral-combined_k141_173\|\|full | Not-determined |
| 18-105-final-viral-combined_k141_17960\|\|full | Low-quality |
| 18-105-final-viral-combined_k141_17965\|\|full | Not-determined |
| 18-105-final-viral-combined_k141_115943\|\|full | Not-determined |
| 18-105-final-viral-combined_k141_107022\|\|full | Not-determined |
| 18-105-final-viral-combined_k141_26858\|\|full | Not-determined |
| 18-105-final-viral-combined_k141_107036\|\|full | Low-quality |
| 18-105-final-viral-combined_k141_71431\|\|full | Low-quality |
| 18-105-final-viral-combined_k141_151631\|\|full | Not-determined |
| 18-105-final-viral-combined_k141_133814\|\|full | Not-determined |
| 18-105-final-viral-combined_k141_18012\|\|full | Not-determined |
| 18-105-final-viral-combined_k141_196155\|\|full | Low-quality |
| 18-105-final-viral-combined_k141_107044\|\|full | Low-quality |
| 18-105-final-viral-combined_k141_71446\|\|full | Low-quality |
| 18-105-final-viral-combined_k141_196156\|\|full | Not-determined |
| 18-105-final-viral-combined_k141_26907\|\|full | Low-quality |
| 18-105-final-viral-combined_k141_237\|\|full | Not-determined |
| 18-105-final-viral-combined_k141_107049\|\|full | Low-quality |
| 18-105-final-viral-combined_k141_107051\|\|full | Low-quality |
| 18-105-final-viral-combined_k141_53567\|\|full | Not-determined |
| 18-105-final-viral-combined_k141_115971\|\|full | Low-quality |
| 18-105-final-viral-combined_k141_245\|\|full | Not-determined |
| 18-105-final-viral-combined_k141_98132\|\|full | Low-quality |
| 18-105-final-viral-combined_k141_35690\|\|full | Low-quality |
| 18-105-final-viral-combined_k141_204958\|\|full | Low-quality |
| 18-105-final-viral-combined_k141_115985\|\|full | Low-quality |
| 18-105-final-viral-combined_k141_80306\|\|full | Low-quality |
| 18-105-final-viral-combined_k141_26926\|\|full | Low-quality |
| 18-105-final-viral-combined_k141_160542\|\|full | Low-quality |
| 18-105-final-viral-combined_k141_62485\|\|full | Not-determined |
| 18-105-final-viral-combined_k141_18046\|\|full | Low-quality |
| 18-105-final-viral-combined_k141_71491\|\|full | Low-quality |
| 18-105-final-viral-combined_k141_98163\|\|full | Not-determined |
| 18-105-final-viral-combined_k141_35709\|\|full | Low-quality |
| 18-105-final-viral-combined_k141_178287\|\|full | Low-quality |
| 18-105-final-viral-combined_k141_53622\|\|full | Not-determined |
| 18-105-final-viral-combined_k141_62496\|\|full | Low-quality |
| 18-105-final-viral-combined_k141_107111\|\|full | Not-determined |
| 18-105-final-viral-combined_k141_53629\|\|full | Not-determined |
| 18-105-final-viral-combined_k141_178304\|\|full | Low-quality |
| 18-105-final-viral-combined_k141_116003\|\|full | Not-determined |
| 18-105-final-viral-combined_k141_71534\|\|full | Not-determined |
| 18-105-final-viral-combined_k141_98179\|\|full | Not-determined |
| 18-105-final-viral-combined_k141_71535\|\|full | Low-quality |
| 18-105-final-viral-combined_k141_116021\|\|full | Low-quality |
| 18-105-final-viral-combined_k141_339\|\|full | Low-quality |
| 18-105-final-viral-combined_k141_116026\|\|full | Low-quality |
| 18-105-final-viral-combined_k141_98189\|\|full | Low-quality |
| 18-105-final-viral-combined_k141_80337\|\|full | Low-quality |
| 18-105-final-viral-combined_k141_346\|\|full | Low-quality |
| 18-105-final-viral-combined_k141_178336\|\|full | Low-quality |
| 18-105-final-viral-combined_k141_80372\|\|full | Not-determined |
| 18-105-final-viral-combined_k141_71542\|\|full | Low-quality |
| 18-105-final-viral-combined_k141_142683\|\|full | Not-determined |
| 18-105-final-viral-combined_k141_53686\|\|full | Not-determined |
| 18-105-final-viral-combined_k141_372\|\|full | Not-determined |
| 18-105-final-viral-combined_k141_89214\|\|full | Not-determined |
| 18-105-final-viral-combined_k141_53700\|\|full | Low-quality |
| 18-105-final-viral-combined_k141_142708\|\|full | Low-quality |
| 18-105-final-viral-combined_k141_400\|\|full | Not-determined |
| 18-105-final-viral-combined_k141_142720\|\|full | Not-determined |
| 18-105-final-viral-combined_k141_196229\|\|full | Low-quality |
| 18-105-final-viral-combined_k141_196231\|\|full | Not-determined |
| 18-105-final-viral-combined_k141_89237\|\|full | Low-quality |
| 18-105-final-viral-combined_k141_419\|\|full | Not-determined |
| 18-105-final-viral-combined_k141_178365\|\|full | Not-determined |
| 18-105-final-viral-combined_k141_187241\|\|full | Not-determined |
| 18-105-final-viral-combined_k141_124950\|\|full | Not-determined |
| 18-105-final-viral-combined_k141_178408\|\|full | Not-determined |
| 18-105-final-viral-combined_k141_169330\|\|full | Low-quality |
| 18-105-final-viral-combined_k141_116109\|\|full | Not-determined |
| 18-105-final-viral-combined_k141_124965\|\|full | Not-determined |
| 18-105-final-viral-combined_k141_71574\|\|full | Not-determined |
| 18-105-final-viral-combined_k141_178431\|\|full | Not-determined |
| 18-105-final-viral-combined_k141_169349\|\|full | Not-determined |
| 18-105-final-viral-combined_k141_124969\|\|full | Not-determined |
| 18-105-final-viral-combined_k141_107242\|\|full | Not-determined |
| 18-105-final-viral-combined_k141_451\|\|full | Low-quality |
| 18-105-final-viral-combined_k141_80426\|\|full | Not-determined |
| 18-105-final-viral-combined_k141_116138\|\|full | Not-determined |
| 18-105-final-viral-combined_k141_160639\|\|full | Not-determined |
| 18-105-final-viral-combined_k141_471\|\|full | Low-quality |
| 18-105-final-viral-combined_k141_124987\|\|full | Not-determined |
| 18-105-final-viral-combined_k141_160665\|\|full | Low-quality |
| 18-105-final-viral-combined_k141_116143\|\|full | Not-determined |
| 18-105-final-viral-combined_k141_116144\|\|full | Not-determined |
| 18-105-final-viral-combined_k141_160666\|\|full | Low-quality |
| 18-105-final-viral-combined_k141_492\|\|full | Low-quality |
| 18-105-final-viral-combined_k141_116176\|\|full | Not-determined |
| 18-105-final-viral-combined_k141_142783\|\|full | Low-quality |
| 18-105-final-viral-combined_k141_53763\|\|full | Low-quality |
| 18-105-final-viral-combined_k141_62591\|\|full | Not-determined |
| 18-105-final-viral-combined_k141_53768\|\|full | Not-determined |
| 18-105-final-viral-combined_k141_160720\|\|full | Low-quality |
| 18-105-final-viral-combined_k141_187294\|\|full | Low-quality |
| 18-105-final-viral-combined_k141_53786\|\|full | Not-determined |
| 18-105-final-viral-combined_k141_18144\|\|full | Not-determined |
| 18-105-final-viral-combined_k141_62598\|\|full | Not-determined |
| 18-105-final-viral-combined_k141_71638\|\|full | Low-quality |
| 18-105-final-viral-combined_k141_62606\|\|full | Not-determined |
| 18-105-final-viral-combined_k141_169403\|\|full | Low-quality |
| 18-105-final-viral-combined_k141_187313\|\|full | Not-determined |
| 18-105-final-viral-combined_k141_187316\|\|full | Low-quality |
| 18-105-final-viral-combined_k141_53806\|\|full | Low-quality |
| 18-105-final-viral-combined_k141_187321\|\|full | Not-determined |
| 18-105-final-viral-combined_k141_196378\|\|full | Not-determined |
| 18-105-final-viral-combined_k141_44631\|\|full | Low-quality |
| 18-105-final-viral-combined_k141_71676\|\|full | Low-quality |
| 18-105-final-viral-combined_k141_116216\|\|full | Low-quality |
| 18-105-final-viral-combined_k141_134047\|\|full | Low-quality |
| 18-105-final-viral-combined_k141_71691\|\|full | Not-determined |
| 18-110-final-viral-combined_k141_169835\|\|full | Medium-quality |
| 18-105-final-viral-combined_k141_160778\|\|full | Low-quality |
| 18-105-final-viral-combined_k141_169427\|\|full | Not-determined |
| 18-105-final-viral-combined_k141_71700\|\|full | Low-quality |
| 18-105-final-viral-combined_k141_205049\|\|full | Not-determined |
| 18-105-final-viral-combined_k141_98224\|\|full | Not-determined |
| 18-105-final-viral-combined_k141_196430\|\|full | Low-quality |
| 18-105-final-viral-combined_k141_134082\|\|full | Not-determined |
| 18-105-final-viral-combined_k141_62641\|\|full | Not-determined |
| 18-105-final-viral-combined_k141_187382\|\|full | Low-quality |
| 18-105-final-viral-combined_k141_205062\|\|full | Not-determined |
| 18-105-final-viral-combined_k141_98243\|\|full | Not-determined |
| 18-105-final-viral-combined_k141_80491\|\|full | Low-quality |
| 18-105-final-viral-combined_k141_596\|\|full | Low-quality |
| 18-105-final-viral-combined_k141_187398\|\|full | Not-determined |
| 18-105-final-viral-combined_k141_612\|\|full | Not-determined |
| 18-105-final-viral-combined_k141_62664\|\|full | Low-quality |
| 18-105-final-viral-combined_k141_160792\|\|full | Not-determined |
| 18-105-final-viral-combined_k141_169468\|\|full | Low-quality |
| 18-105-final-viral-combined_k141_196460\|\|full | Low-quality |
| 18-105-final-viral-combined_k141_9406\|\|full | Not-determined |
| 18-105-final-viral-combined_k141_169476\|\|full | Low-quality |
| 18-105-final-viral-combined_k141_9417\|\|full | Not-determined |
| 18-105-final-viral-combined_k141_187420\|\|full | Low-quality |
| 18-105-final-viral-combined_k141_89252\|\|full | Not-determined |
| 18-105-final-viral-combined_k141_80536\|\|full | Low-quality |
| 18-105-final-viral-combined_k141_205084\|\|full | Low-quality |
| 18-105-final-viral-combined_k141_35825\|\|full | Not-determined |
| 18-105-final-viral-combined_k141_205092\|\|full | Low-quality |
| 18-105-final-viral-combined_k141_98300\|\|full | Low-quality |
| 18-105-final-viral-combined_k141_98305\|\|full | Low-quality |
| 18-105-final-viral-combined_k141_44736\|\|full | Not-determined |
| 18-105-final-viral-combined_k141_35838\|\|full | Low-quality |
| 18-105-final-viral-combined_k141_9452\|\|full | Not-determined |
| 18-105-final-viral-combined_k141_35842\|\|full | Not-determined |
| 18-105-final-viral-combined_k141_169496\|\|full | Not-determined |
| 18-106_k141_132588 | Medium-quality |
| 18-105-final-viral-combined_k141_80558\|\|full | Not-determined |
| 18-105-final-viral-combined_k141_71784\|\|full | Not-determined |
| 18-105-final-viral-combined_k141_187450\|\|full | Not-determined |
| 18-105-final-viral-combined_k141_681\|\|full | Low-quality |
| 18-105-final-viral-combined_k141_9477\|\|full | Not-determined |
| 18-105-final-viral-combined_k141_142903\|\|full | Low-quality |
| 18-105-final-viral-combined_k141_71809\|\|full | Low-quality |
| 18-105-final-viral-combined_k141_169514\|\|full | Low-quality |
| 18-105-final-viral-combined_k141_142906\|\|full | Not-determined |
| 18-105-final-viral-combined_k141_107301\|\|full | Low-quality |
| 18-105-final-viral-combined_k141_71823\|\|full | Not-determined |
| 18-105-final-viral-combined_k141_89346\|\|full | Low-quality |
| 18-105-final-viral-combined_k141_62708\|\|full | Low-quality |
| 18-105-final-viral-combined_k141_107308\|\|full | Low-quality |
| 18-105-final-viral-combined_k141_169526\|\|full | Low-quality |
| 18-105-final-viral-combined_k141_151660\|\|full | Not-determined |
| 18-105-final-viral-combined_k141_89383\|\|full | Not-determined |
| 18-105-final-viral-combined_k141_35861\|\|full | Low-quality |
| 18-105-final-viral-combined_k141_196531\|\|full | Not-determined |
| 18-105-final-viral-combined_k141_80617\|\|full | Not-determined |
| 18-105-final-viral-combined_k141_80625\|\|full | Not-determined |
| 18-105-final-viral-combined_k141_107332\|\|full | Low-quality |
| 18-105-final-viral-combined_k141_35888\|\|full | Not-determined |
| 18-105-final-viral-combined_k141_143002\|\|full | Not-determined |
| 18-105-final-viral-combined_k141_44832\|\|full | Not-determined |
| 18-105-final-viral-combined_k141_35920\|\|full | Low-quality |
| 18-105-final-viral-combined_k141_196567\|\|full | Low-quality |
| 18-105-final-viral-combined_k141_107381\|\|full | Low-quality |
| 18-105-final-viral-combined_k141_44869\|\|full | Not-determined |
| 18-105-final-viral-combined_k141_89459\|\|full | Low-quality |
| 18-105-final-viral-combined_k141_772\|\|full | Not-determined |
| 18-105-final-viral-combined_k141_27043\|\|full | Low-quality |
| 18-105-final-viral-combined_k141_160858\|\|full | Low-quality |
| 18-105-final-viral-combined_k141_44888\|\|full | Low-quality |
| 18-105-final-viral-combined_k141_44898\|\|full | Not-determined |
| 18-105-final-viral-combined_k141_27059\|\|full | Not-determined |
| 18-105-final-viral-combined_k141_89492\|\|full | Low-quality |
| 18-105-final-viral-combined_k141_18290\|\|full | Low-quality |
| 18-105-final-viral-combined_k141_169632\|\|full | Low-quality |
| 18-105-final-viral-combined_k141_80700\|\|full | Low-quality |
| 18-105-final-viral-combined_k141_107415\|\|full | Not-determined |
| 18-105-final-viral-combined_k141_169644\|\|full | Not-determined |
| 18-105-final-viral-combined_k141_160883\|\|full | Low-quality |
| 18-105-final-viral-combined_k141_98390\|\|full | Not-determined |
| 18-105-final-viral-combined_k141_35964\|\|full | Low-quality |
| 18-105-final-viral-combined_k141_9583\|\|full | Not-determined |
| 18-105-final-viral-combined_k141_9588\|\|full | Low-quality |
| 18-105-final-viral-combined_k141_134138\|\|full | Low-quality |
| 18-105-final-viral-combined_k141_80730\|\|full | Not-determined |
| 18-105-final-viral-combined_k141_819\|\|full | Low-quality |
| 18-105-final-viral-combined_k141_44973\|\|full | Not-determined |
| 18-105-final-viral-combined_k141_143100\|\|full | Not-determined |
| 18-105-final-viral-combined_k141_71829\|\|full | Not-determined |
| 18-105-final-viral-combined_k141_71841\|\|full | Low-quality |
| 18-105-final-viral-combined_k141_151816\|\|full | Low-quality |
| 18-105-final-viral-combined_k141_125116\|\|full | Low-quality |
| 18-102-final-viral-combined_k141_143737\|\|0_partial | Medium-quality |
| 18-105-final-viral-combined_k141_18360\|\|full | Low-quality |
| 18-105-final-viral-combined_k141_54011\|\|full | Low-quality |
| 18-105-final-viral-combined_k141_71865\|\|full | Low-quality |
| 18-105-final-viral-combined_k141_169713\|\|full | Not-determined |
| 18-105-final-viral-combined_k141_71869\|\|full | Low-quality |
| 18-105-final-viral-combined_k141_18372\|\|full | Low-quality |
| 18-105-final-viral-combined_k141_160989\|\|full | Low-quality |
| 18-105-final-viral-combined_k141_169735\|\|full | Not-determined |
| 18-105-final-viral-combined_k141_143165\|\|full | Low-quality |
| 18-105-final-viral-combined_k141_143180\|\|full | Low-quality |
| 18-105-final-viral-combined_k141_125162\|\|full | Low-quality |
| 18-105-final-viral-combined_k141_205305\|\|full | Low-quality |
| 18-105-final-viral-combined_k141_909\|\|full | Low-quality |
| 18-105-final-viral-combined_k141_125167\|\|full | Low-quality |
| 18-105-final-viral-combined_k141_18379\|\|full | Low-quality |
| 18-105-final-viral-combined_k141_71900\|\|full | Not-determined |
| 18-105-final-viral-combined_k141_920\|\|full | Low-quality |
| 18-105-final-viral-combined_k141_116289\|\|full | Not-determined |
| 18-105-final-viral-combined_k141_205322\|\|full | Not-determined |
| 18-105-final-viral-combined_k141_125200\|\|full | Low-quality |
| 18-105-final-viral-combined_k141_18402\|\|full | Low-quality |
| 18-105-final-viral-combined_k141_116314\|\|full | Not-determined |
| 18-105-final-viral-combined_k141_125207\|\|full | Low-quality |
| 18-105-final-viral-combined_k141_89621\|\|full | Not-determined |
| 18-105-final-viral-combined_k141_169796\|\|full | Not-determined |
| 18-105-final-viral-combined_k141_196613\|\|full | Low-quality |
| 18-105-final-viral-combined_k141_178485\|\|full | Not-determined |
| 18-105-final-viral-combined_k141_205349\|\|full | Not-determined |
| 18-105-final-viral-combined_k141_89643\|\|full | Not-determined |
| 18-105-final-viral-combined_k141_80788\|\|full | Not-determined |
| 18-105-final-viral-combined_k141_196643\|\|full | Low-quality |
| 18-105-final-viral-combined_k141_116337\|\|full | Low-quality |
| 18-105-final-viral-combined_k141_27127\|\|full | Low-quality |
| 18-105-final-viral-combined_k141_196663\|\|full | Low-quality |
| 18-105-final-viral-combined_k141_98609\|\|full | Low-quality |
| 18-105-final-viral-combined_k141_169826\|\|full | Not-determined |
| 18-105-final-viral-combined_k141_151958\|\|full | Low-quality |
| 18-105-final-viral-combined_k141_18446\|\|full | Low-quality |
| 18-105-final-viral-combined_k141_27141\|\|full | Not-determined |
| 18-105-final-viral-combined_k141_125224\|\|full | Not-determined |
| 18-105-final-viral-combined_k141_125229\|\|full | Low-quality |
| 18-105-final-viral-combined_k141_71959\|\|full | Low-quality |
| 18-105-final-viral-combined_k141_98626\|\|full | Not-determined |
| 18-105-final-viral-combined_k141_62804\|\|full | Low-quality |
| 18-105-final-viral-combined_k141_62806\|\|full | Not-determined |
| 18-105-final-viral-combined_k141_125250\|\|full | Low-quality |
| 18-105-final-viral-combined_k141_98628\|\|full | Low-quality |
| 18-105-final-viral-combined_k141_9780\|\|full | Not-determined |
| 18-105-final-viral-combined_k141_1057\|\|full | Not-determined |
| 18-105-final-viral-combined_k141_89712\|\|full | Not-determined |
| 18-105-final-viral-combined_k141_178518\|\|full | Low-quality |
| 18-105-final-viral-combined_k141_18479\|\|full | Low-quality |
| 18-105-final-viral-combined_k141_89727\|\|full | Not-determined |
| 18-105-final-viral-combined_k141_1074\|\|full | Low-quality |
| 18-105-final-viral-combined_k141_161117\|\|full | Low-quality |
| 18-105-final-viral-combined_k141_169894\|\|full | Not-determined |
| 18-105-final-viral-combined_k141_1084\|\|full | Low-quality |
| 18-105-final-viral-combined_k141_45071\|\|full | Not-determined |
| 18-105-final-viral-combined_k141_125335\|\|full | Low-quality |
| 18-105-final-viral-combined_k141_107589\|\|full | Not-determined |
| 18-105-final-viral-combined_k141_9815\|\|full | Low-quality |
| 18-105-final-viral-combined_k141_161149\|\|full | Low-quality |
| 18-105-final-viral-combined_k141_9825\|\|full | Not-determined |
| 18-105-final-viral-combined_k141_205396\|\|full | Not-determined |
| 18-105-final-viral-combined_k141_205409\|\|full | Not-determined |
| 18-105-final-viral-combined_k141_205415\|\|full | Not-determined |
| 18-105-final-viral-combined_k141_36147\|\|full | Not-determined |
| 18-105-final-viral-combined_k141_152066\|\|full | Not-determined |
| 18-105-final-viral-combined_k141_62852\|\|full | Not-determined |
| 18-105-final-viral-combined_k141_187618\|\|full | Low-quality |
| 18-105-final-viral-combined_k141_89846\|\|full | Not-determined |
| 18-105-final-viral-combined_k141_196751\|\|full | Low-quality |
| 18-105-final-viral-combined_k141_178624\|\|full | Low-quality |
| 18-105-final-viral-combined_k141_107676\|\|full | Low-quality |
| 18-105-final-viral-combined_k141_9832\|\|full | Low-quality |
| 18-105-final-viral-combined_k141_196767\|\|full | Not-determined |
| 18-105-final-viral-combined_k141_196772\|\|full | Low-quality |
| 18-105-final-viral-combined_k141_169978\|\|full | Not-determined |
| 18-105-final-viral-combined_k141_196777\|\|full | Not-determined |
| 18-105-final-viral-combined_k141_62885\|\|full | Not-determined |
| 18-105-final-viral-combined_k141_116409\|\|full | Low-quality |
| 18-105-final-viral-combined_k141_45124\|\|full | Not-determined |
| 18-105-final-viral-combined_k141_98724\|\|full | Low-quality |
| 18-105-final-viral-combined_k141_36182\|\|full | Not-determined |
| 18-105-final-viral-combined_k141_89871\|\|full | Low-quality |
| 18-105-final-viral-combined_k141_178667\|\|full | Not-determined |
| 18-105-final-viral-combined_k141_178670\|\|full | Low-quality |
| 18-105-final-viral-combined_k141_161170\|\|full | Low-quality |
| 18-105-final-viral-combined_k141_107678\|\|full | Low-quality |
| 18-105-final-viral-combined_k141_125347\|\|full | Low-quality |
| 18-105-final-viral-combined_k141_125352\|\|full | Low-quality |
| 18-105-final-viral-combined_k141_98759\|\|full | Low-quality |
| 18-105-final-viral-combined_k141_36228\|\|full | Not-determined |
| 18-105-final-viral-combined_k141_45140\|\|full | Low-quality |
| 18-105-final-viral-combined_k141_1114\|\|full | Not-determined |
| 18-105-final-viral-combined_k141_1129\|\|full | Low-quality |
| 18-105-final-viral-combined_k141_170011\|\|full | Not-determined |
| 18-105-final-viral-combined_k141_170013\|\|full | Low-quality |
| 18-105-final-viral-combined_k141_170017\|\|full | Low-quality |
| 18-105-final-viral-combined_k141_170026\|\|full | Low-quality |
| 18-105-final-viral-combined_k141_178725\|\|full | Low-quality |
| 18-105-final-viral-combined_k141_54081\|\|full | Low-quality |
| 18-105-final-viral-combined_k141_178730\|\|full | Not-determined |
| 18-105-final-viral-combined_k141_36244\|\|full | Low-quality |
| 18-105-final-viral-combined_k141_45179\|\|full | Not-determined |
| 18-105-final-viral-combined_k141_205487\|\|full | Not-determined |
| 18-105-final-viral-combined_k141_36254\|\|full | Not-determined |
| 18-105-final-viral-combined_k141_205496\|\|full | Not-determined |
| 18-105-final-viral-combined_k141_178751\|\|full | Not-determined |
| 18-105-final-viral-combined_k141_36260\|\|full | Low-quality |
| 18-105-final-viral-combined_k141_107758\|\|full | Low-quality |
| 18-105-final-viral-combined_k141_125375\|\|full | Not-determined |
| 18-105-final-viral-combined_k141_196867\|\|full | Low-quality |
| 18-105-final-viral-combined_k141_170073\|\|full | Low-quality |
| 18-105-final-viral-combined_k141_36282\|\|full | Not-determined |
| 18-105-final-viral-combined_k141_27214\|\|full | Low-quality |
| 18-105-final-viral-combined_k141_54116\|\|full | Not-determined |
| 18-105-final-viral-combined_k141_125406\|\|full | Not-determined |
| 18-105-final-viral-combined_k141_1229\|\|full | Not-determined |
| 18-105-final-viral-combined_k141_89982\|\|full | Low-quality |
| 18-105-final-viral-combined_k141_27242\|\|full | Not-determined |
| 18-105-final-viral-combined_k141_89990\|\|full | Not-determined |
| 18-105-final-viral-combined_k141_107779\|\|full | Not-determined |
| 18-105-final-viral-combined_k141_152083\|\|full | Not-determined |
| 18-105-final-viral-combined_k141_178769\|\|full | Low-quality |
| 18-105-final-viral-combined_k141_143301\|\|full | Low-quality |
| 18-105-final-viral-combined_k141_178772\|\|full | Not-determined |
| 18-105-final-viral-combined_k141_1291\|\|full | Not-determined |
| 18-105-final-viral-combined_k141_134340\|\|full | Low-quality |
| 18-105-final-viral-combined_k141_143319\|\|full | Low-quality |
| 18-105-final-viral-combined_k141_90019\|\|full | Low-quality |
| 18-105-final-viral-combined_k141_36319\|\|full | Low-quality |
| 18-105-final-viral-combined_k141_18504\|\|full | Not-determined |
| 18-105-final-viral-combined_k141_143327\|\|full | Not-determined |
| 18-105-final-viral-combined_k141_170154\|\|full | Low-quality |
| 18-105-final-viral-combined_k141_178798\|\|full | Low-quality |
| 18-105-final-viral-combined_k141_62968\|\|full | Low-quality |
| 18-105-final-viral-combined_k141_125492\|\|full | Low-quality |
| 18-105-final-viral-combined_k141_62984\|\|full | Low-quality |
| 18-105-final-viral-combined_k141_45275\|\|full | Low-quality |
| 18-105-final-viral-combined_k141_170188\|\|full | Not-determined |

18-105-final-viral-combined_k141_36377||full Low-quality
18-105-final-viral-combined_k141_125501||full Not-determined
18-105-final-viral-combined_k141_80873||full Not-determined
18-105-final-viral-combined_k141_143341||full Low-quality
18-105-final-viral-combined_k141_170203||full Not-determined
18-105-final-viral-combined_k141_80874||full Low-quality
18-105-final-viral-combined_k141_9928||full Low-quality
18-105-final-viral-combined_k141_80892||full Not-determined
18-105-final-viral-combined_k141_152187||full Not-determined
18-105-final-viral-combined_k141_63004||full Not-determined
18-105-final-viral-combined_k141_134375||full Low-quality
18-105-final-viral-combined_k141_116588||full Low-quality
18-105-final-viral-combined_k141_170231||full Not-determined
18-105-final-viral-combined_k141_54278||full Low-quality
18-105-final-viral-combined_k141_143364||full Not-determined
18-105-final-viral-combined_k141_9937||full Low-quality
18-105-final-viral-combined_k141_80906||full Not-determined
18-105-final-viral-combined_k141_45327||full Low-quality
18-105-final-viral-combined_k141_187757||full Not-determined
18-105-final-viral-combined_k141_54284||full Not-determined
18-105-final-viral-combined_k141_90123||full Low-quality
18-105-final-viral-combined_k141_63036||full Low-quality
18-105-final-viral-combined_k141_18535||full Low-quality
18-105-final-viral-combined_k141_18538||full Low-quality
18-105-final-viral-combined_k141_45351||full Not-determined
18-105-final-viral-combined_k141_1373||full Low-quality
18-106-final-viral-combined_k141_132588||full Medium-quality
18-105-final-viral-combined_k141_152264||full Low-quality
18-105-final-viral-combined_k141_196926||full Low-quality
18-105-final-viral-combined_k141_1382||full Not-determined
18-105-final-viral-combined_k141_90133||full Low-quality
18-105-final-viral-combined_k141_152270||full Low-quality
18-105-final-viral-combined_k141_45379||full Low-quality
18-105-final-viral-combined_k141_27431||full Low-quality
18-105-final-viral-combined_k141_18560||full Low-quality
18-105-final-viral-combined_k141_45398||full Low-quality
18-105-final-viral-combined_k141_63072||full Not-determined
18-105-final-viral-combined_k141_161234||full Low-quality
18-105-final-viral-combined_k141_1414||full Not-determined
18-105-final-viral-combined_k141_143404||full Not-determined
18-105-final-viral-combined_k141_63073||full Low-quality
17-108_k141_14390 Complete
18-105-final-viral-combined_k141_178901||full Low-quality
18-105-final-viral-combined_k141_63075||full Not-determined
18-105-final-viral-combined_k141_63081||full Not-determined
18-105-final-viral-combined_k141_45422||full Low-quality
18-105-final-viral-combined_k141_45425||full Not-determined
18-105-final-viral-combined_k141_1436||full Not-determined
18-105-final-viral-combined_k141_143437||full Not-determined
18-105-final-viral-combined_k141_152297||full Not-determined
18-105-final-viral-combined_k141_152307||full Low-quality
18-105-final-viral-combined_k141_178953||full Not-determined
18-105-final-viral-combined_k141_90209||full Low-quality
18-105-final-viral-combined_k141_27494||full Not-determined
18-105-final-viral-combined_k141_178963||full Not-determined
18-105-final-viral-combined_k141_152322||full Low-quality
18-105-final-viral-combined_k141_170287||full Not-determined
18-105-final-viral-combined_k141_10005||full Low-quality
18-105-final-viral-combined_k141_116688||full Low-quality
18-105-final-viral-combined_k141_54401||full Not-determined
18-105-final-viral-combined_k141_10039||full Low-quality
18-105-final-viral-combined_k141_178998||full Not-determined
18-105-final-viral-combined_k141_45454||full Not-determined
18-105-final-viral-combined_k141_1481||full Low-quality
18-105-final-viral-combined_k141_80941||full Not-determined
18-105-final-viral-combined_k141_80944||full Low-quality
18-105-final-viral-combined_k141_107911||full Low-quality
18-105-final-viral-combined_k141_134467||full Not-determined
18-105-final-viral-combined_k141_179027||full Not-determined
18-105-final-viral-combined_k141_143458||full Low-quality
18-105-final-viral-combined_k141_197003||full Not-determined
18-105-final-viral-combined_k141_1525||full Not-determined
18-105-final-viral-combined_k141_1529||full Not-determined
18-105-final-viral-combined_k141_179069||full Not-determined
18-105-final-viral-combined_k141_116744||full Low-quality
18-105-final-viral-combined_k141_1537||full Low-quality
18-105-final-viral-combined_k141_161271||full Low-quality
18-105-final-viral-combined_k141_90268||full Low-quality
18-105-final-viral-combined_k141_18627||full Low-quality
18-105-final-viral-combined_k141_108018||full Low-quality
18-105-final-viral-combined_k141_90277||full Not-determined
18-105-final-viral-combined_k141_108027||full Not-determined
18-105-final-viral-combined_k141_18653||full Not-determined
18-105-final-viral-combined_k141_1578||full Not-determined
18-105-final-viral-combined_k141_161291||full Not-determined
18-105-final-viral-combined_k141_54450||full Not-determined
18-105-final-viral-combined_k141_54455||full Not-determined
18-105-final-viral-combined_k141_90309||full Low-quality
18-105-final-viral-combined_k141_54473||full Not-determined
18-105-final-viral-combined_k141_27554||full Not-determined
18-105-final-viral-combined_k141_205701||full Not-determined
18-105-final-viral-combined_k141_205703||full Not-determined
18-105-final-viral-combined_k141_27570||full Not-determined
18-105-final-viral-combined_k141_205715||full Low-quality
18-105-final-viral-combined_k141_27571||full Low-quality
18-105-final-viral-combined_k141_10070||full Low-quality
18-105-final-viral-combined_k141_27573||full Low-quality
18-105-final-viral-combined_k141_134539||full Low-quality
18-105-final-viral-combined_k141_205721||full Not-determined
18-105-final-viral-combined_k141_63159||full Not-determined
18-105-final-viral-combined_k141_27580||full Low-quality
18-105-final-viral-combined_k141_143528||full Low-quality
18-105-final-viral-combined_k141_205739||full Not-determined
18-105-final-viral-combined_k141_125717||full Low-quality
18-105-final-viral-combined_k141_116805||full Not-determined
18-105-final-viral-combined_k141_1655||full Low-quality
18-105-final-viral-combined_k141_98955||full Low-quality
18-105-final-viral-combined_k141_63200||full Low-quality
18-105-final-viral-combined_k141_36544||full Not-determined
18-105-final-viral-combined_k141_116843||full Not-determined
18-105-final-viral-combined_k141_98974||full Not-determined
18-105-final-viral-combined_k141_179203||full Not-determined
18-105-final-viral-combined_k141_179218||full Not-determined
18-105-final-viral-combined_k141_187886||full Not-determined
18-105-final-viral-combined_k141_45541||full Not-determined
18-105-final-viral-combined_k141_10127||full Not-determined
18-102_k141_107329 Medium-quality
18-105-final-viral-combined_k141_81050||full Not-determined
18-105-final-viral-combined_k141_143597||full Not-determined
18-105-final-viral-combined_k141_197043||full Low-quality
18-105-final-viral-combined_k141_143599||full Not-determined
18-105-final-viral-combined_k141_81069||full Not-determined
18-105-final-viral-combined_k141_197056||full Not-determined
18-105-final-viral-combined_k141_116864||full Low-quality
18-105-final-viral-combined_k141_134667||full Not-determined
18-105-final-viral-combined_k141_1723||full Low-quality
18-105-final-viral-combined_k141_27682||full Low-quality
18-105-final-viral-combined_k141_72268||full Not-determined
18-105-final-viral-combined_k141_152443||full Low-quality
18-105-final-viral-combined_k141_143649||full Low-quality
18-105-final-viral-combined_k141_81138||full Low-quality
18-105-final-viral-combined_k141_99011||full Low-quality
18-105-final-viral-combined_k141_81143||full Not-determined
18-105-final-viral-combined_k141_72288||full Not-determined
18-105-final-viral-combined_k141_187922||full Low-quality
18-105-final-viral-combined_k141_205761||full Low-quality
18-105-final-viral-combined_k141_1761||full Not-determined
18-105-final-viral-combined_k141_90359||full Low-quality
18-105-final-viral-combined_k141_27739||full Low-quality
18-105-final-viral-combined_k141_81182||full Low-quality
18-105-final-viral-combined_k141_197089||full Low-quality
18-105-final-viral-combined_k141_205773||full Low-quality
18-105-final-viral-combined_k141_197093||full Not-determined
18-105-final-viral-combined_k141_81190||full Not-determined
18-105-final-viral-combined_k141_197099||full Low-quality
18-105-final-viral-combined_k141_205786||full Not-determined
18-105-final-viral-combined_k141_99051||full Not-determined
18-105-final-viral-combined_k141_197102||full Low-quality
18-105-final-viral-combined_k141_90386||full Low-quality
18-105-final-viral-combined_k141_90407||full Low-quality
18-105-final-viral-combined_k141_116950||full Low-quality
18-105-final-viral-combined_k141_108205||full Not-determined
18-105-final-viral-combined_k141_45606||full Low-quality
18-105-final-viral-combined_k141_72373||full Low-quality
18-105-final-viral-combined_k141_45611||full Low-quality
18-105-final-viral-combined_k141_197118||full Low-quality
18-105-final-viral-combined_k141_81267||full Low-quality
18-105-final-viral-combined_k141_205821||full Low-quality
18-105-final-viral-combined_k141_152512||full Not-determined
18-105-final-viral-combined_k141_108219||full Low-quality
18-105-final-viral-combined_k141_81281||full Low-quality
18-105-final-viral-combined_k141_117004||full Low-quality
18-105-final-viral-combined_k141_54548||full Low-quality
18-105-final-viral-combined_k141_45636||full Low-quality
18-105-final-viral-combined_k141_10161||full Not-determined
18-105-final-viral-combined_k141_45645||full Low-quality
18-105-final-viral-combined_k141_18702||full Low-quality
18-105-final-viral-combined_k141_197143||full Not-determined
18-105-final-viral-combined_k141_10168||full Not-determined
18-105-final-viral-combined_k141_36616||full Low-quality
18-105-final-viral-combined_k141_152548||full Low-quality
18-105-final-viral-combined_k141_81301||full Not-determined
18-105-final-viral-combined_k141_152551||full Not-determined
18-105-final-viral-combined_k141_161329||full Not-determined
18-105-final-viral-combined_k141_161331||full Low-quality
18-105-final-viral-combined_k141_143717||full Low-quality
18-105-final-viral-combined_k141_36641||full Low-quality
18-105-final-viral-combined_k141_54561||full Low-quality
18-105-final-viral-combined_k141_205873||full Not-determined
18-105-final-viral-combined_k141_36643||full Not-determined
18-105-final-viral-combined_k141_108267||full Low-quality
18-105-final-viral-combined_k141_27827||full Low-quality
18-105-final-viral-combined_k141_63307||full Low-quality
18-105-final-viral-combined_k141_27837||full Low-quality
18-105-final-viral-combined_k141_1853||full Low-quality
18-105-final-viral-combined_k141_108272||full Not-determined
18-105-final-viral-combined_k141_63315||full Low-quality
18-105-final-viral-combined_k141_54581||full Low-quality
18-105-final-viral-combined_k141_108273||full Not-determined
18-105-final-viral-combined_k141_45678||full Low-quality
18-105-final-viral-combined_k141_99088||full Not-determined
18-105-final-viral-combined_k141_152610||full Low-quality
18-102-final-viral-combined_k141_107329||full Medium-quality
18-105-final-viral-combined_k141_134745||full Low-quality
18-105-final-viral-combined_k141_170493||full Not-determined
18-105-final-viral-combined_k141_108301||full Not-determined
18-105-final-viral-combined_k141_205920||full Low-quality
18-105-final-viral-combined_k141_205934||full Low-quality
18-105-final-viral-combined_k141_54646||full Low-quality
18-105-final-viral-combined_k141_134773||full Not-determined
17-108-final-viral-combined_k141_168048||full Complete
18-105-final-viral-combined_k141_179250||full Low-quality
18-105-final-viral-combined_k141_179263||full Low-quality
18-105-final-viral-combined_k141_36653||full Not-determined
18-105-final-viral-combined_k141_45738||full Not-determined
18-105-final-viral-combined_k141_188063||full Low-quality
18-105-final-viral-combined_k141_134832||full Not-determined
18-105-final-viral-combined_k141_134834||full Low-quality
18-105-final-viral-combined_k141_45754||full Low-quality
18-105-final-viral-combined_k141_36669||full Low-quality
18-105-final-viral-combined_k141_143796||full Low-quality
18-105-final-viral-combined_k141_36684||full Low-quality
18-105-final-viral-combined_k141_205977||full Not-determined
18-105-final-viral-combined_k141_18774||full Not-determined
18-105-final-viral-combined_k141_108312||full Low-quality
18-105-final-viral-combined_k141_54681||full Not-determined
18-105-final-viral-combined_k141_45774||full Not-determined
18-105-final-viral-combined_k141_143829||full Not-determined
18-105-final-viral-combined_k141_205981||full Low-quality
18-105-final-viral-combined_k141_179362||full Low-quality
18-105-final-viral-combined_k141_161434||full Not-determined
18-105-final-viral-combined_k141_108376||full Not-determined
18-105-final-viral-combined_k141_179389||full Low-quality
18-105-final-viral-combined_k141_81424||full Not-determined
18-105-final-viral-combined_k141_179394||full Low-quality
18-105-final-viral-combined_k141_18783||full Low-quality
18-105-final-viral-combined_k141_161464||full Low-quality
18-105-final-viral-combined_k141_179410||full Not-determined
18-105-final-viral-combined_k141_143896||full Not-determined
17-108-final-viral-combined_k141_14390||full High-quality
18-110-final-viral-combined_k141_226076||full High-quality
18-105-final-viral-combined_k141_27865||full Not-determined
18-105-final-viral-combined_k141_18807||full Low-quality
18-105-final-viral-combined_k141_72437||full Not-determined
18-105-final-viral-combined_k141_170708||full Not-determined
18-105-final-viral-combined_k141_99176||full Low-quality
18-105-final-viral-combined_k141_90471||full Not-determined
18-105-final-viral-combined_k141_72447||full Low-quality
18-105-final-viral-combined_k141_197230||full Low-quality
18-105-final-viral-combined_k141_197232||full Low-quality
18-105-final-viral-combined_k141_54755||full Low-quality
18-105-final-viral-combined_k141_134926||full Low-quality
18-105-final-viral-combined_k141_179447||full Low-quality
18-105-final-viral-combined_k141_206012||full Low-quality
18-105-final-viral-combined_k141_179455||full Not-determined
18-105-final-viral-combined_k141_117165||full Not-determined
18-105-final-viral-combined_k141_170731||full Not-determined
18-105-final-viral-combined_k141_206022||full Low-quality
18-105-final-viral-combined_k141_18875||full Low-quality
18-105-final-viral-combined_k141_18879||full Low-quality
18-105-final-viral-combined_k141_170758||full Low-quality
18-105-final-viral-combined_k141_179487||full Not-determined
18-105-final-viral-combined_k141_108478||full Low-quality
18-105-final-viral-combined_k141_179489||full Not-determined
18-105-final-viral-combined_k141_197278||full Low-quality
18-105-final-viral-combined_k141_108488||full Not-determined
18-105-final-viral-combined_k141_117201||full Low-quality
18-105-final-viral-combined_k141_170765||full Not-determined
18-105-final-viral-combined_k141_108517||full Not-determined
18-105-final-viral-combined_k141_108519||full Low-quality
18-105-final-viral-combined_k141_54800||full Low-quality
18-105-final-viral-combined_k141_1995||full Not-determined
18-105-final-viral-combined_k141_54809||full Not-determined
18-105-final-viral-combined_k141_27904||full Low-quality
18-105-final-viral-combined_k141_36777||full Low-quality
18-105-final-viral-combined_k141_179548||full Not-determined
18-105-final-viral-combined_k141_10258||full Low-quality
18-110_k141_226076 High-quality
18-105-final-viral-combined_k141_188288||full Low-quality
18-105-final-viral-combined_k141_152740||full Not-determined
18-105-final-viral-combined_k141_90543||full Low-quality
18-105-final-viral-combined_k141_2043||full Not-determined
18-105-final-viral-combined_k141_152755||full Not-determined
18-105-final-viral-combined_k141_152757||full Low-quality
18-105-final-viral-combined_k141_2063||full Low-quality
18-105-final-viral-combined_k141_197324||full Not-determined
18-105-final-viral-combined_k141_197338||full Low-quality
18-105-final-viral-combined_k141_27948||full Low-quality
18-105-final-viral-combined_k141_72548||full Not-determined
18-105-final-viral-combined_k141_10312||full Not-determined
18-105-final-viral-combined_k141_144044||full Not-determined
18-105-final-viral-combined_k141_170814||full Not-determined
18-105-final-viral-combined_k141_81473||full Low-quality
18-105-final-viral-combined_k141_170816||full Not-determined
18-105-final-viral-combined_k141_2088||full Not-determined
18-105-final-viral-combined_k141_197352||full Not-determined
18-105-final-viral-combined_k141_170829||full Low-quality
18-105-final-viral-combined_k141_188342||full Low-quality
18-105-final-viral-combined_k141_161534||full Low-quality
18-105-final-viral-combined_k141_18907||full Low-quality
18-105-final-viral-combined_k141_179714||full Not-determined
18-105-final-viral-combined_k141_18910||full Not-determined
18-105-final-viral-combined_k141_170848||full Not-determined
18-105-final-viral-combined_k141_27988||full Low-quality
18-105-final-viral-combined_k141_54863||full Not-determined
18-105-final-viral-combined_k141_18931||full Not-determined
18-105-final-viral-combined_k141_161547||full Low-quality
18-105-final-viral-combined_k141_54884||full Low-quality
18-105-final-viral-combined_k141_170885||full Low-quality
18-105-final-viral-combined_k141_125978||full Not-determined
18-105-final-viral-combined_k141_179761||full Not-determined
18-105-final-viral-combined_k141_170906||full Low-quality
18-105-final-viral-combined_k141_90603||full Not-determined
18-105-final-viral-combined_k141_90605||full Not-determined
18-105-final-viral-combined_k141_125999||full Not-determined
18-105-final-viral-combined_k141_81548||full Low-quality
18-105-final-viral-combined_k141_108573||full Not-determined
18-105-final-viral-combined_k141_36894||full Low-quality
18-105-final-viral-combined_k141_126037||full Not-determined
18-105-final-viral-combined_k141_179816||full Low-quality
18-105-final-viral-combined_k141_179830||full Not-determined
18-105-final-viral-combined_k141_63592||full Not-determined
18-105-final-viral-combined_k141_179838||full Low-quality
18-105-final-viral-combined_k141_90668||full Not-determined
18-105-final-viral-combined_k141_179846||full Not-determined
18-105-final-viral-combined_k141_117249||full Not-determined
18-105-final-viral-combined_k141_90681||full Low-quality
18-105-final-viral-combined_k141_2158||full Low-quality
18-105-final-viral-combined_k141_28044||full Low-quality
18-105-final-viral-combined_k141_81600||full Not-determined
18-105-final-viral-combined_k141_19016||full Low-quality
18-105-final-viral-combined_k141_134947||full Not-determined
18-105-final-viral-combined_k141_81615||full Low-quality
18-105-final-viral-combined_k141_134950||full Low-quality
18-105-final-viral-combined_k141_188375||full Not-determined
18-105-final-viral-combined_k141_45993||full Not-determined
18-105-final-viral-combined_k141_54964||full Low-quality
18-105-final-viral-combined_k141_188391||full Low-quality
18-105-final-viral-combined_k141_28074||full Not-determined
18-105-final-viral-combined_k141_72699||full Not-determined
18-105-final-viral-combined_k141_28081||full Not-determined
18-105-final-viral-combined_k141_19070||full Low-quality
18-105-final-viral-combined_k141_197371||full Low-quality
18-105-final-viral-combined_k141_134969||full Not-determined
18-105-final-viral-combined_k141_170959||full Low-quality
18-105-final-viral-combined_k141_10407||full Low-quality
18-105-final-viral-combined_k141_81637||full Not-determined
18-105-final-viral-combined_k141_144227||full Low-quality
18-105-final-viral-combined_k141_10426||full Low-quality
18-105-final-viral-combined_k141_28135||full Low-quality
18-105-final-viral-combined_k141_144235||full Not-determined
18-105-final-viral-combined_k141_10431||full Low-quality
18-105-final-viral-combined_k141_19109||full Low-quality
18-105-final-viral-combined_k141_179852||full Low-quality
18-105-final-viral-combined_k141_81666||full Not-determined
18-105-final-viral-combined_k141_134974||full Not-determined
18-105-final-viral-combined_k141_179862||full Not-determined
18-105-final-viral-combined_k141_152902||full Not-determined
18-105-final-viral-combined_k141_188428||full Low-quality
18-105-final-viral-combined_k141_55010||full Low-quality
18-105-final-viral-combined_k141_55016||full Low-quality
18-105-final-viral-combined_k141_144267||full Low-quality
18-105-final-viral-combined_k141_90785||full Low-quality
18-105-final-viral-combined_k141_171016||full Low-quality
18-105-final-viral-combined_k141_144286||full Low-quality
18-105-final-viral-combined_k141_135015||full Low-quality
18-105-final-viral-combined_k141_179912||full Not-determined
18-105-final-viral-combined_k141_135028||full Low-quality
18-105-final-viral-combined_k141_55054||full Low-quality
18-105-final-viral-combined_k141_55055||full Not-determined
18-105-final-viral-combined_k141_188496||full Low-quality
18-105-final-viral-combined_k141_188500||full Low-quality
18-105-final-viral-combined_k141_126097||full Not-determined
18-105-final-viral-combined_k141_197471||full Not-determined
18-105-final-viral-combined_k141_188503||full Not-determined
18-105-final-viral-combined_k141_197477||full Not-determined
18-105-final-viral-combined_k141_171085||full Not-determined
18-105-final-viral-combined_k141_81734||full Low-quality
18-105-final-viral-combined_k141_171092||full Low-quality
18-105-final-viral-combined_k141_2254||full Not-determined
18-105-final-viral-combined_k141_28141||full Low-quality
18-105-final-viral-combined_k141_161642||full Low-quality
18-105-final-viral-combined_k141_28142||full Low-quality
18-105-final-viral-combined_k141_171127||full Low-quality
18-105-final-viral-combined_k141_2276||full Low-quality
18-105-final-viral-combined_k141_171134||full Not-determined
18-105-final-viral-combined_k141_161660||full Not-determined
18-105-final-viral-combined_k141_197499||full Low-quality
18-105-final-viral-combined_k141_2297||full Low-quality
18-105-final-viral-combined_k141_55097||full Low-quality
18-105-final-viral-combined_k141_37035||full Low-quality
18-105-final-viral-combined_k141_99295||full Low-quality
18-105-final-viral-combined_k141_206150||full Low-quality
18-105-final-viral-combined_k141_171176||full Low-quality
18-105-final-viral-combined_k141_206165||full Low-quality
18-105-final-viral-combined_k141_90830||full Not-determined
18-105-final-viral-combined_k141_126184||full Not-determined
18-105-final-viral-combined_k141_108659||full Not-determined
18-105-final-viral-combined_k141_10597||full Not-determined
18-105-final-viral-combined_k141_135066||full Not-determined
18-105-final-viral-combined_k141_197577||full Low-quality
18-105-final-viral-combined_k141_188585||full Not-determined
18-105-final-viral-combined_k141_37055||full Not-determined
18-105-final-viral-combined_k141_126218||full Not-determined
18-105-final-viral-combined_k141_206204||full Not-determined
18-105-final-viral-combined_k141_2351||full Low-quality
18-101-final-viral-combined_k141_322931||full Medium-quality
18-105-final-viral-combined_k141_63625||full Low-quality
18-105-final-viral-combined_k141_153040||full Not-determined
18-105-final-viral-combined_k141_37093||full Not-determined
18-105-final-viral-combined_k141_188623||full Low-quality
18-105-final-viral-combined_k141_180009||full Not-determined
18-105-final-viral-combined_k141_37094||full Not-determined
18-105-final-viral-combined_k141_72878||full Low-quality
18-105-final-viral-combined_k141_180018||full Not-determined
18-105-final-viral-combined_k141_72884||full Not-determined
18-105-final-viral-combined_k141_46069||full Low-quality
18-105-final-viral-combined_k141_19173||full Not-determined
18-105-final-viral-combined_k141_46071||full Low-quality
18-105-final-viral-combined_k141_19175||full Not-determined
18-105-final-viral-combined_k141_37104||full Low-quality
18-105-final-viral-combined_k141_46073||full Low-quality
18-105-final-viral-combined_k141_126253||full Low-quality
18-105-final-viral-combined_k141_161727||full Not-determined
18-105-final-viral-combined_k141_99403||full Not-determined
18-105-final-viral-combined_k141_37106||full Low-quality
18-105-final-viral-combined_k141_180025||full Not-determined
18-105-final-viral-combined_k141_72904||full Low-quality
18-105-final-viral-combined_k141_81820||full Not-determined
18-105-final-viral-combined_k141_37141||full Not-determined
18-105-final-viral-combined_k141_161736||full Not-determined
18-105-final-viral-combined_k141_63651||full Not-determined
18-105-final-viral-combined_k141_144416||full Not-determined
18-105-final-viral-combined_k141_161744||full Low-quality
18-105-final-viral-combined_k141_81838||full Low-quality
18-105-final-viral-combined_k141_46157||full Not-determined
18-105-final-viral-combined_k141_90916||full Low-quality
18-105-final-viral-combined_k141_72958||full Low-quality
18-105-final-viral-combined_k141_63657||full Low-quality
18-105-final-viral-combined_k141_19208||full Not-determined
18-105-final-viral-combined_k141_197591||full Low-quality
18-105-final-viral-combined_k141_161769||full Not-determined
18-105-final-viral-combined_k141_188639||full Not-determined
18-105-final-viral-combined_k141_63678||full Not-determined
18-105-final-viral-combined_k141_161777||full Not-determined
18-105-final-viral-combined_k141_37177||full Not-determined
18-105-final-viral-combined_k141_90935||full Not-determined
18-105-final-viral-combined_k141_161787||full Low-quality
18-105-final-viral-combined_k141_28166||full Low-quality
18-105-final-viral-combined_k141_171291||full Low-quality
18-105-final-viral-combined_k141_73010||full Low-quality
18-105-final-viral-combined_k141_180070||full Not-determined
18-105-final-viral-combined_k141_188661||full Not-determined
18-105-final-viral-combined_k141_171306||full Not-determined
18-105-final-viral-combined_k141_171313||full Not-determined
18-105-final-viral-combined_k141_73026||full Not-determined
18-105-final-viral-combined_k141_10672||full Low-quality
18-105-final-viral-combined_k141_171330||full Not-determined
18-105-final-viral-combined_k141_90960||full Not-determined
18-105-final-viral-combined_k141_73046||full Low-quality
18-105-final-viral-combined_k141_161844||full Low-quality
18-105-final-viral-combined_k141_46240||full Low-quality
18-105-final-viral-combined_k141_206267||full Low-quality
18-105-final-viral-combined_k141_188693||full Low-quality
18-105-final-viral-combined_k141_188701||full Low-quality
18-105-final-viral-combined_k141_63708||full Not-determined
18-105-final-viral-combined_k141_144463||full Not-determined
18-105-final-viral-combined_k141_99561||full Low-quality
18-105-final-viral-combined_k141_46275||full Low-quality
18-105-final-viral-combined_k141_180190||full Not-determined
18-105-final-viral-combined_k141_117352||full Not-determined
18-105-final-viral-combined_k141_180195||full Low-quality
18-105-final-viral-combined_k141_10686||full Low-quality
18-105-final-viral-combined_k141_206321||full Not-determined
18-105-final-viral-combined_k141_63723||full Low-quality
18-105-final-viral-combined_k141_73102||full Not-determined
18-105-final-viral-combined_k141_73106||full Not-determined
18-105-final-viral-combined_k141_144516||full Not-determined
18-105-final-viral-combined_k141_171418||full Low-quality
18-105-final-viral-combined_k141_180209||full Not-determined
18-105-final-viral-combined_k141_117393||full Low-quality
18-105-final-viral-combined_k141_28208||full Not-determined
18-105-final-viral-combined_k141_206341||full Not-determined
18-105-final-viral-combined_k141_117415||full Low-quality
18-105-final-viral-combined_k141_206344||full Not-determined
18-105-final-viral-combined_k141_37281||full Low-quality
18-105-final-viral-combined_k141_99570||full Not-determined
18-105-final-viral-combined_k141_90994||full Low-quality
18-105-final-viral-combined_k141_73158||full Not-determined
18-105-final-viral-combined_k141_10718||full Low-quality
18-105-final-viral-combined_k141_91008||full Low-quality
18-105-final-viral-combined_k141_10730||full Low-quality
18-105-final-viral-combined_k141_135208||full Low-quality
18-105-final-viral-combined_k141_99589||full Low-quality
18-105-final-viral-combined_k141_144572||full Not-determined
18-105-final-viral-combined_k141_81920||full Low-quality
18-105-final-viral-combined_k141_197671||full Low-quality
18-105-final-viral-combined_k141_144576||full Low-quality
18-105-final-viral-combined_k141_37315||full Not-determined
18-105-final-viral-combined_k141_117428||full Low-quality
18-105-final-viral-combined_k141_99607||full Low-quality
18-105-final-viral-combined_k141_206361||full Low-quality
18-105-final-viral-combined_k141_99623||full Not-determined
18-105-final-viral-combined_k141_180244||full Low-quality
18-105-final-viral-combined_k141_161907||full Low-quality
18-105-final-viral-combined_k141_99641||full Low-quality
18-105-final-viral-combined_k141_19295||full Not-determined
18-105-final-viral-combined_k141_73212||full Not-determined
18-105-final-viral-combined_k141_180255||full Not-determined
18-105-final-viral-combined_k141_55248||full Low-quality
18-105-final-viral-combined_k141_19304||full Low-quality
18-105-final-viral-combined_k141_73229||full Not-determined
18-105-final-viral-combined_k141_117469||full Low-quality
18-105-final-viral-combined_k141_197729||full Not-determined
18-105-final-viral-combined_k141_55274||full Not-determined
18-105-final-viral-combined_k141_171536||full Not-determined
18-105-final-viral-combined_k141_91051||full Not-determined
18-105-final-viral-combined_k141_135257||full Not-determined
18-105-final-viral-combined_k141_126511||full Low-quality
18-105-final-viral-combined_k141_10769||full Not-determined
18-105-final-viral-combined_k141_82000||full Not-determined
18-105-final-viral-combined_k141_82003||full Not-determined
18-105-final-viral-combined_k141_144608||full Low-quality
18-105-final-viral-combined_k141_153236||full Not-determined
18-109-final-viral-combined_k141_21693||full High-quality
18-105-final-viral-combined_k141_82013||full Not-determined
18-105-final-viral-combined_k141_206388||full Low-quality
18-105-final-viral-combined_k141_144628||full Not-determined
18-105-final-viral-combined_k141_19352||full Low-quality
18-105-final-viral-combined_k141_188875||full Not-determined
18-105-final-viral-combined_k141_82022||full Low-quality
18-105-final-viral-combined_k141_206438||full Not-determined
18-105-final-viral-combined_k141_135319||full Low-quality
18-105-final-viral-combined_k141_206459||full Low-quality
18-105-final-viral-combined_k141_19366||full Low-quality
18-105-final-viral-combined_k141_171584||full Low-quality
18-105-final-viral-combined_k141_82052||full Not-determined
18-105-final-viral-combined_k141_144693||full Low-quality
18-105-final-viral-combined_k141_188884||full Not-determined
18-105-final-viral-combined_k141_82056||full Low-quality
18-105-final-viral-combined_k141_188898||full Not-determined
18-105-final-viral-combined_k141_117510||full Not-determined
18-105-final-viral-combined_k141_63859||full Low-quality
18-105-final-viral-combined_k141_188919||full Low-quality
18-105-final-viral-combined_k141_180304||full Low-quality
18-105-final-viral-combined_k141_153266||full Low-quality
18-105-final-viral-combined_k141_2630||full Not-determined
18-101-final-viral-combined_k141_52708||full High-quality
18-105-final-viral-combined_k141_171637||full Not-determined
18-105-final-viral-combined_k141_197834||full Not-determined
18-105-final-viral-combined_k141_171662||full Not-determined
18-105-final-viral-combined_k141_28323||full Not-determined
18-105-final-viral-combined_k141_28331||full Not-determined
18-105-final-viral-combined_k141_63921||full Not-determined
18-105-final-viral-combined_k141_188992||full Low-quality
18-105-final-viral-combined_k141_2661||full Not-determined
18-105-final-viral-combined_k141_28335||full Low-quality
18-105-final-viral-combined_k141_2663||full Not-determined
18-105-final-viral-combined_k141_135348||full Low-quality
18-105-final-viral-combined_k141_189008||full Low-quality
18-105-final-viral-combined_k141_63937||full Low-quality
18-105-final-viral-combined_k141_28347||full Low-quality
18-105-final-viral-combined_k141_197865||full Low-quality
18-105-final-viral-combined_k141_171725||full Low-quality
18-105-final-viral-combined_k141_46462||full Low-quality
18-105-final-viral-combined_k141_135373||full Not-determined
18-105-final-viral-combined_k141_73334||full Low-quality
18-105-final-viral-combined_k141_197876||full Not-determined
18-105-final-viral-combined_k141_46470||full Not-determined
18-105-final-viral-combined_k141_189045||full Low-quality
18-105-final-viral-combined_k141_91194||full Not-determined
18-105-final-viral-combined_k141_135395||full Low-quality
18-105-final-viral-combined_k141_135398||full Not-determined
18-105-final-viral-combined_k141_91200||full Low-quality
18-105-final-viral-combined_k141_153361||full Low-quality
18-105-final-viral-combined_k141_91209||full Low-quality
18-105-final-viral-combined_k141_135406||full Low-quality
18-105-final-viral-combined_k141_91210||full Low-quality
18-105-final-viral-combined_k141_91212||full Low-quality
18-105-final-viral-combined_k141_73368||full Low-quality
18-105-final-viral-combined_k141_2706||full Low-quality
18-105-final-viral-combined_k141_180379||full Low-quality
18-105-final-viral-combined_k141_117598||full Low-quality
18-105-final-viral-combined_k141_135439||full Not-determined
18-105-final-viral-combined_k141_28378||full Not-determined
18-105-final-viral-combined_k141_55357||full Not-determined
18-105-final-viral-combined_k141_99803||full Not-determined
18-105-final-viral-combined_k141_64016||full Low-quality
18-105-final-viral-combined_k141_46534||full Not-determined
18-105-final-viral-combined_k141_99804||full Not-determined
18-105-final-viral-combined_k141_206518||full Not-determined
18-105-final-viral-combined_k141_189071||full Low-quality
18-105-final-viral-combined_k141_153406||full Not-determined
18-105-final-viral-combined_k141_144718||full Not-determined
18-105-final-viral-combined_k141_135448||full Low-quality
18-105-final-viral-combined_k141_108749||full Low-quality
18-105-final-viral-combined_k141_162026||full Low-quality
18-105-final-viral-combined_k141_144735||full Not-determined
18-105-final-viral-combined_k141_135457||full Low-quality

| | |
|---|---|
| 18-105-final-viral-combined_k141_19526\|\|full | Low-quality |
| 18-105-final-viral-combined_k141_2761\|\|full | Not-determined |
| 18-105-final-viral-combined_k141_19537\|\|full | Not-determined |
| 18-105-final-viral-combined_k141_144773\|\|full | Not-determined |
| 18-105-final-viral-combined_k141_64049\|\|full | Not-determined |
| 18-105-final-viral-combined_k141_135479\|\|full | Not-determined |
| 18-105-final-viral-combined_k141_91272\|\|full | Low-quality |
| 18-105-final-viral-combined_k141_37482\|\|full | Low-quality |
| 18-105-final-viral-combined_k141_171780\|\|full | Not-determined |
| 18-105-final-viral-combined_k141_64075\|\|full | Low-quality |
| 18-105-final-viral-combined_k141_28509\|\|full | Low-quality |
| 18-105-final-viral-combined_k141_135537\|\|full | Not-determined |
| 18-105-final-viral-combined_k141_162080\|\|full | Low-quality |
| 18-105-final-viral-combined_k141_153436\|\|full | Low-quality |
| 18-105-final-viral-combined_k141_37516\|\|full | Not-determined |
| 18-105-final-viral-combined_k141_126710\|\|full | Not-determined |
| 18-105-final-viral-combined_k141_171816\|\|full | Low-quality |
| 18-105-final-viral-combined_k141_153442\|\|full | Low-quality |
| 18-105-final-viral-combined_k141_99868\|\|full | Low-quality |
| 18-105-final-viral-combined_k141_126721\|\|full | Not-determined |
| 18-105-final-viral-combined_k141_28559\|\|full | Low-quality |
| 18-105-final-viral-combined_k141_46648\|\|full | Low-quality |
| 18-105-final-viral-combined_k141_73447\|\|full | Low-quality |
| 18-105-final-viral-combined_k141_2856\|\|full | Low-quality |
| 18-105-final-viral-combined_k141_189106\|\|full | Not-determined |
| 18-105-final-viral-combined_k141_206546\|\|full | Not-determined |
| 18-105-final-viral-combined_k141_46663\|\|full | Low-quality |
| 18-105-final-viral-combined_k141_126730\|\|full | Low-quality |
| 18-105-final-viral-combined_k141_153474\|\|full | Low-quality |
| 18-105-final-viral-combined_k141_46687\|\|full | Not-determined |
| 18-105-final-viral-combined_k141_153477\|\|full | Low-quality |
| 18-105-final-viral-combined_k141_162090\|\|full | Low-quality |
| 18-105-final-viral-combined_k141_206564\|\|full | Not-determined |
| 18-105-final-viral-combined_k141_126745\|\|full | Low-quality |
| 18-105-final-viral-combined_k141_206570\|\|full | Low-quality |
| 18-105-final-viral-combined_k141_82132\|\|full | Low-quality |
| 18-105-final-viral-combined_k141_2872\|\|full | Not-determined |
| 18-105-final-viral-combined_k141_37553\|\|full | Not-determined |
| 18-105-final-viral-combined_k141_189164\|\|full | Not-determined |
| 18-105-final-viral-combined_k141_206592\|\|full | Not-determined |
| 18-105-final-viral-combined_k141_99939\|\|full | Not-determined |
| 18-105-final-viral-combined_k141_126779\|\|full | Not-determined |
| 18-105-final-viral-combined_k141_37557\|\|full | Not-determined |
| 18-105-final-viral-combined_k141_171858\|\|full | Low-quality |
| 18-105-final-viral-combined_k141_82150\|\|full | Low-quality |
| 18-105-final-viral-combined_k141_153520\|\|full | Not-determined |
| 18-105-final-viral-combined_k141_99962\|\|full | Low-quality |
| 18-105-final-viral-combined_k141_171863\|\|full | Not-determined |
| 18-105-final-viral-combined_k141_91300\|\|full | Low-quality |
| 18-105-final-viral-combined_k141_126811\|\|full | Low-quality |
| 18-105-final-viral-combined_k141_189189\|\|full | Not-determined |
| 18-105-final-viral-combined_k141_37582\|\|full | Low-quality |
| 18-105-final-viral-combined_k141_189207\|\|full | Low-quality |
| 18-105-final-viral-combined_k141_37593\|\|full | Low-quality |
| 18-105-final-viral-combined_k141_99998\|\|full | Low-quality |
| 18-105-final-viral-combined_k141_144899\|\|full | Not-determined |
| 18-105-final-viral-combined_k141_28592\|\|full | Not-determined |
| 18-105-final-viral-combined_k141_180427\|\|full | Not-determined |
| 18-105-final-viral-combined_k141_153553\|\|full | Low-quality |
| 18-105-final-viral-combined_k141_197921\|\|full | Not-determined |
| 18-105-final-viral-combined_k141_117630\|\|full | Low-quality |
| 18-105-final-viral-combined_k141_117637\|\|full | Not-determined |
| 18-105-final-viral-combined_k141_37647\|\|full | Not-determined |
| 18-105-final-viral-combined_k141_206636\|\|full | Not-determined |
| 18-105-final-viral-combined_k141_206642\|\|full | Not-determined |
| 17-106-final-viral-combined_k141_5526\|\|0_partial | Medium-quality |
| 18-105-final-viral-combined_k141_10843\|\|full | Low-quality |
| 18-105-final-viral-combined_k141_100031\|\|full | Not-determined |
| 18-105-final-viral-combined_k141_37662\|\|full | Not-determined |
| 18-105-final-viral-combined_k141_91362\|\|full | Low-quality |
| 18-105-final-viral-combined_k141_180493\|\|full | Not-determined |
| 18-105-final-viral-combined_k141_28626\|\|full | Not-determined |
| 18-105-final-viral-combined_k141_91366\|\|full | Low-quality |
| 18-105-final-viral-combined_k141_162145\|\|full | Not-determined |
| 18-105-final-viral-combined_k141_171891\|\|full | Not-determined |
| 18-105-final-viral-combined_k141_100048\|\|full | Low-quality |
| 18-105-final-viral-combined_k141_126862\|\|full | Low-quality |
| 18-105-final-viral-combined_k141_180510\|\|full | Low-quality |
| 18-105-final-viral-combined_k141_100062\|\|full | Not-determined |
| 18-105-final-viral-combined_k141_73504\|\|full | Not-determined |
| 18-105-final-viral-combined_k141_73511\|\|full | Not-determined |
| 18-105-final-viral-combined_k141_46798\|\|full | Low-quality |
| 18-105-final-viral-combined_k141_73513\|\|full | Not-determined |
| 18-105-final-viral-combined_k141_198033\|\|full | Low-quality |
| 18-105-final-viral-combined_k141_117707\|\|full | Not-determined |
| 18-105-final-viral-combined_k141_100091\|\|full | Not-determined |
| 18-105-final-viral-combined_k141_46822\|\|full | Not-determined |
| 18-105-final-viral-combined_k141_135569\|\|full | Not-determined |
| 18-105-final-viral-combined_k141_91381\|\|full | Not-determined |
| 18-105-final-viral-combined_k141_198064\|\|full | Not-determined |
| 18-105-final-viral-combined_k141_46826\|\|full | Not-determined |
| 18-105-final-viral-combined_k141_64147\|\|full | Not-determined |
| 18-105-final-viral-combined_k141_144929\|\|full | Low-quality |
| 18-105-final-viral-combined_k141_100111\|\|full | Low-quality |
| 18-105-final-viral-combined_k141_206740\|\|full | Not-determined |
| 18-105-final-viral-combined_k141_46832\|\|full | Not-determined |
| 18-105-final-viral-combined_k141_117729\|\|full | Low-quality |
| 18-105-final-viral-combined_k141_144947\|\|full | Low-quality |
| 18-105-final-viral-combined_k141_162193\|\|full | Low-quality |
| 18-105-final-viral-combined_k141_171975\|\|full | Low-quality |
| 18-105-final-viral-combined_k141_198100\|\|full | Not-determined |
| 18-105-final-viral-combined_k141_64169\|\|full | Low-quality |
| 18-105-final-viral-combined_k141_171987\|\|full | Not-determined |
| 18-105-final-viral-combined_k141_180533\|\|full | Low-quality |
| 18-105-final-viral-combined_k141_55576\|\|full | Not-determined |
| 18-105-final-viral-combined_k141_82230\|\|full | Not-determined |
| 18-105-final-viral-combined_k141_2952\|\|full | Low-quality |
| 18-105-final-viral-combined_k141_37697\|\|full | Not-determined |
| 18-105-final-viral-combined_k141_64198\|\|full | Low-quality |
| 18-105-final-viral-combined_k141_28684\|\|full | Low-quality |
| 18-105-final-viral-combined_k141_162286\|\|full | Low-quality |
| 18-105-final-viral-combined_k141_180583\|\|full | Low-quality |
| 18-105-final-viral-combined_k141_73572\|\|full | Not-determined |
| 18-105-final-viral-combined_k141_10916\|\|full | Low-quality |
| 18-105-final-viral-combined_k141_37717\|\|full | Low-quality |
| 18-105-final-viral-combined_k141_55606\|\|full | Low-quality |
| 18-105-final-viral-combined_k141_64252\|\|full | Low-quality |
| 18-105-final-viral-combined_k141_153728\|\|full | Low-quality |
| 18-105-final-viral-combined_k141_180589\|\|full | Low-quality |
| 18-105-final-viral-combined_k141_19598\|\|full | Not-determined |
| 17-108-final-viral-combined_k141_329493\|\|full | High-quality |
| [illegible] | [illegible] |
| 18-105-final-viral-combined_k141_100184\|\|full | Not-determined |
| 18-105-final-viral-combined_k141_100187\|\|full | Low-quality |
| 18-105-final-viral-combined_k141_100193\|\|full | Low-quality |
| 18-105-final-viral-combined_k141_28721\|\|full | Not-determined |
| 18-105-final-viral-combined_k141_135627\|\|full | Not-determined |
| 18-105-final-viral-combined_k141_189348\|\|full | Not-determined |
| 18-105-final-viral-combined_k141_117838\|\|full | Low-quality |
| 18-105-final-viral-combined_k141_64318\|\|full | Low-quality |
| 18-105-final-viral-combined_k141_189375\|\|full | Not-determined |
| 18-105-final-viral-combined_k141_127055\|\|full | Not-determined |
| 18-105-final-viral-combined_k141_46896\|\|full | Low-quality |
| 18-105-final-viral-combined_k141_91518\|\|full | Low-quality |
| 18-105-final-viral-combined_k141_100267\|\|full | Not-determined |
| 18-105-final-viral-combined_k141_2991\|\|full | Low-quality |
| 18-105-final-viral-combined_k141_91527\|\|full | Not-determined |
| 18-105-final-viral-combined_k141_3003\|\|full | Not-determined |
| 18-105-final-viral-combined_k141_162361\|\|full | Low-quality |
| 18-105-final-viral-combined_k141_28743\|\|full | Low-quality |
| 18-105-final-viral-combined_k141_100312\|\|full | Low-quality |
| 18-105-final-viral-combined_k141_189456\|\|full | Not-determined |
| 18-105-final-viral-combined_k141_135667\|\|full | Low-quality |
| 18-105-final-viral-combined_k141_162395\|\|full | Low-quality |
| 18-105-final-viral-combined_k141_73603\|\|full | Not-determined |
| 18-105-final-viral-combined_k141_28766\|\|full | Low-quality |
| 18-105-final-viral-combined_k141_64358\|\|full | Low-quality |
| 18-105-final-viral-combined_k141_189485\|\|full | Not-determined |
| 18-105-final-viral-combined_k141_100371\|\|full | Low-quality |
| 18-105-final-viral-combined_k141_189498\|\|full | Low-quality |
| 18-105-final-viral-combined_k141_11066\|\|full | Low-quality |
| 18-105-final-viral-combined_k141_3074\|\|full | Not-determined |
| 18-105-final-viral-combined_k141_135705\|\|full | Not-determined |
| 18-105-final-viral-combined_k141_172035\|\|full | Not-determined |
| 18-105-final-viral-combined_k141_135712\|\|full | Low-quality |
| 18-105-final-viral-combined_k141_145048\|\|full | Low-quality |
| 18-105-final-viral-combined_k141_46964\|\|full | Low-quality |
| 18-105-final-viral-combined_k141_73626\|\|full | Low-quality |
| 18-105-final-viral-combined_k141_73627\|\|full | Low-quality |
| 18-105-final-viral-combined_k141_28795\|\|full | Not-determined |
| 18-105-final-viral-combined_k141_64376\|\|full | Low-quality |
| 18-105-final-viral-combined_k141_28796\|\|full | Low-quality |
| 18-105-final-viral-combined_k141_64381\|\|full | Low-quality |
| 18-105-final-viral-combined_k141_100415\|\|full | Not-determined |
| 18-105-final-viral-combined_k141_189527\|\|full | Not-determined |
| 18-105-final-viral-combined_k141_206873\|\|full | Low-quality |
| 18-105-final-viral-combined_k141_206875\|\|full | Low-quality |
| 18-105-final-viral-combined_k141_135715\|\|full | Low-quality |
| 18-105-final-viral-combined_k141_100436\|\|full | Not-determined |
| 18-105-final-viral-combined_k141_3108\|\|full | Low-quality |
| 18-105-final-viral-combined_k141_55608\|\|full | Low-quality |
| 18-110-final-viral-combined_k141_225114\|\|full | High-quality |
| 18-105-final-viral-combined_k141_73671\|\|full | Low-quality |
| 18-105-final-viral-combined_k141_198248\|\|full | Not-determined |
| 18-105-final-viral-combined_k141_127132\|\|full | Low-quality |
| 18-105-final-viral-combined_k141_11120\|\|full | Low-quality |
| 18-105-final-viral-combined_k141_108907\|\|full | Low-quality |
| 18-105-final-viral-combined_k141_73701\|\|full | Not-determined |
| 18-105-final-viral-combined_k141_73706\|\|full | Not-determined |
| 18-105-final-viral-combined_k141_180786\|\|full | Not-determined |
| 18-105-final-viral-combined_k141_198278\|\|full | Not-determined |
| 18-105-final-viral-combined_k141_127174\|\|full | Not-determined |
| 18-105-final-viral-combined_k141_91581\|\|full | Not-determined |
| 18-105-final-viral-combined_k141_64496\|\|full | Not-determined |
| 18-105-final-viral-combined_k141_135776\|\|full | Not-determined |
| 18-105-final-viral-combined_k141_189590\|\|full | Low-quality |
| 18-105-final-viral-combined_k141_189596\|\|full | Low-quality |
| 18-105-final-viral-combined_k141_64525\|\|full | Low-quality |
| 18-105-final-viral-combined_k141_189616\|\|full | Low-quality |
| 18-105-final-viral-combined_k141_180817\|\|full | Not-determined |
| 18-105-final-viral-combined_k141_198339\|\|full | Not-determined |
| 18-105-final-viral-combined_k141_189638\|\|full | Low-quality |
| 18-105-final-viral-combined_k141_91609\|\|full | Not-determined |
| 18-105-final-viral-combined_k141_28890\|\|full | Low-quality |
| 18-105-final-viral-combined_k141_73740\|\|full | Low-quality |
| 18-105-final-viral-combined_k141_91620\|\|full | Low-quality |
| 18-105-final-viral-combined_k141_73744\|\|full | Low-quality |
| 18-105-final-viral-combined_k141_117921\|\|full | Not-determined |
| 18-105-final-viral-combined_k141_73746\|\|full | Not-determined |
| 18-105-final-viral-combined_k141_108936\|\|full | Low-quality |
| 18-105-final-viral-combined_k141_117945\|\|full | Not-determined |
| 18-105-final-viral-combined_k141_189674\|\|full | Not-determined |
| 18-105-final-viral-combined_k141_108949\|\|full | Not-determined |
| 18-105-final-viral-combined_k141_145114\|\|full | Not-determined |
| 18-105-final-viral-combined_k141_180872\|\|full | Low-quality |
| 18-105-final-viral-combined_k141_180881\|\|full | Low-quality |
| 18-105-final-viral-combined_k141_37834\|\|full | Low-quality |
| 18-105-final-viral-combined_k141_100607\|\|full | Low-quality |
| 18-106-final-viral-combined_k141_117756\|\|full | High-quality |
| 18-105-final-viral-combined_k141_172177\|\|full | Not-determined |
| 18-105-final-viral-combined_k141_207005\|\|full | Low-quality |
| 18-105-final-viral-combined_k141_207015\|\|full | Low-quality |
| 18-105-final-viral-combined_k141_64604\|\|full | Not-determined |
| 18-105-final-viral-combined_k141_162499\|\|full | Not-determined |
| 18-105-final-viral-combined_k141_100625\|\|full | Low-quality |
| 18-105-final-viral-combined_k141_207036\|\|full | Not-determined |
| 18-105-final-viral-combined_k141_117991\|\|full | Not-determined |
| 18-105-final-viral-combined_k141_162525\|\|full | Not-determined |
| 18-105-final-viral-combined_k141_100642\|\|full | Not-determined |
| 18-105-final-viral-combined_k141_207058\|\|full | Not-determined |
| 18-105-final-viral-combined_k141_180920\|\|full | Low-quality |
| 18-105-final-viral-combined_k141_55654\|\|full | Low-quality |
| 18-105-final-viral-combined_k141_28927\|\|full | Low-quality |
| 18-105-final-viral-combined_k141_198365\|\|full | Low-quality |
| 18-105-final-viral-combined_k141_118036\|\|full | Low-quality |
| 18-105-final-viral-combined_k141_3165\|\|full | Low-quality |
| 18-105-final-viral-combined_k141_118057\|\|full | Not-determined |
| 18-105-final-viral-combined_k141_198368\|\|full | Low-quality |
| 18-105-final-viral-combined_k141_64623\|\|full | Low-quality |
| 18-105-final-viral-combined_k141_162580\|\|full | Not-determined |
| 18-105-final-viral-combined_k141_19708\|\|full | Not-determined |
| 18-105-final-viral-combined_k141_82291\|\|full | Low-quality |
| 18-105-final-viral-combined_k141_64625\|\|full | Not-determined |
| 18-105-final-viral-combined_k141_145151\|\|full | Low-quality |
| 18-105-final-viral-combined_k141_153749\|\|full | Low-quality |
| 18-105-final-viral-combined_k141_198391\|\|full | Low-quality |
| 18-105-final-viral-combined_k141_28977\|\|full | Not-determined |
| 18-105-final-viral-combined_k141_189735\|\|full | Low-quality |
| 18-105-final-viral-combined_k141_47147\|\|full | Not-determined |
| 18-105-final-viral-combined_k141_145160\|\|full | Not-determined |
| 18-105-final-viral-combined_k141_172276\|\|full | Low-quality |
| 18-105-final-viral-combined_k141_189742\|\|full | Not-determined |
| 18-105-final-viral-combined_k141_162620\|\|full | Not-determined |
| 18-105-final-viral-combined_k141_135895\|\|full | Not-determined |
| 18-105-final-viral-combined_k141_135897\|\|full | Not-determined |
| 18-105-final-viral-combined_k141_3188\|\|full | Low-quality |
| 18-105-final-viral-combined_k141_19732\|\|full | Not-determined |
| 18-105-final-viral-combined_k141_100716\|\|full | Low-quality |
| 18-105-final-viral-combined_k141_180976\|\|full | Not-determined |
| 18-105-final-viral-combined_k141_19743\|\|full | Not-determined |
| 18-105-final-viral-combined_k141_189775\|\|full | Not-determined |
| 18-105-final-viral-combined_k141_180981\|\|full | Not-determined |
| 18-105-final-viral-combined_k141_29045\|\|full | Low-quality |
| 18-105-final-viral-combined_k141_108991\|\|full | Low-quality |
| 18-105-final-viral-combined_k141_153784\|\|full | Low-quality |
| 18-105-final-viral-combined_k141_100722\|\|full | Not-determined |
| 18-105-final-viral-combined_k141_91701\|\|full | Not-determined |
| 18-105-final-viral-combined_k141_100734\|\|full | Low-quality |
| 18-105-final-viral-combined_k141_91703\|\|full | Low-quality |
| 18-105-final-viral-combined_k141_145209\|\|full | Low-quality |
| 18-105-final-viral-combined_k141_47178\|\|full | Low-quality |
| 18-105-final-viral-combined_k141_91715\|\|full | Not-determined |
| 18-101-final-viral-combined_k141_132934\|\|full | Complete |
| 18-105-final-viral-combined_k141_198400\|\|full | Not-determined |
| 18-110-final-viral-combined_k141_161181\|\|full | Medium-quality |
| 18-105-final-viral-combined_k141_172356\|\|full | Low-quality |
| 18-105-final-viral-combined_k141_172358\|\|full | Low-quality |
| 18-105-final-viral-combined_k141_181009\|\|full | Not-determined |
| 18-105-final-viral-combined_k141_91739\|\|full | Not-determined |
| 18-105-final-viral-combined_k141_29060\|\|full | Low-quality |
| 18-105-final-viral-combined_k141_91742\|\|full | Not-determined |
| 18-105-final-viral-combined_k141_91744\|\|full | Low-quality |
| 18-105-final-viral-combined_k141_11275\|\|full | Not-determined |
| 18-105-final-viral-combined_k141_135931\|\|full | Low-quality |
| 18-105-final-viral-combined_k141_11279\|\|full | Low-quality |
| 18-105-final-viral-combined_k141_181026\|\|full | Not-determined |
| 18-105-final-viral-combined_k141_91763\|\|full | Not-determined |
| 18-105-final-viral-combined_k141_135942\|\|full | Low-quality |
| 18-105-final-viral-combined_k141_55675\|\|full | Not-determined |
| 18-105-final-viral-combined_k141_73800\|\|full | Low-quality |
| 18-105-final-viral-combined_k141_198438\|\|full | Low-quality |
| 18-105-final-viral-combined_k141_55699\|\|full | Not-determined |
| 18-105-final-viral-combined_k141_91807\|\|full | Not-determined |
| 18-105-final-viral-combined_k141_135993\|\|full | Not-determined |
| 18-105-final-viral-combined_k141_47296\|\|full | Not-determined |
| 18-105-final-viral-combined_k141_153827\|\|full | Not-determined |
| 18-105-final-viral-combined_k141_37952\|\|full | Not-determined |
| 18-105-final-viral-combined_k141_127404\|\|full | Not-determined |
| 18-105-final-viral-combined_k141_55735\|\|full | Low-quality |
| 18-105-final-viral-combined_k141_136045\|\|full | Low-quality |
| 18-105-final-viral-combined_k141_189879\|\|full | Low-quality |
| 18-105-final-viral-combined_k141_37972\|\|full | Low-quality |
| 18-105-final-viral-combined_k141_127423\|\|full | Not-determined |
| 18-105-final-viral-combined_k141_136062\|\|full | Not-determined |
| 18-105-final-viral-combined_k141_127424\|\|full | Low-quality |
| 18-105-final-viral-combined_k141_47368\|\|full | Low-quality |
| 18-105-final-viral-combined_k141_181085\|\|full | Low-quality |
| 18-105-final-viral-combined_k141_118217\|\|full | Low-quality |
| 18-105-final-viral-combined_k141_47379\|\|full | Low-quality |
| 18-105-final-viral-combined_k141_82453\|\|full | Low-quality |
| 18-105-final-viral-combined_k141_172474\|\|full | Not-determined |
| 18-102_k141_79944 | Medium-quality |
| 18-105-final-viral-combined_k141_172483\|\|full | Not-determined |
| 18-105-final-viral-combined_k141_47405\|\|full | Not-determined |
| 18-105-final-viral-combined_k141_3392\|\|full | Not-determined |
| 18-105-final-viral-combined_k141_47418\|\|full | Not-determined |
| 18-105-final-viral-combined_k141_136148\|\|full | Not-determined |
| 18-105-final-viral-combined_k141_29176\|\|full | Low-quality |
| 18-105-final-viral-combined_k141_73894\|\|full | Low-quality |
| 18-105-final-viral-combined_k141_172522\|\|full | Not-determined |
| 18-105-final-viral-combined_k141_127477\|\|full | Low-quality |
| 18-105-final-viral-combined_k141_73909\|\|full | Not-determined |
| 18-105-final-viral-combined_k141_19935\|\|full | Not-determined |
| 18-105-final-viral-combined_k141_145349\|\|full | Low-quality |
| 18-105-final-viral-combined_k141_55814\|\|full | Low-quality |
| 18-105-final-viral-combined_k141_145354\|\|full | Not-determined |
| 18-105-final-viral-combined_k141_3443\|\|full | Not-determined |
| 18-105-final-viral-combined_k141_189934\|\|full | Not-determined |
| 18-105-final-viral-combined_k141_145367\|\|full | Low-quality |
| 18-105-final-viral-combined_k141_3450\|\|full | Low-quality |
| 18-105-final-viral-combined_k141_181155\|\|full | Low-quality |
| 18-105-final-viral-combined_k141_145385\|\|full | Not-determined |
| 18-105-final-viral-combined_k141_136196\|\|full | Not-determined |
| 18-105-final-viral-combined_k141_172539\|\|full | Not-determined |
| 18-105-final-viral-combined_k141_64704\|\|full | Low-quality |
| 18-105-final-viral-combined_k141_127548\|\|full | Not-determined |
| 18-105-final-viral-combined_k141_172559\|\|full | Not-determined |
| 18-105-final-viral-combined_k141_181161\|\|full | Not-determined |
| 18-105-final-viral-combined_k141_136232\|\|full | Not-determined |
| 18-105-final-viral-combined_k141_198602\|\|full | Low-quality |
| 18-105-final-viral-combined_k141_3483\|\|full | Low-quality |
| 18-105-final-viral-combined_k141_55858\|\|full | Not-determined |
| 18-105-final-viral-combined_k141_172580\|\|full | Low-quality |
| 18-105-final-viral-combined_k141_181183\|\|full | Low-quality |
| 18-105-final-viral-combined_k141_181194\|\|full | Not-determined |
| 18-105-final-viral-combined_k141_118234\|\|full | Low-quality |
| 18-105-final-viral-combined_k141_136274\|\|full | Not-determined |
| 18-105-final-viral-combined_k141_64783\|\|full | Not-determined |
| 18-105-final-viral-combined_k141_181223\|\|full | Not-determined |
| 18-105-final-viral-combined_k141_11331\|\|full | Not-determined |
| 18-105-final-viral-combined_k141_172644\|\|full | Not-determined |
| 18-105-final-viral-combined_k141_127621\|\|full | Not-determined |
| 18-105-final-viral-combined_k141_172649\|\|full | Not-determined |
| 18-105-final-viral-combined_k141_55913\|\|full | Not-determined |
| 18-105-final-viral-combined_k141_64793\|\|full | Low-quality |
| 18-105-final-viral-combined_k141_162835\|\|full | Not-determined |
| 18-105-final-viral-combined_k141_73944\|\|full | Not-determined |
| 18-105-final-viral-combined_k141_109141\|\|full | Low-quality |
| 18-105-final-viral-combined_k141_198613\|\|full | Low-quality |
| 18-105-final-viral-combined_k141_47566\|\|full | Low-quality |
| 18-105-final-viral-combined_k141_64805\|\|full | Low-quality |
| 18-105-final-viral-combined_k141_136313\|\|full | Not-determined |
| 18-105-final-viral-combined_k141_162846\|\|full | Low-quality |
| 18-105-final-viral-combined_k141_55933\|\|full | Low-quality |
| 18-105-final-viral-combined_k141_64812\|\|full | Low-quality |
| 18-105-final-viral-combined_k141_162849\|\|full | Not-determined |
| 18-105-final-viral-combined_k141_3546\|\|full | Not-determined |
| 18-105-final-viral-combined_k141_162859\|\|full | Low-quality |
| 18-105-final-viral-combined_k141_118268\|\|full | Low-quality |
| 18-105-final-viral-combined_k141_162864\|\|full | Low-quality |
| 18-105-final-viral-combined_k141_47599\|\|full | Not-determined |
| 18-105-final-viral-combined_k141_118290\|\|full | Low-quality |
| 18-105-final-viral-combined_k141_3554\|\|full | Low-quality |
| 18-105-final-viral-combined_k141_64866\|\|full | Not-determined |
| 18-105-final-viral-combined_k141_47617\|\|full | Not-determined |
| 18-105-final-viral-combined_k141_162895\|\|full | Low-quality |
| 18-105-final-viral-combined_k141_47642\|\|full | Low-quality |
| 18-105-final-viral-combined_k141_3580\|\|full | Low-quality |
| 18-105-final-viral-combined_k141_38144\|\|full | Not-determined |
| 18-105-final-viral-combined_k141_47653\|\|full | Low-quality |
| 18-105-final-viral-combined_k141_11394\|\|full | Not-determined |
| 18-105-final-viral-combined_k141_118336\|\|full | Low-quality |
| 18-105-final-viral-combined_k141_189993\|\|full | Not-determined |
| 18-105-final-viral-combined_k141_189996\|\|full | Low-quality |
| 18-105-final-viral-combined_k141_47667\|\|full | Low-quality |
| 18-105-final-viral-combined_k141_64904\|\|full | Not-determined |
| 18-105-final-viral-combined_k141_74002\|\|full | Low-quality |
| 18-105-final-viral-combined_k141_189998\|\|full | Not-determined |
| 18-105-final-viral-combined_k141_74021\|\|full | Low-quality |
| 18-105-final-viral-combined_k141_64919\|\|full | Not-determined |
| 18-105-final-viral-combined_k141_47688\|\|full | Not-determined |
| 18-105-final-viral-combined_k141_74037\|\|full | Not-determined |
| 18-105-final-viral-combined_k141_29246\|\|full | Not-determined |
| 18-105-final-viral-combined_k141_127702\|\|full | Not-determined |
| 18-105-final-viral-combined_k141_82549\|\|full | Low-quality |
| 18-105-final-viral-combined_k141_82554\|\|full | Low-quality |
| 18-105-final-viral-combined_k141_198633\|\|full | Low-quality |
| 18-105-final-viral-combined_k141_29255\|\|full | Low-quality |
| 18-105-final-viral-combined_k141_38188\|\|full | Not-determined |
| 18-105-final-viral-combined_k141_198648\|\|full | Not-determined |
| 18-105-final-viral-combined_k141_207427\|\|full | Low-quality |
| 18-105-final-viral-combined_k141_207439\|\|full | Low-quality |
| 18-105-final-viral-combined_k141_11430\|\|full | Not-determined |
| 18-105-final-viral-combined_k141_47739\|\|full | Not-determined |
| 18-105-final-viral-combined_k141_92056\|\|full | Not-determined |
| 18-105-final-viral-combined_k141_38228\|\|full | Not-determined |
| 18-105-final-viral-combined_k141_3625\|\|full | Not-determined |
| 18-105-final-viral-combined_k141_162980\|\|full | Not-determined |
| 18-105-final-viral-combined_k141_172754\|\|full | Not-determined |
| 18-105-final-viral-combined_k141_207484\|\|full | Low-quality |
| 18-105-final-viral-combined_k141_38261\|\|full | Not-determined |
| 18-105-final-viral-combined_k141_198676\|\|full | Not-determined |
| 18-105-final-viral-combined_k141_163011\|\|full | Not-determined |
| 18-105-final-viral-combined_k141_3666\|\|full | Not-determined |
| 18-105-final-viral-combined_k141_82625\|\|full | Low-quality |
| 18-105-final-viral-combined_k141_190116\|\|full | Not-determined |
| 18-105-final-viral-combined_k141_74091\|\|full | Not-determined |
| 18-105-final-viral-combined_k141_207544\|\|full | Not-determined |
| 18-105-final-viral-combined_k141_82655\|\|full | Low-quality |
| 18-105-final-viral-combined_k141_74098\|\|full | Low-quality |
| 18-105-final-viral-combined_k141_127788\|\|full | Not-determined |
| 18-105-final-viral-combined_k141_207545\|\|full | Not-determined |
| 18-105-final-viral-combined_k141_145560\|\|full | Low-quality |
| 18-105-final-viral-combined_k141_47798\|\|full | Not-determined |
| 18-105-final-viral-combined_k141_181265\|\|full | Not-determined |
| 18-105-final-viral-combined_k141_11490\|\|full | Low-quality |
| 18-105-final-viral-combined_k141_207549\|\|full | Low-quality |
| 18-105-final-viral-combined_k141_181273\|\|full | Low-quality |
| 18-105-final-viral-combined_k141_163033\|\|full | Low-quality |
| 18-105-final-viral-combined_k141_207556\|\|full | Low-quality |
| 18-105-final-viral-combined_k141_190157\|\|full | Low-quality |
| 18-105-final-viral-combined_k141_190161\|\|full | Low-quality |
| 18-105-final-viral-combined_k141_109231\|\|full | Not-determined |
| 18-105-final-viral-combined_k141_172786\|\|full | Low-quality |
| 18-105-final-viral-combined_k141_20043\|\|full | Not-determined |
| 18-105-final-viral-combined_k141_181300\|\|full | Not-determined |
| 18-105-final-viral-combined_k141_47841\|\|full | Not-determined |
| 18-105-final-viral-combined_k141_190194\|\|full | Not-determined |
| 18-105-final-viral-combined_k141_11501\|\|full | Not-determined |
| 18-105-final-viral-combined_k141_74132\|\|full | Not-determined |
| 18-105-final-viral-combined_k141_181319\|\|full | Low-quality |
| 18-105-final-viral-combined_k141_163063\|\|full | Low-quality |
| 18-105-final-viral-combined_k141_181326\|\|full | Not-determined |
| 18-105-final-viral-combined_k141_47869\|\|full | Not-determined |
| 18-105-final-viral-combined_k141_181336\|\|full | Not-determined |
| 18-105-final-viral-combined_k141_11531\|\|full | Low-quality |
| 18-105-final-viral-combined_k141_190234\|\|full | Low-quality |
| 18-105-final-viral-combined_k141_11552\|\|full | Low-quality |
| 17-109-final-viral-combined_k141_284122\|\|full | High-quality |
| 18-105-final-viral-combined_k141_190237\|\|full | Low-quality |
| 18-105-final-viral-combined_k141_55990\|\|full | Low-quality |
| 18-105-final-viral-combined_k141_190246\|\|full | Not-determined |
| 18-105-final-viral-combined_k141_190248\|\|full | Low-quality |
| 18-105-final-viral-combined_k141_181375\|\|full | Low-quality |
| 18-105-final-viral-combined_k141_11586\|\|full | Not-determined |
| 18-105-final-viral-combined_k141_74213\|\|full | Not-determined |
| 18-105-final-viral-combined_k141_11598\|\|full | Low-quality |
| 18-105-final-viral-combined_k141_29307\|\|full | Not-determined |
| 18-105-final-viral-combined_k141_190295\|\|full | Not-determined |
| 18-105-final-viral-combined_k141_127866\|\|full | Not-determined |
| 18-105-final-viral-combined_k141_207620\|\|full | Low-quality |
| 18-105-final-viral-combined_k141_127872\|\|full | Low-quality |
| 18-105-final-viral-combined_k141_207630\|\|full | Not-determined |
| 18-105-final-viral-combined_k141_127882\|\|full | Low-quality |
| 18-105-final-viral-combined_k141_181431\|\|full | Not-determined |
| 18-105-final-viral-combined_k141_101070\|\|full | Not-determined |
| 18-105-final-viral-combined_k141_65108\|\|full | Not-determined |
| 18-105-final-viral-combined_k141_207641\|\|full | Low-quality |
| 18-105-final-viral-combined_k141_3899\|\|full | Low-quality |
| 18-105-final-viral-combined_k141_127912\|\|full | Low-quality |
| 18-105-final-viral-combined_k141_154171\|\|full | Not-determined |
| 18-105-final-viral-combined_k141_47958\|\|full | Not-determined |
| 18-105-final-viral-combined_k141_118404\|\|full | Low-quality |
| 18-105-final-viral-combined_k141_190324\|\|full | Low-quality |
| 18-105-final-viral-combined_k141_145769\|\|full | Low-quality |
| 18-105-final-viral-combined_k141_181520\|\|full | Not-determined |
| 18-105-final-viral-combined_k141_47992\|\|full | Not-determined |
| 18-105-final-viral-combined_k141_48017\|\|full | Not-determined |
| 18-105-final-viral-combined_k141_163197\|\|full | Not-determined |
| 18-105-final-viral-combined_k141_3954\|\|full | Not-determined |
| 18-105-final-viral-combined_k141_207746\|\|full | Not-determined |
| 18-105-final-viral-combined_k141_82741\|\|full | Low-quality |
| 18-105-final-viral-combined_k141_154248\|\|full | Not-determined |
| 18-105-final-viral-combined_k141_190346\|\|full | Low-quality |
| 18-105-final-viral-combined_k141_3962\|\|full | Low-quality |
| 18-105-final-viral-combined_k141_127999\|\|full | Not-determined |
| 18-105-final-viral-combined_k141_190353\|\|full | Low-quality |
| 18-105-final-viral-combined_k141_109454\|\|full | Not-determined |
| 18-105-final-viral-combined_k141_48043\|\|full | Low-quality |
| 18-105-final-viral-combined_k141_92198\|\|full | Not-determined |
| 18-105-final-viral-combined_k141_145820\|\|full | Low-quality |
| 18-105-final-viral-combined_k141_48053\|\|full | Not-determined |
| 18-105-final-viral-combined_k141_163215\|\|full | Low-quality |
| 18-105-final-viral-combined_k141_145834\|\|full | Low-quality |
| 18-105-final-viral-combined_k141_145836\|\|full | Not-determined |
| 18-105-final-viral-combined_k141_38370\|\|full | Low-quality |
| 18-105-final-viral-combined_k141_101218\|\|full | Not-determined |
| 18-105-final-viral-combined_k141_29349\|\|full | Low-quality |
| 18-105-final-viral-combined_k141_163230\|\|full | Low-quality |
| 18-105-final-viral-combined_k141_109519\|\|full | Low-quality |
| 18-105-final-viral-combined_k141_190415\|\|full | Not-determined |
| 18-105-final-viral-combined_k141_101249\|\|full | Low-quality |
| 18-105-final-viral-combined_k141_11663\|\|full | Low-quality |
| 18-105-final-viral-combined_k141_109547\|\|full | Low-quality |
| 18-105-final-viral-combined_k141_128094\|\|full | Not-determined |
| 18-105-final-viral-combined_k141_172854\|\|full | Low-quality |
| 18-105-final-viral-combined_k141_29396\|\|full | Not-determined |
| 18-105-final-viral-combined_k141_109553\|\|full | Not-determined |
| 18-105-final-viral-combined_k141_145898\|\|full | Low-quality |
| 18-105-final-viral-combined_k141_74314\|\|full | Not-determined |
| 18-105-final-viral-combined_k141_163286\|\|full | Low-quality |
| 18-105-final-viral-combined_k141_56100\|\|full | Low-quality |
| 18-105-final-viral-combined_k141_128122\|\|full | Not-determined |
| 18-105-final-viral-combined_k141_190433\|\|full | Not-determined |
| 18-105-final-viral-combined_k141_136352\|\|full | Low-quality |
| 18-105-final-viral-combined_k141_136357\|\|full | Low-quality |
| 18-105-final-viral-combined_k141_118543\|\|full | Low-quality |
| 18-105-final-viral-combined_k141_136369\|\|full | Low-quality |
| 18-105-final-viral-combined_k141_101279\|\|full | Low-quality |
| 18-105-final-viral-combined_k141_145965\|\|full | Not-determined |
| 18-105-final-viral-combined_k141_65168\|\|full | Low-quality |
| 18-105-final-viral-combined_k141_11742\|\|full | Low-quality |
| 18-105-final-viral-combined_k141_128162\|\|full | Not-determined |
| 18-105-final-viral-combined_k141_181610\|\|full | Low-quality |
| 18-105-final-viral-combined_k141_109582\|\|full | Low-quality |
| 18-105-final-viral-combined_k141_146001\|\|full | Low-quality |
| 18-105-final-viral-combined_k141_154358\|\|full | Not-determined |
| 18-105-final-viral-combined_k141_128189\|\|full | Low-quality |
| 18-105-final-viral-combined_k141_128193\|\|full | Not-determined |
| 18-105-final-viral-combined_k141_4139\|\|full | Low-quality |
| 18-105-final-viral-combined_k141_29424\|\|full | Low-quality |
| 18-105-final-viral-combined_k141_74353\|\|full | Not-determined |
| 18-105-final-viral-combined_k141_20217\|\|full | Not-determined |
| 18-105-final-viral-combined_k141_163318\|\|full | Low-quality |
| 18-105-final-viral-combined_k141_198810\|\|full | Not-determined |
| 18-105-final-viral-combined_k141_163326\|\|full | Low-quality |
| 18-105-final-viral-combined_k141_65262\|\|full | Low-quality |
| 18-105-final-viral-combined_k141_82886\|\|full | Low-quality |
| 18-105-final-viral-combined_k141_82890\|\|full | Low-quality |
| 18-105-final-viral-combined_k141_74394\|\|full | Not-determined |
| 18-105-final-viral-combined_k141_207812\|\|full | Not-determined |
| 18-105-final-viral-combined_k141_20282\|\|full | Not-determined |
| 18-105-final-viral-combined_k141_163371\|\|full | Not-determined |
| 18-105-final-viral-combined_k141_146055\|\|full | Low-quality |
| 18-105-final-viral-combined_k141_154415\|\|full | Low-quality |
| 18-105-final-viral-combined_k141_4201\|\|full | Low-quality |
| 18-105-final-viral-combined_k141_74437\|\|full | Low-quality |
| 18-105-final-viral-combined_k141_172921\|\|full | Not-determined |
| 18-105-final-viral-combined_k141_4203\|\|full | Not-determined |
| 18-105-final-viral-combined_k141_109667\|\|full | Low-quality |
| 18-105-final-viral-combined_k141_48135\|\|full | Low-quality |
| 18-105-final-viral-combined_k141_172930\|\|full | Not-determined |
| 18-105-final-viral-combined_k141_65310\|\|full | Not-determined |
| 18-105-final-viral-combined_k141_4215\|\|full | Not-determined |
| 18-109-final-viral-combined_k141_137858\|\|full | Medium-quality |
| 18-105-final-viral-combined_k141_163427\|\|full | Low-quality |
| 18-105-final-viral-combined_k141_65317\|\|full | Not-determined |
| 18-105-final-viral-combined_k141_146090\|\|full | Low-quality |
| 18-105-final-viral-combined_k141_146091\|\|full | Not-determined |
| 18-105-final-viral-combined_k141_65323\|\|full | Not-determined |
| 18-105-final-viral-combined_k141_4241\|\|full | Low-quality |
| 18-105-final-viral-combined_k141_172951\|\|full | Low-quality |
| 18-105-final-viral-combined_k141_4246\|\|full | Not-determined |
| 18-105-final-viral-combined_k141_118657\|\|full | Low-quality |
| 18-105-final-viral-combined_k141_4255\|\|full | Low-quality |
| 18-105-final-viral-combined_k141_109687\|\|full | Low-quality |
| 18-105-final-viral-combined_k141_118665\|\|full | Low-quality |
| 18-105-final-viral-combined_k141_4270\|\|full | Not-determined |
| 18-105-final-viral-combined_k141_172978\|\|full | Low-quality |
| 18-105-final-viral-combined_k141_136404\|\|full | Low-quality |
| 18-105-final-viral-combined_k141_118672\|\|full | Not-determined |
| 18-105-final-viral-combined_k141_11807\|\|full | Not-determined |
| 18-105-final-viral-combined_k141_146107\|\|full | Low-quality |
| 18-102-final-viral-combined_k141_79944\|\|full | Medium-quality |
| 18-105-final-viral-combined_k141_38514\|\|full | Not-determined |
| 18-105-final-viral-combined_k141_20371\|\|full | Low-quality |
| 18-105-final-viral-combined_k141_118684\|\|full | Not-determined |
| 18-105-final-viral-combined_k141_38519\|\|full | Not-determined |
| 18-105-final-viral-combined_k141_65380\|\|full | Low-quality |
| 18-105-final-viral-combined_k141_11822\|\|full | Low-quality |
| 18-105-final-viral-combined_k141_118700\|\|full | Not-determined |
| 18-105-final-viral-combined_k141_82987\|\|full | Low-quality |
| 18-105-final-viral-combined_k141_118704\|\|full | Low-quality |
| 18-105-final-viral-combined_k141_136440\|\|full | Low-quality |
| 18-105-final-viral-combined_k141_29462\|\|full | Low-quality |
| 18-105-final-viral-combined_k141_29464\|\|full | Low-quality |
| 18-105-final-viral-combined_k141_11839\|\|full | Not-determined |
| 18-105-final-viral-combined_k141_11842\|\|full | Low-quality |
| 18-105-final-viral-combined_k141_92338\|\|full | Low-quality |
| 18-105-final-viral-combined_k141_173014\|\|full | Low-quality |
| 18-105-final-viral-combined_k141_118735\|\|full | Low-quality |
| 18-105-final-viral-combined_k141_146111\|\|full | Not-determined |
| 18-105-final-viral-combined_k141_198947\|\|full | Low-quality |
| 18-105-final-viral-combined_k141_56158\|\|full | Low-quality |
| 18-105-final-viral-combined_k141_136451\|\|full | Not-determined |
| 18-105-final-viral-combined_k141_38543\|\|full | Low-quality |
| 18-105-final-viral-combined_k141_154442\|\|full | Not-determined |
| 18-105-final-viral-combined_k141_163513\|\|full | Low-quality |
| 18-105-final-viral-combined_k141_29468\|\|full | Low-quality |
| 18-105-final-viral-combined_k141_38555\|\|full | Not-determined |

| | |
|---|---|
| 18-105-final-viral-combined_k141_136473\|\|full | Low-quality |
| 18-105-final-viral-combined_k141_92375\|\|full | Low-quality |
| 18-105-final-viral-combined_k141_190532\|\|full | Low-quality |
| 18-105-final-viral-combined_k141_136478\|\|full | Low-quality |
| 18-105-final-viral-combined_k141_4356\|\|full | Low-quality |
| 18-105-final-viral-combined_k141_11872\|\|full | Not-determined |
| 18-105-final-viral-combined_k141_190536\|\|full | Not-determined |
| 18-105-final-viral-combined_k141_38565\|\|full | Not-determined |
| 18-105-final-viral-combined_k141_92389\|\|full | Low-quality |
| 18-105-final-viral-combined_k141_74574\|\|full | Not-determined |
| 18-105-final-viral-combined_k141_65439\|\|full | Not-determined |
| 18-105-final-viral-combined_k141_163550\|\|full | Low-quality |
| 18-105-final-viral-combined_k141_207864\|\|full | Low-quality |
| 18-105-final-viral-combined_k141_38576\|\|full | Low-quality |
| 18-105-final-viral-combined_k141_190554\|\|full | Low-quality |
| 18-105-final-viral-combined_k141_83043\|\|full | Not-determined |
| 18-105-final-viral-combined_k141_190561\|\|full | Low-quality |
| 18-105-final-viral-combined_k141_136497\|\|full | Not-determined |
| 18-105-final-viral-combined_k141_11897\|\|full | Not-determined |
| 18-105-final-viral-combined_k141_190565\|\|full | Low-quality |
| 18-105-final-viral-combined_k141_109769\|\|full | Low-quality |
| 18-105-final-viral-combined_k141_199027\|\|full | Low-quality |
| 18-105-final-viral-combined_k141_74610\|\|full | Not-determined |
| 18-105-final-viral-combined_k141_83069\|\|full | Low-quality |
| 18-105-final-viral-combined_k141_154523\|\|full | Low-quality |
| 18-105-final-viral-combined_k141_11912\|\|full | Not-determined |
| 18-105-final-viral-combined_k141_56188\|\|full | Not-determined |
| 18-105-final-viral-combined_k141_38601\|\|full | Low-quality |
| 18-105-final-viral-combined_k141_136524\|\|full | Not-determined |
| 18-105-final-viral-combined_k141_154525\|\|full | Not-determined |
| 18-105-final-viral-combined_k141_11926\|\|full | Low-quality |
| 18-105-final-viral-combined_k141_207904\|\|full | Low-quality |
| 18-105-final-viral-combined_k141_74630\|\|full | Not-determined |
| 18-105-final-viral-combined_k141_38613\|\|full | Low-quality |
| 18-105-final-viral-combined_k141_20497\|\|full | Low-quality |
| 18-105-final-viral-combined_k141_56193\|\|full | Not-determined |
| 18-105-final-viral-combined_k141_38619\|\|full | Not-determined |
| 18-105-final-viral-combined_k141_207932\|\|full | Not-determined |
| 18-105-final-viral-combined_k141_154550\|\|full | Low-quality |
| 18-105-final-viral-combined_k141_128392\|\|full | Low-quality |
| 18-105-final-viral-combined_k141_56211\|\|full | Low-quality |
| 18-105-final-viral-combined_k141_48302\|\|full | Not-determined |
| 18-105-final-viral-combined_k141_207951\|\|full | Not-determined |
| 18-105-final-viral-combined_k141_74667\|\|full | Not-determined |
| 18-105-final-viral-combined_k141_38634\|\|full | Not-determined |
| 18-105-final-viral-combined_k141_11964\|\|full | Not-determined |
| 18-105-final-viral-combined_k141_74702\|\|full | Not-determined |
| 18-105-final-viral-combined_k141_48346\|\|full | Low-quality |
| 18-105-final-viral-combined_k141_92448\|\|full | Low-quality |
| 18-105-final-viral-combined_k141_109805\|\|full | Not-determined |
| 18-105-final-viral-combined_k141_4460\|\|full | Low-quality |
| 18-105-final-viral-combined_k141_190577\|\|full | Low-quality |
| 18-105-final-viral-combined_k141_163576\|\|full | Not-determined |
| 18-105-final-viral-combined_k141_38656\|\|full | Low-quality |
| 18-105-final-viral-combined_k141_109819\|\|full | Low-quality |
| 18-105-final-viral-combined_k141_199090\|\|full | Not-determined |
| 18-105-final-viral-combined_k141_92463\|\|full | Not-determined |
| 18-105-final-viral-combined_k141_92468\|\|full | Not-determined |
| 18-105-final-viral-combined_k141_208023\|\|full | Not-determined |
| 18-105-final-viral-combined_k141_56285\|\|full | Low-quality |
| 18-105-final-viral-combined_k141_92477\|\|full | Low-quality |
| 18-105-final-viral-combined_k141_190617\|\|full | Not-determined |
| 18-105-final-viral-combined_k141_199101\|\|full | Not-determined |
| 18-105-final-viral-combined_k141_11978\|\|full | Low-quality |
| 18-105-final-viral-combined_k141_4494\|\|full | Not-determined |
| 18-105-final-viral-combined_k141_190623\|\|full | Not-determined |
| 18-105-final-viral-combined_k141_208039\|\|full | Not-determined |
| 18-105-final-viral-combined_k141_173148\|\|full | Not-determined |
| 18-105-final-viral-combined_k141_11992\|\|full | Not-determined |
| 18-105-final-viral-combined_k141_38691\|\|full | Low-quality |
| 18-105-final-viral-combined_k141_208061\|\|full | Not-determined |
| 18-105-final-viral-combined_k141_56317\|\|full | Not-determined |
| 18-105-final-viral-combined_k141_4537\|\|full | Not-determined |
| 18-105-final-viral-combined_k141_92523\|\|full | Low-quality |
| 18-105-final-viral-combined_k141_20561\|\|full | Not-determined |
| 18-105-final-viral-combined_k141_38697\|\|full | Low-quality |
| 18-105-final-viral-combined_k141_56337\|\|full | Not-determined |
| 18-105-final-viral-combined_k141_56338\|\|full | Not-determined |
| 18-105-final-viral-combined_k141_4541\|\|full | Not-determined |
| 18-105-final-viral-combined_k141_190645\|\|full | Not-determined |
| 18-105-final-viral-combined_k141_38699\|\|full | Low-quality |
| 18-105-final-viral-combined_k141_208077\|\|full | Not-determined |
| 18-105-final-viral-combined_k141_56346\|\|full | Low-quality |
| 18-105-final-viral-combined_k141_38710\|\|full | Not-determined |
| 18-105-final-viral-combined_k141_146219\|\|full | Not-determined |
| 18-105-final-viral-combined_k141_29589\|\|full | Low-quality |
| 18-105-final-viral-combined_k141_154669\|\|full | Low-quality |
| 18-105-final-viral-combined_k141_4552\|\|full | Not-determined |
| 18-105-final-viral-combined_k141_4561\|\|full | Not-determined |
| 18-105-final-viral-combined_k141_48431\|\|full | Not-determined |
| 18-105-final-viral-combined_k141_181741\|\|full | Not-determined |
| 18-105-final-viral-combined_k141_74864\|\|full | Not-determined |
| 18-105-final-viral-combined_k141_208095\|\|full | Not-determined |
| 18-105-final-viral-combined_k141_20617\|\|full | Not-determined |
| 18-105-final-viral-combined_k141_163661\|\|full | Low-quality |
| 18-105-final-viral-combined_k141_136738\|\|full | Not-determined |
| 18-105-final-viral-combined_k141_101606\|\|full | Not-determined |
| 18-105-final-viral-combined_k141_29658\|\|full | Not-determined |
| 18-105-final-viral-combined_k141_154707\|\|full | Low-quality |
| 18-105-final-viral-combined_k141_4599\|\|full | Not-determined |
| 18-105-final-viral-combined_k141_56367\|\|full | Not-determined |
| 18-105-final-viral-combined_k141_56369\|\|full | Not-determined |
| 18-105-final-viral-combined_k141_163693\|\|full | Not-determined |
| 18-105-final-viral-combined_k141_146271\|\|full | Low-quality |
| 18-105-final-viral-combined_k141_154739\|\|full | Low-quality |
| 18-105-final-viral-combined_k141_56391\|\|full | Not-determined |
| 18-105-final-viral-combined_k141_29697\|\|full | Not-determined |
| 18-105-final-viral-combined_k141_154751\|\|full | Low-quality |
| 18-105-final-viral-combined_k141_146297\|\|full | Not-determined |
| 18-105-final-viral-combined_k141_173239\|\|full | Low-quality |
| 18-105-final-viral-combined_k141_29723\|\|full | Low-quality |
| 18-109-final-viral-combined_k141_2122\|\|full | Medium-quality |
| 18-105-final-viral-combined_k141_146308\|\|full | Low-quality |
| 18-105-final-viral-combined_k141_199142\|\|full | Low-quality |
| 18-105-final-viral-combined_k141_109930\|\|full | Low-quality |
| 18-105-final-viral-combined_k141_12116\|\|full | Low-quality |
| 18-105-final-viral-combined_k141_154804\|\|full | Not-determined |
| 18-105-final-viral-combined_k141_4633\|\|full | Low-quality |
| 18-105-final-viral-combined_k141_163756\|\|full | Low-quality |
| 18-105-final-viral-combined_k141_163761\|\|full | Not-determined |
| 18-105-final-viral-combined_k141_136838\|\|full | Not-determined |
| 18-105-final-viral-combined_k141_109944\|\|full | Low-quality |
| 18-105-final-viral-combined_k141_12127\|\|full | Low-quality |
| 18-105-final-viral-combined_k141_173261\|\|full | Low-quality |
| 18-105-final-viral-combined_k141_56475\|\|full | Low-quality |
| 18-105-final-viral-combined_k141_29750\|\|full | Not-determined |
| 18-105-final-viral-combined_k141_65488\|\|full | Not-determined |
| 18-105-final-viral-combined_k141_136845\|\|full | Not-determined |
| 18-105-final-viral-combined_k141_173266\|\|full | Not-determined |
| 18-105-final-viral-combined_k141_136852\|\|full | Low-quality |
| 18-105-final-viral-combined_k141_101638\|\|full | Not-determined |
| 18-105-final-viral-combined_k141_12155\|\|full | Low-quality |
| 18-105-final-viral-combined_k141_146354\|\|full | Not-determined |
| 18-105-final-viral-combined_k141_199164\|\|full | Not-determined |
| 18-105-final-viral-combined_k141_173308\|\|full | Low-quality |
| 18-105-final-viral-combined_k141_199172\|\|full | Low-quality |
| 18-105-final-viral-combined_k141_65518\|\|full | Low-quality |
| 18-105-final-viral-combined_k141_74945\|\|full | Not-determined |
| 18-105-final-viral-combined_k141_173312\|\|full | Not-determined |
| 18-105-final-viral-combined_k141_4687\|\|full | Not-determined |
| 18-105-final-viral-combined_k141_101695\|\|full | Low-quality |
| 18-105-final-viral-combined_k141_199195\|\|full | Not-determined |
| 18-105-final-viral-combined_k141_4698\|\|full | Low-quality |
| 18-105-final-viral-combined_k141_173343\|\|full | Low-quality |
| 18-105-final-viral-combined_k141_190771\|\|full | Not-determined |
| 18-105-final-viral-combined_k141_118985\|\|full | Not-determined |
| 18-105-final-viral-combined_k141_128597\|\|full | Low-quality |
| 18-105-final-viral-combined_k141_101745\|\|full | Not-determined |
| 18-105-final-viral-combined_k141_4725\|\|full | Low-quality |
| 18-105-final-viral-combined_k141_20689\|\|full | Low-quality |
| 18-105-final-viral-combined_k141_92743\|\|full | Not-determined |
| 18-105-final-viral-combined_k141_20699\|\|full | Low-quality |
| 18-105-final-viral-combined_k141_65573\|\|full | Not-determined |
| 18-105-final-viral-combined_k141_128614\|\|full | Low-quality |
| 17-106-final-viral-combined_k141_93232\|\|full | High-quality |
| 18-105-final-viral-combined_k141_56493\|\|full | Not-determined |
| 18-105-final-viral-combined_k141_12249\|\|full | Not-determined |
| 18-105-final-viral-combined_k141_65588\|\|full | Low-quality |
| 18-105-final-viral-combined_k141_20709\|\|full | Not-determined |
| 18-105-final-viral-combined_k141_208205\|\|full | Low-quality |
| 18-105-final-viral-combined_k141_4760\|\|full | Not-determined |
| 17-109-final-viral-combined_k141_329242\|\|full | High-quality |
| 18-105-final-viral-combined_k141_65603\|\|full | Low-quality |
| 18-105-final-viral-combined_k141_83380\|\|full | Not-determined |
| 18-105-final-viral-combined_k141_83389\|\|full | Low-quality |
| 18-105-final-viral-combined_k141_208220\|\|full | Low-quality |
| 18-105-final-viral-combined_k141_38863\|\|full | Low-quality |
| 18-105-final-viral-combined_k141_208229\|\|full | Not-determined |
| 18-105-final-viral-combined_k141_119010\|\|full | Low-quality |
| 18-105-final-viral-combined_k141_136901\|\|full | Low-quality |
| 18-105-final-viral-combined_k141_38875\|\|full | Not-determined |
| [illegible] | [illegible] |
| 18-105-final-viral-combined_k141_209077\|\|full | Low-quality |
| 18-105-final-viral-combined_k141_39696\|\|full | Not-determined |
| 18-105-final-viral-combined_k141_84286\|\|full | Low-quality |
| 18-105-final-viral-combined_k141_155507\|\|full | Low-quality |
| 18-105-final-viral-combined_k141_57564\|\|full | Low-quality |
| 18-105-final-viral-combined_k141_5845\|\|full | Not-determined |
| 18-105-final-viral-combined_k141_147044\|\|full | Not-determined |
| 18-105-final-viral-combined_k141_110966\|\|full | Low-quality |
| 18-105-final-viral-combined_k141_39714\|\|full | Not-determined |
| 18-105-final-viral-combined_k141_129375\|\|full | Low-quality |
| 18-105-final-viral-combined_k141_129380\|\|full | Low-quality |
| 18-105-final-viral-combined_k141_57589\|\|full | Low-quality |
| 18-105-final-viral-combined_k141_182413\|\|full | Low-quality |
| 18-105-final-viral-combined_k141_164549\|\|full | Not-determined |
| 18-105-final-viral-combined_k141_174100\|\|full | Low-quality |
| 18-105-final-viral-combined_k141_147106\|\|full | Not-determined |
| 18-105-final-viral-combined_k141_200192\|\|full | Not-determined |
| 18-105-final-viral-combined_k141_200211\|\|full | Low-quality |
| 18-105-final-viral-combined_k141_129419\|\|full | Low-quality |
| 18-105-final-viral-combined_k141_39767\|\|full | Not-determined |
| 18-105-final-viral-combined_k141_191424\|\|full | Not-determined |
| 18-105-final-viral-combined_k141_147142\|\|full | Not-determined |
| 18-105-final-viral-combined_k141_191434\|\|full | Not-determined |
| 18-105-final-viral-combined_k141_182473\|\|full | Low-quality |
| 18-105-final-viral-combined_k141_164591\|\|full | Low-quality |
| 18-105-final-viral-combined_k141_102869\|\|full | Not-determined |
| 18-105-final-viral-combined_k141_75935\|\|full | Not-determined |
| 18-105-final-viral-combined_k141_57652\|\|full | Low-quality |
| 18-105-final-viral-combined_k141_102894\|\|full | Low-quality |
| 18-105-final-viral-combined_k141_111052\|\|full | Not-determined |
| 18-105-final-viral-combined_k141_84402\|\|full | Not-determined |
| 18-105-final-viral-combined_k141_75981\|\|full | Low-quality |
| 18-105-final-viral-combined_k141_111070\|\|full | Not-determined |
| 18-105-final-viral-combined_k141_129479\|\|full | Low-quality |
| 18-101-final-viral-combined_k141_118167\|\|full | Medium-quality |
| 18-105-final-viral-combined_k141_30464\|\|full | Low-quality |
| 18-105-final-viral-combined_k141_30469\|\|full | Not-determined |
| 18-105-final-viral-combined_k141_147230\|\|full | Not-determined |
| 18-105-final-viral-combined_k141_147234\|\|full | Not-determined |
| 18-105-final-viral-combined_k141_76007\|\|full | Low-quality |
| 18-105-final-viral-combined_k141_111090\|\|full | Low-quality |
| 18-105-final-viral-combined_k141_129521\|\|full | Low-quality |
| 18-105-final-viral-combined_k141_111093\|\|full | Low-quality |
| 18-105-final-viral-combined_k141_147252\|\|full | Not-determined |
| 18-105-final-viral-combined_k141_57709\|\|full | Low-quality |
| 18-105-final-viral-combined_k141_102913\|\|full | Low-quality |
| 18-105-final-viral-combined_k141_39779\|\|full | Not-determined |
| 18-105-final-viral-combined_k141_102923\|\|full | Not-determined |
| 18-105-final-viral-combined_k141_102937\|\|full | Low-quality |
| 18-105-final-viral-combined_k141_66701\|\|full | Low-quality |
| 18-105-final-viral-combined_k141_66714\|\|full | Not-determined |
| 18-105-final-viral-combined_k141_182535\|\|full | Low-quality |
| 18-105-final-viral-combined_k141_6078\|\|full | Low-quality |
| 18-105-final-viral-combined_k141_6081\|\|full | Low-quality |
| 18-105-final-viral-combined_k141_111144\|\|full | Not-determined |
| 18-105-final-viral-combined_k141_200245\|\|full | Low-quality |
| 18-105-final-viral-combined_k141_30515\|\|full | Low-quality |
| 18-105-final-viral-combined_k141_93599\|\|full | Not-determined |
| 18-105-final-viral-combined_k141_93602\|\|full | Not-determined |
| 18-105-final-viral-combined_k141_13443\|\|full | Low-quality |
| 18-105-final-viral-combined_k141_174237\|\|full | Not-determined |
| 18-105-final-viral-combined_k141_66727\|\|full | Not-determined |
| 18-105-final-viral-combined_k141_209279\|\|full | Low-quality |
| 18-105-final-viral-combined_k141_93635\|\|full | Not-determined |
| 18-105-final-viral-combined_k141_111179\|\|full | Low-quality |
| 18-105-final-viral-combined_k141_66766\|\|full | Low-quality |
| 18-105-final-viral-combined_k141_111194\|\|full | Low-quality |
| 18-105-final-viral-combined_k141_84525\|\|full | Not-determined |
| 18-105-final-viral-combined_k141_147370\|\|full | Not-determined |
| 18-105-final-viral-combined_k141_182590\|\|full | Low-quality |
| 18-105-final-viral-combined_k141_147378\|\|full | Not-determined |
| 18-105-final-viral-combined_k141_200287\|\|full | Low-quality |
| 18-105-final-viral-combined_k141_137615\|\|full | Not-determined |
| 18-105-final-viral-combined_k141_119948\|\|full | Not-determined |
| 18-105-final-viral-combined_k141_182611\|\|full | Low-quality |
| 18-105-final-viral-combined_k141_30558\|\|full | Low-quality |
| 18-105-final-viral-combined_k141_119961\|\|full | Not-determined |
| 18-105-final-viral-combined_k141_119971\|\|full | Not-determined |
| 18-105-final-viral-combined_k141_164628\|\|full | Not-determined |
| 18-105-final-viral-combined_k141_6139\|\|full | Low-quality |
| 18-105-final-viral-combined_k141_111255\|\|full | Low-quality |
| 18-105-final-viral-combined_k141_30578\|\|full | Not-determined |
| 18-105-final-viral-combined_k141_39835\|\|full | Not-determined |
| 18-105-final-viral-combined_k141_93718\|\|full | Low-quality |
| 18-105-final-viral-combined_k141_129546\|\|full | Not-determined |
| 18-105-final-viral-combined_k141_182654\|\|full | Not-determined |
| 18-105-final-viral-combined_k141_182660\|\|full | Low-quality |
| 18-105-final-viral-combined_k141_182672\|\|full | Low-quality |

| | |
|---|---|
| 18-105-final-viral-combined_k141_13534\|\|full | Not-determined |
| 18-105-final-viral-combined_k141_200334\|\|full | Low-quality |
| 18-105-final-viral-combined_k141_200339\|\|full | Low-quality |
| 18-105-final-viral-combined_k141_30609\|\|full | Not-determined |
| 18-105-final-viral-combined_k141_76068\|\|full | Low-quality |
| 18-105-final-viral-combined_k141_93754\|\|full | Not-determined |
| 18-105-final-viral-combined_k141_84609\|\|full | Not-determined |
| 18-105-final-viral-combined_k141_164640\|\|full | Low-quality |
| 18-105-final-viral-combined_k141_93762\|\|full | Low-quality |
| 18-105-final-viral-combined_k141_93772\|\|full | Not-determined |
| 18-105-final-viral-combined_k141_111346\|\|full | Low-quality |
| 18-105-final-viral-combined_k141_21370\|\|full | Not-determined |
| 18-105-final-viral-combined_k141_129585\|\|full | Not-determined |
| 18-105-final-viral-combined_k141_111358\|\|full | Low-quality |
| 18-105-final-viral-combined_k141_93787\|\|full | Low-quality |
| 18-105-final-viral-combined_k141_191485\|\|full | Low-quality |
| 18-105-final-viral-combined_k141_209386\|\|full | Not-determined |
| 18-105-final-viral-combined_k141_191497\|\|full | Low-quality |
| 18-105-final-viral-combined_k141_84634\|\|full | Low-quality |
| 18-105-final-viral-combined_k141_76077\|\|full | Low-quality |
| 18-105-final-viral-combined_k141_30638\|\|full | Low-quality |
| 18-105-final-viral-combined_k141_39897\|\|full | Low-quality |
| 18-105-final-viral-combined_k141_209423\|\|full | Not-determined |
| 18-105-final-viral-combined_k141_30651\|\|full | Low-quality |
| 18-105-final-viral-combined_k141_49268\|\|full | Low-quality |
| 18-105-final-viral-combined_k141_191547\|\|full | Low-quality |
| 18-105-final-viral-combined_k141_155593\|\|full | Low-quality |
| 18-105-final-viral-combined_k141_103051\|\|full | Not-determined |
| 18-105-final-viral-combined_k141_13568\|\|full | Low-quality |
| 18-105-final-viral-combined_k141_111414\|\|full | Low-quality |
| 18-105-final-viral-combined_k141_120052\|\|full | Not-determined |
| 18-105-final-viral-combined_k141_111428\|\|full | Not-determined |
| 18-105-final-viral-combined_k141_93857\|\|full | Low-quality |
| 18-105-final-viral-combined_k141_200369\|\|full | Low-quality |
| 18-105-final-viral-combined_k141_6227\|\|full | Low-quality |
| 18-105-final-viral-combined_k141_182832\|\|full | Not-determined |
| 18-105-final-viral-combined_k141_13600\|\|full | Low-quality |
| 18-105-final-viral-combined_k141_93876\|\|full | Low-quality |
| 18-105-final-viral-combined_k141_93880\|\|full | Low-quality |
| 18-105-final-viral-combined_k141_6244\|\|full | Not-determined |
| 18-105-final-viral-combined_k141_49277\|\|full | Not-determined |
| 18-105-final-viral-combined_k141_191607\|\|full | Not-determined |
| 18-105-final-viral-combined_k141_209502\|\|full | Not-determined |
| 18-105-final-viral-combined_k141_191622\|\|full | Low-quality |
| 18-105-final-viral-combined_k141_191629\|\|full | Not-determined |
| 18-105-final-viral-combined_k141_137805\|\|full | Not-determined |
| 18-105-final-viral-combined_k141_84754\|\|full | Not-determined |
| 18-105-final-viral-combined_k141_84762\|\|full | Not-determined |
| 18-105-final-viral-combined_k141_182893\|\|full | Not-determined |
| 18-105-final-viral-combined_k141_21394\|\|full | Low-quality |
| 18-105-final-viral-combined_k141_21403\|\|full | Low-quality |
| 18-105-final-viral-combined_k141_200395\|\|full | Not-determined |
| 18-105-final-viral-combined_k141_30683\|\|full | Low-quality |
| 18-105-final-viral-combined_k141_13662\|\|full | Not-determined |
| 18-105-final-viral-combined_k141_155691\|\|full | Not-determined |
| 18-105-final-viral-combined_k141_191666\|\|full | Not-determined |
| 18-105-final-viral-combined_k141_6314\|\|full | Not-determined |
| 18-105-final-viral-combined_k141_30694\|\|full | Low-quality |
| 18-105-final-viral-combined_k141_209542\|\|full | Low-quality |
| 18-105-final-viral-combined_k141_13679\|\|full | Low-quality |
| 18-105-final-viral-combined_k141_39964\|\|full | Low-quality |
| 18-105-final-viral-combined_k141_39970\|\|full | Not-determined |
| 18-105-final-viral-combined_k141_174359\|\|full | Low-quality |
| 18-105-final-viral-combined_k141_6328\|\|full | Not-determined |
| 18-105-final-viral-combined_k141_57853\|\|full | Low-quality |
| 18-105-final-viral-combined_k141_155705\|\|full | Not-determined |
| 18-105-final-viral-combined_k141_111468\|\|full | Low-quality |
| 18-105-final-viral-combined_k141_84818\|\|full | Not-determined |
| 18-105-final-viral-combined_k141_30713\|\|full | Low-quality |
| 18-105-final-viral-combined_k141_57879\|\|full | Not-determined |
| 18-105-final-viral-combined_k141_155736\|\|full | Low-quality |
| 18-105-final-viral-combined_k141_57886\|\|full | Not-determined |
| 18-105-final-viral-combined_k141_84835\|\|full | Low-quality |
| 18-105-final-viral-combined_k141_6360\|\|full | Low-quality |
| 18-105-final-viral-combined_k141_57891\|\|full | Not-determined |
| 18-105-final-viral-combined_k141_84843\|\|full | Low-quality |
| 18-105-final-viral-combined_k141_155747\|\|full | Not-determined |
| 18-105-final-viral-combined_k141_13701\|\|full | Not-determined |
| 18-105-final-viral-combined_k141_200451\|\|full | Not-determined |
| 18-105-final-viral-combined_k141_120069\|\|full | Not-determined |
| 18-105-final-viral-combined_k141_21514\|\|full | Low-quality |
| 18-109-final-viral-combined_k141_135154\|\|full | Medium-quality |
| 18-105-final-viral-combined_k141_164798\|\|full | Not-determined |
| 18-105-final-viral-combined_k141_164806\|\|full | Not-determined |
| 18-105-final-viral-combined_k141_200471\|\|full | Low-quality |
| 18-105-final-viral-combined_k141_13756\|\|full | Low-quality |
| 18-105-final-viral-combined_k141_111573\|\|full | Low-quality |
| 18-105-final-viral-combined_k141_103165\|\|full | Low-quality |
| 18-105-final-viral-combined_k141_137867\|\|full | Low-quality |
| 18-105-final-viral-combined_k141_155808\|\|full | Low-quality |
| 18-105-final-viral-combined_k141_76262\|\|full | Not-determined |
| 18-105-final-viral-combined_k141_209623\|\|full | Low-quality |
| 18-105-final-viral-combined_k141_155821\|\|full | Not-determined |
| 18-105-final-viral-combined_k141_191771\|\|full | Low-quality |
| 18-105-final-viral-combined_k141_13801\|\|full | Not-determined |
| 18-105-final-viral-combined_k141_13805\|\|full | Not-determined |
| 18-105-final-viral-combined_k141_6393\|\|full | Low-quality |
| 18-105-final-viral-combined_k141_174454\|\|full | Low-quality |
| 18-105-final-viral-combined_k141_40018\|\|full | Not-determined |
| 18-105-final-viral-combined_k141_13820\|\|full | Not-determined |
| 18-105-final-viral-combined_k141_200576\|\|full | Not-determined |
| 18-105-final-viral-combined_k141_40026\|\|full | Not-determined |
| 18-105-final-viral-combined_k141_174460\|\|full | Not-determined |
| 18-105-final-viral-combined_k141_40029\|\|full | Not-determined |
| 18-105-final-viral-combined_k141_200592\|\|full | Low-quality |
| 18-105-final-viral-combined_k141_129685\|\|full | Low-quality |
| 18-105-final-viral-combined_k141_76337\|\|full | Low-quality |
| 18-105-final-viral-combined_k141_164828\|\|full | Not-determined |
| 18-105-final-viral-combined_k141_164832\|\|full | Not-determined |
| 18-105-final-viral-combined_k141_164842\|\|full | Low-quality |
| 18-105-final-viral-combined_k141_200618\|\|full | Low-quality |
| 18-105-final-viral-combined_k141_76356\|\|full | Not-determined |
| 18-105-final-viral-combined_k141_6444\|\|full | Low-quality |
| 18-105-final-viral-combined_k141_147419\|\|full | Low-quality |
| 18-105-final-viral-combined_k141_147422\|\|full | Low-quality |
| 18-105-final-viral-combined_k141_155861\|\|full | Not-determined |
| 18-105-final-viral-combined_k141_120116\|\|full | Not-determined |
| 18-105-final-viral-combined_k141_13871\|\|full | Low-quality |
| 18-105-final-viral-combined_k141_40095\|\|full | Not-determined |
| 18-105-final-viral-combined_k141_137918\|\|full | Not-determined |
| 18-105-final-viral-combined_k141_40106\|\|full | Low-quality |
| 18-105-final-viral-combined_k141_191823\|\|full | Not-determined |
| 18-105-final-viral-combined_k141_40114\|\|full | Not-determined |
| 18-105-final-viral-combined_k141_182950\|\|full | Low-quality |
| 18-105-final-viral-combined_k141_111648\|\|full | Not-determined |
| 18-105-final-viral-combined_k141_103228\|\|full | Not-determined |
| 18-105-final-viral-combined_k141_182955\|\|full | Not-determined |
| 18-105-final-viral-combined_k141_30797\|\|full | Not-determined |
| 18-105-final-viral-combined_k141_13925\|\|full | Not-determined |
| 18-105-final-viral-combined_k141_40149\|\|full | Low-quality |
| 18-105-final-viral-combined_k141_30809\|\|full | Not-determined |
| 18-105-final-viral-combined_k141_164901\|\|full | Low-quality |
| 18-105-final-viral-combined_k141_191866\|\|full | Not-determined |
| 18-105-final-viral-combined_k141_84946\|\|full | Not-determined |
| 18-105-final-viral-combined_k141_191875\|\|full | Not-determined |
| 18-105-final-viral-combined_k141_6562\|\|full | Low-quality |
| 18-105-final-viral-combined_k141_6576\|\|full | Low-quality |
| 18-105-final-viral-combined_k141_111686\|\|full | Low-quality |
| 18-105-final-viral-combined_k141_147447\|\|full | Low-quality |
| 18-105-final-viral-combined_k141_200678\|\|full | Low-quality |
| 18-105-final-viral-combined_k141_200682\|\|full | Not-determined |
| 18-105-final-viral-combined_k141_13969\|\|full | Low-quality |
| 18-105-final-viral-combined_k141_21614\|\|full | Not-determined |
| 18-105-final-viral-combined_k141_120206\|\|full | Low-quality |
| 18-105-final-viral-combined_k141_174601\|\|full | Not-determined |
| 18-105-final-viral-combined_k141_191946\|\|full | Low-quality |
| 18-105-final-viral-combined_k141_120226\|\|full | Not-determined |
| 18-105-final-viral-combined_k141_164925\|\|full | Low-quality |
| 18-105-final-viral-combined_k141_209650\|\|full | Low-quality |
| 18-105-final-viral-combined_k141_84977\|\|full | Low-quality |
| 18-105-final-viral-combined_k141_174635\|\|full | Not-determined |
| 18-105-final-viral-combined_k141_174637\|\|full | Not-determined |
| 18-105-final-viral-combined_k141_174638\|\|full | Not-determined |
| 18-105-final-viral-combined_k141_137964\|\|full | Low-quality |
| 18-105-final-viral-combined_k141_58068\|\|full | Low-quality |
| 18-105-final-viral-combined_k141_103265\|\|full | Low-quality |
| 18-105-final-viral-combined_k141_58094\|\|full | Low-quality |
| 18-105-final-viral-combined_k141_30909\|\|full | Low-quality |
| 18-105-final-viral-combined_k141_58096\|\|full | Not-determined |
| 18-105-final-viral-combined_k141_76462\|\|full | Not-determined |
| 18-105-final-viral-combined_k141_58107\|\|full | Low-quality |
| 18-105-final-viral-combined_k141_40287\|\|full | Not-determined |
| 18-105-final-viral-combined_k141_137979\|\|full | Low-quality |
| 18-105-final-viral-combined_k141_120260\|\|full | Not-determined |
| 18-105-final-viral-combined_k141_137992\|\|full | Low-quality |
| 18-105-final-viral-combined_k141_49338\|\|full | Not-determined |
| 18-105-final-viral-combined_k141_209708\|\|full | Not-determined |
| 18-105-final-viral-combined_k141_85088\|\|full | Low-quality |
| 18-105-final-viral-combined_k141_30994\|\|full | Not-determined |
| 18-105-final-viral-combined_k141_58161\|\|full | Not-determined |
| 18-105-final-viral-combined_k141_85091\|\|full | Low-quality |
| 18-105-final-viral-combined_k141_120299\|\|full | Not-determined |
| 18-105-final-viral-combined_k141_49360\|\|full | Not-determined |
| 18-105-final-viral-combined_k141_58176\|\|full | Low-quality |
| 18-105-final-viral-combined_k141_111783\|\|full | Low-quality |
| 18-105-final-viral-combined_k141_200900\|\|full | Low-quality |
| 18-105-final-viral-combined_k141_40333\|\|full | Low-quality |
| 18-105-final-viral-combined_k141_85112\|\|full | Low-quality |
| 18-105-final-viral-combined_k141_183108\|\|full | Low-quality |
| 18-105-final-viral-combined_k141_49399\|\|full | Not-determined |
| 18-105-final-viral-combined_k141_6591\|\|full | Low-quality |
| 18-105-final-viral-combined_k141_138042\|\|full | Low-quality |
| 18-105-final-viral-combined_k141_76500\|\|full | Low-quality |
| 18-105-final-viral-combined_k141_138048\|\|full | Low-quality |
| 18-105-final-viral-combined_k141_58241\|\|full | Low-quality |
| 18-105-final-viral-combined_k141_31087\|\|full | Not-determined |
| 18-105-final-viral-combined_k141_111849\|\|full | Low-quality |
| 18-105-final-viral-combined_k141_6608\|\|full | Low-quality |
| 18-105-final-viral-combined_k141_76524\|\|full | Not-determined |
| 18-105-final-viral-combined_k141_164977\|\|full | Not-determined |
| 18-105-final-viral-combined_k141_21732\|\|full | Not-determined |
| 18-105-final-viral-combined_k141_58271\|\|full | Low-quality |
| 18-105-final-viral-combined_k141_103279\|\|full | Not-determined |
| 18-105-final-viral-combined_k141_31120\|\|full | Not-determined |
| 18-105-final-viral-combined_k141_14013\|\|full | Not-determined |
| 18-105-final-viral-combined_k141_111877\|\|full | Not-determined |
| 18-105-final-viral-combined_k141_66891\|\|full | Not-determined |
| 18-105-final-viral-combined_k141_58302\|\|full | Not-determined |
| 17-109-final-viral-combined_k141_184338\|\|full | Complete |
| 18-105-final-viral-combined_k141_49422\|\|full | Not-determined |
| 18-105-final-viral-combined_k141_183145\|\|full | Not-determined |
| 18-105-final-viral-combined_k141_209778\|\|full | Low-quality |
| 18-105-final-viral-combined_k141_49437\|\|full | Not-determined |
| 18-105-final-viral-combined_k141_6624\|\|full | Low-quality |
| 18-105-final-viral-combined_k141_138091\|\|full | Not-determined |
| 18-105-final-viral-combined_k141_192020\|\|full | Not-determined |
| 18-105-final-viral-combined_k141_66924\|\|full | Not-determined |
| 18-105-final-viral-combined_k141_147578\|\|full | Not-determined |
| 18-105-final-viral-combined_k141_49454\|\|full | Low-quality |
| 18-105-final-viral-combined_k141_192025\|\|full | Low-quality |
| 18-105-final-viral-combined_k141_6664\|\|full | Low-quality |
| 18-105-final-viral-combined_k141_192038\|\|full | Low-quality |
| 18-105-final-viral-combined_k141_14078\|\|full | Not-determined |
| 18-105-final-viral-combined_k141_21801\|\|full | Low-quality |
| 18-105-final-viral-combined_k141_58369\|\|full | Low-quality |
| 18-105-final-viral-combined_k141_40453\|\|full | Not-determined |
| 18-105-final-viral-combined_k141_183201\|\|full | Not-determined |
| 18-105-final-viral-combined_k141_94026\|\|full | Low-quality |
| 18-105-final-viral-combined_k141_85210\|\|full | Low-quality |
| 18-105-final-viral-combined_k141_14106\|\|full | Not-determined |
| 18-105-final-viral-combined_k141_156005\|\|full | Low-quality |
| 18-105-final-viral-combined_k141_129833\|\|full | Not-determined |
| 18-105-final-viral-combined_k141_147604\|\|full | Low-quality |
| 18-105-final-viral-combined_k141_165051\|\|full | Not-determined |
| 18-105-final-viral-combined_k141_192062\|\|full | Not-determined |
| 18-105-final-viral-combined_k141_94049\|\|full | Not-determined |
| 18-105-final-viral-combined_k141_165064\|\|full | Low-quality |
| 18-105-final-viral-combined_k141_147640\|\|full | Low-quality |
| 18-105-final-viral-combined_k141_94064\|\|full | Low-quality |
| 18-105-final-viral-combined_k141_14151\|\|full | Not-determined |
| 18-105-final-viral-combined_k141_85259\|\|full | Low-quality |
| 18-105-final-viral-combined_k141_6766\|\|full | Not-determined |
| 18-105-final-viral-combined_k141_94077\|\|full | Low-quality |
| 18-105-final-viral-combined_k141_138120\|\|full | Low-quality |
| 18-105-final-viral-combined_k141_138131\|\|full | Low-quality |
| 18-105-final-viral-combined_k141_6801\|\|full | Low-quality |
| 18-105-final-viral-combined_k141_209874\|\|full | Not-determined |
| 18-105-final-viral-combined_k141_209879\|\|full | Not-determined |
| 18-105-final-viral-combined_k141_174694\|\|full | Low-quality |
| 18-105-final-viral-combined_k141_129899\|\|full | Low-quality |
| 18-105-final-viral-combined_k141_6831\|\|full | Low-quality |
| 18-105-final-viral-combined_k141_138157\|\|full | Low-quality |
| 18-105-final-viral-combined_k141_147678\|\|full | Low-quality |
| 18-105-final-viral-combined_k141_165139\|\|full | Not-determined |
| 18-105-final-viral-combined_k141_58429\|\|full | Low-quality |
| 18-105-final-viral-combined_k141_209903\|\|full | Low-quality |
| 18-105-final-viral-combined_k141_103351\|\|full | Low-quality |
| 18-105-final-viral-combined_k141_147692\|\|full | Not-determined |
| 18-105-final-viral-combined_k141_31160\|\|full | Low-quality |
| 18-105-final-viral-combined_k141_85321\|\|full | Low-quality |
| 18-105-final-viral-combined_k141_209914\|\|full | Not-determined |
| 18-105-final-viral-combined_k141_58468\|\|full | Low-quality |
| 18-105-final-viral-combined_k141_6855\|\|full | Low-quality |
| 18-105-final-viral-combined_k141_58489\|\|full | Not-determined |
| 18-105-final-viral-combined_k141_6861\|\|full | Low-quality |
| 18-105-final-viral-combined_k141_103384\|\|full | Not-determined |
| 18-105-final-viral-combined_k141_6870\|\|full | Low-quality |
| 18-105-final-viral-combined_k141_183394\|\|full | Low-quality |
| 18-105-final-viral-combined_k141_209959\|\|full | Not-determined |
| 18-105-final-viral-combined_k141_147723\|\|full | Low-quality |
| 18-105-final-viral-combined_k141_94141\|\|full | Low-quality |
| 18-105-final-viral-combined_k141_6876\|\|full | Not-determined |
| 18-105-final-viral-combined_k141_49585\|\|full | Not-determined |
| 18-105-final-viral-combined_k141_49596\|\|full | Low-quality |
| 18-105-final-viral-combined_k141_174766\|\|full | Low-quality |
| 18-105-final-viral-combined_k141_49600\|\|full | Not-determined |
| 18-105-final-viral-combined_k141_31216\|\|full | Not-determined |
| 18-105-final-viral-combined_k141_183414\|\|full | Low-quality |
| 18-105-final-viral-combined_k141_120358\|\|full | Low-quality |
| 18-105-final-viral-combined_k141_6912\|\|full | Low-quality |
| 18-105-final-viral-combined_k141_156067\|\|full | Low-quality |
| 18-105-final-viral-combined_k141_183421\|\|full | Not-determined |
| 18-105-final-viral-combined_k141_112040\|\|full | Low-quality |
| 18-105-final-viral-combined_k141_31238\|\|full | Low-quality |
| 18-105-final-viral-combined_k141_174827\|\|full | Low-quality |
| 18-105-final-viral-combined_k141_112046\|\|full | Low-quality |
| 18-105-final-viral-combined_k141_200974\|\|full | Low-quality |
| 18-105-final-viral-combined_k141_183458\|\|full | Low-quality |
| 18-105-final-viral-combined_k141_21981\|\|full | Not-determined |
| 18-105-final-viral-combined_k141_67030\|\|full | Low-quality |
| 18-105-final-viral-combined_k141_31265\|\|full | Low-quality |
| 18-105-final-viral-combined_k141_210040\|\|full | Not-determined |
| 18-105-final-viral-combined_k141_192128\|\|full | Not-determined |
| 18-105-final-viral-combined_k141_210046\|\|full | Low-quality |
| 18-105-final-viral-combined_k141_31287\|\|full | Not-determined |
| 18-105-final-viral-combined_k141_156092\|\|full | Not-determined |
| 18-105-final-viral-combined_k141_31308\|\|full | Not-determined |
| 18-105-final-viral-combined_k141_138283\|\|full | Low-quality |
| 18-105-final-viral-combined_k141_112080\|\|full | Not-determined |
| 18-105-final-viral-combined_k141_192161\|\|full | Low-quality |
| 18-105-final-viral-combined_k141_58650\|\|full | Not-determined |
| 18-105-final-viral-combined_k141_200992\|\|full | Low-quality |
| 18-105-final-viral-combined_k141_192195\|\|full | Low-quality |
| 18-105-final-viral-combined_k141_58658\|\|full | Not-determined |
| 18-105-final-viral-combined_k141_58660\|\|full | Low-quality |
| 18-105-final-viral-combined_k141_165196\|\|full | Low-quality |
| 18-105-final-viral-combined_k141_22059\|\|full | Low-quality |
| 18-105-final-viral-combined_k141_58671\|\|full | Low-quality |
| 18-105-final-viral-combined_k141_129972\|\|full | Not-determined |
| 18-105-final-viral-combined_k141_138355\|\|full | Low-quality |
| 18-105-final-viral-combined_k141_138364\|\|full | Low-quality |
| 18-105-final-viral-combined_k141_40700\|\|full | Not-determined |
| 18-105-final-viral-combined_k141_120443\|\|full | Not-determined |
| 18-105-final-viral-combined_k141_165218\|\|full | Low-quality |
| 18-105-final-viral-combined_k141_165230\|\|full | Not-determined |
| 18-105-final-viral-combined_k141_40719\|\|full | Low-quality |
| 18-105-final-viral-combined_k141_22081\|\|full | Low-quality |
| 18-105-final-viral-combined_k141_58696\|\|full | Not-determined |
| 18-105-final-viral-combined_k141_130016\|\|full | Not-determined |
| 18-105-final-viral-combined_k141_156180\|\|full | Low-quality |
| 18-105-final-viral-combined_k141_210140\|\|full | Not-determined |
| 18-105-final-viral-combined_k141_31425\|\|full | Not-determined |
| 18-105-final-viral-combined_k141_156204\|\|full | Low-quality |
| 18-105-final-viral-combined_k141_138406\|\|full | Not-determined |
| 18-105-final-viral-combined_k141_210162\|\|full | Low-quality |
| 18-105-final-viral-combined_k141_67108\|\|full | Low-quality |
| 18-105-final-viral-combined_k141_138420\|\|full | Not-determined |
| 18-105-final-viral-combined_k141_103467\|\|full | Low-quality |
| 18-105-final-viral-combined_k141_40810\|\|full | Low-quality |
| 18-105-final-viral-combined_k141_58757\|\|full | Low-quality |
| 18-105-final-viral-combined_k141_103476\|\|full | Low-quality |
| 18-105-final-viral-combined_k141_183544\|\|full | Low-quality |
| 18-105-final-viral-combined_k141_183550\|\|full | Not-determined |
| 18-105-final-viral-combined_k141_14312\|\|full | Not-determined |
| 18-105-final-viral-combined_k141_130069\|\|full | Low-quality |
| 18-105-final-viral-combined_k141_40858\|\|full | Low-quality |
| 18-105-final-viral-combined_k141_49741\|\|full | Low-quality |
| 18-105-final-viral-combined_k141_58805\|\|full | Low-quality |
| 18-105-final-viral-combined_k141_40859\|\|full | Not-determined |
| 18-105-final-viral-combined_k141_210186\|\|full | Low-quality |
| 18-105-final-viral-combined_k141_165286\|\|full | Not-determined |
| 18-105-final-viral-combined_k141_165290\|\|full | Not-determined |
| 18-105-final-viral-combined_k141_174899\|\|full | Low-quality |
| 18-105-final-viral-combined_k141_58837\|\|full | Low-quality |
| 18-105-final-viral-combined_k141_165309\|\|full | Not-determined |
| 18-105-final-viral-combined_k141_94195\|\|full | Low-quality |
| 18-105-final-viral-combined_k141_174903\|\|full | Not-determined |
| 18-105-final-viral-combined_k141_201112\|\|full | Low-quality |
| 18-105-final-viral-combined_k141_94202\|\|full | Not-determined |
| 18-105-final-viral-combined_k141_7075\|\|full | Not-determined |
| 18-105-final-viral-combined_k141_94213\|\|full | Not-determined |
| 18-105-final-viral-combined_k141_130088\|\|full | Low-quality |
| 18-105-final-viral-combined_k141_183626\|\|full | Not-determined |
| 18-105-final-viral-combined_k141_138443\|\|full | Not-determined |
| 18-105-final-viral-combined_k141_138451\|\|full | Not-determined |
| 18-105-final-viral-combined_k141_192308\|\|full | Not-determined |
| 18-105-final-viral-combined_k141_138457\|\|full | Not-determined |
| 18-105-final-viral-combined_k141_22188\|\|full | Low-quality |
| 18-105-final-viral-combined_k141_130109\|\|full | Not-determined |
| 18-105-final-viral-combined_k141_40893\|\|full | Low-quality |
| 18-105-final-viral-combined_k141_103598\|\|full | Low-quality |
| 18-105-final-viral-combined_k141_94235\|\|full | Low-quality |
| 18-105-final-viral-combined_k141_112096\|\|full | Low-quality |
| 18-105-final-viral-combined_k141_138488\|\|full | Low-quality |
| 18-105-final-viral-combined_k141_31466\|\|full | Not-determined |
| 18-105-final-viral-combined_k141_130132\|\|full | Not-determined |
| 18-105-final-viral-combined_k141_192370\|\|full | Not-determined |
| 18-105-final-viral-combined_k141_58896\|\|full | Not-determined |
| 18-105-final-viral-combined_k141_174931\|\|full | Low-quality |
| 18-105-final-viral-combined_k141_120681\|\|full | Low-quality |
| 18-105-final-viral-combined_k141_14342\|\|full | Not-determined |
| 18-101_k141_296850 | High-quality |
| 18-105-final-viral-combined_k141_58925\|\|full | Low-quality |
| 18-105-final-viral-combined_k141_14360\|\|full | Not-determined |
| 18-105-final-viral-combined_k141_147825\|\|full | Low-quality |
| 18-105-final-viral-combined_k141_112138\|\|full | Not-determined |
| 18-105-final-viral-combined_k141_49794\|\|full | Low-quality |
| 18-105-final-viral-combined_k141_192433\|\|full | Low-quality |
| 18-105-final-viral-combined_k141_138505\|\|full | Low-quality |
| 18-105-final-viral-combined_k141_103666\|\|full | Not-determined |
| 18-105-final-viral-combined_k141_103678\|\|full | Not-determined |
| 18-105-final-viral-combined_k141_130216\|\|full | Low-quality |
| 18-105-final-viral-combined_k141_67259\|\|full | Not-determined |
| 18-105-final-viral-combined_k141_22231\|\|full | Not-determined |
| 18-105-final-viral-combined_k141_31549\|\|full | Not-determined |
| 18-105-final-viral-combined_k141_31558\|\|full | Not-determined |
| 18-105-final-viral-combined_k141_192475\|\|full | Low-quality |
| 18-105-final-viral-combined_k141_59015\|\|full | Not-determined |
| 18-105-final-viral-combined_k141_138575\|\|full | Low-quality |
| 18-105-final-viral-combined_k141_59030\|\|full | Not-determined |
| 18-105-final-viral-combined_k141_138584\|\|full | Low-quality |
| 18-105-final-viral-combined_k141_112198\|\|full | Low-quality |
| 18-101-final-viral-combined_k141_296850\|\|full | High-quality |
| 18-105-final-viral-combined_k141_130269\|\|full | Not-determined |
| 18-105-final-viral-combined_k141_112209\|\|full | Low-quality |
| 18-105-final-viral-combined_k141_130278\|\|full | Not-determined |
| 18-105-final-viral-combined_k141_120718\|\|full | Low-quality |
| 18-105-final-viral-combined_k141_112226\|\|full | Not-determined |
| 18-105-final-viral-combined_k141_103740\|\|full | Not-determined |
| 18-105-final-viral-combined_k141_7180\|\|full | Low-quality |
| 18-105-final-viral-combined_k141_94374\|\|full | Low-quality |
| 18-105-final-viral-combined_k141_31647\|\|full | Low-quality |
| 18-105-final-viral-combined_k141_138611\|\|full | Low-quality |
| 18-105-final-viral-combined_k141_130314\|\|full | Low-quality |
| 18-105-final-viral-combined_k141_156225\|\|full | Not-determined |
| 18-105-final-viral-combined_k141_103771\|\|full | Low-quality |
| 18-105-final-viral-combined_k141_22282\|\|full | Low-quality |
| 18-105-final-viral-combined_k141_175060\|\|full | Not-determined |
| 18-105-final-viral-combined_k141_41007\|\|full | Not-determined |
| 18-105-final-viral-combined_k141_85372\|\|full | Not-determined |
| 18-105-final-viral-combined_k141_192521\|\|full | Not-determined |
| 18-105-final-viral-combined_k141_94393\|\|full | Not-determined |
| 18-105-final-viral-combined_k141_94395\|\|full | Not-determined |
| 18-105-final-viral-combined_k141_156246\|\|full | Low-quality |
| 18-105-final-viral-combined_k141_103790\|\|full | Not-determined |
| 18-105-final-viral-combined_k141_49840\|\|full | Low-quality |
| 18-105-final-viral-combined_k141_138631\|\|full | Low-quality |
| 18-105-final-viral-combined_k141_156266\|\|full | Not-determined |
| 18-105-final-viral-combined_k141_192548\|\|full | Low-quality |
| 18-105-final-viral-combined_k141_201136\|\|full | Not-determined |
| 18-105-final-viral-combined_k141_22328\|\|full | Not-determined |
| 18-105-final-viral-combined_k141_130367\|\|full | Not-determined |
| 18-105-final-viral-combined_k141_192569\|\|full | Low-quality |
| 18-105-final-viral-combined_k141_41053\|\|full | Low-quality |
| 18-105-final-viral-combined_k141_22342\|\|full | Not-determined |
| 18-105-final-viral-combined_k141_192570\|\|full | Low-quality |
| 18-105-final-viral-combined_k141_201140\|\|full | Low-quality |
| 18-105-final-viral-combined_k141_112282\|\|full | Low-quality |
| 18-105-final-viral-combined_k141_183734\|\|full | Not-determined |
| 18-105-final-viral-combined_k141_156316\|\|full | Not-determined |
| 18-105-final-viral-combined_k141_7254\|\|full | Low-quality |
| 18-105-final-viral-combined_k141_94422\|\|full | Low-quality |
| 18-105-final-viral-combined_k141_41102\|\|full | Low-quality |
| 18-105-final-viral-combined_k141_103844\|\|full | Low-quality |
| KM-final-viral-combined_k141_247494\|\|full | Medium-quality |
| 18-105-final-viral-combined_k141_22371\|\|full | Low-quality |
| 18-105-final-viral-combined_k141_7267\|\|full | Low-quality |
| 18-105-final-viral-combined_k141_103851\|\|full | Not-determined |
| 18-105-final-viral-combined_k141_130402\|\|full | Not-determined |
| 18-105-final-viral-combined_k141_103856\|\|full | Low-quality |
| 17-108-final-viral-combined_k141_74983\|\|full | High-quality |
| 18-105-final-viral-combined_k141_85483\|\|full | Not-determined |
| 18-105-final-viral-combined_k141_103877\|\|full | Low-quality |
| 18-105-final-viral-combined_k141_112352\|\|full | Low-quality |
| 18-105-final-viral-combined_k141_156346\|\|full | Low-quality |
| 17-109-final-viral-combined_k141_91231\|\|full | Medium-quality |
| 18-105-final-viral-combined_k141_85510\|\|full | Low-quality |
| 18-105-final-viral-combined_k141_67348\|\|full | Not-determined |
| 18-105-final-viral-combined_k141_14430\|\|full | Low-quality |
| 18-105-final-viral-combined_k141_201161\|\|full | Not-determined |
| 18-105-final-viral-combined_k141_201163\|\|full | Low-quality |
| 18-105-final-viral-combined_k141_183794\|\|full | Not-determined |
| 18-105-final-viral-combined_k141_22407\|\|full | Not-determined |
| 18-105-final-viral-combined_k141_22408\|\|full | Not-determined |
| 18-105-final-viral-combined_k141_130470\|\|full | Low-quality |
| 18-105-final-viral-combined_k141_201203\|\|full | Low-quality |
| 18-102-final-viral-combined_k141_113619\|\|full | High-quality |
| 18-105-final-viral-combined_k141_14477\|\|full | Low-quality |
| 18-105-final-viral-combined_k141_31717\|\|full | Not-determined |
| 18-105-final-viral-combined_k141_14479\|\|full | Low-quality |
| 18-105-final-viral-combined_k141_7351\|\|full | Low-quality |
| 18-105-final-viral-combined_k141_49966\|\|full | Low-quality |
| 18-105-final-viral-combined_k141_41177\|\|full | Low-quality |
| 18-105-final-viral-combined_k141_14487\|\|full | Low-quality |
| 18-105-final-viral-combined_k141_148128\|\|full | Low-quality |
| 18-105-final-viral-combined_k141_31738\|\|full | Not-determined |
| 18-105-final-viral-combined_k141_201240\|\|full | Low-quality |
| 18-105-final-viral-combined_k141_85616\|\|full | Low-quality |
| 18-105-final-viral-combined_k141_31747\|\|full | Low-quality |
| 18-105-final-viral-combined_k141_175117\|\|full | Low-quality |
| 18-105-final-viral-combined_k141_85623\|\|full | Low-quality |
| 18-105-final-viral-combined_k141_31751\|\|full | Low-quality |
| 18-105-final-viral-combined_k141_7375\|\|full | Not-determined |
| 18-105-final-viral-combined_k141_76987\|\|full | Low-quality |
| 18-105-final-viral-combined_k141_31755\|\|full | Low-quality |
| 18-105-final-viral-combined_k141_59092\|\|full | Not-determined |
| 18-105-final-viral-combined_k141_85629\|\|full | Low-quality |
| 18-105-final-viral-combined_k141_201264\|\|full | Low-quality |
| 18-105-final-viral-combined_k141_77000\|\|full | Not-determined |
| 18-105-final-viral-combined_k141_156360\|\|full | Low-quality |
| 18-105-final-viral-combined_k141_148170\|\|full | Not-determined |
| 18-105-final-viral-combined_k141_148179\|\|full | Not-determined |
| 18-105-final-viral-combined_k141_183888\|\|full | Not-determined |
| 18-105-final-viral-combined_k141_120928\|\|full | Low-quality |
| 18-105-final-viral-combined_k141_31776\|\|full | Low-quality |
| 18-105-final-viral-combined_k141_77012\|\|full | Low-quality |
| 18-105-final-viral-combined_k141_183894\|\|full | Low-quality |
| 18-105-final-viral-combined_k141_50042\|\|full | Not-determined |
| 18-105-final-viral-combined_k141_138727\|\|full | Low-quality |
| 18-105-final-viral-combined_k141_41229\|\|full | Not-determined |
| 18-102-final-viral-combined_k141_52568\|\|full | Complete |
| 18-105-final-viral-combined_k141_210217\|\|full | Low-quality |
| 18-105-final-viral-combined_k141_103943\|\|full | Low-quality |
| 18-105-final-viral-combined_k141_77051\|\|full | Low-quality |
| 18-105-final-viral-combined_k141_210219\|\|full | Low-quality |
| 18-105-final-viral-combined_k141_41244\|\|full | Low-quality |
| 18-105-final-viral-combined_k141_183935\|\|full | Not-determined |
| 18-105-final-viral-combined_k141_103958\|\|full | Low-quality |
| 18-105-final-viral-combined_k141_201316\|\|full | Not-determined |
| 18-105-final-viral-combined_k141_7442\|\|full | Not-determined |
| 17-108-final-viral-combined_k141_60498\|\|full | High-quality |
| 18-105-final-viral-combined_k141_156388\|\|full | Not-determined |
| 18-105-final-viral-combined_k141_201331\|\|full | Not-determined |
| 18-105-final-viral-combined_k141_31845\|\|full | Low-quality |
| 18-105-final-viral-combined_k141_103989\|\|full | Low-quality |
| 18-105-final-viral-combined_k141_67484\|\|full | Low-quality |
| 18-105-final-viral-combined_k141_165363\|\|full | Low-quality |
| 18-105-final-viral-combined_k141_59151\|\|full | Low-quality |
| 18-105-final-viral-combined_k141_7475\|\|full | Low-quality |
| 18-105-final-viral-combined_k141_165371\|\|full | Low-quality |
| 18-105-final-viral-combined_k141_31859\|\|full | Not-determined |
| 18-105-final-viral-combined_k141_50090\|\|full | Not-determined |
| 18-105-final-viral-combined_k141_67496\|\|full | Not-determined |
| 18-105-final-viral-combined_k141_31861\|\|full | Low-quality |
| 18-105-final-viral-combined_k141_31866\|\|full | Not-determined |
| 18-105-final-viral-combined_k141_14523\|\|full | Not-determined |
| 18-105-final-viral-combined_k141_104005\|\|full | Not-determined |
| 18-105-final-viral-combined_k141_7484\|\|full | Not-determined |
| 18-105-final-viral-combined_k141_156407\|\|full | Not-determined |
| 18-105-final-viral-combined_k141_50112\|\|full | Not-determined |
| 18-105-final-viral-combined_k141_156411\|\|full | Low-quality |
| 18-105-final-viral-combined_k141_156417\|\|full | Not-determined |
| 18-105-final-viral-combined_k141_165418\|\|full | Not-determined |
| 18-105-final-viral-combined_k141_67520\|\|full | Low-quality |
| 18-105-final-viral-combined_k141_67522\|\|full | Not-determined |
| 18-105-final-viral-combined_k141_121017\|\|full | Not-determined |
| 18-105-final-viral-combined_k141_112413\|\|full | Low-quality |
| 18-105-final-viral-combined_k141_156442\|\|full | Not-determined |
| 18-105-final-viral-combined_k141_104054\|\|full | Not-determined |
| 18-105-final-viral-combined_k141_121042\|\|full | Not-determined |
| 18-105-final-viral-combined_k141_175320\|\|full | Low-quality |
| 18-105-final-viral-combined_k141_67561\|\|full | Not-determined |
| 18-105-final-viral-combined_k141_22504\|\|full | Not-determined |
| 18-105-final-viral-combined_k141_201369\|\|full | Low-quality |
| 18-105-final-viral-combined_k141_22508\|\|full | Low-quality |
| 18-105-final-viral-combined_k141_192742\|\|full | Low-quality |
| 18-105-final-viral-combined_k141_148244\|\|full | Not-determined |
| 18-105-final-viral-combined_k141_112417\|\|full | Low-quality |
| 18-105-final-viral-combined_k141_112418\|\|full | Low-quality |
| 17-108-final-viral-combined_k141_48246\|\|full | Medium-quality |
| 18-105-final-viral-combined_k141_14604\|\|full | Low-quality |
| 18-105-final-viral-combined_k141_94485\|\|full | Not-determined |
| 18-105-final-viral-combined_k141_138839\|\|full | Not-determined |
| 18-105-final-viral-combined_k141_112424\|\|full | Low-quality |
| 18-105-final-viral-combined_k141_50201\|\|full | Low-quality |
| 18-105-final-viral-combined_k141_183940\|\|full | Not-determined |
| 18-105-final-viral-combined_k141_183967\|\|full | Low-quality |
| 18-105-final-viral-combined_k141_210245\|\|full | Not-determined |
| 18-105-final-viral-combined_k141_138888\|\|full | Not-determined |
| 18-105-final-viral-combined_k141_41338\|\|full | Low-quality |
| 18-105-final-viral-combined_k141_183973\|\|full | Not-determined |
| 18-105-final-viral-combined_k141_41341\|\|full | Low-quality |
| 18-105-final-viral-combined_k141_31992\|\|full | Low-quality |
| 18-105-final-viral-combined_k141_165494\|\|full | Not-determined |
| 18-105-final-viral-combined_k141_138903\|\|full | Low-quality |
| 18-105-final-viral-combined_k141_32004\|\|full | Low-quality |
| 18-105-final-viral-combined_k141_175353\|\|full | Low-quality |
| 18-105-final-viral-combined_k141_85727\|\|full | Not-determined |
| 18-105-final-viral-combined_k141_165525\|\|full | Low-quality |
| 18-105-final-viral-combined_k141_50225\|\|full | Not-determined |
| 18-105-final-viral-combined_k141_94495\|\|full | Not-determined |
| 18-105-final-viral-combined_k141_94501\|\|full | Not-determined |
| 18-105-final-viral-combined_k141_112483\|\|full | Not-determined |
| 18-105-final-viral-combined_k141_210309\|\|full | Not-determined |
| 18-105-final-viral-combined_k141_175391\|\|full | Not-determined |
| 18-105-final-viral-combined_k141_32050\|\|full | Not-determined |
| 18-105-final-viral-combined_k141_50254\|\|full | Not-determined |
| 18-105-final-viral-combined_k141_41387\|\|full | Not-determined |
| 18-105-final-viral-combined_k141_77233\|\|full | Not-determined |
| 18-105-final-viral-combined_k141_77236\|\|full | Low-quality |
| 18-105-final-viral-combined_k141_175421\|\|full | Low-quality |
| 18-105-final-viral-combined_k141_7525\|\|full | Not-determined |
| 18-105-final-viral-combined_k141_14663\|\|full | Not-determined |
| 18-105-final-viral-combined_k141_77257\|\|full | Low-quality |
| 18-105-final-viral-combined_k141_210316\|\|full | Low-quality |
| 18-105-final-viral-combined_k141_130729\|\|full | Low-quality |
| 18-105-final-viral-combined_k141_148305\|\|full | Not-determined |
| 18-105-final-viral-combined_k141_156555\|\|full | Not-determined |
| 18-105-final-viral-combined_k141_192791\|\|full | Low-quality |
| 18-105-final-viral-combined_k141_184036\|\|full | Not-determined |
| 18-105-final-viral-combined_k141_112539\|\|full | Not-determined |
| 18-105-final-viral-combined_k141_138998\|\|full | Not-determined |
| 18-105-final-viral-combined_k141_165592\|\|full | Not-determined |
| 18-105-final-viral-combined_k141_148328\|\|full | Not-determined |
| 18-105-final-viral-combined_k141_121134\|\|full | Low-quality |
| 18-105-final-viral-combined_k141_85745\|\|full | Low-quality |
| 18-105-final-viral-combined_k141_14703\|\|full | Low-quality |
| 18-105-final-viral-combined_k141_175449\|\|full | Not-determined |
| 18-105-final-viral-combined_k141_112570\|\|full | Not-determined |
| 18-105-final-viral-combined_k141_210336\|\|full | Low-quality |
| 18-105-final-viral-combined_k141_139013\|\|full | Not-determined |
| 18-105-final-viral-combined_k141_210346\|\|full | Not-determined |
| 18-105-final-viral-combined_k141_22536\|\|full | Low-quality |
| 18-105-final-viral-combined_k141_67649\|\|full | Not-determined |
| 18-105-final-viral-combined_k141_130782\|\|full | Not-determined |
| 18-105-final-viral-combined_k141_112605\|\|full | Not-determined |
| 18-105-final-viral-combined_k141_85750\|\|full | Low-quality |
| 18-105-final-viral-combined_k141_130796\|\|full | Low-quality |
| 18-105-final-viral-combined_k141_112612\|\|full | Low-quality |

18-105-final-viral-combined_k141_7576||full Low-quality
18-105-final-viral-combined_k141_14717||full Not-determined
18-105-final-viral-combined_k141_148381||full Not-determined
18-105-final-viral-combined_k141_77347||full Low-quality
18-105-final-viral-combined_k141_130813||full Not-determined
18-105-final-viral-combined_k141_148385||full Not-determined
18-105-final-viral-combined_k141_59278||full Low-quality
18-105-final-viral-combined_k141_59279||full Low-quality
18-105-final-viral-combined_k141_32112||full Low-quality
18-105-final-viral-combined_k141_104100||full Low-quality
18-105-final-viral-combined_k141_59281||full Low-quality
18-105-final-viral-combined_k141_7601||full Low-quality
18-105-final-viral-combined_k141_77383||full Not-determined
18-105-final-viral-combined_k141_148441||full Low-quality
18-105-final-viral-combined_k141_139079||full Low-quality
18-105-final-viral-combined_k141_77389||full Low-quality
18-105-final-viral-combined_k141_94576||full Low-quality
18-105-final-viral-combined_k141_77393||full Low-quality
18-105-final-viral-combined_k141_32173||full Low-quality
18-105-final-viral-combined_k141_165673||full Not-determined
18-105-final-viral-combined_k141_50341||full Not-determined
18-105-final-viral-combined_k141_7633||full Low-quality
18-105-final-viral-combined_k141_148491||full Low-quality
17-108-final-viral-combined_k141_92114||full Medium-quality
18-105-final-viral-combined_k141_50347||full Low-quality
18-105-final-viral-combined_k141_14816||full Not-determined
18-105-final-viral-combined_k141_184196||full Not-determined
18-105-final-viral-combined_k141_50381||full Low-quality
18-105-final-viral-combined_k141_192940||full Low-quality
18-105-final-viral-combined_k141_14842||full Not-determined
18-105-final-viral-combined_k141_50392||full Not-determined
18-105-final-viral-combined_k141_139118||full Low-quality
18-105-final-viral-combined_k141_148514||full Not-determined
18-105-final-viral-combined_k141_112708||full Low-quality
18-105-final-viral-combined_k141_77455||full Not-determined
18-105-final-viral-combined_k141_184259||full Low-quality
18-105-final-viral-combined_k141_32196||full Not-determined
18-105-final-viral-combined_k141_104159||full Low-quality
18-105-final-viral-combined_k141_201583||full Low-quality
18-105-final-viral-combined_k141_77464||full Low-quality
18-105-final-viral-combined_k141_7693||full Not-determined
18-102_k141_248324 Complete
18-105-final-viral-combined_k141_156719||full Low-quality
18-105-final-viral-combined_k141_41457||full Not-determined
18-105-final-viral-combined_k141_7707||full Not-determined
18-105-final-viral-combined_k141_184297||full Not-determined
18-105-final-viral-combined_k141_104174||full Not-determined
18-105-final-viral-combined_k141_32200||full Not-determined
18-105-final-viral-combined_k141_32205||full Low-quality
18-105-final-viral-combined_k141_104187||full Low-quality
18-105-final-viral-combined_k141_175534||full Not-determined
18-105-final-viral-combined_k141_210474||full Not-determined
18-105-final-viral-combined_k141_7736||full Low-quality
18-105-final-viral-combined_k141_32221||full Low-quality
18-105-final-viral-combined_k141_59394||full Not-determined
18-105-final-viral-combined_k141_156737||full Not-determined
18-105-final-viral-combined_k141_94681||full Low-quality
18-105-final-viral-combined_k141_59403||full Not-determined
18-105-final-viral-combined_k141_184336||full Not-determined
18-105-final-viral-combined_k141_77522||full Not-determined
18-105-final-viral-combined_k141_112779||full Not-determined
18-105-final-viral-combined_k141_77527||full Low-quality
18-105-final-viral-combined_k141_22677||full Not-determined
18-105-final-viral-combined_k141_59423||full Not-determined
18-105-final-viral-combined_k141_32263||full Not-determined
18-105-final-viral-combined_k141_121263||full Low-quality
18-105-final-viral-combined_k141_156754||full Low-quality
18-105-final-viral-combined_k141_112787||full Low-quality
18-105-final-viral-combined_k141_32279||full Not-determined
18-105-final-viral-combined_k141_175574||full Low-quality
18-105-final-viral-combined_k141_121290||full Low-quality
18-105-final-viral-combined_k141_77539||full Low-quality
18-105-final-viral-combined_k141_121305||full Not-determined
18-105-final-viral-combined_k141_94693||full Low-quality
18-105-final-viral-combined_k141_32289||full Not-determined
18-105-final-viral-combined_k141_77551||full Not-determined
18-105-final-viral-combined_k141_156765||full Not-determined
18-105-final-viral-combined_k141_184365||full Low-quality
18-105-final-viral-combined_k141_59431||full Not-determined
18-105-final-viral-combined_k141_156784||full Low-quality
18-105-final-viral-combined_k141_139224||full Low-quality
18-105-final-viral-combined_k141_131107||full Not-determined
18-105-final-viral-combined_k141_201612||full Not-determined
18-105-final-viral-combined_k141_148537||full Low-quality
18-105-final-viral-combined_k141_156791||full Not-determined
18-105-final-viral-combined_k141_131113||full Low-quality
18-105-final-viral-combined_k141_32313||full Low-quality
18-105-final-viral-combined_k141_7786||full Low-quality
18-105-final-viral-combined_k141_148560||full Not-determined
18-105-final-viral-combined_k141_201657||full Low-quality
18-105-final-viral-combined_k141_201658||full Not-determined
18-105-final-viral-combined_k141_112805||full Not-determined
18-105-final-viral-combined_k141_22746||full Not-determined
18-105-final-viral-combined_k141_175602||full Not-determined
18-105-final-viral-combined_k141_67775||full Low-quality
18-105-final-viral-combined_k141_32326||full Not-determined
18-105-final-viral-combined_k141_175617||full Not-determined
18-105-final-viral-combined_k141_184449||full Low-quality
18-105-final-viral-combined_k141_59514||full Low-quality
18-105-final-viral-combined_k141_148626||full Not-determined
18-105-final-viral-combined_k141_32361||full Low-quality
18-105-final-viral-combined_k141_32362||full Not-determined
18-105-final-viral-combined_k141_184484||full Not-determined
18-105-final-viral-combined_k141_112842||full Low-quality
18-105-final-viral-combined_k141_14876||full Not-determined
18-105-final-viral-combined_k141_59546||full Not-determined
18-105-final-viral-combined_k141_156921||full Low-quality
18-105-final-viral-combined_k141_156922||full Low-quality
18-105-final-viral-combined_k141_32393||full Not-determined
18-105-final-viral-combined_k141_184522||full Low-quality
18-105-final-viral-combined_k141_86108||full Not-determined
18-105-final-viral-combined_k141_59565||full Low-quality
18-105-final-viral-combined_k141_148680||full Not-determined
18-105-final-viral-combined_k141_184545||full Low-quality
18-105-final-viral-combined_k141_184548||full Not-determined
18-105-final-viral-combined_k141_175654||full Low-quality
18-105-final-viral-combined_k141_86134||full Low-quality
18-105-final-viral-combined_k141_77674||full Low-quality
18-105-final-viral-combined_k141_165977||full Not-determined
18-106-final-viral-combined_k141_130995||full Medium-quality
18-105-final-viral-combined_k141_50475||full Low-quality
18-105-final-viral-combined_k141_59595||full Not-determined
18-105-final-viral-combined_k141_148741||full Not-determined
18-105-final-viral-combined_k141_86160||full Low-quality
18-105-final-viral-combined_k141_210687||full Not-determined
18-105-final-viral-combined_k141_14956||full Low-quality
18-105-final-viral-combined_k141_86182||full Low-quality
18-105-final-viral-combined_k141_184591||full Low-quality
18-105-final-viral-combined_k141_184608||full Not-determined
18-105-final-viral-combined_k141_22839||full Not-determined
18-105-final-viral-combined_k141_22843||full Low-quality
18-105-final-viral-combined_k141_131244||full Low-quality
18-105-final-viral-combined_k141_22856||full Low-quality
18-105-final-viral-combined_k141_32405||full Low-quality
18-105-final-viral-combined_k141_50516||full Not-determined
18-105-final-viral-combined_k141_86216||full Not-determined
18-105-final-viral-combined_k141_193228||full Low-quality
18-105-final-viral-combined_k141_86227||full Not-determined
18-105-final-viral-combined_k141_193234||full Not-determined
18-105-final-viral-combined_k141_32437||full Not-determined
18-105-final-viral-combined_k141_131291||full Not-determined
18-105-final-viral-combined_k141_7842||full Not-determined
18-105-final-viral-combined_k141_156932||full Low-quality
18-105-final-viral-combined_k141_50546||full Low-quality
18-105-final-viral-combined_k141_156935||full Not-determined
18-105-final-viral-combined_k141_193235||full Low-quality
18-105-final-viral-combined_k141_184689||full Low-quality
18-105-final-viral-combined_k141_193241||full Low-quality
18-105-final-viral-combined_k141_104419||full Not-determined
18-105-final-viral-combined_k141_22918||full Low-quality
18-105-final-viral-combined_k141_166059||full Low-quality
18-105-final-viral-combined_k141_22926||full Low-quality
18-105-final-viral-combined_k141_22930||full Low-quality
18-105-final-viral-combined_k141_166071||full Not-determined
18-105-final-viral-combined_k141_201802||full Not-determined
18-105-final-viral-combined_k141_193288||full Low-quality
18-105-final-viral-combined_k141_86301||full Not-determined
18-105-final-viral-combined_k141_131338||full Not-determined
18-105-final-viral-combined_k141_59689||full Not-determined
18-105-final-viral-combined_k141_156995||full Low-quality
18-105-final-viral-combined_k141_86307||full Not-determined
18-105-final-viral-combined_k141_7867||full Low-quality
18-105-final-viral-combined_k141_121501||full Low-quality
18-105-final-viral-combined_k141_50634||full Not-determined
18-105-final-viral-combined_k141_41563||full Not-determined
18-105-final-viral-combined_k141_201845||full Low-quality
18-105-final-viral-combined_k141_77751||full Not-determined
18-105-final-viral-combined_k141_121526||full Low-quality
18-105-final-viral-combined_k141_7896||full Low-quality
18-105-final-viral-combined_k141_193295||full Not-determined
18-105-final-viral-combined_k141_166114||full Not-determined
18-105-final-viral-combined_k141_157067||full Not-determined
18-105-final-viral-combined_k141_77794||full Not-determined
18-105-final-viral-combined_k141_59720||full Not-determined
18-105-final-viral-combined_k141_32467||full Low-quality
18-105-final-viral-combined_k141_201879||full Not-determined
18-105-final-viral-combined_k141_77806||full Not-determined
18-105-final-viral-combined_k141_22955||full Not-determined
18-105-final-viral-combined_k141_86376||full Not-determined
18-105-final-viral-combined_k141_121569||full Low-quality
18-105-final-viral-combined_k141_22963||full Low-quality
18-105-final-viral-combined_k141_166174||full Not-determined
18-105-final-viral-combined_k141_77824||full Low-quality
18-105-final-viral-combined_k141_112977||full Low-quality
18-105-final-viral-combined_k141_210889||full Not-determined
18-105-final-viral-combined_k141_94836||full Low-quality
18-105-final-viral-combined_k141_50715||full Not-determined
18-105-final-viral-combined_k141_175936||full Low-quality
18-105-final-viral-combined_k141_22994||full Low-quality
18-105-final-viral-combined_k141_32508||full Low-quality
18-105-final-viral-combined_k141_175940||full Not-determined
18-105-final-viral-combined_k141_201915||full Low-quality
18-105-final-viral-combined_k141_59765||full Low-quality
18-105-final-viral-combined_k141_166186||full Not-determined
18-105-final-viral-combined_k141_131354||full Not-determined
18-105-final-viral-combined_k141_50733||full Not-determined
18-105-final-viral-combined_k141_201933||full Not-determined
18-105-final-viral-combined_k141_148888||full Low-quality
18-105-final-viral-combined_k141_148894||full Low-quality
18-105-final-viral-combined_k141_50747||full Not-determined
18-105-final-viral-combined_k141_175963||full Not-determined
18-105-final-viral-combined_k141_94860||full Low-quality
18-105-final-viral-combined_k141_77881||full Low-quality
18-105-final-viral-combined_k141_32538||full Not-determined
18-105-final-viral-combined_k141_32542||full Not-determined
18-105-final-viral-combined_k141_32547||full Not-determined
18-105-final-viral-combined_k141_210958||full Not-determined
18-105-final-viral-combined_k141_15036||full Not-determined
18-105-final-viral-combined_k141_50777||full Not-determined
18-105-final-viral-combined_k141_67901||full Not-determined
18-105-final-viral-combined_k141_59844||full Low-quality
18-105-final-viral-combined_k141_67910||full Low-quality
18-105-final-viral-combined_k141_15066||full Low-quality
18-105-final-viral-combined_k141_176047||full Low-quality
18-105-final-viral-combined_k141_131454||full Not-determined
18-105-final-viral-combined_k141_67929||full Low-quality
18-105-final-viral-combined_k141_32559||full Low-quality
18-105-final-viral-combined_k141_176065||full Low-quality
18-105-final-viral-combined_k141_15084||full Low-quality
18-105-final-viral-combined_k141_15099||full Not-determined
18-105-final-viral-combined_k141_201988||full Not-determined
18-105-final-viral-combined_k141_157187||full Low-quality
18-105-final-viral-combined_k141_148915||full Low-quality
18-105-final-viral-combined_k141_32568||full Not-determined
18-105-final-viral-combined_k141_113072||full Not-determined
18-105-final-viral-combined_k141_77911||full Low-quality
18-105-final-viral-combined_k141_50820||full Not-determined
18-105-final-viral-combined_k141_67957||full Low-quality
18-105-final-viral-combined_k141_184868||full Not-determined
18-105-final-viral-combined_k141_202004||full Low-quality
18-105-final-viral-combined_k141_176074||full Low-quality
18-105-final-viral-combined_k141_67965||full Low-quality
18-105-final-viral-combined_k141_121740||full Not-determined
18-105-final-viral-combined_k141_67977||full Low-quality
18-105-final-viral-combined_k141_50861||full Low-quality
18-105-final-viral-combined_k141_50868||full Low-quality
18-105-final-viral-combined_k141_15121||full Not-determined
18-105-final-viral-combined_k141_15124||full Not-determined
18-105-final-viral-combined_k141_166252||full Not-determined
18-105-final-viral-combined_k141_131545||full Low-quality
18-105-final-viral-combined_k141_113139||full Low-quality
18-105-final-viral-combined_k141_41618||full Low-quality
18-105-final-viral-combined_k141_41626||full Not-determined
18-105-final-viral-combined_k141_166271||full Not-determined
18-105-final-viral-combined_k141_32588||full Low-quality
18-105-final-viral-combined_k141_121780||full Low-quality
18-105-final-viral-combined_k141_104557||full Low-quality
18-105-final-viral-combined_k141_68003||full Not-determined
18-105-final-viral-combined_k141_139537||full Low-quality
18-105-final-viral-combined_k141_32593||full Not-determined
18-105-final-viral-combined_k141_157278||full Low-quality
18-105-final-viral-combined_k141_50904||full Low-quality
18-105-final-viral-combined_k141_15134||full Not-determined
18-105-final-viral-combined_k141_157286||full Not-determined
18-105-final-viral-combined_k141_50915||full Not-determined
18-105-final-viral-combined_k141_50918||full Not-determined
18-105-final-viral-combined_k141_50921||full Not-determined
18-105-final-viral-combined_k141_139568||full Low-quality
18-105-final-viral-combined_k141_78030||full Not-determined
18-105-final-viral-combined_k141_23094||full Low-quality
18-105-final-viral-combined_k141_139573||full Low-quality
18-105-final-viral-combined_k141_68012||full Low-quality
18-105-final-viral-combined_k141_23106||full Not-determined
18-105-final-viral-combined_k141_113189||full Low-quality
18-105-final-viral-combined_k141_68040||full Low-quality
18-105-final-viral-combined_k141_50955||full Not-determined
18-105-final-viral-combined_k141_104587||full Low-quality
18-105-final-viral-combined_k141_41682||full Low-quality
18-105-final-viral-combined_k141_121872||full Low-quality
18-105-final-viral-combined_k141_166318||full Not-determined
18-105-final-viral-combined_k141_166324||full Not-determined
18-105-final-viral-combined_k141_139612||full Low-quality
18-105-final-viral-combined_k141_166331||full Low-quality
18-105-final-viral-combined_k141_51001||full Not-determined
18-105-final-viral-combined_k141_23159||full Low-quality
18-105-final-viral-combined_k141_32652||full Not-determined
18-105-final-viral-combined_k141_32653||full Not-determined
18-105-final-viral-combined_k141_8092||full Low-quality
18-105-final-viral-combined_k141_15165||full Low-quality
18-105-final-viral-combined_k141_68086||full Not-determined
18-105-final-viral-combined_k141_86538||full Low-quality
18-105-final-viral-combined_k141_23177||full Not-determined
18-105-final-viral-combined_k141_113227||full Low-quality
18-105-final-viral-combined_k141_166378||full Not-determined
18-105-final-viral-combined_k141_23188||full Not-determined
18-105-final-viral-combined_k141_185020||full Low-quality
18-105-final-viral-combined_k141_166393||full Low-quality
18-105-final-viral-combined_k141_139674||full Not-determined
18-105-final-viral-combined_k141_51053||full Not-determined
18-105-final-viral-combined_k141_8153||full Not-determined
18-105-final-viral-combined_k141_157409||full Not-determined
18-102-final-viral-combined_k141_170845||full Medium-quality
18-105-final-viral-combined_k141_15228||full Not-determined
18-105-final-viral-combined_k141_185062||full Not-determined
18-105-final-viral-combined_k141_185067||full Not-determined
18-105-final-viral-combined_k141_121980||full Not-determined
18-109_k141_118607 High-quality
18-105-final-viral-combined_k141_15261||full Low-quality
18-105-final-viral-combined_k141_78157||full Low-quality
18-105-final-viral-combined_k141_211010||full Low-quality
18-105-final-viral-combined_k141_166462||full Low-quality
18-105-final-viral-combined_k141_51109||full Low-quality
18-105-final-viral-combined_k141_78162||full Not-determined
18-105-final-viral-combined_k141_32764||full Not-determined
18-105-final-viral-combined_k141_202083||full Low-quality
18-105-final-viral-combined_k141_113342||full Not-determined
18-105-final-viral-combined_k141_104681||full Not-determined
18-105-final-viral-combined_k141_185089||full Not-determined
18-105-final-viral-combined_k141_149080||full Low-quality
18-105-final-viral-combined_k141_122001||full Low-quality
18-105-final-viral-combined_k141_68222||full Low-quality
18-105-final-viral-combined_k141_193470||full Not-determined
18-105-final-viral-combined_k141_104689||full Not-determined
18-105-final-viral-combined_k141_166486||full Not-determined
18-105-final-viral-combined_k141_32780||full Low-quality
18-105-final-viral-combined_k141_211030||full Low-quality
18-105-final-viral-combined_k141_202089||full Low-quality
18-105-final-viral-combined_k141_149097||full Not-determined
18-105-final-viral-combined_k141_113367||full Low-quality
18-105-final-viral-combined_k141_157443||full Low-quality
18-105-final-viral-combined_k141_202101||full Not-determined
18-105-final-viral-combined_k141_193491||full Low-quality
18-105-final-viral-combined_k141_8189||full Low-quality
18-105-final-viral-combined_k141_23259||full Not-determined
18-105-final-viral-combined_k141_185109||full Low-quality
18-105-final-viral-combined_k141_51165||full Low-quality
18-105-final-viral-combined_k141_122038||full Not-determined
18-105-final-viral-combined_k141_193500||full Low-quality
18-105-final-viral-combined_k141_68268||full Not-determined
18-105-final-viral-combined_k141_15300||full Low-quality
18-105-final-viral-combined_k141_41766||full Not-determined
18-105-final-viral-combined_k141_51173||full Low-quality
18-105-final-viral-combined_k141_95083||full Not-determined
18-105-final-viral-combined_k141_185138||full Not-determined
18-105-final-viral-combined_k141_139815||full Low-quality
18-105-final-viral-combined_k141_23288||full Not-determined
18-105-final-viral-combined_k141_149127||full Low-quality
18-105-final-viral-combined_k141_23311||full Low-quality
18-105-final-viral-combined_k141_157512||full Not-determined
18-105-final-viral-combined_k141_157513||full Low-quality
18-105-final-viral-combined_k141_149136||full Not-determined
18-105-final-viral-combined_k141_157522||full Not-determined
18-105-final-viral-combined_k141_23319||full Not-determined
18-105-final-viral-combined_k141_149140||full Not-determined
18-105-final-viral-combined_k141_15371||full Low-quality
18-105-final-viral-combined_k141_78190||full Low-quality
18-105-final-viral-combined_k141_157535||full Not-determined
18-105-final-viral-combined_k141_211072||full Low-quality
18-105-final-viral-combined_k141_157545||full Not-determined
18-105-final-viral-combined_k141_176255||full Low-quality
18-105-final-viral-combined_k141_23351||full Not-determined
18-105-final-viral-combined_k141_8257||full Not-determined
18-105-final-viral-combined_k141_149172||full Not-determined
18-105-final-viral-combined_k141_41801||full Low-quality
18-105-final-viral-combined_k141_176277||full Low-quality
18-105-final-viral-combined_k141_32878||full Not-determined
18-105-final-viral-combined_k141_32882||full Low-quality
18-105-final-viral-combined_k141_185141||full Not-determined
18-105-final-viral-combined_k141_211127||full Low-quality
18-105-final-viral-combined_k141_41830||full Not-determined
18-105-final-viral-combined_k141_139900||full Not-determined
18-105-final-viral-combined_k141_176302||full Low-quality
18-105-final-viral-combined_k141_23412||full Not-determined
18-105-final-viral-combined_k141_23433||full Low-quality
18-105-final-viral-combined_k141_193560||full Not-determined
18-105-final-viral-combined_k141_41835||full Low-quality
18-105-final-viral-combined_k141_113445||full Not-determined
18-105-final-viral-combined_k141_32936||full Low-quality
18-105-final-viral-combined_k141_149203||full Not-determined
18-105-final-viral-combined_k141_86812||full Not-determined
18-105-final-viral-combined_k141_51305||full Not-determined
18-105-final-viral-combined_k141_15526||full Low-quality
18-105-final-viral-combined_k141_41868||full Not-determined
18-105-final-viral-combined_k141_15535||full Not-determined
18-105-final-viral-combined_k141_157595||full Not-determined
18-105-final-viral-combined_k141_131787||full Low-quality
18-105-final-viral-combined_k141_185161||full Low-quality
18-105-final-viral-combined_k141_122100||full Low-quality
18-105-final-viral-combined_k141_95148||full Not-determined
18-105-final-viral-combined_k141_185175||full Low-quality
18-105-final-viral-combined_k141_86846||full Not-determined
18-105-final-viral-combined_k141_166558||full Low-quality
18-105-final-viral-combined_k141_86852||full Low-quality
18-105-final-viral-combined_k141_193632||full Not-determined
18-105-final-viral-combined_k141_104827||full Low-quality
18-105-final-viral-combined_k141_95167||full Not-determined
18-105-final-viral-combined_k141_193654||full Not-determined
18-105-final-viral-combined_k141_193655||full Not-determined
18-105-final-viral-combined_k141_104842||full Not-determined
18-105-final-viral-combined_k141_122117||full Not-determined
18-105-final-viral-combined_k141_113513||full Not-determined
18-105-final-viral-combined_k141_202191||full Not-determined
18-105-final-viral-combined_k141_78405||full Not-determined
18-105-final-viral-combined_k141_157644||full Low-quality
18-105-final-viral-combined_k141_41950||full Not-determined
18-105-final-viral-combined_k141_78419||full Not-determined
18-105-final-viral-combined_k141_202208||full Not-determined
18-105-final-viral-combined_k141_157659||full Low-quality
18-105-final-viral-combined_k141_122165||full Low-quality
18-105-final-viral-combined_k141_140006||full Low-quality
18-105-final-viral-combined_k141_51371||full Not-determined
18-105-final-viral-combined_k141_166594||full Not-determined
18-105-final-viral-combined_k141_166595||full Not-determined
18-105-final-viral-combined_k141_176409||full Low-quality
18-105-final-viral-combined_k141_185242||full Not-determined
18-105-final-viral-combined_k141_113537||full Low-quality
18-105-final-viral-combined_k141_140037||full Low-quality
18-105-final-viral-combined_k141_122199||full Not-determined
18-105-final-viral-combined_k141_193708||full Low-quality
18-105-final-viral-combined_k141_211161||full Low-quality
18-105-final-viral-combined_k141_131875||full Low-quality
18-105-final-viral-combined_k141_86936||full Low-quality
18-105-final-viral-combined_k141_149240||full Low-quality
18-105-final-viral-combined_k141_32974||full Not-determined
18-105-final-viral-combined_k141_193712||full Not-determined
18-105-final-viral-combined_k141_122217||full Not-determined
18-105-final-viral-combined_k141_131899||full Not-determined
18-105-final-viral-combined_k141_86962||full Low-quality
18-105-final-viral-combined_k141_104899||full Low-quality
18-105-final-viral-combined_k141_166674||full Not-determined
18-105-final-viral-combined_k141_33006||full Not-determined
18-105-final-viral-combined_k141_157769||full Low-quality
18-105-final-viral-combined_k141_140102||full Low-quality
18-105-final-viral-combined_k141_68547||full Not-determined
18-105-final-viral-combined_k141_104921||full Not-determined
18-105-final-viral-combined_k141_211203||full Low-quality
18-105-final-viral-combined_k141_33010||full Low-quality
18-105-final-viral-combined_k141_68571||full Low-quality
18-105-final-viral-combined_k141_33025||full Low-quality
18-105-final-viral-combined_k141_185260||full Low-quality
18-105-final-viral-combined_k141_193764||full Low-quality
18-105-final-viral-combined_k141_33037||full Low-quality
18-105-final-viral-combined_k141_122263||full Not-determined
18-105-final-viral-combined_k141_211252||full Low-quality
18-105-final-viral-combined_k141_95280||full Not-determined
18-105-final-viral-combined_k141_8357||full Low-quality
18-105-final-viral-combined_k141_33058||full Not-determined
18-105-final-viral-combined_k141_51421||full Not-determined
18-105-final-viral-combined_k141_51424||full Not-determined
18-105-final-viral-combined_k141_157834||full Not-determined
18-105-final-viral-combined_k141_8368||full Not-determined
18-105-final-viral-combined_k141_60213||full Not-determined
18-105-final-viral-combined_k141_68608||full Not-determined
18-105-final-viral-combined_k141_60216||full Not-determined
18-105-final-viral-combined_k141_33077||full Low-quality
18-105-final-viral-combined_k141_122295||full Not-determined
18-105-final-viral-combined_k141_122300||full Not-determined
18-105-final-viral-combined_k141_193824||full Low-quality
18-105-final-viral-combined_k141_104969||full Low-quality
18-105-final-viral-combined_k141_193833||full Low-quality
18-105-final-viral-combined_k141_41960||full Low-quality
18-105-final-viral-combined_k141_87099||full Low-quality
18-105-final-viral-combined_k141_131999||full Low-quality
18-105-final-viral-combined_k141_166780||full Not-determined
18-105-final-viral-combined_k141_68633||full Low-quality
18-105-final-viral-combined_k141_68646||full Low-quality
18-105-final-viral-combined_k141_211336||full Not-determined
17-106-final-viral-combined_k141_31456||full Medium-quality
18-105-final-viral-combined_k141_105007||full Not-determined
18-105-final-viral-combined_k141_15657||full Low-quality
18-105-final-viral-combined_k141_105010||full Low-quality
18-105-final-viral-combined_k141_8409||full Not-determined
18-105-final-viral-combined_k141_132036||full Low-quality
18-105-final-viral-combined_k141_105028||full Not-determined
18-105-final-viral-combined_k141_140133||full Low-quality
18-105-final-viral-combined_k141_78463||full Low-quality
18-105-final-viral-combined_k141_68698||full Low-quality
18-105-final-viral-combined_k141_157919||full Not-determined
18-105-final-viral-combined_k141_15687||full Not-determined
18-105-final-viral-combined_k141_78485||full Low-quality
18-105-final-viral-combined_k141_122376||full Not-determined
18-105-final-viral-combined_k141_95384||full Low-quality
18-105-final-viral-combined_k141_140194||full Not-determined
18-105-final-viral-combined_k141_68721||full Low-quality
18-105-final-viral-combined_k141_23676||full Not-determined
18-105-final-viral-combined_k141_157965||full Low-quality
18-105-final-viral-combined_k141_78539||full Not-determined
18-105-final-viral-combined_k141_157969||full Low-quality
18-105-final-viral-combined_k141_78543||full Low-quality
18-105-final-viral-combined_k141_166890||full Not-determined
18-105-final-viral-combined_k141_68748||full Not-determined
18-105-final-viral-combined_k141_211421||full Low-quality
18-105-final-viral-combined_k141_166901||full Not-determined
18-105-final-viral-combined_k141_105097||full Low-quality
18-105-final-viral-combined_k141_105104||full Not-determined
18-105-final-viral-combined_k141_140230||full Low-quality
18-105-final-viral-combined_k141_122397||full Low-quality
18-105-final-viral-combined_k141_185495||full Low-quality
18-105-final-viral-combined_k141_78573||full Not-determined
18-105-final-viral-combined_k141_68793||full Low-quality
18-105-final-viral-combined_k141_60337||full Not-determined
18-105-final-viral-combined_k141_60340||full Low-quality
18-105-final-viral-combined_k141_211462||full Low-quality
18-105-final-viral-combined_k141_157993||full Low-quality
18-105-final-viral-combined_k141_23736||full Low-quality
18-105-final-viral-combined_k141_211468||full Not-determined
18-105-final-viral-combined_k141_23745||full Low-quality
18-105-final-viral-combined_k141_51587||full Not-determined
18-105-final-viral-combined_k141_166926||full Low-quality
18-105-final-viral-combined_k141_140300||full Not-determined
18-105-final-viral-combined_k141_87195||full Low-quality
18-105-final-viral-combined_k141_33215||full Low-quality
18-105-final-viral-combined_k141_78615||full Low-quality
18-105-final-viral-combined_k141_105175||full Not-determined
18-105-final-viral-combined_k141_68869||full Low-quality
18-105-final-viral-combined_k141_132134||full Low-quality
18-105-final-viral-combined_k141_132138||full Low-quality
18-105-final-viral-combined_k141_105180||full Not-determined
18-105-final-viral-combined_k141_105181||full Low-quality
18-105-final-viral-combined_k141_41997||full Low-quality
18-105-final-viral-combined_k141_122453||full Low-quality
18-105-final-viral-combined_k141_132154||full Low-quality
18-105-final-viral-combined_k141_23763||full Low-quality
18-105-final-viral-combined_k141_149317||full Low-quality
18-105-final-viral-combined_k141_60396||full Low-quality
18-105-final-viral-combined_k141_140330||full Not-determined
18-105-final-viral-combined_k141_105193||full Low-quality
18-105-final-viral-combined_k141_78645||full Not-determined
18-105-final-viral-combined_k141_8517||full Not-determined
18-105-final-viral-combined_k141_23788||full Low-quality
18-105-final-viral-combined_k141_122476||full Not-determined
18-105-final-viral-combined_k141_105213||full Not-determined
18-105-final-viral-combined_k141_166992||full Not-determined
18-105-final-viral-combined_k141_68928||full Not-determined
18-105-final-viral-combined_k141_122486||full Not-determined
18-105-final-viral-combined_k141_87262||full Low-quality
18-105-final-viral-combined_k141_60429||full Low-quality
18-105-final-viral-combined_k141_68932||full Not-determined
18-105-final-viral-combined_k141_211538||full Not-determined
18-105-final-viral-combined_k141_78667||full Low-quality
18-105-final-viral-combined_k141_78673||full Low-quality
18-105-final-viral-combined_k141_87267||full Not-determined
18-105-final-viral-combined_k141_194098||full Not-determined
18-105-final-viral-combined_k141_122510||full Not-determined
18-105-final-viral-combined_k141_33219||full Low-quality
18-105-final-viral-combined_k141_78692||full Not-determined
18-105-final-viral-combined_k141_149370||full Not-determined
18-105-final-viral-combined_k141_87288||full Not-determined
18-105-final-viral-combined_k141_194125||full Not-determined
18-105-final-viral-combined_k141_194126||full Not-determined
18-105-final-viral-combined_k141_140366||full Not-determined
18-105-final-viral-combined_k141_202409||full Low-quality
18-105-final-viral-combined_k141_8577||full Not-determined
18-105-final-viral-combined_k141_194131||full Low-quality
18-105-final-viral-combined_k141_87311||full Not-determined
18-105-final-viral-combined_k141_105250||full Not-determined
18-105-final-viral-combined_k141_33297||full Not-determined
18-105-final-viral-combined_k141_95580||full Low-quality
18-105-final-viral-combined_k141_194184||full Low-quality
18-105-final-viral-combined_k141_8651||full Low-quality
18-105-final-viral-combined_k141_105292||full Not-determined
18-105-final-viral-combined_k141_33324||full Low-quality
18-105-final-viral-combined_k141_122597||full Not-determined
18-105-final-viral-combined_k141_33330||full Low-quality
18-105-final-viral-combined_k141_33332||full Not-determined
18-105-final-viral-combined_k141_194204||full Low-quality
18-105-final-viral-combined_k141_60566||full Low-quality
18-105-final-viral-combined_k141_194227||full Not-determined
18-105-final-viral-combined_k141_87393||full Not-determined
18-105-final-viral-combined_k141_68979||full Low-quality
18-105-final-viral-combined_k141_8695||full Not-determined
18-105-final-viral-combined_k141_42096||full Low-quality
18-105-final-viral-combined_k141_60597||full Not-determined
18-105-final-viral-combined_k141_23874||full Low-quality
18-105-final-viral-combined_k141_68985||full Not-determined
18-105-final-viral-combined_k141_105346||full Low-quality
18-105-final-viral-combined_k141_23878||full Not-determined
18-105-final-viral-combined_k141_105356||full Not-determined
18-105-final-viral-combined_k141_33375||full Low-quality
18-105-final-viral-combined_k141_87426||full Not-determined
18-105-final-viral-combined_k141_87428||full Low-quality
18-105-final-viral-combined_k141_8717||full Low-quality
18-105-final-viral-combined_k141_42105||full Low-quality
18-105-final-viral-combined_k141_185530||full Not-determined
18-105-final-viral-combined_k141_87441||full Not-determined
18-105-final-viral-combined_k141_42121||full Not-determined
18-105-final-viral-combined_k141_51749||full Not-determined
18-105-final-viral-combined_k141_132270||full Low-quality
18-105-final-viral-combined_k141_185542||full Not-determined
18-105-final-viral-combined_k141_60650||full Not-determined
18-105-final-viral-combined_k141_122708||full Low-quality
18-105-final-viral-combined_k141_8760||full Not-determined
18-105-final-viral-combined_k141_132286||full Low-quality
18-105-final-viral-combined_k141_105370||full Low-quality
18-105-final-viral-combined_k141_87464||full Not-determined
18-105-final-viral-combined_k141_51783||full Low-quality
18-105-final-viral-combined_k141_122724||full Low-quality
18-105-final-viral-combined_k141_167015||full Low-quality
18-105-final-viral-combined_k141_194282||full Not-determined

18-105-final-viral-combined_k141_167023||full Low-quality
18-105-final-viral-combined_k141_167039||full Not-determined
18-105-final-viral-combined_k141_122754||full Low-quality
18-105-final-viral-combined_k141_33482||full Not-determined
18-105-final-viral-combined_k141_60721||full Low-quality
18-105-final-viral-combined_k141_105411||full Low-quality
18-105-final-viral-combined_k141_87517||full Low-quality
18-105-final-viral-combined_k141_42162||full Low-quality
18-105-final-viral-combined_k141_15928||full Low-quality
18-105-final-viral-combined_k141_33487||full Not-determined
18-105-final-viral-combined_k141_149516||full Not-determined
18-105-final-viral-combined_k141_60731||full Not-determined
18-105-final-viral-combined_k141_149530||full Not-determined
18-105-final-viral-combined_k141_15935||full Not-determined
18-105-final-viral-combined_k141_167072||full Low-quality
18-105-final-viral-combined_k141_95763||full Not-determined
18-105-final-viral-combined_k141_132389||full Not-determined
18-105-final-viral-combined_k141_149546||full Low-quality
18-105-final-viral-combined_k141_87521||full Low-quality
18-105-final-viral-combined_k141_15945||full Low-quality
18-105-final-viral-combined_k141_185558||full Not-determined
18-105-final-viral-combined_k141_167111||full Not-determined
18-105-final-viral-combined_k141_87535||full Not-determined
18-105-final-viral-combined_k141_113617||full Low-quality
18-105-final-viral-combined_k141_51904||full Low-quality
18-105-final-viral-combined_k141_33534||full Low-quality
18-105-final-viral-combined_k141_23977||full Low-quality
18-105-final-viral-combined_k141_113629||full Low-quality
18-105-final-viral-combined_k141_60821||full Not-determined
18-105-final-viral-combined_k141_176632||full Low-quality
18-105-final-viral-combined_k141_140567||full Low-quality
18-105-final-viral-combined_k141_140568||full Not-determined
18-105-final-viral-combined_k141_51920||full Not-determined
18-105-final-viral-combined_k141_60829||full Not-determined
18-105-final-viral-combined_k141_8855||full Low-quality
18-105-final-viral-combined_k141_122853||full Not-determined
18-105-final-viral-combined_k141_95841||full Not-determined
18-105-final-viral-combined_k141_149573||full Low-quality
18-105-final-viral-combined_k141_95856||full Low-quality
18-105-final-viral-combined_k141_42211||full Not-determined
18-105-final-viral-combined_k141_176675||full Not-determined
18-105-final-viral-combined_k141_69244||full Not-determined
18-105-final-viral-combined_k141_194404||full Low-quality
18-105-final-viral-combined_k141_8900||full Low-quality
18-105-final-viral-combined_k141_194407||full Low-quality
18-105-final-viral-combined_k141_132456||full Low-quality
18-105-final-viral-combined_k141_122895||full Low-quality
18-105-final-viral-combined_k141_140651||full Not-determined
18-105-final-viral-combined_k141_167189||full Not-determined
18-105-final-viral-combined_k141_140662||full Not-determined
18-105-final-viral-combined_k141_176694||full Low-quality
18-105-final-viral-combined_k141_33669||full Not-determined
18-105-final-viral-combined_k141_15980||full Low-quality
18-105-final-viral-combined_k141_42258||full Not-determined
18-105-final-viral-combined_k141_52039||full Low-quality
18-105-final-viral-combined_k141_87611||full Low-quality
18-105-final-viral-combined_k141_122936||full Not-determined
18-105-final-viral-combined_k141_52064||full Not-determined
18-105-final-viral-combined_k141_24055||full Not-determined
18-105-final-viral-combined_k141_167222||full Low-quality
18-105-final-viral-combined_k141_52073||full Low-quality
18-105-final-viral-combined_k141_105592||full Not-determined
18-105-final-viral-combined_k141_202710||full Not-determined
18-105-final-viral-combined_k141_24110||full Low-quality
18-105-final-viral-combined_k141_105612||full Low-quality
18-105-final-viral-combined_k141_202730||full Not-determined
18-105-final-viral-combined_k141_87708||full Low-quality
18-105-final-viral-combined_k141_149634||full Low-quality
18-105-final-viral-combined_k141_123026||full Low-quality
18-105-final-viral-combined_k141_105620||full Not-determined
18-105-final-viral-combined_k141_185614||full Not-determined
18-105-final-viral-combined_k141_42369||full Low-quality
18-105-final-viral-combined_k141_105634||full Not-determined
18-105-final-viral-combined_k141_149661||full Low-quality
18-105-final-viral-combined_k141_176778||full Not-determined
18-105-final-viral-combined_k141_167266||full Low-quality
18-105-final-viral-combined_k141_33737||full Low-quality
18-105-final-viral-combined_k141_52179||full Low-quality
18-105-final-viral-combined_k141_211802||full Not-determined
18-105-final-viral-combined_k141_149703||full Not-determined
18-105-final-viral-combined_k141_158204||full Not-determined
18-105-final-viral-combined_k141_132583||full Low-quality
18-105-final-viral-combined_k141_149719||full Not-determined
18-105-final-viral-combined_k141_167314||full Not-determined
18-105-final-viral-combined_k141_42419||full Not-determined
18-105-final-viral-combined_k141_33763||full Low-quality
18-105-final-viral-combined_k141_87759||full Low-quality
18-105-final-viral-combined_k141_185708||full Not-determined
18-105-final-viral-combined_k141_33780||full Low-quality
18-105-final-viral-combined_k141_194488||full Low-quality
18-105-final-viral-combined_k141_194497||full Low-quality
18-105-final-viral-combined_k141_24137||full Low-quality
18-105-final-viral-combined_k141_132615||full Low-quality
18-105-final-viral-combined_k141_194502||full Low-quality
18-105-final-viral-combined_k141_24147||full Low-quality
18-105-final-viral-combined_k141_113933||full Not-determined
18-105-final-viral-combined_k141_158243||full Low-quality
18-105-final-viral-combined_k141_158247||full Low-quality
18-105-final-viral-combined_k141_167378||full Low-quality
18-105-final-viral-combined_k141_78976||full Not-determined
18-105-final-viral-combined_k141_176885||full Low-quality
18-105-final-viral-combined_k141_202837||full Not-determined
18-105-final-viral-combined_k141_96000||full Low-quality
18-105-final-viral-combined_k141_52291||full Low-quality
18-105-final-viral-combined_k141_176896||full Low-quality
18-105-final-viral-combined_k141_24194||full Not-determined
18-105-final-viral-combined_k141_87863||full Low-quality
18-105-final-viral-combined_k141_194551||full Not-determined
18-105-final-viral-combined_k141_24199||full Low-quality
18-105-final-viral-combined_k141_194557||full Low-quality
18-105-final-viral-combined_k141_194558||full Not-determined
18-105-final-viral-combined_k141_158278||full Low-quality
18-105-final-viral-combined_k141_158286||full Not-determined
18-105-final-viral-combined_k141_123129||full Low-quality
18-105-final-viral-combined_k141_33855||full Low-quality
18-105-final-viral-combined_k141_176932||full Low-quality
18-105-final-viral-combined_k141_194573||full Not-determined
18-105-final-viral-combined_k141_202878||full Not-determined
18-105-final-viral-combined_k141_96034||full Low-quality
18-105-final-viral-combined_k141_69313||full Low-quality
18-105-final-viral-combined_k141_194587||full Not-determined
18-105-final-viral-combined_k141_149829||full Not-determined
18-105-final-viral-combined_k141_149831||full Not-determined
18-105-final-viral-combined_k141_176981||full Low-quality
18-105-final-viral-combined_k141_79042||full Not-determined
18-105-final-viral-combined_k141_185792||full Low-quality
18-105-final-viral-combined_k141_194619||full Not-determined
18-105-final-viral-combined_k141_33919||full Not-determined
18-105-final-viral-combined_k141_87901||full Not-determined
18-105-final-viral-combined_k141_33927||full Not-determined
18-105-final-viral-combined_k141_105689||full Low-quality
18-105-final-viral-combined_k141_16036||full Not-determined
18-105-final-viral-combined_k141_114085||full Low-quality
18-105-final-viral-combined_k141_211900||full Low-quality
18-105-final-viral-combined_k141_194648||full Not-determined
18-105-final-viral-combined_k141_167550||full Low-quality
18-105-final-viral-combined_k141_123222||full Not-determined
18-105-final-viral-combined_k141_177049||full Low-quality
18-105-final-viral-combined_k141_177056||full Not-determined
18-105-final-viral-combined_k141_167569||full Not-determined
18-105-final-viral-combined_k141_194672||full Not-determined
18-105-final-viral-combined_k141_194677||full Low-quality
18-105-final-viral-combined_k141_96073||full Low-quality
18-105-final-viral-combined_k141_24271||full Not-determined
18-105-final-viral-combined_k141_96078||full Low-quality
18-105-final-viral-combined_k141_96080||full Not-determined
18-105-final-viral-combined_k141_123248||full Low-quality
18-105-final-viral-combined_k141_42602||full Low-quality
18-105-final-viral-combined_k141_24288||full Not-determined
18-105-final-viral-combined_k141_140824||full Low-quality
18-105-final-viral-combined_k141_194708||full Low-quality
18-105-final-viral-combined_k141_34001||full Low-quality
18-105-final-viral-combined_k141_158408||full Not-determined
18-105-final-viral-combined_k141_167612||full Low-quality
18-105-final-viral-combined_k141_61117||full Not-determined
18-105-final-viral-combined_k141_87970||full Not-determined
18-105-final-viral-combined_k141_34008||full Not-determined
18-105-final-viral-combined_k141_194723||full Low-quality
18-105-final-viral-combined_k141_69343||full Low-quality
18-105-final-viral-combined_k141_87984||full Low-quality
18-105-final-viral-combined_k141_185862||full Low-quality
18-105-final-viral-combined_k141_123285||full Not-determined
18-105-final-viral-combined_k141_42633||full Low-quality
18-105-final-viral-combined_k141_34012||full Low-quality
18-105-final-viral-combined_k141_34014||full Low-quality
18-105-final-viral-combined_k141_52499||full Low-quality
18-105-final-viral-combined_k141_211970||full Not-determined
18-105-final-viral-combined_k141_69357||full Not-determined
18-105-final-viral-combined_k141_167643||full Low-quality
18-105-final-viral-combined_k141_211975||full Low-quality
18-105-final-viral-combined_k141_123306||full Not-determined
18-105-final-viral-combined_k141_167650||full Not-determined
18-105-final-viral-combined_k141_123314||full Low-quality
18-105-final-viral-combined_k141_149945||full Low-quality
18-105-final-viral-combined_k141_24357||full Low-quality
18-105-final-viral-combined_k141_96165||full Not-determined
18-105-final-viral-combined_k141_42659||full Low-quality
18-105-final-viral-combined_k141_105757||full Not-determined
18-105-final-viral-combined_k141_177143||full Not-determined
18-105-final-viral-combined_k141_185889||full Low-quality
18-105-final-viral-combined_k141_34046||full Low-quality
18-105-final-viral-combined_k141_158459||full Not-determined
18-105-final-viral-combined_k141_149962||full Not-determined
18-105-final-viral-combined_k141_177153||full Not-determined
18-105-final-viral-combined_k141_52537||full Low-quality
18-105-final-viral-combined_k141_42682||full Not-determined
18-105-final-viral-combined_k141_167690||full Not-determined
18-105-final-viral-combined_k141_34083||full Low-quality
18-105-final-viral-combined_k141_52560||full Low-quality
18-105-final-viral-combined_k141_42707||full Low-quality
18-105-final-viral-combined_k141_52561||full Low-quality
18-105-final-viral-combined_k141_34106||full Low-quality
18-105-final-viral-combined_k141_123334||full Not-determined
18-105-final-viral-combined_k141_140956||full Low-quality
18-105-final-viral-combined_k141_212027||full Not-determined
18-105-final-viral-combined_k141_52572||full Low-quality
18-105-final-viral-combined_k141_177179||full Low-quality
18-105-final-viral-combined_k141_105830||full Low-quality
18-105-final-viral-combined_k141_88014||full Not-determined
18-105-final-viral-combined_k141_202965||full Not-determined
18-105-final-viral-combined_k141_194871||full Low-quality
18-105-final-viral-combined_k141_177208||full Low-quality
18-105-final-viral-combined_k141_79206||full Not-determined
18-105-final-viral-combined_k141_42775||full Not-determined
18-105-final-viral-combined_k141_16110||full Not-determined
KM-final-viral-combined_k141_244708||full High-quality
18-105-final-viral-combined_k141_105898||full Not-determined
18-105-final-viral-combined_k141_16130||full Low-quality
18-105-final-viral-combined_k141_88048||full Not-determined
18-105-final-viral-combined_k141_203034||full Not-determined
18-105-final-viral-combined_k141_16143||full Not-determined
18-105-final-viral-combined_k141_203043||full Not-determined
18-105-final-viral-combined_k141_69447||full Not-determined
18-105-final-viral-combined_k141_88083||full Low-quality
18-105-final-viral-combined_k141_185976||full Not-determined
18-105-final-viral-combined_k141_185984||full Not-determined
18-105-final-viral-combined_k141_69458||full Low-quality
18-105-final-viral-combined_k141_16181||full Not-determined
18-105-final-viral-combined_k141_150100||full Low-quality
18-105-final-viral-combined_k141_185991||full Not-determined
18-105-final-viral-combined_k141_158674||full Low-quality
18-105-final-viral-combined_k141_69462||full Low-quality
18-105-final-viral-combined_k141_88098||full Not-determined
18-105-final-viral-combined_k141_114125||full Not-determined
18-105-final-viral-combined_k141_16192||full Not-determined
18-109-final-viral-combined_k141_118607||full High-quality
18-105-final-viral-combined_k141_79223||full Not-determined
18-105-final-viral-combined_k141_123517||full Low-quality
18-105-final-viral-combined_k141_141089||full Low-quality
18-105-final-viral-combined_k141_194983||full Low-quality
18-105-final-viral-combined_k141_42814||full Low-quality
18-105-final-viral-combined_k141_16226||full Low-quality
18-105-final-viral-combined_k141_79292||full Low-quality
18-105-final-viral-combined_k141_96343||full Low-quality
18-105-final-viral-combined_k141_105995||full Not-determined
18-105-final-viral-combined_k141_194999||full Low-quality
18-105-final-viral-combined_k141_16235||full Not-determined
18-105-final-viral-combined_k141_96354||full Not-determined
18-105-final-viral-combined_k141_34172||full Not-determined
18-105-final-viral-combined_k141_96375||full Low-quality
18-105-final-viral-combined_k141_167855||full Not-determined
18-105-final-viral-combined_k141_24422||full Not-determined
18-105-final-viral-combined_k141_79331||full Low-quality
18-105-final-viral-combined_k141_114191||full Not-determined
18-105-final-viral-combined_k141_106007||full Low-quality
18-105-final-viral-combined_k141_114209||full Low-quality
18-105-final-viral-combined_k141_195069||full Not-determined
18-105-final-viral-combined_k141_34210||full Low-quality
18-105-final-viral-combined_k141_106021||full Not-determined
18-105-final-viral-combined_k141_88204||full Not-determined
18-105-final-viral-combined_k141_114227||full Not-determined
18-105-final-viral-combined_k141_16310||full Low-quality
18-105-final-viral-combined_k141_96447||full Not-determined
18-105-final-viral-combined_k141_16313||full Not-determined
18-105-final-viral-combined_k141_42835||full Not-determined
18-105-final-viral-combined_k141_96453||full Not-determined
18-105-final-viral-combined_k141_106064||full Not-determined
18-105-final-viral-combined_k141_42856||full Low-quality
18-105-final-viral-combined_k141_88243||full Not-determined
18-105-final-viral-combined_k141_24518||full Low-quality
18-105-final-viral-combined_k141_52731||full Low-quality
18-105-final-viral-combined_k141_52734||full Not-determined
18-105-final-viral-combined_k141_96478||full Low-quality
18-105-final-viral-combined_k141_203096||full Not-determined
18-105-final-viral-combined_k141_203097||full Low-quality
18-105-final-viral-combined_k141_186130||full Not-determined
18-105-final-viral-combined_k141_150195||full Not-determined
18-105-final-viral-combined_k141_203113||full Not-determined
18-105-final-viral-combined_k141_195130||full Not-determined
18-105-final-viral-combined_k141_106116||full Low-quality
18-105-final-viral-combined_k141_79420||full Low-quality
18-105-final-viral-combined_k141_123633||full Not-determined
18-105-final-viral-combined_k141_88308||full Low-quality
18-105-final-viral-combined_k141_203131||full Low-quality
18-105-final-viral-combined_k141_114372||full Not-determined
18-105-final-viral-combined_k141_69601||full Low-quality
18-105-final-viral-combined_k141_24621||full Low-quality
18-105-final-viral-combined_k141_79474||full Low-quality
18-105-final-viral-combined_k141_96577||full Not-determined
18-105-final-viral-combined_k141_212258||full Low-quality
18-105-final-viral-combined_k141_158758||full Not-determined
18-105-final-viral-combined_k141_24636||full Low-quality
18-105-final-viral-combined_k141_123688||full Not-determined
18-105-final-viral-combined_k141_203168||full Not-determined
18-105-final-viral-combined_k141_123703||full Low-quality
18-105-final-viral-combined_k141_141165||full Not-determined
18-105-final-viral-combined_k141_195178||full Low-quality
18-105-final-viral-combined_k141_24694||full Not-determined
18-105-final-viral-combined_k141_123742||full Not-determined
18-105-final-viral-combined_k141_88392||full Not-determined
18-105-final-viral-combined_k141_195209||full Not-determined
18-105-final-viral-combined_k141_96664||full Not-determined
18-105-final-viral-combined_k141_69716||full Low-quality
18-105-final-viral-combined_k141_177298||full Not-determined
18-105-final-viral-combined_k141_79497||full Not-determined
18-105-final-viral-combined_k141_88395||full Not-determined
18-105-final-viral-combined_k141_123767||full Low-quality
18-105-final-viral-combined_k141_88411||full Not-determined
18-105-final-viral-combined_k141_132847||full Not-determined
18-105-final-viral-combined_k141_195240||full Not-determined
18-105-final-viral-combined_k141_123788||full Low-quality
18-105-final-viral-combined_k141_79525||full Not-determined
18-105-final-viral-combined_k141_150331||full Not-determined
18-105-final-viral-combined_k141_52762||full Not-determined
18-105-final-viral-combined_k141_186196||full Low-quality
18-105-final-viral-combined_k141_167942||full Low-quality
18-105-final-viral-combined_k141_150349||full Low-quality
18-105-final-viral-combined_k141_52775||full Low-quality
18-105-final-viral-combined_k141_212310||full Low-quality
18-105-final-viral-combined_k141_123823||full Low-quality
18-105-final-viral-combined_k141_79548||full Low-quality
18-105-final-viral-combined_k141_79553||full Low-quality
18-105-final-viral-combined_k141_16569||full Not-determined
18-105-final-viral-combined_k141_158860||full Low-quality
18-105-final-viral-combined_k141_88485||full Low-quality
18-105-final-viral-combined_k141_96747||full Not-determined
18-105-final-viral-combined_k141_34440||full Low-quality
18-105-final-viral-combined_k141_186223||full Not-determined
18-105-final-viral-combined_k141_167983||full Low-quality
18-105-final-viral-combined_k141_16579||full Low-quality
18-105-final-viral-combined_k141_96762||full Low-quality
18-105-final-viral-combined_k141_106239||full Low-quality
18-105-final-viral-combined_k141_158877||full Low-quality
18-105-final-viral-combined_k141_203280||full Not-determined
18-105-final-viral-combined_k141_150382||full Not-determined
18-105-final-viral-combined_k141_212365||full Not-determined
18-105-final-viral-combined_k141_186238||full Not-determined
18-105-final-viral-combined_k141_114481||full Not-determined
18-105-final-viral-combined_k141_195284||full Not-determined
18-105-final-viral-combined_k141_106260||full Low-quality
18-105-final-viral-combined_k141_186246||full Not-determined
18-105-final-viral-combined_k141_106274||full Low-quality
18-105-final-viral-combined_k141_150420||full Low-quality
18-105-final-viral-combined_k141_212374||full Not-determined
18-105-final-viral-combined_k141_158917||full Not-determined
18-105-final-viral-combined_k141_158928||full Not-determined
18-105-final-viral-combined_k141_150440||full Low-quality
18-105-final-viral-combined_k141_168021||full Not-determined
18-105-final-viral-combined_k141_96849||full Low-quality
18-105-final-viral-combined_k141_106309||full Low-quality
18-105-final-viral-combined_k141_88536||full Low-quality
18-105-final-viral-combined_k141_123944||full Not-determined
18-105-final-viral-combined_k141_203352||full Low-quality
18-105-final-viral-combined_k141_132999||full Low-quality
18-105-final-viral-combined_k141_195328||full Low-quality
18-105-final-viral-combined_k141_195337||full Low-quality
18-105-final-viral-combined_k141_123949||full Not-determined
18-105-final-viral-combined_k141_203369||full Not-determined
18-105-final-viral-combined_k141_96888||full Low-quality
18-105-final-viral-combined_k141_79608||full Low-quality
18-105-final-viral-combined_k141_133020||full Low-quality
18-105-final-viral-combined_k141_16690||full Not-determined
18-105-final-viral-combined_k141_186308||full Not-determined
18-105-final-viral-combined_k141_141358||full Low-quality
18-105-final-viral-combined_k141_16694||full Low-quality
18-105-final-viral-combined_k141_34509||full Low-quality
18-105-final-viral-combined_k141_168088||full Not-determined
18-105-final-viral-combined_k141_177349||full Low-quality
18-105-final-viral-combined_k141_124006||full Low-quality
18-105-final-viral-combined_k141_106394||full Not-determined
18-105-final-viral-combined_k141_34528||full Not-determined
18-105-final-viral-combined_k141_24756||full Low-quality
18-105-final-viral-combined_k141_106415||full Low-quality
18-105-final-viral-combined_k141_24770||full Not-determined
18-105-final-viral-combined_k141_34548||full Low-quality
18-105-final-viral-combined_k141_177386||full Low-quality
18-105-final-viral-combined_k141_52879||full Low-quality
18-105-final-viral-combined_k141_159031||full Low-quality
18-105-final-viral-combined_k141_43072||full Not-determined
18-105-final-viral-combined_k141_141390||full Not-determined
18-105-final-viral-combined_k141_43083||full Low-quality
18-105-final-viral-combined_k141_61234||full Not-determined
18-105-final-viral-combined_k141_79694||full Low-quality
18-105-final-viral-combined_k141_34562||full Not-determined
18-105-final-viral-combined_k141_177437||full Low-quality
18-105-final-viral-combined_k141_79710||full Not-determined
18-105-final-viral-combined_k141_24838||full Low-quality
18-105-final-viral-combined_k141_61259||full Low-quality
18-105-final-viral-combined_k141_114605||full Low-quality
18-105-final-viral-combined_k141_79713||full Low-quality
18-105-final-viral-combined_k141_16807||full Low-quality
18-105-final-viral-combined_k141_133109||full Not-determined
18-105-final-viral-combined_k141_34574||full Low-quality
18-105-final-viral-combined_k141_195411||full Low-quality
18-105-final-viral-combined_k141_195413||full Low-quality
18-105-final-viral-combined_k141_69797||full Low-quality
18-105-final-viral-combined_k141_114616||full Low-quality
18-105-final-viral-combined_k141_186422||full Low-quality
18-105-final-viral-combined_k141_106481||full Not-determined
18-105-final-viral-combined_k141_43119||full Not-determined
18-105-final-viral-combined_k141_79731||full Low-quality
18-105-final-viral-combined_k141_212459||full Low-quality
18-105-final-viral-combined_k141_16811||full Low-quality
18-105-final-viral-combined_k141_133140||full Not-determined
18-105-final-viral-combined_k141_97066||full Not-determined
18-105-final-viral-combined_k141_195435||full Not-determined
18-105-final-viral-combined_k141_177496||full Not-determined
18-105-final-viral-combined_k141_168186||full Not-determined
18-105-final-viral-combined_k141_43177||full Low-quality
18-105-final-viral-combined_k141_133172||full Not-determined
18-105-final-viral-combined_k141_168200||full Not-determined
18-105-final-viral-combined_k141_97091||full Low-quality
18-105-final-viral-combined_k141_195486||full Not-determined
18-105-final-viral-combined_k141_43204||full Not-determined
18-105-final-viral-combined_k141_186501||full Low-quality
18-105-final-viral-combined_k141_43221||full Not-determined
18-105-final-viral-combined_k141_203488||full Not-determined
18-105-final-viral-combined_k141_195525||full Not-determined
18-105-final-viral-combined_k141_16840||full Not-determined
18-105-final-viral-combined_k141_106523||full Low-quality
18-105-final-viral-combined_k141_141438||full Not-determined
18-105-final-viral-combined_k141_79823||full Not-determined
18-105-final-viral-combined_k141_141440||full Low-quality
18-105-final-viral-combined_k141_212553||full Not-determined
18-105-final-viral-combined_k141_24966||full Not-determined
18-105-final-viral-combined_k141_212556||full Low-quality
18-105-final-viral-combined_k141_88696||full Low-quality
18-105-final-viral-combined_k141_16866||full Low-quality
18-105-final-viral-combined_k141_159144||full Not-determined
18-105-final-viral-combined_k141_141462||full Low-quality
18-105-final-viral-combined_k141_69912||full Low-quality
18-105-final-viral-combined_k141_203520||full Not-determined
18-105-final-viral-combined_k141_133267||full Not-determined
18-105-final-viral-combined_k141_97105||full Low-quality
18-105-final-viral-combined_k141_133269||full Low-quality
18-105-final-viral-combined_k141_88715||full Low-quality
18-105-final-viral-combined_k141_203529||full Not-determined
18-103_k141_22744 High-quality
18-105-final-viral-combined_k141_150709||full Not-determined
18-105-final-viral-combined_k141_150713||full Low-quality
18-105-final-viral-combined_k141_212603||full Low-quality
18-105-final-viral-combined_k141_61367||full Low-quality
18-105-final-viral-combined_k141_159195||full Not-determined
18-105-final-viral-combined_k141_25028||full Low-quality
18-105-final-viral-combined_k141_159202||full Low-quality
18-105-final-viral-combined_k141_114701||full Not-determined
18-105-final-viral-combined_k141_52943||full Low-quality
18-105-final-viral-combined_k141_168334||full Low-quality
18-105-final-viral-combined_k141_43288||full Not-determined
18-105-final-viral-combined_k141_159212||full Low-quality
18-105-final-viral-combined_k141_168346||full Not-determined
18-105-final-viral-combined_k141_114714||full Not-determined
18-105-final-viral-combined_k141_97162||full Not-determined
18-105-final-viral-combined_k141_52962||full Not-determined
18-105-final-viral-combined_k141_212658||full Not-determined
18-105-final-viral-combined_k141_79882||full Not-determined
18-105-final-viral-combined_k141_159248||full Not-determined
18-105-final-viral-combined_k141_114737||full Low-quality
18-103-final-viral-combined_k141_22744||full High-quality
18-105-final-viral-combined_k141_52982||full Not-determined
18-105-final-viral-combined_k141_212672||full Low-quality
18-105-final-viral-combined_k141_25068||full Not-determined
18-105-final-viral-combined_k141_97208||full Not-determined
18-105-final-viral-combined_k141_25075||full Not-determined
18-105-final-viral-combined_k141_114747||full Low-quality
18-105-final-viral-combined_k141_61432||full Not-determined
18-105-final-viral-combined_k141_124167||full Not-determined
18-105-final-viral-combined_k141_79904||full Not-determined
18-105-final-viral-combined_k141_88802||full Low-quality
18-105-final-viral-combined_k141_25084||full Low-quality
18-105-final-viral-combined_k141_97223||full Low-quality
18-105-final-viral-combined_k141_106635||full Not-determined
18-105-final-viral-combined_k141_141604||full Not-determined
18-105-final-viral-combined_k141_70000||full Low-quality
18-105-final-viral-combined_k141_97249||full Not-determined
18-105-final-viral-combined_k141_195650||full Low-quality
18-105-final-viral-combined_k141_212678||full Not-determined
18-105-final-viral-combined_k141_150798||full Not-determined
18-105-final-viral-combined_k141_70025||full Not-determined
18-105-final-viral-combined_k141_53054||full Not-determined
18-105-final-viral-combined_k141_203673||full Not-determined
18-105-final-viral-combined_k141_141635||full Not-determined
18-105-final-viral-combined_k141_168456||full Low-quality
18-105-final-viral-combined_k141_133403||full Low-quality
18-105-final-viral-combined_k141_61498||full Not-determined
18-105-final-viral-combined_k141_53073||full Low-quality
18-105-final-viral-combined_k141_203702||full Low-quality
18-105-final-viral-combined_k141_168469||full Not-determined
18-105-final-viral-combined_k141_88884||full Not-determined
18-105-final-viral-combined_k141_212716||full Low-quality
18-105-final-viral-combined_k141_70060||full Low-quality
18-105-final-viral-combined_k141_34648||full Not-determined
18-105-final-viral-combined_k141_212718||full Low-quality
18-105-final-viral-combined_k141_141680||full Not-determined
18-105-final-viral-combined_k141_70080||full Low-quality
18-105-final-viral-combined_k141_141693||full Low-quality
18-105-final-viral-combined_k141_141704||full Low-quality
18-105-final-viral-combined_k141_159341||full Not-determined
18-105-final-viral-combined_k141_124219||full Not-determined
18-105-final-viral-combined_k141_124225||full Not-determined
18-105-final-viral-combined_k141_124230||full Not-determined
18-105-final-viral-combined_k141_97290||full Not-determined
18-105-final-viral-combined_k141_61590||full Not-determined
18-105-final-viral-combined_k141_16993||full Not-determined
18-105-final-viral-combined_k141_212805||full Not-determined
18-105-final-viral-combined_k141_34687||full Low-quality
18-105-final-viral-combined_k141_141753||full Low-quality
18-105-final-viral-combined_k141_106668||full Not-determined
18-105-final-viral-combined_k141_80036||full Low-quality
18-105-final-viral-combined_k141_159401||full Not-determined
18-105-final-viral-combined_k141_88938||full Not-determined
18-105-final-viral-combined_k141_61605||full Not-determined
18-105-final-viral-combined_k141_150884||full Not-determined
18-105-final-viral-combined_k141_203745||full Low-quality
18-105-final-viral-combined_k141_106692||full Low-quality
18-105-final-viral-combined_k141_133469||full Low-quality
18-105-final-viral-combined_k141_61615||full Low-quality
18-105-final-viral-combined_k141_61617||full Low-quality
18-105-final-viral-combined_k141_53231||full Not-determined
18-105-final-viral-combined_k141_70163||full Not-determined
18-105-final-viral-combined_k141_97397||full Not-determined
18-105-final-viral-combined_k141_34740||full Not-determined
18-105-final-viral-combined_k141_186696||full Not-determined
18-105-final-viral-combined_k141_88965||full Low-quality
18-105-final-viral-combined_k141_124337||full Not-determined
18-105-final-viral-combined_k141_70187||full Low-quality
18-105-final-viral-combined_k141_80044||full Low-quality
18-105-final-viral-combined_k141_61662||full Not-determined
18-105-final-viral-combined_k141_97438||full Low-quality
18-105-final-viral-combined_k141_34783||full Low-quality
18-105-final-viral-combined_k141_80062||full Low-quality
18-105-final-viral-combined_k141_70215||full Not-determined
18-105-final-viral-combined_k141_53292||full Not-determined
18-105-final-viral-combined_k141_106759||full Not-determined
18-105-final-viral-combined_k141_34793||full Low-quality
18-105-final-viral-combined_k141_106763||full Low-quality
18-105-final-viral-combined_k141_97497||full Not-determined
18-105-final-viral-combined_k141_53333||full Low-quality
18-105-final-viral-combined_k141_177688||full Low-quality
18-105-final-viral-combined_k141_61727||full Low-quality
18-105-final-viral-combined_k141_61730||full Not-determined
18-105-final-viral-combined_k141_80100||full Low-quality
18-105-final-viral-combined_k141_80105||full Low-quality
18-105-final-viral-combined_k141_177729||full Low-quality
18-105-final-viral-combined_k141_61757||full Low-quality
18-105-final-viral-combined_k141_124401||full Low-quality
18-105-final-viral-combined_k141_43559||full Low-quality
18-105-final-viral-combined_k141_133518||full Not-determined
18-105-final-viral-combined_k141_61799||full Low-quality
18-105-final-viral-combined_k141_186746||full Low-quality
18-105-final-viral-combined_k141_150977||full Not-determined
18-105-final-viral-combined_k141_203836||full Not-determined
18-105-final-viral-combined_k141_212925||full Low-quality
18-105-final-viral-combined_k141_212927||full Low-quality
18-105-final-viral-combined_k141_150995||full Not-determined
18-105-final-viral-combined_k141_150999||full Not-determined
18-105-final-viral-combined_k141_212934||full Low-quality
18-105-final-viral-combined_k141_106846||full Not-determined
18-105-final-viral-combined_k141_34955||full Low-quality
18-105-final-viral-combined_k141_17152||full Low-quality
18-105-final-viral-combined_k141_168657||full Low-quality
18-105-final-viral-combined_k141_133589||full Low-quality
18-105-final-viral-combined_k141_61874||full Low-quality
18-105-final-viral-combined_k141_168660||full Not-determined
18-105-final-viral-combined_k141_43651||full Not-determined
18-105-final-viral-combined_k141_17168||full Low-quality
18-105-final-viral-combined_k141_70297||full Not-determined
18-105-final-viral-combined_k141_97636||full Not-determined
18-105-final-viral-combined_k141_97645||full Not-determined
18-105-final-viral-combined_k141_97665||full Low-quality
18-105-final-viral-combined_k141_177800||full Low-quality
18-105-final-viral-combined_k141_213014||full Not-determined
18-105-final-viral-combined_k141_141914||full Low-quality
18-105-final-viral-combined_k141_35021||full Low-quality
18-105-final-viral-combined_k141_115194||full Low-quality
18-105-final-viral-combined_k141_141931||full Low-quality
18-105-final-viral-combined_k141_168680||full Low-quality
18-105-final-viral-combined_k141_97714||full Not-determined
18-105-final-viral-combined_k141_70353||full Low-quality
18-105-final-viral-combined_k141_141934||full Low-quality
18-105-final-viral-combined_k141_70355||full Not-determined
18-105-final-viral-combined_k141_115242||full Low-quality
18-105-final-viral-combined_k141_177862||full Low-quality
18-105-final-viral-combined_k141_61973||full Not-determined
18-105-final-viral-combined_k141_213055||full Not-determined
18-105-final-viral-combined_k141_124605||full Not-determined
18-105-final-viral-combined_k141_141982||full Low-quality
18-105-final-viral-combined_k141_168751||full Not-determined
18-105-final-viral-combined_k141_124613||full Low-quality
18-105-final-viral-combined_k141_115300||full Not-determined
18-105-final-viral-combined_k141_43717||full Not-determined
18-105-final-viral-combined_k141_168774||full Low-quality
18-105-final-viral-combined_k141_70399||full Low-quality
18-105-final-viral-combined_k141_177901||full Low-quality
18-105-final-viral-combined_k141_159567||full Low-quality
18-105-final-viral-combined_k141_213093||full Low-quality
18-105-final-viral-combined_k141_25411||full Low-quality
18-105-final-viral-combined_k141_35148||full Low-quality
18-105-final-viral-combined_k141_17230||full Low-quality

| | |
|---|---|
| 18-105-final-viral-combined_k141_177922\|\|full | Low-quality |
| 18-105-final-viral-combined_k141_35155\|\|full | Not-determined |
| 18-105-final-viral-combined_k141_213099\|\|full | Not-determined |
| 18-105-final-viral-combined_k141_186775\|\|full | Not-determined |
| 18-105-final-viral-combined_k141_177929\|\|full | Not-determined |
| 18-105-final-viral-combined_k141_177931\|\|full | Low-quality |
| 18-105-final-viral-combined_k141_43760\|\|full | Not-determined |
| 18-105-final-viral-combined_k141_17238\|\|full | Low-quality |
| 18-105-final-viral-combined_k141_43778\|\|full | Not-determined |
| 18-105-final-viral-combined_k141_142035\|\|full | Low-quality |
| 18-105-final-viral-combined_k141_168816\|\|full | Low-quality |
| 18-105-final-viral-combined_k141_159614\|\|full | Not-determined |
| 18-105-final-viral-combined_k141_43785\|\|full | Low-quality |
| 18-105-final-viral-combined_k141_168820\|\|full | Low-quality |
| 18-105-final-viral-combined_k141_62005\|\|full | Not-determined |
| 18-105-final-viral-combined_k141_70445\|\|full | Low-quality |
| 18-105-final-viral-combined_k141_159631\|\|full | Not-determined |
| 18-105-final-viral-combined_k141_62037\|\|full | Not-determined |
| 18-105-final-viral-combined_k141_97872\|\|full | Low-quality |
| 18-105-final-viral-combined_k141_151191\|\|full | Low-quality |
| 18-105-final-viral-combined_k141_43863\|\|full | Low-quality |
| 18-105-final-viral-combined_k141_178008\|\|full | Low-quality |
| 18-105-final-viral-combined_k141_43874\|\|full | Low-quality |
| 18-105-final-viral-combined_k141_17310\|\|full | Low-quality |
| 18-105-final-viral-combined_k141_17313\|\|full | Low-quality |
| 18-105-final-viral-combined_k141_178025\|\|full | Not-determined |
| 18-105-final-viral-combined_k141_35280\|\|full | Not-determined |
| 18-105-final-viral-combined_k141_17329\|\|full | Not-determined |
| 18-105-final-viral-combined_k141_70477\|\|full | Not-determined |
| 18-105-final-viral-combined_k141_178052\|\|full | Not-determined |
| 18-105-final-viral-combined_k141_35300\|\|full | Low-quality |
| 18-105-final-viral-combined_k141_178073\|\|full | Not-determined |
| 18-105-final-viral-combined_k141_35318\|\|full | Not-determined |
| 18-105-final-viral-combined_k141_43916\|\|full | Not-determined |
| 18-105-final-viral-combined_k141_186911\|\|full | Low-quality |
| 18-105-final-viral-combined_k141_186913\|\|full | Low-quality |
| 18-105-final-viral-combined_k141_178108\|\|full | Low-quality |
| 18-105-final-viral-combined_k141_115332\|\|full | Low-quality |
| 18-105-final-viral-combined_k141_159809\|\|full | Not-determined |
| 18-105-final-viral-combined_k141_204160\|\|full | Low-quality |
| 18-105-final-viral-combined_k141_17441\|\|full | Not-determined |
| 18-105-final-viral-combined_k141_204183\|\|full | Low-quality |
| 18-105-final-viral-combined_k141_43949\|\|full | Not-determined |
| 18-105-final-viral-combined_k141_186998\|\|full | Low-quality |
| 18-105-final-viral-combined_k141_43980\|\|full | Low-quality |
| 18-105-final-viral-combined_k141_17469\|\|full | Low-quality |
| 18-105-final-viral-combined_k141_213121\|\|full | Not-determined |
| 18-105-final-viral-combined_k141_187031\|\|full | Not-determined |
| 18-105-final-viral-combined_k141_142196\|\|full | Low-quality |
| 18-105-final-viral-combined_k141_151416\|\|full | Not-determined |
| 18-105-final-viral-combined_k141_168988\|\|full | Low-quality |
| 18-105-final-viral-combined_k141_187042\|\|full | Low-quality |
| 18-105-final-viral-combined_k141_159890\|\|full | Not-determined |
| 18-105-final-viral-combined_k141_151429\|\|full | Low-quality |
| 18-105-final-viral-combined_k141_25561\|\|full | Not-determined |
| 18-105-final-viral-combined_k141_213174\|\|full | Not-determined |
| 18-105-final-viral-combined_k141_115446\|\|full | Low-quality |
| 18-105-final-viral-combined_k141_17493\|\|full | Not-determined |
| 18-105-final-viral-combined_k141_44053\|\|full | Not-determined |
| 18-105-final-viral-combined_k141_97925\|\|full | Not-determined |
| 18-105-final-viral-combined_k141_169014\|\|full | Low-quality |
| 18-105-final-viral-combined_k141_17503\|\|full | Low-quality |
| 18-105-final-viral-combined_k141_187048\|\|full | Low-quality |
| 18-105-final-viral-combined_k141_70587\|\|full | Not-determined |
| 18-105-final-viral-combined_k141_204238\|\|full | Low-quality |
| 18-105-final-viral-combined_k141_204259\|\|full | Not-determined |
| 18-105-final-viral-combined_k141_70609\|\|full | Low-quality |
| 18-105-final-viral-combined_k141_25590\|\|full | Not-determined |
| 18-105-final-viral-combined_k141_97932\|\|full | Not-determined |
| 18-105-final-viral-combined_k141_25604\|\|full | Low-quality |
| 18-105-final-viral-combined_k141_213182\|\|full | Low-quality |
| 18-105-final-viral-combined_k141_142271\|\|full | Not-determined |
| 18-105-final-viral-combined_k141_115529\|\|full | Low-quality |
| 18-105-final-viral-combined_k141_62192\|\|full | Low-quality |
| 18-105-final-viral-combined_k141_70659\|\|full | Low-quality |
| 18-105-final-viral-combined_k141_169075\|\|full | Not-determined |
| 18-105-final-viral-combined_k141_169084\|\|full | Not-determined |
| 18-105-final-viral-combined_k141_160015\|\|full | Not-determined |
| 18-105-final-viral-combined_k141_204344\|\|full | Low-quality |
| 18-105-final-viral-combined_k141_35426\|\|full | Low-quality |
| 18-105-final-viral-combined_k141_204359\|\|full | Low-quality |
| 18-105-final-viral-combined_k141_204367\|\|full | Low-quality |
| 18-105-final-viral-combined_k141_115587\|\|full | Not-determined |
| 18-105-final-viral-combined_k141_35455\|\|full | Not-determined |
| 18-105-final-viral-combined_k141_35468\|\|full | Low-quality |
| 18-105-final-viral-combined_k141_213249\|\|full | Not-determined |
| 18-105-final-viral-combined_k141_160077\|\|full | Not-determined |
| 18-105-final-viral-combined_k141_213275\|\|full | Not-determined |
| 18-105-final-viral-combined_k141_204394\|\|full | Not-determined |
| 18-105-final-viral-combined_k141_142423\|\|full | Not-determined |
| 18-105-final-viral-combined_k141_25647\|\|full | Not-determined |
| 18-105-final-viral-combined_k141_115629\|\|full | Low-quality |
| 18-105-final-viral-combined_k141_213287\|\|full | Not-determined |
| 18-105-final-viral-combined_k141_70800\|\|full | Low-quality |
| 18-105-final-viral-combined_k141_204432\|\|full | Not-determined |
| 18-105-final-viral-combined_k141_213297\|\|full | Low-quality |
| 18-105-final-viral-combined_k141_204451\|\|full | Low-quality |
| 18-105-final-viral-combined_k141_115663\|\|full | Low-quality |
| 18-105-final-viral-combined_k141_160156\|\|full | Low-quality |
| 18-105-final-viral-combined_k141_204459\|\|full | Low-quality |
| 18-105-final-viral-combined_k141_169177\|\|full | Low-quality |
| 18-105-final-viral-combined_k141_204475\|\|full | Not-determined |
| 18-105-final-viral-combined_k141_17641\|\|full | Low-quality |
| 18-105-final-viral-combined_k141_204478\|\|full | Low-quality |
| 18-105-final-viral-combined_k141_169196\|\|full | Low-quality |
| 18-105-final-viral-combined_k141_53411\|\|full | Low-quality |
| 18-105-final-viral-combined_k141_204500\|\|full | Low-quality |
| 18-105-final-viral-combined_k141_213360\|\|full | Not-determined |
| 18-105-final-viral-combined_k141_53416\|\|full | Not-determined |
| 18-105-final-viral-combined_k141_115698\|\|full | Low-quality |
| 18-105-final-viral-combined_k141_17673\|\|full | Not-determined |
| 18-105-final-viral-combined_k141_25709\|\|full | Not-determined |
| 18-105-final-viral-combined_k141_17682\|\|full | Not-determined |
| 18-105-final-viral-combined_k141_115728\|\|full | Not-determined |
| 18-105-final-viral-combined_k141_213370\|\|full | Not-determined |
| 18-105-final-viral-combined_k141_44138\|\|full | Low-quality |
| 18-105-final-viral-combined_k141_142489\|\|full | Not-determined |
| 18-105-final-viral-combined_k141_142492\|\|full | Not-determined |
| 18-105-final-viral-combined_k141_169216\|\|full | Low-quality |
| 18-105-final-viral-combined_k141_169223\|\|full | Low-quality |
| 18-105-final-viral-combined_k141_35552\|\|full | Not-determined |
| 18-105-final-viral-combined_k141_160219\|\|full | Not-determined |
| 18-105-final-viral-combined_k141_35559\|\|full | Low-quality |
| 18-105-final-viral-combined_k141_35560\|\|full | Not-determined |
| 18-105-final-viral-combined_k141_169246\|\|full | Low-quality |
| 18-105-final-viral-combined_k141_25766\|\|full | Not-determined |
| 18-105-final-viral-combined_k141_142512\|\|full | Not-determined |
| 18-105-final-viral-combined_k141_53447\|\|full | Low-quality |
| 18-105-final-viral-combined_k141_25798\|\|full | Not-determined |
| 18-105-final-viral-combined_k141_44187\|\|full | Not-determined |
| 18-105-final-viral-combined_k141_115757\|\|full | Low-quality |
| 18-105-final-viral-combined_k141_44191\|\|full | Not-determined |
| 18-105-final-viral-combined_k141_115766\|\|full | Not-determined |
| 18-105-final-viral-combined_k141_44193\|\|full | Not-determined |
| 18-105-final-viral-combined_k141_160274\|\|full | Low-quality |
| 18-105-final-viral-combined_k141_17726\|\|full | Low-quality |
| 18-105-final-viral-combined_k141_25823\|\|full | Low-quality |
| 18-105-final-viral-combined_k141_44216\|\|full | Low-quality |
| 18-105-final-viral-combined_k141_44227\|\|full | Not-determined |
| 18-105-final-viral-combined_k141_213420\|\|full | Low-quality |
| 18-105-final-viral-combined_k141_17763\|\|full | Low-quality |
| 18-105-final-viral-combined_k141_35596\|\|full | Low-quality |
| 18-105-final-viral-combined_k141_17785\|\|full | Not-determined |
| 18-105-final-viral-combined_k141_97987\|\|full | Low-quality |
| 18-105-final-viral-combined_k141_17788\|\|full | Not-determined |
| 18-105-final-viral-combined_k141_204681\|\|full | Not-determined |
| 18-105-final-viral-combined_k141_25902\|\|full | Not-determined |
| 18-105-final-viral-combined_k141_70908\|\|full | Not-determined |
| 18-105-final-viral-combined_k141_70910\|\|full | Low-quality |
| 18-105-final-viral-combined_k141_204736\|\|full | Not-determined |
| 18-105-final-viral-combined_k141_213459\|\|full | Low-quality |
| 18-105-final-viral-combined_k141_44347\|\|full | Not-determined |
| 18-105-final-viral-combined_k141_70929\|\|full | Low-quality |
| 18-105-final-viral-combined_k141_25965\|\|full | Not-determined |
| 18-105-final-viral-combined_k141_213476\|\|full | Low-quality |
| 18-105-final-viral-combined_k141_44359\|\|full | Low-quality |
| 18-105-final-viral-combined_k141_44378\|\|full | Not-determined |
| 18-105-final-viral-combined_k141_26006\|\|full | Low-quality |
| 18-105-final-viral-combined_k141_70940\|\|full | Not-determined |
| KM_k141_29082 | High-quality |
| 18-105-final-viral-combined_k141_26033\|\|full | Not-determined |
| 18-105-final-viral-combined_k141_62347\|\|full | Not-determined |
| 18-105-final-viral-combined_k141_70990\|\|full | Not-determined |
| 18-105-final-viral-combined_k141_26035\|\|full | Not-determined |
| 18-105-final-viral-combined_k141_26058\|\|full | Not-determined |
| 18-105-final-viral-combined_k141_204860\|\|full | Not-determined |
| 18-105-final-viral-combined_k141_204879\|\|full | Not-determined |
| 18-105-final-viral-combined_k141_44405\|\|full | Not-determined |
| 18-105-final-viral-combined_k141_71033\|\|full | Not-determined |
| 18-105-final-viral-combined_k141_44419\|\|full | Low-quality |
| 18-105-final-viral-combined_k141_44421\|\|full | Not-determined |
| 18-105-final-viral-combined_k141_71050\|\|full | Not-determined |
| 18-105-final-viral-combined_k141_26118\|\|full | Low-quality |
| 18-105-final-viral-combined_k141_26120\|\|full | Not-determined |
| 18-105-final-viral-combined_k141_26130\|\|full | Low-quality |
| 18-105-final-viral-combined_k141_26173\|\|full | Low-quality |
| 18-105-final-viral-combined_k141_71212\|\|full | Not-determined |
| 18-105-final-viral-combined_k141_71223\|\|full | Not-determined |
| 18-105-final-viral-combined_k141_71225\|\|full | Not-determined |
| 18-105-final-viral-combined_k141_71249\|\|full | Not-determined |
| 18-105-final-viral-combined_k141_71258\|\|full | Not-determined |
| 18-105-final-viral-combined_k141_71266\|\|full | Low-quality |
| 18-105-final-viral-combined_k141_26412\|\|full | Low-quality |
| 18-105-final-viral-combined_k141_26419\|\|full | Not-determined |
| 18-105-final-viral-combined_k141_26434\|\|full | Not-determined |
| 18-105-final-viral-combined_k141_26467\|\|full | Low-quality |
| 18-105-final-viral-combined_k141_26475\|\|full | Low-quality |
| 18-105-final-viral-combined_k141_26503\|\|full | Not-determined |
| 18-105-final-viral-combined_k141_26531\|\|full | Not-determined |
| 18-105-final-viral-combined_k141_26543\|\|full | Not-determined |
| 18-105-final-viral-combined_k141_26622\|\|full | Low-quality |
| 18-105-final-viral-combined_k141_26647\|\|full | Not-determined |
| 18-105-final-viral-combined_k141_26665\|\|full | Low-quality |
| 18-105-final-viral-combined_k141_26681\|\|full | Not-determined |
| 18-105-final-viral-combined_k141_26702\|\|full | Low-quality |
| 18-105-final-viral-combined_k141_26717\|\|full | Low-quality |
| 18-105-final-viral-combined_k141_26849\|\|0_partial | Low-quality |
| 18-105-final-viral-combined_k141_204940\|\|0_partial | Low-quality |
| 18-105-final-viral-combined_k141_196197\|\|0_partial | Not-determined |
| 18-105-final-viral-combined_k141_124904\|\|0_partial | Low-quality |
| 18-104-final-viral-combined_k141_57123\|\|full | Medium-quality |
| 17-109-final-viral-combined_k141_25459\|\|full | High-quality |
| 18-105-final-viral-combined_k141_142873\|\|0_partial | Not-determined |
| 18-105-final-viral-combined_k141_26983\|\|0_partial | Low-quality |
| 18-105-final-viral-combined_k141_27009\|\|0_partial | Low-quality |
| 18-105-final-viral-combined_k141_98307\|\|0_partial | Low-quality |
| 18-105-final-viral-combined_k141_9528\|\|0_partial | Low-quality |
| 18-105-final-viral-combined_k141_27036\|\|0_partial | Not-determined |
| 18-110_k141_82880 | Medium-quality |
| 18-105-final-viral-combined_k141_179168\|\|0_partial | Low-quality |
| 18-105-final-viral-combined_k141_18679\|\|0_partial | Low-quality |
| 18-105-final-viral-combined_k141_161321\|\|0_partial | Low-quality |
| 18-102-final-viral-combined_k141_248324\|\|full | High-quality |
| 18-105-final-viral-combined_k141_161469\|\|0_partial | Not-determined |
| 18-105-final-viral-combined_k141_63378\|\|0_partial | Low-quality |
| 18-105-final-viral-combined_k141_152631\|\|0_partial | Not-determined |
| 18-105-final-viral-combined_k141_45803\|\|0_partial | Low-quality |
| 18-105-final-viral-combined_k141_45803\|\|1_partial | Low-quality |
| 18-110-final-viral-combined_k141_82880\|\|full | Medium-quality |
| 18-105-final-viral-combined_k141_99254\|\|0_partial | Low-quality |
| 18-105-final-viral-combined_k141_46035\|\|0_partial | Not-determined |
| 18-105-final-viral-combined_k141_197664\|\|0_partial | Low-quality |
| 18-105-final-viral-combined_k141_206359\|\|0_partial | Low-quality |
| 18-105-final-viral-combined_k141_108718\|\|0_partial | Low-quality |
| 18-105-final-viral-combined_k141_180367\|\|0_partial | Low-quality |
| 18-105-final-viral-combined_k141_144698\|\|0_partial | Low-quality |
| 18-105-final-viral-combined_k141_91275\|\|0_partial | Low-quality |
| 18-105-final-viral-combined_k141_180393\|\|0_partial | Low-quality |
| 18-105-final-viral-combined_k141_10819\|\|0_partial | Low-quality |
| 18-105-final-viral-combined_k141_19663\|\|0_partial | Not-determined |
| 18-105-final-viral-combined_k141_162433\|\|0_partial | Not-determined |
| 18-105-final-viral-combined_k141_189786\|\|0_partial | Low-quality |
| 18-105-final-viral-combined_k141_91926\|\|0_partial | Low-quality |
| 18-105-final-viral-combined_k141_73929\|\|0_partial | Low-quality |
| 18-105-final-viral-combined_k141_109133\|\|0_partial | Low-quality |
| 18-105-final-viral-combined_k141_207323\|\|0_partial | Not-determined |
| 18-105-final-viral-combined_k141_172722\|\|0_partial | Low-quality |
| 18-101_k141_225995 | High-quality |
| 18-105-final-viral-combined_k141_172874\|\|0_partial | Not-determined |
| 18-105-final-viral-combined_k141_92221\|\|0_partial | Not-determined |
| 18-105-final-viral-combined_k141_48351\|\|0_partial | Not-determined |
| 18-105-final-viral-combined_k141_163801\|\|0_partial | Low-quality |
| 18-105-final-viral-combined_k141_48525\|\|0_partial | Low-quality |
| 18-105-final-viral-combined_k141_119198\|\|0_partial | Not-determined |
| 18-105-final-viral-combined_k141_20957\|\|0_partial | Low-quality |
| 18-105-final-viral-combined_k141_137171\|\|0_partial | Low-quality |
| 18-105-final-viral-combined_k141_110550\|\|0_partial | Low-quality |
| 18-105-final-viral-combined_k141_182127\|\|0_partial | Low-quality |
| 18-105-final-viral-combined_k141_173869\|\|0_partial | Not-determined |
| 18-105-final-viral-combined_k141_155356\|\|0_partial | Not-determined |
| 18-105-final-viral-combined_k141_30475\|\|0_partial | Not-determined |
| 18-101-final-viral-combined_k141_225995\|\|full | High-quality |
| 18-105-final-viral-combined_k141_164620\|\|0_partial | Low-quality |
| 18-105-final-viral-combined_k141_120070\|\|0_partial | Low-quality |
| 18-105-final-viral-combined_k141_191791\|\|0_partial | Low-quality |
| 18-105-final-viral-combined_k141_21581\|\|0_partial | Not-determined |
| 18-105-final-viral-combined_k141_155902\|\|0_partial | Low-quality |
| 17-108_k141_211679 | High-quality |
| 18-105-final-viral-combined_k141_93953\|\|0_partial | Low-quality |
| 18-105-final-viral-combined_k141_85202\|\|0_partial | Low-quality |
| 18-105-final-viral-combined_k141_200932\|\|0_partial | Not-determined |
| 18-105-final-viral-combined_k141_192105\|\|0_partial | Low-quality |
| 18-105-final-viral-combined_k141_40688\|\|0_partial | Low-quality |
| 18-105-final-viral-combined_k141_156223\|\|0_partial | Not-determined |
| 18-105-final-viral-combined_k141_147938\|\|0_partial | Low-quality |
| 18-105-final-viral-combined_k141_67335\|\|0_partial | Not-determined |
| 18-105-final-viral-combined_k141_112407\|\|0_partial | Not-determined |
| 18-105-final-viral-combined_k141_130617\|\|0_partial | Not-determined |
| 18-105-final-viral-combined_k141_7742\|\|0_partial | Low-quality |
| 18-105-final-viral-combined_k141_104194\|\|0_partial | Low-quality |
| 18-105-final-viral-combined_k141_41485\|\|0_partial | Low-quality |
| 18-105-final-viral-combined_k141_15022\|\|0_partial | Not-determined |
| 18-105-final-viral-combined_k141_193422\|\|0_partial | Not-determined |
| 18-105-final-viral-combined_k141_122056\|\|0_partial | Low-quality |
| 18-105-final-viral-combined_k141_202147\|\|0_partial | Low-quality |
| 18-105-final-viral-combined_k141_41953\|\|0_partial | Low-quality |
| 18-105-final-viral-combined_k141_202237\|\|0_partial | Low-quality |
| 18-105-final-viral-combined_k141_15798\|\|0_partial | Low-quality |
| 18-105-final-viral-combined_k141_194446\|\|0_partial | Not-determined |
| 18-105-final-viral-combined_k141_69272\|\|0_partial | Low-quality |
| 18-105-final-viral-combined_k141_211849\|\|0_partial | Not-determined |
| 18-105-final-viral-combined_k141_52566\|\|0_partial | Low-quality |
| 18-105-final-viral-combined_k141_42829\|\|0_partial | Not-determined |
| 18-105-final-viral-combined_k141_42829\|\|1_partial | Low-quality |
| 18-105-final-viral-combined_k141_203080\|\|0_partial | Low-quality |
| KM-final-viral-combined_k141_29082\|\|full | High-quality |
| 18-105-final-viral-combined_k141_167999\|\|0_partial | Not-determined |
| 18-105-final-viral-combined_k141_69722\|\|0_partial | Not-determined |
| 18-105-final-viral-combined_k141_141803\|\|0_partial | Low-quality |
| 18-105-final-viral-combined_k141_213019\|\|0_partial | Not-determined |
| 18-105-final-viral-combined_k141_35379\|\|0_partial | Low-quality |
| 18-105-final-viral-combined_k141_213553\|\|0_partial | Low-quality |
| 18-105-final-viral-combined_k141_80180\|\|lt2gene | Not-determined |
| 18-105-final-viral-combined_k141_204892\|\|lt2gene | Not-determined |
| 18-105-final-viral-combined_k141_80185\|\|lt2gene | Low-quality |
| 18-105-final-viral-combined_k141_115824\|\|lt2gene | Low-quality |
| 18-105-final-viral-combined_k141_106911\|\|lt2gene | Not-determined |
| 18-105-final-viral-combined_k141_115833\|\|lt2gene | Not-determined |
| 18-105-final-viral-combined_k141_53478\|\|lt2gene | Low-quality |
| 18-105-final-viral-combined_k141_160369\|\|lt2gene | Not-determined |
| 18-105-final-viral-combined_k141_8952\|\|lt2gene | Low-quality |
| 18-105-final-viral-combined_k141_151466\|\|lt2gene | Low-quality |
| 18-105-final-viral-combined_k141_35648\|\|lt2gene | Not-determined |
| 18-105-final-viral-combined_k141_187088\|\|lt2gene | Low-quality |
| 18-105-final-viral-combined_k141_124771\|\|lt2gene | Low-quality |
| 18-105-final-viral-combined_k141_8963\|\|lt2gene | Low-quality |
| 18-105-final-viral-combined_k141_133652\|\|lt2gene | Not-determined |
| 18-105-final-viral-combined_k141_151493\|\|lt2gene | Low-quality |
| 18-105-final-viral-combined_k141_44559\|\|lt2gene | Not-determined |
| 18-105-final-viral-combined_k141_80240\|\|lt2gene | Low-quality |
| 18-105-final-viral-combined_k141_187117\|\|lt2gene | Low-quality |
| 18-105-final-viral-combined_k141_71340\|\|lt2gene | Not-determined |
| 18-105-final-viral-combined_k141_124807\|\|lt2gene | Not-determined |
| 18-105-final-viral-combined_k141_133707\|\|lt2gene | Low-quality |
| 18-105-final-viral-combined_k141_17875\|\|lt2gene | Low-quality |
| 18-105-final-viral-combined_k141_80262\|\|lt2gene | Low-quality |
| 18-105-final-viral-combined_k141_133734\|\|lt2gene | Not-determined |
| 18-105-final-viral-combined_k141_26846\|\|lt2gene | Low-quality |
| 18-105-final-viral-combined_k141_196051\|\|lt2gene | Low-quality |
| 18-105-final-viral-combined_k141_133777\|\|lt2gene | Low-quality |
| 18-105-final-viral-combined_k141_160523\|\|lt2gene | Low-quality |
| 18-105-final-viral-combined_k141_18003\|\|lt2gene | Low-quality |
| 18-105-final-viral-combined_k141_9126\|\|lt2gene | Low-quality |
| 18-105-final-viral-combined_k141_62441\|\|lt2gene | Low-quality |
| 18-105-final-viral-combined_k141_71442\|\|lt2gene | Low-quality |
| 18-105-final-viral-combined_k141_9130\|\|lt2gene | Low-quality |
| 18-105-final-viral-combined_k141_98120\|\|lt2gene | Low-quality |
| 18-105-final-viral-combined_k141_71460\|\|lt2gene | Low-quality |
| 18-105-final-viral-combined_k141_98126\|\|lt2gene | Low-quality |
| 18-105-final-viral-combined_k141_133848\|\|lt2gene | Low-quality |
| 18-105-final-viral-combined_k141_107092\|\|lt2gene | Not-determined |
| 18-105-final-viral-combined_k141_160563\|\|lt2gene | Low-quality |
| 18-105-final-viral-combined_k141_9165\|\|lt2gene | Low-quality |
| 18-105-final-viral-combined_k141_71540\|\|lt2gene | Low-quality |
| 18-105-final-viral-combined_k141_98185\|\|lt2gene | Not-determined |
| 18-105-final-viral-combined_k141_80343\|\|lt2gene | Low-quality |
| 18-105-final-viral-combined_k141_107161\|\|lt2gene | Low-quality |
| 18-105-final-viral-combined_k141_196208\|\|lt2gene | Low-quality |
| 18-105-final-viral-combined_k141_160603\|\|lt2gene | Low-quality |
| 18-105-final-viral-combined_k141_142703\|\|lt2gene | Not-determined |
| 18-105-final-viral-combined_k141_124922\|\|lt2gene | Low-quality |
| 18-105-final-viral-combined_k141_89224\|\|lt2gene | Low-quality |
| 18-105-final-viral-combined_k141_9222\|\|lt2gene | Not-determined |
| 18-105-final-viral-combined_k141_196252\|\|lt2gene | Low-quality |
| 18-105-final-viral-combined_k141_169342\|\|lt2gene | Low-quality |
| 18-105-final-viral-combined_k141_9241\|\|lt2gene | Low-quality |
| 18-105-final-viral-combined_k141_142768\|\|lt2gene | Low-quality |
| 18-105-final-viral-combined_k141_124980\|\|lt2gene | Low-quality |
| 18-105-final-viral-combined_k141_80456\|\|lt2gene | Low-quality |
| 18-105-final-viral-combined_k141_160659\|\|lt2gene | Low-quality |
| 18-105-final-viral-combined_k141_124989\|\|lt2gene | Low-quality |
| 18-105-final-viral-combined_k141_124995\|\|lt2gene | Low-quality |
| 18-105-final-viral-combined_k141_124999\|\|lt2gene | Low-quality |
| 18-105-final-viral-combined_k141_125001\|\|lt2gene | Low-quality |
| 18-105-final-viral-combined_k141_80469\|\|lt2gene | Not-determined |
| 18-105-final-viral-combined_k141_80481\|\|lt2gene | Low-quality |
| 18-105-final-viral-combined_k141_116184\|\|lt2gene | Low-quality |
| 18-105-final-viral-combined_k141_205011\|\|lt2gene | Low-quality |
| 18-105-final-viral-combined_k141_62609\|\|lt2gene | Low-quality |
| 18-105-final-viral-combined_k141_518\|\|lt2gene | Low-quality |
| 18-105-final-viral-combined_k141_62627\|\|lt2gene | Low-quality |
| 18-105-final-viral-combined_k141_9326\|\|lt2gene | Low-quality |
| 18-105-final-viral-combined_k141_9334\|\|lt2gene | Low-quality |
| 18-105-final-viral-combined_k141_71662\|\|lt2gene | Low-quality |
| 18-105-final-viral-combined_k141_35794\|\|lt2gene | Low-quality |
| 18-105-final-viral-combined_k141_142788\|\|lt2gene | Low-quality |
| 18-105-final-viral-combined_k141_71686\|\|lt2gene | Low-quality |
| 18-105-final-viral-combined_k141_98228\|\|lt2gene | Low-quality |
| 18-105-final-viral-combined_k141_98255\|\|lt2gene | Low-quality |
| 18-105-final-viral-combined_k141_9402\|\|lt2gene | Low-quality |
| 18-105-final-viral-combined_k141_35819\|\|lt2gene | Low-quality |
| 18-105-final-viral-combined_k141_205106\|\|lt2gene | Low-quality |
| 18-105-final-viral-combined_k141_196470\|\|lt2gene | Low-quality |
| 18-105-final-viral-combined_k141_196476\|\|lt2gene | Low-quality |
| 18-105-final-viral-combined_k141_169490\|\|lt2gene | Not-determined |
| 18-105-final-viral-combined_k141_151641\|\|lt2gene | Low-quality |
| 18-105-final-viral-combined_k141_169495\|\|lt2gene | Low-quality |
| 18-105-final-viral-combined_k141_196495\|\|lt2gene | Not-determined |
| 18-105-final-viral-combined_k141_71789\|\|lt2gene | Low-quality |
| 18-105-final-viral-combined_k141_142914\|\|lt2gene | Low-quality |
| 18-105-final-viral-combined_k141_9527\|\|lt2gene | Not-determined |
| 18-105-final-viral-combined_k141_35851\|\|lt2gene | Not-determined |
| 18-105-final-viral-combined_k141_35854\|\|lt2gene | Not-determined |
| 18-105-final-viral-combined_k141_35864\|\|lt2gene | Low-quality |
| 18-105-final-viral-combined_k141_169574\|\|lt2gene | Not-determined |
| 18-105-final-viral-combined_k141_80656\|\|lt2gene | Low-quality |
| 18-105-final-viral-combined_k141_18235\|\|lt2gene | Not-determined |
| 18-105-final-viral-combined_k141_35907\|\|lt2gene | Low-quality |
| 18-105-final-viral-combined_k141_44838\|\|lt2gene | Low-quality |
| 18-105-final-viral-combined_k141_89439\|\|lt2gene | Low-quality |
| 18-105-final-viral-combined_k141_98317\|\|lt2gene | Low-quality |
| 18-105-final-viral-combined_k141_53941\|\|lt2gene | Low-quality |
| 18-105-final-viral-combined_k141_205194\|\|lt2gene | Low-quality |
| 18-105-final-viral-combined_k141_169608\|\|lt2gene | Low-quality |
| 18-105-final-viral-combined_k141_89471\|\|lt2gene | Low-quality |
| 18-105-final-viral-combined_k141_789\|\|lt2gene | Low-quality |
| 18-105-final-viral-combined_k141_9566\|\|lt2gene | Low-quality |
| 18-105-final-viral-combined_k141_9574\|\|lt2gene | Low-quality |
| 18-105-final-viral-combined_k141_160882\|\|lt2gene | Not-determined |
| 18-105-final-viral-combined_k141_9616\|\|lt2gene | Low-quality |
| 18-105-final-viral-combined_k141_80754\|\|lt2gene | Low-quality |
| 18-105-final-viral-combined_k141_98460\|\|lt2gene | Low-quality |
| 18-105-final-viral-combined_k141_9672\|\|lt2gene | Not-determined |
| 18-105-final-viral-combined_k141_107461\|\|lt2gene | Not-determined |
| 18-105-final-viral-combined_k141_18362\|\|lt2gene | Not-determined |
| 18-105-final-viral-combined_k141_44999\|\|lt2gene | Low-quality |
| 18-105-final-viral-combined_k141_18365\|\|lt2gene | Low-quality |
| 18-105-final-viral-combined_k141_89600\|\|lt2gene | Not-determined |
| 18-105-final-viral-combined_k141_125168\|\|lt2gene | Low-quality |
| 18-105-final-viral-combined_k141_98495\|\|lt2gene | Low-quality |
| 18-105-final-viral-combined_k141_921\|\|lt2gene | Low-quality |
| 18-105-final-viral-combined_k141_125194\|\|lt2gene | Low-quality |
| 18-105-final-viral-combined_k141_125198\|\|lt2gene | Low-quality |
| 18-105-final-viral-combined_k141_98523\|\|lt2gene | Low-quality |
| 18-105-final-viral-combined_k141_98545\|\|lt2gene | Low-quality |
| 18-105-final-viral-combined_k141_143219\|\|lt2gene | Not-determined |
| 18-105-final-viral-combined_k141_205368\|\|lt2gene | Low-quality |
| 18-105-final-viral-combined_k141_169815\|\|lt2gene | Not-determined |
| 18-105-final-viral-combined_k141_98623\|\|lt2gene | Low-quality |
| 18-105-final-viral-combined_k141_151965\|\|lt2gene | Low-quality |
| 18-105-final-viral-combined_k141_169836\|\|lt2gene | Not-determined |
| 18-105-final-viral-combined_k141_18461\|\|lt2gene | Low-quality |
| 18-105-final-viral-combined_k141_36096\|\|lt2gene | Low-quality |
| 18-105-final-viral-combined_k141_71979\|\|lt2gene | Low-quality |
| 18-105-final-viral-combined_k141_125270\|\|lt2gene | Low-quality |
| 18-105-final-viral-combined_k141_71990\|\|lt2gene | Low-quality |
| 18-105-final-viral-combined_k141_54037\|\|lt2gene | Low-quality |
| 18-105-final-viral-combined_k141_125299\|\|lt2gene | Not-determined |
| 18-105-final-viral-combined_k141_27182\|\|lt2gene | Low-quality |
| 18-105-final-viral-combined_k141_152017\|\|lt2gene | Low-quality |
| 18-105-final-viral-combined_k141_45068\|\|lt2gene | Not-determined |
| 18-105-final-viral-combined_k141_27200\|\|lt2gene | Low-quality |
| 18-105-final-viral-combined_k141_178561\|\|lt2gene | Not-determined |
| 18-105-final-viral-combined_k141_71997\|\|lt2gene | Low-quality |
| 18-105-final-viral-combined_k141_89859\|\|lt2gene | Low-quality |
| 18-105-final-viral-combined_k141_169995\|\|lt2gene | Low-quality |
| 18-105-final-viral-combined_k141_107694\|\|lt2gene | Low-quality |
| 18-105-final-viral-combined_k141_89909\|\|lt2gene | Low-quality |
| 18-105-final-viral-combined_k141_62921\|\|lt2gene | Low-quality |
| 18-105-final-viral-combined_k141_178709\|\|lt2gene | Low-quality |
| 18-105-final-viral-combined_k141_205478\|\|lt2gene | Low-quality |
| 18-105-final-viral-combined_k141_178728\|\|lt2gene | Low-quality |
| 18-105-final-viral-combined_k141_80835\|\|lt2gene | Low-quality |
| 18-105-final-viral-combined_k141_107736\|\|lt2gene | Low-quality |
| 18-105-final-viral-combined_k141_45201\|\|lt2gene | Low-quality |
| 18-105-final-viral-combined_k141_1183\|\|lt2gene | Low-quality |
| 18-105-final-viral-combined_k141_107751\|\|lt2gene | Low-quality |
| 18-105-final-viral-combined_k141_80848\|\|lt2gene | Not-determined |
| 18-105-final-viral-combined_k141_170076\|\|lt2gene | Not-determined |
| 18-105-final-viral-combined_k141_143283\|\|lt2gene | Low-quality |
| 18-105-final-viral-combined_k141_170105\|\|lt2gene | Not-determined |
| 18-105-final-viral-combined_k141_1243\|\|lt2gene | Low-quality |
| 18-105-final-viral-combined_k141_27255\|\|lt2gene | Low-quality |
| 18-105-final-viral-combined_k141_54150\|\|lt2gene | Low-quality |
| 18-105-final-viral-combined_k141_134319\|\|lt2gene | Low-quality |
| 18-105-final-viral-combined_k141_1305\|\|lt2gene | Low-quality |
| 18-105-final-viral-combined_k141_54198\|\|lt2gene | Low-quality |
| 18-105-final-viral-combined_k141_90033\|\|lt2gene | Low-quality |
| 18-105-final-viral-combined_k141_90040\|\|lt2gene | Low-quality |
| 18-105-final-viral-combined_k141_152130\|\|lt2gene | Not-determined |
| 18-105-final-viral-combined_k141_170164\|\|lt2gene | Low-quality |
| 18-105-final-viral-combined_k141_62975\|\|lt2gene | Low-quality |
| 18-105-final-viral-combined_k141_45286\|\|lt2gene | Low-quality |
| 18-105-final-viral-combined_k141_90055\|\|lt2gene | Low-quality |
| 18-105-final-viral-combined_k141_90056\|\|lt2gene | Low-quality |
| 18-105-final-viral-combined_k141_187736\|\|lt2gene | Low-quality |
| 18-105-final-viral-combined_k141_90073\|\|lt2gene | Low-quality |
| 18-105-final-viral-combined_k141_9923\|\|lt2gene | Low-quality |
| 18-105-final-viral-combined_k141_54271\|\|lt2gene | Low-quality |
| 18-105-final-viral-combined_k141_27359\|\|lt2gene | Low-quality |
| 18-105-final-viral-combined_k141_90094\|\|lt2gene | Not-determined |
| 18-105-final-viral-combined_k141_80902\|\|lt2gene | Not-determined |
| 18-105-final-viral-combined_k141_9943\|\|lt2gene | Not-determined |
| 18-105-final-viral-combined_k141_1359\|\|lt2gene | Low-quality |
| 18-105-final-viral-combined_k141_1363\|\|lt2gene | Low-quality |
| 18-105-final-viral-combined_k141_170243\|\|lt2gene | Low-quality |
| 18-105-final-viral-combined_k141_1368\|\|lt2gene | Low-quality |
| 18-105-final-viral-combined_k141_98811\|\|lt2gene | Low-quality |
| 18-105-final-viral-combined_k141_90128\|\|lt2gene | Low-quality |
| 18-105-final-viral-combined_k141_54320\|\|lt2gene | Not-determined |
| 18-105-final-viral-combined_k141_1388\|\|lt2gene | Low-quality |
| 18-105-final-viral-combined_k141_161212\|\|lt2gene | Not-determined |
| 18-105-final-viral-combined_k141_187783\|\|lt2gene | Low-quality |
| 18-105-final-viral-combined_k141_161250\|\|lt2gene | Low-quality |
| 18-105-final-viral-combined_k141_143422\|\|lt2gene | Not-determined |
| 18-105-final-viral-combined_k141_178946\|\|lt2gene | Low-quality |
| 18-105-final-viral-combined_k141_178952\|\|lt2gene | Low-quality |
| 18-105-final-viral-combined_k141_9994\|\|lt2gene | Not-determined |
| 18-105-final-viral-combined_k141_170290\|\|lt2gene | Low-quality |
| 18-105-final-viral-combined_k141_178995\|\|lt2gene | Low-quality |
| 18-105-final-viral-combined_k141_45472\|\|lt2gene | Low-quality |
| 18-105-final-viral-combined_k141_116703\|\|lt2gene | Low-quality |
| 18-105-final-viral-combined_k141_107927\|\|lt2gene | Low-quality |
| 18-105-final-viral-combined_k141_107929\|\|lt2gene | Not-determined |
| 18-105-final-viral-combined_k141_107941\|\|lt2gene | Low-quality |
| 18-105-final-viral-combined_k141_187857\|\|lt2gene | Low-quality |
| 18-105-final-viral-combined_k141_134506\|\|lt2gene | Low-quality |
| 18-105-final-viral-combined_k141_98904\|\|lt2gene | Not-determined |
| 18-105-final-viral-combined_k141_108010\|\|lt2gene | Not-determined |
| 18-105-final-viral-combined_k141_179104\|\|lt2gene | Low-quality |
| 18-105-final-viral-combined_k141_116754\|\|lt2gene | Not-determined |
| 18-105-final-viral-combined_k141_108039\|\|lt2gene | Low-quality |
| 18-105-final-viral-combined_k141_125681\|\|lt2gene | Not-determined |
| 18-105-final-viral-combined_k141_98936\|\|lt2gene | Not-determined |
| 18-105-final-viral-combined_k141_125697\|\|lt2gene | Low-quality |
| 18-105-final-viral-combined_k141_187871\|\|lt2gene | Not-determined |
| 18-105-final-viral-combined_k141_143546\|\|lt2gene | Not-determined |
| 18-105-final-viral-combined_k141_1659\|\|lt2gene | Low-quality |
| 18-105-final-viral-combined_k141_72182\|\|lt2gene | Not-determined |
| 18-105-final-viral-combined_k141_179187\|\|lt2gene | Low-quality |
| 18-105-final-viral-combined_k141_179188\|\|lt2gene | Low-quality |
| 18-105-final-viral-combined_k141_134593\|\|lt2gene | Low-quality |
| 18-105-final-viral-combined_k141_72225\|\|lt2gene | Low-quality |
| 18-105-final-viral-combined_k141_27658\|\|lt2gene | Low-quality |
| 18-105-final-viral-combined_k141_143589\|\|lt2gene | Low-quality |
| 18-105-final-viral-combined_k141_125756\|\|lt2gene | Low-quality |
| 18-105-final-viral-combined_k141_197034\|\|lt2gene | Low-quality |
| 18-105-final-viral-combined_k141_125792\|\|lt2gene | Low-quality |
| 18-105-final-viral-combined_k141_116862\|\|lt2gene | Not-determined |
| 18-105-final-viral-combined_k141_81099\|\|lt2gene | Not-determined |
| 18-105-final-viral-combined_k141_134666\|\|lt2gene | Not-determined |
| 18-105-final-viral-combined_k141_72270\|\|lt2gene | Not-determined |
| 18-105-final-viral-combined_k141_197065\|\|lt2gene | Low-quality |
| 18-105-final-viral-combined_k141_116893\|\|lt2gene | Not-determined |
| 18-105-final-viral-combined_k141_90374\|\|lt2gene | Low-quality |
| 18-105-final-viral-combined_k141_1770\|\|lt2gene | Not-determined |
| 18-105-final-viral-combined_k141_72323\|\|lt2gene | Low-quality |
| 18-105-final-viral-combined_k141_152479\|\|lt2gene | Low-quality |
| 18-105-final-viral-combined_k141_72341\|\|lt2gene | Low-quality |
| 18-105-final-viral-combined_k141_152485\|\|lt2gene | Not-determined |
| 18-105-final-viral-combined_k141_125894\|\|lt2gene | Low-quality |
| 18-105-final-viral-combined_k141_108199\|\|lt2gene | Low-quality |
| 18-105-final-viral-combined_k141_152500\|\|lt2gene | Not-determined |
| 18-105-final-viral-combined_k141_72380\|\|lt2gene | Low-quality |
| 18-105-final-viral-combined_k141_187992\|\|lt2gene | Low-quality |
| 18-105-final-viral-combined_k141_1795\|\|lt2gene | Low-quality |
| 18-105-final-viral-combined_k141_1813\|\|lt2gene | Not-determined |
| 18-105-final-viral-combined_k141_36633\|\|lt2gene | Low-quality |
| 18-105-final-viral-combined_k141_1827\|\|lt2gene | Low-quality |
| 18-105-final-viral-combined_k141_54591\|\|lt2gene | Low-quality |
| 18-105-final-viral-combined_k141_117060\|\|lt2gene | Not-determined |
| 18-105-final-viral-combined_k141_117063\|\|lt2gene | Low-quality |
| 18-105-final-viral-combined_k141_54596\|\|lt2gene | Low-quality |
| 18-105-final-viral-combined_k141_205928\|\|lt2gene | Low-quality |
| 18-105-final-viral-combined_k141_161409\|\|lt2gene | Low-quality |
| 18-105-final-viral-combined_k141_143773\|\|lt2gene | Low-quality |
| 18-105-final-viral-combined_k141_179307\|\|lt2gene | Low-quality |
| 18-105-final-viral-combined_k141_54710\|\|lt2gene | Low-quality |
| 18-105-final-viral-combined_k141_1925\|\|lt2gene | Low-quality |
| 18-105-final-viral-combined_k141_1943\|\|lt2gene | Low-quality |
| 18-105-final-viral-combined_k141_63365\|\|lt2gene | Low-quality |
| 18-105-final-viral-combined_k141_45802\|\|lt2gene | Low-quality |
| 18-105-final-viral-combined_k141_161444\|\|lt2gene | Low-quality |
| 18-105-final-viral-combined_k141_143873\|\|lt2gene | Low-quality |
| 18-105-final-viral-combined_k141_108386\|\|lt2gene | Low-quality |
| 18-105-final-viral-combined_k141_108406\|\|lt2gene | Not-determined |
| 18-105-final-viral-combined_k141_143883\|\|lt2gene | Low-quality |
| 18-105-final-viral-combined_k141_117125\|\|lt2gene | Low-quality |
| 18-105-final-viral-combined_k141_117133\|\|lt2gene | Low-quality |
| 18-105-final-viral-combined_k141_90477\|\|lt2gene | Not-determined |
| 18-105-final-viral-combined_k141_206009\|\|lt2gene | Low-quality |
| 18-105-final-viral-combined_k141_206010\|\|lt2gene | Low-quality |
| 18-105-final-viral-combined_k141_117191\|\|lt2gene | Not-determined |
| 18-105-final-viral-combined_k141_188204\|\|lt2gene | Low-quality |
| 18-105-final-viral-combined_k141_1965\|\|lt2gene | Low-quality |
| 18-105-final-viral-combined_k141_36768\|\|lt2gene | Low-quality |
| 18-105-final-viral-combined_k141_10238\|\|lt2gene | Low-quality |
| 18-105-final-viral-combined_k141_108527\|\|lt2gene | Low-quality |
| 18-105-final-viral-combined_k141_152710\|\|lt2gene | Low-quality |
| 18-105-final-viral-combined_k141_108531\|\|lt2gene | Low-quality |
| 18-105-final-viral-combined_k141_2007\|\|lt2gene | Low-quality |
| 18-105-final-viral-combined_k141_108539\|\|lt2gene | Not-determined |
| 18-105-final-viral-combined_k141_63457\|\|lt2gene | Low-quality |
| 18-105-final-viral-combined_k141_45813\|\|lt2gene | Low-quality |
| 18-105-final-viral-combined_k141_90533\|\|lt2gene | Low-quality |
| 18-105-final-viral-combined_k141_144009\|\|lt2gene | Not-determined |
| 18-105-final-viral-combined_k141_10291\|\|lt2gene | Low-quality |
| 18-105-final-viral-combined_k141_27962\|\|lt2gene | Low-quality |
| 18-105-final-viral-combined_k141_72593\|\|lt2gene | Not-determined |
| 18-105-final-viral-combined_k141_161536\|\|lt2gene | Low-quality |
| 18-105-final-viral-combined_k141_125933\|\|lt2gene | Low-quality |
| 18-105-final-viral-combined_k141_18924\|\|lt2gene | Not-determined |
| 18-105-final-viral-combined_k141_179723\|\|lt2gene | Not-determined |
| 18-105-final-viral-combined_k141_28007\|\|lt2gene | Low-quality |
| 18-105-final-viral-combined_k141_81508\|\|lt2gene | Low-quality |
| 18-105-final-viral-combined_k141_117239\|\|lt2gene | Not-determined |
| 18-105-final-viral-combined_k141_126023\|\|lt2gene | Low-quality |
| 18-105-final-viral-combined_k141_72654\|\|lt2gene | Low-quality |
| 18-105-final-viral-combined_k141_36903\|\|lt2gene | Low-quality |

| | |
|---|---|
| 18-105-final-viral-combined_k141_179828\|\|lt2gene | Low-quality |
| 18-105-final-viral-combined_k141_144189\|\|lt2gene | Low-quality |
| 18-105-final-viral-combined_k141_117250\|\|lt2gene | Not-determined |
| 18-105-final-viral-combined_k141_19007\|\|lt2gene | Not-determined |
| 18-105-final-viral-combined_k141_54946\|\|lt2gene | Low-quality |
| 18-105-final-viral-combined_k141_19015\|\|lt2gene | Not-determined |
| 18-105-final-viral-combined_k141_28057\|\|lt2gene | Low-quality |
| 18-105-final-viral-combined_k141_90695\|\|lt2gene | Low-quality |
| 18-105-final-viral-combined_k141_45975\|\|lt2gene | Low-quality |
| 18-105-final-viral-combined_k141_134955\|\|lt2gene | Low-quality |
| 18-105-final-viral-combined_k141_45986\|\|lt2gene | Not-determined |
| 18-105-final-viral-combined_k141_90717\|\|lt2gene | Low-quality |
| 18-105-final-viral-combined_k141_197378\|\|lt2gene | Not-determined |
| 18-105-final-viral-combined_k141_152892\|\|lt2gene | Not-determined |
| 18-105-final-viral-combined_k141_134985\|\|lt2gene | Not-determined |
| 18-105-final-viral-combined_k141_152915\|\|lt2gene | Low-quality |
| 18-105-final-viral-combined_k141_188475\|\|lt2gene | Not-determined |
| 18-105-final-viral-combined_k141_144302\|\|lt2gene | Low-quality |
| 18-105-final-viral-combined_k141_126091\|\|lt2gene | Not-determined |
| 18-105-final-viral-combined_k141_144326\|\|lt2gene | Low-quality |
| 18-105-final-viral-combined_k141_126109\|\|lt2gene | Not-determined |
| 18-105-final-viral-combined_k141_10488\|\|lt2gene | Low-quality |
| 18-105-final-viral-combined_k141_152926\|\|lt2gene | Not-determined |
| 18-105-final-viral-combined_k141_161636\|\|lt2gene | Low-quality |
| 18-105-final-viral-combined_k141_152961\|\|lt2gene | Low-quality |
| 18-105-final-viral-combined_k141_2283\|\|lt2gene | Low-quality |
| 18-105-final-viral-combined_k141_126123\|\|lt2gene | Low-quality |
| 18-105-final-viral-combined_k141_206143\|\|lt2gene | Low-quality |
| 18-105-final-viral-combined_k141_126157\|\|lt2gene | Not-determined |
| 18-105-final-viral-combined_k141_10556\|\|lt2gene | Low-quality |
| 18-105-final-viral-combined_k141_72805\|\|lt2gene | Not-determined |
| 18-105-final-viral-combined_k141_10595\|\|lt2gene | Low-quality |
| 18-105-final-viral-combined_k141_144379\|\|lt2gene | Low-quality |
| 18-105-final-viral-combined_k141_188612\|\|lt2gene | Not-determined |
| 18-105-final-viral-combined_k141_206210\|\|lt2gene | Low-quality |
| 18-105-final-viral-combined_k141_46055\|\|lt2gene | Low-quality |
| 18-105-final-viral-combined_k141_19162\|\|lt2gene | Low-quality |
| 18-105-final-viral-combined_k141_90843\|\|lt2gene | Low-quality |
| 18-105-final-viral-combined_k141_180026\|\|lt2gene | Low-quality |
| 18-105-final-viral-combined_k141_37128\|\|lt2gene | Not-determined |
| 18-105-final-viral-combined_k141_46122\|\|lt2gene | Low-quality |
| 18-105-final-viral-combined_k141_81860\|\|lt2gene | Low-quality |
| 18-105-final-viral-combined_k141_46171\|\|lt2gene | Not-determined |
| 18-105-final-viral-combined_k141_117291\|\|lt2gene | Not-determined |
| 18-105-final-viral-combined_k141_161829\|\|lt2gene | Not-determined |
| 18-105-final-viral-combined_k141_206288\|\|lt2gene | Low-quality |
| 18-105-final-viral-combined_k141_180156\|\|lt2gene | Low-quality |
| 18-105-final-viral-combined_k141_206297\|\|lt2gene | Low-quality |
| 18-105-final-viral-combined_k141_37239\|\|lt2gene | Not-determined |
| 18-105-final-viral-combined_k141_188724\|\|lt2gene | Low-quality |
| 18-105-final-viral-combined_k141_188730\|\|lt2gene | Low-quality |
| 18-105-final-viral-combined_k141_73111\|\|lt2gene | Low-quality |
| 18-105-final-viral-combined_k141_55229\|\|lt2gene | Not-determined |
| 18-105-final-viral-combined_k141_144522\|\|lt2gene | Low-quality |
| 18-105-final-viral-combined_k141_180205\|\|lt2gene | Not-determined |
| 18-105-final-viral-combined_k141_28199\|\|lt2gene | Low-quality |
| 18-105-final-viral-combined_k141_144533\|\|lt2gene | Low-quality |
| 18-105-final-viral-combined_k141_180216\|\|lt2gene | Low-quality |
| 18-105-final-viral-combined_k141_46330\|\|lt2gene | Low-quality |
| 18-105-final-viral-combined_k141_171500\|\|lt2gene | Low-quality |
| 18-105-final-viral-combined_k141_2456\|\|lt2gene | Not-determined |
| 18-105-final-viral-combined_k141_126482\|\|lt2gene | Low-quality |
| 18-105-final-viral-combined_k141_19312\|\|lt2gene | Low-quality |
| 18-105-final-viral-combined_k141_55262\|\|lt2gene | Low-quality |
| 18-105-final-viral-combined_k141_153217\|\|lt2gene | Not-determined |
| 18-105-final-viral-combined_k141_19329\|\|lt2gene | Low-quality |
| 18-105-final-viral-combined_k141_135253\|\|lt2gene | Low-quality |
| 18-105-final-viral-combined_k141_37359\|\|lt2gene | Low-quality |
| 18-105-final-viral-combined_k141_37365\|\|lt2gene | Not-determined |
| 18-105-final-viral-combined_k141_99693\|\|lt2gene | Not-determined |
| 18-105-final-viral-combined_k141_91088\|\|lt2gene | Not-determined |
| 18-105-final-viral-combined_k141_37383\|\|lt2gene | Low-quality |
| 18-105-final-viral-combined_k141_37387\|\|lt2gene | Not-determined |
| 18-105-final-viral-combined_k141_108736\|\|lt2gene | Low-quality |
| 18-105-final-viral-combined_k141_91096\|\|lt2gene | Low-quality |
| 18-105-final-viral-combined_k141_144634\|\|lt2gene | Low-quality |
| 18-105-final-viral-combined_k141_144673\|\|lt2gene | Low-quality |
| 18-105-final-viral-combined_k141_2565\|\|lt2gene | Low-quality |
| 18-105-final-viral-combined_k141_188899\|\|lt2gene | Low-quality |
| 18-105-final-viral-combined_k141_171624\|\|lt2gene | Not-determined |
| 18-105-final-viral-combined_k141_171647\|\|lt2gene | Not-determined |
| 18-105-final-viral-combined_k141_91136\|\|lt2gene | Low-quality |
| 18-105-final-viral-combined_k141_2648\|\|lt2gene | Not-determined |
| 18-105-final-viral-combined_k141_2664\|\|lt2gene | Not-determined |
| 18-105-final-viral-combined_k141_189023\|\|lt2gene | Low-quality |
| 18-105-final-viral-combined_k141_135377\|\|lt2gene | Low-quality |
| 18-105-final-viral-combined_k141_91179\|\|lt2gene | Low-quality |
| 18-105-final-viral-combined_k141_189043\|\|lt2gene | Low-quality |
| 18-105-final-viral-combined_k141_135389\|\|lt2gene | Low-quality |
| 18-105-final-viral-combined_k141_189065\|\|lt2gene | Not-determined |
| 18-105-final-viral-combined_k141_91213\|\|lt2gene | Low-quality |
| 18-105-final-viral-combined_k141_19502\|\|lt2gene | Low-quality |
| 18-105-final-viral-combined_k141_63982\|\|lt2gene | Low-quality |
| 18-105-final-viral-combined_k141_117603\|\|lt2gene | Low-quality |
| 18-105-final-viral-combined_k141_180389\|\|lt2gene | Not-determined |
| 18-105-final-viral-combined_k141_99795\|\|lt2gene | Low-quality |
| 18-105-final-viral-combined_k141_206492\|\|lt2gene | Not-determined |
| 18-105-final-viral-combined_k141_46528\|\|lt2gene | Low-quality |
| 18-105-final-viral-combined_k141_144726\|\|lt2gene | Low-quality |
| 18-105-final-viral-combined_k141_64030\|\|lt2gene | Low-quality |
| 18-105-final-viral-combined_k141_2765\|\|lt2gene | Low-quality |
| 18-105-final-viral-combined_k141_108761\|\|lt2gene | Not-determined |
| 18-105-final-viral-combined_k141_126686\|\|lt2gene | Not-determined |
| 18-105-final-viral-combined_k141_2824\|\|lt2gene | Low-quality |
| 18-105-final-viral-combined_k141_55392\|\|lt2gene | Low-quality |
| 18-105-final-viral-combined_k141_171793\|\|lt2gene | Low-quality |
| 18-105-final-viral-combined_k141_46624\|\|lt2gene | Not-determined |
| 18-105-final-viral-combined_k141_73424\|\|lt2gene | Low-quality |
| 18-105-final-viral-combined_k141_73426\|\|lt2gene | Low-quality |
| 18-105-final-viral-combined_k141_2841\|\|lt2gene | Low-quality |
| 18-105-final-viral-combined_k141_64100\|\|lt2gene | Not-determined |
| 18-105-final-viral-combined_k141_99911\|\|lt2gene | Low-quality |
| 18-105-final-viral-combined_k141_189142\|\|lt2gene | Low-quality |
| 18-105-final-viral-combined_k141_46700\|\|lt2gene | Low-quality |
| 18-105-final-viral-combined_k141_99958\|\|lt2gene | Low-quality |
| 18-105-final-viral-combined_k141_91294\|\|lt2gene | Not-determined |
| 18-105-final-viral-combined_k141_55443\|\|lt2gene | Not-determined |
| 18-105-final-viral-combined_k141_162102\|\|lt2gene | Low-quality |
| 18-105-final-viral-combined_k141_100033\|\|lt2gene | Low-quality |
| 18-105-final-viral-combined_k141_28630\|\|lt2gene | Low-quality |
| 18-105-final-viral-combined_k141_37684\|\|lt2gene | Low-quality |
| 18-105-final-viral-combined_k141_171929\|\|lt2gene | Low-quality |
| 18-105-final-viral-combined_k141_64131\|\|lt2gene | Low-quality |
| 18-105-final-viral-combined_k141_117726\|\|lt2gene | Low-quality |
| 18-105-final-viral-combined_k141_144945\|\|lt2gene | Low-quality |
| 18-105-final-viral-combined_k141_162207\|\|lt2gene | Low-quality |
| 18-105-final-viral-combined_k141_46864\|\|lt2gene | Not-determined |
| 18-105-final-viral-combined_k141_100165\|\|lt2gene | Low-quality |
| 18-105-final-viral-combined_k141_198104\|\|lt2gene | Not-determined |
| 18-105-final-viral-combined_k141_117776\|\|lt2gene | Low-quality |
| 18-105-final-viral-combined_k141_10873\|\|lt2gene | Low-quality |
| 18-105-final-viral-combined_k141_82225\|\|lt2gene | Low-quality |
| 18-105-final-viral-combined_k141_153695\|\|lt2gene | Low-quality |
| 18-105-final-viral-combined_k141_162288\|\|lt2gene | Low-quality |
| 18-105-final-viral-combined_k141_162296\|\|lt2gene | Low-quality |
| 18-105-final-viral-combined_k141_100179\|\|lt2gene | Low-quality |
| 18-105-final-viral-combined_k141_127001\|\|lt2gene | Low-quality |
| 18-105-final-viral-combined_k141_127007\|\|lt2gene | Low-quality |
| 18-105-final-viral-combined_k141_127026\|\|lt2gene | Low-quality |
| 18-105-final-viral-combined_k141_162350\|\|lt2gene | Low-quality |
| 18-105-final-viral-combined_k141_3026\|\|lt2gene | Low-quality |
| 18-105-final-viral-combined_k141_100332\|\|lt2gene | Not-determined |
| 18-105-final-viral-combined_k141_135702\|\|lt2gene | Low-quality |
| 18-105-final-viral-combined_k141_11070\|\|lt2gene | Not-determined |
| 18-105-final-viral-combined_k141_206867\|\|lt2gene | Not-determined |
| 18-105-final-viral-combined_k141_172078\|\|lt2gene | Low-quality |
| 18-105-final-viral-combined_k141_189541\|\|lt2gene | Low-quality |
| 18-105-final-viral-combined_k141_206882\|\|lt2gene | Low-quality |
| 18-105-final-viral-combined_k141_162400\|\|lt2gene | Not-determined |
| 18-105-final-viral-combined_k141_28805\|\|lt2gene | Low-quality |
| 18-105-final-viral-combined_k141_73675\|\|lt2gene | Low-quality |
| 18-105-final-viral-combined_k141_145079\|\|lt2gene | Not-determined |
| 18-105-final-viral-combined_k141_100523\|\|lt2gene | Low-quality |
| 18-105-final-viral-combined_k141_180797\|\|lt2gene | Low-quality |
| 18-105-final-viral-combined_k141_28870\|\|lt2gene | Low-quality |
| 18-105-final-viral-combined_k141_189619\|\|lt2gene | Low-quality |
| 18-105-final-viral-combined_k141_91618\|\|lt2gene | Low-quality |
| 18-105-final-viral-combined_k141_180844\|\|lt2gene | Low-quality |
| 18-105-final-viral-combined_k141_117976\|\|lt2gene | Low-quality |
| 18-105-final-viral-combined_k141_37862\|\|lt2gene | Low-quality |
| 18-105-final-viral-combined_k141_172205\|\|lt2gene | Low-quality |
| 18-105-final-viral-combined_k141_47039\|\|lt2gene | Low-quality |
| 18-105-final-viral-combined_k141_198360\|\|lt2gene | Low-quality |
| 18-105-final-viral-combined_k141_47061\|\|lt2gene | Low-quality |
| 18-105-final-viral-combined_k141_162565\|\|lt2gene | Low-quality |
| 18-105-final-viral-combined_k141_127249\|\|lt2gene | Not-determined |
| 18-105-final-viral-combined_k141_28951\|\|lt2gene | Low-quality |
| 18-105-final-viral-combined_k141_82270\|\|lt2gene | Not-determined |
| 18-105-final-viral-combined_k141_47094\|\|lt2gene | Not-determined |
| 18-105-final-viral-combined_k141_82276\|\|lt2gene | Low-quality |
| 18-105-final-viral-combined_k141_118092\|\|lt2gene | Low-quality |
| 18-105-final-viral-combined_k141_127284\|\|lt2gene | Not-determined |
| 18-105-final-viral-combined_k141_207159\|\|lt2gene | Not-determined |
| 18-105-final-viral-combined_k141_172285\|\|lt2gene | Not-determined |
| 18-105-final-viral-combined_k141_145191\|\|lt2gene | Low-quality |
| 18-105-final-viral-combined_k141_145194\|\|lt2gene | Low-quality |
| 18-105-final-viral-combined_k141_91696\|\|lt2gene | Low-quality |
| 18-105-final-viral-combined_k141_207210\|\|lt2gene | Low-quality |
| 18-105-final-viral-combined_k141_91704\|\|lt2gene | Low-quality |
| 18-105-final-viral-combined_k141_91722\|\|lt2gene | Low-quality |
| 18-105-final-viral-combined_k141_153809\|\|lt2gene | Low-quality |
| 18-105-final-viral-combined_k141_198417\|\|lt2gene | Low-quality |
| 18-105-final-viral-combined_k141_3227\|\|lt2gene | Not-determined |
| 18-105-final-viral-combined_k141_135930\|\|lt2gene | Low-quality |
| 18-105-final-viral-combined_k141_11289\|\|lt2gene | Low-quality |
| 18-105-final-viral-combined_k141_172368\|\|lt2gene | Not-determined |
| 18-105-final-viral-combined_k141_29085\|\|lt2gene | Low-quality |
| 18-105-final-viral-combined_k141_135968\|\|lt2gene | Not-determined |
| 18-105-final-viral-combined_k141_47249\|\|lt2gene | Low-quality |
| 18-105-final-viral-combined_k141_136011\|\|lt2gene | Low-quality |
| 18-105-final-viral-combined_k141_118152\|\|lt2gene | Low-quality |
| 18-105-final-viral-combined_k141_47319\|\|lt2gene | Low-quality |
| 18-105-final-viral-combined_k141_153841\|\|lt2gene | Low-quality |
| 18-105-final-viral-combined_k141_172405\|\|lt2gene | Low-quality |
| 18-105-final-viral-combined_k141_153854\|\|lt2gene | Not-determined |
| 18-105-final-viral-combined_k141_162659\|\|lt2gene | Low-quality |
| 18-105-final-viral-combined_k141_47342\|\|lt2gene | Low-quality |
| 18-105-final-viral-combined_k141_198471\|\|lt2gene | Low-quality |
| 18-105-final-viral-combined_k141_91868\|\|lt2gene | Low-quality |
| 18-105-final-viral-combined_k141_145264\|\|lt2gene | Low-quality |
| 18-105-final-viral-combined_k141_19877\|\|lt2gene | Low-quality |
| 18-105-final-viral-combined_k141_145265\|\|lt2gene | Not-determined |
| 18-105-final-viral-combined_k141_19882\|\|lt2gene | Low-quality |
| 18-105-final-viral-combined_k141_136080\|\|lt2gene | Not-determined |
| 18-105-final-viral-combined_k141_136118\|\|lt2gene | Not-determined |
| 18-105-final-viral-combined_k141_145308\|\|lt2gene | Not-determined |
| 18-105-final-viral-combined_k141_38048\|\|lt2gene | Low-quality |
| 18-105-final-viral-combined_k141_38053\|\|lt2gene | Low-quality |
| 18-105-final-viral-combined_k141_198522\|\|lt2gene | Low-quality |
| 18-105-final-viral-combined_k141_136151\|\|lt2gene | Not-determined |
| 18-105-final-viral-combined_k141_136162\|\|lt2gene | Low-quality |
| 18-105-final-viral-combined_k141_181119\|\|lt2gene | Low-quality |
| 18-105-final-viral-combined_k141_3448\|\|lt2gene | Low-quality |
| 18-105-final-viral-combined_k141_55822\|\|lt2gene | Low-quality |
| 18-105-final-viral-combined_k141_189959\|\|lt2gene | Low-quality |
| 18-105-final-viral-combined_k141_198575\|\|lt2gene | Low-quality |
| 18-105-final-viral-combined_k141_145406\|\|lt2gene | Not-determined |
| 18-105-final-viral-combined_k141_162759\|\|lt2gene | Low-quality |
| 18-105-final-viral-combined_k141_127601\|\|lt2gene | Not-determined |
| 18-105-final-viral-combined_k141_127602\|\|lt2gene | Low-quality |
| 18-105-final-viral-combined_k141_47515\|\|lt2gene | Low-quality |
| 18-105-final-viral-combined_k141_64776\|\|lt2gene | Low-quality |
| 18-105-final-viral-combined_k141_100903\|\|lt2gene | Not-determined |
| 18-105-final-viral-combined_k141_181235\|\|lt2gene | Low-quality |
| 18-105-final-viral-combined_k141_38102\|\|lt2gene | Not-determined |
| 18-105-final-viral-combined_k141_47581\|\|lt2gene | Low-quality |
| 18-105-final-viral-combined_k141_145422\|\|lt2gene | Not-determined |
| 18-105-final-viral-combined_k141_47596\|\|lt2gene | Not-determined |
| 18-105-final-viral-combined_k141_162886\|\|lt2gene | Low-quality |
| 18-105-final-viral-combined_k141_3559\|\|lt2gene | Low-quality |
| 18-105-final-viral-combined_k141_47616\|\|lt2gene | Low-quality |
| 18-105-final-viral-combined_k141_11375\|\|lt2gene | Low-quality |
| 18-105-final-viral-combined_k141_162922\|\|lt2gene | Not-determined |
| 18-105-final-viral-combined_k141_207392\|\|lt2gene | Low-quality |
| 18-105-final-viral-combined_k141_145463\|\|lt2gene | Low-quality |
| 18-105-final-viral-combined_k141_172703\|\|lt2gene | Low-quality |
| 18-105-final-viral-combined_k141_172710\|\|lt2gene | Not-determined |
| 18-105-final-viral-combined_k141_172721\|\|lt2gene | Not-determined |
| 18-105-final-viral-combined_k141_154091\|\|lt2gene | Low-quality |
| 18-105-final-viral-combined_k141_190079\|\|lt2gene | Low-quality |
| 18-105-final-viral-combined_k141_154100\|\|lt2gene | Not-determined |
| 18-105-final-viral-combined_k141_198660\|\|lt2gene | Low-quality |
| 18-105-final-viral-combined_k141_3641\|\|lt2gene | Low-quality |
| 18-105-final-viral-combined_k141_145529\|\|lt2gene | Low-quality |
| 18-105-final-viral-combined_k141_145537\|\|lt2gene | Low-quality |
| 18-105-final-viral-combined_k141_207541\|\|lt2gene | Low-quality |
| 18-105-final-viral-combined_k141_3710\|\|lt2gene | Low-quality |
| 18-105-final-viral-combined_k141_127828\|\|lt2gene | Low-quality |
| 18-105-final-viral-combined_k141_145589\|\|lt2gene | Low-quality |
| 18-105-final-viral-combined_k141_20039\|\|lt2gene | Low-quality |
| 18-105-final-viral-combined_k141_127845\|\|lt2gene | Low-quality |
| 18-105-final-viral-combined_k141_145608\|\|lt2gene | Low-quality |
| 18-105-final-viral-combined_k141_181312\|\|lt2gene | Not-determined |
| 18-105-final-viral-combined_k141_207596\|\|lt2gene | Not-determined |
| 18-105-final-viral-combined_k141_190215\|\|lt2gene | Low-quality |
| 18-105-final-viral-combined_k141_145629\|\|lt2gene | Low-quality |
| 18-105-final-viral-combined_k141_3769\|\|lt2gene | Not-determined |
| 18-105-final-viral-combined_k141_198705\|\|lt2gene | Not-determined |
| 18-105-final-viral-combined_k141_11546\|\|lt2gene | Low-quality |
| 18-105-final-viral-combined_k141_74184\|\|lt2gene | Not-determined |
| 18-105-final-viral-combined_k141_65080\|\|lt2gene | Not-determined |
| 18-105-final-viral-combined_k141_190256\|\|lt2gene | Low-quality |
| 18-105-final-viral-combined_k141_11580\|\|lt2gene | Low-quality |
| 18-105-final-viral-combined_k141_11581\|\|lt2gene | Low-quality |
| 18-105-final-viral-combined_k141_163150\|\|lt2gene | Low-quality |
| 18-105-final-viral-combined_k141_3851\|\|lt2gene | Low-quality |
| 18-105-final-viral-combined_k141_145685\|\|lt2gene | Not-determined |
| 18-105-final-viral-combined_k141_11606\|\|lt2gene | Low-quality |
| 18-105-final-viral-combined_k141_127917\|\|lt2gene | Not-determined |
| 18-105-final-viral-combined_k141_127923\|\|lt2gene | Not-determined |
| 18-105-final-viral-combined_k141_181491\|\|lt2gene | Low-quality |
| 18-105-final-viral-combined_k141_127958\|\|lt2gene | Low-quality |
| 18-105-final-viral-combined_k141_48002\|\|lt2gene | Not-determined |
| 18-105-final-viral-combined_k141_163192\|\|lt2gene | Low-quality |
| 18-105-final-viral-combined_k141_207736\|\|lt2gene | Low-quality |
| 18-105-final-viral-combined_k141_181546\|\|lt2gene | Low-quality |
| 18-105-final-viral-combined_k141_181552\|\|lt2gene | Low-quality |
| 18-105-final-viral-combined_k141_145816\|\|lt2gene | Not-determined |
| 18-105-final-viral-combined_k141_48046\|\|lt2gene | Not-determined |
| 18-105-final-viral-combined_k141_92205\|\|lt2gene | Low-quality |
| 18-105-final-viral-combined_k141_128016\|\|lt2gene | Not-determined |
| 18-105-final-viral-combined_k141_190410\|\|lt2gene | Low-quality |
| 18-105-final-viral-combined_k141_163236\|\|lt2gene | Low-quality |
| 18-105-final-viral-combined_k141_82784\|\|lt2gene | Low-quality |
| 18-105-final-viral-combined_k141_38387\|\|lt2gene | Low-quality |
| 18-105-final-viral-combined_k141_11641\|\|lt2gene | Not-determined |
| 18-105-final-viral-combined_k141_29370\|\|lt2gene | Not-determined |
| 18-105-final-viral-combined_k141_109528\|\|lt2gene | Low-quality |
| 18-105-final-viral-combined_k141_38407\|\|lt2gene | Low-quality |
| 18-105-final-viral-combined_k141_154277\|\|lt2gene | Low-quality |
| 18-105-final-viral-combined_k141_11682\|\|lt2gene | Low-quality |
| 18-105-final-viral-combined_k141_11686\|\|lt2gene | Low-quality |
| 18-105-final-viral-combined_k141_163296\|\|lt2gene | Low-quality |
| 18-105-final-viral-combined_k141_101262\|\|lt2gene | Not-determined |
| 18-105-final-viral-combined_k141_109573\|\|lt2gene | Low-quality |
| 18-105-final-viral-combined_k141_109575\|\|lt2gene | Low-quality |
| 18-105-final-viral-combined_k141_136385\|\|lt2gene | Not-determined |
| 18-105-final-viral-combined_k141_145961\|\|lt2gene | Low-quality |
| 18-105-final-viral-combined_k141_198792\|\|lt2gene | Not-determined |
| 18-105-final-viral-combined_k141_128175\|\|lt2gene | Low-quality |
| 18-105-final-viral-combined_k141_4106\|\|lt2gene | Low-quality |
| 18-105-final-viral-combined_k141_82820\|\|lt2gene | Low-quality |
| 18-105-final-viral-combined_k141_20205\|\|lt2gene | Low-quality |
| 18-105-final-viral-combined_k141_198836\|\|lt2gene | Not-determined |
| 18-105-final-viral-combined_k141_128228\|\|lt2gene | Not-determined |
| 18-105-final-viral-combined_k141_82896\|\|lt2gene | Not-determined |
| 18-105-final-viral-combined_k141_109631\|\|lt2gene | Low-quality |
| 18-105-final-viral-combined_k141_198871\|\|lt2gene | Low-quality |
| 18-105-final-viral-combined_k141_48149\|\|lt2gene | Low-quality |
| 18-105-final-viral-combined_k141_181668\|\|lt2gene | Not-determined |
| 18-105-final-viral-combined_k141_74459\|\|lt2gene | Low-quality |
| 18-105-final-viral-combined_k141_20345\|\|lt2gene | Low-quality |
| 18-105-final-viral-combined_k141_163471\|\|lt2gene | Not-determined |
| 18-105-final-viral-combined_k141_11798\|\|lt2gene | Low-quality |
| 18-105-final-viral-combined_k141_74486\|\|lt2gene | Low-quality |
| 18-105-final-viral-combined_k141_109701\|\|lt2gene | Not-determined |
| 18-105-final-viral-combined_k141_20376\|\|lt2gene | Low-quality |
| 18-105-final-viral-combined_k141_92302\|\|lt2gene | Low-quality |
| 18-105-final-viral-combined_k141_29458\|\|lt2gene | Not-determined |
| 18-105-final-viral-combined_k141_48194\|\|lt2gene | Not-determined |
| 18-105-final-viral-combined_k141_82989\|\|lt2gene | Low-quality |
| 18-105-final-viral-combined_k141_163488\|\|lt2gene | Not-determined |
| 18-105-final-viral-combined_k141_173012\|\|lt2gene | Low-quality |
| 18-105-final-viral-combined_k141_163497\|\|lt2gene | Not-determined |
| 18-105-final-viral-combined_k141_65408\|\|lt2gene | Not-determined |
| 18-105-final-viral-combined_k141_101393\|\|lt2gene | Low-quality |
| 18-105-final-viral-combined_k141_48221\|\|lt2gene | Low-quality |
| 18-105-final-viral-combined_k141_136458\|\|lt2gene | Not-determined |
| 18-105-final-viral-combined_k141_136460\|\|lt2gene | Low-quality |
| 18-105-final-viral-combined_k141_92346\|\|lt2gene | Low-quality |
| 18-105-final-viral-combined_k141_163516\|\|lt2gene | Not-determined |
| 18-105-final-viral-combined_k141_92354\|\|lt2gene | Low-quality |
| 18-105-final-viral-combined_k141_11857\|\|lt2gene | Low-quality |
| 18-105-final-viral-combined_k141_4337\|\|lt2gene | Low-quality |
| 18-105-final-viral-combined_k141_11866\|\|lt2gene | Low-quality |
| 18-105-final-viral-combined_k141_163538\|\|lt2gene | Low-quality |
| 18-105-final-viral-combined_k141_92386\|\|lt2gene | Low-quality |
| 18-105-final-viral-combined_k141_199006\|\|lt2gene | Not-determined |
| 18-105-final-viral-combined_k141_190539\|\|lt2gene | Low-quality |
| 18-105-final-viral-combined_k141_207857\|\|lt2gene | Low-quality |
| 18-105-final-viral-combined_k141_74597\|\|lt2gene | Low-quality |
| 18-105-final-viral-combined_k141_109757\|\|lt2gene | Low-quality |
| 18-105-final-viral-combined_k141_20454\|\|lt2gene | Low-quality |
| 18-105-final-viral-combined_k141_190562\|\|lt2gene | Low-quality |
| 18-105-final-viral-combined_k141_83049\|\|lt2gene | Low-quality |
| 18-105-final-viral-combined_k141_11899\|\|lt2gene | Not-determined |
| 18-105-final-viral-combined_k141_74606\|\|lt2gene | Low-quality |
| 18-105-final-viral-combined_k141_83067\|\|lt2gene | Low-quality |
| 18-105-final-viral-combined_k141_56190\|\|lt2gene | Not-determined |
| 18-105-final-viral-combined_k141_154543\|\|lt2gene | Low-quality |
| 18-105-final-viral-combined_k141_56213\|\|lt2gene | Not-determined |
| 18-105-final-viral-combined_k141_29541\|\|lt2gene | Low-quality |
| 18-105-final-viral-combined_k141_11939\|\|lt2gene | Low-quality |
| 18-105-final-viral-combined_k141_11947\|\|lt2gene | Not-determined |
| 18-105-final-viral-combined_k141_4419\|\|lt2gene | Not-determined |
| 18-105-final-viral-combined_k141_4431\|\|lt2gene | Not-determined |
| 18-105-final-viral-combined_k141_83148\|\|lt2gene | Low-quality |
| 18-105-final-viral-combined_k141_92427\|\|lt2gene | Low-quality |
| 18-105-final-viral-combined_k141_101491\|\|lt2gene | Low-quality |
| 18-105-final-viral-combined_k141_173082\|\|lt2gene | Low-quality |
| 18-105-final-viral-combined_k141_83154\|\|lt2gene | Low-quality |
| 18-105-final-viral-combined_k141_154594\|\|lt2gene | Not-determined |
| 18-105-final-viral-combined_k141_65468\|\|lt2gene | Low-quality |
| 18-105-final-viral-combined_k141_20509\|\|lt2gene | Low-quality |
| 18-105-final-viral-combined_k141_92474\|\|lt2gene | Not-determined |
| 18-105-final-viral-combined_k141_190618\|\|lt2gene | Low-quality |
| 18-105-final-viral-combined_k141_190621\|\|lt2gene | Not-determined |
| 18-105-final-viral-combined_k141_109838\|\|lt2gene | Low-quality |
| 18-105-final-viral-combined_k141_12000\|\|lt2gene | Not-determined |
| 18-105-final-viral-combined_k141_29568\|\|lt2gene | Low-quality |
| 18-105-final-viral-combined_k141_92501\|\|lt2gene | Low-quality |
| 18-105-final-viral-combined_k141_4518\|\|lt2gene | Low-quality |
| 18-105-final-viral-combined_k141_92506\|\|lt2gene | Not-determined |
| 18-105-final-viral-combined_k141_146189\|\|lt2gene | Low-quality |
| 18-105-final-viral-combined_k141_20562\|\|lt2gene | Low-quality |
| 18-105-final-viral-combined_k141_12050\|\|lt2gene | Low-quality |
| 18-105-final-viral-combined_k141_12068\|\|lt2gene | Low-quality |
| 18-105-final-viral-combined_k141_154662\|\|lt2gene | Low-quality |
| 18-105-final-viral-combined_k141_48405\|\|lt2gene | Low-quality |
| 18-105-final-viral-combined_k141_20609\|\|lt2gene | Low-quality |
| 18-105-final-viral-combined_k141_74848\|\|lt2gene | Low-quality |
| 18-105-final-viral-combined_k141_29651\|\|lt2gene | Not-determined |
| 18-105-final-viral-combined_k141_163665\|\|lt2gene | Not-determined |
| 18-105-final-viral-combined_k141_4600\|\|lt2gene | Not-determined |
| 18-105-final-viral-combined_k141_29676\|\|lt2gene | Not-determined |
| 18-105-final-viral-combined_k141_181804\|\|lt2gene | Low-quality |
| 18-105-final-viral-combined_k141_173240\|\|lt2gene | Low-quality |
| 18-105-final-viral-combined_k141_190732\|\|lt2gene | Low-quality |
| 18-105-final-viral-combined_k141_101636\|\|lt2gene | Low-quality |
| 18-105-final-viral-combined_k141_38775\|\|lt2gene | Low-quality |
| 18-105-final-viral-combined_k141_83343\|\|lt2gene | Low-quality |
| 18-105-final-viral-combined_k141_12154\|\|lt2gene | Not-determined |
| 18-105-final-viral-combined_k141_12188\|\|lt2gene | Not-determined |
| 18-105-final-viral-combined_k141_92672\|\|lt2gene | Not-determined |
| 18-105-final-viral-combined_k141_74953\|\|lt2gene | Low-quality |
| 18-105-final-viral-combined_k141_199193\|\|lt2gene | Low-quality |
| 18-105-final-viral-combined_k141_92711\|\|lt2gene | Low-quality |
| 18-105-final-viral-combined_k141_163790\|\|lt2gene | Low-quality |
| 18-105-final-viral-combined_k141_163792\|\|lt2gene | Low-quality |
| 18-105-final-viral-combined_k141_101735\|\|lt2gene | Low-quality |
| 18-105-final-viral-combined_k141_4752\|\|lt2gene | Low-quality |
| 18-105-final-viral-combined_k141_173360\|\|lt2gene | Low-quality |
| 18-105-final-viral-combined_k141_75032\|\|lt2gene | Low-quality |
| 18-105-final-viral-combined_k141_208260\|\|lt2gene | Low-quality |
| 18-105-final-viral-combined_k141_56564\|\|lt2gene | Low-quality |
| 18-105-final-viral-combined_k141_75060\|\|lt2gene | Low-quality |
| 18-105-final-viral-combined_k141_190807\|\|lt2gene | Low-quality |
| 18-105-final-viral-combined_k141_163824\|\|lt2gene | Low-quality |
| 18-105-final-viral-combined_k141_56588\|\|lt2gene | Low-quality |
| 18-105-final-viral-combined_k141_163828\|\|lt2gene | Low-quality |
| 18-105-final-viral-combined_k141_29789\|\|lt2gene | Low-quality |
| 18-105-final-viral-combined_k141_38954\|\|lt2gene | Not-determined |
| 18-105-final-viral-combined_k141_136923\|\|lt2gene | Low-quality |
| 18-105-final-viral-combined_k141_92833\|\|lt2gene | Low-quality |
| 18-105-final-viral-combined_k141_173397\|\|lt2gene | Low-quality |
| 18-105-final-viral-combined_k141_119137\|\|lt2gene | Low-quality |
| 18-105-final-viral-combined_k141_29812\|\|lt2gene | Not-determined |
| 18-105-final-viral-combined_k141_92891\|\|lt2gene | Low-quality |
| 18-105-final-viral-combined_k141_110008\|\|lt2gene | Low-quality |
| 18-105-final-viral-combined_k141_56651\|\|lt2gene | Low-quality |
| 18-105-final-viral-combined_k141_65722\|\|lt2gene | Low-quality |
| 18-105-final-viral-combined_k141_83522\|\|lt2gene | Low-quality |
| 18-105-final-viral-combined_k141_101830\|\|lt2gene | Not-determined |
| 18-105-final-viral-combined_k141_146439\|\|lt2gene | Not-determined |
| 18-105-final-viral-combined_k141_208329\|\|lt2gene | Low-quality |
| 18-105-final-viral-combined_k141_208361\|\|lt2gene | Not-determined |
| 18-105-final-viral-combined_k141_146460\|\|lt2gene | Low-quality |
| 18-105-final-viral-combined_k141_65727\|\|lt2gene | Low-quality |
| 18-105-final-viral-combined_k141_146461\|\|lt2gene | Not-determined |
| 18-105-final-viral-combined_k141_4967\|\|lt2gene | Low-quality |
| 18-105-final-viral-combined_k141_190910\|\|lt2gene | Low-quality |
| 18-105-final-viral-combined_k141_173522\|\|lt2gene | Low-quality |
| 18-105-final-viral-combined_k141_56721\|\|lt2gene | Low-quality |
| 18-105-final-viral-combined_k141_12457\|\|lt2gene | Low-quality |
| 18-105-final-viral-combined_k141_12465\|\|lt2gene | Not-determined |
| 18-105-final-viral-combined_k141_65779\|\|lt2gene | Low-quality |
| 18-105-final-viral-combined_k141_173587\|\|lt2gene | Low-quality |
| 18-105-final-viral-combined_k141_146545\|\|lt2gene | Not-determined |
| 18-105-final-viral-combined_k141_83612\|\|lt2gene | Low-quality |
| 18-105-final-viral-combined_k141_48677\|\|lt2gene | Not-determined |
| 18-105-final-viral-combined_k141_75196\|\|lt2gene | Low-quality |
| 18-105-final-viral-combined_k141_83639\|\|lt2gene | Low-quality |
| 18-105-final-viral-combined_k141_199364\|\|lt2gene | Not-determined |
| 18-105-final-viral-combined_k141_101910\|\|lt2gene | Low-quality |
| 18-105-final-viral-combined_k141_48715\|\|lt2gene | Low-quality |
| 18-105-final-viral-combined_k141_137060\|\|lt2gene | Low-quality |
| 18-105-final-viral-combined_k141_119293\|\|lt2gene | Not-determined |
| 18-105-final-viral-combined_k141_137064\|\|lt2gene | Not-determined |
| 18-105-final-viral-combined_k141_119308\|\|lt2gene | Low-quality |
| 18-105-final-viral-combined_k141_20835\|\|lt2gene | Not-determined |
| 18-105-final-viral-combined_k141_39050\|\|lt2gene | Not-determined |
| 18-105-final-viral-combined_k141_39064\|\|lt2gene | Low-quality |
| 18-105-final-viral-combined_k141_48742\|\|lt2gene | Not-determined |
| 18-105-final-viral-combined_k141_65855\|\|lt2gene | Low-quality |
| 18-105-final-viral-combined_k141_20873\|\|lt2gene | Low-quality |
| 18-105-final-viral-combined_k141_75333\|\|lt2gene | Not-determined |
| 18-105-final-viral-combined_k141_20907\|\|lt2gene | Not-determined |
| 18-105-final-viral-combined_k141_102066\|\|lt2gene | Not-determined |
| 18-105-final-viral-combined_k141_102114\|\|lt2gene | Low-quality |
| 18-105-final-viral-combined_k141_65974\|\|lt2gene | Low-quality |
| 18-105-final-viral-combined_k141_65977\|\|lt2gene | Low-quality |
| 18-105-final-viral-combined_k141_154941\|\|lt2gene | Low-quality |
| 18-105-final-viral-combined_k141_110159\|\|lt2gene | Not-determined |
| 18-105-final-viral-combined_k141_146709\|\|lt2gene | Low-quality |
| 18-105-final-viral-combined_k141_66001\|\|lt2gene | Low-quality |
| 18-105-final-viral-combined_k141_154951\|\|lt2gene | Low-quality |
| 18-105-final-viral-combined_k141_119474\|\|lt2gene | Not-determined |
| 18-105-final-viral-combined_k141_56981\|\|lt2gene | Low-quality |
| 18-105-final-viral-combined_k141_146752\|\|lt2gene | Low-quality |
| 18-105-final-viral-combined_k141_12557\|\|lt2gene | Low-quality |
| 18-105-final-viral-combined_k141_199570\|\|lt2gene | Low-quality |
| 18-105-final-viral-combined_k141_83785\|\|lt2gene | Low-quality |
| 18-105-final-viral-combined_k141_208557\|\|lt2gene | Low-quality |
| 18-105-final-viral-combined_k141_208563\|\|lt2gene | Not-determined |
| 18-105-final-viral-combined_k141_48904\|\|lt2gene | Low-quality |
| 18-105-final-viral-combined_k141_83819\|\|lt2gene | Low-quality |
| 18-105-final-viral-combined_k141_182042\|\|lt2gene | Low-quality |
| 18-105-final-viral-combined_k141_83858\|\|lt2gene | Low-quality |
| 18-105-final-viral-combined_k141_137207\|\|lt2gene | Not-determined |
| 18-105-final-viral-combined_k141_21088\|\|lt2gene | Not-determined |
| 18-105-final-viral-combined_k141_48978\|\|lt2gene | Not-determined |
| 18-105-final-viral-combined_k141_21092\|\|lt2gene | Low-quality |
| 18-105-final-viral-combined_k141_208622\|\|lt2gene | Low-quality |
| 18-105-final-viral-combined_k141_199773\|\|lt2gene | Low-quality |
| 18-105-final-viral-combined_k141_21138\|\|lt2gene | Not-determined |
| 18-105-final-viral-combined_k141_137251\|\|lt2gene | Low-quality |
| 18-105-final-viral-combined_k141_30000\|\|lt2gene | Low-quality |
| 18-105-final-viral-combined_k141_5378\|\|lt2gene | Low-quality |
| 18-105-final-viral-combined_k141_75498\|\|lt2gene | Not-determined |
| 18-105-final-viral-combined_k141_5385\|\|lt2gene | Low-quality |
| 18-105-final-viral-combined_k141_102267\|\|lt2gene | Low-quality |
| 18-105-final-viral-combined_k141_75500\|\|lt2gene | Low-quality |
| 18-105-final-viral-combined_k141_75501\|\|lt2gene | Low-quality |
| 18-105-final-viral-combined_k141_137278\|\|lt2gene | Not-determined |
| 18-105-final-viral-combined_k141_5393\|\|lt2gene | Low-quality |
| 18-105-final-viral-combined_k141_102279\|\|lt2gene | Not-determined |
| 18-105-final-viral-combined_k141_137300\|\|lt2gene | Low-quality |
| 18-105-final-viral-combined_k141_137303\|\|lt2gene | Low-quality |
| 18-105-final-viral-combined_k141_119668\|\|lt2gene | Not-determined |
| 18-105-final-viral-combined_k141_173805\|\|lt2gene | Low-quality |
| 18-105-final-viral-combined_k141_48989\|\|lt2gene | Low-quality |
| 18-105-final-viral-combined_k141_119676\|\|lt2gene | Low-quality |
| 18-105-final-viral-combined_k141_30013\|\|lt2gene | Not-determined |
| 18-105-final-viral-combined_k141_12818\|\|lt2gene | Not-determined |
| 18-105-final-viral-combined_k141_57164\|\|lt2gene | Low-quality |
| 18-105-final-viral-combined_k141_57175\|\|lt2gene | Low-quality |
| 18-105-final-viral-combined_k141_137352\|\|lt2gene | Not-determined |
| 18-105-final-viral-combined_k141_119720\|\|lt2gene | Not-determined |
| 18-105-final-viral-combined_k141_164144\|\|lt2gene | Not-determined |
| 18-105-final-viral-combined_k141_83941\|\|lt2gene | Low-quality |
| 18-105-final-viral-combined_k141_102343\|\|lt2gene | Low-quality |
| 18-105-final-viral-combined_k141_66211\|\|lt2gene | Low-quality |
| 18-105-final-viral-combined_k141_208768\|\|lt2gene | Low-quality |
| 18-105-final-viral-combined_k141_102414\|\|lt2gene | Low-quality |
| 18-105-final-viral-combined_k141_119804\|\|lt2gene | Low-quality |
| 18-105-final-viral-combined_k141_129091\|\|lt2gene | Low-quality |
| 18-105-final-viral-combined_k141_155252\|\|lt2gene | Low-quality |
| 18-105-final-viral-combined_k141_39424\|\|lt2gene | Not-determined |
| 18-105-final-viral-combined_k141_66303\|\|lt2gene | Low-quality |
| 18-105-final-viral-combined_k141_12951\|\|lt2gene | Low-quality |
| 18-105-final-viral-combined_k141_164289\|\|lt2gene | Low-quality |
| 18-105-final-viral-combined_k141_110625\|\|lt2gene | Not-determined |
| 18-105-final-viral-combined_k141_13003\|\|lt2gene | Not-determined |
| 18-105-final-viral-combined_k141_102509\|\|lt2gene | Low-quality |
| 18-105-final-viral-combined_k141_155342\|\|lt2gene | Low-quality |
| 18-105-final-viral-combined_k141_30182\|\|lt2gene | Low-quality |
| 18-105-final-viral-combined_k141_129123\|\|lt2gene | Low-quality |
| 18-105-final-viral-combined_k141_137513\|\|lt2gene | Low-quality |
| 18-105-final-viral-combined_k141_66344\|\|lt2gene | Low-quality |
| 18-105-final-viral-combined_k141_119869\|\|lt2gene | Low-quality |
| 18-105-final-viral-combined_k141_39535\|\|lt2gene | Not-determined |
| 18-105-final-viral-combined_k141_102568\|\|lt2gene | Low-quality |
| 18-105-final-viral-combined_k141_39548\|\|lt2gene | Low-quality |
| 18-105-final-viral-combined_k141_84093\|\|lt2gene | Not-determined |
| 18-105-final-viral-combined_k141_110709\|\|lt2gene | Low-quality |
| 18-105-final-viral-combined_k141_84100\|\|lt2gene | Not-determined |
| 18-105-final-viral-combined_k141_137563\|\|lt2gene | Low-quality |
| 18-105-final-viral-combined_k141_13084\|\|lt2gene | Low-quality |
| 18-105-final-viral-combined_k141_208913\|\|lt2gene | Low-quality |
| 18-105-final-viral-combined_k141_102599\|\|lt2gene | Low-quality |
| 18-105-final-viral-combined_k141_102608\|\|lt2gene | Low-quality |
| 18-105-final-viral-combined_k141_182239\|\|lt2gene | Low-quality |
| 18-105-final-viral-combined_k141_129168\|\|lt2gene | Not-determined |
| 18-105-final-viral-combined_k141_208971\|\|lt2gene | Low-quality |
| 18-105-final-viral-combined_k141_5642\|\|lt2gene | Low-quality |
| 18-105-final-viral-combined_k141_164425\|\|lt2gene | Low-quality |
| 18-105-final-viral-combined_k141_30325\|\|lt2gene | Low-quality |
| 18-105-final-viral-combined_k141_30326\|\|lt2gene | Low-quality |
| 18-105-final-viral-combined_k141_30331\|\|lt2gene | Low-quality |
| 18-105-final-viral-combined_k141_129284\|\|lt2gene | Low-quality |
| 18-105-final-viral-combined_k141_191337\|\|lt2gene | Not-determined |
| 18-105-final-viral-combined_k141_129325\|\|lt2gene | Low-quality |
| 18-105-final-viral-combined_k141_39687\|\|lt2gene | Low-quality |
| 18-105-final-viral-combined_k141_39700\|\|lt2gene | Low-quality |
| 18-105-final-viral-combined_k141_191374\|\|lt2gene | Low-quality |
| 18-105-final-viral-combined_k141_21274\|\|lt2gene | Not-determined |
| 18-105-final-viral-combined_k141_147035\|\|lt2gene | Low-quality |
| 18-105-final-viral-combined_k141_66650\|\|lt2gene | Low-quality |
| 18-105-final-viral-combined_k141_84295\|\|lt2gene | Not-determined |
| 18-105-final-viral-combined_k141_21292\|\|lt2gene | Not-determined |
| 18-105-final-viral-combined_k141_93492\|\|lt2gene | Low-quality |
| 18-105-final-viral-combined_k141_57571\|\|lt2gene | Not-determined |
| 18-105-final-viral-combined_k141_5857\|\|lt2gene | Not-determined |
| 18-105-final-viral-combined_k141_200166\|\|lt2gene | Low-quality |
| 18-105-final-viral-combined_k141_191392\|\|lt2gene | Low-quality |
| 18-105-final-viral-combined_k141_111005\|\|lt2gene | Low-quality |
| 18-105-final-viral-combined_k141_182447\|\|lt2gene | Low-quality |
| 18-105-final-viral-combined_k141_147131\|\|lt2gene | Low-quality |
| 18-105-final-viral-combined_k141_5908\|\|lt2gene | Low-quality |
| 18-105-final-viral-combined_k141_5925\|\|lt2gene | Low-quality |
| 18-105-final-viral-combined_k141_164598\|\|lt2gene | Low-quality |
| 18-105-final-viral-combined_k141_119937\|\|lt2gene | Low-quality |
| 18-105-final-viral-combined_k141_129447\|\|lt2gene | Low-quality |
| 18-105-final-viral-combined_k141_129449\|\|lt2gene | Not-determined |
| 18-105-final-viral-combined_k141_93543\|\|lt2gene | Low-quality |
| 18-105-final-viral-combined_k141_111116\|\|lt2gene | Not-determined |
| 18-105-final-viral-combined_k141_39781\|\|lt2gene | Low-quality |
| 18-105-final-viral-combined_k141_13449\|\|lt2gene | Not-determined |
| 18-105-final-viral-combined_k141_174248\|\|lt2gene | Low-quality |
| 18-105-final-viral-combined_k141_209292\|\|lt2gene | Low-quality |
| 18-105-final-viral-combined_k141_13476\|\|lt2gene | Not-determined |
| 18-105-final-viral-combined_k141_147358\|\|lt2gene | Low-quality |
| 18-105-final-viral-combined_k141_119947\|\|lt2gene | Low-quality |
| 18-105-final-viral-combined_k141_200317\|\|lt2gene | Not-determined |
| 18-105-final-viral-combined_k141_129539\|\|lt2gene | Low-quality |
| 18-105-final-viral-combined_k141_30598\|\|lt2gene | Low-quality |
| 18-105-final-viral-combined_k141_30607\|\|lt2gene | Not-determined |
| 18-105-final-viral-combined_k141_93805\|\|lt2gene | Not-determined |
| 18-105-final-viral-combined_k141_30636\|\|lt2gene | Not-determined |

| | |
|---|---|
| 18-105-final-viral-combined_k141_182765\|\|lt2gene | Not-determined |
| 18-105-final-viral-combined_k141_39903\|\|lt2gene | Low-quality |
| 18-105-final-viral-combined_k141_174312\|\|lt2gene | Low-quality |
| 18-105-final-viral-combined_k141_174322\|\|lt2gene | Not-determined |
| 18-105-final-viral-combined_k141_57774\|\|lt2gene | Not-determined |
| 18-105-final-viral-combined_k141_84743\|\|lt2gene | Low-quality |
| 18-105-final-viral-combined_k141_13636\|\|lt2gene | Low-quality |
| 18-105-final-viral-combined_k141_76171\|\|lt2gene | Low-quality |
| 18-105-final-viral-combined_k141_76178\|\|lt2gene | Low-quality |
| 18-105-final-viral-combined_k141_155677\|\|lt2gene | Low-quality |
| 18-105-final-viral-combined_k141_129637\|\|lt2gene | Low-quality |
| 18-105-final-viral-combined_k141_174373\|\|lt2gene | Low-quality |
| 18-105-final-viral-combined_k141_57874\|\|lt2gene | Low-quality |
| 18-105-final-viral-combined_k141_209558\|\|lt2gene | Not-determined |
| 18-105-final-viral-combined_k141_21503\|\|lt2gene | Low-quality |
| 18-105-final-viral-combined_k141_209584\|\|lt2gene | Not-determined |
| 18-105-final-viral-combined_k141_164790\|\|lt2gene | Not-determined |
| 18-105-final-viral-combined_k141_57926\|\|lt2gene | Not-determined |
| 18-105-final-viral-combined_k141_111541\|\|lt2gene | Not-determined |
| 18-105-final-viral-combined_k141_200480\|\|lt2gene | Low-quality |
| 18-105-final-viral-combined_k141_84878\|\|lt2gene | Low-quality |
| 18-105-final-viral-combined_k141_200494\|\|lt2gene | Low-quality |
| 18-105-final-viral-combined_k141_191788\|\|lt2gene | Not-determined |
| 18-105-final-viral-combined_k141_13824\|\|lt2gene | Low-quality |
| 18-105-final-viral-combined_k141_76333\|\|lt2gene | Low-quality |
| 18-105-final-viral-combined_k141_76364\|\|lt2gene | Not-determined |
| 18-105-final-viral-combined_k141_137903\|\|lt2gene | Low-quality |
| 18-105-final-viral-combined_k141_40108\|\|lt2gene | Low-quality |
| 18-105-final-viral-combined_k141_6515\|\|lt2gene | Not-determined |
| 18-105-final-viral-combined_k141_30865\|\|lt2gene | Not-determined |
| 18-105-final-viral-combined_k141_57996\|\|lt2gene | Low-quality |
| 18-105-final-viral-combined_k141_200708\|\|lt2gene | Low-quality |
| 18-105-final-viral-combined_k141_200716\|\|lt2gene | Low-quality |
| 18-105-final-viral-combined_k141_164912\|\|lt2gene | Low-quality |
| 18-105-final-viral-combined_k141_200727\|\|lt2gene | Low-quality |
| 18-105-final-viral-combined_k141_200729\|\|lt2gene | Low-quality |
| 18-105-final-viral-combined_k141_137949\|\|lt2gene | Low-quality |
| 18-105-final-viral-combined_k141_40245\|\|lt2gene | Low-quality |
| 18-105-final-viral-combined_k141_58076\|\|lt2gene | Not-determined |
| 18-105-final-viral-combined_k141_147508\|\|lt2gene | Low-quality |
| 18-105-final-viral-combined_k141_120239\|\|lt2gene | Not-determined |
| 18-105-final-viral-combined_k141_200860\|\|lt2gene | Low-quality |
| 18-105-final-viral-combined_k141_30991\|\|lt2gene | Not-determined |
| 18-105-final-viral-combined_k141_111758\|\|lt2gene | Low-quality |
| 18-105-final-viral-combined_k141_49365\|\|lt2gene | Not-determined |
| 18-105-final-viral-combined_k141_58182\|\|lt2gene | Low-quality |
| 18-105-final-viral-combined_k141_49372\|\|lt2gene | Not-determined |
| 18-105-final-viral-combined_k141_191993\|\|lt2gene | Not-determined |
| 18-105-final-viral-combined_k141_58275\|\|lt2gene | Low-quality |
| 18-105-final-viral-combined_k141_21749\|\|lt2gene | Low-quality |
| 18-105-final-viral-combined_k141_66876\|\|lt2gene | Low-quality |
| 18-105-final-viral-combined_k141_138093\|\|lt2gene | Low-quality |
| 18-105-final-viral-combined_k141_49447\|\|lt2gene | Not-determined |
| 18-105-final-viral-combined_k141_40452\|\|lt2gene | Not-determined |
| 18-105-final-viral-combined_k141_129811\|\|lt2gene | Low-quality |
| 18-105-final-viral-combined_k141_183196\|\|lt2gene | Low-quality |
| 18-105-final-viral-combined_k141_103294\|\|lt2gene | Low-quality |
| 18-105-final-viral-combined_k141_129815\|\|lt2gene | Low-quality |
| 18-105-final-viral-combined_k141_6698\|\|lt2gene | Low-quality |
| 18-105-final-viral-combined_k141_94028\|\|lt2gene | Low-quality |
| 18-105-final-viral-combined_k141_94029\|\|lt2gene | Not-determined |
| 18-105-final-viral-combined_k141_165053\|\|lt2gene | Not-determined |
| 18-105-final-viral-combined_k141_85216\|\|lt2gene | Low-quality |
| 18-105-final-viral-combined_k141_85218\|\|lt2gene | Not-determined |
| 18-105-final-viral-combined_k141_40468\|\|lt2gene | Low-quality |
| 18-105-final-viral-combined_k141_156022\|\|lt2gene | Low-quality |
| 18-105-final-viral-combined_k141_85233\|\|lt2gene | Low-quality |
| 18-105-final-viral-combined_k141_94063\|\|lt2gene | Low-quality |
| 18-105-final-viral-combined_k141_6787\|\|lt2gene | Low-quality |
| 18-105-final-viral-combined_k141_183333\|\|lt2gene | Low-quality |
| 18-105-final-viral-combined_k141_183347\|\|lt2gene | Low-quality |
| 18-105-final-viral-combined_k141_183349\|\|lt2gene | Low-quality |
| 18-105-final-viral-combined_k141_21876\|\|lt2gene | Not-determined |
| 18-105-final-viral-combined_k141_183360\|\|lt2gene | Not-determined |
| 18-105-final-viral-combined_k141_174720\|\|lt2gene | Low-quality |
| 18-105-final-viral-combined_k141_6838\|\|lt2gene | Low-quality |
| 18-105-final-viral-combined_k141_147698\|\|lt2gene | Not-determined |
| 18-105-final-viral-combined_k141_58479\|\|lt2gene | Not-determined |
| 18-105-final-viral-combined_k141_6896\|\|lt2gene | Low-quality |
| 18-105-final-viral-combined_k141_94165\|\|lt2gene | Low-quality |
| 18-105-final-viral-combined_k141_21953\|\|lt2gene | Low-quality |
| 18-105-final-viral-combined_k141_21977\|\|lt2gene | Low-quality |
| 18-105-final-viral-combined_k141_67032\|\|lt2gene | Low-quality |
| 18-105-final-viral-combined_k141_31268\|\|lt2gene | Low-quality |
| 18-105-final-viral-combined_k141_156085\|\|lt2gene | Not-determined |
| 18-105-final-viral-combined_k141_31297\|\|lt2gene | Low-quality |
| 18-105-final-viral-combined_k141_192144\|\|lt2gene | Low-quality |
| 18-105-final-viral-combined_k141_192148\|\|lt2gene | Low-quality |
| 18-105-final-viral-combined_k141_120392\|\|lt2gene | Not-determined |
| 18-105-final-viral-combined_k141_156118\|\|lt2gene | Low-quality |
| 18-105-final-viral-combined_k141_156122\|\|lt2gene | Not-determined |
| 18-105-final-viral-combined_k141_120414\|\|lt2gene | Low-quality |
| 18-105-final-viral-combined_k141_165204\|\|lt2gene | Low-quality |
| 18-105-final-viral-combined_k141_31365\|\|lt2gene | Not-determined |
| 18-105-final-viral-combined_k141_138368\|\|lt2gene | Low-quality |
| 18-105-final-viral-combined_k141_129991\|\|lt2gene | Low-quality |
| 18-105-final-viral-combined_k141_14233\|\|lt2gene | Low-quality |
| 18-105-final-viral-combined_k141_58697\|\|lt2gene | Not-determined |
| 18-105-final-viral-combined_k141_210114\|\|lt2gene | Low-quality |
| 18-105-final-viral-combined_k141_40744\|\|lt2gene | Low-quality |
| 18-105-final-viral-combined_k141_14247\|\|lt2gene | Low-quality |
| 18-105-final-viral-combined_k141_6942\|\|lt2gene | Not-determined |
| 18-105-final-viral-combined_k141_120468\|\|lt2gene | Low-quality |
| 18-105-final-viral-combined_k141_49685\|\|lt2gene | Low-quality |
| 18-105-final-viral-combined_k141_183507\|\|lt2gene | Low-quality |
| 18-105-final-viral-combined_k141_103449\|\|lt2gene | Low-quality |
| 18-105-final-viral-combined_k141_201033\|\|lt2gene | Not-determined |
| 18-105-final-viral-combined_k141_40805\|\|lt2gene | Low-quality |
| 18-105-final-viral-combined_k141_120492\|\|lt2gene | Low-quality |
| 18-105-final-viral-combined_k141_58756\|\|lt2gene | Not-determined |
| 18-105-final-viral-combined_k141_40816\|\|lt2gene | Low-quality |
| 18-105-final-viral-combined_k141_49714\|\|lt2gene | Low-quality |
| 18-105-final-viral-combined_k141_201099\|\|lt2gene | Low-quality |
| 18-105-final-viral-combined_k141_192285\|\|lt2gene | Low-quality |
| 18-105-final-viral-combined_k141_94215\|\|lt2gene | Low-quality |
| 18-105-final-viral-combined_k141_130094\|\|lt2gene | Low-quality |
| 18-105-final-viral-combined_k141_67156\|\|lt2gene | Low-quality |
| 18-105-final-viral-combined_k141_103572\|\|lt2gene | Not-determined |
| 18-105-final-viral-combined_k141_31433\|\|lt2gene | Not-determined |
| 18-105-final-viral-combined_k141_76766\|\|lt2gene | Low-quality |
| 18-105-final-viral-combined_k141_103618\|\|lt2gene | Low-quality |
| 18-105-final-viral-combined_k141_147811\|\|lt2gene | Low-quality |
| 18-105-final-viral-combined_k141_130157\|\|lt2gene | Low-quality |
| 18-105-final-viral-combined_k141_174966\|\|lt2gene | Not-determined |
| 18-105-final-viral-combined_k141_67200\|\|lt2gene | Not-determined |
| 18-105-final-viral-combined_k141_58959\|\|lt2gene | Low-quality |
| 18-105-final-viral-combined_k141_22222\|\|lt2gene | Low-quality |
| 18-105-final-viral-combined_k141_112171\|\|lt2gene | Low-quality |
| 18-105-final-viral-combined_k141_31523\|\|lt2gene | Not-determined |
| 18-105-final-viral-combined_k141_31570\|\|lt2gene | Low-quality |
| 18-105-final-viral-combined_k141_31574\|\|lt2gene | Low-quality |
| 18-105-final-viral-combined_k141_130249\|\|lt2gene | Not-determined |
| 18-105-final-viral-combined_k141_138574\|\|lt2gene | Low-quality |
| 18-105-final-viral-combined_k141_40926\|\|lt2gene | Not-determined |
| 18-105-final-viral-combined_k141_120721\|\|lt2gene | Not-determined |
| 18-105-final-viral-combined_k141_138605\|\|lt2gene | Low-quality |
| 18-105-final-viral-combined_k141_138606\|\|lt2gene | Not-determined |
| 18-105-final-viral-combined_k141_112248\|\|lt2gene | Low-quality |
| 18-105-final-viral-combined_k141_156240\|\|lt2gene | Low-quality |
| 18-105-final-viral-combined_k141_138625\|\|lt2gene | Low-quality |
| 18-105-final-viral-combined_k141_175062\|\|lt2gene | Not-determined |
| 18-105-final-viral-combined_k141_156249\|\|lt2gene | Low-quality |
| 18-105-final-viral-combined_k141_94400\|\|lt2gene | Low-quality |
| 18-105-final-viral-combined_k141_192538\|\|lt2gene | Low-quality |
| 18-105-final-viral-combined_k141_59051\|\|lt2gene | Not-determined |
| 18-105-final-viral-combined_k141_76862\|\|lt2gene | Low-quality |
| 18-105-final-viral-combined_k141_7219\|\|lt2gene | Low-quality |
| 18-105-final-viral-combined_k141_120783\|\|lt2gene | Low-quality |
| 18-105-final-viral-combined_k141_147959\|\|lt2gene | Low-quality |
| 18-105-final-viral-combined_k141_120792\|\|lt2gene | Not-determined |
| 18-105-final-viral-combined_k141_130383\|\|lt2gene | Low-quality |
| 18-105-final-viral-combined_k141_41081\|\|lt2gene | Low-quality |
| 18-105-final-viral-combined_k141_183753\|\|lt2gene | Low-quality |
| 18-105-final-viral-combined_k141_130396\|\|lt2gene | Low-quality |
| 18-105-final-viral-combined_k141_85467\|\|lt2gene | Not-determined |
| 18-105-final-viral-combined_k141_41124\|\|lt2gene | Low-quality |
| 18-105-final-viral-combined_k141_41153\|\|lt2gene | Low-quality |
| 18-105-final-viral-combined_k141_67399\|\|lt2gene | Low-quality |
| 18-105-final-viral-combined_k141_31705\|\|lt2gene | Low-quality |
| 18-105-final-viral-combined_k141_67419\|\|lt2gene | Low-quality |
| 18-105-final-viral-combined_k141_130493\|\|lt2gene | Low-quality |
| 18-105-final-viral-combined_k141_76991\|\|lt2gene | Low-quality |
| 18-105-final-viral-combined_k141_175142\|\|lt2gene | Low-quality |
| 18-105-final-viral-combined_k141_50019\|\|lt2gene | Low-quality |
| 18-105-final-viral-combined_k141_130542\|\|lt2gene | Low-quality |
| 18-105-final-viral-combined_k141_201287\|\|lt2gene | Low-quality |
| 18-105-final-viral-combined_k141_201297\|\|lt2gene | Not-determined |
| 18-105-final-viral-combined_k141_130578\|\|lt2gene | Low-quality |
| 18-105-final-viral-combined_k141_210236\|\|lt2gene | Not-determined |
| 18-105-final-viral-combined_k141_165352\|\|lt2gene | Low-quality |
| 18-105-final-viral-combined_k141_85659\|\|lt2gene | Low-quality |
| 18-105-final-viral-combined_k141_59156\|\|lt2gene | Low-quality |
| 18-105-final-viral-combined_k141_175241\|\|lt2gene | Low-quality |
| 18-105-final-viral-combined_k141_7478\|\|lt2gene | Low-quality |
| 18-105-final-viral-combined_k141_165377\|\|lt2gene | Low-quality |
| 18-105-final-viral-combined_k141_175266\|\|lt2gene | Low-quality |
| 18-105-final-viral-combined_k141_50153\|\|lt2gene | Low-quality |
| 18-105-final-viral-combined_k141_138781\|\|lt2gene | Not-determined |
| 18-105-final-viral-combined_k141_50176\|\|lt2gene | Low-quality |
| 18-105-final-viral-combined_k141_31933\|\|lt2gene | Low-quality |
| 18-105-final-viral-combined_k141_77169\|\|lt2gene | Low-quality |
| 18-105-final-viral-combined_k141_165453\|\|lt2gene | Low-quality |
| 18-105-final-viral-combined_k141_138869\|\|lt2gene | Low-quality |
| 18-105-final-viral-combined_k141_201407\|\|lt2gene | Not-determined |
| 18-105-final-viral-combined_k141_175360\|\|lt2gene | Not-determined |
| 18-105-final-viral-combined_k141_148260\|\|lt2gene | Not-determined |
| 18-105-final-viral-combined_k141_156502\|\|lt2gene | Low-quality |
| 18-105-final-viral-combined_k141_41370\|\|lt2gene | Low-quality |
| 18-105-final-viral-combined_k141_130724\|\|lt2gene | Low-quality |
| 18-105-final-viral-combined_k141_165567\|\|lt2gene | Not-determined |
| 18-105-final-viral-combined_k141_156551\|\|lt2gene | Low-quality |
| 18-105-final-viral-combined_k141_156565\|\|lt2gene | Low-quality |
| 18-105-final-viral-combined_k141_130774\|\|lt2gene | Not-determined |
| 18-105-final-viral-combined_k141_130777\|\|lt2gene | Low-quality |
| 18-105-final-viral-combined_k141_130793\|\|lt2gene | Low-quality |
| 18-105-final-viral-combined_k141_148373\|\|lt2gene | Not-determined |
| 18-105-final-viral-combined_k141_175502\|\|lt2gene | Not-determined |
| 18-105-final-viral-combined_k141_112619\|\|lt2gene | Not-determined |
| 18-105-final-viral-combined_k141_130819\|\|lt2gene | Low-quality |
| 18-105-final-viral-combined_k141_201476\|\|lt2gene | Low-quality |
| 18-105-final-viral-combined_k141_130839\|\|lt2gene | Low-quality |
| 18-105-final-viral-combined_k141_59322\|\|lt2gene | Low-quality |
| 18-105-final-viral-combined_k141_184131\|\|lt2gene | Low-quality |
| 18-105-final-viral-combined_k141_130890\|\|lt2gene | Not-determined |
| 18-105-final-viral-combined_k141_148473\|\|lt2gene | Not-determined |
| 18-105-final-viral-combined_k141_104117\|\|lt2gene | Low-quality |
| 18-105-final-viral-combined_k141_210377\|\|lt2gene | Low-quality |
| 18-105-final-viral-combined_k141_7645\|\|lt2gene | Not-determined |
| 18-105-final-viral-combined_k141_50364\|\|lt2gene | Low-quality |
| 18-105-final-viral-combined_k141_50373\|\|lt2gene | Not-determined |
| 18-105-final-viral-combined_k141_210438\|\|lt2gene | Low-quality |
| 18-105-final-viral-combined_k141_7680\|\|lt2gene | Low-quality |
| 18-105-final-viral-combined_k141_131008\|\|lt2gene | Not-determined |
| 18-105-final-viral-combined_k141_165718\|\|lt2gene | Low-quality |
| 18-105-final-viral-combined_k141_50421\|\|lt2gene | Low-quality |
| 18-105-final-viral-combined_k141_156693\|\|lt2gene | Low-quality |
| 18-105-final-viral-combined_k141_22641\|\|lt2gene | Low-quality |
| 18-105-final-viral-combined_k141_77477\|\|lt2gene | Low-quality |
| 18-105-final-viral-combined_k141_112730\|\|lt2gene | Low-quality |
| 18-105-final-viral-combined_k141_121208\|\|lt2gene | Low-quality |
| 18-105-final-viral-combined_k141_165773\|\|lt2gene | Not-determined |
| 18-105-final-viral-combined_k141_104188\|\|lt2gene | Low-quality |
| 18-105-final-viral-combined_k141_32249\|\|lt2gene | Low-quality |
| 18-105-final-viral-combined_k141_139188\|\|lt2gene | Low-quality |
| 18-105-final-viral-combined_k141_94685\|\|lt2gene | Low-quality |
| 18-105-final-viral-combined_k141_32285\|\|lt2gene | Not-determined |
| 18-105-final-viral-combined_k141_121307\|\|lt2gene | Low-quality |
| 18-105-final-viral-combined_k141_156770\|\|lt2gene | Low-quality |
| 18-105-final-viral-combined_k141_32300\|\|lt2gene | Low-quality |
| 18-105-final-viral-combined_k141_32301\|\|lt2gene | Low-quality |
| 18-105-final-viral-combined_k141_184373\|\|lt2gene | Low-quality |
| 18-105-final-viral-combined_k141_32303\|\|lt2gene | Low-quality |
| 18-105-final-viral-combined_k141_165787\|\|lt2gene | Low-quality |
| 18-105-final-viral-combined_k141_94709\|\|lt2gene | Low-quality |
| 18-105-final-viral-combined_k141_175588\|\|lt2gene | Low-quality |
| 18-105-final-viral-combined_k141_67755\|\|lt2gene | Low-quality |
| 18-105-final-viral-combined_k141_7815\|\|lt2gene | Low-quality |
| 18-105-final-viral-combined_k141_193105\|\|lt2gene | Low-quality |
| 18-105-final-viral-combined_k141_156854\|\|lt2gene | Low-quality |
| 18-105-final-viral-combined_k141_22758\|\|lt2gene | Low-quality |
| 18-105-final-viral-combined_k141_104215\|\|lt2gene | Not-determined |
| 18-105-final-viral-combined_k141_156862\|\|lt2gene | Low-quality |
| 18-105-final-viral-combined_k141_22775\|\|lt2gene | Low-quality |
| 18-105-final-viral-combined_k141_184474\|\|lt2gene | Low-quality |
| 18-105-final-viral-combined_k141_32371\|\|lt2gene | Low-quality |
| 18-105-final-viral-combined_k141_22799\|\|lt2gene | Low-quality |
| 18-105-final-viral-combined_k141_59542\|\|lt2gene | Not-determined |
| 18-105-final-viral-combined_k141_201710\|\|lt2gene | Low-quality |
| 18-105-final-viral-combined_k141_210635\|\|lt2gene | Not-determined |
| 18-105-final-viral-combined_k141_86144\|\|lt2gene | Low-quality |
| 18-105-final-viral-combined_k141_184568\|\|lt2gene | Low-quality |
| 18-105-final-viral-combined_k141_14929\|\|lt2gene | Low-quality |
| 18-105-final-viral-combined_k141_86154\|\|lt2gene | Low-quality |
| 18-105-final-viral-combined_k141_104351\|\|lt2gene | Low-quality |
| 18-105-final-viral-combined_k141_14960\|\|lt2gene | Not-determined |
| 18-105-final-viral-combined_k141_175716\|\|lt2gene | Low-quality |
| 18-105-final-viral-combined_k141_14966\|\|lt2gene | Low-quality |
| 18-105-final-viral-combined_k141_139397\|\|lt2gene | Not-determined |
| 18-105-final-viral-combined_k141_121403\|\|lt2gene | Low-quality |
| 18-105-final-viral-combined_k141_139424\|\|lt2gene | Low-quality |
| 18-105-final-viral-combined_k141_121411\|\|lt2gene | Not-determined |
| 18-105-final-viral-combined_k141_104403\|\|lt2gene | Low-quality |
| 18-105-final-viral-combined_k141_86200\|\|lt2gene | Not-determined |
| 18-105-final-viral-combined_k141_32420\|\|lt2gene | Low-quality |
| 18-105-final-viral-combined_k141_121450\|\|lt2gene | Low-quality |
| 18-105-final-viral-combined_k141_175793\|\|lt2gene | Low-quality |
| 18-105-final-viral-combined_k141_121467\|\|lt2gene | Low-quality |
| 18-105-final-viral-combined_k141_121479\|\|lt2gene | Low-quality |
| 18-105-final-viral-combined_k141_22921\|\|lt2gene | Not-determined |
| 18-105-final-viral-combined_k141_22923\|\|lt2gene | Not-determined |
| 18-105-final-viral-combined_k141_175811\|\|lt2gene | Low-quality |
| 18-105-final-viral-combined_k141_50578\|\|lt2gene | Low-quality |
| 18-105-final-viral-combined_k141_175812\|\|lt2gene | Low-quality |
| 18-105-final-viral-combined_k141_156960\|\|lt2gene | Low-quality |
| 18-105-final-viral-combined_k141_121484\|\|lt2gene | Low-quality |
| 18-105-final-viral-combined_k141_86299\|\|lt2gene | Not-determined |
| 18-105-final-viral-combined_k141_184747\|\|lt2gene | Low-quality |
| 18-105-final-viral-combined_k141_156991\|\|lt2gene | Low-quality |
| 18-105-final-viral-combined_k141_41541\|\|lt2gene | Low-quality |
| 18-105-final-viral-combined_k141_166105\|\|lt2gene | Low-quality |
| 18-105-final-viral-combined_k141_7888\|\|lt2gene | Not-determined |
| 18-105-final-viral-combined_k141_175891\|\|lt2gene | Low-quality |
| 18-105-final-viral-combined_k141_22947\|\|lt2gene | Low-quality |
| 18-105-final-viral-combined_k141_59715\|\|lt2gene | Low-quality |
| 18-105-final-viral-combined_k141_193311\|\|lt2gene | Not-determined |
| 18-105-final-viral-combined_k141_193312\|\|lt2gene | Low-quality |
| 18-105-final-viral-combined_k141_77780\|\|lt2gene | Not-determined |
| 18-105-final-viral-combined_k141_59719\|\|lt2gene | Low-quality |
| 18-105-final-viral-combined_k141_131346\|\|lt2gene | Low-quality |
| 18-105-final-viral-combined_k141_184785\|\|lt2gene | Low-quality |
| 18-105-final-viral-combined_k141_77830\|\|lt2gene | Low-quality |
| 18-105-final-viral-combined_k141_210901\|\|lt2gene | Low-quality |
| 18-105-final-viral-combined_k141_59775\|\|lt2gene | Low-quality |
| 18-105-final-viral-combined_k141_121625\|\|lt2gene | Low-quality |
| 18-105-final-viral-combined_k141_131362\|\|lt2gene | Not-determined |
| 18-105-final-viral-combined_k141_59794\|\|lt2gene | Low-quality |
| 18-105-final-viral-combined_k141_131399\|\|lt2gene | Low-quality |
| 18-105-final-viral-combined_k141_210939\|\|lt2gene | Low-quality |
| 18-105-final-viral-combined_k141_67876\|\|lt2gene | Low-quality |
| 18-105-final-viral-combined_k141_131420\|\|lt2gene | Low-quality |
| 18-105-final-viral-combined_k141_121676\|\|lt2gene | Low-quality |
| 18-105-final-viral-combined_k141_176034\|\|lt2gene | Low-quality |
| 18-105-final-viral-combined_k141_23058\|\|lt2gene | Low-quality |
| 18-105-final-viral-combined_k141_176039\|\|lt2gene | Low-quality |
| 18-105-final-viral-combined_k141_166218\|\|lt2gene | Low-quality |
| 18-105-final-viral-combined_k141_121702\|\|lt2gene | Low-quality |
| 18-105-final-viral-combined_k141_176058\|\|lt2gene | Low-quality |
| 18-105-final-viral-combined_k141_131486\|\|lt2gene | Not-determined |
| 18-105-final-viral-combined_k141_131506\|\|lt2gene | Low-quality |
| 18-105-final-viral-combined_k141_157196\|\|lt2gene | Not-determined |
| 18-105-final-viral-combined_k141_77934\|\|lt2gene | Low-quality |
| 18-105-final-viral-combined_k141_104519\|\|lt2gene | Low-quality |
| 18-105-final-viral-combined_k141_113091\|\|lt2gene | Not-determined |
| 18-105-final-viral-combined_k141_104521\|\|lt2gene | Low-quality |
| 18-105-final-viral-combined_k141_7972\|\|lt2gene | Low-quality |
| 18-105-final-viral-combined_k141_202027\|\|lt2gene | Low-quality |
| 18-105-final-viral-combined_k141_131526\|\|lt2gene | Low-quality |
| 18-105-final-viral-combined_k141_121774\|\|lt2gene | Low-quality |
| 18-105-final-viral-combined_k141_41632\|\|lt2gene | Low-quality |
| 18-105-final-viral-combined_k141_121795\|\|lt2gene | Low-quality |
| 18-105-final-viral-combined_k141_131575\|\|lt2gene | Low-quality |
| 18-105-final-viral-combined_k141_121806\|\|lt2gene | Low-quality |
| 18-105-final-viral-combined_k141_32609\|\|lt2gene | Low-quality |
| 18-105-final-viral-combined_k141_50919\|\|lt2gene | Low-quality |
| 18-105-final-viral-combined_k141_8025\|\|lt2gene | Low-quality |
| 18-105-final-viral-combined_k141_139581\|\|lt2gene | Low-quality |
| 18-105-final-viral-combined_k141_50939\|\|lt2gene | Low-quality |
| 18-105-final-viral-combined_k141_32619\|\|lt2gene | Low-quality |
| 18-105-final-viral-combined_k141_157311\|\|lt2gene | Low-quality |
| 18-105-final-viral-combined_k141_86487\|\|lt2gene | Not-determined |
| 18-105-final-viral-combined_k141_50966\|\|lt2gene | Not-determined |
| 18-105-final-viral-combined_k141_176165\|\|lt2gene | Low-quality |
| 18-105-final-viral-combined_k141_23141\|\|lt2gene | Low-quality |
| 18-105-final-viral-combined_k141_166349\|\|lt2gene | Low-quality |
| 18-105-final-viral-combined_k141_8100\|\|lt2gene | Low-quality |
| 18-105-final-viral-combined_k141_113230\|\|lt2gene | Not-determined |
| 18-105-final-viral-combined_k141_51028\|\|lt2gene | Low-quality |
| 18-105-final-viral-combined_k141_51049\|\|lt2gene | Low-quality |
| 18-105-final-viral-combined_k141_185029\|\|lt2gene | Low-quality |
| 18-105-final-viral-combined_k141_51057\|\|lt2gene | Low-quality |
| 18-105-final-viral-combined_k141_166414\|\|lt2gene | Low-quality |
| 18-105-final-viral-combined_k141_139723\|\|lt2gene | Low-quality |
| 18-105-final-viral-combined_k141_210987\|\|lt2gene | Not-determined |
| 18-105-final-viral-combined_k141_193432\|\|lt2gene | Low-quality |
| 18-105-final-viral-combined_k141_32750\|\|lt2gene | Low-quality |
| 18-105-final-viral-combined_k141_185086\|\|lt2gene | Not-determined |
| 18-105-final-viral-combined_k141_149074\|\|lt2gene | Low-quality |
| 18-105-final-viral-combined_k141_60044\|\|lt2gene | Low-quality |
| 18-105-final-viral-combined_k141_166502\|\|lt2gene | Low-quality |
| 18-105-final-viral-combined_k141_131722\|\|lt2gene | Low-quality |
| 18-105-final-viral-combined_k141_113372\|\|lt2gene | Low-quality |
| 18-105-final-viral-combined_k141_149112\|\|lt2gene | Not-determined |
| 18-105-final-viral-combined_k141_15288\|\|lt2gene | Not-determined |
| 18-105-final-viral-combined_k141_185117\|\|lt2gene | Low-quality |
| 18-105-final-viral-combined_k141_122048\|\|lt2gene | Not-determined |
| 18-105-final-viral-combined_k141_157490\|\|lt2gene | Low-quality |
| 18-105-final-viral-combined_k141_23331\|\|lt2gene | Low-quality |
| 18-105-final-viral-combined_k141_32848\|\|lt2gene | Not-determined |
| 18-105-final-viral-combined_k141_15376\|\|lt2gene | Low-quality |
| 18-105-final-viral-combined_k141_104748\|\|lt2gene | Not-determined |
| 18-105-final-viral-combined_k141_51195\|\|lt2gene | Low-quality |
| 18-105-final-viral-combined_k141_78211\|\|lt2gene | Low-quality |
| 18-105-final-viral-combined_k141_32870\|\|lt2gene | Not-determined |
| 18-105-final-viral-combined_k141_8267\|\|lt2gene | Not-determined |
| 18-105-final-viral-combined_k141_202146\|\|lt2gene | Low-quality |
| 18-105-final-viral-combined_k141_23382\|\|lt2gene | Low-quality |
| 18-105-final-viral-combined_k141_23385\|\|lt2gene | Low-quality |
| 18-105-final-viral-combined_k141_51239\|\|lt2gene | Low-quality |
| 18-105-final-viral-combined_k141_51244\|\|lt2gene | Low-quality |
| 18-105-final-viral-combined_k141_193547\|\|lt2gene | Not-determined |
| 18-105-final-viral-combined_k141_15490\|\|lt2gene | Low-quality |
| 18-105-final-viral-combined_k141_68367\|\|lt2gene | Not-determined |
| 18-105-final-viral-combined_k141_113440\|\|lt2gene | Low-quality |
| 18-105-final-viral-combined_k141_104785\|\|lt2gene | Low-quality |
| 18-105-final-viral-combined_k141_41867\|\|lt2gene | Low-quality |
| 18-105-final-viral-combined_k141_41870\|\|lt2gene | Low-quality |
| 18-105-final-viral-combined_k141_41891\|\|lt2gene | Low-quality |
| 18-105-final-viral-combined_k141_157616\|\|lt2gene | Not-determined |
| 18-105-final-viral-combined_k141_113489\|\|lt2gene | Low-quality |
| 18-105-final-viral-combined_k141_15599\|\|lt2gene | Low-quality |
| 18-105-final-viral-combined_k141_104847\|\|lt2gene | Not-determined |
| 18-105-final-viral-combined_k141_15602\|\|lt2gene | Not-determined |
| 18-105-final-viral-combined_k141_60096\|\|lt2gene | Low-quality |
| 18-105-final-viral-combined_k141_95206\|\|lt2gene | Not-determined |
| 18-105-final-viral-combined_k141_41934\|\|lt2gene | Not-determined |
| 18-105-final-viral-combined_k141_78396\|\|lt2gene | Low-quality |
| 18-105-final-viral-combined_k141_60110\|\|lt2gene | Low-quality |
| 18-105-final-viral-combined_k141_78424\|\|lt2gene | Low-quality |
| 18-105-final-viral-combined_k141_157666\|\|lt2gene | Low-quality |
| 18-105-final-viral-combined_k141_157670\|\|lt2gene | Low-quality |
| 18-105-final-viral-combined_k141_157672\|\|lt2gene | Low-quality |
| 18-105-final-viral-combined_k141_122185\|\|lt2gene | Low-quality |
| 18-105-final-viral-combined_k141_176422\|\|lt2gene | Low-quality |
| 18-105-final-viral-combined_k141_185245\|\|lt2gene | Low-quality |
| 18-105-final-viral-combined_k141_166618\|\|lt2gene | Low-quality |
| 18-105-final-viral-combined_k141_176470\|\|lt2gene | Not-determined |
| 18-105-final-viral-combined_k141_86975\|\|lt2gene | Not-determined |
| 18-105-final-viral-combined_k141_68536\|\|lt2gene | Low-quality |
| 18-105-final-viral-combined_k141_104910\|\|lt2gene | Low-quality |
| 18-105-final-viral-combined_k141_211204\|\|lt2gene | Low-quality |
| 18-105-final-viral-combined_k141_166683\|\|lt2gene | Low-quality |
| 18-105-final-viral-combined_k141_211226\|\|lt2gene | Low-quality |
| 18-105-final-viral-combined_k141_122247\|\|lt2gene | Not-determined |
| 18-105-final-viral-combined_k141_87018\|\|lt2gene | Low-quality |
| 18-105-final-viral-combined_k141_193759\|\|lt2gene | Not-determined |
| 18-105-final-viral-combined_k141_131941\|\|lt2gene | Not-determined |
| 18-105-final-viral-combined_k141_87042\|\|lt2gene | Low-quality |
| 18-105-final-viral-combined_k141_131971\|\|lt2gene | Low-quality |
| 18-105-final-viral-combined_k141_122298\|\|lt2gene | Low-quality |
| 18-105-final-viral-combined_k141_33087\|\|lt2gene | Not-determined |
| 18-105-final-viral-combined_k141_8375\|\|lt2gene | Low-quality |
| 18-105-final-viral-combined_k141_87102\|\|lt2gene | Low-quality |
| 18-105-final-viral-combined_k141_60271\|\|lt2gene | Low-quality |
| 18-105-final-viral-combined_k141_8392\|\|lt2gene | Low-quality |
| 18-105-final-viral-combined_k141_132012\|\|lt2gene | Not-determined |
| 18-105-final-viral-combined_k141_193886\|\|lt2gene | Low-quality |
| 18-105-final-viral-combined_k141_105011\|\|lt2gene | Low-quality |
| 18-105-final-viral-combined_k141_140119\|\|lt2gene | Not-determined |
| 18-105-final-viral-combined_k141_68675\|\|lt2gene | Low-quality |
| 18-105-final-viral-combined_k141_132041\|\|lt2gene | Low-quality |
| 18-105-final-viral-combined_k141_157910\|\|lt2gene | Low-quality |
| 18-105-final-viral-combined_k141_122340\|\|lt2gene | Not-determined |
| 18-105-final-viral-combined_k141_87159\|\|lt2gene | Not-determined |
| 18-105-final-viral-combined_k141_140148\|\|lt2gene | Low-quality |
| 18-105-final-viral-combined_k141_23638\|\|lt2gene | Not-determined |
| 18-105-final-viral-combined_k141_15704\|\|lt2gene | Low-quality |
| 18-105-final-viral-combined_k141_105058\|\|lt2gene | Low-quality |
| 18-105-final-viral-combined_k141_132092\|\|lt2gene | Not-determined |
| 18-105-final-viral-combined_k141_15722\|\|lt2gene | Low-quality |
| 18-105-final-viral-combined_k141_78527\|\|lt2gene | Low-quality |
| 18-105-final-viral-combined_k141_68743\|\|lt2gene | Low-quality |
| 18-105-final-viral-combined_k141_185467\|\|lt2gene | Low-quality |
| 18-105-final-viral-combined_k141_15759\|\|lt2gene | Low-quality |
| 18-105-final-viral-combined_k141_95424\|\|lt2gene | Low-quality |
| 18-105-final-viral-combined_k141_68779\|\|lt2gene | Low-quality |
| 18-105-final-viral-combined_k141_122425\|\|lt2gene | Low-quality |
| 18-105-final-viral-combined_k141_166910\|\|lt2gene | Low-quality |
| 18-105-final-viral-combined_k141_140290\|\|lt2gene | Not-determined |
| 18-105-final-viral-combined_k141_87192\|\|lt2gene | Low-quality |
| 18-105-final-viral-combined_k141_132124\|\|lt2gene | Low-quality |
| 18-105-final-viral-combined_k141_193996\|\|lt2gene | Low-quality |
| 18-105-final-viral-combined_k141_140309\|\|lt2gene | Not-determined |
| 18-105-final-viral-combined_k141_68859\|\|lt2gene | Low-quality |
| 18-105-final-viral-combined_k141_78620\|\|lt2gene | Low-quality |
| 18-105-final-viral-combined_k141_132137\|\|lt2gene | Not-determined |
| 18-105-final-viral-combined_k141_202346\|\|lt2gene | Low-quality |
| 18-105-final-viral-combined_k141_132142\|\|lt2gene | Not-determined |
| 18-105-final-viral-combined_k141_42000\|\|lt2gene | Not-determined |
| 18-105-final-viral-combined_k141_60392\|\|lt2gene | Low-quality |
| 18-105-final-viral-combined_k141_194037\|\|lt2gene | Low-quality |
| 18-105-final-viral-combined_k141_87238\|\|lt2gene | Low-quality |
| 18-105-final-viral-combined_k141_105197\|\|lt2gene | Low-quality |
| 18-105-final-viral-combined_k141_211513\|\|lt2gene | Not-determined |
| 18-105-final-viral-combined_k141_194047\|\|lt2gene | Not-determined |
| 18-105-final-viral-combined_k141_68907\|\|lt2gene | Low-quality |
| 18-105-final-viral-combined_k141_68910\|\|lt2gene | Not-determined |
| 18-105-final-viral-combined_k141_158030\|\|lt2gene | Low-quality |
| 18-105-final-viral-combined_k141_194077\|\|lt2gene | Low-quality |
| 18-105-final-viral-combined_k141_194080\|\|lt2gene | Low-quality |
| 18-105-final-viral-combined_k141_78691\|\|lt2gene | Not-determined |
| 18-105-final-viral-combined_k141_105230\|\|lt2gene | Low-quality |
| 18-105-final-viral-combined_k141_23804\|\|lt2gene | Low-quality |
| 18-105-final-viral-combined_k141_95553\|\|lt2gene | Not-determined |
| 18-105-final-viral-combined_k141_78750\|\|lt2gene | Low-quality |
| 18-105-final-viral-combined_k141_105269\|\|lt2gene | Low-quality |
| 18-105-final-viral-combined_k141_122562\|\|lt2gene | Not-determined |
| 18-105-final-viral-combined_k141_122566\|\|lt2gene | Low-quality |
| 18-105-final-viral-combined_k141_105289\|\|lt2gene | Low-quality |
| 18-105-final-viral-combined_k141_87373\|\|lt2gene | Low-quality |
| 18-105-final-viral-combined_k141_42049\|\|lt2gene | Low-quality |
| 18-105-final-viral-combined_k141_211676\|\|lt2gene | Low-quality |
| 18-105-final-viral-combined_k141_122623\|\|lt2gene | Low-quality |
| 18-105-final-viral-combined_k141_51687\|\|lt2gene | Not-determined |
| 18-105-final-viral-combined_k141_95608\|\|lt2gene | Low-quality |
| 18-105-final-viral-combined_k141_87412\|\|lt2gene | Low-quality |
| 18-105-final-viral-combined_k141_60604\|\|lt2gene | Low-quality |
| 18-105-final-viral-combined_k141_8709\|\|lt2gene | Low-quality |
| 18-105-final-viral-combined_k141_68993\|\|lt2gene | Not-determined |
| 18-105-final-viral-combined_k141_15831\|\|lt2gene | Low-quality |
| 18-105-final-viral-combined_k141_105355\|\|lt2gene | Low-quality |
| 18-105-final-viral-combined_k141_15835\|\|lt2gene | Low-quality |
| 18-105-final-viral-combined_k141_122653\|\|lt2gene | Low-quality |
| 18-105-final-viral-combined_k141_15872\|\|lt2gene | Low-quality |
| 18-105-final-viral-combined_k141_42151\|\|lt2gene | Low-quality |
| 18-105-final-viral-combined_k141_15879\|\|lt2gene | Low-quality |
| 18-105-final-viral-combined_k141_23921\|\|lt2gene | Not-determined |
| 18-105-final-viral-combined_k141_132307\|\|lt2gene | Not-determined |
| 18-105-final-viral-combined_k141_202524\|\|lt2gene | Low-quality |
| 18-105-final-viral-combined_k141_105391\|\|lt2gene | Not-determined |
| 18-105-final-viral-combined_k141_69074\|\|lt2gene | Low-quality |
| 18-105-final-viral-combined_k141_78822\|\|lt2gene | Low-quality |
| 18-105-final-viral-combined_k141_23939\|\|lt2gene | Low-quality |
| 18-105-final-viral-combined_k141_78845\|\|lt2gene | Low-quality |
| 18-105-final-viral-combined_k141_176563\|\|lt2gene | Low-quality |
| 18-105-final-viral-combined_k141_149512\|\|lt2gene | Not-determined |
| 18-105-final-viral-combined_k141_158161\|\|lt2gene | Low-quality |
| 18-105-final-viral-combined_k141_23951\|\|lt2gene | Not-determined |
| 18-105-final-viral-combined_k141_132360\|\|lt2gene | Low-quality |
| 18-105-final-viral-combined_k141_60746\|\|lt2gene | Low-quality |
| 18-105-final-viral-combined_k141_8808\|\|lt2gene | Not-determined |
| 18-105-final-viral-combined_k141_95774\|\|lt2gene | Not-determined |
| 18-105-final-viral-combined_k141_185556\|\|lt2gene | Low-quality |
| 18-105-final-viral-combined_k141_60777\|\|lt2gene | Not-determined |
| 18-105-final-viral-combined_k141_87538\|\|lt2gene | Low-quality |
| 18-105-final-viral-combined_k141_95789\|\|lt2gene | Low-quality |
| 18-105-final-viral-combined_k141_95795\|\|lt2gene | Low-quality |
| 18-105-final-viral-combined_k141_69185\|\|lt2gene | Low-quality |
| 18-105-final-viral-combined_k141_23991\|\|lt2gene | Low-quality |
| 18-105-final-viral-combined_k141_176641\|\|lt2gene | Not-determined |
| 18-105-final-viral-combined_k141_69249\|\|lt2gene | Low-quality |
| 18-105-final-viral-combined_k141_202636\|\|lt2gene | Low-quality |
| 18-105-final-viral-combined_k141_211700\|\|lt2gene | Low-quality |
| 18-105-final-viral-combined_k141_113713\|\|lt2gene | Low-quality |
| 18-105-final-viral-combined_k141_176705\|\|lt2gene | Low-quality |
| 18-105-final-viral-combined_k141_167209\|\|lt2gene | Not-determined |
| 18-105-final-viral-combined_k141_176736\|\|lt2gene | Not-determined |
| 18-105-final-viral-combined_k141_42299\|\|lt2gene | Low-quality |
| 18-105-final-viral-combined_k141_176760\|\|lt2gene | Low-quality |
| 18-105-final-viral-combined_k141_132528\|\|lt2gene | Not-determined |
| 18-105-final-viral-combined_k141_42324\|\|lt2gene | Low-quality |
| 18-105-final-viral-combined_k141_87692\|\|lt2gene | Low-quality |
| 18-105-final-viral-combined_k141_42343\|\|lt2gene | Low-quality |
| 18-105-final-viral-combined_k141_42349\|\|lt2gene | Low-quality |
| 18-105-final-viral-combined_k141_185597\|\|lt2gene | Not-determined |
| 18-105-final-viral-combined_k141_185647\|\|lt2gene | Low-quality |
| 18-105-final-viral-combined_k141_176810\|\|lt2gene | Low-quality |
| 18-105-final-viral-combined_k141_113842\|\|lt2gene | Low-quality |
| 18-105-final-viral-combined_k141_185668\|\|lt2gene | Not-determined |
| 18-105-final-viral-combined_k141_113899\|\|lt2gene | Low-quality |
| 18-105-final-viral-combined_k141_78928\|\|lt2gene | Low-quality |
| 18-105-final-viral-combined_k141_60977\|\|lt2gene | Low-quality |
| 18-105-final-viral-combined_k141_167342\|\|lt2gene | Not-determined |
| 18-105-final-viral-combined_k141_95989\|\|lt2gene | Low-quality |
| 18-105-final-viral-combined_k141_132619\|\|lt2gene | Low-quality |
| 18-105-final-viral-combined_k141_87796\|\|lt2gene | Low-quality |
| 18-105-final-viral-combined_k141_52242\|\|lt2gene | Not-determined |
| 18-105-final-viral-combined_k141_78951\|\|lt2gene | Low-quality |
| 18-105-final-viral-combined_k141_87817\|\|lt2gene | Low-quality |
| 18-105-final-viral-combined_k141_149783\|\|lt2gene | Low-quality |
| 18-105-final-viral-combined_k141_33815\|\|lt2gene | Low-quality |
| 18-105-final-viral-combined_k141_149788\|\|lt2gene | Low-quality |
| 18-105-final-viral-combined_k141_149791\|\|lt2gene | Low-quality |
| 18-105-final-viral-combined_k141_167379\|\|lt2gene | Low-quality |
| 18-105-final-viral-combined_k141_87836\|\|lt2gene | Not-determined |
| 18-105-final-viral-combined_k141_87837\|\|lt2gene | Not-determined |
| 18-105-final-viral-combined_k141_78977\|\|lt2gene | Low-quality |
| 18-105-final-viral-combined_k141_87861\|\|lt2gene | Low-quality |
| 18-105-final-viral-combined_k141_194541\|\|lt2gene | Not-determined |
| 18-105-final-viral-combined_k141_79004\|\|lt2gene | Low-quality |
| 18-105-final-viral-combined_k141_194549\|\|lt2gene | Low-quality |
| 18-105-final-viral-combined_k141_52305\|\|lt2gene | Low-quality |
| 18-105-final-viral-combined_k141_113987\|\|lt2gene | Low-quality |
| 18-105-final-viral-combined_k141_52315\|\|lt2gene | Not-determined |
| 18-105-final-viral-combined_k141_52333\|\|lt2gene | Low-quality |
| 18-105-final-viral-combined_k141_194572\|\|lt2gene | Low-quality |
| 18-105-final-viral-combined_k141_24208\|\|lt2gene | Low-quality |
| 18-105-final-viral-combined_k141_24216\|\|lt2gene | Low-quality |
| 18-105-final-viral-combined_k141_52350\|\|lt2gene | Low-quality |
| 18-105-final-viral-combined_k141_123151\|\|lt2gene | Not-determined |
| 18-105-final-viral-combined_k141_194600\|\|lt2gene | Low-quality |
| 18-105-final-viral-combined_k141_194609\|\|lt2gene | Low-quality |
| 18-105-final-viral-combined_k141_177000\|\|lt2gene | Low-quality |
| 18-105-final-viral-combined_k141_105674\|\|lt2gene | Not-determined |
| 18-105-final-viral-combined_k141_123185\|\|lt2gene | Not-determined |
| 18-105-final-viral-combined_k141_211870\|\|lt2gene | Low-quality |
| 18-105-final-viral-combined_k141_149885\|\|lt2gene | Not-determined |
| 18-105-final-viral-combined_k141_167558\|\|lt2gene | Not-determined |
| 18-105-final-viral-combined_k141_167565\|\|lt2gene | Low-quality |
| 18-105-final-viral-combined_k141_194675\|\|lt2gene | Not-determined |
| 18-105-final-viral-combined_k141_87950\|\|lt2gene | Not-determined |
| 18-105-final-viral-combined_k141_194707\|\|lt2gene | Low-quality |
| 18-105-final-viral-combined_k141_105737\|\|lt2gene | Low-quality |
| 18-105-final-viral-combined_k141_16078\|\|lt2gene | Low-quality |
| 18-105-final-viral-combined_k141_158416\|\|lt2gene | Low-quality |
| 18-105-final-viral-combined_k141_69336\|\|lt2gene | Low-quality |
| 18-105-final-viral-combined_k141_87978\|\|lt2gene | Not-determined |
| 18-105-final-viral-combined_k141_24333\|\|lt2gene | Low-quality |
| 18-105-final-viral-combined_k141_42636\|\|lt2gene | Low-quality |
| 18-105-final-viral-combined_k141_194737\|\|lt2gene | Low-quality |
| 18-105-final-viral-combined_k141_96135\|\|lt2gene | Not-determined |
| 18-105-final-viral-combined_k141_52496\|\|lt2gene | Low-quality |
| 18-105-final-viral-combined_k141_149931\|\|lt2gene | Not-determined |
| 18-105-final-viral-combined_k141_194750\|\|lt2gene | Not-determined |
| 18-105-final-viral-combined_k141_211976\|\|lt2gene | Low-quality |
| 18-105-final-viral-combined_k141_140879\|\|lt2gene | Not-determined |
| 18-105-final-viral-combined_k141_177142\|\|lt2gene | Low-quality |
| 18-105-final-viral-combined_k141_149955\|\|lt2gene | Not-determined |
| 18-105-final-viral-combined_k141_105761\|\|lt2gene | Low-quality |
| 18-105-final-viral-combined_k141_149970\|\|lt2gene | Low-quality |

| | |
|---|---|
| 18-105-final-viral-combined_k141_194785\|\|lt2gene | Low-quality |
| 18-105-final-viral-combined_k141_42679\|\|lt2gene | Low-quality |
| 18-105-final-viral-combined_k141_96177\|\|lt2gene | Not-determined |
| 18-105-final-viral-combined_k141_42691\|\|lt2gene | Not-determined |
| 18-105-final-viral-combined_k141_140953\|\|lt2gene | Not-determined |
| 18-105-final-viral-combined_k141_140955\|\|lt2gene | Not-determined |
| 18-105-final-viral-combined_k141_87991\|\|lt2gene | Low-quality |
| 18-105-final-viral-combined_k141_42746\|\|lt2gene | Not-determined |
| 18-105-final-viral-combined_k141_42751\|\|lt2gene | Low-quality |
| 18-105-final-viral-combined_k141_105848\|\|lt2gene | Low-quality |
| 18-105-final-viral-combined_k141_42757\|\|lt2gene | Not-determined |
| 18-105-final-viral-combined_k141_140984\|\|lt2gene | Not-determined |
| 18-105-final-viral-combined_k141_52603\|\|lt2gene | Low-quality |
| 18-105-final-viral-combined_k141_202982\|\|lt2gene | Low-quality |
| 18-105-final-viral-combined_k141_96238\|\|lt2gene | Low-quality |
| 18-105-final-viral-combined_k141_167747\|\|lt2gene | Not-determined |
| 18-105-final-viral-combined_k141_96242\|\|lt2gene | Not-determined |
| 18-105-final-viral-combined_k141_141020\|\|lt2gene | Low-quality |
| 18-105-final-viral-combined_k141_123419\|\|lt2gene | Low-quality |
| 18-105-final-viral-combined_k141_123428\|\|lt2gene | Low-quality |
| 18-105-final-viral-combined_k141_96266\|\|lt2gene | Not-determined |
| 18-105-final-viral-combined_k141_177236\|\|lt2gene | Low-quality |
| 18-105-final-viral-combined_k141_212073\|\|lt2gene | Low-quality |
| 18-105-final-viral-combined_k141_88071\|\|lt2gene | Low-quality |
| 18-105-final-viral-combined_k141_88078\|\|lt2gene | Low-quality |
| 18-105-final-viral-combined_k141_88080\|\|lt2gene | Low-quality |
| 18-105-final-viral-combined_k141_132721\|\|lt2gene | Low-quality |
| 18-105-final-viral-combined_k141_69452\|\|lt2gene | Low-quality |
| 18-105-final-viral-combined_k141_141054\|\|lt2gene | Not-determined |
| 18-105-final-viral-combined_k141_185983\|\|lt2gene | Low-quality |
| 18-105-final-viral-combined_k141_88105\|\|lt2gene | Low-quality |
| 18-105-final-viral-combined_k141_186002\|\|lt2gene | Low-quality |
| 18-105-final-viral-combined_k141_167798\|\|lt2gene | Low-quality |
| 18-105-final-viral-combined_k141_123522\|\|lt2gene | Low-quality |
| 18-105-final-viral-combined_k141_114165\|\|lt2gene | Not-determined |
| 18-105-final-viral-combined_k141_79278\|\|lt2gene | Low-quality |
| 18-105-final-viral-combined_k141_88172\|\|lt2gene | Low-quality |
| 18-105-final-viral-combined_k141_42823\|\|lt2gene | Low-quality |
| 18-105-final-viral-combined_k141_150156\|\|lt2gene | Low-quality |
| 18-105-final-viral-combined_k141_96339\|\|lt2gene | Not-determined |
| 18-105-final-viral-combined_k141_34163\|\|lt2gene | Low-quality |
| 18-105-final-viral-combined_k141_79300\|\|lt2gene | Low-quality |
| 18-105-final-viral-combined_k141_167845\|\|lt2gene | Not-determined |
| 18-105-final-viral-combined_k141_96389\|\|lt2gene | Low-quality |
| 18-105-final-viral-combined_k141_195065\|\|lt2gene | Low-quality |
| [illegible] | [illegible] |

| | |
|---|---|
| 18-105_k141_142765 | Not-determined |
| 18-105_k141_142766 | Not-determined |
| 18-105_k141_160650 | Not-determined |
| 18-105_k141_160653 | Not-determined |
| 18-105_k141_160659 | Low-quality |
| 18-105_k141_107271 | Not-determined |
| 18-105_k141_80469 | Not-determined |
| 18-105_k141_494 | Not-determined |
| 18-105_k141_495 | Not-determined |
| 18-105_k141_496 | Not-determined |
| 18-105_k141_116153 | Not-determined |
| 18-105_k141_80477 | Not-determined |
| 18-105_k141_133946 | Not-determined |
| 18-105_k141_133947 | Not-determined |
| 18-105_k141_53763 | Low-quality |
| 18-105_k141_187285 | Not-determined |
| 18-105_k141_187286 | Low-quality |
| 18-105_k141_160703 | Not-determined |
| 18-105_k141_187289 | Not-determined |
| 18-105_k141_116187 | Not-determined |
| 18-105_k141_9280 | Not-determined |
| 18-105_k141_125026 | Not-determined |
| 18-105_k141_125032 | Low-quality |
| 18-105_k141_133965 | Not-determined |
| 18-105_k141_125036 | Low-quality |
| 18-105_k141_98203 | Low-quality |
| 18-105_k141_18138 | Not-determined |
| 18-105_k141_98204 | Not-determined |
| 18-105_k141_205019 | Not-determined |
| 18-105_k141_18141 | Not-determined |
| 18-105_k141_205020 | Not-determined |
| 18-105_k141_71630 | Low-quality |
| 18-105_k141_187303 | Not-determined |
| 18-105_k141_196349 | Low-quality |
| 18-105_k141_205031 | Not-determined |
| 18-105_k141_196351 | Not-determined |
| 18-105_k141_205037 | Not-determined |
| 18-105_k141_62598 | Not-determined |
| 18-105_k141_196361 | Not-determined |
| 18-105_k141_18156 | Not-determined |
| 18-105_k141_44606 | Not-determined |
| 18-105_k141_18158 | Not-determined |
| 18-105_k141_187308 | Not-determined |
| 18-105_k141_160733 | Not-determined |
| 18-105_k141_507 | Not-determined |
| 18-105_k141_509 | Not-determined |
| 18-105_k141_133990 | Not-determined |
| 18-105_k141_187313 | Not-determined |
| 18-105_k141_71648 | Not-determined |
| 18-105_k141_62615 | Not-determined |
| 18-105_k141_523 | Not-determined |
| 18-105_k141_35779 | Not-determined |
| 18-105_k141_35781 | Not-determined |
| 18-105_k141_53799 | Not-determined |
| 18-105_k141_62624 | Not-determined |
| 18-105_k141_134012 | Not-determined |
| 18-105_k141_44615 | Not-determined |
| 18-105_k141_134014 | Not-determined |
| 18-105_k141_62633 | Not-determined |
| 18-105_k141_18163 | Not-determined |
| 18-105_k141_62635 | Not-determined |
| 18-105_k141_160760 | Not-determined |
| 18-105_k141_71660 | Not-determined |
| 18-105_k141_160761 | Not-determined |
| 18-105_k141_71662 | Low-quality |
| 18-105_k141_134027 | Not-determined |
| 18-105_k141_44630 | Not-determined |
| 18-105_k141_116208 | Not-determined |
| 18-105_k141_160765 | Not-determined |
| 18-105_k141_71669 | Not-determined |
| 18-105_k141_35790 | Not-determined |
| 18-105_k141_187328 | Not-determined |
| 18-105_k141_196384 | Not-determined |
| 18-105_k141_134040 | Not-determined |
| 18-105_k141_196393 | Not-determined |
| 18-105_k141_134041 | Low-quality |
| 18-105_k141_35801 | Not-determined |
| 18-105_k141_71682 | Not-determined |
| 18-105_k141_18182 | Not-determined |
| 18-105_k141_71683 | Not-determined |
| 18-105_k141_35807 | Not-determined |
| 18-105_k141_18188 | Not-determined |
| 18-105_k141_142793 | Low-quality |
| 18-105_k141_196407 | Not-determined |
| 18-105_k141_196410 | Low-quality |
| 18-105_k141_196412 | Not-determined |
| 18-105_k141_554 | Not-determined |
| 18-105_k141_558 | Not-determined |
| 18-105_k141_196414 | Not-determined |
| 18-105_k141_116222 | Not-determined |
| 18-105_k141_187346 | Not-determined |
| 18-105_k141_9367 | Not-determined |
| 18-105_k141_196419 | Not-determined |
| 18-105_k141_98212 | Not-determined |
| 18-105_k141_187355 | Not-determined |
| 18-105_k141_98226 | Not-determined |
| 18-105_k141_116227 | Not-determined |
| 18-105_k141_187367 | Not-determined |
| 18-105_k141_116233 | Not-determined |
| 18-105_k141_44656 | Not-determined |
| 18-105_k141_187374 | Not-determined |
| 18-105_k141_142829 | Not-determined |
| 18-105_k141_187380 | Not-determined |
| 18-105_k141_580 | Not-determined |
| 18-105_k141_62639 | Not-determined |
| 18-105_k141_71703 | Not-determined |
| 18-105_k141_142843 | Not-determined |
| 18-105_k141_589 | Not-determined |
| 18-105_k141_590 | Not-determined |
| 18-105_k141_9377 | Not-determined |
| 18-105_k141_187386 | Not-determined |
| 18-105_k141_134088 | Not-determined |
| 18-105_k141_80490 | Low-quality |
| 18-105_k141_594 | Not-determined |
| 18-105_k141_187395 | Not-determined |
| 18-105_k141_187402 | Not-determined |
| 18-105_k141_205074 | Not-determined |
| 18-105_k141_187404 | Not-determined |
| 18-105_k141_205078 | Low-quality |
| 18-105_k141_196453 | Not-determined |
| 18-105_k141_9397 | Not-determined |
| 18-105_k141_71729 | Not-determined |
| 18-105_k141_196455 | Not-determined |
| 18-105_k141_71737 | Not-determined |
| 18-105_k141_98272 | Not-determined |
| 18-105_k141_9400 | Not-determined |
| 18-105_k141_98289 | Not-determined |
| 18-105_k141_187419 | Not-determined |
| 18-105_k141_89251 | Not-determined |
| 18-105_k141_629 | Low-quality |
| 18-105_k141_89253 | Not-determined |
| 18-105_k141_205084 | Low-quality |
| 18-105_k141_35821 | Not-determined |
| 18-105_k141_53833 | Not-determined |
| 18-105_k141_35824 | Not-determined |
| 18-105_k141_9423 | Not-determined |
| 18-105_k141_187429 | Not-determined |
| 18-105_k141_80541 | Not-determined |
| 18-105_k141_160816 | Not-determined |
| 18-105_k141_160817 | Not-determined |
| 18-105_k141_35833 | Not-determined |
| 18-105_k141_637 | Not-determined |
| 18-105_k141_89271 | Not-determined |
| 18-105_k141_9444 | Not-determined |
| 18-105_k141_205108 | Not-determined |
| 18-105_k141_35838 | Low-quality |
| 18-105_k141_89284 | Not-determined |
| 18-105_k141_205114 | Not-determined |
| 18-105_k141_80547 | Not-determined |
| 18-105_k141_125044 | Not-determined |
| 18-105_k141_89292 | Not-determined |
| 18-105_k141_89293 | Not-determined |
| 18-105_k141_187432 | Not-determined |
| 18-105_k141_647 | Not-determined |
| 18-105_k141_648 | Not-determined |
| 18-105_k141_649 | Not-determined |
| 18-105_k141_205132 | Not-determined |
| 18-105_k141_134104 | Not-determined |
| 18-105_k141_9470 | Not-determined |
| 18-105_k141_9472 | Not-determined |
| 18-105_k141_205143 | Not-determined |
| 18-105_k141_134115 | Not-determined |
| 18-105_k141_71770 | Not-determined |
| 18-105_k141_661 | Not-determined |
| 18-105_k141_142878 | Not-determined |
| 18-102-final-viral-combined_k141_57293\|\|full | High-quality |
| 18-105_k141_116244 | Not-determined |
| 18-105_k141_196485 | Not-determined |
| 18-105_k141_80563 | Not-determined |
| 18-105_k141_26981 | Not-determined |
| 18-105_k141_71781 | Not-determined |
| 18-105_k141_53857 | Not-determined |
| 18-105_k141_53861 | Not-determined |
| 18-105_k141_71788 | Not-determined |
| 18-105_k141_196496 | Not-determined |
| 18-105_k141_18198 | Not-determined |
| 18-105_k141_71802 | Not-determined |
| 18-105_k141_53872 | Not-determined |
| 18-105_k141_9480 | Not-determined |
| 18-105_k141_107298 | Not-determined |
| 18-105_k141_107299 | Not-determined |
| 18-105_k141_9491 | Low-quality |
| 18-105_k141_703 | Not-determined |
| 18-105_k141_44757 | Not-determined |
| 18-105_k141_107304 | Not-determined |
| 18-105_k141_18207 | Not-determined |
| 18-105_k141_26998 | Not-determined |
| 18-105_k141_62701 | Not-determined |
| 18-105_k141_71824 | Not-determined |
| 18-105_k141_142911 | Low-quality |
| 18-105_k141_142913 | Not-determined |
| 18-105_k141_187479 | Not-determined |
| 18-105_k141_62708 | Low-quality |
| 18-105_k141_142916 | Not-determined |
| 18-105_k141_89350 | Not-determined |
| 18-105_k141_196505 | Not-determined |
| 18-105_k141_107312 | Not-determined |
| 18-105_k141_107314 | Not-determined |
| 18-105_k141_107315 | Not-determined |
| 18-105_k141_27003 | Not-determined |
| 18-105_k141_187506 | Not-determined |
| 18-105_k141_89368 | Not-determined |
| 18-105_k141_62731 | Not-determined |
| 18-105_k141_62737 | Not-determined |
| 18-105_k141_169531 | Not-determined |
| 18-105_k141_205166 | Not-determined |
| 18-105_k141_169540 | Not-determined |
| 18-105_k141_35856 | Not-determined |
| 18-105_k141_151672 | Not-determined |
| 18-105_k141_196517 | Not-determined |
| 18-105_k141_169550 | Not-determined |
| 18-105_k141_169551 | Not-determined |
| 18-105_k141_205172 | Not-determined |
| 18-105_k141_169555 | Not-determined |
| 18-105_k141_169559 | Not-determined |
| 18-105_k141_35865 | Not-determined |
| 18-105_k141_151685 | Not-determined |
| 18-105_k141_142977 | Not-determined |
| 18-105_k141_35871 | Not-determined |
| 18-105_k141_142980 | Not-determined |
| 18-105_k141_169562 | Not-determined |
| 18-105_k141_53899 | Not-determined |
| 18-105_k141_107333 | Not-determined |
| 18-105_k141_169572 | Not-determined |
| 18-105_k141_35881 | Not-determined |
| 18-105_k141_196532 | Not-determined |
| 18-105_k141_169575 | Not-determined |
| 18-105_k141_80644 | Not-determined |
| 18-105_k141_53910 | Not-determined |
| 18-105_k141_27016 | Not-determined |
| 18-105_k141_142991 | Not-determined |
| 18-105_k141_151695 | Not-determined |
| 18-105_k141_732 | Not-determined |
| 18-105_k141_80658 | Not-determined |
| 18-105_k141_151704 | Not-determined |
| 18-105_k141_53920 | Not-determined |
| 18-105_k141_27026 | Not-determined |
| 18-105_k141_187547 | Not-determined |
| 18-105_k141_44823 | Not-determined |
| 18-105_k141_53930 | Not-determined |
| 18-105_k141_143006 | Not-determined |
| 18-105_k141_35900 | Not-determined |
| 18-105_k141_107342 | Low-quality |
| 18-105_k141_18241 | Not-determined |
| 18-105_k141_44835 | Not-determined |
| 18-105_k141_18242 | Not-determined |
| 18-105_k141_18243 | Not-determined |
| 18-105_k141_89435 | Not-determined |
| 18-105_k141_151732 | Not-determined |
| 18-105_k141_107353 | Not-determined |
| 18-105_k141_735 | Not-determined |
| 18-105_k141_98314 | Not-determined |
| 18-105_k141_205181 | Low-quality |
| 18-105_k141_53941 | Low-quality |
| 18-105_k141_98323 | Not-determined |
| 18-105_k141_187568 | Not-determined |
| 18-105_k141_196557 | Not-determined |
| 18-105_k141_196560 | Not-determined |
| 18-105_k141_196562 | Not-determined |
| 18-105_k141_53953 | Not-determined |
| 18-105_k141_44856 | Not-determined |
| 18-105_k141_18247 | Not-determined |
| 18-105_k141_761 | Not-determined |
| 18-105_k141_107381 | Low-quality |
| 18-105_k141_53958 | Not-determined |
| 18-105_k141_169598 | Not-determined |
| 18-105_k141_35934 | Not-determined |
| 18-105_k141_62759 | Not-determined |
| 18-105_k141_98346 | Not-determined |
| 18-105_k141_62761 | Not-determined |
| 18-105_k141_62764 | Not-determined |
| 18-105_k141_18262 | Not-determined |
| 18-105_k141_205198 | Not-determined |
| 18-105_k141_9534 | Not-determined |
| 18-105_k141_107397 | Not-determined |
| 18-105_k141_773 | Not-determined |
| 18-105_k141_143026 | Not-determined |
| 18-105_k141_205202 | Not-determined |
| 18-105_k141_160857 | Not-determined |
| 18-105_k141_9537 | Not-determined |
| 18-105_k141_9538 | Not-determined |
| 18-105_k141_9539 | Not-determined |
| 18-105_k141_160860 | Not-determined |
| 18-105_k141_143044 | Not-determined |
| 18-105_k141_89481 | Not-determined |
| 18-105_k141_160868 | Not-determined |
| 18-105_k141_18280 | Low-quality |
| 18-105_k141_107409 | Not-determined |
| 18-105_k141_205239 | Low-quality |
| 18-105_k141_98386 | Not-determined |
| 18-105_k141_169633 | Not-determined |
| 18-105_k141_44909 | Not-determined |
| 18-105_k141_107414 | Not-determined |
| 18-105_k141_44921 | Not-determined |
| 18-105_k141_89518 | Low-quality |
| 18-105_k141_169652 | Low-quality |
| 18-105_k141_44932 | Not-determined |
| 18-105_k141_89528 | Not-determined |
| 18-105_k141_44935 | Not-determined |
| 18-105_k141_89534 | Not-determined |
| 18-105_k141_809 | Not-determined |
| 18-105_k141_205269 | Not-determined |
| 18-105_k141_35964 | Low-quality |
| 18-105_k141_143085 | Not-determined |
| 18-105_k141_143088 | Not-determined |
| 18-105_k141_125058 | Not-determined |
| 18-105_k141_125060 | Not-determined |
| 18-105_k141_98422 | Not-determined |
| 18-105_k141_98424 | Not-determined |
| 18-105_k141_53973 | Low-quality |
| 18-105_k141_98426 | Not-determined |
| 18-105_k141_160934 | Not-determined |
| 18-105_k141_53983 | Not-determined |
| 18-105_k141_205279 | Not-determined |
| 18-105_k141_89567 | Not-determined |
| 18-105_k141_819 | Low-quality |
| 18-105_k141_125090 | Low-quality |
| 18-105_k141_151752 | Not-determined |
| 18-105_k141_18327 | Not-determined |
| 18-105_k141_160952 | Not-determined |
| 18-105_k141_18331 | Not-determined |
| 18-105_k141_160955 | Not-determined |
| 18-105_k141_89578 | Not-determined |
| 18-105_k141_9640 | Not-determined |
| 18-105_k141_151766 | Not-determined |
| 18-105_k141_89585 | Not-determined |
| 18-105_k141_36002 | Not-determined |
| 18-105_k141_151769 | Not-determined |
| 18-105_k141_44971 | Not-determined |
| 18-105_k141_18342 | Not-determined |
| 18-105_k141_18346 | Not-determined |
| 18-105_k141_169679 | Not-determined |
| 18-105_k141_98463 | Not-determined |
| 18-105_k141_44972 | Not-determined |
| 18-105_k141_169685 | Not-determined |
| 18-105_k141_841 | Not-determined |
| 18-105_k141_169686 | Not-determined |
| 18-105_k141_160965 | Not-determined |
| 18-105_k141_842 | Not-determined |
| 18-105_k141_54006 | Not-determined |
| 18-105_k141_160973 | Not-determined |
| 18-105_k141_851 | Not-determined |
| 18-105_k141_854 | Not-determined |
| 18-105_k141_134167 | Not-determined |
| 18-105_k141_71848 | Not-determined |
| 18-105_k141_27085 | Not-determined |
| 18-105_k141_151823 | Not-determined |
| 18-105_k141_27090 | Not-determined |
| 18-105_k141_80761 | Not-determined |
| 18-105_k141_44985 | Not-determined |
| 18-105_k141_44987 | Not-determined |
| 18-105_k141_44991 | Not-determined |
| 18-105_k141_125119 | Low-quality |
| 18-105_k141_125121 | Not-determined |
| 18-105_k141_71862 | Not-determined |
| 18-105_k141_116256 | Not-determined |
| 18-105_k141_71868 | Not-determined |
| 18-105_k141_205284 | Not-determined |
| 18-105_k141_71869 | Low-quality |
| 18-105_k141_894 | Not-determined |
| 18-105_k141_898 | Low-quality |
| 18-105_k141_205298 | Not-determined |
| 18-105_k141_169734 | Not-determined |
| 18-105_k141_205302 | Not-determined |
| 18-105_k141_169757 | Not-determined |
| 18-105_k141_169765 | Low-quality |
| 18-105_k141_908 | Not-determined |
| 18-105_k141_89603 | Not-determined |
| 18-105_k141_98492 | Not-determined |
| 18-105_k141_125172 | Not-determined |
| 18-105_k141_98495 | Low-quality |
| 18-105_k141_125177 | Not-determined |
| 18-105_k141_918 | Not-determined |
| 18-105_k141_71909 | Not-determined |
| 18-105_k141_98503 | Not-determined |
| 18-105_k141_926 | Not-determined |
| 18-105_k141_116297 | Not-determined |
| 18-105_k141_116298 | Not-determined |
| 18-105_k141_178448 | Not-determined |
| 18-105_k141_125191 | Not-determined |
| 18-105_k141_89613 | Not-determined |
| 18-105_k141_116305 | Not-determined |
| 18-105_k141_125198 | Low-quality |
| 18-105_k141_151897 | Not-determined |
| 18-105_k141_178458 | Not-determined |
| 18-105_k141_71922 | Not-determined |
| 18-105_k141_18402 | Low-quality |
| 18-105_k141_71924 | Not-determined |
| 18-105_k141_71935 | Not-determined |
| 18-105_k141_71942 | Not-determined |
| 18-105_k141_949 | Not-determined |
| 18-105_k141_71948 | Not-determined |
| 18-105_k141_954 | Not-determined |
| 18-105_k141_151935 | Not-determined |
| 18-105_k141_143217 | Not-determined |
| 18-105_k141_205331 | Not-determined |
| 18-105_k141_18415 | Not-determined |
| 18-105_k141_98560 | Not-determined |
| 18-105_k141_18422 | Not-determined |
| 18-105_k141_125215 | Not-determined |
| 18-105_k141_98570 | Not-determined |
| 18-105_k141_957 | Not-determined |
| 18-105_k141_36067 | Not-determined |
| 18-105_k141_89638 | Not-determined |
| 18-105_k141_196627 | Not-determined |
| 18-105_k141_107542 | Not-determined |
| 18-105_k141_107545 | Not-determined |
| 18-105_k141_107552 | Not-determined |
| 18-105_k141_178497 | Low-quality |
| 18-105_k141_196637 | Not-determined |
| 18-105_k141_62769 | Not-determined |
| 18-105_k141_80795 | Not-determined |
| 18-105_k141_107564 | Not-determined |
| 18-105_k141_161035 | Not-determined |
| 18-105_k141_974 | Not-determined |
| 18-105_k141_196650 | Not-determined |
| 18-105_k141_27121 | Not-determined |
| 18-105_k141_18431 | Not-determined |
| 18-105_k141_169815 | Not-determined |
| 18-105_k141_116344 | Not-determined |
| 18-105_k141_27131 | Not-determined |
| 18-105_k141_98611 | Not-determined |
| 18-105_k141_62778 | Not-determined |
| 18-105_k141_116347 | Not-determined |
| 18-105_k141_62781 | Not-determined |
| 18-105_k141_62784 | Not-determined |
| 18-105_k141_98620 | Not-determined |
| 18-105_k141_116350 | Not-determined |
| 18-105_k141_98623 | Low-quality |
| 18-105_k141_151959 | Not-determined |
| 18-105_k141_18445 | Not-determined |
| 18-105_k141_27140 | Not-determined |
| 18-105_k141_151965 | Low-quality |
| 18-105_k141_151966 | Not-determined |
| 18-105_k141_1001 | Not-determined |
| 18-105_k141_161052 | Not-determined |
| 18-105_k141_125228 | Not-determined |
| 18-105_k141_196678 | Not-determined |
| 18-105_k141_62795 | Low-quality |
| 18-105_k141_169842 | Not-determined |
| 18-105_k141_116366 | Not-determined |
| 18-105_k141_89684 | Not-determined |
| 18-105_k141_169846 | Not-determined |
| 18-105_k141_1013 | Not-determined |
| 18-105_k141_27157 | Not-determined |
| 18-105_k141_134213 | Not-determined |
| 18-105_k141_125249 | Not-determined |
| 18-105_k141_27163 | Not-determined |
| 18-105_k141_9749 | Not-determined |
| 18-105_k141_169854 | Not-determined |
| 18-105_k141_1029 | Not-determined |
| 18-105_k141_125250 | Low-quality |
| 18-105_k141_134231 | Not-determined |
| 18-105_k141_125253 | Not-determined |
| 18-105_k141_196710 | Not-determined |
| 18-105_k141_1037 | Low-quality |
| 18-105_k141_125257 | Not-determined |
| 18-105_k141_116376 | Not-determined |
| 18-105_k141_71981 | Not-determined |
| 18-105_k141_116378 | Not-determined |
| 18-105_k141_9759 | Not-determined |
| 18-105_k141_125258 | Not-determined |
| 18-105_k141_125265 | Not-determined |
| 18-105_k141_125266 | Not-determined |
| 18-105_k141_54034 | Not-determined |
| 18-105_k141_151977 | Not-determined |
| 18-105_k141_1050 | Not-determined |
| 18-105_k141_161075 | Not-determined |
| 18-105_k141_169869 | Not-determined |
| 18-105_k141_89700 | Not-determined |
| 18-105_k141_125272 | Not-determined |
| 18-105_k141_89703 | Not-determined |
| 18-105_k141_45031 | Not-determined |
| 18-105_k141_125284 | Not-determined |
| 18-105_k141_98644 | Low-quality |
| 18-105_k141_98646 | Low-quality |
| 18-105_k141_1059 | Not-determined |
| 18-105_k141_151991 | Not-determined |
| 18-105_k141_36119 | Not-determined |
| 18-105_k141_125288 | Not-determined |
| 18-105_k141_18469 | Not-determined |
| 18-105_k141_45039 | Not-determined |
| 18-105_k141_178518 | Low-quality |
| 18-105_k141_89716 | Not-determined |
| 18-105_k141_98654 | Not-determined |
| 18-105_k141_98655 | Not-determined |
| 18-105_k141_1073 | Not-determined |
| 18-105_k141_36125 | Not-determined |
| 18-105_k141_161098 | Not-determined |
| 18-105_k141_18486 | Not-determined |
| 18-105_k141_89732 | Not-determined |
| 18-105_k141_178543 | Not-determined |
| 18-105_k141_178545 | Not-determined |
| 18-105_k141_27187 | Not-determined |
| 18-105_k141_161117 | Low-quality |
| 18-105_k141_134248 | Not-determined |
| 18-105_k141_54061 | Not-determined |
| 18-105_k141_134254 | Not-determined |
| 18-105_k141_27197 | Low-quality |
| 18-105_k141_45070 | Not-determined |
| 18-105_k141_205378 | Not-determined |
| 18-105_k141_45073 | Not-determined |
| 18-105_k141_169900 | Not-determined |
| 18-105_k141_134261 | Not-determined |
| 18-105_k141_134264 | Not-determined |
| 18-105_k141_1098 | Not-determined |
| 18-105_k141_125333 | Not-determined |
| 18-105_k141_125335 | Low-quality |
| 18-105_k141_54072 | Not-determined |
| 18-105_k141_89752 | Not-determined |
| 18-105_k141_71993 | Not-determined |
| 18-105_k141_205384 | Not-determined |
| 18-105_k141_169911 | Not-determined |
| 18-105_k141_125337 | Not-determined |
| 18-105_k141_89769 | Not-determined |
| 18-105_k141_134281 | Not-determined |
| 18-105_k141_107599 | Not-determined |
| 18-105_k141_9817 | Not-determined |
| 18-105_k141_72024 | Not-determined |
| 18-105_k141_98703 | Not-determined |
| 18-105_k141_98704 | Not-determined |
| 18-105_k141_134295 | Not-determined |
| 18-105_k141_107623 | Not-determined |
| 18-105_k141_89798 | Not-determined |
| 18-105_k141_178605 | Not-determined |
| 18-105_k141_107643 | Not-determined |
| 18-105_k141_205397 | Not-determined |
| 18-105_k141_205399 | Not-determined |
| 18-105_k141_72040 | Not-determined |
| 18-105_k141_178610 | Not-determined |
| 18-105_k141_152058 | Not-determined |
| 18-105_k141_205400 | Low-quality |
| 18-105_k141_205403 | Not-determined |
| 18-105_k141_187591 | Not-determined |
| 18-105_k141_196732 | Not-determined |
| 18-105_k141_98709 | Not-determined |
| 18-105_k141_205414 | Not-determined |
| 18-105_k141_36146 | Not-determined |
| 18-105_k141_62835 | Not-determined |
| 18-105_k141_169929 | Not-determined |
| 18-105_k141_205422 | Not-determined |
| 18-105_k141_62850 | Not-determined |
| 18-105_k141_169945 | Not-determined |
| 18-105_k141_169950 | Not-determined |
| 18-105_k141_205435 | Not-determined |
| 18-105_k141_205436 | Not-determined |
| 18-105_k141_169960 | Not-determined |
| 18-105_k141_178619 | Not-determined |
| 18-105_k141_196749 | Low-quality |
| 18-105_k141_45106 | Not-determined |
| 18-105_k141_89849 | Not-determined |
| 18-105_k141_205446 | Not-determined |
| 18-105_k141_178628 | Not-determined |
| 18-105_k141_187623 | Not-determined |
| 18-105_k141_89854 | Not-determined |
| 18-105_k141_72052 | Not-determined |
| 18-105_k141_9832 | Low-quality |
| 18-105_k141_196776 | Not-determined |
| 18-105_k141_45120 | Not-determined |
| 18-105_k141_62876 | Not-determined |
| 18-105_k141_196782 | Not-determined |
| 18-105_k141_36172 | Not-determined |
| 18-105_k141_178659 | Not-determined |
| 18-105_k141_98717 | Not-determined |
| 18-105_k141_98722 | Not-determined |
| 18-105_k141_196796 | Not-determined |
| 18-105_k141_196798 | Not-determined |
| 18-105_k141_178667 | Not-determined |
| 18-105_k141_89881 | Not-determined |
| 18-105_k141_125341 | Not-determined |
| 18-105_k141_36203 | Not-determined |
| 18-105_k141_125350 | Not-determined |
| 18-105_k141_36213 | Not-determined |
| 18-105_k141_196810 | Not-determined |
| 18-105_k141_80816 | Not-determined |
| 18-105_k141_125356 | Not-determined |
| 18-105_k141_125357 | Low-quality |
| 18-105_k141_80830 | Not-determined |
| 18-105_k141_196817 | Not-determined |
| 18-105_k141_45131 | Not-determined |
| 18-105_k141_98770 | Not-determined |
| 18-105_k141_98771 | Low-quality |
| 18-105_k141_45134 | Not-determined |
| 18-105_k141_45135 | Not-determined |
| 18-105_k141_125368 | Not-determined |
| 18-105_k141_178694 | Not-determined |
| 18-105_k141_178696 | Not-determined |
| 18-105_k141_178705 | Low-quality |
| 18-105_k141_178709 | Low-quality |

| | |
|---|---|
| 18-105_k141_178711 | Not-determined |
| 18-105_k141_205476 | Not-determined |
| 18-105_k141_116449 | Low-quality |
| 18-105_k141_205478 | Low-quality |
| 18-105_k141_170007 | Not-determined |
| 18-105_k141_1141 | Not-determined |
| 18-105_k141_170014 | Not-determined |
| 18-105_k141_54080 | Not-determined |
| 18-105_k141_107725 | Not-determined |
| 18-105_k141_134309 | Not-determined |
| 18-105_k141_178728 | Low-quality |
| 18-105_k141_54083 | Not-determined |
| 18-105_k141_134314 | Not-determined |
| 18-105_k141_36244 | Low-quality |
| 18-105_k141_187656 | Not-determined |
| 18-105_k141_1167 | Not-determined |
| 18-105_k141_205492 | Not-determined |
| 18-105_k141_205493 | Not-determined |
| 18-105_k141_89957 | Not-determined |
| 18-105_k141_54090 | Not-determined |
| 18-105_k141_36255 | Not-determined |
| 18-105_k141_116462 | Not-determined |
| 18-105_k141_178751 | Not-determined |
| 18-105_k141_205507 | Not-determined |
| 18-105_k141_36265 | Not-determined |
| 18-105_k141_1180 | Not-determined |
| 18-105_k141_170059 | Not-determined |
| 18-105_k141_80838 | Not-determined |
| 18-105_k141_80839 | Not-determined |
| 18-105_k141_80840 | Not-determined |
| 18-105_k141_107750 | Not-determined |
| 18-105_k141_205520 | Not-determined |
| 18-105_k141_107756 | Not-determined |
| 18-105_k141_107759 | Not-determined |
| 18-105_k141_196864 | Not-determined |
| 18-105_k141_80853 | Not-determined |
| 18-105_k141_89968 | Not-determined |
| 18-105_k141_89969 | Not-determined |
| 18-105_k141_125379 | Not-determined |
| 18-105_k141_80856 | Not-determined |
| 18-105_k141_196872 | Not-determined |
| 18-105_k141_27209 | Not-determined |
| 18-105_k141_125383 | Not-determined |
| 18-105_k141_116481 | Not-determined |
| 18-105_k141_125388 | Not-determined |
| 18-105_k141_116492 | Not-determined |
| 18-105_k141_27214 | Low-quality |
| 18-105_k141_205544 | Not-determined |
| 18-105_k141_27222 | Not-determined |
| 18-105_k141_205551 | Not-determined |
| 18-105_k141_1227 | Not-determined |
| 18-105_k141_187681 | Not-determined |
| 18-105_k141_170100 | Not-determined |
| 18-105_k141_205570 | Low-quality |
| 18-105_k141_89982 | Low-quality |
| 18-105_k141_54131 | Not-determined |
| 18-105_k141_1241 | Not-determined |
| 18-105_k141_27251 | Not-determined |
| 18-105_k141_62946 | Not-determined |
| 18-105_k141_27255 | Low-quality |
| 18-105_k141_205584 | Not-determined |
| 18-105_k141_54138 | Not-determined |
| 18-105_k141_89995 | Not-determined |
| 18-105_k141_89996 | Not-determined |
| 18-105_k141_205593 | Not-determined |
| 18-105_k141_1263 | Not-determined |
| 18-105_k141_1266 | Not-determined |
| 18-105_k141_45223 | Not-determined |
| 18-105_k141_90008 | Not-determined |
| 18-105_k141_54149 | Not-determined |
| 18-105_k141_152086 | Not-determined |
| 18-105_k141_9865 | Not-determined |
| 18-105_k141_116536 | Not-determined |
| 18-105_k141_54161 | Not-determined |
| 18-105_k141_134319 | Low-quality |
| 18-105_k141_54162 | Not-determined |
| 18-105_k141_116539 | Not-determined |
| 18-105_k141_116540 | Not-determined |
| 18-105_k141_143302 | Not-determined |
| 18-105_k141_27273 | Not-determined |
| 18-105_k141_27274 | Not-determined |
| 18-105_k141_143304 | Not-determined |
| 18-105_k141_143305 | Not-determined |
| 18-105_k141_170126 | Not-determined |
| 18-105_k141_9868 | Not-determined |
| 18-105_k141_205612 | Not-determined |
| 18-105_k141_27278 | Not-determined |
| 18-105_k141_125451 | Not-determined |
| 18-105_k141_143311 | Not-determined |
| 18-105_k141_90016 | Not-determined |
| 18-105_k141_27289 | Not-determined |
| 18-105_k141_134341 | Not-determined |
| 18-105_k141_116554 | Not-determined |
| 18-105_k141_1301 | Not-determined |
| 18-105_k141_143318 | Not-determined |
| 18-105_k141_90019 | Low-quality |
| 18-105_k141_116557 | Not-determined |
| 18-105_k141_54184 | Not-determined |
| 18-105_k141_143324 | Not-determined |
| 18-105_k141_54186 | Not-determined |
| 18-105_k141_27302 | Not-determined |
| 18-105_k141_170153 | Not-determined |
| 18-105_k141_178789 | Low-quality |
| 18-105_k141_187700 | Not-determined |
| 18-105_k141_90032 | Not-determined |
| 18-105_k141_54204 | Not-determined |
| 18-105_k141_36325 | Not-determined |
| 18-105_k141_90040 | Low-quality |
| 18-105_k141_18510 | Not-determined |
| 18-105_k141_54208 | Not-determined |
| 18-105_k141_152121 | Not-determined |
| 18-105_k141_178798 | Low-quality |
| 18-105_k141_27320 | Not-determined |
| 18-105_k141_178807 | Not-determined |
| 18-105_k141_152136 | Not-determined |
| 18-105_k141_62973 | Not-determined |
| 18-105_k141_187710 | Not-determined |
| 18-105_k141_170173 | Not-determined |
| 18-105_k141_62977 | Not-determined |
| 18-105_k141_18523 | Not-determined |
| 18-105_k141_187718 | Not-determined |
| 18-105_k141_18529 | Not-determined |
| 18-105_k141_187720 | Not-determined |
| 18-105_k141_107812 | Not-determined |
| 18-105_k141_107815 | Not-determined |
| 18-105_k141_45276 | Not-determined |
| 18-105_k141_178826 | Not-determined |
| 18-105_k141_170191 | Not-determined |
| 18-105_k141_152161 | Not-determined |
| 18-105_k141_36373 | Not-determined |
| 18-105_k141_134359 | Not-determined |
| 18-105_k141_62991 | Not-determined |
| 18-105_k141_90056 | Low-quality |
| 18-105_k141_62995 | Not-determined |
| 18-105_k141_63000 | Not-determined |
| 18-105_k141_90068 | Not-determined |
| 18-105_k141_107829 | Not-determined |
| 18-105_k141_125506 | Not-determined |
| 18-105_k141_36385 | Not-determined |
| 18-105_k141_143343 | Not-determined |
| 18-105_k141_1317 | Not-determined |
| 18-105_k141_116567 | Not-determined |
| 18-105_k141_27355 | Not-determined |
| 18-105_k141_80886 | Not-determined |
| 18-105_k141_45304 | Not-determined |
| 18-105_k141_134370 | Low-quality |
| 18-105_k141_90081 | Not-determined |
| 18-105_k141_125517 | Not-determined |
| 18-105_k141_134374 | Not-determined |
| 18-105_k141_196884 | Low-quality |
| 18-105_k141_170229 | Not-determined |
| 18-105_k141_170231 | Not-determined |
| 18-105_k141_54278 | Low-quality |
| 18-105_k141_90103 | Not-determined |
| 18-105_k141_45318 | Not-determined |
| 18-105_k141_45324 | Not-determined |
| 18-105_k141_9939 | Not-determined |
| 18-105_k141_9942 | Not-determined |
| 18-105_k141_9943 | Not-determined |
| 18-105_k141_80909 | Not-determined |
| 18-105_k141_63024 | Low-quality |
| 18-105_k141_45331 | Not-determined |
| 18-105_k141_63033 | Not-determined |
| 18-105_k141_143387 | Not-determined |
| 18-105_k141_27384 | Not-determined |
| 18-105_k141_1352 | Not-determined |
| 18-105_k141_178856 | Not-determined |
| 18-105_k141_54299 | Not-determined |
| 18-105_k141_27401 | Not-determined |
| 18-105_k141_27402 | Not-determined |
| 18-105_k141_27404 | Not-determined |
| 18-105_k141_27407 | Low-quality |
| 18-105_k141_63051 | Not-determined |
| 18-105_k141_27410 | Not-determined |
| 18-105_k141_1361 | Not-determined |
| 18-105_k141_18535 | Low-quality |
| 18-105_k141_1364 | Not-determined |
| 18-105_k141_170243 | Low-quality |
| 18-105_k141_116626 | Not-determined |
| 18-105_k141_170246 | Not-determined |
| 18-105_k141_27417 | Not-determined |
| 18-105_k141_1370 | Not-determined |
| 18-105_k141_170247 | Not-determined |
| 18-105_k141_196911 | Not-determined |
| 18-105_k141_125558 | Not-determined |
| 18-105_k141_116636 | Not-determined |
| 18-105_k141_45356 | Not-determined |
| 18-105_k141_170254 | Not-determined |
| 18-105_k141_196920 | Not-determined |
| 18-105_k141_98817 | Not-determined |
| 18-105_k141_45360 | Not-determined |
| 18-105_k141_1379 | Not-determined |
| 18-105_k141_45363 | Not-determined |
| 18-105_k141_98821 | Not-determined |
| 18-105_k141_45365 | Not-determined |
| 18-105_k141_54316 | Low-quality |
| 18-105_k141_196927 | Not-determined |
| 18-105_k141_125564 | Low-quality |
| 18-105_k141_54320 | Not-determined |
| 18-105_k141_143396 | Not-determined |
| 18-105_k141_54326 | Not-determined |
| 18-105_k141_90138 | Not-determined |
| 18-105_k141_90144 | Not-determined |
| 18-105_k141_45380 | Not-determined |
| 18-105_k141_45381 | Low-quality |
| 18-105_k141_54336 | Not-determined |
| 18-105_k141_1398 | Not-determined |
| 18-105_k141_161221 | Not-determined |
| 18-105_k141_187783 | Low-quality |
| 18-105_k141_187786 | Not-determined |
| 18-105_k141_161227 | Not-determined |
| 18-105_k141_45393 | Not-determined |
| 18-105_k141_9953 | Not-determined |
| 18-105_k141_45394 | Not-determined |
| 18-105_k141_9956 | Not-determined |
| 18-105_k141_90162 | Not-determined |
| 18-105_k141_161233 | Not-determined |
| 18-105_k141_90168 | Not-determined |
| 18-105_k141_1414 | Low-quality |
| 18-105_k141_18574 | Not-determined |
| 18-105_k141_1425 | Not-determined |
| 18-105_k141_18581 | Not-determined |
| 18-105_k141_161239 | Low-quality |
| 18-105_k141_187795 | Not-determined |
| 18-105_k141_45416 | Not-determined |
| 18-105_k141_1430 | Not-determined |
| 18-105_k141_134386 | Not-determined |
| 18-105_k141_187817 | Not-determined |
| 18-105_k141_27438 | Not-determined |
| 18-105_k141_178917 | Not-determined |
| 18-105_k141_134398 | Not-determined |
| 18-105_k141_187822 | Low-quality |
| 18-105_k141_170261 | Not-determined |
| 18-105_k141_107833 | Not-determined |
| 18-105_k141_45426 | Low-quality |
| 18-105_k141_1436 | Not-determined |
| 18-105_k141_45429 | Not-determined |
| 18-105_k141_90182 | Not-determined |
| 18-105_k141_1439 | Not-determined |
| 18-105_k141_27464 | Not-determined |
| 18-105_k141_27469 | Not-determined |
| 18-105_k141_1449 | Not-determined |
| 18-105_k141_27477 | Not-determined |
| 18-105_k141_170270 | Not-determined |
| 18-105_k141_27480 | Not-determined |
| 18-105_k141_178946 | Low-quality |
| 18-105_k141_107853 | Not-determined |
| 18-105_k141_9991 | Not-determined |
| 18-105_k141_54386 | Not-determined |
| 18-105_k141_9993 | Not-determined |
| 18-105_k141_54393 | Not-determined |
| 18-105_k141_54394 | Not-determined |
| 18-105_k141_107865 | Not-determined |
| 18-105_k141_27507 | Not-determined |
| 18-105_k141_10006 | Not-determined |
| 18-105_k141_116687 | Not-determined |
| 18-105_k141_170294 | Not-determined |
| 18-105_k141_10013 | Not-determined |
| 18-105_k141_90230 | Not-determined |
| 18-105_k141_134420 | Not-determined |
| 18-105_k141_10018 | Not-determined |
| 18-105_k141_90234 | Low-quality |
| 18-105_k141_36406 | Not-determined |
| 18-105_k141_134426 | Not-determined |
| 18-105_k141_45444 | Not-determined |
| 18-105_k141_54401 | Not-determined |
| 18-105_k141_45449 | Not-determined |
| 18-105_k141_178985 | Not-determined |
| 18-105_k141_134430 | Not-determined |
| 18-105_k141_10029 | Not-determined |
| 18-105_k141_107887 | Not-determined |
| 18-105_k141_107894 | Not-determined |
| 18-105_k141_10037 | Not-determined |
| 18-105_k141_134451 | Not-determined |
| 18-105_k141_45469 | Not-determined |
| 18-105_k141_196977 | Not-determined |
| 18-105_k141_196981 | Not-determined |
| 18-105_k141_179023 | Not-determined |
| 18-105_k141_80954 | Not-determined |
| 18-105_k141_116695 | Not-determined |
| 18-105_k141_187843 | Not-determined |
| 18-105_k141_116704 | Not-determined |
| 18-105_k141_116705 | Not-determined |
| 18-105_k141_179029 | Not-determined |
| 18-105_k141_170304 | Not-determined |
| 18-105_k141_107922 | Not-determined |
| 18-105_k141_63128 | Not-determined |
| 18-105_k141_196998 | Not-determined |
| 18-105_k141_1500 | Not-determined |
| 18-105_k141_197000 | Not-determined |
| 18-105_k141_116716 | Not-determined |
| 18-105_k141_54421 | Not-determined |
| 18-105_k141_134483 | Not-determined |
| 18-105_k141_98890 | Not-determined |
| 18-105_k141_116724 | Not-determined |
| 18-105_k141_54424 | Not-determined |
| 18-105_k141_179046 | Not-determined |
| 18-105_k141_205619 | Not-determined |
| 18-105_k141_98894 | Not-determined |
| 18-105_k141_107942 | Not-determined |
| 18-105_k141_98897 | Not-determined |
| 18-105_k141_179049 | Not-determined |
| 18-105_k141_107956 | Not-determined |
| 18-105_k141_187853 | Not-determined |
| 18-105_k141_179053 | Not-determined |
| 18-105_k141_187854 | Not-determined |
| 18-105_k141_187861 | Not-determined |
| 18-105_k141_179062 | Not-determined |
| 18-105_k141_98906 | Not-determined |
| 18-105_k141_36478 | Not-determined |
| 18-105_k141_179068 | Not-determined |
| 18-105_k141_116734 | Not-determined |
| 18-105_k141_161262 | Not-determined |
| 18-105_k141_107988 | Not-determined |
| 18-105_k141_125606 | Not-determined |
| 18-105_k141_161273 | Not-determined |
| 18-105_k141_80967 | Not-determined |
| 18-105_k141_90265 | Not-determined |
| 18-105_k141_108008 | Not-determined |
| 18-105_k141_125615 | Not-determined |
| 18-105_k141_179099 | Low-quality |
| 18-105_k141_18631 | Low-quality |
| 18-105_k141_161288 | Not-determined |
| 18-105_k141_125620 | Not-determined |
| 18-105_k141_36504 | Not-determined |
| 18-105_k141_18636 | Not-determined |
| 18-105_k141_179113 | Not-determined |
| 18-105_k141_179115 | Low-quality |
| 18-105_k141_125631 | Not-determined |
| 18-105_k141_36515 | Not-determined |
| 18-105_k141_1560 | Not-determined |
| 18-105_k141_116760 | Not-determined |
| 18-105_k141_125636 | Not-determined |
| 18-105_k141_125637 | Not-determined |
| 18-105_k141_108032 | Not-determined |
| 18-105_k141_179119 | Not-determined |
| 18-105_k141_143493 | Not-determined |
| 18-105_k141_179122 | Not-determined |
| 18-105_k141_90293 | Not-determined |
| 18-105_k141_161291 | Not-determined |
| 18-105_k141_45485 | Not-determined |
| 18-105_k141_170340 | Not-determined |
| 18-105_k141_205658 | Not-determined |
| 18-105_k141_170355 | Not-determined |
| 18-105_k141_45499 | Not-determined |
| 18-105_k141_54459 | Not-determined |
| 18-105_k141_18667 | Not-determined |
| 18-105_k141_205682 | Not-determined |
| 18-105_k141_80989 | Not-determined |
| 18-105_k141_45507 | Not-determined |
| 18-105_k141_80996 | Not-determined |
| 18-105_k141_27538 | Not-determined |
| 18-105_k141_98924 | Not-determined |
| 18-105_k141_54485 | Not-determined |
| 18-105_k141_27545 | Not-determined |
| 18-105_k141_54494 | Not-determined |
| 18-105_k141_134514 | Not-determined |
| 18-105_k141_134518 | Not-determined |
| 18-105_k141_179158 | Low-quality |
| 18-105_k141_98938 | Not-determined |
| 18-105_k141_72138 | Not-determined |
| 18-105_k141_108080 | Not-determined |
| 18-105_k141_98949 | Not-determined |
| 18-105_k141_170376 | Not-determined |
| 18-105_k141_81009 | Not-determined |
| 18-105_k141_54507 | Low-quality |
| 18-105_k141_1611 | Not-determined |
| 18-105_k141_205706 | Low-quality |
| 18-105_k141_27570 | Not-determined |
| 18-105_k141_54514 | Not-determined |
| 18-105_k141_108088 | Not-determined |
| 18-105_k141_10070 | Low-quality |
| 18-105_k141_27573 | Low-quality |
| 18-105_k141_10074 | Not-determined |
| 18-105_k141_81014 | Not-determined |
| 18-105_k141_81017 | Not-determined |
| 18-105_k141_125706 | Not-determined |
| 18-105_k141_63162 | Not-determined |
| 18-105_k141_205726 | Not-determined |
| 18-105_k141_10081 | Low-quality |
| 18-105_k141_205727 | Not-determined |
| 18-105_k141_108099 | Not-determined |
| 18-105_k141_36537 | Not-determined |
| 18-105_k141_134554 | Not-determined |
| 18-105_k141_1645 | Not-determined |
| 18-105_k141_45527 | Not-determined |
| 18-105_k141_134564 | Not-determined |
| 18-105_k141_63187 | Not-determined |
| 18-105_k141_134566 | Not-determined |
| 18-105_k141_63192 | Not-determined |
| 18-105_k141_161304 | Not-determined |
| 18-105_k141_63194 | Not-determined |
| 18-105_k141_161306 | Not-determined |
| 18-105_k141_1655 | Low-quality |
| 18-105_k141_72167 | Not-determined |
| 18-105_k141_63198 | Not-determined |
| 18-105_k141_1659 | Low-quality |
| 18-105_k141_1661 | Not-determined |
| 18-105_k141_63202 | Low-quality |
| 18-105_k141_98958 | Not-determined |
| 18-105_k141_108119 | Not-determined |
| 18-105_k141_27587 | Not-determined |
| 18-105_k141_179169 | Not-determined |
| 18-105_k141_63212 | Not-determined |
| 18-105_k141_116839 | Not-determined |
| 18-105_k141_63215 | Not-determined |
| 18-105_k141_1685 | Not-determined |
| 18-105_k141_116840 | Low-quality |
| 18-105_k141_179184 | Not-determined |
| 18-105_k141_134590 | Not-determined |
| 18-105_k141_179190 | Low-quality |
| 18-105_k141_134592 | Not-determined |
| 18-105_k141_116845 | Not-determined |
| 18-105_k141_27623 | Not-determined |
| 18-105_k141_134608 | Not-determined |
| 18-105_k141_134612 | Not-determined |
| 18-105_k141_152350 | Not-determined |
| 18-105_k141_45532 | Not-determined |
| 18-105_k141_98983 | Not-determined |
| 18-105_k141_45534 | Not-determined |
| 18-105_k141_179207 | Not-determined |
| 18-105_k141_152356 | Not-determined |
| 18-105_k141_98991 | Not-determined |
| 18-105_k141_98993 | Not-determined |
| 18-105_k141_72209 | Not-determined |
| 18-105_k141_179215 | Not-determined |
| 18-105_k141_187885 | Not-determined |
| 18-105_k141_72215 | Not-determined |
| 18-105_k141_143563 | Not-determined |
| 18-105_k141_72219 | Not-determined |
| 18-105_k141_152366 | Not-determined |
| 18-105_k141_81035 | Not-determined |
| 18-105_k141_108140 | Not-determined |
| 18-105_k141_179226 | Not-determined |
| 18-105_k141_179227 | Not-determined |
| 18-105_k141_143579 | Low-quality |
| 18-105_k141_152372 | Not-determined |
| 18-105_k141_197023 | Not-determined |
| 18-105_k141_143583 | Not-determined |
| 18-105_k141_81041 | Not-determined |
| 18-105_k141_143589 | Low-quality |
| 18-105_k141_197031 | Not-determined |
| 18-105_k141_125756 | Low-quality |
| 18-105_k141_125761 | Not-determined |
| 18-105_k141_197035 | Not-determined |
| 18-105_k141_152380 | Low-quality |
| 18-105_k141_197040 | Not-determined |
| 18-105_k141_45567 | Not-determined |
| 18-105_k141_197042 | Not-determined |
| 18-105_k141_81054 | Not-determined |
| 18-105_k141_45570 | Not-determined |
| 18-105_k141_10147 | Not-determined |
| 18-105_k141_45576 | Not-determined |
| 18-105_k141_90340 | Not-determined |
| 18-105_k141_72242 | Not-determined |
| 18-105_k141_134634 | Not-determined |
| 18-105_k141_125766 | Not-determined |
| 18-105_k141_108161 | Not-determined |
| 18-105_k141_125771 | Not-determined |
| 18-105_k141_90342 | Low-quality |
| 18-105_k141_45589 | Not-determined |
| 18-105_k141_108167 | Not-determined |
| 18-105_k141_170429 | Low-quality |
| 18-105_k141_1696 | Low-quality |
| 18-105_k141_134648 | Not-determined |
| 18-105_k141_125784 | Not-determined |
| 18-105_k141_81084 | Not-determined |
| 18-105_k141_90349 | Low-quality |
| 18-105_k141_1703 | Not-determined |
| 18-105_k141_152405 | Not-determined |
| 18-105_k141_134657 | Not-determined |
| 18-105_k141_152410 | Not-determined |
| 18-105_k141_125793 | Not-determined |
| 18-105_k141_125795 | Low-quality |
| 18-105_k141_81095 | Not-determined |
| 18-105_k141_1712 | Not-determined |
| 18-105_k141_170462 | Not-determined |
| 18-105_k141_187904 | Not-determined |
| 18-105_k141_125805 | Low-quality |
| 18-105_k141_116873 | Not-determined |
| 18-105_k141_99002 | Not-determined |
| 18-105_k141_72259 | Not-determined |
| 18-105_k141_99003 | Not-determined |
| 18-105_k141_134673 | Not-determined |
| 18-105_k141_116882 | Not-determined |
| 18-105_k141_125813 | Not-determined |
| 18-105_k141_152432 | Low-quality |
| 18-105_k141_72274 | Not-determined |
| 18-105_k141_143642 | Not-determined |
| 18-105_k141_72281 | Not-determined |
| 18-105_k141_205742 | Not-determined |
| 18-105_k141_1743 | Not-determined |
| 18-105_k141_143657 | Not-determined |
| 18-105_k141_143658 | Not-determined |
| 18-105_k141_27708 | Not-determined |
| 18-105_k141_205750 | Not-determined |
| 18-105_k141_125834 | Not-determined |
| 18-105_k141_197067 | Not-determined |
| 18-105_k141_72288 | Not-determined |
| 18-105_k141_27712 | Not-determined |
| 18-105_k141_1751 | Not-determined |
| 18-105_k141_152450 | Not-determined |
| 18-105_k141_152451 | Not-determined |
| 18-105_k141_125845 | Not-determined |
| 18-105_k141_27718 | Not-determined |
| 18-105_k141_54535 | Not-determined |
| 18-105_k141_54537 | Not-determined |
| 18-105_k141_54539 | Not-determined |
| 18-105_k141_205761 | Low-quality |
| 18-105_k141_152458 | Low-quality |
| 18-105_k141_187930 | Low-quality |
| 18-105_k141_116898 | Not-determined |
| 18-105_k141_116905 | Not-determined |
| 18-105_k141_116908 | Not-determined |
| 18-105_k141_197089 | Low-quality |
| 18-105_k141_72314 | Not-determined |
| 18-105_k141_99031 | Not-determined |
| 18-105_k141_72319 | Low-quality |
| 18-105_k141_72322 | Not-determined |
| 18-105_k141_205784 | Not-determined |
| 18-105_k141_72323 | Low-quality |
| 18-105_k141_116918 | Not-determined |
| 18-105_k141_72325 | Not-determined |
| 18-105_k141_134712 | Not-determined |
| 18-105_k141_197095 | Not-determined |
| 18-105_k141_36573 | Not-determined |
| 18-105_k141_99042 | Not-determined |
| 18-105_k141_99043 | Not-determined |
| 18-105_k141_81196 | Not-determined |
| 18-105_k141_116925 | Not-determined |
| 18-105_k141_81206 | Not-determined |
| 18-105_k141_108189 | Not-determined |
| 18-105_k141_116928 | Not-determined |
| 18-105_k141_99057 | Not-determined |
| 18-105_k141_72344 | Not-determined |
| 18-105_k141_152486 | Not-determined |
| 18-105_k141_187944 | Not-determined |
| 18-105_k141_125888 | Not-determined |
| 18-105_k141_197104 | Not-determined |
| 18-105_k141_36590 | Not-determined |
| 18-105_k141_90386 | Low-quality |
| 18-105_k141_45602 | Not-determined |
| 18-105_k141_197106 | Not-determined |
| 18-105_k141_72354 | Low-quality |
| 18-105_k141_81218 | Not-determined |
| 18-105_k141_125892 | Not-determined |
| 18-105_k141_81219 | Not-determined |
| 18-105_k141_134736 | Not-determined |
| 18-105_k141_108199 | Low-quality |
| 18-105_k141_108200 | Not-determined |
| 18-105_k141_170480 | Not-determined |
| 18-105_k141_108204 | Not-determined |
| 18-105_k141_205796 | Low-quality |
| 18-105_k141_18681 | Low-quality |
| 18-105_k141_45607 | Not-determined |
| 18-105_k141_72375 | Not-determined |
| 18-105_k141_45610 | Not-determined |
| 18-105_k141_152490 | Not-determined |
| 18-105_k141_18688 | Low-quality |
| 18-105_k141_152493 | Not-determined |
| 18-105_k141_143685 | Not-determined |
| 18-105_k141_143689 | Not-determined |
| 18-105_k141_197116 | Not-determined |
| 18-105_k141_116974 | Not-determined |
| 18-105_k141_187976 | Not-determined |
| 18-105_k141_90414 | Not-determined |
| 18-105_k141_90416 | Not-determined |
| 18-105_k141_187994 | Not-determined |
| 18-105_k141_116988 | Not-determined |
| 18-105_k141_187997 | Not-determined |
| 18-105_k141_116994 | Not-determined |
| 18-105_k141_205825 | Not-determined |
| 18-105_k141_99075 | Not-determined |
| 18-105_k141_1788 | Not-determined |
| 18-105_k141_81275 | Not-determined |
| 18-105_k141_72401 | Not-determined |
| 18-105_k141_1790 | Not-determined |
| 18-105_k141_205832 | Low-quality |
| 18-105_k141_99077 | Not-determined |
| 18-105_k141_27777 | Not-determined |
| 18-105_k141_45624 | Not-determined |
| 18-105_k141_117005 | Not-determined |
| 18-105_k141_10160 | Not-determined |
| 18-105_k141_1805 | Not-determined |
| 18-105_k141_45647 | Not-determined |
| 18-105_k141_197146 | Not-determined |
| 18-105_k141_143716 | Low-quality |
| 18-105_k141_36622 | Not-determined |
| 18-105_k141_205854 | Not-determined |

| | |
|---|---|
| 18-105_k141_205857 | Low-quality |
| 18-105_k141_205864 | Not-determined |
| 18-105_k141_27817 | Not-determined |
| 18-105_k141_1827 | Low-quality |
| 18-105_k141_1832 | Not-determined |
| 18-105_k141_1834 | Not-determined |
| 18-105_k141_152564 | Not-determined |
| 18-105_k141_36641 | Low-quality |
| 18-105_k141_54564 | Low-quality |
| 18-105_k141_54567 | Not-determined |
| 18-105_k141_27828 | Not-determined |
| 18-105_k141_27830 | Not-determined |
| 18-105_k141_197152 | Not-determined |
| 18-105_k141_117047 | Not-determined |
| 18-105_k141_143746 | Not-determined |
| 18-105_k141_117049 | Not-determined |
| 18-105_k141_197160 | Not-determined |
| 18-105_k141_125910 | Not-determined |
| 18-105_k141_125911 | Not-determined |
| 18-105_k141_45679 | Not-determined |
| 18-105_k141_117055 | Not-determined |
| 18-105_k141_45682 | Not-determined |
| 18-105_k141_18725 | Not-determined |
| 18-105_k141_117060 | Not-determined |
| 18-105_k141_99085 | Not-determined |
| 18-105_k141_205884 | Not-determined |
| 18-105_k141_117062 | Not-determined |
| 18-105_k141_108286 | Not-determined |
| 18-105_k141_117064 | Not-determined |
| 18-105_k141_45696 | Not-determined |
| 18-105_k141_81340 | Low-quality |
| 18-105_k141_161370 | Not-determined |
| 18-105_k141_45705 | Not-determined |
| 18-105_k141_45708 | Not-determined |
| 18-105_k141_99108 | Not-determined |
| 18-105_k141_152614 | Low-quality |
| 18-105_k141_54614 | Not-determined |
| 18-105_k141_152620 | Low-quality |
| 18-105_k141_108292 | Not-determined |
| 18-105_k141_99116 | Not-determined |
| 18-105_k141_152624 | Not-determined |
| 18-105_k141_99123 | Not-determined |
| 18-105_k141_18748 | Not-determined |
| 18-105_k141_134761 | Not-determined |
| 18-105_k141_188010 | Not-determined |
| 18-105_k141_205928 | Low-quality |
| 18-105_k141_134769 | Not-determined |
| 18-105_k141_205937 | Not-determined |
| 18-105_k141_205938 | Not-determined |
| 18-105_k141_205941 | Not-determined |
| 18-105_k141_170513 | Not-determined |
| 18-105_k141_134775 | Not-determined |
| 18-105_k141_134776 | Low-quality |
| 18-105_k141_170518 | Not-determined |
| 18-105_k141_134783 | Not-determined |
| 18-105_k141_170527 | Not-determined |
| 18-105_k141_63331 | Low-quality |
| 18-105_k141_170531 | Low-quality |
| 18-105_k141_27851 | Not-determined |
| 18-105_k141_143762 | Not-determined |
| 18-105_k141_45716 | Not-determined |
| 18-105_k141_45718 | Not-determined |
| 18-105_k141_143775 | Not-determined |
| 18-105_k141_143777 | Not-determined |
| 18-105_k141_179266 | Not-determined |
| 18-105_k141_45732 | Not-determined |
| 18-105_k141_143783 | Not-determined |
| 18-105_k141_45738 | Not-determined |
| 18-105_k141_45742 | Not-determined |
| 18-105_k141_170575 | Not-determined |
| 18-105_k141_63333 | Not-determined |
| 18-105_k141_179271 | Not-determined |
| 18-105_k141_63337 | Not-determined |
| 18-105_k141_205960 | Not-determined |
| 18-105_k141_143795 | Not-determined |
| 18-105_k141_54661 | Not-determined |
| 18-105_k141_54662 | Not-determined |
| 18-105_k141_1880 | Not-determined |
| 18-105_k141_54667 | Not-determined |
| 18-105_k141_179304 | Not-determined |
| 18-105_k141_18769 | Not-determined |
| 18-105_k141_179310 | Not-determined |
| 18-105_k141_81362 | Low-quality |
| 18-105_k141_179324 | Not-determined |
| 18-105_k141_108327 | Not-determined |
| 18-105_k141_179338 | Not-determined |
| 18-105_k141_36728 | Not-determined |
| 18-105_k141_188086 | Not-determined |
| 18-105_k141_36732 | Not-determined |
| 18-105_k141_205986 | Not-determined |
| 18-105_k141_54709 | Not-determined |
| 18-105_k141_179351 | Not-determined |
| 18-105_k141_188093 | Not-determined |
| 18-105_k141_54714 | Not-determined |
| 18-105_k141_188097 | Not-determined |
| 18-105_k141_63349 | Not-determined |
| 18-105_k141_188101 | Not-determined |
| 18-105_k141_63357 | Not-determined |
| 18-105_k141_170614 | Not-determined |
| 18-105_k141_170615 | Not-determined |
| 18-105_k141_63361 | Not-determined |
| 18-105_k141_45792 | Low-quality |
| 18-105_k141_188107 | Not-determined |
| 18-105_k141_188108 | Not-determined |
| 18-105_k141_205990 | Low-quality |
| 18-105_k141_170628 | Not-determined |
| 18-105_k141_81394 | Not-determined |
| 18-105_k141_161439 | Not-determined |
| 18-105_k141_143866 | Not-determined |
| 18-105_k141_161444 | Low-quality |
| 18-105_k141_188116 | Not-determined |
| 18-105_k141_170650 | Not-determined |
| 18-105_k141_179375 | Not-determined |
| 18-105_k141_161447 | Not-determined |
| 18-105_k141_81418 | Not-determined |
| 18-105_k141_161455 | Not-determined |
| 18-105_k141_117083 | Not-determined |
| 18-105_k141_81425 | Not-determined |
| 18-105_k141_179398 | Not-determined |
| 18-105_k141_108393 | Not-determined |
| 18-105_k141_108406 | Not-determined |
| 18-105_k141_170654 | Not-determined |
| 18-105_k141_18793 | Low-quality |
| 18-105_k141_170663 | Not-determined |
| 18-105_k141_108421 | Not-determined |
| 18-105_k141_179429 | Low-quality |
| 18-105_k141_117096 | Not-determined |
| 18-105_k141_179433 | Not-determined |
| 18-105_k141_170679 | Not-determined |
| 18-105_k141_27861 | Not-determined |
| 18-105_k141_108432 | Not-determined |
| 18-105_k141_170682 | Not-determined |
| 18-105_k141_170683 | Not-determined |
| 18-105_k141_179444 | Not-determined |
| 18-105_k141_72413 | Not-determined |
| 18-105_k141_27865 | Not-determined |
| 18-105_k141_27879 | Low-quality |
| 18-105_k141_161470 | Not-determined |
| 18-105_k141_108447 | Not-determined |
| 18-105_k141_134897 | Not-determined |
| 18-105_k141_27899 | Not-determined |
| 18-105_k141_99167 | Not-determined |
| 18-105_k141_54732 | Not-determined |
| 18-105_k141_197212 | Not-determined |
| 18-105_k141_99176 | Low-quality |
| 18-105_k141_206002 | Not-determined |
| 18-105_k141_188143 | Not-determined |
| 18-105_k141_99189 | Not-determined |
| 18-105_k141_90478 | Not-determined |
| 18-105_k141_99197 | Not-determined |
| 18-105_k141_188150 | Not-determined |
| 18-105_k141_170721 | Not-determined |
| 18-105_k141_170726 | Not-determined |
| 18-105_k141_188151 | Not-determined |
| 18-105_k141_197235 | Low-quality |
| 18-105_k141_18860 | Not-determined |
| 18-105_k141_18864 | Not-determined |
| 18-105_k141_117161 | Not-determined |
| 18-105_k141_197239 | Not-determined |
| 18-105_k141_170727 | Low-quality |
| 18-105_k141_206014 | Not-determined |
| 18-105_k141_179459 | Not-determined |
| 18-105_k141_170736 | Not-determined |
| 18-105_k141_117181 | Not-determined |
| 18-105_k141_197246 | Not-determined |
| 18-105_k141_188185 | Not-determined |
| 18-105_k141_152652 | Not-determined |
| 18-105_k141_18887 | Not-determined |
| 18-105_k141_197251 | Low-quality |
| 18-105_k141_36762 | Not-determined |
| 18-105_k141_99208 | Not-determined |
| 18-105_k141_1951 | Not-determined |
| 18-105_k141_152663 | Not-determined |
| 18-105_k141_197279 | Not-determined |
| 18-105_k141_197281 | Not-determined |
| 18-105_k141_108487 | Not-determined |
| 18-105_k141_188209 | Not-determined |
| 18-105_k141_170767 | Not-determined |
| 18-105_k141_197287 | Not-determined |
| 18-105_k141_72472 | Not-determined |
| 18-105_k141_108495 | Not-determined |
| 18-105_k141_170770 | Not-determined |
| 18-105_k141_179498 | Not-determined |
| 18-105_k141_54785 | Not-determined |
| 18-105_k141_63420 | Low-quality |
| 18-105_k141_170775 | Not-determined |
| 18-105_k141_143925 | Not-determined |
| 18-105_k141_179505 | Low-quality |
| 18-105_k141_152684 | Low-quality |
| 18-105_k141_179508 | Not-determined |
| 18-105_k141_179517 | Not-determined |
| 18-105_k141_108514 | Low-quality |
| 18-105_k141_152694 | Not-determined |
| 18-105_k141_108521 | Not-determined |
| 18-105_k141_143948 | Not-determined |
| 18-105_k141_108523 | Not-determined |
| 18-105_k141_152706 | Low-quality |
| 18-105_k141_188227 | Not-determined |
| 18-105_k141_179534 | Not-determined |
| 18-105_k141_10252 | Not-determined |
| 18-105_k141_108539 | Not-determined |
| 18-105_k141_36779 | Not-determined |
| 18-105_k141_143954 | Not-determined |
| 18-105_k141_36785 | Not-determined |
| 18-105_k141_179555 | Not-determined |
| 18-105_k141_197290 | Not-determined |
| 18-105_k141_152712 | Not-determined |
| 18-105_k141_10258 | Low-quality |
| 18-105_k141_63461 | Not-determined |
| 18-105_k141_36803 | Not-determined |
| 18-105_k141_179565 | Not-determined |
| 18-105_k141_45804 | Not-determined |
| 18-105_k141_143980 | Not-determined |
| 18-105_k141_206054 | Not-determined |
| 18-105_k141_10269 | Not-determined |
| 18-105_k141_36810 | Low-quality |
| 18-105_k141_36815 | Not-determined |
| 18-105_k141_36818 | Not-determined |
| 18-105_k141_63473 | Low-quality |
| 18-105_k141_179571 | Low-quality |
| 18-105_k141_143998 | Low-quality |
| 18-105_k141_179585 | Not-determined |
| 18-105_k141_188287 | Not-determined |
| 18-105_k141_152733 | Not-determined |
| 18-105_k141_188293 | Not-determined |
| 18-105_k141_179602 | Not-determined |
| 18-105_k141_36831 | Not-determined |
| 18-105_k141_54816 | Low-quality |
| 18-105_k141_188297 | Not-determined |
| 18-105_k141_36840 | Not-determined |
| 18-105_k141_90533 | Low-quality |
| 18-105_k141_144010 | Not-determined |
| 18-105_k141_2041 | Not-determined |
| 18-105_k141_144014 | Not-determined |
| 18-105_k141_72518 | Not-determined |
| 18-105_k141_170788 | Not-determined |
| 18-105_k141_72520 | Not-determined |
| 18-105_k141_152755 | Not-determined |
| 18-105_k141_72522 | Not-determined |
| 18-105_k141_206060 | Not-determined |
| 18-105_k141_188313 | Not-determined |
| 18-105_k141_206065 | Not-determined |
| 18-105_k141_179620 | Not-determined |
| 18-105_k141_197314 | Not-determined |
| 18-105_k141_99219 | Not-determined |
| 18-105_k141_161490 | Not-determined |
| 18-105_k141_161491 | Not-determined |
| 18-105_k141_2084 | Not-determined |
| 18-105_k141_81453 | Not-determined |
| 18-105_k141_81456 | Not-determined |
| 18-105_k141_197340 | Not-determined |
| 18-105_k141_10296 | Not-determined |
| 18-105_k141_27946 | Not-determined |
| 18-105_k141_179658 | Low-quality |
| 18-105_k141_63478 | Not-determined |
| 18-105_k141_10299 | Low-quality |
| 18-105_k141_10303 | Not-determined |
| 18-105_k141_197351 | Not-determined |
| 18-105_k141_99245 | Not-determined |
| 18-105_k141_188326 | Not-determined |
| 18-105_k141_72565 | Not-determined |
| 18-105_k141_10324 | Not-determined |
| 18-105_k141_81481 | Not-determined |
| 18-105_k141_188335 | Not-determined |
| 18-105_k141_72575 | Low-quality |
| 18-105_k141_99249 | Not-determined |
| 18-105_k141_179700 | Not-determined |
| 18-105_k141_81482 | Not-determined |
| 18-105_k141_144064 | Not-determined |
| 18-105_k141_72585 | Not-determined |
| 18-105_k141_144072 | Low-quality |
| 18-105_k141_36864 | Low-quality |
| 18-105_k141_54847 | Not-determined |
| 18-105_k141_18900 | Not-determined |
| 18-105_k141_18904 | Not-determined |
| 18-105_k141_161535 | Not-determined |
| 18-105_k141_27977 | Not-determined |
| 18-105_k141_161540 | Not-determined |
| 18-105_k141_161543 | Not-determined |
| 18-105_k141_18908 | Not-determined |
| 18-105_k141_10349 | Not-determined |
| 18-105_k141_27986 | Not-determined |
| 18-105_k141_81490 | Not-determined |
| 18-105_k141_179720 | Not-determined |
| 18-105_k141_125940 | Not-determined |
| 18-105_k141_54867 | Not-determined |
| 18-105_k141_161545 | Not-determined |
| 18-105_k141_125944 | Not-determined |
| 18-105_k141_170859 | Not-determined |
| 18-105_k141_36881 | Not-determined |
| 18-105_k141_170866 | Not-determined |
| 18-105_k141_81497 | Not-determined |
| 18-105_k141_81504 | Not-determined |
| 18-105_k141_170873 | Not-determined |
| 18-105_k141_54881 | Not-determined |
| 18-105_k141_152785 | Not-determined |
| 18-105_k141_152786 | Not-determined |
| 18-105_k141_206092 | Not-determined |
| 18-105_k141_90581 | Not-determined |
| 18-105_k141_179744 | Not-determined |
| 18-105_k141_161554 | Low-quality |
| 18-105_k141_45868 | Not-determined |
| 18-105_k141_179753 | Not-determined |
| 18-105_k141_45874 | Not-determined |
| 18-105_k141_81507 | Not-determined |
| 18-105_k141_125975 | Not-determined |
| 18-105_k141_170889 | Not-determined |
| 18-105_k141_125977 | Not-determined |
| 18-105_k141_18954 | Not-determined |
| 18-105_k141_18958 | Not-determined |
| 18-105_k141_90599 | Not-determined |
| 18-105_k141_90602 | Not-determined |
| 18-105_k141_170909 | Not-determined |
| 18-105_k141_54906 | Not-determined |
| 18-105_k141_54907 | Not-determined |
| 18-105_k141_63554 | Not-determined |
| 18-105_k141_36888 | Low-quality |
| 18-105_k141_152818 | Not-determined |
| 18-105_k141_18966 | Not-determined |
| 18-105_k141_72641 | Not-determined |
| 18-105_k141_126014 | Not-determined |
| 18-105_k141_90615 | Not-determined |
| 18-105_k141_72642 | Not-determined |
| 18-105_k141_170924 | Not-determined |
| 18-105_k141_170925 | Not-determined |
| 18-105_k141_90619 | Not-determined |
| 18-105_k141_81537 | Low-quality |
| 18-105_k141_117239 | Not-determined |
| 18-105_k141_18976 | Not-determined |
| 18-105_k141_144146 | Low-quality |
| 18-105_k141_81539 | Not-determined |
| 18-105_k141_108562 | Not-determined |
| 18-105_k141_126023 | Low-quality |
| 18-105_k141_18985 | Not-determined |
| 18-105_k141_45924 | Not-determined |
| 18-105_k141_81547 | Not-determined |
| 18-105_k141_108572 | Not-determined |
| 18-105_k141_170942 | Not-determined |
| 18-105_k141_36894 | Low-quality |
| 18-105_k141_126037 | Not-determined |
| 18-105_k141_72670 | Not-determined |
| 18-105_k141_179819 | Not-determined |
| 18-105_k141_179820 | Not-determined |
| 18-105_k141_170947 | Not-determined |
| 18-105_k141_36902 | Not-determined |
| 18-105_k141_54911 | Not-determined |
| 18-105_k141_179825 | Not-determined |
| 18-105_k141_2141 | Not-determined |
| 18-105_k141_126051 | Not-determined |
| 18-105_k141_90656 | Low-quality |
| 18-105_k141_36912 | Not-determined |
| 18-105_k141_72673 | Not-determined |
| 18-105_k141_108585 | Not-determined |
| 18-105_k141_161581 | Not-determined |
| 18-105_k141_45952 | Not-determined |
| 18-105_k141_126061 | Not-determined |
| 18-105_k141_144174 | Not-determined |
| 18-105_k141_72686 | Not-determined |
| 18-105_k141_108603 | Not-determined |
| 18-105_k141_179840 | Not-determined |
| 18-105_k141_161588 | Not-determined |
| 18-105_k141_152848 | Not-determined |
| 18-105_k141_81578 | Not-determined |
| 18-105_k141_45963 | Not-determined |
| 18-105_k141_54931 | Not-determined |
| 18-105_k141_63604 | Not-determined |
| 18-105_k141_179849 | Not-determined |
| 18-105_k141_63606 | Low-quality |
| 18-105_k141_72688 | Not-determined |
| 18-105_k141_2155 | Not-determined |
| 18-105_k141_19007 | Not-determined |
| 18-105_k141_152860 | Not-determined |
| 18-105_k141_152868 | Not-determined |
| 18-105_k141_117261 | Not-determined |
| 18-105_k141_28052 | Not-determined |
| 18-105_k141_54944 | Not-determined |
| 18-105_k141_134940 | Not-determined |
| 18-105_k141_81609 | Not-determined |
| 18-105_k141_90691 | Not-determined |
| 18-105_k141_152873 | Low-quality |
| 18-105_k141_90695 | Low-quality |
| 18-105_k141_2174 | Not-determined |
| 18-105_k141_19021 | Low-quality |
| 18-105_k141_19024 | Not-determined |
| 18-105_k141_2179 | Not-determined |
| 18-105_k141_90704 | Not-determined |
| 18-105_k141_19032 | Not-determined |
| 18-105_k141_2185 | Not-determined |
| 18-105_k141_19036 | Not-determined |
| 18-105_k141_134954 | Not-determined |
| 18-105_k141_2187 | Not-determined |
| 18-105_k141_19045 | Not-determined |
| 18-105_k141_45991 | Not-determined |
| 18-105_k141_188388 | Not-determined |
| 18-105_k141_90729 | Not-determined |
| 18-105_k141_19059 | Not-determined |
| 18-105_k141_188402 | Not-determined |
| 18-105_k141_2193 | Not-determined |
| 18-105_k141_28085 | Low-quality |
| 18-105_k141_72701 | Not-determined |
| 18-105_k141_19069 | Not-determined |
| 18-105_k141_54972 | Not-determined |
| 18-105_k141_54976 | Not-determined |
| 18-105_k141_2203 | Not-determined |
| 18-105_k141_28093 | Not-determined |
| 18-105_k141_28096 | Not-determined |
| 18-105_k141_188410 | Not-determined |
| 18-105_k141_170954 | Not-determined |
| 18-105_k141_188414 | Not-determined |
| 18-105_k141_170956 | Not-determined |
| 18-105_k141_197377 | Not-determined |
| 18-105_k141_72716 | Not-determined |
| 18-105_k141_170959 | Low-quality |
| 18-105_k141_72720 | Not-determined |
| 18-105_k141_197386 | Not-determined |
| 18-105_k141_28116 | Not-determined |
| 18-105_k141_19090 | Not-determined |
| 18-105_k141_2218 | Not-determined |
| 18-105_k141_197396 | Not-determined |
| 18-105_k141_10412 | Not-determined |
| 18-105_k141_197398 | Not-determined |
| 18-105_k141_197399 | Not-determined |
| 18-105_k141_10415 | Not-determined |
| 18-105_k141_28128 | Not-determined |
| 18-105_k141_63619 | Low-quality |
| 18-105_k141_63621 | Not-determined |
| 18-105_k141_144230 | Not-determined |
| 18-105_k141_72732 | Not-determined |
| 18-105_k141_197403 | Low-quality |
| 18-105_k141_72733 | Not-determined |
| 18-105_k141_197405 | Not-determined |
| 18-105_k141_170972 | Not-determined |
| 18-105_k141_197406 | Not-determined |
| 18-105_k141_10426 | Not-determined |
| 18-105_k141_197409 | Not-determined |
| 18-105_k141_197410 | Not-determined |
| 18-105_k141_19102 | Not-determined |
| 18-105_k141_170983 | Not-determined |
| 18-105_k141_81663 | Not-determined |
| 18-105_k141_144245 | Not-determined |
| 18-105_k141_10437 | Low-quality |
| 18-105_k141_72747 | Not-determined |
| 18-105_k141_72754 | Not-determined |
| 18-105_k141_188428 | Low-quality |
| 18-105_k141_134982 | Not-determined |
| 18-105_k141_188430 | Not-determined |
| 18-105_k141_134983 | Not-determined |
| 18-105_k141_179869 | Not-determined |
| 18-105_k141_55010 | Low-quality |
| 18-105_k141_197419 | Not-determined |
| 18-105_k141_197420 | Not-determined |
| 18-105_k141_144262 | Not-determined |
| 18-105_k141_10448 | Not-determined |
| 18-105_k141_171008 | Not-determined |
| 18-105_k141_144269 | Not-determined |
| 18-105_k141_179886 | Not-determined |
| 18-105_k141_144275 | Not-determined |
| 18-105_k141_188460 | Not-determined |
| 18-105_k141_55032 | Not-determined |
| 18-105_k141_72788 | Not-determined |
| 18-105_k141_135013 | Not-determined |
| 18-105_k141_135015 | Low-quality |
| 18-105_k141_144289 | Not-determined |
| 18-105_k141_179906 | Not-determined |
| 18-105_k141_206122 | Not-determined |
| 18-105_k141_197450 | Not-determined |
| 18-105_k141_197452 | Not-determined |
| 18-105_k141_197455 | Not-determined |
| 18-105_k141_206133 | Not-determined |
| 18-105_k141_55057 | Not-determined |
| 18-105_k141_188497 | Not-determined |
| 18-105_k141_81701 | Not-determined |
| 18-105_k141_10472 | Not-determined |
| 18-105_k141_36971 | Not-determined |
| 18-105_k141_81710 | Not-determined |
| 18-105_k141_10477 | Not-determined |
| 18-105_k141_81712 | Not-determined |
| 18-105_k141_10479 | Not-determined |
| 18-105_k141_197474 | Not-determined |
| 18-105_k141_144325 | Not-determined |
| 18-105_k141_161622 | Not-determined |
| 18-105_k141_161623 | Not-determined |
| 18-105_k141_126109 | Not-determined |
| 18-105_k141_55070 | Not-determined |
| 18-105_k141_144328 | Not-determined |
| 18-105_k141_197482 | Not-determined |
| 18-105_k141_144329 | Not-determined |
| 18-105_k141_55076 | Not-determined |
| 18-105_k141_197485 | Not-determined |
| 18-105_k141_126113 | Not-determined |
| 18-105_k141_126115 | Not-determined |
| 18-105_k141_171087 | Not-determined |
| 18-105_k141_81734 | Low-quality |
| 18-105_k141_99268 | Not-determined |
| 18-105_k141_81738 | Not-determined |
| 18-105_k141_161635 | Not-determined |
| 18-105_k141_2253 | Not-determined |
| 18-105_k141_46020 | Not-determined |
| 18-105_k141_2256 | Not-determined |
| 18-105_k141_36987 | Not-determined |
| 18-105_k141_161639 | Not-determined |
| 18-105_k141_2263 | Not-determined |
| 18-105_k141_2266 | Not-determined |
| 18-105_k141_179940 | Not-determined |
| 18-105_k141_152951 | Not-determined |
| 18-105_k141_152953 | Not-determined |
| 18-105_k141_152956 | Not-determined |
| 18-105_k141_179942 | Low-quality |
| 18-105_k141_179943 | Low-quality |
| 18-105_k141_179945 | Not-determined |
| 18-105_k141_152963 | Low-quality |
| 18-105_k141_2276 | Low-quality |
| 18-105_k141_72793 | Not-determined |
| 18-105_k141_55080 | Not-determined |
| 18-105_k141_171134 | Not-determined |
| 18-105_k141_152970 | Not-determined |
| 18-105_k141_2283 | Low-quality |
| 18-105_k141_10504 | Not-determined |
| 18-105_k141_161663 | Low-quality |
| 18-105_k141_161664 | Not-determined |
| 18-105_k141_2284 | Not-determined |
| 18-105_k141_171136 | Not-determined |
| 18-105_k141_2291 | Not-determined |
| 18-105_k141_10515 | Not-determined |
| 18-105_k141_37024 | Not-determined |
| 18-105_k141_37028 | Not-determined |
| 18-105_k141_10527 | Not-determined |
| 18-105_k141_152990 | Not-determined |
| 18-105_k141_179968 | Not-determined |
| 18-105_k141_28157 | Not-determined |
| 18-105_k141_108611 | Not-determined |
| 18-105_k141_179971 | Not-determined |
| 18-105_k141_55096 | Not-determined |
| 18-105_k141_126128 | Not-determined |
| 18-105_k141_55101 | Not-determined |
| 18-105_k141_171159 | Not-determined |
| 18-105_k141_188513 | Not-determined |
| 18-105_k141_37036 | Not-determined |
| 18-105_k141_188514 | Not-determined |
| 18-105_k141_188518 | Not-determined |
| 18-105_k141_188519 | Not-determined |
| 18-105_k141_10538 | Not-determined |
| 18-105_k141_161688 | Not-determined |
| 18-105_k141_171168 | Not-determined |
| 18-105_k141_161690 | Not-determined |
| 18-105_k141_206143 | Low-quality |
| 18-105_k141_108629 | Not-determined |
| 18-105_k141_171172 | Not-determined |
| 18-105_k141_126145 | Not-determined |
| 18-105_k141_10546 | Not-determined |
| 18-105_k141_108638 | Not-determined |
| 18-105_k141_108639 | Not-determined |
| 18-105_k141_126151 | Not-determined |
| 18-105_k141_197518 | Not-determined |
| 18-105_k141_126152 | Not-determined |
| 18-105_k141_197519 | Not-determined |
| 18-105_k141_206154 | Not-determined |
| 18-105_k141_108642 | Not-determined |
| 18-105_k141_126155 | Not-determined |
| 18-105_k141_197525 | Not-determined |
| 18-105_k141_10559 | Low-quality |
| 18-105_k141_188530 | Not-determined |
| 18-105_k141_161704 | Not-determined |
| 18-105_k141_108653 | Not-determined |
| 18-105_k141_161705 | Not-determined |
| 18-105_k141_90829 | Not-determined |
| 18-105_k141_188540 | Not-determined |
| 18-105_k141_188541 | Not-determined |
| 18-105_k141_188543 | Not-determined |
| 18-105_k141_161711 | Not-determined |
| 18-105_k141_2323 | Not-determined |
| 18-105_k141_2324 | Not-determined |
| 18-105_k141_144361 | Not-determined |
| 18-105_k141_72830 | Not-determined |
| 18-105_k141_206174 | Not-determined |
| 18-105_k141_153013 | Not-determined |
| 18-105_k141_153017 | Low-quality |
| 18-105_k141_108661 | Not-determined |
| 18-105_k141_10590 | Not-determined |
| 18-105_k141_153019 | Not-determined |
| 18-105_k141_188553 | Not-determined |
| 18-105_k141_2341 | Not-determined |
| 18-105_k141_2342 | Not-determined |
| 18-105_k141_126192 | Not-determined |
| 18-105_k141_197549 | Not-determined |
| 18-105_k141_206185 | Not-determined |
| 18-105_k141_197559 | Not-determined |
| 18-105_k141_126205 | Low-quality |
| 18-105_k141_108678 | Not-determined |

| | |
|---|---|
| 18-105_k141_108679 | Not-determined |
| 18-105_k141_197571 | Not-determined |
| 18-105_k141_126211 | Not-determined |
| 18-105_k141_99330 | Not-determined |
| 18-105_k141_188578 | Not-determined |
| 18-105_k141_99332 | Not-determined |
| 18-105_k141_188582 | Not-determined |
| 18-105_k141_108691 | Not-determined |
| 18-105_k141_135074 | Not-determined |
| 18-105_k141_206201 | Not-determined |
| 18-105_k141_108697 | Not-determined |
| 18-105_k141_135079 | Not-determined |
| 18-105_k141_171213 | Not-determined |
| 18-105_k141_2348 | Not-determined |
| 18-105_k141_188608 | Not-determined |
| 18-105_k141_37066 | Not-determined |
| 18-105_k141_2350 | Low-quality |
| 18-105_k141_108708 | Not-determined |
| 18-105_k141_171217 | Not-determined |
| 18-105_k141_188614 | Not-determined |
| 18-105_k141_99345 | Not-determined |
| 18-105_k141_99350 | Not-determined |
| 18-105_k141_188615 | Not-determined |
| 18-105_k141_144387 | Not-determined |
| 18-105_k141_55153 | Not-determined |
| 18-105_k141_55155 | Low-quality |
| 18-105_k141_144392 | Not-determined |
| 18-105_k141_153031 | Not-determined |
| 18-105_k141_144397 | Not-determined |
| 18-105_k141_46063 | Not-determined |
| 18-105_k141_10644 | Not-determined |
| 18-105_k141_179992 | Not-determined |
| 18-105_k141_55179 | Low-quality |
| 18-105_k141_10653 | Not-determined |
| 18-105_k141_19149 | Not-determined |
| 18-105_k141_19150 | Low-quality |
| 18-105_k141_171238 | Not-determined |
| 18-105_k141_180003 | Low-quality |
| 18-105_k141_72874 | Not-determined |
| 18-105_k141_153049 | Not-determined |
| 18-105_k141_19155 | Low-quality |
| 18-105_k141_180008 | Not-determined |
| 18-105_k141_135103 | Low-quality |
| 18-105_k141_19162 | Low-quality |
| 18-105_k141_180015 | Not-determined |
| 18-105_k141_81772 | Not-determined |
| 18-105_k141_90850 | Not-determined |
| 18-105_k141_99399 | Not-determined |
| 18-105_k141_19177 | Not-determined |
| 18-105_k141_81786 | Not-determined |
| 18-105_k141_99402 | Not-determined |
| 18-105_k141_37106 | Low-quality |
| 18-105_k141_90859 | Not-determined |
| 18-105_k141_90860 | Not-determined |
| 18-105_k141_99408 | Not-determined |
| 18-105_k141_37107 | Not-determined |
| 18-105_k141_37108 | Not-determined |
| 18-105_k141_99409 | Not-determined |
| 18-105_k141_46083 | Not-determined |
| 18-105_k141_180028 | Not-determined |
| 18-105_k141_180029 | Not-determined |
| 18-105_k141_19187 | Not-determined |
| 18-105_k141_81797 | Not-determined |
| 18-105_k141_81798 | Not-determined |
| 18-105_k141_46091 | Not-determined |
| 18-105_k141_81802 | Not-determined |
| 18-105_k141_37119 | Not-determined |
| 18-105_k141_126271 | Not-determined |
| 18-105_k141_126272 | Not-determined |
| 18-105_k141_180043 | Not-determined |
| 18-105_k141_135119 | Not-determined |
| 18-105_k141_126279 | Not-determined |
| 18-105_k141_90874 | Not-determined |
| 18-105_k141_37135 | Not-determined |
| 18-105_k141_180047 | Low-quality |
| 18-105_k141_99424 | Not-determined |
| 18-105_k141_72914 | Not-determined |
| 18-105_k141_19202 | Not-determined |
| 18-105_k141_2404 | Not-determined |
| 18-105_k141_153069 | Not-determined |
| 18-105_k141_63642 | Not-determined |
| 18-105_k141_63644 | Not-determined |
| 18-105_k141_161733 | Not-determined |
| 18-105_k141_90900 | Not-determined |
| 18-105_k141_63646 | Not-determined |
| 18-105_k141_153084 | Not-determined |
| 18-105_k141_99446 | Not-determined |
| 18-105_k141_72918 | Not-determined |
| 18-105_k141_46137 | Not-determined |
| 18-105_k141_153088 | Not-determined |
| 18-105_k141_126310 | Not-determined |
| 18-105_k141_37147 | Not-determined |
| 18-105_k141_72932 | Not-determined |
| 18-105_k141_161745 | Not-determined |
| 18-105_k141_99456 | Not-determined |
| 18-105_k141_37150 | Not-determined |
| 18-105_k141_90907 | Not-determined |
| 18-105_k141_99460 | Not-determined |
| 18-105_k141_197587 | Not-determined |
| 18-105_k141_90910 | Not-determined |
| 18-105_k141_99463 | Not-determined |
| 18-105_k141_197590 | Not-determined |
| 18-105_k141_72951 | Not-determined |
| 18-105_k141_90915 | Not-determined |
| 18-105_k141_135139 | Not-determined |
| 18-105_k141_46158 | Not-determined |
| 18-105_k141_117281 | Not-determined |
| 18-105_k141_72958 | Low-quality |
| 18-105_k141_63659 | Not-determined |
| 18-105_k141_63661 | Not-determined |
| 18-105_k141_46162 | Not-determined |
| 18-105_k141_19211 | Not-determined |
| 18-105_k141_161765 | Not-determined |
| 18-105_k141_126337 | Not-determined |
| 18-105_k141_90923 | Low-quality |
| 18-105_k141_153124 | Not-determined |
| 18-105_k141_37155 | Low-quality |
| 18-105_k141_72969 | Not-determined |
| 18-105_k141_117292 | Not-determined |
| 18-105_k141_153132 | Not-determined |
| 18-105_k141_90930 | Not-determined |
| 18-105_k141_135151 | Not-determined |
| 18-105_k141_117296 | Not-determined |
| 18-105_k141_37170 | Not-determined |
| 18-105_k141_197605 | Low-quality |
| 18-105_k141_153139 | Low-quality |
| 18-105_k141_72985 | Not-determined |
| 18-105_k141_81872 | Not-determined |
| 18-105_k141_19228 | Not-determined |
| 18-105_k141_99495 | Not-determined |
| 18-105_k141_99501 | Not-determined |
| 18-105_k141_153154 | Not-determined |
| 18-105_k141_28162 | Not-determined |
| 18-105_k141_99507 | Not-determined |
| 18-105_k141_46195 | Not-determined |
| 18-105_k141_37191 | Not-determined |
| 18-105_k141_99509 | Not-determined |
| 18-105_k141_206230 | Not-determined |
| 18-105_k141_135199 | Not-determined |
| 18-105_k141_161799 | Not-determined |
| 18-105_k141_188651 | Not-determined |
| 18-105_k141_188652 | Not-determined |
| 18-105_k141_153173 | Not-determined |
| 18-105_k141_28176 | Not-determined |
| 18-105_k141_126376 | Not-determined |
| 18-105_k141_144443 | Not-determined |
| 18-105_k141_28186 | Not-determined |
| 18-105_k141_46224 | Not-determined |
| 18-105_k141_144446 | Not-determined |
| 18-105_k141_19236 | Not-determined |
| 18-105_k141_28192 | Low-quality |
| 18-105_k141_126390 | Not-determined |
| 18-105_k141_180088 | Low-quality |
| 18-105_k141_171316 | Not-determined |
| 18-105_k141_19249 | Not-determined |
| 18-105_k141_161808 | Not-determined |
| 18-105_k141_126405 | Not-determined |
| 18-105_k141_161809 | Not-determined |
| 18-105_k141_19254 | Not-determined |
| 18-105_k141_19258 | Not-determined |
| 18-105_k141_90955 | Not-determined |
| 18-105_k141_171334 | Not-determined |
| 18-105_k141_161835 | Not-determined |
| 18-105_k141_161839 | Not-determined |
| 18-105_k141_161843 | Not-determined |
| 18-105_k141_117334 | Not-determined |
| 18-105_k141_171362 | Not-determined |
| 18-105_k141_188680 | Not-determined |
| 18-105_k141_46244 | Not-determined |
| 18-105_k141_180145 | Not-determined |
| 18-105_k141_180146 | Not-determined |
| 18-105_k141_180149 | Not-determined |
| 18-105_k141_188692 | Not-determined |
| 18-105_k141_37209 | Not-determined |
| 18-105_k141_206288 | Low-quality |
| 18-105_k141_180157 | Not-determined |
| 18-105_k141_46261 | Not-determined |
| 18-105_k141_126426 | Not-determined |
| 18-105_k141_37225 | Not-determined |
| 18-105_k141_188712 | Not-determined |
| 18-105_k141_180186 | Not-determined |
| 18-105_k141_197609 | Not-determined |
| 18-105_k141_81886 | Not-determined |
| 18-105_k141_188729 | Not-determined |
| 18-105_k141_206312 | Not-determined |
| 18-105_k141_37254 | Not-determined |
| 18-105_k141_63713 | Not-determined |
| 18-105_k141_117357 | Not-determined |
| 18-105_k141_197621 | Not-determined |
| 18-105_k141_117360 | Not-determined |
| 18-105_k141_37264 | Not-determined |
| 18-105_k141_63719 | Not-determined |
| 18-105_k141_63721 | Not-determined |
| 18-105_k141_144496 | Not-determined |
| 18-105_k141_55212 | Not-determined |
| 18-105_k141_188747 | Not-determined |
| 18-105_k141_55218 | Not-determined |
| 18-105_k141_63727 | Not-determined |
| 18-105_k141_37269 | Not-determined |
| 18-105_k141_144508 | Not-determined |
| 18-105_k141_2428 | Low-quality |
| 18-105_k141_63733 | Not-determined |
| 18-105_k141_2440 | Not-determined |
| 18-105_k141_2442 | Not-determined |
| 18-105_k141_55231 | Not-determined |
| 18-105_k141_117372 | Not-determined |
| 18-105_k141_73122 | Not-determined |
| 18-105_k141_144527 | Not-determined |
| 18-105_k141_188774 | Not-determined |
| 18-105_k141_2448 | Not-determined |
| 18-105_k141_28199 | Low-quality |
| 18-105_k141_171423 | Not-determined |
| 18-105_k141_197651 | Low-quality |
| 18-105_k141_28216 | Not-determined |
| 18-105_k141_28219 | Not-determined |
| 18-105_k141_73142 | Not-determined |
| 18-105_k141_188786 | Low-quality |
| 18-105_k141_171430 | Not-determined |
| 18-105_k141_117416 | Not-determined |
| 18-105_k141_117418 | Not-determined |
| 18-105_k141_206349 | Low-quality |
| 18-105_k141_90978 | Not-determined |
| 18-105_k141_188799 | Not-determined |
| 18-105_k141_144552 | Not-determined |
| 18-105_k141_206352 | Low-quality |
| 18-105_k141_206354 | Not-determined |
| 18-105_k141_90986 | Low-quality |
| 18-105_k141_206356 | Not-determined |
| 18-105_k141_81911 | Not-determined |
| 18-105_k141_188804 | Not-determined |
| 18-105_k141_171437 | Not-determined |
| 18-105_k141_28233 | Not-determined |
| 18-105_k141_117424 | Not-determined |
| 18-105_k141_46301 | Not-determined |
| 18-105_k141_117425 | Not-determined |
| 18-105_k141_188809 | Low-quality |
| 18-105_k141_90996 | Not-determined |
| 18-105_k141_10709 | Not-determined |
| 18-105_k141_188814 | Not-determined |
| 18-105_k141_10711 | Low-quality |
| 18-105_k141_10712 | Not-determined |
| 18-105_k141_73160 | Not-determined |
| 18-105_k141_171451 | Not-determined |
| 18-105_k141_46310 | Not-determined |
| 18-105_k141_91005 | Not-determined |
| 18-105_k141_73166 | Not-determined |
| 18-105_k141_10718 | Low-quality |
| 18-105_k141_91011 | Not-determined |
| 18-105_k141_73179 | Not-determined |
| 18-105_k141_135204 | Low-quality |
| 18-105_k141_144562 | Not-determined |
| 18-105_k141_10735 | Not-determined |
| 18-105_k141_135208 | Low-quality |
| 18-105_k141_99586 | Not-determined |
| 18-105_k141_99589 | Low-quality |
| 18-105_k141_171471 | Low-quality |
| 18-105_k141_126447 | Not-determined |
| 18-105_k141_161901 | Not-determined |
| 18-105_k141_126450 | Not-determined |
| 18-105_k141_135222 | Not-determined |
| 18-105_k141_28251 | Not-determined |
| 18-105_k141_197678 | Not-determined |
| 18-105_k141_206361 | Low-quality |
| 18-105_k141_99609 | Not-determined |
| 18-105_k141_144584 | Not-determined |
| 18-105_k141_197682 | Not-determined |
| 18-105_k141_206362 | Not-determined |
| 18-105_k141_99614 | Not-determined |
| 18-105_k141_144592 | Not-determined |
| 18-105_k141_99617 | Not-determined |
| 18-105_k141_171490 | Not-determined |
| 18-105_k141_19283 | Not-determined |
| 18-105_k141_81934 | Not-determined |
| 18-105_k141_180242 | Not-determined |
| 18-105_k141_73203 | Not-determined |
| 18-105_k141_37320 | Not-determined |
| 18-105_k141_180248 | Not-determined |
| 18-105_k141_161913 | Not-determined |
| 18-105_k141_117449 | Low-quality |
| 18-105_k141_55244 | Not-determined |
| 18-105_k141_81950 | Not-determined |
| 18-105_k141_99649 | Low-quality |
| 18-105_k141_73217 | Not-determined |
| 18-105_k141_188851 | Not-determined |
| 18-105_k141_55249 | Low-quality |
| 18-105_k141_117460 | Not-determined |
| 18-105_k141_63781 | Not-determined |
| 18-105_k141_153201 | Not-determined |
| 18-105_k141_197706 | Not-determined |
| 18-105_k141_171525 | Low-quality |
| 18-105_k141_63786 | Not-determined |
| 18-105_k141_73228 | Not-determined |
| 18-105_k141_73230 | Not-determined |
| 18-105_k141_171526 | Not-determined |
| 18-105_k141_19312 | Low-quality |
| 18-105_k141_28261 | Low-quality |
| 18-105_k141_81973 | Not-determined |
| 18-105_k141_197719 | Not-determined |
| 18-105_k141_91033 | Not-determined |
| 18-105_k141_135248 | Not-determined |
| 18-105_k141_197724 | Not-determined |
| 18-105_k141_81978 | Low-quality |
| 18-105_k141_10761 | Low-quality |
| 18-105_k141_126489 | Not-determined |
| 18-105_k141_55273 | Not-determined |
| 18-105_k141_10764 | Not-determined |
| 18-105_k141_117472 | Low-quality |
| 18-105_k141_81983 | Not-determined |
| 18-105_k141_2490 | Not-determined |
| 18-105_k141_197738 | Not-determined |
| 18-105_k141_91051 | Not-determined |
| 18-105_k141_99672 | Not-determined |
| 18-105_k141_135256 | Low-quality |
| 18-105_k141_99675 | Not-determined |
| 18-105_k141_63798 | Not-determined |
| 18-105_k141_126511 | Low-quality |
| 18-105_k141_10769 | Not-determined |
| 18-105_k141_10772 | Low-quality |
| 18-105_k141_10778 | Not-determined |
| 18-105_k141_19342 | Not-determined |
| 18-105_k141_19343 | Not-determined |
| 18-105_k141_135266 | Not-determined |
| 18-105_k141_126523 | Not-determined |
| 18-105_k141_82001 | Low-quality |
| 18-105_k141_91070 | Not-determined |
| 18-105_k141_153224 | Not-determined |
| 18-105_k141_63808 | Not-determined |
| 18-105_k141_197763 | Low-quality |
| 18-105_k141_144602 | Not-determined |
| 18-105_k141_63812 | Not-determined |
| 18-105_k141_144605 | Not-determined |
| 18-105_k141_37369 | Not-determined |
| 18-105_k141_153237 | Not-determined |
| 18-105_k141_73248 | Not-determined |
| 18-105_k141_144618 | Not-determined |
| 18-105_k141_91080 | Not-determined |
| 18-105_k141_82010 | Not-determined |
| 18-105_k141_135287 | Low-quality |
| 18-105_k141_73256 | Low-quality |
| 18-102-final-viral-combined_k141_207000\|\|0_partial | High-quality |
| 18-105_k141_144626 | Not-determined |
| 18-105_k141_144627 | Not-determined |
| 18-105_k141_10787 | Not-determined |
| 18-105_k141_206388 | Low-quality |
| 18-105_k141_135296 | Not-determined |
| 18-105_k141_171550 | Not-determined |
| 18-105_k141_171551 | Not-determined |
| 18-105_k141_206397 | Not-determined |
| 18-105_k141_206400 | Not-determined |
| 18-105_k141_2526 | Not-determined |
| 18-105_k141_188872 | Not-determined |
| 18-105_k141_206406 | Not-determined |
| 18-105_k141_91099 | Not-determined |
| 18-105_k141_46378 | Not-determined |
| 18-105_k141_2538 | Not-determined |
| 18-105_k141_2539 | Not-determined |
| 18-105_k141_144646 | Not-determined |
| 18-105_k141_135305 | Not-determined |
| 18-105_k141_37388 | Low-quality |
| 18-105_k141_2546 | Not-determined |
| 18-105_k141_126530 | Not-determined |
| 18-105_k141_126531 | Not-determined |
| 18-105_k141_144664 | Not-determined |
| 18-105_k141_161927 | Not-determined |
| 18-105_k141_10811 | Not-determined |
| 18-105_k141_10812 | Not-determined |
| 18-105_k141_2555 | Low-quality |
| 18-105_k141_28292 | Not-determined |
| 18-105_k141_28293 | Not-determined |
| 18-105_k141_2557 | Not-determined |
| 18-105_k141_161934 | Not-determined |
| 18-105_k141_197773 | Not-determined |
| 18-105_k141_206457 | Not-determined |
| 18-105_k141_171583 | Not-determined |
| 18-105_k141_19366 | Low-quality |
| 18-105_k141_144687 | Not-determined |
| 18-105_k141_2565 | Low-quality |
| 18-105_k141_188882 | Not-determined |
| 18-105_k141_144693 | Low-quality |
| 18-105_k141_188896 | Not-determined |
| 18-105_k141_2589 | Not-determined |
| 18-105_k141_19401 | Not-determined |
| 18-105_k141_197802 | Not-determined |
| 18-105_k141_63845 | Not-determined |
| 18-105_k141_188902 | Not-determined |
| 18-105_k141_117505 | Not-determined |
| 18-105_k141_188904 | Not-determined |
| 18-105_k141_19413 | Not-determined |
| 18-105_k141_188915 | Not-determined |
| 18-105_k141_197816 | Not-determined |
| 18-105_k141_19425 | Not-determined |
| 18-105_k141_63864 | Not-determined |
| 18-105_k141_153254 | Not-determined |
| 18-105_k141_2614 | Not-determined |
| 18-105_k141_153264 | Not-determined |
| 18-105_k141_19443 | Not-determined |
| 18-105_k141_19444 | Not-determined |
| 18-105_k141_171603 | Not-determined |
| 18-105_k141_63866 | Not-determined |
| 18-105_k141_188933 | Not-determined |
| 18-105_k141_46409 | Not-determined |
| 18-105_k141_171614 | Not-determined |
| 18-105_k141_63874 | Not-determined |
| 18-105_k141_63875 | Not-determined |
| 18-105_k141_63881 | Not-determined |
| 18-105_k141_180336 | Not-determined |
| 18-105_k141_171632 | Not-determined |
| 18-105_k141_63897 | Not-determined |
| 18-105_k141_171635 | Not-determined |
| 18-105_k141_19474 | Not-determined |
| 18-105_k141_180354 | Not-determined |
| 18-105_k141_153279 | Low-quality |
| 18-105_k141_180360 | Low-quality |
| 18-105_k141_73307 | Not-determined |
| 18-105_k141_171650 | Not-determined |
| 18-105_k141_171654 | Not-determined |
| 18-105_k141_153284 | Not-determined |
| 18-105_k141_171657 | Not-determined |
| 18-105_k141_28313 | Not-determined |
| 18-105_k141_91125 | Not-determined |
| 18-105_k141_99750 | Not-determined |
| 18-105_k141_28321 | Not-determined |
| 18-105_k141_188955 | Not-determined |
| 18-105_k141_171666 | Not-determined |
| 18-105_k141_188958 | Not-determined |
| 18-105_k141_188960 | Not-determined |
| 18-105_k141_91136 | Low-quality |
| 18-105_k141_46421 | Not-determined |
| 18-105_k141_171671 | Not-determined |
| 18-105_k141_171675 | Not-determined |
| 18-105_k141_91145 | Not-determined |
| 18-105_k141_91146 | Not-determined |
| 18-105_k141_126542 | Low-quality |
| 18-105_k141_91154 | Not-determined |
| 18-105_k141_171684 | Not-determined |
| 18-105_k141_153316 | Not-determined |
| 18-105_k141_171694 | Not-determined |
| 18-105_k141_46441 | Low-quality |
| 18-105_k141_153319 | Not-determined |
| 18-105_k141_171703 | Low-quality |
| 18-105_k141_28341 | Not-determined |
| 18-105_k141_153324 | Not-determined |
| 18-105_k141_28342 | Not-determined |
| 18-105_k141_37413 | Low-quality |
| 18-105_k141_91164 | Not-determined |
| 18-105_k141_126577 | Not-determined |
| 18-105_k141_63942 | Not-determined |
| 18-105_k141_197864 | Not-determined |
| 18-105_k141_126583 | Not-determined |
| 18-105_k141_189032 | Not-determined |
| 18-105_k141_135375 | Not-determined |
| 18-105_k141_171739 | Not-determined |
| 18-105_k141_46469 | Not-determined |
| 18-105_k141_73335 | Not-determined |
| 18-105_k141_153344 | Low-quality |
| 18-105_k141_153345 | Not-determined |
| 18-105_k141_117556 | Not-determined |
| 18-105_k141_153351 | Not-determined |
| 18-105_k141_2685 | Not-determined |
| 18-105_k141_91188 | Not-determined |
| 18-105_k141_73349 | Not-determined |
| 18-105_k141_117557 | Not-determined |
| 18-105_k141_46484 | Not-determined |
| 18-105_k141_135396 | Not-determined |
| 18-105_k141_189063 | Not-determined |
| 18-105_k141_189066 | Not-determined |
| 18-105_k141_99779 | Not-determined |
| 18-105_k141_91216 | Not-determined |
| 18-105_k141_91221 | Not-determined |
| 18-105_k141_46494 | Not-determined |
| 18-105_k141_19502 | Low-quality |
| 18-105_k141_37442 | Not-determined |
| 18-105_k141_180377 | Not-determined |
| 18-105_k141_117585 | Not-determined |
| 18-105_k141_73374 | Not-determined |
| 18-105_k141_135429 | Not-determined |
| 18-105_k141_117593 | Not-determined |
| 18-105_k141_55322 | Not-determined |
| 18-105_k141_135430 | Not-determined |
| 18-105_k141_63987 | Not-determined |
| 18-105_k141_55329 | Not-determined |
| 18-105_k141_91237 | Not-determined |
| 18-105_k141_135437 | Not-determined |
| 18-105_k141_46510 | Not-determined |
| 18-105_k141_180385 | Not-determined |
| 18-105_k141_126601 | Not-determined |
| 18-105_k141_55340 | Not-determined |
| 18-105_k141_180392 | Not-determined |
| 18-105_k141_2716 | Not-determined |
| 18-105_k141_126606 | Not-determined |
| 18-105_k141_144700 | Not-determined |
| 18-105_k141_206490 | Not-determined |
| 18-105_k141_46522 | Not-determined |
| 18-105_k141_206496 | Not-determined |
| 18-105_k141_2729 | Not-determined |
| 18-105_k141_144708 | Not-determined |
| 18-105_k141_64018 | Not-determined |
| 18-105_k141_189069 | Not-determined |
| 18-105_k141_91242 | Not-determined |
| 18-105_k141_91244 | Low-quality |
| 18-105_k141_144716 | Not-determined |
| 18-105_k141_91247 | Not-determined |
| 18-105_k141_28409 | Not-determined |
| 18-105_k141_91248 | Not-determined |
| 18-105_k141_91250 | Not-determined |
| 18-105_k141_28416 | Low-quality |
| 18-105_k141_19512 | Not-determined |
| 18-105_k141_19515 | Not-determined |
| 18-105_k141_135450 | Not-determined |
| 18-105_k141_19518 | Not-determined |
| 18-105_k141_46557 | Not-determined |
| 18-105_k141_144731 | Not-determined |
| 18-105_k141_64031 | Not-determined |
| 18-105_k141_144745 | Not-determined |
| 18-105_k141_162036 | Not-determined |
| 18-105_k141_28447 | Not-determined |
| 18-105_k141_135457 | Low-quality |
| 18-105_k141_19531 | Not-determined |
| 18-105_k141_135464 | Not-determined |
| 18-105_k141_135466 | Not-determined |
| 18-105_k141_144768 | Not-determined |
| 18-105_k141_153422 | Not-determined |
| 18-105_k141_55379 | Low-quality |
| 18-105_k141_55381 | Not-determined |
| 18-105_k141_2783 | Not-determined |
| 18-105_k141_2787 | Low-quality |
| 18-105_k141_108764 | Not-determined |
| 18-105_k141_37466 | Not-determined |
| 18-105_k141_37467 | Not-determined |
| 18-105_k141_144787 | Not-determined |
| 18-105_k141_2804 | Not-determined |
| 18-105_k141_2808 | Not-determined |
| 18-105_k141_46579 | Not-determined |
| 18-105_k141_28473 | Low-quality |
| 18-105_k141_2810 | Not-determined |
| 18-105_k141_108775 | Not-determined |
| 18-105_k141_162057 | Not-determined |
| 18-105_k141_37481 | Not-determined |
| 18-105_k141_28474 | Not-determined |
| 18-105_k141_28475 | Not-determined |
| 18-105_k141_64067 | Not-determined |
| 18-105_k141_73384 | Not-determined |
| 18-105_k141_162062 | Not-determined |
| 18-105_k141_73391 | Not-determined |
| 18-105_k141_162067 | Not-determined |
| 18-105_k141_2818 | Not-determined |
| 18-105_k141_135517 | Not-determined |
| 18-105_k141_64072 | Low-quality |
| 18-105_k141_171780 | Not-determined |
| 18-105_k141_55395 | Low-quality |
| 18-105_k141_64075 | Low-quality |
| 18-105_k141_171791 | Not-determined |
| 18-105_k141_37488 | Not-determined |
| 18-105_k141_28519 | Not-determined |
| 18-105_k141_28521 | Low-quality |
| 18-105_k141_46634 | Not-determined |
| 18-105_k141_28527 | Not-determined |
| 18-105_k141_73427 | Not-determined |
| 18-105_k141_171801 | Low-quality |
| 18-105_k141_171805 | Not-determined |
| 18-105_k141_206537 | Not-determined |
| 18-105_k141_37517 | Not-determined |
| 18-105_k141_28539 | Not-determined |
| 18-105_k141_99864 | Not-determined |
| 18-105_k141_28558 | Not-determined |
| 18-105_k141_144805 | Not-determined |
| 18-105_k141_46643 | Low-quality |
| 18-105_k141_126728 | Not-determined |
| 18-105_k141_189095 | Not-determined |
| 18-105_k141_99874 | Not-determined |
| 18-105_k141_99877 | Not-determined |
| 18-105_k141_99879 | Not-determined |
| 18-105_k141_28570 | Not-determined |
| 18-105_k141_144822 | Not-determined |
| 18-105_k141_46656 | Low-quality |
| 18-105_k141_144832 | Low-quality |
| 18-105_k141_206545 | Not-determined |
| 18-105_k141_206548 | Not-determined |
| 18-105_k141_46666 | Not-determined |
| 18-105_k141_73471 | Not-determined |
| 18-105_k141_64111 | Not-determined |
| 18-105_k141_189115 | Not-determined |
| 18-105_k141_99911 | Low-quality |
| 18-105_k141_153469 | Not-determined |
| 18-105_k141_189123 | Not-determined |
| 18-105_k141_99922 | Not-determined |
| 18-105_k141_99925 | Low-quality |
| 18-105_k141_144849 | Not-determined |
| 18-105_k141_189129 | Not-determined |

| | |
|---|---|
| 18-105_k141_99931 | Not-determined |
| 18-105_k141_171825 | Not-determined |
| 18-105_k141_153479 | Low-quality |
| 18-105_k141_37535 | Not-determined |
| 18-105_k141_206567 | Not-determined |
| 18-105_k141_126750 | Not-determined |
| 18-105_k141_46695 | Not-determined |
| 18-105_k141_189141 | Not-determined |
| 18-105_k141_2870 | Not-determined |
| 18-105_k141_37546 | Not-determined |
| 18-105_k141_189157 | Not-determined |
| 18-105_k141_206587 | Not-determined |
| 18-105_k141_37555 | Not-determined |
| 18-105_k141_99939 | Not-determined |
| 18-105_k141_46700 | Low-quality |
| 18-105_k141_206599 | Not-determined |
| 18-105_k141_153517 | Low-quality |
| 18-105_k141_153520 | Low-quality |
| 18-105_k141_82152 | Not-determined |
| 18-105_k141_46706 | Not-determined |
| 18-105_k141_37568 | Not-determined |
| 18-105_k141_206607 | Not-determined |
| 18-105_k141_126795 | Not-determined |
| 18-105_k141_91295 | Not-determined |
| 18-105_k141_206611 | Not-determined |
| 18-105_k141_46721 | Not-determined |
| 18-105_k141_82168 | Not-determined |
| 18-105_k141_144872 | Not-determined |
| 18-105_k141_126800 | Not-determined |
| 18-105_k141_19550 | Not-determined |
| 18-105_k141_91305 | Not-determined |
| 18-105_k141_126809 | Low-quality |
| 18-105_k141_126810 | Low-quality |
| 18-105_k141_153539 | Not-determined |
| 18-105_k141_153540 | Not-determined |
| 18-105_k141_91313 | Not-determined |
| 18-105_k141_206632 | Not-determined |
| 18-105_k141_189190 | Not-determined |
| 18-105_k141_189197 | Not-determined |
| 18-105_k141_99980 | Not-determined |
| 18-105_k141_153548 | Not-determined |
| 18-105_k141_37591 | Not-determined |
| 18-105_k141_180402 | Not-determined |
| 18-105_k141_189207 | Low-quality |
| 18-105_k141_55434 | Not-determined |
| 18-105_k141_37606 | Low-quality |
| 18-105_k141_55439 | Not-determined |
| 18-105_k141_37610 | Not-determined |
| 18-105_k141_144895 | Not-determined |
| 18-105_k141_55445 | Not-determined |
| 18-105_k141_153551 | Not-determined |
| 18-105_k141_197914 | Not-determined |
| 18-105_k141_37623 | Not-determined |
| 18-105_k141_197917 | Not-determined |
| 18-105_k141_197920 | Not-determined |
| 18-105_k141_126834 | Not-determined |
| 18-105_k141_189231 | Not-determined |
| 18-105_k141_37630 | Not-determined |
| 18-105_k141_55456 | Not-determined |
| 18-105_k141_180449 | Not-determined |
| 18-105_k141_117634 | Low-quality |
| 18-105_k141_117635 | Not-determined |
| 18-105_k141_55464 | Not-determined |
| 18-105_k141_2906 | Low-quality |
| 18-105_k141_126845 | Not-determined |
| 18-105_k141_197932 | Not-determined |
| 18-105_k141_180459 | Not-determined |
| 18-105_k141_28607 | Not-determined |
| 18-105_k141_180461 | Not-determined |
| 18-105_k141_189244 | Not-determined |
| 18-105_k141_180473 | Not-determined |
| 18-105_k141_206643 | Not-determined |
| 18-105_k141_10830 | Not-determined |
| 18-105_k141_206646 | Not-determined |
| 18-105_k141_180482 | Not-determined |
| 18-105_k141_37654 | Not-determined |
| 18-105_k141_37660 | Not-determined |
| 18-105_k141_100029 | Low-quality |
| 18-105_k141_10842 | Not-determined |
| 18-105_k141_37661 | Not-determined |
| 18-105_k141_37667 | Not-determined |
| 18-105_k141_91356 | Not-determined |
| 18-105_k141_100037 | Low-quality |
| 18-105_k141_153611 | Not-determined |
| 18-105_k141_153612 | Not-determined |
| 18-105_k141_73489 | Not-determined |
| 18-105_k141_46741 | Not-determined |
| 18-105_k141_91364 | Not-determined |
| 18-105_k141_37677 | Not-determined |
| 18-105_k141_46743 | Not-determined |
| 18-105_k141_153622 | Not-determined |
| 18-105_k141_46749 | Not-determined |
| 18-105_k141_46754 | Not-determined |
| 18-105_k141_10869 | Not-determined |
| 18-105_k141_162152 | Not-determined |
| 18-105_k141_197984 | Not-determined |
| 18-105_k141_197988 | Not-determined |
| 18-105_k141_37679 | Not-determined |
| 18-105_k141_28645 | Not-determined |
| 18-105_k141_206687 | Not-determined |
| 18-105_k141_100054 | Not-determined |
| 18-105_k141_171902 | Not-determined |
| 18-105_k141_117658 | Not-determined |
| 18-105_k141_117661 | Not-determined |
| 18-105_k141_126874 | Low-quality |
| 18-105_k141_117667 | Not-determined |
| 18-105_k141_126877 | Not-determined |
| 18-105_k141_162178 | Not-determined |
| 18-105_k141_73498 | Not-determined |
| 18-105_k141_126887 | Not-determined |
| 18-105_k141_46793 | Not-determined |
| 18-105_k141_198020 | Not-determined |
| 18-105_k141_126906 | Not-determined |
| 18-105_k141_198022 | Not-determined |
| 18-105_k141_171928 | Not-determined |
| 18-105_k141_117690 | Not-determined |
| 18-105_k141_198030 | Not-determined |
| 18-105_k141_198036 | Not-determined |
| 18-105_k141_135548 | Low-quality |
| 18-105_k141_108854 | Not-determined |
| 18-105_k141_171937 | Not-determined |
| 18-105_k141_55498 | Not-determined |
| 18-105_k141_55501 | Not-determined |
| 18-105_k141_206720 | Not-determined |
| 18-105_k141_91377 | Not-determined |
| 18-105_k141_91379 | Not-determined |
| 18-105_k141_206724 | Low-quality |
| 18-105_k141_100104 | Not-determined |
| 18-105_k141_206731 | Not-determined |
| 18-105_k141_171941 | Not-determined |
| 18-105_k141_91386 | Not-determined |
| 18-105_k141_198066 | Not-determined |
| 18-105_k141_206737 | Not-determined |
| 18-105_k141_64151 | Not-determined |
| 18-105_k141_55524 | Not-determined |
| 18-105_k141_171953 | Not-determined |
| 18-105_k141_91387 | Low-quality |
| 18-105_k141_73518 | Low-quality |
| 18-105_k141_117724 | Not-determined |
| 18-105_k141_73525 | Not-determined |
| 18-105_k141_144936 | Not-determined |
| 18-105_k141_171962 | Not-determined |
| 18-105_k141_198088 | Not-determined |
| 18-105_k141_108873 | Not-determined |
| 18-105_k141_2918 | Not-determined |
| 18-105_k141_46859 | Not-determined |
| 18-105_k141_198092 | Not-determined |
| 18-105_k141_198093 | Not-determined |
| 18-105_k141_108879 | Not-determined |
| 18-105_k141_135574 | Not-determined |
| 18-105_k141_206761 | Not-determined |
| 18-105_k141_206765 | Not-determined |
| 18-105_k141_73529 | Not-determined |
| 18-105_k141_73532 | Not-determined |
| 18-105_k141_117745 | Not-determined |
| 18-105_k141_144966 | Not-determined |
| 18-105_k141_117750 | Not-determined |
| 18-105_k141_162214 | Not-determined |
| 18-105_k141_206776 | Not-determined |
| 18-105_k141_162217 | Not-determined |
| 18-105_k141_117761 | Not-determined |
| 18-105_k141_171978 | Not-determined |
| 18-105_k141_144969 | Not-determined |
| 18-105_k141_126965 | Low-quality |
| 18-105_k141_64161 | Not-determined |
| 18-105_k141_144978 | Low-quality |
| 18-105_k141_144980 | Not-determined |
| 18-105_k141_144984 | Not-determined |
| 18-105_k141_144985 | Not-determined |
| 18-105_k141_126977 | Low-quality |
| 18-105_k141_126980 | Not-determined |
| 18-105_k141_189260 | Not-determined |
| 18-105_k141_82197 | Not-determined |
| 18-105_k141_55547 | Not-determined |
| 18-105_k141_55549 | Not-determined |
| 18-105_k141_82199 | Low-quality |
| 18-105_k141_198112 | Not-determined |
| 18-105_k141_117774 | Not-determined |
| 18-105_k141_126988 | Not-determined |
| 18-105_k141_198116 | Not-determined |
| 18-105_k141_55553 | Not-determined |
| 18-105_k141_126991 | Not-determined |
| 18-105_k141_64176 | Low-quality |
| 18-105_k141_162219 | Not-determined |
| 18-105_k141_55559 | Not-determined |
| 18-105_k141_162220 | Not-determined |
| 18-105_k141_180534 | Not-determined |
| 18-105_k141_64179 | Not-determined |
| 18-105_k141_171993 | Not-determined |
| 18-105_k141_10881 | Not-determined |
| 18-105_k141_162228 | Not-determined |
| 18-105_k141_162231 | Not-determined |
| 18-105_k141_162233 | Low-quality |
| 18-105_k141_55572 | Not-determined |
| 18-105_k141_198129 | Not-determined |
| 18-105_k141_19579 | Not-determined |
| 18-105_k141_55579 | Low-quality |
| 18-105_k141_55583 | Not-determined |
| 18-105_k141_55586 | Not-determined |
| 18-105_k141_82237 | Not-determined |
| 18-105_k141_55592 | Not-determined |
| 18-105_k141_189286 | Not-determined |
| 18-105_k141_162271 | Not-determined |
| 18-105_k141_198155 | Not-determined |
| 18-105_k141_180564 | Not-determined |
| 18-105_k141_73560 | Not-determined |
| 18-105_k141_64205 | Not-determined |
| 18-105_k141_180568 | Not-determined |
| 18-105_k141_10891 | Not-determined |
| 18-105_k141_198162 | Not-determined |
| 18-105_k141_153694 | Not-determined |
| 18-105_k141_153696 | Not-determined |
| 18-105_k141_153697 | Not-determined |
| 18-105_k141_180578 | Not-determined |
| 18-105_k141_10905 | Not-determined |
| 18-105_k141_37711 | Not-determined |
| 18-105_k141_153711 | Not-determined |
| 18-105_k141_91450 | Not-determined |
| 18-105_k141_55598 | Not-determined |
| 18-105_k141_135583 | Not-determined |
| 18-105_k141_55602 | Not-determined |
| 18-105_k141_135586 | Not-determined |
| 18-105_k141_55605 | Not-determined |
| 18-105_k141_28697 | Not-determined |
| 18-105_k141_91461 | Not-determined |
| 18-105_k141_64241 | Not-determined |
| 18-105_k141_10928 | Not-determined |
| 18-105_k141_135601 | Not-determined |
| 18-105_k141_91467 | Not-determined |
| 18-105_k141_145026 | Not-determined |
| 18-105_k141_135610 | Not-determined |
| 18-105_k141_10942 | Low-quality |
| 18-105_k141_10944 | Not-determined |
| 18-105_k141_189299 | Not-determined |
| 18-105_k141_189301 | Not-determined |
| 18-105_k141_162323 | Not-determined |
| 18-105_k141_117823 | Not-determined |
| 18-105_k141_10956 | Not-determined |
| 18-105_k141_162329 | Not-determined |
| 18-105_k141_180613 | Not-determined |
| 18-105_k141_10966 | Not-determined |
| 18-105_k141_127001 | Low-quality |
| 18-105_k141_10969 | Not-determined |
| 18-105_k141_64277 | Not-determined |
| 18-105_k141_127012 | Not-determined |
| 18-105_k141_135628 | Not-determined |
| 18-105_k141_180624 | Not-determined |
| 18-105_k141_180626 | Not-determined |
| 18-105_k141_180631 | Not-determined |
| 18-105_k141_135633 | Not-determined |
| 18-105_k141_46874 | Not-determined |
| 18-105_k141_127019 | Low-quality |
| 18-105_k141_100223 | Not-determined |
| 18-105_k141_100227 | Not-determined |
| 18-105_k141_64300 | Not-determined |
| 18-105_k141_100244 | Not-determined |
| 18-105_k141_127038 | Low-quality |
| 18-105_k141_91497 | Not-determined |
| 18-105_k141_189356 | Not-determined |
| 18-105_k141_117851 | Not-determined |
| 18-105_k141_64318 | Low-quality |
| 18-105_k141_189362 | Not-determined |
| 18-105_k141_198174 | Low-quality |
| 18-105_k141_91503 | Not-determined |
| 18-105_k141_91505 | Not-determined |
| 18-105_k141_10972 | Not-determined |
| 18-105_k141_198189 | Not-determined |
| 18-105_k141_117869 | Not-determined |
| 18-105_k141_180675 | Low-quality |
| 18-105_k141_117871 | Not-determined |
| 18-105_k141_198196 | Not-determined |
| 18-105_k141_46891 | Not-determined |
| 18-105_k141_198199 | Not-determined |
| 18-105_k141_189386 | Not-determined |
| 18-105_k141_189390 | Not-determined |
| 18-105_k141_117881 | Not-determined |
| 18-105_k141_10995 | Not-determined |
| 18-105_k141_91516 | Not-determined |
| 18-105_k141_46899 | Not-determined |
| 18-105_k141_91517 | Not-determined |
| 18-105_k141_11003 | Not-determined |
| 18-105_k141_117892 | Not-determined |
| 18-105_k141_46912 | Not-determined |
| 18-105_k141_37730 | Not-determined |
| 18-105_k141_189416 | Not-determined |
| 18-105_k141_189424 | Not-determined |
| 18-105_k141_19619 | Not-determined |
| 18-105_k141_46916 | Not-determined |
| 18-105_k141_19624 | Not-determined |
| 18-105_k141_162362 | Not-determined |
| 18-105_k141_135641 | Not-determined |
| 18-105_k141_19632 | Not-determined |
| 18-105_k141_19634 | Not-determined |
| 18-105_k141_162367 | Not-determined |
| 18-105_k141_100301 | Not-determined |
| 18-105_k141_37770 | Not-determined |
| 18-105_k141_19637 | Not-determined |
| 18-105_k141_46932 | Not-determined |
| 18-105_k141_37772 | Not-determined |
| 18-105_k141_100310 | Not-determined |
| 18-105_k141_100312 | Low-quality |
| 18-105_k141_46944 | Not-determined |
| 18-105_k141_3026 | Low-quality |
| 18-105_k141_3031 | Not-determined |
| 18-105_k141_46953 | Not-determined |
| 18-105_k141_28749 | Not-determined |
| 18-105_k141_180687 | Not-determined |
| 18-105_k141_11025 | Not-determined |
| 18-105_k141_11026 | Not-determined |
| 18-105_k141_11027 | Low-quality |
| 18-105_k141_127083 | Not-determined |
| 18-105_k141_11031 | Not-determined |
| 18-105_k141_135662 | Not-determined |
| 18-105_k141_127087 | Not-determined |
| 18-105_k141_180703 | Not-determined |
| 18-105_k141_100339 | Low-quality |
| 18-105_k141_73602 | Not-determined |
| 18-105_k141_64344 | Not-determined |
| 18-105_k141_100349 | Not-determined |
| 18-105_k141_189464 | Not-determined |
| 18-105_k141_180722 | Not-determined |
| 18-105_k141_100359 | Not-determined |
| 18-105_k141_180730 | Not-determined |
| 18-105_k141_206840 | Not-determined |
| 18-105_k141_189482 | Not-determined |
| 18-105_k141_100375 | Not-determined |
| 18-105_k141_11050 | Not-determined |
| 18-105_k141_135688 | Not-determined |
| 18-105_k141_189495 | Not-determined |
| 18-105_k141_3064 | Not-determined |
| 18-105_k141_91534 | Not-determined |
| 18-105_k141_172022 | Not-determined |
| 18-105_k141_180765 | Not-determined |
| 18-105_k141_172037 | Not-determined |
| 18-105_k141_145046 | Not-determined |
| 18-105_k141_64365 | Not-determined |
| 18-105_k141_172052 | Not-determined |
| 18-105_k141_46966 | Not-determined |
| 18-105_k141_64369 | Not-determined |
| 18-105_k141_73640 | Not-determined |
| 18-105_k141_189522 | Not-determined |
| 18-105_k141_28800 | Not-determined |
| 18-105_k141_198210 | Low-quality |
| 18-105_k141_64388 | Not-determined |
| 18-105_k141_206868 | Not-determined |
| 18-105_k141_189535 | Not-determined |
| 18-105_k141_206871 | Not-determined |
| 18-105_k141_172078 | Low-quality |
| 18-105_k141_172082 | Not-determined |
| 18-105_k141_64406 | Not-determined |
| 18-105_k141_189546 | Not-determined |
| 18-105_k141_100432 | Low-quality |
| 18-105_k141_206876 | Not-determined |
| 18-105_k141_198233 | Low-quality |
| 18-105_k141_206879 | Not-determined |
| 18-105_k141_55610 | Not-determined |
| 18-105_k141_198240 | Not-determined |
| 18-105_k141_55617 | Not-determined |
| 18-105_k141_135723 | Not-determined |
| 18-105_k141_172112 | Not-determined |
| 18-105_k141_64440 | Not-determined |
| 18-105_k141_172116 | Low-quality |
| 18-105_k141_64441 | Not-determined |
| 18-105_k141_28810 | Low-quality |
| 18-105_k141_28814 | Not-determined |
| 18-105_k141_162414 | Not-determined |
| 18-105_k141_206892 | Not-determined |
| 18-105_k141_206897 | Not-determined |
| 18-105_k141_73671 | Low-quality |
| 18-105_k141_206899 | Not-determined |
| 18-105_k141_28822 | Not-determined |
| 18-105_k141_100451 | Not-determined |
| 18-105_k141_91554 | Not-determined |
| 18-105_k141_100456 | Not-determined |
| 18-105_k141_11104 | Not-determined |
| 18-105_k141_11107 | Low-quality |
| 18-105_k141_127132 | Low-quality |
| 18-105_k141_28828 | Not-determined |
| 18-105_k141_11111 | Not-determined |
| 18-105_k141_172142 | Not-determined |
| 18-105_k141_145081 | Not-determined |
| 18-105_k141_206908 | Not-determined |
| 18-105_k141_127147 | Not-determined |
| 18-105_k141_11120 | Low-quality |
| 18-105_k141_127151 | Not-determined |
| 18-105_k141_100478 | Not-determined |
| 18-105_k141_206917 | Low-quality |
| 18-105_k141_73693 | Not-determined |
| 18-105_k141_180772 | Not-determined |
| 18-105_k141_198269 | Not-determined |
| 18-105_k141_108909 | Not-determined |
| 18-105_k141_64474 | Not-determined |
| 18-105_k141_198276 | Not-determined |
| 18-105_k141_11128 | Not-determined |
| 18-105_k141_28837 | Not-determined |
| 18-105_k141_180784 | Not-determined |
| 18-105_k141_64481 | Not-determined |
| 18-105_k141_11133 | Not-determined |
| 18-105_k141_206933 | Not-determined |
| 18-105_k141_73711 | Not-determined |
| 18-105_k141_206941 | Not-determined |
| 18-105_k141_73719 | Not-determined |
| 18-105_k141_73720 | Not-determined |
| 18-105_k141_64485 | Not-determined |
| 18-105_k141_127176 | Not-determined |
| 18-105_k141_206949 | Not-determined |
| 18-105_k141_91573 | Not-determined |
| 18-105_k141_28844 | Not-determined |
| 18-105_k141_100509 | Not-determined |
| 18-105_k141_28849 | Not-determined |
| 18-105_k141_11149 | Not-determined |
| 18-105_k141_37793 | Not-determined |
| 18-105_k141_11154 | Not-determined |
| 18-105_k141_28856 | Not-determined |
| 18-105_k141_11159 | Not-determined |
| 18-105_k141_189584 | Not-determined |
| 18-105_k141_198303 | Not-determined |
| 18-105_k141_100533 | Not-determined |
| 18-105_k141_135781 | Not-determined |
| 18-105_k141_198319 | Not-determined |
| 18-105_k141_135787 | Not-determined |
| 18-105_k141_189596 | Low-quality |
| 18-105_k141_11178 | Not-determined |
| 18-105_k141_180805 | Not-determined |
| 18-105_k141_11184 | Not-determined |
| 18-105_k141_11188 | Not-determined |
| 18-105_k141_180813 | Not-determined |
| 18-105_k141_100550 | Not-determined |
| 18-105_k141_64535 | Not-determined |
| 18-105_k141_127214 | Not-determined |
| 18-105_k141_145087 | Not-determined |
| 18-105_k141_180833 | Not-determined |
| 18-105_k141_64553 | Not-determined |
| 18-105_k141_145092 | Not-determined |
| 18-105_k141_91618 | Low-quality |
| 18-105_k141_91619 | Not-determined |
| 18-105_k141_19665 | Not-determined |
| 18-105_k141_145098 | Not-determined |
| 18-105_k141_189643 | Not-determined |
| 18-105_k141_100571 | Not-determined |
| 18-105_k141_37810 | Not-determined |
| 18-105_k141_189648 | Not-determined |
| 18-105_k141_117925 | Not-determined |
| 18-105_k141_73746 | Not-determined |
| 18-105_k141_73747 | Not-determined |
| 18-105_k141_73748 | Not-determined |
| 18-105_k141_28903 | Not-determined |
| 18-105_k141_11219 | Not-determined |
| 18-105_k141_11223 | Not-determined |
| 18-105_k141_172161 | Not-determined |
| 18-105_k141_73756 | Not-determined |
| 18-105_k141_206976 | Not-determined |
| 18-105_k141_206977 | Not-determined |
| 18-105_k141_19680 | Not-determined |
| 18-105_k141_180848 | Not-determined |
| 18-105_k141_162455 | Not-determined |
| 18-105_k141_180853 | Not-determined |
| 18-105_k141_117946 | Not-determined |
| 18-105_k141_180857 | Not-determined |
| 18-105_k141_189682 | Low-quality |
| 18-105_k141_162465 | Not-determined |
| 18-105_k141_91638 | Not-determined |
| 18-105_k141_145117 | Not-determined |
| 18-105_k141_108958 | Not-determined |
| 18-105_k141_162472 | Not-determined |
| 18-105_k141_100603 | Not-determined |
| 18-105_k141_91648 | Not-determined |
| 18-105_k141_91649 | Not-determined |
| 18-105_k141_73771 | Not-determined |
| 18-105_k141_64595 | Not-determined |
| 18-105_k141_3132 | Not-determined |
| 18-105_k141_46987 | Low-quality |
| 18-101_k141_29288 | Medium-quality |
| 18-105_k141_73780 | Not-determined |
| 18-105_k141_127220 | Not-determined |
| 18-105_k141_189705 | Not-determined |
| 18-105_k141_117973 | Low-quality |
| 18-105_k141_127225 | Not-determined |
| 18-105_k141_162493 | Not-determined |
| 18-105_k141_127233 | Not-determined |
| 18-105_k141_207017 | Low-quality |
| 18-105_k141_172189 | Not-determined |
| 18-105_k141_73785 | Not-determined |
| 18-105_k141_37843 | Not-determined |
| 18-105_k141_189711 | Not-determined |
| 18-105_k141_47006 | Not-determined |
| 18-105_k141_180896 | Not-determined |
| 18-105_k141_207026 | Not-determined |
| 18-105_k141_100623 | Not-determined |
| 18-105_k141_47010 | Not-determined |
| 18-105_k141_162503 | Low-quality |
| 18-105_k141_47012 | Not-determined |
| 18-105_k141_162506 | Not-determined |
| 18-105_k141_55626 | Not-determined |
| 18-105_k141_55627 | Not-determined |
| 18-105_k141_135807 | Not-determined |
| 18-105_k141_37859 | Not-determined |
| 18-105_k141_135810 | Not-determined |
| 18-105_k141_117992 | Not-determined |
| 18-105_k141_172202 | Not-determined |
| 18-105_k141_37868 | Not-determined |
| 18-105_k141_37871 | Not-determined |
| 18-105_k141_162522 | Not-determined |
| 18-105_k141_118003 | Not-determined |
| 18-105_k141_162529 | Not-determined |
| 18-105_k141_37878 | Not-determined |
| 18-105_k141_100642 | Not-determined |
| 18-105_k141_162534 | Not-determined |
| 18-105_k141_100647 | Not-determined |
| 18-105_k141_162536 | Low-quality |
| 18-105_k141_162537 | Not-determined |
| 18-105_k141_198345 | Not-determined |
| 18-105_k141_55641 | Not-determined |
| 18-105_k141_162540 | Not-determined |
| 18-105_k141_207064 | Not-determined |
| 18-105_k141_55646 | Not-determined |
| 18-105_k141_47040 | Not-determined |
| 18-105_k141_180920 | Low-quality |
| 18-105_k141_100660 | Not-determined |
| 18-105_k141_180929 | Not-determined |
| 18-105_k141_198351 | Not-determined |
| 18-105_k141_55656 | Not-determined |
| 18-105_k141_47048 | Not-determined |
| 18-105_k141_207078 | Not-determined |
| 18-105_k141_135834 | Not-determined |
| 18-105_k141_207081 | Not-determined |
| 18-105_k141_135839 | Not-determined |
| 18-105_k141_118037 | Not-determined |
| 18-105_k141_118039 | Not-determined |
| 18-105_k141_135840 | Low-quality |
| 18-105_k141_135842 | Not-determined |
| 18-105_k141_162565 | Low-quality |
| 18-105_k141_162570 | Not-determined |
| 18-105_k141_118053 | Not-determined |
| 18-105_k141_28939 | Not-determined |
| 18-105_k141_207108 | Not-determined |
| 18-105_k141_82257 | Not-determined |
| 18-105_k141_82258 | Not-determined |
| 18-105_k141_207115 | Not-determined |
| 18-105_k141_82261 | Not-determined |
| 18-105_k141_207118 | Not-determined |
| 18-105_k141_207119 | Not-determined |
| 18-105_k141_47078 | Not-determined |
| 18-105_k141_28954 | Not-determined |
| 18-105_k141_64617 | Not-determined |
| 18-105_k141_198373 | Not-determined |
| 18-105_k141_28958 | Not-determined |
| 18-105_k141_47085 | Not-determined |
| 18-105_k141_207124 | Not-determined |
| 18-105_k141_100681 | Low-quality |
| 18-105_k141_19703 | Not-determined |
| 18-105_k141_82279 | Not-determined |
| 18-105_k141_82281 | Not-determined |
| 18-105_k141_91657 | Not-determined |
| 18-105_k141_82289 | Not-determined |
| 18-105_k141_47109 | Not-determined |
| 18-105_k141_82291 | Low-quality |
| 18-105_k141_47112 | Not-determined |
| 18-105_k141_82294 | Not-determined |
| 18-105_k141_198382 | Not-determined |
| 18-105_k141_47125 | Not-determined |
| 18-105_k141_118091 | Not-determined |
| 18-105_k141_153738 | Not-determined |
| 18-105_k141_64629 | Not-determined |
| 18-105_k141_153742 | Not-determined |
| 18-105_k141_145154 | Not-determined |
| 18-105_k141_19722 | Not-determined |
| 18-105_k141_118105 | Not-determined |
| 18-105_k141_172256 | Not-determined |
| 18-105_k141_189724 | Not-determined |
| 18-105_k141_64637 | Not-determined |
| 18-105_k141_153750 | Not-determined |
| 18-105_k141_100702 | Not-determined |
| 18-105_k141_162615 | Not-determined |
| 18-105_k141_135880 | Not-determined |
| 18-105_k141_162617 | Not-determined |
| 18-105_k141_153759 | Not-determined |
| 18-105_k141_64644 | Not-determined |
| 18-105_k141_189740 | Not-determined |
| 18-105_k141_172275 | Not-determined |
| 18-105_k141_172277 | Not-determined |
| 18-105_k141_47152 | Not-determined |
| 18-105_k141_28990 | Not-determined |
| 18-105_k141_207156 | Not-determined |
| 18-105_k141_172280 | Not-determined |
| 18-105_k141_207157 | Not-determined |
| 18-105_k141_127294 | Not-determined |
| 18-105_k141_64648 | Not-determined |
| 18-105_k141_28996 | Not-determined |

| | |
|---|---|
| 18-105_k141_180944 | Not-determined |
| 18-105_k141_180946 | Not-determined |
| 18-105_k141_47158 | Not-determined |
| 18-105_k141_100712 | Not-determined |
| 18-105_k141_64662 | Not-determined |
| 18-105_k141_91669 | Not-determined |
| 18-105_k141_64670 | Low-quality |
| 18-105_k141_29015 | Not-determined |
| 18-105_k141_118137 | Not-determined |
| 18-105_k141_29019 | Not-determined |
| 18-105_k141_145170 | Not-determined |
| 18-105_k141_3196 | Not-determined |
| 18-105_k141_135910 | Not-determined |
| 18-105_k141_29025 | Not-determined |
| 18-105_k141_180973 | Not-determined |
| 18-105_k141_19742 | Not-determined |
| 18-105_k141_189772 | Low-quality |
| 18-105_k141_207185 | Not-determined |
| 18-105_k141_207186 | Not-determined |
| 18-105_k141_19743 | Not-determined |
| 18-105_k141_172299 | Not-determined |
| 18-105_k141_19755 | Not-determined |
| 18-105_k141_145189 | Not-determined |
| 18-105_k141_180994 | Not-determined |
| 18-105_k141_172307 | Not-determined |
| 18-105_k141_118146 | Not-determined |
| 18-105_k141_109002 | Not-determined |
| 18-105_k141_47175 | Low-quality |
| 18-105_k141_145198 | Not-determined |
| 18-105_k141_172321 | Not-determined |
| 18-105_k141_207218 | Not-determined |
| 18-105_k141_100735 | Not-determined |
| 18-105_k141_91705 | Not-determined |
| 18-105_k141_145212 | Not-determined |
| 18-105_k141_145216 | Not-determined |
| 18-105_k141_172334 | Low-quality |
| 18-105_k141_47191 | Not-determined |
| 18-105_k141_153792 | Not-determined |
| 18-105_k141_91718 | Not-determined |
| 18-105_k141_3203 | Not-determined |
| 18-105_k141_153796 | Not-determined |
| 18-105_k141_145229 | Not-determined |
| 18-105_k141_145230 | Not-determined |
| 18-105_k141_91724 | Not-determined |
| 18-105_k141_3210 | Not-determined |
| 18-105_k141_82331 | Low-quality |
| 18-105_k141_145236 | Not-determined |
| 18-105_k141_109032 | Not-determined |
| 18-105_k141_37935 | Not-determined |
| 18-105_k141_198402 | Not-determined |
| 18-105_k141_100759 | Not-determined |
| 18-105_k141_29054 | Not-determined |
| 18-105_k141_198411 | Not-determined |
| 18-105_k141_198412 | Not-determined |
| 18-105_k141_198420 | Not-determined |
| 18-105_k141_3234 | Not-determined |
| 18-105_k141_127303 | Not-determined |
| 18-105_k141_91748 | Not-determined |
| 18-105_k141_11277 | Not-determined |
| 18-105_k141_135931 | Low-quality |
| 18-105_k141_135935 | Not-determined |
| 18-105_k141_29073 | Not-determined |
| 18-105_k141_47226 | Not-determined |
| 18-105_k141_29074 | Not-determined |
| 18-105_k141_3245 | Not-determined |
| 18-105_k141_73797 | Not-determined |
| 18-105_k141_127324 | Not-determined |
| 18-105_k141_19771 | Not-determined |
| 18-105_k141_181028 | Not-determined |
| 18-105_k141_91774 | Not-determined |
| 18-105_k141_109081 | Not-determined |
| 18-105_k141_172369 | Not-determined |
| 18-105_k141_127333 | Not-determined |
| 18-105_k141_135957 | Not-determined |
| 18-105_k141_29090 | Low-quality |
| 18-105_k141_135964 | Not-determined |
| 18-105_k141_135968 | Not-determined |
| 18-105_k141_11314 | Not-determined |
| 18-105_k141_19795 | Not-determined |
| 18-105_k141_109103 | Not-determined |
| 18-105_k141_91798 | Not-determined |
| 18-105_k141_29098 | Not-determined |
| 18-105_k141_73801 | Not-determined |
| 18-105_k141_3284 | Not-determined |
| 18-105_k141_189791 | Not-determined |
| 18-105_k141_29103 | Not-determined |
| 18-105_k141_198438 | Low-quality |
| 18-105_k141_135982 | Not-determined |
| 18-105_k141_55703 | Not-determined |
| 18-105_k141_73814 | Not-determined |
| 18-105_k141_19808 | Not-determined |
| 18-105_k141_55705 | Not-determined |
| 18-105_k141_135991 | Low-quality |
| 18-105_k141_47273 | Not-determined |
| 18-105_k141_100797 | Not-determined |
| 18-105_k141_198449 | Not-determined |
| 18-105_k141_55715 | Not-determined |
| 18-105_k141_135995 | Not-determined |
| 18-105_k141_100801 | Not-determined |
| 18-105_k141_29116 | Not-determined |
| 18-105_k141_207245 | Not-determined |
| 18-105_k141_127378 | Not-determined |
| 18-105_k141_91823 | Not-determined |
| 18-105_k141_82378 | Not-determined |
| 18-105_k141_3313 | Not-determined |
| 18-105_k141_3314 | Not-determined |
| 18-105_k141_136006 | Not-determined |
| 18-105_k141_172386 | Not-determined |
| 18-105_k141_127386 | Not-determined |
| 18-105_k141_207253 | Not-determined |
| 18-105_k141_136009 | Not-determined |
| 18-105_k141_136011 | Low-quality |
| 18-105_k141_91835 | Not-determined |
| 18-105_k141_136013 | Low-quality |
| 18-105_k141_136014 | Not-determined |
| 18-105_k141_3337 | Not-determined |
| 18-105_k141_189838 | Not-determined |
| 18-105_k141_207263 | Not-determined |
| 18-105_k141_3339 | Not-determined |
| 18-105_k141_172393 | Not-determined |
| 18-105_k141_82399 | Not-determined |
| 18-105_k141_136027 | Low-quality |
| 18-105_k141_82401 | Not-determined |
| 18-105_k141_47306 | Not-determined |
| 18-105_k141_37937 | Not-determined |
| 18-105_k141_189843 | Not-determined |
| 18-105_k141_82404 | Not-determined |
| 18-105_k141_100825 | Not-determined |
| 18-105_k141_100827 | Not-determined |
| 18-105_k141_207274 | Not-determined |
| 18-105_k141_189845 | Not-determined |
| 18-105_k141_136034 | Low-quality |
| 18-105_k141_55721 | Not-determined |
| 18-105_k141_47318 | Not-determined |
| 18-105_k141_3355 | Not-determined |
| 18-105_k141_189852 | Low-quality |
| 18-105_k141_73821 | Not-determined |
| 18-105_k141_37954 | Not-determined |
| 18-105_k141_82415 | Not-determined |
| 18-105_k141_73826 | Not-determined |
| 18-105_k141_118174 | Not-determined |
| 18-105_k141_153843 | Low-quality |
| 18-105_k141_37959 | Not-determined |
| 18-105_k141_3378 | Not-determined |
| 18-105_k141_181067 | Not-determined |
| 18-105_k141_172405 | Low-quality |
| 18-105_k141_64689 | Not-determined |
| 18-105_k141_64691 | Not-determined |
| 18-105_k141_172425 | Not-determined |
| 18-105_k141_198471 | Low-quality |
| 18-105_k141_91856 | Not-determined |
| 18-105_k141_91857 | Not-determined |
| 18-105_k141_73841 | Not-determined |
| 18-105_k141_91868 | Low-quality |
| 18-105_k141_127410 | Not-determined |
| 18-105_k141_47355 | Not-determined |
| 18-105_k141_37972 | Low-quality |
| 18-105_k141_47360 | Not-determined |
| 18-105_k141_47361 | Not-determined |
| 18-105_k141_73850 | Not-determined |
| 18-105_k141_172438 | Not-determined |
| 18-105_k141_118212 | Not-determined |
| 18-105_k141_100843 | Not-determined |
| 18-105_k141_189893 | Not-determined |
| 18-105_k141_37994 | Not-determined |
| 18-105_k141_127430 | Not-determined |
| 18-105_k141_145264 | Low-quality |
| 18-105_k141_82453 | Low-quality |
| 18-105_k141_82455 | Not-determined |
| 18-105_k141_38019 | Not-determined |
| 18-105_k141_172465 | Not-determined |
| 18-105_k141_47392 | Not-determined |
| 18-105_k141_127434 | Not-determined |
| 18-105_k141_172471 | Not-determined |
| 18-105_k141_55755 | Not-determined |
| 18-105_k141_136080 | Not-determined |
| 18-105_k141_172479 | Not-determined |
| 18-105_k141_29142 | Not-determined |
| 18-105_k141_136092 | Not-determined |
| 18-105_k141_136098 | Low-quality |
| 18-105_k141_198500 | Not-determined |
| 18-105_k141_145301 | Not-determined |
| 18-105_k141_3395 | Not-determined |
| 18-105_k141_55793 | Not-determined |
| 18-105_k141_136118 | Not-determined |
| 18-105_k141_136120 | Not-determined |
| 18-105_k141_145307 | Not-determined |
| 18-105_k141_47421 | Not-determined |
| 18-105_k141_73873 | Not-determined |
| 18-105_k141_136127 | Not-determined |
| 18-105_k141_145314 | Not-determined |
| 18-105_k141_100877 | Not-determined |
| 18-105_k141_136134 | Not-determined |
| 18-105_k141_127452 | Not-determined |
| 18-105_k141_127453 | Not-determined |
| 18-105_k141_55810 | Not-determined |
| 18-105_k141_181108 | Not-determined |
| 18-105_k141_127459 | Not-determined |
| 18-105_k141_29170 | Not-determined |
| 18-105_k141_73894 | Low-quality |
| 18-105_k141_3420 | Low-quality |
| 18-105_k141_127473 | Not-determined |
| 18-105_k141_47444 | Not-determined |
| 18-105_k141_47449 | Low-quality |
| 18-105_k141_47450 | Not-determined |
| 18-105_k141_127478 | Not-determined |
| 18-105_k141_136160 | Not-determined |
| 18-105_k141_29197 | Not-determined |
| 18-105_k141_29200 | Not-determined |
| 18-105_k141_172537 | Not-determined |
| 18-105_k141_38069 | Not-determined |
| 18-105_k141_145350 | Not-determined |
| 18-105_k141_127489 | Not-determined |
| 18-105_k141_162721 | Not-determined |
| 18-105_k141_29207 | Not-determined |
| 18-105_k141_91916 | Not-determined |
| 18-105_k141_38078 | Not-determined |
| 18-105_k141_91920 | Not-determined |
| 18-105_k141_73914 | Not-determined |
| 18-105_k141_145360 | Not-determined |
| 18-105_k141_145361 | Not-determined |
| 18-105_k141_127494 | Not-determined |
| 18-105_k141_91922 | Not-determined |
| 18-105_k141_153899 | Not-determined |
| 18-105_k141_145366 | Not-determined |
| 18-105_k141_153905 | Not-determined |
| 18-105_k141_181143 | Not-determined |
| 18-105_k141_3450 | Low-quality |
| 18-105_k141_3455 | Not-determined |
| 18-105_k141_153909 | Not-determined |
| 18-105_k141_189938 | Not-determined |
| 18-105_k141_136181 | Not-determined |
| 18-105_k141_73923 | Not-determined |
| 18-105_k141_136184 | Not-determined |
| 18-105_k141_55828 | Not-determined |
| 18-105_k141_153920 | Not-determined |
| 18-105_k141_55834 | Not-determined |
| 18-105_k141_3464 | Not-determined |
| 18-105_k141_29213 | Not-determined |
| 18-105_k141_19967 | Not-determined |
| 18-105_k141_172540 | Not-determined |
| 18-105_k141_127526 | Not-determined |
| 18-105_k141_47479 | Not-determined |
| 18-105_k141_127527 | Not-determined |
| 18-105_k141_64709 | Not-determined |
| 18-105_k141_145395 | Not-determined |
| 18-105_k141_64710 | Not-determined |
| 18-105_k141_73925 | Not-determined |
| 18-105_k141_127535 | Not-determined |
| 18-105_k141_127537 | Not-determined |
| 18-105_k141_189965 | Not-determined |
| 18-105_k141_73927 | Not-determined |
| 18-105_k141_198564 | Not-determined |
| 18-105_k141_64715 | Not-determined |
| 18-105_k141_127546 | Not-determined |
| 18-105_k141_29231 | Not-determined |
| 18-105_k141_127552 | Not-determined |
| 18-105_k141_127555 | Not-determined |
| 18-105_k141_198581 | Not-determined |
| 18-105_k141_198582 | Not-determined |
| 18-105_k141_198592 | Not-determined |
| 18-105_k141_127561 | Not-determined |
| 18-105_k141_198604 | Not-determined |
| 18-105_k141_19979 | Not-determined |
| 18-105_k141_64740 | Not-determined |
| 18-105_k141_64741 | Not-determined |
| 18-105_k141_127573 | Not-determined |
| 18-105_k141_172576 | Not-determined |
| 18-105_k141_162751 | Not-determined |
| 18-105_k141_198608 | Not-determined |
| 18-105_k141_145410 | Not-determined |
| 18-105_k141_19992 | Not-determined |
| 18-105_k141_172581 | Not-determined |
| 18-105_k141_181182 | Not-determined |
| 18-105_k141_145412 | Not-determined |
| 18-105_k141_55868 | Not-determined |
| 18-105_k141_100889 | Not-determined |
| 18-105_k141_172590 | Not-determined |
| 18-105_k141_172592 | Not-determined |
| 18-105_k141_82480 | Not-determined |
| 18-105_k141_181191 | Not-determined |
| 18-105_k141_172600 | Not-determined |
| 18-105_k141_82483 | Not-determined |
| 18-105_k141_162780 | Not-determined |
| 18-105_k141_172607 | Not-determined |
| 18-105_k141_127591 | Not-determined |
| 18-105_k141_172608 | Not-determined |
| 18-105_k141_64768 | Not-determined |
| 18-105_k141_118227 | Not-determined |
| 18-105_k141_118228 | Not-determined |
| 18-105_k141_82493 | Not-determined |
| 18-105_k141_118233 | Not-determined |
| 18-105_k141_47513 | Not-determined |
| 18-105_k141_47516 | Not-determined |
| 18-105_k141_127605 | Not-determined |
| 18-105_k141_55892 | Not-determined |
| 18-105_k141_162803 | Not-determined |
| 18-105_k141_181213 | Not-determined |
| 18-105_k141_181214 | Not-determined |
| 18-105_k141_127609 | Not-determined |
| 18-105_k141_91930 | Not-determined |
| 18-105_k141_47524 | Not-determined |
| 18-105_k141_136275 | Not-determined |
| 18-105_k141_64781 | Low-quality |
| 18-105_k141_162810 | Not-determined |
| 18-105_k141_162813 | Not-determined |
| 18-105_k141_55905 | Not-determined |
| 18-105_k141_162818 | Not-determined |
| 18-105_k141_47533 | Not-determined |
| 18-105_k141_11331 | Not-determined |
| 18-105_k141_47535 | Not-determined |
| 18-105_k141_153961 | Not-determined |
| 18-105_k141_153962 | Low-quality |
| 18-105_k141_11337 | Not-determined |
| 18-105_k141_11339 | Not-determined |
| 18-105_k141_47545 | Not-determined |
| 18-105_k141_162829 | Not-determined |
| 18-105_k141_127618 | Not-determined |
| 18-105_k141_153971 | Not-determined |
| 18-105_k141_73938 | Not-determined |
| 18-105_k141_127627 | Low-quality |
| 18-105_k141_11348 | Not-determined |
| 18-105_k141_55919 | Not-determined |
| 18-105_k141_64799 | Low-quality |
| 18-105_k141_136307 | Not-determined |
| 18-105_k141_109142 | Not-determined |
| 18-105_k141_82517 | Not-determined |
| 18-105_k141_118248 | Not-determined |
| 18-105_k141_127638 | Low-quality |
| 18-105_k141_82518 | Not-determined |
| 18-105_k141_64805 | Low-quality |
| 18-105_k141_109144 | Not-determined |
| 18-105_k141_198621 | Low-quality |
| 18-105_k141_198624 | Not-determined |
| 18-105_k141_55933 | Low-quality |
| 18-105_k141_38096 | Not-determined |
| 18-105_k141_91940 | Not-determined |
| 18-105_k141_82530 | Not-determined |
| 18-105_k141_162851 | Not-determined |
| 18-105_k141_55940 | Low-quality |
| 18-105_k141_55941 | Not-determined |
| 18-105_k141_11365 | Not-determined |
| 18-105_k141_38100 | Not-determined |
| 18-105_k141_64815 | Not-determined |
| 18-105_k141_118263 | Not-determined |
| 18-105_k141_118265 | Not-determined |
| 18-105_k141_11372 | Not-determined |
| 18-105_k141_162860 | Not-determined |
| 18-105_k141_162861 | Not-determined |
| 18-105_k141_118268 | Low-quality |
| 18-105_k141_47579 | Not-determined |
| 18-105_k141_47587 | Not-determined |
| 18-105_k141_118272 | Low-quality |
| 18-105_k141_91963 | Not-determined |
| 18-105_k141_47595 | Low-quality |
| 18-105_k141_118283 | Low-quality |
| 18-105_k141_118289 | Not-determined |
| 18-105_k141_109178 | Not-determined |
| 18-105_k141_73972 | Not-determined |
| 18-105_k141_3553 | Not-determined |
| 18-105_k141_47611 | Not-determined |
| 18-105_k141_3561 | Not-determined |
| 18-105_k141_55967 | Not-determined |
| 18-105_k141_162901 | Not-determined |
| 18-105_k141_207348 | Not-determined |
| 18-105_k141_38132 | Not-determined |
| 18-105_k141_64880 | Low-quality |
| 18-105_k141_47642 | Low-quality |
| 18-105_k141_118312 | Not-determined |
| 18-105_k141_3582 | Not-determined |
| 18-105_k141_154028 | Low-quality |
| 18-105_k141_162922 | Not-determined |
| 18-105_k141_118320 | Not-determined |
| 18-105_k141_145429 | Not-determined |
| 18-105_k141_162927 | Not-determined |
| 18-105_k141_64889 | Not-determined |
| 18-105_k141_47658 | Not-determined |
| 18-105_k141_118324 | Not-determined |
| 18-105_k141_92007 | Low-quality |
| 18-105_k141_127668 | Not-determined |
| 18-105_k141_118330 | Not-determined |
| 18-105_k141_3601 | Not-determined |
| 18-105_k141_118333 | Not-determined |
| 18-105_k141_118339 | Low-quality |
| 18-105_k141_118341 | Not-determined |
| 18-105_k141_47667 | Low-quality |
| 18-105_k141_73998 | Not-determined |
| 18-105_k141_207380 | Not-determined |
| 18-105_k141_118358 | Not-determined |
| 18-105_k141_172681 | Not-determined |
| 18-105_k141_82543 | Not-determined |
| 18-105_k141_74002 | Low-quality |
| 18-105_k141_38170 | Not-determined |
| 18-105_k141_118364 | Not-determined |
| 18-105_k141_100917 | Not-determined |
| 18-105_k141_74021 | Low-quality |
| 18-105_k141_127688 | Not-determined |
| 18-105_k141_47689 | Not-determined |
| 18-105_k141_127697 | Not-determined |
| 18-105_k141_145453 | Not-determined |
| 18-105_k141_74050 | Not-determined |
| 18-105_k141_74051 | Not-determined |
| 18-105_k141_190029 | Not-determined |
| 18-105_k141_74057 | Not-determined |
| 18-105_k141_172706 | Not-determined |
| 18-105_k141_145472 | Not-determined |
| 18-105_k141_82550 | Not-determined |
| 18-105_k141_190038 | Not-determined |
| 18-105_k141_38181 | Not-determined |
| 18-105_k141_207405 | Not-determined |
| 18-105_k141_127710 | Not-determined |
| 18-105_k141_82561 | Not-determined |
| 18-105_k141_190042 | Not-determined |
| 18-105_k141_190045 | Not-determined |
| 18-105_k141_47720 | Not-determined |
| 18-105_k141_127718 | Low-quality |
| 18-105_k141_154081 | Not-determined |
| 18-105_k141_198649 | Not-determined |
| 18-105_k141_207435 | Not-determined |
| 18-105_k141_38214 | Not-determined |
| 18-105_k141_207441 | Low-quality |
| 18-105_k141_190091 | Not-determined |
| 18-105_k141_11428 | Not-determined |
| 18-105_k141_207449 | Not-determined |
| 18-105_k141_47736 | Not-determined |
| 18-105_k141_190104 | Not-determined |
| 18-105_k141_47739 | Not-determined |
| 18-105_k141_74073 | Not-determined |
| 18-105_k141_172741 | Not-determined |
| 18-105_k141_64931 | Not-determined |
| 18-105_k141_11438 | Not-determined |
| 18-105_k141_64939 | Not-determined |
| 18-105_k141_92059 | Not-determined |
| 18-105_k141_172745 | Not-determined |
| 18-105_k141_92062 | Not-determined |
| 18-105_k141_207475 | Not-determined |
| 18-105_k141_100978 | Not-determined |
| 18-105_k141_3632 | Not-determined |
| 18-105_k141_3633 | Not-determined |
| 18-105_k141_64949 | Low-quality |
| 18-105_k141_92072 | Not-determined |
| 18-105_k141_92076 | Not-determined |
| 18-105_k141_145485 | Not-determined |
| 18-105_k141_109194 | Not-determined |
| 18-105_k141_11442 | Not-determined |
| 18-105_k141_109197 | Not-determined |
| 18-105_k141_109198 | Not-determined |
| 18-105_k141_145487 | Not-determined |
| 18-105_k141_3648 | Not-determined |
| 18-105_k141_207490 | Not-determined |
| 18-105_k141_3650 | Not-determined |
| 18-105_k141_145493 | Not-determined |
| 18-105_k141_198674 | Not-determined |
| 18-105_k141_109209 | Not-determined |
| 18-105_k141_163008 | Not-determined |
| 18-105_k141_11467 | Not-determined |
| 18-105_k141_207498 | Not-determined |
| 18-105_k141_11479 | Not-determined |
| 18-105_k141_47760 | Not-determined |
| 18-105_k141_38273 | Not-determined |
| 18-105_k141_127749 | Not-determined |
| 18-105_k141_127751 | Not-determined |
| 18-105_k141_3677 | Not-determined |
| 18-105_k141_3678 | Not-determined |
| 18-105_k141_82632 | Not-determined |
| 18-105_k141_127775 | Not-determined |
| 18-105_k141_20015 | Not-determined |
| 18-105_k141_145537 | Low-quality |
| 18-105_k141_74083 | Not-determined |
| 18-105_k141_47781 | Not-determined |
| 18-105_k141_145543 | Not-determined |
| 18-105_k141_47785 | Not-determined |
| 18-105_k141_74091 | Not-determined |
| 18-105_k141_3683 | Not-determined |
| 18-105_k141_47795 | Not-determined |
| 18-105_k141_29262 | Not-determined |
| 18-105_k141_64961 | Not-determined |
| 18-105_k141_64962 | Not-determined |
| 18-105_k141_92119 | Not-determined |
| 18-105_k141_145562 | Not-determined |
| 18-105_k141_127798 | Not-determined |
| 18-105_k141_92124 | Not-determined |
| 18-105_k141_127800 | Not-determined |
| 18-105_k141_145565 | Not-determined |
| 18-105_k141_3697 | Not-determined |
| 18-105_k141_92126 | Not-determined |
| 18-105_k141_127804 | Not-determined |
| 18-105_k141_127805 | Not-determined |
| 18-105_k141_3700 | Not-determined |
| 18-105_k141_29271 | Low-quality |
| 18-105_k141_181273 | Low-quality |
| 18-105_k141_190149 | Not-determined |
| 18-105_k141_172761 | Not-determined |
| 18-105_k141_64979 | Not-determined |
| 18-105_k141_92133 | Not-determined |
| 18-105_k141_92135 | Not-determined |
| 18-105_k141_190155 | Not-determined |
| 18-105_k141_47815 | Not-determined |
| 18-105_k141_38295 | Not-determined |
| 18-105_k141_74110 | Not-determined |
| 18-105_k141_145582 | Not-determined |
| 18-105_k141_172784 | Not-determined |
| 18-105_k141_181286 | Not-determined |
| 18-105_k141_47827 | Not-determined |
| 18-105_k141_207565 | Not-determined |
| 18-105_k141_109236 | Not-determined |
| 18-105_k141_190167 | Not-determined |
| 18-105_k141_181291 | Not-determined |
| 18-105_k141_20035 | Not-determined |
| 18-105_k141_38300 | Not-determined |
| 18-105_k141_207568 | Not-determined |
| 18-105_k141_207569 | Not-determined |
| 18-105_k141_38302 | Not-determined |
| 18-105_k141_207570 | Low-quality |
| 18-105_k141_65013 | Not-determined |
| 18-105_k141_65014 | Not-determined |
| 18-105_k141_47829 | Not-determined |
| 18-105_k141_145597 | Not-determined |
| 18-105_k141_207580 | Not-determined |
| 18-105_k141_181296 | Not-determined |
| 18-105_k141_47834 | Not-determined |
| 18-105_k141_65023 | Not-determined |
| 18-105_k141_20053 | Not-determined |
| 18-105_k141_190187 | Low-quality |
| 18-105_k141_20056 | Not-determined |
| 18-105_k141_11502 | Not-determined |
| 18-105_k141_20066 | Not-determined |
| 18-105_k141_47853 | Not-determined |
| 18-105_k141_11512 | Not-determined |
| 18-105_k141_11515 | Not-determined |
| 18-105_k141_74139 | Low-quality |
| 18-105_k141_163060 | Not-determined |
| 18-105_k141_190208 | Not-determined |
| 18-105_k141_20079 | Not-determined |
| 18-105_k141_20081 | Not-determined |
| 18-105_k141_109256 | Not-determined |
| 18-105_k141_11524 | Not-determined |
| 18-105_k141_172793 | Not-determined |
| 18-105_k141_109261 | Low-quality |
| 18-105_k141_11531 | Low-quality |
| 18-105_k141_74168 | Not-determined |
| 18-105_k141_181342 | Low-quality |
| 18-105_k141_198695 | Not-determined |
| 18-105_k141_11534 | Not-determined |
| 18-105_k141_198700 | Not-determined |
| 18-105_k141_163088 | Not-determined |
| 18-105_k141_145644 | Not-determined |
| 18-105_k141_198705 | Not-determined |
| 18-105_k141_172812 | Not-determined |
| 18-105_k141_198712 | Not-determined |
| 18-105_k141_198713 | Not-determined |
| 18-105_k141_3776 | Not-determined |
| 18-105_k141_109284 | Not-determined |
| 18-105_k141_11550 | Not-determined |
| 18-105_k141_181352 | Not-determined |
| 18-105_k141_181353 | Not-determined |
| 18-105_k141_181355 | Not-determined |
| 18-105_k141_3784 | Not-determined |
| 18-105_k141_20091 | Not-determined |
| 18-105_k141_11557 | Not-determined |
| 18-105_k141_65063 | Low-quality |
| 18-105_k141_29281 | Not-determined |
| 18-105_k141_163102 | Not-determined |
| 18-105_k141_163103 | Not-determined |
| 18-105_k141_163106 | Not-determined |
| 18-105_k141_100999 | Not-determined |
| 18-105_k141_109300 | Not-determined |
| 18-105_k141_11566 | Not-determined |
| 18-105_k141_56009 | Not-determined |
| 18-105_k141_101006 | Not-determined |
| 18-105_k141_3797 | Not-determined |
| 18-105_k141_11572 | Not-determined |
| 18-105_k141_65079 | Not-determined |
| 18-105_k141_74189 | Not-determined |
| 18-105_k141_163115 | Not-determined |
| 18-105_k141_3805 | Not-determined |
| 18-105_k141_74195 | Not-determined |
| 18-105_k141_3812 | Not-determined |
| 18-105_k141_3816 | Not-determined |
| 18-105_k141_29294 | Not-determined |
| 18-105_k141_190272 | Not-determined |
| 18-105_k141_3827 | Not-determined |
| 18-105_k141_181390 | Not-determined |
| 18-105_k141_74201 | Low-quality |
| 18-105_k141_190282 | Not-determined |
| 18-105_k141_101033 | Not-determined |
| 18-105_k141_56028 | Not-determined |
| 18-105_k141_56029 | Not-determined |
| 18-105_k141_56030 | Not-determined |

| | |
|---|---|
| 18-105_k141_82664 | Not-determined |
| 18-105_k141_56031 | Low-quality |
| 18-105_k141_82668 | Not-determined |
| 18-105_k141_3841 | Not-determined |
| 18-105_k141_154123 | Not-determined |
| 18-105_k141_20106 | Not-determined |
| 18-105_k141_163155 | Not-determined |
| 18-105_k141_3848 | Low-quality |
| 18-105_k141_82674 | Not-determined |
| 18-105_k141_56050 | Not-determined |
| 18-105_k141_82681 | Not-determined |
| 18-105_k141_3857 | Not-determined |
| 18-105_k141_101052 | Not-determined |
| 18-105_k141_181416 | Not-determined |
| 18-105_k141_65090 | Not-determined |
| 18-105_k141_29312 | Not-determined |
| 18-105_k141_20118 | Not-determined |
| 18-105_k141_29316 | Not-determined |
| 18-105_k141_20122 | Not-determined |
| 18-105_k141_101064 | Not-determined |
| 18-105_k141_65102 | Not-determined |
| 18-105_k141_145704 | Not-determined |
| 18-105_k141_20127 | Not-determined |
| 18-105_k141_29323 | Not-determined |
| 18-105_k141_181426 | Not-determined |
| 18-105_k141_127885 | Not-determined |
| 18-105_k141_145718 | Not-determined |
| 18-105_k141_145719 | Not-determined |
| 18-105_k141_154140 | Not-determined |
| 18-105_k141_47902 | Not-determined |
| 18-105_k141_127895 | Not-determined |
| 18-105_k141_101075 | Not-determined |
| 18-105_k141_20136 | Not-determined |
| 18-105_k141_101076 | Not-determined |
| 18-105_k141_207647 | Low-quality |
| 18-105_k141_207651 | Not-determined |
| 18-105_k141_82706 | Not-determined |
| 18-105_k141_101085 | Not-determined |
| 18-105_k141_65119 | Not-determined |
| 18-105_k141_101086 | Not-determined |
| 18-105_k141_154150 | Not-determined |
| 18-105_k141_207656 | Not-determined |
| 18-105_k141_65130 | Not-determined |
| 18-105_k141_109353 | Not-determined |
| 18-105_k141_109355 | Not-determined |
| 18-105_k141_101100 | Not-determined |
| 18-105_k141_3909 | Not-determined |
| 18-105_k141_101105 | Not-determined |
| 18-105_k141_127908 | Not-determined |
| 18-105_k141_154164 | Not-determined |
| 18-105_k141_127911 | Not-determined |
| 18-105_k141_127912 | Low-quality |
| 18-105_k141_3925 | Not-determined |
| 18-105_k141_207690 | Not-determined |
| 18-105_k141_127923 | Not-determined |
| 18-105_k141_190309 | Low-quality |
| 18-105_k141_65147 | Not-determined |
| 18-105_k141_154186 | Not-determined |
| 18-105_k141_127936 | Not-determined |
| 18-105_k141_207704 | Not-determined |
| 18-105_k141_181498 | Not-determined |
| 18-105_k141_181500 | Not-determined |
| 18-105_k141_127946 | Not-determined |
| 18-105_k141_154203 | Not-determined |
| 18-105_k141_47963 | Low-quality |
| 18-105_k141_181505 | Not-determined |
| 18-105_k141_181508 | Not-determined |
| 18-105_k141_47969 | Not-determined |
| 18-105_k141_101125 | Not-determined |
| 18-105_k141_101130 | Not-determined |
| 18-105_k141_109390 | Low-quality |
| 18-105_k141_127970 | Not-determined |
| 18-105_k141_127976 | Not-determined |
| 18-105_k141_118438 | Low-quality |
| 18-105_k141_109402 | Not-determined |
| 18-105_k141_145775 | Not-determined |
| 18-105_k141_172845 | Not-determined |
| 18-105_k141_48000 | Not-determined |
| 18-105_k141_127991 | Not-determined |
| 18-105_k141_48007 | Low-quality |
| 18-105_k141_92183 | Not-determined |
| 18-105_k141_82722 | Not-determined |
| 18-105_k141_48011 | Not-determined |
| 18-105_k141_190334 | Not-determined |
| 18-105_k141_48014 | Not-determined |
| 18-105_k141_38313 | Not-determined |
| 18-105_k141_163196 | Not-determined |
| 18-105_k141_38319 | Not-determined |
| 18-105_k141_163198 | Not-determined |
| 18-105_k141_38320 | Low-quality |
| 18-105_k141_154238 | Low-quality |
| 18-105_k141_118472 | Not-determined |
| 18-105_k141_101151 | Not-determined |
| 18-105_k141_145800 | Not-determined |
| 18-105_k141_207751 | Not-determined |
| 18-105_k141_163206 | Low-quality |
| 18-105_k141_118474 | Not-determined |
| 18-105_k141_181532 | Not-determined |
| 18-105_k141_181534 | Not-determined |
| 18-105_k141_38334 | Not-determined |
| 18-105_k141_154250 | Not-determined |
| 18-105_k141_163213 | Not-determined |
| 18-105_k141_127998 | Not-determined |
| 18-105_k141_101161 | Not-determined |
| 18-105_k141_190349 | Not-determined |
| 18-105_k141_128001 | Not-determined |
| 18-105_k141_181543 | Not-determined |
| 18-105_k141_29332 | Not-determined |
| 18-105_k141_29333 | Not-determined |
| 18-105_k141_118492 | Not-determined |
| 18-105_k141_11625 | Not-determined |
| 18-105_k141_3973 | Not-determined |
| 18-105_k141_48052 | Not-determined |
| 18-105_k141_48056 | Not-determined |
| 18-105_k141_92213 | Not-determined |
| 18-105_k141_190372 | Low-quality |
| 18-105_k141_29348 | Not-determined |
| 18-105_k141_74228 | Not-determined |
| 18-105_k141_109475 | Not-determined |
| 18-105_k141_163221 | Not-determined |
| 18-105_k141_48074 | Not-determined |
| 18-105_k141_128016 | Not-determined |
| 18-105_k141_109482 | Low-quality |
| 18-105_k141_118520 | Not-determined |
| 18-105_k141_74245 | Not-determined |
| 18-105_k141_145841 | Low-quality |
| 18-105_k141_145844 | Not-determined |
| 18-105_k141_128027 | Not-determined |
| 18-105_k141_101213 | Not-determined |
| 18-105_k141_74257 | Not-determined |
| 18-105_k141_4012 | Not-determined |
| 18-105_k141_145848 | Not-determined |
| 18-105_k141_109495 | Not-determined |
| 18-105_k141_29349 | Low-quality |
| 18-105_k141_190407 | Not-determined |
| 18-105_k141_29354 | Not-determined |
| 18-105_k141_101222 | Not-determined |
| 18-105_k141_128055 | Not-determined |
| 18-105_k141_56089 | Low-quality |
| 18-105_k141_163241 | Not-determined |
| 18-105_k141_82790 | Not-determined |
| 18-105_k141_145865 | Not-determined |
| 18-105_k141_128069 | Not-determined |
| 18-105_k141_82797 | Not-determined |
| 18-105_k141_38396 | Not-determined |
| 18-105_k141_109528 | Low-quality |
| 18-105_k141_38407 | Low-quality |
| 18-105_k141_74270 | Not-determined |
| 18-105_k141_154277 | Low-quality |
| 18-105_k141_4038 | Not-determined |
| 18-105_k141_74275 | Not-determined |
| 18-105_k141_74283 | Not-determined |
| 18-105_k141_190430 | Not-determined |
| 18-105_k141_154284 | Not-determined |
| 18-105_k141_154287 | Not-determined |
| 18-105_k141_11664 | Not-determined |
| 18-105_k141_128091 | Low-quality |
| 18-105_k141_74293 | Low-quality |
| 18-105_k141_145884 | Not-determined |
| 18-105_k141_38422 | Not-determined |
| 18-105_k141_145888 | Not-determined |
| 18-105_k141_154295 | Not-determined |
| 18-105_k141_74299 | Not-determined |
| 18-105_k141_11674 | Low-quality |
| 18-105_k141_163271 | Not-determined |
| 18-105_k141_20194 | Not-determined |
| 18-105_k141_172857 | Not-determined |
| 18-105_k141_74302 | Not-determined |
| 18-105_k141_181576 | Not-determined |
| 18-105_k141_11682 | Low-quality |
| 18-105_k141_145898 | Low-quality |
| 18-105_k141_11684 | Not-determined |
| 18-105_k141_11686 | Low-quality |
| 18-105_k141_145901 | Not-determined |
| 18-105_k141_145902 | Not-determined |
| 18-105_k141_163291 | Not-determined |
| 18-105_k141_181584 | Not-determined |
| 18-105_k141_163292 | Not-determined |
| 18-105_k141_207768 | Not-determined |
| 18-105_k141_181588 | Not-determined |
| 18-105_k141_163306 | Not-determined |
| 18-105_k141_145922 | Not-determined |
| 18-105_k141_128126 | Not-determined |
| 18-105_k141_154321 | Low-quality |
| 18-105_k141_154325 | Not-determined |
| 18-105_k141_207783 | Not-determined |
| 18-105_k141_207784 | Not-determined |
| 18-105_k141_190434 | Not-determined |
| 18-105_k141_38431 | Not-determined |
| 18-105_k141_190444 | Not-determined |
| 18-105_k141_145938 | Not-determined |
| 18-105_k141_145945 | Not-determined |
| 18-105_k141_109574 | Not-determined |
| 18-105_k141_11713 | Not-determined |
| 18-105_k141_118549 | Not-determined |
| 18-105_k141_118551 | Not-determined |
| 18-105_k141_136370 | Not-determined |
| 18-105_k141_145950 | Low-quality |
| 18-105_k141_101269 | Not-determined |
| 18-105_k141_145952 | Not-determined |
| 18-105_k141_145956 | Not-determined |
| 18-105_k141_101281 | Not-determined |
| 18-105_k141_145957 | Not-determined |
| 18-105_k141_101283 | Not-determined |
| 18-105_k141_101284 | Not-determined |
| 18-105_k141_65162 | Not-determined |
| 18-105_k141_109578 | Not-determined |
| 18-105_k141_109580 | Not-determined |
| 18-105_k141_172876 | Not-determined |
| 18-105_k141_4068 | Not-determined |
| 18-105_k141_65175 | Not-determined |
| 18-105_k141_145979 | Low-quality |
| 18-105_k141_181595 | Not-determined |
| 18-105_k141_145983 | Not-determined |
| 18-105_k141_65187 | Not-determined |
| 18-105_k141_145991 | Not-determined |
| 18-105_k141_65196 | Not-determined |
| 18-105_k141_65201 | Low-quality |
| 18-105_k141_198788 | Not-determined |
| 18-105_k141_181611 | Not-determined |
| 18-105_k141_101309 | Not-determined |
| 18-105_k141_101314 | Not-determined |
| 18-105_k141_154365 | Low-quality |
| 18-105_k141_65230 | Not-determined |
| 18-105_k141_118578 | Not-determined |
| 18-105_k141_29420 | Not-determined |
| 18-105_k141_4139 | Low-quality |
| 18-105_k141_82850 | Not-determined |
| 18-105_k141_74349 | Low-quality |
| 18-105_k141_65251 | Not-determined |
| 18-105_k141_181659 | Not-determined |
| 18-105_k141_146025 | Not-determined |
| 18-105_k141_74357 | Low-quality |
| 18-105_k141_82866 | Not-determined |
| 18-105_k141_82868 | Not-determined |
| 18-105_k141_82869 | Not-determined |
| 18-105_k141_92237 | Not-determined |
| 18-105_k141_4167 | Not-determined |
| 18-105_k141_4168 | Low-quality |
| 18-105_k141_4169 | Low-quality |
| 18-105_k141_163334 | Not-determined |
| 18-105_k141_82885 | Not-determined |
| 18-105_k141_74382 | Not-determined |
| 18-105_k141_128222 | Not-determined |
| 18-105_k141_118592 | Not-determined |
| 18-105_k141_65276 | Not-determined |
| 18-105_k141_82890 | Low-quality |
| 18-105_k141_82892 | Not-determined |
| 18-105_k141_82896 | Not-determined |
| 18-105_k141_82898 | Not-determined |
| 18-105_k141_118604 | Low-quality |
| 18-105_k141_118605 | Not-determined |
| 18-105_k141_207801 | Not-determined |
| 18-105_k141_118616 | Not-determined |
| 18-105_k141_118618 | Low-quality |
| 18-105_k141_20267 | Not-determined |
| 18-105_k141_65296 | Not-determined |
| 18-105_k141_198865 | Not-determined |
| 18-105_k141_118631 | Not-determined |
| 18-105_k141_101328 | Not-determined |
| 18-105_k141_190458 | Not-determined |
| 18-105_k141_118634 | Not-determined |
| 18-105_k141_128261 | Not-determined |
| 18-105_k141_128264 | Not-determined |
| 18-105_k141_109651 | Low-quality |
| 18-105_k141_146047 | Not-determined |
| 18-105_k141_20281 | Not-determined |
| 18-105_k141_190466 | Not-determined |
| 18-105_k141_74406 | Not-determined |
| 18-105_k141_163369 | Not-determined |
| 18-105_k141_65299 | Not-determined |
| 18-105_k141_190467 | Low-quality |
| 18-105_k141_172902 | Not-determined |
| 18-105_k141_20290 | Not-determined |
| 18-105_k141_190473 | Not-determined |
| 18-105_k141_48117 | Not-determined |
| 18-105_k141_146063 | Not-determined |
| 18-105_k141_48120 | Not-determined |
| 18-105_k141_74428 | Not-determined |
| 18-105_k141_154413 | Not-determined |
| 18-105_k141_163386 | Not-determined |
| 18-105_k141_48124 | Not-determined |
| 18-105_k141_163388 | Not-determined |
| 18-105_k141_74434 | Not-determined |
| 18-105_k141_4201 | Low-quality |
| 18-105_k141_172918 | Not-determined |
| 18-105_k141_101334 | Not-determined |
| 18-105_k141_172920 | Not-determined |
| 18-105_k141_48134 | Not-determined |
| 18-105_k141_38446 | Not-determined |
| 18-105_k141_109671 | Not-determined |
| 18-105_k141_190479 | Not-determined |
| 18-105_k141_190482 | Not-determined |
| 18-105_k141_38456 | Not-determined |
| 18-105_k141_82918 | Not-determined |
| 18-105_k141_82922 | Not-determined |
| 18-105_k141_4217 | Not-determined |
| 18-105_k141_163426 | Not-determined |
| 18-105_k141_190492 | Not-determined |
| 18-105_k141_101354 | Not-determined |
| 18-105_k141_190495 | Not-determined |
| 18-105_k141_4222 | Not-determined |
| 18-105_k141_11751 | Not-determined |
| 18-105_k141_4225 | Not-determined |
| 18-105_k141_11755 | Not-determined |
| 18-105_k141_20334 | Not-determined |
| 18-105_k141_172945 | Not-determined |
| 18-105_k141_146090 | Low-quality |
| 18-105_k141_4232 | Not-determined |
| 18-105_k141_4235 | Low-quality |
| 18-105_k141_38483 | Not-determined |
| 18-105_k141_128296 | Not-determined |
| 18-105_k141_163449 | Not-determined |
| 18-105_k141_163453 | Not-determined |
| 18-105_k141_11768 | Not-determined |
| 18-105_k141_146101 | Not-determined |
| 18-105_k141_11770 | Not-determined |
| 18-105_k141_65337 | Not-determined |
| 18-105_k141_163461 | Not-determined |
| 18-105_k141_92249 | Not-determined |
| 18-105_k141_48173 | Not-determined |
| 18-105_k141_92252 | Not-determined |
| 18-105_k141_11775 | Not-determined |
| 18-105_k141_82925 | Not-determined |
| 18-105_k141_11780 | Not-determined |
| 18-105_k141_20357 | Not-determined |
| 18-105_k141_172975 | Not-determined |
| 18-105_k141_38498 | Not-determined |
| 18-105_k141_4260 | Not-determined |
| 18-105_k141_20361 | Not-determined |
| 18-105_k141_74464 | Not-determined |
| 18-105_k141_109689 | Not-determined |
| 18-105_k141_20363 | Not-determined |
| 18-105_k141_82927 | Not-determined |
| 18-105_k141_109693 | Not-determined |
| 18-105_k141_74470 | Not-determined |
| 18-105_k141_4266 | Not-determined |
| 18-105_k141_118665 | Low-quality |
| 18-105_k141_11789 | Not-determined |
| 18-105_k141_74473 | Not-determined |
| 18-105_k141_109697 | Not-determined |
| 18-105_k141_74477 | Not-determined |
| 18-105_k141_82939 | Not-determined |
| 18-105_k141_92275 | Not-determined |
| 18-105_k141_11801 | Not-determined |
| 18-105_k141_82951 | Not-determined |
| 18-105_k141_38509 | Not-determined |
| 18-105_k141_38511 | Not-determined |
| 18-105_k141_172988 | Not-determined |
| 18-105_k141_11816 | Not-determined |
| 18-105_k141_20379 | Not-determined |
| 18-105_k141_29456 | Not-determined |
| 18-105_k141_92307 | Not-determined |
| 18-105_k141_48190 | Not-determined |
| 18-105_k141_38519 | Not-determined |
| 18-105_k141_65381 | Not-determined |
| 18-105_k141_92313 | Not-determined |
| 18-105_k141_48197 | Not-determined |
| 18-105_k141_92315 | Not-determined |
| 18-105_k141_4296 | Not-determined |
| 18-105_k141_128303 | Not-determined |
| 18-105_k141_82987 | Low-quality |
| 18-105_k141_82988 | Not-determined |
| 18-105_k141_4299 | Not-determined |
| 18-105_k141_82989 | Low-quality |
| 18-105_k141_163485 | Not-determined |
| 18-105_k141_11832 | Not-determined |
| 18-105_k141_118707 | Not-determined |
| 18-105_k141_163489 | Not-determined |
| 18-105_k141_118710 | Not-determined |
| 18-105_k141_20385 | Not-determined |
| 18-105_k141_118715 | Not-determined |
| 18-105_k141_74522 | Low-quality |
| 18-105_k141_65395 | Not-determined |
| 18-105_k141_4301 | Not-determined |
| 18-105_k141_65403 | Not-determined |
| 18-105_k141_163497 | Not-determined |
| 18-105_k141_20402 | Not-determined |
| 18-105_k141_173020 | Low-quality |
| 18-105_k141_65414 | Not-determined |
| 18-105_k141_101394 | Not-determined |
| 18-105_k141_20404 | Not-determined |
| 18-105_k141_48225 | Not-determined |
| 18-105_k141_48230 | Not-determined |
| 18-105_k141_56153 | Low-quality |
| 18-105_k141_118749 | Not-determined |
| 18-105_k141_48235 | Not-determined |
| 18-105_k141_154437 | Not-determined |
| 18-105_k141_136451 | Not-determined |
| 18-105_k141_20417 | Not-determined |
| 18-105_k141_198955 | Not-determined |
| 18-105_k141_118762 | Not-determined |
| 18-105_k141_207821 | Not-determined |
| 18-105_k141_154442 | Not-determined |
| 18-105_k141_118769 | Not-determined |
| 18-105_k141_136469 | Not-determined |
| 18-105_k141_92346 | Low-quality |
| 18-105_k141_163518 | Not-determined |
| 18-105_k141_92355 | Not-determined |
| 18-105_k141_198975 | Not-determined |
| 18-105_k141_136476 | Not-determined |
| 18-105_k141_82996 | Not-determined |
| 18-105_k141_163524 | Not-determined |
| 18-105_k141_154457 | Not-determined |
| 18-105_k141_4339 | Not-determined |
| 18-105_k141_109726 | Not-determined |
| 18-105_k141_101439 | Not-determined |
| 18-105_k141_163529 | Not-determined |
| 18-105_k141_190528 | Not-determined |
| 18-105_k141_92373 | Not-determined |
| 18-105_k141_11864 | Not-determined |
| 18-105_k141_154470 | Not-determined |
| 18-105_k141_154471 | Not-determined |
| 18-105_k141_11867 | Not-determined |
| 18-105_k141_92380 | Not-determined |
| 18-105_k141_65425 | Not-determined |
| 18-105_k141_4346 | Not-determined |
| 18-105_k141_109732 | Not-determined |
| 18-105_k141_190534 | Low-quality |
| 18-105_k141_207849 | Not-determined |
| 18-105_k141_38573 | Not-determined |
| 18-105_k141_11884 | Not-determined |
| 18-105_k141_74584 | Not-determined |
| 18-105_k141_83030 | Not-determined |
| 18-105_k141_74588 | Not-determined |
| 18-105_k141_109740 | Not-determined |
| 18-105_k141_163550 | Low-quality |
| 18-105_k141_109746 | Not-determined |
| 18-105_k141_163552 | Not-determined |
| 18-105_k141_29485 | Not-determined |
| 18-105_k141_38576 | Low-quality |
| 18-105_k141_74597 | Low-quality |
| 18-105_k141_38581 | Not-determined |
| 18-105_k141_109757 | Low-quality |
| 18-105_k141_190560 | Not-determined |
| 18-105_k141_154493 | Not-determined |
| 18-105_k141_163558 | Not-determined |
| 18-105_k141_83048 | Not-determined |
| 18-105_k141_190566 | Not-determined |
| 18-105_k141_20461 | Not-determined |
| 18-105_k141_20462 | Not-determined |
| 18-105_k141_207879 | Not-determined |
| 18-105_k141_207880 | Not-determined |
| 18-105_k141_83059 | Not-determined |
| 18-105_k141_20467 | Not-determined |
| 18-105_k141_199022 | Not-determined |
| 18-105_k141_20474 | Not-determined |
| 18-105_k141_11907 | Not-determined |
| 18-105_k141_20476 | Not-determined |
| 18-105_k141_29502 | Not-determined |
| 18-105_k141_154522 | Not-determined |
| 18-105_k141_136516 | Low-quality |
| 18-105_k141_11912 | Low-quality |
| 18-105_k141_128369 | Not-determined |
| 18-105_k141_29511 | Not-determined |
| 18-105_k141_207894 | Not-determined |
| 18-105_k141_128371 | Not-determined |
| 18-105_k141_136527 | Not-determined |
| 18-105_k141_128374 | Not-determined |
| 18-105_k141_83081 | Not-determined |
| 18-105_k141_29515 | Not-determined |
| 18-105_k141_20490 | Not-determined |
| 18-105_k141_154536 | Not-determined |
| 18-105_k141_136532 | Not-determined |
| 18-105_k141_128378 | Not-determined |
| 18-105_k141_20504 | Not-determined |
| 18-105_k141_154546 | Not-determined |
| 18-105_k141_136540 | Not-determined |
| 18-105_k141_207926 | Not-determined |
| 18-105_k141_48290 | Not-determined |
| 18-105_k141_207932 | Not-determined |
| 18-105_k141_154552 | Low-quality |
| 18-105_k141_38625 | Not-determined |
| 18-105_k141_74658 | Not-determined |
| 18-105_k141_56212 | Not-determined |
| 18-105_k141_48304 | Not-determined |
| 18-105_k141_118796 | Not-determined |
| 18-105_k141_11939 | Low-quality |
| 18-105_k141_118808 | Not-determined |
| 18-105_k141_83118 | Not-determined |
| 18-105_k141_74683 | Not-determined |
| 18-105_k141_92412 | Not-determined |
| 18-105_k141_11946 | Not-determined |
| 18-105_k141_48322 | Not-determined |
| 18-105_k141_74688 | Not-determined |
| 18-105_k141_56227 | Not-determined |
| 18-105_k141_146154 | Not-determined |
| 18-105_k141_83128 | Not-determined |
| 18-105_k141_118824 | Not-determined |
| 18-105_k141_207967 | Not-determined |
| 18-105_k141_173058 | Not-determined |
| 18-105_k141_118829 | Not-determined |
| 18-105_k141_199035 | Not-determined |
| 18-105_k141_74696 | Not-determined |
| 18-105_k141_207978 | Not-determined |
| 18-105_k141_207979 | Not-determined |
| 18-105_k141_92424 | Not-determined |
| 18-105_k141_83148 | Low-quality |
| 18-105_k141_173079 | Not-determined |
| 18-105_k141_136587 | Not-determined |
| 18-105_k141_38647 | Not-determined |
| 18-105_k141_173086 | Not-determined |
| 18-105_k141_101496 | Low-quality |
| 18-105_k141_199047 | Not-determined |
| 18-105_k141_199048 | Not-determined |
| 18-105_k141_56240 | Not-determined |
| 18-105_k141_128413 | Not-determined |
| 18-105_k141_48344 | Not-determined |
| 18-105_k141_83153 | Not-determined |
| 18-105_k141_83154 | Low-quality |
| 18-105_k141_65445 | Not-determined |
| 18-105_k141_173095 | Not-determined |
| 18-105_k141_74719 | Not-determined |
| 18-105_k141_4454 | Not-determined |
| 18-105_k141_181682 | Not-determined |
| 18-105_k141_109792 | Not-determined |
| 18-105_k141_83165 | Not-determined |
| 18-105_k141_109793 | Not-determined |
| 18-105_k141_56254 | Not-determined |
| 18-105_k141_199059 | Low-quality |
| 18-105_k141_101509 | Not-determined |
| 18-105_k141_128420 | Not-determined |
| 18-105_k141_118840 | Low-quality |
| 18-105_k141_154595 | Not-determined |
| 18-105_k141_83174 | Not-determined |
| 18-105_k141_207991 | Not-determined |
| 18-105_k141_154597 | Not-determined |
| 18-105_k141_92449 | Not-determined |
| 18-105_k141_83182 | Not-determined |
| 18-105_k141_56263 | Not-determined |
| 18-105_k141_83186 | Low-quality |
| 18-105_k141_208001 | Not-determined |
| 18-105_k141_128442 | Not-determined |
| 18-105_k141_173105 | Not-determined |
| 18-105_k141_65466 | Not-determined |
| 18-105_k141_128445 | Not-determined |
| 18-105_k141_146166 | Not-determined |
| 18-105_k141_190590 | Low-quality |
| 18-105_k141_208008 | Not-determined |
| 18-105_k141_92456 | Not-determined |
| 18-105_k141_4472 | Not-determined |
| 18-105_k141_56270 | Not-determined |
| 18-105_k141_146174 | Not-determined |
| 18-105_k141_38662 | Not-determined |
| 18-105_k141_118849 | Not-determined |
| 18-105_k141_20513 | Not-determined |
| 18-105_k141_56275 | Not-determined |
| 18-105_k141_173122 | Not-determined |
| 18-105_k141_74745 | Not-determined |
| 18-105_k141_83208 | Not-determined |
| 18-105_k141_190607 | Not-determined |
| 18-105_k141_109828 | Not-determined |
| 18-105_k141_118860 | Not-determined |
| 18-105_k141_74754 | Not-determined |
| 18-105_k141_4480 | Low-quality |
| 18-105_k141_173129 | Not-determined |
| 18-105_k141_4481 | Low-quality |
| 18-105_k141_20525 | Not-determined |
| 18-105_k141_74763 | Low-quality |
| 18-105_k141_101524 | Not-determined |
| 18-105_k141_199104 | Not-determined |
| 18-105_k141_74773 | Not-determined |
| 18-105_k141_74774 | Not-determined |
| 18-105_k141_83229 | Not-determined |
| 18-105_k141_199108 | Not-determined |
| 18-105_k141_190624 | Not-determined |
| 18-105_k141_208039 | Not-determined |
| 18-105_k141_199110 | Not-determined |
| 18-105_k141_109844 | Not-determined |
| 18-105_k141_4500 | Not-determined |
| 18-105_k141_92488 | Not-determined |
| 18-105_k141_83247 | Not-determined |
| 18-105_k141_190634 | Not-determined |
| 18-105_k141_11996 | Not-determined |
| 18-105_k141_83250 | Not-determined |
| 18-105_k141_83251 | Not-determined |
| 18-105_k141_12002 | Not-determined |
| 18-105_k141_101533 | Not-determined |
| 18-105_k141_48363 | Not-determined |
| 18-105_k141_48364 | Not-determined |
| 18-105_k141_92496 | Not-determined |
| 18-105_k141_48368 | Not-determined |
| 18-105_k141_20540 | Low-quality |
| 18-105_k141_56303 | Not-determined |
| 18-105_k141_92511 | Not-determined |
| 18-105_k141_56307 | Not-determined |
| 18-105_k141_101553 | Not-determined |
| 18-105_k141_20548 | Not-determined |
| 18-105_k141_48388 | Not-determined |

| | |
|---|---|
| 18-105_k141_92514 | Not-determined |
| 18-105_k141_56315 | Not-determined |
| 18-105_k141_163585 | Not-determined |
| 18-105_k141_56318 | Not-determined |
| 18-105_k141_163590 | Not-determined |
| 18-105_k141_118894 | Not-determined |
| 18-105_k141_56327 | Not-determined |
| 18-105_k141_154634 | Not-determined |
| 18-105_k141_163598 | Not-determined |
| 18-105_k141_146189 | Low-quality |
| 18-105_k141_146190 | Not-determined |
| 18-105_k141_12043 | Not-determined |
| 18-105_k141_4542 | Not-determined |
| 18-105_k141_12048 | Not-determined |
| 18-105_k141_208072 | Not-determined |
| 18-105_k141_92526 | Not-determined |
| 18-105_k141_83297 | Not-determined |
| 18-105_k141_190650 | Not-determined |
| 18-105_k141_146204 | Not-determined |
| 18-105_k141_20578 | Not-determined |
| 18-105_k141_4545 | Not-determined |
| 18-105_k141_146209 | Not-determined |
| 18-105_k141_12058 | Not-determined |
| 18-105_k141_20586 | Not-determined |
| 18-105_k141_12062 | Not-determined |
| 18-105_k141_154654 | Not-determined |
| 18-105_k141_74802 | Low-quality |
| 18-105_k141_12073 | Not-determined |
| 18-105_k141_146218 | Not-determined |
| 18-105_k141_12076 | Not-determined |
| 18-105_k141_173187 | Not-determined |
| 18-105_k141_92540 | Not-determined |
| 18-105_k141_48406 | Not-determined |
| 18-105_k141_190667 | Not-determined |
| 18-105_k141_173190 | Not-determined |
| 18-105_k141_20600 | Not-determined |
| 18-105_k141_163626 | Not-determined |
| 18-105_k141_118931 | Not-determined |
| 18-105_k141_154666 | Not-determined |
| 18-105_k141_154667 | Not-determined |
| 18-105_k141_118933 | Not-determined |
| 18-105_k141_173197 | Not-determined |
| 18-105_k141_181702 | Not-determined |
| 18-105_k141_74813 | Not-determined |
| 18-105_k141_146230 | Not-determined |
| 18-105_k141_29590 | Not-determined |
| 18-105_k141_190672 | Not-determined |
| 18-105_k141_163636 | Not-determined |
| 18-105_k141_12085 | Not-determined |
| 18-105_k141_4555 | Low-quality |
| 18-105_k141_38724 | Not-determined |
| 18-105_k141_38725 | Low-quality |
| 18-105_k141_74821 | Not-determined |
| 18-105_k141_163642 | Not-determined |
| 18-105_k141_163648 | Not-determined |
| 18-105_k141_163651 | Not-determined |
| 18-105_k141_173201 | Not-determined |
| 18-105_k141_92557 | Not-determined |
| 18-105_k141_4566 | Not-determined |
| 18-105_k141_118962 | Not-determined |
| 18-105_k141_83312 | Low-quality |
| 18-105_k141_4568 | Low-quality |
| 18-105_k141_136697 | Not-determined |
| 18-105_k141_181716 | Not-determined |
| 18-105_k141_136708 | Not-determined |
| 18-105_k141_101569 | Low-quality |
| 18-105_k141_74855 | Not-determined |
| 18-105_k141_136718 | Not-determined |
| 18-105_k141_173207 | Not-determined |
| 18-105_k141_101578 | Not-determined |
| 18-105_k141_154677 | Low-quality |
| 18-105_k141_154681 | Not-determined |
| 18-105_k141_154684 | Not-determined |
| 18-105_k141_181742 | Not-determined |
| 18-105_k141_208094 | Not-determined |
| 18-105_k141_48442 | Not-determined |
| 18-105_k141_74869 | Low-quality |
| 18-105_k141_181749 | Not-determined |
| 18-105_k141_181750 | Not-determined |
| 18-105_k141_20614 | Not-determined |
| 18-105_k141_20615 | Not-determined |
| 18-105_k141_48446 | Not-determined |
| 18-105_k141_29647 | Not-determined |
| 18-105_k141_173221 | Not-determined |
| 18-105_k141_154698 | Not-determined |
| 18-105_k141_190680 | Not-determined |
| 18-105_k141_4589 | Not-determined |
| 18-105_k141_48451 | Not-determined |
| 18-105_k141_92574 | Not-determined |
| 18-105_k141_4596 | Not-determined |
| 18-105_k141_181762 | Not-determined |
| 18-105_k141_136742 | Not-determined |
| 18-105_k141_173232 | Not-determined |
| 18-105_k141_163668 | Not-determined |
| 18-105_k141_48466 | Not-determined |
| 18-105_k141_56359 | Not-determined |
| 18-105_k141_163670 | Not-determined |
| 18-105_k141_136747 | Not-determined |
| 18-105_k141_4600 | Not-determined |
| 18-105_k141_101613 | Not-determined |
| 18-105_k141_136748 | Not-determined |
| 18-105_k141_136749 | Not-determined |
| 18-105_k141_92581 | Low-quality |
| 18-105_k141_208117 | Not-determined |
| 18-105_k141_181774 | Not-determined |
| 18-105_k141_92586 | Not-determined |
| 18-105_k141_199134 | Not-determined |
| 18-105_k141_29674 | Low-quality |
| 18-105_k141_20632 | Not-determined |
| 18-105_k141_181776 | Low-quality |
| 18-105_k141_154726 | Not-determined |
| 18-105_k141_109880 | Not-determined |
| 18-105_k141_109882 | Not-determined |
| 18-105_k141_109883 | Low-quality |
| 18-105_k141_146261 | Not-determined |
| 18-105_k141_56388 | Not-determined |
| 18-105_k141_20648 | Not-determined |
| 18-105_k141_154735 | Low-quality |
| 18-105_k141_38742 | Not-determined |
| 18-105_k141_56398 | Not-determined |
| 18-105_k141_146279 | Not-determined |
| 18-105_k141_92601 | Not-determined |
| 18-105_k141_56405 | Not-determined |
| 18-105_k141_92602 | Not-determined |
| 18-105_k141_92605 | Not-determined |
| 18-105_k141_136778 | Not-determined |
| 18-105_k141_29705 | Not-determined |
| 18-105_k141_29708 | Not-determined |
| 18-105_k141_163708 | Not-determined |
| 18-105_k141_181803 | Not-determined |
| 18-105_k141_29711 | Not-determined |
| 18-105_k141_181804 | Low-quality |
| 18-105_k141_29717 | Not-determined |
| 18-105_k141_136798 | Not-determined |
| 18-105_k141_173240 | Low-quality |
| 18-105_k141_173242 | Not-determined |
| 18-105_k141_29723 | Low-quality |
| 18-105_k141_173245 | Not-determined |
| 18-105_k141_199137 | Not-determined |
| 18-105_k141_38752 | Not-determined |
| 18-105_k141_163739 | Not-determined |
| 18-105_k141_190708 | Not-determined |
| 18-105_k141_109927 | Not-determined |
| 18-105_k141_109931 | Not-determined |
| 18-105_k141_190714 | Not-determined |
| 18-105_k141_154799 | Not-determined |
| 18-105_k141_56450 | Not-determined |
| 18-105_k141_136823 | Not-determined |
| 18-105_k141_199149 | Not-determined |
| 18-105_k141_163752 | Not-determined |
| 18-105_k141_128478 | Not-determined |
| 18-105_k141_101624 | Not-determined |
| 18-105_k141_163754 | Not-determined |
| 18-105_k141_190726 | Not-determined |
| 18-105_k141_208132 | Not-determined |
| 18-105_k141_56467 | Not-determined |
| 18-105_k141_163756 | Low-quality |
| 18-105_k141_208138 | Not-determined |
| 18-105_k141_154807 | Not-determined |
| 18-105_k141_163762 | Not-determined |
| 18-105_k141_136839 | Not-determined |
| 18-105_k141_163764 | Not-determined |
| 18-105_k141_136841 | Not-determined |
| 18-105_k141_173262 | Not-determined |
| 18-105_k141_190732 | Low-quality |
| 18-105_k141_56474 | Not-determined |
| 18-105_k141_146330 | Not-determined |
| 18-105_k141_101632 | Low-quality |
| 18-105_k141_101635 | Not-determined |
| 18-105_k141_29754 | Not-determined |
| 18-105_k141_128490 | Not-determined |
| 18-105_k141_146339 | Not-determined |
| 18-105_k141_29756 | Not-determined |
| 18-105_k141_92631 | Low-quality |
| 18-105_k141_12135 | Low-quality |
| 18-105_k141_56490 | Not-determined |
| 18-105_k141_29758 | Not-determined |
| 18-105_k141_29760 | Not-determined |
| 18-105_k141_146342 | Not-determined |
| 18-105_k141_128501 | Not-determined |
| 18-105_k141_65505 | Not-determined |
| 18-105_k141_12149 | Not-determined |
| 18-105_k141_128503 | Low-quality |
| 18-105_k141_38788 | Not-determined |
| 18-105_k141_83346 | Low-quality |
| 18-105_k141_74906 | Not-determined |
| 18-105_k141_74907 | Not-determined |
| 18-105_k141_128507 | Not-determined |
| 18-105_k141_101649 | Not-determined |
| 18-105_k141_136873 | Not-determined |
| 18-105_k141_128519 | Low-quality |
| 18-105_k141_173290 | Not-determined |
| 18-105_k141_146357 | Low-quality |
| 18-105_k141_92653 | Not-determined |
| 18-105_k141_92654 | Not-determined |
| 18-105_k141_92655 | Not-determined |
| 18-105_k141_173302 | Not-determined |
| 18-105_k141_208180 | Not-determined |
| 18-105_k141_12178 | Not-determined |
| 18-105_k141_173311 | Not-determined |
| 18-105_k141_190757 | Not-determined |
| 18-105_k141_74941 | Not-determined |
| 18-105_k141_74944 | Not-determined |
| 18-105_k141_92674 | Not-determined |
| 18-105_k141_38799 | Not-determined |
| 18-105_k141_109960 | Not-determined |
| 18-105_k141_101694 | Not-determined |
| 18-105_k141_118971 | Not-determined |
| 18-105_k141_109969 | Not-determined |
| 18-105_k141_118975 | Not-determined |
| 18-105_k141_92691 | Not-determined |
| 18-105_k141_173328 | Not-determined |
| 18-105_k141_199189 | Not-determined |
| 18-105_k141_74956 | Not-determined |
| 18-105_k141_12216 | Not-determined |
| 18-105_k141_173337 | Not-determined |
| 18-105_k141_92702 | Not-determined |
| 18-105_k141_101708 | Not-determined |
| 18-105_k141_173340 | Not-determined |
| 18-105_k141_20667 | Not-determined |
| 18-105_k141_101712 | Not-determined |
| 18-105_k141_199201 | Not-determined |
| 18-105_k141_190770 | Not-determined |
| 18-105_k141_101718 | Not-determined |
| 18-105_k141_12228 | Not-determined |
| 18-105_k141_163787 | Low-quality |
| 18-105_k141_101725 | Not-determined |
| 18-105_k141_190779 | Not-determined |
| 18-105_k141_199212 | Not-determined |
| 18-105_k141_65554 | Not-determined |
| 18-105_k141_92736 | Not-determined |
| 18-105_k141_199228 | Not-determined |
| 18-105_k141_20688 | Not-determined |
| 18-105_k141_4719 | Not-determined |
| 18-105_k141_4731 | Not-determined |
| 18-105_k141_128608 | Not-determined |
| 18-105_k141_92751 | Not-determined |
| 18-105_k141_12234 | Not-determined |
| 18-105_k141_20702 | Not-determined |
| 18-105_k141_38856 | Not-determined |
| 18-105_k141_38859 | Not-determined |
| 18-105_k141_12238 | Not-determined |
| 18-105_k141_4744 | Not-determined |
| 18-105_k141_101753 | Not-determined |
| 18-105_k141_65587 | Not-determined |
| 18-105_k141_128630 | Not-determined |
| 18-105_k141_12256 | Not-determined |
| 18-105_k141_4754 | Not-determined |
| 18-105_k141_173363 | Not-determined |
| 18-105_k141_56499 | Not-determined |
| 18-105_k141_199248 | Not-determined |
| 18-105_k141_12272 | Not-determined |
| 18-105_k141_92763 | Not-determined |
| 18-105_k141_208209 | Not-determined |
| 18-105_k141_48504 | Not-determined |
| 18-105_k141_48508 | Not-determined |
| 18-105_k141_65606 | Not-determined |
| 18-105_k141_92773 | Not-determined |
| 18-105_k141_83382 | Not-determined |
| 18-105_k141_74980 | Not-determined |
| 18-105_k141_83385 | Low-quality |
| 18-105_k141_48514 | Not-determined |
| 18-105_k141_128656 | Not-determined |
| 18-105_k141_208228 | Not-determined |
| 18-105_k141_38867 | Not-determined |
| 18-105_k141_208234 | Not-determined |
| 18-105_k141_75000 | Not-determined |
| 18-105_k141_119010 | Low-quality |
| 18-105_k141_75005 | Not-determined |
| 18-105_k141_65631 | Not-determined |
| 18-105_k141_119015 | Not-determined |
| 18-105_k141_128679 | Not-determined |
| 18-105_k141_75018 | Not-determined |
| 18-105_k141_119022 | Not-determined |
| 18-105_k141_56553 | Not-determined |
| 18-105_k141_75031 | Not-determined |
| 18-105_k141_4811 | Not-determined |
| 18-105_k141_92796 | Not-determined |
| 18-105_k141_208254 | Low-quality |
| 18-105_k141_119029 | Not-determined |
| 18-105_k141_208261 | Not-determined |
| 18-105_k141_29765 | Not-determined |
| 18-105_k141_38896 | Not-determined |
| 18-105_k141_29766 | Not-determined |
| 18-105_k141_38907 | Not-determined |
| 18-105_k141_163808 | Not-determined |
| 18-105_k141_199276 | Not-determined |
| 18-105_k141_65635 | Not-determined |
| 18-105_k141_38913 | Not-determined |
| 18-105_k141_199280 | Not-determined |
| 18-105_k141_190813 | Not-determined |
| 18-105_k141_119064 | Not-determined |
| 18-105_k141_163820 | Low-quality |
| 18-105_k141_163826 | Not-determined |
| 18-105_k141_29777 | Not-determined |
| 18-105_k141_29780 | Not-determined |
| 18-105_k141_119082 | Not-determined |
| 18-105_k141_29783 | Not-determined |
| 18-105_k141_163835 | Not-determined |
| 18-105_k141_146363 | Not-determined |
| 18-105_k141_83457 | Not-determined |
| 18-105_k141_38959 | Not-determined |
| 18-105_k141_119102 | Not-determined |
| 18-105_k141_4844 | Not-determined |
| 18-105_k141_92826 | Not-determined |
| 18-105_k141_75090 | Not-determined |
| 18-105_k141_12377 | Not-determined |
| 18-105_k141_83478 | Low-quality |
| 18-105_k141_199307 | Low-quality |
| 18-105_k141_4848 | Not-determined |
| 18-105_k141_119120 | Not-determined |
| 18-105_k141_136931 | Not-determined |
| 18-105_k141_136937 | Not-determined |
| 18-105_k141_56615 | Not-determined |
| 18-105_k141_119135 | Not-determined |
| 18-105_k141_92847 | Not-determined |
| 18-105_k141_101818 | Not-determined |
| 18-105_k141_136949 | Not-determined |
| 18-105_k141_173406 | Not-determined |
| 18-105_k141_65672 | Not-determined |
| 18-105_k141_29812 | Not-determined |
| 18-105_k141_29814 | Not-determined |
| 18-105_k141_163859 | Not-determined |
| 18-105_k141_4862 | Not-determined |
| 18-105_k141_4864 | Not-determined |
| 18-105_k141_181844 | Not-determined |
| 18-105_k141_4869 | Not-determined |
| 18-105_k141_163868 | Not-determined |
| 18-105_k141_146373 | Not-determined |
| 18-105_k141_109988 | Not-determined |
| 18-105_k141_208267 | Not-determined |
| 18-105_k141_65690 | Not-determined |
| 18-105_k141_136961 | Not-determined |
| 18-105_k141_109997 | Not-determined |
| 18-105_k141_146378 | Low-quality |
| 18-105_k141_65698 | Not-determined |
| 18-105_k141_173420 | Not-determined |
| 18-105_k141_146384 | Not-determined |
| 18-105_k141_146385 | Not-determined |
| 18-105_k141_173428 | Not-determined |
| 18-105_k141_181869 | Low-quality |
| 18-105_k141_48540 | Low-quality |
| 18-105_k141_4877 | Not-determined |
| 18-105_k141_4880 | Not-determined |
| 18-105_k141_119188 | Not-determined |
| 18-105_k141_208302 | Not-determined |
| 18-105_k141_146407 | Not-determined |
| 18-105_k141_119193 | Not-determined |
| 18-105_k141_75145 | Not-determined |
| 18-105_k141_146413 | Not-determined |
| 18-105_k141_4890 | Not-determined |
| 18-105_k141_190851 | Not-determined |
| 18-105_k141_83511 | Not-determined |
| 18-105_k141_83515 | Not-determined |
| 18-105_k141_83518 | Not-determined |
| 18-105_k141_208311 | Not-determined |
| 18-105_k141_92899 | Not-determined |
| 18-105_k141_92906 | Not-determined |
| 18-105_k141_146432 | Not-determined |
| 18-105_k141_12402 | Not-determined |
| 18-105_k141_190861 | Not-determined |
| 18-105_k141_146441 | Not-determined |
| 18-105_k141_92924 | Not-determined |
| 18-105_k141_146447 | Not-determined |
| 18-105_k141_110042 | Low-quality |
| 18-105_k141_4917 | Not-determined |
| 18-105_k141_101840 | Not-determined |
| 18-105_k141_146452 | Not-determined |
| 18-105_k141_208337 | Not-determined |
| 18-105_k141_110051 | Not-determined |
| 18-105_k141_101845 | Not-determined |
| 18-105_k141_190879 | Not-determined |
| 18-105_k141_208342 | Low-quality |
| 18-105_k141_173481 | Not-determined |
| 18-105_k141_173482 | Not-determined |
| 18-105_k141_208348 | Not-determined |
| 18-105_k141_208353 | Not-determined |
| 18-105_k141_92945 | Not-determined |
| 18-105_k141_208360 | Low-quality |
| 18-105_k141_163875 | Not-determined |
| 18-105_k141_4958 | Not-determined |
| 18-105_k141_48597 | Not-determined |
| 18-105_k141_173503 | Not-determined |
| 18-105_k141_190899 | Not-determined |
| 18-105_k141_146461 | Not-determined |
| 18-105_k141_4971 | Not-determined |
| 18-105_k141_4972 | Not-determined |
| 18-105_k141_190905 | Not-determined |
| 18-105_k141_65737 | Not-determined |
| 18-105_k141_190910 | Low-quality |
| 18-105_k141_48606 | Not-determined |
| 18-105_k141_65740 | Low-quality |
| 18-105_k141_65742 | Not-determined |
| 18-105_k141_173514 | Not-determined |
| 18-105_k141_12433 | Not-determined |
| 18-105_k141_119204 | Not-determined |
| 18-105_k141_56710 | Not-determined |
| 18-105_k141_56712 | Not-determined |
| 18-105_k141_48622 | Not-determined |
| 18-105_k141_190917 | Not-determined |
| 18-105_k141_83564 | Not-determined |
| 18-105_k141_75153 | Not-determined |
| 18-105_k141_163890 | Not-determined |
| 18-105_k141_83569 | Not-determined |
| 18-105_k141_83571 | Not-determined |
| 18-105_k141_75156 | Not-determined |
| 18-105_k141_48625 | Low-quality |
| 18-105_k141_146480 | Not-determined |
| 18-105_k141_12444 | Low-quality |
| 18-105_k141_65753 | Not-determined |
| 18-105_k141_56726 | Not-determined |
| 18-105_k141_119219 | Not-determined |
| 18-105_k141_146488 | Not-determined |
| 18-105_k141_110083 | Not-determined |
| 18-105_k141_110090 | Not-determined |
| 18-105_k141_173539 | Low-quality |
| 18-105_k141_39014 | Not-determined |
| 18-105_k141_146504 | Not-determined |
| 18-105_k141_110099 | Not-determined |
| 18-105_k141_190931 | Not-determined |
| 18-105_k141_190934 | Not-determined |
| 18-105_k141_4997 | Not-determined |
| 18-105_k141_12470 | Not-determined |
| 18-105_k141_128692 | Low-quality |
| 18-105_k141_39027 | Not-determined |
| 18-105_k141_29854 | Not-determined |
| 18-105_k141_39032 | Not-determined |
| 18-105_k141_173561 | Not-determined |
| 18-105_k141_29865 | Not-determined |
| 18-105_k141_48652 | Not-determined |
| 18-105_k141_5007 | Not-determined |
| 18-105_k141_29877 | Not-determined |
| 18-105_k141_5017 | Not-determined |
| 18-105_k141_146532 | Not-determined |
| 18-105_k141_48660 | Not-determined |
| 18-105_k141_12483 | Not-determined |
| 18-105_k141_173575 | Not-determined |
| 18-105_k141_146534 | Not-determined |
| 18-105_k141_146539 | Not-determined |
| 18-105_k141_173587 | Low-quality |
| 18-105_k141_75171 | Not-determined |
| 18-105_k141_12492 | Not-determined |
| 18-105_k141_29885 | Not-determined |
| 18-105_k141_5034 | Not-determined |
| 18-105_k141_119262 | Not-determined |
| 18-105_k141_190961 | Not-determined |
| 18-105_k141_56741 | Low-quality |
| 18-105_k141_173604 | Not-determined |
| 18-105_k141_173606 | Not-determined |
| 18-105_k141_83605 | Not-determined |
| 18-105_k141_128697 | Not-determined |
| 18-105_k141_83609 | Not-determined |
| 18-105_k141_29900 | Not-determined |
| 18-105_k141_83611 | Not-determined |
| 18-105_k141_75183 | Not-determined |
| 18-105_k141_48678 | Not-determined |
| 18-105_k141_56765 | Not-determined |
| 18-105_k141_5043 | Not-determined |
| 18-105_k141_83626 | Not-determined |
| 18-105_k141_128724 | Not-determined |
| 18-105_k141_119267 | Not-determined |
| 18-105_k141_119268 | Not-determined |
| 18-105_k141_48690 | Not-determined |
| 18-105_k141_5050 | Not-determined |
| 18-105_k141_12518 | Not-determined |
| 18-105_k141_128733 | Not-determined |
| 18-105_k141_5051 | Low-quality |
| 18-105_k141_5052 | Not-determined |
| 18-105_k141_5053 | Not-determined |
| 18-105_k141_173641 | Low-quality |
| 18-105_k141_83639 | Low-quality |
| 18-105_k141_190997 | Not-determined |
| 18-105_k141_137032 | Not-determined |
| 18-105_k141_191006 | Not-determined |
| 18-105_k141_65819 | Low-quality |
| 18-105_k141_199352 | Not-determined |
| 18-105_k141_48706 | Not-determined |
| 18-105_k141_48707 | Not-determined |
| 18-105_k141_65823 | Not-determined |
| 18-105_k141_75225 | Low-quality |
| 18-105_k141_101910 | Low-quality |
| 18-105_k141_137044 | Low-quality |
| 18-105_k141_208409 | Not-determined |
| 18-105_k141_137049 | Not-determined |
| 18-105_k141_20783 | Not-determined |
| 18-105_k141_101925 | Low-quality |
| 18-105_k141_48720 | Not-determined |
| 18-105_k141_101927 | Low-quality |
| 18-105_k141_119295 | Not-determined |
| 18-105_k141_208429 | Not-determined |
| 18-105_k141_128745 | Not-determined |
| 18-105_k141_75256 | Not-determined |
| 18-105_k141_20820 | Not-determined |
| 18-105_k141_75267 | Not-determined |
| 18-105_k141_20831 | Not-determined |
| 18-105_k141_119320 | Not-determined |
| 18-105_k141_20835 | Not-determined |
| 18-105_k141_39052 | Not-determined |
| 18-105_k141_208454 | Not-determined |
| 18-105_k141_146556 | Not-determined |
| 18-105_k141_119328 | Not-determined |
| 18-105_k141_20845 | Not-determined |
| 18-105_k141_56811 | Not-determined |
| 18-105_k141_56815 | Not-determined |
| 18-105_k141_65836 | Not-determined |
| 18-105_k141_48728 | Not-determined |
| 18-105_k141_146564 | Not-determined |
| 18-105_k141_83670 | Not-determined |
| 18-105_k141_119340 | Not-determined |
| 18-105_k141_39063 | Not-determined |
| 18-105_k141_83675 | Not-determined |
| 18-105_k141_75293 | Not-determined |
| 18-105_k141_83676 | Not-determined |
| 18-105_k141_20857 | Not-determined |
| 18-105_k141_75294 | Not-determined |
| 18-105_k141_101957 | Not-determined |
| 18-105_k141_191051 | Not-determined |
| 18-105_k141_101965 | Not-determined |
| 18-105_k141_119360 | Not-determined |
| 18-105_k141_119362 | Not-determined |
| 18-105_k141_146576 | Not-determined |
| 18-105_k141_119365 | Not-determined |
| 18-105_k141_48744 | Not-determined |
| 18-105_k141_119368 | Not-determined |
| 18-105_k141_48745 | Not-determined |
| 18-105_k141_191063 | Not-determined |
| 18-105_k141_128774 | Not-determined |
| 18-105_k141_5079 | Not-determined |
| 18-105_k141_56836 | Not-determined |
| 18-105_k141_199382 | Not-determined |
| 18-105_k141_39075 | Not-determined |
| 18-105_k141_199391 | Not-determined |
| 18-105_k141_83702 | Not-determined |
| 18-105_k141_110109 | Not-determined |
| 18-105_k141_199393 | Not-determined |
| 18-105_k141_110111 | Not-determined |
| 18-105_k141_39086 | Not-determined |
| 18-105_k141_20866 | Not-determined |
| 18-105_k141_56852 | Not-determined |
| 18-105_k141_20875 | Not-determined |
| 18-105_k141_101992 | Not-determined |
| 18-105_k141_56862 | Not-determined |
| 18-105_k141_65875 | Not-determined |
| 18-105_k141_110132 | Not-determined |
| 18-105_k141_92964 | Not-determined |
| 18-105_k141_110137 | Not-determined |
| 18-105_k141_102006 | Not-determined |
| 18-105_k141_56870 | Low-quality |
| 18-105_k141_173648 | Not-determined |
| 18-105_k141_20887 | Not-determined |
| 18-105_k141_208463 | Not-determined |
| 18-105_k141_102008 | Not-determined |
| 18-105_k141_20889 | Not-determined |
| 18-105_k141_92971 | Not-determined |
| 18-105_k141_110145 | Not-determined |
| 18-105_k141_102016 | Not-determined |
| 18-105_k141_5100 | Not-determined |
| 18-105_k141_65901 | Not-determined |
| 18-105_k141_56888 | Not-determined |
| 18-105_k141_56892 | Not-determined |
| 18-105_k141_199396 | Low-quality |
| 18-105_k141_102036 | Low-quality |
| 18-105_k141_102040 | Not-determined |
| 18-105_k141_173671 | Not-determined |
| 18-105_k141_5121 | Not-determined |
| 18-105_k141_75343 | Not-determined |
| 18-105_k141_5126 | Low-quality |
| 18-105_k141_20907 | Not-determined |
| 18-105_k141_146583 | Not-determined |
| 18-105_k141_5142 | Not-determined |
| 18-105_k141_102052 | Low-quality |
| 18-105_k141_173683 | Not-determined |
| 18-105_k141_173686 | Not-determined |
| 18-105_k141_173687 | Not-determined |
| 18-105_k141_5150 | Low-quality |
| 18-105_k141_137114 | Not-determined |
| 18-105_k141_181935 | Not-determined |
| 18-105_k141_191105 | Low-quality |
| 18-105_k141_191106 | Low-quality |
| 18-105_k141_163922 | Not-determined |
| 18-105_k141_173696 | Not-determined |
| 18-105_k141_5160 | Not-determined |
| 18-105_k141_137119 | Low-quality |
| 18-105_k141_39120 | Low-quality |
| 18-105_k141_146606 | Not-determined |
| 18-105_k141_137124 | Not-determined |
| 18-105_k141_39124 | Low-quality |
| 18-105_k141_146615 | Not-determined |
| 18-105_k141_20953 | Not-determined |

| | |
|---|---|
| 18-105_k141_65957 | Not-determined |
| 18-105_k141_208478 | Not-determined |
| 18-105_k141_56916 | Not-determined |
| 18-105_k141_102088 | Not-determined |
| 18-105_k141_137130 | Not-determined |
| 18-105_k141_56923 | Not-determined |
| 18-105_k141_191117 | Not-determined |
| 18-105_k141_102098 | Not-determined |
| 18-105_k141_208487 | Not-determined |
| 18-105_k141_39148 | Not-determined |
| 18-105_k141_137139 | Not-determined |
| 18-105_k141_199441 | Not-determined |
| 18-105_k141_199442 | Not-determined |
| 18-105_k141_208493 | Low-quality |
| 18-105_k141_199447 | Not-determined |
| 18-105_k141_56946 | Not-determined |
| 18-105_k141_65983 | Not-determined |
| 18-105_k141_5189 | Not-determined |
| 18-105_k141_29908 | Not-determined |
| 18-105_k141_48771 | Not-determined |
| 18-105_k141_163974 | Low-quality |
| 18-105_k141_146667 | Not-determined |
| 18-105_k141_146669 | Not-determined |
| 18-105_k141_146670 | Not-determined |
| 18-105_k141_102126 | Not-determined |
| 18-105_k141_29918 | Not-determined |
| 18-105_k141_29919 | Not-determined |
| 18-105_k141_48778 | Not-determined |
| 18-105_k141_163982 | Not-determined |
| 18-105_k141_199469 | Not-determined |
| 18-105_k141_110152 | Low-quality |
| 18-105_k141_102144 | Not-determined |
| 18-105_k141_56962 | Not-determined |
| 18-105_k141_110164 | Low-quality |
| 18-105_k141_128838 | Not-determined |
| 18-105_k141_39165 | Not-determined |
| 18-105_k141_39174 | Low-quality |
| 18-105_k141_128853 | Not-determined |
| 18-105_k141_20961 | Not-determined |
| 18-105_k141_66001 | Low-quality |
| 18-105_k141_48803 | Not-determined |
| 18-105_k141_102156 | Not-determined |
| 18-105_k141_5218 | Not-determined |
| 18-105_k141_20969 | Not-determined |
| 18-105_k141_154951 | Low-quality |
| 18-105_k141_164000 | Not-determined |
| 18-105_k141_146715 | Not-determined |
| 18-105_k141_199499 | Not-determined |
| 18-105_k141_39191 | Not-determined |
| 18-105_k141_137168 | Not-determined |
| 18-105_k141_154961 | Not-determined |
| 18-105_k141_5230 | Not-determined |
| 18-105_k141_102174 | Not-determined |
| 18-105_k141_146727 | Not-determined |
| 18-105_k141_119475 | Not-determined |
| 18-105_k141_146734 | Not-determined |
| 18-105_k141_164004 | Not-determined |
| 18-105_k141_48820 | Not-determined |
| 18-105_k141_56978 | Low-quality |
| 18-105_k141_56981 | Low-quality |
| 18-105_k141_199523 | Low-quality |
| 18-105_k141_5256 | Not-determined |
| 18-105_k141_56988 | Not-determined |
| 18-105_k141_146744 | Not-determined |
| 18-105_k141_56990 | Not-determined |
| 18-105_k141_56991 | Not-determined |
| 18-105_k141_56992 | Not-determined |
| 18-105_k141_110213 | Not-determined |
| 18-105_k141_146752 | Low-quality |
| 18-105_k141_146753 | Not-determined |
| 18-105_k141_5277 | Not-determined |
| 18-105_k141_48853 | Not-determined |
| 18-105_k141_5291 | Not-determined |
| 18-105_k141_110229 | Low-quality |
| 17-108-final-viral-combined_k141_211679\|\|full | High-quality |
| 18-105_k141_66018 | Not-determined |
| 18-105_k141_110235 | Not-determined |
| 18-105_k141_5313 | Low-quality |
| 18-105_k141_110240 | Low-quality |
| 18-105_k141_5315 | Not-determined |
| 18-105_k141_199552 | Not-determined |
| 18-105_k141_5319 | Not-determined |
| 18-105_k141_83743 | Not-determined |
| 18-105_k141_5323 | Not-determined |
| 18-105_k141_12543 | Low-quality |
| 18-105_k141_5330 | Not-determined |
| 18-105_k141_181988 | Not-determined |
| 18-105_k141_181991 | Not-determined |
| 18-105_k141_48877 | Not-determined |
| 18-105_k141_12564 | Not-determined |
| 18-105_k141_182003 | Not-determined |
| 18-105_k141_199576 | Low-quality |
| 18-105_k141_119504 | Not-determined |
| 18-105_k141_66040 | Not-determined |
| 18-105_k141_83773 | Not-determined |
| 18-105_k141_110260 | Not-determined |
| 18-105_k141_128874 | Not-determined |
| 18-105_k141_128876 | Not-determined |
| 18-105_k141_110266 | Not-determined |
| 18-105_k141_93059 | Not-determined |
| 18-105_k141_102203 | Not-determined |
| 18-105_k141_48898 | Not-determined |
| 18-105_k141_83797 | Not-determined |
| 18-105_k141_83798 | Not-determined |
| 18-105_k141_199605 | Not-determined |
| 18-105_k141_199606 | Not-determined |
| 18-105_k141_21009 | Low-quality |
| 18-105_k141_48910 | Not-determined |
| 18-105_k141_182024 | Not-determined |
| 18-105_k141_119537 | Not-determined |
| 18-105_k141_12582 | Not-determined |
| 18-105_k141_182029 | Not-determined |
| 18-105_k141_57049 | Not-determined |
| 18-105_k141_39241 | Not-determined |
| 18-105_k141_83821 | Not-determined |
| 18-105_k141_83823 | Not-determined |
| 18-105_k141_12595 | Not-determined |
| 18-105_k141_110284 | Not-determined |
| 18-105_k141_102222 | Not-determined |
| 18-105_k141_93074 | Not-determined |
| 18-105_k141_199629 | Not-determined |
| 18-105_k141_12599 | Not-determined |
| 18-105_k141_93076 | Not-determined |
| 18-105_k141_93077 | Not-determined |
| 18-105_k141_48913 | Low-quality |
| 18-105_k141_83831 | Not-determined |
| 18-105_k141_83833 | Not-determined |
| 18-105_k141_12601 | Not-determined |
| 18-105_k141_110299 | Not-determined |
| 18-105_k141_93083 | Not-determined |
| 18-105_k141_93084 | Not-determined |
| 18-105_k141_173719 | Low-quality |
| 18-105_k141_29933 | Low-quality |
| 18-105_k141_93088 | Not-determined |
| 18-105_k141_29938 | Low-quality |
| 18-105_k141_146774 | Not-determined |
| 18-105_k141_39249 | Not-determined |
| 18-105_k141_119556 | Not-determined |
| 18-105_k141_83842 | Not-determined |
| 18-105_k141_21020 | Not-determined |
| 18-105_k141_110313 | Not-determined |
| 18-105_k141_48932 | Not-determined |
| 18-105_k141_75410 | Not-determined |
| 18-105_k141_21033 | Not-determined |
| 18-105_k141_83857 | Not-determined |
| 18-105_k141_110323 | Not-determined |
| 18-105_k141_48939 | Not-determined |
| 18-105_k141_128927 | Not-determined |
| 18-105_k141_199643 | Not-determined |
| 18-105_k141_83863 | Not-determined |
| 18-105_k141_83874 | Not-determined |
| 18-105_k141_154993 | Not-determined |
| 18-105_k141_48951 | Not-determined |
| 18-105_k141_102249 | Not-determined |
| 18-105_k141_12630 | Not-determined |
| 18-105_k141_199668 | Not-determined |
| 18-105_k141_83878 | Low-quality |
| 18-105_k141_164086 | Not-determined |
| 18-105_k141_110337 | Not-determined |
| 18-105_k141_128954 | Not-determined |
| 18-105_k141_173737 | Not-determined |
| 18-105_k141_83884 | Not-determined |
| 18-105_k141_146822 | Low-quality |
| 18-105_k141_12648 | Not-determined |
| 18-105_k141_155027 | Not-determined |
| 18-105_k141_137178 | Not-determined |
| 18-105_k141_12657 | Not-determined |
| 18-105_k141_199690 | Low-quality |
| 18-105_k141_21071 | Not-determined |
| 18-105_k141_182067 | Not-determined |
| 18-105_k141_12658 | Not-determined |
| 18-105_k141_110339 | Not-determined |
| 18-105_k141_83900 | Not-determined |
| 18-105_k141_75452 | Not-determined |
| 18-105_k141_128970 | Not-determined |
| 18-105_k141_137192 | Low-quality |
| 18-105_k141_48964 | Not-determined |
| 18-105_k141_48967 | Not-determined |
| 18-105_k141_137203 | Not-determined |
| 18-105_k141_155042 | Not-determined |
| 18-105_k141_155044 | Not-determined |
| 18-105_k141_199703 | Low-quality |
| 18-105_k141_119585 | Not-determined |
| 18-105_k141_119586 | Not-determined |
| 18-105_k141_199704 | Not-determined |
| 18-105_k141_146836 | Not-determined |
| 18-105_k141_208575 | Not-determined |
| 18-105_k141_208576 | Not-determined |
| 18-105_k141_146842 | Not-determined |
| 18-105_k141_146845 | Not-determined |
| 18-105_k141_93103 | Not-determined |
| 18-105_k141_21087 | Not-determined |
| 18-105_k141_155060 | Not-determined |
| 18-105_k141_12685 | Not-determined |
| 18-105_k141_21092 | Low-quality |
| 18-105_k141_75467 | Not-determined |
| 18-105_k141_146852 | Not-determined |
| 18-105_k141_155063 | Not-determined |
| 18-105_k141_29989 | Not-determined |
| 18-105_k141_75469 | Not-determined |
| 18-105_k141_155071 | Not-determined |
| 18-105_k141_75472 | Not-determined |
| 18-105_k141_75474 | Not-determined |
| 18-105_k141_146866 | Low-quality |
| 18-105_k141_75484 | Not-determined |
| 18-105_k141_155082 | Not-determined |
| 18-105_k141_208592 | Low-quality |
| 18-105_k141_208593 | Not-determined |
| 18-105_k141_66098 | Not-determined |
| 18-105_k141_12716 | Low-quality |
| 18-105_k141_182102 | Not-determined |
| 18-105_k141_199759 | Not-determined |
| 18-105_k141_146877 | Low-quality |
| 18-105_k141_137219 | Low-quality |
| 18-105_k141_199761 | Not-determined |
| 18-105_k141_137220 | Not-determined |
| 18-105_k141_12723 | Not-determined |
| 18-105_k141_199770 | Not-determined |
| 18-105_k141_155102 | Not-determined |
| 18-105_k141_199785 | Not-determined |
| 18-105_k141_199798 | Not-determined |
| 18-105_k141_208644 | Not-determined |
| 18-105_k141_137248 | Not-determined |
| 18-105_k141_66125 | Not-determined |
| 18-105_k141_208648 | Not-determined |
| 18-105_k141_110432 | Not-determined |
| 18-105_k141_137261 | Not-determined |
| 18-105_k141_39278 | Not-determined |
| 18-105_k141_21148 | Not-determined |
| 18-105_k141_5371 | Not-determined |
| 18-105_k141_12761 | Not-determined |
| 18-105_k141_30005 | Low-quality |
| 18-105_k141_5378 | Low-quality |
| 18-105_k141_199819 | Not-determined |
| 18-105_k141_21154 | Not-determined |
| 18-105_k141_39290 | Not-determined |
| 18-105_k141_173773 | Not-determined |
| 18-105_k141_208671 | Not-determined |
| 18-105_k141_75508 | Not-determined |
| 18-105_k141_12784 | Low-quality |
| 18-105_k141_208682 | Not-determined |
| 18-105_k141_102274 | Not-determined |
| 18-105_k141_137289 | Not-determined |
| 18-105_k141_12792 | Not-determined |
| 18-105_k141_119652 | Not-determined |
| 18-105_k141_119654 | Low-quality |
| 18-105_k141_137295 | Not-determined |
| 18-105_k141_208703 | Low-quality |
| 18-105_k141_137300 | Low-quality |
| 18-105_k141_75520 | Not-determined |
| 18-105_k141_119660 | Low-quality |
| 18-105_k141_75524 | Not-determined |
| 18-105_k141_66164 | Not-determined |
| 18-105_k141_208711 | Not-determined |
| 18-105_k141_12802 | Not-determined |
| 18-105_k141_119666 | Low-quality |
| 18-105_k141_5424 | Not-determined |
| 18-105_k141_173811 | Low-quality |
| 18-105_k141_191138 | Not-determined |
| 18-105_k141_12815 | Not-determined |
| 18-105_k141_102304 | Not-determined |
| 18-105_k141_30018 | Not-determined |
| 18-105_k141_173825 | Not-determined |
| 18-105_k141_30022 | Not-determined |
| 18-105_k141_12817 | Not-determined |
| 18-105_k141_110471 | Not-determined |
| 18-105_k141_137329 | Not-determined |
| 18-105_k141_12821 | Not-determined |
| 18-105_k141_110474 | Not-determined |
| 18-105_k141_21197 | Not-determined |
| 18-105_k141_75543 | Not-determined |
| 18-105_k141_102315 | Not-determined |
| 18-105_k141_5442 | Not-determined |
| 18-105_k141_57174 | Not-determined |
| 18-105_k141_75547 | Not-determined |
| 18-105_k141_102326 | Not-determined |
| 18-105_k141_39342 | Not-determined |
| 18-105_k141_110492 | Not-determined |
| 18-105_k141_49007 | Not-determined |
| 18-105_k141_21220 | Not-determined |
| 18-105_k141_39354 | Not-determined |
| 18-105_k141_66178 | Not-determined |
| 18-105_k141_208736 | Not-determined |
| 18-105_k141_21223 | Low-quality |
| 18-105_k141_75561 | Not-determined |
| 18-105_k141_39360 | Low-quality |
| 18-105_k141_208749 | Not-determined |
| 18-105_k141_39366 | Not-determined |
| 18-105_k141_57187 | Not-determined |
| 18-105_k141_49037 | Not-determined |
| 18-105_k141_75571 | Not-determined |
| 18-105_k141_39370 | Not-determined |
| 18-105_k141_5451 | Not-determined |
| 18-105_k141_5452 | Not-determined |
| 18-105_k141_66204 | Not-determined |
| 18-105_k141_66206 | Not-determined |
| 18-105_k141_137352 | Not-determined |
| 18-105_k141_110520 | Not-determined |
| 18-105_k141_164132 | Not-determined |
| 18-105_k141_164134 | Low-quality |
| 18-105_k141_137359 | Not-determined |
| 18-105_k141_57191 | Not-determined |
| 18-105_k141_137362 | Not-determined |
| 18-105_k141_57195 | Not-determined |
| 18-105_k141_5469 | Not-determined |
| 18-105_k141_5470 | Not-determined |
| 18-105_k141_137373 | Not-determined |
| 18-105_k141_57198 | Not-determined |
| 18-105_k141_57199 | Not-determined |
| 18-105_k141_5478 | Not-determined |
| 18-105_k141_5484 | Not-determined |
| 18-105_k141_83913 | Not-determined |
| 18-105_k141_83920 | Not-determined |
| 18-105_k141_57202 | Not-determined |
| 18-105_k141_12841 | Not-determined |
| 18-105_k141_12842 | Not-determined |
| 18-105_k141_83930 | Not-determined |
| 18-105_k141_93121 | Not-determined |
| 18-105_k141_12848 | Not-determined |
| 18-105_k141_49043 | Not-determined |
| 18-105_k141_137387 | Not-determined |
| 18-105_k141_12871 | Not-determined |
| 18-105_k141_5500 | Not-determined |
| 18-105_k141_93150 | Not-determined |
| 18-105_k141_12882 | Not-determined |
| 18-105_k141_93155 | Not-determined |
| 18-105_k141_57215 | Not-determined |
| 18-105_k141_164178 | Not-determined |
| 18-105_k141_12886 | Not-determined |
| 18-105_k141_57220 | Not-determined |
| 18-105_k141_164194 | Not-determined |
| 18-105_k141_155183 | Not-determined |
| 18-105_k141_102347 | Not-determined |
| 18-105_k141_110545 | Low-quality |
| 18-105_k141_30062 | Not-determined |
| 18-105_k141_57229 | Not-determined |
| 18-105_k141_102356 | Not-determined |
| 18-105_k141_199854 | Not-determined |
| 18-105_k141_199857 | Not-determined |
| 18-105_k141_208777 | Not-determined |
| 18-105_k141_129053 | Not-determined |
| 18-105_k141_93169 | Not-determined |
| 18-105_k141_93171 | Not-determined |
| 18-105_k141_93172 | Not-determined |
| 18-105_k141_208788 | Not-determined |
| 18-105_k141_93173 | Not-determined |
| 18-105_k141_66225 | Not-determined |
| 18-105_k141_129054 | Not-determined |
| 18-105_k141_93175 | Not-determined |
| 18-105_k141_93176 | Not-determined |
| 18-105_k141_57257 | Not-determined |
| 18-105_k141_102405 | Not-determined |
| 18-105_k141_155202 | Not-determined |
| 18-105_k141_93191 | Not-determined |
| 18-105_k141_129072 | Not-determined |
| 18-105_k141_129073 | Not-determined |
| 18-105_k141_93193 | Not-determined |
| 18-105_k141_66245 | Not-determined |
| 18-105_k141_12901 | Not-determined |
| 18-105_k141_129085 | Not-determined |
| 18-105_k141_119806 | Not-determined |
| 18-105_k141_164216 | Not-determined |
| 18-105_k141_119814 | Not-determined |
| 18-105_k141_102435 | Not-determined |
| 18-105_k141_164232 | Not-determined |
| 18-105_k141_102437 | Not-determined |
| 18-105_k141_57293 | Not-determined |
| 18-105_k141_110553 | Not-determined |
| 18-105_k141_102443 | Not-determined |
| 18-105_k141_39394 | Not-determined |
| 18-105_k141_164252 | Not-determined |
| 18-105_k141_66277 | Not-determined |
| 18-105_k141_155244 | Low-quality |
| 18-105_k141_164266 | Not-determined |
| 18-105_k141_66284 | Not-determined |
| 18-105_k141_66286 | Not-determined |
| 18-105_k141_39414 | Not-determined |
| 18-105_k141_75586 | Low-quality |
| 18-105_k141_208839 | Not-determined |
| 18-105_k141_75594 | Low-quality |
| 18-105_k141_75596 | Not-determined |
| 18-105_k141_39438 | Not-determined |
| 18-105_k141_208850 | Not-determined |
| 18-105_k141_155276 | Not-determined |
| 18-105_k141_12947 | Not-determined |
| 18-105_k141_110568 | Not-determined |
| 18-105_k141_39444 | Not-determined |
| 18-105_k141_39447 | Not-determined |
| 18-105_k141_39450 | Not-determined |
| 18-105_k141_164293 | Not-determined |
| 18-105_k141_12964 | Not-determined |
| 18-105_k141_30144 | Low-quality |
| 18-105_k141_30153 | Not-determined |
| 18-105_k141_30155 | Not-determined |
| 18-105_k141_84013 | Not-determined |
| 18-105_k141_30157 | Not-determined |
| 18-105_k141_155286 | Not-determined |
| 18-105_k141_110609 | Not-determined |
| 18-105_k141_30164 | Not-determined |
| 18-105_k141_30169 | Not-determined |
| 18-105_k141_30171 | Not-determined |
| 18-105_k141_155292 | Not-determined |
| 18-105_k141_182131 | Not-determined |
| 18-105_k141_155303 | Not-determined |
| 18-105_k141_199889 | Not-determined |
| 18-105_k141_93219 | Not-determined |
| 18-105_k141_110624 | Not-determined |
| 18-105_k141_110625 | Not-determined |
| 18-105_k141_93223 | Not-determined |
| 18-105_k141_13006 | Not-determined |
| 18-105_k141_75615 | Not-determined |
| 18-105_k141_13007 | Not-determined |
| 18-105_k141_199907 | Not-determined |
| 18-105_k141_75620 | Not-determined |
| 18-105_k141_146883 | Not-determined |
| 18-105_k141_49051 | Not-determined |
| 18-105_k141_119836 | Not-determined |
| 18-105_k141_199917 | Not-determined |
| 18-105_k141_199919 | Not-determined |
| 18-105_k141_155324 | Not-determined |
| 18-105_k141_13024 | Not-determined |
| 18-105_k141_137466 | Not-determined |
| 18-105_k141_137467 | Not-determined |
| 18-105_k141_129107 | Not-determined |
| 18-105_k141_199927 | Not-determined |
| 18-105_k141_129108 | Not-determined |
| 18-105_k141_129110 | Not-determined |
| 18-105_k141_199931 | Not-determined |
| 18-105_k141_119844 | Not-determined |
| 18-105_k141_208862 | Not-determined |
| 18-105_k141_199934 | Not-determined |
| 18-105_k141_110655 | Low-quality |
| 18-105_k141_102519 | Not-determined |
| 18-105_k141_137486 | Not-determined |
| 18-105_k141_199941 | Not-determined |
| 18-105_k141_146904 | Low-quality |
| 18-105_k141_66318 | Not-determined |
| 18-105_k141_57383 | Not-determined |
| 18-105_k141_110663 | Not-determined |
| 18-105_k141_66319 | Low-quality |
| 18-105_k141_137499 | Not-determined |
| 18-105_k141_137500 | Not-determined |
| 18-105_k141_110675 | Not-determined |
| 18-105_k141_110682 | Not-determined |
| 18-105_k141_173892 | Not-determined |
| 18-105_k141_102531 | Not-determined |
| 18-105_k141_191179 | Low-quality |
| 18-105_k141_5510 | Not-determined |
| 18-105_k141_66337 | Not-determined |
| 18-105_k141_137519 | Not-determined |
| 18-105_k141_119864 | Not-determined |
| 18-105_k141_129126 | Not-determined |
| 18-105_k141_102537 | Not-determined |
| 18-105_k141_173901 | Not-determined |
| 18-105_k141_110689 | Low-quality |
| 18-105_k141_199948 | Not-determined |
| 18-105_k141_164317 | Low-quality |
| 18-105_k141_84073 | Not-determined |
| 18-105_k141_137526 | Low-quality |
| 18-105_k141_110694 | Not-determined |
| 18-105_k141_191193 | Not-determined |
| 18-105_k141_102549 | Not-determined |
| 18-105_k141_66356 | Not-determined |
| 18-105_k141_182163 | Not-determined |
| 18-105_k141_39533 | Not-determined |
| 18-105_k141_182170 | Not-determined |
| 18-105_k141_57422 | Not-determined |
| 18-105_k141_208888 | Low-quality |
| 18-105_k141_84082 | Not-determined |
| 18-105_k141_146910 | Not-determined |
| 18-105_k141_191204 | Not-determined |
| 18-105_k141_137544 | Not-determined |
| 18-105_k141_66370 | Not-determined |
| 18-105_k141_66371 | Not-determined |
| 18-105_k141_164332 | Not-determined |
| 18-105_k141_39548 | Low-quality |
| 18-105_k141_13058 | Not-determined |
| 18-105_k141_39555 | Not-determined |
| 18-105_k141_110705 | Not-determined |
| 18-105_k141_110707 | Not-determined |
| 18-105_k141_13061 | Not-determined |
| 18-105_k141_13064 | Not-determined |
| 18-105_k141_110712 | Not-determined |
| 18-105_k141_93269 | Not-determined |
| 18-105_k141_30201 | Not-determined |
| 18-105_k141_164351 | Low-quality |
| 18-105_k141_13068 | Not-determined |
| 18-105_k141_191224 | Not-determined |
| 18-105_k141_173918 | Not-determined |
| 18-105_k141_30210 | Not-determined |
| 18-105_k141_30215 | Not-determined |
| 18-105_k141_84118 | Not-determined |
| 18-105_k141_30216 | Not-determined |
| 18-105_k141_84121 | Not-determined |
| 18-105_k141_13081 | Low-quality |
| 18-105_k141_84128 | Not-determined |
| 18-105_k141_164362 | Low-quality |
| 18-105_k141_39573 | Not-determined |
| 18-105_k141_119899 | Not-determined |
| 18-105_k141_39576 | Not-determined |
| 18-105_k141_57455 | Not-determined |
| 18-105_k141_66407 | Low-quality |
| 18-105_k141_182199 | Low-quality |
| 18-105_k141_119910 | Not-determined |
| 18-105_k141_93291 | Low-quality |
| 18-105_k141_102596 | Not-determined |
| 18-105_k141_102597 | Not-determined |
| 18-105_k141_57463 | Not-determined |
| 18-105_k141_49070 | Not-determined |
| 18-105_k141_30223 | Not-determined |
| 18-105_k141_13092 | Not-determined |
| 18-105_k141_5549 | Not-determined |
| 18-105_k141_182204 | Low-quality |
| 18-105_k141_119928 | Not-determined |
| 18-105_k141_13099 | Not-determined |
| 18-105_k141_39588 | Not-determined |
| 18-105_k141_66417 | Not-determined |
| 18-105_k141_66418 | Not-determined |
| 18-105_k141_5560 | Low-quality |
| 18-105_k141_39593 | Not-determined |
| 18-105_k141_39594 | Not-determined |
| 18-105_k141_182220 | Not-determined |
| 18-105_k141_75641 | Not-determined |
| 18-105_k141_102614 | Not-determined |
| 18-105_k141_5570 | Not-determined |
| 18-105_k141_13112 | Not-determined |
| 18-105_k141_84149 | Not-determined |
| 18-105_k141_173964 | Not-determined |
| 18-105_k141_57494 | Not-determined |
| 18-105_k141_66437 | Not-determined |
| 18-105_k141_75662 | Not-determined |
| 18-105_k141_13126 | Not-determined |
| 18-105_k141_208939 | Not-determined |
| 18-105_k141_84166 | Not-determined |
| 18-105_k141_182239 | Low-quality |
| 18-105_k141_39624 | Not-determined |
| 18-105_k141_208942 | Low-quality |
| 18-105_k141_75674 | Not-determined |
| 18-105_k141_102628 | Not-determined |
| 18-105_k141_110756 | Not-determined |
| 18-105_k141_5590 | Not-determined |
| 18-105_k141_155364 | Not-determined |
| 18-105_k141_21237 | Not-determined |
| 18-105_k141_200000 | Not-determined |
| 18-105_k141_5595 | Not-determined |
| 18-105_k141_49115 | Not-determined |
| 18-105_k141_191249 | Not-determined |
| 18-105_k141_75688 | Not-determined |
| 18-105_k141_200003 | Not-determined |
| 18-105_k141_5605 | Not-determined |
| 18-105_k141_191258 | Not-determined |
| 18-105_k141_191259 | Not-determined |
| 18-105_k141_110761 | Not-determined |
| 18-105_k141_200009 | Not-determined |
| 18-105_k141_5612 | Not-determined |
| 18-105_k141_208968 | Not-determined |
| 18-105_k141_57515 | Low-quality |
| 18-105_k141_75704 | Not-determined |
| 18-105_k141_93317 | Not-determined |
| 18-105_k141_200021 | Not-determined |
| 18-105_k141_30281 | Not-determined |
| 18-105_k141_191269 | Not-determined |
| 18-105_k141_191270 | Not-determined |
| 18-105_k141_200029 | Not-determined |
| 18-105_k141_75716 | Not-determined |
| 18-105_k141_75719 | Not-determined |
| 18-105_k141_208982 | Not-determined |
| 18-105_k141_208983 | Not-determined |
| 18-105_k141_110779 | Low-quality |
| 18-105_k141_75723 | Not-determined |
| 18-105_k141_66494 | Not-determined |
| 18-105_k141_182258 | Not-determined |
| 18-105_k141_102689 | Not-determined |
| 18-105_k141_182265 | Not-determined |
| 18-105_k141_174022 | Not-determined |
| 18-105_k141_102696 | Low-quality |
| 18-105_k141_66504 | Not-determined |
| 18-105_k141_102706 | Not-determined |
| 18-105_k141_66508 | Not-determined |
| 18-105_k141_84233 | Not-determined |
| 18-105_k141_13150 | Not-determined |
| 18-105_k141_30291 | Low-quality |
| 18-105_k141_30292 | Not-determined |
| 18-105_k141_13161 | Not-determined |
| 18-105_k141_5670 | Not-determined |
| 18-105_k141_174050 | Not-determined |
| 18-105_k141_30298 | Not-determined |
| 18-105_k141_200047 | Not-determined |
| 18-105_k141_182268 | Not-determined |

| | |
|---|---|
| 18-105_k141_5673 | Not-determined |
| 18-105_k141_191309 | Not-determined |
| 18-105_k141_155402 | Low-quality |
| 18-105_k141_182274 | Not-determined |
| 18-105_k141_102726 | Not-determined |
| 18-105_k141_30307 | Not-determined |
| 18-105_k141_129218 | Not-determined |
| 18-105_k141_13168 | Not-determined |
| 18-105_k141_13172 | Not-determined |
| 18-105_k141_110831 | Low-quality |
| 18-105_k141_129227 | Not-determined |
| 18-105_k141_5696 | Not-determined |
| 18-105_k141_66522 | Not-determined |
| 18-105_k141_93349 | Not-determined |
| 18-105_k141_49122 | Not-determined |
| 18-105_k141_49123 | Not-determined |
| 18-105_k141_174089 | Not-determined |
| 18-105_k141_30330 | Not-determined |
| 18-105_k141_129240 | Not-determined |
| 18-105_k141_129241 | Not-determined |
| 18-105_k141_66541 | Not-determined |
| 18-105_k141_129243 | Not-determined |
| 18-105_k141_5708 | Not-determined |
| 18-105_k141_30336 | Not-determined |
| 18-105_k141_30340 | Not-determined |
| 18-105_k141_102751 | Not-determined |
| 18-105_k141_93363 | Not-determined |
| 18-105_k141_110863 | Not-determined |
| 18-105_k141_5729 | Not-determined |
| 18-105_k141_5730 | Not-determined |
| 18-105_k141_155442 | Not-determined |
| 18-105_k141_30357 | Not-determined |
| 18-105_k141_30359 | Not-determined |
| 18-105_k141_102780 | Not-determined |
| 18-105_k141_75766 | Not-determined |
| 18-105_k141_209008 | Not-determined |
| 18-105_k141_209009 | Not-determined |
| 18-105_k141_155451 | Not-determined |
| 18-105_k141_30363 | Not-determined |
| 18-105_k141_200072 | Not-determined |
| 18-105_k141_209021 | Not-determined |
| 18-105_k141_200074 | Not-determined |
| 18-105_k141_75778 | Not-determined |
| 18-105_k141_5752 | Not-determined |
| 18-105_k141_200077 | Not-determined |
| 18-105_k141_155455 | Low-quality |
| 18-105_k141_155457 | Not-determined |
| 18-105_k141_209030 | Not-determined |
| 18-105_k141_30377 | Not-determined |
| 18-105_k141_5761 | Not-determined |
| 18-105_k141_155459 | Not-determined |
| 18-105_k141_164452 | Not-determined |
| 18-105_k141_146962 | Not-determined |
| 18-105_k141_102801 | Low-quality |
| 18-105_k141_129281 | Not-determined |
| 18-105_k141_110900 | Not-determined |
| 18-105_k141_75790 | Not-determined |
| 18-105_k141_209040 | Not-determined |
| 18-105_k141_129284 | Low-quality |
| 18-105_k141_93406 | Low-quality |
| 18-105_k141_191336 | Not-determined |
| 18-105_k141_146968 | Low-quality |
| 18-105_k141_191338 | Low-quality |
| 18-105_k141_75794 | Not-determined |
| 18-105_k141_13194 | Not-determined |
| 18-105_k141_30400 | Not-determined |
| 18-105_k141_182286 | Not-determined |
| 18-105_k141_5775 | Not-determined |
| 18-105_k141_102814 | Not-determined |
| 18-105_k141_75803 | Not-determined |
| 18-105_k141_129303 | Not-determined |
| 18-105_k141_110912 | Not-determined |
| 18-105_k141_30404 | Not-determined |
| 18-105_k141_93430 | Not-determined |
| 18-105_k141_66589 | Not-determined |
| 18-105_k141_75814 | Low-quality |
| 18-105_k141_84242 | Not-determined |
| 18-105_k141_102832 | Not-determined |
| 18-105_k141_75826 | Not-determined |
| 18-105_k141_146984 | Not-determined |
| 18-105_k141_13218 | Low-quality |
| 18-105_k141_39660 | Not-determined |
| 18-105_k141_49127 | Not-determined |
| 18-105_k141_13224 | Not-determined |
| 18-105_k141_39678 | Not-determined |
| 18-105_k141_182329 | Low-quality |
| 18-105_k141_39683 | Not-determined |
| 18-105_k141_147004 | Not-determined |
| 18-105_k141_49139 | Not-determined |
| 18-105_k141_39687 | Low-quality |
| 18-105_k141_49140 | Not-determined |
| 18-105_k141_147007 | Not-determined |
| 18-105_k141_182332 | Not-determined |
| 18-105_k141_191346 | Low-quality |
| 18-105_k141_164477 | Not-determined |
| 18-105_k141_110920 | Not-determined |
| 18-105_k141_75829 | Not-determined |
| 18-105_k141_5815 | Not-determined |
| 18-105_k141_66616 | Not-determined |
| 18-105_k141_13240 | Not-determined |
| 18-105_k141_66617 | Not-determined |
| 18-105_k141_110924 | Not-determined |
| 18-105_k141_209074 | Not-determined |
| 18-105_k141_66619 | Not-determined |
| 18-105_k141_191358 | Low-quality |
| 18-105_k141_110930 | Low-quality |
| 18-105_k141_191365 | Not-determined |
| 18-105_k141_93446 | Not-determined |
| 18-105_k141_57530 | Not-determined |
| 18-105_k141_129346 | Not-determined |
| 18-105_k141_110940 | Not-determined |
| 18-105_k141_129348 | Not-determined |
| 18-105_k141_164484 | Not-determined |
| 18-105_k141_30429 | Not-determined |
| 18-105_k141_93456 | Not-determined |
| 18-105_k141_84278 | Low-quality |
| 18-105_k141_182350 | Not-determined |
| 18-105_k141_49169 | Not-determined |
| 18-105_k141_5828 | Not-determined |
| 18-105_k141_5829 | Not-determined |
| 18-105_k141_21263 | Not-determined |
| 18-105_k141_191376 | Not-determined |
| 18-105_k141_93464 | Not-determined |
| 18-105_k141_93465 | Not-determined |
| 18-105_k141_191378 | Not-determined |
| 18-105_k141_84283 | Not-determined |
| 18-105_k141_49181 | Not-determined |
| 18-105_k141_57548 | Not-determined |
| 18-105_k141_21273 | Not-determined |
| 18-105_k141_209093 | Not-determined |
| 18-105_k141_147035 | Low-quality |
| 18-105_k141_21281 | Not-determined |
| 18-105_k141_155507 | Low-quality |
| 18-105_k141_182372 | Not-determined |
| 18-105_k141_110958 | Not-determined |
| 18-105_k141_57564 | Low-quality |
| 18-105_k141_75857 | Not-determined |
| 18-105_k141_182379 | Not-determined |
| 18-105_k141_182380 | Not-determined |
| 18-105_k141_129368 | Not-determined |
| 18-105_k141_129369 | Not-determined |
| 18-105_k141_209107 | Not-determined |
| 18-105_k141_164511 | Not-determined |
| 18-105_k141_110969 | Not-determined |
| 18-105_k141_182387 | Not-determined |
| 18-105_k141_209115 | Low-quality |
| 18-105_k141_182388 | Not-determined |
| 18-105_k141_93497 | Not-determined |
| 18-105_k141_30432 | Not-determined |
| 18-105_k141_209119 | Not-determined |
| 18-105_k141_182394 | Not-determined |
| 18-105_k141_5862 | Not-determined |
| 18-105_k141_57578 | Not-determined |
| 18-105_k141_110980 | Not-determined |
| 18-105_k141_209127 | Not-determined |
| 18-105_k141_30442 | Not-determined |
| 18-105_k141_209133 | Not-determined |
| 18-105_k141_57587 | Not-determined |
| 18-105_k141_102853 | Not-determined |
| 18-105_k141_147072 | Not-determined |
| 18-105_k141_13290 | Not-determined |
| 18-105_k141_200160 | Not-determined |
| 18-105_k141_200165 | Not-determined |
| 18-105_k141_182418 | Not-determined |
| 18-105_k141_5895 | Not-determined |
| 18-105_k141_39746 | Not-determined |
| 18-105_k141_75889 | Not-determined |
| 18-105_k141_110989 | Not-determined |
| 18-105_k141_110993 | Not-determined |
| 18-105_k141_147091 | Not-determined |
| 18-105_k141_21311 | Not-determined |
| 18-105_k141_21313 | Not-determined |
| 18-105_k141_147102 | Not-determined |
| 18-105_k141_164561 | Not-determined |
| 18-105_k141_174101 | Not-determined |
| 18-105_k141_13327 | Not-determined |
| 18-105_k141_57598 | Not-determined |
| 18-105_k141_200197 | Not-determined |
| 18-105_k141_84347 | Not-determined |
| 18-105_k141_191405 | Not-determined |
| 18-105_k141_200205 | Not-determined |
| 18-105_k141_84350 | Not-determined |
| 18-105_k141_182438 | Not-determined |
| 18-105_k141_84353 | Not-determined |
| 18-105_k141_174117 | Not-determined |
| 18-105_k141_84362 | Not-determined |
| 18-105_k141_147131 | Low-quality |
| 18-105_k141_200213 | Not-determined |
| 18-105_k141_182455 | Not-determined |
| 18-105_k141_57615 | Not-determined |
| 18-105_k141_39770 | Not-determined |
| 18-105_k141_13355 | Not-determined |
| 18-105_k141_84380 | Not-determined |
| 18-105_k141_111011 | Not-determined |
| 18-105_k141_209173 | Not-determined |
| 18-105_k141_57618 | Not-determined |
| 18-105_k141_191429 | Not-determined |
| 18-105_k141_13364 | Not-determined |
| 18-105_k141_5929 | Not-determined |
| 18-105_k141_57626 | Not-determined |
| 18-105_k141_129431 | Not-determined |
| 18-105_k141_129432 | Not-determined |
| 18-105_k141_147146 | Not-determined |
| 18-105_k141_75935 | Not-determined |
| 18-105_k141_93512 | Not-determined |
| 18-105_k141_13379 | Not-determined |
| 18-105_k141_102870 | Low-quality |
| 18-105_k141_182494 | Not-determined |
| 18-105_k141_75940 | Low-quality |
| 18-105_k141_13383 | Not-determined |
| 18-105_k141_13393 | Not-determined |
| 18-105_k141_75953 | Not-determined |
| 18-105_k141_75956 | Not-determined |
| 18-105_k141_174128 | Not-determined |
| 18-105_k141_174130 | Not-determined |
| 18-105_k141_182518 | Not-determined |
| 18-105_k141_5951 | Not-determined |
| 18-105_k141_147184 | Not-determined |
| 18-105_k141_57670 | Not-determined |
| 18-105_k141_5962 | Not-determined |
| 18-105_k141_147197 | Not-determined |
| 18-105_k141_102906 | Not-determined |
| 18-105_k141_30461 | Not-determined |
| 18-105_k141_209200 | Not-determined |
| 18-105_k141_5975 | Not-determined |
| 18-105_k141_75983 | Not-determined |
| 18-105_k141_93542 | Low-quality |
| 18-105_k141_129479 | Low-quality |
| 18-105_k141_147217 | Not-determined |
| 18-105_k141_30466 | Not-determined |
| 18-105_k141_111072 | Not-determined |
| 18-105_k141_30468 | Not-determined |
| 18-105_k141_30469 | Low-quality |
| 18-105_k141_147225 | Not-determined |
| 18-105_k141_5984 | Not-determined |
| 18-105_k141_147238 | Not-determined |
| 18-105_k141_111078 | Not-determined |
| 18-105_k141_174147 | Not-determined |
| 18-105_k141_66670 | Not-determined |
| 18-105_k141_66673 | Not-determined |
| 18-105_k141_66676 | Not-determined |
| 18-105_k141_66683 | Not-determined |
| 18-105_k141_111089 | Not-determined |
| 18-105_k141_76016 | Not-determined |
| 18-105_k141_111090 | Low-quality |
| 18-105_k141_57697 | Not-determined |
| 18-105_k141_6020 | Not-determined |
| 18-105_k141_57701 | Not-determined |
| 18-105_k141_6025 | Not-determined |
| 18-105_k141_129522 | Low-quality |
| 18-105_k141_6031 | Not-determined |
| 18-105_k141_84436 | Not-determined |
| 18-105_k141_84445 | Not-determined |
| 18-105_k141_174154 | Not-determined |
| 18-105_k141_147263 | Not-determined |
| 18-105_k141_57711 | Not-determined |
| 18-105_k141_174168 | Not-determined |
| 18-105_k141_111114 | Not-determined |
| 18-105_k141_111115 | Not-determined |
| 18-105_k141_84459 | Not-determined |
| 18-105_k141_147275 | Not-determined |
| 18-105_k141_164605 | Not-determined |
| 18-105_k141_102921 | Not-determined |
| 18-105_k141_147278 | Not-determined |
| 18-105_k141_39781 | Low-quality |
| 18-105_k141_93562 | Not-determined |
| 18-105_k141_39788 | Not-determined |
| 18-105_k141_102932 | Not-determined |
| 18-105_k141_93570 | Not-determined |
| 18-105_k141_6055 | Not-determined |
| 18-105_k141_76025 | Not-determined |
| 18-105_k141_147287 | Not-determined |
| 18-105_k141_76028 | Not-determined |
| 18-105_k141_147290 | Not-determined |
| 18-105_k141_66706 | Not-determined |
| 18-105_k141_13409 | Not-determined |
| 18-105_k141_6068 | Not-determined |
| 18-105_k141_174208 | Not-determined |
| 18-105_k141_93583 | Not-determined |
| 18-105_k141_182538 | Not-determined |
| 18-105_k141_182539 | Not-determined |
| 18-105_k141_30510 | Not-determined |
| 18-105_k141_93594 | Low-quality |
| 18-105_k141_200243 | Not-determined |
| 18-105_k141_84472 | Low-quality |
| 18-105_k141_30516 | Not-determined |
| 18-105_k141_84481 | Not-determined |
| 18-105_k141_93601 | Not-determined |
| 18-105_k141_30519 | Not-determined |
| 18-105_k141_6098 | Not-determined |
| 18-105_k141_174242 | Not-determined |
| 18-105_k141_93612 | Not-determined |
| 18-105_k141_147310 | Not-determined |
| 18-105_k141_147311 | Not-determined |
| 18-105_k141_111160 | Not-determined |
| 18-105_k141_93622 | Not-determined |
| 18-105_k141_13457 | Not-determined |
| 18-105_k141_93624 | Not-determined |
| 18-105_k141_93625 | Not-determined |
| 18-105_k141_13461 | Not-determined |
| 18-105_k141_66733 | Not-determined |
| 18-105_k141_200255 | Not-determined |
| 18-105_k141_174247 | Not-determined |
| 18-105_k141_147331 | Not-determined |
| 18-105_k141_147332 | Not-determined |
| 18-105_k141_13469 | Not-determined |
| 18-105_k141_209283 | Not-determined |
| 18-105_k141_174262 | Not-determined |
| 18-105_k141_93644 | Not-determined |
| 18-105_k141_174264 | Not-determined |
| 18-105_k141_174269 | Not-determined |
| 18-105_k141_84483 | Low-quality |
| 18-105_k141_93654 | Not-determined |
| 18-105_k141_147345 | Not-determined |
| 18-105_k141_84491 | Not-determined |
| 18-105_k141_30541 | Not-determined |
| 18-105_k141_84499 | Not-determined |
| 18-105_k141_174284 | Not-determined |
| 18-105_k141_155534 | Not-determined |
| 18-105_k141_155536 | Not-determined |
| 18-105_k141_93671 | Low-quality |
| 18-105_k141_111192 | Not-determined |
| 18-105_k141_111197 | Not-determined |
| 18-105_k141_13485 | Not-determined |
| 18-105_k141_13487 | Not-determined |
| 18-105_k141_111203 | Not-determined |
| 18-105_k141_21366 | Not-determined |
| 18-105_k141_209312 | Not-determined |
| 18-105_k141_84521 | Not-determined |
| 18-105_k141_182575 | Not-determined |
| 18-105_k141_182578 | Not-determined |
| 18-105_k141_182583 | Not-determined |
| 18-105_k141_209325 | Not-determined |
| 18-105_k141_111212 | Not-determined |
| 18-105_k141_66787 | Not-determined |
| 18-105_k141_84537 | Not-determined |
| 18-105_k141_111222 | Not-determined |
| 18-105_k141_84543 | Not-determined |
| 18-105_k141_209336 | Not-determined |
| 18-105_k141_209343 | Not-determined |
| 18-105_k141_39804 | Not-determined |
| 18-105_k141_6102 | Not-determined |
| 18-105_k141_200289 | Not-determined |
| 18-105_k141_182600 | Not-determined |
| 18-105_k141_6109 | Not-determined |
| 18-105_k141_84562 | Not-determined |
| 18-105_k141_182611 | Low-quality |
| 18-105_k141_119952 | Low-quality |
| 18-105_k141_93700 | Not-determined |
| 18-105_k141_182618 | Not-determined |
| 18-105_k141_6128 | Not-determined |
| 18-105_k141_182620 | Not-determined |
| 18-105_k141_111250 | Low-quality |
| 18-105_k141_191470 | Not-determined |
| 18-105_k141_93706 | Not-determined |
| 18-105_k141_147398 | Not-determined |
| 18-105_k141_84570 | Not-determined |
| 18-105_k141_119973 | Not-determined |
| 18-105_k141_200319 | Not-determined |
| 18-105_k141_30574 | Not-determined |
| 18-105_k141_30576 | Not-determined |
| 18-105_k141_39825 | Not-determined |
| 18-105_k141_6146 | Not-determined |
| 18-105_k141_13500 | Not-determined |
| 18-105_k141_129539 | Low-quality |
| 18-105_k141_137657 | Not-determined |
| 18-105_k141_6159 | Not-determined |
| 18-105_k141_6162 | Not-determined |
| 18-105_k141_137666 | Low-quality |
| 18-105_k141_209372 | Not-determined |
| 18-105_k141_111268 | Not-determined |
| 18-105_k141_182657 | Not-determined |
| 18-105_k141_182658 | Not-determined |
| 18-105_k141_200332 | Not-determined |
| 18-105_k141_137678 | Not-determined |
| 18-105_k141_209378 | Not-determined |
| 18-105_k141_30600 | Not-determined |
| 18-105_k141_49204 | Not-determined |
| 18-105_k141_93737 | Not-determined |
| 18-105_k141_111294 | Not-determined |
| 18-105_k141_84600 | Not-determined |
| 18-105_k141_111303 | Not-determined |
| 18-105_k141_200337 | Not-determined |
| 18-105_k141_200338 | Not-determined |
| 18-105_k141_200343 | Not-determined |
| 18-105_k141_200347 | Not-determined |
| 18-105_k141_49224 | Not-determined |
| 18-105_k141_111325 | Not-determined |
| 18-105_k141_200348 | Not-determined |
| 18-105_k141_30618 | Not-determined |
| 18-105_k141_13554 | Not-determined |
| 18-105_k141_49232 | Not-determined |
| 18-105_k141_111334 | Low-quality |
| 18-105_k141_30626 | Not-determined |
| 18-105_k141_49235 | Low-quality |
| 18-105_k141_182688 | Not-determined |
| 18-105_k141_182698 | Not-determined |
| 18-105_k141_137716 | Not-determined |
| 18-105_k141_182703 | Not-determined |
| 18-105_k141_111346 | Low-quality |
| 18-105_k141_137720 | Not-determined |
| 18-105_k141_93774 | Not-determined |
| 18-105_k141_137724 | Not-determined |
| 18-105_k141_21373 | Not-determined |
| 18-105_k141_39853 | Not-determined |
| 18-105_k141_39854 | Not-determined |
| 18-105_k141_39855 | Not-determined |
| 18-105_k141_191484 | Not-determined |
| 18-105_k141_111361 | Not-determined |
| 18-105_k141_155563 | Not-determined |
| 18-105_k141_93790 | Not-determined |
| 18-105_k141_191497 | Low-quality |
| 18-105_k141_39873 | Not-determined |
| 18-105_k141_103031 | Not-determined |
| 18-105_k141_111390 | Low-quality |
| 18-105_k141_30636 | Not-determined |
| 18-105_k141_30639 | Not-determined |
| 18-105_k141_84648 | Not-determined |
| 18-105_k141_209411 | Not-determined |
| 18-105_k141_209412 | Not-determined |
| 18-105_k141_76093 | Not-determined |
| 18-105_k141_182778 | Not-determined |
| 18-105_k141_182779 | Not-determined |
| 18-105_k141_182780 | Not-determined |
| 18-105_k141_39897 | Low-quality |
| 18-105_k141_129605 | Not-determined |
| 18-105_k141_191532 | Not-determined |
| 18-105_k141_39900 | Not-determined |
| 18-105_k141_84663 | Not-determined |
| 18-105_k141_137752 | Not-determined |
| 18-105_k141_30650 | Not-determined |
| 18-105_k141_84668 | Not-determined |
| 18-105_k141_191540 | Not-determined |
| 18-105_k141_39903 | Low-quality |
| 18-105_k141_191547 | Low-quality |
| 18-105_k141_6188 | Not-determined |
| 18-105_k141_39911 | Not-determined |
| 18-105_k141_84676 | Not-determined |
| 18-105_k141_103053 | Not-determined |
| 18-105_k141_103054 | Not-determined |
| 18-105_k141_103055 | Not-determined |
| 18-105_k141_103059 | Not-determined |
| 18-105_k141_209437 | Not-determined |
| 18-105_k141_120039 | Not-determined |
| 18-105_k141_209441 | Not-determined |
| 18-105_k141_120042 | Not-determined |
| 18-105_k141_76108 | Not-determined |
| 18-105_k141_120051 | Low-quality |
| 18-105_k141_111415 | Not-determined |
| 18-105_k141_30669 | Not-determined |
| 18-105_k141_49270 | Not-determined |
| 18-105_k141_103083 | Not-determined |
| 18-105_k141_6211 | Not-determined |
| 18-105_k141_191572 | Not-determined |
| 18-105_k141_155618 | Not-determined |
| 18-105_k141_111423 | Not-determined |
| 18-105_k141_21382 | Not-determined |
| 18-105_k141_84699 | Not-determined |
| 18-105_k141_120060 | Not-determined |
| 18-105_k141_182820 | Not-determined |
| 18-105_k141_137761 | Not-determined |
| 18-105_k141_191583 | Not-determined |
| 18-105_k141_13581 | Not-determined |
| 18-105_k141_6224 | Not-determined |
| 18-105_k141_200373 | Not-determined |
| 18-105_k141_13601 | Not-determined |
| 18-105_k141_137781 | Not-determined |
| 18-105_k141_13603 | Not-determined |
| 18-105_k141_76125 | Not-determined |
| 18-105_k141_93878 | Not-determined |
| 18-105_k141_93879 | Low-quality |
| 18-105_k141_76127 | Not-determined |
| 18-105_k141_209470 | Low-quality |
| 18-105_k141_174312 | Low-quality |
| 18-105_k141_155645 | Low-quality |
| 18-105_k141_155646 | Low-quality |
| 18-105_k141_111437 | Not-determined |
| 18-105_k141_6240 | Not-determined |
| 18-105_k141_209475 | Not-determined |
| 18-105_k141_84719 | Not-determined |
| 18-105_k141_76136 | Not-determined |
| 18-105_k141_182852 | Not-determined |
| 18-105_k141_76147 | Not-determined |
| 18-105_k141_182862 | Not-determined |
| 18-105_k141_174329 | Low-quality |
| 18-105_k141_49283 | Not-determined |
| 18-105_k141_174333 | Not-determined |
| 18-105_k141_209493 | Not-determined |
| 18-105_k141_57773 | Not-determined |
| 18-105_k141_93898 | Not-determined |
| 18-105_k141_182881 | Not-determined |
| 18-105_k141_84740 | Not-determined |
| 18-105_k141_76166 | Not-determined |
| 18-105_k141_209506 | Not-determined |
| 18-105_k141_209507 | Not-determined |
| 18-105_k141_137803 | Not-determined |
| 18-105_k141_191626 | Not-determined |
| 18-105_k141_57796 | Not-determined |
| 18-105_k141_57799 | Not-determined |
| 18-105_k141_209514 | Not-determined |
| 18-105_k141_129623 | Not-determined |
| 18-105_k141_39943 | Not-determined |
| 18-105_k141_84752 | Not-determined |
| 18-105_k141_137813 | Low-quality |
| 18-105_k141_76175 | Not-determined |
| 18-105_k141_182890 | Not-determined |
| 18-105_k141_93917 | Not-determined |
| 18-105_k141_209519 | Low-quality |
| 18-105_k141_30671 | Not-determined |
| 18-105_k141_137822 | Not-determined |
| 18-105_k141_6295 | Not-determined |
| 18-105_k141_21403 | Low-quality |
| 18-105_k141_182902 | Not-determined |
| 18-105_k141_111458 | Low-quality |
| 18-105_k141_76186 | Not-determined |
| 18-105_k141_30683 | Low-quality |
| 18-105_k141_137828 | Not-determined |
| 18-105_k141_209529 | Not-determined |
| 18-105_k141_200402 | Not-determined |
| 18-105_k141_155689 | Not-determined |
| 18-105_k141_6305 | Not-determined |
| 18-105_k141_137841 | Not-determined |
| 18-105_k141_13675 | Not-determined |
| 18-105_k141_30694 | Low-quality |
| 18-105_k141_191673 | Not-determined |
| 18-105_k141_30695 | Not-determined |
| 18-105_k141_39963 | Not-determined |
| 18-105_k141_191677 | Not-determined |
| 18-105_k141_174354 | Not-determined |
| 18-105_k141_57844 | Not-determined |
| 18-105_k141_76206 | Not-determined |
| 18-105_k141_39966 | Not-determined |
| 18-105_k141_76209 | Not-determined |
| 18-105_k141_200421 | Not-determined |
| 18-105_k141_57853 | Low-quality |
| 18-105_k141_30708 | Not-determined |
| 18-105_k141_191689 | Not-determined |
| 18-105_k141_164723 | Not-determined |
| 18-105_k141_13688 | Not-determined |
| 18-105_k141_30711 | Not-determined |
| 18-105_k141_155713 | Not-determined |
| 18-105_k141_155715 | Not-determined |
| 18-105_k141_174373 | Low-quality |
| 18-105_k141_6338 | Not-determined |
| 18-105_k141_111481 | Not-determined |
| 18-105_k141_111483 | Not-determined |
| 18-105_k141_200441 | Not-determined |
| 18-105_k141_155727 | Not-determined |
| 18-105_k141_155729 | Not-determined |
| 18-105_k141_155731 | Not-determined |
| 18-105_k141_164734 | Not-determined |
| 18-105_k141_111497 | Low-quality |
| 18-105_k141_57880 | Not-determined |
| 18-105_k141_49290 | Not-determined |
| 18-105_k141_6357 | Not-determined |
| 18-105_k141_155740 | Not-determined |
| 18-105_k141_164738 | Not-determined |
| 18-105_k141_21475 | Not-determined |
| 18-105_k141_13700 | Not-determined |
| 18-105_k141_21485 | Not-determined |
| 18-105_k141_84845 | Not-determined |
| 18-105_k141_84856 | Not-determined |
| 18-105_k141_174402 | Not-determined |
| 18-105_k141_174403 | Not-determined |
| 18-105_k141_13709 | Not-determined |
| 18-105_k141_200464 | Not-determined |
| 18-105_k141_30716 | Not-determined |
| 18-105_k141_13715 | Not-determined |
| 18-105_k141_21508 | Not-determined |
| 18-105_k141_155753 | Not-determined |
| 18-105_k141_111524 | Not-determined |
| 18-105_k141_21514 | Low-quality |
| 18-105_k141_164791 | Not-determined |
| 18-101-final-viral-combined_k141_29288\|\|full | Medium-quality |
| 18-105_k141_164795 | Not-determined |
| 18-105_k141_21520 | Not-determined |
| 18-105_k141_191716 | Not-determined |
| 18-105_k141_76242 | Not-determined |
| 18-105_k141_209594 | Low-quality |
| 18-105_k141_111545 | Not-determined |
| 18-105_k141_111547 | Not-determined |
| 18-105_k141_39982 | Not-determined |
| 18-105_k141_39983 | Not-determined |
| 18-105_k141_13743 | Not-determined |
| 18-105_k141_21538 | Not-determined |
| 18-105_k141_111564 | Low-quality |
| 18-105_k141_155767 | Not-determined |
| 18-105_k141_155770 | Not-determined |
| 18-105_k141_21548 | Not-determined |

| | |
|---|---|
| 18-105_k141_155773 | Not-determined |
| 18-105_k141_21553 | Low-quality |
| 18-105_k141_21556 | Not-determined |
| 18-105_k141_39998 | Low-quality |
| 18-105_k141_39999 | Not-determined |
| 18-105_k141_84880 | Not-determined |
| 18-105_k141_40003 | Not-determined |
| 18-105_k141_200490 | Not-determined |
| 18-105_k141_191727 | Not-determined |
| 18-105_k141_209605 | Not-determined |
| 18-105_k141_155794 | Not-determined |
| 18-105_k141_155801 | Not-determined |
| 18-105_k141_21571 | Not-determined |
| 18-105_k141_155803 | Not-determined |
| 18-105_k141_191738 | Not-determined |
| 18-105_k141_164808 | Not-determined |
| 18-105_k141_111587 | Not-determined |
| 18-105_k141_209618 | Not-determined |
| 18-105_k141_111595 | Not-determined |
| 18-105_k141_155817 | Not-determined |
| 18-105_k141_76266 | Not-determined |
| 18-105_k141_111602 | Not-determined |
| 18-105_k141_84890 | Not-determined |
| 18-105_k141_174420 | Not-determined |
| 18-105_k141_191758 | Not-determined |
| 18-105_k141_209634 | Not-determined |
| 18-105_k141_57935 | Not-determined |
| 18-105_k141_76276 | Not-determined |
| 18-105_k141_191760 | Not-determined |
| 18-105_k141_200523 | Not-determined |
| 18-105_k141_155831 | Not-determined |
| 18-105_k141_137881 | Not-determined |
| 18-105_k141_200533 | Not-determined |
| 18-105_k141_174428 | Not-determined |
| 18-105_k141_191767 | Not-determined |
| 18-105_k141_200539 | Not-determined |
| 18-105_k141_103189 | Low-quality |
| 18-105_k141_200540 | Not-determined |
| 18-105_k141_111628 | Not-determined |
| 18-105_k141_200554 | Low-quality |
| 18-105_k141_6388 | Not-determined |
| 18-105_k141_129655 | Not-determined |
| 18-105_k141_76305 | Not-determined |
| 18-105_k141_129657 | Low-quality |
| 18-105_k141_200562 | Not-determined |
| 18-105_k141_40015 | Not-determined |
| 18-105_k141_76315 | Not-determined |
| 18-105_k141_200565 | Not-determined |
| 18-105_k141_200569 | Not-determined |
| 18-105_k141_13815 | Not-determined |
| 18-105_k141_40019 | Not-determined |
| 18-105_k141_129664 | Not-determined |
| 18-105_k141_200575 | Not-determined |
| 18-105_k141_13828 | Not-determined |
| 18-105_k141_6415 | Not-determined |
| 18-105_k141_6418 | Not-determined |
| 18-105_k141_6419 | Not-determined |
| 18-105_k141_129682 | Not-determined |
| 18-105_k141_30720 | Low-quality |
| 18-105_k141_120087 | Not-determined |
| 18-105_k141_200590 | Not-determined |
| 18-105_k141_129685 | Low-quality |
| 18-105_k141_30728 | Not-determined |
| 18-105_k141_76337 | Low-quality |
| 18-105_k141_30730 | Not-determined |
| 18-105_k141_76338 | Not-determined |
| 18-105_k141_200601 | Not-determined |
| 18-105_k141_30736 | Not-determined |
| 18-105_k141_164832 | Not-determined |
| 18-105_k141_40046 | Not-determined |
| 18-105_k141_164839 | Not-determined |
| 18-105_k141_40047 | Low-quality |
| 18-105_k141_30742 | Not-determined |
| 18-105_k141_200613 | Not-determined |
| 18-105_k141_129692 | Not-determined |
| 18-105_k141_76350 | Not-determined |
| 18-105_k141_40054 | Not-determined |
| 18-105_k141_76355 | Not-determined |
| 18-105_k141_155836 | Not-determined |
| 18-105_k141_76357 | Not-determined |
| 18-105_k141_13845 | Not-determined |
| 18-105_k141_6457 | Not-determined |
| 18-105_k141_147420 | Not-determined |
| 18-105_k141_13857 | Not-determined |
| 18-105_k141_200624 | Not-determined |
| 18-105_k141_155857 | Not-determined |
| 18-105_k141_191792 | Not-determined |
| 18-105_k141_13870 | Not-determined |
| 18-105_k141_13871 | Low-quality |
| 18-105_k141_137914 | Not-determined |
| 18-105_k141_120126 | Not-determined |
| 18-105_k141_191806 | Not-determined |
| 18-105_k141_84899 | Not-determined |
| 18-105_k141_200635 | Not-determined |
| 18-105_k141_103207 | Not-determined |
| 18-105_k141_191811 | Not-determined |
| 18-105_k141_13880 | Not-determined |
| 18-105_k141_155881 | Not-determined |
| 18-105_k141_40104 | Not-determined |
| 18-105_k141_6487 | Not-determined |
| 18-105_k141_6493 | Not-determined |
| 18-105_k141_103216 | Low-quality |
| 18-105_k141_84911 | Not-determined |
| 18-105_k141_191826 | Not-determined |
| 18-105_k141_191827 | Not-determined |
| 18-105_k141_191833 | Not-determined |
| 18-105_k141_13896 | Low-quality |
| 18-105_k141_174490 | Not-determined |
| 18-105_k141_40117 | Not-determined |
| 18-105_k141_200653 | Not-determined |
| 18-105_k141_40118 | Not-determined |
| 18-105_k141_13903 | Not-determined |
| 18-105_k141_174497 | Low-quality |
| 18-105_k141_40126 | Not-determined |
| 18-105_k141_103242 | Not-determined |
| 18-105_k141_84932 | Not-determined |
| 18-105_k141_191855 | Not-determined |
| 18-105_k141_40147 | Not-determined |
| 18-105_k141_120155 | Not-determined |
| 18-105_k141_6526 | Not-determined |
| 18-105_k141_30812 | Not-determined |
| 18-105_k141_103258 | Not-determined |
| 18-105_k141_57948 | Not-determined |
| 18-105_k141_40168 | Not-determined |
| 18-105_k141_6551 | Not-determined |
| 18-105_k141_200665 | Not-determined |
| 18-105_k141_6558 | Not-determined |
| 18-105_k141_111675 | Not-determined |
| 18-105_k141_76392 | Not-determined |
| 18-105_k141_30837 | Low-quality |
| 18-105_k141_76395 | Not-determined |
| 18-105_k141_6567 | Not-determined |
| 18-105_k141_174547 | Not-determined |
| 18-105_k141_191887 | Not-determined |
| 18-105_k141_30844 | Not-determined |
| 18-105_k141_191892 | Not-determined |
| 18-105_k141_30849 | Not-determined |
| 18-105_k141_120168 | Not-determined |
| 18-105_k141_76415 | Not-determined |
| 18-105_k141_191903 | Not-determined |
| 18-105_k141_13941 | Not-determined |
| 18-105_k141_174569 | Not-determined |
| 18-105_k141_57962 | Not-determined |
| 18-105_k141_191910 | Not-determined |
| 18-105_k141_30868 | Not-determined |
| 18-105_k141_183018 | Not-determined |
| 18-105_k141_21607 | Not-determined |
| 18-105_k141_183022 | Not-determined |
| 18-105_k141_120197 | Not-determined |
| 18-105_k141_155923 | Not-determined |
| 18-105_k141_21610 | Not-determined |
| 18-105_k141_120199 | Not-determined |
| 18-105_k141_155925 | Not-determined |
| 18-105_k141_155927 | Not-determined |
| 18-105_k141_191931 | Not-determined |
| 18-105_k141_21619 | Not-determined |
| 18-105_k141_191933 | Not-determined |
| 18-105_k141_200706 | Not-determined |
| 18-105_k141_191937 | Not-determined |
| 18-105_k141_200707 | Not-determined |
| 18-105_k141_174601 | Not-determined |
| 18-105_k141_21627 | Not-determined |
| 18-105_k141_200715 | Not-determined |
| 18-105_k141_120220 | Not-determined |
| 18-105_k141_58018 | Not-determined |
| 18-105_k141_58021 | Not-determined |
| 18-105_k141_21630 | Not-determined |
| 18-105_k141_21631 | Not-determined |
| 18-105_k141_21632 | Not-determined |
| 18-105_k141_147460 | Low-quality |
| 18-105_k141_164925 | Low-quality |
| 18-105_k141_84962 | Not-determined |
| 18-105_k141_147465 | Low-quality |
| 18-105_k141_84970 | Not-determined |
| 18-105_k141_209648 | Not-determined |
| 18-105_k141_21635 | Not-determined |
| 18-105_k141_84983 | Not-determined |
| 18-105_k141_209655 | Not-determined |
| 18-105_k141_164947 | Low-quality |
| 18-105_k141_58050 | Low-quality |
| 18-105_k141_200740 | Not-determined |
| 18-105_k141_200743 | Not-determined |
| 18-105_k141_200747 | Not-determined |
| 18-105_k141_137966 | Not-determined |
| 18-105_k141_30885 | Not-determined |
| 18-105_k141_85004 | Not-determined |
| 18-105_k141_137969 | Not-determined |
| 18-105_k141_137971 | Not-determined |
| 18-105_k141_191967 | Not-determined |
| 18-105_k141_200756 | Not-determined |
| 18-105_k141_200758 | Not-determined |
| 18-105_k141_30890 | Not-determined |
| 18-105_k141_200766 | Not-determined |
| 18-105_k141_30896 | Not-determined |
| 18-105_k141_30900 | Not-determined |
| 18-105_k141_103265 | Low-quality |
| 18-105_k141_58086 | Not-determined |
| 18-105_k141_85022 | Low-quality |
| 18-105_k141_103268 | Not-determined |
| 18-105_k141_103270 | Not-determined |
| 18-105_k141_21659 | Not-determined |
| 18-105_k141_200780 | Not-determined |
| 18-105_k141_76446 | Not-determined |
| 18-105_k141_76451 | Not-determined |
| 18-105_k141_21665 | Not-determined |
| 18-105_k141_76453 | Not-determined |
| 18-105_k141_200784 | Not-determined |
| 18-105_k141_40265 | Not-determined |
| 18-105_k141_147505 | Not-determined |
| 18-105_k141_174645 | Not-determined |
| 18-105_k141_200809 | Not-determined |
| 18-105_k141_30938 | Not-determined |
| 18-105_k141_200816 | Not-determined |
| 18-105_k141_174654 | Not-determined |
| 18-105_k141_129754 | Not-determined |
| 18-105_k141_147525 | Not-determined |
| 18-105_k141_40281 | Not-determined |
| 18-105_k141_200826 | Not-determined |
| 18-105_k141_58126 | Not-determined |
| 18-105_k141_120244 | Not-determined |
| 18-105_k141_129765 | Not-determined |
| 18-105_k141_209690 | Not-determined |
| 18-105_k141_58129 | Not-determined |
| 18-105_k141_120249 | Not-determined |
| 18-105_k141_58130 | Not-determined |
| 18-105_k141_58137 | Not-determined |
| 18-105_k141_200840 | Not-determined |
| 18-105_k141_120256 | Not-determined |
| 18-105_k141_137985 | Not-determined |
| 18-105_k141_40306 | Low-quality |
| 18-105_k141_209702 | Not-determined |
| 18-105_k141_120261 | Not-determined |
| 18-105_k141_120262 | Not-determined |
| 18-105_k141_21675 | Not-determined |
| 18-105_k141_120265 | Not-determined |
| 18-105_k141_129783 | Not-determined |
| 18-105_k141_30971 | Not-determined |
| 18-105_k141_120272 | Not-determined |
| 18-105_k141_209711 | Not-determined |
| 18-105_k141_49345 | Low-quality |
| 18-105_k141_58151 | Not-determined |
| 18-105_k141_49348 | Not-determined |
| 18-105_k141_30990 | Not-determined |
| 18-105_k141_21687 | Not-determined |
| 18-105_k141_111753 | Low-quality |
| 18-105_k141_30996 | Not-determined |
| 18-105_k141_200878 | Not-determined |
| 18-105_k141_49350 | Not-determined |
| 18-105_k141_200879 | Not-determined |
| 18-105_k141_30999 | Not-determined |
| 18-105_k141_111763 | Not-determined |
| 18-105_k141_85097 | Not-determined |
| 18-105_k141_200885 | Not-determined |
| 18-105_k141_111770 | Not-determined |
| 18-105_k141_111775 | Not-determined |
| 18-105_k141_31009 | Not-determined |
| 18-105_k141_40325 | Not-determined |
| 18-105_k141_200891 | Not-determined |
| 18-105_k141_209729 | Not-determined |
| 18-105_k141_209731 | Not-determined |
| 18-105_k141_31011 | Not-determined |
| 18-105_k141_85099 | Not-determined |
| 18-105_k141_209732 | Not-determined |
| 18-105_k141_49365 | Not-determined |
| 18-105_k141_209737 | Not-determined |
| 18-105_k141_31021 | Not-determined |
| 18-105_k141_164965 | Not-determined |
| 18-105_k141_40327 | Not-determined |
| 18-105_k141_31028 | Not-determined |
| 18-105_k141_209746 | Not-determined |
| 18-105_k141_200912 | Not-determined |
| 18-105_k141_191995 | Not-determined |
| 18-105_k141_49381 | Not-determined |
| 18-105_k141_85116 | Not-determined |
| 18-105_k141_111797 | Not-determined |
| 18-105_k141_209756 | Not-determined |
| 18-105_k141_58206 | Not-determined |
| 18-105_k141_58209 | Not-determined |
| 18-105_k141_138027 | Not-determined |
| 18-105_k141_76493 | Not-determined |
| 18-105_k141_183104 | Not-determined |
| 18-105_k141_58215 | Low-quality |
| 18-105_k141_58216 | Not-determined |
| 18-105_k141_85138 | Not-determined |
| 18-105_k141_183108 | Low-quality |
| 18-105_k141_6589 | Not-determined |
| 18-105_k141_183112 | Not-determined |
| 18-105_k141_21700 | Low-quality |
| 18-105_k141_58226 | Not-determined |
| 18-105_k141_21704 | Not-determined |
| 18-105_k141_6597 | Not-determined |
| 18-105_k141_138047 | Not-determined |
| 18-105_k141_6598 | Not-determined |
| 18-105_k141_58236 | Not-determined |
| 18-105_k141_58238 | Not-determined |
| 18-105_k141_31066 | Low-quality |
| 18-105_k141_31069 | Not-determined |
| 18-105_k141_31072 | Not-determined |
| 18-105_k141_85159 | Not-determined |
| 18-105_k141_21714 | Not-determined |
| 18-105_k141_111842 | Not-determined |
| 18-105_k141_111844 | Low-quality |
| 18-105_k141_164971 | Not-determined |
| 18-105_k141_40350 | Not-determined |
| 18-105_k141_76521 | Not-determined |
| 18-105_k141_58251 | Not-determined |
| 18-105_k141_111849 | Low-quality |
| 18-105_k141_66826 | Not-determined |
| 18-105_k141_21732 | Not-determined |
| 18-105_k141_58258 | Not-determined |
| 18-105_k141_111855 | Not-determined |
| 18-105_k141_21736 | Not-determined |
| 18-105_k141_76530 | Not-determined |
| 18-105_k141_58263 | Not-determined |
| 18-105_k141_13977 | Not-determined |
| 18-105_k141_13978 | Not-determined |
| 18-105_k141_58269 | Not-determined |
| 18-105_k141_76539 | Not-determined |
| 18-105_k141_58273 | Not-determined |
| 18-105_k141_13989 | Not-determined |
| 18-105_k141_13996 | Not-determined |
| 18-105_k141_164986 | Not-determined |
| 18-105_k141_49415 | Not-determined |
| 18-105_k141_164997 | Not-determined |
| 18-105_k141_76549 | Not-determined |
| 18-105_k141_58289 | Not-determined |
| 18-105_k141_58290 | Not-determined |
| 18-105_k141_40388 | Not-determined |
| 18-105_k141_165002 | Not-determined |
| 18-105_k141_58292 | Not-determined |
| 18-105_k141_165003 | Not-determined |
| 18-105_k141_58294 | Not-determined |
| 18-105_k141_31120 | Not-determined |
| 18-105_k141_40392 | Not-determined |
| 18-105_k141_165007 | Not-determined |
| 18-105_k141_40401 | Not-determined |
| 18-105_k141_14011 | Not-determined |
| 18-105_k141_76560 | Not-determined |
| 18-105_k141_165019 | Not-determined |
| 18-105_k141_76568 | Not-determined |
| 18-105_k141_14019 | Not-determined |
| 18-105_k141_14020 | Not-determined |
| 18-105_k141_14021 | Not-determined |
| 18-105_k141_14024 | Not-determined |
| 18-105_k141_58309 | Not-determined |
| 18-105_k141_14032 | Not-determined |
| 18-105_k141_192005 | Not-determined |
| 18-105_k141_14034 | Not-determined |
| 18-105_k141_165033 | Not-determined |
| 18-105_k141_192009 | Low-quality |
| 18-105_k141_209762 | Not-determined |
| 18-105_k141_183140 | Not-determined |
| 18-105_k141_155976 | Not-determined |
| 18-105_k141_155981 | Not-determined |
| 18-105_k141_209771 | Not-determined |
| 18-105_k141_6618 | Not-determined |
| 18-105_k141_6621 | Not-determined |
| 18-105_k141_155985 | Not-determined |
| 18-105_k141_49437 | Not-determined |
| 18-105_k141_66916 | Not-determined |
| 18-105_k141_58328 | Not-determined |
| 18-105_k141_155998 | Not-determined |
| 18-105_k141_49444 | Not-determined |
| 18-105_k141_6631 | Not-determined |
| 18-105_k141_138093 | Low-quality |
| 18-105_k141_49448 | Not-determined |
| 18-105_k141_6638 | Not-determined |
| 18-105_k141_209785 | Not-determined |
| 18-105_k141_209787 | Not-determined |
| 18-105_k141_147584 | Not-determined |
| 18-105_k141_209794 | Not-determined |
| 18-105_k141_49454 | Low-quality |
| 18-105_k141_120311 | Not-determined |
| 18-105_k141_6651 | Not-determined |
| 18-105_k141_49463 | Not-determined |
| 18-105_k141_21782 | Not-determined |
| 18-105_k141_93988 | Not-determined |
| 18-105_k141_21788 | Low-quality |
| 18-105_k141_209817 | Not-determined |
| 18-105_k141_49472 | Not-determined |
| 18-105_k141_49476 | Not-determined |
| 18-105_k141_209825 | Not-determined |
| 18-105_k141_94003 | Not-determined |
| 18-105_k141_129799 | Not-determined |
| 18-105_k141_103289 | Not-determined |
| 18-105_k141_192042 | Not-determined |
| 18-105_k141_40446 | Not-determined |
| 18-105_k141_40448 | Not-determined |
| 18-105_k141_14086 | Not-determined |
| 18-105_k141_94006 | Not-determined |
| 18-105_k141_183186 | Not-determined |
| 18-105_k141_6686 | Not-determined |
| 18-105_k141_14090 | Not-determined |
| 18-105_k141_94010 | Not-determined |
| 18-105_k141_103296 | Not-determined |
| 18-105_k141_21822 | Not-determined |
| 18-105_k141_14096 | Not-determined |
| 18-105_k141_49488 | Not-determined |
| 18-105_k141_49494 | Not-determined |
| 18-105_k141_209839 | Not-determined |
| 18-105_k141_94022 | Not-determined |
| 18-105_k141_209840 | Not-determined |
| 18-105_k141_209841 | Not-determined |
| 18-105_k141_165043 | Not-determined |
| 18-105_k141_209846 | Not-determined |
| 18-105_k141_21834 | Not-determined |
| 18-105_k141_129832 | Not-determined |
| 18-105_k141_209850 | Not-determined |
| 18-105_k141_94028 | Low-quality |
| 18-105_k141_21841 | Not-determined |
| 18-105_k141_40461 | Not-determined |
| 18-105_k141_85215 | Not-determined |
| 18-105_k141_147608 | Not-determined |
| 18-105_k141_40467 | Not-determined |
| 18-105_k141_183228 | Not-determined |
| 18-105_k141_6727 | Not-determined |
| 18-105_k141_111913 | Not-determined |
| 18-105_k141_85224 | Not-determined |
| 18-105_k141_147615 | Not-determined |
| 18-105_k141_183235 | Not-determined |
| 18-105_k141_183238 | Not-determined |
| 18-105_k141_40479 | Not-determined |
| 18-105_k141_111925 | Not-determined |
| 18-105_k141_111929 | Not-determined |
| 18-105_k141_111932 | Not-determined |
| 18-105_k141_40489 | Not-determined |
| 18-105_k141_14149 | Not-determined |
| 18-105_k141_165067 | Not-determined |
| 18-105_k141_165069 | Low-quality |
| 18-105_k141_192079 | Not-determined |
| 18-105_k141_138112 | Not-determined |
| 18-105_k141_6761 | Not-determined |
| 18-105_k141_129856 | Not-determined |
| 18-105_k141_94083 | Not-determined |
| 18-105_k141_6779 | Not-determined |
| 18-105_k141_85285 | Not-determined |
| 18-105_k141_6781 | Not-determined |
| 18-105_k141_85289 | Not-determined |
| 18-105_k141_94098 | Not-determined |
| 18-105_k141_165099 | Not-determined |
| 18-105_k141_85303 | Not-determined |
| 18-105_k141_165103 | Not-determined |
| 18-105_k141_129879 | Low-quality |
| 18-105_k141_120319 | Not-determined |
| 18-105_k141_174685 | Not-determined |
| 18-105_k141_103300 | Not-determined |
| 18-105_k141_192086 | Not-determined |
| 18-105_k141_174693 | Not-determined |
| 18-105_k141_209878 | Low-quality |
| 18-105_k141_66934 | Not-determined |
| 18-105_k141_66942 | Not-determined |
| 18-105_k141_103316 | Not-determined |
| 18-105_k141_183338 | Not-determined |
| 18-105_k141_66947 | Not-determined |
| 18-105_k141_183343 | Not-determined |
| 18-105_k141_21867 | Not-determined |
| 18-105_k141_183349 | Low-quality |
| 18-105_k141_21872 | Not-determined |
| 18-105_k141_183352 | Not-determined |
| 18-105_k141_147675 | Low-quality |
| 18-105_k141_111939 | Not-determined |
| 18-105_k141_21877 | Not-determined |
| 18-105_k141_111942 | Not-determined |
| 18-105_k141_111946 | Not-determined |
| 18-105_k141_138157 | Low-quality |
| 18-105_k141_103337 | Not-determined |
| 18-105_k141_138163 | Not-determined |
| 18-105_k141_138168 | Not-determined |
| 18-105_k141_165145 | Not-determined |
| 18-105_k141_85319 | Not-determined |
| 18-105_k141_183377 | Not-determined |
| 18-105_k141_209903 | Low-quality |
| 18-105_k141_66986 | Not-determined |
| 18-105_k141_174736 | Not-determined |
| 18-105_k141_49524 | Low-quality |
| 18-105_k141_66990 | Not-determined |
| 18-105_k141_103356 | Not-determined |
| 18-105_k141_6837 | Not-determined |
| 18-105_k141_31160 | Low-quality |
| 18-105_k141_209909 | Not-determined |
| 18-105_k141_31165 | Not-determined |
| 18-105_k141_111970 | Not-determined |
| 18-105_k141_31169 | Not-determined |
| 18-105_k141_209923 | Not-determined |
| 18-105_k141_6851 | Not-determined |
| 18-105_k141_58471 | Not-determined |
| 18-105_k141_111979 | Not-determined |
| 18-105_k141_58479 | Not-determined |
| 18-105_k141_129924 | Low-quality |
| 18-105_k141_103371 | Not-determined |
| 18-105_k141_147720 | Not-determined |
| 18-105_k141_94118 | Not-determined |
| 18-105_k141_40595 | Not-determined |
| 18-105_k141_58499 | Not-determined |
| 18-105_k141_103377 | Not-determined |
| 18-105_k141_94123 | Not-determined |
| 18-105_k141_183385 | Not-determined |
| 18-105_k141_94134 | Not-determined |
| 18-105_k141_209955 | Not-determined |
| 18-105_k141_85358 | Not-determined |
| 18-105_k141_209956 | Not-determined |
| 18-105_k141_6871 | Not-determined |
| 18-105_k141_21904 | Not-determined |
| 18-105_k141_147726 | Not-determined |
| 18-105_k141_209967 | Not-determined |
| 18-105_k141_147728 | Low-quality |
| 18-105_k141_40616 | Not-determined |
| 18-105_k141_147735 | Not-determined |
| 18-105_k141_209985 | Not-determined |
| 18-105_k141_165181 | Not-determined |
| 18-105_k141_129927 | Not-determined |
| 18-105_k141_40623 | Not-determined |
| 18-105_k141_120337 | Not-determined |
| 18-105_k141_40632 | Not-determined |
| 18-105_k141_174763 | Low-quality |
| 18-105_k141_6891 | Not-determined |
| 18-105_k141_112012 | Not-determined |
| 18-105_k141_76586 | Low-quality |
| 18-105_k141_21924 | Not-determined |
| 18-105_k141_6899 | Not-determined |
| 18-105_k141_210005 | Not-determined |
| 18-105_k141_31212 | Not-determined |
| 18-105_k141_112018 | Not-determined |
| 18-105_k141_49604 | Not-determined |
| 18-105_k141_21929 | Low-quality |
| 18-105_k141_6904 | Not-determined |
| 18-105_k141_76595 | Not-determined |
| 18-105_k141_76596 | Not-determined |
| 18-105_k141_58555 | Low-quality |
| 18-105_k141_200935 | Not-determined |
| 18-105_k141_21936 | Not-determined |
| 18-105_k141_21938 | Not-determined |
| 18-105_k141_147763 | Not-determined |
| 18-105_k141_94163 | Not-determined |
| 18-105_k141_120358 | Low-quality |
| 18-105_k141_94165 | Low-quality |
| 18-105_k141_112037 | Not-determined |
| 18-105_k141_6914 | Not-determined |
| 18-105_k141_76606 | Not-determined |
| 18-105_k141_138196 | Not-determined |
| 18-105_k141_174795 | Low-quality |
| 18-105_k141_183421 | Not-determined |
| 18-105_k141_58568 | Not-determined |
| 18-105_k141_58578 | Not-determined |
| 18-105_k141_94172 | Not-determined |
| 18-105_k141_76611 | Not-determined |
| 18-105_k141_58592 | Not-determined |
| 18-105_k141_67004 | Not-determined |
| 18-105_k141_40662 | Not-determined |
| 18-105_k141_174829 | Not-determined |
| 18-105_k141_112050 | Not-determined |
| 18-105_k141_112052 | Not-determined |
| 18-105_k141_21966 | Not-determined |
| 18-105_k141_138223 | Not-determined |
| 18-105_k141_174837 | Not-determined |
| 18-105_k141_21977 | Low-quality |
| 18-105_k141_210023 | Not-determined |
| 18-105_k141_174845 | Not-determined |
| 18-105_k141_58604 | Not-determined |
| 18-105_k141_112065 | Not-determined |
| 18-105_k141_120375 | Not-determined |
| 18-105_k141_138236 | Not-determined |
| 18-105_k141_210033 | Low-quality |
| 18-105_k141_112075 | Not-determined |
| 18-105_k141_138253 | Not-determined |
| 18-105_k141_21998 | Not-determined |
| 18-105_k141_138259 | Not-determined |
| 18-105_k141_14175 | Not-determined |
| 18-105_k141_183491 | Not-determined |
| 18-105_k141_210045 | Not-determined |
| 18-105_k141_31283 | Not-determined |
| 18-105_k141_67061 | Not-determined |
| 18-105_k141_183497 | Not-determined |
| 18-105_k141_210063 | Not-determined |
| 18-105_k141_22032 | Not-determined |
| 18-105_k141_120381 | Not-determined |
| 18-105_k141_22034 | Not-determined |
| 18-105_k141_22039 | Not-determined |
| 18-105_k141_14186 | Not-determined |
| 18-105_k141_192148 | Low-quality |
| 18-105_k141_192154 | Not-determined |
| 18-105_k141_14194 | Not-determined |
| 18-105_k141_120382 | Not-determined |
| 18-105_k141_31314 | Not-determined |
| 18-105_k141_120391 | Low-quality |
| 18-105_k141_156112 | Not-determined |
| 18-105_k141_31324 | Not-determined |
| 18-105_k141_120400 | Low-quality |
| 18-105_k141_58639 | Low-quality |

| | |
|---|---|
| 18-105_k141_156133 | Low-quality |
| 18-105_k141_156135 | Not-determined |
| 18-105_k141_200985 | Not-determined |
| 18-105_k141_156140 | Not-determined |
| 18-105_k141_120405 | Not-determined |
| 18-105_k141_138319 | Not-determined |
| 18-105_k141_192202 | Not-determined |
| 18-105_k141_192205 | Low-quality |
| 18-105_k141_138334 | Not-determined |
| 18-105_k141_156168 | Not-determined |
| 18-105_k141_67073 | Not-determined |
| 18-105_k141_138349 | Not-determined |
| 18-105_k141_138350 | Not-determined |
| 18-105_k141_22073 | Low-quality |
| 18-105_k141_120440 | Not-determined |
| 18-105_k141_129979 | Not-determined |
| 18-105_k141_31365 | Not-determined |
| 18-105_k141_31370 | Not-determined |
| 18-105_k141_67094 | Not-determined |
| 18-105_k141_76651 | Low-quality |
| 18-105_k141_165212 | Not-determined |
| 18-105_k141_31379 | Not-determined |
| 18-105_k141_6928 | Not-determined |
| 18-105_k141_49643 | Not-determined |
| 18-105_k141_49645 | Not-determined |
| 18-105_k141_49647 | Not-determined |
| 18-105_k141_129993 | Not-determined |
| 18-105_k141_76663 | Not-determined |
| 18-105_k141_58685 | Not-determined |
| 18-105_k141_129996 | Not-determined |
| 18-105_k141_31396 | Not-determined |
| 18-105_k141_94181 | Not-determined |
| 18-105_k141_192239 | Not-determined |
| 18-105_k141_192242 | Not-determined |
| 18-105_k141_210103 | Not-determined |
| 18-105_k141_40731 | Not-determined |
| 18-105_k141_40735 | Not-determined |
| 18-105_k141_40738 | Not-determined |
| 18-105_k141_210115 | Not-determined |
| 18-105_k141_156178 | Not-determined |
| 18-105_k141_40747 | Not-determined |
| 18-105_k141_210121 | Not-determined |
| 18-105_k141_31420 | Not-determined |
| 18-105_k141_58722 | Not-determined |
| 18-105_k141_14255 | Not-determined |
| 18-105_k141_210130 | Not-determined |
| 18-105_k141_210132 | Not-determined |
| 18-105_k141_22094 | Not-determined |
| 18-105_k141_22095 | Not-determined |
| 18-105_k141_40759 | Not-determined |
| 18-105_k141_67100 | Not-determined |
| 18-105_k141_40765 | Not-determined |
| 18-105_k141_67104 | Not-determined |
| 18-105_k141_22115 | Not-determined |
| 18-105_k141_22117 | Not-determined |
| 18-105_k141_49681 | Low-quality |
| 18-105_k141_201026 | Not-determined |
| 18-105_k141_49684 | Not-determined |
| 18-105_k141_156203 | Not-determined |
| 18-105_k141_201028 | Low-quality |
| 18-105_k141_103451 | Not-determined |
| 18-105_k141_210156 | Not-determined |
| 18-105_k141_6971 | Not-determined |
| 18-105_k141_201036 | Not-determined |
| 18-105_k141_201039 | Not-determined |
| 18-105_k141_183519 | Not-determined |
| 18-105_k141_201040 | Not-determined |
| 18-105_k141_40787 | Not-determined |
| 18-105_k141_76677 | Not-determined |
| 18-105_k141_120477 | Not-determined |
| 18-105_k141_58746 | Not-determined |
| 18-105_k141_120480 | Low-quality |
| 18-105_k141_49693 | Not-determined |
| 18-105_k141_120483 | Not-determined |
| 18-105_k141_22134 | Not-determined |
| 18-105_k141_147767 | Not-determined |
| 18-105_k141_120488 | Low-quality |
| 18-105_k141_138424 | Not-determined |
| 18-105_k141_210178 | Not-determined |
| 18-105_k141_120494 | Low-quality |
| 18-105_k141_103467 | Low-quality |
| 18-105_k141_67113 | Not-determined |
| 18-105_k141_138434 | Not-determined |
| 18-105_k141_40815 | Not-determined |
| 18-105_k141_183540 | Not-determined |
| 18-105_k141_49708 | Not-determined |
| 18-105_k141_165242 | Not-determined |
| 18-105_k141_201067 | Not-determined |
| 18-105_k141_40833 | Low-quality |
| 18-105_k141_14291 | Not-determined |
| 18-105_k141_67129 | Not-determined |
| 18-105_k141_14294 | Not-determined |
| 18-105_k141_58782 | Not-determined |
| 18-105_k141_14299 | Not-determined |
| 18-105_k141_165258 | Not-determined |
| 18-105_k141_76690 | Not-determined |
| 18-105_k141_7023 | Low-quality |
| 18-105_k141_201073 | Not-determined |
| 18-105_k141_201077 | Not-determined |
| 18-105_k141_201078 | Not-determined |
| 18-105_k141_201079 | Not-determined |
| 18-105_k141_183560 | Not-determined |
| 18-105_k141_120543 | Not-determined |
| 18-105_k141_130065 | Not-determined |
| 18-105_k141_174874 | Not-determined |
| 18-105_k141_40858 | Low-quality |
| 18-105_k141_49741 | Low-quality |
| 18-105_k141_103508 | Not-determined |
| 18-105_k141_165283 | Low-quality |
| 18-105_k141_120554 | Not-determined |
| 18-105_k141_165295 | Not-determined |
| 18-105_k141_22170 | Not-determined |
| 18-105_k141_165297 | Not-determined |
| 18-105_k141_22177 | Not-determined |
| 18-105_k141_174894 | Not-determined |
| 18-105_k141_174896 | Not-determined |
| 18-105_k141_49749 | Not-determined |
| 18-105_k141_174898 | Not-determined |
| 18-105_k141_22183 | Not-determined |
| 18-105_k141_7059 | Not-determined |
| 18-105_k141_76724 | Not-determined |
| 18-105_k141_76725 | Not-determined |
| 18-105_k141_7063 | Not-determined |
| 18-105_k141_58833 | Not-determined |
| 18-105_k141_201107 | Not-determined |
| 18-105_k141_183596 | Not-determined |
| 18-105_k141_183600 | Not-determined |
| 18-105_k141_183602 | Not-determined |
| 18-105_k141_94197 | Low-quality |
| 18-105_k141_183605 | Not-determined |
| 18-105_k141_174908 | Not-determined |
| 18-105_k141_130081 | Not-determined |
| 18-105_k141_103551 | Not-determined |
| 18-105_k141_103552 | Not-determined |
| 18-105_k141_103555 | Not-determined |
| 18-105_k141_130086 | Not-determined |
| 18-105_k141_94211 | Not-determined |
| 18-105_k141_120583 | Not-determined |
| 18-105_k141_192284 | Low-quality |
| 18-105_k141_67141 | Not-determined |
| 18-105_k141_130088 | Low-quality |
| 18-105_k141_130089 | Not-determined |
| 18-105_k141_183627 | Not-determined |
| 18-105_k141_192293 | Not-determined |
| 18-105_k141_130092 | Not-determined |
| 18-105_k141_120595 | Not-determined |
| 18-105_k141_192295 | Not-determined |
| 18-105_k141_120603 | Not-determined |
| 18-105_k141_67148 | Not-determined |
| 18-105_k141_183637 | Not-determined |
| 18-105_k141_120605 | Not-determined |
| 18-105_k141_103567 | Not-determined |
| 18-105_k141_183642 | Not-determined |
| 18-105_k141_67157 | Not-determined |
| 18-105_k141_31433 | Not-determined |
| 18-105_k141_192325 | Not-determined |
| 18-105_k141_130106 | Not-determined |
| 18-105_k141_58880 | Not-determined |
| 18-105_k141_130112 | Not-determined |
| 18-105_k141_130119 | Not-determined |
| 18-105_k141_192345 | Not-determined |
| 18-105_k141_130125 | Not-determined |
| 18-105_k141_120644 | Not-determined |
| 18-105_k141_120648 | Not-determined |
| 18-105_k141_192357 | Not-determined |
| 18-105_k141_210203 | Not-determined |
| 18-105_k141_120651 | Not-determined |
| 18-105_k141_14328 | Not-determined |
| 18-105_k141_130137 | Not-determined |
| 18-105_k141_14332 | Not-determined |
| 18-105_k141_31478 | Not-determined |
| 18-105_k141_14339 | Not-determined |
| 18-105_k141_76750 | Not-determined |
| 18-105_k141_112110 | Not-determined |
| 18-105_k141_76754 | Not-determined |
| 18-105_k141_58897 | Not-determined |
| 18-105_k141_76759 | Not-determined |
| 18-105_k141_174936 | Not-determined |
| 18-105_k141_76762 | Not-determined |
| 18-105_k141_192384 | Not-determined |
| 18-105_k141_76765 | Not-determined |
| 18-105_k141_130147 | Not-determined |
| 18-105_k141_120684 | Not-determined |
| 18-105_k141_192388 | Not-determined |
| 18-105_k141_147798 | Not-determined |
| 18-105_k141_14344 | Not-determined |
| 18-105_k141_147802 | Not-determined |
| 18-105_k141_130151 | Not-determined |
| 18-105_k141_192393 | Not-determined |
| 18-105_k141_58918 | Low-quality |
| 18-101-final-viral-combined_k141_394574\|\|full | High-quality |
| 18-105_k141_58919 | Not-determined |
| 18-105_k141_14350 | Not-determined |
| 18-105_k141_147811 | Low-quality |
| 18-105_k141_58922 | Not-determined |
| 18-105_k141_67176 | Low-quality |
| 18-105_k141_192401 | Not-determined |
| 18-105_k141_174957 | Not-determined |
| 18-105_k141_174958 | Not-determined |
| 18-105_k141_192403 | Not-determined |
| 18-105_k141_94255 | Not-determined |
| 18-105_k141_103627 | Not-determined |
| 18-105_k141_94256 | Not-determined |
| 18-105_k141_58932 | Not-determined |
| 18-105_k141_58935 | Not-determined |
| 18-105_k141_192412 | Not-determined |
| 18-105_k141_174963 | Not-determined |
| 18-105_k141_31490 | Not-determined |
| 18-105_k141_192421 | Not-determined |
| 18-105_k141_14377 | Not-determined |
| 18-105_k141_49799 | Not-determined |
| 18-105_k141_31494 | Not-determined |
| 18-105_k141_103652 | Not-determined |
| 18-105_k141_103653 | Not-determined |
| 18-105_k141_58958 | Not-determined |
| 18-105_k141_130186 | Not-determined |
| 18-105_k141_94280 | Not-determined |
| 18-105_k141_31510 | Not-determined |
| 18-105_k141_58965 | Not-determined |
| 18-105_k141_58968 | Not-determined |
| 18-105_k141_94285 | Not-determined |
| 18-105_k141_112165 | Not-determined |
| 18-105_k141_31517 | Not-determined |
| 18-105_k141_22220 | Not-determined |
| 18-105_k141_192435 | Not-determined |
| 18-105_k141_103670 | Not-determined |
| 18-105_k141_22223 | Not-determined |
| 18-105_k141_31528 | Not-determined |
| 18-105_k141_112182 | Not-determined |
| 18-105_k141_22225 | Not-determined |
| 18-105_k141_22226 | Not-determined |
| 18-105_k141_192443 | Not-determined |
| 18-105_k141_67247 | Not-determined |
| 18-105_k141_7143 | Not-determined |
| 18-105_k141_58994 | Not-determined |
| 18-105_k141_103684 | Not-determined |
| 18-105_k141_138528 | Not-determined |
| 18-105_k141_138532 | Low-quality |
| 18-105_k141_103687 | Not-determined |
| 18-105_k141_49819 | Not-determined |
| 18-105_k141_67263 | Not-determined |
| 18-105_k141_94324 | Not-determined |
| 18-105_k141_67266 | Not-determined |
| 18-105_k141_31551 | Not-determined |
| 18-105_k141_76792 | Not-determined |
| 18-105_k141_138538 | Not-determined |
| 18-105_k141_130229 | Not-determined |
| 18-105_k141_130230 | Not-determined |
| 18-105_k141_120696 | Not-determined |
| 18-105_k141_67281 | Not-determined |
| 18-105_k141_174980 | Not-determined |
| 18-105_k141_147884 | Not-determined |
| 18-105_k141_130233 | Not-determined |
| 18-105_k141_31558 | Not-determined |
| 18-105_k141_147888 | Not-determined |
| 18-105_k141_31560 | Not-determined |
| 18-105_k141_67288 | Not-determined |
| 18-105_k141_59007 | Not-determined |
| 18-105_k141_103696 | Not-determined |
| 18-105_k141_94344 | Not-determined |
| 18-105_k141_31570 | Low-quality |
| 18-105_k141_138558 | Not-determined |
| 18-105_k141_59017 | Not-determined |
| 18-105_k141_147897 | Not-determined |
| 18-105_k141_7160 | Not-determined |
| 18-105_k141_138573 | Not-determined |
| 18-105_k141_40919 | Not-determined |
| 18-105_k141_183675 | Not-determined |
| 18-105_k141_103712 | Not-determined |
| 18-105_k141_103713 | Not-determined |
| 18-105_k141_14398 | Not-determined |
| 18-105_k141_14399 | Not-determined |
| 18-105_k141_130265 | Not-determined |
| 18-105_k141_147918 | Not-determined |
| 18-105_k141_120701 | Not-determined |
| 18-105_k141_174997 | Not-determined |
| 18-105_k141_147923 | Not-determined |
| 18-105_k141_112207 | Not-determined |
| 18-105_k141_183690 | Not-determined |
| 18-105_k141_130271 | Low-quality |
| 18-105_k141_130274 | Not-determined |
| 18-105_k141_112218 | Not-determined |
| 18-105_k141_130276 | Not-determined |
| 18-105_k141_130277 | Not-determined |
| 18-105_k141_14412 | Not-determined |
| 18-105_k141_14413 | Not-determined |
| 18-105_k141_103733 | Not-determined |
| 18-105_k141_103735 | Not-determined |
| 18-105_k141_40953 | Not-determined |
| 18-105_k141_31623 | Not-determined |
| 18-105_k141_175014 | Not-determined |
| 18-105_k141_94361 | Low-quality |
| 18-105_k141_120720 | Not-determined |
| 18-105_k141_31628 | Not-determined |
| 18-105_k141_112231 | Not-determined |
| 18-105_k141_40962 | Low-quality |
| 18-105_k141_31635 | Not-determined |
| 18-105_k141_7179 | Not-determined |
| 18-105_k141_112239 | Not-determined |
| 18-105_k141_40970 | Not-determined |
| 18-105_k141_138599 | Not-determined |
| 18-105_k141_175028 | Not-determined |
| 18-105_k141_7185 | Not-determined |
| 18-105_k141_22246 | Not-determined |
| 18-105_k141_22250 | Not-determined |
| 18-105_k141_175034 | Not-determined |
| 18-105_k141_130299 | Not-determined |
| 18-105_k141_22252 | Not-determined |
| 18-105_k141_175035 | Not-determined |
| 18-105_k141_112244 | Not-determined |
| 18-105_k141_112247 | Not-determined |
| 18-105_k141_22264 | Low-quality |
| 18-105_k141_112254 | Low-quality |
| 18-105_k141_175053 | Not-determined |
| 18-105_k141_130316 | Not-determined |
| 18-105_k141_40996 | Not-determined |
| 18-105_k141_22277 | Not-determined |
| 18-105_k141_22278 | Not-determined |
| 18-105_k141_130318 | Low-quality |
| 18-105_k141_85363 | Not-determined |
| 18-105_k141_156233 | Not-determined |
| 18-105_k141_192505 | Not-determined |
| 18-105_k141_76829 | Not-determined |
| 18-105_k141_67304 | Not-determined |
| 18-105_k141_156237 | Not-determined |
| 18-105_k141_130327 | Not-determined |
| 18-105_k141_156240 | Low-quality |
| 18-105_k141_76837 | Not-determined |
| 18-105_k141_156244 | Not-determined |
| 18-105_k141_31661 | Not-determined |
| 18-105_k141_76848 | Not-determined |
| 18-105_k141_94390 | Not-determined |
| 18-105_k141_175073 | Not-determined |
| 18-105_k141_85381 | Not-determined |
| 18-105_k141_192523 | Not-determined |
| 18-105_k141_192524 | Not-determined |
| 18-105_k141_192529 | Not-determined |
| 18-105_k141_76856 | Not-determined |
| 18-105_k141_22325 | Not-determined |
| 18-105_k141_31678 | Not-determined |
| 18-105_k141_41030 | Not-determined |
| 18-105_k141_130341 | Not-determined |
| 18-105_k141_130346 | Not-determined |
| 18-105_k141_76862 | Low-quality |
| 18-105_k141_76863 | Not-determined |
| 18-105_k141_156262 | Not-determined |
| 18-105_k141_156265 | Not-determined |
| 18-105_k141_147945 | Not-determined |
| 18-105_k141_76880 | Not-determined |
| 18-105_k141_138634 | Not-determined |
| 18-105_k141_192554 | Not-determined |
| 18-105_k141_192555 | Not-determined |
| 18-105_k141_138639 | Not-determined |
| 18-105_k141_156281 | Not-determined |
| 18-105_k141_103809 | Not-determined |
| 18-105_k141_147959 | Low-quality |
| 18-105_k141_120789 | Not-determined |
| 18-105_k141_192559 | Not-determined |
| 18-105_k141_192564 | Not-determined |
| 18-105_k141_22339 | Not-determined |
| 18-105_k141_138648 | Not-determined |
| 18-105_k141_41052 | Not-determined |
| 18-105_k141_183722 | Not-determined |
| 18-105_k141_130378 | Not-determined |
| 18-105_k141_120797 | Not-determined |
| 18-105_k141_130383 | Low-quality |
| 18-105_k141_147974 | Not-determined |
| 18-105_k141_49854 | Not-determined |
| 18-105_k141_156300 | Not-determined |
| 18-105_k141_192573 | Not-determined |
| 18-105_k141_147979 | Not-determined |
| 18-105_k141_156304 | Not-determined |
| 18-105_k141_147984 | Not-determined |
| 18-105_k141_147990 | Not-determined |
| 18-105_k141_41076 | Not-determined |
| 18-105_k141_192585 | Not-determined |
| 18-105_k141_41080 | Not-determined |
| 18-105_k141_183746 | Not-determined |
| 18-105_k141_41091 | Not-determined |
| 18-105_k141_183752 | Not-determined |
| 18-105_k141_112303 | Not-determined |
| 18-105_k141_112305 | Not-determined |
| 18-105_k141_76886 | Not-determined |
| 18-105_k141_183763 | Low-quality |
| 18-105_k141_192610 | Not-determined |
| 18-105_k141_183776 | Not-determined |
| 18-105_k141_147994 | Not-determined |
| 18-105_k141_147996 | Not-determined |
| 18-105_k141_120836 | Not-determined |
| 18-105_k141_85454 | Not-determined |
| 18-105_k141_130403 | Not-determined |
| 18-105_k141_130404 | Not-determined |
| 18-105_k141_103858 | Not-determined |
| 18-105_k141_103860 | Not-determined |
| 18-105_k141_130412 | Not-determined |
| 18-105_k141_148023 | Not-determined |
| 18-105_k141_103863 | Not-determined |
| 18-105_k141_120855 | Not-determined |
| 18-105_k141_7284 | Not-determined |
| 18-105_k141_148029 | Low-quality |
| 18-105_k141_7286 | Not-determined |
| 18-105_k141_120858 | Not-determined |
| 18-105_k141_76909 | Not-determined |
| 18-105_k141_85477 | Not-determined |
| 18-105_k141_85480 | Not-determined |
| 18-105_k141_41119 | Not-determined |
| 18-105_k141_112333 | Not-determined |
| 18-105_k141_41123 | Not-determined |
| 18-105_k141_103867 | Not-determined |
| 18-105_k141_156330 | Not-determined |
| 18-105_k141_85495 | Not-determined |
| 18-105_k141_156343 | Not-determined |
| 18-105_k141_112349 | Not-determined |
| 18-105_k141_103876 | Not-determined |
| 18-105_k141_103877 | Low-quality |
| 18-105_k141_85509 | Not-determined |
| 18-105_k141_156351 | Not-determined |
| 18-105_k141_103888 | Not-determined |
| 18-105_k141_103890 | Not-determined |
| 18-105_k141_76934 | Not-determined |
| 18-105_k141_201157 | Not-determined |
| 18-105_k141_148037 | Not-determined |
| 18-105_k141_7300 | Not-determined |
| 18-105_k141_130431 | Not-determined |
| 18-105_k141_85532 | Not-determined |
| 18-105_k141_76953 | Not-determined |
| 18-105_k141_7308 | Not-determined |
| 18-105_k141_67362 | Not-determined |
| 18-105_k141_130436 | Not-determined |
| 18-105_k141_67365 | Not-determined |
| 18-105_k141_49928 | Not-determined |
| 18-105_k141_148059 | Not-determined |
| 18-105_k141_41144 | Not-determined |
| 18-105_k141_67378 | Not-determined |
| 18-105_k141_49932 | Not-determined |
| 18-105_k141_67380 | Not-determined |
| 18-105_k141_201182 | Not-determined |
| 18-105_k141_49936 | Not-determined |
| 18-105_k141_22400 | Not-determined |
| 18-105_k141_22406 | Not-determined |
| 18-105_k141_183795 | Not-determined |
| 18-105_k141_183798 | Not-determined |
| 18-105_k141_85566 | Not-determined |
| 18-105_k141_148086 | Not-determined |
| 18-105_k141_201201 | Not-determined |
| 18-105_k141_67393 | Not-determined |
| 18-105_k141_41155 | Not-determined |
| 18-105_k141_192622 | Not-determined |
| 18-105_k141_183813 | Low-quality |
| 18-105_k141_49952 | Not-determined |
| 18-105_k141_183818 | Not-determined |
| 18-105_k141_201203 | Low-quality |
| 18-105_k141_192634 | Not-determined |
| 18-105_k141_31710 | Not-determined |
| 18-105_k141_31716 | Not-determined |
| 18-105_k141_148102 | Not-determined |
| 18-105_k141_183832 | Not-determined |
| 18-105_k141_22421 | Not-determined |
| 18-105_k141_130484 | Not-determined |
| 18-105_k141_201218 | Not-determined |
| 18-105_k141_49963 | Not-determined |
| 18-105_k141_41166 | Not-determined |
| 18-105_k141_31730 | Not-determined |
| 18-105_k141_183852 | Not-determined |
| 18-105_k141_120865 | Not-determined |
| 18-105_k141_183853 | Not-determined |
| 18-105_k141_85604 | Not-determined |
| 18-105_k141_31735 | Not-determined |
| 18-105_k141_148128 | Low-quality |
| 18-105_k141_59078 | Low-quality |
| 18-105_k141_201240 | Low-quality |
| 18-105_k141_49972 | Not-determined |
| 18-105_k141_59081 | Not-determined |
| 18-105_k141_14492 | Not-determined |
| 18-105_k141_94454 | Not-determined |
| 18-105_k141_49976 | Not-determined |
| 18-105_k141_59084 | Not-determined |
| 18-105_k141_148135 | Not-determined |
| 18-105_k141_41185 | Not-determined |
| 18-105_k141_148139 | Not-determined |
| 18-105_k141_67441 | Not-determined |
| 18-105_k141_41189 | Low-quality |
| 18-105_k141_85619 | Not-determined |
| 18-105_k141_120881 | Not-determined |
| 18-105_k141_183859 | Not-determined |
| 18-105_k141_49985 | Not-determined |
| 18-105_k141_7374 | Not-determined |
| 18-105_k141_7376 | Not-determined |
| 18-105_k141_183873 | Not-determined |
| 18-105_k141_201257 | Not-determined |
| 18-105_k141_175129 | Low-quality |
| 18-105_k141_192673 | Not-determined |
| 18-105_k141_192674 | Not-determined |
| 18-105_k141_85627 | Not-determined |
| 18-105_k141_148156 | Not-determined |
| 18-105_k141_175140 | Not-determined |
| 18-105_k141_175143 | Not-determined |
| 18-105_k141_175144 | Not-determined |
| 18-105_k141_50008 | Not-determined |
| 18-105_k141_7398 | Not-determined |
| 18-105_k141_156360 | Low-quality |
| 18-105_k141_148168 | Not-determined |
| 18-105_k141_192698 | Not-determined |
| 18-105_k141_183877 | Not-determined |
| 18-105_k141_192701 | Not-determined |
| 18-105_k141_77008 | Low-quality |
| 18-105_k141_120917 | Not-determined |
| 18-105_k141_183883 | Not-determined |
| 18-105_k141_183884 | Not-determined |
| 18-105_k141_85649 | Not-determined |
| 18-105_k141_7409 | Not-determined |
| 18-105_k141_41221 | Not-determined |
| 18-105_k141_148182 | Not-determined |
| 18-105_k141_192716 | Not-determined |
| 18-105_k141_201272 | Not-determined |
| 18-105_k141_50029 | Not-determined |
| 18-105_k141_148187 | Not-determined |
| 18-105_k141_183895 | Low-quality |
| 18-105_k141_31782 | Not-determined |
| 18-105_k141_138707 | Not-determined |
| 18-105_k141_130547 | Not-determined |
| 18-105_k141_201287 | Low-quality |
| 18-105_k141_50046 | Not-determined |
| 18-105_k141_103926 | Not-determined |
| 18-105_k141_50049 | Not-determined |
| 18-105_k141_103928 | Not-determined |
| 18-105_k141_103930 | Not-determined |
| 18-105_k141_138717 | Not-determined |
| 18-105_k141_183905 | Not-determined |
| 18-105_k141_183907 | Not-determined |
| 18-105_k141_130565 | Not-determined |
| 18-105_k141_183910 | Not-determined |
| 18-105_k141_201296 | Not-determined |
| 18-105_k141_201297 | Not-determined |
| 18-105_k141_59115 | Not-determined |
| 18-105_k141_41232 | Low-quality |
| 18-105_k141_41239 | Not-determined |
| 18-105_k141_59119 | Not-determined |
| 18-105_k141_59120 | Not-determined |
| 18-105_k141_103945 | Not-determined |
| 18-105_k141_183933 | Not-determined |
| 18-105_k141_59129 | Not-determined |
| 18-105_k141_41246 | Low-quality |
| 18-105_k141_130591 | Not-determined |
| 18-105_k141_77055 | Not-determined |
| 18-105_k141_103948 | Not-determined |
| 18-105_k141_130592 | Not-determined |
| 18-105_k141_31817 | Not-determined |
| 18-105_k141_103950 | Not-determined |
| 18-105_k141_103954 | Not-determined |
| 18-105_k141_103956 | Not-determined |
| 18-105_k141_210237 | Not-determined |
| 18-105_k141_120967 | Not-determined |
| 18-105_k141_175173 | Not-determined |
| 18-105_k141_41261 | Not-determined |
| 18-105_k141_130601 | Not-determined |
| 18-105_k141_31832 | Not-determined |
| 18-105_k141_130602 | Not-determined |
| 18-105_k141_103966 | Not-determined |
| 18-105_k141_165349 | Not-determined |
| 18-105_k141_41277 | Not-determined |
| 18-105_k141_175190 | Not-determined |
| 18-105_k141_175192 | Not-determined |
| 18-105_k141_156367 | Not-determined |
| 18-105_k141_7433 | Not-determined |
| 18-105_k141_156368 | Not-determined |
| 18-105_k141_120993 | Not-determined |
| 18-105_k141_103974 | Not-determined |
| 18-102-final-viral-combined_k141_248316\|\|full | High-quality |
| 18-105_k141_85654 | Not-determined |
| 18-105_k141_7456 | Not-determined |
| 18-105_k141_103982 | Not-determined |
| 18-105_k141_103986 | Not-determined |
| 18-105_k141_31844 | Low-quality |
| 18-105_k141_175207 | Not-determined |
| 18-105_k141_85666 | Not-determined |
| 18-105_k141_156399 | Not-determined |
| 18-105_k141_175216 | Not-determined |
| 18-105_k141_31858 | Not-determined |
| 18-105_k141_14505 | Not-determined |
| 18-105_k141_22431 | Not-determined |
| 18-105_k141_85677 | Not-determined |
| 18-105_k141_77081 | Not-determined |
| 18-105_k141_85680 | Not-determined |
| 18-105_k141_77088 | Not-determined |
| 18-105_k141_175226 | Not-determined |
| 18-105_k141_59158 | Not-determined |
| 18-105_k141_50085 | Not-determined |
| 18-105_k141_7477 | Not-determined |

| | |
|---|---|
| 18-105_k141_175233 | Not-determined |
| 18-105_k141_156401 | Not-determined |
| 18-105_k141_156403 | Not-determined |
| 18-105_k141_31860 | Not-determined |
| 18-105_k141_14520 | Not-determined |
| 18-105_k141_165376 | Not-determined |
| 18-105_k141_175245 | Not-determined |
| 18-105_k141_165377 | Low-quality |
| 18-105_k141_31869 | Not-determined |
| 18-105_k141_165382 | Not-determined |
| 18-105_k141_67502 | Not-determined |
| 18-105_k141_165390 | Not-determined |
| 18-105_k141_175256 | Not-determined |
| 18-105_k141_77105 | Not-determined |
| 18-105_k141_77108 | Not-determined |
| 18-105_k141_104018 | Not-determined |
| 18-105_k141_22440 | Not-determined |
| 18-105_k141_104019 | Not-determined |
| 18-105_k141_59175 | Low-quality |
| 18-105_k141_138743 | Not-determined |
| 18-105_k141_104022 | Not-determined |
| 18-105_k141_50116 | Not-determined |
| 18-105_k141_175272 | Not-determined |
| 18-105_k141_175275 | Not-determined |
| 18-105_k141_201355 | Not-determined |
| 18-105_k141_22461 | Not-determined |
| 18-105_k141_175281 | Not-determined |
| 18-105_k141_50146 | Not-determined |
| 18-105_k141_14548 | Not-determined |
| 18-105_k141_59190 | Not-determined |
| 18-105_k141_104046 | Not-determined |
| 18-105_k141_59197 | Not-determined |
| 18-105_k141_138774 | Not-determined |
| 18-105_k141_59200 | Not-determined |
| 18-105_k141_138777 | Not-determined |
| 18-105_k141_175296 | Low-quality |
| 18-105_k141_77130 | Not-determined |
| 18-105_k141_22483 | Not-determined |
| 18-105_k141_14553 | Not-determined |
| 18-105_k141_50167 | Not-determined |
| 18-105_k141_192720 | Not-determined |
| 18-105_k141_175310 | Not-determined |
| 18-105_k141_121032 | Not-determined |
| 18-105_k141_14573 | Low-quality |
| 18-105_k141_77152 | Not-determined |
| 18-105_k141_59227 | Not-determined |
| 18-105_k141_121046 | Not-determined |
| 18-105_k141_104066 | Not-determined |
| 18-105_k141_31928 | Not-determined |
| 18-105_k141_148230 | Not-determined |
| 18-105_k141_22505 | Not-determined |
| 18-105_k141_104070 | Not-determined |
| 18-105_k141_156452 | Not-determined |
| 18-105_k141_201382 | Not-determined |
| 18-105_k141_50195 | Not-determined |
| 18-105_k141_138839 | Not-determined |
| 18-105_k141_165452 | Not-determined |
| 18-105_k141_22533 | Not-determined |
| 18-105_k141_59256 | Not-determined |
| 18-105_k141_59259 | Not-determined |
| 18-105_k141_156484 | Not-determined |
| 18-105_k141_31950 | Not-determined |
| 18-105_k141_183945 | Not-determined |
| 18-105_k141_165463 | Low-quality |
| 18-105_k141_31958 | Not-determined |
| 18-105_k141_50213 | Not-determined |
| 18-105_k141_31960 | Not-determined |
| 18-105_k141_138856 | Low-quality |
| 18-105_k141_41319 | Not-determined |
| 18-105_k141_121075 | Not-determined |
| 18-105_k141_41322 | Not-determined |
| 18-105_k141_183963 | Not-determined |
| 18-105_k141_41326 | Not-determined |
| 18-105_k141_130652 | Low-quality |
| 18-105_k141_175340 | Not-determined |
| 18-105_k141_183968 | Not-determined |
| 18-105_k141_112438 | Not-determined |
| 18-105_k141_138884 | Not-determined |
| 18-105_k141_41335 | Not-determined |
| 18-105_k141_67600 | Not-determined |
| 18-105_k141_31974 | Not-determined |
| 18-105_k141_31976 | Not-determined |
| 18-105_k141_14627 | Not-determined |
| 18-105_k141_165477 | Not-determined |
| 18-105_k141_138891 | Not-determined |
| 18-105_k141_138893 | Not-determined |
| 18-105_k141_121093 | Not-determined |
| 18-105_k141_138895 | Not-determined |
| 18-105_k141_210253 | Not-determined |
| 18-105_k141_31989 | Not-determined |
| 18-105_k141_31993 | Not-determined |
| 18-105_k141_210270 | Not-determined |
| 18-105_k141_165492 | Not-determined |
| 18-105_k141_67623 | Not-determined |
| 18-105_k141_192761 | Not-determined |
| 18-105_k141_31999 | Not-determined |
| 18-105_k141_67629 | Not-determined |
| 18-105_k141_165501 | Not-determined |
| 18-105_k141_32006 | Not-determined |
| 18-105_k141_85720 | Not-determined |
| 18-105_k141_85722 | Not-determined |
| 18-105_k141_112462 | Not-determined |
| 18-105_k141_165509 | Not-determined |
| 18-105_k141_112463 | Not-determined |
| 18-105_k141_175360 | Not-determined |
| 18-105_k141_165517 | Not-determined |
| 18-105_k141_210278 | Not-determined |
| 18-105_k141_112475 | Not-determined |
| 18-105_k141_112478 | Not-determined |
| 18-105_k141_138927 | Not-determined |
| 18-105_k141_94494 | Not-determined |
| 18-105_k141_50225 | Not-determined |
| 18-105_k141_50226 | Not-determined |
| 18-105_k141_210288 | Not-determined |
| 18-105_k141_165532 | Not-determined |
| 18-105_k141_50231 | Not-determined |
| 18-105_k141_165536 | Not-determined |
| 18-105_k141_7513 | Not-determined |
| 18-105_k141_184001 | Not-determined |
| 18-105_k141_77210 | Low-quality |
| 18-105_k141_130676 | Not-determined |
| 18-105_k141_14629 | Low-quality |
| 18-105_k141_50242 | Not-determined |
| 18-105_k141_175387 | Not-determined |
| 18-105_k141_210300 | Not-determined |
| 18-105_k141_50247 | Not-determined |
| 18-105_k141_77218 | Not-determined |
| 18-105_k141_210308 | Not-determined |
| 18-105_k141_41382 | Not-determined |
| 18-105_k141_32041 | Not-determined |
| 18-105_k141_156516 | Not-determined |
| 18-105_k141_32044 | Not-determined |
| 18-105_k141_112503 | Not-determined |
| 18-105_k141_32048 | Not-determined |
| 18-105_k141_112506 | Not-determined |
| 18-105_k141_175406 | Not-determined |
| 18-105_k141_14645 | Low-quality |
| 18-105_k141_50251 | Not-determined |
| 18-105_k141_138961 | Low-quality |
| 18-105_k141_32058 | Not-determined |
| 18-105_k141_112513 | Low-quality |
| 18-105_k141_156538 | Not-determined |
| 18-105_k141_156541 | Not-determined |
| 18-105_k141_7524 | Not-determined |
| 18-105_k141_7526 | Not-determined |
| 18-105_k141_138967 | Not-determined |
| 18-105_k141_192790 | Not-determined |
| 18-105_k141_112526 | Not-determined |
| 18-105_k141_184016 | Not-determined |
| 18-105_k141_77248 | Not-determined |
| 18-105_k141_77253 | Not-determined |
| 18-105_k141_148292 | Not-determined |
| 18-105_k141_184027 | Not-determined |
| 18-105_k141_77259 | Not-determined |
| 18-105_k141_121115 | Not-determined |
| 18-105_k141_121117 | Not-determined |
| 18-105_k141_165576 | Low-quality |
| 18-105_k141_32065 | Not-determined |
| 18-105_k141_165583 | Not-determined |
| 18-105_k141_14680 | Not-determined |
| 18-105_k141_192793 | Not-determined |
| 18-105_k141_148309 | Not-determined |
| 18-105_k141_192794 | Not-determined |
| 18-105_k141_165585 | Not-determined |
| 18-105_k141_210329 | Not-determined |
| 18-105_k141_184037 | Not-determined |
| 18-105_k141_130734 | Not-determined |
| 18-105_k141_7535 | Not-determined |
| 18-105_k141_7538 | Not-determined |
| 18-105_k141_14689 | Not-determined |
| 18-105_k141_201424 | Not-determined |
| 18-105_k141_7543 | Not-determined |
| 18-105_k141_139001 | Not-determined |
| 18-105_k141_139002 | Not-determined |
| 18-105_k141_77298 | Not-determined |
| 18-105_k141_77304 | Not-determined |
| 18-105_k141_175448 | Not-determined |
| 18-105_k141_210331 | Not-determined |
| 18-105_k141_112557 | Not-determined |
| 18-105_k141_201439 | Not-determined |
| 18-105_k141_14703 | Low-quality |
| 18-105_k141_130752 | Not-determined |
| 18-105_k141_7560 | Not-determined |
| 18-105_k141_210336 | Low-quality |
| 18-105_k141_148331 | Not-determined |
| 18-105_k141_121150 | Not-determined |
| 18-105_k141_210338 | Not-determined |
| 18-105_k141_67643 | Not-determined |
| 18-105_k141_112585 | Not-determined |
| 18-105_k141_67649 | Not-determined |
| 18-105_k141_22537 | Not-determined |
| 18-105_k141_22539 | Not-determined |
| 18-105_k141_184047 | Not-determined |
| 18-105_k141_130780 | Not-determined |
| 18-105_k141_112601 | Low-quality |
| 18-105_k141_130793 | Low-quality |
| 18-105_k141_14716 | Not-determined |
| 18-105_k141_85757 | Not-determined |
| 18-105_k141_175479 | Not-determined |
| 18-105_k141_175482 | Not-determined |
| 18-105_k141_77335 | Not-determined |
| 18-105_k141_201466 | Not-determined |
| 18-105_k141_130812 | Not-determined |
| 18-105_k141_148382 | Not-determined |
| 18-105_k141_175502 | Not-determined |
| 18-105_k141_77347 | Low-quality |
| 18-105_k141_77350 | Not-determined |
| 18-105_k141_156591 | Not-determined |
| 18-105_k141_192799 | Not-determined |
| 18-105_k141_77356 | Not-determined |
| 18-105_k141_14728 | Not-determined |
| 18-105_k141_77359 | Not-determined |
| 18-105_k141_192807 | Not-determined |
| 18-105_k141_156598 | Not-determined |
| 18-105_k141_156599 | Not-determined |
| 18-105_k141_104090 | Not-determined |
| 18-105_k141_59267 | Not-determined |
| 18-105_k141_156611 | Not-determined |
| 18-105_k141_192818 | Not-determined |
| 18-105_k141_139050 | Not-determined |
| 18-105_k141_112621 | Not-determined |
| 18-105_k141_192824 | Not-determined |
| 18-105_k141_130837 | Not-determined |
| 18-105_k141_165622 | Low-quality |
| 18-105_k141_148411 | Not-determined |
| 18-105_k141_32108 | Not-determined |
| 18-105_k141_85796 | Not-determined |
| 18-105_k141_85797 | Not-determined |
| 18-105_k141_85798 | Not-determined |
| 18-105_k141_148415 | Not-determined |
| 18-105_k141_85806 | Not-determined |
| 18-105_k141_156626 | Not-determined |
| 18-105_k141_139074 | Not-determined |
| 18-105_k141_32115 | Not-determined |
| 18-105_k141_130862 | Not-determined |
| 18-105_k141_59283 | Not-determined |
| 18-105_k141_94525 | Not-determined |
| 18-105_k141_94528 | Not-determined |
| 18-105_k141_94530 | Not-determined |
| 18-105_k141_94531 | Not-determined |
| 18-105_k141_85819 | Not-determined |
| 18-105_k141_85823 | Not-determined |
| 18-105_k141_201497 | Not-determined |
| 18-105_k141_148431 | Not-determined |
| 18-105_k141_148432 | Low-quality |
| 18-105_k141_85831 | Not-determined |
| 18-105_k141_94543 | Not-determined |
| 18-105_k141_14755 | Not-determined |
| 18-105_k141_94548 | Not-determined |
| 18-105_k141_192845 | Not-determined |
| 18-105_k141_85843 | Not-determined |
| 18-105_k141_192848 | Not-determined |
| 18-105_k141_139081 | Not-determined |
| 18-105_k141_201510 | Not-determined |
| 18-105_k141_85850 | Not-determined |
| 18-105_k141_50302 | Not-determined |
| 18-105_k141_59310 | Not-determined |
| 18-105_k141_201516 | Not-determined |
| 18-105_k141_192855 | Low-quality |
| 18-105_k141_184117 | Not-determined |
| 18-105_k141_165635 | Not-determined |
| 18-105_k141_14782 | Not-determined |
| 18-105_k141_59315 | Not-determined |
| 18-105_k141_165640 | Low-quality |
| 18-105_k141_210354 | Not-determined |
| 18-105_k141_192864 | Not-determined |
| 18-105_k141_184132 | Not-determined |
| 18-105_k141_156667 | Not-determined |
| 18-105_k141_130888 | Not-determined |
| 18-105_k141_156671 | Not-determined |
| 18-105_k141_59343 | Not-determined |
| 18-105_k141_104110 | Not-determined |
| 18-105_k141_94585 | Not-determined |
| 18-105_k141_165656 | Not-determined |
| 18-105_k141_112649 | Not-determined |
| 18-105_k141_7624 | Not-determined |
| 18-105_k141_165660 | Not-determined |
| 18-105_k141_165662 | Not-determined |
| 18-105_k141_184151 | Not-determined |
| 18-105_k141_104117 | Low-quality |
| 18-105_k141_184153 | Not-determined |
| 18-105_k141_50333 | Not-determined |
| 18-105_k141_210376 | Low-quality |
| 18-105_k141_32175 | Not-determined |
| 18-105_k141_192873 | Not-determined |
| 18-105_k141_77404 | Not-determined |
| 18-105_k141_184160 | Not-determined |
| 18-105_k141_148483 | Not-determined |
| 18-105_k141_148484 | Not-determined |
| 18-105_k141_148486 | Not-determined |
| 18-105_k141_85885 | Not-determined |
| 18-105_k141_201531 | Not-determined |
| 18-105_k141_77410 | Not-determined |
| 18-105_k141_77412 | Not-determined |
| 18-105_k141_192884 | Not-determined |
| 18-105_k141_192889 | Not-determined |
| 18-105_k141_77413 | Low-quality |
| 18-105_k141_210389 | Not-determined |
| 18-105_k141_148498 | Not-determined |
| 18-105_k141_7636 | Not-determined |
| 18-105_k141_184176 | Not-determined |
| 18-105_k141_130941 | Not-determined |
| 18-105_k141_130942 | Not-determined |
| 18-105_k141_85899 | Not-determined |
| 18-105_k141_94619 | Not-determined |
| 18-105_k141_7648 | Not-determined |
| 18-105_k141_192913 | Not-determined |
| 18-105_k141_14800 | Not-determined |
| 18-105_k141_85914 | Not-determined |
| 18-105_k141_184184 | Not-determined |
| 18-105_k141_139099 | Not-determined |
| 18-105_k141_210408 | Not-determined |
| 18-105_k141_14812 | Not-determined |
| 18-105_k141_94632 | Not-determined |
| 18-105_k141_130962 | Not-determined |
| 18-105_k141_50363 | Not-determined |
| 18-105_k141_210415 | Not-determined |
| 18-105_k141_14823 | Not-determined |
| 18-105_k141_50368 | Not-determined |
| 18-105_k141_14825 | Not-determined |
| 18-105_k141_210419 | Not-determined |
| 18-105_k141_184197 | Not-determined |
| 18-105_k141_50379 | Not-determined |
| 18-105_k141_165713 | Not-determined |
| 18-105_k141_50381 | Low-quality |
| 18-105_k141_192931 | Not-determined |
| 18-105_k141_210423 | Low-quality |
| 18-105_k141_22592 | Not-determined |
| 18-105_k141_210424 | Low-quality |
| 18-105_k141_14838 | Not-determined |
| 18-105_k141_184208 | Not-determined |
| 18-105_k141_184211 | Not-determined |
| 18-105_k141_7665 | Not-determined |
| 18-105_k141_210434 | Not-determined |
| 18-105_k141_210436 | Not-determined |
| 18-105_k141_210438 | Low-quality |
| 18-105_k141_184220 | Not-determined |
| 18-105_k141_7672 | Not-determined |
| 18-105_k141_192954 | Not-determined |
| 18-105_k141_50400 | Not-determined |
| 18-105_k141_184227 | Not-determined |
| 18-105_k141_22598 | Not-determined |
| 18-105_k141_201550 | Not-determined |
| 18-105_k141_85969 | Not-determined |
| 18-105_k141_201555 | Not-determined |
| 18-105_k141_77437 | Not-determined |
| 18-105_k141_104134 | Not-determined |
| 18-105_k141_50411 | Not-determined |
| 18-105_k141_210455 | Not-determined |
| 18-105_k141_139124 | Not-determined |
| 18-105_k141_77446 | Not-determined |
| 18-105_k141_104147 | Not-determined |
| 18-105_k141_104151 | Not-determined |
| 18-105_k141_32192 | Not-determined |
| 18-105_k141_50421 | Low-quality |
| 18-105_k141_104155 | Not-determined |
| 18-105_k141_139137 | Not-determined |
| 18-105_k141_131019 | Not-determined |
| 18-105_k141_77460 | Not-determined |
| 18-105_k141_165722 | Not-determined |
| 18-105_k141_77463 | Not-determined |
| 18-105_k141_50430 | Not-determined |
| 18-105_k141_22628 | Not-determined |
| 18-105_k141_192970 | Not-determined |
| 18-105_k141_41455 | Low-quality |
| 18-105_k141_184275 | Not-determined |
| 18-105_k141_156707 | Not-determined |
| 18-105_k141_139156 | Not-determined |
| 18-105_k141_112714 | Not-determined |
| 18-105_k141_22642 | Not-determined |
| 18-105_k141_7696 | Not-determined |
| 17-109-final-viral-combined_k141_264425\|\|full | High-quality |
| 18-105_k141_77478 | Not-determined |
| 18-105_k141_22648 | Not-determined |
| 18-105_k141_165742 | Not-determined |
| 18-105_k141_77479 | Not-determined |
| 18-105_k141_165749 | Not-determined |
| 18-105_k141_165750 | Not-determined |
| 18-105_k141_156729 | Not-determined |
| 18-105_k141_192989 | Not-determined |
| 18-105_k141_192991 | Not-determined |
| 18-105_k141_7709 | Not-determined |
| 18-105_k141_7713 | Not-determined |
| 18-105_k141_104174 | Not-determined |
| 18-105_k141_7714 | Not-determined |
| 18-105_k141_94649 | Not-determined |
| 18-105_k141_112739 | Not-determined |
| 18-105_k141_121210 | Not-determined |
| 18-105_k141_59359 | Not-determined |
| 18-105_k141_7721 | Not-determined |
| 18-105_k141_210466 | Not-determined |
| 18-105_k141_22664 | Not-determined |
| 18-105_k141_32205 | Low-quality |
| 18-105_k141_59371 | Not-determined |
| 18-105_k141_59372 | Not-determined |
| 18-105_k141_165774 | Low-quality |
| 18-105_k141_94661 | Not-determined |
| 18-105_k141_184310 | Not-determined |
| 18-105_k141_112753 | Not-determined |
| 18-105_k141_210477 | Not-determined |
| 18-105_k141_59381 | Not-determined |
| 18-105_k141_59385 | Not-determined |
| 18-105_k141_59387 | Not-determined |
| 18-105_k141_193006 | Not-determined |
| 18-105_k141_32222 | Not-determined |
| 18-105_k141_32223 | Not-determined |
| 18-105_k141_156734 | Not-determined |
| 18-105_k141_86021 | Not-determined |
| 18-105_k141_77511 | Not-determined |
| 18-105_k141_193013 | Not-determined |
| 18-105_k141_139177 | Not-determined |
| 18-105_k141_175555 | Not-determined |
| 18-105_k141_41467 | Not-determined |
| 18-105_k141_41468 | Not-determined |
| 18-105_k141_94676 | Not-determined |
| 18-105_k141_41471 | Not-determined |
| 18-105_k141_193022 | Not-determined |
| 18-105_k141_193023 | Not-determined |
| 18-105_k141_86031 | Not-determined |
| 18-105_k141_156740 | Not-determined |
| 18-105_k141_112773 | Not-determined |
| 18-105_k141_86034 | Not-determined |
| 18-105_k141_94681 | Low-quality |
| 18-105_k141_77515 | Not-determined |
| 18-105_k141_32237 | Not-determined |
| 18-105_k141_77516 | Not-determined |
| 18-105_k141_131073 | Not-determined |
| 18-105_k141_131075 | Not-determined |
| 18-105_k141_32244 | Low-quality |
| 18-105_k141_131077 | Not-determined |
| 18-105_k141_59408 | Not-determined |
| 18-105_k141_131079 | Not-determined |
| 18-105_k141_139183 | Not-determined |
| 18-105_k141_210491 | Not-determined |
| 18-105_k141_139185 | Not-determined |
| 18-105_k141_59413 | Not-determined |
| 18-105_k141_22669 | Not-determined |
| 18-105_k141_184350 | Not-determined |
| 18-105_k141_139186 | Not-determined |
| 18-105_k141_193044 | Not-determined |
| 18-105_k141_139187 | Not-determined |
| 18-105_k141_59418 | Not-determined |
| 18-105_k141_121262 | Not-determined |
| 18-105_k141_22678 | Not-determined |
| 18-105_k141_139196 | Not-determined |
| 18-105_k141_86062 | Not-determined |
| 18-105_k141_22682 | Not-determined |
| 18-105_k141_210502 | Low-quality |
| 18-105_k141_59427 | Low-quality |
| 18-105_k141_59429 | Low-quality |
| 18-105_k141_121276 | Not-determined |
| 18-105_k141_121289 | Not-determined |
| 18-105_k141_131086 | Not-determined |
| 18-105_k141_32286 | Not-determined |
| 18-105_k141_121290 | Low-quality |
| 18-105_k141_77545 | Not-determined |
| 18-105_k141_121307 | Low-quality |
| 18-105_k141_94697 | Not-determined |
| 18-105_k141_201590 | Not-determined |
| 18-105_k141_77554 | Not-determined |
| 18-105_k141_193087 | Not-determined |
| 18-105_k141_156772 | Not-determined |
| 18-105_k141_184369 | Not-determined |
| 18-105_k141_77561 | Not-determined |
| 18-105_k141_7749 | Not-determined |
| 18-105_k141_59435 | Not-determined |
| 18-105_k141_184374 | Not-determined |
| 18-105_k141_67699 | Not-determined |
| 18-105_k141_77567 | Not-determined |
| 18-105_k141_22701 | Not-determined |
| 18-105_k141_67712 | Not-determined |
| 18-105_k141_156791 | Not-determined |
| 18-105_k141_210563 | Not-determined |
| 18-105_k141_201627 | Not-determined |
| 18-105_k141_156798 | Not-determined |
| 18-105_k141_77580 | Not-determined |
| 18-105_k141_59450 | Not-determined |
| 18-105_k141_201636 | Not-determined |
| 18-105_k141_59457 | Not-determined |
| 18-105_k141_7779 | Not-determined |
| 18-105_k141_32311 | Not-determined |
| 18-105_k141_7782 | Not-determined |
| 18-105_k141_148552 | Low-quality |
| 18-105_k141_77589 | Not-determined |
| 18-105_k141_94726 | Not-determined |
| 18-105_k141_77597 | Not-determined |
| 18-105_k141_175588 | Low-quality |
| 18-105_k141_94730 | Not-determined |
| 18-105_k141_77599 | Low-quality |
| 18-105_k141_156822 | Not-determined |
| 18-105_k141_156824 | Not-determined |
| 18-105_k141_131128 | Not-determined |
| 18-105_k141_131131 | Not-determined |
| 18-105_k141_112803 | Not-determined |
| 18-105_k141_148574 | Not-determined |
| 18-105_k141_94744 | Not-determined |
| 18-105_k141_59489 | Low-quality |
| 18-105_k141_131142 | Not-determined |
| 18-105_k141_131143 | Not-determined |
| 18-105_k141_7815 | Low-quality |
| 18-105_k141_67765 | Not-determined |
| 18-105_k141_67766 | Not-determined |
| 18-105_k141_32319 | Not-determined |
| 18-105_k141_175602 | Not-determined |
| 18-105_k141_148587 | Not-determined |
| 18-105_k141_67773 | Not-determined |
| 18-105_k141_156854 | Low-quality |
| 18-105_k141_131164 | Not-determined |
| 18-105_k141_32324 | Not-determined |
| 18-105_k141_59510 | Not-determined |
| 18-105_k141_67778 | Not-determined |
| 18-105_k141_94771 | Not-determined |
| 18-105_k141_94775 | Not-determined |
| 18-105_k141_104217 | Not-determined |
| 18-105_k141_94782 | Not-determined |
| 18-105_k141_193128 | Not-determined |
| 18-105_k141_32336 | Low-quality |
| 18-105_k141_184460 | Not-determined |
| 18-105_k141_104232 | Not-determined |
| 18-105_k141_67790 | Not-determined |
| 18-105_k141_193132 | Not-determined |
| 18-105_k141_139303 | Low-quality |
| 18-105_k141_139304 | Not-determined |
| 18-105_k141_210570 | Not-determined |
| 18-105_k141_156878 | Not-determined |
| 18-105_k141_210571 | Not-determined |
| 18-105_k141_193137 | Not-determined |
| 18-105_k141_175636 | Not-determined |
| 18-105_k141_165878 | Not-determined |
| 18-105_k141_32361 | Low-quality |
| 18-105_k141_104255 | Not-determined |
| 18-105_k141_184481 | Not-determined |
| 18-105_k141_112824 | Not-determined |
| 18-105_k141_104259 | Not-determined |
| 18-105_k141_112828 | Not-determined |
| 18-105_k141_32369 | Not-determined |
| 18-105_k141_32371 | Low-quality |
| 18-105_k141_210587 | Not-determined |
| 18-105_k141_201676 | Not-determined |
| 18-105_k141_210591 | Not-determined |
| 18-105_k141_193144 | Not-determined |
| 18-105_k141_156895 | Not-determined |
| 18-105_k141_104278 | Not-determined |
| 18-105_k141_32386 | Not-determined |
| 18-105_k141_22805 | Not-determined |
| 18-105_k141_210602 | Not-determined |
| 18-105_k141_14858 | Not-determined |
| 18-105_k141_184497 | Not-determined |
| 18-105_k141_201690 | Not-determined |
| 18-105_k141_112847 | Not-determined |
| 18-105_k141_131178 | Not-determined |
| 18-105_k141_14870 | Low-quality |
| 18-105_k141_201698 | Not-determined |
| 18-105_k141_184504 | Not-determined |
| 18-105_k141_184506 | Not-determined |
| 18-105_k141_184509 | Not-determined |
| 18-105_k141_14877 | Not-determined |
| 18-105_k141_77616 | Not-determined |
| 18-105_k141_165925 | Not-determined |
| 18-105_k141_184519 | Not-determined |
| 18-105_k141_77622 | Not-determined |
| 18-105_k141_77627 | Not-determined |
| 18-105_k141_148674 | Not-determined |
| 18-105_k141_59563 | Not-determined |
| 18-105_k141_77636 | Not-determined |
| 18-105_k141_77640 | Not-determined |
| 18-105_k141_77641 | Not-determined |
| 18-105_k141_77642 | Not-determined |
| 18-105_k141_165951 | Low-quality |
| 18-105_k141_210641 | Not-determined |
| 18-105_k141_77655 | Not-determined |
| 18-105_k141_210648 | Not-determined |
| 18-105_k141_210650 | Not-determined |
| 18-105_k141_210654 | Not-determined |
| 18-105_k141_50461 | Not-determined |
| 18-105_k141_14907 | Not-determined |
| 18-105_k141_14910 | Not-determined |
| 18-105_k141_86127 | Not-determined |
| 18-105_k141_86128 | Not-determined |
| 18-105_k141_77668 | Not-determined |
| 18-105_k141_210660 | Not-determined |
| 18-105_k141_210662 | Not-determined |
| 18-105_k141_210663 | Not-determined |
| 18-105_k141_210664 | Not-determined |
| 18-105_k141_148718 | Not-determined |
| 18-105_k141_175669 | Not-determined |
| 18-105_k141_193151 | Not-determined |
| 18-105_k141_112862 | Not-determined |
| 18-105_k141_210673 | Low-quality |

| | |
|---|---|
| 18-105_k141_148729 | Not-determined |
| 18-105_k141_175673 | Not-determined |
| 18-105_k141_14925 | Low-quality |
| 18-105_k141_184567 | Not-determined |
| 18-105_k141_210682 | Not-determined |
| 18-105_k141_121370 | Not-determined |
| 18-105_k141_148747 | Not-determined |
| 18-105_k141_50483 | Not-determined |
| 18-105_k141_94801 | Not-determined |
| 18-105_k141_121384 | Not-determined |
| 18-105_k141_50494 | Low-quality |
| 18-105_k141_94810 | Not-determined |
| 18-105_k141_14947 | Low-quality |
| 18-105_k141_139355 | Not-determined |
| 18-105_k141_14951 | Not-determined |
| 18-105_k141_193167 | Not-determined |
| 18-105_k141_59597 | Not-determined |
| 18-105_k141_139362 | Not-determined |
| 18-105_k141_166005 | Not-determined |
| 18-105_k141_148765 | Not-determined |
| 18-105_k141_59601 | Not-determined |
| 18-105_k141_94826 | Not-determined |
| 18-105_k141_166013 | Not-determined |
| 18-105_k141_77687 | Not-determined |
| 18-105_k141_201737 | Low-quality |
| 18-105_k141_201745 | Not-determined |
| 18-105_k141_139378 | Not-determined |
| 18-105_k141_210706 | Not-determined |
| 18-105_k141_77698 | Not-determined |
| 18-105_k141_14962 | Not-determined |
| 18-105_k141_139385 | Not-determined |
| 18-105_k141_184591 | Low-quality |
| 18-105_k141_14972 | Not-determined |
| 18-105_k141_210715 | Not-determined |
| 18-105_k141_139394 | Not-determined |
| 18-105_k141_104363 | Not-determined |
| 18-105_k141_67794 | Not-determined |
| 18-105_k141_67797 | Not-determined |
| 18-105_k141_104373 | Not-determined |
| 18-105_k141_59616 | Not-determined |
| 18-105_k141_210721 | Not-determined |
| 18-105_k141_175731 | Not-determined |
| 18-105_k141_131207 | Not-determined |
| 18-105_k141_175738 | Not-determined |
| 18-105_k141_14995 | Not-determined |
| 18-105_k141_112903 | Not-determined |
| 18-105_k141_15004 | Not-determined |
| 18-105_k141_139412 | Not-determined |
| 18-105_k141_175754 | Not-determined |
| 18-105_k141_148797 | Low-quality |
| 18-105_k141_193174 | Not-determined |
| 18-105_k141_139423 | Low-quality |
| 18-105_k141_210734 | Not-determined |
| 18-105_k141_131245 | Not-determined |
| 18-105_k141_59652 | Not-determined |
| 18-105_k141_131252 | Not-determined |
| 18-105_k141_32410 | Not-determined |
| 18-105_k141_193203 | Not-determined |
| 18-105_k141_131263 | Low-quality |
| 18-105_k141_139450 | Not-determined |
| 18-105_k141_121416 | Not-determined |
| 18-105_k141_86203 | Not-determined |
| 18-105_k141_166045 | Not-determined |
| 18-105_k141_121423 | Not-determined |
| 18-105_k141_121424 | Not-determined |
| 18-105_k141_210749 | Not-determined |
| 18-105_k141_210752 | Not-determined |
| 18-105_k141_131281 | Not-determined |
| 18-105_k141_210753 | Not-determined |
| 18-105_k141_193227 | Not-determined |
| 18-105_k141_32427 | Not-determined |
| 18-105_k141_32428 | Not-determined |
| 18-105_k141_184651 | Not-determined |
| 18-105_k141_193228 | Low-quality |
| 18-105_k141_22890 | Not-determined |
| 18-105_k141_131282 | Low-quality |
| 18-105_k141_184654 | Not-determined |
| 18-105_k141_112906 | Not-determined |
| 18-105_k141_86240 | Not-determined |
| 18-105_k141_156934 | Not-determined |
| 18-105_k141_7848 | Not-determined |
| 18-105_k141_112909 | Low-quality |
| 18-105_k141_112912 | Not-determined |
| 18-105_k141_210785 | Not-determined |
| 18-105_k141_210799 | Not-determined |
| 18-105_k141_156944 | Not-determined |
| 18-105_k141_131302 | Low-quality |
| 18-105_k141_175804 | Not-determined |
| 18-105_k141_41498 | Not-determined |
| 18-105_k141_22916 | Not-determined |
| 18-105_k141_156948 | Not-determined |
| 18-105_k141_156949 | Not-determined |
| 18-105_k141_22923 | Not-determined |
| 18-105_k141_41508 | Not-determined |
| 18-105_k141_41510 | Not-determined |
| 18-105_k141_175812 | Low-quality |
| 18-105_k141_175818 | Not-determined |
| 18-105_k141_41520 | Not-determined |
| 18-105_k141_156954 | Low-quality |
| 18-105_k141_86269 | Not-determined |
| 18-105_k141_22930 | Low-quality |
| 18-105_k141_41526 | Not-determined |
| 18-105_k141_175830 | Not-determined |
| 18-105_k141_156960 | Low-quality |
| 18-105_k141_50585 | Not-determined |
| 18-105_k141_112920 | Not-determined |
| 18-105_k141_112924 | Not-determined |
| 18-105_k141_184728 | Not-determined |
| 18-105_k141_104433 | Not-determined |
| 18-105_k141_131321 | Not-determined |
| 18-105_k141_156977 | Not-determined |
| 18-105_k141_156979 | Not-determined |
| 18-105_k141_121485 | Not-determined |
| 18-105_k141_86297 | Not-determined |
| 18-105_k141_86299 | Not-determined |
| 18-105_k141_210842 | Not-determined |
| 18-105_k141_50605 | Not-determined |
| 18-105_k141_59683 | Not-determined |
| 18-105_k141_184734 | Not-determined |
| 18-105_k141_50611 | Not-determined |
| 18-105_k141_121488 | Not-determined |
| 18-105_k141_131336 | Not-determined |
| 18-105_k141_131339 | Not-determined |
| 18-105_k141_104445 | Not-determined |
| 18-105_k141_59691 | Not-determined |
| 18-105_k141_139490 | Not-determined |
| 18-105_k141_50622 | Not-determined |
| 18-105_k141_156994 | Low-quality |
| 18-105_k141_201835 | Low-quality |
| 18-105_k141_139502 | Not-determined |
| 18-105_k141_184755 | Not-determined |
| 18-105_k141_86312 | Not-determined |
| 18-105_k141_41545 | Not-determined |
| 18-105_k141_139508 | Not-determined |
| 18-105_k141_77732 | Not-determined |
| 18-105_k141_139510 | Not-determined |
| 18-105_k141_77736 | Not-determined |
| 18-105_k141_86327 | Not-determined |
| 18-105_k141_157013 | Not-determined |
| 18-105_k141_157016 | Not-determined |
| 18-105_k141_50637 | Not-determined |
| 18-105_k141_157017 | Not-determined |
| 18-105_k141_166092 | Not-determined |
| 18-105_k141_157028 | Not-determined |
| 18-105_k141_157030 | Not-determined |
| 18-105_k141_7885 | Not-determined |
| 18-105_k141_157033 | Not-determined |
| 18-105_k141_121519 | Not-determined |
| 18-105_k141_157045 | Not-determined |
| 18-105_k141_166105 | Low-quality |
| 18-105_k141_157050 | Not-determined |
| 18-105_k141_7887 | Not-determined |
| 18-105_k141_121523 | Not-determined |
| 18-105_k141_121524 | Not-determined |
| 18-105_k141_112931 | Not-determined |
| 18-105_k141_201852 | Not-determined |
| 18-105_k141_175891 | Low-quality |
| 18-105_k141_41583 | Not-determined |
| 18-105_k141_7896 | Low-quality |
| 18-105_k141_50666 | Not-determined |
| 18-105_k141_112946 | Not-determined |
| 18-105_k141_7897 | Low-quality |
| 18-105_k141_104460 | Not-determined |
| 18-105_k141_22943 | Not-determined |
| 18-105_k141_86371 | Low-quality |
| 18-105_k141_166115 | Not-determined |
| 18-105_k141_41592 | Not-determined |
| 18-105_k141_77764 | Not-determined |
| 18-105_k141_77769 | Not-determined |
| 18-105_k141_50680 | Not-determined |
| 18-105_k141_77776 | Not-determined |
| 18-105_k141_148838 | Not-determined |
| 18-105_k141_157082 | Not-determined |
| 18-105_k141_104472 | Not-determined |
| 18-105_k141_210883 | Not-determined |
| 18-105_k141_148841 | Not-determined |
| 18-105_k141_166153 | Not-determined |
| 18-105_k141_59720 | Not-determined |
| 18-105_k141_201873 | Not-determined |
| 18-105_k141_148844 | Low-quality |
| 18-105_k141_175907 | Not-determined |
| 18-105_k141_121553 | Low-quality |
| 18-105_k141_112950 | Not-determined |
| 18-105_k141_77804 | Not-determined |
| 18-105_k141_193343 | Not-determined |
| 18-105_k141_184772 | Not-determined |
| 18-105_k141_22957 | Low-quality |
| 18-105_k141_22959 | Not-determined |
| 18-105_k141_121567 | Not-determined |
| 18-105_k141_175925 | Not-determined |
| 18-105_k141_157088 | Not-determined |
| 18-105_k141_7910 | Low-quality |
| 18-105_k141_59742 | Not-determined |
| 18-105_k141_131348 | Not-determined |
| 18-105_k141_131349 | Low-quality |
| 18-105_k141_184787 | Not-determined |
| 18-105_k141_77824 | Low-quality |
| 18-105_k141_184790 | Not-determined |
| 18-105_k141_121589 | Not-determined |
| 18-105_k141_184799 | Low-quality |
| 18-105_k141_32499 | Not-determined |
| 18-105_k141_193356 | Not-determined |
| 18-105_k141_22988 | Not-determined |
| 18-105_k141_175936 | Low-quality |
| 18-105_k141_175937 | Not-determined |
| 18-105_k141_94846 | Not-determined |
| 18-105_k141_148877 | Not-determined |
| 18-105_k141_59765 | Low-quality |
| 18-105_k141_23001 | Not-determined |
| 18-105_k141_67841 | Not-determined |
| 18-105_k141_210907 | Not-determined |
| 18-105_k141_210908 | Not-determined |
| 18-105_k141_32515 | Not-determined |
| 18-105_k141_157112 | Not-determined |
| 18-105_k141_59775 | Low-quality |
| 18-105_k141_67850 | Not-determined |
| 18-105_k141_131361 | Not-determined |
| 18-105_k141_77850 | Not-determined |
| 18-105_k141_77852 | Not-determined |
| 18-105_k141_77853 | Not-determined |
| 18-105_k141_131367 | Not-determined |
| 18-105_k141_184817 | Not-determined |
| 18-105_k141_184820 | Not-determined |
| 18-105_k141_201941 | Not-determined |
| 18-105_k141_148888 | Low-quality |
| 18-105_k141_148889 | Not-determined |
| 18-105_k141_148890 | Not-determined |
| 18-105_k141_121641 | Not-determined |
| 18-105_k141_23007 | Not-determined |
| 18-105_k141_184824 | Not-determined |
| 18-105_k141_184826 | Not-determined |
| 18-105_k141_157133 | Not-determined |
| 18-105_k141_201951 | Not-determined |
| 18-105_k141_113016 | Not-determined |
| 18-105_k141_59795 | Not-determined |
| 18-105_k141_59797 | Not-determined |
| 18-105_k141_59800 | Not-determined |
| 18-105_k141_184839 | Low-quality |
| 18-105_k141_67861 | Not-determined |
| 18-105_k141_77887 | Not-determined |
| 18-105_k141_94873 | Not-determined |
| 18-105_k141_131405 | Not-determined |
| 18-105_k141_32541 | Not-determined |
| 18-105_k141_184860 | Not-determined |
| 18-105_k141_67875 | Not-determined |
| 18-105_k141_184866 | Not-determined |
| 18-105_k141_131420 | Low-quality |
| 18-105_k141_59823 | Not-determined |
| 18-105_k141_193371 | Not-determined |
| 18-105_k141_23032 | Not-determined |
| 18-105_k141_32550 | Not-determined |
| 18-105_k141_210951 | Not-determined |
| 18-105_k141_50768 | Not-determined |
| 18-105_k141_32555 | Not-determined |
| 18-105_k141_67892 | Not-determined |
| 18-105_k141_59830 | Not-determined |
| 18-105_k141_131433 | Not-determined |
| 18-105_k141_113029 | Not-determined |
| 18-105_k141_59838 | Not-determined |
| 18-105_k141_59839 | Not-determined |
| 18-105_k141_59840 | Not-determined |
| 18-105_k141_201976 | Not-determined |
| 18-105_k141_15045 | Not-determined |
| 18-105_k141_113034 | Not-determined |
| 18-105_k141_67907 | Not-determined |
| 18-105_k141_15051 | Not-determined |
| 18-105_k141_121680 | Not-determined |
| 18-105_k141_15056 | Not-determined |
| 18-105_k141_193401 | Not-determined |
| 18-105_k141_67911 | Not-determined |
| 18-105_k141_121691 | Not-determined |
| 18-105_k141_176045 | Not-determined |
| 18-105_k141_131462 | Not-determined |
| 18-105_k141_67933 | Not-determined |
| 18-105_k141_131463 | Not-determined |
| 18-105_k141_15079 | Not-determined |
| 18-105_k141_113048 | Not-determined |
| 18-105_k141_67935 | Low-quality |
| 18-105_k141_193418 | Not-determined |
| 18-105_k141_157164 | Not-determined |
| 18-105_k141_121712 | Not-determined |
| 18-105_k141_50796 | Not-determined |
| 18-105_k141_15088 | Not-determined |
| 18-105_k141_59858 | Not-determined |
| 18-105_k141_67945 | Low-quality |
| 18-105_k141_50808 | Not-determined |
| 18-105_k141_77895 | Not-determined |
| 18-105_k141_94915 | Not-determined |
| 18-105_k141_113068 | Not-determined |
| 18-105_k141_67949 | Low-quality |
| 18-105_k141_131496 | Low-quality |
| 18-105_k141_50816 | Not-determined |
| 18-105_k141_104496 | Not-determined |
| 18-105_k141_202001 | Not-determined |
| 18-105_k141_77920 | Not-determined |
| 18-105_k141_184874 | Not-determined |
| 18-105_k141_50831 | Not-determined |
| 18-105_k141_131506 | Low-quality |
| 18-105_k141_77932 | Not-determined |
| 18-105_k141_94947 | Not-determined |
| 18-105_k141_176077 | Not-determined |
| 18-105_k141_50845 | Not-determined |
| 18-105_k141_67968 | Not-determined |
| 18-105_k141_184887 | Not-determined |
| 18-105_k141_184889 | Not-determined |
| 18-105_k141_67969 | Not-determined |
| 18-105_k141_86394 | Not-determined |
| 18-105_k141_77942 | Not-determined |
| 18-105_k141_77943 | Not-determined |
| 18-105_k141_176090 | Low-quality |
| 18-105_k141_176094 | Not-determined |
| 18-105_k141_121741 | Not-determined |
| 18-105_k141_176096 | Not-determined |
| 18-105_k141_7979 | Not-determined |
| 18-105_k141_121745 | Not-determined |
| 18-105_k141_67984 | Not-determined |
| 18-105_k141_67985 | Not-determined |
| 18-105_k141_131530 | Not-determined |
| 18-105_k141_104539 | Not-determined |
| 18-105_k141_113114 | Low-quality |
| 18-105_k141_176101 | Not-determined |
| 18-105_k141_113118 | Not-determined |
| 18-105_k141_77960 | Not-determined |
| 18-105_k141_7985 | Not-determined |
| 18-105_k141_121756 | Not-determined |
| 18-105_k141_157234 | Not-determined |
| 18-105_k141_148964 | Not-determined |
| 18-105_k141_86419 | Not-determined |
| 18-105_k141_157238 | Not-determined |
| 18-105_k141_7988 | Not-determined |
| 18-105_k141_113134 | Not-determined |
| 18-105_k141_166254 | Not-determined |
| 18-105_k141_50878 | Not-determined |
| 18-105_k141_121763 | Not-determined |
| 18-105_k141_113140 | Not-determined |
| 18-105_k141_77984 | Not-determined |
| 18-105_k141_131549 | Not-determined |
| 18-105_k141_50886 | Not-determined |
| 18-105_k141_41631 | Not-determined |
| 18-105_k141_41632 | Low-quality |
| 18-105_k141_8001 | Not-determined |
| 18-105_k141_139540 | Not-determined |
| 18-105_k141_139541 | Not-determined |
| 18-105_k141_15129 | Low-quality |
| 18-105_k141_68007 | Not-determined |
| 18-105_k141_41644 | Not-determined |
| 18-105_k141_8005 | Not-determined |
| 18-105_k141_50906 | Not-determined |
| 18-105_k141_131575 | Low-quality |
| 18-105_k141_121801 | Not-determined |
| 18-105_k141_113162 | Not-determined |
| 18-105_k141_121804 | Not-determined |
| 18-105_k141_41649 | Not-determined |
| 18-105_k141_50909 | Low-quality |
| 18-105_k141_32610 | Not-determined |
| 18-105_k141_157290 | Not-determined |
| 18-105_k141_59914 | Not-determined |
| 18-105_k141_157292 | Not-determined |
| 18-105_k141_139563 | Not-determined |
| 18-105_k141_131594 | Not-determined |
| 18-105_k141_86456 | Not-determined |
| 18-105_k141_78024 | Not-determined |
| 18-105_k141_78026 | Not-determined |
| 18-105_k141_113185 | Low-quality |
| 18-105_k141_78029 | Not-determined |
| 18-105_k141_139571 | Not-determined |
| 18-105_k141_41667 | Not-determined |
| 18-105_k141_50926 | Not-determined |
| 18-105_k141_41669 | Not-determined |
| 18-105_k141_23096 | Not-determined |
| 18-105_k141_23102 | Not-determined |
| 18-105_k141_139583 | Not-determined |
| 18-105_k141_184900 | Not-determined |
| 18-105_k141_68021 | Not-determined |
| 18-105_k141_68025 | Not-determined |
| 18-105_k141_86465 | Not-determined |
| 18-105_k141_121842 | Not-determined |
| 18-105_k141_184907 | Not-determined |
| 18-105_k141_139593 | Not-determined |
| 18-105_k141_23116 | Not-determined |
| 18-105_k141_86472 | Not-determined |
| 18-105_k141_50952 | Not-determined |
| 18-105_k141_23123 | Not-determined |
| 18-105_k141_41679 | Not-determined |
| 18-105_k141_121864 | Not-determined |
| 18-105_k141_131638 | Not-determined |
| 18-105_k141_68041 | Not-determined |
| 18-105_k141_121869 | Not-determined |
| 18-105_k141_157317 | Not-determined |
| 18-105_k141_166299 | Not-determined |
| 18-105_k141_23130 | Low-quality |
| 18-105_k141_157320 | Not-determined |
| 18-105_k141_59932 | Low-quality |
| 18-105_k141_68051 | Not-determined |
| 18-105_k141_8066 | Not-determined |
| 18-105_k141_166308 | Not-determined |
| 18-105_k141_59942 | Low-quality |
| 18-105_k141_121886 | Not-determined |
| 18-105_k141_121888 | Not-determined |
| 18-105_k141_50981 | Not-determined |
| 18-105_k141_121892 | Not-determined |
| 18-105_k141_139609 | Not-determined |
| 18-105_k141_176180 | Not-determined |
| 18-105_k141_50989 | Not-determined |
| 18-105_k141_184938 | Not-determined |
| 18-105_k141_50995 | Not-determined |
| 18-105_k141_121898 | Not-determined |
| 18-105_k141_23156 | Not-determined |
| 18-105_k141_166333 | Not-determined |
| 18-105_k141_86522 | Not-determined |
| 18-105_k141_32647 | Not-determined |
| 18-105_k141_59953 | Not-determined |
| 18-105_k141_184959 | Not-determined |
| 18-105_k141_139619 | Not-determined |
| 18-105_k141_166345 | Not-determined |
| 18-105_k141_32655 | Not-determined |
| 18-105_k141_8097 | Not-determined |
| 18-105_k141_8101 | Not-determined |
| 18-105_k141_131649 | Not-determined |
| 18-105_k141_8108 | Not-determined |
| 18-105_k141_184979 | Not-determined |
| 18-105_k141_113220 | Not-determined |
| 18-105_k141_86546 | Not-determined |
| 18-105_k141_166362 | Not-determined |
| 18-105_k141_166365 | Not-determined |
| 18-105_k141_166368 | Not-determined |
| 18-105_k141_166370 | Not-determined |
| 18-105_k141_176204 | Not-determined |
| 18-105_k141_148999 | Not-determined |
| 18-105_k141_185004 | Not-determined |
| 18-105_k141_185005 | Not-determined |
| 18-105_k141_139650 | Not-determined |
| 18-105_k141_113241 | Low-quality |
| 18-105_k141_149017 | Not-determined |
| 18-105_k141_149018 | Not-determined |
| 18-105_k141_185020 | Low-quality |
| 18-105_k141_121944 | Not-determined |
| 18-105_k141_157392 | Not-determined |
| 18-105_k141_8135 | Not-determined |
| 18-105_k141_86580 | Not-determined |
| 18-105_k141_23219 | Not-determined |
| 18-105_k141_23220 | Not-determined |
| 18-105_k141_185029 | Low-quality |
| 18-105_k141_185034 | Not-determined |
| 18-105_k141_32693 | Not-determined |
| 18-105_k141_121958 | Not-determined |
| 18-105_k141_78122 | Not-determined |
| 18-105_k141_113265 | Not-determined |
| 18-105_k141_185040 | Not-determined |
| 18-105_k141_185042 | Not-determined |
| 18-105_k141_15221 | Not-determined |
| 18-105_k141_121965 | Not-determined |
| 18-105_k141_68127 | Not-determined |
| 18-105_k141_15223 | Not-determined |
| 18-105_k141_23230 | Not-determined |
| 18-105_k141_113270 | Not-determined |
| 18-105_k141_23236 | Not-determined |
| 18-105_k141_157411 | Not-determined |
| 18-105_k141_185052 | Not-determined |
| 18-105_k141_139690 | Not-determined |
| 18-105_k141_15230 | Not-determined |
| 18-105_k141_8163 | Not-determined |
| 18-105_k141_68143 | Not-determined |
| 18-105_k141_131680 | Not-determined |
| 18-105_k141_210972 | Not-determined |
| 18-105_k141_131684 | Not-determined |
| 18-105_k141_131685 | Not-determined |
| 18-105_k141_131686 | Not-determined |
| 18-105_k141_131692 | Not-determined |
| 18-105_k141_113288 | Not-determined |
| 18-105_k141_15243 | Not-determined |
| 18-105_k141_113292 | Not-determined |
| 18-105_k141_86609 | Low-quality |
| 18-105_k141_139712 | Not-determined |
| 18-105_k141_51071 | Not-determined |
| 18-105_k141_68161 | Not-determined |
| 18-105_k141_121976 | Not-determined |
| 18-105_k141_51073 | Not-determined |
| 18-105_k141_51076 | Not-determined |
| 18-105_k141_68168 | Not-determined |
| 18-105_k141_51078 | Not-determined |
| 18-105_k141_121982 | Not-determined |
| 17-109_k141_264425 | High-quality |
| 18-105_k141_32734 | Not-determined |
| 18-105_k141_166441 | Not-determined |
| 18-105_k141_15260 | Not-determined |
| 18-105_k141_78139 | Not-determined |
| 18-105_k141_51091 | Not-determined |
| 18-105_k141_15262 | Not-determined |
| 18-105_k141_113309 | Not-determined |
| 18-105_k141_60009 | Not-determined |
| 18-105_k141_51095 | Not-determined |
| 18-105_k141_60015 | Not-determined |
| 18-105_k141_210999 | Not-determined |
| 18-105_k141_51098 | Not-determined |
| 18-105_k141_211004 | Not-determined |
| 18-105_k141_68199 | Not-determined |
| 18-105_k141_139746 | Not-determined |
| 18-105_k141_60031 | Not-determined |
| 18-105_k141_139751 | Not-determined |
| 18-105_k141_94992 | Not-determined |
| 18-105_k141_166464 | Not-determined |
| 18-105_k141_104675 | Not-determined |
| 18-105_k141_68203 | Not-determined |
| 18-105_k141_32760 | Not-determined |
| 18-105_k141_149067 | Not-determined |
| 18-105_k141_15277 | Not-determined |
| 18-105_k141_113326 | Not-determined |
| 18-105_k141_149073 | Not-determined |
| 18-105_k141_41714 | Not-determined |
| 18-105_k141_78165 | Not-determined |
| 18-105_k141_51115 | Not-determined |
| 18-105_k141_51116 | Not-determined |
| 18-105_k141_41721 | Not-determined |
| 18-105_k141_41723 | Not-determined |
| 18-105_k141_121997 | Not-determined |
| 18-105_k141_193462 | Not-determined |
| 18-105_k141_121998 | Not-determined |
| 18-105_k141_86643 | Not-determined |
| 18-105_k141_149078 | Not-determined |
| 18-105_k141_95012 | Not-determined |
| 18-105_k141_193467 | Not-determined |
| 18-105_k141_86648 | Not-determined |
| 18-105_k141_193468 | Not-determined |
| 18-105_k141_60066 | Not-determined |
| 18-105_k141_193473 | Not-determined |
| 18-105_k141_166486 | Not-determined |
| 18-105_k141_104692 | Not-determined |
| 18-105_k141_131714 | Not-determined |
| 18-105_k141_122018 | Not-determined |
| 18-105_k141_68232 | Low-quality |
| 18-105_k141_202092 | Not-determined |
| 18-105_k141_193476 | Not-determined |
| 18-105_k141_86668 | Not-determined |
| 18-105_k141_104699 | Not-determined |
| 18-105_k141_139782 | Not-determined |
| 18-105_k141_51135 | Not-determined |
| 18-105_k141_166501 | Not-determined |
| 18-105_k141_139785 | Not-determined |
| 18-105_k141_166502 | Low-quality |
| 18-105_k141_32795 | Not-determined |
| 18-105_k141_193480 | Not-determined |
| 18-105_k141_202097 | Not-determined |
| 18-105_k141_193481 | Not-determined |
| 18-105_k141_211041 | Not-determined |
| 18-105_k141_202102 | Not-determined |
| 18-105_k141_166509 | Not-determined |
| 18-105_k141_131730 | Not-determined |
| 18-105_k141_95041 | Not-determined |
| 18-105_k141_157454 | Not-determined |
| 18-105_k141_131735 | Not-determined |
| 18-105_k141_131737 | Not-determined |
| 18-105_k141_51151 | Not-determined |
| 18-105_k141_113375 | Not-determined |
| 18-105_k141_193492 | Not-determined |
| 18-105_k141_149105 | Not-determined |
| 18-105_k141_113378 | Not-determined |
| 18-105_k141_185102 | Not-determined |
| 18-105_k141_68260 | Low-quality |
| 18-105_k141_122036 | Not-determined |
| 18-105_k141_113381 | Not-determined |
| 18-105_k141_211058 | Not-determined |
| 18-105_k141_32819 | Not-determined |
| 18-105_k141_149115 | Not-determined |
| 18-105_k141_51165 | Low-quality |
| 18-105_k141_51166 | Not-determined |
| 18-105_k141_185117 | Low-quality |
| 18-105_k141_211063 | Not-determined |
| 18-105_k141_157471 | Not-determined |
| 18-105_k141_122041 | Not-determined |
| 18-105_k141_41756 | Not-determined |
| 18-105_k141_68271 | Not-determined |
| 18-105_k141_139802 | Not-determined |
| 18-105_k141_95073 | Not-determined |
| 18-105_k141_122046 | Not-determined |
| 18-105_k141_104733 | Not-determined |
| 18-105_k141_15307 | Not-determined |
| 18-105_k141_51172 | Not-determined |
| 18-105_k141_104735 | Not-determined |
| 18-105_k141_15310 | Not-determined |
| 18-105_k141_193511 | Not-determined |
| 18-105_k141_193512 | Not-determined |
| 18-105_k141_51174 | Not-determined |
| 18-105_k141_193517 | Low-quality |
| 18-105_k141_157492 | Not-determined |
| 18-105_k141_157494 | Not-determined |
| 18-105_k141_139818 | Not-determined |
| 18-105_k141_68295 | Not-determined |
| 18-105_k141_131757 | Not-determined |

| | |
|---|---|
| 18-105_k141_157499 | Not-determined |
| 18-105_k141_157503 | Not-determined |
| 18-105_k141_149126 | Low-quality |
| 18-105_k141_68299 | Not-determined |
| 18-105_k141_23294 | Not-determined |
| 18-105_k141_68302 | Not-determined |
| 18-105_k141_149127 | Low-quality |
| 18-105_k141_149129 | Not-determined |
| 18-105_k141_23298 | Not-determined |
| 18-105_k141_15336 | Not-determined |
| 18-105_k141_157509 | Not-determined |
| 18-105_k141_41782 | Not-determined |
| 18-105_k141_202122 | Not-determined |
| 18-105_k141_15343 | Not-determined |
| 18-105_k141_23310 | Not-determined |
| 18-105_k141_41784 | Not-determined |
| 18-105_k141_41786 | Low-quality |
| 18-105_k141_15347 | Not-determined |
| 18-105_k141_41789 | Not-determined |
| 18-105_k141_157516 | Not-determined |
| 18-105_k141_8226 | Not-determined |
| 18-105_k141_78176 | Not-determined |
| 18-105_k141_131771 | Not-determined |
| 18-105_k141_8234 | Not-determined |
| 18-105_k141_78180 | Not-determined |
| 18-105_k141_78181 | Not-determined |
| 18-105_k141_32830 | Not-determined |
| 18-105_k141_86682 | Not-determined |
| 18-105_k141_23325 | Low-quality |
| 18-105_k141_113389 | Not-determined |
| 18-105_k141_23327 | Not-determined |
| 18-105_k141_113390 | Not-determined |
| 18-105_k141_193542 | Not-determined |
| 18-105_k141_23329 | Not-determined |
| 18-105_k141_23330 | Not-determined |
| 18-105_k141_113394 | Not-determined |
| 18-105_k141_113401 | Not-determined |
| 18-105_k141_113402 | Not-determined |
| 18-105_k141_15371 | Low-quality |
| 18-105_k141_113403 | Not-determined |
| 18-105_k141_78190 | Low-quality |
| 18-105_k141_149150 | Not-determined |
| 18-105_k141_113408 | Not-determined |
| 18-105_k141_104748 | Not-determined |
| 18-105_k141_104749 | Not-determined |
| 18-105_k141_51195 | Low-quality |
| 18-105_k141_23347 | Not-determined |
| 18-105_k141_32861 | Low-quality |
| 18-105_k141_95123 | Not-determined |
| 18-105_k141_95127 | Not-determined |
| 18-105_k141_211086 | Not-determined |
| 18-105_k141_41795 | Not-determined |
| 18-105_k141_15397 | Not-determined |
| 18-105_k141_23354 | Not-determined |
| 18-105_k141_139854 | Not-determined |
| 18-105_k141_86731 | Not-determined |
| 18-105_k141_139856 | Not-determined |
| 18-105_k141_41801 | Low-quality |
| 18-105_k141_8267 | Not-determined |
| 18-105_k141_15417 | Not-determined |
| 18-105_k141_211096 | Not-determined |
| 18-105_k141_23367 | Not-determined |
| 18-105_k141_202133 | Not-determined |
| 18-105_k141_202134 | Not-determined |
| 18-105_k141_202136 | Not-determined |
| 18-105_k141_139867 | Not-determined |
| 18-105_k141_41808 | Not-determined |
| 18-105_k141_202138 | Not-determined |
| 18-105_k141_211099 | Low-quality |
| 18-105_k141_202140 | Not-determined |
| 18-105_k141_211100 | Not-determined |
| 18-105_k141_202141 | Not-determined |
| 18-105_k141_23377 | Not-determined |
| 18-105_k141_51226 | Low-quality |
| 18-105_k141_176285 | Not-determined |
| 18-105_k141_41811 | Not-determined |
| 18-105_k141_51228 | Not-determined |
| 18-105_k141_211108 | Not-determined |
| 18-105_k141_23383 | Not-determined |
| 18-105_k141_149182 | Not-determined |
| 18-105_k141_23387 | Not-determined |
| 18-105_k141_139883 | Not-determined |
| 18-105_k141_23388 | Not-determined |
| 18-105_k141_32883 | Not-determined |
| 18-105_k141_15452 | Not-determined |
| 18-105_k141_51234 | Low-quality |
| 18-105_k141_51235 | Not-determined |
| 18-105_k141_185146 | Low-quality |
| 18-105_k141_51238 | Not-determined |
| 18-105_k141_211133 | Low-quality |
| 18-105_k141_211134 | Not-determined |
| 18-105_k141_78267 | Not-determined |
| 18-105_k141_211135 | Not-determined |
| 18-105_k141_211136 | Not-determined |
| 18-105_k141_157565 | Not-determined |
| 18-105_k141_185150 | Not-determined |
| 18-105_k141_15475 | Not-determined |
| 18-105_k141_78272 | Not-determined |
| 18-105_k141_23401 | Not-determined |
| 18-105_k141_23402 | Not-determined |
| 18-105_k141_68352 | Not-determined |
| 18-105_k141_23408 | Not-determined |
| 18-105_k141_176304 | Not-determined |
| 18-105_k141_139905 | Not-determined |
| 18-105_k141_51244 | Low-quality |
| 18-105_k141_176306 | Not-determined |
| 18-105_k141_32900 | Not-determined |
| 18-105_k141_51252 | Not-determined |
| 18-105_k141_68362 | Not-determined |
| 18-105_k141_104757 | Not-determined |
| 18-105_k141_23423 | Not-determined |
| 18-105_k141_68367 | Not-determined |
| 18-105_k141_32917 | Not-determined |
| 18-105_k141_32919 | Not-determined |
| 18-105_k141_104769 | Not-determined |
| 18-105_k141_149193 | Not-determined |
| 18-105_k141_51265 | Not-determined |
| 18-105_k141_104773 | Not-determined |
| 18-105_k141_51275 | Not-determined |
| 18-105_k141_78302 | Not-determined |
| 18-105_k141_78303 | Not-determined |
| 18-105_k141_78307 | Not-determined |
| 18-105_k141_51283 | Low-quality |
| 18-105_k141_113446 | Not-determined |
| 18-105_k141_51286 | Not-determined |
| 18-105_k141_41843 | Not-determined |
| 18-105_k141_32946 | Low-quality |
| 18-105_k141_104787 | Low-quality |
| 18-105_k141_139943 | Not-determined |
| 18-105_k141_193590 | Not-determined |
| 18-105_k141_157580 | Not-determined |
| 18-105_k141_15522 | Not-determined |
| 18-105_k141_78321 | Not-determined |
| 18-105_k141_15523 | Not-determined |
| 18-105_k141_139946 | Not-determined |
| 18-105_k141_139947 | Not-determined |
| 18-105_k141_185153 | Not-determined |
| 18-105_k141_15529 | Not-determined |
| 18-105_k141_185154 | Not-determined |
| 18-105_k141_185155 | Not-determined |
| 18-105_k141_185156 | Not-determined |
| 18-105_k141_157588 | Not-determined |
| 18-105_k141_185158 | Low-quality |
| 18-105_k141_131785 | Not-determined |
| 18-105_k141_86833 | Not-determined |
| 18-105_k141_176327 | Not-determined |
| 18-105_k141_157595 | Not-determined |
| 18-105_k141_15540 | Not-determined |
| 18-105_k141_176331 | Not-determined |
| 18-105_k141_104796 | Not-determined |
| 18-105_k141_113456 | Not-determined |
| 18-105_k141_32960 | Not-determined |
| 18-105_k141_131793 | Not-determined |
| 18-105_k141_193615 | Not-determined |
| 18-105_k141_131801 | Not-determined |
| 18-105_k141_202149 | Not-determined |
| 18-105_k141_23457 | Not-determined |
| 18-105_k141_104812 | Not-determined |
| 18-105_k141_23458 | Not-determined |
| 18-105_k141_8280 | Not-determined |
| 18-105_k141_23460 | Not-determined |
| 18-105_k141_131802 | Not-determined |
| 18-105_k141_8284 | Not-determined |
| 18-105_k141_23462 | Not-determined |
| 18-105_k141_157626 | Not-determined |
| 18-105_k141_86852 | Low-quality |
| 18-105_k141_157628 | Not-determined |
| 18-105_k141_113474 | Not-determined |
| 18-105_k141_113475 | Not-determined |
| 18-105_k141_193632 | Not-determined |
| 18-105_k141_8290 | Not-determined |
| 18-105_k141_41904 | Not-determined |
| 18-105_k141_78361 | Low-quality |
| 18-105_k141_113485 | Not-determined |
| 18-105_k141_166564 | Not-determined |
| 18-105_k141_193641 | Not-determined |
| 18-105_k141_193645 | Not-determined |
| 18-105_k141_86866 | Not-determined |
| 18-105_k141_51338 | Not-determined |
| 18-105_k141_60074 | Not-determined |
| 18-105_k141_8314 | Not-determined |
| 18-105_k141_122107 | Not-determined |
| 18-105_k141_60080 | Not-determined |
| 18-105_k141_193660 | Not-determined |
| 18-105_k141_113510 | Not-determined |
| 18-105_k141_78374 | Not-determined |
| 18-105_k141_104849 | Not-determined |
| 18-105_k141_15604 | Not-determined |
| 18-105_k141_60093 | Not-determined |
| 18-105_k141_202183 | Not-determined |
| 18-105_k141_113518 | Not-determined |
| 18-105_k141_193664 | Not-determined |
| 18-105_k141_41931 | Not-determined |
| 18-105_k141_60107 | Not-determined |
| 18-105_k141_176360 | Not-determined |
| 18-105_k141_60112 | Not-determined |
| 18-105_k141_104855 | Low-quality |
| 18-105_k141_193672 | Not-determined |
| 18-105_k141_122133 | Not-determined |
| 18-105_k141_176381 | Not-determined |
| 18-105_k141_113523 | Not-determined |
| 18-105_k141_193676 | Not-determined |
| 18-105_k141_211141 | Not-determined |
| 18-105_k141_157645 | Not-determined |
| 18-105_k141_176389 | Not-determined |
| 18-105_k141_86878 | Not-determined |
| 18-105_k141_15611 | Not-determined |
| 18-105_k141_122145 | Low-quality |
| 18-105_k141_139994 | Not-determined |
| 18-105_k141_95227 | Not-determined |
| 18-105_k141_68430 | Low-quality |
| 18-105_k141_185213 | Not-determined |
| 18-105_k141_122151 | Not-determined |
| 18-105_k141_122152 | Not-determined |
| 18-105_k141_68431 | Low-quality |
| 18-105_k141_95233 | Not-determined |
| 18-105_k141_68435 | Not-determined |
| 18-105_k141_68436 | Not-determined |
| 18-105_k141_23487 | Not-determined |
| 18-105_k141_185218 | Not-determined |
| 18-105_k141_104862 | Not-determined |
| 18-105_k141_60138 | Not-determined |
| 18-105_k141_185224 | Not-determined |
| 18-105_k141_211151 | Not-determined |
| 18-105_k141_157660 | Not-determined |
| 18-105_k141_68448 | Low-quality |
| 18-105_k141_122167 | Not-determined |
| 18-105_k141_157666 | Low-quality |
| 18-105_k141_23501 | Not-determined |
| 18-105_k141_211158 | Not-determined |
| 18-105_k141_23504 | Not-determined |
| 18-105_k141_15618 | Not-determined |
| 18-105_k141_185236 | Low-quality |
| 18-105_k141_140020 | Low-quality |
| 18-105_k141_68467 | Not-determined |
| 18-105_k141_185241 | Not-determined |
| 18-105_k141_113537 | Low-quality |
| 18-105_k141_140034 | Not-determined |
| 18-105_k141_140035 | Not-determined |
| 18-105_k141_166604 | Not-determined |
| 18-105_k141_95266 | Not-determined |
| 18-105_k141_166607 | Not-determined |
| 18-105_k141_131862 | Not-determined |
| 18-105_k141_193703 | Not-determined |
| 18-105_k141_68486 | Not-determined |
| 18-105_k141_176427 | Not-determined |
| 18-105_k141_122202 | Not-determined |
| 18-105_k141_157692 | Not-determined |
| 18-105_k141_51376 | Not-determined |
| 18-105_k141_166618 | Low-quality |
| 18-105_k141_86930 | Not-determined |
| 18-105_k141_86931 | Not-determined |
| 18-105_k141_51380 | Not-determined |
| 18-105_k141_211164 | Not-determined |
| 18-105_k141_51383 | Not-determined |
| 18-105_k141_51384 | Not-determined |
| 18-105_k141_176437 | Not-determined |
| 18-105_k141_140049 | Not-determined |
| 18-105_k141_51387 | Not-determined |
| 18-105_k141_51389 | Not-determined |
| 18-105_k141_86941 | Not-determined |
| 18-105_k141_211177 | Not-determined |
| 18-105_k141_51395 | Not-determined |
| 18-105_k141_157708 | Not-determined |
| 18-105_k141_140065 | Not-determined |
| 18-105_k141_140066 | Not-determined |
| 18-105_k141_68499 | Not-determined |
| 18-105_k141_68500 | Not-determined |
| 18-105_k141_86951 | Not-determined |
| 18-105_k141_131891 | Not-determined |
| 18-105_k141_86952 | Not-determined |
| 18-105_k141_140072 | Not-determined |
| 18-105_k141_149260 | Not-determined |
| 18-105_k141_193719 | Not-determined |
| 18-105_k141_157721 | Not-determined |
| 18-105_k141_149262 | Not-determined |
| 18-105_k141_104881 | Not-determined |
| 18-105_k141_166645 | Not-determined |
| 18-105_k141_193724 | Not-determined |
| 18-105_k141_131908 | Not-determined |
| 18-105_k141_86961 | Not-determined |
| 18-105_k141_23518 | Low-quality |
| 18-105_k141_86962 | Low-quality |
| 18-105_k141_140083 | Not-determined |
| 18-105_k141_166654 | Not-determined |
| 18-105_k141_157728 | Not-determined |
| 18-105_k141_23525 | Not-determined |
| 18-105_k141_157739 | Not-determined |
| 18-105_k141_68522 | Not-determined |
| 18-105_k141_32996 | Low-quality |
| 18-105_k141_122230 | Not-determined |
| 18-105_k141_157755 | Not-determined |
| 18-105_k141_211192 | Not-determined |
| 18-105_k141_211196 | Not-determined |
| 18-105_k141_211198 | Not-determined |
| 18-105_k141_68536 | Low-quality |
| 18-105_k141_104910 | Low-quality |
| 18-105_k141_68541 | Not-determined |
| 18-105_k141_104916 | Not-determined |
| 18-105_k141_140098 | Low-quality |
| 18-105_k141_140099 | Not-determined |
| 18-105_k141_86994 | Not-determined |
| 18-105_k141_193746 | Not-determined |
| 18-105_k141_60169 | Not-determined |
| 18-105_k141_140105 | Not-determined |
| 18-105_k141_157775 | Not-determined |
| 18-105_k141_68553 | Not-determined |
| 18-105_k141_104927 | Not-determined |
| 18-105_k141_68554 | Not-determined |
| 18-105_k141_166683 | Low-quality |
| 18-105_k141_166684 | Not-determined |
| 18-105_k141_23548 | Not-determined |
| 18-105_k141_157790 | Not-determined |
| 18-105_k141_60173 | Not-determined |
| 18-105_k141_122241 | Not-determined |
| 18-105_k141_131927 | Not-determined |
| 18-105_k141_131928 | Not-determined |
| 18-105_k141_193757 | Not-determined |
| 18-105_k141_166703 | Not-determined |
| 18-105_k141_87017 | Not-determined |
| 18-105_k141_166706 | Not-determined |
| 18-105_k141_185260 | Low-quality |
| 18-105_k141_23566 | Not-determined |
| 18-105_k141_33030 | Not-determined |
| 18-105_k141_157805 | Not-determined |
| 18-105_k141_87023 | Not-determined |
| 18-105_k141_193765 | Not-determined |
| 18-105_k141_68586 | Not-determined |
| 18-105_k141_211246 | Not-determined |
| 18-105_k141_211250 | Not-determined |
| 18-105_k141_68591 | Not-determined |
| 18-105_k141_60193 | Low-quality |
| 18-105_k141_185275 | Not-determined |
| 18-105_k141_8344 | Not-determined |
| 18-105_k141_211255 | Not-determined |
| 18-105_k141_166723 | Not-determined |
| 18-105_k141_87040 | Not-determined |
| 18-105_k141_87042 | Low-quality |
| 18-105_k141_87043 | Low-quality |
| 18-105_k141_157828 | Not-determined |
| 18-105_k141_211265 | Not-determined |
| 18-105_k141_8366 | Not-determined |
| 18-105_k141_193793 | Not-determined |
| 18-105_k141_87054 | Low-quality |
| 18-105_k141_15624 | Low-quality |
| 18-105_k141_211271 | Low-quality |
| 18-105_k141_193799 | Not-determined |
| 18-105_k141_193802 | Not-determined |
| 18-105_k141_131962 | Low-quality |
| 18-105_k141_157847 | Not-determined |
| 18-105_k141_131966 | Not-determined |
| 18-105_k141_60227 | Not-determined |
| 18-105_k141_104942 | Low-quality |
| 18-105_k141_122284 | Low-quality |
| 18-105_k141_104943 | Not-determined |
| 18-105_k141_193813 | Not-determined |
| 18-105_k141_95306 | Not-determined |
| 18-105_k141_131972 | Not-determined |
| 18-105_k141_122290 | Not-determined |
| 18-105_k141_95309 | Not-determined |
| 18-105_k141_33081 | Not-determined |
| 18-105_k141_87078 | Not-determined |
| 18-105_k141_33083 | Not-determined |
| 18-105_k141_33084 | Not-determined |
| 18-105_k141_87084 | Not-determined |
| 18-105_k141_185327 | Not-determined |
| 18-105_k141_193829 | Low-quality |
| 18-105_k141_131984 | Not-determined |
| 18-105_k141_131989 | Not-determined |
| 18-105_k141_131990 | Not-determined |
| 18-105_k141_95325 | Not-determined |
| 18-105_k141_95327 | Low-quality |
| 18-105_k141_60258 | Not-determined |
| 18-105_k141_193837 | Not-determined |
| 18-105_k141_131997 | Not-determined |
| 18-105_k141_193839 | Not-determined |
| 18-105_k141_8372 | Not-determined |
| 18-105_k141_33111 | Not-determined |
| 18-105_k141_87101 | Not-determined |
| 18-105_k141_95338 | Not-determined |
| 18-105_k141_157856 | Not-determined |
| 18-105_k141_166773 | Not-determined |
| 18-105_k141_23595 | Not-determined |
| 18-105_k141_157859 | Not-determined |
| 18-105_k141_157860 | Not-determined |
| 18-105_k141_68634 | Not-determined |
| 18-105_k141_157865 | Low-quality |
| 18-105_k141_104997 | Not-determined |
| 18-105_k141_15644 | Not-determined |
| 18-105_k141_193866 | Not-determined |
| 18-105_k141_211330 | Not-determined |
| 18-105_k141_211333 | Not-determined |
| 18-105_k141_60274 | Not-determined |
| 18-105_k141_60278 | Not-determined |
| 18-105_k141_211342 | Not-determined |
| 18-105_k141_193878 | Not-determined |
| 18-105_k141_157879 | Not-determined |
| 18-105_k141_132023 | Not-determined |
| 18-105_k141_185348 | Not-determined |
| 18-105_k141_8414 | Low-quality |
| 18-105_k141_60285 | Not-determined |
| 18-105_k141_185354 | Not-determined |
| 18-105_k141_140124 | Not-determined |
| 18-105_k141_105023 | Not-determined |
| 18-105_k141_51482 | Low-quality |
| 18-105_k141_166816 | Not-determined |
| 18-105_k141_185365 | Not-determined |
| 18-105_k141_23610 | Low-quality |
| 18-105_k141_105036 | Not-determined |
| 18-105_k141_122344 | Not-determined |
| 18-105_k141_15669 | Not-determined |
| 18-105_k141_132058 | Not-determined |
| 18-105_k141_185372 | Not-determined |
| 18-105_k141_68695 | Not-determined |
| 18-105_k141_122350 | Not-determined |
| 18-105_k141_33143 | Low-quality |
| 18-105_k141_78473 | Not-determined |
| 18-105_k141_15681 | Not-determined |
| 18-105_k141_157923 | Not-determined |
| 18-105_k141_140159 | Not-determined |
| 18-105_k141_51499 | Not-determined |
| 18-105_k141_23632 | Not-determined |
| 18-105_k141_157926 | Not-determined |
| 18-105_k141_140166 | Not-determined |
| 18-105_k141_95352 | Low-quality |
| 18-105_k141_15692 | Not-determined |
| 18-105_k141_185406 | Not-determined |
| 18-105_k141_166847 | Not-determined |
| 18-105_k141_95358 | Low-quality |
| 18-105_k141_51522 | Not-determined |
| 18-105_k141_166850 | Low-quality |
| 18-105_k141_140181 | Not-determined |
| 18-105_k141_78491 | Not-determined |
| 18-105_k141_15698 | Not-determined |
| 18-105_k141_23648 | Not-determined |
| 18-105_k141_78493 | Not-determined |
| 18-105_k141_95367 | Not-determined |
| 18-105_k141_78495 | Not-determined |
| 18-105_k141_95371 | Not-determined |
| 18-105_k141_23653 | Not-determined |
| 18-105_k141_51541 | Not-determined |
| 18-105_k141_105055 | Not-determined |
| 18-105_k141_185424 | Not-determined |
| 18-105_k141_157950 | Low-quality |
| 18-105_k141_140191 | Not-determined |
| 18-105_k141_132083 | Not-determined |
| 18-105_k141_95379 | Not-determined |
| 18-105_k141_78500 | Low-quality |
| 18-105_k141_211372 | Not-determined |
| 18-105_k141_157951 | Not-determined |
| 18-105_k141_132089 | Not-determined |
| 18-105_k141_23670 | Not-determined |
| 18-105_k141_15720 | Not-determined |
| 18-105_k141_95385 | Not-determined |
| 18-105_k141_132092 | Not-determined |
| 18-105_k141_51557 | Not-determined |
| 18-105_k141_105071 | Not-determined |
| 18-105_k141_78517 | Not-determined |
| 18-105_k141_78526 | Not-determined |
| 18-105_k141_166872 | Not-determined |
| 18-105_k141_105084 | Not-determined |
| 18-105_k141_149268 | Not-determined |
| 18-105_k141_140204 | Not-determined |
| 18-105_k141_166876 | Not-determined |
| 18-105_k141_68732 | Not-determined |
| 18-105_k141_185447 | Not-determined |
| 18-105_k141_68735 | Not-determined |
| 18-105_k141_211409 | Not-determined |
| 18-105_k141_140206 | Not-determined |
| 18-105_k141_60305 | Not-determined |
| 18-105_k141_157965 | Low-quality |
| 18-105_k141_8436 | Not-determined |
| 18-105_k141_166887 | Not-determined |
| 18-105_k141_193960 | Not-determined |
| 18-105_k141_68743 | Low-quality |
| 18-105_k141_68746 | Not-determined |
| 18-105_k141_211421 | Low-quality |
| 18-105_k141_185467 | Low-quality |
| 18-105_k141_193971 | Not-determined |
| 18-105_k141_68753 | Not-determined |
| 18-105_k141_211423 | Not-determined |
| 18-105_k141_95425 | Not-determined |
| 18-105_k141_15769 | Not-determined |
| 18-105_k141_193985 | Not-determined |
| 18-105_k141_105109 | Not-determined |
| 18-105_k141_185486 | Not-determined |
| 18-105_k141_60326 | Not-determined |
| 18-105_k141_78567 | Not-determined |
| 18-105_k141_185491 | Not-determined |
| 18-105_k141_140240 | Not-determined |
| 18-105_k141_202253 | Not-determined |
| 18-105_k141_68785 | Not-determined |
| 18-105_k141_185495 | Low-quality |
| 18-105_k141_95449 | Not-determined |
| 18-105_k141_185498 | Not-determined |
| 18-105_k141_122414 | Not-determined |
| 18-105_k141_78577 | Not-determined |
| 18-105_k141_211443 | Not-determined |
| 18-105_k141_122418 | Not-determined |
| 18-105_k141_140259 | Not-determined |
| 18-105_k141_202273 | Not-determined |
| 18-105_k141_140267 | Not-determined |
| 18-105_k141_95463 | Not-determined |
| 18-105_k141_185514 | Low-quality |
| 18-105_k141_105137 | Not-determined |
| 18-105_k141_105141 | Not-determined |
| 18-105_k141_202293 | Not-determined |
| 18-105_k141_166914 | Not-determined |
| 18-105_k141_23742 | Low-quality |
| 18-105_k141_158001 | Not-determined |
| 18-105_k141_140290 | Not-determined |
| 18-105_k141_78588 | Not-determined |
| 18-105_k141_95490 | Not-determined |
| 18-105_k141_95499 | Not-determined |
| 18-105_k141_8473 | Not-determined |
| 18-105_k141_78595 | Not-determined |
| 18-105_k141_140298 | Not-determined |
| 18-105_k141_33211 | Not-determined |
| 18-105_k141_8477 | Not-determined |
| 18-105_k141_105153 | Not-determined |
| 18-105_k141_87195 | Low-quality |
| 18-105_k141_87196 | Not-determined |
| 18-105_k141_202321 | Not-determined |
| 18-105_k141_140301 | Not-determined |
| 18-105_k141_105160 | Not-determined |
| 18-105_k141_105162 | Low-quality |
| 18-105_k141_41988 | Not-determined |
| 18-105_k141_68853 | Low-quality |
| 18-105_k141_41989 | Not-determined |
| 18-105_k141_87205 | Not-determined |
| 18-105_k141_211484 | Not-determined |
| 18-105_k141_149306 | Not-determined |
| 18-105_k141_78616 | Not-determined |
| 18-105_k141_194003 | Low-quality |
| 18-105_k141_78618 | Not-determined |
| 18-105_k141_60377 | Not-determined |
| 18-105_k141_211489 | Not-determined |
| 18-105_k141_140315 | Not-determined |
| 18-105_k141_140317 | Not-determined |
| 18-105_k141_166957 | Not-determined |
| 18-105_k141_122442 | Not-determined |
| 18-105_k141_87218 | Not-determined |
| 18-105_k141_122443 | Not-determined |
| 18-105_k141_202341 | Not-determined |
| 18-105_k141_78626 | Not-determined |
| 18-105_k141_202345 | Not-determined |
| 18-105_k141_202346 | Low-quality |
| 18-105_k141_68882 | Not-determined |
| 18-105_k141_140322 | Not-determined |
| 18-105_k141_8488 | Not-determined |
| 18-105_k141_122450 | Not-determined |
| 18-105_k141_60384 | Not-determined |
| 18-105_k141_105181 | Low-quality |
| 18-105_k141_132146 | Not-determined |
| 18-105_k141_122453 | Low-quality |
| 18-105_k141_122454 | Not-determined |
| 18-105_k141_122457 | Not-determined |
| 18-105_k141_8491 | Not-determined |
| 18-105_k141_132154 | Low-quality |
| 18-105_k141_87230 | Not-determined |
| 18-105_k141_149316 | Not-determined |
| 18-105_k141_122467 | Not-determined |
| 18-105_k141_149321 | Not-determined |
| 18-105_k141_87234 | Not-determined |
| 18-105_k141_42009 | Not-determined |
| 18-105_k141_87239 | Not-determined |
| 18-105_k141_42012 | Low-quality |
| 18-105_k141_42015 | Not-determined |
| 18-105_k141_68903 | Not-determined |
| 18-105_k141_68904 | Not-determined |
| 18-105_k141_78650 | Not-determined |
| 18-105_k141_211520 | Not-determined |
| 18-105_k141_166981 | Not-determined |
| 18-105_k141_78652 | Not-determined |
| 18-105_k141_78653 | Not-determined |
| 18-105_k141_78654 | Not-determined |
| 18-105_k141_23790 | Not-determined |
| 18-105_k141_60418 | Low-quality |
| 18-105_k141_51594 | Not-determined |
| 18-105_k141_211530 | Not-determined |
| 18-105_k141_149334 | Not-determined |
| 18-105_k141_122483 | Low-quality |
| 18-105_k141_95523 | Not-determined |
| 18-105_k141_8535 | Not-determined |
| 18-105_k141_95525 | Low-quality |
| 18-105_k141_122490 | Not-determined |
| 18-105_k141_60432 | Not-determined |
| 18-105_k141_194074 | Not-determined |
| 18-105_k141_8542 | Not-determined |

| | |
|---|---|
| 18-105_k141_211539 | Not-determined |
| 18-105_k141_95532 | Not-determined |
| 18-105_k141_158037 | Not-determined |
| 18-105_k141_87265 | Not-determined |
| 18-105_k141_60437 | Low-quality |
| 18-105_k141_8553 | Not-determined |
| 18-105_k141_23796 | Not-determined |
| 18-105_k141_194084 | Not-determined |
| 18-105_k141_122501 | Not-determined |
| 18-105_k141_105218 | Not-determined |
| 18-105_k141_122506 | Not-determined |
| 18-105_k141_87276 | Not-determined |
| 18-105_k141_78680 | Low-quality |
| 18-105_k141_78682 | Not-determined |
| 18-105_k141_194101 | Not-determined |
| 18-105_k141_33219 | Low-quality |
| 18-105_k141_8561 | Low-quality |
| 18-105_k141_78694 | Not-determined |
| 18-105_k141_105232 | Not-determined |
| 18-105_k141_78701 | Not-determined |
| 18-105_k141_122521 | Not-determined |
| 18-105_k141_211567 | Not-determined |
| 18-105_k141_194122 | Not-determined |
| 18-105_k141_211576 | Not-determined |
| 18-105_k141_60469 | Not-determined |
| 18-105_k141_149382 | Not-determined |
| 18-105_k141_105243 | Not-determined |
| 18-105_k141_105244 | Not-determined |
| 18-105_k141_194135 | Low-quality |
| 18-105_k141_202419 | Not-determined |
| 18-105_k141_149388 | Not-determined |
| 18-105_k141_33261 | Not-determined |
| 18-105_k141_33263 | Not-determined |
| 18-105_k141_23804 | Low-quality |
| 18-105_k141_33270 | Not-determined |
| 18-105_k141_23809 | Not-determined |
| 18-105_k141_105246 | Not-determined |
| 18-105_k141_95548 | Not-determined |
| 18-105_k141_68954 | Not-determined |
| 18-105_k141_33275 | Not-determined |
| 18-105_k141_60490 | Not-determined |
| 18-105_k141_202429 | Not-determined |
| 18-105_k141_95556 | Not-determined |
| 18-105_k141_68961 | Not-determined |
| 18-105_k141_202436 | Not-determined |
| 18-105_k141_194155 | Not-determined |
| 18-105_k141_8613 | Not-determined |
| 18-105_k141_211606 | Low-quality |
| 18-105_k141_78747 | Not-determined |
| 18-105_k141_105263 | Not-determined |
| 18-105_k141_78749 | Not-determined |
| 18-105_k141_158075 | Not-determined |
| 18-105_k141_87343 | Not-determined |
| 18-105_k141_8623 | Not-determined |
| 18-105_k141_105274 | Not-determined |
| 18-105_k141_78758 | Not-determined |
| 18-105_k141_140394 | Not-determined |
| 18-105_k141_122562 | Not-determined |
| 18-105_k141_8629 | Not-determined |
| 18-105_k141_122565 | Not-determined |
| 18-105_k141_60519 | Not-determined |
| 18-105_k141_122569 | Not-determined |
| 18-105_k141_33301 | Not-determined |
| 18-105_k141_87359 | Low-quality |
| 18-105_k141_87360 | Not-determined |
| 18-105_k141_23826 | Not-determined |
| 18-105_k141_8642 | Not-determined |
| 18-105_k141_60528 | Not-determined |
| 18-105_k141_211630 | Not-determined |
| 18-105_k141_122590 | Not-determined |
| 18-105_k141_33313 | Not-determined |
| 18-105_k141_194189 | Not-determined |
| 18-105_k141_105289 | Low-quality |
| 18-105_k141_140416 | Not-determined |
| 18-105_k141_158095 | Not-determined |
| 18-105_k141_211641 | Not-determined |
| 18-105_k141_176517 | Not-determined |
| 18-105_k141_211645 | Not-determined |
| 18-105_k141_105301 | Not-determined |
| 18-105_k141_105303 | Not-determined |
| 18-105_k141_42031 | Not-determined |
| 18-105_k141_8658 | Not-determined |
| 18-105_k141_211653 | Not-determined |
| 18-105_k141_42036 | Not-determined |
| 18-105_k141_60546 | Not-determined |
| 18-105_k141_8664 | Not-determined |
| 18-105_k141_105314 | Not-determined |
| 18-105_k141_140434 | Not-determined |
| 18-105_k141_51645 | Not-determined |
| 18-105_k141_33330 | Low-quality |
| 18-105_k141_42048 | Not-determined |
| 18-105_k141_149429 | Not-determined |
| 18-105_k141_194211 | Not-determined |
| 18-105_k141_60560 | Not-determined |
| 18-105_k141_33345 | Low-quality |
| 18-105_k141_51664 | Not-determined |
| 18-105_k141_149442 | Not-determined |
| 18-105_k141_194226 | Not-determined |
| 18-105_k141_158112 | Not-determined |
| 18-105_k141_60572 | Not-determined |
| 18-105_k141_15804 | Not-determined |
| 18-105_k141_122623 | Low-quality |
| 18-105_k141_122624 | Not-determined |
| 18-105_k141_140470 | Not-determined |
| 18-105_k141_194232 | Not-determined |
| 18-105_k141_60585 | Not-determined |
| 18-105_k141_23860 | Not-determined |
| 18-105_k141_33362 | Not-determined |
| 18-105_k141_113577 | Not-determined |
| 18-105_k141_8686 | Not-determined |
| 18-105_k141_8688 | Not-determined |
| 18-105_k141_149475 | Not-determined |
| 18-105_k141_8690 | Not-determined |
| 18-105_k141_8693 | Not-determined |
| 18-105_k141_51690 | Not-determined |
| 18-105_k141_42091 | Not-determined |
| 18-105_k141_23872 | Not-determined |
| 18-105_k141_15818 | Low-quality |
| 18-105_k141_132219 | Not-determined |
| 18-105_k141_68981 | Not-determined |
| 18-105_k141_132225 | Not-determined |
| 18-105_k141_23877 | Not-determined |
| 18-105_k141_8704 | Not-determined |
| 18-105_k141_132230 | Not-determined |
| 18-105_k141_68987 | Not-determined |
| 18-105_k141_105348 | Not-determined |
| 18-105_k141_51703 | Not-determined |
| 18-105_k141_51704 | Not-determined |
| 18-105_k141_149493 | Not-determined |
| 18-105_k141_60604 | Low-quality |
| 18-105_k141_149495 | Not-determined |
| 18-105_k141_60606 | Not-determined |
| 18-105_k141_60608 | Not-determined |
| 18-105_k141_194245 | Not-determined |
| 18-105_k141_15830 | Not-determined |
| 18-105_k141_68994 | Not-determined |
| 18-105_k141_122652 | Not-determined |
| 18-105_k141_8711 | Low-quality |
| 18-105_k141_132237 | Not-determined |
| 18-105_k141_68995 | Not-determined |
| 18-105_k141_60610 | Not-determined |
| 18-105_k141_87426 | Not-determined |
| 18-105_k141_95635 | Not-determined |
| 18-105_k141_33380 | Not-determined |
| 18-105_k141_33382 | Not-determined |
| 18-105_k141_60618 | Low-quality |
| 18-105_k141_51723 | Not-determined |
| 18-105_k141_15845 | Not-determined |
| 18-105_k141_51725 | Not-determined |
| 18-105_k141_194262 | Not-determined |
| 18-105_k141_8717 | Low-quality |
| 18-105_k141_23908 | Not-determined |
| 18-105_k141_194271 | Not-determined |
| 18-105_k141_42113 | Not-determined |
| 18-105_k141_23910 | Not-determined |
| 18-105_k141_33397 | Not-determined |
| 18-105_k141_33398 | Not-determined |
| 18-105_k141_69024 | Not-determined |
| 18-105_k141_95663 | Not-determined |
| 18-105_k141_132266 | Not-determined |
| 18-105_k141_51744 | Not-determined |
| 18-105_k141_42125 | Not-determined |
| 18-105_k141_211681 | Not-determined |
| 18-105_k141_8746 | Not-determined |
| 18-105_k141_60636 | Not-determined |
| 18-105_k141_95681 | Not-determined |
| 18-105_k141_60642 | Not-determined |
| 18-105_k141_51755 | Not-determined |
| 18-105_k141_8751 | Not-determined |
| 18-105_k141_8756 | Not-determined |
| 18-105_k141_69048 | Not-determined |
| 18-105_k141_60655 | Not-determined |
| 18-105_k141_15873 | Not-determined |
| 18-105_k141_95703 | Not-determined |
| 18-105_k141_51775 | Not-determined |
| 18-105_k141_69054 | Not-determined |
| 18-105_k141_33430 | Low-quality |
| 18-105_k141_202510 | Not-determined |
| 18-105_k141_69062 | Not-determined |
| 18-105_k141_167001 | Not-determined |
| 18-105_k141_33450 | Not-determined |
| 18-105_k141_202522 | Not-determined |
| 18-105_k141_78808 | Not-determined |
| 18-105_k141_167007 | Not-determined |
| 18-105_k141_78811 | Not-determined |
| 18-105_k141_167010 | Not-determined |
| 18-105_k141_122729 | Not-determined |
| 18-105_k141_33456 | Not-determined |
| 18-105_k141_33462 | Not-determined |
| 18-105_k141_167016 | Not-determined |
| 18-105_k141_78821 | Not-determined |
| 18-105_k141_95720 | Not-determined |
| 18-105_k141_167021 | Not-determined |
| 18-105_k141_60707 | Low-quality |
| 18-105_k141_60709 | Not-determined |
| 18-105_k141_69087 | Not-determined |
| 18-105_k141_15901 | Not-determined |
| 18-105_k141_132325 | Not-determined |
| 18-105_k141_87493 | Low-quality |
| 18-105_k141_15906 | Not-determined |
| 18-105_k141_87494 | Not-determined |
| 18-105_k141_87496 | Not-determined |
| 18-105_k141_140489 | Low-quality |
| 18-105_k141_87500 | Not-determined |
| 18-105_k141_51818 | Not-determined |
| 18-105_k141_140494 | Not-determined |
| 18-105_k141_87502 | Not-determined |
| 18-105_k141_51820 | Not-determined |
| 18-105_k141_140497 | Not-determined |
| 18-105_k141_8777 | Not-determined |
| 18-105_k141_8782 | Not-determined |
| 18-105_k141_78844 | Not-determined |
| 18-105_k141_87512 | Not-determined |
| 18-105_k141_69108 | Not-determined |
| 18-105_k141_69111 | Not-determined |
| 18-105_k141_8786 | Not-determined |
| 18-105_k141_51832 | Not-determined |
| 18-105_k141_140507 | Not-determined |
| 18-105_k141_95734 | Not-determined |
| 18-105_k141_8793 | Not-determined |
| 18-105_k141_132353 | Not-determined |
| 18-105_k141_69116 | Not-determined |
| 18-105_k141_15927 | Not-determined |
| 18-105_k141_176572 | Not-determined |
| 18-105_k141_69123 | Not-determined |
| 18-105_k141_202555 | Not-determined |
| 18-105_k141_78859 | Not-determined |
| 18-105_k141_95749 | Not-determined |
| 18-105_k141_176578 | Not-determined |
| 18-105_k141_149525 | Not-determined |
| 18-105_k141_132362 | Not-determined |
| 18-105_k141_167062 | Not-determined |
| 18-105_k141_51853 | Not-determined |
| 18-105_k141_51858 | Not-determined |
| 18-105_k141_69138 | Not-determined |
| 18-105_k141_69140 | Not-determined |
| 18-105_k141_95759 | Not-determined |
| 18-105_k141_194311 | Not-determined |
| 18-105_k141_69155 | Not-determined |
| 18-105_k141_105415 | Not-determined |
| 18-105_k141_167073 | Not-determined |
| 18-105_k141_51876 | Not-determined |
| 18-105_k141_105419 | Low-quality |
| 18-105_k141_132383 | Not-determined |
| 18-105_k141_149537 | Not-determined |
| 18-105_k141_105423 | Not-determined |
| 18-105_k141_140534 | Not-determined |
| 18-105_k141_105425 | Not-determined |
| 18-105_k141_140544 | Not-determined |
| 18-105_k141_176606 | Not-determined |
| 18-105_k141_87521 | Low-quality |
| 18-105_k141_167094 | Not-determined |
| 18-105_k141_78873 | Not-determined |
| 18-105_k141_33498 | Not-determined |
| 18-105_k141_122795 | Not-determined |
| 18-105_k141_15944 | Not-determined |
| 18-105_k141_60766 | Not-determined |
| 18-105_k141_132400 | Low-quality |
| 18-105_k141_15950 | Not-determined |
| 18-105_k141_33505 | Not-determined |
| 18-105_k141_105449 | Not-determined |
| 18-105_k141_87532 | Not-determined |
| 18-105_k141_132407 | Not-determined |
| 18-105_k141_122809 | Low-quality |
| 18-105_k141_132412 | Not-determined |
| 18-105_k141_87535 | Not-determined |
| 18-105_k141_87536 | Not-determined |
| 18-105_k141_122812 | Low-quality |
| 18-105_k141_122815 | Not-determined |
| 18-105_k141_33525 | Not-determined |
| 18-105_k141_78893 | Not-determined |
| 18-105_k141_51896 | Not-determined |
| 18-105_k141_113620 | Not-determined |
| 18-105_k141_122823 | Not-determined |
| 18-105_k141_87547 | Not-determined |
| 18-105_k141_95791 | Not-determined |
| 18-105_k141_60806 | Not-determined |
| 18-105_k141_23976 | Not-determined |
| 18-105_k141_60811 | Not-determined |
| 18-105_k141_87561 | Not-determined |
| 18-105_k141_51917 | Not-determined |
| 18-105_k141_69182 | Not-determined |
| 18-105_k141_105485 | Not-determined |
| 18-105_k141_42167 | Not-determined |
| 18-105_k141_23991 | Low-quality |
| 18-105_k141_23993 | Not-determined |
| 18-105_k141_8848 | Low-quality |
| 18-105_k141_194329 | Not-determined |
| 18-105_k141_113643 | Not-determined |
| 18-105_k141_140568 | Not-determined |
| 18-105_k141_105498 | Not-determined |
| 18-105_k141_122836 | Not-determined |
| 18-105_k141_51923 | Not-determined |
| 18-105_k141_51924 | Not-determined |
| 18-105_k141_167151 | Not-determined |
| 18-105_k141_51927 | Not-determined |
| 18-105_k141_78921 | Not-determined |
| 18-105_k141_42181 | Not-determined |
| 18-105_k141_51930 | Not-determined |
| 18-105_k141_202574 | Not-determined |
| 18-105_k141_202577 | Not-determined |
| 18-105_k141_24007 | Not-determined |
| 18-105_k141_42187 | Low-quality |
| 18-105_k141_185585 | Not-determined |
| 18-105_k141_202586 | Not-determined |
| 18-105_k141_132429 | Not-determined |
| 18-105_k141_51942 | Not-determined |
| 18-105_k141_202595 | Not-determined |
| 18-105_k141_24019 | Not-determined |
| 18-105_k141_33568 | Low-quality |
| 18-105_k141_95849 | Not-determined |
| 18-105_k141_95850 | Not-determined |
| 18-105_k141_69190 | Not-determined |
| 18-105_k141_8871 | Not-determined |
| 18-105_k141_140602 | Not-determined |
| 18-105_k141_42200 | Not-determined |
| 18-105_k141_60850 | Not-determined |
| 18-105_k141_33575 | Not-determined |
| 18-105_k141_140607 | Not-determined |
| 18-105_k141_122865 | Not-determined |
| 18-105_k141_60858 | Not-determined |
| 18-105_k141_167158 | Not-determined |
| 18-105_k141_51968 | Not-determined |
| 18-105_k141_8877 | Not-determined |
| 18-105_k141_87606 | Not-determined |
| 18-105_k141_60864 | Not-determined |
| 18-105_k141_176660 | Not-determined |
| 18-105_k141_69212 | Not-determined |
| 18-105_k141_176665 | Not-determined |
| 18-105_k141_113681 | Not-determined |
| 18-105_k141_33587 | Not-determined |
| 18-105_k141_60870 | Not-determined |
| 18-105_k141_113683 | Not-determined |
| 18-105_k141_149591 | Not-determined |
| 18-105_k141_113684 | Not-determined |
| 18-105_k141_51981 | Not-determined |
| 18-105_k141_69218 | Not-determined |
| 18-105_k141_122875 | Not-determined |
| 18-105_k141_51987 | Not-determined |
| 18-105_k141_51990 | Not-determined |
| 18-105_k141_51995 | Not-determined |
| 18-105_k141_122885 | Not-determined |
| 18-105_k141_69235 | Not-determined |
| 18-105_k141_194389 | Not-determined |
| 18-105_k141_176676 | Not-determined |
| 18-105_k141_202612 | Not-determined |
| 18-105_k141_132445 | Not-determined |
| 18-105_k141_33624 | Not-determined |
| 18-105_k141_140643 | Not-determined |
| 18-105_k141_149612 | Not-determined |
| 18-105_k141_194405 | Not-determined |
| 18-105_k141_149614 | Not-determined |
| 18-105_k141_132452 | Not-determined |
| 18-105_k141_33635 | Not-determined |
| 18-105_k141_33636 | Not-determined |
| 18-105_k141_140644 | Not-determined |
| 18-105_k141_194408 | Low-quality |
| 18-105_k141_167174 | Not-determined |
| 18-105_k141_105528 | Not-determined |
| 18-105_k141_105532 | Not-determined |
| 18-105_k141_167179 | Not-determined |
| 18-105_k141_42240 | Low-quality |
| 18-105_k141_122895 | Low-quality |
| 18-105_k141_60891 | Not-determined |
| 18-105_k141_95880 | Not-determined |
| 18-105_k141_105538 | Not-determined |
| 18-105_k141_60893 | Not-determined |
| 18-105_k141_33644 | Not-determined |
| 18-105_k141_149629 | Not-determined |
| 18-105_k141_33646 | Not-determined |
| 18-105_k141_176684 | Not-determined |
| 18-105_k141_42251 | Not-determined |
| 18-105_k141_211702 | Not-determined |
| 18-105_k141_113707 | Not-determined |
| 18-105_k141_95888 | Not-determined |
| 18-105_k141_33658 | Not-determined |
| 18-105_k141_202649 | Not-determined |
| 18-105_k141_52030 | Not-determined |
| 18-105_k141_167199 | Not-determined |
| 18-105_k141_202651 | Not-determined |
| 18-105_k141_113713 | Low-quality |
| 18-105_k141_176705 | Low-quality |
| 18-105_k141_52036 | Low-quality |
| 18-105_k141_42262 | Not-determined |
| 18-105_k141_211724 | Low-quality |
| 18-105_k141_167212 | Not-determined |
| 18-105_k141_60912 | Not-determined |
| 18-105_k141_105565 | Low-quality |
| 18-105_k141_113733 | Not-determined |
| 18-105_k141_87610 | Not-determined |
| 18-105_k141_194418 | Not-determined |
| 18-105_k141_52060 | Not-determined |
| 18-105_k141_132486 | Not-determined |
| 18-105_k141_87622 | Not-determined |
| 18-105_k141_132487 | Not-determined |
| 18-105_k141_113752 | Low-quality |
| 18-105_k141_176713 | Not-determined |
| 18-105_k141_60923 | Not-determined |
| 18-105_k141_176719 | Not-determined |
| 18-105_k141_176721 | Not-determined |
| 18-105_k141_132496 | Not-determined |
| 18-105_k141_132499 | Not-determined |
| 18-105_k141_202681 | Not-determined |
| 18-105_k141_167236 | Not-determined |
| 18-105_k141_24076 | Not-determined |
| 18-105_k141_132504 | Not-determined |
| 18-105_k141_69258 | Not-determined |
| 18-105_k141_105595 | Not-determined |
| 18-105_k141_176735 | Low-quality |
| 18-105_k141_95911 | Low-quality |
| 18-105_k141_132511 | Not-determined |
| 18-105_k141_87639 | Not-determined |
| 18-105_k141_24088 | Not-determined |
| 18-105_k141_52089 | Low-quality |
| 18-105_k141_176758 | Not-determined |
| 18-105_k141_176760 | Low-quality |
| 18-105_k141_95916 | Not-determined |
| 18-105_k141_87663 | Not-determined |
| 18-105_k141_95921 | Not-determined |
| 18-105_k141_202695 | Low-quality |
| 18-105_k141_95925 | Not-determined |
| 18-105_k141_113786 | Low-quality |
| 18-105_k141_202702 | Not-determined |
| 18-105_k141_24110 | Low-quality |
| 18-105_k141_24111 | Not-determined |
| 18-105_k141_87683 | Not-determined |
| 18-105_k141_24115 | Not-determined |
| 18-105_k141_202717 | Not-determined |
| 18-105_k141_87687 | Not-determined |
| 18-105_k141_132535 | Not-determined |
| 18-105_k141_42336 | Not-determined |
| 18-105_k141_33702 | Not-determined |
| 18-105_k141_202732 | Low-quality |
| 18-105_k141_132545 | Not-determined |
| 18-105_k141_33712 | Not-determined |
| 18-105_k141_140673 | Not-determined |
| 18-105_k141_33716 | Not-determined |
| 18-105_k141_42363 | Not-determined |
| 18-105_k141_132552 | Not-determined |
| 18-105_k141_132554 | Not-determined |
| 18-105_k141_24130 | Not-determined |
| 18-105_k141_185597 | Not-determined |
| 18-105_k141_123021 | Not-determined |
| 18-105_k141_52117 | Not-determined |
| 18-105_k141_149634 | Low-quality |
| 18-105_k141_149637 | Not-determined |
| 18-105_k141_149641 | Low-quality |
| 18-105_k141_132568 | Not-determined |
| 18-105_k141_149647 | Not-determined |
| 18-105_k141_132582 | Not-determined |
| 18-105_k141_185617 | Not-determined |
| 18-105_k141_149659 | Not-determined |
| 18-105_k141_167249 | Not-determined |
| 18-105_k141_185625 | Not-determined |
| 18-105_k141_52145 | Not-determined |
| 18-105_k141_211759 | Not-determined |
| 18-105_k141_105644 | Not-determined |
| 18-105_k141_176779 | Not-determined |
| 18-105_k141_149674 | Not-determined |
| 18-105_k141_33729 | Not-determined |
| 18-105_k141_52158 | Not-determined |
| 18-105_k141_211763 | Not-determined |
| 18-105_k141_42384 | Not-determined |
| 18-105_k141_176785 | Not-determined |
| 18-105_k141_123065 | Not-determined |
| 18-105_k141_167259 | Not-determined |
| 18-105_k141_167266 | Low-quality |
| 18-105_k141_176799 | Not-determined |
| 18-105_k141_185641 | Not-determined |
| 18-105_k141_123086 | Not-determined |
| 18-105_k141_185644 | Not-determined |
| 18-105_k141_185647 | Low-quality |
| 18-105_k141_52186 | Not-determined |
| 18-105_k141_95937 | Not-determined |
| 18-105_k141_149690 | Not-determined |
| 18-105_k141_113836 | Not-determined |
| 18-105_k141_95945 | Not-determined |
| 18-105_k141_202745 | Not-determined |
| 18-105_k141_52205 | Not-determined |
| 18-105_k141_211810 | Not-determined |
| 18-105_k141_185668 | Not-determined |
| 18-105_k141_202757 | Not-determined |
| 18-105_k141_42396 | Not-determined |
| 18-105_k141_87738 | Not-determined |
| 18-105_k141_185674 | Not-determined |
| 18-105_k141_202761 | Not-determined |
| 18-105_k141_42400 | Not-determined |
| 18-105_k141_87745 | Not-determined |
| 18-105_k141_167305 | Not-determined |
| 18-105_k141_60946 | Not-determined |
| 18-105_k141_132602 | Low-quality |
| 18-105_k141_185692 | Not-determined |
| 18-105_k141_113877 | Not-determined |
| 18-105_k141_149730 | Not-determined |
| 18-105_k141_202773 | Not-determined |
| 18-105_k141_194463 | Not-determined |
| 18-105_k141_60956 | Not-determined |
| 18-105_k141_42429 | Not-determined |
| 18-105_k141_194471 | Not-determined |
| 18-105_k141_60962 | Not-determined |
| 18-105_k141_194473 | Not-determined |
| 18-105_k141_87765 | Not-determined |
| 18-105_k141_167324 | Not-determined |
| 18-105_k141_42441 | Not-determined |
| 18-105_k141_95971 | Not-determined |
| 18-105_k141_42446 | Not-determined |
| 18-105_k141_132609 | Not-determined |
| 18-105_k141_95981 | Not-determined |
| 18-105_k141_167334 | Not-determined |
| 18-105_k141_60977 | Low-quality |
| 18-105_k141_113918 | Not-determined |
| 18-105_k141_42457 | Not-determined |
| 18-105_k141_113919 | Not-determined |
| 18-105_k141_33793 | Not-determined |
| 18-105_k141_176826 | Not-determined |
| 18-105_k141_158220 | Low-quality |
| 18-105_k141_60997 | Not-determined |
| 18-105_k141_52224 | Not-determined |
| 18-105_k141_194502 | Low-quality |
| 18-105_k141_158226 | Not-determined |
| 18-105_k141_176833 | Not-determined |
| 18-105_k141_113929 | Not-determined |
| 18-105_k141_202800 | Not-determined |
| 18-105_k141_176834 | Low-quality |
| 18-105_k141_194503 | Not-determined |
| 18-105_k141_24146 | Not-determined |
| 18-105_k141_167355 | Low-quality |
| 18-105_k141_61007 | Not-determined |
| 18-105_k141_33796 | Not-determined |
| 18-105_k141_202805 | Not-determined |
| 18-105_k141_176847 | Not-determined |
| 18-105_k141_176854 | Not-determined |
| 18-105_k141_176860 | Not-determined |
| 18-105_k141_52248 | Not-determined |
| 18-105_k141_87815 | Not-determined |
| 18-105_k141_87817 | Low-quality |
| 18-105_k141_78956 | Not-determined |
| 18-105_k141_202815 | Not-determined |
| 18-105_k141_167376 | Not-determined |
| 18-105_k141_176869 | Not-determined |
| 18-105_k141_149789 | Not-determined |
| 18-105_k141_52271 | Low-quality |
| 18-105_k141_24173 | Not-determined |
| 18-105_k141_42480 | Not-determined |
| 18-105_k141_78984 | Not-determined |
| 18-105_k141_24181 | Low-quality |
| 18-105_k141_158258 | Not-determined |
| 18-105_k141_42488 | Not-determined |
| 18-105_k141_96001 | Not-determined |
| 18-105_k141_42490 | Not-determined |
| 18-105_k141_24189 | Not-determined |
| 18-105_k141_167403 | Not-determined |
| 18-105_k141_202845 | Not-determined |
| 18-105_k141_78997 | Not-determined |
| 18-105_k141_149813 | Not-determined |
| 18-105_k141_194537 | Not-determined |
| 18-105_k141_42501 | Not-determined |
| 18-105_k141_176898 | Not-determined |
| 18-105_k141_69284 | Not-determined |
| 18-105_k141_176900 | Not-determined |
| 18-105_k141_176901 | Not-determined |
| 18-105_k141_194542 | Not-determined |
| 18-105_k141_52298 | Not-determined |
| 18-105_k141_79007 | Not-determined |
| 18-105_k141_69291 | Not-determined |
| 18-105_k141_113970 | Low-quality |
| 18-105_k141_87863 | Low-quality |
| 18-105_k141_202861 | Not-determined |
| 18-105_k141_113985 | Not-determined |
| 18-105_k141_113986 | Not-determined |
| 18-105_k141_42512 | Not-determined |
| 18-105_k141_42518 | Not-determined |
| 18-105_k141_96013 | Not-determined |
| 18-105_k141_79024 | Not-determined |
| 18-105_k141_113991 | Not-determined |
| 18-105_k141_87882 | Not-determined |
| 18-105_k141_113993 | Not-determined |
| 18-105_k141_42528 | Not-determined |
| 18-105_k141_79030 | Not-determined |
| 18-105_k141_79031 | Not-determined |
| 18-105_k141_158290 | Not-determined |
| 18-105_k141_52328 | Not-determined |
| 18-105_k141_185747 | Not-determined |
| 18-105_k141_158292 | Not-determined |
| 18-105_k141_202874 | Low-quality |
| 18-105_k141_52333 | Low-quality |
| 18-105_k141_140687 | Not-determined |
| 17-109-final-viral-combined_k141_376326\|\|full | Medium-quality |
| 18-105_k141_52340 | Not-determined |

| | |
|---|---|
| 18-105_k141_176938 | Not-determined |
| 18-105_k141_202880 | Not-determined |
| 18-105_k141_194582 | Not-determined |
| 18-105_k141_96040 | Not-determined |
| 18-105_k141_185758 | Not-determined |
| 18-105_k141_167430 | Not-determined |
| 18-105_k141_149827 | Not-determined |
| 18-105_k141_114006 | Not-determined |
| 18-105_k141_33871 | Not-determined |
| 18-105_k141_140704 | Not-determined |
| 18-105_k141_202885 | Not-determined |
| 18-105_k141_96052 | Not-determined |
| 18-105_k141_202886 | Not-determined |
| 18-105_k141_140706 | Not-determined |
| 18-105_k141_96054 | Not-determined |
| 18-105_k141_42556 | Not-determined |
| 18-105_k141_167440 | Not-determined |
| 18-105_k141_69330 | Not-determined |
| 18-105_k141_96058 | Low-quality |
| 18-105_k141_176957 | Not-determined |
| 18-105_k141_114022 | Not-determined |
| 18-105_k141_33881 | Not-determined |
| 18-105_k141_167449 | Not-determined |
| 18-105_k141_167450 | Not-determined |
| 18-105_k141_176964 | Low-quality |
| 18-105_k141_185771 | Not-determined |
| 18-105_k141_185773 | Not-determined |
| 18-105_k141_52364 | Not-determined |
| 18-105_k141_167460 | Not-determined |
| 18-105_k141_33893 | Not-determined |
| 18-105_k141_167462 | Not-determined |
| 18-105_k141_176972 | Not-determined |
| 18-105_k141_52369 | Not-determined |
| 18-105_k141_140724 | Not-determined |
| 18-105_k141_149838 | Not-determined |
| 18-105_k141_79044 | Not-determined |
| 18-105_k141_52379 | Not-determined |
| 18-105_k141_185789 | Not-determined |
| 18-105_k141_123162 | Not-determined |
| 18-105_k141_105652 | Not-determined |
| 18-105_k141_33909 | Low-quality |
| 18-105_k141_105659 | Not-determined |
| 18-105_k141_105660 | Not-determined |
| 18-105_k141_79057 | Not-determined |
| 18-105_k141_123177 | Not-determined |
| 18-105_k141_79061 | Not-determined |
| 18-105_k141_194624 | Not-determined |
| 18-105_k141_16005 | Not-determined |
| 18-105_k141_149856 | Not-determined |
| 18-105_k141_16006 | Not-determined |
| 18-105_k141_105676 | Not-determined |
| 18-105_k141_211856 | Not-determined |
| 18-105_k141_33922 | Not-determined |
| 18-105_k141_16008 | Not-determined |
| 18-105_k141_61020 | Not-determined |
| 18-105_k141_140762 | Not-determined |
| 18-105_k141_105681 | Not-determined |
| 18-105_k141_185802 | Not-determined |
| 18-105_k141_158333 | Not-determined |
| 18-105_k141_158334 | Not-determined |
| 18-105_k141_149866 | Low-quality |
| 18-105_k141_42580 | Not-determined |
| 18-105_k141_52406 | Not-determined |
| 18-105_k141_61030 | Not-determined |
| 18-105_k141_52411 | Not-determined |
| 18-105_k141_185812 | Not-determined |
| 18-105_k141_61033 | Not-determined |
| 18-105_k141_105694 | Not-determined |
| 18-105_k141_167504 | Not-determined |
| 18-105_k141_194629 | Not-determined |
| 18-105_k141_61040 | Not-determined |
| 18-105_k141_87911 | Low-quality |
| 18-105_k141_105701 | Not-determined |
| 18-105_k141_87912 | Not-determined |
| 18-105_k141_87913 | Not-determined |
| 18-105_k141_167512 | Not-determined |
| 18-105_k141_211883 | Not-determined |
| 18-105_k141_105705 | Not-determined |
| 18-105_k141_61047 | Not-determined |
| 18-105_k141_79075 | Not-determined |
| 18-105_k141_16038 | Low-quality |
| 18-105_k141_211888 | Not-determined |
| 18-105_k141_167524 | Not-determined |
| 18-105_k141_194643 | Not-determined |
| 18-105_k141_211894 | Not-determined |
| 18-105_k141_167531 | Not-determined |
| 18-105_k141_177032 | Not-determined |
| 18-105_k141_123209 | Not-determined |
| 18-105_k141_52430 | Not-determined |
| 18-105_k141_61060 | Not-determined |
| 18-105_k141_61062 | Not-determined |
| 18-105_k141_167549 | Low-quality |
| 18-105_k141_52436 | Not-determined |
| 18-105_k141_167550 | Low-quality |
| 18-105_k141_123217 | Not-determined |
| 18-105_k141_87930 | Not-determined |
| 18-105_k141_61066 | Not-determined |
| 18-105_k141_61068 | Not-determined |
| 18-105_k141_24251 | Low-quality |
| 18-105_k141_167565 | Low-quality |
| 18-105_k141_24264 | Not-determined |
| 18-105_k141_211925 | Not-determined |
| 18-105_k141_16057 | Not-determined |
| 18-105_k141_194675 | Not-determined |
| 18-105_k141_24269 | Not-determined |
| 18-105_k141_33964 | Not-determined |
| 18-105_k141_96070 | Not-determined |
| 18-105_k141_96077 | Not-determined |
| 18-105_k141_177073 | Not-determined |
| 18-105_k141_42585 | Not-determined |
| 18-105_k141_140802 | Low-quality |
| 18-105_k141_211949 | Not-determined |
| 18-105_k141_177076 | Not-determined |
| 18-105_k141_123244 | Low-quality |
| 18-105_k141_149899 | Not-determined |
| 18-105_k141_42593 | Not-determined |
| 18-105_k141_211956 | Not-determined |
| 18-105_k141_123249 | Not-determined |
| 18-105_k141_140818 | Not-determined |
| 18-105_k141_33983 | Not-determined |
| 18-105_k141_33985 | Not-determined |
| 18-105_k141_149912 | Not-determined |
| 18-105_k141_52461 | Not-determined |
| 18-105_k141_24299 | Not-determined |
| 18-105_k141_132670 | Not-determined |
| 18-105_k141_167600 | Not-determined |
| 18-105_k141_87947 | Not-determined |
| 18-105_k141_33997 | Low-quality |
| 18-105_k141_105730 | Low-quality |
| 18-105_k141_185858 | Not-determined |
| 18-105_k141_105732 | Not-determined |
| 18-105_k141_194704 | Low-quality |
| 18-105_k141_194705 | Low-quality |
| 18-105_k141_149922 | Not-determined |
| 18-105_k141_132675 | Not-determined |
| 18-105_k141_123267 | Not-determined |
| 18-105_k141_194709 | Not-determined |
| 18-105_k141_87957 | Not-determined |
| 18-105_k141_87960 | Not-determined |
| 18-105_k141_105743 | Not-determined |
| 18-105_k141_42628 | Not-determined |
| 18-105_k141_52465 | Not-determined |
| 18-105_k141_194720 | Not-determined |
| 18-105_k141_52467 | Not-determined |
| 18-105_k141_61119 | Not-determined |
| 18-105_k141_123281 | Not-determined |
| 18-105_k141_177089 | Not-determined |
| 18-105_k141_16079 | Not-determined |
| 18-105_k141_61122 | Not-determined |
| 18-105_k141_87977 | Not-determined |
| 18-105_k141_132696 | Not-determined |
| 18-105_k141_24333 | Low-quality |
| 18-105_k141_167624 | Not-determined |
| 18-105_k141_52478 | Not-determined |
| 18-105_k141_185862 | Low-quality |
| 18-105_k141_185865 | Not-determined |
| 18-105_k141_167630 | Not-determined |
| 18-105_k141_42633 | Low-quality |
| 18-105_k141_42634 | Not-determined |
| 18-105_k141_24341 | Not-determined |
| 18-105_k141_96128 | Not-determined |
| 18-105_k141_34012 | Low-quality |
| 18-105_k141_123289 | Not-determined |
| 18-105_k141_194738 | Not-determined |
| 18-105_k141_140853 | Not-determined |
| 18-105_k141_61138 | Not-determined |
| 18-105_k141_194741 | Not-determined |
| 18-105_k141_52492 | Not-determined |
| 18-105_k141_52493 | Not-determined |
| 18-105_k141_69353 | Not-determined |
| 18-105_k141_34016 | Not-determined |
| 18-105_k141_211967 | Not-determined |
| 18-105_k141_149928 | Not-determined |
| 18-105_k141_149929 | Not-determined |
| 18-105_k141_42642 | Low-quality |
| 18-105_k141_61144 | Not-determined |
| 18-105_k141_52504 | Not-determined |
| 18-105_k141_140869 | Not-determined |
| 18-105_k141_167647 | Not-determined |
| 18-105_k141_61147 | Not-determined |
| 18-105_k141_140879 | Not-determined |
| 18-105_k141_140882 | Not-determined |
| 18-105_k141_34028 | Not-determined |
| 18-105_k141_202930 | Low-quality |
| 18-105_k141_158448 | Not-determined |
| 18-105_k141_34029 | Not-determined |
| 18-105_k141_211983 | Not-determined |
| 18-105_k141_202934 | Not-determined |
| 18-105_k141_34031 | Not-determined |
| 18-105_k141_96163 | Not-determined |
| 18-105_k141_24363 | Not-determined |
| 18-105_k141_194754 | Not-determined |
| 18-105_k141_34037 | Not-determined |
| 18-105_k141_79115 | Not-determined |
| 18-105_k141_42659 | Low-quality |
| 18-105_k141_167658 | Not-determined |
| 18-105_k141_185889 | Low-quality |
| 18-105_k141_42660 | Not-determined |
| 18-105_k141_105761 | Low-quality |
| 18-105_k141_149960 | Low-quality |
| 18-105_k141_79121 | Not-determined |
| 18-105_k141_185898 | Not-determined |
| 18-105_k141_42665 | Not-determined |
| 18-105_k141_140901 | Not-determined |
| 18-105_k141_158469 | Not-determined |
| 18-105_k141_105773 | Not-determined |
| 18-105_k141_149970 | Low-quality |
| 18-105_k141_149973 | Not-determined |
| 18-105_k141_79130 | Not-determined |
| 18-105_k141_140910 | Not-determined |
| 18-105_k141_158485 | Not-determined |
| 18-105_k141_194785 | Low-quality |
| 18-105_k141_79135 | Not-determined |
| 18-105_k141_158489 | Low-quality |
| 18-105_k141_96175 | Not-determined |
| 18-105_k141_149983 | Not-determined |
| 18-105_k141_105784 | Not-determined |
| 18-105_k141_52555 | Not-determined |
| 18-105_k141_212006 | Not-determined |
| 18-105_k141_194806 | Not-determined |
| 18-105_k141_177164 | Not-determined |
| 18-105_k141_158507 | Not-determined |
| 18-105_k141_158508 | Not-determined |
| 18-105_k141_105789 | Not-determined |
| 18-105_k141_194817 | Not-determined |
| 18-105_k141_158512 | Low-quality |
| 18-105_k141_194818 | Not-determined |
| 18-105_k141_34095 | Not-determined |
| 18-105_k141_105794 | Low-quality |
| 18-105_k141_96206 | Not-determined |
| 18-105_k141_105801 | Not-determined |
| 18-105_k141_105802 | Not-determined |
| 18-105_k141_96214 | Not-determined |
| 18-105_k141_79143 | Not-determined |
| 18-105_k141_140946 | Not-determined |
| 18-105_k141_123333 | Not-determined |
| 18-105_k141_158521 | Not-determined |
| 18-105_k141_96226 | Not-determined |
| 18-105_k141_167717 | Not-determined |
| 18-105_k141_167718 | Low-quality |
| 18-105_k141_158530 | Not-determined |
| 18-105_k141_140953 | Not-determined |
| KM-final-viral-combined_k141_260343\|\|full | High-quality |
| 18-105_k141_79159 | Not-determined |
| 18-105_k141_212024 | Not-determined |
| 18-105_k141_167727 | Low-quality |
| 18-105_k141_123346 | Not-determined |
| 18-105_k141_158544 | Not-determined |
| 18-105_k141_69387 | Not-determined |
| 18-105_k141_194851 | Not-determined |
| 18-105_k141_194852 | Not-determined |
| 18-105_k141_105842 | Not-determined |
| 18-105_k141_150020 | Not-determined |
| 18-105_k141_212060 | Not-determined |
| 18-105_k141_123372 | Not-determined |
| 18-105_k141_212071 | Not-determined |
| 18-105_k141_123373 | Not-determined |
| 18-105_k141_123376 | Low-quality |
| 18-105_k141_69403 | Not-determined |
| 18-105_k141_123379 | Not-determined |
| 18-105_k141_69406 | Not-determined |
| 18-105_k141_123381 | Low-quality |
| 18-105_k141_194867 | Not-determined |
| 18-105_k141_202966 | Not-determined |
| 18-105_k141_88025 | Not-determined |
| 18-105_k141_69413 | Not-determined |
| 18-105_k141_158574 | Not-determined |
| 18-105_k141_42766 | Not-determined |
| 18-105_k141_158577 | Not-determined |
| 18-105_k141_52602 | Not-determined |
| 18-105_k141_69419 | Not-determined |
| 18-105_k141_79202 | Not-determined |
| 18-105_k141_177205 | Not-determined |
| 18-105_k141_177210 | Not-determined |
| 18-105_k141_52609 | Not-determined |
| 18-105_k141_141003 | Not-determined |
| 18-105_k141_141004 | Not-determined |
| 18-105_k141_202987 | Not-determined |
| 18-105_k141_185939 | Not-determined |
| 18-105_k141_69429 | Not-determined |
| 18-105_k141_158584 | Not-determined |
| 18-105_k141_105882 | Not-determined |
| 18-105_k141_88035 | Not-determined |
| 18-105_k141_88039 | Not-determined |
| 18-105_k141_69435 | Not-determined |
| 18-105_k141_158593 | Not-determined |
| 18-105_k141_203000 | Not-determined |
| 18-105_k141_96242 | Not-determined |
| 18-105_k141_185947 | Not-determined |
| 18-105_k141_96245 | Not-determined |
| 18-105_k141_16116 | Not-determined |
| 18-105_k141_158603 | Not-determined |
| 18-105_k141_52628 | Not-determined |
| 18-105_k141_105910 | Not-determined |
| 18-105_k141_167753 | Not-determined |
| 18-105_k141_177226 | Not-determined |
| 18-105_k141_16125 | Not-determined |
| 18-105_k141_123418 | Not-determined |
| 18-105_k141_96253 | Not-determined |
| 18-105_k141_203014 | Not-determined |
| 18-105_k141_96255 | Not-determined |
| 18-105_k141_123419 | Low-quality |
| 18-105_k141_158613 | Not-determined |
| 18-105_k141_150030 | Low-quality |
| 18-105_k141_150032 | Not-determined |
| 18-105_k141_203025 | Not-determined |
| 18-105_k141_123431 | Not-determined |
| 18-105_k141_141034 | Not-determined |
| 18-105_k141_141036 | Not-determined |
| 18-105_k141_203026 | Not-determined |
| 18-105_k141_16136 | Not-determined |
| 18-105_k141_203029 | Not-determined |
| 18-105_k141_105940 | Not-determined |
| 18-105_k141_158623 | Not-determined |
| 18-105_k141_177236 | Low-quality |
| 18-105_k141_177238 | Not-determined |
| 18-105_k141_167776 | Not-determined |
| 18-105_k141_96276 | Not-determined |
| 18-105_k141_177242 | Not-determined |
| 18-105_k141_88057 | Not-determined |
| 18-105_k141_16143 | Not-determined |
| 18-105_k141_150055 | Not-determined |
| 18-105_k141_123457 | Not-determined |
| 18-105_k141_185961 | Not-determined |
| 18-105_k141_88069 | Not-determined |
| 18-105_k141_158639 | Not-determined |
| 18-105_k141_105953 | Not-determined |
| 18-105_k141_96281 | Not-determined |
| 18-105_k141_141042 | Not-determined |
| 18-105_k141_212077 | Not-determined |
| 18-105_k141_16156 | Not-determined |
| 18-105_k141_141043 | Not-determined |
| 18-105_k141_194913 | Not-determined |
| 18-105_k141_185967 | Low-quality |
| 18-105_k141_105959 | Not-determined |
| 18-105_k141_141045 | Not-determined |
| 18-105_k141_185969 | Not-determined |
| 18-105_k141_203042 | Not-determined |
| 18-105_k141_88075 | Low-quality |
| 18-105_k141_88076 | Not-determined |
| 18-105_k141_105965 | Not-determined |
| 18-105_k141_96292 | Not-determined |
| 18-105_k141_88080 | Low-quality |
| 18-105_k141_158647 | Not-determined |
| 18-105_k141_114104 | Not-determined |
| 18-105_k141_88083 | Low-quality |
| 18-105_k141_123471 | Not-determined |
| 18-105_k141_69450 | Not-determined |
| 18-105_k141_105975 | Not-determined |
| 18-105_k141_105976 | Not-determined |
| 18-105_k141_16174 | Low-quality |
| 18-105_k141_123492 | Not-determined |
| 18-105_k141_61176 | Not-determined |
| 18-105_k141_42782 | Not-determined |
| 18-105_k141_16188 | Not-determined |
| 18-105_k141_61187 | Not-determined |
| 17-109-final-viral-combined_k141_156234\|\|full | Medium-quality |
| 18-105_k141_69467 | Not-determined |
| 18-105_k141_177267 | Not-determined |
| 18-105_k141_186002 | Low-quality |
| 18-105_k141_194942 | Not-determined |
| 18-105_k141_212097 | Not-determined |
| 18-105_k141_186011 | Not-determined |
| 18-105_k141_212098 | Not-determined |
| 18-105_k141_79237 | Not-determined |
| 18-105_k141_141073 | Not-determined |
| 18-105_k141_79243 | Not-determined |
| 18-105_k141_114147 | Not-determined |
| 18-105_k141_114151 | Not-determined |
| 18-105_k141_114154 | Not-determined |
| 18-105_k141_79255 | Not-determined |
| 18-105_k141_88131 | Not-determined |
| 18-105_k141_150127 | Not-determined |
| 18-105_k141_114157 | Not-determined |
| 18-105_k141_88137 | Not-determined |
| 18-105_k141_186048 | Not-determined |
| 18-105_k141_194967 | Not-determined |
| 18-105_k141_96321 | Not-determined |
| 18-105_k141_186058 | Not-determined |
| 18-105_k141_194974 | Low-quality |
| 18-105_k141_79276 | Not-determined |
| 18-105_k141_194981 | Not-determined |
| 18-105_k141_186066 | Not-determined |
| 18-105_k141_150147 | Not-determined |
| 18-105_k141_88166 | Not-determined |
| 18-105_k141_88169 | Not-determined |
| 18-105_k141_88172 | Low-quality |
| 18-105_k141_34156 | Not-determined |
| 18-105_k141_79287 | Low-quality |
| 18-105_k141_186081 | Not-determined |
| 18-105_k141_96339 | Not-determined |
| 18-105_k141_24396 | Not-determined |
| 18-105_k141_24397 | Not-determined |
| 18-105_k141_88179 | Not-determined |
| 18-105_k141_34161 | Low-quality |
| 18-105_k141_88190 | Not-determined |
| 18-105_k141_79299 | Not-determined |
| 18-105_k141_194997 | Not-determined |
| 18-105_k141_141118 | Not-determined |
| 18-105_k141_88197 | Not-determined |
| 18-105_k141_79307 | Not-determined |
| 18-105_k141_69517 | Not-determined |
| 18-105_k141_212170 | Low-quality |
| 18-105_k141_16249 | Not-determined |
| 18-105_k141_16255 | Not-determined |
| 18-105_k141_79315 | Not-determined |
| 18-105_k141_96374 | Not-determined |
| 18-105_k141_96385 | Not-determined |
| 18-105_k141_52699 | Not-determined |
| 18-105_k141_34186 | Not-determined |
| 18-105_k141_16268 | Not-determined |
| 18-105_k141_114190 | Not-determined |
| 18-105_k141_79334 | Not-determined |
| 18-105_k141_167865 | Not-determined |
| 18-105_k141_16272 | Not-determined |
| 18-105_k141_79337 | Not-determined |
| 18-105_k141_167871 | Not-determined |
| 18-105_k141_52725 | Not-determined |
| 18-105_k141_114206 | Not-determined |
| 18-105_k141_24428 | Not-determined |
| 18-105_k141_24433 | Not-determined |
| 18-105_k141_106015 | Not-determined |
| 18-105_k141_34212 | Not-determined |
| 18-105_k141_96422 | Not-determined |
| 18-105_k141_24455 | Low-quality |
| 18-105_k141_24460 | Not-determined |
| 18-105_k141_16303 | Not-determined |
| 18-105_k141_24467 | Not-determined |
| 18-105_k141_106033 | Not-determined |
| 18-105_k141_106036 | Not-determined |
| 18-105_k141_212180 | Not-determined |
| 18-105_k141_106041 | Not-determined |
| 18-105_k141_96451 | Not-determined |
| 18-105_k141_106046 | Not-determined |
| 18-105_k141_24486 | Not-determined |
| 18-105_k141_114243 | Not-determined |
| 18-105_k141_167931 | Not-determined |
| 18-105_k141_114247 | Not-determined |
| 18-105_k141_114251 | Not-determined |
| 18-105_k141_24497 | Not-determined |
| 18-105_k141_195085 | Not-determined |
| 18-105_k141_24498 | Not-determined |
| 18-105_k141_34239 | Not-determined |
| 18-105_k141_132767 | Not-determined |
| 18-105_k141_42846 | Not-determined |
| 18-105_k141_106060 | Low-quality |
| 18-105_k141_106062 | Not-determined |
| 18-105_k141_79358 | Not-determined |
| 18-105_k141_16331 | Not-determined |
| 18-105_k141_79362 | Not-determined |
| 18-105_k141_88240 | Not-determined |
| 18-105_k141_114265 | Low-quality |
| 18-105_k141_123548 | Not-determined |
| 18-105_k141_24520 | Not-determined |
| 18-105_k141_123549 | Not-determined |
| 18-105_k141_177272 | Not-determined |
| 18-105_k141_123551 | Not-determined |
| 18-105_k141_16348 | Not-determined |
| 18-105_k141_79379 | Not-determined |
| 18-105_k141_24527 | Not-determined |
| 18-105_k141_123554 | Not-determined |
| 18-105_k141_79382 | Not-determined |
| 18-105_k141_114274 | Not-determined |
| 18-105_k141_186098 | Not-determined |
| 18-105_k141_16355 | Not-determined |
| 18-105_k141_42861 | Not-determined |
| 18-105_k141_195089 | Not-determined |
| 18-105_k141_79393 | Not-determined |
| 18-105_k141_88258 | Not-determined |
| 18-105_k141_203085 | Low-quality |
| 18-105_k141_203088 | Not-determined |
| 18-105_k141_132796 | Not-determined |
| 18-105_k141_79406 | Not-determined |
| 18-105_k141_69535 | Not-determined |
| 18-105_k141_114292 | Not-determined |
| 18-105_k141_132803 | Not-determined |
| 18-105_k141_69544 | Not-determined |
| 18-105_k141_150185 | Not-determined |
| 18-105_k141_123576 | Not-determined |
| 18-105_k141_195106 | Not-determined |
| 18-105_k141_69557 | Not-determined |
| 18-105_k141_195108 | Not-determined |
| 18-105_k141_186119 | Not-determined |
| 18-105_k141_24551 | Not-determined |
| 18-105_k141_106113 | Low-quality |
| 18-105_k141_106115 | Not-determined |
| 18-105_k141_123598 | Not-determined |
| 18-105_k141_42909 | Not-determined |
| 18-105_k141_123602 | Not-determined |
| 18-105_k141_186128 | Not-determined |
| 18-105_k141_24555 | Low-quality |
| 18-105_k141_203107 | Not-determined |
| 18-105_k141_212202 | Not-determined |
| 18-105_k141_158739 | Not-determined |
| 18-105_k141_195127 | Low-quality |
| 18-105_k141_16361 | Not-determined |
| 18-105_k141_96518 | Not-determined |
| 18-105_k141_150199 | Not-determined |
| 18-105_k141_24566 | Not-determined |
| 18-105_k141_186139 | Not-determined |
| 18-105_k141_195137 | Not-determined |
| 18-105_k141_114343 | Low-quality |
| 18-105_k141_88288 | Not-determined |
| 18-105_k141_114349 | Not-determined |
| 18-105_k141_24579 | Low-quality |
| 18-105_k141_150210 | Not-determined |
| 18-105_k141_69591 | Not-determined |
| 18-105_k141_24584 | Not-determined |
| 18-105_k141_114354 | Not-determined |
| 18-105_k141_96536 | Not-determined |
| 18-105_k141_96539 | Not-determined |
| 18-105_k141_96542 | Not-determined |
| 18-105_k141_16384 | Not-determined |
| 18-105_k141_123638 | Not-determined |
| 18-105_k141_203123 | Not-determined |
| 18-105_k141_114361 | Not-determined |
| 18-105_k141_96550 | Not-determined |
| 18-105_k141_114366 | Not-determined |
| 18-105_k141_96561 | Not-determined |
| 18-105_k141_203139 | Low-quality |
| 18-105_k141_123653 | Not-determined |
| 18-105_k141_150227 | Not-determined |
| 18-105_k141_79453 | Low-quality |
| 18-105_k141_150230 | Not-determined |
| 18-105_k141_150237 | Not-determined |
| 18-105_k141_79465 | Not-determined |
| 18-105_k141_16413 | Not-determined |
| 18-105_k141_34299 | Not-determined |
| 18-105_k141_88327 | Not-determined |
| 18-105_k141_69607 | Not-determined |
| 18-105_k141_150244 | Not-determined |
| 18-105_k141_88332 | Not-determined |
| 18-105_k141_212255 | Not-determined |
| 18-105_k141_42938 | Not-determined |
| 18-105_k141_203150 | Not-determined |
| 18-105_k141_16425 | Not-determined |
| 18-105_k141_24629 | Not-determined |
| 18-105_k141_212262 | Not-determined |
| 18-105_k141_16428 | Not-determined |
| 18-105_k141_106131 | Not-determined |
| 18-105_k141_123680 | Not-determined |
| 18-105_k141_69627 | Not-determined |
| 18-105_k141_203160 | Not-determined |
| 18-105_k141_16432 | Low-quality |
| 18-105_k141_16433 | Not-determined |
| 18-105_k141_106137 | Not-determined |
| 18-105_k141_106140 | Not-determined |
| 18-105_k141_195170 | Not-determined |
| 18-105_k141_106142 | Not-determined |
| 18-105_k141_24636 | Low-quality |
| 18-105_k141_69632 | Not-determined |
| 18-105_k141_16446 | Low-quality |
| 18-105_k141_212284 | Not-determined |
| 18-105_k141_96598 | Not-determined |
| 18-105_k141_186151 | Not-determined |
| 18-105_k141_42970 | Not-determined |
| 18-105_k141_88336 | Not-determined |
| 18-105_k141_141161 | Not-determined |
| 18-105_k141_96604 | Not-determined |
| 18-105_k141_88339 | Not-determined |
| 18-105_k141_212291 | Not-determined |
| 18-105_k141_16453 | Low-quality |
| 18-105_k141_123696 | Not-determined |
| 18-105_k141_24660 | Not-determined |
| 18-105_k141_24661 | Not-determined |
| 18-105_k141_69644 | Not-determined |
| 18-105_k141_132813 | Not-determined |
| 18-105_k141_16459 | Not-determined |
| 18-105_k141_16463 | Not-determined |
| 18-105_k141_123714 | Not-determined |
| 18-105_k141_150278 | Not-determined |
| 18-105_k141_96619 | Not-determined |
| 18-105_k141_42985 | Not-determined |
| 18-105_k141_150283 | Not-determined |
| 18-105_k141_69665 | Not-determined |
| 18-105_k141_69667 | Not-determined |
| 18-105_k141_150285 | Not-determined |
| 18-105_k141_141165 | Not-determined |
| 18-105_k141_34322 | Not-determined |
| 18-105_k141_24680 | Not-determined |
| 18-105_k141_79481 | Not-determined |
| 18-105_k141_16477 | Not-determined |
| 18-105_k141_79483 | Low-quality |
| 18-105_k141_141171 | Not-determined |
| 18-105_k141_69677 | Not-determined |
| 18-105_k141_69682 | Not-determined |
| 18-105_k141_158774 | Not-determined |
| 18-105_k141_106189 | Not-determined |
| 18-105_k141_195190 | Not-determined |

| | |
|---|---|
| 18-105_k141_69683 | Not-determined |
| 18-105_k141_203220 | Not-determined |
| 18-105_k141_106192 | Not-determined |
| 18-105_k141_123731 | Not-determined |
| 18-105_k141_88377 | Not-determined |
| 18-105_k141_158779 | Not-determined |
| 18-105_k141_24690 | Not-determined |
| 18-105_k141_141182 | Low-quality |
| 18-105_k141_16497 | Not-determined |
| 18-105_k141_16502 | Not-determined |
| 18-105_k141_106206 | Not-determined |
| 18-105_k141_123750 | Not-determined |
| 18-105_k141_203243 | Not-determined |
| 18-105_k141_69716 | Low-quality |
| 18-105_k141_34359 | Not-determined |
| 18-105_k141_195224 | Not-determined |
| 18-105_k141_123757 | Not-determined |
| 18-105_k141_177298 | Not-determined |
| 18-105_k141_114389 | Not-determined |
| 18-105_k141_150304 | Not-determined |
| 18-105_k141_79506 | Not-determined |
| 18-105_k141_177314 | Not-determined |
| 18-105_k141_203259 | Not-determined |
| 18-105_k141_150306 | Not-determined |
| 18-105_k141_203261 | Not-determined |
| 18-105_k141_88415 | Not-determined |
| 18-105_k141_132848 | Not-determined |
| 18-105_k141_34382 | Not-determined |
| 18-105_k141_16523 | Not-determined |
| 18-105_k141_195248 | Not-determined |
| 18-105_k141_195250 | Not-determined |
| 18-105_k141_195254 | Not-determined |
| 18-105_k141_79524 | Not-determined |
| 18-105_k141_52756 | Not-determined |
| 18-105_k141_88439 | Not-determined |
| 18-105_k141_52761 | Low-quality |
| 18-105_k141_203266 | Low-quality |
| 18-105_k141_16534 | Not-determined |
| 18-105_k141_96697 | Not-determined |
| 18-105_k141_16540 | Not-determined |
| 18-105_k141_203273 | Not-determined |
| 18-105_k141_123803 | Not-determined |
| 18-105_k141_96702 | Not-determined |
| 18-105_k141_150346 | Not-determined |
| 18-105_k141_141223 | Low-quality |
| 18-105_k141_96706 | Not-determined |
| 18-105_k141_203275 | Not-determined |
| 18-105_k141_167945 | Low-quality |
| 18-105_k141_195277 | Not-determined |
| 18-105_k141_24737 | Not-determined |
| 18-105_k141_167952 | Not-determined |
| 18-105_k141_123819 | Not-determined |
| 18-105_k141_186202 | Not-determined |
| 18-105_k141_123822 | Not-determined |
| 18-105_k141_106212 | Not-determined |
| 18-105_k141_96718 | Not-determined |
| 18-105_k141_52784 | Not-determined |
| 18-105_k141_123823 | Low-quality |
| 18-105_k141_141235 | Not-determined |
| 18-105_k141_141236 | Not-determined |
| 18-105_k141_114431 | Not-determined |
| 18-105_k141_141238 | Not-determined |
| 18-105_k141_52791 | Not-determined |
| 18-105_k141_106219 | Not-determined |
| 18-105_k141_106221 | Not-determined |
| 18-105_k141_167965 | Not-determined |
| 18-105_k141_79545 | Not-determined |
| 18-105_k141_96727 | Not-determined |
| 18-105_k141_96728 | Not-determined |
| 18-105_k141_167970 | Not-determined |
| 18-105_k141_186213 | Not-determined |
| 18-105_k141_132884 | Not-determined |
| 18-105_k141_114444 | Not-determined |
| 18-105_k141_212326 | Not-determined |
| 18-105_k141_132890 | Low-quality |
| 18-105_k141_158858 | Not-determined |
| 18-105_k141_167981 | Not-determined |
| 18-105_k141_96744 | Not-determined |
| 18-105_k141_79559 | Not-determined |
| 18-105_k141_158864 | Not-determined |
| 18-105_k141_79561 | Not-determined |
| 18-105_k141_88487 | Not-determined |
| 18-105_k141_79564 | Not-determined |
| 18-105_k141_141267 | Low-quality |
| 18-105_k141_141270 | Not-determined |
| 18-105_k141_96756 | Not-determined |
| 18-105_k141_79573 | Not-determined |
| 18-105_k141_96760 | Not-determined |
| 18-105_k141_16578 | Not-determined |
| 18-105_k141_106231 | Not-determined |
| 18-105_k141_114457 | Not-determined |
| 18-105_k141_123847 | Not-determined |
| 18-105_k141_79577 | Not-determined |
| 18-105_k141_88498 | Not-determined |
| 18-105_k141_212347 | Not-determined |
| 18-105_k141_150381 | Not-determined |
| 18-105_k141_96765 | Not-determined |
| 18-105_k141_52815 | Not-determined |
| 18-105_k141_96769 | Not-determined |
| 18-105_k141_158877 | Low-quality |
| 18-105_k141_34451 | Not-determined |
| 18-105_k141_34453 | Not-determined |
| 18-105_k141_88503 | Not-determined |
| 18-105_k141_106244 | Not-determined |
| 18-105_k141_186231 | Not-determined |
| 18-105_k141_203288 | Not-determined |
| 18-105_k141_186238 | Not-determined |
| 18-105_k141_141290 | Not-determined |
| 18-105_k141_114481 | Not-determined |
| 18-105_k141_96789 | Not-determined |
| 18-105_k141_106261 | Not-determined |
| 18-105_k141_195292 | Not-determined |
| 18-105_k141_195293 | Not-determined |
| 18-105_k141_203311 | Not-determined |
| 18-105_k141_132938 | Not-determined |
| 18-105_k141_34481 | Not-determined |
| 18-105_k141_150414 | Not-determined |
| 18-105_k141_106273 | Not-determined |
| 18-105_k141_123879 | Low-quality |
| 18-105_k141_96808 | Not-determined |
| 18-105_k141_186264 | Not-determined |
| 18-105_k141_96810 | Not-determined |
| 18-105_k141_186265 | Not-determined |
| 18-105_k141_186266 | Not-determined |
| 18-105_k141_96816 | Not-determined |
| 18-105_k141_96817 | Not-determined |
| 18-105_k141_96818 | Low-quality |
| 18-105_k141_96829 | Not-determined |
| 18-105_k141_186281 | Not-determined |
| 18-105_k141_132960 | Not-determined |
| 18-105_k141_96836 | Not-determined |
| 18-105_k141_186290 | Not-determined |
| 18-105_k141_79586 | Not-determined |
| 18-105_k141_212399 | Not-determined |
| 18-105_k141_168029 | Not-determined |
| 18-105_k141_195305 | Not-determined |
| 18-105_k141_168031 | Not-determined |
| 18-105_k141_168035 | Not-determined |
| 18-105_k141_114499 | Not-determined |
| 18-105_k141_212404 | Not-determined |
| 18-105_k141_195311 | Low-quality |
| 18-105_k141_96855 | Not-determined |
| 18-105_k141_195319 | Not-determined |
| 18-105_k141_96859 | Not-determined |
| 18-105_k141_16642 | Not-determined |
| 18-105_k141_88536 | Low-quality |
| 18-105_k141_123933 | Not-determined |
| 18-105_k141_177328 | Not-determined |
| 18-105_k141_96865 | Not-determined |
| 18-105_k141_212418 | Not-determined |
| 18-105_k141_177331 | Not-determined |
| 18-105_k141_114520 | Not-determined |
| 18-105_k141_16646 | Not-determined |
| 18-105_k141_150476 | Not-determined |
| 18-105_k141_114523 | Not-determined |
| 18-105_k141_150481 | Not-determined |
| 18-105_k141_16652 | Not-determined |
| 18-105_k141_150483 | Not-determined |
| 18-105_k141_43010 | Not-determined |
| 18-105_k141_133008 | Not-determined |
| 18-105_k141_141322 | Not-determined |
| 18-105_k141_52838 | Not-determined |
| 18-105_k141_114542 | Not-determined |
| 18-105_k141_123959 | Low-quality |
| 18-105_k141_150498 | Not-determined |
| 18-105_k141_114545 | Not-determined |
| 18-105_k141_16676 | Not-determined |
| 18-105_k141_52841 | Not-determined |
| 18-105_k141_88555 | Not-determined |
| 18-105_k141_123961 | Not-determined |
| 18-105_k141_203378 | Not-determined |
| 18-105_k141_123963 | Not-determined |
| 18-105_k141_114550 | Low-quality |
| 18-105_k141_79608 | Low-quality |
| 18-105_k141_43031 | Not-determined |
| 18-105_k141_141339 | Not-determined |
| 18-105_k141_123975 | Not-determined |
| 18-105_k141_168069 | Not-determined |
| 18-105_k141_150506 | Not-determined |
| 18-105_k141_79621 | Not-determined |
| 18-105_k141_133028 | Not-determined |
| 18-105_k141_150517 | Not-determined |
| 18-105_k141_141352 | Not-determined |
| 18-105_k141_150518 | Low-quality |
| 18-105_k141_16693 | Not-determined |
| 18-105_k141_43046 | Not-determined |
| 18-105_k141_34496 | Not-determined |
| 18-105_k141_150524 | Not-determined |
| 18-105_k141_123992 | Not-determined |
| 18-105_k141_141360 | Not-determined |
| 18-105_k141_16697 | Not-determined |
| 18-105_k141_43050 | Not-determined |
| 18-105_k141_123997 | Not-determined |
| 18-105_k141_88588 | Not-determined |
| 18-105_k141_203398 | Not-determined |
| 18-105_k141_124003 | Not-determined |
| 18-105_k141_158993 | Not-determined |
| 18-105_k141_177347 | Not-determined |
| 18-105_k141_186326 | Not-determined |
| 18-105_k141_203402 | Not-determined |
| 18-105_k141_16707 | Not-determined |
| 18-105_k141_34512 | Not-determined |
| 18-105_k141_34513 | Not-determined |
| 18-105_k141_133054 | Not-determined |
| 18-105_k141_52857 | Not-determined |
| 18-105_k141_158997 | Not-determined |
| 18-105_k141_212442 | Not-determined |
| 18-105_k141_79628 | Not-determined |
| 18-105_k141_52860 | Not-determined |
| 18-105_k141_203411 | Low-quality |
| 18-105_k141_16718 | Not-determined |
| 18-105_k141_186337 | Not-determined |
| 18-105_k141_114560 | Not-determined |
| 18-105_k141_52866 | Low-quality |
| 18-105_k141_96929 | Not-determined |
| 18-105_k141_203418 | Not-determined |
| 18-105_k141_124013 | Not-determined |
| 18-105_k141_79641 | Not-determined |
| 18-105_k141_203420 | Not-determined |
| 18-105_k141_106399 | Not-determined |
| 18-105_k141_79643 | Not-determined |
| 18-105_k141_186346 | Not-determined |
| 18-105_k141_186349 | Not-determined |
| 18-105_k141_177361 | Not-determined |
| 18-105_k141_52870 | Not-determined |
| 18-105_k141_96937 | Not-determined |
| 18-105_k141_16731 | Not-determined |
| 18-105_k141_203431 | Not-determined |
| 18-105_k141_106406 | Not-determined |
| 18-105_k141_16737 | Not-determined |
| 18-105_k141_34535 | Not-determined |
| 18-105_k141_16739 | Low-quality |
| 18-105_k141_124036 | Not-determined |
| 18-105_k141_106412 | Low-quality |
| 18-105_k141_69726 | Not-determined |
| 18-105_k141_34544 | Not-determined |
| 18-105_k141_79661 | Not-determined |
| 18-105_k141_114568 | Not-determined |
| 18-105_k141_96952 | Low-quality |
| 18-105_k141_114571 | Not-determined |
| 18-105_k141_79663 | Not-determined |
| 18-105_k141_61215 | Not-determined |
| 18-105_k141_61218 | Not-determined |
| 18-105_k141_159019 | Not-determined |
| 18-105_k141_106424 | Low-quality |
| 18-105_k141_34559 | Low-quality |
| 18-105_k141_106428 | Not-determined |
| 18-105_k141_69739 | Not-determined |
| 18-105_k141_79668 | Low-quality |
| 18-105_k141_52883 | Low-quality |
| 18-105_k141_43061 | Not-determined |
| 18-105_k141_52888 | Not-determined |
| 18-105_k141_24787 | Not-determined |
| 18-105_k141_24788 | Not-determined |
| 18-105_k141_195352 | Not-determined |
| 18-105_k141_177403 | Not-determined |
| 18-105_k141_43078 | Not-determined |
| 18-105_k141_195364 | Not-determined |
| 18-105_k141_133064 | Not-determined |
| 18-105_k141_24800 | Not-determined |
| 18-105_k141_177408 | Not-determined |
| 18-105_k141_195371 | Not-determined |
| 18-105_k141_177410 | Low-quality |
| 18-105_k141_43090 | Low-quality |
| 18-105_k141_24807 | Not-determined |
| 18-105_k141_159043 | Not-determined |
| 18-105_k141_24808 | Not-determined |
| 18-105_k141_79683 | Not-determined |
| 18-105_k141_133075 | Not-determined |
| 18-105_k141_96998 | Not-determined |
| 18-105_k141_69762 | Not-determined |
| 18-105_k141_61239 | Not-determined |
| 18-105_k141_177415 | Not-determined |
| 18-105_k141_69766 | Not-determined |
| 18-105_k141_24816 | Not-determined |
| 18-105_k141_69767 | Low-quality |
| 18-105_k141_69770 | Low-quality |
| 18-105_k141_195384 | Not-determined |
| 18-105_k141_203455 | Not-determined |
| 18-105_k141_69772 | Not-determined |
| 18-105_k141_141410 | Not-determined |
| 18-105_k141_61250 | Not-determined |
| 18-105_k141_159055 | Not-determined |
| 18-105_k141_168115 | Not-determined |
| 18-105_k141_79694 | Low-quality |
| 18-105_k141_203463 | Not-determined |
| 18-105_k141_16791 | Not-determined |
| 18-105_k141_79702 | Not-determined |
| 18-105_k141_114602 | Not-determined |
| 18-105_k141_97025 | Not-determined |
| 18-105_k141_159065 | Not-determined |
| 18-105_k141_34563 | Not-determined |
| 18-105_k141_24833 | Not-determined |
| 18-105_k141_195397 | Low-quality |
| 18-105_k141_195398 | Not-determined |
| 18-105_k141_97031 | Not-determined |
| 18-105_k141_168131 | Not-determined |
| 18-105_k141_177439 | Not-determined |
| 18-105_k141_61257 | Not-determined |
| 18-105_k141_168137 | Not-determined |
| 18-105_k141_24844 | Low-quality |
| 18-105_k141_133109 | Not-determined |
| 18-105_k141_79716 | Not-determined |
| 18-105_k141_97045 | Not-determined |
| 18-105_k141_69797 | Low-quality |
| 18-105_k141_79727 | Not-determined |
| 18-105_k141_34584 | Not-determined |
| 18-105_k141_52890 | Not-determined |
| 18-105_k141_186422 | Low-quality |
| 18-105_k141_97057 | Not-determined |
| 18-105_k141_186424 | Not-determined |
| 18-105_k141_186427 | Not-determined |
| 18-105_k141_133121 | Not-determined |
| 18-105_k141_168156 | Not-determined |
| 18-105_k141_168158 | Not-determined |
| 18-105_k141_61291 | Not-determined |
| 18-105_k141_168160 | Not-determined |
| 18-105_k141_61294 | Not-determined |
| 18-105_k141_114625 | Not-determined |
| 18-105_k141_195426 | Not-determined |
| 18-105_k141_69819 | Not-determined |
| 18-105_k141_61297 | Not-determined |
| 18-105_k141_177463 | Not-determined |
| 18-105_k141_186448 | Not-determined |
| 18-105_k141_43147 | Not-determined |
| 18-105_k141_150559 | Not-determined |
| 18-105_k141_133142 | Not-determined |
| 18-105_k141_150561 | Low-quality |
| 18-105_k141_133144 | Not-determined |
| 18-105_k141_24873 | Not-determined |
| 18-105_k141_61304 | Not-determined |
| 18-105_k141_186455 | Not-determined |
| 18-105_k141_186456 | Not-determined |
| 18-105_k141_97067 | Not-determined |
| 18-105_k141_212487 | Not-determined |
| 18-105_k141_177477 | Not-determined |
| 18-105_k141_212488 | Not-determined |
| 18-105_k141_186463 | Not-determined |
| 18-105_k141_212491 | Not-determined |
| 18-105_k141_150570 | Not-determined |
| 18-105_k141_212492 | Not-determined |
| 18-105_k141_43163 | Not-determined |
| 18-105_k141_212496 | Not-determined |
| 18-105_k141_97078 | Not-determined |
| 18-105_k141_69854 | Not-determined |
| 18-105_k141_61327 | Not-determined |
| 18-105_k141_114648 | Not-determined |
| 18-105_k141_43171 | Not-determined |
| 18-105_k141_186479 | Low-quality |
| 18-105_k141_79766 | Not-determined |
| 18-105_k141_114651 | Not-determined |
| 18-105_k141_133165 | Not-determined |
| 18-105_k141_24903 | Not-determined |
| 18-105_k141_150594 | Not-determined |
| 18-105_k141_177505 | Not-determined |
| 18-105_k141_177508 | Not-determined |
| 18-105_k141_79775 | Low-quality |
| 18-105_k141_79780 | Not-determined |
| 18-105_k141_195476 | Not-determined |
| 18-105_k141_43197 | Not-determined |
| 18-105_k141_159081 | Not-determined |
| 18-105_k141_195481 | Not-determined |
| 18-105_k141_79790 | Not-determined |
| 18-105_k141_159088 | Not-determined |
| 18-105_k141_69861 | Not-determined |
| 18-105_k141_168202 | Not-determined |
| 18-105_k141_43208 | Not-determined |
| 18-105_k141_24917 | Low-quality |
| 18-105_k141_212505 | Not-determined |
| 18-105_k141_69870 | Not-determined |
| 18-105_k141_24919 | Not-determined |
| 18-105_k141_24922 | Not-determined |
| 18-105_k141_195498 | Not-determined |
| 18-105_k141_106499 | Not-determined |
| 18-105_k141_203483 | Not-determined |
| 18-105_k141_203484 | Not-determined |
| 18-105_k141_114679 | Not-determined |
| 18-105_k141_114680 | Not-determined |
| 18-105_k141_79797 | Not-determined |
| 18-105_k141_79799 | Not-determined |
| 18-105_k141_195511 | Not-determined |
| 18-105_k141_159107 | Not-determined |
| 18-105_k141_43222 | Not-determined |
| 18-105_k141_141428 | Not-determined |
| 18-105_k141_159110 | Not-determined |
| 18-105_k141_186515 | Not-determined |
| 18-105_k141_106510 | Low-quality |
| 18-105_k141_195516 | Not-determined |
| 18-105_k141_195521 | Not-determined |
| 18-105_k141_79807 | Not-determined |
| 18-105_k141_150640 | Not-determined |
| 18-105_k141_168223 | Not-determined |
| 18-105_k141_168224 | Not-determined |
| 18-105_k141_150644 | Not-determined |
| 18-105_k141_24944 | Not-determined |
| 18-105_k141_150646 | Not-determined |
| 18-105_k141_203494 | Not-determined |
| 18-105_k141_150651 | Not-determined |
| 18-105_k141_124090 | Not-determined |
| 18-105_k141_133211 | Not-determined |
| 18-105_k141_79817 | Low-quality |
| 18-105_k141_24950 | Not-determined |
| 18-105_k141_133220 | Not-determined |
| 18-105_k141_106527 | Not-determined |
| 18-105_k141_212544 | Not-determined |
| 18-105_k141_16844 | Not-determined |
| 18-105_k141_79824 | Not-determined |
| 18-105_k141_16845 | Not-determined |
| 18-105_k141_195549 | Not-determined |
| 18-105_k141_150667 | Not-determined |
| 18-105_k141_106529 | Not-determined |
| 18-105_k141_150670 | Not-determined |
| 18-105_k141_52916 | Not-determined |
| 18-105_k141_106535 | Not-determined |
| 18-105_k141_150674 | Not-determined |
| 18-105_k141_133239 | Not-determined |
| 18-105_k141_186546 | Not-determined |
| 18-105_k141_16862 | Not-determined |
| 18-105_k141_212560 | Not-determined |
| 18-105_k141_203510 | Low-quality |
| 18-105_k141_212565 | Not-determined |
| 18-105_k141_52921 | Not-determined |
| 18-105_k141_16866 | Low-quality |
| 18-105_k141_88702 | Not-determined |
| 18-105_k141_177549 | Not-determined |
| 18-105_k141_88708 | Not-determined |
| 18-105_k141_159147 | Not-determined |
| 18-105_k141_212574 | Not-determined |
| 18-105_k141_69907 | Not-determined |
| 18-105_k141_69912 | Low-quality |
| 18-105_k141_212587 | Not-determined |
| 18-105_k141_141483 | Not-determined |
| 18-105_k141_212589 | Not-determined |
| 18-105_k141_97102 | Not-determined |
| 18-105_k141_43283 | Not-determined |
| 18-105_k141_69922 | Not-determined |
| 18-105_k141_69923 | Not-determined |
| 18-105_k141_69927 | Not-determined |
| 18-105_k141_150699 | Not-determined |
| 18-105_k141_150701 | Low-quality |
| 18-105_k141_195575 | Low-quality |
| 18-105_k141_195576 | Not-determined |
| 18-105_k141_195580 | Not-determined |
| 18-105_k141_106583 | Not-determined |
| 18-102-final-viral-combined_k141_207832\|\|full | High-quality |
| 18-105_k141_106586 | Not-determined |
| 18-105_k141_186583 | Not-determined |
| 18-105_k141_124114 | Low-quality |
| 18-105_k141_97123 | Not-determined |
| 18-105_k141_133302 | Not-determined |
| 18-105_k141_195589 | Not-determined |
| 18-105_k141_141517 | Not-determined |
| 18-105_k141_124126 | Not-determined |
| 18-105_k141_141520 | Not-determined |
| 18-105_k141_195595 | Low-quality |
| 18-105_k141_25005 | Low-quality |
| 18-105_k141_212626 | Low-quality |
| 18-105_k141_212629 | Not-determined |
| 18-105_k141_25014 | Not-determined |
| 18-105_k141_25015 | Not-determined |
| 18-105_k141_186595 | Not-determined |
| 18-105_k141_34605 | Not-determined |
| 18-105_k141_52935 | Not-determined |
| 18-105_k141_168323 | Not-determined |
| 18-105_k141_133322 | Not-determined |
| 18-105_k141_159202 | Low-quality |
| 18-105_k141_88749 | Not-determined |
| 18-105_k141_114705 | Not-determined |
| 18-105_k141_159211 | Not-determined |
| 18-105_k141_25042 | Not-determined |
| 18-105_k141_34617 | Low-quality |
| 18-105_k141_124133 | Not-determined |
| 18-105_k141_25051 | Not-determined |
| 18-105_k141_141546 | Low-quality |
| 18-105_k141_61381 | Not-determined |
| 18-105_k141_124136 | Not-determined |
| 18-105_k141_212644 | Not-determined |
| 18-105_k141_34624 | Not-determined |
| 18-105_k141_97169 | Not-determined |
| 18-105_k141_97173 | Not-determined |
| 18-105_k141_79862 | Not-determined |
| 18-105_k141_159232 | Not-determined |
| 18-105_k141_79868 | Low-quality |
| 18-105_k141_43296 | Not-determined |
| 18-105_k141_97183 | Not-determined |
| 18-105_k141_141556 | Not-determined |
| 18-105_k141_52963 | Not-determined |
| 18-105_k141_203601 | Not-determined |
| 18-105_k141_114736 | Not-determined |
| 18-105_k141_97194 | Not-determined |
| 18-105_k141_212661 | Not-determined |
| 18-105_k141_97198 | Not-determined |
| 18-105_k141_203615 | Not-determined |
| 18-105_k141_52975 | Not-determined |
| 18-105_k141_150741 | Not-determined |
| 18-105_k141_168383 | Not-determined |
| 18-105_k141_106606 | Not-determined |
| 18-105_k141_97204 | Not-determined |
| 18-105_k141_25067 | Not-determined |
| 18-105_k141_168398 | Not-determined |
| 18-105_k141_124150 | Not-determined |
| 18-105_k141_168399 | Not-determined |
| 18-105_k141_124151 | Not-determined |
| 18-105_k141_133354 | Not-determined |
| 18-105_k141_69963 | Not-determined |
| 18-105_k141_141573 | Not-determined |
| 18-105_k141_133357 | Not-determined |
| 18-105_k141_141574 | Not-determined |
| 18-105_k141_25077 | Not-determined |
| 18-105_k141_141577 | Not-determined |
| 18-105_k141_141579 | Not-determined |
| 18-105_k141_133363 | Not-determined |
| 18-105_k141_43305 | Low-quality |
| 18-105_k141_43306 | Not-determined |
| 18-105_k141_124166 | Not-determined |
| 18-105_k141_114753 | Not-determined |
| 18-105_k141_53013 | Not-determined |
| 18-105_k141_124170 | Not-determined |
| 18-105_k141_43318 | Not-determined |
| 18-105_k141_69976 | Not-determined |
| 18-105_k141_124173 | Low-quality |
| 18-105_k141_159263 | Not-determined |
| 18-105_k141_106625 | Not-determined |
| 18-105_k141_159267 | Not-determined |
| 18-105_k141_25084 | Low-quality |
| 18-105_k141_79910 | Not-determined |
| 18-105_k141_150788 | Low-quality |
| 18-105_k141_159274 | Low-quality |
| 18-105_k141_168414 | Not-determined |
| 18-105_k141_203618 | Low-quality |
| 18-105_k141_195625 | Not-determined |
| 18-105_k141_114762 | Not-determined |
| 18-105_k141_97234 | Low-quality |
| 18-105_k141_53027 | Low-quality |
| 18-105_k141_168423 | Not-determined |
| 18-105_k141_159281 | Not-determined |
| 18-105_k141_203622 | Not-determined |
| 18-105_k141_97237 | Not-determined |
| 18-105_k141_25099 | Not-determined |
| 18-105_k141_70001 | Not-determined |
| 18-105_k141_70002 | Not-determined |
| 18-105_k141_25102 | Not-determined |
| 18-105_k141_53032 | Not-determined |
| 18-105_k141_88817 | Not-determined |
| 18-105_k141_106644 | Not-determined |
| 18-105_k141_159283 | Not-determined |
| 18-105_k141_195648 | Not-determined |
| 18-105_k141_114780 | Not-determined |
| 18-105_k141_114781 | Low-quality |
| 18-105_k141_114784 | Not-determined |
| 18-105_k141_212679 | Not-determined |
| 18-105_k141_168442 | Not-determined |
| 18-105_k141_88827 | Not-determined |
| 18-105_k141_70021 | Not-determined |
| 18-105_k141_88829 | Not-determined |
| 18-105_k141_70022 | Not-determined |
| 18-105_k141_203644 | Not-determined |
| 18-105_k141_150801 | Not-determined |
| 18-105_k141_168449 | Not-determined |
| 18-105_k141_88832 | Not-determined |
| 18-105_k141_106660 | Not-determined |
| 18-105_k141_150806 | Not-determined |
| 18-105_k141_203655 | Not-determined |
| 18-105_k141_212689 | Not-determined |
| 18-105_k141_203668 | Not-determined |
| 18-105_k141_212696 | Not-determined |
| 18-105_k141_203677 | Not-determined |
| 18-105_k141_114802 | Not-determined |
| 18-105_k141_195669 | Low-quality |
| 18-105_k141_61488 | Low-quality |
| 18-105_k141_114811 | Not-determined |
| 18-105_k141_203689 | Not-determined |
| 18-105_k141_203690 | Low-quality |
| 18-105_k141_88856 | Not-determined |
| 18-105_k141_133399 | Not-determined |
| 18-105_k141_88858 | Not-determined |
| 18-105_k141_203695 | Not-determined |
| 18-105_k141_43342 | Low-quality |
| 18-105_k141_88868 | Not-determined |
| 18-105_k141_88872 | Not-determined |
| 18-105_k141_79967 | Not-determined |
| 18-105_k141_195685 | Not-determined |
| 18-105_k141_88878 | Low-quality |
| 18-105_k141_53070 | Not-determined |
| 18-105_k141_70052 | Not-determined |
| 18-105_k141_195689 | Not-determined |
| 18-105_k141_141648 | Not-determined |
| 18-105_k141_195691 | Not-determined |
| 18-105_k141_34646 | Not-determined |

| | |
|---|---|
| 18-105_k141_168469 | Not-determined |
| 18-105_k141_168471 | Not-determined |
| 18-105_k141_203705 | Low-quality |
| 18-105_k141_88886 | Low-quality |
| 18-105_k141_79981 | Not-determined |
| 18-105_k141_195702 | Not-determined |
| 18-105_k141_212716 | Low-quality |
| 18-105_k141_16932 | Not-determined |
| 18-105_k141_203714 | Not-determined |
| 18-105_k141_43351 | Not-determined |
| 18-105_k141_150821 | Not-determined |
| 18-105_k141_195711 | Not-determined |
| 18-105_k141_203718 | Low-quality |
| 18-105_k141_70067 | Not-determined |
| 18-105_k141_114852 | Low-quality |
| 18-105_k141_53083 | Not-determined |
| 18-105_k141_25137 | Not-determined |
| 18-105_k141_212722 | Not-determined |
| 18-105_k141_150829 | Not-determined |
| 18-105_k141_53092 | Not-determined |
| 18-105_k141_150832 | Not-determined |
| 18-105_k141_195721 | Low-quality |
| 18-105_k141_212730 | Not-determined |
| 18-105_k141_70077 | Not-determined |
| 18-105_k141_141685 | Not-determined |
| 18-105_k141_16946 | Not-determined |
| 18-105_k141_25155 | Not-determined |
| 18-105_k141_70082 | Not-determined |
| 18-105_k141_25162 | Not-determined |
| 18-105_k141_150850 | Not-determined |
| 18-105_k141_150852 | Not-determined |
| 18-105_k141_168509 | Not-determined |
| 18-105_k141_212749 | Not-determined |
| 18-105_k141_114878 | Not-determined |
| 18-105_k141_177603 | Not-determined |
| 18-105_k141_177604 | Not-determined |
| 18-105_k141_114893 | Not-determined |
| 18-105_k141_177613 | Not-determined |
| 18-105_k141_195735 | Not-determined |
| 18-105_k141_195736 | Not-determined |
| 18-105_k141_53103 | Not-determined |
| 18-105_k141_124215 | Not-determined |
| 18-105_k141_124216 | Not-determined |
| 18-105_k141_53109 | Not-determined |
| 18-105_k141_53110 | Not-determined |
| 18-105_k141_177626 | Not-determined |
| 18-105_k141_177627 | Not-determined |
| 18-105_k141_177634 | Not-determined |
| 18-105_k141_133434 | Not-determined |
| 18-105_k141_53111 | Not-determined |
| 18-105_k141_53115 | Not-determined |
| 18-105_k141_53117 | Low-quality |
| 18-105_k141_80005 | Not-determined |
| 18-105_k141_114927 | Not-determined |
| 18-105_k141_61554 | Not-determined |
| 18-105_k141_186605 | Not-determined |
| 18-105_k141_61558 | Not-determined |
| 18-105_k141_114940 | Not-determined |
| 18-105_k141_186615 | Not-determined |
| 18-105_k141_168550 | Not-determined |
| 18-105_k141_212779 | Not-determined |
| 18-105_k141_53137 | Low-quality |
| 18-105_k141_212784 | Not-determined |
| 18-105_k141_168557 | Not-determined |
| 18-105_k141_195779 | Not-determined |
| 18-105_k141_88922 | Not-determined |
| 18-105_k141_212793 | Not-determined |
| 18-105_k141_70104 | Not-determined |
| 18-105_k141_16993 | Not-determined |
| 18-105_k141_16994 | Not-determined |
| 18-105_k141_43379 | Low-quality |
| 18-105_k141_70110 | Not-determined |
| 18-105_k141_80009 | Not-determined |
| 18-105_k141_70113 | Not-determined |
| 18-105_k141_53165 | Not-determined |
| 18-105_k141_70119 | Not-determined |
| 18-105_k141_97318 | Not-determined |
| 18-105_k141_80016 | Not-determined |
| 18-105_k141_212814 | Not-determined |
| 18-105_k141_195791 | Low-quality |
| 18-105_k141_80022 | Low-quality |
| 18-105_k141_17013 | Not-determined |
| 18-105_k141_212820 | Not-determined |
| 18-105_k141_168569 | Low-quality |
| 18-105_k141_34694 | Low-quality |
| 18-105_k141_168574 | Not-determined |
| 18-105_k141_17021 | Not-determined |
| 18-105_k141_114979 | Not-determined |
| 18-105_k141_17026 | Not-determined |
| 18-105_k141_124299 | Not-determined |
| 18-105_k141_133453 | Not-determined |
| 18-105_k141_133454 | Not-determined |
| 18-105_k141_124300 | Not-determined |
| 18-105_k141_70133 | Not-determined |
| 18-105_k141_195804 | Not-determined |
| 18-105_k141_114994 | Not-determined |
| 18-105_k141_97358 | Not-determined |
| 18-105_k141_114995 | Not-determined |
| 18-105_k141_97361 | Not-determined |
| 18-105_k141_150876 | Not-determined |
| 18-105_k141_133458 | Not-determined |
| 18-105_k141_61603 | Not-determined |
| 18-105_k141_70147 | Not-determined |
| 18-105_k141_195815 | Not-determined |
| 18-105_k141_150883 | Low-quality |
| 18-105_k141_195819 | Low-quality |
| 18-105_k141_195820 | Not-determined |
| 18-105_k141_97375 | Not-determined |
| 18-105_k141_150888 | Not-determined |
| 18-105_k141_195823 | Not-determined |
| 18-105_k141_43414 | Not-determined |
| 18-105_k141_88943 | Not-determined |
| 18-105_k141_106702 | Not-determined |
| 18-105_k141_195837 | Low-quality |
| 18-105_k141_133471 | Not-determined |
| 18-105_k141_203749 | Low-quality |
| 18-105_k141_61620 | Not-determined |
| 18-105_k141_186685 | Low-quality |
| 18-105_k141_186686 | Not-determined |
| 18-105_k141_34734 | Not-determined |
| 18-105_k141_70166 | Not-determined |
| 18-105_k141_53238 | Not-determined |
| 18-105_k141_34738 | Not-determined |
| 18-105_k141_141794 | Not-determined |
| 18-105_k141_141795 | Not-determined |
| 18-105_k141_97407 | Not-determined |
| 18-105_k141_115034 | Not-determined |
| 18-105_k141_150911 | Not-determined |
| 18-105_k141_61631 | Low-quality |
| 18-105_k141_70175 | Not-determined |
| 18-105_k141_195851 | Not-determined |
| 18-105_k141_150917 | Not-determined |
| 18-105_k141_150921 | Not-determined |
| 18-105_k141_88963 | Not-determined |
| 18-105_k141_195857 | Not-determined |
| 18-105_k141_70183 | Not-determined |
| 18-105_k141_186701 | Not-determined |
| 18-105_k141_186702 | Not-determined |
| 18-105_k141_97425 | Not-determined |
| 18-105_k141_115053 | Not-determined |
| 18-105_k141_177648 | Not-determined |
| 18-105_k141_195864 | Not-determined |
| 18-105_k141_61650 | Not-determined |
| 18-105_k141_61652 | Not-determined |
| 18-105_k141_177655 | Not-determined |
| 18-105_k141_61654 | Not-determined |
| 18-105_k141_124346 | Not-determined |
| 18-105_k141_177660 | Not-determined |
| 18-105_k141_177661 | Not-determined |
| 18-105_k141_88983 | Not-determined |
| 18-105_k141_88989 | Not-determined |
| 18-105_k141_195877 | Not-determined |
| 18-105_k141_34775 | Not-determined |
| 18-105_k141_34777 | Not-determined |
| 18-105_k141_89006 | Not-determined |
| 18-105_k141_70208 | Not-determined |
| 18-105_k141_80058 | Not-determined |
| 18-105_k141_17033 | Not-determined |
| 18-105_k141_186713 | Not-determined |
| 18-105_k141_89012 | Not-determined |
| 18-105_k141_106750 | Not-determined |
| 18-105_k141_186717 | Not-determined |
| 18-105_k141_80062 | Low-quality |
| 18-105_k141_195891 | Low-quality |
| 18-105_k141_195892 | Not-determined |
| 18-105_k141_70224 | Low-quality |
| 18-105_k141_17062 | Not-determined |
| 18-105_k141_97460 | Not-determined |
| 18-105_k141_17063 | Not-determined |
| 18-105_k141_43495 | Not-determined |
| 18-105_k141_17075 | Not-determined |
| 18-105_k141_89038 | Not-determined |
| 18-105_k141_97467 | Not-determined |
| 18-105_k141_17081 | Not-determined |
| 18-105_k141_89050 | Not-determined |
| 18-105_k141_124355 | Not-determined |
| 18-105_k141_97482 | Not-determined |
| 18-105_k141_195914 | Not-determined |
| 18-105_k141_53306 | Not-determined |
| 18-105_k141_212840 | Not-determined |
| 18-105_k141_70230 | Not-determined |
| 18-105_k141_133476 | Not-determined |
| 18-105_k141_34827 | Not-determined |
| 18-105_k141_61695 | Not-determined |
| 18-105_k141_203783 | Not-determined |
| 18-105_k141_70236 | Not-determined |
| 18-105_k141_133486 | Not-determined |
| 18-105_k141_70239 | Not-determined |
| 18-105_k141_212845 | Not-determined |
| 18-105_k141_53330 | Not-determined |
| 18-105_k141_106784 | Not-determined |
| 18-105_k141_177685 | Not-determined |
| 18-105_k141_212856 | Not-determined |
| 18-105_k141_34840 | Not-determined |
| 18-105_k141_25185 | Not-determined |
| 18-105_k141_43531 | Not-determined |
| 18-105_k141_212859 | Not-determined |
| 18-105_k141_177688 | Low-quality |
| 18-105_k141_106794 | Not-determined |
| 18-105_k141_177689 | Not-determined |
| 18-105_k141_212863 | Not-determined |
| 18-105_k141_70252 | Not-determined |
| 18-105_k141_177695 | Not-determined |
| 18-105_k141_203804 | Not-determined |
| 18-105_k141_124366 | Not-determined |
| 18-105_k141_17095 | Not-determined |
| 18-105_k141_124370 | Not-determined |
| 18-105_k141_61729 | Not-determined |
| 18-105_k141_212874 | Not-determined |
| 18-105_k141_80096 | Not-determined |
| 18-105_k141_53341 | Not-determined |
| 18-105_k141_34855 | Low-quality |
| 18-105_k141_124377 | Not-determined |
| 18-105_k141_106810 | Low-quality |
| 18-105_k141_124380 | Not-determined |
| 18-105_k141_212878 | Not-determined |
| 18-105_k141_34861 | Low-quality |
| 18-105_k141_212881 | Not-determined |
| 18-105_k141_212883 | Not-determined |
| 18-105_k141_34865 | Not-determined |
| 18-105_k141_80114 | Not-determined |
| 18-105_k141_25209 | Not-determined |
| 18-105_k141_25211 | Not-determined |
| 18-105_k141_195965 | Not-determined |
| 18-105_k141_124395 | Not-determined |
| 18-105_k141_34874 | Low-quality |
| 18-105_k141_195971 | Low-quality |
| 18-105_k141_124401 | Low-quality |
| 18-105_k141_195977 | Not-determined |
| 18-105_k141_53357 | Not-determined |
| 18-105_k141_53361 | Not-determined |
| 18-105_k141_34890 | Not-determined |
| 18-105_k141_124410 | Not-determined |
| 18-105_k141_168586 | Not-determined |
| 18-105_k141_141805 | Not-determined |
| 18-105_k141_150936 | Not-determined |
| 18-105_k141_53372 | Not-determined |
| 18-105_k141_150939 | Not-determined |
| 18-105_k141_150941 | Not-determined |
| 18-105_k141_53378 | Not-determined |
| 18-105_k141_141813 | Not-determined |
| 18-105_k141_133533 | Not-determined |
| 18-105_k141_133537 | Not-determined |
| 18-105_k141_141823 | Not-determined |
| 18-105_k141_141829 | Not-determined |
| 18-105_k141_124438 | Not-determined |
| 18-105_k141_124439 | Not-determined |
| 18-105_k141_97544 | Not-determined |
| 18-105_k141_177762 | Not-determined |
| 18-105_k141_61795 | Not-determined |
| 18-105_k141_25258 | Not-determined |
| 18-105_k141_141842 | Not-determined |
| 18-105_k141_141844 | Not-determined |
| 18-105_k141_168592 | Not-determined |
| 18-105_k141_61799 | Low-quality |
| 18-105_k141_34935 | Not-determined |
| 18-105_k141_61803 | Not-determined |
| 18-105_k141_124457 | Not-determined |
| 18-105_k141_80158 | Not-determined |
| 18-105_k141_80159 | Not-determined |
| 18-105_k141_203829 | Not-determined |
| 18-105_k141_97568 | Not-determined |
| 18-105_k141_203831 | Low-quality |
| 18-105_k141_203834 | Not-determined |
| 18-105_k141_124468 | Not-determined |
| 18-105_k141_80171 | Not-determined |
| 18-105_k141_80173 | Not-determined |
| 18-105_k141_80174 | Not-determined |
| 18-105_k141_168607 | Not-determined |
| 18-105_k141_159441 | Not-determined |
| 18-105_k141_203844 | Not-determined |
| 18-105_k141_168616 | Not-determined |
| 18-105_k141_61828 | Not-determined |
| 18-105_k141_168620 | Not-determined |
| 18-105_k141_159446 | Not-determined |
| 18-105_k141_150992 | Low-quality |
| 18-105_k141_106828 | Not-determined |
| 18-105_k141_115079 | Not-determined |
| 18-105_k141_133569 | Not-determined |
| 18-105_k141_159454 | Not-determined |
| 18-105_k141_133575 | Not-determined |
| 18-105_k141_159463 | Not-determined |
| 18-105_k141_115090 | Not-determined |
| 18-105_k141_141878 | Not-determined |
| 18-105_k141_212935 | Low-quality |
| 18-105_k141_115092 | Not-determined |
| 18-105_k141_61849 | Not-determined |
| 18-105_k141_61853 | Not-determined |
| 18-105_k141_115094 | Not-determined |
| 18-105_k141_61856 | Not-determined |
| 18-105_k141_124490 | Not-determined |
| 18-105_k141_159474 | Not-determined |
| 18-105_k141_61858 | Not-determined |
| 18-105_k141_89058 | Not-determined |
| 18-105_k141_61860 | Not-determined |
| 18-105_k141_106841 | Not-determined |
| 18-105_k141_141886 | Not-determined |
| 18-105_k141_203872 | Low-quality |
| 18-105_k141_168647 | Not-determined |
| 18-105_k141_124500 | Not-determined |
| 18-105_k141_53398 | Not-determined |
| 18-105_k141_34961 | Not-determined |
| 18-105_k141_212947 | Not-determined |
| 18-105_k141_89084 | Not-determined |
| 18-105_k141_115104 | Low-quality |
| 18-105_k141_124514 | Not-determined |
| 18-105_k141_124515 | Not-determined |
| 18-105_k141_43647 | Not-determined |
| 18-105_k141_34970 | Not-determined |
| 18-105_k141_61875 | Not-determined |
| 18-105_k141_43653 | Not-determined |
| 18-105_k141_106862 | Not-determined |
| 18-105_k141_141898 | Not-determined |
| 18-105_k141_43654 | Not-determined |
| 18-105_k141_124531 | Not-determined |
| 18-105_k141_141902 | Not-determined |
| 18-105_k141_34979 | Not-determined |
| 18-105_k141_212967 | Not-determined |
| 18-105_k141_106871 | Not-determined |
| 18-105_k141_115127 | Not-determined |
| 18-105_k141_17184 | Not-determined |
| 18-105_k141_43672 | Not-determined |
| 18-105_k141_115130 | Not-determined |
| 18-105_k141_43673 | Not-determined |
| 18-105_k141_141905 | Not-determined |
| 18-105_k141_34992 | Not-determined |
| 18-105_k141_203901 | Not-determined |
| 18-105_k141_97643 | Not-determined |
| 18-105_k141_97644 | Not-determined |
| 18-105_k141_212991 | Not-determined |
| 18-105_k141_43680 | Not-determined |
| 18-105_k141_43685 | Not-determined |
| 18-105_k141_124555 | Not-determined |
| 18-105_k141_97649 | Low-quality |
| 18-105_k141_141910 | Low-quality |
| 18-105_k141_115138 | Not-determined |
| 18-105_k141_124558 | Not-determined |
| 18-105_k141_106887 | Low-quality |
| 18-105_k141_97656 | Not-determined |
| 18-105_k141_115142 | Not-determined |
| 18-105_k141_151032 | Not-determined |
| 18-105_k141_106890 | Not-determined |
| 18-105_k141_97664 | Not-determined |
| 18-105_k141_61904 | Not-determined |
| 18-105_k141_213006 | Not-determined |
| 18-105_k141_203920 | Not-determined |
| 18-105_k141_213008 | Not-determined |
| 18-105_k141_70325 | Not-determined |
| 18-105_k141_177798 | Not-determined |
| 18-105_k141_203924 | Not-determined |
| 18-105_k141_177802 | Low-quality |
| 18-105_k141_106902 | Not-determined |
| 18-105_k141_203933 | Not-determined |
| 18-105_k141_70334 | Not-determined |
| 18-105_k141_115161 | Not-determined |
| 18-105_k141_151045 | Not-determined |
| 18-105_k141_141916 | Not-determined |
| 18-105_k141_97686 | Not-determined |
| 18-105_k141_97687 | Low-quality |
| 18-105_k141_115187 | Not-determined |
| 18-105_k141_61919 | Not-determined |
| 18-105_k141_97698 | Not-determined |
| 18-105_k141_97704 | Not-determined |
| 18-105_k141_141932 | Low-quality |
| 18-105_k141_35055 | Not-determined |
| 18-105_k141_168686 | Not-determined |
| 18-105_k141_168687 | Not-determined |
| 18-105_k141_203950 | Not-determined |
| 18-105_k141_203956 | Not-determined |
| 18-105_k141_35071 | Not-determined |
| 18-105_k141_203960 | Not-determined |
| 18-105_k141_151093 | Not-determined |
| 18-105_k141_168691 | Not-determined |
| 18-105_k141_124586 | Not-determined |
| 18-105_k141_97740 | Not-determined |
| 18-105_k141_168697 | Not-determined |
| 18-105_k141_133609 | Not-determined |
| 18-105_k141_177838 | Not-determined |
| 18-105_k141_115219 | Not-determined |
| 18-105_k141_115221 | Not-determined |
| 18-105_k141_168706 | Not-determined |
| 18-105_k141_70371 | Not-determined |
| 18-105_k141_177850 | Not-determined |
| 18-105_k141_151115 | Not-determined |
| 18-105_k141_168715 | Not-determined |
| 18-105_k141_151122 | Not-determined |
| 18-105_k141_35119 | Low-quality |
| 18-105_k141_35123 | Not-determined |
| 18-105_k141_115247 | Not-determined |
| 18-105_k141_115252 | Not-determined |
| 18-105_k141_115254 | Not-determined |
| 18-105_k141_25341 | Not-determined |
| 18-105_k141_203980 | Not-determined |
| 18-105_k141_141967 | Not-determined |
| 18-105_k141_151148 | Not-determined |
| 18-105_k141_61970 | Not-determined |
| 18-105_k141_203988 | Not-determined |
| 18-105_k141_141975 | Not-determined |
| 18-105_k141_159530 | Not-determined |
| 18-105_k141_43709 | Not-determined |
| 18-105_k141_43710 | Low-quality |
| 18-105_k141_159537 | Not-determined |
| 18-105_k141_203999 | Not-determined |
| 18-105_k141_159542 | Not-determined |
| 18-105_k141_43714 | Not-determined |
| 18-105_k141_141984 | Not-determined |
| 18-105_k141_204010 | Not-determined |
| 18-105_k141_168753 | Not-determined |
| 18-105_k141_204011 | Not-determined |
| 18-105_k141_204015 | Not-determined |
| 18-105_k141_159547 | Not-determined |
| 18-105_k141_124622 | Not-determined |
| 18-105_k141_204019 | Not-determined |
| 18-105_k141_177889 | Not-determined |
| 18-105_k141_17214 | Not-determined |
| 18-105_k141_159561 | Not-determined |
| 18-105_k141_43726 | Not-determined |
| 18-105_k141_17222 | Low-quality |
| 18-105_k141_213081 | Not-determined |
| 18-105_k141_142011 | Low-quality |
| 18-105_k141_168787 | Not-determined |
| 18-105_k141_168788 | Low-quality |
| 18-105_k141_159567 | Low-quality |
| 18-105_k141_159569 | Not-determined |
| 18-105_k141_168795 | Not-determined |
| 18-105_k141_204033 | Not-determined |
| 18-105_k141_70414 | Not-determined |
| 18-105_k141_177930 | Not-determined |
| 18-105_k141_204046 | Low-quality |
| 18-105_k141_97828 | Not-determined |
| 18-105_k141_204053 | Not-determined |
| 18-105_k141_97833 | Not-determined |
| 18-105_k141_204057 | Not-determined |
| 18-105_k141_35157 | Not-determined |
| 18-105_k141_177941 | Not-determined |
| 18-105_k141_159591 | Not-determined |
| 18-105_k141_35169 | Not-determined |
| 18-105_k141_142035 | Low-quality |
| 18-105_k141_204066 | Not-determined |
| 18-105_k141_35194 | Not-determined |
| 18-105_k141_17265 | Not-determined |
| 18-105_k141_70428 | Not-determined |
| 18-105_k141_35202 | Not-determined |
| 18-105_k141_17268 | Not-determined |
| 18-105_k141_159623 | Not-determined |
| 18-105_k141_70436 | Not-determined |
| 18-105_k141_35209 | Not-determined |
| 18-105_k141_43803 | Not-determined |
| 18-105_k141_70446 | Not-determined |
| 18-105_k141_168840 | Not-determined |
| 18-105_k141_168841 | Not-determined |
| 18-105_k141_177968 | Not-determined |
| 18-105_k141_97843 | Not-determined |
| 18-105_k141_159627 | Not-determined |
| 18-105_k141_70453 | Not-determined |
| 18-105_k141_168851 | Not-determined |
| 18-105_k141_70460 | Not-determined |
| 18-105_k141_159633 | Not-determined |
| 18-105_k141_70467 | Low-quality |
| 18-105_k141_168862 | Not-determined |
| 18-105_k141_43829 | Not-determined |
| 18-105_k141_97854 | Not-determined |
| 18-105_k141_186794 | Not-determined |
| 18-105_k141_186797 | Not-determined |
| 18-105_k141_35230 | Not-determined |
| 18-105_k141_97863 | Not-determined |
| 18-105_k141_151183 | Not-determined |
| 18-105_k141_35237 | Not-determined |
| 18-105_k141_97869 | Not-determined |
| 18-105_k141_43849 | Not-determined |
| 18-105_k141_62042 | Not-determined |
| 18-105_k141_43853 | Not-determined |
| 18-105_k141_17299 | Not-determined |
| 18-105_k141_62049 | Not-determined |
| 18-105_k141_178001 | Not-determined |
| 18-105_k141_43863 | Low-quality |
| 18-105_k141_97885 | Not-determined |
| 18-105_k141_178003 | Not-determined |
| 18-105_k141_151195 | Not-determined |
| 18-105_k141_97890 | Not-determined |
| 18-105_k141_168886 | Not-determined |
| 18-105_k141_97891 | Low-quality |
| 18-105_k141_17308 | Not-determined |
| 18-105_k141_159669 | Low-quality |
| 18-105_k141_178011 | Not-determined |
| 18-105_k141_142064 | Not-determined |
| 18-105_k141_97898 | Not-determined |
| 18-105_k141_168895 | Not-determined |
| 18-105_k141_186829 | Not-determined |
| 18-105_k141_97903 | Not-determined |
| 18-105_k141_97904 | Not-determined |
| 18-105_k141_43882 | Not-determined |
| 18-105_k141_62063 | Not-determined |
| 18-105_k141_142080 | Not-determined |
| 18-105_k141_70476 | Not-determined |
| 18-105_k141_178029 | Not-determined |
| 18-105_k141_151213 | Not-determined |
| 18-105_k141_178033 | Not-determined |
| 18-105_k141_186847 | Not-determined |
| 18-105_k141_35279 | Not-determined |
| 18-105_k141_17328 | Not-determined |
| 18-105_k141_159695 | Not-determined |
| 18-105_k141_178048 | Not-determined |
| 18-105_k141_62073 | Not-determined |
| 18-105_k141_186860 | Not-determined |
| 18-105_k141_35288 | Not-determined |
| 18-105_k141_151238 | Not-determined |
| 18-105_k141_168921 | Not-determined |
| 18-105_k141_17345 | Not-determined |
| 18-105_k141_35303 | Not-determined |
| 18-105_k141_25440 | Not-determined |
| 18-105_k141_168930 | Not-determined |
| 18-105_k141_142112 | Not-determined |
| 18-105_k141_186886 | Not-determined |
| 18-105_k141_17352 | Not-determined |
| 18-105_k141_151253 | Not-determined |
| 18-105_k141_151254 | Not-determined |
| 18-105_k141_17363 | Not-determined |
| 18-105_k141_35321 | Not-determined |
| 18-105_k141_151261 | Not-determined |
| 18-105_k141_186899 | Not-determined |
| 18-105_k141_17372 | Not-determined |
| 18-105_k141_159739 | Not-determined |
| 18-105_k141_43915 | Not-determined |
| 18-105_k141_151268 | Not-determined |
| 18-105_k141_186907 | Not-determined |
| 18-105_k141_186910 | Not-determined |
| 18-105_k141_17387 | Not-determined |
| 18-105_k141_142118 | Not-determined |
| 18-105_k141_142120 | Not-determined |
| 18-105_k141_159763 | Not-determined |
| 18-105_k141_159764 | Low-quality |
| 18-105_k141_151288 | Not-determined |
| 18-105_k141_178106 | Not-determined |
| 18-105_k141_17389 | Not-determined |
| 18-105_k141_142129 | Low-quality |
| 18-105_k141_142132 | Not-determined |
| 18-105_k141_204112 | Not-determined |
| 18-105_k141_186943 | Not-determined |
| 18-105_k141_178116 | Not-determined |
| 18-105_k141_178117 | Not-determined |
| 18-105_k141_115334 | Not-determined |
| 18-105_k141_159795 | Not-determined |
| 18-105_k141_151308 | Not-determined |
| 18-105_k141_178152 | Not-determined |
| 18-105_k141_204138 | Not-determined |
| 18-105_k141_142141 | Not-determined |
| 18-105_k141_204142 | Not-determined |
| 18-105_k141_70504 | Not-determined |
| 18-105_k141_159805 | Not-determined |
| 18-105_k141_25458 | Not-determined |
| 18-105_k141_151324 | Not-determined |
| 18-105_k141_186964 | Not-determined |
| 18-105_k141_178157 | Not-determined |
| 18-105_k141_204148 | Not-determined |
| 18-105_k141_17430 | Not-determined |
| 18-105_k141_204154 | Not-determined |
| 18-105_k141_186966 | Not-determined |
| 18-105_k141_151338 | Not-determined |
| 18-105_k141_17436 | Not-determined |
| 18-105_k141_115369 | Not-determined |
| 18-105_k141_204160 | Low-quality |
| 18-105_k141_186975 | Not-determined |
| 18-105_k141_115382 | Not-determined |
| 18-105_k141_115390 | Not-determined |
| 18-105_k141_115391 | Not-determined |
| 18-105_k141_43944 | Not-determined |
| 18-105_k141_204180 | Not-determined |
| 18-105_k141_115396 | Not-determined |
| 18-105_k141_115398 | Not-determined |
| 18-105_k141_151359 | Not-determined |
| 18-105_k141_159825 | Not-determined |
| 18-105_k141_17443 | Not-determined |
| 18-105_k141_168966 | Not-determined |
| 18-105_k141_43954 | Not-determined |
| 18-105_k141_151362 | Not-determined |
| 18-105_k141_159834 | Not-determined |
| 18-105_k141_168978 | Not-determined |
| 18-105_k141_124677 | Not-determined |
| 18-105_k141_159838 | Not-determined |
| 18-105_k141_187000 | Not-determined |
| 18-105_k141_168984 | Not-determined |
| 18-105_k141_142156 | Not-determined |
| 18-105_k141_159848 | Not-determined |
| 18-105_k141_159850 | Not-determined |
| 18-105_k141_142165 | Not-determined |
| 18-105_k141_142166 | Not-determined |
| 18-105_k141_115409 | Not-determined |
| 18-105_k141_124699 | Not-determined |

18-105_k141_17470 Not-determined
18-105_k141_213116 Not-determined
18-105_k141_213117 Not-determined
18-105_k141_187021 Not-determined
18-105_k141_213119 Not-determined
18-105_k141_151400 Not-determined
18-105_k141_142182 Not-determined
18-105_k141_124715 Not-determined
18-105_k141_187030 Not-determined
18-105_k141_17481 Not-determined
18-105_k141_187032 Not-determined
18-105_k141_124719 Not-determined
18-105_k141_142197 Not-determined
18-105_k141_142200 Not-determined
18-105_k141_25517 Not-determined
18-105_k141_25519 Not-determined
18-105_k141_25520 Not-determined
18-105_k141_204200 Not-determined
18-105_k141_70525 Low-quality
18-105_k141_151428 Not-determined
18-105_k141_159891 Low-quality
18-105_k141_70539 Not-determined
18-105_k141_169005 Not-determined
18-105_k141_204211 Not-determined
18-105_k141_151442 Not-determined
18-105_k141_213169 Low-quality
18-105_k141_25573 Not-determined
18-105_k141_97913 Not-determined
18-105_k141_97915 Not-determined
18-105_k141_25575 Low-quality
18-105_k141_35357 Not-determined
18-105_k141_204222 Low-quality
18-105_k141_115446 Low-quality
18-105_k141_70559 Not-determined
18-105_k141_44056 Not-determined
18-105_k141_159920 Not-determined
18-105_k141_169020 Not-determined
18-105_k141_204229 Not-determined
18-105_k141_70576 Not-determined
18-105_k141_35377 Low-quality
18-105_k141_70585 Not-determined
18-105_k141_70587 Not-determined
18-105_k141_159943 Not-determined
18-105_k141_44062 Not-determined
18-105_k141_115477 Not-determined
18-105_k141_204254 Not-determined
18-105_k141_70598 Not-determined
18-105_k141_115484 Not-determined
18-105_k141_70605 Not-determined
18-105_k141_44065 Not-determined
18-105_k141_44071 Not-determined
18-105_k141_44075 Not-determined
18-105_k141_97937 Low-quality
18-105_k141_25598 Not-determined
18-105_k141_70619 Not-determined
18-105_k141_97943 Not-determined
18-105_k141_44083 Not-determined
18-105_k141_115494 Not-determined
18-105_k141_62159 Not-determined
18-105_k141_115513 Not-determined
18-105_k141_213185 Not-determined
18-105_k141_204269 Not-determined
18-105_k141_204270 Not-determined
18-105_k141_70643 Not-determined
18-105_k141_213201 Low-quality
18-105_k141_70645 Not-determined
18-105_k141_62169 Low-quality
18-105_k141_142290 Not-determined
18-105_k141_142295 Not-determined
18-105_k141_169053 Not-determined
18-105_k141_142299 Not-determined
18-105_k141_142304 Not-determined
18-105_k141_62193 Not-determined
18-105_k141_115549 Not-determined
18-105_k141_142325 Not-determined
18-105_k141_70659 Low-quality
18-105_k141_159986 Not-determined
18-105_k141_62207 Not-determined
18-105_k141_204299 Not-determined
18-105_k141_97957 Not-determined
18-105_k141_62211 Not-determined
18-105_k141_70671 Not-determined
18-105_k141_70674 Not-determined
18-105_k141_142356 Not-determined
18-105_k141_142360 Low-quality
18-105_k141_142361 Not-determined
18-105_k141_142372 Not-determined
18-105_k141_70701 Not-determined
18-105_k141_204320 Not-determined
18-105_k141_160001 Not-determined
18-105_k141_35392 Not-determined
18-105_k141_160011 Not-determined
18-105_k141_213211 Not-determined
18-105_k141_204340 Not-determined
18-105_k141_115573 Not-determined
18-105_k141_35426 Low-quality
18-105_k141_142388 Not-determined
18-105_k141_115583 Not-determined
18-105_k141_142402 Not-determined
18-105_k141_204367 Low-quality
18-105_k141_213245 Low-quality
18-105_k141_115601 Not-determined
18-105_k141_35469 Not-determined
18-105_k141_115606 Not-determined
18-105_k141_160047 Not-determined
18-105_k141_204374 Not-determined
18-105_k141_17541 Not-determined
18-105_k141_70755 Not-determined
18-105_k141_62281 Not-determined
18-105_k141_169112 Not-determined
18-105_k141_169113 Not-determined
18-105_k141_115615 Not-determined
18-105_k141_35485 Not-determined
18-105_k141_25615 Not-determined
18-105_k141_17565 Not-determined
18-105_k141_142412 Not-determined
18-105_k141_35497 Not-determined
18-105_k141_142416 Not-determined
18-105_k141_213277 Not-determined
18-105_k141_213280 Not-determined
18-105_k141_160082 Not-determined
18-105_k141_62303 Low-quality
18-105_k141_204396 Not-determined
18-105_k141_70772 Low-quality
18-105_k141_70773 Not-determined
18-105_k141_17586 Not-determined
18-105_k141_70779 Not-determined
18-105_k141_160095 Not-determined
18-105_k141_62314 Not-determined
18-105_k141_160101 Low-quality
18-105_k141_70791 Not-determined
18-105_k141_17600 Not-determined
18-105_k141_160108 Not-determined
18-105_k141_169145 Not-determined
18-105_k141_213284 Not-determined
18-105_k141_62316 Not-determined
18-105_k141_115630 Not-determined
18-105_k141_169152 Not-determined
18-105_k141_169154 Not-determined
18-105_k141_160119 Not-determined
18-105_k141_213291 Not-determined
18-105_k141_115639 Not-determined
18-105_k141_213297 Low-quality
18-105_k141_213300 Not-determined
18-105_k141_70809 Not-determined
18-105_k141_160134 Not-determined
18-105_k141_115658 Not-determined
18-105_k141_44090 Not-determined
18-105_k141_160144 Not-determined
18-105_k141_25679 Not-determined
18-105_k141_25680 Not-determined
18-105_k141_169168 Not-determined
18-105_k141_142446 Not-determined
18-105_k141_25686 Not-determined
18-105_k141_17628 Not-determined
18-105_k141_25693 Not-determined
18-105_k141_204466 Not-determined
18-105_k141_142456 Not-determined
18-105_k141_169184 Not-determined
18-105_k141_142461 Not-determined
18-105_k141_204474 Not-determined
18-105_k141_142464 Not-determined
18-105_k141_213336 Not-determined
18-105_k141_17646 Not-determined
18-105_k141_160176 Not-determined
18-105_k141_160183 Not-determined
18-105_k141_44102 Not-determined
18-105_k141_35511 Not-determined
18-105_k141_213357 Not-determined
18-105_k141_35515 Low-quality
18-105_k141_35518 Not-determined
18-105_k141_115698 Low-quality
18-105_k141_115699 Not-determined
18-105_k141_115702 Not-determined
18-105_k141_17675 Not-determined
18-105_k141_142482 Not-determined
18-105_k141_204523 Low-quality
18-105_k141_142485 Not-determined
18-105_k141_25718 Not-determined
18-105_k141_115731 Not-determined
18-105_k141_25727 Not-determined
18-105_k141_25730 Not-determined
18-105_k141_142495 Not-determined
18-105_k141_142498 Not-determined
18-105_k141_35553 Not-determined
18-105_k141_142500 Not-determined
18-105_k141_17692 Not-determined
18-105_k141_44150 Not-determined
18-105_k141_44151 Not-determined
18-105_k141_160215 Not-determined
18-105_k141_160218 Not-determined
18-105_k141_204553 Not-determined
18-105_k141_204556 Not-determined
18-105_k141_35566 Not-determined
18-105_k141_160234 Low-quality
18-105_k141_204562 Not-determined
18-105_k141_204564 Not-determined
18-105_k141_44158 Not-determined
18-105_k141_44167 Not-determined
18-105_k141_53439 Low-quality
18-105_k141_17709 Not-determined
18-105_k141_142506 Low-quality
18-105_k141_44177 Not-determined
18-105_k141_142507 Not-determined
18-105_k141_204575 Not-determined
18-105_k141_169252 Not-determined
18-105_k141_169253 Not-determined
18-105_k141_204579 Not-determined
18-105_k141_213382 Not-determined
18-105_k141_25784 Not-determined
18-105_k141_25793 Not-determined
18-105_k141_25794 Not-determined
18-105_k141_213393 Not-determined
18-105_k141_160248 Not-determined
18-105_k141_213396 Not-determined
18-105_k141_53450 Not-determined
18-105_k141_160252 Not-determined
18-105_k141_204590 Not-determined
18-105_k141_160259 Not-determined
18-105_k141_213406 Not-determined
18-105_k141_142529 Not-determined
18-105_k141_17733 Not-determined
18-105_k141_142533 Low-quality
18-105_k141_44205 Not-determined
18-105_k141_25814 Not-determined
18-105_k141_204617 Not-determined
18-105_k141_25821 Not-determined
18-105_k141_17740 Low-quality
18-105_k141_25822 Not-determined
18-105_k141_70841 Not-determined
18-105_k141_25823 Low-quality
18-105_k141_204621 Not-determined
18-105_k141_35574 Not-determined
18-105_k141_44221 Not-determined
18-105_k141_115784 Not-determined
18-105_k141_17747 Not-determined
18-105_k141_115795 Not-determined
18-105_k141_213416 Not-determined
18-105_k141_25842 Not-determined
18-105_k141_44231 Not-determined
18-105_k141_213421 Not-determined
18-105_k141_160295 Not-determined
18-105_k141_25850 Not-determined
18-105_k141_213430 Low-quality
18-105_k141_213431 Not-determined
18-105_k141_25854 Not-determined
18-105_k141_35587 Not-determined
18-105_k141_160302 Not-determined
18-105_k141_115811 Not-determined
18-105_k141_160304 Not-determined
18-105_k141_17760 Not-determined
18-105_k141_25865 Not-determined
18-105_k141_70875 Not-determined
18-105_k141_70879 Not-determined
18-105_k141_204676 Not-determined
18-105_k141_44272 Not-determined
18-105_k141_35632 Low-quality
18-105_k141_25885 Not-determined
18-105_k141_160328 Not-determined
18-105_k141_70897 Not-determined
18-105_k141_204691 Low-quality
18-105_k141_160333 Not-determined
18-105_k141_44293 Not-determined
18-105_k141_25908 Not-determined
18-105_k141_44297 Not-determined
18-105_k141_44301 Not-determined
18-105_k141_25920 Not-determined
18-105_k141_204732 Not-determined
18-105_k141_25937 Not-determined
18-105_k141_44315 Not-determined
18-105_k141_70923 Not-determined
18-105_k141_204743 Not-determined
18-105_k141_213458 Not-determined
18-105_k141_213459 Low-quality
18-105_k141_25945 Not-determined
18-105_k141_25948 Not-determined
18-105_k141_213465 Low-quality
18-105_k141_204757 Not-determined
18-105_k141_25955 Not-determined
18-105_k141_204772 Not-determined
18-105_k141_204774 Not-determined
18-105_k141_25982 Not-determined
18-105_k141_213492 Not-determined
18-105_k141_25988 Not-determined
18-105_k141_213499 Not-determined
18-105_k141_44385 Not-determined
18-105_k141_213521 Not-determined
18-105_k141_70933 Not-determined
18-105_k141_70938 Not-determined
18-105_k141_70943 Not-determined
18-105_k141_26013 Not-determined
18-105_k141_26015 Not-determined
18-105_k141_26016 Not-determined
18-105_k141_26018 Not-determined
18-105_k141_213544 Not-determined
18-105_k141_204783 Not-determined
18-105_k141_26026 Low-quality
18-105_k141_70978 Not-determined
18-105_k141_70982 Not-determined
18-105_k141_204796 Not-determined
18-105_k141_204807 Not-determined
18-105_k141_204833 Not-determined
18-105_k141_26055 Not-determined
18-105_k141_204838 Not-determined
18-105_k141_26060 Not-determined
18-105_k141_204866 Not-determined
18-105_k141_204878 Not-determined
18-105_k141_204886 Not-determined
18-105_k141_204888 Not-determined
18-105_k141_71028 Not-determined
18-105_k141_71037 Not-determined
18-105_k141_26072 Not-determined
18-105_k141_44436 Low-quality
18-105_k141_71053 Not-determined
18-105_k141_71056 Not-determined
18-105_k141_26090 Not-determined
18-105_k141_44467 Not-determined
18-105_k141_44473 Not-determined
18-105_k141_71081 Not-determined
18-105_k141_71087 Not-determined
18-105_k141_44487 Low-quality
18-105_k141_44489 Not-determined
18-105_k141_44505 Not-determined
18-105_k141_71099 Not-determined
18-105_k141_26141 Not-determined
18-105_k141_26155 Not-determined
18-105_k141_71143 Not-determined
18-105_k141_71145 Low-quality
18-105_k141_26166 Not-determined
18-105_k141_44520 Not-determined
18-105_k141_26174 Not-determined
18-105_k141_44531 Not-determined
18-105_k141_44542 Not-determined
18-105_k141_26201 Not-determined
18-105_k141_71170 Not-determined
18-105_k141_71173 Not-determined
18-105_k141_71175 Not-determined
18-105_k141_71178 Not-determined
18-105_k141_71182 Not-determined
18-105_k141_71183 Low-quality
18-105_k141_26228 Not-determined
18-105_k141_71220 Not-determined
18-105_k141_71222 Low-quality
18-105_k141_71227 Not-determined
18-105_k141_71234 Not-determined
18-105_k141_71255 Not-determined
18-105_k141_26295 Not-determined
18-105_k141_26308 Not-determined
18-105_k141_26320 Not-determined
18-105_k141_26330 Not-determined
18-105_k141_26342 Not-determined
18-105_k141_26351 Not-determined
18-105_k141_26353 Not-determined
18-105_k141_26361 Not-determined
18-105_k141_26370 Not-determined
18-105_k141_26407 Not-determined
18-105_k141_26438 Not-determined
18-105_k141_26453 Not-determined
18-105_k141_26465 Not-determined
18-105_k141_26478 Not-determined
18-105_k141_26492 Not-determined
18-105_k141_26558 Not-determined
18-105_k141_26571 Not-determined
18-105_k141_26576 Not-determined
18-105_k141_26584 Not-determined
18-105_k141_26595 Not-determined
18-105_k141_213582 Not-determined
18-105_k141_213585 Not-determined
18-105_k141_213586 Not-determined
18-105_k141_213587 Not-determined
18-105_k141_213592 Not-determined
18-105_k141_213600 Not-determined
18-105_k141_213604 Not-determined
18-105_k141_213614 Not-determined
18-105_k141_213615 Low-quality
18-105_k141_213621 Not-determined
18-105_k141_213622 Not-determined
18-105_k141_213624 Not-determined
18-105_k141_213626 Not-determined
18-105_k141_213630 Not-determined
18-105_k141_213632 Not-determined
18-105_k141_213634 Not-determined
18-105_k141_213643 Not-determined
18-105_k141_213646 Not-determined
18-105_k141_213652 Not-determined
18-105_k141_213653 Not-determined
18-105_k141_213654 Not-determined
18-105_k141_213656 Not-determined
18-105_k141_213663 Not-determined
18-105_k141_213675 Not-determined
18-105_k141_213680 Not-determined
18-105_k141_213681 Not-determined
18-105_k141_213687 Low-quality
18-105_k141_213690 Not-determined
18-105_k141_213698 Not-determined
18-105_k141_213703 Low-quality
18-105_k141_213709 Not-determined
18-105_k141_213722 Low-quality
18-105_k141_213730 Not-determined
18-105_k141_213739 Not-determined
18-105_k141_213752 Not-determined
18-105_k141_213755 Not-determined
18-105_k141_213758 Not-determined
18-105_k141_213760 Not-determined
18-105_k141_213763 Not-determined
18-105_k141_213764 Not-determined
18-105_k141_213766 Low-quality
18-105_k141_213767 Not-determined
18-105_k141_213769 Not-determined
18-105_k141_213770 Not-determined
18-105_k141_213772 Not-determined
18-105_k141_213777 Not-determined
18-105_k141_213785 Not-determined
18-105_k141_213789 Not-determined
18-105_k141_213790 Not-determined
18-105_k141_213795 Not-determined
18-105_k141_213796 Not-determined
18-105_k141_213798 Not-determined
18-105_k141_26679 Not-determined
18-105_k141_26680 Not-determined
18-105_k141_26701 Not-determined
18-105_k141_26712 Not-determined
18-109-final-viral-combined_k141_112344||full Not-determined
18-109-final-viral-combined_k141_174141||full Low-quality
18-109-final-viral-combined_k141_194825||full Low-quality
18-102-final-viral-combined_k141_166665||full Medium-quality
18-109-final-viral-combined_k141_83411||full Not-determined
17-109_k141_15700 Medium-quality
18-109-final-viral-combined_k141_176909||full Not-determined
17-109-final-viral-combined_k141_15700||full Medium-quality
18-109-final-viral-combined_k141_78243||full Not-determined
18-109-final-viral-combined_k141_232639||full Low-quality
18-109-final-viral-combined_k141_90371||full Not-determined
17-106-final-viral-combined_k141_97755||full Medium-quality
18-106-final-viral-combined_k141_32530||full Medium-quality
18-109-final-viral-combined_k141_244878||full Low-quality
18-109-final-viral-combined_k141_244974||full Not-determined
18-109-final-viral-combined_k141_142965||full Low-quality
18-109-final-viral-combined_k141_204231||full Not-determined
18-109-final-viral-combined_k141_61275||full Not-determined
18-109-final-viral-combined_k141_81707||full Not-determined
18-109-final-viral-combined_k141_204242||full Low-quality
18-109-final-viral-combined_k141_132769||full Not-determined
18-109-final-viral-combined_k141_30649||full Low-quality
18-109-final-viral-combined_k141_173610||full Not-determined
18-109-final-viral-combined_k141_204249||full Not-determined
18-109-final-viral-combined_k141_81712||full Not-determined
18-109-final-viral-combined_k141_153186||full Not-determined
18-109-final-viral-combined_k141_122549||full Not-determined
18-109-final-viral-combined_k141_40873||full Low-quality
18-109-final-viral-combined_k141_204252||full Not-determined
18-109-final-viral-combined_k141_51090||full Not-determined
18-109-final-viral-combined_k141_173637||full Low-quality
18-109-final-viral-combined_k141_214495||full Low-quality
18-109-final-viral-combined_k141_204265||full Not-determined
18-109-final-viral-combined_k141_153205||full Not-determined
18-109-final-viral-combined_k141_91921||full Low-quality
18-109-final-viral-combined_k141_10237||full Low-quality
18-109-final-viral-combined_k141_102157||full Not-determined
18-109-final-viral-combined_k141_112383||full Not-determined
18-109-final-viral-combined_k141_48||full Not-determined
18-109-final-viral-combined_k141_102162||full Not-determined
18-109-final-viral-combined_k141_132808||full Low-quality
18-109-final-viral-combined_k141_91934||full Not-determined
18-109-final-viral-combined_k141_51126||full Low-quality
18-109-final-viral-combined_k141_234918||full Low-quality
18-109-final-viral-combined_k141_173668||full Not-determined
18-109-final-viral-combined_k141_51135||full Not-determined
18-109-final-viral-combined_k141_81744||full Not-determined
18-109-final-viral-combined_k141_234933||full Low-quality
18-109-final-viral-combined_k141_91967||full Low-quality
18-109-final-viral-combined_k141_143040||full Low-quality
18-109-final-viral-combined_k141_10277||full Low-quality
18-109-final-viral-combined_k141_234938||full Low-quality
18-109-final-viral-combined_k141_79||full Low-quality
18-109-final-viral-combined_k141_153268||full Not-determined
18-109-final-viral-combined_k141_173675||full Not-determined
18-109-final-viral-combined_k141_194037||full Low-quality
18-109-final-viral-combined_k141_51158||full Not-determined
18-109-final-viral-combined_k141_90||full Low-quality
18-109-final-viral-combined_k141_112417||full Not-determined
18-109-final-viral-combined_k141_234962||full Low-quality
18-109-final-viral-combined_k141_40912||full Not-determined
18-109-final-viral-combined_k141_163490||full Not-determined
18-109-final-viral-combined_k141_153291||full Not-determined
18-109-final-viral-combined_k141_234985||full Low-quality
18-109-final-viral-combined_k141_118||full Not-determined
18-109-final-viral-combined_k141_214559||full Low-quality
18-109-final-viral-combined_k141_153295||full Not-determined
18-109-final-viral-combined_k141_120||full Not-determined
18-109-final-viral-combined_k141_224741||full Not-determined
18-109-final-viral-combined_k141_122609||full Not-determined
18-109-final-viral-combined_k141_163504||full Low-quality
18-109-final-viral-combined_k141_194085||full Not-determined
18-109-final-viral-combined_k141_153301||full Not-determined
18-109-final-viral-combined_k141_40928||full Low-quality
18-109-final-viral-combined_k141_214573||full Not-determined
18-109-final-viral-combined_k141_102206||full Not-determined
18-109-final-viral-combined_k141_173684||full Low-quality
18-109-final-viral-combined_k141_122630||full Low-quality
18-109-final-viral-combined_k141_235014||full Not-determined
18-109-final-viral-combined_k141_153325||full Not-determined
18-109-final-viral-combined_k141_61355||full Low-quality
18-109-final-viral-combined_k141_40938||full Low-quality
18-109-final-viral-combined_k141_40945||full Not-determined
18-109-final-viral-combined_k141_143106||full Low-quality
18-109-final-viral-combined_k141_40952||full Low-quality
18-109-final-viral-combined_k141_51173||full Not-determined
18-109-final-viral-combined_k141_40956||full Low-quality
18-109-final-viral-combined_k141_61372||full Not-determined
18-109-final-viral-combined_k141_153332||full Low-quality
18-109-final-viral-combined_k141_224779||full Not-determined
18-109-final-viral-combined_k141_194127||full Low-quality
18-109-final-viral-combined_k141_61381||full Low-quality
18-109-final-viral-combined_k141_112485||full Not-determined
18-109-final-viral-combined_k141_92032||full Not-determined
18-109-final-viral-combined_k141_184||full Low-quality
18-109-final-viral-combined_k141_20484||full Not-determined
18-109-final-viral-combined_k141_102260||full Not-determined
18-109-final-viral-combined_k141_112493||full Low-quality
18-109-final-viral-combined_k141_112497||full Not-determined
18-109-final-viral-combined_k141_163551||full Low-quality
18-109-final-viral-combined_k141_163554||full Not-determined
18-109-final-viral-combined_k141_61397||full Low-quality
KM-final-viral-combined_k141_127688||full High-quality
18-109-final-viral-combined_k141_143168||full Low-quality
18-109-final-viral-combined_k141_61406||full Low-quality
18-109-final-viral-combined_k141_214635||full Not-determined
18-109-final-viral-combined_k141_20515||full Not-determined
18-109-final-viral-combined_k141_30757||full Low-quality
18-109-final-viral-combined_k141_229||full Not-determined
18-109-final-viral-combined_k141_153353||full Low-quality
18-109-final-viral-combined_k141_112515||full Low-quality
18-109-final-viral-combined_k141_132826||full Not-determined
18-109-final-viral-combined_k141_143185||full Low-quality
18-109-final-viral-combined_k141_194177||full Low-quality
18-109-final-viral-combined_k141_163593||full Low-quality
18-109-final-viral-combined_k141_92065||full Low-quality
18-109-final-viral-combined_k141_92069||full Not-determined
18-109-final-viral-combined_k141_194188||full Not-determined
18-109-final-viral-combined_k141_204361||full Low-quality
18-109-final-viral-combined_k141_112537||full Not-determined
18-109-final-viral-combined_k141_122675||full Not-determined
18-109-final-viral-combined_k141_153406||full Low-quality
18-109-final-viral-combined_k141_183966||full Low-quality
18-109-final-viral-combined_k141_214665||full Low-quality
18-109-final-viral-combined_k141_132842||full Not-determined
18-109-final-viral-combined_k141_194217||full Low-quality
18-109-final-viral-combined_k141_122692||full Low-quality
KM_k141_127688 High-quality
18-109-final-viral-combined_k141_256||full Not-determined
18-109-final-viral-combined_k141_132857||full Low-quality
18-109-final-viral-combined_k141_153420||full Low-quality
18-109-final-viral-combined_k141_163618||full Not-determined
18-109-final-viral-combined_k141_235055||full Low-quality
18-109-final-viral-combined_k141_112560||full Not-determined
18-109-final-viral-combined_k141_61477||full Not-determined
18-109-final-viral-combined_k141_132882||full Low-quality
18-109-final-viral-combined_k141_224860||full Not-determined
18-109-final-viral-combined_k141_71608||full Low-quality
18-109-final-viral-combined_k141_30793||full Not-determined
18-109-final-viral-combined_k141_10325||full Not-determined
18-109-final-viral-combined_k141_122728||full Not-determined
18-109-final-viral-combined_k141_235091||full Not-determined
18-109-final-viral-combined_k141_30799||full Low-quality
18-109-final-viral-combined_k141_235094||full Low-quality
18-109-final-viral-combined_k141_112578||full Low-quality
18-109-final-viral-combined_k141_132918||full Low-quality
18-109-final-viral-combined_k141_20554||full Not-determined
18-109-final-viral-combined_k141_61501||full Low-quality
18-109-final-viral-combined_k141_10331||full Not-determined
18-109-final-viral-combined_k141_20555||full Not-determined
18-109-final-viral-combined_k141_214721||full Not-determined
18-109-final-viral-combined_k141_235101||full Not-determined
18-109-final-viral-combined_k141_102383||full Low-quality
18-109-final-viral-combined_k141_112627||full Not-determined
18-109-final-viral-combined_k141_163679||full Not-determined
18-109-final-viral-combined_k141_20568||full Not-determined
18-109-final-viral-combined_k141_204405||full Low-quality
18-109-final-viral-combined_k141_51233||full Not-determined
18-109-final-viral-combined_k141_122794||full Low-quality
18-109-final-viral-combined_k141_122798||full Not-determined
18-109-final-viral-combined_k141_30838||full Low-quality
18-109-final-viral-combined_k141_214741||full Not-determined
18-109-final-viral-combined_k141_51257||full Low-quality
18-109-final-viral-combined_k141_132940||full Not-determined
18-109-final-viral-combined_k141_132952||full Not-determined
18-109-final-viral-combined_k141_30855||full Low-quality
18-109-final-viral-combined_k141_204441||full Not-determined

| | |
|---|---|
| 18-109-final-viral-combined_k141_173846\|\|full | Not-determined |
| 18-109-final-viral-combined_k141_224954\|\|full | Not-determined |
| 18-109-final-viral-combined_k141_204446\|\|full | Low-quality |
| 18-109-final-viral-combined_k141_194300\|\|full | Not-determined |
| 18-109-final-viral-combined_k141_313\|\|full | Low-quality |
| 18-109-final-viral-combined_k141_204450\|\|full | Low-quality |
| 18-109-final-viral-combined_k141_112674\|\|full | Low-quality |
| 18-109-final-viral-combined_k141_20605\|\|full | Not-determined |
| 18-109-final-viral-combined_k141_320\|\|full | Not-determined |
| 18-109-final-viral-combined_k141_61543\|\|full | Low-quality |
| 18-109-final-viral-combined_k141_324\|\|full | Not-determined |
| 18-109-final-viral-combined_k141_81801\|\|full | Not-determined |
| 18-109-final-viral-combined_k141_41050\|\|full | Low-quality |
| 18-109-final-viral-combined_k141_184034\|\|full | Low-quality |
| 18-109-final-viral-combined_k141_10440\|\|full | Low-quality |
| 18-109-final-viral-combined_k141_184049\|\|full | Not-determined |
| 18-109-final-viral-combined_k141_204478\|\|full | Low-quality |
| 18-109-final-viral-combined_k141_153434\|\|full | Not-determined |
| 18-109-final-viral-combined_k141_163777\|\|full | Low-quality |
| 18-109-final-viral-combined_k141_354\|\|full | Low-quality |
| 18-101_k141_100010 | High-quality |
| 18-109-final-viral-combined_k141_194325\|\|full | Not-determined |
| 18-109-final-viral-combined_k141_133002\|\|full | Not-determined |
| 18-109-final-viral-combined_k141_20650\|\|full | Not-determined |
| 18-109-final-viral-combined_k141_20654\|\|full | Not-determined |
| 18-109-final-viral-combined_k141_194339\|\|full | Not-determined |
| 18-109-final-viral-combined_k141_133004\|\|full | Low-quality |
| 18-109-final-viral-combined_k141_235185\|\|full | Not-determined |
| 18-109-final-viral-combined_k141_133020\|\|full | Not-determined |
| 18-109-final-viral-combined_k141_92265\|\|full | Low-quality |
| 18-109-final-viral-combined_k141_153508\|\|full | Low-quality |
| 18-109-final-viral-combined_k141_133025\|\|full | Not-determined |
| 18-109-final-viral-combined_k141_235223\|\|full | Low-quality |
| 18-109-final-viral-combined_k141_385\|\|full | Low-quality |
| 18-109-final-viral-combined_k141_173897\|\|full | Not-determined |
| 18-109-final-viral-combined_k141_194406\|\|full | Low-quality |
| 18-109-final-viral-combined_k141_81917\|\|full | Not-determined |
| 18-109-final-viral-combined_k141_173915\|\|full | Low-quality |
| 18-109-final-viral-combined_k141_194421\|\|full | Low-quality |
| 18-109-final-viral-combined_k141_163830\|\|full | Not-determined |
| 18-109-final-viral-combined_k141_112783\|\|full | Not-determined |
| 18-109-final-viral-combined_k141_112789\|\|full | Low-quality |
| 18-109-final-viral-combined_k141_61633\|\|full | Not-determined |
| 18-109-final-viral-combined_k141_102460\|\|full | Low-quality |
| 18-109-final-viral-combined_k141_173938\|\|full | Not-determined |
| 18-109-final-viral-combined_k141_184155\|\|full | Not-determined |
| 18-109-final-viral-combined_k141_225068\|\|full | Not-determined |
| 18-109-final-viral-combined_k141_102462\|\|full | Low-quality |
| 18-109-final-viral-combined_k141_20686\|\|full | Not-determined |
| 18-109-final-viral-combined_k141_61653\|\|full | Low-quality |
| 18-109-final-viral-combined_k141_204503\|\|full | Low-quality |
| 18-109-final-viral-combined_k141_204505\|\|full | Low-quality |
| 18-109-final-viral-combined_k141_10522\|\|full | Not-determined |
| 18-109-final-viral-combined_k141_153593\|\|full | Low-quality |
| 18-109-final-viral-combined_k141_400\|\|full | Low-quality |
| 18-109-final-viral-combined_k141_143342\|\|full | Low-quality |
| 18-109-final-viral-combined_k141_163882\|\|full | Not-determined |
| 18-109-final-viral-combined_k141_415\|\|full | Not-determined |
| 18-109-final-viral-combined_k141_235294\|\|full | Low-quality |
| 18-109-final-viral-combined_k141_61683\|\|full | Low-quality |
| 18-109-final-viral-combined_k141_173981\|\|full | Low-quality |
| 18-109-final-viral-combined_k141_423\|\|full | Low-quality |
| 18-109-final-viral-combined_k141_153611\|\|full | Low-quality |
| 18-109-final-viral-combined_k141_112823\|\|full | Low-quality |
| 18-109-final-viral-combined_k141_163898\|\|full | Not-determined |
| 18-109-final-viral-combined_k141_173995\|\|full | Low-quality |
| 18-109-final-viral-combined_k141_204526\|\|full | Not-determined |
| 18-109-final-viral-combined_k141_61686\|\|full | Low-quality |
| 18-109-final-viral-combined_k141_163922\|\|full | Low-quality |
| 18-109-final-viral-combined_k141_20704\|\|full | Low-quality |
| 18-109-final-viral-combined_k141_20707\|\|full | Not-determined |
| 18-109-final-viral-combined_k141_194511\|\|full | Low-quality |
| 18-109-final-viral-combined_k141_112861\|\|full | Not-determined |
| 18-109-final-viral-combined_k141_31031\|\|full | Low-quality |
| 18-109-final-viral-combined_k141_235314\|\|full | Low-quality |
| 18-109-final-viral-combined_k141_163941\|\|full | Not-determined |
| 18-109-final-viral-combined_k141_41103\|\|full | Low-quality |
| 18-109-final-viral-combined_k141_10585\|\|full | Low-quality |
| 18-109-final-viral-combined_k141_153639\|\|full | Not-determined |
| 18-109-final-viral-combined_k141_453\|\|full | Low-quality |
| 18-109-final-viral-combined_k141_235336\|\|full | Not-determined |
| 18-109-final-viral-combined_k141_225162\|\|full | Low-quality |
| 18-109-final-viral-combined_k141_214802\|\|full | Not-determined |
| 18-109-final-viral-combined_k141_153671\|\|full | Not-determined |
| 18-109-final-viral-combined_k141_225176\|\|full | Not-determined |
| 18-109-final-viral-combined_k141_143416\|\|full | Not-determined |
| 18-109-final-viral-combined_k141_194557\|\|full | Low-quality |
| 18-109-final-viral-combined_k141_214831\|\|full | Not-determined |
| 18-109-final-viral-combined_k141_112927\|\|full | Low-quality |
| 18-109-final-viral-combined_k141_41148\|\|full | Not-determined |
| 18-109-final-viral-combined_k141_133149\|\|full | Low-quality |
| 18-109-final-viral-combined_k141_82043\|\|full | Not-determined |
| 18-109-final-viral-combined_k141_41157\|\|full | Not-determined |
| 18-109-final-viral-combined_k141_506\|\|full | Low-quality |
| 18-109-final-viral-combined_k141_204610\|\|full | Not-determined |
| 18-109-final-viral-combined_k141_153692\|\|full | Not-determined |
| 18-109-final-viral-combined_k141_143429\|\|full | Low-quality |
| 18-109-final-viral-combined_k141_225205\|\|full | Not-determined |
| 18-109-final-viral-combined_k141_133172\|\|full | Not-determined |
| 18-109-final-viral-combined_k141_82062\|\|full | Not-determined |
| 18-109-final-viral-combined_k141_528\|\|full | Not-determined |
| 18-109-final-viral-combined_k141_20751\|\|full | Low-quality |
| 18-109-final-viral-combined_k141_61740\|\|full | Not-determined |
| 18-109-final-viral-combined_k141_184245\|\|full | Not-determined |
| 18-109-final-viral-combined_k141_225219\|\|full | Low-quality |
| 18-109-final-viral-combined_k141_112952\|\|full | Low-quality |
| 18-109-final-viral-combined_k141_102524\|\|full | Not-determined |
| 18-109-final-viral-combined_k141_235402\|\|full | Not-determined |
| 18-109-final-viral-combined_k141_153722\|\|full | Low-quality |
| 18-109-final-viral-combined_k141_71732\|\|full | Low-quality |
| 18-109-final-viral-combined_k141_225238\|\|full | Not-determined |
| 18-109-final-viral-combined_k141_102538\|\|full | Low-quality |
| 18-109-final-viral-combined_k141_41206\|\|full | Low-quality |
| 18-109-final-viral-combined_k141_235413\|\|full | Not-determined |
| 18-109-final-viral-combined_k141_194644\|\|full | Not-determined |
| 18-109-final-viral-combined_k141_122935\|\|full | Low-quality |
| 18-109-final-viral-combined_k141_102559\|\|full | Not-determined |
| 18-109-final-viral-combined_k141_174074\|\|full | Not-determined |
| 18-109-final-viral-combined_k141_214910\|\|full | Low-quality |
| 18-109-final-viral-combined_k141_194656\|\|full | Not-determined |
| 18-109-final-viral-combined_k141_235433\|\|full | Not-determined |
| 18-109-final-viral-combined_k141_556\|\|full | Low-quality |
| 18-109-final-viral-combined_k141_194665\|\|full | Not-determined |
| 18-102_k141_140933 | High-quality |
| 18-109-final-viral-combined_k141_61792\|\|full | Low-quality |
| 18-109-final-viral-combined_k141_143503\|\|full | Low-quality |
| 18-109-final-viral-combined_k141_174085\|\|full | Low-quality |
| 18-109-final-viral-combined_k141_122944\|\|full | Not-determined |
| 18-109-final-viral-combined_k141_71756\|\|full | Low-quality |
| 17-109_k141_179388 | High-quality |
| 18-109-final-viral-combined_k141_102566\|\|full | Low-quality |
| 18-109-final-viral-combined_k141_10642\|\|full | Low-quality |
| 18-109-final-viral-combined_k141_143527\|\|full | Not-determined |
| 18-109-final-viral-combined_k141_184313\|\|full | Low-quality |
| 18-109-final-viral-combined_k141_214967\|\|full | Low-quality |
| 18-109-final-viral-combined_k141_143534\|\|full | Not-determined |
| 18-109-final-viral-combined_k141_204684\|\|full | Low-quality |
| 18-109-final-viral-combined_k141_41216\|\|full | Low-quality |
| 18-109-final-viral-combined_k141_133258\|\|full | Not-determined |
| 18-109-final-viral-combined_k141_10674\|\|full | Not-determined |
| 18-109-final-viral-combined_k141_71784\|\|full | Not-determined |
| 18-109-final-viral-combined_k141_143543\|\|full | Low-quality |
| 18-109-final-viral-combined_k141_92409\|\|full | Not-determined |
| 18-109-final-viral-combined_k141_41223\|\|full | Low-quality |
| 18-109-final-viral-combined_k141_112983\|\|full | Low-quality |
| 18-109-final-viral-combined_k141_102572\|\|full | Low-quality |
| 18-109-final-viral-combined_k141_598\|\|full | Not-determined |
| 18-109-final-viral-combined_k141_204707\|\|full | Low-quality |
| 18-109-final-viral-combined_k141_225300\|\|full | Low-quality |
| 18-109-final-viral-combined_k141_174134\|\|full | Low-quality |
| 18-109-final-viral-combined_k141_71801\|\|full | Low-quality |
| 18-109-final-viral-combined_k141_174138\|\|full | Low-quality |
| 18-109-final-viral-combined_k141_102589\|\|full | Low-quality |
| 18-109-final-viral-combined_k141_82159\|\|full | Not-determined |
| 18-109-final-viral-combined_k141_204714\|\|full | Not-determined |
| 18-109-final-viral-combined_k141_605\|\|full | Not-determined |
| 18-109-final-viral-combined_k141_225326\|\|full | Not-determined |
| 18-109-final-viral-combined_k141_153836\|\|full | Not-determined |
| 18-109-final-viral-combined_k141_20860\|\|full | Not-determined |
| 18-109-final-viral-combined_k141_204726\|\|full | Not-determined |
| 18-109-final-viral-combined_k141_174169\|\|full | Low-quality |
| 18-109-final-viral-combined_k141_20864\|\|full | Low-quality |
| 18-109-final-viral-combined_k141_20865\|\|full | Not-determined |
| 18-109-final-viral-combined_k141_235504\|\|full | Low-quality |
| 18-109-final-viral-combined_k141_630\|\|full | Not-determined |
| 18-109-final-viral-combined_k141_174184\|\|full | Low-quality |
| 18-109-final-viral-combined_k141_225344\|\|full | Not-determined |
| 18-109-final-viral-combined_k141_31129\|\|full | Not-determined |
| 18-109-final-viral-combined_k141_225346\|\|full | Not-determined |
| 18-109-final-viral-combined_k141_174191\|\|full | Low-quality |
| 18-109-final-viral-combined_k141_61874\|\|full | Not-determined |
| 18-109-final-viral-combined_k141_235536\|\|full | Not-determined |
| 18-109-final-viral-combined_k141_225357\|\|full | Not-determined |
| 18-109-final-viral-combined_k141_194783\|\|full | Low-quality |
| 18-109-final-viral-combined_k141_174215\|\|full | Not-determined |
| 18-109-final-viral-combined_k141_194787\|\|full | Low-quality |
| 18-109-final-viral-combined_k141_10710\|\|full | Low-quality |
| 18-109-final-viral-combined_k141_215071\|\|full | Low-quality |
| 18-109-final-viral-combined_k141_41265\|\|full | Not-determined |
| 18-109-final-viral-combined_k141_123018\|\|full | Low-quality |
| 18-109-final-viral-combined_k141_194808\|\|full | Low-quality |
| 18-109-final-viral-combined_k141_215082\|\|full | Not-determined |
| 18-109-final-viral-combined_k141_681\|\|full | Not-determined |
| 18-109-final-viral-combined_k141_194815\|\|full | Low-quality |
| 18-109-final-viral-combined_k141_102655\|\|full | Not-determined |
| 18-109-final-viral-combined_k141_41291\|\|full | Not-determined |
| 18-109-final-viral-combined_k141_215101\|\|full | Not-determined |
| 18-109-final-viral-combined_k141_102666\|\|full | Not-determined |
| 18-109-final-viral-combined_k141_215115\|\|full | Low-quality |
| 18-109-final-viral-combined_k141_153928\|\|full | Low-quality |
| 18-109-final-viral-combined_k141_164020\|\|full | Low-quality |
| 18-109-final-viral-combined_k141_194833\|\|full | Not-determined |
| 18-109-final-viral-combined_k141_20879\|\|full | Not-determined |
| 18-109-final-viral-combined_k141_215134\|\|full | Not-determined |
| 18-109-final-viral-combined_k141_153937\|\|full | Not-determined |
| 18-109-final-viral-combined_k141_153946\|\|full | Low-quality |
| 18-109-final-viral-combined_k141_20904\|\|full | Not-determined |
| 18-109-final-viral-combined_k141_235567\|\|full | Not-determined |
| 18-109-final-viral-combined_k141_51418\|\|full | Not-determined |
| 18-109-final-viral-combined_k141_51439\|\|full | Not-determined |
| 18-109-final-viral-combined_k141_51445\|\|full | Not-determined |
| 18-109-final-viral-combined_k141_10758\|\|full | Low-quality |
| 18-109-final-viral-combined_k141_235605\|\|full | Low-quality |
| 18-109-final-viral-combined_k141_153972\|\|full | Not-determined |
| 18-109-final-viral-combined_k141_215172\|\|full | Low-quality |
| 18-109-final-viral-combined_k141_235609\|\|full | Not-determined |
| 18-109-final-viral-combined_k141_10772\|\|full | Low-quality |
| 18-109-final-viral-combined_k141_102675\|\|full | Not-determined |
| 18-109-final-viral-combined_k141_153985\|\|full | Not-determined |
| 18-102-final-viral-combined_k141_140933\|\|full | High-quality |
| 18-109-final-viral-combined_k141_51463\|\|full | Low-quality |
| 18-109-final-viral-combined_k141_133332\|\|full | Low-quality |
| 18-109-final-viral-combined_k141_123088\|\|full | Low-quality |
| 18-109-final-viral-combined_k141_82237\|\|full | Not-determined |
| 18-109-final-viral-combined_k141_153999\|\|full | Not-determined |
| 18-109-final-viral-combined_k141_71973\|\|full | Not-determined |
| 17-109-final-viral-combined_k141_179388\|\|full | High-quality |
| 18-109-final-viral-combined_k141_164120\|\|full | Not-determined |
| 18-109-final-viral-combined_k141_204853\|\|full | Not-determined |
| 18-109-final-viral-combined_k141_123135\|\|full | Low-quality |
| 18-109-final-viral-combined_k141_215240\|\|full | Not-determined |
| 18-109-final-viral-combined_k141_194939\|\|full | Not-determined |
| 18-109-final-viral-combined_k141_51529\|\|full | Low-quality |
| 18-109-final-viral-combined_k141_61971\|\|full | Not-determined |
| 18-109-final-viral-combined_k141_184416\|\|full | Low-quality |
| 18-109-final-viral-combined_k141_123161\|\|full | Not-determined |
| 18-109-final-viral-combined_k141_215272\|\|full | Not-determined |
| 18-109-final-viral-combined_k141_133408\|\|full | Low-quality |
| 18-109-final-viral-combined_k141_154115\|\|full | Low-quality |
| 18-109-final-viral-combined_k141_51537\|\|full | Low-quality |
| 18-109-final-viral-combined_k141_154138\|\|full | Not-determined |
| 18-109-final-viral-combined_k141_51554\|\|full | Not-determined |
| 18-109-final-viral-combined_k141_51555\|\|full | Low-quality |
| 18-109-final-viral-combined_k141_72015\|\|full | Not-determined |
| 17-109-final-viral-combined_k141_259783\|\|full | High-quality |
| 18-109-final-viral-combined_k141_133449\|\|full | Not-determined |
| 18-109-final-viral-combined_k141_123227\|\|full | Not-determined |
| 18-109-final-viral-combined_k141_72027\|\|full | Not-determined |
| 18-109-final-viral-combined_k141_235716\|\|full | Not-determined |
| 18-109-final-viral-combined_k141_82343\|\|full | Not-determined |
| 18-109-final-viral-combined_k141_215304\|\|full | Low-quality |
| 18-109-final-viral-combined_k141_194979\|\|full | Not-determined |
| 18-109-final-viral-combined_k141_164163\|\|full | Low-quality |
| 18-109-final-viral-combined_k141_41336\|\|full | Low-quality |
| 18-109-final-viral-combined_k141_72038\|\|full | Not-determined |
| 18-109-final-viral-combined_k141_62014\|\|full | Not-determined |
| 18-109-final-viral-combined_k141_174293\|\|full | Low-quality |
| 18-109-final-viral-combined_k141_878\|\|full | Not-determined |
| 18-109-final-viral-combined_k141_113205\|\|full | Low-quality |
| 18-109-final-viral-combined_k141_174302\|\|full | Not-determined |
| 18-109-final-viral-combined_k141_62022\|\|full | Not-determined |
| 18-109-final-viral-combined_k141_215344\|\|full | Not-determined |
| 18-109-final-viral-combined_k141_215351\|\|full | Not-determined |
| 18-109-final-viral-combined_k141_225455\|\|full | Low-quality |
| 18-109-final-viral-combined_k141_154160\|\|full | Low-quality |
| 18-109-final-viral-combined_k141_215362\|\|full | Low-quality |
| 18-109-final-viral-combined_k141_215367\|\|full | Low-quality |
| 18-109-final-viral-combined_k141_225480\|\|full | Not-determined |
| 18-109-final-viral-combined_k141_92531\|\|full | Low-quality |
| 18-109-final-viral-combined_k141_62068\|\|full | Low-quality |
| 18-109-final-viral-combined_k141_225489\|\|full | Low-quality |
| 18-109-final-viral-combined_k141_184499\|\|full | Not-determined |
| 18-109-final-viral-combined_k141_113249\|\|full | Not-determined |
| 18-109-final-viral-combined_k141_10923\|\|full | Not-determined |
| 18-109-final-viral-combined_k141_20971\|\|full | Low-quality |
| 18-109-final-viral-combined_k141_41444\|\|full | Not-determined |
| 18-109-final-viral-combined_k141_41445\|\|full | Not-determined |
| 18-109-final-viral-combined_k141_225510\|\|full | Low-quality |
| 18-109-final-viral-combined_k141_92556\|\|full | Not-determined |
| 18-109-final-viral-combined_k141_31334\|\|full | Low-quality |
| 18-109-final-viral-combined_k141_20975\|\|full | Not-determined |
| 18-109-final-viral-combined_k141_62098\|\|full | Low-quality |
| 18-109-final-viral-combined_k141_51684\|\|full | Not-determined |
| 18-109-final-viral-combined_k141_113290\|\|full | Not-determined |
| 18-109-final-viral-combined_k141_205009\|\|full | Not-determined |
| 18-109-final-viral-combined_k141_92601\|\|full | Low-quality |
| 18-109-final-viral-combined_k141_215380\|\|full | Low-quality |
| 18-109-final-viral-combined_k141_154232\|\|full | Low-quality |
| 18-109-final-viral-combined_k141_41486\|\|full | Low-quality |
| 18-109-final-viral-combined_k141_21028\|\|full | Not-determined |
| 18-109-final-viral-combined_k141_205024\|\|full | Not-determined |
| 18-109-final-viral-combined_k141_72069\|\|full | Not-determined |
| 18-109-final-viral-combined_k141_51724\|\|full | Not-determined |
| 18-109-final-viral-combined_k141_31417\|\|full | Low-quality |
| 18-109-final-viral-combined_k141_31423\|\|full | Not-determined |
| 18-109-final-viral-combined_k141_164291\|\|full | Not-determined |
| 18-109-final-viral-combined_k141_154259\|\|full | Not-determined |
| 18-109-final-viral-combined_k141_31429\|\|full | Not-determined |
| 18-109-final-viral-combined_k141_11020\|\|full | Low-quality |
| 18-109-final-viral-combined_k141_133607\|\|full | Low-quality |
| 18-109-final-viral-combined_k141_164302\|\|full | Not-determined |
| 18-109-final-viral-combined_k141_235737\|\|full | Not-determined |
| 18-109-final-viral-combined_k141_51741\|\|full | Not-determined |
| 18-109-final-viral-combined_k141_184620\|\|full | Not-determined |
| 18-109-final-viral-combined_k141_41519\|\|full | Not-determined |
| 18-109-final-viral-combined_k141_51742\|\|full | Low-quality |
| 18-109-final-viral-combined_k141_92661\|\|full | Low-quality |
| 18-109-final-viral-combined_k141_11036\|\|full | Not-determined |
| 18-109-final-viral-combined_k141_195117\|\|full | Low-quality |
| 18-109-final-viral-combined_k141_164332\|\|full | Not-determined |
| 18-109-final-viral-combined_k141_184632\|\|full | Not-determined |
| 18-109-final-viral-combined_k141_31471\|\|full | Low-quality |
| 18-109-final-viral-combined_k141_235786\|\|full | Not-determined |
| 18-109-final-viral-combined_k141_184655\|\|full | Not-determined |
| 18-109-final-viral-combined_k141_31477\|\|full | Low-quality |
| 18-109-final-viral-combined_k141_215456\|\|full | Low-quality |
| 18-109-final-viral-combined_k141_184657\|\|full | Low-quality |
| 18-109-final-viral-combined_k141_205084\|\|full | Not-determined |
| 18-109-final-viral-combined_k141_215465\|\|full | Low-quality |
| 18-109-final-viral-combined_k141_11068\|\|full | Not-determined |
| 18-109-final-viral-combined_k141_31505\|\|full | Low-quality |
| 18-109-final-viral-combined_k141_82400\|\|full | Not-determined |
| 18-109-final-viral-combined_k141_235803\|\|full | Low-quality |
| 18-109-final-viral-combined_k141_195170\|\|full | Low-quality |
| 18-109-final-viral-combined_k141_205129\|\|full | Low-quality |
| 18-109-final-viral-combined_k141_21069\|\|full | Low-quality |
| 18-109-final-viral-combined_k141_235814\|\|full | Not-determined |
| 18-109-final-viral-combined_k141_154317\|\|full | Low-quality |
| 18-109-final-viral-combined_k141_31532\|\|full | Not-determined |
| 18-109-final-viral-combined_k141_184667\|\|full | Low-quality |
| 18-109-final-viral-combined_k141_215492\|\|full | Not-determined |
| 18-109-final-viral-combined_k141_205132\|\|full | Not-determined |
| 18-109-final-viral-combined_k141_21075\|\|full | Not-determined |
| 18-109-final-viral-combined_k141_154339\|\|full | Not-determined |
| 18-109-final-viral-combined_k141_82438\|\|full | Not-determined |
| 18-109-final-viral-combined_k141_225569\|\|full | Low-quality |
| 18-109-final-viral-combined_k141_41571\|\|full | Not-determined |
| 18-109-final-viral-combined_k141_51849\|\|full | Low-quality |
| 18-109-final-viral-combined_k141_205143\|\|full | Not-determined |
| 18-109-final-viral-combined_k141_235835\|\|full | Not-determined |
| 18-109-final-viral-combined_k141_174331\|\|full | Not-determined |
| 18-109-final-viral-combined_k141_51856\|\|full | Low-quality |
| 18-109-final-viral-combined_k141_154349\|\|full | Low-quality |
| 18-109-final-viral-combined_k141_41582\|\|full | Low-quality |
| 18-109-final-viral-combined_k141_195187\|\|full | Low-quality |
| 18-109-final-viral-combined_k141_225600\|\|full | Low-quality |
| 18-109-final-viral-combined_k141_51866\|\|full | Not-determined |
| 18-109-final-viral-combined_k141_225608\|\|full | Not-determined |
| 18-109-final-viral-combined_k141_235854\|\|full | Not-determined |
| 18-109-final-viral-combined_k141_235859\|\|full | Not-determined |
| 18-109-final-viral-combined_k141_102740\|\|full | Not-determined |
| 18-109-final-viral-combined_k141_164412\|\|full | Not-determined |
| 18-109-final-viral-combined_k141_133687\|\|full | Low-quality |
| 18-109-final-viral-combined_k141_164415\|\|full | Not-determined |
| 18-109-final-viral-combined_k141_143595\|\|full | Low-quality |
| 18-109-final-viral-combined_k141_195218\|\|full | Not-determined |
| 18-109-final-viral-combined_k141_195229\|\|full | Low-quality |
| 18-109-final-viral-combined_k141_143606\|\|full | Not-determined |
| 18-109-final-viral-combined_k141_225629\|\|full | Not-determined |
| 18-109-final-viral-combined_k141_154397\|\|full | Low-quality |
| 18-109-final-viral-combined_k141_143616\|\|full | Not-determined |
| 18-109-final-viral-combined_k141_205164\|\|full | Low-quality |
| 18-109-final-viral-combined_k141_133715\|\|full | Not-determined |
| 18-109-final-viral-combined_k141_235914\|\|full | Low-quality |
| 18-109-final-viral-combined_k141_143620\|\|full | Not-determined |
| 18-109-final-viral-combined_k141_31555\|\|full | Not-determined |
| 18-109-final-viral-combined_k141_225660\|\|full | Not-determined |
| 18-109-final-viral-combined_k141_133736\|\|full | Low-quality |
| 18-109-final-viral-combined_k141_154409\|\|full | Low-quality |
| 18-109-final-viral-combined_k141_123462\|\|full | Low-quality |
| 18-109-final-viral-combined_k141_174355\|\|full | Not-determined |
| 18-109-final-viral-combined_k141_205197\|\|full | Not-determined |
| 18-109-final-viral-combined_k141_123479\|\|full | Low-quality |
| 18-109-final-viral-combined_k141_11147\|\|full | Low-quality |
| 18-109-final-viral-combined_k141_941\|\|full | Low-quality |
| 18-109-final-viral-combined_k141_123484\|\|full | Low-quality |
| 18-109-final-viral-combined_k141_174378\|\|full | Not-determined |
| 18-109-final-viral-combined_k141_102795\|\|full | Low-quality |
| 18-109-final-viral-combined_k141_123497\|\|full | Low-quality |
| 18-109-final-viral-combined_k141_51926\|\|full | Not-determined |
| 18-109-final-viral-combined_k141_164457\|\|full | Low-quality |
| 18-109-final-viral-combined_k141_113390\|\|full | Low-quality |
| 18-109-final-viral-combined_k141_174392\|\|full | Not-determined |
| 18-109-final-viral-combined_k141_123499\|\|full | Not-determined |
| 18-109-final-viral-combined_k141_174398\|\|full | Low-quality |
| 18-109-final-viral-combined_k141_960\|\|full | Low-quality |
| 18-109-final-viral-combined_k141_102796\|\|full | Low-quality |
| 18-109-final-viral-combined_k141_225685\|\|full | Not-determined |
| 18-109-final-viral-combined_k141_133769\|\|full | Low-quality |
| 18-109-final-viral-combined_k141_174418\|\|full | Not-determined |
| 18-109-final-viral-combined_k141_184872\|\|full | Not-determined |
| 18-109-final-viral-combined_k141_113448\|\|full | Not-determined |
| 18-109-final-viral-combined_k141_133794\|\|full | Not-determined |
| 18-109-final-viral-combined_k141_51950\|\|full | Low-quality |
| 18-109-final-viral-combined_k141_133796\|\|full | Not-determined |
| 18-109-final-viral-combined_k141_195275\|\|full | Not-determined |
| 18-109-final-viral-combined_k141_205284\|\|full | Low-quality |
| 18-109-final-viral-combined_k141_205296\|\|full | Low-quality |
| 18-109-final-viral-combined_k141_11235\|\|full | Low-quality |
| 18-109-final-viral-combined_k141_82557\|\|full | Low-quality |
| 18-109-final-viral-combined_k141_102833\|\|full | Not-determined |
| 18-109-final-viral-combined_k141_102840\|\|full | Low-quality |
| 18-109-final-viral-combined_k141_1037\|\|full | Low-quality |
| 18-109-final-viral-combined_k141_41650\|\|full | Not-determined |
| 18-109-final-viral-combined_k141_82567\|\|full | Low-quality |
| 18-101-final-viral-combined_k141_100010\|\|full | High-quality |
| 18-109-final-viral-combined_k141_51992\|\|full | Low-quality |
| 18-109-final-viral-combined_k141_123514\|\|full | Not-determined |
| 18-109-final-viral-combined_k141_133809\|\|full | Not-determined |
| 18-109-final-viral-combined_k141_82588\|\|full | Low-quality |
| 18-109-final-viral-combined_k141_11245\|\|full | Low-quality |
| 18-109-final-viral-combined_k141_82603\|\|full | Not-determined |
| 18-109-final-viral-combined_k141_62155\|\|full | Not-determined |
| 18-109-final-viral-combined_k141_41686\|\|full | Not-determined |
| 18-109-final-viral-combined_k141_62162\|\|full | Not-determined |
| 18-109-final-viral-combined_k141_174489\|\|full | Not-determined |
| 18-109-final-viral-combined_k141_82618\|\|full | Low-quality |
| 18-109-final-viral-combined_k141_113506\|\|full | Not-determined |
| 18-109-final-viral-combined_k141_205355\|\|full | Low-quality |
| 18-109-final-viral-combined_k141_1081\|\|full | Low-quality |
| 18-109-final-viral-combined_k141_225756\|\|full | Not-determined |
| 18-109-final-viral-combined_k141_123550\|\|full | Low-quality |
| 18-109-final-viral-combined_k141_235968\|\|full | Low-quality |
| 18-109-final-viral-combined_k141_123557\|\|full | Low-quality |
| 18-109-final-viral-combined_k141_195355\|\|full | Not-determined |
| 18-109-final-viral-combined_k141_52072\|\|full | Low-quality |
| 18-109-final-viral-combined_k141_205398\|\|full | Low-quality |
| 18-109-final-viral-combined_k141_235970\|\|full | Low-quality |
| 18-109-final-viral-combined_k141_174526\|\|full | Low-quality |
| 18-109-final-viral-combined_k141_11294\|\|full | Low-quality |
| 18-109-final-viral-combined_k141_184985\|\|full | Low-quality |
| 18-109-final-viral-combined_k141_184986\|\|full | Low-quality |
| 18-109-final-viral-combined_k141_133874\|\|full | Not-determined |
| 18-109-final-viral-combined_k141_11305\|\|full | Low-quality |
| 18-109-final-viral-combined_k141_133876\|\|full | Not-determined |
| 18-109-final-viral-combined_k141_113529\|\|full | Low-quality |
| 18-109-final-viral-combined_k141_21159\|\|full | Not-determined |
| 18-109-final-viral-combined_k141_102907\|\|full | Not-determined |
| 18-109-final-viral-combined_k141_82697\|\|full | Not-determined |
| 18-109-final-viral-combined_k141_164517\|\|full | Not-determined |
| 18-109-final-viral-combined_k141_31717\|\|full | Low-quality |
| 18-109-final-viral-combined_k141_235998\|\|full | Not-determined |
| 18-109-final-viral-combined_k141_102919\|\|full | Not-determined |
| 18-109-final-viral-combined_k141_11306\|\|full | Not-determined |
| 18-109-final-viral-combined_k141_31722\|\|full | Low-quality |
| 18-109-final-viral-combined_k141_82701\|\|full | Low-quality |
| 18-109-final-viral-combined_k141_205419\|\|full | Low-quality |
| 18-109-final-viral-combined_k141_195391\|\|full | Low-quality |
| 18-109-final-viral-combined_k141_236010\|\|full | Low-quality |
| 18-109-final-viral-combined_k141_21189\|\|full | Not-determined |
| 18-109-final-viral-combined_k141_21190\|\|full | Low-quality |
| 18-109-final-viral-combined_k141_11325\|\|full | Low-quality |
| 18-109-final-viral-combined_k141_174592\|\|full | Low-quality |
| 18-109-final-viral-combined_k141_113601\|\|full | Low-quality |
| 18-109-final-viral-combined_k141_62204\|\|full | Not-determined |
| 18-109-final-viral-combined_k141_195441\|\|full | Low-quality |
| 18-109-final-viral-combined_k141_205519\|\|full | Not-determined |
| 18-109-final-viral-combined_k141_31774\|\|full | Low-quality |
| 18-109-final-viral-combined_k141_41725\|\|full | Not-determined |
| 18-109-final-viral-combined_k141_52136\|\|full | Low-quality |
| 18-109-final-viral-combined_k141_174634\|\|full | Low-quality |
| 18-109-final-viral-combined_k141_205533\|\|full | Not-determined |
| 18-109-final-viral-combined_k141_185010\|\|full | Not-determined |
| 18-109-final-viral-combined_k141_31796\|\|full | Not-determined |
| 18-109-final-viral-combined_k141_82777\|\|full | Low-quality |
| 18-109-final-viral-combined_k141_133931\|\|full | Low-quality |
| 18-109-final-viral-combined_k141_31803\|\|full | Not-determined |
| 18-109-final-viral-combined_k141_225883\|\|full | Not-determined |
| 18-109-final-viral-combined_k141_103020\|\|full | Low-quality |
| 18-109-final-viral-combined_k141_103023\|\|full | Low-quality |
| 18-109-final-viral-combined_k141_225892\|\|full | Not-determined |
| 18-109-final-viral-combined_k141_62222\|\|full | Not-determined |
| 18-109-final-viral-combined_k141_225897\|\|full | Not-determined |
| 18-109-final-viral-combined_k141_72078\|\|full | Low-quality |
| 18-109-final-viral-combined_k141_62231\|\|full | Not-determined |
| 18-109-final-viral-combined_k141_92746\|\|full | Not-determined |
| 18-109-final-viral-combined_k141_113634\|\|full | Not-determined |
| 18-109-final-viral-combined_k141_195457\|\|full | Low-quality |
| 18-109-final-viral-combined_k141_113636\|\|full | Not-determined |
| 18-109-final-viral-combined_k141_143872\|\|full | Not-determined |
| 18-109-final-viral-combined_k141_62247\|\|full | Not-determined |
| 18-109-final-viral-combined_k141_31822\|\|full | Not-determined |
| 18-109-final-viral-combined_k141_143874\|\|full | Not-determined |
| 18-109-final-viral-combined_k141_62257\|\|full | Low-quality |
| 18-109-final-viral-combined_k141_92759\|\|full | Low-quality |
| 17-108_k141_6203 | Medium-quality |
| 18-109-final-viral-combined_k141_41810\|\|full | Low-quality |
| 18-109-final-viral-combined_k141_21254\|\|full | Not-determined |
| 18-109-final-viral-combined_k141_103052\|\|full | Not-determined |
| 18-109-final-viral-combined_k141_185031\|\|full | Not-determined |
| 18-109-final-viral-combined_k141_185032\|\|full | Low-quality |
| 18-109-final-viral-combined_k141_133970\|\|full | Not-determined |
| 18-109-final-viral-combined_k141_185035\|\|full | Low-quality |
| 18-109-final-viral-combined_k141_41835\|\|full | Low-quality |
| 18-109-final-viral-combined_k141_103087\|\|full | Not-determined |
| 18-109-final-viral-combined_k141_133976\|\|full | Not-determined |
| 18-109-final-viral-combined_k141_103092\|\|full | Low-quality |
| 18-109-final-viral-combined_k141_41848\|\|full | Low-quality |
| 18-109-final-viral-combined_k141_174707\|\|full | Not-determined |
| 18-109-final-viral-combined_k141_195511\|\|full | Low-quality |
| 18-109-final-viral-combined_k141_41853\|\|full | Not-determined |
| 18-109-final-viral-combined_k141_1151\|\|full | Not-determined |
| 18-109-final-viral-combined_k141_225954\|\|full | Not-determined |
| 18-109-final-viral-combined_k141_1156\|\|full | Low-quality |
| 18-109-final-viral-combined_k141_92795\|\|full | Not-determined |
| 18-109-final-viral-combined_k141_41868\|\|full | Low-quality |
| 18-109-final-viral-combined_k141_41875\|\|full | Not-determined |
| 18-109-final-viral-combined_k141_31848\|\|full | Not-determined |
| 18-109-final-viral-combined_k141_225971\|\|full | Not-determined |
| 18-109-final-viral-combined_k141_195564\|\|full | Low-quality |
| 18-109-final-viral-combined_k141_41894\|\|full | Not-determined |
| 18-109-final-viral-combined_k141_103133\|\|full | Low-quality |
| 18-109-final-viral-combined_k141_1213\|\|full | Low-quality |
| 18-109-final-viral-combined_k141_11408\|\|full | Low-quality |
| 18-109-final-viral-combined_k141_21304\|\|full | Not-determined |
| 18-109-final-viral-combined_k141_103144\|\|full | Low-quality |
| 18-109-final-viral-combined_k141_185087\|\|full | Not-determined |
| 18-109-final-viral-combined_k141_62329\|\|full | Not-determined |
| 18-109-final-viral-combined_k141_123777\|\|full | Not-determined |
| 18-109-final-viral-combined_k141_174767\|\|full | Not-determined |
| 18-109-final-viral-combined_k141_21342\|\|full | Low-quality |
| 18-109-final-viral-combined_k141_195606\|\|full | Low-quality |
| 18-109-final-viral-combined_k141_174775\|\|full | Low-quality |
| 18-109-final-viral-combined_k141_154606\|\|full | Low-quality |
| 18-109-final-viral-combined_k141_195615\|\|full | Not-determined |
| 18-109-final-viral-combined_k141_113682\|\|full | Not-determined |
| 18-109-final-viral-combined_k141_31890\|\|full | Low-quality |
| 18-109-final-viral-combined_k141_103195\|\|full | Low-quality |
| 18-109-final-viral-combined_k141_236146\|\|full | Low-quality |
| 18-109-final-viral-combined_k141_164739\|\|full | Not-determined |
| 18-109-final-viral-combined_k141_62358\|\|full | Not-determined |
| 18-109-final-viral-combined_k141_52269\|\|full | Low-quality |
| 18-109-final-viral-combined_k141_31915\|\|full | Not-determined |
| 18-109-final-viral-combined_k141_205715\|\|full | Low-quality |
| 18-109-final-viral-combined_k141_11434\|\|full | Not-determined |
| 18-109-final-viral-combined_k141_236158\|\|full | Not-determined |
| 18-109-final-viral-combined_k141_174821\|\|full | Not-determined |
| 18-109-final-viral-combined_k141_62370\|\|full | Low-quality |
| 18-109-final-viral-combined_k141_185140\|\|full | Low-quality |
| 18-109-final-viral-combined_k141_113695\|\|full | Low-quality |
| 18-109-final-viral-combined_k141_113697\|\|full | Low-quality |
| 18-109-final-viral-combined_k141_123797\|\|full | Not-determined |
| 18-109-final-viral-combined_k141_62397\|\|full | Low-quality |
| 18-109-final-viral-combined_k141_185175\|\|full | Low-quality |
| 18-109-final-viral-combined_k141_215818\|\|full | Not-determined |
| 18-109-final-viral-combined_k141_154651\|\|full | Low-quality |
| 18-109-final-viral-combined_k141_134115\|\|full | Not-determined |
| 18-109-final-viral-combined_k141_72165\|\|full | Not-determined |
| 18-109-final-viral-combined_k141_1275\|\|full | Low-quality |
| 18-109-final-viral-combined_k141_205747\|\|full | Low-quality |
| 18-109-final-viral-combined_k141_185182\|\|full | Not-determined |
| 18-109-final-viral-combined_k141_92810\|\|full | Low-quality |
| 18-109-final-viral-combined_k141_226015\|\|full | Low-quality |
| 18-109-final-viral-combined_k141_164747\|\|full | Not-determined |
| 18-109-final-viral-combined_k141_143936\|\|full | Low-quality |
| 18-109-final-viral-combined_k141_62410\|\|full | Not-determined |
| 18-109-final-viral-combined_k141_92820\|\|full | Not-determined |
| 18-109-final-viral-combined_k141_72176\|\|full | Low-quality |
| 18-109-final-viral-combined_k141_52310\|\|full | Not-determined |
| 18-109-final-viral-combined_k141_92843\|\|full | Not-determined |
| 18-109-final-viral-combined_k141_72196\|\|full | Low-quality |
| 18-109-final-viral-combined_k141_215848\|\|full | Low-quality |
| 18-109-final-viral-combined_k141_62438\|\|full | Low-quality |
| 18-109-final-viral-combined_k141_215858\|\|full | Not-determined |
| 18-109-final-viral-combined_k141_185226\|\|full | Not-determined |
| 18-109-final-viral-combined_k141_236209\|\|full | Not-determined |
| 18-109-final-viral-combined_k141_11473\|\|full | Low-quality |
| 18-109-final-viral-combined_k141_154670\|\|full | Not-determined |
| 18-109-final-viral-combined_k141_185228\|\|full | Low-quality |
| 18-109-final-viral-combined_k141_62448\|\|full | Low-quality |
| 18-109-final-viral-combined_k141_31970\|\|full | Not-determined |
| 18-109-final-viral-combined_k141_143953\|\|full | Low-quality |
| 18-109-final-viral-combined_k141_113747\|\|full | Low-quality |
| 18-109-final-viral-combined_k141_11476\|\|full | Not-determined |
| 18-109-final-viral-combined_k141_52330\|\|full | Not-determined |
| 18-109-final-viral-combined_k141_62455\|\|full | Not-determined |
| 18-109-final-viral-combined_k141_41915\|\|full | Low-quality |
| 18-109-final-viral-combined_k141_72206\|\|full | Not-determined |
| 18-109-final-viral-combined_k141_113762\|\|full | Low-quality |
| 18-109-final-viral-combined_k141_226036\|\|full | Low-quality |
| 18-109-final-viral-combined_k141_123839\|\|full | Not-determined |
| 18-109-final-viral-combined_k141_215888\|\|full | Not-determined |
| 18-109-final-viral-combined_k141_185264\|\|full | Low-quality |
| 18-109-final-viral-combined_k141_62474\|\|full | Low-quality |
| 18-109-final-viral-combined_k141_72240\|\|full | Low-quality |
| 18-109-final-viral-combined_k141_11494\|\|full | Low-quality |
| 18-109-final-viral-combined_k141_103227\|\|full | Low-quality |
| 18-109-final-viral-combined_k141_236219\|\|full | Low-quality |
| 18-109-final-viral-combined_k141_226062\|\|full | Not-determined |
| 18-109-final-viral-combined_k141_185300\|\|full | Low-quality |
| 18-109-final-viral-combined_k141_154718\|\|full | Not-determined |
| 18-109-final-viral-combined_k141_62492\|\|full | Not-determined |
| 18-109-final-viral-combined_k141_1338\|\|full | Not-determined |
| 18-109-final-viral-combined_k141_103239\|\|full | Not-determined |
| 18-109-final-viral-combined_k141_41940\|\|full | Low-quality |
| 18-109-final-viral-combined_k141_236237\|\|full | Low-quality |
| 18-109-final-viral-combined_k141_21386\|\|full | Not-determined |
| 18-109-final-viral-combined_k141_41942\|\|full | Not-determined |
| 18-109-final-viral-combined_k141_226090\|\|full | Not-determined |

| | |
|---|---|
| 18-109-final-viral-combined_k141_103252\|\|full | Not-determined |
| 18-109-final-viral-combined_k141_215915\|\|full | Not-determined |
| 18-109-final-viral-combined_k141_236248\|\|full | Low-quality |
| 18-109-final-viral-combined_k141_236250\|\|full | Low-quality |
| 18-109-final-viral-combined_k141_164821\|\|full | Not-determined |
| 18-109-final-viral-combined_k141_174916\|\|full | Not-determined |
| 18-109-final-viral-combined_k141_195812\|\|full | Not-determined |
| 18-109-final-viral-combined_k141_236251\|\|full | Low-quality |
| 18-109-final-viral-combined_k141_226112\|\|full | Not-determined |
| 18-109-final-viral-combined_k141_143986\|\|full | Not-determined |
| 18-109-final-viral-combined_k141_164830\|\|full | Low-quality |
| 18-109-final-viral-combined_k141_195837\|\|full | Low-quality |
| 18-109-final-viral-combined_k141_164832\|\|full | Low-quality |
| 18-109-final-viral-combined_k141_143996\|\|full | Low-quality |
| 18-109-final-viral-combined_k141_174925\|\|full | Not-determined |
| 18-109-final-viral-combined_k141_134212\|\|full | Low-quality |
| 18-109-final-viral-combined_k141_134215\|\|full | Not-determined |
| 18-109-final-viral-combined_k141_134223\|\|full | Not-determined |
| 18-109-final-viral-combined_k141_144016\|\|full | Low-quality |
| 18-109-final-viral-combined_k141_92885\|\|full | Low-quality |
| 18-109-final-viral-combined_k141_72282\|\|full | Low-quality |
| 18-109-final-viral-combined_k141_52355\|\|full | Not-determined |
| 18-109-final-viral-combined_k141_154790\|\|full | Not-determined |
| 18-109-final-viral-combined_k141_226147\|\|full | Low-quality |
| 18-109-final-viral-combined_k141_154791\|\|full | Not-determined |
| 18-109-final-viral-combined_k141_52370\|\|full | Low-quality |
| 18-109-final-viral-combined_k141_21436\|\|full | Not-determined |
| 18-109-final-viral-combined_k141_21439\|\|full | Not-determined |
| 18-109-final-viral-combined_k141_164889\|\|full | Low-quality |
| 18-109-final-viral-combined_k141_164890\|\|full | Not-determined |
| 18-109-final-viral-combined_k141_226157\|\|full | Not-determined |
| 18-109-final-viral-combined_k141_41950\|\|full | Not-determined |
| 18-109-final-viral-combined_k141_185382\|\|full | Not-determined |
| 18-109-final-viral-combined_k141_62551\|\|full | Not-determined |
| 18-109-final-viral-combined_k141_226165\|\|full | Not-determined |
| 18-109-final-viral-combined_k141_103307\|\|full | Low-quality |
| 18-109-final-viral-combined_k141_185384\|\|full | Not-determined |
| 18-109-final-viral-combined_k141_72290\|\|full | Low-quality |
| 18-109-final-viral-combined_k141_113782\|\|full | Not-determined |
| 18-109-final-viral-combined_k141_154810\|\|full | Not-determined |
| 18-109-final-viral-combined_k141_72295\|\|full | Not-determined |
| 18-109-final-viral-combined_k141_164908\|\|full | Not-determined |
| 18-109-final-viral-combined_k141_72297\|\|full | Not-determined |
| 18-109-final-viral-combined_k141_113795\|\|full | Not-determined |
| 18-109-final-viral-combined_k141_72300\|\|full | Not-determined |
| 18-109-final-viral-combined_k141_123870\|\|full | Not-determined |
| 18-109-final-viral-combined_k141_72303\|\|full | Low-quality |
| 18-109-final-viral-combined_k141_226173\|\|full | Low-quality |
| 18-109-final-viral-combined_k141_185418\|\|full | Not-determined |
| 18-109-final-viral-combined_k141_236302\|\|full | Low-quality |
| 18-109-final-viral-combined_k141_72305\|\|full | Low-quality |
| 18-109-final-viral-combined_k141_123878\|\|full | Low-quality |
| 18-109-final-viral-combined_k141_226195\|\|full | Not-determined |
| 18-109-final-viral-combined_k141_32078\|\|full | Not-determined |
| 18-109-final-viral-combined_k141_134296\|\|full | Low-quality |
| 17-109_k141_401366 | Complete |
| 18-109-final-viral-combined_k141_62583\|\|full | Low-quality |
| 18-109-final-viral-combined_k141_123909\|\|full | Low-quality |
| 18-109-final-viral-combined_k141_236347\|\|full | Low-quality |
| 18-109-final-viral-combined_k141_113827\|\|full | Low-quality |
| 18-109-final-viral-combined_k141_164934\|\|full | Not-determined |
| 18-109-final-viral-combined_k141_32103\|\|full | Low-quality |
| 18-109-final-viral-combined_k141_164950\|\|full | Not-determined |
| 18-109-final-viral-combined_k141_62597\|\|full | Not-determined |
| 18-109-final-viral-combined_k141_205832\|\|full | Not-determined |
| 18-109-final-viral-combined_k141_123922\|\|full | Low-quality |
| 18-109-final-viral-combined_k141_52449\|\|full | Not-determined |
| 18-109-final-viral-combined_k141_195842\|\|full | Low-quality |
| 18-109-final-viral-combined_k141_92921\|\|full | Not-determined |
| 18-109-final-viral-combined_k141_32114\|\|full | Not-determined |
| 18-109-final-viral-combined_k141_154852\|\|full | Not-determined |
| 18-109-final-viral-combined_k141_226227\|\|full | Not-determined |
| 18-109-final-viral-combined_k141_52452\|\|full | Not-determined |
| 18-109-final-viral-combined_k141_195846\|\|full | Low-quality |
| 18-109-final-viral-combined_k141_1407\|\|full | Not-determined |
| 18-109-final-viral-combined_k141_92926\|\|full | Not-determined |
| 18-109-final-viral-combined_k141_52459\|\|full | Low-quality |
| 18-109-final-viral-combined_k141_164969\|\|full | Not-determined |
| 18-109-final-viral-combined_k141_236361\|\|full | Not-determined |
| 18-109-final-viral-combined_k141_144094\|\|full | Low-quality |
| 18-109-final-viral-combined_k141_164979\|\|full | Not-determined |
| 18-109-final-viral-combined_k141_134324\|\|full | Not-determined |
| 18-109-final-viral-combined_k141_52468\|\|full | Not-determined |
| 18-109-final-viral-combined_k141_154862\|\|full | Low-quality |
| 18-109-final-viral-combined_k141_11544\|\|full | Not-determined |
| 18-109-final-viral-combined_k141_123939\|\|full | Not-determined |
| 18-109-final-viral-combined_k141_144112\|\|full | Not-determined |
| 18-109-final-viral-combined_k141_174974\|\|full | Not-determined |
| 18-109-final-viral-combined_k141_154871\|\|full | Low-quality |
| 18-109-final-viral-combined_k141_113847\|\|full | Not-determined |
| 18-109-final-viral-combined_k141_226248\|\|full | Not-determined |
| 18-109-final-viral-combined_k141_52498\|\|full | Not-determined |
| 18-109-final-viral-combined_k141_164983\|\|full | Low-quality |
| 18-109-final-viral-combined_k141_236380\|\|full | Low-quality |
| 18-109-final-viral-combined_k141_113896\|\|full | Not-determined |
| 18-109-final-viral-combined_k141_72382\|\|full | Low-quality |
| 18-109-final-viral-combined_k141_21505\|\|full | Not-determined |
| 18-109-final-viral-combined_k141_82894\|\|full | Not-determined |
| KM-final-viral-combined_k141_253499\|\|full | High-quality |
| 18-109-final-viral-combined_k141_134351\|\|full | Low-quality |
| 18-109-final-viral-combined_k141_123957\|\|full | Not-determined |
| 18-109-final-viral-combined_k141_72393\|\|full | Low-quality |
| 18-109-final-viral-combined_k141_62657\|\|full | Not-determined |
| 18-109-final-viral-combined_k141_134357\|\|full | Not-determined |
| 18-109-final-viral-combined_k141_154922\|\|full | Not-determined |
| 18-109-final-viral-combined_k141_82905\|\|full | Not-determined |
| 18-109-final-viral-combined_k141_52567\|\|full | Low-quality |
| 18-109-final-viral-combined_k141_82909\|\|full | Low-quality |
| 18-109-final-viral-combined_k141_52578\|\|full | Not-determined |
| 18-109-final-viral-combined_k141_62667\|\|full | Not-determined |
| 18-109-final-viral-combined_k141_185576\|\|full | Not-determined |
| 18-109-final-viral-combined_k141_226280\|\|full | Low-quality |
| 18-109-final-viral-combined_k141_205879\|\|full | Low-quality |
| 18-109-final-viral-combined_k141_144198\|\|full | Not-determined |
| 18-109-final-viral-combined_k141_226298\|\|full | Low-quality |
| 18-109-final-viral-combined_k141_113932\|\|full | Low-quality |
| 18-109-final-viral-combined_k141_236427\|\|full | Low-quality |
| 18-109-final-viral-combined_k141_1419\|\|full | Not-determined |
| 18-109-final-viral-combined_k141_103458\|\|full | Not-determined |
| 18-109-final-viral-combined_k141_1431\|\|full | Low-quality |
| 18-109-final-viral-combined_k141_72443\|\|full | Low-quality |
| 18-109-final-viral-combined_k141_82948\|\|full | Not-determined |
| 18-109-final-viral-combined_k141_185613\|\|full | Not-determined |
| 18-109-final-viral-combined_k141_52598\|\|full | Not-determined |
| 18-109-final-viral-combined_k141_123974\|\|full | Not-determined |
| 18-109-final-viral-combined_k141_72454\|\|full | Not-determined |
| 18-109-final-viral-combined_k141_205914\|\|full | Low-quality |
| 18-109-final-viral-combined_k141_123991\|\|full | Low-quality |
| 18-109-final-viral-combined_k141_185643\|\|full | Low-quality |
| 18-109-final-viral-combined_k141_236451\|\|full | Not-determined |
| 18-109-final-viral-combined_k141_165037\|\|full | Low-quality |
| 18-109-final-viral-combined_k141_165039\|\|full | Low-quality |
| 18-109-final-viral-combined_k141_134450\|\|full | Low-quality |
| 18-109-final-viral-combined_k141_154976\|\|full | Not-determined |
| 18-109-final-viral-combined_k141_195911\|\|full | Not-determined |
| 18-109-final-viral-combined_k141_114008\|\|full | Not-determined |
| 18-109-final-viral-combined_k141_42140\|\|full | Low-quality |
| 18-109-final-viral-combined_k141_124018\|\|full | Low-quality |
| 18-109-final-viral-combined_k141_226368\|\|full | Low-quality |
| 18-109-final-viral-combined_k141_226369\|\|full | Low-quality |
| 18-109-final-viral-combined_k141_82953\|\|full | Not-determined |
| 18-109-final-viral-combined_k141_52633\|\|full | Low-quality |
| 18-109-final-viral-combined_k141_1499\|\|full | Not-determined |
| 18-109-final-viral-combined_k141_195955\|\|full | Low-quality |
| 18-109-final-viral-combined_k141_175098\|\|full | Not-determined |
| 18-109-final-viral-combined_k141_124059\|\|full | Not-determined |
| 18-109-final-viral-combined_k141_92971\|\|full | Not-determined |
| 18-109-final-viral-combined_k141_52642\|\|full | Low-quality |
| 18-109-final-viral-combined_k141_92972\|\|full | Not-determined |
| 18-109-final-viral-combined_k141_155024\|\|full | Not-determined |
| 18-109-final-viral-combined_k141_185715\|\|full | Not-determined |
| 18-109-final-viral-combined_k141_236536\|\|full | Not-determined |
| 18-109-final-viral-combined_k141_175115\|\|full | Low-quality |
| 18-109-final-viral-combined_k141_185729\|\|full | Low-quality |
| 18-109-final-viral-combined_k141_42177\|\|full | Not-determined |
| 18-109-final-viral-combined_k141_236563\|\|full | Low-quality |
| 18-109-final-viral-combined_k141_62796\|\|full | Not-determined |
| 18-109-final-viral-combined_k141_236564\|\|full | Low-quality |
| 18-109-final-viral-combined_k141_52680\|\|full | Low-quality |
| 17-109-final-viral-combined_k141_401366\|\|full | High-quality |
| 18-109-final-viral-combined_k141_103566\|\|full | Low-quality |
| 18-109-final-viral-combined_k141_236578\|\|full | Low-quality |
| 18-109-final-viral-combined_k141_175140\|\|full | Low-quality |
| 18-109-final-viral-combined_k141_114092\|\|full | Low-quality |
| 18-109-final-viral-combined_k141_226420\|\|full | Not-determined |
| 18-109-final-viral-combined_k141_32259\|\|full | Not-determined |
| 18-109-final-viral-combined_k141_226427\|\|full | Not-determined |
| 18-109-final-viral-combined_k141_175162\|\|full | Not-determined |
| 18-109-final-viral-combined_k141_165064\|\|full | Not-determined |
| 18-109-final-viral-combined_k141_236602\|\|full | Low-quality |
| 18-109-final-viral-combined_k141_1518\|\|full | Not-determined |
| 18-109-final-viral-combined_k141_236613\|\|full | Low-quality |
| 18-109-final-viral-combined_k141_175184\|\|full | Not-determined |
| 18-109-final-viral-combined_k141_83016\|\|full | Low-quality |
| 18-109-final-viral-combined_k141_93029\|\|full | Low-quality |
| 18-109-final-viral-combined_k141_226465\|\|full | Low-quality |
| 18-109-final-viral-combined_k141_1527\|\|full | Not-determined |
| 18-109-final-viral-combined_k141_103626\|\|full | Not-determined |
| 18-109-final-viral-combined_k141_32311\|\|full | Not-determined |
| 18-109-final-viral-combined_k141_165095\|\|full | Not-determined |
| 18-109-final-viral-combined_k141_175196\|\|full | Low-quality |
| 18-109-final-viral-combined_k141_52783\|\|full | Not-determined |
| KM-final-viral-combined_k141_233761\|\|full | Medium-quality |
| 18-109-final-viral-combined_k141_165100\|\|full | Low-quality |
| 18-109-final-viral-combined_k141_185773\|\|full | Low-quality |
| 18-109-final-viral-combined_k141_93067\|\|full | Not-determined |
| 18-109-final-viral-combined_k141_114146\|\|full | Low-quality |
| 18-109-final-viral-combined_k141_114153\|\|full | Low-quality |
| 18-109-final-viral-combined_k141_155056\|\|full | Not-determined |
| 18-109-final-viral-combined_k141_103660\|\|full | Not-determined |
| 18-109-final-viral-combined_k141_134467\|\|full | Low-quality |
| 18-109-final-viral-combined_k141_124108\|\|full | Low-quality |
| 18-109-final-viral-combined_k141_32340\|\|full | Not-determined |
| 18-109-final-viral-combined_k141_1558\|\|full | Low-quality |
| 18-109-final-viral-combined_k141_124111\|\|full | Not-determined |
| 18-109-final-viral-combined_k141_124115\|\|full | Not-determined |
| 18-109-final-viral-combined_k141_236651\|\|full | Not-determined |
| 18-109-final-viral-combined_k141_226473\|\|full | Low-quality |
| 18-109-final-viral-combined_k141_165138\|\|full | Low-quality |
| 18-109-final-viral-combined_k141_144229\|\|full | Low-quality |
| 18-109-final-viral-combined_k141_114199\|\|full | Low-quality |
| 18-109-final-viral-combined_k141_226484\|\|full | Low-quality |
| 18-109-final-viral-combined_k141_42271\|\|full | Low-quality |
| 18-109-final-viral-combined_k141_1582\|\|full | Low-quality |
| 18-109-final-viral-combined_k141_165147\|\|full | Not-determined |
| 18-109-final-viral-combined_k141_226492\|\|full | Low-quality |
| 18-109-final-viral-combined_k141_32355\|\|full | Not-determined |
| 18-109-final-viral-combined_k141_155093\|\|full | Low-quality |
| 18-109-final-viral-combined_k141_103674\|\|full | Not-determined |
| 18-109-final-viral-combined_k141_226500\|\|full | Low-quality |
| 18-109-final-viral-combined_k141_124145\|\|full | Not-determined |
| 18-109-final-viral-combined_k141_134520\|\|full | Low-quality |
| 18-109-final-viral-combined_k141_165172\|\|full | Not-determined |
| 18-109-final-viral-combined_k141_42307\|\|full | Low-quality |
| 18-109-final-viral-combined_k141_226508\|\|full | Not-determined |
| 18-109-final-viral-combined_k141_236663\|\|full | Low-quality |
| 18-109-final-viral-combined_k141_196022\|\|full | Low-quality |
| 18-109-final-viral-combined_k141_155112\|\|full | Low-quality |
| 18-109-final-viral-combined_k141_144289\|\|full | Low-quality |
| 18-109-final-viral-combined_k141_206166\|\|full | Not-determined |
| 18-109-final-viral-combined_k141_155142\|\|full | Low-quality |
| 18-109-final-viral-combined_k141_83046\|\|full | Not-determined |
| 18-109-final-viral-combined_k141_216103\|\|full | Not-determined |
| 18-109-final-viral-combined_k141_226532\|\|full | Not-determined |
| 18-109-final-viral-combined_k141_206194\|\|full | Not-determined |
| 18-109-final-viral-combined_k141_52860\|\|full | Low-quality |
| 18-109-final-viral-combined_k141_144326\|\|full | Not-determined |
| 18-109-final-viral-combined_k141_52866\|\|full | Low-quality |
| 18-109-final-viral-combined_k141_144328\|\|full | Low-quality |
| 18-109-final-viral-combined_k141_103703\|\|full | Low-quality |
| 18-109-final-viral-combined_k141_185866\|\|full | Not-determined |
| 18-109-final-viral-combined_k141_216126\|\|full | Low-quality |
| 18-109-final-viral-combined_k141_52903\|\|full | Low-quality |
| 18-109-final-viral-combined_k141_216129\|\|full | Low-quality |
| 18-109-final-viral-combined_k141_124203\|\|full | Not-determined |
| 18-109-final-viral-combined_k141_72507\|\|full | Not-determined |
| 18-109-final-viral-combined_k141_236680\|\|full | Not-determined |
| 18-109-final-viral-combined_k141_124215\|\|full | Not-determined |
| 18-109-final-viral-combined_k141_236689\|\|full | Low-quality |
| 18-109-final-viral-combined_k141_226588\|\|full | Not-determined |
| 18-109-final-viral-combined_k141_124227\|\|full | Not-determined |
| 18-109-final-viral-combined_k141_226590\|\|full | Not-determined |
| 18-109-final-viral-combined_k141_216160\|\|full | Low-quality |
| 18-109-final-viral-combined_k141_236715\|\|full | Low-quality |
| 18-109-final-viral-combined_k141_124231\|\|full | Not-determined |
| 18-109-final-viral-combined_k141_236716\|\|full | Not-determined |
| 18-109-final-viral-combined_k141_236722\|\|full | Not-determined |
| 18-109-final-viral-combined_k141_32458\|\|full | Low-quality |
| 18-109-final-viral-combined_k141_21619\|\|full | Low-quality |
| 18-109-final-viral-combined_k141_103754\|\|full | Low-quality |
| 18-109-final-viral-combined_k141_165285\|\|full | Low-quality |
| 18-109-final-viral-combined_k141_196058\|\|full | Not-determined |
| 18-109-final-viral-combined_k141_103764\|\|full | Not-determined |
| 18-109-final-viral-combined_k141_21643\|\|full | Low-quality |
| 18-109-final-viral-combined_k141_114294\|\|full | Not-determined |
| 18-109-final-viral-combined_k141_72514\|\|full | Not-determined |
| 18-109-final-viral-combined_k141_196079\|\|full | Not-determined |
| 18-109-final-viral-combined_k141_11690\|\|full | Low-quality |
| 18-109-final-viral-combined_k141_21661\|\|full | Not-determined |
| 18-109-final-viral-combined_k141_72522\|\|full | Not-determined |
| 18-109-final-viral-combined_k141_1665\|\|full | Low-quality |
| 18-109-final-viral-combined_k141_11700\|\|full | Not-determined |
| 18-109-final-viral-combined_k141_42391\|\|full | Low-quality |
| 18-109-final-viral-combined_k141_42394\|\|full | Not-determined |
| 18-109-final-viral-combined_k141_114306\|\|full | Low-quality |
| 18-109-final-viral-combined_k141_236789\|\|full | Not-determined |
| 18-109-final-viral-combined_k141_1690\|\|full | Low-quality |
| 18-109-final-viral-combined_k141_196091\|\|full | Low-quality |
| 18-109-final-viral-combined_k141_196100\|\|full | Low-quality |
| 18-109-final-viral-combined_k141_103790\|\|full | Not-determined |
| 18-109-final-viral-combined_k141_124308\|\|full | Not-determined |
| 18-109-final-viral-combined_k141_185924\|\|full | Not-determined |
| 18-109-final-viral-combined_k141_1717\|\|full | Low-quality |
| 18-109-final-viral-combined_k141_103808\|\|full | Not-determined |
| 18-109-final-viral-combined_k141_144350\|\|full | Not-determined |
| 18-109-final-viral-combined_k141_124328\|\|full | Not-determined |
| 18-109-final-viral-combined_k141_32505\|\|full | Low-quality |
| 18-109-final-viral-combined_k141_42443\|\|full | Low-quality |
| 18-109-final-viral-combined_k141_206311\|\|full | Not-determined |
| 17-108-final-viral-combined_k141_304239\|\|full | High-quality |
| 18-109-final-viral-combined_k141_206314\|\|full | Not-determined |
| 18-109-final-viral-combined_k141_206317\|\|full | Not-determined |
| 18-109-final-viral-combined_k141_21706\|\|full | Low-quality |
| 18-109-final-viral-combined_k141_21709\|\|full | Low-quality |
| 18-109-final-viral-combined_k141_103832\|\|full | Low-quality |
| 18-109-final-viral-combined_k141_206324\|\|full | Not-determined |
| 18-109-final-viral-combined_k141_236851\|\|full | Low-quality |
| 18-109-final-viral-combined_k141_21711\|\|full | Low-quality |
| 18-109-final-viral-combined_k141_144388\|\|full | Not-determined |
| 18-109-final-viral-combined_k141_226673\|\|full | Low-quality |
| 18-109-final-viral-combined_k141_144401\|\|full | Not-determined |
| 18-109-final-viral-combined_k141_63003\|\|full | Low-quality |
| 18-109-final-viral-combined_k141_32552\|\|full | Not-determined |
| 18-109-final-viral-combined_k141_206342\|\|full | Not-determined |
| 18-109-final-viral-combined_k141_32554\|\|full | Low-quality |
| 18-109-final-viral-combined_k141_32560\|\|full | Not-determined |
| 18-109-final-viral-combined_k141_226704\|\|full | Not-determined |
| 18-109-final-viral-combined_k141_114335\|\|full | Not-determined |
| 18-109-final-viral-combined_k141_114338\|\|full | Not-determined |
| 18-109-final-viral-combined_k141_175378\|\|full | Not-determined |
| 18-109-final-viral-combined_k141_196105\|\|full | Low-quality |
| 18-109-final-viral-combined_k141_226729\|\|full | Not-determined |
| 18-109-final-viral-combined_k141_226730\|\|full | Low-quality |
| 18-109-final-viral-combined_k141_103901\|\|full | Not-determined |
| 18-109-final-viral-combined_k141_83185\|\|full | Low-quality |
| 18-109-final-viral-combined_k141_83187\|\|full | Not-determined |
| 18-109-final-viral-combined_k141_103916\|\|full | Low-quality |
| 18-109-final-viral-combined_k141_53017\|\|full | Not-determined |
| 18-109-final-viral-combined_k141_93265\|\|full | Low-quality |
| 18-109-final-viral-combined_k141_114379\|\|full | Not-determined |
| 18-109-final-viral-combined_k141_83213\|\|full | Not-determined |
| 18-109-final-viral-combined_k141_1795\|\|full | Low-quality |
| 18-109-final-viral-combined_k141_175436\|\|full | Low-quality |
| 18-109-final-viral-combined_k141_114395\|\|full | Not-determined |
| 18-109-final-viral-combined_k141_21762\|\|full | Not-determined |
| 18-109-final-viral-combined_k141_114399\|\|full | Not-determined |
| 18-109-final-viral-combined_k141_42522\|\|full | Not-determined |
| 18-109-final-viral-combined_k141_175465\|\|full | Low-quality |
| 18-109-final-viral-combined_k141_63021\|\|full | Not-determined |
| 18-109-final-viral-combined_k141_11742\|\|full | Low-quality |
| 18-109-final-viral-combined_k141_53072\|\|full | Not-determined |
| 18-109-final-viral-combined_k141_103940\|\|full | Low-quality |
| 18-109-final-viral-combined_k141_144420\|\|full | Low-quality |
| 18-109-final-viral-combined_k141_186027\|\|full | Low-quality |
| 18-109-final-viral-combined_k141_114429\|\|full | Not-determined |
| 18-109-final-viral-combined_k141_1834\|\|full | Not-determined |
| 18-109-final-viral-combined_k141_114432\|\|full | Low-quality |
| 18-109-final-viral-combined_k141_134632\|\|full | Not-determined |
| 18-109-final-viral-combined_k141_124462\|\|full | Not-determined |
| 18-109-final-viral-combined_k141_155198\|\|full | Not-determined |
| 18-109-final-viral-combined_k141_206439\|\|full | Low-quality |
| 18-109-final-viral-combined_k141_72592\|\|full | Not-determined |
| 18-109-final-viral-combined_k141_155208\|\|full | Not-determined |
| 18-109-final-viral-combined_k141_216260\|\|full | Not-determined |
| 18-109-final-viral-combined_k141_186047\|\|full | Low-quality |
| 18-109-final-viral-combined_k141_104007\|\|full | Low-quality |
| 18-109-final-viral-combined_k141_216272\|\|full | Not-determined |
| 18-109-final-viral-combined_k141_11815\|\|full | Low-quality |
| 18-109-final-viral-combined_k141_155223\|\|full | Low-quality |
| 18-109-final-viral-combined_k141_155227\|\|full | Low-quality |
| 18-109-final-viral-combined_k141_104010\|\|full | Not-determined |
| 18-109-final-viral-combined_k141_53124\|\|full | Low-quality |
| 18-109-final-viral-combined_k141_124505\|\|full | Low-quality |
| 18-109-final-viral-combined_k141_32671\|\|full | Not-determined |
| 18-109-final-viral-combined_k141_53148\|\|full | Not-determined |
| 18-109-final-viral-combined_k141_32682\|\|full | Low-quality |
| 18-109-final-viral-combined_k141_155247\|\|full | Not-determined |
| 18-109-final-viral-combined_k141_196228\|\|full | Low-quality |
| 18-109-final-viral-combined_k141_144496\|\|full | Not-determined |
| 18-109-final-viral-combined_k141_206514\|\|full | Not-determined |
| 18-109-final-viral-combined_k141_124567\|\|full | Not-determined |
| 18-109-final-viral-combined_k141_32734\|\|full | Not-determined |
| 18-109-final-viral-combined_k141_114456\|\|full | Not-determined |
| 18-109-final-viral-combined_k141_72627\|\|full | Not-determined |
| 18-109-final-viral-combined_k141_124590\|\|full | Not-determined |
| 18-109-final-viral-combined_k141_83249\|\|full | Not-determined |
| 18-109-final-viral-combined_k141_42665\|\|full | Low-quality |
| 18-109-final-viral-combined_k141_237073\|\|full | Not-determined |
| 18-109-final-viral-combined_k141_155304\|\|full | Not-determined |
| 18-109-final-viral-combined_k141_155305\|\|full | Low-quality |
| 18-109-final-viral-combined_k141_104078\|\|full | Not-determined |
| 18-109-final-viral-combined_k141_216341\|\|full | Low-quality |
| 18-109-final-viral-combined_k141_53197\|\|full | Low-quality |
| 18-109-final-viral-combined_k141_83292\|\|full | Low-quality |
| 18-109-final-viral-combined_k141_83294\|\|full | Low-quality |
| 18-109-final-viral-combined_k141_42685\|\|full | Not-determined |
| 18-109-final-viral-combined_k141_216358\|\|full | Low-quality |
| 18-109-final-viral-combined_k141_216372\|\|full | Low-quality |
| 18-109-final-viral-combined_k141_226848\|\|full | Low-quality |
| 18-109-final-viral-combined_k141_72653\|\|full | Not-determined |
| 18-109-final-viral-combined_k141_134692\|\|full | Not-determined |
| 18-109-final-viral-combined_k141_216381\|\|full | Low-quality |
| 18-109-final-viral-combined_k141_32773\|\|full | Low-quality |
| 18-109-final-viral-combined_k141_104106\|\|full | Not-determined |
| 18-109-final-viral-combined_k141_63112\|\|full | Not-determined |
| 18-109-final-viral-combined_k141_134703\|\|full | Low-quality |
| 18-109-final-viral-combined_k141_104120\|\|full | Low-quality |
| 18-109-final-viral-combined_k141_1923\|\|full | Low-quality |
| 18-109-final-viral-combined_k141_72664\|\|full | Not-determined |
| 18-109-final-viral-combined_k141_104121\|\|full | Low-quality |
| 18-109-final-viral-combined_k141_1925\|\|full | Not-determined |
| 18-109-final-viral-combined_k141_155348\|\|full | Low-quality |
| 18-109-final-viral-combined_k141_216408\|\|full | Low-quality |
| 18-109-final-viral-combined_k141_63124\|\|full | Not-determined |
| 18-109-final-viral-combined_k141_134733\|\|full | Low-quality |
| 18-109-final-viral-combined_k141_32777\|\|full | Low-quality |
| 18-109-final-viral-combined_k141_155360\|\|full | Not-determined |
| 18-109-final-viral-combined_k141_1944\|\|full | Not-determined |
| 18-109-final-viral-combined_k141_144551\|\|full | Low-quality |
| 18-109-final-viral-combined_k141_1951\|\|full | Not-determined |
| 18-109-final-viral-combined_k141_206601\|\|full | Not-determined |
| 18-109-final-viral-combined_k141_165386\|\|full | Low-quality |
| 18-109-final-viral-combined_k141_1952\|\|full | Low-quality |
| 18-109-final-viral-combined_k141_63144\|\|full | Not-determined |
| 18-109-final-viral-combined_k141_21914\|\|full | Low-quality |
| 18-109-final-viral-combined_k141_155383\|\|full | Low-quality |
| 18-109-final-viral-combined_k141_144574\|\|full | Not-determined |
| 18-109-final-viral-combined_k141_11997\|\|full | Low-quality |
| 18-109-final-viral-combined_k141_144586\|\|full | Not-determined |
| 18-109-final-viral-combined_k141_42766\|\|full | Low-quality |
| 18-109-final-viral-combined_k141_134777\|\|full | Low-quality |
| 18-109-final-viral-combined_k141_226924\|\|full | Low-quality |
| 18-109-final-viral-combined_k141_155388\|\|full | Low-quality |
| 18-109-final-viral-combined_k141_155391\|\|full | Not-determined |
| 18-109-final-viral-combined_k141_72692\|\|full | Low-quality |
| 18-109-final-viral-combined_k141_21929\|\|full | Low-quality |
| 18-109-final-viral-combined_k141_72695\|\|full | Low-quality |
| 18-109-final-viral-combined_k141_216438\|\|full | Not-determined |
| 18-109-final-viral-combined_k141_175548\|\|full | Not-determined |
| 18-109-final-viral-combined_k141_196284\|\|full | Low-quality |
| 18-109-final-viral-combined_k141_226940\|\|full | Low-quality |
| 18-109-final-viral-combined_k141_144591\|\|full | Not-determined |
| 18-109-final-viral-combined_k141_12023\|\|full | Low-quality |
| 18-109-final-viral-combined_k141_72709\|\|full | Low-quality |
| 18-109-final-viral-combined_k141_53207\|\|full | Low-quality |
| 18-109-final-viral-combined_k141_53209\|\|full | Not-determined |
| 18-109-final-viral-combined_k141_72731\|\|full | Low-quality |
| 18-109-final-viral-combined_k141_53214\|\|full | Low-quality |
| 18-109-final-viral-combined_k141_12044\|\|full | Low-quality |
| 18-109-final-viral-combined_k141_83366\|\|full | Not-determined |
| 18-109-final-viral-combined_k141_83367\|\|full | Not-determined |
| 18-109-final-viral-combined_k141_72737\|\|full | Low-quality |
| 18-109-final-viral-combined_k141_104147\|\|full | Not-determined |
| 18-109-final-viral-combined_k141_72744\|\|full | Low-quality |
| 18-109-final-viral-combined_k141_12052\|\|full | Low-quality |
| 18-109-final-viral-combined_k141_165438\|\|full | Not-determined |
| 18-109-final-viral-combined_k141_227001\|\|full | Not-determined |
| 18-109-final-viral-combined_k141_237122\|\|full | Not-determined |
| 18-109-final-viral-combined_k141_12066\|\|full | Low-quality |
| 18-109-final-viral-combined_k141_227005\|\|full | Not-determined |
| 18-109-final-viral-combined_k141_206657\|\|full | Not-determined |
| 18-109-final-viral-combined_k141_216453\|\|full | Not-determined |
| 18-109-final-viral-combined_k141_155445\|\|full | Not-determined |
| 18-109-final-viral-combined_k141_114601\|\|full | Not-determined |
| 18-109-final-viral-combined_k141_12084\|\|full | Not-determined |
| 18-109-final-viral-combined_k141_165453\|\|full | Low-quality |
| 18-109-final-viral-combined_k141_22011\|\|full | Low-quality |
| 18-109-final-viral-combined_k141_206670\|\|full | Not-determined |
| 18-109-final-viral-combined_k141_72754\|\|full | Not-determined |
| 18-109-final-viral-combined_k141_155472\|\|full | Not-determined |
| 18-109-final-viral-combined_k141_196358\|\|full | Not-determined |
| 18-109-final-viral-combined_k141_72762\|\|full | Low-quality |
| 18-109-final-viral-combined_k141_196362\|\|full | Low-quality |
| 18-109-final-viral-combined_k141_22029\|\|full | Not-determined |
| 18-109-final-viral-combined_k141_196363\|\|full | Not-determined |
| 18-109-final-viral-combined_k141_104208\|\|full | Not-determined |
| 18-109-final-viral-combined_k141_22030\|\|full | Not-determined |
| 18-109-final-viral-combined_k141_144634\|\|full | Not-determined |
| 18-109-final-viral-combined_k141_12112\|\|full | Low-quality |
| 18-109-final-viral-combined_k141_227033\|\|full | Low-quality |
| 18-109-final-viral-combined_k141_104214\|\|full | Not-determined |
| 18-109-final-viral-combined_k141_32821\|\|full | Low-quality |
| 18-109-final-viral-combined_k141_22047\|\|full | Not-determined |
| 18-109-final-viral-combined_k141_2050\|\|full | Not-determined |
| 18-109-final-viral-combined_k141_104262\|\|full | Low-quality |
| 18-109-final-viral-combined_k141_32841\|\|full | Low-quality |
| 18-109-final-viral-combined_k141_2066\|\|full | Not-determined |
| 18-109-final-viral-combined_k141_124679\|\|full | Low-quality |
| 18-109-final-viral-combined_k141_22049\|\|full | Not-determined |
| 18-109-final-viral-combined_k141_186225\|\|full | Not-determined |
| 18-109-final-viral-combined_k141_2073\|\|full | Not-determined |
| 18-109-final-viral-combined_k141_227065\|\|full | Low-quality |
| 18-109-final-viral-combined_k141_63284\|\|full | Not-determined |
| 18-109-final-viral-combined_k141_124704\|\|full | Low-quality |
| 18-109-final-viral-combined_k141_114637\|\|full | Not-determined |
| 18-109-final-viral-combined_k141_227077\|\|full | Low-quality |
| 18-109-final-viral-combined_k141_186253\|\|full | Low-quality |
| 18-109-final-viral-combined_k141_155484\|\|full | Not-determined |
| 18-109-final-viral-combined_k141_93376\|\|full | Low-quality |
| 18-109-final-viral-combined_k141_53330\|\|full | Low-quality |
| 18-109-final-viral-combined_k141_186271\|\|full | Not-determined |
| 18-109-final-viral-combined_k141_155501\|\|full | Not-determined |
| 18-109-final-viral-combined_k141_63296\|\|full | Not-determined |
| 18-109-final-viral-combined_k141_63297\|\|full | Not-determined |
| 18-109-final-viral-combined_k141_196453\|\|full | Not-determined |
| 18-109-final-viral-combined_k141_155516\|\|full | Low-quality |
| 18-109-final-viral-combined_k141_72815\|\|full | Not-determined |
| 18-109-final-viral-combined_k141_134826\|\|full | Low-quality |
| 18-109-final-viral-combined_k141_63312\|\|full | Low-quality |
| 18-109-final-viral-combined_k141_63320\|\|full | Not-determined |
| 18-109-final-viral-combined_k141_72827\|\|full | Low-quality |
| 18-109-final-viral-combined_k141_63324\|\|full | Not-determined |
| 18-109-final-viral-combined_k141_83431\|\|full | Low-quality |
| 18-109-final-viral-combined_k141_165598\|\|full | Low-quality |
| 18-109-final-viral-combined_k141_63328\|\|full | Low-quality |
| 18-109-final-viral-combined_k141_216542\|\|full | Low-quality |
| 18-109-final-viral-combined_k141_155531\|\|full | Low-quality |
| 18-109-final-viral-combined_k141_83436\|\|full | Not-determined |
| 18-109-final-viral-combined_k141_134848\|\|full | Low-quality |
| 18-109-final-viral-combined_k141_12218\|\|full | Low-quality |
| 18-109-final-viral-combined_k141_63348\|\|full | Low-quality |
| 18-109-final-viral-combined_k141_63351\|\|full | Not-determined |
| 18-109-final-viral-combined_k141_134852\|\|full | Low-quality |
| 18-109-final-viral-combined_k141_72885\|\|full | Low-quality |
| 18-109-final-viral-combined_k141_63353\|\|full | Low-quality |
| 18-109-final-viral-combined_k141_93424\|\|full | Not-determined |
| 18-109-final-viral-combined_k141_104289\|\|full | Not-determined |
| KM-final-viral-combined_k141_72466\|\|full | Medium-quality |
| 18-109-final-viral-combined_k141_104295\|\|full | Low-quality |
| 18-109-final-viral-combined_k141_206830\|\|full | Not-determined |
| 18-109-final-viral-combined_k141_22167\|\|full | Low-quality |
| 18-109-final-viral-combined_k141_12233\|\|full | Low-quality |
| 18-109-final-viral-combined_k141_104301\|\|full | Low-quality |
| 18-109-final-viral-combined_k141_196516\|\|full | Low-quality |
| 18-109-final-viral-combined_k141_93442\|\|full | Low-quality |
| 18-109-final-viral-combined_k141_114641\|\|full | Low-quality |
| 18-109-final-viral-combined_k141_144647\|\|full | Not-determined |
| 18-109-final-viral-combined_k141_114643\|\|full | Not-determined |
| 18-109-final-viral-combined_k141_196537\|\|full | Not-determined |
| 18-109-final-viral-combined_k141_2151\|\|full | Not-determined |
| 18-109-final-viral-combined_k141_124833\|\|full | Low-quality |
| 18-109-final-viral-combined_k141_32900\|\|full | Not-determined |
| 18-109-final-viral-combined_k141_93462\|\|full | Low-quality |
| 18-109-final-viral-combined_k141_2163\|\|full | Not-determined |
| 18-109-final-viral-combined_k141_134906\|\|full | Low-quality |
| 18-109-final-viral-combined_k141_144671\|\|full | Low-quality |
| 18-109-final-viral-combined_k141_237207\|\|full | Low-quality |
| 18-109-final-viral-combined_k141_144674\|\|full | Not-determined |
| 18-109-final-viral-combined_k141_227165\|\|full | Not-determined |
| 18-109-final-viral-combined_k141_216631\|\|full | Low-quality |
| 18-109-final-viral-combined_k141_227167\|\|full | Low-quality |
| 18-109-final-viral-combined_k141_114672\|\|full | Not-determined |
| 18-109-final-viral-combined_k141_227183\|\|full | Low-quality |
| 18-109-final-viral-combined_k141_93489\|\|full | Low-quality |
| 18-109-final-viral-combined_k141_227187\|\|full | Low-quality |
| 18-109-final-viral-combined_k141_165715\|\|full | Low-quality |
| 18-109-final-viral-combined_k141_2186\|\|full | Not-determined |
| 18-109-final-viral-combined_k141_134938\|\|full | Low-quality |
| 18-109-final-viral-combined_k141_216661\|\|full | Low-quality |
| 18-109-final-viral-combined_k141_216666\|\|full | Low-quality |
| 18-109-final-viral-combined_k141_165723\|\|full | Low-quality |
| 18-102-final-viral-combined_k141_64692\|\|full | High-quality |
| 18-109-final-viral-combined_k141_2204\|\|full | Low-quality |
| 18-109-final-viral-combined_k141_22240\|\|full | Not-determined |
| 18-109-final-viral-combined_k141_22241\|\|full | Low-quality |
| 18-109-final-viral-combined_k141_104344\|\|full | Low-quality |
| 18-109-final-viral-combined_k141_104356\|\|full | Low-quality |
| 18-109-final-viral-combined_k141_104360\|\|full | Low-quality |
| 18-109-final-viral-combined_k141_114738\|\|full | Not-determined |
| 18-109-final-viral-combined_k141_22261\|\|full | Low-quality |
| 18-109-final-viral-combined_k141_155633\|\|full | Not-determined |
| 18-109-final-viral-combined_k141_155649\|\|full | Not-determined |
| 18-109-final-viral-combined_k141_237247\|\|full | Not-determined |
| 18-109-final-viral-combined_k141_114773\|\|full | Low-quality |
| 18-109-final-viral-combined_k141_186446\|\|full | Not-determined |
| 18-109-final-viral-combined_k141_227246\|\|full | Not-determined |
| 18-109-final-viral-combined_k141_144746\|\|full | Not-determined |
| 18-109-final-viral-combined_k141_144747\|\|full | Not-determined |
| 18-109-final-viral-combined_k141_53419\|\|full | Not-determined |
| 18-109-final-viral-combined_k141_237268\|\|full | Low-quality |
| 18-109-final-viral-combined_k141_227254\|\|full | Low-quality |
| 18-109-final-viral-combined_k141_43018\|\|full | Low-quality |
| 18-109-final-viral-combined_k141_93578\|\|full | Low-quality |
| 18-109-final-viral-combined_k141_216707\|\|full | Not-determined |
| 18-109-final-viral-combined_k141_63479\|\|full | Not-determined |
| 18-109-final-viral-combined_k141_83578\|\|full | Not-determined |
| 18-109-final-viral-combined_k141_237286\|\|full | Not-determined |
| 18-109-final-viral-combined_k141_144780\|\|full | Not-determined |
| 18-109-final-viral-combined_k141_237287\|\|full | Low-quality |
| 18-109-final-viral-combined_k141_216733\|\|full | Not-determined |
| 18-109-final-viral-combined_k141_2258\|\|full | Not-determined |
| 18-109-final-viral-combined_k141_114816\|\|full | Low-quality |
| 18-109-final-viral-combined_k141_237297\|\|full | Low-quality |
| 18-109-final-viral-combined_k141_2264\|\|full | Not-determined |
| 18-109-final-viral-combined_k141_216747\|\|full | Low-quality |
| 18-109-final-viral-combined_k141_114821\|\|full | Low-quality |
| 18-109-final-viral-combined_k141_12317\|\|full | Low-quality |
| 18-109-final-viral-combined_k141_216751\|\|full | Not-determined |
| 18-109-final-viral-combined_k141_227291\|\|full | Low-quality |
| 18-109-final-viral-combined_k141_72963\|\|full | Low-quality |
| 18-109-final-viral-combined_k141_43034\|\|full | Low-quality |
| 18-109-final-viral-combined_k141_155701\|\|full | Low-quality |
| 18-109-final-viral-combined_k141_12326\|\|full | Not-determined |
| 18-109-final-viral-combined_k141_216761\|\|full | Low-quality |
| 18-109-final-viral-combined_k141_72975\|\|full | Not-determined |
| 18-109-final-viral-combined_k141_2304\|\|full | Not-determined |
| 18-109-final-viral-combined_k141_155719\|\|full | Low-quality |
| 18-109-final-viral-combined_k141_206940\|\|full | Not-determined |
| 18-109-final-viral-combined_k141_165778\|\|full | Low-quality |
| 18-109-final-viral-combined_k141_12344\|\|full | Low-quality |
| 18-109-final-viral-combined_k141_144877\|\|full | Low-quality |
| 18-109-final-viral-combined_k141_144883\|\|full | Not-determined |
| 18-109-final-viral-combined_k141_155732\|\|full | Low-quality |
| 18-109-final-viral-combined_k141_83658\|\|full | Low-quality |
| 18-109-final-viral-combined_k141_155735\|\|full | Not-determined |
| 18-109-final-viral-combined_k141_165806\|\|full | Not-determined |
| 18-109-final-viral-combined_k141_12371\|\|full | Low-quality |
| 18-109-final-viral-combined_k141_83672\|\|full | Not-determined |
| 18-109-final-viral-combined_k141_53481\|\|full | Not-determined |

| | |
|---|---|
| 18-109-final-viral-combined_k141_12275\|\|full | Not-determined |
| 18-109-final-viral-combined_k141_12381\|\|full | Low-quality |
| 18-109-final-viral-combined_k141_144909\|\|full | Not-determined |
| 18-109-final-viral-combined_k141_155754\|\|full | Not-determined |
| 18-109-final-viral-combined_k141_22312\|\|full | Not-determined |
| 18-109-final-viral-combined_k141_206993\|\|full | Low-quality |
| 18-109-final-viral-combined_k141_83689\|\|full | Not-determined |
| 18-109-final-viral-combined_k141_33025\|\|full | Not-determined |
| 18-109-final-viral-combined_k141_206994\|\|full | Low-quality |
| 18-109-final-viral-combined_k141_237370\|\|full | Low-quality |
| 18-109-final-viral-combined_k141_144928\|\|full | Not-determined |
| 18-109-final-viral-combined_k141_237373\|\|full | Low-quality |
| 18-109-final-viral-combined_k141_144930\|\|full | Not-determined |
| 18-109-final-viral-combined_k141_104480\|\|full | Not-determined |
| 18-109-final-viral-combined_k141_196778\|\|full | Not-determined |
| 18-109-final-viral-combined_k141_33047\|\|full | Not-determined |
| 18-109-final-viral-combined_k141_83708\|\|full | Low-quality |
| 18-109-final-viral-combined_k141_63558\|\|full | Not-determined |
| 18-109-final-viral-combined_k141_144940\|\|full | Not-determined |
| 18-109-final-viral-combined_k141_63570\|\|full | Not-determined |
| 18-109-final-viral-combined_k141_73048\|\|full | Not-determined |
| 18-109-final-viral-combined_k141_216814\|\|full | Not-determined |
| 18-109-final-viral-combined_k141_63573\|\|full | Not-determined |
| 18-109-final-viral-combined_k141_155780\|\|full | Low-quality |
| 18-109-final-viral-combined_k141_73051\|\|full | Not-determined |
| 18-109-final-viral-combined_k141_53502\|\|full | Low-quality |
| 18-109-final-viral-combined_k141_114827\|\|full | Not-determined |
| 18-109-final-viral-combined_k141_63582\|\|full | Not-determined |
| 18-109-final-viral-combined_k141_196797\|\|full | Low-quality |
| 18-109-final-viral-combined_k141_114829\|\|full | Not-determined |
| 18-109-final-viral-combined_k141_93684\|\|full | Low-quality |
| 18-109-final-viral-combined_k141_196798\|\|full | Low-quality |
| 18-109-final-viral-combined_k141_124900\|\|full | Not-determined |
| 18-109-final-viral-combined_k141_114838\|\|full | Low-quality |
| 18-109-final-viral-combined_k141_227400\|\|full | Low-quality |
| 18-109-final-viral-combined_k141_73076\|\|full | Low-quality |
| 18-109-final-viral-combined_k141_165859\|\|full | Low-quality |
| 18-109-final-viral-combined_k141_134998\|\|full | Low-quality |
| 18-109-final-viral-combined_k141_53537\|\|full | Not-determined |
| 18-109-final-viral-combined_k141_93711\|\|full | Low-quality |
| 18-109-final-viral-combined_k141_124910\|\|full | Not-determined |
| 18-109-final-viral-combined_k141_135017\|\|full | Low-quality |
| 18-109-final-viral-combined_k141_155809\|\|full | Low-quality |
| 18-109-final-viral-combined_k141_53544\|\|full | Not-determined |
| 18-109-final-viral-combined_k141_196841\|\|full | Not-determined |
| [illegible] | [illegible] |

| | |
|---|---|
| 18-109-final-viral-combined_k141_84726||full | Not-determined |
| 18-109-final-viral-combined_k141_64564||full | Not-determined |
| 18-109-final-viral-combined_k141_166942||full | Low-quality |
| 18-109-final-viral-combined_k141_64570||full | Low-quality |
| 18-109-final-viral-combined_k141_228383||full | Low-quality |
| 18-109-final-viral-combined_k141_145998||full | Low-quality |
| 18-109-final-viral-combined_k141_125962||full | Low-quality |
| 18-109-final-viral-combined_k141_228390||full | Low-quality |
| 18-109-final-viral-combined_k141_74019||full | Low-quality |
| 18-109-final-viral-combined_k141_228400||full | Not-determined |
| 18-109-final-viral-combined_k141_64576||full | Not-determined |
| 18-109-final-viral-combined_k141_228401||full | Not-determined |
| 18-109-final-viral-combined_k141_64582||full | Not-determined |
| 18-109-final-viral-combined_k141_176964||full | Low-quality |
| 18-109-final-viral-combined_k141_146021||full | Not-determined |
| 18-109-final-viral-combined_k141_135925||full | Not-determined |
| 18-109-final-viral-combined_k141_34292||full | Low-quality |
| 18-109-final-viral-combined_k141_156817||full | Not-determined |
| 18-109-final-viral-combined_k141_167021||full | Low-quality |
| 18-109-final-viral-combined_k141_105444||full | Not-determined |
| 18-109-final-viral-combined_k141_84755||full | Not-determined |
| 18-109-final-viral-combined_k141_135953||full | Not-determined |
| 18-109-final-viral-combined_k141_167032||full | Low-quality |
| 18-109-final-viral-combined_k141_105461||full | Low-quality |
| 18-109-final-viral-combined_k141_187835||full | Low-quality |
| 18-109-final-viral-combined_k141_13781||full | Low-quality |
| 18-109-final-viral-combined_k141_126021||full | Low-quality |
| 18-109-final-viral-combined_k141_198074||full | Low-quality |
| 18-109-final-viral-combined_k141_167053||full | Low-quality |
| 18-109-final-viral-combined_k141_44221||full | Low-quality |
| 18-109-final-viral-combined_k141_198085||full | Low-quality |
| 18-109-final-viral-combined_k141_44226||full | Low-quality |
| 18-109-final-viral-combined_k141_94784||full | Not-determined |
| 18-109-final-viral-combined_k141_44231||full | Low-quality |
| 18-109-final-viral-combined_k141_146079||full | Not-determined |
| 18-109-final-viral-combined_k141_64667||full | Not-determined |
| 18-109-final-viral-combined_k141_74071||full | Low-quality |
| 18-109-final-viral-combined_k141_105496||full | Low-quality |
| 18-109-final-viral-combined_k141_84793||full | Not-determined |
| 18-109-final-viral-combined_k141_74087||full | Not-determined |
| 18-109-final-viral-combined_k141_13820||full | Low-quality |
| 18-109-final-viral-combined_k141_64677||full | Not-determined |
| 18-109-final-viral-combined_k141_105506||full | Low-quality |
| 18-109-final-viral-combined_k141_228443||full | Low-quality |
| 18-109-final-viral-combined_k141_34343||full | Low-quality |
| 18-109-final-viral-combined_k141_167095||full | Low-quality |
| 18-109-final-viral-combined_k141_208444||full | Not-determined |
| 18-109-final-viral-combined_k141_187893||full | Not-determined |
| 18-109-final-viral-combined_k141_13838||full | Low-quality |
| 18-109-final-viral-combined_k141_228448||full | Not-determined |
| 18-109-final-viral-combined_k141_13842||full | Low-quality |
| 18-109-final-viral-combined_k141_136018||full | Not-determined |
| 18-109-final-viral-combined_k141_167111||full | Low-quality |
| 18-109-final-viral-combined_k141_74122||full | Low-quality |
| 18-109-final-viral-combined_k141_94809||full | Not-determined |
| 18-109-final-viral-combined_k141_13877||full | Not-determined |
| 18-109-final-viral-combined_k141_136037||full | Not-determined |
| 18-109-final-viral-combined_k141_198124||full | Not-determined |
| 18-109-final-viral-combined_k141_105528||full | Not-determined |
| 18-109-final-viral-combined_k141_198129||full | Not-determined |
| 18-109-final-viral-combined_k141_146126||full | Low-quality |
| 18-109-final-viral-combined_k141_105531||full | Low-quality |
| 18-109-final-viral-combined_k141_126042||full | Not-determined |
| 18-109-final-viral-combined_k141_23194||full | Low-quality |
| 18-109-final-viral-combined_k141_126047||full | Not-determined |
| 18-109-final-viral-combined_k141_3461||full | Not-determined |
| 18-109-final-viral-combined_k141_167154||full | Low-quality |
| 18-109-final-viral-combined_k141_74151||full | Low-quality |
| 18-109-final-viral-combined_k141_187919||full | Low-quality |
| 18-109-final-viral-combined_k141_3469||full | Not-determined |
| 18-109-final-viral-combined_k141_115991||full | Low-quality |
| 18-109-final-viral-combined_k141_217573||full | Not-determined |
| 18-109-final-viral-combined_k141_34377||full | Not-determined |
| 18-109-final-viral-combined_k141_177043||full | Low-quality |
| 18-109-final-viral-combined_k141_74179||full | Not-determined |
| 18-109-final-viral-combined_k141_187946||full | Not-determined |
| 18-109-final-viral-combined_k141_74181||full | Low-quality |
| 18-109-final-viral-combined_k141_84874||full | Low-quality |
| 18-109-final-viral-combined_k141_187954||full | Low-quality |
| 18-109-final-viral-combined_k141_177053||full | Low-quality |
| 18-109-final-viral-combined_k141_208536||full | Low-quality |
| 18-109-final-viral-combined_k141_116013||full | Not-determined |
| 18-109-final-viral-combined_k141_187957||full | Not-determined |
| 18-109-final-viral-combined_k141_198149||full | Low-quality |
| 18-109-final-viral-combined_k141_187958||full | Not-determined |
| 18-109-final-viral-combined_k141_13924||full | Low-quality |
| 18-109-final-viral-combined_k141_64705||full | Low-quality |
| 18-109-final-viral-combined_k141_34393||full | Low-quality |
| 18-109-final-viral-combined_k141_13929||full | Low-quality |
| 18-109-final-viral-combined_k141_228535||full | Not-determined |
| 18-109-final-viral-combined_k141_238642||full | Not-determined |
| 18-109-final-viral-combined_k141_64730||full | Not-determined |
| 18-109-final-viral-combined_k141_116042||full | Low-quality |
| 18-109-final-viral-combined_k141_238678||full | Not-determined |
| 18-109-final-viral-combined_k141_34411||full | Low-quality |
| 18-109-final-viral-combined_k141_105603||full | Low-quality |
| 18-109-final-viral-combined_k141_208542||full | Low-quality |
| 18-109-final-viral-combined_k141_146173||full | Not-determined |
| 18-109-final-viral-combined_k141_3512||full | Low-quality |
| 18-109-final-viral-combined_k141_34421||full | Not-determined |
| 18-109-final-viral-combined_k141_238690||full | Not-determined |
| 18-109-final-viral-combined_k141_64740||full | Low-quality |
| 18-109-final-viral-combined_k141_105621||full | Low-quality |
| 18-109-final-viral-combined_k141_94844||full | Low-quality |
| 18-109-final-viral-combined_k141_116070||full | Low-quality |
| 18-109-final-viral-combined_k141_177077||full | Low-quality |
| 18-109-final-viral-combined_k141_34431||full | Not-determined |
| 18-109-final-viral-combined_k141_44350||full | Low-quality |
| 18-109-final-viral-combined_k141_228579||full | Not-determined |
| 18-109-final-viral-combined_k141_84913||full | Not-determined |
| 18-109-final-viral-combined_k141_44370||full | Not-determined |
| 18-109-final-viral-combined_k141_208577||full | Not-determined |
| 18-109-final-viral-combined_k141_105635||full | Not-determined |
| 18-109-final-viral-combined_k141_228587||full | Low-quality |
| 18-109-final-viral-combined_k141_146205||full | Low-quality |
| 18-109-final-viral-combined_k141_188034||full | Not-determined |
| 18-109-final-viral-combined_k141_74272||full | Not-determined |
| 18-109-final-viral-combined_k141_167206||full | Not-determined |
| 18-109-final-viral-combined_k141_167210||full | Not-determined |
| 18-109-final-viral-combined_k141_74289||full | Low-quality |
| 18-109-final-viral-combined_k141_126166||full | Not-determined |
| 18-109-final-viral-combined_k141_74292||full | Low-quality |
| 18-109-final-viral-combined_k141_146253||full | Not-determined |
| 18-109-final-viral-combined_k141_177152||full | Low-quality |
| 18-109-final-viral-combined_k141_44401||full | Low-quality |
| 18-109-final-viral-combined_k141_177154||full | Not-determined |
| 18-109-final-viral-combined_k141_44406||full | Not-determined |
| 18-109-final-viral-combined_k141_136082||full | Low-quality |
| 18-109-final-viral-combined_k141_126180||full | Low-quality |
| 18-109-final-viral-combined_k141_167246||full | Not-determined |
| 18-109-final-viral-combined_k141_146264||full | Not-determined |
| 18-109-final-viral-combined_k141_94896||full | Low-quality |
| 18-109-final-viral-combined_k141_116162||full | Not-determined |
| 18-109-final-viral-combined_k141_44415||full | Low-quality |
| 18-109-final-viral-combined_k141_94913||full | Low-quality |
| 18-109-final-viral-combined_k141_44421||full | Not-determined |
| 18-109-final-viral-combined_k141_167274||full | Not-determined |
| 18-109-final-viral-combined_k141_167275||full | Low-quality |
| 18-109-final-viral-combined_k141_116181||full | Not-determined |
| 18-109-final-viral-combined_k141_238745||full | Not-determined |
| 18-109-final-viral-combined_k141_136114||full | Low-quality |
| 18-109-final-viral-combined_k141_74356||full | Not-determined |
| 18-109-final-viral-combined_k141_44435||full | Low-quality |
| 18-109-final-viral-combined_k141_74362||full | Not-determined |
| 18-109-final-viral-combined_k141_198229||full | Low-quality |
| 18-109-final-viral-combined_k141_208617||full | Low-quality |
| 18-109-final-viral-combined_k141_44440||full | Low-quality |
| 18-109-final-viral-combined_k141_85051||full | Not-determined |
| 18-109-final-viral-combined_k141_94972||full | Low-quality |
| 18-109-final-viral-combined_k141_198240||full | Low-quality |
| 18-109-final-viral-combined_k141_198251||full | Not-determined |
| 18-109-final-viral-combined_k141_198252||full | Not-determined |
| 18-109-final-viral-combined_k141_94988||full | Low-quality |
| 18-109-final-viral-combined_k141_74399||full | Low-quality |
| 18-109-final-viral-combined_k141_85068||full | Low-quality |
| 18-109-final-viral-combined_k141_74400||full | Low-quality |
| 18-109-final-viral-combined_k141_177213||full | Not-determined |
| 18-109-final-viral-combined_k141_208633||full | Low-quality |
| 18-109-final-viral-combined_k141_198265||full | Low-quality |
| 18-101-final-viral-combined_k141_126027||full | High-quality |
| 18-109-final-viral-combined_k141_44478||full | Low-quality |
| 18-109-final-viral-combined_k141_34517||full | Not-determined |
| 18-109-final-viral-combined_k141_44491||full | Low-quality |
| 18-109-final-viral-combined_k141_146359||full | Not-determined |
| 18-109-final-viral-combined_k141_208644||full | Not-determined |
| 18-109-final-viral-combined_k141_238797||full | Low-quality |
| 18-109-final-viral-combined_k141_126235||full | Not-determined |
| 18-109-final-viral-combined_k141_116289||full | Not-determined |
| 18-109-final-viral-combined_k141_85108||full | Low-quality |
| 18-109-final-viral-combined_k141_74417||full | Not-determined |
| 18-109-final-viral-combined_k141_85113||full | Low-quality |
| 18-109-final-viral-combined_k141_64783||full | Low-quality |
| 18-109-final-viral-combined_k141_146381||full | Not-determined |
| 18-109-final-viral-combined_k141_238833||full | Low-quality |
| 18-109-final-viral-combined_k141_126281||full | Not-determined |
| 18-109-final-viral-combined_k141_74437||full | Low-quality |
| 18-109-final-viral-combined_k141_177255||full | Not-determined |
| 18-109-final-viral-combined_k141_74444||full | Low-quality |
| 18-109-final-viral-combined_k141_198322||full | Low-quality |
| 18-109-final-viral-combined_k141_238852||full | Not-determined |
| 18-109-final-viral-combined_k141_188137||full | Not-determined |
| 18-109-final-viral-combined_k141_64822||full | Not-determined |
| 18-109-final-viral-combined_k141_136220||full | Low-quality |
| 18-109-final-viral-combined_k141_23349||full | Not-determined |
| 18-109-final-viral-combined_k141_74498||full | Not-determined |
| 18-109-final-viral-combined_k141_136232||full | Not-determined |
| 18-109-final-viral-combined_k141_126351||full | Low-quality |
| 18-109-final-viral-combined_k141_23361||full | Low-quality |
| 18-109-final-viral-combined_k141_23365||full | Low-quality |
| 18-109-final-viral-combined_k141_146466||full | Low-quality |
| 18-109-final-viral-combined_k141_188166||full | Low-quality |
| 18-109-final-viral-combined_k141_157107||full | Low-quality |
| 18-109-final-viral-combined_k141_85219||full | Not-determined |
| 18-109-final-viral-combined_k141_126424||full | Low-quality |
| 18-109-final-viral-combined_k141_34621||full | Not-determined |
| 18-109-final-viral-combined_k141_146523||full | Low-quality |
| 18-109-final-viral-combined_k141_136294||full | Not-determined |
| 18-109-final-viral-combined_k141_167452||full | Low-quality |
| 18-109-final-viral-combined_k141_146524||full | Not-determined |
| 18-109-final-viral-combined_k141_157146||full | Not-determined |
| 18-109-final-viral-combined_k141_23438||full | Not-determined |
| 18-109-final-viral-combined_k141_177359||full | Not-determined |
| 18-109-final-viral-combined_k141_177360||full | Not-determined |
| 18-109-final-viral-combined_k141_157165||full | Not-determined |
| 18-109-final-viral-combined_k141_34663||full | Low-quality |
| 18-109-final-viral-combined_k141_34665||full | Not-determined |
| 18-109-final-viral-combined_k141_116369||full | Not-determined |
| 18-109-final-viral-combined_k141_95151||full | Low-quality |
| 18-109-final-viral-combined_k141_95159||full | Low-quality |
| 18-109-final-viral-combined_k141_23450||full | Low-quality |
| 18-109-final-viral-combined_k141_74645||full | Not-determined |
| 18-109-final-viral-combined_k141_23463||full | Not-determined |
| 18-109-final-viral-combined_k141_177383||full | Not-determined |
| 18-109-final-viral-combined_k141_64940||full | Not-determined |
| 18-109-final-viral-combined_k141_85263||full | Low-quality |
| 18-109-final-viral-combined_k141_44637||full | Not-determined |
| 18-109-final-viral-combined_k141_239022||full | Not-determined |
| 18-109-final-viral-combined_k141_74677||full | Not-determined |
| 18-109-final-viral-combined_k141_95205||full | Low-quality |
| 18-109-final-viral-combined_k141_239031||full | Low-quality |
| 18-109-final-viral-combined_k141_177396||full | Not-determined |
| 18-109-final-viral-combined_k141_177401||full | Low-quality |
| 18-109-final-viral-combined_k141_14010||full | Low-quality |
| 18-109-final-viral-combined_k141_95227||full | Not-determined |
| 18-109-final-viral-combined_k141_23484||full | Not-determined |
| 18-109-final-viral-combined_k141_177409||full | Low-quality |
| 18-109-final-viral-combined_k141_217730||full | Low-quality |
| 18-109-final-viral-combined_k141_239068||full | Not-determined |
| 18-109-final-viral-combined_k141_217731||full | Low-quality |
| 18-109-final-viral-combined_k141_44684||full | Not-determined |
| 18-109-final-viral-combined_k141_74721||full | Not-determined |
| 18-109-final-viral-combined_k141_14036||full | Low-quality |
| 18-109-final-viral-combined_k141_64994||full | Not-determined |
| 18-109-final-viral-combined_k141_14038||full | Not-determined |
| 18-109-final-viral-combined_k141_239085||full | Low-quality |
| 18-109-final-viral-combined_k141_85304||full | Not-determined |
| 18-109-final-viral-combined_k141_74739||full | Low-quality |
| 18-109-final-viral-combined_k141_85314||full | Not-determined |
| 18-109-final-viral-combined_k141_85315||full | Not-determined |
| 18-102-final-viral-combined_k141_85381||full | Medium-quality |
| 18-109-final-viral-combined_k141_74742||full | Low-quality |
| 18-109-final-viral-combined_k141_65019||full | Low-quality |
| 18-109-final-viral-combined_k141_14056||full | Not-determined |
| 18-109-final-viral-combined_k141_65033||full | Not-determined |
| 18-109-final-viral-combined_k141_146593||full | Not-determined |
| 18-109-final-viral-combined_k141_54650||full | Not-determined |
| 18-109-final-viral-combined_k141_217783||full | Not-determined |
| 18-109-final-viral-combined_k141_105774||full | Not-determined |
| 18-109-final-viral-combined_k141_126511||full | Not-determined |
| 18-109-final-viral-combined_k141_105782||full | Low-quality |
| 18-109-final-viral-combined_k141_146613||full | Low-quality |
| 18-109-final-viral-combined_k141_198393||full | Low-quality |
| 18-109-final-viral-combined_k141_44748||full | Low-quality |
| 18-109-final-viral-combined_k141_208916||full | Not-determined |
| 18-109-final-viral-combined_k141_167610||full | Low-quality |
| 18-109-final-viral-combined_k141_146615||full | Not-determined |
| 18-109-final-viral-combined_k141_177496||full | Low-quality |
| 18-109-final-viral-combined_k141_85343||full | Not-determined |
| 18-109-final-viral-combined_k141_126514||full | Low-quality |
| 18-109-final-viral-combined_k141_136401||full | Not-determined |
| 18-109-final-viral-combined_k141_146625||full | Not-determined |
| 18-109-final-viral-combined_k141_95299||full | Low-quality |
| 18-109-final-viral-combined_k141_177500||full | Low-quality |
| 18-109-final-viral-combined_k141_188352||full | Low-quality |
| 18-109-final-viral-combined_k141_95316||full | Not-determined |
| 18-109-final-viral-combined_k141_136441||full | Not-determined |
| 18-109-final-viral-combined_k141_177526||full | Not-determined |
| 18-109-final-viral-combined_k141_85361||full | Not-determined |
| 18-109-final-viral-combined_k141_157195||full | Not-determined |
| 18-109-final-viral-combined_k141_228762||full | Low-quality |
| 18-109-final-viral-combined_k141_85363||full | Not-determined |
| 18-109-final-viral-combined_k141_146633||full | Low-quality |
| 18-109-final-viral-combined_k141_85367||full | Not-determined |
| 18-109-final-viral-combined_k141_198455||full | Not-determined |
| 18-109-final-viral-combined_k141_228771||full | Low-quality |
| 18-109-final-viral-combined_k141_177541||full | Low-quality |
| 18-109-final-viral-combined_k141_54716||full | Not-determined |
| 18-109-final-viral-combined_k141_126551||full | Not-determined |
| 18-109-final-viral-combined_k141_177558||full | Not-determined |
| 18-109-final-viral-combined_k141_188387||full | Low-quality |
| 18-109-final-viral-combined_k141_157247||full | Not-determined |
| 18-109-final-viral-combined_k141_136498||full | Low-quality |
| 18-109-final-viral-combined_k141_167642||full | Low-quality |
| 18-109-final-viral-combined_k141_54729||full | Not-determined |
| 18-109-final-viral-combined_k141_95365||full | Low-quality |
| 18-109-final-viral-combined_k141_177566||full | Low-quality |
| 18-109-final-viral-combined_k141_105898||full | Low-quality |
| 18-109-final-viral-combined_k141_126614||full | Not-determined |
| 18-109-final-viral-combined_k141_23605||full | Low-quality |
| 18-109-final-viral-combined_k141_54765||full | Low-quality |
| 18-109-final-viral-combined_k141_136516||full | Not-determined |
| 18-109-final-viral-combined_k141_167670||full | Not-determined |
| 18-109-final-viral-combined_k141_95393||full | Not-determined |
| 18-109-final-viral-combined_k141_228824||full | Low-quality |
| 18-109-final-viral-combined_k141_54773||full | Not-determined |
| 18-109-final-viral-combined_k141_167675||full | Low-quality |
| 18-109-final-viral-combined_k141_116417||full | Low-quality |
| 18-109-final-viral-combined_k141_228835||full | Low-quality |
| 18-109-final-viral-combined_k141_188420||full | Not-determined |
| 18-109-final-viral-combined_k141_188427||full | Low-quality |
| 18-109-final-viral-combined_k141_116427||full | Not-determined |
| 18-109-final-viral-combined_k141_34868||full | Low-quality |
| 18-109-final-viral-combined_k141_65168||full | Low-quality |
| 18-109-final-viral-combined_k141_177622||full | Not-determined |
| 18-109-final-viral-combined_k141_95413||full | Low-quality |
| 18-109-final-viral-combined_k141_177625||full | Not-determined |
| 18-109-final-viral-combined_k141_198524||full | Not-determined |
| 18-109-final-viral-combined_k141_54806||full | Not-determined |
| 18-109-final-viral-combined_k141_239177||full | Not-determined |
| 18-109-final-viral-combined_k141_239182||full | Not-determined |
| 18-109-final-viral-combined_k141_198537||full | Low-quality |
| 18-109-final-viral-combined_k141_177668||full | Low-quality |
| 18-109-final-viral-combined_k141_65220||full | Low-quality |
| 18-109-final-viral-combined_k141_95435||full | Low-quality |
| 18-109-final-viral-combined_k141_14108||full | Low-quality |
| 18-109-final-viral-combined_k141_239211||full | Low-quality |
| 18-109-final-viral-combined_k141_23628||full | Not-determined |
| 18-109-final-viral-combined_k141_209053||full | Low-quality |
| 18-109-final-viral-combined_k141_177679||full | Not-determined |
| 18-109-final-viral-combined_k141_34894||full | Not-determined |
| 18-109-final-viral-combined_k141_136548||full | Not-determined |
| 18-109-final-viral-combined_k141_85482||full | Not-determined |
| 18-109-final-viral-combined_k141_167733||full | Low-quality |
| 18-109-final-viral-combined_k141_177700||full | Not-determined |
| 18-109-final-viral-combined_k141_136575||full | Low-quality |
| 17-108-final-viral-combined_k141_179605||full | Medium-quality |
| 18-109-final-viral-combined_k141_126680||full | Not-determined |
| 18-109-final-viral-combined_k141_105982||full | Low-quality |
| 18-109-final-viral-combined_k141_95464||full | Not-determined |
| 18-109-final-viral-combined_k141_3875||full | Not-determined |
| 18-109-final-viral-combined_k141_228910||full | Not-determined |
| 18-109-final-viral-combined_k141_167741||full | Low-quality |
| 17-109-final-viral-combined_k141_120133||full | Medium-quality |
| 18-109-final-viral-combined_k141_3882||full | Low-quality |
| 18-109-final-viral-combined_k141_23702||full | Not-determined |
| 18-109-final-viral-combined_k141_177730||full | Low-quality |
| 18-109-final-viral-combined_k141_188497||full | Low-quality |
| 18-109-final-viral-combined_k141_136620||full | Not-determined |
| 18-109-final-viral-combined_k141_14165||full | Not-determined |
| 18-109-final-viral-combined_k141_157280||full | Low-quality |
| 18-109-final-viral-combined_k141_95507||full | Low-quality |
| 18-109-final-viral-combined_k141_3895||full | Low-quality |
| 18-109-final-viral-combined_k141_126710||full | Low-quality |
| 18-109-final-viral-combined_k141_136634||full | Not-determined |
| 18-109-final-viral-combined_k141_54857||full | Not-determined |
| 18-109-final-viral-combined_k141_198629||full | Not-determined |
| 18-109-final-viral-combined_k141_54858||full | Low-quality |
| 18-109-final-viral-combined_k141_198635||full | Low-quality |
| 18-109-final-viral-combined_k141_126722||full | Not-determined |
| 18-109-final-viral-combined_k141_167776||full | Low-quality |
| 18-109-final-viral-combined_k141_157284||full | Low-quality |
| 18-109-final-viral-combined_k141_198644||full | Not-determined |
| 18-109-final-viral-combined_k141_106010||full | Not-determined |
| 18-109-final-viral-combined_k141_34924||full | Low-quality |
| 18-109-final-viral-combined_k141_217797||full | Low-quality |
| 18-104-final-viral-combined_k141_14411||full | High-quality |
| 18-109-final-viral-combined_k141_44896||full | Not-determined |
| 18-109-final-viral-combined_k141_167809||full | Low-quality |
| 18-109-final-viral-combined_k141_157296||full | Not-determined |
| 18-109-final-viral-combined_k141_106023||full | Low-quality |
| 18-109-final-viral-combined_k141_198668||full | Low-quality |
| 18-109-final-viral-combined_k141_239282||full | Not-determined |
| 18-109-final-viral-combined_k141_198669||full | Not-determined |
| 18-109-final-viral-combined_k141_146779||full | Not-determined |
| 18-109-final-viral-combined_k141_65373||full | Low-quality |
| 18-109-final-viral-combined_k141_54882||full | Not-determined |
| 18-109-final-viral-combined_k141_228989||full | Low-quality |
| 18-109-final-viral-combined_k141_239305||full | Low-quality |
| 18-109-final-viral-combined_k141_177792||full | Not-determined |
| 18-109-final-viral-combined_k141_54893||full | Not-determined |
| 18-109-final-viral-combined_k141_239314||full | Low-quality |
| 18-109-final-viral-combined_k141_54896||full | Not-determined |
| 18-109-final-viral-combined_k141_239316||full | Not-determined |
| 18-109-final-viral-combined_k141_34963||full | Low-quality |
| 18-109-final-viral-combined_k141_3921||full | Not-determined |
| 18-109-final-viral-combined_k141_54910||full | Not-determined |
| 18-109-final-viral-combined_k141_14181||full | Low-quality |
| 18-109-final-viral-combined_k141_106080||full | Not-determined |
| 18-109-final-viral-combined_k141_14182||full | Not-determined |
| 18-109-final-viral-combined_k141_209073||full | Not-determined |
| 18-109-final-viral-combined_k141_126782||full | Low-quality |
| 18-109-final-viral-combined_k141_3928||full | Low-quality |
| 18-109-final-viral-combined_k141_157350||full | Not-determined |
| 18-109-final-viral-combined_k141_229000||full | Not-determined |
| 18-109-final-viral-combined_k141_23788||full | Not-determined |
| 18-109-final-viral-combined_k141_3936||full | Low-quality |
| 18-109-final-viral-combined_k141_65398||full | Low-quality |
| 18-109-final-viral-combined_k141_23792||full | Low-quality |
| 18-109-final-viral-combined_k141_116546||full | Low-quality |
| 18-109-final-viral-combined_k141_239352||full | Low-quality |
| 18-109-final-viral-combined_k141_23801||full | Not-determined |
| 18-109-final-viral-combined_k141_85533||full | Not-determined |
| 18-109-final-viral-combined_k141_229020||full | Not-determined |
| 18-109-final-viral-combined_k141_126805||full | Not-determined |
| 18-109-final-viral-combined_k141_146841||full | Not-determined |
| 18-109-final-viral-combined_k141_54943||full | Low-quality |
| 18-109-final-viral-combined_k141_14215||full | Low-quality |
| 18-109-final-viral-combined_k141_65429||full | Low-quality |
| 18-109-final-viral-combined_k141_217875||full | Not-determined |
| 18-109-final-viral-combined_k141_126815||full | Not-determined |
| 18-109-final-viral-combined_k141_54961||full | Low-quality |
| 18-109-final-viral-combined_k141_54971||full | Low-quality |
| 18-109-final-viral-combined_k141_14247||full | Low-quality |
| 18-109-final-viral-combined_k141_54984||full | Not-determined |
| 18-109-final-viral-combined_k141_106128||full | Not-determined |
| 18-109-final-viral-combined_k141_85569||full | Not-determined |
| 18-109-final-viral-combined_k141_229060||full | Low-quality |
| 18-109-final-viral-combined_k141_3960||full | Low-quality |
| 18-109-final-viral-combined_k141_157362||full | Low-quality |
| 18-109-final-viral-combined_k141_106140||full | Not-determined |
| 18-109-final-viral-combined_k141_3966||full | Not-determined |
| 18-109-final-viral-combined_k141_126852||full | Low-quality |
| 18-109-final-viral-combined_k141_177859||full | Not-determined |
| 18-109-final-viral-combined_k141_177871||full | Low-quality |
| 18-109-final-viral-combined_k141_229100||full | Low-quality |
| 18-109-final-viral-combined_k141_55026||full | Low-quality |
| 18-109-final-viral-combined_k141_177875||full | Not-determined |
| 18-109-final-viral-combined_k141_3979||full | Low-quality |
| 18-109-final-viral-combined_k141_45014||full | Low-quality |
| 18-109-final-viral-combined_k141_116637||full | Low-quality |
| 18-109-final-viral-combined_k141_198832||full | Low-quality |
| 18-109-final-viral-combined_k141_116645||full | Low-quality |
| 18-109-final-viral-combined_k141_106181||full | Not-determined |
| 18-109-final-viral-combined_k141_85627||full | Not-determined |
| 18-109-final-viral-combined_k141_126870||full | Not-determined |
| 18-109-final-viral-combined_k141_177887||full | Not-determined |
| 18-109-final-viral-combined_k141_55042||full | Not-determined |
| 18-109-final-viral-combined_k141_209123||full | Low-quality |
| 18-109-final-viral-combined_k141_85644||full | Not-determined |
| 18-109-final-viral-combined_k141_106212||full | Not-determined |
| 17-109-final-viral-combined_k141_328554||full | Medium-quality |
| 18-109-final-viral-combined_k141_65554||full | Not-determined |
| 18-109-final-viral-combined_k141_209136||full | Not-determined |
| 18-109-final-viral-combined_k141_45064||full | Low-quality |
| 18-109-final-viral-combined_k141_136655||full | Low-quality |
| 18-109-final-viral-combined_k141_229130||full | Not-determined |
| 18-109-final-viral-combined_k141_55073||full | Not-determined |
| 18-109-final-viral-combined_k141_136660||full | Not-determined |
| 18-110_k141_20528 | High-quality |
| 18-109-final-viral-combined_k141_106218||full | Not-determined |
| 18-109-final-viral-combined_k141_126892||full | Low-quality |
| 18-109-final-viral-combined_k141_74921||full | Low-quality |
| 18-109-final-viral-combined_k141_85688||full | Not-determined |
| 18-109-final-viral-combined_k141_4037||full | Low-quality |
| 18-109-final-viral-combined_k141_217912||full | Not-determined |
| 18-109-final-viral-combined_k141_136682||full | Not-determined |
| 18-109-final-viral-combined_k141_239457||full | Not-determined |
| 18-109-final-viral-combined_k141_229153||full | Not-determined |
| 18-109-final-viral-combined_k141_35092||full | Low-quality |
| 18-110-final-viral-combined_k141_246185||0_partial | High-quality |
| 18-109-final-viral-combined_k141_23851||full | Not-determined |
| 18-109-final-viral-combined_k141_35099||full | Low-quality |
| 18-109-final-viral-combined_k141_209166||full | Not-determined |
| 18-109-final-viral-combined_k141_55094||full | Low-quality |
| 18-109-final-viral-combined_k141_106244||full | Not-determined |
| 18-109-final-viral-combined_k141_239468||full | Not-determined |
| 18-109-final-viral-combined_k141_65609||full | Low-quality |
| 18-109-final-viral-combined_k141_239470||full | Not-determined |
| 18-109-final-viral-combined_k141_209194||full | Not-determined |
| 18-109-final-viral-combined_k141_65629||full | Low-quality |
| 18-109-final-viral-combined_k141_217928||full | Not-determined |
| 18-109-final-viral-combined_k141_95651||full | Not-determined |
| 18-109-final-viral-combined_k141_177959||full | Not-determined |
| 18-109-final-viral-combined_k141_45152||full | Low-quality |
| 18-109-final-viral-combined_k141_55126||full | Low-quality |
| 18-109-final-viral-combined_k141_4090||full | Low-quality |
| 18-109-final-viral-combined_k141_95658||full | Low-quality |
| 18-106-final-viral-combined_k141_78655||full | Medium-quality |
| 18-109-final-viral-combined_k141_126970||full | Not-determined |
| 18-109-final-viral-combined_k141_55148||full | Low-quality |
| 18-109-final-viral-combined_k141_65679||full | Not-determined |
| 18-109-final-viral-combined_k141_55152||full | Not-determined |
| 18-109-final-viral-combined_k141_85718||full | Not-determined |
| 18-109-final-viral-combined_k141_146932||full | Low-quality |
| 18-109-final-viral-combined_k141_95682||full | Not-determined |
| 18-109-final-viral-combined_k141_239555||full | Low-quality |
| 18-109-final-viral-combined_k141_146948||full | Low-quality |
| 18-109-final-viral-combined_k141_157526||full | Low-quality |
| 18-109-final-viral-combined_k141_167881||full | Not-determined |
| 18-109-final-viral-combined_k141_95688||full | Low-quality |
| 18-109-final-viral-combined_k141_23887||full | Not-determined |
| 18-109-final-viral-combined_k141_55176||full | Low-quality |
| 18-109-final-viral-combined_k141_127004||full | Low-quality |
| 18-109-final-viral-combined_k141_239580||full | Not-determined |
| 18-109-final-viral-combined_k141_146971||full | Low-quality |
| 18-109-final-viral-combined_k141_177981||full | Low-quality |
| 18-109-final-viral-combined_k141_209274||full | Low-quality |
| 18-109-final-viral-combined_k141_198945||full | Low-quality |
| 18-109-final-viral-combined_k141_35193||full | Low-quality |
| 18-109-final-viral-combined_k141_157560||full | Low-quality |
| 18-109-final-viral-combined_k141_198960||full | Not-determined |
| 18-109-final-viral-combined_k141_157570||full | Low-quality |
| 18-109-final-viral-combined_k141_127047||full | Not-determined |
| 18-109-final-viral-combined_k141_95727||full | Not-determined |
| 18-109-final-viral-combined_k141_85750||full | Not-determined |
| 18-109-final-viral-combined_k141_55228||full | Not-determined |
| 18-109-final-viral-combined_k141_116793||full | Low-quality |
| 18-109-final-viral-combined_k141_178029||full | Not-determined |
| 18-109-final-viral-combined_k141_95738||full | Not-determined |
| 18-110-final-viral-combined_k141_20528||full | High-quality |
| 18-109-final-viral-combined_k141_65798||full | Not-determined |
| 18-109-final-viral-combined_k141_178035||full | Not-determined |
| 18-109-final-viral-combined_k141_4136||full | Not-determined |
| 18-109-final-viral-combined_k141_116804||full | Not-determined |
| 18-109-final-viral-combined_k141_157585||full | Low-quality |
| 18-109-final-viral-combined_k141_4140||full | Not-determined |
| 18-109-final-viral-combined_k141_167931||full | Low-quality |
| 18-109-final-viral-combined_k141_209318||full | Low-quality |
| 18-109-final-viral-combined_k141_157591||full | Low-quality |
| 18-109-final-viral-combined_k141_106264||full | Low-quality |
| 18-109-final-viral-combined_k141_239648||full | Low-quality |
| 18-109-final-viral-combined_k141_167940||full | Not-determined |
| 18-109-final-viral-combined_k141_4150||full | Not-determined |
| 18-109-final-viral-combined_k141_239658||full | Low-quality |
| 18-109-final-viral-combined_k141_4152||full | Not-determined |
| 18-109-final-viral-combined_k141_35247||full | Not-determined |
| 18-109-final-viral-combined_k141_157623||full | Low-quality |
| 18-109-final-viral-combined_k141_127117||full | Low-quality |
| 18-109-final-viral-combined_k141_188604||full | Low-quality |
| 18-109-final-viral-combined_k141_136803||full | Low-quality |
| 18-109-final-viral-combined_k141_75091||full | Not-determined |
| 18-109-final-viral-combined_k141_4184||full | Low-quality |
| 18-109-final-viral-combined_k141_106293||full | Not-determined |
| 18-109-final-viral-combined_k141_218025||full | Low-quality |
| 18-109-final-viral-combined_k141_127162||full | Low-quality |
| 18-109-final-viral-combined_k141_65825||full | Not-determined |
| 18-109-final-viral-combined_k141_106295||full | Not-determined |
| 18-109-final-viral-combined_k141_106304||full | Not-determined |
| 18-109-final-viral-combined_k141_157644||full | Not-determined |
| 18-109-final-viral-combined_k141_85801||full | Not-determined |
| 18-109-final-viral-combined_k141_95770||full | Low-quality |
| 18-109-final-viral-combined_k141_199038||full | Not-determined |
| 18-109-final-viral-combined_k141_178064||full | Not-determined |
| 18-109-final-viral-combined_k141_75133||full | Not-determined |
| 18-109-final-viral-combined_k141_106310||full | Low-quality |
| 18-109-final-viral-combined_k141_188645||full | Not-determined |
| 18-109-final-viral-combined_k141_157657||full | Low-quality |
| 18-109-final-viral-combined_k141_229252||full | Not-determined |
| 18-109-final-viral-combined_k141_199049||full | Not-determined |
| 18-109-final-viral-combined_k141_218064||full | Not-determined |
| 18-109-final-viral-combined_k141_106321||full | Low-quality |
| 18-109-final-viral-combined_k141_168000||full | Not-determined |
| 18-109-final-viral-combined_k141_14370||full | Low-quality |
| 18-109-final-viral-combined_k141_199059||full | Low-quality |
| 18-109-final-viral-combined_k141_45216||full | Not-determined |
| 18-109-final-viral-combined_k141_188667||full | Not-determined |
| 18-109-final-viral-combined_k141_75156||full | Not-determined |
| 18-109-final-viral-combined_k141_199068||full | Not-determined |
| 18-109-final-viral-combined_k141_218086||full | Not-determined |
| 18-109-final-viral-combined_k141_127222||full | Not-determined |
| 18-109-final-viral-combined_k141_95822||full | Low-quality |
| 18-109-final-viral-combined_k141_85837||full | Not-determined |
| 18-109-final-viral-combined_k141_55309||full | Not-determined |
| 18-109-final-viral-combined_k141_14414||full | Not-determined |
| 18-109-final-viral-combined_k141_14416||full | Low-quality |
| 18-109-final-viral-combined_k141_229326||full | Low-quality |
| 18-109-final-viral-combined_k141_75194||full | Low-quality |
| 18-109-final-viral-combined_k141_116897||full | Low-quality |
| 18-109-final-viral-combined_k141_239773||full | Low-quality |
| 18-109-final-viral-combined_k141_75200||full | Low-quality |
| 18-109-final-viral-combined_k141_116899||full | Not-determined |
| 18-109-final-viral-combined_k141_75201||full | Low-quality |
| 18-109-final-viral-combined_k141_95898||full | Low-quality |
| 18-109-final-viral-combined_k141_116911||full | Not-determined |
| 18-109-final-viral-combined_k141_75213||full | Low-quality |
| 18-109-final-viral-combined_k141_199134||full | Low-quality |
| 18-109-final-viral-combined_k141_157794||full | Low-quality |
| 18-109-final-viral-combined_k141_85857||full | Low-quality |
| 18-109-final-viral-combined_k141_199146||full | Not-determined |
| 18-109-final-viral-combined_k141_4256||full | Not-determined |
| 18-109-final-viral-combined_k141_188746||full | Not-determined |
| 18-109-final-viral-combined_k141_218136||full | Not-determined |
| 18-109-final-viral-combined_k141_188747||full | Low-quality |
| 18-102-final-viral-combined_k141_99446||full | Medium-quality |
| 18-109-final-viral-combined_k141_65892||full | Low-quality |
| 18-109-final-viral-combined_k141_85872||full | Low-quality |
| 18-109-final-viral-combined_k141_239791||full | Not-determined |
| 18-109-final-viral-combined_k141_24072||full | Not-determined |
| 18-109-final-viral-combined_k141_157811||full | Low-quality |
| 18-109-final-viral-combined_k141_85890||full | Not-determined |
| 18-109-final-viral-combined_k141_24074||full | Low-quality |
| 18-109-final-viral-combined_k141_199155||full | Not-determined |
| 18-109-final-viral-combined_k141_65921||full | Not-determined |
| 18-109-final-viral-combined_k141_65922||full | Low-quality |
| 18-109-final-viral-combined_k141_65925||full | Low-quality |
| 18-109-final-viral-combined_k141_35435||full | Not-determined |
| 18-109-final-viral-combined_k141_209538||full | Not-determined |
| 18-109-final-viral-combined_k141_136982||full | Not-determined |
| 18-109-final-viral-combined_k141_45285||full | Low-quality |

| | |
|---|---|
| 18-109-final-viral-combined_k141_209550\|\|full | Low-quality |
| 18-109-final-viral-combined_k141_85940\|\|full | Low-quality |
| KM-final-viral-combined_k141_179771\|\|full | High-quality |
| 18-109-final-viral-combined_k141_136993\|\|full | Not-determined |
| 18-101-final-viral-combined_k141_325646\|\|full | Medium-quality |
| 18-109-final-viral-combined_k141_127260\|\|full | Not-determined |
| 18-109-final-viral-combined_k141_137004\|\|full | Low-quality |
| 18-109-final-viral-combined_k141_239849\|\|full | Low-quality |
| 18-109-final-viral-combined_k141_4324\|\|full | Not-determined |
| 18-109-final-viral-combined_k141_65976\|\|full | Low-quality |
| 18-109-final-viral-combined_k141_35482\|\|full | Not-determined |
| 18-109-final-viral-combined_k141_35486\|\|full | Low-quality |
| 18-109-final-viral-combined_k141_239854\|\|full | Low-quality |
| 18-109-final-viral-combined_k141_35502\|\|full | Not-determined |
| 18-109-final-viral-combined_k141_55337\|\|full | Low-quality |
| 18-109-final-viral-combined_k141_209608\|\|full | Low-quality |
| 18-109-final-viral-combined_k141_157890\|\|full | Low-quality |
| 18-109-final-viral-combined_k141_95985\|\|full | Not-determined |
| 18-109-final-viral-combined_k141_55351\|\|full | Not-determined |
| 18-109-final-viral-combined_k141_4378\|\|full | Low-quality |
| 18-109-final-viral-combined_k141_168158\|\|full | Not-determined |
| 18-109-final-viral-combined_k141_147086\|\|full | Low-quality |
| 18-109-final-viral-combined_k141_55363\|\|full | Low-quality |
| 18-109-final-viral-combined_k141_168165\|\|full | Not-determined |
| 18-109-final-viral-combined_k141_106347\|\|full | Low-quality |
| 18-109-final-viral-combined_k141_178282\|\|full | Not-determined |
| 18-109-final-viral-combined_k141_137033\|\|full | Not-determined |
| 18-109-final-viral-combined_k141_96005\|\|full | Low-quality |
| 18-109-final-viral-combined_k141_147090\|\|full | Low-quality |
| 18-109-final-viral-combined_k141_116974\|\|full | Not-determined |
| 18-109-final-viral-combined_k141_218195\|\|full | Low-quality |
| 18-109-final-viral-combined_k141_45332\|\|full | Low-quality |
| 18-109-final-viral-combined_k141_55368\|\|full | Low-quality |
| 18-109-final-viral-combined_k141_85991\|\|full | Low-quality |
| 18-109-final-viral-combined_k141_218198\|\|full | Not-determined |
| 18-109-final-viral-combined_k141_188756\|\|full | Low-quality |
| 18-109-final-viral-combined_k141_239879\|\|full | Not-determined |
| 18-109-final-viral-combined_k141_35551\|\|full | Low-quality |
| 18-109-final-viral-combined_k141_168181\|\|full | Not-determined |
| 18-109-final-viral-combined_k141_157947\|\|full | Not-determined |
| 18-109-final-viral-combined_k141_157950\|\|full | Low-quality |
| 18-109-final-viral-combined_k141_209627\|\|full | Low-quality |
| 18-109-final-viral-combined_k141_55372\|\|full | Not-determined |
| 18-109-final-viral-combined_k141_147109\|\|full | Low-quality |
| 18-109-final-viral-combined_k141_86003\|\|full | Low-quality |
| 18-109-final-viral-combined_k141_55378\|\|full | Low-quality |
| 18-109-final-viral-combined_k141_209631\|\|full | Low-quality |
| 18-109-final-viral-combined_k141_127306\|\|full | Low-quality |
| 17-106-final-viral-combined_k141_105822\|\|full | High-quality |
| 18-109-final-viral-combined_k141_55387\|\|full | Low-quality |
| 18-109-final-viral-combined_k141_4400\|\|full | Not-determined |
| 18-109-final-viral-combined_k141_168192\|\|full | Low-quality |
| 18-109-final-viral-combined_k141_66008\|\|full | Not-determined |
| 18-109-final-viral-combined_k141_117002\|\|full | Low-quality |
| 18-109-final-viral-combined_k141_127327\|\|full | Not-determined |
| 18-109-final-viral-combined_k141_117017\|\|full | Low-quality |
| 18-109-final-viral-combined_k141_157981\|\|full | Low-quality |
| 18-109-final-viral-combined_k141_137091\|\|full | Not-determined |
| 18-109-final-viral-combined_k141_199237\|\|full | Not-determined |
| 18-109-final-viral-combined_k141_75354\|\|full | Not-determined |
| 18-109-final-viral-combined_k141_137107\|\|full | Low-quality |
| 18-109-final-viral-combined_k141_75365\|\|full | Not-determined |
| 18-109-final-viral-combined_k141_55420\|\|full | Low-quality |
| 18-109-final-viral-combined_k141_4441\|\|full | Not-determined |
| 18-109-final-viral-combined_k141_55421\|\|full | Low-quality |
| 18-109-final-viral-combined_k141_137119\|\|full | Not-determined |
| 18-109-final-viral-combined_k141_117030\|\|full | Low-quality |
| 18-109-final-viral-combined_k141_209657\|\|full | Not-determined |
| 18-109-final-viral-combined_k141_4448\|\|full | Not-determined |
| 18-109-final-viral-combined_k141_178352\|\|full | Low-quality |
| 18-109-final-viral-combined_k141_168224\|\|full | Not-determined |
| 18-109-final-viral-combined_k141_4450\|\|full | Low-quality |
| 18-109-final-viral-combined_k141_75383\|\|full | Low-quality |
| 18-109-final-viral-combined_k141_188798\|\|full | Low-quality |
| 18-109-final-viral-combined_k141_75386\|\|full | Not-determined |
| 18-109-final-viral-combined_k141_137132\|\|full | Low-quality |
| 18-109-final-viral-combined_k141_168240\|\|full | Low-quality |
| 18-109-final-viral-combined_k141_229469\|\|full | Low-quality |
| 18-109-final-viral-combined_k141_14564\|\|full | Low-quality |
| 18-109-final-viral-combined_k141_188808\|\|full | Not-determined |
| 18-109-final-viral-combined_k141_66040\|\|full | Low-quality |
| 18-109-final-viral-combined_k141_158027\|\|full | Low-quality |
| 18-109-final-viral-combined_k141_55462\|\|full | Not-determined |
| 18-109-final-viral-combined_k141_4488\|\|full | Not-determined |
| 18-109-final-viral-combined_k141_209709\|\|full | Low-quality |
| 18-109-final-viral-combined_k141_229486\|\|full | Not-determined |
| 18-109-final-viral-combined_k141_158055\|\|full | Not-determined |
| 18-109-final-viral-combined_k141_158064\|\|full | Low-quality |
| 18-109-final-viral-combined_k141_127384\|\|full | Not-determined |
| 18-109-final-viral-combined_k141_96088\|\|full | Low-quality |
| 18-109-final-viral-combined_k141_127388\|\|full | Low-quality |
| 18-109-final-viral-combined_k141_55489\|\|full | Not-determined |
| 18-109-final-viral-combined_k141_66091\|\|full | Low-quality |
| 18-109-final-viral-combined_k141_55500\|\|full | Not-determined |
| 18-109-final-viral-combined_k141_199264\|\|full | Not-determined |
| 18-109-final-viral-combined_k141_4523\|\|full | Not-determined |
| 18-109-final-viral-combined_k141_158070\|\|full | Low-quality |
| 18-109-final-viral-combined_k141_75429\|\|full | Not-determined |
| 18-109-final-viral-combined_k141_178445\|\|full | Low-quality |
| 18-109-final-viral-combined_k141_239996\|\|full | Low-quality |
| 18-109-final-viral-combined_k141_75443\|\|full | Not-determined |
| 18-109-final-viral-combined_k141_240011\|\|full | Not-determined |
| 18-109-final-viral-combined_k141_127409\|\|full | Low-quality |
| 18-109-final-viral-combined_k141_96108\|\|full | Not-determined |
| 18-109-final-viral-combined_k141_199289\|\|full | Low-quality |
| 18-109-final-viral-combined_k141_14601\|\|full | Low-quality |
| 18-109-final-viral-combined_k141_35673\|\|full | Not-determined |
| 18-109-final-viral-combined_k141_45468\|\|full | Low-quality |
| 18-109-final-viral-combined_k141_45472\|\|full | Not-determined |
| 18-109-final-viral-combined_k141_35684\|\|full | Low-quality |
| 18-109-final-viral-combined_k141_35685\|\|full | Not-determined |
| 18-109-final-viral-combined_k141_86098\|\|full | Low-quality |
| 18-109-final-viral-combined_k141_137216\|\|full | Not-determined |
| 18-109-final-viral-combined_k141_188896\|\|full | Not-determined |
| 18-109-final-viral-combined_k141_199330\|\|full | Low-quality |
| 18-109-final-viral-combined_k141_188897\|\|full | Not-determined |
| 18-109-final-viral-combined_k141_75462\|\|full | Low-quality |
| 18-109-final-viral-combined_k141_127435\|\|full | Low-quality |
| 18-109-final-viral-combined_k141_178495\|\|full | Low-quality |
| 18-109-final-viral-combined_k141_55573\|\|full | Not-determined |
| 18-109-final-viral-combined_k141_4568\|\|full | Not-determined |
| 18-109-final-viral-combined_k141_158113\|\|full | Not-determined |
| 18-109-final-viral-combined_k141_147162\|\|full | Not-determined |
| 18-109-final-viral-combined_k141_66170\|\|full | Not-determined |
| 18-109-final-viral-combined_k141_127453\|\|full | Not-determined |
| 18-109-final-viral-combined_k141_137237\|\|full | Not-determined |
| 18-109-final-viral-combined_k141_4591\|\|full | Not-determined |
| 18-109-final-viral-combined_k141_137239\|\|full | Not-determined |
| 18-109-final-viral-combined_k141_188909\|\|full | Not-determined |
| 18-109-final-viral-combined_k141_240040\|\|full | Low-quality |
| 18-109-final-viral-combined_k141_45526\|\|full | Low-quality |
| 18-109-final-viral-combined_k141_45528\|\|full | Not-determined |
| 18-109-final-viral-combined_k141_178511\|\|full | Low-quality |
| 18-109-final-viral-combined_k141_106446\|\|full | Not-determined |
| 18-109-final-viral-combined_k141_137266\|\|full | Not-determined |
| 18-109-final-viral-combined_k141_229524\|\|full | Not-determined |
| 18-109-final-viral-combined_k141_14684\|\|full | Low-quality |
| 18-109-final-viral-combined_k141_178533\|\|full | Not-determined |
| 18-109-final-viral-combined_k141_45538\|\|full | Not-determined |
| 18-109-final-viral-combined_k141_158189\|\|full | Low-quality |
| 18-109-final-viral-combined_k141_106484\|\|full | Not-determined |
| 18-109-final-viral-combined_k141_96190\|\|full | Low-quality |
| 18-109-final-viral-combined_k141_127523\|\|full | Low-quality |
| 18-109-final-viral-combined_k141_106500\|\|full | Low-quality |
| 18-109-final-viral-combined_k141_147198\|\|full | Low-quality |
| 18-109-final-viral-combined_k141_229600\|\|full | Low-quality |
| 18-109-final-viral-combined_k141_4669\|\|full | Low-quality |
| 18-109-final-viral-combined_k141_55663\|\|full | Not-determined |
| 18-109-final-viral-combined_k141_127566\|\|full | Not-determined |
| 18-109-final-viral-combined_k141_117256\|\|full | Not-determined |
| 18-109-final-viral-combined_k141_35819\|\|full | Low-quality |
| 18-109-final-viral-combined_k141_75492\|\|full | Low-quality |
| 18-109-final-viral-combined_k141_35820\|\|full | Not-determined |
| 18-109-final-viral-combined_k141_199397\|\|full | Low-quality |
| 18-109-final-viral-combined_k141_75494\|\|full | Low-quality |
| 18-109-final-viral-combined_k141_218351\|\|full | Low-quality |
| 18-109-final-viral-combined_k141_168444\|\|full | Not-determined |
| 18-109-final-viral-combined_k141_55675\|\|full | Not-determined |
| 18-109-final-viral-combined_k141_147249\|\|full | Low-quality |
| 18-109-final-viral-combined_k141_55676\|\|full | Low-quality |
| 18-109-final-viral-combined_k141_199406\|\|full | Low-quality |
| 18-109-final-viral-combined_k141_96281\|\|full | Not-determined |
| 18-109-final-viral-combined_k141_158254\|\|full | Not-determined |
| 18-109-final-viral-combined_k141_35849\|\|full | Low-quality |
| 18-109-final-viral-combined_k141_86208\|\|full | Low-quality |
| 18-109-final-viral-combined_k141_35854\|\|full | Low-quality |
| 18-109-final-viral-combined_k141_86222\|\|full | Not-determined |
| 18-109-final-viral-combined_k141_66322\|\|full | Not-determined |
| 18-109-final-viral-combined_k141_137287\|\|full | Not-determined |
| 18-109-final-viral-combined_k141_45599\|\|full | Low-quality |
| 18-109-final-viral-combined_k141_86229\|\|full | Low-quality |
| 18-109-final-viral-combined_k141_117293\|\|full | Not-determined |
| 18-109-final-viral-combined_k141_86231\|\|full | Not-determined |
| 18-109-final-viral-combined_k141_158273\|\|full | Not-determined |
| 18-109-final-viral-combined_k141_75514\|\|full | Low-quality |
| 18-109-final-viral-combined_k141_117305\|\|full | Not-determined |
| 18-109-final-viral-combined_k141_14721\|\|full | Not-determined |
| 18-109-final-viral-combined_k141_55740\|\|full | Low-quality |
| 18-109-final-viral-combined_k141_75538\|\|full | Not-determined |
| 18-109-final-viral-combined_k141_14729\|\|full | Low-quality |
| 18-109-final-viral-combined_k141_66349\|\|full | Not-determined |
| 18-109-final-viral-combined_k141_188996\|\|full | Low-quality |
| 18-109-final-viral-combined_k141_189000\|\|full | Not-determined |
| 18-109-final-viral-combined_k141_55753\|\|full | Low-quality |
| 18-109-final-viral-combined_k141_75554\|\|full | Low-quality |
| 18-109-final-viral-combined_k141_189003\|\|full | Not-determined |
| 18-109-final-viral-combined_k141_14748\|\|full | Not-determined |
| 18-109-final-viral-combined_k141_199420\|\|full | Not-determined |
| 18-109-final-viral-combined_k141_96366\|\|full | Not-determined |
| 18-109-final-viral-combined_k141_96375\|\|full | Low-quality |
| 18-109-final-viral-combined_k141_55777\|\|full | Low-quality |
| 18-109-final-viral-combined_k141_117345\|\|full | Low-quality |
| 18-109-final-viral-combined_k141_178720\|\|full | Not-determined |
| 18-109-final-viral-combined_k141_178729\|\|full | Low-quality |
| 18-109-final-viral-combined_k141_14777\|\|full | Not-determined |
| 18-109-final-viral-combined_k141_168524\|\|full | Low-quality |
| 18-109-final-viral-combined_k141_45644\|\|full | Not-determined |
| 18-109-final-viral-combined_k141_147358\|\|full | Low-quality |
| 18-109-final-viral-combined_k141_189049\|\|full | Low-quality |
| 18-109-final-viral-combined_k141_55822\|\|full | Not-determined |
| 18-109-final-viral-combined_k141_178760\|\|full | Low-quality |
| 18-109-final-viral-combined_k141_45659\|\|full | Not-determined |
| 18-109-final-viral-combined_k141_86304\|\|full | Low-quality |
| 18-109-final-viral-combined_k141_55830\|\|full | Low-quality |
| 18-109-final-viral-combined_k141_240142\|\|full | Low-quality |
| 18-109-final-viral-combined_k141_106608\|\|full | Not-determined |
| 18-109-final-viral-combined_k141_117406\|\|full | Low-quality |
| 18-109-final-viral-combined_k141_96391\|\|full | Low-quality |
| 18-109-final-viral-combined_k141_117412\|\|full | Not-determined |
| 18-109-final-viral-combined_k141_127646\|\|full | Not-determined |
| 18-109-final-viral-combined_k141_4673\|\|full | Not-determined |
| 18-109-final-viral-combined_k141_189089\|\|full | Not-determined |
| 18-109-final-viral-combined_k141_86333\|\|full | Low-quality |
| 18-109-final-viral-combined_k141_96404\|\|full | Not-determined |
| 18-109-final-viral-combined_k141_127666\|\|full | Low-quality |
| 18-109-final-viral-combined_k141_137398\|\|full | Not-determined |
| 18-109-final-viral-combined_k141_168596\|\|full | Not-determined |
| 18-109-final-viral-combined_k141_147385\|\|full | Low-quality |
| 18-109-final-viral-combined_k141_168604\|\|full | Low-quality |
| 18-109-final-viral-combined_k141_199535\|\|full | Not-determined |
| 18-109-final-viral-combined_k141_240188\|\|full | Not-determined |
| 18-109-final-viral-combined_k141_75695\|\|full | Not-determined |
| 18-109-final-viral-combined_k141_14783\|\|full | Low-quality |
| 18-109-final-viral-combined_k141_240193\|\|full | Not-determined |
| 18-109-final-viral-combined_k141_75703\|\|full | Not-determined |
| 18-109-final-viral-combined_k141_229750\|\|full | Not-determined |
| 18-109-final-viral-combined_k141_96450\|\|full | Low-quality |
| 18-109-final-viral-combined_k141_189107\|\|full | Not-determined |
| 18-109-final-viral-combined_k141_229767\|\|full | Not-determined |
| 18-109-final-viral-combined_k141_4737\|\|full | Low-quality |
| 18-109-final-viral-combined_k141_158372\|\|full | Low-quality |
| 18-109-final-viral-combined_k141_189110\|\|full | Not-determined |
| 18-109-final-viral-combined_k141_158378\|\|full | Low-quality |
| 18-109-final-viral-combined_k141_147415\|\|full | Not-determined |
| 18-109-final-viral-combined_k141_66523\|\|full | Not-determined |
| 18-109-final-viral-combined_k141_240253\|\|full | Low-quality |
| 18-109-final-viral-combined_k141_240255\|\|full | Not-determined |
| 18-109-final-viral-combined_k141_199567\|\|full | Low-quality |
| 18-109-final-viral-combined_k141_4769\|\|full | Not-determined |
| 18-109-final-viral-combined_k141_209743\|\|full | Low-quality |
| 18-109-final-viral-combined_k141_45753\|\|full | Not-determined |
| 18-109-final-viral-combined_k141_137414\|\|full | Not-determined |
| 18-109-final-viral-combined_k141_75784\|\|full | Low-quality |
| 18-109-final-viral-combined_k141_147446\|\|full | Not-determined |
| 18-109-final-viral-combined_k141_117472\|\|full | Not-determined |
| 18-109-final-viral-combined_k141_66567\|\|full | Not-determined |
| 18-109-final-viral-combined_k141_147465\|\|full | Low-quality |
| 18-109-final-viral-combined_k141_66579\|\|full | Low-quality |
| 18-109-final-viral-combined_k141_55930\|\|full | Low-quality |
| 18-109-final-viral-combined_k141_55931\|\|full | Low-quality |
| 18-109-final-viral-combined_k141_55944\|\|full | Low-quality |
| 18-109-final-viral-combined_k141_117527\|\|full | Low-quality |
| 18-109-final-viral-combined_k141_199614\|\|full | Low-quality |
| 18-109-final-viral-combined_k141_147498\|\|full | Not-determined |
| 18-109-final-viral-combined_k141_229778\|\|full | Not-determined |
| 18-109-final-viral-combined_k141_14898\|\|full | Not-determined |
| 18-109-final-viral-combined_k141_55953\|\|full | Low-quality |
| 18-109-final-viral-combined_k141_137481\|\|full | Not-determined |
| 18-109-final-viral-combined_k141_229788\|\|full | Not-determined |
| 18-109-final-viral-combined_k141_209798\|\|full | Low-quality |
| 18-109-final-viral-combined_k141_209800\|\|full | Low-quality |
| 18-109-final-viral-combined_k141_229798\|\|full | Not-determined |
| 18-109-final-viral-combined_k141_24233\|\|full | Low-quality |
| 18-109-final-viral-combined_k141_117531\|\|full | Low-quality |
| 18-109-final-viral-combined_k141_199637\|\|full | Low-quality |
| 18-109-final-viral-combined_k141_229803\|\|full | Low-quality |
| 18-109-final-viral-combined_k141_127735\|\|full | Not-determined |
| 18-109-final-viral-combined_k141_24241\|\|full | Not-determined |
| 18-109-final-viral-combined_k141_96499\|\|full | Not-determined |
| 18-109-final-viral-combined_k141_199650\|\|full | Not-determined |
| 18-109-final-viral-combined_k141_199651\|\|full | Low-quality |
| 18-109-final-viral-combined_k141_96500\|\|full | Not-determined |
| 18-109-final-viral-combined_k141_66638\|\|full | Low-quality |
| 18-109-final-viral-combined_k141_96501\|\|full | Not-determined |
| 18-109-final-viral-combined_k141_218391\|\|full | Not-determined |
| 18-109-final-viral-combined_k141_117564\|\|full | Low-quality |
| 18-109-final-viral-combined_k141_218394\|\|full | Not-determined |
| 18-109-final-viral-combined_k141_199661\|\|full | Low-quality |
| 18-109-final-viral-combined_k141_75833\|\|full | Not-determined |
| 18-109-final-viral-combined_k141_178815\|\|full | Low-quality |
| 18-109-final-viral-combined_k141_209817\|\|full | Low-quality |
| 18-109-final-viral-combined_k141_24281\|\|full | Not-determined |
| 18-109-final-viral-combined_k141_56031\|\|full | Low-quality |
| 18-109-final-viral-combined_k141_137512\|\|full | Not-determined |
| 18-109-final-viral-combined_k141_117601\|\|full | Low-quality |
| 18-109-final-viral-combined_k141_199670\|\|full | Not-determined |
| 18-109-final-viral-combined_k141_178829\|\|full | Low-quality |
| 18-109-final-viral-combined_k141_75844\|\|full | Low-quality |
| 18-109-final-viral-combined_k141_127766\|\|full | Low-quality |
| 18-109-final-viral-combined_k141_117605\|\|full | Not-determined |
| 18-109-final-viral-combined_k141_56033\|\|full | Low-quality |
| 18-109-final-viral-combined_k141_199681\|\|full | Low-quality |
| 18-109-final-viral-combined_k141_15009\|\|full | Low-quality |
| 18-109-final-viral-combined_k141_240327\|\|full | Not-determined |
| 18-109-final-viral-combined_k141_199686\|\|full | Low-quality |
| 18-109-final-viral-combined_k141_229851\|\|full | Low-quality |
| 18-109-final-viral-combined_k141_15027\|\|full | Not-determined |
| 18-109-final-viral-combined_k141_4851\|\|full | Low-quality |
| 18-109-final-viral-combined_k141_229862\|\|full | Low-quality |
| 18-109-final-viral-combined_k141_218436\|\|full | Not-determined |
| 18-109-final-viral-combined_k141_66695\|\|full | Low-quality |
| 18-109-final-viral-combined_k141_66698\|\|full | Not-determined |
| 18-109-final-viral-combined_k141_96571\|\|full | Not-determined |
| 18-109-final-viral-combined_k141_96573\|\|full | Low-quality |
| 18-109-final-viral-combined_k141_56057\|\|full | Not-determined |
| 18-109-final-viral-combined_k141_75865\|\|full | Not-determined |
| 18-109-final-viral-combined_k141_117642\|\|full | Not-determined |
| 18-109-final-viral-combined_k141_158475\|\|full | Not-determined |
| 18-109-final-viral-combined_k141_106774\|\|full | Not-determined |
| 18-109-final-viral-combined_k141_209842\|\|full | Low-quality |
| 18-109-final-viral-combined_k141_75884\|\|full | Not-determined |
| 18-109-final-viral-combined_k141_137564\|\|full | Low-quality |
| 18-109-final-viral-combined_k141_75892\|\|full | Low-quality |
| 18-109-final-viral-combined_k141_189202\|\|full | Not-determined |
| 18-109-final-viral-combined_k141_4892\|\|full | Low-quality |
| 18-109-final-viral-combined_k141_75898\|\|full | Not-determined |
| 18-109-final-viral-combined_k141_66752\|\|full | Not-determined |
| 18-109-final-viral-combined_k141_4906\|\|full | Not-determined |
| 18-109-final-viral-combined_k141_66762\|\|full | Low-quality |
| 18-109-final-viral-combined_k141_106824\|\|full | Low-quality |
| 18-109-final-viral-combined_k141_96616\|\|full | Not-determined |
| 18-109-final-viral-combined_k141_218457\|\|full | Low-quality |
| 18-109-final-viral-combined_k141_117675\|\|full | Low-quality |
| 18-109-final-viral-combined_k141_189233\|\|full | Not-determined |
| 18-109-final-viral-combined_k141_117681\|\|full | Low-quality |
| 18-109-final-viral-combined_k141_96626\|\|full | Not-determined |
| 18-109-final-viral-combined_k141_189238\|\|full | Low-quality |
| 17-109_k141_202803 | High-quality |
| 18-109-final-viral-combined_k141_75946\|\|full | Low-quality |
| 18-109-final-viral-combined_k141_4932\|\|full | Low-quality |
| 18-109-final-viral-combined_k141_4939\|\|full | Low-quality |
| 18-109-final-viral-combined_k141_66788\|\|full | Low-quality |
| 18-109-final-viral-combined_k141_199801\|\|full | Low-quality |
| 18-109-final-viral-combined_k141_96669\|\|full | Low-quality |
| 18-109-final-viral-combined_k141_96678\|\|full | Not-determined |
| 18-109-final-viral-combined_k141_218510\|\|full | Low-quality |
| 18-109-final-viral-combined_k141_168841\|\|full | Low-quality |
| 18-109-final-viral-combined_k141_158525\|\|full | Low-quality |
| 18-109-final-viral-combined_k141_147643\|\|full | Not-determined |
| 18-109-final-viral-combined_k141_137603\|\|full | Not-determined |
| 18-109-final-viral-combined_k141_45813\|\|full | Low-quality |
| 18-109-final-viral-combined_k141_4962\|\|full | Not-determined |
| 18-109-final-viral-combined_k141_96700\|\|full | Not-determined |
| 18-109-final-viral-combined_k141_24482\|\|full | Not-determined |
| 18-109-final-viral-combined_k141_168854\|\|full | Low-quality |
| 18-109-final-viral-combined_k141_96701\|\|full | Low-quality |
| 18-109-final-viral-combined_k141_4977\|\|full | Low-quality |
| 18-109-final-viral-combined_k141_127824\|\|full | Low-quality |
| 18-109-final-viral-combined_k141_45818\|\|full | Not-determined |
| 18-109-final-viral-combined_k141_168866\|\|full | Low-quality |
| KM_k141_230221 | High-quality |
| 18-109-final-viral-combined_k141_45821\|\|full | Not-determined |
| 18-109-final-viral-combined_k141_137640\|\|full | Not-determined |
| 18-109-final-viral-combined_k141_158548\|\|full | Not-determined |
| 18-109-final-viral-combined_k141_4987\|\|full | Low-quality |
| 18-109-final-viral-combined_k141_127841\|\|full | Low-quality |
| 18-109-final-viral-combined_k141_137658\|\|full | Low-quality |
| 18-109-final-viral-combined_k141_24503\|\|full | Not-determined |
| 18-109-final-viral-combined_k141_24505\|\|full | Not-determined |
| 18-109-final-viral-combined_k141_35971\|\|full | Not-determined |
| 18-109-final-viral-combined_k141_24508\|\|full | Not-determined |
| 18-109-final-viral-combined_k141_127854\|\|full | Not-determined |
| 18-109-final-viral-combined_k141_158568\|\|full | Not-determined |
| 18-109-final-viral-combined_k141_240421\|\|full | Not-determined |
| 18-109-final-viral-combined_k141_147717\|\|full | Low-quality |
| 18-109-final-viral-combined_k141_96750\|\|full | Not-determined |
| 18-109-final-viral-combined_k141_66849\|\|full | Low-quality |
| 18-109-final-viral-combined_k141_76010\|\|full | Low-quality |
| 18-109-final-viral-combined_k141_189259\|\|full | Low-quality |
| 18-109-final-viral-combined_k141_66860\|\|full | Low-quality |
| 18-109-final-viral-combined_k141_96771\|\|full | Low-quality |
| 18-109-final-viral-combined_k141_240448\|\|full | Not-determined |
| 18-109-final-viral-combined_k141_117795\|\|full | Not-determined |
| 18-109-final-viral-combined_k141_127901\|\|full | Low-quality |
| 18-109-final-viral-combined_k141_218564\|\|full | Not-determined |
| 18-109-final-viral-combined_k141_56109\|\|full | Low-quality |
| 18-109-final-viral-combined_k141_189269\|\|full | Low-quality |
| 18-109-final-viral-combined_k141_240469\|\|full | Not-determined |
| 18-109-final-viral-combined_k141_240470\|\|full | Low-quality |
| 18-109-final-viral-combined_k141_189310\|\|full | Low-quality |
| 18-109-final-viral-combined_k141_45887\|\|full | Not-determined |
| 18-109-final-viral-combined_k141_56126\|\|full | Not-determined |
| 18-109-final-viral-combined_k141_218595\|\|full | Not-determined |
| 18-109-final-viral-combined_k141_117825\|\|full | Low-quality |
| 18-109-final-viral-combined_k141_66888\|\|full | Not-determined |
| 18-109-final-viral-combined_k141_178981\|\|full | Low-quality |
| 18-109-final-viral-combined_k141_189327\|\|full | Low-quality |
| 18-109-final-viral-combined_k141_106863\|\|full | Not-determined |
| 18-109-final-viral-combined_k141_218604\|\|full | Not-determined |
| 18-109-final-viral-combined_k141_15238\|\|full | Not-determined |
| 18-109-final-viral-combined_k141_45912\|\|full | Low-quality |
| 18-109-final-viral-combined_k141_117851\|\|full | Not-determined |
| 18-109-final-viral-combined_k141_117854\|\|full | Not-determined |
| 18-109-final-viral-combined_k141_86430\|\|full | Low-quality |
| 18-109-final-viral-combined_k141_137758\|\|full | Low-quality |
| 18-109-final-viral-combined_k141_189339\|\|full | Low-quality |
| 18-109-final-viral-combined_k141_147737\|\|full | Low-quality |
| 18-109-final-viral-combined_k141_168967\|\|full | Not-determined |
| 18-109-final-viral-combined_k141_5089\|\|full | Not-determined |
| 18-109-final-viral-combined_k141_168976\|\|full | Not-determined |
| 18-109-final-viral-combined_k141_137785\|\|full | Low-quality |
| 18-109-final-viral-combined_k141_179022\|\|full | Low-quality |
| 18-109-final-viral-combined_k141_199970\|\|full | Not-determined |
| 18-109-final-viral-combined_k141_86465\|\|full | Low-quality |
| 18-109-final-viral-combined_k141_15272\|\|full | Not-determined |
| 18-109-final-viral-combined_k141_179026\|\|full | Not-determined |
| 18-109-final-viral-combined_k141_86473\|\|full | Not-determined |
| 18-109-final-viral-combined_k141_56204\|\|full | Not-determined |
| 18-109-final-viral-combined_k141_45970\|\|full | Not-determined |
| 18-109-final-viral-combined_k141_86487\|\|full | Not-determined |
| 18-109-final-viral-combined_k141_137807\|\|full | Not-determined |
| 18-109-final-viral-combined_k141_56210\|\|full | Not-determined |
| 18-109-final-viral-combined_k141_200001\|\|full | Low-quality |
| 18-109-final-viral-combined_k141_66909\|\|full | Low-quality |
| 18-109-final-viral-combined_k141_106932\|\|full | Not-determined |
| 18-109-final-viral-combined_k141_96884\|\|full | Not-determined |
| 18-109-final-viral-combined_k141_127955\|\|full | Low-quality |
| 18-109-final-viral-combined_k141_96886\|\|full | Not-determined |
| 18-109-final-viral-combined_k141_189382\|\|full | Low-quality |
| 18-109-final-viral-combined_k141_86517\|\|full | Low-quality |
| 18-109-final-viral-combined_k141_218641\|\|full | Not-determined |
| 18-109-final-viral-combined_k141_66915\|\|full | Not-determined |
| 18-109-final-viral-combined_k141_15312\|\|full | Low-quality |
| 18-109-final-viral-combined_k141_106934\|\|full | Low-quality |
| 18-109-final-viral-combined_k141_36067\|\|full | Low-quality |
| 18-109-final-viral-combined_k141_189390\|\|full | Not-determined |
| 18-109-final-viral-combined_k141_86528\|\|full | Low-quality |
| 18-109-final-viral-combined_k141_137833\|\|full | Low-quality |
| 18-109-final-viral-combined_k141_179088\|\|full | Low-quality |
| 18-109-final-viral-combined_k141_24679\|\|full | Not-determined |
| 18-109-final-viral-combined_k141_229998\|\|full | Low-quality |
| 18-109-final-viral-combined_k141_189399\|\|full | Low-quality |
| 18-109-final-viral-combined_k141_158667\|\|full | Low-quality |
| 18-109-final-viral-combined_k141_218645\|\|full | Not-determined |
| 18-109-final-viral-combined_k141_86557\|\|full | Low-quality |
| 18-109-final-viral-combined_k141_127990\|\|full | Not-determined |
| 18-109-final-viral-combined_k141_36096\|\|full | Not-determined |
| 18-109-final-viral-combined_k141_46006\|\|full | Not-determined |
| 18-109-final-viral-combined_k141_179124\|\|full | Not-determined |
| 18-109-final-viral-combined_k141_200019\|\|full | Low-quality |
| 18-109-final-viral-combined_k141_76117\|\|full | Low-quality |
| 18-109-final-viral-combined_k141_46015\|\|full | Low-quality |
| 18-109-final-viral-combined_k141_210069\|\|full | Low-quality |
| 18-109-final-viral-combined_k141_24715\|\|full | Low-quality |
| 18-109-final-viral-combined_k141_169058\|\|full | Low-quality |
| 18-109-final-viral-combined_k141_210071\|\|full | Low-quality |
| 18-109-final-viral-combined_k141_66935\|\|full | Low-quality |
| 18-109-final-viral-combined_k141_46031\|\|full | Low-quality |
| 18-109-final-viral-combined_k141_128032\|\|full | Not-determined |
| 18-109-final-viral-combined_k141_106958\|\|full | Low-quality |
| 18-109-final-viral-combined_k141_147772\|\|full | Not-determined |
| 18-109-final-viral-combined_k141_218664\|\|full | Low-quality |
| 18-109-final-viral-combined_k141_106969\|\|full | Low-quality |
| 18-109-final-viral-combined_k141_210084\|\|full | Low-quality |
| 18-109-final-viral-combined_k141_218674\|\|full | Low-quality |
| 18-109-final-viral-combined_k141_117990\|\|full | Not-determined |
| 18-109-final-viral-combined_k141_158695\|\|full | Low-quality |
| 18-109-final-viral-combined_k141_147786\|\|full | Low-quality |
| 18-109-final-viral-combined_k141_179127\|\|full | Low-quality |
| 18-109-final-viral-combined_k141_210102\|\|full | Not-determined |
| 18-109-final-viral-combined_k141_200043\|\|full | Not-determined |
| 18-109-final-viral-combined_k141_118027\|\|full | Low-quality |
| 18-109-final-viral-combined_k141_5181\|\|full | Not-determined |
| 18-109-final-viral-combined_k141_200050\|\|full | Not-determined |
| 18-109-final-viral-combined_k141_158718\|\|full | Not-determined |
| 18-109-final-viral-combined_k141_66997\|\|full | Not-determined |
| 18-109-final-viral-combined_k141_240486\|\|full | Low-quality |
| 18-109-final-viral-combined_k141_240487\|\|full | Not-determined |
| 18-109-final-viral-combined_k141_24739\|\|full | Not-determined |
| KM-final-viral-combined_k141_230221\|\|full | High-quality |
| 18-109-final-viral-combined_k141_210147\|\|full | Low-quality |
| 18-109-final-viral-combined_k141_15357\|\|full | Low-quality |
| 18-109-final-viral-combined_k141_179153\|\|full | Low-quality |
| 18-109-final-viral-combined_k141_147822\|\|full | Not-determined |
| 18-109-final-viral-combined_k141_200090\|\|full | Not-determined |
| 18-109-final-viral-combined_k141_107047\|\|full | Not-determined |
| 18-109-final-viral-combined_k141_86653\|\|full | Low-quality |
| 18-109-final-viral-combined_k141_118057\|\|full | Not-determined |
| 18-109-final-viral-combined_k141_24759\|\|full | Not-determined |
| 18-109-final-viral-combined_k141_118058\|\|full | Not-determined |
| 18-109-final-viral-combined_k141_189530\|\|full | Not-determined |
| 18-109-final-viral-combined_k141_147833\|\|full | Low-quality |
| 18-109-final-viral-combined_k141_46144\|\|full | Low-quality |
| 18-109-final-viral-combined_k141_179165\|\|full | Low-quality |
| 18-109-final-viral-combined_k141_189544\|\|full | Low-quality |
| 18-109-final-viral-combined_k141_76248\|\|full | Not-determined |
| 18-109-final-viral-combined_k141_147857\|\|full | Low-quality |
| 18-109-final-viral-combined_k141_96940\|\|full | Not-determined |
| 18-109-final-viral-combined_k141_169125\|\|full | Low-quality |
| 18-109-final-viral-combined_k141_107088\|\|full | Not-determined |
| 18-109-final-viral-combined_k141_128101\|\|full | Low-quality |
| 18-109-final-viral-combined_k141_96959\|\|full | Not-determined |
| 18-109-final-viral-combined_k141_67056\|\|full | Low-quality |
| 18-109-final-viral-combined_k141_96966\|\|full | Low-quality |
| 18-109-final-viral-combined_k141_96976\|\|full | Not-determined |
| 18-109-final-viral-combined_k141_5258\|\|full | Not-determined |
| 18-109-final-viral-combined_k141_137942\|\|full | Low-quality |
| 18-109-final-viral-combined_k141_200173\|\|full | Not-determined |
| 18-109-final-viral-combined_k141_200178\|\|full | Low-quality |
| 18-109-final-viral-combined_k141_158751\|\|full | Low-quality |
| 18-109-final-viral-combined_k141_128144\|\|full | Low-quality |
| 18-109-final-viral-combined_k141_179219\|\|full | Not-determined |
| 18-109-final-viral-combined_k141_76287\|\|full | Low-quality |
| 18-109-final-viral-combined_k141_218726\|\|full | Low-quality |
| 18-109-final-viral-combined_k141_56248\|\|full | Not-determined |
| 18-109-final-viral-combined_k141_76300\|\|full | Not-determined |
| 18-109-final-viral-combined_k141_5300\|\|full | Low-quality |
| 18-109-final-viral-combined_k141_86699\|\|full | Not-determined |
| 18-109-final-viral-combined_k141_158795\|\|full | Low-quality |
| 18-109-final-viral-combined_k141_107106\|\|full | Low-quality |
| 18-109-final-viral-combined_k141_24814\|\|full | Not-determined |
| 18-109-final-viral-combined_k141_147939\|\|full | Not-determined |
| 18-109-final-viral-combined_k141_5322\|\|full | Low-quality |
| 18-109-final-viral-combined_k141_118185\|\|full | Not-determined |
| 18-109-final-viral-combined_k141_15382\|\|full | Not-determined |
| 18-109-final-viral-combined_k141_230150\|\|full | Low-quality |
| 18-109-final-viral-combined_k141_24835\|\|full | Low-quality |
| 18-109-final-viral-combined_k141_67107\|\|full | Not-determined |
| 18-109-final-viral-combined_k141_179296\|\|full | Not-determined |
| 18-109-final-viral-combined_k141_189616\|\|full | Low-quality |
| 18-109-final-viral-combined_k141_179297\|\|full | Not-determined |
| 18-109-final-viral-combined_k141_147954\|\|full | Low-quality |
| 18-109-final-viral-combined_k141_24837\|\|full | Not-determined |
| 18-109-final-viral-combined_k141_189622\|\|full | Not-determined |
| 18-109-final-viral-combined_k141_46216\|\|full | Low-quality |
| 18-109-final-viral-combined_k141_24845\|\|full | Low-quality |
| 18-109-final-viral-combined_k141_218744\|\|full | Low-quality |
| 18-109-final-viral-combined_k141_169261\|\|full | Low-quality |
| 18-109-final-viral-combined_k141_67150\|\|full | Low-quality |
| 18-109-final-viral-combined_k141_46233\|\|full | Not-determined |
| 18-109-final-viral-combined_k141_218757\|\|full | Low-quality |
| 18-109-final-viral-combined_k141_46238\|\|full | Low-quality |
| 18-109-final-viral-combined_k141_169280\|\|full | Not-determined |
| 18-109-final-viral-combined_k141_107195\|\|full | Low-quality |
| 18-109-final-viral-combined_k141_15443\|\|full | Not-determined |
| 18-109-final-viral-combined_k141_76424\|\|full | Low-quality |
| 18-109-final-viral-combined_k141_36304\|\|full | Low-quality |
| 18-109-final-viral-combined_k141_230213\|\|full | Not-determined |
| 18-109-final-viral-combined_k141_230226\|\|full | Not-determined |
| 18-109-final-viral-combined_k141_46258\|\|full | Not-determined |
| 18-109-final-viral-combined_k141_240588\|\|full | Low-quality |
| 18-109-final-viral-combined_k141_97081\|\|full | Not-determined |
| 18-109-final-viral-combined_k141_76434\|\|full | Not-determined |
| 18-109-final-viral-combined_k141_36333\|\|full | Low-quality |
| 18-109-final-viral-combined_k141_97086\|\|full | Low-quality |
| 18-109-final-viral-combined_k141_169322\|\|full | Low-quality |
| 18-109-final-viral-combined_k141_107229\|\|full | Low-quality |
| 18-109-final-viral-combined_k141_240597\|\|full | Not-determined |
| 18-109-final-viral-combined_k141_36346\|\|full | Not-determined |
| 18-109-final-viral-combined_k141_169329\|\|full | Not-determined |
| 18-109-final-viral-combined_k141_67198\|\|full | Low-quality |
| 18-109-final-viral-combined_k141_148008\|\|full | Not-determined |
| 18-109-final-viral-combined_k141_107239\|\|full | Not-determined |
| 18-109-final-viral-combined_k141_97123\|\|full | Not-determined |
| 18-109-final-viral-combined_k141_97126\|\|full | Low-quality |
| 18-109-final-viral-combined_k141_158879\|\|full | Not-determined |
| 18-109-final-viral-combined_k141_169360\|\|full | Not-determined |
| 18-109-final-viral-combined_k141_15518\|\|full | Not-determined |
| 18-109-final-viral-combined_k141_200296\|\|full | Low-quality |
| 18-109-final-viral-combined_k141_200306\|\|full | Not-determined |
| 18-109-final-viral-combined_k141_118223\|\|full | Not-determined |
| 18-109-final-viral-combined_k141_15534\|\|full | Low-quality |
| 18-109-final-viral-combined_k141_148047\|\|full | Not-determined |
| 18-109-final-viral-combined_k141_46302\|\|full | Not-determined |
| 18-109-final-viral-combined_k141_179398\|\|full | Low-quality |
| 18-109-final-viral-combined_k141_179399\|\|full | Low-quality |
| 18-109-final-viral-combined_k141_46309\|\|full | Low-quality |
| 18-109-final-viral-combined_k141_118243\|\|full | Low-quality |
| 18-109-final-viral-combined_k141_138055\|\|full | Low-quality |
| 18-109-final-viral-combined_k141_36424\|\|full | Not-determined |
| 18-109-final-viral-combined_k141_210309\|\|full | Not-determined |
| 18-109-final-viral-combined_k141_240636\|\|full | Low-quality |
| 18-109-final-viral-combined_k141_46343\|\|full | Not-determined |
| 18-109-final-viral-combined_k141_56462\|\|full | Not-determined |
| 18-109-final-viral-combined_k141_240650\|\|full | Low-quality |
| 18-109-final-viral-combined_k141_97214\|\|full | Not-determined |
| 18-109-final-viral-combined_k141_56469\|\|full | Low-quality |
| 18-109-final-viral-combined_k141_138092\|\|full | Low-quality |
| 18-109-final-viral-combined_k141_5457\|\|full | Low-quality |
| 18-109-final-viral-combined_k141_15599\|\|full | Not-determined |
| 18-109-final-viral-combined_k141_210323\|\|full | Not-determined |
| 18-109-final-viral-combined_k141_210328\|\|full | Low-quality |
| 18-109-final-viral-combined_k141_210330\|\|full | Low-quality |
| 18-109-final-viral-combined_k141_118290\|\|full | Low-quality |
| 18-109-final-viral-combined_k141_200407\|\|full | Low-quality |
| 18-109-final-viral-combined_k141_107265\|\|full | Not-determined |
| 18-109-final-viral-combined_k141_138138\|\|full | Low-quality |
| 18-109-final-viral-combined_k141_25023\|\|full | Low-quality |
| 18-109-final-viral-combined_k141_189772\|\|full | Low-quality |

| | |
|---|---|
| 18-109-final-viral-combined_k141_240707\|\|full | Not-determined |
| 18-109-final-viral-combined_k141_118310\|\|full | Low-quality |
| 18-109-final-viral-combined_k141_46412\|\|full | Not-determined |
| 18-109-final-viral-combined_k141_138150\|\|full | Not-determined |
| 18-109-final-viral-combined_k141_25032\|\|full | Low-quality |
| 18-109-final-viral-combined_k141_189782\|\|full | Low-quality |
| 18-109-final-viral-combined_k141_230337\|\|full | Not-determined |
| 18-109-final-viral-combined_k141_138156\|\|full | Not-determined |
| 18-109-final-viral-combined_k141_189787\|\|full | Low-quality |
| 18-109-final-viral-combined_k141_240728\|\|full | Not-determined |
| 18-109-final-viral-combined_k141_5505\|\|full | Not-determined |
| 18-109-final-viral-combined_k141_36494\|\|full | Low-quality |
| 18-109-final-viral-combined_k141_240739\|\|full | Not-determined |
| 18-109-final-viral-combined_k141_107293\|\|full | Not-determined |
| 18-109-final-viral-combined_k141_200445\|\|full | Low-quality |
| 18-109-final-viral-combined_k141_107307\|\|full | Low-quality |
| 18-109-final-viral-combined_k141_148167\|\|full | Low-quality |
| 18-109-final-viral-combined_k141_128227\|\|full | Low-quality |
| 18-109-final-viral-combined_k141_56557\|\|full | Low-quality |
| 18-109-final-viral-combined_k141_159021\|\|full | Not-determined |
| 18-109-final-viral-combined_k141_46454\|\|full | Not-determined |
| 18-109-final-viral-combined_k141_15685\|\|full | Low-quality |
| 17-109-final-viral-combined_k141_202803\|\|full | High-quality |
| 18-109-final-viral-combined_k141_86822\|\|full | Low-quality |
| 18-109-final-viral-combined_k141_230366\|\|full | Low-quality |
| 18-109-final-viral-combined_k141_128263\|\|full | Low-quality |
| 18-109-final-viral-combined_k141_86831\|\|full | Low-quality |
| 18-109-final-viral-combined_k141_107348\|\|full | Low-quality |
| 18-109-final-viral-combined_k141_97257\|\|full | Not-determined |
| 18-109-final-viral-combined_k141_86846\|\|full | Not-determined |
| 18-109-final-viral-combined_k141_36551\|\|full | Not-determined |
| 18-109-final-viral-combined_k141_25098\|\|full | Not-determined |
| 18-109-final-viral-combined_k141_138206\|\|full | Not-determined |
| 18-109-final-viral-combined_k141_179467\|\|full | Low-quality |
| 18-109-final-viral-combined_k141_218922\|\|full | Low-quality |
| 18-109-final-viral-combined_k141_148205\|\|full | Not-determined |
| 18-109-final-viral-combined_k141_189881\|\|full | Not-determined |
| 18-109-final-viral-combined_k141_179475\|\|full | Low-quality |
| 18-109-final-viral-combined_k141_148215\|\|full | Not-determined |
| 18-109-final-viral-combined_k141_128274\|\|full | Not-determined |
| 18-109-final-viral-combined_k141_76586\|\|full | Not-determined |
| 18-109-final-viral-combined_k141_128275\|\|full | Low-quality |
| 18-109-final-viral-combined_k141_76590\|\|full | Low-quality |
| 18-109-final-viral-combined_k141_107389\|\|full | Low-quality |
| 18-109-final-viral-combined_k141_107393\|\|full | Not-determined |
| 18-109-final-viral-combined_k141_159072\|\|full | Low-quality |
| 18-109-final-viral-combined_k141_5570\|\|full | Low-quality |
| 18-109-final-viral-combined_k141_36605\|\|full | Not-determined |
| 18-109-final-viral-combined_k141_97317\|\|full | Low-quality |
| 18-109-final-viral-combined_k141_67247\|\|full | Low-quality |
| 18-109-final-viral-combined_k141_169414\|\|full | Not-determined |
| 18-109-final-viral-combined_k141_189883\|\|full | Not-determined |
| 18-109-final-viral-combined_k141_5574\|\|full | Low-quality |
| 18-109-final-viral-combined_k141_67262\|\|full | Low-quality |
| 18-109-final-viral-combined_k141_159085\|\|full | Low-quality |
| 18-109-final-viral-combined_k141_15746\|\|full | Not-determined |
| 18-109-final-viral-combined_k141_118347\|\|full | Low-quality |
| 18-109-final-viral-combined_k141_138248\|\|full | Low-quality |
| 18-109-final-viral-combined_k141_97333\|\|full | Low-quality |
| 18-109-final-viral-combined_k141_5584\|\|full | Low-quality |
| 18-109-final-viral-combined_k141_210481\|\|full | Low-quality |
| 18-109-final-viral-combined_k141_138254\|\|full | Not-determined |
| 18-109-final-viral-combined_k141_240805\|\|full | Low-quality |
| 18-109-final-viral-combined_k141_148260\|\|full | Low-quality |
| 18-109-final-viral-combined_k141_138257\|\|full | Low-quality |
| 18-109-final-viral-combined_k141_36639\|\|full | Not-determined |
| 18-109-final-viral-combined_k141_148264\|\|full | Low-quality |
| 18-109-final-viral-combined_k141_107418\|\|full | Not-determined |
| 18-109-final-viral-combined_k141_15768\|\|full | Low-quality |
| 18-109-final-viral-combined_k141_240824\|\|full | Not-determined |
| 18-109-final-viral-combined_k141_138269\|\|full | Not-determined |
| 18-109-final-viral-combined_k141_97356\|\|full | Not-determined |
| 18-109-final-viral-combined_k141_128307\|\|full | Low-quality |
| 18-109-final-viral-combined_k141_56638\|\|full | Low-quality |
| 18-109-final-viral-combined_k141_148266\|\|full | Not-determined |
| 18-109-final-viral-combined_k141_36662\|\|full | Not-determined |
| 18-109-final-viral-combined_k141_118364\|\|full | Not-determined |
| 18-109-final-viral-combined_k141_97378\|\|full | Low-quality |
| 18-109-final-viral-combined_k141_36678\|\|full | Not-determined |
| 18-109-final-viral-combined_k141_169479\|\|full | Not-determined |
| 18-109-final-viral-combined_k141_36681\|\|full | Not-determined |
| 18-109-final-viral-combined_k141_76641\|\|full | Low-quality |
| 18-109-final-viral-combined_k141_76642\|\|full | Not-determined |
| 18-109-final-viral-combined_k141_230470\|\|full | Low-quality |
| 18-109-final-viral-combined_k141_86908\|\|full | Not-determined |
| 18-109-final-viral-combined_k141_15780\|\|full | Not-determined |
| 18-109-final-viral-combined_k141_200556\|\|full | Low-quality |
| 18-109-final-viral-combined_k141_15790\|\|full | Low-quality |
| 18-109-final-viral-combined_k141_97395\|\|full | Not-determined |
| 18-109-final-viral-combined_k141_169503\|\|full | Low-quality |
| 18-109-final-viral-combined_k141_36705\|\|full | Not-determined |
| 18-109-final-viral-combined_k141_56664\|\|full | Not-determined |
| 18-109-final-viral-combined_k141_179549\|\|full | Low-quality |
| 18-109-final-viral-combined_k141_200558\|\|full | Low-quality |
| 18-109-final-viral-combined_k141_107490\|\|full | Not-determined |
| 18-109-final-viral-combined_k141_189969\|\|full | Low-quality |
| 18-109-final-viral-combined_k141_189979\|\|full | Not-determined |
| 18-109-final-viral-combined_k141_76673\|\|full | Not-determined |
| 18-109-final-viral-combined_k141_200581\|\|full | Low-quality |
| 18-109-final-viral-combined_k141_200583\|\|full | Low-quality |
| 18-109-final-viral-combined_k141_128381\|\|full | Low-quality |
| 18-109-final-viral-combined_k141_179564\|\|full | Low-quality |
| 18-109-final-viral-combined_k141_189985\|\|full | Not-determined |
| 18-109-final-viral-combined_k141_46621\|\|full | Low-quality |
| 18-109-final-viral-combined_k141_128385\|\|full | Not-determined |
| 18-109-final-viral-combined_k141_128386\|\|full | Not-determined |
| 18-109-final-viral-combined_k141_56688\|\|full | Low-quality |
| 18-109-final-viral-combined_k141_179571\|\|full | Not-determined |
| 18-109-final-viral-combined_k141_190007\|\|full | Low-quality |
| 18-109-final-viral-combined_k141_179588\|\|full | Low-quality |
| 18-109-final-viral-combined_k141_107523\|\|full | Low-quality |
| 18-109-final-viral-combined_k141_25191\|\|full | Not-determined |
| 18-109-final-viral-combined_k141_118424\|\|full | Low-quality |
| 18-109-final-viral-combined_k141_200609\|\|full | Not-determined |
| 18-109-final-viral-combined_k141_118425\|\|full | Not-determined |
| 18-109-final-viral-combined_k141_36745\|\|full | Low-quality |
| 18-109-final-viral-combined_k141_240944\|\|full | Low-quality |
| 18-109-final-viral-combined_k141_128401\|\|full | Low-quality |
| 18-109-final-viral-combined_k141_210610\|\|full | Low-quality |
| 18-109-final-viral-combined_k141_56733\|\|full | Low-quality |
| 18-109-final-viral-combined_k141_210613\|\|full | Not-determined |
| 18-109-final-viral-combined_k141_179622\|\|full | Not-determined |
| 18-109-final-viral-combined_k141_118429\|\|full | Not-determined |
| 18-109-final-viral-combined_k141_190057\|\|full | Low-quality |
| 18-109-final-viral-combined_k141_76740\|\|full | Not-determined |
| 18-109-final-viral-combined_k141_128445\|\|full | Not-determined |
| 18-109-final-viral-combined_k141_67318\|\|full | Not-determined |
| 18-109-final-viral-combined_k141_190073\|\|full | Not-determined |
| 18-109-final-viral-combined_k141_36770\|\|full | Not-determined |
| 18-109-final-viral-combined_k141_159211\|\|full | Not-determined |
| 18-109-final-viral-combined_k141_210657\|\|full | Low-quality |
| 18-109-final-viral-combined_k141_76759\|\|full | Low-quality |
| 18-109-final-viral-combined_k141_87022\|\|full | Low-quality |
| 17-109-final-viral-combined_k141_107602\|\|full | Medium-quality |
| 18-109-final-viral-combined_k141_230519\|\|full | Not-determined |
| 18-109-final-viral-combined_k141_169582\|\|full | Not-determined |
| 18-109-final-viral-combined_k141_159239\|\|full | Not-determined |
| 18-109-final-viral-combined_k141_107571\|\|full | Not-determined |
| 18-109-final-viral-combined_k141_190118\|\|full | Low-quality |
| 18-109-final-viral-combined_k141_230524\|\|full | Low-quality |
| 18-109-final-viral-combined_k141_200677\|\|full | Low-quality |
| 18-109-final-viral-combined_k141_218972\|\|full | Low-quality |
| 18-109-final-viral-combined_k141_46745\|\|full | Not-determined |
| 18-109-final-viral-combined_k141_87045\|\|full | Low-quality |
| 18-109-final-viral-combined_k141_240982\|\|full | Not-determined |
| 18-109-final-viral-combined_k141_138383\|\|full | Low-quality |
| 18-109-final-viral-combined_k141_240983\|\|full | Low-quality |
| 18-109-final-viral-combined_k141_148336\|\|full | Not-determined |
| 18-109-final-viral-combined_k141_46758\|\|full | Not-determined |
| 18-109-final-viral-combined_k141_138404\|\|full | Not-determined |
| 18-109-final-viral-combined_k141_200700\|\|full | Not-determined |
| 18-109-final-viral-combined_k141_128501\|\|full | Low-quality |
| 18-109-final-viral-combined_k141_148358\|\|full | Low-quality |
| 18-109-final-viral-combined_k141_128506\|\|full | Not-determined |
| 18-109-final-viral-combined_k141_159266\|\|full | Low-quality |
| 18-109-final-viral-combined_k141_97482\|\|full | Low-quality |
| 18-109-final-viral-combined_k141_25200\|\|full | Not-determined |
| 18-109-final-viral-combined_k141_118497\|\|full | Low-quality |
| 18-109-final-viral-combined_k141_87081\|\|full | Low-quality |
| 18-109-final-viral-combined_k141_56810\|\|full | Not-determined |
| 18-109-final-viral-combined_k141_128520\|\|full | Low-quality |
| 18-109-final-viral-combined_k141_128526\|\|full | Not-determined |
| 18-109-final-viral-combined_k141_56816\|\|full | Low-quality |
| 18-109-final-viral-combined_k141_138418\|\|full | Low-quality |
| 18-109-final-viral-combined_k141_25213\|\|full | Not-determined |
| 18-109-final-viral-combined_k141_138439\|\|full | Low-quality |
| 18-109-final-viral-combined_k141_128569\|\|full | Low-quality |
| 18-109-final-viral-combined_k141_5699\|\|full | Not-determined |
| 18-109-final-viral-combined_k141_67446\|\|full | Low-quality |
| 18-109-final-viral-combined_k141_241046\|\|full | Low-quality |
| 18-109-final-viral-combined_k141_190216\|\|full | Not-determined |
| 18-109-final-viral-combined_k141_179781\|\|full | Low-quality |
| 18-109-final-viral-combined_k141_87176\|\|full | Not-determined |
| 18-109-final-viral-combined_k141_179791\|\|full | Not-determined |
| 18-109-final-viral-combined_k141_190235\|\|full | Not-determined |
| 18-109-final-viral-combined_k141_128594\|\|full | Low-quality |
| 18-109-final-viral-combined_k141_67475\|\|full | Not-determined |
| 18-109-final-viral-combined_k141_46805\|\|full | Low-quality |
| 18-109-final-viral-combined_k141_241070\|\|full | Not-determined |
| 18-109-final-viral-combined_k141_138469\|\|full | Not-determined |
| 18-109-final-viral-combined_k141_118606\|\|full | Not-determined |
| 18-109-final-viral-combined_k141_25257\|\|full | Not-determined |
| 18-109-final-viral-combined_k141_219046\|\|full | Not-determined |
| 18-109-final-viral-combined_k141_169723\|\|full | Low-quality |
| 18-109-final-viral-combined_k141_219056\|\|full | Not-determined |
| 18-109-final-viral-combined_k141_67506\|\|full | Not-determined |
| 18-109-final-viral-combined_k141_5711\|\|full | Low-quality |
| 18-109-final-viral-combined_k141_241114\|\|full | Not-determined |
| 18-109-final-viral-combined_k141_169747\|\|full | Not-determined |
| 18-109-final-viral-combined_k141_36944\|\|full | Low-quality |
| 18-109-final-viral-combined_k141_159358\|\|full | Not-determined |
| 18-109-final-viral-combined_k141_219081\|\|full | Not-determined |
| 18-109-final-viral-combined_k141_169753\|\|full | Low-quality |
| 18-109-final-viral-combined_k141_159362\|\|full | Low-quality |
| 18-109-final-viral-combined_k141_67529\|\|full | Not-determined |
| 18-109-final-viral-combined_k141_210723\|\|full | Low-quality |
| 18-109-final-viral-combined_k141_5731\|\|full | Low-quality |
| 18-109-final-viral-combined_k141_76808\|\|full | Low-quality |
| 18-109-final-viral-combined_k141_219099\|\|full | Low-quality |
| 18-109-final-viral-combined_k141_241130\|\|full | Not-determined |
| 18-109-final-viral-combined_k141_5746\|\|full | Low-quality |
| 18-109-final-viral-combined_k141_210738\|\|full | Low-quality |
| 18-109-final-viral-combined_k141_76831\|\|full | Not-determined |
| 18-109-final-viral-combined_k141_138518\|\|full | Low-quality |
| 18-109-final-viral-combined_k141_74829\|\|full | Complete |
| 18-109-final-viral-combined_k141_148475\|\|full | Low-quality |
| 18-109-final-viral-combined_k141_118620\|\|full | Not-determined |
| 18-109-final-viral-combined_k141_159421\|\|full | Not-determined |
| 18-109-final-viral-combined_k141_25305\|\|full | Low-quality |
| 18-109-final-viral-combined_k141_138543\|\|full | Not-determined |
| 18-109-final-viral-combined_k141_159424\|\|full | Low-quality |
| 18-109-final-viral-combined_k141_37013\|\|full | Low-quality |
| 18-109-final-viral-combined_k141_148510\|\|full | Not-determined |
| 18-109-final-viral-combined_k141_138558\|\|full | Not-determined |
| 18-109-final-viral-combined_k141_118634\|\|full | Low-quality |
| 18-109-final-viral-combined_k141_16014\|\|full | Low-quality |
| 18-109-final-viral-combined_k141_210786\|\|full | Not-determined |
| 18-109-final-viral-combined_k141_97590\|\|full | Low-quality |
| 18-109-final-viral-combined_k141_87267\|\|full | Low-quality |
| 18-109-final-viral-combined_k141_138577\|\|full | Not-determined |
| 18-109-final-viral-combined_k141_16022\|\|full | Not-determined |
| 18-109-final-viral-combined_k141_37042\|\|full | Low-quality |
| 18-110-final-viral-combined_k141_25607\|\|full | High-quality |
| 18-109-final-viral-combined_k141_118642\|\|full | Low-quality |
| 18-109-final-viral-combined_k141_169874\|\|full | Not-determined |
| 18-109-final-viral-combined_k141_56910\|\|full | Not-determined |
| 18-109-final-viral-combined_k141_179879\|\|full | Not-determined |
| 18-109-final-viral-combined_k141_97622\|\|full | Low-quality |
| 18-109-final-viral-combined_k141_16034\|\|full | Low-quality |
| 18-109-final-viral-combined_k141_241195\|\|full | Not-determined |
| 18-109-final-viral-combined_k141_97636\|\|full | Low-quality |
| 18-109-final-viral-combined_k141_118662\|\|full | Not-determined |
| 18-109-final-viral-combined_k141_118665\|\|full | Low-quality |
| 18-109-final-viral-combined_k141_219151\|\|full | Low-quality |
| 18-109-final-viral-combined_k141_200750\|\|full | Low-quality |
| 18-109-final-viral-combined_k141_67658\|\|full | Not-determined |
| 18-109-final-viral-combined_k141_159482\|\|full | Low-quality |
| 18-109-final-viral-combined_k141_138606\|\|full | Low-quality |
| 18-109-final-viral-combined_k141_219169\|\|full | Not-determined |
| 18-109-final-viral-combined_k141_230667\|\|full | Not-determined |
| 18-109-final-viral-combined_k141_230674\|\|full | Not-determined |
| 18-109-final-viral-combined_k141_148607\|\|full | Not-determined |
| 18-109-final-viral-combined_k141_241242\|\|full | Not-determined |
| 18-109-final-viral-combined_k141_128737\|\|full | Low-quality |
| 18-109-final-viral-combined_k141_230685\|\|full | Low-quality |
| 18-109-final-viral-combined_k141_241244\|\|full | Not-determined |
| 17-108-final-viral-combined_k141_321938\|\|full | High-quality |
| 18-109-final-viral-combined_k141_56990\|\|full | Low-quality |
| 18-109-final-viral-combined_k141_190288\|\|full | Low-quality |
| 18-109-final-viral-combined_k141_67732\|\|full | Low-quality |
| 18-109-final-viral-combined_k141_210860\|\|full | Not-determined |
| 18-109-final-viral-combined_k141_200797\|\|full | Not-determined |
| 18-109-final-viral-combined_k141_169966\|\|full | Not-determined |
| 18-109-final-viral-combined_k141_76886\|\|full | Not-determined |
| 18-109-final-viral-combined_k141_16100\|\|full | Not-determined |
| 18-109-final-viral-combined_k141_25380\|\|full | Low-quality |
| 18-109-final-viral-combined_k141_169986\|\|full | Low-quality |
| 18-109-final-viral-combined_k141_118724\|\|full | Not-determined |
| 18-109-final-viral-combined_k141_57042\|\|full | Low-quality |
| 18-109-final-viral-combined_k141_219253\|\|full | Low-quality |
| 18-109-final-viral-combined_k141_219255\|\|full | Low-quality |
| 18-109-final-viral-combined_k141_76896\|\|full | Not-determined |
| 18-109-final-viral-combined_k141_138711\|\|full | Not-determined |
| 18-109-final-viral-combined_k141_219269\|\|full | Not-determined |
| 18-109-final-viral-combined_k141_138727\|\|full | Low-quality |
| 18-109-final-viral-combined_k141_16156\|\|full | Not-determined |
| 18-109-final-viral-combined_k141_128754\|\|full | Low-quality |
| 18-109-final-viral-combined_k141_5847\|\|full | Not-determined |
| 18-109-final-viral-combined_k141_25412\|\|full | Not-determined |
| 18-109-final-viral-combined_k141_219294\|\|full | Low-quality |
| 18-109-final-viral-combined_k141_128765\|\|full | Low-quality |
| 18-109-final-viral-combined_k141_25443\|\|full | Low-quality |
| 18-109-final-viral-combined_k141_190338\|\|full | Not-determined |
| 18-109-final-viral-combined_k141_37225\|\|full | Low-quality |
| 18-109-final-viral-combined_k141_67803\|\|full | Low-quality |
| 18-109-final-viral-combined_k141_97766\|\|full | Not-determined |
| 18-109-final-viral-combined_k141_230782\|\|full | Not-determined |
| 18-109-final-viral-combined_k141_170031\|\|full | Not-determined |
| 18-109-final-viral-combined_k141_25474\|\|full | Low-quality |
| 18-109-final-viral-combined_k141_97784\|\|full | Not-determined |
| 18-109-final-viral-combined_k141_210973\|\|full | Not-determined |
| 18-109-final-viral-combined_k141_170044\|\|full | Low-quality |
| 18-109-final-viral-combined_k141_46925\|\|full | Not-determined |
| 18-109-final-viral-combined_k141_210977\|\|full | Low-quality |
| 18-109-final-viral-combined_k141_190398\|\|full | Low-quality |
| 18-109-final-viral-combined_k141_16213\|\|full | Low-quality |
| 18-109-final-viral-combined_k141_230805\|\|full | Low-quality |
| 18-109-final-viral-combined_k141_179993\|\|full | Low-quality |
| 18-109-final-viral-combined_k141_46949\|\|full | Low-quality |
| 18-109-final-viral-combined_k141_67860\|\|full | Low-quality |
| 18-109-final-viral-combined_k141_37278\|\|full | Not-determined |
| 18-109-final-viral-combined_k141_76994\|\|full | Not-determined |
| 18-109-final-viral-combined_k141_57191\|\|full | Not-determined |
| 18-109-final-viral-combined_k141_118799\|\|full | Low-quality |
| 18-109-final-viral-combined_k141_25520\|\|full | Low-quality |
| 18-109-final-viral-combined_k141_5919\|\|full | Low-quality |
| 18-109-final-viral-combined_k141_57209\|\|full | Not-determined |
| 18-109-final-viral-combined_k141_37304\|\|full | Low-quality |
| 18-109-final-viral-combined_k141_118827\|\|full | Not-determined |
| 18-109-final-viral-combined_k141_128899\|\|full | Low-quality |
| 18-109-final-viral-combined_k141_97820\|\|full | Not-determined |
| 18-109-final-viral-combined_k141_128910\|\|full | Not-determined |
| 18-109-final-viral-combined_k141_148766\|\|full | Low-quality |
| 18-109-final-viral-combined_k141_159762\|\|full | Not-determined |
| 18-109-final-viral-combined_k141_77032\|\|full | Not-determined |
| 18-109-final-viral-combined_k141_87473\|\|full | Not-determined |
| 18-109-final-viral-combined_k141_180037\|\|full | Not-determined |
| 18-109-final-viral-combined_k141_128916\|\|full | Not-determined |
| 18-109-final-viral-combined_k141_46983\|\|full | Not-determined |
| 18-109-final-viral-combined_k141_230863\|\|full | Low-quality |
| 18-109-final-viral-combined_k141_148776\|\|full | Low-quality |
| 18-109-final-viral-combined_k141_46985\|\|full | Not-determined |
| 18-109-final-viral-combined_k141_46987\|\|full | Low-quality |
| 18-109-final-viral-combined_k141_148782\|\|full | Not-determined |
| 18-109-final-viral-combined_k141_97845\|\|full | Not-determined |
| 18-109-final-viral-combined_k141_180058\|\|full | Not-determined |
| 18-109-final-viral-combined_k141_170128\|\|full | Low-quality |
| 18-109-final-viral-combined_k141_180068\|\|full | Low-quality |
| 18-109-final-viral-combined_k141_97860\|\|full | Low-quality |
| 18-109-final-viral-combined_k141_16340\|\|full | Not-determined |
| 18-109-final-viral-combined_k141_241346\|\|full | Not-determined |
| 18-109-final-viral-combined_k141_5955\|\|full | Low-quality |
| 18-109-final-viral-combined_k141_219413\|\|full | Low-quality |
| 18-109-final-viral-combined_k141_87502\|\|full | Not-determined |
| 18-109-final-viral-combined_k141_118883\|\|full | Not-determined |
| 18-109-final-viral-combined_k141_148799\|\|full | Low-quality |
| 18-109-final-viral-combined_k141_219420\|\|full | Not-determined |
| 18-109-final-viral-combined_k141_118901\|\|full | Low-quality |
| 18-109-final-viral-combined_k141_170149\|\|full | Low-quality |
| 18-109-final-viral-combined_k141_241356\|\|full | Not-determined |
| 18-109-final-viral-combined_k141_77071\|\|full | Not-determined |
| 18-109-final-viral-combined_k141_16378\|\|full | Not-determined |
| 18-109-final-viral-combined_k141_118922\|\|full | Low-quality |
| 18-109-final-viral-combined_k141_5990\|\|full | Not-determined |
| 18-109-final-viral-combined_k141_97908\|\|full | Low-quality |
| 18-109-final-viral-combined_k141_190558\|\|full | Low-quality |
| 18-109-final-viral-combined_k141_5998\|\|full | Low-quality |
| 18-109-final-viral-combined_k141_148827\|\|full | Not-determined |
| 18-109-final-viral-combined_k141_211047\|\|full | Not-determined |
| 18-109-final-viral-combined_k141_230916\|\|full | Low-quality |
| 18-109-final-viral-combined_k141_107703\|\|full | Not-determined |
| 18-109-final-viral-combined_k141_6007\|\|full | Low-quality |
| 18-109-final-viral-combined_k141_57353\|\|full | Not-determined |
| 18-109-final-viral-combined_k141_211055\|\|full | Low-quality |
| 18-109-final-viral-combined_k141_241412\|\|full | Not-determined |
| 18-109-final-viral-combined_k141_190591\|\|full | Low-quality |
| 18-109-final-viral-combined_k141_77110\|\|full | Not-determined |
| 18-109-final-viral-combined_k141_241416\|\|full | Not-determined |
| 18-109-final-viral-combined_k141_200890\|\|full | Low-quality |
| 18-109-final-viral-combined_k141_190594\|\|full | Not-determined |
| 18-109-final-viral-combined_k141_200891\|\|full | Low-quality |
| 18-109-final-viral-combined_k141_170218\|\|full | Low-quality |
| 18-109-final-viral-combined_k141_6032\|\|full | Not-determined |
| 18-109-final-viral-combined_k141_37344\|\|full | Not-determined |
| 18-109-final-viral-combined_k141_148879\|\|full | Not-determined |
| 18-109-final-viral-combined_k141_87554\|\|full | Not-determined |
| 18-109-final-viral-combined_k141_148886\|\|full | Not-determined |
| 18-109-final-viral-combined_k141_118972\|\|full | Not-determined |
| 18-109-final-viral-combined_k141_170249\|\|full | Low-quality |
| 18-109-final-viral-combined_k141_97955\|\|full | Low-quality |
| 18-109-final-viral-combined_k141_118998\|\|full | Not-determined |
| 18-109-final-viral-combined_k141_87576\|\|full | Not-determined |
| 18-109-final-viral-combined_k141_148923\|\|full | Low-quality |
| 18-109-final-viral-combined_k141_119007\|\|full | Low-quality |
| 18-109-final-viral-combined_k141_170263\|\|full | Not-determined |
| 18-109-final-viral-combined_k141_138912\|\|full | Not-determined |
| 18-109-final-viral-combined_k141_170268\|\|full | Low-quality |
| 18-109-final-viral-combined_k141_200930\|\|full | Low-quality |
| 18-109-final-viral-combined_k141_57407\|\|full | Low-quality |
| 18-109-final-viral-combined_k141_129052\|\|full | Not-determined |
| 18-109-final-viral-combined_k141_47062\|\|full | Not-determined |
| 18-109-final-viral-combined_k141_25764\|\|full | Not-determined |
| 18-109-final-viral-combined_k141_68011\|\|full | Not-determined |
| 18-109-final-viral-combined_k141_87598\|\|full | Low-quality |
| 18-109-final-viral-combined_k141_68012\|\|full | Low-quality |
| 18-109-final-viral-combined_k141_68013\|\|full | Not-determined |
| 18-109-final-viral-combined_k141_77153\|\|full | Not-determined |
| 18-109-final-viral-combined_k141_190659\|\|full | Low-quality |
| 18-109-final-viral-combined_k141_87607\|\|full | Not-determined |
| 18-109-final-viral-combined_k141_47089\|\|full | Low-quality |
| 18-109-final-viral-combined_k141_37384\|\|full | Not-determined |
| 18-109-final-viral-combined_k141_119055\|\|full | Not-determined |
| 18-109-final-viral-combined_k141_119062\|\|full | Not-determined |
| 18-109-final-viral-combined_k141_211161\|\|full | Low-quality |
| 18-109-final-viral-combined_k141_77179\|\|full | Not-determined |
| 17-108-final-viral-combined_k141_64392\|\|full | High-quality |
| 18-109-final-viral-combined_k141_190706\|\|full | Not-determined |
| 18-109-final-viral-combined_k141_148966\|\|full | Low-quality |
| 18-109-final-viral-combined_k141_57426\|\|full | Low-quality |
| 18-109-final-viral-combined_k141_138938\|\|full | Not-determined |
| 18-109-final-viral-combined_k141_148973\|\|full | Low-quality |
| 18-109-final-viral-combined_k141_190720\|\|full | Not-determined |
| 18-109-final-viral-combined_k141_138952\|\|full | Low-quality |
| 18-109-final-viral-combined_k141_6103\|\|full | Low-quality |
| 18-109-final-viral-combined_k141_200976\|\|full | Not-determined |
| 18-109-final-viral-combined_k141_77223\|\|full | Low-quality |
| 18-109-final-viral-combined_k141_190750\|\|full | Low-quality |
| 18-109-final-viral-combined_k141_138981\|\|full | Not-determined |
| 18-109-final-viral-combined_k141_138983\|\|full | Low-quality |
| 18-109-final-viral-combined_k141_190754\|\|full | Low-quality |
| 18-109-final-viral-combined_k141_68043\|\|full | Low-quality |
| 18-109-final-viral-combined_k141_47166\|\|full | Not-determined |
| 18-109-final-viral-combined_k141_231027\|\|full | Low-quality |
| 18-109-final-viral-combined_k141_16509\|\|full | Low-quality |
| 18-109-final-viral-combined_k141_129094\|\|full | Not-determined |
| 18-109-final-viral-combined_k141_47186\|\|full | Not-determined |
| 18-109-final-viral-combined_k141_241461\|\|full | Low-quality |
| 18-109-final-viral-combined_k141_201015\|\|full | Low-quality |
| 18-109-final-viral-combined_k141_98073\|\|full | Low-quality |
| 18-109-final-viral-combined_k141_241466\|\|full | Not-determined |
| 18-109-final-viral-combined_k141_241469\|\|full | Not-determined |
| 18-109-final-viral-combined_k141_77260\|\|full | Low-quality |
| 18-109-final-viral-combined_k141_231075\|\|full | Not-determined |
| 18-109-final-viral-combined_k141_201047\|\|full | Low-quality |
| 18-109-final-viral-combined_k141_231080\|\|full | Low-quality |
| 18-109-final-viral-combined_k141_231091\|\|full | Not-determined |
| 18-109-final-viral-combined_k141_119109\|\|full | Not-determined |
| 18-109-final-viral-combined_k141_129131\|\|full | Low-quality |
| 18-109-final-viral-combined_k141_219501\|\|full | Not-determined |
| 18-109-final-viral-combined_k141_190802\|\|full | Low-quality |
| 18-109-final-viral-combined_k141_231104\|\|full | Not-determined |
| 18-109-final-viral-combined_k141_201074\|\|full | Low-quality |
| 18-109-final-viral-combined_k141_201075\|\|full | Not-determined |
| 18-109-final-viral-combined_k141_6186\|\|full | Low-quality |
| 18-109-final-viral-combined_k141_57492\|\|full | Low-quality |
| 18-109-final-viral-combined_k141_139078\|\|full | Not-determined |
| 18-109-final-viral-combined_k141_180175\|\|full | Not-determined |
| 18-109-final-viral-combined_k141_159815\|\|full | Low-quality |
| 18-109-final-viral-combined_k141_149004\|\|full | Low-quality |
| 18-109-final-viral-combined_k141_170332\|\|full | Low-quality |
| 18-109-final-viral-combined_k141_107775\|\|full | Not-determined |
| 18-109-final-viral-combined_k141_201109\|\|full | Not-determined |
| 18-109-final-viral-combined_k141_201111\|\|full | Low-quality |
| 18-109-final-viral-combined_k141_57515\|\|full | Not-determined |
| 18-109-final-viral-combined_k141_16522\|\|full | Low-quality |
| 18-109-final-viral-combined_k141_57516\|\|full | Not-determined |
| 18-109-final-viral-combined_k141_57522\|\|full | Low-quality |
| 18-109-final-viral-combined_k141_77329\|\|full | Not-determined |
| 18-109-final-viral-combined_k141_16532\|\|full | Not-determined |
| 18-109-final-viral-combined_k141_6201\|\|full | Not-determined |
| 18-109-final-viral-combined_k141_241534\|\|full | Low-quality |
| 18-109-final-viral-combined_k141_6204\|\|full | Low-quality |
| 18-109-final-viral-combined_k141_16549\|\|full | Low-quality |
| 18-109-final-viral-combined_k141_211188\|\|full | Low-quality |
| 18-109-final-viral-combined_k141_231150\|\|full | Low-quality |
| 18-109-final-viral-combined_k141_16555\|\|full | Low-quality |
| 18-109-final-viral-combined_k141_139139\|\|full | Not-determined |
| 18-109-final-viral-combined_k141_119150\|\|full | Low-quality |
| 18-109-final-viral-combined_k141_159831\|\|full | Low-quality |
| 18-109-final-viral-combined_k141_6232\|\|full | Not-determined |
| 18-109-final-viral-combined_k141_47211\|\|full | Not-determined |
| 18-109-final-viral-combined_k141_57547\|\|full | Low-quality |
| 18-109-final-viral-combined_k141_241564\|\|full | Low-quality |
| 18-109-final-viral-combined_k141_57550\|\|full | Low-quality |
| 18-109-final-viral-combined_k141_6235\|\|full | Not-determined |
| 18-109-final-viral-combined_k141_47223\|\|full | Low-quality |
| 18-109-final-viral-combined_k141_6248\|\|full | Not-determined |
| 18-109-final-viral-combined_k141_231202\|\|full | Not-determined |
| 18-109-final-viral-combined_k141_87706\|\|full | Low-quality |
| 18-109-final-viral-combined_k141_57566\|\|full | Not-determined |
| 18-109-final-viral-combined_k141_98199\|\|full | Low-quality |
| 18-109-final-viral-combined_k141_6256\|\|full | Not-determined |
| 18-109-final-viral-combined_k141_16592\|\|full | Low-quality |
| 18-109-final-viral-combined_k141_47247\|\|full | Low-quality |
| 18-109-final-viral-combined_k141_170368\|\|full | Low-quality |
| 18-109-final-viral-combined_k141_6269\|\|full | Not-determined |
| 18-109-final-viral-combined_k141_98222\|\|full | Not-determined |
| 18-109-final-viral-combined_k141_159909\|\|full | Low-quality |
| 18-109-final-viral-combined_k141_6280\|\|full | Low-quality |
| 18-109-final-viral-combined_k141_68248\|\|full | Low-quality |
| 18-109-final-viral-combined_k141_47278\|\|full | Not-determined |
| 18-109-final-viral-combined_k141_139215\|\|full | Not-determined |
| 18-109-final-viral-combined_k141_98249\|\|full | Not-determined |
| 18-109-final-viral-combined_k141_211277\|\|full | Not-determined |
| 18-109-final-viral-combined_k141_87744\|\|full | Not-determined |
| 18-109-final-viral-combined_k141_37572\|\|full | Not-determined |
| 18-109-final-viral-combined_k141_16646\|\|full | Not-determined |
| 18-109-final-viral-combined_k141_77447\|\|full | Not-determined |
| 18-109-final-viral-combined_k141_119188\|\|full | Low-quality |
| 18-109-final-viral-combined_k141_57632\|\|full | Not-determined |
| 18-109-final-viral-combined_k141_37574\|\|full | Low-quality |
| 18-109-final-viral-combined_k141_190896\|\|full | Not-determined |
| 18-109-final-viral-combined_k141_119201\|\|full | Low-quality |
| 18-109-final-viral-combined_k141_37586\|\|full | Low-quality |
| 18-109-final-viral-combined_k141_98303\|\|full | Low-quality |
| 18-109-final-viral-combined_k141_87806\|\|full | Not-determined |
| 18-109-final-viral-combined_k141_190915\|\|full | Not-determined |
| 18-109-final-viral-combined_k141_37604\|\|full | Not-determined |
| 18-109-final-viral-combined_k141_170450\|\|full | Low-quality |
| 18-109-final-viral-combined_k141_149121\|\|full | Not-determined |
| 18-109-final-viral-combined_k141_201318\|\|full | Not-determined |
| 18-109-final-viral-combined_k141_170460\|\|full | Low-quality |
| 18-109-final-viral-combined_k141_219617\|\|full | Not-determined |
| 18-109-final-viral-combined_k141_47365\|\|full | Not-determined |
| 18-109-final-viral-combined_k141_190927\|\|full | Not-determined |
| 18-109-final-viral-combined_k141_47370\|\|full | Low-quality |
| 18-109-final-viral-combined_k141_129238\|\|full | Not-determined |
| 18-109-final-viral-combined_k141_47376\|\|full | Not-determined |
| 18-109-final-viral-combined_k141_190937\|\|full | Low-quality |
| 18-109-final-viral-combined_k141_6396\|\|full | Low-quality |
| 18-109-final-viral-combined_k141_26076\|\|full | Not-determined |
| 18-109-final-viral-combined_k141_26078\|\|full | Low-quality |
| 18-109-final-viral-combined_k141_57695\|\|full | Low-quality |
| 18-109-final-viral-combined_k141_119285\|\|full | Low-quality |
| 18-109-final-viral-combined_k141_211302\|\|full | Low-quality |
| 18-109-final-viral-combined_k141_180260\|\|full | Low-quality |
| 18-109-final-viral-combined_k141_57699\|\|full | Not-determined |
| 18-109-final-viral-combined_k141_190956\|\|full | Not-determined |
| 18-109-final-viral-combined_k141_37678\|\|full | Low-quality |
| 18-109-final-viral-combined_k141_170531\|\|full | Not-determined |
| 18-109-final-viral-combined_k141_190960\|\|full | Not-determined |
| 18-109-final-viral-combined_k141_190962\|\|full | Low-quality |
| 18-109-final-viral-combined_k141_201362\|\|full | Low-quality |
| 18-109-final-viral-combined_k141_37685\|\|full | Low-quality |
| 18-109-final-viral-combined_k141_16711\|\|full | Not-determined |
| 18-109-final-viral-combined_k141_47423\|\|full | Low-quality |
| 18-109-final-viral-combined_k141_107823\|\|full | Low-quality |
| 18-109-final-viral-combined_k141_37692\|\|full | Not-determined |
| 18-109-final-viral-combined_k141_170544\|\|full | Not-determined |
| 18-109-final-viral-combined_k141_57701\|\|full | Not-determined |
| 18-109-final-viral-combined_k141_37697\|\|full | Low-quality |
| 18-109-final-viral-combined_k141_219673\|\|full | Not-determined |
| 18-109-final-viral-combined_k141_57702\|\|full | Not-determined |
| 18-109-final-viral-combined_k141_37703\|\|full | Low-quality |
| 18-109-final-viral-combined_k141_6451\|\|full | Not-determined |
| 18-109-final-viral-combined_k141_6456\|\|full | Not-determined |
| 18-109-final-viral-combined_k141_201379\|\|full | Low-quality |
| 18-109-final-viral-combined_k141_68437\|\|full | Not-determined |
| 18-109-final-viral-combined_k141_107848\|\|full | Not-determined |
| 18-109-final-viral-combined_k141_201382\|\|full | Not-determined |
| 18-109-final-viral-combined_k141_231225\|\|full | Not-determined |
| 18-109-final-viral-combined_k141_37708\|\|full | Not-determined |
| 18-109-final-viral-combined_k141_149187\|\|full | Low-quality |
| 18-109-final-viral-combined_k141_87889\|\|full | Not-determined |
| 18-109-final-viral-combined_k141_98382\|\|full | Low-quality |
| 18-109-final-viral-combined_k141_98383\|\|full | Low-quality |
| 18-109-final-viral-combined_k141_6471\|\|full | Not-determined |
| 18-109-final-viral-combined_k141_6472\|\|full | Low-quality |
| 18-109-final-viral-combined_k141_211309\|\|full | Low-quality |
| 18-109-final-viral-combined_k141_16742\|\|full | Not-determined |
| 18-109-final-viral-combined_k141_87904\|\|full | Not-determined |
| 18-109-final-viral-combined_k141_98421\|\|full | Low-quality |
| 18-109-final-viral-combined_k141_119312\|\|full | Low-quality |
| 18-109-final-viral-combined_k141_26123\|\|full | Not-determined |
| 18-109-final-viral-combined_k141_211333\|\|full | Low-quality |
| 18-109-final-viral-combined_k141_107891\|\|full | Not-determined |
| 18-109-final-viral-combined_k141_170581\|\|full | Low-quality |
| 18-109-final-viral-combined_k141_87930\|\|full | Not-determined |
| 18-109-final-viral-combined_k141_160109\|\|full | Low-quality |
| 18-109-final-viral-combined_k141_170584\|\|full | Low-quality |
| 18-109-final-viral-combined_k141_107906\|\|full | Low-quality |
| 18-109-final-viral-combined_k141_231281\|\|full | Not-determined |
| 18-109-final-viral-combined_k141_180384\|\|full | Low-quality |
| 18-109-final-viral-combined_k141_231286\|\|full | Not-determined |
| 18-109-final-viral-combined_k141_219691\|\|full | Low-quality |
| 18-109-final-viral-combined_k141_231290\|\|full | Low-quality |
| 18-109-final-viral-combined_k141_47522\|\|full | Low-quality |
| 18-109-final-viral-combined_k141_68493\|\|full | Not-determined |
| 18-109-final-viral-combined_k141_211359\|\|full | Not-determined |
| 18-109-final-viral-combined_k141_57787\|\|full | Low-quality |
| 18-109-final-viral-combined_k141_139258\|\|full | Low-quality |
| 18-109-final-viral-combined_k141_139262\|\|full | Not-determined |
| 18-109-final-viral-combined_k141_47539\|\|full | Low-quality |
| 18-109-final-viral-combined_k141_201431\|\|full | Not-determined |
| 18-109-final-viral-combined_k141_37752\|\|full | Low-quality |
| 18-109-final-viral-combined_k141_211376\|\|full | Not-determined |
| 18-109-final-viral-combined_k141_139279\|\|full | Low-quality |
| 18-109-final-viral-combined_k141_231311\|\|full | Low-quality |
| 18-109-final-viral-combined_k141_211380\|\|full | Low-quality |
| 18-109-final-viral-combined_k141_139285\|\|full | Not-determined |
| 18-109-final-viral-combined_k141_57812\|\|full | Low-quality |
| 18-109-final-viral-combined_k141_160166\|\|full | Not-determined |
| 18-109-final-viral-combined_k141_6557\|\|full | Not-determined |
| 18-109-final-viral-combined_k141_47559\|\|full | Not-determined |
| 18-109-final-viral-combined_k141_88004\|\|full | Low-quality |
| 18-109-final-viral-combined_k141_6564\|\|full | Not-determined |
| 18-109-final-viral-combined_k141_170608\|\|full | Not-determined |
| 18-109-final-viral-combined_k141_160179\|\|full | Not-determined |
| 18-109-final-viral-combined_k141_68539\|\|full | Not-determined |
| 18-109-final-viral-combined_k141_231346\|\|full | Not-determined |
| 18-109-final-viral-combined_k141_219709\|\|full | Not-determined |
| 18-109-final-viral-combined_k141_57831\|\|full | Low-quality |
| 18-109-final-viral-combined_k141_47574\|\|full | Not-determined |
| 18-109-final-viral-combined_k141_107981\|\|full | Not-determined |
| 18-109-final-viral-combined_k141_160206\|\|full | Low-quality |
| 18-109-final-viral-combined_k141_6585\|\|full | Low-quality |
| 18-109-final-viral-combined_k141_88053\|\|full | Not-determined |
| 18-109-final-viral-combined_k141_219734\|\|full | Low-quality |
| 18-109-final-viral-combined_k141_108001\|\|full | Low-quality |

| | |
|---|---|
| 18-109-final-viral-combined_k141_98490\|\|full | Not-determined |
| 18-109-final-viral-combined_k141_241751\|\|full | Low-quality |
| 18-109-final-viral-combined_k141_6588\|\|full | Not-determined |
| 18-109-final-viral-combined_k141_108004\|\|full | Low-quality |
| 18-109-final-viral-combined_k141_6598\|\|full | Low-quality |
| 18-109-final-viral-combined_k141_241761\|\|full | Not-determined |
| 18-109-final-viral-combined_k141_57869\|\|full | Low-quality |
| 18-109-final-viral-combined_k141_129416\|\|full | Not-determined |
| 18-109-final-viral-combined_k141_16822\|\|full | Low-quality |
| 18-109-final-viral-combined_k141_241783\|\|full | Low-quality |
| 18-109-final-viral-combined_k141_108011\|\|full | Low-quality |
| 18-109-final-viral-combined_k141_77622\|\|full | Low-quality |
| 18-109-final-viral-combined_k141_57878\|\|full | Not-determined |
| 18-109-final-viral-combined_k141_6625\|\|full | Low-quality |
| 18-109-final-viral-combined_k141_6646\|\|full | Low-quality |
| 18-109-final-viral-combined_k141_16863\|\|full | Not-determined |
| 18-109-final-viral-combined_k141_37846\|\|full | Low-quality |
| 18-109-final-viral-combined_k141_160275\|\|full | Not-determined |
| 18-109-final-viral-combined_k141_108076\|\|full | Not-determined |
| 18-109-final-viral-combined_k141_180474\|\|full | Low-quality |
| 18-109-final-viral-combined_k141_37852\|\|full | Low-quality |
| 18-109-final-viral-combined_k141_160286\|\|full | Low-quality |
| 18-109-final-viral-combined_k141_129496\|\|full | Low-quality |
| 18-109-final-viral-combined_k141_6679\|\|full | Low-quality |
| 18-109-final-viral-combined_k141_180484\|\|full | Low-quality |
| 18-109-final-viral-combined_k141_211444\|\|full | Low-quality |
| 18-109-final-viral-combined_k141_98560\|\|full | Low-quality |
| 18-109-final-viral-combined_k141_108108\|\|full | Not-determined |
| 18-109-final-viral-combined_k141_88106\|\|full | Not-determined |
| 18-109-final-viral-combined_k141_219758\|\|full | Low-quality |
| 18-109-final-viral-combined_k141_180495\|\|full | Low-quality |
| 18-109-final-viral-combined_k141_211464\|\|full | Not-determined |
| 18-109-final-viral-combined_k141_108139\|\|full | Low-quality |
| 18-110-final-viral-combined_k141_276211\|\|full | High-quality |
| 18-109-final-viral-combined_k141_191249\|\|full | Low-quality |
| 18-109-final-viral-combined_k141_26220\|\|full | Low-quality |
| 18-109-final-viral-combined_k141_211473\|\|full | Not-determined |
| 18-109-final-viral-combined_k141_98581\|\|full | Not-determined |
| 18-109-final-viral-combined_k141_26221\|\|full | Not-determined |
| 18-109-final-viral-combined_k141_191273\|\|full | Low-quality |
| 18-109-final-viral-combined_k141_211483\|\|full | Not-determined |
| 18-109-final-viral-combined_k141_77703\|\|full | Low-quality |
| 18-109-final-viral-combined_k141_149283\|\|full | Low-quality |
| 18-109-final-viral-combined_k141_77710\|\|full | Not-determined |
| 18-109-final-viral-combined_k141_219782\|\|full | Not-determined |
| 18-109-final-viral-combined_k141_98585\|\|full | Not-determined |
| 18-109-final-viral-combined_k141_88148\|\|full | Not-determined |
| 18-109-final-viral-combined_k141_160337\|\|full | Low-quality |
| 18-109-final-viral-combined_k141_98599\|\|full | Low-quality |
| 18-109-final-viral-combined_k141_68609\|\|full | Low-quality |
| 18-109-final-viral-combined_k141_231493\|\|full | Not-determined |
| 18-109-final-viral-combined_k141_16938\|\|full | Not-determined |
| 18-109-final-viral-combined_k141_231502\|\|full | Low-quality |
| 18-109-final-viral-combined_k141_16947\|\|full | Low-quality |
| 18-109-final-viral-combined_k141_77732\|\|full | Not-determined |
| 18-109-final-viral-combined_k141_119417\|\|full | Not-determined |
| 18-109-final-viral-combined_k141_98641\|\|full | Low-quality |
| 18-109-final-viral-combined_k141_211524\|\|full | Low-quality |
| 18-109-final-viral-combined_k141_47799\|\|full | Low-quality |
| 18-109-final-viral-combined_k141_129560\|\|full | Low-quality |
| 18-109-final-viral-combined_k141_219828\|\|full | Low-quality |
| 18-109-final-viral-combined_k141_139342\|\|full | Not-determined |
| 18-109-final-viral-combined_k141_160388\|\|full | Not-determined |
| 18-109-final-viral-combined_k141_139346\|\|full | Not-determined |
| 18-109-final-viral-combined_k141_98685\|\|full | Low-quality |
| 18-109-final-viral-combined_k141_37947\|\|full | Low-quality |
| 18-109-final-viral-combined_k141_129586\|\|full | Not-determined |
| 18-109-final-viral-combined_k141_47813\|\|full | Not-determined |
| 18-109-final-viral-combined_k141_219846\|\|full | Not-determined |
| 18-109-final-viral-combined_k141_26306\|\|full | Low-quality |
| 18-109-final-viral-combined_k141_191294\|\|full | Low-quality |
| 18-109-final-viral-combined_k141_6827\|\|full | Not-determined |
| 18-109-final-viral-combined_k141_26327\|\|full | Not-determined |
| 18-109-final-viral-combined_k141_68727\|\|full | Not-determined |
| 18-109-final-viral-combined_k141_160434\|\|full | Not-determined |
| 18-109-final-viral-combined_k141_211579\|\|full | Not-determined |
| 18-109-final-viral-combined_k141_139371\|\|full | Low-quality |
| 18-109-final-viral-combined_k141_119483\|\|full | Not-determined |
| 18-109-final-viral-combined_k141_88246\|\|full | Low-quality |
| 18-109-final-viral-combined_k141_129638\|\|full | Low-quality |
| 18-109-final-viral-combined_k141_139384\|\|full | Low-quality |
| 18-109-final-viral-combined_k141_119486\|\|full | Low-quality |
| 18-109-final-viral-combined_k141_26353\|\|full | Not-determined |
| 18-109-final-viral-combined_k141_191343\|\|full | Not-determined |
| 18-109-final-viral-combined_k141_149348\|\|full | Not-determined |
| 18-109-final-viral-combined_k141_149349\|\|full | Not-determined |
| 18-109-final-viral-combined_k141_77868\|\|full | Low-quality |
| 18-109-final-viral-combined_k141_47866\|\|full | Low-quality |
| 18-109-final-viral-combined_k141_58114\|\|full | Low-quality |
| 18-109-final-viral-combined_k141_191360\|\|full | Low-quality |
| 18-109-final-viral-combined_k141_58120\|\|full | Low-quality |
| 18-109-final-viral-combined_k141_160461\|\|full | Not-determined |
| 18-109-final-viral-combined_k141_191363\|\|full | Not-determined |
| 18-109-final-viral-combined_k141_191384\|\|full | Low-quality |
| 18-109-final-viral-combined_k141_139456\|\|full | Not-determined |
| 18-109-final-viral-combined_k141_6910\|\|full | Not-determined |
| 18-109-final-viral-combined_k141_108202\|\|full | Not-determined |
| 18-109-final-viral-combined_k141_129681\|\|full | Not-determined |
| 18-109-final-viral-combined_k141_26397\|\|full | Not-determined |
| 18-109-final-viral-combined_k141_180673\|\|full | Not-determined |
| 18-109-final-viral-combined_k141_119524\|\|full | Not-determined |
| 18-109-final-viral-combined_k141_180677\|\|full | Not-determined |
| 18-109-final-viral-combined_k141_211657\|\|full | Low-quality |
| 18-109-final-viral-combined_k141_160501\|\|full | Low-quality |
| 18-109-final-viral-combined_k141_219868\|\|full | Low-quality |
| 18-109-final-viral-combined_k141_160513\|\|full | Low-quality |
| 18-109-final-viral-combined_k141_219876\|\|full | Low-quality |
| 18-109-final-viral-combined_k141_170634\|\|full | Low-quality |
| 18-109-final-viral-combined_k141_88367\|\|full | Low-quality |
| 18-109-final-viral-combined_k141_180710\|\|full | Low-quality |
| 18-109-final-viral-combined_k141_219905\|\|full | Not-determined |
| 18-109-final-viral-combined_k141_231631\|\|full | Not-determined |
| 18-109-final-viral-combined_k141_180720\|\|full | Low-quality |
| 18-109-final-viral-combined_k141_211685\|\|full | Low-quality |
| 18-109-final-viral-combined_k141_129716\|\|full | Low-quality |
| 18-109-final-viral-combined_k141_17150\|\|full | Low-quality |
| 18-109-final-viral-combined_k141_231638\|\|full | Not-determined |
| 18-109-final-viral-combined_k141_170669\|\|full | Not-determined |
| 18-109-final-viral-combined_k141_17153\|\|full | Not-determined |
| 18-109-final-viral-combined_k141_119601\|\|full | Low-quality |
| 18-109-final-viral-combined_k141_201634\|\|full | Not-determined |
| 18-109-final-viral-combined_k141_98767\|\|full | Not-determined |
| 18-109-final-viral-combined_k141_149502\|\|full | Low-quality |
| 18-109-final-viral-combined_k141_149506\|\|full | Not-determined |
| 18-109-final-viral-combined_k141_17189\|\|full | Not-determined |
| 18-109-final-viral-combined_k141_119653\|\|full | Not-determined |
| 18-109-final-viral-combined_k141_26440\|\|full | Not-determined |
| 18-109-final-viral-combined_k141_68829\|\|full | Low-quality |
| 18-109-final-viral-combined_k141_191471\|\|full | Low-quality |
| 18-109-final-viral-combined_k141_68846\|\|full | Low-quality |
| 18-109-final-viral-combined_k141_149544\|\|full | Not-determined |
| 18-109-final-viral-combined_k141_201697\|\|full | Low-quality |
| 18-109-final-viral-combined_k141_98834\|\|full | Low-quality |
| 18-109-final-viral-combined_k141_211710\|\|full | Low-quality |
| 18-109-final-viral-combined_k141_139510\|\|full | Low-quality |
| 18-109-final-viral-combined_k141_149557\|\|full | Not-determined |
| 18-109-final-viral-combined_k141_98860\|\|full | Low-quality |
| 18-109-final-viral-combined_k141_160674\|\|full | Not-determined |
| 18-109-final-viral-combined_k141_98872\|\|full | Not-determined |
| 18-109-final-viral-combined_k141_68886\|\|full | Not-determined |
| 18-109-final-viral-combined_k141_231742\|\|full | Low-quality |
| 18-109-final-viral-combined_k141_17265\|\|full | Not-determined |
| 18-109-final-viral-combined_k141_201729\|\|full | Not-determined |
| 18-109-final-viral-combined_k141_77948\|\|full | Not-determined |
| 18-109-final-viral-combined_k141_77951\|\|full | Not-determined |
| 18-109-final-viral-combined_k141_108330\|\|full | Low-quality |
| 18-109-final-viral-combined_k141_139558\|\|full | Low-quality |
| 18-109-final-viral-combined_k141_231785\|\|full | Not-determined |
| 18-109-final-viral-combined_k141_77953\|\|full | Not-determined |
| 18-109-final-viral-combined_k141_180841\|\|full | Low-quality |
| 18-109-final-viral-combined_k141_6997\|\|full | Low-quality |
| 18-109-final-viral-combined_k141_68961\|\|full | Not-determined |
| 18-109-final-viral-combined_k141_38105\|\|full | Not-determined |
| 18-109-final-viral-combined_k141_26503\|\|full | Low-quality |
| 18-109-final-viral-combined_k141_220032\|\|full | Low-quality |
| 18-109-final-viral-combined_k141_88405\|\|full | Low-quality |
| 18-109-final-viral-combined_k141_211795\|\|full | Not-determined |
| 18-109-final-viral-combined_k141_88414\|\|full | Not-determined |
| 18-109-final-viral-combined_k141_88417\|\|full | Low-quality |
| 18-109-final-viral-combined_k141_119770\|\|full | Not-determined |
| 18-109-final-viral-combined_k141_129858\|\|full | Not-determined |
| 18-109-final-viral-combined_k141_17345\|\|full | Not-determined |
| 18-109-final-viral-combined_k141_108376\|\|full | Low-quality |
| 18-109-final-viral-combined_k141_78001\|\|full | Low-quality |
| 18-109-final-viral-combined_k141_17352\|\|full | Low-quality |
| 18-109-final-viral-combined_k141_98899\|\|full | Low-quality |
| 18-109-final-viral-combined_k141_58259\|\|full | Low-quality |
| 18-109-final-viral-combined_k141_191579\|\|full | Not-determined |
| 18-109-final-viral-combined_k141_98921\|\|full | Low-quality |
| 18-109-final-viral-combined_k141_119806\|\|full | Not-determined |
| 18-109-final-viral-combined_k141_220053\|\|full | Not-determined |
| 18-109-final-viral-combined_k141_129898\|\|full | Low-quality |
| 18-109-final-viral-combined_k141_119820\|\|full | Low-quality |
| 18-109-final-viral-combined_k141_170676\|\|full | Not-determined |
| 18-109-final-viral-combined_k141_160785\|\|full | Low-quality |
| 18-102-final-viral-combined_k141_238929\|\|full | High-quality |
| 18-109-final-viral-combined_k141_119825\|\|full | Low-quality |
| 18-109-final-viral-combined_k141_38149\|\|full | Not-determined |
| 18-109-final-viral-combined_k141_191633\|\|full | Not-determined |
| 18-109-final-viral-combined_k141_78033\|\|full | Not-determined |
| 18-109-final-viral-combined_k141_180939\|\|full | Not-determined |
| 18-109-final-viral-combined_k141_48066\|\|full | Not-determined |
| 18-109-final-viral-combined_k141_7029\|\|full | Not-determined |
| 18-109-final-viral-combined_k141_48069\|\|full | Low-quality |
| 18-109-final-viral-combined_k141_201858\|\|full | Low-quality |
| 18-109-final-viral-combined_k141_231893\|\|full | Not-determined |
| 18-109-final-viral-combined_k141_149700\|\|full | Low-quality |
| 18-109-final-viral-combined_k141_170724\|\|full | Not-determined |
| 18-109-final-viral-combined_k141_58301\|\|full | Low-quality |
| 18-109-final-viral-combined_k141_38175\|\|full | Not-determined |
| 18-109-final-viral-combined_k141_170732\|\|full | Not-determined |
| 18-109-final-viral-combined_k141_48103\|\|full | Low-quality |
| 18-109-final-viral-combined_k141_119877\|\|full | Not-determined |
| 18-109-final-viral-combined_k141_88451\|\|full | Low-quality |
| 18-109-final-viral-combined_k141_26537\|\|full | Low-quality |
| 18-109-final-viral-combined_k141_119893\|\|full | Low-quality |
| 18-109-final-viral-combined_k141_108476\|\|full | Not-determined |
| 18-109-final-viral-combined_k141_231931\|\|full | Not-determined |
| 18-109-final-viral-combined_k141_129947\|\|full | Low-quality |
| 18-109-final-viral-combined_k141_69093\|\|full | Not-determined |
| 18-109-final-viral-combined_k141_69094\|\|full | Not-determined |
| 18-109-final-viral-combined_k141_7091\|\|full | Low-quality |
| 18-109-final-viral-combined_k141_17387\|\|full | Not-determined |
| 18-109-final-viral-combined_k141_17395\|\|full | Low-quality |
| 18-109-final-viral-combined_k141_88471\|\|full | Not-determined |
| 18-109-final-viral-combined_k141_149771\|\|full | Not-determined |
| 18-109-final-viral-combined_k141_26571\|\|full | Low-quality |
| 18-109-final-viral-combined_k141_26575\|\|full | Low-quality |
| 18-109-final-viral-combined_k141_170797\|\|full | Low-quality |
| 18-109-final-viral-combined_k141_48183\|\|full | Low-quality |
| 18-109-final-viral-combined_k141_181083\|\|full | Not-determined |
| 18-109-final-viral-combined_k141_149806\|\|full | Not-determined |
| 18-109-final-viral-combined_k141_149809\|\|full | Low-quality |
| 18-109-final-viral-combined_k141_78129\|\|full | Low-quality |
| 18-109-final-viral-combined_k141_201925\|\|full | Low-quality |
| 18-109-final-viral-combined_k141_160925\|\|full | Not-determined |
| 18-109-final-viral-combined_k141_170810\|\|full | Not-determined |
| 18-109-final-viral-combined_k141_119959\|\|full | Not-determined |
| 18-109-final-viral-combined_k141_181098\|\|full | Low-quality |
| 18-109-final-viral-combined_k141_211942\|\|full | Low-quality |
| 18-109-final-viral-combined_k141_69129\|\|full | Low-quality |
| 18-109-final-viral-combined_k141_201957\|\|full | Low-quality |
| 18-109-final-viral-combined_k141_38217\|\|full | Not-determined |
| 18-109-final-viral-combined_k141_232010\|\|full | Low-quality |
| 18-109-final-viral-combined_k141_119980\|\|full | Not-determined |
| 18-109-final-viral-combined_k141_99046\|\|full | Low-quality |
| 18-109-final-viral-combined_k141_191694\|\|full | Low-quality |
| 18-109-final-viral-combined_k141_119993\|\|full | Low-quality |
| 18-109-final-viral-combined_k141_38229\|\|full | Not-determined |
| 18-109-final-viral-combined_k141_38230\|\|full | Not-determined |
| 18-109-final-viral-combined_k141_191709\|\|full | Low-quality |
| 18-109-final-viral-combined_k141_69178\|\|full | Not-determined |
| 18-109-final-viral-combined_k141_17500\|\|full | Not-determined |
| 18-109-final-viral-combined_k141_201995\|\|full | Not-determined |
| 18-109-final-viral-combined_k141_191718\|\|full | Not-determined |
| 18-109-final-viral-combined_k141_202001\|\|full | Not-determined |
| 18-109-final-viral-combined_k141_232049\|\|full | Low-quality |
| 18-109-final-viral-combined_k141_139727\|\|full | Low-quality |
| 18-109-final-viral-combined_k141_191727\|\|full | Low-quality |
| 18-109-final-viral-combined_k141_78168\|\|full | Low-quality |
| 18-109-final-viral-combined_k141_170835\|\|full | Low-quality |
| 18-109-final-viral-combined_k141_26613\|\|full | Not-determined |
| 18-109-final-viral-combined_k141_69225\|\|full | Low-quality |
| 18-109-final-viral-combined_k141_78176\|\|full | Not-determined |
| 18-109-final-viral-combined_k141_202007\|\|full | Low-quality |
| 18-109-final-viral-combined_k141_232064\|\|full | Low-quality |
| 18-109-final-viral-combined_k141_191758\|\|full | Not-determined |
| 18-109-final-viral-combined_k141_161030\|\|full | Not-determined |
| 18-109-final-viral-combined_k141_88587\|\|full | Low-quality |
| 18-109-final-viral-combined_k141_17513\|\|full | Low-quality |
| 18-109-final-viral-combined_k141_181215\|\|full | Not-determined |
| 18-109-final-viral-combined_k141_130068\|\|full | Not-determined |
| 18-109-final-viral-combined_k141_69252\|\|full | Not-determined |
| 18-109-final-viral-combined_k141_202033\|\|full | Not-determined |
| 18-109-final-viral-combined_k141_161041\|\|full | Not-determined |
| 18-109-final-viral-combined_k141_26649\|\|full | Low-quality |
| 18-109-final-viral-combined_k141_139764\|\|full | Low-quality |
| 18-109-final-viral-combined_k141_88600\|\|full | Low-quality |
| 18-109-final-viral-combined_k141_191767\|\|full | Not-determined |
| 18-109-final-viral-combined_k141_202039\|\|full | Low-quality |
| 18-109-final-viral-combined_k141_139767\|\|full | Not-determined |
| 18-109-final-viral-combined_k141_120049\|\|full | Low-quality |
| 18-109-final-viral-combined_k141_58487\|\|full | Not-determined |
| 18-109-final-viral-combined_k141_220178\|\|full | Low-quality |
| 18-109-final-viral-combined_k141_191781\|\|full | Low-quality |
| 18-109-final-viral-combined_k141_38311\|\|full | Not-determined |
| 18-109-final-viral-combined_k141_181246\|\|full | Low-quality |
| 18-109-final-viral-combined_k141_17553\|\|full | Low-quality |
| 18-109-final-viral-combined_k141_88611\|\|full | Low-quality |
| 18-109-final-viral-combined_k141_191788\|\|full | Not-determined |
| 18-109-final-viral-combined_k141_161073\|\|full | Not-determined |
| 18-109-final-viral-combined_k141_191794\|\|full | Low-quality |
| 18-109-final-viral-combined_k141_120086\|\|full | Low-quality |
| 18-109-final-viral-combined_k141_88624\|\|full | Low-quality |
| 18-109-final-viral-combined_k141_38326\|\|full | Not-determined |
| 18-109-final-viral-combined_k141_120088\|\|full | Not-determined |
| 18-109-final-viral-combined_k141_58518\|\|full | Not-determined |
| 18-109-final-viral-combined_k141_191807\|\|full | Not-determined |
| 18-109-final-viral-combined_k141_38342\|\|full | Low-quality |
| 18-109-final-viral-combined_k141_170868\|\|full | Not-determined |
| 18-109-final-viral-combined_k141_88645\|\|full | Low-quality |
| 18-109-final-viral-combined_k141_99185\|\|full | Not-determined |
| 18-109-final-viral-combined_k141_220206\|\|full | Not-determined |
| 18-109-final-viral-combined_k141_232128\|\|full | Low-quality |
| 18-109-final-viral-combined_k141_211999\|\|full | Low-quality |
| 18-109-final-viral-combined_k141_88654\|\|full | Low-quality |
| 18-109-final-viral-combined_k141_69274\|\|full | Low-quality |
| 18-109-final-viral-combined_k141_181284\|\|full | Low-quality |
| 18-109-final-viral-combined_k141_48337\|\|full | Not-determined |
| 18-109-final-viral-combined_k141_26677\|\|full | Not-determined |
| 18-109-final-viral-combined_k141_242019\|\|full | Not-determined |
| 18-109-final-viral-combined_k141_170918\|\|full | Not-determined |
| 18-109-final-viral-combined_k141_149941\|\|full | Not-determined |
| 18-109-final-viral-combined_k141_191834\|\|full | Not-determined |
| 17-109-final-viral-combined_k141_385270\|\|full | Medium-quality |
| 18-109-final-viral-combined_k141_88696\|\|full | Not-determined |
| 18-109-final-viral-combined_k141_120161\|\|full | Low-quality |
| 18-109-final-viral-combined_k141_130141\|\|full | Low-quality |
| 18-109-final-viral-combined_k141_108687\|\|full | Not-determined |
| 18-109-final-viral-combined_k141_58554\|\|full | Not-determined |
| 18-109-final-viral-combined_k141_232189\|\|full | Low-quality |
| 18-109-final-viral-combined_k141_149979\|\|full | Not-determined |
| 18-109-final-viral-combined_k141_69291\|\|full | Low-quality |
| 18-109-final-viral-combined_k141_58578\|\|full | Low-quality |
| 18-109-final-viral-combined_k141_78292\|\|full | Low-quality |
| 18-109-final-viral-combined_k141_181364\|\|full | Not-determined |
| 18-109-final-viral-combined_k141_161180\|\|full | Low-quality |
| 18-109-final-viral-combined_k141_220285\|\|full | Not-determined |
| 18-109-final-viral-combined_k141_48400\|\|full | Not-determined |
| 18-109-final-viral-combined_k141_69319\|\|full | Not-determined |
| 18-109-final-viral-combined_k141_220287\|\|full | Not-determined |
| 18-109-final-viral-combined_k141_78306\|\|full | Low-quality |
| 18-109-final-viral-combined_k141_130162\|\|full | Low-quality |
| 18-109-final-viral-combined_k141_108735\|\|full | Low-quality |
| 18-109-final-viral-combined_k141_232216\|\|full | Low-quality |
| 18-109-final-viral-combined_k141_232229\|\|full | Low-quality |
| 18-109-final-viral-combined_k141_232238\|\|full | Not-determined |
| 18-109-final-viral-combined_k141_161224\|\|full | Low-quality |
| 18-109-final-viral-combined_k141_120200\|\|full | Not-determined |
| 18-109-final-viral-combined_k141_120207\|\|full | Not-determined |
| 18-109-final-viral-combined_k141_69347\|\|full | Low-quality |
| 18-109-final-viral-combined_k141_38478\|\|full | Low-quality |
| 18-109-final-viral-combined_k141_108747\|\|full | Low-quality |
| 18-109-final-viral-combined_k141_130204\|\|full | Not-determined |
| 18-109-final-viral-combined_k141_120217\|\|full | Not-determined |
| 18-109-final-viral-combined_k141_181446\|\|full | Low-quality |
| 18-109-final-viral-combined_k141_78335\|\|full | Not-determined |
| 18-109-final-viral-combined_k141_191925\|\|full | Not-determined |
| 18-109-final-viral-combined_k141_232282\|\|full | Low-quality |
| 18-109-final-viral-combined_k141_202248\|\|full | Low-quality |
| 18-109-final-viral-combined_k141_212124\|\|full | Low-quality |
| 18-109-final-viral-combined_k141_88778\|\|full | Low-quality |
| 18-109-final-viral-combined_k141_191938\|\|full | Low-quality |
| 18-109-final-viral-combined_k141_202258\|\|full | Not-determined |
| 18-109-final-viral-combined_k141_38518\|\|full | Not-determined |
| 18-109-final-viral-combined_k141_212131\|\|full | Not-determined |
| 18-109-final-viral-combined_k141_7298\|\|full | Not-determined |
| 18-109-final-viral-combined_k141_38530\|\|full | Low-quality |
| 18-109-final-viral-combined_k141_7308\|\|full | Not-determined |
| 18-109-final-viral-combined_k141_171009\|\|full | Low-quality |
| 18-109-final-viral-combined_k141_171010\|\|full | Not-determined |
| 18-109-final-viral-combined_k141_171015\|\|full | Low-quality |
| 18-109-final-viral-combined_k141_232300\|\|full | Not-determined |
| 18-109-final-viral-combined_k141_88823\|\|full | Low-quality |
| 18-109-final-viral-combined_k141_48505\|\|full | Not-determined |
| 18-109-final-viral-combined_k141_17607\|\|full | Not-determined |
| 18-109-final-viral-combined_k141_99362\|\|full | Low-quality |
| 18-109-final-viral-combined_k141_150072\|\|full | Low-quality |
| 18-109-final-viral-combined_k141_78382\|\|full | Not-determined |
| 18-109-final-viral-combined_k141_232321\|\|full | Not-determined |
| 18-109-final-viral-combined_k141_108829\|\|full | Not-determined |
| 18-109-final-viral-combined_k141_191966\|\|full | Low-quality |
| 18-109-final-viral-combined_k141_78394\|\|full | Not-determined |
| 18-109-final-viral-combined_k141_38561\|\|full | Not-determined |
| 18-109-final-viral-combined_k141_26723\|\|full | Not-determined |
| 18-109-final-viral-combined_k141_58719\|\|full | Low-quality |
| 18-109-final-viral-combined_k141_17634\|\|full | Not-determined |
| 18-109-final-viral-combined_k141_161343\|\|full | Low-quality |
| 18-109-final-viral-combined_k141_38577\|\|full | Not-determined |
| 18-109-final-viral-combined_k141_99382\|\|full | Low-quality |
| 18-109-final-viral-combined_k141_130291\|\|full | Low-quality |
| 18-109-final-viral-combined_k141_212175\|\|full | Not-determined |
| 18-109-final-viral-combined_k141_150114\|\|full | Not-determined |
| 18-109-final-viral-combined_k141_88839\|\|full | Not-determined |
| 18-109-final-viral-combined_k141_181539\|\|full | Low-quality |
| 18-109-final-viral-combined_k141_150115\|\|full | Not-determined |
| 18-109-final-viral-combined_k141_38595\|\|full | Low-quality |
| 18-109-final-viral-combined_k141_130292\|\|full | Low-quality |
| 18-109-final-viral-combined_k141_99395\|\|full | Not-determined |
| 18-109-final-viral-combined_k141_130296\|\|full | Low-quality |
| 18-109-final-viral-combined_k141_58769\|\|full | Not-determined |
| 18-109-final-viral-combined_k141_108846\|\|full | Low-quality |
| 18-109-final-viral-combined_k141_88860\|\|full | Low-quality |
| 18-109-final-viral-combined_k141_130309\|\|full | Low-quality |
| 18-109-final-viral-combined_k141_171106\|\|full | Low-quality |
| 18-109-final-viral-combined_k141_192003\|\|full | Low-quality |
| 18-109-final-viral-combined_k141_17698\|\|full | Not-determined |
| 18-109-final-viral-combined_k141_212226\|\|full | Not-determined |
| 18-109-final-viral-combined_k141_7402\|\|full | Low-quality |
| 18-109-final-viral-combined_k141_17707\|\|full | Not-determined |
| 18-109-final-viral-combined_k141_38617\|\|full | Low-quality |
| 18-109-final-viral-combined_k141_7411\|\|full | Not-determined |
| 18-109-final-viral-combined_k141_212242\|\|full | Low-quality |
| 18-109-final-viral-combined_k141_7412\|\|full | Not-determined |
| 18-109-final-viral-combined_k141_58830\|\|full | Not-determined |
| 18-109-final-viral-combined_k141_69437\|\|full | Not-determined |
| 18-109-final-viral-combined_k141_26796\|\|full | Not-determined |
| 18-109-final-viral-combined_k141_202391\|\|full | Not-determined |
| 18-109-final-viral-combined_k141_130372\|\|full | Not-determined |
| 18-109-final-viral-combined_k141_212263\|\|full | Low-quality |
| 18-109-final-viral-combined_k141_181609\|\|full | Not-determined |
| 18-109-final-viral-combined_k141_150174\|\|full | Low-quality |
| 18-109-final-viral-combined_k141_17747\|\|full | Not-determined |
| 18-109-final-viral-combined_k141_69457\|\|full | Not-determined |
| 18-109-final-viral-combined_k141_88925\|\|full | Low-quality |
| 18-109-final-viral-combined_k141_99438\|\|full | Not-determined |
| 18-109-final-viral-combined_k141_88929\|\|full | Not-determined |
| 18-109-final-viral-combined_k141_130389\|\|full | Not-determined |
| 18-109-final-viral-combined_k141_69467\|\|full | Low-quality |
| 18-109-final-viral-combined_k141_171189\|\|full | Low-quality |
| 18-109-final-viral-combined_k141_99458\|\|full | Low-quality |
| 18-109-final-viral-combined_k141_202430\|\|full | Low-quality |
| 18-109-final-viral-combined_k141_202447\|\|full | Low-quality |
| 18-109-final-viral-combined_k141_48677\|\|full | Low-quality |
| 18-109-final-viral-combined_k141_130439\|\|full | Low-quality |
| 18-109-final-viral-combined_k141_99474\|\|full | Not-determined |
| 18-109-final-viral-combined_k141_202482\|\|full | Not-determined |
| 18-109-final-viral-combined_k141_130465\|\|full | Not-determined |
| 18-109-final-viral-combined_k141_220417\|\|full | Not-determined |
| 18-109-final-viral-combined_k141_58931\|\|full | Low-quality |
| 18-109-final-viral-combined_k141_242143\|\|full | Not-determined |
| 18-109-final-viral-combined_k141_202503\|\|full | Low-quality |
| 18-109-final-viral-combined_k141_58953\|\|full | Not-determined |
| 18-109-final-viral-combined_k141_212340\|\|full | Not-determined |
| 18-109-final-viral-combined_k141_161435\|\|full | Low-quality |
| 18-109-final-viral-combined_k141_220450\|\|full | Low-quality |
| 18-109-final-viral-combined_k141_161440\|\|full | Not-determined |
| 18-109-final-viral-combined_k141_220457\|\|full | Low-quality |
| 18-109-final-viral-combined_k141_220460\|\|full | Low-quality |
| 18-109-final-viral-combined_k141_26839\|\|full | Not-determined |
| 18-109-final-viral-combined_k141_17879\|\|full | Not-determined |
| 18-109-final-viral-combined_k141_78567\|\|full | Low-quality |
| 18-109-final-viral-combined_k141_150263\|\|full | Low-quality |
| 18-109-final-viral-combined_k141_78576\|\|full | Low-quality |
| 18-109-final-viral-combined_k141_242166\|\|full | Not-determined |
| 18-109-final-viral-combined_k141_17886\|\|full | Not-determined |
| 18-109-final-viral-combined_k141_38810\|\|full | Not-determined |
| 18-109-final-viral-combined_k141_26857\|\|full | Not-determined |
| 18-109-final-viral-combined_k141_38819\|\|full | Not-determined |
| 18-109-final-viral-combined_k141_130522\|\|full | Low-quality |
| 18-109-final-viral-combined_k141_88985\|\|full | Not-determined |
| 18-109-final-viral-combined_k141_48717\|\|full | Low-quality |
| 18-109-final-viral-combined_k141_7577\|\|full | Not-determined |
| 18-109-final-viral-combined_k141_220505\|\|full | Not-determined |
| 18-109-final-viral-combined_k141_192086\|\|full | Low-quality |
| 18-109-final-viral-combined_k141_161485\|\|full | Not-determined |
| 18-109-final-viral-combined_k141_59029\|\|full | Not-determined |
| 18-109-final-viral-combined_k141_26873\|\|full | Low-quality |
| 18-109-final-viral-combined_k141_26874\|\|full | Low-quality |
| 18-109-final-viral-combined_k141_171275\|\|full | Not-determined |
| 18-109-final-viral-combined_k141_48740\|\|full | Low-quality |
| 18-109-final-viral-combined_k141_59031\|\|full | Low-quality |
| 18-109-final-viral-combined_k141_26887\|\|full | Not-determined |
| 18-109-final-viral-combined_k141_99543\|\|full | Not-determined |
| 18-109-final-viral-combined_k141_17917\|\|full | Low-quality |
| 18-109-final-viral-combined_k141_59039\|\|full | Low-quality |
| 18-109-final-viral-combined_k141_242223\|\|full | Not-determined |
| 18-109-final-viral-combined_k141_78646\|\|full | Low-quality |
| 18-109-final-viral-combined_k141_130565\|\|full | Low-quality |
| 18-109-final-viral-combined_k141_69537\|\|full | Low-quality |
| 18-109-final-viral-combined_k141_26896\|\|full | Low-quality |
| 18-109-final-viral-combined_k141_88997\|\|full | Low-quality |
| 18-109-final-viral-combined_k141_192119\|\|full | Not-determined |
| 18-109-final-viral-combined_k141_130567\|\|full | Not-determined |
| 18-109-final-viral-combined_k141_17943\|\|full | Low-quality |
| 18-109-final-viral-combined_k141_120303\|\|full | Low-quality |
| 18-109-final-viral-combined_k141_161545\|\|full | Not-determined |
| 18-109-final-viral-combined_k141_232508\|\|full | Low-quality |
| 18-109-final-viral-combined_k141_202621\|\|full | Low-quality |
| 18-109-final-viral-combined_k141_99592\|\|full | Low-quality |
| 18-109-final-viral-combined_k141_232514\|\|full | Low-quality |
| 18-109-final-viral-combined_k141_120326\|\|full | Low-quality |
| 18-109-final-viral-combined_k141_120327\|\|full | Not-determined |
| 18-109-final-viral-combined_k141_99605\|\|full | Low-quality |
| 18-109-final-viral-combined_k141_78703\|\|full | Low-quality |
| 18-109-final-viral-combined_k141_99609\|\|full | Low-quality |
| 18-109-final-viral-combined_k141_139907\|\|full | Low-quality |
| 18-109-final-viral-combined_k141_48832\|\|full | Not-determined |
| 18-109-final-viral-combined_k141_48833\|\|full | Low-quality |
| 18-109-final-viral-combined_k141_192166\|\|full | Not-determined |
| 18-109-final-viral-combined_k141_171335\|\|full | Low-quality |
| 18-109-final-viral-combined_k141_130624\|\|full | Not-determined |
| 18-109-final-viral-combined_k141_242278\|\|full | Low-quality |
| 18-109-final-viral-combined_k141_78732\|\|full | Not-determined |
| 18-109-final-viral-combined_k141_232542\|\|full | Low-quality |
| 18-109-final-viral-combined_k141_69614\|\|full | Low-quality |
| 18-109-final-viral-combined_k141_78748\|\|full | Not-determined |
| 18-109-final-viral-combined_k141_139943\|\|full | Low-quality |
| 18-109-final-viral-combined_k141_232556\|\|full | Low-quality |
| 18-109-final-viral-combined_k141_120375\|\|full | Low-quality |
| 18-109-final-viral-combined_k141_69630\|\|full | Low-quality |
| 18-109-final-viral-combined_k141_26930\|\|full | Low-quality |
| 18-109-final-viral-combined_k141_161618\|\|full | Not-determined |
| 18-109-final-viral-combined_k141_161631\|\|full | Low-quality |
| 18-109-final-viral-combined_k141_232575\|\|full | Low-quality |
| 18-109-final-viral-combined_k141_26944\|\|full | Not-determined |
| 18-109-final-viral-combined_k141_26946\|\|full | Not-determined |
| 18-109-final-viral-combined_k141_202676\|\|full | Low-quality |
| 18-109-final-viral-combined_k141_232581\|\|full | Low-quality |
| 18-109-final-viral-combined_k141_48896\|\|full | Low-quality |
| 18-109-final-viral-combined_k141_18011\|\|full | Not-determined |
| 18-109-final-viral-combined_k141_242340\|\|full | Low-quality |
| 18-109-final-viral-combined_k141_78807\|\|full | Low-quality |
| 18-109-final-viral-combined_k141_192252\|\|full | Not-determined |
| 18-109-final-viral-combined_k141_109164\|\|full | Not-determined |
| 18-109-final-viral-combined_k141_109165\|\|full | Low-quality |
| 18-109-final-viral-combined_k141_242356\|\|full | Low-quality |
| 18-109-final-viral-combined_k141_69667\|\|full | Low-quality |
| 18-109-final-viral-combined_k141_99678\|\|full | Not-determined |
| 18-109-final-viral-combined_k141_161659\|\|full | Low-quality |
| 18-109-final-viral-combined_k141_212428\|\|full | Low-quality |
| 18-109-final-viral-combined_k141_232595\|\|full | Low-quality |
| 18-109-final-viral-combined_k141_212430\|\|full | Not-determined |
| 18-109-final-viral-combined_k141_99695\|\|full | Not-determined |
| 18-109-final-viral-combined_k141_120411\|\|full | Low-quality |
| 18-109-final-viral-combined_k141_192277\|\|full | Not-determined |
| 18-109-final-viral-combined_k141_202722\|\|full | Low-quality |
| 18-109-final-viral-combined_k141_48930\|\|full | Low-quality |
| 18-109-final-viral-combined_k141_38987\|\|full | Low-quality |
| 18-109-final-viral-combined_k141_181863\|\|full | Not-determined |
| 18-109-final-viral-combined_k141_48937\|\|full | Low-quality |
| 18-109-final-viral-combined_k141_48938\|\|full | Low-quality |
| 18-109-final-viral-combined_k141_150445\|\|full | Not-determined |
| 18-109-final-viral-combined_k141_140040\|\|full | Not-determined |
| 18-109-final-viral-combined_k141_48983\|\|full | Not-determined |
| 18-109-final-viral-combined_k141_181899\|\|full | Low-quality |
| 18-109-final-viral-combined_k141_232657\|\|full | Not-determined |
| 18-109-final-viral-combined_k141_109212\|\|full | Low-quality |
| 18-109-final-viral-combined_k141_232674\|\|full | Low-quality |
| 18-109-final-viral-combined_k141_120485\|\|full | Low-quality |
| 18-109-final-viral-combined_k141_181914\|\|full | Low-quality |
| 18-109-final-viral-combined_k141_212490\|\|full | Low-quality |
| 18-109-final-viral-combined_k141_130751\|\|full | Not-determined |
| 18-109-final-viral-combined_k141_242461\|\|full | Low-quality |
| 18-109-final-viral-combined_k141_78837\|\|full | Not-determined |
| 18-109-final-viral-combined_k141_109223\|\|full | Not-determined |
| 18-109-final-viral-combined_k141_161769\|\|full | Not-determined |
| 18-109-final-viral-combined_k141_109227\|\|full | Low-quality |
| 18-109-final-viral-combined_k141_99822\|\|full | Low-quality |
| 18-109-final-viral-combined_k141_59220\|\|full | Not-determined |
| 18-109-final-viral-combined_k141_120526\|\|full | Not-determined |
| 18-109-final-viral-combined_k141_220631\|\|full | Not-determined |
| 18-109-final-viral-combined_k141_212529\|\|full | Not-determined |
| 18-109-final-viral-combined_k141_220638\|\|full | Low-quality |
| 18-109-final-viral-combined_k141_39053\|\|full | Low-quality |
| 18-109-final-viral-combined_k141_109244\|\|full | Low-quality |
| 18-109-final-viral-combined_k141_140089\|\|full | Low-quality |
| 18-109-final-viral-combined_k141_220646\|\|full | Low-quality |
| 18-109-final-viral-combined_k141_78864\|\|full | Low-quality |
| 18-109-final-viral-combined_k141_27027\|\|full | Low-quality |
| 18-109-final-viral-combined_k141_120538\|\|full | Not-determined |
| 18-109-final-viral-combined_k141_220653\|\|full | Low-quality |
| 18-109-final-viral-combined_k141_220654\|\|full | Not-determined |
| 18-109-final-viral-combined_k141_232745\|\|full | Low-quality |
| 18-109-final-viral-combined_k141_109254\|\|full | Low-quality |
| 18-109-final-viral-combined_k141_171484\|\|full | Low-quality |
| 18-109-final-viral-combined_k141_69735\|\|full | Not-determined |
| 18-109-final-viral-combined_k141_220672\|\|full | Low-quality |
| 18-109-final-viral-combined_k141_89120\|\|full | Low-quality |
| 18-109-final-viral-combined_k141_7716\|\|full | Low-quality |
| 18-109-final-viral-combined_k141_192346\|\|full | Not-determined |
| 18-109-final-viral-combined_k141_202876\|\|full | Not-determined |
| 18-109-final-viral-combined_k141_78892\|\|full | Low-quality |
| 18-109-final-viral-combined_k141_99853\|\|full | Low-quality |
| 18-109-final-viral-combined_k141_232772\|\|full | Low-quality |
| 18-109-final-viral-combined_k141_140116\|\|full | Low-quality |
| 18-109-final-viral-combined_k141_182033\|\|full | Not-determined |
| 18-109-final-viral-combined_k141_171504\|\|full | Not-determined |
| 18-109-final-viral-combined_k141_130850\|\|full | Not-determined |
| 18-109-final-viral-combined_k141_242528\|\|full | Low-quality |
| 18-109-final-viral-combined_k141_99877\|\|full | Not-determined |
| 18-109-final-viral-combined_k141_78914\|\|full | Low-quality |
| 18-109-final-viral-combined_k141_59280\|\|full | Not-determined |
| 18-109-final-viral-combined_k141_69767\|\|full | Low-quality |
| 18-109-final-viral-combined_k141_109302\|\|full | Not-determined |
| 18-109-final-viral-combined_k141_232799\|\|full | Not-determined |
| 18-109-final-viral-combined_k141_109305\|\|full | Low-quality |
| 18-109-final-viral-combined_k141_99885\|\|full | Low-quality |
| 18-109-final-viral-combined_k141_7787\|\|full | Not-determined |
| 18-109-final-viral-combined_k141_89160\|\|full | Not-determined |
| 18-109-final-viral-combined_k141_89171\|\|full | Low-quality |
| 18-109-final-viral-combined_k141_192409\|\|full | Low-quality |
| 18-109-final-viral-combined_k141_89177\|\|full | Not-determined |
| 18-109-final-viral-combined_k141_212550\|\|full | Not-determined |
| 18-109-final-viral-combined_k141_192417\|\|full | Not-determined |
| 18-109-final-viral-combined_k141_161830\|\|full | Not-determined |
| 18-109-final-viral-combined_k141_89181\|\|full | Not-determined |
| 18-109-final-viral-combined_k141_182082\|\|full | Not-determined |
| 18-109-final-viral-combined_k141_109333\|\|full | Low-quality |
| 18-109-final-viral-combined_k141_232826\|\|full | Low-quality |
| 18-109-final-viral-combined_k141_99907\|\|full | Low-quality |
| 18-109-final-viral-combined_k141_109337\|\|full | Not-determined |
| 18-109-final-viral-combined_k141_182091\|\|full | Not-determined |
| 18-109-final-viral-combined_k141_99928\|\|full | Low-quality |
| 18-109-final-viral-combined_k141_49101\|\|full | Not-determined |
| 18-109-final-viral-combined_k141_182093\|\|full | Low-quality |
| 18-109-final-viral-combined_k141_109343\|\|full | Low-quality |
| 18-109-final-viral-combined_k141_109350\|\|full | Not-determined |
| 18-109-final-viral-combined_k141_89218\|\|full | Low-quality |

18-109-final-viral-combined_k141_150575||full Low-quality
18-109-final-viral-combined_k141_212608||full Not-determined
18-109-final-viral-combined_k141_49138||full Not-determined
18-109-final-viral-combined_k141_150578||full Low-quality
18-109-final-viral-combined_k141_59296||full Low-quality
18-109-final-viral-combined_k141_59297||full Low-quality
18-109-final-viral-combined_k141_150582||full Low-quality
18-109-final-viral-combined_k141_192444||full Not-determined
18-109-final-viral-combined_k141_150583||full Not-determined
18-109-final-viral-combined_k141_182128||full Not-determined
18-109-final-viral-combined_k141_140234||full Not-determined
18-109-final-viral-combined_k141_109399||full Not-determined
18-109-final-viral-combined_k141_49163||full Not-determined
18-109-final-viral-combined_k141_182147||full Low-quality
18-109-final-viral-combined_k141_78973||full Not-determined
18-109-final-viral-combined_k141_99942||full Low-quality
18-109-final-viral-combined_k141_89279||full Not-determined
18-109-final-viral-combined_k141_78991||full Low-quality
18-109-final-viral-combined_k141_203058||full Not-determined
18-109-final-viral-combined_k141_212648||full Low-quality
18-109-final-viral-combined_k141_212652||full Low-quality
18-109-final-viral-combined_k141_49197||full Not-determined
18-109-final-viral-combined_k141_79002||full Not-determined
18-109-final-viral-combined_k141_39176||full Not-determined
18-109-final-viral-combined_k141_18253||full Not-determined
18-109-final-viral-combined_k141_99973||full Not-determined
18-109-final-viral-combined_k141_203104||full Low-quality
18-109-final-viral-combined_k141_220853||full Not-determined
18-109-final-viral-combined_k141_7907||full Low-quality
18-109-final-viral-combined_k141_220856||full Low-quality
18-109-final-viral-combined_k141_120610||full Low-quality
18-109-final-viral-combined_k141_161889||full Low-quality
18-109-final-viral-combined_k141_203118||full Low-quality
18-109-final-viral-combined_k141_39180||full Low-quality
18-109-final-viral-combined_k141_192514||full Not-determined
18-109-final-viral-combined_k141_7924||full Low-quality
18-109-final-viral-combined_k141_203124||full Not-determined
18-109-final-viral-combined_k141_182186||full Not-determined
18-109-final-viral-combined_k141_39191||full Low-quality
18-109-final-viral-combined_k141_140288||full Low-quality
18-109-final-viral-combined_k141_192544||full Low-quality
18-109-final-viral-combined_k141_7942||full Low-quality
18-109-final-viral-combined_k141_39209||full Not-determined
18-109-final-viral-combined_k141_79055||full Low-quality
18-109-final-viral-combined_k141_39211||full Not-determined
18-109-final-viral-combined_k141_192546||full Low-quality
18-109-final-viral-combined_k141_89306||full Not-determined
18-109-final-viral-combined_k141_109498||full Not-determined
18-109-final-viral-combined_k141_161936||full Low-quality
18-109-final-viral-combined_k141_212730||full Low-quality
18-109-final-viral-combined_k141_7961||full Not-determined
18-109-final-viral-combined_k141_220880||full Not-determined
18-109-final-viral-combined_k141_233006||full Not-determined
18-109-final-viral-combined_k141_233007||full Not-determined
18-109-final-viral-combined_k141_49253||full Not-determined
18-109-final-viral-combined_k141_89331||full Low-quality
18-109-final-viral-combined_k141_212744||full Low-quality
18-109-final-viral-combined_k141_59394||full Not-determined
18-109-final-viral-combined_k141_212753||full Not-determined
18-109-final-viral-combined_k141_59407||full Low-quality
18-109-final-viral-combined_k141_220938||full Low-quality
18-109-final-viral-combined_k141_49275||full Low-quality
18-109-final-viral-combined_k141_59416||full Low-quality
18-109-final-viral-combined_k141_150701||full Not-determined
18-109-final-viral-combined_k141_100089||full Not-determined
18-109-final-viral-combined_k141_69851||full Low-quality
18-109-final-viral-combined_k141_203239||full Low-quality
18-109-final-viral-combined_k141_120748||full Low-quality
18-109-final-viral-combined_k141_203251||full Low-quality
18-109-final-viral-combined_k141_59441||full Not-determined
18-109-final-viral-combined_k141_39290||full Not-determined
18-109-final-viral-combined_k141_18276||full Not-determined
18-109-final-viral-combined_k141_18277||full Not-determined
18-109-final-viral-combined_k141_69887||full Not-determined
18-109-final-viral-combined_k141_233051||full Not-determined
18-109-final-viral-combined_k141_120762||full Low-quality
18-109-final-viral-combined_k141_39328||full Low-quality
18-109-final-viral-combined_k141_18302||full Low-quality
18-109-final-viral-combined_k141_18321||full Not-determined
18-109-final-viral-combined_k141_192654||full Low-quality
18-109-final-viral-combined_k141_79113||full Not-determined
18-109-final-viral-combined_k141_18326||full Not-determined
18-109-final-viral-combined_k141_59447||full Not-determined
18-109-final-viral-combined_k141_192670||full Not-determined
18-109-final-viral-combined_k141_18334||full Low-quality
18-109-final-viral-combined_k141_8047||full Low-quality
18-109-final-viral-combined_k141_109604||full Not-determined
18-109-final-viral-combined_k141_79159||full Not-determined
18-109-final-viral-combined_k141_8074||full Low-quality
18-109-final-viral-combined_k141_49363||full Low-quality
18-109-final-viral-combined_k141_233127||full Low-quality
18-109-final-viral-combined_k141_120823||full Not-determined
18-109-final-viral-combined_k141_182284||full Not-determined
18-109-final-viral-combined_k141_18373||full Not-determined
18-109-final-viral-combined_k141_233130||full Low-quality
18-109-final-viral-combined_k141_59492||full Not-determined
18-109-final-viral-combined_k141_182292||full Not-determined
18-109-final-viral-combined_k141_192726||full Low-quality
18-109-final-viral-combined_k141_150781||full Low-quality
18-109-final-viral-combined_k141_171838||full Low-quality
18-109-final-viral-combined_k141_150782||full Not-determined
18-109-final-viral-combined_k141_162010||full Low-quality
18-109-final-viral-combined_k141_220984||full Low-quality
18-109-final-viral-combined_k141_8090||full Not-determined
18-109-final-viral-combined_k141_242650||full Not-determined
18-109-final-viral-combined_k141_8098||full Low-quality
18-109-final-viral-combined_k141_242659||full Not-determined
18-109-final-viral-combined_k141_79197||full Not-determined
18-109-final-viral-combined_k141_233162||full Low-quality
18-109-final-viral-combined_k141_100181||full Low-quality
18-109-final-viral-combined_k141_233169||full Low-quality
18-109-final-viral-combined_k141_192776||full Low-quality
18-109-final-viral-combined_k141_182320||full Low-quality
18-109-final-viral-combined_k141_69972||full Low-quality
18-109-final-viral-combined_k141_162044||full Low-quality
18-109-final-viral-combined_k141_69973||full Low-quality
18-109-final-viral-combined_k141_130863||full Low-quality
18-109-final-viral-combined_k141_100222||full Not-determined
18-109-final-viral-combined_k141_242714||full Not-determined
18-109-final-viral-combined_k141_89418||full Not-determined
18-109-final-viral-combined_k141_182341||full Not-determined
18-109-final-viral-combined_k141_120925||full Not-determined
18-109-final-viral-combined_k141_100233||full Not-determined
18-109-final-viral-combined_k141_233235||full Low-quality
18-109-final-viral-combined_k141_27100||full Not-determined
18-109-final-viral-combined_k141_242742||full Not-determined
18-109-final-viral-combined_k141_109751||full Not-determined
18-109-final-viral-combined_k141_59537||full Low-quality
18-109-final-viral-combined_k141_140318||full Not-determined
18-109-final-viral-combined_k141_140325||full Low-quality
18-109-final-viral-combined_k141_162078||full Not-determined
18-109-final-viral-combined_k141_27129||full Low-quality
18-109-final-viral-combined_k141_162083||full Not-determined
18-109-final-viral-combined_k141_120982||full Not-determined
18-109-final-viral-combined_k141_162098||full Not-determined
18-109-final-viral-combined_k141_39496||full Low-quality
18-109-final-viral-combined_k141_59585||full Not-determined
18-109-final-viral-combined_k141_140359||full Not-determined
18-109-final-viral-combined_k141_182407||full Low-quality
18-109-final-viral-combined_k141_233270||full Not-determined
17-106-final-viral-combined_k141_33230||full Medium-quality
18-109-final-viral-combined_k141_121004||full Low-quality
18-109-final-viral-combined_k141_70027||full Not-determined
18-109-final-viral-combined_k141_130903||full Not-determined
18-109-final-viral-combined_k141_121013||full Not-determined
18-109-final-viral-combined_k141_70029||full Not-determined
18-109-final-viral-combined_k141_130918||full Low-quality
18-109-final-viral-combined_k141_203503||full Low-quality
18-109-final-viral-combined_k141_49539||full Not-determined
18-109-final-viral-combined_k141_221131||full Not-determined
18-109-final-viral-combined_k141_121028||full Not-determined
18-109-final-viral-combined_k141_192846||full Not-determined
18-109-final-viral-combined_k141_109828||full Low-quality
18-109-final-viral-combined_k141_70052||full Not-determined
18-109-final-viral-combined_k141_121040||full Low-quality
18-109-final-viral-combined_k141_140412||full Not-determined
18-109-final-viral-combined_k141_182456||full Not-determined
18-109-final-viral-combined_k141_130931||full Not-determined
18-109-final-viral-combined_k141_182464||full Low-quality
18-109-final-viral-combined_k141_221134||full Low-quality
18-109-final-viral-combined_k141_59656||full Not-determined
18-109-final-viral-combined_k141_140429||full Not-determined
18-109-final-viral-combined_k141_8226||full Not-determined
18-109-final-viral-combined_k141_192871||full Not-determined
18-109-final-viral-combined_k141_182492||full Not-determined
18-109-final-viral-combined_k141_203569||full Not-determined
18-109-final-viral-combined_k141_182495||full Not-determined
18-109-final-viral-combined_k141_70070||full Not-determined
18-109-final-viral-combined_k141_130947||full Low-quality
18-109-final-viral-combined_k141_221161||full Not-determined
18-109-final-viral-combined_k141_242894||full Low-quality
18-109-final-viral-combined_k141_70078||full Not-determined
18-109-final-viral-combined_k141_203604||full Low-quality
18-109-final-viral-combined_k141_233347||full Low-quality
18-109-final-viral-combined_k141_130965||full Low-quality
18-109-final-viral-combined_k141_203611||full Not-determined
18-102-final-viral-combined_k141_176376||full Medium-quality
18-109-final-viral-combined_k141_59693||full Not-determined
18-109-final-viral-combined_k141_192925||full Low-quality
18-109-final-viral-combined_k141_221182||full Not-determined
18-109-final-viral-combined_k141_100386||full Low-quality
18-109-final-viral-combined_k141_233365||full Low-quality
18-109-final-viral-combined_k141_100388||full Not-determined
18-109-final-viral-combined_k141_8282||full Low-quality
18-109-final-viral-combined_k141_192931||full Not-determined
18-109-final-viral-combined_k141_121093||full Not-determined
18-109-final-viral-combined_k141_221213||full Low-quality
18-109-final-viral-combined_k141_89496||full Not-determined
18-109-final-viral-combined_k141_49678||full Not-determined
18-109-final-viral-combined_k141_70121||full Not-determined
18-109-final-viral-combined_k141_192956||full Low-quality
18-109-final-viral-combined_k141_212916||full Low-quality
18-109-final-viral-combined_k141_150918||full Not-determined
18-109-final-viral-combined_k141_49701||full Not-determined
18-109-final-viral-combined_k141_89511||full Low-quality
18-109-final-viral-combined_k141_27236||full Not-determined
18-109-final-viral-combined_k141_70133||full Not-determined
18-109-final-viral-combined_k141_27244||full Not-determined
18-109-final-viral-combined_k141_203658||full Low-quality
18-109-final-viral-combined_k141_192976||full Low-quality
18-109-final-viral-combined_k141_131036||full Low-quality
18-109-final-viral-combined_k141_70145||full Not-determined
18-109-final-viral-combined_k141_203664||full Low-quality
18-109-final-viral-combined_k141_192988||full Low-quality
18-109-final-viral-combined_k141_212949||full Not-determined
18-109-final-viral-combined_k141_182554||full Not-determined
18-109-final-viral-combined_k141_212952||full Low-quality
18-109-final-viral-combined_k141_242954||full Not-determined
18-109-final-viral-combined_k141_192992||full Not-determined
18-109-final-viral-combined_k141_49722||full Not-determined
18-109-final-viral-combined_k141_70168||full Low-quality
18-109-final-viral-combined_k141_162167||full Not-determined
18-109-final-viral-combined_k141_18432||full Not-determined
18-109-final-viral-combined_k141_79304||full Low-quality
18-109-final-viral-combined_k141_140570||full Not-determined
18-109-final-viral-combined_k141_27256||full Low-quality
18-109-final-viral-combined_k141_203719||full Low-quality
18-109-final-viral-combined_k141_121178||full Low-quality
18-109-final-viral-combined_k141_162187||full Not-determined
18-109-final-viral-combined_k141_89578||full Low-quality
18-109-final-viral-combined_k141_233435||full Low-quality
18-109-final-viral-combined_k141_151009||full Not-determined
18-109-final-viral-combined_k141_70210||full Low-quality
18-109-final-viral-combined_k141_182595||full Not-determined
18-109-final-viral-combined_k141_121194||full Low-quality
18-109-final-viral-combined_k141_213014||full Not-determined
18-109-final-viral-combined_k141_193071||full Not-determined
18-109-final-viral-combined_k141_182599||full Low-quality
18-109-final-viral-combined_k141_39681||full Not-determined
18-109-final-viral-combined_k141_70227||full Not-determined
18-109-final-viral-combined_k141_203754||full Low-quality
18-109-final-viral-combined_k141_172094||full Low-quality
18-109-final-viral-combined_k141_100491||full Not-determined
18-109-final-viral-combined_k141_70231||full Low-quality
18-109-final-viral-combined_k141_233473||full Not-determined
18-109-final-viral-combined_k141_59804||full Not-determined
18-109-final-viral-combined_k141_233475||full Not-determined
18-109-final-viral-combined_k141_233476||full Low-quality
18-109-final-viral-combined_k141_243050||full Low-quality
18-109-final-viral-combined_k141_89604||full Not-determined
18-109-final-viral-combined_k141_70236||full Not-determined
18-109-final-viral-combined_k141_233481||full Low-quality
18-109-final-viral-combined_k141_213031||full Not-determined
18-109-final-viral-combined_k141_193090||full Not-determined
18-109-final-viral-combined_k141_213043||full Not-determined
18-109-final-viral-combined_k141_70258||full Low-quality
18-109-final-viral-combined_k141_121247||full Low-quality
18-109-final-viral-combined_k141_8392||full Low-quality
18-109-final-viral-combined_k141_100522||full Low-quality
18-109-final-viral-combined_k141_140657||full Not-determined
18-109-final-viral-combined_k141_140662||full Low-quality
18-109-final-viral-combined_k141_100543||full Not-determined
18-109-final-viral-combined_k141_39707||full Low-quality
18-109-final-viral-combined_k141_172128||full Not-determined
18-109-final-viral-combined_k141_18453||full Low-quality
18-109-final-viral-combined_k141_131127||full Low-quality
18-109-final-viral-combined_k141_49830||full Low-quality
18-109-final-viral-combined_k141_70314||full Not-determined
18-109-final-viral-combined_k141_49831||full Not-determined
18-109-final-viral-combined_k141_172136||full Not-determined
18-109-final-viral-combined_k141_59832||full Not-determined
18-109-final-viral-combined_k141_121285||full Low-quality
18-109-final-viral-combined_k141_49838||full Low-quality
18-109-final-viral-combined_k141_8453||full Low-quality
18-109-final-viral-combined_k141_213104||full Not-determined
18-109-final-viral-combined_k141_203772||full Not-determined
18-109-final-viral-combined_k141_110042||full Not-determined
18-109-final-viral-combined_k141_8463||full Low-quality
18-109-final-viral-combined_k141_233550||full Not-determined
18-109-final-viral-combined_k141_59851||full Low-quality
18-109-final-viral-combined_k141_100585||full Low-quality
18-109-final-viral-combined_k141_49888||full Not-determined
18-109-final-viral-combined_k141_59857||full Not-determined
18-109-final-viral-combined_k141_59858||full Low-quality
18-109-final-viral-combined_k141_243108||full Low-quality
18-109-final-viral-combined_k141_59864||full Low-quality
18-109-final-viral-combined_k141_18531||full Not-determined
18-109-final-viral-combined_k141_121365||full Not-determined
18-109-final-viral-combined_k141_89682||full Low-quality
18-109-final-viral-combined_k141_203823||full Low-quality
18-109-final-viral-combined_k141_27312||full Low-quality
18-109-final-viral-combined_k141_49903||full Not-determined
18-109-final-viral-combined_k141_233580||full Low-quality
18-109-final-viral-combined_k141_182657||full Not-determined
18-109-final-viral-combined_k141_203842||full Low-quality
18-109-final-viral-combined_k141_89691||full Not-determined
18-109-final-viral-combined_k141_110077||full Not-determined
18-109-final-viral-combined_k141_162342||full Not-determined
18-109-final-viral-combined_k141_233589||full Low-quality
18-109-final-viral-combined_k141_193156||full Low-quality
18-109-final-viral-combined_k141_49933||full Not-determined
17-109-final-viral-combined_k141_358797||full Medium-quality
18-109-final-viral-combined_k141_79354||full Not-determined
18-109-final-viral-combined_k141_49938||full Not-determined
18-109-final-viral-combined_k141_151115||full Low-quality
18-109-final-viral-combined_k141_70365||full Low-quality
18-109-final-viral-combined_k141_151117||full Low-quality
18-109-final-viral-combined_k141_140739||full Low-quality
18-109-final-viral-combined_k141_110105||full Not-determined
18-109-final-viral-combined_k141_70393||full Low-quality
18-109-final-viral-combined_k141_131236||full Not-determined
18-109-final-viral-combined_k141_243138||full Low-quality
18-109-final-viral-combined_k141_193198||full Low-quality
18-109-final-viral-combined_k141_213151||full Not-determined
18-109-final-viral-combined_k141_8554||full Low-quality
18-109-final-viral-combined_k141_213154||full Not-determined
18-109-final-viral-combined_k141_182732||full Low-quality
18-109-final-viral-combined_k141_151172||full Low-quality
18-109-final-viral-combined_k141_70420||full Not-determined
18-109-final-viral-combined_k141_203906||full Not-determined
18-109-final-viral-combined_k141_79412||full Low-quality
18-109-final-viral-combined_k141_193215||full Not-determined
18-109-final-viral-combined_k141_18587||full Not-determined
18-109-final-viral-combined_k141_182744||full Low-quality
18-109-final-viral-combined_k141_49977||full Low-quality
18-109-final-viral-combined_k141_18594||full Not-determined
18-109-final-viral-combined_k141_59934||full Not-determined
18-109-final-viral-combined_k141_79416||full Not-determined
18-109-final-viral-combined_k141_8574||full Low-quality
18-109-final-viral-combined_k141_221402||full Not-determined
18-109-final-viral-combined_k141_140765||full Low-quality
18-109-final-viral-combined_k141_140767||full Not-determined
18-109-final-viral-combined_k141_221404||full Not-determined
18-109-final-viral-combined_k141_172256||full Low-quality
18-109-final-viral-combined_k141_59949||full Low-quality
18-109-final-viral-combined_k141_203936||full Not-determined
18-109-final-viral-combined_k141_203942||full Low-quality
18-109-final-viral-combined_k141_243164||full Low-quality
18-109-final-viral-combined_k141_79436||full Low-quality
18-109-final-viral-combined_k141_27337||full Low-quality
18-109-final-viral-combined_k141_89791||full Low-quality
18-109-final-viral-combined_k141_162401||full Low-quality
18-109-final-viral-combined_k141_221446||full Not-determined
18-109-final-viral-combined_k141_213219||full Not-determined
18-109-final-viral-combined_k141_89796||full Not-determined
18-109-final-viral-combined_k141_162408||full Low-quality
18-109-final-viral-combined_k141_151233||full Not-determined
18-109-final-viral-combined_k141_27352||full Not-determined
18-109-final-viral-combined_k141_59991||full Not-determined
18-109-final-viral-combined_k141_27355||full Low-quality
18-109-final-viral-combined_k141_100602||full Not-determined
18-109-final-viral-combined_k141_110169||full Not-determined
18-109-final-viral-combined_k141_131285||full Not-determined
18-109-final-viral-combined_k141_121474||full Low-quality
18-109-final-viral-combined_k141_243186||full Low-quality
18-109-final-viral-combined_k141_110182||full Low-quality
18-109-final-viral-combined_k141_151242||full Low-quality
18-109-final-viral-combined_k141_110191||full Not-determined
18-109-final-viral-combined_k141_110193||full Not-determined
18-109-final-viral-combined_k141_60064||full Low-quality
18-109-final-viral-combined_k141_131325||full Low-quality
18-109-final-viral-combined_k141_110201||full Not-determined
18-109-final-viral-combined_k141_50050||full Low-quality
18-109-final-viral-combined_k141_182843||full Low-quality
18-109-final-viral-combined_k141_243228||full Low-quality
18-109-final-viral-combined_k141_70521||full Low-quality
18-109-final-viral-combined_k141_89867||full Not-determined
18-109-final-viral-combined_k141_79527||full Not-determined
18-109-final-viral-combined_k141_213293||full Low-quality
18-109-final-viral-combined_k141_50070||full Not-determined
18-109-final-viral-combined_k141_193340||full Not-determined
18-109-final-viral-combined_k141_89878||full Not-determined
18-109-final-viral-combined_k141_213315||full Not-determined
18-109-final-viral-combined_k141_243236||full Low-quality
18-109-final-viral-combined_k141_233829||full Not-determined
18-109-final-viral-combined_k141_60089||full Not-determined
18-109-final-viral-combined_k141_110230||full Not-determined
18-109-final-viral-combined_k141_140812||full Not-determined
18-109-final-viral-combined_k141_233839||full Not-determined
18-109-final-viral-combined_k141_39895||full Low-quality
18-109-final-viral-combined_k141_221529||full Low-quality
18-109-final-viral-combined_k141_131370||full Low-quality
18-109-final-viral-combined_k141_50113||full Low-quality
18-109-final-viral-combined_k141_8677||full Not-determined
18-109-final-viral-combined_k141_193386||full Low-quality
18-109-final-viral-combined_k141_172366||full Low-quality
18-109-final-viral-combined_k141_110254||full Not-determined
18-109-final-viral-combined_k141_221549||full Low-quality
18-109-final-viral-combined_k141_172383||full Low-quality
18-109-final-viral-combined_k141_8706||full Low-quality
18-109-final-viral-combined_k141_172388||full Low-quality
18-109-final-viral-combined_k141_131406||full Low-quality
18-109-final-viral-combined_k141_110263||full Low-quality
18-109-final-viral-combined_k141_233871||full Not-determined
18-109-final-viral-combined_k141_233886||full Not-determined
18-109-final-viral-combined_k141_50153||full Not-determined
18-109-final-viral-combined_k141_89938||full Low-quality
18-109-final-viral-combined_k141_60109||full Not-determined
18-109-final-viral-combined_k141_193415||full Low-quality
18-109-final-viral-combined_k141_233893||full Low-quality
18-109-final-viral-combined_k141_121616||full Not-determined
18-109-final-viral-combined_k141_172401||full Low-quality
18-109-final-viral-combined_k141_151312||full Low-quality
18-109-final-viral-combined_k141_8725||full Low-quality
18-109-final-viral-combined_k141_233897||full Low-quality
18-109-final-viral-combined_k141_60113||full Low-quality
18-109-final-viral-combined_k141_233904||full Low-quality
18-109-final-viral-combined_k141_233907||full Low-quality
18-109-final-viral-combined_k141_8743||full Not-determined
18-109-final-viral-combined_k141_172424||full Not-determined
18-109-final-viral-combined_k141_8749||full Low-quality
18-109-final-viral-combined_k141_193445||full Low-quality
18-109-final-viral-combined_k141_8752||full Low-quality
18-109-final-viral-combined_k141_213393||full Not-determined
18-109-final-viral-combined_k141_60160||full Not-determined
18-109-final-viral-combined_k141_151383||full Not-determined
18-109-final-viral-combined_k141_60168||full Low-quality
18-109-final-viral-combined_k141_243268||full Low-quality
18-109-final-viral-combined_k141_204204||full Not-determined
18-109-final-viral-combined_k141_233970||full Not-determined
18-109-final-viral-combined_k141_18652||full Low-quality
18-109-final-viral-combined_k141_140846||full Low-quality
18-109-final-viral-combined_k141_100728||full Not-determined
18-109-final-viral-combined_k141_60173||full Low-quality
18-109-final-viral-combined_k141_50217||full Low-quality
18-109-final-viral-combined_k141_221648||full Not-determined
18-109-final-viral-combined_k141_221651||full Not-determined
18-109-final-viral-combined_k141_140867||full Not-determined
18-109-final-viral-combined_k141_100752||full Not-determined
18-109-final-viral-combined_k141_110327||full Not-determined
18-109-final-viral-combined_k141_89967||full Not-determined
18-109-final-viral-combined_k141_50236||full Low-quality
18-109-final-viral-combined_k141_131492||full Not-determined
18-109-final-viral-combined_k141_50240||full Low-quality
18-109-final-viral-combined_k141_18688||full Not-determined
18-109-final-viral-combined_k141_70618||full Not-determined
18-109-final-viral-combined_k141_110340||full Low-quality
18-109-final-viral-combined_k141_151432||full Low-quality
18-109-final-viral-combined_k141_40020||full Low-quality
18-109-final-viral-combined_k141_131518||full Low-quality
18-109-final-viral-combined_k141_213432||full Not-determined
18-109-final-viral-combined_k141_131520||full Low-quality
18-109-final-viral-combined_k141_243290||full Not-determined
18-109-final-viral-combined_k141_193486||full Not-determined
18-109-final-viral-combined_k141_100812||full Not-determined
18-109-final-viral-combined_k141_131529||full Not-determined
18-109-final-viral-combined_k141_121704||full Not-determined
18-109-final-viral-combined_k141_151444||full Low-quality
18-109-final-viral-combined_k141_40043||full Not-determined
18-109-final-viral-combined_k141_8771||full Not-determined
18-109-final-viral-combined_k141_40054||full Not-determined
18-109-final-viral-combined_k141_131548||full Not-determined
18-109-final-viral-combined_k141_213451||full Not-determined
18-109-final-viral-combined_k141_234021||full Not-determined
18-109-final-viral-combined_k141_40069||full Not-determined
18-109-final-viral-combined_k141_213463||full Low-quality
18-109-final-viral-combined_k141_40077||full Not-determined
18-109-final-viral-combined_k141_90035||full Not-determined
18-109-final-viral-combined_k141_40078||full Low-quality
18-109-final-viral-combined_k141_131570||full Low-quality
18-109-final-viral-combined_k141_213466||full Not-determined
18-109-final-viral-combined_k141_50308||full Not-determined
18-109-final-viral-combined_k141_60264||full Low-quality
18-109-final-viral-combined_k141_213472||full Low-quality
18-109-final-viral-combined_k141_27469||full Low-quality
18-109-final-viral-combined_k141_60269||full Not-determined
18-109-final-viral-combined_k141_79635||full Not-determined
18-109-final-viral-combined_k141_172484||full Not-determined
18-109-final-viral-combined_k141_70723||full Not-determined
18-109-final-viral-combined_k141_193534||full Low-quality
18-109-final-viral-combined_k141_131586||full Not-determined
18-109-final-viral-combined_k141_172498||full Not-determined
18-109-final-viral-combined_k141_131596||full Not-determined
18-109-final-viral-combined_k141_70736||full Low-quality
18-109-final-viral-combined_k141_60291||full Low-quality
18-109-final-viral-combined_k141_18726||full Low-quality
18-109-final-viral-combined_k141_243361||full Not-determined
18-109-final-viral-combined_k141_110426||full Low-quality
18-109-final-viral-combined_k141_140998||full Not-determined
18-109-final-viral-combined_k141_182965||full Not-determined
18-109-final-viral-combined_k141_141006||full Not-determined
18-109-final-viral-combined_k141_50336||full Low-quality
18-109-final-viral-combined_k141_213525||full Not-determined
18-109-final-viral-combined_k141_79694||full Not-determined
18-109-final-viral-combined_k141_110433||full Low-quality
18-109-final-viral-combined_k141_182976||full Not-determined
18-109-final-viral-combined_k141_110434||full Not-determined
18-109-final-viral-combined_k141_100923||full Not-determined
18-109-final-viral-combined_k141_50349||full Not-determined
18-109-final-viral-combined_k141_172546||full Low-quality
18-109-final-viral-combined_k141_213526||full Not-determined
18-109-final-viral-combined_k141_141012||full Not-determined
18-109-final-viral-combined_k141_193592||full Low-quality
18-109-final-viral-combined_k141_141024||full Not-determined
18-109-final-viral-combined_k141_110449||full Low-quality
18-109-final-viral-combined_k141_193599||full Not-determined
18-109-final-viral-combined_k141_151504||full Not-determined
18-109-final-viral-combined_k141_131652||full Not-determined
18-109-final-viral-combined_k141_172565||full Low-quality
18-109-final-viral-combined_k141_79714||full Not-determined
18-109-final-viral-combined_k141_234079||full Low-quality
18-109-final-viral-combined_k141_40169||full Not-determined
18-109-final-viral-combined_k141_110472||full Low-quality
18-109-final-viral-combined_k141_90127||full Not-determined
18-109-final-viral-combined_k141_183020||full Low-quality
18-109-final-viral-combined_k141_121836||full Low-quality
18-109-final-viral-combined_k141_100964||full Low-quality
18-109-final-viral-combined_k141_121841||full Low-quality
18-109-final-viral-combined_k141_40177||full Low-quality
18-109-final-viral-combined_k141_70805||full Not-determined
18-109-final-viral-combined_k141_183029||full Low-quality
18-109-final-viral-combined_k141_100972||full Not-determined
18-109-final-viral-combined_k141_50400||full Low-quality
18-109-final-viral-combined_k141_131685||full Not-determined
18-109-final-viral-combined_k141_151557||full Low-quality
18-109-final-viral-combined_k141_213612||full Not-determined
18-109-final-viral-combined_k141_121856||full Low-quality
18-109-final-viral-combined_k141_213614||full Low-quality
18-109-final-viral-combined_k141_50425||full Not-determined
18-109-final-viral-combined_k141_70850||full Low-quality
18-109-final-viral-combined_k141_193693||full Not-determined
18-109-final-viral-combined_k141_141078||full Low-quality
18-109-final-viral-combined_k141_110552||full Low-quality
18-109-final-viral-combined_k141_70870||full Not-determined
18-109-final-viral-combined_k141_193714||full Low-quality
18-109-final-viral-combined_k141_162708||full Not-determined
18-109-final-viral-combined_k141_151593||full Not-determined
18-109-final-viral-combined_k141_234138||full Not-determined
18-109-final-viral-combined_k141_221808||full Not-determined
18-109-final-viral-combined_k141_193730||full Low-quality
18-109-final-viral-combined_k141_90183||full Low-quality
18-109-final-viral-combined_k141_243467||full Low-quality
18-109-final-viral-combined_k141_162729||full Low-quality
18-109-final-viral-combined_k141_60314||full Low-quality
18-109-final-viral-combined_k141_243484||full Not-determined
18-109-final-viral-combined_k141_60317||full Not-determined
18-109-final-viral-combined_k141_221814||full Not-determined
18-109-final-viral-combined_k141_193773||full Not-determined
18-109-final-viral-combined_k141_70922||full Not-determined
18-109-final-viral-combined_k141_90231||full Not-determined
18-109-final-viral-combined_k141_193801||full Not-determined
18-109-final-viral-combined_k141_70939||full Low-quality
18-109-final-viral-combined_k141_50517||full Not-determined
18-109-final-viral-combined_k141_60353||full Not-determined
18-109-final-viral-combined_k141_221855||full Low-quality
18-109-final-viral-combined_k141_183134||full Low-quality
18-109-final-viral-combined_k141_110634||full Not-determined
18-109-final-viral-combined_k141_121957||full Not-determined
18-109-final-viral-combined_k141_60377||full Low-quality
18-109-final-viral-combined_k141_162765||full Low-quality
18-109-final-viral-combined_k141_8789||full Low-quality
18-109-final-viral-combined_k141_213732||full Not-determined
18-109-final-viral-combined_k141_8797||full Low-quality
18-109-final-viral-combined_k141_221875||full Not-determined
18-109-final-viral-combined_k141_90243||full Low-quality
18-109-final-viral-combined_k141_8801||full Low-quality
18-109-final-viral-combined_k141_101056||full Low-quality
18-109-final-viral-combined_k141_70993||full Low-quality
18-109-final-viral-combined_k141_60424||full Not-determined
18-109-final-viral-combined_k141_50586||full Not-determined
18-109-final-viral-combined_k141_221890||full Low-quality
18-109-final-viral-combined_k141_71005||full Low-quality
18-109-final-viral-combined_k141_18894||full Low-quality
18-109-final-viral-combined_k141_234248||full Not-determined
18-109-final-viral-combined_k141_101073||full Not-determined
18-109-final-viral-combined_k141_141176||full Not-determined
18-109-final-viral-combined_k141_243564||full Low-quality
18-109-final-viral-combined_k141_18905||full Not-determined
18-109-final-viral-combined_k141_151611||full Not-determined
18-109-final-viral-combined_k141_90301||full Not-determined
18-109-final-viral-combined_k141_151612||full Low-quality
18-109-final-viral-combined_k141_221904||full Not-determined
18-109-final-viral-combined_k141_8849||full Low-quality
18-109-final-viral-combined_k141_122002||full Not-determined
18-109-final-viral-combined_k141_79748||full Low-quality
18-109-final-viral-combined_k141_234268||full Not-determined
18-109-final-viral-combined_k141_162840||full Not-determined
18-109-final-viral-combined_k141_60484||full Not-determined
18-109-final-viral-combined_k141_8870||full Not-determined
18-109-final-viral-combined_k141_234288||full Not-determined
18-109-final-viral-combined_k141_79760||full Low-quality
18-109-final-viral-combined_k141_18942||full Not-determined
18-109-final-viral-combined_k141_243623||full Low-quality
18-109-final-viral-combined_k141_193893||full Not-determined
18-109-final-viral-combined_k141_183237||full Not-determined
18-109-final-viral-combined_k141_50704||full Not-determined
18-109-final-viral-combined_k141_50712||full Not-determined
18-109-final-viral-combined_k141_71084||full Low-quality
18-109-final-viral-combined_k141_141247||full Not-determined
18-109-final-viral-combined_k141_110756||full Low-quality
18-109-final-viral-combined_k141_60512||full Not-determined
18-109-final-viral-combined_k141_40206||full Low-quality
18-109-final-viral-combined_k141_213797||full Not-determined
18-109-final-viral-combined_k141_234311||full Low-quality
18-109-final-viral-combined_k141_18983||full Not-determined
18-109-final-viral-combined_k141_193915||full Not-determined
18-109-final-viral-combined_k141_172799||full Low-quality
18-109-final-viral-combined_k141_234312||full Low-quality
18-109-final-viral-combined_k141_79772||full Not-determined

18-109-final-viral-combined_k141_234328||full Not-determined
18-109-final-viral-combined_k141_141293||full Low-quality
18-109-final-viral-combined_k141_122095||full Not-determined
18-109-final-viral-combined_k141_234345||full Not-determined
18-109-final-viral-combined_k141_60527||full Not-determined
18-109-final-viral-combined_k141_40254||full Not-determined
18-109-final-viral-combined_k141_193945||full Not-determined
18-109-final-viral-combined_k141_162917||full Not-determined
18-109-final-viral-combined_k141_8916||full Low-quality
18-109-final-viral-combined_k141_222017||full Low-quality
18-109-final-viral-combined_k141_183295||full Low-quality
18-109-final-viral-combined_k141_19022||full Not-determined
18-109-final-viral-combined_k141_151635||full Not-determined
18-109-final-viral-combined_k141_183304||full Low-quality
18-109-final-viral-combined_k141_193964||full Low-quality
18-109-final-viral-combined_k141_8935||full Not-determined
18-109-final-viral-combined_k141_151641||full Low-quality
18-109-final-viral-combined_k141_122108||full Low-quality
18-109-final-viral-combined_k141_8954||full Not-determined
18-109-final-viral-combined_k141_141318||full Low-quality
18-109-final-viral-combined_k141_122120||full Not-determined
18-109-final-viral-combined_k141_90385||full Low-quality
18-109-final-viral-combined_k141_8959||full Not-determined
18-109-final-viral-combined_k141_19058||full Not-determined
18-109-final-viral-combined_k141_193982||full Not-determined
18-109-final-viral-combined_k141_183362||full Low-quality
18-109-final-viral-combined_k141_50765||full Low-quality
18-109-final-viral-combined_k141_8982||full Not-determined
18-109-final-viral-combined_k141_50770||full Not-determined
18-109-final-viral-combined_k141_60596||full Not-determined
KM-final-viral-combined_k141_158354||full Medium-quality
18-109-final-viral-combined_k141_40333||full Low-quality
18-109-final-viral-combined_k141_163001||full Not-determined
18-109-final-viral-combined_k141_79832||full Low-quality
18-109-final-viral-combined_k141_183408||full Not-determined
18-109-final-viral-combined_k141_8998||full Not-determined
18-109-final-viral-combined_k141_193996||full Low-quality
18-109-final-viral-combined_k141_40345||full Low-quality
18-109-final-viral-combined_k141_90435||full Not-determined
18-109-final-viral-combined_k141_9009||full Not-determined
18-109-final-viral-combined_k141_183425||full Not-determined
18-109-final-viral-combined_k141_50814||full Low-quality
18-109-final-viral-combined_k141_40354||full Not-determined
18-109-final-viral-combined_k141_234447||full Low-quality
18-109-final-viral-combined_k141_9014||full Not-determined
18-109-final-viral-combined_k141_9015||full Not-determined
18-109-final-viral-combined_k141_222096||full Low-quality
18-109-final-viral-combined_k141_163035||full Low-quality
18-109-final-viral-combined_k141_50826||full Low-quality
18-109-final-viral-combined_k141_60638||full Not-determined
18-109-final-viral-combined_k141_222100||full Low-quality
18-109-final-viral-combined_k141_101174||full Not-determined
18-109-final-viral-combined_k141_90480||full Low-quality
18-109-final-viral-combined_k141_79865||full Low-quality
18-109-final-viral-combined_k141_101186||full Not-determined
18-109-final-viral-combined_k141_101189||full Low-quality
18-109-final-viral-combined_k141_40399||full Low-quality
18-109-final-viral-combined_k141_183470||full Not-determined
18-109-final-viral-combined_k141_213969||full Not-determined
18-109-final-viral-combined_k141_40412||full Low-quality
18-109-final-viral-combined_k141_110974||full Not-determined
18-109-final-viral-combined_k141_151756||full Low-quality
18-109-final-viral-combined_k141_71162||full Not-determined
18-109-final-viral-combined_k141_90526||full Not-determined
18-109-final-viral-combined_k141_71171||full Low-quality
18-109-final-viral-combined_k141_151767||full Low-quality
18-109-final-viral-combined_k141_213982||full Not-determined
18-109-final-viral-combined_k141_172967||full Not-determined
18-109-final-viral-combined_k141_183501||full Not-determined
18-109-final-viral-combined_k141_71181||full Low-quality
18-109-final-viral-combined_k141_90535||full Not-determined
18-109-final-viral-combined_k141_79929||full Low-quality
18-109-final-viral-combined_k141_71184||full Low-quality
18-109-final-viral-combined_k141_9110||full Low-quality
18-109-final-viral-combined_k141_79937||full Low-quality
18-109-final-viral-combined_k141_234476||full Low-quality
18-109-final-viral-combined_k141_60704||full Low-quality
18-109-final-viral-combined_k141_79970||full Low-quality
18-109-final-viral-combined_k141_71196||full Not-determined
18-109-final-viral-combined_k141_131776||full Low-quality
18-109-final-viral-combined_k141_131788||full Not-determined
18-109-final-viral-combined_k141_101246||full Not-determined
18-109-final-viral-combined_k141_163147||full Low-quality
18-109-final-viral-combined_k141_90599||full Low-quality
18-109-final-viral-combined_k141_151865||full Not-determined
18-109-final-viral-combined_k141_151873||full Low-quality
18-109-final-viral-combined_k141_71229||full Low-quality
18-109-final-viral-combined_k141_90619||full Low-quality
18-109-final-viral-combined_k141_9188||full Low-quality
18-109-final-viral-combined_k141_101256||full Not-determined
18-109-final-viral-combined_k141_243737||full Low-quality
18-109-final-viral-combined_k141_173052||full Low-quality
18-109-final-viral-combined_k141_173053||full Low-quality
18-109-final-viral-combined_k141_163154||full Not-determined
17-106-final-viral-combined_k141_3461||full High-quality
18-109-final-viral-combined_k141_173056||full Not-determined
18-109-final-viral-combined_k141_111007||full Not-determined
18-109-final-viral-combined_k141_40545||full Not-determined
18-109-final-viral-combined_k141_27659||full Low-quality
18-109-final-viral-combined_k141_151939||full Not-determined
18-109-final-viral-combined_k141_90648||full Not-determined
18-109-final-viral-combined_k141_40561||full Not-determined
18-109-final-viral-combined_k141_80046||full Not-determined
18-109-final-viral-combined_k141_71300||full Not-determined
18-109-final-viral-combined_k141_80049||full Not-determined
18-109-final-viral-combined_k141_214066||full Low-quality
18-109-final-viral-combined_k141_101332||full Low-quality
18-109-final-viral-combined_k141_90684||full Not-determined
18-109-final-viral-combined_k141_151991||full Low-quality
18-109-final-viral-combined_k141_9297||full Not-determined
18-109-final-viral-combined_k141_71324||full Low-quality
18-109-final-viral-combined_k141_80070||full Low-quality
18-109-final-viral-combined_k141_19217||full Low-quality
18-109-final-viral-combined_k141_141384||full Not-determined
18-109-final-viral-combined_k141_122185||full Low-quality
18-109-final-viral-combined_k141_40589||full Low-quality
18-109-final-viral-combined_k141_243753||full Not-determined
18-109-final-viral-combined_k141_163253||full Not-determined
18-109-final-viral-combined_k141_80085||full Low-quality
18-109-final-viral-combined_k141_60793||full Not-determined
18-109-final-viral-combined_k141_141398||full Not-determined
18-109-final-viral-combined_k141_183638||full Not-determined
18-109-final-viral-combined_k141_141399||full Not-determined
18-109-final-viral-combined_k141_40592||full Low-quality
18-109-final-viral-combined_k141_141409||full Not-determined
18-109-final-viral-combined_k141_40595||full Low-quality
18-109-final-viral-combined_k141_71343||full Low-quality
18-109-final-viral-combined_k141_152048||full Low-quality
18-109-final-viral-combined_k141_152049||full Not-determined
18-109-final-viral-combined_k141_80111||full Low-quality
18-109-final-viral-combined_k141_222183||full Not-determined
18-109-final-viral-combined_k141_19264||full Not-determined
18-109-final-viral-combined_k141_131833||full Low-quality
18-109-final-viral-combined_k141_90717||full Not-determined
18-109-final-viral-combined_k141_183683||full Low-quality
18-109-final-viral-combined_k141_122254||full Not-determined
18-109-final-viral-combined_k141_152066||full Low-quality
18-109-final-viral-combined_k141_163304||full Low-quality
18-109-final-viral-combined_k141_71380||full Low-quality
18-109-final-viral-combined_k141_131839||full Low-quality
18-109-final-viral-combined_k141_71384||full Low-quality
18-109-final-viral-combined_k141_111106||full Not-determined
18-109-final-viral-combined_k141_152070||full Not-determined
18-109-final-viral-combined_k141_122262||full Not-determined
18-109-final-viral-combined_k141_111116||full Low-quality
18-109-final-viral-combined_k141_71397||full Low-quality
18-109-final-viral-combined_k141_9309||full Low-quality
18-109-final-viral-combined_k141_71402||full Not-determined
18-109-final-viral-combined_k141_222214||full Low-quality
18-109-final-viral-combined_k141_40654||full Not-determined
18-109-final-viral-combined_k141_101383||full Not-determined
18-109-final-viral-combined_k141_19315||full Low-quality
18-109-final-viral-combined_k141_122278||full Low-quality
18-109-final-viral-combined_k141_90719||full Low-quality
18-109-final-viral-combined_k141_141459||full Low-quality
18-109-final-viral-combined_k141_19327||full Low-quality
18-109-final-viral-combined_k141_111137||full Low-quality
18-109-final-viral-combined_k141_40664||full Not-determined
18-109-final-viral-combined_k141_9328||full Low-quality
18-109-final-viral-combined_k141_141468||full Low-quality
18-109-final-viral-combined_k141_71428||full Low-quality
18-109-final-viral-combined_k141_27748||full Low-quality
18-109-final-viral-combined_k141_27752||full Low-quality
18-109-final-viral-combined_k141_183715||full Not-determined
18-109-final-viral-combined_k141_19343||full Not-determined
18-109-final-viral-combined_k141_141473||full Not-determined
18-109-final-viral-combined_k141_71438||full Low-quality
18-109-final-viral-combined_k141_111152||full Not-determined
18-109-final-viral-combined_k141_40687||full Low-quality
18-109-final-viral-combined_k141_173177||full Low-quality
18-109-final-viral-combined_k141_152139||full Not-determined
KM-final-viral-combined_k141_185239||full Medium-quality
18-109-final-viral-combined_k141_183732||full Not-determined
18-109-final-viral-combined_k141_222241||full Low-quality
18-109-final-viral-combined_k141_122336||full Not-determined
18-109-final-viral-combined_k141_234696||full Not-determined
18-109-final-viral-combined_k141_60893||full Low-quality
18-109-final-viral-combined_k141_152167||full Low-quality
18-109-final-viral-combined_k141_234715||full Low-quality
18-109-final-viral-combined_k141_183765||full Low-quality
18-109-final-viral-combined_k141_183771||full Not-determined
18-109-final-viral-combined_k141_80219||full Not-determined
18-109-final-viral-combined_k141_152200||full Not-determined
18-109-final-viral-combined_k141_173234||full Low-quality
17-109-final-viral-combined_k141_250551||full High-quality
18-109-final-viral-combined_k141_40737||full Low-quality
18-109-final-viral-combined_k141_222304||full Not-determined
18-109-final-viral-combined_k141_40742||full Not-determined
18-109-final-viral-combined_k141_163338||full Low-quality
18-109-final-viral-combined_k141_122374||full Not-determined
18-109-final-viral-combined_k141_60986||full Not-determined
18-109-final-viral-combined_k141_80251||full Not-determined
18-109-final-viral-combined_k141_9488||full Not-determined
18-109-final-viral-combined_k141_163345||full Low-quality
18-109-final-viral-combined_k141_141573||full Low-quality
18-109-final-viral-combined_k141_9489||full Not-determined
18-109-final-viral-combined_k141_185016||full High-quality
18-109-final-viral-combined_k141_90788||full Low-quality
18-109-final-viral-combined_k141_222379||full Not-determined
18-109-final-viral-combined_k141_101451||full Not-determined
18-109-final-viral-combined_k141_152294||full Low-quality
18-109-final-viral-combined_k141_19521||full Not-determined
18-109-final-viral-combined_k141_141593||full Not-determined
18-109-final-viral-combined_k141_61029||full Not-determined
18-109-final-viral-combined_k141_222402||full Low-quality
18-109-final-viral-combined_k141_19535||full Not-determined
18-109-final-viral-combined_k141_122415||full Not-determined
18-109-final-viral-combined_k141_9550||full Low-quality
18-109-final-viral-combined_k141_101480||full Not-determined
18-109-final-viral-combined_k141_152312||full Low-quality
18-109-final-viral-combined_k141_80307||full Low-quality
18-109-final-viral-combined_k141_152322||full Low-quality
18-109-final-viral-combined_k141_173304||full Low-quality
18-109-final-viral-combined_k141_101509||full Low-quality
18-109-final-viral-combined_k141_9565||full Low-quality
18-109-final-viral-combined_k141_90855||full Low-quality
18-109-final-viral-combined_k141_19571||full Not-determined
18-109-final-viral-combined_k141_111235||full Not-determined
18-109-final-viral-combined_k141_122444||full Not-determined
18-109-final-viral-combined_k141_50960||full Not-determined
18-109-final-viral-combined_k141_111241||full Not-determined
17-108-final-viral-combined_k141_198512||full Medium-quality
18-109-final-viral-combined_k141_152325||full Not-determined
18-109-final-viral-combined_k141_222433||full Not-determined
18-109-final-viral-combined_k141_90881||full Low-quality
18-109-final-viral-combined_k141_61116||full Not-determined
18-109-final-viral-combined_k141_90886||full Low-quality
18-109-final-viral-combined_k141_50991||full Not-determined
18-109-final-viral-combined_k141_111264||full Not-determined
18-109-final-viral-combined_k141_27877||full Low-quality
18-109-final-viral-combined_k141_141626||full Not-determined
18-109-final-viral-combined_k141_61142||full Not-determined
18-109-final-viral-combined_k141_111288||full Low-quality
18-109-final-viral-combined_k141_80329||full Low-quality
18-109-final-viral-combined_k141_111303||full Not-determined
18-109-final-viral-combined_k141_214235||full Low-quality
18-109-final-viral-combined_k141_141661||full Low-quality
18-109-final-viral-combined_k141_9620||full Low-quality
18-109-final-viral-combined_k141_27907||full Not-determined
18-109-final-viral-combined_k141_27914||full Not-determined
18-109-final-viral-combined_k141_141690||full Not-determined
18-109-final-viral-combined_k141_131940||full Not-determined
18-109-final-viral-combined_k141_152389||full Low-quality
18-109-final-viral-combined_k141_173441||full Low-quality
18-109-final-viral-combined_k141_141704||full Not-determined
18-109-final-viral-combined_k141_101581||full Not-determined
18-109-final-viral-combined_k141_214266||full Low-quality
18-109-final-viral-combined_k141_131955||full Low-quality
18-109-final-viral-combined_k141_80334||full Low-quality
18-109-final-viral-combined_k141_131957||full Not-determined
18-109-final-viral-combined_k141_51000||full Not-determined
18-109-final-viral-combined_k141_101595||full Not-determined
18-109-final-viral-combined_k141_27956||full Low-quality
18-109-final-viral-combined_k141_222506||full Not-determined
18-109-final-viral-combined_k141_173471||full Not-determined
18-109-final-viral-combined_k141_51011||full Low-quality
18-109-final-viral-combined_k141_131983||full Low-quality
18-109-final-viral-combined_k141_111364||full Not-determined
18-109-final-viral-combined_k141_9721||full Low-quality
18-109-final-viral-combined_k141_131984||full Low-quality
18-109-final-viral-combined_k141_131995||full Not-determined
18-109-final-viral-combined_k141_101630||full Low-quality
18-109-final-viral-combined_k141_19667||full Low-quality
18-109-final-viral-combined_k141_222573||full Not-determined
18-109-final-viral-combined_k141_111391||full Low-quality
18-109-final-viral-combined_k141_173503||full Not-determined
18-109-final-viral-combined_k141_173504||full Low-quality
18-109-final-viral-combined_k141_173507||full Low-quality
18-109-final-viral-combined_k141_141731||full Low-quality
18-109-final-viral-combined_k141_27986||full Low-quality
18-109-final-viral-combined_k141_132075||full Low-quality
18-109-final-viral-combined_k141_222628||full Low-quality
18-109-final-viral-combined_k141_19695||full Not-determined
18-109-final-viral-combined_k141_141763||full Not-determined
18-109-final-viral-combined_k141_173544||full Not-determined
18-109-final-viral-combined_k141_214307||full Low-quality
18-109-final-viral-combined_k141_111426||full Not-determined
18-109-final-viral-combined_k141_152458||full Low-quality
18-109-final-viral-combined_k141_19707||full Not-determined
18-109-final-viral-combined_k141_132130||full Low-quality
18-109-final-viral-combined_k141_111435||full Not-determined
18-109-final-viral-combined_k141_28024||full Not-determined
18-109-final-viral-combined_k141_243774||full Not-determined
18-109-final-viral-combined_k141_19728||full Low-quality
18-109-final-viral-combined_k141_28026||full Low-quality
18-109-final-viral-combined_k141_132132||full Low-quality
18-109-final-viral-combined_k141_111456||full Not-determined
18-109-final-viral-combined_k141_91099||full Low-quality
18-109-final-viral-combined_k141_19744||full Not-determined
18-109-final-viral-combined_k141_243810||full Low-quality
18-109-final-viral-combined_k141_111487||full Low-quality
18-109-final-viral-combined_k141_141821||full Low-quality
18-109-final-viral-combined_k141_101758||full Not-determined
18-109-final-viral-combined_k141_214389||full Low-quality
18-109-final-viral-combined_k141_111501||full Not-determined
18-109-final-viral-combined_k141_28064||full Not-determined
18-109-final-viral-combined_k141_19784||full Low-quality
18-109-final-viral-combined_k141_222689||full Low-quality
18-109-final-viral-combined_k141_28069||full Not-determined
18-109-final-viral-combined_k141_152511||full Low-quality
18-109-final-viral-combined_k141_222701||full Not-determined
18-109-final-viral-combined_k141_9828||full Low-quality
18-109-final-viral-combined_k141_19794||full Low-quality
18-109-final-viral-combined_k141_222710||full Not-determined
18-109-final-viral-combined_k141_9849||full Not-determined
18-109-final-viral-combined_k141_9856||full Low-quality
18-109-final-viral-combined_k141_132200||full Not-determined
18-109-final-viral-combined_k141_80541||full Low-quality
18-109-final-viral-combined_k141_132205||full Not-determined
18-109-final-viral-combined_k141_111548||full Low-quality
18-109-final-viral-combined_k141_141939||full Low-quality
18-109-final-viral-combined_k141_111563||full Low-quality
18-109-final-viral-combined_k141_111564||full Not-determined
18-109-final-viral-combined_k141_28129||full Low-quality
18-109-final-viral-combined_k141_9901||full Low-quality
18-109-final-viral-combined_k141_141991||full Not-determined
18-109-final-viral-combined_k141_9915||full Not-determined
18-109-final-viral-combined_k141_132255||full Not-determined
18-109-final-viral-combined_k141_19888||full Not-determined
18-109-final-viral-combined_k141_152573||full Low-quality
18-105_k141_94965 High-quality
18-109-final-viral-combined_k141_80611||full Low-quality
18-109-final-viral-combined_k141_91209||full Not-determined
18-109-final-viral-combined_k141_111650||full Low-quality
18-109-final-viral-combined_k141_19917||full Low-quality
18-109-final-viral-combined_k141_28174||full Low-quality
18-109-final-viral-combined_k141_101967||full Not-determined
18-109-final-viral-combined_k141_152605||full Low-quality
18-109-final-viral-combined_k141_142038||full Low-quality
18-109-final-viral-combined_k141_9982||full Not-determined
18-109-final-viral-combined_k141_152609||full Low-quality
18-109-final-viral-combined_k141_111691||full Low-quality
18-109-final-viral-combined_k141_243828||full Not-determined
18-109-final-viral-combined_k141_152624||full Low-quality
18-109-final-viral-combined_k141_28219||full Low-quality
18-109-final-viral-combined_k141_222751||full Not-determined
18-109-final-viral-combined_k141_222775||full Not-determined
18-109-final-viral-combined_k141_152644||full Not-determined
18-109-final-viral-combined_k141_222783||full Low-quality
18-109-final-viral-combined_k141_222785||full Low-quality
18-109-final-viral-combined_k141_111751||full Low-quality
18-109-final-viral-combined_k141_19976||full Not-determined
18-109-final-viral-combined_k141_111759||full Low-quality
18-109-final-viral-combined_k141_102021||full Low-quality
18-109-final-viral-combined_k141_91295||full Low-quality
18-109-final-viral-combined_k141_243892||full Low-quality
18-109-final-viral-combined_k141_243896||full Not-determined
18-109-final-viral-combined_k141_91347||full Not-determined
18-109-final-viral-combined_k141_102064||full Low-quality
18-109-final-viral-combined_k141_102069||full Low-quality
18-109-final-viral-combined_k141_28330||full Low-quality
18-109-final-viral-combined_k141_80743||full Not-determined
18-109-final-viral-combined_k141_111872||full Not-determined
18-109-final-viral-combined_k141_243942||full Not-determined
18-109-final-viral-combined_k141_222901||full Low-quality
18-109-final-viral-combined_k141_91415||full Not-determined
18-109-final-viral-combined_k141_152753||full Low-quality
18-109-final-viral-combined_k141_132363||full Not-determined
18-109-final-viral-combined_k141_243979||full Low-quality
18-109-final-viral-combined_k141_132381||full Not-determined
18-109-final-viral-combined_k141_222913||full Not-determined
18-109-final-viral-combined_k141_132387||full Low-quality
18-109-final-viral-combined_k141_80796||full Not-determined
18-109-final-viral-combined_k141_28377||full Not-determined
18-109-final-viral-combined_k141_244000||full Not-determined
18-109-final-viral-combined_k141_244005||full Low-quality
18-109-final-viral-combined_k141_132426||full Low-quality
18-109-final-viral-combined_k141_244016||full Not-determined
18-109-final-viral-combined_k141_244018||full Low-quality
18-109-final-viral-combined_k141_222945||full Not-determined
18-109-final-viral-combined_k141_91434||full Not-determined
18-109-final-viral-combined_k141_152798||full Not-determined
18-109-final-viral-combined_k141_222964||full Not-determined
18-109-final-viral-combined_k141_244049||full Not-determined
18-109-final-viral-combined_k141_91465||full Low-quality
18-109-final-viral-combined_k141_28473||full Not-determined
18-109-final-viral-combined_k141_142204||full Not-determined
18-109-final-viral-combined_k141_152866||full Not-determined
18-109-final-viral-combined_k141_142220||full Low-quality
18-109-final-viral-combined_k141_152869||full Not-determined
18-109-final-viral-combined_k141_80833||full Low-quality
18-109-final-viral-combined_k141_142224||full Not-determined
18-109-final-viral-combined_k141_132539||full Not-determined
18-109-final-viral-combined_k141_244078||full Low-quality
18-109-final-viral-combined_k141_28532||full Low-quality
18-109-final-viral-combined_k141_142228||full Not-determined
18-109-final-viral-combined_k141_152900||full Low-quality
18-109-final-viral-combined_k141_111944||full Low-quality
18-109-final-viral-combined_k141_20143||full Not-determined
18-109-final-viral-combined_k141_20145||full Not-determined
18-109-final-viral-combined_k141_111989||full Low-quality
18-109-final-viral-combined_k141_142262||full Low-quality
18-109-final-viral-combined_k141_112011||full Not-determined
18-109-final-viral-combined_k141_132590||full Low-quality
18-109-final-viral-combined_k141_142331||full Low-quality
18-109-final-viral-combined_k141_112029||full Low-quality
18-109-final-viral-combined_k141_244160||full Low-quality
18-109-final-viral-combined_k141_80946||full Not-determined
18-109-final-viral-combined_k141_244166||full Not-determined
18-109-final-viral-combined_k141_10161||full Low-quality
18-109-final-viral-combined_k141_244194||full Low-quality
18-109-final-viral-combined_k141_152958||full Not-determined
18-109-final-viral-combined_k141_28624||full Low-quality
18-109-final-viral-combined_k141_223107||full Not-determined
18-109-final-viral-combined_k141_20174||full Low-quality
18-109-final-viral-combined_k141_223110||full Low-quality
18-109-final-viral-combined_k141_10184||full Not-determined
18-109-final-viral-combined_k141_28640||full Low-quality
18-109-final-viral-combined_k141_152991||full Not-determined
18-109-final-viral-combined_k141_153006||full Not-determined
18-109-final-viral-combined_k141_153029||full Not-determined
18-109-final-viral-combined_k141_28709||full Not-determined
18-109-final-viral-combined_k141_223129||full Low-quality
18-109-final-viral-combined_k141_223149||full Not-determined
18-109-final-viral-combined_k141_142455||full Low-quality
18-109-final-viral-combined_k141_28721||full Low-quality
18-109-final-viral-combined_k141_28729||full Low-quality
18-109-final-viral-combined_k141_153079||full Low-quality
18-109-final-viral-combined_k141_112200||full Not-determined
18-109-final-viral-combined_k141_28739||full Not-determined
18-109-final-viral-combined_k141_223165||full Not-determined
18-109-final-viral-combined_k141_153096||full Not-determined
18-109-final-viral-combined_k141_153097||full Low-quality
18-109-final-viral-combined_k141_244215||full Not-determined
18-109-final-viral-combined_k141_28790||full Not-determined
18-109-final-viral-combined_k141_20328||full Not-determined
18-109-final-viral-combined_k141_20334||full Low-quality
18-109-final-viral-combined_k141_28834||full Not-determined
18-109-final-viral-combined_k141_132699||full Not-determined
18-109-final-viral-combined_k141_112269||full Not-determined
18-109-final-viral-combined_k141_132717||full Not-determined
18-109-final-viral-combined_k141_28858||full Low-quality
18-109-final-viral-combined_k141_112303||full Low-quality
18-109-final-viral-combined_k141_142477||full Low-quality
18-109-final-viral-combined_k141_20382||full Not-determined
18-109-final-viral-combined_k141_28885||full Low-quality
18-109-final-viral-combined_k141_153151||full Low-quality
18-109-final-viral-combined_k141_142495||full Low-quality
18-109-final-viral-combined_k141_132746||full Not-determined
18-109-final-viral-combined_k141_20396||full Not-determined
18-109-final-viral-combined_k141_142501||full Not-determined
18-109-final-viral-combined_k141_91644||full Low-quality
18-109-final-viral-combined_k141_132749||full Low-quality
18-109-final-viral-combined_k141_91650||full Not-determined
18-109-final-viral-combined_k141_142517||full Not-determined
18-109-final-viral-combined_k141_244263||full Low-quality
18-109-final-viral-combined_k141_81145||full Low-quality
18-109-final-viral-combined_k141_81154||full Low-quality
18-109-final-viral-combined_k141_142564||full Low-quality
18-109-final-viral-combined_k141_91759||full Low-quality
18-109-final-viral-combined_k141_91761||full Not-determined
18-109-final-viral-combined_k141_91767||full Not-determined
18-109-final-viral-combined_k141_142620||full Low-quality
18-109-final-viral-combined_k141_28998||full Low-quality
18-109-final-viral-combined_k141_244282||full Not-determined
18-109-final-viral-combined_k141_142657||full Not-determined
18-109-final-viral-combined_k141_29046||full Low-quality
18-109-final-viral-combined_k141_142664||full Low-quality
18-109-final-viral-combined_k141_223339||full Low-quality
18-109-final-viral-combined_k141_244328||full Low-quality
18-109-final-viral-combined_k141_244332||full Not-determined
18-109-final-viral-combined_k141_29107||full Not-determined
18-109-final-viral-combined_k141_244339||full Not-determined
18-109-final-viral-combined_k141_142706||full Not-determined
18-109-final-viral-combined_k141_29147||full Low-quality
18-109-final-viral-combined_k141_29152||full Not-determined
18-109-final-viral-combined_k141_81297||full Low-quality
18-109-final-viral-combined_k141_81325||full Low-quality
18-109-final-viral-combined_k141_81326||full Low-quality
18-109-final-viral-combined_k141_81330||full Not-determined
18-109-final-viral-combined_k141_29172||full Low-quality
18-109-final-viral-combined_k141_142781||full Not-determined
18-109-final-viral-combined_k141_81344||full Low-quality
18-109-final-viral-combined_k141_81370||full Not-determined
18-109-final-viral-combined_k141_81379||full Low-quality
18-109-final-viral-combined_k141_29211||full Not-determined
18-109-final-viral-combined_k141_91820||full Not-determined
18-109-final-viral-combined_k141_81414||full Low-quality
18-109-final-viral-combined_k141_244396||full Not-determined
18-109-final-viral-combined_k141_142804||full Low-quality
18-109-final-viral-combined_k141_81466||full Low-quality
18-109-final-viral-combined_k141_244423||full Low-quality
18-109-final-viral-combined_k141_223452||full Not-determined
18-109-final-viral-combined_k141_29307||full Low-quality
18-109-final-viral-combined_k141_91892||full Not-determined
18-109-final-viral-combined_k141_29336||full Low-quality
18-109-final-viral-combined_k141_29385||full Low-quality
18-101-final-viral-combined_k141_324053||full High-quality
18-109-final-viral-combined_k141_244433||full Not-determined
18-109-final-viral-combined_k141_244444||full Low-quality
18-109-final-viral-combined_k141_244456||full Not-determined
18-109-final-viral-combined_k141_223609||full Low-quality
18-109-final-viral-combined_k141_142891||full Low-quality
18-109-final-viral-combined_k141_244459||full Low-quality
18-109-final-viral-combined_k141_223650||full Low-quality
18-109-final-viral-combined_k141_142945||full Not-determined
18-109-final-viral-combined_k141_244481||full Low-quality
18-109-final-viral-combined_k141_142957||full Low-quality
18-109-final-viral-combined_k141_81559||full Not-determined
18-109-final-viral-combined_k141_223677||full Not-determined
18-109-final-viral-combined_k141_244512||full Not-determined
18-109-final-viral-combined_k141_223752||full Not-determined
18-109-final-viral-combined_k141_223781||full Not-determined
18-109-final-viral-combined_k141_223850||full Low-quality
18-109-final-viral-combined_k141_244561||full Low-quality
18-109-final-viral-combined_k141_223893||full Low-quality
18-109-final-viral-combined_k141_223905||full Not-determined
18-109-final-viral-combined_k141_223938||full Not-determined
18-109-final-viral-combined_k141_223950||full Low-quality
18-109-final-viral-combined_k141_29525||full Not-determined
18-109-final-viral-combined_k141_244624||full Low-quality
18-109-final-viral-combined_k141_29530||full Low-quality
18-109-final-viral-combined_k141_29580||full Not-determined
18-109-final-viral-combined_k141_244638||full Low-quality
18-109-final-viral-combined_k141_224010||full Not-determined
18-109-final-viral-combined_k141_224066||full Low-quality
18-109-final-viral-combined_k141_244653||full Not-determined
18-109-final-viral-combined_k141_29808||full Low-quality
18-109-final-viral-combined_k141_244713||full Low-quality
18-109-final-viral-combined_k141_224150||full Low-quality
18-109-final-viral-combined_k141_224189||full Not-determined
18-109-final-viral-combined_k141_29945||full Not-determined
18-109-final-viral-combined_k141_224199||full Not-determined
18-109-final-viral-combined_k141_224211||full Low-quality
18-109-final-viral-combined_k141_224227||full Low-quality
18-109-final-viral-combined_k141_29968||full Not-determined
18-109-final-viral-combined_k141_224251||full Not-determined
18-109-final-viral-combined_k141_30024||full Not-determined
18-109-final-viral-combined_k141_244746||full Not-determined
18-109-final-viral-combined_k141_224339||full Low-quality
18-109-final-viral-combined_k141_244760||full Not-determined
18-109-final-viral-combined_k141_224373||full Low-quality
18-109-final-viral-combined_k141_30049||full Low-quality
18-109-final-viral-combined_k141_224375||full Not-determined
18-109-final-viral-combined_k141_244765||full Not-determined
18-109-final-viral-combined_k141_224381||full Not-determined
18-109-final-viral-combined_k141_224410||full Not-determined
18-109-final-viral-combined_k141_30113||full Low-quality
18-109-final-viral-combined_k141_224439||full Low-quality
18-109-final-viral-combined_k141_30127||full Low-quality
18-109-final-viral-combined_k141_30140||full Low-quality
18-109-final-viral-combined_k141_244803||full Not-determined
18-109-final-viral-combined_k141_224494||full Not-determined
18-109-final-viral-combined_k141_244821||full Not-determined
18-109-final-viral-combined_k141_30186||full Not-determined
18-105-final-viral-combined_k141_94965||full High-quality
18-109-final-viral-combined_k141_30197||full Low-quality
18-109-final-viral-combined_k141_224557||full Not-determined
18-109-final-viral-combined_k141_30240||full Not-determined
18-109-final-viral-combined_k141_30276||full Not-determined
18-109-final-viral-combined_k141_30299||full Not-determined
18-109-final-viral-combined_k141_30389||full Not-determined
18-109-final-viral-combined_k141_30463||full Not-determined
18-109-final-viral-combined_k141_30497||full Low-quality
18-109-final-viral-combined_k141_30548||full Low-quality
18-109-final-viral-combined_k141_30576||full Not-determined
18-109-final-viral-combined_k141_224688||0_partial Low-quality
17-109-final-viral-combined_k141_132907||full High-quality
18-109-final-viral-combined_k141_163781||0_partial Low-quality
18-109-final-viral-combined_k141_184132||0_partial Not-determined
18-109-final-viral-combined_k141_41228||0_partial Not-determined
18-110-final-viral-combined_k141_246389||full High-quality
18-109-final-viral-combined_k141_102731||0_partial Low-quality
18-109-final-viral-combined_k141_174309||0_partial Not-determined
18-109-final-viral-combined_k141_174309||1_partial Not-determined
18-101_k141_152617 High-quality
18-109-final-viral-combined_k141_113668||0_partial Not-determined
18-109-final-viral-combined_k141_103212||0_partial Low-quality
18-109-final-viral-combined_k141_123861||0_partial Low-quality
18-109-final-viral-combined_k141_92909||0_partial Low-quality
18-109-final-viral-combined_k141_32346||0_partial Low-quality
18-109-final-viral-combined_k141_52797||0_partial Low-quality
18-109-final-viral-combined_k141_114254||0_partial Not-determined
18-109-final-viral-combined_k141_144442||0_partial Not-determined
18-109-final-viral-combined_k141_165331||0_partial Low-quality
18-109-final-viral-combined_k141_165331||1_partial Low-quality
18-109-final-viral-combined_k141_63256||0_partial Low-quality
18-109-final-viral-combined_k141_22291||0_partial Low-quality
18-109-final-viral-combined_k141_72947||0_partial Low-quality
18-109-final-viral-combined_k141_93627||0_partial Not-determined
18-109-final-viral-combined_k141_134974||0_partial Low-quality
18-109-final-viral-combined_k141_134974||1_partial Low-quality
18-109-final-viral-combined_k141_135032||0_partial Not-determined
18-109-final-viral-combined_k141_53754||0_partial Low-quality
18-109-final-viral-combined_k141_237995||0_partial Not-determined
18-109-final-viral-combined_k141_13200||0_partial Low-quality
18-109-final-viral-combined_k141_198152||0_partial Low-quality
18-109-final-viral-combined_k141_54439||0_partial Low-quality

18-109-final-viral-combined_k141_95028||0_partial Low-quality
18-109-final-viral-combined_k141_74802||0_partial Low-quality
18-101-final-viral-combined_k141_152617||full High-quality
18-109-final-viral-combined_k141_23834||0_partial Not-determined
18-109-final-viral-combined_k141_209717||0_partial Low-quality
18-104_k141_45128 Medium-quality
18-109-final-viral-combined_k141_96390||0_partial Not-determined
18-104-final-viral-combined_k141_45128||full Medium-quality
18-109-final-viral-combined_k141_86413||0_partial Not-determined
18-109-final-viral-combined_k141_56372||0_partial Low-quality
18-109-final-viral-combined_k141_97800||0_partial Low-quality
17-109-final-viral-combined_k141_148741||full High-quality
17-109-final-viral-combined_k141_31761||full High-quality
18-109-final-viral-combined_k141_180144||0_partial Low-quality
18-109-final-viral-combined_k141_119356||0_partial Low-quality
18-109-final-viral-combined_k141_139306||0_partial Low-quality
18-109-final-viral-combined_k141_108165||0_partial Not-determined
18-109-final-viral-combined_k141_211691||0_partial Not-determined
18-109-final-viral-combined_k141_119732||0_partial Low-quality
18-109-final-viral-combined_k141_191688||0_partial Not-determined
18-109-final-viral-combined_k141_232465||0_partial Not-determined
18-109-final-viral-combined_k141_242241||0_partial Low-quality
18-109-final-viral-combined_k141_212403||0_partial Low-quality
18-101_k141_130214 Complete
18-109-final-viral-combined_k141_243145||0_partial Low-quality
18-109-final-viral-combined_k141_131244||0_partial Low-quality
18-109-final-viral-combined_k141_162565||0_partial Low-quality
18-109-final-viral-combined_k141_8773||0_partial Low-quality
18-109-final-viral-combined_k141_27553||0_partial Low-quality
18-109-final-viral-combined_k141_19139||0_partial Low-quality
18-109-final-viral-combined_k141_243745||0_partial Not-determined
18-109-final-viral-combined_k141_173587||0_partial Low-quality
18-109-final-viral-combined_k141_81262||0_partial Low-quality
18-109-final-viral-combined_k141_204233||lt2gene Not-determined
18-109-final-viral-combined_k141_10225||lt2gene Low-quality
18-109-final-viral-combined_k141_214453||lt2gene Low-quality
18-109-final-viral-combined_k141_81708||lt2gene Low-quality
18-109-final-viral-combined_k141_30650||lt2gene Low-quality
18-109-final-viral-combined_k141_40871||lt2gene Low-quality
18-109-final-viral-combined_k141_20448||lt2gene Low-quality
18-109-final-viral-combined_k141_214497||lt2gene Low-quality
18-109-final-viral-combined_k141_51106||lt2gene Not-determined
18-109-final-viral-combined_k141_91930||lt2gene Low-quality
18-109-final-viral-combined_k141_143016||lt2gene Low-quality
18-109-final-viral-combined_k141_81742||lt2gene Low-quality
18-109-final-viral-combined_k141_81747||lt2gene Low-quality
18-109-final-viral-combined_k141_183848||lt2gene Low-quality
18-109-final-viral-combined_k141_234950||lt2gene Low-quality
18-109-final-viral-combined_k141_183868||lt2gene Low-quality
18-109-final-viral-combined_k141_234966||lt2gene Low-quality
18-109-final-viral-combined_k141_122595||lt2gene Low-quality
18-109-final-viral-combined_k141_61333||lt2gene Low-quality
18-109-final-viral-combined_k141_153302||lt2gene Not-determined
18-109-final-viral-combined_k141_92011||lt2gene Low-quality
18-109-final-viral-combined_k141_112480||lt2gene Low-quality
18-109-final-viral-combined_k141_224775||lt2gene Low-quality
18-109-final-viral-combined_k141_143138||lt2gene Not-determined
18-109-final-viral-combined_k141_30740||lt2gene Not-determined
18-109-final-viral-combined_k141_143183||lt2gene Low-quality
18-109-final-viral-combined_k141_239||lt2gene Low-quality
18-109-final-viral-combined_k141_112535||lt2gene Low-quality
18-109-final-viral-combined_k141_61466||lt2gene Low-quality
18-109-final-viral-combined_k141_257||lt2gene Low-quality
18-109-final-viral-combined_k141_122703||lt2gene Not-determined
18-109-final-viral-combined_k141_132895||lt2gene Not-determined
18-109-final-viral-combined_k141_214699||lt2gene Low-quality
18-109-final-viral-combined_k141_296||lt2gene Not-determined
18-109-final-viral-combined_k141_163671||lt2gene Not-determined
18-109-final-viral-combined_k141_173830||lt2gene Low-quality
18-109-final-viral-combined_k141_30824||lt2gene Not-determined
18-109-final-viral-combined_k141_304||lt2gene Not-determined
18-109-final-viral-combined_k141_173857||lt2gene Low-quality
18-109-final-viral-combined_k141_204465||lt2gene Low-quality
18-109-final-viral-combined_k141_61539||lt2gene Low-quality
18-109-final-viral-combined_k141_194311||lt2gene Not-determined
18-109-final-viral-combined_k141_153443||lt2gene Low-quality
18-109-final-viral-combined_k141_143293||lt2gene Low-quality
18-109-final-viral-combined_k141_143296||lt2gene Low-quality
18-109-final-viral-combined_k141_20661||lt2gene Low-quality
18-109-final-viral-combined_k141_81885||lt2gene Low-quality
18-109-final-viral-combined_k141_194393||lt2gene Not-determined
18-109-final-viral-combined_k141_122862||lt2gene Low-quality
18-109-final-viral-combined_k141_163818||lt2gene Low-quality
18-109-final-viral-combined_k141_225056||lt2gene Low-quality
18-109-final-viral-combined_k141_173911||lt2gene Low-quality
18-109-final-viral-combined_k141_30977||lt2gene Low-quality
18-109-final-viral-combined_k141_163842||lt2gene Low-quality
18-109-final-viral-combined_k141_225078||lt2gene Low-quality
18-109-final-viral-combined_k141_163862||lt2gene Low-quality
18-109-final-viral-combined_k141_163863||lt2gene Low-quality
18-109-final-viral-combined_k141_173993||lt2gene Not-determined
18-109-final-viral-combined_k141_133135||lt2gene Low-quality
18-109-final-viral-combined_k141_10571||lt2gene Low-quality
18-109-final-viral-combined_k141_204566||lt2gene Low-quality
18-109-final-viral-combined_k141_71688||lt2gene Low-quality
18-109-final-viral-combined_k141_214814||lt2gene Low-quality
18-109-final-viral-combined_k141_143417||lt2gene Low-quality
18-109-final-viral-combined_k141_41146||lt2gene Not-determined
18-109-final-viral-combined_k141_112933||lt2gene Low-quality
18-109-final-viral-combined_k141_20746||lt2gene Not-determined
18-109-final-viral-combined_k141_204615||lt2gene Low-quality
18-109-final-viral-combined_k141_153706||lt2gene Low-quality
18-109-final-viral-combined_k141_235400||lt2gene Low-quality
18-109-final-viral-combined_k141_112969||lt2gene Low-quality
18-109-final-viral-combined_k141_41198||lt2gene Low-quality
18-109-final-viral-combined_k141_204656||lt2gene Low-quality
18-109-final-viral-combined_k141_214931||lt2gene Low-quality
18-109-final-viral-combined_k141_235462||lt2gene Low-quality
18-109-final-viral-combined_k141_204679||lt2gene Low-quality
18-109-final-viral-combined_k141_61839||lt2gene Not-determined
18-109-final-viral-combined_k141_10686||lt2gene Low-quality
18-109-final-viral-combined_k141_112989||lt2gene Low-quality
18-109-final-viral-combined_k141_194729||lt2gene Low-quality
18-109-final-viral-combined_k141_31101||lt2gene Low-quality
18-109-final-viral-combined_k141_215007||lt2gene Low-quality
18-109-final-viral-combined_k141_31110||lt2gene Low-quality
18-109-final-viral-combined_k141_31118||lt2gene Low-quality
18-109-final-viral-combined_k141_174195||lt2gene Not-determined
18-109-final-viral-combined_k141_61900||lt2gene Not-determined
18-109-final-viral-combined_k141_61907||lt2gene Low-quality
18-109-final-viral-combined_k141_669||lt2gene Low-quality
18-109-final-viral-combined_k141_41284||lt2gene Low-quality
18-109-final-viral-combined_k141_204776||lt2gene Low-quality
18-109-final-viral-combined_k141_51383||lt2gene Low-quality
18-109-final-viral-combined_k141_71925||lt2gene Low-quality
18-109-final-viral-combined_k141_82214||lt2gene Not-determined
18-109-final-viral-combined_k141_113135||lt2gene Low-quality
18-109-final-viral-combined_k141_215185||lt2gene Low-quality
18-109-final-viral-combined_k141_71950||lt2gene Low-quality
18-109-final-viral-combined_k141_215216||lt2gene Low-quality
18-109-final-viral-combined_k141_123108||lt2gene Low-quality
18-109-final-viral-combined_k141_113146||lt2gene Low-quality
18-109-final-viral-combined_k141_133355||lt2gene Low-quality
18-109-final-viral-combined_k141_194925||lt2gene Not-determined
18-109-final-viral-combined_k141_61955||lt2gene Low-quality
18-109-final-viral-combined_k141_133381||lt2gene Not-determined
18-109-final-viral-combined_k141_51532||lt2gene Low-quality
18-109-final-viral-combined_k141_113177||lt2gene Low-quality
18-109-final-viral-combined_k141_31213||lt2gene Low-quality
18-109-final-viral-combined_k141_82298||lt2gene Low-quality
18-109-final-viral-combined_k141_82323||lt2gene Low-quality
18-109-final-viral-combined_k141_51563||lt2gene Low-quality
18-109-final-viral-combined_k141_859||lt2gene Low-quality
18-109-final-viral-combined_k141_215319||lt2gene Low-quality
18-109-final-viral-combined_k141_215331||lt2gene Low-quality
18-109-final-viral-combined_k141_31299||lt2gene Low-quality
18-109-final-viral-combined_k141_164199||lt2gene Low-quality
18-109-final-viral-combined_k141_123292||lt2gene Low-quality
18-109-final-viral-combined_k141_31311||lt2gene Low-quality
18-109-final-viral-combined_k141_113280||lt2gene Not-determined
18-109-final-viral-combined_k141_133531||lt2gene Low-quality
18-109-final-viral-combined_k141_205014||lt2gene Not-determined
18-109-final-viral-combined_k141_215398||lt2gene Low-quality
18-109-final-viral-combined_k141_31385||lt2gene Low-quality
18-109-final-viral-combined_k141_92617||lt2gene Not-determined
18-109-final-viral-combined_k141_195103||lt2gene Low-quality
18-109-final-viral-combined_k141_51745||lt2gene Low-quality
18-109-final-viral-combined_k141_41537||lt2gene Low-quality
18-109-final-viral-combined_k141_41538||lt2gene Low-quality
18-109-final-viral-combined_k141_235794||lt2gene Not-determined
18-109-final-viral-combined_k141_164359||lt2gene Low-quality
18-109-final-viral-combined_k141_11083||lt2gene Low-quality
18-109-final-viral-combined_k141_82426||lt2gene Low-quality
18-109-final-viral-combined_k141_154340||lt2gene Low-quality
18-109-final-viral-combined_k141_21081||lt2gene Low-quality
18-109-final-viral-combined_k141_215518||lt2gene Low-quality
18-109-final-viral-combined_k141_184724||lt2gene Not-determined
18-109-final-viral-combined_k141_184732||lt2gene Low-quality
18-109-final-viral-combined_k141_133692||lt2gene Low-quality
18-109-final-viral-combined_k141_154379||lt2gene Low-quality
18-109-final-viral-combined_k141_215538||lt2gene Low-quality
18-109-final-viral-combined_k141_235915||lt2gene Low-quality
18-109-final-viral-combined_k141_102775||lt2gene Low-quality
18-109-final-viral-combined_k141_235931||lt2gene Not-determined
18-109-final-viral-combined_k141_164429||lt2gene Not-determined
18-109-final-viral-combined_k141_184831||lt2gene Low-quality
18-109-final-viral-combined_k141_184842||lt2gene Not-determined
18-109-final-viral-combined_k141_205220||lt2gene Not-determined
18-109-final-viral-combined_k141_113414||lt2gene Low-quality
18-109-final-viral-combined_k141_967||lt2gene Low-quality
18-109-final-viral-combined_k141_113420||lt2gene Low-quality
18-109-final-viral-combined_k141_113437||lt2gene Low-quality
18-109-final-viral-combined_k141_184877||lt2gene Not-determined
18-109-final-viral-combined_k141_51952||lt2gene Not-determined
18-109-final-viral-combined_k141_195271||lt2gene Low-quality
18-109-final-viral-combined_k141_11208||lt2gene Low-quality
18-109-final-viral-combined_k141_143724||lt2gene Low-quality
18-109-final-viral-combined_k141_1036||lt2gene Low-quality
18-109-final-viral-combined_k141_51989||lt2gene Not-determined
18-109-final-viral-combined_k141_1050||lt2gene Low-quality
18-109-final-viral-combined_k141_143757||lt2gene Low-quality
18-109-final-viral-combined_k141_41676||lt2gene Low-quality
18-109-final-viral-combined_k141_195295||lt2gene Low-quality
18-109-final-viral-combined_k141_82600||lt2gene Low-quality
18-109-final-viral-combined_k141_215691||lt2gene Low-quality
18-109-final-viral-combined_k141_102858||lt2gene Low-quality
18-109-final-viral-combined_k141_102861||lt2gene Low-quality
18-109-final-viral-combined_k141_174506||lt2gene Low-quality
18-109-final-viral-combined_k141_195354||lt2gene Not-determined
18-109-final-viral-combined_k141_195365||lt2gene Not-determined
18-109-final-viral-combined_k141_82671||lt2gene Not-determined
18-109-final-viral-combined_k141_82692||lt2gene Not-determined
18-109-final-viral-combined_k141_21160||lt2gene Not-determined
18-109-final-viral-combined_k141_31728||lt2gene Low-quality
18-109-final-viral-combined_k141_164535||lt2gene Low-quality
18-109-final-viral-combined_k141_195402||lt2gene Not-determined
18-109-final-viral-combined_k141_41731||lt2gene Low-quality
18-109-final-viral-combined_k141_52145||lt2gene Low-quality
18-109-final-viral-combined_k141_123670||lt2gene Low-quality
18-109-final-viral-combined_k141_225874||lt2gene Not-determined
18-109-final-viral-combined_k141_133934||lt2gene Low-quality
18-109-final-viral-combined_k141_205565||lt2gene Not-determined
18-109-final-viral-combined_k141_1109||lt2gene Low-quality
18-109-final-viral-combined_k141_31836||lt2gene Low-quality
18-109-final-viral-combined_k141_123736||lt2gene Low-quality
18-109-final-viral-combined_k141_62277||lt2gene Low-quality
18-109-final-viral-combined_k141_11385||lt2gene Low-quality
18-109-final-viral-combined_k141_1166||lt2gene Low-quality
18-109-final-viral-combined_k141_195546||lt2gene Not-determined
18-109-final-viral-combined_k141_174719||lt2gene Low-quality
18-109-final-viral-combined_k141_205641||lt2gene Low-quality
18-109-final-viral-combined_k141_103157||lt2gene Not-determined
18-109-final-viral-combined_k141_21327||lt2gene Low-quality
18-109-final-viral-combined_k141_195620||lt2gene Not-determined
18-109-final-viral-combined_k141_31912||lt2gene Low-quality
18-109-final-viral-combined_k141_205724||lt2gene Low-quality
18-109-final-viral-combined_k141_31933||lt2gene Low-quality
18-109-final-viral-combined_k141_82866||lt2gene Not-determined
18-109-final-viral-combined_k141_215820||lt2gene Not-determined
18-109-final-viral-combined_k141_134121||lt2gene Low-quality
18-109-final-viral-combined_k141_185202||lt2gene Low-quality
18-109-final-viral-combined_k141_11465||lt2gene Low-quality
18-109-final-viral-combined_k141_174836||lt2gene Not-determined
18-109-final-viral-combined_k141_154680||lt2gene Low-quality
18-109-final-viral-combined_k141_195713||lt2gene Low-quality
18-109-final-viral-combined_k141_154691||lt2gene Low-quality
18-109-final-viral-combined_k141_195742||lt2gene Low-quality
18-109-final-viral-combined_k141_11490||lt2gene Low-quality
18-109-final-viral-combined_k141_11505||lt2gene Low-quality
18-109-final-viral-combined_k141_185302||lt2gene Low-quality
18-109-final-viral-combined_k141_185308||lt2gene Low-quality
18-109-final-viral-combined_k141_11526||lt2gene Low-quality
18-109-final-viral-combined_k141_215909||lt2gene Low-quality
18-109-final-viral-combined_k141_226124||lt2gene Low-quality
18-109-final-viral-combined_k141_226127||lt2gene Low-quality
18-109-final-viral-combined_k141_21412||lt2gene Not-determined
18-109-final-viral-combined_k141_62521||lt2gene Low-quality
18-109-final-viral-combined_k141_21424||lt2gene Low-quality
18-109-final-viral-combined_k141_62545||lt2gene Low-quality
18-109-final-viral-combined_k141_226158||lt2gene Low-quality
18-109-final-viral-combined_k141_52383||lt2gene Not-determined
18-109-final-viral-combined_k141_154809||lt2gene Not-determined
18-109-final-viral-combined_k141_154811||lt2gene Not-determined
18-109-final-viral-combined_k141_32057||lt2gene Low-quality
18-109-final-viral-combined_k141_113805||lt2gene Low-quality
18-109-final-viral-combined_k141_123871||lt2gene Low-quality
18-109-final-viral-combined_k141_41988||lt2gene Low-quality
18-109-final-viral-combined_k141_103381||lt2gene Low-quality
18-109-final-viral-combined_k141_103384||lt2gene Low-quality
18-109-final-viral-combined_k141_144092||lt2gene Low-quality
18-109-final-viral-combined_k141_134316||lt2gene Low-quality
18-109-final-viral-combined_k141_52475||lt2gene Not-determined
18-109-final-viral-combined_k141_134339||lt2gene Not-determined
18-109-final-viral-combined_k141_123938||lt2gene Low-quality
18-109-final-viral-combined_k141_185473||lt2gene Low-quality
18-109-final-viral-combined_k141_185508||lt2gene Not-determined
18-109-final-viral-combined_k141_113881||lt2gene Low-quality
18-109-final-viral-combined_k141_21486||lt2gene Not-determined
18-109-final-viral-combined_k141_52541||lt2gene Not-determined
18-109-final-viral-combined_k141_42067||lt2gene Low-quality
18-109-final-viral-combined_k141_185566||lt2gene Not-determined
18-109-final-viral-combined_k141_62678||lt2gene Not-determined
18-109-final-viral-combined_k141_205894||lt2gene Not-determined
18-109-final-viral-combined_k141_103452||lt2gene Not-determined
18-109-final-viral-combined_k141_62700||lt2gene Low-quality
18-109-final-viral-combined_k141_62701||lt2gene Low-quality
18-109-final-viral-combined_k141_113960||lt2gene Low-quality
18-109-final-viral-combined_k141_185611||lt2gene Not-determined
18-109-final-viral-combined_k141_103480||lt2gene Low-quality
18-109-final-viral-combined_k141_185635||lt2gene Low-quality
18-109-final-viral-combined_k141_134436||lt2gene Low-quality
18-109-final-viral-combined_k141_195899||lt2gene Not-determined
18-109-final-viral-combined_k141_123996||lt2gene Low-quality
18-109-final-viral-combined_k141_185649||lt2gene Low-quality
18-109-final-viral-combined_k141_42151||lt2gene Not-determined
18-109-final-viral-combined_k141_226379||lt2gene Low-quality
18-109-final-viral-combined_k141_175082||lt2gene Low-quality
18-109-final-viral-combined_k141_92956||lt2gene Low-quality
18-109-final-viral-combined_k141_236513||lt2gene Not-determined
18-109-final-viral-combined_k141_124071||lt2gene Not-determined
18-109-final-viral-combined_k141_165051||lt2gene Not-determined
18-109-final-viral-combined_k141_114079||lt2gene Low-quality
18-109-final-viral-combined_k141_114083||lt2gene Low-quality
18-109-final-viral-combined_k141_175158||lt2gene Low-quality
18-109-final-viral-combined_k141_175171||lt2gene Low-quality
18-109-final-viral-combined_k141_185762||lt2gene Low-quality
18-109-final-viral-combined_k141_103600||lt2gene Not-determined
18-109-final-viral-combined_k141_155026||lt2gene Low-quality
18-109-final-viral-combined_k141_1540||lt2gene Low-quality
18-109-final-viral-combined_k141_62861||lt2gene Low-quality
18-109-final-viral-combined_k141_206103||lt2gene Low-quality
18-109-final-viral-combined_k141_236659||lt2gene Low-quality
18-109-final-viral-combined_k141_206116||lt2gene Low-quality
18-109-final-viral-combined_k141_32371||lt2gene Not-determined
18-109-final-viral-combined_k141_83031||lt2gene Low-quality
18-109-final-viral-combined_k141_1609||lt2gene Not-determined
18-109-final-viral-combined_k141_103696||lt2gene Low-quality
18-109-final-viral-combined_k141_226552||lt2gene Low-quality
18-109-final-viral-combined_k141_62901||lt2gene Low-quality
18-109-final-viral-combined_k141_32423||lt2gene Not-determined
18-109-final-viral-combined_k141_42370||lt2gene Not-determined
18-109-final-viral-combined_k141_175244||lt2gene Low-quality
18-109-final-viral-combined_k141_93138||lt2gene Low-quality
18-109-final-viral-combined_k141_103718||lt2gene Low-quality
18-109-final-viral-combined_k141_62937||lt2gene Low-quality
18-109-final-viral-combined_k141_124230||lt2gene Low-quality
18-109-final-viral-combined_k141_236706||lt2gene Not-determined
18-109-final-viral-combined_k141_62942||lt2gene Not-determined
18-109-final-viral-combined_k141_93160||lt2gene Low-quality
18-109-final-viral-combined_k141_196095||lt2gene Not-determined
18-109-final-viral-combined_k141_236802||lt2gene Low-quality
18-109-final-viral-combined_k141_72564||lt2gene Not-determined
18-109-final-viral-combined_k141_114370||lt2gene Low-quality
18-109-final-viral-combined_k141_185987||lt2gene Low-quality
18-109-final-viral-combined_k141_175422||lt2gene Low-quality
18-109-final-viral-combined_k141_1800||lt2gene Not-determined
18-109-final-viral-combined_k141_114415||lt2gene Low-quality
18-109-final-viral-combined_k141_175473||lt2gene Low-quality
18-109-final-viral-combined_k141_53075||lt2gene Low-quality
18-109-final-viral-combined_k141_196163||lt2gene Low-quality
18-109-final-viral-combined_k141_196175||lt2gene Low-quality
18-109-final-viral-combined_k141_216279||lt2gene Low-quality
18-109-final-viral-combined_k141_53137||lt2gene Low-quality
18-109-final-viral-combined_k141_134652||lt2gene Low-quality
18-109-final-viral-combined_k141_155241||lt2gene Not-determined
18-109-final-viral-combined_k141_32691||lt2gene Low-quality
18-109-final-viral-combined_k141_144500||lt2gene Low-quality
18-109-final-viral-combined_k141_237031||lt2gene Low-quality
18-109-final-viral-combined_k141_144507||lt2gene Low-quality
18-109-final-viral-combined_k141_237096||lt2gene Low-quality
18-109-final-viral-combined_k141_155327||lt2gene Not-determined
18-109-final-viral-combined_k141_42703||lt2gene Not-determined
18-109-final-viral-combined_k141_206554||lt2gene Not-determined
18-109-final-viral-combined_k141_226879||lt2gene Not-determined
18-109-final-viral-combined_k141_144564||lt2gene Low-quality
18-109-final-viral-combined_k141_42758||lt2gene Low-quality
18-109-final-viral-combined_k141_83303||lt2gene Low-quality
18-109-final-viral-combined_k141_42785||lt2gene Low-quality
18-109-final-viral-combined_k141_63204||lt2gene Not-determined
18-109-final-viral-combined_k141_83368||lt2gene Not-determined
18-109-final-viral-combined_k141_196312||lt2gene Low-quality
18-109-final-viral-combined_k141_104166||lt2gene Low-quality
18-109-final-viral-combined_k141_32789||lt2gene Low-quality
18-109-final-viral-combined_k141_165424||lt2gene Low-quality
18-109-final-viral-combined_k141_186179||lt2gene Low-quality
18-109-final-viral-combined_k141_165446||lt2gene Low-quality
18-109-final-viral-combined_k141_42926||lt2gene Low-quality
18-109-final-viral-combined_k141_206691||lt2gene Low-quality
18-109-final-viral-combined_k141_134799||lt2gene Low-quality
18-109-final-viral-combined_k141_32819||lt2gene Low-quality
18-109-final-viral-combined_k141_227074||lt2gene Not-determined
18-109-final-viral-combined_k141_196415||lt2gene Low-quality
18-109-final-viral-combined_k141_175644||lt2gene Not-determined
18-109-final-viral-combined_k141_165567||lt2gene Low-quality
18-109-final-viral-combined_k141_63313||lt2gene Low-quality
18-109-final-viral-combined_k141_72843||lt2gene Not-determined
18-109-final-viral-combined_k141_104286||lt2gene Not-determined
18-109-final-viral-combined_k141_155553||lt2gene Low-quality
18-109-final-viral-combined_k141_134888||lt2gene Not-determined
18-109-final-viral-combined_k141_206842||lt2gene Low-quality
18-109-final-viral-combined_k141_93443||lt2gene Low-quality
18-109-final-viral-combined_k141_12247||lt2gene Low-quality
18-109-final-viral-combined_k141_206846||lt2gene Not-determined
18-109-final-viral-combined_k141_227172||lt2gene Not-determined
18-109-final-viral-combined_k141_196583||lt2gene Not-determined
18-109-final-viral-combined_k141_114689||lt2gene Low-quality
18-109-final-viral-combined_k141_83464||lt2gene Low-quality
18-109-final-viral-combined_k141_155601||lt2gene Low-quality
18-109-final-viral-combined_k141_227202||lt2gene Low-quality
18-109-final-viral-combined_k141_2202||lt2gene Low-quality
18-109-final-viral-combined_k141_114724||lt2gene Low-quality
18-109-final-viral-combined_k141_83489||lt2gene Low-quality
18-109-final-viral-combined_k141_83503||lt2gene Low-quality
18-109-final-viral-combined_k141_72937||lt2gene Low-quality
18-109-final-viral-combined_k141_53395||lt2gene Not-determined
18-109-final-viral-combined_k141_227264||lt2gene Low-quality
18-109-final-viral-combined_k141_216705||lt2gene Low-quality
18-109-final-viral-combined_k141_2256||lt2gene Low-quality
18-109-final-viral-combined_k141_144821||lt2gene Low-quality
18-109-final-viral-combined_k141_155723||lt2gene Not-determined
18-109-final-viral-combined_k141_12334||lt2gene Low-quality
18-109-final-viral-combined_k141_2341||lt2gene Not-determined
18-109-final-viral-combined_k141_73016||lt2gene Not-determined
18-109-final-viral-combined_k141_124858||lt2gene Low-quality
18-109-final-viral-combined_k141_22322||lt2gene Low-quality
18-109-final-viral-combined_k141_33037||lt2gene Low-quality
18-109-final-viral-combined_k141_227356||lt2gene Low-quality
18-109-final-viral-combined_k141_33053||lt2gene Low-quality
18-109-final-viral-combined_k141_227379||lt2gene Low-quality
18-109-final-viral-combined_k141_22335||lt2gene Low-quality
18-109-final-viral-combined_k141_196788||lt2gene Low-quality
18-109-final-viral-combined_k141_33082||lt2gene Low-quality
18-109-final-viral-combined_k141_207044||lt2gene Low-quality
18-109-final-viral-combined_k141_165858||lt2gene Low-quality
18-109-final-viral-combined_k141_53527||lt2gene Not-determined
18-109-final-viral-combined_k141_43126||lt2gene Low-quality
18-109-final-viral-combined_k141_207068||lt2gene Low-quality
18-109-final-viral-combined_k141_155800||lt2gene Low-quality
18-109-final-viral-combined_k141_186626||lt2gene Low-quality
18-109-final-viral-combined_k141_144964||lt2gene Not-determined
18-109-final-viral-combined_k141_83747||lt2gene Not-determined
18-109-final-viral-combined_k141_114876||lt2gene Low-quality
18-109-final-viral-combined_k141_83833||lt2gene Not-determined
18-109-final-viral-combined_k141_155858||lt2gene Low-quality
18-109-final-viral-combined_k141_53630||lt2gene Not-determined
18-109-final-viral-combined_k141_175723||lt2gene Low-quality
18-109-final-viral-combined_k141_155875||lt2gene Low-quality
18-109-final-viral-combined_k141_155887||lt2gene Low-quality
18-109-final-viral-combined_k141_227491||lt2gene Not-determined
18-109-final-viral-combined_k141_83875||lt2gene Low-quality
18-109-final-viral-combined_k141_124972||lt2gene Low-quality
18-109-final-viral-combined_k141_135108||lt2gene Not-determined
18-109-final-viral-combined_k141_207215||lt2gene Not-determined
18-109-final-viral-combined_k141_186777||lt2gene Not-determined
18-109-final-viral-combined_k141_114893||lt2gene Low-quality
18-109-final-viral-combined_k141_12577||lt2gene Low-quality
18-109-final-viral-combined_k141_73212||lt2gene Not-determined
18-109-final-viral-combined_k141_197006||lt2gene Low-quality
18-109-final-viral-combined_k141_22440||lt2gene Low-quality
18-109-final-viral-combined_k141_43339||lt2gene Low-quality
18-109-final-viral-combined_k141_237644||lt2gene Low-quality
18-109-final-viral-combined_k141_227626||lt2gene Low-quality
18-109-final-viral-combined_k141_73224||lt2gene Low-quality
18-109-final-viral-combined_k141_145098||lt2gene Low-quality
18-109-final-viral-combined_k141_166011||lt2gene Low-quality
18-109-final-viral-combined_k141_12670||lt2gene Low-quality
18-109-final-viral-combined_k141_156050||lt2gene Low-quality
18-109-final-viral-combined_k141_207309||lt2gene Low-quality
18-109-final-viral-combined_k141_2675||lt2gene Low-quality
18-109-final-viral-combined_k141_197078||lt2gene Not-determined
18-109-final-viral-combined_k141_237728||lt2gene Low-quality
18-109-final-viral-combined_k141_33222||lt2gene Low-quality
18-109-final-viral-combined_k141_73265||lt2gene Low-quality
18-109-final-viral-combined_k141_125055||lt2gene Not-determined
18-109-final-viral-combined_k141_186913||lt2gene Not-determined
18-109-final-viral-combined_k141_186927||lt2gene Low-quality
18-109-final-viral-combined_k141_63904||lt2gene Low-quality
18-109-final-viral-combined_k141_93882||lt2gene Not-determined
18-109-final-viral-combined_k141_115077||lt2gene Low-quality
18-109-final-viral-combined_k141_33304||lt2gene Low-quality
18-109-final-viral-combined_k141_237757||lt2gene Low-quality
18-109-final-viral-combined_k141_43495||lt2gene Low-quality
18-109-final-viral-combined_k141_207391||lt2gene Not-determined
18-109-final-viral-combined_k141_175949||lt2gene Not-determined
18-109-final-viral-combined_k141_186987||lt2gene Not-determined
18-109-final-viral-combined_k141_207451||lt2gene Low-quality
18-109-final-viral-combined_k141_115154||lt2gene Not-determined
18-109-final-viral-combined_k141_237896||lt2gene Low-quality
18-109-final-viral-combined_k141_115187||lt2gene Low-quality
18-109-final-viral-combined_k141_43562||lt2gene Low-quality
18-109-final-viral-combined_k141_156205||lt2gene Low-quality
18-109-final-viral-combined_k141_104705||lt2gene Low-quality
18-109-final-viral-combined_k141_84131||lt2gene Low-quality
18-109-final-viral-combined_k141_22580||lt2gene Low-quality
18-109-final-viral-combined_k141_166178||lt2gene Low-quality
18-109-final-viral-combined_k141_22592||lt2gene Low-quality
18-109-final-viral-combined_k141_176073||lt2gene Low-quality
18-109-final-viral-combined_k141_227722||lt2gene Low-quality
18-109-final-viral-combined_k141_94034||lt2gene Low-quality
18-109-final-viral-combined_k141_84175||lt2gene Low-quality
18-109-final-viral-combined_k141_207565||lt2gene Low-quality
18-109-final-viral-combined_k141_207605||lt2gene Not-determined
18-109-final-viral-combined_k141_135286||lt2gene Low-quality
18-109-final-viral-combined_k141_94084||lt2gene Low-quality
18-109-final-viral-combined_k141_187040||lt2gene Not-determined
18-109-final-viral-combined_k141_73481||lt2gene Not-determined
18-109-final-viral-combined_k141_125265||lt2gene Low-quality
18-109-final-viral-combined_k141_135359||lt2gene Low-quality
18-109-final-viral-combined_k141_84294||lt2gene Low-quality
18-109-final-viral-combined_k141_156371||lt2gene Not-determined
18-109-final-viral-combined_k141_94225||lt2gene Not-determined
18-109-final-viral-combined_k141_217121||lt2gene Low-quality
18-109-final-viral-combined_k141_238033||lt2gene Low-quality
18-109-final-viral-combined_k141_176264||lt2gene Low-quality
18-109-final-viral-combined_k141_125318||lt2gene Low-quality
18-109-final-viral-combined_k141_238047||lt2gene Low-quality
18-109-final-viral-combined_k141_217170||lt2gene Low-quality
18-109-final-viral-combined_k141_22758||lt2gene Low-quality
18-109-final-viral-combined_k141_197427||lt2gene Not-determined
18-109-final-viral-combined_k141_125406||lt2gene Low-quality
18-109-final-viral-combined_k141_207789||lt2gene Low-quality
18-109-final-viral-combined_k141_43754||lt2gene Not-determined
18-109-final-viral-combined_k141_94361||lt2gene Low-quality
18-109-final-viral-combined_k141_145517||lt2gene Not-determined
18-109-final-viral-combined_k141_73640||lt2gene Low-quality
18-109-final-viral-combined_k141_94379||lt2gene Low-quality
18-109-final-viral-combined_k141_104976||lt2gene Low-quality
18-109-final-viral-combined_k141_33705||lt2gene Low-quality
18-109-final-viral-combined_k141_43792||lt2gene Low-quality
18-109-final-viral-combined_k141_94420||lt2gene Low-quality
18-109-final-viral-combined_k141_145579||lt2gene Not-determined
18-109-final-viral-combined_k141_43810||lt2gene Low-quality
18-109-final-viral-combined_k141_176378||lt2gene Low-quality
18-109-final-viral-combined_k141_33733||lt2gene Not-determined
18-109-final-viral-combined_k141_207875||lt2gene Low-quality
18-109-final-viral-combined_k141_84366||lt2gene Low-quality
18-109-final-viral-combined_k141_2982||lt2gene Low-quality
18-109-final-viral-combined_k141_135538||lt2gene Low-quality
18-109-final-viral-combined_k141_176429||lt2gene Not-determined
18-109-final-viral-combined_k141_125612||lt2gene Low-quality
18-109-final-viral-combined_k141_54046||lt2gene Low-quality
18-109-final-viral-combined_k141_43867||lt2gene Low-quality
18-109-final-viral-combined_k141_54053||lt2gene Low-quality
18-109-final-viral-combined_k141_197624||lt2gene Not-determined
18-109-final-viral-combined_k141_166470||lt2gene Not-determined
18-109-final-viral-combined_k141_176497||lt2gene Low-quality
18-109-final-viral-combined_k141_166495||lt2gene Not-determined
18-109-final-viral-combined_k141_94522||lt2gene Low-quality
18-109-final-viral-combined_k141_54092||lt2gene Low-quality
18-109-final-viral-combined_k141_187289||lt2gene Low-quality
18-109-final-viral-combined_k141_3029||lt2gene Not-determined
18-109-final-viral-combined_k141_166523||lt2gene Low-quality
18-109-final-viral-combined_k141_187342||lt2gene Low-quality
18-109-final-viral-combined_k141_187344||lt2gene Not-determined
18-109-final-viral-combined_k141_187346||lt2gene Low-quality
18-109-final-viral-combined_k141_13253||lt2gene Not-determined
18-109-final-viral-combined_k141_33798||lt2gene Low-quality
18-109-final-viral-combined_k141_115492||lt2gene Low-quality
18-109-final-viral-combined_k141_13260||lt2gene Low-quality
18-109-final-viral-combined_k141_217262||lt2gene Low-quality
18-109-final-viral-combined_k141_166628||lt2gene Low-quality
18-109-final-viral-combined_k141_228218||lt2gene Low-quality
18-109-final-viral-combined_k141_187427||lt2gene Low-quality
18-109-final-viral-combined_k141_166670||lt2gene Low-quality
18-109-final-viral-combined_k141_135733||lt2gene Low-quality
18-109-final-viral-combined_k141_135734||lt2gene Low-quality
18-109-final-viral-combined_k141_13378||lt2gene Not-determined
18-109-final-viral-combined_k141_135741||lt2gene Low-quality
18-109-final-viral-combined_k141_22927||lt2gene Low-quality
18-109-final-viral-combined_k141_84562||lt2gene Low-quality
18-109-final-viral-combined_k141_197808||lt2gene Low-quality
18-109-final-viral-combined_k141_217392||lt2gene Low-quality
18-109-final-viral-combined_k141_64306||lt2gene Not-determined
18-109-final-viral-combined_k141_115670||lt2gene Low-quality
18-109-final-viral-combined_k141_34001||lt2gene Low-quality
18-109-final-viral-combined_k141_145784||lt2gene Low-quality
18-109-final-viral-combined_k141_54251||lt2gene Low-quality
18-109-final-viral-combined_k141_187566||lt2gene Low-quality
18-109-final-viral-combined_k141_187601||lt2gene Low-quality
18-109-final-viral-combined_k141_187606||lt2gene Low-quality
18-109-final-viral-combined_k141_44076||lt2gene Low-quality
18-109-final-viral-combined_k141_187619||lt2gene Low-quality
18-109-final-viral-combined_k141_84642||lt2gene Low-quality
18-109-final-viral-combined_k141_94694||lt2gene Not-determined
18-109-final-viral-combined_k141_44093||lt2gene Low-quality
18-109-final-viral-combined_k141_176796||lt2gene Low-quality
18-109-final-viral-combined_k141_94716||lt2gene Low-quality
18-109-final-viral-combined_k141_176809||lt2gene Not-determined
18-109-final-viral-combined_k141_156652||lt2gene Low-quality
18-109-final-viral-combined_k141_197891||lt2gene Low-quality
18-109-final-viral-combined_k141_238409||lt2gene Low-quality
18-109-final-viral-combined_k141_34163||lt2gene Low-quality
18-109-final-viral-combined_k141_208227||lt2gene Not-determined
18-109-final-viral-combined_k141_64414||lt2gene Low-quality
18-109-final-viral-combined_k141_115794||lt2gene Low-quality
18-109-final-viral-combined_k141_135839||lt2gene Low-quality
18-109-final-viral-combined_k141_3301||lt2gene Not-determined
18-109-final-viral-combined_k141_34182||lt2gene Low-quality
18-109-final-viral-combined_k141_64436||lt2gene Low-quality
18-109-final-viral-combined_k141_197923||lt2gene Low-quality
18-109-final-viral-combined_k141_187699||lt2gene Low-quality
18-109-final-viral-combined_k141_197933||lt2gene Low-quality
18-109-final-viral-combined_k141_197959||lt2gene Low-quality
18-109-final-viral-combined_k141_13657||lt2gene Low-quality
18-109-final-viral-combined_k141_23125||lt2gene Low-quality
18-109-final-viral-combined_k141_238494||lt2gene Low-quality
18-109-final-viral-combined_k141_217466||lt2gene Not-determined
18-109-final-viral-combined_k141_166900||lt2gene Not-determined
18-109-final-viral-combined_k141_198007||lt2gene Low-quality
18-109-final-viral-combined_k141_125938||lt2gene Low-quality
18-109-final-viral-combined_k141_208312||lt2gene Not-determined
18-109-final-viral-combined_k141_44183||lt2gene Low-quality
18-109-final-viral-combined_k141_44204||lt2gene Not-determined
18-109-final-viral-combined_k141_238535||lt2gene Not-determined
18-109-final-viral-combined_k141_238541||lt2gene Low-quality
18-109-final-viral-combined_k141_167027||lt2gene Low-quality
18-109-final-viral-combined_k141_105457||lt2gene Low-quality
18-109-final-viral-combined_k141_84770||lt2gene Low-quality
18-109-final-viral-combined_k141_105467||lt2gene Low-quality

18-109-final-viral-combined_k141_105478||lt2gene Low-quality
18-109-final-viral-combined_k141_126030||lt2gene Not-determined
18-109-final-viral-combined_k141_156837||lt2gene Low-quality
18-109-final-viral-combined_k141_146110||lt2gene Low-quality
18-109-final-viral-combined_k141_146116||lt2gene Low-quality
18-109-final-viral-combined_k141_228484||lt2gene Not-determined
18-109-final-viral-combined_k141_167142||lt2gene Not-determined
18-109-final-viral-combined_k141_105539||lt2gene Not-determined
18-109-final-viral-combined_k141_167151||lt2gene Low-quality
18-109-final-viral-combined_k141_126051||lt2gene Low-quality
18-109-final-viral-combined_k141_167162||lt2gene Low-quality
18-109-final-viral-combined_k141_187936||lt2gene Not-determined
18-109-final-viral-combined_k141_94836||lt2gene Low-quality
18-109-final-viral-combined_k141_44323||lt2gene Low-quality
18-109-final-viral-combined_k141_228523||lt2gene Low-quality
18-109-final-viral-combined_k141_217606||lt2gene Low-quality
18-109-final-viral-combined_k141_34419||lt2gene Low-quality
18-109-final-viral-combined_k141_146177||lt2gene Low-quality
18-109-final-viral-combined_k141_3527||lt2gene Not-determined
18-109-final-viral-combined_k141_84900||lt2gene Low-quality
18-109-final-viral-combined_k141_84905||lt2gene Low-quality
18-109-final-viral-combined_k141_228583||lt2gene Low-quality
18-109-final-viral-combined_k141_116105||lt2gene Not-determined
18-109-final-viral-combined_k141_228597||lt2gene Low-quality
18-109-final-viral-combined_k141_116118||lt2gene Low-quality
18-109-final-viral-combined_k141_167207||lt2gene Not-determined
18-109-final-viral-combined_k141_167209||lt2gene Not-determined
18-109-final-viral-combined_k141_116137||lt2gene Low-quality
18-109-final-viral-combined_k141_136080||lt2gene Not-determined
18-109-final-viral-combined_k141_84998||lt2gene Not-determined
18-109-final-viral-combined_k141_44431||lt2gene Not-determined
18-109-final-viral-combined_k141_23267||lt2gene Low-quality
18-109-final-viral-combined_k141_177195||lt2gene Low-quality
18-109-final-viral-combined_k141_146321||lt2gene Low-quality
18-109-final-viral-combined_k141_188081||lt2gene Low-quality
18-109-final-viral-combined_k141_116287||lt2gene Low-quality
18-109-final-viral-combined_k141_177234||lt2gene Low-quality
18-109-final-viral-combined_k141_64799||lt2gene Low-quality
18-109-final-viral-combined_k141_85136||lt2gene Low-quality
18-109-final-viral-combined_k141_95038||lt2gene Not-determined
18-109-final-viral-combined_k141_95066||lt2gene Low-quality
18-109-final-viral-combined_k141_146442||lt2gene Low-quality
18-109-final-viral-combined_k141_238870||lt2gene Not-determined
18-109-final-viral-combined_k141_146448||lt2gene Low-quality
18-109-final-viral-combined_k141_23366||lt2gene Low-quality
18-109-final-viral-combined_k141_208728||lt2gene Low-quality
18-109-final-viral-combined_k141_74532||lt2gene Not-determined
18-109-final-viral-combined_k141_208767||lt2gene Low-quality
18-109-final-viral-combined_k141_64846||lt2gene Not-determined
18-109-final-viral-combined_k141_136289||lt2gene Low-quality
18-109-final-viral-combined_k141_177343||lt2gene Not-determined
18-109-final-viral-combined_k141_74679||lt2gene Low-quality
18-109-final-viral-combined_k141_64993||lt2gene Low-quality
18-109-final-viral-combined_k141_228665||lt2gene Low-quality
18-109-final-viral-combined_k141_136349||lt2gene Low-quality
18-109-final-viral-combined_k141_34812||lt2gene Not-determined
18-109-final-viral-combined_k141_14075||lt2gene Not-determined
18-109-final-viral-combined_k141_198384||lt2gene Not-determined
18-109-final-viral-combined_k141_228707||lt2gene Not-determined
18-109-final-viral-combined_k141_14096||lt2gene Low-quality
18-109-final-viral-combined_k141_146619||lt2gene Low-quality
18-109-final-viral-combined_k141_239126||lt2gene Low-quality
18-109-final-viral-combined_k141_177519||lt2gene Low-quality
18-109-final-viral-combined_k141_54676||lt2gene Low-quality
18-109-final-viral-combined_k141_157204||lt2gene Low-quality
18-109-final-viral-combined_k141_54712||lt2gene Low-quality
18-109-final-viral-combined_k141_167641||lt2gene Not-determined
18-109-final-viral-combined_k141_105882||lt2gene Low-quality
18-109-final-viral-combined_k141_146699||lt2gene Not-determined
18-109-final-viral-combined_k141_85431||lt2gene Low-quality
18-109-final-viral-combined_k141_54755||lt2gene Low-quality
18-109-final-viral-combined_k141_54760||lt2gene Low-quality
18-109-final-viral-combined_k141_188412||lt2gene Low-quality
18-109-final-viral-combined_k141_167668||lt2gene Low-quality
18-109-final-viral-combined_k141_23614||lt2gene Low-quality
18-109-final-viral-combined_k141_65161||lt2gene Not-determined
18-109-final-viral-combined_k141_208995||lt2gene Not-determined
18-109-final-viral-combined_k141_198508||lt2gene Low-quality
18-109-final-viral-combined_k141_188446||lt2gene Not-determined
18-109-final-viral-combined_k141_188456||lt2gene Low-quality
18-109-final-viral-combined_k141_228866||lt2gene Low-quality
18-109-final-viral-combined_k141_14130||lt2gene Not-determined
18-109-final-viral-combined_k141_228878||lt2gene Not-determined
18-109-final-viral-combined_k141_3869||lt2gene Low-quality
18-109-final-viral-combined_k141_95473||lt2gene Low-quality
18-109-final-viral-combined_k141_34921||lt2gene Low-quality
18-109-final-viral-combined_k141_65305||lt2gene Low-quality
18-109-final-viral-combined_k141_228921||lt2gene Low-quality
18-109-final-viral-combined_k141_177738||lt2gene Low-quality
18-109-final-viral-combined_k141_74839||lt2gene Not-determined
18-109-final-viral-combined_k141_239285||lt2gene Low-quality
18-109-final-viral-combined_k141_146808||lt2gene Low-quality
18-109-final-viral-combined_k141_34972||lt2gene Low-quality
18-109-final-viral-combined_k141_23786||lt2gene Not-determined
18-109-final-viral-combined_k141_23790||lt2gene Low-quality
18-109-final-viral-combined_k141_85538||lt2gene Low-quality
18-109-final-viral-combined_k141_3954||lt2gene Not-determined
18-109-final-viral-combined_k141_3956||lt2gene Low-quality
18-109-final-viral-combined_k141_106131||lt2gene Low-quality
18-109-final-viral-combined_k141_198765||lt2gene Low-quality
18-109-final-viral-combined_k141_229066||lt2gene Not-determined
18-109-final-viral-combined_k141_23825||lt2gene Low-quality
18-109-final-viral-combined_k141_85593||lt2gene Low-quality
18-109-final-viral-combined_k141_157411||lt2gene Low-quality
18-109-final-viral-combined_k141_146858||lt2gene Low-quality
18-109-final-viral-combined_k141_217905||lt2gene Low-quality
18-109-final-viral-combined_k141_209145||lt2gene Low-quality
18-109-final-viral-combined_k141_177914||lt2gene Low-quality
18-109-final-viral-combined_k141_95611||lt2gene Low-quality
18-109-final-viral-combined_k141_209171||lt2gene Low-quality
18-109-final-viral-combined_k141_45125||lt2gene Low-quality
18-109-final-viral-combined_k141_55111||lt2gene Low-quality
18-109-final-viral-combined_k141_116722||lt2gene Low-quality
18-109-final-viral-combined_k141_65670||lt2gene Low-quality
18-109-final-viral-combined_k141_95666||lt2gene Not-determined
18-109-final-viral-combined_k141_136737||lt2gene Low-quality
18-109-final-viral-combined_k141_198908||lt2gene Low-quality
18-109-final-viral-combined_k141_95722||lt2gene Not-determined
18-109-final-viral-combined_k141_14315||lt2gene Low-quality
18-109-final-viral-combined_k141_167898||lt2gene Low-quality
18-109-final-viral-combined_k141_4133||lt2gene Not-determined
18-109-final-viral-combined_k141_65776||lt2gene Low-quality
18-109-final-viral-combined_k141_127067||lt2gene Not-determined
18-109-final-viral-combined_k141_65797||lt2gene Low-quality
18-109-final-viral-combined_k141_4180||lt2gene Low-quality
18-109-final-viral-combined_k141_106278||lt2gene Low-quality
18-109-final-viral-combined_k141_106284||lt2gene Low-quality
18-109-final-viral-combined_k141_127197||lt2gene Low-quality
18-109-final-viral-combined_k141_157669||lt2gene Low-quality
18-109-final-viral-combined_k141_14364||lt2gene Low-quality
18-109-final-viral-combined_k141_45210||lt2gene Low-quality
18-109-final-viral-combined_k141_157692||lt2gene Not-determined
18-109-final-viral-combined_k141_35268||lt2gene Low-quality
18-109-final-viral-combined_k141_24006||lt2gene Not-determined
18-109-final-viral-combined_k141_209419||lt2gene Not-determined
18-109-final-viral-combined_k141_116858||lt2gene Not-determined
18-109-final-viral-combined_k141_35280||lt2gene Low-quality
18-109-final-viral-combined_k141_229290||lt2gene Low-quality
18-109-final-viral-combined_k141_218092||lt2gene Low-quality
18-109-final-viral-combined_k141_209449||lt2gene Not-determined
18-109-final-viral-combined_k141_45251||lt2gene Not-determined
18-109-final-viral-combined_k141_35305||lt2gene Low-quality
18-109-final-viral-combined_k141_95904||lt2gene Low-quality
18-109-final-viral-combined_k141_85862||lt2gene Low-quality
18-109-final-viral-combined_k141_4261||lt2gene Low-quality
18-109-final-viral-combined_k141_199168||lt2gene Low-quality
18-109-final-viral-combined_k141_157829||lt2gene Low-quality
18-109-final-viral-combined_k141_178200||lt2gene Low-quality
18-109-final-viral-combined_k141_209531||lt2gene Low-quality
18-109-final-viral-combined_k141_168121||lt2gene Not-determined
18-109-final-viral-combined_k141_209552||lt2gene Not-determined
18-109-final-viral-combined_k141_178238||lt2gene Low-quality
18-109-final-viral-combined_k141_137014||lt2gene Low-quality
18-109-final-viral-combined_k141_55338||lt2gene Low-quality
18-109-final-viral-combined_k141_147075||lt2gene Not-determined
18-109-final-viral-combined_k141_85967||lt2gene Not-determined
18-109-final-viral-combined_k141_65987||lt2gene Not-determined
18-109-final-viral-combined_k141_116979||lt2gene Not-determined
18-109-final-viral-combined_k141_168186||lt2gene Not-determined
18-109-final-viral-combined_k141_45361||lt2gene Not-determined
18-109-final-viral-combined_k141_55397||lt2gene Low-quality
18-109-final-viral-combined_k141_229471||lt2gene Not-determined
18-109-final-viral-combined_k141_188809||lt2gene Low-quality
18-109-final-viral-combined_k141_14576||lt2gene Low-quality
18-109-final-viral-combined_k141_45451||lt2gene Not-determined
18-109-final-viral-combined_k141_75440||lt2gene Low-quality
18-109-final-viral-combined_k141_127408||lt2gene Low-quality
18-109-final-viral-combined_k141_158098||lt2gene Low-quality
18-109-final-viral-combined_k141_45479||lt2gene Low-quality
18-109-final-viral-combined_k141_209720||lt2gene Low-quality
18-109-final-viral-combined_k141_96156||lt2gene Low-quality
18-109-final-viral-combined_k141_96162||lt2gene Not-determined
18-109-final-viral-combined_k141_4571||lt2gene Low-quality
18-109-final-viral-combined_k141_35713||lt2gene Low-quality
18-109-final-viral-combined_k141_178507||lt2gene Low-quality
18-109-final-viral-combined_k141_127462||lt2gene Low-quality
18-109-final-viral-combined_k141_137255||lt2gene Low-quality
18-109-final-viral-combined_k141_178513||lt2gene Low-quality
18-109-final-viral-combined_k141_4641||lt2gene Low-quality
18-109-final-viral-combined_k141_106478||lt2gene Low-quality
18-109-final-viral-combined_k141_147186||lt2gene Not-determined
18-109-final-viral-combined_k141_66264||lt2gene Low-quality
18-109-final-viral-combined_k141_24157||lt2gene Low-quality
18-109-final-viral-combined_k141_199369||lt2gene Low-quality
18-109-final-viral-combined_k141_117231||lt2gene Low-quality
18-109-final-viral-combined_k141_4666||lt2gene Low-quality
18-109-final-viral-combined_k141_117246||lt2gene Low-quality
18-109-final-viral-combined_k141_127548||lt2gene Low-quality
18-109-final-viral-combined_k141_86182||lt2gene Low-quality
18-109-final-viral-combined_k141_86250||lt2gene Not-determined
18-109-final-viral-combined_k141_96329||lt2gene Low-quality
18-109-final-viral-combined_k141_14766||lt2gene Low-quality
18-109-final-viral-combined_k141_189020||lt2gene Low-quality
18-109-final-viral-combined_k141_117351||lt2gene Low-quality
18-109-final-viral-combined_k141_35907||lt2gene Low-quality
18-109-final-viral-combined_k141_75603||lt2gene Low-quality
18-109-final-viral-combined_k141_178745||lt2gene Not-determined
18-109-final-viral-combined_k141_178768||lt2gene Low-quality
18-109-final-viral-combined_k141_117387||lt2gene Low-quality
18-109-final-viral-combined_k141_86310||lt2gene Low-quality
18-109-final-viral-combined_k141_96393||lt2gene Low-quality
18-109-final-viral-combined_k141_75646||lt2gene Low-quality
18-109-final-viral-combined_k141_127648||lt2gene Not-determined
18-109-final-viral-combined_k141_127675||lt2gene Low-quality
18-109-final-viral-combined_k141_199528||lt2gene Low-quality
18-109-final-viral-combined_k141_158354||lt2gene Low-quality
18-109-final-viral-combined_k141_4700||lt2gene Low-quality
18-109-final-viral-combined_k141_75700||lt2gene Low-quality
18-109-final-viral-combined_k141_55856||lt2gene Low-quality
18-109-final-viral-combined_k141_229766||lt2gene Not-determined
18-109-final-viral-combined_k141_147435||lt2gene Low-quality
18-109-final-viral-combined_k141_240261||lt2gene Low-quality
18-109-final-viral-combined_k141_147445||lt2gene Low-quality
18-109-final-viral-combined_k141_55914||lt2gene Low-quality
18-109-final-viral-combined_k141_117525||lt2gene Not-determined
18-109-final-viral-combined_k141_127740||lt2gene Not-determined
18-109-final-viral-combined_k141_229817||lt2gene Low-quality
18-109-final-viral-combined_k141_56018||lt2gene Not-determined
18-109-final-viral-combined_k141_35944||lt2gene Low-quality
18-109-final-viral-combined_k141_178824||lt2gene Low-quality
18-109-final-viral-combined_k141_106710||lt2gene Not-determined
18-109-final-viral-combined_k141_4826||lt2gene Low-quality
18-109-final-viral-combined_k141_127767||lt2gene Low-quality
18-109-final-viral-combined_k141_106722||lt2gene Low-quality
18-109-final-viral-combined_k141_24326||lt2gene Not-determined
18-109-final-viral-combined_k141_147582||lt2gene Low-quality
18-109-final-viral-combined_k141_117631||lt2gene Low-quality
18-109-final-viral-combined_k141_168753||lt2gene Not-determined
18-109-final-viral-combined_k141_4897||lt2gene Low-quality
18-109-final-viral-combined_k141_4903||lt2gene Not-determined
18-109-final-viral-combined_k141_96611||lt2gene Low-quality
18-109-final-viral-combined_k141_189219||lt2gene Not-determined
18-109-final-viral-combined_k141_189235||lt2gene Low-quality
18-109-final-viral-combined_k141_199770||lt2gene Low-quality
18-109-final-viral-combined_k141_199772||lt2gene Not-determined
18-109-final-viral-combined_k141_209892||lt2gene Low-quality
18-109-final-viral-combined_k141_240357||lt2gene Low-quality
18-109-final-viral-combined_k141_4940||lt2gene Not-determined
18-109-final-viral-combined_k141_66790||lt2gene Not-determined
18-109-final-viral-combined_k141_96682||lt2gene Not-determined
18-109-final-viral-combined_k141_15128||lt2gene Not-determined
18-109-final-viral-combined_k141_209925||lt2gene Low-quality
18-109-final-viral-combined_k141_240394||lt2gene Low-quality
18-109-final-viral-combined_k141_117733||lt2gene Not-determined
18-109-final-viral-combined_k141_45824||lt2gene Low-quality
18-109-final-viral-combined_k141_137645||lt2gene Not-determined
18-109-final-viral-combined_k141_117753||lt2gene Not-determined
18-109-final-viral-combined_k141_209969||lt2gene Not-determined
18-109-final-viral-combined_k141_15187||lt2gene Low-quality
18-109-final-viral-combined_k141_199902||lt2gene Not-determined
18-109-final-viral-combined_k141_66877||lt2gene Low-quality
18-109-final-viral-combined_k141_24540||lt2gene Low-quality
18-109-final-viral-combined_k141_189301||lt2gene Low-quality
18-109-final-viral-combined_k141_189306||lt2gene Low-quality
18-109-final-viral-combined_k141_96814||lt2gene Low-quality
18-109-final-viral-combined_k141_66882||lt2gene Low-quality
18-109-final-viral-combined_k141_106860||lt2gene Not-determined
18-109-final-viral-combined_k141_168947||lt2gene Low-quality
18-109-final-viral-combined_k141_45915||lt2gene Not-determined
18-109-final-viral-combined_k141_5078||lt2gene Not-determined
18-109-final-viral-combined_k141_179007||lt2gene Low-quality
18-109-final-viral-combined_k141_168980||lt2gene Low-quality
18-109-final-viral-combined_k141_179012||lt2gene Low-quality
18-109-final-viral-combined_k141_106887||lt2gene Low-quality
18-109-final-viral-combined_k141_106901||lt2gene Low-quality
18-109-final-viral-combined_k141_106902||lt2gene Not-determined
18-109-final-viral-combined_k141_179032||lt2gene Not-determined
18-109-final-viral-combined_k141_24663||lt2gene Not-determined
18-109-final-viral-combined_k141_127958||lt2gene Low-quality
18-109-final-viral-combined_k141_127959||lt2gene Low-quality
18-109-final-viral-combined_k141_24676||lt2gene Low-quality
18-109-final-viral-combined_k141_137834||lt2gene Low-quality
18-109-final-viral-combined_k141_76099||lt2gene Low-quality
18-109-final-viral-combined_k141_56236||lt2gene Low-quality
18-109-final-viral-combined_k141_189407||lt2gene Low-quality
18-109-final-viral-combined_k141_158668||lt2gene Low-quality
18-109-final-viral-combined_k141_127997||lt2gene Not-determined
18-109-final-viral-combined_k141_189425||lt2gene Low-quality
18-109-final-viral-combined_k141_128011||lt2gene Low-quality
18-109-final-viral-combined_k141_147749||lt2gene Low-quality
18-109-final-viral-combined_k141_24719||lt2gene Low-quality
18-109-final-viral-combined_k141_76154||lt2gene Low-quality
18-109-final-viral-combined_k141_117995||lt2gene Not-determined
18-109-final-viral-combined_k141_230066||lt2gene Low-quality
18-109-final-viral-combined_k141_230071||lt2gene Not-determined
18-109-final-viral-combined_k141_118008||lt2gene Low-quality
18-109-final-viral-combined_k141_230081||lt2gene Low-quality
18-109-final-viral-combined_k141_15341||lt2gene Low-quality
18-109-final-viral-combined_k141_76200||lt2gene Not-determined
18-109-final-viral-combined_k141_169085||lt2gene Low-quality
18-109-final-viral-combined_k141_67014||lt2gene Not-determined
18-109-final-viral-combined_k141_118042||lt2gene Low-quality
18-109-final-viral-combined_k141_86676||lt2gene Not-determined
18-109-final-viral-combined_k141_179198||lt2gene Low-quality
18-109-final-viral-combined_k141_200177||lt2gene Low-quality
18-109-final-viral-combined_k141_240531||lt2gene Low-quality
18-109-final-viral-combined_k141_46181||lt2gene Low-quality
18-109-final-viral-combined_k141_76298||lt2gene Low-quality
18-109-final-viral-combined_k141_189590||lt2gene Low-quality
18-109-final-viral-combined_k141_147922||lt2gene Low-quality
18-109-final-viral-combined_k141_107105||lt2gene Not-determined
18-109-final-viral-combined_k141_56313||lt2gene Low-quality
18-109-final-viral-combined_k141_200259||lt2gene Low-quality
18-109-final-viral-combined_k141_169246||lt2gene Low-quality
18-109-final-viral-combined_k141_118214||lt2gene Not-determined
18-109-final-viral-combined_k141_56356||lt2gene Low-quality
18-109-final-viral-combined_k141_15458||lt2gene Low-quality
18-109-final-viral-combined_k141_147972||lt2gene Low-quality
18-109-final-viral-combined_k141_169305||lt2gene Not-determined
18-109-final-viral-combined_k141_107216||lt2gene Low-quality
18-109-final-viral-combined_k141_240594||lt2gene Low-quality
18-109-final-viral-combined_k141_158849||lt2gene Low-quality
18-109-final-viral-combined_k141_107242||lt2gene Not-determined
18-109-final-viral-combined_k141_169347||lt2gene Low-quality
18-109-final-viral-combined_k141_56395||lt2gene Low-quality
18-109-final-viral-combined_k141_169348||lt2gene Low-quality
18-109-final-viral-combined_k141_56414||lt2gene Low-quality
18-109-final-viral-combined_k141_56434||lt2gene Low-quality
18-109-final-viral-combined_k141_15565||lt2gene Low-quality
18-109-final-viral-combined_k141_36417||lt2gene Low-quality
18-109-final-viral-combined_k141_24957||lt2gene Not-determined
18-109-final-viral-combined_k141_76455||lt2gene Low-quality
18-109-final-viral-combined_k141_200374||lt2gene Low-quality
18-109-final-viral-combined_k141_118273||lt2gene Low-quality
18-109-final-viral-combined_k141_148146||lt2gene Low-quality
18-109-final-viral-combined_k141_46402||lt2gene Not-determined
18-109-final-viral-combined_k141_240693||lt2gene Not-determined
18-109-final-viral-combined_k141_210349||lt2gene Low-quality
18-109-final-viral-combined_k141_46416||lt2gene Not-determined
18-109-final-viral-combined_k141_118327||lt2gene Low-quality
18-109-final-viral-combined_k141_107283||lt2gene Not-determined
18-109-final-viral-combined_k141_138154||lt2gene Low-quality
18-109-final-viral-combined_k141_189811||lt2gene Low-quality
18-109-final-viral-combined_k141_76534||lt2gene Not-determined
18-109-final-viral-combined_k141_189848||lt2gene Low-quality
18-109-final-viral-combined_k141_179466||lt2gene Not-determined
18-109-final-viral-combined_k141_97282||lt2gene Low-quality
18-109-final-viral-combined_k141_36566||lt2gene Low-quality
18-109-final-viral-combined_k141_179470||lt2gene Low-quality
18-109-final-viral-combined_k141_76573||lt2gene Low-quality
18-109-final-viral-combined_k141_25115||lt2gene Low-quality
18-109-final-viral-combined_k141_56611||lt2gene Low-quality
18-109-final-viral-combined_k141_148217||lt2gene Low-quality
18-109-final-viral-combined_k141_230412||lt2gene Low-quality
18-109-final-viral-combined_k141_148228||lt2gene Not-determined
18-109-final-viral-combined_k141_159077||lt2gene Low-quality
18-109-final-viral-combined_k141_240797||lt2gene Low-quality
18-109-final-viral-combined_k141_200503||lt2gene Low-quality
18-109-final-viral-combined_k141_5611||lt2gene Low-quality
18-109-final-viral-combined_k141_210512||lt2gene Not-determined
18-109-final-viral-combined_k141_86889||lt2gene Low-quality
18-109-final-viral-combined_k141_169478||lt2gene Not-determined
18-109-final-viral-combined_k141_107475||lt2gene Low-quality
18-109-final-viral-combined_k141_210563||lt2gene Low-quality
18-109-final-viral-combined_k141_76717||lt2gene Not-determined
18-109-final-viral-combined_k141_107528||lt2gene Low-quality
18-109-final-viral-combined_k141_210597||lt2gene Low-quality
18-109-final-viral-combined_k141_240942||lt2gene Low-quality
18-109-final-viral-combined_k141_159194||lt2gene Not-determined
18-109-final-viral-combined_k141_87017||lt2gene Low-quality
18-109-final-viral-combined_k141_87019||lt2gene Not-determined
18-109-final-viral-combined_k141_128450||lt2gene Low-quality
18-109-final-viral-combined_k141_46713||lt2gene Low-quality
18-109-final-viral-combined_k141_128459||lt2gene Not-determined
18-109-final-viral-combined_k141_87037||lt2gene Low-quality
18-109-final-viral-combined_k141_138395||lt2gene Not-determined
18-109-final-viral-combined_k141_159253||lt2gene Low-quality
18-109-final-viral-combined_k141_67373||lt2gene Not-determined
18-109-final-viral-combined_k141_67383||lt2gene Not-determined
18-109-final-viral-combined_k141_200721||lt2gene Low-quality
18-109-final-viral-combined_k141_169708||lt2gene Not-determined
18-109-final-viral-combined_k141_118553||lt2gene Not-determined
18-109-final-viral-combined_k141_97513||lt2gene Low-quality
18-109-final-viral-combined_k141_67453||lt2gene Low-quality
18-109-final-viral-combined_k141_36919||lt2gene Not-determined
18-109-final-viral-combined_k141_241104||lt2gene Not-determined
18-109-final-viral-combined_k141_76803||lt2gene Low-quality
18-109-final-viral-combined_k141_107597||lt2gene Low-quality
18-109-final-viral-combined_k141_67526||lt2gene Low-quality
18-109-final-viral-combined_k141_15961||lt2gene Low-quality
18-109-final-viral-combined_k141_5730||lt2gene Low-quality
18-109-final-viral-combined_k141_67590||lt2gene Low-quality
18-109-final-viral-combined_k141_25339||lt2gene Low-quality
18-109-final-viral-combined_k141_118639||lt2gene Low-quality
18-109-final-viral-combined_k141_67617||lt2gene Low-quality
18-109-final-viral-combined_k141_46864||lt2gene Low-quality
18-109-final-viral-combined_k141_179880||lt2gene Low-quality
18-109-final-viral-combined_k141_97631||lt2gene Low-quality
18-109-final-viral-combined_k141_241202||lt2gene Not-determined
18-109-final-viral-combined_k141_219160||lt2gene Low-quality
18-109-final-viral-combined_k141_37126||lt2gene Low-quality
18-109-final-viral-combined_k141_67708||lt2gene Low-quality
18-109-final-viral-combined_k141_219231||lt2gene Low-quality
18-109-final-viral-combined_k141_16095||lt2gene Low-quality
18-109-final-viral-combined_k141_16099||lt2gene Low-quality
18-109-final-viral-combined_k141_169976||lt2gene Not-determined
18-109-final-viral-combined_k141_138687||lt2gene Low-quality
18-109-final-viral-combined_k141_241300||lt2gene Low-quality
18-109-final-viral-combined_k141_190321||lt2gene Low-quality
18-109-final-viral-combined_k141_179909||lt2gene Not-determined
18-109-final-viral-combined_k141_230698||lt2gene Low-quality
18-109-final-viral-combined_k141_219261||lt2gene Low-quality
18-109-final-viral-combined_k141_210922||lt2gene Low-quality
18-109-final-viral-combined_k141_159586||lt2gene Low-quality
18-109-final-viral-combined_k141_5840||lt2gene Low-quality
18-109-final-viral-combined_k141_179963||lt2gene Not-determined
18-109-final-viral-combined_k141_190330||lt2gene Not-determined
18-109-final-viral-combined_k141_97726||lt2gene Low-quality
18-109-final-viral-combined_k141_97727||lt2gene Not-determined
18-109-final-viral-combined_k141_25450||lt2gene Low-quality
18-109-final-viral-combined_k141_159618||lt2gene Not-determined
18-109-final-viral-combined_k141_230783||lt2gene Low-quality
18-109-final-viral-combined_k141_67833||lt2gene Low-quality
18-109-final-viral-combined_k141_159667||lt2gene Low-quality
18-109-final-viral-combined_k141_148706||lt2gene Low-quality
18-109-final-viral-combined_k141_25479||lt2gene Not-determined
18-109-final-viral-combined_k141_16228||lt2gene Not-determined
18-109-final-viral-combined_k141_230816||lt2gene Low-quality
18-109-final-viral-combined_k141_46959||lt2gene Not-determined
18-109-final-viral-combined_k141_180019||lt2gene Low-quality
18-109-final-viral-combined_k141_16266||lt2gene Low-quality
18-109-final-viral-combined_k141_190441||lt2gene Low-quality
18-109-final-viral-combined_k141_118818||lt2gene Low-quality
18-109-final-viral-combined_k141_180028||lt2gene Low-quality
18-109-final-viral-combined_k141_118830||lt2gene Low-quality
18-109-final-viral-combined_k141_210994||lt2gene Low-quality
18-109-final-viral-combined_k141_77025||lt2gene Low-quality
18-109-final-viral-combined_k141_159769||lt2gene Low-quality
18-109-final-viral-combined_k141_25584||lt2gene Low-quality
18-109-final-viral-combined_k141_180066||lt2gene Low-quality
18-109-final-viral-combined_k141_170141||lt2gene Not-determined
18-109-final-viral-combined_k141_87499||lt2gene Low-quality
18-109-final-viral-combined_k141_5962||lt2gene Low-quality
18-109-final-viral-combined_k141_118891||lt2gene Low-quality
18-109-final-viral-combined_k141_190516||lt2gene Low-quality
18-109-final-viral-combined_k141_241365||lt2gene Not-determined
18-109-final-viral-combined_k141_57301||lt2gene Low-quality
18-109-final-viral-combined_k141_190538||lt2gene Low-quality
18-109-final-viral-combined_k141_57308||lt2gene Low-quality
18-109-final-viral-combined_k141_77078||lt2gene Low-quality
18-109-final-viral-combined_k141_5997||lt2gene Low-quality
18-109-final-viral-combined_k141_77095||lt2gene Low-quality
18-109-final-viral-combined_k141_97929||lt2gene Not-determined
18-109-final-viral-combined_k141_138846||lt2gene Not-determined
18-109-final-viral-combined_k141_200908||lt2gene Low-quality
18-109-final-viral-combined_k141_170229||lt2gene Not-determined
18-109-final-viral-combined_k141_190627||lt2gene Not-determined
18-109-final-viral-combined_k141_170275||lt2gene Low-quality
18-109-final-viral-combined_k141_87602||lt2gene Low-quality
18-109-final-viral-combined_k141_119051||lt2gene Low-quality
18-109-final-viral-combined_k141_47091||lt2gene Low-quality
18-109-final-viral-combined_k141_98018||lt2gene Low-quality
18-109-final-viral-combined_k141_6072||lt2gene Not-determined
18-109-final-viral-combined_k141_47130||lt2gene Low-quality
18-109-final-viral-combined_k141_190708||lt2gene Not-determined
18-109-final-viral-combined_k141_129079||lt2gene Low-quality
18-109-final-viral-combined_k141_170306||lt2gene Low-quality
18-109-final-viral-combined_k141_98069||lt2gene Not-determined
18-109-final-viral-combined_k141_16518||lt2gene Low-quality
18-109-final-viral-combined_k141_201038||lt2gene Low-quality
18-109-final-viral-combined_k141_98081||lt2gene Low-quality
18-109-final-viral-combined_k141_139029||lt2gene Low-quality
18-109-final-viral-combined_k141_180149||lt2gene Not-determined
18-109-final-viral-combined_k141_139077||lt2gene Not-determined
18-109-final-viral-combined_k141_180178||lt2gene Low-quality
18-109-final-viral-combined_k141_180194||lt2gene Not-determined
18-109-final-viral-combined_k141_77311||lt2gene Not-determined
18-109-final-viral-combined_k141_219538||lt2gene Low-quality
18-109-final-viral-combined_k141_16543||lt2gene Low-quality
18-109-final-viral-combined_k141_47219||lt2gene Not-determined
18-109-final-viral-combined_k141_6241||lt2gene Low-quality
18-109-final-viral-combined_k141_211199||lt2gene Low-quality
18-109-final-viral-combined_k141_201187||lt2gene Low-quality
18-109-final-viral-combined_k141_57588||lt2gene Low-quality
18-109-final-viral-combined_k141_211231||lt2gene Not-determined
18-109-final-viral-combined_k141_68264||lt2gene Low-quality
18-109-final-viral-combined_k141_25992||lt2gene Not-determined
18-109-final-viral-combined_k141_37547||lt2gene Low-quality
18-109-final-viral-combined_k141_68276||lt2gene Not-determined
18-109-final-viral-combined_k141_170421||lt2gene Not-determined
18-109-final-viral-combined_k141_159946||lt2gene Low-quality
18-109-final-viral-combined_k141_139233||lt2gene Low-quality
18-109-final-viral-combined_k141_87773||lt2gene Not-determined
18-109-final-viral-combined_k141_129183||lt2gene Not-determined
18-109-final-viral-combined_k141_190900||lt2gene Not-determined
18-109-final-viral-combined_k141_201278||lt2gene Low-quality
18-109-final-viral-combined_k141_201290||lt2gene Not-determined
18-109-final-viral-combined_k141_190910||lt2gene Not-determined
18-109-final-viral-combined_k141_170438||lt2gene Not-determined
18-109-final-viral-combined_k141_149122||lt2gene Low-quality
18-109-final-viral-combined_k141_190922||lt2gene Not-determined
18-109-final-viral-combined_k141_219609||lt2gene Low-quality
18-109-final-viral-combined_k141_159998||lt2gene Low-quality
18-109-final-viral-combined_k141_26064||lt2gene Low-quality
18-109-final-viral-combined_k141_68354||lt2gene Low-quality
18-109-final-viral-combined_k141_47377||lt2gene Low-quality
18-109-final-viral-combined_k141_77529||lt2gene Not-determined
18-109-final-viral-combined_k141_180285||lt2gene Low-quality
18-109-final-viral-combined_k141_47411||lt2gene Low-quality
18-109-final-viral-combined_k141_190975||lt2gene Not-determined
18-109-final-viral-combined_k141_6445||lt2gene Low-quality
18-109-final-viral-combined_k141_57713||lt2gene Low-quality
18-109-final-viral-combined_k141_231221||lt2gene Not-determined
18-109-final-viral-combined_k141_201384||lt2gene Not-determined
18-109-final-viral-combined_k141_160085||lt2gene Low-quality
18-109-final-viral-combined_k141_6496||lt2gene Low-quality
18-109-final-viral-combined_k141_77602||lt2gene Low-quality
18-109-final-viral-combined_k141_87935||lt2gene Not-determined
18-109-final-viral-combined_k141_119348||lt2gene Low-quality
18-109-final-viral-combined_k141_241676||lt2gene Low-quality
18-109-final-viral-combined_k141_47544||lt2gene Low-quality
18-109-final-viral-combined_k141_16767||lt2gene Not-determined
18-109-final-viral-combined_k141_47557||lt2gene Not-determined
18-109-final-viral-combined_k141_211396||lt2gene Low-quality
18-109-final-viral-combined_k141_170609||lt2gene Low-quality
18-109-final-viral-combined_k141_107974||lt2gene Low-quality
18-109-final-viral-combined_k141_88042||lt2gene Low-quality
18-109-final-viral-combined_k141_180429||lt2gene Not-determined
18-109-final-viral-combined_k141_47595||lt2gene Low-quality
18-109-final-viral-combined_k141_211419||lt2gene Not-determined
18-109-final-viral-combined_k141_57856||lt2gene Low-quality
18-109-final-viral-combined_k141_47624||lt2gene Low-quality
18-109-final-viral-combined_k141_149202||lt2gene Not-determined
18-109-final-viral-combined_k141_47628||lt2gene Not-determined
18-109-final-viral-combined_k141_16823||lt2gene Low-quality
18-109-final-viral-combined_k141_180444||lt2gene Low-quality
18-109-final-viral-combined_k141_108015||lt2gene Not-determined
18-109-final-viral-combined_k141_108020||lt2gene Low-quality
18-109-final-viral-combined_k141_47672||lt2gene Low-quality
18-109-final-viral-combined_k141_47689||lt2gene Low-quality
18-109-final-viral-combined_k141_98554||lt2gene Low-quality
18-109-final-viral-combined_k141_77673||lt2gene Not-determined
18-109-final-viral-combined_k141_108104||lt2gene Low-quality
18-109-final-viral-combined_k141_37878||lt2gene Not-determined
18-109-final-viral-combined_k141_180497||lt2gene Low-quality
18-109-final-viral-combined_k141_108142||lt2gene Not-determined
18-109-final-viral-combined_k141_180512||lt2gene Low-quality
18-109-final-viral-combined_k141_180513||lt2gene Not-determined
18-109-final-viral-combined_k141_231472||lt2gene Low-quality
18-109-final-viral-combined_k141_98578||lt2gene Not-determined
18-109-final-viral-combined_k141_57948||lt2gene Low-quality
18-109-final-viral-combined_k141_57955||lt2gene Low-quality
18-109-final-viral-combined_k141_180526||lt2gene Low-quality
18-109-final-viral-combined_k141_211484||lt2gene Not-determined
18-109-final-viral-combined_k141_219783||lt2gene Not-determined
18-109-final-viral-combined_k141_88152||lt2gene Low-quality
18-109-final-viral-combined_k141_6742||lt2gene Low-quality
18-109-final-viral-combined_k141_77730||lt2gene Low-quality
18-109-final-viral-combined_k141_180560||lt2gene Not-determined
18-109-final-viral-combined_k141_58010||lt2gene Not-determined
18-109-final-viral-combined_k141_6771||lt2gene Not-determined
18-109-final-viral-combined_k141_241871||lt2gene Low-quality
18-109-final-viral-combined_k141_219822||lt2gene Low-quality
18-109-final-viral-combined_k141_47804||lt2gene Low-quality
18-109-final-viral-combined_k141_58043||lt2gene Low-quality
18-109-final-viral-combined_k141_68675||lt2gene Low-quality
18-109-final-viral-combined_k141_37956||lt2gene Low-quality
18-109-final-viral-combined_k141_241888||lt2gene Low-quality
18-109-final-viral-combined_k141_98717||lt2gene Low-quality
18-109-final-viral-combined_k141_58059||lt2gene Low-quality
18-109-final-viral-combined_k141_180634||lt2gene Low-quality
18-109-final-viral-combined_k141_211623||lt2gene Not-determined
18-109-final-viral-combined_k141_26415||lt2gene Low-quality
18-109-final-viral-combined_k141_191430||lt2gene Low-quality
18-109-final-viral-combined_k141_231593||lt2gene Not-determined
18-109-final-viral-combined_k141_201594||lt2gene Low-quality
18-109-final-viral-combined_k141_170638||lt2gene Low-quality
18-109-final-viral-combined_k141_88363||lt2gene Low-quality
18-109-final-viral-combined_k141_17132||lt2gene Low-quality
18-109-final-viral-combined_k141_17163||lt2gene Low-quality
18-109-final-viral-combined_k141_108219||lt2gene Not-determined
18-109-final-viral-combined_k141_17167||lt2gene Not-determined
18-109-final-viral-combined_k141_17177||lt2gene Low-quality
18-109-final-viral-combined_k141_129787||lt2gene Low-quality
18-109-final-viral-combined_k141_211714||lt2gene Low-quality
18-109-final-viral-combined_k141_26455||lt2gene Low-quality
18-109-final-viral-combined_k141_211717||lt2gene Not-determined
18-109-final-viral-combined_k141_98849||lt2gene Low-quality
18-109-final-viral-combined_k141_98871||lt2gene Low-quality
18-109-final-viral-combined_k141_77928||lt2gene Low-quality
18-109-final-viral-combined_k141_68907||lt2gene Low-quality
18-109-final-viral-combined_k141_68917||lt2gene Low-quality
18-109-final-viral-combined_k141_108319||lt2gene Not-determined
18-109-final-viral-combined_k141_77956||lt2gene Not-determined
18-109-final-viral-combined_k141_17308||lt2gene Not-determined
18-109-final-viral-combined_k141_211779||lt2gene Low-quality
18-109-final-viral-combined_k141_119736||lt2gene Low-quality
18-109-final-viral-combined_k141_38113||lt2gene Low-quality
18-109-final-viral-combined_k141_17362||lt2gene Low-quality

| | |
|---|---|
| 18-109-final-viral-combined_k141_191596\|\|lt2gene | Low-quality |
| 18-109-final-viral-combined_k141_38154\|\|lt2gene | Low-quality |
| 18-109-final-viral-combined_k141_7030\|\|lt2gene | Low-quality |
| 18-109-final-viral-combined_k141_211840\|\|lt2gene | Low-quality |
| 18-109-final-viral-combined_k141_98963\|\|lt2gene | Not-determined |
| 18-109-final-viral-combined_k141_211846\|\|lt2gene | Low-quality |
| 18-109-final-viral-combined_k141_201887\|\|lt2gene | Low-quality |
| 18-109-final-viral-combined_k141_201890\|\|lt2gene | Low-quality |
| 18-109-final-viral-combined_k141_160821\|\|lt2gene | Low-quality |
| 18-109-final-viral-combined_k141_201901\|\|lt2gene | Not-determined |
| 18-109-final-viral-combined_k141_160845\|\|lt2gene | Low-quality |
| 18-109-final-viral-combined_k141_26542\|\|lt2gene | Low-quality |
| 18-109-final-viral-combined_k141_48147\|\|lt2gene | Low-quality |
| 18-109-final-viral-combined_k141_231933\|\|lt2gene | Low-quality |
| 18-109-final-viral-combined_k141_108482\|\|lt2gene | Low-quality |
| 18-109-final-viral-combined_k141_119915\|\|lt2gene | Low-quality |
| 18-109-final-viral-combined_k141_231956\|\|lt2gene | Not-determined |
| 18-109-final-viral-combined_k141_58361\|\|lt2gene | Not-determined |
| 18-109-final-viral-combined_k141_129961\|\|lt2gene | Low-quality |
| 18-109-final-viral-combined_k141_17435\|\|lt2gene | Low-quality |
| 18-109-final-viral-combined_k141_26594\|\|lt2gene | Not-determined |
| 18-109-final-viral-combined_k141_48201\|\|lt2gene | Low-quality |
| 18-109-final-viral-combined_k141_160921\|\|lt2gene | Low-quality |
| 18-109-final-viral-combined_k141_48205\|\|lt2gene | Not-determined |
| 18-109-final-viral-combined_k141_181104\|\|lt2gene | Low-quality |
| 18-109-final-viral-combined_k141_78151\|\|lt2gene | Low-quality |
| 18-109-final-viral-combined_k141_130021\|\|lt2gene | Not-determined |
| 18-109-final-viral-combined_k141_119987\|\|lt2gene | Not-determined |
| 18-109-final-viral-combined_k141_119990\|\|lt2gene | Low-quality |
| 18-109-final-viral-combined_k141_191698\|\|lt2gene | Low-quality |
| 18-109-final-viral-combined_k141_69172\|\|lt2gene | Low-quality |
| 18-109-final-viral-combined_k141_120003\|\|lt2gene | Low-quality |
| 18-109-final-viral-combined_k141_232037\|\|lt2gene | Low-quality |
| 18-109-final-viral-combined_k141_58449\|\|lt2gene | Low-quality |
| 18-109-final-viral-combined_k141_99100\|\|lt2gene | Low-quality |
| 18-109-final-viral-combined_k141_48293\|\|lt2gene | Not-determined |
| 18-109-final-viral-combined_k141_38295\|\|lt2gene | Not-determined |
| 18-109-final-viral-combined_k141_181226\|\|lt2gene | Low-quality |
| 18-109-final-viral-combined_k141_48321\|\|lt2gene | Not-determined |
| 18-109-final-viral-combined_k141_78207\|\|lt2gene | Low-quality |
| 18-109-final-viral-combined_k141_120067\|\|lt2gene | Low-quality |
| 18-109-final-viral-combined_k141_170858\|\|lt2gene | Low-quality |
| 18-109-final-viral-combined_k141_139786\|\|lt2gene | Low-quality |
| 18-109-final-viral-combined_k141_191799\|\|lt2gene | Not-determined |
| 18-109-final-viral-combined_k141_78222\|\|lt2gene | Not-determined |
| 18-109-final-viral-combined_k141_202110\|\|lt2gene | Low-quality |
| 18-109-final-viral-combined_k141_232142\|\|lt2gene | Not-determined |
| 18-109-final-viral-combined_k141_181288\|\|lt2gene | Low-quality |
| 18-109-final-viral-combined_k141_88677\|\|lt2gene | Low-quality |
| 18-109-final-viral-combined_k141_181304\|\|lt2gene | Low-quality |
| 18-109-final-viral-combined_k141_149966\|\|lt2gene | Low-quality |
| 18-109-final-viral-combined_k141_120168\|\|lt2gene | Not-determined |
| 18-109-final-viral-combined_k141_130145\|\|lt2gene | Not-determined |
| 18-109-final-viral-combined_k141_202153\|\|lt2gene | Low-quality |
| 18-109-final-viral-combined_k141_212072\|\|lt2gene | Low-quality |
| 18-109-final-viral-combined_k141_78290\|\|lt2gene | Not-determined |
| 18-109-final-viral-combined_k141_220265\|\|lt2gene | Low-quality |
| 18-109-final-viral-combined_k141_212089\|\|lt2gene | Low-quality |
| 18-109-final-viral-combined_k141_99272\|\|lt2gene | Not-determined |
| 18-109-final-viral-combined_k141_88746\|\|lt2gene | Low-quality |
| 18-109-final-viral-combined_k141_38428\|\|lt2gene | Low-quality |
| 18-109-final-viral-combined_k141_130160\|\|lt2gene | Not-determined |
| 18-109-final-viral-combined_k141_191877\|\|lt2gene | Low-quality |
| 18-109-final-viral-combined_k141_88751\|\|lt2gene | Low-quality |
| 18-109-final-viral-combined_k141_108739\|\|lt2gene | Not-determined |
| 18-109-final-viral-combined_k141_38488\|\|lt2gene | Low-quality |
| 18-109-final-viral-combined_k141_69381\|\|lt2gene | Low-quality |
| 18-109-final-viral-combined_k141_191937\|\|lt2gene | Low-quality |
| 18-109-final-viral-combined_k141_202262\|\|lt2gene | Not-determined |
| 18-109-final-viral-combined_k141_161261\|\|lt2gene | Low-quality |
| 18-109-final-viral-combined_k141_191955\|\|lt2gene | Low-quality |
| 18-109-final-viral-combined_k141_181496\|\|lt2gene | Low-quality |
| 18-109-final-viral-combined_k141_202285\|\|lt2gene | Low-quality |
| 18-109-final-viral-combined_k141_161299\|\|lt2gene | Low-quality |
| 18-109-final-viral-combined_k141_38569\|\|lt2gene | Not-determined |
| 18-109-final-viral-combined_k141_58733\|\|lt2gene | Low-quality |
| 18-109-final-viral-combined_k141_212186\|\|lt2gene | Low-quality |
| 18-109-final-viral-combined_k141_58760\|\|lt2gene | Low-quality |
| 18-109-final-viral-combined_k141_17679\|\|lt2gene | Low-quality |
| 18-109-final-viral-combined_k141_108867\|\|lt2gene | Low-quality |
| 18-109-final-viral-combined_k141_130341\|\|lt2gene | Low-quality |
| 18-109-final-viral-combined_k141_192016\|\|lt2gene | Low-quality |
| 18-109-final-viral-combined_k141_48573\|\|lt2gene | Low-quality |
| 18-109-final-viral-combined_k141_38611\|\|lt2gene | Low-quality |
| 18-109-final-viral-combined_k141_232387\|\|lt2gene | Low-quality |
| 18-109-final-viral-combined_k141_7465\|\|lt2gene | Low-quality |
| 18-109-final-viral-combined_k141_7471\|\|lt2gene | Low-quality |
| 18-109-final-viral-combined_k141_130441\|\|lt2gene | Low-quality |
| 18-109-final-viral-combined_k141_48687\|\|lt2gene | Low-quality |
| 18-109-final-viral-combined_k141_181672\|\|lt2gene | Not-determined |
| 18-109-final-viral-combined_k141_220414\|\|lt2gene | Low-quality |
| 18-109-final-viral-combined_k141_181691\|\|lt2gene | Not-determined |
| 18-109-final-viral-combined_k141_17854\|\|lt2gene | Low-quality |
| 18-109-final-viral-combined_k141_202505\|\|lt2gene | Low-quality |
| 18-109-final-viral-combined_k141_17863\|\|lt2gene | Low-quality |
| 18-109-final-viral-combined_k141_99510\|\|lt2gene | Not-determined |
| 18-109-final-viral-combined_k141_171258\|\|lt2gene | Low-quality |
| 18-109-final-viral-combined_k141_242185\|\|lt2gene | Low-quality |
| 18-109-final-viral-combined_k141_26878\|\|lt2gene | Low-quality |
| 18-109-final-viral-combined_k141_161502\|\|lt2gene | Low-quality |
| 18-109-final-viral-combined_k141_26884\|\|lt2gene | Low-quality |
| 18-109-final-viral-combined_k141_78634\|\|lt2gene | Not-determined |
| 18-109-final-viral-combined_k141_202583\|\|lt2gene | Low-quality |
| 18-109-final-viral-combined_k141_171321\|\|lt2gene | Not-determined |
| 18-109-final-viral-combined_k141_99587\|\|lt2gene | Low-quality |
| 18-109-final-viral-combined_k141_150336\|\|lt2gene | Not-determined |
| 18-109-final-viral-combined_k141_26908\|\|lt2gene | Low-quality |
| 18-109-final-viral-combined_k141_192181\|\|lt2gene | Not-determined |
| 18-109-final-viral-combined_k141_192204\|\|lt2gene | Low-quality |
| 18-109-final-viral-combined_k141_26957\|\|lt2gene | Low-quality |
| 18-109-final-viral-combined_k141_120391\|\|lt2gene | Not-determined |
| 18-109-final-viral-combined_k141_59126\|\|lt2gene | Low-quality |
| 18-109-final-viral-combined_k141_48902\|\|lt2gene | Low-quality |
| 18-109-final-viral-combined_k141_26982\|\|lt2gene | Low-quality |
| 18-109-final-viral-combined_k141_212433\|\|lt2gene | Not-determined |
| 18-109-final-viral-combined_k141_59158\|\|lt2gene | Low-quality |
| 18-109-final-viral-combined_k141_7710\|\|lt2gene | Low-quality |
| 18-109-final-viral-combined_k141_192297\|\|lt2gene | Not-determined |
| 18-109-final-viral-combined_k141_130718\|\|lt2gene | Not-determined |
| 18-109-final-viral-combined_k141_48954\|\|lt2gene | Low-quality |
| 18-109-final-viral-combined_k141_202785\|\|lt2gene | Low-quality |
| 18-109-final-viral-combined_k141_150446\|\|lt2gene | Not-determined |
| 18-109-final-viral-combined_k141_202797\|\|lt2gene | Not-determined |
| 18-109-final-viral-combined_k141_48985\|\|lt2gene | Not-determined |
| 18-109-final-viral-combined_k141_150466\|\|lt2gene | Not-determined |
| 18-109-final-viral-combined_k141_120489\|\|lt2gene | Low-quality |
| 18-109-final-viral-combined_k141_181921\|\|lt2gene | Low-quality |
| 18-109-final-viral-combined_k141_69706\|\|lt2gene | Low-quality |
| 18-109-final-viral-combined_k141_49041\|\|lt2gene | Low-quality |
| 18-109-final-viral-combined_k141_140082\|\|lt2gene | Low-quality |
| 18-109-final-viral-combined_k141_181979\|\|lt2gene | Not-determined |
| 18-109-final-viral-combined_k141_171471\|\|lt2gene | Low-quality |
| 18-109-final-viral-combined_k141_78867\|\|lt2gene | Low-quality |
| 18-109-final-viral-combined_k141_120571\|\|lt2gene | Not-determined |
| 18-109-final-viral-combined_k141_49071\|\|lt2gene | Low-quality |
| 18-109-final-viral-combined_k141_59261\|\|lt2gene | Low-quality |
| 18-109-final-viral-combined_k141_232788\|\|lt2gene | Low-quality |
| 18-109-final-viral-combined_k141_69783\|\|lt2gene | Not-determined |
| 18-109-final-viral-combined_k141_202945\|\|lt2gene | Low-quality |
| 18-109-final-viral-combined_k141_109330\|\|lt2gene | Not-determined |
| 18-109-final-viral-combined_k141_89183\|\|lt2gene | Low-quality |
| 18-109-final-viral-combined_k141_49091\|\|lt2gene | Not-determined |
| 18-109-final-viral-combined_k141_202994\|\|lt2gene | Low-quality |
| 18-109-final-viral-combined_k141_161852\|\|lt2gene | Not-determined |
| 18-109-final-viral-combined_k141_212618\|\|lt2gene | Low-quality |
| 18-109-final-viral-combined_k141_109423\|\|lt2gene | Low-quality |
| 18-109-final-viral-combined_k141_109441\|\|lt2gene | Low-quality |
| 18-109-final-viral-combined_k141_203061\|\|lt2gene | Low-quality |
| 18-109-final-viral-combined_k141_161878\|\|lt2gene | Low-quality |
| 18-109-final-viral-combined_k141_212693\|\|lt2gene | Low-quality |
| 18-109-final-viral-combined_k141_212705\|\|lt2gene | Not-determined |
| 18-109-final-viral-combined_k141_89297\|\|lt2gene | Low-quality |
| 18-109-final-viral-combined_k141_161913\|\|lt2gene | Low-quality |
| 18-109-final-viral-combined_k141_7949\|\|lt2gene | Low-quality |
| 18-109-final-viral-combined_k141_140298\|\|lt2gene | Low-quality |
| 18-109-final-viral-combined_k141_182194\|\|lt2gene | Low-quality |
| 18-109-final-viral-combined_k141_7972\|\|lt2gene | Low-quality |
| 18-109-final-viral-combined_k141_220921\|\|lt2gene | Low-quality |
| 18-109-final-viral-combined_k141_150715\|\|lt2gene | Not-determined |
| 18-109-final-viral-combined_k141_150721\|\|lt2gene | Low-quality |
| 18-109-final-viral-combined_k141_39329\|\|lt2gene | Low-quality |
| 18-109-final-viral-combined_k141_233068\|\|lt2gene | Not-determined |
| 18-109-final-viral-combined_k141_8048\|\|lt2gene | Low-quality |
| 18-109-final-viral-combined_k141_161960\|\|lt2gene | Not-determined |
| 18-109-final-viral-combined_k141_69960\|\|lt2gene | Low-quality |
| 18-109-final-viral-combined_k141_150777\|\|lt2gene | Not-determined |
| 18-109-final-viral-combined_k141_242632\|\|lt2gene | Not-determined |
| 18-109-final-viral-combined_k141_49391\|\|lt2gene | Low-quality |
| 18-109-final-viral-combined_k141_171861\|\|lt2gene | Not-determined |
| 18-109-final-viral-combined_k141_8109\|\|lt2gene | Not-determined |
| 18-109-final-viral-combined_k141_109718\|\|lt2gene | Low-quality |
| 18-109-final-viral-combined_k141_79222\|\|lt2gene | Low-quality |
| 18-109-final-viral-combined_k141_162060\|\|lt2gene | Low-quality |
| 18-109-final-viral-combined_k141_120959\|\|lt2gene | Low-quality |
| 18-109-final-viral-combined_k141_233249\|\|lt2gene | Low-quality |
| 18-109-final-viral-combined_k141_39488\|\|lt2gene | Low-quality |
| 18-109-final-viral-combined_k141_59572\|\|lt2gene | Not-determined |
| 18-109-final-viral-combined_k141_192826\|\|lt2gene | Low-quality |
| 18-109-final-viral-combined_k141_49550\|\|lt2gene | Low-quality |
| 18-109-final-viral-combined_k141_109822\|\|lt2gene | Low-quality |
| 18-109-final-viral-combined_k141_140403\|\|lt2gene | Low-quality |
| 18-109-final-viral-combined_k141_49575\|\|lt2gene | Not-determined |
| 18-109-final-viral-combined_k141_49578\|\|lt2gene | Not-determined |
| 18-109-final-viral-combined_k141_192860\|\|lt2gene | Low-quality |
| 18-109-final-viral-combined_k141_8239\|\|lt2gene | Low-quality |
| 18-109-final-viral-combined_k141_203591\|\|lt2gene | Low-quality |
| 18-109-final-viral-combined_k141_192901\|\|lt2gene | Low-quality |
| 18-109-final-viral-combined_k141_100358\|\|lt2gene | Not-determined |
| 18-109-final-viral-combined_k141_140478\|\|lt2gene | Low-quality |
| 18-109-final-viral-combined_k141_8271\|\|lt2gene | Low-quality |
| 18-109-final-viral-combined_k141_100372\|\|lt2gene | Low-quality |
| 18-109-final-viral-combined_k141_39594\|\|lt2gene | Not-determined |
| 18-109-final-viral-combined_k141_192936\|\|lt2gene | Low-quality |
| 18-109-final-viral-combined_k141_130990\|\|lt2gene | Low-quality |
| 18-109-final-viral-combined_k141_27224\|\|lt2gene | Low-quality |
| 18-109-final-viral-combined_k141_89513\|\|lt2gene | Not-determined |
| 18-109-final-viral-combined_k141_79275\|\|lt2gene | Not-determined |
| 18-109-final-viral-combined_k141_49708\|\|lt2gene | Low-quality |
| 18-109-final-viral-combined_k141_182542\|\|lt2gene | Low-quality |
| 18-109-final-viral-combined_k141_162143\|\|lt2gene | Low-quality |
| 18-109-final-viral-combined_k141_150954\|\|lt2gene | Low-quality |
| 18-109-final-viral-combined_k141_100421\|\|lt2gene | Low-quality |
| 18-109-final-viral-combined_k141_89538\|\|lt2gene | Low-quality |
| 18-109-final-viral-combined_k141_162175\|\|lt2gene | Low-quality |
| 18-109-final-viral-combined_k141_89558\|\|lt2gene | Low-quality |
| 18-109-final-viral-combined_k141_8343\|\|lt2gene | Not-determined |
| 18-109-final-viral-combined_k141_8344\|\|lt2gene | Low-quality |
| 18-109-final-viral-combined_k141_79311\|\|lt2gene | Low-quality |
| 18-109-final-viral-combined_k141_89563\|\|lt2gene | Low-quality |
| 18-109-final-viral-combined_k141_172077\|\|lt2gene | Low-quality |
| 18-109-final-viral-combined_k141_193065\|\|lt2gene | Low-quality |
| 18-109-final-viral-combined_k141_233446\|\|lt2gene | Not-determined |
| 18-109-final-viral-combined_k141_70245\|\|lt2gene | Low-quality |
| 18-109-final-viral-combined_k141_213070\|\|lt2gene | Low-quality |
| 18-109-final-viral-combined_k141_8439\|\|lt2gene | Not-determined |
| 18-109-final-viral-combined_k141_140684\|\|lt2gene | Low-quality |
| 18-109-final-viral-combined_k141_162279\|\|lt2gene | Low-quality |
| 18-109-final-viral-combined_k141_162285\|\|lt2gene | Low-quality |
| 18-109-final-viral-combined_k141_49861\|\|lt2gene | Low-quality |
| 18-109-final-viral-combined_k141_193105\|\|lt2gene | Low-quality |
| 18-109-final-viral-combined_k141_59847\|\|lt2gene | Low-quality |
| 18-109-final-viral-combined_k141_182617\|\|lt2gene | Not-determined |
| 18-109-final-viral-combined_k141_203796\|\|lt2gene | Not-determined |
| 18-109-final-viral-combined_k141_121347\|\|lt2gene | Low-quality |
| 18-109-final-viral-combined_k141_131197\|\|lt2gene | Low-quality |
| 18-109-final-viral-combined_k141_131204\|\|lt2gene | Low-quality |
| 18-109-final-viral-combined_k141_203845\|\|lt2gene | Not-determined |
| 18-109-final-viral-combined_k141_151147\|\|lt2gene | Low-quality |
| 18-109-final-viral-combined_k141_70394\|\|lt2gene | Low-quality |
| 18-109-final-viral-combined_k141_89731\|\|lt2gene | Low-quality |
| 18-109-final-viral-combined_k141_121389\|\|lt2gene | Low-quality |
| 18-109-final-viral-combined_k141_79409\|\|lt2gene | Low-quality |
| 18-109-final-viral-combined_k141_121397\|\|lt2gene | Low-quality |
| 18-109-final-viral-combined_k141_193210\|\|lt2gene | Not-determined |
| 18-109-final-viral-combined_k141_59941\|\|lt2gene | Low-quality |
| 18-109-final-viral-combined_k141_39806\|\|lt2gene | Low-quality |
| 18-109-final-viral-combined_k141_151201\|\|lt2gene | Low-quality |
| 18-109-final-viral-combined_k141_172248\|\|lt2gene | Low-quality |
| 18-109-final-viral-combined_k141_233690\|\|lt2gene | Low-quality |
| 18-109-final-viral-combined_k141_221433\|\|lt2gene | Not-determined |
| 18-109-final-viral-combined_k141_8593\|\|lt2gene | Low-quality |
| 18-109-final-viral-combined_k141_131259\|\|lt2gene | Not-determined |
| 18-109-final-viral-combined_k141_79446\|\|lt2gene | Not-determined |
| 18-109-final-viral-combined_k141_193256\|\|lt2gene | Low-quality |
| 18-109-final-viral-combined_k141_121472\|\|lt2gene | Low-quality |
| 18-109-final-viral-combined_k141_172304\|\|lt2gene | Low-quality |
| 18-109-final-viral-combined_k141_193288\|\|lt2gene | Low-quality |
| 18-109-final-viral-combined_k141_89835\|\|lt2gene | Low-quality |
| 18-109-final-viral-combined_k141_243202\|\|lt2gene | Low-quality |
| 18-109-final-viral-combined_k141_213277\|\|lt2gene | Low-quality |
| 18-109-final-viral-combined_k141_60072\|\|lt2gene | Not-determined |
| 18-109-final-viral-combined_k141_162491\|\|lt2gene | Not-determined |
| 18-109-final-viral-combined_k141_89879\|\|lt2gene | Low-quality |
| 18-109-final-viral-combined_k141_213316\|\|lt2gene | Low-quality |
| 18-109-final-viral-combined_k141_8667\|\|lt2gene | Low-quality |
| 18-109-final-viral-combined_k141_50087\|\|lt2gene | Low-quality |
| 18-109-final-viral-combined_k141_233844\|\|lt2gene | Low-quality |
| 18-109-final-viral-combined_k141_151266\|\|lt2gene | Not-determined |
| 18-109-final-viral-combined_k141_193375\|\|lt2gene | Low-quality |
| 18-109-final-viral-combined_k141_131378\|\|lt2gene | Low-quality |
| 18-109-final-viral-combined_k141_70582\|\|lt2gene | Low-quality |
| 18-109-final-viral-combined_k141_50129\|\|lt2gene | Not-determined |
| 18-109-final-viral-combined_k141_151299\|\|lt2gene | Low-quality |
| 18-109-final-viral-combined_k141_233879\|\|lt2gene | Low-quality |
| 18-109-final-viral-combined_k141_60124\|\|lt2gene | Low-quality |
| 18-109-final-viral-combined_k141_121641\|\|lt2gene | Low-quality |
| 18-109-final-viral-combined_k141_8762\|\|lt2gene | Low-quality |
| 18-109-final-viral-combined_k141_151379\|\|lt2gene | Low-quality |
| 18-109-final-viral-combined_k141_100746\|\|lt2gene | Low-quality |
| 18-109-final-viral-combined_k141_70617\|\|lt2gene | Low-quality |
| 18-109-final-viral-combined_k141_100794\|\|lt2gene | Low-quality |
| 18-109-final-viral-combined_k141_70631\|\|lt2gene | Not-determined |
| 18-109-final-viral-combined_k141_131528\|\|lt2gene | Low-quality |
| 18-109-final-viral-combined_k141_90018\|\|lt2gene | Low-quality |
| 18-109-final-viral-combined_k141_70702\|\|lt2gene | Low-quality |
| 18-109-final-viral-combined_k141_182923\|\|lt2gene | Low-quality |
| 18-109-final-viral-combined_k141_79647\|\|lt2gene | Not-determined |
| 18-109-final-viral-combined_k141_131597\|\|lt2gene | Low-quality |
| 18-109-final-viral-combined_k141_90093\|\|lt2gene | Low-quality |
| 18-109-final-viral-combined_k141_172556\|\|lt2gene | Not-determined |
| 18-109-final-viral-combined_k141_162617\|\|lt2gene | Low-quality |
| 18-109-final-viral-combined_k141_162621\|\|lt2gene | Low-quality |
| 18-109-final-viral-combined_k141_100936\|\|lt2gene | Low-quality |
| 18-109-final-viral-combined_k141_50380\|\|lt2gene | Not-determined |
| 18-109-final-viral-combined_k141_234080\|\|lt2gene | Low-quality |
| 18-109-final-viral-combined_k141_110480\|\|lt2gene | Not-determined |
| 18-109-final-viral-combined_k141_79721\|\|lt2gene | Low-quality |
| 18-109-final-viral-combined_k141_234088\|\|lt2gene | Low-quality |
| 18-109-final-viral-combined_k141_193651\|\|lt2gene | Low-quality |
| 18-109-final-viral-combined_k141_183034\|\|lt2gene | Low-quality |
| 18-109-final-viral-combined_k141_100988\|\|lt2gene | Low-quality |
| 18-109-final-viral-combined_k141_131692\|\|lt2gene | Low-quality |
| 18-109-final-viral-combined_k141_183071\|\|lt2gene | Not-determined |
| 18-109-final-viral-combined_k141_141112\|\|lt2gene | Low-quality |
| 18-109-final-viral-combined_k141_121924\|\|lt2gene | Low-quality |
| 18-109-final-viral-combined_k141_50503\|\|lt2gene | Not-determined |
| 18-109-final-viral-combined_k141_213707\|\|lt2gene | Low-quality |
| 18-109-final-viral-combined_k141_70970\|\|lt2gene | Low-quality |
| 18-109-final-viral-combined_k141_110639\|\|lt2gene | Low-quality |
| 18-109-final-viral-combined_k141_50568\|\|lt2gene | Not-determined |
| 18-109-final-viral-combined_k141_183140\|\|lt2gene | Low-quality |
| 18-109-final-viral-combined_k141_60396\|\|lt2gene | Low-quality |
| 18-109-final-viral-combined_k141_221874\|\|lt2gene | Low-quality |
| 18-109-final-viral-combined_k141_234223\|\|lt2gene | Low-quality |
| 18-109-final-viral-combined_k141_110670\|\|lt2gene | Low-quality |
| 18-109-final-viral-combined_k141_172680\|\|lt2gene | Not-determined |
| 18-109-final-viral-combined_k141_8825\|\|lt2gene | Low-quality |
| 18-109-final-viral-combined_k141_71027\|\|lt2gene | Low-quality |
| 18-109-final-viral-combined_k141_151610\|\|lt2gene | Low-quality |
| 18-109-final-viral-combined_k141_193855\|\|lt2gene | Not-determined |
| 18-109-final-viral-combined_k141_243594\|\|lt2gene | Low-quality |
| 18-109-final-viral-combined_k141_50632\|\|lt2gene | Low-quality |
| 18-109-final-viral-combined_k141_71042\|\|lt2gene | Low-quality |
| 18-109-final-viral-combined_k141_193882\|\|lt2gene | Low-quality |
| 18-109-final-viral-combined_k141_18964\|\|lt2gene | Low-quality |
| 18-109-final-viral-combined_k141_18980\|\|lt2gene | Low-quality |
| 18-109-final-viral-combined_k141_40215\|\|lt2gene | Not-determined |
| 18-109-final-viral-combined_k141_122079\|\|lt2gene | Low-quality |
| 18-109-final-viral-combined_k141_50728\|\|lt2gene | Not-determined |
| 18-109-final-viral-combined_k141_71106\|\|lt2gene | Low-quality |
| 18-109-final-viral-combined_k141_151625\|\|lt2gene | Not-determined |
| 18-109-final-viral-combined_k141_79795\|\|lt2gene | Low-quality |
| 18-109-final-viral-combined_k141_110846\|\|lt2gene | Low-quality |
| 18-109-final-viral-combined_k141_122135\|\|lt2gene | Low-quality |
| 18-109-final-viral-combined_k141_40314\|\|lt2gene | Not-determined |
| 18-109-final-viral-combined_k141_8976\|\|lt2gene | Not-determined |
| 18-109-final-viral-combined_k141_193990\|\|lt2gene | Low-quality |
| 18-109-final-viral-combined_k141_110883\|\|lt2gene | Not-determined |
| 18-109-final-viral-combined_k141_243708\|\|lt2gene | Low-quality |
| 18-109-final-viral-combined_k141_183393\|\|lt2gene | Low-quality |
| 18-109-final-viral-combined_k141_50784\|\|lt2gene | Not-determined |
| 18-109-final-viral-combined_k141_234430\|\|lt2gene | Not-determined |
| 18-109-final-viral-combined_k141_101156\|\|lt2gene | Low-quality |
| 18-109-final-viral-combined_k141_50821\|\|lt2gene | Low-quality |
| 18-109-final-viral-combined_k141_9026\|\|lt2gene | Not-determined |
| 18-109-final-viral-combined_k141_163041\|\|lt2gene | Low-quality |
| 18-109-final-viral-combined_k141_90475\|\|lt2gene | Low-quality |
| 18-109-final-viral-combined_k141_50839\|\|lt2gene | Low-quality |
| 18-109-final-viral-combined_k141_40373\|\|lt2gene | Low-quality |
| 18-109-final-viral-combined_k141_19106\|\|lt2gene | Not-determined |
| 18-109-final-viral-combined_k141_71164\|\|lt2gene | Low-quality |
| 18-109-final-viral-combined_k141_234468\|\|lt2gene | Low-quality |
| 18-109-final-viral-combined_k141_163141\|\|lt2gene | Low-quality |
| 18-109-final-viral-combined_k141_183516\|\|lt2gene | Not-determined |
| 18-109-final-viral-combined_k141_9169\|\|lt2gene | Not-determined |
| 18-109-final-viral-combined_k141_40480\|\|lt2gene | Low-quality |
| 18-109-final-viral-combined_k141_40483\|\|lt2gene | Not-determined |
| 18-109-final-viral-combined_k141_9200\|\|lt2gene | Not-determined |
| 18-109-final-viral-combined_k141_214024\|\|lt2gene | Low-quality |
| 18-109-final-viral-combined_k141_101263\|\|lt2gene | Low-quality |
| 18-109-final-viral-combined_k141_101276\|\|lt2gene | Not-determined |
| 18-109-final-viral-combined_k141_163164\|\|lt2gene | Low-quality |
| 18-109-final-viral-combined_k141_214039\|\|lt2gene | Low-quality |
| 18-109-final-viral-combined_k141_101306\|\|lt2gene | Low-quality |
| 18-109-final-viral-combined_k141_80063\|\|lt2gene | Not-determined |
| 18-109-final-viral-combined_k141_141389\|\|lt2gene | Low-quality |
| 18-109-final-viral-combined_k141_163262\|\|lt2gene | Low-quality |
| 18-109-final-viral-combined_k141_60816\|\|lt2gene | Low-quality |
| 18-109-final-viral-combined_k141_40605\|\|lt2gene | Low-quality |
| 18-109-final-viral-combined_k141_152040\|\|lt2gene | Low-quality |
| 18-109-final-viral-combined_k141_173101\|\|lt2gene | Not-determined |
| 18-109-final-viral-combined_k141_71347\|\|lt2gene | Low-quality |
| 18-109-final-viral-combined_k141_80103\|\|lt2gene | Low-quality |
| 18-109-final-viral-combined_k141_71359\|\|lt2gene | Not-determined |
| 18-109-final-viral-combined_k141_131831\|\|lt2gene | Low-quality |
| 18-109-final-viral-combined_k141_19275\|\|lt2gene | Low-quality |
| 18-109-final-viral-combined_k141_101390\|\|lt2gene | Not-determined |
| 18-109-final-viral-combined_k141_131862\|\|lt2gene | Low-quality |
| 18-109-final-viral-combined_k141_152138\|\|lt2gene | Low-quality |
| 18-109-final-viral-combined_k141_131905\|\|lt2gene | Low-quality |
| 18-109-final-viral-combined_k141_19381\|\|lt2gene | Not-determined |
| 18-109-final-viral-combined_k141_60910\|\|lt2gene | Low-quality |
| 18-109-final-viral-combined_k141_152183\|\|lt2gene | Low-quality |
| 18-109-final-viral-combined_k141_19430\|\|lt2gene | Low-quality |
| 18-109-final-viral-combined_k141_60979\|\|lt2gene | Low-quality |
| 18-109-final-viral-combined_k141_80241\|\|lt2gene | Not-determined |
| 18-109-final-viral-combined_k141_9485\|\|lt2gene | Low-quality |
| 18-109-final-viral-combined_k141_19466\|\|lt2gene | Low-quality |
| 18-109-final-viral-combined_k141_101446\|\|lt2gene | Low-quality |
| 18-109-final-viral-combined_k141_90799\|\|lt2gene | Low-quality |
| 18-109-final-viral-combined_k141_234785\|\|lt2gene | Not-determined |
| 18-109-final-viral-combined_k141_152315\|\|lt2gene | Low-quality |
| 18-109-final-viral-combined_k141_173345\|\|lt2gene | Low-quality |
| 18-109-final-viral-combined_k141_152333\|\|lt2gene | Low-quality |
| 18-109-final-viral-combined_k141_50989\|\|lt2gene | Not-determined |
| 18-109-final-viral-combined_k141_111277\|\|lt2gene | Low-quality |
| 18-109-final-viral-combined_k141_61157\|\|lt2gene | Low-quality |
| 18-109-final-viral-combined_k141_9628\|\|lt2gene | Low-quality |
| 18-109-final-viral-combined_k141_152376\|\|lt2gene | Low-quality |
| 18-109-final-viral-combined_k141_173443\|\|lt2gene | Low-quality |
| 18-109-final-viral-combined_k141_141710\|\|lt2gene | Low-quality |
| 18-109-final-viral-combined_k141_173458\|\|lt2gene | Low-quality |
| 18-109-final-viral-combined_k141_173489\|\|lt2gene | Low-quality |
| 18-109-final-viral-combined_k141_222524\|\|lt2gene | Low-quality |
| 18-109-final-viral-combined_k141_222533\|\|lt2gene | Not-determined |
| 18-109-final-viral-combined_k141_19662\|\|lt2gene | Low-quality |
| 18-109-final-viral-combined_k141_122492\|\|lt2gene | Low-quality |
| 18-109-final-viral-combined_k141_222571\|\|lt2gene | Low-quality |
| 18-109-final-viral-combined_k141_173519\|\|lt2gene | Low-quality |
| 18-109-final-viral-combined_k141_173521\|\|lt2gene | Not-determined |
| 18-109-final-viral-combined_k141_132089\|\|lt2gene | Low-quality |
| 18-109-final-viral-combined_k141_111462\|\|lt2gene | Low-quality |
| 18-109-final-viral-combined_k141_214356\|\|lt2gene | Low-quality |
| 18-109-final-viral-combined_k141_19765\|\|lt2gene | Not-determined |
| 18-109-final-viral-combined_k141_101812\|\|lt2gene | Low-quality |
| 18-109-final-viral-combined_k141_101825\|\|lt2gene | Low-quality |
| 18-109-final-viral-combined_k141_132203\|\|lt2gene | Low-quality |
| 18-109-final-viral-combined_k141_141944\|\|lt2gene | Low-quality |
| 18-109-final-viral-combined_k141_80600\|\|lt2gene | Low-quality |
| 18-109-final-viral-combined_k141_80627\|\|lt2gene | Low-quality |
| 18-109-final-viral-combined_k141_152594\|\|lt2gene | Not-determined |
| 18-109-final-viral-combined_k141_28190\|\|lt2gene | Low-quality |
| 18-109-final-viral-combined_k141_111688\|\|lt2gene | Low-quality |
| 18-109-final-viral-combined_k141_111699\|\|lt2gene | Low-quality |
| 18-109-final-viral-combined_k141_142076\|\|lt2gene | Low-quality |
| 18-109-final-viral-combined_k141_9992\|\|lt2gene | Low-quality |
| 18-109-final-viral-combined_k141_111754\|\|lt2gene | Not-determined |
| 18-109-final-viral-combined_k141_28295\|\|lt2gene | Low-quality |
| 18-109-final-viral-combined_k141_91301\|\|lt2gene | Low-quality |
| 18-109-final-viral-combined_k141_243887\|\|lt2gene | Low-quality |
| 18-109-final-viral-combined_k141_111783\|\|lt2gene | Low-quality |
| 18-109-final-viral-combined_k141_91314\|\|lt2gene | Not-determined |
| 18-109-final-viral-combined_k141_111794\|\|lt2gene | Low-quality |
| 18-109-final-viral-combined_k141_80709\|\|lt2gene | Not-determined |
| 18-109-final-viral-combined_k141_102051\|\|lt2gene | Low-quality |
| 18-109-final-viral-combined_k141_91350\|\|lt2gene | Not-determined |
| 18-109-final-viral-combined_k141_111857\|\|lt2gene | Low-quality |
| 18-109-final-viral-combined_k141_222898\|\|lt2gene | Low-quality |
| 18-109-final-viral-combined_k141_152764\|\|lt2gene | Low-quality |
| 18-109-final-viral-combined_k141_152765\|\|lt2gene | Low-quality |
| 18-109-final-viral-combined_k141_20100\|\|lt2gene | Not-determined |
| 18-109-final-viral-combined_k141_80779\|\|lt2gene | Not-determined |
| 18-109-final-viral-combined_k141_132432\|\|lt2gene | Low-quality |
| 18-109-final-viral-combined_k141_222953\|\|lt2gene | Low-quality |
| 18-109-final-viral-combined_k141_222956\|\|lt2gene | Low-quality |
| 18-109-final-viral-combined_k141_111885\|\|lt2gene | Low-quality |
| 18-109-final-viral-combined_k141_111890\|\|lt2gene | Low-quality |
| 18-109-final-viral-combined_k141_244046\|\|lt2gene | Low-quality |
| 18-109-final-viral-combined_k141_244063\|\|lt2gene | Low-quality |
| 18-109-final-viral-combined_k141_244068\|\|lt2gene | Low-quality |
| 18-109-final-viral-combined_k141_10120\|\|lt2gene | Low-quality |
| 18-109-final-viral-combined_k141_152868\|\|lt2gene | Not-determined |
| 18-109-final-viral-combined_k141_80837\|\|lt2gene | Low-quality |
| 18-109-final-viral-combined_k141_132540\|\|lt2gene | Low-quality |
| 18-109-final-viral-combined_k141_132544\|\|lt2gene | Low-quality |
| 18-109-final-viral-combined_k141_223052\|\|lt2gene | Not-determined |
| 18-109-final-viral-combined_k141_20147\|\|lt2gene | Not-determined |
| 18-109-final-viral-combined_k141_112001\|\|lt2gene | Low-quality |
| 18-109-final-viral-combined_k141_10153\|\|lt2gene | Not-determined |
| 18-109-final-viral-combined_k141_152942\|\|lt2gene | Not-determined |
| 18-109-final-viral-combined_k141_142327\|\|lt2gene | Not-determined |
| 18-109-final-viral-combined_k141_142354\|\|lt2gene | Low-quality |
| 18-109-final-viral-combined_k141_80959\|\|lt2gene | Not-determined |
| 18-109-final-viral-combined_k141_112058\|\|lt2gene | Not-determined |
| 18-109-final-viral-combined_k141_80980\|\|lt2gene | Low-quality |
| 18-109-final-viral-combined_k141_28681\|\|lt2gene | Low-quality |
| 18-109-final-viral-combined_k141_20222\|\|lt2gene | Not-determined |
| 18-109-final-viral-combined_k141_153032\|\|lt2gene | Low-quality |
| 18-109-final-viral-combined_k141_153042\|\|lt2gene | Low-quality |
| 18-109-final-viral-combined_k141_28714\|\|lt2gene | Low-quality |
| 18-109-final-viral-combined_k141_153076\|\|lt2gene | Low-quality |
| 18-109-final-viral-combined_k141_112207\|\|lt2gene | Low-quality |
| 18-109-final-viral-combined_k141_153108\|\|lt2gene | Low-quality |
| 18-109-final-viral-combined_k141_20331\|\|lt2gene | Low-quality |
| 18-109-final-viral-combined_k141_20374\|\|lt2gene | Not-determined |
| 18-109-final-viral-combined_k141_91604\|\|lt2gene | Low-quality |
| 18-109-final-viral-combined_k141_142519\|\|lt2gene | Not-determined |
| 18-109-final-viral-combined_k141_81241\|\|lt2gene | Low-quality |
| 18-109-final-viral-combined_k141_29104\|\|lt2gene | Low-quality |
| 18-109-final-viral-combined_k141_142732\|\|lt2gene | Not-determined |
| 18-109-final-viral-combined_k141_142752\|\|lt2gene | Low-quality |
| 18-109-final-viral-combined_k141_81387\|\|lt2gene | Low-quality |
| 18-109-final-viral-combined_k141_29218\|\|lt2gene | Low-quality |
| 18-109-final-viral-combined_k141_244385\|\|lt2gene | Low-quality |
| 18-109-final-viral-combined_k141_244388\|\|lt2gene | Low-quality |
| 18-109-final-viral-combined_k141_29245\|\|lt2gene | Low-quality |
| 18-109-final-viral-combined_k141_29263\|\|lt2gene | Low-quality |
| 18-109-final-viral-combined_k141_142799\|\|lt2gene | Low-quality |
| 18-109-final-viral-combined_k141_223484\|\|lt2gene | Low-quality |
| 18-109-final-viral-combined_k141_223508\|\|lt2gene | Low-quality |
| 18-109-final-viral-combined_k141_223556\|\|lt2gene | Low-quality |
| 18-109-final-viral-combined_k141_223581\|\|lt2gene | Not-determined |
| 18-109-final-viral-combined_k141_81657\|\|lt2gene | Not-determined |
| 18-109-final-viral-combined_k141_223838\|\|lt2gene | Low-quality |
| 18-109-final-viral-combined_k141_29664\|\|lt2gene | Low-quality |
| 18-109-final-viral-combined_k141_244652\|\|lt2gene | Low-quality |
| 18-109-final-viral-combined_k141_29908\|\|lt2gene | Low-quality |
| 18-109-final-viral-combined_k141_224221\|\|lt2gene | Low-quality |
| 18-109-final-viral-combined_k141_30000\|\|lt2gene | Low-quality |
| 18-109-final-viral-combined_k141_244728\|\|lt2gene | Low-quality |
| 18-109-final-viral-combined_k141_224327\|\|lt2gene | Low-quality |
| 18-109-final-viral-combined_k141_244769\|\|lt2gene | Low-quality |
| 18-109-final-viral-combined_k141_244792\|\|lt2gene | Low-quality |
| 18-109-final-viral-combined_k141_224517\|\|lt2gene | Low-quality |
| 18-109-final-viral-combined_k141_244845\|\|lt2gene | Low-quality |
| 18-109-final-viral-combined_k141_30194\|\|lt2gene | Low-quality |
| 18-109-final-viral-combined_k141_30283\|\|lt2gene | Low-quality |
| 18-109-final-viral-combined_k141_30318\|\|lt2gene | Low-quality |
| 18-109-final-viral-combined_k141_30527\|\|lt2gene | Not-determined |
| 18-109_k141_194020 | Not-determined |
| 18-109_k141_234864 | Not-determined |
| 18-109_k141_194022 | Not-determined |
| 18-109_k141_183808 | Not-determined |
| 18-109_k141_183810 | Not-determined |
| 18-109_k141_234869 | Not-determined |
| 18-109_k141_5 | Not-determined |
| 18-109_k141_8 | Not-determined |
| 18-109_k141_20426 | Not-determined |
| 18-109_k141_224652 | Low-quality |
| 18-109_k141_11 | Not-determined |
| 18-109_k141_132759 | Not-determined |
| 18-109_k141_173604 | Not-determined |
| 18-109_k141_224654 | Not-determined |
| 18-109_k141_10222 | Not-determined |
| 18-109_k141_16 | Not-determined |
| 18-109_k141_122547 | Not-determined |
| 18-109_k141_163395 | Not-determined |
| 18-109_k141_214455 | Not-determined |
| 18-109_k141_30643 | Not-determined |
| 18-109_k141_61280 | Not-determined |
| 18-109_k141_81709 | Not-determined |
| 18-109_k141_224662 | Not-determined |
| 18-109_k141_183817 | Not-determined |
| 18-109_k141_163401 | Not-determined |
| 18-109_k141_214463 | Not-determined |
| 18-109_k141_163405 | Not-determined |
| 18-109_k141_224673 | Not-determined |
| 18-109_k141_112344 | Not-determined |
| 18-109_k141_61285 | Not-determined |
| 18-109_k141_40864 | Not-determined |
| 18-109_k141_142978 | Not-determined |
| 18-109_k141_173612 | Low-quality |
| 18-109_k141_234873 | Not-determined |
| 18-109_k141_142984 | Not-determined |
| 18-109_k141_173618 | Not-determined |
| 18-109_k141_51078 | Not-determined |
| 18-109_k141_81714 | Not-determined |
| 18-109_k141_173619 | Not-determined |
| 18-109_k141_173620 | Not-determined |
| 18-109_k141_163414 | Not-determined |
| 18-109_k141_30661 | Low-quality |
| 18-109_k141_234879 | Not-determined |
| 18-109_k141_173625 | Not-determined |
| 18-109_k141_234882 | Not-determined |
| 18-109_k141_153190 | Low-quality |
| 18-109_k141_163418 | Not-determined |
| 18-109_k141_122559 | Not-determined |
| 18-109_k141_132786 | Not-determined |
| 18-109_k141_30666 | Not-determined |
| 18-109_k141_71497 | Not-determined |
| 18-109_k141_204257 | Not-determined |
| 18-109_k141_204259 | Not-determined |
| 18-109_k141_143000 | Not-determined |
| 18-109_k141_20452 | Not-determined |
| 18-109_k141_143002 | Not-determined |
| 18-109_k141_30670 | Not-determined |
| 18-109_k141_20454 | Not-determined |
| 18-109_k141_173637 | Low-quality |
| 18-109_k141_143003 | Not-determined |
| 18-109_k141_20459 | Not-determined |
| 18-109_k141_51099 | Not-determined |
| 18-109_k141_71509 | Low-quality |
| 18-109_k141_234902 | Not-determined |
| 18-109_k141_102153 | Not-determined |
| 18-109_k141_71512 | Not-determined |
| 18-109_k141_112364 | Not-determined |
| 18-109_k141_153206 | Not-determined |
| 18-109_k141_102156 | Not-determined |
| 18-109_k141_39 | Not-determined |
| 18-109_k141_71524 | Low-quality |
| 18-109_k141_30678 | Not-determined |
| 18-109_k141_30681 | Not-determined |
| 18-109_k141_163454 | Not-determined |
| 18-109_k141_183826 | Not-determined |
| 18-109_k141_163457 | Not-determined |
| 18-109_k141_163458 | Not-determined |
| 18-109_k141_153228 | Not-determined |
| 18-109_k141_30692 | Not-determined |
| 18-109_k141_91935 | Not-determined |
| 18-109_k141_132810 | Not-determined |
| 18-109_k141_54 | Not-determined |
| 18-109_k141_132812 | Not-determined |
| 18-109_k141_30697 | Not-determined |
| 18-109_k141_40886 | Not-determined |
| 18-109_k141_102174 | Not-determined |
| 18-109_k141_234919 | Not-determined |
| 18-109_k141_102178 | Not-determined |
| 18-109_k141_163460 | Not-determined |
| 18-109_k141_214507 | Not-determined |

| | |
|---|---|
| 18-109_k141_143025 | Not-determined |
| 18-109_k141_30712 | Not-determined |
| 18-109_k141_102180 | Not-determined |
| 18-109_k141_102182 | Not-determined |
| 18-109_k141_30716 | Not-determined |
| 18-109_k141_66 | Not-determined |
| 18-109_k141_234930 | Not-determined |
| 18-109_k141_112401 | Not-determined |
| 18-109_k141_91960 | Not-determined |
| 18-109_k141_183839 | Not-determined |
| 18-109_k141_112407 | Not-determined |
| 18-109_k141_234936 | Not-determined |
| 18-109_k141_73 | Not-determined |
| 18-109_k141_91967 | Low-quality |
| 18-109_k141_214517 | Not-determined |
| 18-109_k141_40897 | Not-determined |
| 18-109_k141_153265 | Not-determined |
| 18-109_k141_10273 | Not-determined |
| 18-109_k141_91972 | Not-determined |
| 18-109_k141_224691 | Not-determined |
| 18-109_k141_183848 | Low-quality |
| 18-109_k141_183851 | Not-determined |
| 18-109_k141_81757 | Not-determined |
| 18-109_k141_224696 | Not-determined |
| 18-109_k141_122583 | Not-determined |
| 18-109_k141_183860 | Not-determined |
| 18-109_k141_163480 | Not-determined |
| 18-109_k141_153279 | Not-determined |
| 18-109_k141_224714 | Not-determined |
| 18-109_k141_122587 | Not-determined |
| 18-109_k141_143059 | Not-determined |
| 18-109_k141_143060 | Not-determined |
| 18-109_k141_214548 | Not-determined |
| 18-109_k141_234964 | Not-determined |
| 18-109_k141_153285 | Low-quality |
| 18-109_k141_40911 | Not-determined |
| 18-109_k141_99 | Not-determined |
| 18-109_k141_122592 | Low-quality |
| 18-109_k141_183883 | Not-determined |
| 18-109_k141_40921 | Low-quality |
| 18-109_k141_194064 | Not-determined |
| 18-109_k141_183889 | Not-determined |
| 18-109_k141_163494 | Low-quality |
| 18-109_k141_214558 | Not-determined |
| 18-109_k141_143069 | Not-determined |
| 18-109_k141_183892 | Not-determined |
| 18-109_k141_91983 | Low-quality |
| 18-109_k141_112435 | Not-determined |
| 18-109_k141_112436 | Not-determined |
| 18-109_k141_143074 | Not-determined |
| 18-109_k141_214565 | Not-determined |
| 18-109_k141_143077 | Not-determined |
| 18-109_k141_91989 | Not-determined |
| 18-109_k141_91994 | Not-determined |
| 18-109_k141_112441 | Not-determined |
| 18-109_k141_163506 | Not-determined |
| 18-109_k141_40928 | Low-quality |
| 18-109_k141_40929 | Not-determined |
| 18-109_k141_163513 | Not-determined |
| 18-109_k141_40932 | Not-determined |
| 18-109_k141_153303 | Not-determined |
| 18-109_k141_235010 | Not-determined |
| 18-109_k141_122619 | Not-determined |
| 18-109_k141_112444 | Not-determined |
| 18-109_k141_102213 | Not-determined |
| 18-109_k141_183919 | Not-determined |
| 18-109_k141_183920 | Not-determined |
| 18-109_k141_20464 | Not-determined |
| 18-109_k141_235012 | Not-determined |
| 18-109_k141_183921 | Not-determined |
| 18-109_k141_122624 | Not-determined |
| 18-109_k141_122625 | Not-determined |
| 18-109_k141_122626 | Not-determined |
| 18-109_k141_122627 | Not-determined |
| 18-109_k141_61343 | Not-determined |
| 18-109_k141_71570 | Not-determined |
| 18-109_k141_153312 | Not-determined |
| 18-109_k141_71571 | Not-determined |
| 18-109_k141_10300 | Not-determined |
| 18-109_k141_153322 | Not-determined |
| 18-109_k141_235019 | Not-determined |
| 18-109_k141_112460 | Not-determined |
| 18-109_k141_204303 | Not-determined |
| 18-109_k141_204304 | Not-determined |
| 18-109_k141_40940 | Not-determined |
| 18-109_k141_163524 | Not-determined |
| 18-109_k141_102239 | Not-determined |
| 18-109_k141_224762 | Not-determined |
| 18-109_k141_204306 | Not-determined |
| 18-109_k141_20469 | Not-determined |
| 18-109_k141_143109 | Not-determined |
| 18-109_k141_40953 | Not-determined |
| 18-109_k141_102249 | Not-determined |
| 18-109_k141_162 | Not-determined |
| 18-109_k141_143120 | Not-determined |
| 18-109_k141_51181 | Not-determined |
| 18-109_k141_153329 | Low-quality |
| 18-109_k141_194117 | Not-determined |
| 18-109_k141_224775 | Low-quality |
| 18-109_k141_224777 | Not-determined |
| 18-109_k141_204317 | Not-determined |
| 18-109_k141_179 | Low-quality |
| 18-109_k141_61380 | Low-quality |
| 18-109_k141_92028 | Not-determined |
| 18-109_k141_194130 | Not-determined |
| 18-109_k141_224784 | Not-determined |
| 18-109_k141_102256 | Not-determined |
| 18-109_k141_194135 | Not-determined |
| 18-109_k141_184 | Low-quality |
| 18-109_k141_185 | Not-determined |
| 18-109_k141_163547 | Not-determined |
| 18-109_k141_143150 | Not-determined |
| 18-109_k141_194142 | Not-determined |
| 18-109_k141_102262 | Not-determined |
| 18-109_k141_92042 | Not-determined |
| 18-109_k141_102265 | Not-determined |
| 18-109_k141_30745 | Not-determined |
| 18-109_k141_92046 | Not-determined |
| 18-109_k141_102267 | Not-determined |
| 18-109_k141_204332 | Not-determined |
| 18-109_k141_196 | Not-determined |
| 18-109_k141_30749 | Not-determined |
| 18-109_k141_112497 | Not-determined |
| 18-109_k141_40979 | Not-determined |
| 18-109_k141_204 | Not-determined |
| 18-109_k141_194146 | Not-determined |
| 18-109_k141_194147 | Not-determined |
| 18-109_k141_40985 | Not-determined |
| 18-109_k141_163555 | Not-determined |
| 18-109_k141_143162 | Not-determined |
| 18-109_k141_143163 | Low-quality |
| 18-109_k141_153349 | Not-determined |
| 18-109_k141_132816 | Not-determined |
| 18-109_k141_173717 | Not-determined |
| 18-109_k141_20506 | Low-quality |
| 18-109_k141_220 | Not-determined |
| 18-109_k141_224818 | Not-determined |
| 18-109_k141_173723 | Not-determined |
| 18-109_k141_214633 | Not-determined |
| 18-109_k141_228 | Not-determined |
| 18-109_k141_194156 | Not-determined |
| 18-109_k141_173725 | Not-determined |
| 18-109_k141_194163 | Not-determined |
| 18-109_k141_20521 | Not-determined |
| 18-109_k141_224832 | Not-determined |
| 18-109_k141_41014 | Not-determined |
| 18-109_k141_20531 | Not-determined |
| 18-109_k141_163578 | Not-determined |
| 18-109_k141_194174 | Not-determined |
| 18-109_k141_163586 | Not-determined |
| 18-109_k141_214653 | Not-determined |
| 18-109_k141_20533 | Not-determined |
| 18-109_k141_143190 | Not-determined |
| 18-109_k141_194179 | Not-determined |
| 18-109_k141_20535 | Not-determined |
| 18-109_k141_204359 | Not-determined |
| 18-109_k141_92068 | Not-determined |
| 18-109_k141_194185 | Not-determined |
| 18-109_k141_30779 | Not-determined |
| 18-109_k141_102307 | Not-determined |
| 18-109_k141_143198 | Low-quality |
| 18-109_k141_143199 | Not-determined |
| 18-109_k141_153378 | Not-determined |
| 18-109_k141_204361 | Low-quality |
| 18-109_k141_143201 | Low-quality |
| 18-109_k141_153381 | Not-determined |
| 18-109_k141_61426 | Not-determined |
| 18-109_k141_61427 | Not-determined |
| 18-109_k141_153386 | Not-determined |
| 18-109_k141_214658 | Not-determined |
| 18-109_k141_173741 | Not-determined |
| 18-109_k141_183944 | Not-determined |
| 18-109_k141_143208 | Not-determined |
| 18-109_k141_143209 | Not-determined |
| 18-109_k141_143210 | Not-determined |
| 18-109_k141_112525 | Not-determined |
| 18-109_k141_153397 | Not-determined |
| 18-109_k141_153398 | Not-determined |
| 18-109_k141_153399 | Not-determined |
| 18-109_k141_92073 | Low-quality |
| 18-109_k141_143216 | Not-determined |
| 18-109_k141_183951 | Not-determined |
| 18-109_k141_92078 | Not-determined |
| 18-109_k141_173753 | Not-determined |
| 18-109_k141_20542 | Not-determined |
| 18-109_k141_61463 | Not-determined |
| 18-109_k141_194211 | Not-determined |
| 18-109_k141_250 | Not-determined |
| 18-109_k141_112545 | Not-determined |
| 18-109_k141_61465 | Not-determined |
| 18-109_k141_214666 | Not-determined |
| 18-109_k141_132850 | Not-determined |
| 18-109_k141_92092 | Not-determined |
| 18-109_k141_71593 | Not-determined |
| 18-109_k141_194222 | Not-determined |
| 18-109_k141_235036 | Not-determined |
| 18-109_k141_122703 | Not-determined |
| 18-109_k141_112555 | Not-determined |
| 18-109_k141_92103 | Not-determined |
| 18-109_k141_122708 | Not-determined |
| 18-109_k141_214670 | Not-determined |
| 18-109_k141_214671 | Not-determined |
| 18-109_k141_173778 | Not-determined |
| 18-109_k141_235055 | Low-quality |
| 18-109_k141_224855 | Not-determined |
| 18-109_k141_132882 | Low-quality |
| 18-109_k141_71605 | Low-quality |
| 18-109_k141_122716 | Not-determined |
| 18-109_k141_132885 | Not-determined |
| 18-109_k141_102323 | Not-determined |
| 18-109_k141_163623 | Not-determined |
| 18-109_k141_61483 | Not-determined |
| 18-109_k141_214683 | Not-determined |
| 18-109_k141_163631 | Not-determined |
| 18-109_k141_214686 | Not-determined |
| 18-109_k141_163632 | Not-determined |
| 18-109_k141_61487 | Not-determined |
| 18-109_k141_235077 | Not-determined |
| 18-109_k141_235081 | Not-determined |
| 18-109_k141_122723 | Not-determined |
| 18-109_k141_224868 | Not-determined |
| 18-109_k141_173790 | Not-determined |
| 18-109_k141_122726 | Low-quality |
| 18-109_k141_71623 | Not-determined |
| 18-109_k141_214699 | Low-quality |
| 18-109_k141_122729 | Not-determined |
| 18-109_k141_283 | Not-determined |
| 18-109_k141_235089 | Low-quality |
| 18-109_k141_214705 | Low-quality |
| 18-109_k141_143234 | Not-determined |
| 18-109_k141_235093 | Not-determined |
| 18-109_k141_132911 | Not-determined |
| 18-109_k141_122737 | Not-determined |
| 18-109_k141_143240 | Not-determined |
| 18-109_k141_122742 | Not-determined |
| 18-109_k141_122743 | Not-determined |
| 18-109_k141_143244 | Not-determined |
| 18-109_k141_10329 | Not-determined |
| 18-109_k141_143245 | Not-determined |
| 18-109_k141_92126 | Not-determined |
| 18-109_k141_92127 | Not-determined |
| 18-109_k141_61508 | Not-determined |
| 18-109_k141_112599 | Not-determined |
| 18-109_k141_122753 | Not-determined |
| 18-109_k141_10333 | Not-determined |
| 18-109_k141_204395 | Not-determined |
| 18-109_k141_20555 | Not-determined |
| 18-109_k141_10335 | Not-determined |
| 18-109_k141_214719 | Not-determined |
| 18-109_k141_112606 | Not-determined |
| 18-109_k141_184022 | Not-determined |
| 18-109_k141_173818 | Not-determined |
| 18-109_k141_184025 | Not-determined |
| 18-109_k141_194247 | Not-determined |
| 18-109_k141_163664 | Not-determined |
| 18-109_k141_20561 | Not-determined |
| 18-109_k141_112615 | Not-determined |
| 18-109_k141_112617 | Not-determined |
| 18-109_k141_112623 | Not-determined |
| 18-109_k141_20563 | Low-quality |
| 18-109_k141_224902 | Not-determined |
| 18-109_k141_163672 | Not-determined |
| 18-109_k141_163674 | Not-determined |
| 18-109_k141_224906 | Not-determined |
| 18-109_k141_61530 | Not-determined |
| 18-109_k141_173833 | Not-determined |
| 18-109_k141_214728 | Not-determined |
| 18-109_k141_122786 | Not-determined |
| 18-109_k141_92138 | Not-determined |
| 18-109_k141_122787 | Not-determined |
| 18-109_k141_112638 | Not-determined |
| 18-109_k141_30825 | Low-quality |
| 18-109_k141_122789 | Not-determined |
| 18-109_k141_10368 | Not-determined |
| 18-109_k141_112645 | Not-determined |
| 18-109_k141_92150 | Not-determined |
| 18-109_k141_10370 | Not-determined |
| 18-109_k141_112654 | Not-determined |
| 18-109_k141_194259 | Not-determined |
| 18-109_k141_92163 | Not-determined |
| 18-109_k141_30838 | Low-quality |
| 18-109_k141_224929 | Not-determined |
| 18-109_k141_122812 | Not-determined |
| 18-109_k141_51258 | Not-determined |
| 18-109_k141_204431 | Not-determined |
| 18-109_k141_214745 | Not-determined |
| 18-109_k141_122825 | Not-determined |
| 18-109_k141_10385 | Not-determined |
| 18-109_k141_10388 | Low-quality |
| 18-109_k141_132947 | Not-determined |
| 18-109_k141_122832 | Not-determined |
| 18-109_k141_112662 | Not-determined |
| 18-109_k141_102428 | Not-determined |
| 18-109_k141_92184 | Not-determined |
| 18-109_k141_132959 | Not-determined |
| 18-109_k141_143267 | Not-determined |
| 18-109_k141_10401 | Not-determined |
| 18-109_k141_30869 | Not-determined |
| 18-109_k141_30871 | Not-determined |
| 18-109_k141_102436 | Not-determined |
| 18-109_k141_204446 | Low-quality |
| 18-109_k141_163722 | Not-determined |
| 18-109_k141_194299 | Not-determined |
| 18-109_k141_204448 | Not-determined |
| 18-109_k141_194301 | Not-determined |
| 18-109_k141_224966 | Low-quality |
| 18-109_k141_224969 | Not-determined |
| 18-109_k141_30881 | Not-determined |
| 18-109_k141_30882 | Not-determined |
| 18-109_k141_224973 | Not-determined |
| 18-109_k141_81778 | Not-determined |
| 18-109_k141_235128 | Not-determined |
| 18-109_k141_92203 | Not-determined |
| 18-109_k141_102452 | Not-determined |
| 18-109_k141_81789 | Low-quality |
| 18-109_k141_81791 | Not-determined |
| 18-109_k141_81792 | Not-determined |
| 18-109_k141_323 | Not-determined |
| 18-109_k141_132983 | Not-determined |
| 18-109_k141_41046 | Not-determined |
| 18-109_k141_81802 | Not-determined |
| 18-109_k141_92219 | Not-determined |
| 18-109_k141_10425 | Not-determined |
| 18-109_k141_10428 | Not-determined |
| 18-109_k141_184034 | Low-quality |
| 18-109_k141_112692 | Not-determined |
| 18-109_k141_20616 | Not-determined |
| 18-109_k141_20618 | Not-determined |
| 18-109_k141_163742 | Not-determined |
| 18-109_k141_132995 | Not-determined |
| 18-109_k141_10443 | Not-determined |
| 18-109_k141_224984 | Not-determined |
| 18-109_k141_10447 | Not-determined |
| 18-109_k141_112700 | Not-determined |
| 18-109_k141_10451 | Not-determined |
| 18-109_k141_10455 | Not-determined |
| 18-109_k141_81823 | Not-determined |
| 18-109_k141_10456 | Not-determined |
| 18-109_k141_112708 | Not-determined |
| 18-109_k141_184070 | Not-determined |
| 18-109_k141_92245 | Not-determined |
| 18-109_k141_61564 | Not-determined |
| 18-109_k141_350 | Not-determined |
| 18-109_k141_153442 | Low-quality |
| 18-109_k141_81838 | Not-determined |
| 18-109_k141_30920 | Not-determined |
| 18-109_k141_122838 | Not-determined |
| 18-109_k141_153452 | Not-determined |
| 18-109_k141_30935 | Not-determined |
| 18-109_k141_30942 | Not-determined |
| 18-109_k141_235166 | Not-determined |
| 18-109_k141_235172 | Not-determined |
| 18-109_k141_225024 | Not-determined |
| 18-109_k141_225029 | Not-determined |
| 18-109_k141_133005 | Not-determined |
| 18-109_k141_225037 | Not-determined |
| 18-109_k141_225043 | Not-determined |
| 18-109_k141_143287 | Not-determined |
| 18-109_k141_143290 | Not-determined |
| 18-109_k141_81851 | Not-determined |
| 18-109_k141_133018 | Not-determined |
| 18-109_k141_235189 | Not-determined |
| 18-109_k141_92258 | Not-determined |
| 18-109_k141_153490 | Not-determined |
| 18-109_k141_235195 | Not-determined |
| 18-109_k141_376 | Not-determined |
| 18-109_k141_153497 | Not-determined |
| 18-109_k141_81866 | Not-determined |
| 18-109_k141_51267 | Not-determined |
| 18-109_k141_122846 | Not-determined |
| 18-109_k141_163797 | Not-determined |
| 18-109_k141_92278 | Not-determined |
| 18-109_k141_235211 | Not-determined |
| 18-109_k141_81874 | Not-determined |
| 18-109_k141_81876 | Not-determined |
| 18-109_k141_163802 | Not-determined |
| 18-109_k141_173881 | Not-determined |
| 18-109_k141_235222 | Not-determined |
| 18-109_k141_194386 | Not-determined |
| 18-109_k141_184124 | Not-determined |
| 18-109_k141_153521 | Not-determined |
| 18-109_k141_92289 | Not-determined |
| 18-109_k141_153524 | Not-determined |
| 18-109_k141_81893 | Not-determined |
| 18-109_k141_81894 | Not-determined |
| 18-109_k141_92295 | Not-determined |
| 18-109_k141_81901 | Not-determined |
| 18-109_k141_92296 | Not-determined |
| 18-109_k141_20673 | Not-determined |
| 18-109_k141_235232 | Not-determined |
| 18-109_k141_81906 | Not-determined |
| 18-109_k141_385 | Low-quality |
| 18-109_k141_386 | Not-determined |
| 18-109_k141_81912 | Not-determined |
| 18-109_k141_92305 | Not-determined |
| 18-109_k141_194410 | Not-determined |
| 18-109_k141_112764 | Not-determined |
| 18-109_k141_112767 | Not-determined |
| 18-109_k141_194421 | Low-quality |
| 18-109_k141_81934 | Not-determined |
| 18-109_k141_235251 | Not-determined |
| 18-109_k141_163832 | Not-determined |
| 18-109_k141_61615 | Low-quality |
| 18-109_k141_61619 | Not-determined |
| 18-109_k141_173923 | Not-determined |
| 18-109_k141_235262 | Not-determined |
| 18-109_k141_235263 | Not-determined |
| 18-109_k141_194433 | Not-determined |
| 18-109_k141_30987 | Not-determined |
| 18-109_k141_204485 | Low-quality |
| 18-109_k141_81945 | Not-determined |
| 18-109_k141_10485 | Not-determined |
| 18-109_k141_173939 | Not-determined |
| 18-109_k141_184160 | Not-determined |
| 18-109_k141_102462 | Low-quality |
| 18-109_k141_61648 | Not-determined |
| 18-109_k141_225070 | Not-determined |
| 18-109_k141_20693 | Not-determined |
| 18-109_k141_235270 | Not-determined |
| 18-109_k141_10496 | Not-determined |
| 18-109_k141_112812 | Not-determined |
| 18-109_k141_10506 | Not-determined |
| 18-109_k141_163854 | Not-determined |
| 18-109_k141_225083 | Not-determined |
| 18-109_k141_194466 | Not-determined |
| 18-109_k141_10515 | Not-determined |
| 18-109_k141_143329 | Not-determined |
| 18-109_k141_10519 | Not-determined |
| 18-109_k141_225093 | Not-determined |
| 18-109_k141_133081 | Not-determined |
| 18-109_k141_133083 | Not-determined |
| 18-109_k141_133087 | Not-determined |
| 18-109_k141_225103 | Not-determined |
| 18-109_k141_143340 | Low-quality |
| 18-109_k141_10528 | Not-determined |
| 18-109_k141_143342 | Low-quality |
| 18-109_k141_204516 | Not-determined |
| 18-109_k141_404 | Not-determined |
| 18-109_k141_225113 | Not-determined |
| 18-109_k141_143347 | Not-determined |
| 18-109_k141_133111 | Not-determined |
| 18-109_k141_413 | Not-determined |
| 18-109_k141_133112 | Not-determined |
| 18-109_k141_184165 | Not-determined |
| 18-109_k141_184168 | Not-determined |
| 18-109_k141_194484 | Not-determined |
| 18-109_k141_225118 | Not-determined |
| 18-109_k141_194489 | Not-determined |
| 18-109_k141_163898 | Not-determined |
| 18-109_k141_133126 | Not-determined |
| 18-109_k141_81959 | Not-determined |
| 18-109_k141_112836 | Not-determined |
| 18-109_k141_71641 | Not-determined |
| 18-109_k141_112838 | Not-determined |
| 18-109_k141_122878 | Not-determined |
| 18-109_k141_10556 | Not-determined |
| 18-109_k141_112842 | Not-determined |
| 18-109_k141_133134 | Not-determined |
| 18-109_k141_445 | Not-determined |
| 18-109_k141_194505 | Not-determined |
| 18-109_k141_143357 | Not-determined |
| 18-109_k141_10565 | Not-determined |
| 18-109_k141_163925 | Not-determined |
| 18-109_k141_214780 | Not-determined |
| 18-109_k141_10575 | Not-determined |
| 18-109_k141_20717 | Not-determined |
| 18-109_k141_225149 | Not-determined |
| 18-109_k141_31029 | Low-quality |
| 18-109_k141_143378 | Not-determined |
| 18-109_k141_184183 | Not-determined |
| 18-109_k141_81985 | Not-determined |
| 18-109_k141_204567 | Not-determined |
| 18-109_k141_204568 | Low-quality |
| 18-109_k141_204570 | Not-determined |
| 18-109_k141_112879 | Not-determined |
| 18-109_k141_194528 | Low-quality |
| 18-109_k141_184192 | Not-determined |
| 18-109_k141_102476 | Not-determined |
| 18-109_k141_204585 | Not-determined |
| 18-109_k141_163948 | Not-determined |
| 18-109_k141_81994 | Not-determined |
| 18-109_k141_225158 | Not-determined |
| 18-109_k141_71669 | Not-determined |
| 18-109_k141_10597 | Not-determined |
| 18-109_k141_153646 | Not-determined |
| 18-109_k141_462 | Low-quality |
| 18-109_k141_143405 | Not-determined |
| 18-109_k141_41116 | Not-determined |
| 18-109_k141_71685 | Not-determined |
| 18-109_k141_71686 | Not-determined |
| 18-109_k141_163960 | Not-determined |
| 18-109_k141_112906 | Not-determined |
| 18-109_k141_61698 | Not-determined |
| 18-109_k141_214814 | Low-quality |
| 18-109_k141_163969 | Not-determined |
| 18-109_k141_112908 | Low-quality |
| 18-109_k141_143412 | Not-determined |
| 18-109_k141_112911 | Not-determined |
| 18-109_k141_235348 | Not-determined |
| 18-109_k141_41126 | Low-quality |
| 18-109_k141_225182 | Not-determined |
| 18-109_k141_235352 | Not-determined |
| 18-109_k141_41131 | Not-determined |
| 18-109_k141_194553 | Low-quality |
| 18-109_k141_112924 | Low-quality |
| 18-109_k141_41136 | Not-determined |
| 18-109_k141_112926 | Not-determined |
| 18-109_k141_82021 | Not-determined |
| 18-109_k141_194558 | Low-quality |
| 18-109_k141_235355 | Not-determined |
| 18-109_k141_184218 | Low-quality |
| 18-109_k141_41146 | Not-determined |
| 18-109_k141_41147 | Not-determined |
| 18-109_k141_112930 | Not-determined |
| 18-109_k141_153679 | Not-determined |
| 18-109_k141_82031 | Not-determined |
| 18-109_k141_184223 | Not-determined |
| 18-109_k141_498 | Not-determined |
| 18-109_k141_204606 | Not-determined |
| 18-109_k141_184237 | Not-determined |
| 18-109_k141_153686 | Not-determined |
| 18-109_k141_214842 | Not-determined |
| 18-109_k141_509 | Not-determined |
| 18-109_k141_194581 | Not-determined |
| 18-109_k141_184241 | Low-quality |
| 18-109_k141_71699 | Not-determined |
| 18-109_k141_143427 | Not-determined |
| 18-109_k141_204611 | Not-determined |
| 18-109_k141_20745 | Not-determined |
| 18-109_k141_225199 | Not-determined |
| 18-109_k141_71706 | Not-determined |
| 18-109_k141_102502 | Low-quality |
| 18-109_k141_225203 | Not-determined |
| 18-109_k141_520 | Not-determined |
| 18-109_k141_153707 | Not-determined |
| 18-109_k141_528 | Not-determined |
| 18-109_k141_235375 | Low-quality |
| 18-109_k141_133180 | Not-determined |
| 18-109_k141_153710 | Low-quality |
| 18-109_k141_122904 | Not-determined |
| 18-109_k141_61744 | Not-determined |
| 18-109_k141_214857 | Not-determined |
| 18-109_k141_143439 | Not-determined |
| 18-109_k141_214859 | Not-determined |
| 18-109_k141_235390 | Not-determined |
| 18-109_k141_20762 | Low-quality |
| 18-109_k141_133193 | Not-determined |
| 18-109_k141_51282 | Low-quality |
| 18-109_k141_112962 | Not-determined |
| 18-109_k141_112965 | Not-determined |
| 18-109_k141_112966 | Not-determined |
| 18-109_k141_194611 | Not-determined |
| 18-109_k141_174056 | Not-determined |
| 18-109_k141_235405 | Not-determined |
| 18-109_k141_102531 | Not-determined |
| 18-109_k141_71719 | Not-determined |
| 18-109_k141_61775 | Not-determined |
| 18-109_k141_71726 | Low-quality |
| 18-109_k141_194624 | Not-determined |
| 18-109_k141_194628 | Not-determined |
| 18-109_k141_71732 | Low-quality |
| 18-109_k141_194636 | Not-determined |
| 18-109_k141_194639 | Not-determined |
| 18-109_k141_31051 | Not-determined |
| 18-109_k141_225241 | Not-determined |
| 18-109_k141_20794 | Not-determined |
| 18-109_k141_102543 | Not-determined |
| 18-109_k141_214875 | Not-determined |
| 18-109_k141_31060 | Not-determined |
| 18-109_k141_225248 | Not-determined |
| 18-109_k141_143480 | Not-determined |
| 18-109_k141_122917 | Not-determined |
| 18-109_k141_214886 | Not-determined |
| 18-109_k141_225256 | Not-determined |
| 18-109_k141_20799 | Not-determined |
| 18-109_k141_31064 | Not-determined |
| 18-109_k141_214890 | Not-determined |
| 18-109_k141_51321 | Not-determined |
| 18-109_k141_235426 | Not-determined |
| 18-109_k141_51329 | Not-determined |
| 18-109_k141_51331 | Not-determined |
| 18-109_k141_51333 | Not-determined |
| 18-109_k141_194649 | Not-determined |
| 18-109_k141_214898 | Not-determined |
| 18-109_k141_51335 | Not-determined |
| 18-109_k141_133208 | Not-determined |
| 18-109_k141_122932 | Not-determined |
| 18-109_k141_174068 | Not-determined |
| 18-109_k141_153744 | Not-determined |

| | |
|---|---|
| 18-109_k141_133211 | Not-determined |
| 18-109_k141_102559 | Not-determined |
| 18-109_k141_143492 | Not-determined |
| 18-109_k141_174076 | Not-determined |
| 18-109_k141_214914 | Not-determined |
| 18-109_k141_174080 | Not-determined |
| 18-109_k141_235433 | Not-determined |
| 18-109_k141_143501 | Not-determined |
| 18-109_k141_204656 | Low-quality |
| 18-109_k141_61792 | Low-quality |
| 18-109_k141_174088 | Not-determined |
| 18-109_k141_174094 | Not-determined |
| 18-109_k141_61801 | Low-quality |
| 18-109_k141_71754 | Not-determined |
| 18-109_k141_204664 | Not-determined |
| 18-109_k141_122947 | Not-determined |
| 18-109_k141_122952 | Not-determined |
| 18-109_k141_31070 | Not-determined |
| 18-109_k141_214943 | Not-determined |
| 18-109_k141_10631 | Not-determined |
| 18-109_k141_102568 | Not-determined |
| 18-109_k141_235448 | Low-quality |
| 18-109_k141_214951 | Not-determined |
| 18-109_k141_92385 | Not-determined |
| 18-109_k141_82100 | Not-determined |
| 18-109_k141_133223 | Not-determined |
| 18-109_k141_194697 | Not-determined |
| 18-109_k141_122972 | Not-determined |
| 18-109_k141_92387 | Not-determined |
| 18-109_k141_10644 | Not-determined |
| 18-109_k141_82113 | Not-determined |
| 18-109_k141_82114 | Not-determined |
| 18-109_k141_82120 | Not-determined |
| 18-109_k141_153775 | Not-determined |
| 18-109_k141_143532 | Not-determined |
| 18-109_k141_61833 | Not-determined |
| 18-109_k141_204679 | Low-quality |
| 18-109_k141_153781 | Not-determined |
| 18-109_k141_214968 | Not-determined |
| 18-109_k141_92390 | Not-determined |
| 18-109_k141_235471 | Not-determined |
| 18-109_k141_214972 | Not-determined |
| 18-109_k141_61844 | Not-determined |
| 18-109_k141_214976 | Not-determined |
| 18-109_k141_184320 | Not-determined |
| 18-109_k141_143541 | Low-quality |
| 18-109_k141_577 | Not-determined |
| 18-109_k141_214980 | Not-determined |
| 18-109_k141_61850 | Not-determined |
| 18-109_k141_214982 | Not-determined |
| 18-109_k141_61854 | Not-determined |
| 18-109_k141_204687 | Not-determined |
| 18-109_k141_153797 | Not-determined |
| 18-109_k141_10674 | Not-determined |
| 18-109_k141_184333 | Not-determined |
| 18-109_k141_214986 | Not-determined |
| 18-109_k141_71785 | Not-determined |
| 18-109_k141_174122 | Not-determined |
| 18-109_k141_41222 | Not-determined |
| 18-109_k141_590 | Not-determined |
| 18-109_k141_10680 | Not-determined |
| 18-109_k141_10681 | Not-determined |
| 18-109_k141_235477 | Not-determined |
| 18-109_k141_174127 | Not-determined |
| 18-109_k141_112985 | Low-quality |
| 18-109_k141_92414 | Not-determined |
| 18-109_k141_194723 | Not-determined |
| 18-109_k141_92416 | Not-determined |
| 18-109_k141_31098 | Not-determined |
| 18-109_k141_112992 | Not-determined |
| 18-109_k141_204707 | Low-quality |
| 18-109_k141_225300 | Not-determined |
| 18-109_k141_112994 | Not-determined |
| 18-109_k141_194733 | Not-determined |
| 18-109_k141_143570 | Not-determined |
| 18-109_k141_194749 | Not-determined |
| 18-109_k141_133272 | Low-quality |
| 18-109_k141_204713 | Not-determined |
| 18-109_k141_102589 | Low-quality |
| 18-109_k141_71808 | Not-determined |
| 18-109_k141_174149 | Not-determined |
| 18-109_k141_133278 | Low-quality |
| 18-109_k141_153828 | Not-determined |
| 18-109_k141_610 | Not-determined |
| 18-109_k141_41232 | Not-determined |
| 18-109_k141_174159 | Not-determined |
| 18-109_k141_20857 | Not-determined |
| 18-109_k141_41237 | Not-determined |
| 18-109_k141_225333 | Not-determined |
| 18-109_k141_133295 | Not-determined |
| 18-109_k141_215007 | Low-quality |
| 18-109_k141_235504 | Low-quality |
| 18-109_k141_102613 | Not-determined |
| 18-109_k141_102617 | Not-determined |
| 18-109_k141_153862 | Not-determined |
| 18-109_k141_235514 | Not-determined |
| 18-109_k141_153863 | Not-determined |
| 18-109_k141_31118 | Low-quality |
| 18-109_k141_31120 | Not-determined |
| 18-109_k141_92433 | Not-determined |
| 18-109_k141_92438 | Not-determined |
| 18-109_k141_41258 | Not-determined |
| 18-109_k141_133321 | Not-determined |
| 18-109_k141_637 | Low-quality |
| 18-109_k141_82177 | Not-determined |
| 18-109_k141_92447 | Not-determined |
| 18-109_k141_194768 | Not-determined |
| 18-109_k141_92449 | Not-determined |
| 18-109_k141_51347 | Not-determined |
| 18-109_k141_225349 | Not-determined |
| 18-109_k141_194777 | Not-determined |
| 18-109_k141_61880 | Not-determined |
| 18-109_k141_225362 | Not-determined |
| 18-109_k141_61884 | Not-determined |
| 18-109_k141_92464 | Not-determined |
| 18-109_k141_215051 | Not-determined |
| 18-109_k141_225366 | Not-determined |
| 18-109_k141_225368 | Not-determined |
| 18-109_k141_31147 | Not-determined |
| 18-109_k141_174210 | Not-determined |
| 18-109_k141_82184 | Not-determined |
| 18-109_k141_31151 | Not-determined |
| 18-109_k141_153904 | Not-determined |
| 18-109_k141_71848 | Not-determined |
| 18-109_k141_215071 | Low-quality |
| 18-109_k141_123013 | Not-determined |
| 18-109_k141_31159 | Not-determined |
| 18-109_k141_225382 | Not-determined |
| 18-109_k141_215075 | Not-determined |
| 18-109_k141_61909 | Not-determined |
| 18-109_k141_153915 | Not-determined |
| 18-109_k141_668 | Not-determined |
| 18-109_k141_41271 | Not-determined |
| 18-109_k141_10729 | Not-determined |
| 18-109_k141_677 | Not-determined |
| 18-109_k141_215085 | Not-determined |
| 18-109_k141_113047 | Not-determined |
| 18-109_k141_51363 | Not-determined |
| 18-109_k141_51367 | Not-determined |
| 18-109_k141_113059 | Not-determined |
| 18-109_k141_102659 | Not-determined |
| 18-109_k141_102660 | Not-determined |
| 18-109_k141_113061 | Low-quality |
| 18-109_k141_215102 | Not-determined |
| 18-109_k141_61927 | Not-determined |
| 18-109_k141_703 | Not-determined |
| 18-109_k141_71870 | Not-determined |
| 18-109_k141_41299 | Not-determined |
| 18-109_k141_705 | Low-quality |
| 18-109_k141_706 | Not-determined |
| 18-109_k141_41307 | Not-determined |
| 18-109_k141_51382 | Not-determined |
| 18-109_k141_41318 | Not-determined |
| 18-109_k141_10756 | Not-determined |
| 18-109_k141_225420 | Low-quality |
| 18-109_k141_215123 | Not-determined |
| 18-109_k141_235540 | Not-determined |
| 18-109_k141_720 | Not-determined |
| 18-109_k141_215128 | Low-quality |
| 18-109_k141_71892 | Not-determined |
| 18-109_k141_153935 | Not-determined |
| 18-109_k141_153936 | Not-determined |
| 18-109_k141_41329 | Not-determined |
| 18-109_k141_41330 | Not-determined |
| 18-109_k141_92471 | Not-determined |
| 18-109_k141_731 | Not-determined |
| 18-109_k141_20891 | Not-determined |
| 18-109_k141_123032 | Not-determined |
| 18-109_k141_123036 | Low-quality |
| 18-109_k141_235559 | Not-determined |
| 18-109_k141_51405 | Not-determined |
| 18-109_k141_235564 | Not-determined |
| 18-109_k141_174245 | Not-determined |
| 18-109_k141_71906 | Low-quality |
| 18-109_k141_235569 | Not-determined |
| 18-109_k141_164041 | Not-determined |
| 18-109_k141_235578 | Not-determined |
| 18-109_k141_113095 | Not-determined |
| 18-109_k141_71915 | Low-quality |
| 18-109_k141_51422 | Not-determined |
| 18-109_k141_113100 | Not-determined |
| 18-109_k141_184360 | Not-determined |
| 18-109_k141_174271 | Not-determined |
| 18-109_k141_174272 | Not-determined |
| 18-109_k141_235588 | Not-determined |
| 18-109_k141_82197 | Not-determined |
| 18-109_k141_194856 | Not-determined |
| 18-109_k141_194858 | Not-determined |
| 18-109_k141_82201 | Not-determined |
| 18-109_k141_235598 | Not-determined |
| 18-109_k141_184368 | Not-determined |
| 18-109_k141_71931 | Not-determined |
| 18-109_k141_31171 | Not-determined |
| 18-109_k141_184377 | Not-determined |
| 18-109_k141_113115 | Not-determined |
| 18-109_k141_235606 | Not-determined |
| 18-109_k141_71932 | Not-determined |
| 18-109_k141_153974 | Not-determined |
| 18-109_k141_184386 | Not-determined |
| 18-109_k141_71938 | Not-determined |
| 18-109_k141_71943 | Not-determined |
| 18-109_k141_153981 | Not-determined |
| 18-109_k141_123082 | Not-determined |
| 18-109_k141_113134 | Not-determined |
| 18-109_k141_10768 | Low-quality |
| 18-109_k141_184396 | Not-determined |
| 18-109_k141_71948 | Not-determined |
| 18-109_k141_194877 | Not-determined |
| 18-109_k141_194878 | Not-determined |
| 18-109_k141_235625 | Not-determined |
| 18-109_k141_153985 | Not-determined |
| 18-109_k141_204815 | Not-determined |
| 18-109_k141_153990 | Not-determined |
| 18-109_k141_215194 | Not-determined |
| 18-109_k141_82227 | Not-determined |
| 18-109_k141_82228 | Not-determined |
| 18-109_k141_235643 | Not-determined |
| 18-109_k141_82229 | Not-determined |
| 18-109_k141_71957 | Not-determined |
| 18-109_k141_215200 | Not-determined |
| 18-109_k141_215204 | Not-determined |
| 18-109_k141_215206 | Low-quality |
| 18-109_k141_133344 | Not-determined |
| 18-109_k141_154000 | Not-determined |
| 18-109_k141_215216 | Low-quality |
| 18-109_k141_194910 | Not-determined |
| 18-109_k141_133345 | Not-determined |
| 18-109_k141_71963 | Not-determined |
| 18-109_k141_164091 | Not-determined |
| 18-109_k141_164100 | Not-determined |
| 18-109_k141_92498 | Not-determined |
| 18-109_k141_10787 | Not-determined |
| 18-109_k141_82245 | Not-determined |
| 18-109_k141_102707 | Not-determined |
| 18-109_k141_782 | Not-determined |
| 18-109_k141_235654 | Not-determined |
| 18-109_k141_164101 | Not-determined |
| 18-109_k141_235655 | Not-determined |
| 18-109_k141_92504 | Not-determined |
| 18-101-final-viral-combined_k141_130214\|\|full | High-quality |
| 18-109_k141_123118 | Not-determined |
| 18-109_k141_92510 | Not-determined |
| 18-109_k141_215220 | Not-determined |
| 18-109_k141_133357 | Not-determined |
| 18-109_k141_235672 | Not-determined |
| 18-109_k141_154012 | Not-determined |
| 18-109_k141_235675 | Not-determined |
| 18-109_k141_204842 | Not-determined |
| 18-109_k141_10797 | Not-determined |
| 18-109_k141_154019 | Not-determined |
| 18-109_k141_10802 | Not-determined |
| 18-109_k141_10803 | Not-determined |
| 18-109_k141_10808 | Not-determined |
| 18-109_k141_123130 | Not-determined |
| 18-109_k141_10810 | Not-determined |
| 18-109_k141_235683 | Not-determined |
| 18-109_k141_194924 | Not-determined |
| 18-109_k141_123134 | Not-determined |
| 18-109_k141_10815 | Not-determined |
| 18-109_k141_154026 | Not-determined |
| 18-109_k141_123136 | Not-determined |
| 18-109_k141_123139 | Not-determined |
| 18-109_k141_154034 | Not-determined |
| 18-109_k141_164132 | Not-determined |
| 18-109_k141_51518 | Not-determined |
| 18-109_k141_215250 | Not-determined |
| 18-109_k141_133361 | Not-determined |
| 18-109_k141_51525 | Not-determined |
| 18-109_k141_164137 | Not-determined |
| 18-109_k141_235684 | Low-quality |
| 18-109_k141_31180 | Not-determined |
| 18-109_k141_801 | Not-determined |
| 18-109_k141_133373 | Low-quality |
| 18-109_k141_154044 | Not-determined |
| 18-109_k141_31184 | Not-determined |
| 18-109_k141_164150 | Not-determined |
| 18-109_k141_204878 | Not-determined |
| 18-109_k141_154050 | Not-determined |
| 18-109_k141_154051 | Not-determined |
| 18-109_k141_184406 | Not-determined |
| 18-109_k141_204888 | Not-determined |
| 18-109_k141_61979 | Not-determined |
| 18-109_k141_815 | Not-determined |
| 18-109_k141_154061 | Not-determined |
| 18-109_k141_61984 | Not-determined |
| 18-109_k141_154064 | Not-determined |
| 18-109_k141_184419 | Not-determined |
| 18-109_k141_184420 | Not-determined |
| 18-109_k141_215266 | Not-determined |
| 18-109_k141_31191 | Not-determined |
| 18-109_k141_215273 | Not-determined |
| 18-109_k141_113177 | Low-quality |
| 18-109_k141_31194 | Not-determined |
| 18-109_k141_154087 | Not-determined |
| 18-109_k141_225441 | Not-determined |
| 18-109_k141_184430 | Low-quality |
| 18-109_k141_31202 | Not-determined |
| 18-109_k141_62004 | Not-determined |
| 18-109_k141_204900 | Not-determined |
| 18-109_k141_62006 | Not-determined |
| 18-109_k141_31205 | Not-determined |
| 18-109_k141_10857 | Not-determined |
| 18-109_k141_133402 | Not-determined |
| 18-109_k141_82282 | Not-determined |
| 18-109_k141_194948 | Not-determined |
| 18-109_k141_133408 | Low-quality |
| 18-109_k141_133413 | Not-determined |
| 18-109_k141_51537 | Low-quality |
| 18-109_k141_235695 | Not-determined |
| 18-109_k141_235696 | Not-determined |
| 18-109_k141_235701 | Not-determined |
| 18-109_k141_51544 | Not-determined |
| 18-109_k141_113189 | Not-determined |
| 18-109_k141_31242 | Not-determined |
| 18-109_k141_154141 | Not-determined |
| 18-109_k141_10894 | Low-quality |
| 18-109_k141_113194 | Not-determined |
| 18-109_k141_31247 | Not-determined |
| 18-109_k141_113197 | Not-determined |
| 18-109_k141_123203 | Not-determined |
| 18-109_k141_31252 | Not-determined |
| 18-109_k141_204934 | Not-determined |
| 18-109_k141_31256 | Not-determined |
| 18-109_k141_31258 | Not-determined |
| 18-109_k141_31259 | Low-quality |
| 18-109_k141_51554 | Not-determined |
| 18-109_k141_194974 | Not-determined |
| 18-109_k141_194976 | Not-determined |
| 18-109_k141_133442 | Not-determined |
| 18-109_k141_113198 | Not-determined |
| 18-109_k141_51557 | Not-determined |
| 18-109_k141_72017 | Not-determined |
| 18-109_k141_72020 | Not-determined |
| 18-109_k141_72022 | Not-determined |
| 18-109_k141_20925 | Low-quality |
| 18-109_k141_72028 | Not-determined |
| 18-109_k141_72031 | Not-determined |
| 18-109_k141_164162 | Not-determined |
| 18-109_k141_51577 | Not-determined |
| 18-109_k141_82349 | Low-quality |
| 18-109_k141_215308 | Not-determined |
| 18-109_k141_862 | Not-determined |
| 18-109_k141_235723 | Low-quality |
| 18-109_k141_72040 | Not-determined |
| 18-109_k141_235730 | Not-determined |
| 18-109_k141_874 | Not-determined |
| 18-109_k141_215324 | Not-determined |
| 18-109_k141_174296 | Not-determined |
| 18-109_k141_174300 | Low-quality |
| 18-109_k141_41355 | Not-determined |
| 18-109_k141_82356 | Not-determined |
| 18-109_k141_41360 | Low-quality |
| 18-109_k141_72058 | Not-determined |
| 18-109_k141_215343 | Not-determined |
| 18-109_k141_51593 | Not-determined |
| 18-109_k141_123233 | Not-determined |
| 18-109_k141_164181 | Not-determined |
| 18-109_k141_82364 | Not-determined |
| 18-109_k141_41370 | Not-determined |
| 18-109_k141_195004 | Low-quality |
| 18-109_k141_41376 | Not-determined |
| 18-109_k141_92518 | Not-determined |
| 18-109_k141_82387 | Not-determined |
| 18-109_k141_31271 | Not-determined |
| 18-109_k141_225465 | Not-determined |
| 18-109_k141_92529 | Not-determined |
| 18-109_k141_154159 | Not-determined |
| 18-109_k141_51615 | Not-determined |
| 18-109_k141_31281 | Not-determined |
| 18-109_k141_20942 | Not-determined |
| 18-109_k141_225477 | Not-determined |
| 18-109_k141_215367 | Low-quality |
| 18-109_k141_164187 | Low-quality |
| 18-109_k141_113231 | Not-determined |
| 18-109_k141_62058 | Not-determined |
| 18-109_k141_62061 | Not-determined |
| 18-109_k141_10918 | Not-determined |
| 18-109_k141_215370 | Not-determined |
| 18-109_k141_92533 | Not-determined |
| 18-109_k141_164192 | Not-determined |
| 18-109_k141_195029 | Not-determined |
| 18-109_k141_92542 | Low-quality |
| 18-109_k141_92543 | Not-determined |
| 18-109_k141_113241 | Not-determined |
| 18-109_k141_51634 | Not-determined |
| 18-109_k141_184503 | Not-determined |
| 18-109_k141_184516 | Low-quality |
| 18-109_k141_41405 | Not-determined |
| 18-109_k141_51653 | Not-determined |
| 18-109_k141_195041 | Not-determined |
| 18-109_k141_51666 | Not-determined |
| 18-109_k141_164224 | Not-determined |
| 18-109_k141_164225 | Low-quality |
| 18-109_k141_20952 | Not-determined |
| 18-109_k141_133488 | Not-determined |
| 18-109_k141_204971 | Not-determined |
| 18-109_k141_20958 | Not-determined |
| 18-109_k141_10934 | Not-determined |
| 18-109_k141_195072 | Not-determined |
| 18-109_k141_10945 | Not-determined |
| 18-109_k141_10948 | Not-determined |
| 18-109_k141_184542 | Not-determined |
| 18-109_k141_133502 | Low-quality |
| 18-109_k141_184546 | Not-determined |
| 18-109_k141_184550 | Not-determined |
| 18-109_k141_225499 | Low-quality |
| 18-109_k141_92551 | Not-determined |
| 18-109_k141_133508 | Not-determined |
| 18-109_k141_154185 | Not-determined |
| 18-109_k141_113259 | Not-determined |
| 18-109_k141_62079 | Low-quality |
| 18-109_k141_62081 | Not-determined |
| 18-109_k141_184567 | Not-determined |
| 18-109_k141_123307 | Low-quality |
| 18-109_k141_113281 | Low-quality |
| 18-109_k141_113282 | Not-determined |
| 18-109_k141_62092 | Not-determined |
| 18-109_k141_92569 | Not-determined |
| 18-109_k141_51680 | Not-determined |
| 18-109_k141_92572 | Not-determined |
| 18-109_k141_41465 | Not-determined |
| 18-109_k141_215378 | Not-determined |
| 18-109_k141_154219 | Not-determined |
| 18-109_k141_113290 | Not-determined |
| 18-109_k141_31368 | Not-determined |
| 18-109_k141_154225 | Not-determined |
| 18-109_k141_62109 | Not-determined |
| 18-109_k141_154226 | Not-determined |
| 18-109_k141_31370 | Not-determined |
| 18-109_k141_123320 | Not-determined |
| 18-109_k141_164259 | Not-determined |
| 18-109_k141_164260 | Not-determined |
| 18-109_k141_31378 | Not-determined |
| 18-109_k141_21019 | Not-determined |
| 18-109_k141_10982 | Not-determined |
| 18-109_k141_31379 | Not-determined |
| 18-109_k141_72066 | Not-determined |
| 18-109_k141_133568 | Not-determined |
| 18-109_k141_184593 | Low-quality |
| 18-109_k141_21026 | Not-determined |
| 18-109_k141_164272 | Low-quality |
| 18-109_k141_215401 | Not-determined |
| 18-109_k141_133572 | Not-determined |
| 18-109_k141_10987 | Not-determined |
| 18-109_k141_184599 | Not-determined |
| 18-109_k141_10988 | Not-determined |
| 18-109_k141_21030 | Not-determined |
| 18-109_k141_10992 | Not-determined |
| 18-109_k141_123325 | Not-determined |
| 18-109_k141_51716 | Not-determined |
| 18-109_k141_92620 | Not-determined |
| 18-109_k141_31403 | Not-determined |
| 18-109_k141_51719 | Not-determined |
| 18-109_k141_205038 | Not-determined |
| 18-109_k141_41491 | Not-determined |
| 18-109_k141_225513 | Not-determined |
| 18-109_k141_205051 | Not-determined |
| 18-109_k141_154241 | Not-determined |
| 18-109_k141_31412 | Not-determined |
| 18-109_k141_31414 | Not-determined |
| 18-109_k141_92640 | Low-quality |
| 18-109_k141_225521 | Not-determined |
| 18-109_k141_11012 | Low-quality |
| 18-109_k141_154260 | Not-determined |
| 18-109_k141_164295 | Not-determined |
| 18-109_k141_41508 | Not-determined |
| 18-109_k141_235737 | Not-determined |
| 18-109_k141_154267 | Not-determined |
| 18-109_k141_154268 | Not-determined |
| 18-109_k141_51747 | Not-determined |
| 18-109_k141_184624 | Not-determined |
| 18-109_k141_11030 | Not-determined |
| 18-109_k141_225527 | Not-determined |
| 18-109_k141_215410 | Not-determined |
| 18-109_k141_215413 | Not-determined |
| 18-109_k141_164319 | Not-determined |
| 18-109_k141_235752 | Not-determined |
| 18-109_k141_235760 | Not-determined |
| 18-109_k141_31447 | Low-quality |
| 18-109_k141_41525 | Not-determined |
| 18-109_k141_205059 | Not-determined |
| 18-109_k141_123348 | Not-determined |
| 18-109_k141_51773 | Not-determined |
| 18-109_k141_164335 | Not-determined |
| 18-109_k141_184634 | Not-determined |
| 18-109_k141_113342 | Not-determined |
| 18-109_k141_235783 | Not-determined |
| 18-109_k141_11054 | Not-determined |
| 18-109_k141_215434 | Not-determined |
| 18-109_k141_215435 | Not-determined |
| 18-109_k141_235787 | Not-determined |
| 18-109_k141_195138 | Not-determined |
| 18-109_k141_184654 | Not-determined |
| 18-109_k141_235796 | Not-determined |
| 18-109_k141_113354 | Not-determined |
| 18-109_k141_113357 | Not-determined |
| 18-109_k141_215452 | Not-determined |
| 18-109_k141_205080 | Not-determined |
| 18-109_k141_31477 | Low-quality |
| 18-109_k141_205083 | Not-determined |
| 18-109_k141_41545 | Not-determined |
| 18-109_k141_184657 | Low-quality |
| 18-109_k141_31481 | Not-determined |
| 18-109_k141_195145 | Not-determined |
| 18-109_k141_51816 | Not-determined |
| 18-109_k141_215462 | Not-determined |
| 18-109_k141_21047 | Not-determined |
| 18-109_k141_215473 | Not-determined |
| 18-109_k141_21050 | Not-determined |
| 18-109_k141_31511 | Not-determined |
| 18-109_k141_11075 | Not-determined |
| 18-109_k141_21057 | Not-determined |
| 18-109_k141_21058 | Not-determined |
| 18-109_k141_92679 | Not-determined |
| 18-109_k141_133650 | Not-determined |
| 18-109_k141_31518 | Not-determined |
| 18-109_k141_235807 | Low-quality |
| 18-109_k141_154304 | Not-determined |
| 18-109_k141_215487 | Not-determined |
| 18-109_k141_154305 | Not-determined |
| 18-109_k141_92687 | Not-determined |
| 18-109_k141_31526 | Not-determined |
| 18-109_k141_11088 | Low-quality |
| 18-109_k141_82421 | Not-determined |
| 18-109_k141_154312 | Not-determined |
| 18-109_k141_154317 | Low-quality |
| 18-109_k141_31535 | Not-determined |
| 18-109_k141_225558 | Not-determined |
| 18-109_k141_184672 | Not-determined |
| 18-109_k141_92697 | Low-quality |
| 18-109_k141_41567 | Not-determined |
| 18-109_k141_164375 | Not-determined |
| 18-109_k141_51845 | Not-determined |
| 18-109_k141_184686 | Not-determined |
| 18-109_k141_184692 | Not-determined |
| 18-109_k141_82445 | Not-determined |
| 18-109_k141_82450 | Not-determined |
| 18-109_k141_82452 | Not-determined |
| 18-109_k141_31541 | Not-determined |
| 18-109_k141_225584 | Not-determined |
| 18-109_k141_31545 | Not-determined |
| 18-109_k141_21080 | Not-determined |
| 18-109_k141_225585 | Not-determined |
| 18-109_k141_41575 | Not-determined |
| 18-109_k141_174331 | Not-determined |
| 18-109_k141_164390 | Not-determined |
| 18-109_k141_51856 | Low-quality |
| 18-109_k141_123382 | Not-determined |
| 18-109_k141_184701 | Not-determined |
| 18-109_k141_225592 | Not-determined |
| 18-109_k141_215515 | Not-determined |
| 18-109_k141_235840 | Low-quality |
| 18-109_k141_225596 | Not-determined |
| 18-109_k141_154351 | Low-quality |
| 18-109_k141_184709 | Low-quality |
| 18-109_k141_154357 | Not-determined |
| 18-109_k141_154359 | Not-determined |
| 18-109_k141_11094 | Not-determined |
| 18-109_k141_154365 | Not-determined |
| 18-109_k141_184713 | Not-determined |
| 18-109_k141_123390 | Not-determined |
| 18-109_k141_225611 | Not-determined |
| 18-109_k141_123393 | Not-determined |
| 18-109_k141_235855 | Not-determined |
| 18-109_k141_225613 | Low-quality |
| 18-109_k141_184718 | Not-determined |
| 18-109_k141_154373 | Not-determined |
| 18-109_k141_133673 | Not-determined |
| 18-109_k141_235866 | Not-determined |
| 18-109_k141_164409 | Low-quality |
| 18-109_k141_133686 | Not-determined |
| 18-109_k141_102740 | Not-determined |
| 18-109_k141_164411 | Not-determined |
| 18-109_k141_225618 | Not-determined |
| 18-109_k141_82481 | Not-determined |
| 18-109_k141_102741 | Not-determined |
| 18-109_k141_174341 | Not-determined |
| 18-109_k141_164413 | Not-determined |
| 18-109_k141_133687 | Low-quality |
| 18-109_k141_123409 | Not-determined |
| 18-109_k141_235882 | Not-determined |
| 18-109_k141_174351 | Not-determined |
| 18-109_k141_143595 | Low-quality |
| 18-109_k141_184744 | Not-determined |
| 18-109_k141_154379 | Low-quality |
| 18-109_k141_235891 | Not-determined |
| 18-109_k141_143602 | Not-determined |
| 18-109_k141_887 | Not-determined |
| 18-109_k141_154385 | Not-determined |

| | |
|---|---|
| 18-109_k141_205155 | Not-determined |
| 18-109_k141_11116 | Low-quality |
| 18-109_k141_11117 | Not-determined |
| 18-109_k141_133703 | Not-determined |
| 18-109_k141_215534 | Not-determined |
| 18-109_k141_893 | Not-determined |
| 18-109_k141_133708 | Not-determined |
| 18-109_k141_184760 | Not-determined |
| 18-109_k141_225634 | Not-determined |
| 18-109_k141_215538 | Low-quality |
| 18-109_k141_123427 | Not-determined |
| 18-109_k141_102760 | Not-determined |
| 18-109_k141_184762 | Not-determined |
| 18-109_k141_51914 | Not-determined |
| 18-109_k141_225638 | Not-determined |
| 18-109_k141_82497 | Not-determined |
| 18-109_k141_184769 | Not-determined |
| 18-109_k141_113369 | Not-determined |
| 18-109_k141_205175 | Not-determined |
| 18-109_k141_82504 | Low-quality |
| 18-109_k141_123440 | Not-determined |
| 18-109_k141_133720 | Not-determined |
| 18-109_k141_11123 | Not-determined |
| 18-109_k141_904 | Not-determined |
| 18-109_k141_133722 | Not-determined |
| 18-109_k141_225645 | Not-determined |
| 18-109_k141_102771 | Not-determined |
| 18-109_k141_205176 | Not-determined |
| 18-109_k141_11133 | Low-quality |
| 18-109_k141_919 | Not-determined |
| 18-109_k141_184796 | Not-determined |
| 18-109_k141_154409 | Low-quality |
| 18-109_k141_154410 | Not-determined |
| 18-109_k141_174362 | Not-determined |
| 18-109_k141_174363 | Not-determined |
| 18-109_k141_940 | Not-determined |
| 18-109_k141_184809 | Not-determined |
| 18-109_k141_133752 | Not-determined |
| 18-109_k141_215567 | Not-determined |
| 18-109_k141_154421 | Not-determined |
| 18-109_k141_31575 | Not-determined |
| 18-109_k141_143648 | Not-determined |
| 18-109_k141_143652 | Not-determined |
| 18-109_k141_143653 | Not-determined |
| 18-109_k141_143655 | Not-determined |
| 18-109_k141_154429 | Low-quality |
| 18-109_k141_154431 | Low-quality |
| 18-109_k141_164453 | Not-determined |
| 18-109_k141_164454 | Not-determined |
| 18-109_k141_184822 | Not-determined |
| 18-109_k141_123497 | Low-quality |
| 18-109_k141_31590 | Not-determined |
| 18-109_k141_113390 | Low-quality |
| 18-109_k141_133755 | Not-determined |
| 18-109_k141_133759 | Not-determined |
| 18-109_k141_164464 | Not-determined |
| 18-109_k141_954 | Not-determined |
| 18-109_k141_154440 | Not-determined |
| 18-109_k141_174398 | Low-quality |
| 18-109_k141_11166 | Not-determined |
| 18-109_k141_133762 | Not-determined |
| 18-109_k141_962 | Not-determined |
| 18-109_k141_102798 | Not-determined |
| 18-109_k141_174413 | Not-determined |
| 18-109_k141_102803 | Not-determined |
| 18-109_k141_143690 | Not-determined |
| 18-109_k141_205230 | Not-determined |
| 18-109_k141_133766 | Not-determined |
| 18-109_k141_215584 | Not-determined |
| 18-109_k141_31602 | Not-determined |
| 18-109_k141_133778 | Not-determined |
| 18-109_k141_113437 | Low-quality |
| 18-109_k141_133780 | Not-determined |
| 18-109_k141_205247 | Not-determined |
| 18-109_k141_143693 | Not-determined |
| 18-109_k141_174433 | Low-quality |
| 18-109_k141_143699 | Not-determined |
| 18-109_k141_184878 | Not-determined |
| 18-109_k141_205263 | Not-determined |
| 18-109_k141_1012 | Not-determined |
| 18-109_k141_41621 | Not-determined |
| 18-109_k141_133792 | Not-determined |
| 18-109_k141_174442 | Low-quality |
| 18-109_k141_215612 | Not-determined |
| 18-109_k141_205271 | Not-determined |
| 18-109_k141_143704 | Not-determined |
| 18-109_k141_215613 | Not-determined |
| 18-109_k141_174444 | Not-determined |
| 18-109_k141_215616 | Low-quality |
| 18-109_k141_41627 | Not-determined |
| 18-109_k141_195265 | Not-determined |
| 18-109_k141_164477 | Not-determined |
| 18-109_k141_184893 | Not-determined |
| 18-109_k141_205278 | Not-determined |
| 18-109_k141_11207 | Not-determined |
| 18-109_k141_11215 | Not-determined |
| 18-109_k141_11218 | Not-determined |
| 18-109_k141_164490 | Low-quality |
| 18-109_k141_31622 | Not-determined |
| 18-109_k141_164493 | Low-quality |
| 18-109_k141_215627 | Not-determined |
| 18-109_k141_143724 | Low-quality |
| 18-109_k141_215631 | Not-determined |
| 18-109_k141_82531 | Not-determined |
| 18-109_k141_31626 | Low-quality |
| 18-109_k141_1016 | Not-determined |
| 18-109_k141_205282 | Low-quality |
| 18-109_k141_195282 | Not-determined |
| 18-109_k141_215643 | Not-determined |
| 18-109_k141_82541 | Not-determined |
| 18-109_k141_205284 | Low-quality |
| 18-109_k141_21092 | Not-determined |
| 18-109_k141_21096 | Not-determined |
| 18-109_k141_1025 | Not-determined |
| 18-109_k141_82550 | Not-determined |
| 18-109_k141_1026 | Not-determined |
| 18-109_k141_102827 | Not-determined |
| 18-109_k141_31635 | Not-determined |
| 18-109_k141_41641 | Not-determined |
| 18-109_k141_143734 | Not-determined |
| 18-109_k141_11236 | Not-determined |
| 18-109_k141_41644 | Not-determined |
| 18-109_k141_205298 | Not-determined |
| 18-109_k141_31644 | Not-determined |
| 18-109_k141_21113 | Not-determined |
| 18-109_k141_174463 | Not-determined |
| 18-109_k141_215666 | Not-determined |
| 18-109_k141_215667 | Not-determined |
| 18-109_k141_41655 | Not-determined |
| 18-109_k141_62121 | Not-determined |
| 18-109_k141_143751 | Not-determined |
| 18-109_k141_51994 | Not-determined |
| 18-109_k141_41667 | Not-determined |
| 18-109_k141_82575 | Not-determined |
| 18-109_k141_41668 | Not-determined |
| 18-109_k141_82577 | Not-determined |
| 18-109_k141_184938 | Not-determined |
| 18-109_k141_184940 | Not-determined |
| 18-109_k141_41674 | Low-quality |
| 18-109_k141_62149 | Not-determined |
| 18-109_k141_62151 | Not-determined |
| 18-109_k141_11245 | Low-quality |
| 18-109_k141_205322 | Not-determined |
| 18-109_k141_82600 | Low-quality |
| 18-109_k141_11250 | Not-determined |
| 18-109_k141_41682 | Not-determined |
| 18-109_k141_52006 | Not-determined |
| 18-109_k141_52009 | Not-determined |
| 18-109_k141_11257 | Not-determined |
| 18-109_k141_11265 | Not-determined |
| 18-109_k141_215683 | Not-determined |
| 18-109_k141_205337 | Not-determined |
| 18-109_k141_225727 | Not-determined |
| 18-109_k141_62168 | Not-determined |
| 18-109_k141_62169 | Not-determined |
| 18-109_k141_62170 | Not-determined |
| 18-109_k141_62171 | Not-determined |
| 18-109_k141_205339 | Not-determined |
| 18-109_k141_133834 | Not-determined |
| 18-109_k141_52015 | Not-determined |
| 18-109_k141_11267 | Not-determined |
| 18-109_k141_62177 | Not-determined |
| 18-109_k141_205347 | Not-determined |
| 18-109_k141_195338 | Not-determined |
| 18-109_k141_113506 | Not-determined |
| 18-109_k141_205353 | Not-determined |
| 18-109_k141_123539 | Not-determined |
| 18-109_k141_215701 | Not-determined |
| 18-109_k141_113509 | Not-determined |
| 18-109_k141_113510 | Not-determined |
| 18-109_k141_52040 | Not-determined |
| 18-109_k141_205364 | Not-determined |
| 18-109_k141_225751 | Low-quality |
| 18-109_k141_225755 | Not-determined |
| 18-109_k141_102856 | Not-determined |
| 18-109_k141_21124 | Not-determined |
| 18-109_k141_154489 | Not-determined |
| 18-109_k141_154494 | Not-determined |
| 18-109_k141_113512 | Not-determined |
| 18-109_k141_123559 | Not-determined |
| 18-109_k141_195355 | Not-determined |
| 18-109_k141_184966 | Not-determined |
| 18-109_k141_21136 | Not-determined |
| 18-109_k141_195356 | Not-determined |
| 18-109_k141_184968 | Not-determined |
| 18-109_k141_123570 | Not-determined |
| 18-109_k141_113520 | Not-determined |
| 18-109_k141_123575 | Low-quality |
| 18-109_k141_52080 | Not-determined |
| 18-109_k141_31690 | Not-determined |
| 18-109_k141_154512 | Not-determined |
| 18-109_k141_184975 | Not-determined |
| 18-109_k141_235970 | Low-quality |
| 18-109_k141_41710 | Not-determined |
| 18-109_k141_82668 | Not-determined |
| 18-109_k141_174526 | Low-quality |
| 18-109_k141_195366 | Not-determined |
| 18-109_k141_41712 | Low-quality |
| 18-109_k141_195370 | Not-determined |
| 18-109_k141_235979 | Not-determined |
| 18-109_k141_52102 | Not-determined |
| 18-109_k141_133864 | Not-determined |
| 18-109_k141_52106 | Not-determined |
| 18-109_k141_52107 | Not-determined |
| 18-109_k141_102894 | Not-determined |
| 18-109_k141_164505 | Low-quality |
| 18-109_k141_184986 | Low-quality |
| 18-109_k141_133870 | Not-determined |
| 18-109_k141_82686 | Not-determined |
| 18-109_k141_82687 | Not-determined |
| 18-109_k141_154516 | Not-determined |
| 18-109_k141_11305 | Low-quality |
| 18-109_k141_225780 | Not-determined |
| 18-109_k141_21154 | Not-determined |
| 18-109_k141_102904 | Not-determined |
| 18-109_k141_195377 | Not-determined |
| 18-109_k141_82696 | Not-determined |
| 18-109_k141_21168 | Not-determined |
| 18-109_k141_235998 | Not-determined |
| 18-109_k141_102918 | Not-determined |
| 18-109_k141_113547 | Not-determined |
| 18-109_k141_123595 | Not-determined |
| 18-109_k141_82701 | Low-quality |
| 18-109_k141_236008 | Not-determined |
| 18-109_k141_164525 | Not-determined |
| 18-109_k141_82705 | Not-determined |
| 18-109_k141_31726 | Not-determined |
| 18-109_k141_11316 | Low-quality |
| 18-109_k141_82711 | Not-determined |
| 18-109_k141_174575 | Not-determined |
| 18-109_k141_174577 | Not-determined |
| 18-109_k141_31732 | Not-determined |
| 18-109_k141_11320 | Low-quality |
| 18-109_k141_164538 | Not-determined |
| 18-109_k141_113564 | Not-determined |
| 18-109_k141_31734 | Not-determined |
| 18-109_k141_164540 | Not-determined |
| 18-109_k141_123604 | Not-determined |
| 18-109_k141_123605 | Not-determined |
| 18-109_k141_164541 | Not-determined |
| 18-109_k141_113570 | Not-determined |
| 18-109_k141_164544 | Not-determined |
| 18-109_k141_174589 | Not-determined |
| 18-109_k141_174590 | Not-determined |
| 18-109_k141_174591 | Not-determined |
| 18-109_k141_205437 | Not-determined |
| 18-109_k141_174597 | Not-determined |
| 18-109_k141_205445 | Not-determined |
| 18-109_k141_123618 | Not-determined |
| 18-109_k141_11336 | Not-determined |
| 18-109_k141_31759 | Not-determined |
| 18-109_k141_113588 | Not-determined |
| 18-109_k141_195409 | Not-determined |
| 18-109_k141_236038 | Not-determined |
| 18-109_k141_102951 | Low-quality |
| 18-109_k141_123633 | Not-determined |
| 18-109_k141_123637 | Not-determined |
| 18-109_k141_225807 | Not-determined |
| 18-109_k141_82721 | Not-determined |
| 18-109_k141_225810 | Not-determined |
| 18-109_k141_236044 | Not-determined |
| 18-109_k141_225817 | Not-determined |
| 18-109_k141_205482 | Low-quality |
| 18-109_k141_102959 | Not-determined |
| 18-109_k141_133905 | Not-determined |
| 18-109_k141_195428 | Not-determined |
| 18-109_k141_143848 | Not-determined |
| 18-109_k141_82759 | Not-determined |
| 18-109_k141_205504 | Not-determined |
| 18-109_k141_82761 | Not-determined |
| 18-109_k141_164582 | Not-determined |
| 18-109_k141_82762 | Not-determined |
| 18-109_k141_174627 | Not-determined |
| 18-109_k141_164583 | Not-determined |
| 18-109_k141_102975 | Not-determined |
| 18-109_k141_113616 | Not-determined |
| 18-109_k141_174633 | Not-determined |
| 18-109_k141_41717 | Low-quality |
| 18-109_k141_21217 | Not-determined |
| 18-109_k141_164607 | Low-quality |
| 18-109_k141_184999 | Not-determined |
| 18-109_k141_21224 | Not-determined |
| 18-109_k141_31778 | Not-determined |
| 18-109_k141_205527 | Not-determined |
| 18-109_k141_164613 | Low-quality |
| 18-109_k141_205531 | Not-determined |
| 18-109_k141_225859 | Not-determined |
| 18-109_k141_164618 | Low-quality |
| 18-109_k141_41732 | Not-determined |
| 18-109_k141_205538 | Not-determined |
| 18-109_k141_82775 | Not-determined |
| 18-109_k141_123658 | Not-determined |
| 18-109_k141_123659 | Not-determined |
| 18-109_k141_31791 | Not-determined |
| 18-109_k141_123662 | Not-determined |
| 18-109_k141_21229 | Not-determined |
| 18-109_k141_41751 | Not-determined |
| 18-109_k141_133926 | Not-determined |
| 18-109_k141_82777 | Low-quality |
| 18-109_k141_133928 | Not-determined |
| 18-109_k141_215729 | Not-determined |
| 18-109_k141_41753 | Not-determined |
| 18-109_k141_215730 | Low-quality |
| 18-109_k141_225870 | Low-quality |
| 18-109_k141_205548 | Not-determined |
| 18-109_k141_52151 | Not-determined |
| 18-109_k141_164634 | Not-determined |
| 18-109_k141_164639 | Not-determined |
| 18-109_k141_103017 | Not-determined |
| 18-109_k141_215742 | Not-determined |
| 18-109_k141_123692 | Not-determined |
| 18-109_k141_143856 | Not-determined |
| 18-109_k141_225890 | Not-determined |
| 18-109_k141_103021 | Not-determined |
| 18-109_k141_103024 | Not-determined |
| 18-109_k141_21239 | Not-determined |
| 18-109_k141_103027 | Low-quality |
| 18-109_k141_62227 | Not-determined |
| 18-109_k141_103030 | Not-determined |
| 18-109_k141_133948 | Not-determined |
| 18-109_k141_1087 | Not-determined |
| 18-109_k141_113631 | Not-determined |
| 18-109_k141_154529 | Not-determined |
| 18-109_k141_123693 | Not-determined |
| 18-109_k141_82789 | Not-determined |
| 18-109_k141_205577 | Not-determined |
| 18-109_k141_154533 | Not-determined |
| 18-109_k141_41774 | Not-determined |
| 18-109_k141_1095 | Not-determined |
| 18-109_k141_72090 | Not-determined |
| 18-109_k141_31819 | Low-quality |
| 18-109_k141_225903 | Not-determined |
| 18-109_k141_123702 | Not-determined |
| 18-109_k141_1099 | Low-quality |
| 18-109_k141_195463 | Not-determined |
| 18-109_k141_82805 | Not-determined |
| 18-109_k141_174676 | Not-determined |
| 18-109_k141_62252 | Not-determined |
| 18-109_k141_195469 | Not-determined |
| 18-109_k141_174680 | Low-quality |
| 18-109_k141_31827 | Not-determined |
| 18-109_k141_154544 | Not-determined |
| 18-109_k141_174688 | Not-determined |
| 18-109_k141_164672 | Not-determined |
| 18-109_k141_174690 | Not-determined |
| 18-109_k141_154546 | Not-determined |
| 18-109_k141_195472 | Low-quality |
| 18-109_k141_31832 | Not-determined |
| 18-109_k141_225908 | Not-determined |
| 18-109_k141_143895 | Not-determined |
| 18-109_k141_143897 | Not-determined |
| 18-109_k141_174695 | Not-determined |
| 18-109_k141_143898 | Not-determined |
| 18-109_k141_11339 | Not-determined |
| 18-109_k141_185028 | Not-determined |
| 18-109_k141_72098 | Not-determined |
| 18-109_k141_41818 | Not-determined |
| 18-109_k141_103055 | Not-determined |
| 18-109_k141_11341 | Low-quality |
| 18-109_k141_154572 | Not-determined |
| 18-109_k141_195486 | Not-determined |
| 18-109_k141_143905 | Low-quality |
| 18-109_k141_1114 | Not-determined |
| 18-109_k141_113661 | Low-quality |
| 18-109_k141_103066 | Not-determined |
| 18-109_k141_113664 | Not-determined |
| 18-109_k141_103067 | Low-quality |
| 18-109_k141_225920 | Not-determined |
| 18-109_k141_154583 | Not-determined |
| 18-109_k141_195492 | Not-determined |
| 18-109_k141_225924 | Not-determined |
| 18-109_k141_164689 | Not-determined |
| 18-109_k141_21260 | Not-determined |
| 18-109_k141_11347 | Not-determined |
| 18-109_k141_143915 | Not-determined |
| 18-109_k141_164692 | Not-determined |
| 18-109_k141_154584 | Not-determined |
| 18-109_k141_195498 | Not-determined |
| 18-109_k141_103073 | Not-determined |
| 18-109_k141_164699 | Not-determined |
| 18-109_k141_92768 | Not-determined |
| 18-109_k141_143924 | Not-determined |
| 18-109_k141_1137 | Not-determined |
| 18-109_k141_82822 | Not-determined |
| 18-109_k141_41848 | Low-quality |
| 18-109_k141_133981 | Not-determined |
| 18-109_k141_133982 | Not-determined |
| 18-109_k141_11368 | Not-determined |
| 18-109_k141_185048 | Not-determined |
| 18-109_k141_195513 | Not-determined |
| 18-109_k141_11372 | Not-determined |
| 18-109_k141_174712 | Not-determined |
| 18-109_k141_225948 | Not-determined |
| 18-109_k141_225950 | Not-determined |
| 18-109_k141_92783 | Not-determined |
| 18-109_k141_123734 | Not-determined |
| 18-109_k141_236054 | Not-determined |
| 18-109_k141_11381 | Not-determined |
| 18-109_k141_21272 | Not-determined |
| 18-109_k141_236056 | Not-determined |
| 18-109_k141_41861 | Not-determined |
| 18-109_k141_236057 | Not-determined |
| 18-109_k141_103107 | Not-determined |
| 18-109_k141_195536 | Not-determined |
| 18-109_k141_103109 | Not-determined |
| 18-109_k141_11383 | Not-determined |
| 18-109_k141_11385 | Low-quality |
| 18-109_k141_103113 | Not-determined |
| 18-109_k141_195540 | Not-determined |
| 18-109_k141_195545 | Not-determined |
| 18-109_k141_1169 | Not-determined |
| 18-109_k141_31842 | Not-determined |
| 18-109_k141_62294 | Not-determined |
| 18-109_k141_31847 | Not-determined |
| 18-109_k141_1178 | Not-determined |
| 18-109_k141_215758 | Not-determined |
| 18-109_k141_41882 | Low-quality |
| 18-109_k141_123749 | Not-determined |
| 18-109_k141_236091 | Not-determined |
| 18-109_k141_205636 | Low-quality |
| 18-109_k141_1181 | Not-determined |
| 18-109_k141_215762 | Not-determined |
| 18-109_k141_11397 | Not-determined |
| 18-109_k141_215765 | Not-determined |
| 18-109_k141_236093 | Not-determined |
| 18-109_k141_11399 | Not-determined |
| 18-109_k141_134002 | Not-determined |
| 18-109_k141_123753 | Not-determined |
| 18-109_k141_185058 | Not-determined |
| 18-109_k141_215775 | Not-determined |
| 18-109_k141_21287 | Not-determined |
| 18-109_k141_205643 | Not-determined |
| 18-109_k141_185067 | Not-determined |
| 18-109_k141_205650 | Not-determined |
| 18-109_k141_52195 | Not-determined |
| 18-109_k141_103123 | Not-determined |
| 18-109_k141_31870 | Not-determined |
| 18-109_k141_103126 | Not-determined |
| 18-109_k141_195568 | Not-determined |
| 18-109_k141_31877 | Low-quality |
| 18-109_k141_236120 | Not-determined |
| 18-109_k141_236121 | Not-determined |
| 18-109_k141_236125 | Not-determined |
| 18-109_k141_236126 | Not-determined |
| 18-109_k141_174739 | Not-determined |
| 18-109_k141_154595 | Low-quality |
| 18-109_k141_174745 | Not-determined |
| 18-109_k141_205665 | Not-determined |
| 18-109_k141_21314 | Not-determined |
| 18-109_k141_103157 | Not-determined |
| 18-109_k141_215801 | Not-determined |
| 18-109_k141_103160 | Not-determined |
| 18-109_k141_123766 | Not-determined |
| 18-109_k141_72134 | Not-determined |
| 18-109_k141_123769 | Not-determined |
| 18-109_k141_174757 | Not-determined |
| 18-109_k141_123771 | Not-determined |
| 18-109_k141_195593 | Low-quality |
| 18-109_k141_195599 | Not-determined |
| 18-109_k141_195600 | Not-determined |
| 18-109_k141_52206 | Not-determined |
| 18-109_k141_174766 | Not-determined |
| 18-109_k141_205680 | Not-determined |
| 18-109_k141_174767 | Not-determined |
| 18-109_k141_195602 | Not-determined |
| 18-109_k141_103177 | Not-determined |
| 18-109_k141_52217 | Not-determined |
| 18-109_k141_174769 | Not-determined |
| 18-109_k141_21343 | Not-determined |
| 18-109_k141_103181 | Not-determined |
| 18-109_k141_164723 | Not-determined |
| 18-109_k141_225981 | Not-determined |
| 18-109_k141_225982 | Not-determined |
| 18-109_k141_103189 | Not-determined |
| 18-109_k141_113680 | Low-quality |
| 18-109_k141_174784 | Not-determined |
| 18-109_k141_134067 | Not-determined |
| 18-109_k141_134069 | Not-determined |
| 18-109_k141_134077 | Low-quality |
| 18-109_k141_205688 | Not-determined |
| 18-109_k141_226002 | Not-determined |
| 18-109_k141_31890 | Low-quality |
| 18-109_k141_174794 | Not-determined |
| 18-109_k141_185108 | Not-determined |
| 18-109_k141_31894 | Not-determined |
| 18-109_k141_52245 | Not-determined |
| 18-109_k141_185113 | Not-determined |
| 18-109_k141_164738 | Not-determined |
| 18-109_k141_1223 | Not-determined |
| 18-109_k141_205709 | Not-determined |
| 18-109_k141_52257 | Not-determined |
| 18-109_k141_195653 | Not-determined |
| 18-109_k141_103210 | Low-quality |
| 18-109_k141_103211 | Not-determined |
| 18-109_k141_52262 | Not-determined |
| 18-109_k141_174812 | Not-determined |
| 18-109_k141_154626 | Not-determined |
| 18-109_k141_185130 | Not-determined |
| 18-109_k141_123785 | Not-determined |
| 18-109_k141_11435 | Not-determined |
| 18-109_k141_82849 | Not-determined |
| 18-109_k141_123788 | Not-determined |
| 18-109_k141_154636 | Not-determined |
| 18-109_k141_82852 | Not-determined |
| 18-109_k141_123791 | Not-determined |
| 18-109_k141_185133 | Not-determined |
| 18-109_k141_185135 | Not-determined |
| 18-109_k141_52279 | Not-determined |
| 18-109_k141_205722 | Not-determined |
| 18-109_k141_31932 | Not-determined |
| 18-109_k141_185141 | Not-determined |
| 18-109_k141_185142 | Not-determined |
| 18-109_k141_52281 | Not-determined |
| 18-109_k141_134097 | Not-determined |
| 18-109_k141_205726 | Not-determined |
| 18-109_k141_62384 | Not-determined |
| 18-109_k141_82864 | Not-determined |
| 18-109_k141_185150 | Not-determined |
| 18-109_k141_62389 | Not-determined |
| 18-109_k141_123808 | Not-determined |
| 18-109_k141_31939 | Not-determined |
| 18-109_k141_31941 | Not-determined |
| 18-109_k141_154650 | Not-determined |
| 18-109_k141_82868 | Low-quality |
| 18-109_k141_215819 | Not-determined |
| 18-109_k141_134118 | Not-determined |
| 18-109_k141_72167 | Not-determined |
| 18-109_k141_215822 | Not-determined |
| 18-109_k141_205744 | Not-determined |
| 18-109_k141_1280 | Not-determined |
| 18-109_k141_62401 | Not-determined |
| 18-109_k141_195666 | Not-determined |
| 18-109_k141_143931 | Not-determined |
| 18-109_k141_31952 | Not-determined |
| 18-109_k141_52294 | Not-determined |
| 18-109_k141_236180 | Not-determined |
| 18-109_k141_164748 | Not-determined |
| 18-109_k141_134134 | Not-determined |
| 18-109_k141_52297 | Not-determined |
| 18-109_k141_215831 | Not-determined |
| 18-109_k141_92822 | Not-determined |
| 18-109_k141_143942 | Not-determined |
| 18-109_k141_72176 | Low-quality |
| 18-109_k141_72178 | Not-determined |
| 18-109_k141_72180 | Not-determined |
| 18-109_k141_92834 | Low-quality |
| 18-109_k141_11460 | Not-determined |
| 18-109_k141_92837 | Not-determined |
| 18-109_k141_11464 | Not-determined |
| 18-109_k141_11465 | Low-quality |
| 18-109_k141_92842 | Not-determined |
| 18-109_k141_11469 | Not-determined |
| 18-109_k141_215849 | Not-determined |
| 18-109_k141_72197 | Low-quality |
| 18-109_k141_62433 | Not-determined |
| 18-109_k141_72204 | Not-determined |
| 18-109_k141_185220 | Not-determined |
| 18-109_k141_154669 | Not-determined |
| 18-109_k141_41899 | Not-determined |
| 18-109_k141_143949 | Not-determined |
| 18-109_k141_185232 | Low-quality |
| 18-109_k141_185233 | Not-determined |
| 18-109_k141_185234 | Not-determined |
| 18-109_k141_185237 | Not-determined |
| 18-109_k141_195708 | Not-determined |
| 18-109_k141_154684 | Not-determined |
| 18-109_k141_174846 | Not-determined |
| 18-109_k141_123829 | Not-determined |
| 18-109_k141_103216 | Not-determined |
| 18-109_k141_92852 | Not-determined |
| 18-109_k141_62457 | Not-determined |
| 18-109_k141_62458 | Not-determined |
| 18-109_k141_41920 | Not-determined |
| 18-109_k141_72206 | Not-determined |
| 18-109_k141_143968 | Not-determined |
| 18-109_k141_154690 | Not-determined |
| 18-109_k141_92859 | Not-determined |
| 18-109_k141_72214 | Not-determined |
| 18-109_k141_185254 | Low-quality |
| 18-109_k141_72215 | Not-determined |
| 18-109_k141_164774 | Not-determined |
| 18-109_k141_226033 | Not-determined |
| 18-109_k141_226035 | Not-determined |
| 18-109_k141_174863 | Not-determined |
| 18-109_k141_195737 | Not-determined |
| 18-109_k141_92865 | Not-determined |
| 18-109_k141_113762 | Low-quality |
| 18-109_k141_154701 | Not-determined |
| 18-109_k141_134167 | Low-quality |

| | |
|---|---|
| 18-109_k141_185256 | Low-quality |
| 18-109_k141_123841 | Not-determined |
| 18-109_k141_134171 | Not-determined |
| 18-109_k141_11488 | Not-determined |
| 18-109_k141_185262 | Not-determined |
| 18-109_k141_205779 | Not-determined |
| 18-109_k141_185264 | Low-quality |
| 18-109_k141_185267 | Not-determined |
| 18-109_k141_195748 | Not-determined |
| 18-109_k141_21363 | Not-determined |
| 18-109_k141_21367 | Not-determined |
| 18-109_k141_164780 | Not-determined |
| 18-109_k141_185285 | Not-determined |
| 18-109_k141_185287 | Not-determined |
| 18-109_k141_21369 | Not-determined |
| 18-109_k141_236218 | Not-determined |
| 18-109_k141_195762 | Not-determined |
| 18-109_k141_174885 | Not-determined |
| 18-109_k141_195768 | Not-determined |
| 18-109_k141_62489 | Not-determined |
| 18-109_k141_103231 | Low-quality |
| 18-109_k141_226061 | Not-determined |
| 18-109_k141_195773 | Not-determined |
| 18-109_k141_226067 | Not-determined |
| 18-109_k141_11515 | Not-determined |
| 18-109_k141_195778 | Not-determined |
| 18-109_k141_195780 | Not-determined |
| 18-109_k141_195782 | Not-determined |
| 18-109_k141_11522 | Not-determined |
| 18-109_k141_62494 | Not-determined |
| 18-109_k141_185308 | Low-quality |
| 18-109_k141_226079 | Not-determined |
| 18-109_k141_236235 | Not-determined |
| 18-109_k141_215898 | Low-quality |
| 18-109_k141_11527 | Not-determined |
| 18-109_k141_31976 | Not-determined |
| 18-109_k141_154734 | Not-determined |
| 18-109_k141_11535 | Not-determined |
| 18-109_k141_52340 | Not-determined |
| 18-109_k141_31982 | Not-determined |
| 18-109_k141_11538 | Not-determined |
| 18-109_k141_164815 | Not-determined |
| 18-109_k141_164817 | Not-determined |
| 18-109_k141_1341 | Not-determined |
| 18-109_k141_52348 | Not-determined |
| 18-109_k141_195805 | Not-determined |
| 18-109_k141_1347 | Not-determined |
| 18-109_k141_1349 | Not-determined |
| 18-109_k141_103255 | Low-quality |
| 18-109_k141_1350 | Not-determined |
| 18-109_k141_52350 | Not-determined |
| 18-109_k141_52351 | Not-determined |
| 18-109_k141_123852 | Not-determined |
| 18-109_k141_226100 | Not-determined |
| 18-109_k141_143977 | Not-determined |
| 18-109_k141_215928 | Not-determined |
| 18-109_k141_215929 | Not-determined |
| 18-109_k141_103260 | Not-determined |
| 18-109_k141_226115 | Not-determined |
| 18-109_k141_134198 | Not-determined |
| 18-109_k141_226120 | Low-quality |
| 18-109_k141_134203 | Not-determined |
| 18-109_k141_134205 | Low-quality |
| 18-109_k141_174922 | Not-determined |
| 18-109_k141_154748 | Low-quality |
| 18-109_k141_154752 | Not-determined |
| 18-109_k141_226127 | Low-quality |
| 18-109_k141_143992 | Not-determined |
| 18-109_k141_195839 | Low-quality |
| 18-109_k141_143994 | Not-determined |
| 18-109_k141_62521 | Low-quality |
| 18-109_k141_32001 | Not-determined |
| 18-109_k141_72276 | Not-determined |
| 18-109_k141_215941 | Not-determined |
| 18-109_k141_154768 | Not-determined |
| 18-109_k141_32009 | Not-determined |
| 18-109_k141_144006 | Low-quality |
| 18-109_k141_134217 | Not-determined |
| 18-109_k141_21419 | Not-determined |
| 18-109_k141_32013 | Not-determined |
| 18-109_k141_21426 | Not-determined |
| 18-109_k141_164859 | Not-determined |
| 18-109_k141_103281 | Not-determined |
| 18-109_k141_62540 | Not-determined |
| 18-109_k141_72283 | Not-determined |
| 18-109_k141_226143 | Not-determined |
| 18-109_k141_52360 | Not-determined |
| 18-109_k141_174931 | Not-determined |
| 18-109_k141_154792 | Not-determined |
| 18-109_k141_92903 | Not-determined |
| 18-109_k141_52365 | Not-determined |
| 18-109_k141_92905 | Not-determined |
| 18-109_k141_144036 | Not-determined |
| 18-109_k141_92907 | Not-determined |
| 18-109_k141_21438 | Not-determined |
| 18-109_k141_144045 | Low-quality |
| 18-109_k141_226153 | Not-determined |
| 18-109_k141_52375 | Not-determined |
| 18-109_k141_215969 | Not-determined |
| 18-109_k141_185373 | Not-determined |
| 18-109_k141_41949 | Not-determined |
| 18-109_k141_185374 | Not-determined |
| 18-109_k141_226161 | Not-determined |
| 18-109_k141_72286 | Not-determined |
| 18-109_k141_62553 | Not-determined |
| 18-109_k141_164903 | Not-determined |
| 18-109_k141_52379 | Not-determined |
| 18-109_k141_134264 | Not-determined |
| 18-109_k141_32050 | Not-determined |
| 18-109_k141_215976 | Not-determined |
| 18-109_k141_226169 | Not-determined |
| 18-109_k141_226170 | Not-determined |
| 18-109_k141_236278 | Not-determined |
| 18-109_k141_52392 | Not-determined |
| 18-109_k141_62562 | Not-determined |
| 18-109_k141_103316 | Not-determined |
| 18-109_k141_154815 | Not-determined |
| 18-109_k141_154816 | Not-determined |
| 18-109_k141_41966 | Not-determined |
| 18-109_k141_72298 | Not-determined |
| 18-109_k141_236292 | Not-determined |
| 18-109_k141_103325 | Not-determined |
| 18-109_k141_164916 | Not-determined |
| 18-109_k141_123863 | Not-determined |
| 18-109_k141_113803 | Not-determined |
| 18-109_k141_236300 | Not-determined |
| 18-109_k141_226173 | Low-quality |
| 18-109_k141_134278 | Not-determined |
| 18-109_k141_113812 | Not-determined |
| 18-109_k141_185420 | Not-determined |
| 18-109_k141_123875 | Not-determined |
| 18-109_k141_134289 | Not-determined |
| 18-109_k141_113816 | Not-determined |
| 18-109_k141_226183 | Not-determined |
| 18-109_k141_113823 | Low-quality |
| 18-109_k141_226191 | Not-determined |
| 18-109_k141_154827 | Not-determined |
| 18-109_k141_123878 | Low-quality |
| 18-109_k141_52423 | Not-determined |
| 18-109_k141_62568 | Not-determined |
| 18-109_k141_226202 | Not-determined |
| 18-109_k141_123890 | Not-determined |
| 18-109_k141_226203 | Not-determined |
| 18-109_k141_226207 | Not-determined |
| 18-109_k141_154838 | Not-determined |
| 18-109_k141_103361 | Not-determined |
| 18-109_k141_92911 | Not-determined |
| 18-109_k141_134305 | Not-determined |
| 18-109_k141_103368 | Not-determined |
| 18-109_k141_52436 | Low-quality |
| 18-109_k141_1381 | Not-determined |
| 18-109_k141_174940 | Not-determined |
| 18-109_k141_72319 | Not-determined |
| 18-109_k141_174946 | Not-determined |
| 18-109_k141_103381 | Low-quality |
| 18-109_k141_32103 | Low-quality |
| 18-109_k141_32107 | Low-quality |
| 18-109_k141_32109 | Low-quality |
| 18-109_k141_72334 | Not-determined |
| 18-109_k141_134310 | Not-determined |
| 18-109_k141_226221 | Not-determined |
| 18-109_k141_62610 | Not-determined |
| 18-109_k141_154859 | Not-determined |
| 18-109_k141_1407 | Not-determined |
| 18-109_k141_103408 | Not-determined |
| 18-109_k141_236358 | Not-determined |
| 18-109_k141_103420 | Not-determined |
| 18-109_k141_21451 | Not-determined |
| 18-109_k141_164975 | Not-determined |
| 18-109_k141_32126 | Not-determined |
| 18-109_k141_205860 | Not-determined |
| 18-109_k141_134326 | Low-quality |
| 18-109_k141_226237 | Not-determined |
| 18-109_k141_32133 | Low-quality |
| 18-109_k141_52479 | Not-determined |
| 18-109_k141_144106 | Not-determined |
| 18-109_k141_52491 | Not-determined |
| 18-109_k141_113837 | Not-determined |
| 18-109_k141_113838 | Low-quality |
| 18-109_k141_134342 | Not-determined |
| 18-109_k141_32147 | Low-quality |
| 18-109_k141_32148 | Not-determined |
| 18-109_k141_174975 | Not-determined |
| 18-109_k141_113847 | Not-determined |
| 18-109_k141_174978 | Not-determined |
| 18-109_k141_52499 | Not-determined |
| 18-109_k141_154875 | Not-determined |
| 18-109_k141_52501 | Not-determined |
| 18-109_k141_52503 | Not-determined |
| 18-109_k141_11552 | Not-determined |
| 18-109_k141_11555 | Not-determined |
| 18-109_k141_11557 | Not-determined |
| 18-109_k141_11558 | Not-determined |
| 18-109_k141_164995 | Not-determined |
| 18-109_k141_185486 | Not-determined |
| 18-109_k141_62615 | Not-determined |
| 18-109_k141_144139 | Not-determined |
| 18-109_k141_62621 | Low-quality |
| 18-109_k141_42003 | Low-quality |
| 18-109_k141_21473 | Not-determined |
| 18-109_k141_113873 | Not-determined |
| 18-109_k141_113874 | Not-determined |
| 18-109_k141_52514 | Not-determined |
| 18-109_k141_42010 | Not-determined |
| 18-109_k141_185508 | Not-determined |
| 18-109_k141_62642 | Not-determined |
| 18-109_k141_62644 | Not-determined |
| 18-109_k141_82886 | Not-determined |
| 18-109_k141_52525 | Low-quality |
| 18-109_k141_21493 | Not-determined |
| 18-109_k141_21496 | Not-determined |
| 18-109_k141_21501 | Not-determined |
| 18-109_k141_52534 | Low-quality |
| 18-102-final-viral-combined_k141_32748\|\|0_partial | Medium-quality |
| 18-109_k141_185528 | Not-determined |
| 18-109_k141_123955 | Not-determined |
| 18-109_k141_42034 | Not-determined |
| 18-109_k141_154898 | Not-determined |
| 18-109_k141_236390 | Low-quality |
| 18-109_k141_174990 | Not-determined |
| 18-109_k141_134356 | Not-determined |
| 18-109_k141_185540 | Not-determined |
| 18-109_k141_103431 | Not-determined |
| 18-109_k141_226264 | Not-determined |
| 18-109_k141_72395 | Not-determined |
| 18-109_k141_72397 | Not-determined |
| 18-109_k141_174995 | Not-determined |
| 18-109_k141_52543 | Not-determined |
| 18-109_k141_52548 | Not-determined |
| 18-109_k141_72407 | Not-determined |
| 18-109_k141_72408 | Not-determined |
| 18-109_k141_42057 | Not-determined |
| 18-109_k141_134365 | Low-quality |
| 18-109_k141_134368 | Not-determined |
| 18-109_k141_175002 | Not-determined |
| 18-109_k141_185559 | Not-determined |
| 18-109_k141_226267 | Not-determined |
| 18-109_k141_42065 | Not-determined |
| 18-109_k141_154924 | Low-quality |
| 18-109_k141_154929 | Not-determined |
| 18-109_k141_195875 | Low-quality |
| 18-109_k141_62662 | Not-determined |
| 18-109_k141_134380 | Not-determined |
| 18-109_k141_52571 | Not-determined |
| 18-109_k141_195880 | Not-determined |
| 18-109_k141_52576 | Not-determined |
| 18-109_k141_62663 | Not-determined |
| 18-109_k141_205868 | Not-determined |
| 18-109_k141_134386 | Not-determined |
| 18-109_k141_144192 | Not-determined |
| 18-109_k141_205876 | Not-determined |
| 18-109_k141_226280 | Low-quality |
| 18-109_k141_205880 | Not-determined |
| 18-109_k141_185577 | Not-determined |
| 18-109_k141_62679 | Not-determined |
| 18-109_k141_195883 | Not-determined |
| 18-109_k141_144202 | Not-determined |
| 18-109_k141_205889 | Not-determined |
| 18-109_k141_42070 | Not-determined |
| 18-109_k141_62688 | Not-determined |
| 18-109_k141_185583 | Not-determined |
| 18-109_k141_103442 | Not-determined |
| 18-109_k141_205894 | Not-determined |
| 18-109_k141_226303 | Not-determined |
| 18-109_k141_154942 | Not-determined |
| 18-109_k141_113939 | Not-determined |
| 18-109_k141_32177 | Not-determined |
| 18-109_k141_42078 | Not-determined |
| 18-109_k141_144215 | Not-determined |
| 18-109_k141_236429 | Not-determined |
| 18-109_k141_82929 | Not-determined |
| 18-109_k141_82931 | Not-determined |
| 18-109_k141_144216 | Not-determined |
| 18-109_k141_1421 | Not-determined |
| 18-109_k141_1422 | Not-determined |
| 18-109_k141_226320 | Not-determined |
| 18-109_k141_134401 | Not-determined |
| 18-109_k141_236437 | Not-determined |
| 18-109_k141_154949 | Not-determined |
| 18-109_k141_134403 | Low-quality |
| 18-109_k141_42087 | Not-determined |
| 18-109_k141_72444 | Not-determined |
| 18-109_k141_226331 | Not-determined |
| 18-109_k141_185610 | Not-determined |
| 18-109_k141_113969 | Not-determined |
| 18-109_k141_52599 | Not-determined |
| 18-109_k141_185619 | Not-determined |
| 18-109_k141_226339 | Not-determined |
| 18-109_k141_123975 | Not-determined |
| 18-109_k141_185627 | Not-determined |
| 18-109_k141_205913 | Not-determined |
| 18-109_k141_195889 | Low-quality |
| 18-109_k141_123979 | Not-determined |
| 18-109_k141_72455 | Not-determined |
| 18-109_k141_134425 | Low-quality |
| 18-109_k141_113995 | Not-determined |
| 18-109_k141_113996 | Not-determined |
| 18-109_k141_134429 | Not-determined |
| 18-109_k141_205916 | Not-determined |
| 18-109_k141_205918 | Not-determined |
| 18-109_k141_195893 | Low-quality |
| 18-109_k141_205922 | Not-determined |
| 18-109_k141_62734 | Not-determined |
| 18-109_k141_62736 | Not-determined |
| 18-109_k141_72458 | Not-determined |
| 18-109_k141_42130 | Not-determined |
| 18-109_k141_62740 | Not-determined |
| 18-109_k141_62743 | Not-determined |
| 18-109_k141_72468 | Not-determined |
| 18-109_k141_32196 | Low-quality |
| 18-109_k141_205942 | Not-determined |
| 18-109_k141_72471 | Not-determined |
| 18-109_k141_103502 | Low-quality |
| 18-109_k141_185662 | Not-determined |
| 18-109_k141_32201 | Not-determined |
| 18-109_k141_236458 | Not-determined |
| 18-109_k141_185665 | Not-determined |
| 18-109_k141_32207 | Not-determined |
| 18-109_k141_154974 | Not-determined |
| 18-109_k141_32210 | Not-determined |
| 18-109_k141_114002 | Not-determined |
| 18-109_k141_195910 | Not-determined |
| 18-109_k141_114006 | Not-determined |
| 18-109_k141_236469 | Not-determined |
| 18-109_k141_1459 | Not-determined |
| 18-109_k141_205959 | Not-determined |
| 18-109_k141_195913 | Not-determined |
| 18-109_k141_195921 | Not-determined |
| 18-109_k141_195923 | Not-determined |
| 18-109_k141_236478 | Not-determined |
| 18-109_k141_185673 | Not-determined |
| 18-109_k141_195924 | Not-determined |
| 18-109_k141_175067 | Not-determined |
| 18-109_k141_236481 | Not-determined |
| 18-109_k141_226374 | Not-determined |
| 18-109_k141_32218 | Not-determined |
| 18-109_k141_124031 | Not-determined |
| 18-109_k141_195929 | Not-determined |
| 18-109_k141_226376 | Not-determined |
| 18-109_k141_236488 | Not-determined |
| 18-109_k141_226378 | Not-determined |
| 18-109_k141_175071 | Not-determined |
| 18-109_k141_124038 | Not-determined |
| 18-109_k141_226385 | Not-determined |
| 18-109_k141_175081 | Not-determined |
| 18-109_k141_124040 | Not-determined |
| 18-109_k141_82953 | Not-determined |
| 18-109_k141_185691 | Not-determined |
| 18-109_k141_32222 | Not-determined |
| 18-109_k141_175086 | Low-quality |
| 18-109_k141_32225 | Not-determined |
| 18-109_k141_154988 | Not-determined |
| 18-109_k141_114026 | Not-determined |
| 18-109_k141_195945 | Not-determined |
| 18-109_k141_21529 | Not-determined |
| 18-109_k141_1497 | Not-determined |
| 18-109_k141_226398 | Not-determined |
| 18-109_k141_124050 | Low-quality |
| 18-109_k141_236507 | Not-determined |
| 18-109_k141_1501 | Low-quality |
| 18-109_k141_21530 | Not-determined |
| 18-109_k141_21531 | Not-determined |
| 18-109_k141_195958 | Not-determined |
| 18-109_k141_114035 | Not-determined |
| 18-109_k141_155004 | Not-determined |
| 18-109_k141_236511 | Not-determined |
| 18-109_k141_195964 | Not-determined |
| 18-109_k141_195965 | Not-determined |
| 18-109_k141_236522 | Not-determined |
| 18-109_k141_195967 | Not-determined |
| 18-109_k141_21540 | Not-determined |
| 18-109_k141_21541 | Not-determined |
| 18-109_k141_236524 | Not-determined |
| 18-109_k141_155020 | Not-determined |
| 18-109_k141_52651 | Not-determined |
| 18-109_k141_114051 | Not-determined |
| 18-109_k141_236543 | Not-determined |
| 18-109_k141_42166 | Not-determined |
| 18-109_k141_42175 | Not-determined |
| 18-109_k141_32247 | Not-determined |
| 18-109_k141_195986 | Not-determined |
| 18-109_k141_226402 | Not-determined |
| 18-109_k141_185733 | Not-determined |
| 18-109_k141_32251 | Not-determined |
| 18-109_k141_226404 | Not-determined |
| 18-109_k141_103552 | Not-determined |
| 18-109_k141_52670 | Not-determined |
| 18-109_k141_103554 | Not-determined |
| 18-109_k141_82998 | Not-determined |
| 18-109_k141_83001 | Not-determined |
| 18-109_k141_124079 | Not-determined |
| 18-109_k141_124081 | Not-determined |
| 18-109_k141_83006 | Not-determined |
| 18-109_k141_236563 | Low-quality |
| 18-109_k141_11601 | Not-determined |
| 18-109_k141_62806 | Not-determined |
| 18-109_k141_103566 | Low-quality |
| 18-109_k141_11612 | Not-determined |
| 18-109_k141_114066 | Not-determined |
| 18-109_k141_42194 | Not-determined |
| 18-109_k141_134456 | Low-quality |
| 18-109_k141_103568 | Not-determined |
| 18-109_k141_103571 | Not-determined |
| 18-109_k141_175136 | Not-determined |
| 18-109_k141_1513 | Not-determined |
| 18-109_k141_226412 | Not-determined |
| 18-109_k141_42204 | Not-determined |
| 18-109_k141_52696 | Not-determined |
| 18-109_k141_175138 | Not-determined |
| 18-109_k141_114090 | Not-determined |
| 18-109_k141_175141 | Not-determined |
| 18-109_k141_175144 | Not-determined |
| 18-109_k141_92987 | Not-determined |
| 18-109_k141_114093 | Not-determined |
| 18-109_k141_175148 | Not-determined |
| 18-109_k141_226425 | Not-determined |
| 18-109_k141_175157 | Not-determined |
| 18-109_k141_165060 | Not-determined |
| 18-109_k141_11622 | Not-determined |
| 18-109_k141_52729 | Not-determined |
| 18-109_k141_11623 | Low-quality |
| 18-109_k141_93004 | Not-determined |
| 18-109_k141_11624 | Not-determined |
| 18-109_k141_216006 | Not-determined |
| 18-109_k141_11627 | Not-determined |
| 18-109_k141_42232 | Not-determined |
| 18-109_k141_52738 | Not-determined |
| 18-109_k141_236597 | Not-determined |
| 18-109_k141_175175 | Not-determined |
| 18-109_k141_11637 | Not-determined |
| 18-109_k141_226446 | Not-determined |
| 18-109_k141_226448 | Not-determined |
| 18-109_k141_11640 | Low-quality |
| 18-109_k141_236602 | Low-quality |
| 18-109_k141_103600 | Not-determined |
| 18-109_k141_236608 | Not-determined |
| 18-109_k141_32275 | Not-determined |
| 18-109_k141_32281 | Not-determined |
| 18-109_k141_103614 | Not-determined |
| 18-109_k141_206016 | Not-determined |
| 18-109_k141_114131 | Not-determined |
| 18-109_k141_93034 | Not-determined |
| 18-109_k141_165079 | Not-determined |
| 18-109_k141_226471 | Not-determined |
| 18-109_k141_165082 | Not-determined |
| 18-109_k141_103620 | Not-determined |
| 18-109_k141_11645 | Not-determined |
| 18-109_k141_206030 | Not-determined |
| 18-109_k141_32312 | Not-determined |
| 18-109_k141_52779 | Not-determined |
| 18-109_k141_42245 | Not-determined |
| 18-109_k141_103640 | Not-determined |
| 18-109_k141_32318 | Not-determined |
| 18-109_k141_93061 | Not-determined |
| 18-109_k141_32321 | Not-determined |
| 18-109_k141_1543 | Not-determined |
| 18-109_k141_236621 | Not-determined |
| 18-109_k141_32331 | Not-determined |
| 18-109_k141_206051 | Not-determined |
| 18-109_k141_206056 | Low-quality |
| 18-109_k141_1550 | Not-determined |
| 18-109_k141_62854 | Not-determined |
| 18-109_k141_103663 | Not-determined |
| 18-109_k141_155060 | Low-quality |
| 18-109_k141_62863 | Not-determined |
| 18-109_k141_42258 | Low-quality |
| 18-109_k141_134473 | Not-determined |
| 18-109_k141_236642 | Not-determined |
| 18-109_k141_185817 | Not-determined |
| 18-109_k141_1575 | Not-determined |
| 18-109_k141_226482 | Not-determined |
| 18-109_k141_155079 | Not-determined |
| 18-109_k141_144234 | Not-determined |
| 18-109_k141_144235 | Not-determined |
| 18-109_k141_144237 | Low-quality |
| 18-109_k141_144238 | Not-determined |
| 18-109_k141_134496 | Not-determined |
| 18-109_k141_165147 | Not-determined |
| 18-109_k141_226490 | Not-determined |
| 18-109_k141_114208 | Not-determined |
| 18-109_k141_52807 | Not-determined |
| 18-109_k141_206130 | Not-determined |
| 18-109_k141_155102 | Not-determined |
| 18-109_k141_206134 | Not-determined |
| 18-109_k141_216072 | Not-determined |
| 18-109_k141_226498 | Not-determined |
| 18-109_k141_144261 | Not-determined |
| 18-109_k141_124136 | Not-determined |
| 18-109_k141_32362 | Not-determined |
| 18-109_k141_134519 | Not-determined |
| 18-109_k141_52834 | Not-determined |
| 18-109_k141_124151 | Low-quality |
| 18-109_k141_206148 | Not-determined |
| 18-109_k141_1599 | Not-determined |
| 18-109_k141_32385 | Low-quality |
| 18-109_k141_32386 | Low-quality |
| 18-109_k141_206163 | Not-determined |
| 18-109_k141_226511 | Low-quality |
| 18-109_k141_155123 | Not-determined |
| 18-109_k141_155129 | Not-determined |
| 18-109_k141_144290 | Not-determined |
| 18-109_k141_155132 | Not-determined |
| 18-109_k141_165193 | Not-determined |
| 18-109_k141_144293 | Not-determined |
| 18-109_k141_114241 | Not-determined |
| 18-109_k141_226518 | Not-determined |
| 18-109_k141_62880 | Not-determined |
| 18-109_k141_144306 | Not-determined |
| 18-109_k141_226524 | Not-determined |
| 18-109_k141_42331 | Not-determined |
| 18-109_k141_226530 | Not-determined |
| 18-109_k141_206191 | Low-quality |
| 18-109_k141_42335 | Not-determined |
| 18-109_k141_165204 | Low-quality |
| 18-109_k141_226537 | Low-quality |
| 18-109_k141_1622 | Not-determined |
| 18-109_k141_144321 | Not-determined |
| 18-109_k141_216107 | Not-determined |
| 18-109_k141_93081 | Not-determined |
| 18-109_k141_206201 | Not-determined |
| 18-109_k141_62892 | Not-determined |
| 18-109_k141_124172 | Not-determined |
| 18-109_k141_52863 | Not-determined |
| 18-109_k141_175221 | Not-determined |
| 18-109_k141_83053 | Not-determined |
| 18-109_k141_42351 | Not-determined |
| 18-109_k141_175223 | Not-determined |
| 18-109_k141_52872 | Not-determined |
| 18-109_k141_226556 | Not-determined |
| 18-109_k141_175224 | Not-determined |
| 18-109_k141_124188 | Not-determined |
| 18-109_k141_226561 | Not-determined |
| 18-109_k141_216127 | Not-determined |
| 18-109_k141_52892 | Not-determined |
| 18-109_k141_52896 | Not-determined |
| 18-109_k141_52900 | Not-determined |
| 18-109_k141_52901 | Not-determined |
| 18-109_k141_175229 | Not-determined |
| 18-109_k141_165222 | Not-determined |
| 18-109_k141_62907 | Low-quality |
| 18-109_k141_124204 | Not-determined |
| 18-109_k141_216133 | Not-determined |
| 18-109_k141_144339 | Not-determined |
| 18-109_k141_185870 | Not-determined |
| 18-109_k141_216140 | Not-determined |
| 18-109_k141_42362 | Not-determined |
| 18-109_k141_236678 | Not-determined |
| 18-109_k141_206225 | Not-determined |
| 18-109_k141_93120 | Not-determined |
| 18-109_k141_124214 | Not-determined |
| 18-109_k141_206227 | Not-determined |
| 18-109_k141_32423 | Not-determined |
| 18-109_k141_62919 | Not-determined |
| 18-109_k141_236681 | Not-determined |
| 18-109_k141_62920 | Not-determined |
| 18-109_k141_93128 | Not-determined |
| 18-109_k141_165249 | Not-determined |
| 18-109_k141_93132 | Low-quality |
| 18-109_k141_216156 | Not-determined |
| 18-109_k141_103717 | Not-determined |
| 18-109_k141_236692 | Not-determined |
| 18-109_k141_42380 | Not-determined |
| 18-109_k141_52919 | Not-determined |
| 18-109_k141_103729 | Not-determined |
| 18-109_k141_236705 | Not-determined |
| 18-109_k141_11654 | Not-determined |
| 18-109_k141_196033 | Low-quality |
| 18-109_k141_226599 | Not-determined |
| 18-109_k141_196035 | Not-determined |
| 18-109_k141_165266 | Not-determined |
| 18-109_k141_11656 | Not-determined |
| 18-109_k141_83085 | Not-determined |
| 18-109_k141_62942 | Not-determined |
| 18-109_k141_124231 | Not-determined |
| 18-109_k141_124233 | Not-determined |
| 18-109_k141_32452 | Not-determined |
| 18-109_k141_236720 | Not-determined |
| 18-109_k141_124242 | Not-determined |
| 18-109_k141_103749 | Not-determined |
| 18-109_k141_196056 | Not-determined |
| 18-109_k141_11674 | Not-determined |
| 18-109_k141_165281 | Not-determined |
| 18-109_k141_62960 | Not-determined |
| 18-109_k141_155161 | Not-determined |
| 18-109_k141_236732 | Not-determined |
| 18-109_k141_103762 | Not-determined |
| 18-109_k141_236737 | Not-determined |
| 18-109_k141_1652 | Not-determined |
| 18-109_k141_21640 | Not-determined |

| | |
|---|---|
| 18-109_k141_165291 | Low-quality |
| 18-109_k141_134543 | Not-determined |
| 18-109_k141_236741 | Not-determined |
| 18-109_k141_236744 | Not-determined |
| 18-109_k141_206294 | Not-determined |
| 18-109_k141_21649 | Not-determined |
| 18-109_k141_196073 | Not-determined |
| 18-109_k141_165297 | Not-determined |
| 18-109_k141_236754 | Not-determined |
| 18-109_k141_236759 | Not-determined |
| 18-109_k141_175275 | Not-determined |
| 18-109_k141_52948 | Not-determined |
| 18-109_k141_52951 | Not-determined |
| 18-109_k141_165301 | Not-determined |
| 18-109_k141_165302 | Not-determined |
| 18-109_k141_206299 | Not-determined |
| 18-109_k141_175284 | Not-determined |
| 18-109_k141_103770 | Not-determined |
| 18-109_k141_124264 | Not-determined |
| 18-109_k141_72535 | Not-determined |
| 18-109_k141_93154 | Low-quality |
| 18-109_k141_134577 | Not-determined |
| 18-109_k141_175287 | Low-quality |
| 18-109_k141_1682 | Not-determined |
| 18-109_k141_114300 | Not-determined |
| 18-109_k141_114304 | Not-determined |
| 18-109_k141_124287 | Low-quality |
| 18-109_k141_114306 | Low-quality |
| 18-109_k141_1688 | Low-quality |
| 18-109_k141_1689 | Not-determined |
| 18-109_k141_236789 | Not-determined |
| 18-109_k141_185893 | Not-determined |
| 18-109_k141_236799 | Not-determined |
| 18-109_k141_42408 | Not-determined |
| 18-109_k141_42409 | Not-determined |
| 18-109_k141_103780 | Not-determined |
| 18-109_k141_236807 | Not-determined |
| 18-109_k141_72560 | Not-determined |
| 18-109_k141_226610 | Low-quality |
| 18-109_k141_1699 | Not-determined |
| 18-109_k141_32484 | Not-determined |
| 18-109_k141_42415 | Not-determined |
| 18-109_k141_124316 | Not-determined |
| 18-109_k141_175311 | Not-determined |
| 18-109_k141_1713 | Not-determined |
| 18-109_k141_124326 | Not-determined |
| 18-109_k141_236837 | Not-determined |
| 18-109_k141_42431 | Not-determined |
| 18-109_k141_226624 | Not-determined |
| 18-109_k141_226625 | Not-determined |
| 18-109_k141_124329 | Low-quality |
| 18-109_k141_226634 | Not-determined |
| 18-109_k141_42443 | Low-quality |
| 18-109_k141_93176 | Low-quality |
| 18-109_k141_124341 | Not-determined |
| 18-109_k141_124342 | Not-determined |
| 18-109_k141_62996 | Not-determined |
| 18-109_k141_144360 | Not-determined |
| 18-109_k141_185938 | Low-quality |
| 18-109_k141_206320 | Not-determined |
| 18-109_k141_124350 | Not-determined |
| 18-109_k141_83118 | Not-determined |
| 18-109_k141_185947 | Not-determined |
| 18-109_k141_103832 | Low-quality |
| 18-109_k141_226651 | Not-determined |
| 18-109_k141_226652 | Not-determined |
| 18-109_k141_226655 | Not-determined |
| 18-109_k141_83122 | Not-determined |
| 18-109_k141_226662 | Not-determined |
| 18-109_k141_185965 | Not-determined |
| 18-109_k141_144380 | Not-determined |
| 18-109_k141_185969 | Not-determined |
| 18-109_k141_93208 | Not-determined |
| 18-109_k141_1757 | Not-determined |
| 18-109_k141_236865 | Not-determined |
| 18-109_k141_21713 | Not-determined |
| 18-109_k141_93214 | Not-determined |
| 18-109_k141_93216 | Not-determined |
| 18-109_k141_226673 | Low-quality |
| 18-109_k141_134603 | Low-quality |
| 18-109_k141_206328 | Not-determined |
| 18-109_k141_93223 | Not-determined |
| 18-109_k141_32529 | Low-quality |
| 18-109_k141_134612 | Not-determined |
| 18-109_k141_93224 | Not-determined |
| 18-109_k141_144399 | Not-determined |
| 18-109_k141_32534 | Not-determined |
| 18-109_k141_226683 | Not-determined |
| 18-109_k141_32544 | Not-determined |
| 18-109_k141_144401 | Not-determined |
| 18-109_k141_175359 | Not-determined |
| 18-109_k141_1784 | Not-determined |
| 18-109_k141_1786 | Not-determined |
| 18-109_k141_226696 | Not-determined |
| 18-109_k141_42480 | Not-determined |
| 18-109_k141_32559 | Not-determined |
| 18-109_k141_83151 | Not-determined |
| 18-109_k141_32563 | Not-determined |
| 18-109_k141_42487 | Not-determined |
| 18-109_k141_206356 | Not-determined |
| 18-109_k141_32569 | Not-determined |
| 18-109_k141_1794 | Not-determined |
| 18-109_k141_144408 | Not-determined |
| 18-109_k141_83164 | Low-quality |
| 18-109_k141_206361 | Not-determined |
| 18-109_k141_226714 | Not-determined |
| 18-109_k141_42490 | Not-determined |
| 18-109_k141_175382 | Not-determined |
| 18-109_k141_114349 | Not-determined |
| 18-109_k141_114350 | Not-determined |
| 18-109_k141_175385 | Not-determined |
| 18-109_k141_226726 | Low-quality |
| 18-109_k141_175386 | Not-determined |
| 18-109_k141_52982 | Low-quality |
| 18-109_k141_175388 | Not-determined |
| 18-109_k141_42502 | Not-determined |
| 18-109_k141_175389 | Not-determined |
| 18-109_k141_144417 | Not-determined |
| 18-109_k141_52986 | Not-determined |
| 18-109_k141_83178 | Not-determined |
| 18-109_k141_196110 | Low-quality |
| 18-109_k141_236873 | Not-determined |
| 18-109_k141_114366 | Not-determined |
| 18-109_k141_93251 | Not-determined |
| 18-109_k141_114374 | Not-determined |
| 18-109_k141_124384 | Not-determined |
| 18-109_k141_93258 | Not-determined |
| 18-109_k141_124388 | Low-quality |
| 18-109_k141_175406 | Not-determined |
| 18-109_k141_93260 | Not-determined |
| 18-109_k141_32595 | Not-determined |
| 18-109_k141_93262 | Not-determined |
| 18-109_k141_32598 | Not-determined |
| 18-109_k141_93265 | Low-quality |
| 18-109_k141_83208 | Not-determined |
| 18-109_k141_206410 | Not-determined |
| 18-109_k141_236918 | Not-determined |
| 18-109_k141_185989 | Not-determined |
| 18-109_k141_236922 | Not-determined |
| 18-109_k141_124401 | Not-determined |
| 18-109_k141_32616 | Not-determined |
| 18-109_k141_155175 | Not-determined |
| 18-109_k141_175442 | Not-determined |
| 18-109_k141_21763 | Not-determined |
| 18-109_k141_21765 | Not-determined |
| 18-109_k141_42514 | Not-determined |
| 18-109_k141_185997 | Not-determined |
| 18-109_k141_11730 | Not-determined |
| 18-109_k141_11732 | Not-determined |
| 18-109_k141_21771 | Not-determined |
| 18-109_k141_63020 | Not-determined |
| 18-109_k141_124411 | Not-determined |
| 18-109_k141_186012 | Not-determined |
| 18-109_k141_42532 | Not-determined |
| 18-109_k141_1830 | Not-determined |
| 18-109_k141_186016 | Not-determined |
| 18-109_k141_11737 | Not-determined |
| 18-109_k141_42533 | Not-determined |
| 18-109_k141_103936 | Low-quality |
| 18-109_k141_114419 | Not-determined |
| 18-109_k141_124421 | Not-determined |
| 18-109_k141_226762 | Low-quality |
| 18-109_k141_124422 | Not-determined |
| 18-109_k141_226763 | Not-determined |
| 18-109_k141_11742 | Low-quality |
| 18-109_k141_53073 | Not-determined |
| 18-109_k141_186022 | Not-determined |
| 18-109_k141_175469 | Low-quality |
| 18-109_k141_124429 | Not-determined |
| 18-109_k141_226765 | Not-determined |
| 18-109_k141_226766 | Not-determined |
| 18-109_k141_226767 | Not-determined |
| 18-109_k141_42540 | Not-determined |
| 18-109_k141_175474 | Not-determined |
| 18-109_k141_21791 | Not-determined |
| 18-109_k141_53076 | Not-determined |
| 18-109_k141_144422 | Not-determined |
| 18-109_k141_124435 | Not-determined |
| 18-109_k141_11753 | Not-determined |
| 18-109_k141_226776 | Not-determined |
| 18-109_k141_21801 | Not-determined |
| 18-109_k141_103947 | Not-determined |
| 18-109_k141_11762 | Not-determined |
| 18-109_k141_216235 | Not-determined |
| 18-109_k141_236952 | Not-determined |
| 18-109_k141_1838 | Low-quality |
| 18-109_k141_144425 | Not-determined |
| 18-109_k141_155190 | Not-determined |
| 18-109_k141_11767 | Not-determined |
| 18-109_k141_155196 | Not-determined |
| 18-109_k141_103960 | Not-determined |
| 18-109_k141_103961 | Not-determined |
| 18-109_k141_186036 | Low-quality |
| 18-109_k141_226791 | Not-determined |
| 18-109_k141_103963 | Not-determined |
| 18-109_k141_32652 | Not-determined |
| 18-109_k141_93287 | Not-determined |
| 18-109_k141_124461 | Low-quality |
| 18-109_k141_11776 | Not-determined |
| 18-109_k141_42576 | Not-determined |
| 18-109_k141_32654 | Low-quality |
| 18-109_k141_124470 | Not-determined |
| 18-109_k141_42577 | Not-determined |
| 18-109_k141_196180 | Not-determined |
| 18-109_k141_42587 | Not-determined |
| 18-109_k141_236986 | Not-determined |
| 18-109_k141_103984 | Not-determined |
| 18-109_k141_21803 | Not-determined |
| 18-109_k141_42591 | Not-determined |
| 18-109_k141_216257 | Not-determined |
| 18-109_k141_103995 | Not-determined |
| 18-109_k141_42609 | Low-quality |
| 18-109_k141_206452 | Not-determined |
| 18-109_k141_124487 | Not-determined |
| 18-109_k141_124492 | Not-determined |
| 18-109_k141_21830 | Not-determined |
| 18-109_k141_134639 | Not-determined |
| 18-109_k141_42625 | Not-determined |
| 18-109_k141_186067 | Not-determined |
| 18-109_k141_196204 | Not-determined |
| 18-109_k141_144443 | Not-determined |
| 18-109_k141_53127 | Not-determined |
| 18-109_k141_144445 | Not-determined |
| 18-109_k141_144449 | Not-determined |
| 18-109_k141_53141 | Not-determined |
| 18-109_k141_124504 | Low-quality |
| 18-109_k141_53142 | Not-determined |
| 18-109_k141_144451 | Low-quality |
| 18-109_k141_196216 | Not-determined |
| 18-109_k141_216290 | Not-determined |
| 18-109_k141_32683 | Low-quality |
| 18-109_k141_206469 | Not-determined |
| 18-109_k141_206473 | Low-quality |
| 18-109_k141_32691 | Low-quality |
| 18-109_k141_1851 | Not-determined |
| 18-109_k141_124519 | Not-determined |
| 18-109_k141_237003 | Not-determined |
| 18-109_k141_83224 | Not-determined |
| 18-109_k141_21838 | Not-determined |
| 18-109_k141_32696 | Not-determined |
| 18-109_k141_21840 | Not-determined |
| 18-109_k141_11855 | Not-determined |
| 18-109_k141_124523 | Not-determined |
| 18-109_k141_53164 | Not-determined |
| 18-109_k141_32701 | Low-quality |
| 18-109_k141_83231 | Not-determined |
| 18-109_k141_32704 | Not-determined |
| 18-109_k141_53169 | Not-determined |
| 18-109_k141_42630 | Not-determined |
| 18-109_k141_21845 | Not-determined |
| 18-109_k141_42634 | Not-determined |
| 18-109_k141_216302 | Not-determined |
| 18-109_k141_72603 | Not-determined |
| 18-109_k141_206490 | Not-determined |
| 18-109_k141_11867 | Not-determined |
| 18-109_k141_206492 | Not-determined |
| 18-109_k141_226799 | Not-determined |
| 18-109_k141_216305 | Not-determined |
| 18-109_k141_206495 | Not-determined |
| 18-109_k141_124544 | Not-determined |
| 18-109_k141_1871 | Not-determined |
| 18-109_k141_186089 | Low-quality |
| 18-109_k141_124547 | Low-quality |
| 18-109_k141_104041 | Not-determined |
| 18-109_k141_206500 | Not-determined |
| 18-109_k141_206502 | Not-determined |
| 18-109_k141_1878 | Low-quality |
| 18-109_k141_104045 | Not-determined |
| 18-109_k141_206505 | Not-determined |
| 18-109_k141_216315 | Low-quality |
| 18-109_k141_144491 | Not-determined |
| 18-109_k141_237030 | Not-determined |
| 18-109_k141_237031 | Low-quality |
| 18-109_k141_144505 | Not-determined |
| 18-109_k141_124570 | Not-determined |
| 18-109_k141_32728 | Not-determined |
| 18-109_k141_11879 | Not-determined |
| 18-109_k141_21864 | Not-determined |
| 18-109_k141_11882 | Not-determined |
| 18-109_k141_155294 | Not-determined |
| 18-109_k141_53185 | Not-determined |
| 18-109_k141_206531 | Not-determined |
| 18-109_k141_114456 | Not-determined |
| 18-109_k141_124584 | Not-determined |
| 18-109_k141_21871 | Not-determined |
| 18-109_k141_237061 | Not-determined |
| 18-109_k141_237065 | Not-determined |
| 18-109_k141_114465 | Not-determined |
| 18-109_k141_32738 | Low-quality |
| 18-109_k141_11916 | Not-determined |
| 18-109_k141_42662 | Not-determined |
| 18-109_k141_11918 | Not-determined |
| 18-109_k141_11921 | Not-determined |
| 18-109_k141_11923 | Not-determined |
| 18-109_k141_237073 | Not-determined |
| 18-109_k141_83265 | Low-quality |
| 18-109_k141_83267 | Not-determined |
| 18-109_k141_226833 | Not-determined |
| 18-109_k141_42672 | Not-determined |
| 18-109_k141_63093 | Not-determined |
| 18-109_k141_83276 | Not-determined |
| 18-109_k141_114475 | Not-determined |
| 18-109_k141_42679 | Not-determined |
| 18-109_k141_32744 | Not-determined |
| 18-109_k141_83285 | Not-determined |
| 18-109_k141_237094 | Not-determined |
| 18-109_k141_114486 | Not-determined |
| 18-109_k141_11941 | Not-determined |
| 18-109_k141_72642 | Not-determined |
| 18-109_k141_11948 | Not-determined |
| 18-109_k141_83295 | Not-determined |
| 18-109_k141_21885 | Low-quality |
| 18-109_k141_11951 | Not-determined |
| 18-109_k141_72651 | Not-determined |
| 18-109_k141_21888 | Not-determined |
| 18-109_k141_134685 | Not-determined |
| 18-109_k141_216367 | Not-determined |
| 18-109_k141_72653 | Not-determined |
| 18-109_k141_186133 | Not-determined |
| 18-109_k141_226853 | Not-determined |
| 18-109_k141_155323 | Not-determined |
| 18-109_k141_134695 | Not-determined |
| 18-109_k141_114508 | Not-determined |
| 18-109_k141_206549 | Low-quality |
| 18-109_k141_216382 | Not-determined |
| 18-109_k141_93340 | Not-determined |
| 18-109_k141_114511 | Low-quality |
| 18-109_k141_216386 | Not-determined |
| 18-109_k141_206566 | Not-determined |
| 18-109_k141_165348 | Not-determined |
| 18-109_k141_63105 | Not-determined |
| 18-109_k141_93351 | Not-determined |
| 18-109_k141_1914 | Not-determined |
| 18-109_k141_114524 | Low-quality |
| 18-109_k141_165365 | Not-determined |
| 18-109_k141_226865 | Low-quality |
| 18-109_k141_42726 | Not-determined |
| 18-109_k141_72668 | Not-determined |
| 18-109_k141_134716 | Not-determined |
| 18-109_k141_72673 | Not-determined |
| 18-109_k141_165370 | Not-determined |
| 18-109_k141_155349 | Not-determined |
| 18-109_k141_1933 | Not-determined |
| 18-109_k141_11971 | Not-determined |
| 18-109_k141_124617 | Not-determined |
| 18-109_k141_226871 | Not-determined |
| 18-109_k141_216411 | Not-determined |
| 18-109_k141_134728 | Not-determined |
| 18-109_k141_216417 | Not-determined |
| 18-109_k141_114540 | Not-determined |
| 18-109_k141_124620 | Low-quality |
| 18-109_k141_206584 | Not-determined |
| 18-109_k141_226880 | Not-determined |
| 18-109_k141_32778 | Not-determined |
| 18-109_k141_134738 | Not-determined |
| 18-109_k141_32783 | Not-determined |
| 18-109_k141_114543 | Not-determined |
| 18-109_k141_11978 | Not-determined |
| 18-109_k141_206600 | Not-determined |
| 18-109_k141_134749 | Not-determined |
| 18-109_k141_165389 | Not-determined |
| 18-109_k141_1956 | Low-quality |
| 18-109_k141_63147 | Not-determined |
| 18-109_k141_175522 | Not-determined |
| 18-109_k141_11989 | Not-determined |
| 18-109_k141_206614 | Low-quality |
| 18-109_k141_226899 | Not-determined |
| 18-109_k141_11991 | Not-determined |
| 18-109_k141_11994 | Not-determined |
| 18-109_k141_226911 | Not-determined |
| 18-109_k141_104132 | Not-determined |
| 18-109_k141_226914 | Not-determined |
| 18-109_k141_226916 | Not-determined |
| 18-109_k141_134770 | Not-determined |
| 18-109_k141_63178 | Low-quality |
| 18-109_k141_216423 | Not-determined |
| 18-109_k141_1978 | Not-determined |
| 18-109_k141_226922 | Not-determined |
| 18-109_k141_1985 | Low-quality |
| 18-109_k141_83309 | Not-determined |
| 18-109_k141_134784 | Not-determined |
| 18-109_k141_175540 | Not-determined |
| 18-109_k141_134787 | Not-determined |
| 18-109_k141_104138 | Not-determined |
| 18-109_k141_83315 | Not-determined |
| 18-109_k141_72692 | Low-quality |
| 18-109_k141_226927 | Not-determined |
| 18-109_k141_2001 | Not-determined |
| 18-109_k141_134791 | Not-determined |
| 18-109_k141_2008 | Not-determined |
| 18-109_k141_42782 | Not-determined |
| 18-109_k141_2011 | Low-quality |
| 18-109_k141_226933 | Low-quality |
| 18-109_k141_83324 | Not-determined |
| 18-109_k141_196282 | Not-determined |
| 18-109_k141_226935 | Not-determined |
| 18-109_k141_216446 | Not-determined |
| 18-109_k141_2015 | Not-determined |
| 18-109_k141_72708 | Not-determined |
| 18-109_k141_72709 | Low-quality |
| 18-109_k141_72712 | Not-determined |
| 18-109_k141_144604 | Not-determined |
| 18-109_k141_72720 | Not-determined |
| 18-109_k141_144605 | Not-determined |
| 18-109_k141_63193 | Not-determined |
| 18-109_k141_72726 | Not-determined |
| 18-109_k141_175559 | Low-quality |
| 18-109_k141_175560 | Not-determined |
| 18-109_k141_63203 | Not-determined |
| 18-109_k141_42815 | Not-determined |
| 18-109_k141_226946 | Not-determined |
| 18-109_k141_226949 | Low-quality |
| 18-109_k141_83370 | Not-determined |
| 18-109_k141_42832 | Not-determined |
| 18-109_k141_83376 | Not-determined |
| 18-109_k141_42834 | Not-determined |
| 18-109_k141_104147 | Not-determined |
| 18-109_k141_72742 | Not-determined |
| 18-109_k141_226971 | Not-determined |
| 18-109_k141_63232 | Not-determined |
| 18-109_k141_72744 | Low-quality |
| 18-109_k141_104157 | Not-determined |
| 18-109_k141_21961 | Not-determined |
| 18-109_k141_175594 | Not-determined |
| 18-109_k141_165424 | Low-quality |
| 18-109_k141_32792 | Not-determined |
| 18-109_k141_114565 | Not-determined |
| 18-109_k141_237114 | Not-determined |
| 18-109_k141_165432 | Not-determined |
| 18-109_k141_21975 | Not-determined |
| 18-109_k141_165433 | Not-determined |
| 18-109_k141_175607 | Not-determined |
| 18-109_k141_114575 | Not-determined |
| 18-109_k141_186187 | Not-determined |
| 18-109_k141_42881 | Not-determined |
| 18-109_k141_2039 | Not-determined |
| 18-109_k141_12056 | Not-determined |
| 18-109_k141_53250 | Not-determined |
| 18-109_k141_21981 | Not-determined |
| 18-109_k141_114588 | Not-determined |
| 18-109_k141_53255 | Not-determined |
| 18-109_k141_12063 | Not-determined |
| 18-109_k141_63255 | Not-determined |
| 18-109_k141_206655 | Not-determined |
| 18-109_k141_165441 | Low-quality |
| 18-109_k141_237131 | Low-quality |
| 18-109_k141_227004 | Not-determined |
| 18-109_k141_165444 | Not-determined |
| 18-109_k141_42896 | Not-determined |
| 18-109_k141_186198 | Not-determined |
| 18-109_k141_237142 | Not-determined |
| 18-109_k141_227020 | Not-determined |
| 18-109_k141_227021 | Not-determined |
| 18-109_k141_206667 | Not-determined |
| 18-109_k141_206669 | Not-determined |
| 18-109_k141_12090 | Not-determined |
| 18-109_k141_134794 | Not-determined |
| 18-109_k141_72753 | Not-determined |
| 18-109_k141_42930 | Not-determined |
| 18-109_k141_22023 | Not-determined |
| 18-109_k141_165460 | Not-determined |
| 18-109_k141_114612 | Low-quality |
| 18-109_k141_186205 | Not-determined |
| 18-109_k141_155469 | Not-determined |
| 18-109_k141_144628 | Low-quality |
| 18-109_k141_186217 | Not-determined |
| 18-109_k141_206687 | Not-determined |
| 18-109_k141_114615 | Not-determined |
| 18-109_k141_216487 | Not-determined |
| 18-109_k141_227031 | Not-determined |
| 18-109_k141_196366 | Not-determined |
| 18-109_k141_196369 | Not-determined |
| 18-109_k141_114628 | Not-determined |
| 18-109_k141_227044 | Not-determined |
| 18-109_k141_206697 | Not-determined |
| 18-109_k141_32814 | Not-determined |
| 18-109_k141_32816 | Not-determined |
| 18-109_k141_104220 | Not-determined |
| 18-109_k141_104221 | Not-determined |
| 18-109_k141_165493 | Not-determined |
| 18-109_k141_196385 | Not-determined |
| 18-109_k141_227054 | Not-determined |
| 18-109_k141_22037 | Not-determined |
| 18-109_k141_216509 | Not-determined |
| 18-109_k141_22043 | Not-determined |
| 18-109_k141_165498 | Low-quality |
| 18-109_k141_196395 | Not-determined |
| 18-109_k141_83397 | Low-quality |
| 18-109_k141_104239 | Not-determined |
| 18-109_k141_227062 | Not-determined |
| 18-109_k141_124667 | Not-determined |
| 18-109_k141_104245 | Not-determined |
| 18-109_k141_206713 | Not-determined |
| 18-109_k141_104252 | Not-determined |
| 18-109_k141_83409 | Not-determined |
| 18-109_k141_175628 | Not-determined |
| 18-109_k141_216514 | Not-determined |
| 18-109_k141_175632 | Not-determined |
| 18-109_k141_124670 | Not-determined |
| 18-109_k141_32841 | Not-determined |
| 18-109_k141_12132 | Not-determined |
| 18-109_k141_124677 | Not-determined |
| 18-109_k141_124688 | Not-determined |
| 18-109_k141_237158 | Not-determined |
| 18-109_k141_53285 | Not-determined |
| 18-109_k141_206726 | Not-determined |
| 18-109_k141_53287 | Low-quality |
| 18-109_k141_12154 | Not-determined |
| 18-109_k141_63278 | Not-determined |
| 18-109_k141_22072 | Not-determined |
| 18-109_k141_53303 | Not-determined |
| 18-109_k141_165513 | Not-determined |
| 18-109_k141_186252 | Not-determined |
| 18-109_k141_186256 | Not-determined |
| 18-109_k141_165522 | Not-determined |
| 18-109_k141_165523 | Not-determined |
| 18-109_k141_155487 | Not-determined |
| 18-109_k141_72782 | Not-determined |
| 18-109_k141_196419 | Low-quality |
| 18-109_k141_165525 | Not-determined |
| 18-109_k141_227086 | Not-determined |
| 18-109_k141_165526 | Not-determined |
| 18-109_k141_53312 | Not-determined |
| 18-109_k141_227096 | Not-determined |
| 18-109_k141_165533 | Not-determined |
| 18-109_k141_134812 | Not-determined |
| 18-109_k141_175647 | Not-determined |
| 18-109_k141_186264 | Not-determined |
| 18-109_k141_206758 | Not-determined |
| 18-109_k141_196428 | Low-quality |
| 18-109_k141_124720 | Not-determined |
| 18-109_k141_53328 | Not-determined |
| 18-109_k141_93378 | Not-determined |
| 18-109_k141_155496 | Not-determined |
| 18-109_k141_134822 | Not-determined |
| 18-109_k141_72801 | Not-determined |
| 18-109_k141_124730 | Not-determined |
| 18-109_k141_227110 | Not-determined |
| 18-109_k141_134823 | Not-determined |
| 18-109_k141_134824 | Not-determined |
| 18-109_k141_72806 | Not-determined |
| 18-109_k141_165546 | Not-determined |
| 18-109_k141_186277 | Not-determined |
| 18-109_k141_155504 | Not-determined |
| 18-109_k141_165550 | Not-determined |
| 18-109_k141_42969 | Not-determined |
| 18-109_k141_155508 | Not-determined |
| 18-109_k141_175661 | Not-determined |
| 18-109_k141_12186 | Not-determined |
| 18-109_k141_12188 | Not-determined |
| 18-109_k141_206782 | Not-determined |
| 18-109_k141_83429 | Not-determined |
| 18-109_k141_196449 | Not-determined |
| 18-109_k141_227115 | Not-determined |
| 18-109_k141_2111 | Not-determined |
| 18-109_k141_93398 | Not-determined |
| 18-109_k141_134826 | Low-quality |
| 18-109_k141_206800 | Not-determined |
| 18-109_k141_155523 | Not-determined |
| 18-109_k141_196469 | Not-determined |
| 18-109_k141_165560 | Not-determined |
| 18-109_k141_63311 | Not-determined |
| 18-109_k141_175664 | Not-determined |
| 18-109_k141_72821 | Not-determined |
| 18-109_k141_196476 | Not-determined |
| 18-109_k141_196477 | Not-determined |
| 18-109_k141_72823 | Not-determined |
| 18-109_k141_196478 | Not-determined |
| 18-109_k141_134830 | Low-quality |
| 18-109_k141_175671 | Not-determined |
| 18-109_k141_63314 | Not-determined |
| 18-109_k141_196483 | Not-determined |
| 18-109_k141_124769 | Not-determined |
| 18-109_k141_93412 | Low-quality |
| 18-109_k141_124779 | Not-determined |
| 18-109_k141_72838 | Not-determined |
| 18-109_k141_83432 | Low-quality |
| 18-109_k141_83433 | Low-quality |
| 18-109_k141_22118 | Not-determined |
| 18-109_k141_165592 | Not-determined |
| 18-109_k141_186306 | Not-determined |
| 18-109_k141_186308 | Not-determined |
| 18-109_k141_72849 | Not-determined |
| 18-109_k141_22129 | Not-determined |
| 18-109_k141_155535 | Not-determined |
| 18-109_k141_165599 | Not-determined |
| 18-109_k141_134839 | Not-determined |
| 18-109_k141_134840 | Not-determined |
| 18-109_k141_83435 | Not-determined |

| | |
|---|---|
| 18-109_k141_216543 | Not-determined |
| 18-109_k141_63331 | Not-determined |
| 18-109_k141_155533 | Low-quality |
| 18-109_k141_104279 | Not-determined |
| 18-109_k141_155537 | Not-determined |
| 18-109_k141_63336 | Not-determined |
| 18-109_k141_165606 | Not-determined |
| 18-109_k141_22137 | Low-quality |
| 18-109_k141_72863 | Not-determined |
| 18-109_k141_216553 | Not-determined |
| 18-109_k141_12216 | Not-determined |
| 18-109_k141_83441 | Not-determined |
| 18-109_k141_72876 | Not-determined |
| 18-109_k141_63348 | Low-quality |
| 18-109_k141_165625 | Not-determined |
| 18-109_k141_22154 | Not-determined |
| 18-109_k141_134857 | Not-determined |
| 18-109_k141_165629 | Not-determined |
| 18-109_k141_22157 | Not-determined |
| 18-109_k141_12226 | Low-quality |
| 18-109_k141_124803 | Not-determined |
| 18-109_k141_104284 | Not-determined |
| 18-109_k141_104286 | Not-determined |
| 18-109_k141_63362 | Not-determined |
| 18-109_k141_165637 | Low-quality |
| 18-109_k141_216566 | Not-determined |
| 18-109_k141_124808 | Not-determined |
| 18-109_k141_134870 | Not-determined |
| 18-109_k141_186343 | Low-quality |
| 18-109_k141_216577 | Not-determined |
| 18-109_k141_63370 | Low-quality |
| 18-109_k141_196499 | Not-determined |
| 18-109_k141_186350 | Not-determined |
| 18-109_k141_196500 | Not-determined |
| 18-109_k141_134882 | Not-determined |
| 18-109_k141_83448 | Not-determined |
| 18-109_k141_196503 | Not-determined |
| 18-109_k141_196504 | Not-determined |
| 18-109_k141_216583 | Not-determined |
| 18-109_k141_63382 | Not-determined |
| 18-109_k141_22173 | Low-quality |
| 18-109_k141_134900 | Low-quality |
| 18-109_k141_72904 | Not-determined |
| 18-109_k141_206841 | Not-determined |
| 18-109_k141_93445 | Low-quality |
| 18-109_k141_22188 | Not-determined |
| 18-109_k141_165697 | Not-determined |
| 18-109_k141_186375 | Not-determined |
| 18-109_k141_186376 | Not-determined |
| 18-109_k141_22193 | Not-determined |
| 18-109_k141_216608 | Not-determined |
| 18-109_k141_196545 | Not-determined |
| 18-109_k141_144661 | Not-determined |
| 18-109_k141_12269 | Low-quality |
| 18-109_k141_124840 | Not-determined |
| 18-109_k141_196553 | Not-determined |
| 18-109_k141_237194 | Not-determined |
| 18-109_k141_114662 | Not-determined |
| 18-109_k141_196557 | Not-determined |
| 18-109_k141_216616 | Not-determined |
| 18-109_k141_216617 | Not-determined |
| 18-109_k141_12277 | Not-determined |
| 18-109_k141_237197 | Not-determined |
| 18-109_k141_227152 | Not-determined |
| 18-109_k141_155561 | Not-determined |
| 18-109_k141_237203 | Low-quality |
| 18-109_k141_2166 | Not-determined |
| 18-109_k141_186406 | Not-determined |
| 18-109_k141_155566 | Not-determined |
| 18-109_k141_155567 | Not-determined |
| 18-109_k141_227165 | Not-determined |
| 18-109_k141_216631 | Low-quality |
| 18-109_k141_155574 | Not-determined |
| 18-109_k141_2173 | Not-determined |
| 18-109_k141_155580 | Not-determined |
| 18-109_k141_144693 | Not-determined |
| 18-109_k141_165706 | Not-determined |
| 18-109_k141_227183 | Low-quality |
| 18-109_k141_43002 | Not-determined |
| 18-109_k141_227185 | Not-determined |
| 18-109_k141_216651 | Low-quality |
| 18-109_k141_22223 | Not-determined |
| 18-109_k141_165714 | Not-determined |
| 18-109_k141_134926 | Not-determined |
| 18-109_k141_134929 | Not-determined |
| 18-109_k141_114687 | Not-determined |
| 18-109_k141_83455 | Not-determined |
| 18-109_k141_83457 | Not-determined |
| 18-109_k141_2188 | Low-quality |
| 18-109_k141_93510 | Low-quality |
| 18-109_k141_114696 | Not-determined |
| 18-109_k141_196585 | Low-quality |
| 18-109_k141_83466 | Not-determined |
| 18-109_k141_155601 | Low-quality |
| 18-109_k141_114705 | Not-determined |
| 18-109_k141_2198 | Not-determined |
| 18-109_k141_2200 | Not-determined |
| 18-109_k141_227206 | Low-quality |
| 18-109_k141_216674 | Not-determined |
| 18-102_k141_245232 | High-quality |
| 18-109_k141_196594 | Low-quality |
| 18-109_k141_227213 | Not-determined |
| 18-109_k141_216675 | Not-determined |
| 18-109_k141_32922 | Low-quality |
| 18-109_k141_196596 | Not-determined |
| 18-109_k141_227216 | Not-determined |
| 18-109_k141_22238 | Not-determined |
| 18-109_k141_83480 | Not-determined |
| 18-109_k141_22242 | Not-determined |
| 18-109_k141_186427 | Not-determined |
| 18-109_k141_155613 | Not-determined |
| 18-109_k141_83494 | Not-determined |
| 18-109_k141_104349 | Not-determined |
| 18-109_k141_83501 | Low-quality |
| 18-109_k141_216692 | Not-determined |
| 18-109_k141_22255 | Not-determined |
| 18-109_k141_93531 | Not-determined |
| 18-109_k141_134970 | Not-determined |
| 18-109_k141_83511 | Not-determined |
| 18-109_k141_144723 | Not-determined |
| 18-109_k141_237236 | Not-determined |
| 18-109_k141_114743 | Not-determined |
| 18-109_k141_114744 | Not-determined |
| 18-109_k141_53361 | Not-determined |
| 18-109_k141_196608 | Not-determined |
| 18-109_k141_83523 | Not-determined |
| 18-109_k141_114750 | Not-determined |
| 18-109_k141_83526 | Not-determined |
| 18-109_k141_53364 | Not-determined |
| 18-109_k141_83530 | Not-determined |
| 18-109_k141_144739 | Not-determined |
| 18-109_k141_72924 | Not-determined |
| 18-109_k141_104383 | Not-determined |
| 18-109_k141_83544 | Not-determined |
| 18-109_k141_155640 | Not-determined |
| 18-109_k141_155644 | Not-determined |
| 18-109_k141_155646 | Not-determined |
| 18-109_k141_114766 | Not-determined |
| 18-109_k141_196650 | Not-determined |
| 18-109_k141_32942 | Not-determined |
| 18-109_k141_196657 | Not-determined |
| 18-109_k141_155652 | Not-determined |
| 18-109_k141_83555 | Not-determined |
| 18-109_k141_32948 | Low-quality |
| 18-109_k141_53382 | Not-determined |
| 18-109_k141_83563 | Not-determined |
| 18-109_k141_53390 | Not-determined |
| 18-109_k141_93557 | Not-determined |
| 18-109_k141_93561 | Not-determined |
| 18-109_k141_93563 | Not-determined |
| 18-109_k141_63440 | Not-determined |
| 18-109_k141_206875 | Not-determined |
| 18-109_k141_53409 | Not-determined |
| 18-109_k141_63448 | Low-quality |
| 18-109_k141_206890 | Not-determined |
| 18-109_k141_63460 | Not-determined |
| 18-109_k141_63461 | Not-determined |
| 18-109_k141_53426 | Not-determined |
| 18-109_k141_227254 | Low-quality |
| 18-109_k141_237279 | Not-determined |
| 18-109_k141_216703 | Low-quality |
| 18-109_k141_114797 | Not-determined |
| 18-109_k141_2240 | Not-determined |
| 18-109_k141_2246 | Not-determined |
| 18-109_k141_206904 | Not-determined |
| 18-109_k141_216711 | Not-determined |
| 18-109_k141_227282 | Not-determined |
| 18-109_k141_32977 | Not-determined |
| 18-109_k141_155669 | Not-determined |
| 18-109_k141_227289 | Not-determined |
| 18-109_k141_104418 | Not-determined |
| 18-109_k141_104427 | Not-determined |
| 18-109_k141_144786 | Not-determined |
| 18-109_k141_22299 | Not-determined |
| 18-109_k141_53442 | Not-determined |
| 18-109_k141_237292 | Not-determined |
| 18-109_k141_155676 | Not-determined |
| 18-109_k141_2258 | Not-determined |
| 18-109_k141_114814 | Not-determined |
| 18-109_k141_22302 | Not-determined |
| 18-109_k141_63504 | Not-determined |
| 18-109_k141_2259 | Not-determined |
| 18-109_k141_237297 | Low-quality |
| 18-109_k141_155680 | Not-determined |
| 18-109_k141_12316 | Not-determined |
| 18-109_k141_104438 | Not-determined |
| 18-109_k141_53445 | Not-determined |
| 18-109_k141_83607 | Not-determined |
| 18-109_k141_2271 | Not-determined |
| 18-109_k141_12321 | Not-determined |
| 18-109_k141_93612 | Not-determined |
| 18-109_k141_216756 | Not-determined |
| 18-109_k141_43034 | Low-quality |
| 18-109_k141_93616 | Low-quality |
| 18-109_k141_227292 | Not-determined |
| 18-109_k141_144826 | Not-determined |
| 18-109_k141_165760 | Not-determined |
| 18-109_k141_2291 | Not-determined |
| 18-109_k141_155714 | Not-determined |
| 18-109_k141_83611 | Low-quality |
| 18-109_k141_63541 | Not-determined |
| 18-109_k141_2296 | Not-determined |
| 18-109_k141_216762 | Not-determined |
| 18-109_k141_83615 | Low-quality |
| 18-109_k141_83616 | Not-determined |
| 18-109_k141_83617 | Not-determined |
| 18-109_k141_12329 | Not-determined |
| 18-109_k141_216769 | Not-determined |
| 18-109_k141_144842 | Not-determined |
| 18-109_k141_186508 | Not-determined |
| 18-109_k141_72979 | Not-determined |
| 18-109_k141_2306 | Not-determined |
| 18-109_k141_206938 | Low-quality |
| 18-109_k141_155722 | Not-determined |
| 18-109_k141_165769 | Not-determined |
| 18-109_k141_186517 | Not-determined |
| 18-109_k141_186520 | Not-determined |
| 18-109_k141_43059 | Not-determined |
| 18-109_k141_186521 | Not-determined |
| 18-109_k141_165775 | Not-determined |
| 18-109_k141_12339 | Not-determined |
| 18-109_k141_227306 | Not-determined |
| 18-109_k141_206951 | Not-determined |
| 18-109_k141_186529 | Not-determined |
| 18-109_k141_206961 | Not-determined |
| 18-109_k141_43072 | Not-determined |
| 18-109_k141_83652 | Not-determined |
| 18-109_k141_2333 | Not-determined |
| 18-109_k141_155732 | Low-quality |
| 18-109_k141_206972 | Not-determined |
| 18-109_k141_83658 | Low-quality |
| 18-109_k141_186544 | Low-quality |
| 18-109_k141_2342 | Low-quality |
| 18-109_k141_186549 | Low-quality |
| 18-109_k141_2348 | Not-determined |
| 18-109_k141_12356 | Not-determined |
| 18-109_k141_237338 | Not-determined |
| 18-109_k141_237339 | Not-determined |
| 18-109_k141_155739 | Not-determined |
| 18-109_k141_196732 | Not-determined |
| 18-109_k141_227310 | Not-determined |
| 18-109_k141_12370 | Low-quality |
| 18-109_k141_2367 | Low-quality |
| 18-109_k141_33010 | Low-quality |
| 18-109_k141_196747 | Not-determined |
| 18-109_k141_196752 | Not-determined |
| 18-109_k141_2378 | Not-determined |
| 18-109_k141_124855 | Not-determined |
| 18-109_k141_124859 | Not-determined |
| 18-109_k141_33015 | Not-determined |
| 18-109_k141_206988 | Not-determined |
| 18-109_k141_2402 | Not-determined |
| 18-109_k141_22325 | Not-determined |
| 18-109_k141_93646 | Low-quality |
| 18-109_k141_216801 | Not-determined |
| 18-109_k141_124880 | Not-determined |
| 18-109_k141_206998 | Not-determined |
| 18-109_k141_33036 | Not-determined |
| 18-109_k141_207004 | Not-determined |
| 18-109_k141_186574 | Not-determined |
| 18-109_k141_207010 | Not-determined |
| 18-109_k141_196770 | Not-determined |
| 18-109_k141_237373 | Low-quality |
| 18-109_k141_155775 | Not-determined |
| 18-109_k141_227366 | Not-determined |
| 18-109_k141_237383 | Not-determined |
| 18-109_k141_186585 | Not-determined |
| 18-109_k141_144932 | Not-determined |
| 18-109_k141_227371 | Not-determined |
| 18-109_k141_63557 | Not-determined |
| 18-109_k141_33050 | Not-determined |
| 18-109_k141_207025 | Not-determined |
| 18-109_k141_2410 | Not-determined |
| 18-109_k141_144938 | Not-determined |
| 18-109_k141_104484 | Not-determined |
| 18-109_k141_93670 | Not-determined |
| 18-109_k141_237393 | Not-determined |
| 18-109_k141_104486 | Not-determined |
| 18-109_k141_186595 | Not-determined |
| 18-109_k141_186596 | Not-determined |
| 18-109_k141_144942 | Not-determined |
| 18-109_k141_165813 | Not-determined |
| 18-109_k141_73057 | Low-quality |
| 18-109_k141_93680 | Not-determined |
| 18-109_k141_186604 | Not-determined |
| 18-109_k141_186605 | Not-determined |
| 18-109_k141_196797 | Low-quality |
| 18-109_k141_22348 | Not-determined |
| 18-109_k141_22350 | Not-determined |
| 18-109_k141_124898 | Low-quality |
| 18-109_k141_93684 | Low-quality |
| 18-109_k141_73068 | Not-determined |
| 18-109_k141_216836 | Not-determined |
| 18-109_k141_196799 | Not-determined |
| 18-109_k141_43109 | Low-quality |
| 18-109_k141_196802 | Low-quality |
| 18-109_k141_93688 | Not-determined |
| 18-109_k141_124902 | Not-determined |
| 18-109_k141_165843 | Not-determined |
| 18-109_k141_165845 | Not-determined |
| 18-109_k141_93692 | Not-determined |
| 18-109_k141_196817 | Not-determined |
| 18-109_k141_165853 | Not-determined |
| 18-109_k141_165855 | Not-determined |
| 18-109_k141_33089 | Not-determined |
| 18-109_k141_33090 | Not-determined |
| 18-109_k141_53525 | Not-determined |
| 18-109_k141_237429 | Not-determined |
| 18-109_k141_22372 | Not-determined |
| 18-109_k141_134994 | Not-determined |
| 18-109_k141_155782 | Not-determined |
| 18-109_k141_63606 | Not-determined |
| 18-109_k141_165863 | Not-determined |
| 18-109_k141_53534 | Not-determined |
| 18-109_k141_155789 | Not-determined |
| 18-109_k141_207060 | Not-determined |
| 18-109_k141_155792 | Not-determined |
| 18-109_k141_135010 | Not-determined |
| 18-109_k141_165869 | Not-determined |
| 18-109_k141_135013 | Not-determined |
| 18-109_k141_207068 | Low-quality |
| 18-109_k141_165870 | Not-determined |
| 18-109_k141_165871 | Not-determined |
| 18-109_k141_43136 | Not-determined |
| 18-109_k141_43138 | Not-determined |
| 18-109_k141_237437 | Not-determined |
| 18-109_k141_124913 | Not-determined |
| 18-109_k141_53542 | Not-determined |
| 18-109_k141_63637 | Not-determined |
| 18-109_k141_63640 | Not-determined |
| 18-109_k141_135022 | Not-determined |
| 18-109_k141_135030 | Not-determined |
| 18-109_k141_12446 | Not-determined |
| 18-109_k141_12447 | Not-determined |
| 18-109_k141_53550 | Not-determined |
| 18-109_k141_83746 | Not-determined |
| 18-109_k141_12454 | Not-determined |
| 18-109_k141_93725 | Not-determined |
| 18-109_k141_43150 | Not-determined |
| 18-109_k141_144970 | Not-determined |
| 18-109_k141_83752 | Not-determined |
| 18-109_k141_63667 | Not-determined |
| 18-109_k141_83757 | Not-determined |
| 18-109_k141_63672 | Not-determined |
| 18-109_k141_63673 | Not-determined |
| 18-109_k141_83762 | Not-determined |
| 18-109_k141_237456 | Not-determined |
| 18-109_k141_196861 | Not-determined |
| 18-109_k141_124945 | Not-determined |
| 18-109_k141_186674 | Not-determined |
| 18-109_k141_196876 | Not-determined |
| 18-109_k141_186679 | Not-determined |
| 18-109_k141_2433 | Not-determined |
| 18-109_k141_124954 | Low-quality |
| 18-109_k141_2444 | Not-determined |
| 18-109_k141_196889 | Not-determined |
| 18-109_k141_114856 | Not-determined |
| 18-109_k141_114858 | Not-determined |
| 18-109_k141_114859 | Not-determined |
| 18-109_k141_63696 | Not-determined |
| 18-109_k141_83792 | Low-quality |
| 18-109_k141_216852 | Not-determined |
| 18-109_k141_216856 | Not-determined |
| 18-109_k141_237477 | Not-determined |
| 18-109_k141_207081 | Not-determined |
| 18-109_k141_63705 | Not-determined |
| 18-109_k141_43169 | Not-determined |
| 18-109_k141_196904 | Not-determined |
| 18-109_k141_186695 | Not-determined |
| 18-109_k141_63710 | Not-determined |
| 18-109_k141_43172 | Not-determined |
| 18-109_k141_186700 | Not-determined |
| 18-109_k141_12490 | Not-determined |
| 18-109_k141_53610 | Low-quality |
| 18-109_k141_237485 | Not-determined |
| 18-109_k141_135037 | Not-determined |
| 18-109_k141_175704 | Not-determined |
| 18-109_k141_207099 | Not-determined |
| 18-109_k141_186706 | Not-determined |
| 18-109_k141_135043 | Not-determined |
| 18-109_k141_207102 | Low-quality |
| 18-109_k141_135046 | Not-determined |
| 18-109_k141_207105 | Not-determined |
| 18-109_k141_114885 | Not-determined |
| 18-109_k141_155846 | Not-determined |
| 18-109_k141_175709 | Not-determined |
| 18-109_k141_175711 | Not-determined |
| 18-109_k141_114887 | Not-determined |
| 18-109_k141_83830 | Not-determined |
| 18-109_k141_155853 | Low-quality |
| 18-109_k141_227448 | Not-determined |
| 18-109_k141_145014 | Not-determined |
| 18-109_k141_135053 | Not-determined |
| 18-109_k141_227449 | Not-determined |
| 18-109_k141_33136 | Low-quality |
| 18-109_k141_145019 | Not-determined |
| 18-109_k141_237509 | Not-determined |
| 18-109_k141_207138 | Not-determined |
| 18-109_k141_237519 | Not-determined |
| 18-109_k141_196961 | Not-determined |
| 18-109_k141_53626 | Not-determined |
| 18-109_k141_73125 | Not-determined |
| 18-109_k141_73131 | Not-determined |
| 18-109_k141_165886 | Not-determined |
| 18-109_k141_175719 | Not-determined |
| 18-109_k141_12538 | Low-quality |
| 18-109_k141_43206 | Not-determined |
| 18-109_k141_175723 | Low-quality |
| 18-109_k141_83843 | Not-determined |
| 18-109_k141_43213 | Not-determined |
| 18-109_k141_43214 | Not-determined |
| 18-109_k141_83854 | Not-determined |
| 18-109_k141_33152 | Not-determined |
| 18-109_k141_227466 | Not-determined |
| 18-109_k141_33153 | Not-determined |
| 18-109_k141_186726 | Not-determined |
| 18-109_k141_2493 | Not-determined |
| 18-109_k141_175731 | Not-determined |
| 18-109_k141_73135 | Not-determined |
| 18-109_k141_227470 | Not-determined |
| 18-109_k141_186728 | Not-determined |
| 18-109_k141_155892 | Not-determined |
| 18-109_k141_155896 | Low-quality |
| 18-109_k141_186740 | Not-determined |
| 18-109_k141_53668 | Not-determined |
| 18-109_k141_186743 | Not-determined |
| 18-109_k141_186744 | Not-determined |
| 18-109_k141_186749 | Not-determined |
| 18-109_k141_83872 | Not-determined |
| 18-109_k141_83873 | Not-determined |
| 18-109_k141_124969 | Not-determined |
| 18-109_k141_63760 | Not-determined |
| 18-109_k141_124972 | Low-quality |
| 18-109_k141_43235 | Not-determined |
| 18-109_k141_135073 | Not-determined |
| 18-109_k141_63763 | Not-determined |
| 18-109_k141_155915 | Not-determined |
| 18-109_k141_73170 | Not-determined |
| 18-109_k141_155916 | Not-determined |
| 18-109_k141_43245 | Not-determined |
| 18-109_k141_83882 | Not-determined |
| 18-109_k141_63770 | Not-determined |
| 18-109_k141_135088 | Not-determined |
| 18-109_k141_175745 | Not-determined |
| 18-109_k141_73179 | Not-determined |
| 18-109_k141_135095 | Low-quality |
| 18-109_k141_186774 | Not-determined |
| 18-109_k141_155942 | Low-quality |
| 18-109_k141_125011 | Low-quality |
| 18-109_k141_12563 | Low-quality |
| 18-109_k141_207211 | Not-determined |
| 18-109_k141_83918 | Not-determined |
| 18-109_k141_22414 | Not-determined |
| 18-109_k141_22417 | Not-determined |
| 18-109_k141_207221 | Not-determined |
| 18-109_k141_227541 | Not-determined |
| 18-109_k141_73187 | Low-quality |
| 18-109_k141_196989 | Not-determined |
| 18-109_k141_186789 | Not-determined |
| 18-109_k141_73195 | Low-quality |
| 18-109_k141_196993 | Not-determined |
| 18-109_k141_145068 | Not-determined |
| 18-109_k141_165921 | Not-determined |
| 18-109_k141_12583 | Not-determined |
| 18-109_k141_22423 | Not-determined |
| 18-109_k141_237581 | Low-quality |
| 18-109_k141_165925 | Low-quality |
| 18-109_k141_53704 | Not-determined |
| 18-109_k141_12585 | Low-quality |
| 18-109_k141_63809 | Not-determined |
| 18-109_k141_237587 | Not-determined |
| 18-109_k141_12591 | Low-quality |
| 18-109_k141_207233 | Not-determined |
| 18-109_k141_155958 | Not-determined |
| 18-109_k141_22433 | Not-determined |
| 18-109_k141_165930 | Low-quality |
| 18-109_k141_22435 | Not-determined |
| 18-109_k141_83925 | Not-determined |
| 18-109_k141_2574 | Not-determined |
| 18-109_k141_175776 | Not-determined |
| 18-109_k141_43275 | Not-determined |
| 18-109_k141_237602 | Not-determined |
| 18-109_k141_43279 | Not-determined |
| 18-109_k141_73217 | Not-determined |
| 18-109_k141_197004 | Not-determined |
| 18-109_k141_155969 | Not-determined |
| 18-109_k141_207241 | Low-quality |
| 18-109_k141_43283 | Not-determined |
| 18-109_k141_237609 | Not-determined |
| 18-109_k141_93741 | Not-determined |
| 18-109_k141_155974 | Not-determined |
| 18-109_k141_227561 | Not-determined |
| 18-109_k141_155980 | Not-determined |
| 18-109_k141_43298 | Not-determined |
| 18-109_k141_114934 | Not-determined |
| 18-109_k141_227570 | Not-determined |
| 18-109_k141_197032 | Not-determined |
| 18-109_k141_197033 | Low-quality |
| 18-109_k141_12638 | Not-determined |
| 18-109_k141_175807 | Low-quality |
| 18-109_k141_12640 | Not-determined |
| 18-109_k141_114955 | Low-quality |
| 18-109_k141_2602 | Not-determined |
| 18-109_k141_63822 | Not-determined |
| 18-109_k141_2603 | Not-determined |
| 18-109_k141_145082 | Not-determined |
| 18-109_k141_114958 | Not-determined |
| 18-109_k141_207267 | Low-quality |
| 18-109_k141_175821 | Not-determined |
| 18-109_k141_2607 | Not-determined |
| 18-109_k141_114962 | Not-determined |
| 18-109_k141_2608 | Not-determined |
| 18-109_k141_104515 | Low-quality |
| 18-109_k141_237629 | Not-determined |
| 18-109_k141_104516 | Not-determined |
| 18-109_k141_216871 | Low-quality |
| 18-109_k141_2613 | Not-determined |
| 18-109_k141_43325 | Not-determined |
| 18-109_k141_2614 | Not-determined |
| 18-109_k141_227600 | Not-determined |
| 18-109_k141_63846 | Not-determined |
| 18-109_k141_227607 | Not-determined |
| 18-109_k141_216877 | Not-determined |
| 18-109_k141_83935 | Not-determined |
| 18-109_k141_227615 | Not-determined |
| 18-109_k141_12659 | Not-determined |
| 18-109_k141_175832 | Low-quality |
| 18-109_k141_175833 | Not-determined |
| 18-109_k141_175842 | Not-determined |
| 18-109_k141_227620 | Not-determined |
| 18-109_k141_197052 | Not-determined |
| 18-109_k141_93766 | Low-quality |
| 18-109_k141_135137 | Not-determined |
| 18-109_k141_135142 | Not-determined |
| 18-109_k141_115001 | Not-determined |
| 18-109_k141_237671 | Not-determined |
| 18-109_k141_104547 | Low-quality |
| 18-109_k141_156015 | Not-determined |
| 18-109_k141_216894 | Not-determined |
| 18-109_k141_165982 | Not-determined |
| 18-109_k141_227631 | Not-determined |
| 18-109_k141_63859 | Not-determined |
| 18-109_k141_104558 | Not-determined |
| 18-109_k141_165985 | Not-determined |
| 18-109_k141_216895 | Low-quality |
| 18-109_k141_227635 | Low-quality |
| 18-109_k141_237684 | Low-quality |
| 18-109_k141_33181 | Not-determined |
| 18-109_k141_165998 | Not-determined |
| 18-109_k141_43369 | Not-determined |
| 18-109_k141_175852 | Not-determined |
| 18-109_k141_237690 | Not-determined |
| 18-109_k141_216908 | Not-determined |
| 18-109_k141_93799 | Not-determined |
| 18-109_k141_2651 | Not-determined |
| 18-109_k141_93802 | Low-quality |
| 18-109_k141_166007 | Not-determined |
| 18-109_k141_63868 | Not-determined |
| 18-109_k141_63869 | Not-determined |
| 18-109_k141_63871 | Not-determined |
| 18-109_k141_2665 | Low-quality |
| 18-109_k141_237698 | Not-determined |
| 18-109_k141_237701 | Not-determined |
| 18-109_k141_104576 | Not-determined |
| 18-109_k141_175877 | Not-determined |
| 18-109_k141_207317 | Not-determined |
| 18-109_k141_33195 | Not-determined |
| 18-109_k141_197077 | Not-determined |
| 18-109_k141_186842 | Not-determined |
| 18-109_k141_43397 | Not-determined |
| 18-109_k141_43399 | Not-determined |
| 18-109_k141_2678 | Not-determined |
| 18-109_k141_175884 | Low-quality |
| 18-109_k141_53736 | Not-determined |
| 18-109_k141_175885 | Not-determined |
| 18-109_k141_156058 | Not-determined |
| 18-109_k141_186854 | Not-determined |
| 18-109_k141_43405 | Not-determined |
| 18-109_k141_207335 | Not-determined |
| 18-109_k141_93832 | Not-determined |
| 18-109_k141_53753 | Not-determined |
| 18-109_k141_43413 | Not-determined |
| 18-109_k141_12695 | Not-determined |
| 18-109_k141_12698 | Not-determined |
| 18-109_k141_186871 | Not-determined |
| 18-109_k141_197099 | Not-determined |
| 18-109_k141_186874 | Not-determined |
| 18-109_k141_33222 | Low-quality |
| 18-109_k141_207353 | Not-determined |

| | |
|---|---|
| 18-109_k141_93861 | Not-determined |
| 18-109_k141_186886 | Not-determined |
| 18-109_k141_104607 | Not-determined |
| 18-109_k141_156068 | Not-determined |
| 18-109_k141_12717 | Not-determined |
| 18-109_k141_33258 | Not-determined |
| 18-109_k141_166031 | Not-determined |
| 18-109_k141_186908 | Not-determined |
| 18-109_k141_175917 | Not-determined |
| 18-109_k141_175919 | Not-determined |
| 18-109_k141_33272 | Not-determined |
| 18-109_k141_104617 | Not-determined |
| 18-109_k141_125060 | Not-determined |
| 18-109_k141_125064 | Not-determined |
| 18-109_k141_166052 | Not-determined |
| 18-109_k141_175936 | Not-determined |
| 18-109_k141_207366 | Not-determined |
| 18-109_k141_207367 | Low-quality |
| 18-109_k141_83990 | Not-determined |
| 18-109_k141_197107 | Not-determined |
| 18-109_k141_63900 | Not-determined |
| 18-109_k141_135167 | Not-determined |
| 18-109_k141_156084 | Not-determined |
| 18-109_k141_93876 | Not-determined |
| 18-109_k141_135169 | Not-determined |
| 18-109_k141_84003 | Not-determined |
| 18-109_k141_156088 | Not-determined |
| 18-109_k141_135176 | Not-determined |
| 18-109_k141_84011 | Not-determined |
| 18-109_k141_237746 | Not-determined |
| 18-109_k141_104625 | Not-determined |
| 18-109_k141_125078 | Not-determined |
| 18-109_k141_43444 | Not-determined |
| 18-109_k141_166077 | Not-determined |
| 18-109_k141_33300 | Low-quality |
| 18-109_k141_237752 | Not-determined |
| 18-109_k141_115086 | Not-determined |
| 18-109_k141_156098 | Not-determined |
| 18-109_k141_84018 | Not-determined |
| 18-109_k141_84019 | Not-determined |
| 18-109_k141_84021 | Not-determined |
| 18-109_k141_166091 | Not-determined |
| 18-109_k141_186955 | Not-determined |
| 18-109_k141_135194 | Not-determined |
| 18-109_k141_84023 | Not-determined |
| 18-109_k141_22467 | Not-determined |
| 18-109_k141_186961 | Not-determined |
| 18-109_k141_237767 | Not-determined |
| 18-109_k141_12740 | Not-determined |
| 18-109_k141_156112 | Not-determined |
| 18-109_k141_237768 | Not-determined |
| 18-109_k141_43479 | Not-determined |
| 18-109_k141_12743 | Not-determined |
| 18-109_k141_63914 | Not-determined |
| 18-109_k141_115099 | Not-determined |
| 18-109_k141_12746 | Not-determined |
| 18-109_k141_186969 | Not-determined |
| 18-109_k141_156116 | Not-determined |
| 18-109_k141_135208 | Not-determined |
| 18-109_k141_135209 | Not-determined |
| 18-109_k141_135211 | Not-determined |
| 18-109_k141_12752 | Not-determined |
| 18-109_k141_227648 | Not-determined |
| 18-109_k141_145133 | Not-determined |
| 18-109_k141_237786 | Not-determined |
| 18-109_k141_63926 | Not-determined |
| 18-109_k141_207381 | Not-determined |
| 18-109_k141_237791 | Not-determined |
| 18-109_k141_207382 | Not-determined |
| 18-109_k141_135217 | Not-determined |
| 18-109_k141_207383 | Not-determined |
| 18-109_k141_43499 | Not-determined |
| 18-109_k141_135224 | Not-determined |
| 18-109_k141_104654 | Not-determined |
| 18-109_k141_207388 | Not-determined |
| 18-109_k141_115108 | Not-determined |
| 18-109_k141_104663 | Not-determined |
| 18-109_k141_12761 | Not-determined |
| 18-109_k141_237812 | Not-determined |
| 18-109_k141_73332 | Not-determined |
| 18-109_k141_104674 | Not-determined |
| 18-109_k141_216923 | Not-determined |
| 18-109_k141_216926 | Not-determined |
| 18-109_k141_84047 | Not-determined |
| 18-109_k141_43519 | Not-determined |
| 18-109_k141_115124 | Not-determined |
| 18-109_k141_43526 | Not-determined |
| 18-109_k141_73338 | Not-determined |
| 18-109_k141_73339 | Not-determined |
| 18-109_k141_63939 | Not-determined |
| 18-109_k141_216935 | Not-determined |
| 18-109_k141_63943 | Not-determined |
| 18-109_k141_175968 | Not-determined |
| 18-109_k141_175970 | Low-quality |
| 18-109_k141_22490 | Not-determined |
| 18-109_k141_145161 | Low-quality |
| 18-109_k141_237828 | Not-determined |
| 18-109_k141_12794 | Not-determined |
| 18-109_k141_237831 | Not-determined |
| 18-109_k141_207425 | Not-determined |
| 18-109_k141_33345 | Low-quality |
| 18-109_k141_12809 | Not-determined |
| 18-109_k141_207430 | Not-determined |
| 18-109_k141_84091 | Not-determined |
| 18-109_k141_115144 | Not-determined |
| 18-109_k141_207434 | Not-determined |
| 18-109_k141_63955 | Not-determined |
| 18-109_k141_84097 | Not-determined |
| 18-109_k141_63956 | Not-determined |
| 18-109_k141_22499 | Not-determined |
| 18-109_k141_63957 | Not-determined |
| 18-109_k141_43538 | Not-determined |
| 18-109_k141_176009 | Not-determined |
| 18-109_k141_33356 | Low-quality |
| 18-109_k141_43543 | Not-determined |
| 18-109_k141_12836 | Not-determined |
| 18-109_k141_207454 | Low-quality |
| 18-109_k141_84108 | Not-determined |
| 18-109_k141_63972 | Not-determined |
| 18-109_k141_22515 | Low-quality |
| 18-109_k141_216960 | Not-determined |
| 18-109_k141_176036 | Not-determined |
| 18-109_k141_33377 | Not-determined |
| 18-109_k141_93964 | Not-determined |
| 18-109_k141_22524 | Not-determined |
| 18-109_k141_125097 | Not-determined |
| 18-109_k141_33379 | Low-quality |
| 18-109_k141_115152 | Not-determined |
| 18-109_k141_197167 | Not-determined |
| 18-109_k141_33383 | Not-determined |
| 18-109_k141_93972 | Not-determined |
| 18-109_k141_237871 | Low-quality |
| 18-109_k141_43557 | Not-determined |
| 18-109_k141_197169 | Not-determined |
| 18-109_k141_166115 | Not-determined |
| 18-109_k141_93977 | Low-quality |
| 18-109_k141_176050 | Not-determined |
| 18-109_k141_207467 | Not-determined |
| 18-109_k141_197174 | Not-determined |
| 18-109_k141_93986 | Not-determined |
| 18-109_k141_227653 | Not-determined |
| 18-109_k141_93989 | Not-determined |
| 18-109_k141_33389 | Not-determined |
| 18-109_k141_12861 | Not-determined |
| 18-109_k141_125121 | Not-determined |
| 18-109_k141_135232 | Not-determined |
| 18-109_k141_156182 | Not-determined |
| 18-109_k141_33397 | Not-determined |
| 18-109_k141_115181 | Not-determined |
| 18-109_k141_227666 | Not-determined |
| 18-109_k141_207492 | Not-determined |
| 18-109_k141_156186 | Not-determined |
| 18-109_k141_12877 | Not-determined |
| 18-109_k141_125128 | Not-determined |
| 18-109_k141_227668 | Not-determined |
| 18-109_k141_33401 | Not-determined |
| 18-109_k141_63997 | Not-determined |
| 18-109_k141_115187 | Low-quality |
| 18-109_k141_104687 | Not-determined |
| 18-109_k141_12885 | Not-determined |
| 18-109_k141_64002 | Not-determined |
| 18-109_k141_33405 | Not-determined |
| 18-109_k141_12897 | Low-quality |
| 18-109_k141_166141 | Low-quality |
| 18-109_k141_104696 | Not-determined |
| 18-109_k141_166145 | Not-determined |
| 18-109_k141_156205 | Low-quality |
| 18-109_k141_166157 | Not-determined |
| 18-109_k141_84132 | Low-quality |
| 18-109_k141_237915 | Not-determined |
| 18-109_k141_166165 | Not-determined |
| 18-109_k141_43583 | Not-determined |
| 18-109_k141_22575 | Not-determined |
| 18-109_k141_73357 | Not-determined |
| 18-109_k141_43587 | Not-determined |
| 18-109_k141_237919 | Not-determined |
| 18-109_k141_217005 | Not-determined |
| 18-109_k141_43589 | Not-determined |
| 18-109_k141_207523 | Low-quality |
| 18-109_k141_43591 | Not-determined |
| 18-109_k141_207524 | Not-determined |
| 18-109_k141_176062 | Not-determined |
| 18-109_k141_237924 | Not-determined |
| 18-109_k141_227688 | Not-determined |
| 18-109_k141_84144 | Not-determined |
| 18-109_k141_125154 | Not-determined |
| 18-109_k141_217011 | Not-determined |
| 18-109_k141_22582 | Not-determined |
| 18-109_k141_217013 | Not-determined |
| 18-109_k141_43602 | Not-determined |
| 18-109_k141_43603 | Not-determined |
| 18-109_k141_115195 | Not-determined |
| 18-109_k141_176070 | Not-determined |
| 18-109_k141_2759 | Low-quality |
| 18-109_k141_207539 | Not-determined |
| 18-109_k141_217020 | Not-determined |
| 18-109_k141_12922 | Low-quality |
| 18-109_k141_125170 | Not-determined |
| 18-109_k141_125171 | Low-quality |
| 18-109_k141_227712 | Not-determined |
| 18-109_k141_73370 | Not-determined |
| 18-109_k141_156223 | Not-determined |
| 18-109_k141_125183 | Low-quality |
| 18-109_k141_115205 | Not-determined |
| 18-109_k141_237931 | Not-determined |
| 18-109_k141_237932 | Not-determined |
| 18-109_k141_156232 | Not-determined |
| 18-109_k141_12936 | Not-determined |
| 18-109_k141_33448 | Not-determined |
| 18-109_k141_84149 | Not-determined |
| 18-109_k141_73382 | Not-determined |
| 18-109_k141_84151 | Not-determined |
| 18-109_k141_237944 | Not-determined |
| 18-109_k141_237945 | Not-determined |
| 18-109_k141_115213 | Not-determined |
| 18-109_k141_73387 | Not-determined |
| 18-109_k141_94027 | Not-determined |
| 18-109_k141_12944 | Not-determined |
| 18-109_k141_84155 | Not-determined |
| 18-109_k141_176085 | Not-determined |
| 18-109_k141_166192 | Not-determined |
| 18-109_k141_94029 | Not-determined |
| 18-109_k141_115225 | Not-determined |
| 18-109_k141_115226 | Low-quality |
| 18-109_k141_43647 | Not-determined |
| 18-109_k141_135243 | Not-determined |
| 18-109_k141_43649 | Not-determined |
| 18-109_k141_227741 | Not-determined |
| 18-109_k141_135251 | Not-determined |
| 18-109_k141_43661 | Not-determined |
| 18-109_k141_197203 | Low-quality |
| 18-109_k141_22615 | Not-determined |
| 18-109_k141_73411 | Not-determined |
| 18-109_k141_237963 | Not-determined |
| 18-109_k141_156248 | Not-determined |
| 18-109_k141_156253 | Not-determined |
| 18-109_k141_145245 | Not-determined |
| 18-109_k141_156254 | Not-determined |
| 18-109_k141_207572 | Not-determined |
| 18-109_k141_237965 | Not-determined |
| 18-109_k141_33489 | Not-determined |
| 18-109_k141_43675 | Not-determined |
| 18-109_k141_187009 | Not-determined |
| 18-109_k141_176108 | Not-determined |
| 18-109_k141_176109 | Not-determined |
| 18-109_k141_197213 | Not-determined |
| 18-109_k141_43678 | Not-determined |
| 18-109_k141_156259 | Not-determined |
| 18-109_k141_176113 | Not-determined |
| 18-109_k141_115253 | Low-quality |
| 18-109_k141_2782 | Not-determined |
| 18-109_k141_207582 | Not-determined |
| 18-109_k141_104747 | Not-determined |
| 18-109_k141_125217 | Not-determined |
| 18-109_k141_53826 | Not-determined |
| 18-109_k141_135275 | Not-determined |
| 18-109_k141_207589 | Not-determined |
| 18-109_k141_115262 | Low-quality |
| 18-109_k141_33508 | Not-determined |
| 18-109_k141_43694 | Not-determined |
| 18-109_k141_84180 | Not-determined |
| 18-109_k141_104757 | Low-quality |
| 18-109_k141_73424 | Low-quality |
| 18-109_k141_33515 | Not-determined |
| 18-109_k141_104762 | Not-determined |
| 18-109_k141_115274 | Low-quality |
| 18-109_k141_73431 | Not-determined |
| 18-109_k141_33527 | Not-determined |
| 18-109_k141_33529 | Not-determined |
| 18-109_k141_22646 | Not-determined |
| 18-109_k141_237984 | Not-determined |
| 18-109_k141_104779 | Not-determined |
| 18-109_k141_73444 | Not-determined |
| 18-109_k141_176147 | Not-determined |
| 18-109_k141_176148 | Not-determined |
| 18-109_k141_43710 | Not-determined |
| 18-109_k141_187029 | Not-determined |
| 18-109_k141_237990 | Not-determined |
| 18-109_k141_2809 | Not-determined |
| 18-109_k141_104786 | Not-determined |
| 18-109_k141_176153 | Not-determined |
| 18-109_k141_104789 | Not-determined |
| 18-109_k141_207615 | Not-determined |
| 18-109_k141_207618 | Low-quality |
| 18-109_k141_135291 | Not-determined |
| 18-109_k141_22675 | Not-determined |
| 18-109_k141_135297 | Not-determined |
| 18-109_k141_33564 | Not-determined |
| 18-109_k141_22676 | Not-determined |
| 18-109_k141_22677 | Not-determined |
| 18-109_k141_22678 | Not-determined |
| 18-109_k141_217066 | Not-determined |
| 18-109_k141_187042 | Not-determined |
| 18-109_k141_2823 | Not-determined |
| 18-109_k141_217073 | Not-determined |
| 18-109_k141_135305 | Not-determined |
| 18-109_k141_227752 | Not-determined |
| 18-109_k141_53875 | Not-determined |
| 18-109_k141_33584 | Not-determined |
| 18-109_k141_22691 | Not-determined |
| 18-109_k141_187054 | Not-determined |
| 18-109_k141_197273 | Not-determined |
| 18-109_k141_2845 | Not-determined |
| 18-109_k141_135327 | Not-determined |
| 18-109_k141_187064 | Not-determined |
| 18-109_k141_2847 | Not-determined |
| 18-109_k141_145312 | Not-determined |
| 18-109_k141_135329 | Not-determined |
| 18-109_k141_197283 | Not-determined |
| 18-109_k141_197287 | Not-determined |
| 18-109_k141_176171 | Not-determined |
| 18-109_k141_145317 | Not-determined |
| 18-109_k141_12991 | Not-determined |
| 18-109_k141_145321 | Not-determined |
| 18-109_k141_197297 | Not-determined |
| 18-109_k141_33594 | Not-determined |
| 18-109_k141_84223 | Not-determined |
| 18-109_k141_33599 | Not-determined |
| 18-109_k141_135342 | Not-determined |
| 18-109_k141_84230 | Not-determined |
| 18-109_k141_207628 | Not-determined |
| 18-109_k141_94149 | Not-determined |
| 18-109_k141_156325 | Not-determined |
| 18-109_k141_135345 | Not-determined |
| 18-109_k141_73504 | Not-determined |
| 18-109_k141_217085 | Not-determined |
| 18-109_k141_125264 | Not-determined |
| 18-109_k141_207635 | Not-determined |
| 18-109_k141_145339 | Low-quality |
| 18-109_k141_145341 | Not-determined |
| 18-109_k141_145343 | Not-determined |
| 18-109_k141_22698 | Low-quality |
| 18-109_k141_73517 | Not-determined |
| 18-109_k141_156336 | Low-quality |
| 18-109_k141_217091 | Not-determined |
| 18-109_k141_187116 | Low-quality |
| 18-109_k141_207649 | Not-determined |
| 18-109_k141_156342 | Not-determined |
| 18-109_k141_207652 | Not-determined |
| 18-109_k141_207654 | Not-determined |
| 18-109_k141_217099 | Not-determined |
| 18-109_k141_73523 | Not-determined |
| 18-109_k141_166217 | Not-determined |
| 18-109_k141_166218 | Not-determined |
| 18-109_k141_135364 | Not-determined |
| 18-109_k141_104821 | Not-determined |
| 18-109_k141_217108 | Not-determined |
| 18-109_k141_104822 | Low-quality |
| 18-109_k141_145355 | Not-determined |
| 18-109_k141_145357 | Not-determined |
| 18-109_k141_166232 | Not-determined |
| 18-109_k141_176227 | Not-determined |
| 18-109_k141_84269 | Not-determined |
| 18-109_k141_13042 | Not-determined |
| 18-109_k141_176230 | Not-determined |
| 18-109_k141_104833 | Not-determined |
| 18-109_k141_125276 | Not-determined |
| 18-109_k141_84276 | Not-determined |
| 18-109_k141_145367 | Not-determined |
| 18-109_k141_104840 | Not-determined |
| 18-109_k141_94183 | Not-determined |
| 18-109_k141_176238 | Not-determined |
| 18-109_k141_166250 | Not-determined |
| 18-109_k141_33656 | Not-determined |
| 18-109_k141_237999 | Not-determined |
| 18-109_k141_84292 | Not-determined |
| 18-109_k141_84294 | Low-quality |
| 18-109_k141_227786 | Not-determined |
| 18-109_k141_84295 | Not-determined |
| 18-109_k141_156357 | Not-determined |
| 18-109_k141_145375 | Low-quality |
| 18-109_k141_22710 | Not-determined |
| 18-109_k141_238005 | Not-determined |
| 18-109_k141_227793 | Not-determined |
| 18-109_k141_145381 | Not-determined |
| 18-109_k141_156378 | Not-determined |
| 18-109_k141_227797 | Not-determined |
| 18-109_k141_94211 | Not-determined |
| 18-109_k141_156383 | Not-determined |
| 18-109_k141_94214 | Not-determined |
| 18-109_k141_197343 | Not-determined |
| 18-109_k141_217118 | Not-determined |
| 18-109_k141_156392 | Not-determined |
| 18-109_k141_94220 | Not-determined |
| 18-109_k141_13066 | Not-determined |
| 18-109_k141_94231 | Not-determined |
| 18-109_k141_217131 | Not-determined |
| 18-109_k141_125303 | Not-determined |
| 18-109_k141_13073 | Not-determined |
| 18-109_k141_227813 | Not-determined |
| 18-109_k141_125305 | Not-determined |
| 18-109_k141_104850 | Not-determined |
| 18-109_k141_176263 | Not-determined |
| 18-109_k141_115297 | Not-determined |
| 18-109_k141_156405 | Not-determined |
| 18-109_k141_33667 | Not-determined |
| 18-109_k141_33669 | Not-determined |
| 18-109_k141_176274 | Not-determined |
| 18-109_k141_156410 | Not-determined |
| 18-109_k141_33677 | Not-determined |
| 18-109_k141_13081 | Not-determined |
| 18-109_k141_145434 | Not-determined |
| 18-109_k141_176280 | Low-quality |
| 18-109_k141_13090 | Not-determined |
| 18-109_k141_94239 | Not-determined |
| 18-109_k141_238058 | Not-determined |
| 18-109_k141_115317 | Not-determined |
| 18-109_k141_13099 | Not-determined |
| 18-109_k141_197361 | Not-determined |
| 18-109_k141_94245 | Not-determined |
| 18-109_k141_125335 | Not-determined |
| 18-109_k141_115322 | Not-determined |
| 18-109_k141_197363 | Not-determined |
| 18-109_k141_135402 | Not-determined |
| 18-109_k141_197373 | Not-determined |
| 18-109_k141_166276 | Not-determined |
| 18-109_k141_197375 | Not-determined |
| 18-109_k141_125348 | Not-determined |
| 18-109_k141_217164 | Not-determined |
| 18-109_k141_217166 | Not-determined |
| 18-109_k141_166285 | Not-determined |
| 18-109_k141_22750 | Not-determined |
| 18-109_k141_227843 | Not-determined |
| 18-109_k141_207728 | Low-quality |
| 18-109_k141_135443 | Not-determined |
| 18-109_k141_135444 | Low-quality |
| 18-109_k141_73565 | Not-determined |
| 18-109_k141_207738 | Not-determined |
| 18-109_k141_197402 | Low-quality |
| 18-109_k141_125359 | Not-determined |
| 18-109_k141_94266 | Not-determined |
| 18-109_k141_94269 | Low-quality |
| 18-109_k141_125364 | Not-determined |
| 18-109_k141_104887 | Not-determined |
| 18-109_k141_227845 | Low-quality |
| 18-109_k141_227850 | Not-determined |
| 18-109_k141_207765 | Not-determined |
| 18-109_k141_207766 | Not-determined |
| 18-109_k141_207767 | Not-determined |
| 18-109_k141_125371 | Not-determined |
| 18-109_k141_217176 | Not-determined |
| 18-109_k141_94284 | Not-determined |
| 18-109_k141_176303 | Not-determined |
| 18-109_k141_135465 | Not-determined |
| 18-109_k141_176311 | Not-determined |
| 18-109_k141_135473 | Not-determined |
| 18-109_k141_135476 | Not-determined |
| 18-109_k141_94302 | Not-determined |
| 18-109_k141_227857 | Not-determined |
| 18-109_k141_13126 | Low-quality |
| 18-109_k141_125412 | Not-determined |
| 18-109_k141_238101 | Not-determined |
| 18-109_k141_145470 | Low-quality |
| 18-109_k141_13132 | Not-determined |
| 18-109_k141_125423 | Not-determined |
| 18-109_k141_166311 | Not-determined |
| 18-109_k141_176319 | Not-determined |
| 18-109_k141_166313 | Not-determined |
| 18-109_k141_166317 | Not-determined |
| 18-109_k141_94322 | Not-determined |
| 18-109_k141_43722 | Not-determined |
| 18-109_k141_43729 | Not-determined |
| 18-109_k141_176336 | Not-determined |
| 18-109_k141_166334 | Not-determined |
| 18-109_k141_166335 | Not-determined |
| 18-109_k141_166336 | Not-determined |
| 18-109_k141_207785 | Not-determined |
| 18-109_k141_2875 | Low-quality |
| 18-109_k141_207794 | Not-determined |
| 18-109_k141_64050 | Not-determined |
| 18-109_k141_207795 | Not-determined |
| 18-109_k141_145482 | Not-determined |
| 18-109_k141_135498 | Not-determined |
| 18-109_k141_207799 | Not-determined |
| 18-109_k141_166355 | Not-determined |
| 18-109_k141_217201 | Not-determined |
| 18-109_k141_125446 | Not-determined |
| 18-109_k141_145495 | Not-determined |
| 18-109_k141_207809 | Not-determined |
| 18-109_k141_197509 | Not-determined |
| 18-109_k141_43744 | Not-determined |
| 18-109_k141_207811 | Not-determined |
| 18-109_k141_125452 | Low-quality |
| 18-109_k141_166366 | Not-determined |
| 18-109_k141_166368 | Not-determined |
| 18-109_k141_166369 | Not-determined |
| 18-109_k141_166370 | Not-determined |
| 18-109_k141_115353 | Not-determined |
| 18-109_k141_104922 | Not-determined |
| 18-109_k141_2902 | Not-determined |
| 18-109_k141_43748 | Not-determined |
| 18-109_k141_94354 | Not-determined |
| 18-109_k141_207819 | Low-quality |
| 18-109_k141_73607 | Not-determined |
| 18-109_k141_2908 | Not-determined |
| 18-109_k141_2913 | Not-determined |
| 18-109_k141_197529 | Not-determined |
| 18-109_k141_207829 | Not-determined |
| 18-109_k141_125473 | Not-determined |
| 18-109_k141_104939 | Low-quality |
| 18-109_k141_94362 | Not-determined |
| 18-109_k141_125475 | Low-quality |
| 18-109_k141_238144 | Not-determined |
| 18-109_k141_73631 | Not-determined |
| 18-109_k141_125479 | Not-determined |
| 18-109_k141_2928 | Not-determined |
| 18-109_k141_73634 | Not-determined |
| 18-109_k141_2933 | Not-determined |
| 18-109_k141_43759 | Not-determined |
| 18-109_k141_145524 | Not-determined |
| 18-109_k141_2941 | Not-determined |
| 18-109_k141_104964 | Not-determined |
| 18-109_k141_145533 | Not-determined |
| 18-109_k141_104970 | Not-determined |
| 18-109_k141_207840 | Not-determined |
| 18-109_k141_145539 | Not-determined |
| 18-109_k141_197547 | Not-determined |
| 18-109_k141_64072 | Not-determined |
| 18-109_k141_166388 | Not-determined |
| 18-109_k141_64073 | Not-determined |
| 18-109_k141_227943 | Not-determined |
| 18-109_k141_125498 | Not-determined |
| 18-109_k141_238195 | Not-determined |
| 18-109_k141_145565 | Not-determined |
| 18-109_k141_166395 | Low-quality |
| 18-109_k141_125509 | Not-determined |
| 18-109_k141_2946 | Not-determined |
| 18-109_k141_84324 | Not-determined |
| 18-109_k141_115377 | Not-determined |
| 18-109_k141_94409 | Low-quality |
| 18-109_k141_53946 | Not-determined |
| 18-109_k141_33718 | Not-determined |
| 18-109_k141_115384 | Low-quality |
| 18-109_k141_53956 | Not-determined |
| 18-109_k141_104984 | Not-determined |
| 18-109_k141_115390 | Low-quality |
| 18-109_k141_145579 | Not-determined |
| 18-109_k141_94427 | Not-determined |
| 18-109_k141_197568 | Not-determined |
| 18-109_k141_125528 | Not-determined |
| 18-109_k141_197570 | Not-determined |
| 18-109_k141_125532 | Not-determined |
| 18-109_k141_145589 | Not-determined |
| 18-109_k141_187175 | Not-determined |
| 18-109_k141_94436 | Not-determined |
| 18-109_k141_145595 | Not-determined |
| 18-109_k141_115400 | Not-determined |
| 18-109_k141_115401 | Not-determined |
| 18-109_k141_187183 | Low-quality |
| 18-109_k141_197583 | Low-quality |
| 18-109_k141_115409 | Not-determined |
| 18-109_k141_145603 | Not-determined |
| 18-109_k141_84329 | Not-determined |
| 18-109_k141_227952 | Not-determined |
| 18-109_k141_115414 | Not-determined |
| 18-109_k141_94449 | Not-determined |
| 18-109_k141_94450 | Not-determined |
| 18-109_k141_207857 | Not-determined |
| 18-109_k141_187197 | Not-determined |
| 18-109_k141_2958 | Not-determined |
| 18-109_k141_207862 | Not-determined |
| 18-109_k141_105000 | Not-determined |
| 18-109_k141_125571 | Not-determined |
| 18-109_k141_176373 | Not-determined |
| 18-109_k141_53989 | Not-determined |
| 18-109_k141_125580 | Not-determined |
| 18-109_k141_43829 | Not-determined |
| 18-109_k141_84357 | Not-determined |
| 18-109_k141_33734 | Not-determined |
| 18-109_k141_166438 | Not-determined |
| 18-109_k141_53996 | Not-determined |
| 18-109_k141_135518 | Not-determined |
| 18-109_k141_176388 | Not-determined |
| 18-109_k141_84364 | Not-determined |
| 18-109_k141_227979 | Not-determined |
| 18-109_k141_135523 | Not-determined |
| 18-109_k141_176392 | Not-determined |
| 18-109_k141_176402 | Not-determined |
| 18-109_k141_135534 | Not-determined |
| 18-109_k141_94476 | Not-determined |
| 18-109_k141_145641 | Not-determined |
| 18-109_k141_227987 | Not-determined |
| 18-109_k141_125589 | Not-determined |
| 18-109_k141_125590 | Not-determined |
| 18-109_k141_135542 | Not-determined |
| 18-109_k141_227992 | Not-determined |
| 18-109_k141_54022 | Not-determined |
| 18-109_k141_84384 | Low-quality |
| 18-109_k141_145646 | Low-quality |

| | |
|---|---|
| 18-109_k141_94481 | Not-determined |
| 18-109_k141_115419 | Low-quality |
| 18-109_k141_228002 | Not-determined |
| 18-109_k141_145656 | Not-determined |
| 18-109_k141_207905 | Not-determined |
| 18-109_k141_176447 | Not-determined |
| 18-109_k141_207915 | Low-quality |
| 18-109_k141_115423 | Not-determined |
| 18-109_k141_176460 | Not-determined |
| 18-109_k141_207931 | Not-determined |
| 18-109_k141_187244 | Not-determined |
| 18-109_k141_84399 | Not-determined |
| 18-109_k141_217211 | Not-determined |
| 18-109_k141_187250 | Not-determined |
| 18-109_k141_54046 | Low-quality |
| 18-109_k141_217213 | Not-determined |
| 18-109_k141_64140 | Not-determined |
| 18-109_k141_115432 | Not-determined |
| 18-109_k141_3005 | Not-determined |
| 18-109_k141_238211 | Not-determined |
| 18-109_k141_238212 | Not-determined |
| 18-109_k141_64142 | Not-determined |
| 18-109_k141_115438 | Not-determined |
| 18-109_k141_228011 | Not-determined |
| 18-109_k141_238216 | Not-determined |
| 18-109_k141_176479 | Not-determined |
| 18-109_k141_238220 | Not-determined |
| 18-109_k141_228014 | Not-determined |
| 18-109_k141_187264 | Not-determined |
| 18-109_k141_54062 | Not-determined |
| 18-109_k141_84425 | Not-determined |
| 18-109_k141_43886 | Not-determined |
| 18-109_k141_125629 | Low-quality |
| 18-109_k141_125632 | Not-determined |
| 18-109_k141_125633 | Not-determined |
| 18-109_k141_176493 | Not-determined |
| 18-109_k141_64144 | Not-determined |
| 18-109_k141_125637 | Not-determined |
| 18-109_k141_176507 | Not-determined |
| 18-109_k141_187270 | Low-quality |
| 18-109_k141_105033 | Not-determined |
| 18-109_k141_135585 | Not-determined |
| 18-109_k141_166486 | Not-determined |
| 18-109_k141_187273 | Not-determined |
| 18-109_k141_228043 | Not-determined |
| 18-109_k141_187280 | Not-determined |
| 18-109_k141_105052 | Not-determined |
| 18-109_k141_43921 | Not-determined |
| 18-109_k141_13210 | Low-quality |
| 18-109_k141_43926 | Not-determined |
| 18-109_k141_43927 | Not-determined |
| 18-109_k141_187295 | Not-determined |
| 18-109_k141_176531 | Not-determined |
| 18-109_k141_228058 | Low-quality |
| 18-109_k141_13220 | Not-determined |
| 18-109_k141_43941 | Not-determined |
| 18-109_k141_54102 | Not-determined |
| 18-109_k141_73671 | Not-determined |
| 18-109_k141_73672 | Not-determined |
| 18-109_k141_176543 | Not-determined |
| 18-109_k141_64186 | Not-determined |
| 18-109_k141_156448 | Not-determined |
| 18-109_k141_228083 | Not-determined |
| 18-109_k141_54109 | Not-determined |
| 18-109_k141_145684 | Not-determined |
| 18-109_k141_105068 | Not-determined |
| 18-109_k141_125665 | Not-determined |
| 18-109_k141_84463 | Not-determined |
| 18-109_k141_228090 | Not-determined |
| 18-109_k141_54118 | Not-determined |
| 18-109_k141_156456 | Not-determined |
| 18-109_k141_197654 | Not-determined |
| 18-109_k141_228098 | Not-determined |
| 18-109_k141_115462 | Not-determined |
| 18-109_k141_156464 | Not-determined |
| 18-109_k141_64198 | Not-determined |
| 18-109_k141_176554 | Not-determined |
| 18-109_k141_54127 | Not-determined |
| 18-109_k141_197662 | Low-quality |
| 18-109_k141_187334 | Not-determined |
| 18-109_k141_22794 | Not-determined |
| 18-109_k141_166521 | Not-determined |
| 18-109_k141_125669 | Not-determined |
| 18-109_k141_115470 | Not-determined |
| 18-109_k141_84472 | Not-determined |
| 18-109_k141_115472 | Not-determined |
| 18-109_k141_187342 | Low-quality |
| 18-109_k141_73695 | Not-determined |
| 18-109_k141_22805 | Not-determined |
| 18-109_k141_94545 | Not-determined |
| 18-109_k141_94547 | Not-determined |
| 18-109_k141_228126 | Low-quality |
| 18-109_k141_13253 | Not-determined |
| 18-109_k141_125684 | Not-determined |
| 18-109_k141_228132 | Not-determined |
| 18-109_k141_13254 | Not-determined |
| 18-109_k141_22807 | Low-quality |
| 18-109_k141_13255 | Not-determined |
| 18-109_k141_3043 | Not-determined |
| 18-109_k141_105103 | Not-determined |
| 18-109_k141_33807 | Not-determined |
| 18-109_k141_54146 | Not-determined |
| 18-109_k141_125696 | Not-determined |
| 18-109_k141_105114 | Not-determined |
| 18-109_k141_13269 | Not-determined |
| 18-109_k141_176574 | Not-determined |
| 18-109_k141_176575 | Not-determined |
| 18-109_k141_22825 | Not-determined |
| 18-109_k141_125704 | Not-determined |
| 18-109_k141_228141 | Not-determined |
| 18-109_k141_105126 | Low-quality |
| 18-109_k141_187370 | Low-quality |
| 18-109_k141_228147 | Not-determined |
| 18-109_k141_135652 | Not-determined |
| 18-109_k141_33829 | Not-determined |
| 18-109_k141_187379 | Not-determined |
| 18-109_k141_197688 | Not-determined |
| 18-109_k141_73735 | Not-determined |
| 18-109_k141_22844 | Not-determined |
| 18-109_k141_166579 | Not-determined |
| 18-109_k141_217233 | Not-determined |
| 18-109_k141_217235 | Not-determined |
| 18-109_k141_64220 | Low-quality |
| 18-109_k141_238249 | Not-determined |
| 18-109_k141_197705 | Not-determined |
| 18-109_k141_197707 | Not-determined |
| 18-109_k141_13294 | Not-determined |
| 18-109_k141_105152 | Not-determined |
| 18-109_k141_187394 | Not-determined |
| 18-109_k141_3075 | Not-determined |
| 18-109_k141_187396 | Not-determined |
| 18-109_k141_166597 | Not-determined |
| 18-109_k141_176605 | Not-determined |
| 18-109_k141_22862 | Not-determined |
| 18-109_k141_115510 | Not-determined |
| 18-109_k141_228170 | Not-determined |
| 18-109_k141_176610 | Not-determined |
| 18-109_k141_176613 | Not-determined |
| 18-109_k141_166610 | Not-determined |
| 18-109_k141_187407 | Not-determined |
| 18-109_k141_197723 | Not-determined |
| 18-109_k141_228178 | Low-quality |
| 18-109_k141_135690 | Not-determined |
| 18-109_k141_156518 | Not-determined |
| 18-109_k141_73763 | Not-determined |
| 18-109_k141_33845 | Not-determined |
| 18-109_k141_105166 | Not-determined |
| 18-109_k141_187412 | Not-determined |
| 18-109_k141_176623 | Not-determined |
| 18-109_k141_94553 | Not-determined |
| 18-109_k141_73766 | Not-determined |
| 18-109_k141_187416 | Not-determined |
| 18-109_k141_197743 | Not-determined |
| 18-109_k141_217262 | Low-quality |
| 18-109_k141_33850 | Not-determined |
| 18-109_k141_228192 | Not-determined |
| 18-109_k141_228193 | Not-determined |
| 18-109_k141_228194 | Not-determined |
| 18-109_k141_105172 | Not-determined |
| 18-109_k141_166630 | Not-determined |
| 18-109_k141_217268 | Not-determined |
| 18-109_k141_13326 | Not-determined |
| 18-109_k141_13330 | Not-determined |
| 18-109_k141_13332 | Not-determined |
| 18-109_k141_166640 | Not-determined |
| 18-109_k141_105177 | Not-determined |
| 18-109_k141_197751 | Not-determined |
| 18-109_k141_125715 | Not-determined |
| 18-109_k141_64258 | Not-determined |
| 18-109_k141_64259 | Not-determined |
| 18-109_k141_84527 | Not-determined |
| 18-109_k141_3096 | Not-determined |
| 18-109_k141_33863 | Not-determined |
| 18-109_k141_105189 | Not-determined |
| 18-109_k141_166651 | Not-determined |
| 18-109_k141_228222 | Not-determined |
| 18-109_k141_166654 | Not-determined |
| 18-109_k141_197766 | Not-determined |
| 18-109_k141_166656 | Not-determined |
| 18-109_k141_64264 | Not-determined |
| 18-109_k141_105193 | Not-determined |
| 18-109_k141_197770 | Not-determined |
| 18-109_k141_156529 | Not-determined |
| 18-109_k141_3102 | Not-determined |
| 18-109_k141_207959 | Not-determined |
| 18-109_k141_33875 | Not-determined |
| 18-109_k141_207961 | Not-determined |
| 18-109_k141_228232 | Not-determined |
| 18-109_k141_156534 | Not-determined |
| 18-109_k141_238291 | Low-quality |
| 18-109_k141_13353 | Low-quality |
| 18-109_k141_105203 | Not-determined |
| 18-109_k141_166661 | Not-determined |
| 18-109_k141_197772 | Not-determined |
| 18-109_k141_105206 | Not-determined |
| 18-109_k141_166664 | Not-determined |
| 18-109_k141_3107 | Not-determined |
| 18-109_k141_156546 | Not-determined |
| 18-109_k141_3113 | Not-determined |
| 18-109_k141_238306 | Not-determined |
| 18-109_k141_145737 | Not-determined |
| 18-109_k141_197792 | Not-determined |
| 18-109_k141_43989 | Not-determined |
| 18-109_k141_238315 | Not-determined |
| 18-109_k141_135721 | Low-quality |
| 18-109_k141_33901 | Not-determined |
| 18-109_k141_187439 | Not-determined |
| 18-109_k141_13373 | Not-determined |
| 18-109_k141_207992 | Not-determined |
| 18-109_k141_13374 | Not-determined |
| 18-109_k141_33911 | Low-quality |
| 18-109_k141_115550 | Not-determined |
| 18-109_k141_73792 | Not-determined |
| 18-109_k141_207997 | Not-determined |
| 18-109_k141_156565 | Not-determined |
| 18-109_k141_115557 | Not-determined |
| 18-109_k141_156568 | Low-quality |
| 18-109_k141_105216 | Not-determined |
| 18-109_k141_73806 | Low-quality |
| 18-109_k141_228256 | Not-determined |
| 18-109_k141_238329 | Not-determined |
| 18-109_k141_217324 | Low-quality |
| 18-109_k141_208030 | Not-determined |
| 18-109_k141_125732 | Not-determined |
| 18-109_k141_208031 | Not-determined |
| 18-109_k141_176702 | Low-quality |
| 18-109_k141_73830 | Not-determined |
| 18-109_k141_238335 | Not-determined |
| 18-109_k141_135765 | Not-determined |
| 18-109_k141_3116 | Not-determined |
| 18-109_k141_105237 | Not-determined |
| 18-109_k141_3118 | Low-quality |
| 18-109_k141_22921 | Not-determined |
| 18-109_k141_187451 | Not-determined |
| 18-109_k141_238342 | Not-determined |
| 18-109_k141_84542 | Low-quality |
| 18-109_k141_54154 | Not-determined |
| 18-109_k141_217335 | Not-determined |
| 18-109_k141_217336 | Low-quality |
| 18-109_k141_22929 | Not-determined |
| 18-109_k141_105252 | Low-quality |
| 18-109_k141_176720 | Not-determined |
| 18-109_k141_187453 | Not-determined |
| 18-109_k141_64290 | Not-determined |
| 18-109_k141_115596 | Not-determined |
| 18-109_k141_54167 | Not-determined |
| 18-109_k141_54170 | Not-determined |
| 18-109_k141_94582 | Not-determined |
| 18-109_k141_54172 | Not-determined |
| 18-109_k141_13441 | Not-determined |
| 18-109_k141_54179 | Not-determined |
| 18-109_k141_187466 | Not-determined |
| 18-109_k141_217356 | Not-determined |
| 18-109_k141_84564 | Not-determined |
| 18-109_k141_54185 | Not-determined |
| 18-109_k141_208064 | Low-quality |
| 18-109_k141_228267 | Not-determined |
| 18-109_k141_217362 | Not-determined |
| 18-109_k141_228269 | Not-determined |
| 18-109_k141_228271 | Not-determined |
| 18-109_k141_125776 | Not-determined |
| 18-109_k141_54189 | Not-determined |
| 18-109_k141_54194 | Not-determined |
| 18-109_k141_84572 | Not-determined |
| 18-109_k141_3160 | Not-determined |
| 18-109_k141_3173 | Not-determined |
| 18-109_k141_73847 | Not-determined |
| 18-109_k141_105288 | Low-quality |
| 18-109_k141_166704 | Not-determined |
| 18-109_k141_13463 | Low-quality |
| 18-109_k141_105293 | Not-determined |
| 18-109_k141_238367 | Not-determined |
| 18-109_k141_105296 | Not-determined |
| 18-109_k141_166709 | Not-determined |
| 18-109_k141_3178 | Not-determined |
| 18-109_k141_3180 | Not-determined |
| 18-109_k141_166718 | Not-determined |
| 18-109_k141_3182 | Not-determined |
| 18-109_k141_13476 | Not-determined |
| 18-109_k141_197810 | Not-determined |
| 18-109_k141_208098 | Not-determined |
| 18-109_k141_33985 | Not-determined |
| 18-109_k141_208103 | Not-determined |
| 18-109_k141_208106 | Not-determined |
| 18-109_k141_73862 | Not-determined |
| 18-109_k141_64303 | Low-quality |
| 18-109_k141_13487 | Not-determined |
| 18-109_k141_94620 | Not-determined |
| 18-109_k141_44003 | Not-determined |
| 18-109_k141_208116 | Not-determined |
| 18-109_k141_13490 | Not-determined |
| 18-109_k141_33996 | Not-determined |
| 18-109_k141_208117 | Not-determined |
| 18-109_k141_13493 | Not-determined |
| 18-109_k141_208119 | Not-determined |
| 18-109_k141_73874 | Not-determined |
| 18-109_k141_197821 | Not-determined |
| 18-109_k141_187523 | Not-determined |
| 18-109_k141_3198 | Low-quality |
| 18-109_k141_13500 | Not-determined |
| 18-109_k141_13503 | Not-determined |
| 18-109_k141_94629 | Low-quality |
| 18-109_k141_64328 | Not-determined |
| 18-109_k141_44006 | Not-determined |
| 18-109_k141_44008 | Not-determined |
| 18-109_k141_3204 | Not-determined |
| 18-109_k141_166744 | Not-determined |
| 18-109_k141_187534 | Not-determined |
| 18-109_k141_208145 | Not-determined |
| 18-109_k141_54236 | Not-determined |
| 18-109_k141_208147 | Not-determined |
| 18-109_k141_187543 | Not-determined |
| 18-109_k141_217409 | Not-determined |
| 18-109_k141_187548 | Low-quality |
| 18-109_k141_217418 | Not-determined |
| 18-109_k141_145778 | Not-determined |
| 18-109_k141_54238 | Low-quality |
| 18-109_k141_34035 | Not-determined |
| 18-109_k141_84609 | Not-determined |
| 18-109_k141_22981 | Not-determined |
| 18-109_k141_54242 | Not-determined |
| 18-109_k141_73912 | Not-determined |
| 18-109_k141_54244 | Low-quality |
| 18-109_k141_22988 | Not-determined |
| 18-109_k141_22989 | Not-determined |
| 18-109_k141_22991 | Low-quality |
| 18-109_k141_156578 | Not-determined |
| 18-109_k141_54248 | Low-quality |
| 18-109_k141_208175 | Not-determined |
| 18-109_k141_54252 | Not-determined |
| 18-109_k141_187566 | Low-quality |
| 18-109_k141_176755 | Not-determined |
| 18-109_k141_197845 | Not-determined |
| 18-109_k141_145813 | Not-determined |
| 18-109_k141_197849 | Not-determined |
| 18-109_k141_34054 | Not-determined |
| 18-109_k141_145821 | Not-determined |
| 18-109_k141_34062 | Not-determined |
| 18-109_k141_187576 | Not-determined |
| 18-109_k141_145825 | Not-determined |
| 18-109_k141_3216 | Not-determined |
| 18-109_k141_73934 | Not-determined |
| 18-109_k141_23024 | Not-determined |
| 18-109_k141_176773 | Not-determined |
| 18-109_k141_176774 | Not-determined |
| 18-109_k141_145833 | Not-determined |
| 18-109_k141_145834 | Not-determined |
| 18-109_k141_23028 | Not-determined |
| 18-109_k141_94659 | Not-determined |
| 18-109_k141_187601 | Low-quality |
| 18-109_k141_73955 | Low-quality |
| 18-109_k141_187606 | Low-quality |
| 18-109_k141_94665 | Low-quality |
| 18-109_k141_166795 | Low-quality |
| 18-109_k141_13521 | Not-determined |
| 18-109_k141_94679 | Not-determined |
| 18-109_k141_3223 | Not-determined |
| 18-109_k141_84623 | Not-determined |
| 18-109_k141_64356 | Not-determined |
| 18-109_k141_34088 | Not-determined |
| 18-109_k141_13526 | Not-determined |
| 18-109_k141_13532 | Not-determined |
| 18-109_k141_125855 | Not-determined |
| 18-109_k141_64362 | Not-determined |
| 18-109_k141_176782 | Not-determined |
| 18-109_k141_34105 | Not-determined |
| 18-109_k141_84641 | Not-determined |
| 18-109_k141_156623 | Not-determined |
| 18-109_k141_94694 | Not-determined |
| 18-109_k141_94696 | Not-determined |
| 18-109_k141_64371 | Not-determined |
| 18-109_k141_156632 | Not-determined |
| 18-109_k141_176789 | Not-determined |
| 18-109_k141_84655 | Not-determined |
| 18-109_k141_156638 | Low-quality |
| 18-109_k141_3233 | Not-determined |
| 18-109_k141_176796 | Low-quality |
| 18-109_k141_23047 | Not-determined |
| 18-109_k141_23051 | Not-determined |
| 18-109_k141_84666 | Not-determined |
| 18-109_k141_187629 | Not-determined |
| 18-109_k141_23054 | Not-determined |
| 18-109_k141_166801 | Low-quality |
| 18-109_k141_145875 | Low-quality |
| 18-109_k141_238383 | Not-determined |
| 18-109_k141_115759 | Not-determined |
| 18-109_k141_84679 | Not-determined |
| 18-109_k141_187642 | Not-determined |
| 18-109_k141_54310 | Not-determined |
| 18-109_k141_54312 | Not-determined |
| 18-109_k141_156658 | Not-determined |
| 18-109_k141_135828 | Low-quality |
| 18-109_k141_156660 | Not-determined |
| 18-109_k141_54313 | Low-quality |
| 18-109_k141_187648 | Not-determined |
| 18-109_k141_34128 | Not-determined |
| 18-109_k141_187650 | Not-determined |
| 18-109_k141_94724 | Not-determined |
| 18-109_k141_197893 | Not-determined |
| 18-109_k141_156671 | Not-determined |
| 18-109_k141_115777 | Not-determined |
| 18-109_k141_197896 | Not-determined |
| 18-109_k141_156675 | Not-determined |
| 18-109_k141_197900 | Low-quality |
| 18-109_k141_34143 | Not-determined |
| 18-109_k141_13588 | Low-quality |
| 18-109_k141_176834 | Not-determined |
| 18-109_k141_34144 | Not-determined |
| 18-109_k141_13589 | Not-determined |
| 18-109_k141_64393 | Not-determined |
| 18-109_k141_34151 | Not-determined |
| 18-109_k141_238406 | Not-determined |
| 18-109_k141_176837 | Not-determined |
| 18-109_k141_44112 | Not-determined |
| 18-109_k141_156686 | Not-determined |
| 18-109_k141_34157 | Not-determined |
| 18-109_k141_3266 | Not-determined |
| 18-109_k141_23064 | Not-determined |
| 18-109_k141_176842 | Not-determined |
| 18-109_k141_156695 | Not-determined |
| 18-109_k141_105363 | Not-determined |
| 18-109_k141_3272 | Not-determined |
| 18-109_k141_3276 | Not-determined |
| 18-109_k141_187668 | Not-determined |
| 18-109_k141_208225 | Not-determined |
| 18-109_k141_187670 | Not-determined |
| 18-109_k141_105368 | Not-determined |
| 18-109_k141_115788 | Not-determined |
| 18-109_k141_94755 | Not-determined |
| 18-109_k141_176854 | Not-determined |
| 18-109_k141_176856 | Low-quality |
| 18-109_k141_187673 | Not-determined |
| 18-109_k141_197918 | Not-determined |
| 18-109_k141_197919 | Not-determined |
| 18-109_k141_3283 | Low-quality |
| 18-109_k141_105383 | Not-determined |
| 18-109_k141_13618 | Not-determined |
| 18-109_k141_228303 | Not-determined |
| 18-109_k141_176873 | Not-determined |
| 18-109_k141_54338 | Low-quality |
| 18-109_k141_208239 | Not-determined |
| 18-109_k141_228304 | Low-quality |
| 18-109_k141_166861 | Not-determined |
| 18-109_k141_156711 | Not-determined |
| 18-109_k141_238427 | Not-determined |
| 18-109_k141_166866 | Not-determined |
| 18-109_k141_238428 | Not-determined |
| 18-109_k141_34189 | Not-determined |
| 18-109_k141_64444 | Not-determined |
| 18-109_k141_197924 | Not-determined |
| 18-109_k141_34195 | Not-determined |
| 18-109_k141_125877 | Not-determined |
| 18-109_k141_228317 | Not-determined |
| 18-109_k141_228321 | Not-determined |
| 18-109_k141_166876 | Not-determined |
| 18-109_k141_197928 | Low-quality |
| 18-109_k141_44121 | Not-determined |
| 18-109_k141_197939 | Not-determined |
| 18-109_k141_44130 | Not-determined |
| 18-109_k141_197941 | Not-determined |
| 18-109_k141_238455 | Not-determined |
| 18-109_k141_238457 | Not-determined |
| 18-109_k141_54367 | Not-determined |
| 18-109_k141_187714 | Not-determined |
| 18-109_k141_197950 | Not-determined |
| 18-109_k141_197955 | Not-determined |
| 18-109_k141_23110 | Not-determined |
| 18-109_k141_176880 | Not-determined |
| 18-109_k141_145919 | Not-determined |
| 18-109_k141_187721 | Not-determined |
| 18-109_k141_145921 | Low-quality |
| 18-109_k141_238469 | Not-determined |
| 18-109_k141_13654 | Not-determined |
| 18-109_k141_197959 | Low-quality |
| 18-109_k141_238472 | Not-determined |
| 18-109_k141_54381 | Not-determined |
| 18-109_k141_13667 | Not-determined |
| 18-109_k141_64489 | Not-determined |
| 18-109_k141_54387 | Not-determined |
| 18-109_k141_54400 | Not-determined |
| 18-109_k141_208277 | Not-determined |
| 18-109_k141_208278 | Not-determined |
| 18-109_k141_217441 | Not-determined |
| 18-109_k141_238490 | Not-determined |
| 18-109_k141_125892 | Not-determined |
| 18-109_k141_54407 | Not-determined |
| 18-109_k141_145939 | Not-determined |
| 18-109_k141_125899 | Not-determined |
| 18-109_k141_145942 | Not-determined |
| 18-109_k141_73993 | Not-determined |
| 18-109_k141_176903 | Low-quality |
| 18-109_k141_125909 | Not-determined |
| 18-109_k141_198000 | Not-determined |
| 18-109_k141_208291 | Not-determined |
| 18-109_k141_64516 | Not-determined |
| 18-109_k141_208294 | Low-quality |
| 18-109_k141_217466 | Not-determined |
| 18-109_k141_156735 | Not-determined |
| 18-109_k141_156736 | Not-determined |
| 18-109_k141_156738 | Not-determined |
| 18-109_k141_198008 | Not-determined |
| 18-109_k141_115832 | Not-determined |
| 18-109_k141_228337 | Not-determined |
| 18-109_k141_13678 | Not-determined |
| 18-109_k141_166907 | Not-determined |
| 18-109_k141_176909 | Not-determined |
| 18-109_k141_198012 | Low-quality |
| 18-109_k141_176912 | Not-determined |
| 18-109_k141_54412 | Not-determined |
| 18-109_k141_64526 | Not-determined |
| 18-109_k141_64527 | Not-determined |
| 18-109_k141_23146 | Not-determined |
| 18-109_k141_13698 | Not-determined |
| 18-109_k141_125951 | Not-determined |
| 18-109_k141_208312 | Not-determined |
| 18-109_k141_156758 | Not-determined |
| 18-109_k141_217502 | Not-determined |
| 18-109_k141_176925 | Not-determined |
| 18-109_k141_198034 | Not-determined |
| 18-109_k141_208317 | Not-determined |
| 18-109_k141_13709 | Not-determined |
| 18-109_k141_34258 | Low-quality |
| 18-109_k141_44166 | Not-determined |
| 18-109_k141_228367 | Not-determined |
| 18-109_k141_228368 | Low-quality |
| 18-109_k141_13714 | Not-determined |
| 18-109_k141_34267 | Not-determined |
| 18-109_k141_208331 | Not-determined |
| 18-109_k141_84698 | Not-determined |
| 18-109_k141_84706 | Not-determined |
| 18-109_k141_135860 | Not-determined |
| 18-109_k141_135863 | Not-determined |
| 18-109_k141_156766 | Not-determined |
| 18-109_k141_135866 | Not-determined |
| 18-109_k141_34281 | Not-determined |
| 18-109_k141_3358 | Not-determined |
| 18-109_k141_135871 | Not-determined |
| 18-109_k141_166924 | Not-determined |
| 18-109_k141_166926 | Not-determined |
| 18-109_k141_64559 | Low-quality |
| 18-109_k141_13732 | Not-determined |
| 18-109_k141_135885 | Not-determined |
| 18-109_k141_74002 | Low-quality |
| 18-109_k141_84731 | Not-determined |
| 18-109_k141_74006 | Not-determined |
| 18-109_k141_44205 | Not-determined |
| 18-109_k141_176945 | Not-determined |
| 18-109_k141_156789 | Not-determined |
| 18-109_k141_74012 | Not-determined |
| 18-109_k141_135904 | Not-determined |
| 18-109_k141_105413 | Not-determined |
| 18-109_k141_125962 | Low-quality |
| 18-109_k141_228390 | Low-quality |
| 18-109_k141_74015 | Not-determined |
| 18-109_k141_166956 | Not-determined |
| 18-109_k141_74022 | Not-determined |
| 18-109_k141_105421 | Not-determined |
| 18-109_k141_74024 | Not-determined |
| 18-109_k141_187779 | Not-determined |
| 18-109_k141_166965 | Not-determined |
| 18-109_k141_176954 | Not-determined |
| 18-109_k141_74035 | Not-determined |
| 18-109_k141_125977 | Not-determined |
| 18-109_k141_208357 | Not-determined |
| 18-109_k141_166981 | Not-determined |
| 18-109_k141_125980 | Not-determined |
| 18-109_k141_187806 | Not-determined |
| 18-109_k141_156811 | Not-determined |
| 18-109_k141_135920 | Not-determined |
| 18-109_k141_115888 | Not-determined |
| 18-109_k141_146022 | Not-determined |
| 18-109_k141_166998 | Not-determined |
| 18-109_k141_198059 | Not-determined |
| 18-109_k141_146030 | Not-determined |
| 18-109_k141_135929 | Not-determined |
| 18-109_k141_135930 | Not-determined |
| 18-109_k141_23169 | Not-determined |
| 18-109_k141_105428 | Not-determined |
| 18-109_k141_167011 | Not-determined |
| 18-109_k141_135942 | Not-determined |
| 18-109_k141_84741 | Not-determined |
| 18-109_k141_146042 | Not-determined |
| 18-109_k141_84751 | Not-determined |
| 18-109_k141_146053 | Not-determined |
| 18-109_k141_84754 | Not-determined |
| 18-109_k141_167024 | Low-quality |
| 18-109_k141_126005 | Not-determined |
| 18-109_k141_115900 | Not-determined |
| 18-109_k141_126008 | Not-determined |

| | |
|---|---|
| 18-109_k141_94772 | Not-determined |
| 18-109_k141_105454 | Not-determined |
| 18-109_k141_167035 | Not-determined |
| 18-109_k141_105461 | Low-quality |
| 18-109_k141_228431 | Not-determined |
| 18-109_k141_135965 | Not-determined |
| 18-109_k141_74041 | Low-quality |
| 18-109_k141_13782 | Not-determined |
| 18-109_k141_74043 | Not-determined |
| 18-109_k141_146067 | Not-determined |
| 18-109_k141_176992 | Not-determined |
| 18-109_k141_24313 | Not-determined |
| 18-109_k141_198074 | Low-quality |
| 18-109_k141_187844 | Not-determined |
| 18-109_k141_198075 | Not-determined |
| 18-109_k141_135973 | Not-determined |
| 18-109_k141_94782 | Not-determined |
| 18-109_k141_115911 | Not-determined |
| 18-109_k141_208411 | Not-determined |
| 18-109_k141_105485 | Not-determined |
| 18-109_k141_3366 | Not-determined |
| 18-109_k141_135990 | Low-quality |
| 18-109_k141_64662 | Not-determined |
| 18-109_k141_3368 | Not-determined |
| 18-109_k141_208414 | Not-determined |
| 18-109_k141_64667 | Not-determined |
| 18-109_k141_187875 | Not-determined |
| 18-109_k141_208426 | Not-determined |
| 18-109_k141_84792 | Not-determined |
| 18-109_k141_156832 | Not-determined |
| 18-109_k141_135998 | Not-determined |
| 18-109_k141_23185 | Not-determined |
| 18-109_k141_23190 | Not-determined |
| 18-109_k141_105499 | Not-determined |
| 18-109_k141_167078 | Not-determined |
| 18-109_k141_198087 | Not-determined |
| 18-109_k141_84795 | Not-determined |
| 18-109_k141_198090 | Not-determined |
| 18-109_k141_13820 | Low-quality |
| 18-109_k141_167084 | Not-determined |
| 18-109_k141_84804 | Not-determined |
| 18-109_k141_44249 | Not-determined |
| 18-109_k141_44250 | Not-determined |
| 18-109_k141_105516 | Not-determined |
| 18-109_k141_105519 | Not-determined |
| 18-109_k141_208441 | Low-quality |
| 18-109_k141_208442 | Not-determined |
| 18-109_k141_74106 | Not-determined |
| 18-109_k141_74107 | Not-determined |
| 18-109_k141_198107 | Not-determined |
| 18-109_k141_187898 | Low-quality |
| 18-109_k141_187902 | Not-determined |
| 18-109_k141_198108 | Not-determined |
| 18-109_k141_187907 | Not-determined |
| 18-109_k141_167111 | Low-quality |
| 18-109_k141_208460 | Not-determined |
| 18-109_k141_13859 | Not-determined |
| 18-109_k141_156850 | Not-determined |
| 18-109_k141_167124 | Not-determined |
| 18-109_k141_167130 | Not-determined |
| 18-109_k141_74115 | Not-determined |
| 18-109_k141_156856 | Not-determined |
| 18-109_k141_44275 | Not-determined |
| 18-109_k141_217539 | Low-quality |
| 18-109_k141_3414 | Not-determined |
| 18-109_k141_126040 | Not-determined |
| 18-109_k141_208482 | Not-determined |
| 18-109_k141_228476 | Not-determined |
| 18-109_k141_136036 | Not-determined |
| 18-109_k141_208485 | Not-determined |
| 18-109_k141_156869 | Not-determined |
| 18-109_k141_13881 | Not-determined |
| 18-109_k141_115952 | Not-determined |
| 18-109_k141_208490 | Not-determined |
| 18-109_k141_208491 | Not-determined |
| 18-109_k141_217543 | Not-determined |
| 18-109_k141_136040 | Not-determined |
| 18-109_k141_198126 | Not-determined |
| 18-109_k141_217546 | Not-determined |
| 18-109_k141_94812 | Not-determined |
| 18-109_k141_198132 | Not-determined |
| 18-109_k141_136054 | Not-determined |
| 18-109_k141_3450 | Not-determined |
| 18-109_k141_228481 | Not-determined |
| 18-109_k141_3452 | Not-determined |
| 18-109_k141_228483 | Not-determined |
| 18-109_k141_208510 | Not-determined |
| 18-109_k141_146139 | Not-determined |
| 18-109_k141_146141 | Not-determined |
| 18-109_k141_238587 | Low-quality |
| 18-109_k141_105539 | Not-determined |
| 18-109_k141_105542 | Not-determined |
| 18-109_k141_84852 | Not-determined |
| 18-109_k141_13894 | Not-determined |
| 18-109_k141_115979 | Not-determined |
| 18-109_k141_238602 | Low-quality |
| 18-109_k141_177027 | Not-determined |
| 18-109_k141_156874 | Not-determined |
| 18-109_k141_74156 | Not-determined |
| 18-109_k141_177035 | Low-quality |
| 18-109_k141_228497 | Not-determined |
| 18-109_k141_228500 | Low-quality |
| 18-109_k141_115992 | Not-determined |
| 18-109_k141_115993 | Not-determined |
| 18-109_k141_177040 | Not-determined |
| 18-109_k141_74165 | Not-determined |
| 18-109_k141_217575 | Not-determined |
| 18-109_k141_228507 | Not-determined |
| 18-109_k141_44299 | Not-determined |
| 18-109_k141_198139 | Not-determined |
| 18-109_k141_44308 | Not-determined |
| 18-109_k141_187949 | Not-determined |
| 18-109_k141_177046 | Not-determined |
| 18-109_k141_187953 | Not-determined |
| 18-109_k141_74184 | Not-determined |
| 18-109_k141_228516 | Not-determined |
| 18-109_k141_228517 | Not-determined |
| 18-109_k141_208536 | Low-quality |
| 18-109_k141_217595 | Not-determined |
| 18-109_k141_238617 | Not-determined |
| 18-109_k141_3502 | Not-determined |
| 18-109_k141_217599 | Not-determined |
| 18-109_k141_208540 | Not-determined |
| 18-109_k141_84883 | Not-determined |
| 18-109_k141_238621 | Not-determined |
| 18-109_k141_217602 | Not-determined |
| 18-109_k141_217603 | Not-determined |
| 18-109_k141_187959 | Not-determined |
| 18-109_k141_116026 | Not-determined |
| 18-109_k141_228530 | Not-determined |
| 18-109_k141_167191 | Not-determined |
| 18-109_k141_187965 | Not-determined |
| 18-109_k141_34396 | Not-determined |
| 18-109_k141_187966 | Not-determined |
| 18-109_k141_167194 | Not-determined |
| 18-109_k141_167200 | Not-determined |
| 18-109_k141_217612 | Not-determined |
| 18-109_k141_238641 | Not-determined |
| 18-109_k141_228536 | Not-determined |
| 18-109_k141_74201 | Not-determined |
| 18-109_k141_228538 | Not-determined |
| 18-109_k141_105588 | Not-determined |
| 18-109_k141_238643 | Low-quality |
| 18-109_k141_187980 | Not-determined |
| 18-109_k141_64727 | Not-determined |
| 18-109_k141_228558 | Not-determined |
| 18-109_k141_3508 | Not-determined |
| 18-109_k141_74223 | Not-determined |
| 18-109_k141_74228 | Not-determined |
| 18-109_k141_34410 | Not-determined |
| 18-109_k141_126093 | Low-quality |
| 18-109_k141_156928 | Not-determined |
| 18-109_k141_156929 | Not-determined |
| 18-109_k141_238679 | Not-determined |
| 18-109_k141_188001 | Low-quality |
| 18-109_k141_188002 | Not-determined |
| 18-109_k141_34414 | Not-determined |
| 18-109_k141_177061 | Not-determined |
| 18-109_k141_34417 | Not-determined |
| 18-109_k141_146172 | Not-determined |
| 18-109_k141_177065 | Not-determined |
| 18-109_k141_84897 | Low-quality |
| 18-109_k141_146177 | Low-quality |
| 18-109_k141_74232 | Not-determined |
| 18-109_k141_208554 | Not-determined |
| 18-109_k141_177072 | Not-determined |
| 18-109_k141_74235 | Not-determined |
| 18-109_k141_188011 | Not-determined |
| 18-109_k141_238691 | Not-determined |
| 18-109_k141_94841 | Not-determined |
| 18-109_k141_84906 | Not-determined |
| 18-109_k141_238702 | Not-determined |
| 18-109_k141_177077 | Low-quality |
| 18-109_k141_116078 | Not-determined |
| 18-109_k141_177084 | Not-determined |
| 18-109_k141_177092 | Not-determined |
| 18-109_k141_146193 | Not-determined |
| 18-109_k141_116087 | Not-determined |
| 18-109_k141_3541 | Not-determined |
| 18-109_k141_44374 | Low-quality |
| 18-109_k141_94862 | Low-quality |
| 18-109_k141_44376 | Not-determined |
| 18-109_k141_146209 | Not-determined |
| 18-109_k141_126123 | Not-determined |
| 18-109_k141_3553 | Not-determined |
| 18-109_k141_3554 | Not-determined |
| 18-109_k141_34465 | Not-determined |
| 18-109_k141_74268 | Not-determined |
| 18-109_k141_105639 | Not-determined |
| 18-109_k141_84926 | Not-determined |
| 18-109_k141_3560 | Not-determined |
| 18-109_k141_105643 | Not-determined |
| 18-109_k141_116112 | Not-determined |
| 18-109_k141_3564 | Not-determined |
| 18-109_k141_13986 | Not-determined |
| 18-109_k141_188042 | Not-determined |
| 18-109_k141_74272 | Low-quality |
| 18-109_k141_105652 | Not-determined |
| 18-109_k141_84933 | Not-determined |
| 18-109_k141_238722 | Not-determined |
| 18-109_k141_13996 | Not-determined |
| 18-109_k141_126152 | Not-determined |
| 18-109_k141_167208 | Not-determined |
| 18-109_k141_228615 | Not-determined |
| 18-109_k141_105663 | Not-determined |
| 18-109_k141_105666 | Not-determined |
| 18-109_k141_156939 | Not-determined |
| 18-109_k141_105672 | Not-determined |
| 18-109_k141_167223 | Not-determined |
| 18-109_k141_105675 | Not-determined |
| 18-109_k141_156946 | Not-determined |
| 18-109_k141_146256 | Low-quality |
| 18-109_k141_116128 | Not-determined |
| 18-109_k141_84968 | Not-determined |
| 18-109_k141_74296 | Low-quality |
| 18-109_k141_3591 | Not-determined |
| 18-109_k141_156971 | Not-determined |
| 18-109_k141_116141 | Not-determined |
| 18-109_k141_217645 | Not-determined |
| 18-109_k141_136080 | Not-determined |
| 18-109_k141_198210 | Not-determined |
| 18-109_k141_105694 | Not-determined |
| 18-109_k141_105701 | Not-determined |
| 18-109_k141_126187 | Not-determined |
| 18-109_k141_44412 | Not-determined |
| 18-109_k141_167252 | Low-quality |
| 18-109_k141_94904 | Not-determined |
| 18-109_k141_126195 | Not-determined |
| 18-109_k141_126196 | Not-determined |
| 18-109_k141_116162 | Not-determined |
| 18-109_k141_156990 | Not-determined |
| 18-109_k141_44415 | Low-quality |
| 18-109_k141_3612 | Not-determined |
| 18-109_k141_156998 | Not-determined |
| 18-109_k141_23241 | Not-determined |
| 18-109_k141_116168 | Not-determined |
| 18-109_k141_23246 | Not-determined |
| 18-109_k141_94921 | Not-determined |
| 18-109_k141_44428 | Not-determined |
| 18-109_k141_116173 | Not-determined |
| 18-109_k141_208603 | Not-determined |
| 18-109_k141_23267 | Low-quality |
| 18-109_k141_146270 | Not-determined |
| 18-109_k141_198217 | Not-determined |
| 18-109_k141_54463 | Not-determined |
| 18-109_k141_23274 | Not-determined |
| 18-109_k141_94958 | Not-determined |
| 18-109_k141_146283 | Not-determined |
| 18-109_k141_198227 | Not-determined |
| 18-109_k141_23279 | Not-determined |
| 18-109_k141_74366 | Not-determined |
| 18-109_k141_136130 | Not-determined |
| 18-109_k141_146288 | Low-quality |
| 18-109_k141_44437 | Not-determined |
| 18-109_k141_146289 | Not-determined |
| 18-109_k141_85047 | Not-determined |
| 18-109_k141_44441 | Not-determined |
| 18-109_k141_177187 | Not-determined |
| 18-109_k141_44443 | Not-determined |
| 18-109_k141_198234 | Not-determined |
| 18-109_k141_198240 | Low-quality |
| 18-109_k141_3647 | Not-determined |
| 18-109_k141_44453 | Not-determined |
| 18-109_k141_94977 | Not-determined |
| 18-109_k141_146308 | Not-determined |
| 18-109_k141_198248 | Not-determined |
| 18-109_k141_3651 | Not-determined |
| 18-109_k141_44459 | Not-determined |
| 18-109_k141_146316 | Not-determined |
| 18-109_k141_94988 | Low-quality |
| 18-109_k141_146324 | Not-determined |
| 18-109_k141_167307 | Not-determined |
| 18-109_k141_198254 | Not-determined |
| 18-109_k141_85071 | Not-determined |
| 18-109_k141_74401 | Not-determined |
| 18-109_k141_167309 | Not-determined |
| 18-109_k141_167310 | Not-determined |
| 18-109_k141_34508 | Not-determined |
| 18-109_k141_116255 | Not-determined |
| 18-109_k141_95007 | Not-determined |
| 18-109_k141_146343 | Not-determined |
| 18-109_k141_217664 | Not-determined |
| 18-109_k141_44482 | Not-determined |
| 18-109_k141_85085 | Not-determined |
| 18-109_k141_167325 | Not-determined |
| 18-109_k141_116270 | Not-determined |
| 18-109_k141_198277 | Not-determined |
| 18-109_k141_3678 | Not-determined |
| 18-109_k141_34522 | Not-determined |
| 18-109_k141_116274 | Not-determined |
| 18-109_k141_44491 | Low-quality |
| 18-109_k141_116277 | Not-determined |
| 18-109_k141_238777 | Not-determined |
| 18-109_k141_136148 | Not-determined |
| 18-109_k141_217682 | Not-determined |
| 18-109_k141_238792 | Not-determined |
| 18-109_k141_95022 | Not-determined |
| 18-109_k141_146365 | Not-determined |
| 18-109_k141_34530 | Not-determined |
| 18-109_k141_217696 | Low-quality |
| 18-109_k141_208651 | Not-determined |
| 18-109_k141_177223 | Not-determined |
| 18-109_k141_34535 | Not-determined |
| 18-109_k141_34536 | Not-determined |
| 18-109_k141_85101 | Low-quality |
| 18-109_k141_126244 | Not-determined |
| 18-109_k141_126245 | Not-determined |
| 18-109_k141_3695 | Not-determined |
| 18-109_k141_177238 | Not-determined |
| 18-109_k141_74418 | Not-determined |
| 18-109_k141_126251 | Not-determined |
| 18-109_k141_146372 | Not-determined |
| 18-109_k141_157003 | Not-determined |
| 18-109_k141_198290 | Not-determined |
| 18-109_k141_198292 | Not-determined |
| 18-109_k141_146374 | Not-determined |
| 18-109_k141_198293 | Low-quality |
| 18-109_k141_34554 | Not-determined |
| 18-109_k141_34555 | Not-determined |
| 18-109_k141_157008 | Not-determined |
| 18-109_k141_126264 | Not-determined |
| 18-109_k141_177244 | Not-determined |
| 18-109_k141_34560 | Not-determined |
| 18-109_k141_74432 | Not-determined |
| 18-109_k141_34569 | Not-determined |
| 18-109_k141_157030 | Not-determined |
| 18-109_k141_64792 | Not-determined |
| 18-109_k141_157031 | Not-determined |
| 18-109_k141_146386 | Not-determined |
| 18-109_k141_136170 | Not-determined |
| 18-109_k141_136171 | Not-determined |
| 18-109_k141_157033 | Not-determined |
| 18-109_k141_157038 | Not-determined |
| 18-109_k141_177251 | Not-determined |
| 18-109_k141_177252 | Not-determined |
| 18-109_k141_177253 | Not-determined |
| 18-109_k141_34578 | Not-determined |
| 18-109_k141_238845 | Not-determined |
| 18-109_k141_95033 | Not-determined |
| 18-109_k141_157046 | Not-determined |
| 18-109_k141_126288 | Not-determined |
| 18-109_k141_167359 | Low-quality |
| 18-109_k141_198324 | Not-determined |
| 18-109_k141_64820 | Not-determined |
| 18-109_k141_157056 | Not-determined |
| 18-109_k141_95041 | Not-determined |
| 18-109_k141_188139 | Not-determined |
| 18-109_k141_208681 | Not-determined |
| 18-109_k141_64825 | Not-determined |
| 18-109_k141_44510 | Not-determined |
| 18-109_k141_126295 | Low-quality |
| 18-109_k141_95051 | Not-determined |
| 18-109_k141_167380 | Not-determined |
| 18-109_k141_95058 | Not-determined |
| 18-109_k141_44520 | Not-determined |
| 18-109_k141_146420 | Not-determined |
| 18-109_k141_64833 | Not-determined |
| 18-109_k141_116307 | Not-determined |
| 18-109_k141_116309 | Low-quality |
| 18-109_k141_116310 | Not-determined |
| 18-109_k141_95065 | Not-determined |
| 18-109_k141_23325 | Low-quality |
| 18-109_k141_116314 | Low-quality |
| 18-109_k141_126320 | Low-quality |
| 18-109_k141_157086 | Not-determined |
| 18-109_k141_23345 | Not-determined |
| 18-109_k141_157087 | Not-determined |
| 18-109_k141_157088 | Not-determined |
| 18-109_k141_238868 | Not-determined |
| 18-109_k141_136214 | Not-determined |
| 18-109_k141_74481 | Not-determined |
| 18-109_k141_34594 | Not-determined |
| 18-109_k141_208699 | Not-determined |
| 18-109_k141_167385 | Not-determined |
| 18-109_k141_146444 | Not-determined |
| 18-109_k141_126335 | Not-determined |
| 18-109_k141_23354 | Not-determined |
| 18-109_k141_167393 | Not-determined |
| 18-109_k141_126344 | Not-determined |
| 18-109_k141_188143 | Not-determined |
| 18-109_k141_188146 | Not-determined |
| 18-109_k141_74505 | Low-quality |
| 18-109_k141_146463 | Not-determined |
| 18-109_k141_23360 | Not-determined |
| 18-109_k141_85170 | Not-determined |
| 18-109_k141_177270 | Not-determined |
| 18-109_k141_126354 | Not-determined |
| 18-109_k141_238891 | Not-determined |
| 18-109_k141_167414 | Not-determined |
| 18-109_k141_136259 | Not-determined |
| 18-109_k141_198349 | Not-determined |
| 18-109_k141_188159 | Low-quality |
| 18-109_k141_188163 | Not-determined |
| 18-109_k141_54469 | Not-determined |
| 18-109_k141_238906 | Not-determined |
| 18-109_k141_188173 | Not-determined |
| 18-109_k141_74536 | Not-determined |
| 18-109_k141_167434 | Not-determined |
| 18-109_k141_74544 | Not-determined |
| 18-109_k141_208740 | Not-determined |
| 18-109_k141_54482 | Not-determined |
| 18-109_k141_146485 | Not-determined |
| 18-109_k141_54492 | Not-determined |
| 18-109_k141_126384 | Not-determined |
| 18-109_k141_188201 | Not-determined |
| 18-109_k141_188202 | Not-determined |
| 18-109_k141_208750 | Low-quality |
| 18-109_k141_74562 | Not-determined |
| 18-109_k141_177324 | Not-determined |
| 18-109_k141_85206 | Not-determined |
| 18-109_k141_85213 | Not-determined |
| 18-109_k141_188215 | Not-determined |
| 18-109_k141_74577 | Low-quality |
| 18-109_k141_34612 | Not-determined |
| 18-109_k141_188219 | Not-determined |
| 18-109_k141_95126 | Not-determined |
| 18-109_k141_85220 | Not-determined |
| 18-109_k141_126414 | Not-determined |
| 18-109_k141_157119 | Not-determined |
| 18-109_k141_64851 | Not-determined |
| 18-109_k141_126421 | Not-determined |
| 18-109_k141_74597 | Not-determined |
| 18-109_k141_74601 | Not-determined |
| 18-109_k141_136292 | Not-determined |
| 18-109_k141_238927 | Not-determined |
| 18-109_k141_64867 | Not-determined |
| 18-109_k141_34637 | Not-determined |
| 18-109_k141_136311 | Not-determined |
| 18-109_k141_23434 | Not-determined |
| 18-109_k141_23437 | Not-determined |
| 18-109_k141_44578 | Not-determined |
| 18-109_k141_167472 | Not-determined |
| 18-109_k141_146540 | Not-determined |
| 18-109_k141_34646 | Low-quality |
| 18-109_k141_167474 | Not-determined |
| 18-109_k141_167475 | Not-determined |
| 18-109_k141_64888 | Not-determined |
| 18-109_k141_116355 | Not-determined |
| 18-109_k141_44583 | Not-determined |
| 18-109_k141_238955 | Not-determined |
| 18-109_k141_146546 | Not-determined |
| 18-109_k141_177363 | Not-determined |
| 18-109_k141_34652 | Not-determined |
| 18-109_k141_177364 | Not-determined |
| 18-109_k141_74626 | Not-determined |
| 18-109_k141_116363 | Not-determined |
| 18-109_k141_34657 | Not-determined |
| 18-109_k141_34659 | Not-determined |
| 18-109_k141_167495 | Not-determined |
| 18-109_k141_64905 | Not-determined |
| 18-109_k141_208775 | Not-determined |
| 18-109_k141_95158 | Not-determined |
| 18-109_k141_64918 | Low-quality |
| 18-109_k141_23447 | Not-determined |
| 18-109_k141_238985 | Not-determined |
| 18-109_k141_64923 | Not-determined |
| 18-109_k141_208789 | Not-determined |
| 18-109_k141_238990 | Not-determined |
| 18-109_k141_177377 | Not-determined |
| 18-109_k141_95172 | Not-determined |
| 18-109_k141_239000 | Not-determined |
| 18-109_k141_44623 | Low-quality |
| 18-109_k141_116372 | Not-determined |
| 18-109_k141_239007 | Not-determined |
| 18-109_k141_239011 | Not-determined |
| 18-109_k141_95185 | Not-determined |
| 18-109_k141_126450 | Low-quality |
| 18-109_k141_74655 | Not-determined |
| 18-109_k141_136332 | Not-determined |
| 18-109_k141_177387 | Not-determined |
| 18-109_k141_74665 | Not-determined |
| 18-109_k141_167520 | Not-determined |
| 18-109_k141_208813 | Not-determined |
| 18-109_k141_44644 | Not-determined |
| 18-109_k141_239030 | Not-determined |
| 18-109_k141_64955 | Not-determined |
| 18-109_k141_105720 | Not-determined |
| 18-109_k141_116389 | Not-determined |
| 18-109_k141_95210 | Not-determined |
| 18-109_k141_64967 | Not-determined |
| 18-109_k141_95211 | Not-determined |
| 18-109_k141_188249 | Not-determined |
| 18-109_k141_95213 | Not-determined |
| 18-109_k141_116397 | Not-determined |
| 18-109_k141_116398 | Not-determined |
| 18-109_k141_105729 | Not-determined |
| 18-109_k141_105730 | Not-determined |
| 18-109_k141_44661 | Not-determined |
| 18-109_k141_208817 | Not-determined |
| 18-109_k141_105741 | Not-determined |
| 18-109_k141_217707 | Not-determined |
| 18-109_k141_85270 | Not-determined |
| 18-109_k141_54552 | Not-determined |
| 18-109_k141_85272 | Not-determined |
| 18-109_k141_95220 | Not-determined |
| 18-109_k141_105745 | Not-determined |
| 18-109_k141_105748 | Not-determined |
| 18-109_k141_74694 | Not-determined |
| 18-109_k141_14004 | Not-determined |
| 18-109_k141_188268 | Low-quality |
| 18-109_k141_14006 | Not-determined |
| 18-109_k141_208825 | Not-determined |
| 18-109_k141_14017 | Not-determined |
| 18-109_k141_188284 | Not-determined |
| 18-109_k141_167563 | Not-determined |
| 18-109_k141_54573 | Not-determined |
| 18-109_k141_177410 | Not-determined |
| 18-109_k141_167564 | Low-quality |
| 18-109_k141_95230 | Not-determined |
| 18-109_k141_23489 | Not-determined |
| 18-109_k141_239067 | Not-determined |
| 18-109_k141_167568 | Not-determined |
| 18-109_k141_95237 | Not-determined |
| 18-109_k141_95238 | Not-determined |
| 18-109_k141_95240 | Not-determined |
| 18-109_k141_105753 | Not-determined |
| 18-109_k141_34715 | Not-determined |
| 18-109_k141_74723 | Low-quality |
| 18-109_k141_217739 | Not-determined |
| 18-109_k141_64993 | Low-quality |
| 18-109_k141_34730 | Not-determined |
| 18-109_k141_74729 | Not-determined |
| 18-109_k141_239076 | Not-determined |
| 18-109_k141_85303 | Not-determined |
| 18-109_k141_34743 | Not-determined |
| 18-109_k141_74741 | Not-determined |
| 18-109_k141_217758 | Not-determined |
| 18-109_k141_85309 | Not-determined |
| 18-109_k141_95253 | Not-determined |
| 18-109_k141_177414 | Not-determined |
| 18-109_k141_95260 | Not-determined |
| 18-109_k141_95263 | Not-determined |
| 18-109_k141_34764 | Not-determined |
| 18-109_k141_44699 | Not-determined |
| 18-109_k141_95269 | Not-determined |
| 18-109_k141_65012 | Not-determined |
| 18-109_k141_146590 | Not-determined |
| 18-109_k141_44710 | Not-determined |
| 18-109_k141_3715 | Not-determined |
| 18-109_k141_177431 | Not-determined |
| 18-109_k141_44718 | Not-determined |
| 18-109_k141_188310 | Not-determined |
| 18-109_k141_3718 | Not-determined |
| 18-109_k141_3722 | Not-determined |
| 18-109_k141_228677 | Not-determined |
| 18-109_k141_95291 | Not-determined |
| 18-109_k141_188316 | Not-determined |
| 18-109_k141_228682 | Not-determined |
| 18-109_k141_228683 | Not-determined |
| 18-109_k141_188318 | Not-determined |
| 18-109_k141_208870 | Not-determined |
| 18-109_k141_146591 | Not-determined |
| 18-109_k141_85325 | Not-determined |
| 18-109_k141_3738 | Low-quality |
| 18-109_k141_3739 | Not-determined |
| 18-109_k141_34805 | Not-determined |
| 18-109_k141_85329 | Not-determined |
| 18-109_k141_3751 | Not-determined |
| 18-109_k141_3752 | Not-determined |
| 18-109_k141_136365 | Not-determined |
| 18-109_k141_217780 | Not-determined |
| 18-109_k141_126509 | Not-determined |
| 18-109_k141_54651 | Not-determined |
| 18-109_k141_177464 | Not-determined |
| 18-109_k141_85333 | Not-determined |
| 18-109_k141_34809 | Not-determined |
| 18-109_k141_146599 | Not-determined |
| 18-109_k141_105775 | Not-determined |
| 18-109_k141_65051 | Not-determined |
| 18-109_k141_105779 | Not-determined |
| 18-109_k141_208902 | Not-determined |
| 18-109_k141_34824 | Not-determined |
| 18-109_k141_105785 | Not-determined |
| 18-109_k141_34828 | Not-determined |
| 18-109_k141_177479 | Low-quality |
| 18-109_k141_34829 | Not-determined |
| 18-109_k141_208907 | Not-determined |
| 18-109_k141_198389 | Not-determined |
| 18-109_k141_198391 | Not-determined |
| 18-109_k141_198393 | Low-quality |
| 18-109_k141_177486 | Not-determined |
| 18-109_k141_44744 | Not-determined |
| 18-109_k141_177488 | Not-determined |
| 18-109_k141_177489 | Low-quality |
| 18-109_k141_177491 | Not-determined |
| 18-109_k141_44748 | Low-quality |
| 18-109_k141_188327 | Not-determined |
| 18-109_k141_198404 | Not-determined |
| 18-109_k141_146615 | Not-determined |

| | |
|---|---|
| 18-109_k141_136399 | Not-determined |
| 18-109_k141_167618 | Not-determined |
| 18-109_k141_228723 | Not-determined |
| 18-109_k141_105812 | Not-determined |
| 18-109_k141_239108 | Low-quality |
| 18-109_k141_188345 | Not-determined |
| 18-109_k141_239114 | Not-determined |
| 18-109_k141_85358 | Not-determined |
| 18-109_k141_157175 | Low-quality |
| 18-109_k141_188352 | Low-quality |
| 18-109_k141_3762 | Not-determined |
| 18-109_k141_105821 | Not-determined |
| 18-109_k141_105823 | Not-determined |
| 18-109_k141_157181 | Not-determined |
| 18-109_k141_228733 | Not-determined |
| 18-109_k141_105828 | Not-determined |
| 18-109_k141_228736 | Not-determined |
| 18-109_k141_239127 | Not-determined |
| 18-109_k141_44762 | Not-determined |
| 18-109_k141_105835 | Low-quality |
| 18-109_k141_136424 | Not-determined |
| 18-109_k141_239137 | Not-determined |
| 18-109_k141_177509 | Not-determined |
| 18-109_k141_54667 | Not-determined |
| 18-109_k141_95319 | Not-determined |
| 18-109_k141_198432 | Not-determined |
| 18-109_k141_54673 | Not-determined |
| 18-109_k141_208946 | Not-determined |
| 18-109_k141_54678 | Not-determined |
| 18-109_k141_74760 | Not-determined |
| 18-109_k141_23548 | Not-determined |
| 18-109_k141_188361 | Not-determined |
| 18-109_k141_105858 | Not-determined |
| 18-109_k141_136441 | Not-determined |
| 18-109_k141_188363 | Not-determined |
| 18-109_k141_3766 | Not-determined |
| 18-109_k141_105863 | Not-determined |
| 18-109_k141_136446 | Not-determined |
| 18-109_k141_208955 | Not-determined |
| 18-109_k141_23551 | Not-determined |
| 18-109_k141_44784 | Not-determined |
| 18-109_k141_44792 | Not-determined |
| 18-109_k141_157197 | Not-determined |
| 18-109_k141_177539 | Not-determined |
| 18-109_k141_126536 | Not-determined |
| 18-109_k141_188368 | Not-determined |
| 18-109_k141_65076 | Low-quality |
| 18-109_k141_54704 | Not-determined |
| 18-109_k141_198446 | Not-determined |
| 18-109_k141_146640 | Not-determined |
| 18-109_k141_157201 | Not-determined |
| 18-109_k141_54705 | Low-quality |
| 18-109_k141_208971 | Not-determined |
| 18-109_k141_126546 | Low-quality |
| 18-109_k141_85373 | Not-determined |
| 18-109_k141_228770 | Not-determined |
| 18-109_k141_54713 | Not-determined |
| 18-109_k141_146660 | Not-determined |
| 18-109_k141_198462 | Low-quality |
| 18-109_k141_177546 | Low-quality |
| 18-109_k141_23577 | Low-quality |
| 18-109_k141_198468 | Not-determined |
| 18-109_k141_146669 | Not-determined |
| 18-109_k141_198470 | Not-determined |
| 18-109_k141_23579 | Not-determined |
| 18-109_k141_157231 | Not-determined |
| 18-109_k141_74798 | Not-determined |
| 18-109_k141_198479 | Not-determined |
| 18-109_k141_228783 | Low-quality |
| 18-109_k141_65118 | Not-determined |
| 18-109_k141_198481 | Not-determined |
| 18-109_k141_23591 | Not-determined |
| 18-109_k141_95352 | Not-determined |
| 18-109_k141_95354 | Not-determined |
| 18-109_k141_136494 | Not-determined |
| 18-109_k141_157245 | Not-determined |
| 18-109_k141_198492 | Not-determined |
| 18-109_k141_167635 | Not-determined |
| 18-109_k141_157247 | Not-determined |
| 18-109_k141_198496 | Not-determined |
| 18-109_k141_167642 | Low-quality |
| 18-109_k141_188389 | Not-determined |
| 18-109_k141_54728 | Not-determined |
| 18-109_k141_167643 | Not-determined |
| 18-109_k141_105876 | Low-quality |
| 18-109_k141_54730 | Not-determined |
| 18-109_k141_157251 | Not-determined |
| 18-109_k141_146683 | Not-determined |
| 18-109_k141_65130 | Not-determined |
| 18-109_k141_157256 | Not-determined |
| 18-109_k141_116410 | Not-determined |
| 18-109_k141_228796 | Not-determined |
| 18-109_k141_177577 | Not-determined |
| 18-109_k141_177582 | Not-determined |
| 18-109_k141_105898 | Low-quality |
| 18-109_k141_188406 | Not-determined |
| 18-109_k141_146696 | Not-determined |
| 18-109_k141_126595 | Not-determined |
| 18-109_k141_146699 | Not-determined |
| 18-109_k141_126602 | Not-determined |
| 18-109_k141_85430 | Not-determined |
| 18-109_k141_85431 | Low-quality |
| 18-109_k141_239150 | Not-determined |
| 18-109_k141_239151 | Not-determined |
| 18-109_k141_85434 | Not-determined |
| 18-109_k141_54765 | Low-quality |
| 18-109_k141_177593 | Low-quality |
| 18-109_k141_23608 | Not-determined |
| 18-109_k141_136517 | Not-determined |
| 18-109_k141_177604 | Not-determined |
| 18-109_k141_65151 | Not-determined |
| 18-109_k141_34856 | Not-determined |
| 18-109_k141_208984 | Not-determined |
| 18-109_k141_228833 | Not-determined |
| 18-109_k141_177616 | Low-quality |
| 18-109_k141_85442 | Not-determined |
| 18-109_k141_54781 | Not-determined |
| 18-109_k141_54783 | Not-determined |
| 18-109_k141_23622 | Not-determined |
| 18-109_k141_116422 | Not-determined |
| 18-109_k141_136529 | Not-determined |
| 18-109_k141_198507 | Not-determined |
| 18-109_k141_208996 | Not-determined |
| 18-109_k141_198509 | Not-determined |
| 18-109_k141_65168 | Low-quality |
| 18-109_k141_198514 | Not-determined |
| 18-109_k141_54791 | Not-determined |
| 18-109_k141_188434 | Not-determined |
| 18-109_k141_34870 | Not-determined |
| 18-109_k141_34872 | Not-determined |
| 18-109_k141_239166 | Low-quality |
| 18-109_k141_239172 | Not-determined |
| 18-109_k141_95430 | Not-determined |
| 18-109_k141_188441 | Not-determined |
| 18-109_k141_54812 | Not-determined |
| 18-109_k141_95432 | Not-determined |
| 18-109_k141_54822 | Not-determined |
| 18-109_k141_188454 | Not-determined |
| 18-109_k141_65206 | Not-determined |
| 18-109_k141_239195 | Low-quality |
| 18-109_k141_239196 | Not-determined |
| 18-109_k141_209046 | Not-determined |
| 18-109_k141_105910 | Not-determined |
| 18-109_k141_239207 | Not-determined |
| 18-109_k141_188467 | Not-determined |
| 18-109_k141_239211 | Low-quality |
| 18-109_k141_198556 | Not-determined |
| 18-109_k141_209053 | Low-quality |
| 18-109_k141_209062 | Not-determined |
| 18-109_k141_198563 | Not-determined |
| 18-109_k141_228845 | Not-determined |
| 18-109_k141_65240 | Not-determined |
| 18-109_k141_239219 | Not-determined |
| 18-109_k141_198572 | Not-determined |
| 18-109_k141_177685 | Not-determined |
| 18-109_k141_177689 | Not-determined |
| 18-109_k141_177690 | Not-determined |
| 18-109_k141_14121 | Not-determined |
| 18-109_k141_188476 | Not-determined |
| 18-109_k141_14128 | Not-determined |
| 18-109_k141_167715 | Not-determined |
| 18-109_k141_228858 | Low-quality |
| 18-109_k141_167716 | Not-determined |
| 18-109_k141_167717 | Not-determined |
| 18-109_k141_105926 | Not-determined |
| 18-109_k141_3808 | Low-quality |
| 18-109_k141_188490 | Not-determined |
| 18-109_k141_14129 | Not-determined |
| 18-109_k141_14132 | Not-determined |
| 18-109_k141_14134 | Not-determined |
| 18-109_k141_3816 | Not-determined |
| 18-109_k141_239237 | Not-determined |
| 18-109_k141_167731 | Not-determined |
| 18-109_k141_167732 | Not-determined |
| 18-109_k141_198589 | Not-determined |
| 18-109_k141_239248 | Not-determined |
| 18-109_k141_177696 | Not-determined |
| 18-109_k141_228873 | Not-determined |
| 18-109_k141_177699 | Not-determined |
| 18-109_k141_136557 | Low-quality |
| 18-109_k141_228878 | Not-determined |
| 18-109_k141_14144 | Not-determined |
| 18-109_k141_126663 | Not-determined |
| 18-109_k141_23652 | Not-determined |
| 18-109_k141_23657 | Not-determined |
| 18-109_k141_65265 | Not-determined |
| 18-109_k141_126670 | Not-determined |
| 18-109_k141_228885 | Not-determined |
| 18-109_k141_126672 | Not-determined |
| 18-109_k141_228890 | Not-determined |
| 18-109_k141_95450 | Not-determined |
| 18-109_k141_136586 | Not-determined |
| 18-109_k141_34906 | Not-determined |
| 18-109_k141_177717 | Not-determined |
| 18-109_k141_65285 | Not-determined |
| 18-109_k141_105975 | Not-determined |
| 18-109_k141_105979 | Not-determined |
| 18-109_k141_157262 | Not-determined |
| 18-109_k141_3867 | Not-determined |
| 18-109_k141_105986 | Not-determined |
| 18-109_k141_85508 | Not-determined |
| 18-109_k141_23676 | Not-determined |
| 18-109_k141_136592 | Not-determined |
| 18-109_k141_74808 | Not-determined |
| 18-109_k141_3874 | Not-determined |
| 18-109_k141_23692 | Not-determined |
| 18-109_k141_95483 | Not-determined |
| 18-109_k141_228904 | Low-quality |
| 18-109_k141_146719 | Not-determined |
| 18-109_k141_23697 | Not-determined |
| 18-109_k141_228921 | Low-quality |
| KM-final-viral-combined_k141_146050\|\|full | Medium-quality |
| 18-109_k141_23702 | Not-determined |
| 18-109_k141_136622 | Not-determined |
| 18-109_k141_65324 | Not-determined |
| 18-109_k141_157280 | Low-quality |
| 18-109_k141_167760 | Not-determined |
| 18-109_k141_146732 | Not-determined |
| 18-109_k141_54852 | Not-determined |
| 18-109_k141_65333 | Low-quality |
| 18-109_k141_14174 | Not-determined |
| 18-109_k141_126713 | Not-determined |
| 18-109_k141_126714 | Not-determined |
| 18-109_k141_3901 | Not-determined |
| 18-109_k141_228938 | Not-determined |
| 18-109_k141_65335 | Low-quality |
| 18-109_k141_65336 | Not-determined |
| 18-109_k141_177747 | Not-determined |
| 18-109_k141_65343 | Not-determined |
| 18-109_k141_228946 | Not-determined |
| 18-109_k141_44868 | Not-determined |
| 18-109_k141_126721 | Not-determined |
| 18-109_k141_198638 | Low-quality |
| 18-109_k141_188525 | Not-determined |
| 18-109_k141_198639 | Not-determined |
| 18-109_k141_44876 | Not-determined |
| 18-109_k141_188528 | Not-determined |
| 18-109_k141_167784 | Not-determined |
| 18-109_k141_198645 | Not-determined |
| 18-109_k141_167791 | Not-determined |
| 18-109_k141_44880 | Not-determined |
| 18-109_k141_177763 | Not-determined |
| 18-109_k141_228964 | Not-determined |
| 18-109_k141_44891 | Not-determined |
| 18-109_k141_126746 | Not-determined |
| 18-109_k141_44901 | Not-determined |
| 18-109_k141_157301 | Not-determined |
| 18-109_k141_116471 | Not-determined |
| 18-109_k141_74839 | Not-determined |
| 18-109_k141_65358 | Not-determined |
| 18-109_k141_34930 | Not-determined |
| 18-109_k141_126766 | Not-determined |
| 18-109_k141_65366 | Not-determined |
| 18-109_k141_65368 | Not-determined |
| 18-109_k141_65371 | Not-determined |
| 18-109_k141_34940 | Not-determined |
| 18-109_k141_65372 | Not-determined |
| 18-109_k141_116483 | Not-determined |
| 18-109_k141_34944 | Not-determined |
| 18-109_k141_34945 | Not-determined |
| 18-109_k141_23731 | Not-determined |
| 18-109_k141_177779 | Not-determined |
| 18-109_k141_74859 | Not-determined |
| 18-109_k141_74861 | Not-determined |
| 18-109_k141_54887 | Not-determined |
| 18-109_k141_198684 | Not-determined |
| 18-109_k141_239303 | Not-determined |
| 18-109_k141_65380 | Not-determined |
| 18-109_k141_54888 | Not-determined |
| 18-109_k141_198685 | Not-determined |
| 18-109_k141_23740 | Not-determined |
| 18-109_k141_116489 | Not-determined |
| 18-109_k141_177785 | Not-determined |
| 18-109_k141_65384 | Not-determined |
| 18-109_k141_54892 | Not-determined |
| 18-109_k141_65387 | Not-determined |
| 18-109_k141_23749 | Not-determined |
| 18-109_k141_74870 | Not-determined |
| 18-109_k141_23750 | Not-determined |
| 18-109_k141_177792 | Not-determined |
| 18-109_k141_65389 | Not-determined |
| 18-109_k141_23758 | Not-determined |
| 18-109_k141_34962 | Not-determined |
| 18-109_k141_23765 | Not-determined |
| 18-109_k141_157322 | Not-determined |
| 18-109_k141_198705 | Not-determined |
| 18-109_k141_3922 | Not-determined |
| 18-109_k141_23773 | Not-determined |
| 18-109_k141_126771 | Not-determined |
| 18-109_k141_239323 | Not-determined |
| 18-109_k141_74884 | Not-determined |
| 18-109_k141_126775 | Not-determined |
| 18-109_k141_54903 | Not-determined |
| 18-109_k141_23780 | Not-determined |
| 18-109_k141_239330 | Not-determined |
| 18-109_k141_44957 | Not-determined |
| 18-109_k141_217845 | Not-determined |
| 18-109_k141_116518 | Not-determined |
| 18-109_k141_116524 | Not-determined |
| 18-109_k141_217854 | Low-quality |
| 18-109_k141_228994 | Not-determined |
| 18-109_k141_106087 | Not-determined |
| 18-109_k141_54923 | Not-determined |
| 18-109_k141_217857 | Not-determined |
| 18-109_k141_34974 | Not-determined |
| 18-109_k141_44970 | Low-quality |
| 18-109_k141_106091 | Not-determined |
| 18-109_k141_126783 | Not-determined |
| 18-109_k141_209079 | Not-determined |
| 18-109_k141_126785 | Low-quality |
| 18-109_k141_177809 | Not-determined |
| 18-109_k141_217861 | Not-determined |
| 18-109_k141_177813 | Not-determined |
| 18-109_k141_54934 | Not-determined |
| 18-109_k141_14198 | Low-quality |
| 18-109_k141_65402 | Not-determined |
| 18-109_k141_65405 | Not-determined |
| 18-109_k141_229006 | Low-quality |
| 18-109_k141_54937 | Not-determined |
| 18-109_k141_54938 | Not-determined |
| 18-109_k141_34996 | Not-determined |
| 18-109_k141_23798 | Not-determined |
| 18-109_k141_14205 | Low-quality |
| 18-109_k141_45001 | Not-determined |
| 18-109_k141_65414 | Not-determined |
| 18-109_k141_126804 | Low-quality |
| 18-109_k141_146839 | Not-determined |
| 18-109_k141_146842 | Not-determined |
| 18-109_k141_54941 | Not-determined |
| 18-109_k141_177829 | Not-determined |
| 18-109_k141_65427 | Not-determined |
| 18-109_k141_229032 | Not-determined |
| 18-109_k141_217874 | Low-quality |
| 18-109_k141_95551 | Not-determined |
| 18-109_k141_54952 | Low-quality |
| 18-109_k141_14228 | Not-determined |
| 18-109_k141_54954 | Not-determined |
| 18-109_k141_95556 | Not-determined |
| 18-109_k141_54955 | Not-determined |
| 18-109_k141_106105 | Not-determined |
| 18-109_k141_95561 | Not-determined |
| 18-109_k141_198739 | Not-determined |
| 18-109_k141_95564 | Not-determined |
| 18-109_k141_106112 | Not-determined |
| 18-109_k141_106115 | Not-determined |
| 18-109_k141_85567 | Not-determined |
| 18-109_k141_116556 | Not-determined |
| 18-109_k141_229053 | Low-quality |
| 18-109_k141_116557 | Not-determined |
| 18-109_k141_126827 | Not-determined |
| 18-109_k141_126832 | Not-determined |
| 18-109_k141_198760 | Not-determined |
| 18-109_k141_3962 | Not-determined |
| 18-109_k141_126846 | Not-determined |
| 18-109_k141_198771 | Not-determined |
| 18-109_k141_157374 | Not-determined |
| 18-109_k141_3969 | Not-determined |
| 18-109_k141_177861 | Not-determined |
| 18-109_k141_116588 | Not-determined |
| 18-109_k141_106154 | Not-determined |
| 18-109_k141_23814 | Not-determined |
| 18-109_k141_198790 | Not-determined |
| 18-109_k141_85584 | Not-determined |
| 18-109_k141_3975 | Not-determined |
| 18-109_k141_116598 | Low-quality |
| 18-109_k141_95586 | Not-determined |
| 18-109_k141_23820 | Not-determined |
| 18-109_k141_198800 | Not-determined |
| 18-109_k141_157383 | Not-determined |
| 18-109_k141_35023 | Not-determined |
| 18-109_k141_177865 | Not-determined |
| 18-109_k141_116608 | Not-determined |
| 18-109_k141_85603 | Not-determined |
| 18-109_k141_157398 | Not-determined |
| 18-109_k141_85608 | Not-determined |
| 18-109_k141_116620 | Not-determined |
| 18-109_k141_55026 | Low-quality |
| 18-109_k141_209098 | Not-determined |
| 18-109_k141_116624 | Low-quality |
| 18-109_k141_3980 | Not-determined |
| 18-109_k141_3983 | Not-determined |
| 18-109_k141_85615 | Not-determined |
| 18-109_k141_85616 | Not-determined |
| 18-109_k141_198821 | Not-determined |
| 18-109_k141_239375 | Not-determined |
| 18-109_k141_35038 | Not-determined |
| 18-109_k141_239376 | Not-determined |
| 18-109_k141_35041 | Not-determined |
| 18-109_k141_65511 | Not-determined |
| 18-109_k141_106174 | Not-determined |
| 18-109_k141_55030 | Not-determined |
| 18-109_k141_209108 | Low-quality |
| 18-109_k141_85623 | Not-determined |
| 18-109_k141_85625 | Not-determined |
| 18-109_k141_116646 | Not-determined |
| 18-109_k141_116647 | Not-determined |
| 18-109_k141_106183 | Not-determined |
| 18-109_k141_239396 | Not-determined |
| 18-109_k141_106187 | Not-determined |
| 18-109_k141_3993 | Not-determined |
| 18-109_k141_126880 | Not-determined |
| 18-109_k141_106195 | Not-determined |
| 18-109_k141_198850 | Not-determined |
| 18-109_k141_126885 | Low-quality |
| 18-109_k141_85640 | Not-determined |
| 18-109_k141_3999 | Not-determined |
| 18-109_k141_146845 | Not-determined |
| 18-109_k141_146848 | Not-determined |
| 18-109_k141_45035 | Not-determined |
| 18-109_k141_45037 | Not-determined |
| 18-109_k141_35045 | Not-determined |
| 18-109_k141_146858 | Low-quality |
| 18-109_k141_116669 | Not-determined |
| 18-109_k141_116675 | Not-determined |
| 18-109_k141_65556 | Not-determined |
| 18-109_k141_45047 | Not-determined |
| 18-109_k141_85667 | Not-determined |
| 18-109_k141_65563 | Not-determined |
| 18-109_k141_14270 | Not-determined |
| 18-109_k141_229125 | Not-determined |
| 18-109_k141_74900 | Not-determined |
| 18-109_k141_116693 | Not-determined |
| 18-109_k141_85674 | Not-determined |
| 18-109_k141_85677 | Not-determined |
| 18-109_k141_74910 | Not-determined |
| 18-109_k141_146887 | Not-determined |
| 18-109_k141_55070 | Not-determined |
| 18-109_k141_85685 | Not-determined |
| 18-109_k141_239426 | Not-determined |
| 18-109_k141_55073 | Not-determined |
| 18-109_k141_4021 | Not-determined |
| 18-109_k141_35073 | Low-quality |
| 18-109_k141_177896 | Not-determined |
| 18-109_k141_239433 | Not-determined |
| 18-109_k141_229138 | Not-determined |
| 18-109_k141_65581 | Not-determined |
| 18-109_k141_217897 | Not-determined |
| 18-109_k141_157435 | Not-determined |
| 18-109_k141_157437 | Not-determined |
| 18-109_k141_239444 | Not-determined |
| 18-109_k141_217908 | Not-determined |
| 18-109_k141_74932 | Not-determined |
| 18-109_k141_45090 | Not-determined |
| 18-109_k141_74937 | Not-determined |
| 18-109_k141_4042 | Low-quality |
| 18-109_k141_239460 | Low-quality |
| 18-109_k141_45099 | Not-determined |
| 18-109_k141_146895 | Not-determined |
| 18-109_k141_177907 | Not-determined |
| 18-109_k141_229155 | Not-determined |
| 18-109_k141_45105 | Not-determined |
| 18-109_k141_4048 | Not-determined |
| 18-109_k141_198858 | Not-determined |
| 18-109_k141_126903 | Not-determined |
| 18-109_k141_177918 | Not-determined |
| 18-109_k141_35100 | Not-determined |
| 18-109_k141_239462 | Not-determined |
| 18-109_k141_106243 | Not-determined |
| 18-109_k141_209172 | Not-determined |
| 18-109_k141_146917 | Not-determined |
| 18-109_k141_65612 | Not-determined |
| 18-109_k141_126918 | Not-determined |
| 18-109_k141_136696 | Not-determined |
| 18-109_k141_177939 | Not-determined |
| 18-109_k141_136698 | Not-determined |
| 18-109_k141_217914 | Not-determined |
| 18-109_k141_74968 | Not-determined |
| 18-109_k141_74969 | Not-determined |
| 18-109_k141_35109 | Not-determined |
| 18-109_k141_177948 | Not-determined |
| 18-109_k141_106251 | Not-determined |
| 18-109_k141_4065 | Not-determined |
| 18-109_k141_74979 | Not-determined |
| 18-109_k141_198866 | Low-quality |
| 18-109_k141_136709 | Not-determined |
| 18-109_k141_239496 | Not-determined |
| 18-109_k141_35124 | Not-determined |
| 18-109_k141_209197 | Not-determined |
| 18-109_k141_198871 | Not-determined |
| 18-109_k141_209199 | Not-determined |
| 18-109_k141_198874 | Low-quality |
| 18-109_k141_198875 | Not-determined |
| 18-109_k141_65637 | Not-determined |
| 18-109_k141_74981 | Not-determined |
| 18-109_k141_239506 | Not-determined |
| 18-109_k141_198883 | Not-determined |
| 18-109_k141_239510 | Not-determined |
| 18-109_k141_177963 | Not-determined |
| 18-109_k141_116719 | Not-determined |
| 18-109_k141_14280 | Not-determined |
| 18-109_k141_4086 | Not-determined |
| 18-109_k141_74992 | Not-determined |
| 18-109_k141_126952 | Not-determined |
| 18-109_k141_55134 | Not-determined |
| 18-109_k141_45168 | Not-determined |
| 18-109_k141_167871 | Not-determined |
| 18-109_k141_157480 | Not-determined |
| 18-109_k141_209225 | Not-determined |
| 18-109_k141_209226 | Not-determined |
| 18-109_k141_116736 | Not-determined |
| 18-109_k141_126967 | Not-determined |
| 18-109_k141_116740 | Not-determined |
| 18-109_k141_116741 | Not-determined |
| 18-109_k141_95658 | Low-quality |
| 18-109_k141_217937 | Not-determined |
| 18-109_k141_85715 | Not-determined |
| 18-109_k141_229166 | Not-determined |
| 18-109_k141_65672 | Not-determined |
| 18-109_k141_35156 | Not-determined |
| 18-109_k141_95666 | Not-determined |
| 18-109_k141_116748 | Not-determined |
| 18-109_k141_116749 | Not-determined |
| 18-109_k141_95670 | Not-determined |
| 18-109_k141_75013 | Not-determined |
| 18-109_k141_4113 | Not-determined |
| 18-109_k141_95675 | Not-determined |
| 18-109_k141_146928 | Not-determined |
| 18-109_k141_136720 | Not-determined |
| 18-109_k141_146930 | Not-determined |
| 18-109_k141_177973 | Not-determined |
| 18-109_k141_85717 | Not-determined |
| 18-109_k141_126985 | Not-determined |
| 18-109_k141_75023 | Not-determined |
| 18-109_k141_136725 | Not-determined |
| 18-109_k141_146933 | Not-determined |
| 18-109_k141_146935 | Not-determined |
| 18-109_k141_116757 | Not-determined |
| 18-109_k141_65694 | Not-determined |
| 18-109_k141_55161 | Not-determined |
| 18-109_k141_146944 | Not-determined |
| 18-109_k141_23878 | Not-determined |
| 18-109_k141_239562 | Not-determined |
| 18-109_k141_136744 | Not-determined |
| 18-109_k141_167874 | Low-quality |
| 18-109_k141_35175 | Not-determined |
| 18-109_k141_146954 | Not-determined |
| 18-109_k141_146956 | Not-determined |
| 18-109_k141_198915 | Not-determined |
| 18-109_k141_198920 | Not-determined |
| 18-109_k141_136751 | Not-determined |
| 18-109_k141_209268 | Not-determined |
| 18-109_k141_157538 | Not-determined |
| 18-109_k141_146967 | Not-determined |
| 18-109_k141_14298 | Low-quality |
| 18-109_k141_157540 | Not-determined |
| 18-109_k141_95701 | Not-determined |
| 18-109_k141_146973 | Not-determined |
| 18-109_k141_239586 | Not-determined |
| 18-109_k141_127012 | Not-determined |
| 18-109_k141_65726 | Not-determined |
| 18-109_k141_95714 | Not-determined |
| 18-109_k141_35186 | Not-determined |
| 18-109_k141_239597 | Not-determined |
| 18-109_k141_217975 | Not-determined |
| 18-109_k141_55187 | Low-quality |
| 18-109_k141_95720 | Not-determined |
| 18-109_k141_136756 | Not-determined |
| 18-109_k141_55195 | Not-determined |
| 18-109_k141_4126 | Not-determined |
| 18-109_k141_65740 | Low-quality |
| 18-109_k141_65743 | Not-determined |
| 18-109_k141_127036 | Not-determined |
| 18-109_k141_157560 | Low-quality |
| 18-109_k141_65749 | Low-quality |
| 18-109_k141_198964 | Not-determined |
| 18-109_k141_157566 | Not-determined |
| 18-109_k141_177996 | Not-determined |
| 18-109_k141_198967 | Not-determined |
| 18-109_k141_178001 | Not-determined |
| 18-109_k141_178004 | Not-determined |
| 18-109_k141_116763 | Not-determined |
| 18-109_k141_55211 | Low-quality |
| 18-109_k141_85747 | Not-determined |
| 18-109_k141_55225 | Not-determined |
| 18-109_k141_85749 | Not-determined |
| 18-109_k141_178020 | Low-quality |
| 18-109_k141_209288 | Not-determined |
| 18-109_k141_65772 | Not-determined |
| 18-109_k141_127060 | Not-determined |
| 18-109_k141_65776 | Low-quality |
| 18-109_k141_229191 | Not-determined |
| 18-109_k141_209295 | Not-determined |
| 18-109_k141_209303 | Not-determined |
| 18-109_k141_35217 | Not-determined |
| 18-109_k141_75042 | Not-determined |
| 18-109_k141_167917 | Not-determined |
| 18-109_k141_75053 | Low-quality |
| 18-109_k141_167920 | Not-determined |
| 18-109_k141_75055 | Not-determined |

| | |
|---|---|
| 18-109_k141_178034 | Not-determined |
| 18-109_k141_35226 | Not-determined |
| 18-109_k141_55244 | Not-determined |
| 18-109_k141_167929 | Not-determined |
| 18-109_k141_75066 | Not-determined |
| 18-109_k141_106261 | Not-determined |
| 18-109_k141_239642 | Not-determined |
| 18-109_k141_127084 | Low-quality |
| 18-109_k141_116815 | Not-determined |
| 18-109_k141_209319 | Not-determined |
| 18-109_k141_127087 | Not-determined |
| 18-109_k141_116817 | Not-determined |
| 18-109_k141_106264 | Low-quality |
| 18-109_k141_116818 | Not-determined |
| 18-109_k141_217998 | Not-determined |
| 18-109_k141_239647 | Not-determined |
| 18-109_k141_55253 | Not-determined |
| 18-109_k141_65812 | Not-determined |
| 18-109_k141_4149 | Not-determined |
| 18-109_k141_239655 | Not-determined |
| 18-109_k141_239658 | Low-quality |
| 18-109_k141_106273 | Not-determined |
| 18-109_k141_95761 | Not-determined |
| 18-109_k141_136776 | Not-determined |
| 18-109_k141_239663 | Not-determined |
| 18-109_k141_95767 | Not-determined |
| 18-109_k141_127103 | Not-determined |
| 18-109_k141_23969 | Not-determined |
| 18-109_k141_4162 | Not-determined |
| 18-109_k141_157623 | Low-quality |
| 18-109_k141_188597 | Not-determined |
| 18-109_k141_136801 | Not-determined |
| 18-109_k141_209360 | Not-determined |
| 18-109_k141_127119 | Not-determined |
| 18-109_k141_127123 | Not-determined |
| 18-109_k141_136803 | Low-quality |
| 18-109_k141_127127 | Not-determined |
| 18-109_k141_209376 | Not-determined |
| 18-109_k141_136814 | Not-determined |
| 18-109_k141_136818 | Low-quality |
| 18-109_k141_127139 | Not-determined |
| 18-109_k141_209384 | Not-determined |
| 18-109_k141_209385 | Not-determined |
| 18-109_k141_23985 | Not-determined |
| 18-109_k141_23987 | Not-determined |
| 18-109_k141_188618 | Not-determined |
| 18-109_k141_198996 | Not-determined |
| 18-109_k141_188621 | Not-determined |
| 18-109_k141_127150 | Not-determined |
| 18-109_k141_85768 | Not-determined |
| 18-109_k141_136835 | Not-determined |
| 18-109_k141_188626 | Low-quality |
| 18-109_k141_218025 | Low-quality |
| 18-109_k141_239698 | Not-determined |
| 18-109_k141_136837 | Not-determined |
| 18-109_k141_127157 | Low-quality |
| 18-109_k141_209402 | Not-determined |
| 18-109_k141_218034 | Not-determined |
| 18-109_k141_136850 | Not-determined |
| 18-109_k141_218040 | Not-determined |
| 18-109_k141_218041 | Not-determined |
| 18-109_k141_14322 | Not-determined |
| 18-109_k141_85786 | Not-determined |
| 18-109_k141_229228 | Not-determined |
| 18-109_k141_127177 | Not-determined |
| 18-109_k141_178054 | Low-quality |
| 18-109_k141_136866 | Not-determined |
| 18-109_k141_85798 | Not-determined |
| 18-109_k141_136870 | Not-determined |
| 18-109_k141_178061 | Not-determined |
| 18-109_k141_136872 | Not-determined |
| 18-109_k141_239720 | Not-determined |
| 18-109_k141_157653 | Not-determined |
| 18-109_k141_167993 | Not-determined |
| 18-109_k141_167994 | Not-determined |
| 18-109_k141_45185 | Not-determined |
| 18-109_k141_85811 | Low-quality |
| 18-109_k141_106316 | Not-determined |
| 18-109_k141_229249 | Not-determined |
| 18-109_k141_157664 | Not-determined |
| 18-109_k141_106318 | Low-quality |
| 18-109_k141_218062 | Not-determined |
| 18-109_k141_65844 | Low-quality |
| 18-109_k141_95795 | Not-determined |
| 18-109_k141_168001 | Not-determined |
| 18-109_k141_116833 | Not-determined |
| 18-109_k141_45209 | Not-determined |
| 18-109_k141_4209 | Not-determined |
| 18-109_k141_106330 | Not-determined |
| 18-109_k141_14373 | Not-determined |
| 18-109_k141_75155 | Not-determined |
| 18-109_k141_127210 | Not-determined |
| 18-109_k141_24001 | Not-determined |
| 18-109_k141_229261 | Not-determined |
| 18-109_k141_116842 | Not-determined |
| 18-109_k141_188668 | Not-determined |
| 18-109_k141_45223 | Not-determined |
| 18-109_k141_188669 | Not-determined |
| 18-109_k141_146984 | Not-determined |
| 18-109_k141_218081 | Low-quality |
| 18-109_k141_136886 | Not-determined |
| 18-109_k141_239744 | Low-quality |
| 18-109_k141_116851 | Not-determined |
| 18-109_k141_55275 | Not-determined |
| 18-109_k141_14385 | Not-determined |
| 18-109_k141_178086 | Low-quality |
| 18-109_k141_55280 | Not-determined |
| 18-109_k141_209424 | Not-determined |
| 18-109_k141_209425 | Not-determined |
| 18-109_k141_168042 | Not-determined |
| 18-109_k141_136903 | Not-determined |
| 18-109_k141_168043 | Not-determined |
| 18-109_k141_157717 | Low-quality |
| 18-109_k141_199099 | Not-determined |
| 18-109_k141_146999 | Not-determined |
| 18-109_k141_35285 | Not-determined |
| 18-109_k141_199106 | Low-quality |
| 18-109_k141_65871 | Not-determined |
| 18-109_k141_95836 | Not-determined |
| 18-109_k141_14400 | Not-determined |
| 18-109_k141_65877 | Not-determined |
| 18-109_k141_188710 | Not-determined |
| 18-109_k141_218092 | Low-quality |
| 18-109_k141_14411 | Not-determined |
| 18-109_k141_45253 | Not-determined |
| 18-109_k141_35290 | Not-determined |
| 18-109_k141_95862 | Low-quality |
| 18-109_k141_95870 | Not-determined |
| 18-109_k141_229321 | Not-determined |
| 18-109_k141_168081 | Low-quality |
| 18-109_k141_95877 | Not-determined |
| 18-109_k141_95881 | Not-determined |
| 18-109_k141_168088 | Not-determined |
| 18-109_k141_35301 | Not-determined |
| 18-109_k141_116888 | Not-determined |
| 18-109_k141_116891 | Not-determined |
| 18-109_k141_157770 | Low-quality |
| 18-109_k141_239773 | Low-quality |
| 18-109_k141_14444 | Not-determined |
| 18-109_k141_218105 | Not-determined |
| 18-109_k141_239776 | Not-determined |
| 18-109_k141_239778 | Not-determined |
| 18-109_k141_35311 | Not-determined |
| 18-109_k141_239782 | Not-determined |
| 18-109_k141_75208 | Not-determined |
| 18-109_k141_95894 | Not-determined |
| 18-109_k141_116905 | Not-determined |
| 18-109_k141_178127 | Not-determined |
| 18-109_k141_209460 | Not-determined |
| 18-109_k141_95898 | Low-quality |
| 18-109_k141_178133 | Not-determined |
| 18-109_k141_85846 | Not-determined |
| 18-109_k141_116911 | Not-determined |
| 18-109_k141_75211 | Not-determined |
| 18-109_k141_95904 | Low-quality |
| 18-109_k141_157784 | Not-determined |
| 18-109_k141_178141 | Not-determined |
| 18-109_k141_157785 | Not-determined |
| 18-109_k141_199133 | Not-determined |
| 18-109_k141_218130 | Not-determined |
| 18-109_k141_35329 | Not-determined |
| 18-109_k141_229365 | Not-determined |
| 18-109_k141_24033 | Not-determined |
| 18-109_k141_75229 | Not-determined |
| 18-109_k141_229370 | Not-determined |
| 18-109_k141_116919 | Not-determined |
| 18-109_k141_116920 | Not-determined |
| 18-109_k141_178148 | Not-determined |
| 18-109_k141_157800 | Not-determined |
| 18-109_k141_75236 | Low-quality |
| 18-109_k141_188730 | Not-determined |
| 18-109_k141_199137 | Not-determined |
| 18-109_k141_14460 | Not-determined |
| 18-109_k141_188738 | Not-determined |
| 18-109_k141_178161 | Not-determined |
| 18-109_k141_229389 | Not-determined |
| 18-109_k141_178163 | Not-determined |
| 18-109_k141_199145 | Not-determined |
| 18-109_k141_209492 | Not-determined |
| 18-109_k141_24040 | Not-determined |
| 18-109_k141_24044 | Not-determined |
| 18-109_k141_218138 | Not-determined |
| 18-109_k141_218139 | Low-quality |
| 18-109_k141_239789 | Not-determined |
| 18-109_k141_95921 | Not-determined |
| 18-109_k141_85871 | Not-determined |
| 18-109_k141_75254 | Not-determined |
| 18-109_k141_24052 | Not-determined |
| 18-109_k141_95927 | Not-determined |
| 18-109_k141_116935 | Not-determined |
| 18-109_k141_218143 | Not-determined |
| 18-109_k141_199152 | Not-determined |
| 18-109_k141_4267 | Not-determined |
| 18-109_k141_178178 | Not-determined |
| 18-109_k141_116939 | Not-determined |
| 18-109_k141_24068 | Not-determined |
| 18-109_k141_75270 | Not-determined |
| 18-109_k141_178188 | Not-determined |
| 18-109_k141_85891 | Low-quality |
| 18-109_k141_35389 | Not-determined |
| 18-109_k141_65915 | Not-determined |
| 18-109_k141_209514 | Low-quality |
| 18-109_k141_85903 | Not-determined |
| 18-109_k141_209517 | Not-determined |
| 18-109_k141_65919 | Not-determined |
| 18-109_k141_35415 | Not-determined |
| 18-109_k141_178200 | Low-quality |
| 18-109_k141_209528 | Not-determined |
| 18-109_k141_157831 | Low-quality |
| 18-109_k141_239823 | Not-determined |
| 18-109_k141_157843 | Not-determined |
| 18-109_k141_209539 | Not-determined |
| 18-109_k141_75311 | Not-determined |
| 18-109_k141_85931 | Not-determined |
| 18-109_k141_168115 | Not-determined |
| 18-109_k141_209543 | Not-determined |
| 18-109_k141_178210 | Not-determined |
| 18-109_k141_209553 | Not-determined |
| 18-109_k141_127240 | Not-determined |
| 18-109_k141_178218 | Not-determined |
| 18-109_k141_14509 | Not-determined |
| 18-109_k141_14510 | Not-determined |
| 17-108-final-viral-combined_k141_83571\|\|full | Medium-quality |
| 18-109_k141_35462 | Not-determined |
| 18-109_k141_178225 | Not-determined |
| 18-109_k141_178226 | Not-determined |
| 18-109_k141_65953 | Low-quality |
| 18-109_k141_157861 | Not-determined |
| 18-109_k141_168136 | Not-determined |
| 18-109_k141_75317 | Not-determined |
| 18-106-final-viral-combined_k141_41246\|\|full | High-quality |
| 18-109_k141_127259 | Not-determined |
| 18-109_k141_157873 | Not-determined |
| 18-109_k141_127265 | Not-determined |
| 18-109_k141_65962 | Not-determined |
| 18-109_k141_137008 | Not-determined |
| 18-109_k141_24097 | Not-determined |
| 18-109_k141_209577 | Not-determined |
| 18-109_k141_65969 | Not-determined |
| 18-109_k141_95940 | Low-quality |
| 18-109_k141_14526 | Not-determined |
| 18-109_k141_178247 | Not-determined |
| 18-109_k141_147046 | Not-determined |
| 18-109_k141_95944 | Not-determined |
| 18-109_k141_178248 | Not-determined |
| 18-109_k141_209584 | Not-determined |
| 18-109_k141_116966 | Not-determined |
| 18-109_k141_95951 | Not-determined |
| 18-109_k141_95953 | Not-determined |
| 18-109_k141_35477 | Not-determined |
| 18-109_k141_209593 | Not-determined |
| 18-109_k141_4353 | Not-determined |
| 18-109_k141_35484 | Not-determined |
| 18-109_k141_209599 | Not-determined |
| 18-109_k141_95960 | Not-determined |
| 18-109_k141_127280 | Not-determined |
| 18-109_k141_239853 | Not-determined |
| 18-109_k141_4367 | Not-determined |
| 18-109_k141_239856 | Not-determined |
| 18-109_k141_157885 | Not-determined |
| 18-109_k141_147072 | Not-determined |
| 18-109_k141_95977 | Not-determined |
| 18-109_k141_55341 | Not-determined |
| 18-109_k141_85967 | Not-determined |
| 18-109_k141_106334 | Not-determined |
| 18-109_k141_4376 | Not-determined |
| 18-109_k141_95991 | Low-quality |
| 18-109_k141_85975 | Not-determined |
| 18-109_k141_55353 | Not-determined |
| 18-109_k141_95996 | Not-determined |
| 18-109_k141_55357 | Not-determined |
| 18-109_k141_85979 | Low-quality |
| 18-109_k141_218178 | Not-determined |
| 18-109_k141_55362 | Not-determined |
| 18-109_k141_106352 | Not-determined |
| 18-109_k141_157913 | Not-determined |
| 18-109_k141_116973 | Low-quality |
| 18-109_k141_178287 | Not-determined |
| 18-109_k141_96007 | Not-determined |
| 18-109_k141_147091 | Not-determined |
| 18-109_k141_96009 | Not-determined |
| 18-109_k141_85988 | Not-determined |
| 18-109_k141_178289 | Low-quality |
| 18-109_k141_188754 | Not-determined |
| 18-109_k141_45333 | Not-determined |
| 18-109_k141_96016 | Not-determined |
| 18-109_k141_65987 | Not-determined |
| 18-109_k141_106369 | Not-determined |
| 18-109_k141_147101 | Not-determined |
| 18-109_k141_35543 | Not-determined |
| 18-109_k141_127283 | Not-determined |
| 18-109_k141_157933 | Not-determined |
| 18-109_k141_116982 | Low-quality |
| 18-109_k141_157934 | Not-determined |
| 18-109_k141_127290 | Not-determined |
| 18-109_k141_45347 | Not-determined |
| 18-109_k141_209624 | Not-determined |
| 18-109_k141_168185 | Not-determined |
| 18-109_k141_178306 | Not-determined |
| 18-109_k141_55371 | Not-determined |
| 18-109_k141_127301 | Not-determined |
| 18-109_k141_106389 | Not-determined |
| 18-109_k141_4394 | Not-determined |
| 18-109_k141_106395 | Not-determined |
| 18-109_k141_147118 | Not-determined |
| 18-109_k141_14531 | Not-determined |
| 18-109_k141_239889 | Low-quality |
| 18-109_k141_117000 | Not-determined |
| 18-109_k141_137071 | Not-determined |
| 18-109_k141_127326 | Not-determined |
| 18-109_k141_147135 | Not-determined |
| 18-109_k141_66019 | Not-determined |
| 18-109_k141_66020 | Not-determined |
| 18-109_k141_117011 | Not-determined |
| 18-109_k141_55393 | Not-determined |
| 18-109_k141_55399 | Not-determined |
| 18-109_k141_75336 | Low-quality |
| 18-109_k141_239908 | Not-determined |
| 18-109_k141_168200 | Not-determined |
| 18-109_k141_157981 | Low-quality |
| 18-109_k141_229450 | Not-determined |
| 18-109_k141_239928 | Not-determined |
| 18-109_k141_137110 | Not-determined |
| 18-109_k141_14547 | Not-determined |
| 18-109_k141_168214 | Not-determined |
| 18-109_k141_4438 | Not-determined |
| 18-109_k141_75368 | Not-determined |
| 18-109_k141_178338 | Not-determined |
| 18-109_k141_66025 | Not-determined |
| 18-109_k141_209654 | Not-determined |
| 18-109_k141_66029 | Not-determined |
| 18-109_k141_45379 | Not-determined |
| 18-109_k141_75376 | Not-determined |
| 18-109_k141_178350 | Not-determined |
| 18-109_k141_55432 | Not-determined |
| 18-109_k141_45393 | Not-determined |
| 18-109_k141_66037 | Not-determined |
| 18-109_k141_55436 | Not-determined |
| 18-109_k141_4454 | Not-determined |
| 18-109_k141_86008 | Not-determined |
| 18-109_k141_239953 | Not-determined |
| 18-109_k141_4460 | Not-determined |
| 18-109_k141_178365 | Not-determined |
| 18-109_k141_45401 | Not-determined |
| 18-109_k141_4465 | Low-quality |
| 18-109_k141_229459 | Not-determined |
| 18-109_k141_45408 | Not-determined |
| 18-109_k141_45409 | Not-determined |
| 18-109_k141_4468 | Not-determined |
| 18-109_k141_86013 | Not-determined |
| 18-109_k141_75389 | Not-determined |
| 18-109_k141_188805 | Low-quality |
| 18-109_k141_209673 | Not-determined |
| 18-109_k141_4473 | Not-determined |
| 18-109_k141_188810 | Not-determined |
| 18-109_k141_158022 | Not-determined |
| 18-109_k141_178378 | Not-determined |
| 18-109_k141_55442 | Not-determined |
| 18-109_k141_117063 | Not-determined |
| 18-109_k141_55444 | Low-quality |
| 18-109_k141_158026 | Not-determined |
| 18-109_k141_86026 | Not-determined |
| 18-109_k141_209687 | Not-determined |
| 18-109_k141_35571 | Not-determined |
| 18-109_k141_209690 | Not-determined |
| 18-109_k141_188818 | Not-determined |
| 18-109_k141_229484 | Not-determined |
| 18-109_k141_86028 | Not-determined |
| 18-109_k141_229485 | Low-quality |
| 18-109_k141_96053 | Not-determined |
| 18-109_k141_218206 | Not-determined |
| 18-109_k141_209708 | Not-determined |
| 18-109_k141_218210 | Not-determined |
| 18-109_k141_158050 | Not-determined |
| 18-109_k141_178403 | Not-determined |
| 18-109_k141_35586 | Not-determined |
| 18-109_k141_66067 | Not-determined |
| 18-109_k141_229488 | Not-determined |
| 18-109_k141_4494 | Not-determined |
| 18-109_k141_127372 | Not-determined |
| 18-109_k141_66077 | Not-determined |
| 18-109_k141_35594 | Not-determined |
| 18-109_k141_4498 | Not-determined |
| 18-109_k141_127381 | Not-determined |
| 18-109_k141_117091 | Not-determined |
| 18-109_k141_137152 | Not-determined |
| 18-109_k141_218228 | Not-determined |
| 18-109_k141_86038 | Not-determined |
| 18-109_k141_178424 | Not-determined |
| 18-109_k141_96078 | Not-determined |
| 18-109_k141_218231 | Not-determined |
| 18-109_k141_55487 | Not-determined |
| 18-109_k141_127384 | Not-determined |
| 18-109_k141_35609 | Not-determined |
| 18-109_k141_35610 | Not-determined |
| 18-109_k141_218237 | Not-determined |
| 18-109_k141_4517 | Not-determined |
| 18-109_k141_117095 | Low-quality |
| 18-109_k141_55492 | Not-determined |
| 18-109_k141_66092 | Not-determined |
| 18-109_k141_55501 | Not-determined |
| 18-109_k141_137182 | Not-determined |
| 18-109_k141_66095 | Not-determined |
| 18-109_k141_239989 | Not-determined |
| 18-109_k141_35635 | Not-determined |
| 18-109_k141_35638 | Not-determined |
| 18-109_k141_86046 | Not-determined |
| 18-109_k141_75432 | Not-determined |
| 18-109_k141_158077 | Not-determined |
| 18-109_k141_240001 | Not-determined |
| 18-109_k141_199272 | Low-quality |
| 18-109_k141_35646 | Not-determined |
| 18-109_k141_158084 | Not-determined |
| 18-109_k141_218244 | Not-determined |
| 18-109_k141_86054 | Not-determined |
| 18-109_k141_158087 | Not-determined |
| 18-109_k141_86056 | Not-determined |
| 18-109_k141_158089 | Not-determined |
| 18-109_k141_199286 | Not-determined |
| 18-109_k141_86066 | Not-determined |
| 18-109_k141_188892 | Not-determined |
| 18-109_k141_86071 | Not-determined |
| 18-109_k141_66115 | Not-determined |
| 18-109_k141_127415 | Not-determined |
| 18-109_k141_55528 | Not-determined |
| 18-109_k141_66119 | Not-determined |
| 18-109_k141_45463 | Not-determined |
| 18-109_k141_55530 | Not-determined |
| 18-109_k141_127420 | Not-determined |
| 18-109_k141_14609 | Not-determined |
| 18-109_k141_199302 | Not-determined |
| 18-109_k141_96120 | Not-determined |
| 18-109_k141_4552 | Not-determined |
| 18-109_k141_45469 | Not-determined |
| 18-109_k141_86089 | Not-determined |
| 18-109_k141_66125 | Not-determined |
| 18-109_k141_178476 | Not-determined |
| 18-109_k141_4553 | Not-determined |
| 18-109_k141_168275 | Not-determined |
| 18-109_k141_137201 | Not-determined |
| 18-109_k141_209720 | Low-quality |
| 18-109_k141_4558 | Not-determined |
| 18-109_k141_178489 | Not-determined |
| 18-109_k141_168288 | Not-determined |
| 18-109_k141_168291 | Not-determined |
| 18-109_k141_158108 | Not-determined |
| 18-109_k141_96154 | Not-determined |
| 18-109_k141_35699 | Not-determined |
| 18-109_k141_14626 | Not-determined |
| 18-109_k141_127439 | Not-determined |
| 18-109_k141_178496 | Not-determined |
| 18-109_k141_96162 | Not-determined |
| 18-109_k141_45499 | Not-determined |
| 18-109_k141_45500 | Not-determined |
| 18-109_k141_4568 | Not-determined |
| 18-109_k141_96163 | Not-determined |
| 18-109_k141_168302 | Not-determined |
| 18-109_k141_66161 | Not-determined |
| 18-109_k141_147163 | Not-determined |
| 18-109_k141_158119 | Not-determined |
| 18-109_k141_14640 | Not-determined |
| 18-109_k141_4577 | Not-determined |
| 18-109_k141_218285 | Not-determined |
| 18-109_k141_168317 | Not-determined |
| 18-109_k141_199349 | Not-determined |
| 18-109_k141_24120 | Not-determined |
| 18-109_k141_24121 | Not-determined |
| 18-109_k141_14652 | Not-determined |
| 18-109_k141_106432 | Not-determined |
| 18-109_k141_168325 | Not-determined |
| 18-109_k141_168326 | Not-determined |
| 18-109_k141_188907 | Not-determined |
| 18-109_k141_240035 | Not-determined |
| 18-109_k141_240036 | Not-determined |
| 18-109_k141_127458 | Not-determined |
| 18-109_k141_168332 | Not-determined |
| 18-109_k141_55593 | Not-determined |
| 18-109_k141_137247 | Not-determined |
| 18-109_k141_188919 | Not-determined |
| 18-109_k141_106437 | Not-determined |
| 18-109_k141_240039 | Not-determined |
| 18-109_k141_127467 | Not-determined |
| 18-109_k141_106438 | Not-determined |
| 18-109_k141_4604 | Not-determined |
| 18-109_k141_14663 | Not-determined |
| 18-109_k141_4607 | Not-determined |
| 18-109_k141_106445 | Low-quality |
| 18-109_k141_240046 | Not-determined |
| 18-109_k141_127481 | Not-determined |
| 18-109_k141_127483 | Not-determined |
| 18-109_k141_218293 | Not-determined |
| 18-109_k141_168347 | Not-determined |
| 18-109_k141_35718 | Not-determined |
| 18-109_k141_86116 | Not-determined |
| 18-109_k141_218294 | Not-determined |
| 18-109_k141_229520 | Low-quality |
| 18-109_k141_117174 | Not-determined |
| 18-109_k141_218298 | Not-determined |
| 18-109_k141_218302 | Not-determined |
| 18-109_k141_158179 | Not-determined |
| 18-109_k141_240072 | Not-determined |
| 18-109_k141_35741 | Not-determined |
| 18-109_k141_66229 | Low-quality |
| 18-109_k141_168369 | Not-determined |
| 18-109_k141_168371 | Not-determined |
| 18-109_k141_218306 | Not-determined |
| 18-109_k141_86122 | Not-determined |
| 18-109_k141_106480 | Not-determined |
| 18-109_k141_66237 | Not-determined |
| 18-109_k141_127504 | Not-determined |
| 18-109_k141_218311 | Not-determined |
| 18-109_k141_66239 | Not-determined |
| 18-109_k141_55618 | Not-determined |
| 18-109_k141_96182 | Not-determined |
| 18-109_k141_158193 | Not-determined |
| 18-109_k141_55620 | Not-determined |
| 18-109_k141_66247 | Not-determined |
| 18-109_k141_127510 | Not-determined |
| 18-109_k141_158196 | Not-determined |
| 18-109_k141_55623 | Not-determined |
| 18-109_k141_66254 | Not-determined |
| 18-109_k141_127518 | Low-quality |
| 18-109_k141_127520 | Not-determined |
| 18-109_k141_106499 | Not-determined |
| 18-109_k141_147183 | Not-determined |
| 18-109_k141_147185 | Not-determined |
| 18-109_k141_106508 | Not-determined |
| 18-109_k141_127531 | Low-quality |
| 18-109_k141_240094 | Not-determined |
| 18-109_k141_106512 | Not-determined |
| 18-109_k141_199363 | Not-determined |
| 18-109_k141_66267 | Not-determined |
| 18-109_k141_96210 | Not-determined |
| 18-109_k141_168400 | Low-quality |
| 18-109_k141_86138 | Not-determined |
| 18-109_k141_96214 | Not-determined |
| 18-109_k141_147196 | Not-determined |
| 18-109_k141_218323 | Not-determined |
| 18-109_k141_178561 | Not-determined |
| 18-109_k141_178562 | Not-determined |
| 18-109_k141_168410 | Not-determined |
| 18-109_k141_229565 | Not-determined |
| 18-109_k141_147198 | Low-quality |
| 18-109_k141_106522 | Not-determined |
| 18-109_k141_24163 | Not-determined |
| 18-109_k141_229567 | Low-quality |
| 18-109_k141_147202 | Not-determined |
| 18-109_k141_86154 | Not-determined |
| 18-109_k141_4660 | Not-determined |
| 18-109_k141_45582 | Not-determined |
| 18-109_k141_178582 | Not-determined |
| 18-109_k141_147208 | Not-determined |
| 18-109_k141_86160 | Low-quality |
| 18-109_k141_55645 | Not-determined |
| 18-109_k141_147214 | Not-determined |
| 18-109_k141_229580 | Not-determined |
| 18-109_k141_199370 | Not-determined |
| 18-109_k141_229583 | Not-determined |
| 18-109_k141_24176 | Not-determined |
| 18-109_k141_199373 | Low-quality |
| 18-109_k141_229586 | Not-determined |
| 18-109_k141_147221 | Not-determined |
| 18-109_k141_117231 | Low-quality |
| 18-109_k141_229589 | Not-determined |
| 18-109_k141_117234 | Not-determined |
| 18-109_k141_199378 | Not-determined |
| 18-109_k141_168423 | Not-determined |
| 18-109_k141_4664 | Not-determined |
| 18-109_k141_229604 | Not-determined |
| 18-109_k141_147233 | Not-determined |
| 18-109_k141_147234 | Not-determined |
| 18-109_k141_229607 | Not-determined |
| 18-109_k141_127547 | Not-determined |
| 18-109_k141_240106 | Not-determined |
| 18-109_k141_199387 | Not-determined |
| 18-109_k141_24192 | Not-determined |
| 18-109_k141_35808 | Low-quality |
| 18-109_k141_96246 | Not-determined |
| 18-109_k141_75487 | Not-determined |
| 18-109_k141_178623 | Not-determined |
| 18-109_k141_75490 | Not-determined |
| 18-109_k141_96249 | Not-determined |
| 18-109_k141_96255 | Not-determined |
| 18-109_k141_127563 | Not-determined |
| 18-109_k141_24210 | Not-determined |
| 18-109_k141_178631 | Not-determined |
| 18-109_k141_96260 | Low-quality |
| 18-109_k141_168438 | Not-determined |
| 18-109_k141_75493 | Not-determined |

| | |
|---|---|
| 18-109_k141_168443 | Not-determined |
| 18-109_k141_199397 | Low-quality |
| 18-109_k141_168444 | Not-determined |
| 18-109_k141_168445 | Not-determined |
| 18-109_k141_75502 | Not-determined |
| 18-109_k141_86181 | Low-quality |
| 18-109_k141_35832 | Not-determined |
| 18-109_k141_96273 | Not-determined |
| 18-109_k141_127580 | Not-determined |
| 18-109_k141_178634 | Not-determined |
| 18-109_k141_96278 | Not-determined |
| 18-109_k141_35836 | Not-determined |
| 18-109_k141_14697 | Low-quality |
| 18-109_k141_35838 | Not-determined |
| 18-109_k141_66306 | Not-determined |
| 18-109_k141_35844 | Not-determined |
| 18-109_k141_158252 | Not-determined |
| 18-109_k141_24226 | Not-determined |
| 18-109_k141_147258 | Not-determined |
| 18-109_k141_147262 | Low-quality |
| 18-109_k141_127582 | Not-determined |
| 18-109_k141_199412 | Not-determined |
| 18-109_k141_168466 | Not-determined |
| 18-109_k141_168472 | Not-determined |
| 18-109_k141_96290 | Not-determined |
| 18-109_k141_35852 | Not-determined |
| 18-109_k141_218363 | Not-determined |
| 18-109_k141_35854 | Low-quality |
| 18-109_k141_188944 | Low-quality |
| 18-109_k141_96296 | Not-determined |
| 18-109_k141_188945 | Not-determined |
| 18-109_k141_168481 | Not-determined |
| 18-109_k141_66328 | Not-determined |
| 18-109_k141_45598 | Not-determined |
| 18-109_k141_86228 | Low-quality |
| 18-109_k141_55720 | Not-determined |
| 18-109_k141_188961 | Not-determined |
| 18-109_k141_137295 | Not-determined |
| 18-109_k141_188963 | Not-determined |
| 18-109_k141_14714 | Not-determined |
| 18-109_k141_117295 | Not-determined |
| 18-109_k141_137297 | Not-determined |
| 18-109_k141_86232 | Not-determined |
| 18-109_k141_106534 | Not-determined |
| 18-109_k141_75517 | Not-determined |
| 18-109_k141_127601 | Not-determined |
| 18-109_k141_158283 | Not-determined |
| 18-109_k141_168494 | Not-determined |
| 18-109_k141_158288 | Not-determined |
| 18-109_k141_96317 | Not-determined |
| 18-109_k141_137307 | Not-determined |
| 18-109_k141_45604 | Not-determined |
| 18-109_k141_45605 | Not-determined |
| 18-109_k141_106539 | Not-determined |
| 18-109_k141_158295 | Not-determined |
| 18-109_k141_158299 | Not-determined |
| 18-109_k141_66341 | Not-determined |
| 18-109_k141_188981 | Low-quality |
| 18-109_k141_35880 | Not-determined |
| 18-109_k141_127613 | Not-determined |
| 18-109_k141_66352 | Not-determined |
| 18-109_k141_127616 | Low-quality |
| 18-109_k141_14735 | Low-quality |
| 18-109_k141_178681 | Not-determined |
| 18-109_k141_66357 | Not-determined |
| 18-109_k141_158303 | Low-quality |
| 18-109_k141_189000 | Not-determined |
| 18-109_k141_96356 | Low-quality |
| 18-109_k141_66368 | Not-determined |
| 18-109_k141_55765 | Not-determined |
| 18-109_k141_66374 | Not-determined |
| 18-109_k141_127625 | Not-determined |
| 18-109_k141_75561 | Not-determined |
| 18-109_k141_127628 | Not-determined |
| 18-109_k141_14751 | Not-determined |
| 18-109_k141_127632 | Not-determined |
| 18-109_k141_189008 | Not-determined |
| 18-109_k141_75578 | Not-determined |
| 18-109_k141_96375 | Low-quality |
| 18-109_k141_45615 | Not-determined |
| 18-109_k141_45616 | Not-determined |
| 18-109_k141_14756 | Not-determined |
| 18-109_k141_229654 | Not-determined |
| 18-109_k141_35898 | Not-determined |
| 18-109_k141_75586 | Not-determined |
| 18-109_k141_35902 | Not-determined |
| 18-109_k141_168517 | Not-determined |
| 18-109_k141_35904 | Not-determined |
| 18-109_k141_229664 | Low-quality |
| 18-109_k141_14767 | Not-determined |
| 18-109_k141_45628 | Not-determined |
| 18-109_k141_229667 | Not-determined |
| 18-109_k141_147328 | Not-determined |
| 18-109_k141_199429 | Not-determined |
| 18-109_k141_199434 | Not-determined |
| 18-109_k141_117357 | Not-determined |
| 18-109_k141_199436 | Not-determined |
| 18-109_k141_106578 | Not-determined |
| 18-109_k141_147345 | Not-determined |
| 18-109_k141_55800 | Not-determined |
| 18-109_k141_147348 | Not-determined |
| 18-109_k141_117373 | Not-determined |
| 18-109_k141_75604 | Not-determined |
| 18-109_k141_147351 | Not-determined |
| 18-109_k141_75606 | Not-determined |
| 18-109_k141_75607 | Not-determined |
| 18-109_k141_45643 | Not-determined |
| 18-109_k141_117377 | Not-determined |
| 18-109_k141_45655 | Not-determined |
| 18-109_k141_45656 | Not-determined |
| 18-109_k141_106593 | Not-determined |
| 18-109_k141_178759 | Not-determined |
| 18-109_k141_199466 | Not-determined |
| 18-109_k141_45660 | Not-determined |
| 18-109_k141_127639 | Not-determined |
| 18-109_k141_168551 | Not-determined |
| 18-109_k141_86300 | Not-determined |
| 18-109_k141_117394 | Not-determined |
| 18-109_k141_147364 | Not-determined |
| 18-109_k141_189070 | Not-determined |
| 18-109_k141_117399 | Not-determined |
| 18-109_k141_229688 | Not-determined |
| 18-109_k141_86308 | Not-determined |
| 18-109_k141_158313 | Not-determined |
| 18-109_k141_229692 | Not-determined |
| 18-109_k141_178772 | Not-determined |
| 18-109_k141_86315 | Not-determined |
| 18-109_k141_86317 | Not-determined |
| 18-109_k141_240154 | Not-determined |
| 18-109_k141_96393 | Low-quality |
| 18-109_k141_66450 | Not-determined |
| 18-109_k141_178779 | Not-determined |
| 18-109_k141_75648 | Not-determined |
| 18-109_k141_178780 | Not-determined |
| 18-109_k141_86321 | Not-determined |
| 18-109_k141_55839 | Not-determined |
| 18-109_k141_117417 | Not-determined |
| 18-109_k141_86324 | Not-determined |
| 18-109_k141_229711 | Not-determined |
| 18-109_k141_229715 | Not-determined |
| 18-109_k141_4682 | Not-determined |
| 18-109_k141_75665 | Not-determined |
| 18-109_k141_240161 | Not-determined |
| 18-109_k141_147372 | Low-quality |
| 18-109_k141_240163 | Not-determined |
| 18-109_k141_199498 | Not-determined |
| 18-109_k141_137390 | Not-determined |
| 18-109_k141_168568 | Not-determined |
| 18-109_k141_240174 | Not-determined |
| 18-109_k141_127660 | Not-determined |
| 18-109_k141_86345 | Not-determined |
| 18-109_k141_137402 | Not-determined |
| 18-109_k141_199508 | Not-determined |
| 18-109_k141_168580 | Low-quality |
| 18-109_k141_45684 | Not-determined |
| 18-109_k141_158335 | Low-quality |
| 18-109_k141_45692 | Not-determined |
| 18-109_k141_127668 | Not-determined |
| 18-109_k141_86356 | Not-determined |
| 18-109_k141_96423 | Not-determined |
| 18-109_k141_117429 | Not-determined |
| 18-109_k141_199524 | Not-determined |
| 18-109_k141_158352 | Not-determined |
| 18-109_k141_96435 | Not-determined |
| 18-109_k141_86365 | Not-determined |
| 18-109_k141_106627 | Not-determined |
| 18-109_k141_45707 | Not-determined |
| 18-109_k141_117440 | Not-determined |
| 18-109_k141_86369 | Not-determined |
| 18-109_k141_14786 | Not-determined |
| 18-109_k141_66465 | Low-quality |
| 18-109_k141_199545 | Not-determined |
| 18-109_k141_240197 | Low-quality |
| 18-109_k141_168627 | Low-quality |
| 18-109_k141_45717 | Not-determined |
| 18-109_k141_14791 | Not-determined |
| 18-109_k141_86377 | Not-determined |
| 18-109_k141_55868 | Not-determined |
| 18-109_k141_55869 | Not-determined |
| 18-109_k141_96448 | Not-determined |
| 18-109_k141_240219 | Not-determined |
| 18-109_k141_240220 | Not-determined |
| 18-109_k141_240222 | Not-determined |
| 18-109_k141_229764 | Not-determined |
| 18-109_k141_55887 | Not-determined |
| 18-109_k141_168636 | Not-determined |
| 18-109_k141_168638 | Not-determined |
| 18-109_k141_86386 | Not-determined |
| 18-109_k141_199562 | Not-determined |
| 18-109_k141_75731 | Not-determined |
| 18-109_k141_199565 | Not-determined |
| 18-109_k141_4743 | Not-determined |
| 18-109_k141_4744 | Not-determined |
| 18-109_k141_168644 | Not-determined |
| 18-109_k141_75742 | Not-determined |
| 18-109_k141_45724 | Not-determined |
| 18-109_k141_240241 | Not-determined |
| 18-109_k141_158372 | Low-quality |
| 18-109_k141_86403 | Low-quality |
| 18-109_k141_14831 | Not-determined |
| 18-109_k141_168646 | Not-determined |
| 18-109_k141_75746 | Not-determined |
| 18-109_k141_127703 | Not-determined |
| 18-109_k141_86407 | Not-determined |
| 18-109_k141_66520 | Not-determined |
| 18-109_k141_147415 | Not-determined |
| 18-109_k141_189111 | Not-determined |
| 18-109_k141_55892 | Not-determined |
| 18-109_k141_147423 | Not-determined |
| 18-109_k141_199568 | Not-determined |
| 18-109_k141_4764 | Not-determined |
| 18-109_k141_106653 | Not-determined |
| 18-109_k141_4767 | Not-determined |
| 18-109_k141_14842 | Low-quality |
| 18-109_k141_199589 | Not-determined |
| 18-109_k141_14847 | Low-quality |
| 18-109_k141_14850 | Not-determined |
| 18-109_k141_55901 | Not-determined |
| 18-109_k141_147435 | Low-quality |
| 18-109_k141_14854 | Not-determined |
| 18-109_k141_66550 | Not-determined |
| 18-109_k141_199601 | Not-determined |
| 18-109_k141_209752 | Not-determined |
| 18-109_k141_66553 | Not-determined |
| 18-109_k141_66554 | Not-determined |
| 18-109_k141_45755 | Not-determined |
| 18-109_k141_45756 | Low-quality |
| 18-109_k141_209755 | Not-determined |
| 18-109_k141_147439 | Not-determined |
| 18-109_k141_106666 | Not-determined |
| 18-109_k141_209763 | Not-determined |
| 18-109_k141_240276 | Not-determined |
| 18-109_k141_178789 | Not-determined |
| 18-109_k141_168673 | Not-determined |
| 18-109_k141_66569 | Not-determined |
| 18-109_k141_14876 | Low-quality |
| 18-109_k141_55916 | Low-quality |
| 18-109_k141_66574 | Not-determined |
| 18-109_k141_117491 | Not-determined |
| 18-109_k141_147472 | Not-determined |
| 18-109_k141_117497 | Not-determined |
| 18-109_k141_137442 | Not-determined |
| 18-109_k141_137444 | Not-determined |
| 18-109_k141_14885 | Not-determined |
| 18-109_k141_14886 | Not-determined |
| 18-109_k141_117513 | Not-determined |
| 18-109_k141_240306 | Not-determined |
| 18-109_k141_209783 | Not-determined |
| 18-109_k141_127712 | Not-determined |
| 18-109_k141_240309 | Not-determined |
| 18-109_k141_147495 | Low-quality |
| 18-109_k141_137471 | Not-determined |
| 18-109_k141_147497 | Not-determined |
| 18-109_k141_55952 | Not-determined |
| 18-109_k141_209786 | Not-determined |
| 18-109_k141_229778 | Not-determined |
| 18-109_k141_199617 | Not-determined |
| 18-109_k141_75791 | Not-determined |
| 18-109_k141_147502 | Not-determined |
| 18-109_k141_137482 | Not-determined |
| 18-109_k141_14909 | Not-determined |
| 18-109_k141_147506 | Not-determined |
| 18-109_k141_106684 | Not-determined |
| 18-109_k141_14913 | Not-determined |
| 18-109_k141_75796 | Not-determined |
| 18-109_k141_147509 | Not-determined |
| 18-109_k141_147512 | Not-determined |
| 18-109_k141_147514 | Not-determined |
| 18-109_k141_209798 | Low-quality |
| 18-109_k141_199634 | Not-determined |
| 18-109_k141_209801 | Not-determined |
| 18-109_k141_14923 | Low-quality |
| 18-109_k141_209802 | Not-determined |
| 18-109_k141_75803 | Not-determined |
| 18-109_k141_229801 | Not-determined |
| 18-109_k141_24235 | Not-determined |
| 18-109_k141_55972 | Not-determined |
| 18-109_k141_14934 | Low-quality |
| 18-109_k141_14935 | Low-quality |
| 18-109_k141_75807 | Not-determined |
| 18-109_k141_199639 | Not-determined |
| 18-109_k141_75810 | Not-determined |
| 18-109_k141_14940 | Not-determined |
| 18-109_k141_96494 | Not-determined |
| 18-109_k141_199646 | Not-determined |
| 18-109_k141_75820 | Not-determined |
| 18-109_k141_66635 | Not-determined |
| 18-109_k141_218381 | Not-determined |
| 18-109_k141_127752 | Not-determined |
| 18-109_k141_117554 | Not-determined |
| 18-109_k141_178794 | Not-determined |
| 18-109_k141_127758 | Not-determined |
| 18-109_k141_158401 | Not-determined |
| 18-109_k141_199654 | Not-determined |
| 18-109_k141_66641 | Not-determined |
| 18-109_k141_14962 | Not-determined |
| 18-109_k141_178806 | Not-determined |
| 18-109_k141_55997 | Not-determined |
| 18-109_k141_24256 | Not-determined |
| 18-109_k141_14972 | Not-determined |
| 18-109_k141_66645 | Low-quality |
| 18-109_k141_117577 | Not-determined |
| 18-109_k141_158416 | Not-determined |
| 18-109_k141_96520 | Not-determined |
| 18-109_k141_35932 | Not-determined |
| 18-109_k141_66651 | Not-determined |
| 18-109_k141_56006 | Not-determined |
| 18-109_k141_178817 | Not-determined |
| 18-109_k141_96529 | Not-determined |
| 18-109_k141_14981 | Not-determined |
| 18-109_k141_66660 | Not-determined |
| 18-109_k141_56016 | Not-determined |
| 18-109_k141_45787 | Low-quality |
| 18-109_k141_96533 | Not-determined |
| 18-109_k141_178822 | Not-determined |
| 18-109_k141_117588 | Not-determined |
| 18-109_k141_24274 | Not-determined |
| 18-109_k141_106702 | Not-determined |
| 18-109_k141_4821 | Not-determined |
| 18-109_k141_209827 | Not-determined |
| 18-109_k141_106707 | Not-determined |
| 18-109_k141_178829 | Low-quality |
| 18-109_k141_117604 | Not-determined |
| 18-109_k141_189137 | Not-determined |
| 18-109_k141_229839 | Not-determined |
| 18-109_k141_189142 | Not-determined |
| 18-109_k141_75849 | Not-determined |
| 18-109_k141_4832 | Low-quality |
| 18-109_k141_147553 | Not-determined |
| 18-109_k141_127771 | Not-determined |
| 18-109_k141_14998 | Not-determined |
| 18-109_k141_168697 | Not-determined |
| 18-109_k141_127779 | Not-determined |
| 18-109_k141_178852 | Not-determined |
| 18-109_k141_96558 | Not-determined |
| 18-109_k141_218409 | Not-determined |
| 18-109_k141_117622 | Not-determined |
| 18-109_k141_218411 | Not-determined |
| 18-109_k141_66671 | Not-determined |
| 18-109_k141_24325 | Not-determined |
| 18-109_k141_75858 | Not-determined |
| 18-109_k141_75859 | Not-determined |
| 18-109_k141_15025 | Not-determined |
| 18-109_k141_240335 | Not-determined |
| 18-109_k141_66688 | Not-determined |
| 18-109_k141_218434 | Not-determined |
| 18-109_k141_24344 | Not-determined |
| 18-109_k141_56047 | Not-determined |
| 18-109_k141_218449 | Not-determined |
| 18-109_k141_96571 | Not-determined |
| 18-109_k141_168714 | Low-quality |
| 18-109_k141_147594 | Not-determined |
| 18-109_k141_24364 | Not-determined |
| 18-109_k141_56059 | Not-determined |
| 18-109_k141_24367 | Not-determined |
| 18-109_k141_147599 | Not-determined |
| 18-109_k141_117635 | Not-determined |
| 18-109_k141_189158 | Low-quality |
| 18-109_k141_229878 | Not-determined |
| 18-109_k141_117638 | Not-determined |
| 18-109_k141_147600 | Not-determined |
| 18-109_k141_75866 | Low-quality |
| 18-109_k141_75869 | Low-quality |
| 18-109_k141_209830 | Not-determined |
| 18-109_k141_189173 | Not-determined |
| 18-109_k141_137550 | Not-determined |
| 18-109_k141_4875 | Not-determined |
| 18-109_k141_96582 | Not-determined |
| 18-109_k141_189177 | Not-determined |
| 18-109_k141_96585 | Not-determined |
| 18-109_k141_106771 | Not-determined |
| 18-109_k141_158476 | Not-determined |
| 18-109_k141_229885 | Not-determined |
| 18-109_k141_209841 | Not-determined |
| 18-109_k141_66725 | Not-determined |
| 18-109_k141_4888 | Not-determined |
| 18-109_k141_229895 | Not-determined |
| 18-109_k141_229896 | Not-determined |
| 18-109_k141_158487 | Not-determined |
| 18-109_k141_137565 | Not-determined |
| 18-109_k141_24398 | Not-determined |
| 18-109_k141_96602 | Low-quality |
| 18-109_k141_96606 | Not-determined |
| 18-109_k141_106803 | Not-determined |
| 18-109_k141_4900 | Not-determined |
| 18-109_k141_96611 | Low-quality |
| 18-109_k141_24404 | Not-determined |
| 18-109_k141_106806 | Not-determined |
| 18-109_k141_4905 | Not-determined |
| 18-109_k141_4906 | Not-determined |
| 18-109_k141_15064 | Not-determined |
| 18-109_k141_209864 | Not-determined |
| 18-109_k141_229910 | Not-determined |
| 18-109_k141_4915 | Not-determined |
| 18-109_k141_106821 | Low-quality |
| 18-109_k141_15071 | Not-determined |
| 18-109_k141_106822 | Not-determined |
| 18-109_k141_168773 | Not-determined |
| 18-109_k141_106824 | Low-quality |
| 18-109_k141_4921 | Not-determined |
| 18-109_k141_178868 | Not-determined |
| 18-109_k141_4922 | Not-determined |
| 18-109_k141_168779 | Not-determined |
| 18-109_k141_168785 | Not-determined |
| 18-109_k141_218461 | Not-determined |
| 18-109_k141_189228 | Not-determined |
| 18-109_k141_96622 | Not-determined |
| 18-109_k141_189230 | Not-determined |
| 18-109_k141_218469 | Not-determined |
| 18-109_k141_209869 | Not-determined |
| 18-109_k141_96630 | Not-determined |
| 18-110-final-viral-combined_k141_97848\|\|full | Medium-quality |
| 18-109_k141_189242 | Not-determined |
| 18-109_k141_218474 | Not-determined |
| 18-109_k141_24429 | Not-determined |
| 18-109_k141_168801 | Not-determined |
| 18-109_k141_96635 | Not-determined |
| 18-109_k141_229925 | Not-determined |
| 18-109_k141_15087 | Not-determined |
| 18-109_k141_209876 | Not-determined |
| 18-109_k141_117698 | Not-determined |
| 18-109_k141_209892 | Low-quality |
| 18-109_k141_117705 | Low-quality |
| 18-109_k141_24439 | Not-determined |
| 18-109_k141_117707 | Not-determined |
| 18-109_k141_168816 | Not-determined |
| 18-109_k141_75944 | Not-determined |
| 18-109_k141_96644 | Not-determined |
| 18-109_k141_168822 | Low-quality |
| 18-109_k141_168825 | Not-determined |
| 18-109_k141_158507 | Not-determined |
| 18-109_k141_15111 | Not-determined |
| 18-109_k141_199795 | Not-determined |
| 18-109_k141_96660 | Not-determined |
| 18-109_k141_15117 | Not-determined |
| 18-109_k141_96663 | Not-determined |
| 18-109_k141_218493 | Not-determined |
| 18-109_k141_229948 | Not-determined |
| 18-109_k141_66788 | Low-quality |
| 18-109_k141_218498 | Not-determined |
| 18-109_k141_240375 | Not-determined |
| 18-109_k141_178893 | Low-quality |
| 18-109_k141_240380 | Not-determined |
| 18-109_k141_15120 | Not-determined |
| 18-109_k141_218503 | Not-determined |
| 18-109_k141_4945 | Not-determined |
| 18-109_k141_24469 | Not-determined |
| 18-109_k141_199819 | Not-determined |
| 18-109_k141_96687 | Not-determined |
| 18-109_k141_218512 | Not-determined |
| 18-109_k141_75960 | Not-determined |
| 18-109_k141_218513 | Not-determined |
| 18-109_k141_209916 | Not-determined |
| 18-109_k141_117722 | Not-determined |
| 18-109_k141_96693 | Not-determined |
| 18-109_k141_117724 | Not-determined |
| 18-109_k141_218517 | Not-determined |
| 18-109_k141_218518 | Not-determined |
| 18-109_k141_209924 | Not-determined |
| 18-109_k141_209925 | Low-quality |
| 18-109_k141_96699 | Not-determined |
| 18-109_k141_218522 | Low-quality |
| 18-109_k141_218525 | Not-determined |
| 18-109_k141_199829 | Not-determined |
| 18-109_k141_45814 | Not-determined |
| 18-109_k141_137618 | Not-determined |
| 18-109_k141_4965 | Not-determined |
| 18-109_k141_147653 | Not-determined |
| 18-109_k141_158534 | Not-determined |
| 18-109_k141_137623 | Not-determined |
| 18-109_k141_158536 | Not-determined |
| 18-109_k141_158537 | Not-determined |
| 18-109_k141_240387 | Not-determined |
| 18-109_k141_240394 | Low-quality |
| 18-109_k141_158547 | Not-determined |
| 18-109_k141_96709 | Not-determined |
| 18-109_k141_168866 | Low-quality |
| 18-109_k141_240402 | Not-determined |
| 18-109_k141_117736 | Not-determined |
| 18-109_k141_45822 | Not-determined |
| 18-109_k141_168872 | Not-determined |
| 18-109_k141_4985 | Not-determined |
| 18-109_k141_24493 | Not-determined |
| 18-109_k141_127836 | Not-determined |
| 18-109_k141_147669 | Not-determined |
| 18-109_k141_66812 | Not-determined |
| 18-109_k141_75997 | Not-determined |
| 18-109_k141_117741 | Not-determined |
| 18-109_k141_199845 | Not-determined |
| 18-109_k141_15169 | Not-determined |
| 18-109_k141_209933 | Not-determined |
| 18-109_k141_117747 | Not-determined |
| 18-109_k141_147684 | Not-determined |
| 18-109_k141_168888 | Not-determined |
| 18-109_k141_178935 | Not-determined |
| 18-109_k141_4989 | Not-determined |
| 18-109_k141_117756 | Not-determined |
| 18-109_k141_178939 | Not-determined |
| 18-109_k141_117758 | Not-determined |
| 18-109_k141_117759 | Not-determined |
| 18-109_k141_117761 | Not-determined |
| 18-109_k141_199859 | Not-determined |
| 18-109_k141_56081 | Not-determined |
| 18-109_k141_147702 | Not-determined |
| 18-109_k141_209953 | Not-determined |
| 18-109_k141_137663 | Not-determined |
| 18-109_k141_209955 | Not-determined |
| 18-109_k141_240416 | Low-quality |
| 18-109_k141_127860 | Not-determined |
| 18-109_k141_199874 | Low-quality |
| 18-109_k141_96737 | Not-determined |
| 18-109_k141_209971 | Not-determined |
| 18-109_k141_209975 | Not-determined |
| 18-109_k141_4994 | Not-determined |
| 18-109_k141_147718 | Not-determined |
| 18-109_k141_5000 | Not-determined |
| 18-109_k141_66849 | Low-quality |
| 18-109_k141_127874 | Low-quality |
| 18-109_k141_76008 | Not-determined |
| 18-109_k141_35983 | Not-determined |
| 18-109_k141_137685 | Not-determined |
| 18-109_k141_96765 | Low-quality |
| 18-109_k141_35988 | Not-determined |
| 18-109_k141_76020 | Not-determined |
| 18-109_k141_76021 | Not-determined |
| 18-109_k141_96767 | Not-determined |
| 18-109_k141_35993 | Not-determined |
| 18-109_k141_35994 | Not-determined |
| 18-109_k141_66864 | Not-determined |
| 18-109_k141_199894 | Not-determined |
| 18-109_k141_199895 | Not-determined |
| 18-109_k141_137694 | Not-determined |
| 18-109_k141_56098 | Low-quality |
| 18-109_k141_117790 | Not-determined |
| 18-109_k141_240449 | Not-determined |
| 18-109_k141_218559 | Not-determined |
| 18-109_k141_35999 | Not-determined |
| 18-109_k141_240451 | Not-determined |
| 18-109_k141_36002 | Not-determined |
| 18-109_k141_117794 | Not-determined |
| 18-109_k141_240458 | Not-determined |
| 18-109_k141_15194 | Low-quality |
| 18-109_k141_189268 | Not-determined |
| 18-109_k141_15199 | Not-determined |
| 18-109_k141_189272 | Low-quality |
| 18-109_k141_96777 | Not-determined |
| 18-109_k141_24518 | Low-quality |
| 18-109_k141_96779 | Not-determined |
| 18-109_k141_45840 | Low-quality |
| 18-109_k141_189277 | Not-determined |
| 18-109_k141_24526 | Not-determined |
| 18-109_k141_24528 | Not-determined |
| 18-109_k141_240461 | Not-determined |
| 18-109_k141_15213 | Not-determined |
| 18-109_k141_106838 | Not-determined |
| 18-109_k141_15215 | Not-determined |
| 18-109_k141_24542 | Not-determined |
| 18-109_k141_189290 | Not-determined |
| 18-109_k141_137721 | Not-determined |
| 18-109_k141_45861 | Not-determined |
| 18-109_k141_218573 | Not-determined |
| 18-109_k141_189299 | Not-determined |
| 18-109_k141_45871 | Not-determined |
| 18-109_k141_158588 | Not-determined |
| 18-109_k141_158590 | Not-determined |
| 18-109_k141_24555 | Not-determined |
| 18-109_k141_189304 | Not-determined |
| 18-109_k141_178967 | Low-quality |
| 18-109_k141_45881 | Not-determined |
| 18-109_k141_45884 | Not-determined |
| 18-109_k141_56116 | Not-determined |
| 18-109_k141_45886 | Not-determined |
| 18-109_k141_137725 | Not-determined |
| 18-109_k141_218588 | Not-determined |
| 18-109_k141_168928 | Not-determined |
| 18-109_k141_218591 | Not-determined |
| 18-109_k141_218592 | Low-quality |
| 18-109_k141_96816 | Not-determined |
| 18-109_k141_24568 | Not-determined |
| 18-109_k141_117825 | Low-quality |
| 18-109_k141_96830 | Not-determined |
| 18-109_k141_45900 | Low-quality |
| 18-109_k141_45901 | Not-determined |
| 18-109_k141_218601 | Low-quality |
| 18-109_k141_137740 | Low-quality |
| 18-109_k141_45903 | Not-determined |
| 18-109_k141_168939 | Not-determined |
| 18-109_k141_106862 | Not-determined |
| 18-109_k141_5065 | Low-quality |

| | |
|---|---|
| 18-109_k141_189327 | Low-quality |
| 18-109_k141_189329 | Not-determined |
| 18-109_k141_178985 | Not-determined |
| 18-109_k141_147726 | Not-determined |
| 18-109_k141_168949 | Not-determined |
| 18-109_k141_178990 | Not-determined |
| 18-109_k141_158599 | Not-determined |
| 18-109_k141_189335 | Not-determined |
| 18-109_k141_218605 | Not-determined |
| 18-109_k141_117848 | Not-determined |
| 18-109_k141_147731 | Not-determined |
| 18-109_k141_117852 | Not-determined |
| 18-109_k141_137754 | Not-determined |
| 18-109_k141_147732 | Not-determined |
| 18-109_k141_117856 | Not-determined |
| 18-109_k141_45918 | Not-determined |
| 18-109_k141_209993 | Not-determined |
| 18-109_k141_209995 | Not-determined |
| 18-109_k141_56161 | Not-determined |
| 18-109_k141_15244 | Not-determined |
| 18-109_k141_15246 | Not-determined |
| 18-109_k141_56170 | Not-determined |
| 18-109_k141_168959 | Not-determined |
| 18-109_k141_45931 | Not-determined |
| 18-109_k141_199943 | Not-determined |
| 18-109_k141_127917 | Not-determined |
| 18-109_k141_56173 | Low-quality |
| 18-109_k141_15260 | Not-determined |
| 18-109_k141_127922 | Not-determined |
| 18-109_k141_106873 | Not-determined |
| 18-109_k141_45940 | Not-determined |
| 18-109_k141_179007 | Low-quality |
| 18-109_k141_168972 | Not-determined |
| 18-109_k141_127926 | Low-quality |
| 18-109_k141_137781 | Not-determined |
| 18-109_k141_45949 | Not-determined |
| 18-109_k141_45952 | Low-quality |
| 18-109_k141_45953 | Not-determined |
| 18-109_k141_106883 | Not-determined |
| 18-109_k141_56188 | Not-determined |
| 18-109_k141_76054 | Not-determined |
| 18-109_k141_86458 | Not-determined |
| 18-109_k141_5105 | Not-determined |
| 18-109_k141_86459 | Not-determined |
| 18-109_k141_5108 | Not-determined |
| 18-109_k141_86461 | Not-determined |
| 18-109_k141_45963 | Not-determined |
| 18-109_k141_5113 | Not-determined |
| 18-109_k141_137788 | Not-determined |
| 18-109_k141_168999 | Not-determined |
| 18-109_k141_169000 | Not-determined |
| 18-109_k141_56197 | Not-determined |
| 18-109_k141_86469 | Not-determined |
| 18-109_k141_117897 | Not-determined |
| 18-109_k141_137800 | Not-determined |
| 18-109_k141_106903 | Not-determined |
| 18-109_k141_169011 | Not-determined |
| 18-109_k141_137802 | Not-determined |
| 18-109_k141_106908 | Not-determined |
| 18-109_k141_106913 | Not-determined |
| 18-109_k141_117900 | Not-determined |
| 18-109_k141_117903 | Not-determined |
| 18-109_k141_86485 | Not-determined |
| 18-109_k141_24637 | Low-quality |
| 18-109_k141_45972 | Not-determined |
| 18-109_k141_36038 | Not-determined |
| 18-109_k141_36039 | Not-determined |
| 18-109_k141_96860 | Not-determined |
| 18-109_k141_56212 | Not-determined |
| 18-109_k141_5136 | Not-determined |
| 18-109_k141_45985 | Not-determined |
| 18-109_k141_45986 | Not-determined |
| 18-109_k141_45989 | Not-determined |
| 18-109_k141_56216 | Not-determined |
| 18-109_k141_106930 | Not-determined |
| 18-109_k141_86494 | Not-determined |
| 18-109_k141_66897 | Not-determined |
| 18-109_k141_56221 | Not-determined |
| 18-109_k141_15287 | Not-determined |
| 18-109_k141_189373 | Low-quality |
| 18-109_k141_117909 | Not-determined |
| 18-109_k141_86500 | Not-determined |
| 18-109_k141_5147 | Not-determined |
| 18-109_k141_96880 | Not-determined |
| 18-109_k141_96881 | Not-determined |
| 18-109_k141_137819 | Not-determined |
| 18-109_k141_96884 | Not-determined |
| 18-109_k141_15299 | Not-determined |
| 18-109_k141_36062 | Not-determined |
| 18-109_k141_96888 | Not-determined |
| 18-109_k141_127965 | Low-quality |
| 18-109_k141_189382 | Low-quality |
| 18-109_k141_45996 | Not-determined |
| 18-109_k141_86526 | Not-determined |
| 18-109_k141_106942 | Not-determined |
| 18-109_k141_189393 | Low-quality |
| 18-109_k141_15321 | Not-determined |
| 18-109_k141_117939 | Not-determined |
| 18-109_k141_86541 | Not-determined |
| 18-109_k141_158654 | Not-determined |
| 18-109_k141_86544 | Not-determined |
| 18-109_k141_210060 | Not-determined |
| 18-109_k141_24688 | Not-determined |
| 18-109_k141_36084 | Not-determined |
| 18-109_k141_189406 | Not-determined |
| 18-109_k141_86554 | Not-determined |
| 18-109_k141_127988 | Low-quality |
| 18-109_k141_158664 | Not-determined |
| 18-109_k141_117958 | Not-determined |
| 18-109_k141_189415 | Not-determined |
| 18-109_k141_179123 | Not-determined |
| 18-109_k141_127997 | Not-determined |
| 18-109_k141_230017 | Not-determined |
| 18-109_k141_46008 | Low-quality |
| 18-109_k141_46012 | Not-determined |
| 18-109_k141_179125 | Not-determined |
| 18-109_k141_46014 | Not-determined |
| 18-109_k141_76117 | Low-quality |
| 18-109_k141_158679 | Not-determined |
| 18-109_k141_76122 | Not-determined |
| 18-109_k141_230037 | Not-determined |
| 18-109_k141_117971 | Not-determined |
| 18-109_k141_117973 | Not-determined |
| 18-109_k141_66927 | Not-determined |
| 18-109_k141_210067 | Not-determined |
| 18-109_k141_24711 | Not-determined |
| 18-109_k141_218655 | Not-determined |
| 18-109_k141_128023 | Not-determined |
| 18-109_k141_128024 | Not-determined |
| 18-109_k141_24715 | Low-quality |
| 18-109_k141_46032 | Not-determined |
| 18-109_k141_36128 | Not-determined |
| 18-109_k141_218663 | Not-determined |
| 18-109_k141_147765 | Not-determined |
| 18-109_k141_76137 | Not-determined |
| 18-109_k141_158692 | Not-determined |
| 18-109_k141_86567 | Not-determined |
| 18-109_k141_76143 | Not-determined |
| 18-109_k141_106965 | Not-determined |
| 18-109_k141_169075 | Not-determined |
| 18-109_k141_218667 | Not-determined |
| 18-109_k141_76148 | Low-quality |
| 18-109_k141_76151 | Not-determined |
| 18-109_k141_106972 | Not-determined |
| 18-109_k141_76156 | Not-determined |
| 18-109_k141_210091 | Not-determined |
| 18-109_k141_106980 | Not-determined |
| 18-109_k141_147785 | Not-determined |
| 18-109_k141_230071 | Not-determined |
| 18-109_k141_66955 | Not-determined |
| 18-109_k141_230072 | Not-determined |
| 18-109_k141_66957 | Not-determined |
| 18-109_k141_118004 | Low-quality |
| 18-109_k141_46055 | Not-determined |
| 18-109_k141_66964 | Not-determined |
| 18-109_k141_218689 | Not-determined |
| 18-109_k141_118016 | Not-determined |
| 18-109_k141_46059 | Not-determined |
| 18-109_k141_118020 | Not-determined |
| 18-109_k141_210111 | Not-determined |
| 18-109_k141_147801 | Not-determined |
| 18-109_k141_118025 | Not-determined |
| 18-109_k141_230080 | Not-determined |
| 18-109_k141_118026 | Not-determined |
| 18-109_k141_189474 | Not-determined |
| 18-109_k141_118027 | Low-quality |
| 18-109_k141_210119 | Not-determined |
| 18-109_k141_200046 | Not-determined |
| 18-109_k141_15336 | Not-determined |
| 18-109_k141_118029 | Not-determined |
| 18-109_k141_5188 | Not-determined |
| 18-109_k141_66984 | Not-determined |
| 18-109_k141_200056 | Not-determined |
| 18-109_k141_86626 | Not-determined |
| 18-109_k141_210134 | Not-determined |
| 18-109_k141_218695 | Not-determined |
| 18-109_k141_240475 | Not-determined |
| 18-109_k141_15345 | Not-determined |
| 18-109_k141_240478 | Not-determined |
| 18-109_k141_15347 | Not-determined |
| 18-109_k141_76209 | Not-determined |
| 18-109_k141_107036 | Not-determined |
| 18-109_k141_24740 | Not-determined |
| 18-109_k141_200072 | Not-determined |
| 18-109_k141_189495 | Not-determined |
| 18-109_k141_240494 | Not-determined |
| 18-109_k141_67011 | Not-determined |
| 18-109_k141_210152 | Not-determined |
| 18-109_k141_24742 | Not-determined |
| 18-109_k141_189502 | Not-determined |
| 18-109_k141_76218 | Not-determined |
| 18-109_k141_218711 | Not-determined |
| 18-109_k141_210157 | Not-determined |
| 18-109_k141_67018 | Not-determined |
| 18-109_k141_46106 | Low-quality |
| 18-109_k141_218715 | Not-determined |
| 18-109_k141_67022 | Not-determined |
| 18-109_k141_86643 | Not-determined |
| 18-109_k141_118050 | Not-determined |
| 18-109_k141_179153 | Low-quality |
| 18-109_k141_200087 | Not-determined |
| 18-109_k141_46108 | Low-quality |
| 18-109_k141_189510 | Not-determined |
| 18-109_k141_179160 | Not-determined |
| 18-109_k141_137872 | Not-determined |
| 18-109_k141_200092 | Not-determined |
| 18-109_k141_210163 | Not-determined |
| 18-109_k141_230097 | Not-determined |
| 18-109_k141_107047 | Not-determined |
| 18-109_k141_200096 | Not-determined |
| 18-109_k141_147828 | Not-determined |
| 18-109_k141_46122 | Not-determined |
| 18-109_k141_118058 | Not-determined |
| 18-109_k141_107055 | Not-determined |
| 18-109_k141_118060 | Not-determined |
| 18-109_k141_189534 | Not-determined |
| 18-109_k141_189536 | Not-determined |
| 18-109_k141_210182 | Not-determined |
| 18-109_k141_107060 | Not-determined |
| 18-109_k141_76235 | Not-determined |
| 18-109_k141_240503 | Not-determined |
| 18-109_k141_96920 | Not-determined |
| 18-109_k141_169106 | Not-determined |
| 18-109_k141_118070 | Not-determined |
| 18-109_k141_137907 | Not-determined |
| 18-109_k141_96932 | Not-determined |
| 18-109_k141_96933 | Not-determined |
| 18-109_k141_76244 | Not-determined |
| 18-109_k141_118074 | Not-determined |
| 18-109_k141_128093 | Not-determined |
| 18-109_k141_67052 | Not-determined |
| 18-109_k141_67053 | Not-determined |
| 18-109_k141_210186 | Not-determined |
| 18-109_k141_118080 | Not-determined |
| 18-109_k141_76249 | Not-determined |
| 18-109_k141_128098 | Not-determined |
| 18-109_k141_107083 | Not-determined |
| 18-109_k141_200143 | Low-quality |
| 18-109_k141_169130 | Not-determined |
| 18-109_k141_36163 | Not-determined |
| 18-109_k141_86664 | Not-determined |
| 18-109_k141_210197 | Not-determined |
| 18-109_k141_210200 | Low-quality |
| 18-109_k141_36166 | Not-determined |
| 18-109_k141_36168 | Not-determined |
| 18-109_k141_179190 | Not-determined |
| 18-109_k141_76267 | Not-determined |
| 18-109_k141_179192 | Not-determined |
| 18-109_k141_128122 | Not-determined |
| 18-109_k141_67056 | Low-quality |
| 18-109_k141_137923 | Not-determined |
| 18-109_k141_96971 | Not-determined |
| 18-109_k141_5241 | Not-determined |
| 18-109_k141_76279 | Low-quality |
| 18-109_k141_67067 | Not-determined |
| 18-109_k141_179199 | Not-determined |
| 18-109_k141_179200 | Not-determined |
| 18-109_k141_118115 | Not-determined |
| 18-109_k141_24764 | Not-determined |
| 18-109_k141_5251 | Not-determined |
| 18-109_k141_36178 | Low-quality |
| 18-109_k141_200167 | Not-determined |
| 18-109_k141_179205 | Not-determined |
| 18-109_k141_36181 | Not-determined |
| 18-109_k141_137937 | Not-determined |
| 18-109_k141_169143 | Not-determined |
| 18-109_k141_36183 | Not-determined |
| 18-109_k141_67074 | Not-determined |
| 18-109_k141_96988 | Not-determined |
| 18-109_k141_24776 | Not-determined |
| 18-109_k141_137942 | Low-quality |
| 18-109_k141_200173 | Not-determined |
| 18-109_k141_200175 | Not-determined |
| 18-109_k141_147879 | Not-determined |
| 18-109_k141_46175 | Not-determined |
| 18-109_k141_96998 | Not-determined |
| 18-109_k141_67078 | Not-determined |
| 18-109_k141_158751 | Low-quality |
| 18-109_k141_97001 | Not-determined |
| 18-109_k141_200183 | Not-determined |
| 18-109_k141_169153 | Not-determined |
| 18-109_k141_5268 | Not-determined |
| 18-109_k141_76288 | Not-determined |
| 18-109_k141_46183 | Not-determined |
| 18-109_k141_158761 | Not-determined |
| 18-109_k141_118142 | Not-determined |
| 18-109_k141_97008 | Low-quality |
| 18-109_k141_179225 | Low-quality |
| 18-109_k141_200190 | Not-determined |
| 18-109_k141_147895 | Not-determined |
| 18-109_k141_169158 | Low-quality |
| 18-109_k141_200194 | Not-determined |
| 18-109_k141_76298 | Low-quality |
| 18-109_k141_147898 | Not-determined |
| 18-109_k141_56248 | Not-determined |
| 18-109_k141_179233 | Not-determined |
| 18-109_k141_24783 | Not-determined |
| 18-109_k141_137964 | Not-determined |
| 18-109_k141_200206 | Low-quality |
| 18-109_k141_169168 | Not-determined |
| 18-109_k141_5279 | Low-quality |
| 18-109_k141_97015 | Not-determined |
| 18-109_k141_86687 | Not-determined |
| 18-109_k141_5286 | Not-determined |
| 18-109_k141_5287 | Not-determined |
| 18-109_k141_56266 | Not-determined |
| 18-109_k141_118150 | Not-determined |
| 18-109_k141_200227 | Not-determined |
| 18-109_k141_169193 | Not-determined |
| 18-109_k141_158782 | Not-determined |
| 18-109_k141_56274 | Not-determined |
| 18-109_k141_169196 | Not-determined |
| 18-109_k141_230142 | Not-determined |
| 18-109_k141_118160 | Not-determined |
| 18-109_k141_128190 | Not-determined |
| 18-109_k141_24794 | Not-determined |
| 18-109_k141_56278 | Not-determined |
| 18-109_k141_169202 | Not-determined |
| 18-109_k141_158786 | Not-determined |
| 18-109_k141_158788 | Not-determined |
| 18-109_k141_36206 | Not-determined |
| 18-109_k141_76318 | Not-determined |
| 18-109_k141_36211 | Not-determined |
| 18-109_k141_76325 | Not-determined |
| 18-109_k141_56295 | Low-quality |
| 18-109_k141_24809 | Not-determined |
| 18-109_k141_86704 | Not-determined |
| 18-109_k141_36224 | Low-quality |
| 18-109_k141_24813 | Not-determined |
| 18-109_k141_128212 | Not-determined |
| 18-109_k141_158805 | Not-determined |
| 18-109_k141_76340 | Not-determined |
| 18-109_k141_200233 | Not-determined |
| 18-109_k141_36228 | Not-determined |
| 18-109_k141_86710 | Not-determined |
| 18-109_k141_36231 | Not-determined |
| 18-109_k141_147948 | Not-determined |
| 18-109_k141_107114 | Not-determined |
| 18-109_k141_97040 | Not-determined |
| 18-109_k141_200247 | Low-quality |
| 18-109_k141_86723 | Not-determined |
| 18-109_k141_46192 | Not-determined |
| 18-109_k141_169230 | Not-determined |
| 18-109_k141_158809 | Not-determined |
| 18-109_k141_15385 | Not-determined |
| 18-109_k141_86727 | Not-determined |
| 18-109_k141_76368 | Not-determined |
| 18-109_k141_158814 | Not-determined |
| 18-109_k141_107123 | Not-determined |
| 18-109_k141_56326 | Not-determined |
| 18-109_k141_86731 | Not-determined |
| 18-109_k141_86732 | Not-determined |
| 18-109_k141_189601 | Not-determined |
| 18-109_k141_189602 | Not-determined |
| 18-109_k141_56328 | Low-quality |
| 18-109_k141_76371 | Low-quality |
| 18-109_k141_36248 | Not-determined |
| 18-109_k141_86736 | Not-determined |
| 18-109_k141_118195 | Not-determined |
| 18-109_k141_86738 | Not-determined |
| 18-109_k141_189608 | Not-determined |
| 18-109_k141_189609 | Not-determined |
| 18-109_k141_118208 | Not-determined |
| 18-109_k141_107131 | Not-determined |
| 18-109_k141_5337 | Low-quality |
| 18-109_k141_169244 | Not-determined |
| 18-109_k141_107133 | Not-determined |
| 18-109_k141_5338 | Not-determined |
| 18-109_k141_230153 | Not-determined |
| 18-109_k141_107137 | Not-determined |
| 18-109_k141_189614 | Not-determined |
| 18-109_k141_189616 | Low-quality |
| 18-109_k141_67118 | Not-determined |
| 18-109_k141_179303 | Not-determined |
| 18-109_k141_15401 | Not-determined |
| 18-109_k141_56352 | Low-quality |
| 18-109_k141_56353 | Not-determined |
| 18-109_k141_118220 | Not-determined |
| 18-109_k141_56356 | Low-quality |
| 18-109_k141_230171 | Not-determined |
| 18-109_k141_107155 | Not-determined |
| 18-109_k141_147959 | Not-determined |
| 18-109_k141_218746 | Not-determined |
| 18-109_k141_218748 | Not-determined |
| 18-109_k141_15419 | Not-determined |
| 18-109_k141_107167 | Not-determined |
| 18-109_k141_46222 | Low-quality |
| 18-109_k141_179327 | Not-determined |
| 18-109_k141_169265 | Not-determined |
| 18-109_k141_46228 | Not-determined |
| 18-109_k141_36285 | Not-determined |
| 18-109_k141_5348 | Not-determined |
| 18-109_k141_107184 | Not-determined |
| 18-109_k141_86756 | Low-quality |
| 18-109_k141_5353 | Not-determined |
| 18-109_k141_107186 | Not-determined |
| 18-109_k141_67162 | Not-determined |
| 18-109_k141_107205 | Not-determined |
| 18-109_k141_36304 | Low-quality |
| 18-109_k141_230213 | Not-determined |
| 18-109_k141_36307 | Not-determined |
| 18-109_k141_169293 | Not-determined |
| 18-109_k141_169296 | Not-determined |
| 18-109_k141_36313 | Not-determined |
| 18-109_k141_169301 | Low-quality |
| 18-109_k141_5386 | Not-determined |
| 18-109_k141_46259 | Not-determined |
| 18-109_k141_97072 | Not-determined |
| 18-109_k141_97073 | Not-determined |
| 18-109_k141_230229 | Not-determined |
| 18-109_k141_169303 | Not-determined |
| 18-109_k141_240588 | Low-quality |
| 18-109_k141_107216 | Low-quality |
| 18-109_k141_15473 | Not-determined |
| 18-109_k141_15475 | Not-determined |
| 18-109_k141_97082 | Not-determined |
| 18-109_k141_169309 | Not-determined |
| 18-109_k141_67174 | Not-determined |
| 18-109_k141_189644 | Not-determined |
| 18-109_k141_240592 | Not-determined |
| 18-109_k141_210219 | Not-determined |
| 18-109_k141_36339 | Not-determined |
| 18-109_k141_210222 | Not-determined |
| 18-109_k141_210224 | Not-determined |
| 18-109_k141_210226 | Not-determined |
| 18-109_k141_137972 | Low-quality |
| 18-109_k141_147991 | Not-determined |
| 18-109_k141_230253 | Not-determined |
| 18-109_k141_46272 | Low-quality |
| 18-109_k141_107230 | Not-determined |
| 18-109_k141_218795 | Not-determined |
| 18-109_k141_97101 | Not-determined |
| 18-109_k141_137975 | Not-determined |
| 18-109_k141_67195 | Not-determined |
| 18-109_k141_36351 | Not-determined |
| 18-109_k141_107235 | Not-determined |
| 18-109_k141_179335 | Low-quality |
| 18-109_k141_218803 | Not-determined |
| 18-109_k141_46279 | Not-determined |
| 18-109_k141_148004 | Not-determined |
| 18-109_k141_169332 | Not-determined |
| 18-109_k141_240608 | Not-determined |
| 18-109_k141_240610 | Not-determined |
| 18-109_k141_240613 | Not-determined |
| 18-109_k141_36363 | Not-determined |
| 18-109_k141_15495 | Not-determined |
| 18-109_k141_15499 | Not-determined |
| 18-109_k141_169343 | Not-determined |
| 18-109_k141_97121 | Not-determined |
| 18-109_k141_24921 | Not-determined |
| 18-109_k141_169345 | Not-determined |
| 18-109_k141_240617 | Low-quality |
| 18-109_k141_158869 | Not-determined |
| 18-109_k141_210261 | Low-quality |
| 18-109_k141_200272 | Not-determined |
| 18-109_k141_97123 | Not-determined |
| 18-109_k141_148026 | Not-determined |
| 18-109_k141_56402 | Not-determined |
| 18-109_k141_24931 | Not-determined |
| 18-109_k141_179353 | Not-determined |
| 18-109_k141_189683 | Not-determined |
| 18-109_k141_189684 | Not-determined |
| 18-109_k141_200284 | Not-determined |
| 18-109_k141_56414 | Low-quality |
| 18-109_k141_179362 | Not-determined |
| 18-109_k141_200289 | Not-determined |
| 18-109_k141_67214 | Not-determined |
| 18-109_k141_138011 | Not-determined |
| 18-109_k141_56422 | Not-determined |
| 18-109_k141_138015 | Low-quality |
| 18-109_k141_189685 | Not-determined |
| 18-109_k141_67221 | Not-determined |
| 18-109_k141_67222 | Not-determined |
| 18-109_k141_36380 | Not-determined |
| 18-109_k141_97147 | Not-determined |
| 18-109_k141_36385 | Not-determined |
| 18-109_k141_148037 | Not-determined |
| 18-109_k141_36388 | Not-determined |
| 18-109_k141_169368 | Not-determined |
| 18-109_k141_179386 | Not-determined |
| 18-109_k141_158900 | Not-determined |
| 18-109_k141_169373 | Not-determined |
| 18-109_k141_46298 | Not-determined |
| 18-109_k141_179390 | Not-determined |
| 18-109_k141_56428 | Not-determined |
| 18-109_k141_97160 | Low-quality |
| 18-109_k141_158910 | Not-determined |
| 18-109_k141_189705 | Not-determined |
| 18-109_k141_15540 | Not-determined |
| 18-109_k141_15544 | Not-determined |
| 18-109_k141_56442 | Not-determined |
| 18-109_k141_179408 | Not-determined |
| 18-109_k141_97170 | Not-determined |
| 18-109_k141_189715 | Not-determined |
| 18-109_k141_36407 | Low-quality |
| 18-109_k141_46319 | Not-determined |
| 18-109_k141_118244 | Not-determined |
| 18-109_k141_118245 | Not-determined |
| 18-109_k141_46324 | Not-determined |
| 18-109_k141_36413 | Not-determined |
| 18-109_k141_148077 | Not-determined |
| 18-109_k141_118256 | Not-determined |
| 18-109_k141_138055 | Low-quality |
| 18-109_k141_118259 | Not-determined |
| 18-109_k141_56457 | Not-determined |
| 18-109_k141_218834 | Not-determined |
| 18-109_k141_230320 | Not-determined |
| 18-109_k141_218836 | Not-determined |
| 18-109_k141_189736 | Not-determined |
| 18-109_k141_230324 | Not-determined |
| 18-109_k141_240637 | Not-determined |
| 18-109_k141_15579 | Not-determined |
| 18-109_k141_210313 | Not-determined |
| 18-109_k141_218842 | Not-determined |
| 18-109_k141_179428 | Not-determined |
| 18-109_k141_189745 | Not-determined |
| 18-109_k141_230329 | Not-determined |
| 18-109_k141_179431 | Not-determined |
| 18-109_k141_200319 | Not-determined |
| 18-109_k141_240645 | Not-determined |
| 18-109_k141_46347 | Not-determined |
| 18-109_k141_189752 | Not-determined |
| 18-109_k141_97212 | Not-determined |
| 18-109_k141_240647 | Not-determined |
| 18-102_k141_161986 | High-quality |
| 18-109_k141_210316 | Not-determined |
| 18-109_k141_56466 | Not-determined |
| 18-109_k141_200332 | Not-determined |
| 18-109_k141_5439 | Not-determined |
| 18-109_k141_200338 | Not-determined |
| 18-109_k141_15593 | Not-determined |
| 18-109_k141_200340 | Not-determined |
| 18-109_k141_158938 | Not-determined |
| 18-109_k141_158942 | Not-determined |
| 18-109_k141_5451 | Not-determined |
| 18-109_k141_138080 | Not-determined |
| 18-109_k141_46355 | Not-determined |
| 18-109_k141_138081 | Not-determined |
| 18-109_k141_56487 | Not-determined |
| 18-109_k141_138087 | Not-determined |
| 18-109_k141_240666 | Not-determined |
| 18-109_k141_86763 | Not-determined |
| 18-109_k141_218877 | Not-determined |
| 18-109_k141_76456 | Not-determined |
| 18-109_k141_97234 | Not-determined |
| 18-109_k141_5456 | Not-determined |
| 18-109_k141_56495 | Not-determined |
| 18-109_k141_138092 | Low-quality |
| 18-109_k141_5458 | Low-quality |
| 18-109_k141_56500 | Not-determined |
| 18-109_k141_200361 | Low-quality |
| 18-109_k141_158952 | Not-determined |
| 18-109_k141_5462 | Not-determined |
| 18-109_k141_148125 | Not-determined |
| 18-109_k141_36462 | Not-determined |
| 18-109_k141_5469 | Low-quality |
| 18-109_k141_148131 | Not-determined |
| 18-109_k141_36468 | Not-determined |
| 18-109_k141_15600 | Low-quality |
| 18-109_k141_25004 | Not-determined |
| 18-109_k141_200386 | Not-determined |
| 18-109_k141_25006 | Not-determined |
| 18-109_k141_200387 | Not-determined |
| 18-109_k141_118281 | Not-determined |
| 18-109_k141_138105 | Not-determined |
| 18-109_k141_86775 | Not-determined |
| 18-109_k141_86776 | Not-determined |
| 18-109_k141_138111 | Not-determined |
| 18-109_k141_25010 | Low-quality |
| 18-109_k141_240686 | Not-determined |
| 18-109_k141_118288 | Not-determined |
| 18-109_k141_76469 | Not-determined |
| 18-109_k141_5474 | Not-determined |
| 18-109_k141_200399 | Not-determined |
| 18-109_k141_76475 | Not-determined |
| 18-109_k141_56523 | Not-determined |
| 18-109_k141_210338 | Not-determined |
| 18-109_k141_76477 | Not-determined |
| 18-109_k141_118299 | Low-quality |
| 18-109_k141_158966 | Not-determined |
| 18-109_k141_56535 | Not-determined |
| 18-109_k141_158973 | Not-determined |
| 18-109_k141_138139 | Not-determined |
| 18-109_k141_210360 | Not-determined |
| 18-109_k141_76500 | Low-quality |
| 18-109_k141_158980 | Not-determined |
| 18-109_k141_36484 | Not-determined |
| 18-109_k141_240713 | Not-determined |
| 18-109_k141_138145 | Not-determined |

| | |
|---|---|
| 18-109_k141_189780 | Not-determined |
| 18-109_k141_240718 | Not-determined |
| 18-109_k141_200425 | Low-quality |
| 18-109_k141_210376 | Not-determined |
| 18-109_k141_15634 | Not-determined |
| 18-109_k141_240720 | Not-determined |
| 18-109_k141_240722 | Not-determined |
| 18-109_k141_210379 | Not-determined |
| 18-109_k141_36490 | Not-determined |
| 18-109_k141_240725 | Not-determined |
| 18-109_k141_118329 | Not-determined |
| 18-109_k141_107284 | Not-determined |
| 18-109_k141_107285 | Not-determined |
| 18-109_k141_76513 | Not-determined |
| 18-109_k141_25044 | Low-quality |
| 18-109_k141_25047 | Not-determined |
| 18-109_k141_25051 | Not-determined |
| 18-109_k141_230344 | Not-determined |
| 18-109_k141_15647 | Not-determined |
| 18-109_k141_5512 | Not-determined |
| 18-109_k141_25059 | Not-determined |
| 18-109_k141_15658 | Not-determined |
| 18-109_k141_240741 | Not-determined |
| 18-109_k141_210392 | Not-determined |
| 18-109_k141_15661 | Not-determined |
| 18-109_k141_107305 | Not-determined |
| 18-109_k141_46445 | Not-determined |
| 18-109_k141_240748 | Not-determined |
| 18-109_k141_159011 | Not-determined |
| 18-109_k141_76521 | Not-determined |
| 18-109_k141_15667 | Low-quality |
| 18-109_k141_230358 | Not-determined |
| 18-109_k141_189810 | Low-quality |
| 18-109_k141_36505 | Not-determined |
| 18-109_k141_15671 | Not-determined |
| 18-109_k141_200446 | Not-determined |
| 18-109_k141_56558 | Not-determined |
| 18-109_k141_56562 | Not-determined |
| 18-109_k141_86813 | Low-quality |
| 18-109_k141_218893 | Not-determined |
| 18-109_k141_86814 | Not-determined |
| 18-109_k141_138166 | Low-quality |
| 18-109_k141_138167 | Not-determined |
| 18-109_k141_128249 | Not-determined |
| 18-109_k141_138170 | Not-determined |
| 18-109_k141_56564 | Not-determined |
| 18-109_k141_86818 | Not-determined |
| 18-109_k141_36522 | Not-determined |
| 18-109_k141_107320 | Not-determined |
| 18-109_k141_169376 | Not-determined |
| 18-109_k141_148180 | Not-determined |
| 18-109_k141_200450 | Not-determined |
| 18-109_k141_148181 | Not-determined |
| 18-109_k141_189842 | Not-determined |
| 18-109_k141_200454 | Not-determined |
| 18-109_k141_159043 | Not-determined |
| 18-109_k141_128263 | Low-quality |
| 18-109_k141_107334 | Not-determined |
| 18-109_k141_46477 | Not-determined |
| 18-109_k141_107340 | Low-quality |
| 18-109_k141_107342 | Low-quality |
| 18-109_k141_36542 | Not-determined |
| 18-109_k141_107343 | Not-determined |
| 18-109_k141_56581 | Not-determined |
| 18-109_k141_210414 | Not-determined |
| 18-109_k141_210415 | Not-determined |
| 18-109_k141_200459 | Not-determined |
| 18-109_k141_210420 | Not-determined |
| 18-109_k141_15705 | Not-determined |
| 18-109_k141_107353 | Not-determined |
| 18-109_k141_15708 | Not-determined |
| 18-109_k141_97263 | Not-determined |
| 18-109_k141_189855 | Not-determined |
| 18-109_k141_218912 | Not-determined |
| 18-109_k141_189859 | Not-determined |
| 18-109_k141_86847 | Not-determined |
| 18-109_k141_25095 | Not-determined |
| 18-109_k141_36551 | Not-determined |
| 18-109_k141_25100 | Not-determined |
| 18-109_k141_25101 | Not-determined |
| 18-109_k141_5532 | Not-determined |
| 18-109_k141_148196 | Not-determined |
| 18-109_k141_189869 | Not-determined |
| 18-109_k141_179464 | Not-determined |
| 18-109_k141_76565 | Low-quality |
| 18-109_k141_76566 | Not-determined |
| 18-109_k141_36560 | Not-determined |
| 18-109_k141_15713 | Not-determined |
| 18-109_k141_159054 | Not-determined |
| 18-109_k141_97281 | Not-determined |
| 18-109_k141_97284 | Not-determined |
| 18-109_k141_230393 | Not-determined |
| 18-109_k141_148206 | Not-determined |
| 18-109_k141_189874 | Not-determined |
| 18-109_k141_86863 | Not-determined |
| 18-109_k141_189876 | Not-determined |
| 18-109_k141_230401 | Not-determined |
| 18-109_k141_36574 | Not-determined |
| 18-109_k141_230402 | Not-determined |
| 18-109_k141_159063 | Low-quality |
| 18-109_k141_148215 | Not-determined |
| 18-109_k141_36580 | Not-determined |
| 18-109_k141_86872 | Not-determined |
| 18-109_k141_25119 | Not-determined |
| 18-109_k141_36584 | Not-determined |
| 18-109_k141_179480 | Not-determined |
| 18-109_k141_86876 | Not-determined |
| 18-109_k141_200485 | Not-determined |
| 18-109_k141_25125 | Not-determined |
| 18-109_k141_56613 | Not-determined |
| 18-109_k141_36597 | Not-determined |
| 18-109_k141_240763 | Not-determined |
| 18-109_k141_36602 | Not-determined |
| 18-109_k141_46507 | Not-determined |
| 18-109_k141_46508 | Not-determined |
| 18-109_k141_107399 | Not-determined |
| 18-109_k141_67246 | Not-determined |
| 18-109_k141_67250 | Low-quality |
| 18-109_k141_189884 | Not-determined |
| 18-109_k141_5574 | Low-quality |
| 18-109_k141_5575 | Not-determined |
| 18-109_k141_138237 | Not-determined |
| 18-109_k141_36621 | Not-determined |
| 18-109_k141_210466 | Not-determined |
| 18-109_k141_128303 | Not-determined |
| 18-109_k141_138243 | Not-determined |
| 18-109_k141_169430 | Not-determined |
| 18-109_k141_210474 | Not-determined |
| 18-109_k141_189895 | Not-determined |
| 18-109_k141_15745 | Not-determined |
| 18-109_k141_148252 | Not-determined |
| 18-109_k141_118345 | Not-determined |
| 18-109_k141_189903 | Not-determined |
| 18-109_k141_15752 | Not-determined |
| 18-109_k141_97334 | Not-determined |
| 18-109_k141_240801 | Not-determined |
| 18-109_k141_159088 | Not-determined |
| 18-109_k141_5588 | Not-determined |
| 18-109_k141_210486 | Not-determined |
| 18-109_k141_210490 | Not-determined |
| 18-109_k141_240809 | Not-determined |
| 18-109_k141_200503 | Low-quality |
| 18-109_k141_15764 | Not-determined |
| 18-109_k141_138262 | Not-determined |
| 18-109_k141_36640 | Not-determined |
| 18-109_k141_15766 | Not-determined |
| 18-109_k141_138266 | Not-determined |
| 18-109_k141_76638 | Not-determined |
| 18-109_k141_56630 | Not-determined |
| 18-109_k141_25162 | Not-determined |
| 18-109_k141_15772 | Not-determined |
| 18-109_k141_36652 | Not-determined |
| 18-109_k141_169447 | Not-determined |
| 18-109_k141_210511 | Not-determined |
| 18-109_k141_210518 | Not-determined |
| 18-109_k141_36661 | Not-determined |
| 18-109_k141_169453 | Low-quality |
| 18-109_k141_210522 | Low-quality |
| 18-109_k141_240839 | Not-determined |
| 18-109_k141_128316 | Not-determined |
| 18-109_k141_36665 | Not-determined |
| 18-109_k141_148278 | Not-determined |
| 18-109_k141_169462 | Not-determined |
| 18-109_k141_240851 | Not-determined |
| 18-109_k141_230433 | Not-determined |
| 18-109_k141_189935 | Not-determined |
| 18-109_k141_36672 | Not-determined |
| 18-109_k141_128324 | Low-quality |
| 18-109_k141_230435 | Low-quality |
| 18-109_k141_210535 | Not-determined |
| 18-109_k141_118361 | Not-determined |
| 18-109_k141_46537 | Not-determined |
| 18-109_k141_67275 | Not-determined |
| 18-109_k141_169473 | Not-determined |
| 18-109_k141_169477 | Not-determined |
| 18-109_k141_86899 | Low-quality |
| 18-109_k141_118366 | Not-determined |
| 18-109_k141_97379 | Low-quality |
| 18-109_k141_67283 | Not-determined |
| 18-109_k141_179531 | Not-determined |
| 18-109_k141_169481 | Not-determined |
| 18-109_k141_46555 | Not-determined |
| 18-109_k141_230458 | Not-determined |
| 18-109_k141_36681 | Not-determined |
| 18-109_k141_46557 | Not-determined |
| 18-109_k141_189943 | Not-determined |
| 18-109_k141_118369 | Not-determined |
| 18-109_k141_36686 | Not-determined |
| 18-109_k141_76641 | Low-quality |
| 18-109_k141_46561 | Not-determined |
| 18-109_k141_76650 | Not-determined |
| 18-109_k141_179536 | Not-determined |
| 18-109_k141_46569 | Not-determined |
| 18-109_k141_56648 | Not-determined |
| 18-109_k141_230478 | Not-determined |
| 18-109_k141_230479 | Low-quality |
| 18-109_k141_107473 | Low-quality |
| 18-109_k141_200546 | Not-determined |
| 18-109_k141_159112 | Not-determined |
| 18-109_k141_189952 | Not-determined |
| 18-109_k141_46580 | Not-determined |
| 18-109_k141_169494 | Not-determined |
| 18-109_k141_36697 | Low-quality |
| 18-109_k141_86924 | Not-determined |
| 18-109_k141_210561 | Not-determined |
| 18-109_k141_76657 | Not-determined |
| 18-109_k141_56662 | Not-determined |
| 18-109_k141_210565 | Not-determined |
| 18-109_k141_159118 | Not-determined |
| 18-109_k141_240893 | Not-determined |
| 18-109_k141_240895 | Not-determined |
| 18-109_k141_86930 | Not-determined |
| 18-109_k141_210573 | Not-determined |
| 18-109_k141_15801 | Not-determined |
| 18-109_k141_240902 | Not-determined |
| 18-109_k141_179552 | Not-determined |
| 18-109_k141_86942 | Not-determined |
| 18-109_k141_107490 | Not-determined |
| 18-109_k141_46596 | Not-determined |
| 18-109_k141_97405 | Not-determined |
| 18-109_k141_97406 | Not-determined |
| 18-109_k141_210578 | Not-determined |
| 18-109_k141_159140 | Not-determined |
| 18-109_k141_240904 | Not-determined |
| 18-109_k141_107501 | Not-determined |
| 18-109_k141_46603 | Not-determined |
| 18-109_k141_56678 | Not-determined |
| 18-109_k141_56679 | Not-determined |
| 18-109_k141_159142 | Not-determined |
| 18-109_k141_169511 | Not-determined |
| 18-109_k141_76682 | Low-quality |
| 18-109_k141_86962 | Not-determined |
| 18-109_k141_97413 | Not-determined |
| 18-109_k141_189983 | Low-quality |
| 18-109_k141_46627 | Not-determined |
| 18-109_k141_46629 | Not-determined |
| 18-109_k141_128391 | Not-determined |
| 18-109_k141_128394 | Not-determined |
| 18-109_k141_56691 | Not-determined |
| 18-109_k141_76702 | Not-determined |
| 18-109_k141_179573 | Not-determined |
| 18-109_k141_15804 | Low-quality |
| 18-109_k141_15805 | Not-determined |
| 18-109_k141_169522 | Not-determined |
| 18-109_k141_118410 | Not-determined |
| 18-109_k141_190007 | Low-quality |
| 18-109_k141_118412 | Not-determined |
| 18-109_k141_15818 | Not-determined |
| 18-109_k141_107523 | Low-quality |
| 18-109_k141_118416 | Not-determined |
| 18-109_k141_107525 | Not-determined |
| 18-109_k141_107528 | Low-quality |
| 18-109_k141_25187 | Not-determined |
| 18-109_k141_56708 | Not-determined |
| 18-109_k141_159161 | Not-determined |
| 18-109_k141_56712 | Not-determined |
| 18-109_k141_179596 | Not-determined |
| 18-109_k141_36742 | Not-determined |
| 18-109_k141_107534 | Not-determined |
| 18-109_k141_159166 | Not-determined |
| 18-109_k141_159168 | Not-determined |
| 18-109_k141_118428 | Not-determined |
| 18-109_k141_210605 | Not-determined |
| 18-109_k141_159173 | Low-quality |
| 18-109_k141_46671 | Not-determined |
| 18-109_k141_179617 | Not-determined |
| 18-109_k141_76722 | Not-determined |
| 18-109_k141_179627 | Not-determined |
| 18-109_k141_56741 | Not-determined |
| 18-109_k141_118429 | Not-determined |
| 18-109_k141_210620 | Not-determined |
| 18-109_k141_56744 | Not-determined |
| 18-109_k141_76731 | Not-determined |
| 18-109_k141_210630 | Not-determined |
| 18-109_k141_56751 | Not-determined |
| 18-109_k141_46681 | Not-determined |
| 18-109_k141_87007 | Not-determined |
| 18-109_k141_200643 | Not-determined |
| 18-109_k141_76744 | Not-determined |
| 18-109_k141_128434 | Low-quality |
| 18-109_k141_46691 | Not-determined |
| 18-109_k141_128436 | Not-determined |
| 18-109_k141_128439 | Not-determined |
| 18-109_k141_87017 | Low-quality |
| 18-109_k141_190070 | Not-determined |
| 18-109_k141_36761 | Not-determined |
| 18-109_k141_210645 | Not-determined |
| 18-109_k141_169555 | Not-determined |
| 18-109_k141_210647 | Not-determined |
| 18-109_k141_200655 | Not-determined |
| 18-109_k141_169561 | Not-determined |
| 18-109_k141_128449 | Not-determined |
| 18-109_k141_128451 | Not-determined |
| 18-109_k141_200659 | Not-determined |
| 18-109_k141_36775 | Not-determined |
| 18-109_k141_169574 | Not-determined |
| 18-109_k141_210657 | Low-quality |
| 18-109_k141_138322 | Not-determined |
| 18-109_k141_138323 | Not-determined |
| 18-109_k141_76754 | Not-determined |
| 18-109_k141_76755 | Not-determined |
| 18-109_k141_240949 | Not-determined |
| 18-109_k141_230514 | Not-determined |
| 18-109_k141_46730 | Not-determined |
| 18-109_k141_46732 | Not-determined |
| 18-109_k141_118459 | Not-determined |
| 18-109_k141_190084 | Not-determined |
| 18-109_k141_138331 | Not-determined |
| 18-109_k141_190089 | Not-determined |
| 18-109_k141_200673 | Low-quality |
| 18-109_k141_240950 | Low-quality |
| 18-109_k141_190096 | Not-determined |
| 18-109_k141_138343 | Not-determined |
| 18-109_k141_169583 | Not-determined |
| 18-109_k141_159238 | Not-determined |
| 18-109_k141_138347 | Not-determined |
| 18-109_k141_67329 | Not-determined |
| 18-109_k141_190105 | Not-determined |
| 18-109_k141_5643 | Not-determined |
| 18-109_k141_138351 | Not-determined |
| 18-109_k141_5645 | Low-quality |
| 18-109_k141_240960 | Not-determined |
| 18-109_k141_240962 | Not-determined |
| 18-109_k141_107571 | Not-determined |
| 18-109_k141_97439 | Not-determined |
| 18-109_k141_138358 | Not-determined |
| 18-109_k141_148321 | Not-determined |
| 18-109_k141_179687 | Not-determined |
| 18-109_k141_240971 | Not-determined |
| 18-109_k141_148330 | Not-determined |
| 18-109_k141_107583 | Not-determined |
| 18-109_k141_107584 | Not-determined |
| 18-109_k141_169593 | Not-determined |
| 18-109_k141_179697 | Low-quality |
| 18-109_k141_97450 | Not-determined |
| 18-109_k141_240975 | Not-determined |
| 18-109_k141_230530 | Not-determined |
| 18-109_k141_138374 | Not-determined |
| 18-109_k141_56765 | Not-determined |
| 18-109_k141_200678 | Not-determined |
| 18-109_k141_128469 | Not-determined |
| 18-109_k141_169607 | Not-determined |
| 18-109_k141_240983 | Low-quality |
| 18-109_k141_15842 | Not-determined |
| 18-109_k141_46748 | Not-determined |
| 18-109_k141_148338 | Not-determined |
| 18-109_k141_97462 | Not-determined |
| 18-109_k141_230532 | Not-determined |
| 18-109_k141_179715 | Not-determined |
| 18-109_k141_118466 | Not-determined |
| 18-109_k141_128480 | Not-determined |
| 18-109_k141_118467 | Not-determined |
| 18-109_k141_138392 | Not-determined |
| 18-109_k141_67357 | Not-determined |
| 18-109_k141_159250 | Not-determined |
| 18-109_k141_87059 | Not-determined |
| 18-109_k141_118472 | Low-quality |
| 18-109_k141_97473 | Not-determined |
| 18-109_k141_200699 | Not-determined |
| 18-109_k141_148356 | Not-determined |
| 18-109_k141_118479 | Not-determined |
| 18-109_k141_5679 | Not-determined |
| 18-109_k141_97478 | Not-determined |
| 18-109_k141_200703 | Not-determined |
| 18-109_k141_241002 | Not-determined |
| 18-109_k141_128507 | Low-quality |
| 18-109_k141_67376 | Not-determined |
| 18-109_k141_169650 | Not-determined |
| 18-109_k141_67378 | Not-determined |
| 18-109_k141_190148 | Not-determined |
| 18-109_k141_67383 | Not-determined |
| 18-109_k141_25202 | Not-determined |
| 18-109_k141_190157 | Not-determined |
| 18-109_k141_36831 | Not-determined |
| 18-109_k141_87079 | Low-quality |
| 18-109_k141_67389 | Not-determined |
| 18-109_k141_118497 | Low-quality |
| 18-109_k141_190165 | Low-quality |
| 18-109_k141_97491 | Not-determined |
| 18-109_k141_219009 | Not-determined |
| 18-109_k141_169669 | Not-determined |
| 18-109_k141_87086 | Not-determined |
| 18-109_k141_87093 | Low-quality |
| 18-109_k141_118509 | Not-determined |
| 18-109_k141_169673 | Not-determined |
| 18-109_k141_118514 | Not-determined |
| 18-109_k141_87097 | Not-determined |
| 18-109_k141_118520 | Not-determined |
| 18-109_k141_67407 | Not-determined |
| 18-109_k141_190177 | Not-determined |
| 18-109_k141_67409 | Not-determined |
| 18-109_k141_241037 | Not-determined |
| 18-109_k141_56822 | Not-determined |
| 18-109_k141_138419 | Not-determined |
| 18-109_k141_200724 | Not-determined |
| 18-109_k141_25217 | Low-quality |
| 18-109_k141_179741 | Not-determined |
| 18-109_k141_15904 | Not-determined |
| 18-109_k141_25222 | Not-determined |
| 18-109_k141_15907 | Not-determined |
| 18-109_k141_159273 | Low-quality |
| 18-109_k141_200728 | Low-quality |
| 18-109_k141_25224 | Not-determined |
| 18-109_k141_87126 | Not-determined |
| 18-109_k141_97495 | Not-determined |
| 18-109_k141_5691 | Not-determined |
| 18-109_k141_190196 | Not-determined |
| 18-109_k141_36860 | Not-determined |
| 18-109_k141_15916 | Not-determined |
| 18-109_k141_118552 | Not-determined |
| 18-109_k141_97502 | Not-determined |
| 18-109_k141_138444 | Not-determined |
| 18-109_k141_5700 | Not-determined |
| 18-109_k141_118558 | Not-determined |
| 18-109_k141_179755 | Not-determined |
| 18-109_k141_148370 | Not-determined |
| 18-109_k141_5705 | Not-determined |
| 18-109_k141_5708 | Not-determined |
| 18-109_k141_159308 | Not-determined |
| 18-109_k141_36876 | Low-quality |
| 18-109_k141_36878 | Not-determined |
| 18-109_k141_159313 | Not-determined |
| 18-109_k141_219042 | Not-determined |
| 18-109_k141_159318 | Not-determined |
| 18-109_k141_179775 | Not-determined |
| 18-109_k141_15927 | Not-determined |
| 18-109_k141_56845 | Not-determined |
| 18-109_k141_97519 | Not-determined |
| 18-109_k141_190221 | Not-determined |
| 18-109_k141_159334 | Not-determined |
| 18-109_k141_190227 | Not-determined |
| 18-109_k141_148397 | Not-determined |
| 18-109_k141_25243 | Not-determined |
| 18-109_k141_190231 | Not-determined |
| 18-109_k141_190233 | Not-determined |
| 18-109_k141_230562 | Not-determined |
| 18-109_k141_179787 | Not-determined |
| 18-109_k141_148407 | Low-quality |
| 18-109_k141_241067 | Not-determined |
| 18-109_k141_46797 | Not-determined |
| 18-109_k141_148408 | Not-determined |
| 18-109_k141_87178 | Not-determined |
| 18-109_k141_230573 | Not-determined |
| 18-109_k141_148416 | Not-determined |
| 18-109_k141_36905 | Not-determined |
| 18-109_k141_179791 | Not-determined |
| 18-109_k141_138466 | Not-determined |
| 18-109_k141_67473 | Not-determined |
| 18-109_k141_46802 | Not-determined |
| 18-109_k141_230576 | Not-determined |
| 18-109_k141_118596 | Not-determined |
| 18-109_k141_67475 | Not-determined |
| 18-109_k141_210665 | Not-determined |
| 18-109_k141_210666 | Not-determined |
| 18-109_k141_118602 | Not-determined |
| 18-109_k141_148430 | Not-determined |
| 18-109_k141_210682 | Not-determined |
| 18-109_k141_87198 | Not-determined |
| 18-109_k141_87200 | Not-determined |
| 18-109_k141_219046 | Not-determined |
| 18-109_k141_25262 | Not-determined |
| 18-109_k141_219053 | Not-determined |
| 18-109_k141_15946 | Not-determined |
| 18-109_k141_67500 | Not-determined |
| 18-109_k141_148434 | Not-determined |
| 18-109_k141_36928 | Not-determined |
| 18-109_k141_107593 | Not-determined |
| 18-109_k141_67509 | Not-determined |
| 18-109_k141_67511 | Not-determined |
| 18-109_k141_169742 | Not-determined |
| 18-109_k141_67517 | Not-determined |
| 18-109_k141_241113 | Not-determined |
| 18-109_k141_148440 | Not-determined |
| 18-109_k141_210712 | Not-determined |
| 18-109_k141_76800 | Not-determined |
| 18-109_k141_87218 | Not-determined |
| 18-109_k141_5719 | Not-determined |
| 18-109_k141_5724 | Not-determined |
| 18-109_k141_138498 | Not-determined |
| 18-109_k141_138502 | Not-determined |
| 18-109_k141_67539 | Not-determined |
| 18-109_k141_107606 | Not-determined |
| 18-109_k141_219096 | Not-determined |
| 18-109_k141_36969 | Not-determined |
| 18-109_k141_5744 | Not-determined |
| 18-109_k141_219101 | Not-determined |
| 18-109_k141_241136 | Not-determined |
| 18-109_k141_159380 | Not-determined |
| 18-109_k141_128607 | Not-determined |
| 18-109_k141_128616 | Not-determined |
| 18-109_k141_138518 | Low-quality |
| 18-109_k141_169782 | Not-determined |
| 18-109_k141_179843 | Not-determined |
| 18-109_k141_67563 | Not-determined |
| 18-109_k141_25282 | Not-determined |
| 18-109_k141_5757 | Not-determined |
| 18-102-final-viral-combined_k141_245232\|\|full | Medium-quality |
| 18-109_k141_169792 | Not-determined |
| 18-109_k141_159405 | Not-determined |
| 18-109_k141_36994 | Not-determined |
| 18-109_k141_128629 | Not-determined |
| 18-109_k141_159408 | Not-determined |
| 18-109_k141_128634 | Not-determined |
| 18-109_k141_118616 | Not-determined |
| 18-109_k141_210764 | Not-determined |
| 18-109_k141_107617 | Not-determined |
| 18-109_k141_210767 | Not-determined |
| 18-109_k141_76843 | Not-determined |
| 18-109_k141_138546 | Not-determined |
| 18-109_k141_37003 | Not-determined |
| 18-109_k141_15976 | Not-determined |
| 18-109_k141_169813 | Not-determined |
| 18-109_k141_219116 | Not-determined |
| 18-109_k141_118622 | Not-determined |
| 18-109_k141_128659 | Not-determined |
| 18-109_k141_128661 | Not-determined |
| 18-109_k141_5790 | Not-determined |
| 18-109_k141_67590 | Low-quality |
| 18-109_k141_128665 | Not-determined |
| 18-109_k141_148493 | Not-determined |
| 18-109_k141_230594 | Not-determined |
| 18-109_k141_118629 | Not-determined |
| 18-109_k141_67594 | Not-determined |
| 18-109_k141_169828 | Not-determined |
| 18-109_k141_15991 | Not-determined |
| 18-109_k141_210771 | Not-determined |
| 18-109_k141_148505 | Not-determined |
| 18-109_k141_87253 | Not-determined |
| 18-109_k141_230607 | Not-determined |
| 18-109_k141_15999 | Not-determined |
| 18-109_k141_56878 | Not-determined |
| 18-109_k141_97573 | Not-determined |
| 18-109_k141_46827 | Not-determined |
| 18-109_k141_37021 | Not-determined |
| 18-109_k141_87266 | Not-determined |
| 18-109_k141_37026 | Not-determined |
| 18-109_k141_179853 | Not-determined |
| 18-109_k141_128680 | Not-determined |
| 18-109_k141_16014 | Low-quality |
| 18-109_k141_230620 | Not-determined |
| 18-109_k141_148527 | Not-determined |
| 18-109_k141_230626 | Not-determined |
| 18-109_k141_138576 | Not-determined |
| 18-109_k141_37037 | Not-determined |
| 18-109_k141_87267 | Low-quality |
| 18-109_k141_97595 | Not-determined |
| 18-109_k141_107671 | Not-determined |
| 18-109_k141_230628 | Not-determined |
| 18-109_k141_148540 | Not-determined |
| 18-109_k141_179865 | Not-determined |
| 18-109_k141_118640 | Not-determined |
| 18-109_k141_190270 | Not-determined |
| 18-109_k141_107678 | Not-determined |
| 18-109_k141_128690 | Not-determined |
| 18-109_k141_230630 | Not-determined |
| 18-101-final-viral-combined_k141_360285\|\|full | Medium-quality |
| 18-109_k141_148547 | Not-determined |
| 18-109_k141_37048 | Not-determined |
| 18-109_k141_97602 | Not-determined |
| 18-109_k141_16028 | Not-determined |
| 18-109_k141_148555 | Not-determined |
| 18-109_k141_230636 | Not-determined |
| 18-109_k141_56905 | Not-determined |
| 18-109_k141_76868 | Not-determined |
| 18-109_k141_118644 | Not-determined |
| 18-109_k141_190278 | Not-determined |
| 18-109_k141_230646 | Not-determined |
| 18-109_k141_97621 | Not-determined |
| 18-109_k141_219130 | Not-determined |
| 18-109_k141_219132 | Not-determined |
| 18-109_k141_128703 | Not-determined |
| 18-109_k141_46878 | Not-determined |
| 18-109_k141_230659 | Not-determined |
| 18-109_k141_67632 | Not-determined |
| 18-109_k141_37082 | Not-determined |
| 18-109_k141_67640 | Not-determined |
| 18-109_k141_25359 | Not-determined |
| 18-109_k141_25362 | Not-determined |
| 18-109_k141_159464 | Not-determined |
| 18-109_k141_67646 | Low-quality |
| 18-109_k141_219146 | Not-determined |
| 18-109_k141_37092 | Not-determined |
| 18-109_k141_118662 | Not-determined |
| 18-109_k141_87304 | Not-determined |
| 18-109_k141_67653 | Not-determined |
| 18-109_k141_97657 | Not-determined |
| 18-109_k141_67655 | Not-determined |
| 18-109_k141_159470 | Not-determined |
| 18-109_k141_148588 | Not-determined |

| | |
|---|---|
| 18-109_k141_159472 | Low-quality |
| 18-109_k141_118671 | Low-quality |
| 18-109_k141_200751 | Not-determined |
| 18-109_k141_210799 | Not-determined |
| 18-109_k141_159482 | Low-quality |
| 18-109_k141_159484 | Not-determined |
| 18-109_k141_5798 | Not-determined |
| 18-109_k141_46907 | Not-determined |
| 18-109_k141_200763 | Not-determined |
| 18-109_k141_37112 | Not-determined |
| 18-109_k141_241230 | Not-determined |
| 18-109_k141_37113 | Low-quality |
| 18-109_k141_138609 | Not-determined |
| 18-109_k141_5804 | Not-determined |
| 18-109_k141_200766 | Low-quality |
| 18-109_k141_169897 | Low-quality |
| 18-109_k141_16041 | Not-determined |
| 18-109_k141_5806 | Not-determined |
| 18-109_k141_148599 | Not-determined |
| 18-109_k141_5811 | Not-determined |
| 18-109_k141_16048 | Not-determined |
| 18-109_k141_67675 | Not-determined |
| 18-109_k141_138627 | Not-determined |
| 18-109_k141_200775 | Not-determined |
| 18-109_k141_128736 | Not-determined |
| 18-109_k141_230682 | Not-determined |
| 18-109_k141_179902 | Not-determined |
| 18-109_k141_219191 | Not-determined |
| 18-109_k141_67695 | Not-determined |
| 18-109_k141_67700 | Not-determined |
| 18-109_k141_128744 | Not-determined |
| 18-109_k141_37144 | Low-quality |
| 18-109_k141_87347 | Not-determined |
| 18-109_k141_169950 | Not-determined |
| 18-109_k141_118674 | Not-determined |
| 18-109_k141_87357 | Not-determined |
| 18-109_k141_56979 | Not-determined |
| 18-109_k141_219206 | Not-determined |
| 18-109_k141_118678 | Not-determined |
| 18-109_k141_56981 | Not-determined |
| 18-109_k141_219207 | Not-determined |
| 18-109_k141_118680 | Not-determined |
| 18-109_k141_67710 | Not-determined |
| 18-109_k141_16068 | Not-determined |
| 18-109_k141_190282 | Not-determined |
| 18-109_k141_56990 | Low-quality |
| 18-109_k141_190287 | Not-determined |
| 18-109_k141_67718 | Not-determined |
| 18-109_k141_169953 | Not-determined |
| 18-109_k141_56996 | Not-determined |
| 18-109_k141_56997 | Not-determined |
| 18-109_k141_241260 | Not-determined |
| 18-109_k141_190291 | Not-determined |
| 18-109_k141_210854 | Not-determined |
| 18-109_k141_210856 | Not-determined |
| 18-109_k141_169965 | Not-determined |
| 18-109_k141_241266 | Not-determined |
| 18-109_k141_118706 | Not-determined |
| 18-109_k141_16094 | Not-determined |
| 18-109_k141_16096 | Not-determined |
| 18-109_k141_76885 | Not-determined |
| 18-109_k141_118718 | Not-determined |
| 18-109_k141_5820 | Not-determined |
| 18-109_k141_200820 | Not-determined |
| 18-109_k141_169977 | Not-determined |
| 18-109_k141_219244 | Not-determined |
| 18-109_k141_57039 | Not-determined |
| 18-109_k141_241293 | Not-determined |
| 18-109_k141_25380 | Low-quality |
| 18-109_k141_57041 | Not-determined |
| 18-109_k141_200828 | Not-determined |
| 18-109_k141_138687 | Low-quality |
| 18-109_k141_169989 | Not-determined |
| 18-109_k141_241300 | Low-quality |
| 18-109_k141_138691 | Not-determined |
| 18-109_k141_169994 | Not-determined |
| 18-109_k141_159530 | Low-quality |
| 18-109_k141_118728 | Not-determined |
| 18-109_k141_241310 | Not-determined |
| 18-109_k141_159533 | Low-quality |
| 18-109_k141_159535 | Not-determined |
| 18-109_k141_5825 | Not-determined |
| 18-109_k141_76889 | Not-determined |
| 18-109_k141_25392 | Not-determined |
| 18-109_k141_25395 | Not-determined |
| 18-109_k141_57056 | Not-determined |
| 18-109_k141_200834 | Not-determined |
| 18-109_k141_76893 | Not-determined |
| 18-109_k141_138705 | Not-determined |
| 18-109_k141_159549 | Not-determined |
| 18-109_k141_210907 | Not-determined |
| 18-109_k141_25411 | Not-determined |
| 18-109_k141_148644 | Not-determined |
| 18-109_k141_219265 | Not-determined |
| 18-109_k141_76906 | Not-determined |
| 18-109_k141_76907 | Not-determined |
| 18-109_k141_67747 | Not-determined |
| 18-109_k141_138724 | Not-determined |
| 18-109_k141_219271 | Not-determined |
| 18-109_k141_230725 | Not-determined |
| 18-109_k141_76918 | Not-determined |
| 18-109_k141_219275 | Not-determined |
| 18-109_k141_179944 | Not-determined |
| 18-109_k141_76922 | Not-determined |
| 18-109_k141_138732 | Not-determined |
| 18-109_k141_67752 | Not-determined |
| 18-109_k141_76924 | Not-determined |
| 18-109_k141_97713 | Low-quality |
| 18-109_k141_128754 | Low-quality |
| 18-109_k141_159589 | Not-determined |
| 18-109_k141_159591 | Not-determined |
| 18-109_k141_138744 | Not-determined |
| 18-109_k141_230747 | Not-determined |
| 18-109_k141_57100 | Not-determined |
| 18-109_k141_37188 | Not-determined |
| 18-109_k141_25413 | Low-quality |
| 18-109_k141_37190 | Not-determined |
| 18-109_k141_219299 | Not-determined |
| 18-109_k141_25426 | Not-determined |
| 18-109_k141_219300 | Not-determined |
| 18-109_k141_118753 | Not-determined |
| 18-109_k141_16172 | Not-determined |
| 18-109_k141_37193 | Low-quality |
| 18-109_k141_219307 | Not-determined |
| 18-109_k141_5859 | Not-determined |
| 18-109_k141_148665 | Not-determined |
| 18-109_k141_97729 | Not-determined |
| 18-109_k141_97731 | Not-determined |
| 18-109_k141_16175 | Not-determined |
| 18-109_k141_57106 | Not-determined |
| 18-109_k141_230754 | Not-determined |
| 18-109_k141_128774 | Not-determined |
| 18-109_k141_25439 | Not-determined |
| 18-109_k141_57111 | Not-determined |
| 18-109_k141_16181 | Not-determined |
| 18-109_k141_97739 | Not-determined |
| 18-109_k141_159609 | Not-determined |
| 18-109_k141_97741 | Not-determined |
| 18-109_k141_219311 | Low-quality |
| 18-109_k141_219315 | Not-determined |
| 18-109_k141_230762 | Not-determined |
| 18-109_k141_230763 | Not-determined |
| 18-109_k141_210950 | Not-determined |
| 18-109_k141_219317 | Not-determined |
| 18-109_k141_57124 | Not-determined |
| 18-109_k141_57126 | Not-determined |
| 18-109_k141_57130 | Not-determined |
| 18-109_k141_159627 | Not-determined |
| 18-109_k141_190356 | Not-determined |
| 18-109_k141_210958 | Not-determined |
| 18-109_k141_37229 | Not-determined |
| 18-109_k141_57144 | Not-determined |
| 18-109_k141_219338 | Not-determined |
| 18-109_k141_37234 | Not-determined |
| 18-109_k141_67806 | Not-determined |
| 18-109_k141_37243 | Low-quality |
| 18-109_k141_159638 | Not-determined |
| 18-109_k141_16201 | Not-determined |
| 18-109_k141_170029 | Not-determined |
| 18-109_k141_87376 | Not-determined |
| 18-109_k141_128820 | Not-determined |
| 18-109_k141_219345 | Not-determined |
| 18-109_k141_219346 | Not-determined |
| 18-109_k141_118774 | Not-determined |
| 18-109_k141_210965 | Not-determined |
| 18-109_k141_190386 | Not-determined |
| 18-109_k141_57158 | Not-determined |
| 18-109_k141_170039 | Not-determined |
| 18-109_k141_87399 | Not-determined |
| 18-109_k141_179967 | Not-determined |
| 18-109_k141_46917 | Not-determined |
| 18-109_k141_16208 | Not-determined |
| 18-109_k141_190389 | Not-determined |
| 18-109_k141_5900 | Not-determined |
| 18-109_k141_159669 | Not-determined |
| 18-109_k141_159670 | Low-quality |
| 18-109_k141_16211 | Not-determined |
| 18-109_k141_159672 | Not-determined |
| 18-109_k141_210976 | Not-determined |
| 18-109_k141_118784 | Not-determined |
| 18-109_k141_87412 | Not-determined |
| 18-109_k141_179981 | Not-determined |
| 18-109_k141_210979 | Not-determined |
| 18-109_k141_5905 | Not-determined |
| 18-109_k141_190400 | Low-quality |
| 18-109_k141_37255 | Not-determined |
| 18-109_k141_210982 | Not-determined |
| 18-109_k141_37259 | Low-quality |
| 18-109_k141_37261 | Not-determined |
| 18-109_k141_37263 | Not-determined |
| 18-109_k141_25482 | Not-determined |
| 18-109_k141_159685 | Not-determined |
| 18-109_k141_37267 | Not-determined |
| 18-109_k141_159687 | Not-determined |
| 18-109_k141_25486 | Not-determined |
| 18-109_k141_148722 | Not-determined |
| 18-109_k141_25487 | Not-determined |
| 18-109_k141_148723 | Not-determined |
| 18-109_k141_190418 | Not-determined |
| 18-109_k141_16235 | Not-determined |
| 18-109_k141_25490 | Not-determined |
| 18-109_k141_25493 | Not-determined |
| 18-109_k141_148733 | Not-determined |
| 18-109_k141_128840 | Not-determined |
| 18-109_k141_128842 | Not-determined |
| 18-109_k141_159700 | Not-determined |
| 18-109_k141_25502 | Not-determined |
| 18-109_k141_148738 | Not-determined |
| 18-109_k141_230814 | Not-determined |
| 18-109_k141_37280 | Not-determined |
| 18-109_k141_159708 | Not-determined |
| 18-109_k141_16254 | Not-determined |
| 18-109_k141_170089 | Not-determined |
| 18-109_k141_180018 | Not-determined |
| 18-109_k141_67870 | Not-determined |
| 18-109_k141_76999 | Not-determined |
| 18-109_k141_37290 | Not-determined |
| 18-109_k141_87431 | Low-quality |
| 18-109_k141_46967 | Not-determined |
| 18-109_k141_118794 | Not-determined |
| 18-109_k141_87435 | Not-determined |
| 18-109_k141_230823 | Not-determined |
| 18-109_k141_219372 | Not-determined |
| 18-109_k141_67871 | Not-determined |
| 18-109_k141_67872 | Not-determined |
| 18-109_k141_16269 | Not-determined |
| 18-109_k141_25511 | Not-determined |
| 18-109_k141_118799 | Low-quality |
| 18-109_k141_159716 | Not-determined |
| 18-109_k141_148750 | Not-determined |
| 18-109_k141_159720 | Not-determined |
| 18-109_k141_67879 | Not-determined |
| 18-109_k141_148755 | Not-determined |
| 18-109_k141_190448 | Not-determined |
| 18-109_k141_87453 | Not-determined |
| 18-109_k141_148758 | Not-determined |
| 18-109_k141_57203 | Not-determined |
| 18-109_k141_230836 | Not-determined |
| 18-109_k141_230837 | Not-determined |
| 18-109_k141_46982 | Not-determined |
| 18-109_k141_57204 | Not-determined |
| 18-109_k141_118814 | Not-determined |
| 18-109_k141_97805 | Not-determined |
| 18-109_k141_159746 | Not-determined |
| 18-109_k141_128882 | Not-determined |
| 18-109_k141_170099 | Not-determined |
| 18-109_k141_159748 | Not-determined |
| 18-109_k141_159749 | Not-determined |
| 18-109_k141_77017 | Not-determined |
| 18-109_k141_87460 | Not-determined |
| 18-109_k141_25543 | Not-determined |
| 18-109_k141_159753 | Not-determined |
| 18-109_k141_25546 | Not-determined |
| 18-109_k141_67897 | Not-determined |
| 18-109_k141_25550 | Not-determined |
| 18-109_k141_118836 | Not-determined |
| 18-109_k141_37318 | Not-determined |
| 18-109_k141_57234 | Not-determined |
| 18-109_k141_211001 | Not-determined |
| 18-109_k141_211002 | Not-determined |
| 18-109_k141_97824 | Not-determined |
| 18-109_k141_211007 | Not-determined |
| 18-109_k141_97832 | Not-determined |
| 18-109_k141_67913 | Not-determined |
| 18-109_k141_118850 | Not-determined |
| 18-109_k141_5929 | Not-determined |
| 18-109_k141_118852 | Not-determined |
| 18-109_k141_67923 | Not-determined |
| 18-109_k141_25571 | Not-determined |
| 18-109_k141_211019 | Not-determined |
| 18-109_k141_25574 | Not-determined |
| 18-109_k141_219388 | Low-quality |
| 18-109_k141_211022 | Not-determined |
| 18-109_k141_16327 | Not-determined |
| 18-109_k141_128936 | Not-determined |
| 18-109_k141_180059 | Not-determined |
| 18-109_k141_97849 | Not-determined |
| 18-109_k141_180065 | Not-determined |
| 18-109_k141_87487 | Not-determined |
| 18-109_k141_25596 | Not-determined |
| 18-109_k141_77053 | Low-quality |
| 18-109_k141_25600 | Not-determined |
| 18-109_k141_148787 | Not-determined |
| 18-109_k141_170141 | Not-determined |
| 18-109_k141_118873 | Not-determined |
| 18-109_k141_230876 | Not-determined |
| 18-109_k141_180078 | Not-determined |
| 18-109_k141_241346 | Not-determined |
| 18-109_k141_5955 | Low-quality |
| 18-109_k141_241349 | Not-determined |
| 18-109_k141_241350 | Not-determined |
| 18-109_k141_57265 | Not-determined |
| 18-109_k141_180085 | Not-determined |
| 18-109_k141_16349 | Not-determined |
| 18-109_k141_77065 | Not-determined |
| 18-109_k141_118881 | Not-determined |
| 18-109_k141_25618 | Not-determined |
| 18-109_k141_219413 | Low-quality |
| 18-109_k141_5959 | Not-determined |
| 18-109_k141_57277 | Not-determined |
| 18-109_k141_180092 | Low-quality |
| 18-109_k141_97876 | Not-determined |
| 18-109_k141_180096 | Not-determined |
| 18-109_k141_57285 | Not-determined |
| 18-109_k141_138793 | Not-determined |
| 18-109_k141_97882 | Not-determined |
| 18-109_k141_118900 | Not-determined |
| 18-109_k141_97885 | Not-determined |
| 18-109_k141_97886 | Not-determined |
| 18-109_k141_190526 | Not-determined |
| 18-109_k141_241357 | Not-determined |
| 18-109_k141_57293 | Low-quality |
| 18-109_k141_118905 | Not-determined |
| 18-109_k141_47004 | Not-determined |
| 18-109_k141_138804 | Not-determined |
| 18-109_k141_219428 | Not-determined |
| 18-109_k141_219430 | Not-determined |
| 18-109_k141_118910 | Not-determined |
| 18-109_k141_241365 | Not-determined |
| 18-109_k141_5976 | Not-determined |
| 18-109_k141_200843 | Not-determined |
| 18-109_k141_241366 | Not-determined |
| 18-109_k141_97900 | Low-quality |
| 18-109_k141_87512 | Not-determined |
| 18-109_k141_57307 | Not-determined |
| 18-109_k141_118917 | Not-determined |
| 18-109_k141_180115 | Not-determined |
| 18-109_k141_87517 | Not-determined |
| 18-109_k141_241367 | Not-determined |
| 18-109_k141_118920 | Not-determined |
| 18-109_k141_5986 | Not-determined |
| 18-109_k141_138818 | Not-determined |
| 18-109_k141_16378 | Not-determined |
| 18-109_k141_148808 | Not-determined |
| 18-109_k141_87523 | Not-determined |
| 18-109_k141_170163 | Not-determined |
| 18-109_k141_97908 | Low-quality |
| 18-109_k141_241373 | Low-quality |
| 18-109_k141_5997 | Low-quality |
| 18-109_k141_190558 | Low-quality |
| 18-109_k141_47019 | Not-determined |
| 18-109_k141_77080 | Low-quality |
| 18-109_k141_190561 | Not-determined |
| 18-109_k141_107693 | Low-quality |
| 18-109_k141_87526 | Low-quality |
| 18-109_k141_138826 | Low-quality |
| 18-109_k141_170177 | Not-determined |
| 18-109_k141_77082 | Low-quality |
| 18-109_k141_87530 | Not-determined |
| 18-109_k141_57333 | Low-quality |
| 18-109_k141_77087 | Not-determined |
| 18-109_k141_170185 | Not-determined |
| 18-109_k141_107697 | Not-determined |
| 18-109_k141_148828 | Not-determined |
| 18-109_k141_128979 | Not-determined |
| 18-109_k141_6003 | Not-determined |
| 18-109_k141_25652 | Not-determined |
| 18-109_k141_128981 | Not-determined |
| 18-109_k141_57344 | Not-determined |
| 18-109_k141_97927 | Not-determined |
| 18-109_k141_128982 | Not-determined |
| 18-109_k141_230921 | Low-quality |
| 18-109_k141_97929 | Not-determined |
| 18-109_k141_138836 | Not-determined |
| 18-109_k141_57351 | Not-determined |
| 18-109_k141_6013 | Not-determined |
| 18-109_k141_241402 | Not-determined |
| 18-109_k141_25665 | Low-quality |
| 18-109_k141_6015 | Not-determined |
| 18-109_k141_148848 | Not-determined |
| 18-109_k141_170197 | Not-determined |
| 18-109_k141_138850 | Not-determined |
| 18-109_k141_190582 | Not-determined |
| 18-109_k141_211054 | Not-determined |
| 18-109_k141_170200 | Not-determined |
| 18-109_k141_211056 | Not-determined |
| 18-109_k141_148860 | Not-determined |
| 18-109_k141_200875 | Not-determined |
| 18-109_k141_230926 | Not-determined |
| 18-109_k141_241414 | Low-quality |
| 18-109_k141_77108 | Not-determined |
| 18-109_k141_37338 | Not-determined |
| 18-109_k141_6026 | Not-determined |
| 18-109_k141_200888 | Not-determined |
| 18-109_k141_211064 | Not-determined |
| 18-109_k141_25690 | Low-quality |
| 18-109_k141_25693 | Not-determined |
| 18-109_k141_107742 | Not-determined |
| 18-109_k141_230930 | Not-determined |
| 18-109_k141_47029 | Low-quality |
| 18-109_k141_200898 | Not-determined |
| 18-109_k141_77119 | Not-determined |
| 18-109_k141_138863 | Low-quality |
| 18-109_k141_138865 | Not-determined |
| 18-109_k141_118954 | Not-determined |
| 18-109_k141_87548 | Not-determined |
| 18-109_k141_230935 | Not-determined |
| 18-109_k141_118961 | Not-determined |
| 18-109_k141_230938 | Not-determined |
| 18-109_k141_87552 | Not-determined |
| 18-109_k141_170222 | Not-determined |
| 18-109_k141_170225 | Not-determined |
| 18-109_k141_57380 | Low-quality |
| 18-109_k141_47047 | Not-determined |
| 18-109_k141_148884 | Not-determined |
| 18-109_k141_87557 | Not-determined |
| 18-109_k141_57382 | Not-determined |
| 18-109_k141_230945 | Not-determined |
| 18-109_k141_67956 | Not-determined |
| 18-109_k141_230946 | Not-determined |
| 18-109_k141_47053 | Not-determined |
| 18-109_k141_230947 | Not-determined |
| 18-109_k141_129011 | Not-determined |
| 18-109_k141_170231 | Not-determined |
| 18-109_k141_47059 | Low-quality |
| 18-109_k141_170235 | Not-determined |
| 18-109_k141_129021 | Not-determined |
| 18-109_k141_190627 | Not-determined |
| 18-109_k141_170236 | Not-determined |
| 18-109_k141_148903 | Not-determined |
| 18-109_k141_87571 | Not-determined |
| 18-109_k141_190630 | Not-determined |
| 18-109_k141_190631 | Not-determined |
| 18-109_k141_190632 | Not-determined |
| 18-109_k141_190633 | Not-determined |
| 18-109_k141_241440 | Not-determined |
| 18-109_k141_170249 | Low-quality |
| 18-109_k141_57401 | Not-determined |
| 18-109_k141_67971 | Not-determined |
| 18-109_k141_211108 | Not-determined |
| 18-109_k141_67975 | Not-determined |
| 18-109_k141_148916 | Not-determined |
| 18-109_k141_25720 | Not-determined |
| 18-109_k141_25722 | Not-determined |
| 18-109_k141_67982 | Not-determined |
| 18-109_k141_211119 | Not-determined |
| 18-109_k141_67985 | Not-determined |
| 18-109_k141_170258 | Not-determined |
| 18-109_k141_129045 | Low-quality |
| 18-109_k141_37359 | Not-determined |
| 18-109_k141_170260 | Not-determined |
| 18-109_k141_25736 | Not-determined |
| 18-109_k141_138903 | Not-determined |
| 18-109_k141_97967 | Not-determined |
| 18-109_k141_119006 | Not-determined |
| 18-109_k141_37368 | Not-determined |
| 18-109_k141_119008 | Not-determined |
| 18-109_k141_148927 | Not-determined |
| 18-109_k141_119010 | Not-determined |
| 18-109_k141_16429 | Not-determined |
| 18-109_k141_57407 | Low-quality |
| 18-109_k141_6037 | Not-determined |
| 18-109_k141_211127 | Not-determined |
| 18-109_k141_6040 | Not-determined |
| 18-109_k141_87588 | Not-determined |
| 18-109_k141_6042 | Low-quality |
| 18-109_k141_211130 | Not-determined |
| 18-109_k141_119022 | Not-determined |
| 18-109_k141_190653 | Not-determined |
| 18-109_k141_148944 | Not-determined |
| 18-109_k141_119023 | Not-determined |
| 18-109_k141_47067 | Not-determined |
| 18-109_k141_97989 | Not-determined |
| 18-109_k141_68009 | Not-determined |
| 18-109_k141_211133 | Not-determined |
| 18-109_k141_87593 | Not-determined |
| 18-109_k141_77149 | Not-determined |
| 18-109_k141_87595 | Low-quality |
| 18-109_k141_119034 | Not-determined |
| 18-109_k141_77150 | Not-determined |
| 18-109_k141_211139 | Not-determined |
| 18-109_k141_77152 | Not-determined |
| 18-109_k141_119040 | Not-determined |
| 18-109_k141_25775 | Not-determined |
| 18-109_k141_119042 | Not-determined |
| 18-109_k141_119043 | Not-determined |
| 18-109_k141_190660 | Not-determined |
| 18-109_k141_170285 | Not-determined |
| 18-109_k141_6062 | Not-determined |
| 18-109_k141_211145 | Not-determined |
| 18-109_k141_119053 | Not-determined |
| 18-109_k141_119054 | Not-determined |
| 18-109_k141_87610 | Not-determined |
| 18-109_k141_211150 | Low-quality |
| 18-109_k141_98022 | Not-determined |
| 18-109_k141_190685 | Not-determined |
| 18-109_k141_6070 | Low-quality |
| 18-109_k141_47112 | Not-determined |
| 18-109_k141_87619 | Not-determined |
| 18-109_k141_16459 | Not-determined |
| 18-109_k141_129063 | Not-determined |
| 18-109_k141_47124 | Not-determined |
| KM-final-viral-combined_k141_71626\|\|full | Medium-quality |
| 18-109_k141_190692 | Not-determined |
| 18-109_k141_129069 | Not-determined |
| 18-109_k141_77190 | Not-determined |
| 18-109_k141_190697 | Not-determined |
| 18-109_k141_170302 | Not-determined |
| 18-109_k141_77199 | Not-determined |
| 18-109_k141_57417 | Not-determined |
| 18-109_k141_16474 | Not-determined |
| 18-109_k141_190711 | Not-determined |
| 18-109_k141_57428 | Not-determined |
| 18-109_k141_138939 | Not-determined |
| 18-109_k141_57429 | Not-determined |
| 18-109_k141_25806 | Low-quality |
| 18-109_k141_16484 | Not-determined |
| 18-109_k141_25808 | Not-determined |
| 18-109_k141_230982 | Not-determined |
| 18-109_k141_230984 | Not-determined |
| 18-109_k141_230985 | Not-determined |
| 18-109_k141_77213 | Not-determined |
| 18-109_k141_77214 | Not-determined |
| 18-109_k141_190722 | Not-determined |
| 18-109_k141_190724 | Not-determined |
| 18-109_k141_6102 | Not-determined |
| 18-109_k141_230990 | Not-determined |
| 18-109_k141_68038 | Not-determined |
| 18-109_k141_57432 | Not-determined |
| 18-109_k141_200964 | Not-determined |
| 18-109_k141_200967 | Not-determined |
| 18-109_k141_200971 | Low-quality |
| 18-109_k141_200972 | Not-determined |
| 18-109_k141_138959 | Not-determined |
| 18-109_k141_190733 | Low-quality |
| 18-109_k141_57441 | Not-determined |
| 18-109_k141_138968 | Not-determined |
| 18-109_k141_190741 | Not-determined |
| 18-109_k141_231010 | Not-determined |
| 18-109_k141_200981 | Not-determined |
| 18-109_k141_231015 | Not-determined |
| 18-109_k141_98050 | Not-determined |
| 18-109_k141_98051 | Not-determined |
| 18-109_k141_6114 | Not-determined |
| 18-109_k141_190754 | Low-quality |
| 18-109_k141_16501 | Not-determined |
| 18-109_k141_200986 | Not-determined |
| 18-109_k141_138991 | Low-quality |
| 18-109_k141_25853 | Not-determined |
| 18-109_k141_57469 | Not-determined |
| 18-109_k141_190761 | Not-determined |
| 18-109_k141_241454 | Low-quality |
| 18-109_k141_77233 | Not-determined |
| 18-109_k141_98069 | Not-determined |
| 18-109_k141_68064 | Not-determined |
| 18-109_k141_47183 | Not-determined |
| 18-109_k141_25864 | Not-determined |
| 18-109_k141_25869 | Not-determined |
| 18-109_k141_16520 | Not-determined |
| 18-109_k141_231033 | Not-determined |
| 18-109_k141_25878 | Not-determined |
| 18-109_k141_68078 | Not-determined |
| 18-109_k141_129110 | Not-determined |
| 18-109_k141_37429 | Not-determined |
| 18-109_k141_57478 | Not-determined |
| 18-109_k141_98081 | Low-quality |
| 18-109_k141_159773 | Not-determined |
| 18-109_k141_77253 | Low-quality |
| 18-109_k141_68090 | Not-determined |
| 18-109_k141_129117 | Not-determined |
| 18-109_k141_219489 | Not-determined |
| 18-109_k141_190775 | Not-determined |
| 18-109_k141_25885 | Not-determined |
| 18-109_k141_37440 | Not-determined |
| 18-109_k141_129120 | Not-determined |
| 18-109_k141_139028 | Not-determined |
| 18-109_k141_219494 | Not-determined |
| 18-109_k141_231083 | Not-determined |
| 18-109_k141_129122 | Not-determined |
| 18-109_k141_57482 | Not-determined |
| 18-109_k141_98099 | Not-determined |
| 18-109_k141_139044 | Not-determined |
| 18-109_k141_201054 | Not-determined |
| 18-109_k141_77281 | Not-determined |
| 18-109_k141_201058 | Not-determined |
| 18-109_k141_68108 | Not-determined |
| 18-109_k141_68110 | Not-determined |
| 18-109_k141_77283 | Not-determined |
| 18-109_k141_139049 | Not-determined |
| 18-109_k141_201067 | Not-determined |
| 18-109_k141_159789 | Not-determined |
| 18-109_k141_159790 | Not-determined |
| 18-109_k141_107745 | Not-determined |
| 18-109_k141_6168 | Not-determined |
| 18-109_k141_180150 | Not-determined |
| 18-109_k141_6169 | Not-determined |
| 18-109_k141_107747 | Not-determined |
| 18-109_k141_129149 | Not-determined |
| 18-109_k141_170313 | Not-determined |

| | |
|---|---|
| 18-109_k141_170320 | Not-determined |
| 18-109_k141_107760 | Not-determined |
| 18-109_k141_107761 | Not-determined |
| 18-109_k141_119114 | Not-determined |
| 18-109_k141_6182 | Not-determined |
| 18-109_k141_159808 | Not-determined |
| 18-109_k141_159809 | Not-determined |
| 18-109_k141_77295 | Not-determined |
| 18-109_k141_98132 | Not-determined |
| 18-109_k141_211165 | Not-determined |
| 18-109_k141_219524 | Not-determined |
| 18-109_k141_219526 | Low-quality |
| 18-109_k141_180177 | Not-determined |
| 18-109_k141_211169 | Not-determined |
| 18-109_k141_68131 | Not-determined |
| 18-109_k141_241507 | Low-quality |
| 18-109_k141_77300 | Not-determined |
| 18-109_k141_77304 | Not-determined |
| 18-109_k141_201096 | Not-determined |
| 18-109_k141_201098 | Not-determined |
| 18-109_k141_170333 | Not-determined |
| 18-109_k141_201100 | Not-determined |
| 18-109_k141_25911 | Not-determined |
| 18-109_k141_57510 | Not-determined |
| 18-109_k141_68152 | Not-determined |
| 18-109_k141_139094 | Not-determined |
| 18-109_k141_201109 | Not-determined |
| 18-109_k141_231128 | Not-determined |
| 18-109_k141_16522 | Low-quality |
| 18-109_k141_170352 | Not-determined |
| 18-109_k141_119123 | Not-determined |
| 18-109_k141_119129 | Low-quality |
| 18-109_k141_201120 | Not-determined |
| 18-109_k141_98170 | Not-determined |
| 18-109_k141_57523 | Not-determined |
| 18-109_k141_201126 | Not-determined |
| 18-109_k141_219549 | Not-determined |
| 18-109_k141_98176 | Not-determined |
| 18-109_k141_241525 | Not-determined |
| 18-109_k141_98177 | Not-determined |
| 18-109_k141_201130 | Not-determined |
| 18-109_k141_219554 | Not-determined |
| 18-109_k141_77336 | Not-determined |
| 18-109_k141_139120 | Not-determined |
| 18-109_k141_16539 | Not-determined |
| 18-109_k141_77343 | Not-determined |
| 18-109_k141_16544 | Not-determined |
| 18-109_k141_107796 | Not-determined |
| 18-109_k141_241538 | Not-determined |
| 18-109_k141_68172 | Not-determined |
| 18-109_k141_25929 | Not-determined |
| 18-109_k141_149029 | Not-determined |
| 18-109_k141_68176 | Not-determined |
| 18-109_k141_159826 | Not-determined |
| 18-109_k141_77351 | Not-determined |
| 18-109_k141_68180 | Not-determined |
| 18-109_k141_231172 | Not-determined |
| 18-109_k141_159829 | Not-determined |
| 18-109_k141_201146 | Not-determined |
| 18-109_k141_149036 | Not-determined |
| 18-109_k141_87668 | Not-determined |
| 18-109_k141_241559 | Not-determined |
| 18-109_k141_149044 | Not-determined |
| 18-109_k141_25944 | Not-determined |
| 18-109_k141_87676 | Not-determined |
| 18-109_k141_87677 | Low-quality |
| 18-109_k141_159840 | Low-quality |
| 18-109_k141_87682 | Not-determined |
| 18-109_k141_149047 | Not-determined |
| 18-109_k141_87684 | Not-determined |
| 18-109_k141_201160 | Not-determined |
| 18-109_k141_47216 | Not-determined |
| 18-109_k141_119156 | Not-determined |
| 18-109_k141_231198 | Not-determined |
| 18-109_k141_37489 | Not-determined |
| 18-109_k141_25951 | Not-determined |
| 18-109_k141_87694 | Not-determined |
| 18-109_k141_37493 | Low-quality |
| 18-109_k141_6241 | Low-quality |
| 18-109_k141_6242 | Not-determined |
| 18-109_k141_57558 | Not-determined |
| 18-109_k141_201176 | Not-determined |
| 18-109_k141_37501 | Not-determined |
| 18-109_k141_241578 | Not-determined |
| 18-109_k141_241579 | Not-determined |
| 18-109_k141_57565 | Not-determined |
| 18-109_k141_16575 | Not-determined |
| 18-109_k141_139155 | Not-determined |
| 18-109_k141_98195 | Low-quality |
| 18-109_k141_211211 | Not-determined |
| 18-109_k141_37506 | Not-determined |
| 18-109_k141_139157 | Not-determined |
| 18-109_k141_77392 | Not-determined |
| 18-109_k141_77393 | Not-determined |
| 18-109_k141_37509 | Not-determined |
| 18-109_k141_47237 | Not-determined |
| 18-109_k141_16587 | Not-determined |
| 18-109_k141_170362 | Not-determined |
| 18-109_k141_159883 | Not-determined |
| 18-109_k141_47247 | Low-quality |
| 18-109_k141_16596 | Not-determined |
| 18-109_k141_87719 | Not-determined |
| 18-109_k141_87724 | Not-determined |
| 18-109_k141_98212 | Not-determined |
| 18-109_k141_201204 | Not-determined |
| 18-109_k141_98213 | Not-determined |
| 18-109_k141_211224 | Not-determined |
| 18-109_k141_47257 | Not-determined |
| 18-109_k141_68224 | Not-determined |
| 18-109_k141_201206 | Not-determined |
| 18-109_k141_159895 | Not-determined |
| 18-109_k141_25961 | Not-determined |
| 18-109_k141_170377 | Not-determined |
| 18-109_k141_159898 | Low-quality |
| 18-109_k141_139179 | Not-determined |
| 18-109_k141_98222 | Not-determined |
| 18-109_k141_170386 | Not-determined |
| 18-109_k141_47264 | Low-quality |
| 18-109_k141_68246 | Not-determined |
| 18-109_k141_47271 | Not-determined |
| 18-109_k141_68248 | Low-quality |
| 18-109_k141_16615 | Not-determined |
| 18-109_k141_139201 | Not-determined |
| 18-109_k141_139202 | Not-determined |
| 18-109_k141_159929 | Not-determined |
| 18-109_k141_68258 | Not-determined |
| 18-109_k141_98241 | Not-determined |
| 18-109_k141_211263 | Low-quality |
| 18-109_k141_68260 | Not-determined |
| 18-109_k141_68265 | Not-determined |
| 18-109_k141_68266 | Not-determined |
| 18-109_k141_57611 | Not-determined |
| 18-109_k141_190852 | Not-determined |
| 18-109_k141_139216 | Not-determined |
| 18-109_k141_190856 | Low-quality |
| 18-109_k141_37546 | Not-determined |
| 18-109_k141_47291 | Not-determined |
| 18-109_k141_190863 | Not-determined |
| 18-109_k141_149084 | Not-determined |
| 18-109_k141_98257 | Low-quality |
| 18-109_k141_47298 | Not-determined |
| 18-109_k141_47299 | Not-determined |
| 18-109_k141_6328 | Not-determined |
| 18-109_k141_98264 | Not-determined |
| 18-109_k141_170428 | Not-determined |
| 18-109_k141_98267 | Not-determined |
| 18-109_k141_47302 | Not-determined |
| 18-109_k141_211289 | Not-determined |
| 18-109_k141_6331 | Low-quality |
| 18-109_k141_37570 | Not-determined |
| 18-109_k141_87768 | Not-determined |
| 18-109_k141_87770 | Not-determined |
| 18-109_k141_87773 | Not-determined |
| 18-109_k141_129177 | Not-determined |
| 18-109_k141_190886 | Not-determined |
| 18-109_k141_129179 | Not-determined |
| 18-109_k141_47317 | Not-determined |
| 18-109_k141_6348 | Not-determined |
| 18-109_k141_87782 | Not-determined |
| 18-109_k141_98286 | Not-determined |
| 18-109_k141_37577 | Not-determined |
| 18-109_k141_37581 | Low-quality |
| 18-109_k141_190897 | Low-quality |
| 18-109_k141_119196 | Low-quality |
| 18-109_k141_119200 | Low-quality |
| 18-109_k141_129197 | Not-determined |
| 18-109_k141_6359 | Not-determined |
| 18-109_k141_16669 | Not-determined |
| 18-109_k141_119214 | Not-determined |
| 18-109_k141_87798 | Low-quality |
| 18-109_k141_98303 | Low-quality |
| 18-109_k141_57643 | Low-quality |
| 18-109_k141_68321 | Not-determined |
| 18-109_k141_119221 | Not-determined |
| 18-109_k141_119222 | Not-determined |
| 18-109_k141_201297 | Not-determined |
| 18-109_k141_77465 | Low-quality |
| 18-109_k141_6362 | Not-determined |
| 18-109_k141_170449 | Not-determined |
| 18-109_k141_190919 | Not-determined |
| 18-109_k141_219592 | Not-determined |
| 18-109_k141_37614 | Not-determined |
| 18-109_k141_26029 | Not-determined |
| 18-109_k141_149124 | Not-determined |
| 18-109_k141_47361 | Not-determined |
| 18-109_k141_16687 | Not-determined |
| 18-109_k141_26039 | Not-determined |
| 18-109_k141_149130 | Not-determined |
| 18-109_k141_149131 | Not-determined |
| 18-109_k141_219602 | Low-quality |
| 18-109_k141_159981 | Not-determined |
| 18-109_k141_219604 | Not-determined |
| 18-109_k141_180231 | Not-determined |
| 18-109_k141_57661 | Not-determined |
| 18-109_k141_180236 | Not-determined |
| 18-109_k141_219613 | Not-determined |
| 18-109_k141_6385 | Not-determined |
| 18-109_k141_98313 | Not-determined |
| 18-109_k141_170465 | Not-determined |
| 18-109_k141_37638 | Not-determined |
| 18-109_k141_129224 | Not-determined |
| 18-109_k141_149140 | Not-determined |
| 18-109_k141_37642 | Not-determined |
| 18-109_k141_77503 | Not-determined |
| 18-109_k141_47369 | Not-determined |
| 18-109_k141_26062 | Low-quality |
| 18-109_k141_129240 | Not-determined |
| 18-109_k141_87846 | Not-determined |
| 18-109_k141_6398 | Not-determined |
| 18-109_k141_129243 | Not-determined |
| 18-109_k141_87853 | Not-determined |
| 18-109_k141_129244 | Not-determined |
| 18-109_k141_170497 | Not-determined |
| 18-109_k141_190944 | Not-determined |
| 18-109_k141_119268 | Not-determined |
| 18-109_k141_87858 | Not-determined |
| 18-109_k141_149166 | Not-determined |
| 18-109_k141_47383 | Not-determined |
| 18-109_k141_241597 | Not-determined |
| 18-109_k141_37659 | Not-determined |
| 18-109_k141_129249 | Low-quality |
| 18-109_k141_180257 | Not-determined |
| 18-109_k141_160013 | Not-determined |
| 18-109_k141_37661 | Not-determined |
| 18-109_k141_119275 | Not-determined |
| 18-109_k141_87866 | Not-determined |
| 18-109_k141_16691 | Not-determined |
| 18-109_k141_170507 | Not-determined |
| 18-109_k141_26082 | Not-determined |
| 18-109_k141_129256 | Not-determined |
| 18-109_k141_241609 | Not-determined |
| 18-109_k141_87869 | Not-determined |
| 18-109_k141_149177 | Not-determined |
| 18-109_k141_149179 | Not-determined |
| 18-109_k141_219637 | Not-determined |
| 18-109_k141_180262 | Not-determined |
| 18-109_k141_219641 | Not-determined |
| 18-109_k141_180265 | Not-determined |
| 18-109_k141_219642 | Not-determined |
| 18-109_k141_6414 | Not-determined |
| 18-109_k141_37668 | Not-determined |
| 18-109_k141_47398 | Low-quality |
| 18-109_k141_119294 | Not-determined |
| 18-109_k141_241618 | Not-determined |
| 18-109_k141_6418 | Not-determined |
| 18-109_k141_6419 | Not-determined |
| 18-109_k141_37671 | Not-determined |
| 18-109_k141_219648 | Not-determined |
| 18-109_k141_87879 | Not-determined |
| 18-109_k141_37672 | Not-determined |
| 18-109_k141_201357 | Not-determined |
| 18-109_k141_129262 | Low-quality |
| 18-109_k141_170528 | Low-quality |
| 18-109_k141_16696 | Not-determined |
| 18-109_k141_16697 | Not-determined |
| 18-109_k141_180283 | Not-determined |
| 18-109_k141_68384 | Not-determined |
| 18-109_k141_16703 | Not-determined |
| 18-109_k141_170533 | Low-quality |
| 18-109_k141_180286 | Low-quality |
| 18-109_k141_241634 | Not-determined |
| 18-109_k141_68397 | Not-determined |
| 18-109_k141_37686 | Not-determined |
| 18-109_k141_190965 | Not-determined |
| 18-109_k141_201365 | Not-determined |
| 18-109_k141_170539 | Not-determined |
| 18-109_k141_98355 | Not-determined |
| 18-109_k141_107825 | Not-determined |
| 18-109_k141_37696 | Not-determined |
| 18-109_k141_68411 | Not-determined |
| 18-109_k141_170550 | Not-determined |
| 18-109_k141_201376 | Not-determined |
| 18-109_k141_160029 | Not-determined |
| 18-109_k141_190991 | Not-determined |
| 18-109_k141_16717 | Not-determined |
| 18-109_k141_16722 | Not-determined |
| 18-109_k141_241640 | Not-determined |
| 18-109_k141_57704 | Not-determined |
| 18-109_k141_190999 | Not-determined |
| 18-109_k141_68424 | Not-determined |
| 18-109_k141_107841 | Low-quality |
| 18-109_k141_160039 | Not-determined |
| 18-109_k141_170554 | Low-quality |
| 18-109_k141_191005 | Not-determined |
| 18-109_k141_107846 | Low-quality |
| 18-109_k141_107847 | Not-determined |
| 18-109_k141_160046 | Not-determined |
| 18-109_k141_180326 | Not-determined |
| 18-109_k141_191021 | Not-determined |
| 18-109_k141_57720 | Not-determined |
| 18-109_k141_68442 | Not-determined |
| 18-109_k141_201383 | Not-determined |
| 18-109_k141_201384 | Not-determined |
| 18-109_k141_57725 | Not-determined |
| 18-109_k141_160056 | Not-determined |
| 18-109_k141_180337 | Not-determined |
| 18-109_k141_241654 | Low-quality |
| 18-109_k141_107864 | Not-determined |
| 18-109_k141_231233 | Not-determined |
| 18-109_k141_231234 | Not-determined |
| 18-109_k141_160074 | Not-determined |
| 18-109_k141_68462 | Not-determined |
| 18-109_k141_6469 | Not-determined |
| 18-109_k141_180348 | Not-determined |
| 18-109_k141_160086 | Not-determined |
| 18-109_k141_98411 | Not-determined |
| 18-109_k141_47459 | Not-determined |
| 18-109_k141_57757 | Not-determined |
| 18-109_k141_211309 | Low-quality |
| 18-109_k141_211315 | Not-determined |
| 18-109_k141_37734 | Not-determined |
| 18-109_k141_191065 | Not-determined |
| 18-109_k141_87905 | Not-determined |
| 18-109_k141_98417 | Not-determined |
| 18-109_k141_87906 | Not-determined |
| 18-109_k141_98418 | Not-determined |
| 18-109_k141_47472 | Not-determined |
| 18-109_k141_139249 | Low-quality |
| 18-109_k141_107871 | Not-determined |
| 18-109_k141_47474 | Not-determined |
| 18-109_k141_107873 | Not-determined |
| 18-109_k141_26122 | Not-determined |
| 18-109_k141_107877 | Not-determined |
| 18-109_k141_211327 | Not-determined |
| 18-109_k141_211329 | Not-determined |
| 18-109_k141_180352 | Not-determined |
| 18-109_k141_6499 | Low-quality |
| 18-109_k141_180355 | Not-determined |
| 18-109_k141_119320 | Not-determined |
| 18-109_k141_47485 | Not-determined |
| 18-109_k141_47488 | Not-determined |
| 18-109_k141_107886 | Not-determined |
| 18-109_k141_211335 | Not-determined |
| 18-109_k141_191080 | Not-determined |
| 18-109_k141_47495 | Not-determined |
| 18-109_k141_77589 | Not-determined |
| 18-109_k141_26136 | Not-determined |
| 18-109_k141_107892 | Not-determined |
| 18-109_k141_170575 | Not-determined |
| 18-109_k141_211341 | Not-determined |
| 18-109_k141_107896 | Not-determined |
| 18-109_k141_26138 | Not-determined |
| 18-109_k141_98434 | Not-determined |
| 18-109_k141_107902 | Not-determined |
| 18-109_k141_160109 | Low-quality |
| 18-109_k141_191084 | Not-determined |
| 18-109_k141_87934 | Low-quality |
| 18-109_k141_57769 | Not-determined |
| 18-109_k141_129301 | Not-determined |
| 18-109_k141_77608 | Not-determined |
| 18-109_k141_47503 | Not-determined |
| 18-109_k141_47505 | Low-quality |
| 18-109_k141_129306 | Not-determined |
| 18-109_k141_231277 | Not-determined |
| 18-109_k141_87939 | Not-determined |
| 18-109_k141_119347 | Not-determined |
| 18-109_k141_219679 | Not-determined |
| 18-109_k141_47515 | Not-determined |
| 18-109_k141_231283 | Not-determined |
| 18-109_k141_191094 | Not-determined |
| 18-109_k141_57785 | Not-determined |
| 18-109_k141_160128 | Not-determined |
| 18-109_k141_211343 | Not-determined |
| 18-109_k141_68491 | Not-determined |
| 18-109_k141_47522 | Low-quality |
| 18-109_k141_26152 | Not-determined |
| 18-109_k141_191103 | Not-determined |
| 18-109_k141_180391 | Not-determined |
| 18-109_k141_201417 | Low-quality |
| 18-109_k141_26161 | Not-determined |
| 18-109_k141_26162 | Not-determined |
| 18-109_k141_6521 | Not-determined |
| 18-109_k141_231305 | Not-determined |
| 18-109_k141_201421 | Not-determined |
| 18-109_k141_98448 | Low-quality |
| 18-109_k141_16747 | Not-determined |
| 18-109_k141_129321 | Not-determined |
| 18-109_k141_47532 | Not-determined |
| 18-109_k141_201424 | Not-determined |
| 18-109_k141_87968 | Not-determined |
| 18-109_k141_98452 | Not-determined |
| 18-109_k141_87970 | Not-determined |
| 18-109_k141_6526 | Not-determined |
| 18-109_k141_26177 | Not-determined |
| 18-109_k141_160150 | Not-determined |
| 18-109_k141_160152 | Not-determined |
| 18-109_k141_87983 | Not-determined |
| 18-109_k141_47542 | Not-determined |
| 18-109_k141_6536 | Not-determined |
| 18-109_k141_211377 | Not-determined |
| 18-109_k141_57800 | Not-determined |
| 18-109_k141_57804 | Not-determined |
| 18-109_k141_129332 | Not-determined |
| 18-109_k141_241697 | Not-determined |
| 18-109_k141_57809 | Not-determined |
| 18-109_k141_231312 | Not-determined |
| 18-109_k141_211381 | Not-determined |
| 18-109_k141_191121 | Not-determined |
| 18-109_k141_6548 | Not-determined |
| 18-109_k141_68513 | Not-determined |
| 18-109_k141_139289 | Not-determined |
| 18-109_k141_231321 | Low-quality |
| 18-109_k141_47557 | Not-determined |
| 18-109_k141_26205 | Not-determined |
| 18-109_k141_231325 | Not-determined |
| 18-109_k141_129364 | Not-determined |
| 18-109_k141_241717 | Not-determined |
| 18-109_k141_170602 | Not-determined |
| 18-109_k141_68524 | Not-determined |
| 18-109_k141_191128 | Not-determined |
| 18-109_k141_16772 | Not-determined |
| 18-109_k141_47563 | Not-determined |
| 18-109_k141_68530 | Not-determined |
| 18-109_k141_129371 | Not-determined |
| 18-109_k141_68536 | Not-determined |
| 18-109_k141_88010 | Not-determined |
| 18-109_k141_37786 | Not-determined |
| 18-109_k141_129377 | Not-determined |
| 18-109_k141_37787 | Not-determined |
| 18-109_k141_68544 | Not-determined |
| 18-109_k141_231350 | Low-quality |
| 18-109_k141_129385 | Not-determined |
| 18-109_k141_88025 | Not-determined |
| 18-109_k141_88026 | Not-determined |
| 18-109_k141_160189 | Not-determined |
| 18-109_k141_211413 | Not-determined |
| 18-109_k141_57832 | Not-determined |
| 18-109_k141_98476 | Not-determined |
| 18-109_k141_160201 | Not-determined |
| 18-109_k141_68563 | Not-determined |
| 18-109_k141_68565 | Not-determined |
| 18-109_k141_180418 | Not-determined |
| 18-109_k141_241739 | Not-determined |
| 18-109_k141_180419 | Low-quality |
| 18-109_k141_160207 | Not-determined |
| 18-109_k141_16786 | Not-determined |
| 18-109_k141_88049 | Not-determined |
| 18-109_k141_98483 | Not-determined |
| 18-109_k141_88052 | Not-determined |
| 18-109_k141_68567 | Not-determined |
| 18-109_k141_98485 | Not-determined |
| 18-109_k141_180429 | Not-determined |
| 18-109_k141_241747 | Not-determined |
| 18-109_k141_129403 | Not-determined |
| 18-109_k141_108000 | Low-quality |
| 18-109_k141_129406 | Not-determined |
| 18-109_k141_57841 | Not-determined |
| 18-109_k141_241750 | Low-quality |
| 18-109_k141_231376 | Not-determined |
| 18-109_k141_98497 | Not-determined |
| 18-109_k141_68578 | Not-determined |
| 18-109_k141_57850 | Not-determined |
| 18-109_k141_231377 | Not-determined |
| 18-109_k141_241759 | Not-determined |
| 18-109_k141_211424 | Not-determined |
| 18-109_k141_98502 | Not-determined |
| 18-109_k141_108007 | Not-determined |
| 18-109_k141_6597 | Not-determined |
| 18-109_k141_68583 | Not-determined |
| 18-109_k141_68585 | Not-determined |
| 18-109_k141_47609 | Not-determined |
| 18-109_k141_6598 | Low-quality |
| 18-109_k141_68588 | Not-determined |
| 18-109_k141_6600 | Not-determined |
| 18-109_k141_191167 | Not-determined |
| 18-109_k141_16795 | Not-determined |
| 18-109_k141_16797 | Not-determined |
| 18-109_k141_98518 | Not-determined |
| 18-109_k141_149200 | Not-determined |
| 18-109_k141_16813 | Not-determined |
| 18-109_k141_160245 | Low-quality |
| 18-109_k141_47634 | Not-determined |
| 18-109_k141_98536 | Not-determined |
| 18-109_k141_160251 | Not-determined |
| 18-109_k141_160252 | Not-determined |
| 18-109_k141_16822 | Low-quality |
| 18-109_k141_129436 | Low-quality |
| 18-109_k141_77611 | Not-determined |
| 18-109_k141_16825 | Not-determined |
| 18-109_k141_77612 | Not-determined |
| 18-109_k141_191190 | Not-determined |
| 18-109_k141_180444 | Low-quality |
| 18-109_k141_16829 | Not-determined |
| 18-109_k141_129446 | Not-determined |
| 18-109_k141_129448 | Not-determined |
| 18-109_k141_191195 | Not-determined |
| 18-109_k141_129449 | Not-determined |
| 18-109_k141_231402 | Low-quality |
| 18-109_k141_108011 | Low-quality |
| 18-109_k141_201511 | Not-determined |
| 18-109_k141_108019 | Low-quality |
| 18-109_k141_108024 | Low-quality |
| 18-109_k141_108026 | Not-determined |
| 18-109_k141_191208 | Not-determined |
| 18-109_k141_160260 | Low-quality |
| 18-109_k141_160261 | Not-determined |
| 18-109_k141_160262 | Not-determined |
| 18-109_k141_6639 | Not-determined |
| 18-109_k141_57889 | Low-quality |
| 18-109_k141_149239 | Not-determined |
| 18-109_k141_149240 | Low-quality |
| 18-109_k141_57891 | Not-determined |
| 18-109_k141_191217 | Not-determined |
| 18-109_k141_16855 | Not-determined |
| 18-109_k141_77647 | Not-determined |
| 18-109_k141_129476 | Not-determined |
| 18-109_k141_57898 | Not-determined |
| 18-109_k141_129481 | Not-determined |
| 18-109_k141_191229 | Low-quality |
| 18-109_k141_16862 | Not-determined |
| 18-109_k141_88081 | Not-determined |
| 18-109_k141_57905 | Not-determined |
| 18-109_k141_57907 | Not-determined |
| 18-109_k141_129485 | Not-determined |
| 18-109_k141_241822 | Not-determined |
| 18-109_k141_108072 | Not-determined |
| 18-109_k141_37850 | Not-determined |
| 18-109_k141_241834 | Not-determined |
| 18-109_k141_129489 | Not-determined |
| 18-109_k141_108077 | Low-quality |
| 18-109_k141_108084 | Not-determined |
| 18-109_k141_231438 | Not-determined |
| 18-109_k141_6677 | Not-determined |
| 18-109_k141_98549 | Not-determined |
| 18-109_k141_108094 | Not-determined |
| 18-109_k141_6680 | Low-quality |
| 18-109_k141_16890 | Not-determined |
| 18-109_k141_37865 | Not-determined |
| 18-109_k141_88096 | Not-determined |
| 18-109_k141_16895 | Not-determined |
| 18-109_k141_6690 | Not-determined |
| 18-109_k141_108114 | Not-determined |
| 18-109_k141_108116 | Not-determined |
| 18-109_k141_77682 | Not-determined |
| 18-109_k141_77684 | Low-quality |
| 18-109_k141_77685 | Not-determined |
| 18-109_k141_88111 | Not-determined |
| 18-109_k141_108132 | Not-determined |
| 18-109_k141_88113 | Not-determined |
| 18-109_k141_88117 | Not-determined |
| 18-109_k141_47736 | Not-determined |
| 18-109_k141_180496 | Not-determined |
| 18-109_k141_47740 | Not-determined |
| 18-109_k141_191246 | Not-determined |
| 18-109_k141_98571 | Not-determined |
| 18-109_k141_211462 | Not-determined |
| 18-109_k141_57930 | Not-determined |
| 18-109_k141_219765 | Not-determined |
| 18-109_k141_108139 | Low-quality |
| 18-109_k141_47752 | Not-determined |
| 18-109_k141_119362 | Not-determined |
| 18-109_k141_108142 | Not-determined |
| 18-109_k141_88122 | Not-determined |
| 18-109_k141_180507 | Not-determined |
| 18-109_k141_191253 | Not-determined |
| 18-109_k141_231469 | Not-determined |
| 18-109_k141_88131 | Not-determined |
| 18-109_k141_16918 | Not-determined |
| 18-109_k141_191259 | Not-determined |
| 18-109_k141_211476 | Not-determined |
| 18-109_k141_6723 | Not-determined |
| 18-109_k141_108148 | Not-determined |
| 18-109_k141_57948 | Low-quality |
| 18-109_k141_219771 | Not-determined |
| 18-109_k141_47755 | Not-determined |
| 18-109_k141_191266 | Not-determined |
| 18-109_k141_160325 | Not-determined |
| 18-109_k141_231476 | Not-determined |
| 18-109_k141_47767 | Low-quality |
| 18-109_k141_231478 | Not-determined |
| 18-109_k141_149283 | Low-quality |
| 18-109_k141_88145 | Not-determined |
| 18-109_k141_119389 | Low-quality |
| 18-109_k141_98587 | Not-determined |
| 18-109_k141_231487 | Not-determined |
| 18-109_k141_77715 | Not-determined |
| 18-109_k141_231491 | Not-determined |
| 18-109_k141_219788 | Not-determined |
| 18-109_k141_77723 | Not-determined |
| 18-109_k141_26232 | Not-determined |
| 18-109_k141_26233 | Not-determined |
| 18-109_k141_26234 | Not-determined |
| 18-109_k141_6742 | Low-quality |
| 18-109_k141_47789 | Not-determined |
| 18-109_k141_98608 | Low-quality |
| 18-109_k141_68604 | Not-determined |
| 18-109_k141_37893 | Not-determined |
| 18-109_k141_68608 | Not-determined |
| 18-109_k141_37897 | Not-determined |

| | |
|---|---|
| 18-109_k141_37898 | Not-determined |
| 18-109_k141_68609 | Low-quality |
| 18-109_k141_180548 | Not-determined |
| 18-109_k141_6749 | Not-determined |
| 18-109_k141_68611 | Not-determined |
| 18-109_k141_231497 | Not-determined |
| 18-109_k141_68612 | Not-determined |
| 18-109_k141_6752 | Not-determined |
| 18-109_k141_119407 | Not-determined |
| 18-109_k141_219802 | Not-determined |
| 18-109_k141_211507 | Not-determined |
| 18-109_k141_160352 | Not-determined |
| 18-109_k141_180555 | Not-determined |
| 18-109_k141_98626 | Not-determined |
| 18-109_k141_219805 | Not-determined |
| 18-109_k141_88174 | Low-quality |
| 18-109_k141_98630 | Not-determined |
| 18-109_k141_58001 | Not-determined |
| 18-109_k141_68633 | Not-determined |
| 18-109_k141_129527 | Not-determined |
| 18-109_k141_98637 | Not-determined |
| 18-109_k141_98638 | Not-determined |
| 18-109_k141_77745 | Not-determined |
| 18-109_k141_26259 | Not-determined |
| 18-109_k141_77748 | Low-quality |
| 18-109_k141_119420 | Not-determined |
| 18-109_k141_26263 | Not-determined |
| 18-109_k141_180574 | Not-determined |
| 18-109_k141_6769 | Not-determined |
| 18-109_k141_119427 | Not-determined |
| 18-109_k141_180581 | Not-determined |
| 18-109_k141_88193 | Not-determined |
| 18-109_k141_6779 | Low-quality |
| 18-109_k141_129559 | Not-determined |
| 18-109_k141_139318 | Not-determined |
| 18-109_k141_6781 | Not-determined |
| 18-109_k141_68654 | Not-determined |
| 18-109_k141_58034 | Not-determined |
| 18-109_k141_68656 | Not-determined |
| 18-109_k141_139327 | Not-determined |
| 18-109_k141_129563 | Not-determined |
| 18-109_k141_211535 | Not-determined |
| 18-109_k141_139329 | Low-quality |
| 18-109_k141_139330 | Low-quality |
| 18-109_k141_219827 | Not-determined |
| 18-109_k141_6791 | Not-determined |
| 18-109_k141_16980 | Not-determined |
| 18-109_k141_139334 | Low-quality |
| 18-109_k141_37933 | Not-determined |
| 18-109_k141_160386 | Not-determined |
| 18-109_k141_219840 | Not-determined |
| 18-109_k141_58038 | Not-determined |
| 18-109_k141_16994 | Not-determined |
| 18-109_k141_211552 | Not-determined |
| 18-109_k141_160389 | Not-determined |
| 18-109_k141_160390 | Not-determined |
| 18-109_k141_231535 | Not-determined |
| 18-109_k141_68675 | Low-quality |
| 18-109_k141_77778 | Not-determined |
| 18-109_k141_211559 | Not-determined |
| 18-109_k141_88222 | Not-determined |
| 18-109_k141_6813 | Not-determined |
| 18-109_k141_119447 | Not-determined |
| 18-109_k141_211562 | Not-determined |
| 18-109_k141_37956 | Low-quality |
| 18-109_k141_241878 | Low-quality |
| 18-109_k141_77793 | Not-determined |
| 18-109_k141_6814 | Not-determined |
| 18-109_k141_139364 | Not-determined |
| 18-109_k141_241884 | Not-determined |
| 18-109_k141_160395 | Low-quality |
| 18-109_k141_58048 | Not-determined |
| 18-109_k141_6822 | Not-determined |
| 18-109_k141_129600 | Not-determined |
| 18-109_k141_58052 | Not-determined |
| 18-109_k141_98716 | Not-determined |
| 18-109_k141_58058 | Not-determined |
| 18-109_k141_26306 | Low-quality |
| 18-109_k141_160410 | Not-determined |
| 18-109_k141_68699 | Not-determined |
| 18-109_k141_180618 | Not-determined |
| 18-109_k141_77804 | Not-determined |
| 18-109_k141_191299 | Not-determined |
| 18-109_k141_231558 | Not-determined |
| 18-109_k141_68705 | Not-determined |
| 18-109_k141_191302 | Not-determined |
| 18-109_k141_88236 | Not-determined |
| 18-109_k141_231562 | Not-determined |
| 18-109_k141_47836 | Not-determined |
| 18-109_k141_180627 | Not-determined |
| 18-109_k141_68715 | Not-determined |
| 18-109_k141_211575 | Not-determined |
| 18-109_k141_231566 | Not-determined |
| 18-109_k141_241897 | Not-determined |
| 18-109_k141_129621 | Not-determined |
| 18-109_k141_139369 | Not-determined |
| 18-109_k141_191311 | Not-determined |
| 18-109_k141_68724 | Not-determined |
| 18-109_k141_201561 | Not-determined |
| 18-109_k141_77826 | Not-determined |
| 18-109_k141_129626 | Not-determined |
| 18-109_k141_26328 | Not-determined |
| 18-109_k141_68727 | Not-determined |
| 18-109_k141_68731 | Not-determined |
| 18-109_k141_231583 | Not-determined |
| 18-109_k141_17020 | Not-determined |
| 18-109_k141_129638 | Low-quality |
| 18-109_k141_68733 | Not-determined |
| 18-109_k141_191325 | Not-determined |
| 18-109_k141_26341 | Not-determined |
| 18-109_k141_211594 | Not-determined |
| 18-109_k141_26345 | Not-determined |
| 18-109_k141_139382 | Not-determined |
| 18-109_k141_88257 | Not-determined |
| 18-109_k141_241910 | Not-determined |
| 18-109_k141_88260 | Not-determined |
| 18-109_k141_88262 | Not-determined |
| 18-109_k141_17035 | Not-determined |
| 18-109_k141_139394 | Not-determined |
| 18-109_k141_26356 | Not-determined |
| 18-109_k141_37969 | Not-determined |
| 18-109_k141_191346 | Not-determined |
| 18-109_k141_6859 | Not-determined |
| 18-109_k141_37973 | Not-determined |
| 18-109_k141_17042 | Not-determined |
| 18-109_k141_17049 | Not-determined |
| 18-109_k141_77867 | Not-determined |
| 18-109_k141_191354 | Not-determined |
| 18-109_k141_6871 | Not-determined |
| 18-109_k141_17058 | Low-quality |
| 18-109_k141_37989 | Low-quality |
| 18-109_k141_77868 | Low-quality |
| 18-109_k141_160450 | Not-determined |
| 18-109_k141_6873 | Not-determined |
| 18-109_k141_77870 | Not-determined |
| 18-109_k141_160454 | Not-determined |
| 18-109_k141_17066 | Not-determined |
| 18-109_k141_58114 | Low-quality |
| 18-109_k141_241929 | Not-determined |
| 18-109_k141_38004 | Not-determined |
| 18-109_k141_58120 | Low-quality |
| 18-109_k141_88274 | Not-determined |
| 18-109_k141_211617 | Not-determined |
| 18-109_k141_17081 | Not-determined |
| 18-109_k141_160468 | Not-determined |
| 18-109_k141_129651 | Not-determined |
| 18-109_k141_58139 | Not-determined |
| 18-109_k141_108182 | Not-determined |
| 18-109_k141_211627 | Not-determined |
| 18-109_k141_17089 | Not-determined |
| 18-109_k141_77882 | Not-determined |
| 18-109_k141_77883 | Not-determined |
| 18-109_k141_180660 | Not-determined |
| 18-109_k141_119514 | Not-determined |
| 18-109_k141_191386 | Not-determined |
| 18-109_k141_6898 | Not-determined |
| 18-109_k141_38020 | Not-determined |
| 18-109_k141_160478 | Not-determined |
| 18-109_k141_77886 | Not-determined |
| 18-109_k141_6905 | Not-determined |
| 18-109_k141_160482 | Not-determined |
| 18-109_k141_241956 | Not-determined |
| 18-109_k141_211648 | Not-determined |
| 18-109_k141_241964 | Not-determined |
| 18-109_k141_129675 | Not-determined |
| 18-109_k141_129680 | Not-determined |
| 18-109_k141_88304 | Not-determined |
| 18-109_k141_149410 | Not-determined |
| 18-109_k141_180671 | Not-determined |
| 18-109_k141_119521 | Not-determined |
| 18-109_k141_149423 | Not-determined |
| 18-109_k141_191413 | Not-determined |
| 18-109_k141_119533 | Not-determined |
| 18-109_k141_119535 | Not-determined |
| 18-109_k141_149429 | Not-determined |
| 18-109_k141_26405 | Not-determined |
| 18-109_k141_160500 | Not-determined |
| 18-109_k141_38033 | Not-determined |
| 18-109_k141_160502 | Not-determined |
| 18-109_k141_108203 | Low-quality |
| 18-109_k141_108204 | Not-determined |
| 18-109_k141_211665 | Not-determined |
| 18-109_k141_211666 | Not-determined |
| 18-109_k141_180687 | Not-determined |
| 18-109_k141_149438 | Not-determined |
| 18-109_k141_201580 | Low-quality |
| 18-109_k141_191430 | Low-quality |
| 18-109_k141_119550 | Not-determined |
| 18-109_k141_88336 | Not-determined |
| 18-109_k141_149449 | Not-determined |
| 18-109_k141_88342 | Not-determined |
| 18-109_k141_231588 | Not-determined |
| 18-109_k141_201595 | Not-determined |
| 18-109_k141_98723 | Not-determined |
| 18-109_k141_170636 | Not-determined |
| 18-109_k141_88357 | Not-determined |
| 18-109_k141_38040 | Not-determined |
| 18-109_k141_231611 | Not-determined |
| 18-109_k141_17127 | Not-determined |
| 18-109_k141_47904 | Not-determined |
| 18-109_k141_38048 | Not-determined |
| 18-109_k141_231621 | Not-determined |
| 18-109_k141_241992 | Not-determined |
| 18-109_k141_98733 | Not-determined |
| 18-109_k141_98734 | Not-determined |
| 18-109_k141_38056 | Not-determined |
| 18-109_k141_180718 | Not-determined |
| 18-109_k141_219909 | Not-determined |
| 18-109_k141_98741 | Not-determined |
| 18-109_k141_219912 | Not-determined |
| 18-109_k141_119578 | Not-determined |
| 18-109_k141_119582 | Not-determined |
| 18-109_k141_170658 | Not-determined |
| 18-109_k141_219917 | Not-determined |
| 18-109_k141_68812 | Not-determined |
| 18-109_k141_211687 | Low-quality |
| 18-109_k141_68815 | Low-quality |
| 18-109_k141_219924 | Not-determined |
| 18-109_k141_242004 | Not-determined |
| 18-109_k141_201618 | Not-determined |
| 18-109_k141_17150 | Low-quality |
| 18-109_k141_242009 | Not-determined |
| 18-109_k141_17151 | Low-quality |
| 18-109_k141_149457 | Not-determined |
| 18-109_k141_149458 | Not-determined |
| 18-109_k141_149459 | Not-determined |
| 18-109_k141_231643 | Not-determined |
| 18-109_k141_98755 | Not-determined |
| 18-109_k141_108209 | Not-determined |
| 18-109_k141_149466 | Not-determined |
| 18-109_k141_231652 | Not-determined |
| 18-109_k141_149468 | Not-determined |
| 18-109_k141_219936 | Not-determined |
| 18-109_k141_47924 | Not-determined |
| 18-109_k141_201633 | Not-determined |
| 18-109_k141_47930 | Not-determined |
| 18-109_k141_219939 | Low-quality |
| 18-109_k141_47937 | Not-determined |
| 18-109_k141_119607 | Not-determined |
| 18-109_k141_47941 | Not-determined |
| 18-109_k141_47944 | Not-determined |
| 18-109_k141_47946 | Not-determined |
| 18-109_k141_108230 | Not-determined |
| 18-109_k141_119610 | Not-determined |
| 18-109_k141_108236 | Not-determined |
| 18-109_k141_160582 | Not-determined |
| 18-109_k141_6922 | Not-determined |
| 18-109_k141_98776 | Low-quality |
| 18-109_k141_108242 | Not-determined |
| 18-109_k141_98777 | Not-determined |
| 18-109_k141_149498 | Not-determined |
| 18-109_k141_108245 | Not-determined |
| 18-109_k141_17183 | Not-determined |
| 18-109_k141_180749 | Not-determined |
| 18-109_k141_149504 | Not-determined |
| 18-109_k141_180751 | Not-determined |
| 18-109_k141_149505 | Not-determined |
| 18-109_k141_160596 | Not-determined |
| 18-109_k141_160600 | Not-determined |
| 18-109_k141_219942 | Low-quality |
| 18-109_k141_160603 | Not-determined |
| 18-109_k141_160604 | Not-determined |
| 18-109_k141_26426 | Low-quality |
| 18-109_k141_98792 | Not-determined |
| 18-109_k141_108252 | Not-determined |
| 18-109_k141_129764 | Not-determined |
| 18-109_k141_119633 | Not-determined |
| 18-109_k141_108261 | Not-determined |
| 18-109_k141_108264 | Not-determined |
| 18-109_k141_47958 | Not-determined |
| 18-109_k141_119641 | Not-determined |
| 18-109_k141_129776 | Not-determined |
| 18-109_k141_17198 | Not-determined |
| 18-109_k141_160629 | Not-determined |
| 18-109_k141_219947 | Not-determined |
| 18-109_k141_68827 | Not-determined |
| 18-109_k141_129784 | Not-determined |
| 18-109_k141_231688 | Not-determined |
| 18-109_k141_119654 | Not-determined |
| 18-109_k141_219948 | Not-determined |
| 18-109_k141_17204 | Not-determined |
| 18-109_k141_231696 | Not-determined |
| 18-109_k141_129790 | Not-determined |
| 18-109_k141_17209 | Not-determined |
| 18-109_k141_17211 | Not-determined |
| 18-109_k141_211692 | Not-determined |
| 18-109_k141_211695 | Not-determined |
| 18-109_k141_26446 | Not-determined |
| 18-109_k141_47978 | Not-determined |
| 18-109_k141_191486 | Not-determined |
| 18-109_k141_149544 | Not-determined |
| 18-109_k141_211703 | Not-determined |
| 18-109_k141_58179 | Not-determined |
| 18-109_k141_119672 | Not-determined |
| 18-109_k141_129804 | Not-determined |
| 18-109_k141_129805 | Not-determined |
| 18-109_k141_201702 | Not-determined |
| 18-109_k141_191496 | Not-determined |
| 18-109_k141_191498 | Not-determined |
| 18-109_k141_98836 | Not-determined |
| 18-109_k141_47989 | Not-determined |
| 18-109_k141_98839 | Not-determined |
| 18-109_k141_119686 | Not-determined |
| 18-109_k141_68852 | Not-determined |
| 18-109_k141_17238 | Not-determined |
| 18-109_k141_26456 | Not-determined |
| 18-109_k141_47996 | Not-determined |
| 18-109_k141_191501 | Not-determined |
| 18-109_k141_201714 | Not-determined |
| 18-109_k141_58190 | Not-determined |
| 18-109_k141_180789 | Not-determined |
| 18-109_k141_211722 | Not-determined |
| 18-109_k141_191505 | Not-determined |
| 18-109_k141_48005 | Not-determined |
| 18-109_k141_98853 | Not-determined |
| 18-109_k141_180791 | Low-quality |
| 18-109_k141_211724 | Not-determined |
| 18-109_k141_58194 | Not-determined |
| 18-109_k141_129817 | Not-determined |
| 18-109_k141_191511 | Not-determined |
| 18-109_k141_58199 | Not-determined |
| 18-109_k141_6949 | Not-determined |
| 18-109_k141_68862 | Not-determined |
| 18-109_k141_191513 | Not-determined |
| 18-109_k141_48015 | Not-determined |
| 18-109_k141_231727 | Not-determined |
| 18-109_k141_48025 | Not-determined |
| 18-109_k141_191525 | Not-determined |
| 18-109_k141_58209 | Not-determined |
| 18-109_k141_77916 | Not-determined |
| 18-109_k141_6958 | Not-determined |
| 18-109_k141_17252 | Not-determined |
| 18-109_k141_98867 | Not-determined |
| 18-109_k141_231737 | Not-determined |
| 18-109_k141_48032 | Not-determined |
| 18-109_k141_98872 | Not-determined |
| 18-109_k141_129835 | Not-determined |
| 18-109_k141_38066 | Not-determined |
| 18-109_k141_17263 | Not-determined |
| 18-109_k141_108311 | Not-determined |
| 18-109_k141_211751 | Low-quality |
| 18-109_k141_108312 | Not-determined |
| 18-109_k141_149581 | Not-determined |
| 18-109_k141_38068 | Not-determined |
| 18-109_k141_219990 | Low-quality |
| 18-109_k141_48047 | Not-determined |
| 18-109_k141_119731 | Not-determined |
| 18-109_k141_139522 | Not-determined |
| 18-109_k141_48050 | Not-determined |
| 18-109_k141_139525 | Not-determined |
| 18-109_k141_201733 | Not-determined |
| 18-109_k141_68896 | Not-determined |
| 18-109_k141_58229 | Not-determined |
| 18-109_k141_68900 | Not-determined |
| 18-109_k141_58231 | Not-determined |
| 18-109_k141_220005 | Not-determined |
| 18-109_k141_17282 | Not-determined |
| 18-109_k141_149605 | Not-determined |
| 18-109_k141_180814 | Not-determined |
| 18-109_k141_149609 | Not-determined |
| 18-109_k141_201758 | Not-determined |
| 18-109_k141_139545 | Not-determined |
| 18-109_k141_68920 | Not-determined |
| 18-109_k141_231768 | Not-determined |
| 18-109_k141_38081 | Not-determined |
| 18-109_k141_68925 | Not-determined |
| 18-109_k141_58244 | Low-quality |
| 18-109_k141_68928 | Not-determined |
| 18-109_k141_108325 | Not-determined |
| 18-109_k141_231774 | Not-determined |
| 18-109_k141_68939 | Not-determined |
| 18-109_k141_191544 | Not-determined |
| 18-109_k141_139557 | Not-determined |
| 18-109_k141_139558 | Low-quality |
| 18-109_k141_108340 | Low-quality |
| 18-109_k141_77955 | Not-determined |
| 18-109_k141_180841 | Low-quality |
| 18-109_k141_180842 | Not-determined |
| 18-109_k141_17298 | Not-determined |
| 18-109_k141_17301 | Not-determined |
| 18-109_k141_220017 | Not-determined |
| 18-109_k141_77963 | Not-determined |
| 18-109_k141_220018 | Not-determined |
| 18-109_k141_160722 | Not-determined |
| 18-109_k141_220019 | Not-determined |
| 18-109_k141_180852 | Not-determined |
| 18-109_k141_191559 | Not-determined |
| 18-109_k141_26503 | Low-quality |
| 18-109_k141_160734 | Not-determined |
| 18-109_k141_108347 | Not-determined |
| 18-109_k141_88400 | Not-determined |
| 18-109_k141_88404 | Not-determined |
| 18-109_k141_68986 | Not-determined |
| 18-109_k141_68987 | Not-determined |
| 18-109_k141_231796 | Not-determined |
| 18-109_k141_211801 | Not-determined |
| 18-109_k141_201780 | Not-determined |
| 18-109_k141_129843 | Not-determined |
| 18-109_k141_129845 | Not-determined |
| 18-109_k141_77995 | Not-determined |
| 18-109_k141_88420 | Not-determined |
| 18-109_k141_26521 | Not-determined |
| 18-109_k141_119776 | Low-quality |
| 18-109_k141_17346 | Not-determined |
| 18-109_k141_17347 | Not-determined |
| 18-109_k141_38107 | Not-determined |
| 18-109_k141_88423 | Low-quality |
| 18-109_k141_17351 | Not-determined |
| 18-109_k141_38116 | Not-determined |
| 18-109_k141_17354 | Not-determined |
| 18-109_k141_180891 | Not-determined |
| 18-109_k141_201813 | Not-determined |
| 18-109_k141_98902 | Not-determined |
| 18-109_k141_201817 | Not-determined |
| 18-109_k141_69013 | Not-determined |
| 18-109_k141_149630 | Low-quality |
| 18-109_k141_180902 | Not-determined |
| 18-109_k141_149638 | Not-determined |
| 18-109_k141_98916 | Not-determined |
| 18-109_k141_139623 | Not-determined |
| 18-109_k141_180913 | Not-determined |
| 18-109_k141_129878 | Not-determined |
| 18-109_k141_139630 | Not-determined |
| 18-109_k141_119804 | Not-determined |
| 18-109_k141_119810 | Not-determined |
| 18-109_k141_129893 | Low-quality |
| 18-109_k141_160771 | Not-determined |
| 18-109_k141_211814 | Not-determined |
| 18-109_k141_220047 | Not-determined |
| 18-109_k141_98934 | Not-determined |
| 18-109_k141_119814 | Not-determined |
| 18-109_k141_220051 | Not-determined |
| 18-109_k141_149654 | Not-determined |
| 18-109_k141_149656 | Low-quality |
| 18-109_k141_7000 | Not-determined |
| 18-109_k141_180919 | Not-determined |
| 18-109_k141_149660 | Not-determined |
| 18-109_k141_129898 | Low-quality |
| 18-109_k141_7002 | Not-determined |
| 18-109_k141_38137 | Not-determined |
| 18-109_k141_108415 | Not-determined |
| 18-109_k141_191612 | Not-determined |
| 18-109_k141_220058 | Low-quality |
| 18-109_k141_119824 | Not-determined |
| 18-109_k141_7007 | Not-determined |
| 18-109_k141_191621 | Not-determined |
| 18-109_k141_149676 | Not-determined |
| 18-109_k141_129906 | Not-determined |
| 18-109_k141_69030 | Low-quality |
| 18-109_k141_191626 | Not-determined |
| 18-109_k141_170700 | Not-determined |
| 18-109_k141_38154 | Low-quality |
| 18-109_k141_149683 | Not-determined |
| 18-109_k141_170706 | Not-determined |
| 18-109_k141_48064 | Not-determined |
| 18-109_k141_139667 | Not-determined |
| 18-109_k141_108431 | Not-determined |
| 18-109_k141_58286 | Not-determined |
| 18-109_k141_58287 | Not-determined |
| 18-109_k141_58288 | Low-quality |
| 18-109_k141_48067 | Not-determined |
| 18-109_k141_78038 | Low-quality |
| 18-109_k141_191636 | Low-quality |
| 18-109_k141_7027 | Not-determined |
| 18-109_k141_201857 | Not-determined |
| 18-109_k141_108445 | Not-determined |
| 18-109_k141_180943 | Not-determined |
| 18-109_k141_119852 | Not-determined |
| 18-109_k141_48069 | Low-quality |
| 18-109_k141_220068 | Low-quality |
| 18-109_k141_7034 | Low-quality |
| 18-109_k141_69043 | Not-determined |
| 18-109_k141_26530 | Not-determined |
| 18-109_k141_149702 | Not-determined |
| 18-109_k141_170718 | Not-determined |
| 18-109_k141_149707 | Not-determined |
| 18-109_k141_220075 | Not-determined |
| 18-109_k141_78063 | Not-determined |
| 18-109_k141_58293 | Low-quality |
| 18-109_k141_211846 | Low-quality |
| 18-109_k141_211849 | Not-determined |
| 18-109_k141_211851 | Not-determined |
| 18-109_k141_48091 | Not-determined |
| 18-109_k141_201875 | Not-determined |
| 18-109_k141_191652 | Low-quality |
| 18-109_k141_58300 | Not-determined |
| 18-109_k141_129931 | Not-determined |
| 18-109_k141_180963 | Not-determined |
| 18-109_k141_220083 | Not-determined |
| 18-109_k141_180965 | Not-determined |
| 18-109_k141_98987 | Not-determined |
| 18-109_k141_180968 | Not-determined |
| 18-109_k141_231908 | Not-determined |
| 18-109_k141_170736 | Low-quality |
| 18-109_k141_98991 | Not-determined |
| 18-109_k141_211859 | Not-determined |
| 18-109_k141_160812 | Not-determined |
| 18-109_k141_7048 | Low-quality |
| 18-109_k141_201890 | Low-quality |
| 18-109_k141_191656 | Not-determined |
| 18-109_k141_160816 | Not-determined |
| 18-109_k141_69052 | Not-determined |
| 18-109_k141_48106 | Not-determined |
| 18-109_k141_160821 | Low-quality |
| 18-109_k141_170741 | Not-determined |
| 18-109_k141_160824 | Not-determined |
| 18-109_k141_160825 | Not-determined |
| 18-109_k141_108466 | Low-quality |
| 18-109_k141_231919 | Not-determined |
| 18-109_k141_38197 | Not-determined |
| 18-109_k141_191681 | Not-determined |
| 18-109_k141_48126 | Not-determined |
| 18-109_k141_98994 | Not-determined |
| 18-109_k141_160834 | Not-determined |
| 18-109_k141_160837 | Not-determined |
| 18-109_k141_160841 | Not-determined |
| 18-109_k141_69058 | Not-determined |
| 18-109_k141_160842 | Not-determined |
| 18-109_k141_48136 | Low-quality |
| 18-109_k141_69062 | Not-determined |
| 18-109_k141_160844 | Not-determined |
| 18-109_k141_181007 | Not-determined |
| 18-109_k141_48146 | Not-determined |
| 18-109_k141_48148 | Not-determined |
| 18-109_k141_139671 | Not-determined |
| 18-109_k141_139673 | Not-determined |
| 18-109_k141_69078 | Not-determined |
| 18-109_k141_7075 | Not-determined |
| 18-109_k141_48153 | Not-determined |
| 18-109_k141_99010 | Not-determined |
| 18-109_k141_119905 | Not-determined |
| 18-109_k141_129942 | Not-determined |
| 18-109_k141_181032 | Not-determined |
| 18-109_k141_69092 | Not-determined |
| 18-109_k141_58320 | Low-quality |
| 18-109_k141_181033 | Not-determined |
| 18-109_k141_58324 | Not-determined |
| 18-109_k141_7086 | Not-determined |
| 18-109_k141_181038 | Not-determined |
| 18-109_k141_7087 | Not-determined |
| 18-109_k141_69098 | Not-determined |
| 18-109_k141_119920 | Not-determined |
| 18-109_k141_211890 | Not-determined |
| 18-109_k141_88467 | Not-determined |
| 18-109_k141_211894 | Not-determined |
| 18-109_k141_26559 | Not-determined |
| 18-109_k141_108502 | Not-determined |
| 18-109_k141_181049 | Not-determined |
| 18-109_k141_149767 | Not-determined |
| 18-109_k141_88469 | Not-determined |
| 18-109_k141_26560 | Not-determined |
| 18-109_k141_17399 | Not-determined |
| 18-109_k141_88472 | Not-determined |
| 18-109_k141_58339 | Not-determined |
| 18-109_k141_78088 | Not-determined |
| 18-109_k141_88475 | Not-determined |
| 18-109_k141_211912 | Not-determined |
| 18-109_k141_108518 | Not-determined |
| 18-109_k141_17407 | Not-determined |
| 18-109_k141_78096 | Low-quality |
| 18-109_k141_108523 | Not-determined |
| 18-109_k141_160884 | Not-determined |
| 18-109_k141_160887 | Not-determined |
| 18-109_k141_88488 | Not-determined |
| 18-109_k141_26572 | Not-determined |
| 18-109_k141_48181 | Not-determined |
| 18-109_k141_78106 | Not-determined |
| 18-109_k141_149788 | Not-determined |
| 18-109_k141_129954 | Not-determined |
| 18-109_k141_119931 | Not-determined |
| 18-109_k141_108531 | Low-quality |
| 18-109_k141_149797 | Not-determined |
| 18-109_k141_58361 | Not-determined |
| 18-109_k141_220093 | Not-determined |
| 18-109_k141_231969 | Not-determined |
| 18-109_k141_48186 | Not-determined |
| 18-109_k141_48188 | Not-determined |
| 18-109_k141_170803 | Not-determined |
| 18-109_k141_48192 | Low-quality |
| 18-109_k141_181083 | Not-determined |
| 18-109_k141_78117 | Not-determined |
| 18-109_k141_78119 | Not-determined |
| 18-109_k141_48197 | Not-determined |
| 18-109_k141_108545 | Not-determined |
| 18-109_k141_231983 | Not-determined |
| 18-109_k141_211930 | Not-determined |
| 18-109_k141_88515 | Not-determined |
| 18-109_k141_201917 | Not-determined |
| 18-109_k141_26594 | Not-determined |

| | |
|---|---|
| 18-109_k141_58376 | Not-determined |
| 18-109_k141_119958 | Not-determined |
| 18-109_k141_129969 | Low-quality |
| 18-109_k141_78129 | Low-quality |
| 18-109_k141_181097 | Not-determined |
| 18-109_k141_88523 | Not-determined |
| 18-109_k141_58383 | Not-determined |
| 18-109_k141_17455 | Low-quality |
| 18-109_k141_26602 | Not-determined |
| 18-109_k141_58385 | Not-determined |
| 18-109_k141_78139 | Not-determined |
| 18-109_k141_26605 | Not-determined |
| 18-109_k141_139695 | Not-determined |
| 18-109_k141_160929 | Not-determined |
| 18-109_k141_170811 | Not-determined |
| 18-109_k141_7123 | Low-quality |
| 18-109_k141_139698 | Not-determined |
| 18-109_k141_201935 | Not-determined |
| 18-109_k141_58392 | Not-determined |
| 18-109_k141_88532 | Not-determined |
| 18-109_k141_201938 | Not-determined |
| 18-109_k141_58393 | Not-determined |
| 18-109_k141_160938 | Not-determined |
| 18-109_k141_38213 | Not-determined |
| 18-109_k141_69131 | Not-determined |
| 18-109_k141_139701 | Not-determined |
| 18-109_k141_181107 | Not-determined |
| 18-109_k141_129995 | Not-determined |
| 18-109_k141_108561 | Not-determined |
| 18-109_k141_211949 | Not-determined |
| 18-109_k141_17474 | Not-determined |
| 18-109_k141_211950 | Not-determined |
| 18-109_k141_201950 | Low-quality |
| 18-109_k141_160941 | Not-determined |
| 18-109_k141_220103 | Not-determined |
| 18-109_k141_181114 | Not-determined |
| 18-109_k141_119976 | Not-determined |
| 18-109_k141_130000 | Not-determined |
| 18-109_k141_58409 | Not-determined |
| 18-109_k141_130006 | Low-quality |
| 18-109_k141_99038 | Not-determined |
| 18-109_k141_7142 | Not-determined |
| 18-109_k141_130009 | Not-determined |
| 18-109_k141_232010 | Low-quality |
| 18-109_k141_108573 | Not-determined |
| 18-109_k141_201970 | Not-determined |
| 18-109_k141_220122 | Not-determined |
| 18-109_k141_130014 | Not-determined |
| 18-109_k141_88547 | Not-determined |
| 18-109_k141_48224 | Not-determined |
| 18-109_k141_130019 | Not-determined |
| 18-109_k141_99046 | Low-quality |
| 18-109_k141_130026 | Not-determined |
| 18-109_k141_99048 | Not-determined |
| 18-109_k141_119991 | Not-determined |
| 18-109_k141_130031 | Not-determined |
| 18-109_k141_69167 | Not-determined |
| 18-109_k141_181144 | Not-determined |
| 18-109_k141_139720 | Not-determined |
| 18-109_k141_38234 | Not-determined |
| 18-109_k141_191709 | Low-quality |
| 18-109_k141_48241 | Not-determined |
| 18-109_k141_120008 | Not-determined |
| 18-109_k141_38237 | Not-determined |
| 18-109_k141_160972 | Not-determined |
| 18-109_k141_130034 | Not-determined |
| 18-109_k141_130037 | Not-determined |
| 18-109_k141_120011 | Not-determined |
| 18-109_k141_160976 | Not-determined |
| 18-109_k141_17501 | Not-determined |
| 18-109_k141_99066 | Not-determined |
| 18-109_k141_58428 | Not-determined |
| 18-109_k141_120014 | Low-quality |
| 18-109_k141_120016 | Not-determined |
| 18-109_k141_220139 | Not-determined |
| 18-109_k141_120027 | Not-determined |
| 18-109_k141_48256 | Not-determined |
| 18-109_k141_48259 | Not-determined |
| 18-109_k141_211969 | Not-determined |
| 18-109_k141_48263 | Not-determined |
| 18-109_k141_7168 | Not-determined |
| 18-109_k141_78169 | Not-determined |
| 18-109_k141_181179 | Not-determined |
| 18-109_k141_220147 | Not-determined |
| 18-109_k141_69208 | Not-determined |
| 18-109_k141_160996 | Not-determined |
| 18-109_k141_99084 | Low-quality |
| 18-109_k141_139736 | Not-determined |
| 18-109_k141_220150 | Not-determined |
| 18-109_k141_38265 | Not-determined |
| 18-109_k141_69215 | Not-determined |
| 18-109_k141_99092 | Not-determined |
| 18-109_k141_7173 | Not-determined |
| 18-109_k141_220158 | Not-determined |
| 18-109_k141_69225 | Low-quality |
| 18-109_k141_191745 | Not-determined |
| 18-109_k141_108599 | Not-determined |
| 18-109_k141_181188 | Low-quality |
| 18-109_k141_191748 | Not-determined |
| 18-109_k141_99102 | Not-determined |
| 18-109_k141_139752 | Not-determined |
| 18-109_k141_130057 | Not-determined |
| 18-109_k141_181195 | Not-determined |
| 18-109_k141_38282 | Not-determined |
| 18-109_k141_202010 | Not-determined |
| 18-109_k141_181197 | Not-determined |
| 18-109_k141_202015 | Not-determined |
| 18-109_k141_139760 | Not-determined |
| 18-109_k141_181204 | Not-determined |
| 18-109_k141_58463 | Not-determined |
| 18-109_k141_191758 | Not-determined |
| 18-109_k141_130067 | Low-quality |
| 18-109_k141_48292 | Not-determined |
| 18-109_k141_38290 | Not-determined |
| 18-109_k141_191760 | Not-determined |
| 18-109_k141_38293 | Low-quality |
| 18-109_k141_130068 | Not-determined |
| 18-109_k141_26639 | Not-determined |
| 18-109_k141_202030 | Not-determined |
| 18-109_k141_7189 | Not-determined |
| 18-109_k141_120044 | Not-determined |
| 18-109_k141_202041 | Not-determined |
| 18-109_k141_191773 | Not-determined |
| 18-109_k141_202048 | Not-determined |
| 18-109_k141_130084 | Not-determined |
| 18-109_k141_69271 | Low-quality |
| 18-109_k141_191781 | Low-quality |
| 18-109_k141_17542 | Not-determined |
| 18-109_k141_161054 | Not-determined |
| 18-109_k141_161057 | Low-quality |
| 18-109_k141_161059 | Not-determined |
| 18-109_k141_202053 | Not-determined |
| 18-109_k141_120062 | Not-determined |
| 18-109_k141_120063 | Not-determined |
| 18-109_k141_78205 | Not-determined |
| 18-109_k141_191784 | Not-determined |
| 18-109_k141_170842 | Not-determined |
| 18-109_k141_232093 | Not-determined |
| 18-109_k141_220189 | Not-determined |
| 18-109_k141_161068 | Not-determined |
| 18-109_k141_139781 | Not-determined |
| 18-109_k141_149878 | Not-determined |
| 18-109_k141_17556 | Low-quality |
| 18-109_k141_220191 | Not-determined |
| 18-109_k141_232104 | Not-determined |
| 18-109_k141_232108 | Not-determined |
| 18-109_k141_26659 | Not-determined |
| 18-109_k141_170858 | Low-quality |
| 18-109_k141_26660 | Not-determined |
| 18-109_k141_99173 | Low-quality |
| 18-109_k141_149892 | Not-determined |
| 18-109_k141_139795 | Not-determined |
| 18-109_k141_232116 | Not-determined |
| 18-109_k141_181259 | Not-determined |
| 18-109_k141_181260 | Not-determined |
| 18-109_k141_170869 | Not-determined |
| 18-109_k141_181264 | Not-determined |
| 18-109_k141_202092 | Not-determined |
| 18-109_k141_120096 | Not-determined |
| 18-109_k141_232129 | Not-determined |
| 18-109_k141_232131 | Not-determined |
| 18-109_k141_58533 | Not-determined |
| 18-109_k141_78232 | Not-determined |
| 18-109_k141_58534 | Not-determined |
| 18-109_k141_232133 | Low-quality |
| 18-109_k141_78238 | Not-determined |
| 18-109_k141_181282 | Not-determined |
| 18-109_k141_99200 | Not-determined |
| 18-109_k141_202122 | Not-determined |
| 18-109_k141_108624 | Not-determined |
| 18-109_k141_232142 | Not-determined |
| 18-109_k141_99206 | Not-determined |
| 18-109_k141_108629 | Not-determined |
| 18-109_k141_161120 | Not-determined |
| 18-109_k141_232147 | Not-determined |
| 18-109_k141_88671 | Not-determined |
| 18-109_k141_78242 | Not-determined |
| 18-109_k141_212021 | Not-determined |
| 18-109_k141_88676 | Not-determined |
| 18-109_k141_191821 | Not-determined |
| 18-109_k141_88690 | Not-determined |
| 18-109_k141_242028 | Low-quality |
| 18-109_k141_108659 | Not-determined |
| 18-109_k141_108667 | Not-determined |
| 18-109_k141_78260 | Not-determined |
| 18-109_k141_191835 | Not-determined |
| 18-109_k141_99215 | Not-determined |
| 18-109_k141_242039 | Not-determined |
| 18-109_k141_99222 | Not-determined |
| 18-109_k141_88703 | Low-quality |
| 18-109_k141_88704 | Not-determined |
| 18-109_k141_120158 | Not-determined |
| 18-109_k141_191846 | Not-determined |
| 18-109_k141_232174 | Not-determined |
| 18-109_k141_38384 | Not-determined |
| 18-109_k141_38386 | Low-quality |
| 18-109_k141_242059 | Not-determined |
| 18-109_k141_38387 | Not-determined |
| 18-109_k141_139826 | Low-quality |
| 18-109_k141_99235 | Not-determined |
| 18-109_k141_130141 | Low-quality |
| 18-109_k141_99238 | Not-determined |
| 18-109_k141_232182 | Not-determined |
| 18-109_k141_120169 | Not-determined |
| 18-109_k141_120170 | Not-determined |
| 18-109_k141_78280 | Not-determined |
| 18-109_k141_149971 | Not-determined |
| 18-109_k141_202150 | Not-determined |
| 18-109_k141_149975 | Not-determined |
| 18-109_k141_69276 | Not-determined |
| 18-109_k141_202152 | Not-determined |
| 18-109_k141_58555 | Not-determined |
| 18-109_k141_48360 | Not-determined |
| 18-109_k141_212069 | Not-determined |
| 18-109_k141_38395 | Not-determined |
| 18-109_k141_149978 | Not-determined |
| 18-109_k141_108701 | Not-determined |
| 18-109_k141_212074 | Not-determined |
| 18-109_k141_161159 | Not-determined |
| 18-109_k141_212076 | Not-determined |
| 18-109_k141_108704 | Not-determined |
| 18-109_k141_38400 | Not-determined |
| 18-109_k141_38402 | Not-determined |
| 18-109_k141_220253 | Not-determined |
| 18-109_k141_161163 | Low-quality |
| 18-109_k141_139857 | Not-determined |
| 18-109_k141_202165 | Not-determined |
| 18-109_k141_78283 | Not-determined |
| 18-109_k141_202166 | Not-determined |
| 18-109_k141_38407 | Not-determined |
| 18-109_k141_202173 | Not-determined |
| 18-109_k141_88726 | Not-determined |
| 18-109_k141_88728 | Not-determined |
| 18-109_k141_69298 | Not-determined |
| 18-109_k141_78291 | Not-determined |
| 18-109_k141_181352 | Not-determined |
| 18-109_k141_161172 | Not-determined |
| 18-109_k141_220265 | Low-quality |
| 18-109_k141_17585 | Not-determined |
| 18-109_k141_48384 | Not-determined |
| 18-109_k141_99269 | Low-quality |
| 18-109_k141_69302 | Not-determined |
| 18-109_k141_88737 | Not-determined |
| 18-109_k141_58586 | Low-quality |
| 18-109_k141_161179 | Not-determined |
| 18-109_k141_170958 | Not-determined |
| 18-109_k141_120175 | Not-determined |
| 18-109_k141_170961 | Not-determined |
| 18-109_k141_99276 | Not-determined |
| 18-109_k141_58589 | Not-determined |
| 18-109_k141_232207 | Not-determined |
| 18-109_k141_38417 | Not-determined |
| 18-109_k141_161182 | Not-determined |
| 18-109_k141_58593 | Not-determined |
| 18-109_k141_202193 | Not-determined |
| 18-109_k141_181377 | Not-determined |
| 18-109_k141_48404 | Not-determined |
| 18-109_k141_78304 | Not-determined |
| 18-109_k141_88748 | Not-determined |
| 18-109_k141_191879 | Not-determined |
| 18-109_k141_181386 | Not-determined |
| 18-109_k141_161198 | Not-determined |
| 18-109_k141_181392 | Not-determined |
| 18-109_k141_181393 | Not-determined |
| 18-109_k141_58614 | Not-determined |
| 18-109_k141_38441 | Not-determined |
| 18-109_k141_202213 | Not-determined |
| 18-109_k141_38443 | Not-determined |
| 18-109_k141_58615 | Not-determined |
| 18-109_k141_161204 | Not-determined |
| 18-109_k141_130172 | Not-determined |
| 18-109_k141_130173 | Not-determined |
| 18-109_k141_232223 | Not-determined |
| 18-109_k141_181398 | Not-determined |
| 18-109_k141_69328 | Not-determined |
| 18-109_k141_220295 | Not-determined |
| 18-109_k141_38453 | Not-determined |
| 18-109_k141_181405 | Not-determined |
| 18-109_k141_7240 | Not-determined |
| 18-109_k141_130187 | Not-determined |
| 18-109_k141_220303 | Low-quality |
| 18-109_k141_212108 | Not-determined |
| 18-109_k141_161221 | Not-determined |
| 18-109_k141_202224 | Low-quality |
| 18-109_k141_69338 | Not-determined |
| 18-109_k141_191893 | Not-determined |
| 18-109_k141_161224 | Low-quality |
| 18-109_k141_181418 | Not-determined |
| 18-109_k141_191902 | Not-determined |
| 18-109_k141_7254 | Not-determined |
| 18-109_k141_220316 | Not-determined |
| 18-109_k141_232253 | Not-determined |
| 18-109_k141_232254 | Not-determined |
| 18-109_k141_69346 | Not-determined |
| 18-109_k141_48430 | Low-quality |
| 18-109_k141_48432 | Not-determined |
| 18-109_k141_232260 | Not-determined |
| 18-109_k141_7265 | Not-determined |
| 18-109_k141_69356 | Not-determined |
| 18-109_k141_232269 | Not-determined |
| 18-109_k141_7269 | Low-quality |
| 18-109_k141_69359 | Not-determined |
| 18-109_k141_38490 | Not-determined |
| 18-109_k141_58636 | Not-determined |
| 18-109_k141_191915 | Not-determined |
| 18-109_k141_242103 | Not-determined |
| 18-109_k141_242105 | Not-determined |
| 18-109_k141_78332 | Not-determined |
| 18-109_k141_181441 | Not-determined |
| 18-109_k141_58640 | Not-determined |
| 18-109_k141_181443 | Not-determined |
| 18-109_k141_78335 | Not-determined |
| 18-109_k141_7275 | Not-determined |
| 18-109_k141_48446 | Not-determined |
| 18-109_k141_181448 | Not-determined |
| 18-109_k141_58646 | Not-determined |
| 18-109_k141_161240 | Not-determined |
| 18-109_k141_120222 | Not-determined |
| 18-109_k141_191926 | Not-determined |
| 18-109_k141_181453 | Not-determined |
| 18-109_k141_120226 | Not-determined |
| 18-109_k141_120231 | Low-quality |
| 18-109_k141_181458 | Not-determined |
| 18-109_k141_120232 | Not-determined |
| 18-109_k141_232285 | Not-determined |
| 18-109_k141_202248 | Low-quality |
| 18-109_k141_232286 | Not-determined |
| 18-109_k141_78343 | Not-determined |
| 18-109_k141_38502 | Not-determined |
| 18-109_k141_108766 | Low-quality |
| 18-109_k141_120237 | Not-determined |
| 18-109_k141_108768 | Not-determined |
| 18-109_k141_232292 | Not-determined |
| 18-109_k141_48455 | Not-determined |
| 18-109_k141_150026 | Not-determined |
| 18-109_k141_181470 | Not-determined |
| 18-109_k141_48462 | Not-determined |
| 18-109_k141_88786 | Low-quality |
| 18-109_k141_120249 | Not-determined |
| 18-109_k141_48465 | Not-determined |
| 18-109_k141_99329 | Not-determined |
| 18-109_k141_202262 | Not-determined |
| 18-109_k141_202267 | Not-determined |
| 18-109_k141_130239 | Not-determined |
| 18-109_k141_150040 | Low-quality |
| 18-109_k141_202270 | Not-determined |
| 18-109_k141_130243 | Not-determined |
| 18-109_k141_161258 | Not-determined |
| 18-109_k141_171000 | Not-determined |
| 18-109_k141_99340 | Not-determined |
| 18-109_k141_191955 | Low-quality |
| 18-109_k141_17599 | Not-determined |
| 18-109_k141_108791 | Not-determined |
| 18-109_k141_108794 | Not-determined |
| 18-109_k141_88801 | Not-determined |
| 18-109_k141_7315 | Not-determined |
| 18-109_k141_171009 | Low-quality |
| 18-109_k141_161270 | Not-determined |
| 18-109_k141_150050 | Not-determined |
| 18-109_k141_212141 | Not-determined |
| 18-109_k141_38548 | Not-determined |
| 18-109_k141_48493 | Not-determined |
| 18-109_k141_202284 | Not-determined |
| 18-109_k141_7325 | Not-determined |
| 18-109_k141_58677 | Not-determined |
| 18-109_k141_150060 | Not-determined |
| 18-109_k141_171023 | Not-determined |
| 18-109_k141_150062 | Not-determined |
| 18-109_k141_150063 | Not-determined |
| 18-109_k141_108801 | Not-determined |
| 18-109_k141_7334 | Not-determined |
| 18-109_k141_108803 | Low-quality |
| 18-109_k141_108804 | Low-quality |
| 18-109_k141_78376 | Not-determined |
| 18-109_k141_108807 | Not-determined |
| 18-109_k141_17610 | Not-determined |
| 18-109_k141_17611 | Not-determined |
| 18-109_k141_7339 | Not-determined |
| 18-109_k141_181511 | Not-determined |
| 18-109_k141_130258 | Not-determined |
| 18-109_k141_99367 | Not-determined |
| 18-109_k141_38556 | Not-determined |
| 18-109_k141_17617 | Not-determined |
| 18-109_k141_17620 | Not-determined |
| 18-109_k141_161314 | Not-determined |
| 18-109_k141_181522 | Not-determined |
| 18-109_k141_78394 | Not-determined |
| 18-109_k141_17624 | Not-determined |
| 18-109_k141_181527 | Not-determined |
| 18-109_k141_99374 | Not-determined |
| 18-109_k141_202303 | Not-determined |
| 18-109_k141_26726 | Not-determined |
| 18-109_k141_78407 | Low-quality |
| 18-109_k141_150083 | Not-determined |
| 18-109_k141_78410 | Not-determined |
| 18-109_k141_58723 | Not-determined |
| 18-109_k141_161337 | Not-determined |
| 18-109_k141_38571 | Not-determined |
| 18-109_k141_161339 | Not-determined |
| 18-109_k141_7371 | Low-quality |
| 18-109_k141_78421 | Not-determined |
| 18-109_k141_212158 | Not-determined |
| 18-109_k141_161347 | Low-quality |
| 18-109_k141_58728 | Not-determined |
| 18-109_k141_212167 | Not-determined |
| 18-109_k141_171074 | Not-determined |
| 18-109_k141_130285 | Not-determined |
| 18-109_k141_171081 | Not-determined |
| 18-109_k141_38590 | Low-quality |
| 18-109_k141_212178 | Not-determined |
| 18-109_k141_48530 | Low-quality |
| 18-109_k141_212181 | Not-determined |
| 18-109_k141_38595 | Low-quality |
| 18-109_k141_130292 | Low-quality |
| 18-109_k141_69402 | Not-determined |
| 18-109_k141_26731 | Low-quality |
| 18-109_k141_161361 | Not-determined |
| 18-109_k141_212191 | Not-determined |
| 18-109_k141_78452 | Not-determined |
| 18-109_k141_26733 | Not-determined |
| 18-109_k141_202357 | Not-determined |
| 18-109_k141_181541 | Low-quality |
| 18-109_k141_78454 | Not-determined |
| 18-109_k141_161371 | Not-determined |
| 18-109_k141_191985 | Not-determined |
| 18-109_k141_130299 | Not-determined |
| 18-109_k141_26739 | Not-determined |
| 18-109_k141_58770 | Not-determined |
| 18-109_k141_130300 | Not-determined |
| 18-109_k141_17668 | Not-determined |
| 18-109_k141_108846 | Low-quality |
| 18-109_k141_17673 | Low-quality |
| 18-109_k141_130308 | Not-determined |
| 18-109_k141_232357 | Not-determined |
| 18-109_k141_171103 | Not-determined |
| 18-109_k141_108848 | Not-determined |
| 18-109_k141_58778 | Not-determined |
| 18-109_k141_212201 | Not-determined |
| 18-109_k141_26752 | Low-quality |
| 18-109_k141_17683 | Not-determined |
| 18-109_k141_58785 | Not-determined |
| 18-109_k141_108856 | Not-determined |
| 18-109_k141_58786 | Not-determined |
| 18-109_k141_220344 | Not-determined |
| 18-109_k141_69416 | Low-quality |
| 18-109_k141_150133 | Not-determined |
| 18-109_k141_88865 | Not-determined |
| 18-109_k141_7396 | Not-determined |
| 18-109_k141_7399 | Not-determined |
| 18-109_k141_17697 | Not-determined |
| 18-109_k141_212218 | Not-determined |
| 18-109_k141_48569 | Not-determined |
| 18-109_k141_192016 | Low-quality |
| 18-109_k141_192020 | Not-determined |
| 18-109_k141_17700 | Not-determined |
| 18-109_k141_212227 | Not-determined |
| 18-109_k141_48575 | Not-determined |
| 18-109_k141_88875 | Not-determined |
| 18-109_k141_48576 | Not-determined |
| 18-109_k141_48577 | Not-determined |
| 18-109_k141_88881 | Not-determined |
| 18-109_k141_88882 | Not-determined |
| 18-109_k141_88884 | Not-determined |
| 18-109_k141_150150 | Not-determined |
| 18-109_k141_7406 | Not-determined |
| 18-109_k141_171139 | Not-determined |
| 18-109_k141_7407 | Low-quality |
| 18-109_k141_48584 | Not-determined |
| 18-109_k141_38614 | Not-determined |
| 18-109_k141_17718 | Not-determined |
| 18-109_k141_69428 | Not-determined |
| 18-109_k141_171146 | Not-determined |
| 18-109_k141_139883 | Not-determined |
| 18-109_k141_26779 | Not-determined |
| 18-109_k141_88902 | Not-determined |
| 18-109_k141_232381 | Not-determined |
| 18-109_k141_7413 | Not-determined |
| 18-109_k141_58836 | Not-determined |
| 18-109_k141_7414 | Not-determined |
| 18-109_k141_58838 | Not-determined |
| 18-109_k141_150173 | Not-determined |
| 18-109_k141_17737 | Not-determined |
| 18-109_k141_130360 | Not-determined |
| 18-109_k141_99419 | Low-quality |
| 18-109_k141_171156 | Not-determined |
| 18-109_k141_130362 | Not-determined |
| 18-109_k141_7424 | Not-determined |
| 18-109_k141_48618 | Not-determined |
| 18-109_k141_38637 | Not-determined |
| 18-109_k141_130370 | Not-determined |
| 18-109_k141_17742 | Not-determined |
| 18-109_k141_181601 | Low-quality |
| 18-109_k141_99424 | Not-determined |
| 18-109_k141_192048 | Not-determined |
| 18-109_k141_108892 | Not-determined |
| 18-109_k141_192052 | Not-determined |
| 18-109_k141_192059 | Not-determined |
| 18-109_k141_48623 | Not-determined |
| 18-109_k141_192064 | Low-quality |
| 18-109_k141_108903 | Not-determined |
| 18-109_k141_171171 | Not-determined |
| 18-109_k141_58849 | Not-determined |
| 18-109_k141_17757 | Not-determined |
| 18-109_k141_232422 | Not-determined |
| 18-109_k141_171176 | Not-determined |
| 18-109_k141_212293 | Not-determined |
| 18-109_k141_17766 | Not-determined |
| 18-109_k141_7454 | Not-determined |
| 18-109_k141_232424 | Not-determined |
| 18-109_k141_26816 | Not-determined |
| 18-109_k141_232428 | Low-quality |
| 18-109_k141_48648 | Not-determined |
| 18-109_k141_232433 | Not-determined |
| 18-109_k141_88931 | Not-determined |
| 18-109_k141_99446 | Low-quality |
| 18-109_k141_69468 | Not-determined |
| 18-109_k141_7469 | Not-determined |
| 18-109_k141_17781 | Not-determined |
| 18-109_k141_181624 | Not-determined |
| 18-109_k141_88933 | Not-determined |
| 18-109_k141_99460 | Not-determined |
| 18-109_k141_58860 | Not-determined |
| 18-109_k141_38681 | Not-determined |
| 18-109_k141_88938 | Not-determined |
| 18-109_k141_38684 | Not-determined |
| 18-109_k141_88939 | Not-determined |
| 18-109_k141_88940 | Not-determined |
| 18-109_k141_171195 | Not-determined |
| 18-109_k141_58861 | Not-determined |
| 18-109_k141_171197 | Not-determined |
| 18-109_k141_171199 | Not-determined |
| 18-109_k141_181635 | Not-determined |
| 18-109_k141_108953 | Not-determined |
| 18-109_k141_108954 | Not-determined |
| 18-109_k141_48672 | Not-determined |
| 18-109_k141_130437 | Not-determined |
| 18-109_k141_181640 | Not-determined |
| 18-109_k141_48676 | Not-determined |
| 18-109_k141_48679 | Not-determined |
| 18-109_k141_58885 | Not-determined |
| 18-109_k141_130441 | Low-quality |
| 18-109_k141_99468 | Not-determined |
| 18-109_k141_171212 | Not-determined |
| 18-109_k141_38711 | Not-determined |
| 18-109_k141_181649 | Not-determined |
| 18-109_k141_38712 | Low-quality |
| 18-109_k141_17807 | Not-determined |
| 18-109_k141_58896 | Not-determined |
| 18-109_k141_161384 | Not-determined |
| 18-109_k141_78505 | Not-determined |
| 18-109_k141_78506 | Not-determined |
| 18-109_k141_7497 | Not-determined |
| 18-109_k141_202474 | Not-determined |
| 18-109_k141_161390 | Not-determined |
| 18-109_k141_202479 | Not-determined |
| 18-109_k141_150185 | Not-determined |
| 18-109_k141_150188 | Low-quality |
| 18-109_k141_150189 | Not-determined |
| 18-109_k141_171227 | Not-determined |
| 18-109_k141_17832 | Not-determined |
| 18-109_k141_48699 | Not-determined |
| 18-109_k141_58914 | Not-determined |
| 18-109_k141_212324 | Not-determined |
| 18-109_k141_161418 | Not-determined |
| 18-109_k141_78528 | Not-determined |
| 18-109_k141_181688 | Not-determined |
| 18-109_k141_212331 | Not-determined |
| 18-109_k141_212332 | Not-determined |
| 18-109_k141_181689 | Not-determined |
| 18-109_k141_220424 | Not-determined |
| 18-109_k141_181694 | Not-determined |
| 18-109_k141_78536 | Not-determined |
| 18-109_k141_7524 | Low-quality |
| 18-109_k141_17854 | Low-quality |
| 18-109_k141_58942 | Not-determined |
| 18-109_k141_242138 | Not-determined |
| 18-109_k141_171239 | Not-determined |
| 18-109_k141_242142 | Not-determined |
| 18-109_k141_58949 | Not-determined |
| 18-109_k141_58951 | Not-determined |
| 18-109_k141_130488 | Low-quality |
| 18-109_k141_161431 | Not-determined |
| 18-109_k141_171244 | Not-determined |
| 18-109_k141_220441 | Not-determined |
| 18-109_k141_171246 | Not-determined |
| 18-109_k141_242151 | Not-determined |
| 18-109_k141_171249 | Not-determined |
| 18-109_k141_78557 | Not-determined |
| 18-109_k141_212344 | Not-determined |

| | |
|---|---|
| 18-109_k141_212352 | Not-determined |
| 18-109_k141_220450 | Low-quality |
| 18-109_k141_99501 | Not-determined |
| 18-109_k141_181724 | Not-determined |
| 18-109_k141_181725 | Not-determined |
| 18-109_k141_108964 | Not-determined |
| 18-109_k141_181734 | Not-determined |
| 18-109_k141_58972 | Not-determined |
| 18-109_k141_150235 | Not-determined |
| 18-109_k141_220459 | Not-determined |
| 18-109_k141_181737 | Not-determined |
| 18-109_k141_220460 | Low-quality |
| 18-109_k141_220464 | Not-determined |
| 18-109_k141_108979 | Not-determined |
| 18-109_k141_17865 | Not-determined |
| 18-109_k141_38791 | Not-determined |
| 18-109_k141_181745 | Not-determined |
| 18-109_k141_220472 | Not-determined |
| 18-109_k141_150249 | Not-determined |
| 18-109_k141_38796 | Not-determined |
| 18-109_k141_108987 | Not-determined |
| 18-109_k141_26843 | Not-determined |
| 18-109_k141_78570 | Not-determined |
| 18-109_k141_181754 | Not-determined |
| 18-109_k141_78572 | Not-determined |
| 18-109_k141_26847 | Low-quality |
| 18-109_k141_150267 | Not-determined |
| 18-109_k141_150268 | Not-determined |
| 18-109_k141_26849 | Not-determined |
| 18-109_k141_150269 | Not-determined |
| 18-109_k141_7556 | Not-determined |
| 18-109_k141_130504 | Not-determined |
| 18-109_k141_181755 | Low-quality |
| 18-109_k141_58996 | Not-determined |
| 18-109_k141_150276 | Not-determined |
| 18-109_k141_69480 | Not-determined |
| 18-109_k141_212362 | Not-determined |
| 18-109_k141_220491 | Not-determined |
| 18-109_k141_59002 | Not-determined |
| 18-109_k141_108991 | Not-determined |
| 18-109_k141_69482 | Not-determined |
| 18-109_k141_78588 | Not-determined |
| 18-109_k141_7569 | Not-determined |
| 18-109_k141_212370 | Not-determined |
| 18-109_k141_99511 | Not-determined |
| 18-109_k141_38818 | Not-determined |
| 18-109_k141_78595 | Not-determined |
| 18-109_k141_48715 | Not-determined |
| 18-109_k141_38824 | Not-determined |
| 18-109_k141_202543 | Not-determined |
| 18-109_k141_59015 | Not-determined |
| 18-109_k141_59016 | Not-determined |
| 18-109_k141_59017 | Not-determined |
| 18-109_k141_7578 | Not-determined |
| 18-109_k141_38827 | Not-determined |
| 18-109_k141_69492 | Not-determined |
| 18-109_k141_78606 | Not-determined |
| 18-109_k141_220508 | Not-determined |
| 18-109_k141_171261 | Not-determined |
| 18-109_k141_202545 | Not-determined |
| 18-109_k141_202547 | Not-determined |
| 18-109_k141_109007 | Not-determined |
| 18-109_k141_130529 | Not-determined |
| 18-109_k141_202553 | Not-determined |
| 18-109_k141_242183 | Not-determined |
| 18-109_k141_99529 | Not-determined |
| 18-109_k141_17913 | Not-determined |
| 18-109_k141_38834 | Not-determined |
| 18-109_k141_220520 | Not-determined |
| 18-109_k141_130533 | Not-determined |
| 18-109_k141_130535 | Not-determined |
| 18-109_k141_99538 | Not-determined |
| 18-109_k141_38837 | Not-determined |
| 18-109_k141_38838 | Not-determined |
| 18-109_k141_192091 | Not-determined |
| 18-109_k141_212394 | Low-quality |
| 18-109_k141_242196 | Not-determined |
| 18-109_k141_69506 | Low-quality |
| 18-109_k141_242208 | Not-determined |
| 18-109_k141_38850 | Not-determined |
| 18-109_k141_38852 | Not-determined |
| 18-109_k141_38853 | Not-determined |
| 18-109_k141_109028 | Not-determined |
| 18-109_k141_242212 | Low-quality |
| 18-109_k141_150290 | Not-determined |
| 18-109_k141_48754 | Not-determined |
| 18-109_k141_69521 | Not-determined |
| 18-109_k141_212402 | Low-quality |
| 18-109_k141_38861 | Not-determined |
| 18-109_k141_120284 | Not-determined |
| 18-109_k141_109033 | Not-determined |
| 18-109_k141_181765 | Not-determined |
| 18-109_k141_48761 | Not-determined |
| 18-109_k141_150297 | Not-determined |
| 18-109_k141_202566 | Not-determined |
| 18-109_k141_69526 | Not-determined |
| 18-109_k141_181772 | Not-determined |
| 18-109_k141_48767 | Not-determined |
| 18-109_k141_17917 | Low-quality |
| 18-109_k141_48769 | Not-determined |
| 18-109_k141_242222 | Not-determined |
| 18-109_k141_232473 | Not-determined |
| 18-109_k141_48775 | Not-determined |
| 18-109_k141_48776 | Not-determined |
| 18-109_k141_109048 | Not-determined |
| 18-109_k141_48778 | Not-determined |
| 18-109_k141_109051 | Not-determined |
| 18-109_k141_171299 | Not-determined |
| 18-109_k141_78646 | Low-quality |
| 18-109_k141_232483 | Not-determined |
| 18-109_k141_59045 | Not-determined |
| 18-109_k141_59046 | Not-determined |
| 18-109_k141_78649 | Not-determined |
| 18-109_k141_48783 | Not-determined |
| 18-109_k141_59048 | Not-determined |
| 18-109_k141_202580 | Not-determined |
| 18-109_k141_181787 | Not-determined |
| 18-109_k141_78654 | Not-determined |
| 18-109_k141_161525 | Not-determined |
| 18-109_k141_38872 | Not-determined |
| 18-109_k141_78659 | Not-determined |
| 18-109_k141_59055 | Not-determined |
| 18-109_k141_88997 | Low-quality |
| 18-109_k141_88998 | Not-determined |
| 18-109_k141_99563 | Not-determined |
| 18-109_k141_99566 | Not-determined |
| 18-109_k141_192123 | Not-determined |
| 18-109_k141_69545 | Not-determined |
| 18-109_k141_89005 | Not-determined |
| 18-109_k141_202592 | Not-determined |
| 18-109_k141_38884 | Not-determined |
| 18-109_k141_120294 | Not-determined |
| 18-109_k141_59062 | Not-determined |
| 18-109_k141_120299 | Not-determined |
| 18-109_k141_181804 | Not-determined |
| 18-109_k141_89012 | Not-determined |
| 18-109_k141_59063 | Low-quality |
| 18-109_k141_181805 | Not-determined |
| 18-109_k141_89015 | Not-determined |
| 18-109_k141_220544 | Not-determined |
| 18-109_k141_99578 | Not-determined |
| 18-109_k141_38894 | Not-determined |
| 18-109_k141_130577 | Not-determined |
| 18-109_k141_99580 | Not-determined |
| 18-109_k141_161538 | Not-determined |
| 18-109_k141_69561 | Low-quality |
| 18-109_k141_89022 | Not-determined |
| 18-109_k141_202609 | Not-determined |
| 18-109_k141_192133 | Not-determined |
| 18-109_k141_130586 | Not-determined |
| 18-109_k141_120309 | Not-determined |
| 18-109_k141_120310 | Not-determined |
| 18-109_k141_232508 | Low-quality |
| 18-109_k141_220554 | Not-determined |
| 18-109_k141_38903 | Not-determined |
| 18-109_k141_99586 | Not-determined |
| 18-109_k141_17953 | Not-determined |
| 18-109_k141_78684 | Low-quality |
| 18-109_k141_232513 | Not-determined |
| 18-109_k141_220555 | Not-determined |
| 18-109_k141_69585 | Not-determined |
| 18-109_k141_232525 | Not-determined |
| 18-109_k141_17961 | Not-determined |
| 18-109_k141_150330 | Not-determined |
| 18-109_k141_242250 | Not-determined |
| 18-109_k141_99609 | Low-quality |
| 18-109_k141_150336 | Not-determined |
| 18-109_k141_232535 | Not-determined |
| 18-109_k141_99616 | Not-determined |
| 18-109_k141_78721 | Not-determined |
| 18-109_k141_99621 | Not-determined |
| 18-109_k141_220568 | Not-determined |
| 18-109_k141_150346 | Not-determined |
| 18-109_k141_120356 | Not-determined |
| 18-109_k141_202633 | Not-determined |
| 18-109_k141_139909 | Not-determined |
| 18-109_k141_99629 | Not-determined |
| 18-109_k141_99630 | Not-determined |
| 18-109_k141_139911 | Not-determined |
| 18-109_k141_17988 | Not-determined |
| 18-109_k141_242274 | Low-quality |
| 18-109_k141_192175 | Not-determined |
| 18-109_k141_99637 | Not-determined |
| 18-109_k141_120360 | Not-determined |
| 18-109_k141_120361 | Not-determined |
| 18-109_k141_192177 | Not-determined |
| 18-109_k141_26907 | Not-determined |
| 18-109_k141_220577 | Not-determined |
| 18-109_k141_242278 | Low-quality |
| 18-109_k141_38913 | Not-determined |
| 18-109_k141_181810 | Not-determined |
| 18-109_k141_69613 | Not-determined |
| 18-109_k141_48844 | Not-determined |
| 18-109_k141_109089 | Low-quality |
| 18-109_k141_202653 | Not-determined |
| 18-109_k141_48846 | Not-determined |
| 18-109_k141_109095 | Not-determined |
| 18-109_k141_150362 | Not-determined |
| 18-109_k141_78742 | Not-determined |
| 18-109_k141_7601 | Not-determined |
| 18-109_k141_161598 | Not-determined |
| 18-109_k141_130637 | Not-determined |
| 18-109_k141_69617 | Not-determined |
| 18-109_k141_139935 | Not-determined |
| 18-109_k141_161604 | Not-determined |
| 18-109_k141_69620 | Not-determined |
| 18-109_k141_181828 | Not-determined |
| 18-109_k141_232550 | Not-determined |
| 18-109_k141_161607 | Not-determined |
| 18-109_k141_78749 | Not-determined |
| 18-109_k141_48859 | Not-determined |
| 18-109_k141_26922 | Not-determined |
| 18-109_k141_192197 | Not-determined |
| 18-109_k141_78761 | Not-determined |
| 18-109_k141_130654 | Not-determined |
| 18-109_k141_192207 | Not-determined |
| 18-109_k141_78765 | Not-determined |
| 18-109_k141_202657 | Not-determined |
| 18-109_k141_109126 | Not-determined |
| 18-109_k141_109128 | Not-determined |
| 18-109_k141_48875 | Not-determined |
| 18-109_k141_242316 | Not-determined |
| 18-109_k141_59094 | Not-determined |
| 18-109_k141_139959 | Not-determined |
| 18-109_k141_78769 | Not-determined |
| 18-109_k141_130670 | Not-determined |
| 18-109_k141_192216 | Not-determined |
| 18-109_k141_69631 | Not-determined |
| 18-109_k141_202660 | Not-determined |
| 18-109_k141_109135 | Not-determined |
| 18-109_k141_130675 | Not-determined |
| 18-109_k141_242321 | Not-determined |
| 18-109_k141_130677 | Not-determined |
| 18-109_k141_161622 | Not-determined |
| 18-109_k141_69637 | Not-determined |
| 18-109_k141_99649 | Not-determined |
| 18-109_k141_202665 | Not-determined |
| 18-109_k141_161628 | Not-determined |
| 18-109_k141_59098 | Not-determined |
| 18-109_k141_38930 | Not-determined |
| 18-109_k141_78791 | Not-determined |
| 18-109_k141_69656 | Not-determined |
| 18-109_k141_202683 | Not-determined |
| 18-109_k141_232578 | Not-determined |
| 18-109_k141_7658 | Not-determined |
| 18-109_k141_120381 | Not-determined |
| 18-109_k141_38944 | Not-determined |
| 18-109_k141_59114 | Low-quality |
| 18-109_k141_202689 | Not-determined |
| 18-109_k141_161642 | Not-determined |
| 18-109_k141_59117 | Not-determined |
| 18-109_k141_161643 | Not-determined |
| 18-109_k141_78805 | Not-determined |
| 18-109_k141_59128 | Not-determined |
| 18-109_k141_161648 | Not-determined |
| 18-109_k141_109157 | Not-determined |
| 18-109_k141_26966 | Not-determined |
| 18-109_k141_139977 | Not-determined |
| 18-109_k141_78809 | Not-determined |
| 18-109_k141_220612 | Not-determined |
| 18-109_k141_99668 | Not-determined |
| 18-109_k141_7672 | Not-determined |
| 18-109_k141_202699 | Not-determined |
| 18-109_k141_7673 | Not-determined |
| 18-109_k141_99671 | Not-determined |
| 18-109_k141_109163 | Not-determined |
| 18-109_k141_48905 | Low-quality |
| 18-109_k141_242349 | Not-determined |
| 18-109_k141_109164 | Not-determined |
| 18-109_k141_212404 | Not-determined |
| 18-109_k141_7678 | Low-quality |
| 18-109_k141_7680 | Not-determined |
| 18-109_k141_99676 | Not-determined |
| 18-109_k141_7683 | Not-determined |
| 18-109_k141_242357 | Not-determined |
| 18-109_k141_212416 | Not-determined |
| 18-109_k141_26976 | Not-determined |
| 18-109_k141_232592 | Not-determined |
| 18-109_k141_120403 | Not-determined |
| 18-109_k141_99688 | Not-determined |
| 18-109_k141_7691 | Not-determined |
| 18-109_k141_202713 | Low-quality |
| 18-109_k141_59139 | Not-determined |
| 18-109_k141_212428 | Low-quality |
| 18-109_k141_242367 | Not-determined |
| 18-109_k141_232595 | Low-quality |
| 18-109_k141_181853 | Low-quality |
| 18-109_k141_99704 | Not-determined |
| 18-109_k141_242373 | Not-determined |
| 18-109_k141_120417 | Not-determined |
| 18-109_k141_48921 | Not-determined |
| 18-109_k141_171368 | Not-determined |
| 18-109_k141_171373 | Not-determined |
| 18-109_k141_171377 | Not-determined |
| 18-109_k141_171384 | Not-determined |
| 18-109_k141_26997 | Not-determined |
| 18-109_k141_109179 | Not-determined |
| 18-109_k141_161694 | Not-determined |
| 18-109_k141_202729 | Not-determined |
| 18-109_k141_150403 | Not-determined |
| 18-109_k141_109182 | Not-determined |
| 18-109_k141_242401 | Not-determined |
| 18-109_k141_202730 | Not-determined |
| 18-109_k141_27000 | Not-determined |
| 18-109_k141_192292 | Not-determined |
| 18-109_k141_27003 | Not-determined |
| 18-109_k141_181863 | Not-determined |
| 18-109_k141_192297 | Not-determined |
| 18-109_k141_99731 | Low-quality |
| 18-109_k141_99732 | Not-determined |
| 18-109_k141_140006 | Not-determined |
| 18-109_k141_140007 | Low-quality |
| 18-109_k141_89053 | Not-determined |
| 18-109_k141_140011 | Not-determined |
| 18-109_k141_192301 | Not-determined |
| 18-109_k141_140012 | Not-determined |
| 18-109_k141_99739 | Not-determined |
| 18-109_k141_150422 | Not-determined |
| 18-109_k141_202757 | Low-quality |
| 18-109_k141_171398 | Not-determined |
| 18-109_k141_232631 | Not-determined |
| 18-109_k141_120440 | Not-determined |
| 18-109_k141_212459 | Not-determined |
| 18-109_k141_109196 | Not-determined |
| 18-109_k141_181878 | Not-determined |
| 18-109_k141_109197 | Not-determined |
| 18-109_k141_130730 | Not-determined |
| 18-109_k141_130732 | Not-determined |
| 18-109_k141_130733 | Not-determined |
| 18-109_k141_212464 | Not-determined |
| 18-109_k141_69694 | Not-determined |
| 18-109_k141_212475 | Not-determined |
| 18-109_k141_202776 | Not-determined |
| 18-109_k141_48956 | Not-determined |
| 18-109_k141_212476 | Not-determined |
| 18-109_k141_18031 | Not-determined |
| 18-109_k141_120463 | Not-determined |
| 18-109_k141_212481 | Not-determined |
| 18-109_k141_120467 | Not-determined |
| 18-109_k141_140035 | Not-determined |
| 18-109_k141_242413 | Not-determined |
| 18-109_k141_48974 | Not-determined |
| 18-109_k141_140039 | Not-determined |
| 18-109_k141_109203 | Not-determined |
| 18-109_k141_109209 | Not-determined |
| 18-109_k141_99776 | Not-determined |
| 18-109_k141_150454 | Not-determined |
| 18-109_k141_140046 | Not-determined |
| 18-109_k141_89069 | Not-determined |
| 18-109_k141_89071 | Not-determined |
| 18-109_k141_140051 | Not-determined |
| 18-109_k141_171423 | Not-determined |
| 18-109_k141_39028 | Not-determined |
| 18-109_k141_232657 | Not-determined |
| 18-109_k141_171429 | Not-determined |
| 18-109_k141_120482 | Not-determined |
| 18-109_k141_39036 | Not-determined |
| 18-109_k141_232669 | Not-determined |
| 18-109_k141_232671 | Not-determined |
| 18-109_k141_78816 | Not-determined |
| 18-109_k141_150477 | Not-determined |
| 18-109_k141_18049 | Low-quality |
| 18-109_k141_78823 | Not-determined |
| 18-109_k141_120493 | Not-determined |
| 18-109_k141_99799 | Not-determined |
| 18-109_k141_99801 | Not-determined |
| 18-109_k141_242453 | Not-determined |
| 18-109_k141_120503 | Not-determined |
| 18-109_k141_242461 | Low-quality |
| 18-109_k141_120511 | Not-determined |
| 18-109_k141_242462 | Not-determined |
| 18-109_k141_49022 | Not-determined |
| 18-109_k141_130765 | Not-determined |
| 18-109_k141_202815 | Not-determined |
| 18-109_k141_99816 | Not-determined |
| 18-109_k141_181948 | Not-determined |
| 18-109_k141_99818 | Not-determined |
| 18-109_k141_192318 | Low-quality |
| 18-109_k141_120521 | Not-determined |
| 18-109_k141_242477 | Not-determined |
| 18-109_k141_161772 | Not-determined |
| 18-109_k141_181953 | Not-determined |
| 18-109_k141_18073 | Not-determined |
| 18-109_k141_181959 | Low-quality |
| 18-109_k141_242485 | Not-determined |
| 18-109_k141_192329 | Not-determined |
| 18-109_k141_59222 | Not-determined |
| 18-109_k141_49043 | Not-determined |
| 18-109_k141_18077 | Not-determined |
| 18-109_k141_49045 | Not-determined |
| 18-109_k141_220632 | Not-determined |
| 18-109_k141_27021 | Not-determined |
| 18-109_k141_202837 | Not-determined |
| 18-109_k141_109241 | Not-determined |
| 18-109_k141_212531 | Not-determined |
| 18-109_k141_181971 | Not-determined |
| 18-109_k141_18082 | Not-determined |
| 18-109_k141_130789 | Not-determined |
| 18-109_k141_49056 | Not-determined |
| 18-109_k141_59242 | Not-determined |
| 18-109_k141_130790 | Not-determined |
| 18-109_k141_109250 | Not-determined |
| 18-109_k141_120538 | Not-determined |
| 18-109_k141_181981 | Not-determined |
| 18-109_k141_140095 | Not-determined |
| 18-109_k141_140097 | Not-determined |
| 18-109_k141_140099 | Not-determined |
| 18-109_k141_242506 | Not-determined |
| 18-109_k141_220655 | Not-determined |
| 18-109_k141_171476 | Not-determined |
| 18-109_k141_120545 | Not-determined |
| 18-109_k141_78871 | Not-determined |
| 18-109_k141_232745 | Low-quality |
| 18-109_k141_202861 | Not-determined |
| 18-109_k141_220660 | Not-determined |
| 18-109_k141_192339 | Low-quality |
| 18-109_k141_192342 | Not-determined |
| 18-109_k141_27034 | Not-determined |
| 18-109_k141_130805 | Not-determined |
| 18-109_k141_27037 | Not-determined |
| 18-109_k141_18103 | Not-determined |
| 18-109_k141_192344 | Not-determined |
| 18-109_k141_120554 | Not-determined |
| 18-109_k141_109260 | Not-determined |
| 18-109_k141_202874 | Not-determined |
| 18-109_k141_49060 | Not-determined |
| 18-109_k141_212543 | Not-determined |
| 18-109_k141_18110 | Not-determined |
| 18-109_k141_39096 | Not-determined |
| 18-109_k141_120561 | Not-determined |
| 18-109_k141_232757 | Not-determined |
| 18-109_k141_192348 | Not-determined |
| 18-109_k141_120564 | Not-determined |
| 18-109_k141_89130 | Not-determined |
| 18-109_k141_220678 | Not-determined |
| 18-109_k141_7723 | Not-determined |
| 18-109_k141_130831 | Not-determined |
| 18-109_k141_78893 | Not-determined |
| 18-109_k141_192359 | Not-determined |
| 18-109_k141_89142 | Not-determined |
| 18-109_k141_232766 | Low-quality |
| 18-109_k141_171494 | Not-determined |
| 18-109_k141_7733 | Not-determined |
| 18-109_k141_18125 | Not-determined |
| 18-109_k141_78900 | Not-determined |
| 18-109_k141_130832 | Not-determined |
| 18-109_k141_192365 | Not-determined |
| 18-109_k141_232772 | Low-quality |
| 18-109_k141_202881 | Not-determined |
| 18-109_k141_99860 | Not-determined |
| 18-109_k141_89152 | Not-determined |
| 18-109_k141_7744 | Not-determined |
| 18-109_k141_99870 | Low-quality |
| 18-109_k141_130847 | Not-determined |
| 18-109_k141_49071 | Low-quality |
| 18-109_k141_171506 | Not-determined |
| 18-109_k141_232781 | Not-determined |
| 18-109_k141_232783 | Not-determined |
| 18-109_k141_109286 | Not-determined |
| 18-109_k141_202900 | Not-determined |
| 18-109_k141_161794 | Not-determined |
| 18-109_k141_192382 | Low-quality |
| 18-109_k141_59267 | Not-determined |
| 18-109_k141_7760 | Not-determined |
| 18-109_k141_7761 | Not-determined |
| 18-109_k141_59271 | Not-determined |
| 18-109_k141_232791 | Not-determined |
| 18-109_k141_232796 | Not-determined |
| 18-109_k141_99882 | Not-determined |
| 18-109_k141_150507 | Not-determined |
| 18-109_k141_69768 | Low-quality |
| 18-109_k141_78925 | Low-quality |
| 18-109_k141_182054 | Not-determined |
| 18-109_k141_150513 | Not-determined |
| 18-109_k141_69788 | Not-determined |
| 18-109_k141_140169 | Not-determined |
| 18-109_k141_192404 | Not-determined |
| 18-109_k141_7793 | Not-determined |
| 18-109_k141_220719 | Not-determined |
| 18-109_k141_140171 | Not-determined |
| 18-109_k141_192407 | Not-determined |
| 18-109_k141_49079 | Not-determined |
| 18-109_k141_232802 | Not-determined |
| 18-109_k141_89167 | Not-determined |
| 18-109_k141_49082 | Not-determined |
| 18-109_k141_18130 | Not-determined |
| 18-109_k141_182070 | Not-determined |
| 18-109_k141_99894 | Not-determined |
| 18-109_k141_109319 | Not-determined |
| 18-109_k141_89178 | Not-determined |
| 18-109_k141_150521 | Not-determined |
| 18-109_k141_109327 | Not-determined |
| 18-109_k141_242536 | Not-determined |
| 18-109_k141_99906 | Not-determined |
| 18-109_k141_242537 | Not-determined |
| 18-109_k141_49091 | Not-determined |
| 18-109_k141_89186 | Not-determined |
| 18-109_k141_171524 | Not-determined |
| 18-109_k141_18143 | Not-determined |
| 18-109_k141_89190 | Not-determined |
| 18-109_k141_7817 | Low-quality |
| 18-109_k141_171530 | Not-determined |
| 18-109_k141_150534 | Not-determined |
| 18-109_k141_182088 | Not-determined |
| 18-109_k141_150535 | Not-determined |
| 18-109_k141_150537 | Not-determined |
| 18-109_k141_18148 | Not-determined |
| 18-109_k141_18149 | Not-determined |
| 18-109_k141_89192 | Not-determined |
| 18-109_k141_99923 | Not-determined |
| 18-109_k141_212575 | Not-determined |
| 18-109_k141_18162 | Not-determined |
| 18-109_k141_202977 | Not-determined |
| 18-109_k141_150548 | Not-determined |
| 18-109_k141_150551 | Not-determined |
| 18-109_k141_18176 | Not-determined |
| 18-109_k141_171551 | Not-determined |
| 18-109_k141_109342 | Not-determined |
| 18-109_k141_150556 | Low-quality |
| 18-109_k141_109344 | Not-determined |
| 18-109_k141_150560 | Not-determined |
| 18-109_k141_182109 | Not-determined |
| 18-109_k141_140216 | Not-determined |
| 18-109_k141_49116 | Not-determined |
| 18-109_k141_212598 | Not-determined |
| 18-109_k141_140219 | Not-determined |
| 18-109_k141_49123 | Not-determined |
| 18-109_k141_150562 | Not-determined |
| 18-109_k141_49126 | Not-determined |
| 18-109_k141_182118 | Not-determined |
| 18-109_k141_232850 | Not-determined |
| 18-109_k141_220747 | Not-determined |
| 18-109_k141_220751 | Not-determined |
| 18-109_k141_150567 | Not-determined |
| 18-109_k141_109364 | Not-determined |
| 18-109_k141_78958 | Not-determined |
| 18-109_k141_232861 | Not-determined |
| 18-109_k141_232863 | Not-determined |
| 18-109_k141_7853 | Not-determined |
| 18-109_k141_150582 | Low-quality |
| 18-109_k141_7858 | Not-determined |
| 18-109_k141_49145 | Not-determined |
| 18-109_k141_192445 | Not-determined |
| 18-109_k141_232875 | Not-determined |
| 18-109_k141_109387 | Not-determined |
| 18-109_k141_182130 | Low-quality |
| 18-109_k141_192448 | Not-determined |
| 18-109_k141_192450 | Not-determined |
| 18-109_k141_109396 | Not-determined |
| 18-109_k141_192454 | Not-determined |
| 18-109_k141_59310 | Not-determined |
| 18-109_k141_89241 | Low-quality |
| 18-109_k141_232892 | Not-determined |
| 18-109_k141_7875 | Not-determined |
| 18-109_k141_89243 | Not-determined |
| 18-109_k141_232893 | Not-determined |
| 18-109_k141_150599 | Not-determined |
| 18-109_k141_203037 | Not-determined |
| 18-109_k141_109400 | Not-determined |
| 18-109_k141_109401 | Not-determined |
| 18-109_k141_192463 | Not-determined |
| 18-109_k141_192464 | Not-determined |
| 18-109_k141_232901 | Not-determined |
| 18-109_k141_220788 | Not-determined |
| 18-109_k141_220789 | Not-determined |
| 18-109_k141_203043 | Low-quality |
| 18-109_k141_171600 | Not-determined |
| 18-109_k141_49162 | Not-determined |
| 18-109_k141_182147 | Low-quality |
| 18-109_k141_182157 | Not-determined |
| 18-109_k141_7898 | Not-determined |
| 18-109_k141_109423 | Low-quality |
| 18-109_k141_49172 | Not-determined |
| 18-109_k141_59331 | Not-determined |
| 18-109_k141_220798 | Not-determined |
| 18-109_k141_220799 | Not-determined |
| 18-109_k141_78975 | Low-quality |
| 18-109_k141_192476 | Not-determined |
| 18-109_k141_59334 | Not-determined |
| 18-109_k141_220803 | Not-determined |
| 18-109_k141_49180 | Not-determined |
| 18-109_k141_89269 | Not-determined |
| 18-109_k141_109433 | Not-determined |
| 18-109_k141_39149 | Not-determined |
| 18-109_k141_18209 | Not-determined |
| 18-109_k141_203051 | Not-determined |
| 18-109_k141_220807 | Not-determined |
| 18-109_k141_203055 | Not-determined |
| 18-109_k141_192486 | Not-determined |

| | |
|---|---|
| 18-109_k141_232932 | Not-determined |
| 18-109_k141_89283 | Not-determined |
| 18-109_k141_59344 | Not-determined |
| 18-109_k141_232935 | Not-determined |
| 18-109_k141_18223 | Not-determined |
| 18-109_k141_18225 | Low-quality |
| 18-109_k141_150617 | Not-determined |
| 18-109_k141_220823 | Not-determined |
| 18-109_k141_232942 | Not-determined |
| 18-109_k141_150619 | Not-determined |
| 18-109_k141_120578 | Low-quality |
| 18-109_k141_203074 | Not-determined |
| 18-109_k141_150635 | Not-determined |
| 18-109_k141_150638 | Not-determined |
| 18-109_k141_220835 | Not-determined |
| 18-109_k141_79007 | Not-determined |
| 18-109_k141_212668 | Not-determined |
| 18-109_k141_150640 | Not-determined |
| 18-109_k141_220842 | Not-determined |
| 18-109_k141_18248 | Not-determined |
| 18-109_k141_99963 | Low-quality |
| 18-109_k141_99973 | Not-determined |
| 18-109_k141_140240 | Not-determined |
| 18-109_k141_203104 | Low-quality |
| 18-109_k141_120598 | Not-determined |
| 18-109_k141_49216 | Not-determined |
| 18-109_k141_49217 | Not-determined |
| 18-109_k141_120604 | Not-determined |
| 18-109_k141_49219 | Low-quality |
| 18-109_k141_99982 | Not-determined |
| 18-109_k141_203119 | Not-determined |
| 18-109_k141_39180 | Low-quality |
| 18-109_k141_161892 | Not-determined |
| 18-109_k141_99988 | Not-determined |
| 18-109_k141_140265 | Not-determined |
| 18-109_k141_120617 | Not-determined |
| 18-109_k141_220861 | Not-determined |
| 18-109_k141_182176 | Not-determined |
| 18-109_k141_192514 | Not-determined |
| 18-109_k141_7919 | Not-determined |
| 18-109_k141_212694 | Not-determined |
| 18-109_k141_89285 | Not-determined |
| 18-109_k141_171668 | Not-determined |
| 18-109_k141_120639 | Not-determined |
| 18-109_k141_171671 | Not-determined |
| 18-109_k141_171673 | Not-determined |
| 18-109_k141_39188 | Low-quality |
| 18-109_k141_212705 | Not-determined |
| 18-109_k141_7924 | Low-quality |
| 18-109_k141_89289 | Not-determined |
| 18-109_k141_7927 | Not-determined |
| 18-109_k141_7928 | Not-determined |
| 18-109_k141_203129 | Not-determined |
| 18-109_k141_161900 | Not-determined |
| 18-109_k141_192534 | Not-determined |
| 18-109_k141_39197 | Not-determined |
| 18-109_k141_171692 | Not-determined |
| 18-109_k141_171697 | Not-determined |
| 18-109_k141_232992 | Low-quality |
| 18-109_k141_161915 | Low-quality |
| 18-109_k141_39207 | Not-determined |
| 18-109_k141_79049 | Not-determined |
| 18-109_k141_161918 | Not-determined |
| 18-109_k141_7949 | Low-quality |
| 18-109_k141_150665 | Not-determined |
| 18-109_k141_79058 | Not-determined |
| 18-109_k141_120655 | Not-determined |
| 18-109_k141_100017 | Not-determined |
| 18-109_k141_192546 | Low-quality |
| 18-109_k141_161928 | Not-determined |
| 18-109_k141_161929 | Not-determined |
| 18-109_k141_79062 | Not-determined |
| 18-109_k141_161933 | Not-determined |
| 18-109_k141_109492 | Not-determined |
| 18-109_k141_39219 | Not-determined |
| 18-109_k141_150672 | Not-determined |
| 18-109_k141_220874 | Not-determined |
| 18-109_k141_49237 | Not-determined |
| 18-109_k141_69799 | Not-determined |
| 18-109_k141_49241 | Not-determined |
| 18-109_k141_182202 | Not-determined |
| 18-109_k141_69806 | Not-determined |
| 18-109_k141_220884 | Not-determined |
| 18-109_k141_120680 | Not-determined |
| 18-109_k141_220887 | Not-determined |
| 18-109_k141_7969 | Not-determined |
| 18-109_k141_233008 | Not-determined |
| 18-109_k141_49247 | Not-determined |
| 18-109_k141_120682 | Not-determined |
| 18-109_k141_220894 | Not-determined |
| 18-109_k141_120684 | Not-determined |
| 18-109_k141_203176 | Not-determined |
| 18-109_k141_49250 | Not-determined |
| 18-109_k141_182204 | Not-determined |
| 18-109_k141_109517 | Not-determined |
| 18-109_k141_49253 | Not-determined |
| 18-109_k141_120688 | Not-determined |
| 18-109_k141_192553 | Not-determined |
| 18-109_k141_109521 | Not-determined |
| 18-109_k141_79093 | Not-determined |
| 18-109_k141_212742 | Not-determined |
| 18-109_k141_100048 | Not-determined |
| 18-109_k141_192558 | Not-determined |
| 18-109_k141_49261 | Not-determined |
| 18-109_k141_89345 | Not-determined |
| 18-109_k141_120698 | Not-determined |
| 18-109_k141_39238 | Not-determined |
| 18-109_k141_192562 | Not-determined |
| 18-109_k141_150682 | Not-determined |
| 18-109_k141_69828 | Not-determined |
| 18-109_k141_220922 | Not-determined |
| 18-109_k141_171722 | Not-determined |
| 18-109_k141_49271 | Not-determined |
| 18-109_k141_203208 | Not-determined |
| 18-109_k141_49272 | Not-determined |
| 18-109_k141_182229 | Not-determined |
| 18-109_k141_212758 | Low-quality |
| 18-109_k141_220931 | Not-determined |
| 18-109_k141_89364 | Not-determined |
| 18-109_k141_182233 | Not-determined |
| 18-109_k141_100067 | Not-determined |
| 18-109_k141_171726 | Not-determined |
| 18-109_k141_100070 | Not-determined |
| 18-109_k141_182234 | Not-determined |
| 18-109_k141_182236 | Not-determined |
| 18-109_k141_203219 | Not-determined |
| 18-109_k141_100076 | Not-determined |
| 18-109_k141_39261 | Not-determined |
| 18-109_k141_171739 | Not-determined |
| 18-109_k141_192588 | Not-determined |
| 18-109_k141_39266 | Not-determined |
| 18-109_k141_212777 | Not-determined |
| 18-109_k141_69843 | Not-determined |
| 18-109_k141_59428 | Low-quality |
| 18-109_k141_192598 | Not-determined |
| 18-109_k141_69847 | Not-determined |
| 18-109_k141_49300 | Not-determined |
| 18-109_k141_192603 | Not-determined |
| 18-109_k141_69854 | Not-determined |
| 18-109_k141_39279 | Not-determined |
| 18-109_k141_89388 | Not-determined |
| 18-109_k141_120743 | Not-determined |
| 18-109_k141_39286 | Low-quality |
| 18-109_k141_18271 | Not-determined |
| 18-109_k141_203251 | Low-quality |
| 18-109_k141_171764 | Not-determined |
| 18-109_k141_69859 | Not-determined |
| 18-109_k141_69860 | Not-determined |
| 18-109_k141_120750 | Not-determined |
| 18-109_k141_18276 | Not-determined |
| 18-109_k141_69866 | Not-determined |
| 18-109_k141_69867 | Not-determined |
| 18-109_k141_100111 | Not-determined |
| 18-109_k141_69868 | Not-determined |
| 18-109_k141_69870 | Not-determined |
| 18-109_k141_203275 | Not-determined |
| 18-109_k141_39291 | Not-determined |
| 18-109_k141_39298 | Low-quality |
| 18-109_k141_109553 | Not-determined |
| 18-109_k141_212807 | Not-determined |
| 18-109_k141_212809 | Not-determined |
| 18-109_k141_192634 | Not-determined |
| 18-109_k141_233050 | Not-determined |
| 18-109_k141_39309 | Not-determined |
| 18-109_k141_18285 | Not-determined |
| 18-109_k141_192646 | Not-determined |
| 18-109_k141_120768 | Not-determined |
| 18-109_k141_203282 | Not-determined |
| 18-109_k141_8010 | Not-determined |
| 18-109_k141_8013 | Not-determined |
| 18-109_k141_69896 | Not-determined |
| 18-109_k141_49331 | Not-determined |
| 18-109_k141_39317 | Not-determined |
| 18-109_k141_39319 | Not-determined |
| 18-109_k141_69898 | Not-determined |
| 18-109_k141_150728 | Not-determined |
| 18-109_k141_69901 | Not-determined |
| 18-109_k141_212831 | Not-determined |
| 18-109_k141_39329 | Low-quality |
| 18-109_k141_8019 | Not-determined |
| 18-109_k141_69909 | Not-determined |
| 18-109_k141_79106 | Low-quality |
| 18-109_k141_8029 | Not-determined |
| 18-109_k141_150742 | Not-determined |
| 18-109_k141_18319 | Not-determined |
| 18-109_k141_39336 | Not-determined |
| 18-109_k141_79115 | Not-determined |
| 18-109_k141_192660 | Not-determined |
| 18-109_k141_79119 | Not-determined |
| 18-109_k141_242567 | Not-determined |
| 18-109_k141_18338 | Not-determined |
| 18-109_k141_39352 | Not-determined |
| 18-109_k141_79139 | Not-determined |
| 18-109_k141_59461 | Not-determined |
| 18-109_k141_233094 | Low-quality |
| 18-109_k141_109602 | Not-determined |
| 18-109_k141_8047 | Low-quality |
| 18-109_k141_203305 | Not-determined |
| 18-109_k141_242579 | Not-determined |
| 18-109_k141_39364 | Not-determined |
| 18-109_k141_39367 | Not-determined |
| 18-109_k141_109609 | Not-determined |
| 18-109_k141_59465 | Not-determined |
| 18-109_k141_39370 | Not-determined |
| 18-109_k141_8062 | Not-determined |
| 18-109_k141_69926 | Not-determined |
| 18-109_k141_59467 | Not-determined |
| 18-109_k141_242589 | Not-determined |
| 18-109_k141_233118 | Not-determined |
| 18-109_k141_100133 | Not-determined |
| 18-109_k141_49355 | Not-determined |
| 18-109_k141_242596 | Not-determined |
| 18-109_k141_69942 | Not-determined |
| 18-109_k141_182266 | Not-determined |
| 18-109_k141_8074 | Low-quality |
| 18-109_k141_220940 | Not-determined |
| 18-109_k141_182269 | Not-determined |
| 18-109_k141_49369 | Not-determined |
| 18-109_k141_59478 | Not-determined |
| 18-109_k141_212864 | Not-determined |
| 18-109_k141_212865 | Not-determined |
| 18-109_k141_120819 | Not-determined |
| 18-109_k141_109627 | Not-determined |
| 18-109_k141_79165 | Not-determined |
| 18-109_k141_59485 | Not-determined |
| 18-109_k141_8083 | Low-quality |
| 18-109_k141_100166 | Not-determined |
| 18-109_k141_192711 | Not-determined |
| 18-109_k141_182287 | Not-determined |
| 18-109_k141_220952 | Not-determined |
| 18-109_k141_39401 | Not-determined |
| 18-109_k141_120835 | Not-determined |
| 18-109_k141_120839 | Not-determined |
| 18-109_k141_161978 | Not-determined |
| 18-109_k141_69957 | Not-determined |
| 18-109_k141_242627 | Not-determined |
| 18-109_k141_120843 | Not-determined |
| 18-109_k141_203357 | Not-determined |
| 18-109_k141_233135 | Not-determined |
| 18-109_k141_59499 | Not-determined |
| 18-109_k141_171828 | Not-determined |
| 18-109_k141_150781 | Low-quality |
| 18-109_k141_120849 | Not-determined |
| 18-109_k141_162001 | Not-determined |
| 18-109_k141_8086 | Not-determined |
| 18-109_k141_233148 | Not-determined |
| 18-109_k141_171841 | Not-determined |
| 18-109_k141_162003 | Not-determined |
| 18-109_k141_8089 | Not-determined |
| 18-109_k141_192732 | Low-quality |
| 18-109_k141_162005 | Not-determined |
| 18-109_k141_171845 | Not-determined |
| 18-109_k141_120854 | Not-determined |
| 18-109_k141_120855 | Not-determined |
| 18-109_k141_109650 | Not-determined |
| 18-109_k141_233157 | Low-quality |
| 18-109_k141_8091 | Not-determined |
| 18-109_k141_233161 | Not-determined |
| 18-109_k141_242651 | Not-determined |
| 18-109_k141_242653 | Not-determined |
| 18-109_k141_192746 | Not-determined |
| 18-109_k141_109658 | Not-determined |
| 18-109_k141_242661 | Not-determined |
| 18-109_k141_162026 | Not-determined |
| 18-109_k141_203362 | Not-determined |
| 18-109_k141_242671 | Not-determined |
| 18-109_k141_49406 | Not-determined |
| 18-109_k141_221007 | Not-determined |
| 18-109_k141_79200 | Not-determined |
| 18-109_k141_171874 | Not-determined |
| 18-109_k141_109665 | Not-determined |
| 18-109_k141_171877 | Not-determined |
| 18-109_k141_100187 | Not-determined |
| 18-109_k141_100188 | Not-determined |
| 18-109_k141_109669 | Low-quality |
| 18-109_k141_100190 | Not-determined |
| 18-109_k141_203374 | Not-determined |
| 18-109_k141_203377 | Not-determined |
| 18-109_k141_171887 | Not-determined |
| 18-109_k141_49414 | Not-determined |
| 18-109_k141_109679 | Not-determined |
| 18-109_k141_27049 | Not-determined |
| 18-109_k141_171892 | Not-determined |
| 18-109_k141_203385 | Not-determined |
| 18-109_k141_27052 | Not-determined |
| 18-109_k141_192771 | Not-determined |
| 18-109_k141_109685 | Not-determined |
| 18-109_k141_233181 | Not-determined |
| 18-109_k141_59517 | Low-quality |
| 18-109_k141_192784 | Not-determined |
| 18-109_k141_100212 | Not-determined |
| 18-109_k141_233190 | Not-determined |
| 18-109_k141_171903 | Not-determined |
| 18-109_k141_120910 | Not-determined |
| 18-109_k141_27067 | Not-determined |
| 18-109_k141_27069 | Not-determined |
| 18-109_k141_162043 | Low-quality |
| 18-109_k141_233203 | Not-determined |
| 18-109_k141_120917 | Not-determined |
| 18-109_k141_49433 | Not-determined |
| 18-109_k141_100220 | Not-determined |
| 18-109_k141_221043 | Not-determined |
| 18-109_k141_89417 | Not-determined |
| 18-109_k141_100223 | Not-determined |
| 18-109_k141_109716 | Not-determined |
| 18-109_k141_89418 | Not-determined |
| 18-109_k141_182342 | Not-determined |
| 18-109_k141_100233 | Not-determined |
| 18-109_k141_100235 | Not-determined |
| 18-109_k141_233231 | Not-determined |
| 18-109_k141_100244 | Not-determined |
| 18-109_k141_233235 | Low-quality |
| 18-109_k141_109743 | Not-determined |
| 18-109_k141_242734 | Low-quality |
| 18-109_k141_182364 | Not-determined |
| 18-109_k141_8154 | Not-determined |
| 18-109_k141_182369 | Not-determined |
| 18-109_k141_109754 | Not-determined |
| 18-109_k141_150795 | Not-determined |
| 18-109_k141_59537 | Low-quality |
| 18-109_k141_39451 | Not-determined |
| 18-109_k141_120941 | Not-determined |
| 18-109_k141_242746 | Not-determined |
| 18-109_k141_89452 | Not-determined |
| 18-109_k141_150798 | Not-determined |
| 18-109_k141_130882 | Not-determined |
| 18-109_k141_130884 | Not-determined |
| 18-109_k141_39458 | Not-determined |
| 18-109_k141_69991 | Not-determined |
| 18-109_k141_130887 | Not-determined |
| 18-109_k141_150802 | Not-determined |
| 18-109_k141_120952 | Not-determined |
| 18-109_k141_79220 | Not-determined |
| 18-109_k141_130889 | Not-determined |
| 18-109_k141_39461 | Not-determined |
| 18-109_k141_120955 | Not-determined |
| 18-109_k141_140316 | Not-determined |
| 18-109_k141_8165 | Not-determined |
| 18-109_k141_150809 | Not-determined |
| 18-109_k141_221098 | Not-determined |
| 18-109_k141_242758 | Not-determined |
| 18-109_k141_242759 | Not-determined |
| 18-109_k141_140319 | Not-determined |
| 18-109_k141_192798 | Not-determined |
| 18-109_k141_171919 | Not-determined |
| 18-109_k141_70002 | Not-determined |
| 18-109_k141_192802 | Not-determined |
| 18-109_k141_79223 | Not-determined |
| 18-109_k141_242765 | Not-determined |
| 18-109_k141_109776 | Not-determined |
| 18-109_k141_203452 | Not-determined |
| 18-109_k141_203454 | Not-determined |
| 18-109_k141_140329 | Not-determined |
| 18-109_k141_182393 | Not-determined |
| 18-109_k141_100267 | Not-determined |
| 18-109_k141_59551 | Not-determined |
| 18-109_k141_242788 | Not-determined |
| 18-109_k141_221112 | Not-determined |
| 18-109_k141_49494 | Not-determined |
| 18-109_k141_100278 | Not-determined |
| 18-109_k141_8183 | Not-determined |
| 18-109_k141_49499 | Not-determined |
| 18-109_k141_49500 | Not-determined |
| 18-109_k141_59564 | Not-determined |
| 18-109_k141_89461 | Not-determined |
| 18-109_k141_59568 | Not-determined |
| 18-109_k141_39489 | Not-determined |
| 18-109_k141_27138 | Not-determined |
| 18-109_k141_221129 | Not-determined |
| 18-109_k141_49506 | Not-determined |
| 18-109_k141_27144 | Not-determined |
| 18-109_k141_242811 | Not-determined |
| 18-109_k141_242813 | Not-determined |
| 18-109_k141_49512 | Not-determined |
| 18-109_k141_49513 | Not-determined |
| 18-109_k141_120992 | Not-determined |
| 18-109_k141_49514 | Not-determined |
| 18-109_k141_70017 | Not-determined |
| 18-109_k141_233265 | Not-determined |
| 18-109_k141_39502 | Not-determined |
| 18-109_k141_49522 | Not-determined |
| 18-109_k141_233267 | Not-determined |
| 18-109_k141_39507 | Not-determined |
| 18-109_k141_242824 | Low-quality |
| 18-109_k141_121005 | Not-determined |
| 18-109_k141_39514 | Not-determined |
| 18-109_k141_121008 | Not-determined |
| 18-109_k141_130903 | Not-determined |
| 18-109_k141_8195 | Not-determined |
| 18-109_k141_130909 | Not-determined |
| 18-109_k141_109806 | Not-determined |
| 18-109_k141_49535 | Not-determined |
| 18-109_k141_121014 | Not-determined |
| 18-109_k141_140378 | Not-determined |
| 18-109_k141_109809 | Not-determined |
| 18-109_k141_130914 | Not-determined |
| 18-109_k141_39525 | Not-determined |
| 18-109_k141_49538 | Not-determined |
| 18-109_k141_39527 | Not-determined |
| 18-109_k141_59599 | Not-determined |
| 18-109_k141_182420 | Not-determined |
| 18-109_k141_140387 | Not-determined |
| 18-109_k141_203508 | Not-determined |
| 18-109_k141_8203 | Not-determined |
| 18-109_k141_100295 | Not-determined |
| 18-109_k141_8205 | Not-determined |
| 18-109_k141_59607 | Low-quality |
| 18-109_k141_49550 | Low-quality |
| 18-109_k141_39538 | Not-determined |
| 18-109_k141_130922 | Not-determined |
| 18-109_k141_242853 | Not-determined |
| 18-109_k141_140403 | Low-quality |
| 18-109_k141_59618 | Not-determined |
| 18-109_k141_140405 | Low-quality |
| 18-109_k141_70044 | Not-determined |
| 18-109_k141_70045 | Low-quality |
| 18-109_k141_70046 | Not-determined |
| 18-109_k141_140409 | Not-determined |
| 18-109_k141_121029 | Low-quality |
| 18-109_k141_203529 | Not-determined |
| 18-109_k141_192847 | Not-determined |
| 18-109_k141_233293 | Not-determined |
| 18-109_k141_49577 | Not-determined |
| 18-109_k141_233296 | Low-quality |
| 18-109_k141_233298 | Not-determined |
| 18-109_k141_70055 | Low-quality |
| 18-109_k141_192850 | Not-determined |
| 18-109_k141_49584 | Not-determined |
| 18-109_k141_130927 | Not-determined |
| 18-109_k141_203544 | Not-determined |
| 18-109_k141_233309 | Not-determined |
| 18-109_k141_89477 | Not-determined |
| 18-109_k141_233311 | Not-determined |
| 18-109_k141_171943 | Not-determined |
| 18-109_k141_130932 | Not-determined |
| 18-109_k141_59648 | Low-quality |
| 18-109_k141_39560 | Not-determined |
| 18-109_k141_182464 | Low-quality |
| 18-109_k141_233315 | Low-quality |
| 18-109_k141_49600 | Not-determined |
| 18-109_k141_100320 | Not-determined |
| 18-109_k141_100321 | Not-determined |
| 18-109_k141_233321 | Not-determined |
| 18-109_k141_140429 | Not-determined |
| 18-109_k141_39568 | Not-determined |
| 18-109_k141_39569 | Not-determined |
| 18-109_k141_100333 | Not-determined |
| 18-109_k141_39572 | Not-determined |
| 18-109_k141_182488 | Not-determined |
| 18-109_k141_182491 | Not-determined |
| 18-109_k141_242877 | Not-determined |
| 18-109_k141_203579 | Not-determined |
| 18-109_k141_140447 | Not-determined |
| 18-109_k141_192883 | Not-determined |
| 18-109_k141_221150 | Low-quality |
| 18-109_k141_203582 | Not-determined |
| 18-109_k141_242886 | Not-determined |
| 18-109_k141_171975 | Not-determined |
| 18-109_k141_182505 | Not-determined |
| 18-109_k141_203588 | Not-determined |
| 18-109_k141_171980 | Not-determined |
| 18-109_k141_171981 | Not-determined |
| 18-109_k141_203592 | Not-determined |
| 18-109_k141_70067 | Not-determined |
| 18-109_k141_171983 | Not-determined |
| 18-109_k141_100355 | Not-determined |
| 18-109_k141_130944 | Not-determined |
| 18-109_k141_100358 | Not-determined |
| 18-109_k141_233339 | Not-determined |
| 18-109_k141_70070 | Not-determined |
| 18-109_k141_59679 | Low-quality |
| 18-109_k141_100361 | Not-determined |
| 18-109_k141_192906 | Not-determined |
| 18-109_k141_8252 | Not-determined |
| 18-109_k141_221167 | Not-determined |
| 18-109_k141_130955 | Not-determined |
| 18-109_k141_140473 | Not-determined |
| 18-109_k141_130956 | Not-determined |
| 18-109_k141_221172 | Not-determined |
| 18-109_k141_49626 | Not-determined |
| 18-109_k141_192910 | Not-determined |
| 18-109_k141_203604 | Low-quality |
| 18-109_k141_192912 | Not-determined |
| 18-109_k141_8263 | Not-determined |
| 18-109_k141_130958 | Not-determined |
| 18-109_k141_221175 | Not-determined |
| 18-109_k141_8266 | Not-determined |
| 18-109_k141_8270 | Not-determined |
| 18-109_k141_140487 | Not-determined |
| 18-109_k141_150866 | Not-determined |
| 18-109_k141_172003 | Not-determined |
| 18-109_k141_172004 | Not-determined |
| 18-109_k141_172005 | Not-determined |
| 18-109_k141_49644 | Not-determined |
| 18-109_k141_100376 | Not-determined |
| 18-109_k141_49648 | Not-determined |
| 18-109_k141_59691 | Not-determined |
| 18-109_k141_203623 | Not-determined |
| 18-109_k141_27180 | Not-determined |
| 18-109_k141_8280 | Not-determined |
| 18-109_k141_59696 | Not-determined |
| 18-109_k141_203631 | Not-determined |
| 18-109_k141_233362 | Not-determined |
| 18-109_k141_233363 | Not-determined |
| 18-109_k141_150881 | Not-determined |
| 18-109_k141_233364 | Not-determined |
| 18-109_k141_27185 | Not-determined |
| 18-109_k141_140512 | Not-determined |
| 18-109_k141_150889 | Not-determined |
| 18-109_k141_121076 | Not-determined |
| 18-109_k141_150895 | Not-determined |
| 18-109_k141_192937 | Not-determined |
| 18-109_k141_221201 | Not-determined |
| 18-109_k141_192943 | Not-determined |
| 18-109_k141_27204 | Low-quality |
| 18-109_k141_49677 | Not-determined |
| 18-109_k141_59725 | Not-determined |
| 18-109_k141_89500 | Low-quality |
| 18-109_k141_192953 | Not-determined |
| 18-109_k141_27210 | Not-determined |
| 18-109_k141_121100 | Not-determined |
| 18-109_k141_221225 | Not-determined |
| 18-109_k141_79240 | Not-determined |
| 18-109_k141_70115 | Not-determined |
| 18-109_k141_121114 | Low-quality |
| 18-109_k141_131006 | Not-determined |
| 18-109_k141_79253 | Not-determined |
| 18-109_k141_192962 | Not-determined |
| 18-109_k141_192966 | Not-determined |
| 18-109_k141_203653 | Low-quality |
| 18-109_k141_27234 | Not-determined |
| 18-109_k141_79268 | Not-determined |
| 18-109_k141_233379 | Not-determined |
| 18-109_k141_27241 | Low-quality |
| 18-109_k141_150932 | Not-determined |
| 18-109_k141_79275 | Not-determined |
| 18-109_k141_109898 | Not-determined |
| 18-109_k141_109899 | Not-determined |
| 18-109_k141_212934 | Not-determined |
| 18-109_k141_59753 | Not-determined |
| 18-109_k141_162135 | Not-determined |
| 18-109_k141_212936 | Not-determined |
| 18-109_k141_182551 | Not-determined |
| 18-109_k141_192976 | Low-quality |
| 18-109_k141_49720 | Not-determined |
| 18-109_k141_109919 | Not-determined |
| 18-109_k141_162141 | Not-determined |
| 18-109_k141_70145 | Not-determined |
| 18-109_k141_109920 | Not-determined |
| 18-109_k141_192982 | Not-determined |
| 18-109_k141_150949 | Not-determined |
| 18-109_k141_100403 | Low-quality |
| 18-109_k141_8330 | Not-determined |
| 18-109_k141_140537 | Not-determined |
| 18-109_k141_150956 | Not-determined |
| 18-109_k141_18426 | Not-determined |
| 18-109_k141_212952 | Low-quality |
| 18-109_k141_70156 | Low-quality |
| 18-109_k141_242963 | Not-determined |
| 18-109_k141_109933 | Not-determined |
| 18-109_k141_131053 | Not-determined |
| 18-109_k141_221255 | Not-determined |
| 18-109_k141_212972 | Low-quality |
| 18-109_k141_131059 | Not-determined |
| 18-109_k141_70166 | Not-determined |
| 18-109_k141_131071 | Not-determined |
| 18-109_k141_203698 | Not-determined |
| 18-109_k141_100435 | Not-determined |
| 18-109_k141_89544 | Not-determined |
| 18-109_k141_242991 | Not-determined |
| 18-109_k141_172038 | Not-determined |
| 18-109_k141_121155 | Not-determined |
| 18-109_k141_121158 | Not-determined |
| 18-109_k141_121161 | Not-determined |
| 18-109_k141_193022 | Not-determined |
| 18-109_k141_49737 | Not-determined |
| 18-109_k141_131084 | Not-determined |
| 18-109_k141_140570 | Not-determined |
| 18-109_k141_221264 | Not-determined |
| 18-109_k141_243003 | Not-determined |
| 18-109_k141_150969 | Not-determined |
| 18-109_k141_121169 | Not-determined |
| 18-109_k141_140573 | Not-determined |
| 18-109_k141_233413 | Not-determined |
| 18-109_k141_140577 | Not-determined |
| 18-109_k141_193033 | Low-quality |
| 18-109_k141_70191 | Not-determined |
| 18-109_k141_100451 | Not-determined |
| 18-109_k141_100455 | Not-determined |

| | |
|---|---|
| 18-109_k141_150980 | Not-determined |
| 18-109_k141_59791 | Not-determined |
| 18-109_k141_233430 | Not-determined |
| 18-109_k141_89565 | Not-determined |
| 18-109_k141_79320 | Not-determined |
| 18-109_k141_109978 | Not-determined |
| 18-109_k141_79321 | Not-determined |
| 18-109_k141_243021 | Not-determined |
| 18-109_k141_121179 | Not-determined |
| 18-109_k141_131094 | Not-determined |
| 18-109_k141_131095 | Not-determined |
| 18-109_k141_150987 | Low-quality |
| 18-109_k141_49754 | Not-determined |
| 18-109_k141_162185 | Not-determined |
| 18-109_k141_131099 | Not-determined |
| 18-109_k141_49758 | Low-quality |
| 18-109_k141_221280 | Not-determined |
| 18-109_k141_150996 | Not-determined |
| 18-109_k141_89577 | Not-determined |
| 18-109_k141_182584 | Not-determined |
| 18-109_k141_140601 | Not-determined |
| 18-109_k141_151002 | Not-determined |
| 18-109_k141_121192 | Not-determined |
| 18-109_k141_172069 | Not-determined |
| 18-109_k141_121193 | Not-determined |
| 18-109_k141_39669 | Not-determined |
| 18-109_k141_221288 | Not-determined |
| 18-109_k141_100475 | Not-determined |
| 18-109_k141_172079 | Not-determined |
| 18-109_k141_39673 | Not-determined |
| 18-109_k141_193070 | Not-determined |
| 18-109_k141_233446 | Not-determined |
| 18-109_k141_162194 | Low-quality |
| 18-109_k141_70217 | Not-determined |
| 18-109_k141_70219 | Not-determined |
| 18-109_k141_100478 | Not-determined |
| 18-109_k141_100479 | Not-determined |
| 18-109_k141_233452 | Low-quality |
| 18-109_k141_121197 | Not-determined |
| 18-109_k141_121199 | Not-determined |
| 18-109_k141_233455 | Not-determined |
| 18-109_k141_121203 | Not-determined |
| 18-109_k141_39681 | Not-determined |
| 18-109_k141_121205 | Not-determined |
| 18-109_k141_89596 | Not-determined |
| 18-109_k141_121207 | Not-determined |
| 18-109_k141_151018 | Not-determined |
| 18-109_k141_162211 | Not-determined |
| 18-109_k141_140622 | Not-determined |
| 18-109_k141_243035 | Not-determined |
| 18-109_k141_49769 | Not-determined |
| 18-109_k141_59802 | Not-determined |
| 18-109_k141_100503 | Not-determined |
| 18-109_k141_131104 | Not-determined |
| 18-109_k141_121219 | Not-determined |
| 18-109_k141_233480 | Not-determined |
| 18-109_k141_121223 | Not-determined |
| 18-109_k141_140631 | Not-determined |
| 18-109_k141_162233 | Not-determined |
| 18-109_k141_233484 | Low-quality |
| 18-109_k141_109998 | Not-determined |
| 18-109_k141_121239 | Not-determined |
| 18-109_k141_8391 | Not-determined |
| 18-109_k141_193085 | Not-determined |
| 18-109_k141_213031 | Not-determined |
| 18-109_k141_49775 | Low-quality |
| 18-109_k141_162243 | Low-quality |
| 18-109_k141_213038 | Low-quality |
| 18-109_k141_89617 | Not-determined |
| 18-109_k141_110000 | Not-determined |
| 18-109_k141_213044 | Not-determined |
| 18-109_k141_140651 | Not-determined |
| 18-109_k141_213049 | Not-determined |
| 18-109_k141_110013 | Not-determined |
| 18-109_k141_49795 | Not-determined |
| 18-109_k141_131119 | Not-determined |
| 18-109_k141_49796 | Not-determined |
| 18-109_k141_89630 | Not-determined |
| 18-109_k141_49798 | Not-determined |
| 18-109_k141_49799 | Not-determined |
| 18-109_k141_49802 | Not-determined |
| 18-109_k141_151029 | Not-determined |
| 18-109_k141_8397 | Not-determined |
| 18-109_k141_151032 | Not-determined |
| 18-109_k141_8399 | Low-quality |
| 18-109_k141_140668 | Not-determined |
| 18-109_k141_213063 | Not-determined |
| 18-109_k141_243071 | Not-determined |
| 18-109_k141_121267 | Not-determined |
| 18-109_k141_243074 | Not-determined |
| 18-109_k141_162254 | Not-determined |
| 18-109_k141_100551 | Not-determined |
| 18-109_k141_8418 | Not-determined |
| 18-109_k141_213082 | Not-determined |
| 18-109_k141_131132 | Not-determined |
| 18-109_k141_172132 | Not-determined |
| 18-109_k141_70305 | Not-determined |
| 18-109_k141_8430 | Not-determined |
| 18-109_k141_162264 | Not-determined |
| 18-109_k141_70308 | Not-determined |
| 18-109_k141_233527 | Not-determined |
| 18-109_k141_70314 | Not-determined |
| 18-109_k141_18476 | Not-determined |
| 18-109_k141_8438 | Not-determined |
| 18-109_k141_233540 | Not-determined |
| 18-109_k141_243098 | Not-determined |
| 18-109_k141_162273 | Not-determined |
| 18-109_k141_162275 | Not-determined |
| 18-109_k141_110020 | Not-determined |
| 18-109_k141_100564 | Not-determined |
| 18-109_k141_233545 | Not-determined |
| 18-109_k141_131140 | Not-determined |
| 18-109_k141_8442 | Not-determined |
| 18-109_k141_140681 | Not-determined |
| 18-109_k141_151070 | Not-determined |
| 18-109_k141_121290 | Not-determined |
| 18-109_k141_121294 | Not-determined |
| 18-109_k141_8450 | Not-determined |
| 18-109_k141_110031 | Not-determined |
| 18-109_k141_49846 | Low-quality |
| 18-109_k141_121303 | Not-determined |
| 18-109_k141_151081 | Not-determined |
| 18-109_k141_121308 | Not-determined |
| 18-109_k141_213107 | Low-quality |
| 18-109_k141_39741 | Not-determined |
| 18-109_k141_39744 | Not-determined |
| 18-109_k141_121312 | Not-determined |
| 18-109_k141_203773 | Not-determined |
| 18-109_k141_89644 | Not-determined |
| 18-109_k141_70321 | Not-determined |
| 18-109_k141_79328 | Not-determined |
| 18-109_k141_140712 | Not-determined |
| 18-109_k141_79330 | Not-determined |
| 18-109_k141_151100 | Not-determined |
| 18-109_k141_121328 | Not-determined |
| 18-109_k141_151101 | Not-determined |
| 18-109_k141_193111 | Not-determined |
| 18-109_k141_89657 | Not-determined |
| 18-109_k141_89659 | Not-determined |
| 18-109_k141_221332 | Not-determined |
| 18-109_k141_59851 | Low-quality |
| 18-109_k141_89665 | Not-determined |
| 18-109_k141_27300 | Not-determined |
| 18-109_k141_221341 | Low-quality |
| 18-109_k141_59855 | Not-determined |
| 18-109_k141_100590 | Not-determined |
| 18-109_k141_110055 | Not-determined |
| 18-109_k141_110062 | Low-quality |
| 18-109_k141_49893 | Not-determined |
| 18-109_k141_151111 | Not-determined |
| 18-109_k141_203813 | Not-determined |
| 18-109_k141_233569 | Not-determined |
| 18-109_k141_8489 | Not-determined |
| 18-109_k141_18532 | Not-determined |
| 18-109_k141_221361 | Not-determined |
| 18-109_k141_59868 | Not-determined |
| 18-109_k141_59869 | Not-determined |
| 18-109_k141_243122 | Not-determined |
| 18-109_k141_89682 | Low-quality |
| 18-109_k141_182642 | Not-determined |
| 18-109_k141_221366 | Not-determined |
| 18-109_k141_243123 | Not-determined |
| 18-109_k141_89683 | Not-determined |
| 18-109_k141_8496 | Not-determined |
| 18-109_k141_49900 | Not-determined |
| 18-109_k141_8497 | Not-determined |
| 18-109_k141_70351 | Not-determined |
| 18-109_k141_162327 | Not-determined |
| 18-109_k141_49904 | Not-determined |
| 18-109_k141_18535 | Not-determined |
| 18-109_k141_221378 | Not-determined |
| 18-109_k141_8498 | Not-determined |
| 18-109_k141_131193 | Not-determined |
| 18-109_k141_131197 | Low-quality |
| 18-109_k141_193150 | Not-determined |
| 18-109_k141_131201 | Not-determined |
| 18-109_k141_110075 | Low-quality |
| 18-109_k141_70359 | Not-determined |
| 18-109_k141_70362 | Not-determined |
| 18-109_k141_49928 | Not-determined |
| 18-109_k141_89696 | Not-determined |
| 18-109_k141_89699 | Not-determined |
| 18-109_k141_203846 | Not-determined |
| 18-109_k141_110082 | Not-determined |
| 18-102-final-viral-combined_k141_161986\|\|full | High-quality |
| 18-109_k141_8528 | Not-determined |
| 18-109_k141_193166 | Not-determined |
| 18-109_k141_59886 | Not-determined |
| 18-109_k141_79354 | Not-determined |
| 18-109_k141_79363 | Not-determined |
| 18-109_k141_172177 | Not-determined |
| 18-109_k141_213114 | Not-determined |
| 18-109_k141_79364 | Low-quality |
| 18-109_k141_140732 | Not-determined |
| 18-109_k141_140734 | Not-determined |
| 18-109_k141_49945 | Not-determined |
| 18-109_k141_39766 | Not-determined |
| 18-109_k141_59892 | Not-determined |
| 18-109_k141_213123 | Not-determined |
| 18-109_k141_59894 | Not-determined |
| 18-109_k141_213126 | Not-determined |
| 18-109_k141_233613 | Not-determined |
| 18-109_k141_213128 | Not-determined |
| 18-109_k141_182688 | Not-determined |
| 18-109_k141_59903 | Not-determined |
| 18-109_k141_182689 | Not-determined |
| 18-109_k141_79384 | Not-determined |
| 18-109_k141_79385 | Not-determined |
| 18-109_k141_59908 | Not-determined |
| 18-109_k141_110102 | Not-determined |
| 18-109_k141_193182 | Not-determined |
| 18-109_k141_70385 | Not-determined |
| 18-109_k141_89719 | Not-determined |
| 18-109_k141_213137 | Not-determined |
| 18-109_k141_151151 | Low-quality |
| 18-109_k141_131221 | Not-determined |
| 18-109_k141_151153 | Not-determined |
| 18-109_k141_213141 | Not-determined |
| 18-109_k141_193191 | Not-determined |
| 18-109_k141_213143 | Not-determined |
| 18-109_k141_70397 | Not-determined |
| 18-109_k141_18558 | Not-determined |
| 18-109_k141_39776 | Not-determined |
| 18-109_k141_131236 | Not-determined |
| 18-109_k141_18566 | Not-determined |
| 18-109_k141_213150 | Not-determined |
| 18-109_k141_70408 | Not-determined |
| 18-109_k141_203893 | Not-determined |
| 18-109_k141_151167 | Not-determined |
| 18-109_k141_70413 | Not-determined |
| 18-109_k141_193203 | Not-determined |
| 18-109_k141_89737 | Not-determined |
| 18-109_k141_89738 | Not-determined |
| 18-109_k141_121393 | Not-determined |
| 18-109_k141_193206 | Not-determined |
| 18-109_k141_39793 | Not-determined |
| 18-109_k141_121397 | Low-quality |
| 18-109_k141_203905 | Not-determined |
| 18-109_k141_193210 | Not-determined |
| 18-109_k141_193211 | Not-determined |
| 18-109_k141_172217 | Not-determined |
| 18-109_k141_203906 | Not-determined |
| 18-109_k141_172222 | Not-determined |
| 18-109_k141_203911 | Not-determined |
| 18-109_k141_182744 | Low-quality |
| 18-109_k141_233645 | Not-determined |
| 18-109_k141_39797 | Not-determined |
| 18-109_k141_182745 | Not-determined |
| 18-109_k141_140752 | Low-quality |
| 18-109_k141_151189 | Not-determined |
| 18-109_k141_182749 | Not-determined |
| 18-109_k141_203916 | Not-determined |
| 18-109_k141_70439 | Not-determined |
| 18-109_k141_182753 | Not-determined |
| 18-109_k141_151195 | Not-determined |
| 18-109_k141_213174 | Not-determined |
| 18-109_k141_89753 | Not-determined |
| 18-109_k141_203921 | Not-determined |
| 18-109_k141_233657 | Not-determined |
| 18-109_k141_70443 | Not-determined |
| 18-109_k141_121414 | Not-determined |
| 18-109_k141_172242 | Not-determined |
| 18-109_k141_79419 | Not-determined |
| 18-109_k141_203927 | Not-determined |
| 18-109_k141_172248 | Low-quality |
| 18-109_k141_8576 | Not-determined |
| 18-109_k141_221403 | Not-determined |
| 18-109_k141_121422 | Not-determined |
| 18-109_k141_233667 | Not-determined |
| 18-109_k141_121427 | Not-determined |
| 18-109_k141_110154 | Not-determined |
| 18-109_k141_59949 | Low-quality |
| 18-109_k141_8579 | Not-determined |
| 18-109_k141_182777 | Not-determined |
| 18-109_k141_243150 | Not-determined |
| 18-109_k141_243151 | Not-determined |
| 18-109_k141_213198 | Not-determined |
| 18-109_k141_203944 | Not-determined |
| 18-109_k141_243156 | Not-determined |
| 18-109_k141_70461 | Not-determined |
| 18-109_k141_151211 | Not-determined |
| 18-109_k141_39841 | Low-quality |
| 18-109_k141_121444 | Not-determined |
| 18-109_k141_193244 | Not-determined |
| 18-109_k141_193245 | Not-determined |
| 18-109_k141_89782 | Not-determined |
| 18-109_k141_8593 | Low-quality |
| 18-109_k141_121447 | Not-determined |
| 18-109_k141_59965 | Not-determined |
| 18-109_k141_50019 | Not-determined |
| 18-109_k141_131253 | Not-determined |
| 18-109_k141_182806 | Not-determined |
| 18-109_k141_233697 | Not-determined |
| 18-109_k141_79443 | Not-determined |
| 18-109_k141_131260 | Not-determined |
| 18-109_k141_233700 | Not-determined |
| 18-109_k141_18602 | Not-determined |
| 18-109_k141_193254 | Not-determined |
| 18-109_k141_140805 | Not-determined |
| 18-109_k141_89798 | Not-determined |
| 18-109_k141_233710 | Low-quality |
| 18-109_k141_59982 | Not-determined |
| 18-109_k141_27345 | Not-determined |
| 18-109_k141_203967 | Not-determined |
| 18-109_k141_151234 | Not-determined |
| 18-109_k141_233717 | Not-determined |
| 18-109_k141_59984 | Low-quality |
| 18-109_k141_59986 | Not-determined |
| 18-109_k141_213229 | Not-determined |
| 18-109_k141_79461 | Not-determined |
| 18-109_k141_79462 | Not-determined |
| 18-109_k141_193264 | Not-determined |
| 18-109_k141_193265 | Not-determined |
| 18-109_k141_193269 | Not-determined |
| 18-109_k141_193270 | Not-determined |
| 18-109_k141_203984 | Not-determined |
| 18-109_k141_172295 | Not-determined |
| 18-109_k141_27360 | Not-determined |
| 18-109_k141_59994 | Not-determined |
| 18-109_k141_172300 | Not-determined |
| 18-109_k141_79471 | Not-determined |
| 18-109_k141_60002 | Not-determined |
| 18-109_k141_172306 | Not-determined |
| 18-109_k141_221466 | Not-determined |
| 18-109_k141_110170 | Low-quality |
| 18-109_k141_100606 | Not-determined |
| 18-109_k141_100609 | Not-determined |
| 18-109_k141_243186 | Low-quality |
| 18-109_k141_213244 | Not-determined |
| 18-109_k141_193296 | Not-determined |
| 18-109_k141_213245 | Not-determined |
| 18-109_k141_243187 | Not-determined |
| 18-109_k141_172315 | Not-determined |
| 18-109_k141_162435 | Not-determined |
| 18-109_k141_162436 | Not-determined |
| 18-109_k141_60027 | Not-determined |
| 18-109_k141_172317 | Not-determined |
| 18-109_k141_233746 | Not-determined |
| 18-109_k141_233748 | Not-determined |
| 18-109_k141_89847 | Not-determined |
| 18-109_k141_193312 | Not-determined |
| 18-109_k141_60037 | Not-determined |
| 18-109_k141_233756 | Not-determined |
| 18-109_k141_243202 | Low-quality |
| 18-109_k141_243203 | Not-determined |
| 18-109_k141_243206 | Not-determined |
| 18-109_k141_60047 | Not-determined |
| 18-109_k141_213261 | Not-determined |
| 18-109_k141_213262 | Not-determined |
| 18-109_k141_204037 | Not-determined |
| 18-109_k141_182835 | Not-determined |
| 18-109_k141_221514 | Not-determined |
| 18-109_k141_27366 | Not-determined |
| 18-109_k141_162447 | Not-determined |
| 18-109_k141_79505 | Low-quality |
| 18-109_k141_233764 | Not-determined |
| 18-109_k141_110182 | Not-determined |
| 18-109_k141_204040 | Not-determined |
| 18-109_k141_8616 | Not-determined |
| 18-109_k141_60057 | Not-determined |
| 18-109_k141_233777 | Not-determined |
| 18-109_k141_89864 | Not-determined |
| 18-109_k141_60064 | Low-quality |
| 18-109_k141_60065 | Not-determined |
| 18-109_k141_233784 | Not-determined |
| 18-109_k141_79513 | Not-determined |
| 18-109_k141_131325 | Low-quality |
| 18-109_k141_121520 | Not-determined |
| 18-109_k141_162476 | Not-determined |
| 18-109_k141_70520 | Not-determined |
| 18-109_k141_193329 | Not-determined |
| 18-109_k141_213285 | Not-determined |
| 18-109_k141_110202 | Not-determined |
| 18-109_k141_193330 | Not-determined |
| 18-109_k141_233792 | Not-determined |
| 18-109_k141_70528 | Not-determined |
| 18-109_k141_243232 | Not-determined |
| 18-109_k141_79520 | Not-determined |
| 18-109_k141_172329 | Low-quality |
| 18-109_k141_89869 | Not-determined |
| 18-109_k141_162488 | Not-determined |
| 18-109_k141_89872 | Low-quality |
| 18-109_k141_213296 | Not-determined |
| 18-109_k141_121534 | Not-determined |
| 18-109_k141_162491 | Not-determined |
| 18-109_k141_193334 | Not-determined |
| 18-109_k141_182852 | Not-determined |
| 18-109_k141_182862 | Not-determined |
| 18-109_k141_213308 | Not-determined |
| 18-109_k141_131340 | Not-determined |
| 18-109_k141_70546 | Not-determined |
| 18-109_k141_89879 | Low-quality |
| 18-109_k141_233819 | Not-determined |
| 18-109_k141_100649 | Not-determined |
| 18-109_k141_162498 | Not-determined |
| 18-109_k141_213316 | Low-quality |
| 18-109_k141_100651 | Not-determined |
| 18-109_k141_27405 | Not-determined |
| 18-109_k141_110222 | Not-determined |
| 18-109_k141_162506 | Not-determined |
| 18-109_k141_89888 | Not-determined |
| 18-109_k141_100655 | Not-determined |
| 18-109_k141_193360 | Not-determined |
| 18-109_k141_243246 | Not-determined |
| 18-109_k141_233833 | Not-determined |
| 18-109_k141_18606 | Low-quality |
| 18-109_k141_110229 | Not-determined |
| 18-109_k141_100658 | Not-determined |
| 18-109_k141_50087 | Low-quality |
| 18-109_k141_110238 | Not-determined |
| 18-109_k141_233840 | Low-quality |
| 18-109_k141_110241 | Not-determined |
| 18-109_k141_140821 | Not-determined |
| 18-109_k141_50097 | Not-determined |
| 18-109_k141_131360 | Not-determined |
| 18-109_k141_221540 | Not-determined |
| 18-109_k141_79564 | Not-determined |
| 18-109_k141_213343 | Not-determined |
| 18-109_k141_221542 | Not-determined |
| 18-109_k141_121588 | Not-determined |
| 18-109_k141_233861 | Low-quality |
| 18-109_k141_89913 | Not-determined |
| 18-109_k141_79571 | Not-determined |
| 18-109_k141_204107 | Not-determined |
| 18-109_k141_89916 | Not-determined |
| 18-109_k141_100661 | Not-determined |
| 18-109_k141_172370 | Not-determined |
| 18-109_k141_110247 | Not-determined |
| 18-109_k141_70584 | Not-determined |
| 18-109_k141_70585 | Not-determined |
| 18-109_k141_70586 | Not-determined |
| 18-109_k141_131384 | Not-determined |
| 18-109_k141_50126 | Not-determined |
| 18-109_k141_8697 | Not-determined |
| 18-109_k141_172374 | Not-determined |
| 18-109_k141_60102 | Not-determined |
| 18-109_k141_60104 | Not-determined |
| 18-109_k141_204123 | Not-determined |
| 18-109_k141_221553 | Not-determined |
| 18-109_k141_193398 | Not-determined |
| 18-109_k141_172383 | Low-quality |
| 18-109_k141_131401 | Not-determined |
| 18-109_k141_221558 | Not-determined |
| 18-109_k141_162536 | Not-determined |
| 18-109_k141_50135 | Not-determined |
| 18-109_k141_131403 | Not-determined |
| 18-109_k141_131404 | Low-quality |
| 18-109_k141_8707 | Not-determined |
| 18-109_k141_151287 | Not-determined |
| 18-109_k141_39937 | Not-determined |
| 18-109_k141_162542 | Not-determined |
| 18-109_k141_131417 | Not-determined |
| 18-109_k141_193409 | Not-determined |
| 18-109_k141_131422 | Not-determined |
| 18-109_k141_151299 | Low-quality |
| 18-109_k141_151300 | Not-determined |
| 18-109_k141_100683 | Not-determined |
| 18-109_k141_221566 | Not-determined |
| 18-109_k141_233885 | Not-determined |
| 18-109_k141_221569 | Not-determined |
| 18-109_k141_204152 | Not-determined |
| 18-109_k141_100690 | Not-determined |
| 18-109_k141_89943 | Not-determined |
| 18-109_k141_89945 | Not-determined |
| 18-109_k141_172402 | Not-determined |
| 18-109_k141_50166 | Not-determined |
| 18-109_k141_172410 | Not-determined |
| 18-109_k141_233902 | Not-determined |
| 18-109_k141_151319 | Not-determined |
| 18-109_k141_50168 | Not-determined |
| 18-109_k141_100700 | Not-determined |
| 18-109_k141_221573 | Low-quality |
| 18-109_k141_221574 | Not-determined |
| 18-109_k141_193432 | Not-determined |
| 18-109_k141_193433 | Not-determined |
| 18-109_k141_100708 | Not-determined |
| 18-109_k141_8735 | Not-determined |
| 18-109_k141_233911 | Not-determined |
| 18-109_k141_60128 | Not-determined |
| 18-109_k141_233913 | Not-determined |
| 18-109_k141_89953 | Not-determined |
| 18-109_k141_39949 | Not-determined |
| 18-109_k141_8740 | Low-quality |
| 18-109_k141_151345 | Not-determined |
| 18-109_k141_151346 | Not-determined |
| 18-109_k141_213380 | Not-determined |
| 18-109_k141_27408 | Not-determined |
| 18-109_k141_110280 | Not-determined |
| 18-109_k141_60131 | Not-determined |
| 18-109_k141_151357 | Not-determined |
| 18-109_k141_39964 | Not-determined |
| 18-109_k141_172429 | Not-determined |
| 18-109_k141_172432 | Not-determined |
| 18-109_k141_39967 | Low-quality |
| 18-109_k141_151363 | Not-determined |
| 18-109_k141_39969 | Not-determined |
| 18-109_k141_172435 | Not-determined |
| 18-109_k141_50187 | Not-determined |
| 18-109_k141_151368 | Not-determined |
| 18-109_k141_131445 | Not-determined |
| 18-109_k141_50191 | Not-determined |
| 18-109_k141_50192 | Not-determined |
| 18-109_k141_121640 | Not-determined |
| 18-109_k141_121643 | Not-determined |
| 18-109_k141_204191 | Not-determined |
| 18-109_k141_172452 | Not-determined |
| 18-109_k141_60161 | Not-determined |
| 18-109_k141_233954 | Not-determined |
| 18-109_k141_221627 | Not-determined |
| 18-109_k141_233964 | Not-determined |
| 18-109_k141_110309 | Not-determined |
| 18-109_k141_221630 | Not-determined |
| 18-109_k141_131470 | Not-determined |
| 18-109_k141_131474 | Not-determined |
| 18-109_k141_100727 | Not-determined |
| 18-109_k141_140845 | Not-determined |
| 18-109_k141_18653 | Low-quality |
| 18-109_k141_18655 | Not-determined |
| 18-109_k141_204218 | Not-determined |
| 18-109_k141_221647 | Not-determined |
| 18-109_k141_151401 | Not-determined |
| 18-109_k141_151402 | Not-determined |
| 18-109_k141_100739 | Not-determined |
| 18-109_k141_110316 | Low-quality |
| 18-109_k141_110321 | Low-quality |
| 18-109_k141_121664 | Not-determined |
| 18-109_k141_100746 | Low-quality |
| 18-109_k141_18669 | Not-determined |
| 18-109_k141_27431 | Not-determined |
| 18-109_k141_50233 | Not-determined |
| 18-109_k141_100763 | Not-determined |
| 18-109_k141_27440 | Not-determined |
| 18-109_k141_50238 | Not-determined |
| 18-109_k141_243279 | Not-determined |
| 18-109_k141_89975 | Not-determined |
| 18-109_k141_60193 | Not-determined |
| 18-109_k141_89976 | Not-determined |
| 18-109_k141_100768 | Not-determined |
| 18-109_k141_110337 | Not-determined |
| 18-109_k141_131493 | Low-quality |
| 18-109_k141_193459 | Not-determined |
| 18-109_k141_193460 | Not-determined |
| 18-109_k141_18686 | Not-determined |
| 18-109_k141_121688 | Not-determined |
| 18-109_k141_193466 | Not-determined |
| 18-109_k141_100780 | Not-determined |
| 18-109_k141_140885 | Not-determined |
| 18-109_k141_121694 | Not-determined |
| 18-109_k141_70623 | Not-determined |
| 18-109_k141_131511 | Not-determined |
| 18-109_k141_100796 | Not-determined |
| 18-109_k141_151429 | Not-determined |
| 18-109_k141_50252 | Not-determined |
| 18-109_k141_70633 | Low-quality |
| 18-109_k141_233989 | Not-determined |
| 18-109_k141_131515 | Not-determined |
| 18-109_k141_131516 | Not-determined |
| 18-109_k141_151432 | Low-quality |
| 18-109_k141_151433 | Not-determined |
| 18-109_k141_50261 | Not-determined |
| 18-109_k141_213431 | Not-determined |
| 18-109_k141_70638 | Not-determined |
| 18-109_k141_70640 | Not-determined |
| 18-109_k141_131525 | Not-determined |
| 18-109_k141_110355 | Not-determined |
| 18-109_k141_213437 | Not-determined |
| 18-109_k141_172470 | Low-quality |
| 18-109_k141_243299 | Not-determined |
| 18-109_k141_151438 | Not-determined |
| 18-109_k141_140920 | Low-quality |
| 18-109_k141_151439 | Low-quality |
| 18-109_k141_110360 | Not-determined |
| 18-109_k141_50275 | Not-determined |
| 18-109_k141_151444 | Low-quality |
| 18-109_k141_90010 | Not-determined |
| 18-109_k141_60241 | Not-determined |
| 18-109_k141_90011 | Not-determined |
| 18-109_k141_40040 | Not-determined |
| 18-109_k141_151448 | Not-determined |
| 18-109_k141_100824 | Not-determined |
| 18-109_k141_70661 | Not-determined |
| 18-109_k141_60243 | Low-quality |
| 18-109_k141_100834 | Not-determined |
| 18-109_k141_131541 | Not-determined |
| 18-109_k141_110384 | Not-determined |
| 18-109_k141_50289 | Not-determined |
| 18-109_k141_100838 | Not-determined |
| 18-109_k141_90015 | Not-determined |
| 18-109_k141_131549 | Not-determined |

| | |
|---|---|
| 18-109_k141_131555 | Not-determined |
| 18-109_k141_110394 | Not-determined |
| 18-109_k141_110395 | Not-determined |
| 18-109_k141_110396 | Not-determined |
| 18-109_k141_40063 | Not-determined |
| 18-109_k141_234016 | Not-determined |
| 18-109_k141_70686 | Not-determined |
| 18-109_k141_40065 | Not-determined |
| 18-109_k141_40067 | Not-determined |
| 18-109_k141_110401 | Not-determined |
| 18-109_k141_100854 | Low-quality |
| 18-109_k141_140947 | Not-determined |
| 18-109_k141_90031 | Not-determined |
| 18-109_k141_70692 | Not-determined |
| 18-109_k141_131563 | Not-determined |
| 18-109_k141_131567 | Not-determined |
| 18-109_k141_140962 | Not-determined |
| 18-109_k141_40084 | Not-determined |
| 18-109_k141_79626 | Not-determined |
| 18-109_k141_121745 | Not-determined |
| 18-109_k141_234047 | Not-determined |
| 18-109_k141_234048 | Not-determined |
| 18-109_k141_60267 | Not-determined |
| 18-109_k141_100864 | Not-determined |
| 18-109_k141_40087 | Not-determined |
| 18-109_k141_79636 | Not-determined |
| 18-109_k141_79637 | Not-determined |
| 18-109_k141_172485 | Not-determined |
| 18-109_k141_110417 | Not-determined |
| 18-109_k141_27475 | Not-determined |
| 18-109_k141_182931 | Not-determined |
| 18-109_k141_100873 | Not-determined |
| 18-109_k141_40094 | Not-determined |
| 18-109_k141_182933 | Not-determined |
| 18-109_k141_90053 | Not-determined |
| 18-109_k141_193528 | Not-determined |
| 18-109_k141_182937 | Not-determined |
| 18-109_k141_243336 | Low-quality |
| 18-109_k141_193535 | Not-determined |
| 18-109_k141_131586 | Not-determined |
| 18-109_k141_131589 | Not-determined |
| 18-109_k141_243339 | Not-determined |
| 18-109_k141_213486 | Not-determined |
| 18-109_k141_40101 | Not-determined |
| 18-109_k141_70734 | Not-determined |
| 18-109_k141_79657 | Not-determined |
| 18-109_k141_40102 | Not-determined |
| 18-109_k141_213489 | Not-determined |
| 18-109_k141_243346 | Not-determined |
| 18-109_k141_213491 | Not-determined |
| 18-109_k141_90065 | Not-determined |
| 18-109_k141_100893 | Not-determined |
| 18-109_k141_60291 | Low-quality |
| 18-109_k141_90073 | Not-determined |
| 18-109_k141_121767 | Low-quality |
| 18-109_k141_213502 | Not-determined |
| 18-109_k141_70739 | Not-determined |
| 18-109_k141_234069 | Not-determined |
| 18-109_k141_162566 | Not-determined |
| 18-109_k141_79673 | Not-determined |
| 18-109_k141_193559 | Not-determined |
| 18-109_k141_70748 | Not-determined |
| 18-109_k141_162577 | Not-determined |
| 18-109_k141_243362 | Not-determined |
| 18-109_k141_40120 | Not-determined |
| 18-109_k141_221688 | Not-determined |
| 18-109_k141_221692 | Not-determined |
| 18-109_k141_193569 | Low-quality |
| 18-109_k141_131606 | Not-determined |
| 18-109_k141_70759 | Low-quality |
| 18-109_k141_18743 | Not-determined |
| 18-109_k141_40130 | Not-determined |
| 18-109_k141_18744 | Not-determined |
| 18-109_k141_40131 | Not-determined |
| 18-109_k141_182968 | Not-determined |
| 18-109_k141_40133 | Not-determined |
| 18-109_k141_193573 | Not-determined |
| 18-109_k141_90093 | Low-quality |
| 18-109_k141_100913 | Not-determined |
| 18-109_k141_213525 | Not-determined |
| 18-109_k141_50338 | Not-determined |
| 18-109_k141_50339 | Not-determined |
| 18-109_k141_90102 | Not-determined |
| 18-109_k141_193584 | Not-determined |
| 18-109_k141_221695 | Not-determined |
| 18-109_k141_50349 | Not-determined |
| 18-109_k141_50354 | Not-determined |
| 18-109_k141_110438 | Not-determined |
| 18-109_k141_193590 | Not-determined |
| 18-109_k141_110441 | Not-determined |
| 18-109_k141_193594 | Not-determined |
| 18-109_k141_151494 | Low-quality |
| 18-109_k141_141028 | Not-determined |
| 18-109_k141_182986 | Not-determined |
| 18-109_k141_70787 | Not-determined |
| 18-109_k141_213544 | Not-determined |
| 18-109_k141_193599 | Not-determined |
| 18-109_k141_50362 | Not-determined |
| 18-109_k141_193601 | Not-determined |
| 18-109_k141_151501 | Not-determined |
| 18-109_k141_40162 | Not-determined |
| 18-109_k141_141041 | Not-determined |
| 18-109_k141_50367 | Not-determined |
| 18-109_k141_151503 | Not-determined |
| 18-109_k141_182995 | Not-determined |
| 18-109_k141_193605 | Not-determined |
| 18-109_k141_70794 | Not-determined |
| 18-109_k141_121814 | Not-determined |
| 18-109_k141_50386 | Not-determined |
| 18-109_k141_141047 | Not-determined |
| 18-109_k141_193614 | Not-determined |
| 18-109_k141_110469 | Not-determined |
| 18-109_k141_193619 | Not-determined |
| 18-109_k141_100946 | Low-quality |
| 18-109_k141_151522 | Not-determined |
| 18-109_k141_121828 | Not-determined |
| 18-109_k141_234075 | Not-determined |
| 18-109_k141_193626 | Not-determined |
| 18-109_k141_162645 | Not-determined |
| 18-109_k141_162647 | Not-determined |
| 18-109_k141_151527 | Not-determined |
| 18-109_k141_213582 | Not-determined |
| 18-109_k141_243404 | Not-determined |
| 18-109_k141_162653 | Not-determined |
| 18-109_k141_100958 | Not-determined |
| 18-109_k141_193635 | Low-quality |
| 18-109_k141_110483 | Not-determined |
| 18-109_k141_162656 | Not-determined |
| 18-109_k141_110484 | Low-quality |
| 18-109_k141_172583 | Not-determined |
| 18-109_k141_172585 | Not-determined |
| 18-109_k141_162663 | Not-determined |
| 18-109_k141_40179 | Not-determined |
| 18-109_k141_243414 | Not-determined |
| 18-109_k141_40180 | Not-determined |
| 18-109_k141_193646 | Not-determined |
| 18-109_k141_221732 | Not-determined |
| 18-109_k141_221735 | Not-determined |
| 18-109_k141_40181 | Low-quality |
| 18-109_k141_193651 | Low-quality |
| 18-109_k141_193652 | Not-determined |
| 18-109_k141_70815 | Not-determined |
| 18-109_k141_183038 | Low-quality |
| 18-109_k141_27483 | Not-determined |
| 18-109_k141_70822 | Not-determined |
| 18-109_k141_70823 | Not-determined |
| 18-109_k141_100981 | Not-determined |
| 18-109_k141_70831 | Not-determined |
| 18-109_k141_162688 | Not-determined |
| 18-109_k141_27502 | Not-determined |
| 18-109_k141_234111 | Not-determined |
| 18-109_k141_100990 | Not-determined |
| 18-109_k141_70845 | Not-determined |
| 18-109_k141_27514 | Not-determined |
| 18-109_k141_100993 | Not-determined |
| 18-109_k141_70847 | Not-determined |
| 18-109_k141_110522 | Not-determined |
| 18-109_k141_183060 | Not-determined |
| 18-109_k141_90136 | Not-determined |
| 18-109_k141_141059 | Not-determined |
| 18-109_k141_90142 | Not-determined |
| 18-109_k141_70849 | Not-determined |
| 18-109_k141_90143 | Not-determined |
| 18-109_k141_213620 | Not-determined |
| 18-109_k141_110532 | Not-determined |
| 18-109_k141_70853 | Not-determined |
| 18-109_k141_141066 | Low-quality |
| 18-109_k141_90148 | Not-determined |
| 18-109_k141_70855 | Not-determined |
| 18-109_k141_70861 | Not-determined |
| 18-109_k141_141077 | Not-determined |
| 18-109_k141_183068 | Not-determined |
| 18-109_k141_213637 | Low-quality |
| 18-109_k141_121874 | Not-determined |
| 18-109_k141_213641 | Low-quality |
| 18-109_k141_162702 | Not-determined |
| 18-109_k141_151581 | Low-quality |
| 18-109_k141_213645 | Not-determined |
| 18-109_k141_70877 | Not-determined |
| 18-109_k141_193714 | Low-quality |
| 18-109_k141_234120 | Not-determined |
| 18-109_k141_193720 | Low-quality |
| 18-109_k141_234124 | Not-determined |
| 18-109_k141_234125 | Not-determined |
| 18-109_k141_162706 | Low-quality |
| 18-109_k141_90163 | Not-determined |
| 18-109_k141_27542 | Not-determined |
| 18-109_k141_234134 | Not-determined |
| 18-109_k141_213660 | Not-determined |
| 18-109_k141_243447 | Not-determined |
| 18-109_k141_121886 | Not-determined |
| 18-109_k141_27549 | Not-determined |
| 18-109_k141_221806 | Low-quality |
| 18-109_k141_162719 | Not-determined |
| 18-109_k141_243458 | Not-determined |
| 18-109_k141_162720 | Not-determined |
| 18-109_k141_243460 | Not-determined |
| 18-109_k141_162723 | Not-determined |
| 18-109_k141_243464 | Not-determined |
| 18-109_k141_60299 | Not-determined |
| 18-109_k141_121893 | Not-determined |
| 18-109_k141_50448 | Not-determined |
| 18-109_k141_90188 | Not-determined |
| 18-109_k141_141103 | Not-determined |
| 18-109_k141_172612 | Not-determined |
| 18-109_k141_50449 | Not-determined |
| 18-109_k141_90189 | Not-determined |
| 18-109_k141_90192 | Not-determined |
| 18-109_k141_162731 | Not-determined |
| 18-109_k141_243476 | Not-determined |
| 18-109_k141_172619 | Not-determined |
| 18-109_k141_90205 | Not-determined |
| 18-109_k141_162736 | Not-determined |
| 18-109_k141_70902 | Not-determined |
| 18-109_k141_243479 | Not-determined |
| 18-109_k141_193756 | Not-determined |
| 18-109_k141_243482 | Not-determined |
| 18-109_k141_50478 | Not-determined |
| 18-109_k141_243487 | Not-determined |
| 18-109_k141_141124 | Not-determined |
| 18-109_k141_70906 | Not-determined |
| 18-109_k141_60323 | Not-determined |
| 18-109_k141_121930 | Not-determined |
| 18-109_k141_110574 | Not-determined |
| 18-109_k141_121932 | Not-determined |
| 18-109_k141_90217 | Not-determined |
| 18-109_k141_110575 | Not-determined |
| 18-109_k141_70912 | Not-determined |
| 18-109_k141_243494 | Low-quality |
| 18-109_k141_243495 | Low-quality |
| 18-109_k141_213686 | Not-determined |
| 18-109_k141_90226 | Low-quality |
| 18-109_k141_110588 | Not-determined |
| 18-109_k141_221830 | Not-determined |
| 18-109_k141_50495 | Low-quality |
| 18-109_k141_193791 | Not-determined |
| 18-109_k141_234169 | Not-determined |
| 18-109_k141_60341 | Not-determined |
| 18-109_k141_193800 | Low-quality |
| 18-109_k141_234173 | Low-quality |
| 18-109_k141_121942 | Not-determined |
| 18-109_k141_110602 | Not-determined |
| 18-109_k141_193803 | Not-determined |
| 18-109_k141_70938 | Low-quality |
| 18-109_k141_221837 | Not-determined |
| 18-109_k141_70939 | Low-quality |
| 18-109_k141_60347 | Not-determined |
| 18-109_k141_183111 | Low-quality |
| 18-109_k141_110608 | Not-determined |
| 18-109_k141_162755 | Not-determined |
| 18-109_k141_110610 | Not-determined |
| 18-109_k141_110612 | Not-determined |
| 18-109_k141_183120 | Not-determined |
| 18-109_k141_221846 | Not-determined |
| 18-109_k141_221847 | Low-quality |
| 18-109_k141_101005 | Not-determined |
| 18-109_k141_18821 | Not-determined |
| 18-109_k141_60356 | Not-determined |
| 18-109_k141_193818 | Not-determined |
| 18-109_k141_213712 | Not-determined |
| 18-109_k141_234192 | Not-determined |
| 18-109_k141_221855 | Low-quality |
| 18-109_k141_50527 | Low-quality |
| 18-109_k141_110625 | Low-quality |
| 18-109_k141_60367 | Not-determined |
| 18-109_k141_18838 | Not-determined |
| 18-109_k141_121953 | Not-determined |
| 18-109_k141_60371 | Not-determined |
| 18-109_k141_183136 | Not-determined |
| 18-109_k141_110633 | Not-determined |
| 18-109_k141_60374 | Low-quality |
| 18-109_k141_101018 | Not-determined |
| 18-109_k141_141131 | Not-determined |
| 18-109_k141_243516 | Low-quality |
| 18-109_k141_18851 | Not-determined |
| 18-109_k141_221860 | Not-determined |
| 18-109_k141_110647 | Not-determined |
| 18-109_k141_50562 | Not-determined |
| 18-109_k141_60383 | Not-determined |
| 18-109_k141_221866 | Not-determined |
| 18-109_k141_60387 | Not-determined |
| 18-109_k141_121961 | Not-determined |
| 18-109_k141_221871 | Low-quality |
| 18-109_k141_243519 | Not-determined |
| 18-109_k141_243520 | Not-determined |
| 18-109_k141_60394 | Not-determined |
| 18-109_k141_90242 | Not-determined |
| 18-109_k141_213732 | Not-determined |
| 18-109_k141_234217 | Not-determined |
| 18-109_k141_213733 | Not-determined |
| 18-109_k141_213735 | Not-determined |
| 18-109_k141_221874 | Low-quality |
| 18-109_k141_101037 | Not-determined |
| 18-109_k141_221882 | Not-determined |
| 18-109_k141_243526 | Not-determined |
| 18-109_k141_162780 | Not-determined |
| 18-109_k141_162781 | Not-determined |
| 18-109_k141_183153 | Not-determined |
| 18-109_k141_162782 | Not-determined |
| 18-109_k141_121968 | Not-determined |
| 18-109_k141_18879 | Not-determined |
| 18-109_k141_141147 | Not-determined |
| 18-109_k141_50581 | Not-determined |
| 18-109_k141_183157 | Not-determined |
| 18-109_k141_18888 | Not-determined |
| 18-109_k141_183160 | Not-determined |
| 18-109_k141_70989 | Not-determined |
| 18-109_k141_70990 | Not-determined |
| 18-109_k141_243540 | Not-determined |
| 18-109_k141_70991 | Not-determined |
| 18-109_k141_243544 | Not-determined |
| 18-109_k141_101061 | Not-determined |
| 18-109_k141_8803 | Not-determined |
| 18-109_k141_50585 | Not-determined |
| 18-109_k141_183169 | Not-determined |
| 18-109_k141_90275 | Not-determined |
| 18-109_k141_110678 | Not-determined |
| 18-109_k141_121977 | Not-determined |
| 18-109_k141_121978 | Not-determined |
| 18-109_k141_243558 | Not-determined |
| 18-109_k141_183181 | Not-determined |
| 18-109_k141_71015 | Not-determined |
| 18-109_k141_172680 | Not-determined |
| 18-109_k141_18904 | Not-determined |
| 18-109_k141_8825 | Low-quality |
| 18-109_k141_71027 | Low-quality |
| 18-109_k141_141176 | Not-determined |
| 18-109_k141_71028 | Not-determined |
| 18-109_k141_234258 | Not-determined |
| 18-109_k141_172696 | Not-determined |
| 18-109_k141_141185 | Not-determined |
| 18-109_k141_50608 | Not-determined |
| 18-109_k141_90300 | Not-determined |
| 18-109_k141_162814 | Not-determined |
| 18-109_k141_121988 | Not-determined |
| 18-109_k141_50612 | Not-determined |
| 18-109_k141_90301 | Not-determined |
| 18-109_k141_50616 | Not-determined |
| 18-109_k141_151612 | Low-quality |
| 18-109_k141_60452 | Low-quality |
| 18-109_k141_141196 | Not-determined |
| 18-109_k141_172705 | Not-determined |
| 18-109_k141_151614 | Not-determined |
| 18-109_k141_172707 | Not-determined |
| 18-109_k141_71029 | Not-determined |
| 18-109_k141_141203 | Not-determined |
| 18-109_k141_60459 | Not-determined |
| 18-109_k141_79735 | Not-determined |
| 18-109_k141_162832 | Not-determined |
| 18-109_k141_131704 | Not-determined |
| 18-109_k141_60469 | Not-determined |
| 18-109_k141_18923 | Not-determined |
| 18-109_k141_60472 | Not-determined |
| 18-109_k141_79745 | Not-determined |
| 18-109_k141_71036 | Not-determined |
| 18-109_k141_243600 | Not-determined |
| 18-109_k141_183213 | Not-determined |
| 18-109_k141_50648 | Not-determined |
| 18-109_k141_8864 | Not-determined |
| 18-109_k141_183224 | Low-quality |
| 18-109_k141_50664 | Not-determined |
| 18-109_k141_172744 | Not-determined |
| 18-109_k141_110712 | Not-determined |
| 18-109_k141_18938 | Not-determined |
| 18-109_k141_8867 | Not-determined |
| 18-109_k141_50668 | Not-determined |
| 18-109_k141_141217 | Not-determined |
| 18-109_k141_90322 | Not-determined |
| 18-109_k141_90325 | Not-determined |
| 18-109_k141_110726 | Not-determined |
| 18-109_k141_183231 | Not-determined |
| 18-109_k141_8871 | Low-quality |
| 18-109_k141_79755 | Not-determined |
| 18-109_k141_122019 | Not-determined |
| 18-109_k141_243620 | Not-determined |
| 18-109_k141_213782 | Not-determined |
| 18-109_k141_110733 | Not-determined |
| 18-109_k141_213783 | Not-determined |
| 18-109_k141_50687 | Not-determined |
| 18-109_k141_172771 | Not-determined |
| 18-109_k141_193890 | Not-determined |
| 18-109_k141_79763 | Not-determined |
| 18-109_k141_8876 | Not-determined |
| 18-109_k141_221964 | Not-determined |
| 18-109_k141_141226 | Not-determined |
| 18-109_k141_221971 | Not-determined |
| 18-109_k141_141230 | Not-determined |
| 18-109_k141_71069 | Not-determined |
| 18-109_k141_234304 | Not-determined |
| 18-109_k141_18963 | Not-determined |
| 18-109_k141_71074 | Not-determined |
| 18-109_k141_18964 | Low-quality |
| 18-109_k141_141245 | Not-determined |
| 18-109_k141_162885 | Not-determined |
| 18-109_k141_18967 | Not-determined |
| 18-109_k141_18969 | Not-determined |
| 18-109_k141_18970 | Not-determined |
| 18-109_k141_18971 | Not-determined |
| 18-109_k141_193904 | Not-determined |
| 18-109_k141_110750 | Not-determined |
| 18-109_k141_122050 | Low-quality |
| 18-109_k141_141250 | Not-determined |
| 18-109_k141_122054 | Not-determined |
| 18-109_k141_60512 | Not-determined |
| 18-109_k141_110763 | Not-determined |
| 18-109_k141_162891 | Not-determined |
| 18-109_k141_40212 | Not-determined |
| 18-109_k141_40214 | Low-quality |
| 18-109_k141_221995 | Not-determined |
| 18-109_k141_131717 | Low-quality |
| 18-109_k141_221999 | Not-determined |
| 18-109_k141_71089 | Not-determined |
| 18-109_k141_172804 | Not-determined |
| 18-109_k141_79778 | Not-determined |
| 18-109_k141_141265 | Not-determined |
| 18-109_k141_110772 | Not-determined |
| 18-109_k141_50718 | Not-determined |
| 18-109_k141_193926 | Not-determined |
| 18-109_k141_40224 | Not-determined |
| 18-109_k141_40225 | Not-determined |
| 18-109_k141_40226 | Not-determined |
| 18-109_k141_162911 | Not-determined |
| 18-109_k141_50729 | Not-determined |
| 18-109_k141_141286 | Not-determined |
| 18-109_k141_141287 | Not-determined |
| 18-109_k141_172813 | Not-determined |
| 18-109_k141_172814 | Not-determined |
| 18-109_k141_172815 | Not-determined |
| 18-109_k141_79790 | Not-determined |
| 18-109_k141_183270 | Not-determined |
| 18-109_k141_243645 | Not-determined |
| 18-109_k141_141293 | Low-quality |
| 18-109_k141_234335 | Not-determined |
| 18-109_k141_172821 | Not-determined |
| 18-109_k141_183277 | Not-determined |
| 18-109_k141_213840 | Not-determined |
| 18-109_k141_122090 | Not-determined |
| 18-109_k141_60523 | Not-determined |
| 18-109_k141_193943 | Not-determined |
| 18-109_k141_243647 | Not-determined |
| 18-109_k141_141302 | Not-determined |
| 18-109_k141_141305 | Not-determined |
| 18-109_k141_122096 | Not-determined |
| 18-109_k141_151620 | Not-determined |
| 18-109_k141_151621 | Not-determined |
| 18-109_k141_172826 | Not-determined |
| 18-109_k141_172827 | Not-determined |
| 18-109_k141_71113 | Not-determined |
| 18-109_k141_141310 | Not-determined |
| 18-109_k141_71115 | Not-determined |
| 18-109_k141_60530 | Not-determined |
| 18-109_k141_222005 | Not-determined |
| 18-109_k141_222006 | Not-determined |
| 18-109_k141_234354 | Not-determined |
| 18-109_k141_40261 | Not-determined |
| 18-109_k141_243651 | Not-determined |
| 18-109_k141_8919 | Not-determined |
| 18-109_k141_8920 | Not-determined |
| 18-109_k141_213850 | Not-determined |
| 18-109_k141_60547 | Not-determined |
| 18-109_k141_8926 | Not-determined |
| 18-109_k141_151635 | Not-determined |
| 18-109_k141_8928 | Not-determined |
| 18-109_k141_162932 | Not-determined |
| 18-109_k141_193969 | Not-determined |
| 18-109_k141_60560 | Not-determined |
| 18-109_k141_8944 | Not-determined |
| 18-109_k141_162935 | Not-determined |
| 18-109_k141_101087 | Low-quality |
| 18-109_k141_8950 | Not-determined |
| 18-109_k141_40269 | Not-determined |
| 18-109_k141_122108 | Low-quality |
| 18-109_k141_162946 | Not-determined |
| 18-109_k141_40280 | Not-determined |
| 18-109_k141_101097 | Low-quality |
| 18-109_k141_151656 | Not-determined |
| 18-109_k141_101107 | Not-determined |
| 18-109_k141_243676 | Not-determined |
| 18-109_k141_183339 | Not-determined |
| 18-109_k141_8958 | Not-determined |
| 18-109_k141_40298 | Not-determined |
| 18-109_k141_110837 | Not-determined |
| 18-109_k141_101110 | Not-determined |
| 18-109_k141_151670 | Not-determined |
| 18-109_k141_234375 | Not-determined |
| 18-109_k141_101111 | Not-determined |
| 18-109_k141_234376 | Not-determined |
| 18-109_k141_222041 | Not-determined |
| 18-109_k141_110846 | Low-quality |
| 18-109_k141_222044 | Not-determined |
| 18-109_k141_122126 | Not-determined |
| 18-109_k141_110852 | Not-determined |
| 18-109_k141_243691 | Not-determined |
| 18-109_k141_193983 | Not-determined |
| 18-109_k141_50760 | Not-determined |
| 18-109_k141_183360 | Not-determined |
| 18-109_k141_40315 | Not-determined |
| 18-109_k141_8976 | Not-determined |
| 18-109_k141_8977 | Not-determined |
| 18-109_k141_172853 | Not-determined |
| 18-109_k141_183366 | Not-determined |
| 18-109_k141_183369 | Not-determined |
| 18-109_k141_183374 | Not-determined |
| 18-109_k141_162977 | Not-determined |
| 18-109_k141_172859 | Not-determined |
| 18-109_k141_213881 | Not-determined |
| 18-109_k141_172862 | Not-determined |
| 18-109_k141_234393 | Not-determined |
| 18-109_k141_19084 | Not-determined |
| 18-109_k141_19085 | Not-determined |
| 18-109_k141_213884 | Not-determined |
| 18-109_k141_60597 | Not-determined |
| 18-109_k141_243707 | Not-determined |
| 18-109_k141_243708 | Low-quality |
| 18-109_k141_172866 | Not-determined |
| 18-109_k141_162987 | Not-determined |
| 18-109_k141_243712 | Not-determined |
| 18-109_k141_234407 | Not-determined |
| 18-109_k141_222059 | Not-determined |
| 18-109_k141_141329 | Low-quality |
| 18-109_k141_234411 | Not-determined |
| 18-109_k141_101125 | Not-determined |
| 18-109_k141_101126 | Not-determined |
| 18-109_k141_50778 | Not-determined |
| 18-109_k141_101127 | Not-determined |
| 18-109_k141_222064 | Not-determined |
| 18-109_k141_8985 | Not-determined |
| 18-109_k141_183397 | Not-determined |
| 18-109_k141_162996 | Not-determined |
| 18-109_k141_8986 | Not-determined |
| 18-109_k141_90417 | Not-determined |
| 18-109_k141_172877 | Low-quality |
| 18-109_k141_213903 | Not-determined |
| 18-109_k141_50794 | Not-determined |
| 18-109_k141_163001 | Not-determined |
| 18-109_k141_60605 | Not-determined |
| 18-109_k141_79834 | Not-determined |
| 18-109_k141_183412 | Not-determined |
| 18-109_k141_110911 | Not-determined |
| 18-109_k141_40342 | Not-determined |
| 18-109_k141_60616 | Not-determined |
| 18-109_k141_141356 | Not-determined |
| 18-109_k141_141357 | Low-quality |
| 18-109_k141_110917 | Not-determined |
| 18-109_k141_194001 | Not-determined |
| 18-109_k141_183427 | Low-quality |
| 18-109_k141_101158 | Not-determined |
| 18-109_k141_9013 | Not-determined |
| 18-109_k141_79851 | Not-determined |
| 18-109_k141_110926 | Not-determined |
| 18-109_k141_50820 | Not-determined |
| 18-109_k141_213928 | Not-determined |
| 18-109_k141_60630 | Not-determined |
| 18-109_k141_172916 | Not-determined |
| 18-109_k141_110937 | Not-determined |
| 18-109_k141_60635 | Not-determined |
| 18-109_k141_79864 | Not-determined |
| 18-109_k141_9026 | Not-determined |
| 18-109_k141_183444 | Not-determined |
| 18-109_k141_90467 | Not-determined |
| 18-109_k141_163042 | Not-determined |
| 18-109_k141_163044 | Not-determined |
| 18-109_k141_60647 | Not-determined |
| 18-109_k141_213939 | Not-determined |
| 18-109_k141_40371 | Not-determined |
| 18-109_k141_50841 | Not-determined |
| 18-109_k141_40373 | Low-quality |
| 18-109_k141_19094 | Not-determined |
| 18-109_k141_79865 | Low-quality |
| 18-109_k141_79869 | Not-determined |
| 18-109_k141_40377 | Not-determined |
| 18-109_k141_19097 | Not-determined |
| 18-109_k141_19099 | Not-determined |
| 18-109_k141_101188 | Not-determined |
| 18-109_k141_9051 | Not-determined |
| 18-109_k141_60666 | Not-determined |
| 18-109_k141_19105 | Not-determined |
| 18-109_k141_50853 | Not-determined |
| 18-109_k141_71145 | Not-determined |
| 18-109_k141_163050 | Not-determined |
| 18-109_k141_163055 | Not-determined |
| 18-109_k141_163056 | Not-determined |
| 18-109_k141_9058 | Not-determined |
| 18-109_k141_79876 | Low-quality |
| 18-109_k141_50858 | Not-determined |
| 18-109_k141_40399 | Low-quality |
| 18-109_k141_183469 | Not-determined |
| 18-109_k141_163065 | Not-determined |
| 18-109_k141_71156 | Not-determined |

| | |
|---|---|
| 18-109_k141_79904 | Not-determined |
| 18-109_k141_163074 | Not-determined |
| 18-109_k141_19116 | Not-determined |
| 18-109_k141_19125 | Not-determined |
| 18-109_k141_79914 | Not-determined |
| 18-109_k141_213981 | Not-determined |
| 18-109_k141_40429 | Not-determined |
| 18-109_k141_71170 | Not-determined |
| 18-109_k141_90526 | Not-determined |
| 18-109_k141_19137 | Not-determined |
| 18-109_k141_71171 | Low-quality |
| 18-109_k141_163090 | Not-determined |
| 18-109_k141_40433 | Not-determined |
| 18-109_k141_172958 | Not-determined |
| 18-109_k141_79920 | Not-determined |
| 18-109_k141_71179 | Not-determined |
| 18-109_k141_213995 | Not-determined |
| 18-109_k141_213999 | Not-determined |
| 18-109_k141_9101 | Not-determined |
| 18-109_k141_79928 | Not-determined |
| 18-109_k141_131738 | Not-determined |
| 18-109_k141_131739 | Not-determined |
| 18-109_k141_79937 | Low-quality |
| 18-109_k141_101208 | Not-determined |
| 18-109_k141_9120 | Not-determined |
| 18-109_k141_40468 | Not-determined |
| 18-109_k141_163128 | Not-determined |
| 18-109_k141_151806 | Not-determined |
| 18-109_k141_234476 | Low-quality |
| 18-109_k141_27575 | Not-determined |
| 18-109_k141_79961 | Low-quality |
| 18-109_k141_101229 | Not-determined |
| 18-109_k141_234489 | Not-determined |
| 18-109_k141_234491 | Not-determined |
| 18-109_k141_60710 | Not-determined |
| 18-109_k141_9140 | Not-determined |
| 18-109_k141_79971 | Not-determined |
| 18-109_k141_60717 | Not-determined |
| 18-109_k141_60719 | Not-determined |
| 18-109_k141_79978 | Not-determined |
| 18-109_k141_151835 | Not-determined |
| 18-109_k141_151840 | Not-determined |
| 18-109_k141_71213 | Not-determined |
| 18-109_k141_173012 | Not-determined |
| 18-109_k141_151842 | Not-determined |
| 18-109_k141_131787 | Not-determined |
| 18-109_k141_79987 | Not-determined |
| 18-109_k141_151843 | Not-determined |
| 18-109_k141_173015 | Not-determined |
| 18-109_k141_79995 | Not-determined |
| 18-109_k141_27606 | Not-determined |
| 18-109_k141_151851 | Not-determined |
| 18-109_k141_151855 | Low-quality |
| 18-109_k141_151857 | Not-determined |
| 18-109_k141_19150 | Not-determined |
| 18-109_k141_79998 | Not-determined |
| 18-109_k141_183510 | Not-determined |
| 18-109_k141_27612 | Not-determined |
| 18-109_k141_183515 | Not-determined |
| 18-109_k141_234511 | Not-determined |
| 18-109_k141_163147 | Low-quality |
| 18-109_k141_90599 | Low-quality |
| 18-109_k141_173031 | Not-determined |
| 18-109_k141_234514 | Not-determined |
| 18-109_k141_173034 | Not-determined |
| 18-109_k141_234522 | Not-determined |
| 18-109_k141_234523 | Not-determined |
| 18-109_k141_183524 | Not-determined |
| 18-109_k141_183530 | Not-determined |
| 18-109_k141_19170 | Not-determined |
| 18-109_k141_151879 | Not-determined |
| 18-109_k141_40482 | Low-quality |
| 18-109_k141_40486 | Not-determined |
| 18-109_k141_90620 | Low-quality |
| 18-109_k141_151889 | Not-determined |
| 18-109_k141_151892 | Not-determined |
| 18-109_k141_173049 | Not-determined |
| 18-109_k141_40493 | Not-determined |
| 18-109_k141_151893 | Not-determined |
| 18-109_k141_183563 | Not-determined |
| 18-109_k141_183568 | Not-determined |
| 18-109_k141_90632 | Not-determined |
| 18-109_k141_27646 | Not-determined |
| 18-109_k141_90633 | Not-determined |
| 18-109_k141_71246 | Not-determined |
| 18-109_k141_40498 | Not-determined |
| 18-109_k141_71249 | Not-determined |
| 18-109_k141_27652 | Not-determined |
| 18-109_k141_234533 | Not-determined |
| 18-109_k141_71259 | Not-determined |
| 18-109_k141_163161 | Not-determined |
| 18-109_k141_71270 | Not-determined |
| 18-109_k141_40526 | Not-determined |
| 18-109_k141_40528 | Not-determined |
| 18-109_k141_40531 | Not-determined |
| 18-109_k141_40534 | Not-determined |
| 18-109_k141_101285 | Not-determined |
| 18-109_k141_173059 | Not-determined |
| 18-109_k141_234557 | Not-determined |
| 18-109_k141_111014 | Low-quality |
| 18-109_k141_173064 | Low-quality |
| 18-109_k141_111019 | Not-determined |
| 18-109_k141_101293 | Not-determined |
| 18-109_k141_101296 | Not-determined |
| 18-109_k141_214031 | Not-determined |
| 18-109_k141_234578 | Not-determined |
| 18-109_k141_234580 | Not-determined |
| 18-109_k141_234582 | Low-quality |
| 18-109_k141_101303 | Not-determined |
| 18-109_k141_163188 | Not-determined |
| 18-109_k141_183577 | Not-determined |
| 18-109_k141_101307 | Not-determined |
| 18-109_k141_214047 | Not-determined |
| 18-109_k141_101309 | Not-determined |
| 18-109_k141_80031 | Not-determined |
| 18-109_k141_151942 | Not-determined |
| 18-109_k141_214050 | Not-determined |
| 18-109_k141_151945 | Not-determined |
| 18-109_k141_19208 | Not-determined |
| 18-109_k141_183581 | Not-determined |
| 18-109_k141_151952 | Not-determined |
| 18-109_k141_90654 | Not-determined |
| 18-109_k141_122151 | Not-determined |
| 18-109_k141_9271 | Low-quality |
| 18-109_k141_151963 | Not-determined |
| 18-109_k141_9275 | Not-determined |
| 18-109_k141_183596 | Not-determined |
| 18-109_k141_101328 | Not-determined |
| 18-109_k141_163221 | Not-determined |
| 18-109_k141_90672 | Not-determined |
| 18-109_k141_141374 | Not-determined |
| 18-109_k141_50893 | Not-determined |
| 18-109_k141_80054 | Not-determined |
| 18-109_k141_151981 | Not-determined |
| 18-109_k141_111041 | Not-determined |
| 18-109_k141_173089 | Not-determined |
| 18-109_k141_222119 | Not-determined |
| 18-109_k141_80061 | Not-determined |
| 18-109_k141_111050 | Not-determined |
| 18-109_k141_111052 | Low-quality |
| 18-109_k141_151995 | Not-determined |
| 18-109_k141_183611 | Not-determined |
| 18-109_k141_71322 | Not-determined |
| 18-109_k141_71323 | Not-determined |
| 18-109_k141_111073 | Not-determined |
| 18-109_k141_163242 | Not-determined |
| 18-109_k141_71338 | Not-determined |
| 18-109_k141_243750 | Low-quality |
| 18-109_k141_19224 | Not-determined |
| 18-109_k141_40589 | Low-quality |
| 18-109_k141_152007 | Not-determined |
| 18-109_k141_122192 | Not-determined |
| 18-109_k141_152008 | Not-determined |
| 18-109_k141_183631 | Not-determined |
| 18-109_k141_19226 | Not-determined |
| 18-109_k141_222141 | Not-determined |
| 18-109_k141_122193 | Low-quality |
| 18-109_k141_122199 | Not-determined |
| 18-109_k141_122206 | Not-determined |
| 18-109_k141_183634 | Not-determined |
| 18-109_k141_60794 | Not-determined |
| 18-109_k141_80090 | Not-determined |
| 18-109_k141_183639 | Not-determined |
| 18-109_k141_80095 | Not-determined |
| 18-109_k141_27703 | Not-determined |
| 18-109_k141_27705 | Not-determined |
| 18-109_k141_80098 | Not-determined |
| 18-109_k141_80100 | Not-determined |
| 18-109_k141_40596 | Not-determined |
| 18-109_k141_60812 | Not-determined |
| 18-109_k141_122235 | Not-determined |
| 18-109_k141_141414 | Not-determined |
| 18-109_k141_163273 | Not-determined |
| 18-109_k141_122241 | Low-quality |
| 18-109_k141_141425 | Not-determined |
| 18-109_k141_163279 | Not-determined |
| 18-109_k141_40609 | Not-determined |
| 18-109_k141_163281 | Not-determined |
| 18-109_k141_183662 | Not-determined |
| 18-109_k141_222169 | Not-determined |
| 18-109_k141_60823 | Low-quality |
| 18-109_k141_222172 | Not-determined |
| 18-109_k141_27718 | Not-determined |
| 18-109_k141_173116 | Not-determined |
| 18-109_k141_131826 | Not-determined |
| 18-109_k141_222177 | Not-determined |
| 18-109_k141_101344 | Not-determined |
| 18-109_k141_101345 | Not-determined |
| 18-109_k141_183675 | Not-determined |
| 18-109_k141_80112 | Not-determined |
| 18-109_k141_60832 | Not-determined |
| 18-109_k141_19262 | Not-determined |
| 18-109_k141_71371 | Not-determined |
| 18-109_k141_60834 | Low-quality |
| 18-109_k141_60835 | Not-determined |
| 18-109_k141_234606 | Not-determined |
| 18-109_k141_234607 | Not-determined |
| 18-109_k141_101353 | Not-determined |
| 18-109_k141_80121 | Not-determined |
| 18-109_k141_80124 | Not-determined |
| 18-109_k141_60847 | Not-determined |
| 18-109_k141_40633 | Not-determined |
| 18-109_k141_19281 | Not-determined |
| 18-109_k141_173137 | Not-determined |
| 18-109_k141_222192 | Not-determined |
| 18-109_k141_163304 | Low-quality |
| 18-109_k141_122258 | Not-determined |
| 18-109_k141_60854 | Not-determined |
| 18-109_k141_60857 | Not-determined |
| 18-109_k141_131843 | Not-determined |
| 18-109_k141_234623 | Low-quality |
| 18-109_k141_40644 | Not-determined |
| 18-109_k141_131849 | Not-determined |
| 18-109_k141_19301 | Not-determined |
| 18-109_k141_234626 | Not-determined |
| 18-109_k141_111109 | Not-determined |
| 18-109_k141_152075 | Not-determined |
| 18-109_k141_111113 | Low-quality |
| 18-109_k141_111123 | Not-determined |
| 18-109_k141_19314 | Not-determined |
| 18-109_k141_80150 | Not-determined |
| 18-109_k141_71404 | Not-determined |
| 18-109_k141_173145 | Not-determined |
| 18-109_k141_183696 | Not-determined |
| 18-109_k141_27743 | Not-determined |
| 18-109_k141_19325 | Low-quality |
| 18-109_k141_71421 | Not-determined |
| 18-109_k141_122294 | Not-determined |
| 18-109_k141_71422 | Not-determined |
| 18-109_k141_122295 | Not-determined |
| 18-109_k141_122296 | Not-determined |
| 18-109_k141_234645 | Not-determined |
| 18-109_k141_9331 | Not-determined |
| 18-109_k141_141472 | Not-determined |
| 18-109_k141_111147 | Not-determined |
| 18-109_k141_9343 | Not-determined |
| 18-109_k141_183720 | Not-determined |
| 18-109_k141_131880 | Low-quality |
| 18-109_k141_234671 | Not-determined |
| 18-109_k141_71448 | Not-determined |
| 18-109_k141_40681 | Not-determined |
| 18-109_k141_152123 | Not-determined |
| 18-109_k141_173169 | Not-determined |
| 18-109_k141_9353 | Not-determined |
| 18-109_k141_152129 | Not-determined |
| 18-109_k141_152130 | Not-determined |
| 18-109_k141_152135 | Not-determined |
| 18-109_k141_19365 | Not-determined |
| 18-109_k141_9361 | Not-determined |
| 18-109_k141_60879 | Not-determined |
| 18-109_k141_40694 | Not-determined |
| 18-109_k141_9364 | Not-determined |
| 18-109_k141_9366 | Not-determined |
| 18-109_k141_90732 | Not-determined |
| 18-109_k141_122328 | Not-determined |
| 18-109_k141_122329 | Not-determined |
| 18-109_k141_9370 | Not-determined |
| 18-109_k141_141501 | Not-determined |
| 18-109_k141_80167 | Not-determined |
| 18-109_k141_122331 | Not-determined |
| 18-109_k141_152148 | Not-determined |
| 18-109_k141_80168 | Not-determined |
| 18-109_k141_19374 | Not-determined |
| 18-109_k141_222242 | Not-determined |
| 18-109_k141_19377 | Not-determined |
| 18-109_k141_60884 | Not-determined |
| 18-109_k141_234689 | Not-determined |
| 18-109_k141_27783 | Not-determined |
| 18-109_k141_9376 | Not-determined |
| 18-109_k141_40705 | Not-determined |
| 18-109_k141_111177 | Not-determined |
| 18-109_k141_60893 | Low-quality |
| 18-109_k141_9380 | Not-determined |
| 18-109_k141_183744 | Not-determined |
| 18-109_k141_183748 | Not-determined |
| 18-109_k141_80182 | Not-determined |
| 18-109_k141_131912 | Not-determined |
| 18-109_k141_27796 | Not-determined |
| 18-109_k141_222248 | Not-determined |
| 18-109_k141_80184 | Not-determined |
| 18-109_k141_27801 | Not-determined |
| 18-109_k141_80187 | Not-determined |
| 18-109_k141_152168 | Not-determined |
| 18-109_k141_9396 | Not-determined |
| 18-109_k141_122357 | Not-determined |
| 18-109_k141_9398 | Not-determined |
| 18-109_k141_122363 | Not-determined |
| 18-109_k141_222258 | Not-determined |
| 18-109_k141_122367 | Not-determined |
| 18-109_k141_122368 | Not-determined |
| 18-109_k141_122370 | Not-determined |
| 18-109_k141_80195 | Not-determined |
| 18-109_k141_40716 | Not-determined |
| 18-109_k141_19399 | Not-determined |
| 18-109_k141_173212 | Not-determined |
| 18-109_k141_141519 | Not-determined |
| 18-109_k141_9423 | Not-determined |
| 18-109_k141_40726 | Not-determined |
| 18-109_k141_152194 | Low-quality |
| 18-109_k141_214089 | Not-determined |
| 18-109_k141_163330 | Low-quality |
| 18-109_k141_152198 | Not-determined |
| 18-109_k141_141533 | Not-determined |
| 18-109_k141_173231 | Not-determined |
| 18-109_k141_19433 | Not-determined |
| 18-109_k141_141542 | Not-determined |
| 18-109_k141_234721 | Low-quality |
| 18-109_k141_152215 | Not-determined |
| 18-109_k141_141550 | Not-determined |
| 18-109_k141_141552 | Not-determined |
| 18-109_k141_80236 | Not-determined |
| 18-109_k141_27820 | Not-determined |
| 18-109_k141_9447 | Not-determined |
| 18-109_k141_222313 | Not-determined |
| 18-109_k141_173254 | Not-determined |
| 18-109_k141_222318 | Not-determined |
| 18-109_k141_122376 | Not-determined |
| 18-109_k141_152224 | Not-determined |
| 18-109_k141_152225 | Not-determined |
| 18-109_k141_9461 | Not-determined |
| 18-109_k141_60976 | Low-quality |
| 18-109_k141_234742 | Not-determined |
| 18-109_k141_9467 | Not-determined |
| 18-109_k141_152239 | Not-determined |
| 18-109_k141_152243 | Not-determined |
| 18-109_k141_152244 | Not-determined |
| 18-109_k141_183804 | Not-determined |
| 18-109_k141_9477 | Not-determined |
| 18-109_k141_234752 | Not-determined |
| 18-109_k141_101394 | Not-determined |
| 18-109_k141_9481 | Not-determined |
| 18-109_k141_122387 | Not-determined |
| 18-109_k141_183806 | Not-determined |
| 18-109_k141_141569 | Not-determined |
| 18-109_k141_19466 | Low-quality |
| 18-109_k141_214115 | Not-determined |
| 18-109_k141_214117 | Not-determined |
| 18-109_k141_173272 | Not-determined |
| 18-109_k141_222342 | Not-determined |
| 18-109_k141_173278 | Not-determined |
| 18-109_k141_163353 | Not-determined |
| 18-109_k141_19481 | Not-determined |
| 18-109_k141_222353 | Low-quality |
| 18-109_k141_152259 | Not-determined |
| 18-109_k141_19489 | Not-determined |
| 18-109_k141_101423 | Not-determined |
| 18-109_k141_101424 | Not-determined |
| 18-109_k141_111188 | Not-determined |
| 18-109_k141_163366 | Not-determined |
| 18-109_k141_101429 | Not-determined |
| 18-109_k141_9509 | Not-determined |
| 18-109_k141_101431 | Not-determined |
| 18-109_k141_19498 | Not-determined |
| 18-109_k141_19500 | Not-determined |
| 18-109_k141_90783 | Not-determined |
| 17-109-final-viral-combined_k141_371541\|\|full | Medium-quality |
| 18-109_k141_90784 | Not-determined |
| 18-109_k141_61020 | Not-determined |
| 18-109_k141_141580 | Not-determined |
| 18-109_k141_80259 | Not-determined |
| 18-109_k141_90790 | Not-determined |
| 18-109_k141_80262 | Low-quality |
| 18-109_k141_61027 | Not-determined |
| 18-109_k141_122393 | Not-determined |
| 18-109_k141_122394 | Not-determined |
| 18-109_k141_80268 | Not-determined |
| 18-109_k141_122395 | Not-determined |
| 18-109_k141_222382 | Not-determined |
| 18-109_k141_152291 | Low-quality |
| 18-109_k141_9535 | Not-determined |
| 18-109_k141_222395 | Low-quality |
| 18-109_k141_122402 | Not-determined |
| 18-109_k141_101462 | Not-determined |
| 18-109_k141_80288 | Not-determined |
| 18-109_k141_122407 | Not-determined |
| 18-109_k141_40754 | Low-quality |
| 18-109_k141_152307 | Not-determined |
| 18-109_k141_141610 | Not-determined |
| 18-109_k141_222406 | Not-determined |
| 18-109_k141_234784 | Not-determined |
| 18-109_k141_61042 | Not-determined |
| 18-109_k141_111209 | Not-determined |
| 18-109_k141_234789 | Not-determined |
| 18-109_k141_19544 | Not-determined |
| 18-109_k141_61044 | Not-determined |
| 18-109_k141_111212 | Not-determined |
| 18-109_k141_234794 | Not-determined |
| 18-109_k141_40771 | Not-determined |
| 18-109_k141_173295 | Not-determined |
| 18-109_k141_80317 | Not-determined |
| 18-109_k141_234801 | Not-determined |
| 18-109_k141_9561 | Not-determined |
| 18-109_k141_40778 | Not-determined |
| 18-109_k141_234804 | Low-quality |
| 18-109_k141_80322 | Not-determined |
| 18-109_k141_173304 | Low-quality |
| 18-109_k141_19553 | Not-determined |
| 18-109_k141_50941 | Not-determined |
| 18-109_k141_173310 | Not-determined |
| 18-109_k141_173313 | Not-determined |
| 18-109_k141_90843 | Not-determined |
| 18-109_k141_90844 | Not-determined |
| 18-109_k141_27845 | Not-determined |
| 18-109_k141_50953 | Not-determined |
| 18-109_k141_9569 | Not-determined |
| 18-109_k141_101523 | Not-determined |
| 18-109_k141_9572 | Not-determined |
| 18-109_k141_19571 | Not-determined |
| 18-109_k141_111229 | Low-quality |
| 18-109_k141_234831 | Not-determined |
| 18-109_k141_9586 | Not-determined |
| 18-109_k141_61082 | Not-determined |
| 18-109_k141_234834 | Not-determined |
| 18-109_k141_122451 | Not-determined |
| 18-109_k141_222430 | Not-determined |
| 18-109_k141_61086 | Not-determined |
| 18-109_k141_9592 | Not-determined |
| 18-109_k141_173348 | Not-determined |
| 18-109_k141_214150 | Not-determined |
| 18-109_k141_122454 | Not-determined |
| 18-109_k141_111248 | Not-determined |
| 18-109_k141_111251 | Not-determined |
| 18-109_k141_234852 | Not-determined |
| 18-109_k141_111253 | Not-determined |
| 18-109_k141_111255 | Not-determined |
| 18-109_k141_122462 | Not-determined |
| 18-109_k141_40790 | Not-determined |
| 18-109_k141_152329 | Not-determined |
| 18-109_k141_214163 | Not-determined |
| 18-109_k141_122467 | Not-determined |
| 18-109_k141_50988 | Low-quality |
| 18-109_k141_214167 | Not-determined |
| 18-109_k141_111257 | Not-determined |
| 18-109_k141_19593 | Not-determined |
| 18-109_k141_111263 | Not-determined |
| 18-109_k141_61112 | Not-determined |
| 18-109_k141_214188 | Not-determined |
| 18-109_k141_90886 | Low-quality |
| 18-109_k141_90889 | Not-determined |
| 18-109_k141_19609 | Not-determined |
| 18-109_k141_214198 | Not-determined |
| 18-109_k141_90896 | Not-determined |
| 18-109_k141_111267 | Not-determined |
| 18-109_k141_214207 | Low-quality |
| 18-109_k141_27875 | Not-determined |
| 18-109_k141_27876 | Not-determined |
| 18-109_k141_90904 | Not-determined |
| 18-109_k141_222450 | Low-quality |
| 18-109_k141_173379 | Not-determined |
| 18-109_k141_90912 | Not-determined |
| 18-109_k141_61137 | Not-determined |
| 18-109_k141_111276 | Not-determined |
| 18-109_k141_141619 | Not-determined |
| 18-109_k141_141627 | Not-determined |
| 18-109_k141_61146 | Not-determined |
| 18-109_k141_173393 | Not-determined |
| 18-109_k141_173396 | Not-determined |
| 18-109_k141_101528 | Not-determined |
| 18-109_k141_27883 | Not-determined |
| 18-109_k141_27884 | Not-determined |
| 18-109_k141_152357 | Not-determined |
| 18-109_k141_27889 | Not-determined |
| 18-109_k141_152359 | Not-determined |
| 18-109_k141_9602 | Low-quality |
| 18-109_k141_111302 | Not-determined |
| 18-109_k141_214224 | Not-determined |
| 18-109_k141_141648 | Not-determined |
| 18-109_k141_27903 | Low-quality |
| 18-109_k141_214232 | Not-determined |
| 18-109_k141_9619 | Not-determined |
| 18-109_k141_173413 | Not-determined |
| 18-109_k141_61180 | Not-determined |
| 18-109_k141_173423 | Not-determined |
| 18-109_k141_90931 | Not-determined |
| 18-109_k141_222464 | Not-determined |
| 18-109_k141_173429 | Not-determined |
| 18-109_k141_141679 | Not-determined |
| 18-109_k141_152376 | Low-quality |
| 18-109_k141_90944 | Not-determined |
| 18-109_k141_101560 | Not-determined |
| 18-109_k141_90948 | Not-determined |
| 18-109_k141_101566 | Not-determined |
| 18-109_k141_131942 | Low-quality |
| 18-109_k141_131943 | Not-determined |
| 18-109_k141_141693 | Not-determined |
| 18-109_k141_222475 | Not-determined |
| 18-109_k141_27922 | Not-determined |
| 18-109_k141_141696 | Not-determined |
| 18-109_k141_111325 | Not-determined |
| 18-109_k141_214259 | Not-determined |
| 18-109_k141_9661 | Not-determined |
| 18-109_k141_131955 | Low-quality |
| 18-109_k141_131956 | Not-determined |
| 18-109_k141_9664 | Not-determined |
| 18-109_k141_80343 | Not-determined |
| 18-109_k141_101594 | Not-determined |
| 18-109_k141_9675 | Not-determined |
| 18-109_k141_141716 | Not-determined |
| 18-109_k141_80351 | Not-determined |
| 18-109_k141_214270 | Not-determined |
| 18-109_k141_173464 | Not-determined |
| 18-109_k141_27954 | Not-determined |
| 18-109_k141_90965 | Not-determined |
| 18-109_k141_27957 | Not-determined |
| 18-109_k141_131976 | Not-determined |
| 18-109_k141_152408 | Not-determined |
| 18-109_k141_101609 | Not-determined |
| 18-109_k141_90972 | Not-determined |
| 18-109_k141_90973 | Not-determined |
| 18-109_k141_51007 | Not-determined |
| 18-109_k141_111342 | Not-determined |
| 18-109_k141_90976 | Not-determined |
| 18-109_k141_80365 | Not-determined |
| 18-109_k141_173477 | Not-determined |
| 18-109_k141_173480 | Not-determined |
| 18-109_k141_80367 | Not-determined |
| 18-109_k141_173481 | Not-determined |
| 18-109_k141_152426 | Not-determined |
| 18-109_k141_9696 | Not-determined |
| 18-109_k141_214294 | Not-determined |
| 18-109_k141_9700 | Not-determined |
| 18-109_k141_152429 | Not-determined |
| 18-109_k141_9704 | Not-determined |
| 18-109_k141_80379 | Not-determined |
| 18-109_k141_152436 | Not-determined |
| 18-109_k141_152439 | Not-determined |
| 18-109_k141_152443 | Not-determined |
| 18-109_k141_9716 | Not-determined |
| 18-109_k141_9724 | Not-determined |
| 18-109_k141_19644 | Not-determined |
| 18-109_k141_90988 | Not-determined |
| 18-109_k141_111365 | Low-quality |
| 18-109_k141_80398 | Not-determined |
| 18-109_k141_51043 | Not-determined |
| 18-109_k141_80400 | Not-determined |
| 18-109_k141_40810 | Not-determined |
| 18-109_k141_222527 | Not-determined |
| 18-109_k141_90997 | Not-determined |
| 18-109_k141_90998 | Not-determined |
| 18-109_k141_40821 | Not-determined |
| 18-109_k141_51059 | Not-determined |
| 18-109_k141_101622 | Not-determined |
| 18-109_k141_61190 | Not-determined |
| 18-109_k141_9736 | Not-determined |
| 18-109_k141_222537 | Not-determined |
| 18-109_k141_122489 | Not-determined |
| 18-109_k141_101628 | Not-determined |
| 18-109_k141_222544 | Not-determined |
| 18-109_k141_122493 | Not-determined |
| 18-109_k141_61206 | Not-determined |
| 18-109_k141_122497 | Not-determined |
| 18-109_k141_101643 | Not-determined |
| 18-109_k141_101646 | Not-determined |
| 18-109_k141_122501 | Not-determined |
| 18-109_k141_91010 | Not-determined |
| 18-109_k141_91011 | Not-determined |
| 18-109_k141_101652 | Not-determined |
| 18-109_k141_132033 | Not-determined |
| 18-109_k141_101662 | Not-determined |
| 18-109_k141_132036 | Not-determined |
| 18-109_k141_222571 | Low-quality |
| 18-109_k141_122513 | Not-determined |
| 18-109_k141_91022 | Not-determined |
| 18-109_k141_222580 | Not-determined |
| 18-109_k141_61226 | Not-determined |
| 18-109_k141_111399 | Not-determined |
| 18-109_k141_122529 | Low-quality |
| 18-109_k141_111408 | Not-determined |
| 18-109_k141_91030 | Not-determined |
| 18-109_k141_122536 | Not-determined |
| 18-109_k141_27980 | Not-determined |
| 18-109_k141_91040 | Not-determined |
| 18-109_k141_222598 | Not-determined |
| 18-109_k141_173510 | Not-determined |
| 18-109_k141_222615 | Not-determined |
| 18-109_k141_19690 | Not-determined |
| 18-109_k141_101719 | Not-determined |
| 18-109_k141_91052 | Not-determined |
| 18-109_k141_132071 | Not-determined |
| 18-109_k141_132073 | Low-quality |
| 18-109_k141_173515 | Not-determined |
| 18-109_k141_222624 | Not-determined |
| 18-109_k141_173520 | Not-determined |
| 18-109_k141_173522 | Not-determined |
| 18-109_k141_132093 | Not-determined |
| 18-109_k141_27995 | Not-determined |
| 18-109_k141_61268 | Not-determined |
| 18-109_k141_28005 | Not-determined |
| 18-109_k141_214298 | Not-determined |

| | |
|---|---|
| 18-109_k141_28010 | Not-determined |
| 18-109_k141_91066 | Not-determined |
| 18-109_k141_222638 | Not-determined |
| 18-109_k141_91071 | Not-determined |
| 18-109_k141_222650 | Not-determined |
| 18-109_k141_80434 | Not-determined |
| 18-109_k141_80435 | Not-determined |
| 18-109_k141_173562 | Not-determined |
| 18-109_k141_91087 | Not-determined |
| 18-109_k141_111428 | Not-determined |
| 18-109_k141_91089 | Not-determined |
| 18-109_k141_91091 | Not-determined |
| 18-109_k141_152458 | Low-quality |
| 18-109_k141_91093 | Not-determined |
| 18-109_k141_214325 | Not-determined |
| 18-109_k141_243770 | Not-determined |
| 18-109_k141_132131 | Not-determined |
| 18-109_k141_19726 | Not-determined |
| 18-109_k141_243775 | Not-determined |
| 18-109_k141_152472 | Not-determined |
| 18-109_k141_28026 | Low-quality |
| 18-109_k141_214344 | Not-determined |
| 18-109_k141_9787 | Not-determined |
| 18-109_k141_214347 | Not-determined |
| 18-109_k141_214349 | Not-determined |
| 18-109_k141_91098 | Low-quality |
| 18-109_k141_111468 | Not-determined |
| 18-109_k141_91106 | Not-determined |
| 18-109_k141_214373 | Not-determined |
| 18-109_k141_173565 | Not-determined |
| 18-109_k141_91111 | Not-determined |
| 18-109_k141_173570 | Not-determined |
| 18-109_k141_152475 | Not-determined |
| 18-109_k141_101747 | Not-determined |
| 18-109_k141_243810 | Low-quality |
| 18-109_k141_28037 | Not-determined |
| 18-109_k141_243813 | Not-determined |
| 18-109_k141_91120 | Low-quality |
| 18-109_k141_222674 | Not-determined |
| 18-109_k141_141823 | Not-determined |
| 18-109_k141_152491 | Low-quality |
| 18-109_k141_214391 | Low-quality |
| 18-109_k141_132165 | Not-determined |
| 18-109_k141_132166 | Not-determined |
| 18-109_k141_132169 | Not-determined |
| 18-109_k141_152504 | Not-determined |
| 18-109_k141_152505 | Not-determined |
| 18-109_k141_101796 | Not-determined |
| 18-109_k141_19761 | Not-determined |
| 18-109_k141_28051 | Not-determined |
| 18-109_k141_141864 | Not-determined |
| 18-109_k141_28052 | Not-determined |
| 18-109_k141_28053 | Not-determined |
| 18-109_k141_80440 | Not-determined |
| 18-109_k141_9797 | Not-determined |
| 18-109_k141_9801 | Not-determined |
| 18-109_k141_80447 | Not-determined |
| 18-109_k141_80455 | Not-determined |
| 18-109_k141_80456 | Not-determined |
| 18-109_k141_80462 | Not-determined |
| 18-109_k141_28068 | Not-determined |
| 18-109_k141_9811 | Not-determined |
| 18-109_k141_222691 | Not-determined |
| 18-109_k141_101805 | Not-determined |
| 18-109_k141_28069 | Not-determined |
| 18-109_k141_80472 | Not-determined |
| 18-109_k141_222696 | Not-determined |
| 18-109_k141_222698 | Not-determined |
| 18-109_k141_80476 | Not-determined |
| 18-109_k141_152514 | Not-determined |
| 18-109_k141_152515 | Low-quality |
| 18-109_k141_152517 | Not-determined |
| 18-109_k141_111515 | Not-determined |
| 18-109_k141_101812 | Low-quality |
| 18-109_k141_91143 | Not-determined |
| 18-109_k141_80485 | Not-determined |
| 18-109_k141_80487 | Not-determined |
| 18-109_k141_28086 | Not-determined |
| 18-109_k141_19793 | Low-quality |
| 18-109_k141_19796 | Not-determined |
| 18-109_k141_111527 | Not-determined |
| 18-109_k141_141909 | Not-determined |
| 18-109_k141_222719 | Low-quality |
| 18-109_k141_111536 | Not-determined |
| 18-109_k141_19809 | Not-determined |
| 18-109_k141_80518 | Not-determined |
| 18-109_k141_28095 | Not-determined |
| 18-109_k141_80524 | Not-determined |
| 18-109_k141_101847 | Not-determined |
| 18-109_k141_132187 | Not-determined |
| 18-109_k141_9857 | Not-determined |
| 18-109_k141_9859 | Not-determined |
| 18-109_k141_9862 | Not-determined |
| 18-109_k141_9863 | Not-determined |
| 18-109_k141_141914 | Not-determined |
| 18-109_k141_132197 | Not-determined |
| 18-109_k141_141925 | Not-determined |
| 18-109_k141_9871 | Low-quality |
| 18-109_k141_132204 | Not-determined |
| 18-109_k141_214410 | Not-determined |
| 18-109_k141_111553 | Not-determined |
| 18-109_k141_132214 | Not-determined |
| 18-109_k141_101862 | Not-determined |
| 18-109_k141_141944 | Low-quality |
| 18-109_k141_141945 | Not-determined |
| 18-109_k141_80554 | Not-determined |
| 18-109_k141_28117 | Not-determined |
| 18-109_k141_101872 | Not-determined |
| 18-109_k141_141949 | Not-determined |
| 18-109_k141_91175 | Low-quality |
| 18-109_k141_141957 | Not-determined |
| 18-109_k141_80561 | Not-determined |
| 18-109_k141_80564 | Not-determined |
| 18-109_k141_80568 | Not-determined |
| 18-109_k141_111571 | Not-determined |
| 18-109_k141_101889 | Not-determined |
| 18-109_k141_80575 | Not-determined |
| 18-109_k141_111582 | Not-determined |
| 18-109_k141_80582 | Low-quality |
| 18-109_k141_101905 | Not-determined |
| 18-109_k141_19858 | Not-determined |
| 18-109_k141_19860 | Not-determined |
| 18-109_k141_214438 | Not-determined |
| 18-109_k141_101910 | Not-determined |
| 18-109_k141_80590 | Not-determined |
| 18-109_k141_132263 | Not-determined |
| 18-109_k141_91187 | Not-determined |
| 18-109_k141_132265 | Not-determined |
| 18-109_k141_91190 | Not-determined |
| 18-109_k141_91191 | Not-determined |
| 18-109_k141_132270 | Not-determined |
| 18-109_k141_111607 | Not-determined |
| 18-109_k141_101923 | Not-determined |
| 18-109_k141_111613 | Not-determined |
| 18-109_k141_9928 | Not-determined |
| 18-109_k141_111618 | Not-determined |
| 18-109_k141_9936 | Not-determined |
| 18-109_k141_19894 | Not-determined |
| 18-109_k141_19895 | Not-determined |
| 18-109_k141_19897 | Not-determined |
| 18-109_k141_19901 | Not-determined |
| 18-109_k141_132287 | Not-determined |
| 18-109_k141_222729 | Not-determined |
| 18-109_k141_152575 | Not-determined |
| 18-109_k141_111642 | Not-determined |
| 18-109_k141_101951 | Not-determined |
| 18-109_k141_142012 | Not-determined |
| 18-109_k141_80618 | Not-determined |
| 18-109_k141_28157 | Not-determined |
| 18-109_k141_152585 | Not-determined |
| 18-109_k141_152586 | Not-determined |
| 18-109_k141_101954 | Not-determined |
| 18-109_k141_28158 | Not-determined |
| 18-109_k141_132306 | Not-determined |
| 18-109_k141_9969 | Not-determined |
| 18-109_k141_91216 | Not-determined |
| 18-109_k141_152591 | Not-determined |
| 18-109_k141_111652 | Not-determined |
| 18-109_k141_19923 | Not-determined |
| 18-109_k141_28171 | Not-determined |
| 18-109_k141_152596 | Not-determined |
| 18-109_k141_80632 | Not-determined |
| 18-109_k141_152603 | Not-determined |
| 18-109_k141_28182 | Not-determined |
| 18-109_k141_91237 | Not-determined |
| 18-109_k141_91242 | Not-determined |
| 18-109_k141_111677 | Not-determined |
| 18-109_k141_91244 | Not-determined |
| 18-109_k141_19939 | Not-determined |
| 18-109_k141_152605 | Low-quality |
| 18-109_k141_101977 | Not-determined |
| 18-109_k141_111679 | Not-determined |
| 18-109_k141_101982 | Not-determined |
| 18-109_k141_111687 | Not-determined |
| 18-109_k141_19951 | Not-determined |
| 18-109_k141_111692 | Not-determined |
| 18-109_k141_243835 | Not-determined |
| 18-109_k141_28222 | Not-determined |
| 18-109_k141_28223 | Not-determined |
| 18-109_k141_91256 | Not-determined |
| 18-109_k141_19960 | Not-determined |
| 18-109_k141_243837 | Not-determined |
| 18-109_k141_111711 | Not-determined |
| 18-109_k141_91260 | Not-determined |
| 18-109_k141_111717 | Not-determined |
| 18-109_k141_111721 | Not-determined |
| 18-109_k141_222768 | Not-determined |
| 18-109_k141_222773 | Not-determined |
| 18-109_k141_101993 | Not-determined |
| 18-109_k141_101999 | Low-quality |
| 18-109_k141_142078 | Not-determined |
| 18-109_k141_111732 | Not-determined |
| 18-109_k141_243855 | Not-determined |
| 18-109_k141_102002 | Not-determined |
| 18-109_k141_9996 | Low-quality |
| 18-109_k141_80638 | Not-determined |
| 18-109_k141_19970 | Not-determined |
| 18-109_k141_142082 | Not-determined |
| 18-109_k141_111757 | Not-determined |
| 18-109_k141_152671 | Not-determined |
| 18-109_k141_152672 | Not-determined |
| 18-109_k141_243863 | Not-determined |
| 18-109_k141_91287 | Not-determined |
| 18-109_k141_243866 | Not-determined |
| 18-109_k141_222808 | Not-determined |
| 18-109_k141_222809 | Not-determined |
| 18-109_k141_19983 | Not-determined |
| 18-109_k141_19986 | Not-determined |
| 18-109_k141_10019 | Not-determined |
| 18-109_k141_80657 | Not-determined |
| 18-109_k141_28294 | Not-determined |
| 18-109_k141_19992 | Not-determined |
| 18-109_k141_142096 | Not-determined |
| 18-109_k141_19995 | Not-determined |
| 18-109_k141_222816 | Not-determined |
| 18-109_k141_142098 | Not-determined |
| 18-109_k141_142106 | Not-determined |
| 18-109_k141_142107 | Not-determined |
| 18-109_k141_111789 | Not-determined |
| 18-109_k141_111791 | Not-determined |
| 18-109_k141_102034 | Low-quality |
| 18-109_k141_142113 | Not-determined |
| 18-109_k141_243899 | Not-determined |
| 18-109_k141_222833 | Not-determined |
| 18-109_k141_80690 | Not-determined |
| 18-109_k141_91331 | Low-quality |
| 18-109_k141_80694 | Not-determined |
| 18-109_k141_111807 | Not-determined |
| 18-109_k141_20011 | Not-determined |
| 18-109_k141_152701 | Not-determined |
| 18-109_k141_142120 | Not-determined |
| 18-109_k141_142125 | Not-determined |
| 18-109_k141_111821 | Not-determined |
| 18-109_k141_142130 | Not-determined |
| 18-109_k141_243926 | Not-determined |
| 18-109_k141_111829 | Not-determined |
| 18-109_k141_102048 | Not-determined |
| 18-109_k141_10049 | Not-determined |
| 18-109_k141_111836 | Not-determined |
| 18-109_k141_222867 | Not-determined |
| 18-109_k141_102052 | Not-determined |
| 18-109_k141_111843 | Not-determined |
| 18-109_k141_243929 | Not-determined |
| 18-109_k141_10061 | Not-determined |
| 18-109_k141_111847 | Not-determined |
| 18-109_k141_10065 | Not-determined |
| 18-109_k141_10069 | Not-determined |
| 18-109_k141_20047 | Not-determined |
| 18-109_k141_152719 | Not-determined |
| 18-109_k141_80729 | Not-determined |
| 18-109_k141_91374 | Not-determined |
| 18-109_k141_91381 | Not-determined |
| 18-109_k141_91383 | Not-determined |
| 18-109_k141_91384 | Not-determined |
| 18-109_k141_132327 | Not-determined |
| 18-109_k141_20061 | Not-determined |
| 18-109_k141_243946 | Not-determined |
| 18-109_k141_20063 | Not-determined |
| 18-109_k141_91389 | Not-determined |
| 18-109_k141_20069 | Not-determined |
| 18-109_k141_20070 | Not-determined |
| 18-109_k141_20072 | Not-determined |
| 18-109_k141_222890 | Not-determined |
| 18-109_k141_91405 | Not-determined |
| 18-109_k141_132357 | Not-determined |
| 18-109_k141_80767 | Not-determined |
| 18-109_k141_152759 | Not-determined |
| 18-109_k141_10084 | Not-determined |
| 18-109_k141_152760 | Not-determined |
| 18-109_k141_20094 | Low-quality |
| 18-109_k141_20095 | Not-determined |
| 18-109_k141_10085 | Not-determined |
| 18-109_k141_20100 | Not-determined |
| 18-109_k141_10087 | Not-determined |
| 18-109_k141_243965 | Not-determined |
| 18-109_k141_102098 | Not-determined |
| 18-109_k141_132370 | Not-determined |
| 18-109_k141_132372 | Low-quality |
| 18-109_k141_102103 | Not-determined |
| 18-109_k141_102104 | Not-determined |
| 18-109_k141_80786 | Not-determined |
| 18-109_k141_80789 | Not-determined |
| 18-109_k141_243988 | Not-determined |
| 18-109_k141_243990 | Not-determined |
| 18-109_k141_132386 | Not-determined |
| 18-109_k141_243993 | Not-determined |
| 18-109_k141_132410 | Not-determined |
| 18-109_k141_142177 | Not-determined |
| 18-109_k141_244011 | Not-determined |
| 18-109_k141_244014 | Not-determined |
| 18-109_k141_152788 | Not-determined |
| 18-109_k141_80825 | Not-determined |
| 18-109_k141_152790 | Not-determined |
| 18-109_k141_132445 | Not-determined |
| 18-109_k141_152791 | Not-determined |
| 18-109_k141_244020 | Not-determined |
| 18-109_k141_222946 | Not-determined |
| 18-109_k141_244028 | Not-determined |
| 18-109_k141_111879 | Not-determined |
| 18-109_k141_152798 | Not-determined |
| 18-109_k141_111882 | Not-determined |
| 18-109_k141_111892 | Not-determined |
| 18-109_k141_244043 | Not-determined |
| 18-109_k141_244054 | Not-determined |
| 18-109_k141_152813 | Not-determined |
| 18-109_k141_244062 | Not-determined |
| 18-109_k141_152820 | Not-determined |
| 18-109_k141_111899 | Not-determined |
| 18-109_k141_91464 | Low-quality |
| 18-109_k141_28435 | Not-determined |
| 18-109_k141_91466 | Not-determined |
| 18-109_k141_111914 | Not-determined |
| 18-109_k141_222990 | Not-determined |
| 18-109_k141_222992 | Not-determined |
| 18-109_k141_28464 | Not-determined |
| 18-109_k141_132493 | Not-determined |
| 18-109_k141_28470 | Not-determined |
| 18-109_k141_111927 | Not-determined |
| 18-109_k141_132502 | Low-quality |
| 18-109_k141_223003 | Not-determined |
| 18-109_k141_91491 | Not-determined |
| 18-109_k141_152860 | Not-determined |
| 18-109_k141_132517 | Not-determined |
| 18-109_k141_142221 | Not-determined |
| 18-109_k141_152882 | Not-determined |
| 18-109_k141_28503 | Not-determined |
| 18-109_k141_28506 | Not-determined |
| 18-109_k141_111938 | Not-determined |
| 18-109_k141_80845 | Low-quality |
| 18-109_k141_132533 | Not-determined |
| 18-109_k141_132540 | Low-quality |
| 18-109_k141_132541 | Not-determined |
| 18-109_k141_223023 | Not-determined |
| 18-109_k141_80860 | Not-determined |
| 18-109_k141_132544 | Low-quality |
| 18-109_k141_28532 | Low-quality |
| 18-109_k141_244082 | Not-determined |
| 18-109_k141_80868 | Not-determined |
| 18-109_k141_80873 | Not-determined |
| 18-109_k141_152900 | Low-quality |
| 18-109_k141_152906 | Not-determined |
| 18-109_k141_80883 | Not-determined |
| 18-109_k141_80884 | Not-determined |
| 18-109_k141_223045 | Not-determined |
| 18-109_k141_80893 | Not-determined |
| 18-109_k141_223050 | Not-determined |
| 18-109_k141_111957 | Low-quality |
| 18-109_k141_132578 | Not-determined |
| 18-109_k141_223070 | Not-determined |
| 18-109_k141_80909 | Not-determined |
| 18-109_k141_80916 | Not-determined |
| 18-109_k141_142259 | Not-determined |
| 18-109_k141_111989 | Low-quality |
| 18-109_k141_111992 | Not-determined |
| 18-109_k141_244104 | Not-determined |
| 18-109_k141_112003 | Not-determined |
| 18-109_k141_152932 | Not-determined |
| 18-109_k141_80936 | Not-determined |
| 18-109_k141_142284 | Not-determined |
| 18-109_k141_28572 | Not-determined |
| 18-109_k141_28576 | Not-determined |
| 18-109_k141_142289 | Not-determined |
| 18-109_k141_10149 | Not-determined |
| 18-109_k141_112023 | Not-determined |
| 18-109_k141_132592 | Not-determined |
| 18-109_k141_142318 | Not-determined |
| 18-109_k141_152938 | Not-determined |
| 18-109_k141_142319 | Not-determined |
| 18-109_k141_142321 | Not-determined |
| 18-109_k141_28584 | Not-determined |
| 18-109_k141_28591 | Not-determined |
| 18-109_k141_142332 | Not-determined |
| 18-109_k141_244165 | Not-determined |
| 18-109_k141_112034 | Not-determined |
| 18-109_k141_142348 | Not-determined |
| 18-109_k141_112043 | Not-determined |
| 18-109_k141_142352 | Not-determined |
| 18-109_k141_244177 | Not-determined |
| 18-109_k141_80962 | Not-determined |
| 18-109_k141_244183 | Not-determined |
| 18-109_k141_223090 | Not-determined |
| 18-109_k141_244184 | Low-quality |
| 18-109_k141_112061 | Not-determined |
| 18-109_k141_10174 | Not-determined |
| 18-109_k141_223101 | Not-determined |
| 18-109_k141_152953 | Not-determined |
| 18-109_k141_223109 | Not-determined |
| 18-109_k141_80987 | Not-determined |
| 18-109_k141_80990 | Low-quality |
| 18-109_k141_28634 | Not-determined |
| 18-109_k141_80993 | Not-determined |
| 18-109_k141_28636 | Not-determined |
| 18-109_k141_223118 | Not-determined |
| 18-109_k141_28643 | Not-determined |
| 18-109_k141_20186 | Low-quality |
| 18-109_k141_142373 | Not-determined |
| 18-109_k141_81013 | Not-determined |
| 18-109_k141_81014 | Not-determined |
| 18-109_k141_10195 | Not-determined |
| 18-109_k141_81016 | Not-determined |
| 18-109_k141_20192 | Not-determined |
| 18-109_k141_81028 | Not-determined |
| 18-109_k141_10209 | Not-determined |
| 18-109_k141_28683 | Not-determined |
| 18-109_k141_132630 | Not-determined |
| 18-109_k141_142375 | Not-determined |
| 18-109_k141_28693 | Not-determined |
| 18-109_k141_81049 | Not-determined |
| 18-109_k141_81054 | Not-determined |
| 18-109_k141_112098 | Not-determined |
| 18-109_k141_112100 | Not-determined |
| 18-109_k141_28707 | Not-determined |
| 18-109_k141_20233 | Not-determined |
| 18-109_k141_112120 | Not-determined |
| 18-109_k141_153031 | Low-quality |
| 18-109_k141_153033 | Not-determined |
| 18-109_k141_28711 | Not-determined |
| 18-109_k141_28712 | Not-determined |
| 18-109_k141_142403 | Not-determined |
| 18-109_k141_244205 | Not-determined |
| 18-109_k141_132654 | Not-determined |
| 18-109_k141_20254 | Not-determined |
| 18-109_k141_142409 | Not-determined |
| 18-109_k141_142426 | Not-determined |
| 18-109_k141_28720 | Not-determined |
| 18-109_k141_112144 | Not-determined |
| 18-109_k141_112146 | Not-determined |
| 18-109_k141_132677 | Not-determined |
| 18-109_k141_142437 | Not-determined |
| 18-109_k141_112170 | Not-determined |
| 18-109_k141_112173 | Low-quality |
| 18-109_k141_132690 | Not-determined |
| 18-109_k141_112191 | Not-determined |
| 18-109_k141_153079 | Low-quality |
| 18-109_k141_153097 | Low-quality |
| 18-109_k141_153100 | Not-determined |
| 18-109_k141_28760 | Not-determined |
| 18-109_k141_112229 | Not-determined |
| 18-109_k141_28762 | Not-determined |
| 18-109_k141_244221 | Not-determined |
| 18-109_k141_244224 | Not-determined |
| 18-109_k141_223198 | Not-determined |
| 18-109_k141_223200 | Not-determined |
| 18-109_k141_28781 | Not-determined |
| 18-109_k141_244234 | Not-determined |
| 18-109_k141_28791 | Not-determined |
| 18-109_k141_28822 | Not-determined |
| 18-109_k141_112244 | Not-determined |
| 18-109_k141_81078 | Not-determined |
| 18-109_k141_28839 | Not-determined |
| 18-109_k141_91537 | Low-quality |
| 18-109_k141_153118 | Not-determined |
| 18-109_k141_153133 | Not-determined |
| 18-109_k141_91555 | Not-determined |
| 18-109_k141_132721 | Low-quality |
| 18-109_k141_91562 | Not-determined |
| 18-109_k141_91563 | Not-determined |
| 18-109_k141_28848 | Not-determined |
| 18-109_k141_28853 | Not-determined |
| 18-109_k141_223222 | Low-quality |
| 18-109_k141_153147 | Not-determined |
| 18-109_k141_142466 | Not-determined |
| 18-109_k141_28865 | Not-determined |
| 18-109_k141_112308 | Not-determined |
| 18-109_k141_142480 | Not-determined |
| 18-109_k141_91603 | Not-determined |
| 18-109_k141_223257 | Not-determined |
| 18-109_k141_91609 | Not-determined |
| 18-109_k141_223261 | Not-determined |
| 18-109_k141_223263 | Not-determined |
| 18-109_k141_223264 | Not-determined |
| 18-109_k141_20382 | Not-determined |
| 18-109_k141_28892 | Not-determined |
| 18-109_k141_153153 | Not-determined |
| 18-109_k141_153155 | Not-determined |
| 18-109_k141_223281 | Not-determined |
| 18-109_k141_132749 | Low-quality |
| 18-109_k141_91646 | Not-determined |
| 18-109_k141_223294 | Not-determined |
| 18-109_k141_28912 | Not-determined |
| 18-109_k141_91661 | Not-determined |
| 18-109_k141_142525 | Not-determined |
| 18-109_k141_91673 | Not-determined |
| 18-109_k141_91675 | Not-determined |
| 18-109_k141_244242 | Not-determined |
| 18-109_k141_153158 | Low-quality |
| 18-109_k141_244250 | Not-determined |
| 18-109_k141_244256 | Not-determined |
| 18-109_k141_244259 | Not-determined |
| 18-109_k141_91703 | Not-determined |
| 18-109_k141_91704 | Not-determined |
| 18-109_k141_91706 | Not-determined |
| 18-109_k141_153173 | Not-determined |
| 18-109_k141_20401 | Not-determined |
| 18-109_k141_81154 | Low-quality |
| 18-109_k141_28930 | Not-determined |
| 18-109_k141_28932 | Not-determined |
| 18-109_k141_91734 | Not-determined |
| 18-109_k141_81161 | Not-determined |
| 18-109_k141_81171 | Not-determined |
| 18-109_k141_81175 | Low-quality |
| 18-109_k141_20409 | Not-determined |
| 18-109_k141_142556 | Not-determined |
| 18-109_k141_28950 | Not-determined |
| 18-109_k141_28954 | Not-determined |
| 18-109_k141_91758 | Not-determined |
| 18-109_k141_142562 | Not-determined |
| 18-109_k141_81183 | Not-determined |
| 18-109_k141_142566 | Not-determined |
| 18-109_k141_91762 | Not-determined |
| 18-109_k141_91764 | Not-determined |
| 18-109_k141_142572 | Not-determined |
| 18-109_k141_28978 | Not-determined |
| 18-109_k141_142586 | Not-determined |
| 18-109_k141_81195 | Low-quality |
| 18-109_k141_91776 | Not-determined |
| 18-109_k141_142616 | Not-determined |
| 18-109_k141_142627 | Not-determined |
| 18-109_k141_81220 | Not-determined |
| 18-109_k141_81221 | Not-determined |
| 18-109_k141_81224 | Not-determined |
| 18-109_k141_81228 | Not-determined |
| 18-109_k141_81233 | Not-determined |
| 18-109_k141_29001 | Not-determined |
| 18-109_k141_81239 | Not-determined |
| 18-109_k141_81246 | Not-determined |
| 18-109_k141_81247 | Not-determined |
| 18-109_k141_142648 | Not-determined |
| 18-109_k141_29019 | Not-determined |
| 18-109_k141_91792 | Not-determined |
| 18-109_k141_223328 | Not-determined |
| 18-109_k141_29045 | Not-determined |
| 18-109_k141_142666 | Not-determined |
| 18-109_k141_29048 | Not-determined |
| 18-109_k141_142673 | Low-quality |
| 18-109_k141_29062 | Low-quality |
| 18-109_k141_142680 | Not-determined |
| 18-109_k141_29064 | Not-determined |
| 18-109_k141_244323 | Low-quality |
| 18-109_k141_142694 | Not-determined |
| 18-109_k141_29088 | Not-determined |
| 18-109_k141_29095 | Not-determined |
| 18-109_k141_142704 | Not-determined |
| 18-109_k141_244337 | Not-determined |
| 18-109_k141_29112 | Not-determined |
| 18-109_k141_244342 | Not-determined |
| 18-109_k141_29123 | Not-determined |
| 18-109_k141_142729 | Not-determined |
| 18-109_k141_81281 | Not-determined |
| 18-109_k141_81287 | Low-quality |
| 18-109_k141_81296 | Not-determined |
| 18-109_k141_29157 | Not-determined |
| 18-109_k141_142746 | Not-determined |
| 18-109_k141_142747 | Not-determined |
| 18-109_k141_142756 | Not-determined |
| 18-109_k141_142758 | Not-determined |
| 18-109_k141_81327 | Low-quality |
| 18-109_k141_29167 | Not-determined |
| 18-109_k141_81347 | Not-determined |
| 18-109_k141_223363 | Not-determined |
| 18-109_k141_29198 | Not-determined |
| 18-109_k141_223373 | Not-determined |
| 18-109_k141_223374 | Not-determined |
| 18-109_k141_223376 | Low-quality |
| 18-109_k141_223381 | Low-quality |
| 18-109_k141_81378 | Not-determined |
| 18-109_k141_223387 | Not-determined |
| 18-109_k141_81408 | Not-determined |
| 18-109_k141_223403 | Not-determined |
| 18-109_k141_244366 | Not-determined |
| 18-109_k141_244369 | Not-determined |
| 18-109_k141_223410 | Not-determined |
| 18-109_k141_29234 | Low-quality |
| 18-109_k141_244373 | Not-determined |
| 18-109_k141_244378 | Not-determined |
| 18-109_k141_223421 | Not-determined |
| 18-109_k141_91837 | Not-determined |
| 18-109_k141_29242 | Not-determined |
| 18-109_k141_223432 | Not-determined |
| 18-109_k141_29249 | Not-determined |
| 18-109_k141_29251 | Not-determined |
| 18-109_k141_91850 | Not-determined |
| 18-109_k141_244398 | Not-determined |
| 18-109_k141_91858 | Not-determined |
| 18-109_k141_244402 | Not-determined |
| 18-109_k141_29263 | Low-quality |
| 18-109_k141_223437 | Not-determined |
| 18-109_k141_81467 | Not-determined |
| 18-109_k141_29281 | Not-determined |

| | |
|---|---|
| 18-109_k141_29290 | Not-determined |
| 18-109_k141_142826 | Not-determined |
| 18-109_k141_223463 | Not-determined |
| 18-109_k141_223464 | Not-determined |
| 18-109_k141_91869 | Low-quality |
| 18-109_k141_223465 | Low-quality |
| 18-109_k141_91888 | Not-determined |
| 18-109_k141_29311 | Not-determined |
| 18-109_k141_29313 | Not-determined |
| 18-109_k141_142855 | Not-determined |
| 18-109_k141_142856 | Not-determined |
| 18-109_k141_29327 | Low-quality |
| 18-109_k141_29332 | Not-determined |
| 18-109_k141_29336 | Low-quality |
| 18-109_k141_29370 | Not-determined |
| 18-109_k141_223510 | Not-determined |
| 18-109_k141_142877 | Low-quality |
| 18-109_k141_29373 | Not-determined |
| 18-109_k141_223531 | Not-determined |
| 18-110-final-viral-combined_k141_276204\|\|full | High-quality |
| 18-109_k141_29389 | Not-determined |
| 18-109_k141_244426 | Not-determined |
| 18-109_k141_29396 | Not-determined |
| 18-109_k141_223536 | Not-determined |
| 18-109_k141_223546 | Not-determined |
| 18-109_k141_223564 | Not-determined |
| 18-109_k141_223573 | Not-determined |
| 18-109_k141_244455 | Not-determined |
| 18-109_k141_29415 | Not-determined |
| 18-109_k141_29417 | Not-determined |
| 18-109_k141_81510 | Low-quality |
| 18-109_k141_223601 | Low-quality |
| 18-109_k141_223605 | Not-determined |
| 18-109_k141_223610 | Not-determined |
| 18-109_k141_81523 | Not-determined |
| 18-109_k141_81526 | Not-determined |
| 18-109_k141_81527 | Not-determined |
| 18-109_k141_142885 | Not-determined |
| 18-109_k141_142895 | Not-determined |
| 18-109_k141_29425 | Not-determined |
| 18-109_k141_81540 | Not-determined |
| 18-109_k141_142905 | Not-determined |
| 18-109_k141_142910 | Low-quality |
| 18-109_k141_142922 | Not-determined |
| 18-109_k141_29437 | Not-determined |
| 18-109_k141_81545 | Not-determined |
| 18-109_k141_29454 | Not-determined |
| 18-109_k141_142957 | Low-quality |
| 18-109_k141_223662 | Not-determined |
| 18-109_k141_29459 | Not-determined |
| 18-109_k141_244487 | Not-determined |
| 18-109_k141_244488 | Not-determined |
| 18-109_k141_223673 | Not-determined |
| 18-109_k141_81559 | Not-determined |
| 18-109_k141_223676 | Not-determined |
| 18-109_k141_223677 | Not-determined |
| 18-109_k141_223681 | Not-determined |
| 18-109_k141_29473 | Not-determined |
| 18-109_k141_244507 | Not-determined |
| 18-109_k141_223708 | Not-determined |
| 18-109_k141_81587 | Not-determined |
| 18-109_k141_223717 | Not-determined |
| 18-109_k141_223725 | Low-quality |
| 18-109_k141_223731 | Not-determined |
| 18-109_k141_244513 | Not-determined |
| 18-109_k141_244517 | Not-determined |
| 18-109_k141_244521 | Not-determined |
| 18-109_k141_81611 | Not-determined |
| 18-109_k141_81617 | Not-determined |
| 18-109_k141_81627 | Not-determined |
| 18-109_k141_244530 | Not-determined |
| 18-109_k141_81644 | Not-determined |
| 18-109_k141_223791 | Not-determined |
| 18-109_k141_223794 | Not-determined |
| 18-109_k141_81679 | Not-determined |
| 18-109_k141_81683 | Not-determined |
| 18-109_k141_81685 | Not-determined |
| 18-109_k141_81693 | Not-determined |
| 18-109_k141_223829 | Not-determined |
| 18-109_k141_223838 | Low-quality |
| 18-109_k141_223842 | Not-determined |
| 18-109_k141_223844 | Not-determined |
| 18-109_k141_223856 | Not-determined |
| 18-109_k141_244562 | Not-determined |
| 18-109_k141_244581 | Not-determined |
| 18-109_k141_244585 | Not-determined |
| 18-109_k141_244589 | Low-quality |
| 18-109_k141_223898 | Not-determined |
| 18-109_k141_223903 | Not-determined |
| 18-109_k141_244598 | Not-determined |
| 18-109_k141_29497 | Not-determined |
| 18-109_k141_223952 | Not-determined |
| 18-109_k141_29557 | Not-determined |
| 18-109_k141_29580 | Not-determined |
| 18-109_k141_29582 | Low-quality |
| 18-109_k141_29622 | Not-determined |
| 18-109_k141_29634 | Not-determined |
| 18-109_k141_224000 | Low-quality |
| 18-109_k141_29639 | Not-determined |
| 18-109_k141_29652 | Not-determined |
| 18-109_k141_244636 | Low-quality |
| 18-109_k141_224026 | Not-determined |
| 18-109_k141_29701 | Not-determined |
| 18-109_k141_29712 | Not-determined |
| 18-109_k141_29716 | Not-determined |
| 18-109_k141_244647 | Not-determined |
| 18-109_k141_29721 | Low-quality |
| 18-109_k141_224048 | Not-determined |
| 18-109_k141_224053 | Not-determined |
| 18-109_k141_29735 | Not-determined |
| 18-109_k141_224060 | Not-determined |
| 18-109_k141_29739 | Not-determined |
| 18-109_k141_29752 | Not-determined |
| 18-109_k141_224067 | Not-determined |
| 18-109_k141_29757 | Not-determined |
| 18-109_k141_29772 | Not-determined |
| 18-109_k141_29775 | Not-determined |
| 18-109_k141_244667 | Not-determined |
| 18-109_k141_244674 | Not-determined |
| 18-109_k141_244696 | Not-determined |
| 18-109_k141_244700 | Not-determined |
| 18-109_k141_244702 | Low-quality |
| 18-109_k141_29821 | Not-determined |
| 18-109_k141_29837 | Not-determined |
| 18-109_k141_244713 | Not-determined |
| 18-109_k141_224123 | Not-determined |
| 18-109_k141_224129 | Not-determined |
| 18-109_k141_224133 | Not-determined |
| 18-109_k141_224147 | Not-determined |
| 18-109_k141_224148 | Not-determined |
| 18-109_k141_29900 | Not-determined |
| 18-109_k141_29920 | Not-determined |
| 18-109_k141_29922 | Not-determined |
| 18-109_k141_224182 | Not-determined |
| 18-109_k141_29928 | Not-determined |
| 18-109_k141_29933 | Not-determined |
| 18-109_k141_224203 | Not-determined |
| 18-109_k141_29961 | Low-quality |
| 18-109_k141_29964 | Low-quality |
| 18-109_k141_224213 | Not-determined |
| 18-109_k141_224227 | Low-quality |
| 18-109_k141_224229 | Not-determined |
| 18-109_k141_224230 | Not-determined |
| 18-109_k141_224241 | Not-determined |
| 18-109_k141_224255 | Not-determined |
| 18-109_k141_224259 | Not-determined |
| 18-109_k141_224266 | Not-determined |
| 18-109_k141_224273 | Not-determined |
| 18-109_k141_29983 | Not-determined |
| 18-109_k141_224276 | Not-determined |
| 18-109_k141_29987 | Not-determined |
| 18-109_k141_30009 | Not-determined |
| 18-109_k141_224290 | Not-determined |
| 18-109_k141_244731 | Not-determined |
| 18-109_k141_224291 | Not-determined |
| 18-109_k141_30027 | Not-determined |
| 18-109_k141_30039 | Not-determined |
| 18-109_k141_224310 | Not-determined |
| 18-109_k141_244741 | Not-determined |
| 18-109_k141_244745 | Not-determined |
| 18-109_k141_224321 | Not-determined |
| 18-109_k141_224326 | Not-determined |
| 18-109_k141_224327 | Low-quality |
| 18-109_k141_224329 | Not-determined |
| 18-109_k141_244754 | Not-determined |
| 18-109_k141_224376 | Not-determined |
| 18-109_k141_244766 | Not-determined |
| 18-109_k141_244773 | Not-determined |
| 18-109_k141_244775 | Not-determined |
| 18-109_k141_244776 | Not-determined |
| 18-109_k141_30073 | Not-determined |
| 18-109_k141_30074 | Low-quality |
| 18-109_k141_244792 | Low-quality |
| 18-109_k141_244797 | Not-determined |
| 18-109_k141_30091 | Not-determined |
| 18-109_k141_30096 | Not-determined |
| 18-109_k141_30101 | Low-quality |
| 18-109_k141_224418 | Not-determined |
| 18-109_k141_30105 | Not-determined |
| 18-109_k141_224424 | Not-determined |
| 18-109_k141_30109 | Not-determined |
| 18-109_k141_224431 | Not-determined |
| 18-109_k141_30133 | Not-determined |
| 18-109_k141_224461 | Low-quality |
| 18-109_k141_244821 | Not-determined |
| 18-109_k141_30150 | Not-determined |
| 18-109_k141_30151 | Not-determined |
| 18-109_k141_30159 | Not-determined |
| 18-109_k141_244845 | Low-quality |
| 18-109_k141_30166 | Not-determined |
| 18-109_k141_30185 | Not-determined |
| 18-109_k141_224528 | Not-determined |
| 18-109_k141_30191 | Not-determined |
| 18-109_k141_224540 | Not-determined |
| 18-109_k141_224554 | Not-determined |
| 18-109_k141_224562 | Not-determined |
| 18-109_k141_224572 | Not-determined |
| 18-109_k141_224587 | Not-determined |
| 18-109_k141_30226 | Not-determined |
| 18-109_k141_30231 | Not-determined |
| 18-109_k141_30232 | Not-determined |
| 18-109_k141_244861 | Not-determined |
| 18-109_k141_30247 | Not-determined |
| 18-109_k141_30288 | Not-determined |
| 18-109_k141_30289 | Not-determined |
| 18-109_k141_30302 | Not-determined |
| 18-109_k141_30327 | Not-determined |
| 17-109_k141_386692 | Complete |
| 18-109_k141_30341 | Low-quality |
| 18-109_k141_30352 | Not-determined |
| 18-109_k141_30362 | Not-determined |
| 18-109_k141_30391 | Not-determined |
| 18-109_k141_30397 | Not-determined |
| 18-109_k141_244874 | Not-determined |
| 18-109_k141_244882 | Not-determined |
| 18-109_k141_244893 | Not-determined |
| 18-109_k141_244915 | Not-determined |
| 18-109_k141_244920 | Not-determined |
| 18-109_k141_244921 | Not-determined |
| 18-109_k141_244924 | Not-determined |
| 18-109_k141_244935 | Not-determined |
| 18-109_k141_244937 | Not-determined |
| 18-109_k141_244938 | Not-determined |
| 18-109_k141_30446 | Not-determined |
| 18-109_k141_244943 | Not-determined |
| 18-109_k141_244946 | Not-determined |
| 18-109_k141_244947 | Not-determined |
| 18-109_k141_244953 | Not-determined |
| 18-109_k141_244958 | Not-determined |
| 18-109_k141_244964 | Not-determined |
| 18-109_k141_244965 | Not-determined |
| 18-109_k141_244967 | Not-determined |
| 18-109_k141_244968 | Not-determined |
| 18-109_k141_244971 | Not-determined |
| 18-109_k141_30468 | Low-quality |
| 18-109_k141_244974 | Not-determined |
| 18-109_k141_244976 | Not-determined |
| 18-109_k141_244986 | Not-determined |
| 18-109_k141_244988 | Not-determined |
| 18-109_k141_30473 | Not-determined |
| 18-109_k141_30481 | Not-determined |
| 18-109_k141_245003 | Not-determined |
| 18-109_k141_30486 | Not-determined |
| 18-109_k141_30487 | Not-determined |
| 18-109_k141_245018 | Not-determined |
| 18-109_k141_245019 | Not-determined |
| 18-109_k141_245020 | Not-determined |
| 18-109_k141_30489 | Not-determined |
| 18-109_k141_245025 | Not-determined |
| 18-109_k141_245027 | Not-determined |
| 18-109_k141_245036 | Not-determined |
| 18-109_k141_245045 | Not-determined |
| 18-109_k141_245046 | Not-determined |
| 18-109_k141_245050 | Not-determined |
| 18-109_k141_245059 | Not-determined |
| 18-109_k141_245060 | Not-determined |
| 18-109_k141_245061 | Not-determined |
| 18-109_k141_245068 | Not-determined |
| 18-109_k141_245073 | Not-determined |
| 18-109_k141_30503 | Low-quality |
| 18-109_k141_30516 | Not-determined |
| 18-109_k141_30545 | Low-quality |
| 18-109_k141_30549 | Not-determined |
| 18-109_k141_30583 | Not-determined |
| 18-109_k141_30633 | Not-determined |
| KM-final-viral-combined_k141_167045\|\|full | Not-determined |
| 18-101_k141_170115 | High-quality |
| KM-final-viral-combined_k141_69879\|\|full | Not-determined |
| KM-final-viral-combined_k141_159480\|\|full | Low-quality |
| KM-final-viral-combined_k141_138473\|\|full | Not-determined |
| KM-final-viral-combined_k141_239963\|\|full | Low-quality |
| KM-final-viral-combined_k141_253476\|\|full | Not-determined |
| KM-final-viral-combined_k141_266507\|\|full | Not-determined |
| 17-109-final-viral-combined_k141_293354\|\|full | Medium-quality |
| KM-final-viral-combined_k141_266513\|\|full | Not-determined |
| KM-final-viral-combined_k141_266517\|\|full | Not-determined |
| KM-final-viral-combined_k141_266526\|\|full | Not-determined |
| KM-final-viral-combined_k141_266544\|\|full | Not-determined |
| KM-final-viral-combined_k141_266550\|\|full | Not-determined |
| KM-final-viral-combined_k141_177808\|\|full | Low-quality |
| KM-final-viral-combined_k141_11117\|\|full | Not-determined |
| KM-final-viral-combined_k141_88904\|\|full | Not-determined |
| KM-final-viral-combined_k141_111131\|\|full | Low-quality |
| KM-final-viral-combined_k141_66685\|\|full | Not-determined |
| KM-final-viral-combined_k141_77806\|\|full | Low-quality |
| KM-final-viral-combined_k141_188930\|\|full | Not-determined |
| KM-final-viral-combined_k141_44464\|\|full | Not-determined |
| KM-final-viral-combined_k141_33348\|\|full | Low-quality |
| KM-final-viral-combined_k141_144485\|\|full | Low-quality |
| KM-final-viral-combined_k141_244516\|\|full | Not-determined |
| KM-final-viral-combined_k141_244517\|\|full | Not-determined |
| KM-final-viral-combined_k141_111148\|\|full | Not-determined |
| KM-final-viral-combined_k141_188948\|\|full | Low-quality |
| KM-final-viral-combined_k141_77807\|\|full | Low-quality |
| KM-final-viral-combined_k141_133379\|\|full | Not-determined |
| KM-final-viral-combined_k141_255625\|\|full | Not-determined |
| KM-final-viral-combined_k141_200077\|\|full | Not-determined |
| KM-final-viral-combined_k141_88932\|\|full | Not-determined |
| KM-final-viral-combined_k141_111172\|\|full | Low-quality |
| KM-final-viral-combined_k141_200089\|\|full | Not-determined |
| KM-final-viral-combined_k141_211169\|\|full | Not-determined |
| KM-final-viral-combined_k141_144514\|\|full | Not-determined |
| KM-final-viral-combined_k141_188998\|\|full | Low-quality |
| KM-final-viral-combined_k141_200117\|\|full | Low-quality |
| KM-final-viral-combined_k141_53\|\|full | Not-determined |
| KM-final-viral-combined_k141_222325\|\|full | Not-determined |
| KM-final-viral-combined_k141_77829\|\|full | Not-determined |
| KM-final-viral-combined_k141_100057\|\|full | Not-determined |
| KM-final-viral-combined_k141_211195\|\|full | Not-determined |
| KM-final-viral-combined_k141_100059\|\|full | Not-determined |
| KM-final-viral-combined_k141_100062\|\|full | Low-quality |
| KM-final-viral-combined_k141_66705\|\|full | Not-determined |
| KM-final-viral-combined_k141_77849\|\|full | Low-quality |
| KM-final-viral-combined_k141_200144\|\|full | Not-determined |
| KM-final-viral-combined_k141_66718\|\|full | Low-quality |
| KM-final-viral-combined_k141_122285\|\|full | Low-quality |
| KM-final-viral-combined_k141_33424\|\|full | Not-determined |
| KM-final-viral-combined_k141_155664\|\|full | Low-quality |
| KM-final-viral-combined_k141_122291\|\|full | Not-determined |
| KM-final-viral-combined_k141_80\|\|full | Low-quality |
| KM-final-viral-combined_k141_66729\|\|full | Not-determined |
| KM-final-viral-combined_k141_255689\|\|full | Low-quality |
| KM-final-viral-combined_k141_11133\|\|full | Not-determined |
| KM-final-viral-combined_k141_88970\|\|full | Not-determined |
| KM-final-viral-combined_k141_66751\|\|full | Not-determined |
| KM-final-viral-combined_k141_166775\|\|full | Not-determined |
| KM-final-viral-combined_k141_177817\|\|full | Low-quality |
| KM-final-viral-combined_k141_244577\|\|full | Low-quality |
| KM-final-viral-combined_k141_177843\|\|full | Low-quality |
| KM-final-viral-combined_k141_177849\|\|full | Not-determined |
| KM-final-viral-combined_k141_211268\|\|full | Not-determined |
| KM-final-viral-combined_k141_122331\|\|full | Not-determined |
| KM-final-viral-combined_k141_22282\|\|full | Low-quality |
| KM-final-viral-combined_k141_189051\|\|full | Not-determined |
| KM-final-viral-combined_k141_222385\|\|full | Low-quality |
| KM-final-viral-combined_k141_233470\|\|full | Not-determined |
| KM-final-viral-combined_k141_200224\|\|full | Low-quality |
| KM-final-viral-combined_k141_189060\|\|full | Low-quality |
| KM-final-viral-combined_k141_111245\|\|full | Not-determined |
| KM-final-viral-combined_k141_222392\|\|full | Not-determined |
| KM-final-viral-combined_k141_189075\|\|full | Low-quality |
| KM-final-viral-combined_k141_255751\|\|full | Low-quality |
| KM-final-viral-combined_k141_189081\|\|full | Not-determined |
| KM-final-viral-combined_k141_111257\|\|full | Low-quality |
| KM-final-viral-combined_k141_211299\|\|full | Not-determined |
| KM-final-viral-combined_k141_200246\|\|full | Low-quality |
| KM-final-viral-combined_k141_89000\|\|full | Low-quality |
| KM-final-viral-combined_k141_91\|\|full | Not-determined |
| KM-final-viral-combined_k141_255759\|\|full | Low-quality |
| KM-final-viral-combined_k141_122398\|\|full | Low-quality |
| KM-final-viral-combined_k141_177889\|\|full | Low-quality |
| KM-final-viral-combined_k141_11184\|\|full | Not-determined |
| KM-final-viral-combined_k141_89019\|\|full | Low-quality |
| KM-final-viral-combined_k141_144690\|\|full | Low-quality |
| KM-final-viral-combined_k141_255778\|\|full | Low-quality |
| KM-final-viral-combined_k141_22329\|\|full | Not-determined |
| KM-final-viral-combined_k141_122402\|\|full | Not-determined |
| KM-final-viral-combined_k141_109\|\|full | Not-determined |
| KM-final-viral-combined_k141_11208\|\|full | Low-quality |
| KM-final-viral-combined_k141_255781\|\|full | Not-determined |
| KM-final-viral-combined_k141_55676\|\|full | Low-quality |
| KM-final-viral-combined_k141_222442\|\|full | Low-quality |
| KM-final-viral-combined_k141_55680\|\|full | Not-determined |
| KM-final-viral-combined_k141_55683\|\|full | Low-quality |
| KM-final-viral-combined_k141_200284\|\|full | Not-determined |
| KM-final-viral-combined_k141_22340\|\|full | Low-quality |
| KM-final-viral-combined_k141_222459\|\|full | Not-determined |
| KM-final-viral-combined_k141_222464\|\|full | Not-determined |
| KM-final-viral-combined_k141_189132\|\|full | Not-determined |
| KM-final-viral-combined_k141_255794\|\|full | Not-determined |
| KM-final-viral-combined_k141_133544\|\|full | Low-quality |
| KM-final-viral-combined_k141_189137\|\|full | Not-determined |
| KM-final-viral-combined_k141_189138\|\|full | Not-determined |
| KM-final-viral-combined_k141_122424\|\|full | Not-determined |
| KM-final-viral-combined_k141_189140\|\|full | Low-quality |
| KM-final-viral-combined_k141_233549\|\|full | Low-quality |
| KM-final-viral-combined_k141_55728\|\|full | Not-determined |
| KM-final-viral-combined_k141_78042\|\|full | Low-quality |
| KM-final-viral-combined_k141_200335\|\|full | Not-determined |
| KM-final-viral-combined_k141_100117\|\|full | Low-quality |
| KM-final-viral-combined_k141_55739\|\|full | Low-quality |
| KM-final-viral-combined_k141_211369\|\|full | Not-determined |
| KM-final-viral-combined_k141_222488\|\|full | Not-determined |
| KM-final-viral-combined_k141_177937\|\|full | Low-quality |
| KM-final-viral-combined_k141_177940\|\|full | Not-determined |
| KM-final-viral-combined_k141_22375\|\|full | Not-determined |
| KM-final-viral-combined_k141_122443\|\|full | Not-determined |
| KM-final-viral-combined_k141_222503\|\|full | Not-determined |
| KM-final-viral-combined_k141_66866\|\|full | Low-quality |
| KM-final-viral-combined_k141_122469\|\|full | Low-quality |
| KM-final-viral-combined_k141_255856\|\|full | Low-quality |
| KM-final-viral-combined_k141_222527\|\|full | Not-determined |
| KM-final-viral-combined_k141_144770\|\|full | Not-determined |
| KM-final-viral-combined_k141_155760\|\|full | Not-determined |
| KM-final-viral-combined_k141_144775\|\|full | Low-quality |
| KM-final-viral-combined_k141_111398\|\|full | Not-determined |
| KM-final-viral-combined_k141_33676\|\|full | Low-quality |
| KM-final-viral-combined_k141_11283\|\|full | Not-determined |
| KM-final-viral-combined_k141_89084\|\|full | Not-determined |
| KM-final-viral-combined_k141_178000\|\|full | Not-determined |
| KM-final-viral-combined_k141_100146\|\|full | Low-quality |
| KM-final-viral-combined_k141_55769\|\|full | Low-quality |
| KM_k141_266511 | Complete |
| KM-final-viral-combined_k141_166838\|\|full | Not-determined |
| KM-final-viral-combined_k141_211420\|\|full | Low-quality |
| KM-final-viral-combined_k141_211423\|\|full | Not-determined |
| KM-final-viral-combined_k141_255887\|\|full | Not-determined |
| KM-final-viral-combined_k141_200355\|\|full | Low-quality |
| KM-final-viral-combined_k141_122511\|\|full | Low-quality |
| KM-final-viral-combined_k141_78158\|\|full | Not-determined |
| KM-final-viral-combined_k141_133575\|\|full | Low-quality |
| KM-final-viral-combined_k141_122518\|\|full | Low-quality |
| KM-final-viral-combined_k141_133586\|\|full | Not-determined |
| KM-final-viral-combined_k141_111403\|\|full | Not-determined |
| KM-final-viral-combined_k141_155795\|\|full | Low-quality |
| KM-final-viral-combined_k141_111405\|\|full | Not-determined |
| KM-final-viral-combined_k141_22469\|\|full | Not-determined |
| KM-final-viral-combined_k141_155797\|\|full | Not-determined |
| KM-final-viral-combined_k141_155799\|\|full | Not-determined |
| KM-final-viral-combined_k141_200386\|\|full | Not-determined |
| KM-final-viral-combined_k141_89125\|\|full | Not-determined |
| KM-final-viral-combined_k141_11317\|\|full | Low-quality |
| KM-final-viral-combined_k141_211487\|\|full | Low-quality |
| KM-final-viral-combined_k141_144815\|\|full | Low-quality |
| KM-final-viral-combined_k141_33740\|\|full | Low-quality |
| KM-final-viral-combined_k141_66969\|\|full | Not-determined |
| KM-final-viral-combined_k141_222606\|\|full | Low-quality |
| KM-final-viral-combined_k141_211494\|\|full | Not-determined |
| KM-final-viral-combined_k141_255926\|\|full | Not-determined |
| KM-final-viral-combined_k141_89129\|\|full | Not-determined |
| KM-final-viral-combined_k141_233648\|\|full | Not-determined |
| KM-final-viral-combined_k141_11332\|\|full | Low-quality |
| KM-final-viral-combined_k141_166894\|\|full | Low-quality |
| KM-final-viral-combined_k141_122543\|\|full | Low-quality |
| KM-final-viral-combined_k141_78198\|\|full | Low-quality |
| KM-final-viral-combined_k141_211514\|\|full | Low-quality |
| KM-final-viral-combined_k141_122554\|\|full | Not-determined |
| KM-final-viral-combined_k141_178014\|\|full | Not-determined |
| KM-final-viral-combined_k141_89137\|\|full | Low-quality |
| KM-final-viral-combined_k141_122572\|\|full | Low-quality |
| KM-final-viral-combined_k141_67037\|\|full | Not-determined |
| KM-final-viral-combined_k141_55864\|\|full | Low-quality |
| KM-final-viral-combined_k141_44645\|\|full | Low-quality |
| KM-final-viral-combined_k141_233705\|\|full | Not-determined |
| KM-final-viral-combined_k141_244805\|\|full | Low-quality |
| KM-final-viral-combined_k141_67056\|\|full | Not-determined |
| KM-final-viral-combined_k141_133662\|\|full | Low-quality |
| KM-final-viral-combined_k141_100234\|\|full | Low-quality |
| KM-final-viral-combined_k141_244820\|\|full | Not-determined |
| KM-final-viral-combined_k141_233726\|\|full | Not-determined |
| KM-final-viral-combined_k141_189227\|\|full | Low-quality |
| KM-final-viral-combined_k141_111461\|\|full | Not-determined |
| KM-final-viral-combined_k141_111462\|\|full | Low-quality |
| KM-final-viral-combined_k141_303\|\|full | Not-determined |
| KM-final-viral-combined_k141_244832\|\|full | Not-determined |
| KM-final-viral-combined_k141_155832\|\|full | Low-quality |
| KM-final-viral-combined_k141_22517\|\|full | Not-determined |
| KM-final-viral-combined_k141_222655\|\|full | Not-determined |
| KM-final-viral-combined_k141_233742\|\|full | Not-determined |
| KM-final-viral-combined_k141_22519\|\|full | Not-determined |
| KM-final-viral-combined_k141_222661\|\|full | Not-determined |
| KM-final-viral-combined_k141_122596\|\|full | Low-quality |
| KM-final-viral-combined_k141_100266\|\|full | Low-quality |
| KM-final-viral-combined_k141_89187\|\|full | Not-determined |
| KM-final-viral-combined_k141_200439\|\|full | Low-quality |
| KM-final-viral-combined_k141_211542\|\|full | Low-quality |
| KM-final-viral-combined_k141_89193\|\|full | Not-determined |
| KM-final-viral-combined_k141_144935\|\|full | Not-determined |
| KM-final-viral-combined_k141_55918\|\|full | Low-quality |
| KM-final-viral-combined_k141_350\|\|full | Not-determined |
| KM-final-viral-combined_k141_111521\|\|full | Low-quality |
| KM-final-viral-combined_k141_44705\|\|full | Low-quality |
| KM-final-viral-combined_k141_22549\|\|full | Low-quality |
| KM-final-viral-combined_k141_111542\|\|full | Low-quality |
| KM-final-viral-combined_k141_155873\|\|full | Not-determined |
| KM-final-viral-combined_k141_78314\|\|full | Not-determined |
| KM-final-viral-combined_k141_44707\|\|full | Low-quality |
| KM-final-viral-combined_k141_211568\|\|full | Not-determined |
| KM-final-viral-combined_k141_244862\|\|full | Low-quality |
| KM-final-viral-combined_k141_11387\|\|full | Not-determined |
| KM-final-viral-combined_k141_222686\|\|full | Not-determined |
| KM-final-viral-combined_k141_11390\|\|full | Low-quality |
| 17-109-final-viral-combined_k141_168259\|\|0_partial | High-quality |
| KM-final-viral-combined_k141_111554\|\|full | Not-determined |
| KM-final-viral-combined_k141_122637\|\|full | Not-determined |
| KM-final-viral-combined_k141_244869\|\|full | Low-quality |
| KM-final-viral-combined_k141_111561\|\|full | Not-determined |
| KM-final-viral-combined_k141_67101\|\|full | Not-determined |
| KM-final-viral-combined_k141_67105\|\|full | Low-quality |
| KM-final-viral-combined_k141_244884\|\|full | Not-determined |
| KM-final-viral-combined_k141_122656\|\|full | Not-determined |
| KM-final-viral-combined_k141_111574\|\|full | Not-determined |
| KM-final-viral-combined_k141_389\|\|full | Not-determined |
| KM-final-viral-combined_k141_111583\|\|full | Not-determined |
| KM-final-viral-combined_k141_133702\|\|full | Not-determined |
| KM-final-viral-combined_k141_67132\|\|full | Low-quality |
| KM-final-viral-combined_k141_100345\|\|full | Low-quality |
| KM-final-viral-combined_k141_200455\|\|full | Not-determined |
| KM-final-viral-combined_k141_89247\|\|full | Not-determined |
| KM-final-viral-combined_k141_111595\|\|full | Not-determined |
| KM-final-viral-combined_k141_111596\|\|full | Low-quality |
| 18-110-final-viral-combined_k141_74275\|\|full | Medium-quality |
| KM-final-viral-combined_k141_67138\|\|full | Low-quality |
| KM-final-viral-combined_k141_222717\|\|full | Low-quality |
| KM-final-viral-combined_k141_166973\|\|full | Low-quality |
| KM-final-viral-combined_k141_89259\|\|full | Not-determined |
| KM-final-viral-combined_k141_427\|\|full | Low-quality |
| KM-final-viral-combined_k141_166993\|\|full | Low-quality |
| KM-final-viral-combined_k141_111610\|\|full | Not-determined |
| KM-final-viral-combined_k141_200493\|\|full | Low-quality |
| KM-final-viral-combined_k141_440\|\|full | Not-determined |
| KM-final-viral-combined_k141_200499\|\|full | Not-determined |
| KM-final-viral-combined_k141_89276\|\|full | Not-determined |
| KM-final-viral-combined_k141_44750\|\|full | Low-quality |
| KM-final-viral-combined_k141_441\|\|full | Low-quality |
| KM-final-viral-combined_k141_22610\|\|full | Not-determined |
| KM-final-viral-combined_k141_22612\|\|full | Not-determined |
| KM-final-viral-combined_k141_122708\|\|full | Low-quality |
| KM-final-viral-combined_k141_122710\|\|full | Low-quality |
| KM-final-viral-combined_k141_222788\|\|full | Not-determined |
| KM-final-viral-combined_k141_133731\|\|full | Not-determined |
| KM-final-viral-combined_k141_67211\|\|full | Not-determined |
| KM-final-viral-combined_k141_200527\|\|full | Low-quality |
| KM-final-viral-combined_k141_133746\|\|full | Not-determined |
| KM-final-viral-combined_k141_200530\|\|full | Low-quality |
| KM-final-viral-combined_k141_470\|\|full | Low-quality |
| KM-final-viral-combined_k141_155958\|\|full | Not-determined |
| KM-final-viral-combined_k141_256032\|\|full | Low-quality |
| KM-final-viral-combined_k141_200536\|\|full | Not-determined |
| KM-final-viral-combined_k141_155963\|\|full | Low-quality |
| KM-final-viral-combined_k141_476\|\|full | Low-quality |
| KM-final-viral-combined_k141_145080\|\|full | Low-quality |
| KM-final-viral-combined_k141_111634\|\|full | Low-quality |
| KM-final-viral-combined_k141_155965\|\|full | Not-determined |
| KM-final-viral-combined_k141_244984\|\|full | Not-determined |
| KM_k141_205587 | High-quality |
| KM-final-viral-combined_k141_256045\|\|full | Low-quality |
| KM-final-viral-combined_k141_211644\|\|full | Low-quality |
| KM-final-viral-combined_k141_145085\|\|full | Not-determined |
| KM-final-viral-combined_k141_78377\|\|full | Low-quality |
| KM-final-viral-combined_k141_167032\|\|full | Not-determined |
| KM-final-viral-combined_k141_486\|\|full | Not-determined |
| KM-final-viral-combined_k141_78380\|\|full | Not-determined |
| KM-final-viral-combined_k141_211653\|\|full | Low-quality |
| KM-final-viral-combined_k141_67259\|\|full | Not-determined |
| KM-final-viral-combined_k141_499\|\|full | Not-determined |
| KM-final-viral-combined_k141_233799\|\|full | Not-determined |
| KM-final-viral-combined_k141_155997\|\|full | Not-determined |
| KM-final-viral-combined_k141_167055\|\|full | Not-determined |
| KM-final-viral-combined_k141_167062\|\|full | Low-quality |
| KM-final-viral-combined_k141_122744\|\|full | Low-quality |
| KM-final-viral-combined_k141_233803\|\|full | Low-quality |
| KM-final-viral-combined_k141_67292\|\|full | Not-determined |
| KM-final-viral-combined_k141_167086\|\|full | Low-quality |
| KM-final-viral-combined_k141_111697\|\|full | Not-determined |
| KM-final-viral-combined_k141_211701\|\|full | Low-quality |
| KM-final-viral-combined_k141_167090\|\|full | Not-determined |
| KM-final-viral-combined_k141_33843\|\|full | Not-determined |
| KM-final-viral-combined_k141_67305\|\|full | Low-quality |
| KM-final-viral-combined_k141_122764\|\|full | Low-quality |
| KM-final-viral-combined_k141_11481\|\|full | Low-quality |
| KM-final-viral-combined_k141_78447\|\|full | Low-quality |
| KM-final-viral-combined_k141_122785\|\|full | Low-quality |
| KM-final-viral-combined_k141_22650\|\|full | Low-quality |
| KM-final-viral-combined_k141_521\|\|full | Low-quality |
| KM-final-viral-combined_k141_33854\|\|full | Low-quality |
| KM-final-viral-combined_k141_67338\|\|full | Low-quality |
| KM-final-viral-combined_k141_200620\|\|full | Not-determined |
| KM-final-viral-combined_k141_200622\|\|full | Not-determined |
| KM-final-viral-combined_k141_256069\|\|full | Low-quality |
| KM-final-viral-combined_k141_78476\|\|full | Not-determined |
| KM-final-viral-combined_k141_156045\|\|full | Not-determined |
| KM-final-viral-combined_k141_222863\|\|full | Not-determined |
| KM-final-viral-combined_k141_200634\|\|full | Not-determined |
| KM-final-viral-combined_k141_156048\|\|full | Not-determined |
| KM-final-viral-combined_k141_256074\|\|full | Not-determined |
| KM-final-viral-combined_k141_178113\|\|full | Not-determined |
| KM-final-viral-combined_k141_33868\|\|full | Not-determined |
| KM-final-viral-combined_k141_211718\|\|full | Not-determined |
| KM-final-viral-combined_k141_122816\|\|full | Low-quality |
| KM-final-viral-combined_k141_100445\|\|full | Low-quality |
| KM-final-viral-combined_k141_133816\|\|full | Low-quality |
| KM-final-viral-combined_k141_89315\|\|full | Low-quality |
| KM-final-viral-combined_k141_200651\|\|full | Low-quality |
| KM-final-viral-combined_k141_200652\|\|full | Not-determined |
| KM-final-viral-combined_k141_67371\|\|full | Low-quality |
| KM-final-viral-combined_k141_178149\|\|full | Low-quality |
| KM-final-viral-combined_k141_67373\|\|full | Low-quality |
| KM-final-viral-combined_k141_67376\|\|full | Low-quality |

| | |
|---|---|
| KM-final-viral-combined_k141_122833||full | Low-quality |
| KM-final-viral-combined_k141_44822||full | Low-quality |
| KM-final-viral-combined_k141_78491||full | Low-quality |
| KM-final-viral-combined_k141_222893||full | Low-quality |
| KM-final-viral-combined_k141_222894||full | Low-quality |
| KM-final-viral-combined_k141_222896||full | Low-quality |
| KM-final-viral-combined_k141_222901||full | Low-quality |
| KM-final-viral-combined_k141_78507||full | Not-determined |
| KM-final-viral-combined_k141_256133||full | Not-determined |
| KM-final-viral-combined_k141_89341||full | Not-determined |
| KM-final-viral-combined_k141_156069||full | Low-quality |
| KM-final-viral-combined_k141_256143||full | Not-determined |
| KM-final-viral-combined_k141_100459||full | Low-quality |
| KM-final-viral-combined_k141_89358||full | Not-determined |
| KM-final-viral-combined_k141_200684||full | Low-quality |
| KM-final-viral-combined_k141_566||full | Low-quality |
| KM-final-viral-combined_k141_178193||full | Not-determined |
| KM-final-viral-combined_k141_44870||full | Low-quality |
| KM-final-viral-combined_k141_89365||full | Not-determined |
| KM-final-viral-combined_k141_67414||full | Not-determined |
| KM-final-viral-combined_k141_233852||full | Low-quality |
| KM-final-viral-combined_k141_211753||full | Low-quality |
| KM-final-viral-combined_k141_11542||full | Not-determined |
| KM-final-viral-combined_k141_44875||full | Low-quality |
| KM-final-viral-combined_k141_44881||full | Low-quality |
| KM-final-viral-combined_k141_200716||full | Low-quality |
| KM-final-viral-combined_k141_111807||full | Not-determined |
| KM-final-viral-combined_k141_167154||full | Low-quality |
| KM-final-viral-combined_k141_200726||full | Low-quality |
| KM-final-viral-combined_k141_256175||full | Not-determined |
| KM-final-viral-combined_k141_100484||full | Low-quality |
| KM-final-viral-combined_k141_200730||full | Not-determined |
| KM-final-viral-combined_k141_222925||full | Low-quality |
| KM-final-viral-combined_k141_606||full | Low-quality |
| KM-final-viral-combined_k141_233855||full | Low-quality |
| KM-final-viral-combined_k141_167162||full | Low-quality |
| KM-final-viral-combined_k141_178210||full | Not-determined |
| KM-final-viral-combined_k141_100497||full | Not-determined |
| KM-final-viral-combined_k141_67443||full | Not-determined |
| KM-final-viral-combined_k141_122923||full | Not-determined |
| KM-final-viral-combined_k141_145156||full | Not-determined |
| KM-final-viral-combined_k141_205587||full | High-quality |
| KM-final-viral-combined_k141_200767||full | Low-quality |
| KM-final-viral-combined_k141_33978||full | Not-determined |
| KM-final-viral-combined_k141_145166||full | Not-determined |
| KM-final-viral-combined_k141_56077||full | Not-determined |
| KM-final-viral-combined_k141_619||full | Low-quality |
| KM-final-viral-combined_k141_111858||full | Low-quality |
| KM-final-viral-combined_k141_156129||full | Low-quality |
| KM-final-viral-combined_k141_211838||full | Not-determined |
| KM-final-viral-combined_k141_200796||full | Not-determined |
| KM-final-viral-combined_k141_133974||full | Low-quality |
| KM-final-viral-combined_k141_256281||full | Not-determined |
| KM-final-viral-combined_k141_178244||full | Low-quality |
| KM-final-viral-combined_k141_56110||full | Not-determined |
| KM-final-viral-combined_k141_211885||full | Not-determined |
| KM-final-viral-combined_k141_100509||full | Not-determined |
| KM-final-viral-combined_k141_178250||full | Not-determined |
| KM-final-viral-combined_k141_67475||full | Low-quality |
| KM-final-viral-combined_k141_211891||full | Not-determined |
| KM-final-viral-combined_k141_660||full | Not-determined |
| KM-final-viral-combined_k141_189489||full | Low-quality |
| KM-final-viral-combined_k141_200836||full | Not-determined |
| KM-final-viral-combined_k141_156175||full | Not-determined |
| KM-final-viral-combined_k141_111913||full | Low-quality |
| KM-final-viral-combined_k141_89420||full | Low-quality |
| KM-final-viral-combined_k141_67500||full | Low-quality |
| KM-final-viral-combined_k141_156178||full | Low-quality |
| KM-final-viral-combined_k141_222984||full | Low-quality |
| KM-final-viral-combined_k141_133997||full | Low-quality |
| KM-final-viral-combined_k141_56142||full | Not-determined |
| KM-final-viral-combined_k141_156193||full | Low-quality |
| KM-final-viral-combined_k141_56159||full | Not-determined |
| KM-final-viral-combined_k141_78587||full | Low-quality |
| KM-final-viral-combined_k141_189511||full | Not-determined |
| KM-final-viral-combined_k141_67538||full | Not-determined |
| KM-final-viral-combined_k141_167231||full | Low-quality |
| KM-final-viral-combined_k141_33998||full | Not-determined |
| KM-final-viral-combined_k141_78612||full | Low-quality |
| KM-final-viral-combined_k141_34004||full | Not-determined |
| KM-final-viral-combined_k141_178315||full | Low-quality |
| KM-final-viral-combined_k141_89462||full | Not-determined |
| KM-final-viral-combined_k141_67562||full | Not-determined |
| KM-final-viral-combined_k141_11647||full | Low-quality |
| KM-final-viral-combined_k141_189540||full | Not-determined |
| KM-final-viral-combined_k141_78638||full | Low-quality |
| KM-final-viral-combined_k141_200872||full | Not-determined |
| KM-final-viral-combined_k141_178340||full | Low-quality |
| KM-final-viral-combined_k141_200874||full | Not-determined |
| KM-final-viral-combined_k141_67579||full | Not-determined |
| KM-final-viral-combined_k141_89480||full | Low-quality |
| KM-final-viral-combined_k141_111953||full | Low-quality |
| KM-final-viral-combined_k141_134052||full | Not-determined |
| KM-final-viral-combined_k141_111978||full | Not-determined |
| KM-final-viral-combined_k141_256378||full | Not-determined |
| KM-final-viral-combined_k141_233946||full | Not-determined |
| KM-final-viral-combined_k141_189555||full | Low-quality |
| KM-final-viral-combined_k141_734||full | Low-quality |
| KM-final-viral-combined_k141_145284||full | Not-determined |
| KM-final-viral-combined_k141_178382||full | Not-determined |
| KM-final-viral-combined_k141_245240||full | Not-determined |
| KM-final-viral-combined_k141_56186||full | Low-quality |
| KM-final-viral-combined_k141_245241||full | Not-determined |
| KM-final-viral-combined_k141_178388||full | Low-quality |
| KM-final-viral-combined_k141_200948||full | Not-determined |
| KM-final-viral-combined_k141_760||full | Not-determined |
| KM-final-viral-combined_k141_189597||full | Not-determined |
| KM-final-viral-combined_k141_178402||full | Low-quality |
| KM-final-viral-combined_k141_78701||full | Not-determined |
| KM-final-viral-combined_k141_189605||full | Not-determined |
| KM-final-viral-combined_k141_112002||full | Not-determined |
| KM-final-viral-combined_k141_245271||full | Low-quality |
| KM-final-viral-combined_k141_189614||full | Low-quality |
| KM-final-viral-combined_k141_78711||full | Not-determined |
| KM-final-viral-combined_k141_787||full | Not-determined |
| KM-final-viral-combined_k141_112013||full | Not-determined |
| KM-final-viral-combined_k141_145341||full | Low-quality |
| KM-final-viral-combined_k141_34119||full | Not-determined |
| KM-final-viral-combined_k141_134075||full | Low-quality |
| KM-final-viral-combined_k141_189633||full | Low-quality |
| KM-final-viral-combined_k141_800||full | Low-quality |
| KM-final-viral-combined_k141_34137||full | Low-quality |
| KM-final-viral-combined_k141_212052||full | Low-quality |
| KM-final-viral-combined_k141_112028||full | Not-determined |
| KM-final-viral-combined_k141_56263||full | Not-determined |
| KM-final-viral-combined_k141_112050||full | Low-quality |
| KM-final-viral-combined_k141_78740||full | Not-determined |
| KM-final-viral-combined_k141_178426||full | Not-determined |
| KM-final-viral-combined_k141_201001||full | Low-quality |
| KM-final-viral-combined_k141_178441||full | Low-quality |
| KM-final-viral-combined_k141_100598||full | Low-quality |
| KM-final-viral-combined_k141_201021||full | Low-quality |
| KM-final-viral-combined_k141_233982||full | Not-determined |
| KM-final-viral-combined_k141_145398||full | Not-determined |
| KM-final-viral-combined_k141_34155||full | Low-quality |
| KM-final-viral-combined_k141_112085||full | Not-determined |
| KM-final-viral-combined_k141_201040||full | Not-determined |
| KM-final-viral-combined_k141_100635||full | Not-determined |
| KM-final-viral-combined_k141_112107||full | Not-determined |
| KM-final-viral-combined_k141_145409||full | Low-quality |
| KM-final-viral-combined_k141_78796||full | Low-quality |
| KM-final-viral-combined_k141_201062||full | Low-quality |
| KM-final-viral-combined_k141_178477||full | Not-determined |
| KM-final-viral-combined_k141_167303||full | Not-determined |
| KM-final-viral-combined_k141_22787||full | Low-quality |
| KM-final-viral-combined_k141_212108||full | Not-determined |
| KM-final-viral-combined_k141_201078||full | Low-quality |
| KM-final-viral-combined_k141_212123||full | Not-determined |
| KM-final-viral-combined_k141_233995||full | Not-determined |
| KM-final-viral-combined_k141_45035||full | Not-determined |
| KM-final-viral-combined_k141_56321||full | Low-quality |
| KM-final-viral-combined_k141_11787||full | Not-determined |
| KM-final-viral-combined_k141_22808||full | Low-quality |
| KM-final-viral-combined_k141_223186||full | Low-quality |
| KM-final-viral-combined_k141_78825||full | Not-determined |
| KM-final-viral-combined_k141_56326||full | Not-determined |
| KM-final-viral-combined_k141_22814||full | Low-quality |
| KM-final-viral-combined_k141_212155||full | Low-quality |
| KM-final-viral-combined_k141_234001||full | Not-determined |
| KM-final-viral-combined_k141_123183||full | Not-determined |
| KM-final-viral-combined_k141_223200||full | Low-quality |
| KM-final-viral-combined_k141_22818||full | Not-determined |
| KM-final-viral-combined_k141_145461||full | Not-determined |
| KM-final-viral-combined_k141_156354||full | Not-determined |
| KM-final-viral-combined_k141_11842||full | Low-quality |
| KM-final-viral-combined_k141_45076||full | Not-determined |
| KM-final-viral-combined_k141_156362||full | Not-determined |
| KM-final-viral-combined_k141_189741||full | Low-quality |
| KM-final-viral-combined_k141_89666||full | Low-quality |
| KM-final-viral-combined_k141_11850||full | Not-determined |
| KM-final-viral-combined_k141_212188||full | Low-quality |
| KM-final-viral-combined_k141_245372||full | Not-determined |
| KM-final-viral-combined_k141_223226||full | Low-quality |
| KM-final-viral-combined_k141_245376||full | Not-determined |
| KM-final-viral-combined_k141_178560||full | Not-determined |
| KM-final-viral-combined_k141_11855||full | Not-determined |
| KM-final-viral-combined_k141_223229||full | Low-quality |
| KM-final-viral-combined_k141_45099||full | Not-determined |
| KM-final-viral-combined_k141_167389||full | Not-determined |
| KM-final-viral-combined_k141_34187||full | Not-determined |
| KM-final-viral-combined_k141_167397||full | Not-determined |
| KM-final-viral-combined_k141_78886||full | Not-determined |
| KM-final-viral-combined_k141_178605||full | Low-quality |
| KM-final-viral-combined_k141_245398||full | Low-quality |
| KM-final-viral-combined_k141_78898||full | Not-determined |
| KM-final-viral-combined_k141_67729||full | Not-determined |
| KM-final-viral-combined_k141_145530||full | Low-quality |
| KM-final-viral-combined_k141_67735||full | Not-determined |
| KM-final-viral-combined_k141_34211||full | Low-quality |
| KM-final-viral-combined_k141_100685||full | Not-determined |
| KM-final-viral-combined_k141_234067||full | Low-quality |
| KM-final-viral-combined_k141_34212||full | Low-quality |
| KM-final-viral-combined_k141_11887||full | Low-quality |
| KM-final-viral-combined_k141_34217||full | Low-quality |
| KM-final-viral-combined_k141_11896||full | Not-determined |
| KM-final-viral-combined_k141_67771||full | Not-determined |
| KM-final-viral-combined_k141_22853||full | Not-determined |
| KM-final-viral-combined_k141_22855||full | Not-determined |
| KM-final-viral-combined_k141_145564||full | Not-determined |
| KM-final-viral-combined_k141_56461||full | Low-quality |
| KM-final-viral-combined_k141_11924||full | Low-quality |
| KM-final-viral-combined_k141_56476||full | Not-determined |
| KM-final-viral-combined_k141_67817||full | Not-determined |
| KM-final-viral-combined_k141_34276||full | Not-determined |
| KM-final-viral-combined_k141_78984||full | Low-quality |
| KM-final-viral-combined_k141_100733||full | Low-quality |
| KM-final-viral-combined_k141_22881||full | Not-determined |
| KM-final-viral-combined_k141_256540||full | Not-determined |
| KM-final-viral-combined_k141_100738||full | Not-determined |
| KM-final-viral-combined_k141_256545||full | Low-quality |
| KM-final-viral-combined_k141_201189||full | Not-determined |
| KM-final-viral-combined_k141_56483||full | Not-determined |
| KM-final-viral-combined_k141_156475||full | Not-determined |
| KM-final-viral-combined_k141_245422||full | Not-determined |
| KM-final-viral-combined_k141_256547||full | Not-determined |
| KM-final-viral-combined_k141_123273||full | Low-quality |
| KM-final-viral-combined_k141_34287||full | Not-determined |
| KM-final-viral-combined_k141_100759||full | Not-determined |
| KM-final-viral-combined_k141_56504||full | Not-determined |
| KM-final-viral-combined_k141_100763||full | Low-quality |
| KM-final-viral-combined_k141_45110||full | Low-quality |
| KM-final-viral-combined_k141_892||full | Low-quality |
| KM-final-viral-combined_k141_79023||full | Not-determined |
| KM-final-viral-combined_k141_178706||full | Not-determined |
| KM-final-viral-combined_k141_223346||full | Low-quality |
| KM-final-viral-combined_k141_123295||full | Not-determined |
| KM-final-viral-combined_k141_201203||full | Low-quality |
| KM-final-viral-combined_k141_178721||full | Not-determined |
| KM-final-viral-combined_k141_223349||full | Low-quality |
| KM-final-viral-combined_k141_56513||full | Low-quality |
| KM-final-viral-combined_k141_201211||full | Not-determined |
| KM-final-viral-combined_k141_178728||full | Not-determined |
| KM-final-viral-combined_k141_212243||full | Low-quality |
| KM-final-viral-combined_k141_902||full | Low-quality |
| KM-final-viral-combined_k141_201212||full | Low-quality |
| KM-final-viral-combined_k141_223365||full | Low-quality |
| KM-final-viral-combined_k141_22943||full | Low-quality |
| KM-final-viral-combined_k141_256574||full | Not-determined |
| KM-final-viral-combined_k141_256575||full | Low-quality |
| KM-final-viral-combined_k141_11980||full | Low-quality |
| KM-final-viral-combined_k141_79064||full | Low-quality |
| KM-final-viral-combined_k141_189832||full | Not-determined |
| KM-final-viral-combined_k141_212268||full | Low-quality |
| KM-final-viral-combined_k141_123329||full | Not-determined |
| KM-final-viral-combined_k141_112294||full | Low-quality |
| KM-final-viral-combined_k141_56555||full | Not-determined |
| KM-final-viral-combined_k141_145626||full | Low-quality |
| KM-final-viral-combined_k141_123339||full | Low-quality |
| KM-final-viral-combined_k141_123342||full | Low-quality |
| KM-final-viral-combined_k141_112295||full | Low-quality |
| KM-final-viral-combined_k141_234163||full | Low-quality |
| KM-final-viral-combined_k141_134188||full | Not-determined |
| KM-final-viral-combined_k141_167540||full | Low-quality |
| KM-final-viral-combined_k141_223404||full | Low-quality |
| KM-final-viral-combined_k141_945||full | Not-determined |
| KM-final-viral-combined_k141_156553||full | Low-quality |
| KM-final-viral-combined_k141_223433||full | Low-quality |
| KM-final-viral-combined_k141_112304||full | Not-determined |
| KM-final-viral-combined_k141_123366||full | Low-quality |
| KM-final-viral-combined_k141_234183||full | Not-determined |
| KM-final-viral-combined_k141_100809||full | Low-quality |
| KM-final-viral-combined_k141_234185||full | Not-determined |
| KM-final-viral-combined_k141_145636||full | Not-determined |
| KM-final-viral-combined_k141_112318||full | Low-quality |
| KM-final-viral-combined_k141_145641||full | Not-determined |
| KM-final-viral-combined_k141_56620||full | Low-quality |
| KM-final-viral-combined_k141_145643||full | Not-determined |
| KM-final-viral-combined_k141_234197||full | Not-determined |
| KM-final-viral-combined_k141_201293||full | Not-determined |
| KM-final-viral-combined_k141_12004||full | Not-determined |
| KM-final-viral-combined_k141_123393||full | Low-quality |
| KM-final-viral-combined_k141_12005||full | Not-determined |
| KM-final-viral-combined_k141_167568||full | Low-quality |
| KM-final-viral-combined_k141_201298||full | Low-quality |
| KM-final-viral-combined_k141_45159||full | Low-quality |
| KM-final-viral-combined_k141_1007||full | Low-quality |
| KM-final-viral-combined_k141_123401||full | Not-determined |
| KM-final-viral-combined_k141_156618||full | Not-determined |
| KM-final-viral-combined_k141_89722||full | Not-determined |
| KM-final-viral-combined_k141_123413||full | Low-quality |
| KM-final-viral-combined_k141_212351||full | Low-quality |
| KM-final-viral-combined_k141_1027||full | Not-determined |
| KM-final-viral-combined_k141_178771||full | Not-determined |
| KM-final-viral-combined_k141_212363||full | Not-determined |
| KM-final-viral-combined_k141_89747||full | Low-quality |
| KM-final-viral-combined_k141_34389||full | Not-determined |
| KM-final-viral-combined_k141_145665||full | Low-quality |
| KM-final-viral-combined_k141_212373||full | Low-quality |
| KM-final-viral-combined_k141_212375||full | Not-determined |
| KM-final-viral-combined_k141_56660||full | Not-determined |
| KM-final-viral-combined_k141_112385||full | Low-quality |
| KM-final-viral-combined_k141_123460||full | Low-quality |
| KM-final-viral-combined_k141_34419||full | Low-quality |
| KM-final-viral-combined_k141_45183||full | Not-determined |
| KM-final-viral-combined_k141_123483||full | Low-quality |
| KM-final-viral-combined_k141_45192||full | Low-quality |
| KM-final-viral-combined_k141_56677||full | Low-quality |
| KM-final-viral-combined_k141_201332||full | Low-quality |
| KM-final-viral-combined_k141_56682||full | Low-quality |
| KM-final-viral-combined_k141_123509||full | Not-determined |
| KM-final-viral-combined_k141_34436||full | Not-determined |
| KM-final-viral-combined_k141_189929||full | Not-determined |
| KM-final-viral-combined_k141_223535||full | Not-determined |
| KM-final-viral-combined_k141_256650||full | Not-determined |
| KM-final-viral-combined_k141_167692||full | Low-quality |
| KM-final-viral-combined_k141_167695||full | Not-determined |
| KM-final-viral-combined_k141_201345||full | Low-quality |
| KM-final-viral-combined_k141_134360||full | Not-determined |
| KM-final-viral-combined_k141_67838||full | Low-quality |
| KM-final-viral-combined_k141_145687||full | Not-determined |
| KM-final-viral-combined_k141_223554||full | Low-quality |
| KM-final-viral-combined_k141_167710||full | Low-quality |
| KM-final-viral-combined_k141_67839||full | Not-determined |
| KM-final-viral-combined_k141_223560||full | Low-quality |
| KM-final-viral-combined_k141_201365||full | Not-determined |
| KM-final-viral-combined_k141_12076||full | Not-determined |
| KM-final-viral-combined_k141_112443||full | Not-determined |
| KM-final-viral-combined_k141_145707||full | Not-determined |
| KM-final-viral-combined_k141_212390||full | Low-quality |
| KM-final-viral-combined_k141_67864||full | Not-determined |
| KM-final-viral-combined_k141_112446||full | Not-determined |
| KM-final-viral-combined_k141_201383||full | Low-quality |
| KM-final-viral-combined_k141_201389||full | Low-quality |
| KM-final-viral-combined_k141_12087||full | Not-determined |
| KM-final-viral-combined_k141_178959||full | Low-quality |
| KM-final-viral-combined_k141_212412||full | Not-determined |
| KM-final-viral-combined_k141_1138||full | Not-determined |
| KM-final-viral-combined_k141_201399||full | Low-quality |
| KM-final-viral-combined_k141_212417||full | Not-determined |
| KM-final-viral-combined_k141_89787||full | Not-determined |
| KM-final-viral-combined_k141_67895||full | Not-determined |
| KM-final-viral-combined_k141_134395||full | Not-determined |
| KM-final-viral-combined_k141_178974||full | Not-determined |
| KM-final-viral-combined_k141_145733||full | Low-quality |
| KM-final-viral-combined_k141_178984||full | Not-determined |
| KM-final-viral-combined_k141_201413||full | Low-quality |
| KM-final-viral-combined_k141_245589||full | Low-quality |
| KM-final-viral-combined_k141_112476||full | Low-quality |
| KM-final-viral-combined_k141_234376||full | Low-quality |
| KM-final-viral-combined_k141_134408||full | Not-determined |
| KM-final-viral-combined_k141_79266||full | Not-determined |
| KM-final-viral-combined_k141_134411||full | Not-determined |
| KM-final-viral-combined_k141_156640||full | Low-quality |
| KM-final-viral-combined_k141_212435||full | Low-quality |
| KM-final-viral-combined_k141_23144||full | Not-determined |
| KM-final-viral-combined_k141_212437||full | Low-quality |
| KM-final-viral-combined_k141_245604||full | Low-quality |
| KM-final-viral-combined_k141_212440||full | Low-quality |
| KM-final-viral-combined_k141_179024||full | Low-quality |
| KM-final-viral-combined_k141_12118||full | Not-determined |
| KM-final-viral-combined_k141_89808||full | Not-determined |
| KM-final-viral-combined_k141_156650||full | Low-quality |
| KM-final-viral-combined_k141_100917||full | Low-quality |
| KM-final-viral-combined_k141_34508||full | Not-determined |
| KM-final-viral-combined_k141_100922||full | Low-quality |
| KM-final-viral-combined_k141_79300||full | Not-determined |
| KM-final-viral-combined_k141_67960||full | Not-determined |
| KM-final-viral-combined_k141_234412||full | Not-determined |
| KM-final-viral-combined_k141_223678||full | Not-determined |
| KM-final-viral-combined_k141_212457||full | Not-determined |
| KM-final-viral-combined_k141_89822||full | Not-determined |
| KM-final-viral-combined_k141_134417||full | Not-determined |
| KM-final-viral-combined_k141_256713||full | Low-quality |
| KM-final-viral-combined_k141_100937||full | Not-determined |
| KM-final-viral-combined_k141_234424||full | Not-determined |
| KM-final-viral-combined_k141_179061||full | Low-quality |
| KM-final-viral-combined_k141_212470||full | Not-determined |
| KM-final-viral-combined_k141_123580||full | Not-determined |
| KM-final-viral-combined_k141_23185||full | Low-quality |
| KM-final-viral-combined_k141_12172||full | Low-quality |
| KM-final-viral-combined_k141_212472||full | Not-determined |
| KM-final-viral-combined_k141_45348||full | Low-quality |
| KM-final-viral-combined_k141_212479||full | Low-quality |
| KM-final-viral-combined_k141_23193||full | Not-determined |
| KM-final-viral-combined_k141_45357||full | Not-determined |
| KM-final-viral-combined_k141_223719||full | Not-determined |
| KM-final-viral-combined_k141_23201||full | Not-determined |
| KM-final-viral-combined_k141_179084||full | Not-determined |
| KM-final-viral-combined_k141_223727||full | Not-determined |
| KM-final-viral-combined_k141_68018||full | Not-determined |
| KM-final-viral-combined_k141_23213||full | Not-determined |
| KM-final-viral-combined_k141_89875||full | Low-quality |
| KM-final-viral-combined_k141_212516||full | Not-determined |
| KM-final-viral-combined_k141_23216||full | Not-determined |
| KM-final-viral-combined_k141_112486||full | Not-determined |
| KM-final-viral-combined_k141_223731||full | Not-determined |
| KM-final-viral-combined_k141_123600||full | Low-quality |
| KM-final-viral-combined_k141_79364||full | Low-quality |
| KM-final-viral-combined_k141_123607||full | Low-quality |
| KM-final-viral-combined_k141_212533||full | Low-quality |
| KM-final-viral-combined_k141_68064||full | Low-quality |
| KM-final-viral-combined_k141_123621||full | Not-determined |
| KM-final-viral-combined_k141_101007||full | Low-quality |
| KM-final-viral-combined_k141_101010||full | Not-determined |
| KM-final-viral-combined_k141_212554||full | Not-determined |
| KM-final-viral-combined_k141_201522||full | Low-quality |
| KM-final-viral-combined_k141_266511||full | High-quality |
| KM-final-viral-combined_k141_23233||full | Low-quality |
| KM-final-viral-combined_k141_145846||full | Not-determined |
| KM-final-viral-combined_k141_167794||full | Low-quality |
| KM-final-viral-combined_k141_212571||full | Low-quality |
| KM-final-viral-combined_k141_201536||full | Low-quality |
| KM-final-viral-combined_k141_212573||full | Not-determined |
| KM-final-viral-combined_k141_79391||full | Low-quality |
| KM-final-viral-combined_k141_201542||full | Not-determined |
| KM-final-viral-combined_k141_167804||full | Low-quality |
| KM-final-viral-combined_k141_68081||full | Not-determined |
| KM-final-viral-combined_k141_89914||full | Low-quality |
| KM-final-viral-combined_k141_1270||full | Not-determined |
| KM-final-viral-combined_k141_223757||full | Low-quality |
| KM-final-viral-combined_k141_34611||full | Low-quality |
| KM-final-viral-combined_k141_68100||full | Low-quality |
| KM-final-viral-combined_k141_34621||full | Not-determined |
| KM-final-viral-combined_k141_256782||full | Not-determined |
| KM-final-viral-combined_k141_145887||full | Not-determined |
| KM-final-viral-combined_k141_245671||full | Low-quality |
| KM-final-viral-combined_k141_223773||full | Not-determined |
| KM-final-viral-combined_k141_123692||full | Not-determined |
| KM-final-viral-combined_k141_167857||full | Not-determined |
| KM-final-viral-combined_k141_167859||full | Not-determined |
| KM-final-viral-combined_k141_68131||full | Not-determined |
| KM-final-viral-combined_k141_134549||full | Not-determined |
| KM-final-viral-combined_k141_101094||full | Low-quality |
| KM-final-viral-combined_k141_256800||full | Low-quality |
| KM-final-viral-combined_k141_156704||full | Not-determined |
| KM-final-viral-combined_k141_56796||full | Not-determined |
| KM-final-viral-combined_k141_89952||full | Low-quality |
| KM-final-viral-combined_k141_245703||full | Low-quality |
| KM-final-viral-combined_k141_56811||full | Low-quality |
| KM-final-viral-combined_k141_179160||full | Low-quality |
| KM-final-viral-combined_k141_223800||full | Not-determined |
| KM-final-viral-combined_k141_79491||full | Not-determined |
| KM-final-viral-combined_k141_56819||full | Not-determined |
| KM-final-viral-combined_k141_190116||full | Not-determined |
| KM-final-viral-combined_k141_89979||full | Not-determined |
| KM-final-viral-combined_k141_156754||full | Low-quality |
| KM-final-viral-combined_k141_201609||full | Low-quality |
| KM-final-viral-combined_k141_79514||full | Not-determined |
| KM-final-viral-combined_k141_89983||full | Low-quality |
| KM-final-viral-combined_k141_123768||full | Not-determined |
| KM-final-viral-combined_k141_212648||full | Low-quality |
| KM-final-viral-combined_k141_101111||full | Not-determined |
| KM-final-viral-combined_k141_89994||full | Not-determined |
| KM-final-viral-combined_k141_245755||full | Not-determined |
| KM-final-viral-combined_k141_45377||full | Low-quality |
| KM-final-viral-combined_k141_223836||full | Not-determined |
| KM-final-viral-combined_k141_212670||full | Not-determined |
| KM-final-viral-combined_k141_201651||full | Not-determined |
| KM-final-viral-combined_k141_68196||full | Low-quality |
| KM-final-viral-combined_k141_68198||full | Low-quality |
| KM-final-viral-combined_k141_1326||full | Low-quality |
| KM-final-viral-combined_k141_90005||full | Not-determined |
| KM-final-viral-combined_k141_101128||full | Low-quality |
| KM-final-viral-combined_k141_190187||full | Not-determined |
| KM-final-viral-combined_k141_1337||full | Low-quality |
| KM-final-viral-combined_k141_34624||full | Not-determined |
| KM-final-viral-combined_k141_45413||full | Not-determined |
| KM-final-viral-combined_k141_245795||full | Low-quality |
| KM-final-viral-combined_k141_1342||full | Not-determined |
| KM-final-viral-combined_k141_34627||full | Low-quality |
| KM-final-viral-combined_k141_23317||full | Low-quality |
| KM-final-viral-combined_k141_190203||full | Low-quality |
| KM-final-viral-combined_k141_34632||full | Low-quality |
| KM-final-viral-combined_k141_212705||full | Not-determined |
| KM-final-viral-combined_k141_23330||full | Low-quality |
| KM-final-viral-combined_k141_156787||full | Not-determined |
| KM-final-viral-combined_k141_90033||full | Not-determined |
| KM-final-viral-combined_k141_23338||full | Not-determined |
| KM-final-viral-combined_k141_23340||full | Low-quality |
| KM-final-viral-combined_k141_79573||full | Not-determined |
| KM-final-viral-combined_k141_167905||full | Low-quality |
| KM-final-viral-combined_k141_123862||full | Low-quality |
| KM-final-viral-combined_k141_34668||full | Not-determined |
| KM-final-viral-combined_k141_123866||full | Low-quality |
| KM-final-viral-combined_k141_79580||full | Not-determined |
| KM-final-viral-combined_k141_256895||full | Low-quality |
| KM-final-viral-combined_k141_167908||full | Low-quality |
| KM-final-viral-combined_k141_90047||full | Not-determined |
| KM-final-viral-combined_k141_34680||full | Not-determined |
| KM-final-viral-combined_k141_79582||full | Low-quality |
| KM-final-viral-combined_k141_1373||full | Not-determined |
| KM-final-viral-combined_k141_112600||full | Low-quality |
| KM-final-viral-combined_k141_201716||full | Low-quality |
| KM-final-viral-combined_k141_56878||full | Low-quality |
| KM-final-viral-combined_k141_1386||full | Low-quality |
| KM-final-viral-combined_k141_201723||full | Not-determined |
| KM-final-viral-combined_k141_179235||full | Low-quality |
| KM-final-viral-combined_k141_167920||full | Not-determined |
| KM-final-viral-combined_k141_212783||full | Not-determined |
| KM-final-viral-combined_k141_167926||full | Low-quality |
| KM-final-viral-combined_k141_145970||full | Not-determined |
| KM-final-viral-combined_k141_45462||full | Not-determined |
| KM-final-viral-combined_k141_156854||full | Low-quality |
| KM-final-viral-combined_k141_234515||full | Low-quality |
| KM-final-viral-combined_k141_45469||full | Low-quality |
| KM-final-viral-combined_k141_201753||full | Low-quality |
| KM-final-viral-combined_k141_223919||full | Low-quality |
| KM-final-viral-combined_k141_212801||full | Low-quality |
| KM-final-viral-combined_k141_79643||full | Low-quality |
| KM-final-viral-combined_k141_234546||full | Low-quality |
| KM-final-viral-combined_k141_201780||full | Low-quality |
| KM-final-viral-combined_k141_256942||full | Not-determined |
| KM-final-viral-combined_k141_212836||full | Not-determined |
| KM-final-viral-combined_k141_256949||full | Low-quality |
| KM-final-viral-combined_k141_56937||full | Low-quality |
| KM-final-viral-combined_k141_201806||full | Not-determined |
| KM-final-viral-combined_k141_134752||full | Not-determined |
| KM-final-viral-combined_k141_256973||full | Not-determined |
| KM-final-viral-combined_k141_12288||full | Not-determined |
| KM-final-viral-combined_k141_201813||full | Not-determined |
| KM-final-viral-combined_k141_56950||full | Low-quality |
| KM-final-viral-combined_k141_223960||full | Low-quality |
| KM-final-viral-combined_k141_45520||full | Low-quality |
| KM-final-viral-combined_k141_79706||full | Low-quality |
| KM-final-viral-combined_k141_12312||full | Not-determined |
| KM-final-viral-combined_k141_146018||full | Low-quality |
| KM-final-viral-combined_k141_212854||full | Not-determined |
| KM-final-viral-combined_k141_223984||full | Not-determined |
| KM-final-viral-combined_k141_223987||full | Low-quality |
| KM-final-viral-combined_k141_101255||full | Not-determined |
| KM-final-viral-combined_k141_23378||full | Not-determined |
| 17-109-final-viral-combined_k141_386692||full | High-quality |
| KM-final-viral-combined_k141_257006||full | Low-quality |
| KM-final-viral-combined_k141_201837||full | Low-quality |
| KM-final-viral-combined_k141_90065||full | Low-quality |
| KM-final-viral-combined_k141_156946||full | Not-determined |
| KM-final-viral-combined_k141_90069||full | Low-quality |
| KM-final-viral-combined_k141_1462||full | Low-quality |
| KM-final-viral-combined_k141_257015||full | Not-determined |
| KM-final-viral-combined_k141_90077||full | Low-quality |
| KM-final-viral-combined_k141_79729||full | Low-quality |
| KM-final-viral-combined_k141_257028||full | Low-quality |
| KM-final-viral-combined_k141_234626||full | Not-determined |
| KM-final-viral-combined_k141_112719||full | Not-determined |
| KM-final-viral-combined_k141_257034||full | Not-determined |
| KM-final-viral-combined_k141_112729||full | Not-determined |
| KM-final-viral-combined_k141_56973||full | Low-quality |
| KM-final-viral-combined_k141_257055||full | Low-quality |
| KM-final-viral-combined_k141_12359||full | Low-quality |
| KM-final-viral-combined_k141_257058||full | Low-quality |
| KM-final-viral-combined_k141_123979||full | Not-determined |
| KM-final-viral-combined_k141_68315||full | Low-quality |
| KM-final-viral-combined_k141_224051||full | Low-quality |
| KM-final-viral-combined_k141_257072||full | Not-determined |
| KM-final-viral-combined_k141_34804||full | Not-determined |
| KM-final-viral-combined_k141_190365||full | Not-determined |
| 18-105_k141_67169 | High-quality |
| KM-final-viral-combined_k141_201863||full | Low-quality |
| KM-final-viral-combined_k141_156994||full | Low-quality |
| KM-final-viral-combined_k141_201868||full | Not-determined |
| KM-final-viral-combined_k141_146061||full | Low-quality |
| KM-final-viral-combined_k141_146063||full | Not-determined |
| KM-final-viral-combined_k141_23465||full | Not-determined |
| KM-final-viral-combined_k141_190390||full | Low-quality |
| KM-final-viral-combined_k141_157033||full | Low-quality |
| KM-final-viral-combined_k141_201904||full | Not-determined |
| KM-final-viral-combined_k141_234664||full | Low-quality |
| KM-final-viral-combined_k141_168104||full | Low-quality |
| KM-final-viral-combined_k141_179325||full | Not-determined |
| KM-final-viral-combined_k141_168111||full | Not-determined |
| KM-final-viral-combined_k141_146109||full | Not-determined |
| KM-final-viral-combined_k141_124011||full | Not-determined |
| KM-final-viral-combined_k141_179329||full | Low-quality |
| KM-final-viral-combined_k141_212884||full | Not-determined |
| KM-final-viral-combined_k141_34835||full | Not-determined |
| KM-final-viral-combined_k141_134846||full | Not-determined |
| KM-final-viral-combined_k141_90127||full | Low-quality |
| KM-final-viral-combined_k141_146117||full | Low-quality |
| KM-final-viral-combined_k141_134859||full | Not-determined |
| KM-final-viral-combined_k141_1541||full | Low-quality |
| KM-final-viral-combined_k141_157036||full | Low-quality |
| KM-final-viral-combined_k141_90143||full | Low-quality |
| KM-final-viral-combined_k141_257147||full | Not-determined |
| KM-final-viral-combined_k141_224121||full | Low-quality |
| KM-final-viral-combined_k141_12393||full | Not-determined |
| KM-final-viral-combined_k141_124043||full | Low-quality |
| KM-final-viral-combined_k141_90155||full | Not-determined |
| 18-105-final-viral-combined_k141_67169||full | High-quality |
| KM-final-viral-combined_k141_68419||full | Not-determined |
| KM-final-viral-combined_k141_68421||full | Low-quality |
| KM-final-viral-combined_k141_201943||full | Not-determined |
| KM-final-viral-combined_k141_224142||full | Low-quality |
| KM-final-viral-combined_k141_124065||full | Low-quality |
| KM-final-viral-combined_k141_190496||full | Not-determined |
| KM-final-viral-combined_k141_190497||full | Not-determined |
| KM-final-viral-combined_k141_23545||full | Not-determined |
| KM-final-viral-combined_k141_190527||full | Low-quality |
| KM-final-viral-combined_k141_146139||full | Low-quality |
| KM-final-viral-combined_k141_257221||full | Low-quality |

| | |
|---|---|
| KM-final-viral-combined_k141_190540\|\|full | Low-quality |
| KM-final-viral-combined_k141_257222\|\|full | Not-determined |
| KM-final-viral-combined_k141_1617\|\|full | Not-determined |
| KM-final-viral-combined_k141_146149\|\|full | Not-determined |
| KM-final-viral-combined_k141_23558\|\|full | Not-determined |
| KM-final-viral-combined_k141_112910\|\|full | Low-quality |
| KM-final-viral-combined_k141_179400\|\|full | Not-determined |
| KM-final-viral-combined_k141_112912\|\|full | Low-quality |
| KM-final-viral-combined_k141_90229\|\|full | Low-quality |
| KM-final-viral-combined_k141_146168\|\|full | Low-quality |
| KM-final-viral-combined_k141_45708\|\|full | Not-determined |
| KM-final-viral-combined_k141_134948\|\|full | Low-quality |
| KM-final-viral-combined_k141_101400\|\|full | Low-quality |
| KM-final-viral-combined_k141_23584\|\|full | Low-quality |
| KM-final-viral-combined_k141_79902\|\|full | Not-determined |
| KM-final-viral-combined_k141_146178\|\|full | Not-determined |
| KM-final-viral-combined_k141_1657\|\|full | Low-quality |
| KM-final-viral-combined_k141_134956\|\|full | Low-quality |
| KM-final-viral-combined_k141_1662\|\|full | Not-determined |
| KM-final-viral-combined_k141_23592\|\|full | Not-determined |
| KM-final-viral-combined_k141_134960\|\|full | Not-determined |
| KM-final-viral-combined_k141_124148\|\|full | Low-quality |
| KM-final-viral-combined_k141_224228\|\|full | Not-determined |
| KM-final-viral-combined_k141_68541\|\|full | Low-quality |
| KM-final-viral-combined_k141_79952\|\|full | Low-quality |
| KM-final-viral-combined_k141_79958\|\|full | Low-quality |
| KM-final-viral-combined_k141_1686\|\|full | Low-quality |
| KM-final-viral-combined_k141_202024\|\|full | Low-quality |
| KM-final-viral-combined_k141_1693\|\|full | Low-quality |
| KM-final-viral-combined_k141_202041\|\|full | Not-determined |
| KM-final-viral-combined_k141_135001\|\|full | Not-determined |
| KM-final-viral-combined_k141_224251\|\|full | Low-quality |
| KM-final-viral-combined_k141_124170\|\|full | Not-determined |
| KM-final-viral-combined_k141_101445\|\|full | Not-determined |
| KM-final-viral-combined_k141_1719\|\|full | Low-quality |
| KM-final-viral-combined_k141_168254\|\|full | Not-determined |
| KM-final-viral-combined_k141_1721\|\|full | Not-determined |
| KM-final-viral-combined_k141_1722\|\|full | Not-determined |
| KM-final-viral-combined_k141_257322\|\|full | Not-determined |
| KM-final-viral-combined_k141_179456\|\|full | Low-quality |
| KM-final-viral-combined_k141_101474\|\|full | Not-determined |
| KM-final-viral-combined_k141_190622\|\|full | Not-determined |
| KM-final-viral-combined_k141_113016\|\|full | Low-quality |
| KM-final-viral-combined_k141_90314\|\|full | Low-quality |
| KM-final-viral-combined_k141_212980\|\|full | Low-quality |
| KM-final-viral-combined_k141_224281\|\|full | Low-quality |
| KM-final-viral-combined_k141_90317\|\|full | Low-quality |
| KM-final-viral-combined_k141_68583\|\|full | Low-quality |
| KM-final-viral-combined_k141_168283\|\|full | Not-determined |
| KM-final-viral-combined_k141_113041\|\|full | Not-determined |
| KM-final-viral-combined_k141_23688\|\|full | Not-determined |
| KM-final-viral-combined_k141_213008\|\|full | Not-determined |
| KM-final-viral-combined_k141_1762\|\|full | Not-determined |
| KM-final-viral-combined_k141_35045\|\|full | Low-quality |
| KM-final-viral-combined_k141_23700\|\|full | Low-quality |
| KM-final-viral-combined_k141_257342\|\|full | Low-quality |
| KM-final-viral-combined_k141_113062\|\|full | Low-quality |
| KM-final-viral-combined_k141_113066\|\|full | Not-determined |
| KM-final-viral-combined_k141_124232\|\|full | Not-determined |
| KM-final-viral-combined_k141_202105\|\|full | Low-quality |
| KM-final-viral-combined_k141_224342\|\|full | Not-determined |
| KM-final-viral-combined_k141_202108\|\|full | Not-determined |
| KM-final-viral-combined_k141_190683\|\|full | Not-determined |
| KM-final-viral-combined_k141_179527\|\|full | Low-quality |
| KM-final-viral-combined_k141_68618\|\|full | Not-determined |
| KM-final-viral-combined_k141_257351\|\|full | Low-quality |
| KM-final-viral-combined_k141_224353\|\|full | Low-quality |
| KM-final-viral-combined_k141_45737\|\|full | Low-quality |
| KM-final-viral-combined_k141_135068\|\|full | Low-quality |
| KM-final-viral-combined_k141_90353\|\|full | Low-quality |
| KM-final-viral-combined_k141_190694\|\|full | Not-determined |
| KM-final-viral-combined_k141_168351\|\|full | Low-quality |
| KM-final-viral-combined_k141_135094\|\|full | Low-quality |
| KM-final-viral-combined_k141_213065\|\|full | Not-determined |
| KM-final-viral-combined_k141_90362\|\|full | Not-determined |
| KM-final-viral-combined_k141_157124\|\|full | Low-quality |
| KM-final-viral-combined_k141_23727\|\|full | Low-quality |
| KM-final-viral-combined_k141_179572\|\|full | Not-determined |
| KM-final-viral-combined_k141_168376\|\|full | Not-determined |
| KM-final-viral-combined_k141_224379\|\|full | Not-determined |
| KM-final-viral-combined_k141_101525\|\|full | Not-determined |
| KM-final-viral-combined_k141_213117\|\|full | Low-quality |
| KM-final-viral-combined_k141_45772\|\|full | Not-determined |
| KM-final-viral-combined_k141_213123\|\|full | Low-quality |
| KM-final-viral-combined_k141_257399\|\|full | Not-determined |
| KM-final-viral-combined_k141_245912\|\|full | Not-determined |
| KM-final-viral-combined_k141_146276\|\|full | Low-quality |
| KM-final-viral-combined_k141_79987\|\|full | Not-determined |
| 18-106-final-viral-combined_k141_138710\|\|full | High-quality |
| KM-final-viral-combined_k141_57242\|\|full | Not-determined |
| KM-final-viral-combined_k141_124281\|\|full | Low-quality |
| KM-final-viral-combined_k141_190763\|\|full | Low-quality |
| KM-final-viral-combined_k141_90382\|\|full | Low-quality |
| KM-final-viral-combined_k141_124294\|\|full | Not-determined |
| KM-final-viral-combined_k141_68635\|\|full | Low-quality |
| KM-final-viral-combined_k141_245938\|\|full | Not-determined |
| KM-final-viral-combined_k141_79998\|\|full | Not-determined |
| KM-final-viral-combined_k141_68638\|\|full | Not-determined |
| KM-final-viral-combined_k141_213143\|\|full | Low-quality |
| KM-final-viral-combined_k141_35106\|\|full | Not-determined |
| KM-final-viral-combined_k141_157167\|\|full | Low-quality |
| KM-final-viral-combined_k141_135192\|\|full | Low-quality |
| KM-final-viral-combined_k141_113156\|\|full | Not-determined |
| KM-final-viral-combined_k141_179619\|\|full | Not-determined |
| 18-101-final-viral-combined_k141_170115\|\|full | High-quality |
| KM-final-viral-combined_k141_68679\|\|full | Not-determined |
| KM-final-viral-combined_k141_190805\|\|full | Not-determined |
| KM-final-viral-combined_k141_35134\|\|full | Not-determined |
| KM-final-viral-combined_k141_190807\|\|full | Low-quality |
| KM-final-viral-combined_k141_57290\|\|full | Not-determined |
| KM-final-viral-combined_k141_35142\|\|full | Not-determined |
| KM-final-viral-combined_k141_224386\|\|full | Low-quality |
| KM-final-viral-combined_k141_1818\|\|full | Not-determined |
| KM-final-viral-combined_k141_68705\|\|full | Low-quality |
| KM-final-viral-combined_k141_213192\|\|full | Not-determined |
| KM-final-viral-combined_k141_224402\|\|full | Not-determined |
| KM-final-viral-combined_k141_213194\|\|full | Not-determined |
| KM-final-viral-combined_k141_90443\|\|full | Low-quality |
| KM-final-viral-combined_k141_179667\|\|full | Low-quality |
| KM-final-viral-combined_k141_234984\|\|full | Low-quality |
| KM-final-viral-combined_k141_202328\|\|full | Low-quality |
| KM-final-viral-combined_k141_213239\|\|full | Not-determined |
| KM-final-viral-combined_k141_12473\|\|full | Low-quality |
| KM-final-viral-combined_k141_113234\|\|full | Not-determined |
| KM-final-viral-combined_k141_124393\|\|full | Low-quality |
| KM-final-viral-combined_k141_135279\|\|full | Not-determined |
| KM-final-viral-combined_k141_80068\|\|full | Not-determined |
| KM-final-viral-combined_k141_45857\|\|full | Not-determined |
| KM-final-viral-combined_k141_113239\|\|full | Low-quality |
| KM-final-viral-combined_k141_168523\|\|full | Not-determined |
| KM-final-viral-combined_k141_113240\|\|full | Not-determined |
| KM-final-viral-combined_k141_224415\|\|full | Low-quality |
| KM-final-viral-combined_k141_80069\|\|full | Not-determined |
| KM-final-viral-combined_k141_23836\|\|full | Not-determined |
| KM-final-viral-combined_k141_124411\|\|full | Low-quality |
| KM-final-viral-combined_k141_235013\|\|full | Not-determined |
| KM-final-viral-combined_k141_213265\|\|full | Not-determined |
| KM-final-viral-combined_k141_124415\|\|full | Low-quality |
| KM-final-viral-combined_k141_235018\|\|full | Low-quality |
| KM-final-viral-combined_k141_80090\|\|full | Not-determined |
| KM-final-viral-combined_k141_168556\|\|full | Not-determined |
| KM-final-viral-combined_k141_80102\|\|full | Low-quality |
| KM-final-viral-combined_k141_113284\|\|full | Low-quality |
| KM-final-viral-combined_k141_1931\|\|full | Not-determined |
| KM-final-viral-combined_k141_157224\|\|full | Low-quality |
| KM-final-viral-combined_k141_90494\|\|full | Low-quality |
| KM-final-viral-combined_k141_135315\|\|full | Not-determined |
| KM-final-viral-combined_k141_213306\|\|full | Low-quality |
| KM-final-viral-combined_k141_68761\|\|full | Not-determined |
| KM-final-viral-combined_k141_23864\|\|full | Not-determined |
| KM-final-viral-combined_k141_113303\|\|full | Low-quality |
| KM-final-viral-combined_k141_90496\|\|full | Not-determined |
| KM-final-viral-combined_k141_80127\|\|full | Not-determined |
| KM-final-viral-combined_k141_101673\|\|full | Low-quality |
| KM-final-viral-combined_k141_113311\|\|full | Not-determined |
| KM-final-viral-combined_k141_1950\|\|full | Not-determined |
| KM-final-viral-combined_k141_213318\|\|full | Low-quality |
| KM-final-viral-combined_k141_23886\|\|full | Low-quality |
| KM-final-viral-combined_k141_157241\|\|full | Low-quality |
| KM-final-viral-combined_k141_45893\|\|full | Not-determined |
| KM-final-viral-combined_k141_202424\|\|full | Not-determined |
| KM-final-viral-combined_k141_23899\|\|full | Low-quality |
| KM-final-viral-combined_k141_1962\|\|full | Low-quality |
| KM-final-viral-combined_k141_90533\|\|full | Not-determined |
| KM-final-viral-combined_k141_45898\|\|full | Not-determined |
| KM-final-viral-combined_k141_235038\|\|full | Not-determined |
| KM-final-viral-combined_k141_101703\|\|full | Not-determined |
| KM-final-viral-combined_k141_90535\|\|full | Not-determined |
| KM-final-viral-combined_k141_213329\|\|full | Low-quality |
| KM-final-viral-combined_k141_213332\|\|full | Low-quality |
| KM-final-viral-combined_k141_257534\|\|full | Low-quality |
| KM-final-viral-combined_k141_90547\|\|full | Not-determined |
| KM-final-viral-combined_k141_101721\|\|full | Low-quality |
| KM-final-viral-combined_k141_190898\|\|full | Low-quality |
| KM-final-viral-combined_k141_246020\|\|full | Not-determined |
| KM-final-viral-combined_k141_146406\|\|full | Low-quality |
| KM-final-viral-combined_k141_235056\|\|full | Not-determined |
| KM-final-viral-combined_k141_80138\|\|full | Low-quality |
| KM-final-viral-combined_k141_202480\|\|full | Low-quality |
| KM-final-viral-combined_k141_68781\|\|full | Low-quality |
| KM-final-viral-combined_k141_80152\|\|full | Low-quality |
| KM-final-viral-combined_k141_213355\|\|full | Low-quality |
| KM-final-viral-combined_k141_168634\|\|full | Not-determined |
| KM-final-viral-combined_k141_213357\|\|full | Low-quality |
| KM-final-viral-combined_k141_224499\|\|full | Not-determined |
| KM-final-viral-combined_k141_35285\|\|full | Not-determined |
| KM-final-viral-combined_k141_202506\|\|full | Not-determined |
| KM-final-viral-combined_k141_246054\|\|full | Low-quality |
| KM-final-viral-combined_k141_57395\|\|full | Low-quality |
| KM-final-viral-combined_k141_235099\|\|full | Not-determined |
| KM-final-viral-combined_k141_202536\|\|full | Low-quality |
| KM-final-viral-combined_k141_101761\|\|full | Not-determined |
| KM-final-viral-combined_k141_101766\|\|full | Not-determined |
| KM-final-viral-combined_k141_246070\|\|full | Not-determined |
| 18-101-final-viral-combined_k141_94988\|\|0_partial | Medium-quality |
| KM-final-viral-combined_k141_157305\|\|full | Low-quality |
| KM-final-viral-combined_k141_2015\|\|full | Low-quality |
| KM-final-viral-combined_k141_202558\|\|full | Not-determined |
| KM-final-viral-combined_k141_146479\|\|full | Not-determined |
| KM-final-viral-combined_k141_190961\|\|full | Low-quality |
| KM-final-viral-combined_k141_235110\|\|full | Not-determined |
| KM-final-viral-combined_k141_35307\|\|full | Low-quality |
| KM-final-viral-combined_k141_57426\|\|full | Low-quality |
| KM-final-viral-combined_k141_2033\|\|full | Low-quality |
| KM-final-viral-combined_k141_113412\|\|full | Low-quality |
| KM-final-viral-combined_k141_12560\|\|full | Low-quality |
| KM-final-viral-combined_k141_35318\|\|full | Low-quality |
| KM-final-viral-combined_k141_2052\|\|full | Not-determined |
| KM-final-viral-combined_k141_146504\|\|full | Low-quality |
| KM-final-viral-combined_k141_157338\|\|full | Not-determined |
| KM-final-viral-combined_k141_35327\|\|full | Not-determined |
| KM-final-viral-combined_k141_190984\|\|full | Low-quality |
| KM-final-viral-combined_k141_2067\|\|full | Not-determined |
| KM-final-viral-combined_k141_57443\|\|full | Not-determined |
| KM-final-viral-combined_k141_113460\|\|full | Low-quality |
| KM-final-viral-combined_k141_213443\|\|full | Low-quality |
| KM-final-viral-combined_k141_90608\|\|full | Not-determined |
| KM-final-viral-combined_k141_179826\|\|full | Not-determined |
| KM-final-viral-combined_k141_213444\|\|full | Low-quality |
| KM-final-viral-combined_k141_179834\|\|full | Low-quality |
| KM-final-viral-combined_k141_80244\|\|full | Low-quality |
| KM-final-viral-combined_k141_257608\|\|full | Not-determined |
| KM-final-viral-combined_k141_191041\|\|full | Not-determined |
| KM-final-viral-combined_k141_35347\|\|full | Low-quality |
| KM-final-viral-combined_k141_202566\|\|full | Not-determined |
| KM-final-viral-combined_k141_257609\|\|full | Low-quality |
| KM-final-viral-combined_k141_235182\|\|full | Not-determined |
| KM-final-viral-combined_k141_168764\|\|full | Low-quality |
| KM-final-viral-combined_k141_202572\|\|full | Not-determined |
| KM-final-viral-combined_k141_146535\|\|full | Not-determined |
| KM-final-viral-combined_k141_113523\|\|full | Not-determined |
| KM-final-viral-combined_k141_246097\|\|full | Low-quality |
| KM-final-viral-combined_k141_191076\|\|full | Not-determined |
| KM-final-viral-combined_k141_168787\|\|full | Not-determined |
| KM-final-viral-combined_k141_68903\|\|full | Low-quality |
| KM-final-viral-combined_k141_213484\|\|full | Not-determined |
| KM-final-viral-combined_k141_80273\|\|full | Not-determined |
| KM-final-viral-combined_k141_135524\|\|full | Low-quality |
| KM-final-viral-combined_k141_12610\|\|full | Low-quality |
| KM-final-viral-combined_k141_235214\|\|full | Not-determined |
| KM-final-viral-combined_k141_179882\|\|full | Not-determined |
| KM-final-viral-combined_k141_146590\|\|full | Not-determined |
| KM-final-viral-combined_k141_135571\|\|full | Not-determined |
| KM-final-viral-combined_k141_12641\|\|full | Not-determined |
| KM-final-viral-combined_k141_57534\|\|full | Not-determined |
| KM-final-viral-combined_k141_113552\|\|full | Low-quality |
| KM-final-viral-combined_k141_2127\|\|full | Not-determined |
| KM-final-viral-combined_k141_146600\|\|full | Not-determined |
| KM-final-viral-combined_k141_68957\|\|full | Not-determined |
| KM-final-viral-combined_k141_257628\|\|full | Low-quality |
| KM-final-viral-combined_k141_90644\|\|full | Not-determined |
| KM-final-viral-combined_k141_146613\|\|full | Not-determined |
| KM-final-viral-combined_k141_246140\|\|full | Not-determined |
| KM-final-viral-combined_k141_257642\|\|full | Not-determined |
| KM-final-viral-combined_k141_35394\|\|full | Low-quality |
| KM-final-viral-combined_k141_57559\|\|full | Not-determined |
| KM-final-viral-combined_k141_213501\|\|full | Not-determined |
| KM-final-viral-combined_k141_179911\|\|full | Not-determined |
| KM-final-viral-combined_k141_35402\|\|full | Not-determined |
| KM-final-viral-combined_k141_235234\|\|full | Not-determined |
| KM-final-viral-combined_k141_191110\|\|full | Not-determined |
| KM-final-viral-combined_k141_246162\|\|full | Low-quality |
| KM-final-viral-combined_k141_146640\|\|full | Low-quality |
| KM-final-viral-combined_k141_191124\|\|full | Not-determined |
| KM-final-viral-combined_k141_202629\|\|full | Low-quality |
| KM-final-viral-combined_k141_113585\|\|full | Low-quality |
| KM-final-viral-combined_k141_101837\|\|full | Not-determined |
| KM-final-viral-combined_k141_202635\|\|full | Low-quality |
| KM-final-viral-combined_k141_124512\|\|full | Not-determined |
| KM-final-viral-combined_k141_135615\|\|full | Low-quality |
| KM-final-viral-combined_k141_80347\|\|full | Low-quality |
| KM-final-viral-combined_k141_191144\|\|full | Low-quality |
| KM-final-viral-combined_k141_113601\|\|full | Not-determined |
| KM-final-viral-combined_k141_12677\|\|full | Low-quality |
| KM-final-viral-combined_k141_124521\|\|full | Not-determined |
| KM-final-viral-combined_k141_213547\|\|full | Low-quality |
| KM-final-viral-combined_k141_224612\|\|full | Not-determined |
| KM-final-viral-combined_k141_168815\|\|full | Low-quality |
| KM-final-viral-combined_k141_168818\|\|full | Low-quality |
| KM-final-viral-combined_k141_113623\|\|full | Low-quality |
| KM-final-viral-combined_k141_124531\|\|full | Not-determined |
| KM-final-viral-combined_k141_168821\|\|full | Not-determined |
| KM-final-viral-combined_k141_57578\|\|full | Low-quality |
| KM-final-viral-combined_k141_124537\|\|full | Not-determined |
| KM-final-viral-combined_k141_224633\|\|full | Low-quality |
| KM-final-viral-combined_k141_213570\|\|full | Low-quality |
| KM-final-viral-combined_k141_146671\|\|full | Not-determined |
| KM-final-viral-combined_k141_135631\|\|full | Low-quality |
| KM-final-viral-combined_k141_12708\|\|full | Not-determined |
| KM-final-viral-combined_k141_224651\|\|full | Low-quality |
| KM-final-viral-combined_k141_213582\|\|full | Not-determined |
| KM-final-viral-combined_k141_57619\|\|full | Low-quality |
| KM-final-viral-combined_k141_35467\|\|full | Low-quality |
| KM-final-viral-combined_k141_12734\|\|full | Low-quality |
| KM-final-viral-combined_k141_2159\|\|full | Low-quality |
| KM-final-viral-combined_k141_257688\|\|full | Not-determined |
| KM-final-viral-combined_k141_124591\|\|full | Low-quality |
| KM-final-viral-combined_k141_2164\|\|full | Not-determined |
| KM-final-viral-combined_k141_246202\|\|full | Not-determined |
| KM-final-viral-combined_k141_45992\|\|full | Low-quality |
| KM-final-viral-combined_k141_124595\|\|full | Low-quality |
| KM-final-viral-combined_k141_90698\|\|full | Not-determined |
| KM-final-viral-combined_k141_202680\|\|full | Not-determined |
| KM-final-viral-combined_k141_168849\|\|full | Not-determined |
| KM-final-viral-combined_k141_235297\|\|full | Low-quality |
| KM-final-viral-combined_k141_202684\|\|full | Low-quality |
| KM-final-viral-combined_k141_90722\|\|full | Low-quality |
| KM-final-viral-combined_k141_124611\|\|full | Not-determined |
| KM-final-viral-combined_k141_213609\|\|full | Low-quality |
| KM-final-viral-combined_k141_157501\|\|full | Not-determined |
| KM-final-viral-combined_k141_57682\|\|full | Low-quality |
| KM-final-viral-combined_k141_113710\|\|full | Low-quality |
| KM-final-viral-combined_k141_235326\|\|full | Low-quality |
| KM-final-viral-combined_k141_80442\|\|full | Not-determined |
| KM-final-viral-combined_k141_101896\|\|full | Low-quality |
| KM-final-viral-combined_k141_146700\|\|full | Not-determined |
| KM-final-viral-combined_k141_101903\|\|full | Not-determined |
| KM-final-viral-combined_k141_24068\|\|full | Not-determined |
| KM-final-viral-combined_k141_146711\|\|full | Not-determined |
| KM-final-viral-combined_k141_24079\|\|full | Not-determined |
| KM-final-viral-combined_k141_12762\|\|full | Low-quality |
| KM-final-viral-combined_k141_202704\|\|full | Not-determined |
| KM-final-viral-combined_k141_191263\|\|full | Not-determined |
| KM-final-viral-combined_k141_168877\|\|full | Not-determined |
| KM-final-viral-combined_k141_180062\|\|full | Low-quality |
| KM-final-viral-combined_k141_257743\|\|full | Not-determined |
| KM-final-viral-combined_k141_257749\|\|full | Not-determined |
| KM-final-viral-combined_k141_246271\|\|full | Low-quality |
| KM-final-viral-combined_k141_46087\|\|full | Low-quality |
| KM-final-viral-combined_k141_124690\|\|full | Low-quality |
| KM-final-viral-combined_k141_57712\|\|full | Low-quality |
| KM-final-viral-combined_k141_213646\|\|full | Not-determined |
| KM-final-viral-combined_k141_113744\|\|full | Not-determined |
| KM-final-viral-combined_k141_213651\|\|full | Low-quality |
| KM-final-viral-combined_k141_57715\|\|full | Low-quality |
| KM-final-viral-combined_k141_35555\|\|full | Not-determined |
| KM-final-viral-combined_k141_12785\|\|full | Low-quality |
| KM-final-viral-combined_k141_202741\|\|full | Not-determined |
| KM-final-viral-combined_k141_191266\|\|full | Low-quality |
| KM-final-viral-combined_k141_135731\|\|full | Not-determined |
| KM-final-viral-combined_k141_46108\|\|full | Not-determined |
| KM-final-viral-combined_k141_124725\|\|full | Low-quality |
| KM-final-viral-combined_k141_46115\|\|full | Not-determined |
| KM-final-viral-combined_k141_69099\|\|full | Not-determined |
| KM-final-viral-combined_k141_213668\|\|full | Not-determined |
| KM-final-viral-combined_k141_235374\|\|full | Not-determined |
| KM-final-viral-combined_k141_168942\|\|full | Low-quality |
| KM-final-viral-combined_k141_157526\|\|full | Not-determined |
| KM-final-viral-combined_k141_157531\|\|full | Low-quality |
| KM-final-viral-combined_k141_257777\|\|full | Not-determined |
| KM-final-viral-combined_k141_235381\|\|full | Not-determined |
| KM-final-viral-combined_k141_213677\|\|full | Not-determined |
| KM-final-viral-combined_k141_101923\|\|full | Not-determined |
| KM-final-viral-combined_k141_146753\|\|full | Not-determined |
| KM-final-viral-combined_k141_202770\|\|full | Not-determined |
| KM-final-viral-combined_k141_202775\|\|full | Low-quality |
| KM-final-viral-combined_k141_202776\|\|full | Low-quality |
| KM-final-viral-combined_k141_113777\|\|full | Not-determined |
| KM-final-viral-combined_k141_57722\|\|full | Not-determined |
| KM-final-viral-combined_k141_113779\|\|full | Low-quality |
| KM-final-viral-combined_k141_191299\|\|full | Low-quality |
| KM-final-viral-combined_k141_124746\|\|full | Not-determined |
| KM-final-viral-combined_k141_69125\|\|full | Not-determined |
| KM-final-viral-combined_k141_46145\|\|full | Not-determined |
| KM-final-viral-combined_k141_135767\|\|full | Not-determined |
| KM-final-viral-combined_k141_257798\|\|full | Low-quality |
| KM-final-viral-combined_k141_191307\|\|full | Low-quality |
| KM-final-viral-combined_k141_69127\|\|full | Low-quality |
| KM-final-viral-combined_k141_35604\|\|full | Not-determined |
| KM-final-viral-combined_k141_124750\|\|full | Low-quality |
| KM-final-viral-combined_k141_202790\|\|full | Low-quality |
| KM-final-viral-combined_k141_224722\|\|full | Not-determined |
| KM-final-viral-combined_k141_202791\|\|full | Not-determined |
| KM-final-viral-combined_k141_202796\|\|full | Low-quality |
| KM-final-viral-combined_k141_157565\|\|full | Low-quality |
| KM-final-viral-combined_k141_124772\|\|full | Low-quality |
| KM-final-viral-combined_k141_90823\|\|full | Not-determined |
| KM-final-viral-combined_k141_124786\|\|full | Low-quality |
| KM-final-viral-combined_k141_224762\|\|full | Not-determined |
| KM-final-viral-combined_k141_46180\|\|full | Low-quality |
| KM-final-viral-combined_k141_113837\|\|full | Not-determined |
| KM-final-viral-combined_k141_24158\|\|full | Low-quality |
| KM-final-viral-combined_k141_235451\|\|full | Not-determined |
| KM-final-viral-combined_k141_224791\|\|full | Not-determined |
| 18-101-final-viral-combined_k141_191622\|\|full | High-quality |
| KM-final-viral-combined_k141_169026\|\|full | Not-determined |
| KM-final-viral-combined_k141_213700\|\|full | Low-quality |
| KM-final-viral-combined_k141_24181\|\|full | Low-quality |
| KM-final-viral-combined_k141_246314\|\|full | Not-determined |
| KM-final-viral-combined_k141_135826\|\|full | Low-quality |
| KM-final-viral-combined_k141_191357\|\|full | Not-determined |
| KM-final-viral-combined_k141_69158\|\|full | Not-determined |
| KM-final-viral-combined_k141_169064\|\|full | Low-quality |
| KM-final-viral-combined_k141_213724\|\|full | Not-determined |
| KM-final-viral-combined_k141_146820\|\|full | Not-determined |
| KM-final-viral-combined_k141_146824\|\|full | Not-determined |
| KM-final-viral-combined_k141_80567\|\|full | Not-determined |
| KM-final-viral-combined_k141_113892\|\|full | Low-quality |
| KM-final-viral-combined_k141_224840\|\|full | Not-determined |
| KM-final-viral-combined_k141_224844\|\|full | Low-quality |
| KM-final-viral-combined_k141_191382\|\|full | Not-determined |
| KM-final-viral-combined_k141_213743\|\|full | Not-determined |
| KM-final-viral-combined_k141_257825\|\|full | Not-determined |
| KM-final-viral-combined_k141_180225\|\|full | Low-quality |
| KM-final-viral-combined_k141_102034\|\|full | Not-determined |
| KM-final-viral-combined_k141_202847\|\|full | Not-determined |
| KM-final-viral-combined_k141_57815\|\|full | Low-quality |
| KM-final-viral-combined_k141_235527\|\|full | Low-quality |
| KM-final-viral-combined_k141_180242\|\|full | Not-determined |
| KM-final-viral-combined_k141_246368\|\|full | Not-determined |
| KM-final-viral-combined_k141_90886\|\|full | Low-quality |
| KM-final-viral-combined_k141_157610\|\|full | Not-determined |
| KM-final-viral-combined_k141_113931\|\|full | Not-determined |
| KM-final-viral-combined_k141_69204\|\|full | Low-quality |
| 18-102_k141_23749 | Medium-quality |
| KM-final-viral-combined_k141_146853\|\|full | Low-quality |
| KM-final-viral-combined_k141_257869\|\|full | Low-quality |
| KM-final-viral-combined_k141_24262\|\|full | Low-quality |
| KM-final-viral-combined_k141_191429\|\|full | Not-determined |
| KM-final-viral-combined_k141_124958\|\|full | Low-quality |
| KM-final-viral-combined_k141_257896\|\|full | Low-quality |
| KM-final-viral-combined_k141_124963\|\|full | Not-determined |
| KM-final-viral-combined_k141_102080\|\|full | Not-determined |
| KM-final-viral-combined_k141_235543\|\|full | Not-determined |
| KM-final-viral-combined_k141_57851\|\|full | Not-determined |
| KM-final-viral-combined_k141_24294\|\|full | Not-determined |
| KM-final-viral-combined_k141_2283\|\|full | Not-determined |
| KM-final-viral-combined_k141_169129\|\|full | Low-quality |
| KM-final-viral-combined_k141_90912\|\|full | Not-determined |
| KM-final-viral-combined_k141_113971\|\|full | Low-quality |
| KM-final-viral-combined_k141_113974\|\|full | Not-determined |
| KM-final-viral-combined_k141_69257\|\|full | Low-quality |
| KM-final-viral-combined_k141_224914\|\|full | Low-quality |
| KM-final-viral-combined_k141_69263\|\|full | Not-determined |
| KM-final-viral-combined_k141_224926\|\|full | Low-quality |
| KM-final-viral-combined_k141_224927\|\|full | Not-determined |
| KM-final-viral-combined_k141_113989\|\|full | Not-determined |
| KM-final-viral-combined_k141_113990\|\|full | Not-determined |
| KM-final-viral-combined_k141_235587\|\|full | Not-determined |
| KM-final-viral-combined_k141_224938\|\|full | Not-determined |
| KM-final-viral-combined_k141_257936\|\|full | Low-quality |
| KM-final-viral-combined_k141_113998\|\|full | Low-quality |
| KM-final-viral-combined_k141_146906\|\|full | Low-quality |
| KM-final-viral-combined_k141_12953\|\|full | Not-determined |
| KM-final-viral-combined_k141_257940\|\|full | Low-quality |
| KM-final-viral-combined_k141_69297\|\|full | Not-determined |
| KM-final-viral-combined_k141_46218\|\|full | Not-determined |
| KM-final-viral-combined_k141_257955\|\|full | Low-quality |
| KM-final-viral-combined_k141_246447\|\|full | Not-determined |
| KM-final-viral-combined_k141_246450\|\|full | Low-quality |
| KM-final-viral-combined_k141_202941\|\|full | Low-quality |
| KM-final-viral-combined_k141_246451\|\|full | Not-determined |
| KM-final-viral-combined_k141_191529\|\|full | Low-quality |
| KM-final-viral-combined_k141_169200\|\|full | Low-quality |
| KM-final-viral-combined_k141_35690\|\|full | Not-determined |
| KM-final-viral-combined_k141_2327\|\|full | Not-determined |
| KM-final-viral-combined_k141_224988\|\|full | Not-determined |
| KM-final-viral-combined_k141_257986\|\|full | Low-quality |
| KM-final-viral-combined_k141_35700\|\|full | Low-quality |
| KM-final-viral-combined_k141_69336\|\|full | Low-quality |
| KM-final-viral-combined_k141_169219\|\|full | Low-quality |
| KM-final-viral-combined_k141_257999\|\|full | Not-determined |
| KM-final-viral-combined_k141_2345\|\|full | Low-quality |
| KM-final-viral-combined_k141_2347\|\|full | Low-quality |
| KM-final-viral-combined_k141_90990\|\|full | Not-determined |
| KM-final-viral-combined_k141_90993\|\|full | Low-quality |
| KM-final-viral-combined_k141_2348\|\|full | Not-determined |
| KM-final-viral-combined_k141_157676\|\|full | Low-quality |
| KM-final-viral-combined_k141_235632\|\|full | Low-quality |
| KM-final-viral-combined_k141_213887\|\|full | Not-determined |
| KM-final-viral-combined_k141_46260\|\|full | Not-determined |
| KM-final-viral-combined_k141_135938\|\|full | Not-determined |
| KM-final-viral-combined_k141_157685\|\|full | Not-determined |
| KM-final-viral-combined_k141_235644\|\|full | Low-quality |
| KM-final-viral-combined_k141_102133\|\|full | Low-quality |
| KM-final-viral-combined_k141_157693\|\|full | Not-determined |
| KM-final-viral-combined_k141_258027\|\|full | Not-determined |
| KM-final-viral-combined_k141_114065\|\|full | Not-determined |
| KM-final-viral-combined_k141_114070\|\|full | Not-determined |
| KM-final-viral-combined_k141_135993\|\|full | Low-quality |
| KM-final-viral-combined_k141_180309\|\|full | Not-determined |
| KM-final-viral-combined_k141_191555\|\|full | Not-determined |
| KM-final-viral-combined_k141_169246\|\|full | Low-quality |
| KM-final-viral-combined_k141_114094\|\|full | Not-determined |
| KM-final-viral-combined_k141_136009\|\|full | Not-determined |
| KM-final-viral-combined_k141_191558\|\|full | Low-quality |
| KM-final-viral-combined_k141_213914\|\|full | Low-quality |
| KM-final-viral-combined_k141_136015\|\|full | Not-determined |
| KM-final-viral-combined_k141_180330\|\|full | Low-quality |
| KM-final-viral-combined_k141_35762\|\|full | Not-determined |
| KM-final-viral-combined_k141_125057\|\|full | Not-determined |
| KM-final-viral-combined_k141_46295\|\|full | Low-quality |
| KM-final-viral-combined_k141_258086\|\|full | Not-determined |
| KM-final-viral-combined_k141_125063\|\|full | Not-determined |
| KM-final-viral-combined_k141_114127\|\|full | Not-determined |
| KM-final-viral-combined_k141_35780\|\|full | Not-determined |
| KM-final-viral-combined_k141_147060\|\|full | Low-quality |
| KM-final-viral-combined_k141_147061\|\|full | Not-determined |
| KM-final-viral-combined_k141_91067\|\|full | Low-quality |
| KM-final-viral-combined_k141_91071\|\|full | Not-determined |
| KM-final-viral-combined_k141_46301\|\|full | Low-quality |
| KM-final-viral-combined_k141_13035\|\|full | Not-determined |
| KM-final-viral-combined_k141_246521\|\|full | Not-determined |
| KM-final-viral-combined_k141_80617\|\|full | Low-quality |
| KM-final-viral-combined_k141_213958\|\|full | Low-quality |
| KM-final-viral-combined_k141_180347\|\|full | Low-quality |
| KM-final-viral-combined_k141_171305\|\|full | Medium-quality |
| KM-final-viral-combined_k141_147098\|\|full | Not-determined |
| KM-final-viral-combined_k141_246525\|\|full | Low-quality |
| KM-final-viral-combined_k141_169290\|\|full | Not-determined |
| KM-final-viral-combined_k141_80629\|\|full | Not-determined |
| KM-final-viral-combined_k141_13074\|\|full | Not-determined |
| KM-final-viral-combined_k141_57957\|\|full | Not-determined |
| KM-final-viral-combined_k141_46338\|\|full | Low-quality |
| KM-final-viral-combined_k141_213988\|\|full | Not-determined |
| KM-final-viral-combined_k141_102252\|\|full | Not-determined |
| KM-final-viral-combined_k141_80637\|\|full | Low-quality |
| KM-final-viral-combined_k141_225116\|\|full | Not-determined |
| KM-final-viral-combined_k141_225118\|\|full | Not-determined |
| KM-final-viral-combined_k141_80641\|\|full | Not-determined |
| KM-final-viral-combined_k141_102262\|\|full | Not-determined |
| KM-final-viral-combined_k141_57980\|\|full | Low-quality |
| KM-final-viral-combined_k141_114254\|\|full | Low-quality |
| KM-final-viral-combined_k141_114262\|\|full | Low-quality |
| KM-final-viral-combined_k141_80689\|\|full | Not-determined |
| KM-final-viral-combined_k141_69466\|\|full | Low-quality |
| KM-final-viral-combined_k141_147166\|\|full | Not-determined |
| KM-final-viral-combined_k141_258108\|\|full | Low-quality |
| KM-final-viral-combined_k141_191662\|\|full | Not-determined |
| KM-final-viral-combined_k141_2447\|\|full | Low-quality |
| 17-108-final-viral-combined_k141_193148\|\|full | Medium-quality |
| KM-final-viral-combined_k141_203145\|\|full | Low-quality |
| KM-final-viral-combined_k141_169354\|\|full | Not-determined |
| KM-final-viral-combined_k141_180420\|\|full | Not-determined |
| KM-final-viral-combined_k141_80704\|\|full | Not-determined |
| KM-final-viral-combined_k141_147176\|\|full | Low-quality |
| KM-final-viral-combined_k141_35846\|\|full | Low-quality |
| KM-final-viral-combined_k141_180435\|\|full | Low-quality |
| KM-final-viral-combined_k141_69471\|\|full | Low-quality |
| KM-final-viral-combined_k141_147191\|\|full | Not-determined |
| KM-final-viral-combined_k141_13117\|\|full | Not-determined |
| KM-final-viral-combined_k141_157770\|\|full | Not-determined |
| KM-final-viral-combined_k141_2463\|\|full | Not-determined |
| KM-final-viral-combined_k141_191680\|\|full | Not-determined |
| KM-final-viral-combined_k141_203175\|\|full | Not-determined |
| KM-final-viral-combined_k141_136055\|\|full | Not-determined |
| KM-final-viral-combined_k141_69488\|\|full | Not-determined |
| KM-final-viral-combined_k141_2482\|\|full | Not-determined |
| KM-final-viral-combined_k141_235838\|\|full | Not-determined |
| KM-final-viral-combined_k141_258128\|\|full | Not-determined |
| KM-final-viral-combined_k141_136066\|\|full | Low-quality |
| KM-final-viral-combined_k141_69499\|\|full | Low-quality |
| KM-final-viral-combined_k141_191698\|\|full | Low-quality |
| KM-final-viral-combined_k141_125105\|\|full | Low-quality |
| KM-final-viral-combined_k141_235851\|\|full | Not-determined |
| KM-final-viral-combined_k141_258153\|\|full | Not-determined |
| KM-final-viral-combined_k141_114333\|\|full | Low-quality |
| KM-final-viral-combined_k141_69514\|\|full | Not-determined |
| KM-final-viral-combined_k141_125117\|\|full | Not-determined |
| KM-final-viral-combined_k141_235874\|\|full | Low-quality |
| KM-final-viral-combined_k141_258162\|\|full | Low-quality |
| KM-final-viral-combined_k141_125118\|\|full | Not-determined |
| KM-final-viral-combined_k141_114349\|\|full | Not-determined |
| KM-final-viral-combined_k141_214109\|\|full | Not-determined |
| KM-final-viral-combined_k141_69531\|\|full | Low-quality |
| KM-final-viral-combined_k141_169428\|\|full | Not-determined |
| KM-final-viral-combined_k141_24403\|\|full | Not-determined |
| KM-final-viral-combined_k141_136107\|\|full | Not-determined |
| KM-final-viral-combined_k141_258185\|\|full | Not-determined |
| KM-final-viral-combined_k141_157809\|\|full | Low-quality |
| KM-final-viral-combined_k141_225183\|\|full | Low-quality |
| KM-final-viral-combined_k141_258192\|\|full | Not-determined |
| KM-final-viral-combined_k141_235889\|\|full | Not-determined |
| KM-final-viral-combined_k141_102365\|\|full | Not-determined |
| KM-final-viral-combined_k141_35910\|\|full | Not-determined |
| KM-final-viral-combined_k141_246657\|\|full | Not-determined |
| KM-final-viral-combined_k141_147245\|\|full | Not-determined |
| KM-final-viral-combined_k141_2511\|\|full | Low-quality |
| KM-final-viral-combined_k141_157821\|\|full | Low-quality |
| KM-final-viral-combined_k141_235906\|\|full | Low-quality |
| KM-final-viral-combined_k141_114388\|\|full | Not-determined |
| KM-final-viral-combined_k141_191732\|\|full | Low-quality |
| KM-final-viral-combined_k141_246671\|\|full | Not-determined |
| KM-final-viral-combined_k141_169481\|\|full | Not-determined |
| KM-final-viral-combined_k141_180504\|\|full | Not-determined |
| KM-final-viral-combined_k141_214129\|\|full | Not-determined |
| KM-final-viral-combined_k141_69583\|\|full | Low-quality |
| KM-final-viral-combined_k141_169492\|\|full | Not-determined |
| KM-final-viral-combined_k141_180519\|\|full | Low-quality |

| | |
|---|---|
| KM-final-viral-combined_k141_24459||full | Not-determined |
| KM-final-viral-combined_k141_80796||full | Low-quality |
| KM-final-viral-combined_k141_125171||full | Low-quality |
| KM-final-viral-combined_k141_214158||full | Not-determined |
| KM-final-viral-combined_k141_225215||full | Low-quality |
| KM-final-viral-combined_k141_35937||full | Not-determined |
| KM-final-viral-combined_k141_80800||full | Not-determined |
| KM-final-viral-combined_k141_235942||full | Low-quality |
| KM-final-viral-combined_k141_225219||full | Not-determined |
| KM-final-viral-combined_k141_46406||full | Not-determined |
| KM-final-viral-combined_k141_203274||full | Low-quality |
| KM-final-viral-combined_k141_214190||full | Low-quality |
| KM-final-viral-combined_k141_180545||full | Low-quality |
| KM-final-viral-combined_k141_246728||full | Not-determined |
| KM-final-viral-combined_k141_191743||full | Not-determined |
| KM-final-viral-combined_k141_35944||full | Low-quality |
| KM-final-viral-combined_k141_157842||full | Low-quality |
| KM-final-viral-combined_k141_13267||full | Not-determined |
| KM-final-viral-combined_k141_58063||full | Low-quality |
| KM-final-viral-combined_k141_35955||full | Not-determined |
| KM-final-viral-combined_k141_24508||full | Not-determined |
| KM-final-viral-combined_k141_80826||full | Low-quality |
| KM-final-viral-combined_k141_258258||full | Low-quality |
| KM-final-viral-combined_k141_157844||full | Not-determined |
| KM-final-viral-combined_k141_214239||full | Not-determined |
| KM-final-viral-combined_k141_80837||full | Not-determined |
| KM-final-viral-combined_k141_13280||full | Low-quality |
| KM-final-viral-combined_k141_180572||full | Not-determined |
| KM-final-viral-combined_k141_102408||full | Low-quality |
| KM-final-viral-combined_k141_58082||full | Not-determined |
| KM-final-viral-combined_k141_258267||full | Not-determined |
| KM-final-viral-combined_k141_180606||full | Not-determined |
| KM-final-viral-combined_k141_102410||full | Not-determined |
| KM-final-viral-combined_k141_2599||full | Not-determined |
| KM-final-viral-combined_k141_114489||full | Low-quality |
| KM-final-viral-combined_k141_235977||full | Low-quality |
| KM-final-viral-combined_k141_147336||full | Low-quality |
| KM-final-viral-combined_k141_13308||full | Not-determined |
| KM-final-viral-combined_k141_13312||full | Low-quality |
| KM-final-viral-combined_k141_2612||full | Low-quality |
| KM-final-viral-combined_k141_157873||full | Not-determined |
| KM-final-viral-combined_k141_235985||full | Not-determined |
| KM-final-viral-combined_k141_180630||full | Not-determined |
| KM-final-viral-combined_k141_58120||full | Low-quality |
| KM-final-viral-combined_k141_102431||full | Not-determined |
| [illegible] | [illegible] |

| | |
|---|---|
| KM-final-viral-combined_k141_215578\|\|full | Low-quality |
| KM-final-viral-combined_k141_116034\|\|full | Not-determined |
| KM-final-viral-combined_k141_3644\|\|full | Low-quality |
| KM-final-viral-combined_k141_103683\|\|full | Low-quality |
| KM-final-viral-combined_k141_237002\|\|full | Not-determined |
| KM-final-viral-combined_k141_158830\|\|full | Not-determined |
| KM-final-viral-combined_k141_215611\|\|full | Low-quality |
| KM-final-viral-combined_k141_59157\|\|full | Low-quality |
| KM-final-viral-combined_k141_47009\|\|full | Low-quality |
| KM-final-viral-combined_k141_59161\|\|full | Not-determined |
| KM-final-viral-combined_k141_193187\|\|full | Low-quality |
| KM-final-viral-combined_k141_204465\|\|full | Not-determined |
| KM-final-viral-combined_k141_237008\|\|full | Not-determined |
| KM-final-viral-combined_k141_137476\|\|full | Not-determined |
| KM-final-viral-combined_k141_137487\|\|full | Not-determined |
| KM-final-viral-combined_k141_3687\|\|full | Low-quality |
| KM-final-viral-combined_k141_25771\|\|full | Low-quality |
| KM-final-viral-combined_k141_126411\|\|full | Not-determined |
| KM-final-viral-combined_k141_237034\|\|full | Not-determined |
| KM-final-viral-combined_k141_70921\|\|full | Not-determined |
| KM-final-viral-combined_k141_182180\|\|full | Low-quality |
| KM-final-viral-combined_k141_237039\|\|full | Not-determined |
| KM-final-viral-combined_k141_47025\|\|full | Low-quality |
| KM-final-viral-combined_k141_47031\|\|full | Low-quality |
| KM-final-viral-combined_k141_70936\|\|full | Not-determined |
| KM-final-viral-combined_k141_82307\|\|full | Low-quality |
| KM-final-viral-combined_k141_137521\|\|full | Low-quality |
| KM-final-viral-combined_k141_137522\|\|full | Not-determined |
| KM-final-viral-combined_k141_116139\|\|full | Not-determined |
| KM-final-viral-combined_k141_3722\|\|full | Not-determined |
| KM-final-viral-combined_k141_204502\|\|full | Not-determined |
| KM-final-viral-combined_k141_193249\|\|full | Low-quality |
| KM-final-viral-combined_k141_215651\|\|full | Not-determined |
| KM-final-viral-combined_k141_126460\|\|full | Not-determined |
| KM-final-viral-combined_k141_59192\|\|full | Not-determined |
| KM-final-viral-combined_k141_3742\|\|full | Not-determined |
| KM-final-viral-combined_k141_59195\|\|full | Not-determined |
| KM-final-viral-combined_k141_103752\|\|full | Low-quality |
| KM-final-viral-combined_k141_14362\|\|full | Not-determined |
| KM-final-viral-combined_k141_148546\|\|full | Low-quality |
| KM-final-viral-combined_k141_148555\|\|full | Low-quality |
| KM-final-viral-combined_k141_226537\|\|full | Not-determined |
| KM-final-viral-combined_k141_59226\|\|full | Not-determined |
| KM-final-viral-combined_k141_126492\|\|full | Not-determined |
| KM-final-viral-combined_k141_237091\|\|full | Not-determined |
| KM-final-viral-combined_k141_126494\|\|full | Not-determined |
| KM-final-viral-combined_k141_170578\|\|full | Low-quality |
| KM-final-viral-combined_k141_37078\|\|full | Low-quality |
| KM-final-viral-combined_k141_193287\|\|full | Low-quality |
| KM-final-viral-combined_k141_158877\|\|full | Low-quality |
| KM-final-viral-combined_k141_215723\|\|full | Not-determined |
| KM-final-viral-combined_k141_237113\|\|full | Low-quality |
| KM-final-viral-combined_k141_126508\|\|full | Low-quality |
| 17-106-final-viral-combined_k141_2042\|\|full | Medium-quality |
| KM-final-viral-combined_k141_158884\|\|full | Not-determined |
| KM-final-viral-combined_k141_37093\|\|full | Low-quality |
| KM-final-viral-combined_k141_103782\|\|full | Not-determined |
| KM-final-viral-combined_k141_170607\|\|full | Low-quality |
| KM-final-viral-combined_k141_59269\|\|full | Not-determined |
| KM-final-viral-combined_k141_71002\|\|full | Not-determined |
| KM-final-viral-combined_k141_137588\|\|full | Low-quality |
| KM-final-viral-combined_k141_237131\|\|full | Low-quality |
| KM-final-viral-combined_k141_82370\|\|full | Low-quality |
| KM-final-viral-combined_k141_137591\|\|full | Not-determined |
| KM-final-viral-combined_k141_103839\|\|full | Not-determined |
| KM-final-viral-combined_k141_182290\|\|full | Not-determined |
| KM-final-viral-combined_k141_215753\|\|full | Low-quality |
| KM-final-viral-combined_k141_237172\|\|full | Not-determined |
| KM-final-viral-combined_k141_3832\|\|full | Low-quality |
| KM-final-viral-combined_k141_37150\|\|full | Not-determined |
| KM-final-viral-combined_k141_126588\|\|full | Not-determined |
| KM-final-viral-combined_k141_182322\|\|full | Not-determined |
| KM-final-viral-combined_k141_25890\|\|full | Low-quality |
| KM-final-viral-combined_k141_82436\|\|full | Not-determined |
| KM-final-viral-combined_k141_72837\|\|full | High-quality |
| KM-final-viral-combined_k141_259111\|\|full | Not-determined |
| KM-final-viral-combined_k141_137631\|\|full | Low-quality |
| KM-final-viral-combined_k141_82443\|\|full | Low-quality |
| KM-final-viral-combined_k141_148647\|\|full | Low-quality |
| KM-final-viral-combined_k141_170653\|\|full | Low-quality |
| KM-final-viral-combined_k141_148654\|\|full | Not-determined |
| KM-final-viral-combined_k141_116230\|\|full | Not-determined |
| KM-final-viral-combined_k141_158947\|\|full | Low-quality |
| KM-final-viral-combined_k141_158949\|\|full | Not-determined |
| KM-final-viral-combined_k141_71028\|\|full | Not-determined |
| KM-final-viral-combined_k141_182347\|\|full | Low-quality |
| KM-final-viral-combined_k141_116237\|\|full | Low-quality |
| KM-final-viral-combined_k141_137654\|\|full | Not-determined |
| KM-final-viral-combined_k141_37171\|\|full | Not-determined |
| KM-final-viral-combined_k141_71030\|\|full | Low-quality |
| KM-final-viral-combined_k141_237191\|\|full | Low-quality |
| KM-final-viral-combined_k141_91969\|\|full | Low-quality |
| KM-final-viral-combined_k141_59309\|\|full | Not-determined |
| KM-final-viral-combined_k141_71045\|\|full | Low-quality |
| KM-final-viral-combined_k141_91976\|\|full | Not-determined |
| KM-final-viral-combined_k141_126612\|\|full | Not-determined |
| KM-final-viral-combined_k141_259128\|\|full | Low-quality |
| KM-final-viral-combined_k141_25943\|\|full | Low-quality |
| KM-final-viral-combined_k141_3865\|\|full | Low-quality |
| KM-final-viral-combined_k141_148681\|\|full | Not-determined |
| KM-final-viral-combined_k141_3866\|\|full | Not-determined |
| KM-final-viral-combined_k141_59314\|\|full | Not-determined |
| KM-final-viral-combined_k141_71049\|\|full | Low-quality |
| KM-final-viral-combined_k141_247820\|\|full | Low-quality |
| KM-final-viral-combined_k141_25954\|\|full | Low-quality |
| KM-final-viral-combined_k141_237217\|\|full | Not-determined |
| KM-final-viral-combined_k141_182395\|\|full | Low-quality |
| KM-final-viral-combined_k141_91993\|\|full | Low-quality |
| KM-final-viral-combined_k141_59332\|\|full | Not-determined |
| 18-109-final-viral-combined_k141_71592\|\|full | Medium-quality |
| KM-final-viral-combined_k141_259146\|\|full | Not-determined |
| KM-final-viral-combined_k141_237232\|\|full | Not-determined |
| KM-final-viral-combined_k141_182417\|\|full | Not-determined |
| KM-final-viral-combined_k141_226757\|\|full | Low-quality |
| KM-final-viral-combined_k141_215866\|\|full | Not-determined |
| KM-final-viral-combined_k141_170675\|\|full | Low-quality |
| KM-final-viral-combined_k141_71090\|\|full | Low-quality |
| KM-final-viral-combined_k141_47100\|\|full | Not-determined |
| KM-final-viral-combined_k141_193360\|\|full | Not-determined |
| KM-final-viral-combined_k141_237251\|\|full | Not-determined |
| KM-final-viral-combined_k141_71113\|\|full | Not-determined |
| KM-final-viral-combined_k141_59368\|\|full | Low-quality |
| KM-final-viral-combined_k141_116312\|\|full | Not-determined |
| KM-final-viral-combined_k141_82548\|\|full | Not-determined |
| KM-final-viral-combined_k141_148737\|\|full | Not-determined |
| KM-final-viral-combined_k141_14447\|\|full | Not-determined |
| KM-final-viral-combined_k141_47128\|\|full | Not-determined |
| KM-final-viral-combined_k141_26016\|\|full | Not-determined |
| KM-final-viral-combined_k141_59387\|\|full | Not-determined |
| KM-final-viral-combined_k141_116333\|\|full | Low-quality |
| KM-final-viral-combined_k141_170743\|\|full | Not-determined |
| KM-final-viral-combined_k141_148755\|\|full | Not-determined |
| KM-final-viral-combined_k141_14476\|\|full | Not-determined |
| KM-final-viral-combined_k141_193408\|\|full | Not-determined |
| KM-final-viral-combined_k141_193410\|\|full | Low-quality |
| KM-final-viral-combined_k141_47138\|\|full | Not-determined |
| KM-final-viral-combined_k141_170758\|\|full | Low-quality |
| KM-final-viral-combined_k141_71157\|\|full | Not-determined |
| KM-final-viral-combined_k141_82604\|\|full | Not-determined |
| KM-final-viral-combined_k141_103988\|\|full | Low-quality |
| KM-final-viral-combined_k141_59438\|\|full | Low-quality |
| KM-final-viral-combined_k141_37254\|\|full | Not-determined |
| KM-final-viral-combined_k141_193466\|\|full | Not-determined |
| KM-final-viral-combined_k141_37257\|\|full | Low-quality |
| KM-final-viral-combined_k141_92112\|\|full | Low-quality |
| KM-final-viral-combined_k141_170769\|\|full | Not-determined |
| KM-final-viral-combined_k141_148797\|\|full | Low-quality |
| KM-final-viral-combined_k141_193482\|\|full | Low-quality |
| KM-final-viral-combined_k141_148801\|\|full | Low-quality |
| KM-final-viral-combined_k141_116382\|\|full | Not-determined |
| KM-final-viral-combined_k141_170774\|\|full | Not-determined |
| KM-final-viral-combined_k141_182496\|\|full | Low-quality |
| KM-final-viral-combined_k141_92121\|\|full | Low-quality |
| KM-final-viral-combined_k141_226828\|\|full | Not-determined |
| KM-final-viral-combined_k141_92132\|\|full | Not-determined |
| KM-final-viral-combined_k141_148817\|\|full | Not-determined |
| KM-final-viral-combined_k141_116411\|\|full | Low-quality |
| KM-final-viral-combined_k141_104013\|\|full | Not-determined |
| KM-final-viral-combined_k141_126659\|\|full | Low-quality |
| KM-final-viral-combined_k141_148821\|\|full | Low-quality |
| KM-final-viral-combined_k141_170786\|\|full | Low-quality |
| KM-final-viral-combined_k141_116420\|\|full | Low-quality |
| KM-final-viral-combined_k141_104028\|\|full | Low-quality |
| KM-final-viral-combined_k141_204594\|\|full | Low-quality |
| KM-final-viral-combined_k141_170812\|\|full | Low-quality |
| KM-final-viral-combined_k141_37291\|\|full | Low-quality |
| KM-final-viral-combined_k141_215929\|\|full | Low-quality |
| KM-final-viral-combined_k141_116432\|\|full | Not-determined |
| KM-final-viral-combined_k141_71221\|\|full | Not-determined |
| KM-final-viral-combined_k141_226883\|\|full | Low-quality |
| KM-final-viral-combined_k141_170828\|\|full | Low-quality |
| KM-final-viral-combined_k141_159034\|\|full | Low-quality |
| KM-final-viral-combined_k141_126688\|\|full | Not-determined |
| KM-final-viral-combined_k141_3934\|\|full | Not-determined |
| KM-final-viral-combined_k141_82687\|\|full | Not-determined |
| KM-final-viral-combined_k141_82688\|\|full | Low-quality |
| KM-final-viral-combined_k141_148866\|\|full | Low-quality |
| KM-final-viral-combined_k141_126713\|\|full | Not-determined |
| KM-final-viral-combined_k141_170860\|\|full | Not-determined |
| KM-final-viral-combined_k141_170865\|\|full | Not-determined |
| KM-final-viral-combined_k141_104087\|\|full | Low-quality |
| KM-final-viral-combined_k141_237336\|\|full | Not-determined |
| KM-final-viral-combined_k141_226917\|\|full | Not-determined |
| KM-final-viral-combined_k141_37312\|\|full | Not-determined |
| KM-final-viral-combined_k141_182543\|\|full | Not-determined |
| KM-final-viral-combined_k141_71269\|\|full | Low-quality |
| KM-final-viral-combined_k141_104098\|\|full | Not-determined |
| KM-final-viral-combined_k141_226928\|\|full | Not-determined |
| KM-final-viral-combined_k141_82737\|\|full | Not-determined |
| KM-final-viral-combined_k141_104099\|\|full | Low-quality |
| KM-final-viral-combined_k141_14504\|\|full | Not-determined |
| KM-final-viral-combined_k141_37316\|\|full | Low-quality |
| KM-final-viral-combined_k141_226930\|\|full | Not-determined |
| KM-final-viral-combined_k141_104108\|\|full | Not-determined |
| KM-final-viral-combined_k141_226938\|\|full | Low-quality |
| KM-final-viral-combined_k141_247962\|\|full | Not-determined |
| KM-final-viral-combined_k141_82754\|\|full | Not-determined |
| KM-final-viral-combined_k141_148886\|\|full | Not-determined |
| KM-final-viral-combined_k141_71281\|\|full | Low-quality |
| KM-final-viral-combined_k141_215974\|\|full | Low-quality |
| KM-final-viral-combined_k141_215981\|\|full | Low-quality |
| KM-final-viral-combined_k141_137797\|\|full | Not-determined |
| KM-final-viral-combined_k141_82763\|\|full | Not-determined |
| KM-final-viral-combined_k141_37360\|\|full | Not-determined |
| KM-final-viral-combined_k141_137802\|\|full | Not-determined |
| KM-final-viral-combined_k141_104131\|\|full | Not-determined |
| KM-final-viral-combined_k141_82779\|\|full | Not-determined |
| KM-final-viral-combined_k141_204674\|\|full | Low-quality |
| KM-final-viral-combined_k141_59476\|\|full | Low-quality |
| KM-final-viral-combined_k141_104156\|\|full | Not-determined |
| KM-final-viral-combined_k141_216010\|\|full | Not-determined |
| KM-final-viral-combined_k141_237357\|\|full | Low-quality |
| KM-final-viral-combined_k141_182580\|\|full | Not-determined |
| KM-final-viral-combined_k141_237359\|\|full | Not-determined |
| KM-final-viral-combined_k141_116518\|\|full | Not-determined |
| KM-final-viral-combined_k141_259221\|\|full | Low-quality |
| KM-final-viral-combined_k141_71332\|\|full | Not-determined |
| KM-final-viral-combined_k141_14536\|\|full | Low-quality |
| KM-final-viral-combined_k141_59498\|\|full | Low-quality |
| KM-final-viral-combined_k141_82797\|\|full | Not-determined |
| KM-final-viral-combined_k141_92164\|\|full | Not-determined |
| KM-final-viral-combined_k141_59509\|\|full | Low-quality |
| KM-final-viral-combined_k141_4013\|\|full | Low-quality |
| KM-final-viral-combined_k141_116546\|\|full | Low-quality |
| KM-final-viral-combined_k141_216051\|\|full | Not-determined |
| KM-final-viral-combined_k141_26175\|\|full | Low-quality |
| KM-final-viral-combined_k141_26176\|\|full | Not-determined |
| KM-final-viral-combined_k141_92180\|\|full | Low-quality |
| KM-final-viral-combined_k141_14555\|\|full | Low-quality |
| KM-final-viral-combined_k141_4022\|\|full | Not-determined |
| KM-final-viral-combined_k141_182610\|\|full | Low-quality |
| KM-final-viral-combined_k141_47191\|\|full | Not-determined |
| KM-final-viral-combined_k141_237391\|\|full | Not-determined |
| KM-final-viral-combined_k141_4026\|\|full | Not-determined |
| KM-final-viral-combined_k141_227002\|\|full | Low-quality |
| KM-final-viral-combined_k141_247968\|\|full | Not-determined |
| KM-final-viral-combined_k141_182613\|\|full | Not-determined |
| KM-final-viral-combined_k141_182615\|\|full | Low-quality |
| KM-final-viral-combined_k141_204734\|\|full | Not-determined |
| KM-final-viral-combined_k141_227022\|\|full | Low-quality |
| KM-final-viral-combined_k141_182626\|\|full | Low-quality |
| KM-final-viral-combined_k141_237416\|\|full | Not-determined |
| KM-final-viral-combined_k141_227028\|\|full | Low-quality |
| KM-final-viral-combined_k141_237417\|\|full | Not-determined |
| KM-final-viral-combined_k141_204759\|\|full | Not-determined |
| KM-final-viral-combined_k141_92220\|\|full | Low-quality |
| KM-final-viral-combined_k141_4069\|\|full | Low-quality |
| KM-final-viral-combined_k141_159143\|\|full | Low-quality |
| KM-final-viral-combined_k141_92222\|\|full | Low-quality |
| KM-final-viral-combined_k141_193598\|\|full | Low-quality |
| KM-final-viral-combined_k141_182649\|\|full | Not-determined |
| 18-101-final-viral-combined_k141_332194\|\|full | Medium-quality |
| KM-final-viral-combined_k141_104233\|\|full | Not-determined |
| KM-final-viral-combined_k141_47244\|\|full | Low-quality |
| 18-104-final-viral-combined_k141_42647\|\|full | High-quality |
| KM-final-viral-combined_k141_26240\|\|full | Low-quality |
| KM-final-viral-combined_k141_14570\|\|full | Low-quality |
| KM-final-viral-combined_k141_237447\|\|full | Not-determined |
| KM-final-viral-combined_k141_193620\|\|full | Not-determined |
| KM-final-viral-combined_k141_126782\|\|full | Not-determined |
| KM-final-viral-combined_k141_116621\|\|full | Not-determined |
| KM-final-viral-combined_k141_104243\|\|full | Low-quality |
| KM-final-viral-combined_k141_14571\|\|full | Low-quality |
| KM-final-viral-combined_k141_14576\|\|full | Not-determined |
| KM-final-viral-combined_k141_116637\|\|full | Not-determined |
| KM-final-viral-combined_k141_14579\|\|full | Low-quality |
| KM-final-viral-combined_k141_104256\|\|full | Low-quality |
| KM-final-viral-combined_k141_116641\|\|full | Not-determined |
| KM-final-viral-combined_k141_26244\|\|full | Low-quality |
| KM-final-viral-combined_k141_259278\|\|full | Low-quality |
| KM-final-viral-combined_k141_237474\|\|full | Not-determined |
| KM-final-viral-combined_k141_26252\|\|full | Low-quality |
| KM-final-viral-combined_k141_259285\|\|full | Low-quality |
| KM-final-viral-combined_k141_82930\|\|full | Low-quality |
| KM-final-viral-combined_k141_126820\|\|full | Low-quality |
| KM-final-viral-combined_k141_47281\|\|full | Low-quality |
| KM-final-viral-combined_k141_216111\|\|full | Low-quality |
| KM-final-viral-combined_k141_104266\|\|full | Low-quality |
| KM-final-viral-combined_k141_126822\|\|full | Not-determined |
| KM-final-viral-combined_k141_182681\|\|full | Low-quality |
| KM-final-viral-combined_k141_248042\|\|full | Not-determined |
| KM-final-viral-combined_k141_116675\|\|full | Low-quality |
| KM-final-viral-combined_k141_227101\|\|full | Low-quality |
| KM-final-viral-combined_k141_82947\|\|full | Not-determined |
| KM-final-viral-combined_k141_104279\|\|full | Not-determined |
| KM-final-viral-combined_k141_171025\|\|full | Not-determined |
| KM-final-viral-combined_k141_204793\|\|full | Not-determined |
| KM-final-viral-combined_k141_4155\|\|full | Not-determined |
| KM-final-viral-combined_k141_59556\|\|full | Low-quality |
| KM-final-viral-combined_k141_82965\|\|full | Low-quality |
| KM-final-viral-combined_k141_82966\|\|full | Not-determined |
| KM-final-viral-combined_k141_47304\|\|full | Not-determined |
| KM-final-viral-combined_k141_237531\|\|full | Low-quality |
| KM-final-viral-combined_k141_216147\|\|full | Low-quality |
| KM-final-viral-combined_k141_237547\|\|full | Low-quality |
| KM-final-viral-combined_k141_92287\|\|full | Low-quality |
| KM-final-viral-combined_k141_204819\|\|full | Low-quality |
| KM-final-viral-combined_k141_26300\|\|full | Low-quality |
| KM-final-viral-combined_k141_148946\|\|full | Low-quality |
| KM-final-viral-combined_k141_204836\|\|full | Not-determined |
| KM-final-viral-combined_k141_4196\|\|full | Low-quality |
| KM-final-viral-combined_k141_83019\|\|full | Not-determined |
| KM-final-viral-combined_k141_71414\|\|full | Low-quality |
| KM-final-viral-combined_k141_26334\|\|full | Not-determined |
| KM-final-viral-combined_k141_83030\|\|full | Not-determined |
| KM-final-viral-combined_k141_26337\|\|full | Low-quality |
| KM-final-viral-combined_k141_104313\|\|full | Not-determined |
| KM-final-viral-combined_k141_59593\|\|full | Not-determined |
| KM-final-viral-combined_k141_4214\|\|full | Not-determined |
| KM-final-viral-combined_k141_37497\|\|full | Low-quality |
| KM-final-viral-combined_k141_204860\|\|full | Low-quality |
| KM-final-viral-combined_k141_237582\|\|full | Not-determined |
| KM-final-viral-combined_k141_37510\|\|full | Low-quality |
| KM-final-viral-combined_k141_26353\|\|full | Low-quality |
| KM-final-viral-combined_k141_248109\|\|full | Not-determined |
| KM-final-viral-combined_k141_193695\|\|full | Not-determined |
| KM-final-viral-combined_k141_14661\|\|full | Not-determined |
| KM-final-viral-combined_k141_83051\|\|full | Low-quality |
| KM-final-viral-combined_k141_204869\|\|full | Not-determined |
| KM-final-viral-combined_k141_248123\|\|full | Low-quality |
| KM-final-viral-combined_k141_237606\|\|full | Not-determined |
| KM-final-viral-combined_k141_14682\|\|full | Not-determined |
| KM-final-viral-combined_k141_159220\|\|full | Not-determined |
| KM-final-viral-combined_k141_26368\|\|full | Low-quality |
| KM-final-viral-combined_k141_59669\|\|full | Low-quality |
| KM-final-viral-combined_k141_204885\|\|full | Low-quality |
| KM-final-viral-combined_k141_116772\|\|full | Not-determined |
| KM-final-viral-combined_k141_171058\|\|full | Not-determined |
| KM-final-viral-combined_k141_26376\|\|full | Low-quality |
| KM-final-viral-combined_k141_204889\|\|full | Not-determined |
| KM-final-viral-combined_k141_248136\|\|full | Low-quality |
| KM-final-viral-combined_k141_26381\|\|full | Low-quality |
| KM-final-viral-combined_k141_59688\|\|full | Not-determined |
| KM-final-viral-combined_k141_259352\|\|full | Not-determined |
| KM-final-viral-combined_k141_92349\|\|full | Low-quality |
| KM-final-viral-combined_k141_248153\|\|full | Not-determined |
| KM-final-viral-combined_k141_149035\|\|full | Not-determined |
| KM-final-viral-combined_k141_193758\|\|full | Low-quality |
| KM-final-viral-combined_k141_92354\|\|full | Not-determined |
| KM-final-viral-combined_k141_137945\|\|full | Not-determined |
| KM-final-viral-combined_k141_71454\|\|full | Not-determined |
| KM-final-viral-combined_k141_237650\|\|full | Low-quality |
| KM-final-viral-combined_k141_47383\|\|full | Not-determined |
| KM-final-viral-combined_k141_259368\|\|full | Low-quality |
| KM-final-viral-combined_k141_92378\|\|full | Low-quality |
| KM-final-viral-combined_k141_227251\|\|full | Low-quality |
| KM-final-viral-combined_k141_227258\|\|full | Not-determined |
| KM-final-viral-combined_k141_182844\|\|full | Not-determined |
| KM-final-viral-combined_k141_47412\|\|full | Low-quality |
| KM-final-viral-combined_k141_248207\|\|full | Low-quality |
| KM-final-viral-combined_k141_116817\|\|full | Low-quality |
| KM-final-viral-combined_k141_92388\|\|full | Low-quality |
| KM-final-viral-combined_k141_92390\|\|full | Low-quality |
| KM-final-viral-combined_k141_159314\|\|full | Not-determined |
| KM-final-viral-combined_k141_149074\|\|full | Low-quality |
| KM-final-viral-combined_k141_149078\|\|full | Low-quality |
| KM-final-viral-combined_k141_37608\|\|full | Low-quality |
| KM-final-viral-combined_k141_248229\|\|full | Not-determined |
| KM-final-viral-combined_k141_149084\|\|full | Low-quality |
| KM-final-viral-combined_k141_37610\|\|full | Low-quality |
| KM-final-viral-combined_k141_216255\|\|full | Not-determined |
| KM-final-viral-combined_k141_227311\|\|full | Low-quality |
| KM-final-viral-combined_k141_216264\|\|full | Not-determined |
| KM-final-viral-combined_k141_83105\|\|full | Low-quality |
| KM-final-viral-combined_k141_248251\|\|full | Not-determined |
| KM-final-viral-combined_k141_171149\|\|full | Low-quality |
| KM-final-viral-combined_k141_171151\|\|full | Low-quality |
| KM-final-viral-combined_k141_92416\|\|full | Low-quality |
| KM-final-viral-combined_k141_182899\|\|full | Not-determined |
| KM-final-viral-combined_k141_227357\|\|full | Low-quality |
| KM-final-viral-combined_k141_116926\|\|full | Low-quality |
| KM-final-viral-combined_k141_14748\|\|full | Low-quality |
| KM-final-viral-combined_k141_159372\|\|full | Not-determined |
| KM-final-viral-combined_k141_116928\|\|full | Low-quality |
| KM-final-viral-combined_k141_116929\|\|full | Low-quality |
| KM-final-viral-combined_k141_83133\|\|full | Low-quality |
| KM-final-viral-combined_k141_92433\|\|full | Not-determined |
| KM-final-viral-combined_k141_59779\|\|full | Low-quality |
| KM-final-viral-combined_k141_37654\|\|full | Not-determined |
| KM-final-viral-combined_k141_216296\|\|full | Not-determined |
| KM-final-viral-combined_k141_104405\|\|full | Low-quality |
| KM-final-viral-combined_k141_126981\|\|full | Not-determined |
| KM-final-viral-combined_k141_204977\|\|full | Low-quality |
| KM-final-viral-combined_k141_59801\|\|full | Not-determined |
| KM-final-viral-combined_k141_37674\|\|full | Low-quality |
| KM-final-viral-combined_k141_26438\|\|full | Not-determined |
| KM-final-viral-combined_k141_248283\|\|full | Low-quality |
| KM-final-viral-combined_k141_237692\|\|full | Not-determined |
| KM-final-viral-combined_k141_149168\|\|full | Not-determined |
| KM-final-viral-combined_k141_227394\|\|full | Not-determined |
| KM-final-viral-combined_k141_37689\|\|full | Low-quality |
| KM-final-viral-combined_k141_182945\|\|full | Low-quality |
| KM-final-viral-combined_k141_149186\|\|full | Low-quality |
| KM-final-viral-combined_k141_182957\|\|full | Low-quality |
| KM-final-viral-combined_k141_237702\|\|full | Not-determined |
| KM-final-viral-combined_k141_171229\|\|full | Low-quality |
| KM-final-viral-combined_k141_92468\|\|full | Low-quality |
| KM-final-viral-combined_k141_171237\|\|full | Low-quality |
| KM-final-viral-combined_k141_138042\|\|full | Not-determined |
| KM-final-viral-combined_k141_116998\|\|full | Low-quality |
| KM-final-viral-combined_k141_193896\|\|full | Not-determined |
| KM-final-viral-combined_k141_59855\|\|full | Low-quality |
| KM-final-viral-combined_k141_47494\|\|full | Not-determined |
| KM-final-viral-combined_k141_59858\|\|full | Low-quality |
| KM-final-viral-combined_k141_216353\|\|full | Low-quality |
| KM-final-viral-combined_k141_37724\|\|full | Not-determined |
| KM-final-viral-combined_k141_237720\|\|full | Low-quality |
| KM-final-viral-combined_k141_216359\|\|full | Low-quality |
| KM-final-viral-combined_k141_216361\|\|full | Low-quality |
| KM-final-viral-combined_k141_117023\|\|full | Not-determined |
| KM-final-viral-combined_k141_47495\|\|full | Not-determined |
| KM-final-viral-combined_k141_193920\|\|full | Low-quality |
| KM-final-viral-combined_k141_183018\|\|full | Not-determined |
| KM-final-viral-combined_k141_149262\|\|full | Not-determined |
| KM-final-viral-combined_k141_138075\|\|full | Low-quality |
| KM-final-viral-combined_k141_14811\|\|full | Not-determined |
| KM-final-viral-combined_k141_205076\|\|full | Low-quality |
| KM-final-viral-combined_k141_216380\|\|full | Low-quality |
| KM-final-viral-combined_k141_138080\|\|full | Low-quality |
| KM-final-viral-combined_k141_26521\|\|full | Not-determined |
| KM-final-viral-combined_k141_127022\|\|full | Not-determined |
| KM-final-viral-combined_k141_227491\|\|full | Not-determined |
| KM-final-viral-combined_k141_47522\|\|full | Not-determined |
| KM-final-viral-combined_k141_149286\|\|full | Low-quality |
| KM-final-viral-combined_k141_227518\|\|full | Not-determined |
| KM-final-viral-combined_k141_138106\|\|full | Low-quality |
| KM-final-viral-combined_k141_205102\|\|full | Low-quality |
| KM-final-viral-combined_k141_117088\|\|full | Not-determined |
| KM-final-viral-combined_k141_259395\|\|full | Low-quality |
| KM-final-viral-combined_k141_4313\|\|full | Low-quality |
| KM-final-viral-combined_k141_71523\|\|full | Not-determined |
| KM-final-viral-combined_k141_83278\|\|full | Not-determined |
| KM-final-viral-combined_k141_237757\|\|full | Low-quality |
| KM-final-viral-combined_k141_14838\|\|full | Low-quality |
| KM-final-viral-combined_k141_259398\|\|full | Low-quality |
| KM-final-viral-combined_k141_138120\|\|full | Low-quality |
| KM-final-viral-combined_k141_138139\|\|full | Low-quality |
| KM-final-viral-combined_k141_205139\|\|full | Not-determined |
| KM-final-viral-combined_k141_216389\|\|full | Low-quality |
| KM-final-viral-combined_k141_183064\|\|full | Not-determined |
| KM-final-viral-combined_k141_205144\|\|full | Not-determined |
| KM-final-viral-combined_k141_104478\|\|full | Low-quality |
| KM-final-viral-combined_k141_71565\|\|full | Not-determined |
| KM-final-viral-combined_k141_183065\|\|full | Low-quality |
| KM-final-viral-combined_k141_149322\|\|full | Not-determined |
| KM-final-viral-combined_k141_4350\|\|full | Not-determined |
| KM-final-viral-combined_k141_183071\|\|full | Low-quality |
| KM-final-viral-combined_k141_237785\|\|full | Low-quality |
| KM-final-viral-combined_k141_216413\|\|full | Low-quality |
| KM-final-viral-combined_k141_92559\|\|full | Not-determined |
| KM-final-viral-combined_k141_205172\|\|full | Not-determined |
| KM-final-viral-combined_k141_216415\|\|full | Not-determined |
| KM-final-viral-combined_k141_4369\|\|full | Low-quality |
| KM-final-viral-combined_k141_83307\|\|full | Low-quality |
| KM-final-viral-combined_k141_104518\|\|full | Not-determined |
| KM-final-viral-combined_k141_216420\|\|full | Low-quality |
| KM-final-viral-combined_k141_92565\|\|full | Low-quality |
| KM-final-viral-combined_k141_83321\|\|full | Low-quality |
| KM-final-viral-combined_k141_227579\|\|full | Not-determined |
| KM-final-viral-combined_k141_47580\|\|full | Low-quality |
| KM-final-viral-combined_k141_83329\|\|full | Low-quality |
| 17-108_k141_3116 | Medium-quality |
| KM-final-viral-combined_k141_14867\|\|full | Not-determined |
| KM-final-viral-combined_k141_183098\|\|full | Low-quality |
| KM-final-viral-combined_k141_104535\|\|full | Low-quality |
| 17-108-final-viral-combined_k141_3116\|\|full | Medium-quality |
| KM-final-viral-combined_k141_216449\|\|full | Low-quality |
| KM-final-viral-combined_k141_149353\|\|full | Low-quality |
| KM-final-viral-combined_k141_171331\|\|full | Not-determined |
| KM-final-viral-combined_k141_127058\|\|full | Not-determined |
| KM-final-viral-combined_k141_171333\|\|full | Low-quality |
| KM-final-viral-combined_k141_227634\|\|full | Low-quality |
| KM-final-viral-combined_k141_205184\|\|full | Not-determined |
| KM-final-viral-combined_k141_59972\|\|full | Low-quality |
| KM-final-viral-combined_k141_248340\|\|full | Not-determined |
| KM-final-viral-combined_k141_227640\|\|full | Low-quality |
| KM-final-viral-combined_k141_248347\|\|full | Not-determined |
| KM-final-viral-combined_k141_227643\|\|full | Not-determined |
| KM-final-viral-combined_k141_37767\|\|full | Low-quality |
| KM-final-viral-combined_k141_37775\|\|full | Low-quality |
| KM-final-viral-combined_k141_83359\|\|full | Low-quality |
| KM-final-viral-combined_k141_37786\|\|full | Low-quality |
| KM-final-viral-combined_k141_117244\|\|full | Low-quality |
| KM-final-viral-combined_k141_26668\|\|full | Not-determined |
| KM-final-viral-combined_k141_37798\|\|full | Low-quality |
| KM-final-viral-combined_k141_183188\|\|full | Low-quality |
| KM-final-viral-combined_k141_14934\|\|full | Not-determined |
| KM-final-viral-combined_k141_104590\|\|full | Low-quality |
| KM-final-viral-combined_k141_127114\|\|full | Low-quality |
| KM-final-viral-combined_k141_205240\|\|full | Low-quality |
| KM-final-viral-combined_k141_83400\|\|full | Low-quality |
| KM-final-viral-combined_k141_37815\|\|full | Not-determined |
| KM-final-viral-combined_k141_104601\|\|full | Not-determined |
| 17-108-final-viral-combined_k141_200084\|\|full | Medium-quality |
| KM-final-viral-combined_k141_83418\|\|full | Not-determined |
| KM-final-viral-combined_k141_205271\|\|full | Not-determined |
| KM-final-viral-combined_k141_37840\|\|full | Not-determined |
| KM-final-viral-combined_k141_60011\|\|full | Low-quality |
| KM-final-viral-combined_k141_127154\|\|full | Not-determined |
| KM-final-viral-combined_k141_159577\|\|full | Low-quality |
| KM-final-viral-combined_k141_216543\|\|full | Not-determined |
| KM-final-viral-combined_k141_127160\|\|full | Low-quality |
| KM-final-viral-combined_k141_216549\|\|full | Not-determined |
| KM-final-viral-combined_k141_183235\|\|full | Not-determined |
| KM-final-viral-combined_k141_171381\|\|full | Not-determined |
| KM-final-viral-combined_k141_183237\|\|full | Not-determined |
| KM-final-viral-combined_k141_104641\|\|full | Not-determined |
| KM-final-viral-combined_k141_26750\|\|full | Not-determined |
| KM-final-viral-combined_k141_71684\|\|full | Not-determined |
| KM-final-viral-combined_k141_37889\|\|full | Low-quality |
| KM-final-viral-combined_k141_37890\|\|full | Low-quality |
| KM-final-viral-combined_k141_71690\|\|full | Not-determined |
| KM-final-viral-combined_k141_47654\|\|full | Low-quality |
| KM-final-viral-combined_k141_83470\|\|full | Not-determined |
| KM-final-viral-combined_k141_71694\|\|full | Low-quality |
| KM-final-viral-combined_k141_4450\|\|full | Not-determined |
| KM-final-viral-combined_k141_205319\|\|full | Not-determined |
| KM-final-viral-combined_k141_227749\|\|full | Low-quality |
| KM-final-viral-combined_k141_71697\|\|full | Low-quality |
| KM-final-viral-combined_k141_248446\|\|full | Low-quality |
| KM-final-viral-combined_k141_216594\|\|full | Not-determined |
| KM-final-viral-combined_k141_175924\|\|full | High-quality |
| KM-final-viral-combined_k141_4470\|\|full | Not-determined |
| KM-final-viral-combined_k141_71725\|\|full | Not-determined |
| KM-final-viral-combined_k141_193978\|\|full | Low-quality |
| KM-final-viral-combined_k141_83505\|\|full | Low-quality |
| KM-final-viral-combined_k141_47675\|\|full | Low-quality |
| KM-final-viral-combined_k141_83516\|\|full | Low-quality |
| KM-final-viral-combined_k141_159685\|\|full | Not-determined |
| KM-final-viral-combined_k141_92665\|\|full | Not-determined |
| KM-final-viral-combined_k141_37908\|\|full | Not-determined |
| KM-final-viral-combined_k141_104648\|\|full | Low-quality |
| KM-final-viral-combined_k141_216630\|\|full | Not-determined |
| KM-final-viral-combined_k141_237856\|\|full | Not-determined |
| KM-final-viral-combined_k141_92668\|\|full | Low-quality |
| KM-final-viral-combined_k141_37910\|\|full | Low-quality |
| KM-final-viral-combined_k141_159695\|\|full | Not-determined |
| KM-final-viral-combined_k141_104657\|\|full | Not-determined |
| KM-final-viral-combined_k141_14950\|\|full | Low-quality |
| KM-final-viral-combined_k141_159706\|\|full | Not-determined |
| KM-final-viral-combined_k141_183309\|\|full | Low-quality |
| KM-final-viral-combined_k141_83549\|\|full | Low-quality |
| KM-final-viral-combined_k141_149468\|\|full | Low-quality |
| KM-final-viral-combined_k141_104679\|\|full | Not-determined |
| KM-final-viral-combined_k141_227813\|\|full | Not-determined |
| KM-final-viral-combined_k141_37962\|\|full | Low-quality |
| KM-final-viral-combined_k141_149479\|\|full | Not-determined |
| KM-final-viral-combined_k141_138390\|\|full | Low-quality |
| KM-final-viral-combined_k141_138393\|\|full | Not-determined |
| KM-final-viral-combined_k141_104691\|\|full | Not-determined |
| KM-final-viral-combined_k141_4514\|\|full | Low-quality |
| KM-final-viral-combined_k141_37969\|\|full | Not-determined |
| KM-final-viral-combined_k141_117339\|\|full | Low-quality |
| KM-final-viral-combined_k141_47697\|\|full | Not-determined |
| KM-final-viral-combined_k141_4521\|\|full | Low-quality |
| KM-final-viral-combined_k141_227825\|\|full | Not-determined |
| KM-final-viral-combined_k141_71761\|\|full | Not-determined |
| KM-final-viral-combined_k141_259627\|\|full | Low-quality |
| KM-final-viral-combined_k141_248504\|\|full | Low-quality |
| KM-final-viral-combined_k141_127300\|\|full | Low-quality |
| KM-final-viral-combined_k141_92726\|\|full | Low-quality |
| KM-final-viral-combined_k141_159738\|\|full | Not-determined |
| KM-final-viral-combined_k141_205434\|\|full | Not-determined |
| KM-final-viral-combined_k141_4550\|\|full | Not-determined |
| KM-final-viral-combined_k141_37999\|\|full | Not-determined |
| KM-final-viral-combined_k141_138428\|\|full | Low-quality |
| KM-final-viral-combined_k141_92730\|\|full | Low-quality |
| KM-final-viral-combined_k141_71783\|\|full | Not-determined |
| KM-final-viral-combined_k141_26863\|\|full | Not-determined |
| KM-final-viral-combined_k141_26866\|\|full | Not-determined |
| KM-final-viral-combined_k141_248546\|\|full | Not-determined |
| KM-final-viral-combined_k141_47737\|\|full | Not-determined |
| KM-final-viral-combined_k141_171582\|\|full | Low-quality |
| KM-final-viral-combined_k141_183386\|\|full | Not-determined |
| KM-final-viral-combined_k141_4582\|\|full | Low-quality |
| KM-final-viral-combined_k141_149544\|\|full | Not-determined |
| KM-final-viral-combined_k141_4592\|\|full | Low-quality |
| KM-final-viral-combined_k141_127364\|\|full | Not-determined |
| KM-final-viral-combined_k141_83579\|\|full | Low-quality |
| KM-final-viral-combined_k141_117412\|\|full | Low-quality |
| KM-final-viral-combined_k141_183426\|\|full | Low-quality |
| KM-final-viral-combined_k141_83583\|\|full | Not-determined |
| KM-final-viral-combined_k141_127400\|\|full | Low-quality |
| KM-final-viral-combined_k141_194040\|\|full | Not-determined |
| KM-final-viral-combined_k141_127402\|\|full | Low-quality |
| KM-final-viral-combined_k141_83597\|\|full | Not-determined |
| KM-final-viral-combined_k141_71843\|\|full | Low-quality |
| KM-final-viral-combined_k141_205462\|\|full | Not-determined |
| KM-final-viral-combined_k141_205466\|\|full | Low-quality |

| | |
|---|---|
| KM-final-viral-combined_k141_127404||full | Not-determined |
| KM-final-viral-combined_k141_259683||full | Low-quality |
| KM-final-viral-combined_k141_149555||full | Not-determined |
| KM-final-viral-combined_k141_60168||full | Low-quality |
| KM-final-viral-combined_k141_83623||full | Low-quality |
| KM-final-viral-combined_k141_194071||full | Not-determined |
| KM-final-viral-combined_k141_47801||full | Not-determined |
| KM-final-viral-combined_k141_83624||full | Not-determined |
| KM-final-viral-combined_k141_227935||full | Not-determined |
| KM-final-viral-combined_k141_15104||full | Not-determined |
| KM-final-viral-combined_k141_171645||full | Low-quality |
| KM-final-viral-combined_k141_171657||full | Low-quality |
| KM-final-viral-combined_k141_183473||full | Not-determined |
| KM-final-viral-combined_k141_237971||full | Low-quality |
| KM-final-viral-combined_k141_15132||full | Low-quality |
| KM-final-viral-combined_k141_47820||full | Not-determined |
| KM-final-viral-combined_k141_149607||full | Not-determined |
| KM-final-viral-combined_k141_47824||full | Not-determined |
| KM-final-viral-combined_k141_183483||full | Low-quality |
| KM-final-viral-combined_k141_227962||full | Not-determined |
| KM-final-viral-combined_k141_237988||full | Not-determined |
| KM-final-viral-combined_k141_237991||full | Low-quality |
| KM-final-viral-combined_k141_227974||full | Not-determined |
| KM-final-viral-combined_k141_159848||full | Not-determined |
| KM-final-viral-combined_k141_60246||full | Low-quality |
| KM-final-viral-combined_k141_248634||full | Low-quality |
| KM-final-viral-combined_k141_194178||full | Not-determined |
| KM-final-viral-combined_k141_127503||full | Not-determined |
| KM-final-viral-combined_k141_227999||full | Low-quality |
| KM-final-viral-combined_k141_4710||full | Low-quality |
| KM-final-viral-combined_k141_15164||full | Low-quality |
| KM-final-viral-combined_k141_92788||full | Not-determined |
| KM-final-viral-combined_k141_138583||full | Low-quality |
| KM-final-viral-combined_k141_26999||full | Low-quality |
| KM-final-viral-combined_k141_117446||full | Not-determined |
| KM-final-viral-combined_k141_83718||full | Not-determined |
| KM-final-viral-combined_k141_159885||full | Low-quality |
| KM-final-viral-combined_k141_259693||full | Not-determined |
| KM-final-viral-combined_k141_127505||full | Low-quality |
| KM-final-viral-combined_k141_248661||full | Low-quality |
| KM-final-viral-combined_k141_38130||full | Not-determined |
| KM-final-viral-combined_k141_194216||full | Low-quality |
| KM-final-viral-combined_k141_248665||full | Low-quality |
| KM-final-viral-combined_k141_92804||full | Low-quality |
| KM-final-viral-combined_k141_83744||full | Low-quality |
| KM-final-viral-combined_k141_171744||full | Not-determined |
| KM-final-viral-combined_k141_171746||full | Not-determined |
| KM-final-viral-combined_k141_104906||full | Not-determined |
| KM-final-viral-combined_k141_171751||full | Not-determined |
| KM-final-viral-combined_k141_194242||full | Not-determined |
| KM-final-viral-combined_k141_238038||full | Low-quality |
| KM-final-viral-combined_k141_194250||full | Not-determined |
| KM-final-viral-combined_k141_83747||full | Low-quality |
| KM-final-viral-combined_k141_127551||full | Not-determined |
| KM-final-viral-combined_k141_183580||full | Not-determined |
| KM-final-viral-combined_k141_38158||full | Low-quality |
| KM-final-viral-combined_k141_194262||full | Not-determined |
| KM-final-viral-combined_k141_149684||full | Low-quality |
| KM-final-viral-combined_k141_194264||full | Not-determined |
| 18-104-final-viral-combined_k141_83492||full | Medium-quality |
| KM-final-viral-combined_k141_38166||full | Low-quality |
| KM-final-viral-combined_k141_71998||full | Low-quality |
| KM-final-viral-combined_k141_83764||full | Not-determined |
| KM-final-viral-combined_k141_104931||full | Low-quality |
| KM-final-viral-combined_k141_194274||full | Low-quality |
| KM-final-viral-combined_k141_127564||full | Not-determined |
| KM-final-viral-combined_k141_92810||full | Not-determined |
| KM-final-viral-combined_k141_83792||full | Low-quality |
| KM-final-viral-combined_k141_248699||full | Not-determined |
| KM-final-viral-combined_k141_194299||full | Not-determined |
| KM-final-viral-combined_k141_38196||full | Low-quality |
| KM-final-viral-combined_k141_83797||full | Not-determined |
| KM-final-viral-combined_k141_216884||full | Not-determined |
| KM-final-viral-combined_k141_228015||full | Not-determined |
| KM-final-viral-combined_k141_92819||full | Not-determined |
| KM-final-viral-combined_k141_117536||full | Low-quality |
| KM-final-viral-combined_k141_205635||full | Not-determined |
| KM-final-viral-combined_k141_138690||full | Low-quality |
| KM-final-viral-combined_k141_38219||full | Low-quality |
| KM-final-viral-combined_k141_205645||full | Low-quality |
| KM-final-viral-combined_k141_127600||full | Not-determined |
| KM-final-viral-combined_k141_159924||full | Not-determined |
| KM-final-viral-combined_k141_248708||full | Not-determined |
| 17-109-final-viral-combined_k141_320262||full | Medium-quality |
| KM-final-viral-combined_k141_47862||full | Low-quality |
| KM-final-viral-combined_k141_92852||full | Low-quality |
| KM-final-viral-combined_k141_38239||full | Not-determined |
| KM-final-viral-combined_k141_47873||full | Low-quality |
| KM-final-viral-combined_k141_228060||full | Not-determined |
| KM-final-viral-combined_k141_38250||full | Low-quality |
| KM-final-viral-combined_k141_138744||full | Low-quality |
| KM-final-viral-combined_k141_248732||full | Low-quality |
| KM-final-viral-combined_k141_183693||full | Not-determined |
| KM-final-viral-combined_k141_127625||full | Low-quality |
| KM-final-viral-combined_k141_149755||full | Not-determined |
| KM-final-viral-combined_k141_127628||full | Not-determined |
| KM-final-viral-combined_k141_60339||full | Not-determined |
| KM-final-viral-combined_k141_38277||full | Low-quality |
| KM-final-viral-combined_k141_72087||full | Low-quality |
| KM-final-viral-combined_k141_228107||full | Low-quality |
| KM-final-viral-combined_k141_238172||full | Low-quality |
| KM-final-viral-combined_k141_248761||full | Not-determined |
| KM-final-viral-combined_k141_47899||full | Not-determined |
| KM-final-viral-combined_k141_149789||full | Not-determined |
| KM-final-viral-combined_k141_15291||full | Not-determined |
| KM-final-viral-combined_k141_160006||full | Low-quality |
| KM-final-viral-combined_k141_205710||full | Low-quality |
| KM-final-viral-combined_k141_194404||full | Low-quality |
| KM-final-viral-combined_k141_149802||full | Not-determined |
| KM-final-viral-combined_k141_15297||full | Not-determined |
| KM-final-viral-combined_k141_205725||full | Not-determined |
| KM-final-viral-combined_k141_15299||full | Not-determined |
| KM-final-viral-combined_k141_47928||full | Low-quality |
| KM-final-viral-combined_k141_60400||full | Not-determined |
| KM-final-viral-combined_k141_38308||full | Not-determined |
| KM-final-viral-combined_k141_138806||full | Not-determined |
| KM-final-viral-combined_k141_4818||full | Not-determined |
| KM-final-viral-combined_k141_216965||full | Low-quality |
| KM-final-viral-combined_k141_160051||full | Not-determined |
| 18-104_k141_118690 | Medium-quality |
| KM-final-viral-combined_k141_92888||full | Not-determined |
| KM-final-viral-combined_k141_228178||full | Not-determined |
| KM-final-viral-combined_k141_149839||full | Low-quality |
| KM-final-viral-combined_k141_171932||full | Low-quality |
| KM_k141_116248 | High-quality |
| KM-final-viral-combined_k141_171938||full | Low-quality |
| KM-final-viral-combined_k141_216990||full | Low-quality |
| KM-final-viral-combined_k141_171939||full | Low-quality |
| KM-final-viral-combined_k141_83946||full | Low-quality |
| KM-final-viral-combined_k141_117684||full | Low-quality |
| KM-final-viral-combined_k141_105096||full | Low-quality |
| KM-final-viral-combined_k141_47972||full | Not-determined |
| KM-final-viral-combined_k141_183798||full | Low-quality |
| KM-final-viral-combined_k141_259837||full | Low-quality |
| KM-final-viral-combined_k141_171967||full | Low-quality |
| KM-final-viral-combined_k141_27172||full | Low-quality |
| KM-final-viral-combined_k141_83981||full | Low-quality |
| KM-final-viral-combined_k141_60519||full | Low-quality |
| KM-final-viral-combined_k141_83987||full | Low-quality |
| KM-final-viral-combined_k141_60524||full | Not-determined |
| KM-final-viral-combined_k141_117724||full | Not-determined |
| KM-final-viral-combined_k141_228216||full | Not-determined |
| KM-final-viral-combined_k141_160115||full | Low-quality |
| KM-final-viral-combined_k141_194497||full | Not-determined |
| KM-final-viral-combined_k141_84004||full | Not-determined |
| KM-final-viral-combined_k141_72163||full | Not-determined |
| KM-final-viral-combined_k141_248903||full | Low-quality |
| KM-final-viral-combined_k141_160122||full | Low-quality |
| KM-final-viral-combined_k141_117737||full | Not-determined |
| KM-final-viral-combined_k141_259851||full | Not-determined |
| KM-final-viral-combined_k141_72173||full | Low-quality |
| KM-final-viral-combined_k141_48012||full | Low-quality |
| KM-final-viral-combined_k141_194506||full | Not-determined |
| KM-final-viral-combined_k141_48016||full | Not-determined |
| KM-final-viral-combined_k141_238277||full | Not-determined |
| KM-final-viral-combined_k141_60551||full | Not-determined |
| KM-final-viral-combined_k141_127722||full | Not-determined |
| KM-final-viral-combined_k141_149913||full | Low-quality |
| KM-final-viral-combined_k141_149916||full | Not-determined |
| KM-final-viral-combined_k141_4901||full | Not-determined |
| KM-final-viral-combined_k141_228287||full | Low-quality |
| KM-final-viral-combined_k141_205864||full | Not-determined |
| KM-final-viral-combined_k141_48029||full | Low-quality |
| KM-final-viral-combined_k141_105192||full | Low-quality |
| KM-final-viral-combined_k141_238287||full | Not-determined |
| KM-final-viral-combined_k141_84074||full | Not-determined |
| KM-final-viral-combined_k141_15351||full | Not-determined |
| KM-final-viral-combined_k141_149930||full | Low-quality |
| KM-final-viral-combined_k141_138877||full | Low-quality |
| KM-final-viral-combined_k141_27200||full | Low-quality |
| KM-final-viral-combined_k141_27209||full | Low-quality |
| KM-final-viral-combined_k141_27212||full | Low-quality |
| KM-final-viral-combined_k141_160159||full | Not-determined |
| KM-final-viral-combined_k141_248957||full | Not-determined |
| KM-final-viral-combined_k141_138899||full | Low-quality |
| KM-final-viral-combined_k141_4934||full | Not-determined |
| KM-final-viral-combined_k141_117795||full | Not-determined |
| KM-final-viral-combined_k141_27220||full | Low-quality |
| KM-final-viral-combined_k141_160175||full | Low-quality |
| KM-final-viral-combined_k141_149959||full | Not-determined |
| KM-final-viral-combined_k141_72250||full | Not-determined |
| KM-final-viral-combined_k141_38389||full | Not-determined |
| KM-final-viral-combined_k141_217130||full | Not-determined |
| KM-final-viral-combined_k141_160188||full | Low-quality |
| KM-final-viral-combined_k141_84126||full | Low-quality |
| KM-final-viral-combined_k141_84130||full | Low-quality |
| KM-final-viral-combined_k141_84134||full | Low-quality |
| KM-final-viral-combined_k141_194626||full | Not-determined |
| KM-final-viral-combined_k141_15401||full | Low-quality |
| KM-final-viral-combined_k141_105250||full | Not-determined |
| KM-final-viral-combined_k141_160214||full | Not-determined |
| KM-final-viral-combined_k141_127772||full | Not-determined |
| KM-final-viral-combined_k141_259947||full | Low-quality |
| KM-final-viral-combined_k141_60637||full | Not-determined |
| KM-final-viral-combined_k141_160226||full | Low-quality |
| KM-final-viral-combined_k141_27262||full | Low-quality |
| KM-final-viral-combined_k141_194637||full | Not-determined |
| KM-final-viral-combined_k141_228356||full | Not-determined |
| KM-final-viral-combined_k141_228362||full | Low-quality |
| KM-final-viral-combined_k141_72296||full | Not-determined |
| KM-final-viral-combined_k141_194653||full | Not-determined |
| KM-final-viral-combined_k141_60660||full | Low-quality |
| KM-final-viral-combined_k141_150027||full | Low-quality |
| KM-final-viral-combined_k141_117860||full | Low-quality |
| KM-final-viral-combined_k141_27283||full | Low-quality |
| KM-final-viral-combined_k141_228381||full | Low-quality |
| KM-final-viral-combined_k141_127777||full | Not-determined |
| KM-final-viral-combined_k141_138962||full | Not-determined |
| KM-final-viral-combined_k141_238378||full | Low-quality |
| KM-final-viral-combined_k141_205943||full | Not-determined |
| KM-final-viral-combined_k141_93005||full | Low-quality |
| KM-final-viral-combined_k141_183932||full | Not-determined |
| KM-final-viral-combined_k141_138984||full | Not-determined |
| KM-final-viral-combined_k141_194665||full | Not-determined |
| KM-final-viral-combined_k141_259997||full | Low-quality |
| KM-final-viral-combined_k141_172044||full | Not-determined |
| KM-final-viral-combined_k141_15468||full | Not-determined |
| KM-final-viral-combined_k141_172050||full | Not-determined |
| KM-final-viral-combined_k141_60686||full | Not-determined |
| KM-final-viral-combined_k141_117919||full | Low-quality |
| KM-final-viral-combined_k141_15478||full | Not-determined |
| KM-final-viral-combined_k141_5055||full | Low-quality |
| KM-final-viral-combined_k141_194684||full | Not-determined |
| KM-final-viral-combined_k141_117921||full | Low-quality |
| KM-final-viral-combined_k141_105318||full | Low-quality |
| KM-final-viral-combined_k141_117931||full | Low-quality |
| KM-final-viral-combined_k141_139027||full | Not-determined |
| KM-final-viral-combined_k141_183958||full | Not-determined |
| KM-final-viral-combined_k141_60706||full | Low-quality |
| KM-final-viral-combined_k141_38427||full | Not-determined |
| KM-final-viral-combined_k141_27338||full | Low-quality |
| KM-final-viral-combined_k141_260027||full | Not-determined |
| KM-final-viral-combined_k141_172091||full | Not-determined |
| KM-final-viral-combined_k141_183963||full | Low-quality |
| KM-final-viral-combined_k141_60715||full | Low-quality |
| KM-final-viral-combined_k141_127849||full | Not-determined |
| KM-final-viral-combined_k141_206029||full | Not-determined |
| KM-final-viral-combined_k141_160283||full | Low-quality |
| KM-final-viral-combined_k141_217232||full | Not-determined |
| KM-final-viral-combined_k141_15517||full | Low-quality |
| KM-final-viral-combined_k141_127863||full | Low-quality |
| KM-final-viral-combined_k141_260043||full | Not-determined |
| KM-final-viral-combined_k141_139077||full | Low-quality |
| KM-final-viral-combined_k141_150083||full | Not-determined |
| KM-final-viral-combined_k141_249003||full | Low-quality |
| KM-final-viral-combined_k141_117949||full | Not-determined |
| KM-final-viral-combined_k141_194720||full | Not-determined |
| KM-final-viral-combined_k141_249006||full | Low-quality |
| KM-final-viral-combined_k141_183988||full | Not-determined |
| KM-final-viral-combined_k141_15524||full | Not-determined |
| KM-final-viral-combined_k141_117954||full | Low-quality |
| KM-final-viral-combined_k141_38454||full | Low-quality |
| KM-final-viral-combined_k141_194728||full | Not-determined |
| KM-final-viral-combined_k141_260059||full | Low-quality |
| KM-final-viral-combined_k141_93068||full | Low-quality |
| KM-final-viral-combined_k141_38460||full | Not-determined |
| KM-final-viral-combined_k141_72412||full | Low-quality |
| KM-final-viral-combined_k141_249035||full | Not-determined |
| KM-final-viral-combined_k141_238467||full | Low-quality |
| KM-final-viral-combined_k141_249036||full | Not-determined |
| KM-final-viral-combined_k141_127910||full | Low-quality |
| KM-final-viral-combined_k141_38478||full | Not-determined |
| KM-final-viral-combined_k141_27382||full | Not-determined |
| KM-final-viral-combined_k141_228466||full | Low-quality |
| KM-final-viral-combined_k141_72422||full | Low-quality |
| KM-final-viral-combined_k141_93112||full | Low-quality |
| KM-final-viral-combined_k141_48094||full | Not-determined |
| KM-final-viral-combined_k141_15555||full | Low-quality |
| KM-final-viral-combined_k141_194743||full | Not-determined |
| KM-final-viral-combined_k141_72437||full | Not-determined |
| KM-final-viral-combined_k141_249041||full | Not-determined |
| KM-final-viral-combined_k141_249049||full | Not-determined |
| KM-final-viral-combined_k141_105422||full | Low-quality |
| KM-final-viral-combined_k141_194772||full | Not-determined |
| KM-final-viral-combined_k141_206096||full | Not-determined |
| KM-final-viral-combined_k141_93132||full | Not-determined |
| KM-final-viral-combined_k141_172105||full | Not-determined |
| KM-final-viral-combined_k141_48111||full | Low-quality |
| KM-final-viral-combined_k141_184050||full | Not-determined |
| KM-final-viral-combined_k141_48116||full | Not-determined |
| KM-final-viral-combined_k141_217311||full | Not-determined |
| KM-final-viral-combined_k141_206105||full | Low-quality |
| KM-final-viral-combined_k141_139169||full | Not-determined |
| KM-final-viral-combined_k141_48119||full | Not-determined |
| KM-final-viral-combined_k141_160387||full | Low-quality |
| KM-final-viral-combined_k141_48121||full | Not-determined |
| KM-final-viral-combined_k141_118057||full | Low-quality |
| KM-final-viral-combined_k141_105476||full | Not-determined |
| KM-final-viral-combined_k141_118065||full | Not-determined |
| KM-final-viral-combined_k141_116248||full | High-quality |
| KM-final-viral-combined_k141_160405||full | Low-quality |
| KM-final-viral-combined_k141_48130||full | Not-determined |
| KM-final-viral-combined_k141_5177||full | Low-quality |
| KM-final-viral-combined_k141_206167||full | Not-determined |
| KM-final-viral-combined_k141_160420||full | Low-quality |
| KM-final-viral-combined_k141_184084||full | Not-determined |
| KM-final-viral-combined_k141_160425||full | Low-quality |
| KM-final-viral-combined_k141_48164||full | Not-determined |
| KM-final-viral-combined_k141_60820||full | Not-determined |
| KM-final-viral-combined_k141_27470||full | Low-quality |
| KM-final-viral-combined_k141_84228||full | Not-determined |
| KM-final-viral-combined_k141_139207||full | Not-determined |
| KM-final-viral-combined_k141_38594||full | Not-determined |
| KM-final-viral-combined_k141_194872||full | Not-determined |
| KM-final-viral-combined_k141_127955||full | Low-quality |
| KM-final-viral-combined_k141_93177||full | Not-determined |
| KM-final-viral-combined_k141_194874||full | Low-quality |
| KM-final-viral-combined_k141_127960||full | Not-determined |
| KM-final-viral-combined_k141_5203||full | Low-quality |
| KM-final-viral-combined_k141_48189||full | Low-quality |
| KM-final-viral-combined_k141_160437||full | Not-determined |
| KM-final-viral-combined_k141_105517||full | Low-quality |
| KM-final-viral-combined_k141_84255||full | Low-quality |
| KM-final-viral-combined_k141_93201||full | Not-determined |
| KM-final-viral-combined_k141_5221||full | Low-quality |
| KM-final-viral-combined_k141_60863||full | Not-determined |
| KM-final-viral-combined_k141_194893||full | Not-determined |
| KM-final-viral-combined_k141_38628||full | Not-determined |
| KM-final-viral-combined_k141_84264||full | Not-determined |
| KM-final-viral-combined_k141_228577||full | Low-quality |
| KM-final-viral-combined_k141_238602||full | Not-determined |
| KM-final-viral-combined_k141_228583||full | Not-determined |
| KM-final-viral-combined_k141_238612||full | Not-determined |
| KM-final-viral-combined_k141_48214||full | Not-determined |
| KM-final-viral-combined_k141_93219||full | Not-determined |
| KM-final-viral-combined_k141_228594||full | Low-quality |
| KM-final-viral-combined_k141_249075||full | Low-quality |
| KM-final-viral-combined_k141_15659||full | Not-determined |
| KM-final-viral-combined_k141_118136||full | Not-determined |
| KM-final-viral-combined_k141_105563||full | Not-determined |
| KM-final-viral-combined_k141_139274||full | Low-quality |
| KM-final-viral-combined_k141_228604||full | Low-quality |
| KM-final-viral-combined_k141_172154||full | Low-quality |
| KM-final-viral-combined_k141_217442||full | Low-quality |
| KM-final-viral-combined_k141_84302||full | Low-quality |
| KM-final-viral-combined_k141_238637||full | Low-quality |
| KM-final-viral-combined_k141_184109||full | Not-determined |
| KM-final-viral-combined_k141_249103||full | Low-quality |
| KM-final-viral-combined_k141_160496||full | Low-quality |
| KM-final-viral-combined_k141_206298||full | Low-quality |
| KM-final-viral-combined_k141_118144||full | Not-determined |
| KM-final-viral-combined_k141_72562||full | Low-quality |
| KM-final-viral-combined_k141_228633||full | Not-determined |
| KM-final-viral-combined_k141_249118||full | Low-quality |
| KM-final-viral-combined_k141_5265||full | Not-determined |
| KM-final-viral-combined_k141_238657||full | Not-determined |
| KM-final-viral-combined_k141_84316||full | Low-quality |
| KM-final-viral-combined_k141_38662||full | Not-determined |
| KM-final-viral-combined_k141_228639||full | Low-quality |
| KM-final-viral-combined_k141_48257||full | Not-determined |
| KM-final-viral-combined_k141_84324||full | Low-quality |
| KM-final-viral-combined_k141_38671||full | Not-determined |
| KM-final-viral-combined_k141_128066||full | Not-determined |
| KM-final-viral-combined_k141_72587||full | Low-quality |
| KM-final-viral-combined_k141_72592||full | Not-determined |
| KM-final-viral-combined_k141_27581||full | Not-determined |
| KM-final-viral-combined_k141_48265||full | Low-quality |
| KM-final-viral-combined_k141_249132||full | Not-determined |
| KM-final-viral-combined_k141_184151||full | Not-determined |
| KM-final-viral-combined_k141_249135||full | Not-determined |
| KM-final-viral-combined_k141_38686||full | Not-determined |
| KM-final-viral-combined_k141_38687||full | Low-quality |
| KM-final-viral-combined_k141_128071||full | Low-quality |
| KM-final-viral-combined_k141_38692||full | Not-determined |
| KM-final-viral-combined_k141_84337||full | Low-quality |
| KM-final-viral-combined_k141_128075||full | Not-determined |
| KM-final-viral-combined_k141_15700||full | Low-quality |
| KM-final-viral-combined_k141_72625||full | Not-determined |
| KM-final-viral-combined_k141_260238||full | Not-determined |
| KM-final-viral-combined_k141_15702||full | Not-determined |
| KM-final-viral-combined_k141_5294||full | Not-determined |
| KM-final-viral-combined_k141_160569||full | Not-determined |
| KM-final-viral-combined_k141_228664||full | Not-determined |
| KM-final-viral-combined_k141_172211||full | Not-determined |
| KM-final-viral-combined_k141_60981||full | Low-quality |
| KM-final-viral-combined_k141_27621||full | Not-determined |
| KM-final-viral-combined_k141_217540||full | Not-determined |
| KM-final-viral-combined_k141_238722||full | Low-quality |
| KM-final-viral-combined_k141_72650||full | Low-quality |
| KM-final-viral-combined_k141_260253||full | Not-determined |
| KM-final-viral-combined_k141_160582||full | Low-quality |
| KM-final-viral-combined_k141_5320||full | Not-determined |
| KM-final-viral-combined_k141_184195||full | Not-determined |
| KM-final-viral-combined_k141_228680||full | Not-determined |
| KM-final-viral-combined_k141_139362||full | Low-quality |
| KM-final-viral-combined_k141_139364||full | Low-quality |
| KM-final-viral-combined_k141_105651||full | Not-determined |
| KM-final-viral-combined_k141_84384||full | Not-determined |
| KM-final-viral-combined_k141_228698||full | Not-determined |
| KM-final-viral-combined_k141_194993||full | Low-quality |
| KM-final-viral-combined_k141_118195||full | Low-quality |
| KM-final-viral-combined_k141_206409||full | Low-quality |
| KM-final-viral-combined_k141_5344||full | Not-determined |
| KM-final-viral-combined_k141_184223||full | Low-quality |
| KM-final-viral-combined_k141_217566||full | Low-quality |
| KM-final-viral-combined_k141_172266||full | Not-determined |
| KM-final-viral-combined_k141_238766||full | Not-determined |
| KM-final-viral-combined_k141_184245||full | Low-quality |
| KM-final-viral-combined_k141_48340||full | Low-quality |
| KM-final-viral-combined_k141_238776||full | Low-quality |
| KM-final-viral-combined_k141_195056||full | Low-quality |
| KM-final-viral-combined_k141_15761||full | Not-determined |
| KM-final-viral-combined_k141_84422||full | Low-quality |
| KM-final-viral-combined_k141_128129||full | Not-determined |
| KM-final-viral-combined_k141_84431||full | Low-quality |
| KM-final-viral-combined_k141_217594||full | Low-quality |
| KM-final-viral-combined_k141_72725||full | Not-determined |
| KM-final-viral-combined_k141_150322||full | Low-quality |
| KM-final-viral-combined_k141_238804||full | Not-determined |
| KM-final-viral-combined_k141_238806||full | Not-determined |
| KM-final-viral-combined_k141_5417||full | Low-quality |
| KM-final-viral-combined_k141_206482||full | Not-determined |
| KM-final-viral-combined_k141_195105||full | Not-determined |
| KM-final-viral-combined_k141_160656||full | Low-quality |
| KM-final-viral-combined_k141_48381||full | Not-determined |
| KM-final-viral-combined_k141_105720||full | Low-quality |
| KM-final-viral-combined_k141_150338||full | Not-determined |
| KM-final-viral-combined_k141_93328||full | Low-quality |
| KM-final-viral-combined_k141_5425||full | Not-determined |
| KM-final-viral-combined_k141_105726||full | Not-determined |
| KM-final-viral-combined_k141_228804||full | Low-quality |
| KM-final-viral-combined_k141_217636||full | Not-determined |
| KM-final-viral-combined_k141_238843||full | Low-quality |
| KM-final-viral-combined_k141_206531||full | Not-determined |
| KM-final-viral-combined_k141_228823||full | Not-determined |
| KM-final-viral-combined_k141_105740||full | Low-quality |
| KM-final-viral-combined_k141_118327||full | Low-quality |
| KM-final-viral-combined_k141_260336||full | Not-determined |
| KM-final-viral-combined_k141_128220||full | Not-determined |
| KM-final-viral-combined_k141_160697||full | Not-determined |
| KM-final-viral-combined_k141_118359||full | Low-quality |
| KM-final-viral-combined_k141_160713||full | Not-determined |
| KM-final-viral-combined_k141_195156||full | Not-determined |
| KM-final-viral-combined_k141_84455||full | Low-quality |
| KM-final-viral-combined_k141_128232||full | Not-determined |
| KM-final-viral-combined_k141_72766||full | Not-determined |
| KM-final-viral-combined_k141_72780||full | Low-quality |
| KM-final-viral-combined_k141_184374||full | Low-quality |
| KM-final-viral-combined_k141_72784||full | Low-quality |
| KM-final-viral-combined_k141_195169||full | Not-determined |
| KM-final-viral-combined_k141_150400||full | Not-determined |
| KM-final-viral-combined_k141_48424||full | Not-determined |
| KM-final-viral-combined_k141_184383||full | Not-determined |
| KM-final-viral-combined_k141_150410||full | Low-quality |
| KM-final-viral-combined_k141_38764||full | Low-quality |
| KM-final-viral-combined_k141_61073||full | Low-quality |
| KM-final-viral-combined_k141_93398||full | Not-determined |
| KM-final-viral-combined_k141_38767||full | Not-determined |
| KM-final-viral-combined_k141_38771||full | Low-quality |
| KM-final-viral-combined_k141_72821||full | Not-determined |
| KM-final-viral-combined_k141_38772||full | Not-determined |
| 17-108_k141_87293 | Complete |
| KM-final-viral-combined_k141_5493||full | Not-determined |
| KM-final-viral-combined_k141_195188||full | Low-quality |
| KM-final-viral-combined_k141_172365||full | Low-quality |
| KM-final-viral-combined_k141_260367||full | Low-quality |
| KM-final-viral-combined_k141_184427||full | Low-quality |
| KM-final-viral-combined_k141_206616||full | Not-determined |
| KM-final-viral-combined_k141_238908||full | Low-quality |
| KM-final-viral-combined_k141_48445||full | Low-quality |
| KM-final-viral-combined_k141_238909||full | Low-quality |
| KM-final-viral-combined_k141_195228||full | Not-determined |
| KM-final-viral-combined_k141_195231||full | Not-determined |
| KM-final-viral-combined_k141_15827||full | Low-quality |
| KM-final-viral-combined_k141_260386||full | Low-quality |
| 18-104-final-viral-combined_k141_118690||full | Medium-quality |
| KM-final-viral-combined_k141_93453||full | Low-quality |
| KM-final-viral-combined_k141_93454||full | Low-quality |
| KM-final-viral-combined_k141_184484||full | Low-quality |
| KM-final-viral-combined_k141_84512||full | Low-quality |
| KM-final-viral-combined_k141_128356||full | Not-determined |
| KM-final-viral-combined_k141_128357||full | Not-determined |
| KM-final-viral-combined_k141_84524||full | Low-quality |
| KM-final-viral-combined_k141_217794||full | Not-determined |
| KM-final-viral-combined_k141_118471||full | Low-quality |
| KM-final-viral-combined_k141_72852||full | Low-quality |
| KM-final-viral-combined_k141_217806||full | Low-quality |
| KM-final-viral-combined_k141_206659||full | Low-quality |
| KM-final-viral-combined_k141_38834||full | Low-quality |
| KM-final-viral-combined_k141_238966||full | Not-determined |
| KM-final-viral-combined_k141_184531||full | Low-quality |
| KM-final-viral-combined_k141_38838||full | Low-quality |
| KM-final-viral-combined_k141_84540||full | Not-determined |
| KM-final-viral-combined_k141_118498||full | Not-determined |
| KM-final-viral-combined_k141_238986||full | Low-quality |
| KM-final-viral-combined_k141_217819||full | Not-determined |
| KM-final-viral-combined_k141_195277||full | Not-determined |
| KM-final-viral-combined_k141_195280||full | Low-quality |
| KM-final-viral-combined_k141_195285||full | Not-determined |
| KM-final-viral-combined_k141_61197||full | Not-determined |
| KM-final-viral-combined_k141_184547||full | Low-quality |
| KM-final-viral-combined_k141_206691||full | Not-determined |
| KM-final-viral-combined_k141_217843||full | Low-quality |
| KM-final-viral-combined_k141_150547||full | Low-quality |
| KM-final-viral-combined_k141_128404||full | Low-quality |
| KM-final-viral-combined_k141_15892||full | Low-quality |
| KM-final-viral-combined_k141_184559||full | Not-determined |
| KM-final-viral-combined_k141_206700||full | Not-determined |
| KM-final-viral-combined_k141_150554||full | Not-determined |
| KM-final-viral-combined_k141_249456||full | Low-quality |
| KM-final-viral-combined_k141_184568||full | Not-determined |
| KM-final-viral-combined_k141_118526||full | Not-determined |
| KM-final-viral-combined_k141_61250||full | Low-quality |
| KM-final-viral-combined_k141_93504||full | Not-determined |
| KM-final-viral-combined_k141_260469||full | Not-determined |
| KM-final-viral-combined_k141_195302||full | Not-determined |
| KM-final-viral-combined_k141_206716||full | Low-quality |
| KM-final-viral-combined_k141_260489||full | Low-quality |
| KM-final-viral-combined_k141_195319||full | Not-determined |
| KM-final-viral-combined_k141_5570||full | Low-quality |
| KM-final-viral-combined_k141_184643||full | Low-quality |
| KM-final-viral-combined_k141_228991||full | Not-determined |
| KM-final-viral-combined_k141_38903||full | Not-determined |
| KM-final-viral-combined_k141_139679||full | Low-quality |
| KM-final-viral-combined_k141_249501||full | Not-determined |
| KM-final-viral-combined_k141_15932||full | Not-determined |
| KM-final-viral-combined_k141_260505||full | Not-determined |
| KM-final-viral-combined_k141_206762||full | Low-quality |
| KM-final-viral-combined_k141_139696||full | Low-quality |
| KM-final-viral-combined_k141_195348||full | Not-determined |
| KM-final-viral-combined_k141_139701||full | Low-quality |
| KM-final-viral-combined_k141_229010||full | Low-quality |
| KM-final-viral-combined_k141_139703||full | Low-quality |
| KM-final-viral-combined_k141_128423||full | Low-quality |
| KM-final-viral-combined_k141_105837||full | Not-determined |
| KM-final-viral-combined_k141_172489||full | Low-quality |
| KM-final-viral-combined_k141_139720||full | Low-quality |
| KM-final-viral-combined_k141_84605||full | Low-quality |
| KM-final-viral-combined_k141_160895||full | Not-determined |
| KM-final-viral-combined_k141_239082||full | Not-determined |
| KM-final-viral-combined_k141_217916||full | Low-quality |
| KM-final-viral-combined_k141_38930||full | Not-determined |
| KM-final-viral-combined_k141_239089||full | Low-quality |
| KM-final-viral-combined_k141_160911||full | Low-quality |
| KM-final-viral-combined_k141_195368||full | Not-determined |
| KM-final-viral-combined_k141_72990||full | Not-determined |
| KM-final-viral-combined_k141_160915||full | Not-determined |
| KM-final-viral-combined_k141_184683||full | Low-quality |
| KM-final-viral-combined_k141_139759||full | Not-determined |
| KM-final-viral-combined_k141_38939||full | Low-quality |
| KM-final-viral-combined_k141_239103||full | Not-determined |
| KM-final-viral-combined_k141_195378||full | Low-quality |
| KM-final-viral-combined_k141_128478||full | Low-quality |
| KM-final-viral-combined_k141_61313||full | Not-determined |
| KM-final-viral-combined_k141_206819||full | Low-quality |
| KM-final-viral-combined_k141_93597||full | Not-determined |
| KM-final-viral-combined_k141_105876||full | Not-determined |
| KM-final-viral-combined_k141_93601||full | Not-determined |
| KM-final-viral-combined_k141_195388||full | Not-determined |
| KM-final-viral-combined_k141_260557||full | Not-determined |
| KM-final-viral-combined_k141_184705||full | Low-quality |
| KM-final-viral-combined_k141_229111||full | Not-determined |
| KM-final-viral-combined_k141_105895||full | Low-quality |
| KM-final-viral-combined_k141_260590||full | Low-quality |
| KM-final-viral-combined_k141_61369||full | Not-determined |
| KM-final-viral-combined_k141_93625||full | Not-determined |
| KM-final-viral-combined_k141_27744||full | Not-determined |
| KM-final-viral-combined_k141_217989||full | Not-determined |
| KM-final-viral-combined_k141_93626||full | Low-quality |
| KM-final-viral-combined_k141_260601||full | Not-determined |
| KM-final-viral-combined_k141_195469||full | Low-quality |
| KM-final-viral-combined_k141_105932||full | Not-determined |
| KM-final-viral-combined_k141_260606||full | Low-quality |
| KM-final-viral-combined_k141_229164||full | Low-quality |
| KM-final-viral-combined_k141_172610||full | Not-determined |
| KM-final-viral-combined_k141_48656||full | Low-quality |
| KM-final-viral-combined_k141_93640||full | Not-determined |
| KM-final-viral-combined_k141_16042||full | Low-quality |
| KM-final-viral-combined_k141_172633||full | Low-quality |
| KM-final-viral-combined_k141_249597||full | Not-determined |
| KM-final-viral-combined_k141_184765||full | Not-determined |
| KM-final-viral-combined_k141_172669||full | Low-quality |
| KM-final-viral-combined_k141_229203||full | Not-determined |
| KM-final-viral-combined_k141_27792||full | Not-determined |
| KM-final-viral-combined_k141_16079||full | Low-quality |
| KM-final-viral-combined_k141_206929||full | Not-determined |
| KM-final-viral-combined_k141_84699||full | Not-determined |
| KM-final-viral-combined_k141_229212||full | Low-quality |
| KM-final-viral-combined_k141_139835||full | Not-determined |
| KM-final-viral-combined_k141_260652||full | Not-determined |
| KM-final-viral-combined_k141_218026||full | Not-determined |
| KM-final-viral-combined_k141_5665||full | Not-determined |
| KM-final-viral-combined_k141_249608||full | Low-quality |
| KM-final-viral-combined_k141_38993||full | Low-quality |
| KM-final-viral-combined_k141_229230||full | Not-determined |
| KM-final-viral-combined_k141_249612||full | Not-determined |
| KM-final-viral-combined_k141_195560||full | Not-determined |
| KM-final-viral-combined_k141_128576||full | Not-determined |
| KM-final-viral-combined_k141_5677||full | Low-quality |

| | |
|---|---|
| KM-final-viral-combined_k141_195565\|\|full | Not-determined |
| KM-final-viral-combined_k141_184831\|\|full | Low-quality |
| KM-final-viral-combined_k141_218043\|\|full | Not-determined |
| KM-final-viral-combined_k141_106024\|\|full | Not-determined |
| KM-final-viral-combined_k141_150683\|\|full | Low-quality |
| KM-final-viral-combined_k141_128588\|\|full | Not-determined |
| KM-final-viral-combined_k141_48698\|\|full | Low-quality |
| KM-final-viral-combined_k141_61420\|\|full | Low-quality |
| KM-final-viral-combined_k141_48701\|\|full | Not-determined |
| KM-final-viral-combined_k141_139859\|\|full | Not-determined |
| KM-final-viral-combined_k141_184861\|\|full | Low-quality |
| KM-final-viral-combined_k141_260689\|\|full | Low-quality |
| KM-final-viral-combined_k141_184867\|\|full | Not-determined |
| KM-final-viral-combined_k141_128605\|\|full | Not-determined |
| KM-final-viral-combined_k141_150697\|\|full | Low-quality |
| KM-final-viral-combined_k141_93712\|\|full | Low-quality |
| KM-final-viral-combined_k141_61437\|\|full | Low-quality |
| KM-final-viral-combined_k141_61438\|\|full | Not-determined |
| KM-final-viral-combined_k141_93719\|\|full | Low-quality |
| KM-final-viral-combined_k141_249639\|\|full | Not-determined |
| KM-final-viral-combined_k141_239233\|\|full | Low-quality |
| KM-final-viral-combined_k141_61454\|\|full | Not-determined |
| KM-final-viral-combined_k141_27815\|\|full | Not-determined |
| KM-final-viral-combined_k141_39032\|\|full | Not-determined |
| KM-final-viral-combined_k141_161057\|\|full | Low-quality |
| KM-final-viral-combined_k141_161065\|\|full | Low-quality |
| KM-final-viral-combined_k141_27834\|\|full | Not-determined |
| KM-final-viral-combined_k141_128656\|\|full | Low-quality |
| KM-final-viral-combined_k141_139928\|\|full | Not-determined |
| KM-final-viral-combined_k141_27848\|\|full | Low-quality |
| KM-final-viral-combined_k141_207007\|\|full | Low-quality |
| KM-final-viral-combined_k141_73141\|\|full | Low-quality |
| KM-final-viral-combined_k141_73143\|\|full | Not-determined |
| KM-final-viral-combined_k141_218130\|\|full | Not-determined |
| KM-final-viral-combined_k141_139946\|\|full | Low-quality |
| KM-final-viral-combined_k141_5742\|\|full | Low-quality |
| KM-final-viral-combined_k141_128681\|\|full | Not-determined |
| KM-final-viral-combined_k141_218132\|\|full | Low-quality |
| KM-final-viral-combined_k141_229317\|\|full | Low-quality |
| KM-final-viral-combined_k141_260744\|\|full | Not-determined |
| KM-final-viral-combined_k141_218146\|\|full | Low-quality |
| KM-final-viral-combined_k141_207028\|\|full | Not-determined |
| KM-final-viral-combined_k141_150739\|\|full | Low-quality |
| KM-final-viral-combined_k141_39093\|\|full | Low-quality |
| KM-final-viral-combined_k141_139976\|\|full | Not-determined |
| KM-final-viral-combined_k141_260775\|\|full | Not-determined |
| KM-final-viral-combined_k141_184906\|\|full | Low-quality |
| KM-final-viral-combined_k141_84847\|\|full | Not-determined |
| KM-final-viral-combined_k141_61495\|\|full | Not-determined |
| KM-final-viral-combined_k141_84850\|\|full | Low-quality |
| KM-final-viral-combined_k141_184913\|\|full | Low-quality |
| KM-final-viral-combined_k141_106087\|\|full | Not-determined |
| KM-final-viral-combined_k141_39129\|\|full | Low-quality |
| KM-final-viral-combined_k141_118744\|\|full | Not-determined |
| KM-final-viral-combined_k141_128697\|\|full | Low-quality |
| KM-final-viral-combined_k141_16143\|\|full | Not-determined |
| KM-final-viral-combined_k141_118747\|\|full | Not-determined |
| KM-final-viral-combined_k141_218218\|\|full | Not-determined |
| KM-final-viral-combined_k141_172848\|\|full | Low-quality |
| KM-final-viral-combined_k141_84883\|\|full | Low-quality |
| KM-final-viral-combined_k141_139990\|\|full | Not-determined |
| KM-final-viral-combined_k141_195731\|\|full | Not-determined |
| KM-final-viral-combined_k141_5802\|\|full | Not-determined |
| KM-final-viral-combined_k141_140012\|\|full | Low-quality |
| KM-final-viral-combined_k141_207058\|\|full | Not-determined |
| KM-final-viral-combined_k141_260821\|\|full | Not-determined |
| KM-final-viral-combined_k141_106127\|\|full | Not-determined |
| KM-final-viral-combined_k141_239341\|\|full | Low-quality |
| KM-final-viral-combined_k141_229399\|\|full | Not-determined |
| KM-final-viral-combined_k141_249734\|\|full | Low-quality |
| KM-final-viral-combined_k141_218227\|\|full | Not-determined |
| KM-final-viral-combined_k141_48836\|\|full | Low-quality |
| KM-final-viral-combined_k141_106138\|\|full | Not-determined |
| KM-final-viral-combined_k141_73251\|\|full | Low-quality |
| KM-final-viral-combined_k141_106139\|\|full | Not-determined |
| KM-final-viral-combined_k141_84903\|\|full | Not-determined |
| KM-final-viral-combined_k141_218247\|\|full | Low-quality |
| KM-final-viral-combined_k141_239357\|\|full | Not-determined |
| KM-final-viral-combined_k141_172896\|\|full | Low-quality |
| KM-final-viral-combined_k141_218266\|\|full | Not-determined |
| KM-final-viral-combined_k141_207101\|\|full | Not-determined |
| KM-final-viral-combined_k141_260864\|\|full | Not-determined |
| KM-final-viral-combined_k141_140089\|\|full | Low-quality |
| KM-final-viral-combined_k141_218268\|\|full | Low-quality |
| KM-final-viral-combined_k141_229424\|\|full | Not-determined |
| KM-final-viral-combined_k141_184951\|\|full | Low-quality |
| KM-final-viral-combined_k141_260866\|\|full | Low-quality |
| KM-final-viral-combined_k141_106175\|\|full | Not-determined |
| KM-final-viral-combined_k141_207116\|\|full | Not-determined |
| KM-final-viral-combined_k141_239395\|\|full | Not-determined |
| KM-final-viral-combined_k141_260869\|\|full | Low-quality |
| KM-final-viral-combined_k141_140102\|\|full | Low-quality |
| KM-final-viral-combined_k141_172906\|\|full | Not-determined |
| KM-final-viral-combined_k141_218281\|\|full | Not-determined |
| KM-final-viral-combined_k141_16167\|\|full | Not-determined |
| KM-final-viral-combined_k141_73312\|\|full | Not-determined |
| KM-final-viral-combined_k141_73315\|\|full | Low-quality |
| KM-final-viral-combined_k141_73320\|\|full | Not-determined |
| KM-final-viral-combined_k141_106190\|\|full | Low-quality |
| KM-final-viral-combined_k141_16176\|\|full | Low-quality |
| KM-final-viral-combined_k141_150798\|\|full | Low-quality |
| KM-final-viral-combined_k141_172921\|\|full | Not-determined |
| KM-final-viral-combined_k141_172922\|\|full | Low-quality |
| KM-final-viral-combined_k141_106203\|\|full | Low-quality |
| KM-final-viral-combined_k141_16188\|\|full | Not-determined |
| KM-final-viral-combined_k141_5879\|\|full | Low-quality |
| KM-final-viral-combined_k141_239429\|\|full | Not-determined |
| KM-final-viral-combined_k141_5889\|\|full | Not-determined |
| KM-final-viral-combined_k141_93802\|\|full | Low-quality |
| KM-final-viral-combined_k141_239432\|\|full | Not-determined |
| KM-final-viral-combined_k141_195766\|\|full | Low-quality |
| KM-final-viral-combined_k141_229469\|\|full | Low-quality |
| KM-final-viral-combined_k141_39241\|\|full | Not-determined |
| KM-final-viral-combined_k141_28011\|\|full | Low-quality |
| KM-final-viral-combined_k141_207194\|\|full | Low-quality |
| KM-final-viral-combined_k141_73391\|\|full | Not-determined |
| KM-final-viral-combined_k141_61625\|\|full | Low-quality |
| KM-final-viral-combined_k141_239462\|\|full | Not-determined |
| KM-final-viral-combined_k141_73393\|\|full | Low-quality |
| KM-final-viral-combined_k141_207210\|\|full | Low-quality |
| KM-final-viral-combined_k141_48924\|\|full | Low-quality |
| KM-final-viral-combined_k141_5923\|\|full | Not-determined |
| KM-final-viral-combined_k141_195792\|\|full | Low-quality |
| KM-final-viral-combined_k141_184974\|\|full | Not-determined |
| KM-final-viral-combined_k141_39295\|\|full | Low-quality |
| KM-final-viral-combined_k141_16257\|\|full | Not-determined |
| KM-final-viral-combined_k141_173001\|\|full | Low-quality |
| KM-final-viral-combined_k141_249778\|\|full | Low-quality |
| KM-final-viral-combined_k141_195804\|\|full | Not-determined |
| KM-final-viral-combined_k141_195811\|\|full | Low-quality |
| KM-final-viral-combined_k141_195815\|\|full | Low-quality |
| KM-final-viral-combined_k141_185008\|\|full | Not-determined |
| KM-final-viral-combined_k141_48929\|\|full | Not-determined |
| KM-final-viral-combined_k141_195822\|\|full | Low-quality |
| KM-final-viral-combined_k141_5955\|\|full | Not-determined |
| KM-final-viral-combined_k141_229541\|\|full | Low-quality |
| KM-final-viral-combined_k141_150869\|\|full | Low-quality |
| KM-final-viral-combined_k141_39347\|\|full | Low-quality |
| KM-final-viral-combined_k141_28068\|\|full | Low-quality |
| KM-final-viral-combined_k141_16304\|\|full | Not-determined |
| KM-final-viral-combined_k141_5963\|\|full | Low-quality |
| KM-final-viral-combined_k141_185049\|\|full | Not-determined |
| KM-final-viral-combined_k141_173024\|\|full | Low-quality |
| KM-final-viral-combined_k141_185058\|\|full | Not-determined |
| KM-final-viral-combined_k141_218406\|\|full | Low-quality |
| KM-final-viral-combined_k141_261017\|\|full | Low-quality |
| KM-final-viral-combined_k141_140201\|\|full | Not-determined |
| KM-final-viral-combined_k141_161183\|\|full | Low-quality |
| KM-final-viral-combined_k141_39369\|\|full | Not-determined |
| KM-final-viral-combined_k141_229556\|\|full | Not-determined |
| KM-final-viral-combined_k141_195858\|\|full | Not-determined |
| KM-final-viral-combined_k141_150874\|\|full | Low-quality |
| KM-final-viral-combined_k141_239553\|\|full | Not-determined |
| KM-final-viral-combined_k141_85094\|\|full | Not-determined |
| 17-108-final-viral-combined_k141_87293\|\|full | High-quality |
| KM-final-viral-combined_k141_218427\|\|full | Low-quality |
| KM-final-viral-combined_k141_140212\|\|full | Not-determined |
| 18-109_k141_113137 | High-quality |
| KM-final-viral-combined_k141_218430\|\|full | Not-determined |
| KM-final-viral-combined_k141_207246\|\|full | Not-determined |
| KM-final-viral-combined_k141_173028\|\|full | Not-determined |
| KM-final-viral-combined_k141_218438\|\|full | Low-quality |
| KM-final-viral-combined_k141_150900\|\|full | Low-quality |
| KM-final-viral-combined_k141_5982\|\|full | Low-quality |
| KM-final-viral-combined_k141_140227\|\|full | Not-determined |
| KM-final-viral-combined_k141_261054\|\|full | Low-quality |
| KM-final-viral-combined_k141_150906\|\|full | Not-determined |
| KM-final-viral-combined_k141_249822\|\|full | Low-quality |
| KM-final-viral-combined_k141_207265\|\|full | Low-quality |
| KM-final-viral-combined_k141_61717\|\|full | Low-quality |
| KM-final-viral-combined_k141_229612\|\|full | Low-quality |
| KM-final-viral-combined_k141_49008\|\|full | Not-determined |
| KM-final-viral-combined_k141_185096\|\|full | Not-determined |
| KM-final-viral-combined_k141_249833\|\|full | Not-determined |
| KM-final-viral-combined_k141_16340\|\|full | Not-determined |
| KM-final-viral-combined_k141_150937\|\|full | Low-quality |
| KM-final-viral-combined_k141_85114\|\|full | Not-determined |
| KM-final-viral-combined_k141_161223\|\|full | Not-determined |
| KM-final-viral-combined_k141_185121\|\|full | Low-quality |
| KM-final-viral-combined_k141_261090\|\|full | Not-determined |
| KM-final-viral-combined_k141_229619\|\|full | Low-quality |
| KM-final-viral-combined_k141_150947\|\|full | Not-determined |
| KM-final-viral-combined_k141_218496\|\|full | Low-quality |
| KM-final-viral-combined_k141_195924\|\|full | Not-determined |
| KM-final-viral-combined_k141_39387\|\|full | Low-quality |
| KM-final-viral-combined_k141_39388\|\|full | Not-determined |
| KM-final-viral-combined_k141_239611\|\|full | Low-quality |
| KM-final-viral-combined_k141_185130\|\|full | Not-determined |
| KM-final-viral-combined_k141_173086\|\|full | Not-determined |
| KM-final-viral-combined_k141_73479\|\|full | Not-determined |
| KM-final-viral-combined_k141_128829\|\|full | Not-determined |
| KM-final-viral-combined_k141_185144\|\|full | Low-quality |
| KM-final-viral-combined_k141_261117\|\|full | Not-determined |
| KM-final-viral-combined_k141_6027\|\|full | Low-quality |
| KM-final-viral-combined_k141_185153\|\|full | Low-quality |
| KM-final-viral-combined_k141_118984\|\|full | Not-determined |
| KM-final-viral-combined_k141_185157\|\|full | Not-determined |
| KM-final-viral-combined_k141_6034\|\|full | Low-quality |
| KM-final-viral-combined_k141_195949\|\|full | Low-quality |
| KM-final-viral-combined_k141_140268\|\|full | Low-quality |
| KM-final-viral-combined_k141_93915\|\|full | Low-quality |
| KM-final-viral-combined_k141_119009\|\|full | Low-quality |
| KM-final-viral-combined_k141_49036\|\|full | Not-determined |
| KM-final-viral-combined_k141_93926\|\|full | Low-quality |
| KM-final-viral-combined_k141_207358\|\|full | Low-quality |
| KM-final-viral-combined_k141_119017\|\|full | Low-quality |
| KM-final-viral-combined_k141_39425\|\|full | Low-quality |
| KM-final-viral-combined_k141_106310\|\|full | Low-quality |
| KM-final-viral-combined_k141_218568\|\|full | Not-determined |
| KM-final-viral-combined_k141_73517\|\|full | Low-quality |
| KM-final-viral-combined_k141_106311\|\|full | Not-determined |
| KM-final-viral-combined_k141_249900\|\|full | Not-determined |
| KM-final-viral-combined_k141_28138\|\|full | Not-determined |
| KM-final-viral-combined_k141_61852\|\|full | Not-determined |
| KM-final-viral-combined_k141_161284\|\|full | Not-determined |
| KM-final-viral-combined_k141_28140\|\|full | Not-determined |
| KM-final-viral-combined_k141_161285\|\|full | Low-quality |
| KM-final-viral-combined_k141_261183\|\|full | Not-determined |
| KM-final-viral-combined_k141_28141\|\|full | Low-quality |
| KM-final-viral-combined_k141_173178\|\|full | Low-quality |
| KM-final-viral-combined_k141_161302\|\|full | Not-determined |
| KM-final-viral-combined_k141_249914\|\|full | Not-determined |
| KM-final-viral-combined_k141_207410\|\|full | Not-determined |
| KM-final-viral-combined_k141_185169\|\|full | Low-quality |
| KM-final-viral-combined_k141_16443\|\|full | Low-quality |
| KM-final-viral-combined_k141_173192\|\|full | Not-determined |
| KM-final-viral-combined_k141_196068\|\|full | Not-determined |
| KM-final-viral-combined_k141_16448\|\|full | Not-determined |
| KM-final-viral-combined_k141_249936\|\|full | Not-determined |
| KM-final-viral-combined_k141_229686\|\|full | Not-determined |
| KM-final-viral-combined_k141_161348\|\|full | Not-determined |
| KM-final-viral-combined_k141_39510\|\|full | Not-determined |
| KM-final-viral-combined_k141_85210\|\|full | Not-determined |
| KM-final-viral-combined_k141_39513\|\|full | Low-quality |
| KM-final-viral-combined_k141_6119\|\|full | Low-quality |
| KM-final-viral-combined_k141_218601\|\|full | Not-determined |
| KM-final-viral-combined_k141_61930\|\|full | Not-determined |
| KM-final-viral-combined_k141_218606\|\|full | Low-quality |
| KM-final-viral-combined_k141_6128\|\|full | Not-determined |
| KM-final-viral-combined_k141_151021\|\|full | Low-quality |
| KM-final-viral-combined_k141_239624\|\|full | Low-quality |
| KM-final-viral-combined_k141_61947\|\|full | Low-quality |
| KM-final-viral-combined_k141_151026\|\|full | Low-quality |
| KM-final-viral-combined_k141_6129\|\|full | Not-determined |
| KM-final-viral-combined_k141_106398\|\|full | Low-quality |
| KM-final-viral-combined_k141_6136\|\|full | Low-quality |
| KM-final-viral-combined_k141_28222\|\|full | Low-quality |
| KM-final-viral-combined_k141_249961\|\|full | Low-quality |
| KM-final-viral-combined_k141_6140\|\|full | Low-quality |
| KM-final-viral-combined_k141_119050\|\|full | Low-quality |
| KM-final-viral-combined_k141_218650\|\|full | Not-determined |
| KM-final-viral-combined_k141_229733\|\|full | Not-determined |
| KM-final-viral-combined_k141_28242\|\|full | Low-quality |
| KM-final-viral-combined_k141_249984\|\|full | Low-quality |
| KM-final-viral-combined_k141_229736\|\|full | Not-determined |
| KM-final-viral-combined_k141_49146\|\|full | Not-determined |
| KM-final-viral-combined_k141_28249\|\|full | Low-quality |
| KM-final-viral-combined_k141_218693\|\|full | Not-determined |
| KM-final-viral-combined_k141_61992\|\|full | Low-quality |
| KM-final-viral-combined_k141_218694\|\|full | Low-quality |
| KM-final-viral-combined_k141_119078\|\|full | Not-determined |
| KM-final-viral-combined_k141_173249\|\|full | Not-determined |
| KM-final-viral-combined_k141_61994\|\|full | Not-determined |
| KM-final-viral-combined_k141_249997\|\|full | Not-determined |
| KM-final-viral-combined_k141_119083\|\|full | Not-determined |
| KM-final-viral-combined_k141_6163\|\|full | Low-quality |
| KM-final-viral-combined_k141_196171\|\|full | Low-quality |
| KM-final-viral-combined_k141_49153\|\|full | Low-quality |
| KM-final-viral-combined_k141_85241\|\|full | Low-quality |
| KM-final-viral-combined_k141_119097\|\|full | Low-quality |
| KM-final-viral-combined_k141_28259\|\|full | Not-determined |
| KM-final-viral-combined_k141_128904\|\|full | Not-determined |
| KM-final-viral-combined_k141_239690\|\|full | Not-determined |
| KM-final-viral-combined_k141_151093\|\|full | Low-quality |
| KM-final-viral-combined_k141_49168\|\|full | Not-determined |
| KM-final-viral-combined_k141_250014\|\|full | Not-determined |
| KM-final-viral-combined_k141_16537\|\|full | Not-determined |
| KM-final-viral-combined_k141_106438\|\|full | Not-determined |
| KM-final-viral-combined_k141_94015\|\|full | Not-determined |
| KM-final-viral-combined_k141_207471\|\|full | Low-quality |
| KM-final-viral-combined_k141_49179\|\|full | Low-quality |
| KM-final-viral-combined_k141_49191\|\|full | Not-determined |
| KM-final-viral-combined_k141_207492\|\|full | Low-quality |
| KM-final-viral-combined_k141_218728\|\|full | Not-determined |
| KM-final-viral-combined_k141_128943\|\|full | Low-quality |
| KM-final-viral-combined_k141_62039\|\|full | Not-determined |
| KM-final-viral-combined_k141_173283\|\|full | Not-determined |
| KM-final-viral-combined_k141_128948\|\|full | Low-quality |
| KM-final-viral-combined_k141_39597\|\|full | Low-quality |
| KM-final-viral-combined_k141_6181\|\|full | Low-quality |
| KM-final-viral-combined_k141_106459\|\|full | Not-determined |
| KM-final-viral-combined_k141_161467\|\|full | Low-quality |
| KM-final-viral-combined_k141_239717\|\|full | Not-determined |
| KM-final-viral-combined_k141_73676\|\|full | Not-determined |
| KM-final-viral-combined_k141_39612\|\|full | Low-quality |
| 17-109-final-viral-combined_k141_21504\|\|full | Medium-quality |
| KM-final-viral-combined_k141_128964\|\|full | Low-quality |
| KM-final-viral-combined_k141_151148\|\|full | Low-quality |
| KM-final-viral-combined_k141_128968\|\|full | Not-determined |
| KM-final-viral-combined_k141_62067\|\|full | Not-determined |
| KM-final-viral-combined_k141_151156\|\|full | Not-determined |
| KM-final-viral-combined_k141_196248\|\|full | Low-quality |
| KM-final-viral-combined_k141_250059\|\|full | Low-quality |
| KM-final-viral-combined_k141_39638\|\|full | Not-determined |
| KM-final-viral-combined_k141_161498\|\|full | Not-determined |
| KM-final-viral-combined_k141_94064\|\|full | Not-determined |
| KM-final-viral-combined_k141_161500\|\|full | Not-determined |
| KM-final-viral-combined_k141_106499\|\|full | Low-quality |
| KM-final-viral-combined_k141_261331\|\|full | Low-quality |
| KM-final-viral-combined_k141_185264\|\|full | Low-quality |
| KM-final-viral-combined_k141_151179\|\|full | Not-determined |
| KM-final-viral-combined_k141_173328\|\|full | Low-quality |
| KM-final-viral-combined_k141_85265\|\|full | Not-determined |
| KM-final-viral-combined_k141_28334\|\|full | Not-determined |
| KM-final-viral-combined_k141_161521\|\|full | Low-quality |
| KM-final-viral-combined_k141_119172\|\|full | Low-quality |
| KM-final-viral-combined_k141_39662\|\|full | Not-determined |
| KM-final-viral-combined_k141_161528\|\|full | Low-quality |
| KM-final-viral-combined_k141_28336\|\|full | Low-quality |
| KM-final-viral-combined_k141_207532\|\|full | Not-determined |
| KM-final-viral-combined_k141_261343\|\|full | Low-quality |
| KM-final-viral-combined_k141_62122\|\|full | Not-determined |
| KM-final-viral-combined_k141_151211\|\|full | Low-quality |
| KM-final-viral-combined_k141_73732\|\|full | Not-determined |
| KM-final-viral-combined_k141_229843\|\|full | Not-determined |
| KM-final-viral-combined_k141_119180\|\|full | Not-determined |
| KM-final-viral-combined_k141_73744\|\|full | Not-determined |
| KM-final-viral-combined_k141_129024\|\|full | Low-quality |
| KM-final-viral-combined_k141_73751\|\|full | Low-quality |
| KM-final-viral-combined_k141_28373\|\|full | Low-quality |
| KM-final-viral-combined_k141_261382\|\|full | Not-determined |
| KM-final-viral-combined_k141_261386\|\|full | Not-determined |
| KM-final-viral-combined_k141_28384\|\|full | Low-quality |
| KM-final-viral-combined_k141_151241\|\|full | Low-quality |
| 17-109_k141_386815 | High-quality |
| KM-final-viral-combined_k141_151254\|\|full | Not-determined |
| KM-final-viral-combined_k141_185338\|\|full | Low-quality |
| KM-final-viral-combined_k141_6276\|\|full | Not-determined |
| KM-final-viral-combined_k141_16547\|\|full | Low-quality |
| KM-final-viral-combined_k141_173397\|\|full | Low-quality |
| KM-final-viral-combined_k141_85356\|\|full | Not-determined |
| KM-final-viral-combined_k141_207580\|\|full | Low-quality |
| KM-final-viral-combined_k141_185368\|\|full | Low-quality |
| KM-final-viral-combined_k141_73788\|\|full | Low-quality |
| KM-final-viral-combined_k141_16559\|\|full | Not-determined |
| KM-final-viral-combined_k141_16561\|\|full | Low-quality |
| KM-final-viral-combined_k141_62178\|\|full | Low-quality |
| KM-final-viral-combined_k141_250115\|\|full | Low-quality |
| KM-final-viral-combined_k141_16565\|\|full | Low-quality |
| KM-final-viral-combined_k141_106605\|\|full | Not-determined |
| KM-final-viral-combined_k141_119244\|\|full | Not-determined |
| KM-final-viral-combined_k141_28427\|\|full | Low-quality |
| KM-final-viral-combined_k141_49270\|\|full | Not-determined |
| KM-final-viral-combined_k141_261419\|\|full | Low-quality |
| KM-final-viral-combined_k141_185396\|\|full | Low-quality |
| KM-final-viral-combined_k141_49271\|\|full | Not-determined |
| KM-final-viral-combined_k141_151315\|\|full | Low-quality |
| KM-final-viral-combined_k141_62199\|\|full | Not-determined |
| KM-final-viral-combined_k141_85401\|\|full | Not-determined |
| KM-final-viral-combined_k141_129129\|\|full | Not-determined |
| KM-final-viral-combined_k141_218889\|\|full | Low-quality |
| KM-final-viral-combined_k141_62216\|\|full | Not-determined |
| KM-final-viral-combined_k141_218900\|\|full | Not-determined |
| KM-final-viral-combined_k141_185427\|\|full | Not-determined |
| KM-final-viral-combined_k141_106618\|\|full | Not-determined |
| KM-final-viral-combined_k141_94140\|\|full | Low-quality |
| KM-final-viral-combined_k141_62233\|\|full | Low-quality |
| KM-final-viral-combined_k141_173449\|\|full | Not-determined |
| KM-final-viral-combined_k141_6294\|\|full | Not-determined |
| KM-final-viral-combined_k141_140405\|\|full | Not-determined |
| KM-final-viral-combined_k141_49295\|\|full | Not-determined |
| KM-final-viral-combined_k141_129167\|\|full | Low-quality |
| KM-final-viral-combined_k141_85450\|\|full | Not-determined |
| KM-final-viral-combined_k141_119273\|\|full | Low-quality |
| KM-final-viral-combined_k141_119274\|\|full | Low-quality |
| KM-final-viral-combined_k141_119280\|\|full | Not-determined |
| KM-final-viral-combined_k141_196379\|\|full | Low-quality |
| KM-final-viral-combined_k141_173472\|\|full | Low-quality |
| KM-final-viral-combined_k141_39717\|\|full | Low-quality |
| KM-final-viral-combined_k141_94158\|\|full | Not-determined |
| KM-final-viral-combined_k141_229969\|\|full | Low-quality |
| KM-final-viral-combined_k141_129176\|\|full | Low-quality |
| KM-final-viral-combined_k141_218952\|\|full | Not-determined |
| KM-final-viral-combined_k141_106649\|\|full | Not-determined |
| KM-final-viral-combined_k141_229984\|\|full | Not-determined |
| KM-final-viral-combined_k141_6326\|\|full | Low-quality |
| KM-final-viral-combined_k141_39724\|\|full | Low-quality |
| KM-final-viral-combined_k141_173487\|\|full | Not-determined |
| KM-final-viral-combined_k141_49317\|\|full | Not-determined |
| KM-final-viral-combined_k141_239961\|\|full | Not-determined |
| KM-final-viral-combined_k141_73887\|\|full | Not-determined |
| KM-final-viral-combined_k141_140454\|\|full | Not-determined |
| KM-final-viral-combined_k141_73896\|\|full | Low-quality |
| KM-final-viral-combined_k141_230004\|\|full | Low-quality |
| KM-final-viral-combined_k141_85515\|\|full | Low-quality |
| KM-final-viral-combined_k141_94180\|\|full | Low-quality |
| KM-final-viral-combined_k141_85517\|\|full | Low-quality |
| KM-final-viral-combined_k141_239966\|\|full | Low-quality |
| KM-final-viral-combined_k141_73904\|\|full | Not-determined |
| KM-final-viral-combined_k141_261494\|\|full | Not-determined |
| KM-final-viral-combined_k141_151388\|\|full | Low-quality |
| KM-final-viral-combined_k141_6342\|\|full | Not-determined |
| KM-final-viral-combined_k141_140470\|\|full | Low-quality |
| KM-final-viral-combined_k141_119322\|\|full | Low-quality |
| KM-final-viral-combined_k141_230021\|\|full | Low-quality |
| KM-final-viral-combined_k141_62275\|\|full | Low-quality |
| KM-final-viral-combined_k141_218987\|\|full | Not-determined |
| KM-final-viral-combined_k141_218990\|\|full | Not-determined |
| KM-final-viral-combined_k141_62277\|\|full | Not-determined |
| KM-final-viral-combined_k141_119338\|\|full | Not-determined |
| 18-110-final-viral-combined_k141_18481\|\|full | Medium-quality |
| KM-final-viral-combined_k141_49364\|\|full | Low-quality |
| KM-final-viral-combined_k141_39774\|\|full | Low-quality |
| KM-final-viral-combined_k141_62293\|\|full | Low-quality |
| KM-final-viral-combined_k141_230025\|\|full | Not-determined |
| KM-final-viral-combined_k141_140490\|\|full | Low-quality |
| KM-final-viral-combined_k141_240018\|\|full | Low-quality |
| KM-final-viral-combined_k141_151428\|\|full | Low-quality |
| KM-final-viral-combined_k141_240022\|\|full | Not-determined |
| KM-final-viral-combined_k141_250241\|\|full | Low-quality |
| KM-final-viral-combined_k141_250242\|\|full | Not-determined |
| KM-final-viral-combined_k141_119359\|\|full | Not-determined |
| KM-final-viral-combined_k141_62305\|\|full | Not-determined |
| KM-final-viral-combined_k141_140497\|\|full | Not-determined |
| KM-final-viral-combined_k141_119361\|\|full | Low-quality |
| KM-final-viral-combined_k141_230041\|\|full | Low-quality |
| KM-final-viral-combined_k141_151448\|\|full | Not-determined |
| KM-final-viral-combined_k141_62306\|\|full | Low-quality |
| KM-final-viral-combined_k141_39792\|\|full | Not-determined |
| KM-final-viral-combined_k141_161615\|\|full | Low-quality |
| KM-final-viral-combined_k141_196430\|\|full | Not-determined |
| KM-final-viral-combined_k141_39807\|\|full | Low-quality |
| KM-final-viral-combined_k141_240045\|\|full | Low-quality |
| KM-final-viral-combined_k141_230077\|\|full | Not-determined |
| KM-final-viral-combined_k141_185512\|\|full | Not-determined |
| KM-final-viral-combined_k141_261547\|\|full | Low-quality |
| KM-final-viral-combined_k141_250269\|\|full | Low-quality |
| KM-final-viral-combined_k141_28586\|\|full | Low-quality |
| KM-final-viral-combined_k141_85598\|\|full | Not-determined |
| 18-106-final-viral-combined_k141_745\|\|full | High-quality |
| KM-final-viral-combined_k141_219045\|\|full | Not-determined |
| KM-final-viral-combined_k141_207717\|\|full | Low-quality |
| KM-final-viral-combined_k141_207718\|\|full | Not-determined |
| KM-final-viral-combined_k141_140522\|\|full | Not-determined |
| KM-final-viral-combined_k141_151483\|\|full | Not-determined |
| KM-final-viral-combined_k141_207728\|\|full | Low-quality |
| KM-final-viral-combined_k141_129345\|\|full | Low-quality |
| KM-final-viral-combined_k141_49456\|\|full | Not-determined |
| KM-final-viral-combined_k141_261595\|\|full | Not-determined |
| KM-final-viral-combined_k141_85636\|\|full | Not-determined |
| KM-final-viral-combined_k141_140572\|\|full | Not-determined |
| KM-final-viral-combined_k141_196545\|\|full | Low-quality |
| KM-final-viral-combined_k141_185565\|\|full | Low-quality |
| KM-final-viral-combined_k141_85675\|\|full | Low-quality |
| KM-final-viral-combined_k141_173643\|\|full | Not-determined |
| KM-final-viral-combined_k141_230156\|\|full | Not-determined |
| KM-final-viral-combined_k141_6437\|\|full | Low-quality |
| KM-final-viral-combined_k141_106761\|\|full | Low-quality |
| KM-final-viral-combined_k141_173653\|\|full | Not-determined |
| KM-final-viral-combined_k141_140587\|\|full | Not-determined |
| KM-final-viral-combined_k141_250340\|\|full | Low-quality |
| KM-final-viral-combined_k141_85689\|\|full | Low-quality |
| KM-final-viral-combined_k141_62415\|\|full | Low-quality |
| KM-final-viral-combined_k141_250348\|\|full | Low-quality |
| KM-final-viral-combined_k141_250349\|\|full | Not-determined |
| KM-final-viral-combined_k141_250368\|\|full | Low-quality |
| KM-final-viral-combined_k141_185622\|\|full | Not-determined |
| KM-final-viral-combined_k141_119415\|\|full | Not-determined |
| KM-final-viral-combined_k141_230212\|\|full | Not-determined |
| KM-final-viral-combined_k141_173708\|\|full | Low-quality |
| KM-final-viral-combined_k141_173709\|\|full | Not-determined |
| KM-final-viral-combined_k141_119427\|\|full | Not-determined |
| KM-final-viral-combined_k141_207827\|\|full | Not-determined |
| KM-final-viral-combined_k141_140628\|\|full | Low-quality |
| KM-final-viral-combined_k141_28661\|\|full | Low-quality |
| KM-final-viral-combined_k141_129435\|\|full | Low-quality |
| KM-final-viral-combined_k141_119445\|\|full | Not-determined |
| KM-final-viral-combined_k141_129438\|\|full | Low-quality |
| KM-final-viral-combined_k141_129448\|\|full | Low-quality |
| KM-final-viral-combined_k141_240176\|\|full | Low-quality |
| KM-final-viral-combined_k141_94351\|\|full | Low-quality |
| KM-final-viral-combined_k141_39939\|\|full | Low-quality |
| KM-final-viral-combined_k141_207866\|\|full | Not-determined |
| KM-final-viral-combined_k141_207869\|\|full | Not-determined |
| KM-final-viral-combined_k141_250389\|\|full | Low-quality |
| KM-final-viral-combined_k141_106809\|\|full | Not-determined |
| KM-final-viral-combined_k141_185683\|\|full | Not-determined |
| KM-final-viral-combined_k141_74019\|\|full | Not-determined |
| KM-final-viral-combined_k141_161742\|\|full | Low-quality |
| KM-final-viral-combined_k141_129474\|\|full | Not-determined |
| 18-102-final-viral-combined_k141_159670\|\|full | Medium-quality |
| KM-final-viral-combined_k141_106825\|\|full | Not-determined |
| KM-final-viral-combined_k141_240195\|\|full | Not-determined |
| KM-final-viral-combined_k141_85713\|\|full | Not-determined |
| KM-final-viral-combined_k141_250403\|\|full | Not-determined |
| KM-final-viral-combined_k141_119512\|\|full | Not-determined |
| KM-final-viral-combined_k141_6542\|\|full | Not-determined |
| KM-final-viral-combined_k141_129508\|\|full | Low-quality |
| KM-final-viral-combined_k141_250404\|\|full | Low-quality |
| KM-final-viral-combined_k141_16830\|\|full | Low-quality |
| KM-final-viral-combined_k141_85731\|\|full | Not-determined |
| KM-final-viral-combined_k141_6547\|\|full | Not-determined |
| KM-final-viral-combined_k141_74067\|\|full | Not-determined |
| KM-final-viral-combined_k141_250408\|\|full | Low-quality |
| KM-final-viral-combined_k141_39989\|\|full | Not-determined |
| KM-final-viral-combined_k141_49554\|\|full | Low-quality |
| KM-final-viral-combined_k141_219110\|\|full | Low-quality |
| KM-final-viral-combined_k141_74072\|\|full | Low-quality |
| KM-final-viral-combined_k141_173796\|\|full | Not-determined |
| KM-final-viral-combined_k141_85743\|\|full | Low-quality |
| KM-final-viral-combined_k141_94387\|\|full | Low-quality |
| KM-final-viral-combined_k141_39998\|\|full | Not-determined |
| KM-final-viral-combined_k141_261758\|\|full | Not-determined |
| KM-final-viral-combined_k141_151655\|\|full | Not-determined |
| KM-final-viral-combined_k141_240230\|\|full | Low-quality |
| KM-final-viral-combined_k141_151661\|\|full | Low-quality |
| KM-final-viral-combined_k141_62539\|\|full | Not-determined |
| KM-final-viral-combined_k141_85759\|\|full | Not-determined |
| KM-final-viral-combined_k141_6584\|\|full | Not-determined |
| KM-final-viral-combined_k141_119539\|\|full | Low-quality |
| KM-final-viral-combined_k141_230238\|\|full | Not-determined |
| KM-final-viral-combined_k141_129553\|\|full | Not-determined |
| KM-final-viral-combined_k141_129557\|\|full | Low-quality |
| KM-final-viral-combined_k141_40044\|\|full | Not-determined |
| KM-final-viral-combined_k141_196693\|\|full | Low-quality |
| KM-final-viral-combined_k141_196696\|\|full | Low-quality |
| KM-final-viral-combined_k141_250448\|\|full | Low-quality |
| KM-final-viral-combined_k141_219168\|\|full | Low-quality |
| KM-final-viral-combined_k141_207989\|\|full | Low-quality |
| KM-final-viral-combined_k141_151732\|\|full | Not-determined |
| KM-final-viral-combined_k141_16904\|\|full | Not-determined |
| KM-final-viral-combined_k141_106929\|\|full | Low-quality |
| KM-final-viral-combined_k141_151748\|\|full | Low-quality |
| KM-final-viral-combined_k141_49585\|\|full | Not-determined |
| KM-final-viral-combined_k141_49593\|\|full | Not-determined |
| KM-final-viral-combined_k141_230287\|\|full | Low-quality |
| KM-final-viral-combined_k141_250483\|\|full | Low-quality |
| KM-final-viral-combined_k141_173897\|\|full | Not-determined |
| KM-final-viral-combined_k141_161873\|\|full | Low-quality |
| KM-final-viral-combined_k141_261845\|\|full | Not-determined |
| KM-final-viral-combined_k141_219213\|\|full | Not-determined |
| KM-final-viral-combined_k141_119635\|\|full | Low-quality |
| KM-final-viral-combined_k141_119638\|\|full | Not-determined |
| KM-final-viral-combined_k141_196740\|\|full | Low-quality |
| KM-final-viral-combined_k141_74201\|\|full | Low-quality |
| KM-final-viral-combined_k141_62595\|\|full | Not-determined |
| KM-final-viral-combined_k141_49631\|\|full | Low-quality |
| KM-final-viral-combined_k141_161898\|\|full | Not-determined |
| KM-final-viral-combined_k141_173913\|\|full | Low-quality |
| KM-final-viral-combined_k141_161901\|\|full | Not-determined |
| KM-final-viral-combined_k141_219253\|\|full | Not-determined |
| KM-final-viral-combined_k141_6613\|\|full | Not-determined |
| KM-final-viral-combined_k141_161917\|\|full | Not-determined |
| KM-final-viral-combined_k141_250536\|\|full | Not-determined |
| KM-final-viral-combined_k141_173942\|\|full | Low-quality |
| KM-final-viral-combined_k141_161918\|\|full | Not-determined |
| KM-final-viral-combined_k141_49649\|\|full | Low-quality |
| KM-final-viral-combined_k141_261902\|\|full | Not-determined |
| KM-final-viral-combined_k141_230361\|\|full | Not-determined |
| KM-final-viral-combined_k141_90378\|\|full | High-quality |
| KM-final-viral-combined_k141_94511\|\|full | Not-determined |
| KM-final-viral-combined_k141_261923\|\|full | Not-determined |
| KM-final-viral-combined_k141_161941\|\|full | Not-determined |
| KM-final-viral-combined_k141_49661\|\|full | Not-determined |
| KM-final-viral-combined_k141_261926\|\|full | Low-quality |
| KM-final-viral-combined_k141_85869\|\|full | Not-determined |
| KM-final-viral-combined_k141_173968\|\|full | Low-quality |
| KM-final-viral-combined_k141_129649\|\|full | Not-determined |
| KM-final-viral-combined_k141_240309\|\|full | Low-quality |
| KM-final-viral-combined_k141_196787\|\|full | Not-determined |
| KM-final-viral-combined_k141_106947\|\|full | Low-quality |
| KM-final-viral-combined_k141_106949\|\|full | Low-quality |
| KM-final-viral-combined_k141_40186\|\|full | Not-determined |
| KM-final-viral-combined_k141_173978\|\|full | Not-determined |
| KM-final-viral-combined_k141_161953\|\|full | Low-quality |
| KM-final-viral-combined_k141_28895\|\|full | Not-determined |
| KM-final-viral-combined_k141_240310\|\|full | Low-quality |
| KM-final-viral-combined_k141_94524\|\|full | Low-quality |
| KM-final-viral-combined_k141_185774\|\|full | Not-determined |
| KM-final-viral-combined_k141_28913\|\|full | Low-quality |
| KM-final-viral-combined_k141_119765\|\|full | Low-quality |
| KM-final-viral-combined_k141_40206\|\|full | Not-determined |
| KM-final-viral-combined_k141_140827\|\|full | Not-determined |
| KM-final-viral-combined_k141_40213\|\|full | Low-quality |
| KM-final-viral-combined_k141_174031\|\|full | Low-quality |
| KM-final-viral-combined_k141_219349\|\|full | Low-quality |
| KM-final-viral-combined_k141_230442\|\|full | Not-determined |
| KM-final-viral-combined_k141_219352\|\|full | Low-quality |
| KM-final-viral-combined_k141_185816\|\|full | Low-quality |

KM-final-viral-combined_k141_208108||full Low-quality
KM-final-viral-combined_k141_119806||full Low-quality
KM-final-viral-combined_k141_62731||full Low-quality
KM-final-viral-combined_k141_140852||full Low-quality
KM-final-viral-combined_k141_151826||full Not-determined
KM-final-viral-combined_k141_196813||full Low-quality
KM-final-viral-combined_k141_161995||full Low-quality
KM-final-viral-combined_k141_6698||full Low-quality
KM-final-viral-combined_k141_107036||full Not-determined
KM-final-viral-combined_k141_140869||full Not-determined
KM-final-viral-combined_k141_219373||full Low-quality
KM-final-viral-combined_k141_219374||full Low-quality
KM-final-viral-combined_k141_74276||full Not-determined
KM-final-viral-combined_k141_62766||full Not-determined
KM-final-viral-combined_k141_119850||full Low-quality
KM-final-viral-combined_k141_107057||full Low-quality
KM-final-viral-combined_k141_196841||full Low-quality
KM-final-viral-combined_k141_17061||full Low-quality
KM-final-viral-combined_k141_28922||full Low-quality
KM-final-viral-combined_k141_140907||full Low-quality
KM-final-viral-combined_k141_28923||full Not-determined
KM-final-viral-combined_k141_208150||full Not-determined
KM-final-viral-combined_k141_196855||full Low-quality
KM-final-viral-combined_k141_250570||full Not-determined
KM-final-viral-combined_k141_17066||full Low-quality
KM-final-viral-combined_k141_49759||full Not-determined
KM-final-viral-combined_k141_185894||full Low-quality
KM-final-viral-combined_k141_49763||full Low-quality
KM-final-viral-combined_k141_119875||full Not-determined
KM-final-viral-combined_k141_250594||full Not-determined
KM-final-viral-combined_k141_208175||full Low-quality
KM-final-viral-combined_k141_151902||full Low-quality
KM-final-viral-combined_k141_185903||full Not-determined
KM-final-viral-combined_k141_219423||full Not-determined
KM-final-viral-combined_k141_230575||full Low-quality
KM-final-viral-combined_k141_140950||full Low-quality
KM-final-viral-combined_k141_174054||full Low-quality
KM-final-viral-combined_k141_140951||full Low-quality
KM-final-viral-combined_k141_40292||full Low-quality
KM-final-viral-combined_k141_262044||full Not-determined
KM-final-viral-combined_k141_174059||full Low-quality
KM-final-viral-combined_k141_174061||full Low-quality
KM-final-viral-combined_k141_219441||full Not-determined
KM-final-viral-combined_k141_17097||full Not-determined
KM-final-viral-combined_k141_86006||full Not-determined
KM-final-viral-combined_k141_49808||full Not-determined
KM-final-viral-combined_k141_174083||full Low-quality
KM-final-viral-combined_k141_86025||full Low-quality
KM-final-viral-combined_k141_208229||full Not-determined
KM-final-viral-combined_k141_219461||full Not-determined
KM-final-viral-combined_k141_94665||full Not-determined
KM-final-viral-combined_k141_141003||full Not-determined
KM-final-viral-combined_k141_62890||full Not-determined
KM-final-viral-combined_k141_230617||full Not-determined
KM-final-viral-combined_k141_141013||full Not-determined
KM-final-viral-combined_k141_86053||full Not-determined
KM-final-viral-combined_k141_174094||full Low-quality
KM-final-viral-combined_k141_250629||full Low-quality
KM-final-viral-combined_k141_94678||full Low-quality
KM-final-viral-combined_k141_129781||full Low-quality
KM-final-viral-combined_k141_28964||full Not-determined
KM-final-viral-combined_k141_107110||full Not-determined
KM-final-viral-combined_k141_86073||full Not-determined
KM-final-viral-combined_k141_107115||full Not-determined
KM-final-viral-combined_k141_219517||full Low-quality
KM-final-viral-combined_k141_94704||full Not-determined
KM-final-viral-combined_k141_151933||full Not-determined
KM-final-viral-combined_k141_74417||full Low-quality
KM-final-viral-combined_k141_6773||full Low-quality
KM-final-viral-combined_k141_17113||full Low-quality
KM-final-viral-combined_k141_107134||full Not-determined
KM-final-viral-combined_k141_141068||full Not-determined
KM-final-viral-combined_k141_250663||full Low-quality
KM-final-viral-combined_k141_74430||full Not-determined
KM-final-viral-combined_k141_174157||full Low-quality
KM-final-viral-combined_k141_151952||full Low-quality
KM-final-viral-combined_k141_129795||full Low-quality
KM-final-viral-combined_k141_40374||full Not-determined
KM-final-viral-combined_k141_86107||full Not-determined
KM-final-viral-combined_k141_62926||full Low-quality
KM-final-viral-combined_k141_196986||full Low-quality
KM-final-viral-combined_k141_219544||full Low-quality
KM-final-viral-combined_k141_240370||full Low-quality
KM-final-viral-combined_k141_119929||full Not-determined
KM-final-viral-combined_k141_62933||full Not-determined
KM-final-viral-combined_k141_151960||full Low-quality
KM-final-viral-combined_k141_162058||full Not-determined
KM-final-viral-combined_k141_119935||full Low-quality
KM-final-viral-combined_k141_129807||full Low-quality
KM-final-viral-combined_k141_40391||full Low-quality
KM-final-viral-combined_k141_162075||full Not-determined
KM-final-viral-combined_k141_262099||full Not-determined
KM-final-viral-combined_k141_107159||full Not-determined
KM-final-viral-combined_k141_129812||full Low-quality
KM-final-viral-combined_k141_250695||full Not-determined
KM-final-viral-combined_k141_219560||full Not-determined
KM-final-viral-combined_k141_162089||full Not-determined
KM-final-viral-combined_k141_17152||full Not-determined
KM-final-viral-combined_k141_230742||full Low-quality
KM-final-viral-combined_k141_186018||full Low-quality
KM-final-viral-combined_k141_197025||full Not-determined
KM-final-viral-combined_k141_219575||full Not-determined
KM-final-viral-combined_k141_151985||full Low-quality
KM-final-viral-combined_k141_186042||full Low-quality
KM-final-viral-combined_k141_162139||full Low-quality
KM-final-viral-combined_k141_74446||full Low-quality
KM-final-viral-combined_k141_262151||full Not-determined
KM-final-viral-combined_k141_107215||full Not-determined
KM-final-viral-combined_k141_86136||full Low-quality
KM-final-viral-combined_k141_17213||full Low-quality
KM-final-viral-combined_k141_230779||full Low-quality
KM-final-viral-combined_k141_119983||full Low-quality
KM-final-viral-combined_k141_174254||full Not-determined
KM-final-viral-combined_k141_74460||full Not-determined
KM-final-viral-combined_k141_162159||full Not-determined
KM-final-viral-combined_k141_197067||full Low-quality
KM-final-viral-combined_k141_86171||full Not-determined
KM-final-viral-combined_k141_141181||full Not-determined
KM-final-viral-combined_k141_197090||full Low-quality
KM-final-viral-combined_k141_141184||full Not-determined
KM-final-viral-combined_k141_208385||full Not-determined
KM-final-viral-combined_k141_86184||full Not-determined
KM-final-viral-combined_k141_197100||full Low-quality
KM-final-viral-combined_k141_29054||full Not-determined
KM-final-viral-combined_k141_141199||full Not-determined
KM-final-viral-combined_k141_141200||full Low-quality
KM-final-viral-combined_k141_86201||full Low-quality
KM-final-viral-combined_k141_162194||full Not-determined
KM-final-viral-combined_k141_174296||full Not-determined
KM-final-viral-combined_k141_86207||full Not-determined
KM-final-viral-combined_k141_141218||full Not-determined
KM-final-viral-combined_k141_230826||full Low-quality
KM-final-viral-combined_k141_219583||full Low-quality
KM-final-viral-combined_k141_197131||full Low-quality
KM-final-viral-combined_k141_262192||full Low-quality
KM-final-viral-combined_k141_141231||full Low-quality
KM-final-viral-combined_k141_17296||full Low-quality
KM-final-viral-combined_k141_62993||full Not-determined
KM-final-viral-combined_k141_208416||full Not-determined
KM-final-viral-combined_k141_120076||full Not-determined
KM-final-viral-combined_k141_186141||full Low-quality
KM-final-viral-combined_k141_29080||full Not-determined
KM-final-viral-combined_k141_86217||full Not-determined
KM-final-viral-combined_k141_240493||full Low-quality
KM-final-viral-combined_k141_74563||full Not-determined
KM-final-viral-combined_k141_74577||full Low-quality
KM-final-viral-combined_k141_230847||full Low-quality
KM-final-viral-combined_k141_186164||full Not-determined
KM-final-viral-combined_k141_17313||full Low-quality
KM-final-viral-combined_k141_49916||full Low-quality
KM-final-viral-combined_k141_74586||full Not-determined
KM-final-viral-combined_k141_240538||full Low-quality
KM-final-viral-combined_k141_240539||full Not-determined
KM-final-viral-combined_k141_186183||full Not-determined
KM-final-viral-combined_k141_63057||full Not-determined
KM-final-viral-combined_k141_174388||full Not-determined
KM-final-viral-combined_k141_86241||full Not-determined
KM-final-viral-combined_k141_174390||full Low-quality
KM-final-viral-combined_k141_120117||full Low-quality
KM-final-viral-combined_k141_230875||full Low-quality
KM-final-viral-combined_k141_240555||full Low-quality
KM-final-viral-combined_k141_49931||full Low-quality
KM-final-viral-combined_k141_141277||full Low-quality
KM-final-viral-combined_k141_141281||full Low-quality
KM-final-viral-combined_k141_208507||full Low-quality
KM-final-viral-combined_k141_186233||full Not-determined
KM-final-viral-combined_k141_240584||full Not-determined
KM-final-viral-combined_k141_219670||full Low-quality
KM-final-viral-combined_k141_94826||full Low-quality
17-109-final-viral-combined_k141_386815||full High-quality
KM-final-viral-combined_k141_17359||full Not-determined
KM-final-viral-combined_k141_141329||full Low-quality
KM-final-viral-combined_k141_74664||full Not-determined
KM-final-viral-combined_k141_208533||full Low-quality
KM-final-viral-combined_k141_240602||full Not-determined
KM-final-viral-combined_k141_74676||full Low-quality
KM-final-viral-combined_k141_186265||full Low-quality
KM-final-viral-combined_k141_240614||full Not-determined
KM-final-viral-combined_k141_94840||full Low-quality
KM-final-viral-combined_k141_197240||full Low-quality
KM-final-viral-combined_k141_152173||full Not-determined
KM-final-viral-combined_k141_86345||full Low-quality
KM-final-viral-combined_k141_152179||full Not-determined
KM-final-viral-combined_k141_29127||full Not-determined
KM-final-viral-combined_k141_230943||full Low-quality
KM-final-viral-combined_k141_174475||full Low-quality
KM-final-viral-combined_k141_152192||full Not-determined
KM-final-viral-combined_k141_230950||full Low-quality
KM-final-viral-combined_k141_186283||full Not-determined
KM-final-viral-combined_k141_240658||full Not-determined
KM-final-viral-combined_k141_17441||full Low-quality
KM-final-viral-combined_k141_197274||full Not-determined
KM-final-viral-combined_k141_152231||full Not-determined
KM-final-viral-combined_k141_107384||full Low-quality
KM-final-viral-combined_k141_174545||full Not-determined
KM-final-viral-combined_k141_86412||full Low-quality
KM-final-viral-combined_k141_262361||full Not-determined
KM-final-viral-combined_k141_17455||full Low-quality
KM-final-viral-combined_k141_94885||full Not-determined
KM-final-viral-combined_k141_262370||full Low-quality
KM-final-viral-combined_k141_120196||full Low-quality
KM-final-viral-combined_k141_197319||full Low-quality
KM-final-viral-combined_k141_120197||full Not-determined
KM-final-viral-combined_k141_141473||full Low-quality
KM-final-viral-combined_k141_208670||full Not-determined
KM-final-viral-combined_k141_29202||full Not-determined
KM-final-viral-combined_k141_86437||full Low-quality
KM-final-viral-combined_k141_152278||full Low-quality
KM-final-viral-combined_k141_74766||full Not-determined
KM-final-viral-combined_k141_17479||full Not-determined
KM-final-viral-combined_k141_208712||full Low-quality
KM-final-viral-combined_k141_29209||full Not-determined
KM-final-viral-combined_k141_152288||full Low-quality
KM-final-viral-combined_k141_174631||full Not-determined
KM-final-viral-combined_k141_174635||full Low-quality
KM-final-viral-combined_k141_174638||full Low-quality
KM-final-viral-combined_k141_231047||full Not-determined
KM-final-viral-combined_k141_63233||full Low-quality
KM-final-viral-combined_k141_29227||full Low-quality
KM-final-viral-combined_k141_174648||full Not-determined
KM-final-viral-combined_k141_86449||full Low-quality
KM-final-viral-combined_k141_262449||full Low-quality
KM-final-viral-combined_k141_94908||full Not-determined
KM-final-viral-combined_k141_120268||full Low-quality
KM-final-viral-combined_k141_120270||full Low-quality
KM-final-viral-combined_k141_208742||full Not-determined
KM-final-viral-combined_k141_231082||full Low-quality
KM-final-viral-combined_k141_250936||full Low-quality
KM-final-viral-combined_k141_94922||full Low-quality
KM-final-viral-combined_k141_17530||full Not-determined
KM-final-viral-combined_k141_74804||full Not-determined
18-105-final-viral-combined_k141_204782||full High-quality
KM-final-viral-combined_k141_231112||full Not-determined
KM-final-viral-combined_k141_208764||full Low-quality
KM-final-viral-combined_k141_186315||full Not-determined
KM-final-viral-combined_k141_141501||full Not-determined
KM-final-viral-combined_k141_141515||full Low-quality
KM-final-viral-combined_k141_120296||full Not-determined
KM-final-viral-combined_k141_63264||full Low-quality
KM-final-viral-combined_k141_197468||full Low-quality
KM-final-viral-combined_k141_219906||full Not-determined
KM-final-viral-combined_k141_6953||full Low-quality
KM-final-viral-combined_k141_208784||full Not-determined
KM-final-viral-combined_k141_262515||full Low-quality
KM-final-viral-combined_k141_74855||full Low-quality
KM-final-viral-combined_k141_174723||full Low-quality
KM-final-viral-combined_k141_6960||full Not-determined
KM-final-viral-combined_k141_74862||full Low-quality
KM-final-viral-combined_k141_29314||full Not-determined
KM-final-viral-combined_k141_262531||full Not-determined
KM-final-viral-combined_k141_107563||full Not-determined
KM-final-viral-combined_k141_120306||full Not-determined
KM-final-viral-combined_k141_141539||full Low-quality
KM-final-viral-combined_k141_162492||full Not-determined
KM-final-viral-combined_k141_208814||full Not-determined
KM-final-viral-combined_k141_86531||full Low-quality
KM-final-viral-combined_k141_186375||full Not-determined
KM-final-viral-combined_k141_186376||full Low-quality
KM-final-viral-combined_k141_6989||full Low-quality
KM-final-viral-combined_k141_120331||full Not-determined
KM-final-viral-combined_k141_63314||full Low-quality
KM-final-viral-combined_k141_50140||full Not-determined
KM-final-viral-combined_k141_208849||full Low-quality
KM-final-viral-combined_k141_29354||full Low-quality
KM-final-viral-combined_k141_74914||full Low-quality
KM-final-viral-combined_k141_74916||full Low-quality
KM-final-viral-combined_k141_186391||full Not-determined
KM-final-viral-combined_k141_240861||full Low-quality
KM-final-viral-combined_k141_240868||full Not-determined
KM-final-viral-combined_k141_107621||full Low-quality
KM-final-viral-combined_k141_219979||full Low-quality
KM-final-viral-combined_k141_231184||full Low-quality
KM-final-viral-combined_k141_86577||full Not-determined
KM-final-viral-combined_k141_86579||full Not-determined
KM-final-viral-combined_k141_63361||full Not-determined
KM-final-viral-combined_k141_231192||full Not-determined
KM-final-viral-combined_k141_152393||full Low-quality
KM-final-viral-combined_k141_17599||full Low-quality
KM-final-viral-combined_k141_197564||full Not-determined
KM-final-viral-combined_k141_240886||full Not-determined
KM-final-viral-combined_k141_86605||full Low-quality
KM-final-viral-combined_k141_29385||full Low-quality
KM-final-viral-combined_k141_231213||full Not-determined
KM-final-viral-combined_k141_186437||full Low-quality
KM-final-viral-combined_k141_86609||full Not-determined
KM-final-viral-combined_k141_240892||full Low-quality
KM-final-viral-combined_k141_162561||full Not-determined
KM-final-viral-combined_k141_186460||full Not-determined
KM-final-viral-combined_k141_17629||full Low-quality
KM-final-viral-combined_k141_17645||full Low-quality
KM-final-viral-combined_k141_231246||full Low-quality
KM-final-viral-combined_k141_95043||full Low-quality
KM-final-viral-combined_k141_208913||full Not-determined
KM-final-viral-combined_k141_95054||full Low-quality
KM-final-viral-combined_k141_120431||full Not-determined
KM-final-viral-combined_k141_152452||full Low-quality
KM-final-viral-combined_k141_141676||full Not-determined
KM-final-viral-combined_k141_240939||full Not-determined
KM-final-viral-combined_k141_262590||full Low-quality
KM-final-viral-combined_k141_186509||full Low-quality
KM-final-viral-combined_k141_152455||full Low-quality
KM-final-viral-combined_k141_240950||full Not-determined
KM-final-viral-combined_k141_186523||full Not-determined
KM-final-viral-combined_k141_152459||full Not-determined
KM-final-viral-combined_k141_86670||full Not-determined
KM-final-viral-combined_k141_63447||full Low-quality
KM-final-viral-combined_k141_86676||full Low-quality
KM-final-viral-combined_k141_152468||full Low-quality
KM-final-viral-combined_k141_95099||full Not-determined
KM-final-viral-combined_k141_197657||full Low-quality
KM-final-viral-combined_k141_197666||full Not-determined
KM-final-viral-combined_k141_162629||full Not-determined
KM-final-viral-combined_k141_186565||full Not-determined
KM-final-viral-combined_k141_40560||full Not-determined
KM-final-viral-combined_k141_197677||full Not-determined
KM-final-viral-combined_k141_208998||full Not-determined
KM-final-viral-combined_k141_186580||full Not-determined
KM-final-viral-combined_k141_186583||full Low-quality
KM-final-viral-combined_k141_50281||full Not-determined
KM-final-viral-combined_k141_241007||full Low-quality
KM-final-viral-combined_k141_107702||full Not-determined
KM-final-viral-combined_k141_86733||full Not-determined
KM-final-viral-combined_k141_7106||full Not-determined
KM-final-viral-combined_k141_186600||full Not-determined
KM-final-viral-combined_k141_129986||full Low-quality
KM-final-viral-combined_k141_162669||full Low-quality
KM-final-viral-combined_k141_186608||full Low-quality
KM-final-viral-combined_k141_152554||full Low-quality
KM-final-viral-combined_k141_7112||full Not-determined
KM-final-viral-combined_k141_40581||full Not-determined
KM-final-viral-combined_k141_40583||full Not-determined
KM-final-viral-combined_k141_141726||full Not-determined
KM-final-viral-combined_k141_186624||full Low-quality
KM-final-viral-combined_k141_17721||full Not-determined
KM-final-viral-combined_k141_130017||full Not-determined
KM-final-viral-combined_k141_241043||full Not-determined
KM-final-viral-combined_k141_162681||full Not-determined
KM-final-viral-combined_k141_220171||full Not-determined
KM-final-viral-combined_k141_186641||full Not-determined
KM-final-viral-combined_k141_130026||full Not-determined
KM-final-viral-combined_k141_220176||full Not-determined
KM-final-viral-combined_k141_251050||full Not-determined
KM-final-viral-combined_k141_174905||full Low-quality
KM-final-viral-combined_k141_17736||full Low-quality
KM-final-viral-combined_k141_220185||full Not-determined
KM-final-viral-combined_k141_162696||full Not-determined
KM-final-viral-combined_k141_107744||full Not-determined
KM-final-viral-combined_k141_241071||full Low-quality
KM-final-viral-combined_k141_29580||full Not-determined
KM-final-viral-combined_k141_162717||full Not-determined
KM-final-viral-combined_k141_141773||full Low-quality
KM-final-viral-combined_k141_120541||full Low-quality
KM-final-viral-combined_k141_231407||full Not-determined
KM-final-viral-combined_k141_7167||full Low-quality
KM-final-viral-combined_k141_186659||full Not-determined
KM-final-viral-combined_k141_197730||full Low-quality
KM-final-viral-combined_k141_17780||full Not-determined
KM-final-viral-combined_k141_63588||full Low-quality
KM-final-viral-combined_k141_95124||full Low-quality
KM-final-viral-combined_k141_174976||full Not-determined
KM-final-viral-combined_k141_241082||full Low-quality
KM-final-viral-combined_k141_251114||full Not-determined
KM-final-viral-combined_k141_174980||full Low-quality
KM-final-viral-combined_k141_231437||full Low-quality
KM-final-viral-combined_k141_63602||full Not-determined
KM-final-viral-combined_k141_241094||full Not-determined
KM-final-viral-combined_k141_174988||full Low-quality
KM-final-viral-combined_k141_251127||full Not-determined
KM-final-viral-combined_k141_209112||full Not-determined
KM-final-viral-combined_k141_231452||full Low-quality
KM-final-viral-combined_k141_209115||full Not-determined
KM-final-viral-combined_k141_95155||full Not-determined
KM-final-viral-combined_k141_251143||full Not-determined
KM-final-viral-combined_k141_241120||full Not-determined
KM-final-viral-combined_k141_251156||full Not-determined
KM-final-viral-combined_k141_220279||full Low-quality
KM-final-viral-combined_k141_241127||full Not-determined
KM-final-viral-combined_k141_17823||full Low-quality
KM-final-viral-combined_k141_7244||full Low-quality
KM-final-viral-combined_k141_7245||full Low-quality
KM-final-viral-combined_k141_7247||full Not-determined
KM-final-viral-combined_k141_95173||full Low-quality
KM-final-viral-combined_k141_241130||full Not-determined
KM-final-viral-combined_k141_197807||full Low-quality
KM-final-viral-combined_k141_29659||full Low-quality
KM-final-viral-combined_k141_141819||full Not-determined
KM-final-viral-combined_k141_251183||full Not-determined
KM-final-viral-combined_k141_162779||full Low-quality
KM-final-viral-combined_k141_251195||full Not-determined
KM-final-viral-combined_k141_241149||full Low-quality
KM-final-viral-combined_k141_40704||full Not-determined
KM-final-viral-combined_k141_152578||full Low-quality
KM-final-viral-combined_k141_29678||full Not-determined
KM-final-viral-combined_k141_197824||full Not-determined
KM-final-viral-combined_k141_120680||full Low-quality
KM-final-viral-combined_k141_152587||full Low-quality
KM-final-viral-combined_k141_220295||full Not-determined
KM-final-viral-combined_k141_209140||full Low-quality
KM-final-viral-combined_k141_40713||full Not-determined
KM-final-viral-combined_k141_209147||full Not-determined
KM-final-viral-combined_k141_220311||full Low-quality
KM-final-viral-combined_k141_209162||full Not-determined
KM-final-viral-combined_k141_63668||full Low-quality
KM-final-viral-combined_k141_107813||full Low-quality
KM-final-viral-combined_k141_17871||full Low-quality
KM-final-viral-combined_k141_75306||full Not-determined
KM-final-viral-combined_k141_220319||full Low-quality
KM-final-viral-combined_k141_186730||full Not-determined
KM-final-viral-combined_k141_120731||full Low-quality
KM-final-viral-combined_k141_7330||full Low-quality
KM-final-viral-combined_k141_231579||full Not-determined
KM-final-viral-combined_k141_197837||full Not-determined
KM-final-viral-combined_k141_86903||full Low-quality
KM-final-viral-combined_k141_186744||full Low-quality
KM-final-viral-combined_k141_175049||full Not-determined
KM-final-viral-combined_k141_7348||full Low-quality
KM-final-viral-combined_k141_7349||full Low-quality
KM-final-viral-combined_k141_186763||full Not-determined
KM-final-viral-combined_k141_40755||full Low-quality
KM-final-viral-combined_k141_95250||full Low-quality
KM-final-viral-combined_k141_231611||full Not-determined
KM-final-viral-combined_k141_141841||full Low-quality
KM-final-viral-combined_k141_40756||full Not-determined
KM-final-viral-combined_k141_152658||full Low-quality
KM-final-viral-combined_k141_17907||full Not-determined
KM-final-viral-combined_k141_7380||full Not-determined
KM-final-viral-combined_k141_175074||full Not-determined
KM-final-viral-combined_k141_17920||full Not-determined
KM-final-viral-combined_k141_86936||full Not-determined
KM-final-viral-combined_k141_7394||full Low-quality
KM-final-viral-combined_k141_120805||full Low-quality
KM-final-viral-combined_k141_95277||full Not-determined
KM-final-viral-combined_k141_86966||full Low-quality
KM-final-viral-combined_k141_120822||full Low-quality
KM-final-viral-combined_k141_95289||full Not-determined
KM-final-viral-combined_k141_241289||full Not-determined
KM-final-viral-combined_k141_231621||full Not-determined
KM-final-viral-combined_k141_75356||full Not-determined
KM-final-viral-combined_k141_86990||full Not-determined
KM-final-viral-combined_k141_152713||full Low-quality
KM-final-viral-combined_k141_209272||full Low-quality
KM-final-viral-combined_k141_197927||full Low-quality
KM-final-viral-combined_k141_186805||full Low-quality
KM-final-viral-combined_k141_75407||full Not-determined
KM-final-viral-combined_k141_186809||full Not-determined
KM-final-viral-combined_k141_40832||full Not-determined
KM-final-viral-combined_k141_186824||full Not-determined
KM-final-viral-combined_k141_241317||full Low-quality
KM-final-viral-combined_k141_141927||full Low-quality
17-108_k141_330479 Complete
KM-final-viral-combined_k141_95349||full Not-determined
KM-final-viral-combined_k141_175164||full Low-quality
KM-final-viral-combined_k141_262761||full Low-quality
KM-final-viral-combined_k141_220457||full Not-determined
KM-final-viral-combined_k141_162798||full Low-quality
KM-final-viral-combined_k141_7437||full Not-determined
KM-final-viral-combined_k141_75443||full Low-quality
KM-final-viral-combined_k141_175197||full Low-quality
KM-final-viral-combined_k141_87087||full Not-determined
KM-final-viral-combined_k141_75444||full Not-determined
KM-final-viral-combined_k141_107973||full Low-quality
KM-final-viral-combined_k141_152772||full Low-quality
KM-final-viral-combined_k141_162827||full Not-determined
KM-final-viral-combined_k141_152774||full Low-quality
KM-final-viral-combined_k141_231679||full Not-determined
KM-final-viral-combined_k141_241353||full Low-quality
KM-final-viral-combined_k141_63838||full Low-quality
KM-final-viral-combined_k141_241362||full Low-quality
KM-final-viral-combined_k141_50525||full Not-determined
KM-final-viral-combined_k141_152793||full Low-quality
KM-final-viral-combined_k141_262782||full Not-determined
KM-final-viral-combined_k141_120914||full Low-quality
KM-final-viral-combined_k141_50530||full Not-determined
KM-final-viral-combined_k141_175210||full Not-determined
KM-final-viral-combined_k141_63846||full Not-determined
KM-final-viral-combined_k141_50540||full Not-determined
KM-final-viral-combined_k141_50541||full Low-quality
KM-final-viral-combined_k141_186910||full Not-determined
KM-final-viral-combined_k141_162866||full Low-quality
KM-final-viral-combined_k141_50548||full Not-determined
KM-final-viral-combined_k141_75533||full Not-determined
KM-final-viral-combined_k141_209386||full Not-determined
KM-final-viral-combined_k141_162878||full Not-determined
KM-final-viral-combined_k141_29765||full Low-quality
KM-final-viral-combined_k141_63884||full Low-quality
KM-final-viral-combined_k141_162884||full Low-quality
KM-final-viral-combined_k141_142022||full Not-determined
KM-final-viral-combined_k141_162888||full Not-determined
KM-final-viral-combined_k141_75542||full Low-quality
KM-final-viral-combined_k141_63887||full Not-determined
KM-final-viral-combined_k141_231690||full Low-quality
KM-final-viral-combined_k141_220566||full Low-quality
KM-final-viral-combined_k141_75557||full Not-determined
KM-final-viral-combined_k141_50564||full Not-determined
KM-final-viral-combined_k141_75566||full Not-determined
KM-final-viral-combined_k141_130160||full Not-determined
KM-final-viral-combined_k141_198071||full Low-quality
KM-final-viral-combined_k141_75574||full Not-determined
KM-final-viral-combined_k141_251356||full Low-quality
KM-final-viral-combined_k141_152838||full Not-determined
KM-final-viral-combined_k141_7516||full Not-determined
KM-final-viral-combined_k141_120971||full Not-determined
KM-final-viral-combined_k141_262839||full Not-determined
KM-final-viral-combined_k141_241416||full Not-determined
KM-final-viral-combined_k141_175274||full Low-quality
KM-final-viral-combined_k141_142076||full Low-quality
KM-final-viral-combined_k141_50590||full Low-quality
KM-final-viral-combined_k141_120981||full Not-determined
KM-final-viral-combined_k141_29847||full Low-quality
KM-final-viral-combined_k141_175283||full Not-determined
KM-final-viral-combined_k141_63956||full Not-determined
KM-final-viral-combined_k141_120987||full Not-determined
KM-final-viral-combined_k141_162978||full Not-determined
KM-final-viral-combined_k141_75619||full Low-quality
KM-final-viral-combined_k141_120989||full Not-determined
KM-final-viral-combined_k141_186998||full Not-determined
KM-final-viral-combined_k141_50598||full Low-quality
KM-final-viral-combined_k141_87227||full Not-determined
18-109-final-viral-combined_k141_113137||full High-quality
KM-final-viral-combined_k141_121007||full Low-quality
KM-final-viral-combined_k141_75652||full Low-quality
KM-final-viral-combined_k141_75653||full Low-quality
KM-final-viral-combined_k141_75657||full Not-determined
KM-final-viral-combined_k141_63996||full Not-determined
KM-final-viral-combined_k141_251436||full Low-quality
KM-final-viral-combined_k141_64001||full Low-quality
KM-final-viral-combined_k141_95388||full Not-determined
KM-final-viral-combined_k141_142133||full Not-determined
KM-final-viral-combined_k141_262902||full Low-quality
KM-final-viral-combined_k141_187056||full Low-quality
KM-final-viral-combined_k141_87267||full Low-quality
KM-final-viral-combined_k141_75668||full Not-determined
KM-final-viral-combined_k141_198157||full Low-quality
KM-final-viral-combined_k141_108072||full Not-determined
17-108-final-viral-combined_k141_330479||full Medium-quality
KM-final-viral-combined_k141_75673||full Low-quality
KM-final-viral-combined_k141_121037||full Not-determined
KM-final-viral-combined_k141_87281||full Not-determined
KM-final-viral-combined_k141_220750||full Not-determined
KM-final-viral-combined_k141_187076||full Low-quality
KM-final-viral-combined_k141_142171||full Not-determined
KM-final-viral-combined_k141_163013||full Not-determined
KM-final-viral-combined_k141_108097||full Low-quality
KM-final-viral-combined_k141_262930||full Low-quality
KM-final-viral-combined_k141_121064||full Not-determined
KM-final-viral-combined_k141_198195||full Not-determined
KM-final-viral-combined_k141_7618||full Not-determined
KM-final-viral-combined_k141_108107||full Not-determined
KM-final-viral-combined_k141_75683||full Low-quality
KM-final-viral-combined_k141_50716||full Not-determined
KM-final-viral-combined_k141_29909||full Not-determined
KM-final-viral-combined_k141_262940||full Low-quality
KM-final-viral-combined_k141_108127||full Low-quality
KM-final-viral-combined_k141_121098||full Low-quality
KM-final-viral-combined_k141_87287||full Not-determined
KM-final-viral-combined_k141_95443||full Not-determined
KM-final-viral-combined_k141_220789||full Not-determined
KM-final-viral-combined_k141_262951||full Not-determined
KM-final-viral-combined_k141_130310||full Low-quality
KM-final-viral-combined_k141_87300||full Not-determined
KM-final-viral-combined_k141_18175||full Not-determined
KM-final-viral-combined_k141_142203||full Low-quality
KM-final-viral-combined_k141_130321||full Low-quality
KM-final-viral-combined_k141_75704||full Not-determined
KM-final-viral-combined_k141_50764||full Not-determined
KM-final-viral-combined_k141_121124||full Low-quality
KM-final-viral-combined_k141_198228||full Low-quality
KM-final-viral-combined_k141_251507||full Not-determined
KM-final-viral-combined_k141_95460||full Not-determined
KM-final-viral-combined_k141_130351||full Low-quality
KM-final-viral-combined_k141_75735||full Not-determined
KM-final-viral-combined_k141_142240||full Low-quality
KM-final-viral-combined_k141_95484||full Not-determined
KM-final-viral-combined_k141_18213||full Low-quality
KM-final-viral-combined_k141_142252||full Not-determined
KM-final-viral-combined_k141_18226||full Not-determined
KM-final-viral-combined_k141_231842||full Not-determined
KM-final-viral-combined_k141_108192||full Low-quality
KM-final-viral-combined_k141_153056||full Not-determined
KM-final-viral-combined_k141_175464||full Low-quality
KM-final-viral-combined_k141_163119||full Low-quality
KM-final-viral-combined_k141_231865||full Not-determined
KM-final-viral-combined_k141_50793||full Low-quality
KM-final-viral-combined_k141_95511||full Low-quality
KM-final-viral-combined_k141_95516||full Not-determined
KM-final-viral-combined_k141_163126||full Low-quality
KM-final-viral-combined_k141_75780||full Not-determined
KM-final-viral-combined_k141_263028||full Low-quality
KM-final-viral-combined_k141_231881||full Not-determined

| | |
|---|---|
| KM-final-viral-combined_k141_163136\|\|full | Not-determined |
| KM-final-viral-combined_k141_241609\|\|full | Not-determined |
| KM-final-viral-combined_k141_187145\|\|full | Not-determined |
| KM-final-viral-combined_k141_7724\|\|full | Not-determined |
| KM-final-viral-combined_k141_163139\|\|full | Low-quality |
| KM-final-viral-combined_k141_18259\|\|full | Not-determined |
| KM-final-viral-combined_k141_50810\|\|full | Low-quality |
| KM-final-viral-combined_k141_220859\|\|full | Not-determined |
| KM-final-viral-combined_k141_87404\|\|full | Not-determined |
| KM-final-viral-combined_k141_87408\|\|full | Not-determined |
| KM-final-viral-combined_k141_75836\|\|full | Low-quality |
| KM-final-viral-combined_k141_263066\|\|full | Not-determined |
| KM-final-viral-combined_k141_187173\|\|full | Not-determined |
| KM-final-viral-combined_k141_241647\|\|full | Not-determined |
| KM-final-viral-combined_k141_231908\|\|full | Low-quality |
| KM-final-viral-combined_k141_187182\|\|full | Low-quality |
| KM-final-viral-combined_k141_231931\|\|full | Low-quality |
| KM-final-viral-combined_k141_163171\|\|full | Not-determined |
| KM-final-viral-combined_k141_50834\|\|full | Low-quality |
| KM-final-viral-combined_k141_209490\|\|full | Low-quality |
| KM-final-viral-combined_k141_142374\|\|full | Not-determined |
| KM-final-viral-combined_k141_108255\|\|full | Not-determined |
| KM-final-viral-combined_k141_50848\|\|full | Low-quality |
| KM-final-viral-combined_k141_30035\|\|full | Not-determined |
| KM-final-viral-combined_k141_50853\|\|full | Low-quality |
| KM-final-viral-combined_k141_241675\|\|full | Low-quality |
| KM-final-viral-combined_k141_87450\|\|full | Low-quality |
| KM-final-viral-combined_k141_175516\|\|full | Low-quality |
| KM-final-viral-combined_k141_209512\|\|full | Not-determined |
| KM-final-viral-combined_k141_187214\|\|full | Not-determined |
| KM-final-viral-combined_k141_153154\|\|full | Low-quality |
| KM-final-viral-combined_k141_251599\|\|full | Low-quality |
| KM-final-viral-combined_k141_41051\|\|full | Low-quality |
| KM-final-viral-combined_k141_209526\|\|full | Not-determined |
| KM-final-viral-combined_k141_30060\|\|full | Not-determined |
| KM-final-viral-combined_k141_251608\|\|full | Low-quality |
| KM-final-viral-combined_k141_87456\|\|full | Low-quality |
| KM-final-viral-combined_k141_87459\|\|full | Low-quality |
| KM-final-viral-combined_k141_18326\|\|full | Low-quality |
| KM-final-viral-combined_k141_64112\|\|full | Low-quality |
| 17-109-final-viral-combined_k141_190114\|\|full | Medium-quality |
| KM-final-viral-combined_k141_30077\|\|full | Not-determined |
| KM-final-viral-combined_k141_251624\|\|full | Not-determined |
| KM-final-viral-combined_k141_95611\|\|full | Not-determined |
| KM-final-viral-combined_k141_41075\|\|full | Low-quality |
| KM-final-viral-combined_k141_175566\|\|full | Not-determined |
| KM-final-viral-combined_k141_241697\|\|full | Low-quality |
| KM-final-viral-combined_k141_187272\|\|full | Not-determined |
| KM-final-viral-combined_k141_41093\|\|full | Not-determined |
| KM-final-viral-combined_k141_232004\|\|full | Low-quality |
| KM-final-viral-combined_k141_163217\|\|full | Not-determined |
| KM-final-viral-combined_k141_187293\|\|full | Low-quality |
| KM-final-viral-combined_k141_220968\|\|full | Not-determined |
| KM-final-viral-combined_k141_175579\|\|full | Not-determined |
| KM-final-viral-combined_k141_87505\|\|full | Low-quality |
| KM-final-viral-combined_k141_198350\|\|full | Low-quality |
| KM-final-viral-combined_k141_163237\|\|full | Not-determined |
| KM-final-viral-combined_k141_121259\|\|full | Low-quality |
| KM-final-viral-combined_k141_87515\|\|full | Not-determined |
| KM-final-viral-combined_k141_18364\|\|full | Not-determined |
| KM-final-viral-combined_k141_7838\|\|full | Low-quality |
| KM-final-viral-combined_k141_220994\|\|full | Not-determined |
| KM-final-viral-combined_k141_220998\|\|full | Not-determined |
| KM-final-viral-combined_k141_209588\|\|full | Not-determined |
| KM-final-viral-combined_k141_187324\|\|full | Not-determined |
| KM-final-viral-combined_k141_232048\|\|full | Not-determined |
| KM-final-viral-combined_k141_121286\|\|full | Not-determined |
| KM-final-viral-combined_k141_121287\|\|full | Low-quality |
| KM-final-viral-combined_k141_30104\|\|full | Not-determined |
| KM-final-viral-combined_k141_198371\|\|full | Low-quality |
| KM-final-viral-combined_k141_87525\|\|full | Low-quality |
| KM-final-viral-combined_k141_142479\|\|full | Not-determined |
| KM-final-viral-combined_k141_87540\|\|full | Not-determined |
| KM-final-viral-combined_k141_75957\|\|full | Not-determined |
| KM-final-viral-combined_k141_221012\|\|full | Not-determined |
| KM-final-viral-combined_k141_75959\|\|full | Not-determined |
| KM-final-viral-combined_k141_130471\|\|full | Not-determined |
| KM-final-viral-combined_k141_209617\|\|full | Not-determined |
| KM-final-viral-combined_k141_263230\|\|full | Low-quality |
| KM-final-viral-combined_k141_30136\|\|full | Not-determined |
| KM-final-viral-combined_k141_95653\|\|full | Not-determined |
| KM-final-viral-combined_k141_7899\|\|full | Low-quality |
| KM-final-viral-combined_k141_75971\|\|full | Low-quality |
| KM-final-viral-combined_k141_87575\|\|full | Not-determined |
| KM-final-viral-combined_k141_163289\|\|full | Low-quality |
| KM-final-viral-combined_k141_241804\|\|full | Low-quality |
| KM-final-viral-combined_k141_198411\|\|full | Low-quality |
| KM-final-viral-combined_k141_187330\|\|full | Low-quality |
| KM-final-viral-combined_k141_251709\|\|full | Not-determined |
| KM-final-viral-combined_k141_153206\|\|full | Not-determined |
| KM-final-viral-combined_k141_64203\|\|full | Low-quality |
| KM-final-viral-combined_k141_50952\|\|full | Not-determined |
| KM-final-viral-combined_k141_64213\|\|full | Low-quality |
| KM-final-viral-combined_k141_263254\|\|full | Not-determined |
| KM-final-viral-combined_k141_108423\|\|full | Not-determined |
| KM-final-viral-combined_k141_153230\|\|full | Low-quality |
| KM-final-viral-combined_k141_87622\|\|full | Not-determined |
| KM-final-viral-combined_k141_263255\|\|full | Not-determined |
| KM-final-viral-combined_k141_76028\|\|full | Low-quality |
| KM-final-viral-combined_k141_87627\|\|full | Not-determined |
| KM-final-viral-combined_k141_142573\|\|full | Low-quality |
| KM-final-viral-combined_k141_95700\|\|full | Not-determined |
| KM-final-viral-combined_k141_209668\|\|full | Low-quality |
| KM-final-viral-combined_k141_76031\|\|full | Not-determined |
| KM-final-viral-combined_k141_108439\|\|full | Low-quality |
| KM-final-viral-combined_k141_187367\|\|full | Low-quality |
| KM-final-viral-combined_k141_209670\|\|full | Not-determined |
| KM-final-viral-combined_k141_209671\|\|full | Low-quality |
| KM-final-viral-combined_k141_221085\|\|full | Low-quality |
| KM-final-viral-combined_k141_209673\|\|full | Low-quality |
| KM-final-viral-combined_k141_130542\|\|full | Not-determined |
| KM-final-viral-combined_k141_50982\|\|full | Low-quality |
| KM-final-viral-combined_k141_30210\|\|full | Low-quality |
| KM-final-viral-combined_k141_209681\|\|full | Low-quality |
| KM-final-viral-combined_k141_142603\|\|full | Not-determined |
| KM-final-viral-combined_k141_87664\|\|full | Low-quality |
| KM-final-viral-combined_k141_41239\|\|full | Not-determined |
| KM-final-viral-combined_k141_95719\|\|full | Not-determined |
| KM-final-viral-combined_k141_121376\|\|full | Not-determined |
| KM-final-viral-combined_k141_51002\|\|full | Not-determined |
| KM-final-viral-combined_k141_251780\|\|full | Not-determined |
| KM-final-viral-combined_k141_209708\|\|full | Not-determined |
| KM-final-viral-combined_k141_251790\|\|full | Not-determined |
| KM-final-viral-combined_k141_130547\|\|full | Low-quality |
| KM-final-viral-combined_k141_108484\|\|full | Not-determined |
| KM-final-viral-combined_k141_7961\|\|full | Not-determined |
| KM-final-viral-combined_k141_187408\|\|full | Not-determined |
| KM-final-viral-combined_k141_198507\|\|full | Not-determined |
| KM-final-viral-combined_k141_108496\|\|full | Not-determined |
| KM-final-viral-combined_k141_87690\|\|full | Not-determined |
| KM-final-viral-combined_k141_232059\|\|full | Not-determined |
| KM-final-viral-combined_k141_41259\|\|full | Low-quality |
| KM-final-viral-combined_k141_251830\|\|full | Not-determined |
| KM-final-viral-combined_k141_41270\|\|full | Low-quality |
| KM-final-viral-combined_k141_251834\|\|full | Not-determined |
| KM-final-viral-combined_k141_209743\|\|full | Low-quality |
| KM-final-viral-combined_k141_130580\|\|full | Not-determined |
| KM-final-viral-combined_k141_232097\|\|full | Not-determined |
| KM-final-viral-combined_k141_142654\|\|full | Low-quality |
| KM-final-viral-combined_k141_263303\|\|full | Low-quality |
| KM-final-viral-combined_k141_121431\|\|full | Not-determined |
| KM-final-viral-combined_k141_198551\|\|full | Not-determined |
| KM-final-viral-combined_k141_153338\|\|full | Not-determined |
| KM-final-viral-combined_k141_30290\|\|full | Not-determined |
| KM-final-viral-combined_k141_18491\|\|full | Low-quality |
| KM-final-viral-combined_k141_209767\|\|full | Low-quality |
| KM-final-viral-combined_k141_251866\|\|full | Not-determined |
| KM-final-viral-combined_k141_64299\|\|full | Not-determined |
| KM-final-viral-combined_k141_175695\|\|full | Not-determined |
| KM-final-viral-combined_k141_95786\|\|full | Not-determined |
| KM-final-viral-combined_k141_8044\|\|full | Not-determined |
| KM-final-viral-combined_k141_175705\|\|full | Low-quality |
| KM-final-viral-combined_k141_198593\|\|full | Low-quality |
| KM-final-viral-combined_k141_209807\|\|full | Not-determined |
| KM-final-viral-combined_k141_76145\|\|full | Not-determined |
| KM-final-viral-combined_k141_251901\|\|full | Not-determined |
| KM-final-viral-combined_k141_95819\|\|full | Low-quality |
| KM-final-viral-combined_k141_64346\|\|full | Low-quality |
| KM-final-viral-combined_k141_130651\|\|full | Low-quality |
| KM-final-viral-combined_k141_108588\|\|full | Low-quality |
| KM-final-viral-combined_k141_8094\|\|full | Not-determined |
| KM-final-viral-combined_k141_64350\|\|full | Not-determined |
| KM-final-viral-combined_k141_198626\|\|full | Not-determined |
| 18-109-final-viral-combined_k141_107679\|\|1_partial | Medium-quality |
| KM-final-viral-combined_k141_130666\|\|full | Low-quality |
| KM-final-viral-combined_k141_198628\|\|full | Not-determined |
| KM-final-viral-combined_k141_8108\|\|full | Not-determined |
| KM-final-viral-combined_k141_87721\|\|full | Not-determined |
| KM-final-viral-combined_k141_130682\|\|full | Low-quality |
| KM-final-viral-combined_k141_30320\|\|full | Low-quality |
| KM-final-viral-combined_k141_209851\|\|full | Not-determined |
| KM-final-viral-combined_k141_108615\|\|full | Not-determined |
| KM-final-viral-combined_k141_87734\|\|full | Low-quality |
| KM-final-viral-combined_k141_121456\|\|full | Low-quality |
| KM-final-viral-combined_k141_8118\|\|full | Low-quality |
| KM-final-viral-combined_k141_232205\|\|full | Low-quality |
| KM-final-viral-combined_k141_241993\|\|full | Not-determined |
| KM-final-viral-combined_k141_76183\|\|full | Not-determined |
| KM-final-viral-combined_k141_18552\|\|full | Not-determined |
| KM-final-viral-combined_k141_51128\|\|full | Low-quality |
| KM-final-viral-combined_k141_209884\|\|full | Low-quality |
| KM-final-viral-combined_k141_30341\|\|full | Not-determined |
| KM-final-viral-combined_k141_108624\|\|full | Not-determined |
| KM-final-viral-combined_k141_130732\|\|full | Low-quality |
| KM-final-viral-combined_k141_108628\|\|full | Not-determined |
| KM-final-viral-combined_k141_232243\|\|full | Not-determined |
| KM-final-viral-combined_k141_175742\|\|full | Low-quality |
| KM-final-viral-combined_k141_163406\|\|full | Not-determined |
| KM-final-viral-combined_k141_64369\|\|full | Not-determined |
| KM-final-viral-combined_k141_30370\|\|full | Not-determined |
| KM-final-viral-combined_k141_198687\|\|full | Low-quality |
| KM-final-viral-combined_k141_76219\|\|full | Low-quality |
| KM-final-viral-combined_k141_51166\|\|full | Not-determined |
| KM-final-viral-combined_k141_142770\|\|full | Low-quality |
| KM-final-viral-combined_k141_251959\|\|full | Not-determined |
| KM-final-viral-combined_k141_198697\|\|full | Low-quality |
| KM-final-viral-combined_k141_8180\|\|full | Low-quality |
| KM-final-viral-combined_k141_221153\|\|full | Low-quality |
| KM-final-viral-combined_k141_30386\|\|full | Low-quality |
| KM-final-viral-combined_k141_8181\|\|full | Low-quality |
| KM-final-viral-combined_k141_130765\|\|full | Not-determined |
| KM-final-viral-combined_k141_87783\|\|full | Not-determined |
| KM-final-viral-combined_k141_153489\|\|full | Not-determined |
| KM-final-viral-combined_k141_221171\|\|full | Not-determined |
| KM-final-viral-combined_k141_251966\|\|full | Low-quality |
| KM-final-viral-combined_k141_221185\|\|full | Not-determined |
| KM-final-viral-combined_k141_142809\|\|full | Low-quality |
| KM-final-viral-combined_k141_108699\|\|full | Not-determined |
| KM-final-viral-combined_k141_64418\|\|full | Low-quality |
| KM-final-viral-combined_k141_198728\|\|full | Not-determined |
| KM-final-viral-combined_k141_263344\|\|full | Not-determined |
| KM-final-viral-combined_k141_232291\|\|full | Not-determined |
| KM-final-viral-combined_k141_41360\|\|full | Not-determined |
| KM-final-viral-combined_k141_130818\|\|full | Low-quality |
| KM-final-viral-combined_k141_187490\|\|full | Not-determined |
| KM-final-viral-combined_k141_64439\|\|full | Not-determined |
| KM-final-viral-combined_k141_121531\|\|full | Not-determined |
| KM-final-viral-combined_k141_8278\|\|full | Low-quality |
| KM-final-viral-combined_k141_163489\|\|full | Not-determined |
| KM-final-viral-combined_k141_121537\|\|full | Not-determined |
| KM-final-viral-combined_k141_221245\|\|full | Low-quality |
| KM-final-viral-combined_k141_76292\|\|full | Not-determined |
| KM-final-viral-combined_k141_142841\|\|full | Not-determined |
| KM-final-viral-combined_k141_95930\|\|full | Low-quality |
| KM-final-viral-combined_k141_76297\|\|full | Low-quality |
| KM-final-viral-combined_k141_64471\|\|full | Not-determined |
| KM-final-viral-combined_k141_18692\|\|full | Not-determined |
| KM-final-viral-combined_k141_175862\|\|full | Not-determined |
| KM-final-viral-combined_k141_51250\|\|full | Low-quality |
| KM-final-viral-combined_k141_163516\|\|full | Not-determined |
| KM-final-viral-combined_k141_263398\|\|full | Low-quality |
| KM-final-viral-combined_k141_142858\|\|full | Not-determined |
| KM-final-viral-combined_k141_242067\|\|full | Not-determined |
| KM-final-viral-combined_k141_187533\|\|full | Not-determined |
| KM-final-viral-combined_k141_8315\|\|full | Not-determined |
| KM-final-viral-combined_k141_51263\|\|full | Not-determined |
| KM-final-viral-combined_k141_232360\|\|full | Not-determined |
| KM-final-viral-combined_k141_18707\|\|full | Not-determined |
| KM-final-viral-combined_k141_163524\|\|full | Not-determined |
| KM-final-viral-combined_k141_51277\|\|full | Low-quality |
| KM-final-viral-combined_k141_64515\|\|full | Not-determined |
| KM-final-viral-combined_k141_142893\|\|full | Not-determined |
| KM-final-viral-combined_k141_163526\|\|full | Not-determined |
| KM-final-viral-combined_k141_242090\|\|full | Not-determined |
| KM-final-viral-combined_k141_198800\|\|full | Low-quality |
| KM-final-viral-combined_k141_64524\|\|full | Not-determined |
| KM-final-viral-combined_k141_121605\|\|full | Low-quality |
| KM-final-viral-combined_k141_87902\|\|full | Not-determined |
| KM-final-viral-combined_k141_263420\|\|full | Not-determined |
| KM-final-viral-combined_k141_163542\|\|full | Low-quality |
| KM-final-viral-combined_k141_108825\|\|full | Not-determined |
| KM-final-viral-combined_k141_142918\|\|full | Not-determined |
| KM-final-viral-combined_k141_175913\|\|full | Low-quality |
| KM-final-viral-combined_k141_64554\|\|full | Low-quality |
| KM-final-viral-combined_k141_242109\|\|full | Low-quality |
| KM-final-viral-combined_k141_175919\|\|full | Not-determined |
| KM-final-viral-combined_k141_221322\|\|full | Low-quality |
| KM-final-viral-combined_k141_76397\|\|full | Not-determined |
| KM-final-viral-combined_k141_187601\|\|full | Not-determined |
| KM-final-viral-combined_k141_108860\|\|full | Low-quality |
| KM-final-viral-combined_k141_8364\|\|full | Low-quality |
| KM-final-viral-combined_k141_187613\|\|full | Low-quality |
| KM-final-viral-combined_k141_41462\|\|full | Low-quality |
| KM-final-viral-combined_k141_51345\|\|full | Not-determined |
| KM-final-viral-combined_k141_187616\|\|full | Not-determined |
| KM-final-viral-combined_k141_163594\|\|full | Low-quality |
| KM-final-viral-combined_k141_96019\|\|full | Not-determined |
| KM-final-viral-combined_k141_163596\|\|full | Low-quality |
| KM-final-viral-combined_k141_232394\|\|full | Not-determined |
| 18-102-final-viral-combined_k141_227800\|\|full | Medium-quality |
| KM-final-viral-combined_k141_41476\|\|full | Not-determined |
| KM-final-viral-combined_k141_153606\|\|full | Not-determined |
| KM-final-viral-combined_k141_198878\|\|full | Not-determined |
| KM-final-viral-combined_k141_96028\|\|full | Low-quality |
| KM-final-viral-combined_k141_18755\|\|full | Low-quality |
| KM-final-viral-combined_k141_30569\|\|full | Low-quality |
| KM-final-viral-combined_k141_232408\|\|full | Not-determined |
| KM-final-viral-combined_k141_8418\|\|full | Not-determined |
| KM-final-viral-combined_k141_143004\|\|full | Low-quality |
| KM-final-viral-combined_k141_198899\|\|full | Not-determined |
| KM-final-viral-combined_k141_209945\|\|full | Not-determined |
| KM-final-viral-combined_k141_96062\|\|full | Low-quality |
| KM-final-viral-combined_k141_121718\|\|full | Low-quality |
| KM-final-viral-combined_k141_232422\|\|full | Low-quality |
| KM-final-viral-combined_k141_187670\|\|full | Low-quality |
| KM-final-viral-combined_k141_41511\|\|full | Not-determined |
| KM-final-viral-combined_k141_163623\|\|full | Low-quality |
| KM-final-viral-combined_k141_153648\|\|full | Not-determined |
| KM-final-viral-combined_k141_51389\|\|full | Low-quality |
| KM-final-viral-combined_k141_153649\|\|full | Low-quality |
| KM-final-viral-combined_k141_18770\|\|full | Not-determined |
| KM-final-viral-combined_k141_130993\|\|full | Not-determined |
| KM-final-viral-combined_k141_41517\|\|full | Low-quality |
| KM-final-viral-combined_k141_130995\|\|full | Low-quality |
| KM-final-viral-combined_k141_30609\|\|full | Not-determined |
| KM-final-viral-combined_k141_18798\|\|full | Low-quality |
| KM-final-viral-combined_k141_175998\|\|full | Low-quality |
| KM-final-viral-combined_k141_221484\|\|full | Not-determined |
| KM-final-viral-combined_k141_251996\|\|full | Not-determined |
| KM-final-viral-combined_k141_108939\|\|full | Not-determined |
| KM-final-viral-combined_k141_199006\|\|full | Low-quality |
| KM-final-viral-combined_k141_41596\|\|full | Low-quality |
| KM-final-viral-combined_k141_221534\|\|full | Low-quality |
| KM-final-viral-combined_k141_88049\|\|full | Not-determined |
| KM-final-viral-combined_k141_221545\|\|full | Low-quality |
| KM-final-viral-combined_k141_143110\|\|full | Not-determined |
| KM-final-viral-combined_k141_41615\|\|full | Low-quality |
| KM-final-viral-combined_k141_64797\|\|full | Not-determined |
| KM-final-viral-combined_k141_51466\|\|full | Low-quality |
| KM-final-viral-combined_k141_64812\|\|full | Not-determined |
| KM-final-viral-combined_k141_131069\|\|full | Not-determined |
| KM-final-viral-combined_k141_143135\|\|full | Low-quality |
| KM-final-viral-combined_k141_18819\|\|full | Not-determined |
| KM-final-viral-combined_k141_64837\|\|full | Low-quality |
| KM-final-viral-combined_k141_88082\|\|full | Low-quality |
| KM-final-viral-combined_k141_131088\|\|full | Not-determined |
| KM-final-viral-combined_k141_109005\|\|full | Low-quality |
| KM-final-viral-combined_k141_30678\|\|full | Low-quality |
| KM-final-viral-combined_k141_242256\|\|full | Low-quality |
| KM-final-viral-combined_k141_18835\|\|full | Not-determined |
| KM-final-viral-combined_k141_64848\|\|full | Low-quality |
| KM-final-viral-combined_k141_18838\|\|full | Not-determined |
| KM-final-viral-combined_k141_41658\|\|full | Not-determined |
| KM-final-viral-combined_k141_221568\|\|full | Low-quality |
| KM-final-viral-combined_k141_64855\|\|full | Low-quality |
| KM-final-viral-combined_k141_176075\|\|full | Not-determined |
| KM-final-viral-combined_k141_263593\|\|full | Not-determined |
| KM-final-viral-combined_k141_187778\|\|full | Not-determined |
| KM-final-viral-combined_k141_252102\|\|full | Not-determined |
| KM-final-viral-combined_k141_18855\|\|full | Low-quality |
| KM-final-viral-combined_k141_121843\|\|full | Not-determined |
| KM-final-viral-combined_k141_109046\|\|full | Low-quality |
| KM-final-viral-combined_k141_96139\|\|full | Low-quality |
| KM-final-viral-combined_k141_163784\|\|full | Not-determined |
| KM-final-viral-combined_k141_199030\|\|full | Not-determined |
| KM-final-viral-combined_k141_263624\|\|full | Low-quality |
| KM-final-viral-combined_k141_210106\|\|full | Low-quality |
| KM-final-viral-combined_k141_199036\|\|full | Not-determined |
| KM-final-viral-combined_k141_30720\|\|full | Low-quality |
| KM-final-viral-combined_k141_210112\|\|full | Not-determined |
| KM-final-viral-combined_k141_96150\|\|full | Not-determined |
| KM-final-viral-combined_k141_30728\|\|full | Low-quality |
| KM-final-viral-combined_k141_18900\|\|full | Not-determined |
| KM-final-viral-combined_k141_210126\|\|full | Not-determined |
| KM-final-viral-combined_k141_64870\|\|full | Not-determined |
| KM-final-viral-combined_k141_96175\|\|full | Low-quality |
| KM-final-viral-combined_k141_263664\|\|full | Low-quality |
| KM-final-viral-combined_k141_143206\|\|full | Not-determined |
| KM-final-viral-combined_k141_18916\|\|full | Not-determined |
| KM-final-viral-combined_k141_210155\|\|full | Not-determined |
| KM-final-viral-combined_k141_242341\|\|full | Not-determined |
| KM-final-viral-combined_k141_88217\|\|full | Not-determined |
| KM-final-viral-combined_k141_252168\|\|full | Not-determined |
| KM-final-viral-combined_k141_131175\|\|full | Not-determined |
| KM-final-viral-combined_k141_153888\|\|full | Not-determined |
| KM-final-viral-combined_k141_121904\|\|full | Not-determined |
| KM-final-viral-combined_k141_163860\|\|full | Not-determined |
| KM-final-viral-combined_k141_121909\|\|full | Low-quality |
| KM-final-viral-combined_k141_18932\|\|full | Low-quality |
| KM-final-viral-combined_k141_242362\|\|full | Not-determined |
| KM-final-viral-combined_k141_153903\|\|full | Not-determined |
| KM-final-viral-combined_k141_18940\|\|full | Low-quality |
| KM-final-viral-combined_k141_232684\|\|full | Not-determined |
| KM-final-viral-combined_k141_88241\|\|full | Not-determined |
| KM-final-viral-combined_k141_41735\|\|full | Not-determined |
| KM-final-viral-combined_k141_8495\|\|full | Not-determined |
| KM-final-viral-combined_k141_163872\|\|full | Low-quality |
| KM-final-viral-combined_k141_51532\|\|full | Low-quality |
| KM-final-viral-combined_k141_263710\|\|full | Not-determined |
| KM-final-viral-combined_k141_221700\|\|full | Not-determined |
| KM-final-viral-combined_k141_232695\|\|full | Low-quality |
| KM-final-viral-combined_k141_121931\|\|full | Low-quality |
| KM-final-viral-combined_k141_232699\|\|full | Low-quality |
| KM-final-viral-combined_k141_88261\|\|full | Not-determined |
| KM-final-viral-combined_k141_232703\|\|full | Not-determined |
| KM-final-viral-combined_k141_163894\|\|full | Not-determined |
| KM-final-viral-combined_k141_187909\|\|full | Not-determined |
| KM-final-viral-combined_k141_41768\|\|full | Not-determined |
| KM-final-viral-combined_k141_199172\|\|full | Low-quality |
| KM-final-viral-combined_k141_143272\|\|full | Not-determined |
| KM-final-viral-combined_k141_163900\|\|full | Low-quality |
| KM-final-viral-combined_k141_76751\|\|full | Low-quality |
| KM-final-viral-combined_k141_64943\|\|full | Not-determined |
| KM-final-viral-combined_k141_8526\|\|full | Low-quality |
| KM-final-viral-combined_k141_163902\|\|full | Low-quality |
| KM-final-viral-combined_k141_232740\|\|full | Not-determined |
| KM-final-viral-combined_k141_221730\|\|full | Low-quality |
| KM-final-viral-combined_k141_232753\|\|full | Not-determined |
| KM-final-viral-combined_k141_76772\|\|full | Low-quality |
| KM-final-viral-combined_k141_76773\|\|full | Not-determined |
| KM-final-viral-combined_k141_153950\|\|full | Not-determined |
| KM-final-viral-combined_k141_163925\|\|full | Not-determined |
| KM-final-viral-combined_k141_19002\|\|full | Not-determined |
| KM-final-viral-combined_k141_19003\|\|full | Not-determined |
| KM-final-viral-combined_k141_131205\|\|full | Low-quality |
| KM-final-viral-combined_k141_88323\|\|full | Low-quality |
| KM-final-viral-combined_k141_96261\|\|full | Not-determined |
| KM-final-viral-combined_k141_76790\|\|full | Not-determined |
| KM-final-viral-combined_k141_199221\|\|full | Not-determined |
| KM-final-viral-combined_k141_143330\|\|full | Not-determined |
| KM-final-viral-combined_k141_153993\|\|full | Not-determined |
| KM-final-viral-combined_k141_30857\|\|full | Not-determined |
| KM-final-viral-combined_k141_199236\|\|full | Not-determined |
| KM-final-viral-combined_k141_88356\|\|full | Low-quality |
| KM-final-viral-combined_k141_232815\|\|full | Low-quality |
| KM-final-viral-combined_k141_131244\|\|full | Low-quality |
| KM-final-viral-combined_k141_176248\|\|full | Not-determined |
| KM-final-viral-combined_k141_76814\|\|full | Low-quality |
| KM-final-viral-combined_k141_242388\|\|full | Not-determined |
| KM-final-viral-combined_k141_163950\|\|full | Not-determined |
| KM-final-viral-combined_k141_232832\|\|full | Not-determined |
| KM-final-viral-combined_k141_131252\|\|full | Not-determined |
| KM-final-viral-combined_k141_51627\|\|full | Low-quality |
| KM-final-viral-combined_k141_51629\|\|full | Low-quality |
| KM-final-viral-combined_k141_199251\|\|full | Not-determined |
| KM-final-viral-combined_k141_8585\|\|full | Not-determined |
| KM-final-viral-combined_k141_199256\|\|full | Not-determined |
| KM-final-viral-combined_k141_88375\|\|full | Low-quality |
| KM-final-viral-combined_k141_187981\|\|full | Low-quality |
| KM-final-viral-combined_k141_163963\|\|full | Not-determined |
| KM-final-viral-combined_k141_131266\|\|full | Not-determined |
| KM-final-viral-combined_k141_109151\|\|full | Low-quality |
| KM-final-viral-combined_k141_210276\|\|full | Not-determined |
| KM-final-viral-combined_k141_19025\|\|full | Not-determined |
| KM-final-viral-combined_k141_163974\|\|full | Not-determined |
| KM-final-viral-combined_k141_163977\|\|full | Low-quality |
| KM-final-viral-combined_k141_65098\|\|full | Not-determined |
| KM-final-viral-combined_k141_221862\|\|full | Not-determined |
| KM-final-viral-combined_k141_109164\|\|full | Not-determined |
| KM-final-viral-combined_k141_242463\|\|full | Not-determined |
| KM-final-viral-combined_k141_30884\|\|full | Not-determined |
| KM-final-viral-combined_k141_263802\|\|full | Low-quality |
| KM-final-viral-combined_k141_65129\|\|full | Not-determined |
| KM-final-viral-combined_k141_88462\|\|full | Not-determined |
| KM-final-viral-combined_k141_96316\|\|full | Not-determined |
| KM-final-viral-combined_k141_30909\|\|full | Not-determined |
| KM-final-viral-combined_k141_210309\|\|full | Not-determined |
| KM-final-viral-combined_k141_188033\|\|full | Low-quality |
| KM-final-viral-combined_k141_88476\|\|full | Low-quality |
| KM-final-viral-combined_k141_51685\|\|full | Not-determined |
| KM-final-viral-combined_k141_131326\|\|full | Low-quality |
| KM-final-viral-combined_k141_109193\|\|full | Low-quality |
| KM-final-viral-combined_k141_88505\|\|full | Not-determined |
| KM-final-viral-combined_k141_164026\|\|full | Low-quality |
| KM-final-viral-combined_k141_176353\|\|full | Low-quality |
| KM-final-viral-combined_k141_263850\|\|full | Not-determined |
| KM-final-viral-combined_k141_252322\|\|full | Low-quality |
| KM-final-viral-combined_k141_109199\|\|full | Low-quality |
| KM-final-viral-combined_k141_210346\|\|full | Not-determined |
| KM-final-viral-combined_k141_109210\|\|full | Low-quality |
| KM-final-viral-combined_k141_30924\|\|full | Not-determined |
| KM-final-viral-combined_k141_188074\|\|full | Low-quality |
| KM-final-viral-combined_k141_199342\|\|full | Low-quality |
| KM-final-viral-combined_k141_122044\|\|full | Low-quality |
| KM-final-viral-combined_k141_263867\|\|full | Not-determined |
| KM-final-viral-combined_k141_164054\|\|full | Low-quality |
| KM-final-viral-combined_k141_8682\|\|full | Not-determined |
| KM-final-viral-combined_k141_263869\|\|full | Low-quality |
| KM-final-viral-combined_k141_188097\|\|full | Not-determined |
| KM-final-viral-combined_k141_210381\|\|full | Not-determined |
| KM-final-viral-combined_k141_188106\|\|full | Low-quality |
| KM-final-viral-combined_k141_76903\|\|full | Not-determined |
| KM-final-viral-combined_k141_252371\|\|full | Not-determined |
| KM-final-viral-combined_k141_96378\|\|full | Low-quality |
| KM-final-viral-combined_k141_199368\|\|full | Low-quality |
| KM-final-viral-combined_k141_176401\|\|full | Not-determined |
| KM-final-viral-combined_k141_242541\|\|full | Not-determined |
| KM-final-viral-combined_k141_76911\|\|full | Not-determined |
| KM-final-viral-combined_k141_65196\|\|full | Low-quality |
| KM-final-viral-combined_k141_263886\|\|full | Low-quality |
| KM-final-viral-combined_k141_30984\|\|full | Not-determined |
| KM-final-viral-combined_k141_210410\|\|full | Low-quality |
| KM-final-viral-combined_k141_76921\|\|full | Not-determined |
| KM-final-viral-combined_k141_30988\|\|full | Not-determined |
| KM-final-viral-combined_k141_242549\|\|full | Low-quality |
| KM-final-viral-combined_k141_242550\|\|full | Not-determined |
| KM-final-viral-combined_k141_41932\|\|full | Low-quality |
| KM-final-viral-combined_k141_131405\|\|full | Not-determined |
| KM-final-viral-combined_k141_19140\|\|full | Low-quality |
| KM-final-viral-combined_k141_199379\|\|full | Low-quality |
| KM-final-viral-combined_k141_65219\|\|full | Low-quality |
| KM-final-viral-combined_k141_176437\|\|full | Not-determined |
| KM-final-viral-combined_k141_41938\|\|full | Low-quality |
| KM-final-viral-combined_k141_143505\|\|full | Not-determined |
| KM-final-viral-combined_k141_252406\|\|full | Low-quality |
| KM-final-viral-combined_k141_221963\|\|full | Not-determined |
| KM-final-viral-combined_k141_143518\|\|full | Low-quality |
| KM-final-viral-combined_k141_233024\|\|full | Low-quality |
| KM-final-viral-combined_k141_122078\|\|full | Low-quality |
| KM-final-viral-combined_k141_210470\|\|full | Not-determined |
| KM-final-viral-combined_k141_233031\|\|full | Not-determined |
| KM-final-viral-combined_k141_19169\|\|full | Low-quality |
| KM-final-viral-combined_k141_154207\|\|full | Not-determined |
| KM-final-viral-combined_k141_8791\|\|full | Not-determined |
| KM-final-viral-combined_k141_31036\|\|full | Not-determined |
| KM-final-viral-combined_k141_131444\|\|full | Not-determined |
| KM-final-viral-combined_k141_221990\|\|full | Not-determined |
| KM-final-viral-combined_k141_51741\|\|full | Low-quality |
| KM-final-viral-combined_k141_8795\|\|full | Not-determined |
| KM-final-viral-combined_k141_154228\|\|full | Low-quality |
| KM-final-viral-combined_k141_88574\|\|full | Not-determined |
| KM-final-viral-combined_k141_164152\|\|full | Low-quality |
| KM-final-viral-combined_k141_222001\|\|full | Low-quality |
| KM-final-viral-combined_k141_76985\|\|full | Low-quality |
| KM-final-viral-combined_k141_176498\|\|full | Not-determined |
| KM-final-viral-combined_k141_76988\|\|full | Not-determined |
| KM-final-viral-combined_k141_88584\|\|full | Not-determined |
| KM-final-viral-combined_k141_154240\|\|full | Not-determined |
| KM-final-viral-combined_k141_109310\|\|full | Not-determined |
| KM-final-viral-combined_k141_233033\|\|full | Not-determined |
| KM-final-viral-combined_k141_164167\|\|full | Low-quality |
| KM-final-viral-combined_k141_176500\|\|full | Not-determined |
| KM-final-viral-combined_k141_154242\|\|full | Low-quality |
| KM-final-viral-combined_k141_252454\|\|full | Not-determined |
| KM-final-viral-combined_k141_188207\|\|full | Low-quality |
| KM-final-viral-combined_k141_154248\|\|full | Low-quality |
| KM-final-viral-combined_k141_109319\|\|full | Not-determined |
| 18-105-final-viral-combined_k141_179241\|\|0_partial | Medium-quality |
| KM-final-viral-combined_k141_199451\|\|full | Low-quality |
| KM-final-viral-combined_k141_233056\|\|full | Low-quality |
| KM-final-viral-combined_k141_31098\|\|full | Not-determined |
| KM-final-viral-combined_k141_164185\|\|full | Low-quality |
| KM-final-viral-combined_k141_8847\|\|full | Not-determined |
| KM-final-viral-combined_k141_122146\|\|full | Not-determined |
| KM-final-viral-combined_k141_8869\|\|full | Low-quality |
| KM-final-viral-combined_k141_233078\|\|full | Not-determined |
| KM-final-viral-combined_k141_122164\|\|full | Not-determined |
| KM-final-viral-combined_k141_210567\|\|full | Not-determined |
| KM-final-viral-combined_k141_222056\|\|full | Low-quality |
| KM-final-viral-combined_k141_77005\|\|full | Low-quality |
| KM-final-viral-combined_k141_143636\|\|full | Not-determined |
| KM-final-viral-combined_k141_31120\|\|full | Not-determined |
| KM-final-viral-combined_k141_77009\|\|full | Low-quality |
| KM-final-viral-combined_k141_222066\|\|full | Not-determined |
| KM-final-viral-combined_k141_222067\|\|full | Not-determined |
| KM-final-viral-combined_k141_42079\|\|full | Not-determined |
| KM-final-viral-combined_k141_131504\|\|full | Not-determined |
| KM-final-viral-combined_k141_164220\|\|full | Not-determined |
| KM-final-viral-combined_k141_88670\|\|full | Not-determined |
| KM-final-viral-combined_k141_210606\|\|full | Not-determined |
| KM-final-viral-combined_k141_222077\|\|full | Not-determined |
| KM-final-viral-combined_k141_131512\|\|full | Not-determined |
| KM-final-viral-combined_k141_19241\|\|full | Low-quality |
| KM-final-viral-combined_k141_263981\|\|full | Low-quality |
| KM-final-viral-combined_k141_252537\|\|full | Not-determined |
| KM-final-viral-combined_k141_176601\|\|full | Low-quality |
| KM-final-viral-combined_k141_233141\|\|full | Low-quality |
| KM-final-viral-combined_k141_109394\|\|full | Not-determined |
| KM-final-viral-combined_k141_131538\|\|full | Not-determined |
| KM-final-viral-combined_k141_188299\|\|full | Not-determined |
| KM-final-viral-combined_k141_31169\|\|full | Not-determined |
| KM-final-viral-combined_k141_154356\|\|full | Not-determined |
| KM-final-viral-combined_k141_8955\|\|full | Not-determined |
| KM-final-viral-combined_k141_88705\|\|full | Not-determined |
| KM-final-viral-combined_k141_109414\|\|full | Low-quality |
| KM-final-viral-combined_k141_131553\|\|full | Not-determined |
| KM-final-viral-combined_k141_188321\|\|full | Low-quality |
| KM-final-viral-combined_k141_222138\|\|full | Not-determined |
| KM-final-viral-combined_k141_77050\|\|full | Low-quality |
| KM-final-viral-combined_k141_210687\|\|full | Low-quality |
| KM-final-viral-combined_k141_42139\|\|full | Low-quality |
| KM-final-viral-combined_k141_188330\|\|full | Not-determined |
| KM-final-viral-combined_k141_199532\|\|full | Not-determined |
| KM-final-viral-combined_k141_77063\|\|full | Not-determined |
| KM-final-viral-combined_k141_264041\|\|full | Not-determined |
| KM-final-viral-combined_k141_199536\|\|full | Low-quality |
| KM-final-viral-combined_k141_210711\|\|full | Not-determined |
| KM-final-viral-combined_k141_19297\|\|full | Low-quality |
| KM-final-viral-combined_k141_42157\|\|full | Low-quality |
| KM-final-viral-combined_k141_51751\|\|full | Low-quality |
| KM-final-viral-combined_k141_210718\|\|full | Low-quality |
| KM-final-viral-combined_k141_9025\|\|full | Not-determined |
| KM-final-viral-combined_k141_77104\|\|full | Low-quality |
| KM-final-viral-combined_k141_210739\|\|full | Not-determined |
| KM-final-viral-combined_k141_51773\|\|full | Low-quality |
| KM-final-viral-combined_k141_154398\|\|full | Not-determined |
| KM-final-viral-combined_k141_51778\|\|full | Not-determined |
| KM-final-viral-combined_k141_131580\|\|full | Low-quality |
| KM-final-viral-combined_k141_77111\|\|full | Low-quality |
| KM-final-viral-combined_k141_252590\|\|full | Low-quality |
| KM-final-viral-combined_k141_210759\|\|full | Not-determined |
| KM-final-viral-combined_k141_252592\|\|full | Not-determined |
| KM-final-viral-combined_k141_176691\|\|full | Not-determined |
| KM-final-viral-combined_k141_19336\|\|full | Not-determined |
| KM-final-viral-combined_k141_210775\|\|full | Low-quality |
| KM-final-viral-combined_k141_131592\|\|full | Not-determined |
| KM-final-viral-combined_k141_176695\|\|full | Not-determined |
| KM-final-viral-combined_k141_122197\|\|full | Not-determined |
| KM-final-viral-combined_k141_9073\|\|full | Low-quality |

| | |
|---|---|
| KM-final-viral-combined_k141_96623\|\|full | Low-quality |
| KM-final-viral-combined_k141_233228\|\|full | Not-determined |
| KM-final-viral-combined_k141_143779\|\|full | Not-determined |
| KM-final-viral-combined_k141_96633\|\|full | Not-determined |
| KM-final-viral-combined_k141_42218\|\|full | Not-determined |
| KM-final-viral-combined_k141_143791\|\|full | Low-quality |
| KM-final-viral-combined_k141_19346\|\|full | Not-determined |
| KM-final-viral-combined_k141_222197\|\|full | Not-determined |
| KM-final-viral-combined_k141_77172\|\|full | Low-quality |
| KM-final-viral-combined_k141_19358\|\|full | Low-quality |
| KM-final-viral-combined_k141_154448\|\|full | Low-quality |
| KM-final-viral-combined_k141_199611\|\|full | Not-determined |
| KM-final-viral-combined_k141_210818\|\|full | Not-determined |
| KM-final-viral-combined_k141_210822\|\|full | Not-determined |
| KM-final-viral-combined_k141_210827\|\|full | Not-determined |
| KM-final-viral-combined_k141_42240\|\|full | Low-quality |
| KM-final-viral-combined_k141_199615\|\|full | Not-determined |
| KM-final-viral-combined_k141_188397\|\|full | Low-quality |
| KM-final-viral-combined_k141_42245\|\|full | Not-determined |
| KM-final-viral-combined_k141_9119\|\|full | Not-determined |
| KM-final-viral-combined_k141_164306\|\|full | Not-determined |
| KM-final-viral-combined_k141_131663\|\|full | Not-determined |
| KM-final-viral-combined_k141_164309\|\|full | Low-quality |
| KM-final-viral-combined_k141_188410\|\|full | Low-quality |
| KM-final-viral-combined_k141_188411\|\|full | Low-quality |
| KM-final-viral-combined_k141_199621\|\|full | Low-quality |
| KM-final-viral-combined_k141_88806\|\|full | Not-determined |
| KM-final-viral-combined_k141_242723\|\|full | Not-determined |
| KM-final-viral-combined_k141_154496\|\|full | Not-determined |
| KM-final-viral-combined_k141_154499\|\|full | Not-determined |
| KM-final-viral-combined_k141_264129\|\|full | Not-determined |
| KM-final-viral-combined_k141_233299\|\|full | Not-determined |
| KM-final-viral-combined_k141_199657\|\|full | Low-quality |
| KM-final-viral-combined_k141_122221\|\|full | Not-determined |
| KM-final-viral-combined_k141_9123\|\|full | Low-quality |
| KM-final-viral-combined_k141_199659\|\|full | Not-determined |
| KM-final-viral-combined_k141_143866\|\|full | Not-determined |
| KM-final-viral-combined_k141_164346\|\|full | Not-determined |
| KM-final-viral-combined_k141_176801\|\|full | Low-quality |
| KM-final-viral-combined_k141_176807\|\|full | Low-quality |
| KM-final-viral-combined_k141_210887\|\|full | Low-quality |
| KM-final-viral-combined_k141_264146\|\|full | Low-quality |
| KM-final-viral-combined_k141_176816\|\|full | Not-determined |
| KM-final-viral-combined_k141_42285\|\|full | Not-determined |
| KM-final-viral-combined_k141_264162\|\|full | Low-quality |
| KM-final-viral-combined_k141_199686\|\|full | Not-determined |
| KM-final-viral-combined_k141_131757\|\|full | Not-determined |
| KM-final-viral-combined_k141_210921\|\|full | Low-quality |
| KM-final-viral-combined_k141_199707\|\|full | Not-determined |
| KM-final-viral-combined_k141_65518\|\|full | Low-quality |
| KM-final-viral-combined_k141_242772\|\|full | Not-determined |
| KM-final-viral-combined_k141_65520\|\|full | Not-determined |
| KM-final-viral-combined_k141_242773\|\|full | Low-quality |
| KM-final-viral-combined_k141_131776\|\|full | Not-determined |
| KM-final-viral-combined_k141_65528\|\|full | Not-determined |
| KM-final-viral-combined_k141_210942\|\|full | Low-quality |
| KM-final-viral-combined_k141_154536\|\|full | Low-quality |
| KM-final-viral-combined_k141_176866\|\|full | Low-quality |
| KM-final-viral-combined_k141_154555\|\|full | Not-determined |
| KM-final-viral-combined_k141_222249\|\|full | Not-determined |
| KM-final-viral-combined_k141_31301\|\|full | Not-determined |
| KM-final-viral-combined_k141_42335\|\|full | Low-quality |
| KM-final-viral-combined_k141_222255\|\|full | Not-determined |
| KM-final-viral-combined_k141_65558\|\|full | Low-quality |
| KM-final-viral-combined_k141_242828\|\|full | Not-determined |
| KM-final-viral-combined_k141_42350\|\|full | Low-quality |
| KM-final-viral-combined_k141_9220\|\|full | Not-determined |
| KM-final-viral-combined_k141_31313\|\|full | Not-determined |
| KM-final-viral-combined_k141_88886\|\|full | Not-determined |
| KM-final-viral-combined_k141_51901\|\|full | Not-determined |
| KM-final-viral-combined_k141_77276\|\|full | Not-determined |
| KM-final-viral-combined_k141_42353\|\|full | Not-determined |
| KM-final-viral-combined_k141_96731\|\|full | Low-quality |
| KM-final-viral-combined_k141_19487\|\|full | Not-determined |
| KM-final-viral-combined_k141_65603\|\|full | Not-determined |
| KM-final-viral-combined_k141_65609\|\|full | Low-quality |
| KM-final-viral-combined_k141_131836\|\|full | Not-determined |
| KM-final-viral-combined_k141_77302\|\|full | Low-quality |
| KM-final-viral-combined_k141_65621\|\|full | Low-quality |
| KM-final-viral-combined_k141_176928\|\|full | Not-determined |
| KM-final-viral-combined_k141_131850\|\|full | Low-quality |
| KM-final-viral-combined_k141_51940\|\|full | Not-determined |
| KM-final-viral-combined_k141_19515\|\|full | Not-determined |
| KM-final-viral-combined_k141_210953\|\|full | Low-quality |
| KM-final-viral-combined_k141_109700\|\|full | Not-determined |
| KM-final-viral-combined_k141_42414\|\|full | Low-quality |
| KM-final-viral-combined_k141_65658\|\|full | Low-quality |
| KM-final-viral-combined_k141_143904\|\|full | Not-determined |
| KM-final-viral-combined_k141_188486\|\|full | Not-determined |
| KM-final-viral-combined_k141_109709\|\|full | Not-determined |
| KM-final-viral-combined_k141_252727\|\|full | Low-quality |
| KM-final-viral-combined_k141_96796\|\|full | Low-quality |
| KM-final-viral-combined_k141_242885\|\|full | Not-determined |
| KM-final-viral-combined_k141_42430\|\|full | Not-determined |
| KM-final-viral-combined_k141_176977\|\|full | Not-determined |
| KM-final-viral-combined_k141_77366\|\|full | Low-quality |
| KM-final-viral-combined_k141_176982\|\|full | Not-determined |
| KM-final-viral-combined_k141_51975\|\|full | Not-determined |
| KM-final-viral-combined_k141_264251\|\|full | Not-determined |
| KM-final-viral-combined_k141_51997\|\|full | Not-determined |
| KM-final-viral-combined_k141_164489\|\|full | Low-quality |
| KM-final-viral-combined_k141_96837\|\|full | Not-determined |
| KM-final-viral-combined_k141_242920\|\|full | Not-determined |
| KM-final-viral-combined_k141_77406\|\|full | Low-quality |
| KM-final-viral-combined_k141_52018\|\|full | Low-quality |
| KM-final-viral-combined_k141_131973\|\|full | Low-quality |
| KM-final-viral-combined_k141_164501\|\|full | Low-quality |
| KM-final-viral-combined_k141_264269\|\|full | Not-determined |
| KM-final-viral-combined_k141_143938\|\|full | Not-determined |
| KM-final-viral-combined_k141_143946\|\|full | Low-quality |
| KM-final-viral-combined_k141_52021\|\|full | Low-quality |
| 18-101-final-viral-combined_k141_41369\|\|full | Medium-quality |
| KM-final-viral-combined_k141_77427\|\|full | Not-determined |
| KM-final-viral-combined_k141_31485\|\|full | Low-quality |
| KM-final-viral-combined_k141_188530\|\|full | Not-determined |
| KM-final-viral-combined_k141_77437\|\|full | Not-determined |
| KM-final-viral-combined_k141_9346\|\|full | Low-quality |
| KM-final-viral-combined_k141_177044\|\|full | Not-determined |
| KM-final-viral-combined_k141_242968\|\|full | Not-determined |
| KM-final-viral-combined_k141_132032\|\|full | Not-determined |
| KM-final-viral-combined_k141_9363\|\|full | Low-quality |
| KM-final-viral-combined_k141_164544\|\|full | Not-determined |
| KM-final-viral-combined_k141_31510\|\|full | Low-quality |
| KM-final-viral-combined_k141_252805\|\|full | Not-determined |
| KM-final-viral-combined_k141_31511\|\|full | Not-determined |
| KM-final-viral-combined_k141_132048\|\|full | Low-quality |
| KM-final-viral-combined_k141_199782\|\|full | Low-quality |
| KM-final-viral-combined_k141_199786\|\|full | Low-quality |
| KM-final-viral-combined_k141_188597\|\|full | Not-determined |
| KM-final-viral-combined_k141_199794\|\|full | Low-quality |
| KM-final-viral-combined_k141_132062\|\|full | Low-quality |
| KM-final-viral-combined_k141_96913\|\|full | Not-determined |
| KM-final-viral-combined_k141_9386\|\|full | Low-quality |
| KM-final-viral-combined_k141_65785\|\|full | Not-determined |
| KM-final-viral-combined_k141_199812\|\|full | Not-determined |
| KM-final-viral-combined_k141_211045\|\|full | Not-determined |
| KM-final-viral-combined_k141_31573\|\|full | Low-quality |
| KM-final-viral-combined_k141_243018\|\|full | Not-determined |
| KM-final-viral-combined_k141_243019\|\|full | Low-quality |
| KM-final-viral-combined_k141_199825\|\|full | Low-quality |
| KM-final-viral-combined_k141_188626\|\|full | Low-quality |
| KM-final-viral-combined_k141_65800\|\|full | Low-quality |
| KM-final-viral-combined_k141_264322\|\|full | Not-determined |
| KM-final-viral-combined_k141_9416\|\|full | Not-determined |
| KM-final-viral-combined_k141_31591\|\|full | Low-quality |
| KM-final-viral-combined_k141_77538\|\|full | Low-quality |
| KM-final-viral-combined_k141_154784\|\|full | Not-determined |
| KM-final-viral-combined_k141_264335\|\|full | Not-determined |
| KM-final-viral-combined_k141_188638\|\|full | Low-quality |
| KM-final-viral-combined_k141_19618\|\|full | Not-determined |
| KM-final-viral-combined_k141_77559\|\|full | Low-quality |
| KM-final-viral-combined_k141_164655\|\|full | Not-determined |
| KM-final-viral-combined_k141_144009\|\|full | Not-determined |
| KM-final-viral-combined_k141_19625\|\|full | Not-determined |
| KM-final-viral-combined_k141_211076\|\|full | Low-quality |
| KM-final-viral-combined_k141_31621\|\|full | Not-determined |
| KM-final-viral-combined_k141_144030\|\|full | Low-quality |
| KM-final-viral-combined_k141_9468\|\|full | Low-quality |
| KM-final-viral-combined_k141_9470\|\|full | Not-determined |
| KM-final-viral-combined_k141_31629\|\|full | Not-determined |
| KM-final-viral-combined_k141_77571\|\|full | Low-quality |
| KM-final-viral-combined_k141_31632\|\|full | Not-determined |
| KM-final-viral-combined_k141_211096\|\|full | Low-quality |
| KM-final-viral-combined_k141_177149\|\|full | Low-quality |
| KM-final-viral-combined_k141_164678\|\|full | Not-determined |
| KM-final-viral-combined_k141_65835\|\|full | Low-quality |
| KM-final-viral-combined_k141_65836\|\|full | Low-quality |
| KM-final-viral-combined_k141_164683\|\|full | Not-determined |
| KM-final-viral-combined_k141_177169\|\|full | Low-quality |
| KM-final-viral-combined_k141_211122\|\|full | Low-quality |
| KM-final-viral-combined_k141_188683\|\|full | Low-quality |
| KM-final-viral-combined_k141_188684\|\|full | Low-quality |
| KM-final-viral-combined_k141_31667\|\|full | Low-quality |
| KM-final-viral-combined_k141_188686\|\|full | Not-determined |
| KM-final-viral-combined_k141_65855\|\|full | Not-determined |
| KM-final-viral-combined_k141_252910\|\|full | Not-determined |
| KM-final-viral-combined_k141_9528\|\|full | Not-determined |
| KM-final-viral-combined_k141_211139\|\|full | Low-quality |
| KM-final-viral-combined_k141_188704\|\|full | Low-quality |
| KM-final-viral-combined_k141_264414\|\|full | Not-determined |
| KM-final-viral-combined_k141_211141\|\|full | Not-determined |
| KM-final-viral-combined_k141_31691\|\|full | Not-determined |
| KM-final-viral-combined_k141_154831\|\|full | Not-determined |
| KM-final-viral-combined_k141_97011\|\|full | Low-quality |
| KM-final-viral-combined_k141_164717\|\|full | Not-determined |
| KM-final-viral-combined_k141_52190\|\|full | Low-quality |
| KM-final-viral-combined_k141_188761\|\|full | Not-determined |
| KM-final-viral-combined_k141_188786\|\|full | Low-quality |
| KM-final-viral-combined_k141_97057\|\|full | Not-determined |
| KM-final-viral-combined_k141_132250\|\|full | Low-quality |
| KM-final-viral-combined_k141_177267\|\|full | Not-determined |
| KM-final-viral-combined_k141_188801\|\|full | Low-quality |
| KM-final-viral-combined_k141_154894\|\|full | Not-determined |
| KM-final-viral-combined_k141_164773\|\|full | Not-determined |
| KM-final-viral-combined_k141_77663\|\|full | Low-quality |
| KM-final-viral-combined_k141_243072\|\|full | Not-determined |
| KM-final-viral-combined_k141_109863\|\|full | Not-determined |
| KM-final-viral-combined_k141_132264\|\|full | Not-determined |
| KM-final-viral-combined_k141_31713\|\|full | Low-quality |
| KM-final-viral-combined_k141_31718\|\|full | Not-determined |
| KM-final-viral-combined_k141_42743\|\|full | Not-determined |
| KM-final-viral-combined_k141_31727\|\|full | Not-determined |
| KM-final-viral-combined_k141_199904\|\|full | Not-determined |
| KM-final-viral-combined_k141_199907\|\|full | Not-determined |
| KM-final-viral-combined_k141_164840\|\|full | Not-determined |
| KM-final-viral-combined_k141_188853\|\|full | Low-quality |
| KM-final-viral-combined_k141_199924\|\|full | Not-determined |
| KM-final-viral-combined_k141_9614\|\|full | Not-determined |
| KM-final-viral-combined_k141_243110\|\|full | Not-determined |
| KM-final-viral-combined_k141_253044\|\|full | Not-determined |
| KM-final-viral-combined_k141_243121\|\|full | Not-determined |
| KM-final-viral-combined_k141_77687\|\|full | Not-determined |
| KM-final-viral-combined_k141_177319\|\|full | Low-quality |
| KM-final-viral-combined_k141_9638\|\|full | Low-quality |
| KM-final-viral-combined_k141_9650\|\|full | Low-quality |
| KM-final-viral-combined_k141_31777\|\|full | Low-quality |
| KM-final-viral-combined_k141_9665\|\|full | Low-quality |
| KM-final-viral-combined_k141_132308\|\|full | Not-determined |
| KM-final-viral-combined_k141_199959\|\|full | Not-determined |
| KM-final-viral-combined_k141_164896\|\|full | Low-quality |
| KM-final-viral-combined_k141_9676\|\|full | Low-quality |
| KM-final-viral-combined_k141_97155\|\|full | Low-quality |
| KM-final-viral-combined_k141_264487\|\|full | Low-quality |
| KM-final-viral-combined_k141_132331\|\|full | Not-determined |
| KM-final-viral-combined_k141_253099\|\|full | Not-determined |
| KM-final-viral-combined_k141_66087\|\|full | Not-determined |
| KM-final-viral-combined_k141_199985\|\|full | Not-determined |
| KM-final-viral-combined_k141_109946\|\|full | Low-quality |
| KM-final-viral-combined_k141_199998\|\|full | Low-quality |
| KM-final-viral-combined_k141_66115\|\|full | Not-determined |
| KM-final-viral-combined_k141_9707\|\|full | Low-quality |
| KM-final-viral-combined_k141_177376\|\|full | Not-determined |
| KM-final-viral-combined_k141_154948\|\|full | Low-quality |
| KM-final-viral-combined_k141_154954\|\|full | Not-determined |
| KM-final-viral-combined_k141_9722\|\|full | Low-quality |
| KM-final-viral-combined_k141_264520\|\|full | Not-determined |
| KM-final-viral-combined_k141_109987\|\|full | Low-quality |
| KM-final-viral-combined_k141_177381\|\|full | Not-determined |
| KM-final-viral-combined_k141_164984\|\|full | Not-determined |
| KM-final-viral-combined_k141_264523\|\|full | Low-quality |
| 18-110-final-viral-combined_k141_185611\|\|full | Medium-quality |
| KM-final-viral-combined_k141_144060\|\|full | Low-quality |
| KM-final-viral-combined_k141_243215\|\|full | Not-determined |
| KM-final-viral-combined_k141_19814\|\|full | Not-determined |
| KM-final-viral-combined_k141_110008\|\|full | Not-determined |
| KM-final-viral-combined_k141_264552\|\|full | Not-determined |
| KM-final-viral-combined_k141_19831\|\|full | Not-determined |
| KM-final-viral-combined_k141_9750\|\|full | Low-quality |
| KM-final-viral-combined_k141_243257\|\|full | Low-quality |
| KM-final-viral-combined_k141_31801\|\|full | Low-quality |
| KM-final-viral-combined_k141_177443\|\|full | Not-determined |
| KM-final-viral-combined_k141_155012\|\|full | Low-quality |
| KM-final-viral-combined_k141_177447\|\|full | Low-quality |
| KM-final-viral-combined_k141_110061\|\|full | Not-determined |
| KM-final-viral-combined_k141_253163\|\|full | Not-determined |
| KM-final-viral-combined_k141_264571\|\|full | Not-determined |
| KM-final-viral-combined_k141_19851\|\|full | Not-determined |
| KM-final-viral-combined_k141_110080\|\|full | Low-quality |
| KM-final-viral-combined_k141_42920\|\|full | Not-determined |
| KM-final-viral-combined_k141_253191\|\|full | Not-determined |
| KM-final-viral-combined_k141_66185\|\|full | Not-determined |
| KM-final-viral-combined_k141_97288\|\|full | Low-quality |
| KM-final-viral-combined_k141_66193\|\|full | Not-determined |
| KM-final-viral-combined_k141_52362\|\|full | Not-determined |
| KM-final-viral-combined_k141_42932\|\|full | Not-determined |
| KM-final-viral-combined_k141_31860\|\|full | Not-determined |
| KM-final-viral-combined_k141_253212\|\|full | Not-determined |
| KM-final-viral-combined_k141_264614\|\|full | Low-quality |
| KM-final-viral-combined_k141_177502\|\|full | Low-quality |
| KM-final-viral-combined_k141_42955\|\|full | Low-quality |
| KM-final-viral-combined_k141_19889\|\|full | Low-quality |
| KM-final-viral-combined_k141_9808\|\|full | Low-quality |
| KM-final-viral-combined_k141_253220\|\|full | Low-quality |
| KM-final-viral-combined_k141_110125\|\|full | Low-quality |
| KM-final-viral-combined_k141_66229\|\|full | Low-quality |
| KM-final-viral-combined_k141_19897\|\|full | Not-determined |
| KM-final-viral-combined_k141_132430\|\|full | Low-quality |
| KM-final-viral-combined_k141_110133\|\|full | Not-determined |
| KM-final-viral-combined_k141_42986\|\|full | Low-quality |
| KM-final-viral-combined_k141_177536\|\|full | Low-quality |
| KM-final-viral-combined_k141_253237\|\|full | Low-quality |
| KM-final-viral-combined_k141_165158\|\|full | Not-determined |
| KM-final-viral-combined_k141_264664\|\|full | Not-determined |
| KM-final-viral-combined_k141_97364\|\|full | Not-determined |
| KM-final-viral-combined_k141_177561\|\|full | Low-quality |
| KM-final-viral-combined_k141_97371\|\|full | Low-quality |
| KM-final-viral-combined_k141_97377\|\|full | Not-determined |
| KM-final-viral-combined_k141_177575\|\|full | Not-determined |
| KM-final-viral-combined_k141_264690\|\|full | Low-quality |
| KM-final-viral-combined_k141_165188\|\|full | Not-determined |
| KM-final-viral-combined_k141_243312\|\|full | Low-quality |
| KM-final-viral-combined_k141_144139\|\|full | Not-determined |
| KM-final-viral-combined_k141_19979\|\|full | Not-determined |
| KM-final-viral-combined_k141_144145\|\|full | Not-determined |
| KM-final-viral-combined_k141_97398\|\|full | Low-quality |
| KM-final-viral-combined_k141_43028\|\|full | Not-determined |
| KM-final-viral-combined_k141_97410\|\|full | Not-determined |
| 18-101-final-viral-combined_k141_143459\|\|full | Medium-quality |
| KM-final-viral-combined_k141_77758\|\|full | Not-determined |
| KM-final-viral-combined_k141_177636\|\|full | Low-quality |
| KM-final-viral-combined_k141_253315\|\|full | Not-determined |
| KM-final-viral-combined_k141_253316\|\|full | Low-quality |
| KM-final-viral-combined_k141_97424\|\|full | Not-determined |
| KM-final-viral-combined_k141_9880\|\|full | Not-determined |
| KM-final-viral-combined_k141_144175\|\|full | Not-determined |
| KM-final-viral-combined_k141_77760\|\|full | Not-determined |
| KM-final-viral-combined_k141_165243\|\|full | Not-determined |
| KM-final-viral-combined_k141_43072\|\|full | Not-determined |
| KM-final-viral-combined_k141_155076\|\|full | Not-determined |
| KM-final-viral-combined_k141_155077\|\|full | Not-determined |
| KM-final-viral-combined_k141_66285\|\|full | Low-quality |
| KM-final-viral-combined_k141_155089\|\|full | Not-determined |
| KM-final-viral-combined_k141_155099\|\|full | Low-quality |
| KM-final-viral-combined_k141_243351\|\|full | Low-quality |
| KM-final-viral-combined_k141_264765\|\|full | Low-quality |
| KM-final-viral-combined_k141_253340\|\|full | Low-quality |
| KM-final-viral-combined_k141_243364\|\|full | Low-quality |
| KM-final-viral-combined_k141_243365\|\|full | Low-quality |
| KM-final-viral-combined_k141_253354\|\|full | Not-determined |
| KM-final-viral-combined_k141_132577\|\|full | Not-determined |
| KM-final-viral-combined_k141_43180\|\|full | Not-determined |
| KM-final-viral-combined_k141_144240\|\|full | Low-quality |
| KM-final-viral-combined_k141_43182\|\|full | Not-determined |
| KM-final-viral-combined_k141_264801\|\|full | Low-quality |
| KM-final-viral-combined_k141_97512\|\|full | Not-determined |
| KM-final-viral-combined_k141_43194\|\|full | Not-determined |
| KM-final-viral-combined_k141_132603\|\|full | Low-quality |
| KM-final-viral-combined_k141_97526\|\|full | Low-quality |
| KM-final-viral-combined_k141_19999\|\|full | Not-determined |
| KM-final-viral-combined_k141_165334\|\|full | Not-determined |
| KM-final-viral-combined_k141_9985\|\|full | Low-quality |
| KM-final-viral-combined_k141_155169\|\|full | Low-quality |
| KM-final-viral-combined_k141_132611\|\|full | Not-determined |
| KM-final-viral-combined_k141_132615\|\|full | Not-determined |
| KM-final-viral-combined_k141_253417\|\|full | Not-determined |
| KM-final-viral-combined_k141_43221\|\|full | Low-quality |
| KM-final-viral-combined_k141_66419\|\|full | Not-determined |
| KM-final-viral-combined_k141_144277\|\|full | Low-quality |
| KM-final-viral-combined_k141_97554\|\|full | Not-determined |
| KM-final-viral-combined_k141_43224\|\|full | Not-determined |
| KM-final-viral-combined_k141_32009\|\|full | Low-quality |
| KM-final-viral-combined_k141_97556\|\|full | Not-determined |
| KM-final-viral-combined_k141_144284\|\|full | Not-determined |
| KM-final-viral-combined_k141_66431\|\|full | Not-determined |
| KM-final-viral-combined_k141_155192\|\|full | Not-determined |
| KM-final-viral-combined_k141_132655\|\|full | Not-determined |
| KM-final-viral-combined_k141_10029\|\|full | Low-quality |
| KM-final-viral-combined_k141_32040\|\|full | Low-quality |
| KM-final-viral-combined_k141_110306\|\|full | Not-determined |
| KM-final-viral-combined_k141_155214\|\|full | Not-determined |
| KM-final-viral-combined_k141_243411\|\|full | Not-determined |
| KM-final-viral-combined_k141_264857\|\|full | Not-determined |
| KM-final-viral-combined_k141_66469\|\|full | Not-determined |
| KM-final-viral-combined_k141_97582\|\|full | Low-quality |
| KM-final-viral-combined_k141_144335\|\|full | Not-determined |
| KM-final-viral-combined_k141_97589\|\|full | Low-quality |
| KM-final-viral-combined_k141_165425\|\|full | Not-determined |
| KM-final-viral-combined_k141_253477\|\|full | Low-quality |
| KM-final-viral-combined_k141_52703\|\|full | Low-quality |
| KM-final-viral-combined_k141_264878\|\|full | Not-determined |
| KM-final-viral-combined_k141_66492\|\|full | Low-quality |
| KM-final-viral-combined_k141_110337\|\|full | Not-determined |
| KM-final-viral-combined_k141_132727\|\|full | Low-quality |
| KM-final-viral-combined_k141_43305\|\|full | Low-quality |
| KM-final-viral-combined_k141_165472\|\|full | Not-determined |
| KM-final-viral-combined_k141_132744\|\|full | Not-determined |
| KM-final-viral-combined_k141_165509\|\|full | Low-quality |
| KM-final-viral-combined_k141_264920\|\|full | Low-quality |
| KM-final-viral-combined_k141_165510\|\|full | Not-determined |
| KM-final-viral-combined_k141_66512\|\|full | Not-determined |
| KM-final-viral-combined_k141_32135\|\|full | Not-determined |
| KM-final-viral-combined_k141_165515\|\|full | Not-determined |
| KM-final-viral-combined_k141_144419\|\|full | Low-quality |
| 17-108-final-viral-combined_k141_14636\|\|full | High-quality |
| KM-final-viral-combined_k141_253504\|\|full | Not-determined |
| KM-final-viral-combined_k141_264938\|\|full | Low-quality |
| KM-final-viral-combined_k141_165525\|\|full | Low-quality |
| KM-final-viral-combined_k141_253512\|\|full | Low-quality |
| KM-final-viral-combined_k141_20089\|\|full | Low-quality |
| KM-final-viral-combined_k141_144428\|\|full | Not-determined |
| KM-final-viral-combined_k141_144430\|\|full | Not-determined |
| KM-final-viral-combined_k141_32153\|\|full | Low-quality |
| KM-final-viral-combined_k141_66522\|\|full | Not-determined |
| KM-final-viral-combined_k141_43397\|\|full | Not-determined |
| KM-final-viral-combined_k141_43398\|\|full | Low-quality |
| KM-final-viral-combined_k141_253535\|\|full | Low-quality |
| KM-final-viral-combined_k141_144450\|\|full | Not-determined |
| KM-final-viral-combined_k141_97657\|\|full | Low-quality |
| KM-final-viral-combined_k141_110434\|\|full | Low-quality |
| KM-final-viral-combined_k141_97681\|\|full | Not-determined |
| KM-final-viral-combined_k141_144462\|\|full | Not-determined |
| KM-final-viral-combined_k141_144465\|\|full | Low-quality |
| KM-final-viral-combined_k141_43441\|\|full | Not-determined |
| KM-final-viral-combined_k141_20178\|\|full | Low-quality |
| KM-final-viral-combined_k141_132809\|\|full | Not-determined |
| KM-final-viral-combined_k141_97705\|\|full | Not-determined |
| KM-final-viral-combined_k141_43458\|\|full | Not-determined |
| KM-final-viral-combined_k141_20191\|\|full | Low-quality |
| KM-final-viral-combined_k141_264993\|\|full | Not-determined |
| KM-final-viral-combined_k141_43470\|\|full | Not-determined |
| KM-final-viral-combined_k141_43481\|\|full | Not-determined |
| KM-final-viral-combined_k141_20208\|\|full | Not-determined |
| KM-final-viral-combined_k141_165666\|\|full | Not-determined |
| KM-final-viral-combined_k141_265009\|\|full | Not-determined |
| KM-final-viral-combined_k141_110476\|\|full | Not-determined |
| KM-final-viral-combined_k141_52897\|\|full | Not-determined |
| KM-final-viral-combined_k141_132833\|\|full | Not-determined |
| KM-final-viral-combined_k141_97765\|\|full | Low-quality |
| KM-final-viral-combined_k141_165682\|\|full | Low-quality |
| KM-final-viral-combined_k141_253647\|\|full | Low-quality |
| KM-final-viral-combined_k141_43532\|\|full | Not-determined |
| KM-final-viral-combined_k141_265045\|\|full | Low-quality |
| KM-final-viral-combined_k141_155369\|\|full | Low-quality |
| KM-final-viral-combined_k141_10280\|\|full | Not-determined |
| KM-final-viral-combined_k141_253679\|\|full | Low-quality |
| KM-final-viral-combined_k141_165711\|\|full | Not-determined |
| KM-final-viral-combined_k141_243733\|\|full | Not-determined |
| KM-final-viral-combined_k141_10292\|\|full | Not-determined |
| KM-final-viral-combined_k141_20297\|\|full | Low-quality |
| KM-final-viral-combined_k141_253698\|\|full | Not-determined |
| KM-final-viral-combined_k141_253699\|\|full | Low-quality |
| KM-final-viral-combined_k141_32208\|\|full | Low-quality |
| KM-final-viral-combined_k141_132862\|\|full | Not-determined |
| KM-final-viral-combined_k141_110518\|\|full | Not-determined |
| KM-final-viral-combined_k141_155400\|\|full | Not-determined |
| KM-final-viral-combined_k141_132870\|\|full | Low-quality |
| KM-final-viral-combined_k141_165765\|\|full | Not-determined |
| KM-final-viral-combined_k141_132872\|\|full | Low-quality |
| KM-final-viral-combined_k141_20347\|\|full | Low-quality |
| KM-final-viral-combined_k141_20350\|\|full | Not-determined |
| KM-final-viral-combined_k141_132879\|\|full | Low-quality |
| KM-final-viral-combined_k141_155431\|\|full | Low-quality |
| KM-final-viral-combined_k141_43557\|\|full | Low-quality |
| KM-final-viral-combined_k141_155440\|\|full | Low-quality |
| KM-final-viral-combined_k141_43559\|\|full | Low-quality |
| KM-final-viral-combined_k141_10354\|\|full | Not-determined |
| KM-final-viral-combined_k141_32272\|\|full | Low-quality |
| KM-final-viral-combined_k141_243803\|\|full | Low-quality |
| KM-final-viral-combined_k141_132894\|\|full | Not-determined |
| KM-final-viral-combined_k141_32277\|\|full | Low-quality |
| KM-final-viral-combined_k141_110560\|\|full | Low-quality |
| KM-final-viral-combined_k141_97863\|\|full | Low-quality |
| KM-final-viral-combined_k141_20413\|\|full | Not-determined |
| KM-final-viral-combined_k141_110565\|\|full | Low-quality |
| KM-final-viral-combined_k141_132913\|\|full | Not-determined |
| KM-final-viral-combined_k141_32301\|\|full | Low-quality |
| KM-final-viral-combined_k141_20424\|\|full | Low-quality |
| KM-final-viral-combined_k141_32313\|\|full | Low-quality |
| KM-final-viral-combined_k141_243866\|\|full | Low-quality |
| KM-final-viral-combined_k141_97896\|\|full | Not-determined |
| KM-final-viral-combined_k141_110588\|\|full | Not-determined |
| KM-final-viral-combined_k141_97902\|\|full | Not-determined |
| KM-final-viral-combined_k141_20477\|\|full | Low-quality |
| KM-final-viral-combined_k141_243877\|\|full | Not-determined |
| KM-final-viral-combined_k141_243879\|\|full | Not-determined |
| KM-final-viral-combined_k141_43669\|\|full | Not-determined |
| KM-final-viral-combined_k141_20487\|\|full | Low-quality |
| KM-final-viral-combined_k141_132966\|\|full | Not-determined |
| KM-final-viral-combined_k141_155507\|\|full | Not-determined |
| KM-final-viral-combined_k141_43681\|\|full | Not-determined |
| KM-final-viral-combined_k141_97939\|\|full | Not-determined |
| KM-final-viral-combined_k141_20534\|\|full | Low-quality |
| KM-final-viral-combined_k141_133000\|\|full | Not-determined |
| KM-final-viral-combined_k141_155543\|\|full | Low-quality |
| KM-final-viral-combined_k141_243971\|\|full | Not-determined |
| KM-final-viral-combined_k141_97972\|\|full | Not-determined |
| KM-final-viral-combined_k141_43746\|\|full | Low-quality |
| KM-final-viral-combined_k141_165932\|\|full | Not-determined |
| KM-final-viral-combined_k141_32480\|\|full | Not-determined |
| KM-final-viral-combined_k141_165945\|\|full | Not-determined |
| KM-final-viral-combined_k141_97990\|\|full | Not-determined |
| KM-final-viral-combined_k141_43786\|\|full | Not-determined |
| KM-final-viral-combined_k141_265180\|\|full | Low-quality |
| KM-final-viral-combined_k141_97996\|\|full | Not-determined |
| KM-final-viral-combined_k141_244027\|\|full | Not-determined |
| KM-final-viral-combined_k141_133075\|\|full | Low-quality |
| KM-final-viral-combined_k141_43791\|\|full | Low-quality |
| KM-final-viral-combined_k141_97999\|\|full | Not-determined |
| KM-final-viral-combined_k141_53018\|\|full | Not-determined |
| KM-final-viral-combined_k141_244032\|\|full | Low-quality |
| KM-final-viral-combined_k141_265194\|\|full | Not-determined |
| KM-final-viral-combined_k141_133090\|\|full | Not-determined |
| KM-final-viral-combined_k141_165989\|\|full | Not-determined |
| KM-final-viral-combined_k141_110605\|\|full | Not-determined |
| KM-final-viral-combined_k141_20615\|\|full | Low-quality |
| KM-final-viral-combined_k141_43807\|\|full | Not-determined |
| KM-final-viral-combined_k141_43813\|\|full | Not-determined |
| KM-final-viral-combined_k141_244040\|\|full | Not-determined |
| KM-final-viral-combined_k141_110615\|\|full | Low-quality |
| KM-final-viral-combined_k141_166018\|\|full | Not-determined |
| KM-final-viral-combined_k141_110623\|\|full | Low-quality |
| KM-final-viral-combined_k141_244053\|\|full | Low-quality |
| KM-final-viral-combined_k141_20639\|\|full | Low-quality |
| KM-final-viral-combined_k141_53060\|\|full | Low-quality |
| KM-final-viral-combined_k141_265215\|\|full | Low-quality |
| KM-final-viral-combined_k141_20656\|\|full | Not-determined |
| KM-final-viral-combined_k141_32543\|\|full | Low-quality |
| KM-final-viral-combined_k141_133121\|\|full | Low-quality |
| KM-final-viral-combined_k141_53072\|\|full | Low-quality |
| KM-final-viral-combined_k141_20678\|\|full | Not-determined |
| KM-final-viral-combined_k141_265263\|\|full | Low-quality |
| KM-final-viral-combined_k141_133168\|\|full | Not-determined |
| KM-final-viral-combined_k141_166076\|\|full | Low-quality |
| KM-final-viral-combined_k141_133175\|\|full | Not-determined |
| KM-final-viral-combined_k141_43883\|\|full | Low-quality |
| KM-final-viral-combined_k141_53104\|\|full | Low-quality |
| KM-final-viral-combined_k141_254094\|\|full | Not-determined |
| KM-final-viral-combined_k141_254097\|\|full | Low-quality |
| KM-final-viral-combined_k141_43909\|\|full | Not-determined |
| KM-final-viral-combined_k141_244073\|\|full | Low-quality |
| KM-final-viral-combined_k141_110680\|\|full | Not-determined |
| KM-final-viral-combined_k141_133180\|\|full | Low-quality |
| KM-final-viral-combined_k141_244090\|\|full | Not-determined |
| KM-final-viral-combined_k141_166124\|\|full | Not-determined |
| KM-final-viral-combined_k141_43956\|\|full | Low-quality |
| KM-final-viral-combined_k141_20694\|\|full | Not-determined |
| KM-final-viral-combined_k141_43969\|\|full | Not-determined |
| KM-final-viral-combined_k141_20713\|\|full | Low-quality |
| KM-final-viral-combined_k141_53193\|\|full | Not-determined |
| KM-final-viral-combined_k141_98187\|\|full | Low-quality |
| KM-final-viral-combined_k141_32685\|\|full | Not-determined |
| KM-final-viral-combined_k141_20741\|\|full | Not-determined |
| KM-final-viral-combined_k141_44008\|\|full | Low-quality |
| KM-final-viral-combined_k141_20749\|\|full | Not-determined |
| KM-final-viral-combined_k141_166154\|\|full | Not-determined |
| KM-final-viral-combined_k141_98229\|\|full | Not-determined |
| KM-final-viral-combined_k141_265399\|\|full | Not-determined |
| KM-final-viral-combined_k141_20774\|\|full | Not-determined |
| KM-final-viral-combined_k141_166161\|\|full | Not-determined |
| KM-final-viral-combined_k141_98256\|\|full | Not-determined |
| KM-final-viral-combined_k141_265432\|\|full | Low-quality |
| KM-final-viral-combined_k141_44049\|\|full | Not-determined |
| KM-final-viral-combined_k141_133273\|\|full | Not-determined |
| KM-final-viral-combined_k141_44057\|\|full | Not-determined |
| KM-final-viral-combined_k141_98288\|\|full | Not-determined |
| KM-final-viral-combined_k141_166178\|\|full | Not-determined |
| KM-final-viral-combined_k141_98303\|\|full | Low-quality |
| KM-final-viral-combined_k141_32792\|\|full | Not-determined |
| KM-final-viral-combined_k141_44078\|\|full | Not-determined |
| KM-final-viral-combined_k141_44082\|\|full | Not-determined |
| KM-final-viral-combined_k141_133301\|\|full | Not-determined |
| KM-final-viral-combined_k141_44119\|\|full | Low-quality |
| KM-final-viral-combined_k141_254248\|\|full | Low-quality |
| KM-final-viral-combined_k141_32818\|\|full | Not-determined |
| KM-final-viral-combined_k141_254250\|\|full | Low-quality |
| KM-final-viral-combined_k141_254261\|\|full | Low-quality |
| KM-final-viral-combined_k141_32825\|\|full | Low-quality |
| KM-final-viral-combined_k141_133327\|\|full | Low-quality |
| KM-final-viral-combined_k141_110764\|\|full | Not-determined |
| KM-final-viral-combined_k141_244286\|\|full | Not-determined |
| KM-final-viral-combined_k141_44142\|\|full | Low-quality |
| KM-final-viral-combined_k141_110775\|\|full | Not-determined |
| KM-final-viral-combined_k141_265469\|\|full | Low-quality |
| KM-final-viral-combined_k141_98369\|\|full | Low-quality |
| KM-final-viral-combined_k141_265475\|\|full | Not-determined |
| KM-final-viral-combined_k141_244306\|\|full | Not-determined |
| KM-final-viral-combined_k141_32872\|\|full | Low-quality |
| KM-final-viral-combined_k141_10749\|\|full | Low-quality |
| KM-final-viral-combined_k141_166190\|\|full | Low-quality |
| KM-final-viral-combined_k141_244315\|\|full | Not-determined |
| KM-final-viral-combined_k141_166192\|\|full | Not-determined |
| KM-final-viral-combined_k141_32886\|\|full | Not-determined |
| KM-final-viral-combined_k141_265501\|\|full | Low-quality |
| KM-final-viral-combined_k141_265503\|\|full | Low-quality |
| KM-final-viral-combined_k141_20954\|\|full | Low-quality |
| KM-final-viral-combined_k141_53333\|\|full | Not-determined |
| KM-final-viral-combined_k141_265510\|\|full | Not-determined |
| KM-final-viral-combined_k141_44169\|\|full | Not-determined |
| KM-final-viral-combined_k141_10770\|\|full | Not-determined |
| KM-final-viral-combined_k141_254285\|\|full | Not-determined |
| KM-final-viral-combined_k141_265542\|\|full | Low-quality |
| KM-final-viral-combined_k141_98460\|\|full | Not-determined |
| KM-final-viral-combined_k141_21000\|\|full | Low-quality |
| KM-final-viral-combined_k141_254299\|\|full | Not-determined |
| KM-final-viral-combined_k141_244363\|\|full | Low-quality |
| KM-final-viral-combined_k141_10794\|\|full | Not-determined |
| KM-final-viral-combined_k141_44241\|\|full | Not-determined |
| KM-final-viral-combined_k141_244371\|\|full | Not-determined |
| KM-final-viral-combined_k141_21035\|\|full | Not-determined |
| KM-final-viral-combined_k141_254342\|\|full | Low-quality |
| KM-final-viral-combined_k141_244379\|\|full | Low-quality |
| KM-final-viral-combined_k141_21063\|\|full | Low-quality |
| KM-final-viral-combined_k141_32946\|\|full | Low-quality |
| KM-final-viral-combined_k141_265594\|\|full | Low-quality |
| KM-final-viral-combined_k141_254372\|\|full | Low-quality |
| KM-final-viral-combined_k141_44287\|\|full | Not-determined |
| KM-final-viral-combined_k141_166246\|\|full | Low-quality |
| KM-final-viral-combined_k141_254409\|\|full | Low-quality |
| KM-final-viral-combined_k141_21089\|\|full | Not-determined |

| | |
|---|---|
| KM-final-viral-combined_k141_166273||full | Not-determined |
| KM-final-viral-combined_k141_32988||full | Low-quality |
| KM-final-viral-combined_k141_254433||full | Not-determined |
| KM-final-viral-combined_k141_244430||full | Not-determined |
| KM-final-viral-combined_k141_244432||full | Not-determined |
| KM-final-viral-combined_k141_44329||full | Low-quality |
| KM-final-viral-combined_k141_110875||full | Not-determined |
| KM-final-viral-combined_k141_110883||full | Low-quality |
| KM-final-viral-combined_k141_98567||full | Not-determined |
| KM-final-viral-combined_k141_33027||full | Not-determined |
| KM-final-viral-combined_k141_53393||full | Not-determined |
| KM-final-viral-combined_k141_110900||full | Low-quality |
| KM-final-viral-combined_k141_53414||full | Not-determined |
| KM-final-viral-combined_k141_244480||full | Low-quality |
| KM-final-viral-combined_k141_10961||full | Not-determined |
| KM-final-viral-combined_k141_265659||full | Low-quality |
| KM-final-viral-combined_k141_44402||full | Not-determined |
| KM-final-viral-combined_k141_10995||full | Low-quality |
| KM-final-viral-combined_k141_21215||full | Not-determined |
| KM-final-viral-combined_k141_98680||full | Low-quality |
| KM-final-viral-combined_k141_53460||full | Low-quality |
| KM-final-viral-combined_k141_53473||full | Not-determined |
| KM-final-viral-combined_k141_33105||full | Not-determined |
| KM-final-viral-combined_k141_265702||full | Not-determined |
| KM-final-viral-combined_k141_11041||full | Not-determined |
| KM-final-viral-combined_k141_98756||full | Low-quality |
| KM-final-viral-combined_k141_53494||full | Low-quality |
| KM-final-viral-combined_k141_110958||full | Low-quality |
| KM-final-viral-combined_k141_98757||full | Not-determined |
| KM-final-viral-combined_k141_33140||full | Not-determined |
| KM-final-viral-combined_k141_166321||full | Low-quality |
| KM-final-viral-combined_k141_21281||full | Low-quality |
| KM-final-viral-combined_k141_265756||full | Not-determined |
| KM-final-viral-combined_k141_11071||full | Low-quality |
| KM-final-viral-combined_k141_265767||full | Not-determined |
| KM-final-viral-combined_k141_11093||full | Low-quality |
| KM-final-viral-combined_k141_11109||full | Low-quality |
| KM-final-viral-combined_k141_111037||full | Not-determined |
| KM-final-viral-combined_k141_21314||full | Not-determined |
| KM-final-viral-combined_k141_53526||full | Not-determined |
| KM-final-viral-combined_k141_33173||full | Not-determined |
| KM-final-viral-combined_k141_33175||full | Not-determined |
| KM-final-viral-combined_k141_254575||full | Not-determined |
| KM-final-viral-combined_k141_33192||full | Low-quality |
| KM-final-viral-combined_k141_166391||full | Not-determined |
| KM-final-viral-combined_k141_53572||full | Low-quality |
| KM-final-viral-combined_k141_21358||full | Low-quality |
| KM-final-viral-combined_k141_254591||full | Not-determined |
| KM-final-viral-combined_k141_254596||full | Not-determined |
| KM-final-viral-combined_k141_98916||full | Not-determined |
| KM-final-viral-combined_k141_33245||full | Low-quality |
| KM-final-viral-combined_k141_265885||full | Low-quality |
| KM-final-viral-combined_k141_254618||full | Low-quality |
| KM-final-viral-combined_k141_254627||full | Not-determined |
| KM-final-viral-combined_k141_33259||full | Not-determined |
| KM-final-viral-combined_k141_21416||full | Not-determined |
| KM-final-viral-combined_k141_265941||full | Not-determined |
| KM-final-viral-combined_k141_21434||full | Not-determined |
| KM-final-viral-combined_k141_21440||full | Not-determined |
| KM-final-viral-combined_k141_111104||full | Not-determined |
| KM-final-viral-combined_k141_33293||full | Not-determined |
| KM-final-viral-combined_k141_166410||full | Not-determined |
| KM-final-viral-combined_k141_265982||full | Not-determined |
| KM-final-viral-combined_k141_33323||full | Low-quality |
| KM-final-viral-combined_k141_53794||full | Low-quality |
| KM-final-viral-combined_k141_99075||full | Low-quality |
| KM-final-viral-combined_k141_266149||full | Low-quality |
| KM-final-viral-combined_k141_99077||full | Low-quality |
| KM-final-viral-combined_k141_166512||full | Low-quality |
| KM-final-viral-combined_k141_254689||full | Low-quality |
| KM-final-viral-combined_k141_266170||full | Not-determined |
| KM-final-viral-combined_k141_166539||full | Low-quality |
| KM-final-viral-combined_k141_254710||full | Not-determined |
| KM-final-viral-combined_k141_166560||full | Low-quality |
| KM-final-viral-combined_k141_266257||full | Low-quality |
| KM-final-viral-combined_k141_99141||full | Low-quality |
| KM-final-viral-combined_k141_254747||full | Low-quality |
| KM-final-viral-combined_k141_266279||full | Not-determined |
| KM-final-viral-combined_k141_254754||full | Low-quality |
| KM-final-viral-combined_k141_254758||full | Low-quality |
| KM-final-viral-combined_k141_166650||full | Not-determined |
| KM-final-viral-combined_k141_166657||full | Not-determined |
| KM-final-viral-combined_k141_21733||full | Not-determined |
| KM-final-viral-combined_k141_266335||full | Low-quality |
| KM-final-viral-combined_k141_266339||full | Low-quality |
| KM-final-viral-combined_k141_266381||full | Not-determined |
| KM-final-viral-combined_k141_254861||full | Low-quality |
| KM-final-viral-combined_k141_21781||full | Low-quality |
| 18-101-final-viral-combined_k141_353757||full | High-quality |
| KM-final-viral-combined_k141_99271||full | Low-quality |
| KM-final-viral-combined_k141_254877||full | Not-determined |
| KM-final-viral-combined_k141_254900||full | Low-quality |
| KM-final-viral-combined_k141_254904||full | Not-determined |
| KM-final-viral-combined_k141_254911||full | Low-quality |
| KM-final-viral-combined_k141_99306||full | Not-determined |
| KM-final-viral-combined_k141_21821||full | Low-quality |
| KM-final-viral-combined_k141_21890||full | Low-quality |
| KM-final-viral-combined_k141_54059||full | Low-quality |
| KM-final-viral-combined_k141_266464||full | Low-quality |
| KM-final-viral-combined_k141_266479||full | Not-determined |
| KM-final-viral-combined_k141_21908||full | Not-determined |
| KM-final-viral-combined_k141_99399||full | Low-quality |
| KM-final-viral-combined_k141_99430||full | Low-quality |
| KM-final-viral-combined_k141_254977||full | Not-determined |
| KM-final-viral-combined_k141_99522||full | Not-determined |
| KM-final-viral-combined_k141_21983||full | Low-quality |
| KM-final-viral-combined_k141_21986||full | Not-determined |
| KM-final-viral-combined_k141_99543||full | Low-quality |
| KM-final-viral-combined_k141_54226||full | Not-determined |
| KM-final-viral-combined_k141_22005||full | Low-quality |
| KM-final-viral-combined_k141_22030||full | Low-quality |
| KM-final-viral-combined_k141_22031||full | Low-quality |
| KM-final-viral-combined_k141_54290||full | Low-quality |
| KM-final-viral-combined_k141_99577||full | Not-determined |
| KM-final-viral-combined_k141_54354||full | Low-quality |
| KM-final-viral-combined_k141_54368||full | Not-determined |
| KM-final-viral-combined_k141_54376||full | Low-quality |
| KM-final-viral-combined_k141_255279||full | Not-determined |
| KM-final-viral-combined_k141_255298||full | Low-quality |
| KM-final-viral-combined_k141_99667||full | Not-determined |
| KM-final-viral-combined_k141_255321||full | Low-quality |
| KM-final-viral-combined_k141_99687||full | Not-determined |
| KM-final-viral-combined_k141_99694||full | Not-determined |
| KM-final-viral-combined_k141_255337||full | Low-quality |
| KM-final-viral-combined_k141_99708||full | Low-quality |
| KM-final-viral-combined_k141_99722||full | Low-quality |
| KM-final-viral-combined_k141_54506||full | Low-quality |
| KM-final-viral-combined_k141_22213||full | Not-determined |
| KM-final-viral-combined_k141_255435||full | Low-quality |
| KM-final-viral-combined_k141_54592||full | Low-quality |
| KM-final-viral-combined_k141_54603||full | Low-quality |
| KM-final-viral-combined_k141_54632||full | Not-determined |
| KM-final-viral-combined_k141_54638||full | Low-quality |
| KM-final-viral-combined_k141_99780||full | Not-determined |
| KM-final-viral-combined_k141_255509||full | Low-quality |
| KM-final-viral-combined_k141_54677||full | Low-quality |
| KM-final-viral-combined_k141_255537||full | Not-determined |
| KM-final-viral-combined_k141_255563||full | Low-quality |
| KM-final-viral-combined_k141_54743||full | Not-determined |
| KM-final-viral-combined_k141_99861||full | Low-quality |
| KM-final-viral-combined_k141_54816||full | Not-determined |
| KM-final-viral-combined_k141_54873||full | Not-determined |
| KM-final-viral-combined_k141_54877||full | Not-determined |
| KM-final-viral-combined_k141_99949||full | Low-quality |
| KM-final-viral-combined_k141_99966||full | Not-determined |
| KM-final-viral-combined_k141_99973||full | Not-determined |
| KM-final-viral-combined_k141_99992||full | Low-quality |
| KM-final-viral-combined_k141_55018||full | Low-quality |
| KM-final-viral-combined_k141_55103||full | Low-quality |
| KM-final-viral-combined_k141_55239||full | Not-determined |
| KM-final-viral-combined_k141_55277||full | Low-quality |
| KM-final-viral-combined_k141_55332||full | Low-quality |
| KM-final-viral-combined_k141_55443||full | Low-quality |
| KM-final-viral-combined_k141_55459||full | Not-determined |
| KM-final-viral-combined_k141_55532||full | Low-quality |
| KM-final-viral-combined_k141_55550||full | Not-determined |
| KM-final-viral-combined_k141_55554||full | Not-determined |
| KM-final-viral-combined_k141_122280||0_partial | Low-quality |
| KM-final-viral-combined_k141_22268||0_partial | Not-determined |
| KM-final-viral-combined_k141_167117||0_partial | Low-quality |
| KM-final-viral-combined_k141_178208||0_partial | Low-quality |
| KM-final-viral-combined_k141_156231||0_partial | Low-quality |
| KM-final-viral-combined_k141_56179||0_partial | Low-quality |
| KM-final-viral-combined_k141_112180||0_partial | Low-quality |
| KM-final-viral-combined_k141_189865||0_partial | Low-quality |
| KM-final-viral-combined_k141_179183||0_partial | Low-quality |
| KM-final-viral-combined_k141_234491||0_partial | Low-quality |
| KM-final-viral-combined_k141_212874||0_partial | Not-determined |
| KM-final-viral-combined_k141_203353||0_partial | Not-determined |
| KM-final-viral-combined_k141_91111||0_partial | Low-quality |
| KM-final-viral-combined_k141_115233||0_partial | Low-quality |
| KM-final-viral-combined_k141_13918||0_partial | Not-determined |
| KM-final-viral-combined_k141_158852||0_partial | Low-quality |
| KM-final-viral-combined_k141_47162||0_partial | Low-quality |
| KM-final-viral-combined_k141_138002||0_partial | Not-determined |
| KM-final-viral-combined_k141_248622||0_partial | Low-quality |
| KM-final-viral-combined_k141_84190||0_partial | Not-determined |
| KM-final-viral-combined_k141_249059||0_partial | Not-determined |
| KM-final-viral-combined_k141_73126||0_partial | Low-quality |
| KM-final-viral-combined_k141_16146||0_partial | Low-quality |
| KM-final-viral-combined_k141_128756||0_partial | Low-quality |
| KM-final-viral-combined_k141_140294||0_partial | Low-quality |
| KM-final-viral-combined_k141_74016||0_partial | Low-quality |
| KM-final-viral-combined_k141_162054||0_partial | Low-quality |
| KM-final-viral-combined_k141_120194||0_partial | Low-quality |
| KM-final-viral-combined_k141_50505||0_partial | Low-quality |
| KM-final-viral-combined_k141_221567||0_partial | Not-determined |
| KM-final-viral-combined_k141_242634||0_partial | Low-quality |
| KM-final-viral-combined_k141_122180||0_partial | Low-quality |
| KM-final-viral-combined_k141_122229||0_partial | Not-determined |
| KM-final-viral-combined_k141_109840||0_partial | Low-quality |
| KM-final-viral-combined_k141_166407||0_partial | Not-determined |
| KM-final-viral-combined_k141_22234||lt2gene | Low-quality |
| KM-final-viral-combined_k141_88924||lt2gene | Low-quality |
| KM-final-viral-combined_k141_255616||lt2gene | Low-quality |
| KM-final-viral-combined_k141_188977||lt2gene | Not-determined |
| KM-final-viral-combined_k141_100065||lt2gene | Low-quality |
| KM-final-viral-combined_k141_66707||lt2gene | Low-quality |
| KM-final-viral-combined_k141_244561||lt2gene | Low-quality |
| KM-final-viral-combined_k141_166732||lt2gene | Low-quality |
| KM-final-viral-combined_k141_33419||lt2gene | Low-quality |
| KM-final-viral-combined_k141_77873||lt2gene | Low-quality |
| KM-final-viral-combined_k141_88964||lt2gene | Not-determined |
| KM-final-viral-combined_k141_200192||lt2gene | Low-quality |
| KM-final-viral-combined_k141_66756||lt2gene | Low-quality |
| KM-final-viral-combined_k141_111233||lt2gene | Low-quality |
| KM-final-viral-combined_k141_233493||lt2gene | Low-quality |
| KM-final-viral-combined_k141_222413||lt2gene | Low-quality |
| KM-final-viral-combined_k141_211305||lt2gene | Low-quality |
| KM-final-viral-combined_k141_189093||lt2gene | Low-quality |
| KM-final-viral-combined_k141_144666||lt2gene | Low-quality |
| KM-final-viral-combined_k141_111262||lt2gene | Low-quality |
| KM-final-viral-combined_k141_77944||lt2gene | Not-determined |
| KM-final-viral-combined_k141_200273||lt2gene | Low-quality |
| KM-final-viral-combined_k141_55647||lt2gene | Not-determined |
| KM-final-viral-combined_k141_211326||lt2gene | Not-determined |
| KM-final-viral-combined_k141_211339||lt2gene | Not-determined |
| KM-final-viral-combined_k141_77993||lt2gene | Low-quality |
| KM-final-viral-combined_k141_111302||lt2gene | Low-quality |
| KM-final-viral-combined_k141_11241||lt2gene | Low-quality |
| KM-final-viral-combined_k141_11244||lt2gene | Low-quality |
| KM-final-viral-combined_k141_155738||lt2gene | Low-quality |
| KM-final-viral-combined_k141_222477||lt2gene | Not-determined |
| KM-final-viral-combined_k141_89036||lt2gene | Low-quality |
| KM-final-viral-combined_k141_189176||lt2gene | Low-quality |
| KM-final-viral-combined_k141_66859||lt2gene | Low-quality |
| KM-final-viral-combined_k141_111375||lt2gene | Low-quality |
| KM-final-viral-combined_k141_100137||lt2gene | Not-determined |
| KM-final-viral-combined_k141_166828||lt2gene | Low-quality |
| KM-final-viral-combined_k141_233602||lt2gene | Not-determined |
| KM-final-viral-combined_k141_33695||lt2gene | Low-quality |
| KM-final-viral-combined_k141_122506||lt2gene | Low-quality |
| KM-final-viral-combined_k141_200364||lt2gene | Low-quality |
| KM-final-viral-combined_k141_155785||lt2gene | Not-determined |
| KM-final-viral-combined_k141_166863||lt2gene | Low-quality |
| KM-final-viral-combined_k141_100184||lt2gene | Low-quality |
| KM-final-viral-combined_k141_55841||lt2gene | Low-quality |
| KM-final-viral-combined_k141_22485||lt2gene | Not-determined |
| KM-final-viral-combined_k141_22492||lt2gene | Low-quality |
| KM-final-viral-combined_k141_222647||lt2gene | Low-quality |
| KM-final-viral-combined_k141_100238||lt2gene | Low-quality |
| KM-final-viral-combined_k141_233749||lt2gene | Low-quality |
| KM-final-viral-combined_k141_189275||lt2gene | Low-quality |
| KM-final-viral-combined_k141_44688||lt2gene | Low-quality |
| KM-final-viral-combined_k141_44698||lt2gene | Low-quality |
| KM-final-viral-combined_k141_44704||lt2gene | Low-quality |
| KM-final-viral-combined_k141_22561||lt2gene | Low-quality |
| KM-final-viral-combined_k141_211563||lt2gene | Low-quality |
| KM-final-viral-combined_k141_144983||lt2gene | Low-quality |
| KM-final-viral-combined_k141_11392||lt2gene | Not-determined |
| KM-final-viral-combined_k141_22577||lt2gene | Low-quality |
| KM-final-viral-combined_k141_211587||lt2gene | Not-determined |
| KM-final-viral-combined_k141_55990||lt2gene | Low-quality |
| KM-final-viral-combined_k141_89249||lt2gene | Low-quality |
| KM-final-viral-combined_k141_200466||lt2gene | Low-quality |
| KM-final-viral-combined_k141_166972||lt2gene | Not-determined |
| KM-final-viral-combined_k141_200489||lt2gene | Not-determined |
| KM-final-viral-combined_k141_145027||lt2gene | Not-determined |
| KM-final-viral-combined_k141_67205||lt2gene | Low-quality |
| KM-final-viral-combined_k141_145084||lt2gene | Low-quality |
| KM-final-viral-combined_k141_133767||lt2gene | Low-quality |
| KM-final-viral-combined_k141_111650||lt2gene | Not-determined |
| KM-final-viral-combined_k141_78403||lt2gene | Not-determined |
| KM-final-viral-combined_k141_122748||lt2gene | Low-quality |
| KM-final-viral-combined_k141_22629||lt2gene | Low-quality |
| KM-final-viral-combined_k141_520||lt2gene | Low-quality |
| KM-final-viral-combined_k141_111716||lt2gene | Low-quality |
| KM-final-viral-combined_k141_89332||lt2gene | Not-determined |
| KM-final-viral-combined_k141_200693||lt2gene | Not-determined |
| KM-final-viral-combined_k141_100471||lt2gene | Low-quality |
| KM-final-viral-combined_k141_178202||lt2gene | Low-quality |
| KM-final-viral-combined_k141_211792||lt2gene | Low-quality |
| KM-final-viral-combined_k141_233871||lt2gene | Low-quality |
| KM-final-viral-combined_k141_156115||lt2gene | Low-quality |
| KM-final-viral-combined_k141_133932||lt2gene | Not-determined |
| KM-final-viral-combined_k141_33981||lt2gene | Not-determined |
| KM-final-viral-combined_k141_111874||lt2gene | Not-determined |
| KM-final-viral-combined_k141_167183||lt2gene | Low-quality |
| KM-final-viral-combined_k141_67491||lt2gene | Low-quality |
| KM-final-viral-combined_k141_674||lt2gene | Low-quality |
| KM-final-viral-combined_k141_679||lt2gene | Low-quality |
| KM-final-viral-combined_k141_167243||lt2gene | Low-quality |
| KM-final-viral-combined_k141_67580||lt2gene | Not-determined |
| KM-final-viral-combined_k141_200910||lt2gene | Not-determined |
| KM-final-viral-combined_k141_200913||lt2gene | Not-determined |
| KM-final-viral-combined_k141_67599||lt2gene | Low-quality |
| KM-final-viral-combined_k141_178375||lt2gene | Low-quality |
| KM-final-viral-combined_k141_245249||lt2gene | Not-determined |
| KM-final-viral-combined_k141_189598||lt2gene | Low-quality |
| KM-final-viral-combined_k141_34092||lt2gene | Low-quality |
| KM-final-viral-combined_k141_776||lt2gene | Low-quality |
| KM-final-viral-combined_k141_212021||lt2gene | Not-determined |
| KM-final-viral-combined_k141_89558||lt2gene | Low-quality |
| KM-final-viral-combined_k141_100564||lt2gene | Not-determined |
| KM-final-viral-combined_k141_89562||lt2gene | Not-determined |
| KM-final-viral-combined_k141_167278||lt2gene | Low-quality |
| KM-final-viral-combined_k141_156303||lt2gene | Low-quality |
| KM-final-viral-combined_k141_100584||lt2gene | Low-quality |
| KM-final-viral-combined_k141_245322||lt2gene | Low-quality |
| KM-final-viral-combined_k141_212081||lt2gene | Low-quality |
| KM-final-viral-combined_k141_156327||lt2gene | Low-quality |
| KM-final-viral-combined_k141_34164||lt2gene | Not-determined |
| KM-final-viral-combined_k141_78795||lt2gene | Not-determined |
| KM-final-viral-combined_k141_189702||lt2gene | Not-determined |
| KM-final-viral-combined_k141_11768||lt2gene | Low-quality |
| KM-final-viral-combined_k141_22809||lt2gene | Low-quality |
| KM-final-viral-combined_k141_67668||lt2gene | Not-determined |
| KM-final-viral-combined_k141_178515||lt2gene | Low-quality |
| KM-final-viral-combined_k141_145500||lt2gene | Not-determined |
| KM-final-viral-combined_k141_78862||lt2gene | Low-quality |
| KM-final-viral-combined_k141_178578||lt2gene | Low-quality |
| KM-final-viral-combined_k141_223242||lt2gene | Low-quality |
| KM-final-viral-combined_k141_123237||lt2gene | Not-determined |
| KM-final-viral-combined_k141_178635||lt2gene | Low-quality |
| KM-final-viral-combined_k141_156429||lt2gene | Not-determined |
| KM-final-viral-combined_k141_859||lt2gene | Not-determined |
| KM-final-viral-combined_k141_78993||lt2gene | Low-quality |
| KM-final-viral-combined_k141_123277||lt2gene | Not-determined |
| KM-final-viral-combined_k141_123288||lt2gene | Low-quality |
| KM-final-viral-combined_k141_123296||lt2gene | Low-quality |
| KM-final-viral-combined_k141_189799||lt2gene | Not-determined |
| KM-final-viral-combined_k141_156528||lt2gene | Low-quality |
| KM-final-viral-combined_k141_234141||lt2gene | Low-quality |
| KM-final-viral-combined_k141_245482||lt2gene | Not-determined |
| KM-final-viral-combined_k141_212276||lt2gene | Low-quality |
| KM-final-viral-combined_k141_134205||lt2gene | Low-quality |
| KM-final-viral-combined_k141_245501||lt2gene | Not-determined |
| KM-final-viral-combined_k141_34343||lt2gene | Not-determined |
| KM-final-viral-combined_k141_22983||lt2gene | Not-determined |
| KM-final-viral-combined_k141_256614||lt2gene | Low-quality |
| KM-final-viral-combined_k141_156592||lt2gene | Low-quality |
| KM-final-viral-combined_k141_112349||lt2gene | Low-quality |
| KM-final-viral-combined_k141_100846||lt2gene | Low-quality |
| KM-final-viral-combined_k141_178811||lt2gene | Low-quality |
| KM-final-viral-combined_k141_56661||lt2gene | Low-quality |
| KM-final-viral-combined_k141_45180||lt2gene | Not-determined |
| KM-final-viral-combined_k141_123482||lt2gene | Low-quality |
| KM-final-viral-combined_k141_189919||lt2gene | Not-determined |
| KM-final-viral-combined_k141_201350||lt2gene | Not-determined |
| KM-final-viral-combined_k141_123526||lt2gene | Low-quality |
| KM-final-viral-combined_k141_67907||lt2gene | Not-determined |
| KM-final-viral-combined_k141_100929||lt2gene | Low-quality |
| KM-final-viral-combined_k141_234413||lt2gene | Low-quality |
| KM-final-viral-combined_k141_134424||lt2gene | Low-quality |
| KM-final-viral-combined_k141_201453||lt2gene | Not-determined |
| KM-final-viral-combined_k141_134472||lt2gene | Low-quality |
| KM-final-viral-combined_k141_68021||lt2gene | Low-quality |
| KM-final-viral-combined_k141_134483||lt2gene | Not-determined |
| KM-final-viral-combined_k141_79365||lt2gene | Low-quality |
| KM-final-viral-combined_k141_145829||lt2gene | Not-determined |
| KM-final-viral-combined_k141_212572||lt2gene | Low-quality |
| KM-final-viral-combined_k141_145854||lt2gene | Not-determined |
| KM-final-viral-combined_k141_34607||lt2gene | Low-quality |
| KM-final-viral-combined_k141_68082||lt2gene | Low-quality |
| KM-final-viral-combined_k141_79409||lt2gene | Low-quality |
| KM-final-viral-combined_k141_1274||lt2gene | Not-determined |
| KM-final-viral-combined_k141_89927||lt2gene | Low-quality |
| KM-final-viral-combined_k141_1291||lt2gene | Low-quality |
| KM-final-viral-combined_k141_245675||lt2gene | Low-quality |
| KM-final-viral-combined_k141_1308||lt2gene | Not-determined |
| KM-final-viral-combined_k141_145935||lt2gene | Low-quality |
| KM-final-viral-combined_k141_223815||lt2gene | Low-quality |
| KM-final-viral-combined_k141_201635||lt2gene | Not-determined |
| KM-final-viral-combined_k141_45384||lt2gene | Not-determined |
| KM-final-viral-combined_k141_212700||lt2gene | Low-quality |
| KM-final-viral-combined_k141_167878||lt2gene | Not-determined |
| KM-final-viral-combined_k141_212711||lt2gene | Low-quality |
| KM-final-viral-combined_k141_1353||lt2gene | Low-quality |
| KM-final-viral-combined_k141_79578||lt2gene | Not-determined |
| KM-final-viral-combined_k141_201707||lt2gene | Low-quality |
| KM-final-viral-combined_k141_201712||lt2gene | Not-determined |
| KM-final-viral-combined_k141_79610||lt2gene | Low-quality |
| KM-final-viral-combined_k141_79615||lt2gene | Low-quality |
| KM-final-viral-combined_k141_79630||lt2gene | Not-determined |
| KM-final-viral-combined_k141_212799||lt2gene | Low-quality |
| KM-final-viral-combined_k141_1422||lt2gene | Low-quality |
| KM-final-viral-combined_k141_156882||lt2gene | Not-determined |
| KM-final-viral-combined_k141_12271||lt2gene | Low-quality |
| KM-final-viral-combined_k141_212827||lt2gene | Not-determined |
| KM-final-viral-combined_k141_256960||lt2gene | Low-quality |
| KM-final-viral-combined_k141_112644||lt2gene | Low-quality |
| KM-final-viral-combined_k141_34758||lt2gene | Low-quality |
| KM-final-viral-combined_k141_56946||lt2gene | Not-determined |
| KM-final-viral-combined_k141_146013||lt2gene | Low-quality |
| KM-final-viral-combined_k141_134763||lt2gene | Low-quality |
| KM-final-viral-combined_k141_223992||lt2gene | Low-quality |
| KM-final-viral-combined_k141_224002||lt2gene | Not-determined |
| KM-final-viral-combined_k141_190317||lt2gene | Low-quality |
| KM-final-viral-combined_k141_112696||lt2gene | Low-quality |
| KM-final-viral-combined_k141_79732||lt2gene | Not-determined |
| KM-final-viral-combined_k141_134790||lt2gene | Low-quality |
| KM-final-viral-combined_k141_168026||lt2gene | Low-quality |
| KM-final-viral-combined_k141_134815||lt2gene | Low-quality |
| KM-final-viral-combined_k141_112812||lt2gene | Not-determined |
| KM-final-viral-combined_k141_1515||lt2gene | Low-quality |
| KM-final-viral-combined_k141_101317||lt2gene | Low-quality |
| KM-final-viral-combined_k141_90146||lt2gene | Not-determined |
| KM-final-viral-combined_k141_34853||lt2gene | Low-quality |
| KM-final-viral-combined_k141_190456||lt2gene | Low-quality |
| KM-final-viral-combined_k141_79819||lt2gene | Low-quality |
| KM-final-viral-combined_k141_201950||lt2gene | Low-quality |
| KM-final-viral-combined_k141_224199||lt2gene | Low-quality |
| KM-final-viral-combined_k141_112925||lt2gene | Not-determined |
| KM-final-viral-combined_k141_234792||lt2gene | Low-quality |
| KM-final-viral-combined_k141_1661||lt2gene | Not-determined |
| KM-final-viral-combined_k141_202009||lt2gene | Not-determined |
| KM-final-viral-combined_k141_124143||lt2gene | Low-quality |
| KM-final-viral-combined_k141_257281||lt2gene | Low-quality |
| KM-final-viral-combined_k141_79956||lt2gene | Low-quality |
| KM-final-viral-combined_k141_1691||lt2gene | Low-quality |
| KM-final-viral-combined_k141_135034||lt2gene | Low-quality |
| KM-final-viral-combined_k141_1733||lt2gene | Low-quality |
| KM-final-viral-combined_k141_124199||lt2gene | Low-quality |
| KM-final-viral-combined_k141_213010||lt2gene | Low-quality |
| KM-final-viral-combined_k141_245839||lt2gene | Low-quality |
| KM-final-viral-combined_k141_135063||lt2gene | Not-determined |
| KM-final-viral-combined_k141_179561||lt2gene | Low-quality |
| KM-final-viral-combined_k141_202165||lt2gene | Low-quality |
| KM-final-viral-combined_k141_146254||lt2gene | Low-quality |
| KM-final-viral-combined_k141_135138||lt2gene | Not-determined |
| KM-final-viral-combined_k141_213127||lt2gene | Low-quality |
| KM-final-viral-combined_k141_1797||lt2gene | Low-quality |
| KM-final-viral-combined_k141_202228||lt2gene | Low-quality |
| KM-final-viral-combined_k141_57281||lt2gene | Low-quality |
| KM-final-viral-combined_k141_245967||lt2gene | Low-quality |
| KM-final-viral-combined_k141_234945||lt2gene | Low-quality |
| KM-final-viral-combined_k141_124336||lt2gene | Low-quality |
| KM-final-viral-combined_k141_234962||lt2gene | Low-quality |
| KM-final-viral-combined_k141_90445||lt2gene | Low-quality |
| KM-final-viral-combined_k141_90455||lt2gene | Low-quality |
| KM-final-viral-combined_k141_23835||lt2gene | Not-determined |
| KM-final-viral-combined_k141_146358||lt2gene | Low-quality |
| KM-final-viral-combined_k141_57326||lt2gene | Low-quality |
| KM-final-viral-combined_k141_157223||lt2gene | Low-quality |
| KM-final-viral-combined_k141_113308||lt2gene | Low-quality |
| KM-final-viral-combined_k141_213309||lt2gene | Low-quality |
| KM-final-viral-combined_k141_202402||lt2gene | Not-determined |
| KM-final-viral-combined_k141_179755||lt2gene | Not-determined |
| KM-final-viral-combined_k141_1978||lt2gene | Not-determined |
| KM-final-viral-combined_k141_12501||lt2gene | Not-determined |
| KM-final-viral-combined_k141_213351||lt2gene | Not-determined |
| KM-final-viral-combined_k141_146420||lt2gene | Not-determined |
| KM-final-viral-combined_k141_157287||lt2gene | Low-quality |
| KM-final-viral-combined_k141_45963||lt2gene | Low-quality |
| KM-final-viral-combined_k141_35305||lt2gene | Low-quality |
| KM-final-viral-combined_k141_179782||lt2gene | Low-quality |
| KM-final-viral-combined_k141_80220||lt2gene | Low-quality |
| KM-final-viral-combined_k141_146507||lt2gene | Low-quality |
| KM-final-viral-combined_k141_257577||lt2gene | Low-quality |
| KM-final-viral-combined_k141_168728||lt2gene | Low-quality |
| KM-final-viral-combined_k141_135479||lt2gene | Low-quality |
| KM-final-viral-combined_k141_101798||lt2gene | Low-quality |
| KM-final-viral-combined_k141_213468||lt2gene | Not-determined |
| KM-final-viral-combined_k141_113518||lt2gene | Not-determined |
| KM-final-viral-combined_k141_12619||lt2gene | Low-quality |
| KM-final-viral-combined_k141_80298||lt2gene | Low-quality |
| KM-final-viral-combined_k141_224571||lt2gene | Not-determined |
| KM-final-viral-combined_k141_202637||lt2gene | Low-quality |
| KM-final-viral-combined_k141_101862||lt2gene | Low-quality |
| KM-final-viral-combined_k141_191170||lt2gene | Low-quality |
| KM-final-viral-combined_k141_124571||lt2gene | Low-quality |
| KM-final-viral-combined_k141_113652||lt2gene | Low-quality |
| KM-final-viral-combined_k141_80380||lt2gene | Low-quality |
| KM-final-viral-combined_k141_80391||lt2gene | Not-determined |
| KM-final-viral-combined_k141_246218||lt2gene | Low-quality |
| KM-final-viral-combined_k141_224688||lt2gene | Not-determined |
| KM-final-viral-combined_k141_24088||lt2gene | Low-quality |
| KM-final-viral-combined_k141_246253||lt2gene | Low-quality |
| KM-final-viral-combined_k141_24098||lt2gene | Low-quality |
| KM-final-viral-combined_k141_180065||lt2gene | Not-determined |
| KM-final-viral-combined_k141_46089||lt2gene | Low-quality |
| KM-final-viral-combined_k141_35560||lt2gene | Not-determined |
| KM-final-viral-combined_k141_12795||lt2gene | Low-quality |
| KM-final-viral-combined_k141_90779||lt2gene | Low-quality |
| KM-final-viral-combined_k141_235363||lt2gene | Low-quality |
| KM-final-viral-combined_k141_146765||lt2gene | Not-determined |
| KM-final-viral-combined_k141_135777||lt2gene | Low-quality |
| KM-final-viral-combined_k141_80503||lt2gene | Not-determined |
| KM-final-viral-combined_k141_57753||lt2gene | Low-quality |
| KM-final-viral-combined_k141_235448||lt2gene | Low-quality |
| KM-final-viral-combined_k141_135813||lt2gene | Not-determined |
| KM-final-viral-combined_k141_169051||lt2gene | Low-quality |
| KM-final-viral-combined_k141_246319||lt2gene | Not-determined |
| KM-final-viral-combined_k141_202836||lt2gene | Low-quality |
| KM-final-viral-combined_k141_246356||lt2gene | Low-quality |
| KM-final-viral-combined_k141_124912||lt2gene | Low-quality |
| KM-final-viral-combined_k141_69188||lt2gene | Low-quality |
| KM-final-viral-combined_k141_35615||lt2gene | Low-quality |
| KM-final-viral-combined_k141_224884||lt2gene | Not-determined |
| KM-final-viral-combined_k141_69214||lt2gene | Not-determined |
| KM-final-viral-combined_k141_124953||lt2gene | Not-determined |
| KM-final-viral-combined_k141_257915||lt2gene | Not-determined |
| KM-final-viral-combined_k141_169133||lt2gene | Low-quality |
| KM-final-viral-combined_k141_113973||lt2gene | Not-determined |
| KM-final-viral-combined_k141_180279||lt2gene | Low-quality |
| KM-final-viral-combined_k141_135887||lt2gene | Low-quality |
| KM-final-viral-combined_k141_125036||lt2gene | Low-quality |
| KM-final-viral-combined_k141_2349||lt2gene | Low-quality |
| KM-final-viral-combined_k141_235637||lt2gene | Not-determined |
| KM-final-viral-combined_k141_225018||lt2gene | Low-quality |
| KM-final-viral-combined_k141_147000||lt2gene | Low-quality |
| KM-final-viral-combined_k141_202998||lt2gene | Low-quality |
| KM-final-viral-combined_k141_147069||lt2gene | Low-quality |
| KM-final-viral-combined_k141_180364||lt2gene | Low-quality |
| KM-final-viral-combined_k141_191597||lt2gene | Not-determined |
| KM-final-viral-combined_k141_35817||lt2gene | Low-quality |
| KM-final-viral-combined_k141_180410||lt2gene | Low-quality |
| KM-final-viral-combined_k141_114252||lt2gene | Low-quality |
| KM-final-viral-combined_k141_203139||lt2gene | Low-quality |
| KM-final-viral-combined_k141_203140||lt2gene | Not-determined |
| KM-final-viral-combined_k141_246593||lt2gene | Not-determined |
| KM-final-viral-combined_k141_180425||lt2gene | Low-quality |
| KM-final-viral-combined_k141_58003||lt2gene | Low-quality |
| KM-final-viral-combined_k141_125078||lt2gene | Low-quality |
| KM-final-viral-combined_k141_169382||lt2gene | Low-quality |
| KM-final-viral-combined_k141_180449||lt2gene | Low-quality |
| KM-final-viral-combined_k141_102311||lt2gene | Low-quality |
| KM-final-viral-combined_k141_136059||lt2gene | Not-determined |
| KM-final-viral-combined_k141_13131||lt2gene | Low-quality |
| KM-final-viral-combined_k141_235855||lt2gene | Low-quality |
| KM-final-viral-combined_k141_246612||lt2gene | Not-determined |
| KM-final-viral-combined_k141_246621||lt2gene | Low-quality |
| KM-final-viral-combined_k141_35895||lt2gene | Low-quality |
| KM-final-viral-combined_k141_24399||lt2gene | Low-quality |
| KM-final-viral-combined_k141_69543||lt2gene | Low-quality |
| KM-final-viral-combined_k141_235891||lt2gene | Low-quality |
| KM-final-viral-combined_k141_235898||lt2gene | Low-quality |
| KM-final-viral-combined_k141_24428||lt2gene | Low-quality |
| KM-final-viral-combined_k141_147259||lt2gene | Low-quality |
| KM-final-viral-combined_k141_13212||lt2gene | Low-quality |
| KM-final-viral-combined_k141_147269||lt2gene | Low-quality |
| KM-final-viral-combined_k141_214145||lt2gene | Low-quality |
| KM-final-viral-combined_k141_180515||lt2gene | Low-quality |
| KM-final-viral-combined_k141_180530||lt2gene | Not-determined |
| KM-final-viral-combined_k141_157845||lt2gene | Not-determined |
| KM-final-viral-combined_k141_136167||lt2gene | Low-quality |
| KM-final-viral-combined_k141_80831||lt2gene | Not-determined |
| KM-final-viral-combined_k141_147334||lt2gene | Low-quality |
| KM-final-viral-combined_k141_258272||lt2gene | Low-quality |
| KM-final-viral-combined_k141_214271||lt2gene | Low-quality |
| KM-final-viral-combined_k141_214278||lt2gene | Low-quality |
| KM-final-viral-combined_k141_258281||lt2gene | Not-determined |
| KM-final-viral-combined_k141_13344||lt2gene | Low-quality |
| KM-final-viral-combined_k141_58144||lt2gene | Not-determined |
| KM-final-viral-combined_k141_169573||lt2gene | Not-determined |
| KM-final-viral-combined_k141_36029||lt2gene | Low-quality |
| KM-final-viral-combined_k141_102499||lt2gene | Not-determined |
| KM-final-viral-combined_k141_2678||lt2gene | Low-quality |
| KM-final-viral-combined_k141_36061||lt2gene | Not-determined |
| KM-final-viral-combined_k141_102513||lt2gene | Not-determined |
| KM-final-viral-combined_k141_80944||lt2gene | Not-determined |
| KM-final-viral-combined_k141_13416||lt2gene | Low-quality |
| KM-final-viral-combined_k141_225384||lt2gene | Not-determined |
| KM-final-viral-combined_k141_125367||lt2gene | Low-quality |
| KM-final-viral-combined_k141_102585||lt2gene | Low-quality |
| KM-final-viral-combined_k141_24661||lt2gene | Low-quality |
| KM-final-viral-combined_k141_180878||lt2gene | Low-quality |
| KM-final-viral-combined_k141_102639||lt2gene | Not-determined |
| KM-final-viral-combined_k141_203391||lt2gene | Not-determined |
| KM-final-viral-combined_k141_36179||lt2gene | Low-quality |
| KM-final-viral-combined_k141_69863||lt2gene | Low-quality |
| KM-final-viral-combined_k141_203421||lt2gene | Not-determined |
| KM-final-viral-combined_k141_114672||lt2gene | Low-quality |
| KM-final-viral-combined_k141_81177||lt2gene | Low-quality |
| KM-final-viral-combined_k141_203435||lt2gene | Low-quality |
| KM-final-viral-combined_k141_24759||lt2gene | Low-quality |
| KM-final-viral-combined_k141_114743||lt2gene | Low-quality |
| KM-final-viral-combined_k141_13547||lt2gene | Low-quality |
| KM-final-viral-combined_k141_203482||lt2gene | Not-determined |
| KM-final-viral-combined_k141_13562||lt2gene | Low-quality |
| KM-final-viral-combined_k141_2844||lt2gene | Low-quality |
| KM-final-viral-combined_k141_114799||lt2gene | Low-quality |
| KM-final-viral-combined_k141_203518||lt2gene | Low-quality |
| KM-final-viral-combined_k141_24819||lt2gene | Low-quality |
| KM-final-viral-combined_k141_214619||lt2gene | Low-quality |
| KM-final-viral-combined_k141_214623||lt2gene | Low-quality |
| KM-final-viral-combined_k141_125590||lt2gene | Low-quality |
| KM-final-viral-combined_k141_258552||lt2gene | Not-determined |
| KM-final-viral-combined_k141_147623||lt2gene | Low-quality |
| KM-final-viral-combined_k141_236282||lt2gene | Not-determined |
| KM-final-viral-combined_k141_58415||lt2gene | Low-quality |
| KM-final-viral-combined_k141_58419||lt2gene | Low-quality |
| KM-final-viral-combined_k141_192183||lt2gene | Low-quality |
| KM-final-viral-combined_k141_114957||lt2gene | Low-quality |
| KM-final-viral-combined_k141_125690||lt2gene | Low-quality |
| KM-final-viral-combined_k141_36359||lt2gene | Low-quality |
| KM-final-viral-combined_k141_81365||lt2gene | Not-determined |
| KM-final-viral-combined_k141_69999||lt2gene | Not-determined |
| KM-final-viral-combined_k141_102951||lt2gene | Low-quality |

KM-final-viral-combined_k141_36416||lt2gene Low-quality
KM-final-viral-combined_k141_13677||lt2gene Not-determined
KM-final-viral-combined_k141_225710||lt2gene Low-quality
KM-final-viral-combined_k141_81461||lt2gene Not-determined
KM-final-viral-combined_k141_147759||lt2gene Low-quality
KM-final-viral-combined_k141_115124||lt2gene Not-determined
KM-final-viral-combined_k141_214800||lt2gene Low-quality
KM-final-viral-combined_k141_147824||lt2gene Low-quality
KM-final-viral-combined_k141_192395||lt2gene Low-quality
KM-final-viral-combined_k141_58596||lt2gene Not-determined
KM-final-viral-combined_k141_192472||lt2gene Low-quality
KM-final-viral-combined_k141_25099||lt2gene Low-quality
KM-final-viral-combined_k141_70130||lt2gene Not-determined
KM-final-viral-combined_k141_36561||lt2gene Not-determined
KM-final-viral-combined_k141_103121||lt2gene Not-determined
KM-final-viral-combined_k141_103123||lt2gene Not-determined
KM-final-viral-combined_k141_81565||lt2gene Not-determined
KM-final-viral-combined_k141_91481||lt2gene Not-determined
KM-final-viral-combined_k141_181391||lt2gene Low-quality
KM-final-viral-combined_k141_158318||lt2gene Low-quality
KM-final-viral-combined_k141_115238||lt2gene Not-determined
KM-final-viral-combined_k141_258708||lt2gene Low-quality
KM-final-viral-combined_k141_3275||lt2gene Low-quality
KM-final-viral-combined_k141_203860||lt2gene Low-quality
KM-final-viral-combined_k141_236569||lt2gene Low-quality
KM-final-viral-combined_k141_91502||lt2gene Low-quality
KM-final-viral-combined_k141_46870||lt2gene Low-quality
KM-final-viral-combined_k141_225881||lt2gene Low-quality
KM-final-viral-combined_k141_225887||lt2gene Not-determined
KM-final-viral-combined_k141_170038||lt2gene Low-quality
KM-final-viral-combined_k141_170043||lt2gene Low-quality
KM-final-viral-combined_k141_103228||lt2gene Low-quality
KM-final-viral-combined_k141_203923||lt2gene Not-determined
KM-final-viral-combined_k141_247273||lt2gene Low-quality
KM-final-viral-combined_k141_225934||lt2gene Low-quality
KM-final-viral-combined_k141_158421||lt2gene Low-quality
KM-final-viral-combined_k141_3402||lt2gene Low-quality
KM-final-viral-combined_k141_192628||lt2gene Not-determined
KM-final-viral-combined_k141_115349||lt2gene Low-quality
KM-final-viral-combined_k141_137032||lt2gene Low-quality
KM-final-viral-combined_k141_137036||lt2gene Low-quality
KM-final-viral-combined_k141_115404||lt2gene Not-determined
KM-final-viral-combined_k141_103261||lt2gene Not-determined
KM-final-viral-combined_k141_81806||lt2gene Low-quality
KM-final-viral-combined_k141_236684||lt2gene Low-quality
KM-final-viral-combined_k141_247397||lt2gene Low-quality
KM-final-viral-combined_k141_81859||lt2gene Low-quality
KM-final-viral-combined_k141_247404||lt2gene Low-quality
KM-final-viral-combined_k141_247406||lt2gene Low-quality
KM-final-viral-combined_k141_115529||lt2gene Low-quality
KM-final-viral-combined_k141_103309||lt2gene Not-determined
KM-final-viral-combined_k141_103324||lt2gene Low-quality
KM-final-viral-combined_k141_158530||lt2gene Low-quality
KM-final-viral-combined_k141_13998||lt2gene Not-determined
KM-final-viral-combined_k141_158544||lt2gene Low-quality
KM-final-viral-combined_k141_247483||lt2gene Low-quality
KM-final-viral-combined_k141_215245||lt2gene Low-quality
KM-final-viral-combined_k141_148217||lt2gene Low-quality
KM-final-viral-combined_k141_115650||lt2gene Not-determined
KM-final-viral-combined_k141_170269||lt2gene Low-quality
KM-final-viral-combined_k141_115685||lt2gene Low-quality
KM-final-viral-combined_k141_81955||lt2gene Low-quality
KM-final-viral-combined_k141_215348||lt2gene Low-quality
KM-final-viral-combined_k141_215355||lt2gene Low-quality
KM-final-viral-combined_k141_236752||lt2gene Not-determined
KM-final-viral-combined_k141_103484||lt2gene Low-quality
KM-final-viral-combined_k141_226177||lt2gene Low-quality
KM-final-viral-combined_k141_115802||lt2gene Low-quality
KM-final-viral-combined_k141_36871||lt2gene Low-quality
KM-final-viral-combined_k141_70814||lt2gene Low-quality
KM-final-viral-combined_k141_82162||lt2gene Low-quality
KM-final-viral-combined_k141_170433||lt2gene Not-determined
KM-final-viral-combined_k141_14169||lt2gene Not-determined
KM-final-viral-combined_k141_259038||lt2gene Not-determined
KM-final-viral-combined_k141_25678||lt2gene Low-quality
KM-final-viral-combined_k141_91904||lt2gene Low-quality
KM-final-viral-combined_k141_25702||lt2gene Not-determined
KM-final-viral-combined_k141_204368||lt2gene Low-quality
KM-final-viral-combined_k141_59110||lt2gene Low-quality
KM-final-viral-combined_k141_204407||lt2gene Low-quality
KM-final-viral-combined_k141_236994||lt2gene Not-determined
KM-final-viral-combined_k141_182148||lt2gene Not-determined
KM-final-viral-combined_k141_59165||lt2gene Not-determined
KM-final-viral-combined_k141_182161||lt2gene Low-quality
KM-final-viral-combined_k141_3673||lt2gene Low-quality
KM-final-viral-combined_k141_3677||lt2gene Low-quality
KM-final-viral-combined_k141_126407||lt2gene Not-determined
KM-final-viral-combined_k141_37012||lt2gene Not-determined
KM-final-viral-combined_k141_47039||lt2gene Low-quality
KM-final-viral-combined_k141_226504||lt2gene Low-quality
KM-final-viral-combined_k141_82313||lt2gene Not-determined
KM-final-viral-combined_k141_59193||lt2gene Not-determined
KM-final-viral-combined_k141_215684||lt2gene Not-determined
KM-final-viral-combined_k141_25848||lt2gene Not-determined
KM-final-viral-combined_k141_148583||lt2gene Not-determined
KM-final-viral-combined_k141_37104||lt2gene Not-determined
KM-final-viral-combined_k141_82361||lt2gene Low-quality
KM-final-viral-combined_k141_82373||lt2gene Low-quality
KM-final-viral-combined_k141_182284||lt2gene Low-quality
KM-final-viral-combined_k141_215754||lt2gene Not-determined
KM-final-viral-combined_k141_37147||lt2gene Low-quality
KM-final-viral-combined_k141_82427||lt2gene Low-quality
KM-final-viral-combined_k141_25900||lt2gene Low-quality
KM-final-viral-combined_k141_226675||lt2gene Low-quality
KM-final-viral-combined_k141_116221||lt2gene Low-quality
KM-final-viral-combined_k141_25944||lt2gene Low-quality
KM-final-viral-combined_k141_103895||lt2gene Not-determined
KM-final-viral-combined_k141_259138||lt2gene Low-quality
KM-final-viral-combined_k141_71068||lt2gene Low-quality
KM-final-viral-combined_k141_37191||lt2gene Not-determined
KM-final-viral-combined_k141_103916||lt2gene Low-quality
KM-final-viral-combined_k141_37202||lt2gene Low-quality
KM-final-viral-combined_k141_182420||lt2gene Not-determined
KM-final-viral-combined_k141_103935||lt2gene Not-determined
KM-final-viral-combined_k141_137679||lt2gene Not-determined
KM-final-viral-combined_k141_116282||lt2gene Low-quality
KM-final-viral-combined_k141_59367||lt2gene Low-quality
KM-final-viral-combined_k141_148741||lt2gene Low-quality
KM-final-viral-combined_k141_82577||lt2gene Low-quality
KM-final-viral-combined_k141_82612||lt2gene Low-quality
KM-final-viral-combined_k141_226807||lt2gene Low-quality
KM-final-viral-combined_k141_26083||lt2gene Low-quality
KM-final-viral-combined_k141_26086||lt2gene Not-determined
KM-final-viral-combined_k141_226840||lt2gene Low-quality
KM-final-viral-combined_k141_170794||lt2gene Low-quality
KM-final-viral-combined_k141_226864||lt2gene Low-quality
KM-final-viral-combined_k141_182505||lt2gene Low-quality
KM-final-viral-combined_k141_182512||lt2gene Not-determined
KM-final-viral-combined_k141_71244||lt2gene Low-quality
KM-final-viral-combined_k141_126721||lt2gene Low-quality
KM-final-viral-combined_k141_116467||lt2gene Low-quality
KM-final-viral-combined_k141_3958||lt2gene Low-quality
KM-final-viral-combined_k141_226922||lt2gene Low-quality
KM-final-viral-combined_k141_14501||lt2gene Not-determined
KM-final-viral-combined_k141_82762||lt2gene Low-quality
KM-final-viral-combined_k141_159086||lt2gene Low-quality
KM-final-viral-combined_k141_116570||lt2gene Low-quality
KM-final-viral-combined_k141_26208||lt2gene Not-determined
KM-final-viral-combined_k141_159133||lt2gene Not-determined
KM-final-viral-combined_k141_137848||lt2gene Low-quality
KM-final-viral-combined_k141_159141||lt2gene Low-quality
KM-final-viral-combined_k141_4072||lt2gene Not-determined
KM-final-viral-combined_k141_47247||lt2gene Not-determined
KM-final-viral-combined_k141_259282||lt2gene Low-quality
KM-final-viral-combined_k141_137856||lt2gene Low-quality
KM-final-viral-combined_k141_14614||lt2gene Low-quality
KM-final-viral-combined_k141_14617||lt2gene Low-quality
KM-final-viral-combined_k141_137878||lt2gene Low-quality
KM-final-viral-combined_k141_116689||lt2gene Low-quality
KM-final-viral-combined_k141_83007||lt2gene Not-determined
KM-final-viral-combined_k141_216155||lt2gene Not-determined
KM-final-viral-combined_k141_227178||lt2gene Low-quality
KM-final-viral-combined_k141_104318||lt2gene Low-quality
KM-final-viral-combined_k141_37511||lt2gene Low-quality
KM-final-viral-combined_k141_237597||lt2gene Low-quality
KM-final-viral-combined_k141_159214||lt2gene Low-quality
KM-final-viral-combined_k141_171054||lt2gene Not-determined
KM-final-viral-combined_k141_71419||lt2gene Not-determined
KM-final-viral-combined_k141_171072||lt2gene Low-quality
KM-final-viral-combined_k141_237634||lt2gene Low-quality
KM-final-viral-combined_k141_104328||lt2gene Low-quality
KM-final-viral-combined_k141_182812||lt2gene Low-quality
KM-final-viral-combined_k141_216241||lt2gene Low-quality
KM-final-viral-combined_k141_116802||lt2gene Low-quality
KM-final-viral-combined_k141_92362||lt2gene Low-quality
KM-final-viral-combined_k141_37597||lt2gene Low-quality
KM-final-viral-combined_k141_149093||lt2gene Low-quality
KM-final-viral-combined_k141_204944||lt2gene Low-quality
KM-final-viral-combined_k141_37618||lt2gene Low-quality
KM-final-viral-combined_k141_116917||lt2gene Low-quality
KM-final-viral-combined_k141_171173||lt2gene Low-quality
KM-final-viral-combined_k141_216298||lt2gene Not-determined
KM-final-viral-combined_k141_204979||lt2gene Low-quality
KM-final-viral-combined_k141_4271||lt2gene Not-determined
KM-final-viral-combined_k141_92469||lt2gene Low-quality
KM-final-viral-combined_k141_205025||lt2gene Not-determined
KM-final-viral-combined_k141_117005||lt2gene Low-quality
KM-final-viral-combined_k141_159446||lt2gene Low-quality
KM-final-viral-combined_k141_159450||lt2gene Low-quality
KM-final-viral-combined_k141_159470||lt2gene Not-determined
KM-final-viral-combined_k141_117093||lt2gene Low-quality
KM-final-viral-combined_k141_4358||lt2gene Not-determined
KM-final-viral-combined_k141_4363||lt2gene Not-determined
KM-final-viral-combined_k141_159491||lt2gene Not-determined
KM-final-viral-combined_k141_259447||lt2gene Low-quality
KM-final-viral-combined_k141_183106||lt2gene Not-determined
KM-final-viral-combined_k141_159513||lt2gene Low-quality
KM-final-viral-combined_k141_171334||lt2gene Low-quality
KM-final-viral-combined_k141_71588||lt2gene Low-quality
KM-final-viral-combined_k141_47645||lt2gene Low-quality
KM-final-viral-combined_k141_59998||lt2gene Low-quality
KM-final-viral-combined_k141_193957||lt2gene Low-quality
KM-final-viral-combined_k141_37822||lt2gene Low-quality
KM-final-viral-combined_k141_117295||lt2gene Low-quality
KM-final-viral-combined_k141_216541||lt2gene Not-determined
KM-final-viral-combined_k141_183236||lt2gene Low-quality
KM-final-viral-combined_k141_216559||lt2gene Low-quality
KM-final-viral-combined_k141_183251||lt2gene Low-quality
KM-final-viral-combined_k141_4445||lt2gene Low-quality
KM-final-viral-combined_k141_83500||lt2gene Low-quality
KM-final-viral-combined_k141_4497||lt2gene Not-determined
KM-final-viral-combined_k141_104650||lt2gene Low-quality
KM-final-viral-combined_k141_71732||lt2gene Low-quality
KM-final-viral-combined_k141_159707||lt2gene Low-quality
KM-final-viral-combined_k141_159710||lt2gene Low-quality
KM-final-viral-combined_k141_37974||lt2gene Not-determined
KM-final-viral-combined_k141_216708||lt2gene Low-quality
KM-final-viral-combined_k141_237893||lt2gene Not-determined
KM-final-viral-combined_k141_227837||lt2gene Low-quality
KM-final-viral-combined_k141_227839||lt2gene Low-quality
KM-final-viral-combined_k141_38001||lt2gene Low-quality
KM-final-viral-combined_k141_71796||lt2gene Not-determined
KM-final-viral-combined_k141_127366||lt2gene Not-determined
KM-final-viral-combined_k141_171604||lt2gene Low-quality
KM-final-viral-combined_k141_117413||lt2gene Low-quality
KM-final-viral-combined_k141_38063||lt2gene Low-quality
KM-final-viral-combined_k141_159751||lt2gene Not-determined
KM-final-viral-combined_k141_171632||lt2gene Not-determined
KM-final-viral-combined_k141_4678||lt2gene Not-determined
KM-final-viral-combined_k141_38110||lt2gene Low-quality
KM-final-viral-combined_k141_4713||lt2gene Low-quality
KM-final-viral-combined_k141_104854||lt2gene Low-quality
KM-final-viral-combined_k141_117455||lt2gene Low-quality
KM-final-viral-combined_k141_194185||lt2gene Low-quality
KM-final-viral-combined_k141_38175||lt2gene Low-quality
KM-final-viral-combined_k141_205603||lt2gene Not-determined
KM-final-viral-combined_k141_248691||lt2gene Not-determined
KM-final-viral-combined_k141_27046||lt2gene Low-quality
KM-final-viral-combined_k141_27056||lt2gene Low-quality
KM-final-viral-combined_k141_38217||lt2gene Low-quality
KM-final-viral-combined_k141_138701||lt2gene Low-quality
KM-final-viral-combined_k141_117557||lt2gene Low-quality
KM-final-viral-combined_k141_149724||lt2gene Low-quality
KM-final-viral-combined_k141_194357||lt2gene Not-determined
KM-final-viral-combined_k141_105040||lt2gene Low-quality
KM-final-viral-combined_k141_171835||lt2gene Low-quality
KM-final-viral-combined_k141_127634||lt2gene Low-quality
KM-final-viral-combined_k141_159988||lt2gene Low-quality
KM-final-viral-combined_k141_105049||lt2gene Low-quality
KM-final-viral-combined_k141_47909||lt2gene Low-quality
KM-final-viral-combined_k141_248775||lt2gene Low-quality
KM-final-viral-combined_k141_216944||lt2gene Low-quality
KM-final-viral-combined_k141_117679||lt2gene Low-quality
KM-final-viral-combined_k141_205776||lt2gene Not-determined
KM-final-viral-combined_k141_47975||lt2gene Low-quality
KM-final-viral-combined_k141_228230||lt2gene Low-quality
KM-final-viral-combined_k141_4891||lt2gene Low-quality
KM-final-viral-combined_k141_127719||lt2gene Low-quality
KM-final-viral-combined_k141_149918||lt2gene Low-quality
KM-final-viral-combined_k141_117773||lt2gene Low-quality
KM-final-viral-combined_k141_92943||lt2gene Not-determined
KM-final-viral-combined_k141_84113||lt2gene Low-quality
KM-final-viral-combined_k141_248967||lt2gene Low-quality
KM-final-viral-combined_k141_84120||lt2gene Not-determined
KM-final-viral-combined_k141_15394||lt2gene Low-quality
KM-final-viral-combined_k141_5020||lt2gene Not-determined
KM-final-viral-combined_k141_194659||lt2gene Low-quality
KM-final-viral-combined_k141_205939||lt2gene Low-quality
KM-final-viral-combined_k141_205983||lt2gene Not-determined
KM-final-viral-combined_k141_160258||lt2gene Low-quality
KM-final-viral-combined_k141_160269||lt2gene Low-quality
KM-final-viral-combined_k141_150068||lt2gene Low-quality
KM-final-viral-combined_k141_93059||lt2gene Low-quality
KM-final-viral-combined_k141_249012||lt2gene Not-determined
KM-final-viral-combined_k141_206043||lt2gene Low-quality
KM-final-viral-combined_k141_105400||lt2gene Low-quality
KM-final-viral-combined_k141_93109||lt2gene Low-quality
KM-final-viral-combined_k141_150121||lt2gene Not-determined
KM-final-viral-combined_k141_260095||lt2gene Low-quality
KM-final-viral-combined_k141_206059||lt2gene Low-quality
KM-final-viral-combined_k141_93117||lt2gene Low-quality
KM-final-viral-combined_k141_15556||lt2gene Low-quality
KM-final-viral-combined_k141_105417||lt2gene Low-quality
KM-final-viral-combined_k141_206088||lt2gene Not-determined
KM-final-viral-combined_k141_194780||lt2gene Not-determined
KM-final-viral-combined_k141_194800||lt2gene Low-quality
KM-final-viral-combined_k141_48115||lt2gene Low-quality
KM-final-viral-combined_k141_60793||lt2gene Low-quality
KM-final-viral-combined_k141_194857||lt2gene Low-quality
KM-final-viral-combined_k141_72467||lt2gene Not-determined
KM-final-viral-combined_k141_48134||lt2gene Low-quality
KM-final-viral-combined_k141_238561||lt2gene Low-quality
KM-final-viral-combined_k141_93194||lt2gene Low-quality
KM-final-viral-combined_k141_206238||lt2gene Low-quality
KM-final-viral-combined_k141_105535||lt2gene Low-quality
KM-final-viral-combined_k141_5239||lt2gene Low-quality
KM-final-viral-combined_k141_139255||lt2gene Low-quality
KM-final-viral-combined_k141_48222||lt2gene Low-quality
KM-final-viral-combined_k141_60890||lt2gene Low-quality
KM-final-viral-combined_k141_27529||lt2gene Low-quality
KM-final-viral-combined_k141_206283||lt2gene Low-quality
KM-final-viral-combined_k141_72547||lt2gene Low-quality
KM-final-viral-combined_k141_249104||lt2gene Low-quality
KM-final-viral-combined_k141_27552||lt2gene Low-quality
KM-final-viral-combined_k141_84342||lt2gene Low-quality
KM-final-viral-combined_k141_238721||lt2gene Low-quality
KM-final-viral-combined_k141_172215||lt2gene Low-quality
KM-final-viral-combined_k141_139365||lt2gene Low-quality
KM-final-viral-combined_k141_228696||lt2gene Low-quality
KM-final-viral-combined_k141_5343||lt2gene Not-determined
KM-final-viral-combined_k141_249202||lt2gene Low-quality
KM-final-viral-combined_k141_228739||lt2gene Not-determined
KM-final-viral-combined_k141_84424||lt2gene Low-quality
KM-final-viral-combined_k141_195073||lt2gene Low-quality
KM-final-viral-combined_k141_118271||lt2gene Low-quality
KM-final-viral-combined_k141_150319||lt2gene Not-determined
KM-final-viral-combined_k141_128200||lt2gene Low-quality
KM-final-viral-combined_k141_48401||lt2gene Low-quality
KM-final-viral-combined_k141_184343||lt2gene Low-quality
KM-final-viral-combined_k141_128227||lt2gene Low-quality
KM-final-viral-combined_k141_38727||lt2gene Low-quality
KM-final-viral-combined_k141_38754||lt2gene Low-quality
KM-final-viral-combined_k141_195176||lt2gene Low-quality
KM-final-viral-combined_k141_118409||lt2gene Low-quality
KM-final-viral-combined_k141_228846||lt2gene Low-quality
KM-final-viral-combined_k141_184419||lt2gene Low-quality
KM-final-viral-combined_k141_139532||lt2gene Not-determined
KM-final-viral-combined_k141_184420||lt2gene Not-determined
KM-final-viral-combined_k141_48457||lt2gene Not-determined
KM-final-viral-combined_k141_15825||lt2gene Not-determined
KM-final-viral-combined_k141_5520||lt2gene Not-determined
KM-final-viral-combined_k141_206636||lt2gene Low-quality
KM-final-viral-combined_k141_150482||lt2gene Low-quality
KM-final-viral-combined_k141_238921||lt2gene Not-determined
KM-final-viral-combined_k141_93456||lt2gene Low-quality
KM-final-viral-combined_k141_139566||lt2gene Low-quality
KM-final-viral-combined_k141_260420||lt2gene Not-determined
KM-final-viral-combined_k141_238955||lt2gene Low-quality
KM-final-viral-combined_k141_206667||lt2gene Not-determined
KM-final-viral-combined_k141_228941||lt2gene Low-quality
KM-final-viral-combined_k141_84553||lt2gene Low-quality
KM-final-viral-combined_k141_38866||lt2gene Low-quality
KM-final-viral-combined_k141_61226||lt2gene Low-quality
KM-final-viral-combined_k141_72911||lt2gene Not-determined
KM-final-viral-combined_k141_15918||lt2gene Not-determined
KM-final-viral-combined_k141_15925||lt2gene Not-determined
KM-final-viral-combined_k141_139684||lt2gene Low-quality
KM-final-viral-combined_k141_139692||lt2gene Low-quality
KM-final-viral-combined_k141_239066||lt2gene Low-quality
KM-final-viral-combined_k141_61289||lt2gene Low-quality
KM-final-viral-combined_k141_139739||lt2gene Low-quality
KM-final-viral-combined_k141_195374||lt2gene Low-quality
KM-final-viral-combined_k141_195384||lt2gene Low-quality
KM-final-viral-combined_k141_206825||lt2gene Low-quality
KM-final-viral-combined_k141_38960||lt2gene Low-quality
KM-final-viral-combined_k141_61342||lt2gene Low-quality
KM-final-viral-combined_k141_239122||lt2gene Low-quality
KM-final-viral-combined_k141_118618||lt2gene Low-quality
KM-final-viral-combined_k141_184725||lt2gene Not-determined
KM-final-viral-combined_k141_239127||lt2gene Low-quality
KM-final-viral-combined_k141_260592||lt2gene Low-quality
KM-final-viral-combined_k141_61366||lt2gene Not-determined
KM-final-viral-combined_k141_206889||lt2gene Low-quality
KM-final-viral-combined_k141_128532||lt2gene Low-quality
KM-final-viral-combined_k141_16047||lt2gene Low-quality
KM-final-viral-combined_k141_93642||lt2gene Not-determined
KM-final-viral-combined_k141_218010||lt2gene Low-quality
KM-final-viral-combined_k141_184807||lt2gene Low-quality
KM-final-viral-combined_k141_206937||lt2gene Low-quality
KM-final-viral-combined_k141_139865||lt2gene Low-quality
KM-final-viral-combined_k141_118680||lt2gene Low-quality
KM-final-viral-combined_k141_218116||lt2gene Low-quality
KM-final-viral-combined_k141_39057||lt2gene Low-quality
KM-final-viral-combined_k141_172767||lt2gene Not-determined
KM-final-viral-combined_k141_84807||lt2gene Low-quality
KM-final-viral-combined_k141_84820||lt2gene Low-quality
KM-final-viral-combined_k141_16128||lt2gene Low-quality
KM-final-viral-combined_k141_249671||lt2gene Low-quality
KM-final-viral-combined_k141_106077||lt2gene Not-determined
KM-final-viral-combined_k141_195719||lt2gene Low-quality
KM-final-viral-combined_k141_229363||lt2gene Low-quality
KM-final-viral-combined_k141_106122||lt2gene Low-quality
KM-final-viral-combined_k141_229388||lt2gene Not-determined
KM-final-viral-combined_k141_140036||lt2gene Low-quality
KM-final-viral-combined_k141_118772||lt2gene Low-quality
KM-final-viral-combined_k141_48839||lt2gene Not-determined
KM-final-viral-combined_k141_106171||lt2gene Low-quality
KM-final-viral-combined_k141_140097||lt2gene Low-quality
KM-final-viral-combined_k141_207112||lt2gene Low-quality
KM-final-viral-combined_k141_106187||lt2gene Not-determined
KM-final-viral-combined_k141_207127||lt2gene Not-determined
KM-final-viral-combined_k141_239409||lt2gene Not-determined
KM-final-viral-combined_k141_73341||lt2gene Low-quality
KM-final-viral-combined_k141_106227||lt2gene Low-quality
KM-final-viral-combined_k141_207185||lt2gene Low-quality
KM-final-viral-combined_k141_106237||lt2gene Low-quality
KM-final-viral-combined_k141_218325||lt2gene Low-quality
KM-final-viral-combined_k141_184989||lt2gene Not-determined
KM-final-viral-combined_k141_73420||lt2gene Low-quality
KM-final-viral-combined_k141_195825||lt2gene Low-quality
KM-final-viral-combined_k141_16299||lt2gene Low-quality
KM-final-viral-combined_k141_5974||lt2gene Low-quality
KM-final-viral-combined_k141_140208||lt2gene Not-determined
KM-final-viral-combined_k141_249814||lt2gene Not-determined
KM-final-viral-combined_k141_49014||lt2gene Not-determined
KM-final-viral-combined_k141_61748||lt2gene Low-quality
KM-final-viral-combined_k141_261098||lt2gene Low-quality
KM-final-viral-combined_k141_39406||lt2gene Low-quality
KM-final-viral-combined_k141_207336||lt2gene Low-quality
KM-final-viral-combined_k141_207339||lt2gene Low-quality
KM-final-viral-combined_k141_261137||lt2gene Not-determined
KM-final-viral-combined_k141_49048||lt2gene Not-determined
KM-final-viral-combined_k141_16408||lt2gene Low-quality
KM-final-viral-combined_k141_207382||lt2gene Low-quality
KM-final-viral-combined_k141_6064||lt2gene Low-quality
KM-final-viral-combined_k141_28149||lt2gene Low-quality
KM-final-viral-combined_k141_73536||lt2gene Low-quality
KM-final-viral-combined_k141_93976||lt2gene Low-quality
KM-final-viral-combined_k141_196092||lt2gene Low-quality
KM-final-viral-combined_k141_185196||lt2gene Not-determined
KM-final-viral-combined_k141_85207||lt2gene Not-determined
KM-final-viral-combined_k141_28198||lt2gene Low-quality
KM-final-viral-combined_k141_218626||lt2gene Low-quality
KM-final-viral-combined_k141_119096||lt2gene Low-quality
KM-final-viral-combined_k141_28270||lt2gene Low-quality
KM-final-viral-combined_k141_261281||lt2gene Low-quality
KM-final-viral-combined_k141_151135||lt2gene Not-determined
KM-final-viral-combined_k141_73680||lt2gene Low-quality
KM-final-viral-combined_k141_229776||lt2gene Not-determined
KM-final-viral-combined_k141_196257||lt2gene Not-determined
KM-final-viral-combined_k141_173327||lt2gene Low-quality
KM-final-viral-combined_k141_73728||lt2gene Low-quality
KM-final-viral-combined_k141_185304||lt2gene Low-quality
KM-final-viral-combined_k141_106551||lt2gene Low-quality
KM-final-viral-combined_k141_106588||lt2gene Not-determined
KM-final-viral-combined_k141_151276||lt2gene Low-quality
KM-final-viral-combined_k141_207592||lt2gene Low-quality
KM-final-viral-combined_k141_173416||lt2gene Low-quality
KM-final-viral-combined_k141_119247||lt2gene Low-quality
KM-final-viral-combined_k141_239908||lt2gene Low-quality
KM-final-viral-combined_k141_62195||lt2gene Low-quality
KM-final-viral-combined_k141_151337||lt2gene Not-determined
KM-final-viral-combined_k141_173465||lt2gene Low-quality
KM-final-viral-combined_k141_261453||lt2gene Low-quality
KM-final-viral-combined_k141_151370||lt2gene Not-determined
KM-final-viral-combined_k141_49345||lt2gene Low-quality
KM-final-viral-combined_k141_140464||lt2gene Low-quality
KM-final-viral-combined_k141_185457||lt2gene Low-quality
KM-final-viral-combined_k141_119334||lt2gene Not-determined
KM-final-viral-combined_k141_73947||lt2gene Low-quality
KM-final-viral-combined_k141_129239||lt2gene Low-quality
KM-final-viral-combined_k141_28550||lt2gene Not-determined
KM-final-viral-combined_k141_230047||lt2gene Low-quality
KM-final-viral-combined_k141_185501||lt2gene Not-determined
KM-final-viral-combined_k141_16650||lt2gene Not-determined
KM-final-viral-combined_k141_196452||lt2gene Low-quality
KM-final-viral-combined_k141_106734||lt2gene Low-quality
KM-final-viral-combined_k141_16671||lt2gene Not-determined
KM-final-viral-combined_k141_151500||lt2gene Low-quality
KM-final-viral-combined_k141_140571||lt2gene Low-quality
KM-final-viral-combined_k141_6447||lt2gene Not-determined
KM-final-viral-combined_k141_106773||lt2gene Low-quality
KM-final-viral-combined_k141_119392||lt2gene Low-quality
KM-final-viral-combined_k141_185608||lt2gene Low-quality
KM-final-viral-combined_k141_161656||lt2gene Low-quality
KM-final-viral-combined_k141_140623||lt2gene Not-determined
KM-final-viral-combined_k141_173755||lt2gene Low-quality
KM-final-viral-combined_k141_106826||lt2gene Low-quality
KM-final-viral-combined_k141_207914||lt2gene Not-determined
KM-final-viral-combined_k141_74061||lt2gene Not-determined
KM-final-viral-combined_k141_161765||lt2gene Not-determined
KM-final-viral-combined_k141_74069||lt2gene Low-quality
KM-final-viral-combined_k141_85746||lt2gene Low-quality
KM-final-viral-combined_k141_40026||lt2gene Low-quality
KM-final-viral-combined_k141_230229||lt2gene Low-quality
KM-final-viral-combined_k141_119530||lt2gene Low-quality
KM-final-viral-combined_k141_6586||lt2gene Low-quality
KM-final-viral-combined_k141_140722||lt2gene Not-determined
KM-final-viral-combined_k141_62557||lt2gene Not-determined
KM-final-viral-combined_k141_6596||lt2gene Low-quality
KM-final-viral-combined_k141_250437||lt2gene Low-quality
KM-final-viral-combined_k141_94432||lt2gene Not-determined
KM-final-viral-combined_k141_119550||lt2gene Low-quality
KM-final-viral-combined_k141_261791||lt2gene Low-quality
KM-final-viral-combined_k141_261802||lt2gene Low-quality
KM-final-viral-combined_k141_230255||lt2gene Not-determined
KM-final-viral-combined_k141_151738||lt2gene Low-quality
KM-final-viral-combined_k141_62616||lt2gene Low-quality
KM-final-viral-combined_k141_119688||lt2gene Not-determined
KM-final-viral-combined_k141_261870||lt2gene Not-determined
KM-final-viral-combined_k141_208057||lt2gene Low-quality
KM-final-viral-combined_k141_28873||lt2gene Low-quality
KM-final-viral-combined_k141_85868||lt2gene Not-determined
KM-final-viral-combined_k141_6645||lt2gene Low-quality
KM-final-viral-combined_k141_28903||lt2gene Low-quality
KM-final-viral-combined_k141_230406||lt2gene Not-determined
KM-final-viral-combined_k141_174012||lt2gene Low-quality
KM-final-viral-combined_k141_129722||lt2gene Low-quality
KM-final-viral-combined_k141_106991||lt2gene Not-determined
KM-final-viral-combined_k141_219360||lt2gene Low-quality
KM-final-viral-combined_k141_17014||lt2gene Low-quality
KM-final-viral-combined_k141_17045||lt2gene Low-quality
KM-final-viral-combined_k141_17046||lt2gene Low-quality
KM-final-viral-combined_k141_119846||lt2gene Not-determined
KM-final-viral-combined_k141_230499||lt2gene Low-quality
KM-final-viral-combined_k141_250588||lt2gene Not-determined
KM-final-viral-combined_k141_151904||lt2gene Not-determined
KM-final-viral-combined_k141_230567||lt2gene Low-quality
KM-final-viral-combined_k141_62841||lt2gene Low-quality
KM-final-viral-combined_k141_94661||lt2gene Low-quality
KM-final-viral-combined_k141_174092||lt2gene Low-quality
KM-final-viral-combined_k141_28950||lt2gene Not-determined
KM-final-viral-combined_k141_119903||lt2gene Low-quality
KM-final-viral-combined_k141_219498||lt2gene Low-quality
KM-final-viral-combined_k141_208287||lt2gene Low-quality
KM-final-viral-combined_k141_208292||lt2gene Not-determined
KM-final-viral-combined_k141_162055||lt2gene Low-quality
KM-final-viral-combined_k141_219520||lt2gene Low-quality
KM-final-viral-combined_k141_141079||lt2gene Not-determined
KM-final-viral-combined_k141_6793||lt2gene Low-quality
KM-final-viral-combined_k141_186000||lt2gene Low-quality
KM-final-viral-combined_k141_186005||lt2gene Low-quality
KM-final-viral-combined_k141_230726||lt2gene Low-quality
KM-final-viral-combined_k141_151992||lt2gene Low-quality
KM-final-viral-combined_k141_6867||lt2gene Not-determined
KM-final-viral-combined_k141_174234||lt2gene Low-quality
KM-final-viral-combined_k141_29021||lt2gene Low-quality
KM-final-viral-combined_k141_162157||lt2gene Low-quality
KM-final-viral-combined_k141_162160||lt2gene Low-quality
KM-final-viral-combined_k141_120010||lt2gene Low-quality
KM-final-viral-combined_k141_197082||lt2gene Low-quality
KM-final-viral-combined_k141_240458||lt2gene Low-quality
KM-final-viral-combined_k141_74550||lt2gene Low-quality
KM-final-viral-combined_k141_107262||lt2gene Low-quality
KM-final-viral-combined_k141_29073||lt2gene Not-determined
KM-final-viral-combined_k141_186140||lt2gene Low-quality
KM-final-viral-combined_k141_120078||lt2gene Not-determined
KM-final-viral-combined_k141_120081||lt2gene Not-determined
KM-final-viral-combined_k141_86233||lt2gene Low-quality
KM-final-viral-combined_k141_63053||lt2gene Low-quality
KM-final-viral-combined_k141_17334||lt2gene Low-quality
KM-final-viral-combined_k141_262260||lt2gene Low-quality
KM-final-viral-combined_k141_186189||lt2gene Not-determined
KM-final-viral-combined_k141_120122||lt2gene Not-determined
KM-final-viral-combined_k141_17356||lt2gene Not-determined
KM-final-viral-combined_k141_250787||lt2gene Low-quality
KM-final-viral-combined_k141_94819||lt2gene Low-quality
KM-final-viral-combined_k141_186226||lt2gene Low-quality
KM-final-viral-combined_k141_120191||lt2gene Low-quality
KM-final-viral-combined_k141_94828||lt2gene Low-quality
KM-final-viral-combined_k141_240608||lt2gene Low-quality
KM-final-viral-combined_k141_29143||lt2gene Low-quality
KM-final-viral-combined_k141_219757||lt2gene Not-determined
KM-final-viral-combined_k141_186296||lt2gene Low-quality
KM-final-viral-combined_k141_107386||lt2gene Low-quality
KM-final-viral-combined_k141_250894||lt2gene Low-quality
KM-final-viral-combined_k141_120236||lt2gene Low-quality
KM-final-viral-combined_k141_197371||lt2gene Not-determined
KM-final-viral-combined_k141_240744||lt2gene Low-quality
KM-final-viral-combined_k141_240773||lt2gene Not-determined
KM-final-viral-combined_k141_174672||lt2gene Low-quality
KM-final-viral-combined_k141_174680||lt2gene Low-quality
KM-final-viral-combined_k141_262497||lt2gene Low-quality
KM-final-viral-combined_k141_63281||lt2gene Low-quality
KM-final-viral-combined_k141_17557||lt2gene Not-determined
KM-final-viral-combined_k141_63298||lt2gene Low-quality
KM-final-viral-combined_k141_86528||lt2gene Low-quality
KM-final-viral-combined_k141_50133||lt2gene Low-quality
KM-final-viral-combined_k141_50137||lt2gene Low-quality
KM-final-viral-combined_k141_63315||lt2gene Low-quality
KM-final-viral-combined_k141_141577||lt2gene Low-quality
KM-final-viral-combined_k141_63332||lt2gene Low-quality
KM-final-viral-combined_k141_29368||lt2gene Low-quality
KM-final-viral-combined_k141_162536||lt2gene Low-quality
KM-final-viral-combined_k141_50184||lt2gene Low-quality
KM-final-viral-combined_k141_17638||lt2gene Low-quality
KM-final-viral-combined_k141_86630||lt2gene Low-quality
KM-final-viral-combined_k141_17653||lt2gene Not-determined
KM-final-viral-combined_k141_95049||lt2gene Not-determined
KM-final-viral-combined_k141_220022||lt2gene Low-quality
KM-final-viral-combined_k141_7026||lt2gene Not-determined
KM-final-viral-combined_k141_220058||lt2gene Low-quality
KM-final-viral-combined_k141_162615||lt2gene Low-quality
KM-final-viral-combined_k141_197638||lt2gene Low-quality
KM-final-viral-combined_k141_220080||lt2gene Not-determined
KM-final-viral-combined_k141_29492||lt2gene Not-determined
KM-final-viral-combined_k141_29510||lt2gene Low-quality
KM-final-viral-combined_k141_186571||lt2gene Low-quality
KM-final-viral-combined_k141_186588||lt2gene Low-quality
KM-final-viral-combined_k141_152522||lt2gene Not-determined
KM-final-viral-combined_k141_120503||lt2gene Low-quality
KM-final-viral-combined_k141_17720||lt2gene Low-quality
KM-final-viral-combined_k141_174898||lt2gene Low-quality
KM-final-viral-combined_k141_75099||lt2gene Low-quality
KM-final-viral-combined_k141_17741||lt2gene Low-quality
KM-final-viral-combined_k141_29578||lt2gene Not-determined
KM-final-viral-combined_k141_50342||lt2gene Low-quality
KM-final-viral-combined_k141_251095||lt2gene Low-quality
KM-final-viral-combined_k141_220240||lt2gene Not-determined

| | |
|---|---|
| KM-final-viral-combined_k141_174981\|\|lt2gene | Low-quality |
| KM-final-viral-combined_k141_231443\|\|lt2gene | Not-determined |
| KM-final-viral-combined_k141_209108\|\|lt2gene | Not-determined |
| KM-final-viral-combined_k141_174990\|\|lt2gene | Not-determined |
| KM-final-viral-combined_k141_209120\|\|lt2gene | Low-quality |
| KM-final-viral-combined_k141_75189\|\|lt2gene | Low-quality |
| KM-final-viral-combined_k141_141794\|\|lt2gene | Low-quality |
| KM-final-viral-combined_k141_17827\|\|lt2gene | Not-determined |
| KM-final-viral-combined_k141_231486\|\|lt2gene | Low-quality |
| KM-final-viral-combined_k141_17840\|\|lt2gene | Not-determined |
| KM-final-viral-combined_k141_251185\|\|lt2gene | Low-quality |
| KM-final-viral-combined_k141_152588\|\|lt2gene | Low-quality |
| KM-final-viral-combined_k141_7316\|\|lt2gene | Not-determined |
| KM-final-viral-combined_k141_86902\|\|lt2gene | Low-quality |
| KM-final-viral-combined_k141_50453\|\|lt2gene | Low-quality |
| KM-final-viral-combined_k141_17893\|\|lt2gene | Low-quality |
| KM-final-viral-combined_k141_231597\|\|lt2gene | Not-determined |
| KM-final-viral-combined_k141_241244\|\|lt2gene | Low-quality |
| KM-final-viral-combined_k141_95247\|\|lt2gene | Low-quality |
| KM-final-viral-combined_k141_120767\|\|lt2gene | Low-quality |
| KM-final-viral-combined_k141_251264\|\|lt2gene | Low-quality |
| KM-final-viral-combined_k141_107876\|\|lt2gene | Low-quality |
| KM-final-viral-combined_k141_63764\|\|lt2gene | Low-quality |
| KM-final-viral-combined_k141_75368\|\|lt2gene | Low-quality |
| KM-final-viral-combined_k141_209271\|\|lt2gene | Not-determined |
| KM-final-viral-combined_k141_63806\|\|lt2gene | Low-quality |
| KM-final-viral-combined_k141_152733\|\|lt2gene | Low-quality |
| KM-final-viral-combined_k141_220437\|\|lt2gene | Low-quality |
| KM-final-viral-combined_k141_209319\|\|lt2gene | Not-determined |
| KM-final-viral-combined_k141_107946\|\|lt2gene | Low-quality |
| KM-final-viral-combined_k141_40840\|\|lt2gene | Not-determined |
| KM-final-viral-combined_k141_209331\|\|lt2gene | Low-quality |
| KM-final-viral-combined_k141_209332\|\|lt2gene | Low-quality |
| KM-final-viral-combined_k141_152752\|\|lt2gene | Low-quality |
| KM-final-viral-combined_k141_251323\|\|lt2gene | Low-quality |
| KM-final-viral-combined_k141_63841\|\|lt2gene | Low-quality |
| KM-final-viral-combined_k141_87136\|\|lt2gene | Low-quality |
| KM-final-viral-combined_k141_120917\|\|lt2gene | Low-quality |
| KM-final-viral-combined_k141_75523\|\|lt2gene | Low-quality |
| KM-final-viral-combined_k141_63866\|\|lt2gene | Low-quality |
| KM-final-viral-combined_k141_209401\|\|lt2gene | Not-determined |
| KM-final-viral-combined_k141_87157\|\|lt2gene | Low-quality |
| KM-final-viral-combined_k141_75580\|\|lt2gene | Low-quality |
| KM-final-viral-combined_k141_175255\|\|lt2gene | Not-determined |
| KM-final-viral-combined_k141_152846\|\|lt2gene | Low-quality |
| KM-final-viral-combined_k141_262843\|\|lt2gene | Low-quality |
| KM-final-viral-combined_k141_63940\|\|lt2gene | Low-quality |
| KM-final-viral-combined_k141_209451\|\|lt2gene | Not-determined |
| KM-final-viral-combined_k141_87223\|\|lt2gene | Low-quality |
| KM-final-viral-combined_k141_63987\|\|lt2gene | Low-quality |
| KM-final-viral-combined_k141_251429\|\|lt2gene | Low-quality |
| KM-final-viral-combined_k141_187023\|\|lt2gene | Not-determined |
| KM-final-viral-combined_k141_40929\|\|lt2gene | Low-quality |
| KM-final-viral-combined_k141_251450\|\|lt2gene | Low-quality |
| KM-final-viral-combined_k141_7604\|\|lt2gene | Low-quality |
| KM-final-viral-combined_k141_18109\|\|lt2gene | Low-quality |
| KM-final-viral-combined_k141_198173\|\|lt2gene | Not-determined |
| KM-final-viral-combined_k141_251474\|\|lt2gene | Low-quality |
| KM-final-viral-combined_k141_198206\|\|lt2gene | Not-determined |
| KM-final-viral-combined_k141_231783\|\|lt2gene | Low-quality |
| KM-final-viral-combined_k141_241532\|\|lt2gene | Low-quality |
| KM-final-viral-combined_k141_153002\|\|lt2gene | Not-determined |
| KM-final-viral-combined_k141_175416\|\|lt2gene | Low-quality |
| KM-final-viral-combined_k141_231820\|\|lt2gene | Low-quality |
| KM-final-viral-combined_k141_75723\|\|lt2gene | Low-quality |
| KM-final-viral-combined_k141_262963\|\|lt2gene | Not-determined |
| KM-final-viral-combined_k141_75745\|\|lt2gene | Not-determined |
| KM-final-viral-combined_k141_241588\|\|lt2gene | Low-quality |
| KM-final-viral-combined_k141_241589\|\|lt2gene | Low-quality |
| KM-final-viral-combined_k141_7703\|\|lt2gene | Low-quality |
| KM-final-viral-combined_k141_142318\|\|lt2gene | Low-quality |
| KM-final-viral-combined_k141_263052\|\|lt2gene | Not-determined |
| KM-final-viral-combined_k141_75851\|\|lt2gene | Low-quality |
| KM-final-viral-combined_k141_231926\|\|lt2gene | Low-quality |
| KM-final-viral-combined_k141_18277\|\|lt2gene | Low-quality |
| KM-final-viral-combined_k141_108263\|\|lt2gene | Low-quality |
| KM-final-viral-combined_k141_163185\|\|lt2gene | Low-quality |
| KM-final-viral-combined_k141_251597\|\|lt2gene | Low-quality |
| KM-final-viral-combined_k141_198301\|\|lt2gene | Not-determined |
| KM-final-viral-combined_k141_7797\|\|lt2gene | Low-quality |
| KM-final-viral-combined_k141_18341\|\|lt2gene | Low-quality |
| KM-final-viral-combined_k141_263144\|\|lt2gene | Low-quality |
| KM-final-viral-combined_k141_7804\|\|lt2gene | Low-quality |
| KM-final-viral-combined_k141_108312\|\|lt2gene | Low-quality |
| KM-final-viral-combined_k141_220946\|\|lt2gene | Low-quality |
| KM-final-viral-combined_k141_121275\|\|lt2gene | Not-determined |
| KM-final-viral-combined_k141_263218\|\|lt2gene | Low-quality |
| KM-final-viral-combined_k141_41159\|\|lt2gene | Low-quality |
| KM-final-viral-combined_k141_198417\|\|lt2gene | Not-determined |
| KM-final-viral-combined_k141_64208\|\|lt2gene | Low-quality |
| KM-final-viral-combined_k141_198459\|\|lt2gene | Not-determined |
| KM-final-viral-combined_k141_241837\|\|lt2gene | Low-quality |
| KM-final-viral-combined_k141_95701\|\|lt2gene | Low-quality |
| KM-final-viral-combined_k141_7931\|\|lt2gene | Low-quality |
| KM-final-viral-combined_k141_87643\|\|lt2gene | Low-quality |
| KM-final-viral-combined_k141_263280\|\|lt2gene | Low-quality |
| KM-final-viral-combined_k141_142588\|\|lt2gene | Not-determined |
| KM-final-viral-combined_k141_221092\|\|lt2gene | Low-quality |
| KM-final-viral-combined_k141_7940\|\|lt2gene | Low-quality |
| KM-final-viral-combined_k141_241881\|\|lt2gene | Low-quality |
| KM-final-viral-combined_k141_7951\|\|lt2gene | Low-quality |
| KM-final-viral-combined_k141_142629\|\|lt2gene | Not-determined |
| KM-final-viral-combined_k141_251789\|\|lt2gene | Not-determined |
| KM-final-viral-combined_k141_51013\|\|lt2gene | Low-quality |
| KM-final-viral-combined_k141_108520\|\|lt2gene | Low-quality |
| KM-final-viral-combined_k141_251836\|\|lt2gene | Not-determined |
| KM-final-viral-combined_k141_121425\|\|lt2gene | Low-quality |
| KM-final-viral-combined_k141_153333\|\|lt2gene | Not-determined |
| KM-final-viral-combined_k141_64266\|\|lt2gene | Low-quality |
| KM-final-viral-combined_k141_232102\|\|lt2gene | Low-quality |
| KM-final-viral-combined_k141_108560\|\|lt2gene | Low-quality |
| KM-final-viral-combined_k141_241927\|\|lt2gene | Low-quality |
| KM-final-viral-combined_k141_95795\|\|lt2gene | Low-quality |
| KM-final-viral-combined_k141_51088\|\|lt2gene | Low-quality |
| KM-final-viral-combined_k141_130635\|\|lt2gene | Not-determined |
| KM-final-viral-combined_k141_232159\|\|lt2gene | Low-quality |
| KM-final-viral-combined_k141_251939\|\|lt2gene | Low-quality |
| KM-final-viral-combined_k141_95830\|\|lt2gene | Not-determined |
| KM-final-viral-combined_k141_30374\|\|lt2gene | Low-quality |
| KM-final-viral-combined_k141_251961\|\|lt2gene | Low-quality |
| KM-final-viral-combined_k141_76241\|\|lt2gene | Low-quality |
| KM-final-viral-combined_k141_153508\|\|lt2gene | Low-quality |
| KM-final-viral-combined_k141_153512\|\|lt2gene | Low-quality |
| KM-final-viral-combined_k141_41365\|\|lt2gene | Not-determined |
| KM-final-viral-combined_k141_163481\|\|lt2gene | Low-quality |
| KM-final-viral-combined_k141_198760\|\|lt2gene | Low-quality |
| KM-final-viral-combined_k141_232319\|\|lt2gene | Low-quality |
| KM-final-viral-combined_k141_198770\|\|lt2gene | Not-determined |
| KM-final-viral-combined_k141_130848\|\|lt2gene | Low-quality |
| KM-final-viral-combined_k141_142842\|\|lt2gene | Low-quality |
| KM-final-viral-combined_k141_41388\|\|lt2gene | Low-quality |
| KM-final-viral-combined_k141_121546\|\|lt2gene | Low-quality |
| KM-final-viral-combined_k141_95960\|\|lt2gene | Not-determined |
| KM-final-viral-combined_k141_121586\|\|lt2gene | Low-quality |
| KM-final-viral-combined_k141_41412\|\|lt2gene | Low-quality |
| KM-final-viral-combined_k141_108788\|\|lt2gene | Low-quality |
| KM-final-viral-combined_k141_51280\|\|lt2gene | Not-determined |
| KM-final-viral-combined_k141_18721\|\|lt2gene | Not-determined |
| KM-final-viral-combined_k141_95992\|\|lt2gene | Low-quality |
| KM-final-viral-combined_k141_87930\|\|lt2gene | Low-quality |
| KM-final-viral-combined_k141_242130\|\|lt2gene | Low-quality |
| KM-final-viral-combined_k141_30524\|\|lt2gene | Low-quality |
| KM-final-viral-combined_k141_30536\|\|lt2gene | Not-determined |
| KM-final-viral-combined_k141_30548\|\|lt2gene | Low-quality |
| KM-final-viral-combined_k141_143008\|\|lt2gene | Low-quality |
| KM-final-viral-combined_k141_209971\|\|lt2gene | Low-quality |
| KM-final-viral-combined_k141_143031\|\|lt2gene | Not-determined |
| KM-final-viral-combined_k141_143035\|\|lt2gene | Low-quality |
| KM-final-viral-combined_k141_30621\|\|lt2gene | Low-quality |
| KM-final-viral-combined_k141_252001\|\|lt2gene | Low-quality |
| KM-final-viral-combined_k141_163670\|\|lt2gene | Low-quality |
| KM-final-viral-combined_k141_252021\|\|lt2gene | Low-quality |
| KM-final-viral-combined_k141_176025\|\|lt2gene | Not-determined |
| KM-final-viral-combined_k141_88046\|\|lt2gene | Low-quality |
| KM-final-viral-combined_k141_41611\|\|lt2gene | Low-quality |
| KM-final-viral-combined_k141_131057\|\|lt2gene | Low-quality |
| KM-final-viral-combined_k141_18822\|\|lt2gene | Low-quality |
| KM-final-viral-combined_k141_131086\|\|lt2gene | Low-quality |
| KM-final-viral-combined_k141_109002\|\|lt2gene | Low-quality |
| KM-final-viral-combined_k141_263632\|\|lt2gene | Low-quality |
| KM-final-viral-combined_k141_210111\|\|lt2gene | Low-quality |
| KM-final-viral-combined_k141_252133\|\|lt2gene | Low-quality |
| KM-final-viral-combined_k141_176134\|\|lt2gene | Not-determined |
| KM-final-viral-combined_k141_252135\|\|lt2gene | Not-determined |
| KM-final-viral-combined_k141_176138\|\|lt2gene | Not-determined |
| KM-final-viral-combined_k141_232622\|\|lt2gene | Not-determined |
| KM-final-viral-combined_k141_30754\|\|lt2gene | Low-quality |
| KM-final-viral-combined_k141_8473\|\|lt2gene | Low-quality |
| KM-final-viral-combined_k141_187856\|\|lt2gene | Low-quality |
| KM-final-viral-combined_k141_76672\|\|lt2gene | Low-quality |
| KM-final-viral-combined_k141_30770\|\|lt2gene | Low-quality |
| KM-final-viral-combined_k141_96202\|\|lt2gene | Low-quality |
| KM-final-viral-combined_k141_121922\|\|lt2gene | Low-quality |
| KM-final-viral-combined_k141_221691\|\|lt2gene | Low-quality |
| KM-final-viral-combined_k141_41741\|\|lt2gene | Low-quality |
| KM-final-viral-combined_k141_109095\|\|lt2gene | Not-determined |
| KM-final-viral-combined_k141_41769\|\|lt2gene | Low-quality |
| KM-final-viral-combined_k141_109109\|\|lt2gene | Not-determined |
| KM-final-viral-combined_k141_8527\|\|lt2gene | Low-quality |
| KM-final-viral-combined_k141_64976\|\|lt2gene | Low-quality |
| KM-final-viral-combined_k141_176212\|\|lt2gene | Low-quality |
| KM-final-viral-combined_k141_221761\|\|lt2gene | Not-determined |
| KM-final-viral-combined_k141_76793\|\|lt2gene | Not-determined |
| KM-final-viral-combined_k141_76797\|\|lt2gene | Not-determined |
| KM-final-viral-combined_k141_153988\|\|lt2gene | Not-determined |
| KM-final-viral-combined_k141_242387\|\|lt2gene | Not-determined |
| KM-final-viral-combined_k141_65081\|\|lt2gene | Low-quality |
| KM-final-viral-combined_k141_8610\|\|lt2gene | Not-determined |
| KM-final-viral-combined_k141_199272\|\|lt2gene | Low-quality |
| KM-final-viral-combined_k141_8613\|\|lt2gene | Low-quality |
| KM-final-viral-combined_k141_65097\|\|lt2gene | Low-quality |
| KM-final-viral-combined_k141_109158\|\|lt2gene | Low-quality |
| KM-final-viral-combined_k141_88455\|\|lt2gene | Low-quality |
| KM-final-viral-combined_k141_88460\|\|lt2gene | Low-quality |
| KM-final-viral-combined_k141_263817\|\|lt2gene | Not-determined |
| KM-final-viral-combined_k141_176318\|\|lt2gene | Not-determined |
| KM-final-viral-combined_k141_8656\|\|lt2gene | Low-quality |
| KM-final-viral-combined_k141_51675\|\|lt2gene | Low-quality |
| KM-final-viral-combined_k141_109191\|\|lt2gene | Low-quality |
| KM-final-viral-combined_k141_221915\|\|lt2gene | Not-determined |
| KM-final-viral-combined_k141_131342\|\|lt2gene | Low-quality |
| KM-final-viral-combined_k141_154104\|\|lt2gene | Low-quality |
| KM-final-viral-combined_k141_51715\|\|lt2gene | Low-quality |
| KM-final-viral-combined_k141_154147\|\|lt2gene | Low-quality |
| KM-final-viral-combined_k141_143485\|\|lt2gene | Not-determined |
| KM-final-viral-combined_k141_199380\|\|lt2gene | Low-quality |
| KM-final-viral-combined_k141_131421\|\|lt2gene | Not-determined |
| KM-final-viral-combined_k141_8753\|\|lt2gene | Not-determined |
| KM-final-viral-combined_k141_8756\|\|lt2gene | Low-quality |
| KM-final-viral-combined_k141_221951\|\|lt2gene | Low-quality |
| KM-final-viral-combined_k141_122071\|\|lt2gene | Low-quality |
| KM-final-viral-combined_k141_154206\|\|lt2gene | Low-quality |
| KM-final-viral-combined_k141_154213\|\|lt2gene | Low-quality |
| KM-final-viral-combined_k141_263908\|\|lt2gene | Low-quality |
| KM-final-viral-combined_k141_41978\|\|lt2gene | Not-determined |
| KM-final-viral-combined_k141_96486\|\|lt2gene | Low-quality |
| KM-final-viral-combined_k141_96495\|\|lt2gene | Low-quality |
| KM-final-viral-combined_k141_19190\|\|lt2gene | Low-quality |
| KM-final-viral-combined_k141_252455\|\|lt2gene | Low-quality |
| KM-final-viral-combined_k141_143588\|\|lt2gene | Low-quality |
| KM-final-viral-combined_k141_164179\|\|lt2gene | Low-quality |
| KM-final-viral-combined_k141_154260\|\|lt2gene | Low-quality |
| KM-final-viral-combined_k141_42027\|\|lt2gene | Low-quality |
| KM-final-viral-combined_k141_222039\|\|lt2gene | Low-quality |
| KM-final-viral-combined_k141_8872\|\|lt2gene | Low-quality |
| KM-final-viral-combined_k141_176570\|\|lt2gene | Not-determined |
| KM-final-viral-combined_k141_31131\|\|lt2gene | Low-quality |
| KM-final-viral-combined_k141_19219\|\|lt2gene | Low-quality |
| KM-final-viral-combined_k141_176573\|\|lt2gene | Low-quality |
| KM-final-viral-combined_k141_263960\|\|lt2gene | Low-quality |
| KM-final-viral-combined_k141_188274\|\|lt2gene | Not-determined |
| KM-final-viral-combined_k141_77033\|\|lt2gene | Low-quality |
| KM-final-viral-combined_k141_88700\|\|lt2gene | Low-quality |
| KM-final-viral-combined_k141_31188\|\|lt2gene | Low-quality |
| KM-final-viral-combined_k141_199509\|\|lt2gene | Low-quality |
| KM-final-viral-combined_k141_199510\|\|lt2gene | Low-quality |
| KM-final-viral-combined_k141_210681\|\|lt2gene | Low-quality |
| KM-final-viral-combined_k141_65381\|\|lt2gene | Not-determined |
| KM-final-viral-combined_k141_8991\|\|lt2gene | Low-quality |
| KM-final-viral-combined_k141_42149\|\|lt2gene | Not-determined |
| KM-final-viral-combined_k141_264047\|\|lt2gene | Not-determined |
| KM-final-viral-combined_k141_51762\|\|lt2gene | Not-determined |
| KM-final-viral-combined_k141_131578\|\|lt2gene | Low-quality |
| KM-final-viral-combined_k141_96593\|\|lt2gene | Not-determined |
| KM-final-viral-combined_k141_199587\|\|lt2gene | Low-quality |
| KM-final-viral-combined_k141_131598\|\|lt2gene | Not-determined |
| KM-final-viral-combined_k141_9104\|\|lt2gene | Not-determined |
| KM-final-viral-combined_k141_210843\|\|lt2gene | Low-quality |
| KM-final-viral-combined_k141_154483\|\|lt2gene | Not-determined |
| KM-final-viral-combined_k141_65470\|\|lt2gene | Low-quality |
| KM-final-viral-combined_k141_65476\|\|lt2gene | Low-quality |
| KM-final-viral-combined_k141_31279\|\|lt2gene | Low-quality |
| KM-final-viral-combined_k141_51877\|\|lt2gene | Low-quality |
| KM-final-viral-combined_k141_42262\|\|lt2gene | Low-quality |
| KM-final-viral-combined_k141_131740\|\|lt2gene | Not-determined |
| KM-final-viral-combined_k141_42282\|\|lt2gene | Not-determined |
| KM-final-viral-combined_k141_242766\|\|lt2gene | Low-quality |
| KM-final-viral-combined_k141_242782\|\|lt2gene | Low-quality |
| KM-final-viral-combined_k141_242834\|\|lt2gene | Low-quality |
| KM-final-viral-combined_k141_77311\|\|lt2gene | Low-quality |
| KM-final-viral-combined_k141_252701\|\|lt2gene | Not-determined |
| KM-final-viral-combined_k141_242867\|\|lt2gene | Low-quality |
| KM-final-viral-combined_k141_264240\|\|lt2gene | Not-determined |
| KM-final-viral-combined_k141_77349\|\|lt2gene | Not-determined |
| KM-final-viral-combined_k141_164458\|\|lt2gene | Low-quality |
| KM-final-viral-combined_k141_176980\|\|lt2gene | Low-quality |
| KM-final-viral-combined_k141_252732\|\|lt2gene | Low-quality |
| KM-final-viral-combined_k141_109727\|\|lt2gene | Low-quality |
| KM-final-viral-combined_k141_264255\|\|lt2gene | Not-determined |
| KM-final-viral-combined_k141_131934\|\|lt2gene | Low-quality |
| KM-final-viral-combined_k141_252760\|\|lt2gene | Low-quality |
| KM-final-viral-combined_k141_77386\|\|lt2gene | Not-determined |
| KM-final-viral-combined_k141_96838\|\|lt2gene | Low-quality |
| KM-final-viral-combined_k141_31474\|\|lt2gene | Not-determined |
| KM-final-viral-combined_k141_143943\|\|lt2gene | Low-quality |
| KM-final-viral-combined_k141_131985\|\|lt2gene | Low-quality |
| KM-final-viral-combined_k141_96849\|\|lt2gene | Low-quality |
| KM-final-viral-combined_k141_96851\|\|lt2gene | Low-quality |
| KM-final-viral-combined_k141_52028\|\|lt2gene | Low-quality |
| KM-final-viral-combined_k141_96862\|\|lt2gene | Low-quality |
| KM-final-viral-combined_k141_199760\|\|lt2gene | Low-quality |
| KM-final-viral-combined_k141_252803\|\|lt2gene | Low-quality |
| KM-final-viral-combined_k141_96895\|\|lt2gene | Low-quality |
| KM-final-viral-combined_k141_77472\|\|lt2gene | Low-quality |
| KM-final-viral-combined_k141_77476\|\|lt2gene | Low-quality |
| KM-final-viral-combined_k141_143993\|\|lt2gene | Low-quality |
| KM-final-viral-combined_k141_188652\|\|lt2gene | Not-determined |
| KM-final-viral-combined_k141_164670\|\|lt2gene | Low-quality |
| KM-final-viral-combined_k141_9529\|\|lt2gene | Low-quality |
| KM-final-viral-combined_k141_65892\|\|lt2gene | Low-quality |
| KM-final-viral-combined_k141_252958\|\|lt2gene | Low-quality |
| KM-final-viral-combined_k141_154856\|\|lt2gene | Low-quality |
| KM-final-viral-combined_k141_177221\|\|lt2gene | Low-quality |
| KM-final-viral-combined_k141_177251\|\|lt2gene | Low-quality |
| KM-final-viral-combined_k141_188794\|\|lt2gene | Low-quality |
| KM-final-viral-combined_k141_97067\|\|lt2gene | Low-quality |
| KM-final-viral-combined_k141_164826\|\|lt2gene | Low-quality |
| KM-final-viral-combined_k141_243100\|\|lt2gene | Low-quality |
| KM-final-viral-combined_k141_243115\|\|lt2gene | Low-quality |
| KM-final-viral-combined_k141_188874\|\|lt2gene | Low-quality |
| KM-final-viral-combined_k141_199948\|\|lt2gene | Low-quality |
| KM-final-viral-combined_k141_77709\|\|lt2gene | Not-determined |
| KM-final-viral-combined_k141_97152\|\|lt2gene | Not-determined |
| KM-final-viral-combined_k141_66068\|\|lt2gene | Low-quality |
| KM-final-viral-combined_k141_177342\|\|lt2gene | Not-determined |
| KM-final-viral-combined_k141_109955\|\|lt2gene | Low-quality |
| KM-final-viral-combined_k141_42846\|\|lt2gene | Low-quality |
| KM-final-viral-combined_k141_52288\|\|lt2gene | Low-quality |
| KM-final-viral-combined_k141_144071\|\|lt2gene | Low-quality |
| KM-final-viral-combined_k141_154991\|\|lt2gene | Low-quality |
| KM-final-viral-combined_k141_9757\|\|lt2gene | Low-quality |
| KM-final-viral-combined_k141_97266\|\|lt2gene | Low-quality |
| KM-final-viral-combined_k141_177474\|\|lt2gene | Not-determined |
| KM-final-viral-combined_k141_97315\|\|lt2gene | Low-quality |
| KM-final-viral-combined_k141_165113\|\|lt2gene | Not-determined |
| KM-final-viral-combined_k141_165144\|\|lt2gene | Not-determined |
| KM-final-viral-combined_k141_253243\|\|lt2gene | Not-determined |
| KM-final-viral-combined_k141_155078\|\|lt2gene | Not-determined |
| KM-final-viral-combined_k141_253326\|\|lt2gene | Not-determined |
| KM-final-viral-combined_k141_110201\|\|lt2gene | Not-determined |
| KM-final-viral-combined_k141_144186\|\|lt2gene | Not-determined |
| KM-final-viral-combined_k141_264728\|\|lt2gene | Not-determined |
| KM-final-viral-combined_k141_132571\|\|lt2gene | Low-quality |
| KM-final-viral-combined_k141_43142\|\|lt2gene | Low-quality |
| KM-final-viral-combined_k141_177719\|\|lt2gene | Not-determined |
| KM-final-viral-combined_k141_66351\|\|lt2gene | Low-quality |
| KM-final-viral-combined_k141_144245\|\|lt2gene | Low-quality |
| KM-final-viral-combined_k141_110268\|\|lt2gene | Low-quality |
| KM-final-viral-combined_k141_66376\|\|lt2gene | Low-quality |
| KM-final-viral-combined_k141_264820\|\|lt2gene | Low-quality |
| KM-final-viral-combined_k141_177777\|\|lt2gene | Not-determined |
| KM-final-viral-combined_k141_9998\|\|lt2gene | Not-determined |
| KM-final-viral-combined_k141_32042\|\|lt2gene | Not-determined |
| KM-final-viral-combined_k141_66457\|\|lt2gene | Low-quality |
| KM-final-viral-combined_k141_110317\|\|lt2gene | Low-quality |
| KM-final-viral-combined_k141_20051\|\|lt2gene | Not-determined |
| KM-final-viral-combined_k141_144392\|\|lt2gene | Low-quality |
| KM-final-viral-combined_k141_243509\|\|lt2gene | Low-quality |
| KM-final-viral-combined_k141_97636\|\|lt2gene | Not-determined |
| KM-final-viral-combined_k141_20077\|\|lt2gene | Low-quality |
| KM-final-viral-combined_k141_20096\|\|lt2gene | Low-quality |
| KM-final-viral-combined_k141_110426\|\|lt2gene | Not-determined |
| KM-final-viral-combined_k141_264971\|\|lt2gene | Not-determined |
| KM-final-viral-combined_k141_43444\|\|lt2gene | Low-quality |
| KM-final-viral-combined_k141_253567\|\|lt2gene | Low-quality |
| KM-final-viral-combined_k141_52862\|\|lt2gene | Low-quality |
| KM-final-viral-combined_k141_10223\|\|lt2gene | Low-quality |
| KM-final-viral-combined_k141_243689\|\|lt2gene | Low-quality |
| KM-final-viral-combined_k141_155344\|\|lt2gene | Low-quality |
| KM-final-viral-combined_k141_110508\|\|lt2gene | Low-quality |
| KM-final-viral-combined_k141_20264\|\|lt2gene | Low-quality |
| KM-final-viral-combined_k141_66610\|\|lt2gene | Not-determined |
| KM-final-viral-combined_k141_20280\|\|lt2gene | Low-quality |
| KM-final-viral-combined_k141_10376\|\|lt2gene | Not-determined |
| KM-final-viral-combined_k141_243820\|\|lt2gene | Low-quality |
| KM-final-viral-combined_k141_20422\|\|lt2gene | Low-quality |
| KM-final-viral-combined_k141_253815\|\|lt2gene | Low-quality |
| KM-final-viral-combined_k141_132983\|\|lt2gene | Not-determined |
| KM-final-viral-combined_k141_97949\|\|lt2gene | Not-determined |
| KM-final-viral-combined_k141_10511\|\|lt2gene | Not-determined |
| KM-final-viral-combined_k141_43731\|\|lt2gene | Low-quality |
| KM-final-viral-combined_k141_243969\|\|lt2gene | Low-quality |
| KM-final-viral-combined_k141_20561\|\|lt2gene | Not-determined |
| KM-final-viral-combined_k141_155553\|\|lt2gene | Low-quality |
| KM-final-viral-combined_k141_10578\|\|lt2gene | Low-quality |
| KM-final-viral-combined_k141_20597\|\|lt2gene | Low-quality |
| KM-final-viral-combined_k141_20604\|\|lt2gene | Not-determined |
| KM-final-viral-combined_k141_53049\|\|lt2gene | Not-determined |
| KM-final-viral-combined_k141_20642\|\|lt2gene | Not-determined |
| KM-final-viral-combined_k141_20658\|\|lt2gene | Not-determined |
| KM-final-viral-combined_k141_110662\|\|lt2gene | Low-quality |
| KM-final-viral-combined_k141_244101\|\|lt2gene | Low-quality |
| KM-final-viral-combined_k141_244105\|\|lt2gene | Not-determined |
| KM-final-viral-combined_k141_133241\|\|lt2gene | Not-determined |
| KM-final-viral-combined_k141_32714\|\|lt2gene | Low-quality |
| KM-final-viral-combined_k141_32721\|\|lt2gene | Low-quality |
| KM-final-viral-combined_k141_166168\|\|lt2gene | Not-determined |
| KM-final-viral-combined_k141_133263\|\|lt2gene | Not-determined |
| KM-final-viral-combined_k141_20800\|\|lt2gene | Not-determined |
| KM-final-viral-combined_k141_20805\|\|lt2gene | Not-determined |
| KM-final-viral-combined_k141_20948\|\|lt2gene | Low-quality |
| KM-final-viral-combined_k141_265544\|\|lt2gene | Low-quality |
| KM-final-viral-combined_k141_265584\|\|lt2gene | Low-quality |
| KM-final-viral-combined_k141_244378\|\|lt2gene | Low-quality |
| KM-final-viral-combined_k141_265642\|\|lt2gene | Low-quality |
| KM-final-viral-combined_k141_53378\|\|lt2gene | Low-quality |
| KM-final-viral-combined_k141_244453\|\|lt2gene | Low-quality |
| KM-final-viral-combined_k141_33034\|\|lt2gene | Low-quality |
| KM-final-viral-combined_k141_244479\|\|lt2gene | Not-determined |
| KM-final-viral-combined_k141_98610\|\|lt2gene | Low-quality |
| KM-final-viral-combined_k141_254494\|\|lt2gene | Low-quality |
| KM-final-viral-combined_k141_265730\|\|lt2gene | Low-quality |
| KM-final-viral-combined_k141_98766\|\|lt2gene | Low-quality |
| KM-final-viral-combined_k141_53502\|\|lt2gene | Low-quality |
| KM-final-viral-combined_k141_98835\|\|lt2gene | Low-quality |
| KM-final-viral-combined_k141_53547\|\|lt2gene | Not-determined |
| KM-final-viral-combined_k141_33211\|\|lt2gene | Low-quality |
| KM-final-viral-combined_k141_98937\|\|lt2gene | Low-quality |
| KM-final-viral-combined_k141_21412\|\|lt2gene | Low-quality |
| KM-final-viral-combined_k141_33286\|\|lt2gene | Low-quality |
| KM-final-viral-combined_k141_53734\|\|lt2gene | Not-determined |
| KM-final-viral-combined_k141_53741\|\|lt2gene | Not-determined |
| KM-final-viral-combined_k141_53791\|\|lt2gene | Low-quality |
| KM-final-viral-combined_k141_266103\|\|lt2gene | Not-determined |
| KM-final-viral-combined_k141_266115\|\|lt2gene | Not-determined |
| KM-final-viral-combined_k141_254676\|\|lt2gene | Low-quality |
| KM-final-viral-combined_k141_21587\|\|lt2gene | Not-determined |
| KM-final-viral-combined_k141_266271\|\|lt2gene | Low-quality |
| KM-final-viral-combined_k141_21725\|\|lt2gene | Not-determined |
| KM-final-viral-combined_k141_99227\|\|lt2gene | Low-quality |
| KM-final-viral-combined_k141_266467\|\|lt2gene | Not-determined |
| KM-final-viral-combined_k141_254947\|\|lt2gene | Low-quality |
| KM-final-viral-combined_k141_54072\|\|lt2gene | Not-determined |
| KM-final-viral-combined_k141_254980\|\|lt2gene | Not-determined |
| KM-final-viral-combined_k141_21949\|\|lt2gene | Low-quality |
| KM-final-viral-combined_k141_21957\|\|lt2gene | Low-quality |
| KM-final-viral-combined_k141_99526\|\|lt2gene | Not-determined |
| KM-final-viral-combined_k141_22042\|\|lt2gene | Not-determined |
| KM-final-viral-combined_k141_22069\|\|lt2gene | Not-determined |
| KM-final-viral-combined_k141_255166\|\|lt2gene | Low-quality |
| KM-final-viral-combined_k141_54395\|\|lt2gene | Low-quality |
| KM-final-viral-combined_k141_255271\|\|lt2gene | Low-quality |
| KM-final-viral-combined_k141_99682\|\|lt2gene | Low-quality |
| KM-final-viral-combined_k141_99836\|\|lt2gene | Low-quality |
| KM-final-viral-combined_k141_54756\|\|lt2gene | Low-quality |
| KM-final-viral-combined_k141_54828\|\|lt2gene | Low-quality |
| KM-final-viral-combined_k141_55089\|\|lt2gene | Low-quality |
| KM-final-viral-combined_k141_55393\|\|lt2gene | Not-determined |
| KM-final-viral-combined_k141_55519\|\|lt2gene | Low-quality |
| KM_k141_211149 | Not-determined |
| KM_k141_88907 | Not-determined |
| KM_k141_77802 | Not-determined |
| KM_k141_100022 | Not-determined |
| KM_k141_133366 | Not-determined |
| KM_k141_244501 | Not-determined |
| KM_k141_55576 | Not-determined |
| KM_k141_111146 | Not-determined |
| KM_k141_244508 | Not-determined |
| KM_k141_144485 | Low-quality |
| KM_k141_255599 | Not-determined |
| KM_k141_88920 | Not-determined |
| KM_k141_244513 | Not-determined |
| KM_k141_100030 | Not-determined |
| KM_k141_255606 | Not-determined |
| KM_k141_188943 | Not-determined |
| KM_k141_100034 | Not-determined |
| KM_k141_88929 | Not-determined |
| KM_k141_222268 | Not-determined |
| KM_k141_222270 | Not-determined |
| KM_k141_111148 | Not-determined |
| KM_k141_188948 | Low-quality |
| KM_k141_200058 | Not-determined |
| KM_k141_3 | Not-determined |
| KM_k141_155595 | Not-determined |
| KM_k141_44480 | Low-quality |
| KM_k141_255618 | Not-determined |
| KM_k141_233393 | Not-determined |
| KM_k141_244540 | Not-determined |
| KM_k141_222277 | Not-determined |
| KM_k141_133385 | Not-determined |
| KM_k141_44492 | Not-determined |
| KM_k141_18 | Not-determined |
| KM_k141_33383 | Not-determined |
| KM_k141_31 | Not-determined |
| KM_k141_222299 | Low-quality |
| KM_k141_32 | Not-determined |
| KM_k141_222304 | Not-determined |
| KM_k141_211179 | Not-determined |
| KM_k141_211182 | Not-determined |
| KM_k141_33391 | Not-determined |
| KM_k141_188989 | Not-determined |
| KM_k141_111193 | Not-determined |
| KM_k141_255655 | Not-determined |
| KM_k141_255656 | Not-determined |
| KM_k141_188995 | Not-determined |
| KM_k141_133405 | Not-determined |
| KM_k141_33406 | Not-determined |
| KM_k141_77827 | Not-determined |
| KM_k141_189007 | Not-determined |
| KM_k141_33409 | Not-determined |
| KM_k141_200129 | Not-determined |
| KM_k141_144537 | Not-determined |
| KM_k141_55608 | Not-determined |
| KM_k141_55616 | Not-determined |
| KM_k141_55617 | Not-determined |
| KM_k141_55618 | Not-determined |
| KM_k141_55620 | Not-determined |
| KM_k141_155630 | Not-determined |
| KM_k141_55622 | Not-determined |
| KM_k141_200139 | Not-determined |
| KM_k141_255665 | Not-determined |
| KM_k141_155643 | Not-determined |
| KM_k141_66708 | Not-determined |
| KM_k141_77848 | Not-determined |
| KM_k141_244563 | Not-determined |
| KM_k141_77854 | Not-determined |
| KM_k141_77855 | Not-determined |
| KM_k141_33420 | Not-determined |
| KM_k141_66717 | Not-determined |
| KM_k141_200150 | Not-determined |
| KM_k141_200151 | Not-determined |
| KM_k141_255673 | Not-determined |
| KM_k141_133421 | Not-determined |
| KM_k141_222338 | Not-determined |
| KM_k141_222340 | Not-determined |
| KM_k141_222342 | Not-determined |
| KM_k141_33425 | Not-determined |
| KM_k141_77870 | Not-determined |
| KM_k141_200172 | Not-determined |
| KM_k141_33430 | Not-determined |
| KM_k141_11126 | Not-determined |
| KM_k141_33432 | Not-determined |
| KM_k141_233412 | Not-determined |
| KM_k141_77875 | Not-determined |
| KM_k141_144564 | Not-determined |
| KM_k141_189009 | Not-determined |
| KM_k141_81 | Not-determined |
| KM_k141_11130 | Not-determined |
| KM_k141_166760 | Not-determined |
| KM_k141_33441 | Not-determined |
| KM_k141_255689 | Low-quality |
| KM_k141_200180 | Low-quality |
| KM_k141_200182 | Not-determined |
| KM_k141_166767 | Not-determined |
| KM_k141_211220 | Not-determined |
| KM_k141_77889 | Not-determined |
| KM_k141_33450 | Not-determined |
| KM_k141_66739 | Not-determined |
| KM_k141_22250 | Not-determined |
| KM_k141_66742 | Not-determined |
| KM_k141_44530 | Not-determined |
| KM_k141_133428 | Not-determined |
| KM_k141_133434 | Not-determined |
| KM_k141_66749 | Not-determined |
| KM_k141_77899 | Not-determined |
| KM_k141_144583 | Not-determined |
| KM_k141_211228 | Low-quality |
| KM_k141_22256 | Not-determined |
| KM_k141_88974 | Not-determined |
| KM_k141_144595 | Not-determined |
| KM_k141_33471 | Not-determined |
| KM_k141_189025 | Not-determined |
| KM_k141_66762 | Not-determined |
| KM_k141_122311 | Not-determined |
| KM_k141_177819 | Not-determined |
| KM_k141_189032 | Low-quality |
| KM_k141_144601 | Not-determined |
| KM_k141_66768 | Not-determined |
| KM_k141_88984 | Not-determined |
| KM_k141_11157 | Not-determined |
| KM_k141_177828 | Not-determined |
| KM_k141_177831 | Not-determined |
| KM_k141_33481 | Not-determined |
| KM_k141_144608 | Not-determined |
| KM_k141_66773 | Not-determined |
| KM_k141_177837 | Not-determined |
| KM_k141_177840 | Not-determined |
| KM_k141_11166 | Not-determined |
| KM_k141_177843 | Low-quality |
| KM_k141_177845 | Not-determined |
| KM_k141_111233 | Low-quality |
| KM_k141_244588 | Not-determined |
| KM_k141_222376 | Not-determined |
| KM_k141_122331 | Not-determined |
| KM_k141_144622 | Not-determined |
| KM_k141_244591 | Not-determined |
| KM_k141_111238 | Not-determined |
| KM_k141_122340 | Not-determined |
| KM_k141_177855 | Not-determined |
| KM_k141_133458 | Not-determined |
| KM_k141_66783 | Not-determined |
| KM_k141_122355 | Not-determined |
| KM_k141_255729 | Not-determined |
| KM_k141_133464 | Not-determined |
| KM_k141_222395 | Low-quality |
| KM_k141_133469 | Not-determined |
| KM_k141_144635 | Not-determined |
| KM_k141_255739 | Not-determined |
| KM_k141_166800 | Not-determined |
| KM_k141_189072 | Not-determined |
| KM_k141_255743 | Not-determined |
| KM_k141_255748 | Not-determined |
| KM_k141_211288 | Not-determined |
| KM_k141_111251 | Not-determined |
| KM_k141_233483 | Not-determined |
| KM_k141_144647 | Not-determined |
| KM_k141_22296 | Not-determined |
| KM_k141_144650 | Not-determined |
| KM_k141_133479 | Not-determined |

| | |
|---|---|
| KM_k141_111257 | Low-quality |
| KM_k141_233493 | Low-quality |
| KM_k141_133482 | Not-determined |
| KM_k141_33513 | Not-determined |
| KM_k141_144661 | Not-determined |
| KM_k141_222416 | Not-determined |
| KM_k141_222418 | Not-determined |
| KM_k141_200255 | Not-determined |
| KM_k141_189095 | Not-determined |
| KM_k141_144666 | Low-quality |
| KM_k141_33520 | Not-determined |
| KM_k141_122395 | Low-quality |
| KM_k141_133493 | Not-determined |
| KM_k141_233498 | Not-determined |
| KM_k141_244629 | Not-determined |
| KM_k141_77944 | Not-determined |
| KM_k141_66791 | Not-determined |
| KM_k141_77948 | Not-determined |
| KM_k141_189108 | Not-determined |
| KM_k141_55634 | Not-determined |
| KM_k141_33541 | Not-determined |
| KM_k141_255763 | Not-determined |
| KM_k141_155694 | Not-determined |
| KM_k141_77971 | Not-determined |
| KM_k141_155699 | Not-determined |
| KM_k141_177889 | Low-quality |
| KM_k141_177890 | Not-determined |
| KM_k141_11180 | Not-determined |
| KM_k141_177893 | Not-determined |
| KM_k141_55656 | Not-determined |
| KM_k141_98 | Low-quality |
| KM_k141_99 | Not-determined |
| KM_k141_89020 | Not-determined |
| KM_k141_244634 | Not-determined |
| KM_k141_22328 | Not-determined |
| KM_k141_11197 | Not-determined |
| KM_k141_33547 | Not-determined |
| KM_k141_177905 | Not-determined |
| KM_k141_133519 | Not-determined |
| KM_k141_112 | Not-determined |
| KM_k141_77978 | Not-determined |
| KM_k141_55676 | Low-quality |
| KM_k141_144699 | Not-determined |
| KM_k141_222444 | Not-determined |
| KM_k141_111282 | Not-determined |
| KM_k141_11215 | Not-determined |
| KM_k141_11216 | Not-determined |
| KM_k141_11217 | Not-determined |
| KM_k141_122416 | Not-determined |
| KM_k141_122 | Not-determined |
| KM_k141_55685 | Not-determined |
| KM_k141_222451 | Not-determined |
| KM_k141_100102 | Not-determined |
| KM_k141_126 | Not-determined |
| KM_k141_144702 | Not-determined |
| KM_k141_55690 | Not-determined |
| KM_k141_22335 | Not-determined |
| KM_k141_200289 | Not-determined |
| KM_k141_144704 | Not-determined |
| KM_k141_77987 | Not-determined |
| KM_k141_200294 | Not-determined |
| KM_k141_77990 | Not-determined |
| KM_k141_200297 | Not-determined |
| KM_k141_111295 | Not-determined |
| KM_k141_200299 | Not-determined |
| KM_k141_111297 | Not-determined |
| KM_k141_22341 | Not-determined |
| KM_k141_134 | Not-determined |
| KM_k141_33577 | Not-determined |
| KM_k141_22349 | Not-determined |
| KM_k141_144715 | Not-determined |
| KM_k141_145 | Low-quality |
| KM_k141_233515 | Not-determined |
| KM_k141_144716 | Not-determined |
| KM_k141_144720 | Not-determined |
| KM_k141_153 | Not-determined |
| KM_k141_78005 | Not-determined |
| KM_k141_255795 | Not-determined |
| KM_k141_78007 | Not-determined |
| KM_k141_133545 | Not-determined |
| KM_k141_11249 | Not-determined |
| KM_k141_133550 | Not-determined |
| KM_k141_122424 | Not-determined |
| KM_k141_33596 | Not-determined |
| KM_k141_78034 | Not-determined |
| KM_k141_177924 | Not-determined |
| KM_k141_78039 | Not-determined |
| KM_k141_144728 | Not-determined |
| KM_k141_233549 | Low-quality |
| KM_k141_100116 | Not-determined |
| KM_k141_200333 | Not-determined |
| KM_k141_174 | Not-determined |
| KM_k141_55732 | Not-determined |
| KM_k141_122430 | Not-determined |
| KM_k141_144741 | Not-determined |
| KM_k141_100118 | Not-determined |
| KM_k141_244692 | Not-determined |
| KM_k141_166814 | Not-determined |
| KM_k141_78049 | Low-quality |
| KM_k141_244694 | Not-determined |
| KM_k141_200339 | Not-determined |
| KM_k141_211361 | Not-determined |
| KM_k141_233557 | Not-determined |
| KM_k141_100121 | Not-determined |
| KM_k141_33612 | Low-quality |
| KM_k141_33613 | Not-determined |
| KM_k141_44577 | Low-quality |
| KM_k141_122440 | Low-quality |
| KM_k141_33622 | Low-quality |
| KM_k141_233572 | Not-determined |
| KM_k141_11278 | Not-determined |
| KM_k141_33630 | Not-determined |
| KM_k141_44588 | Not-determined |
| KM_k141_255808 | Not-determined |
| KM_k141_22370 | Not-determined |
| KM_k141_222492 | Not-determined |
| KM_k141_188 | Not-determined |
| KM_k141_177942 | Not-determined |
| KM_k141_33637 | Not-determined |
| KM_k141_222494 | Not-determined |
| KM_k141_222495 | Not-determined |
| KM_k141_22381 | Not-determined |
| KM_k141_255825 | Not-determined |
| KM_k141_111356 | Not-determined |
| KM_k141_22386 | Not-determined |
| KM_k141_78068 | Not-determined |
| KM_k141_78071 | Not-determined |
| KM_k141_197 | Not-determined |
| KM_k141_66861 | Not-determined |
| KM_k141_111363 | Not-determined |
| KM_k141_66864 | Not-determined |
| KM_k141_89044 | Low-quality |
| KM_k141_144758 | Not-determined |
| KM_k141_144760 | Not-determined |
| KM_k141_222508 | Not-determined |
| KM_k141_255834 | Not-determined |
| KM_k141_78084 | Not-determined |
| KM_k141_155749 | Not-determined |
| KM_k141_255843 | Not-determined |
| KM_k141_33659 | Not-determined |
| KM_k141_255844 | Not-determined |
| KM_k141_222516 | Not-determined |
| KM_k141_33664 | Not-determined |
| KM_k141_78091 | Not-determined |
| KM_k141_255850 | Not-determined |
| KM_k141_78094 | Not-determined |
| KM_k141_255852 | Not-determined |
| KM_k141_177979 | Not-determined |
| KM_k141_111394 | Not-determined |
| KM_k141_255856 | Low-quality |
| KM_k141_189183 | Not-determined |
| KM_k141_66893 | Not-determined |
| KM_k141_89077 | Not-determined |
| KM_k141_222528 | Low-quality |
| KM_k141_33669 | Not-determined |
| KM_k141_222529 | Not-determined |
| KM_k141_144775 | Low-quality |
| KM_k141_144776 | Not-determined |
| KM_k141_255873 | Not-determined |
| KM_k141_211404 | Not-determined |
| KM_k141_122487 | Not-determined |
| KM_k141_255876 | Not-determined |
| KM_k141_177993 | Not-determined |
| KM_k141_177995 | Not-determined |
| KM_k141_144784 | Not-determined |
| KM_k141_211411 | Not-determined |
| KM_k141_11284 | Not-determined |
| KM_k141_166831 | Not-determined |
| KM_k141_33681 | Low-quality |
| KM_k141_89087 | Not-determined |
| KM_k141_233600 | Not-determined |
| KM_k141_55766 | Not-determined |
| KM_k141_144788 | Not-determined |
| KM_k141_78125 | Not-determined |
| KM_k141_33687 | Not-determined |
| KM_k141_55773 | Not-determined |
| KM_k141_33691 | Not-determined |
| KM_k141_78131 | Not-determined |
| KM_k141_78138 | Not-determined |
| KM_k141_211430 | Not-determined |
| KM_k141_100152 | Not-determined |
| KM_k141_244719 | Not-determined |
| KM_k141_11294 | Not-determined |
| KM_k141_255888 | Not-determined |
| KM_k141_22428 | Not-determined |
| KM_k141_55791 | Not-determined |
| KM_k141_155770 | Not-determined |
| KM_k141_89102 | Not-determined |
| KM_k141_122503 | Not-determined |
| KM_k141_244730 | Not-determined |
| KM_k141_66932 | Low-quality |
| KM_k141_211441 | Not-determined |
| KM_k141_66934 | Not-determined |
| KM_k141_78154 | Not-determined |
| KM_k141_78156 | Not-determined |
| KM_k141_244735 | Not-determined |
| KM_k141_78157 | Not-determined |
| KM_k141_166857 | Not-determined |
| KM_k141_33703 | Not-determined |
| KM_k141_200367 | Not-determined |
| KM_k141_66941 | Not-determined |
| KM_k141_222575 | Low-quality |
| KM_k141_211452 | Not-determined |
| KM_k141_222581 | Not-determined |
| KM_k141_89113 | Not-determined |
| KM_k141_133579 | Not-determined |
| KM_k141_33713 | Not-determined |
| KM_k141_100167 | Not-determined |
| KM_k141_133584 | Not-determined |
| KM_k141_133585 | Not-determined |
| KM_k141_122527 | Not-determined |
| KM_k141_144802 | Not-determined |
| KM_k141_144803 | Not-determined |
| KM_k141_33721 | Not-determined |
| KM_k141_22467 | Not-determined |
| KM_k141_22468 | Not-determined |
| KM_k141_255904 | Not-determined |
| KM_k141_133589 | Not-determined |
| KM_k141_55817 | Not-determined |
| KM_k141_55818 | Low-quality |
| KM_k141_155801 | Low-quality |
| KM_k141_55827 | Not-determined |
| KM_k141_22473 | Not-determined |
| KM_k141_211480 | Low-quality |
| KM_k141_144810 | Not-determined |
| KM_k141_66963 | Not-determined |
| KM_k141_33734 | Not-determined |
| KM_k141_200389 | Not-determined |
| KM_k141_200392 | Not-determined |
| KM_k141_100188 | Not-determined |
| KM_k141_211486 | Not-determined |
| KM_k141_133602 | Not-determined |
| KM_k141_111422 | Low-quality |
| KM_k141_222605 | Not-determined |
| KM_k141_89128 | Not-determined |
| KM_k141_78179 | Not-determined |
| KM_k141_22485 | Not-determined |
| KM_k141_100195 | Not-determined |
| KM_k141_255923 | Not-determined |
| KM_k141_44615 | Not-determined |
| KM_k141_100197 | Not-determined |
| KM_k141_255925 | Not-determined |
| KM_k141_222608 | Not-determined |
| KM_k141_22489 | Not-determined |
| KM_k141_255926 | Not-determined |
| KM_k141_22492 | Low-quality |
| KM_k141_233645 | Low-quality |
| KM_k141_233646 | Not-determined |
| KM_k141_66985 | Not-determined |
| KM_k141_200403 | Low-quality |
| KM_k141_200407 | Not-determined |
| KM_k141_22499 | Not-determined |
| KM_k141_78189 | Not-determined |
| KM_k141_144817 | Not-determined |
| KM_k141_100203 | Not-determined |
| KM_k141_211498 | Not-determined |
| KM_k141_55851 | Not-determined |
| KM_k141_100208 | Not-determined |
| KM_k141_166892 | Not-determined |
| KM_k141_242 | Low-quality |
| KM_k141_233658 | Not-determined |
| KM_k141_11339 | Not-determined |
| KM_k141_44627 | Not-determined |
| KM_k141_222623 | Not-determined |
| KM_k141_211513 | Not-determined |
| KM_k141_122542 | Not-determined |
| KM_k141_78198 | Low-quality |
| KM_k141_256 | Not-determined |
| KM_k141_259 | Low-quality |
| KM_k141_222637 | Not-determined |
| KM_k141_44635 | Not-determined |
| KM_k141_264 | Not-determined |
| KM_k141_133627 | Not-determined |
| KM_k141_266 | Not-determined |
| KM_k141_122562 | Not-determined |
| KM_k141_189196 | Not-determined |
| KM_k141_122565 | Not-determined |
| KM_k141_89141 | Low-quality |
| KM_k141_55859 | Not-determined |
| KM_k141_268 | Not-determined |
| KM_k141_67033 | Not-determined |
| KM_k141_144851 | Low-quality |
| KM_k141_274 | Not-determined |
| KM_k141_122573 | Not-determined |
| KM_k141_133647 | Not-determined |
| KM_k141_67039 | Low-quality |
| KM_k141_55872 | Not-determined |
| KM_k141_55881 | Not-determined |
| KM_k141_155812 | Low-quality |
| KM_k141_133662 | Low-quality |
| KM_k141_55886 | Not-determined |
| KM_k141_55887 | Not-determined |
| KM_k141_111446 | Not-determined |
| KM_k141_67062 | Not-determined |
| KM_k141_111450 | Not-determined |
| KM_k141_22504 | Not-determined |
| KM_k141_189225 | Not-determined |
| KM_k141_55898 | Low-quality |
| KM_k141_189230 | Not-determined |
| KM_k141_111462 | Low-quality |
| KM_k141_233732 | Not-determined |
| KM_k141_178023 | Not-determined |
| KM_k141_133666 | Not-determined |
| KM_k141_233736 | Not-determined |
| KM_k141_133671 | Not-determined |
| KM_k141_78228 | Not-determined |
| KM_k141_255950 | Not-determined |
| KM_k141_255954 | Not-determined |
| KM_k141_233739 | Low-quality |
| KM_k141_255956 | Not-determined |
| KM_k141_255957 | Not-determined |
| KM_k141_155843 | Not-determined |
| KM_k141_189254 | Not-determined |
| KM_k141_189259 | Low-quality |
| KM_k141_122593 | Not-determined |
| KM_k141_22527 | Not-determined |
| KM_k141_189262 | Not-determined |
| KM_k141_189265 | Not-determined |
| KM_k141_233754 | Not-determined |
| KM_k141_233755 | Not-determined |
| KM_k141_233758 | Not-determined |
| KM_k141_328 | Not-determined |
| KM_k141_255988 | Not-determined |
| KM_k141_78258 | Not-determined |
| KM_k141_178026 | Low-quality |
| KM_k141_200436 | Low-quality |
| KM_k141_44669 | Not-determined |
| KM_k141_200438 | Not-determined |
| KM_k141_78263 | Not-determined |
| KM_k141_78265 | Not-determined |
| KM_k141_22542 | Not-determined |
| KM_k141_233760 | Not-determined |
| KM_k141_22546 | Not-determined |
| KM_k141_44674 | Not-determined |
| KM_k141_189269 | Low-quality |
| KM_k141_78274 | Not-determined |
| KM_k141_211547 | Not-determined |
| KM_k141_55917 | Not-determined |
| KM_k141_44690 | Not-determined |
| KM_k141_144945 | Not-determined |
| KM_k141_155864 | Not-determined |
| KM_k141_166916 | Not-determined |
| KM_k141_144957 | Not-determined |
| KM_k141_11358 | Low-quality |
| KM_k141_111522 | Not-determined |
| KM_k141_189289 | Not-determined |
| KM_k141_144969 | Not-determined |
| KM_k141_55935 | Not-determined |
| KM_k141_44702 | Not-determined |
| KM_k141_55943 | Not-determined |
| KM_k141_100296 | Not-determined |
| KM_k141_89201 | Not-determined |
| KM_k141_67088 | Not-determined |
| KM_k141_67096 | Not-determined |
| KM_k141_144985 | Low-quality |
| KM_k141_155883 | Not-determined |
| KM_k141_122634 | Not-determined |
| KM_k141_244862 | Low-quality |
| KM_k141_11386 | Not-determined |
| KM_k141_155885 | Not-determined |
| KM_k141_44713 | Not-determined |
| KM_k141_155889 | Not-determined |
| KM_k141_244868 | Not-determined |
| KM_k141_155897 | Not-determined |
| KM_k141_89226 | Not-determined |
| KM_k141_89229 | Not-determined |
| KM_k141_78330 | Not-determined |
| KM_k141_244879 | Not-determined |
| KM_k141_122645 | Not-determined |
| KM_k141_122648 | Not-determined |
| KM_k141_222701 | Not-determined |
| KM_k141_233765 | Not-determined |
| KM_k141_67108 | Not-determined |
| KM_k141_233768 | Not-determined |
| KM_k141_122658 | Not-determined |
| KM_k141_55958 | Not-determined |
| KM_k141_244892 | Not-determined |
| KM_k141_55959 | Not-determined |
| KM_k141_222711 | Not-determined |
| KM_k141_11411 | Not-determined |
| KM_k141_155909 | Not-determined |
| KM_k141_166929 | Not-determined |
| KM_k141_11418 | Not-determined |
| KM_k141_55970 | Not-determined |
| KM_k141_100336 | Not-determined |
| KM_k141_122673 | Not-determined |
| KM_k141_78349 | Not-determined |
| KM_k141_122678 | Not-determined |
| KM_k141_145004 | Not-determined |
| KM_k141_55983 | Not-determined |
| KM_k141_100342 | Not-determined |
| KM_k141_145007 | Not-determined |
| KM_k141_55986 | Not-determined |
| KM_k141_166951 | Not-determined |
| KM_k141_111585 | Low-quality |
| KM_k141_55990 | Low-quality |
| KM_k141_11433 | Not-determined |
| KM_k141_166957 | Not-determined |
| KM_k141_403 | Not-determined |
| KM_k141_11438 | Not-determined |
| KM_k141_11440 | Not-determined |
| KM_k141_407 | Not-determined |
| KM_k141_166962 | Not-determined |
| KM_k141_412 | Not-determined |
| KM_k141_166967 | Not-determined |
| KM_k141_100358 | Not-determined |
| KM_k141_89253 | Not-determined |
| KM_k141_200468 | Not-determined |
| KM_k141_414 | Not-determined |
| KM_k141_22591 | Not-determined |
| KM_k141_200474 | Not-determined |
| KM_k141_166974 | Not-determined |
| KM_k141_22595 | Not-determined |
| KM_k141_67152 | Not-determined |
| KM_k141_200479 | Not-determined |
| KM_k141_256005 | Not-determined |
| KM_k141_222749 | Not-determined |
| KM_k141_211614 | Not-determined |
| KM_k141_67171 | Not-determined |
| KM_k141_256014 | Low-quality |
| KM_k141_211620 | Not-determined |
| KM_k141_256017 | Not-determined |
| KM_k141_67179 | Not-determined |
| KM_k141_200495 | Not-determined |
| KM_k141_200497 | Not-determined |
| KM_k141_178047 | Not-determined |
| KM_k141_44748 | Not-determined |
| KM_k141_256022 | Not-determined |
| KM_k141_178050 | Not-determined |
| KM_k141_178051 | Not-determined |
| KM_k141_122702 | Not-determined |
| KM_k141_22600 | Low-quality |
| KM_k141_67190 | Not-determined |
| KM_k141_145035 | Not-determined |
| KM_k141_178064 | Not-determined |
| KM_k141_447 | Not-determined |
| KM_k141_100387 | Low-quality |
| KM_k141_155925 | Low-quality |
| KM_k141_22613 | Not-determined |
| KM_k141_22617 | Not-determined |
| KM_k141_122708 | Low-quality |
| KM_k141_122714 | Not-determined |
| KM_k141_145053 | Not-determined |
| KM_k141_222789 | Not-determined |
| KM_k141_178080 | Not-determined |
| KM_k141_200518 | Not-determined |
| KM_k141_244962 | Not-determined |
| KM_k141_178088 | Not-determined |
| KM_k141_222801 | Not-determined |
| KM_k141_100408 | Not-determined |
| KM_k141_44774 | Not-determined |
| KM_k141_145061 | Low-quality |
| KM_k141_44783 | Not-determined |
| KM_k141_464 | Not-determined |
| KM_k141_155947 | Not-determined |
| KM_k141_111615 | Not-determined |
| KM_k141_155950 | Not-determined |
| KM_k141_111618 | Not-determined |
| KM_k141_155954 | Not-determined |
| KM_k141_133747 | Not-determined |
| KM_k141_100421 | Not-determined |
| KM_k141_111622 | Low-quality |
| KM_k141_133751 | Not-determined |
| KM_k141_133753 | Not-determined |
| KM_k141_244976 | Not-determined |
| KM_k141_145073 | Not-determined |
| KM_k141_233782 | Not-determined |
| KM_k141_475 | Not-determined |
| KM_k141_100424 | Low-quality |
| KM_k141_145079 | Not-determined |
| KM_k141_111635 | Not-determined |
| KM_k141_167017 | Not-determined |
| KM_k141_111639 | Not-determined |
| KM_k141_78369 | Not-determined |
| KM_k141_256044 | Not-determined |
| KM_k141_111645 | Not-determined |
| KM_k141_155974 | Not-determined |
| KM_k141_167027 | Not-determined |
| KM_k141_56011 | Not-determined |
| KM_k141_256051 | Not-determined |
| KM_k141_67222 | Not-determined |
| KM_k141_256054 | Not-determined |
| KM_k141_200567 | Not-determined |
| KM_k141_167033 | Not-determined |
| KM_k141_167035 | Not-determined |
| KM_k141_244998 | Not-determined |
| KM_k141_33784 | Not-determined |
| KM_k141_211653 | Low-quality |
| KM_k141_245007 | Not-determined |
| KM_k141_200579 | Not-determined |
| KM_k141_211659 | Not-determined |
| KM_k141_111661 | Not-determined |
| KM_k141_211664 | Not-determined |
| KM_k141_111667 | Not-determined |
| KM_k141_111669 | Not-determined |
| KM_k141_122726 | Not-determined |
| KM_k141_167046 | Not-determined |
| KM_k141_122728 | Not-determined |
| KM_k141_503 | Not-determined |
| KM_k141_222830 | Not-determined |
| KM_k141_155997 | Not-determined |
| KM_k141_67270 | Not-determined |
| KM_k141_222832 | Not-determined |
| KM_k141_67273 | Not-determined |
| KM_k141_133789 | Not-determined |
| KM_k141_155999 | Not-determined |
| KM_k141_156000 | Not-determined |
| KM_k141_22624 | Not-determined |
| KM_k141_211674 | Not-determined |
| KM_k141_211675 | Not-determined |
| KM_k141_122739 | Low-quality |
| KM_k141_211678 | Not-determined |
| KM_k141_167068 | Not-determined |
| KM_k141_233801 | Not-determined |
| KM_k141_33820 | Not-determined |
| KM_k141_167071 | Not-determined |
| KM_k141_111680 | Not-determined |
| KM_k141_167074 | Low-quality |
| KM_k141_145091 | Low-quality |
| KM_k141_33825 | Not-determined |
| KM_k141_133808 | Not-determined |
| KM_k141_245019 | Not-determined |
| KM_k141_122745 | Low-quality |
| KM_k141_33827 | Not-determined |
| KM_k141_11457 | Not-determined |
| KM_k141_11459 | Not-determined |
| KM_k141_156017 | Not-determined |
| KM_k141_245023 | Not-determined |
| KM_k141_111690 | Not-determined |
| KM_k141_78416 | Not-determined |
| KM_k141_211694 | Not-determined |
| KM_k141_222839 | Low-quality |
| KM_k141_11463 | Not-determined |
| KM_k141_78419 | Not-determined |
| KM_k141_211698 | Not-determined |
| KM_k141_211699 | Not-determined |
| KM_k141_11467 | Not-determined |
| KM_k141_33836 | Not-determined |
| KM_k141_122754 | Low-quality |
| KM_k141_245029 | Not-determined |
| KM_k141_33838 | Not-determined |
| KM_k141_56037 | Not-determined |
| KM_k141_167095 | Not-determined |
| KM_k141_156026 | Not-determined |
| KM_k141_67305 | Low-quality |
| KM_k141_67307 | Not-determined |
| KM_k141_222848 | Low-quality |
| KM_k141_122765 | Not-determined |
| KM_k141_56041 | Not-determined |
| KM_k141_122767 | Not-determined |
| KM_k141_11476 | Not-determined |
| KM_k141_11477 | Not-determined |
| KM_k141_11479 | Not-determined |
| KM_k141_167100 | Not-determined |
| KM_k141_122778 | Not-determined |
| KM_k141_189332 | Not-determined |
| KM_k141_11489 | Not-determined |
| KM_k141_145121 | Not-determined |
| KM_k141_22648 | Not-determined |
| KM_k141_222855 | Not-determined |
| KM_k141_11493 | Not-determined |
| KM_k141_167114 | Not-determined |
| KM_k141_167115 | Not-determined |
| KM_k141_245048 | Not-determined |
| KM_k141_122791 | Not-determined |
| KM_k141_78458 | Not-determined |
| KM_k141_11498 | Low-quality |
| KM_k141_56044 | Low-quality |
| KM_k141_122796 | Not-determined |
| KM_k141_89297 | Not-determined |
| KM_k141_245054 | Not-determined |
| KM_k141_122800 | Not-determined |
| KM_k141_33855 | Not-determined |
| KM_k141_189348 | Not-determined |
| KM_k141_78470 | Not-determined |
| KM_k141_156041 | Not-determined |
| KM_k141_256069 | Low-quality |
| KM_k141_78473 | Not-determined |
| KM_k141_78479 | Not-determined |
| KM_k141_78483 | Not-determined |
| KM_k141_256078 | Not-determined |
| KM_k141_211717 | Not-determined |
| KM_k141_189358 | Not-determined |
| KM_k141_111725 | Not-determined |
| KM_k141_22657 | Not-determined |
| KM_k141_33869 | Not-determined |
| KM_k141_211720 | Not-determined |
| KM_k141_122816 | Low-quality |
| KM_k141_56054 | Not-determined |

| | |
|---|---|
| KM_k141_122824 | Not-determined |
| KM_k141_245068 | Not-determined |
| KM_k141_156061 | Not-determined |
| KM_k141_89308 | Not-determined |
| KM_k141_67359 | Not-determined |
| KM_k141_145136 | Low-quality |
| KM_k141_156064 | Not-determined |
| KM_k141_156065 | Not-determined |
| KM_k141_256095 | Not-determined |
| KM_k141_178131 | Not-determined |
| KM_k141_133820 | Not-determined |
| KM_k141_133822 | Not-determined |
| KM_k141_89315 | Low-quality |
| KM_k141_200651 | Low-quality |
| KM_k141_245077 | Not-determined |
| KM_k141_67364 | Not-determined |
| KM_k141_11531 | Not-determined |
| KM_k141_100450 | Not-determined |
| KM_k141_111744 | Not-determined |
| KM_k141_11536 | Not-determined |
| KM_k141_67378 | Not-determined |
| KM_k141_200668 | Not-determined |
| KM_k141_89331 | Not-determined |
| KM_k141_222889 | Not-determined |
| KM_k141_89333 | Not-determined |
| KM_k141_256117 | Not-determined |
| KM_k141_111748 | Not-determined |
| KM_k141_111750 | Not-determined |
| KM_k141_44836 | Not-determined |
| KM_k141_111753 | Not-determined |
| KM_k141_256128 | Not-determined |
| KM_k141_133841 | Not-determined |
| KM_k141_133844 | Not-determined |
| KM_k141_67391 | Not-determined |
| KM_k141_133848 | Not-determined |
| KM_k141_222901 | Low-quality |
| KM_k141_133851 | Not-determined |
| KM_k141_89336 | Not-determined |
| KM_k141_67393 | Not-determined |
| KM_k141_89338 | Not-determined |
| KM_k141_67396 | Not-determined |
| KM_k141_133859 | Not-determined |
| KM_k141_111765 | Not-determined |
| KM_k141_222909 | Not-determined |
| KM_k141_256134 | Not-determined |
| KM_k141_78509 | Low-quality |
| KM_k141_67402 | Not-determined |
| KM_k141_111772 | Not-determined |
| KM_k141_122863 | Not-determined |
| KM_k141_156071 | Not-determined |
| KM_k141_562 | Low-quality |
| KM_k141_133869 | Not-determined |
| KM_k141_563 | Not-determined |
| KM_k141_89352 | Not-determined |
| KM_k141_67407 | Not-determined |
| KM_k141_67408 | Not-determined |
| KM_k141_189390 | Low-quality |
| KM_k141_111783 | Not-determined |
| KM_k141_200688 | Not-determined |
| KM_k141_100467 | Not-determined |
| KM_k141_211747 | Not-determined |
| KM_k141_189394 | Not-determined |
| KM_k141_574 | Not-determined |
| KM_k141_33917 | Not-determined |
| KM_k141_200701 | Not-determined |
| KM_k141_100474 | Not-determined |
| KM_k141_178203 | Not-determined |
| KM_k141_111790 | Not-determined |
| KM_k141_44873 | Not-determined |
| KM_k141_11540 | Not-determined |
| KM_k141_200711 | Not-determined |
| KM_k141_189406 | Not-determined |
| KM_k141_78524 | Not-determined |
| KM_k141_122890 | Low-quality |
| KM_k141_11545 | Low-quality |
| KM_k141_211759 | Not-determined |
| KM_k141_167138 | Not-determined |
| KM_k141_11548 | Not-determined |
| KM_k141_584 | Not-determined |
| KM_k141_167144 | Not-determined |
| KM_k141_200720 | Not-determined |
| KM_k141_111806 | Not-determined |
| KM_k141_145155 | Not-determined |
| KM_k141_11556 | Not-determined |
| KM_k141_67429 | Not-determined |
| KM_k141_33937 | Not-determined |
| KM_k141_156087 | Not-determined |
| KM_k141_167156 | Low-quality |
| KM_k141_111813 | Not-determined |
| KM_k141_167158 | Not-determined |
| KM_k141_11562 | Not-determined |
| KM_k141_133890 | Low-quality |
| KM_k141_211791 | Not-determined |
| KM_k141_11565 | Not-determined |
| KM_k141_598 | Not-determined |
| KM_k141_189415 | Not-determined |
| KM_k141_200729 | Not-determined |
| KM_k141_122903 | Not-determined |
| KM_k141_211802 | Not-determined |
| KM_k141_256187 | Low-quality |
| KM_k141_133908 | Not-determined |
| KM_k141_133909 | Not-determined |
| KM_k141_133913 | Not-determined |
| KM_k141_256194 | Not-determined |
| KM_k141_200734 | Not-determined |
| KM_k141_33953 | Not-determined |
| KM_k141_200739 | Not-determined |
| KM_k141_156110 | Not-determined |
| KM_k141_222951 | Not-determined |
| KM_k141_122926 | Not-determined |
| KM_k141_122927 | Not-determined |
| KM_k141_222954 | Not-determined |
| KM_k141_11603 | Not-determined |
| KM_k141_245101 | Not-determined |
| KM_k141_11607 | Not-determined |
| KM_k141_89381 | Not-determined |
| KM_k141_67450 | Not-determined |
| KM_k141_145163 | Not-determined |
| KM_k141_122936 | Not-determined |
| KM_k141_122937 | Not-determined |
| KM_k141_200767 | Low-quality |
| KM_k141_156116 | Not-determined |
| KM_k141_122941 | Not-determined |
| KM_k141_33981 | Not-determined |
| KM_k141_111850 | Not-determined |
| KM_k141_245124 | Not-determined |
| KM_k141_145175 | Not-determined |
| KM_k141_111862 | Not-determined |
| KM_k141_624 | Not-determined |
| KM_k141_145181 | Not-determined |
| KM_k141_627 | Low-quality |
| KM_k141_133959 | Not-determined |
| KM_k141_189448 | Not-determined |
| KM_k141_156129 | Low-quality |
| KM_k141_189454 | Not-determined |
| KM_k141_189455 | Low-quality |
| KM_k141_256245 | Not-determined |
| KM_k141_200786 | Not-determined |
| KM_k141_256253 | Not-determined |
| KM_k141_200799 | Not-determined |
| KM_k141_78535 | Not-determined |
| KM_k141_78536 | Not-determined |
| KM_k141_200805 | Not-determined |
| KM_k141_111870 | Not-determined |
| KM_k141_245132 | Not-determined |
| KM_k141_111872 | Not-determined |
| KM_k141_245134 | Not-determined |
| KM_k141_78546 | Not-determined |
| KM_k141_78547 | Not-determined |
| KM_k141_122973 | Not-determined |
| KM_k141_167178 | Not-determined |
| KM_k141_111881 | Not-determined |
| KM_k141_178233 | Not-determined |
| KM_k141_200811 | Not-determined |
| KM_k141_211869 | Low-quality |
| KM_k141_78560 | Not-determined |
| KM_k141_122983 | Not-determined |
| KM_k141_211872 | Not-determined |
| KM_k141_211874 | Not-determined |
| KM_k141_256288 | Not-determined |
| KM_k141_211878 | Not-determined |
| KM_k141_89392 | Low-quality |
| KM_k141_256291 | Low-quality |
| KM_k141_89394 | Not-determined |
| KM_k141_156169 | Not-determined |
| KM_k141_211884 | Not-determined |
| KM_k141_56111 | Not-determined |
| KM_k141_256303 | Not-determined |
| KM_k141_56114 | Not-determined |
| KM_k141_100511 | Not-determined |
| KM_k141_167196 | Not-determined |
| KM_k141_655 | Not-determined |
| KM_k141_656 | Not-determined |
| KM_k141_189473 | Not-determined |
| KM_k141_222969 | Not-determined |
| KM_k141_167200 | Not-determined |
| KM_k141_67476 | Not-determined |
| KM_k141_89405 | Not-determined |
| KM_k141_111905 | Low-quality |
| KM_k141_111907 | Not-determined |
| KM_k141_67482 | Not-determined |
| KM_k141_663 | Not-determined |
| KM_k141_67485 | Not-determined |
| KM_k141_56131 | Not-determined |
| KM_k141_67490 | Not-determined |
| KM_k141_67491 | Low-quality |
| KM_k141_211899 | Not-determined |
| KM_k141_189492 | Not-determined |
| KM_k141_222979 | Not-determined |
| KM_k141_178272 | Not-determined |
| KM_k141_670 | Not-determined |
| KM_k141_671 | Not-determined |
| KM_k141_233893 | Not-determined |
| KM_k141_222985 | Not-determined |
| KM_k141_89425 | Not-determined |
| KM_k141_200846 | Low-quality |
| KM_k141_200847 | Not-determined |
| KM_k141_245148 | Not-determined |
| KM_k141_145232 | Not-determined |
| KM_k141_233906 | Not-determined |
| KM_k141_256336 | Not-determined |
| KM_k141_67515 | Not-determined |
| KM_k141_67518 | Not-determined |
| KM_k141_89440 | Not-determined |
| KM_k141_702 | Low-quality |
| KM_k141_44899 | Not-determined |
| KM_k141_233920 | Low-quality |
| KM_k141_167222 | Not-determined |
| KM_k141_44906 | Not-determined |
| KM_k141_707 | Not-determined |
| KM_k141_145249 | Not-determined |
| KM_k141_167225 | Low-quality |
| KM_k141_167229 | Not-determined |
| KM_k141_245159 | Not-determined |
| KM_k141_56177 | Not-determined |
| KM_k141_34003 | Not-determined |
| KM_k141_211956 | Low-quality |
| KM_k141_67549 | Not-determined |
| KM_k141_78612 | Low-quality |
| KM_k141_211960 | Not-determined |
| KM_k141_223022 | Not-determined |
| KM_k141_256367 | Not-determined |
| KM_k141_256369 | Not-determined |
| KM_k141_89460 | Not-determined |
| KM_k141_256372 | Not-determined |
| KM_k141_78622 | Not-determined |
| KM_k141_200861 | Not-determined |
| KM_k141_89465 | Not-determined |
| KM_k141_189537 | Not-determined |
| KM_k141_67557 | Not-determined |
| KM_k141_78629 | Not-determined |
| KM_k141_245177 | Not-determined |
| KM_k141_178323 | Not-determined |
| KM_k141_189541 | Not-determined |
| KM_k141_11649 | Not-determined |
| KM_k141_78640 | Not-determined |
| KM_k141_167249 | Not-determined |
| KM_k141_134037 | Not-determined |
| KM_k141_245189 | Not-determined |
| KM_k141_178347 | Not-determined |
| KM_k141_223035 | Not-determined |
| KM_k141_78645 | Not-determined |
| KM_k141_245194 | Not-determined |
| KM_k141_89479 | Not-determined |
| KM_k141_67580 | Not-determined |
| KM_k141_78647 | Not-determined |
| KM_k141_67582 | Not-determined |
| KM_k141_111941 | Not-determined |
| KM_k141_34045 | Not-determined |
| KM_k141_111945 | Not-determined |
| KM_k141_111948 | Not-determined |
| KM_k141_78651 | Not-determined |
| KM_k141_156238 | Not-determined |
| KM_k141_223049 | Not-determined |
| KM_k141_223051 | Not-determined |
| KM_k141_123010 | Not-determined |
| KM_k141_200893 | Not-determined |
| KM_k141_111961 | Not-determined |
| KM_k141_200901 | Not-determined |
| KM_k141_245213 | Not-determined |
| KM_k141_178355 | Not-determined |
| KM_k141_156251 | Not-determined |
| KM_k141_111972 | Not-determined |
| KM_k141_111976 | Not-determined |
| KM_k141_34055 | Not-determined |
| KM_k141_123028 | Not-determined |
| KM_k141_156258 | Not-determined |
| KM_k141_156263 | Not-determined |
| KM_k141_256380 | Not-determined |
| KM_k141_78676 | Not-determined |
| KM_k141_145270 | Not-determined |
| KM_k141_89501 | Not-determined |
| KM_k141_245223 | Not-determined |
| KM_k141_256390 | Not-determined |
| KM_k141_11673 | Not-determined |
| KM_k141_211963 | Low-quality |
| KM_k141_211965 | Not-determined |
| KM_k141_145277 | Not-determined |
| KM_k141_11677 | Not-determined |
| KM_k141_178373 | Not-determined |
| KM_k141_145280 | Not-determined |
| KM_k141_245230 | Not-determined |
| KM_k141_178375 | Low-quality |
| KM_k141_200925 | Not-determined |
| KM_k141_123043 | Not-determined |
| KM_k141_89514 | Not-determined |
| KM_k141_178380 | Not-determined |
| KM_k141_200933 | Not-determined |
| KM_k141_200934 | Not-determined |
| KM_k141_741 | Not-determined |
| KM_k141_44909 | Not-determined |
| KM_k141_211979 | Low-quality |
| KM_k141_89522 | Not-determined |
| KM_k141_211981 | Not-determined |
| KM_k141_145292 | Not-determined |
| KM_k141_89524 | Not-determined |
| KM_k141_89526 | Not-determined |
| KM_k141_123049 | Not-determined |
| KM_k141_145299 | Not-determined |
| KM_k141_200944 | Low-quality |
| KM_k141_178390 | Not-determined |
| KM_k141_123054 | Not-determined |
| KM_k141_256406 | Not-determined |
| KM_k141_89530 | Not-determined |
| KM_k141_145303 | Not-determined |
| KM_k141_145304 | Not-determined |
| KM_k141_189584 | Not-determined |
| KM_k141_134066 | Not-determined |
| KM_k141_189586 | Not-determined |
| KM_k141_89534 | Not-determined |
| KM_k141_78697 | Not-determined |
| KM_k141_56211 | Not-determined |
| KM_k141_200952 | Not-determined |
| KM_k141_212004 | Low-quality |
| KM_k141_245257 | Not-determined |
| KM_k141_56214 | Not-determined |
| KM_k141_245261 | Not-determined |
| KM_k141_44931 | Not-determined |
| KM_k141_200958 | Not-determined |
| KM_k141_123069 | Not-determined |
| KM_k141_200961 | Not-determined |
| KM_k141_34102 | Low-quality |
| KM_k141_123074 | Not-determined |
| KM_k141_22704 | Not-determined |
| KM_k141_233955 | Not-determined |
| KM_k141_123080 | Not-determined |
| KM_k141_145322 | Not-determined |
| KM_k141_145327 | Low-quality |
| KM_k141_44940 | Not-determined |
| KM_k141_178407 | Not-determined |
| KM_k141_178411 | Not-determined |
| KM_k141_233960 | Not-determined |
| KM_k141_78713 | Not-determined |
| KM_k141_145332 | Low-quality |
| KM_k141_123086 | Not-determined |
| KM_k141_245280 | Not-determined |
| KM_k141_34113 | Not-determined |
| KM_k141_245281 | Not-determined |
| KM_k141_78716 | Not-determined |
| KM_k141_34115 | Not-determined |
| KM_k141_34117 | Not-determined |
| KM_k141_123092 | Not-determined |
| KM_k141_256429 | Not-determined |
| KM_k141_123100 | Not-determined |
| KM_k141_134077 | Not-determined |
| KM_k141_134078 | Not-determined |
| KM_k141_245288 | Not-determined |
| KM_k141_212033 | Not-determined |
| KM_k141_100557 | Not-determined |
| KM_k141_34122 | Not-determined |
| KM_k141_134083 | Low-quality |
| KM_k141_22723 | Not-determined |
| KM_k141_34128 | Not-determined |
| KM_k141_134084 | Low-quality |
| KM_k141_134085 | Not-determined |
| KM_k141_134087 | Not-determined |
| KM_k141_795 | Not-determined |
| KM_k141_145347 | Not-determined |
| KM_k141_44957 | Not-determined |
| KM_k141_56248 | Not-determined |
| KM_k141_189632 | Not-determined |
| KM_k141_78721 | Not-determined |
| KM_k141_145351 | Not-determined |
| KM_k141_11707 | Not-determined |
| KM_k141_145353 | Not-determined |
| KM_k141_799 | Not-determined |
| KM_k141_200982 | Not-determined |
| KM_k141_200985 | Not-determined |
| KM_k141_11712 | Not-determined |
| KM_k141_200987 | Not-determined |
| KM_k141_189643 | Not-determined |
| KM_k141_189644 | Not-determined |
| KM_k141_145361 | Not-determined |
| KM_k141_112020 | Not-determined |
| KM_k141_145363 | Not-determined |
| KM_k141_223119 | Not-determined |
| KM_k141_89566 | Not-determined |
| KM_k141_223127 | Not-determined |
| KM_k141_189647 | Not-determined |
| KM_k141_22752 | Not-determined |
| KM_k141_134101 | Not-determined |
| KM_k141_223138 | Not-determined |
| KM_k141_123134 | Not-determined |
| KM_k141_178415 | Not-determined |
| KM_k141_123139 | Not-determined |
| KM_k141_112047 | Not-determined |
| KM_k141_145378 | Not-determined |
| KM_k141_11751 | Not-determined |
| KM_k141_223150 | Not-determined |
| KM_k141_56272 | Not-determined |
| KM_k141_44998 | Not-determined |
| KM_k141_89582 | Not-determined |
| KM_k141_156299 | Not-determined |
| KM_k141_45001 | Not-determined |
| KM_k141_112056 | Not-determined |
| KM_k141_189660 | Not-determined |
| KM_k141_201000 | Low-quality |
| KM_k141_189667 | Not-determined |
| KM_k141_100584 | Low-quality |
| KM_k141_100594 | Not-determined |
| KM_k141_201008 | Not-determined |
| KM_k141_201011 | Not-determined |
| KM_k141_156308 | Low-quality |
| KM_k141_223164 | Not-determined |
| KM_k141_156312 | Low-quality |
| KM_k141_178440 | Not-determined |
| KM_k141_223167 | Not-determined |
| KM_k141_201017 | Not-determined |
| KM_k141_123152 | Not-determined |
| KM_k141_245316 | Not-determined |
| KM_k141_145396 | Not-determined |
| KM_k141_89593 | Low-quality |
| KM_k141_100603 | Not-determined |
| KM_k141_245322 | Low-quality |
| KM_k141_201023 | Not-determined |
| KM_k141_22761 | Not-determined |
| KM_k141_212069 | Not-determined |
| KM_k141_212081 | Low-quality |
| KM_k141_201024 | Not-determined |
| KM_k141_201025 | Not-determined |
| KM_k141_178459 | Low-quality |
| KM_k141_34166 | Low-quality |
| KM_k141_212092 | Not-determined |
| KM_k141_156333 | Not-determined |
| KM_k141_178465 | Not-determined |
| KM_k141_100635 | Not-determined |
| KM_k141_56289 | Not-determined |
| KM_k141_56290 | Not-determined |
| KM_k141_67646 | Not-determined |
| KM_k141_100637 | Not-determined |
| KM_k141_112109 | Not-determined |
| KM_k141_134151 | Not-determined |
| KM_k141_189693 | Not-determined |
| KM_k141_201049 | Not-determined |
| KM_k141_201054 | Not-determined |
| KM_k141_178472 | Not-determined |
| KM_k141_112116 | Not-determined |
| KM_k141_22781 | Not-determined |
| KM_k141_178475 | Not-determined |
| KM_k141_112123 | Not-determined |
| KM_k141_56302 | Not-determined |
| KM_k141_11761 | Not-determined |
| KM_k141_78803 | Not-determined |
| KM_k141_56312 | Not-determined |
| KM_k141_11768 | Low-quality |
| KM_k141_45026 | Not-determined |
| KM_k141_11770 | Not-determined |
| KM_k141_145418 | Not-determined |
| KM_k141_100646 | Not-determined |
| KM_k141_89609 | Not-determined |
| KM_k141_201075 | Not-determined |
| KM_k141_201076 | Not-determined |
| KM_k141_212116 | Not-determined |
| KM_k141_212117 | Not-determined |
| KM_k141_212119 | Not-determined |
| KM_k141_167318 | Not-determined |
| KM_k141_145427 | Not-determined |
| KM_k141_123162 | Not-determined |
| KM_k141_178486 | Not-determined |
| KM_k141_178487 | Not-determined |
| KM_k141_201088 | Not-determined |
| KM_k141_212135 | Not-determined |
| KM_k141_22812 | Not-determined |
| KM_k141_89618 | Not-determined |
| KM_k141_11793 | Not-determined |
| KM_k141_78827 | Not-determined |
| KM_k141_256445 | Not-determined |
| KM_k141_223192 | Not-determined |
| KM_k141_223194 | Not-determined |
| KM_k141_201102 | Not-determined |
| KM_k141_167334 | Not-determined |
| KM_k141_45045 | Low-quality |
| KM_k141_233999 | Not-determined |
| KM_k141_212151 | Not-determined |
| KM_k141_11805 | Not-determined |
| KM_k141_212153 | Not-determined |
| KM_k141_212155 | Low-quality |
| KM_k141_112165 | Not-determined |
| KM_k141_145436 | Not-determined |
| KM_k141_100662 | Not-determined |
| KM_k141_67664 | Not-determined |
| KM_k141_100665 | Not-determined |
| KM_k141_67667 | Not-determined |
| KM_k141_11818 | Low-quality |
| KM_k141_123184 | Low-quality |
| KM_k141_234010 | Not-determined |
| KM_k141_45055 | Not-determined |
| KM_k141_234011 | Not-determined |
| KM_k141_34173 | Not-determined |
| KM_k141_67675 | Not-determined |
| KM_k141_178500 | Not-determined |
| KM_k141_67678 | Not-determined |
| KM_k141_123189 | Not-determined |
| KM_k141_234017 | Not-determined |
| KM_k141_256450 | Not-determined |
| KM_k141_178509 | Not-determined |
| KM_k141_212171 | Not-determined |
| KM_k141_178514 | Not-determined |
| KM_k141_56341 | Low-quality |
| KM_k141_123196 | Not-determined |
| KM_k141_145460 | Not-determined |
| KM_k141_89634 | Not-determined |
| KM_k141_123202 | Not-determined |
| KM_k141_89636 | Not-determined |
| KM_k141_223203 | Low-quality |
| KM_k141_234020 | Not-determined |
| KM_k141_89647 | Not-determined |
| KM_k141_189721 | Not-determined |
| KM_k141_56355 | Not-determined |
| KM_k141_156359 | Not-determined |
| KM_k141_223214 | Not-determined |
| KM_k141_123207 | Not-determined |
| KM_k141_67685 | Not-determined |
| KM_k141_89659 | Not-determined |
| KM_k141_256456 | Low-quality |
| KM_k141_156364 | Not-determined |
| KM_k141_67692 | Not-determined |
| KM_k141_56361 | Not-determined |
| KM_k141_245364 | Not-determined |
| KM_k141_245368 | Not-determined |
| KM_k141_45085 | Low-quality |
| KM_k141_178539 | Not-determined |
| KM_k141_256459 | Low-quality |
| KM_k141_234036 | Not-determined |
| KM_k141_167375 | Low-quality |
| KM_k141_145490 | Low-quality |
| KM_k141_245371 | Not-determined |
| KM_k141_234045 | Not-determined |
| KM_k141_234048 | Not-determined |
| KM_k141_833 | Not-determined |
| KM_k141_167380 | Not-determined |
| KM_k141_134170 | Not-determined |
| KM_k141_100672 | Not-determined |
| KM_k141_100673 | Not-determined |
| KM_k141_223228 | Not-determined |
| KM_k141_34184 | Not-determined |
| KM_k141_145502 | Not-determined |
| KM_k141_178565 | Not-determined |
| KM_k141_78862 | Low-quality |
| KM_k141_145506 | Not-determined |
| KM_k141_839 | Not-determined |
| KM_k141_178574 | Not-determined |
| KM_k141_178577 | Not-determined |
| KM_k141_223240 | Not-determined |
| KM_k141_223243 | Not-determined |
| KM_k141_156390 | Not-determined |
| KM_k141_167389 | Not-determined |
| KM_k141_178583 | Not-determined |
| KM_k141_145521 | Not-determined |
| KM_k141_67715 | Not-determined |
| KM_k141_11874 | Not-determined |
| KM_k141_167399 | Not-determined |
| KM_k141_178599 | Not-determined |
| KM_k141_189778 | Not-determined |
| KM_k141_223249 | Not-determined |
| KM_k141_78891 | Not-determined |
| KM_k141_234063 | Not-determined |
| KM_k141_245396 | Not-determined |
| KM_k141_256484 | Not-determined |
| KM_k141_145529 | Not-determined |
| KM_k141_56414 | Not-determined |
| KM_k141_56416 | Not-determined |
| KM_k141_256498 | Not-determined |
| KM_k141_256504 | Not-determined |
| KM_k141_78908 | Not-determined |
| KM_k141_201133 | Not-determined |
| KM_k141_201137 | Not-determined |
| KM_k141_34220 | Not-determined |
| KM_k141_78917 | Not-determined |
| KM_k141_223275 | Not-determined |
| KM_k141_223277 | Not-determined |
| KM_k141_11899 | Not-determined |
| KM_k141_78926 | Not-determined |
| KM_k141_34225 | Not-determined |
| KM_k141_234087 | Not-determined |
| KM_k141_234088 | Not-determined |
| KM_k141_89680 | Not-determined |
| KM_k141_234094 | Not-determined |
| KM_k141_78948 | Not-determined |
| KM_k141_167445 | Not-determined |
| KM_k141_167446 | Not-determined |
| KM_k141_234100 | Not-determined |
| KM_k141_167451 | Not-determined |
| KM_k141_22847 | Not-determined |
| KM_k141_167453 | Not-determined |
| KM_k141_34228 | Not-determined |
| KM_k141_178634 | Not-determined |
| KM_k141_34232 | Not-determined |
| KM_k141_56446 | Not-determined |

| | |
|---|---|
| KM_k141_56448 | Not-determined |
| KM_k141_89698 | Not-determined |
| KM_k141_34239 | Not-determined |
| KM_k141_123265 | Not-determined |
| KM_k141_167462 | Not-determined |
| KM_k141_67777 | Not-determined |
| KM_k141_11912 | Not-determined |
| KM_k141_145551 | Low-quality |
| KM_k141_67780 | Not-determined |
| KM_k141_201153 | Not-determined |
| KM_k141_167469 | Not-determined |
| KM_k141_78968 | Not-determined |
| KM_k141_145558 | Not-determined |
| KM_k141_22857 | Not-determined |
| KM_k141_100724 | Not-determined |
| KM_k141_234108 | Not-determined |
| KM_k141_156441 | Not-determined |
| KM_k141_67800 | Not-determined |
| KM_k141_112183 | Not-determined |
| KM_k141_56463 | Not-determined |
| KM_k141_234112 | Not-determined |
| KM_k141_78976 | Not-determined |
| KM_k141_178654 | Not-determined |
| KM_k141_156443 | Not-determined |
| KM_k141_22871 | Not-determined |
| KM_k141_100730 | Not-determined |
| KM_k141_145585 | Not-determined |
| KM_k141_223297 | Not-determined |
| KM_k141_145588 | Not-determined |
| KM_k141_245411 | Not-determined |
| KM_k141_201171 | Not-determined |
| KM_k141_67818 | Low-quality |
| KM_k141_223306 | Not-determined |
| KM_k141_245414 | Not-determined |
| KM_k141_78989 | Not-determined |
| KM_k141_256535 | Not-determined |
| KM_k141_56482 | Not-determined |
| KM_k141_201186 | Not-determined |
| KM_k141_22893 | Not-determined |
| KM_k141_156472 | Not-determined |
| KM_k141_178672 | Not-determined |
| KM_k141_78999 | Not-determined |
| KM_k141_112224 | Not-determined |
| KM_k141_201190 | Not-determined |
| KM_k141_876 | Not-determined |
| KM_k141_178676 | Not-determined |
| KM_k141_100741 | Not-determined |
| KM_k141_245421 | Not-determined |
| KM_k141_79005 | Not-determined |
| KM_k141_100746 | Not-determined |
| KM_k141_178682 | Not-determined |
| KM_k141_178684 | Not-determined |
| KM_k141_112235 | Low-quality |
| KM_k141_245424 | Not-determined |
| KM_k141_11935 | Low-quality |
| KM_k141_112240 | Not-determined |
| KM_k141_223331 | Not-determined |
| KM_k141_123269 | Not-determined |
| KM_k141_22904 | Not-determined |
| KM_k141_100755 | Not-determined |
| KM_k141_56495 | Not-determined |
| KM_k141_156493 | Not-determined |
| KM_k141_178692 | Not-determined |
| KM_k141_223338 | Not-determined |
| KM_k141_11947 | Not-determined |
| KM_k141_11949 | Not-determined |
| KM_k141_245434 | Not-determined |
| KM_k141_56504 | Not-determined |
| KM_k141_45112 | Not-determined |
| KM_k141_123286 | Not-determined |
| KM_k141_245439 | Not-determined |
| KM_k141_245441 | Not-determined |
| KM_k141_201204 | Not-determined |
| KM_k141_178720 | Not-determined |
| KM_k141_189808 | Not-determined |
| KM_k141_112249 | Not-determined |
| KM_k141_45130 | Not-determined |
| KM_k141_178727 | Not-determined |
| KM_k141_178731 | Not-determined |
| KM_k141_123315 | Not-determined |
| KM_k141_112265 | Not-determined |
| KM_k141_223360 | Not-determined |
| KM_k141_112267 | Not-determined |
| KM_k141_256560 | Not-determined |
| KM_k141_56518 | Not-determined |
| KM_k141_79048 | Not-determined |
| KM_k141_212253 | Not-determined |
| KM_k141_22942 | Low-quality |
| KM_k141_256568 | Not-determined |
| KM_k141_79052 | Not-determined |
| KM_k141_223374 | Not-determined |
| KM_k141_223375 | Not-determined |
| KM_k141_145617 | Low-quality |
| KM_k141_79059 | Not-determined |
| KM_k141_189824 | Not-determined |
| KM_k141_256584 | Not-determined |
| KM_k141_123321 | Not-determined |
| KM_k141_223387 | Not-determined |
| KM_k141_245460 | Not-determined |
| KM_k141_178739 | Not-determined |
| KM_k141_11989 | Not-determined |
| KM_k141_22953 | Not-determined |
| KM_k141_234147 | Not-determined |
| KM_k141_112294 | Low-quality |
| KM_k141_922 | Low-quality |
| KM_k141_123334 | Not-determined |
| KM_k141_123337 | Not-determined |
| KM_k141_925 | Not-determined |
| KM_k141_189851 | Not-determined |
| KM_k141_234158 | Not-determined |
| KM_k141_134176 | Not-determined |
| KM_k141_56566 | Not-determined |
| KM_k141_933 | Low-quality |
| KM_k141_112295 | Low-quality |
| KM_k141_134191 | Not-determined |
| KM_k141_178761 | Not-determined |
| KM_k141_134194 | Not-determined |
| KM_k141_189859 | Not-determined |
| KM_k141_100778 | Not-determined |
| KM_k141_167540 | Low-quality |
| KM_k141_123355 | Low-quality |
| KM_k141_56574 | Not-determined |
| KM_k141_56576 | Not-determined |
| KM_k141_56581 | Not-determined |
| KM_k141_79078 | Not-determined |
| KM_k141_245500 | Not-determined |
| KM_k141_79082 | Not-determined |
| KM_k141_223422 | Not-determined |
| KM_k141_156552 | Not-determined |
| KM_k141_234171 | Not-determined |
| KM_k141_100792 | Not-determined |
| KM_k141_223427 | Not-determined |
| KM_k141_223432 | Not-determined |
| KM_k141_201254 | Not-determined |
| KM_k141_223433 | Low-quality |
| KM_k141_156556 | Not-determined |
| KM_k141_949 | Not-determined |
| KM_k141_112305 | Not-determined |
| KM_k141_79090 | Not-determined |
| KM_k141_245514 | Not-determined |
| KM_k141_79091 | Not-determined |
| KM_k141_201259 | Not-determined |
| KM_k141_123367 | Not-determined |
| KM_k141_201261 | Not-determined |
| KM_k141_156562 | Not-determined |
| KM_k141_245522 | Not-determined |
| KM_k141_145634 | Not-determined |
| KM_k141_201271 | Not-determined |
| KM_k141_965 | Not-determined |
| KM_k141_245526 | Not-determined |
| KM_k141_145640 | Not-determined |
| KM_k141_134236 | Not-determined |
| KM_k141_79106 | Not-determined |
| KM_k141_79108 | Not-determined |
| KM_k141_112325 | Not-determined |
| KM_k141_201276 | Not-determined |
| KM_k141_223444 | Not-determined |
| KM_k141_223446 | Not-determined |
| KM_k141_134249 | Not-determined |
| KM_k141_56626 | Low-quality |
| KM_k141_256621 | Not-determined |
| KM_k141_11995 | Not-determined |
| KM_k141_145649 | Not-determined |
| KM_k141_201279 | Not-determined |
| KM_k141_134253 | Not-determined |
| KM_k141_45145 | Not-determined |
| KM_k141_56627 | Not-determined |
| KM_k141_45148 | Not-determined |
| KM_k141_234204 | Not-determined |
| KM_k141_156594 | Not-determined |
| KM_k141_123387 | Not-determined |
| KM_k141_123390 | Low-quality |
| KM_k141_167547 | Not-determined |
| KM_k141_167548 | Not-determined |
| KM_k141_989 | Not-determined |
| KM_k141_134262 | Not-determined |
| KM_k141_79124 | Not-determined |
| KM_k141_156602 | Not-determined |
| KM_k141_56641 | Not-determined |
| KM_k141_156605 | Not-determined |
| KM_k141_34357 | Not-determined |
| KM_k141_79133 | Not-determined |
| KM_k141_156607 | Not-determined |
| KM_k141_1004 | Not-determined |
| KM_k141_223459 | Not-determined |
| KM_k141_112349 | Low-quality |
| KM_k141_212339 | Not-determined |
| KM_k141_1017 | Not-determined |
| KM_k141_1019 | Not-determined |
| KM_k141_245539 | Not-determined |
| KM_k141_100820 | Low-quality |
| KM_k141_79148 | Not-determined |
| KM_k141_156626 | Not-determined |
| KM_k141_100824 | Not-determined |
| KM_k141_12032 | Low-quality |
| KM_k141_167586 | Low-quality |
| KM_k141_167589 | Not-determined |
| KM_k141_201309 | Not-determined |
| KM_k141_212349 | Not-determined |
| KM_k141_123419 | Not-determined |
| KM_k141_223471 | Not-determined |
| KM_k141_89734 | Not-determined |
| KM_k141_223475 | Not-determined |
| KM_k141_167603 | Low-quality |
| KM_k141_178771 | Not-determined |
| KM_k141_56644 | Not-determined |
| KM_k141_12044 | Not-determined |
| KM_k141_189868 | Not-determined |
| KM_k141_123439 | Not-determined |
| KM_k141_212366 | Not-determined |
| KM_k141_234235 | Not-determined |
| KM_k141_89758 | Not-determined |
| KM_k141_167615 | Not-determined |
| KM_k141_79170 | Not-determined |
| KM_k141_167624 | Not-determined |
| KM_k141_145667 | Not-determined |
| KM_k141_112373 | Not-determined |
| KM_k141_212368 | Not-determined |
| KM_k141_145671 | Not-determined |
| KM_k141_79177 | Not-determined |
| KM_k141_167628 | Not-determined |
| KM_k141_189891 | Not-determined |
| KM_k141_189894 | Not-determined |
| KM_k141_79187 | Not-determined |
| KM_k141_100863 | Not-determined |
| KM_k141_167629 | Not-determined |
| KM_k141_234239 | Not-determined |
| KM_k141_234241 | Not-determined |
| KM_k141_1033 | Not-determined |
| KM_k141_167635 | Not-determined |
| KM_k141_234242 | Not-determined |
| KM_k141_234243 | Not-determined |
| KM_k141_34418 | Not-determined |
| KM_k141_178829 | Not-determined |
| KM_k141_134305 | Not-determined |
| KM_k141_123469 | Not-determined |
| KM_k141_34425 | Not-determined |
| KM_k141_167650 | Not-determined |
| KM_k141_178844 | Not-determined |
| KM_k141_1062 | Not-determined |
| KM_k141_234264 | Not-determined |
| KM_k141_23067 | Not-determined |
| KM_k141_45194 | Not-determined |
| KM_k141_189908 | Not-determined |
| KM_k141_234275 | Not-determined |
| KM_k141_112413 | Not-determined |
| KM_k141_234278 | Not-determined |
| KM_k141_100905 | Not-determined |
| KM_k141_234286 | Not-determined |
| KM_k141_234287 | Not-determined |
| KM_k141_256638 | Not-determined |
| KM_k141_134334 | Not-determined |
| KM_k141_56694 | Not-determined |
| KM_k141_23087 | Low-quality |
| KM_k141_256644 | Not-determined |
| KM_k141_189925 | Not-determined |
| KM_k141_167682 | Not-determined |
| KM_k141_56699 | Not-determined |
| KM_k141_178875 | Not-determined |
| KM_k141_234310 | Not-determined |
| KM_k141_256657 | Not-determined |
| KM_k141_123518 | Not-determined |
| KM_k141_167693 | Not-determined |
| KM_k141_234316 | Not-determined |
| KM_k141_34446 | Not-determined |
| KM_k141_23103 | Not-determined |
| KM_k141_234318 | Not-determined |
| KM_k141_134349 | Not-determined |
| KM_k141_234324 | Not-determined |
| KM_k141_234325 | Not-determined |
| KM_k141_34453 | Not-determined |
| KM_k141_256667 | Not-determined |
| KM_k141_45217 | Not-determined |
| KM_k141_145684 | Not-determined |
| KM_k141_256674 | Not-determined |
| KM_k141_256675 | Not-determined |
| KM_k141_256676 | Not-determined |
| KM_k141_67835 | Not-determined |
| KM_k141_1097 | Not-determined |
| KM_k141_12071 | Not-determined |
| KM_k141_56722 | Not-determined |
| KM_k141_134374 | Not-determined |
| KM_k141_12075 | Not-determined |
| KM_k141_234333 | Not-determined |
| KM_k141_223560 | Low-quality |
| KM_k141_223563 | Not-determined |
| KM_k141_234336 | Not-determined |
| KM_k141_45236 | Not-determined |
| KM_k141_245556 | Not-determined |
| KM_k141_223578 | Not-determined |
| KM_k141_256684 | Not-determined |
| KM_k141_145694 | Not-determined |
| KM_k141_245559 | Not-determined |
| KM_k141_256687 | Not-determined |
| KM_k141_123532 | Not-determined |
| KM_k141_178926 | Not-determined |
| KM_k141_123534 | Not-determined |
| KM_k141_212385 | Not-determined |
| KM_k141_145701 | Low-quality |
| KM_k141_112442 | Not-determined |
| KM_k141_1116 | Not-determined |
| KM_k141_178937 | Not-determined |
| KM_k141_45250 | Low-quality |
| KM_k141_1120 | Not-determined |
| KM_k141_201379 | Not-determined |
| KM_k141_212392 | Not-determined |
| KM_k141_79240 | Not-determined |
| KM_k141_145717 | Low-quality |
| KM_k141_201382 | Not-determined |
| KM_k141_145718 | Not-determined |
| KM_k141_178953 | Not-determined |
| KM_k141_112451 | Not-determined |
| KM_k141_67875 | Not-determined |
| KM_k141_145724 | Not-determined |
| KM_k141_212401 | Not-determined |
| KM_k141_112457 | Not-determined |
| KM_k141_201389 | Low-quality |
| KM_k141_45271 | Not-determined |
| KM_k141_156637 | Not-determined |
| KM_k141_212411 | Not-determined |
| KM_k141_178963 | Not-determined |
| KM_k141_67885 | Not-determined |
| KM_k141_145727 | Not-determined |
| KM_k141_178966 | Not-determined |
| KM_k141_45275 | Not-determined |
| KM_k141_112468 | Not-determined |
| KM_k141_112470 | Not-determined |
| KM_k141_67892 | Not-determined |
| KM_k141_256694 | Low-quality |
| KM_k141_89788 | Not-determined |
| KM_k141_234349 | Low-quality |
| KM_k141_56731 | Low-quality |
| KM_k141_67902 | Not-determined |
| KM_k141_67903 | Not-determined |
| KM_k141_145742 | Not-determined |
| KM_k141_212424 | Not-determined |
| KM_k141_234371 | Not-determined |
| KM_k141_201416 | Not-determined |
| KM_k141_201417 | Not-determined |
| KM_k141_67907 | Not-determined |
| KM_k141_112478 | Not-determined |
| KM_k141_234377 | Not-determined |
| KM_k141_178994 | Not-determined |
| KM_k141_167762 | Not-determined |
| KM_k141_201425 | Low-quality |
| KM_k141_223636 | Not-determined |
| KM_k141_245595 | Not-determined |
| KM_k141_179003 | Not-determined |
| KM_k141_45302 | Not-determined |
| KM_k141_201427 | Not-determined |
| KM_k141_145750 | Not-determined |
| KM_k141_179007 | Not-determined |
| KM_k141_112482 | Not-determined |
| KM_k141_156648 | Not-determined |
| KM_k141_89795 | Not-determined |
| KM_k141_179014 | Not-determined |
| KM_k141_245604 | Low-quality |
| KM_k141_89802 | Not-determined |
| KM_k141_167769 | Not-determined |
| KM_k141_123549 | Low-quality |
| KM_k141_34486 | Not-determined |
| KM_k141_212442 | Low-quality |
| KM_k141_145769 | Not-determined |
| KM_k141_179028 | Not-determined |
| KM_k141_67927 | Low-quality |
| KM_k141_34495 | Not-determined |
| KM_k141_34496 | Not-determined |
| KM_k141_156650 | Low-quality |
| KM_k141_23155 | Not-determined |
| KM_k141_179041 | Not-determined |
| KM_k141_45322 | Not-determined |
| KM_k141_67948 | Not-determined |
| KM_k141_67953 | Low-quality |
| KM_k141_179050 | Not-determined |
| KM_k141_67959 | Not-determined |
| KM_k141_12138 | Not-determined |
| KM_k141_89816 | Not-determined |
| KM_k141_234408 | Not-determined |
| KM_k141_245620 | Not-determined |
| KM_k141_1179 | Not-determined |
| KM_k141_12143 | Not-determined |
| KM_k141_79304 | Not-determined |
| KM_k141_245627 | Not-determined |
| KM_k141_256709 | Not-determined |
| KM_k141_45334 | Not-determined |
| KM_k141_212459 | Not-determined |
| KM_k141_245635 | Not-determined |
| KM_k141_12151 | Not-determined |
| KM_k141_212461 | Low-quality |
| KM_k141_89829 | Not-determined |
| KM_k141_79315 | Not-determined |
| KM_k141_79316 | Not-determined |
| KM_k141_145793 | Not-determined |
| KM_k141_134429 | Low-quality |
| KM_k141_100941 | Not-determined |
| KM_k141_100942 | Not-determined |
| KM_k141_245646 | Not-determined |
| KM_k141_179061 | Low-quality |
| KM_k141_234426 | Not-determined |
| KM_k141_189960 | Not-determined |
| KM_k141_145798 | Not-determined |
| KM_k141_179063 | Low-quality |
| KM_k141_79326 | Not-determined |
| KM_k141_23181 | Not-determined |
| KM_k141_179066 | Not-determined |
| KM_k141_145803 | Not-determined |
| KM_k141_201433 | Not-determined |
| KM_k141_134432 | Not-determined |
| KM_k141_134434 | Not-determined |
| KM_k141_256725 | Not-determined |
| KM_k141_201436 | Not-determined |
| KM_k141_145805 | Not-determined |
| KM_k141_189976 | Not-determined |
| KM_k141_89840 | Not-determined |
| KM_k141_201442 | Not-determined |
| KM_k141_45353 | Not-determined |
| KM_k141_100952 | Not-determined |
| KM_k141_89846 | Not-determined |
| KM_k141_223703 | Not-determined |
| KM_k141_223704 | Not-determined |
| KM_k141_100954 | Not-determined |
| KM_k141_1209 | Not-determined |
| KM_k141_1210 | Not-determined |
| KM_k141_89849 | Not-determined |
| KM_k141_34544 | Not-determined |
| KM_k141_145814 | Not-determined |
| KM_k141_89854 | Low-quality |
| KM_k141_123585 | Not-determined |
| KM_k141_245655 | Not-determined |
| KM_k141_223720 | Not-determined |
| KM_k141_79346 | Low-quality |
| KM_k141_134455 | Not-determined |
| KM_k141_223722 | Not-determined |
| KM_k141_134458 | Not-determined |
| KM_k141_245658 | Not-determined |
| KM_k141_68003 | Not-determined |
| KM_k141_68004 | Not-determined |
| KM_k141_68012 | Not-determined |
| KM_k141_1234 | Not-determined |
| KM_k141_68015 | Not-determined |
| KM_k141_256740 | Not-determined |
| KM_k141_100969 | Low-quality |
| KM_k141_68021 | Low-quality |
| KM_k141_134482 | Not-determined |
| KM_k141_12202 | Not-determined |
| KM_k141_190003 | Not-determined |
| KM_k141_201474 | Low-quality |
| KM_k141_68031 | Not-determined |
| KM_k141_100976 | Not-determined |
| KM_k141_223731 | Not-determined |
| KM_k141_68040 | Not-determined |
| KM_k141_223733 | Low-quality |
| KM_k141_134497 | Not-determined |
| KM_k141_201488 | Not-determined |
| KM_k141_156660 | Not-determined |
| KM_k141_100985 | Not-determined |
| KM_k141_134500 | Not-determined |
| KM_k141_100994 | Not-determined |
| KM_k141_212528 | Not-determined |
| KM_k141_190021 | Not-determined |
| KM_k141_201500 | Low-quality |
| KM_k141_234456 | Not-determined |
| KM_k141_68062 | Not-determined |
| KM_k141_68063 | Not-determined |
| KM_k141_145833 | Not-determined |
| KM_k141_79371 | Not-determined |
| KM_k141_12213 | Not-determined |
| KM_k141_123613 | Not-determined |
| KM_k141_123614 | Not-determined |
| KM_k141_212540 | Not-determined |
| KM_k141_234459 | Not-determined |
| KM_k141_234460 | Not-determined |
| KM_k141_79379 | Not-determined |
| KM_k141_212543 | Not-determined |
| KM_k141_101002 | Not-determined |
| KM_k141_79387 | Not-determined |
| KM_k141_123629 | Not-determined |
| KM_k141_89901 | Not-determined |
| KM_k141_68079 | Not-determined |
| KM_k141_1243 | Not-determined |
| KM_k141_201521 | Not-determined |
| KM_k141_134534 | Not-determined |
| KM_k141_101012 | Not-determined |
| KM_k141_234465 | Low-quality |
| KM_k141_112489 | Not-determined |
| KM_k141_101019 | Not-determined |
| KM_k141_101020 | Not-determined |
| KM_k141_134543 | Not-determined |
| KM_k141_134544 | Low-quality |
| KM_k141_234472 | Not-determined |
| KM_k141_145845 | Not-determined |
| KM_k141_167788 | Not-determined |
| KM_k141_1257 | Not-determined |
| KM_k141_212569 | Not-determined |
| KM_k141_112505 | Not-determined |
| KM_k141_112506 | Not-determined |
| KM_k141_145849 | Not-determined |
| KM_k141_190043 | Not-determined |
| KM_k141_112510 | Not-determined |
| KM_k141_23246 | Not-determined |
| KM_k141_123638 | Low-quality |
| KM_k141_123640 | Not-determined |
| KM_k141_79396 | Not-determined |
| KM_k141_179110 | Not-determined |
| KM_k141_145862 | Not-determined |
| KM_k141_179115 | Low-quality |
| KM_k141_179118 | Not-determined |
| KM_k141_112520 | Not-determined |
| KM_k141_179119 | Not-determined |
| KM_k141_68085 | Not-determined |
| KM_k141_68086 | Low-quality |
| KM_k141_167814 | Not-determined |
| KM_k141_212596 | Not-determined |
| KM_k141_79404 | Not-determined |
| KM_k141_112526 | Not-determined |
| KM_k141_223760 | Not-determined |
| KM_k141_190073 | Not-determined |
| KM_k141_89920 | Not-determined |
| KM_k141_167827 | Not-determined |
| KM_k141_190079 | Not-determined |
| KM_k141_89922 | Low-quality |
| KM_k141_123652 | Not-determined |
| KM_k141_101052 | Not-determined |
| KM_k141_101054 | Not-determined |
| KM_k141_23267 | Not-determined |
| KM_k141_179129 | Low-quality |
| KM_k141_167837 | Not-determined |
| KM_k141_145886 | Not-determined |
| KM_k141_89940 | Not-determined |
| KM_k141_167842 | Not-determined |
| KM_k141_223772 | Not-determined |
| KM_k141_101071 | Low-quality |
| KM_k141_245667 | Not-determined |
| KM_k141_101075 | Not-determined |
| KM_k141_156688 | Not-determined |
| KM_k141_1297 | Not-determined |
| KM_k141_167850 | Not-determined |
| KM_k141_156692 | Not-determined |
| KM_k141_12249 | Not-determined |
| KM_k141_123686 | Not-determined |
| KM_k141_23300 | Not-determined |
| KM_k141_79449 | Not-determined |
| KM_k141_123689 | Not-determined |
| KM_k141_179136 | Not-determined |
| KM_k141_156696 | Not-determined |
| KM_k141_179139 | Not-determined |
| KM_k141_79456 | Not-determined |
| KM_k141_245683 | Not-determined |
| KM_k141_201551 | Not-determined |
| KM_k141_156699 | Not-determined |
| KM_k141_68127 | Not-determined |
| KM_k141_1304 | Not-determined |
| KM_k141_145897 | Low-quality |
| KM_k141_179148 | Not-determined |
| KM_k141_23309 | Not-determined |
| KM_k141_68132 | Low-quality |
| KM_k141_79470 | Not-determined |
| KM_k141_134553 | Not-determined |
| KM_k141_123705 | Low-quality |
| KM_k141_256802 | Not-determined |
| KM_k141_123712 | Low-quality |
| KM_k141_145911 | Not-determined |
| KM_k141_245695 | Not-determined |
| KM_k141_256808 | Not-determined |
| KM_k141_156712 | Not-determined |
| KM_k141_79478 | Low-quality |
| KM_k141_134567 | Not-determined |
| KM_k141_223783 | Not-determined |
| KM_k141_134572 | Not-determined |
| KM_k141_56790 | Not-determined |
| KM_k141_256815 | Not-determined |
| KM_k141_56797 | Not-determined |
| KM_k141_123736 | Not-determined |
| KM_k141_134575 | Not-determined |
| KM_k141_245707 | Not-determined |
| KM_k141_245708 | Not-determined |
| KM_k141_89958 | Not-determined |
| KM_k141_256819 | Not-determined |
| KM_k141_245711 | Low-quality |
| KM_k141_89963 | Not-determined |
| KM_k141_56809 | Not-determined |
| KM_k141_56810 | Not-determined |
| KM_k141_201586 | Not-determined |
| KM_k141_245717 | Not-determined |
| KM_k141_245724 | Not-determined |
| KM_k141_223801 | Not-determined |
| KM_k141_56818 | Not-determined |
| KM_k141_245731 | Not-determined |
| KM_k141_256836 | Not-determined |

| | |
|---|---|
| KM_k141_201595 | Not-determined |
| KM_k141_201597 | Not-determined |
| KM_k141_156739 | Not-determined |
| KM_k141_201600 | Not-determined |
| KM_k141_101098 | Not-determined |
| KM_k141_156744 | Not-determined |
| KM_k141_212634 | Not-determined |
| KM_k141_79511 | Not-determined |
| KM_k141_245735 | Not-determined |
| KM_k141_101101 | Not-determined |
| KM_k141_101102 | Not-determined |
| KM_k141_123759 | Not-determined |
| KM_k141_201613 | Not-determined |
| KM_k141_145931 | Not-determined |
| KM_k141_112560 | Not-determined |
| KM_k141_212655 | Not-determined |
| KM_k141_223811 | Not-determined |
| KM_k141_201624 | Not-determined |
| KM_k141_89987 | Not-determined |
| KM_k141_145947 | Not-determined |
| KM_k141_223818 | Not-determined |
| KM_k141_190142 | Not-determined |
| KM_k141_145952 | Not-determined |
| KM_k141_223822 | Not-determined |
| KM_k141_190150 | Not-determined |
| KM_k141_89997 | Not-determined |
| KM_k141_201639 | Not-determined |
| KM_k141_56826 | Not-determined |
| KM_k141_56829 | Not-determined |
| KM_k141_245760 | Not-determined |
| KM_k141_190157 | Not-determined |
| KM_k141_190165 | Not-determined |
| KM_k141_223836 | Not-determined |
| KM_k141_212675 | Not-determined |
| KM_k141_256852 | Not-determined |
| KM_k141_112591 | Not-determined |
| KM_k141_223837 | Not-determined |
| KM_k141_212686 | Not-determined |
| KM_k141_68227 | Not-determined |
| KM_k141_123782 | Not-determined |
| KM_k141_212690 | Not-determined |
| KM_k141_101133 | Not-determined |
| KM_k141_68236 | Not-determined |
| KM_k141_68237 | Not-determined |
| KM_k141_68240 | Not-determined |
| KM_k141_45411 | Not-determined |
| KM_k141_1337 | Low-quality |
| KM_k141_68246 | Not-determined |
| KM_k141_1340 | Not-determined |
| KM_k141_90015 | Not-determined |
| KM_k141_223859 | Not-determined |
| KM_k141_56848 | Not-determined |
| KM_k141_245795 | Low-quality |
| KM_k141_45416 | Not-determined |
| KM_k141_23315 | Not-determined |
| KM_k141_90022 | Not-determined |
| KM_k141_1342 | Not-determined |
| KM_k141_12259 | Not-determined |
| KM_k141_245801 | Not-determined |
| KM_k141_123816 | Not-determined |
| KM_k141_190207 | Not-determined |
| KM_k141_45422 | Not-determined |
| KM_k141_201658 | Not-determined |
| KM_k141_256874 | Not-determined |
| KM_k141_123820 | Not-determined |
| KM_k141_134655 | Not-determined |
| KM_k141_167878 | Not-determined |
| KM_k141_90028 | Low-quality |
| KM_k141_34635 | Not-determined |
| KM_k141_90029 | Not-determined |
| KM_k141_245806 | Not-determined |
| KM_k141_79548 | Not-determined |
| KM_k141_34641 | Not-determined |
| KM_k141_212707 | Not-determined |
| KM_k141_79552 | Not-determined |
| KM_k141_223871 | Not-determined |
| KM_k141_167889 | Not-determined |
| KM_k141_23332 | Not-determined |
| KM_k141_212711 | Low-quality |
| KM_k141_223872 | Not-determined |
| KM_k141_201670 | Not-determined |
| KM_k141_167892 | Not-determined |
| KM_k141_245815 | Not-determined |
| KM_k141_45439 | Not-determined |
| KM_k141_245819 | Not-determined |
| KM_k141_79566 | Not-determined |
| KM_k141_256883 | Not-determined |
| KM_k141_201681 | Not-determined |
| KM_k141_134680 | Not-determined |
| KM_k141_34663 | Not-determined |
| KM_k141_23344 | Not-determined |
| KM_k141_256892 | Not-determined |
| KM_k141_212731 | Not-determined |
| KM_k141_201691 | Not-determined |
| KM_k141_23347 | Not-determined |
| KM_k141_123872 | Not-determined |
| KM_k141_223885 | Not-determined |
| KM_k141_201704 | Not-determined |
| KM_k141_1371 | Not-determined |
| KM_k141_201710 | Not-determined |
| KM_k141_201712 | Not-determined |
| KM_k141_134688 | Not-determined |
| KM_k141_101181 | Not-determined |
| KM_k141_79592 | Not-determined |
| KM_k141_79597 | Not-determined |
| KM_k141_34690 | Not-determined |
| KM_k141_256902 | Not-determined |
| KM_k141_1385 | Not-determined |
| KM_k141_123883 | Not-determined |
| KM_k141_34696 | Not-determined |
| KM_k141_156831 | Not-determined |
| KM_k141_223901 | Not-determined |
| KM_k141_156833 | Not-determined |
| KM_k141_79609 | Not-determined |
| KM_k141_112614 | Not-determined |
| KM_k141_101186 | Not-determined |
| KM_k141_201724 | Not-determined |
| KM_k141_179236 | Not-determined |
| KM_k141_179237 | Not-determined |
| KM_k141_23353 | Not-determined |
| KM_k141_167921 | Low-quality |
| KM_k141_34703 | Not-determined |
| KM_k141_201729 | Not-determined |
| KM_k141_56888 | Low-quality |
| KM_k141_179244 | Not-determined |
| KM_k141_212785 | Not-determined |
| KM_k141_167925 | Not-determined |
| KM_k141_145968 | Not-determined |
| KM_k141_145969 | Not-determined |
| KM_k141_101194 | Not-determined |
| KM_k141_234510 | Not-determined |
| KM_k141_1400 | Not-determined |
| KM_k141_123894 | Not-determined |
| KM_k141_201733 | Not-determined |
| KM_k141_1401 | Not-determined |
| KM_k141_190252 | Not-determined |
| KM_k141_23364 | Not-determined |
| KM_k141_34720 | Not-determined |
| KM_k141_201741 | Not-determined |
| KM_k141_101201 | Not-determined |
| KM_k141_79623 | Not-determined |
| KM_k141_34726 | Not-determined |
| KM_k141_223915 | Not-determined |
| KM_k141_167938 | Not-determined |
| KM_k141_212795 | Not-determined |
| KM_k141_167939 | Not-determined |
| KM_k141_179275 | Not-determined |
| KM_k141_56900 | Not-determined |
| KM_k141_45475 | Not-determined |
| KM_k141_123913 | Not-determined |
| KM_k141_123914 | Not-determined |
| KM_k141_256927 | Not-determined |
| KM_k141_223928 | Not-determined |
| KM_k141_79635 | Low-quality |
| KM_k141_223933 | Not-determined |
| KM_k141_1431 | Low-quality |
| KM_k141_79643 | Low-quality |
| KM_k141_201768 | Not-determined |
| KM_k141_56912 | Not-determined |
| KM_k141_156883 | Not-determined |
| KM_k141_79649 | Not-determined |
| KM_k141_101205 | Low-quality |
| KM_k141_45507 | Low-quality |
| KM_k141_156888 | Not-determined |
| KM_k141_156890 | Not-determined |
| KM_k141_156891 | Not-determined |
| KM_k141_212817 | Not-determined |
| KM_k141_145988 | Not-determined |
| KM_k141_212823 | Not-determined |
| KM_k141_212827 | Not-determined |
| KM_k141_145990 | Low-quality |
| KM_k141_134739 | Low-quality |
| KM_k141_101229 | Not-determined |
| KM_k141_79675 | Not-determined |
| KM_k141_134744 | Not-determined |
| KM_k141_256949 | Low-quality |
| KM_k141_145996 | Low-quality |
| KM_k141_167963 | Not-determined |
| KM_k141_56938 | Not-determined |
| KM_k141_256957 | Not-determined |
| KM_k141_12274 | Not-determined |
| KM_k141_79694 | Low-quality |
| KM_k141_12276 | Not-determined |
| KM_k141_101234 | Not-determined |
| KM_k141_12283 | Low-quality |
| KM_k141_190292 | Not-determined |
| KM_k141_212845 | Not-determined |
| KM_k141_112644 | Low-quality |
| KM_k141_79703 | Not-determined |
| KM_k141_156897 | Not-determined |
| KM_k141_45515 | Not-determined |
| KM_k141_45517 | Not-determined |
| KM_k141_201820 | Low-quality |
| KM_k141_234577 | Not-determined |
| KM_k141_223960 | Low-quality |
| KM_k141_256986 | Not-determined |
| KM_k141_256990 | Not-determined |
| KM_k141_256994 | Not-determined |
| KM_k141_101246 | Not-determined |
| KM_k141_101249 | Not-determined |
| KM_k141_256997 | Not-determined |
| KM_k141_223977 | Low-quality |
| KM_k141_190313 | Not-determined |
| KM_k141_12317 | Not-determined |
| KM_k141_257003 | Not-determined |
| KM_k141_68295 | Not-determined |
| KM_k141_234610 | Not-determined |
| KM_k141_23377 | Not-determined |
| KM_k141_167992 | Not-determined |
| KM_k141_201825 | Low-quality |
| KM_k141_12321 | Not-determined |
| KM_k141_56956 | Not-determined |
| KM_k141_212861 | Not-determined |
| KM_k141_212862 | Not-determined |
| KM_k141_101258 | Not-determined |
| KM_k141_45554 | Not-determined |
| KM_k141_45556 | Not-determined |
| KM_k141_146034 | Not-determined |
| KM_k141_201828 | Not-determined |
| KM_k141_223993 | Not-determined |
| KM_k141_201833 | Not-determined |
| KM_k141_224001 | Not-determined |
| KM_k141_23381 | Not-determined |
| KM_k141_156940 | Not-determined |
| KM_k141_79713 | Low-quality |
| KM_k141_212871 | Not-determined |
| KM_k141_101270 | Not-determined |
| KM_k141_146044 | Not-determined |
| KM_k141_156954 | Not-determined |
| KM_k141_1470 | Not-determined |
| KM_k141_56963 | Not-determined |
| KM_k141_257018 | Low-quality |
| KM_k141_112704 | Not-determined |
| KM_k141_146048 | Not-determined |
| KM_k141_156966 | Not-determined |
| KM_k141_23409 | Not-determined |
| KM_k141_234624 | Not-determined |
| KM_k141_168014 | Not-determined |
| KM_k141_224031 | Not-determined |
| KM_k141_234631 | Not-determined |
| KM_k141_23422 | Not-determined |
| KM_k141_201842 | Not-determined |
| KM_k141_201843 | Not-determined |
| KM_k141_234634 | Not-determined |
| KM_k141_234635 | Not-determined |
| KM_k141_123948 | Not-determined |
| KM_k141_168028 | Not-determined |
| KM_k141_12347 | Not-determined |
| KM_k141_79755 | Not-determined |
| KM_k141_34782 | Not-determined |
| KM_k141_134806 | Not-determined |
| KM_k141_123959 | Not-determined |
| KM_k141_224040 | Low-quality |
| KM_k141_190341 | Not-determined |
| KM_k141_112738 | Not-determined |
| KM_k141_112740 | Not-determined |
| KM_k141_56992 | Not-determined |
| KM_k141_112746 | Not-determined |
| KM_k141_123973 | Not-determined |
| KM_k141_112753 | Not-determined |
| KM_k141_112757 | Not-determined |
| KM_k141_12366 | Not-determined |
| KM_k141_90088 | Not-determined |
| KM_k141_57005 | Not-determined |
| KM_k141_112772 | Not-determined |
| KM_k141_90093 | Not-determined |
| KM_k141_257071 | Not-determined |
| KM_k141_34807 | Not-determined |
| KM_k141_34808 | Not-determined |
| KM_k141_179286 | Not-determined |
| KM_k141_156991 | Low-quality |
| KM_k141_179288 | Not-determined |
| KM_k141_90097 | Not-determined |
| KM_k141_190366 | Not-determined |
| KM_k141_179295 | Not-determined |
| KM_k141_257080 | Not-determined |
| KM_k141_45597 | Not-determined |
| KM_k141_201864 | Not-determined |
| KM_k141_146056 | Not-determined |
| KM_k141_57016 | Not-determined |
| KM_k141_179299 | Not-determined |
| KM_k141_257090 | Low-quality |
| KM_k141_57022 | Not-determined |
| KM_k141_146064 | Low-quality |
| KM_k141_57025 | Not-determined |
| KM_k141_45606 | Not-determined |
| KM_k141_146075 | Not-determined |
| KM_k141_68332 | Not-determined |
| KM_k141_257097 | Not-determined |
| KM_k141_23474 | Not-determined |
| KM_k141_201889 | Not-determined |
| KM_k141_257104 | Not-determined |
| KM_k141_23479 | Not-determined |
| KM_k141_157014 | Not-determined |
| KM_k141_79758 | Not-determined |
| KM_k141_257116 | Not-determined |
| KM_k141_234652 | Low-quality |
| KM_k141_68352 | Not-determined |
| KM_k141_79762 | Not-determined |
| KM_k141_23486 | Not-determined |
| KM_k141_257119 | Not-determined |
| KM_k141_146092 | Not-determined |
| KM_k141_68358 | Not-determined |
| KM_k141_201902 | Not-determined |
| KM_k141_201903 | Not-determined |
| KM_k141_23497 | Low-quality |
| KM_k141_168099 | Not-determined |
| KM_k141_257123 | Not-determined |
| KM_k141_190410 | Not-determined |
| KM_k141_68367 | Not-determined |
| KM_k141_190413 | Not-determined |
| KM_k141_179326 | Not-determined |
| KM_k141_201910 | Not-determined |
| KM_k141_79784 | Not-determined |
| KM_k141_112814 | Not-determined |
| KM_k141_234669 | Low-quality |
| KM_k141_34829 | Not-determined |
| KM_k141_234672 | Not-determined |
| KM_k141_23507 | Not-determined |
| KM_k141_34831 | Not-determined |
| KM_k141_101313 | Not-determined |
| KM_k141_201920 | Not-determined |
| KM_k141_224073 | Not-determined |
| KM_k141_1520 | Not-determined |
| KM_k141_224074 | Not-determined |
| KM_k141_201924 | Not-determined |
| KM_k141_124017 | Not-determined |
| KM_k141_257137 | Not-determined |
| KM_k141_112830 | Not-determined |
| KM_k141_224083 | Not-determined |
| KM_k141_1526 | Not-determined |
| KM_k141_257144 | Not-determined |
| KM_k141_190426 | Not-determined |
| KM_k141_234683 | Not-determined |
| KM_k141_45640 | Low-quality |
| KM_k141_224088 | Not-determined |
| KM_k141_112839 | Not-determined |
| KM_k141_134855 | Not-determined |
| KM_k141_224098 | Not-determined |
| KM_k141_45648 | Low-quality |
| KM_k141_179349 | Not-determined |
| KM_k141_112847 | Not-determined |
| KM_k141_224107 | Not-determined |
| KM_k141_146125 | Not-determined |
| KM_k141_134864 | Not-determined |
| KM_k141_157036 | Low-quality |
| KM_k141_112858 | Not-determined |
| KM_k141_90144 | Not-determined |
| KM_k141_257150 | Not-determined |
| KM_k141_134866 | Not-determined |
| KM_k141_134869 | Not-determined |
| KM_k141_68416 | Not-determined |
| KM_k141_68418 | Not-determined |
| KM_k141_34856 | Not-determined |
| KM_k141_190456 | Low-quality |
| KM_k141_57083 | Not-determined |
| KM_k141_224136 | Not-determined |
| KM_k141_224138 | Not-determined |
| KM_k141_157060 | Not-determined |
| KM_k141_257166 | Not-determined |
| KM_k141_201933 | Not-determined |
| KM_k141_157063 | Not-determined |
| KM_k141_190463 | Not-determined |
| KM_k141_157066 | Not-determined |
| KM_k141_34864 | Not-determined |
| KM_k141_168130 | Not-determined |
| KM_k141_68420 | Not-determined |
| KM_k141_90164 | Not-determined |
| KM_k141_12406 | Not-determined |
| KM_k141_190474 | Not-determined |
| KM_k141_12408 | Not-determined |
| KM_k141_134901 | Low-quality |
| KM_k141_168143 | Not-determined |
| KM_k141_190478 | Not-determined |
| KM_k141_34873 | Not-determined |
| KM_k141_257189 | Not-determined |
| KM_k141_201955 | Not-determined |
| KM_k141_68436 | Not-determined |
| KM_k141_201959 | Not-determined |
| KM_k141_68437 | Not-determined |
| KM_k141_1569 | Not-determined |
| KM_k141_90178 | Not-determined |
| KM_k141_34876 | Not-determined |
| KM_k141_90182 | Not-determined |
| KM_k141_134914 | Not-determined |
| KM_k141_179378 | Not-determined |
| KM_k141_68454 | Not-determined |
| KM_k141_68458 | Not-determined |
| KM_k141_68462 | Not-determined |
| KM_k141_23534 | Not-determined |
| KM_k141_1596 | Not-determined |
| KM_k141_23537 | Low-quality |
| KM_k141_68471 | Not-determined |
| KM_k141_101334 | Not-determined |
| KM_k141_201983 | Not-determined |
| KM_k141_23543 | Not-determined |
| KM_k141_134920 | Not-determined |
| KM_k141_168183 | Not-determined |
| KM_k141_224167 | Not-determined |
| KM_k141_234735 | Not-determined |
| KM_k141_190520 | Not-determined |
| KM_k141_201994 | Not-determined |
| KM_k141_190522 | Not-determined |
| KM_k141_79829 | Not-determined |
| KM_k141_157099 | Not-determined |
| KM_k141_190523 | Not-determined |
| KM_k141_146138 | Not-determined |
| KM_k141_157104 | Not-determined |
| KM_k141_23550 | Not-determined |
| KM_k141_124100 | Not-determined |
| KM_k141_157108 | Not-determined |
| KM_k141_124102 | Not-determined |
| KM_k141_146141 | Not-determined |
| KM_k141_124103 | Not-determined |
| KM_k141_146143 | Not-determined |
| KM_k141_112872 | Not-determined |
| KM_k141_79857 | Not-determined |
| KM_k141_234743 | Not-determined |
| KM_k141_57141 | Not-determined |
| KM_k141_68486 | Not-determined |
| KM_k141_68489 | Not-determined |
| KM_k141_234745 | Not-determined |
| KM_k141_257223 | Not-determined |
| KM_k141_101356 | Not-determined |
| KM_k141_112895 | Not-determined |
| KM_k141_112896 | Not-determined |
| KM_k141_34883 | Not-determined |
| KM_k141_257224 | Not-determined |
| KM_k141_90195 | Not-determined |
| KM_k141_112898 | Not-determined |
| KM_k141_112900 | Not-determined |
| KM_k141_1619 | Not-determined |
| KM_k141_45673 | Not-determined |
| KM_k141_257226 | Not-determined |
| KM_k141_1621 | Not-determined |
| KM_k141_101359 | Not-determined |
| KM_k141_224196 | Not-determined |
| KM_k141_212908 | Not-determined |
| KM_k141_146152 | Not-determined |
| KM_k141_1624 | Not-determined |
| KM_k141_179396 | Not-determined |
| KM_k141_179398 | Not-determined |
| KM_k141_101368 | Not-determined |
| KM_k141_112910 | Low-quality |
| KM_k141_146155 | Not-determined |
| KM_k141_168224 | Not-determined |
| KM_k141_224206 | Not-determined |
| KM_k141_179407 | Not-determined |
| KM_k141_112914 | Not-determined |
| KM_k141_146162 | Not-determined |
| KM_k141_112920 | Not-determined |
| KM_k141_112925 | Not-determined |
| KM_k141_112926 | Not-determined |
| KM_k141_79876 | Not-determined |
| KM_k141_146165 | Not-determined |
| KM_k141_101382 | Not-determined |
| KM_k141_101384 | Not-determined |
| KM_k141_257233 | Not-determined |
| KM_k141_112940 | Not-determined |
| KM_k141_234779 | Not-determined |
| KM_k141_190557 | Not-determined |
| KM_k141_79895 | Not-determined |
| KM_k141_124116 | Not-determined |
| KM_k141_101396 | Not-determined |
| KM_k141_34925 | Not-determined |
| KM_k141_68501 | Not-determined |
| KM_k141_146170 | Not-determined |
| KM_k141_45711 | Not-determined |
| KM_k141_190562 | Not-determined |
| KM_k141_1644 | Not-determined |
| KM_k141_101401 | Not-determined |
| KM_k141_257245 | Not-determined |
| KM_k141_112949 | Not-determined |
| KM_k141_112952 | Not-determined |
| KM_k141_179426 | Low-quality |
| KM_k141_1655 | Not-determined |
| KM_k141_23587 | Not-determined |
| KM_k141_79911 | Not-determined |
| KM_k141_134951 | Not-determined |
| KM_k141_190580 | Not-determined |
| KM_k141_190581 | Not-determined |
| KM_k141_190584 | Not-determined |
| KM_k141_212914 | Not-determined |
| KM_k141_124136 | Not-determined |
| KM_k141_1668 | Not-determined |
| KM_k141_168238 | Not-determined |
| KM_k141_202019 | Not-determined |
| KM_k141_79932 | Not-determined |
| KM_k141_79936 | Not-determined |
| KM_k141_23599 | Low-quality |
| KM_k141_112971 | Not-determined |
| KM_k141_124152 | Not-determined |
| KM_k141_79940 | Not-determined |
| KM_k141_68535 | Not-determined |
| KM_k141_112974 | Not-determined |
| KM_k141_90249 | Not-determined |
| KM_k141_168248 | Not-determined |
| KM_k141_224232 | Low-quality |
| KM_k141_146187 | Not-determined |
| KM_k141_90253 | Not-determined |
| KM_k141_34950 | Not-determined |
| KM_k141_134979 | Not-determined |
| KM_k141_79947 | Not-determined |
| KM_k141_224233 | Not-determined |
| KM_k141_1680 | Not-determined |
| KM_k141_1683 | Not-determined |
| KM_k141_79957 | Not-determined |
| KM_k141_68544 | Not-determined |
| KM_k141_90264 | Low-quality |
| KM_k141_134998 | Not-determined |
| KM_k141_134999 | Not-determined |
| KM_k141_146196 | Low-quality |
| KM_k141_224250 | Not-determined |
| KM_k141_68555 | Not-determined |
| KM_k141_146199 | Not-determined |
| KM_k141_146201 | Not-determined |
| KM_k141_124165 | Not-determined |
| KM_k141_146204 | Not-determined |
| KM_k141_90281 | Not-determined |
| KM_k141_234843 | Not-determined |
| KM_k141_257305 | Not-determined |
| KM_k141_124171 | Not-determined |
| KM_k141_34984 | Not-determined |
| KM_k141_101449 | Not-determined |
| KM_k141_1719 | Low-quality |
| KM_k141_34988 | Not-determined |
| KM_k141_135008 | Not-determined |
| KM_k141_224258 | Not-determined |
| KM_k141_1721 | Low-quality |
| KM_k141_179440 | Not-determined |
| KM_k141_101461 | Not-determined |
| KM_k141_202052 | Not-determined |
| KM_k141_202055 | Not-determined |
| KM_k141_101473 | Not-determined |
| KM_k141_224262 | Not-determined |
| KM_k141_224263 | Not-determined |
| KM_k141_124189 | Not-determined |
| KM_k141_202059 | Not-determined |
| KM_k141_234870 | Not-determined |
| KM_k141_124194 | Not-determined |
| KM_k141_224270 | Not-determined |
| KM_k141_190630 | Not-determined |
| KM_k141_68562 | Not-determined |
| KM_k141_68564 | Low-quality |
| KM_k141_113017 | Not-determined |
| KM_k141_35017 | Not-determined |
| KM_k141_168275 | Not-determined |
| KM_k141_168277 | Not-determined |
| KM_k141_68571 | Not-determined |
| KM_k141_1737 | Not-determined |
| KM_k141_190640 | Not-determined |
| KM_k141_224273 | Not-determined |
| KM_k141_224275 | Not-determined |
| KM_k141_68579 | Not-determined |
| KM_k141_1742 | Not-determined |
| KM_k141_124212 | Not-determined |
| KM_k141_124215 | Not-determined |
| KM_k141_212988 | Not-determined |
| KM_k141_212994 | Not-determined |
| KM_k141_202069 | Not-determined |
| KM_k141_168284 | Not-determined |
| KM_k141_212999 | Not-determined |
| KM_k141_68586 | Not-determined |
| KM_k141_23676 | Not-determined |
| KM_k141_23677 | Not-determined |
| KM_k141_124224 | Not-determined |
| KM_k141_179477 | Not-determined |
| KM_k141_35037 | Not-determined |
| KM_k141_1761 | Not-determined |
| KM_k141_113047 | Not-determined |
| KM_k141_168290 | Not-determined |
| KM_k141_23691 | Not-determined |
| KM_k141_90326 | Not-determined |
| KM_k141_1766 | Not-determined |
| KM_k141_245824 | Not-determined |
| KM_k141_245828 | Not-determined |
| KM_k141_257339 | Low-quality |
| KM_k141_257341 | Not-determined |
| KM_k141_113061 | Not-determined |
| KM_k141_257345 | Not-determined |
| KM_k141_213018 | Not-determined |
| KM_k141_168296 | Not-determined |
| KM_k141_113069 | Not-determined |
| KM_k141_202093 | Not-determined |
| KM_k141_68602 | Not-determined |
| KM_k141_202099 | Not-determined |
| KM_k141_224340 | Not-determined |
| KM_k141_179528 | Not-determined |
| KM_k141_124242 | Not-determined |

| | |
|---|---|
| KM_k141_90348 | Low-quality |
| KM_k141_135049 | Not-determined |
| KM_k141_135051 | Not-determined |
| KM_k141_45723 | Not-determined |
| KM_k141_213036 | Not-determined |
| KM_k141_135054 | Not-determined |
| KM_k141_45726 | Not-determined |
| KM_k141_23705 | Not-determined |
| KM_k141_68620 | Not-determined |
| KM_k141_213045 | Not-determined |
| KM_k141_68626 | Not-determined |
| KM_k141_179540 | Low-quality |
| KM_k141_202126 | Not-determined |
| KM_k141_135063 | Not-determined |
| KM_k141_168337 | Not-determined |
| KM_k141_202130 | Not-determined |
| KM_k141_113075 | Not-determined |
| KM_k141_135077 | Not-determined |
| KM_k141_257359 | Not-determined |
| KM_k141_124256 | Not-determined |
| KM_k141_245876 | Not-determined |
| KM_k141_124259 | Not-determined |
| KM_k141_179556 | Not-determined |
| KM_k141_135084 | Not-determined |
| KM_k141_202143 | Low-quality |
| KM_k141_179557 | Not-determined |
| KM_k141_224366 | Not-determined |
| KM_k141_245880 | Not-determined |
| KM_k141_135088 | Not-determined |
| KM_k141_135090 | Not-determined |
| KM_k141_168350 | Not-determined |
| KM_k141_213060 | Not-determined |
| KM_k141_257373 | Not-determined |
| KM_k141_202146 | Not-determined |
| KM_k141_202147 | Not-determined |
| KM_k141_202148 | Not-determined |
| KM_k141_202151 | Not-determined |
| KM_k141_213068 | Not-determined |
| KM_k141_234903 | Not-determined |
| KM_k141_135121 | Not-determined |
| KM_k141_135123 | Not-determined |
| KM_k141_245893 | Not-determined |
| KM_k141_245897 | Not-determined |
| KM_k141_168388 | Not-determined |
| KM_k141_179586 | Not-determined |
| KM_k141_190734 | Not-determined |
| KM_k141_190740 | Not-determined |
| KM_k141_135145 | Not-determined |
| KM_k141_35073 | Not-determined |
| KM_k141_202178 | Not-determined |
| KM_k141_35078 | Not-determined |
| KM_k141_190749 | Low-quality |
| KM_k141_35090 | Not-determined |
| KM_k141_190750 | Not-determined |
| KM_k141_257396 | Not-determined |
| KM_k141_213118 | Low-quality |
| KM_k141_245911 | Not-determined |
| KM_k141_202193 | Not-determined |
| KM_k141_45779 | Not-determined |
| KM_k141_190758 | Not-determined |
| KM_k141_202203 | Not-determined |
| KM_k141_245914 | Not-determined |
| KM_k141_224384 | Not-determined |
| KM_k141_146277 | Not-determined |
| KM_k141_190762 | Not-determined |
| KM_k141_213132 | Not-determined |
| KM_k141_23766 | Not-determined |
| KM_k141_113095 | Not-determined |
| KM_k141_257408 | Not-determined |
| KM_k141_168407 | Not-determined |
| KM_k141_23773 | Not-determined |
| KM_k141_113100 | Not-determined |
| KM_k141_113107 | Not-determined |
| KM_k141_135165 | Not-determined |
| KM_k141_57242 | Not-determined |
| KM_k141_124281 | Low-quality |
| KM_k141_23778 | Not-determined |
| KM_k141_113114 | Not-determined |
| KM_k141_79995 | Not-determined |
| KM_k141_135171 | Not-determined |
| KM_k141_168424 | Not-determined |
| KM_k141_190766 | Not-determined |
| KM_k141_68635 | Low-quality |
| KM_k141_245936 | Not-determined |
| KM_k141_23782 | Not-determined |
| KM_k141_113131 | Not-determined |
| KM_k141_157158 | Not-determined |
| KM_k141_57255 | Not-determined |
| KM_k141_202228 | Low-quality |
| KM_k141_213136 | Not-determined |
| KM_k141_202229 | Not-determined |
| KM_k141_68641 | Not-determined |
| KM_k141_23786 | Not-determined |
| KM_k141_202232 | Not-determined |
| KM_k141_68643 | Low-quality |
| KM_k141_35104 | Not-determined |
| KM_k141_113140 | Not-determined |
| KM_k141_68653 | Not-determined |
| KM_k141_190790 | Not-determined |
| KM_k141_57267 | Not-determined |
| KM_k141_190793 | Not-determined |
| KM_k141_113145 | Not-determined |
| KM_k141_113148 | Low-quality |
| KM_k141_135185 | Not-determined |
| KM_k141_68659 | Not-determined |
| KM_k141_57278 | Not-determined |
| KM_k141_157172 | Low-quality |
| KM_k141_45791 | Not-determined |
| KM_k141_45794 | Not-determined |
| KM_k141_157177 | Not-determined |
| KM_k141_80009 | Not-determined |
| KM_k141_245954 | Not-determined |
| KM_k141_157181 | Not-determined |
| KM_k141_190796 | Not-determined |
| 18-103_k141_23958 | High-quality |
| KM_k141_101543 | Not-determined |
| KM_k141_179626 | Not-determined |
| KM_k141_190802 | Not-determined |
| KM_k141_80020 | Not-determined |
| KM_k141_124315 | Not-determined |
| KM_k141_135204 | Not-determined |
| KM_k141_190804 | Not-determined |
| KM_k141_80026 | Not-determined |
| KM_k141_90424 | Not-determined |
| KM_k141_257432 | Not-determined |
| KM_k141_135212 | Not-determined |
| KM_k141_23796 | Not-determined |
| KM_k141_190809 | Not-determined |
| KM_k141_257439 | Not-determined |
| KM_k141_168452 | Not-determined |
| KM_k141_135218 | Low-quality |
| KM_k141_245970 | Low-quality |
| KM_k141_179642 | Not-determined |
| KM_k141_213177 | Not-determined |
| KM_k141_257446 | Not-determined |
| KM_k141_190822 | Not-determined |
| KM_k141_68698 | Not-determined |
| KM_k141_1819 | Not-determined |
| KM_k141_68705 | Low-quality |
| KM_k141_190826 | Not-determined |
| KM_k141_90441 | Not-determined |
| KM_k141_245986 | Not-determined |
| KM_k141_179649 | Low-quality |
| KM_k141_245987 | Not-determined |
| KM_k141_23802 | Not-determined |
| KM_k141_80044 | Not-determined |
| KM_k141_12426 | Not-determined |
| KM_k141_80047 | Not-determined |
| KM_k141_157203 | Not-determined |
| KM_k141_135223 | Not-determined |
| KM_k141_257471 | Not-determined |
| KM_k141_146315 | Not-determined |
| KM_k141_234966 | Low-quality |
| KM_k141_12435 | Not-determined |
| KM_k141_90450 | Not-determined |
| KM_k141_213208 | Not-determined |
| KM_k141_45829 | Not-determined |
| KM_k141_135232 | Not-determined |
| KM_k141_1842 | Not-determined |
| KM_k141_45833 | Not-determined |
| KM_k141_202301 | Not-determined |
| KM_k141_113193 | Not-determined |
| KM_k141_202304 | Not-determined |
| KM_k141_213220 | Not-determined |
| KM_k141_12443 | Not-determined |
| KM_k141_101591 | Not-determined |
| KM_k141_12446 | Not-determined |
| KM_k141_12448 | Not-determined |
| KM_k141_45840 | Not-determined |
| KM_k141_124368 | Not-determined |
| KM_k141_234982 | Not-determined |
| KM_k141_202325 | Not-determined |
| KM_k141_135256 | Not-determined |
| KM_k141_113213 | Not-determined |
| KM_k141_113214 | Not-determined |
| KM_k141_1860 | Not-determined |
| KM_k141_12469 | Not-determined |
| KM_k141_1864 | Not-determined |
| KM_k141_113225 | Not-determined |
| KM_k141_23823 | Not-determined |
| KM_k141_35171 | Not-determined |
| KM_k141_179678 | Not-determined |
| KM_k141_202335 | Not-determined |
| KM_k141_179680 | Low-quality |
| KM_k141_135278 | Not-determined |
| KM_k141_12474 | Not-determined |
| KM_k141_135283 | Not-determined |
| KM_k141_234998 | Not-determined |
| KM_k141_234999 | Not-determined |
| KM_k141_168507 | Not-determined |
| KM_k141_35178 | Not-determined |
| KM_k141_35179 | Not-determined |
| KM_k141_12476 | Not-determined |
| KM_k141_179693 | Not-determined |
| KM_k141_179694 | Not-determined |
| KM_k141_179696 | Not-determined |
| KM_k141_213249 | Not-determined |
| KM_k141_113241 | Not-determined |
| KM_k141_168536 | Not-determined |
| KM_k141_168538 | Not-determined |
| KM_k141_35199 | Not-determined |
| KM_k141_80073 | Not-determined |
| KM_k141_179710 | Not-determined |
| KM_k141_101648 | Not-determined |
| KM_k141_23838 | Not-determined |
| KM_k141_101650 | Not-determined |
| KM_k141_146356 | Not-determined |
| KM_k141_146358 | Low-quality |
| KM_k141_23844 | Not-determined |
| KM_k141_1879 | Not-determined |
| KM_k141_80076 | Not-determined |
| KM_k141_146362 | Not-determined |
| KM_k141_35208 | Not-determined |
| KM_k141_135297 | Not-determined |
| KM_k141_146364 | Not-determined |
| KM_k141_35209 | Low-quality |
| KM_k141_168546 | Not-determined |
| KM_k141_235018 | Low-quality |
| KM_k141_80092 | Not-determined |
| KM_k141_213284 | Low-quality |
| KM_k141_113259 | Not-determined |
| KM_k141_90484 | Not-determined |
| KM_k141_224425 | Not-determined |
| KM_k141_1907 | Not-determined |
| KM_k141_235025 | Not-determined |
| KM_k141_190852 | Not-determined |
| KM_k141_202361 | Not-determined |
| KM_k141_224428 | Not-determined |
| KM_k141_57310 | Not-determined |
| KM_k141_213298 | Not-determined |
| KM_k141_57316 | Not-determined |
| KM_k141_80098 | Not-determined |
| KM_k141_202366 | Not-determined |
| KM_k141_68743 | Not-determined |
| KM_k141_57328 | Not-determined |
| KM_k141_80108 | Low-quality |
| KM_k141_68756 | Not-determined |
| KM_k141_113291 | Not-determined |
| KM_k141_157225 | Not-determined |
| KM_k141_101664 | Not-determined |
| KM_k141_101665 | Not-determined |
| KM_k141_257491 | Not-determined |
| KM_k141_68761 | Not-determined |
| KM_k141_68764 | Not-determined |
| KM_k141_101668 | Not-determined |
| KM_k141_202397 | Not-determined |
| KM_k141_80129 | Not-determined |
| KM_k141_190866 | Not-determined |
| KM_k141_101673 | Low-quality |
| KM_k141_257497 | Not-determined |
| KM_k141_135333 | Not-determined |
| KM_k141_202404 | Not-determined |
| KM_k141_146386 | Not-determined |
| KM_k141_257504 | Not-determined |
| KM_k141_45888 | Not-determined |
| KM_k141_90499 | Not-determined |
| KM_k141_1954 | Not-determined |
| KM_k141_157235 | Low-quality |
| KM_k141_190874 | Not-determined |
| KM_k141_213322 | Not-determined |
| KM_k141_202414 | Low-quality |
| KM_k141_202416 | Low-quality |
| KM_k141_202417 | Not-determined |
| KM_k141_224457 | Not-determined |
| KM_k141_90511 | Not-determined |
| KM_k141_113325 | Low-quality |
| KM_k141_157245 | Low-quality |
| KM_k141_157249 | Low-quality |
| KM_k141_257519 | Not-determined |
| KM_k141_235028 | Not-determined |
| KM_k141_135347 | Not-determined |
| KM_k141_101687 | Not-determined |
| KM_k141_35227 | Not-determined |
| KM_k141_101688 | Not-determined |
| KM_k141_101690 | Not-determined |
| KM_k141_179748 | Not-determined |
| KM_k141_202447 | Not-determined |
| KM_k141_1970 | Not-determined |
| KM_k141_235041 | Not-determined |
| KM_k141_135374 | Not-determined |
| KM_k141_202454 | Not-determined |
| KM_k141_179754 | Not-determined |
| KM_k141_246016 | Not-determined |
| KM_k141_1973 | Not-determined |
| KM_k141_213331 | Not-determined |
| KM_k141_12496 | Low-quality |
| KM_k141_179761 | Not-determined |
| KM_k141_35242 | Not-determined |
| KM_k141_101718 | Not-determined |
| KM_k141_124440 | Not-determined |
| KM_k141_101726 | Not-determined |
| KM_k141_213340 | Not-determined |
| KM_k141_190901 | Not-determined |
| KM_k141_12498 | Not-determined |
| KM_k141_124445 | Not-determined |
| KM_k141_168619 | Not-determined |
| KM_k141_157265 | Not-determined |
| KM_k141_235055 | Not-determined |
| KM_k141_12504 | Not-determined |
| KM_k141_35251 | Low-quality |
| KM_k141_202471 | Not-determined |
| KM_k141_35258 | Not-determined |
| KM_k141_1987 | Not-determined |
| KM_k141_146409 | Not-determined |
| KM_k141_35265 | Not-determined |
| KM_k141_213346 | Not-determined |
| KM_k141_235069 | Not-determined |
| KM_k141_80145 | Not-determined |
| KM_k141_113341 | Not-determined |
| KM_k141_202484 | Low-quality |
| KM_k141_213351 | Not-determined |
| KM_k141_135417 | Not-determined |
| KM_k141_224489 | Not-determined |
| KM_k141_190919 | Not-determined |
| KM_k141_213355 | Low-quality |
| KM_k141_146422 | Not-determined |
| KM_k141_190926 | Not-determined |
| KM_k141_246045 | Not-determined |
| KM_k141_224497 | Not-determined |
| KM_k141_246046 | Not-determined |
| KM_k141_12525 | Not-determined |
| KM_k141_168638 | Not-determined |
| KM_k141_213363 | Not-determined |
| KM_k141_113364 | Not-determined |
| KM_k141_190937 | Not-determined |
| KM_k141_146433 | Not-determined |
| KM_k141_113368 | Not-determined |
| KM_k141_113371 | Not-determined |
| KM_k141_68806 | Not-determined |
| KM_k141_68810 | Not-determined |
| KM_k141_146438 | Not-determined |
| KM_k141_68811 | Not-determined |
| KM_k141_113376 | Not-determined |
| KM_k141_113377 | Not-determined |
| KM_k141_68813 | Not-determined |
| KM_k141_213377 | Not-determined |
| KM_k141_168669 | Not-determined |
| KM_k141_246059 | Not-determined |
| KM_k141_168674 | Not-determined |
| KM_k141_45937 | Not-determined |
| KM_k141_146452 | Not-determined |
| KM_k141_224501 | Not-determined |
| KM_k141_168676 | Not-determined |
| KM_k141_168677 | Not-determined |
| KM_k141_235093 | Not-determined |
| KM_k141_101758 | Not-determined |
| KM_k141_45946 | Not-determined |
| KM_k141_157285 | Not-determined |
| KM_k141_113394 | Not-determined |
| KM_k141_157292 | Not-determined |
| KM_k141_23928 | Not-determined |
| KM_k141_68819 | Not-determined |
| KM_k141_146465 | Low-quality |
| KM_k141_257570 | Not-determined |
| KM_k141_80194 | Not-determined |
| KM_k141_146468 | Not-determined |
| KM_k141_80197 | Not-determined |
| KM_k141_23932 | Not-determined |
| KM_k141_45960 | Not-determined |
| KM_k141_202552 | Not-determined |
| KM_k141_202554 | Not-determined |
| KM_k141_80204 | Not-determined |
| KM_k141_68830 | Not-determined |
| KM_k141_157310 | Low-quality |
| KM_k141_45970 | Not-determined |
| KM_k141_57413 | Not-determined |
| KM_k141_135439 | Not-determined |
| KM_k141_213396 | Low-quality |
| KM_k141_157314 | Not-determined |
| KM_k141_80207 | Not-determined |
| KM_k141_2025 | Not-determined |
| KM_k141_35298 | Not-determined |
| KM_k141_135443 | Not-determined |
| KM_k141_135444 | Not-determined |
| KM_k141_135445 | Not-determined |
| KM_k141_213401 | Not-determined |
| KM_k141_45976 | Low-quality |
| KM_k141_35304 | Not-determined |
| KM_k141_235126 | Not-determined |
| KM_k141_57426 | Low-quality |
| KM_k141_179783 | Not-determined |
| KM_k141_12549 | Not-determined |
| KM_k141_23957 | Not-determined |
| KM_k141_12551 | Not-determined |
| KM_k141_80215 | Low-quality |
| KM_k141_12552 | Not-determined |
| KM_k141_135453 | Not-determined |
| KM_k141_113423 | Not-determined |
| KM_k141_12561 | Low-quality |
| KM_k141_80221 | Not-determined |
| KM_k141_146503 | Not-determined |
| KM_k141_179792 | Not-determined |
| KM_k141_35323 | Not-determined |
| KM_k141_35324 | Not-determined |
| KM_k141_135457 | Not-determined |
| KM_k141_146506 | Not-determined |
| KM_k141_135465 | Not-determined |
| KM_k141_101774 | Not-determined |
| KM_k141_101778 | Not-determined |
| KM_k141_68842 | Not-determined |
| KM_k141_35327 | Not-determined |
| KM_k141_190982 | Not-determined |
| KM_k141_213411 | Not-determined |
| KM_k141_213415 | Not-determined |
| KM_k141_246076 | Not-determined |
| KM_k141_246078 | Not-determined |
| KM_k141_168704 | Not-determined |
| KM_k141_90596 | Low-quality |
| KM_k141_213421 | Low-quality |
| KM_k141_257587 | Not-determined |
| KM_k141_235153 | Not-determined |
| KM_k141_235154 | Not-determined |
| KM_k141_57447 | Not-determined |
| KM_k141_191004 | Not-determined |
| KM_k141_57454 | Not-determined |
| KM_k141_179816 | Not-determined |
| KM_k141_191008 | Not-determined |
| KM_k141_213437 | Not-determined |
| KM_k141_113466 | Not-determined |
| KM_k141_113467 | Not-determined |
| KM_k141_68852 | Not-determined |
| KM_k141_191024 | Not-determined |
| KM_k141_68856 | Not-determined |
| KM_k141_57467 | Not-determined |
| KM_k141_168730 | Low-quality |
| KM_k141_168734 | Low-quality |
| KM_k141_57469 | Not-determined |
| KM_k141_191029 | Not-determined |
| KM_k141_113482 | Not-determined |
| KM_k141_80239 | Not-determined |
| KM_k141_179841 | Not-determined |
| KM_k141_168745 | Not-determined |
| KM_k141_68859 | Not-determined |
| KM_k141_113492 | Not-determined |
| KM_k141_213455 | Not-determined |
| KM_k141_113495 | Not-determined |
| KM_k141_257607 | Not-determined |
| KM_k141_68868 | Not-determined |
| KM_k141_68869 | Not-determined |
| KM_k141_146525 | Not-determined |
| KM_k141_168758 | Not-determined |
| KM_k141_68871 | Not-determined |
| KM_k141_235177 | Not-determined |
| KM_k141_101803 | Not-determined |
| KM_k141_57496 | Not-determined |
| KM_k141_213468 | Not-determined |
| KM_k141_191043 | Not-determined |
| KM_k141_68874 | Not-determined |
| KM_k141_35348 | Not-determined |
| KM_k141_80263 | Not-determined |
| KM_k141_146534 | Not-determined |
| KM_k141_57506 | Not-determined |
| KM_k141_12579 | Not-determined |
| KM_k141_57509 | Not-determined |
| KM_k141_35360 | Not-determined |
| KM_k141_191065 | Not-determined |
| KM_k141_35365 | Low-quality |
| KM_k141_246089 | Not-determined |
| KM_k141_246090 | Not-determined |
| KM_k141_146544 | Low-quality |
| KM_k141_246093 | Not-determined |
| KM_k141_12586 | Not-determined |
| KM_k141_12587 | Not-determined |
| KM_k141_179866 | Not-determined |
| KM_k141_224521 | Not-determined |
| KM_k141_12591 | Not-determined |
| KM_k141_157343 | Not-determined |
| KM_k141_191073 | Not-determined |
| KM_k141_179874 | Not-determined |
| KM_k141_157348 | Not-determined |
| KM_k141_157351 | Not-determined |
| KM_k141_235204 | Not-determined |
| KM_k141_179877 | Not-determined |
| KM_k141_157354 | Not-determined |
| KM_k141_68901 | Not-determined |
| KM_k141_12608 | Not-determined |
| KM_k141_23998 | Not-determined |
| KM_k141_135528 | Not-determined |
| KM_k141_146552 | Not-determined |
| KM_k141_35378 | Low-quality |
| KM_k141_113535 | Not-determined |
| KM_k141_113539 | Not-determined |
| KM_k141_113540 | Not-determined |
| KM_k141_157373 | Not-determined |
| KM_k141_80281 | Not-determined |
| KM_k141_135544 | Not-determined |
| KM_k141_235212 | Not-determined |
| KM_k141_224529 | Not-determined |
| KM_k141_135545 | Not-determined |
| KM_k141_235214 | Not-determined |
| KM_k141_146565 | Not-determined |
| KM_k141_80288 | Not-determined |
| KM_k141_146567 | Not-determined |
| KM_k141_146574 | Not-determined |
| KM_k141_80295 | Not-determined |
| KM_k141_12635 | Not-determined |
| KM_k141_146584 | Not-determined |
| KM_k141_35386 | Not-determined |
| KM_k141_135564 | Not-determined |
| KM_k141_57530 | Not-determined |
| KM_k141_80301 | Not-determined |
| KM_k141_80302 | Not-determined |
| KM_k141_213487 | Not-determined |
| KM_k141_213488 | Not-determined |
| KM_k141_135569 | Not-determined |
| KM_k141_213493 | Not-determined |
| KM_k141_124459 | Not-determined |
| KM_k141_213494 | Not-determined |
| KM_k141_124462 | Not-determined |
| KM_k141_146594 | Not-determined |
| KM_k141_68947 | Not-determined |
| KM_k141_146596 | Not-determined |
| KM_k141_224554 | Not-determined |
| KM_k141_90634 | Not-determined |
| KM_k141_113553 | Not-determined |
| KM_k141_80310 | Not-determined |
| KM_k141_179891 | Not-determined |
| KM_k141_90638 | Not-determined |
| KM_k141_146601 | Not-determined |
| KM_k141_135587 | Not-determined |
| KM_k141_80312 | Low-quality |
| KM_k141_157378 | Not-determined |
| KM_k141_80321 | Not-determined |
| KM_k141_2137 | Not-determined |
| KM_k141_257633 | Not-determined |
| KM_k141_191095 | Not-determined |
| KM_k141_191097 | Not-determined |
| KM_k141_68964 | Not-determined |
| KM_k141_179905 | Not-determined |
| KM_k141_157390 | Not-determined |
| KM_k141_224579 | Not-determined |
| KM_k141_157393 | Not-determined |
| KM_k141_12647 | Not-determined |
| KM_k141_135589 | Not-determined |
| KM_k141_157405 | Not-determined |
| KM_k141_12650 | Not-determined |
| KM_k141_257645 | Not-determined |
| KM_k141_12654 | Not-determined |
| KM_k141_12657 | Low-quality |
| KM_k141_157413 | Not-determined |
| KM_k141_146627 | Not-determined |
| KM_k141_246156 | Low-quality |
| KM_k141_2151 | Low-quality |
| KM_k141_157418 | Not-determined |
| KM_k141_202611 | Not-determined |
| KM_k141_191120 | Not-determined |
| KM_k141_246162 | Low-quality |
| KM_k141_157422 | Not-determined |
| KM_k141_213524 | Not-determined |
| KM_k141_135600 | Not-determined |
| KM_k141_202624 | Not-determined |
| KM_k141_146645 | Not-determined |
| KM_k141_146648 | Not-determined |
| KM_k141_113587 | Not-determined |
| KM_k141_202635 | Low-quality |
| KM_k141_257670 | Not-determined |
| KM_k141_157438 | Low-quality |
| KM_k141_101842 | Not-determined |
| KM_k141_213537 | Not-determined |
| KM_k141_113597 | Not-determined |
| KM_k141_124515 | Not-determined |
| KM_k141_124518 | Not-determined |
| KM_k141_246186 | Not-determined |
| KM_k141_101853 | Not-determined |
| KM_k141_113602 | Not-determined |
| KM_k141_246190 | Not-determined |
| KM_k141_80353 | Not-determined |
| KM_k141_168819 | Not-determined |
| KM_k141_57569 | Not-determined |
| KM_k141_90662 | Not-determined |
| KM_k141_57574 | Not-determined |
| KM_k141_146663 | Not-determined |
| KM_k141_101868 | Not-determined |
| KM_k141_113626 | Not-determined |
| KM_k141_146668 | Not-determined |
| KM_k141_101882 | Not-determined |
| KM_k141_35442 | Not-determined |
| KM_k141_113629 | Not-determined |
| KM_k141_113630 | Not-determined |
| KM_k141_168829 | Not-determined |
| KM_k141_57589 | Not-determined |
| KM_k141_57590 | Not-determined |
| KM_k141_213569 | Not-determined |
| KM_k141_179938 | Not-determined |
| KM_k141_179939 | Low-quality |
| KM_k141_235262 | Not-determined |
| KM_k141_124555 | Not-determined |
| KM_k141_179942 | Low-quality |
| KM_k141_35451 | Not-determined |

| | |
|---|---|
| KM_k141_57596 | Not-determined |
| KM_k141_179945 | Not-determined |
| KM_k141_146675 | Not-determined |
| KM_k141_12702 | Not-determined |
| KM_k141_12703 | Not-determined |
| KM_k141_202644 | Not-determined |
| KM_k141_12706 | Not-determined |
| KM_k141_12709 | Not-determined |
| KM_k141_57610 | Not-determined |
| KM_k141_124577 | Not-determined |
| KM_k141_57613 | Not-determined |
| KM_k141_35463 | Not-determined |
| KM_k141_57617 | Low-quality |
| KM_k141_80379 | Not-determined |
| KM_k141_113657 | Not-determined |
| KM_k141_12733 | Not-determined |
| KM_k141_80380 | Low-quality |
| KM_k141_257683 | Low-quality |
| KM_k141_235280 | Not-determined |
| KM_k141_257684 | Not-determined |
| KM_k141_57625 | Not-determined |
| KM_k141_191183 | Not-determined |
| KM_k141_124587 | Not-determined |
| KM_k141_2159 | Low-quality |
| KM_k141_202651 | Not-determined |
| KM_k141_80382 | Not-determined |
| KM_k141_191185 | Not-determined |
| KM_k141_235287 | Not-determined |
| KM_k141_12744 | Not-determined |
| KM_k141_257697 | Not-determined |
| KM_k141_135644 | Not-determined |
| KM_k141_246202 | Low-quality |
| KM_k141_135645 | Not-determined |
| KM_k141_257698 | Not-determined |
| KM_k141_80390 | Not-determined |
| KM_k141_12751 | Not-determined |
| KM_k141_80392 | Not-determined |
| KM_k141_90685 | Not-determined |
| KM_k141_90686 | Not-determined |
| KM_k141_57638 | Not-determined |
| KM_k141_46000 | Not-determined |
| KM_k141_246216 | Not-determined |
| KM_k141_191193 | Low-quality |
| KM_k141_80398 | Low-quality |
| KM_k141_168847 | Not-determined |
| KM_k141_80401 | Not-determined |
| KM_k141_80402 | Not-determined |
| KM_k141_90702 | Not-determined |
| KM_k141_235294 | Not-determined |
| KM_k141_191198 | Not-determined |
| KM_k141_24041 | Not-determined |
| KM_k141_135661 | Not-determined |
| KM_k141_157479 | Not-determined |
| KM_k141_257727 | Not-determined |
| KM_k141_135669 | Not-determined |
| KM_k141_213602 | Not-determined |
| KM_k141_157486 | Not-determined |
| KM_k141_257729 | Not-determined |
| KM_k141_168861 | Not-determined |
| KM_k141_90720 | Not-determined |
| KM_k141_113681 | Not-determined |
| KM_k141_46014 | Not-determined |
| KM_k141_191220 | Not-determined |
| KM_k141_90722 | Low-quality |
| KM_k141_224661 | Not-determined |
| KM_k141_57669 | Not-determined |
| KM_k141_179995 | Not-determined |
| KM_k141_224668 | Not-determined |
| KM_k141_90734 | Not-determined |
| KM_k141_124621 | Not-determined |
| KM_k141_35500 | Not-determined |
| KM_k141_90736 | Not-determined |
| KM_k141_80426 | Not-determined |
| KM_k141_191239 | Not-determined |
| KM_k141_57677 | Not-determined |
| KM_k141_46023 | Not-determined |
| KM_k141_57679 | Not-determined |
| KM_k141_69014 | Not-determined |
| KM_k141_80434 | Low-quality |
| KM_k141_224684 | Not-determined |
| KM_k141_224687 | Not-determined |
| KM_k141_246245 | Not-determined |
| KM_k141_124628 | Not-determined |
| KM_k141_246248 | Not-determined |
| KM_k141_90749 | Not-determined |
| KM_k141_90751 | Not-determined |
| KM_k141_235331 | Not-determined |
| KM_k141_124635 | Not-determined |
| KM_k141_69027 | Not-determined |
| KM_k141_69028 | Not-determined |
| KM_k141_113724 | Not-determined |
| KM_k141_101893 | Not-determined |
| KM_k141_24062 | Not-determined |
| KM_k141_146696 | Not-determined |
| KM_k141_46035 | Not-determined |
| KM_k141_180023 | Not-determined |
| KM_k141_191250 | Not-determined |
| KM_k141_124643 | Not-determined |
| KM_k141_46039 | Not-determined |
| KM_k141_124645 | Not-determined |
| KM_k141_180029 | Not-determined |
| KM_k141_213627 | Not-determined |
| KM_k141_146705 | Not-determined |
| KM_k141_180034 | Not-determined |
| KM_k141_213630 | Low-quality |
| KM_k141_46043 | Not-determined |
| KM_k141_24075 | Not-determined |
| KM_k141_113729 | Not-determined |
| KM_k141_46050 | Not-determined |
| KM_k141_202698 | Not-determined |
| KM_k141_46052 | Not-determined |
| KM_k141_180042 | Not-determined |
| KM_k141_124659 | Not-determined |
| KM_k141_213632 | Not-determined |
| KM_k141_46060 | Not-determined |
| KM_k141_202707 | Not-determined |
| KM_k141_24090 | Not-determined |
| KM_k141_69042 | Not-determined |
| KM_k141_202715 | Not-determined |
| KM_k141_124666 | Not-determined |
| KM_k141_235341 | Not-determined |
| KM_k141_202718 | Not-determined |
| KM_k141_257742 | Not-determined |
| KM_k141_213641 | Not-determined |
| KM_k141_246259 | Not-determined |
| KM_k141_2190 | Not-determined |
| KM_k141_246265 | Not-determined |
| KM_k141_90758 | Not-determined |
| KM_k141_46076 | Not-determined |
| KM_k141_2193 | Not-determined |
| KM_k141_202725 | Not-determined |
| KM_k141_257751 | Not-determined |
| KM_k141_57703 | Not-determined |
| KM_k141_57707 | Not-determined |
| KM_k141_180084 | Not-determined |
| KM_k141_180086 | Not-determined |
| KM_k141_57711 | Not-determined |
| KM_k141_46089 | Low-quality |
| KM_k141_35546 | Not-determined |
| KM_k141_35547 | Not-determined |
| KM_k141_124698 | Not-determined |
| KM_k141_180097 | Not-determined |
| KM_k141_113744 | Not-determined |
| KM_k141_213650 | Not-determined |
| KM_k141_90773 | Not-determined |
| KM_k141_12781 | Not-determined |
| KM_k141_202736 | Not-determined |
| KM_k141_24112 | Not-determined |
| KM_k141_146733 | Not-determined |
| KM_k141_168893 | Not-determined |
| KM_k141_202740 | Not-determined |
| KM_k141_113752 | Not-determined |
| KM_k141_124707 | Not-determined |
| KM_k141_113753 | Not-determined |
| KM_k141_12791 | Not-determined |
| KM_k141_24117 | Not-determined |
| KM_k141_213652 | Not-determined |
| KM_k141_12795 | Low-quality |
| KM_k141_69083 | Not-determined |
| KM_k141_191268 | Not-determined |
| KM_k141_24119 | Not-determined |
| KM_k141_168907 | Not-determined |
| KM_k141_124719 | Not-determined |
| KM_k141_46100 | Not-determined |
| KM_k141_235360 | Not-determined |
| KM_k141_80462 | Not-determined |
| KM_k141_135726 | Not-determined |
| KM_k141_69092 | Not-determined |
| KM_k141_146742 | Not-determined |
| KM_k141_69097 | Not-determined |
| KM_k141_191278 | Not-determined |
| KM_k141_124732 | Low-quality |
| KM_k141_146746 | Not-determined |
| KM_k141_202764 | Not-determined |
| KM_k141_80477 | Low-quality |
| KM_k141_157531 | Low-quality |
| KM_k141_35587 | Not-determined |
| KM_k141_135750 | Not-determined |
| KM_k141_191296 | Not-determined |
| KM_k141_235386 | Not-determined |
| KM_k141_191298 | Not-determined |
| KM_k141_46132 | Not-determined |
| KM_k141_157535 | Not-determined |
| KM_k141_80479 | Not-determined |
| KM_k141_12817 | Not-determined |
| KM_k141_12819 | Not-determined |
| KM_k141_202775 | Low-quality |
| KM_k141_146759 | Not-determined |
| KM_k141_257787 | Not-determined |
| KM_k141_202779 | Not-determined |
| KM_k141_202781 | Not-determined |
| KM_k141_213679 | Not-determined |
| KM_k141_12820 | Not-determined |
| KM_k141_191299 | Low-quality |
| KM_k141_35598 | Not-determined |
| KM_k141_101940 | Not-determined |
| KM_k141_168965 | Not-determined |
| KM_k141_101946 | Not-determined |
| KM_k141_35605 | Not-determined |
| KM_k141_168973 | Not-determined |
| KM_k141_46155 | Not-determined |
| KM_k141_235413 | Not-determined |
| KM_k141_180129 | Not-determined |
| KM_k141_101953 | Not-determined |
| KM_k141_46159 | Low-quality |
| KM_k141_157558 | Not-determined |
| KM_k141_46166 | Not-determined |
| KM_k141_101955 | Not-determined |
| KM_k141_113800 | Not-determined |
| KM_k141_101957 | Not-determined |
| KM_k141_113804 | Not-determined |
| KM_k141_80499 | Not-determined |
| KM_k141_235420 | Not-determined |
| KM_k141_80503 | Not-determined |
| KM_k141_157565 | Low-quality |
| KM_k141_224748 | Not-determined |
| KM_k141_168999 | Not-determined |
| KM_k141_69137 | Not-determined |
| KM_k141_57741 | Not-determined |
| KM_k141_57743 | Not-determined |
| KM_k141_69146 | Not-determined |
| KM_k141_24129 | Not-determined |
| KM_k141_235429 | Low-quality |
| KM_k141_146779 | Not-determined |
| KM_k141_46176 | Not-determined |
| KM_k141_80521 | Not-determined |
| KM_k141_224763 | Not-determined |
| KM_k141_224764 | Low-quality |
| KM_k141_80529 | Not-determined |
| KM_k141_80530 | Not-determined |
| KM_k141_157579 | Not-determined |
| KM_k141_235447 | Not-determined |
| KM_k141_169014 | Low-quality |
| KM_k141_113838 | Not-determined |
| KM_k141_135805 | Not-determined |
| KM_k141_224777 | Not-determined |
| KM_k141_146794 | Not-determined |
| KM_k141_101981 | Not-determined |
| KM_k141_135811 | Not-determined |
| KM_k141_113846 | Not-determined |
| KM_k141_24160 | Low-quality |
| KM_k141_191333 | Not-determined |
| KM_k141_191339 | Not-determined |
| KM_k141_224796 | Not-determined |
| KM_k141_113857 | Not-determined |
| KM_k141_202824 | Not-determined |
| KM_k141_113860 | Low-quality |
| KM_k141_2241 | Not-determined |
| KM_k141_2242 | Not-determined |
| KM_k141_180166 | Not-determined |
| KM_k141_180169 | Not-determined |
| KM_k141_180171 | Not-determined |
| KM_k141_180173 | Not-determined |
| KM_k141_235468 | Not-determined |
| KM_k141_80539 | Not-determined |
| KM_k141_180181 | Not-determined |
| KM_k141_124847 | Not-determined |
| KM_k141_224810 | Not-determined |
| KM_k141_180186 | Not-determined |
| KM_k141_102003 | Not-determined |
| KM_k141_224812 | Not-determined |
| KM_k141_69157 | Not-determined |
| KM_k141_24196 | Not-determined |
| KM_k141_146813 | Not-determined |
| KM_k141_135833 | Not-determined |
| KM_k141_146815 | Not-determined |
| KM_k141_224818 | Not-determined |
| KM_k141_235485 | Not-determined |
| KM_k141_80555 | Not-determined |
| KM_k141_246328 | Not-determined |
| KM_k141_224825 | Low-quality |
| KM_k141_191369 | Not-determined |
| KM_k141_69164 | Not-determined |
| KM_k141_124855 | Not-determined |
| KM_k141_169065 | Not-determined |
| KM_k141_213728 | Not-determined |
| KM_k141_235490 | Not-determined |
| KM_k141_124861 | Not-determined |
| KM_k141_169075 | Not-determined |
| KM_k141_80566 | Not-determined |
| KM_k141_213738 | Not-determined |
| KM_k141_24201 | Not-determined |
| KM_k141_146832 | Not-determined |
| KM_k141_113877 | Not-determined |
| KM_k141_191378 | Not-determined |
| KM_k141_191379 | Not-determined |
| KM_k141_24208 | Low-quality |
| KM_k141_180196 | Not-determined |
| KM_k141_235509 | Not-determined |
| KM_k141_57776 | Not-determined |
| KM_k141_90839 | Not-determined |
| KM_k141_46200 | Not-determined |
| KM_k141_24218 | Not-determined |
| KM_k141_12848 | Not-determined |
| KM_k141_246349 | Not-determined |
| KM_k141_124882 | Not-determined |
| KM_k141_24231 | Not-determined |
| KM_k141_180213 | Not-determined |
| KM_k141_12855 | Not-determined |
| KM_k141_180221 | Not-determined |
| KM_k141_180222 | Not-determined |
| KM_k141_213745 | Not-determined |
| KM_k141_213748 | Not-determined |
| KM_k141_12865 | Not-determined |
| KM_k141_12866 | Not-determined |
| KM_k141_124894 | Not-determined |
| KM_k141_124897 | Not-determined |
| KM_k141_12870 | Not-determined |
| KM_k141_124902 | Low-quality |
| KM_k141_102032 | Not-determined |
| KM_k141_90861 | Not-determined |
| KM_k141_102033 | Not-determined |
| KM_k141_57796 | Not-determined |
| KM_k141_57799 | Not-determined |
| KM_k141_113911 | Not-determined |
| KM_k141_57810 | Not-determined |
| KM_k141_102041 | Not-determined |
| KM_k141_257846 | Not-determined |
| KM_k141_24251 | Not-determined |
| KM_k141_157600 | Not-determined |
| KM_k141_157607 | Not-determined |
| KM_k141_124917 | Not-determined |
| KM_k141_124919 | Not-determined |
| KM_k141_57824 | Not-determined |
| KM_k141_90877 | Not-determined |
| KM_k141_124922 | Not-determined |
| KM_k141_235532 | Low-quality |
| KM_k141_146849 | Not-determined |
| KM_k141_57830 | Not-determined |
| KM_k141_257860 | Not-determined |
| KM_k141_57833 | Not-determined |
| KM_k141_202853 | Not-determined |
| KM_k141_69187 | Low-quality |
| KM_k141_57837 | Not-determined |
| KM_k141_235534 | Low-quality |
| KM_k141_224873 | Not-determined |
| KM_k141_213758 | Not-determined |
| KM_k141_213759 | Not-determined |
| KM_k141_113931 | Not-determined |
| KM_k141_69196 | Not-determined |
| KM_k141_69198 | Not-determined |
| KM_k141_213763 | Not-determined |
| KM_k141_12890 | Not-determined |
| KM_k141_213765 | Not-determined |
| KM_k141_213766 | Not-determined |
| KM_k141_202864 | Not-determined |
| KM_k141_213768 | Not-determined |
| KM_k141_35610 | Not-determined |
| KM_k141_35611 | Not-determined |
| KM_k141_57841 | Not-determined |
| KM_k141_102068 | Not-determined |
| KM_k141_69212 | Not-determined |
| KM_k141_246391 | Not-determined |
| KM_k141_57845 | Not-determined |
| KM_k141_257869 | Low-quality |
| KM_k141_35619 | Not-determined |
| KM_k141_35621 | Not-determined |
| KM_k141_224893 | Low-quality |
| KM_k141_246397 | Not-determined |
| KM_k141_35624 | Not-determined |
| KM_k141_124941 | Not-determined |
| KM_k141_246398 | Not-determined |
| KM_k141_257871 | Not-determined |
| KM_k141_257874 | Not-determined |
| KM_k141_2263 | Not-determined |
| KM_k141_202882 | Not-determined |
| KM_k141_90893 | Not-determined |
| KM_k141_90894 | Not-determined |
| KM_k141_246404 | Not-determined |
| KM_k141_113950 | Not-determined |
| KM_k141_57848 | Not-determined |
| KM_k141_90896 | Not-determined |
| KM_k141_191432 | Not-determined |
| KM_k141_202889 | Not-determined |
| 18-103-final-viral-combined_k141_23958\|\|full | High-quality |
| KM_k141_169104 | Not-determined |
| KM_k141_169106 | Not-determined |
| KM_k141_235537 | Not-determined |
| KM_k141_257892 | Not-determined |
| KM_k141_102075 | Not-determined |
| KM_k141_102076 | Not-determined |
| KM_k141_146864 | Not-determined |
| KM_k141_213794 | Not-determined |
| KM_k141_24285 | Low-quality |
| KM_k141_102077 | Not-determined |
| KM_k141_24288 | Not-determined |
| KM_k141_124965 | Not-determined |
| KM_k141_202897 | Low-quality |
| KM_k141_246424 | Not-determined |
| KM_k141_213798 | Not-determined |
| KM_k141_169121 | Not-determined |
| KM_k141_124976 | Low-quality |
| KM_k141_169125 | Not-determined |
| KM_k141_180273 | Not-determined |
| KM_k141_213804 | Not-determined |
| KM_k141_213807 | Not-determined |
| KM_k141_257915 | Not-determined |
| KM_k141_69249 | Not-determined |
| KM_k141_24300 | Not-determined |
| KM_k141_113973 | Not-determined |
| KM_k141_24305 | Not-determined |
| KM_k141_235568 | Not-determined |
| KM_k141_69256 | Not-determined |
| KM_k141_69257 | Low-quality |
| KM_k141_169134 | Not-determined |
| KM_k141_35667 | Not-determined |
| KM_k141_57873 | Low-quality |
| KM_k141_213830 | Not-determined |
| KM_k141_213831 | Not-determined |
| KM_k141_69269 | Not-determined |
| KM_k141_125003 | Not-determined |
| KM_k141_213833 | Not-determined |
| KM_k141_69276 | Not-determined |
| KM_k141_113994 | Not-determined |
| KM_k141_224937 | Not-determined |
| KM_k141_180276 | Not-determined |
| KM_k141_235589 | Not-determined |
| KM_k141_180277 | Not-determined |
| KM_k141_12944 | Not-determined |
| KM_k141_125020 | Not-determined |
| KM_k141_125021 | Not-determined |
| KM_k141_125022 | Not-determined |
| KM_k141_180287 | Not-determined |
| KM_k141_235592 | Not-determined |
| KM_k141_69292 | Not-determined |
| KM_k141_191495 | Not-determined |
| KM_k141_257948 | Low-quality |
| KM_k141_69299 | Not-determined |
| KM_k141_169176 | Not-determined |
| KM_k141_90942 | Not-determined |
| KM_k141_135892 | Not-determined |
| KM_k141_114012 | Not-determined |
| KM_k141_114013 | Not-determined |
| KM_k141_114014 | Not-determined |
| KM_k141_146918 | Not-determined |
| KM_k141_2309 | Not-determined |
| KM_k141_235610 | Not-determined |
| KM_k141_135898 | Not-determined |
| KM_k141_46224 | Not-determined |
| KM_k141_146923 | Not-determined |
| KM_k141_135899 | Not-determined |
| KM_k141_146924 | Not-determined |
| KM_k141_135901 | Not-determined |
| KM_k141_12970 | Not-determined |
| KM_k141_191516 | Not-determined |
| KM_k141_224962 | Not-determined |
| KM_k141_235618 | Not-determined |
| KM_k141_46231 | Not-determined |
| KM_k141_146940 | Not-determined |
| KM_k141_46233 | Low-quality |
| KM_k141_146945 | Not-determined |
| KM_k141_90957 | Not-determined |
| KM_k141_169189 | Not-determined |
| KM_k141_235624 | Not-determined |
| KM_k141_69314 | Not-determined |
| KM_k141_202948 | Not-determined |
| KM_k141_69319 | Not-determined |
| KM_k141_35685 | Not-determined |
| KM_k141_202953 | Not-determined |
| KM_k141_246455 | Not-determined |
| KM_k141_257978 | Not-determined |
| KM_k141_2326 | Not-determined |
| KM_k141_213842 | Not-determined |
| KM_k141_191535 | Not-determined |
| KM_k141_169202 | Not-determined |
| KM_k141_202959 | Not-determined |
| KM_k141_169205 | Not-determined |
| KM_k141_213844 | Low-quality |
| KM_k141_35695 | Not-determined |
| KM_k141_69335 | Not-determined |
| KM_k141_146963 | Not-determined |
| KM_k141_213862 | Not-determined |
| KM_k141_213864 | Not-determined |
| KM_k141_146966 | Not-determined |
| KM_k141_90984 | Not-determined |
| KM_k141_157671 | Not-determined |
| KM_k141_90987 | Not-determined |
| KM_k141_125034 | Not-determined |
| KM_k141_2344 | Not-determined |
| KM_k141_202963 | Not-determined |
| KM_k141_69348 | Not-determined |
| KM_k141_157675 | Not-determined |
| KM_k141_146983 | Low-quality |
| KM_k141_213882 | Not-determined |
| KM_k141_235629 | Not-determined |
| KM_k141_202974 | Not-determined |
| KM_k141_146991 | Not-determined |
| KM_k141_46260 | Not-determined |
| KM_k141_225020 | Not-determined |
| KM_k141_146993 | Not-determined |
| KM_k141_46266 | Not-determined |
| KM_k141_46268 | Not-determined |
| KM_k141_2354 | Not-determined |
| KM_k141_146996 | Not-determined |
| KM_k141_102126 | Not-determined |
| KM_k141_12987 | Not-determined |
| KM_k141_235653 | Not-determined |
| KM_k141_213911 | Not-determined |
| KM_k141_191542 | Not-determined |
| KM_k141_258024 | Not-determined |
| KM_k141_2363 | Not-determined |
| KM_k141_35737 | Not-determined |
| KM_k141_135965 | Not-determined |
| KM_k141_258028 | Not-determined |
| KM_k141_157694 | Not-determined |
| KM_k141_135968 | Not-determined |
| KM_k141_69379 | Not-determined |
| KM_k141_225051 | Not-determined |
| KM_k141_102142 | Not-determined |
| KM_k141_235668 | Not-determined |
| KM_k141_191554 | Not-determined |
| KM_k141_46284 | Not-determined |
| KM_k141_180302 | Not-determined |
| KM_k141_91018 | Not-determined |
| KM_k141_258041 | Not-determined |
| KM_k141_35741 | Not-determined |
| KM_k141_135997 | Not-determined |
| KM_k141_258042 | Not-determined |
| KM_k141_135998 | Not-determined |
| KM_k141_203012 | Not-determined |
| KM_k141_203015 | Not-determined |
| KM_k141_2371 | Not-determined |
| KM_k141_258047 | Not-determined |
| KM_k141_57888 | Not-determined |
| KM_k141_91033 | Not-determined |
| KM_k141_91034 | Not-determined |
| KM_k141_114095 | Not-determined |
| KM_k141_2376 | Not-determined |
| KM_k141_2380 | Not-determined |
| KM_k141_258066 | Not-determined |
| KM_k141_225079 | Not-determined |
| KM_k141_169259 | Not-determined |
| KM_k141_225081 | Not-determined |
| KM_k141_225084 | Not-determined |
| KM_k141_114100 | Not-determined |
| KM_k141_203028 | Not-determined |
| KM_k141_258074 | Not-determined |
| KM_k141_213918 | Not-determined |
| KM_k141_258075 | Not-determined |
| KM_k141_147039 | Not-determined |
| KM_k141_203033 | Not-determined |
| KM_k141_91052 | Not-determined |
| KM_k141_114114 | Not-determined |
| KM_k141_169269 | Not-determined |
| KM_k141_2393 | Low-quality |
| KM_k141_136028 | Not-determined |
| KM_k141_147051 | Not-determined |
| KM_k141_147053 | Not-determined |
| KM_k141_213928 | Not-determined |
| KM_k141_80596 | Not-determined |
| KM_k141_125059 | Not-determined |
| KM_k141_203051 | Low-quality |
| KM_k141_213930 | Low-quality |
| KM_k141_246505 | Not-determined |
| KM_k141_80599 | Not-determined |
| KM_k141_91061 | Not-determined |
| KM_k141_13021 | Not-determined |
| KM_k141_246510 | Not-determined |
| KM_k141_147062 | Not-determined |
| KM_k141_57913 | Low-quality |
| KM_k141_235692 | Not-determined |
| KM_k141_46298 | Not-determined |
| KM_k141_235695 | Not-determined |
| KM_k141_102185 | Not-determined |
| KM_k141_102186 | Not-determined |
| KM_k141_246515 | Not-determined |
| KM_k141_57926 | Low-quality |
| KM_k141_169281 | Not-determined |
| KM_k141_114141 | Not-determined |
| KM_k141_114142 | Not-determined |
| KM_k141_80613 | Not-determined |
| KM_k141_114143 | Not-determined |
| KM_k141_235712 | Not-determined |
| KM_k141_46314 | Not-determined |
| KM_k141_180346 | Not-determined |
| KM_k141_102204 | Not-determined |
| KM_k141_13048 | Not-determined |
| KM_k141_180350 | Not-determined |
| KM_k141_102213 | Low-quality |
| KM_k141_102215 | Not-determined |
| KM_k141_213964 | Not-determined |
| KM_k141_147095 | Not-determined |
| KM_k141_114167 | Not-determined |
| KM_k141_258091 | Not-determined |
| KM_k141_235728 | Not-determined |
| KM_k141_35782 | Not-determined |
| KM_k141_258093 | Not-determined |
| KM_k141_57942 | Not-determined |
| KM_k141_258095 | Not-determined |

| | |
|---|---|
| KM_k141_180364 | Low-quality |
| KM_k141_213977 | Not-determined |
| KM_k141_24351 | Not-determined |
| KM_k141_180366 | Not-determined |
| KM_k141_80625 | Not-determined |
| KM_k141_13069 | Not-determined |
| KM_k141_2405 | Not-determined |
| KM_k141_169292 | Not-determined |
| KM_k141_57948 | Not-determined |
| KM_k141_102235 | Not-determined |
| KM_k141_114186 | Not-determined |
| KM_k141_203070 | Not-determined |
| KM_k141_46338 | Low-quality |
| KM_k141_203076 | Not-determined |
| KM_k141_114208 | Not-determined |
| KM_k141_180384 | Not-determined |
| KM_k141_35798 | Not-determined |
| KM_k141_225107 | Not-determined |
| KM_k141_13089 | Not-determined |
| KM_k141_225109 | Not-determined |
| KM_k141_13093 | Not-determined |
| KM_k141_157742 | Not-determined |
| KM_k141_214002 | Not-determined |
| KM_k141_246536 | Not-determined |
| KM_k141_114216 | Not-determined |
| KM_k141_80642 | Not-determined |
| KM_k141_57974 | Not-determined |
| KM_k141_80652 | Not-determined |
| KM_k141_102263 | Not-determined |
| KM_k141_246546 | Not-determined |
| KM_k141_246549 | Not-determined |
| KM_k141_102272 | Low-quality |
| KM_k141_80663 | Not-determined |
| KM_k141_35815 | Not-determined |
| KM_k141_235775 | Not-determined |
| KM_k141_191622 | Not-determined |
| KM_k141_114232 | Not-determined |
| KM_k141_114233 | Low-quality |
| KM_k141_102274 | Not-determined |
| KM_k141_80668 | Not-determined |
| KM_k141_214015 | Not-determined |
| KM_k141_80671 | Not-determined |
| KM_k141_2429 | Not-determined |
| KM_k141_191639 | Not-determined |
| KM_k141_203115 | Not-determined |
| KM_k141_13099 | Not-determined |
| KM_k141_114263 | Not-determined |
| KM_k141_214034 | Not-determined |
| KM_k141_235780 | Not-determined |
| KM_k141_191660 | Not-determined |
| KM_k141_246574 | Not-determined |
| KM_k141_225133 | Low-quality |
| KM_k141_214037 | Not-determined |
| KM_k141_214043 | Not-determined |
| KM_k141_2447 | Low-quality |
| KM_k141_13106 | Not-determined |
| KM_k141_13107 | Not-determined |
| KM_k141_258117 | Not-determined |
| KM_k141_57991 | Not-determined |
| KM_k141_203147 | Not-determined |
| KM_k141_46354 | Not-determined |
| KM_k141_225143 | Not-determined |
| KM_k141_2454 | Not-determined |
| KM_k141_225148 | Not-determined |
| KM_k141_46361 | Not-determined |
| KM_k141_24354 | Not-determined |
| KM_k141_35844 | Not-determined |
| KM_k141_147181 | Not-determined |
| KM_k141_91094 | Not-determined |
| KM_k141_24359 | Not-determined |
| KM_k141_169368 | Not-determined |
| KM_k141_203152 | Not-determined |
| KM_k141_58003 | Low-quality |
| KM_k141_191668 | Not-determined |
| KM_k141_235817 | Not-determined |
| KM_k141_235818 | Not-determined |
| KM_k141_35849 | Not-determined |
| KM_k141_180439 | Low-quality |
| KM_k141_235823 | Not-determined |
| KM_k141_35852 | Not-determined |
| KM_k141_157768 | Not-determined |
| KM_k141_80720 | Not-determined |
| KM_k141_58008 | Not-determined |
| KM_k141_102306 | Not-determined |
| KM_k141_80722 | Not-determined |
| KM_k141_35860 | Not-determined |
| KM_k141_125079 | Low-quality |
| KM_k141_2463 | Not-determined |
| KM_k141_180449 | Low-quality |
| KM_k141_102308 | Low-quality |
| KM_k141_203167 | Not-determined |
| KM_k141_169388 | Low-quality |
| KM_k141_102311 | Low-quality |
| KM_k141_235835 | Not-determined |
| KM_k141_180460 | Not-determined |
| KM_k141_102313 | Not-determined |
| KM_k141_214061 | Not-determined |
| KM_k141_214062 | Not-determined |
| KM_k141_191683 | Not-determined |
| KM_k141_258124 | Not-determined |
| KM_k141_214072 | Not-determined |
| KM_k141_136057 | Not-determined |
| KM_k141_214076 | Not-determined |
| KM_k141_214078 | Not-determined |
| KM_k141_80739 | Not-determined |
| KM_k141_69492 | Not-determined |
| KM_k141_235839 | Not-determined |
| KM_k141_180472 | Not-determined |
| KM_k141_125089 | Not-determined |
| KM_k141_58027 | Not-determined |
| KM_k141_2493 | Not-determined |
| KM_k141_235844 | Low-quality |
| KM_k141_125099 | Not-determined |
| KM_k141_69508 | Not-determined |
| KM_k141_13140 | Not-determined |
| KM_k141_125104 | Not-determined |
| KM_k141_169404 | Not-determined |
| KM_k141_258144 | Not-determined |
| KM_k141_246604 | Not-determined |
| KM_k141_114314 | Not-determined |
| KM_k141_35878 | Not-determined |
| KM_k141_125106 | Low-quality |
| KM_k141_13146 | Not-determined |
| KM_k141_2499 | Not-determined |
| KM_k141_169406 | Not-determined |
| KM_k141_58033 | Not-determined |
| KM_k141_125111 | Not-determined |
| KM_k141_235858 | Not-determined |
| KM_k141_2507 | Not-determined |
| KM_k141_147211 | Not-determined |
| KM_k141_258158 | Not-determined |
| KM_k141_246619 | Not-determined |
| KM_k141_246621 | Low-quality |
| KM_k141_258162 | Low-quality |
| KM_k141_191709 | Low-quality |
| KM_k141_246627 | Not-determined |
| KM_k141_80773 | Not-determined |
| KM_k141_157798 | Not-determined |
| KM_k141_114349 | Not-determined |
| KM_k141_13157 | Not-determined |
| KM_k141_258170 | Not-determined |
| KM_k141_258173 | Not-determined |
| KM_k141_35899 | Not-determined |
| KM_k141_147225 | Not-determined |
| KM_k141_13166 | Not-determined |
| KM_k141_258178 | Not-determined |
| KM_k141_125130 | Not-determined |
| KM_k141_24397 | Not-determined |
| KM_k141_246640 | Not-determined |
| KM_k141_13174 | Not-determined |
| KM_k141_180490 | Not-determined |
| KM_k141_258188 | Not-determined |
| KM_k141_246647 | Not-determined |
| KM_k141_169448 | Not-determined |
| KM_k141_136112 | Not-determined |
| KM_k141_35907 | Not-determined |
| KM_k141_203214 | Not-determined |
| KM_k141_180495 | Not-determined |
| KM_k141_180497 | Not-determined |
| KM_k141_125140 | Not-determined |
| KM_k141_114362 | Not-determined |
| KM_k141_225187 | Not-determined |
| KM_k141_147236 | Not-determined |
| KM_k141_246654 | Not-determined |
| KM_k141_114369 | Not-determined |
| KM_k141_246656 | Not-determined |
| KM_k141_102369 | Not-determined |
| KM_k141_169463 | Not-determined |
| KM_k141_102371 | Not-determined |
| KM_k141_13185 | Not-determined |
| KM_k141_2510 | Not-determined |
| KM_k141_246660 | Low-quality |
| KM_k141_13193 | Not-determined |
| KM_k141_203229 | Low-quality |
| KM_k141_246662 | Not-determined |
| KM_k141_258208 | Not-determined |
| KM_k141_203234 | Not-determined |
| KM_k141_13204 | Not-determined |
| KM_k141_235906 | Low-quality |
| KM_k141_235907 | Not-determined |
| KM_k141_246667 | Not-determined |
| KM_k141_147251 | Not-determined |
| KM_k141_147252 | Low-quality |
| KM_k141_2522 | Not-determined |
| KM_k141_147259 | Low-quality |
| KM_k141_24437 | Not-determined |
| KM_k141_13212 | Low-quality |
| KM_k141_169481 | Not-determined |
| KM_k141_114393 | Not-determined |
| KM_k141_169483 | Not-determined |
| KM_k141_235913 | Not-determined |
| KM_k141_125154 | Not-determined |
| KM_k141_13215 | Not-determined |
| KM_k141_246672 | Not-determined |
| KM_k141_69575 | Not-determined |
| KM_k141_180508 | Not-determined |
| KM_k141_214130 | Not-determined |
| KM_k141_13224 | Not-determined |
| KM_k141_24443 | Not-determined |
| KM_k141_102390 | Not-determined |
| KM_k141_102393 | Not-determined |
| KM_k141_24447 | Not-determined |
| KM_k141_24448 | Not-determined |
| KM_k141_169494 | Not-determined |
| KM_k141_169495 | Not-determined |
| KM_k141_69597 | Not-determined |
| KM_k141_235920 | Not-determined |
| KM_k141_180514 | Not-determined |
| KM_k141_180520 | Not-determined |
| KM_k141_246685 | Not-determined |
| KM_k141_203273 | Not-determined |
| KM_k141_69616 | Not-determined |
| KM_k141_147278 | Not-determined |
| KM_k141_58042 | Not-determined |
| KM_k141_2550 | Not-determined |
| KM_k141_80798 | Not-determined |
| KM_k141_180531 | Not-determined |
| KM_k141_125176 | Not-determined |
| KM_k141_35936 | Not-determined |
| KM_k141_235936 | Not-determined |
| KM_k141_246698 | Low-quality |
| KM_k141_258228 | Not-determined |
| KM_k141_46396 | Not-determined |
| KM_k141_258229 | Not-determined |
| KM_k141_225223 | Not-determined |
| KM_k141_246702 | Not-determined |
| KM_k141_24480 | Not-determined |
| KM_k141_114434 | Not-determined |
| KM_k141_24482 | Not-determined |
| KM_k141_69637 | Not-determined |
| KM_k141_214173 | Not-determined |
| KM_k141_24487 | Not-determined |
| KM_k141_214175 | Not-determined |
| KM_k141_214177 | Not-determined |
| KM_k141_46407 | Not-determined |
| KM_k141_180535 | Not-determined |
| KM_k141_80816 | Not-determined |
| KM_k141_169521 | Not-determined |
| KM_k141_80819 | Not-determined |
| KM_k141_58049 | Not-determined |
| KM_k141_24495 | Not-determined |
| KM_k141_136130 | Not-determined |
| KM_k141_258243 | Not-determined |
| KM_k141_35942 | Not-determined |
| KM_k141_147305 | Not-determined |
| KM_k141_147307 | Not-determined |
| KM_k141_191742 | Not-determined |
| KM_k141_169528 | Not-determined |
| KM_k141_13255 | Not-determined |
| KM_k141_58058 | Low-quality |
| KM_k141_191753 | Not-determined |
| KM_k141_114455 | Not-determined |
| KM_k141_69648 | Not-determined |
| KM_k141_35947 | Not-determined |
| KM_k141_69653 | Not-determined |
| KM_k141_69654 | Not-determined |
| KM_k141_258250 | Not-determined |
| KM_k141_69655 | Not-determined |
| KM_k141_246745 | Not-determined |
| KM_k141_169544 | Not-determined |
| KM_k141_214220 | Not-determined |
| KM_k141_35960 | Not-determined |
| KM_k141_58064 | Low-quality |
| KM_k141_114469 | Not-determined |
| KM_k141_80826 | Low-quality |
| KM_k141_191773 | Not-determined |
| KM_k141_191774 | Not-determined |
| KM_k141_246757 | Low-quality |
| KM_k141_136167 | Low-quality |
| KM_k141_180561 | Not-determined |
| KM_k141_80835 | Not-determined |
| KM_k141_246758 | Not-determined |
| KM_k141_180567 | Not-determined |
| KM_k141_80838 | Not-determined |
| KM_k141_246763 | Not-determined |
| KM_k141_102401 | Not-determined |
| KM_k141_203309 | Not-determined |
| KM_k141_180575 | Not-determined |
| KM_k141_180576 | Not-determined |
| KM_k141_136184 | Not-determined |
| KM_k141_203317 | Not-determined |
| KM_k141_80845 | Low-quality |
| KM_k141_235959 | Not-determined |
| KM_k141_235962 | Not-determined |
| KM_k141_180595 | Not-determined |
| KM_k141_203323 | Not-determined |
| KM_k141_235966 | Not-determined |
| KM_k141_214258 | Not-determined |
| KM_k141_80852 | Not-determined |
| KM_k141_2597 | Low-quality |
| KM_k141_180604 | Not-determined |
| KM_k141_246779 | Not-determined |
| KM_k141_169552 | Not-determined |
| KM_k141_80858 | Not-determined |
| KM_k141_80862 | Not-determined |
| KM_k141_136212 | Not-determined |
| KM_k141_180616 | Not-determined |
| KM_k141_136217 | Not-determined |
| KM_k141_58106 | Not-determined |
| KM_k141_80876 | Not-determined |
| KM_k141_214277 | Not-determined |
| KM_k141_157878 | Not-determined |
| KM_k141_102428 | Not-determined |
| KM_k141_58121 | Not-determined |
| KM_k141_36007 | Low-quality |
| KM_k141_191792 | Not-determined |
| KM_k141_102436 | Not-determined |
| KM_k141_258285 | Not-determined |
| KM_k141_191795 | Not-determined |
| KM_k141_58132 | Not-determined |
| KM_k141_13339 | Not-determined |
| KM_k141_147353 | Not-determined |
| KM_k141_147357 | Not-determined |
| KM_k141_236001 | Not-determined |
| KM_k141_214288 | Not-determined |
| KM_k141_147362 | Not-determined |
| KM_k141_180662 | Not-determined |
| KM_k141_114501 | Not-determined |
| KM_k141_24543 | Not-determined |
| KM_k141_24545 | Not-determined |
| KM_k141_169565 | Not-determined |
| KM_k141_102447 | Not-determined |
| KM_k141_236003 | Not-determined |
| KM_k141_147376 | Not-determined |
| KM_k141_236004 | Not-determined |
| KM_k141_191824 | Not-determined |
| KM_k141_258301 | Low-quality |
| KM_k141_203344 | Not-determined |
| KM_k141_69688 | Not-determined |
| KM_k141_69689 | Not-determined |
| KM_k141_214304 | Not-determined |
| KM_k141_114515 | Not-determined |
| KM_k141_236014 | Not-determined |
| KM_k141_114517 | Not-determined |
| KM_k141_169573 | Not-determined |
| KM_k141_36014 | Not-determined |
| KM_k141_246814 | Not-determined |
| KM_k141_180680 | Not-determined |
| KM_k141_147383 | Not-determined |
| KM_k141_236023 | Not-determined |
| KM_k141_58159 | Not-determined |
| KM_k141_58161 | Not-determined |
| KM_k141_191847 | Not-determined |
| KM_k141_102463 | Not-determined |
| KM_k141_114534 | Not-determined |
| KM_k141_46415 | Not-determined |
| KM_k141_147397 | Not-determined |
| KM_k141_180696 | Not-determined |
| KM_k141_157903 | Low-quality |
| KM_k141_157905 | Not-determined |
| KM_k141_58166 | Not-determined |
| KM_k141_180700 | Not-determined |
| KM_k141_114542 | Not-determined |
| KM_k141_147399 | Not-determined |
| KM_k141_246823 | Not-determined |
| KM_k141_46420 | Not-determined |
| KM_k141_214311 | Not-determined |
| KM_k141_147403 | Not-determined |
| KM_k141_147404 | Not-determined |
| KM_k141_214314 | Not-determined |
| KM_k141_136241 | Not-determined |
| KM_k141_214316 | Not-determined |
| KM_k141_102477 | Not-determined |
| KM_k141_225316 | Not-determined |
| KM_k141_225318 | Not-determined |
| KM_k141_214323 | Not-determined |
| KM_k141_157913 | Not-determined |
| KM_k141_58180 | Not-determined |
| KM_k141_58183 | Not-determined |
| KM_k141_180713 | Not-determined |
| KM_k141_125209 | Not-determined |
| KM_k141_46432 | Not-determined |
| KM_k141_136257 | Not-determined |
| KM_k141_136261 | Not-determined |
| KM_k141_214339 | Not-determined |
| KM_k141_46444 | Not-determined |
| KM_k141_46446 | Not-determined |
| KM_k141_114556 | Not-determined |
| KM_k141_36043 | Not-determined |
| KM_k141_36047 | Not-determined |
| KM_k141_36049 | Not-determined |
| KM_k141_58199 | Not-determined |
| KM_k141_225326 | Not-determined |
| KM_k141_258305 | Not-determined |
| KM_k141_125234 | Not-determined |
| KM_k141_258311 | Not-determined |
| KM_k141_24561 | Not-determined |
| KM_k141_69709 | Not-determined |
| KM_k141_136281 | Not-determined |
| KM_k141_24569 | Not-determined |
| KM_k141_136286 | Not-determined |
| KM_k141_2683 | Not-determined |
| KM_k141_36057 | Not-determined |
| KM_k141_102511 | Not-determined |
| KM_k141_46463 | Not-determined |
| KM_k141_36061 | Not-determined |
| KM_k141_236081 | Not-determined |
| KM_k141_125253 | Not-determined |
| KM_k141_58229 | Not-determined |
| KM_k141_214374 | Not-determined |
| KM_k141_136296 | Not-determined |
| KM_k141_24587 | Not-determined |
| KM_k141_180770 | Not-determined |
| KM_k141_180772 | Not-determined |
| KM_k141_236092 | Not-determined |
| KM_k141_69725 | Not-determined |
| KM_k141_157961 | Not-determined |
| KM_k141_136304 | Not-determined |
| KM_k141_258331 | Low-quality |
| KM_k141_2697 | Not-determined |
| KM_k141_2699 | Not-determined |
| KM_k141_147408 | Not-determined |
| KM_k141_157964 | Not-determined |
| KM_k141_80939 | Not-determined |
| KM_k141_180787 | Not-determined |
| KM_k141_125265 | Low-quality |
| KM_k141_236105 | Not-determined |
| KM_k141_136321 | Not-determined |
| KM_k141_136324 | Not-determined |
| KM_k141_46477 | Not-determined |
| KM_k141_114589 | Not-determined |
| KM_k141_180803 | Not-determined |
| KM_k141_46483 | Not-determined |
| KM_k141_69736 | Not-determined |
| KM_k141_69737 | Low-quality |
| KM_k141_258352 | Not-determined |
| KM_k141_157981 | Not-determined |
| KM_k141_191860 | Not-determined |
| KM_k141_58274 | Not-determined |
| KM_k141_258361 | Not-determined |
| KM_k141_46497 | Not-determined |
| KM_k141_191865 | Not-determined |
| KM_k141_157983 | Not-determined |
| KM_k141_46500 | Not-determined |
| KM_k141_13415 | Not-determined |
| KM_k141_191870 | Not-determined |
| KM_k141_157988 | Not-determined |
| KM_k141_191873 | Not-determined |
| KM_k141_136333 | Not-determined |
| KM_k141_225362 | Not-determined |
| KM_k141_236115 | Not-determined |
| KM_k141_46504 | Not-determined |
| KM_k141_258380 | Not-determined |
| KM_k141_58296 | Not-determined |
| KM_k141_46507 | Not-determined |
| KM_k141_225366 | Not-determined |
| KM_k141_36077 | Not-determined |
| KM_k141_13418 | Low-quality |
| KM_k141_58300 | Not-determined |
| KM_k141_125310 | Not-determined |
| KM_k141_136344 | Not-determined |
| KM_k141_69761 | Not-determined |
| KM_k141_225370 | Low-quality |
| KM_k141_147421 | Not-determined |
| KM_k141_58304 | Not-determined |
| KM_k141_46516 | Not-determined |
| KM_k141_125320 | Not-determined |
| KM_k141_2736 | Not-determined |
| KM_k141_69766 | Low-quality |
| KM_k141_158000 | Not-determined |
| KM_k141_147428 | Not-determined |
| KM_k141_158001 | Not-determined |
| KM_k141_147433 | Not-determined |
| KM_k141_158011 | Not-determined |
| KM_k141_13427 | Not-determined |
| KM_k141_158015 | Not-determined |
| KM_k141_147439 | Not-determined |
| KM_k141_125338 | Not-determined |
| KM_k141_225378 | Not-determined |
| KM_k141_36101 | Low-quality |
| KM_k141_69782 | Not-determined |
| KM_k141_13438 | Not-determined |
| KM_k141_147445 | Not-determined |
| KM_k141_136376 | Not-determined |
| KM_k141_214402 | Low-quality |
| KM_k141_214413 | Not-determined |
| KM_k141_125356 | Not-determined |
| KM_k141_191905 | Not-determined |
| KM_k141_191909 | Not-determined |
| KM_k141_36112 | Not-determined |
| KM_k141_36113 | Not-determined |
| KM_k141_214421 | Not-determined |
| KM_k141_225387 | Not-determined |
| KM_k141_214430 | Not-determined |
| KM_k141_225400 | Not-determined |
| KM_k141_158028 | Not-determined |
| KM_k141_203374 | Not-determined |
| KM_k141_214436 | Low-quality |
| KM_k141_102596 | Not-determined |
| KM_k141_13447 | Not-determined |
| KM_k141_136391 | Not-determined |
| KM_k141_125386 | Not-determined |
| KM_k141_214445 | Not-determined |
| KM_k141_191919 | Not-determined |
| KM_k141_136393 | Low-quality |
| KM_k141_24634 | Not-determined |
| KM_k141_125394 | Not-determined |
| KM_k141_125395 | Not-determined |
| KM_k141_24640 | Not-determined |
| KM_k141_246830 | Not-determined |
| KM_k141_2774 | Not-determined |
| KM_k141_24642 | Not-determined |
| KM_k141_136399 | Not-determined |
| KM_k141_81064 | Low-quality |
| KM_k141_24652 | Not-determined |
| KM_k141_214473 | Not-determined |
| KM_k141_180835 | Not-determined |
| KM_k141_246841 | Not-determined |
| KM_k141_69811 | Not-determined |
| KM_k141_180836 | Not-determined |
| KM_k141_125407 | Not-determined |
| KM_k141_180842 | Not-determined |
| KM_k141_169630 | Not-determined |
| KM_k141_58344 | Not-determined |
| KM_k141_214478 | Not-determined |
| KM_k141_69820 | Low-quality |
| KM_k141_214479 | Not-determined |
| KM_k141_125411 | Not-determined |
| KM_k141_180863 | Not-determined |
| KM_k141_69829 | Not-determined |
| KM_k141_69830 | Not-determined |
| KM_k141_81083 | Not-determined |
| KM_k141_102631 | Not-determined |
| KM_k141_24673 | Not-determined |
| KM_k141_214491 | Not-determined |
| KM_k141_180878 | Low-quality |
| KM_k141_114634 | Not-determined |
| KM_k141_114635 | Not-determined |
| KM_k141_125434 | Not-determined |
| KM_k141_136432 | Not-determined |
| KM_k141_114636 | Not-determined |
| KM_k141_136433 | Not-determined |
| KM_k141_191949 | Low-quality |
| KM_k141_24686 | Not-determined |
| KM_k141_58352 | Not-determined |
| KM_k141_125440 | Not-determined |
| KM_k141_246859 | Not-determined |
| KM_k141_191955 | Not-determined |
| KM_k141_158039 | Not-determined |
| KM_k141_169645 | Low-quality |
| KM_k141_13480 | Not-determined |
| KM_k141_158049 | Not-determined |
| KM_k141_125444 | Not-determined |
| KM_k141_36152 | Not-determined |
| KM_k141_24703 | Not-determined |
| KM_k141_125457 | Not-determined |
| KM_k141_13491 | Not-determined |
| KM_k141_36161 | Low-quality |
| KM_k141_24709 | Not-determined |
| KM_k141_158060 | Not-determined |
| KM_k141_169665 | Low-quality |
| KM_k141_136458 | Not-determined |
| KM_k141_36168 | Low-quality |
| KM_k141_225459 | Not-determined |
| KM_k141_24721 | Not-determined |
| KM_k141_236131 | Not-determined |
| KM_k141_225461 | Not-determined |
| KM_k141_24723 | Not-determined |
| KM_k141_203397 | Not-determined |
| KM_k141_236134 | Not-determined |
| KM_k141_91168 | Low-quality |
| KM_k141_102674 | Not-determined |
| KM_k141_81123 | Not-determined |
| KM_k141_81124 | Not-determined |
| KM_k141_46567 | Not-determined |
| KM_k141_69834 | Not-determined |
| KM_k141_24730 | Not-determined |
| KM_k141_36183 | Not-determined |
| KM_k141_81131 | Not-determined |
| KM_k141_81143 | Not-determined |
| KM_k141_158069 | Not-determined |
| KM_k141_91173 | Not-determined |
| KM_k141_125486 | Not-determined |
| KM_k141_203405 | Not-determined |
| KM_k141_158072 | Not-determined |
| KM_k141_136478 | Not-determined |
| KM_k141_158073 | Low-quality |
| KM_k141_246906 | Not-determined |
| KM_k141_102683 | Not-determined |
| KM_k141_91181 | Low-quality |
| KM_k141_246910 | Not-determined |
| KM_k141_203417 | Low-quality |
| KM_k141_158077 | Not-determined |
| KM_k141_158078 | Not-determined |
| KM_k141_114668 | Not-determined |
| KM_k141_169684 | Not-determined |
| KM_k141_114673 | Not-determined |
| KM_k141_136500 | Not-determined |
| KM_k141_225469 | Not-determined |

| | |
|---|---|
| KM_k141_225470 | Not-determined |
| KM_k141_246922 | Not-determined |
| KM_k141_36213 | Not-determined |
| KM_k141_246925 | Not-determined |
| KM_k141_246926 | Not-determined |
| KM_k141_81172 | Not-determined |
| KM_k141_102697 | Not-determined |
| KM_k141_36221 | Not-determined |
| KM_k141_214528 | Not-determined |
| KM_k141_102712 | Not-determined |
| KM_k141_114705 | Not-determined |
| KM_k141_169707 | Not-determined |
| KM_k141_225491 | Low-quality |
| KM_k141_225493 | Not-determined |
| KM_k141_258403 | Not-determined |
| KM_k141_136519 | Not-determined |
| KM_k141_225497 | Not-determined |
| KM_k141_225498 | Low-quality |
| KM_k141_136526 | Not-determined |
| KM_k141_191987 | Not-determined |
| KM_k141_81194 | Not-determined |
| KM_k141_147495 | Not-determined |
| KM_k141_81198 | Not-determined |
| KM_k141_147500 | Low-quality |
| KM_k141_214540 | Not-determined |
| KM_k141_102745 | Not-determined |
| KM_k141_24753 | Not-determined |
| KM_k141_225512 | Not-determined |
| KM_k141_81208 | Not-determined |
| KM_k141_258425 | Not-determined |
| KM_k141_24759 | Low-quality |
| KM_k141_236161 | Not-determined |
| KM_k141_147507 | Not-determined |
| KM_k141_2789 | Not-determined |
| KM_k141_81217 | Low-quality |
| KM_k141_203455 | Not-determined |
| KM_k141_114738 | Not-determined |
| KM_k141_13532 | Not-determined |
| KM_k141_13533 | Not-determined |
| KM_k141_2802 | Not-determined |
| KM_k141_91232 | Not-determined |
| KM_k141_2805 | Not-determined |
| KM_k141_46603 | Not-determined |
| KM_k141_2807 | Not-determined |
| KM_k141_136569 | Not-determined |
| KM_k141_81237 | Not-determined |
| KM_k141_136586 | Not-determined |
| KM_k141_136589 | Not-determined |
| KM_k141_125528 | Not-determined |
| KM_k141_246952 | Not-determined |
| KM_k141_2827 | Not-determined |
| KM_k141_125536 | Low-quality |
| KM_k141_2829 | Not-determined |
| KM_k141_2831 | Not-determined |
| KM_k141_114767 | Not-determined |
| KM_k141_13553 | Not-determined |
| KM_k141_125541 | Not-determined |
| KM_k141_2838 | Not-determined |
| KM_k141_91246 | Not-determined |
| KM_k141_258460 | Not-determined |
| KM_k141_81260 | Not-determined |
| KM_k141_91248 | Not-determined |
| KM_k141_125546 | Low-quality |
| KM_k141_191998 | Not-determined |
| KM_k141_246959 | Not-determined |
| KM_k141_13558 | Not-determined |
| KM_k141_46615 | Not-determined |
| KM_k141_136608 | Not-determined |
| KM_k141_125551 | Not-determined |
| KM_k141_13562 | Low-quality |
| KM_k141_91252 | Not-determined |
| KM_k141_46624 | Not-determined |
| KM_k141_2848 | Not-determined |
| KM_k141_203490 | Not-determined |
| KM_k141_2853 | Not-determined |
| KM_k141_181003 | Not-determined |
| KM_k141_81264 | Not-determined |
| KM_k141_214561 | Not-determined |
| KM_k141_24781 | Not-determined |
| KM_k141_24783 | Not-determined |
| KM_k141_236214 | Not-determined |
| KM_k141_102785 | Not-determined |
| KM_k141_81267 | Not-determined |
| KM_k141_236216 | Not-determined |
| KM_k141_258476 | Not-determined |
| KM_k141_102790 | Not-determined |
| KM_k141_258478 | Not-determined |
| KM_k141_258479 | Not-determined |
| KM_k141_258487 | Not-determined |
| KM_k141_114815 | Not-determined |
| KM_k141_246987 | Not-determined |
| KM_k141_246988 | Not-determined |
| KM_k141_236219 | Not-determined |
| KM_k141_91281 | Not-determined |
| KM_k141_192044 | Not-determined |
| KM_k141_192047 | Not-determined |
| KM_k141_258492 | Not-determined |
| KM_k141_13592 | Not-determined |
| KM_k141_181039 | Not-determined |
| KM_k141_181040 | Not-determined |
| KM_k141_225527 | Not-determined |
| KM_k141_36243 | Not-determined |
| KM_k141_91285 | Not-determined |
| KM_k141_246996 | Not-determined |
| KM_k141_258496 | Not-determined |
| KM_k141_246997 | Not-determined |
| KM_k141_192052 | Not-determined |
| KM_k141_36249 | Not-determined |
| KM_k141_181048 | Not-determined |
| KM_k141_192053 | Not-determined |
| KM_k141_214572 | Not-determined |
| KM_k141_114829 | Low-quality |
| KM_k141_181055 | Not-determined |
| KM_k141_36254 | Not-determined |
| KM_k141_225535 | Not-determined |
| KM_k141_114834 | Not-determined |
| KM_k141_236237 | Not-determined |
| KM_k141_236239 | Not-determined |
| KM_k141_2895 | Not-determined |
| KM_k141_225545 | Not-determined |
| KM_k141_203512 | Not-determined |
| KM_k141_236240 | Not-determined |
| KM_k141_247011 | Not-determined |
| KM_k141_258506 | Not-determined |
| KM_k141_203517 | Not-determined |
| KM_k141_181080 | Low-quality |
| KM_k141_158096 | Not-determined |
| KM_k141_81283 | Not-determined |
| KM_k141_203529 | Not-determined |
| KM_k141_81286 | Not-determined |
| KM_k141_114853 | Not-determined |
| KM_k141_69905 | Not-determined |
| KM_k141_114855 | Not-determined |
| KM_k141_69907 | Not-determined |
| KM_k141_214591 | Not-determined |
| KM_k141_69911 | Not-determined |
| KM_k141_181088 | Not-determined |
| KM_k141_203544 | Not-determined |
| KM_k141_147578 | Not-determined |
| KM_k141_114871 | Not-determined |
| KM_k141_225574 | Not-determined |
| KM_k141_24834 | Not-determined |
| KM_k141_81301 | Not-determined |
| KM_k141_203548 | Not-determined |
| KM_k141_258534 | Not-determined |
| KM_k141_2924 | Not-determined |
| KM_k141_2930 | Not-determined |
| KM_k141_125570 | Not-determined |
| KM_k141_114883 | Not-determined |
| KM_k141_81313 | Not-determined |
| KM_k141_81315 | Not-determined |
| KM_k141_214620 | Not-determined |
| KM_k141_24854 | Not-determined |
| KM_k141_125576 | Not-determined |
| KM_k141_81324 | Not-determined |
| KM_k141_102827 | Not-determined |
| KM_k141_225591 | Not-determined |
| KM_k141_114898 | Not-determined |
| KM_k141_247028 | Not-determined |
| KM_k141_46648 | Not-determined |
| KM_k141_125584 | Low-quality |
| KM_k141_36283 | Not-determined |
| KM_k141_102835 | Not-determined |
| KM_k141_46652 | Not-determined |
| KM_k141_247033 | Not-determined |
| KM_k141_46653 | Not-determined |
| KM_k141_258537 | Not-determined |
| KM_k141_214630 | Not-determined |
| KM_k141_114909 | Not-determined |
| KM_k141_102842 | Not-determined |
| KM_k141_158111 | Not-determined |
| KM_k141_247035 | Low-quality |
| KM_k141_102846 | Not-determined |
| KM_k141_13651 | Not-determined |
| KM_k141_13652 | Not-determined |
| KM_k141_69920 | Not-determined |
| KM_k141_225605 | Not-determined |
| KM_k141_258548 | Not-determined |
| KM_k141_24864 | Not-determined |
| KM_k141_181120 | Not-determined |
| KM_k141_192099 | Not-determined |
| KM_k141_125600 | Not-determined |
| KM_k141_91319 | Not-determined |
| KM_k141_258551 | Not-determined |
| KM_k141_91320 | Not-determined |
| KM_k141_169792 | Not-determined |
| KM_k141_36295 | Not-determined |
| KM_k141_2945 | Not-determined |
| KM_k141_114929 | Not-determined |
| KM_k141_203568 | Low-quality |
| KM_k141_24876 | Not-determined |
| KM_k141_203570 | Not-determined |
| KM_k141_102860 | Not-determined |
| KM_k141_125616 | Not-determined |
| KM_k141_192121 | Not-determined |
| KM_k141_258565 | Not-determined |
| KM_k141_214668 | Not-determined |
| KM_k141_102870 | Not-determined |
| KM_k141_46661 | Not-determined |
| KM_k141_147622 | Not-determined |
| KM_k141_2958 | Not-determined |
| KM_k141_258570 | Not-determined |
| KM_k141_147625 | Not-determined |
| KM_k141_147626 | Not-determined |
| KM_k141_169816 | Not-determined |
| KM_k141_102882 | Low-quality |
| KM_k141_225634 | Not-determined |
| KM_k141_58380 | Not-determined |
| KM_k141_236255 | Not-determined |
| KM_k141_258580 | Not-determined |
| KM_k141_58381 | Not-determined |
| KM_k141_58382 | Not-determined |
| KM_k141_236258 | Not-determined |
| KM_k141_125642 | Not-determined |
| KM_k141_102888 | Not-determined |
| KM_k141_169824 | Not-determined |
| KM_k141_125646 | Not-determined |
| KM_k141_36317 | Not-determined |
| KM_k141_2971 | Not-determined |
| KM_k141_236270 | Low-quality |
| KM_k141_147637 | Not-determined |
| KM_k141_181136 | Not-determined |
| KM_k141_58389 | Not-determined |
| KM_k141_2974 | Not-determined |
| KM_k141_181143 | Not-determined |
| KM_k141_2979 | Not-determined |
| KM_k141_181144 | Not-determined |
| KM_k141_136630 | Not-determined |
| KM_k141_136633 | Not-determined |
| KM_k141_58399 | Not-determined |
| KM_k141_147642 | Low-quality |
| KM_k141_136637 | Not-determined |
| KM_k141_236282 | Not-determined |
| KM_k141_125671 | Not-determined |
| KM_k141_181152 | Not-determined |
| KM_k141_203610 | Low-quality |
| KM_k141_203613 | Not-determined |
| KM_k141_203614 | Low-quality |
| KM_k141_136646 | Not-determined |
| KM_k141_125680 | Not-determined |
| KM_k141_136651 | Not-determined |
| KM_k141_158157 | Not-determined |
| KM_k141_24919 | Not-determined |
| KM_k141_192165 | Not-determined |
| KM_k141_2995 | Not-determined |
| KM_k141_192170 | Not-determined |
| KM_k141_114950 | Not-determined |
| KM_k141_46677 | Low-quality |
| KM_k141_192179 | Not-determined |
| KM_k141_46680 | Not-determined |
| KM_k141_69937 | Not-determined |
| KM_k141_36345 | Not-determined |
| KM_k141_3000 | Not-determined |
| KM_k141_192183 | Low-quality |
| KM_k141_225666 | Not-determined |
| KM_k141_181160 | Not-determined |
| KM_k141_158172 | Low-quality |
| KM_k141_203627 | Not-determined |
| KM_k141_203628 | Not-determined |
| KM_k141_24929 | Not-determined |
| KM_k141_69943 | Not-determined |
| KM_k141_214691 | Low-quality |
| KM_k141_3011 | Not-determined |
| KM_k141_81368 | Not-determined |
| KM_k141_81371 | Not-determined |
| KM_k141_69951 | Not-determined |
| KM_k141_114983 | Low-quality |
| KM_k141_114989 | Not-determined |
| KM_k141_91326 | Not-determined |
| KM_k141_125703 | Not-determined |
| KM_k141_147675 | Not-determined |
| KM_k141_81377 | Not-determined |
| KM_k141_69959 | Not-determined |
| KM_k141_192212 | Low-quality |
| KM_k141_236289 | Not-determined |
| KM_k141_36382 | Not-determined |
| KM_k141_236295 | Not-determined |
| KM_k141_203649 | Not-determined |
| KM_k141_203651 | Not-determined |
| KM_k141_236297 | Not-determined |
| KM_k141_192223 | Not-determined |
| KM_k141_136657 | Not-determined |
| KM_k141_91339 | Not-determined |
| KM_k141_3034 | Not-determined |
| KM_k141_203657 | Low-quality |
| KM_k141_102913 | Not-determined |
| KM_k141_136669 | Not-determined |
| KM_k141_236303 | Not-determined |
| KM_k141_214707 | Not-determined |
| KM_k141_91351 | Not-determined |
| KM_k141_102923 | Not-determined |
| KM_k141_236309 | Not-determined |
| KM_k141_36396 | Not-determined |
| KM_k141_214716 | Not-determined |
| KM_k141_203676 | Not-determined |
| KM_k141_102933 | Not-determined |
| KM_k141_147696 | Not-determined |
| KM_k141_125720 | Not-determined |
| KM_k141_24948 | Not-determined |
| KM_k141_115022 | Not-determined |
| KM_k141_125727 | Not-determined |
| KM_k141_125730 | Not-determined |
| KM_k141_69997 | Not-determined |
| KM_k141_236325 | Not-determined |
| KM_k141_69999 | Not-determined |
| KM_k141_236327 | Not-determined |
| KM_k141_236328 | Not-determined |
| KM_k141_147706 | Not-determined |
| KM_k141_70004 | Not-determined |
| KM_k141_192244 | Not-determined |
| KM_k141_214737 | Not-determined |
| KM_k141_91375 | Not-determined |
| KM_k141_181232 | Not-determined |
| KM_k141_169857 | Not-determined |
| KM_k141_70013 | Not-determined |
| KM_k141_158185 | Not-determined |
| KM_k141_214741 | Low-quality |
| KM_k141_192253 | Not-determined |
| KM_k141_91381 | Low-quality |
| KM_k141_214744 | Not-determined |
| KM_k141_169865 | Low-quality |
| KM_k141_3063 | Not-determined |
| KM_k141_192260 | Not-determined |
| KM_k141_158194 | Low-quality |
| KM_k141_24974 | Not-determined |
| KM_k141_24975 | Not-determined |
| KM_k141_102959 | Not-determined |
| KM_k141_192269 | Not-determined |
| KM_k141_3069 | Not-determined |
| KM_k141_102960 | Not-determined |
| KM_k141_136713 | Not-determined |
| KM_k141_36430 | Not-determined |
| KM_k141_258606 | Low-quality |
| KM_k141_225688 | Not-determined |
| KM_k141_3074 | Not-determined |
| KM_k141_58460 | Low-quality |
| KM_k141_169878 | Not-determined |
| KM_k141_236372 | Not-determined |
| KM_k141_81418 | Not-determined |
| KM_k141_46781 | Not-determined |
| KM_k141_36439 | Not-determined |
| KM_k141_70042 | Not-determined |
| KM_k141_236384 | Not-determined |
| KM_k141_258625 | Not-determined |
| KM_k141_136743 | Not-determined |
| KM_k141_136746 | Not-determined |
| KM_k141_147721 | Not-determined |
| KM_k141_115059 | Low-quality |
| KM_k141_102967 | Not-determined |
| KM_k141_147723 | Not-determined |
| KM_k141_3095 | Low-quality |
| KM_k141_115065 | Not-determined |
| KM_k141_3096 | Not-determined |
| KM_k141_125742 | Low-quality |
| KM_k141_46794 | Not-determined |
| KM_k141_3101 | Not-determined |
| KM_k141_91413 | Not-determined |
| KM_k141_81445 | Not-determined |
| KM_k141_46799 | Not-determined |
| KM_k141_70056 | Not-determined |
| KM_k141_102979 | Not-determined |
| KM_k141_136761 | Low-quality |
| KM_k141_136766 | Low-quality |
| KM_k141_125745 | Not-determined |
| KM_k141_236412 | Not-determined |
| KM_k141_225717 | Not-determined |
| KM_k141_115086 | Not-determined |
| KM_k141_192307 | Not-determined |
| KM_k141_236416 | Not-determined |
| KM_k141_91421 | Not-determined |
| KM_k141_136774 | Not-determined |
| KM_k141_3115 | Not-determined |
| KM_k141_58487 | Not-determined |
| KM_k141_58488 | Not-determined |
| KM_k141_58490 | Not-determined |
| KM_k141_236422 | Not-determined |
| KM_k141_225733 | Low-quality |
| KM_k141_81471 | Not-determined |
| KM_k141_81474 | Not-determined |
| KM_k141_181261 | Not-determined |
| KM_k141_147746 | Not-determined |
| KM_k141_115102 | Not-determined |
| KM_k141_147758 | Low-quality |
| KM_k141_236442 | Not-determined |
| KM_k141_24989 | Not-determined |
| KM_k141_214783 | Not-determined |
| KM_k141_247111 | Not-determined |
| KM_k141_147767 | Not-determined |
| KM_k141_24997 | Not-determined |
| KM_k141_192347 | Not-determined |
| KM_k141_24998 | Not-determined |
| KM_k141_181272 | Not-determined |
| KM_k141_25002 | Not-determined |
| KM_k141_147774 | Not-determined |
| KM_k141_147776 | Not-determined |
| KM_k141_125776 | Not-determined |
| KM_k141_25007 | Not-determined |
| KM_k141_3162 | Not-determined |
| KM_k141_70071 | Not-determined |
| KM_k141_13701 | Low-quality |
| KM_k141_147785 | Not-determined |
| KM_k141_136791 | Not-determined |
| KM_k141_125785 | Not-determined |
| KM_k141_70079 | Not-determined |
| KM_k141_192360 | Not-determined |
| KM_k141_25024 | Not-determined |
| KM_k141_115126 | Not-determined |
| KM_k141_81505 | Not-determined |
| KM_k141_125791 | Not-determined |
| KM_k141_192366 | Not-determined |
| KM_k141_3165 | Not-determined |
| KM_k141_181292 | Not-determined |
| KM_k141_81510 | Low-quality |
| KM_k141_3168 | Not-determined |
| KM_k141_125800 | Not-determined |
| KM_k141_203715 | Not-determined |
| KM_k141_203716 | Not-determined |
| KM_k141_203722 | Not-determined |
| KM_k141_25045 | Not-determined |
| KM_k141_36455 | Not-determined |
| KM_k141_70097 | Not-determined |
| KM_k141_125809 | Not-determined |
| KM_k141_81520 | Not-determined |
| KM_k141_115137 | Not-determined |
| KM_k141_147831 | Not-determined |
| KM_k141_103028 | Not-determined |
| KM_k141_36461 | Not-determined |
| KM_k141_136836 | Not-determined |
| KM_k141_125821 | Not-determined |
| KM_k141_225751 | Not-determined |
| KM_k141_81527 | Not-determined |
| KM_k141_58537 | Not-determined |
| KM_k141_136847 | Not-determined |
| KM_k141_58539 | Not-determined |
| KM_k141_58540 | Not-determined |
| KM_k141_25059 | Low-quality |
| KM_k141_58543 | Low-quality |
| KM_k141_169930 | Not-determined |
| KM_k141_225759 | Not-determined |
| KM_k141_58551 | Not-determined |
| KM_k141_25064 | Not-determined |
| KM_k141_103041 | Not-determined |
| KM_k141_25067 | Not-determined |
| KM_k141_147855 | Not-determined |
| KM_k141_258651 | Not-determined |
| KM_k141_258652 | Not-determined |
| KM_k141_247139 | Not-determined |
| KM_k141_36484 | Not-determined |
| KM_k141_247146 | Not-determined |
| KM_k141_258657 | Not-determined |
| KM_k141_247147 | Not-determined |
| KM_k141_258658 | Not-determined |
| KM_k141_36491 | Not-determined |
| KM_k141_58578 | Low-quality |
| KM_k141_25078 | Not-determined |
| KM_k141_192414 | Not-determined |
| KM_k141_58586 | Not-determined |
| KM_k141_58587 | Not-determined |
| KM_k141_147870 | Not-determined |
| KM_k141_192420 | Not-determined |
| KM_k141_115175 | Not-determined |
| KM_k141_192427 | Not-determined |
| KM_k141_115182 | Not-determined |
| KM_k141_147881 | Not-determined |
| KM_k141_13725 | Not-determined |
| KM_k141_115183 | Not-determined |
| KM_k141_70106 | Not-determined |
| KM_k141_13726 | Not-determined |
| KM_k141_192435 | Not-determined |
| KM_k141_125838 | Not-determined |
| KM_k141_192439 | Not-determined |
| KM_k141_203771 | Not-determined |
| KM_k141_192443 | Not-determined |
| KM_k141_147889 | Not-determined |
| KM_k141_181360 | Not-determined |
| KM_k141_247159 | Not-determined |
| KM_k141_36510 | Not-determined |
| KM_k141_203778 | Not-determined |
| KM_k141_192461 | Not-determined |
| KM_k141_192462 | Not-determined |
| KM_k141_158250 | Not-determined |
| KM_k141_25091 | Not-determined |
| KM_k141_115209 | Not-determined |
| KM_k141_103085 | Not-determined |
| KM_k141_236490 | Not-determined |
| KM_k141_225783 | Not-determined |
| KM_k141_225784 | Not-determined |
| KM_k141_103092 | Not-determined |
| KM_k141_25099 | Low-quality |
| KM_k141_103094 | Not-determined |
| KM_k141_25102 | Not-determined |
| KM_k141_125858 | Not-determined |
| KM_k141_236494 | Not-determined |
| KM_k141_125861 | Not-determined |
| KM_k141_36530 | Not-determined |
| KM_k141_181367 | Not-determined |
| KM_k141_147904 | Not-determined |
| KM_k141_225798 | Not-determined |
| KM_k141_181371 | Low-quality |
| KM_k141_70133 | Not-determined |
| KM_k141_225801 | Not-determined |
| KM_k141_225802 | Not-determined |
| KM_k141_125873 | Not-determined |
| KM_k141_236500 | Not-determined |
| KM_k141_115232 | Not-determined |
| KM_k141_103100 | Not-determined |
| KM_k141_136877 | Not-determined |
| KM_k141_236504 | Not-determined |
| KM_k141_214841 | Not-determined |
| KM_k141_203813 | Not-determined |
| KM_k141_136878 | Not-determined |
| KM_k141_58626 | Low-quality |
| KM_k141_225817 | Not-determined |
| KM_k141_25128 | Not-determined |
| KM_k141_91456 | Not-determined |
| KM_k141_70148 | Not-determined |
| KM_k141_158283 | Not-determined |
| KM_k141_13765 | Not-determined |
| KM_k141_214850 | Not-determined |
| KM_k141_58633 | Not-determined |
| KM_k141_36552 | Not-determined |
| KM_k141_103111 | Not-determined |
| KM_k141_169964 | Not-determined |
| KM_k141_25133 | Not-determined |
| KM_k141_81556 | Not-determined |
| KM_k141_70160 | Not-determined |
| KM_k141_36557 | Not-determined |
| KM_k141_247175 | Low-quality |
| KM_k141_25139 | Not-determined |
| KM_k141_91468 | Not-determined |
| KM_k141_91469 | Not-determined |
| KM_k141_181375 | Not-determined |
| KM_k141_13771 | Not-determined |
| KM_k141_181377 | Not-determined |
| KM_k141_225836 | Not-determined |
| KM_k141_13774 | Not-determined |
| KM_k141_225840 | Not-determined |
| KM_k141_125885 | Not-determined |
| KM_k141_158300 | Not-determined |
| KM_k141_136888 | Not-determined |
| KM_k141_58641 | Not-determined |
| KM_k141_181384 | Not-determined |
| KM_k141_225851 | Low-quality |
| KM_k141_25152 | Not-determined |
| KM_k141_103129 | Not-determined |
| KM_k141_192501 | Not-determined |
| KM_k141_258673 | Not-determined |
| KM_k141_247184 | Not-determined |
| KM_k141_36574 | Not-determined |
| KM_k141_91480 | Not-determined |
| KM_k141_192503 | Not-determined |
| KM_k141_81579 | Not-determined |
| KM_k141_181395 | Not-determined |
| KM_k141_58664 | Low-quality |
| KM_k141_169983 | Low-quality |
| KM_k141_36577 | Not-determined |
| KM_k141_181403 | Not-determined |
| KM_k141_258681 | Not-determined |
| KM_k141_203828 | Not-determined |
| KM_k141_258684 | Not-determined |
| KM_k141_192509 | Not-determined |
| KM_k141_3258 | Not-determined |
| KM_k141_258686 | Not-determined |
| KM_k141_3265 | Not-determined |
| KM_k141_258694 | Not-determined |
| KM_k141_70200 | Not-determined |
| KM_k141_70201 | Not-determined |
| KM_k141_115238 | Not-determined |
| KM_k141_115239 | Not-determined |
| KM_k141_36598 | Not-determined |
| KM_k141_258697 | Not-determined |
| KM_k141_136904 | Not-determined |
| KM_k141_70204 | Not-determined |
| KM_k141_258699 | Not-determined |
| KM_k141_169996 | Not-determined |
| KM_k141_115241 | Not-determined |
| KM_k141_115244 | Not-determined |
| KM_k141_115246 | Low-quality |
| KM_k141_192518 | Not-determined |
| KM_k141_181423 | Not-determined |
| KM_k141_136928 | Not-determined |
| KM_k141_192526 | Not-determined |
| KM_k141_203859 | Not-determined |
| KM_k141_70217 | Not-determined |
| KM_k141_247194 | Not-determined |
| KM_k141_3287 | Not-determined |
| KM_k141_91491 | Not-determined |
| KM_k141_3294 | Not-determined |
| KM_k141_91496 | Not-determined |

| | |
|---|---|
| KM_k141_247207 | Not-determined |
| KM_k141_70227 | Not-determined |
| KM_k141_236551 | Not-determined |
| KM_k141_236553 | Not-determined |
| KM_k141_81602 | Not-determined |
| KM_k141_3299 | Low-quality |
| KM_k141_247214 | Low-quality |
| KM_k141_236565 | Not-determined |
| KM_k141_58706 | Not-determined |
| KM_k141_103180 | Not-determined |
| KM_k141_203869 | Not-determined |
| KM_k141_58710 | Not-determined |
| KM_k141_58712 | Not-determined |
| KM_k141_203872 | Not-determined |
| KM_k141_147915 | Not-determined |
| KM_k141_203879 | Low-quality |
| KM_k141_236575 | Not-determined |
| KM_k141_147922 | Not-determined |
| KM_k141_192561 | Not-determined |
| KM_k141_3324 | Not-determined |
| KM_k141_147925 | Low-quality |
| KM_k141_147927 | Not-determined |
| KM_k141_181470 | Low-quality |
| KM_k141_36608 | Not-determined |
| KM_k141_36612 | Not-determined |
| KM_k141_136948 | Not-determined |
| KM_k141_25199 | Not-determined |
| KM_k141_192585 | Not-determined |
| KM_k141_36617 | Not-determined |
| KM_k141_136958 | Not-determined |
| KM_k141_147951 | Not-determined |
| KM_k141_25207 | Not-determined |
| KM_k141_158364 | Not-determined |
| KM_k141_91516 | Not-determined |
| KM_k141_192593 | Not-determined |
| KM_k141_192594 | Not-determined |
| KM_k141_115264 | Not-determined |
| KM_k141_181489 | Low-quality |
| KM_k141_192595 | Low-quality |
| KM_k141_81635 | Not-determined |
| KM_k141_192596 | Not-determined |
| KM_k141_13833 | Not-determined |
| KM_k141_115267 | Not-determined |
| KM_k141_91522 | Not-determined |
| KM_k141_236595 | Not-determined |
| KM_k141_170028 | Not-determined |
| KM_k141_25216 | Not-determined |
| KM_k141_170030 | Not-determined |
| KM_k141_25218 | Not-determined |
| KM_k141_91527 | Not-determined |
| KM_k141_170034 | Not-determined |
| KM_k141_225889 | Low-quality |
| KM_k141_247236 | Not-determined |
| KM_k141_203910 | Not-determined |
| KM_k141_225893 | Not-determined |
| KM_k141_103215 | Low-quality |
| KM_k141_25226 | Not-determined |
| KM_k141_81655 | Not-determined |
| KM_k141_247245 | Not-determined |
| KM_k141_225901 | Not-determined |
| KM_k141_81657 | Not-determined |
| KM_k141_158385 | Not-determined |
| KM_k141_81658 | Not-determined |
| KM_k141_158386 | Not-determined |
| KM_k141_225902 | Low-quality |
| KM_k141_203917 | Not-determined |
| KM_k141_81662 | Not-determined |
| KM_k141_115286 | Not-determined |
| KM_k141_115287 | Not-determined |
| KM_k141_147971 | Not-determined |
| KM_k141_70251 | Low-quality |
| KM_k141_225911 | Not-determined |
| KM_k141_58726 | Not-determined |
| KM_k141_58728 | Not-determined |
| KM_k141_91544 | Not-determined |
| KM_k141_103233 | Not-determined |
| KM_k141_203925 | Not-determined |
| KM_k141_147973 | Low-quality |
| KM_k141_158398 | Not-determined |
| KM_k141_147977 | Not-determined |
| KM_k141_147979 | Not-determined |
| KM_k141_125977 | Low-quality |
| KM_k141_203933 | Not-determined |
| KM_k141_3369 | Not-determined |
| KM_k141_25250 | Low-quality |
| KM_k141_203936 | Not-determined |
| KM_k141_170048 | Not-determined |
| KM_k141_225925 | Not-determined |
| KM_k141_225928 | Not-determined |
| KM_k141_81681 | Not-determined |
| KM_k141_258726 | Not-determined |
| KM_k141_247264 | Not-determined |
| KM_k141_70267 | Not-determined |
| KM_k141_147984 | Not-determined |
| KM_k141_136991 | Not-determined |
| KM_k141_247268 | Not-determined |
| KM_k141_181563 | Not-determined |
| KM_k141_214956 | Not-determined |
| KM_k141_214961 | Not-determined |
| KM_k141_3387 | Not-determined |
| KM_k141_70276 | Low-quality |
| KM_k141_181571 | Not-determined |
| KM_k141_3389 | Not-determined |
| KM_k141_25262 | Not-determined |
| KM_k141_247272 | Not-determined |
| KM_k141_81686 | Not-determined |
| KM_k141_225934 | Low-quality |
| KM_k141_115314 | Not-determined |
| KM_k141_170065 | Not-determined |
| KM_k141_258737 | Low-quality |
| KM_k141_247285 | Not-determined |
| KM_k141_91571 | Not-determined |
| KM_k141_115321 | Not-determined |
| KM_k141_81688 | Not-determined |
| KM_k141_158416 | Not-determined |
| KM_k141_170077 | Not-determined |
| KM_k141_13864 | Not-determined |
| KM_k141_115326 | Low-quality |
| KM_k141_192620 | Not-determined |
| KM_k141_203958 | Not-determined |
| KM_k141_13868 | Not-determined |
| KM_k141_170089 | Not-determined |
| KM_k141_148014 | Not-determined |
| KM_k141_126019 | Not-determined |
| KM_k141_126020 | Not-determined |
| KM_k141_126021 | Low-quality |
| KM_k141_181611 | Low-quality |
| KM_k141_70321 | Not-determined |
| KM_k141_192631 | Not-determined |
| KM_k141_158436 | Not-determined |
| KM_k141_170095 | Not-determined |
| KM_k141_126029 | Low-quality |
| KM_k141_225957 | Not-determined |
| KM_k141_158442 | Not-determined |
| KM_k141_158443 | Not-determined |
| KM_k141_115336 | Not-determined |
| KM_k141_81698 | Not-determined |
| KM_k141_158448 | Not-determined |
| KM_k141_181625 | Not-determined |
| KM_k141_203977 | Not-determined |
| KM_k141_115345 | Not-determined |
| KM_k141_115349 | Low-quality |
| KM_k141_3419 | Not-determined |
| KM_k141_81712 | Not-determined |
| KM_k141_81713 | Not-determined |
| KM_k141_215009 | Not-determined |
| KM_k141_215010 | Not-determined |
| KM_k141_3430 | Not-determined |
| KM_k141_236609 | Not-determined |
| KM_k141_215014 | Not-determined |
| KM_k141_225968 | Not-determined |
| KM_k141_215016 | Not-determined |
| KM_k141_115359 | Not-determined |
| KM_k141_215017 | Not-determined |
| KM_k141_158457 | Not-determined |
| KM_k141_148033 | Not-determined |
| KM_k141_126039 | Not-determined |
| KM_k141_158460 | Not-determined |
| KM_k141_158465 | Not-determined |
| KM_k141_36683 | Not-determined |
| KM_k141_91591 | Not-determined |
| KM_k141_115366 | Low-quality |
| KM_k141_58762 | Not-determined |
| KM_k141_225974 | Not-determined |
| KM_k141_137022 | Not-determined |
| KM_k141_81731 | Not-determined |
| KM_k141_215037 | Not-determined |
| KM_k141_115367 | Low-quality |
| KM_k141_247310 | Not-determined |
| KM_k141_70363 | Not-determined |
| KM_k141_215040 | Not-determined |
| KM_k141_103242 | Not-determined |
| KM_k141_103243 | Not-determined |
| KM_k141_70372 | Low-quality |
| KM_k141_103245 | Not-determined |
| KM_k141_115375 | Not-determined |
| KM_k141_13912 | Low-quality |
| KM_k141_81745 | Not-determined |
| KM_k141_192648 | Low-quality |
| KM_k141_115390 | Not-determined |
| KM_k141_192651 | Not-determined |
| KM_k141_115403 | Not-determined |
| KM_k141_192654 | Not-determined |
| KM_k141_91612 | Not-determined |
| KM_k141_115405 | Not-determined |
| KM_k141_81754 | Not-determined |
| KM_k141_225996 | Not-determined |
| KM_k141_204004 | Not-determined |
| KM_k141_91620 | Not-determined |
| KM_k141_170111 | Not-determined |
| KM_k141_58792 | Not-determined |
| KM_k141_91627 | Not-determined |
| KM_k141_170115 | Not-determined |
| KM_k141_126069 | Not-determined |
| KM_k141_81765 | Not-determined |
| KM_k141_192660 | Low-quality |
| KM_k141_137054 | Low-quality |
| KM_k141_58804 | Not-determined |
| KM_k141_170128 | Not-determined |
| KM_k141_58812 | Not-determined |
| KM_k141_115436 | Not-determined |
| KM_k141_170130 | Not-determined |
| KM_k141_126085 | Not-determined |
| KM_k141_192673 | Not-determined |
| KM_k141_192677 | Not-determined |
| KM_k141_192682 | Low-quality |
| KM_k141_103273 | Not-determined |
| KM_k141_170139 | Not-determined |
| KM_k141_3435 | Not-determined |
| KM_k141_115451 | Not-determined |
| KM_k141_126092 | Not-determined |
| KM_k141_192690 | Not-determined |
| KM_k141_81796 | Not-determined |
| KM_k141_70375 | Not-determined |
| KM_k141_226017 | Not-determined |
| KM_k141_236674 | Not-determined |
| KM_k141_103279 | Not-determined |
| KM_k141_226023 | Not-determined |
| KM_k141_204016 | Not-determined |
| KM_k141_181692 | Not-determined |
| KM_k141_3445 | Not-determined |
| KM_k141_81811 | Not-determined |
| KM_k141_70384 | Not-determined |
| KM_k141_170160 | Not-determined |
| KM_k141_137068 | Not-determined |
| KM_k141_70391 | Low-quality |
| KM_k141_236681 | Not-determined |
| KM_k141_91648 | Not-determined |
| KM_k141_36713 | Low-quality |
| KM_k141_25319 | Not-determined |
| KM_k141_36722 | Not-determined |
| KM_k141_215066 | Not-determined |
| KM_k141_103289 | Not-determined |
| KM_k141_247374 | Not-determined |
| KM_k141_103292 | Low-quality |
| KM_k141_170186 | Not-determined |
| KM_k141_70422 | Not-determined |
| KM_k141_148066 | Not-determined |
| KM_k141_192699 | Not-determined |
| KM_k141_70426 | Not-determined |
| KM_k141_25334 | Not-determined |
| KM_k141_158480 | Not-determined |
| KM_k141_226033 | Not-determined |
| KM_k141_158485 | Not-determined |
| KM_k141_25343 | Not-determined |
| KM_k141_25344 | Not-determined |
| KM_k141_25345 | Not-determined |
| KM_k141_126120 | Not-determined |
| KM_k141_81850 | Not-determined |
| KM_k141_13939 | Not-determined |
| KM_k141_137085 | Not-determined |
| KM_k141_126124 | Not-determined |
| KM_k141_137094 | Not-determined |
| KM_k141_226043 | Not-determined |
| KM_k141_226044 | Not-determined |
| KM_k141_170203 | Not-determined |
| KM_k141_126138 | Not-determined |
| KM_k141_81861 | Not-determined |
| KM_k141_247405 | Not-determined |
| KM_k141_236703 | Not-determined |
| KM_k141_3478 | Not-determined |
| KM_k141_103299 | Not-determined |
| KM_k141_91651 | Not-determined |
| KM_k141_158496 | Not-determined |
| KM_k141_137104 | Not-determined |
| KM_k141_36727 | Not-determined |
| KM_k141_148084 | Not-determined |
| KM_k141_115528 | Not-determined |
| KM_k141_36731 | Not-determined |
| KM_k141_115529 | Low-quality |
| KM_k141_103301 | Not-determined |
| KM_k141_170215 | Not-determined |
| KM_k141_126156 | Not-determined |
| KM_k141_192742 | Not-determined |
| KM_k141_103306 | Not-determined |
| KM_k141_158515 | Not-determined |
| KM_k141_148096 | Not-determined |
| KM_k141_192749 | Not-determined |
| KM_k141_148111 | Not-determined |
| KM_k141_3503 | Not-determined |
| KM_k141_148113 | Not-determined |
| KM_k141_103322 | Not-determined |
| KM_k141_103324 | Low-quality |
| KM_k141_137141 | Not-determined |
| KM_k141_226067 | Low-quality |
| KM_k141_3521 | Not-determined |
| KM_k141_36757 | Not-determined |
| KM_k141_103334 | Not-determined |
| KM_k141_91673 | Not-determined |
| KM_k141_215164 | Not-determined |
| KM_k141_91676 | Low-quality |
| KM_k141_148138 | Not-determined |
| KM_k141_148139 | Not-determined |
| KM_k141_103343 | Not-determined |
| KM_k141_226072 | Not-determined |
| KM_k141_36763 | Not-determined |
| KM_k141_170230 | Not-determined |
| KM_k141_247460 | Not-determined |
| KM_k141_192770 | Not-determined |
| KM_k141_148142 | Not-determined |
| KM_k141_25407 | Low-quality |
| KM_k141_3532 | Not-determined |
| KM_k141_3538 | Not-determined |
| KM_k141_3540 | Not-determined |
| KM_k141_25417 | Not-determined |
| KM_k141_158537 | Not-determined |
| KM_k141_226090 | Low-quality |
| KM_k141_192786 | Not-determined |
| KM_k141_215184 | Not-determined |
| KM_k141_226094 | Not-determined |
| KM_k141_215185 | Not-determined |
| KM_k141_158541 | Low-quality |
| KM_k141_215189 | Not-determined |
| KM_k141_70473 | Not-determined |
| KM_k141_103367 | Not-determined |
| KM_k141_36774 | Not-determined |
| KM_k141_70481 | Not-determined |
| KM_k141_148159 | Not-determined |
| KM_k141_215209 | Not-determined |
| KM_k141_58886 | Not-determined |
| KM_k141_36784 | Not-determined |
| KM_k141_192798 | Not-determined |
| KM_k141_148178 | Not-determined |
| KM_k141_58890 | Not-determined |
| KM_k141_192803 | Not-determined |
| KM_k141_204057 | Not-determined |
| KM_k141_204058 | Not-determined |
| KM_k141_70495 | Not-determined |
| KM_k141_14015 | Not-determined |
| KM_k141_115605 | Not-determined |
| KM_k141_25441 | Not-determined |
| KM_k141_158564 | Not-determined |
| KM_k141_126185 | Not-determined |
| KM_k141_226117 | Not-determined |
| KM_k141_115610 | Not-determined |
| KM_k141_115613 | Not-determined |
| KM_k141_115614 | Not-determined |
| KM_k141_91703 | Not-determined |
| KM_k141_192814 | Not-determined |
| KM_k141_215233 | Not-determined |
| KM_k141_215238 | Not-determined |
| KM_k141_25455 | Not-determined |
| KM_k141_158575 | Not-determined |
| KM_k141_115619 | Low-quality |
| KM_k141_204077 | Not-determined |
| KM_k141_148194 | Not-determined |
| KM_k141_58921 | Not-determined |
| KM_k141_215250 | Not-determined |
| KM_k141_91717 | Not-determined |
| KM_k141_58929 | Not-determined |
| KM_k141_91721 | Not-determined |
| KM_k141_192825 | Not-determined |
| KM_k141_192827 | Not-determined |
| KM_k141_192828 | Not-determined |
| KM_k141_181808 | Not-determined |
| KM_k141_192833 | Not-determined |
| KM_k141_115639 | Not-determined |
| KM_k141_204094 | Not-determined |
| KM_k141_192839 | Not-determined |
| KM_k141_192840 | Not-determined |
| KM_k141_91735 | Not-determined |
| KM_k141_115652 | Not-determined |
| KM_k141_103409 | Not-determined |
| KM_k141_81916 | Low-quality |
| KM_k141_70530 | Not-determined |
| KM_k141_215271 | Not-determined |
| KM_k141_204111 | Not-determined |
| KM_k141_81921 | Low-quality |
| KM_k141_236723 | Low-quality |
| KM_k141_236724 | Not-determined |
| KM_k141_258876 | Not-determined |
| KM_k141_81924 | Not-determined |
| KM_k141_81928 | Not-determined |
| KM_k141_204117 | Not-determined |
| KM_k141_103437 | Not-determined |
| KM_k141_70559 | Not-determined |
| KM_k141_215296 | Not-determined |
| KM_k141_215300 | Not-determined |
| KM_k141_81938 | Not-determined |
| KM_k141_70563 | Not-determined |
| KM_k141_148227 | Not-determined |
| KM_k141_70573 | Not-determined |
| KM_k141_192861 | Not-determined |
| KM_k141_126190 | Low-quality |
| KM_k141_215310 | Not-determined |
| KM_k141_70578 | Not-determined |
| KM_k141_192867 | Low-quality |
| KM_k141_204132 | Not-determined |
| KM_k141_204133 | Not-determined |
| KM_k141_170276 | Low-quality |
| KM_k141_115695 | Not-determined |
| KM_k141_204135 | Not-determined |
| KM_k141_25490 | Not-determined |
| KM_k141_115697 | Not-determined |
| KM_k141_215317 | Not-determined |
| KM_k141_215319 | Not-determined |
| KM_k141_258893 | Not-determined |
| KM_k141_115699 | Not-determined |
| KM_k141_115703 | Not-determined |
| KM_k141_170285 | Not-determined |
| KM_k141_103466 | Not-determined |
| KM_k141_103467 | Not-determined |
| KM_k141_70599 | Not-determined |
| KM_k141_70601 | Not-determined |
| KM_k141_115716 | Not-determined |
| KM_k141_170289 | Not-determined |
| KM_k141_115719 | Not-determined |
| KM_k141_3579 | Not-determined |
| KM_k141_215340 | Not-determined |
| KM_k141_3582 | Not-determined |
| KM_k141_170294 | Not-determined |
| KM_k141_115726 | Low-quality |
| KM_k141_148270 | Not-determined |
| KM_k141_215359 | Not-determined |
| KM_k141_115731 | Not-determined |
| KM_k141_58954 | Not-determined |
| KM_k141_58956 | Not-determined |
| KM_k141_226164 | Not-determined |
| KM_k141_70625 | Not-determined |
| KM_k141_70627 | Not-determined |
| KM_k141_236757 | Not-determined |
| KM_k141_25525 | Not-determined |
| KM_k141_226165 | Low-quality |
| KM_k141_247521 | Not-determined |
| KM_k141_103484 | Low-quality |
| KM_k141_103485 | Not-determined |
| KM_k141_103487 | Not-determined |
| KM_k141_58976 | Not-determined |
| KM_k141_25533 | Not-determined |
| KM_k141_258933 | Not-determined |
| KM_k141_215364 | Not-determined |
| KM_k141_170308 | Not-determined |
| KM_k141_215370 | Not-determined |
| KM_k141_236768 | Not-determined |
| KM_k141_70672 | Low-quality |
| KM_k141_247545 | Not-determined |
| KM_k141_25550 | Not-determined |
| KM_k141_236769 | Not-determined |
| KM_k141_247546 | Low-quality |
| KM_k141_236772 | Low-quality |
| KM_k141_36796 | Not-determined |
| KM_k141_70676 | Not-determined |
| KM_k141_170315 | Not-determined |
| KM_k141_25561 | Not-determined |
| KM_k141_126230 | Low-quality |
| KM_k141_204167 | Not-determined |
| KM_k141_236780 | Not-determined |
| KM_k141_126235 | Not-determined |
| KM_k141_204171 | Low-quality |
| KM_k141_137169 | Not-determined |
| KM_k141_181834 | Not-determined |
| KM_k141_81990 | Not-determined |
| KM_k141_236788 | Not-determined |
| KM_k141_70686 | Not-determined |
| KM_k141_148299 | Not-determined |
| KM_k141_25575 | Not-determined |
| KM_k141_258954 | Not-determined |
| KM_k141_14049 | Not-determined |
| KM_k141_247570 | Not-determined |
| KM_k141_14054 | Not-determined |
| KM_k141_36810 | Low-quality |
| KM_k141_91823 | Low-quality |
| KM_k141_204186 | Not-determined |
| KM_k141_247578 | Not-determined |
| KM_k141_82001 | Not-determined |
| KM_k141_126256 | Not-determined |
| KM_k141_103514 | Not-determined |
| KM_k141_14057 | Not-determined |
| KM_k141_115776 | Not-determined |
| KM_k141_236797 | Not-determined |
| KM_k141_148309 | Not-determined |
| KM_k141_158594 | Low-quality |
| KM_k141_70711 | Not-determined |
| KM_k141_158596 | Not-determined |
| KM_k141_103525 | Not-determined |
| KM_k141_236809 | Not-determined |
| KM_k141_158601 | Not-determined |
| KM_k141_36833 | Not-determined |
| KM_k141_137177 | Not-determined |
| KM_k141_126270 | Not-determined |
| KM_k141_236816 | Not-determined |
| KM_k141_58996 | Not-determined |
| KM_k141_258961 | Not-determined |
| KM_k141_192920 | Low-quality |
| KM_k141_70719 | Not-determined |
| KM_k141_82006 | Not-determined |
| KM_k141_181848 | Low-quality |
| KM_k141_192925 | Not-determined |
| KM_k141_70722 | Low-quality |
| KM_k141_236826 | Not-determined |
| KM_k141_158608 | Not-determined |
| KM_k141_236830 | Not-determined |
| KM_k141_158610 | Not-determined |
| KM_k141_258967 | Not-determined |
| KM_k141_14081 | Not-determined |
| KM_k141_204197 | Not-determined |
| KM_k141_14082 | Not-determined |
| KM_k141_14083 | Not-determined |
| KM_k141_103544 | Low-quality |
| KM_k141_82018 | Not-determined |
| KM_k141_236839 | Not-determined |
| KM_k141_181875 | Not-determined |
| KM_k141_215398 | Low-quality |
| KM_k141_236845 | Not-determined |
| KM_k141_236848 | Not-determined |
| KM_k141_137201 | Not-determined |
| KM_k141_103556 | Low-quality |
| KM_k141_82034 | Not-determined |
| KM_k141_258975 | Not-determined |
| KM_k141_247580 | Not-determined |
| KM_k141_181892 | Not-determined |
| KM_k141_70749 | Not-determined |
| KM_k141_181894 | Not-determined |
| KM_k141_82044 | Not-determined |
| KM_k141_247585 | Not-determined |
| KM_k141_82052 | Not-determined |
| KM_k141_70760 | Not-determined |
| KM_k141_158645 | Low-quality |
| KM_k141_103568 | Not-determined |
| KM_k141_192952 | Not-determined |
| KM_k141_148365 | Not-determined |
| KM_k141_82058 | Low-quality |
| KM_k141_70774 | Not-determined |
| KM_k141_3601 | Not-determined |
| KM_k141_36867 | Not-determined |
| KM_k141_215437 | Not-determined |
| KM_k141_103577 | Not-determined |
| KM_k141_70778 | Not-determined |
| KM_k141_181920 | Not-determined |
| KM_k141_36869 | Not-determined |
| KM_k141_14106 | Not-determined |
| KM_k141_148375 | Not-determined |
| KM_k141_170374 | Not-determined |
| KM_k141_148377 | Not-determined |
| KM_k141_258988 | Not-determined |
| KM_k141_36880 | Not-determined |
| KM_k141_25600 | Not-determined |
| KM_k141_181930 | Not-determined |
| KM_k141_215448 | Not-determined |
| KM_k141_258998 | Not-determined |
| KM_k141_226243 | Not-determined |
| KM_k141_158674 | Not-determined |
| KM_k141_25607 | Not-determined |
| KM_k141_259006 | Not-determined |
| KM_k141_204233 | Not-determined |
| KM_k141_204234 | Not-determined |
| KM_k141_204235 | Not-determined |
| KM_k141_215456 | Not-determined |
| KM_k141_115843 | Not-determined |
| KM_k141_115844 | Not-determined |
| KM_k141_170386 | Not-determined |
| KM_k141_170388 | Not-determined |
| KM_k141_226261 | Not-determined |
| KM_k141_70803 | Not-determined |
| KM_k141_70804 | Not-determined |
| KM_k141_236897 | Not-determined |
| KM_k141_236899 | Not-determined |
| KM_k141_192996 | Not-determined |
| KM_k141_170394 | Not-determined |
| KM_k141_259022 | Low-quality |
| KM_k141_193000 | Not-determined |
| KM_k141_25628 | Not-determined |
| KM_k141_193003 | Not-determined |
| KM_k141_46889 | Not-determined |
| KM_k141_82130 | Not-determined |
| KM_k141_70819 | Not-determined |
| KM_k141_70820 | Not-determined |
| KM_k141_91825 | Not-determined |
| KM_k141_158691 | Not-determined |
| KM_k141_170400 | Not-determined |
| KM_k141_236905 | Not-determined |
| KM_k141_91829 | Not-determined |
| KM_k141_91830 | Not-determined |
| KM_k141_193014 | Not-determined |
| KM_k141_204271 | Not-determined |
| KM_k141_236907 | Not-determined |
| KM_k141_46904 | Low-quality |
| KM_k141_226302 | Not-determined |
| KM_k141_82146 | Not-determined |
| KM_k141_103601 | Not-determined |
| KM_k141_46911 | Not-determined |
| KM_k141_25644 | Not-determined |
| KM_k141_25647 | Not-determined |
| KM_k141_204277 | Not-determined |
| KM_k141_46918 | Not-determined |
| KM_k141_14148 | Not-determined |
| KM_k141_181989 | Not-determined |
| KM_k141_91848 | Not-determined |
| KM_k141_193023 | Not-determined |

| | |
|---|---|
| KM_k141_82164 | Not-determined |
| KM_k141_204290 | Not-determined |
| KM_k141_82167 | Not-determined |
| KM_k141_82169 | Not-determined |
| KM_k141_204297 | Not-determined |
| KM_k141_170428 | Not-determined |
| KM_k141_236914 | Low-quality |
| KM_k141_236916 | Low-quality |
| KM_k141_59017 | Not-determined |
| KM_k141_36900 | Not-determined |
| KM_k141_14169 | Not-determined |
| KM_k141_115910 | Not-determined |
| KM_k141_226305 | Not-determined |
| KM_k141_226307 | Not-determined |
| KM_k141_158742 | Not-determined |
| KM_k141_59025 | Not-determined |
| KM_k141_182008 | Not-determined |
| KM_k141_236932 | Not-determined |
| KM_k141_182009 | Not-determined |
| KM_k141_91878 | Not-determined |
| KM_k141_215503 | Not-determined |
| KM_k141_236933 | Low-quality |
| KM_k141_215507 | Not-determined |
| KM_k141_259046 | Not-determined |
| KM_k141_204324 | Not-determined |
| KM_k141_215512 | Not-determined |
| KM_k141_137325 | Not-determined |
| KM_k141_170457 | Not-determined |
| KM_k141_236939 | Not-determined |
| KM_k141_215519 | Low-quality |
| KM_k141_182020 | Not-determined |
| KM_k141_170460 | Not-determined |
| KM_k141_158762 | Not-determined |
| KM_k141_14199 | Not-determined |
| KM_k141_226322 | Not-determined |
| KM_k141_215524 | Not-determined |
| KM_k141_14200 | Not-determined |
| KM_k141_148430 | Not-determined |
| KM_k141_215528 | Not-determined |
| KM_k141_14206 | Not-determined |
| KM_k141_226331 | Low-quality |
| KM_k141_36903 | Not-determined |
| KM_k141_70844 | Low-quality |
| KM_k141_137340 | Not-determined |
| KM_k141_193058 | Low-quality |
| KM_k141_25678 | Low-quality |
| KM_k141_25681 | Not-determined |
| KM_k141_182029 | Not-determined |
| KM_k141_126327 | Not-determined |
| KM_k141_91890 | Not-determined |
| KM_k141_25689 | Low-quality |
| KM_k141_215555 | Not-determined |
| KM_k141_70860 | Not-determined |
| KM_k141_25693 | Not-determined |
| KM_k141_91896 | Not-determined |
| KM_k141_14228 | Not-determined |
| KM_k141_115938 | Not-determined |
| KM_k141_25697 | Not-determined |
| KM_k141_193071 | Not-determined |
| KM_k141_115941 | Not-determined |
| KM_k141_14233 | Not-determined |
| KM_k141_59090 | Not-determined |
| KM_k141_204367 | Not-determined |
| KM_k141_82185 | Not-determined |
| KM_k141_148462 | Not-determined |
| KM_k141_137362 | Not-determined |
| KM_k141_36923 | Not-determined |
| KM_k141_182054 | Not-determined |
| KM_k141_59101 | Not-determined |
| KM_k141_59102 | Low-quality |
| KM_k141_226359 | Not-determined |
| KM_k141_193081 | Not-determined |
| KM_k141_115963 | Not-determined |
| KM_k141_259069 | Not-determined |
| KM_k141_182073 | Not-determined |
| KM_k141_46946 | Not-determined |
| KM_k141_182076 | Low-quality |
| KM_k141_158782 | Not-determined |
| KM_k141_70882 | Not-determined |
| KM_k141_182081 | Not-determined |
| KM_k141_182086 | Not-determined |
| KM_k141_193093 | Not-determined |
| KM_k141_46950 | Not-determined |
| KM_k141_193098 | Not-determined |
| KM_k141_82208 | Not-determined |
| KM_k141_170506 | Low-quality |
| KM_k141_14260 | Not-determined |
| KM_k141_193103 | Not-determined |
| KM_k141_126337 | Low-quality |
| KM_k141_193108 | Not-determined |
| KM_k141_126338 | Not-determined |
| KM_k141_204393 | Not-determined |
| KM_k141_226374 | Low-quality |
| KM_k141_226375 | Not-determined |
| KM_k141_182091 | Not-determined |
| KM_k141_103646 | Not-determined |
| KM_k141_25746 | Low-quality |
| KM_k141_126346 | Not-determined |
| KM_k141_126347 | Not-determined |
| KM_k141_82218 | Not-determined |
| KM_k141_115974 | Not-determined |
| KM_k141_182104 | Not-determined |
| KM_k141_115976 | Low-quality |
| KM_k141_137414 | Not-determined |
| KM_k141_126362 | Not-determined |
| KM_k141_137419 | Not-determined |
| KM_k141_14282 | Not-determined |
| KM_k141_226387 | Not-determined |
| KM_k141_59138 | Not-determined |
| KM_k141_204412 | Not-determined |
| KM_k141_70901 | Not-determined |
| KM_k141_46974 | Not-determined |
| KM_k141_137432 | Not-determined |
| KM_k141_115994 | Not-determined |
| KM_k141_247624 | Not-determined |
| KM_k141_14293 | Not-determined |
| KM_k141_126366 | Not-determined |
| KM_k141_116006 | Not-determined |
| KM_k141_247632 | Not-determined |
| KM_k141_247633 | Low-quality |
| KM_k141_14297 | Not-determined |
| KM_k141_103651 | Not-determined |
| KM_k141_158815 | Not-determined |
| KM_k141_247642 | Not-determined |
| KM_k141_116016 | Not-determined |
| KM_k141_158821 | Not-determined |
| KM_k141_204421 | Not-determined |
| KM_k141_59143 | Not-determined |
| KM_k141_226410 | Not-determined |
| KM_k141_14302 | Not-determined |
| KM_k141_148486 | Not-determined |
| KM_k141_137442 | Not-determined |
| KM_k141_3643 | Not-determined |
| KM_k141_126384 | Not-determined |
| KM_k141_103676 | Not-determined |
| KM_k141_3644 | Low-quality |
| KM_k141_47001 | Not-determined |
| KM_k141_215606 | Not-determined |
| KM_k141_158828 | Not-determined |
| KM_k141_126389 | Not-determined |
| KM_k141_116042 | Not-determined |
| KM_k141_116046 | Not-determined |
| KM_k141_14316 | Not-determined |
| KM_k141_3648 | Not-determined |
| KM_k141_116053 | Not-determined |
| KM_k141_47003 | Not-determined |
| KM_k141_82249 | Not-determined |
| KM_k141_126398 | Not-determined |
| KM_k141_158843 | Not-determined |
| KM_k141_226449 | Not-determined |
| KM_k141_182149 | Not-determined |
| KM_k141_116066 | Not-determined |
| KM_k141_59157 | Low-quality |
| KM_k141_116071 | Not-determined |
| KM_k141_158845 | Not-determined |
| KM_k141_148503 | Not-determined |
| KM_k141_70915 | Low-quality |
| KM_k141_204456 | Not-determined |
| KM_k141_36982 | Not-determined |
| KM_k141_116074 | Not-determined |
| KM_k141_59164 | Not-determined |
| KM_k141_116077 | Not-determined |
| KM_k141_170569 | Not-determined |
| KM_k141_247663 | Not-determined |
| KM_k141_116081 | Not-determined |
| KM_k141_14339 | Not-determined |
| KM_k141_193192 | Not-determined |
| KM_k141_182163 | Not-determined |
| KM_k141_204466 | Not-determined |
| KM_k141_237007 | Not-determined |
| KM_k141_36993 | Not-determined |
| KM_k141_137470 | Not-determined |
| KM_k141_247672 | Not-determined |
| KM_k141_137472 | Not-determined |
| KM_k141_193200 | Not-determined |
| KM_k141_237019 | Low-quality |
| KM_k141_247678 | Not-determined |
| KM_k141_116095 | Not-determined |
| KM_k141_137479 | Not-determined |
| KM_k141_116101 | Not-determined |
| KM_k141_59185 | Not-determined |
| KM_k141_193208 | Not-determined |
| KM_k141_148518 | Not-determined |
| KM_k141_116118 | Not-determined |
| KM_k141_204496 | Not-determined |
| KM_k141_25773 | Not-determined |
| KM_k141_182177 | Low-quality |
| KM_k141_59187 | Not-determined |
| KM_k141_82286 | Not-determined |
| KM_k141_59189 | Low-quality |
| KM_k141_126416 | Not-determined |
| KM_k141_25780 | Not-determined |
| KM_k141_137499 | Not-determined |
| KM_k141_47020 | Not-determined |
| KM_k141_3700 | Not-determined |
| KM_k141_82290 | Not-determined |
| KM_k141_3703 | Not-determined |
| KM_k141_193217 | Not-determined |
| KM_k141_3705 | Not-determined |
| KM_k141_25790 | Not-determined |
| KM_k141_182189 | Not-determined |
| KM_k141_193223 | Not-determined |
| KM_k141_193224 | Not-determined |
| KM_k141_193225 | Not-determined |
| KM_k141_193226 | Not-determined |
| KM_k141_47028 | Not-determined |
| KM_k141_3713 | Not-determined |
| KM_k141_137512 | Not-determined |
| KM_k141_37014 | Low-quality |
| KM_k141_182193 | Not-determined |
| KM_k141_116132 | Not-determined |
| KM_k141_116133 | Not-determined |
| KM_k141_137524 | Not-determined |
| KM_k141_116137 | Not-determined |
| KM_k141_226504 | Low-quality |
| KM_k141_116138 | Not-determined |
| KM_k141_226513 | Low-quality |
| KM_k141_103742 | Not-determined |
| KM_k141_103745 | Not-determined |
| KM_k141_103747 | Not-determined |
| KM_k141_25799 | Not-determined |
| KM_k141_193249 | Low-quality |
| KM_k141_25801 | Not-determined |
| KM_k141_182210 | Not-determined |
| KM_k141_82322 | Not-determined |
| KM_k141_237071 | Not-determined |
| KM_k141_47057 | Not-determined |
| KM_k141_82329 | Not-determined |
| KM_k141_182220 | Not-determined |
| KM_k141_14352 | Not-determined |
| KM_k141_59194 | Not-determined |
| KM_k141_126469 | Not-determined |
| KM_k141_182222 | Not-determined |
| KM_k141_59199 | Not-determined |
| KM_k141_70972 | Not-determined |
| KM_k141_126475 | Not-determined |
| KM_k141_82339 | Not-determined |
| KM_k141_103752 | Low-quality |
| KM_k141_247700 | Not-determined |
| KM_k141_103757 | Not-determined |
| KM_k141_137535 | Not-determined |
| KM_k141_215682 | Not-determined |
| KM_k141_247705 | Low-quality |
| KM_k141_37048 | Not-determined |
| KM_k141_193255 | Not-determined |
| KM_k141_137549 | Not-determined |
| KM_k141_59215 | Not-determined |
| KM_k141_137553 | Not-determined |
| KM_k141_82348 | Not-determined |
| KM_k141_14377 | Not-determined |
| KM_k141_14378 | Not-determined |
| KM_k141_204521 | Not-determined |
| KM_k141_37065 | Not-determined |
| KM_k141_204524 | Not-determined |
| KM_k141_226544 | Not-determined |
| KM_k141_148562 | Not-determined |
| KM_k141_126484 | Not-determined |
| KM_k141_25839 | Not-determined |
| KM_k141_226564 | Not-determined |
| KM_k141_37072 | Not-determined |
| KM_k141_37073 | Not-determined |
| KM_k141_25842 | Low-quality |
| KM_k141_215713 | Not-determined |
| KM_k141_59226 | Low-quality |
| KM_k141_91941 | Not-determined |
| KM_k141_247730 | Not-determined |
| KM_k141_126496 | Not-determined |
| KM_k141_59243 | Not-determined |
| KM_k141_126503 | Not-determined |
| KM_k141_226580 | Not-determined |
| KM_k141_116199 | Not-determined |
| KM_k141_25855 | Not-determined |
| KM_k141_182262 | Not-determined |
| KM_k141_182264 | Not-determined |
| KM_k141_137572 | Not-determined |
| KM_k141_70988 | Low-quality |
| KM_k141_103788 | Not-determined |
| KM_k141_70989 | Not-determined |
| KM_k141_148598 | Not-determined |
| KM_k141_126522 | Not-determined |
| KM_k141_91945 | Not-determined |
| KM_k141_182271 | Not-determined |
| KM_k141_137582 | Not-determined |
| KM_k141_126525 | Not-determined |
| KM_k141_247751 | Not-determined |
| KM_k141_3800 | Not-determined |
| KM_k141_237128 | Not-determined |
| KM_k141_170615 | Not-determined |
| KM_k141_82365 | Not-determined |
| KM_k141_103813 | Not-determined |
| KM_k141_37118 | Low-quality |
| KM_k141_158903 | Not-determined |
| KM_k141_237133 | Not-determined |
| KM_k141_170622 | Not-determined |
| KM_k141_37120 | Not-determined |
| KM_k141_82367 | Not-determined |
| KM_k141_37124 | Not-determined |
| KM_k141_82372 | Not-determined |
| KM_k141_182281 | Not-determined |
| KM_k141_182284 | Low-quality |
| KM_k141_116207 | Not-determined |
| KM_k141_170636 | Not-determined |
| KM_k141_82381 | Not-determined |
| KM_k141_59281 | Not-determined |
| KM_k141_82388 | Not-determined |
| KM_k141_37135 | Not-determined |
| KM_k141_47071 | Not-determined |
| KM_k141_25873 | Not-determined |
| KM_k141_47073 | Not-determined |
| KM_k141_47077 | Not-determined |
| KM_k141_158927 | Not-determined |
| KM_k141_25881 | Not-determined |
| KM_k141_226631 | Not-determined |
| KM_k141_71023 | Not-determined |
| KM_k141_215749 | Not-determined |
| KM_k141_158929 | Not-determined |
| KM_k141_37139 | Not-determined |
| KM_k141_158932 | Not-determined |
| KM_k141_215753 | Low-quality |
| KM_k141_3825 | Not-determined |
| KM_k141_82405 | Not-determined |
| KM_k141_148623 | Not-determined |
| KM_k141_237174 | Low-quality |
| KM_k141_215764 | Not-determined |
| KM_k141_37148 | Not-determined |
| KM_k141_226645 | Not-determined |
| KM_k141_226646 | Not-determined |
| KM_k141_126583 | Low-quality |
| KM_k141_247783 | Not-determined |
| KM_k141_182312 | Not-determined |
| KM_k141_182316 | Not-determined |
| KM_k141_247787 | Not-determined |
| KM_k141_226656 | Not-determined |
| KM_k141_82421 | Not-determined |
| KM_k141_215779 | Not-determined |
| KM_k141_148626 | Not-determined |
| KM_k141_148627 | Not-determined |
| KM_k141_82428 | Low-quality |
| KM_k141_103857 | Not-determined |
| KM_k141_148636 | Not-determined |
| KM_k141_247793 | Not-determined |
| KM_k141_259105 | Low-quality |
| KM_k141_259109 | Not-determined |
| KM_k141_215788 | Low-quality |
| KM_k141_226673 | Low-quality |
| KM_k141_247795 | Not-determined |
| KM_k141_116221 | Low-quality |
| KM_k141_215796 | Not-determined |
| KM_k141_182345 | Not-determined |
| KM_k141_25911 | Not-determined |
| KM_k141_226681 | Not-determined |
| KM_k141_259118 | Not-determined |
| KM_k141_226686 | Not-determined |
| KM_k141_116233 | Low-quality |
| KM_k141_170661 | Not-determined |
| KM_k141_215813 | Not-determined |
| KM_k141_116237 | Low-quality |
| KM_k141_182351 | Not-determined |
| KM_k141_158953 | Not-determined |
| KM_k141_215822 | Not-determined |
| KM_k141_37163 | Not-determined |
| KM_k141_25920 | Low-quality |
| KM_k141_37167 | Not-determined |
| KM_k141_182362 | Not-determined |
| KM_k141_215830 | Not-determined |
| KM_k141_3847 | Not-determined |
| KM_k141_158965 | Not-determined |
| KM_k141_59295 | Not-determined |
| KM_k141_59297 | Not-determined |
| KM_k141_59298 | Not-determined |
| KM_k141_59299 | Not-determined |
| KM_k141_158968 | Not-determined |
| KM_k141_103881 | Not-determined |
| KM_k141_71033 | Not-determined |
| KM_k141_37173 | Not-determined |
| KM_k141_182373 | Not-determined |
| KM_k141_237193 | Not-determined |
| KM_k141_71037 | Not-determined |
| KM_k141_237196 | Not-determined |
| KM_k141_226710 | Not-determined |
| KM_k141_158971 | Not-determined |
| KM_k141_59310 | Not-determined |
| KM_k141_3859 | Low-quality |
| KM_k141_247814 | Not-determined |
| KM_k141_59313 | Low-quality |
| KM_k141_37180 | Not-determined |
| KM_k141_82481 | Not-determined |
| KM_k141_3863 | Not-determined |
| KM_k141_259131 | Not-determined |
| KM_k141_148681 | Not-determined |
| KM_k141_148683 | Not-determined |
| KM_k141_226729 | Not-determined |
| KM_k141_226730 | Not-determined |
| KM_k141_71052 | Not-determined |
| KM_k141_71053 | Not-determined |
| KM_k141_259138 | Low-quality |
| KM_k141_158973 | Low-quality |
| KM_k141_215848 | Not-determined |
| KM_k141_103905 | Low-quality |
| KM_k141_59325 | Low-quality |
| KM_k141_59328 | Not-determined |
| KM_k141_103907 | Not-determined |
| KM_k141_215854 | Not-determined |
| 18-106-final-viral-combined_k141_97548\|\|full | Medium-quality |
| KM_k141_148707 | Low-quality |
| KM_k141_215855 | Not-determined |
| KM_k141_71077 | Not-determined |
| KM_k141_37190 | Not-determined |
| KM_k141_71078 | Not-determined |
| KM_k141_215862 | Not-determined |
| KM_k141_116255 | Not-determined |
| KM_k141_148715 | Not-determined |
| KM_k141_37194 | Low-quality |
| KM_k141_148716 | Not-determined |
| KM_k141_182416 | Not-determined |
| KM_k141_14409 | Not-determined |
| KM_k141_116256 | Low-quality |
| KM_k141_14413 | Not-determined |
| KM_k141_103928 | Not-determined |
| KM_k141_182420 | Not-determined |
| KM_k141_170673 | Not-determined |
| KM_k141_116264 | Low-quality |
| KM_k141_116265 | Not-determined |
| KM_k141_137674 | Not-determined |
| KM_k141_193334 | Not-determined |
| KM_k141_82509 | Not-determined |
| KM_k141_137678 | Not-determined |
| KM_k141_59346 | Low-quality |
| KM_k141_92023 | Low-quality |
| KM_k141_237242 | Not-determined |
| KM_k141_59349 | Not-determined |
| KM_k141_82513 | Not-determined |
| KM_k141_116276 | Not-determined |
| KM_k141_47103 | Not-determined |
| KM_k141_237249 | Not-determined |
| KM_k141_59354 | Not-determined |
| KM_k141_59357 | Not-determined |
| KM_k141_92030 | Not-determined |
| KM_k141_25992 | Low-quality |
| KM_k141_47113 | Not-determined |
| KM_k141_247838 | Not-determined |
| KM_k141_182430 | Not-determined |
| KM_k141_237251 | Not-determined |
| KM_k141_116292 | Not-determined |
| KM_k141_170688 | Not-determined |
| KM_k141_82533 | Not-determined |
| KM_k141_71116 | Not-determined |
| KM_k141_59372 | Not-determined |
| KM_k141_103955 | Not-determined |
| KM_k141_92038 | Not-determined |
| KM_k141_37216 | Not-determined |
| KM_k141_103960 | Not-determined |
| KM_k141_116313 | Not-determined |
| KM_k141_226780 | Not-determined |
| KM_k141_170711 | Not-determined |
| KM_k141_148739 | Not-determined |
| KM_k141_170716 | Not-determined |
| KM_k141_204534 | Not-determined |
| KM_k141_71135 | Not-determined |
| KM_k141_37223 | Not-determined |
| KM_k141_14452 | Not-determined |
| KM_k141_82552 | Not-determined |
| KM_k141_37226 | Not-determined |
| KM_k141_170728 | Not-determined |
| KM_k141_247848 | Not-determined |
| KM_k141_247850 | Not-determined |
| KM_k141_82553 | Not-determined |
| KM_k141_71140 | Not-determined |
| KM_k141_14464 | Not-determined |
| KM_k141_148744 | Not-determined |
| KM_k141_170732 | Not-determined |
| KM_k141_26023 | Not-determined |
| KM_k141_148746 | Not-determined |
| KM_k141_170737 | Not-determined |
| KM_k141_14468 | Not-determined |
| KM_k141_193396 | Not-determined |
| KM_k141_126630 | Not-determined |
| KM_k141_82565 | Not-determined |
| KM_k141_247855 | Not-determined |
| KM_k141_92054 | Not-determined |
| KM_k141_92055 | Not-determined |
| KM_k141_247860 | Not-determined |
| KM_k141_116334 | Not-determined |
| KM_k141_116336 | Not-determined |
| KM_k141_82572 | Not-determined |
| KM_k141_82574 | Not-determined |
| KM_k141_14477 | Not-determined |
| KM_k141_71148 | Not-determined |
| KM_k141_193417 | Low-quality |
| KM_k141_215917 | Not-determined |
| KM_k141_47141 | Not-determined |
| KM_k141_193419 | Not-determined |
| KM_k141_59413 | Not-determined |
| KM_k141_193420 | Not-determined |
| KM_k141_59417 | Not-determined |
| KM_k141_193424 | Not-determined |
| KM_k141_92084 | Not-determined |
| KM_k141_92085 | Not-determined |
| KM_k141_215918 | Low-quality |
| KM_k141_103980 | Not-determined |
| KM_k141_47150 | Not-determined |
| KM_k141_92091 | Not-determined |
| KM_k141_82596 | Not-determined |
| KM_k141_103984 | Not-determined |
| KM_k141_204552 | Not-determined |
| KM_k141_59421 | Not-determined |
| KM_k141_47152 | Not-determined |
| KM_k141_82600 | Not-determined |
| KM_k141_82602 | Not-determined |
| KM_k141_204556 | Not-determined |
| KM_k141_204557 | Not-determined |
| KM_k141_116353 | Not-determined |
| KM_k141_47159 | Low-quality |
| KM_k141_193436 | Not-determined |
| KM_k141_82610 | Not-determined |
| KM_k141_193439 | Not-determined |
| KM_k141_237314 | Not-determined |
| KM_k141_103992 | Not-determined |
| KM_k141_82620 | Not-determined |
| KM_k141_82621 | Not-determined |
| KM_k141_158988 | Not-determined |
| KM_k141_182469 | Not-determined |
| KM_k141_204565 | Low-quality |
| KM_k141_237323 | Not-determined |
| KM_k141_3898 | Not-determined |
| KM_k141_247881 | Low-quality |
| KM_k141_193456 | Not-determined |
| KM_k141_182474 | Not-determined |
| KM_k141_193459 | Not-determined |
| KM_k141_158999 | Not-determined |
| KM_k141_103997 | Not-determined |
| KM_k141_82643 | Not-determined |
| KM_k141_226815 | Not-determined |
| KM_k141_193468 | Not-determined |
| KM_k141_193469 | Not-determined |
| KM_k141_247889 | Not-determined |
| KM_k141_37260 | Not-determined |
| KM_k141_226818 | Not-determined |
| KM_k141_193470 | Not-determined |
| KM_k141_226820 | Not-determined |
| KM_k141_193472 | Not-determined |
| KM_k141_37262 | Not-determined |
| KM_k141_3910 | Not-determined |
| KM_k141_26067 | Not-determined |
| KM_k141_193476 | Not-determined |
| KM_k141_193479 | Not-determined |
| KM_k141_148799 | Not-determined |
| KM_k141_148800 | Not-determined |
| KM_k141_104003 | Not-determined |
| KM_k141_182495 | Not-determined |
| KM_k141_126639 | Not-determined |
| KM_k141_193484 | Not-determined |
| KM_k141_247904 | Not-determined |
| KM_k141_159013 | Not-determined |
| KM_k141_159016 | Not-determined |
| KM_k141_92125 | Not-determined |
| KM_k141_116399 | Not-determined |
| KM_k141_137692 | Low-quality |
| KM_k141_116409 | Not-determined |
| KM_k141_116413 | Not-determined |
| KM_k141_104014 | Not-determined |
| KM_k141_193499 | Not-determined |
| KM_k141_226830 | Not-determined |
| KM_k141_26095 | Not-determined |
| KM_k141_148824 | Not-determined |
| KM_k141_26098 | Low-quality |
| KM_k141_26099 | Low-quality |
| KM_k141_226841 | Not-determined |
| KM_k141_126667 | Not-determined |
| KM_k141_71191 | Not-determined |
| KM_k141_226843 | Not-determined |
| KM_k141_170794 | Low-quality |
| KM_k141_116420 | Low-quality |
| KM_k141_82661 | Not-determined |
| KM_k141_170798 | Not-determined |
| KM_k141_104032 | Not-determined |
| KM_k141_170800 | Not-determined |
| KM_k141_193514 | Not-determined |
| KM_k141_126678 | Not-determined |
| KM_k141_71206 | Not-determined |
| KM_k141_170807 | Not-determined |
| KM_k141_204611 | Not-determined |
| KM_k141_37278 | Not-determined |
| KM_k141_226870 | Not-determined |
| KM_k141_170814 | Not-determined |
| KM_k141_104049 | Not-determined |
| KM_k141_71216 | Not-determined |
| KM_k141_71218 | Not-determined |
| KM_k141_215929 | Low-quality |

| | |
|---|---|
| KM_k141_170826 | Not-determined |
| KM_k141_116434 | Not-determined |
| KM_k141_82684 | Not-determined |
| KM_k141_82685 | Not-determined |
| KM_k141_37297 | Not-determined |
| KM_k141_170830 | Not-determined |
| KM_k141_237329 | Not-determined |
| KM_k141_104072 | Not-determined |
| KM_k141_3929 | Not-determined |
| KM_k141_137712 | Not-determined |
| KM_k141_137713 | Not-determined |
| KM_k141_37304 | Not-determined |
| KM_k141_182511 | Not-determined |
| KM_k141_182512 | Not-determined |
| KM_k141_92159 | Not-determined |
| KM_k141_3934 | Not-determined |
| KM_k141_159036 | Not-determined |
| KM_k141_170838 | Not-determined |
| KM_k141_159037 | Not-determined |
| KM_k141_71229 | Not-determined |
| KM_k141_137721 | Not-determined |
| KM_k141_26113 | Not-determined |
| KM_k141_247940 | Not-determined |
| KM_k141_126703 | Not-determined |
| KM_k141_3941 | Not-determined |
| KM_k141_82699 | Not-determined |
| KM_k141_259176 | Not-determined |
| KM_k141_159041 | Not-determined |
| KM_k141_82703 | Not-determined |
| KM_k141_137735 | Not-determined |
| KM_k141_137737 | Not-determined |
| KM_k141_3947 | Not-determined |
| KM_k141_226896 | Not-determined |
| KM_k141_148869 | Not-determined |
| KM_k141_71247 | Not-determined |
| KM_k141_159047 | Not-determined |
| KM_k141_82716 | Not-determined |
| KM_k141_116468 | Not-determined |
| KM_k141_26119 | Not-determined |
| KM_k141_26121 | Not-determined |
| KM_k141_170869 | Not-determined |
| KM_k141_116477 | Not-determined |
| KM_k141_226914 | Not-determined |
| KM_k141_116481 | Low-quality |
| KM_k141_126736 | Not-determined |
| KM_k141_82730 | Not-determined |
| KM_k141_37315 | Not-determined |
| KM_k141_137755 | Not-determined |
| KM_k141_14502 | Low-quality |
| KM_k141_82743 | Not-determined |
| KM_k141_148877 | Not-determined |
| KM_k141_137771 | Not-determined |
| KM_k141_37328 | Not-determined |
| KM_k141_247953 | Not-determined |
| KM_k141_137783 | Not-determined |
| KM_k141_104117 | Not-determined |
| KM_k141_137785 | Not-determined |
| KM_k141_137786 | Not-determined |
| KM_k141_26162 | Low-quality |
| KM_k141_259197 | Not-determined |
| KM_k141_71282 | Not-determined |
| KM_k141_82759 | Not-determined |
| KM_k141_148891 | Not-determined |
| KM_k141_137792 | Not-determined |
| KM_k141_3987 | Not-determined |
| KM_k141_14520 | Not-determined |
| KM_k141_3995 | Not-determined |
| KM_k141_159063 | Not-determined |
| KM_k141_82762 | Low-quality |
| KM_k141_159065 | Not-determined |
| KM_k141_215981 | Not-determined |
| KM_k141_137797 | Not-determined |
| KM_k141_71292 | Not-determined |
| KM_k141_104126 | Not-determined |
| KM_k141_37356 | Not-determined |
| KM_k141_37361 | Not-determined |
| KM_k141_170921 | Low-quality |
| KM_k141_170923 | Not-determined |
| KM_k141_226955 | Low-quality |
| KM_k141_215995 | Low-quality |
| KM_k141_71304 | Not-determined |
| KM_k141_104144 | Not-determined |
| KM_k141_116489 | Not-determined |
| KM_k141_204674 | Low-quality |
| KM_k141_37366 | Not-determined |
| KM_k141_71309 | Not-determined |
| KM_k141_259210 | Low-quality |
| KM_k141_204679 | Not-determined |
| KM_k141_182576 | Not-determined |
| KM_k141_204681 | Not-determined |
| KM_k141_159084 | Not-determined |
| KM_k141_71314 | Not-determined |
| KM_k141_116501 | Not-determined |
| KM_k141_116503 | Not-determined |
| KM_k141_82792 | Not-determined |
| KM_k141_159094 | Not-determined |
| KM_k141_237357 | Low-quality |
| KM_k141_259222 | Not-determined |
| KM_k141_104171 | Not-determined |
| KM_k141_259224 | Not-determined |
| KM_k141_259227 | Not-determined |
| KM_k141_226968 | Low-quality |
| KM_k141_14539 | Not-determined |
| KM_k141_137811 | Not-determined |
| KM_k141_59506 | Not-determined |
| KM_k141_137813 | Not-determined |
| KM_k141_237375 | Not-determined |
| KM_k141_193532 | Not-determined |
| KM_k141_4011 | Not-determined |
| KM_k141_26172 | Not-determined |
| KM_k141_116544 | Not-determined |
| KM_k141_59510 | Not-determined |
| KM_k141_4013 | Low-quality |
| KM_k141_26174 | Not-determined |
| KM_k141_14548 | Not-determined |
| KM_k141_116550 | Not-determined |
| KM_k141_71341 | Not-determined |
| KM_k141_104186 | Not-determined |
| KM_k141_226981 | Not-determined |
| KM_k141_226982 | Not-determined |
| KM_k141_237387 | Not-determined |
| KM_k141_237388 | Not-determined |
| KM_k141_4017 | Not-determined |
| KM_k141_4019 | Not-determined |
| KM_k141_182598 | Not-determined |
| KM_k141_193547 | Not-determined |
| KM_k141_59526 | Not-determined |
| KM_k141_59531 | Not-determined |
| KM_k141_4023 | Low-quality |
| KM_k141_182608 | Not-determined |
| KM_k141_116569 | Not-determined |
| KM_k141_116570 | Low-quality |
| KM_k141_47190 | Not-determined |
| KM_k141_59534 | Not-determined |
| KM_k141_92189 | Not-determined |
| KM_k141_82821 | Not-determined |
| KM_k141_82822 | Not-determined |
| KM_k141_204712 | Not-determined |
| KM_k141_14562 | Not-determined |
| KM_k141_247968 | Not-determined |
| KM_k141_204720 | Not-determined |
| KM_k141_204722 | Not-determined |
| KM_k141_204724 | Not-determined |
| KM_k141_193559 | Not-determined |
| KM_k141_170969 | Low-quality |
| KM_k141_4032 | Not-determined |
| KM_k141_26200 | Not-determined |
| KM_k141_137823 | Not-determined |
| KM_k141_204727 | Not-determined |
| KM_k141_193561 | Low-quality |
| KM_k141_137827 | Not-determined |
| KM_k141_204730 | Low-quality |
| KM_k141_182623 | Not-determined |
| KM_k141_227018 | Not-determined |
| KM_k141_4046 | Not-determined |
| KM_k141_193575 | Not-determined |
| KM_k141_47195 | Not-determined |
| KM_k141_47196 | Not-determined |
| KM_k141_204738 | Not-determined |
| KM_k141_137836 | Not-determined |
| KM_k141_182626 | Low-quality |
| KM_k141_227025 | Not-determined |
| KM_k141_204742 | Not-determined |
| KM_k141_204745 | Not-determined |
| KM_k141_92214 | Not-determined |
| KM_k141_259261 | Not-determined |
| KM_k141_104199 | Not-determined |
| KM_k141_227026 | Not-determined |
| KM_k141_82853 | Not-determined |
| KM_k141_4064 | Low-quality |
| KM_k141_104210 | Not-determined |
| KM_k141_204766 | Not-determined |
| KM_k141_26229 | Not-determined |
| KM_k141_159145 | Not-determined |
| KM_k141_227050 | Not-determined |
| KM_k141_227052 | Not-determined |
| KM_k141_4079 | Not-determined |
| KM_k141_104219 | Not-determined |
| KM_k141_47222 | Not-determined |
| KM_k141_104220 | Not-determined |
| KM_k141_204773 | Low-quality |
| KM_k141_204774 | Not-determined |
| KM_k141_47227 | Not-determined |
| KM_k141_148928 | Not-determined |
| KM_k141_247989 | Not-determined |
| KM_k141_182654 | Not-determined |
| KM_k141_4087 | Not-determined |
| KM_k141_82872 | Not-determined |
| KM_k141_193606 | Low-quality |
| KM_k141_159150 | Not-determined |
| 17-108-final-viral-combined_k141_58538\|\|full | Medium-quality |
| KM_k141_82877 | Not-determined |
| KM_k141_104231 | Not-determined |
| KM_k141_4095 | Not-determined |
| KM_k141_82884 | Not-determined |
| KM_k141_82891 | Not-determined |
| KM_k141_47245 | Not-determined |
| KM_k141_82892 | Low-quality |
| KM_k141_116613 | Not-determined |
| KM_k141_82895 | Not-determined |
| KM_k141_126776 | Not-determined |
| KM_k141_227064 | Not-determined |
| KM_k141_92232 | Low-quality |
| KM_k141_92233 | Not-determined |
| KM_k141_116619 | Not-determined |
| KM_k141_14573 | Not-determined |
| KM_k141_82920 | Not-determined |
| KM_k141_237466 | Not-determined |
| KM_k141_37382 | Not-determined |
| KM_k141_47265 | Not-determined |
| KM_k141_14582 | Not-determined |
| KM_k141_14585 | Not-determined |
| KM_k141_14587 | Not-determined |
| KM_k141_259277 | Not-determined |
| KM_k141_37390 | Not-determined |
| KM_k141_170983 | Not-determined |
| KM_k141_37392 | Not-determined |
| KM_k141_14596 | Not-determined |
| KM_k141_259283 | Not-determined |
| KM_k141_37396 | Not-determined |
| KM_k141_170990 | Not-determined |
| KM_k141_126815 | Not-determined |
| KM_k141_193649 | Not-determined |
| KM_k141_47275 | Not-determined |
| KM_k141_26252 | Low-quality |
| KM_k141_137853 | Not-determined |
| KM_k141_237480 | Not-determined |
| KM_k141_126818 | Not-determined |
| KM_k141_137856 | Low-quality |
| KM_k141_14603 | Not-determined |
| KM_k141_82930 | Low-quality |
| KM_k141_104260 | Low-quality |
| KM_k141_14605 | Not-determined |
| KM_k141_4118 | Not-determined |
| KM_k141_104266 | Low-quality |
| KM_k141_237490 | Not-determined |
| KM_k141_182681 | Low-quality |
| KM_k141_137861 | Not-determined |
| KM_k141_116660 | Not-determined |
| KM_k141_71382 | Not-determined |
| KM_k141_4125 | Not-determined |
| KM_k141_71385 | Not-determined |
| KM_k141_137867 | Not-determined |
| KM_k141_14617 | Low-quality |
| KM_k141_92263 | Low-quality |
| KM_k141_116668 | Not-determined |
| KM_k141_182692 | Low-quality |
| KM_k141_171016 | Not-determined |
| KM_k141_204784 | Not-determined |
| KM_k141_216123 | Not-determined |
| KM_k141_204785 | Not-determined |
| KM_k141_171018 | Not-determined |
| KM_k141_59539 | Not-determined |
| KM_k141_26259 | Not-determined |
| KM_k141_59543 | Not-determined |
| KM_k141_116686 | Not-determined |
| KM_k141_4142 | Not-determined |
| KM_k141_47298 | Not-determined |
| KM_k141_237510 | Low-quality |
| KM_k141_4144 | Not-determined |
| KM_k141_159164 | Not-determined |
| KM_k141_116698 | Not-determined |
| KM_k141_59555 | Not-determined |
| KM_k141_4155 | Not-determined |
| KM_k141_237527 | Not-determined |
| KM_k141_137882 | Not-determined |
| KM_k141_237528 | Not-determined |
| KM_k141_248064 | Not-determined |
| KM_k141_159179 | Not-determined |
| KM_k141_116709 | Not-determined |
| KM_k141_47310 | Not-determined |
| KM_k141_227124 | Not-determined |
| KM_k141_237536 | Not-determined |
| KM_k141_137889 | Not-determined |
| KM_k141_82977 | Not-determined |
| KM_k141_116720 | Not-determined |
| KM_k141_4170 | Not-determined |
| KM_k141_82983 | Not-determined |
| KM_k141_37457 | Not-determined |
| KM_k141_116724 | Not-determined |
| KM_k141_237539 | Not-determined |
| KM_k141_104290 | Low-quality |
| KM_k141_182726 | Not-determined |
| KM_k141_116735 | Not-determined |
| KM_k141_237542 | Not-determined |
| KM_k141_82995 | Not-determined |
| KM_k141_237548 | Not-determined |
| KM_k141_83001 | Not-determined |
| KM_k141_83002 | Not-determined |
| KM_k141_4181 | Not-determined |
| KM_k141_83005 | Not-determined |
| KM_k141_204818 | Not-determined |
| KM_k141_204820 | Not-determined |
| KM_k141_227142 | Not-determined |
| KM_k141_227145 | Not-determined |
| KM_k141_182748 | Not-determined |
| KM_k141_148946 | Low-quality |
| KM_k141_237571 | Not-determined |
| KM_k141_71404 | Not-determined |
| KM_k141_4193 | Not-determined |
| KM_k141_148953 | Not-determined |
| KM_k141_92306 | Not-determined |
| KM_k141_148957 | Not-determined |
| KM_k141_227163 | Not-determined |
| KM_k141_83014 | Not-determined |
| KM_k141_26313 | Not-determined |
| KM_k141_83016 | Low-quality |
| KM_k141_26315 | Not-determined |
| KM_k141_171040 | Not-determined |
| KM_k141_137930 | Not-determined |
| KM_k141_4207 | Low-quality |
| KM_k141_59592 | Not-determined |
| KM_k141_83026 | Not-determined |
| KM_k141_216190 | Low-quality |
| KM_k141_83028 | Low-quality |
| KM_k141_248086 | Not-determined |
| KM_k141_83033 | Not-determined |
| KM_k141_26337 | Low-quality |
| KM_k141_126866 | Not-determined |
| KM_k141_4211 | Not-determined |
| KM_k141_216196 | Not-determined |
| KM_k141_204846 | Not-determined |
| KM_k141_104317 | Low-quality |
| KM_k141_26341 | Not-determined |
| KM_k141_59596 | Not-determined |
| KM_k141_83041 | Not-determined |
| KM_k141_83042 | Not-determined |
| KM_k141_14642 | Not-determined |
| KM_k141_83045 | Not-determined |
| KM_k141_37506 | Not-determined |
| KM_k141_14645 | Low-quality |
| KM_k141_116746 | Not-determined |
| KM_k141_59612 | Not-determined |
| KM_k141_59616 | Not-determined |
| KM_k141_59618 | Not-determined |
| KM_k141_248105 | Not-determined |
| KM_k141_259319 | Not-determined |
| KM_k141_148988 | Low-quality |
| KM_k141_37526 | Not-determined |
| KM_k141_26353 | Low-quality |
| KM_k141_193695 | Not-determined |
| KM_k141_59632 | Low-quality |
| KM_k141_126894 | Not-determined |
| KM_k141_237589 | Not-determined |
| KM_k141_59641 | Not-determined |
| KM_k141_83053 | Not-determined |
| KM_k141_193701 | Not-determined |
| KM_k141_193703 | Not-determined |
| KM_k141_204873 | Not-determined |
| KM_k141_59650 | Low-quality |
| KM_k141_92320 | Not-determined |
| KM_k141_116761 | Not-determined |
| KM_k141_116763 | Not-determined |
| KM_k141_126917 | Not-determined |
| KM_k141_259343 | Not-determined |
| KM_k141_237603 | Not-determined |
| KM_k141_259344 | Not-determined |
| KM_k141_259345 | Not-determined |
| KM_k141_204879 | Not-determined |
| KM_k141_193714 | Low-quality |
| KM_k141_193715 | Not-determined |
| KM_k141_171051 | Not-determined |
| KM_k141_47361 | Not-determined |
| KM_k141_182776 | Low-quality |
| KM_k141_149014 | Not-determined |
| KM_k141_71421 | Not-determined |
| KM_k141_26373 | Not-determined |
| KM_k141_37538 | Not-determined |
| KM_k141_37540 | Not-determined |
| KM_k141_193726 | Not-determined |
| KM_k141_193729 | Not-determined |
| KM_k141_182782 | Not-determined |
| KM_k141_59677 | Not-determined |
| KM_k141_37548 | Not-determined |
| KM_k141_92332 | Not-determined |
| KM_k141_204895 | Not-determined |
| KM_k141_159246 | Not-determined |
| KM_k141_26379 | Not-determined |
| KM_k141_14688 | Not-determined |
| KM_k141_204899 | Not-determined |
| KM_k141_14689 | Not-determined |
| KM_k141_193736 | Not-determined |
| KM_k141_193737 | Not-determined |
| KM_k141_204904 | Not-determined |
| KM_k141_171073 | Not-determined |
| KM_k141_227198 | Low-quality |
| KM_k141_227200 | Not-determined |
| KM_k141_237627 | Not-determined |
| KM_k141_159265 | Not-determined |
| KM_k141_237629 | Not-determined |
| KM_k141_216222 | Not-determined |
| KM_k141_137938 | Not-determined |
| KM_k141_149032 | Not-determined |
| KM_k141_227213 | Not-determined |
| KM_k141_159271 | Not-determined |
| KM_k141_248159 | Not-determined |
| KM_k141_71438 | Low-quality |
| KM_k141_216238 | Low-quality |
| KM_k141_116795 | Not-determined |
| KM_k141_171080 | Not-determined |
| KM_k141_26386 | Not-determined |
| KM_k141_248164 | Not-determined |
| KM_k141_193759 | Low-quality |
| KM_k141_116798 | Low-quality |
| KM_k141_182816 | Not-determined |
| KM_k141_248173 | Not-determined |
| KM_k141_227224 | Not-determined |
| KM_k141_104343 | Not-determined |
| KM_k141_259365 | Not-determined |
| KM_k141_137951 | Not-determined |
| KM_k141_47376 | Not-determined |
| KM_k141_193768 | Not-determined |
| KM_k141_26394 | Not-determined |
| KM_k141_149040 | Not-determined |
| KM_k141_149043 | Not-determined |
| KM_k141_149045 | Not-determined |
| KM_k141_47383 | Not-determined |
| KM_k141_227237 | Not-determined |
| KM_k141_259368 | Low-quality |
| KM_k141_137961 | Not-determined |
| KM_k141_26396 | Not-determined |
| KM_k141_259371 | Not-determined |
| KM_k141_92371 | Not-determined |
| KM_k141_248195 | Not-determined |
| KM_k141_59700 | Not-determined |
| KM_k141_137968 | Not-determined |
| KM_k141_159291 | Not-determined |
| KM_k141_182823 | Not-determined |
| KM_k141_248197 | Not-determined |
| KM_k141_182828 | Not-determined |
| KM_k141_193784 | Not-determined |
| KM_k141_216247 | Not-determined |
| KM_k141_104365 | Not-determined |
| KM_k141_182833 | Not-determined |
| KM_k141_248201 | Not-determined |
| KM_k141_47402 | Not-determined |
| KM_k141_149061 | Not-determined |
| KM_k141_227253 | Not-determined |
| KM_k141_227254 | Not-determined |
| KM_k141_104371 | Not-determined |
| KM_k141_126925 | Not-determined |
| KM_k141_104375 | Not-determined |
| KM_k141_137981 | Low-quality |
| KM_k141_4240 | Not-determined |
| KM_k141_126933 | Not-determined |
| KM_k141_149067 | Not-determined |
| KM_k141_126935 | Not-determined |
| KM_k141_159308 | Not-determined |
| KM_k141_237656 | Low-quality |
| KM_k141_126938 | Not-determined |
| KM_k141_116817 | Low-quality |
| KM_k141_159312 | Not-determined |
| KM_k141_149073 | Not-determined |
| KM_k141_83057 | Not-determined |
| KM_k141_116824 | Not-determined |
| KM_k141_59715 | Not-determined |
| KM_k141_37587 | Not-determined |
| KM_k141_193805 | Not-determined |
| KM_k141_126940 | Not-determined |
| KM_k141_227278 | Not-determined |
| KM_k141_193807 | Not-determined |
| KM_k141_193808 | Not-determined |
| KM_k141_83064 | Not-determined |
| KM_k141_193810 | Not-determined |
| KM_k141_47425 | Not-determined |
| KM_k141_47426 | Not-determined |
| KM_k141_47429 | Not-determined |
| KM_k141_171115 | Not-determined |
| KM_k141_37601 | Not-determined |
| KM_k141_248222 | Not-determined |
| KM_k141_227280 | Not-determined |
| KM_k141_227282 | Not-determined |
| KM_k141_227284 | Not-determined |
| KM_k141_4247 | Not-determined |
| KM_k141_171123 | Not-determined |
| KM_k141_204933 | Low-quality |
| KM_k141_116848 | Not-determined |
| KM_k141_37607 | Not-determined |
| KM_k141_182866 | Not-determined |
| KM_k141_59733 | Not-determined |
| KM_k141_116856 | Not-determined |
| KM_k141_116858 | Not-determined |
| KM_k141_149092 | Not-determined |
| KM_k141_216253 | Low-quality |
| KM_k141_159335 | Not-determined |
| KM_k141_149095 | Not-determined |
| KM_k141_227304 | Not-determined |
| KM_k141_182875 | Not-determined |
| KM_k141_149100 | Not-determined |
| KM_k141_83099 | Not-determined |
| KM_k141_59752 | Not-determined |
| KM_k141_216259 | Not-determined |
| KM_k141_216260 | Not-determined |
| KM_k141_14723 | Not-determined |
| KM_k141_137985 | Not-determined |
| KM_k141_26431 | Not-determined |
| KM_k141_248254 | Not-determined |
| KM_k141_227330 | Not-determined |
| KM_k141_182888 | Not-determined |
| KM_k141_14727 | Not-determined |
| KM_k141_216270 | Not-determined |
| KM_k141_182890 | Not-determined |
| KM_k141_116890 | Not-determined |
| KM_k141_47438 | Not-determined |
| KM_k141_116896 | Not-determined |
| KM_k141_92411 | Not-determined |
| KM_k141_193819 | Not-determined |
| KM_k141_137995 | Not-determined |
| KM_k141_83120 | Not-determined |
| KM_k141_37612 | Not-determined |
| KM_k141_116910 | Not-determined |
| KM_k141_14740 | Not-determined |
| KM_k141_159365 | Not-determined |
| KM_k141_182900 | Not-determined |
| KM_k141_237662 | Not-determined |
| KM_k141_216277 | Not-determined |
| KM_k141_159373 | Not-determined |
| KM_k141_149133 | Not-determined |
| KM_k141_159378 | Not-determined |
| KM_k141_237668 | Low-quality |
| KM_k141_216288 | Not-determined |
| KM_k141_92434 | Low-quality |
| KM_k141_92438 | Not-determined |
| KM_k141_116950 | Not-determined |
| KM_k141_59772 | Not-determined |
| KM_k141_171172 | Not-determined |
| KM_k141_104397 | Not-determined |
| KM_k141_126970 | Not-determined |
| KM_k141_171175 | Not-determined |
| KM_k141_216292 | Not-determined |
| KM_k141_171176 | Not-determined |
| KM_k141_248269 | Not-determined |
| KM_k141_92445 | Not-determined |
| KM_k141_37656 | Not-determined |
| KM_k141_204970 | Low-quality |
| KM_k141_59791 | Not-determined |
| KM_k141_14773 | Not-determined |
| KM_k141_248276 | Not-determined |
| KM_k141_237683 | Not-determined |
| KM_k141_104412 | Not-determined |
| KM_k141_149160 | Not-determined |
| KM_k141_83164 | Not-determined |
| KM_k141_37668 | Low-quality |
| KM_k141_216314 | Not-determined |
| KM_k141_83170 | Not-determined |
| KM_k141_159411 | Not-determined |
| KM_k141_4272 | Not-determined |
| KM_k141_149161 | Not-determined |
| KM_k141_26437 | Not-determined |
| KM_k141_126992 | Not-determined |
| KM_k141_193861 | Not-determined |
| KM_k141_83181 | Not-determined |
| KM_k141_26446 | Not-determined |
| KM_k141_193867 | Not-determined |
| KM_k141_83187 | Not-determined |
| KM_k141_37685 | Not-determined |
| KM_k141_248288 | Not-determined |
| KM_k141_193873 | Not-determined |
| KM_k141_83199 | Not-determined |
| KM_k141_193874 | Not-determined |
| KM_k141_59824 | Not-determined |
| KM_k141_83201 | Not-determined |
| KM_k141_193877 | Not-determined |
| KM_k141_159419 | Not-determined |
| KM_k141_227396 | Not-determined |
| KM_k141_205000 | Not-determined |
| KM_k141_138038 | Not-determined |
| KM_k141_205004 | Not-determined |
| KM_k141_83207 | Not-determined |
| KM_k141_248300 | Not-determined |
| KM_k141_37691 | Not-determined |
| KM_k141_182948 | Not-determined |
| KM_k141_159425 | Not-determined |
| KM_k141_26461 | Not-determined |
| KM_k141_171215 | Not-determined |
| KM_k141_26464 | Not-determined |
| KM_k141_171218 | Not-determined |
| KM_k141_227406 | Not-determined |
| KM_k141_26466 | Not-determined |
| KM_k141_59835 | Not-determined |
| KM_k141_59836 | Not-determined |
| KM_k141_59837 | Not-determined |
| KM_k141_71487 | Not-determined |
| KM_k141_59838 | Not-determined |
| KM_k141_171226 | Not-determined |
| KM_k141_59839 | Not-determined |
| KM_k141_104446 | Not-determined |
| KM_k141_205015 | Not-determined |
| KM_k141_248302 | Not-determined |

| | |
|---|---|
| KM_k141_92462 | Not-determined |
| KM_k141_171229 | Low-quality |
| KM_k141_248305 | Not-determined |
| KM_k141_116976 | Not-determined |
| KM_k141_104447 | Not-determined |
| KM_k141_227422 | Not-determined |
| KM_k141_227425 | Low-quality |
| KM_k141_47462 | Not-determined |
| KM_k141_205028 | Not-determined |
| KM_k141_37699 | Not-determined |
| KM_k141_182969 | Low-quality |
| KM_k141_227431 | Not-determined |
| KM_k141_205036 | Not-determined |
| KM_k141_92474 | Not-determined |
| KM_k141_205042 | Not-determined |
| KM_k141_216342 | Not-determined |
| KM_k141_182987 | Not-determined |
| KM_k141_205050 | Not-determined |
| KM_k141_216344 | Not-determined |
| KM_k141_26469 | Not-determined |
| KM_k141_47486 | Not-determined |
| KM_k141_149218 | Not-determined |
| KM_k141_47487 | Not-determined |
| KM_k141_149220 | Not-determined |
| KM_k141_59851 | Not-determined |
| KM_k141_193898 | Not-determined |
| KM_k141_227456 | Not-determined |
| KM_k141_193902 | Not-determined |
| KM_k141_83236 | Not-determined |
| KM_k141_83238 | Not-determined |
| KM_k141_138073 | Not-determined |
| KM_k141_183005 | Not-determined |
| KM_k141_117019 | Not-determined |
| KM_k141_237720 | Low-quality |
| KM_k141_171259 | Low-quality |
| KM_k141_171260 | Not-determined |
| KM_k141_193906 | Not-determined |
| KM_k141_117028 | Not-determined |
| KM_k141_159454 | Not-determined |
| KM_k141_216372 | Not-determined |
| KM_k141_14794 | Not-determined |
| KM_k141_26510 | Not-determined |
| KM_k141_193920 | Low-quality |
| KM_k141_259375 | Not-determined |
| KM_k141_183018 | Not-determined |
| KM_k141_14805 | Not-determined |
| KM_k141_259380 | Low-quality |
| KM_k141_259383 | Not-determined |
| KM_k141_171279 | Not-determined |
| KM_k141_117047 | Not-determined |
| KM_k141_183024 | Not-determined |
| KM_k141_59873 | Not-determined |
| KM_k141_138078 | Not-determined |
| KM_k141_4298 | Not-determined |
| KM_k141_216380 | Low-quality |
| KM_k141_59881 | Not-determined |
| KM_k141_205082 | Not-determined |
| KM_k141_205084 | Not-determined |
| KM_k141_227494 | Not-determined |
| KM_k141_138092 | Not-determined |
| KM_k141_237730 | Not-determined |
| KM_k141_14825 | Not-determined |
| KM_k141_14826 | Not-determined |
| KM_k141_26535 | Not-determined |
| KM_k141_14827 | Not-determined |
| KM_k141_183039 | Not-determined |
| KM_k141_227509 | Not-determined |
| KM_k141_205097 | Not-determined |
| KM_k141_26539 | Not-determined |
| KM_k141_138099 | Not-determined |
| KM_k141_259386 | Not-determined |
| KM_k141_159466 | Not-determined |
| KM_k141_159470 | Not-determined |
| KM_k141_83268 | Not-determined |
| KM_k141_149286 | Low-quality |
| KM_k141_83271 | Not-determined |
| KM_k141_83273 | Not-determined |
| KM_k141_237748 | Not-determined |
| KM_k141_47532 | Not-determined |
| KM_k141_104460 | Not-determined |
| KM_k141_159477 | Not-determined |
| KM_k141_117088 | Not-determined |
| KM_k141_83276 | Not-determined |
| KM_k141_205103 | Not-determined |
| KM_k141_83277 | Not-determined |
| KM_k141_47537 | Not-determined |
| KM_k141_71531 | Not-determined |
| KM_k141_237756 | Not-determined |
| KM_k141_149305 | Not-determined |
| KM_k141_127027 | Not-determined |
| KM_k141_227531 | Not-determined |
| KM_k141_47542 | Not-determined |
| KM_k141_59912 | Not-determined |
| KM_k141_47543 | Not-determined |
| KM_k141_227538 | Not-determined |
| KM_k141_4325 | Not-determined |
| KM_k141_259405 | Not-determined |
| KM_k141_205136 | Not-determined |
| KM_k141_237762 | Not-determined |
| KM_k141_138140 | Not-determined |
| KM_k141_205139 | Not-determined |
| KM_k141_92532 | Not-determined |
| KM_k141_117115 | Not-determined |
| KM_k141_227548 | Low-quality |
| KM_k141_171297 | Not-determined |
| KM_k141_205141 | Not-determined |
| KM_k141_117120 | Not-determined |
| KM_k141_92536 | Not-determined |
| KM_k141_4337 | Not-determined |
| KM_k141_104479 | Not-determined |
| KM_k141_171302 | Not-determined |
| KM_k141_4339 | Low-quality |
| KM_k141_171304 | Not-determined |
| KM_k141_26558 | Not-determined |
| KM_k141_47555 | Not-determined |
| KM_k141_104486 | Not-determined |
| KM_k141_4343 | Not-determined |
| KM_k141_183072 | Not-determined |
| KM_k141_83298 | Not-determined |
| KM_k141_183076 | Not-determined |
| KM_k141_259423 | Not-determined |
| KM_k141_92559 | Not-determined |
| KM_k141_205169 | Not-determined |
| KM_k141_14843 | Not-determined |
| KM_k141_4366 | Not-determined |
| KM_k141_159491 | Not-determined |
| KM_k141_14847 | Not-determined |
| KM_k141_259430 | Not-determined |
| KM_k141_237789 | Not-determined |
| KM_k141_117167 | Not-determined |
| KM_k141_104518 | Not-determined |
| KM_k141_183081 | Not-determined |
| KM_k141_138187 | Not-determined |
| KM_k141_26581 | Not-determined |
| KM_k141_216420 | Low-quality |
| KM_k141_183085 | Not-determined |
| KM_k141_83318 | Not-determined |
| KM_k141_117175 | Not-determined |
| KM_k141_227574 | Not-determined |
| KM_k141_159497 | Not-determined |
| KM_k141_216427 | Not-determined |
| KM_k141_227582 | Not-determined |
| KM_k141_159503 | Not-determined |
| KM_k141_159504 | Not-determined |
| KM_k141_104524 | Not-determined |
| KM_k141_259449 | Not-determined |
| KM_k141_104531 | Not-determined |
| KM_k141_183097 | Not-determined |
| KM_k141_216428 | Not-determined |
| KM_k141_259456 | Not-determined |
| KM_k141_149339 | Not-determined |
| KM_k141_227598 | Not-determined |
| KM_k141_216436 | Not-determined |
| KM_k141_216439 | Not-determined |
| KM_k141_59950 | Not-determined |
| KM_k141_14875 | Not-determined |
| KM_k141_117202 | Not-determined |
| KM_k141_117204 | Not-determined |
| KM_k141_104537 | Not-determined |
| KM_k141_227610 | Not-determined |
| KM_k141_237807 | Not-determined |
| KM_k141_227613 | Not-determined |
| KM_k141_237808 | Not-determined |
| KM_k141_237809 | Not-determined |
| KM_k141_117211 | Not-determined |
| KM_k141_227619 | Not-determined |
| KM_k141_127050 | Not-determined |
| KM_k141_92576 | Not-determined |
| KM_k141_26595 | Not-determined |
| KM_k141_59956 | Not-determined |
| KM_k141_127056 | Not-determined |
| KM_k141_59958 | Not-determined |
| KM_k141_183121 | Not-determined |
| KM_k141_127064 | Not-determined |
| KM_k141_14880 | Not-determined |
| KM_k141_104548 | Not-determined |
| KM_k141_159523 | Not-determined |
| KM_k141_127066 | Not-determined |
| KM_k141_216457 | Not-determined |
| KM_k141_14882 | Not-determined |
| KM_k141_117223 | Not-determined |
| KM_k141_117225 | Not-determined |
| KM_k141_47607 | Not-determined |
| KM_k141_14884 | Not-determined |
| KM_k141_59964 | Not-determined |
| KM_k141_59966 | Not-determined |
| KM_k141_59967 | Not-determined |
| KM_k141_205188 | Not-determined |
| KM_k141_59971 | Not-determined |
| KM_k141_216468 | Not-determined |
| KM_k141_71593 | Not-determined |
| KM_k141_71595 | Not-determined |
| KM_k141_59975 | Not-determined |
| KM_k141_205198 | Not-determined |
| KM_k141_59978 | Not-determined |
| KM_k141_127073 | Not-determined |
| KM_k141_248338 | Not-determined |
| KM_k141_216479 | Not-determined |
| KM_k141_37766 | Not-determined |
| KM_k141_248341 | Not-determined |
| KM_k141_159536 | Not-determined |
| KM_k141_71615 | Not-determined |
| KM_k141_183148 | Not-determined |
| KM_k141_259466 | Not-determined |
| KM_k141_183150 | Not-determined |
| KM_k141_159542 | Not-determined |
| KM_k141_227644 | Low-quality |
| KM_k141_127091 | Not-determined |
| KM_k141_37767 | Low-quality |
| KM_k141_14906 | Not-determined |
| KM_k141_83343 | Not-determined |
| KM_k141_26639 | Not-determined |
| KM_k141_259473 | Not-determined |
| KM_k141_171356 | Low-quality |
| KM_k141_248358 | Not-determined |
| KM_k141_183166 | Not-determined |
| KM_k141_83351 | Not-determined |
| KM_k141_37776 | Not-determined |
| KM_k141_183170 | Not-determined |
| KM_k141_47632 | Not-determined |
| KM_k141_4414 | Not-determined |
| KM_k141_138215 | Not-determined |
| KM_k141_259479 | Not-determined |
| KM_k141_14916 | Not-determined |
| KM_k141_47639 | Not-determined |
| KM_k141_248364 | Not-determined |
| KM_k141_4416 | Not-determined |
| KM_k141_71620 | Not-determined |
| KM_k141_138222 | Low-quality |
| KM_k141_83365 | Not-determined |
| KM_k141_149368 | Not-determined |
| KM_k141_193953 | Not-determined |
| KM_k141_227671 | Not-determined |
| KM_k141_37788 | Not-determined |
| KM_k141_83373 | Not-determined |
| KM_k141_59982 | Not-determined |
| KM_k141_47645 | Low-quality |
| KM_k141_59983 | Not-determined |
| KM_k141_47648 | Not-determined |
| KM_k141_183182 | Not-determined |
| KM_k141_37797 | Not-determined |
| KM_k141_149379 | Not-determined |
| KM_k141_59991 | Not-determined |
| KM_k141_59993 | Not-determined |
| KM_k141_117253 | Not-determined |
| KM_k141_37802 | Not-determined |
| KM_k141_14934 | Not-determined |
| KM_k141_104586 | Not-determined |
| KM_k141_26677 | Not-determined |
| KM_k141_205236 | Not-determined |
| KM_k141_14941 | Not-determined |
| KM_k141_117265 | Low-quality |
| KM_k141_259511 | Not-determined |
| KM_k141_127114 | Low-quality |
| KM_k141_205238 | Low-quality |
| KM_k141_183202 | Not-determined |
| KM_k141_183209 | Not-determined |
| KM_k141_205249 | Not-determined |
| KM_k141_138229 | Not-determined |
| KM_k141_26686 | Not-determined |
| KM_k141_227710 | Not-determined |
| KM_k141_183222 | Low-quality |
| KM_k141_227713 | Not-determined |
| KM_k141_127131 | Not-determined |
| KM_k141_138244 | Not-determined |
| KM_k141_127138 | Not-determined |
| KM_k141_138248 | Not-determined |
| KM_k141_216516 | Not-determined |
| KM_k141_117293 | Not-determined |
| KM_k141_127145 | Not-determined |
| KM_k141_37839 | Not-determined |
| KM_k141_205279 | Not-determined |
| KM_k141_37844 | Not-determined |
| KM_k141_216530 | Not-determined |
| KM_k141_71640 | Not-determined |
| KM_k141_205285 | Not-determined |
| KM_k141_83436 | Not-determined |
| KM_k141_127154 | Not-determined |
| KM_k141_104626 | Not-determined |
| KM_k141_71644 | Not-determined |
| KM_k141_127155 | Not-determined |
| KM_k141_26711 | Not-determined |
| KM_k141_248374 | Not-determined |
| KM_k141_205293 | Not-determined |
| KM_k141_71652 | Not-determined |
| KM_k141_26718 | Not-determined |
| KM_k141_248377 | Not-determined |
| KM_k141_37862 | Not-determined |
| KM_k141_60021 | Not-determined |
| KM_k141_248381 | Not-determined |
| KM_k141_104634 | Not-determined |
| KM_k141_37865 | Not-determined |
| KM_k141_159584 | Not-determined |
| KM_k141_159596 | Low-quality |
| KM_k141_37876 | Not-determined |
| KM_k141_248396 | Not-determined |
| KM_k141_216559 | Low-quality |
| KM_k141_60035 | Not-determined |
| KM_k141_171389 | Not-determined |
| KM_k141_259530 | Not-determined |
| KM_k141_71669 | Not-determined |
| KM_k141_259531 | Not-determined |
| KM_k141_26741 | Low-quality |
| KM_k141_104640 | Not-determined |
| KM_k141_171391 | Not-determined |
| KM_k141_138291 | Not-determined |
| KM_k141_248403 | Not-determined |
| KM_k141_183251 | Low-quality |
| KM_k141_26750 | Not-determined |
| KM_k141_159620 | Not-determined |
| KM_k141_26755 | Not-determined |
| KM_k141_92619 | Low-quality |
| KM_k141_92621 | Not-determined |
| KM_k141_227736 | Not-determined |
| KM_k141_117303 | Low-quality |
| KM_k141_138310 | Not-determined |
| KM_k141_117305 | Not-determined |
| KM_k141_37885 | Not-determined |
| KM_k141_205306 | Not-determined |
| KM_k141_183263 | Not-determined |
| KM_k141_183264 | Not-determined |
| KM_k141_248424 | Not-determined |
| KM_k141_4442 | Not-determined |
| KM_k141_248425 | Not-determined |
| KM_k141_4444 | Not-determined |
| KM_k141_259547 | Not-determined |
| KM_k141_159636 | Not-determined |
| KM_k141_60040 | Not-determined |
| KM_k141_149398 | Not-determined |
| KM_k141_83468 | Not-determined |
| KM_k141_4451 | Not-determined |
| KM_k141_4454 | Not-determined |
| KM_k141_47660 | Not-determined |
| KM_k141_47663 | Not-determined |
| KM_k141_83476 | Not-determined |
| KM_k141_138316 | Not-determined |
| KM_k141_60052 | Not-determined |
| KM_k141_248440 | Not-determined |
| KM_k141_4460 | Not-determined |
| KM_k141_71700 | Not-determined |
| KM_k141_4463 | Not-determined |
| KM_k141_149411 | Not-determined |
| KM_k141_83481 | Not-determined |
| KM_k141_205323 | Low-quality |
| KM_k141_127220 | Not-determined |
| KM_k141_60057 | Not-determined |
| KM_k141_159651 | Not-determined |
| KM_k141_159652 | Not-determined |
| KM_k141_259557 | Not-determined |
| KM_k141_71707 | Not-determined |
| KM_k141_259562 | Not-determined |
| KM_k141_259564 | Not-determined |
| KM_k141_216595 | Not-determined |
| KM_k141_237828 | Not-determined |
| KM_k141_193962 | Not-determined |
| KM_k141_193964 | Not-determined |
| KM_k141_60066 | Not-determined |
| KM_k141_83497 | Not-determined |
| KM_k141_4480 | Low-quality |
| KM_k141_183290 | Not-determined |
| KM_k141_216612 | Low-quality |
| KM_k141_71723 | Not-determined |
| KM_k141_183292 | Not-determined |
| KM_k141_127243 | Not-determined |
| KM_k141_26797 | Not-determined |
| KM_k141_83506 | Not-determined |
| KM_k141_149456 | Not-determined |
| KM_k141_205352 | Not-determined |
| KM_k141_4483 | Not-determined |
| KM_k141_216616 | Not-determined |
| KM_k141_92655 | Not-determined |
| KM_k141_216617 | Not-determined |
| KM_k141_92660 | Not-determined |
| KM_k141_37901 | Not-determined |
| KM_k141_227771 | Not-determined |
| KM_k141_216625 | Not-determined |
| KM_k141_83517 | Not-determined |
| KM_k141_205354 | Not-determined |
| KM_k141_92663 | Not-determined |
| KM_k141_47688 | Not-determined |
| KM_k141_227779 | Not-determined |
| KM_k141_26809 | Not-determined |
| KM_k141_248458 | Not-determined |
| KM_k141_248462 | Not-determined |
| KM_k141_248463 | Not-determined |
| KM_k141_237860 | Not-determined |
| KM_k141_216636 | Not-determined |
| KM_k141_248467 | Not-determined |
| KM_k141_248468 | Not-determined |
| KM_k141_138343 | Not-determined |
| KM_k141_14949 | Not-determined |
| KM_k141_248474 | Not-determined |
| KM_k141_227798 | Not-determined |
| KM_k141_259582 | Not-determined |
| KM_k141_205368 | Not-determined |
| KM_k141_14952 | Not-determined |
| KM_k141_227809 | Not-determined |
| KM_k141_259586 | Not-determined |
| KM_k141_171473 | Not-determined |
| KM_k141_71731 | Not-determined |
| KM_k141_104666 | Not-determined |
| KM_k141_216664 | Low-quality |
| KM_k141_216665 | Not-determined |
| KM_k141_104672 | Not-determined |
| KM_k141_216676 | Not-determined |
| KM_k141_71746 | Not-determined |
| KM_k141_183312 | Not-determined |
| KM_k141_127287 | Not-determined |
| KM_k141_171483 | Not-determined |
| KM_k141_237869 | Not-determined |
| KM_k141_138374 | Not-determined |
| KM_k141_127293 | Not-determined |
| KM_k141_127298 | Not-determined |
| KM_k141_237873 | Not-determined |
| KM_k141_205385 | Not-determined |
| KM_k141_205388 | Low-quality |
| KM_k141_183322 | Not-determined |
| KM_k141_83559 | Low-quality |
| KM_k141_4509 | Not-determined |
| KM_k141_4510 | Not-determined |
| KM_k141_37958 | Not-determined |
| KM_k141_183328 | Not-determined |
| KM_k141_183329 | Not-determined |
| KM_k141_92704 | Not-determined |
| KM_k141_37963 | Not-determined |
| KM_k141_227819 | Not-determined |
| KM_k141_138392 | Not-determined |
| KM_k141_104691 | Not-determined |
| KM_k141_183334 | Not-determined |
| KM_k141_205396 | Not-determined |
| KM_k141_14980 | Not-determined |
| KM_k141_104695 | Not-determined |
| KM_k141_60116 | Not-determined |
| KM_k141_171514 | Not-determined |
| KM_k141_171518 | Not-determined |
| KM_k141_4521 | Low-quality |
| KM_k141_37975 | Not-determined |
| KM_k141_171521 | Not-determined |
| KM_k141_14991 | Not-determined |
| KM_k141_259620 | Not-determined |
| KM_k141_159731 | Not-determined |
| KM_k141_37980 | Not-determined |
| KM_k141_83575 | Not-determined |
| KM_k141_216706 | Not-determined |
| KM_k141_171532 | Not-determined |
| KM_k141_183347 | Not-determined |
| KM_k141_138407 | Not-determined |
| KM_k141_171538 | Not-determined |
| KM_k141_205415 | Not-determined |
| KM_k141_4540 | Not-determined |
| KM_k141_71768 | Not-determined |
| KM_k141_4543 | Not-determined |
| KM_k141_248514 | Not-determined |
| KM_k141_227846 | Not-determined |
| KM_k141_149505 | Not-determined |
| KM_k141_104717 | Not-determined |
| KM_k141_183355 | Not-determined |
| KM_k141_4551 | Not-determined |
| KM_k141_60122 | Not-determined |
| KM_k141_4554 | Not-determined |
| KM_k141_60124 | Not-determined |
| KM_k141_138425 | Not-determined |
| KM_k141_26858 | Not-determined |
| KM_k141_4558 | Not-determined |
| KM_k141_47722 | Not-determined |
| KM_k141_37999 | Not-determined |
| KM_k141_127320 | Not-determined |
| KM_k141_38001 | Low-quality |
| KM_k141_138429 | Not-determined |
| KM_k141_117365 | Not-determined |
| KM_k141_47727 | Not-determined |
| KM_k141_183362 | Not-determined |
| KM_k141_15009 | Low-quality |
| KM_k141_138436 | Not-determined |
| KM_k141_4574 | Not-determined |
| KM_k141_138440 | Not-determined |
| KM_k141_171558 | Not-determined |
| KM_k141_138441 | Not-determined |
| KM_k141_138443 | Not-determined |
| KM_k141_171560 | Not-determined |
| KM_k141_15014 | Not-determined |
| KM_k141_171563 | Not-determined |
| KM_k141_127333 | Not-determined |
| KM_k141_248544 | Not-determined |
| KM_k141_15018 | Not-determined |
| KM_k141_71794 | Not-determined |
| KM_k141_183374 | Not-determined |
| KM_k141_38007 | Not-determined |
| KM_k141_149532 | Not-determined |
| KM_k141_92742 | Not-determined |
| KM_k141_183377 | Not-determined |
| KM_k141_26881 | Not-determined |
| KM_k141_92745 | Not-determined |
| KM_k141_38012 | Not-determined |
| KM_k141_117374 | Not-determined |
| KM_k141_248553 | Not-determined |
| KM_k141_183386 | Not-determined |
| KM_k141_15026 | Not-determined |
| KM_k141_15027 | Not-determined |
| KM_k141_4584 | Not-determined |
| KM_k141_4587 | Not-determined |
| KM_k141_248557 | Low-quality |
| KM_k141_183398 | Not-determined |
| KM_k141_47756 | Not-determined |
| KM_k141_237929 | Not-determined |
| KM_k141_248564 | Not-determined |
| KM_k141_4593 | Not-determined |
| KM_k141_38027 | Not-determined |
| KM_k141_138464 | Not-determined |
| KM_k141_183402 | Not-determined |
| KM_k141_47763 | Low-quality |
| KM_k141_216758 | Low-quality |
| KM_k141_117402 | Not-determined |
| KM_k141_183411 | Not-determined |
| KM_k141_117406 | Not-determined |
| KM_k141_38036 | Not-determined |
| KM_k141_171604 | Low-quality |
| KM_k141_15043 | Not-determined |
| KM_k141_117408 | Not-determined |
| KM_k141_71814 | Not-determined |
| KM_k141_227878 | Low-quality |
| KM_k141_171609 | Not-determined |
| KM_k141_259673 | Not-determined |
| KM_k141_71820 | Not-determined |
| KM_k141_205455 | Not-determined |
| KM_k141_38049 | Not-determined |
| KM_k141_227891 | Not-determined |
| KM_k141_26916 | Not-determined |
| KM_k141_26917 | Not-determined |
| KM_k141_183422 | Not-determined |
| KM_k141_138488 | Not-determined |
| KM_k141_227900 | Not-determined |
| KM_k141_127392 | Not-determined |
| KM_k141_248582 | Not-determined |
| KM_k141_38062 | Not-determined |
| KM_k141_83587 | Not-determined |
| KM_k141_127397 | Not-determined |
| KM_k141_26928 | Not-determined |
| KM_k141_104745 | Not-determined |
| KM_k141_4606 | Not-determined |
| KM_k141_26936 | Low-quality |
| KM_k141_26938 | Not-determined |
| KM_k141_194049 | Not-determined |
| KM_k141_194050 | Not-determined |
| KM_k141_194052 | Not-determined |
| KM_k141_104758 | Not-determined |
| KM_k141_71843 | Low-quality |
| KM_k141_138506 | Not-determined |
| KM_k141_60155 | Not-determined |
| KM_k141_60156 | Low-quality |
| KM_k141_159754 | Not-determined |
| KM_k141_83606 | Not-determined |
| KM_k141_71848 | Not-determined |
| KM_k141_60160 | Not-determined |
| KM_k141_227908 | Not-determined |
| KM_k141_138518 | Not-determined |
| KM_k141_47791 | Not-determined |
| KM_k141_138520 | Not-determined |
| KM_k141_149552 | Not-determined |
| KM_k141_47792 | Not-determined |
| KM_k141_117429 | Not-determined |
| KM_k141_227917 | Not-determined |
| KM_k141_205475 | Not-determined |
| KM_k141_194069 | Not-determined |
| KM_k141_60168 | Low-quality |
| KM_k141_47798 | Not-determined |
| KM_k141_60169 | Not-determined |
| KM_k141_47799 | Not-determined |
| KM_k141_117432 | Not-determined |
| KM_k141_194073 | Not-determined |
| KM_k141_117434 | Not-determined |
| KM_k141_194080 | Not-determined |
| KM_k141_194081 | Not-determined |
| KM_k141_183438 | Not-determined |
| KM_k141_47806 | Not-determined |
| KM_k141_83628 | Not-determined |
| KM_k141_71868 | Not-determined |
| KM_k141_60175 | Not-determined |
| KM_k141_248595 | Not-determined |
| KM_k141_171622 | Not-determined |
| KM_k141_237949 | Not-determined |
| KM_k141_171625 | Not-determined |
| KM_k141_171628 | Not-determined |
| KM_k141_127426 | Not-determined |
| KM_k141_15104 | Not-determined |
| KM_k141_149568 | Not-determined |
| KM_k141_171631 | Not-determined |
| KM_k141_4646 | Not-determined |
| KM_k141_92760 | Not-determined |

| | |
|---|---|
| KM_k141_205484 | Not-determined |
| KM_k141_71880 | Not-determined |
| KM_k141_194101 | Not-determined |
| KM_k141_159780 | Not-determined |
| KM_k141_194105 | Not-determined |
| KM_k141_248616 | Not-determined |
| KM_k141_127433 | Not-determined |
| KM_k141_248617 | Not-determined |
| KM_k141_149578 | Not-determined |
| KM_k141_248618 | Not-determined |
| KM_k141_216772 | Not-determined |
| KM_k141_149581 | Not-determined |
| KM_k141_149582 | Not-determined |
| KM_k141_159788 | Not-determined |
| KM_k141_216774 | Not-determined |
| KM_k141_60193 | Not-determined |
| KM_k141_205497 | Not-determined |
| KM_k141_60199 | Not-determined |
| KM_k141_4660 | Not-determined |
| KM_k141_149592 | Not-determined |
| KM_k141_227955 | Not-determined |
| KM_k141_38085 | Not-determined |
| KM_k141_26956 | Not-determined |
| KM_k141_26957 | Not-determined |
| KM_k141_149597 | Not-determined |
| KM_k141_159801 | Not-determined |
| KM_k141_237963 | Not-determined |
| KM_k141_183468 | Not-determined |
| KM_k141_15123 | Not-determined |
| KM_k141_15124 | Not-determined |
| KM_k141_171662 | Not-determined |
| KM_k141_237968 | Not-determined |
| KM_k141_127452 | Not-determined |
| KM_k141_71894 | Not-determined |
| KM_k141_171665 | Not-determined |
| KM_k141_205516 | Not-determined |
| KM_k141_92766 | Not-determined |
| KM_k141_205519 | Not-determined |
| KM_k141_60220 | Low-quality |
| KM_k141_92769 | Not-determined |
| KM_k141_194141 | Not-determined |
| KM_k141_183476 | Not-determined |
| KM_k141_38099 | Not-determined |
| KM_k141_83670 | Not-determined |
| KM_k141_149606 | Not-determined |
| KM_k141_4684 | Not-determined |
| KM_k141_47820 | Not-determined |
| KM_k141_4687 | Not-determined |
| KM_k141_205541 | Not-determined |
| KM_k141_237983 | Not-determined |
| KM_k141_104811 | Not-determined |
| KM_k141_4695 | Low-quality |
| KM_k141_171676 | Not-determined |
| KM_k141_4696 | Not-determined |
| KM_k141_83691 | Not-determined |
| KM_k141_4698 | Not-determined |
| KM_k141_104814 | Not-determined |
| KM_k141_248623 | Not-determined |
| KM_k141_104816 | Not-determined |
| KM_k141_227966 | Not-determined |
| KM_k141_159829 | Not-determined |
| KM_k141_248627 | Not-determined |
| KM_k141_127487 | Not-determined |
| KM_k141_149613 | Not-determined |
| KM_k141_171691 | Not-determined |
| KM_k141_237993 | Not-determined |
| KM_k141_159836 | Not-determined |
| KM_k141_104821 | Not-determined |
| KM_k141_26974 | Low-quality |
| KM_k141_60234 | Not-determined |
| KM_k141_149622 | Not-determined |
| KM_k141_104826 | Not-determined |
| KM_k141_149624 | Not-determined |
| KM_k141_60241 | Not-determined |
| KM_k141_205566 | Not-determined |
| KM_k141_183493 | Not-determined |
| KM_k141_227977 | Not-determined |
| KM_k141_149629 | Not-determined |
| KM_k141_149630 | Not-determined |
| KM_k141_60244 | Low-quality |
| KM_k141_159850 | Not-determined |
| KM_k141_92784 | Not-determined |
| KM_k141_216795 | Not-determined |
| KM_k141_227986 | Not-determined |
| KM_k141_38115 | Not-determined |
| KM_k141_15157 | Not-determined |
| KM_k141_127498 | Not-determined |
| KM_k141_60250 | Not-determined |
| KM_k141_60251 | Not-determined |
| KM_k141_227990 | Not-determined |
| KM_k141_248640 | Not-determined |
| KM_k141_227997 | Not-determined |
| KM_k141_117442 | Not-determined |
| KM_k141_117443 | Not-determined |
| KM_k141_15163 | Low-quality |
| KM_k141_171706 | Not-determined |
| KM_k141_4720 | Not-determined |
| KM_k141_104846 | Not-determined |
| KM_k141_47834 | Low-quality |
| KM_k141_27006 | Not-determined |
| KM_k141_183528 | Not-determined |
| KM_k141_92798 | Not-determined |
| KM_k141_104854 | Low-quality |
| KM_k141_47841 | Not-determined |
| KM_k141_83717 | Not-determined |
| KM_k141_83718 | Not-determined |
| KM_k141_138593 | Not-determined |
| KM_k141_104862 | Not-determined |
| KM_k141_104863 | Not-determined |
| KM_k141_47849 | Not-determined |
| KM_k141_117455 | Low-quality |
| KM_k141_83725 | Not-determined |
| KM_k141_216817 | Not-determined |
| KM_k141_194194 | Not-determined |
| KM_k141_205579 | Not-determined |
| KM_k141_183538 | Not-determined |
| KM_k141_205581 | Not-determined |
| KM_k141_127509 | Not-determined |
| KM_k141_71938 | Not-determined |
| KM_k141_171719 | Not-determined |
| KM_k141_38125 | Not-determined |
| KM_k141_194199 | Not-determined |
| KM_k141_171725 | Not-determined |
| KM_k141_117474 | Not-determined |
| KM_k141_71946 | Not-determined |
| KM_k141_83741 | Not-determined |
| KM_k141_117475 | Not-determined |
| KM_k141_194209 | Not-determined |
| KM_k141_205585 | Not-determined |
| KM_k141_127519 | Not-determined |
| KM_k141_183547 | Not-determined |
| KM_k141_149662 | Low-quality |
| KM_k141_171738 | Not-determined |
| KM_k141_127527 | Not-determined |
| KM_k141_117489 | Not-determined |
| KM_k141_138627 | Not-determined |
| KM_k141_92808 | Not-determined |
| KM_k141_71968 | Not-determined |
| KM_k141_127530 | Not-determined |
| KM_k141_194233 | Not-determined |
| KM_k141_104908 | Not-determined |
| KM_k141_171751 | Not-determined |
| KM_k141_183565 | Not-determined |
| KM_k141_117503 | Not-determined |
| KM_k141_104912 | Not-determined |
| KM_k141_183567 | Not-determined |
| KM_k141_117506 | Not-determined |
| KM_k141_238038 | Low-quality |
| KM_k141_238040 | Not-determined |
| KM_k141_138640 | Not-determined |
| KM_k141_117511 | Not-determined |
| KM_k141_194254 | Not-determined |
| KM_k141_138644 | Not-determined |
| KM_k141_171763 | Not-determined |
| KM_k141_194257 | Not-determined |
| KM_k141_117517 | Not-determined |
| KM_k141_138646 | Not-determined |
| KM_k141_71984 | Not-determined |
| KM_k141_149677 | Not-determined |
| KM_k141_138655 | Not-determined |
| KM_k141_71991 | Not-determined |
| KM_k141_4739 | Not-determined |
| KM_k141_104926 | Not-determined |
| KM_k141_183590 | Not-determined |
| 18-109-final-viral-combined_k141_157434\|\|full | Medium-quality |
| KM_k141_83758 | Not-determined |
| KM_k141_149693 | Not-determined |
| KM_k141_4742 | Not-determined |
| KM_k141_205593 | Not-determined |
| KM_k141_183601 | Not-determined |
| KM_k141_27027 | Not-determined |
| KM_k141_205595 | Not-determined |
| KM_k141_60283 | Not-determined |
| KM_k141_171783 | Not-determined |
| KM_k141_194280 | Not-determined |
| KM_k141_248682 | Not-determined |
| KM_k141_60287 | Not-determined |
| KM_k141_216864 | Not-determined |
| KM_k141_83772 | Not-determined |
| KM_k141_15220 | Not-determined |
| KM_k141_194295 | Not-determined |
| KM_k141_216873 | Not-determined |
| KM_k141_138660 | Not-determined |
| KM_k141_183620 | Not-determined |
| KM_k141_183621 | Not-determined |
| KM_k141_228007 | Not-determined |
| KM_k141_183624 | Not-determined |
| KM_k141_38192 | Not-determined |
| KM_k141_228011 | Not-determined |
| KM_k141_83788 | Not-determined |
| KM_k141_83789 | Not-determined |
| KM_k141_205613 | Not-determined |
| KM_k141_238072 | Not-determined |
| KM_k141_238073 | Not-determined |
| KM_k141_205618 | Not-determined |
| KM_k141_127571 | Not-determined |
| KM_k141_104937 | Not-determined |
| KM_k141_259738 | Not-determined |
| KM_k141_259739 | Not-determined |
| KM_k141_259740 | Not-determined |
| KM_k141_228017 | Not-determined |
| KM_k141_205625 | Not-determined |
| KM_k141_159913 | Not-determined |
| KM_k141_159914 | Not-determined |
| KM_k141_171793 | Not-determined |
| KM_k141_38202 | Not-determined |
| KM_k141_171795 | Not-determined |
| KM_k141_92825 | Not-determined |
| KM_k141_27056 | Low-quality |
| KM_k141_183640 | Not-determined |
| KM_k141_183642 | Not-determined |
| KM_k141_92827 | Not-determined |
| KM_k141_238080 | Not-determined |
| KM_k141_60293 | Not-determined |
| KM_k141_238084 | Not-determined |
| KM_k141_117534 | Not-determined |
| KM_k141_92829 | Not-determined |
| KM_k141_60296 | Not-determined |
| KM_k141_216899 | Low-quality |
| KM_k141_228030 | Not-determined |
| KM_k141_183648 | Not-determined |
| KM_k141_83815 | Not-determined |
| KM_k141_117541 | Not-determined |
| KM_k141_216903 | Not-determined |
| KM_k141_149707 | Not-determined |
| KM_k141_259762 | Not-determined |
| KM_k141_127603 | Not-determined |
| KM_k141_117557 | Low-quality |
| KM_k141_228038 | Not-determined |
| KM_k141_117561 | Not-determined |
| KM_k141_183659 | Not-determined |
| KM_k141_92837 | Not-determined |
| KM_k141_228045 | Not-determined |
| KM_k141_92841 | Not-determined |
| KM_k141_38231 | Not-determined |
| KM_k141_238099 | Not-determined |
| KM_k141_149730 | Low-quality |
| KM_k141_72029 | Low-quality |
| KM_k141_138713 | Not-determined |
| KM_k141_47865 | Not-determined |
| KM_k141_228055 | Not-determined |
| KM_k141_138721 | Not-determined |
| KM_k141_138722 | Not-determined |
| KM_k141_149736 | Low-quality |
| KM_k141_259788 | Not-determined |
| KM_k141_38240 | Not-determined |
| KM_k141_138727 | Not-determined |
| KM_k141_183672 | Not-determined |
| KM_k141_194330 | Not-determined |
| KM_k141_149744 | Not-determined |
| KM_k141_104984 | Not-determined |
| KM_k141_27080 | Not-determined |
| KM_k141_183682 | Not-determined |
| KM_k141_238123 | Not-determined |
| KM_k141_117583 | Not-determined |
| KM_k141_15256 | Not-determined |
| KM_k141_138740 | Not-determined |
| KM_k141_38252 | Not-determined |
| KM_k141_27096 | Not-determined |
| KM_k141_238144 | Not-determined |
| KM_k141_15266 | Not-determined |
| KM_k141_15267 | Not-determined |
| KM_k141_38254 | Not-determined |
| KM_k141_159948 | Not-determined |
| KM_k141_194359 | Not-determined |
| KM_k141_194361 | Not-determined |
| KM_k141_38258 | Low-quality |
| KM_k141_15273 | Not-determined |
| KM_k141_117605 | Not-determined |
| KM_k141_72065 | Not-determined |
| KM_k141_238157 | Not-determined |
| KM_k141_228079 | Not-determined |
| KM_k141_105016 | Not-determined |
| KM_k141_159965 | Not-determined |
| KM_k141_105017 | Not-determined |
| KM_k141_138767 | Not-determined |
| KM_k141_47889 | Not-determined |
| KM_k141_105021 | Not-determined |
| KM_k141_194370 | Not-determined |
| KM_k141_259810 | Not-determined |
| KM_k141_72077 | Not-determined |
| KM_k141_138776 | Not-determined |
| KM_k141_92865 | Not-determined |
| KM_k141_194374 | Not-determined |
| KM_k141_83852 | Not-determined |
| KM_k141_117623 | Not-determined |
| KM_k141_60343 | Low-quality |
| KM_k141_159976 | Low-quality |
| KM_k141_183715 | Not-determined |
| KM_k141_38275 | Not-determined |
| KM_k141_72084 | Not-determined |
| KM_k141_183717 | Not-determined |
| KM_k141_248753 | Not-determined |
| KM_k141_15285 | Not-determined |
| KM_k141_105039 | Not-determined |
| KM_k141_171835 | Low-quality |
| KM_k141_171836 | Not-determined |
| KM_k141_228107 | Low-quality |
| KM_k141_72095 | Not-determined |
| KM_k141_15289 | Not-determined |
| KM_k141_159988 | Low-quality |
| KM_k141_15290 | Not-determined |
| KM_k141_205692 | Not-determined |
| KM_k141_117635 | Not-determined |
| KM_k141_205697 | Not-determined |
| KM_k141_159995 | Not-determined |
| KM_k141_228117 | Not-determined |
| KM_k141_72106 | Low-quality |
| KM_k141_72109 | Not-determined |
| KM_k141_248769 | Not-determined |
| KM_k141_228120 | Not-determined |
| KM_k141_117643 | Not-determined |
| KM_k141_194394 | Not-determined |
| KM_k141_47905 | Not-determined |
| KM_k141_4770 | Not-determined |
| KM_k141_117647 | Not-determined |
| KM_k141_160002 | Not-determined |
| KM_k141_228126 | Low-quality |
| KM_k141_60367 | Not-determined |
| KM_k141_238180 | Not-determined |
| KM_k141_15293 | Low-quality |
| KM_k141_15294 | Not-determined |
| KM_k141_83877 | Not-determined |
| KM_k141_171862 | Not-determined |
| KM_k141_38292 | Not-determined |
| KM_k141_238185 | Low-quality |
| KM_k141_248773 | Not-determined |
| KM_k141_160012 | Not-determined |
| KM_k141_138782 | Not-determined |
| KM_k141_205726 | Not-determined |
| KM_k141_171874 | Not-determined |
| KM_k141_117657 | Not-determined |
| KM_k141_238196 | Not-determined |
| KM_k141_15316 | Not-determined |
| KM_k141_149813 | Not-determined |
| KM_k141_160022 | Not-determined |
| KM_k141_160026 | Not-determined |
| KM_k141_4786 | Not-determined |
| KM_k141_47950 | Not-determined |
| KM_k141_205734 | Not-determined |
| KM_k141_60408 | Not-determined |
| KM_k141_194428 | Not-determined |
| KM_k141_149828 | Not-determined |
| KM_k141_138809 | Low-quality |
| KM_k141_60430 | Not-determined |
| KM_k141_194441 | Not-determined |
| KM_k141_216951 | Not-determined |
| KM_k141_238215 | Not-determined |
| KM_k141_92873 | Not-determined |
| KM_k141_92874 | Not-determined |
| KM_k141_248822 | Low-quality |
| KM_k141_60438 | Not-determined |
| KM_k141_238218 | Not-determined |
| KM_k141_248827 | Not-determined |
| KM_k141_92878 | Not-determined |
| KM_k141_248833 | Not-determined |
| KM_k141_83923 | Low-quality |
| KM_k141_248837 | Not-determined |
| KM_k141_38342 | Not-determined |
| KM_k141_83927 | Not-determined |
| KM_k141_83928 | Not-determined |
| KM_k141_27141 | Not-determined |
| KM_k141_149837 | Not-determined |
| KM_k141_183758 | Not-determined |
| KM_k141_216975 | Not-determined |
| KM_k141_15322 | Not-determined |
| KM_k141_183759 | Not-determined |
| KM_k141_83940 | Not-determined |
| KM_k141_205745 | Low-quality |
| KM_k141_15325 | Not-determined |
| KM_k141_238227 | Not-determined |
| KM_k141_160066 | Not-determined |
| KM_k141_72130 | Not-determined |
| KM_k141_248844 | Not-determined |
| 17-108-final-viral-combined_k141_194205\|\|full | Medium-quality |
| KM_k141_92892 | Not-determined |
| KM_k141_160067 | Low-quality |
| KM_k141_27159 | Not-determined |
| KM_k141_205760 | Not-determined |
| KM_k141_60475 | Not-determined |
| KM_k141_205762 | Not-determined |
| KM_k141_72139 | Not-determined |
| KM_k141_72141 | Not-determined |
| KM_k141_60478 | Low-quality |
| KM_k141_60479 | Not-determined |
| KM_k141_105092 | Not-determined |
| KM_k141_38357 | Not-determined |
| KM_k141_228187 | Not-determined |
| KM_k141_92906 | Not-determined |
| KM_k141_205778 | Not-determined |
| KM_k141_228192 | Not-determined |
| KM_k141_92909 | Not-determined |
| KM_k141_60483 | Not-determined |
| KM_k141_259830 | Not-determined |
| KM_k141_194470 | Not-determined |
| KM_k141_183788 | Not-determined |
| KM_k141_160095 | Not-determined |
| KM_k141_47976 | Not-determined |
| KM_k141_47977 | Not-determined |
| KM_k141_205790 | Not-determined |
| KM_k141_160100 | Not-determined |
| KM_k141_4860 | Not-determined |
| KM_k141_248873 | Low-quality |
| KM_k141_117705 | Not-determined |
| KM_k141_259837 | Low-quality |
| KM_k141_259841 | Not-determined |
| KM_k141_149867 | Low-quality |
| KM_k141_117720 | Not-determined |
| KM_k141_105118 | Not-determined |
| KM_k141_47990 | Low-quality |
| KM_k141_72146 | Not-determined |
| KM_k141_60527 | Not-determined |
| KM_k141_149873 | Not-determined |
| KM_k141_194482 | Not-determined |
| KM_k141_92939 | Not-determined |
| KM_k141_83999 | Not-determined |
| KM_k141_228218 | Not-determined |
| KM_k141_72156 | Not-determined |
| KM_k141_47998 | Not-determined |
| KM_k141_48001 | Not-determined |
| KM_k141_248897 | Not-determined |
| KM_k141_217014 | Not-determined |
| KM_k141_205831 | Not-determined |
| KM_k141_160116 | Not-determined |
| KM_k141_183821 | Not-determined |
| KM_k141_217015 | Not-determined |
| KM_k141_194495 | Not-determined |
| KM_k141_183824 | Not-determined |
| KM_k141_194496 | Not-determined |
| KM_k141_84005 | Not-determined |
| KM_k141_117733 | Not-determined |
| KM_k141_259850 | Not-determined |
| KM_k141_172001 | Not-determined |
| KM_k141_238250 | Not-determined |
| KM_k141_149889 | Not-determined |
| KM_k141_72167 | Not-determined |
| KM_k141_238255 | Not-determined |
| KM_k141_105154 | Not-determined |
| KM_k141_84014 | Not-determined |
| KM_k141_238259 | Not-determined |
| KM_k141_105160 | Not-determined |
| KM_k141_238261 | Not-determined |
| KM_k141_238262 | Not-determined |
| KM_k141_172015 | Not-determined |
| KM_k141_4880 | Low-quality |
| KM_k141_238263 | Not-determined |
| KM_k141_238267 | Not-determined |
| KM_k141_4882 | Not-determined |
| KM_k141_238268 | Not-determined |
| KM_k141_4884 | Not-determined |
| KM_k141_205848 | Not-determined |
| KM_k141_84026 | Not-determined |
| KM_k141_4888 | Not-determined |
| KM_k141_194511 | Not-determined |
| KM_k141_127711 | Not-determined |
| KM_k141_4891 | Low-quality |
| KM_k141_27187 | Not-determined |
| KM_k141_105174 | Not-determined |
| KM_k141_72186 | Not-determined |
| KM_k141_238278 | Not-determined |
| KM_k141_183832 | Not-determined |
| KM_k141_105178 | Not-determined |
| KM_k141_194520 | Not-determined |
| KM_k141_259860 | Not-determined |
| KM_k141_228270 | Not-determined |
| KM_k141_183839 | Not-determined |
| KM_k141_60554 | Not-determined |
| KM_k141_149915 | Not-determined |
| KM_k141_217041 | Not-determined |
| KM_k141_4901 | Not-determined |
| KM_k141_105185 | Not-determined |
| KM_k141_194528 | Not-determined |
| KM_k141_105188 | Not-determined |
| KM_k141_194535 | Not-determined |
| KM_k141_217052 | Not-determined |
| KM_k141_105191 | Not-determined |
| KM_k141_248927 | Not-determined |
| KM_k141_72200 | Not-determined |
| KM_k141_183852 | Not-determined |
| KM_k141_138862 | Not-determined |
| KM_k141_84064 | Not-determined |
| KM_k141_149925 | Not-determined |
| KM_k141_149926 | Not-determined |
| KM_k141_48034 | Not-determined |
| KM_k141_248935 | Not-determined |
| KM_k141_183861 | Not-determined |
| KM_k141_27194 | Not-determined |
| KM_k141_183864 | Not-determined |
| KM_k141_84079 | Not-determined |
| KM_k141_84081 | Not-determined |
| KM_k141_138870 | Low-quality |
| KM_k141_238294 | Not-determined |
| KM_k141_217071 | Not-determined |
| KM_k141_228314 | Not-determined |
| KM_k141_127731 | Low-quality |
| KM_k141_238300 | Not-determined |
| KM_k141_248945 | Low-quality |
| KM_k141_117781 | Not-determined |
| KM_k141_105210 | Not-determined |
| KM_k141_27201 | Not-determined |
| KM_k141_60571 | Not-determined |
| KM_k141_15358 | Not-determined |
| KM_k141_105211 | Not-determined |
| KM_k141_238309 | Not-determined |
| KM_k141_138886 | Not-determined |
| KM_k141_60578 | Not-determined |
| KM_k141_138889 | Not-determined |
| KM_k141_105222 | Not-determined |
| KM_k141_72231 | Not-determined |
| KM_k141_138890 | Not-determined |
| KM_k141_127739 | Not-determined |
| KM_k141_117793 | Low-quality |
| KM_k141_72233 | Not-determined |
| KM_k141_60585 | Not-determined |
| KM_k141_72236 | Not-determined |
| KM_k141_27214 | Not-determined |
| KM_k141_160163 | Not-determined |
| KM_k141_60586 | Not-determined |
| KM_k141_138896 | Not-determined |
| KM_k141_183880 | Not-determined |
| KM_k141_105226 | Not-determined |
| KM_k141_117795 | Not-determined |
| KM_k141_4938 | Not-determined |
| KM_k141_60596 | Not-determined |
| KM_k141_183885 | Not-determined |
| KM_k141_38375 | Low-quality |
| KM_k141_48061 | Not-determined |
| KM_k141_48063 | Not-determined |
| KM_k141_84113 | Low-quality |
| KM_k141_92949 | Not-determined |
| KM_k141_72247 | Not-determined |
| KM_k141_117813 | Not-determined |
| KM_k141_238330 | Not-determined |
| KM_k141_4953 | Not-determined |
| KM_k141_138908 | Not-determined |
| KM_k141_138913 | Not-determined |
| KM_k141_4959 | Not-determined |
| KM_k141_160188 | Low-quality |
| KM_k141_84125 | Not-determined |
| KM_k141_138920 | Not-determined |
| KM_k141_127757 | Not-determined |
| KM_k141_38393 | Not-determined |
| KM_k141_38394 | Not-determined |
| KM_k141_160192 | Not-determined |
| KM_k141_138925 | Not-determined |
| KM_k141_149972 | Not-determined |
| KM_k141_194598 | Not-determined |
| KM_k141_205870 | Not-determined |
| KM_k141_38401 | Not-determined |
| KM_k141_183905 | Not-determined |
| KM_k141_183906 | Low-quality |
| KM_k141_259911 | Not-determined |
| KM_k141_60609 | Not-determined |
| KM_k141_15392 | Not-determined |
| KM_k141_138933 | Not-determined |
| KM_k141_259914 | Not-determined |
| KM_k141_194614 | Low-quality |
| KM_k141_194615 | Not-determined |
| KM_k141_259916 | Not-determined |
| KM_k141_15394 | Low-quality |
| KM_k141_248974 | Not-determined |
| KM_k141_15395 | Not-determined |
| KM_k141_60613 | Low-quality |
| KM_k141_248975 | Not-determined |
| KM_k141_138937 | Not-determined |
| KM_k141_127765 | Low-quality |
| KM_k141_4987 | Not-determined |
| KM_k141_4989 | Not-determined |
| KM_k141_149990 | Not-determined |
| KM_k141_84141 | Not-determined |
| KM_k141_160210 | Not-determined |
| KM_k141_27241 | Not-determined |
| KM_k141_15408 | Not-determined |
| KM_k141_92974 | Not-determined |
| KM_k141_92976 | Not-determined |
| KM_k141_117823 | Low-quality |
| KM_k141_117825 | Not-determined |
| KM_k141_127772 | Low-quality |
| KM_k141_259942 | Not-determined |
| KM_k141_217145 | Not-determined |
| KM_k141_217146 | Not-determined |
| KM_k141_160223 | Not-determined |
| KM_k141_217148 | Not-determined |
| KM_k141_259948 | Not-determined |
| KM_k141_5008 | Not-determined |
| KM_k141_238348 | Not-determined |
| KM_k141_60639 | Low-quality |
| KM_k141_84159 | Not-determined |
| KM_k141_117839 | Not-determined |

| | |
|---|---|
| KM_k141_160227 | Not-determined |
| KM_k141_228355 | Not-determined |
| KM_k141_205912 | Not-determined |
| KM_k141_217164 | Not-determined |
| KM_k141_228360 | Not-determined |
| KM_k141_228362 | Low-quality |
| KM_k141_194650 | Not-determined |
| KM_k141_217170 | Not-determined |
| KM_k141_15429 | Not-determined |
| KM_k141_228368 | Not-determined |
| KM_k141_5014 | Not-determined |
| KM_k141_205928 | Not-determined |
| KM_k141_217174 | Not-determined |
| KM_k141_117852 | Not-determined |
| KM_k141_150027 | Low-quality |
| KM_k141_15436 | Not-determined |
| KM_k141_172021 | Not-determined |
| KM_k141_5021 | Not-determined |
| KM_k141_72304 | Not-determined |
| KM_k141_72318 | Not-determined |
| KM_k141_117873 | Not-determined |
| KM_k141_93005 | Low-quality |
| KM_k141_228394 | Not-determined |
| KM_k141_15457 | Not-determined |
| KM_k141_93014 | Not-determined |
| KM_k141_183927 | Not-determined |
| KM_k141_93017 | Not-determined |
| KM_k141_259979 | Not-determined |
| KM_k141_84185 | Not-determined |
| KM_k141_138975 | Not-determined |
| KM_k141_5025 | Not-determined |
| KM_k141_72333 | Not-determined |
| KM_k141_27293 | Not-determined |
| KM_k141_5027 | Not-determined |
| KM_k141_93024 | Not-determined |
| KM_k141_238395 | Not-determined |
| KM_k141_93027 | Not-determined |
| KM_k141_93029 | Not-determined |
| KM_k141_138991 | Not-determined |
| KM_k141_105285 | Not-determined |
| KM_k141_238402 | Not-determined |
| KM_k141_172045 | Not-determined |
| KM_k141_105290 | Not-determined |
| KM_k141_105291 | Not-determined |
| KM_k141_205969 | Not-determined |
| KM_k141_172048 | Low-quality |
| KM_k141_5038 | Not-determined |
| KM_k141_228407 | Not-determined |
| KM_k141_60683 | Not-determined |
| KM_k141_228408 | Not-determined |
| KM_k141_160252 | Not-determined |
| KM_k141_238406 | Not-determined |
| KM_k141_172054 | Not-determined |
| KM_k141_194674 | Not-determined |
| KM_k141_72346 | Not-determined |
| KM_k141_205981 | Not-determined |
| KM_k141_27314 | Not-determined |
| KM_k141_5050 | Not-determined |
| KM_k141_127815 | Not-determined |
| KM_k141_150037 | Not-determined |
| KM_k141_217217 | Not-determined |
| KM_k141_72356 | Not-determined |
| KM_k141_105299 | Not-determined |
| KM_k141_105303 | Not-determined |
| KM_k141_105306 | Not-determined |
| KM_k141_72359 | Not-determined |
| KM_k141_205993 | Not-determined |
| KM_k141_72360 | Not-determined |
| KM_k141_127826 | Not-determined |
| KM_k141_5061 | Not-determined |
| KM_k141_15485 | Not-determined |
| KM_k141_5064 | Not-determined |
| KM_k141_93042 | Not-determined |
| KM_k141_150045 | Low-quality |
| KM_k141_172076 | Not-determined |
| KM_k141_139020 | Low-quality |
| KM_k141_228419 | Not-determined |
| KM_k141_105311 | Not-determined |
| KM_k141_194694 | Not-determined |
| KM_k141_60695 | Not-determined |
| KM_k141_117928 | Not-determined |
| KM_k141_27328 | Not-determined |
| KM_k141_206010 | Not-determined |
| KM_k141_105317 | Not-determined |
| KM_k141_183954 | Not-determined |
| KM_k141_38423 | Not-determined |
| KM_k141_38425 | Not-determined |
| KM_k141_206017 | Not-determined |
| KM_k141_160269 | Low-quality |
| KM_k141_139040 | Not-determined |
| KM_k141_260027 | Not-determined |
| KM_k141_60709 | Not-determined |
| KM_k141_172092 | Not-determined |
| KM_k141_183964 | Not-determined |
| KM_k141_260030 | Not-determined |
| KM_k141_72379 | Not-determined |
| KM_k141_15507 | Not-determined |
| KM_k141_127850 | Not-determined |
| KM_k141_127852 | Not-determined |
| KM_k141_105343 | Not-determined |
| KM_k141_160283 | Low-quality |
| KM_k141_260032 | Low-quality |
| KM_k141_160285 | Not-determined |
| KM_k141_105347 | Not-determined |
| KM_k141_127858 | Not-determined |
| KM_k141_5080 | Not-determined |
| KM_k141_206032 | Not-determined |
| KM_k141_5086 | Not-determined |
| KM_k141_5087 | Not-determined |
| KM_k141_139068 | Not-determined |
| KM_k141_260040 | Not-determined |
| KM_k141_150075 | Not-determined |
| KM_k141_117941 | Not-determined |
| KM_k141_160295 | Low-quality |
| KM_k141_60730 | Not-determined |
| KM_k141_139074 | Not-determined |
| KM_k141_127870 | Low-quality |
| KM_k141_5099 | Not-determined |
| KM_k141_127871 | Not-determined |
| KM_k141_160302 | Not-determined |
| KM_k141_139091 | Not-determined |
| KM_k141_93062 | Not-determined |
| KM_k141_260055 | Not-determined |
| KM_k141_249010 | Not-determined |
| KM_k141_15524 | Not-determined |
| KM_k141_38451 | Not-determined |
| KM_k141_127889 | Not-determined |
| KM_k141_38454 | Low-quality |
| KM_k141_260059 | Low-quality |
| KM_k141_206038 | Not-determined |
| KM_k141_139107 | Not-determined |
| KM_k141_206043 | Low-quality |
| KM_k141_217247 | Not-determined |
| KM_k141_105386 | Not-determined |
| KM_k141_260066 | Not-determined |
| KM_k141_160325 | Not-determined |
| KM_k141_238452 | Not-determined |
| KM_k141_238458 | Not-determined |
| KM_k141_105390 | Not-determined |
| KM_k141_38466 | Not-determined |
| KM_k141_249034 | Not-determined |
| KM_k141_184005 | Not-determined |
| KM_k141_117988 | Not-determined |
| KM_k141_117990 | Not-determined |
| KM_k141_127907 | Not-determined |
| KM_k141_93100 | Not-determined |
| KM_k141_27380 | Not-determined |
| KM_k141_72420 | Not-determined |
| KM_k141_105395 | Not-determined |
| KM_k141_105396 | Not-determined |
| KM_k141_118002 | Low-quality |
| KM_k141_118005 | Not-determined |
| KM_k141_127914 | Low-quality |
| KM_k141_60759 | Not-determined |
| KM_k141_238477 | Not-determined |
| KM_k141_238479 | Not-determined |
| KM_k141_118011 | Not-determined |
| KM_k141_238480 | Not-determined |
| KM_k141_60766 | Not-determined |
| KM_k141_184009 | Not-determined |
| KM_k141_238483 | Not-determined |
| KM_k141_118016 | Not-determined |
| KM_k141_118018 | Not-determined |
| KM_k141_184018 | Not-determined |
| KM_k141_206056 | Not-determined |
| KM_k141_38481 | Not-determined |
| KM_k141_118027 | Not-determined |
| KM_k141_38482 | Not-determined |
| KM_k141_72436 | Not-determined |
| KM_k141_15555 | Low-quality |
| KM_k141_38486 | Not-determined |
| KM_k141_15557 | Not-determined |
| KM_k141_38495 | Not-determined |
| KM_k141_72443 | Not-determined |
| KM_k141_217291 | Not-determined |
| KM_k141_238493 | Not-determined |
| KM_k141_15567 | Low-quality |
| KM_k141_194759 | Not-determined |
| KM_k141_15569 | Not-determined |
| KM_k141_139130 | Not-determined |
| KM_k141_238498 | Not-determined |
| KM_k141_249054 | Not-determined |
| KM_k141_72453 | Not-determined |
| KM_k141_160331 | Not-determined |
| KM_k141_194771 | Not-determined |
| KM_k141_194772 | Not-determined |
| KM_k141_38516 | Not-determined |
| KM_k141_38517 | Not-determined |
| KM_k141_105428 | Not-determined |
| KM_k141_5146 | Not-determined |
| KM_k141_194781 | Not-determined |
| KM_k141_38522 | Low-quality |
| KM_k141_38523 | Not-determined |
| KM_k141_217302 | Not-determined |
| KM_k141_260127 | Not-determined |
| KM_k141_194805 | Low-quality |
| KM_k141_127932 | Not-determined |
| KM_k141_206103 | Not-determined |
| KM_k141_228503 | Not-determined |
| KM_k141_27401 | Not-determined |
| KM_k141_217312 | Not-determined |
| KM_k141_228506 | Not-determined |
| KM_k141_27412 | Not-determined |
| KM_k141_228513 | Not-determined |
| KM_k141_27421 | Not-determined |
| KM_k141_194840 | Not-determined |
| KM_k141_194842 | Not-determined |
| KM_k141_194844 | Not-determined |
| KM_k141_160381 | Not-determined |
| KM_k141_160382 | Not-determined |
| KM_k141_238518 | Not-determined |
| KM_k141_160390 | Not-determined |
| KM_k141_206125 | Not-determined |
| KM_k141_48128 | Not-determined |
| KM_k141_118066 | Not-determined |
| KM_k141_217349 | Not-determined |
| KM_k141_217353 | Not-determined |
| KM_k141_238527 | Not-determined |
| KM_k141_260150 | Not-determined |
| KM_k141_72473 | Low-quality |
| KM_k141_48130 | Not-determined |
| KM_k141_194861 | Not-determined |
| KM_k141_118077 | Not-determined |
| KM_k141_206150 | Not-determined |
| KM_k141_172131 | Not-determined |
| KM_k141_206152 | Not-determined |
| KM_k141_184066 | Not-determined |
| KM_k141_172136 | Not-determined |
| KM_k141_260155 | Not-determined |
| KM_k141_27450 | Not-determined |
| KM_k141_27451 | Not-determined |
| KM_k141_93149 | Not-determined |
| KM_k141_127936 | Not-determined |
| KM_k141_160412 | Not-determined |
| KM_k141_260163 | Not-determined |
| KM_k141_84211 | Not-determined |
| KM_k141_93151 | Not-determined |
| KM_k141_206167 | Not-determined |
| KM_k141_38562 | Not-determined |
| KM_k141_184079 | Not-determined |
| KM_k141_118093 | Not-determined |
| KM_k141_260170 | Not-determined |
| KM_k141_118096 | Not-determined |
| KM_k141_260171 | Not-determined |
| KM_k141_38571 | Not-determined |
| KM_k141_139193 | Low-quality |
| KM_k141_27465 | Not-determined |
| KM_k141_38574 | Not-determined |
| KM_k141_38575 | Not-determined |
| KM_k141_127943 | Low-quality |
| KM_k141_160429 | Not-determined |
| KM_k141_127946 | Low-quality |
| KM_k141_48168 | Not-determined |
| KM_k141_38580 | Not-determined |
| KM_k141_206181 | Not-determined |
| KM_k141_118104 | Not-determined |
| KM_k141_84226 | Not-determined |
| KM_k141_48173 | Not-determined |
| KM_k141_105503 | Not-determined |
| KM_k141_150161 | Not-determined |
| KM_k141_206185 | Not-determined |
| KM_k141_93168 | Not-determined |
| KM_k141_48177 | Not-determined |
| KM_k141_27481 | Not-determined |
| KM_k141_38592 | Not-determined |
| KM_k141_15585 | Low-quality |
| KM_k141_27484 | Not-determined |
| KM_k141_84233 | Not-determined |
| KM_k141_194873 | Not-determined |
| KM_k141_60843 | Not-determined |
| KM_k141_139212 | Not-determined |
| KM_k141_194874 | Low-quality |
| KM_k141_27493 | Not-determined |
| KM_k141_118120 | Not-determined |
| KM_k141_93184 | Not-determined |
| KM_k141_194876 | Not-determined |
| KM_k141_5206 | Not-determined |
| KM_k141_84246 | Not-determined |
| KM_k141_118121 | Not-determined |
| KM_k141_48189 | Low-quality |
| KM_k141_238569 | Not-determined |
| KM_k141_15597 | Not-determined |
| KM_k141_93188 | Not-determined |
| KM_k141_260186 | Not-determined |
| KM_k141_84252 | Not-determined |
| KM_k141_38607 | Not-determined |
| KM_k141_206213 | Not-determined |
| KM_k141_194880 | Not-determined |
| KM_k141_5213 | Not-determined |
| KM_k141_206217 | Not-determined |
| KM_k141_194883 | Not-determined |
| KM_k141_105525 | Not-determined |
| KM_k141_48199 | Not-determined |
| KM_k141_228564 | Low-quality |
| KM_k141_60860 | Not-determined |
| KM_k141_105528 | Not-determined |
| KM_k141_238585 | Not-determined |
| KM_k141_105530 | Not-determined |
| KM_k141_5224 | Not-determined |
| KM_k141_160448 | Not-determined |
| KM_k141_38628 | Not-determined |
| KM_k141_48209 | Not-determined |
| KM_k141_48210 | Not-determined |
| KM_k141_139243 | Not-determined |
| KM_k141_15623 | Not-determined |
| KM_k141_238594 | Low-quality |
| KM_k141_160456 | Not-determined |
| KM_k141_84268 | Not-determined |
| KM_k141_84270 | Not-determined |
| KM_k141_5237 | Not-determined |
| KM_k141_84271 | Not-determined |
| KM_k141_206248 | Not-determined |
| KM_k141_206249 | Not-determined |
| KM_k141_84274 | Not-determined |
| KM_k141_217397 | Not-determined |
| KM_k141_84276 | Not-determined |
| KM_k141_60878 | Not-determined |
| KM_k141_228582 | Not-determined |
| KM_k141_72500 | Not-determined |
| KM_k141_105539 | Not-determined |
| KM_k141_60884 | Not-determined |
| KM_k141_206252 | Not-determined |
| KM_k141_60886 | Not-determined |
| KM_k141_160463 | Not-determined |
| KM_k141_160464 | Not-determined |
| KM_k141_105542 | Not-determined |
| KM_k141_217408 | Not-determined |
| KM_k141_72514 | Not-determined |
| KM_k141_72516 | Not-determined |
| KM_k141_48216 | Not-determined |
| KM_k141_15646 | Low-quality |
| KM_k141_217417 | Not-determined |
| KM_k141_217420 | Not-determined |
| KM_k141_27512 | Not-determined |
| KM_k141_249075 | Low-quality |
| KM_k141_128004 | Not-determined |
| KM_k141_160473 | Not-determined |
| KM_k141_38643 | Not-determined |
| KM_k141_238623 | Not-determined |
| KM_k141_27519 | Not-determined |
| KM_k141_72527 | Not-determined |
| KM_k141_48222 | Low-quality |
| KM_k141_249076 | Not-determined |
| KM_k141_238627 | Not-determined |
| KM_k141_217435 | Not-determined |
| KM_k141_15661 | Not-determined |
| KM_k141_128013 | Not-determined |
| KM_k141_128015 | Not-determined |
| KM_k141_93224 | Low-quality |
| KM_k141_206284 | Not-determined |
| KM_k141_27537 | Low-quality |
| KM_k141_139281 | Not-determined |
| KM_k141_217445 | Not-determined |
| KM_k141_105571 | Not-determined |
| KM_k141_249087 | Not-determined |
| KM_k141_27540 | Not-determined |
| KM_k141_249090 | Not-determined |
| KM_k141_238632 | Not-determined |
| KM_k141_249094 | Not-determined |
| KM_k141_60897 | Not-determined |
| KM_k141_60900 | Not-determined |
| KM_k141_5255 | Not-determined |
| KM_k141_172163 | Not-determined |
| KM_k141_217455 | Not-determined |
| KM_k141_118140 | Not-determined |
| KM_k141_172166 | Not-determined |
| KM_k141_150203 | Not-determined |
| KM_k141_128035 | Not-determined |
| KM_k141_60908 | Not-determined |
| KM_k141_128037 | Not-determined |
| KM_k141_139285 | Not-determined |
| KM_k141_38652 | Low-quality |
| KM_k141_160498 | Not-determined |
| KM_k141_38654 | Not-determined |
| KM_k141_72562 | Low-quality |
| KM_k141_72563 | Not-determined |
| KM_k141_249114 | Not-determined |
| KM_k141_160506 | Not-determined |
| KM_k141_172172 | Not-determined |
| KM_k141_27556 | Not-determined |
| KM_k141_249116 | Not-determined |
| KM_k141_27559 | Not-determined |
| KM_k141_260224 | Not-determined |
| KM_k141_249118 | Low-quality |
| KM_k141_206309 | Not-determined |
| KM_k141_238658 | Not-determined |
| KM_k141_217476 | Not-determined |
| KM_k141_194925 | Not-determined |
| KM_k141_150217 | Not-determined |
| KM_k141_217479 | Not-determined |
| KM_k141_118165 | Not-determined |
| KM_k141_93258 | Not-determined |
| KM_k141_5272 | Not-determined |
| KM_k141_60925 | Not-determined |
| KM_k141_60926 | Not-determined |
| KM_k141_60931 | Not-determined |
| KM_k141_118174 | Not-determined |
| KM_k141_206324 | Not-determined |
| KM_k141_118181 | Not-determined |
| KM_k141_72585 | Not-determined |
| KM_k141_160525 | Not-determined |
| KM_k141_238673 | Not-determined |
| KM_k141_5280 | Not-determined |
| KM_k141_5282 | Not-determined |
| KM_k141_93265 | Low-quality |
| KM_k141_249123 | Low-quality |
| KM_k141_194942 | Low-quality |
| KM_k141_206333 | Not-determined |
| KM_k141_15681 | Not-determined |
| KM_k141_160533 | Not-determined |
| KM_k141_93268 | Not-determined |
| KM_k141_48262 | Not-determined |
| KM_k141_84332 | Not-determined |
| KM_k141_93270 | Not-determined |
| KM_k141_249129 | Not-determined |
| KM_k141_194947 | Not-determined |
| KM_k141_217499 | Not-determined |
| KM_k141_249133 | Not-determined |
| KM_k141_172191 | Low-quality |
| KM_k141_48271 | Not-determined |
| KM_k141_27585 | Not-determined |
| KM_k141_228652 | Not-determined |
| KM_k141_139340 | Not-determined |
| KM_k141_27588 | Not-determined |
| KM_k141_27591 | Not-determined |
| KM_k141_228657 | Not-determined |
| KM_k141_228658 | Not-determined |
| KM_k141_172197 | Not-determined |
| KM_k141_206343 | Not-determined |
| KM_k141_150243 | Low-quality |
| KM_k141_15688 | Not-determined |
| KM_k141_105607 | Not-determined |
| KM_k141_105610 | Not-determined |
| KM_k141_160554 | Not-determined |
| KM_k141_93284 | Not-determined |
| KM_k141_206350 | Not-determined |
| KM_k141_184165 | Not-determined |
| KM_k141_128074 | Not-determined |
| KM_k141_184168 | Not-determined |
| KM_k141_238697 | Not-determined |
| KM_k141_72614 | Not-determined |
| KM_k141_72615 | Not-determined |
| KM_k141_48291 | Not-determined |
| KM_k141_249150 | Not-determined |
| KM_k141_93288 | Not-determined |
| KM_k141_27602 | Not-determined |
| KM_k141_105620 | Not-determined |
| KM_k141_249165 | Not-determined |
| KM_k141_217528 | Not-determined |
| KM_k141_184170 | Not-determined |
| KM_k141_93299 | Not-determined |
| KM_k141_172206 | Not-determined |
| KM_k141_238715 | Not-determined |
| KM_k141_249172 | Not-determined |
| KM_k141_206355 | Not-determined |
| KM_k141_238720 | Low-quality |
| KM_k141_27613 | Not-determined |
| KM_k141_260248 | Not-determined |
| KM_k141_60979 | Not-determined |
| KM_k141_5310 | Not-determined |
| KM_k141_228669 | Low-quality |
| KM_k141_27622 | Not-determined |
| KM_k141_228672 | Not-determined |
| KM_k141_27628 | Not-determined |
| KM_k141_93308 | Not-determined |
| KM_k141_238723 | Not-determined |
| KM_k141_105636 | Not-determined |
| KM_k141_194981 | Not-determined |
| KM_k141_160586 | Not-determined |
| KM_k141_5320 | Not-determined |
| KM_k141_206369 | Not-determined |
| KM_k141_172224 | Not-determined |
| KM_k141_5323 | Not-determined |
| KM_k141_160595 | Not-determined |
| KM_k141_184201 | Not-determined |
| KM_k141_206378 | Not-determined |
| KM_k141_160598 | Not-determined |
| KM_k141_84373 | Not-determined |
| KM_k141_160602 | Not-determined |
| KM_k141_228683 | Not-determined |
| KM_k141_139364 | Low-quality |
| KM_k141_217552 | Not-determined |
| KM_k141_160608 | Not-determined |
| KM_k141_5334 | Not-determined |
| KM_k141_184212 | Not-determined |
| KM_k141_206402 | Not-determined |
| KM_k141_172245 | Low-quality |
| KM_k141_184216 | Not-determined |
| KM_k141_48308 | Not-determined |
| KM_k141_61008 | Not-determined |
| KM_k141_172251 | Not-determined |
| KM_k141_105661 | Not-determined |
| KM_k141_48311 | Not-determined |
| KM_k141_84389 | Not-determined |
| KM_k141_105664 | Not-determined |
| KM_k141_48313 | Not-determined |
| KM_k141_228710 | Not-determined |
| KM_k141_184222 | Low-quality |
| KM_k141_128085 | Low-quality |
| KM_k141_195002 | Not-determined |
| KM_k141_61016 | Not-determined |
| KM_k141_72667 | Not-determined |
| KM_k141_139390 | Not-determined |
| KM_k141_195008 | Low-quality |
| KM_k141_139397 | Not-determined |
| KM_k141_206417 | Not-determined |
| KM_k141_238755 | Not-determined |
| KM_k141_15722 | Not-determined |
| KM_k141_84409 | Not-determined |
| KM_k141_84412 | Not-determined |
| KM_k141_228731 | Not-determined |
| KM_k141_5348 | Low-quality |
| KM_k141_48327 | Not-determined |
| KM_k141_48330 | Not-determined |
| KM_k141_206431 | Not-determined |
| KM_k141_195031 | Not-determined |
| KM_k141_5350 | Not-determined |
| KM_k141_15731 | Not-determined |
| KM_k141_118214 | Not-determined |
| KM_k141_72691 | Not-determined |
| KM_k141_184241 | Not-determined |
| KM_k141_184242 | Not-determined |
| KM_k141_195038 | Not-determined |
| KM_k141_128109 | Not-determined |
| KM_k141_217571 | Not-determined |
| KM_k141_217574 | Not-determined |
| KM_k141_217576 | Not-determined |
| KM_k141_228744 | Not-determined |
| KM_k141_217579 | Not-determined |
| KM_k141_139413 | Not-determined |
| KM_k141_5364 | Not-determined |
| KM_k141_118230 | Not-determined |
| KM_k141_139415 | Not-determined |
| KM_k141_72700 | Not-determined |
| KM_k141_195052 | Not-determined |
| KM_k141_72703 | Not-determined |
| KM_k141_15750 | Not-determined |
| KM_k141_72710 | Not-determined |
| KM_k141_128121 | Not-determined |
| KM_k141_206453 | Not-determined |
| KM_k141_118243 | Not-determined |
| KM_k141_238778 | Low-quality |
| KM_k141_5374 | Low-quality |
| KM_k141_139429 | Not-determined |
| KM_k141_48344 | Not-determined |
| KM_k141_118248 | Not-determined |
| KM_k141_150293 | Not-determined |
| KM_k141_150294 | Not-determined |
| KM_k141_128131 | Not-determined |
| KM_k141_27643 | Not-determined |
| KM_k141_238788 | Low-quality |
| KM_k141_5383 | Not-determined |
| KM_k141_84434 | Not-determined |
| KM_k141_84435 | Not-determined |
| KM_k141_228755 | Not-determined |
| KM_k141_172292 | Low-quality |
| KM_k141_238797 | Not-determined |
| KM_k141_118273 | Not-determined |
| KM_k141_249244 | Not-determined |
| KM_k141_5405 | Not-determined |
| KM_k141_217595 | Not-determined |
| KM_k141_150319 | Not-determined |
| KM_k141_118278 | Not-determined |
| KM_k141_184269 | Not-determined |
| KM_k141_128145 | Not-determined |
| KM_k141_206470 | Not-determined |
| KM_k141_150322 | Low-quality |
| KM_k141_206474 | Not-determined |
| KM_k141_128152 | Not-determined |
| KM_k141_228770 | Not-determined |
| KM_k141_238806 | Not-determined |
| KM_k141_249253 | Not-determined |
| KM_k141_105702 | Not-determined |
| KM_k141_72731 | Not-determined |
| KM_k141_72732 | Not-determined |
| KM_k141_206481 | Not-determined |
| KM_k141_84445 | Not-determined |
| KM_k141_217604 | Not-determined |
| KM_k141_228776 | Not-determined |
| KM_k141_72735 | Not-determined |
| KM_k141_238813 | Not-determined |
| KM_k141_206484 | Not-determined |
| KM_k141_160655 | Not-determined |
| KM_k141_228778 | Not-determined |
| KM_k141_238818 | Not-determined |
| KM_k141_150331 | Not-determined |
| KM_k141_48380 | Not-determined |
| KM_k141_118293 | Not-determined |
| KM_k141_238822 | Not-determined |

| | |
|---|---|
| KM_k141_160657 | Not-determined |
| KM_k141_260317 | Not-determined |
| KM_k141_217619 | Not-determined |
| KM_k141_195118 | Low-quality |
| KM_k141_5426 | Not-determined |
| KM_k141_184298 | Not-determined |
| KM_k141_228797 | Not-determined |
| KM_k141_150349 | Not-determined |
| KM_k141_150350 | Not-determined |
| KM_k141_249275 | Not-determined |
| KM_k141_105729 | Not-determined |
| KM_k141_61032 | Not-determined |
| KM_k141_206513 | Not-determined |
| KM_k141_150354 | Not-determined |
| KM_k141_128188 | Not-determined |
| KM_k141_72743 | Not-determined |
| KM_k141_217632 | Not-determined |
| KM_k141_228811 | Not-determined |
| KM_k141_228812 | Low-quality |
| KM_k141_150365 | Not-determined |
| KM_k141_160681 | Low-quality |
| KM_k141_249284 | Not-determined |
| KM_k141_228819 | Not-determined |
| KM_k141_228820 | Not-determined |
| KM_k141_238843 | Low-quality |
| KM_k141_206526 | Not-determined |
| KM_k141_184318 | Not-determined |
| KM_k141_61040 | Not-determined |
| KM_k141_172330 | Not-determined |
| KM_k141_5445 | Not-determined |
| KM_k141_61042 | Not-determined |
| KM_k141_184325 | Low-quality |
| KM_k141_61045 | Not-determined |
| KM_k141_228823 | Low-quality |
| KM_k141_48392 | Not-determined |
| KM_k141_160688 | Not-determined |
| KM_k141_195134 | Not-determined |
| KM_k141_118331 | Not-determined |
| KM_k141_160693 | Not-determined |
| KM_k141_195141 | Not-determined |
| KM_k141_93347 | Not-determined |
| KM_k141_184337 | Not-determined |
| KM_k141_260339 | Not-determined |
| KM_k141_128223 | Not-determined |
| KM_k141_5451 | Not-determined |
| KM_k141_150380 | Not-determined |
| KM_k141_139474 | Low-quality |
| KM_k141_139478 | Low-quality |
| KM_k141_139482 | Not-determined |
| KM_k141_118350 | Not-determined |
| KM_k141_139487 | Not-determined |
| KM_k141_238863 | Not-determined |
| KM_k141_184350 | Not-determined |
| KM_k141_217663 | Not-determined |
| KM_k141_84450 | Not-determined |
| KM_k141_195158 | Not-determined |
| KM_k141_238876 | Not-determined |
| KM_k141_72780 | Low-quality |
| KM_k141_38743 | Not-determined |
| KM_k141_15780 | Not-determined |
| KM_k141_128243 | Not-determined |
| KM_k141_105762 | Not-determined |
| KM_k141_128245 | Not-determined |
| KM_k141_105766 | Not-determined |
| KM_k141_217689 | Not-determined |
| KM_k141_105768 | Not-determined |
| KM_k141_184377 | Not-determined |
| KM_k141_195172 | Not-determined |
| KM_k141_128251 | Not-determined |
| KM_k141_206591 | Not-determined |
| KM_k141_118382 | Not-determined |
| KM_k141_5484 | Not-determined |
| KM_k141_15790 | Not-determined |
| KM_k141_61058 | Not-determined |
| KM_k141_48422 | Low-quality |
| KM_k141_206598 | Not-determined |
| KM_k141_48428 | Not-determined |
| KM_k141_128261 | Not-determined |
| KM_k141_128263 | Low-quality |
| KM_k141_72803 | Not-determined |
| KM_k141_118400 | Not-determined |
| KM_k141_249316 | Not-determined |
| KM_k141_184406 | Low-quality |
| KM_k141_72809 | Not-determined |
| KM_k141_72810 | Not-determined |
| KM_k141_48438 | Not-determined |
| KM_k141_238885 | Not-determined |
| KM_k141_238886 | Not-determined |
| KM_k141_118406 | Not-determined |
| KM_k141_195181 | Low-quality |
| KM_k141_72823 | Not-determined |
| KM_k141_238889 | Not-determined |
| KM_k141_93412 | Not-determined |
| KM_k141_5492 | Not-determined |
| KM_k141_72828 | Not-determined |
| KM_k141_93417 | Not-determined |
| KM_k141_150419 | Not-determined |
| KM_k141_249326 | Not-determined |
| KM_k141_128279 | Not-determined |
| KM_k141_118419 | Not-determined |
| KM_k141_118420 | Not-determined |
| KM_k141_93422 | Not-determined |
| KM_k141_128288 | Not-determined |
| KM_k141_128295 | Not-determined |
| KM_k141_128296 | Not-determined |
| KM_k141_228844 | Not-determined |
| KM_k141_260360 | Not-determined |
| KM_k141_260361 | Not-determined |
| KM_k141_260363 | Not-determined |
| KM_k141_160744 | Not-determined |
| KM_k141_238900 | Not-determined |
| KM_k141_260365 | Not-determined |
| KM_k141_238901 | Not-determined |
| KM_k141_128298 | Not-determined |
| KM_k141_260370 | Not-determined |
| KM_k141_184422 | Not-determined |
| KM_k141_160753 | Not-determined |
| KM_k141_38784 | Not-determined |
| KM_k141_249340 | Low-quality |
| KM_k141_15806 | Not-determined |
| KM_k141_61095 | Not-determined |
| KM_k141_249346 | Not-determined |
| KM_k141_150436 | Not-determined |
| KM_k141_61099 | Not-determined |
| KM_k141_61103 | Not-determined |
| KM_k141_217729 | Not-determined |
| KM_k141_160768 | Not-determined |
| KM_k141_184445 | Not-determined |
| KM_k141_249356 | Not-determined |
| KM_k141_184446 | Not-determined |
| KM_k141_61109 | Low-quality |
| KM_k141_84485 | Not-determined |
| KM_k141_217739 | Not-determined |
| KM_k141_217740 | Not-determined |
| KM_k141_217741 | Not-determined |
| KM_k141_15819 | Not-determined |
| KM_k141_84493 | Not-determined |
| KM_k141_172388 | Not-determined |
| KM_k141_184455 | Not-determined |
| KM_k141_93432 | Not-determined |
| KM_k141_238915 | Not-determined |
| KM_k141_84496 | Not-determined |
| KM_k141_184458 | Not-determined |
| KM_k141_260382 | Not-determined |
| KM_k141_150471 | Not-determined |
| KM_k141_38804 | Not-determined |
| KM_k141_48457 | Not-determined |
| KM_k141_15824 | Not-determined |
| KM_k141_48459 | Not-determined |
| KM_k141_260385 | Not-determined |
| KM_k141_260386 | Low-quality |
| KM_k141_206632 | Not-determined |
| KM_k141_48468 | Not-determined |
| KM_k141_206635 | Not-determined |
| KM_k141_48470 | Not-determined |
| KM_k141_249379 | Not-determined |
| KM_k141_118435 | Not-determined |
| KM_k141_84504 | Not-determined |
| KM_k141_15839 | Not-determined |
| KM_k141_217765 | Not-determined |
| KM_k141_206641 | Not-determined |
| KM_k141_84506 | Not-determined |
| KM_k141_228891 | Not-determined |
| KM_k141_260395 | Not-determined |
| KM_k141_228894 | Not-determined |
| KM_k141_150485 | Not-determined |
| KM_k141_38822 | Not-determined |
| KM_k141_184478 | Not-determined |
| KM_k141_15848 | Not-determined |
| KM_k141_260404 | Not-determined |
| KM_k141_118446 | Low-quality |
| KM_k141_160811 | Not-determined |
| KM_k141_15858 | Not-determined |
| KM_k141_128349 | Not-determined |
| KM_k141_184492 | Not-determined |
| KM_k141_118453 | Not-determined |
| KM_k141_217785 | Not-determined |
| KM_k141_61133 | Not-determined |
| KM_k141_118457 | Not-determined |
| KM_k141_139574 | Not-determined |
| KM_k141_184499 | Not-determined |
| KM_k141_160818 | Not-determined |
| KM_k141_118462 | Not-determined |
| KM_k141_217793 | Not-determined |
| KM_k141_84525 | Not-determined |
| KM_k141_238943 | Not-determined |
| KM_k141_5542 | Not-determined |
| KM_k141_228925 | Not-determined |
| KM_k141_238952 | Not-determined |
| KM_k141_238953 | Not-determined |
| KM_k141_184515 | Not-determined |
| KM_k141_118474 | Not-determined |
| KM_k141_249430 | Not-determined |
| KM_k141_217803 | Not-determined |
| KM_k141_38827 | Not-determined |
| KM_k141_184521 | Not-determined |
| KM_k141_38829 | Not-determined |
| KM_k141_48514 | Not-determined |
| KM_k141_238960 | Not-determined |
| KM_k141_48516 | Not-determined |
| KM_k141_217806 | Low-quality |
| KM_k141_128370 | Not-determined |
| KM_k141_184527 | Not-determined |
| KM_k141_128371 | Not-determined |
| KM_k141_249437 | Not-determined |
| KM_k141_48526 | Not-determined |
| KM_k141_84539 | Not-determined |
| KM_k141_61169 | Not-determined |
| KM_k141_48532 | Not-determined |
| KM_k141_217812 | Not-determined |
| KM_k141_128382 | Not-determined |
| KM_k141_150530 | Not-determined |
| KM_k141_139594 | Low-quality |
| KM_k141_118496 | Not-determined |
| KM_k141_195269 | Not-determined |
| KM_k141_61175 | Not-determined |
| KM_k141_150537 | Not-determined |
| KM_k141_195275 | Not-determined |
| KM_k141_206682 | Not-determined |
| KM_k141_238989 | Not-determined |
| KM_k141_206686 | Not-determined |
| KM_k141_15865 | Not-determined |
| KM_k141_238995 | Not-determined |
| KM_k141_128388 | Low-quality |
| KM_k141_15869 | Not-determined |
| KM_k141_93481 | Not-determined |
| KM_k141_249447 | Not-determined |
| KM_k141_84550 | Not-determined |
| KM_k141_118508 | Not-determined |
| KM_k141_160838 | Not-determined |
| KM_k141_15877 | Not-determined |
| KM_k141_217833 | Not-determined |
| KM_k141_128397 | Not-determined |
| KM_k141_150544 | Low-quality |
| KM_k141_118509 | Not-determined |
| KM_k141_118510 | Not-determined |
| KM_k141_139616 | Not-determined |
| KM_k141_72886 | Low-quality |
| KM_k141_118515 | Not-determined |
| KM_k141_184549 | Not-determined |
| KM_k141_206692 | Not-determined |
| KM_k141_93493 | Not-determined |
| KM_k141_84561 | Not-determined |
| KM_k141_249455 | Not-determined |
| KM_k141_61218 | Not-determined |
| KM_k141_61220 | Not-determined |
| KM_k141_139638 | Not-determined |
| KM_k141_184563 | Not-determined |
| KM_k141_38865 | Not-determined |
| KM_k141_118520 | Not-determined |
| KM_k141_38868 | Not-determined |
| KM_k141_61229 | Not-determined |
| KM_k141_239027 | Low-quality |
| KM_k141_260459 | Not-determined |
| KM_k141_139655 | Not-determined |
| KM_k141_61241 | Not-determined |
| KM_k141_61244 | Not-determined |
| KM_k141_184582 | Not-determined |
| KM_k141_184583 | Not-determined |
| KM_k141_249468 | Not-determined |
| KM_k141_249470 | Not-determined |
| KM_k141_72900 | Not-determined |
| KM_k141_139665 | Not-determined |
| KM_k141_184592 | Not-determined |
| KM_k141_260474 | Not-determined |
| KM_k141_184602 | Not-determined |
| KM_k141_260475 | Low-quality |
| KM_k141_72912 | Not-determined |
| KM_k141_206719 | Not-determined |
| KM_k141_172419 | Not-determined |
| KM_k141_72913 | Not-determined |
| KM_k141_239043 | Not-determined |
| KM_k141_150579 | Not-determined |
| KM_k141_105793 | Not-determined |
| KM_k141_105794 | Not-determined |
| KM_k141_93507 | Not-determined |
| KM_k141_260487 | Not-determined |
| KM_k141_228972 | Not-determined |
| KM_k141_93508 | Not-determined |
| KM_k141_118545 | Not-determined |
| KM_k141_239052 | Not-determined |
| KM_k141_184612 | Not-determined |
| KM_k141_228979 | Not-determined |
| KM_k141_15911 | Not-determined |
| KM_k141_206732 | Not-determined |
| KM_k141_228983 | Not-determined |
| KM_k141_105802 | Not-determined |
| KM_k141_105804 | Not-determined |
| KM_k141_93521 | Not-determined |
| KM_k141_118554 | Not-determined |
| KM_k141_72929 | Not-determined |
| KM_k141_184633 | Not-determined |
| KM_k141_249485 | Not-determined |
| KM_k141_5587 | Low-quality |
| KM_k141_15919 | Low-quality |
| KM_k141_150603 | Not-determined |
| KM_k141_206748 | Not-determined |
| KM_k141_5596 | Not-determined |
| KM_k141_5598 | Not-determined |
| KM_k141_118574 | Not-determined |
| KM_k141_184653 | Not-determined |
| KM_k141_195333 | Not-determined |
| KM_k141_195335 | Not-determined |
| KM_k141_195336 | Not-determined |
| KM_k141_139682 | Not-determined |
| KM_k141_172458 | Not-determined |
| KM_k141_15930 | Not-determined |
| KM_k141_139688 | Not-determined |
| KM_k141_93541 | Not-determined |
| KM_k141_15934 | Not-determined |
| KM_k141_249510 | Not-determined |
| KM_k141_93546 | Not-determined |
| KM_k141_229003 | Not-determined |
| KM_k141_172463 | Not-determined |
| KM_k141_229005 | Not-determined |
| KM_k141_139697 | Not-determined |
| KM_k141_260518 | Not-determined |
| KM_k141_128415 | Not-determined |
| KM_k141_239062 | Not-determined |
| KM_k141_15945 | Not-determined |
| KM_k141_128418 | Not-determined |
| KM_k141_128421 | Not-determined |
| KM_k141_150604 | Not-determined |
| KM_k141_93556 | Low-quality |
| KM_k141_160875 | Not-determined |
| KM_k141_172483 | Not-determined |
| KM_k141_84595 | Not-determined |
| KM_k141_105835 | Not-determined |
| KM_k141_217898 | Not-determined |
| KM_k141_160880 | Not-determined |
| KM_k141_105837 | Not-determined |
| KM_k141_128428 | Not-determined |
| KM_k141_260529 | Not-determined |
| KM_k141_15955 | Low-quality |
| KM_k141_229021 | Not-determined |
| KM_k141_217900 | Not-determined |
| KM_k141_118592 | Not-determined |
| KM_k141_38911 | Not-determined |
| KM_k141_5618 | Not-determined |
| KM_k141_206776 | Not-determined |
| KM_k141_260536 | Not-determined |
| KM_k141_38920 | Not-determined |
| KM_k141_229030 | Not-determined |
| KM_k141_105850 | Not-determined |
| KM_k141_206785 | Not-determined |
| KM_k141_15957 | Not-determined |
| KM_k141_229034 | Not-determined |
| KM_k141_229036 | Not-determined |
| KM_k141_61298 | Not-determined |
| KM_k141_128445 | Not-determined |
| KM_k141_184672 | Not-determined |
| KM_k141_206789 | Low-quality |
| KM_k141_172518 | Not-determined |
| KM_k141_172519 | Not-determined |
| KM_k141_206792 | Not-determined |
| KM_k141_229044 | Not-determined |
| KM_k141_260549 | Not-determined |
| KM_k141_184678 | Not-determined |
| KM_k141_105860 | Not-determined |
| KM_k141_229052 | Not-determined |
| KM_k141_195364 | Not-determined |
| KM_k141_195366 | Not-determined |
| KM_k141_206802 | Not-determined |
| KM_k141_93585 | Not-determined |
| KM_k141_84611 | Not-determined |
| KM_k141_72989 | Not-determined |
| KM_k141_239100 | Not-determined |
| KM_k141_84616 | Not-determined |
| KM_k141_195374 | Low-quality |
| KM_k141_206814 | Not-determined |
| KM_k141_48578 | Not-determined |
| KM_k141_217937 | Not-determined |
| KM_k141_139761 | Not-determined |
| KM_k141_128477 | Not-determined |
| KM_k141_172531 | Not-determined |
| KM_k141_15990 | Not-determined |
| KM_k141_48585 | Low-quality |
| KM_k141_206822 | Not-determined |
| KM_k141_84630 | Not-determined |
| KM_k141_229078 | Not-determined |
| KM_k141_48591 | Not-determined |
| KM_k141_84632 | Not-determined |
| KM_k141_84634 | Not-determined |
| KM_k141_160934 | Not-determined |
| KM_k141_73002 | Not-determined |
| KM_k141_229083 | Not-determined |
| KM_k141_206829 | Not-determined |
| KM_k141_206834 | Not-determined |
| KM_k141_150648 | Not-determined |
| KM_k141_38949 | Not-determined |
| KM_k141_229088 | Not-determined |
| KM_k141_38950 | Not-determined |
| KM_k141_229091 | Not-determined |
| KM_k141_105886 | Not-determined |
| KM_k141_73010 | Not-determined |
| KM_k141_61328 | Not-determined |
| KM_k141_229095 | Not-determined |
| KM_k141_229098 | Low-quality |
| KM_k141_229099 | Not-determined |
| KM_k141_229102 | Low-quality |
| KM_k141_73014 | Not-determined |
| KM_k141_73021 | Not-determined |
| KM_k141_172550 | Not-determined |
| KM_k141_61341 | Low-quality |
| KM_k141_93615 | Not-determined |
| KM_k141_184714 | Not-determined |
| KM_k141_260571 | Not-determined |
| KM_k141_217957 | Not-determined |
| KM_k141_229122 | Not-determined |
| KM_k141_105904 | Not-determined |
| KM_k141_239123 | Not-determined |
| KM_k141_260576 | Not-determined |
| KM_k141_61357 | Not-determined |
| KM_k141_184721 | Not-determined |
| KM_k141_48610 | Not-determined |
| KM_k141_27732 | Not-determined |
| KM_k141_229127 | Not-determined |
| KM_k141_172580 | Not-determined |
| KM_k141_73040 | Not-determined |
| KM_k141_206853 | Not-determined |
| KM_k141_118623 | Not-determined |
| KM_k141_229131 | Not-determined |
| KM_k141_195438 | Not-determined |
| KM_k141_118630 | Not-determined |
| KM_k141_118639 | Not-determined |
| KM_k141_48625 | Not-determined |
| KM_k141_160960 | Not-determined |
| KM_k141_260596 | Not-determined |
| KM_k141_172590 | Not-determined |
| KM_k141_160962 | Not-determined |
| KM_k141_93623 | Not-determined |
| KM_k141_260598 | Not-determined |
| KM_k141_160967 | Not-determined |
| KM_k141_118649 | Not-determined |
| KM_k141_118651 | Not-determined |
| KM_k141_16020 | Not-determined |
| KM_k141_118657 | Not-determined |
| KM_k141_217990 | Not-determined |
| KM_k141_128511 | Not-determined |
| KM_k141_61378 | Not-determined |
| KM_k141_93632 | Not-determined |
| KM_k141_184734 | Not-determined |
| KM_k141_217993 | Not-determined |
| KM_k141_239152 | Not-determined |
| KM_k141_172595 | Not-determined |
| KM_k141_172597 | Not-determined |
| KM_k141_84641 | Not-determined |
| KM_k141_16028 | Not-determined |
| KM_k141_160979 | Not-determined |
| KM_k141_27759 | Not-determined |
| KM_k141_249567 | Not-determined |
| KM_k141_249571 | Not-determined |
| KM_k141_160985 | Not-determined |
| KM_k141_260606 | Low-quality |
| KM_k141_160989 | Not-determined |
| KM_k141_172610 | Not-determined |
| KM_k141_260614 | Not-determined |
| KM_k141_206892 | Not-determined |
| KM_k141_249584 | Not-determined |
| KM_k141_160992 | Not-determined |
| KM_k141_84657 | Not-determined |
| KM_k141_128532 | Low-quality |
| KM_k141_84661 | Not-determined |
| KM_k141_16047 | Low-quality |
| KM_k141_84663 | Not-determined |
| KM_k141_105956 | Not-determined |
| KM_k141_195479 | Not-determined |
| KM_k141_48666 | Not-determined |
| KM_k141_249586 | Not-determined |
| KM_k141_229177 | Not-determined |
| KM_k141_195484 | Not-determined |
| KM_k141_48669 | Not-determined |
| KM_k141_184751 | Not-determined |
| KM_k141_16067 | Not-determined |
| KM_k141_260631 | Not-determined |
| KM_k141_16071 | Not-determined |
| KM_k141_128545 | Not-determined |
| KM_k141_61385 | Not-determined |
| KM_k141_184756 | Not-determined |
| KM_k141_172640 | Not-determined |
| KM_k141_139807 | Not-determined |
| KM_k141_139809 | Not-determined |
| KM_k141_172650 | Low-quality |
| KM_k141_218008 | Not-determined |
| KM_k141_27788 | Low-quality |
| KM_k141_218015 | Not-determined |
| KM_k141_128555 | Not-determined |
| KM_k141_218017 | Not-determined |
| KM_k141_93666 | Not-determined |
| KM_k141_260640 | Not-determined |
| KM_k141_161012 | Not-determined |
| KM_k141_93667 | Not-determined |
| KM_k141_150664 | Not-determined |
| KM_k141_206921 | Not-determined |
| KM_k141_48677 | Not-determined |
| KM_k141_206922 | Low-quality |
| KM_k141_16079 | Low-quality |
| KM_k141_206923 | Not-determined |
| KM_k141_150670 | Not-determined |
| KM_k141_218019 | Not-determined |
| KM_k141_184792 | Not-determined |
| KM_k141_184796 | Not-determined |
| KM_k141_161025 | Not-determined |
| KM_k141_172675 | Not-determined |
| KM_k141_161029 | Not-determined |
| KM_k141_172678 | Not-determined |
| KM_k141_105998 | Not-determined |
| KM_k141_184803 | Not-determined |
| KM_k141_161035 | Not-determined |
| KM_k141_229220 | Not-determined |
| KM_k141_93675 | Not-determined |
| KM_k141_239172 | Low-quality |
| KM_k141_184813 | Not-determined |
| KM_k141_229223 | Not-determined |
| KM_k141_239173 | Not-determined |
| KM_k141_218031 | Not-determined |
| KM_k141_260664 | Not-determined |
| KM_k141_184817 | Not-determined |
| KM_k141_184819 | Not-determined |
| KM_k141_218034 | Not-determined |
| KM_k141_5672 | Not-determined |
| KM_k141_260668 | Not-determined |
| KM_k141_229239 | Not-determined |
| KM_k141_239186 | Not-determined |
| KM_k141_229243 | Not-determined |
| KM_k141_84732 | Not-determined |
| KM_k141_5678 | Not-determined |
| KM_k141_106011 | Not-determined |
| KM_k141_260671 | Not-determined |
| KM_k141_172697 | Not-determined |
| KM_k141_16109 | Not-determined |
| KM_k141_218047 | Not-determined |
| KM_k141_218052 | Low-quality |
| KM_k141_48696 | Not-determined |
| KM_k141_184851 | Not-determined |
| KM_k141_218058 | Not-determined |
| KM_k141_184854 | Not-determined |
| KM_k141_84744 | Not-determined |
| KM_k141_118666 | Not-determined |
| KM_k141_128592 | Not-determined |
| KM_k141_84749 | Not-determined |
| KM_k141_195599 | Not-determined |
| KM_k141_5698 | Not-determined |
| KM_k141_128597 | Not-determined |
| KM_k141_128598 | Not-determined |
| KM_k141_84757 | Not-determined |
| KM_k141_128599 | Not-determined |
| KM_k141_206969 | Not-determined |
| KM_k141_184861 | Low-quality |
| KM_k141_239210 | Not-determined |
| KM_k141_139865 | Low-quality |
| KM_k141_218082 | Not-determined |
| KM_k141_206981 | Not-determined |
| KM_k141_206983 | Not-determined |
| KM_k141_128614 | Not-determined |
| KM_k141_161044 | Not-determined |
| KM_k141_161047 | Low-quality |
| KM_k141_249635 | Not-determined |
| KM_k141_218093 | Not-determined |
| KM_k141_93718 | Not-determined |
| KM_k141_229265 | Not-determined |
| KM_k141_150713 | Not-determined |
| KM_k141_139892 | Not-determined |
| KM_k141_239232 | Not-determined |
| KM_k141_118692 | Not-determined |
| KM_k141_93725 | Not-determined |
| KM_k141_195625 | Not-determined |
| KM_k141_184893 | Not-determined |
| KM_k141_239233 | Low-quality |
| KM_k141_139899 | Not-determined |
| KM_k141_239236 | Not-determined |
| KM_k141_184897 | Low-quality |
| KM_k141_239238 | Not-determined |
| KM_k141_5720 | Not-determined |
| KM_k141_93735 | Not-determined |
| KM_k141_195629 | Not-determined |
| KM_k141_218114 | Not-determined |
| KM_k141_218116 | Low-quality |
| KM_k141_93740 | Not-determined |
| KM_k141_27822 | Not-determined |
| KM_k141_239247 | Not-determined |
| KM_k141_139906 | Not-determined |
| KM_k141_229276 | Not-determined |
| KM_k141_239250 | Not-determined |
| KM_k141_128638 | Not-determined |
| KM_k141_27829 | Not-determined |
| KM_k141_93744 | Not-determined |
| KM_k141_239255 | Not-determined |
| KM_k141_93745 | Not-determined |

| | |
|---|---|
| KM_k141_206990 | Not-determined |
| KM_k141_161060 | Not-determined |
| KM_k141_249648 | Not-determined |
| KM_k141_229284 | Not-determined |
| KM_k141_48742 | Not-determined |
| KM_k141_207001 | Not-determined |
| KM_k141_195653 | Not-determined |
| KM_k141_161066 | Not-determined |
| KM_k141_150729 | Low-quality |
| KM_k141_260717 | Not-determined |
| KM_k141_128657 | Not-determined |
| KM_k141_229300 | Not-determined |
| KM_k141_84779 | Not-determined |
| KM_k141_139930 | Not-determined |
| KM_k141_39052 | Not-determined |
| KM_k141_48751 | Not-determined |
| KM_k141_39054 | Not-determined |
| KM_k141_84791 | Low-quality |
| KM_k141_39060 | Not-determined |
| KM_k141_139939 | Not-determined |
| KM_k141_39063 | Low-quality |
| KM_k141_73147 | Not-determined |
| KM_k141_93762 | Not-determined |
| KM_k141_73151 | Not-determined |
| KM_k141_73152 | Not-determined |
| KM_k141_260730 | Not-determined |
| KM_k141_139946 | Low-quality |
| KM_k141_48769 | Not-determined |
| KM_k141_260733 | Low-quality |
| KM_k141_195670 | Not-determined |
| KM_k141_195671 | Not-determined |
| KM_k141_239293 | Not-determined |
| KM_k141_229317 | Low-quality |
| KM_k141_48776 | Not-determined |
| KM_k141_172773 | Not-determined |
| KM_k141_39079 | Not-determined |
| KM_k141_84810 | Not-determined |
| KM_k141_161076 | Not-determined |
| KM_k141_73159 | Not-determined |
| KM_k141_172783 | Not-determined |
| KM_k141_139965 | Not-determined |
| KM_k141_73165 | Not-determined |
| KM_k141_229328 | Not-determined |
| KM_k141_218149 | Not-determined |
| KM_k141_39084 | Not-determined |
| KM_k141_73171 | Not-determined |
| KM_k141_27874 | Not-determined |
| KM_k141_48789 | Not-determined |
| KM_k141_139970 | Not-determined |
| KM_k141_260759 | Low-quality |
| KM_k141_260760 | Not-determined |
| KM_k141_260762 | Low-quality |
| KM_k141_218166 | Not-determined |
| KM_k141_172804 | Not-determined |
| KM_k141_27890 | Not-determined |
| KM_k141_260771 | Not-determined |
| KM_k141_84842 | Not-determined |
| KM_k141_73178 | Not-determined |
| KM_k141_84844 | Not-determined |
| KM_k141_195694 | Not-determined |
| KM_k141_249672 | Not-determined |
| KM_k141_249678 | Not-determined |
| KM_k141_249679 | Not-determined |
| KM_k141_218180 | Not-determined |
| KM_k141_260778 | Not-determined |
| KM_k141_260780 | Not-determined |
| KM_k141_172816 | Not-determined |
| KM_k141_27897 | Not-determined |
| KM_k141_61492 | Not-determined |
| KM_k141_218185 | Low-quality |
| KM_k141_161094 | Not-determined |
| KM_k141_161095 | Not-determined |
| KM_k141_161096 | Not-determined |
| KM_k141_207039 | Not-determined |
| KM_k141_106080 | Not-determined |
| KM_k141_172830 | Not-determined |
| KM_k141_73197 | Not-determined |
| KM_k141_218187 | Not-determined |
| KM_k141_106084 | Not-determined |
| KM_k141_73200 | Not-determined |
| KM_k141_5775 | Not-determined |
| KM_k141_61498 | Not-determined |
| KM_k141_207046 | Not-determined |
| KM_k141_207047 | Not-determined |
| KM_k141_161102 | Not-determined |
| KM_k141_249692 | Not-determined |
| KM_k141_239330 | Not-determined |
| KM_k141_150765 | Not-determined |
| KM_k141_73203 | Not-determined |
| KM_k141_207051 | Not-determined |
| KM_k141_106090 | Not-determined |
| KM_k141_84857 | Not-determined |
| KM_k141_106094 | Not-determined |
| KM_k141_249698 | Not-determined |
| KM_k141_195720 | Low-quality |
| KM_k141_218211 | Not-determined |
| KM_k141_16138 | Not-determined |
| KM_k141_118745 | Not-determined |
| KM_k141_260790 | Not-determined |
| KM_k141_184918 | Not-determined |
| KM_k141_128694 | Not-determined |
| KM_k141_73217 | Not-determined |
| KM_k141_260794 | Not-determined |
| KM_k141_27917 | Not-determined |
| KM_k141_16145 | Not-determined |
| KM_k141_84871 | Not-determined |
| KM_k141_128704 | Not-determined |
| KM_k141_195726 | Not-determined |
| KM_k141_128705 | Not-determined |
| KM_k141_195728 | Not-determined |
| KM_k141_229366 | Not-determined |
| KM_k141_5799 | Not-determined |
| KM_k141_39147 | Not-determined |
| KM_k141_128709 | Not-determined |
| KM_k141_61516 | Not-determined |
| KM_k141_106118 | Not-determined |
| KM_k141_184925 | Low-quality |
| KM_k141_27935 | Not-determined |
| KM_k141_106119 | Not-determined |
| KM_k141_106120 | Not-determined |
| KM_k141_61519 | Low-quality |
| KM_k141_84889 | Not-determined |
| KM_k141_260809 | Low-quality |
| KM_k141_61526 | Not-determined |
| KM_k141_195741 | Not-determined |
| KM_k141_128719 | Not-determined |
| KM_k141_73247 | Not-determined |
| KM_k141_260817 | Not-determined |
| KM_k141_229382 | Not-determined |
| KM_k141_260819 | Not-determined |
| KM_k141_207058 | Not-determined |
| KM_k141_140018 | Not-determined |
| KM_k141_39155 | Not-determined |
| KM_k141_61539 | Not-determined |
| KM_k141_229396 | Not-determined |
| KM_k141_195743 | Not-determined |
| KM_k141_61541 | Not-determined |
| KM_k141_260824 | Not-determined |
| KM_k141_172871 | Not-determined |
| KM_k141_39163 | Not-determined |
| KM_k141_5812 | Low-quality |
| KM_k141_260828 | Not-determined |
| KM_k141_140038 | Not-determined |
| KM_k141_172883 | Not-determined |
| KM_k141_48839 | Not-determined |
| KM_k141_249738 | Not-determined |
| KM_k141_106139 | Not-determined |
| KM_k141_140051 | Not-determined |
| KM_k141_140052 | Not-determined |
| KM_k141_73259 | Not-determined |
| KM_k141_84905 | Not-determined |
| KM_k141_260835 | Not-determined |
| KM_k141_260836 | Not-determined |
| KM_k141_118788 | Low-quality |
| KM_k141_140056 | Low-quality |
| KM_k141_229417 | Not-determined |
| KM_k141_84910 | Not-determined |
| KM_k141_118793 | Not-determined |
| KM_k141_207073 | Not-determined |
| KM_k141_118797 | Not-determined |
| KM_k141_27952 | Not-determined |
| KM_k141_260846 | Not-determined |
| KM_k141_118799 | Not-determined |
| KM_k141_207077 | Low-quality |
| KM_k141_260850 | Not-determined |
| KM_k141_27958 | Not-determined |
| KM_k141_207083 | Not-determined |
| KM_k141_61580 | Not-determined |
| KM_k141_260855 | Low-quality |
| KM_k141_207088 | Not-determined |
| KM_k141_207089 | Not-determined |
| KM_k141_48857 | Not-determined |
| KM_k141_5830 | Not-determined |
| KM_k141_118805 | Not-determined |
| KM_k141_106154 | Not-determined |
| KM_k141_128728 | Not-determined |
| KM_k141_106155 | Low-quality |
| KM_k141_118806 | Not-determined |
| KM_k141_84928 | Not-determined |
| KM_k141_128731 | Not-determined |
| KM_k141_5836 | Not-determined |
| KM_k141_39185 | Not-determined |
| KM_k141_184943 | Not-determined |
| KM_k141_106160 | Not-determined |
| KM_k141_239377 | Not-determined |
| KM_k141_16150 | Not-determined |
| KM_k141_239383 | Not-determined |
| KM_k141_39192 | Not-determined |
| KM_k141_128734 | Not-determined |
| KM_k141_239384 | Low-quality |
| KM_k141_239385 | Not-determined |
| KM_k141_207104 | Not-determined |
| KM_k141_218273 | Not-determined |
| KM_k141_140095 | Not-determined |
| KM_k141_128741 | Not-determined |
| KM_k141_161165 | Low-quality |
| KM_k141_184953 | Not-determined |
| KM_k141_161168 | Not-determined |
| KM_k141_207111 | Not-determined |
| KM_k141_207112 | Low-quality |
| KM_k141_84936 | Not-determined |
| KM_k141_5848 | Not-determined |
| KM_k141_140107 | Not-determined |
| KM_k141_84945 | Not-determined |
| KM_k141_140108 | Not-determined |
| KM_k141_218281 | Not-determined |
| KM_k141_172907 | Low-quality |
| KM_k141_218282 | Not-determined |
| KM_k141_93774 | Not-determined |
| KM_k141_150785 | Not-determined |
| KM_k141_229442 | Not-determined |
| KM_k141_207125 | Not-determined |
| KM_k141_249763 | Not-determined |
| KM_k141_61602 | Not-determined |
| KM_k141_93782 | Not-determined |
| KM_k141_5864 | Not-determined |
| KM_k141_84952 | Not-determined |
| KM_k141_207129 | Low-quality |
| KM_k141_73318 | Not-determined |
| KM_k141_218299 | Not-determined |
| KM_k141_39216 | Not-determined |
| KM_k141_150791 | Not-determined |
| KM_k141_239415 | Not-determined |
| KM_k141_218301 | Not-determined |
| KM_k141_150793 | Not-determined |
| KM_k141_229457 | Not-determined |
| KM_k141_239417 | Not-determined |
| KM_k141_239418 | Not-determined |
| KM_k141_106191 | Not-determined |
| KM_k141_150795 | Not-determined |
| KM_k141_229460 | Not-determined |
| KM_k141_5873 | Not-determined |
| KM_k141_207133 | Low-quality |
| KM_k141_93786 | Not-determined |
| KM_k141_207135 | Not-determined |
| KM_k141_106195 | Not-determined |
| KM_k141_39225 | Not-determined |
| KM_k141_207140 | Not-determined |
| KM_k141_106200 | Low-quality |
| KM_k141_16181 | Low-quality |
| KM_k141_16182 | Not-determined |
| KM_k141_260885 | Not-determined |
| KM_k141_207142 | Not-determined |
| KM_k141_73335 | Not-determined |
| KM_k141_218311 | Not-determined |
| KM_k141_61609 | Not-determined |
| KM_k141_195748 | Not-determined |
| KM_k141_48895 | Not-determined |
| KM_k141_16187 | Not-determined |
| KM_k141_48896 | Not-determined |
| KM_k141_61612 | Not-determined |
| KM_k141_48899 | Not-determined |
| KM_k141_195757 | Not-determined |
| KM_k141_48903 | Not-determined |
| KM_k141_61618 | Not-determined |
| KM_k141_39230 | Not-determined |
| KM_k141_84983 | Not-determined |
| KM_k141_84986 | Not-determined |
| KM_k141_5886 | Not-determined |
| KM_k141_39232 | Not-determined |
| KM_k141_106213 | Not-determined |
| KM_k141_106214 | Not-determined |
| KM_k141_195760 | Not-determined |
| KM_k141_140149 | Not-determined |
| KM_k141_195762 | Not-determined |
| KM_k141_229467 | Not-determined |
| KM_k141_140151 | Not-determined |
| KM_k141_5894 | Not-determined |
| KM_k141_73367 | Not-determined |
| KM_k141_207174 | Not-determined |
| KM_k141_39245 | Not-determined |
| KM_k141_239438 | Not-determined |
| KM_k141_106223 | Not-determined |
| KM_k141_207179 | Not-determined |
| KM_k141_93809 | Not-determined |
| KM_k141_28006 | Not-determined |
| KM_k141_207181 | Not-determined |
| KM_k141_73378 | Low-quality |
| KM_k141_85001 | Not-determined |
| KM_k141_5902 | Not-determined |
| KM_k141_39249 | Not-determined |
| KM_k141_28011 | Low-quality |
| KM_k141_85006 | Not-determined |
| KM_k141_150809 | Not-determined |
| KM_k141_106230 | Not-determined |
| KM_k141_150810 | Not-determined |
| KM_k141_150811 | Not-determined |
| KM_k141_260928 | Not-determined |
| KM_k141_207191 | Not-determined |
| KM_k141_16222 | Not-determined |
| KM_k141_239456 | Not-determined |
| KM_k141_73379 | Not-determined |
| KM_k141_93813 | Not-determined |
| KM_k141_106237 | Low-quality |
| KM_k141_150821 | Not-determined |
| KM_k141_93817 | Not-determined |
| KM_k141_16231 | Not-determined |
| KM_k141_106241 | Not-determined |
| KM_k141_172981 | Not-determined |
| KM_k141_16234 | Not-determined |
| KM_k141_218322 | Not-determined |
| KM_k141_229499 | Not-determined |
| KM_k141_93828 | Not-determined |
| KM_k141_39277 | Not-determined |
| KM_k141_16243 | Low-quality |
| KM_k141_39284 | Not-determined |
| KM_k141_16251 | Not-determined |
| KM_k141_195795 | Not-determined |
| KM_k141_260950 | Not-determined |
| KM_k141_106259 | Not-determined |
| KM_k141_184982 | Not-determined |
| KM_k141_218351 | Not-determined |
| KM_k141_184987 | Not-determined |
| KM_k141_73414 | Not-determined |
| KM_k141_28036 | Not-determined |
| KM_k141_218354 | Not-determined |
| KM_k141_184990 | Not-determined |
| KM_k141_16264 | Not-determined |
| KM_k141_61651 | Not-determined |
| KM_k141_150850 | Not-determined |
| KM_k141_195804 | Not-determined |
| KM_k141_195806 | Not-determined |
| KM_k141_39311 | Not-determined |
| KM_k141_229526 | Low-quality |
| KM_k141_195807 | Not-determined |
| KM_k141_195813 | Not-determined |
| KM_k141_28053 | Not-determined |
| KM_k141_118874 | Not-determined |
| KM_k141_260959 | Low-quality |
| KM_k141_195819 | Not-determined |
| KM_k141_73421 | Not-determined |
| KM_k141_218365 | Not-determined |
| KM_k141_28063 | Not-determined |
| KM_k141_48936 | Not-determined |
| KM_k141_185017 | Not-determined |
| KM_k141_16287 | Not-determined |
| KM_k141_185019 | Not-determined |
| KM_k141_260971 | Not-determined |
| KM_k141_229540 | Not-determined |
| KM_k141_249785 | Not-determined |
| KM_k141_185029 | Low-quality |
| KM_k141_118879 | Not-determined |
| KM_k141_61675 | Not-determined |
| KM_k141_195825 | Low-quality |
| KM_k141_229543 | Not-determined |
| KM_k141_39337 | Not-determined |
| KM_k141_85056 | Not-determined |
| KM_k141_39345 | Not-determined |
| KM_k141_85062 | Not-determined |
| KM_k141_185034 | Not-determined |
| KM_k141_195837 | Not-determined |
| KM_k141_260998 | Not-determined |
| KM_k141_161174 | Not-determined |
| KM_k141_185043 | Not-determined |
| KM_k141_16307 | Not-determined |
| KM_k141_195841 | Not-determined |
| KM_k141_73428 | Not-determined |
| KM_k141_5961 | Not-determined |
| KM_k141_195842 | Not-determined |
| KM_k141_39358 | Not-determined |
| KM_k141_85076 | Not-determined |
| KM_k141_61682 | Not-determined |
| KM_k141_28075 | Not-determined |
| KM_k141_249799 | Not-determined |
| KM_k141_185053 | Not-determined |
| KM_k141_173026 | Not-determined |
| KM_k141_39366 | Not-determined |
| KM_k141_61690 | Not-determined |
| KM_k141_261015 | Not-determined |
| KM_k141_140198 | Not-determined |
| KM_k141_218413 | Not-determined |
| KM_k141_140199 | Not-determined |
| KM_k141_48960 | Not-determined |
| KM_k141_118914 | Not-determined |
| KM_k141_261023 | Not-determined |
| KM_k141_61699 | Not-determined |
| KM_k141_229558 | Not-determined |
| KM_k141_61701 | Not-determined |
| KM_k141_61704 | Not-determined |
| KM_k141_61707 | Not-determined |
| KM_k141_150880 | Not-determined |
| KM_k141_106263 | Low-quality |
| KM_k141_48988 | Not-determined |
| KM_k141_261043 | Not-determined |
| KM_k141_229583 | Not-determined |
| KM_k141_118924 | Not-determined |
| KM_k141_161209 | Not-determined |
| KM_k141_207255 | Not-determined |
| KM_k141_229589 | Not-determined |
| KM_k141_195875 | Not-determined |
| KM_k141_128769 | Not-determined |
| KM_k141_261056 | Not-determined |
| KM_k141_140229 | Not-determined |
| KM_k141_28095 | Not-determined |
| KM_k141_128776 | Not-determined |
| KM_k141_140234 | Low-quality |
| KM_k141_48994 | Not-determined |
| KM_k141_239559 | Not-determined |
| KM_k141_28098 | Not-determined |
| KM_k141_229602 | Not-determined |
| KM_k141_118935 | Low-quality |
| KM_k141_229605 | Not-determined |
| KM_k141_118937 | Not-determined |
| KM_k141_185089 | Not-determined |
| KM_k141_185090 | Not-determined |
| KM_k141_218441 | Not-determined |
| KM_k141_239564 | Low-quality |
| KM_k141_128787 | Not-determined |
| KM_k141_150914 | Not-determined |
| KM_k141_195890 | Not-determined |
| KM_k141_218444 | Not-determined |
| KM_k141_218447 | Not-determined |
| KM_k141_61719 | Not-determined |
| KM_k141_185091 | Not-determined |
| KM_k141_218451 | Not-determined |
| KM_k141_173054 | Not-determined |
| KM_k141_49007 | Not-determined |
| KM_k141_249830 | Not-determined |
| KM_k141_61726 | Not-determined |
| KM_k141_61727 | Not-determined |
| KM_k141_249831 | Low-quality |
| KM_k141_61734 | Not-determined |
| KM_k141_49012 | Not-determined |
| KM_k141_140253 | Not-determined |
| KM_k141_49015 | Not-determined |
| KM_k141_218462 | Not-determined |
| KM_k141_249836 | Not-determined |
| KM_k141_261067 | Not-determined |
| KM_k141_207279 | Not-determined |
| KM_k141_195903 | Not-determined |
| KM_k141_185106 | Not-determined |
| KM_k141_128807 | Not-determined |
| KM_k141_239583 | Not-determined |
| KM_k141_118947 | Not-determined |
| KM_k141_185113 | Not-determined |
| KM_k141_118949 | Not-determined |
| KM_k141_239592 | Not-determined |
| KM_k141_218484 | Not-determined |
| KM_k141_261085 | Not-determined |
| KM_k141_85120 | Not-determined |
| KM_k141_195918 | Not-determined |
| KM_k141_195919 | Not-determined |
| KM_k141_173070 | Low-quality |
| KM_k141_218489 | Not-determined |
| KM_k141_249847 | Not-determined |
| KM_k141_218490 | Not-determined |
| KM_k141_39384 | Not-determined |
| KM_k141_195925 | Not-determined |
| KM_k141_150950 | Not-determined |
| KM_k141_261098 | Low-quality |
| KM_k141_185129 | Not-determined |
| KM_k141_118964 | Not-determined |
| KM_k141_261101 | Not-determined |
| KM_k141_61759 | Not-determined |
| KM_k141_150956 | Not-determined |
| KM_k141_207303 | Not-determined |
| KM_k141_61764 | Not-determined |
| KM_k141_239611 | Low-quality |
| KM_k141_73477 | Not-determined |
| KM_k141_128814 | Low-quality |
| KM_k141_73479 | Not-determined |
| KM_k141_118971 | Not-determined |
| KM_k141_118975 | Not-determined |
| KM_k141_16367 | Not-determined |
| KM_k141_106285 | Not-determined |
| KM_k141_185140 | Not-determined |
| KM_k141_218519 | Not-determined |
| KM_k141_118981 | Not-determined |
| KM_k141_229629 | Not-determined |
| KM_k141_73493 | Not-determined |
| KM_k141_173107 | Not-determined |
| KM_k141_218524 | Low-quality |
| KM_k141_73495 | Not-determined |
| KM_k141_185146 | Not-determined |
| KM_k141_218529 | Not-determined |
| KM_k141_128843 | Low-quality |
| KM_k141_73499 | Not-determined |
| KM_k141_261124 | Low-quality |
| KM_k141_207322 | Low-quality |
| KM_k141_106299 | Not-determined |
| KM_k141_128858 | Not-determined |
| KM_k141_161231 | Not-determined |
| KM_k141_93914 | Not-determined |
| KM_k141_218542 | Not-determined |
| KM_k141_128863 | Not-determined |
| KM_k141_150982 | Not-determined |
| KM_k141_249864 | Not-determined |
| KM_k141_118991 | Not-determined |
| KM_k141_195952 | Not-determined |
| KM_k141_249865 | Not-determined |
| KM_k141_118994 | Not-determined |
| KM_k141_106309 | Not-determined |
| KM_k141_249867 | Not-determined |
| KM_k141_207344 | Not-determined |
| KM_k141_6037 | Not-determined |
| KM_k141_85127 | Not-determined |
| KM_k141_6038 | Not-determined |
| KM_k141_93920 | Not-determined |
| KM_k141_119004 | Not-determined |
| KM_k141_195957 | Not-determined |
| KM_k141_195958 | Not-determined |
| KM_k141_161241 | Not-determined |
| KM_k141_16388 | Not-determined |
| KM_k141_119011 | Not-determined |
| KM_k141_119012 | Low-quality |
| KM_k141_161244 | Not-determined |
| KM_k141_207358 | Low-quality |
| KM_k141_150994 | Not-determined |
| KM_k141_119019 | Not-determined |
| KM_k141_16393 | Not-determined |
| KM_k141_218557 | Low-quality |
| KM_k141_195967 | Not-determined |
| KM_k141_61813 | Not-determined |
| KM_k141_73510 | Not-determined |
| KM_k141_85142 | Not-determined |
| KM_k141_49048 | Not-determined |
| KM_k141_49049 | Not-determined |
| KM_k141_161256 | Not-determined |
| KM_k141_140290 | Not-determined |
| KM_k141_161258 | Not-determined |
| KM_k141_85143 | Not-determined |
| KM_k141_161259 | Not-determined |
| KM_k141_49057 | Low-quality |
| KM_k141_73517 | Low-quality |
| KM_k141_119022 | Low-quality |
| KM_k141_85148 | Not-determined |
| KM_k141_106312 | Not-determined |
| KM_k141_195980 | Not-determined |
| KM_k141_16408 | Low-quality |
| KM_k141_195983 | Not-determined |
| KM_k141_229652 | Not-determined |
| KM_k141_261152 | Not-determined |
| KM_k141_261154 | Not-determined |
| KM_k141_161274 | Not-determined |
| KM_k141_61829 | Not-determined |
| KM_k141_93956 | Not-determined |
| KM_k141_261159 | Not-determined |
| KM_k141_229659 | Not-determined |
| KM_k141_207383 | Low-quality |
| KM_k141_106331 | Not-determined |
| KM_k141_196003 | Low-quality |
| KM_k141_28128 | Not-determined |
| KM_k141_61838 | Not-determined |
| KM_k141_229668 | Not-determined |
| KM_k141_229669 | Not-determined |
| KM_k141_61844 | Not-determined |
| KM_k141_61845 | Not-determined |
| KM_k141_28137 | Not-determined |
| KM_k141_173160 | Low-quality |
| KM_k141_173165 | Not-determined |
| KM_k141_196023 | Not-determined |
| KM_k141_173169 | Not-determined |
| KM_k141_173170 | Not-determined |
| KM_k141_173171 | Not-determined |
| KM_k141_28142 | Not-determined |
| KM_k141_61859 | Not-determined |
| KM_k141_196032 | Not-determined |
| KM_k141_173177 | Not-determined |
| KM_k141_39449 | Not-determined |
| KM_k141_106340 | Not-determined |
| KM_k141_161295 | Low-quality |
| KM_k141_106343 | Not-determined |
| KM_k141_161299 | Not-determined |
| KM_k141_61872 | Not-determined |
| KM_k141_39453 | Not-determined |
| KM_k141_106348 | Not-determined |
| KM_k141_85167 | Not-determined |
| KM_k141_28154 | Not-determined |
| KM_k141_61883 | Not-determined |
| KM_k141_6090 | Not-determined |
| KM_k141_161315 | Not-determined |
| KM_k141_93974 | Low-quality |
| KM_k141_207419 | Not-determined |
| KM_k141_161321 | Not-determined |
| KM_k141_6100 | Not-determined |
| KM_k141_106356 | Not-determined |
| KM_k141_185175 | Not-determined |
| KM_k141_49085 | Low-quality |
| KM_k141_49086 | Not-determined |
| KM_k141_106366 | Not-determined |
| KM_k141_39487 | Not-determined |
| KM_k141_140298 | Not-determined |
| KM_k141_106368 | Not-determined |
| KM_k141_207434 | Not-determined |
| KM_k141_106371 | Not-determined |
| KM_k141_16448 | Not-determined |
| KM_k141_16456 | Not-determined |
| KM_k141_261199 | Not-determined |
| KM_k141_106376 | Not-determined |

| | |
|---|---|
| KM_k141_28175 | Not-determined |
| KM_k141_229682 | Not-determined |
| KM_k141_207449 | Not-determined |
| KM_k141_73563 | Not-determined |
| KM_k141_16461 | Not-determined |
| KM_k141_28182 | Not-determined |
| KM_k141_28183 | Not-determined |
| KM_k141_73564 | Not-determined |
| KM_k141_161348 | Not-determined |
| KM_k141_39506 | Not-determined |
| KM_k141_16469 | Not-determined |
| KM_k141_185202 | Not-determined |
| KM_k141_229700 | Not-determined |
| KM_k141_161359 | Not-determined |
| KM_k141_218593 | Not-determined |
| KM_k141_106386 | Not-determined |
| KM_k141_106388 | Not-determined |
| KM_k141_261206 | Not-determined |
| KM_k141_16481 | Not-determined |
| KM_k141_261209 | Not-determined |
| KM_k141_6122 | Not-determined |
| KM_k141_229709 | Not-determined |
| KM_k141_6125 | Not-determined |
| KM_k141_28199 | Not-determined |
| KM_k141_239623 | Not-determined |
| KM_k141_6127 | Not-determined |
| KM_k141_239625 | Not-determined |
| KM_k141_218619 | Not-determined |
| KM_k141_239626 | Not-determined |
| KM_k141_196125 | Not-determined |
| KM_k141_39537 | Not-determined |
| KM_k141_161369 | Not-determined |
| KM_k141_261231 | Not-determined |
| KM_k141_173222 | Not-determined |
| KM_k141_106401 | Not-determined |
| KM_k141_106402 | Not-determined |
| KM_k141_173225 | Not-determined |
| KM_k141_16493 | Not-determined |
| KM_k141_16497 | Low-quality |
| KM_k141_28230 | Not-determined |
| KM_k141_16500 | Not-determined |
| KM_k141_173238 | Not-determined |
| KM_k141_28234 | Not-determined |
| KM_k141_196150 | Not-determined |
| KM_k141_28237 | Low-quality |
| KM_k141_39563 | Not-determined |
| KM_k141_6147 | Not-determined |
| KM_k141_261241 | Not-determined |
| KM_k141_196156 | Not-determined |
| KM_k141_161393 | Not-determined |
| KM_k141_151049 | Not-determined |
| KM_k141_218670 | Not-determined |
| KM_k141_161395 | Not-determined |
| KM_k141_161397 | Not-determined |
| KM_k141_218674 | Not-determined |
| KM_k141_218676 | Not-determined |
| KM_k141_151058 | Not-determined |
| KM_k141_249981 | Not-determined |
| KM_k141_261248 | Not-determined |
| KM_k141_28242 | Low-quality |
| KM_k141_185217 | Not-determined |
| KM_k141_119062 | Not-determined |
| KM_k141_16531 | Not-determined |
| KM_k141_239660 | Not-determined |
| KM_k141_73614 | Not-determined |
| KM_k141_49146 | Not-determined |
| KM_k141_161415 | Not-determined |
| KM_k141_185226 | Not-determined |
| KM_k141_185227 | Not-determined |
| KM_k141_28254 | Not-determined |
| KM_k141_61989 | Not-determined |
| KM_k141_73626 | Not-determined |
| KM_k141_161416 | Not-determined |
| KM_k141_106412 | Not-determined |
| KM_k141_239669 | Low-quality |
| KM_k141_6157 | Not-determined |
| KM_k141_119079 | Not-determined |
| KM_k141_218696 | Not-determined |
| KM_k141_239674 | Not-determined |
| KM_k141_196167 | Not-determined |
| KM_k141_151077 | Not-determined |
| KM_k141_151079 | Not-determined |
| KM_k141_196170 | Not-determined |
| KM_k141_229753 | Not-determined |
| KM_k141_218704 | Not-determined |
| KM_k141_173260 | Low-quality |
| KM_k141_196174 | Not-determined |
| KM_k141_85240 | Not-determined |
| KM_k141_94000 | Not-determined |
| KM_k141_173263 | Not-determined |
| KM_k141_161438 | Low-quality |
| KM_k141_119097 | Low-quality |
| KM_k141_250009 | Not-determined |
| KM_k141_207461 | Not-determined |
| KM_k141_119103 | Not-determined |
| KM_k141_218715 | Not-determined |
| KM_k141_106437 | Not-determined |
| KM_k141_128914 | Not-determined |
| KM_k141_94016 | Not-determined |
| KM_k141_261266 | Not-determined |
| KM_k141_261267 | Not-determined |
| KM_k141_151103 | Not-determined |
| KM_k141_196190 | Not-determined |
| KM_k141_207480 | Not-determined |
| KM_k141_94031 | Not-determined |
| KM_k141_161454 | Not-determined |
| KM_k141_128932 | Not-determined |
| KM_k141_128933 | Low-quality |
| KM_k141_161456 | Not-determined |
| KM_k141_196202 | Low-quality |
| KM_k141_128941 | Not-determined |
| KM_k141_128942 | Low-quality |
| KM_k141_62033 | Not-determined |
| KM_k141_39591 | Not-determined |
| KM_k141_39593 | Not-determined |
| KM_k141_261275 | Not-determined |
| KM_k141_62049 | Not-determined |
| KM_k141_39600 | Not-determined |
| KM_k141_261282 | Low-quality |
| KM_k141_49204 | Not-determined |
| KM_k141_28291 | Not-determined |
| KM_k141_106462 | Not-determined |
| KM_k141_128950 | Not-determined |
| KM_k141_261288 | Not-determined |
| KM_k141_119121 | Not-determined |
| KM_k141_94054 | Not-determined |
| KM_k141_196234 | Not-determined |
| KM_k141_151142 | Not-determined |
| KM_k141_119127 | Not-determined |
| KM_k141_73685 | Not-determined |
| KM_k141_173299 | Not-determined |
| KM_k141_119131 | Not-determined |
| KM_k141_173303 | Not-determined |
| KM_k141_239737 | Not-determined |
| KM_k141_196243 | Not-determined |
| KM_k141_239743 | Not-determined |
| KM_k141_173309 | Not-determined |
| KM_k141_229782 | Not-determined |
| KM_k141_119143 | Not-determined |
| KM_k141_73706 | Not-determined |
| KM_k141_28306 | Not-determined |
| KM_k141_161495 | Not-determined |
| KM_k141_119150 | Not-determined |
| KM_k141_62084 | Low-quality |
| KM_k141_119152 | Not-determined |
| KM_k141_229786 | Not-determined |
| KM_k141_229789 | Not-determined |
| KM_k141_39640 | Not-determined |
| KM_k141_119157 | Not-determined |
| KM_k141_39642 | Not-determined |
| KM_k141_49210 | Not-determined |
| KM_k141_196255 | Not-determined |
| KM_k141_28325 | Not-determined |
| KM_k141_207524 | Not-determined |
| KM_k141_6206 | Not-determined |
| KM_k141_261328 | Not-determined |
| KM_k141_207528 | Not-determined |
| KM_k141_218753 | Not-determined |
| KM_k141_161509 | Not-determined |
| KM_k141_94069 | Not-determined |
| KM_k141_239762 | Not-determined |
| KM_k141_28333 | Not-determined |
| KM_k141_196265 | Not-determined |
| KM_k141_6215 | Not-determined |
| KM_k141_106506 | Not-determined |
| KM_k141_185274 | Not-determined |
| KM_k141_6220 | Not-determined |
| KM_k141_94079 | Not-determined |
| KM_k141_185276 | Not-determined |
| KM_k141_106509 | Not-determined |
| KM_k141_229803 | Not-determined |
| KM_k141_6223 | Not-determined |
| KM_k141_94084 | Not-determined |
| KM_k141_119172 | Low-quality |
| KM_k141_239778 | Not-determined |
| KM_k141_85278 | Not-determined |
| KM_k141_229809 | Not-determined |
| KM_k141_39662 | Not-determined |
| KM_k141_196280 | Not-determined |
| KM_k141_229814 | Not-determined |
| KM_k141_85284 | Not-determined |
| KM_k141_106521 | Not-determined |
| KM_k141_28336 | Low-quality |
| KM_k141_207532 | Not-determined |
| KM_k141_173334 | Low-quality |
| KM_k141_28339 | Not-determined |
| KM_k141_6227 | Not-determined |
| KM_k141_85293 | Not-determined |
| KM_k141_128993 | Not-determined |
| KM_k141_73723 | Not-determined |
| KM_k141_261351 | Not-determined |
| KM_k141_129000 | Not-determined |
| KM_k141_229829 | Not-determined |
| KM_k141_207542 | Not-determined |
| KM_k141_129005 | Not-determined |
| KM_k141_28343 | Low-quality |
| KM_k141_196298 | Not-determined |
| KM_k141_85300 | Not-determined |
| KM_k141_196305 | Not-determined |
| KM_k141_85308 | Not-determined |
| KM_k141_73733 | Not-determined |
| KM_k141_173343 | Not-determined |
| KM_k141_261362 | Not-determined |
| KM_k141_85311 | Not-determined |
| KM_k141_119179 | Not-determined |
| KM_k141_73738 | Low-quality |
| KM_k141_129015 | Not-determined |
| KM_k141_119180 | Not-determined |
| KM_k141_119181 | Low-quality |
| KM_k141_229849 | Not-determined |
| KM_k141_207564 | Not-determined |
| KM_k141_129019 | Not-determined |
| KM_k141_185293 | Low-quality |
| KM_k141_73748 | Not-determined |
| KM_k141_229857 | Not-determined |
| KM_k141_239826 | Not-determined |
| KM_k141_85319 | Not-determined |
| KM_k141_119190 | Low-quality |
| KM_k141_239831 | Low-quality |
| KM_k141_62139 | Not-determined |
| KM_k141_161564 | Not-determined |
| KM_k141_161565 | Not-determined |
| KM_k141_140329 | Not-determined |
| KM_k141_106549 | Not-determined |
| KM_k141_49227 | Not-determined |
| KM_k141_49228 | Not-determined |
| KM_k141_85327 | Not-determined |
| KM_k141_161566 | Not-determined |
| KM_k141_129047 | Not-determined |
| KM_k141_239837 | Not-determined |
| KM_k141_151224 | Not-determined |
| KM_k141_6267 | Not-determined |
| KM_k141_140339 | Not-determined |
| KM_k141_73761 | Not-determined |
| KM_k141_161569 | Not-determined |
| KM_k141_73762 | Not-determined |
| KM_k141_119209 | Not-determined |
| KM_k141_173380 | Not-determined |
| KM_k141_85331 | Not-determined |
| KM_k141_85335 | Not-determined |
| KM_k141_250081 | Not-determined |
| KM_k141_119220 | Not-determined |
| KM_k141_28390 | Not-determined |
| KM_k141_250087 | Low-quality |
| KM_k141_207571 | Not-determined |
| KM_k141_218798 | Not-determined |
| KM_k141_239857 | Not-determined |
| KM_k141_106562 | Low-quality |
| KM_k141_185330 | Not-determined |
| KM_k141_151251 | Not-determined |
| KM_k141_218807 | Not-determined |
| KM_k141_140355 | Not-determined |
| KM_k141_39690 | Not-determined |
| KM_k141_185338 | Low-quality |
| KM_k141_173392 | Not-determined |
| KM_k141_16544 | Not-determined |
| KM_k141_229877 | Low-quality |
| KM_k141_62160 | Not-determined |
| KM_k141_140361 | Not-determined |
| KM_k141_218814 | Not-determined |
| KM_k141_73775 | Not-determined |
| KM_k141_218815 | Not-determined |
| KM_k141_28403 | Not-determined |
| KM_k141_106571 | Not-determined |
| KM_k141_106572 | Not-determined |
| KM_k141_218818 | Not-determined |
| KM_k141_185348 | Not-determined |
| KM_k141_185349 | Not-determined |
| KM_k141_106573 | Not-determined |
| KM_k141_185352 | Not-determined |
| KM_k141_106578 | Not-determined |
| KM_k141_129079 | Not-determined |
| KM_k141_173398 | Not-determined |
| KM_k141_207584 | Low-quality |
| KM_k141_151268 | Not-determined |
| KM_k141_185359 | Not-determined |
| KM_k141_218833 | Not-determined |
| KM_k141_49255 | Low-quality |
| KM_k141_106586 | Not-determined |
| KM_k141_218838 | Not-determined |
| KM_k141_185365 | Not-determined |
| KM_k141_173400 | Not-determined |
| KM_k141_173401 | Not-determined |
| KM_k141_196309 | Not-determined |
| KM_k141_6283 | Not-determined |
| KM_k141_16559 | Not-determined |
| KM_k141_85382 | Not-determined |
| KM_k141_49256 | Not-determined |
| KM_k141_207595 | Not-determined |
| KM_k141_229893 | Not-determined |
| KM_k141_239891 | Not-determined |
| KM_k141_73793 | Low-quality |
| KM_k141_151293 | Not-determined |
| KM_k141_196318 | Not-determined |
| KM_k141_119241 | Not-determined |
| KM_k141_250123 | Not-determined |
| KM_k141_173421 | Not-determined |
| KM_k141_119245 | Not-determined |
| KM_k141_185385 | Not-determined |
| KM_k141_250130 | Not-determined |
| KM_k141_28431 | Not-determined |
| KM_k141_239906 | Not-determined |
| KM_k141_28436 | Not-determined |
| KM_k141_250135 | Not-determined |
| KM_k141_261413 | Not-determined |
| KM_k141_207600 | Not-determined |
| KM_k141_140391 | Not-determined |
| KM_k141_239913 | Not-determined |
| KM_k141_218874 | Not-determined |
| KM_k141_129123 | Low-quality |
| KM_k141_261424 | Not-determined |
| KM_k141_218881 | Not-determined |
| KM_k141_218885 | Not-determined |
| KM_k141_94124 | Not-determined |
| KM_k141_94127 | Not-determined |
| KM_k141_218890 | Not-determined |
| KM_k141_239920 | Not-determined |
| KM_k141_39710 | Not-determined |
| KM_k141_173426 | Not-determined |
| KM_k141_151325 | Not-determined |
| KM_k141_62209 | Not-determined |
| KM_k141_106615 | Not-determined |
| KM_k141_85420 | Low-quality |
| KM_k141_151327 | Not-determined |
| KM_k141_229926 | Not-determined |
| KM_k141_62218 | Not-determined |
| KM_k141_73828 | Not-determined |
| KM_k141_73829 | Not-determined |
| KM_k141_229933 | Low-quality |
| KM_k141_229936 | Not-determined |
| KM_k141_106618 | Not-determined |
| KM_k141_229940 | Not-determined |
| KM_k141_229941 | Not-determined |
| KM_k141_62229 | Not-determined |
| KM_k141_261436 | Not-determined |
| KM_k141_151337 | Not-determined |
| KM_k141_85435 | Not-determined |
| KM_k141_173448 | Not-determined |
| KM_k141_28458 | Low-quality |
| KM_k141_129163 | Not-determined |
| KM_k141_85445 | Low-quality |
| KM_k141_173454 | Not-determined |
| KM_k141_196366 | Not-determined |
| KM_k141_218937 | Not-determined |
| KM_k141_85449 | Not-determined |
| KM_k141_151354 | Not-determined |
| KM_k141_207623 | Not-determined |
| KM_k141_207625 | Not-determined |
| KM_k141_6300 | Not-determined |
| KM_k141_140412 | Not-determined |
| KM_k141_151358 | Not-determined |
| KM_k141_207630 | Not-determined |
| KM_k141_6302 | Not-determined |
| KM_k141_173460 | Not-determined |
| KM_k141_119277 | Not-determined |
| KM_k141_6305 | Not-determined |
| KM_k141_6306 | Not-determined |
| KM_k141_28466 | Not-determined |
| KM_k141_28468 | Not-determined |
| KM_k141_85458 | Not-determined |
| KM_k141_73855 | Not-determined |
| KM_k141_85459 | Low-quality |
| KM_k141_73856 | Not-determined |
| KM_k141_151364 | Not-determined |
| KM_k141_207640 | Not-determined |
| KM_k141_207641 | Not-determined |
| KM_k141_6310 | Not-determined |
| KM_k141_62250 | Not-determined |
| KM_k141_49304 | Not-determined |
| KM_k141_261456 | Not-determined |
| KM_k141_261458 | Not-determined |
| KM_k141_261466 | Not-determined |
| KM_k141_85475 | Not-determined |
| KM_k141_49307 | Not-determined |
| KM_k141_85477 | Not-determined |
| KM_k141_119293 | Not-determined |
| KM_k141_229980 | Not-determined |
| KM_k141_94161 | Not-determined |
| KM_k141_73867 | Not-determined |
| KM_k141_94170 | Not-determined |
| KM_k141_229984 | Not-determined |
| KM_k141_49315 | Not-determined |
| KM_k141_261483 | Not-determined |
| KM_k141_85486 | Low-quality |
| KM_k141_106653 | Not-determined |
| KM_k141_85490 | Not-determined |
| KM_k141_106656 | Not-determined |
| KM_k141_218959 | Not-determined |
| KM_k141_49318 | Not-determined |
| KM_k141_62255 | Not-determined |
| KM_k141_173497 | Not-determined |
| KM_k141_229997 | Not-determined |
| KM_k141_250180 | Not-determined |
| KM_k141_173505 | Not-determined |
| KM_k141_28488 | Not-determined |
| KM_k141_73897 | Not-determined |
| KM_k141_49326 | Not-determined |
| KM_k141_173511 | Not-determined |
| KM_k141_119307 | Not-determined |
| KM_k141_28491 | Not-determined |
| KM_k141_119309 | Not-determined |
| KM_k141_140461 | Not-determined |
| KM_k141_129202 | Not-determined |
| KM_k141_94180 | Low-quality |
| KM_k141_49334 | Not-determined |
| KM_k141_85526 | Not-determined |
| KM_k141_151388 | Low-quality |
| KM_k141_49342 | Not-determined |
| KM_k141_151392 | Not-determined |
| KM_k141_28512 | Not-determined |
| KM_k141_239970 | Not-determined |
| KM_k141_250199 | Not-determined |
| KM_k141_85530 | Not-determined |
| KM_k141_140464 | Low-quality |
| KM_k141_85534 | Not-determined |
| KM_k141_85536 | Not-determined |
| KM_k141_39743 | Low-quality |
| KM_k141_6342 | Not-determined |
| KM_k141_73923 | Not-determined |
| KM_k141_49352 | Not-determined |
| KM_k141_119324 | Low-quality |
| KM_k141_230021 | Low-quality |
| KM_k141_151404 | Not-determined |
| KM_k141_230022 | Not-determined |
| KM_k141_119328 | Not-determined |
| KM_k141_39758 | Not-determined |
| KM_k141_73934 | Not-determined |
| KM_k141_185462 | Not-determined |
| KM_k141_16593 | Not-determined |
| KM_k141_49360 | Not-determined |
| KM_k141_73940 | Not-determined |
| KM_k141_6358 | Not-determined |
| KM_k141_129223 | Not-determined |
| 17-108-final-viral-combined_k141_120257\|\|full | Medium-quality |
| KM_k141_218998 | Not-determined |
| KM_k141_85542 | Not-determined |
| KM_k141_140476 | Not-determined |
| KM_k141_62284 | Not-determined |
| KM_k141_73947 | Low-quality |
| KM_k141_28531 | Not-determined |
| KM_k141_6367 | Not-determined |
| KM_k141_106677 | Not-determined |
| KM_k141_173544 | Not-determined |
| KM_k141_49368 | Not-determined |
| KM_k141_16606 | Not-determined |
| KM_k141_73960 | Not-determined |
| KM_k141_161595 | Not-determined |
| KM_k141_185477 | Not-determined |
| KM_k141_39777 | Not-determined |
| KM_k141_28538 | Not-determined |
| KM_k141_261513 | Not-determined |
| KM_k141_129239 | Low-quality |
| KM_k141_49374 | Not-determined |
| KM_k141_230030 | Not-determined |
| KM_k141_28548 | Not-determined |
| KM_k141_151433 | Not-determined |
| KM_k141_250240 | Not-determined |
| KM_k141_151437 | Not-determined |
| KM_k141_129257 | Not-determined |
| KM_k141_28555 | Not-determined |
| KM_k141_261517 | Not-determined |
| KM_k141_73980 | Not-determined |
| KM_k141_62305 | Not-determined |
| KM_k141_73983 | Not-determined |
| KM_k141_16627 | Not-determined |
| KM_k141_151447 | Not-determined |
| KM_k141_28562 | Not-determined |
| KM_k141_106696 | Not-determined |
| KM_k141_129265 | Not-determined |
| KM_k141_230042 | Not-determined |
| KM_k141_261522 | Not-determined |
| KM_k141_28564 | Not-determined |
| KM_k141_140510 | Not-determined |
| KM_k141_39795 | Not-determined |
| KM_k141_240040 | Not-determined |
| KM_k141_129273 | Not-determined |
| KM_k141_151458 | Not-determined |
| KM_k141_230048 | Low-quality |
| KM_k141_129281 | Not-determined |
| KM_k141_230051 | Not-determined |
| KM_k141_94210 | Low-quality |
| KM_k141_173586 | Not-determined |
| KM_k141_185500 | Low-quality |
| KM_k141_106714 | Not-determined |
| KM_k141_49395 | Not-determined |
| KM_k141_250253 | Not-determined |
| KM_k141_230068 | Not-determined |
| KM_k141_49397 | Not-determined |
| KM_k141_62335 | Not-determined |
| KM_k141_185506 | Not-determined |
| KM_k141_173598 | Not-determined |
| KM_k141_28576 | Not-determined |
| KM_k141_219022 | Low-quality |
| KM_k141_39814 | Not-determined |
| KM_k141_230073 | Not-determined |
| KM_k141_230075 | Not-determined |
| KM_k141_49406 | Not-determined |
| KM_k141_240049 | Not-determined |
| KM_k141_16655 | Low-quality |
| KM_k141_16657 | Low-quality |
| KM_k141_16658 | Not-determined |
| KM_k141_28581 | Not-determined |
| KM_k141_62344 | Not-determined |
| KM_k141_240054 | Low-quality |
| KM_k141_196444 | Not-determined |
| KM_k141_49420 | Not-determined |
| KM_k141_240058 | Not-determined |
| KM_k141_196453 | Not-determined |
| KM_k141_230086 | Not-determined |
| KM_k141_219039 | Not-determined |
| KM_k141_49426 | Not-determined |
| KM_k141_219043 | Not-determined |
| KM_k141_85599 | Not-determined |
| KM_k141_173611 | Not-determined |
| KM_k141_140521 | Not-determined |
| KM_k141_129332 | Not-determined |
| KM_k141_6398 | Not-determined |
| KM_k141_151484 | Not-determined |
| KM_k141_85608 | Not-determined |
| KM_k141_85609 | Not-determined |
| KM_k141_62368 | Not-determined |
| KM_k141_207728 | Low-quality |
| KM_k141_261578 | Not-determined |
| KM_k141_151486 | Not-determined |
| KM_k141_74001 | Low-quality |
| KM_k141_207731 | Not-determined |
| KM_k141_74004 | Not-determined |
| KM_k141_219055 | Not-determined |
| KM_k141_6410 | Not-determined |
| KM_k141_74005 | Not-determined |
| KM_k141_140531 | Low-quality |
| KM_k141_6412 | Not-determined |
| KM_k141_196480 | Not-determined |
| KM_k141_85616 | Not-determined |
| KM_k141_151492 | Not-determined |
| KM_k141_85617 | Not-determined |
| KM_k141_106747 | Not-determined |
| KM_k141_85619 | Not-determined |
| KM_k141_250286 | Not-determined |
| KM_k141_230095 | Not-determined |
| KM_k141_207740 | Not-determined |
| KM_k141_196486 | Low-quality |
| KM_k141_16682 | Not-determined |
| KM_k141_240080 | Not-determined |
| KM_k141_219057 | Not-determined |
| KM_k141_219058 | Not-determined |
| KM_k141_62374 | Not-determined |
| KM_k141_28595 | Not-determined |
| KM_k141_219065 | Not-determined |
| KM_k141_207743 | Not-determined |
| KM_k141_62379 | Not-determined |
| KM_k141_240090 | Not-determined |
| KM_k141_196493 | Not-determined |
| KM_k141_196494 | Not-determined |
| KM_k141_28602 | Not-determined |
| KM_k141_85629 | Not-determined |
| KM_k141_94250 | Not-determined |
| KM_k141_62386 | Not-determined |
| KM_k141_196512 | Not-determined |
| KM_k141_129365 | Not-determined |
| KM_k141_85649 | Not-determined |
| KM_k141_230137 | Not-determined |
| KM_k141_140554 | Not-determined |
| KM_k141_129370 | Not-determined |
| KM_k141_16699 | Not-determined |
| KM_k141_196526 | Not-determined |
| KM_k141_140559 | Not-determined |
| KM_k141_240126 | Not-determined |
| KM_k141_39837 | Not-determined |
| KM_k141_161624 | Not-determined |
| KM_k141_62399 | Not-determined |
| KM_k141_196542 | Not-determined |
| KM_k141_161628 | Not-determined |
| KM_k141_62401 | Not-determined |
| KM_k141_16714 | Not-determined |
| KM_k141_151557 | Low-quality |
| KM_k141_207776 | Not-determined |
| KM_k141_39846 | Not-determined |
| KM_k141_250319 | Low-quality |
| KM_k141_207778 | Low-quality |
| KM_k141_230159 | Not-determined |
| KM_k141_16723 | Not-determined |
| KM_k141_185586 | Not-determined |
| KM_k141_185591 | Not-determined |
| KM_k141_261624 | Not-determined |
| KM_k141_261625 | Low-quality |
| KM_k141_173656 | Not-determined |
| KM_k141_230170 | Not-determined |
| KM_k141_85679 | Not-determined |

| | |
|---|---|
| KM_k141_250332 | Not-determined |
| KM_k141_140591 | Not-determined |
| KM_k141_185597 | Not-determined |
| KM_k141_161630 | Not-determined |
| KM_k141_250334 | Not-determined |
| KM_k141_6452 | Not-determined |
| KM_k141_6454 | Not-determined |
| KM_k141_49467 | Not-determined |
| KM_k141_6457 | Low-quality |
| KM_k141_6458 | Low-quality |
| KM_k141_230181 | Not-determined |
| KM_k141_185599 | Not-determined |
| KM_k141_230184 | Not-determined |
| KM_k141_250340 | Low-quality |
| KM_k141_261639 | Not-determined |
| KM_k141_39881 | Not-determined |
| KM_k141_6465 | Not-determined |
| KM_k141_173665 | Not-determined |
| KM_k141_173666 | Not-determined |
| KM_k141_140603 | Not-determined |
| KM_k141_119383 | Not-determined |
| KM_k141_106773 | Low-quality |
| KM_k141_129415 | Not-determined |
| KM_k141_119390 | Not-determined |
| KM_k141_119391 | Not-determined |
| KM_k141_261650 | Not-determined |
| KM_k141_119397 | Not-determined |
| KM_k141_49480 | Not-determined |
| KM_k141_261651 | Low-quality |
| KM_k141_16750 | Not-determined |
| KM_k141_173680 | Low-quality |
| KM_k141_173682 | Not-determined |
| KM_k141_261654 | Not-determined |
| KM_k141_230198 | Not-determined |
| KM_k141_261655 | Not-determined |
| KM_k141_119404 | Not-determined |
| KM_k141_250354 | Not-determined |
| KM_k141_173692 | Not-determined |
| KM_k141_230204 | Not-determined |
| KM_k141_173695 | Not-determined |
| KM_k141_49494 | Not-determined |
| KM_k141_49495 | Not-determined |
| KM_k141_49496 | Not-determined |
| KM_k141_49497 | Not-determined |
| KM_k141_161661 | Not-determined |
| KM_k141_230207 | Not-determined |
| KM_k141_250370 | Low-quality |
| KM_k141_151614 | Not-determined |
| KM_k141_161671 | Not-determined |
| KM_k141_39912 | Low-quality |
| KM_k141_173704 | Not-determined |
| KM_k141_161674 | Not-determined |
| KM_k141_261666 | Low-quality |
| KM_k141_49505 | Not-determined |
| KM_k141_119420 | Not-determined |
| KM_k141_261671 | Not-determined |
| KM_k141_49511 | Not-determined |
| KM_k141_207822 | Not-determined |
| KM_k141_185629 | Not-determined |
| KM_k141_119424 | Not-determined |
| KM_k141_39924 | Not-determined |
| KM_k141_161681 | Not-determined |
| KM_k141_119428 | Not-determined |
| KM_k141_161686 | Not-determined |
| KM_k141_161688 | Not-determined |
| KM_k141_196578 | Not-determined |
| KM_k141_94323 | Low-quality |
| KM_k141_207833 | Not-determined |
| KM_k141_39930 | Not-determined |
| KM_k141_119436 | Not-determined |
| KM_k141_94327 | Not-determined |
| KM_k141_62439 | Not-determined |
| KM_k141_140630 | Not-determined |
| KM_k141_62443 | Not-determined |
| KM_k141_140632 | Not-determined |
| KM_k141_261685 | Not-determined |
| KM_k141_16791 | Not-determined |
| KM_k141_16794 | Not-determined |
| KM_k141_49528 | Low-quality |
| KM_k141_185648 | Low-quality |
| KM_k141_49530 | Not-determined |
| KM_k141_16799 | Not-determined |
| KM_k141_261700 | Not-determined |
| KM_k141_185652 | Not-determined |
| KM_k141_16801 | Not-determined |
| KM_k141_119459 | Not-determined |
| KM_k141_140649 | Not-determined |
| KM_k141_129453 | Not-determined |
| KM_k141_28681 | Not-determined |
| KM_k141_6490 | Not-determined |
| KM_k141_185667 | Not-determined |
| KM_k141_173732 | Not-determined |
| KM_k141_129460 | Not-determined |
| KM_k141_49537 | Not-determined |
| KM_k141_250389 | Low-quality |
| KM_k141_161725 | Not-determined |
| KM_k141_161726 | Not-determined |
| KM_k141_196616 | Not-determined |
| KM_k141_196618 | Not-determined |
| KM_k141_196619 | Not-determined |
| KM_k141_119465 | Not-determined |
| KM_k141_161728 | Not-determined |
| KM_k141_106810 | Not-determined |
| KM_k141_119475 | Not-determined |
| KM_k141_62487 | Not-determined |
| KM_k141_185684 | Not-determined |
| KM_k141_161740 | Not-determined |
| KM_k141_185689 | Low-quality |
| KM_k141_74024 | Not-determined |
| KM_k141_219084 | Not-determined |
| KM_k141_74027 | Not-determined |
| KM_k141_207887 | Not-determined |
| KM_k141_151630 | Not-determined |
| KM_k141_151631 | Not-determined |
| KM_k141_74033 | Not-determined |
| KM_k141_261725 | Not-determined |
| KM_k141_6520 | Not-determined |
| 18-101-final-viral-combined_k141_158905\|\|full | Medium-quality |
| KM_k141_161744 | Not-determined |
| KM_k141_6522 | Low-quality |
| KM_k141_39951 | Not-determined |
| KM_k141_173761 | Not-determined |
| KM_k141_173762 | Not-determined |
| KM_k141_196640 | Low-quality |
| KM_k141_161749 | Not-determined |
| KM_k141_39959 | Not-determined |
| KM_k141_161753 | Not-determined |
| KM_k141_106832 | Not-determined |
| KM_k141_106833 | Not-determined |
| KM_k141_261731 | Not-determined |
| KM_k141_219104 | Not-determined |
| KM_k141_250402 | Not-determined |
| KM_k141_185719 | Not-determined |
| KM_k141_106841 | Not-determined |
| KM_k141_119505 | Not-determined |
| KM_k141_185721 | Not-determined |
| KM_k141_74055 | Not-determined |
| KM_k141_16825 | Not-determined |
| KM_k141_161760 | Not-determined |
| KM_k141_62503 | Not-determined |
| KM_k141_16827 | Not-determined |
| KM_k141_39970 | Not-determined |
| KM_k141_207911 | Not-determined |
| KM_k141_185726 | Not-determined |
| KM_k141_28720 | Not-determined |
| KM_k141_106847 | Not-determined |
| KM_k141_94380 | Not-determined |
| KM_k141_250404 | Low-quality |
| KM_k141_28725 | Not-determined |
| KM_k141_74063 | Not-determined |
| KM_k141_49546 | Not-determined |
| KM_k141_161766 | Not-determined |
| KM_k141_196660 | Not-determined |
| KM_k141_161770 | Not-determined |
| KM_k141_151644 | Not-determined |
| KM_k141_16839 | Not-determined |
| KM_k141_94383 | Not-determined |
| KM_k141_261751 | Not-determined |
| KM_k141_6555 | Not-determined |
| KM_k141_16843 | Not-determined |
| KM_k141_6556 | Not-determined |
| KM_k141_28736 | Not-determined |
| KM_k141_49557 | Not-determined |
| KM_k141_6563 | Not-determined |
| KM_k141_129530 | Low-quality |
| KM_k141_6565 | Not-determined |
| KM_k141_94391 | Not-determined |
| KM_k141_94394 | Not-determined |
| KM_k141_219125 | Not-determined |
| KM_k141_6570 | Not-determined |
| KM_k141_94398 | Low-quality |
| KM_k141_28745 | Not-determined |
| KM_k141_74080 | Low-quality |
| KM_k141_250421 | Not-determined |
| KM_k141_74082 | Not-determined |
| KM_k141_16861 | Low-quality |
| KM_k141_129534 | Low-quality |
| KM_k141_40015 | Not-determined |
| KM_k141_196673 | Low-quality |
| KM_k141_16866 | Low-quality |
| KM_k141_16869 | Not-determined |
| KM_k141_151665 | Not-determined |
| KM_k141_16870 | Not-determined |
| KM_k141_230229 | Low-quality |
| KM_k141_94408 | Not-determined |
| KM_k141_207941 | Not-determined |
| KM_k141_85761 | Not-determined |
| KM_k141_16882 | Not-determined |
| KM_k141_161799 | Not-determined |
| KM_k141_94412 | Not-determined |
| KM_k141_119534 | Not-determined |
| KM_k141_94416 | Not-determined |
| KM_k141_74105 | Not-determined |
| KM_k141_74109 | Not-determined |
| KM_k141_230238 | Not-determined |
| KM_k141_85773 | Not-determined |
| KM_k141_6588 | Not-determined |
| KM_k141_129554 | Not-determined |
| KM_k141_230242 | Not-determined |
| KM_k141_230243 | Not-determined |
| KM_k141_40036 | Not-determined |
| KM_k141_40038 | Not-determined |
| KM_k141_230246 | Not-determined |
| KM_k141_16889 | Not-determined |
| KM_k141_62556 | Not-determined |
| KM_k141_250435 | Not-determined |
| KM_k141_6595 | Not-determined |
| KM_k141_129557 | Low-quality |
| KM_k141_16892 | Not-determined |
| KM_k141_85784 | Not-determined |
| KM_k141_161815 | Not-determined |
| KM_k141_6601 | Not-determined |
| KM_k141_129563 | Not-determined |
| KM_k141_161818 | Low-quality |
| KM_k141_173829 | Not-determined |
| KM_k141_94436 | Not-determined |
| KM_k141_74132 | Not-determined |
| KM_k141_16900 | Not-determined |
| KM_k141_250443 | Not-determined |
| KM_k141_151716 | Not-determined |
| KM_k141_196694 | Not-determined |
| KM_k141_230252 | Not-determined |
| KM_k141_106903 | Low-quality |
| KM_k141_140735 | Not-determined |
| KM_k141_250448 | Low-quality |
| KM_k141_240249 | Not-determined |
| KM_k141_261792 | Not-determined |
| KM_k141_151722 | Not-determined |
| KM_k141_207983 | Not-determined |
| KM_k141_106907 | Not-determined |
| KM_k141_261795 | Not-determined |
| KM_k141_49565 | Not-determined |
| KM_k141_196704 | Not-determined |
| KM_k141_106913 | Not-determined |
| KM_k141_207989 | Low-quality |
| KM_k141_261802 | Low-quality |
| KM_k141_261804 | Not-determined |
| KM_k141_230255 | Not-determined |
| KM_k141_196711 | Not-determined |
| KM_k141_140745 | Not-determined |
| KM_k141_151739 | Not-determined |
| KM_k141_240265 | Not-determined |
| KM_k141_219185 | Not-determined |
| KM_k141_16909 | Not-determined |
| KM_k141_140750 | Not-determined |
| KM_k141_40077 | Not-determined |
| KM_k141_16912 | Not-determined |
| KM_k141_119592 | Not-determined |
| KM_k141_240274 | Not-determined |
| KM_k141_161858 | Not-determined |
| KM_k141_250454 | Low-quality |
| KM_k141_161859 | Not-determined |
| KM_k141_250458 | Not-determined |
| KM_k141_49581 | Not-determined |
| KM_k141_28785 | Not-determined |
| KM_k141_261821 | Not-determined |
| KM_k141_240275 | Not-determined |
| KM_k141_250463 | Not-determined |
| KM_k141_261823 | Not-determined |
| KM_k141_119617 | Not-determined |
| KM_k141_250464 | Not-determined |
| KM_k141_173874 | Not-determined |
| KM_k141_74140 | Not-determined |
| KM_k141_161865 | Not-determined |
| KM_k141_49593 | Not-determined |
| KM_k141_28798 | Not-determined |
| KM_k141_49595 | Not-determined |
| KM_k141_16920 | Not-determined |
| KM_k141_85814 | Not-determined |
| KM_k141_219207 | Not-determined |
| KM_k141_173891 | Not-determined |
| KM_k141_94442 | Not-determined |
| KM_k141_94444 | Not-determined |
| KM_k141_74159 | Not-determined |
| KM_k141_261840 | Not-determined |
| KM_k141_74162 | Low-quality |
| KM_k141_74164 | Not-determined |
| KM_k141_28813 | Not-determined |
| KM_k141_16933 | Not-determined |
| KM_k141_161872 | Not-determined |
| KM_k141_250485 | Not-determined |
| KM_k141_85825 | Not-determined |
| KM_k141_161873 | Low-quality |
| KM_k141_85829 | Not-determined |
| KM_k141_230291 | Not-determined |
| KM_k141_28818 | Not-determined |
| KM_k141_250491 | Not-determined |
| KM_k141_74171 | Not-determined |
| KM_k141_250493 | Not-determined |
| KM_k141_161877 | Not-determined |
| KM_k141_119638 | Not-determined |
| KM_k141_16949 | Low-quality |
| KM_k141_140786 | Not-determined |
| KM_k141_28830 | Not-determined |
| KM_k141_62591 | Not-determined |
| KM_k141_49623 | Not-determined |
| KM_k141_49624 | Not-determined |
| KM_k141_40102 | Not-determined |
| KM_k141_196744 | Not-determined |
| KM_k141_28838 | Not-determined |
| KM_k141_119649 | Not-determined |
| KM_k141_219231 | Not-determined |
| KM_k141_74202 | Not-determined |
| KM_k141_40114 | Not-determined |
| KM_k141_74215 | Not-determined |
| KM_k141_161895 | Not-determined |
| KM_k141_119667 | Not-determined |
| KM_k141_219239 | Not-determined |
| KM_k141_40121 | Not-determined |
| KM_k141_261858 | Not-determined |
| KM_k141_94471 | Not-determined |
| KM_k141_250519 | Not-determined |
| KM_k141_219246 | Not-determined |
| KM_k141_119677 | Not-determined |
| KM_k141_208050 | Not-determined |
| KM_k141_161905 | Not-determined |
| KM_k141_62616 | Low-quality |
| KM_k141_40132 | Not-determined |
| KM_k141_129611 | Not-determined |
| KM_k141_119695 | Not-determined |
| KM_k141_119696 | Low-quality |
| KM_k141_261877 | Not-determined |
| KM_k141_230345 | Not-determined |
| KM_k141_62636 | Not-determined |
| KM_k141_173937 | Not-determined |
| KM_k141_6617 | Not-determined |
| KM_k141_49640 | Not-determined |
| KM_k141_230347 | Not-determined |
| KM_k141_129615 | Not-determined |
| KM_k141_119706 | Not-determined |
| KM_k141_219273 | Not-determined |
| KM_k141_40152 | Not-determined |
| KM_k141_49650 | Not-determined |
| KM_k141_250551 | Not-determined |
| KM_k141_119722 | Not-determined |
| KM_k141_161928 | Not-determined |
| KM_k141_250553 | Not-determined |
| KM_k141_161934 | Not-determined |
| KM_k141_240285 | Not-determined |
| KM_k141_230365 | Not-determined |
| KM_k141_129629 | Not-determined |
| KM_k141_240292 | Not-determined |
| KM_k141_129630 | Not-determined |
| KM_k141_230371 | Not-determined |
| KM_k141_250562 | Low-quality |
| KM_k141_62669 | Not-determined |
| KM_k141_173948 | Not-determined |
| KM_k141_240300 | Not-determined |
| KM_k141_230380 | Not-determined |
| KM_k141_173952 | Not-determined |
| KM_k141_85860 | Not-determined |
| KM_k141_240305 | Not-determined |
| KM_k141_94513 | Not-determined |
| KM_k141_94514 | Not-determined |
| KM_k141_173956 | Not-determined |
| KM_k141_196772 | Not-determined |
| KM_k141_173959 | Not-determined |
| KM_k141_196773 | Not-determined |
| KM_k141_196777 | Not-determined |
| KM_k141_173965 | Not-determined |
| KM_k141_28884 | Not-determined |
| KM_k141_62672 | Low-quality |
| KM_k141_40184 | Not-determined |
| KM_k141_129657 | Not-determined |
| KM_k141_62675 | Not-determined |
| KM_k141_49682 | Not-determined |
| KM_k141_106951 | Low-quality |
| KM_k141_119738 | Not-determined |
| KM_k141_129667 | Not-determined |
| KM_k141_219309 | Low-quality |
| KM_k141_28893 | Not-determined |
| KM_k141_106956 | Not-determined |
| KM_k141_240310 | Low-quality |
| KM_k141_85891 | Not-determined |
| KM_k141_185761 | Not-determined |
| KM_k141_85892 | Not-determined |
| KM_k141_161956 | Not-determined |
| KM_k141_208083 | Not-determined |
| KM_k141_62690 | Not-determined |
| KM_k141_230401 | Not-determined |
| KM_k141_85895 | Not-determined |
| KM_k141_62692 | Not-determined |
| KM_k141_119752 | Not-determined |
| KM_k141_106964 | Not-determined |
| KM_k141_49695 | Not-determined |
| KM_k141_85899 | Not-determined |
| KM_k141_62695 | Not-determined |
| KM_k141_40201 | Not-determined |
| KM_k141_74226 | Not-determined |
| KM_k141_74230 | Not-determined |
| KM_k141_161967 | Not-determined |
| KM_k141_173989 | Not-determined |
| KM_k141_140809 | Not-determined |
| KM_k141_28915 | Not-determined |
| KM_k141_62704 | Not-determined |
| KM_k141_161971 | Not-determined |
| KM_k141_28918 | Not-determined |
| KM_k141_161972 | Not-determined |
| KM_k141_74237 | Not-determined |
| KM_k141_140812 | Not-determined |
| KM_k141_185797 | Low-quality |
| KM_k141_261937 | Not-determined |
| KM_k141_119773 | Not-determined |
| KM_k141_106983 | Not-determined |
| KM_k141_261945 | Not-determined |
| KM_k141_85918 | Not-determined |
| KM_k141_174009 | Not-determined |
| KM_k141_129717 | Not-determined |
| KM_k141_106992 | Not-determined |
| KM_k141_40213 | Low-quality |
| KM_k141_106997 | Not-determined |
| KM_k141_119784 | Low-quality |
| KM_k141_219348 | Not-determined |
| KM_k141_119788 | Not-determined |
| KM_k141_230437 | Not-determined |
| KM_k141_151803 | Not-determined |
| KM_k141_40223 | Not-determined |
| KM_k141_174040 | Not-determined |
| KM_k141_140835 | Not-determined |
| KM_k141_174042 | Not-determined |
| KM_k141_230442 | Not-determined |
| KM_k141_151809 | Not-determined |
| KM_k141_40229 | Not-determined |
| KM_k141_230446 | Not-determined |
| KM_k141_94559 | Not-determined |
| KM_k141_107012 | Low-quality |
| KM_k141_230451 | Not-determined |
| KM_k141_17012 | Not-determined |
| KM_k141_85954 | Not-determined |
| KM_k141_94564 | Not-determined |
| KM_k141_140847 | Not-determined |
| KM_k141_62732 | Not-determined |
| KM_k141_140848 | Not-determined |
| KM_k141_208113 | Not-determined |
| KM_k141_17019 | Not-determined |
| KM_k141_40244 | Not-determined |
| KM_k141_119811 | Not-determined |
| KM_k141_94573 | Not-determined |
| KM_k141_161992 | Not-determined |
| KM_k141_196804 | Not-determined |
| KM_k141_119812 | Not-determined |
| KM_k141_219365 | Low-quality |
| KM_k141_40253 | Not-determined |
| KM_k141_196807 | Not-determined |
| KM_k141_140858 | Not-determined |
| KM_k141_85962 | Not-determined |
| KM_k141_107026 | Not-determined |
| KM_k141_107027 | Not-determined |
| KM_k141_107028 | Not-determined |
| KM_k141_230468 | Not-determined |
| KM_k141_119819 | Not-determined |
| KM_k141_185829 | Not-determined |
| KM_k141_119821 | Not-determined |
| KM_k141_119824 | Not-determined |
| KM_k141_129742 | Not-determined |
| KM_k141_119830 | Not-determined |
| KM_k141_107038 | Not-determined |
| KM_k141_230480 | Not-determined |
| KM_k141_140879 | Not-determined |
| KM_k141_74272 | Not-determined |
| KM_k141_74274 | Not-determined |
| KM_k141_74276 | Not-determined |
| KM_k141_107047 | Not-determined |
| KM_k141_162007 | Not-determined |
| KM_k141_208122 | Not-determined |
| KM_k141_151849 | Not-determined |
| KM_k141_119845 | Not-determined |
| KM_k141_17053 | Not-determined |
| KM_k141_208124 | Not-determined |
| KM_k141_119847 | Not-determined |
| KM_k141_17054 | Not-determined |
| KM_k141_151855 | Low-quality |
| KM_k141_17055 | Not-determined |
| KM_k141_219379 | Not-determined |
| KM_k141_151858 | Not-determined |
| KM_k141_151861 | Not-determined |
| KM_k141_74285 | Not-determined |
| KM_k141_208134 | Not-determined |
| KM_k141_230507 | Not-determined |
| KM_k141_240314 | Not-determined |
| KM_k141_230509 | Not-determined |
| KM_k141_151866 | Not-determined |
| KM_k141_94603 | Not-determined |
| KM_k141_94605 | Not-determined |
| KM_k141_94607 | Not-determined |
| KM_k141_208140 | Not-determined |
| KM_k141_230514 | Not-determined |
| KM_k141_6710 | Not-determined |
| KM_k141_151871 | Not-determined |
| KM_k141_17064 | Not-determined |
| KM_k141_196846 | Not-determined |
| KM_k141_240326 | Not-determined |
| KM_k141_62783 | Not-determined |
| KM_k141_240327 | Not-determined |
| KM_k141_230523 | Not-determined |
| KM_k141_230527 | Not-determined |
| KM_k141_185872 | Not-determined |
| KM_k141_107069 | Not-determined |
| KM_k141_62791 | Not-determined |
| KM_k141_262011 | Not-determined |
| KM_k141_208150 | Not-determined |
| KM_k141_208153 | Not-determined |
| KM_k141_62799 | Not-determined |
| KM_k141_49750 | Not-determined |
| KM_k141_196859 | Not-determined |
| KM_k141_17068 | Not-determined |
| KM_k141_17069 | Not-determined |
| KM_k141_230538 | Not-determined |
| KM_k141_62807 | Not-determined |
| KM_k141_17070 | Not-determined |
| KM_k141_74302 | Not-determined |
| KM_k141_74305 | Not-determined |
| KM_k141_185890 | Not-determined |
| KM_k141_94630 | Not-determined |
| KM_k141_230547 | Not-determined |
| KM_k141_151890 | Not-determined |
| KM_k141_151896 | Not-determined |
| KM_k141_140927 | Not-determined |
| KM_k141_49763 | Low-quality |
| KM_k141_140930 | Not-determined |
| KM_k141_219414 | Not-determined |
| KM_k141_49771 | Not-determined |
| KM_k141_85974 | Not-determined |
| KM_k141_219422 | Not-determined |
| KM_k141_196885 | Not-determined |
| KM_k141_49774 | Not-determined |
| KM_k141_250602 | Not-determined |
| KM_k141_6745 | Not-determined |
| KM_k141_262033 | Not-determined |
| KM_k141_49775 | Not-determined |
| KM_k141_250606 | Not-determined |
| KM_k141_49782 | Not-determined |
| KM_k141_185907 | Not-determined |
| KM_k141_49783 | Not-determined |
| KM_k141_162035 | Not-determined |
| KM_k141_107086 | Not-determined |
| KM_k141_28938 | Not-determined |
| KM_k141_62834 | Not-determined |
| KM_k141_174050 | Not-determined |
| KM_k141_74335 | Not-determined |
| KM_k141_94656 | Not-determined |
| KM_k141_119885 | Not-determined |
| KM_k141_219436 | Not-determined |
| KM_k141_6754 | Not-determined |
| KM_k141_208192 | Not-determined |
| KM_k141_17086 | Not-determined |
| KM_k141_262043 | Not-determined |
| KM_k141_62846 | Low-quality |
| KM_k141_17090 | Not-determined |
| KM_k141_230579 | Not-determined |
| KM_k141_162044 | Not-determined |
| KM_k141_208205 | Not-determined |
| KM_k141_62855 | Not-determined |
| KM_k141_262049 | Low-quality |
| KM_k141_140963 | Not-determined |
| KM_k141_196905 | Low-quality |
| KM_k141_185920 | Not-determined |
| KM_k141_49800 | Not-determined |
| KM_k141_240350 | Low-quality |
| KM_k141_208212 | Not-determined |
| KM_k141_185923 | Not-determined |
| KM_k141_219446 | Not-determined |
| KM_k141_174075 | Not-determined |
| KM_k141_86018 | Not-determined |
| KM_k141_129767 | Not-determined |
| KM_k141_219456 | Not-determined |
| KM_k141_196916 | Not-determined |
| KM_k141_219458 | Not-determined |
| KM_k141_151918 | Not-determined |
| KM_k141_174087 | Low-quality |
| KM_k141_174089 | Not-determined |
| KM_k141_219459 | Low-quality |
| KM_k141_86032 | Not-determined |
| KM_k141_140991 | Not-determined |
| KM_k141_40325 | Low-quality |
| KM_k141_86039 | Not-determined |
| KM_k141_208247 | Not-determined |
| KM_k141_185954 | Not-determined |
| KM_k141_208252 | Not-determined |
| KM_k141_185961 | Not-determined |
| KM_k141_219465 | Not-determined |
| KM_k141_208253 | Not-determined |
| KM_k141_230603 | Not-determined |
| KM_k141_28941 | Not-determined |
| KM_k141_141009 | Not-determined |
| KM_k141_219482 | Not-determined |
| KM_k141_74379 | Not-determined |
| KM_k141_86052 | Not-determined |
| KM_k141_74381 | Low-quality |

| | |
|---|---|
| KM_k141_262072 | Not-determined |
| KM_k141_219490 | Not-determined |
| KM_k141_208265 | Not-determined |
| KM_k141_74386 | Not-determined |
| KM_k141_94677 | Not-determined |
| KM_k141_86054 | Not-determined |
| KM_k141_86055 | Not-determined |
| KM_k141_262080 | Not-determined |
| KM_k141_262081 | Not-determined |
| KM_k141_174101 | Not-determined |
| KM_k141_28959 | Not-determined |
| KM_k141_40342 | Not-determined |
| KM_k141_174108 | Not-determined |
| KM_k141_49835 | Not-determined |
| KM_k141_141035 | Not-determined |
| KM_k141_40345 | Not-determined |
| KM_k141_119912 | Not-determined |
| KM_k141_196931 | Not-determined |
| KM_k141_196932 | Not-determined |
| KM_k141_17103 | Not-determined |
| KM_k141_230634 | Not-determined |
| KM_k141_174114 | Not-determined |
| KM_k141_107114 | Not-determined |
| KM_k141_94697 | Not-determined |
| KM_k141_174118 | Not-determined |
| KM_k141_28966 | Not-determined |
| KM_k141_196942 | Not-determined |
| KM_k141_174121 | Not-determined |
| KM_k141_196944 | Not-determined |
| KM_k141_250647 | Not-determined |
| KM_k141_262087 | Not-determined |
| KM_k141_230645 | Not-determined |
| KM_k141_230646 | Not-determined |
| KM_k141_162055 | Low-quality |
| KM_k141_94707 | Not-determined |
| KM_k141_28977 | Not-determined |
| KM_k141_74411 | Not-determined |
| KM_k141_219525 | Not-determined |
| KM_k141_250659 | Not-determined |
| KM_k141_74413 | Not-determined |
| KM_k141_74415 | Not-determined |
| KM_k141_240359 | Not-determined |
| KM_k141_74416 | Not-determined |
| KM_k141_240362 | Not-determined |
| KM_k141_196964 | Not-determined |
| KM_k141_230657 | Not-determined |
| KM_k141_196967 | Not-determined |
| KM_k141_250662 | Not-determined |
| KM_k141_219529 | Not-determined |
| KM_k141_74421 | Not-determined |
| KM_k141_94714 | Not-determined |
| KM_k141_129789 | Not-determined |
| KM_k141_230670 | Not-determined |
| KM_k141_250663 | Low-quality |
| KM_k141_174156 | Not-determined |
| KM_k141_230675 | Not-determined |
| KM_k141_86103 | Not-determined |
| KM_k141_230677 | Not-determined |
| KM_k141_174160 | Not-determined |
| KM_k141_250669 | Not-determined |
| KM_k141_250670 | Not-determined |
| KM_k141_94723 | Not-determined |
| KM_k141_141075 | Not-determined |
| KM_k141_151954 | Not-determined |
| KM_k141_94727 | Not-determined |
| KM_k141_230686 | Not-determined |
| KM_k141_250681 | Not-determined |
| KM_k141_62930 | Not-determined |
| KM_k141_6792 | Not-determined |
| KM_k141_186000 | Low-quality |
| KM_k141_94731 | Not-determined |
| KM_k141_62932 | Not-determined |
| KM_k141_219550 | Not-determined |
| KM_k141_119935 | Low-quality |
| KM_k141_119937 | Not-determined |
| KM_k141_119938 | Not-determined |
| KM_k141_129806 | Low-quality |
| KM_k141_129807 | Low-quality |
| KM_k141_196994 | Not-determined |
| KM_k141_230694 | Not-determined |
| KM_k141_230695 | Not-determined |
| KM_k141_162071 | Not-determined |
| KM_k141_240382 | Not-determined |
| KM_k141_141095 | Not-determined |
| KM_k141_162077 | Not-determined |
| KM_k141_208303 | Not-determined |
| KM_k141_230709 | Not-determined |
| KM_k141_62957 | Not-determined |
| KM_k141_240390 | Not-determined |
| KM_k141_162081 | Not-determined |
| KM_k141_129812 | Low-quality |
| KM_k141_262105 | Not-determined |
| KM_k141_208314 | Not-determined |
| KM_k141_174194 | Not-determined |
| KM_k141_197007 | Not-determined |
| KM_k141_174196 | Not-determined |
| KM_k141_219564 | Not-determined |
| KM_k141_186008 | Not-determined |
| KM_k141_250705 | Not-determined |
| KM_k141_129826 | Not-determined |
| KM_k141_17153 | Not-determined |
| KM_k141_230734 | Not-determined |
| KM_k141_162100 | Not-determined |
| KM_k141_262121 | Not-determined |
| KM_k141_208334 | Not-determined |
| KM_k141_141117 | Not-determined |
| KM_k141_141118 | Not-determined |
| KM_k141_17160 | Not-determined |
| KM_k141_219570 | Not-determined |
| KM_k141_186023 | Not-determined |
| KM_k141_186025 | Not-determined |
| KM_k141_17162 | Not-determined |
| KM_k141_107183 | Not-determined |
| KM_k141_6844 | Not-determined |
| KM_k141_262136 | Not-determined |
| KM_k141_174215 | Not-determined |
| KM_k141_107192 | Not-determined |
| KM_k141_197032 | Not-determined |
| KM_k141_107196 | Not-determined |
| KM_k141_174221 | Low-quality |
| KM_k141_162136 | Not-determined |
| KM_k141_6857 | Not-determined |
| KM_k141_262140 | Not-determined |
| KM_k141_240423 | Low-quality |
| KM_k141_197042 | Not-determined |
| KM_k141_240426 | Not-determined |
| KM_k141_230753 | Not-determined |
| KM_k141_230755 | Not-determined |
| KM_k141_240428 | Not-determined |
| KM_k141_262145 | Not-determined |
| KM_k141_174228 | Not-determined |
| KM_k141_262147 | Not-determined |
| KM_k141_107209 | Not-determined |
| KM_k141_119970 | Low-quality |
| KM_k141_186048 | Not-determined |
| KM_k141_141155 | Low-quality |
| KM_k141_74450 | Not-determined |
| KM_k141_186052 | Not-determined |
| KM_k141_28997 | Not-determined |
| KM_k141_40416 | Not-determined |
| KM_k141_29000 | Not-determined |
| KM_k141_186059 | Not-determined |
| KM_k141_6881 | Not-determined |
| KM_k141_29004 | Not-determined |
| KM_k141_119980 | Low-quality |
| KM_k141_17214 | Low-quality |
| KM_k141_29007 | Not-determined |
| KM_k141_29008 | Not-determined |
| KM_k141_230779 | Low-quality |
| KM_k141_186070 | Not-determined |
| KM_k141_230781 | Not-determined |
| KM_k141_86145 | Not-determined |
| KM_k141_186073 | Not-determined |
| KM_k141_129876 | Not-determined |
| KM_k141_186079 | Not-determined |
| KM_k141_186080 | Not-determined |
| KM_k141_86151 | Not-determined |
| KM_k141_40426 | Not-determined |
| KM_k141_186083 | Not-determined |
| KM_k141_174257 | Not-determined |
| KM_k141_94767 | Not-determined |
| KM_k141_129880 | Not-determined |
| KM_k141_40430 | Not-determined |
| KM_k141_152015 | Not-determined |
| KM_k141_40432 | Not-determined |
| KM_k141_86161 | Not-determined |
| KM_k141_17220 | Not-determined |
| KM_k141_86162 | Not-determined |
| KM_k141_152020 | Not-determined |
| KM_k141_94773 | Low-quality |
| KM_k141_162160 | Low-quality |
| KM_k141_74469 | Not-determined |
| KM_k141_17225 | Low-quality |
| KM_k141_141170 | Not-determined |
| KM_k141_86170 | Low-quality |
| KM_k141_262152 | Not-determined |
| KM_k141_40435 | Not-determined |
| KM_k141_107223 | Low-quality |
| KM_k141_174274 | Not-determined |
| KM_k141_107225 | Not-determined |
| KM_k141_120009 | Not-determined |
| KM_k141_186100 | Not-determined |
| KM_k141_240453 | Not-determined |
| KM_k141_162177 | Not-determined |
| KM_k141_162179 | Not-determined |
| KM_k141_86179 | Not-determined |
| KM_k141_29039 | Not-determined |
| KM_k141_74490 | Not-determined |
| KM_k141_197088 | Not-determined |
| KM_k141_129909 | Not-determined |
| KM_k141_262158 | Not-determined |
| KM_k141_29041 | Not-determined |
| KM_k141_141182 | Not-determined |
| KM_k141_86183 | Not-determined |
| KM_k141_74492 | Not-determined |
| KM_k141_120020 | Not-determined |
| KM_k141_174283 | Low-quality |
| KM_k141_129916 | Not-determined |
| KM_k141_29048 | Not-determined |
| KM_k141_141192 | Not-determined |
| KM_k141_40441 | Not-determined |
| KM_k141_120026 | Not-determined |
| KM_k141_62968 | Not-determined |
| KM_k141_162187 | Not-determined |
| KM_k141_6889 | Not-determined |
| KM_k141_186119 | Not-determined |
| KM_k141_6896 | Not-determined |
| KM_k141_186122 | Low-quality |
| KM_k141_262166 | Not-determined |
| KM_k141_230811 | Not-determined |
| KM_k141_62978 | Not-determined |
| KM_k141_208399 | Not-determined |
| KM_k141_141203 | Not-determined |
| KM_k141_174288 | Low-quality |
| KM_k141_120047 | Not-determined |
| KM_k141_74519 | Not-determined |
| KM_k141_230818 | Not-determined |
| KM_k141_49884 | Not-determined |
| KM_k141_141212 | Low-quality |
| KM_k141_120050 | Not-determined |
| KM_k141_197126 | Not-determined |
| KM_k141_120052 | Not-determined |
| KM_k141_120053 | Not-determined |
| KM_k141_40462 | Not-determined |
| KM_k141_208407 | Not-determined |
| KM_k141_152058 | Not-determined |
| KM_k141_262181 | Not-determined |
| KM_k141_141215 | Not-determined |
| KM_k141_49889 | Not-determined |
| KM_k141_49891 | Not-determined |
| KM_k141_120060 | Not-determined |
| KM_k141_262183 | Not-determined |
| KM_k141_174302 | Not-determined |
| KM_k141_174307 | Not-determined |
| KM_k141_219590 | Not-determined |
| KM_k141_17286 | Not-determined |
| KM_k141_62989 | Not-determined |
| KM_k141_74539 | Not-determined |
| KM_k141_49901 | Not-determined |
| KM_k141_219597 | Not-determined |
| KM_k141_197136 | Low-quality |
| KM_k141_74545 | Not-determined |
| KM_k141_174313 | Not-determined |
| KM_k141_74547 | Not-determined |
| KM_k141_120073 | Not-determined |
| KM_k141_208415 | Not-determined |
| KM_k141_120074 | Not-determined |
| KM_k141_74550 | Low-quality |
| KM_k141_17296 | Low-quality |
| KM_k141_74554 | Not-determined |
| KM_k141_107262 | Low-quality |
| KM_k141_29067 | Not-determined |
| KM_k141_29068 | Not-determined |
| KM_k141_129963 | Not-determined |
| KM_k141_107266 | Not-determined |
| KM_k141_174316 | Not-determined |
| KM_k141_174319 | Not-determined |
| KM_k141_208419 | Not-determined |
| KM_k141_262203 | Not-determined |
| KM_k141_17299 | Low-quality |
| KM_k141_262205 | Not-determined |
| KM_k141_107275 | Not-determined |
| KM_k141_63003 | Not-determined |
| KM_k141_240491 | Not-determined |
| KM_k141_63008 | Not-determined |
| KM_k141_262209 | Not-determined |
| KM_k141_74562 | Not-determined |
| KM_k141_107282 | Not-determined |
| KM_k141_129966 | Not-determined |
| KM_k141_174337 | Not-determined |
| KM_k141_208428 | Not-determined |
| KM_k141_197139 | Not-determined |
| KM_k141_107288 | Not-determined |
| KM_k141_262214 | Not-determined |
| KM_k141_219611 | Not-determined |
| KM_k141_162228 | Not-determined |
| KM_k141_174349 | Not-determined |
| KM_k141_94795 | Not-determined |
| KM_k141_219622 | Low-quality |
| KM_k141_74579 | Low-quality |
| KM_k141_262225 | Not-determined |
| KM_k141_86234 | Not-determined |
| KM_k141_94804 | Not-determined |
| KM_k141_174362 | Not-determined |
| KM_k141_174363 | Not-determined |
| KM_k141_63036 | Not-determined |
| KM_k141_63041 | Not-determined |
| KM_k141_49920 | Low-quality |
| KM_k141_141258 | Not-determined |
| KM_k141_240535 | Not-determined |
| KM_k141_17323 | Not-determined |
| KM_k141_208462 | Not-determined |
| KM_k141_162264 | Not-determined |
| KM_k141_174382 | Not-determined |
| KM_k141_63053 | Low-quality |
| KM_k141_120111 | Not-determined |
| KM_k141_17326 | Low-quality |
| KM_k141_250770 | Not-determined |
| KM_k141_17330 | Not-determined |
| KM_k141_219635 | Not-determined |
| KM_k141_17332 | Not-determined |
| KM_k141_250771 | Not-determined |
| KM_k141_230875 | Low-quality |
| KM_k141_230877 | Not-determined |
| KM_k141_17341 | Not-determined |
| KM_k141_208471 | Not-determined |
| KM_k141_186192 | Not-determined |
| KM_k141_186193 | Not-determined |
| KM_k141_240556 | Not-determined |
| KM_k141_208475 | Not-determined |
| KM_k141_186197 | Not-determined |
| KM_k141_230890 | Not-determined |
| KM_k141_86259 | Low-quality |
| KM_k141_230891 | Not-determined |
| KM_k141_208487 | Low-quality |
| KM_k141_186210 | Not-determined |
| KM_k141_208488 | Not-determined |
| KM_k141_186214 | Not-determined |
| KM_k141_250781 | Not-determined |
| KM_k141_94809 | Not-determined |
| KM_k141_219647 | Not-determined |
| KM_k141_94813 | Not-determined |
| KM_k141_250787 | Low-quality |
| KM_k141_230895 | Not-determined |
| KM_k141_120167 | Not-determined |
| KM_k141_120169 | Not-determined |
| KM_k141_174425 | Not-determined |
| KM_k141_141282 | Not-determined |
| KM_k141_86274 | Not-determined |
| KM_k141_120178 | Not-determined |
| KM_k141_197175 | Not-determined |
| KM_k141_197176 | Not-determined |
| KM_k141_186242 | Not-determined |
| KM_k141_186243 | Not-determined |
| KM_k141_120182 | Not-determined |
| KM_k141_74646 | Not-determined |
| KM_k141_74648 | Not-determined |
| KM_k141_197182 | Not-determined |
| KM_k141_250820 | Not-determined |
| KM_k141_86302 | Not-determined |
| KM_k141_74651 | Not-determined |
| KM_k141_120189 | Not-determined |
| KM_k141_230922 | Not-determined |
| KM_k141_197186 | Not-determined |
| KM_k141_230924 | Not-determined |
| KM_k141_86309 | Not-determined |
| KM_k141_120193 | Not-determined |
| KM_k141_174437 | Not-determined |
| KM_k141_162302 | Not-determined |
| KM_k141_107308 | Not-determined |
| KM_k141_141321 | Not-determined |
| 18-104_k141_141607 | Medium-quality |
| KM_k141_29083 | Not-determined |
| KM_k141_197195 | Not-determined |
| KM_k141_74656 | Not-determined |
| KM_k141_186257 | Not-determined |
| KM_k141_141329 | Low-quality |
| KM_k141_29090 | Not-determined |
| KM_k141_262280 | Not-determined |
| KM_k141_250837 | Not-determined |
| KM_k141_197201 | Not-determined |
| KM_k141_230929 | Not-determined |
| KM_k141_29092 | Not-determined |
| KM_k141_230931 | Low-quality |
| KM_k141_186264 | Not-determined |
| KM_k141_219690 | Not-determined |
| KM_k141_49981 | Not-determined |
| KM_k141_49984 | Not-determined |
| KM_k141_74672 | Not-determined |
| KM_k141_29103 | Not-determined |
| KM_k141_74675 | Not-determined |
| KM_k141_240605 | Not-determined |
| KM_k141_74678 | Not-determined |
| KM_k141_240606 | Not-determined |
| KM_k141_63101 | Not-determined |
| KM_k141_17379 | Not-determined |
| KM_k141_74679 | Not-determined |
| KM_k141_197211 | Not-determined |
| KM_k141_74680 | Low-quality |
| KM_k141_197213 | Not-determined |
| KM_k141_240608 | Low-quality |
| KM_k141_74684 | Not-determined |
| KM_k141_240610 | Not-determined |
| KM_k141_86331 | Not-determined |
| KM_k141_29115 | Not-determined |
| KM_k141_240616 | Not-determined |
| KM_k141_141365 | Not-determined |
| KM_k141_49993 | Not-determined |
| KM_k141_94840 | Low-quality |
| KM_k141_63114 | Not-determined |
| KM_k141_208548 | Not-determined |
| KM_k141_50000 | Low-quality |
| KM_k141_107329 | Not-determined |
| KM_k141_262302 | Not-determined |
| KM_k141_208556 | Not-determined |
| KM_k141_262303 | Not-determined |
| KM_k141_219708 | Not-determined |
| KM_k141_219709 | Not-determined |
| KM_k141_162338 | Not-determined |
| KM_k141_6910 | Not-determined |
| KM_k141_162340 | Not-determined |
| KM_k141_17400 | Not-determined |
| KM_k141_162344 | Not-determined |
| KM_k141_29124 | Not-determined |
| KM_k141_17407 | Not-determined |
| KM_k141_208562 | Not-determined |
| KM_k141_17409 | Not-determined |
| KM_k141_17410 | Not-determined |
| KM_k141_240633 | Not-determined |
| KM_k141_174471 | Not-determined |
| KM_k141_208567 | Not-determined |
| KM_k141_219727 | Not-determined |
| KM_k141_152185 | Not-determined |
| KM_k141_208569 | Not-determined |
| KM_k141_152186 | Not-determined |
| KM_k141_152187 | Not-determined |
| KM_k141_219728 | Not-determined |
| KM_k141_107357 | Not-determined |
| KM_k141_240642 | Not-determined |
| KM_k141_174475 | Low-quality |
| KM_k141_174479 | Not-determined |
| KM_k141_230954 | Not-determined |
| KM_k141_186275 | Not-determined |
| KM_k141_162362 | Not-determined |
| KM_k141_86364 | Not-determined |
| KM_k141_152200 | Not-determined |
| KM_k141_152201 | Not-determined |
| KM_k141_29143 | Low-quality |
| KM_k141_50016 | Not-determined |
| KM_k141_174492 | Not-determined |
| KM_k141_141411 | Not-determined |
| KM_k141_63133 | Not-determined |
| KM_k141_174496 | Not-determined |
| KM_k141_152212 | Not-determined |
| KM_k141_152215 | Not-determined |
| KM_k141_262335 | Not-determined |
| KM_k141_230964 | Not-determined |
| KM_k141_17437 | Not-determined |
| KM_k141_94858 | Not-determined |
| KM_k141_250867 | Not-determined |
| KM_k141_250871 | Not-determined |
| KM_k141_240665 | Not-determined |
| KM_k141_174509 | Not-determined |
| KM_k141_262344 | Not-determined |
| KM_k141_50031 | Not-determined |
| KM_k141_186298 | Not-determined |
| KM_k141_107373 | Not-determined |
| KM_k141_174512 | Not-determined |
| KM_k141_197275 | Not-determined |
| KM_k141_17447 | Not-determined |
| KM_k141_262349 | Not-determined |
| KM_k141_174517 | Not-determined |
| KM_k141_86386 | Not-determined |
| KM_k141_29153 | Not-determined |
| KM_k141_230975 | Not-determined |
| KM_k141_262352 | Not-determined |
| KM_k141_240671 | Not-determined |
| KM_k141_107383 | Not-determined |
| KM_k141_208614 | Not-determined |
| KM_k141_86401 | Low-quality |
| KM_k141_141440 | Not-determined |
| KM_k141_74693 | Not-determined |
| KM_k141_107384 | Low-quality |
| KM_k141_50045 | Not-determined |
| KM_k141_208621 | Not-determined |
| KM_k141_208622 | Not-determined |
| KM_k141_94873 | Not-determined |
| KM_k141_74702 | Not-determined |
| KM_k141_107391 | Low-quality |
| KM_k141_250896 | Not-determined |
| KM_k141_50051 | Not-determined |
| KM_k141_219792 | Not-determined |
| KM_k141_152240 | Not-determined |
| KM_k141_262360 | Low-quality |
| KM_k141_107399 | Not-determined |
| KM_k141_17456 | Not-determined |
| KM_k141_86417 | Not-determined |
| KM_k141_240690 | Not-determined |
| KM_k141_219802 | Not-determined |
| KM_k141_240691 | Not-determined |
| KM_k141_208645 | Not-determined |
| KM_k141_262365 | Not-determined |
| KM_k141_86422 | Not-determined |
| KM_k141_152247 | Not-determined |
| KM_k141_50064 | Not-determined |
| KM_k141_231002 | Not-determined |
| KM_k141_174562 | Not-determined |
| KM_k141_74730 | Not-determined |
| KM_k141_74734 | Not-determined |
| KM_k141_174568 | Not-determined |
| KM_k141_94890 | Low-quality |
| KM_k141_240703 | Not-determined |
| KM_k141_74737 | Not-determined |
| KM_k141_29162 | Not-determined |
| KM_k141_152252 | Not-determined |
| KM_k141_29164 | Not-determined |
| KM_k141_50070 | Not-determined |
| KM_k141_120196 | Low-quality |
| KM_k141_197314 | Not-determined |
| KM_k141_219821 | Not-determined |
| KM_k141_197318 | Not-determined |
| KM_k141_174578 | Not-determined |
| KM_k141_197322 | Not-determined |
| KM_k141_120202 | Not-determined |
| KM_k141_120203 | Not-determined |
| KM_k141_29172 | Not-determined |
| KM_k141_50079 | Not-determined |
| KM_k141_120206 | Not-determined |
| KM_k141_262385 | Not-determined |
| KM_k141_197337 | Not-determined |
| KM_k141_29180 | Not-determined |
| KM_k141_120215 | Low-quality |
| KM_k141_63203 | Not-determined |
| KM_k141_197345 | Not-determined |
| KM_k141_197350 | Not-determined |
| KM_k141_197352 | Not-determined |
| KM_k141_231032 | Not-determined |
| KM_k141_29194 | Not-determined |
| KM_k141_17469 | Not-determined |
| KM_k141_231038 | Not-determined |
| KM_k141_29195 | Not-determined |
| KM_k141_231039 | Not-determined |
| KM_k141_250903 | Not-determined |
| KM_k141_219852 | Not-determined |
| KM_k141_174617 | Not-determined |
| KM_k141_120238 | Not-determined |
| KM_k141_208700 | Not-determined |
| KM_k141_120239 | Not-determined |
| KM_k141_86440 | Not-determined |
| KM_k141_74766 | Not-determined |
| KM_k141_240720 | Not-determined |
| KM_k141_208705 | Not-determined |
| KM_k141_17478 | Not-determined |
| KM_k141_240724 | Not-determined |
| KM_k141_250918 | Not-determined |
| KM_k141_240725 | Not-determined |
| KM_k141_107474 | Low-quality |
| KM_k141_174636 | Not-determined |
| KM_k141_262425 | Not-determined |
| KM_k141_262426 | Not-determined |
| KM_k141_208720 | Not-determined |
| KM_k141_74783 | Not-determined |
| KM_k141_250928 | Not-determined |
| KM_k141_17486 | Not-determined |
| KM_k141_231051 | Low-quality |
| KM_k141_6931 | Not-determined |
| KM_k141_262433 | Not-determined |
| KM_k141_29223 | Not-determined |
| KM_k141_29228 | Not-determined |
| KM_k141_231055 | Not-determined |
| KM_k141_141484 | Not-determined |
| KM_k141_86449 | Low-quality |
| KM_k141_208727 | Not-determined |
| KM_k141_240747 | Not-determined |
| KM_k141_17494 | Not-determined |
| KM_k141_240749 | Not-determined |
| KM_k141_240750 | Not-determined |
| KM_k141_17495 | Not-determined |
| KM_k141_29236 | Not-determined |
| KM_k141_262450 | Low-quality |
| KM_k141_17501 | Not-determined |
| KM_k141_94911 | Not-determined |
| KM_k141_120267 | Not-determined |
| KM_k141_94913 | Not-determined |
| KM_k141_63246 | Not-determined |
| KM_k141_152318 | Not-determined |
| KM_k141_17506 | Not-determined |
| KM_k141_152319 | Not-determined |
| KM_k141_197422 | Not-determined |
| KM_k141_94919 | Not-determined |
| KM_k141_86462 | Not-determined |
| KM_k141_86463 | Not-determined |
| KM_k141_197427 | Not-determined |
| KM_k141_63254 | Not-determined |
| KM_k141_231085 | Not-determined |
| KM_k141_208747 | Low-quality |
| KM_k141_240768 | Not-determined |
| KM_k141_231091 | Not-determined |
| KM_k141_107505 | Not-determined |
| KM_k141_262467 | Low-quality |
| KM_k141_17526 | Not-determined |
| KM_k141_107509 | Not-determined |
| KM_k141_197434 | Not-determined |
| KM_k141_74802 | Not-determined |
| KM_k141_86485 | Not-determined |
| KM_k141_231097 | Not-determined |
| KM_k141_29254 | Not-determined |
| KM_k141_152336 | Not-determined |
| KM_k141_29257 | Not-determined |

| | |
|---|---|
| KM_k141_162407 | Not-determined |
| KM_k141_262475 | Not-determined |
| KM_k141_197440 | Not-determined |
| KM_k141_152340 | Not-determined |
| KM_k141_174680 | Low-quality |
| KM_k141_174684 | Not-determined |
| KM_k141_250950 | Not-determined |
| KM_k141_86491 | Low-quality |
| KM_k141_219866 | Not-determined |
| KM_k141_141501 | Not-determined |
| KM_k141_197459 | Not-determined |
| KM_k141_94949 | Not-determined |
| KM_k141_197463 | Not-determined |
| KM_k141_186319 | Low-quality |
| KM_k141_120297 | Not-determined |
| KM_k141_208772 | Not-determined |
| KM_k141_6938 | Not-determined |
| KM_k141_6940 | Not-determined |
| KM_k141_63270 | Not-determined |
| KM_k141_29290 | Not-determined |
| KM_k141_250974 | Not-determined |
| KM_k141_219896 | Not-determined |
| KM_k141_29294 | Not-determined |
| KM_k141_141525 | Low-quality |
| KM_k141_208780 | Not-determined |
| KM_k141_197475 | Low-quality |
| KM_k141_50101 | Not-determined |
| KM_k141_162448 | Not-determined |
| KM_k141_29300 | Not-determined |
| KM_k141_94956 | Not-determined |
| KM_k141_94958 | Not-determined |
| KM_k141_74841 | Not-determined |
| KM_k141_219904 | Not-determined |
| KM_k141_74843 | Not-determined |
| KM_k141_63282 | Not-determined |
| KM_k141_29303 | Not-determined |
| KM_k141_240803 | Not-determined |
| KM_k141_174715 | Not-determined |
| KM_k141_74850 | Not-determined |
| KM_k141_50102 | Not-determined |
| KM_k141_197488 | Not-determined |
| KM_k141_174720 | Not-determined |
| KM_k141_6953 | Low-quality |
| KM_k141_197490 | Not-determined |
| KM_k141_262515 | Low-quality |
| KM_k141_208790 | Not-determined |
| KM_k141_162463 | Not-determined |
| KM_k141_17549 | Not-determined |
| KM_k141_94968 | Not-determined |
| KM_k141_186339 | Not-determined |
| KM_k141_231120 | Not-determined |
| KM_k141_219910 | Not-determined |
| KM_k141_17554 | Not-determined |
| KM_k141_262529 | Not-determined |
| KM_k141_17556 | Not-determined |
| KM_k141_17557 | Not-determined |
| KM_k141_197506 | Not-determined |
| KM_k141_94976 | Not-determined |
| KM_k141_162478 | Low-quality |
| KM_k141_74866 | Not-determined |
| KM_k141_74868 | Not-determined |
| KM_k141_231130 | Not-determined |
| KM_k141_262534 | Not-determined |
| KM_k141_107558 | Low-quality |
| KM_k141_29327 | Not-determined |
| KM_k141_86519 | Not-determined |
| KM_k141_17566 | Not-determined |
| KM_k141_197514 | Not-determined |
| KM_k141_162484 | Not-determined |
| KM_k141_174733 | Not-determined |
| KM_k141_74877 | Not-determined |
| KM_k141_152368 | Not-determined |
| KM_k141_107567 | Not-determined |
| KM_k141_107569 | Not-determined |
| KM_k141_240826 | Not-determined |
| KM_k141_107571 | Not-determined |
| KM_k141_141540 | Not-determined |
| KM_k141_120310 | Not-determined |
| KM_k141_174744 | Not-determined |
| KM_k141_17578 | Not-determined |
| KM_k141_186373 | Not-determined |
| KM_k141_86534 | Not-determined |
| KM_k141_208815 | Not-determined |
| KM_k141_86537 | Not-determined |
| KM_k141_120318 | Not-determined |
| KM_k141_50130 | Not-determined |
| KM_k141_197518 | Not-determined |
| KM_k141_141551 | Not-determined |
| KM_k141_141555 | Not-determined |
| KM_k141_6991 | Not-determined |
| KM_k141_86548 | Not-determined |
| KM_k141_6995 | Not-determined |
| KM_k141_50138 | Not-determined |
| KM_k141_208840 | Not-determined |
| KM_k141_74895 | Not-determined |
| KM_k141_107591 | Not-determined |
| KM_k141_208842 | Low-quality |
| KM_k141_74898 | Not-determined |
| KM_k141_74901 | Not-determined |
| KM_k141_120348 | Not-determined |
| KM_k141_120352 | Not-determined |
| KM_k141_231158 | Not-determined |
| KM_k141_50145 | Low-quality |
| KM_k141_120358 | Not-determined |
| KM_k141_231167 | Low-quality |
| KM_k141_63330 | Not-determined |
| KM_k141_29349 | Not-determined |
| KM_k141_7000 | Not-determined |
| KM_k141_197537 | Not-determined |
| KM_k141_94990 | Not-determined |
| KM_k141_219966 | Not-determined |
| KM_k141_219968 | Not-determined |
| KM_k141_63340 | Not-determined |
| KM_k141_240855 | Not-determined |
| KM_k141_120369 | Not-determined |
| KM_k141_162516 | Not-determined |
| KM_k141_141581 | Not-determined |
| KM_k141_208857 | Not-determined |
| KM_k141_186391 | Not-determined |
| KM_k141_141583 | Not-determined |
| KM_k141_86565 | Not-determined |
| KM_k141_29360 | Not-determined |
| KM_k141_29362 | Not-determined |
| KM_k141_231173 | Not-determined |
| KM_k141_186395 | Not-determined |
| KM_k141_174780 | Not-determined |
| KM_k141_74923 | Low-quality |
| KM_k141_50159 | Low-quality |
| KM_k141_63354 | Not-determined |
| KM_k141_240868 | Not-determined |
| KM_k141_29370 | Not-determined |
| KM_k141_186407 | Not-determined |
| KM_k141_240871 | Not-determined |
| KM_k141_63360 | Not-determined |
| KM_k141_86581 | Not-determined |
| KM_k141_120384 | Not-determined |
| KM_k141_86582 | Not-determined |
| KM_k141_50166 | Not-determined |
| KM_k141_107626 | Not-determined |
| KM_k141_162538 | Not-determined |
| KM_k141_50168 | Not-determined |
| KM_k141_63366 | Low-quality |
| KM_k141_240879 | Not-determined |
| KM_k141_162544 | Not-determined |
| KM_k141_197565 | Not-determined |
| KM_k141_86590 | Not-determined |
| KM_k141_120387 | Low-quality |
| KM_k141_107631 | Not-determined |
| KM_k141_197566 | Not-determined |
| KM_k141_120392 | Low-quality |
| KM_k141_208867 | Not-determined |
| KM_k141_141600 | Not-determined |
| KM_k141_208868 | Not-determined |
| KM_k141_50183 | Not-determined |
| KM_k141_86604 | Not-determined |
| KM_k141_74951 | Not-determined |
| KM_k141_74952 | Not-determined |
| KM_k141_174803 | Not-determined |
| KM_k141_152418 | Not-determined |
| KM_k141_74959 | Not-determined |
| KM_k141_107643 | Not-determined |
| KM_k141_63393 | Not-determined |
| KM_k141_29396 | Not-determined |
| KM_k141_186445 | Not-determined |
| KM_k141_174809 | Not-determined |
| KM_k141_107647 | Not-determined |
| KM_k141_152426 | Not-determined |
| KM_k141_174815 | Not-determined |
| KM_k141_186449 | Not-determined |
| KM_k141_141617 | Not-determined |
| KM_k141_186450 | Not-determined |
| KM_k141_120402 | Not-determined |
| KM_k141_152432 | Low-quality |
| KM_k141_152433 | Not-determined |
| KM_k141_120406 | Not-determined |
| KM_k141_107650 | Not-determined |
| KM_k141_141622 | Not-determined |
| KM_k141_186456 | Not-determined |
| KM_k141_74968 | Not-determined |
| KM_k141_231226 | Not-determined |
| KM_k141_208887 | Not-determined |
| KM_k141_208890 | Not-determined |
| KM_k141_63395 | Not-determined |
| KM_k141_174823 | Low-quality |
| KM_k141_197593 | Not-determined |
| KM_k141_220000 | Not-determined |
| KM_k141_220002 | Not-determined |
| KM_k141_186467 | Not-determined |
| KM_k141_208893 | Not-determined |
| KM_k141_197605 | Not-determined |
| KM_k141_220009 | Not-determined |
| KM_k141_141636 | Not-determined |
| KM_k141_29424 | Not-determined |
| KM_k141_251016 | Not-determined |
| KM_k141_17639 | Not-determined |
| KM_k141_197607 | Not-determined |
| KM_k141_17645 | Low-quality |
| KM_k141_240914 | Not-determined |
| KM_k141_95035 | Not-determined |
| KM_k141_74977 | Not-determined |
| KM_k141_174834 | Not-determined |
| KM_k141_95038 | Not-determined |
| KM_k141_208907 | Not-determined |
| KM_k141_95041 | Not-determined |
| KM_k141_50218 | Not-determined |
| KM_k141_162574 | Not-determined |
| KM_k141_74988 | Not-determined |
| KM_k141_107665 | Not-determined |
| KM_k141_86639 | Not-determined |
| KM_k141_95048 | Not-determined |
| KM_k141_50227 | Not-determined |
| KM_k141_107666 | Not-determined |
| KM_k141_141651 | Not-determined |
| KM_k141_120428 | Not-determined |
| KM_k141_231258 | Not-determined |
| KM_k141_220022 | Low-quality |
| KM_k141_231264 | Not-determined |
| KM_k141_29439 | Not-determined |
| KM_k141_231266 | Not-determined |
| KM_k141_50231 | Not-determined |
| KM_k141_152438 | Not-determined |
| KM_k141_50232 | Not-determined |
| KM_k141_86650 | Not-determined |
| KM_k141_120429 | Low-quality |
| KM_k141_95056 | Not-determined |
| KM_k141_50237 | Not-determined |
| KM_k141_231270 | Not-determined |
| KM_k141_186498 | Not-determined |
| KM_k141_141663 | Not-determined |
| KM_k141_141667 | Not-determined |
| KM_k141_86651 | Low-quality |
| KM_k141_120433 | Not-determined |
| KM_k141_75004 | Not-determined |
| KM_k141_220044 | Not-determined |
| KM_k141_162593 | Not-determined |
| KM_k141_240931 | Not-determined |
| KM_k141_29453 | Not-determined |
| KM_k141_7028 | Not-determined |
| KM_k141_86656 | Low-quality |
| KM_k141_141671 | Not-determined |
| KM_k141_7030 | Not-determined |
| KM_k141_95060 | Not-determined |
| KM_k141_141672 | Not-determined |
| KM_k141_7034 | Not-determined |
| KM_k141_231284 | Not-determined |
| KM_k141_162597 | Not-determined |
| KM_k141_107686 | Not-determined |
| KM_k141_220050 | Not-determined |
| KM_k141_40493 | Not-determined |
| KM_k141_107688 | Not-determined |
| KM_k141_197613 | Not-determined |
| KM_k141_152454 | Low-quality |
| KM_k141_197618 | Not-determined |
| KM_k141_29462 | Not-determined |
| KM_k141_75013 | Low-quality |
| KM_k141_7039 | Low-quality |
| KM_k141_231290 | Not-determined |
| KM_k141_63413 | Not-determined |
| KM_k141_208931 | Low-quality |
| KM_k141_174868 | Not-determined |
| KM_k141_162606 | Not-determined |
| KM_k141_162609 | Not-determined |
| KM_k141_220064 | Not-determined |
| KM_k141_262594 | Not-determined |
| KM_k141_95071 | Not-determined |
| KM_k141_40513 | Not-determined |
| KM_k141_63425 | Not-determined |
| KM_k141_162615 | Low-quality |
| KM_k141_29477 | Not-determined |
| KM_k141_40517 | Not-determined |
| KM_k141_162618 | Not-determined |
| KM_k141_63429 | Not-determined |
| KM_k141_162619 | Not-determined |
| KM_k141_262598 | Not-determined |
| KM_k141_75017 | Not-determined |
| KM_k141_262600 | Not-determined |
| KM_k141_86665 | Not-determined |
| KM_k141_63432 | Not-determined |
| KM_k141_95085 | Not-determined |
| KM_k141_208951 | Not-determined |
| KM_k141_197644 | Not-determined |
| KM_k141_197648 | Not-determined |
| KM_k141_95091 | Low-quality |
| KM_k141_208959 | Not-determined |
| KM_k141_208961 | Not-determined |
| KM_k141_220089 | Not-determined |
| KM_k141_95095 | Not-determined |
| KM_k141_220091 | Not-determined |
| KM_k141_197654 | Not-determined |
| KM_k141_120484 | Not-determined |
| KM_k141_186534 | Not-determined |
| KM_k141_152470 | Not-determined |
| KM_k141_63453 | Not-determined |
| KM_k141_29507 | Not-determined |
| KM_k141_120486 | Not-determined |
| KM_k141_220095 | Not-determined |
| KM_k141_120489 | Not-determined |
| KM_k141_63459 | Not-determined |
| KM_k141_162626 | Not-determined |
| KM_k141_197662 | Not-determined |
| KM_k141_186548 | Not-determined |
| KM_k141_208982 | Not-determined |
| KM_k141_240978 | Not-determined |
| KM_k141_7062 | Not-determined |
| KM_k141_141706 | Not-determined |
| KM_k141_86689 | Not-determined |
| KM_k141_63468 | Not-determined |
| KM_k141_152485 | Low-quality |
| KM_k141_86696 | Not-determined |
| KM_k141_220110 | Not-determined |
| KM_k141_7074 | Not-determined |
| KM_k141_63479 | Not-determined |
| KM_k141_86703 | Not-determined |
| KM_k141_262610 | Not-determined |
| KM_k141_240987 | Not-determined |
| KM_k141_220120 | Not-determined |
| KM_k141_40559 | Not-determined |
| KM_k141_63486 | Not-determined |
| KM_k141_186571 | Low-quality |
| KM_k141_262618 | Not-determined |
| KM_k141_7081 | Not-determined |
| KM_k141_152503 | Not-determined |
| KM_k141_152507 | Not-determined |
| KM_k141_209004 | Not-determined |
| KM_k141_209006 | Not-determined |
| KM_k141_186583 | Low-quality |
| KM_k141_262625 | Not-determined |
| KM_k141_75059 | Not-determined |
| KM_k141_7091 | Not-determined |
| KM_k141_197682 | Low-quality |
| KM_k141_231307 | Low-quality |
| KM_k141_197685 | Not-determined |
| KM_k141_186592 | Not-determined |
| KM_k141_50281 | Not-determined |
| KM_k141_17698 | Not-determined |
| KM_k141_86723 | Not-determined |
| KM_k141_129974 | Not-determined |
| KM_k141_162658 | Not-determined |
| KM_k141_231325 | Not-determined |
| KM_k141_241016 | Not-determined |
| KM_k141_241021 | Not-determined |
| KM_k141_17712 | Not-determined |
| KM_k141_220151 | Not-determined |
| KM_k141_220153 | Not-determined |
| KM_k141_220156 | Not-determined |
| KM_k141_220159 | Not-determined |
| KM_k141_152538 | Not-determined |
| KM_k141_220162 | Not-determined |
| KM_k141_129986 | Low-quality |
| KM_k141_152542 | Not-determined |
| KM_k141_186603 | Not-determined |
| KM_k141_162669 | Low-quality |
| KM_k141_186604 | Not-determined |
| KM_k141_50290 | Not-determined |
| KM_k141_107710 | Not-determined |
| KM_k141_107711 | Not-determined |
| KM_k141_129994 | Not-determined |
| KM_k141_241034 | Not-determined |
| KM_k141_197713 | Not-determined |
| KM_k141_29543 | Not-determined |
| KM_k141_129996 | Not-determined |
| KM_k141_186616 | Not-determined |
| KM_k141_197718 | Not-determined |
| KM_k141_63513 | Not-determined |
| KM_k141_174894 | Not-determined |
| KM_k141_40582 | Not-determined |
| KM_k141_129998 | Not-determined |
| KM_k141_174901 | Not-determined |
| KM_k141_231350 | Not-determined |
| KM_k141_174904 | Not-determined |
| KM_k141_231351 | Not-determined |
| KM_k141_162677 | Not-determined |
| KM_k141_251039 | Not-determined |
| KM_k141_17722 | Not-determined |
| KM_k141_86755 | Not-determined |
| KM_k141_17724 | Not-determined |
| KM_k141_231357 | Not-determined |
| KM_k141_29567 | Not-determined |
| KM_k141_130019 | Not-determined |
| KM_k141_186635 | Not-determined |
| KM_k141_186636 | Not-determined |
| KM_k141_29569 | Not-determined |
| KM_k141_17731 | Not-determined |
| KM_k141_63533 | Not-determined |
| KM_k141_29572 | Not-determined |
| KM_k141_130031 | Not-determined |
| KM_k141_141749 | Not-determined |
| KM_k141_141752 | Not-determined |
| KM_k141_241048 | Not-determined |
| KM_k141_86766 | Not-determined |
| KM_k141_220181 | Not-determined |
| KM_k141_40601 | Not-determined |
| KM_k141_107740 | Not-determined |
| KM_k141_120514 | Not-determined |
| KM_k141_7143 | Not-determined |
| KM_k141_40607 | Not-determined |
| KM_k141_209028 | Not-determined |
| KM_k141_251062 | Not-determined |
| KM_k141_174917 | Not-determined |
| KM_k141_209038 | Not-determined |
| KM_k141_251072 | Not-determined |
| KM_k141_40613 | Not-determined |
| KM_k141_7152 | Not-determined |
| KM_k141_231381 | Not-determined |
| KM_k141_50326 | Not-determined |
| KM_k141_95108 | Not-determined |
| KM_k141_174939 | Not-determined |
| KM_k141_220208 | Low-quality |
| KM_k141_141768 | Not-determined |
| KM_k141_162719 | Not-determined |
| KM_k141_141770 | Not-determined |
| KM_k141_50333 | Not-determined |
| KM_k141_231397 | Not-determined |
| KM_k141_120544 | Not-determined |
| KM_k141_251088 | Not-determined |
| KM_k141_231401 | Not-determined |
| KM_k141_231402 | Not-determined |
| KM_k141_7164 | Not-determined |
| KM_k141_50342 | Low-quality |
| KM_k141_75142 | Not-determined |
| KM_k141_251096 | Not-determined |
| KM_k141_29600 | Not-determined |
| KM_k141_75148 | Not-determined |
| KM_k141_75152 | Not-determined |
| KM_k141_63583 | Not-determined |
| KM_k141_63586 | Not-determined |
| KM_k141_209089 | Not-determined |
| KM_k141_197730 | Low-quality |
| KM_k141_120562 | Low-quality |
| KM_k141_40618 | Not-determined |
| KM_k141_120564 | Not-determined |
| KM_k141_209096 | Not-determined |
| KM_k141_107764 | Not-determined |
| KM_k141_120569 | Not-determined |
| KM_k141_63588 | Low-quality |
| KM_k141_29612 | Not-determined |
| KM_k141_75173 | Low-quality |
| KM_k141_63591 | Not-determined |
| KM_k141_75174 | Not-determined |
| KM_k141_220229 | Not-determined |
| KM_k141_50365 | Low-quality |
| KM_k141_220230 | Not-determined |
| KM_k141_95130 | Not-determined |
| KM_k141_162729 | Not-determined |
| KM_k141_75181 | Not-determined |
| KM_k141_75183 | Not-determined |
| KM_k141_220235 | Not-determined |
| KM_k141_63596 | Not-determined |
| KM_k141_220237 | Not-determined |
| KM_k141_241082 | Low-quality |
| KM_k141_241083 | Not-determined |
| KM_k141_231435 | Not-determined |
| KM_k141_120585 | Not-determined |
| KM_k141_220240 | Not-determined |
| KM_k141_95142 | Not-determined |
| KM_k141_86841 | Not-determined |
| KM_k141_95144 | Not-determined |
| KM_k141_86842 | Not-determined |
| KM_k141_63601 | Low-quality |
| KM_k141_7200 | Not-determined |
| KM_k141_120593 | Not-determined |
| KM_k141_231441 | Not-determined |
| KM_k141_107788 | Not-determined |
| KM_k141_162743 | Not-determined |
| KM_k141_231443 | Not-determined |
| KM_k141_220248 | Not-determined |
| KM_k141_63609 | Not-determined |
| KM_k141_63614 | Not-determined |
| KM_k141_251127 | Not-determined |
| KM_k141_209111 | Not-determined |
| KM_k141_120607 | Not-determined |
| KM_k141_197756 | Not-determined |
| KM_k141_7212 | Not-determined |
| KM_k141_40642 | Not-determined |
| KM_k141_7214 | Not-determined |
| KM_k141_174999 | Not-determined |
| KM_k141_220256 | Not-determined |
| KM_k141_175000 | Not-determined |
| KM_k141_241115 | Not-determined |
| KM_k141_50370 | Not-determined |
| KM_k141_40650 | Not-determined |
| KM_k141_186691 | Not-determined |
| KM_k141_186692 | Not-determined |
| KM_k141_95159 | Not-determined |
| KM_k141_231466 | Not-determined |
| KM_k141_17813 | Not-determined |
| KM_k141_197782 | Not-determined |
| KM_k141_197784 | Not-determined |
| KM_k141_141788 | Not-determined |
| KM_k141_251153 | Not-determined |
| KM_k141_220271 | Not-determined |
| KM_k141_162757 | Not-determined |
| KM_k141_209126 | Not-determined |
| KM_k141_50389 | Not-determined |
| KM_k141_141793 | Low-quality |
| KM_k141_241125 | Not-determined |
| KM_k141_241126 | Not-determined |
| KM_k141_75211 | Not-determined |
| KM_k141_162761 | Not-determined |
| KM_k141_29649 | Not-determined |
| KM_k141_251170 | Not-determined |
| KM_k141_141809 | Not-determined |
| KM_k141_175018 | Not-determined |
| KM_k141_63643 | Not-determined |
| KM_k141_251174 | Not-determined |
| KM_k141_120634 | Not-determined |
| KM_k141_95174 | Not-determined |
| KM_k141_29653 | Not-determined |
| KM_k141_29654 | Not-determined |
| KM_k141_17831 | Not-determined |
| KM_k141_17835 | Not-determined |
| KM_k141_120646 | Not-determined |
| KM_k141_75238 | Not-determined |
| KM_k141_241132 | Not-determined |
| KM_k141_162775 | Not-determined |
| KM_k141_152563 | Not-determined |
| KM_k141_231508 | Low-quality |
| KM_k141_75245 | Not-determined |
| KM_k141_152567 | Not-determined |
| KM_k141_120661 | Not-determined |
| KM_k141_7269 | Not-determined |
| KM_k141_241145 | Not-determined |
| KM_k141_7271 | Not-determined |
| KM_k141_209134 | Not-determined |
| KM_k141_29667 | Not-determined |
| KM_k141_29668 | Low-quality |
| KM_k141_197820 | Not-determined |
| KM_k141_241150 | Not-determined |
| KM_k141_220290 | Not-determined |
| KM_k141_251198 | Not-determined |
| KM_k141_120668 | Not-determined |
| KM_k141_120669 | Not-determined |
| KM_k141_120670 | Not-determined |
| KM_k141_29676 | Not-determined |
| KM_k141_75257 | Not-determined |
| KM_k141_29679 | Not-determined |
| KM_k141_75263 | Not-determined |
| KM_k141_231523 | Not-determined |
| KM_k141_231524 | Not-determined |
| KM_k141_7290 | Not-determined |
| KM_k141_241163 | Low-quality |
| KM_k141_40710 | Not-determined |
| KM_k141_120683 | Not-determined |
| KM_k141_186699 | Not-determined |
| KM_k141_40713 | Not-determined |
| KM_k141_40718 | Not-determined |
| KM_k141_75280 | Not-determined |
| KM_k141_120698 | Not-determined |
| KM_k141_29693 | Not-determined |
| KM_k141_209151 | Not-determined |
| KM_k141_120710 | Not-determined |
| KM_k141_7298 | Not-determined |
| KM_k141_241191 | Not-determined |
| KM_k141_75291 | Not-determined |
| KM_k141_63659 | Low-quality |
| KM_k141_86864 | Not-determined |
| KM_k141_63662 | Not-determined |
| KM_k141_29698 | Not-determined |
| KM_k141_209161 | Not-determined |
| KM_k141_40728 | Not-determined |
| KM_k141_241195 | Low-quality |
| KM_k141_241197 | Not-determined |
| KM_k141_29705 | Not-determined |
| KM_k141_40733 | Not-determined |
| KM_k141_50415 | Not-determined |
| KM_k141_40738 | Not-determined |
| KM_k141_107815 | Not-determined |
| KM_k141_7315 | Not-determined |
| KM_k141_231561 | Not-determined |
| KM_k141_107820 | Not-determined |
| KM_k141_120725 | Not-determined |
| KM_k141_209166 | Not-determined |
| KM_k141_86878 | Not-determined |
| KM_k141_17871 | Low-quality |
| KM_k141_29711 | Not-determined |
| KM_k141_40744 | Not-determined |
| KM_k141_75307 | Not-determined |
| KM_k141_209171 | Not-determined |
| KM_k141_107825 | Not-determined |
| KM_k141_175029 | Not-determined |
| KM_k141_107828 | Not-determined |
| KM_k141_86887 | Not-determined |
| KM_k141_152625 | Not-determined |
| KM_k141_120737 | Not-determined |
| KM_k141_17876 | Not-determined |
| KM_k141_63687 | Not-determined |
| KM_k141_7333 | Not-determined |
| KM_k141_152631 | Not-determined |
| KM_k141_231581 | Low-quality |

| | |
|---|---|
| KM_k141_17881 | Not-determined |
| KM_k141_63693 | Not-determined |
| KM_k141_241225 | Not-determined |
| KM_k141_7338 | Not-determined |
| KM_k141_86907 | Not-determined |
| KM_k141_220340 | Not-determined |
| KM_k141_241229 | Not-determined |
| KM_k141_175043 | Not-determined |
| KM_k141_7342 | Not-determined |
| KM_k141_75323 | Not-determined |
| KM_k141_220347 | Not-determined |
| KM_k141_175047 | Low-quality |
| KM_k141_251222 | Not-determined |
| KM_k141_231592 | Not-determined |
| KM_k141_17890 | Not-determined |
| KM_k141_17891 | Not-determined |
| KM_k141_17893 | Low-quality |
| KM_k141_186760 | Not-determined |
| KM_k141_175054 | Not-determined |
| KM_k141_17896 | Not-determined |
| KM_k141_107851 | Not-determined |
| KM_k141_175056 | Not-determined |
| KM_k141_17898 | Not-determined |
| KM_k141_17899 | Low-quality |
| KM_k141_107854 | Not-determined |
| KM_k141_251229 | Not-determined |
| KM_k141_120755 | Not-determined |
| KM_k141_50459 | Not-determined |
| KM_k141_86918 | Low-quality |
| KM_k141_197856 | Not-determined |
| KM_k141_251236 | Not-determined |
| KM_k141_40755 | Low-quality |
| KM_k141_63722 | Not-determined |
| KM_k141_251241 | Not-determined |
| KM_k141_120767 | Low-quality |
| KM_k141_86926 | Not-determined |
| KM_k141_120776 | Not-determined |
| KM_k141_231615 | Not-determined |
| KM_k141_120778 | Low-quality |
| KM_k141_40756 | Not-determined |
| KM_k141_241257 | Not-determined |
| KM_k141_141849 | Low-quality |
| KM_k141_40763 | Not-determined |
| KM_k141_262684 | Not-determined |
| KM_k141_86932 | Not-determined |
| KM_k141_63735 | Not-determined |
| KM_k141_17925 | Not-determined |
| KM_k141_63742 | Not-determined |
| KM_k141_63745 | Not-determined |
| KM_k141_86945 | Not-determined |
| KM_k141_86947 | Not-determined |
| KM_k141_40773 | Not-determined |
| KM_k141_86951 | Not-determined |
| KM_k141_86953 | Not-determined |
| KM_k141_241272 | Not-determined |
| KM_k141_17935 | Not-determined |
| KM_k141_152679 | Not-determined |
| KM_k141_251273 | Not-determined |
| KM_k141_241280 | Low-quality |
| KM_k141_95284 | Not-determined |
| KM_k141_120826 | Not-determined |
| KM_k141_197892 | Not-determined |
| KM_k141_95291 | Not-determined |
| KM_k141_175095 | Not-determined |
| KM_k141_175097 | Not-determined |
| KM_k141_220381 | Not-determined |
| KM_k141_251286 | Not-determined |
| KM_k141_40805 | Not-determined |
| KM_k141_17952 | Not-determined |
| KM_k141_17953 | Not-determined |
| KM_k141_75350 | Not-determined |
| KM_k141_197900 | Not-determined |
| KM_k141_40810 | Not-determined |
| KM_k141_220402 | Not-determined |
| KM_k141_17964 | Not-determined |
| KM_k141_197911 | Not-determined |
| KM_k141_63762 | Not-determined |
| KM_k141_75359 | Not-determined |
| KM_k141_95309 | Not-determined |
| KM_k141_63763 | Low-quality |
| KM_k141_63764 | Low-quality |
| KM_k141_220411 | Not-determined |
| KM_k141_120846 | Not-determined |
| KM_k141_17971 | Not-determined |
| KM_k141_75374 | Not-determined |
| KM_k141_175117 | Not-determined |
| KM_k141_95319 | Not-determined |
| KM_k141_75378 | Not-determined |
| KM_k141_197923 | Not-determined |
| KM_k141_209275 | Not-determined |
| KM_k141_175121 | Not-determined |
| KM_k141_209278 | Not-determined |
| KM_k141_107905 | Low-quality |
| KM_k141_120869 | Not-determined |
| KM_k141_120870 | Low-quality |
| KM_k141_209292 | Not-determined |
| KM_k141_120872 | Not-determined |
| KM_k141_75401 | Low-quality |
| KM_k141_209299 | Not-determined |
| KM_k141_262721 | Not-determined |
| KM_k141_186806 | Not-determined |
| KM_k141_107914 | Not-determined |
| KM_k141_262726 | Not-determined |
| KM_k141_63798 | Not-determined |
| KM_k141_87024 | Not-determined |
| KM_k141_120889 | Not-determined |
| KM_k141_186810 | Not-determined |
| KM_k141_262731 | Not-determined |
| KM_k141_220428 | Not-determined |
| KM_k141_107920 | Not-determined |
| KM_k141_120896 | Not-determined |
| KM_k141_175140 | Not-determined |
| KM_k141_95330 | Not-determined |
| KM_k141_152732 | Not-determined |
| KM_k141_152737 | Not-determined |
| KM_k141_95338 | Not-determined |
| KM_k141_63816 | Not-determined |
| KM_k141_87038 | Not-determined |
| KM_k141_186823 | Not-determined |
| KM_k141_107929 | Not-determined |
| KM_k141_241316 | Not-determined |
| KM_k141_87041 | Not-determined |
| KM_k141_107931 | Not-determined |
| KM_k141_197955 | Not-determined |
| KM_k141_231641 | Not-determined |
| KM_k141_18004 | Not-determined |
| KM_k141_197961 | Not-determined |
| KM_k141_220441 | Not-determined |
| KM_k141_220443 | Not-determined |
| KM_k141_141928 | Not-determined |
| KM_k141_63829 | Not-determined |
| KM_k141_209318 | Low-quality |
| KM_k141_107943 | Not-determined |
| KM_k141_241325 | Not-determined |
| KM_k141_107944 | Not-determined |
| KM_k141_241327 | Not-determined |
| KM_k141_186835 | Low-quality |
| KM_k141_162790 | Not-determined |
| KM_k141_197976 | Low-quality |
| KM_k141_186837 | Not-determined |
| KM_k141_141939 | Not-determined |
| KM_k141_175173 | Not-determined |
| KM_k141_7431 | Not-determined |
| KM_k141_251305 | Not-determined |
| KM_k141_162805 | Not-determined |
| KM_k141_220460 | Not-determined |
| KM_k141_75439 | Not-determined |
| KM_k141_175181 | Not-determined |
| KM_k141_87068 | Not-determined |
| KM_k141_162806 | Not-determined |
| KM_k141_186848 | Not-determined |
| KM_k141_251310 | Not-determined |
| KM_k141_7442 | Not-determined |
| KM_k141_29726 | Not-determined |
| KM_k141_262772 | Not-determined |
| KM_k141_162810 | Not-determined |
| KM_k141_152752 | Low-quality |
| KM_k141_197998 | Not-determined |
| KM_k141_198000 | Not-determined |
| KM_k141_175194 | Not-determined |
| KM_k141_40853 | Not-determined |
| KM_k141_107967 | Not-determined |
| KM_k141_241338 | Not-determined |
| KM_k141_107970 | Not-determined |
| KM_k141_152757 | Not-determined |
| KM_k141_220473 | Not-determined |
| KM_k141_152764 | Not-determined |
| KM_k141_186865 | Not-determined |
| KM_k141_130076 | Not-determined |
| KM_k141_241349 | Not-determined |
| KM_k141_186871 | Not-determined |
| KM_k141_241351 | Not-determined |
| KM_k141_162825 | Not-determined |
| KM_k141_186874 | Not-determined |
| KM_k141_231674 | Not-determined |
| KM_k141_87109 | Not-determined |
| KM_k141_186880 | Not-determined |
| KM_k141_75470 | Not-determined |
| KM_k141_75471 | Not-determined |
| KM_k141_152781 | Not-determined |
| KM_k141_40864 | Not-determined |
| KM_k141_141976 | Not-determined |
| KM_k141_220499 | Not-determined |
| KM_k141_198007 | Not-determined |
| KM_k141_87127 | Not-determined |
| KM_k141_120907 | Not-determined |
| KM_k141_18036 | Not-determined |
| KM_k141_87139 | Not-determined |
| KM_k141_141984 | Not-determined |
| KM_k141_75492 | Low-quality |
| KM_k141_29749 | Not-determined |
| KM_k141_75494 | Low-quality |
| KM_k141_152799 | Not-determined |
| KM_k141_186902 | Not-determined |
| KM_k141_220531 | Not-determined |
| KM_k141_120925 | Not-determined |
| KM_k141_141999 | Not-determined |
| KM_k141_75510 | Not-determined |
| KM_k141_175221 | Not-determined |
| KM_k141_262794 | Not-determined |
| KM_k141_40876 | Not-determined |
| KM_k141_75518 | Not-determined |
| KM_k141_175234 | Not-determined |
| KM_k141_29759 | Not-determined |
| KM_k141_75528 | Not-determined |
| KM_k141_75529 | Not-determined |
| KM_k141_198026 | Not-determined |
| KM_k141_50547 | Not-determined |
| KM_k141_162869 | Not-determined |
| KM_k141_18051 | Not-determined |
| KM_k141_18052 | Not-determined |
| KM_k141_198034 | Not-determined |
| KM_k141_63871 | Not-determined |
| KM_k141_162876 | Not-determined |
| KM_k141_220545 | Not-determined |
| KM_k141_40900 | Not-determined |
| KM_k141_209393 | Not-determined |
| KM_k141_220548 | Not-determined |
| KM_k141_262809 | Not-determined |
| KM_k141_262811 | Not-determined |
| KM_k141_186942 | Not-determined |
| KM_k141_262812 | Not-determined |
| KM_k141_130123 | Not-determined |
| KM_k141_209398 | Not-determined |
| KM_k141_262816 | Low-quality |
| KM_k141_29765 | Low-quality |
| KM_k141_162884 | Low-quality |
| KM_k141_29769 | Not-determined |
| KM_k141_209406 | Not-determined |
| KM_k141_220557 | Not-determined |
| KM_k141_29775 | Not-determined |
| KM_k141_198045 | Not-determined |
| KM_k141_186950 | Not-determined |
| KM_k141_162890 | Not-determined |
| KM_k141_29784 | Not-determined |
| KM_k141_162892 | Not-determined |
| KM_k141_75543 | Not-determined |
| KM_k141_262821 | Not-determined |
| KM_k141_50561 | Low-quality |
| KM_k141_162901 | Not-determined |
| KM_k141_130146 | Not-determined |
| KM_k141_130149 | Not-determined |
| KM_k141_130150 | Not-determined |
| KM_k141_87166 | Not-determined |
| KM_k141_7498 | Not-determined |
| KM_k141_75562 | Not-determined |
| KM_k141_75563 | Not-determined |
| KM_k141_7499 | Not-determined |
| KM_k141_152822 | Not-determined |
| KM_k141_130155 | Not-determined |
| KM_k141_50565 | Not-determined |
| KM_k141_231708 | Not-determined |
| KM_k141_209421 | Not-determined |
| KM_k141_7505 | Not-determined |
| KM_k141_120953 | Low-quality |
| KM_k141_209425 | Not-determined |
| KM_k141_120956 | Not-determined |
| KM_k141_198075 | Not-determined |
| KM_k141_152832 | Not-determined |
| KM_k141_75573 | Not-determined |
| KM_k141_87167 | Low-quality |
| KM_k141_87168 | Not-determined |
| KM_k141_75577 | Not-determined |
| KM_k141_63918 | Not-determined |
| KM_k141_251361 | Not-determined |
| KM_k141_220610 | Not-determined |
| KM_k141_209432 | Not-determined |
| KM_k141_75582 | Not-determined |
| KM_k141_198089 | Not-determined |
| KM_k141_120975 | Not-determined |
| KM_k141_7524 | Not-determined |
| KM_k141_75586 | Not-determined |
| KM_k141_198090 | Not-determined |
| KM_k141_220619 | Not-determined |
| KM_k141_107997 | Not-determined |
| KM_k141_29817 | Not-determined |
| KM_k141_220620 | Not-determined |
| KM_k141_162930 | Not-determined |
| KM_k141_152847 | Not-determined |
| KM_k141_198091 | Not-determined |
| KM_k141_220623 | Not-determined |
| KM_k141_152849 | Low-quality |
| KM_k141_130164 | Not-determined |
| KM_k141_209438 | Not-determined |
| KM_k141_186966 | Not-determined |
| KM_k141_262846 | Not-determined |
| KM_k141_87179 | Not-determined |
| KM_k141_162939 | Not-determined |
| KM_k141_241402 | Not-determined |
| KM_k141_130170 | Not-determined |
| KM_k141_29823 | Not-determined |
| KM_k141_251379 | Not-determined |
| KM_k141_241405 | Not-determined |
| KM_k141_29825 | Not-determined |
| KM_k141_241410 | Not-determined |
| KM_k141_142065 | Not-determined |
| KM_k141_186979 | Not-determined |
| KM_k141_29830 | Not-determined |
| KM_k141_186982 | Not-determined |
| KM_k141_142066 | Not-determined |
| KM_k141_152857 | Not-determined |
| KM_k141_186985 | Not-determined |
| KM_k141_63943 | Not-determined |
| KM_k141_220643 | Not-determined |
| KM_k141_209450 | Not-determined |
| KM_k141_220646 | Not-determined |
| KM_k141_63947 | Not-determined |
| KM_k141_175274 | Low-quality |
| KM_k141_262855 | Not-determined |
| KM_k141_262858 | Not-determined |
| KM_k141_142076 | Low-quality |
| KM_k141_87196 | Not-determined |
| KM_k141_130195 | Not-determined |
| KM_k141_63949 | Not-determined |
| KM_k141_152877 | Low-quality |
| KM_k141_50594 | Not-determined |
| KM_k141_75598 | Not-determined |
| KM_k141_75602 | Not-determined |
| KM_k141_175282 | Not-determined |
| KM_k141_130205 | Not-determined |
| KM_k141_241433 | Not-determined |
| KM_k141_75606 | Not-determined |
| KM_k141_220675 | Not-determined |
| KM_k141_175284 | Low-quality |
| KM_k141_75614 | Not-determined |
| KM_k141_108013 | Not-determined |
| KM_k141_251402 | Not-determined |
| KM_k141_50597 | Not-determined |
| KM_k141_152891 | Not-determined |
| KM_k141_63964 | Not-determined |
| KM_k141_251409 | Not-determined |
| KM_k141_251410 | Not-determined |
| KM_k141_152899 | Not-determined |
| KM_k141_241442 | Low-quality |
| KM_k141_241444 | Not-determined |
| KM_k141_108025 | Not-determined |
| KM_k141_186996 | Not-determined |
| KM_k141_231714 | Not-determined |
| KM_k141_220690 | Not-determined |
| KM_k141_108027 | Not-determined |
| KM_k141_231716 | Not-determined |
| KM_k141_186998 | Not-determined |
| KM_k141_251417 | Not-determined |
| KM_k141_108033 | Not-determined |
| KM_k141_75630 | Not-determined |
| KM_k141_241455 | Not-determined |
| KM_k141_108040 | Not-determined |
| KM_k141_187005 | Not-determined |
| KM_k141_50604 | Not-determined |
| KM_k141_231722 | Not-determined |
| KM_k141_75639 | Low-quality |
| KM_k141_87229 | Not-determined |
| KM_k141_251424 | Not-determined |
| KM_k141_121008 | Not-determined |
| KM_k141_75645 | Not-determined |
| KM_k141_187014 | Not-determined |
| KM_k141_63981 | Not-determined |
| KM_k141_50612 | Low-quality |
| KM_k141_63984 | Not-determined |
| KM_k141_241461 | Not-determined |
| KM_k141_262891 | Not-determined |
| KM_k141_187020 | Not-determined |
| KM_k141_152919 | Not-determined |
| KM_k141_18070 | Not-determined |
| KM_k141_18073 | Not-determined |
| KM_k141_187023 | Not-determined |
| KM_k141_40920 | Not-determined |
| KM_k141_50628 | Not-determined |
| KM_k141_40927 | Not-determined |
| KM_k141_241466 | Not-determined |
| KM_k141_241467 | Not-determined |
| KM_k141_220708 | Not-determined |
| KM_k141_220711 | Not-determined |
| KM_k141_251443 | Low-quality |
| KM_k141_7577 | Low-quality |
| KM_k141_108048 | Not-determined |
| KM_k141_50636 | Not-determined |
| KM_k141_220714 | Not-determined |
| KM_k141_198134 | Not-determined |
| KM_k141_187038 | Not-determined |
| KM_k141_50641 | Not-determined |
| KM_k141_18085 | Not-determined |
| KM_k141_142129 | Not-determined |
| KM_k141_142131 | Not-determined |
| KM_k141_87260 | Not-determined |
| KM_k141_162988 | Not-determined |
| KM_k141_40939 | Not-determined |
| KM_k141_175318 | Low-quality |
| KM_k141_7583 | Not-determined |
| KM_k141_198140 | Not-determined |
| KM_k141_95392 | Not-determined |
| KM_k141_7588 | Not-determined |
| KM_k141_187048 | Not-determined |
| KM_k141_251462 | Not-determined |
| KM_k141_251463 | Not-determined |
| KM_k141_18093 | Not-determined |
| KM_k141_142140 | Not-determined |
| KM_k141_40950 | Not-determined |
| KM_k141_50655 | Not-determined |
| KM_k141_262906 | Not-determined |
| KM_k141_95408 | Not-determined |
| KM_k141_7602 | Not-determined |
| KM_k141_64025 | Not-determined |
| KM_k141_87271 | Not-determined |
| KM_k141_142154 | Not-determined |
| KM_k141_142158 | Not-determined |
| KM_k141_7609 | Not-determined |
| KM_k141_87277 | Low-quality |
| KM_k141_220745 | Low-quality |
| KM_k141_50669 | Not-determined |
| KM_k141_130216 | Not-determined |
| KM_k141_18111 | Not-determined |
| KM_k141_75673 | Low-quality |
| KM_k141_187066 | Not-determined |
| KM_k141_95412 | Not-determined |
| KM_k141_7612 | Not-determined |
| KM_k141_29877 | Not-determined |
| KM_k141_40965 | Not-determined |
| KM_k141_40966 | Not-determined |
| KM_k141_87282 | Not-determined |
| KM_k141_187072 | Not-determined |
| KM_k141_198174 | Not-determined |
| KM_k141_121050 | Not-determined |
| KM_k141_241494 | Not-determined |
| KM_k141_18117 | Not-determined |
| KM_k141_18119 | Not-determined |
| KM_k141_198179 | Not-determined |
| KM_k141_152954 | Not-determined |
| KM_k141_142177 | Not-determined |
| KM_k141_175345 | Not-determined |
| KM_k141_152963 | Low-quality |
| KM_k141_163013 | Not-determined |
| KM_k141_262928 | Not-determined |
| KM_k141_29899 | Not-determined |
| KM_k141_108097 | Low-quality |
| KM_k141_241502 | Not-determined |
| KM_k141_241503 | Not-determined |
| KM_k141_241505 | Not-determined |
| KM_k141_108101 | Not-determined |
| KM_k141_18131 | Not-determined |
| KM_k141_121065 | Not-determined |
| KM_k141_241509 | Not-determined |
| KM_k141_198194 | Not-determined |
| KM_k141_18135 | Not-determined |
| KM_k141_29906 | Not-determined |
| KM_k141_50697 | Not-determined |
| KM_k141_152977 | Not-determined |
| KM_k141_175370 | Not-determined |
| KM_k141_130267 | Not-determined |
| KM_k141_64039 | Not-determined |
| KM_k141_251474 | Low-quality |
| KM_k141_231775 | Low-quality |
| KM_k141_130272 | Not-determined |
| KM_k141_187100 | Not-determined |
| KM_k141_175378 | Not-determined |
| KM_k141_220767 | Low-quality |
| KM_k141_18148 | Not-determined |
| KM_k141_198206 | Not-determined |
| KM_k141_220771 | Not-determined |
| KM_k141_64051 | Not-determined |
| KM_k141_7622 | Not-determined |
| KM_k141_121086 | Not-determined |
| KM_k141_130282 | Not-determined |
| KM_k141_108120 | Not-determined |
| KM_k141_198213 | Not-determined |
| KM_k141_152996 | Not-determined |
| KM_k141_7625 | Not-determined |
| KM_k141_87287 | Not-determined |
| KM_k141_95441 | Not-determined |
| KM_k141_29915 | Not-determined |
| KM_k141_29916 | Not-determined |
| KM_k141_251488 | Not-determined |
| KM_k141_163050 | Not-determined |
| KM_k141_198222 | Not-determined |
| KM_k141_18160 | Not-determined |
| KM_k141_231800 | Not-determined |
| KM_k141_130301 | Not-determined |
| KM_k141_75693 | Not-determined |
| KM_k141_87298 | Not-determined |
| KM_k141_18168 | Not-determined |
| KM_k141_7647 | Not-determined |
| KM_k141_7649 | Low-quality |
| KM_k141_142198 | Not-determined |
| KM_k141_121101 | Not-determined |
| KM_k141_7650 | Not-determined |
| KM_k141_231806 | Not-determined |
| KM_k141_87302 | Not-determined |
| KM_k141_87305 | Not-determined |
| KM_k141_175419 | Not-determined |
| KM_k141_198225 | Not-determined |
| KM_k141_175421 | Not-determined |
| KM_k141_130317 | Not-determined |
| KM_k141_121109 | Not-determined |
| KM_k141_108151 | Not-determined |
| KM_k141_87311 | Not-determined |
| KM_k141_231811 | Not-determined |
| KM_k141_121111 | Not-determined |
| KM_k141_142204 | Not-determined |
| KM_k141_29926 | Not-determined |
| KM_k141_50754 | Not-determined |
| KM_k141_64077 | Not-determined |
| KM_k141_29931 | Not-determined |
| KM_k141_130331 | Not-determined |
| KM_k141_231818 | Not-determined |
| KM_k141_95455 | Not-determined |
| KM_k141_130336 | Not-determined |
| KM_k141_142219 | Not-determined |
| KM_k141_75710 | Not-determined |
| KM_k141_175433 | Not-determined |
| KM_k141_130339 | Not-determined |
| KM_k141_231821 | Not-determined |
| KM_k141_75717 | Not-determined |
| KM_k141_251506 | Not-determined |
| KM_k141_251509 | Not-determined |
| KM_k141_18193 | Not-determined |
| KM_k141_29951 | Not-determined |
| KM_k141_7666 | Not-determined |
| KM_k141_251517 | Not-determined |
| KM_k141_153030 | Not-determined |
| KM_k141_130351 | Low-quality |
| KM_k141_262963 | Not-determined |
| KM_k141_95465 | Not-determined |
| KM_k141_75735 | Not-determined |
| KM_k141_153033 | Not-determined |
| KM_k141_18202 | Not-determined |
| KM_k141_29960 | Not-determined |
| KM_k141_7671 | Not-determined |
| KM_k141_241549 | Not-determined |
| KM_k141_241550 | Not-determined |
| KM_k141_251531 | Not-determined |
| KM_k141_142240 | Low-quality |
| KM_k141_95472 | Not-determined |
| KM_k141_130362 | Low-quality |
| KM_k141_262976 | Not-determined |
| KM_k141_163090 | Not-determined |
| KM_k141_175453 | Not-determined |
| KM_k141_241556 | Not-determined |
| KM_k141_130370 | Not-determined |
| KM_k141_87353 | Not-determined |
| KM_k141_153048 | Not-determined |
| KM_k141_29971 | Not-determined |
| KM_k141_130377 | Not-determined |
| KM_k141_29974 | Not-determined |
| KM_k141_241570 | Not-determined |
| KM_k141_50771 | Not-determined |
| KM_k141_262988 | Low-quality |
| KM_k141_231843 | Not-determined |
| KM_k141_50775 | Not-determined |
| KM_k141_153056 | Not-determined |
| KM_k141_153057 | Not-determined |
| KM_k141_75753 | Not-determined |
| KM_k141_142274 | Not-determined |
| KM_k141_231852 | Not-determined |
| KM_k141_50787 | Not-determined |
| KM_k141_231854 | Not-determined |
| KM_k141_130392 | Not-determined |
| KM_k141_263004 | Not-determined |
| KM_k141_75764 | Not-determined |
| KM_k141_18240 | Not-determined |
| KM_k141_187122 | Not-determined |
| KM_k141_7680 | Not-determined |
| KM_k141_198251 | Not-determined |
| KM_k141_95506 | Not-determined |
| KM_k141_251548 | Not-determined |
| KM_k141_130399 | Not-determined |
| KM_k141_263009 | Not-determined |
| KM_k141_163124 | Low-quality |
| KM_k141_7691 | Not-determined |
| KM_k141_95517 | Not-determined |
| KM_k141_87367 | Not-determined |
| KM_k141_95518 | Not-determined |
| KM_k141_87369 | Not-determined |
| KM_k141_29987 | Not-determined |
| KM_k141_75781 | Not-determined |
| KM_k141_231875 | Not-determined |
| KM_k141_75783 | Not-determined |
| KM_k141_231876 | Not-determined |
| KM_k141_108226 | Not-determined |
| KM_k141_263027 | Not-determined |
| KM_k141_7703 | Low-quality |
| KM_k141_220843 | Not-determined |
| KM_k141_251554 | Low-quality |
| KM_k141_263028 | Low-quality |
| KM_k141_108229 | Not-determined |
| KM_k141_75795 | Not-determined |
| KM_k141_142307 | Not-determined |
| KM_k141_142308 | Not-determined |
| KM_k141_18251 | Not-determined |
| KM_k141_87388 | Not-determined |

| | |
|---|---|
| KM_k141_87390 | Not-determined |
| KM_k141_121172 | Not-determined |
| KM_k141_87393 | Low-quality |
| KM_k141_241612 | Not-determined |
| KM_k141_95536 | Not-determined |
| KM_k141_75813 | Low-quality |
| KM_k141_18259 | Not-determined |
| KM_k141_142322 | Not-determined |
| KM_k141_209461 | Not-determined |
| KM_k141_50810 | Low-quality |
| KM_k141_50811 | Not-determined |
| KM_k141_121178 | Not-determined |
| KM_k141_75822 | Not-determined |
| KM_k141_209469 | Not-determined |
| KM_k141_95546 | Not-determined |
| KM_k141_187154 | Low-quality |
| KM_k141_263055 | Not-determined |
| KM_k141_263057 | Not-determined |
| KM_k141_231891 | Not-determined |
| KM_k141_95550 | Not-determined |
| KM_k141_50816 | Not-determined |
| KM_k141_121188 | Not-determined |
| KM_k141_50817 | Not-determined |
| KM_k141_187167 | Not-determined |
| KM_k141_231900 | Not-determined |
| KM_k141_75839 | Low-quality |
| KM_k141_175502 | Not-determined |
| KM_k141_231902 | Not-determined |
| KM_k141_153084 | Not-determined |
| KM_k141_153087 | Not-determined |
| KM_k141_231906 | Not-determined |
| KM_k141_241645 | Not-determined |
| KM_k141_263067 | Not-determined |
| KM_k141_95558 | Low-quality |
| KM_k141_198270 | Not-determined |
| KM_k141_130420 | Not-determined |
| KM_k141_231908 | Low-quality |
| KM_k141_50821 | Not-determined |
| KM_k141_87414 | Not-determined |
| KM_k141_231919 | Not-determined |
| KM_k141_241656 | Not-determined |
| KM_k141_130427 | Not-determined |
| KM_k141_231921 | Not-determined |
| KM_k141_163166 | Not-determined |
| KM_k141_209479 | Not-determined |
| KM_k141_41009 | Not-determined |
| KM_k141_263083 | Not-determined |
| KM_k141_231930 | Not-determined |
| KM_k141_7739 | Not-determined |
| KM_k141_187188 | Not-determined |
| KM_k141_241671 | Not-determined |
| KM_k141_7743 | Not-determined |
| KM_k141_187190 | Not-determined |
| KM_k141_95584 | Not-determined |
| KM_k141_18279 | Not-determined |
| KM_k141_153115 | Not-determined |
| KM_k141_153116 | Not-determined |
| KM_k141_41023 | Low-quality |
| KM_k141_7746 | Not-determined |
| KM_k141_142373 | Not-determined |
| KM_k141_50840 | Not-determined |
| KM_k141_7751 | Not-determined |
| KM_k141_50843 | Not-determined |
| KM_k141_142377 | Not-determined |
| KM_k141_50844 | Not-determined |
| KM_k141_263105 | Not-determined |
| KM_k141_50845 | Not-determined |
| KM_k141_18285 | Not-determined |
| KM_k141_220873 | Not-determined |
| KM_k141_87435 | Not-determined |
| KM_k141_263108 | Not-determined |
| KM_k141_163172 | Low-quality |
| KM_k141_209497 | Not-determined |
| KM_k141_41030 | Not-determined |
| KM_k141_7756 | Not-determined |
| KM_k141_163176 | Not-determined |
| KM_k141_220874 | Low-quality |
| KM_k141_231954 | Not-determined |
| KM_k141_50848 | Low-quality |
| KM_k141_108260 | Not-determined |
| KM_k141_18293 | Not-determined |
| KM_k141_231961 | Not-determined |
| KM_k141_251587 | Not-determined |
| KM_k141_18301 | Not-determined |
| KM_k141_241675 | Low-quality |
| KM_k141_209511 | Not-determined |
| KM_k141_153138 | Not-determined |
| KM_k141_142391 | Low-quality |
| KM_k141_153140 | Not-determined |
| KM_k141_153141 | Not-determined |
| KM_k141_220886 | Low-quality |
| KM_k141_30046 | Not-determined |
| KM_k141_153143 | Not-determined |
| KM_k141_142399 | Not-determined |
| KM_k141_30049 | Not-determined |
| KM_k141_187214 | Not-determined |
| KM_k141_30050 | Not-determined |
| KM_k141_251597 | Low-quality |
| KM_k141_220894 | Not-determined |
| KM_k141_142404 | Not-determined |
| KM_k141_163197 | Not-determined |
| KM_k141_41048 | Not-determined |
| KM_k141_187222 | Not-determined |
| KM_k141_163199 | Not-determined |
| KM_k141_18319 | Low-quality |
| KM_k141_153157 | Not-determined |
| KM_k141_220903 | Not-determined |
| KM_k141_41051 | Low-quality |
| KM_k141_142417 | Low-quality |
| KM_k141_231984 | Low-quality |
| KM_k141_41058 | Not-determined |
| KM_k141_121206 | Not-determined |
| KM_k141_187239 | Not-determined |
| KM_k141_198286 | Not-determined |
| KM_k141_198288 | Not-determined |
| KM_k141_30068 | Not-determined |
| KM_k141_231994 | Not-determined |
| KM_k141_30069 | Not-determined |
| KM_k141_64111 | Not-determined |
| KM_k141_142425 | Not-determined |
| KM_k141_7788 | Not-determined |
| KM_k141_50884 | Not-determined |
| KM_k141_142428 | Not-determined |
| KM_k141_121218 | Not-determined |
| KM_k141_87469 | Not-determined |
| KM_k141_87472 | Not-determined |
| KM_k141_30078 | Not-determined |
| KM_k141_187261 | Not-determined |
| KM_k141_209529 | Not-determined |
| KM_k141_108291 | Not-determined |
| KM_k141_220927 | Not-determined |
| KM_k141_175563 | Not-determined |
| KM_k141_64124 | Not-determined |
| KM_k141_198310 | Not-determined |
| KM_k141_75905 | Not-determined |
| KM_k141_175564 | Not-determined |
| KM_k141_209539 | Not-determined |
| KM_k141_251629 | Not-determined |
| KM_k141_198313 | Not-determined |
| KM_k141_153182 | Not-determined |
| KM_k141_41079 | Low-quality |
| KM_k141_75908 | Not-determined |
| KM_k141_263124 | Low-quality |
| KM_k141_142438 | Not-determined |
| KM_k141_220934 | Not-determined |
| KM_k141_241698 | Not-determined |
| KM_k141_108304 | Not-determined |
| KM_k141_263138 | Not-determined |
| KM_k141_187272 | Not-determined |
| KM_k141_108310 | Not-determined |
| KM_k141_263145 | Not-determined |
| KM_k141_187285 | Not-determined |
| KM_k141_50907 | Not-determined |
| KM_k141_187292 | Not-determined |
| KM_k141_232020 | Not-determined |
| KM_k141_121234 | Not-determined |
| KM_k141_241713 | Not-determined |
| KM_k141_50915 | Not-determined |
| KM_k141_18347 | Not-determined |
| KM_k141_241719 | Not-determined |
| KM_k141_175573 | Low-quality |
| KM_k141_87498 | Not-determined |
| KM_k141_87499 | Not-determined |
| KM_k141_50917 | Low-quality |
| KM_k141_64142 | Not-determined |
| KM_k141_50918 | Not-determined |
| KM_k141_198341 | Not-determined |
| KM_k141_163235 | Not-determined |
| KM_k141_198348 | Not-determined |
| KM_k141_64146 | Not-determined |
| KM_k141_198351 | Low-quality |
| KM_k141_263174 | Not-determined |
| KM_k141_87510 | Not-determined |
| KM_k141_209570 | Not-determined |
| KM_k141_121255 | Low-quality |
| KM_k141_121256 | Not-determined |
| KM_k141_263179 | Not-determined |
| KM_k141_18358 | Not-determined |
| KM_k141_7832 | Not-determined |
| KM_k141_121259 | Low-quality |
| KM_k141_232033 | Not-determined |
| KM_k141_232034 | Not-determined |
| KM_k141_75938 | Not-determined |
| KM_k141_198354 | Not-determined |
| KM_k141_108347 | Not-determined |
| KM_k141_41105 | Not-determined |
| KM_k141_220993 | Not-determined |
| KM_k141_121276 | Not-determined |
| KM_k141_121277 | Not-determined |
| KM_k141_142469 | Not-determined |
| KM_k141_187319 | Not-determined |
| KM_k141_263189 | Not-determined |
| KM_k141_187321 | Not-determined |
| KM_k141_232046 | Not-determined |
| KM_k141_18371 | Not-determined |
| KM_k141_209593 | Not-determined |
| KM_k141_232050 | Not-determined |
| KM_k141_121288 | Not-determined |
| KM_k141_7854 | Not-determined |
| KM_k141_30106 | Not-determined |
| KM_k141_163262 | Not-determined |
| KM_k141_87528 | Not-determined |
| KM_k141_263200 | Not-determined |
| KM_k141_87534 | Not-determined |
| KM_k141_87537 | Not-determined |
| KM_k141_251682 | Not-determined |
| KM_k141_187326 | Not-determined |
| KM_k141_95628 | Not-determined |
| KM_k141_50929 | Not-determined |
| KM_k141_251684 | Not-determined |
| KM_k141_130463 | Not-determined |
| KM_k141_108371 | Not-determined |
| KM_k141_30113 | Not-determined |
| KM_k141_41126 | Not-determined |
| KM_k141_50932 | Not-determined |
| KM_k141_121308 | Not-determined |
| KM_k141_142490 | Not-determined |
| KM_k141_130465 | Not-determined |
| KM_k141_87546 | Not-determined |
| KM_k141_64163 | Not-determined |
| KM_k141_241762 | Not-determined |
| KM_k141_30123 | Not-determined |
| KM_k141_263218 | Low-quality |
| KM_k141_108380 | Not-determined |
| KM_k141_41143 | Not-determined |
| KM_k141_18373 | Not-determined |
| KM_k141_263230 | Low-quality |
| KM_k141_7888 | Not-determined |
| KM_k141_209628 | Not-determined |
| KM_k141_87565 | Not-determined |
| KM_k141_263233 | Not-determined |
| KM_k141_209636 | Not-determined |
| KM_k141_198403 | Not-determined |
| KM_k141_108394 | Low-quality |
| KM_k141_175626 | Not-determined |
| KM_k141_263245 | Not-determined |
| KM_k141_87581 | Low-quality |
| KM_k141_121348 | Not-determined |
| KM_k141_41171 | Not-determined |
| KM_k141_130480 | Not-determined |
| KM_k141_64194 | Low-quality |
| KM_k141_142511 | Not-determined |
| KM_k141_209648 | Not-determined |
| KM_k141_18389 | Not-determined |
| KM_k141_209653 | Not-determined |
| KM_k141_41179 | Not-determined |
| KM_k141_130488 | Not-determined |
| KM_k141_163294 | Not-determined |
| KM_k141_121355 | Not-determined |
| KM_k141_209659 | Not-determined |
| KM_k141_87596 | Not-determined |
| KM_k141_241814 | Not-determined |
| KM_k141_50950 | Not-determined |
| KM_k141_64205 | Not-determined |
| KM_k141_18400 | Not-determined |
| KM_k141_41188 | Not-determined |
| KM_k141_187340 | Low-quality |
| KM_k141_142523 | Not-determined |
| KM_k141_163301 | Not-determined |
| KM_k141_153220 | Not-determined |
| KM_k141_251722 | Not-determined |
| KM_k141_198438 | Not-determined |
| KM_k141_130508 | Not-determined |
| KM_k141_198442 | Not-determined |
| KM_k141_41199 | Not-determined |
| KM_k141_76013 | Not-determined |
| KM_k141_87608 | Not-determined |
| KM_k141_142531 | Not-determined |
| KM_k141_198445 | Not-determined |
| KM_k141_30167 | Not-determined |
| KM_k141_95674 | Not-determined |
| KM_k141_198448 | Not-determined |
| KM_k141_251734 | Not-determined |
| KM_k141_198453 | Not-determined |
| KM_k141_95684 | Not-determined |
| KM_k141_130516 | Low-quality |
| KM_k141_187343 | Not-determined |
| KM_k141_251740 | Not-determined |
| KM_k141_153235 | Not-determined |
| KM_k141_142552 | Not-determined |
| KM_k141_41209 | Not-determined |
| KM_k141_95690 | Not-determined |
| KM_k141_95693 | Not-determined |
| KM_k141_187345 | Low-quality |
| KM_k141_95694 | Not-determined |
| KM_k141_163306 | Low-quality |
| KM_k141_187351 | Not-determined |
| KM_k141_130531 | Not-determined |
| KM_k141_87627 | Not-determined |
| KM_k141_163312 | Not-determined |
| KM_k141_142563 | Not-determined |
| KM_k141_241832 | Not-determined |
| KM_k141_108430 | Not-determined |
| KM_k141_18435 | Low-quality |
| KM_k141_142566 | Not-determined |
| KM_k141_251755 | Not-determined |
| KM_k141_50970 | Not-determined |
| KM_k141_221070 | Not-determined |
| KM_k141_241840 | Not-determined |
| KM_k141_221074 | Not-determined |
| KM_k141_108440 | Not-determined |
| KM_k141_142577 | Not-determined |
| KM_k141_142579 | Not-determined |
| KM_k141_142582 | Not-determined |
| KM_k141_142584 | Not-determined |
| KM_k141_76036 | Not-determined |
| KM_k141_87643 | Low-quality |
| KM_k141_108441 | Not-determined |
| KM_k141_263280 | Low-quality |
| KM_k141_209677 | Not-determined |
| KM_k141_251775 | Not-determined |
| KM_k141_41236 | Not-determined |
| KM_k141_221092 | Low-quality |
| KM_k141_76051 | Not-determined |
| KM_k141_142595 | Not-determined |
| KM_k141_187387 | Not-determined |
| KM_k141_30209 | Not-determined |
| KM_k141_30214 | Not-determined |
| KM_k141_187400 | Not-determined |
| KM_k141_153263 | Not-determined |
| KM_k141_198465 | Not-determined |
| KM_k141_163320 | Not-determined |
| KM_k141_209681 | Low-quality |
| KM_k141_153264 | Not-determined |
| KM_k141_7940 | Low-quality |
| KM_k141_108460 | Not-determined |
| KM_k141_87667 | Not-determined |
| KM_k141_108466 | Not-determined |
| KM_k141_142612 | Not-determined |
| KM_k141_142614 | Not-determined |
| KM_k141_7948 | Low-quality |
| KM_k141_50995 | Not-determined |
| KM_k141_95724 | Not-determined |
| KM_k141_50998 | Not-determined |
| KM_k141_153279 | Low-quality |
| KM_k141_108472 | Not-determined |
| KM_k141_108474 | Not-determined |
| KM_k141_30228 | Not-determined |
| KM_k141_87681 | Not-determined |
| KM_k141_142631 | Not-determined |
| KM_k141_198491 | Not-determined |
| KM_k141_51005 | Not-determined |
| KM_k141_198496 | Not-determined |
| KM_k141_187405 | Not-determined |
| KM_k141_251791 | Not-determined |
| KM_k141_187407 | Low-quality |
| KM_k141_209713 | Not-determined |
| KM_k141_130547 | Low-quality |
| KM_k141_198497 | Not-determined |
| KM_k141_130553 | Not-determined |
| KM_k141_7964 | Not-determined |
| KM_k141_108489 | Not-determined |
| KM_k141_163344 | Not-determined |
| KM_k141_163345 | Not-determined |
| KM_k141_121381 | Not-determined |
| KM_k141_163347 | Not-determined |
| KM_k141_209720 | Not-determined |
| KM_k141_95748 | Not-determined |
| KM_k141_232054 | Not-determined |
| KM_k141_30244 | Not-determined |
| KM_k141_87690 | Not-determined |
| KM_k141_130565 | Not-determined |
| KM_k141_51024 | Not-determined |
| KM_k141_108499 | Not-determined |
| KM_k141_187422 | Not-determined |
| KM_k141_232061 | Not-determined |
| KM_k141_198515 | Not-determined |
| KM_k141_263287 | Not-determined |
| KM_k141_232073 | Not-determined |
| KM_k141_263292 | Not-determined |
| KM_k141_263294 | Not-determined |
| KM_k141_251822 | Not-determined |
| KM_k141_18476 | Not-determined |
| KM_k141_7987 | Not-determined |
| KM_k141_51035 | Not-determined |
| KM_k141_263301 | Not-determined |
| KM_k141_232082 | Not-determined |
| KM_k141_209729 | Not-determined |
| KM_k141_209731 | Not-determined |
| KM_k141_18483 | Not-determined |
| KM_k141_108529 | Not-determined |
| KM_k141_251837 | Not-determined |
| KM_k141_95772 | Not-determined |
| KM_k141_121422 | Not-determined |
| KM_k141_142650 | Not-determined |
| KM_k141_142651 | Not-determined |
| KM_k141_8015 | Not-determined |
| KM_k141_121426 | Not-determined |
| KM_k141_130586 | Not-determined |
| KM_k141_64263 | Not-determined |
| KM_k141_142655 | Not-determined |
| KM_k141_64266 | Low-quality |
| KM_k141_251846 | Not-determined |
| KM_k141_175662 | Not-determined |
| KM_k141_175663 | Not-determined |
| KM_k141_8025 | Not-determined |
| KM_k141_121434 | Not-determined |
| KM_k141_41279 | Not-determined |
| KM_k141_251857 | Not-determined |
| KM_k141_153341 | Not-determined |
| KM_k141_18490 | Not-determined |
| KM_k141_51045 | Not-determined |
| KM_k141_41283 | Not-determined |
| KM_k141_18493 | Not-determined |
| KM_k141_198564 | Not-determined |
| KM_k141_142661 | Not-determined |
| KM_k141_142664 | Not-determined |
| KM_k141_51053 | Not-determined |
| KM_k141_142665 | Not-determined |
| KM_k141_8032 | Low-quality |
| KM_k141_51057 | Not-determined |
| KM_k141_8034 | Not-determined |
| KM_k141_142667 | Not-determined |
| KM_k141_64292 | Not-determined |
| KM_k141_251874 | Low-quality |
| KM_k141_209776 | Not-determined |
| KM_k141_232113 | Not-determined |
| KM_k141_76100 | Not-determined |
| KM_k141_232115 | Not-determined |
| KM_k141_130602 | Not-determined |
| KM_k141_51062 | Not-determined |
| KM_k141_142670 | Not-determined |
| KM_k141_130604 | Not-determined |
| KM_k141_232119 | Not-determined |
| KM_k141_41297 | Not-determined |
| KM_k141_108558 | Not-determined |
| KM_k141_108560 | Low-quality |
| KM_k141_64302 | Not-determined |
| KM_k141_41302 | Not-determined |
| KM_k141_209786 | Not-determined |
| KM_k141_41305 | Not-determined |
| KM_k141_41306 | Not-determined |
| KM_k141_130623 | Not-determined |
| KM_k141_251893 | Not-determined |
| KM_k141_18511 | Not-determined |
| KM_k141_76124 | Not-determined |
| KM_k141_76125 | Not-determined |
| KM_k141_18513 | Not-determined |
| KM_k141_95795 | Low-quality |
| KM_k141_64318 | Not-determined |
| KM_k141_263337 | Not-determined |
| KM_k141_142685 | Not-determined |
| KM_k141_95804 | Not-determined |
| KM_k141_209808 | Not-determined |
| KM_k141_95807 | Not-determined |
| KM_k141_76137 | Not-determined |
| KM_k141_198601 | Not-determined |
| KM_k141_153384 | Not-determined |
| KM_k141_130637 | Low-quality |
| KM_k141_241941 | Not-determined |
| KM_k141_153387 | Not-determined |
| KM_k141_64337 | Not-determined |
| KM_k141_130643 | Not-determined |
| KM_k141_232160 | Not-determined |
| KM_k141_153394 | Not-determined |
| KM_k141_130648 | Not-determined |
| KM_k141_64346 | Low-quality |
| KM_k141_198615 | Not-determined |
| KM_k141_142705 | Not-determined |
| KM_k141_251909 | Not-determined |
| KM_k141_76154 | Not-determined |
| KM_k141_142710 | Not-determined |
| KM_k141_108590 | Not-determined |
| KM_k141_87710 | Not-determined |
| KM_k141_76156 | Not-determined |
| KM_k141_87712 | Not-determined |
| KM_k141_251925 | Not-determined |
| KM_k141_51108 | Not-determined |
| KM_k141_130670 | Not-determined |
| KM_k141_8102 | Not-determined |
| KM_k141_251932 | Not-determined |
| KM_k141_30314 | Not-determined |
| KM_k141_232184 | Not-determined |
| KM_k141_30317 | Not-determined |
| KM_k141_209843 | Not-determined |
| KM_k141_108608 | Not-determined |
| KM_k141_153427 | Not-determined |
| KM_k141_130682 | Low-quality |
| KM_k141_76170 | Not-determined |
| KM_k141_221120 | Not-determined |
| KM_k141_8110 | Not-determined |
| KM_k141_153430 | Not-determined |
| KM_k141_232193 | Not-determined |
| KM_k141_209851 | Not-determined |
| KM_k141_130691 | Not-determined |
| KM_k141_51115 | Not-determined |
| KM_k141_163366 | Not-determined |
| KM_k141_209859 | Low-quality |
| KM_k141_209860 | Not-determined |
| KM_k141_18547 | Not-determined |
| KM_k141_18548 | Not-determined |
| KM_k141_30330 | Not-determined |
| KM_k141_163373 | Not-determined |
| KM_k141_209866 | Not-determined |
| KM_k141_153442 | Not-determined |
| KM_k141_153451 | Not-determined |
| KM_k141_8126 | Not-determined |
| KM_k141_18555 | Not-determined |
| KM_k141_130711 | Not-determined |
| KM_k141_121461 | Not-determined |
| KM_k141_198660 | Not-determined |
| KM_k141_76189 | Not-determined |
| KM_k141_130716 | Not-determined |
| KM_k141_130718 | Not-determined |
| KM_k141_108617 | Not-determined |
| KM_k141_76193 | Not-determined |
| KM_k141_198666 | Not-determined |
| KM_k141_8134 | Not-determined |
| KM_k141_130728 | Not-determined |
| KM_k141_87744 | Not-determined |
| KM_k141_76197 | Low-quality |
| KM_k141_87748 | Not-determined |
| KM_k141_232225 | Not-determined |
| KM_k141_163390 | Not-determined |
| KM_k141_18559 | Not-determined |
| KM_k141_142763 | Not-determined |
| KM_k141_30343 | Not-determined |
| KM_k141_76199 | Not-determined |
| KM_k141_8143 | Not-determined |
| KM_k141_242007 | Not-determined |
| KM_k141_8148 | Not-determined |
| KM_k141_209901 | Not-determined |
| KM_k141_8150 | Not-determined |
| KM_k141_242011 | Not-determined |
| KM_k141_8153 | Not-determined |
| KM_k141_153460 | Not-determined |
| KM_k141_153461 | Not-determined |
| KM_k141_18574 | Not-determined |
| KM_k141_76210 | Not-determined |
| KM_k141_153463 | Not-determined |
| KM_k141_87760 | Not-determined |
| KM_k141_8159 | Not-determined |
| KM_k141_51161 | Not-determined |
| KM_k141_163399 | Not-determined |
| KM_k141_187436 | Not-determined |
| KM_k141_18580 | Not-determined |
| KM_k141_163401 | Not-determined |
| KM_k141_108645 | Not-determined |
| KM_k141_175749 | Not-determined |
| KM_k141_95834 | Not-determined |
| KM_k141_187443 | Low-quality |
| KM_k141_8167 | Not-determined |
| KM_k141_242024 | Not-determined |
| KM_k141_130743 | Not-determined |
| KM_k141_130747 | Not-determined |
| KM_k141_175757 | Not-determined |
| KM_k141_64372 | Not-determined |
| KM_k141_175759 | Not-determined |
| KM_k141_175760 | Not-determined |
| KM_k141_221145 | Not-determined |
| KM_k141_187456 | Not-determined |
| KM_k141_64381 | Not-determined |
| KM_k141_198698 | Not-determined |
| KM_k141_18598 | Not-determined |
| KM_k141_30385 | Not-determined |
| KM_k141_175769 | Not-determined |
| KM_k141_76231 | Not-determined |
| KM_k141_198703 | Low-quality |
| KM_k141_130768 | Not-determined |
| KM_k141_198705 | Not-determined |
| KM_k141_232249 | Not-determined |
| KM_k141_87790 | Low-quality |
| KM_k141_76234 | Not-determined |
| KM_k141_221166 | Not-determined |
| KM_k141_76235 | Not-determined |
| KM_k141_232251 | Not-determined |
| KM_k141_153495 | Not-determined |
| KM_k141_8193 | Not-determined |
| KM_k141_130779 | Not-determined |
| KM_k141_163430 | Low-quality |
| KM_k141_163431 | Not-determined |
| KM_k141_95863 | Not-determined |
| KM_k141_163434 | Not-determined |
| KM_k141_30396 | Not-determined |
| KM_k141_232258 | Not-determined |
| KM_k141_163439 | Not-determined |
| KM_k141_87807 | Not-determined |
| KM_k141_130789 | Not-determined |
| KM_k141_130790 | Not-determined |
| KM_k141_130791 | Not-determined |
| KM_k141_121495 | Not-determined |
| KM_k141_142803 | Not-determined |
| KM_k141_51178 | Not-determined |
| KM_k141_108682 | Not-determined |
| KM_k141_232268 | Not-determined |
| KM_k141_108688 | Not-determined |
| KM_k141_121507 | Low-quality |
| KM_k141_198716 | Not-determined |
| KM_k141_95878 | Not-determined |

| | |
|---|---|
| KM_k141_95879 | Not-determined |
| KM_k141_163453 | Not-determined |
| KM_k141_221190 | Not-determined |
| KM_k141_8229 | Not-determined |
| KM_k141_8232 | Not-determined |
| KM_k141_142817 | Not-determined |
| KM_k141_163456 | Not-determined |
| KM_k141_95889 | Not-determined |
| KM_k141_221192 | Not-determined |
| KM_k141_18640 | Not-determined |
| KM_k141_142825 | Not-determined |
| KM_k141_95894 | Not-determined |
| KM_k141_108725 | Not-determined |
| KM_k141_8245 | Not-determined |
| KM_k141_221200 | Not-determined |
| KM_k141_221201 | Not-determined |
| KM_k141_95903 | Not-determined |
| KM_k141_198737 | Not-determined |
| KM_k141_153515 | Not-determined |
| KM_k141_175809 | Not-determined |
| KM_k141_142828 | Not-determined |
| KM_k141_18652 | Not-determined |
| KM_k141_51211 | Not-determined |
| KM_k141_8258 | Not-determined |
| KM_k141_51213 | Not-determined |
| KM_k141_87838 | Not-determined |
| KM_k141_232302 | Not-determined |
| KM_k141_198742 | Not-determined |
| KM_k141_18660 | Not-determined |
| KM_k141_18662 | Not-determined |
| KM_k141_41369 | Not-determined |
| KM_k141_153531 | Not-determined |
| KM_k141_198745 | Not-determined |
| KM_k141_209919 | Not-determined |
| KM_k141_41372 | Not-determined |
| KM_k141_163485 | Not-determined |
| KM_k141_41375 | Not-determined |
| KM_k141_18672 | Not-determined |
| KM_k141_142835 | Not-determined |
| KM_k141_64450 | Not-determined |
| KM_k141_142837 | Not-determined |
| KM_k141_251987 | Not-determined |
| KM_k141_198768 | Not-determined |
| KM_k141_153544 | Not-determined |
| KM_k141_64457 | Not-determined |
| KM_k141_51220 | Not-determined |
| KM_k141_198770 | Not-determined |
| KM_k141_41383 | Not-determined |
| KM_k141_130845 | Not-determined |
| KM_k141_130846 | Not-determined |
| KM_k141_108743 | Not-determined |
| KM_k141_130847 | Not-determined |
| KM_k141_153551 | Not-determined |
| KM_k141_108747 | Not-determined |
| KM_k141_187507 | Not-determined |
| KM_k141_263383 | Not-determined |
| KM_k141_221250 | Not-determined |
| KM_k141_263384 | Not-determined |
| KM_k141_121548 | Not-determined |
| KM_k141_41391 | Not-determined |
| KM_k141_121549 | Not-determined |
| KM_k141_95930 | Low-quality |
| KM_k141_8299 | Not-determined |
| KM_k141_175852 | Not-determined |
| KM_k141_221257 | Not-determined |
| KM_k141_64466 | Not-determined |
| KM_k141_41392 | Not-determined |
| KM_k141_41394 | Not-determined |
| KM_k141_51241 | Not-determined |
| KM_k141_163498 | Not-determined |
| KM_k141_142847 | Not-determined |
| KM_k141_153567 | Not-determined |
| KM_k141_87860 | Not-determined |
| KM_k141_108764 | Not-determined |
| KM_k141_87866 | Not-determined |
| KM_k141_130870 | Not-determined |
| KM_k141_163510 | Not-determined |
| KM_k141_41401 | Not-determined |
| KM_k141_163513 | Not-determined |
| KM_k141_232352 | Not-determined |
| KM_k141_163514 | Not-determined |
| KM_k141_232355 | Not-determined |
| KM_k141_130885 | Not-determined |
| KM_k141_232358 | Not-determined |
| KM_k141_51257 | Not-determined |
| KM_k141_8313 | Not-determined |
| KM_k141_121567 | Not-determined |
| KM_k141_263401 | Not-determined |
| KM_k141_121575 | Not-determined |
| KM_k141_76326 | Not-determined |
| KM_k141_263406 | Not-determined |
| KM_k141_87889 | Not-determined |
| KM_k141_76330 | Not-determined |
| KM_k141_242074 | Not-determined |
| KM_k141_221275 | Not-determined |
| KM_k141_95955 | Low-quality |
| KM_k141_121582 | Not-determined |
| KM_k141_41409 | Not-determined |
| KM_k141_121584 | Not-determined |
| KM_k141_51272 | Not-determined |
| KM_k141_41411 | Not-determined |
| KM_k141_51274 | Not-determined |
| KM_k141_142885 | Not-determined |
| KM_k141_121590 | Not-determined |
| KM_k141_175870 | Not-determined |
| KM_k141_121591 | Not-determined |
| KM_k141_108786 | Not-determined |
| KM_k141_108787 | Not-determined |
| KM_k141_41419 | Not-determined |
| KM_k141_232366 | Not-determined |
| KM_k141_76351 | Not-determined |
| KM_k141_121595 | Not-determined |
| KM_k141_142895 | Not-determined |
| KM_k141_242090 | Not-determined |
| KM_k141_142897 | Not-determined |
| KM_k141_108792 | Not-determined |
| KM_k141_108793 | Not-determined |
| KM_k141_130899 | Not-determined |
| KM_k141_95966 | Not-determined |
| KM_k141_108796 | Not-determined |
| KM_k141_121601 | Not-determined |
| KM_k141_242095 | Not-determined |
| KM_k141_142904 | Not-determined |
| KM_k141_76356 | Not-determined |
| KM_k141_242099 | Not-determined |
| KM_k141_232377 | Not-determined |
| KM_k141_232378 | Not-determined |
| KM_k141_64529 | Not-determined |
| KM_k141_95973 | Not-determined |
| KM_k141_198814 | Not-determined |
| KM_k141_121614 | Low-quality |
| KM_k141_198817 | Not-determined |
| KM_k141_64534 | Not-determined |
| KM_k141_221301 | Not-determined |
| KM_k141_108806 | Not-determined |
| KM_k141_18716 | Not-determined |
| KM_k141_232382 | Not-determined |
| KM_k141_18717 | Not-determined |
| KM_k141_30475 | Not-determined |
| KM_k141_51294 | Not-determined |
| KM_k141_130909 | Not-determined |
| KM_k141_18724 | Not-determined |
| KM_k141_263423 | Not-determined |
| KM_k141_232389 | Not-determined |
| KM_k141_95987 | Not-determined |
| KM_k141_263427 | Not-determined |
| KM_k141_108824 | Not-determined |
| KM_k141_263428 | Not-determined |
| KM_k141_130913 | Not-determined |
| KM_k141_263429 | Not-determined |
| KM_k141_163547 | Not-determined |
| KM_k141_130919 | Low-quality |
| KM_k141_198835 | Not-determined |
| KM_k141_87918 | Not-determined |
| KM_k141_175913 | Low-quality |
| KM_k141_30486 | Not-determined |
| KM_k141_175914 | Not-determined |
| KM_k141_64555 | Not-determined |
| KM_k141_242110 | Not-determined |
| KM_k141_242111 | Not-determined |
| KM_k141_263437 | Low-quality |
| KM_k141_108832 | Low-quality |
| KM_k141_187590 | Not-determined |
| KM_k141_51311 | Not-determined |
| KM_k141_121629 | Not-determined |
| KM_k141_221334 | Not-determined |
| KM_k141_51327 | Not-determined |
| KM_k141_30502 | Not-determined |
| KM_k141_221340 | Not-determined |
| KM_k141_130926 | Not-determined |
| KM_k141_41445 | Not-determined |
| KM_k141_242130 | Low-quality |
| KM_k141_142948 | Not-determined |
| KM_k141_51330 | Not-determined |
| KM_k141_163572 | Not-determined |
| KM_k141_108854 | Not-determined |
| KM_k141_263453 | Not-determined |
| KM_k141_242149 | Not-determined |
| KM_k141_142960 | Low-quality |
| KM_k141_51340 | Not-determined |
| KM_k141_242151 | Not-determined |
| KM_k141_8363 | Not-determined |
| KM_k141_108860 | Low-quality |
| KM_k141_8364 | Low-quality |
| KM_k141_198861 | Not-determined |
| KM_k141_30536 | Not-determined |
| KM_k141_163588 | Low-quality |
| KM_k141_30539 | Not-determined |
| KM_k141_242155 | Not-determined |
| KM_k141_142965 | Not-determined |
| KM_k141_87952 | Not-determined |
| KM_k141_142973 | Not-determined |
| KM_k141_242165 | Not-determined |
| KM_k141_263462 | Not-determined |
| KM_k141_18730 | Not-determined |
| KM_k141_242166 | Not-determined |
| KM_k141_64597 | Not-determined |
| KM_k141_187622 | Not-determined |
| KM_k141_51356 | Not-determined |
| KM_k141_175926 | Not-determined |
| KM_k141_8386 | Not-determined |
| KM_k141_108867 | Not-determined |
| KM_k141_163606 | Not-determined |
| KM_k141_8393 | Not-determined |
| KM_k141_108869 | Not-determined |
| KM_k141_64623 | Low-quality |
| KM_k141_87977 | Not-determined |
| KM_k141_221392 | Not-determined |
| KM_k141_263483 | Not-determined |
| KM_k141_175933 | Not-determined |
| KM_k141_142987 | Not-determined |
| KM_k141_263484 | Low-quality |
| KM_k141_51363 | Not-determined |
| KM_k141_153610 | Not-determined |
| KM_k141_121679 | Not-determined |
| KM_k141_130957 | Not-determined |
| KM_k141_263486 | Low-quality |
| KM_k141_51371 | Not-determined |
| KM_k141_51372 | Not-determined |
| KM_k141_96036 | Low-quality |
| KM_k141_187648 | Not-determined |
| KM_k141_153616 | Not-determined |
| KM_k141_175935 | Not-determined |
| KM_k141_8414 | Not-determined |
| KM_k141_221401 | Not-determined |
| KM_k141_8416 | Not-determined |
| KM_k141_263490 | Not-determined |
| KM_k141_221404 | Not-determined |
| KM_k141_175944 | Not-determined |
| KM_k141_175945 | Not-determined |
| KM_k141_96049 | Not-determined |
| KM_k141_198890 | Not-determined |
| KM_k141_30578 | Not-determined |
| KM_k141_143001 | Low-quality |
| KM_k141_221407 | Not-determined |
| KM_k141_41503 | Not-determined |
| KM_k141_198896 | Not-determined |
| KM_k141_175956 | Not-determined |
| KM_k141_163618 | Not-determined |
| KM_k141_163622 | Not-determined |
| KM_k141_130981 | Not-determined |
| KM_k141_242183 | Not-determined |
| KM_k141_175958 | Not-determined |
| KM_k141_198902 | Not-determined |
| KM_k141_198903 | Not-determined |
| KM_k141_143012 | Low-quality |
| KM_k141_232417 | Not-determined |
| KM_k141_187664 | Not-determined |
| KM_k141_263499 | Not-determined |
| KM_k141_242191 | Not-determined |
| KM_k141_108895 | Not-determined |
| KM_k141_64649 | Not-determined |
| KM_k141_30590 | Not-determined |
| KM_k141_232426 | Not-determined |
| KM_k141_163626 | Not-determined |
| KM_k141_221428 | Not-determined |
| KM_k141_209960 | Not-determined |
| KM_k141_30594 | Not-determined |
| KM_k141_130989 | Not-determined |
| KM_k141_187678 | Not-determined |
| KM_k141_163629 | Not-determined |
| KM_k141_8445 | Not-determined |
| KM_k141_187679 | Not-determined |
| KM_k141_153650 | Not-determined |
| KM_k141_121726 | Not-determined |
| KM_k141_198920 | Not-determined |
| KM_k141_18771 | Not-determined |
| KM_k141_153653 | Not-determined |
| KM_k141_18774 | Not-determined |
| KM_k141_51398 | Not-determined |
| KM_k141_263507 | Low-quality |
| KM_k141_30601 | Not-determined |
| KM_k141_163638 | Not-determined |
| KM_k141_18778 | Not-determined |
| KM_k141_64669 | Not-determined |
| KM_k141_121739 | Not-determined |
| KM_k141_221445 | Not-determined |
| KM_k141_121740 | Not-determined |
| KM_k141_18783 | Not-determined |
| KM_k141_64677 | Not-determined |
| KM_k141_209977 | Not-determined |
| KM_k141_221455 | Not-determined |
| KM_k141_18795 | Not-determined |
| KM_k141_64685 | Not-determined |
| KM_k141_263526 | Not-determined |
| KM_k141_64686 | Not-determined |
| KM_k141_198937 | Not-determined |
| KM_k141_131007 | Not-determined |
| KM_k141_64694 | Not-determined |
| KM_k141_263529 | Not-determined |
| KM_k141_131012 | Not-determined |
| KM_k141_108921 | Not-determined |
| KM_k141_198942 | Not-determined |
| KM_k141_41542 | Not-determined |
| KM_k141_198947 | Not-determined |
| KM_k141_175991 | Not-determined |
| KM_k141_108931 | Not-determined |
| KM_k141_163650 | Not-determined |
| KM_k141_163651 | Not-determined |
| KM_k141_263541 | Not-determined |
| KM_k141_30621 | Low-quality |
| KM_k141_221489 | Not-determined |
| KM_k141_221490 | Not-determined |
| KM_k141_76461 | Not-determined |
| KM_k141_64723 | Not-determined |
| KM_k141_64724 | Not-determined |
| KM_k141_76462 | Not-determined |
| KM_k141_108936 | Not-determined |
| KM_k141_88018 | Not-determined |
| KM_k141_163657 | Low-quality |
| KM_k141_263551 | Not-determined |
| KM_k141_88020 | Not-determined |
| KM_k141_30627 | Not-determined |
| KM_k141_30630 | Not-determined |
| KM_k141_153702 | Not-determined |
| KM_k141_187695 | Not-determined |
| KM_k141_76467 | Not-determined |
| KM_k141_163658 | Not-determined |
| KM_k141_30633 | Not-determined |
| KM_k141_153706 | Not-determined |
| KM_k141_263564 | Low-quality |
| KM_k141_51413 | Not-determined |
| KM_k141_51414 | Not-determined |
| KM_k141_232462 | Not-determined |
| KM_k141_64738 | Not-determined |
| KM_k141_176015 | Not-determined |
| KM_k141_187706 | Not-determined |
| KM_k141_108944 | Not-determined |
| KM_k141_30642 | Not-determined |
| KM_k141_108948 | Not-determined |
| KM_k141_76473 | Not-determined |
| KM_k141_163672 | Not-determined |
| KM_k141_64750 | Not-determined |
| KM_k141_210004 | Not-determined |
| KM_k141_51421 | Not-determined |
| KM_k141_88026 | Low-quality |
| KM_k141_163684 | Not-determined |
| KM_k141_198983 | Not-determined |
| KM_k141_64758 | Low-quality |
| KM_k141_51430 | Not-determined |
| KM_k141_163688 | Not-determined |
| KM_k141_232480 | Not-determined |
| KM_k141_108959 | Not-determined |
| KM_k141_64760 | Low-quality |
| KM_k141_76492 | Not-determined |
| KM_k141_108963 | Not-determined |
| KM_k141_210012 | Not-determined |
| KM_k141_30662 | Not-determined |
| KM_k141_51436 | Not-determined |
| KM_k141_198992 | Not-determined |
| KM_k141_210017 | Not-determined |
| KM_k141_76499 | Not-determined |
| KM_k141_121774 | Not-determined |
| KM_k141_76508 | Not-determined |
| KM_k141_96076 | Not-determined |
| KM_k141_121777 | Not-determined |
| KM_k141_88032 | Not-determined |
| KM_k141_41592 | Not-determined |
| KM_k141_153735 | Not-determined |
| KM_k141_96080 | Not-determined |
| KM_k141_153741 | Not-determined |
| KM_k141_143096 | Not-determined |
| KM_k141_76518 | Not-determined |
| KM_k141_143102 | Not-determined |
| KM_k141_88044 | Not-determined |
| KM_k141_64787 | Not-determined |
| KM_k141_41609 | Not-determined |
| KM_k141_143106 | Not-determined |
| KM_k141_252038 | Not-determined |
| KM_k141_108974 | Low-quality |
| KM_k141_51451 | Not-determined |
| KM_k141_88052 | Not-determined |
| KM_k141_76525 | Not-determined |
| KM_k141_176051 | Not-determined |
| KM_k141_252043 | Not-determined |
| KM_k141_163712 | Not-determined |
| KM_k141_41615 | Low-quality |
| KM_k141_163714 | Not-determined |
| KM_k141_163715 | Not-determined |
| KM_k141_51463 | Not-determined |
| KM_k141_121791 | Not-determined |
| KM_k141_242210 | Not-determined |
| KM_k141_41617 | Not-determined |
| KM_k141_143130 | Not-determined |
| KM_k141_176056 | Low-quality |
| KM_k141_18807 | Not-determined |
| KM_k141_96088 | Not-determined |
| KM_k141_76529 | Not-determined |
| KM_k141_51466 | Low-quality |
| KM_k141_41625 | Not-determined |
| KM_k141_18815 | Not-determined |
| KM_k141_76537 | Not-determined |
| KM_k141_76538 | Not-determined |
| KM_k141_76541 | Not-determined |
| KM_k141_121803 | Low-quality |
| KM_k141_131072 | Not-determined |
| KM_k141_187757 | Not-determined |
| KM_k141_121811 | Not-determined |
| KM_k141_232533 | Not-determined |
| KM_k141_96104 | Not-determined |
| KM_k141_210063 | Not-determined |
| KM_k141_242235 | Low-quality |
| KM_k141_242237 | Not-determined |
| KM_k141_242239 | Not-determined |
| KM_k141_232542 | Not-determined |
| KM_k141_121822 | Not-determined |
| KM_k141_242243 | Not-determined |
| KM_k141_18832 | Not-determined |
| KM_k141_210067 | Not-determined |
| KM_k141_64837 | Low-quality |
| KM_k141_242245 | Not-determined |
| KM_k141_210069 | Not-determined |
| KM_k141_96118 | Not-determined |
| KM_k141_153800 | Not-determined |
| KM_k141_64844 | Not-determined |
| KM_k141_30678 | Low-quality |
| KM_k141_252084 | Not-determined |
| KM_k141_163748 | Not-determined |
| KM_k141_252086 | Not-determined |
| KM_k141_30681 | Not-determined |
| KM_k141_41652 | Low-quality |
| KM_k141_232559 | Not-determined |
| KM_k141_41658 | Not-determined |
| KM_k141_41660 | Not-determined |
| KM_k141_221570 | Not-determined |
| KM_k141_163755 | Not-determined |
| KM_k141_41662 | Not-determined |
| KM_k141_210082 | Not-determined |
| KM_k141_210086 | Not-determined |
| KM_k141_252095 | Not-determined |
| KM_k141_242268 | Not-determined |
| KM_k141_143179 | Not-determined |
| KM_k141_41669 | Not-determined |
| KM_k141_18847 | Not-determined |
| KM_k141_76577 | Not-determined |
| KM_k141_131105 | Not-determined |
| KM_k141_263594 | Not-determined |
| KM_k141_18850 | Not-determined |
| KM_k141_153825 | Not-determined |
| KM_k141_176078 | Not-determined |
| KM_k141_232581 | Not-determined |
| KM_k141_176079 | Not-determined |
| KM_k141_131111 | Not-determined |
| KM_k141_76580 | Not-determined |
| KM_k141_263604 | Not-determined |
| KM_k141_187782 | Not-determined |
| KM_k141_252102 | Not-determined |
| KM_k141_153833 | Not-determined |
| KM_k141_131123 | Not-determined |
| KM_k141_176090 | Not-determined |
| KM_k141_232585 | Not-determined |
| KM_k141_221597 | Low-quality |
| KM_k141_242275 | Not-determined |
| KM_k141_176098 | Low-quality |
| KM_k141_252110 | Not-determined |
| KM_k141_242276 | Not-determined |
| KM_k141_176100 | Not-determined |
| KM_k141_88121 | Not-determined |
| KM_k141_263618 | Not-determined |
| KM_k141_199027 | Not-determined |
| KM_k141_176110 | Not-determined |
| KM_k141_242290 | Not-determined |
| KM_k141_131138 | Not-determined |
| KM_k141_30711 | Not-determined |
| KM_k141_187799 | Not-determined |
| KM_k141_76601 | Not-determined |
| KM_k141_18879 | Not-determined |
| KM_k141_88137 | Not-determined |
| KM_k141_187804 | Not-determined |
| KM_k141_121858 | Low-quality |
| KM_k141_163786 | Not-determined |
| KM_k141_242294 | Not-determined |
| KM_k141_76605 | Not-determined |
| KM_k141_242298 | Not-determined |
| KM_k141_176115 | Not-determined |
| KM_k141_109060 | Not-determined |
| KM_k141_109061 | Not-determined |
| KM_k141_76614 | Not-determined |
| KM_k141_76615 | Not-determined |
| KM_k141_242303 | Not-determined |
| KM_k141_41686 | Low-quality |
| KM_k141_252124 | Not-determined |
| KM_k141_163796 | Not-determined |
| KM_k141_96146 | Not-determined |
| KM_k141_163797 | Not-determined |
| KM_k141_199041 | Not-determined |
| KM_k141_96148 | Not-determined |
| KM_k141_163799 | Not-determined |
| KM_k141_242313 | Not-determined |
| KM_k141_131145 | Low-quality |
| KM_k141_153860 | Not-determined |
| KM_k141_221616 | Not-determined |
| KM_k141_210109 | Low-quality |
| KM_k141_41691 | Not-determined |
| KM_k141_88161 | Not-determined |
| KM_k141_88166 | Not-determined |
| KM_k141_187817 | Not-determined |
| KM_k141_252140 | Not-determined |
| KM_k141_30728 | Low-quality |
| KM_k141_41701 | Not-determined |
| KM_k141_121887 | Not-determined |
| KM_k141_30732 | Not-determined |
| KM_k141_88176 | Not-determined |
| KM_k141_210123 | Not-determined |
| KM_k141_51497 | Not-determined |
| KM_k141_76634 | Low-quality |
| KM_k141_30736 | Not-determined |
| KM_k141_187830 | Not-determined |
| KM_k141_76639 | Not-determined |
| KM_k141_252144 | Not-determined |
| KM_k141_121892 | Not-determined |
| KM_k141_163827 | Not-determined |
| KM_k141_187835 | Low-quality |
| KM_k141_30745 | Not-determined |
| KM_k141_96162 | Not-determined |
| KM_k141_163831 | Not-determined |
| KM_k141_199064 | Not-determined |
| KM_k141_242331 | Not-determined |
| KM_k141_121894 | Not-determined |
| KM_k141_187847 | Not-determined |
| KM_k141_64886 | Not-determined |
| KM_k141_163842 | Low-quality |
| KM_k141_232626 | Not-determined |
| KM_k141_76652 | Not-determined |
| KM_k141_163846 | Not-determined |
| KM_k141_232630 | Not-determined |
| KM_k141_232631 | Not-determined |
| KM_k141_187851 | Not-determined |
| KM_k141_263664 | Low-quality |
| KM_k141_199083 | Not-determined |
| KM_k141_232636 | Not-determined |
| KM_k141_263666 | Not-determined |
| KM_k141_64905 | Not-determined |
| KM_k141_232640 | Not-determined |
| KM_k141_252165 | Not-determined |
| KM_k141_76660 | Low-quality |
| KM_k141_121897 | Not-determined |
| KM_k141_76663 | Not-determined |
| KM_k141_143209 | Not-determined |
| KM_k141_96186 | Not-determined |
| KM_k141_88221 | Not-determined |
| KM_k141_64913 | Not-determined |
| KM_k141_263679 | Not-determined |
| KM_k141_18922 | Not-determined |
| KM_k141_88224 | Not-determined |
| KM_k141_109074 | Not-determined |
| KM_k141_153885 | Not-determined |
| KM_k141_232652 | Not-determined |
| KM_k141_18926 | Not-determined |
| KM_k141_143215 | Low-quality |
| KM_k141_153888 | Not-determined |
| KM_k141_153889 | Not-determined |
| KM_k141_176158 | Not-determined |
| KM_k141_64917 | Not-determined |
| KM_k141_143219 | Not-determined |
| KM_k141_153894 | Not-determined |
| KM_k141_199103 | Not-determined |
| KM_k141_76678 | Not-determined |
| KM_k141_76679 | Not-determined |
| KM_k141_252173 | Not-determined |
| KM_k141_143222 | Not-determined |
| KM_k141_252174 | Not-determined |
| KM_k141_252175 | Not-determined |
| KM_k141_121914 | Not-determined |
| KM_k141_153901 | Not-determined |
| KM_k141_232675 | Not-determined |
| KM_k141_143231 | Not-determined |
| KM_k141_121917 | Not-determined |
| KM_k141_30778 | Not-determined |
| KM_k141_199109 | Not-determined |
| KM_k141_221673 | Not-determined |
| KM_k141_88235 | Not-determined |
| KM_k141_96212 | Not-determined |
| KM_k141_96214 | Not-determined |
| KM_k141_88238 | Not-determined |
| KM_k141_8488 | Low-quality |
| KM_k141_88239 | Not-determined |
| KM_k141_221680 | Not-determined |
| KM_k141_199117 | Not-determined |
| KM_k141_30787 | Not-determined |
| KM_k141_221681 | Not-determined |
| KM_k141_199122 | Not-determined |
| KM_k141_18944 | Not-determined |
| KM_k141_153910 | Not-determined |
| KM_k141_30792 | Not-determined |
| KM_k141_76710 | Not-determined |
| KM_k141_187882 | Not-determined |
| KM_k141_8495 | Not-determined |
| KM_k141_41740 | Not-determined |

| | |
|---|---|
| KM_k141_163872 | Low-quality |
| KM_k141_41741 | Low-quality |
| KM_k141_88249 | Low-quality |
| KM_k141_263712 | Not-determined |
| KM_k141_8504 | Not-determined |
| KM_k141_8507 | Low-quality |
| KM_k141_143247 | Not-determined |
| KM_k141_176173 | Low-quality |
| KM_k141_109096 | Not-determined |
| KM_k141_187897 | Low-quality |
| KM_k141_88256 | Not-determined |
| KM_k141_163881 | Not-determined |
| KM_k141_96226 | Not-determined |
| KM_k141_41748 | Not-determined |
| KM_k141_41750 | Not-determined |
| KM_k141_96229 | Not-determined |
| KM_k141_210201 | Not-determined |
| KM_k141_30810 | Not-determined |
| KM_k141_18969 | Not-determined |
| KM_k141_18970 | Not-determined |
| KM_k141_41756 | Not-determined |
| KM_k141_153922 | Not-determined |
| KM_k141_199153 | Not-determined |
| KM_k141_163891 | Not-determined |
| KM_k141_199157 | Not-determined |
| KM_k141_232713 | Not-determined |
| KM_k141_199161 | Not-determined |
| KM_k141_18980 | Not-determined |
| KM_k141_163895 | Not-determined |
| KM_k141_143253 | Low-quality |
| KM_k141_263727 | Not-determined |
| KM_k141_8519 | Not-determined |
| KM_k141_18984 | Not-determined |
| KM_k141_121944 | Not-determined |
| KM_k141_143260 | Not-determined |
| KM_k141_187917 | Not-determined |
| KM_k141_176190 | Not-determined |
| KM_k141_153926 | Not-determined |
| KM_k141_51560 | Not-determined |
| KM_k141_153928 | Not-determined |
| KM_k141_30825 | Not-determined |
| KM_k141_263737 | Not-determined |
| KM_k141_76743 | Not-determined |
| KM_k141_76747 | Not-determined |
| KM_k141_232733 | Not-determined |
| KM_k141_64943 | Not-determined |
| KM_k141_41779 | Not-determined |
| KM_k141_143283 | Not-determined |
| KM_k141_143287 | Not-determined |
| KM_k141_64955 | Not-determined |
| KM_k141_64956 | Not-determined |
| KM_k141_76770 | Not-determined |
| KM_k141_163903 | Not-determined |
| KM_k141_41793 | Not-determined |
| KM_k141_232743 | Not-determined |
| KM_k141_163912 | Not-determined |
| KM_k141_199190 | Not-determined |
| KM_k141_232752 | Not-determined |
| KM_k141_232753 | Not-determined |
| KM_k141_143298 | Not-determined |
| KM_k141_41804 | Not-determined |
| KM_k141_41807 | Not-determined |
| KM_k141_88296 | Not-determined |
| KM_k141_221740 | Not-determined |
| KM_k141_143302 | Not-determined |
| KM_k141_41808 | Not-determined |
| KM_k141_76775 | Not-determined |
| KM_k141_199202 | Not-determined |
| KM_k141_30838 | Not-determined |
| KM_k141_210236 | Not-determined |
| KM_k141_143305 | Not-determined |
| KM_k141_8551 | Not-determined |
| KM_k141_252210 | Not-determined |
| KM_k141_176210 | Not-determined |
| KM_k141_8559 | Not-determined |
| KM_k141_242371 | Not-determined |
| KM_k141_30844 | Not-determined |
| KM_k141_8562 | Not-determined |
| KM_k141_88323 | Low-quality |
| KM_k141_187948 | Not-determined |
| KM_k141_121950 | Not-determined |
| KM_k141_65014 | Not-determined |
| KM_k141_121953 | Not-determined |
| KM_k141_221776 | Not-determined |
| KM_k141_121954 | Not-determined |
| KM_k141_263770 | Not-determined |
| KM_k141_153977 | Not-determined |
| KM_k141_221779 | Not-determined |
| KM_k141_76797 | Not-determined |
| KM_k141_88331 | Not-determined |
| KM_k141_143327 | Not-determined |
| KM_k141_51598 | Not-determined |
| KM_k141_121959 | Not-determined |
| KM_k141_199225 | Not-determined |
| KM_k141_121961 | Not-determined |
| KM_k141_232790 | Not-determined |
| KM_k141_263776 | Not-determined |
| KM_k141_30854 | Not-determined |
| KM_k141_76808 | Not-determined |
| KM_k141_153986 | Not-determined |
| KM_k141_65026 | Not-determined |
| KM_k141_263780 | Not-determined |
| KM_k141_143332 | Not-determined |
| KM_k141_242381 | Not-determined |
| KM_k141_88347 | Low-quality |
| KM_k141_210258 | Not-determined |
| KM_k141_65032 | Not-determined |
| KM_k141_65033 | Not-determined |
| KM_k141_51614 | Not-determined |
| KM_k141_163935 | Not-determined |
| KM_k141_143335 | Not-determined |
| KM_k141_131239 | Not-determined |
| KM_k141_65044 | Not-determined |
| KM_k141_210272 | Not-determined |
| KM_k141_88358 | Not-determined |
| KM_k141_210273 | Not-determined |
| KM_k141_163946 | Not-determined |
| KM_k141_88360 | Low-quality |
| KM_k141_252248 | Not-determined |
| KM_k141_76814 | Low-quality |
| KM_k141_65055 | Not-determined |
| KM_k141_143350 | Not-determined |
| KM_k141_252259 | Not-determined |
| KM_k141_242402 | Not-determined |
| KM_k141_65060 | Not-determined |
| KM_k141_221813 | Low-quality |
| KM_k141_76833 | Not-determined |
| KM_k141_121984 | Low-quality |
| KM_k141_154004 | Not-determined |
| KM_k141_121991 | Not-determined |
| KM_k141_96265 | Not-determined |
| KM_k141_88376 | Not-determined |
| KM_k141_88378 | Not-determined |
| KM_k141_143355 | Not-determined |
| KM_k141_187981 | Low-quality |
| KM_k141_41820 | Not-determined |
| KM_k141_187985 | Not-determined |
| KM_k141_187986 | Not-determined |
| KM_k141_242420 | Not-determined |
| KM_k141_30865 | Not-determined |
| KM_k141_242424 | Not-determined |
| KM_k141_30866 | Not-determined |
| KM_k141_176279 | Not-determined |
| KM_k141_143369 | Not-determined |
| KM_k141_154019 | Not-determined |
| KM_k141_41827 | Not-determined |
| KM_k141_41828 | Not-determined |
| KM_k141_51635 | Not-determined |
| KM_k141_143373 | Not-determined |
| KM_k141_176282 | Not-determined |
| KM_k141_76847 | Not-determined |
| KM_k141_232868 | Not-determined |
| KM_k141_252274 | Not-determined |
| KM_k141_96282 | Not-determined |
| KM_k141_8608 | Not-determined |
| KM_k141_19026 | Not-determined |
| KM_k141_88416 | Low-quality |
| KM_k141_210282 | Not-determined |
| KM_k141_51639 | Not-determined |
| KM_k141_51642 | Not-determined |
| KM_k141_8616 | Not-determined |
| KM_k141_51645 | Not-determined |
| KM_k141_96287 | Not-determined |
| KM_k141_96288 | Not-determined |
| KM_k141_88433 | Not-determined |
| KM_k141_143380 | Not-determined |
| KM_k141_210290 | Not-determined |
| KM_k141_242454 | Not-determined |
| KM_k141_109161 | Not-determined |
| KM_k141_65110 | Not-determined |
| KM_k141_76866 | Not-determined |
| KM_k141_242461 | Not-determined |
| KM_k141_199283 | Not-determined |
| KM_k141_252295 | Not-determined |
| KM_k141_131280 | Not-determined |
| KM_k141_221867 | Low-quality |
| KM_k141_221868 | Not-determined |
| KM_k141_263801 | Not-determined |
| KM_k141_19048 | Not-determined |
| KM_k141_51664 | Not-determined |
| KM_k141_19052 | Not-determined |
| KM_k141_88457 | Not-determined |
| KM_k141_96302 | Not-determined |
| KM_k141_30893 | Not-determined |
| KM_k141_199292 | Not-determined |
| KM_k141_109177 | Not-determined |
| KM_k141_163992 | Not-determined |
| KM_k141_30895 | Not-determined |
| KM_k141_30896 | Not-determined |
| KM_k141_154053 | Not-determined |
| KM_k141_88463 | Not-determined |
| KM_k141_51672 | Not-determined |
| KM_k141_30900 | Not-determined |
| KM_k141_163996 | Not-determined |
| KM_k141_232884 | Not-determined |
| KM_k141_199301 | Not-determined |
| KM_k141_263824 | Not-determined |
| KM_k141_154064 | Not-determined |
| KM_k141_41864 | Not-determined |
| KM_k141_30908 | Not-determined |
| KM_k141_19087 | Not-determined |
| KM_k141_8659 | Not-determined |
| KM_k141_51678 | Not-determined |
| KM_k141_88476 | Low-quality |
| KM_k141_96322 | Not-determined |
| KM_k141_210313 | Not-determined |
| KM_k141_41877 | Not-determined |
| KM_k141_109191 | Low-quality |
| KM_k141_51687 | Not-determined |
| KM_k141_188042 | Not-determined |
| KM_k141_131326 | Low-quality |
| KM_k141_188044 | Not-determined |
| KM_k141_252305 | Not-determined |
| KM_k141_109195 | Not-determined |
| KM_k141_154080 | Not-determined |
| KM_k141_30922 | Not-determined |
| KM_k141_221888 | Not-determined |
| KM_k141_210326 | Not-determined |
| KM_k141_252313 | Not-determined |
| KM_k141_131334 | Not-determined |
| KM_k141_164022 | Not-determined |
| KM_k141_199312 | Not-determined |
| KM_k141_51693 | Not-determined |
| KM_k141_263841 | Not-determined |
| KM_k141_164023 | Not-determined |
| KM_k141_122019 | Not-determined |
| KM_k141_96341 | Not-determined |
| KM_k141_199317 | Not-determined |
| KM_k141_210338 | Low-quality |
| KM_k141_232921 | Not-determined |
| KM_k141_221895 | Not-determined |
| KM_k141_41882 | Not-determined |
| KM_k141_88505 | Not-determined |
| KM_k141_210342 | Not-determined |
| KM_k141_232923 | Not-determined |
| KM_k141_242484 | Not-determined |
| KM_k141_164032 | Not-determined |
| KM_k141_176355 | Not-determined |
| KM_k141_41887 | Not-determined |
| KM_k141_242485 | Low-quality |
| KM_k141_252323 | Not-determined |
| KM_k141_88512 | Not-determined |
| KM_k141_88514 | Not-determined |
| KM_k141_242491 | Not-determined |
| KM_k141_188068 | Not-determined |
| KM_k141_164040 | Not-determined |
| KM_k141_109207 | Not-determined |
| KM_k141_188069 | Not-determined |
| KM_k141_199330 | Not-determined |
| KM_k141_188071 | Not-determined |
| KM_k141_88527 | Not-determined |
| KM_k141_154101 | Not-determined |
| KM_k141_252336 | Not-determined |
| KM_k141_164042 | Not-determined |
| KM_k141_199336 | Not-determined |
| KM_k141_131349 | Not-determined |
| KM_k141_176376 | Not-determined |
| KM_k141_51716 | Not-determined |
| KM_k141_30932 | Not-determined |
| KM_k141_188082 | Not-determined |
| KM_k141_131358 | Not-determined |
| KM_k141_242507 | Not-determined |
| KM_k141_164055 | Not-determined |
| KM_k141_242510 | Not-determined |
| KM_k141_188089 | Not-determined |
| KM_k141_154119 | Not-determined |
| KM_k141_242512 | Not-determined |
| KM_k141_263869 | Low-quality |
| KM_k141_210372 | Not-determined |
| KM_k141_154125 | Not-determined |
| KM_k141_263870 | Not-determined |
| KM_k141_210374 | Not-determined |
| KM_k141_188095 | Low-quality |
| KM_k141_210377 | Not-determined |
| KM_k141_252359 | Not-determined |
| KM_k141_19100 | Not-determined |
| KM_k141_41917 | Not-determined |
| KM_k141_30951 | Not-determined |
| KM_k141_154132 | Not-determined |
| KM_k141_109221 | Not-determined |
| KM_k141_122055 | Not-determined |
| KM_k141_76899 | Not-determined |
| KM_k141_41931 | Not-determined |
| KM_k141_199358 | Not-determined |
| KM_k141_232967 | Not-determined |
| KM_k141_30959 | Not-determined |
| KM_k141_30961 | Not-determined |
| KM_k141_199364 | Not-determined |
| KM_k141_30962 | Low-quality |
| KM_k141_232974 | Not-determined |
| KM_k141_199366 | Not-determined |
| KM_k141_19112 | Not-determined |
| KM_k141_210388 | Not-determined |
| KM_k141_242535 | Low-quality |
| KM_k141_164075 | Low-quality |
| KM_k141_176403 | Not-determined |
| KM_k141_19120 | Not-determined |
| KM_k141_143477 | Not-determined |
| KM_k141_210403 | Not-determined |
| KM_k141_96388 | Not-determined |
| KM_k141_76911 | Not-determined |
| KM_k141_19125 | Not-determined |
| KM_k141_65196 | Low-quality |
| KM_k141_30982 | Not-determined |
| KM_k141_176406 | Not-determined |
| KM_k141_210410 | Low-quality |
| KM_k141_8723 | Not-determined |
| KM_k141_221923 | Not-determined |
| KM_k141_143486 | Not-determined |
| KM_k141_221925 | Not-determined |
| KM_k141_176416 | Not-determined |
| KM_k141_30991 | Not-determined |
| KM_k141_188134 | Not-determined |
| KM_k141_154154 | Not-determined |
| KM_k141_65204 | Not-determined |
| KM_k141_154156 | Not-determined |
| KM_k141_164090 | Not-determined |
| KM_k141_164091 | Not-determined |
| KM_k141_210427 | Not-determined |
| KM_k141_242557 | Not-determined |
| KM_k141_154173 | Not-determined |
| KM_k141_19140 | Low-quality |
| KM_k141_154175 | Not-determined |
| KM_k141_164098 | Not-determined |
| KM_k141_199380 | Low-quality |
| KM_k141_242563 | Not-determined |
| KM_k141_199381 | Not-determined |
| KM_k141_242564 | Not-determined |
| KM_k141_176432 | Low-quality |
| KM_k141_96425 | Not-determined |
| KM_k141_164104 | Not-determined |
| KM_k141_76940 | Not-determined |
| KM_k141_19151 | Low-quality |
| KM_k141_164107 | Not-determined |
| KM_k141_65225 | Not-determined |
| KM_k141_143502 | Not-determined |
| KM_k141_31018 | Not-determined |
| KM_k141_232995 | Not-determined |
| KM_k141_242571 | Not-determined |
| KM_k141_232996 | Not-determined |
| KM_k141_8767 | Not-determined |
| KM_k141_210445 | Not-determined |
| KM_k141_164114 | Not-determined |
| KM_k141_252397 | Not-determined |
| KM_k141_122074 | Not-determined |
| KM_k141_233005 | Not-determined |
| KM_k141_41941 | Not-determined |
| KM_k141_252399 | Not-determined |
| KM_k141_252401 | Not-determined |
| KM_k141_41942 | Not-determined |
| KM_k141_65234 | Not-determined |
| KM_k141_164119 | Not-determined |
| KM_k141_164120 | Not-determined |
| KM_k141_233014 | Not-determined |
| KM_k141_233015 | Not-determined |
| KM_k141_154189 | Not-determined |
| KM_k141_252404 | Not-determined |
| KM_k141_242579 | Not-determined |
| KM_k141_109268 | Not-determined |
| KM_k141_210456 | Not-determined |
| KM_k141_109270 | Not-determined |
| KM_k141_199403 | Not-determined |
| KM_k141_199406 | Not-determined |
| KM_k141_252411 | Not-determined |
| KM_k141_242588 | Not-determined |
| KM_k141_252415 | Not-determined |
| KM_k141_242593 | Not-determined |
| KM_k141_154206 | Low-quality |
| KM_k141_188165 | Not-determined |
| KM_k141_122081 | Not-determined |
| KM_k141_188168 | Not-determined |
| KM_k141_31025 | Not-determined |
| KM_k141_188169 | Not-determined |
| KM_k141_96456 | Not-determined |
| KM_k141_31027 | Not-determined |
| KM_k141_210478 | Not-determined |
| KM_k141_96458 | Not-determined |
| KM_k141_242602 | Not-determined |
| KM_k141_263904 | Not-determined |
| KM_k141_188176 | Not-determined |
| KM_k141_96465 | Not-determined |
| KM_k141_176463 | Not-determined |
| KM_k141_96469 | Not-determined |
| KM_k141_188184 | Not-determined |
| KM_k141_176467 | Not-determined |
| KM_k141_210494 | Not-determined |
| KM_k141_154220 | Not-determined |
| KM_k141_143538 | Not-determined |
| KM_k141_109293 | Not-determined |
| KM_k141_221995 | Not-determined |
| KM_k141_154228 | Low-quality |
| KM_k141_122099 | Not-determined |
| KM_k141_41991 | Not-determined |
| KM_k141_176483 | Not-determined |
| KM_k141_252437 | Not-determined |
| KM_k141_41992 | Not-determined |
| KM_k141_8807 | Not-determined |
| KM_k141_252441 | Not-determined |
| KM_k141_96487 | Not-determined |
| KM_k141_8809 | Not-determined |
| KM_k141_252443 | Not-determined |
| KM_k141_42001 | Not-determined |
| KM_k141_8811 | Not-determined |
| KM_k141_154235 | Not-determined |
| KM_k141_96495 | Low-quality |
| KM_k141_242626 | Not-determined |
| KM_k141_242628 | Not-determined |
| KM_k141_19189 | Not-determined |
| KM_k141_122108 | Not-determined |
| KM_k141_88585 | Not-determined |
| KM_k141_143567 | Not-determined |
| KM_k141_31069 | Not-determined |
| KM_k141_65256 | Not-determined |
| KM_k141_233040 | Not-determined |
| KM_k141_42008 | Low-quality |
| KM_k141_188208 | Not-determined |
| KM_k141_154248 | Low-quality |
| KM_k141_210526 | Not-determined |
| KM_k141_210527 | Not-determined |
| KM_k141_176510 | Not-determined |
| KM_k141_222018 | Low-quality |
| KM_k141_65267 | Not-determined |
| KM_k141_188215 | Not-determined |
| KM_k141_188219 | Not-determined |
| KM_k141_154257 | Not-determined |
| KM_k141_88602 | Not-determined |
| KM_k141_42015 | Not-determined |
| KM_k141_222029 | Not-determined |
| KM_k141_210531 | Not-determined |
| KM_k141_42021 | Not-determined |
| KM_k141_143597 | Not-determined |
| KM_k141_131476 | Not-determined |
| KM_k141_109335 | Not-determined |
| KM_k141_199449 | Not-determined |
| KM_k141_65282 | Not-determined |
| KM_k141_109339 | Not-determined |
| KM_k141_131487 | Low-quality |
| KM_k141_65288 | Low-quality |
| KM_k141_252475 | Not-determined |
| KM_k141_31098 | Not-determined |
| KM_k141_176536 | Not-determined |
| KM_k141_8840 | Not-determined |
| KM_k141_131493 | Not-determined |
| KM_k141_143611 | Not-determined |
| KM_k141_131499 | Not-determined |
| KM_k141_88603 | Low-quality |
| KM_k141_154274 | Not-determined |
| KM_k141_8850 | Not-determined |
| KM_k141_8853 | Not-determined |
| KM_k141_88608 | Not-determined |
| KM_k141_263929 | Not-determined |
| KM_k141_176546 | Not-determined |
| KM_k141_263931 | Not-determined |
| KM_k141_122147 | Not-determined |
| KM_k141_88612 | Not-determined |
| KM_k141_122156 | Not-determined |
| KM_k141_8866 | Not-determined |
| KM_k141_88617 | Not-determined |
| KM_k141_252491 | Not-determined |
| KM_k141_176555 | Not-determined |
| KM_k141_176556 | Not-determined |
| KM_k141_176558 | Not-determined |
| KM_k141_176561 | Not-determined |
| KM_k141_176562 | Not-determined |
| KM_k141_263933 | Low-quality |
| KM_k141_8872 | Low-quality |
| KM_k141_96515 | Not-determined |
| KM_k141_88626 | Not-determined |
| KM_k141_42063 | Not-determined |
| KM_k141_263939 | Not-determined |
| KM_k141_164193 | Not-determined |
| KM_k141_19205 | Not-determined |
| KM_k141_88629 | Not-determined |
| KM_k141_96519 | Not-determined |
| KM_k141_143637 | Not-determined |
| KM_k141_88631 | Not-determined |
| KM_k141_31120 | Not-determined |
| KM_k141_154292 | Not-determined |
| KM_k141_65307 | Not-determined |
| KM_k141_263948 | Not-determined |
| KM_k141_88639 | Low-quality |
| KM_k141_31126 | Not-determined |
| KM_k141_154297 | Low-quality |
| KM_k141_222066 | Not-determined |
| KM_k141_154299 | Not-determined |
| KM_k141_42074 | Not-determined |
| KM_k141_31130 | Not-determined |
| KM_k141_31131 | Low-quality |
| KM_k141_143646 | Not-determined |
| KM_k141_164207 | Low-quality |
| KM_k141_143649 | Not-determined |
| KM_k141_88659 | Not-determined |
| KM_k141_65320 | Not-determined |
| KM_k141_109370 | Not-determined |
| KM_k141_65321 | Low-quality |
| KM_k141_242636 | Not-determined |
| KM_k141_263959 | Not-determined |
| KM_k141_143657 | Not-determined |
| KM_k141_88665 | Low-quality |
| KM_k141_19226 | Not-determined |
| KM_k141_19227 | Not-determined |
| KM_k141_42080 | Not-determined |
| KM_k141_8898 | Not-determined |
| KM_k141_8900 | Not-determined |
| KM_k141_164217 | Not-determined |
| KM_k141_242642 | Not-determined |
| KM_k141_252527 | Not-determined |
| KM_k141_233112 | Not-determined |
| KM_k141_96525 | Not-determined |
| KM_k141_131509 | Not-determined |
| KM_k141_242644 | Not-determined |
| KM_k141_8910 | Not-determined |
| KM_k141_188253 | Not-determined |
| KM_k141_176585 | Not-determined |
| KM_k141_88673 | Low-quality |
| KM_k141_88674 | Not-determined |
| KM_k141_8915 | Not-determined |
| KM_k141_96531 | Low-quality |
| KM_k141_19240 | Not-determined |
| KM_k141_77021 | Not-determined |
| KM_k141_42102 | Not-determined |
| KM_k141_77028 | Not-determined |
| KM_k141_154335 | Not-determined |
| KM_k141_263982 | Not-determined |
| KM_k141_8924 | Not-determined |
| KM_k141_263983 | Not-determined |
| KM_k141_263985 | Not-determined |
| KM_k141_263987 | Not-determined |
| KM_k141_188278 | Not-determined |
| KM_k141_143684 | Not-determined |
| KM_k141_176597 | Not-determined |
| KM_k141_222082 | Not-determined |
| KM_k141_222083 | Not-determined |
| KM_k141_263992 | Not-determined |
| KM_k141_242656 | Not-determined |
| KM_k141_65347 | Not-determined |
| KM_k141_8939 | Not-determined |
| KM_k141_42116 | Not-determined |
| KM_k141_96548 | Not-determined |
| KM_k141_42117 | Not-determined |
| KM_k141_42118 | Not-determined |
| KM_k141_233134 | Not-determined |
| KM_k141_264000 | Not-determined |
| KM_k141_264001 | Not-determined |
| KM_k141_8944 | Not-determined |
| KM_k141_222092 | Not-determined |
| KM_k141_176608 | Not-determined |
| KM_k141_8948 | Not-determined |
| KM_k141_31165 | Not-determined |
| KM_k141_109393 | Not-determined |
| KM_k141_88692 | Not-determined |
| KM_k141_222094 | Not-determined |
| KM_k141_154353 | Not-determined |
| KM_k141_222095 | Not-determined |
| KM_k141_199483 | Not-determined |
| KM_k141_109396 | Not-determined |
| KM_k141_31169 | Not-determined |
| KM_k141_65363 | Not-determined |
| KM_k141_210645 | Not-determined |
| KM_k141_109399 | Not-determined |
| KM_k141_109400 | Not-determined |
| KM_k141_176619 | Not-determined |
| KM_k141_109401 | Not-determined |
| KM_k141_222102 | Not-determined |
| KM_k141_31187 | Not-determined |
| KM_k141_31188 | Low-quality |
| KM_k141_222111 | Not-determined |
| KM_k141_109412 | Not-determined |
| KM_k141_154369 | Not-determined |
| KM_k141_233151 | Not-determined |
| KM_k141_88709 | Not-determined |
| KM_k141_42134 | Not-determined |
| KM_k141_42137 | Not-determined |
| KM_k141_65376 | Not-determined |
| KM_k141_199505 | Not-determined |
| KM_k141_199506 | Not-determined |
| KM_k141_222119 | Not-determined |
| KM_k141_42138 | Not-determined |
| KM_k141_210671 | Not-determined |
| KM_k141_199509 | Low-quality |
| KM_k141_109416 | Not-determined |
| KM_k141_109417 | Low-quality |
| KM_k141_109418 | Not-determined |
| KM_k141_131552 | Not-determined |
| KM_k141_199516 | Low-quality |
| KM_k141_242666 | Not-determined |
| KM_k141_109423 | Not-determined |
| KM_k141_188319 | Not-determined |
| KM_k141_233163 | Not-determined |

| | |
|---|---|
| KM_k141_233165 | Not-determined |
| KM_k141_109432 | Not-determined |
| KM_k141_77048 | Not-determined |
| KM_k141_8988 | Not-determined |
| KM_k141_77052 | Not-determined |
| KM_k141_222143 | Not-determined |
| KM_k141_233176 | Not-determined |
| KM_k141_252555 | Not-determined |
| KM_k141_199536 | Low-quality |
| KM_k141_199538 | Not-determined |
| KM_k141_77067 | Not-determined |
| KM_k141_42146 | Low-quality |
| KM_k141_188337 | Not-determined |
| KM_k141_96577 | Not-determined |
| KM_k141_143712 | Not-determined |
| KM_k141_252566 | Not-determined |
| KM_k141_222160 | Not-determined |
| KM_k141_42155 | Not-determined |
| KM_k141_222161 | Not-determined |
| KM_k141_164234 | Not-determined |
| KM_k141_222165 | Not-determined |
| KM_k141_143718 | Not-determined |
| KM_k141_210718 | Low-quality |
| KM_k141_210719 | Low-quality |
| KM_k141_19304 | Not-determined |
| KM_k141_210721 | Not-determined |
| KM_k141_51754 | Not-determined |
| KM_k141_233184 | Not-determined |
| KM_k141_264050 | Not-determined |
| KM_k141_233186 | Not-determined |
| KM_k141_233188 | Not-determined |
| KM_k141_77088 | Not-determined |
| KM_k141_88751 | Not-determined |
| KM_k141_154395 | Not-determined |
| KM_k141_96583 | Not-determined |
| KM_k141_9032 | Not-determined |
| KM_k141_65399 | Not-determined |
| KM_k141_143731 | Not-determined |
| KM_k141_264055 | Not-determined |
| KM_k141_199548 | Not-determined |
| KM_k141_252574 | Not-determined |
| KM_k141_199549 | Not-determined |
| KM_k141_264060 | Not-determined |
| KM_k141_19319 | Not-determined |
| KM_k141_233198 | Not-determined |
| KM_k141_96589 | Not-determined |
| KM_k141_210743 | Low-quality |
| KM_k141_96591 | Not-determined |
| KM_k141_42174 | Not-determined |
| KM_k141_96593 | Not-determined |
| KM_k141_51781 | Not-determined |
| KM_k141_176667 | Not-determined |
| KM_k141_264068 | Not-determined |
| KM_k141_96604 | Not-determined |
| KM_k141_51786 | Not-determined |
| KM_k141_122184 | Low-quality |
| KM_k141_77119 | Not-determined |
| KM_k141_96606 | Not-determined |
| KM_k141_210759 | Not-determined |
| KM_k141_176676 | Not-determined |
| KM_k141_210761 | Not-determined |
| KM_k141_210762 | Not-determined |
| KM_k141_77125 | Not-determined |
| KM_k141_176681 | Not-determined |
| KM_k141_199570 | Not-determined |
| KM_k141_65413 | Not-determined |
| KM_k141_19326 | Not-determined |
| KM_k141_9058 | Not-determined |
| KM_k141_252599 | Not-determined |
| KM_k141_9061 | Not-determined |
| KM_k141_19327 | Low-quality |
| KM_k141_199578 | Not-determined |
| KM_k141_19333 | Not-determined |
| KM_k141_51796 | Not-determined |
| KM_k141_51798 | Not-determined |
| KM_k141_143770 | Not-determined |
| KM_k141_252605 | Not-determined |
| KM_k141_176693 | Low-quality |
| KM_k141_51805 | Not-determined |
| KM_k141_77146 | Not-determined |
| KM_k141_199589 | Not-determined |
| KM_k141_131598 | Not-determined |
| KM_k141_19339 | Not-determined |
| KM_k141_96617 | Not-determined |
| KM_k141_131602 | Not-determined |
| KM_k141_96620 | Not-determined |
| KM_k141_109475 | Not-determined |
| KM_k141_51810 | Not-determined |
| KM_k141_154431 | Not-determined |
| KM_k141_176708 | Not-determined |
| KM_k141_42213 | Not-determined |
| KM_k141_109489 | Not-determined |
| KM_k141_77167 | Not-determined |
| KM_k141_164280 | Low-quality |
| KM_k141_109493 | Not-determined |
| KM_k141_88773 | Not-determined |
| KM_k141_9081 | Not-determined |
| KM_k141_210790 | Not-determined |
| KM_k141_176716 | Not-determined |
| KM_k141_109501 | Not-determined |
| KM_k141_42222 | Not-determined |
| KM_k141_88775 | Not-determined |
| KM_k141_222196 | Not-determined |
| KM_k141_131617 | Not-determined |
| KM_k141_19346 | Not-determined |
| KM_k141_233234 | Not-determined |
| KM_k141_233235 | Not-determined |
| KM_k141_210795 | Not-determined |
| KM_k141_143797 | Not-determined |
| KM_k141_131621 | Not-determined |
| KM_k141_252623 | Not-determined |
| KM_k141_143803 | Not-determined |
| KM_k141_31203 | Not-determined |
| KM_k141_143808 | Not-determined |
| KM_k141_222205 | Low-quality |
| KM_k141_154448 | Low-quality |
| KM_k141_199608 | Not-determined |
| KM_k141_154451 | Not-determined |
| KM_k141_210806 | Not-determined |
| KM_k141_9099 | Not-determined |
| KM_k141_122218 | Not-determined |
| KM_k141_252629 | Not-determined |
| KM_k141_42232 | Not-determined |
| KM_k141_210813 | Not-determined |
| KM_k141_96650 | Not-determined |
| KM_k141_88781 | Not-determined |
| KM_k141_131636 | Not-determined |
| KM_k141_19374 | Not-determined |
| KM_k141_65430 | Not-determined |
| KM_k141_9104 | Not-determined |
| KM_k141_88792 | Not-determined |
| KM_k141_77191 | Low-quality |
| KM_k141_210833 | Not-determined |
| KM_k141_131654 | Not-determined |
| KM_k141_65438 | Not-determined |
| KM_k141_31228 | Low-quality |
| KM_k141_42245 | Not-determined |
| KM_k141_96668 | Not-determined |
| KM_k141_210837 | Not-determined |
| KM_k141_188402 | Not-determined |
| KM_k141_264099 | Not-determined |
| KM_k141_42251 | Not-determined |
| KM_k141_42252 | Not-determined |
| KM_k141_210843 | Low-quality |
| KM_k141_264101 | Not-determined |
| KM_k141_65443 | Not-determined |
| KM_k141_264103 | Not-determined |
| KM_k141_210847 | Not-determined |
| KM_k141_210849 | Not-determined |
| KM_k141_65450 | Not-determined |
| KM_k141_176757 | Not-determined |
| KM_k141_88798 | Not-determined |
| KM_k141_88799 | Not-determined |
| KM_k141_143834 | Low-quality |
| KM_k141_31243 | Not-determined |
| KM_k141_88801 | Not-determined |
| KM_k141_131677 | Not-determined |
| KM_k141_188420 | Not-determined |
| KM_k141_154474 | Not-determined |
| KM_k141_131682 | Not-determined |
| KM_k141_65453 | Low-quality |
| KM_k141_65457 | Not-determined |
| KM_k141_131684 | Not-determined |
| KM_k141_51853 | Low-quality |
| KM_k141_143840 | Not-determined |
| KM_k141_143845 | Not-determined |
| KM_k141_51857 | Not-determined |
| KM_k141_164326 | Not-determined |
| KM_k141_199644 | Not-determined |
| KM_k141_65465 | Not-determined |
| KM_k141_176771 | Not-determined |
| KM_k141_210854 | Low-quality |
| KM_k141_65468 | Not-determined |
| KM_k141_154493 | Not-determined |
| KM_k141_143850 | Not-determined |
| KM_k141_176777 | Not-determined |
| KM_k141_264121 | Not-determined |
| KM_k141_176779 | Not-determined |
| KM_k141_176782 | Not-determined |
| KM_k141_88810 | Not-determined |
| KM_k141_88812 | Not-determined |
| KM_k141_164337 | Not-determined |
| KM_k141_109574 | Not-determined |
| KM_k141_131711 | Not-determined |
| KM_k141_109575 | Not-determined |
| KM_k141_31274 | Not-determined |
| KM_k141_176796 | Low-quality |
| KM_k141_188436 | Not-determined |
| KM_k141_65487 | Low-quality |
| KM_k141_143865 | Not-determined |
| KM_k141_154503 | Not-determined |
| KM_k141_109585 | Not-determined |
| KM_k141_154504 | Not-determined |
| KM_k141_31281 | Low-quality |
| KM_k141_42265 | Not-determined |
| KM_k141_31285 | Not-determined |
| KM_k141_199673 | Not-determined |
| KM_k141_210889 | Low-quality |
| KM_k141_77215 | Not-determined |
| KM_k141_131732 | Not-determined |
| KM_k141_131736 | Not-determined |
| KM_k141_210899 | Not-determined |
| KM_k141_131739 | Not-determined |
| KM_k141_176817 | Not-determined |
| KM_k141_176820 | Not-determined |
| KM_k141_154520 | Not-determined |
| KM_k141_9135 | Not-determined |
| KM_k141_164374 | Not-determined |
| KM_k141_42285 | Not-determined |
| KM_k141_9143 | Not-determined |
| KM_k141_242750 | Not-determined |
| KM_k141_131753 | Not-determined |
| KM_k141_210904 | Not-determined |
| KM_k141_109623 | Not-determined |
| KM_k141_210906 | Not-determined |
| KM_k141_264164 | Not-determined |
| KM_k141_77223 | Not-determined |
| KM_k141_42290 | Not-determined |
| KM_k141_96705 | Not-determined |
| KM_k141_77227 | Not-determined |
| KM_k141_96706 | Not-determined |
| KM_k141_88827 | Not-determined |
| KM_k141_242767 | Not-determined |
| KM_k141_164386 | Not-determined |
| KM_k141_222209 | Not-determined |
| KM_k141_222210 | Not-determined |
| KM_k141_164390 | Not-determined |
| KM_k141_264173 | Not-determined |
| KM_k141_199708 | Not-determined |
| KM_k141_242772 | Not-determined |
| KM_k141_164394 | Not-determined |
| KM_k141_210927 | Not-determined |
| KM_k141_210928 | Not-determined |
| KM_k141_199714 | Not-determined |
| KM_k141_42302 | Not-determined |
| KM_k141_88840 | Not-determined |
| KM_k141_252678 | Not-determined |
| KM_k141_65524 | Not-determined |
| KM_k141_9179 | Not-determined |
| KM_k141_242786 | Not-determined |
| KM_k141_77249 | Not-determined |
| KM_k141_109638 | Not-determined |
| KM_k141_19432 | Not-determined |
| KM_k141_88851 | Not-determined |
| KM_k141_9182 | Not-determined |
| KM_k141_42316 | Not-determined |
| KM_k141_88856 | Not-determined |
| KM_k141_131785 | Not-determined |
| KM_k141_154536 | Low-quality |
| KM_k141_242800 | Not-determined |
| KM_k141_242802 | Not-determined |
| KM_k141_77260 | Not-determined |
| KM_k141_264199 | Not-determined |
| KM_k141_131789 | Not-determined |
| KM_k141_65544 | Not-determined |
| KM_k141_176863 | Not-determined |
| KM_k141_176865 | Not-determined |
| KM_k141_154543 | Not-determined |
| KM_k141_131790 | Not-determined |
| KM_k141_109658 | Not-determined |
| KM_k141_176866 | Low-quality |
| KM_k141_131792 | Not-determined |
| KM_k141_9191 | Not-determined |
| KM_k141_242812 | Not-determined |
| KM_k141_222246 | Not-determined |
| KM_k141_88867 | Not-determined |
| KM_k141_131795 | Not-determined |
| KM_k141_164424 | Not-determined |
| KM_k141_9199 | Not-determined |
| KM_k141_109669 | Not-determined |
| KM_k141_176881 | Not-determined |
| KM_k141_31315 | Not-determined |
| KM_k141_42352 | Low-quality |
| KM_k141_65567 | Not-determined |
| KM_k141_65569 | Not-determined |
| KM_k141_176891 | Not-determined |
| KM_k141_176896 | Not-determined |
| KM_k141_9234 | Not-determined |
| KM_k141_131813 | Not-determined |
| KM_k141_19488 | Not-determined |
| KM_k141_51909 | Not-determined |
| KM_k141_65587 | Not-determined |
| KM_k141_65588 | Not-determined |
| KM_k141_77288 | Not-determined |
| KM_k141_176904 | Not-determined |
| KM_k141_51914 | Not-determined |
| KM_k141_31331 | Not-determined |
| KM_k141_51916 | Not-determined |
| KM_k141_42373 | Not-determined |
| KM_k141_264212 | Low-quality |
| KM_k141_42374 | Not-determined |
| KM_k141_176912 | Not-determined |
| KM_k141_264219 | Not-determined |
| KM_k141_19500 | Not-determined |
| KM_k141_65609 | Low-quality |
| KM_k141_176917 | Not-determined |
| KM_k141_31349 | Not-determined |
| KM_k141_19503 | Not-determined |
| KM_k141_77298 | Not-determined |
| KM_k141_65616 | Not-determined |
| KM_k141_42387 | Not-determined |
| KM_k141_109680 | Not-determined |
| KM_k141_96757 | Not-determined |
| KM_k141_77302 | Low-quality |
| KM_k141_42389 | Not-determined |
| KM_k141_51930 | Not-determined |
| KM_k141_188460 | Low-quality |
| KM_k141_65631 | Not-determined |
| KM_k141_65632 | Not-determined |
| KM_k141_96764 | Not-determined |
| KM_k141_176927 | Not-determined |
| KM_k141_9254 | Not-determined |
| KM_k141_9258 | Not-determined |
| KM_k141_176941 | Not-determined |
| KM_k141_242851 | Not-determined |
| KM_k141_96782 | Not-determined |
| KM_k141_31378 | Not-determined |
| KM_k141_9267 | Not-determined |
| KM_k141_19524 | Not-determined |
| KM_k141_188484 | Not-determined |
| KM_k141_31387 | Not-determined |
| KM_k141_154608 | Not-determined |
| KM_k141_31388 | Not-determined |
| KM_k141_51961 | Not-determined |
| KM_k141_131858 | Not-determined |
| KM_k141_264233 | Not-determined |
| KM_k141_210969 | Not-determined |
| KM_k141_65676 | Not-determined |
| KM_k141_154620 | Not-determined |
| KM_k141_42420 | Not-determined |
| KM_k141_199742 | Not-determined |
| KM_k141_19543 | Not-determined |
| KM_k141_31403 | Not-determined |
| KM_k141_131866 | Not-determined |
| KM_k141_264238 | Not-determined |
| KM_k141_42425 | Not-determined |
| KM_k141_143915 | Not-determined |
| KM_k141_210980 | Not-determined |
| KM_k141_176960 | Not-determined |
| KM_k141_143924 | Not-determined |
| KM_k141_77356 | Not-determined |
| KM_k141_210988 | Not-determined |
| KM_k141_9299 | Not-determined |
| KM_k141_19552 | Not-determined |
| KM_k141_9301 | Not-determined |
| KM_k141_131887 | Not-determined |
| KM_k141_131889 | Not-determined |
| KM_k141_9302 | Not-determined |
| KM_k141_264245 | Not-determined |
| KM_k141_211000 | Not-determined |
| KM_k141_164462 | Not-determined |
| KM_k141_77367 | Not-determined |
| KM_k141_31417 | Not-determined |
| KM_k141_51970 | Not-determined |
| KM_k141_77370 | Not-determined |
| KM_k141_31419 | Not-determined |
| KM_k141_51971 | Not-determined |
| KM_k141_19560 | Not-determined |
| KM_k141_65696 | Not-determined |
| KM_k141_65698 | Not-determined |
| KM_k141_154636 | Not-determined |
| KM_k141_154639 | Not-determined |
| KM_k141_154644 | Not-determined |
| KM_k141_252742 | Not-determined |
| KM_k141_242896 | Not-determined |
| KM_k141_143928 | Not-determined |
| KM_k141_51983 | Not-determined |
| KM_k141_176994 | Not-determined |
| KM_k141_154647 | Not-determined |
| KM_k141_96816 | Not-determined |
| KM_k141_65711 | Not-determined |
| KM_k141_51992 | Not-determined |
| KM_k141_65713 | Not-determined |
| KM_k141_19576 | Not-determined |
| KM_k141_19577 | Not-determined |
| KM_k141_31434 | Not-determined |
| KM_k141_51995 | Not-determined |
| KM_k141_31437 | Not-determined |
| KM_k141_176999 | Low-quality |
| KM_k141_51998 | Not-determined |
| KM_k141_154660 | Not-determined |
| KM_k141_131927 | Not-determined |
| KM_k141_31445 | Not-determined |
| KM_k141_19585 | Low-quality |
| KM_k141_252760 | Low-quality |
| KM_k141_252763 | Not-determined |
| KM_k141_131960 | Not-determined |
| KM_k141_252771 | Not-determined |
| KM_k141_242915 | Not-determined |
| KM_k141_131963 | Not-determined |
| KM_k141_42458 | Not-determined |
| KM_k141_242919 | Not-determined |
| KM_k141_177018 | Not-determined |
| KM_k141_31466 | Not-determined |
| KM_k141_264263 | Not-determined |
| KM_k141_242922 | Not-determined |
| KM_k141_264264 | Not-determined |
| KM_k141_131971 | Not-determined |
| KM_k141_242928 | Not-determined |
| KM_k141_42477 | Not-determined |
| KM_k141_131982 | Not-determined |
| KM_k141_164511 | Not-determined |
| KM_k141_143946 | Low-quality |
| KM_k141_143948 | Not-determined |
| KM_k141_188517 | Not-determined |
| KM_k141_131990 | Not-determined |
| KM_k141_42492 | Not-determined |
| KM_k141_164519 | Not-determined |
| KM_k141_9322 | Not-determined |
| KM_k141_31483 | Not-determined |
| KM_k141_31484 | Not-determined |
| KM_k141_242936 | Not-determined |
| KM_k141_31485 | Low-quality |
| KM_k141_109743 | Low-quality |
| KM_k141_242940 | Not-determined |
| KM_k141_132010 | Not-determined |
| KM_k141_109747 | Low-quality |
| KM_k141_177032 | Not-determined |
| KM_k141_143960 | Not-determined |
| KM_k141_42506 | Not-determined |
| KM_k141_188538 | Not-determined |
| KM_k141_42510 | Not-determined |
| KM_k141_52039 | Not-determined |
| KM_k141_52040 | Not-determined |
| KM_k141_52041 | Not-determined |
| KM_k141_188542 | Not-determined |
| KM_k141_177039 | Not-determined |
| KM_k141_164540 | Not-determined |
| KM_k141_52048 | Not-determined |
| KM_k141_143964 | Not-determined |
| KM_k141_77439 | Not-determined |
| KM_k141_164541 | Not-determined |
| KM_k141_199760 | Low-quality |
| KM_k141_96871 | Not-determined |
| KM_k141_264296 | Not-determined |
| KM_k141_242964 | Not-determined |
| KM_k141_132027 | Not-determined |
| KM_k141_65727 | Not-determined |
| KM_k141_242968 | Not-determined |
| KM_k141_154686 | Not-determined |
| KM_k141_154687 | Not-determined |
| KM_k141_132033 | Not-determined |
| KM_k141_77450 | Not-determined |
| KM_k141_65736 | Not-determined |
| KM_k141_65737 | Not-determined |
| KM_k141_177064 | Not-determined |
| KM_k141_154695 | Not-determined |
| KM_k141_252796 | Not-determined |
| KM_k141_65742 | Not-determined |
| KM_k141_154698 | Low-quality |
| KM_k141_109785 | Not-determined |
| KM_k141_65746 | Low-quality |
| KM_k141_252803 | Low-quality |
| KM_k141_242974 | Not-determined |
| KM_k141_154705 | Not-determined |
| KM_k141_77461 | Not-determined |
| KM_k141_96898 | Not-determined |
| KM_k141_252813 | Not-determined |
| KM_k141_96902 | Not-determined |
| KM_k141_132044 | Low-quality |
| KM_k141_132047 | Not-determined |
| KM_k141_77471 | Not-determined |
| KM_k141_154713 | Not-determined |
| KM_k141_188586 | Not-determined |
| KM_k141_77481 | Not-determined |
| KM_k141_132052 | Not-determined |
| KM_k141_188590 | Not-determined |
| KM_k141_109805 | Not-determined |
| KM_k141_154722 | Not-determined |
| KM_k141_42529 | Not-determined |
| KM_k141_52058 | Not-determined |
| KM_k141_31531 | Not-determined |
| KM_k141_109808 | Not-determined |
| KM_k141_96907 | Low-quality |
| KM_k141_211027 | Not-determined |
| KM_k141_154727 | Not-determined |
| KM_k141_143973 | Not-determined |
| KM_k141_52068 | Not-determined |
| KM_k141_52070 | Not-determined |
| KM_k141_143976 | Low-quality |
| KM_k141_65760 | Not-determined |
| KM_k141_9376 | Not-determined |
| KM_k141_242995 | Low-quality |
| KM_k141_9379 | Not-determined |
| KM_k141_9381 | Not-determined |
| KM_k141_164582 | Not-determined |
| KM_k141_96919 | Not-determined |
| KM_k141_252835 | Low-quality |
| KM_k141_199809 | Not-determined |
| KM_k141_9385 | Not-determined |
| KM_k141_77495 | Not-determined |
| KM_k141_252837 | Not-determined |
| KM_k141_143981 | Not-determined |
| KM_k141_9390 | Not-determined |
| KM_k141_52089 | Not-determined |
| KM_k141_264308 | Not-determined |
| KM_k141_243004 | Not-determined |
| KM_k141_132078 | Not-determined |
| KM_k141_154748 | Not-determined |
| KM_k141_42570 | Not-determined |
| KM_k141_31563 | Not-determined |
| KM_k141_19590 | Not-determined |
| KM_k141_264320 | Not-determined |
| KM_k141_211052 | Not-determined |
| KM_k141_31573 | Low-quality |
| KM_k141_9403 | Not-determined |
| KM_k141_243019 | Low-quality |
| KM_k141_9404 | Not-determined |
| KM_k141_243022 | Not-determined |
| KM_k141_132088 | Not-determined |
| KM_k141_77525 | Not-determined |
| KM_k141_31583 | Not-determined |
| KM_k141_243025 | Not-determined |
| KM_k141_132092 | Not-determined |
| KM_k141_199828 | Not-determined |
| KM_k141_77529 | Not-determined |
| KM_k141_164615 | Not-determined |
| KM_k141_77532 | Not-determined |
| KM_k141_164623 | Not-determined |
| KM_k141_9419 | Not-determined |
| KM_k141_77536 | Not-determined |
| KM_k141_42585 | Not-determined |
| KM_k141_132106 | Not-determined |
| KM_k141_252873 | Not-determined |
| KM_k141_132111 | Not-determined |
| KM_k141_9434 | Not-determined |
| KM_k141_31601 | Not-determined |
| KM_k141_143997 | Not-determined |
| KM_k141_211067 | Not-determined |
| KM_k141_9439 | Not-determined |
| KM_k141_164642 | Not-determined |
| KM_k141_264335 | Not-determined |
| KM_k141_42603 | Not-determined |
| KM_k141_77556 | Not-determined |
| KM_k141_264340 | Not-determined |
| KM_k141_264342 | Not-determined |
| KM_k141_211069 | Low-quality |
| KM_k141_154793 | Not-determined |
| KM_k141_264346 | Not-determined |
| KM_k141_31609 | Not-determined |
| KM_k141_211074 | Not-determined |
| KM_k141_19620 | Not-determined |
| KM_k141_96926 | Not-determined |
| KM_k141_9453 | Not-determined |
| KM_k141_96932 | Not-determined |
| KM_k141_164657 | Not-determined |
| KM_k141_42620 | Not-determined |
| KM_k141_177118 | Not-determined |
| KM_k141_177122 | Not-determined |
| KM_k141_96941 | Not-determined |
| KM_k141_19628 | Not-determined |
| KM_k141_154802 | Not-determined |
| KM_k141_154804 | Not-determined |
| KM_k141_243041 | Not-determined |
| KM_k141_154806 | Not-determined |
| KM_k141_144026 | Not-determined |
| KM_k141_132144 | Not-determined |
| KM_k141_132147 | Not-determined |
| KM_k141_211088 | Not-determined |
| KM_k141_264358 | Not-determined |
| KM_k141_31633 | Not-determined |
| KM_k141_177148 | Not-determined |
| KM_k141_77576 | Not-determined |
| KM_k141_188659 | Not-determined |
| KM_k141_164670 | Low-quality |
| KM_k141_264373 | Not-determined |
| KM_k141_154811 | Not-determined |
| KM_k141_31647 | Not-determined |
| KM_k141_96973 | Not-determined |
| KM_k141_177163 | Not-determined |
| KM_k141_96975 | Not-determined |
| KM_k141_188668 | Low-quality |
| KM_k141_211113 | Not-determined |
| KM_k141_264378 | Not-determined |
| KM_k141_52162 | Not-determined |
| KM_k141_77589 | Not-determined |
| KM_k141_31650 | Not-determined |
| KM_k141_144054 | Not-determined |
| KM_k141_252896 | Not-determined |
| KM_k141_96984 | Not-determined |
| KM_k141_9491 | Not-determined |
| KM_k141_154818 | Not-determined |
| KM_k141_264389 | Not-determined |
| KM_k141_42641 | Not-determined |
| KM_k141_42642 | Not-determined |
| KM_k141_42643 | Not-determined |
| KM_k141_264394 | Not-determined |
| KM_k141_31661 | Not-determined |
| KM_k141_9506 | Not-determined |

| | |
|---|---|
| KM_k141_188677 | Not-determined |
| KM_k141_42651 | Not-determined |
| KM_k141_164690 | Not-determined |
| KM_k141_9513 | Not-determined |
| KM_k141_9519 | Not-determined |
| KM_k141_132189 | Not-determined |
| KM_k141_65856 | Not-determined |
| KM_k141_65861 | Not-determined |
| KM_k141_164697 | Not-determined |
| KM_k141_31675 | Not-determined |
| KM_k141_42667 | Not-determined |
| KM_k141_9527 | Not-determined |
| KM_k141_19644 | Not-determined |
| KM_k141_42675 | Not-determined |
| KM_k141_252924 | Not-determined |
| KM_k141_52173 | Not-determined |
| KM_k141_19651 | Not-determined |
| KM_k141_65871 | Not-determined |
| KM_k141_264413 | Not-determined |
| KM_k141_177192 | Not-determined |
| KM_k141_31688 | Not-determined |
| KM_k141_97001 | Not-determined |
| KM_k141_97004 | Not-determined |
| KM_k141_177200 | Low-quality |
| KM_k141_252935 | Not-determined |
| KM_k141_264421 | Not-determined |
| KM_k141_264423 | Not-determined |
| KM_k141_31699 | Not-determined |
| KM_k141_97010 | Not-determined |
| KM_k141_31704 | Not-determined |
| KM_k141_77603 | Not-determined |
| KM_k141_77604 | Not-determined |
| KM_k141_164713 | Not-determined |
| KM_k141_188739 | Not-determined |
| KM_k141_199865 | Not-determined |
| KM_k141_97016 | Not-determined |
| KM_k141_97020 | Not-determined |
| KM_k141_164720 | Not-determined |
| KM_k141_77609 | Not-determined |
| KM_k141_177214 | Not-determined |
| KM_k141_77613 | Not-determined |
| KM_k141_264437 | Low-quality |
| KM_k141_9540 | Not-determined |
| KM_k141_65901 | Low-quality |
| KM_k141_65904 | Low-quality |
| KM_k141_19670 | Not-determined |
| KM_k141_164747 | Not-determined |
| KM_k141_19673 | Not-determined |
| KM_k141_188759 | Not-determined |
| KM_k141_177226 | Low-quality |
| KM_k141_52203 | Low-quality |
| KM_k141_77616 | Not-determined |
| KM_k141_77620 | Not-determined |
| KM_k141_9544 | Not-determined |
| KM_k141_164758 | Not-determined |
| KM_k141_19688 | Not-determined |
| KM_k141_42699 | Not-determined |
| KM_k141_77629 | Not-determined |
| KM_k141_199888 | Not-determined |
| KM_k141_77633 | Not-determined |
| KM_k141_154875 | Low-quality |
| KM_k141_77636 | Not-determined |
| KM_k141_252988 | Not-determined |
| KM_k141_52217 | Not-determined |
| KM_k141_65926 | Not-determined |
| KM_k141_52218 | Low-quality |
| KM_k141_97051 | Not-determined |
| KM_k141_19697 | Not-determined |
| KM_k141_132239 | Not-determined |
| KM_k141_253000 | Not-determined |
| KM_k141_132241 | Not-determined |
| KM_k141_177249 | Not-determined |
| KM_k141_132247 | Not-determined |
| KM_k141_52230 | Not-determined |
| KM_k141_52231 | Not-determined |
| KM_k141_42715 | Not-determined |
| KM_k141_109843 | Not-determined |
| KM_k141_188794 | Low-quality |
| KM_k141_109847 | Not-determined |
| KM_k141_97066 | Not-determined |
| KM_k141_97067 | Low-quality |
| KM_k141_42719 | Not-determined |
| KM_k141_177271 | Not-determined |
| KM_k141_188804 | Not-determined |
| KM_k141_109861 | Not-determined |
| KM_k141_19705 | Not-determined |
| KM_k141_97076 | Not-determined |
| KM_k141_65951 | Not-determined |
| KM_k141_154899 | Low-quality |
| KM_k141_164779 | Not-determined |
| KM_k141_154905 | Not-determined |
| KM_k141_65958 | Not-determined |
| KM_k141_109873 | Not-determined |
| KM_k141_264453 | Not-determined |
| KM_k141_154908 | Not-determined |
| KM_k141_42738 | Not-determined |
| KM_k141_42741 | Low-quality |
| KM_k141_9591 | Not-determined |
| KM_k141_9592 | Not-determined |
| KM_k141_188827 | Not-determined |
| KM_k141_264459 | Not-determined |
| KM_k141_264460 | Not-determined |
| KM_k141_31725 | Not-determined |
| KM_k141_264462 | Not-determined |
| KM_k141_31728 | Not-determined |
| KM_k141_19728 | Not-determined |
| KM_k141_19730 | Not-determined |
| KM_k141_164809 | Not-determined |
| KM_k141_19732 | Not-determined |
| KM_k141_177286 | Not-determined |
| KM_k141_264470 | Not-determined |
| KM_k141_31730 | Not-determined |
| KM_k141_154916 | Not-determined |
| KM_k141_31732 | Not-determined |
| KM_k141_253028 | Not-determined |
| KM_k141_97097 | Not-determined |
| KM_k141_264474 | Not-determined |
| KM_k141_9603 | Not-determined |
| KM_k141_243100 | Low-quality |
| KM_k141_109881 | Low-quality |
| KM_k141_19749 | Not-determined |
| KM_k141_164844 | Not-determined |
| KM_k141_97102 | Not-determined |
| KM_k141_164846 | Not-determined |
| KM_k141_97103 | Not-determined |
| KM_k141_31741 | Not-determined |
| KM_k141_97105 | Not-determined |
| KM_k141_243115 | Low-quality |
| KM_k141_132274 | Not-determined |
| KM_k141_132275 | Not-determined |
| KM_k141_243117 | Not-determined |
| KM_k141_132277 | Not-determined |
| KM_k141_177315 | Not-determined |
| KM_k141_132282 | Not-determined |
| KM_k141_164868 | Not-determined |
| KM_k141_253052 | Not-determined |
| KM_k141_164874 | Not-determined |
| KM_k141_66012 | Not-determined |
| KM_k141_243124 | Not-determined |
| KM_k141_66018 | Not-determined |
| KM_k141_199934 | Not-determined |
| KM_k141_253061 | Not-determined |
| KM_k141_199939 | Not-determined |
| KM_k141_42782 | Not-determined |
| KM_k141_243134 | Not-determined |
| KM_k141_132294 | Not-determined |
| KM_k141_77700 | Low-quality |
| KM_k141_97127 | Not-determined |
| KM_k141_97132 | Not-determined |
| KM_k141_9645 | Not-determined |
| KM_k141_243150 | Not-determined |
| KM_k141_188882 | Low-quality |
| KM_k141_31772 | Low-quality |
| KM_k141_154924 | Not-determined |
| KM_k141_164878 | Low-quality |
| KM_k141_77713 | Not-determined |
| KM_k141_66048 | Low-quality |
| KM_k141_253084 | Not-determined |
| KM_k141_66052 | Not-determined |
| KM_k141_77714 | Not-determined |
| KM_k141_66060 | Not-determined |
| KM_k141_164892 | Not-determined |
| KM_k141_66069 | Not-determined |
| KM_k141_264486 | Not-determined |
| KM_k141_199967 | Low-quality |
| KM_k141_66071 | Not-determined |
| KM_k141_164905 | Not-determined |
| KM_k141_66080 | Not-determined |
| KM_k141_109927 | Not-determined |
| KM_k141_164906 | Not-determined |
| KM_k141_66085 | Not-determined |
| KM_k141_66086 | Not-determined |
| KM_k141_164912 | Not-determined |
| KM_k141_9687 | Not-determined |
| KM_k141_253099 | Not-determined |
| KM_k141_264493 | Not-determined |
| KM_k141_132336 | Not-determined |
| KM_k141_109932 | Not-determined |
| KM_k141_52235 | Not-determined |
| KM_k141_177346 | Not-determined |
| KM_k141_52239 | Not-determined |
| KM_k141_177349 | Not-determined |
| KM_k141_164924 | Not-determined |
| KM_k141_109934 | Not-determined |
| KM_k141_9691 | Not-determined |
| KM_k141_97169 | Not-determined |
| KM_k141_109941 | Not-determined |
| KM_k141_66103 | Not-determined |
| KM_k141_253113 | Not-determined |
| KM_k141_164931 | Not-determined |
| KM_k141_253114 | Not-determined |
| KM_k141_199985 | Not-determined |
| KM_k141_52257 | Not-determined |
| KM_k141_42824 | Not-determined |
| KM_k141_42825 | Not-determined |
| KM_k141_97185 | Low-quality |
| KM_k141_97187 | Not-determined |
| KM_k141_177358 | Low-quality |
| KM_k141_66111 | Not-determined |
| KM_k141_97188 | Not-determined |
| KM_k141_132358 | Not-determined |
| KM_k141_109958 | Not-determined |
| KM_k141_9701 | Not-determined |
| KM_k141_109961 | Not-determined |
| KM_k141_97194 | Not-determined |
| KM_k141_177372 | Low-quality |
| KM_k141_243162 | Not-determined |
| KM_k141_177375 | Not-determined |
| KM_k141_9707 | Low-quality |
| KM_k141_243165 | Not-determined |
| KM_k141_52274 | Not-determined |
| KM_k141_52275 | Not-determined |
| KM_k141_177376 | Not-determined |
| KM_k141_154948 | Low-quality |
| KM_k141_109974 | Not-determined |
| KM_k141_243171 | Not-determined |
| KM_k141_264508 | Not-determined |
| KM_k141_264510 | Not-determined |
| KM_k141_264518 | Not-determined |
| KM_k141_264522 | Not-determined |
| KM_k141_243197 | Not-determined |
| KM_k141_154965 | Not-determined |
| KM_k141_109989 | Not-determined |
| KM_k141_200022 | Not-determined |
| KM_k141_200024 | Not-determined |
| KM_k141_52299 | Not-determined |
| KM_k141_52301 | Low-quality |
| KM_k141_154969 | Not-determined |
| KM_k141_164988 | Not-determined |
| KM_k141_52307 | Not-determined |
| KM_k141_42854 | Not-determined |
| KM_k141_144058 | Not-determined |
| KM_k141_264528 | Not-determined |
| KM_k141_264529 | Not-determined |
| KM_k141_19810 | Not-determined |
| KM_k141_243210 | Not-determined |
| KM_k141_109996 | Not-determined |
| KM_k141_264532 | Not-determined |
| KM_k141_264537 | Not-determined |
| KM_k141_264538 | Not-determined |
| KM_k141_52317 | Not-determined |
| KM_k141_264541 | Not-determined |
| KM_k141_144076 | Not-determined |
| KM_k141_9735 | Not-determined |
| KM_k141_154996 | Not-determined |
| KM_k141_42859 | Not-determined |
| KM_k141_154999 | Not-determined |
| KM_k141_264552 | Not-determined |
| KM_k141_42860 | Not-determined |
| KM_k141_9739 | Not-determined |
| KM_k141_42865 | Not-determined |
| KM_k141_19826 | Not-determined |
| KM_k141_42866 | Not-determined |
| KM_k141_155006 | Not-determined |
| KM_k141_155007 | Not-determined |
| KM_k141_264558 | Low-quality |
| KM_k141_42872 | Not-determined |
| KM_k141_165007 | Not-determined |
| KM_k141_97245 | Not-determined |
| KM_k141_110027 | Not-determined |
| KM_k141_9748 | Not-determined |
| KM_k141_177423 | Not-determined |
| KM_k141_42875 | Not-determined |
| KM_k141_177427 | Not-determined |
| KM_k141_110041 | Not-determined |
| KM_k141_165027 | Not-determined |
| KM_k141_253151 | Not-determined |
| KM_k141_243263 | Not-determined |
| KM_k141_31807 | Not-determined |
| KM_k141_264565 | Not-determined |
| KM_k141_9764 | Not-determined |
| KM_k141_243277 | Not-determined |
| KM_k141_110061 | Not-determined |
| KM_k141_42895 | Not-determined |
| KM_k141_97266 | Low-quality |
| KM_k141_177451 | Not-determined |
| KM_k141_253164 | Low-quality |
| KM_k141_264571 | Not-determined |
| KM_k141_264572 | Not-determined |
| KM_k141_42906 | Not-determined |
| KM_k141_264578 | Not-determined |
| KM_k141_253179 | Not-determined |
| KM_k141_42909 | Not-determined |
| KM_k141_19853 | Not-determined |
| KM_k141_42911 | Not-determined |
| KM_k141_97274 | Not-determined |
| KM_k141_42913 | Not-determined |
| KM_k141_264585 | Low-quality |
| KM_k141_31838 | Not-determined |
| KM_k141_31840 | Not-determined |
| KM_k141_264588 | Not-determined |
| KM_k141_97282 | Not-determined |
| KM_k141_19865 | Not-determined |
| KM_k141_253194 | Not-determined |
| KM_k141_42926 | Not-determined |
| KM_k141_177483 | Not-determined |
| KM_k141_110095 | Not-determined |
| KM_k141_110097 | Not-determined |
| KM_k141_31855 | Not-determined |
| KM_k141_42931 | Not-determined |
| KM_k141_52362 | Not-determined |
| KM_k141_253208 | Not-determined |
| KM_k141_66198 | Not-determined |
| KM_k141_9788 | Not-determined |
| KM_k141_42944 | Not-determined |
| KM_k141_165065 | Low-quality |
| KM_k141_165066 | Not-determined |
| KM_k141_42947 | Low-quality |
| KM_k141_110110 | Not-determined |
| KM_k141_110111 | Not-determined |
| KM_k141_155035 | Not-determined |
| KM_k141_165083 | Not-determined |
| KM_k141_165084 | Not-determined |
| KM_k141_165090 | Not-determined |
| KM_k141_52386 | Low-quality |
| KM_k141_66210 | Not-determined |
| KM_k141_132417 | Not-determined |
| KM_k141_52390 | Not-determined |
| KM_k141_177504 | Not-determined |
| KM_k141_132421 | Not-determined |
| KM_k141_177505 | Not-determined |
| KM_k141_52396 | Not-determined |
| KM_k141_165099 | Not-determined |
| KM_k141_97318 | Not-determined |
| KM_k141_9805 | Not-determined |
| KM_k141_132429 | Not-determined |
| KM_k141_97320 | Not-determined |
| KM_k141_31887 | Not-determined |
| KM_k141_52403 | Not-determined |
| KM_k141_66222 | Not-determined |
| KM_k141_165110 | Not-determined |
| KM_k141_253220 | Low-quality |
| KM_k141_110124 | Not-determined |
| KM_k141_144098 | Not-determined |
| KM_k141_97329 | Not-determined |
| KM_k141_97332 | Not-determined |
| KM_k141_155048 | Low-quality |
| KM_k141_42971 | Not-determined |
| KM_k141_97336 | Not-determined |
| KM_k141_9819 | Not-determined |
| KM_k141_19898 | Not-determined |
| KM_k141_9820 | Not-determined |
| KM_k141_97339 | Not-determined |
| KM_k141_9824 | Not-determined |
| KM_k141_110136 | Not-determined |
| KM_k141_110137 | Low-quality |
| KM_k141_165136 | Not-determined |
| KM_k141_177542 | Not-determined |
| KM_k141_42996 | Not-determined |
| KM_k141_110140 | Not-determined |
| KM_k141_264634 | Not-determined |
| KM_k141_253236 | Low-quality |
| KM_k141_97342 | Not-determined |
| KM_k141_9848 | Not-determined |
| KM_k141_132452 | Not-determined |
| KM_k141_19906 | Not-determined |
| KM_k141_253239 | Not-determined |
| KM_k141_66258 | Not-determined |
| KM_k141_97349 | Not-determined |
| KM_k141_66264 | Not-determined |
| KM_k141_52437 | Low-quality |
| KM_k141_144115 | Not-determined |
| KM_k141_132459 | Not-determined |
| KM_k141_144118 | Low-quality |
| KM_k141_165165 | Not-determined |
| KM_k141_97358 | Not-determined |
| KM_k141_165169 | Not-determined |
| KM_k141_132473 | Not-determined |
| KM_k141_52446 | Not-determined |
| KM_k141_19935 | Not-determined |
| KM_k141_66278 | Not-determined |
| KM_k141_52450 | Not-determined |
| KM_k141_264674 | Not-determined |
| KM_k141_52454 | Not-determined |
| KM_k141_132493 | Not-determined |
| KM_k141_132500 | Not-determined |
| KM_k141_264689 | Low-quality |
| KM_k141_19963 | Not-determined |
| KM_k141_253271 | Not-determined |
| KM_k141_19966 | Not-determined |
| KM_k141_253277 | Not-determined |
| KM_k141_253279 | Not-determined |
| KM_k141_43015 | Not-determined |
| KM_k141_243312 | Low-quality |
| KM_k141_97393 | Not-determined |
| KM_k141_177588 | Not-determined |
| KM_k141_177596 | Not-determined |
| KM_k141_52481 | Not-determined |
| KM_k141_52482 | Not-determined |
| KM_k141_97397 | Not-determined |
| KM_k141_132507 | Not-determined |
| KM_k141_144149 | Not-determined |
| KM_k141_97400 | Not-determined |
| KM_k141_132511 | Not-determined |
| KM_k141_52496 | Not-determined |
| KM_k141_177613 | Not-determined |
| KM_k141_77741 | Not-determined |
| KM_k141_264700 | Not-determined |
| KM_k141_243325 | Not-determined |
| KM_k141_43048 | Not-determined |
| KM_k141_77756 | Not-determined |
| KM_k141_144156 | Not-determined |
| KM_k141_177624 | Not-determined |
| KM_k141_264711 | Not-determined |
| KM_k141_243336 | Not-determined |
| KM_k141_43053 | Not-determined |
| KM_k141_144165 | Not-determined |
| KM_k141_253314 | Not-determined |
| KM_k141_110184 | Not-determined |
| KM_k141_52524 | Not-determined |
| KM_k141_52525 | Not-determined |
| KM_k141_43062 | Not-determined |
| KM_k141_52534 | Not-determined |
| KM_k141_177647 | Not-determined |
| KM_k141_43065 | Not-determined |
| KM_k141_177648 | Not-determined |
| KM_k141_9885 | Not-determined |
| KM_k141_243339 | Not-determined |
| KM_k141_31951 | Not-determined |
| KM_k141_144181 | Not-determined |
| KM_k141_31955 | Not-determined |
| KM_k141_177659 | Not-determined |
| KM_k141_177660 | Not-determined |
| KM_k141_253327 | Not-determined |
| KM_k141_43081 | Not-determined |
| KM_k141_110207 | Low-quality |
| KM_k141_9905 | Not-determined |
| KM_k141_97445 | Not-determined |
| KM_k141_9906 | Not-determined |
| KM_k141_253335 | Not-determined |
| KM_k141_155085 | Not-determined |
| KM_k141_264746 | Not-determined |
| KM_k141_155088 | Not-determined |
| KM_k141_52557 | Not-determined |
| KM_k141_155096 | Not-determined |
| KM_k141_264756 | Not-determined |
| KM_k141_9929 | Not-determined |
| KM_k141_43119 | Not-determined |
| KM_k141_264760 | Low-quality |
| KM_k141_264763 | Not-determined |
| KM_k141_243351 | Low-quality |
| KM_k141_155102 | Not-determined |
| KM_k141_52563 | Not-determined |
| KM_k141_144201 | Not-determined |
| KM_k141_97459 | Not-determined |
| KM_k141_177679 | Not-determined |
| KM_k141_97460 | Not-determined |
| KM_k141_97461 | Not-determined |
| KM_k141_43124 | Not-determined |
| KM_k141_177684 | Not-determined |
| KM_k141_43128 | Not-determined |
| KM_k141_144209 | Not-determined |
| KM_k141_43133 | Not-determined |
| KM_k141_43135 | Low-quality |
| KM_k141_52573 | Not-determined |
| KM_k141_132570 | Not-determined |
| KM_k141_97465 | Not-determined |
| KM_k141_264773 | Not-determined |
| KM_k141_264777 | Not-determined |
| KM_k141_97474 | Not-determined |
| KM_k141_165287 | Not-determined |
| KM_k141_243373 | Not-determined |
| KM_k141_165289 | Not-determined |
| KM_k141_43145 | Not-determined |
| KM_k141_66337 | Low-quality |
| KM_k141_165296 | Not-determined |
| KM_k141_43147 | Not-determined |
| KM_k141_177719 | Not-determined |
| KM_k141_52583 | Not-determined |
| KM_k141_66346 | Not-determined |
| KM_k141_264794 | Not-determined |
| KM_k141_253362 | Not-determined |
| KM_k141_243385 | Low-quality |
| KM_k141_155126 | Not-determined |
| KM_k141_97488 | Low-quality |
| KM_k141_43179 | Not-determined |
| KM_k141_177733 | Not-determined |
| KM_k141_155131 | Not-determined |
| KM_k141_253373 | Low-quality |
| KM_k141_253374 | Not-determined |
| KM_k141_253376 | Not-determined |
| KM_k141_253377 | Not-determined |
| KM_k141_144251 | Not-determined |
| KM_k141_97506 | Not-determined |
| KM_k141_155140 | Not-determined |
| KM_k141_264801 | Low-quality |
| KM_k141_66376 | Low-quality |
| KM_k141_19997 | Not-determined |
| KM_k141_9976 | Not-determined |
| KM_k141_165326 | Not-determined |
| KM_k141_97524 | Not-determined |
| KM_k141_155156 | Not-determined |
| KM_k141_165331 | Not-determined |
| KM_k141_177757 | Not-determined |
| KM_k141_31969 | Not-determined |
| KM_k141_31975 | Not-determined |
| KM_k141_43200 | Not-determined |
| KM_k141_155165 | Not-determined |
| KM_k141_9985 | Low-quality |
| KM_k141_43206 | Not-determined |
| KM_k141_66398 | Not-determined |
| KM_k141_144265 | Not-determined |
| KM_k141_20012 | Not-determined |
| KM_k141_52625 | Not-determined |
| KM_k141_43207 | Not-determined |
| KM_k141_43211 | Not-determined |
| KM_k141_110282 | Not-determined |
| KM_k141_264822 | Not-determined |
| KM_k141_253410 | Not-determined |
| KM_k141_31990 | Not-determined |
| KM_k141_144274 | Not-determined |
| KM_k141_264825 | Low-quality |
| KM_k141_165352 | Not-determined |
| KM_k141_97545 | Not-determined |
| KM_k141_97546 | Not-determined |
| KM_k141_52640 | Not-determined |
| KM_k141_253420 | Not-determined |
| KM_k141_165363 | Not-determined |
| KM_k141_253424 | Not-determined |
| KM_k141_132628 | Not-determined |
| KM_k141_66428 | Not-determined |
| KM_k141_132631 | Not-determined |
| KM_k141_97554 | Not-determined |
| KM_k141_155188 | Not-determined |
| KM_k141_144285 | Not-determined |
| KM_k141_144286 | Not-determined |
| KM_k141_32016 | Not-determined |
| KM_k141_52657 | Not-determined |
| KM_k141_144292 | Not-determined |
| KM_k141_155194 | Low-quality |
| KM_k141_155195 | Not-determined |
| KM_k141_43253 | Not-determined |
| KM_k141_253438 | Not-determined |
| KM_k141_144298 | Low-quality |
| KM_k141_144299 | Not-determined |
| KM_k141_43255 | Low-quality |
| KM_k141_43256 | Not-determined |
| KM_k141_43268 | Not-determined |
| KM_k141_32041 | Not-determined |
| KM_k141_66459 | Not-determined |
| KM_k141_155207 | Not-determined |
| KM_k141_43274 | Not-determined |
| KM_k141_155209 | Not-determined |
| KM_k141_110305 | Not-determined |
| KM_k141_155216 | Not-determined |
| KM_k141_32051 | Not-determined |
| KM_k141_43285 | Low-quality |
| KM_k141_52696 | Not-determined |
| KM_k141_52698 | Not-determined |
| KM_k141_243420 | Not-determined |
| KM_k141_32064 | Not-determined |
| KM_k141_66475 | Not-determined |
| KM_k141_144323 | Not-determined |
| KM_k141_144329 | Not-determined |
| KM_k141_43297 | Not-determined |
| KM_k141_165420 | Not-determined |
| KM_k141_165424 | Not-determined |
| KM_k141_253476 | Not-determined |
| KM_k141_10042 | Not-determined |
| KM_k141_52707 | Low-quality |
| KM_k141_243443 | Not-determined |
| KM_k141_10048 | Not-determined |
| KM_k141_52716 | Not-determined |
| KM_k141_66501 | Not-determined |
| KM_k141_243456 | Not-determined |
| KM_k141_165452 | Low-quality |
| KM_k141_253498 | Not-determined |
| KM_k141_144358 | Not-determined |
| KM_k141_110342 | Not-determined |
| KM_k141_20056 | Not-determined |
| KM_k141_97606 | Not-determined |
| KM_k141_144367 | Low-quality |
| KM_k141_20060 | Not-determined |
| KM_k141_43313 | Not-determined |
| KM_k141_10080 | Not-determined |
| KM_k141_132742 | Not-determined |
| KM_k141_32105 | Not-determined |
| KM_k141_144391 | Not-determined |
| KM_k141_165477 | Not-determined |
| KM_k141_165478 | Not-determined |
| KM_k141_132751 | Not-determined |
| KM_k141_243476 | Not-determined |
| KM_k141_165484 | Not-determined |
| KM_k141_97630 | Not-determined |
| KM_k141_10098 | Not-determined |
| KM_k141_243480 | Not-determined |
| KM_k141_243482 | Not-determined |
| KM_k141_243483 | Not-determined |
| KM_k141_264900 | Not-determined |

| | |
|---|---|
| KM_k141_243489 | Not-determined |
| KM_k141_43337 | Low-quality |
| KM_k141_264906 | Not-determined |
| KM_k141_264911 | Low-quality |
| KM_k141_43352 | Not-determined |
| KM_k141_110391 | Not-determined |
| KM_k141_132767 | Not-determined |
| KM_k141_243507 | Not-determined |
| KM_k141_155244 | Not-determined |
| KM_k141_52757 | Not-determined |
| KM_k141_165510 | Not-determined |
| KM_k141_52761 | Not-determined |
| KM_k141_264921 | Not-determined |
| KM_k141_66513 | Not-determined |
| KM_k141_66515 | Not-determined |
| KM_k141_66517 | Not-determined |
| KM_k141_264925 | Not-determined |
| KM_k141_110399 | Not-determined |
| KM_k141_32139 | Not-determined |
| KM_k141_144418 | Not-determined |
| KM_k141_264932 | Not-determined |
| KM_k141_10112 | Not-determined |
| KM_k141_165522 | Not-determined |
| KM_k141_165527 | Not-determined |
| KM_k141_253515 | Not-determined |
| KM_k141_264943 | Not-determined |
| KM_k141_155259 | Not-determined |
| KM_k141_43387 | Not-determined |
| KM_k141_264949 | Not-determined |
| KM_k141_66521 | Low-quality |
| KM_k141_264950 | Not-determined |
| KM_k141_32153 | Low-quality |
| KM_k141_43390 | Low-quality |
| KM_k141_253521 | Not-determined |
| KM_k141_155268 | Not-determined |
| KM_k141_52789 | Not-determined |
| KM_k141_52791 | Not-determined |
| KM_k141_52794 | Not-determined |
| KM_k141_20111 | Not-determined |
| KM_k141_132796 | Not-determined |
| KM_k141_97649 | Not-determined |
| KM_k141_52797 | Not-determined |
| KM_k141_66532 | Low-quality |
| KM_k141_97650 | Not-determined |
| KM_k141_66536 | Not-determined |
| KM_k141_165542 | Low-quality |
| KM_k141_144449 | Not-determined |
| KM_k141_20121 | Not-determined |
| KM_k141_144452 | Low-quality |
| KM_k141_43414 | Not-determined |
| KM_k141_52813 | Not-determined |
| KM_k141_253540 | Low-quality |
| KM_k141_43418 | Low-quality |
| KM_k141_52814 | Not-determined |
| KM_k141_165553 | Not-determined |
| KM_k141_253541 | Not-determined |
| KM_k141_20125 | Not-determined |
| KM_k141_10144 | Not-determined |
| KM_k141_97668 | Not-determined |
| KM_k141_10146 | Not-determined |
| KM_k141_165563 | Not-determined |
| KM_k141_52821 | Not-determined |
| KM_k141_43433 | Not-determined |
| KM_k141_144460 | Not-determined |
| KM_k141_10153 | Not-determined |
| KM_k141_43436 | Not-determined |
| KM_k141_97673 | Not-determined |
| KM_k141_165575 | Not-determined |
| KM_k141_264970 | Not-determined |
| KM_k141_32197 | Not-determined |
| KM_k141_97676 | Not-determined |
| KM_k141_20139 | Not-determined |
| KM_k141_66547 | Not-determined |
| KM_k141_165580 | Not-determined |
| KM_k141_20143 | Not-determined |
| KM_k141_20145 | Not-determined |
| KM_k141_243586 | Not-determined |
| KM_k141_243587 | Not-determined |
| KM_k141_52827 | Not-determined |
| KM_k141_52829 | Low-quality |
| KM_k141_20159 | Not-determined |
| KM_k141_253557 | Not-determined |
| KM_k141_264974 | Not-determined |
| KM_k141_97691 | Not-determined |
| KM_k141_110442 | Not-determined |
| KM_k141_165592 | Low-quality |
| KM_k141_253562 | Not-determined |
| KM_k141_165596 | Not-determined |
| KM_k141_52835 | Not-determined |
| KM_k141_10167 | Not-determined |
| KM_k141_155304 | Not-determined |
| KM_k141_43444 | Low-quality |
| KM_k141_253565 | Low-quality |
| KM_k141_10170 | Not-determined |
| KM_k141_52839 | Not-determined |
| KM_k141_66568 | Not-determined |
| KM_k141_243600 | Not-determined |
| KM_k141_253568 | Not-determined |
| KM_k141_43450 | Not-determined |
| KM_k141_155312 | Not-determined |
| KM_k141_97706 | Not-determined |
| KM_k141_243614 | Low-quality |
| KM_k141_243617 | Not-determined |
| KM_k141_264986 | Not-determined |
| KM_k141_243626 | Not-determined |
| KM_k141_20187 | Not-determined |
| KM_k141_20197 | Not-determined |
| KM_k141_265001 | Not-determined |
| KM_k141_52857 | Not-determined |
| KM_k141_52858 | Not-determined |
| KM_k141_43472 | Not-determined |
| KM_k141_97724 | Not-determined |
| KM_k141_97728 | Not-determined |
| KM_k141_52862 | Low-quality |
| KM_k141_110468 | Not-determined |
| KM_k141_243647 | Not-determined |
| KM_k141_52878 | Not-determined |
| KM_k141_20209 | Not-determined |
| KM_k141_97750 | Not-determined |
| KM_k141_97751 | Not-determined |
| KM_k141_243657 | Not-determined |
| KM_k141_97752 | Not-determined |
| KM_k141_97755 | Not-determined |
| KM_k141_97756 | Not-determined |
| KM_k141_253619 | Not-determined |
| KM_k141_20222 | Not-determined |
| KM_k141_20224 | Not-determined |
| KM_k141_52892 | Not-determined |
| KM_k141_10233 | Not-determined |
| KM_k141_66584 | Not-determined |
| KM_k141_66593 | Not-determined |
| KM_k141_97764 | Not-determined |
| KM_k141_155328 | Not-determined |
| KM_k141_155329 | Not-determined |
| KM_k141_43511 | Not-determined |
| KM_k141_155332 | Not-determined |
| KM_k141_243695 | Not-determined |
| KM_k141_165676 | Not-determined |
| KM_k141_243698 | Not-determined |
| KM_k141_97767 | Not-determined |
| KM_k141_110491 | Not-determined |
| KM_k141_10246 | Not-determined |
| KM_k141_110499 | Not-determined |
| KM_k141_265036 | Not-determined |
| KM_k141_155344 | Low-quality |
| KM_k141_10251 | Not-determined |
| KM_k141_165690 | Not-determined |
| KM_k141_165692 | Not-determined |
| KM_k141_43524 | Not-determined |
| KM_k141_243714 | Not-determined |
| KM_k141_265042 | Not-determined |
| KM_k141_10257 | Not-determined |
| KM_k141_43529 | Not-determined |
| KM_k141_132847 | Not-determined |
| KM_k141_20258 | Not-determined |
| KM_k141_155361 | Not-determined |
| KM_k141_10267 | Not-determined |
| KM_k141_10272 | Not-determined |
| KM_k141_66610 | Not-determined |
| KM_k141_265063 | Not-determined |
| KM_k141_265067 | Not-determined |
| KM_k141_52929 | Not-determined |
| KM_k141_243730 | Not-determined |
| KM_k141_243731 | Not-determined |
| KM_k141_20284 | Low-quality |
| KM_k141_155376 | Not-determined |
| KM_k141_165719 | Not-determined |
| KM_k141_243738 | Not-determined |
| KM_k141_265083 | Not-determined |
| KM_k141_20295 | Not-determined |
| KM_k141_20298 | Not-determined |
| KM_k141_253699 | Low-quality |
| KM_k141_10300 | Not-determined |
| KM_k141_265090 | Not-determined |
| KM_k141_32209 | Not-determined |
| KM_k141_32214 | Not-determined |
| KM_k141_97779 | Not-determined |
| KM_k141_110513 | Low-quality |
| KM_k141_97780 | Not-determined |
| KM_k141_110514 | Not-determined |
| KM_k141_165741 | Not-determined |
| KM_k141_10316 | Not-determined |
| KM_k141_20317 | Not-determined |
| KM_k141_20320 | Not-determined |
| KM_k141_10321 | Not-determined |
| KM_k141_32225 | Low-quality |
| KM_k141_97798 | Not-determined |
| KM_k141_243764 | Not-determined |
| KM_k141_32233 | Not-determined |
| KM_k141_20338 | Not-determined |
| KM_k141_97806 | Not-determined |
| KM_k141_110534 | Not-determined |
| KM_k141_20341 | Not-determined |
| KM_k141_155415 | Not-determined |
| KM_k141_20344 | Not-determined |
| KM_k141_20345 | Not-determined |
| KM_k141_110540 | Not-determined |
| KM_k141_253723 | Not-determined |
| KM_k141_155422 | Not-determined |
| KM_k141_97811 | Not-determined |
| KM_k141_132878 | Not-determined |
| KM_k141_20351 | Not-determined |
| KM_k141_97814 | Not-determined |
| KM_k141_97816 | Low-quality |
| KM_k141_110546 | Low-quality |
| KM_k141_110547 | Not-determined |
| KM_k141_155427 | Not-determined |
| KM_k141_165770 | Not-determined |
| KM_k141_165771 | Low-quality |
| KM_k141_155430 | Not-determined |
| KM_k141_66653 | Not-determined |
| KM_k141_155431 | Low-quality |
| KM_k141_20358 | Not-determined |
| KM_k141_243777 | Not-determined |
| KM_k141_253734 | Not-determined |
| KM_k141_20363 | Not-determined |
| KM_k141_97826 | Not-determined |
| KM_k141_243781 | Not-determined |
| KM_k141_97828 | Not-determined |
| KM_k141_155438 | Not-determined |
| KM_k141_155439 | Not-determined |
| KM_k141_52970 | Not-determined |
| KM_k141_20374 | Not-determined |
| KM_k141_243784 | Not-determined |
| KM_k141_253748 | Not-determined |
| KM_k141_253749 | Not-determined |
| KM_k141_155446 | Not-determined |
| KM_k141_243788 | Low-quality |
| KM_k141_32267 | Not-determined |
| KM_k141_253758 | Not-determined |
| KM_k141_43576 | Not-determined |
| KM_k141_243800 | Not-determined |
| KM_k141_20396 | Not-determined |
| KM_k141_43581 | Not-determined |
| KM_k141_10365 | Not-determined |
| KM_k141_253760 | Low-quality |
| KM_k141_32275 | Not-determined |
| KM_k141_10377 | Not-determined |
| KM_k141_132897 | Not-determined |
| KM_k141_20404 | Not-determined |
| KM_k141_43587 | Low-quality |
| KM_k141_165805 | Low-quality |
| KM_k141_165806 | Not-determined |
| KM_k141_20411 | Not-determined |
| KM_k141_165807 | Not-determined |
| KM_k141_32294 | Not-determined |
| KM_k141_20416 | Not-determined |
| KM_k141_32297 | Not-determined |
| KM_k141_43601 | Not-determined |
| KM_k141_43606 | Not-determined |
| KM_k141_243835 | Not-determined |
| KM_k141_10395 | Low-quality |
| KM_k141_243838 | Not-determined |
| KM_k141_243839 | Not-determined |
| KM_k141_10396 | Not-determined |
| KM_k141_243841 | Not-determined |
| KM_k141_20427 | Low-quality |
| KM_k141_32313 | Low-quality |
| KM_k141_110574 | Not-determined |
| KM_k141_265103 | Not-determined |
| KM_k141_165828 | Not-determined |
| KM_k141_20444 | Not-determined |
| KM_k141_243862 | Not-determined |
| KM_k141_20445 | Not-determined |
| KM_k141_155484 | Low-quality |
| KM_k141_20448 | Not-determined |
| KM_k141_243865 | Not-determined |
| KM_k141_165835 | Low-quality |
| KM_k141_243869 | Not-determined |
| KM_k141_132946 | Not-determined |
| KM_k141_20460 | Not-determined |
| KM_k141_165843 | Low-quality |
| KM_k141_110586 | Not-determined |
| KM_k141_253813 | Not-determined |
| KM_k141_97896 | Not-determined |
| KM_k141_10426 | Not-determined |
| KM_k141_20462 | Not-determined |
| KM_k141_155494 | Not-determined |
| KM_k141_20467 | Not-determined |
| KM_k141_165857 | Not-determined |
| KM_k141_20475 | Not-determined |
| KM_k141_155498 | Not-determined |
| KM_k141_43659 | Low-quality |
| KM_k141_32362 | Not-determined |
| KM_k141_10442 | Not-determined |
| KM_k141_265116 | Not-determined |
| KM_k141_10445 | Not-determined |
| KM_k141_97915 | Not-determined |
| KM_k141_43673 | Not-determined |
| KM_k141_43674 | Not-determined |
| KM_k141_20488 | Not-determined |
| KM_k141_265121 | Not-determined |
| KM_k141_43677 | Not-determined |
| KM_k141_10456 | Not-determined |
| KM_k141_155510 | Not-determined |
| KM_k141_253829 | Not-determined |
| KM_k141_265128 | Low-quality |
| KM_k141_20503 | Not-determined |
| KM_k141_165891 | Not-determined |
| KM_k141_20504 | Not-determined |
| KM_k141_10469 | Not-determined |
| KM_k141_265137 | Not-determined |
| KM_k141_20513 | Not-determined |
| KM_k141_265139 | Low-quality |
| KM_k141_32408 | Not-determined |
| KM_k141_97934 | Not-determined |
| KM_k141_32413 | Not-determined |
| KM_k141_43686 | Not-determined |
| KM_k141_243910 | Not-determined |
| KM_k141_10483 | Not-determined |
| KM_k141_97939 | Not-determined |
| KM_k141_20533 | Not-determined |
| KM_k141_132997 | Not-determined |
| KM_k141_243922 | Not-determined |
| KM_k141_43700 | Not-determined |
| KM_k141_155535 | Not-determined |
| KM_k141_243928 | Not-determined |
| KM_k141_155540 | Not-determined |
| KM_k141_155542 | Not-determined |
| KM_k141_243935 | Low-quality |
| KM_k141_155547 | Not-determined |
| KM_k141_243944 | Not-determined |
| KM_k141_32442 | Not-determined |
| KM_k141_243946 | Not-determined |
| KM_k141_32448 | Not-determined |
| KM_k141_243948 | Not-determined |
| KM_k141_10510 | Low-quality |
| KM_k141_32452 | Not-determined |
| KM_k141_32454 | Not-determined |
| KM_k141_10512 | Not-determined |
| KM_k141_133019 | Not-determined |
| KM_k141_97957 | Not-determined |
| KM_k141_253858 | Not-determined |
| KM_k141_253859 | Not-determined |
| KM_k141_32462 | Not-determined |
| KM_k141_133032 | Not-determined |
| KM_k141_253870 | Low-quality |
| KM_k141_253872 | Not-determined |
| KM_k141_253875 | Not-determined |
| KM_k141_10527 | Not-determined |
| KM_k141_10530 | Not-determined |
| KM_k141_243984 | Low-quality |
| KM_k141_265154 | Not-determined |
| KM_k141_10537 | Not-determined |
| KM_k141_43755 | Not-determined |
| KM_k141_20566 | Not-determined |
| KM_k141_133050 | Not-determined |
| KM_k141_133051 | Not-determined |
| KM_k141_32477 | Not-determined |
| KM_k141_165937 | Not-determined |
| KM_k141_155553 | Low-quality |
| KM_k141_52991 | Not-determined |
| KM_k141_244002 | Not-determined |
| KM_k141_265167 | Not-determined |
| KM_k141_10547 | Not-determined |
| KM_k141_52997 | Not-determined |
| KM_k141_10551 | Not-determined |
| KM_k141_53004 | Not-determined |
| KM_k141_253896 | Not-determined |
| KM_k141_133064 | Not-determined |
| KM_k141_155565 | Not-determined |
| KM_k141_10559 | Not-determined |
| KM_k141_253900 | Not-determined |
| KM_k141_253908 | Not-determined |
| KM_k141_32492 | Not-determined |
| KM_k141_32494 | Low-quality |
| KM_k141_43791 | Low-quality |
| KM_k141_43792 | Not-determined |
| KM_k141_10579 | Not-determined |
| KM_k141_253917 | Not-determined |
| KM_k141_53020 | Not-determined |
| KM_k141_265191 | Low-quality |
| KM_k141_253925 | Not-determined |
| KM_k141_265194 | Not-determined |
| KM_k141_98010 | Not-determined |
| KM_k141_20597 | Low-quality |
| KM_k141_98011 | Not-determined |
| KM_k141_20598 | Not-determined |
| KM_k141_133091 | Not-determined |
| KM_k141_20601 | Not-determined |
| KM_k141_20604 | Not-determined |
| KM_k141_32517 | Not-determined |
| KM_k141_53032 | Not-determined |
| KM_k141_32519 | Not-determined |
| KM_k141_166006 | Not-determined |
| KM_k141_20619 | Not-determined |
| KM_k141_133096 | Not-determined |
| KM_k141_53039 | Not-determined |
| KM_k141_32527 | Not-determined |
| KM_k141_155580 | Not-determined |
| KM_k141_166017 | Not-determined |
| KM_k141_10596 | Not-determined |
| KM_k141_98031 | Low-quality |
| KM_k141_53051 | Not-determined |
| KM_k141_244052 | Not-determined |
| KM_k141_98032 | Not-determined |
| KM_k141_110623 | Low-quality |
| KM_k141_253957 | Not-determined |
| KM_k141_244056 | Not-determined |
| KM_k141_253959 | Not-determined |
| KM_k141_20641 | Not-determined |
| KM_k141_20643 | Not-determined |
| KM_k141_265214 | Not-determined |
| KM_k141_110629 | Low-quality |
| KM_k141_20654 | Not-determined |
| KM_k141_32534 | Not-determined |
| KM_k141_10625 | Not-determined |
| KM_k141_133120 | Not-determined |
| KM_k141_20658 | Not-determined |
| KM_k141_43845 | Not-determined |
| KM_k141_265230 | Low-quality |
| KM_k141_10629 | Not-determined |
| KM_k141_166039 | Not-determined |
| KM_k141_254005 | Not-determined |
| KM_k141_265241 | Not-determined |
| KM_k141_98068 | Not-determined |
| KM_k141_265244 | Not-determined |
| KM_k141_254017 | Not-determined |
| KM_k141_98074 | Not-determined |
| KM_k141_254019 | Not-determined |
| KM_k141_43848 | Not-determined |
| KM_k141_265253 | Not-determined |
| KM_k141_43856 | Not-determined |
| KM_k141_254029 | Low-quality |
| KM_k141_265256 | Not-determined |
| KM_k141_254032 | Not-determined |
| KM_k141_32577 | Not-determined |
| KM_k141_98086 | Low-quality |
| KM_k141_43868 | Not-determined |
| KM_k141_265262 | Low-quality |
| KM_k141_166070 | Not-determined |
| KM_k141_265263 | Low-quality |
| KM_k141_133160 | Not-determined |
| KM_k141_98089 | Not-determined |
| KM_k141_10656 | Not-determined |
| KM_k141_10657 | Not-determined |
| KM_k141_133176 | Not-determined |
| KM_k141_43877 | Not-determined |
| KM_k141_43879 | Not-determined |
| KM_k141_98099 | Not-determined |
| KM_k141_254067 | Not-determined |
| KM_k141_43883 | Low-quality |
| KM_k141_244067 | Not-determined |
| KM_k141_98107 | Not-determined |
| KM_k141_98108 | Not-determined |
| KM_k141_265275 | Not-determined |
| KM_k141_265276 | Not-determined |
| KM_k141_265278 | Not-determined |
| KM_k141_10667 | Not-determined |
| KM_k141_43891 | Not-determined |
| KM_k141_32608 | Not-determined |
| KM_k141_43895 | Not-determined |
| KM_k141_10669 | Not-determined |
| KM_k141_244071 | Not-determined |
| KM_k141_98122 | Not-determined |
| KM_k141_265304 | Not-determined |
| KM_k141_254104 | Not-determined |
| KM_k141_244075 | Low-quality |
| KM_k141_166096 | Not-determined |
| KM_k141_244081 | Not-determined |
| KM_k141_98129 | Not-determined |
| KM_k141_43924 | Low-quality |
| KM_k141_110685 | Not-determined |
| KM_k141_254115 | Not-determined |
| KM_k141_254117 | Not-determined |
| KM_k141_133187 | Not-determined |
| KM_k141_166110 | Not-determined |
| KM_k141_43933 | Not-determined |
| KM_k141_244105 | Not-determined |
| KM_k141_53136 | Not-determined |
| KM_k141_32629 | Low-quality |
| KM_k141_254135 | Low-quality |
| KM_k141_265333 | Not-determined |
| KM_k141_166127 | Low-quality |
| KM_k141_254147 | Not-determined |
| KM_k141_98162 | Not-determined |
| KM_k141_43945 | Not-determined |
| KM_k141_166132 | Not-determined |
| KM_k141_20685 | Low-quality |
| KM_k141_43952 | Not-determined |
| KM_k141_166139 | Not-determined |
| KM_k141_254151 | Not-determined |
| KM_k141_244129 | Not-determined |
| KM_k141_32642 | Not-determined |
| KM_k141_53150 | Low-quality |
| KM_k141_20691 | Not-determined |
| KM_k141_32644 | Not-determined |
| KM_k141_244134 | Not-determined |
| KM_k141_32650 | Not-determined |
| KM_k141_20697 | Not-determined |
| KM_k141_20699 | Not-determined |
| KM_k141_53158 | Not-determined |
| KM_k141_20701 | Not-determined |
| KM_k141_244148 | Not-determined |
| KM_k141_53161 | Not-determined |
| KM_k141_244152 | Not-determined |
| KM_k141_43980 | Not-determined |
| KM_k141_53181 | Not-determined |
| KM_k141_244168 | Not-determined |
| KM_k141_244173 | Not-determined |
| KM_k141_32677 | Not-determined |
| KM_k141_53200 | Not-determined |
| KM_k141_20725 | Not-determined |
| [illegible] | [illegible] |
| KM_k141_53211 | Not-determined |
| KM_k141_53212 | Not-determined |
| KM_k141_133211 | Not-determined |
| KM_k141_244194 | Not-determined |
| KM_k141_98195 | Low-quality |
| KM_k141_32684 | Not-determined |
| KM_k141_166151 | Not-determined |
| KM_k141_98207 | Not-determined |
| KM_k141_133229 | Not-determined |
| KM_k141_32689 | Low-quality |
| KM_k141_133230 | Not-determined |
| KM_k141_133234 | Not-determined |
| KM_k141_32695 | Not-determined |
| KM_k141_265394 | Not-determined |
| KM_k141_32705 | Not-determined |
| KM_k141_98230 | Not-determined |
| KM_k141_44021 | Not-determined |
| KM_k141_265400 | Not-determined |
| KM_k141_98240 | Not-determined |
| KM_k141_32714 | Low-quality |
| KM_k141_20767 | Not-determined |
| KM_k141_265403 | Not-determined |
| KM_k141_254207 | Low-quality |
| KM_k141_32721 | Low-quality |
| KM_k141_44034 | Not-determined |
| KM_k141_166162 | Not-determined |
| KM_k141_98251 | Not-determined |
| KM_k141_32728 | Low-quality |
| KM_k141_244235 | Not-determined |
| KM_k141_32732 | Not-determined |
| KM_k141_133253 | Not-determined |
| KM_k141_32737 | Not-determined |
| KM_k141_32738 | Not-determined |
| KM_k141_265423 | Not-determined |
| KM_k141_244241 | Not-determined |
| KM_k141_53226 | Not-determined |
| KM_k141_244243 | Not-determined |
| KM_k141_244245 | Not-determined |
| KM_k141_265437 | Not-determined |
| KM_k141_98269 | Low-quality |
| KM_k141_44048 | Not-determined |
| KM_k141_244258 | Not-determined |
| KM_k141_133272 | Not-determined |
| KM_k141_133273 | Not-determined |
| KM_k141_98275 | Not-determined |
| KM_k141_32750 | Not-determined |
| KM_k141_265443 | Low-quality |
| KM_k141_32752 | Not-determined |
| KM_k141_53240 | Not-determined |
| KM_k141_32754 | Not-determined |
| KM_k141_32756 | Not-determined |
| KM_k141_20797 | Not-determined |
| KM_k141_133285 | Low-quality |
| KM_k141_53247 | Not-determined |
| KM_k141_166172 | Not-determined |
| KM_k141_44067 | Not-determined |
| KM_k141_32760 | Not-determined |
| KM_k141_133286 | Not-determined |
| KM_k141_265450 | Not-determined |
| KM_k141_53250 | Not-determined |
| KM_k141_32775 | Not-determined |
| KM_k141_32776 | Not-determined |
| KM_k141_20823 | Not-determined |
| KM_k141_98292 | Not-determined |
| KM_k141_32783 | Not-determined |
| KM_k141_44076 | Not-determined |
| KM_k141_44088 | Not-determined |
| KM_k141_10682 | Not-determined |
| KM_k141_44092 | Not-determined |
| KM_k141_98317 | Not-determined |
| KM_k141_133307 | Not-determined |
| KM_k141_254241 | Not-determined |
| KM_k141_44103 | Not-determined |
| KM_k141_44107 | Low-quality |
| KM_k141_254246 | Not-determined |
| KM_k141_254247 | Not-determined |
| KM_k141_98326 | Not-determined |
| KM_k141_110742 | Low-quality |
| KM_k141_53281 | Not-determined |
| KM_k141_98333 | Not-determined |
| KM_k141_53287 | Not-determined |
| KM_k141_20876 | Low-quality |

| | |
|---|---|
| KM_k141_110754 | Not-determined |
| KM_k141_98338 | Not-determined |
| KM_k141_254260 | Not-determined |
| KM_k141_98343 | Not-determined |
| KM_k141_265459 | Not-determined |
| KM_k141_10706 | Not-determined |
| KM_k141_133325 | Not-determined |
| KM_k141_98349 | Not-determined |
| KM_k141_20890 | Not-determined |
| KM_k141_32844 | Low-quality |
| KM_k141_10726 | Not-determined |
| KM_k141_53306 | Not-determined |
| KM_k141_98361 | Not-determined |
| KM_k141_20904 | Not-determined |
| KM_k141_244298 | Not-determined |
| KM_k141_44155 | Not-determined |
| KM_k141_110777 | Not-determined |
| KM_k141_44159 | Low-quality |
| KM_k141_98371 | Not-determined |
| KM_k141_44164 | Not-determined |
| KM_k141_32864 | Not-determined |
| KM_k141_32866 | Not-determined |
| KM_k141_10742 | Not-determined |
| KM_k141_265476 | Not-determined |
| KM_k141_32872 | Low-quality |
| KM_k141_20935 | Not-determined |
| KM_k141_20936 | Not-determined |
| KM_k141_98407 | Not-determined |
| KM_k141_98409 | Not-determined |
| KM_k141_53338 | Not-determined |
| KM_k141_53343 | Not-determined |
| KM_k141_265518 | Not-determined |
| KM_k141_254271 | Not-determined |
| KM_k141_44173 | Not-determined |
| KM_k141_265524 | Not-determined |
| KM_k141_110808 | Not-determined |
| KM_k141_53356 | Not-determined |
| KM_k141_98426 | Not-determined |
| KM_k141_254282 | Not-determined |
| KM_k141_265531 | Not-determined |
| KM_k141_133333 | Not-determined |
| KM_k141_10782 | Not-determined |
| KM_k141_53365 | Not-determined |
| KM_k141_265538 | Low-quality |
| KM_k141_244331 | Not-determined |
| KM_k141_44197 | Not-determined |
| KM_k141_44204 | Not-determined |
| KM_k141_20990 | Not-determined |
| KM_k141_110822 | Not-determined |
| KM_k141_265544 | Low-quality |
| KM_k141_44213 | Not-determined |
| KM_k141_133345 | Not-determined |
| KM_k141_98456 | Not-determined |
| KM_k141_98459 | Not-determined |
| KM_k141_20995 | Not-determined |
| KM_k141_20998 | Not-determined |
| KM_k141_133352 | Not-determined |
| KM_k141_32905 | Not-determined |
| KM_k141_254304 | Not-determined |
| KM_k141_244356 | Not-determined |
| KM_k141_32914 | Not-determined |
| KM_k141_21015 | Low-quality |
| KM_k141_244364 | Not-determined |
| KM_k141_21022 | Not-determined |
| KM_k141_265562 | Not-determined |
| KM_k141_98484 | Not-determined |
| KM_k141_265575 | Not-determined |
| KM_k141_32921 | Low-quality |
| KM_k141_10800 | Not-determined |
| KM_k141_265579 | Not-determined |
| KM_k141_10803 | Not-determined |
| KM_k141_44248 | Not-determined |
| KM_k141_166213 | Not-determined |
| KM_k141_166221 | Not-determined |
| KM_k141_265593 | Not-determined |
| KM_k141_244389 | Not-determined |
| KM_k141_21054 | Not-determined |
| KM_k141_98507 | Not-determined |
| KM_k141_44264 | Not-determined |
| KM_k141_32942 | Not-determined |
| KM_k141_21060 | Not-determined |
| KM_k141_166232 | Not-determined |
| KM_k141_98513 | Not-determined |
| KM_k141_44267 | Not-determined |
| KM_k141_21066 | Not-determined |
| KM_k141_21069 | Not-determined |
| KM_k141_44270 | Not-determined |
| KM_k141_110832 | Not-determined |
| KM_k141_10828 | Not-determined |
| KM_k141_32949 | Not-determined |
| KM_k141_21079 | Not-determined |
| KM_k141_44281 | Not-determined |
| KM_k141_244403 | Not-determined |
| KM_k141_265607 | Not-determined |
| KM_k141_44287 | Not-determined |
| KM_k141_254385 | Not-determined |
| KM_k141_244411 | Not-determined |
| KM_k141_10846 | Not-determined |
| KM_k141_110840 | Not-determined |
| KM_k141_98537 | Not-determined |
| KM_k141_244419 | Not-determined |
| KM_k141_110843 | Not-determined |
| KM_k141_32973 | Not-determined |
| KM_k141_244422 | Not-determined |
| KM_k141_166253 | Not-determined |
| KM_k141_32977 | Not-determined |
| KM_k141_110849 | Low-quality |
| KM_k141_265615 | Not-determined |
| KM_k141_110851 | Not-determined |
| KM_k141_44316 | Not-determined |
| KM_k141_44319 | Not-determined |
| KM_k141_166273 | Not-determined |
| KM_k141_166275 | Not-determined |
| KM_k141_10898 | Not-determined |
| KM_k141_21098 | Not-determined |
| KM_k141_21099 | Not-determined |
| KM_k141_21101 | Low-quality |
| KM_k141_33007 | Not-determined |
| KM_k141_244438 | Not-determined |
| KM_k141_21109 | Not-determined |
| KM_k141_21111 | Not-determined |
| KM_k141_10909 | Not-determined |
| KM_k141_265646 | Not-determined |
| KM_k141_10912 | Not-determined |
| KM_k141_166283 | Not-determined |
| KM_k141_166295 | Not-determined |
| KM_k141_244449 | Not-determined |
| KM_k141_33022 | Not-determined |
| KM_k141_21133 | Not-determined |
| KM_k141_244458 | Not-determined |
| KM_k141_244459 | Not-determined |
| KM_k141_53396 | Not-determined |
| KM_k141_244463 | Not-determined |
| KM_k141_166299 | Not-determined |
| KM_k141_265650 | Not-determined |
| KM_k141_33041 | Not-determined |
| KM_k141_21166 | Not-determined |
| KM_k141_98606 | Not-determined |
| KM_k141_33055 | Not-determined |
| KM_k141_10944 | Not-determined |
| KM_k141_10945 | Not-determined |
| KM_k141_21191 | Low-quality |
| KM_k141_21192 | Not-determined |
| KM_k141_21195 | Not-determined |
| KM_k141_44388 | Low-quality |
| KM_k141_98624 | Not-determined |
| KM_k141_44395 | Not-determined |
| KM_k141_53421 | Not-determined |
| KM_k141_44397 | Not-determined |
| KM_k141_44401 | Not-determined |
| KM_k141_265671 | Not-determined |
| KM_k141_21207 | Not-determined |
| KM_k141_98648 | Not-determined |
| KM_k141_98659 | Not-determined |
| KM_k141_11012 | Not-determined |
| KM_k141_98670 | Not-determined |
| KM_k141_110925 | Not-determined |
| KM_k141_11017 | Not-determined |
| KM_k141_11029 | Not-determined |
| KM_k141_33075 | Not-determined |
| KM_k141_98700 | Not-determined |
| KM_k141_53481 | Not-determined |
| KM_k141_44436 | Not-determined |
| KM_k141_98710 | Not-determined |
| KM_k141_265699 | Not-determined |
| KM_k141_265701 | Not-determined |
| KM_k141_265709 | Not-determined |
| KM_k141_11035 | Not-determined |
| KM_k141_11036 | Not-determined |
| KM_k141_33126 | Not-determined |
| KM_k141_98746 | Not-determined |
| KM_k141_33131 | Not-determined |
| KM_k141_265730 | Low-quality |
| KM_k141_265732 | Not-determined |
| KM_k141_265736 | Not-determined |
| KM_k141_98760 | Not-determined |
| KM_k141_98779 | Not-determined |
| KM_k141_98784 | Not-determined |
| KM_k141_21265 | Not-determined |
| KM_k141_21267 | Not-determined |
| KM_k141_166318 | Not-determined |
| KM_k141_110987 | Not-determined |
| KM_k141_33140 | Not-determined |
| KM_k141_265752 | Not-determined |
| KM_k141_11067 | Not-determined |
| KM_k141_166325 | Not-determined |
| KM_k141_166329 | Not-determined |
| KM_k141_110996 | Not-determined |
| KM_k141_21284 | Low-quality |
| KM_k141_111000 | Not-determined |
| KM_k141_265754 | Not-determined |
| KM_k141_111013 | Not-determined |
| KM_k141_53503 | Not-determined |
| KM_k141_111016 | Not-determined |
| KM_k141_21300 | Not-determined |
| KM_k141_11084 | Not-determined |
| KM_k141_53511 | Not-determined |
| KM_k141_265774 | Not-determined |
| KM_k141_265775 | Not-determined |
| KM_k141_53514 | Not-determined |
| KM_k141_11093 | Low-quality |
| KM_k141_265778 | Not-determined |
| KM_k141_11101 | Not-determined |
| KM_k141_11105 | Not-determined |
| KM_k141_265792 | Not-determined |
| KM_k141_33162 | Not-determined |
| KM_k141_265801 | Not-determined |
| KM_k141_53520 | Not-determined |
| KM_k141_98812 | Not-determined |
| KM_k141_265802 | Not-determined |
| KM_k141_166347 | Not-determined |
| KM_k141_53522 | Low-quality |
| KM_k141_98818 | Not-determined |
| KM_k141_98828 | Not-determined |
| KM_k141_21323 | Not-determined |
| KM_k141_33173 | Not-determined |
| KM_k141_166356 | Not-determined |
| KM_k141_166358 | Not-determined |
| KM_k141_98837 | Not-determined |
| KM_k141_33177 | Not-determined |
| KM_k141_33184 | Not-determined |
| KM_k141_53545 | Not-determined |
| KM_k141_166384 | Not-determined |
| KM_k141_254571 | Not-determined |
| KM_k141_265820 | Not-determined |
| KM_k141_33192 | Low-quality |
| KM_k141_98853 | Low-quality |
| KM_k141_265825 | Not-determined |
| KM_k141_21344 | Low-quality |
| KM_k141_21351 | Not-determined |
| KM_k141_53570 | Not-determined |
| KM_k141_265839 | Not-determined |
| KM_k141_33211 | Low-quality |
| KM_k141_21358 | Low-quality |
| KM_k141_21359 | Not-determined |
| KM_k141_21360 | Not-determined |
| KM_k141_53594 | Not-determined |
| KM_k141_254587 | Not-determined |
| KM_k141_254588 | Not-determined |
| KM_k141_265853 | Not-determined |
| KM_k141_98881 | Not-determined |
| KM_k141_111062 | Not-determined |
| KM_k141_98884 | Not-determined |
| KM_k141_98887 | Not-determined |
| KM_k141_111064 | Not-determined |
| KM_k141_111065 | Not-determined |
| KM_k141_53627 | Not-determined |
| KM_k141_33237 | Not-determined |
| KM_k141_21375 | Low-quality |
| KM_k141_265860 | Not-determined |
| KM_k141_265861 | Not-determined |
| KM_k141_98909 | Not-determined |
| KM_k141_98910 | Low-quality |
| KM_k141_53645 | Not-determined |
| KM_k141_53647 | Not-determined |
| KM_k141_53649 | Not-determined |
| KM_k141_98920 | Not-determined |
| KM_k141_265884 | Not-determined |
| KM_k141_111070 | Not-determined |
| KM_k141_98923 | Not-determined |
| KM_k141_111071 | Not-determined |
| KM_k141_33249 | Not-determined |
| KM_k141_254618 | Low-quality |
| KM_k141_265895 | Not-determined |
| KM_k141_254631 | Not-determined |
| KM_k141_265899 | Not-determined |
| KM_k141_33254 | Not-determined |
| KM_k141_53661 | Low-quality |
| KM_k141_53663 | Not-determined |
| KM_k141_265910 | Not-determined |
| KM_k141_33258 | Not-determined |
| KM_k141_98941 | Not-determined |
| KM_k141_98947 | Not-determined |
| KM_k141_33264 | Not-determined |
| KM_k141_33265 | Not-determined |
| KM_k141_265936 | Low-quality |
| KM_k141_33283 | Not-determined |
| KM_k141_33286 | Low-quality |
| KM_k141_21416 | Not-determined |
| KM_k141_111088 | Low-quality |
| KM_k141_21424 | Not-determined |
| KM_k141_21439 | Not-determined |
| KM_k141_265944 | Not-determined |
| KM_k141_265948 | Not-determined |
| KM_k141_53703 | Not-determined |
| KM_k141_53707 | Not-determined |
| KM_k141_53711 | Not-determined |
| KM_k141_53719 | Not-determined |
| KM_k141_53723 | Not-determined |
| KM_k141_21461 | Not-determined |
| KM_k141_265966 | Not-determined |
| KM_k141_111117 | Not-determined |
| KM_k141_166418 | Not-determined |
| KM_k141_265993 | Not-determined |
| KM_k141_265994 | Not-determined |
| KM_k141_21470 | Not-determined |
| KM_k141_111120 | Not-determined |
| KM_k141_265999 | Not-determined |
| KM_k141_98991 | Not-determined |
| KM_k141_266008 | Not-determined |
| KM_k141_266011 | Not-determined |
| KM_k141_33313 | Not-determined |
| KM_k141_33315 | Not-determined |
| KM_k141_166436 | Not-determined |
| KM_k141_266022 | Low-quality |
| KM_k141_33322 | Not-determined |
| KM_k141_166440 | Not-determined |
| KM_k141_266026 | Not-determined |
| KM_k141_33334 | Not-determined |
| KM_k141_266048 | Not-determined |
| KM_k141_266053 | Not-determined |
| KM_k141_21498 | Not-determined |
| KM_k141_21504 | Not-determined |
| KM_k141_99039 | Not-determined |
| KM_k141_99048 | Not-determined |
| KM_k141_166463 | Not-determined |
| KM_k141_53793 | Not-determined |
| KM_k141_254659 | Not-determined |
| KM_k141_254661 | Not-determined |
| KM_k141_266110 | Low-quality |
| KM_k141_254668 | Not-determined |
| KM_k141_53802 | Not-determined |
| KM_k141_254671 | Not-determined |
| KM_k141_266112 | Not-determined |
| KM_k141_99068 | Not-determined |
| KM_k141_21541 | Not-determined |
| KM_k141_266120 | Not-determined |
| KM_k141_53824 | Not-determined |
| KM_k141_166496 | Not-determined |
| KM_k141_166499 | Not-determined |
| KM_k141_21548 | Not-determined |
| KM_k141_21560 | Not-determined |
| KM_k141_166503 | Not-determined |
| KM_k141_21585 | Not-determined |
| KM_k141_254689 | Low-quality |
| KM_k141_53866 | Not-determined |
| KM_k141_53867 | Not-determined |
| KM_k141_53871 | Not-determined |
| KM_k141_266173 | Not-determined |
| KM_k141_266178 | Not-determined |
| KM_k141_266182 | Not-determined |
| KM_k141_21624 | Not-determined |
| KM_k141_53889 | Not-determined |
| KM_k141_53890 | Not-determined |
| KM_k141_21641 | Not-determined |
| KM_k141_21644 | Not-determined |
| KM_k141_21649 | Not-determined |
| KM_k141_266219 | Not-determined |
| KM_k141_266227 | Not-determined |
| KM_k141_266228 | Not-determined |
| KM_k141_166581 | Not-determined |
| KM_k141_166588 | Not-determined |
| KM_k141_166590 | Not-determined |
| KM_k141_166594 | Not-determined |
| KM_k141_254716 | Not-determined |
| KM_k141_21681 | Not-determined |
| KM_k141_254726 | Not-determined |
| KM_k141_254730 | Low-quality |
| KM_k141_21693 | Not-determined |
| KM_k141_166601 | Not-determined |
| KM_k141_166611 | Low-quality |
| KM_k141_254740 | Not-determined |
| KM_k141_266268 | Not-determined |
| KM_k141_99147 | Not-determined |
| KM_k141_166620 | Not-determined |
| KM_k141_21708 | Not-determined |
| KM_k141_266283 | Not-determined |
| KM_k141_99151 | Not-determined |
| KM_k141_53911 | Not-determined |
| KM_k141_21722 | Not-determined |
| KM_k141_99159 | Not-determined |
| KM_k141_21728 | Not-determined |
| KM_k141_266287 | Not-determined |
| KM_k141_166642 | Not-determined |
| KM_k141_166646 | Not-determined |
| KM_k141_254773 | Not-determined |
| KM_k141_266296 | Not-determined |
| KM_k141_99180 | Not-determined |
| KM_k141_266301 | Not-determined |
| KM_k141_266302 | Not-determined |
| KM_k141_254790 | Not-determined |
| KM_k141_254792 | Not-determined |
| KM_k141_166670 | Not-determined |
| KM_k141_99193 | Not-determined |
| KM_k141_99195 | Not-determined |
| KM_k141_53927 | Not-determined |
| KM_k141_99199 | Not-determined |
| KM_k141_166681 | Not-determined |
| KM_k141_266333 | Not-determined |
| KM_k141_266339 | Low-quality |
| KM_k141_266340 | Not-determined |
| KM_k141_266342 | Not-determined |
| KM_k141_266349 | Not-determined |
| KM_k141_266355 | Not-determined |
| KM_k141_266357 | Not-determined |
| KM_k141_266360 | Not-determined |
| KM_k141_266368 | Not-determined |
| KM_k141_254837 | Not-determined |
| KM_k141_254861 | Low-quality |
| KM_k141_53968 | Not-determined |
| KM_k141_21768 | Not-determined |
| KM_k141_21769 | Not-determined |
| KM_k141_99262 | Not-determined |
| 18-106-final-viral-combined_k141_56464\|\|full | Medium-quality |
| KM_k141_99272 | Low-quality |
| KM_k141_266393 | Not-determined |
| KM_k141_254890 | Not-determined |
| KM_k141_254893 | Not-determined |
| KM_k141_254902 | Not-determined |
| KM_k141_21808 | Not-determined |
| KM_k141_266403 | Not-determined |
| KM_k141_21813 | Not-determined |
| KM_k141_99300 | Not-determined |
| KM_k141_53992 | Not-determined |
| KM_k141_21817 | Not-determined |
| KM_k141_21822 | Not-determined |
| KM_k141_99312 | Not-determined |
| KM_k141_21828 | Not-determined |
| KM_k141_266424 | Not-determined |
| KM_k141_21837 | Not-determined |
| KM_k141_54002 | Not-determined |
| KM_k141_54008 | Not-determined |
| KM_k141_21840 | Not-determined |
| KM_k141_54015 | Not-determined |
| KM_k141_21863 | Not-determined |
| KM_k141_21865 | Not-determined |
| KM_k141_21867 | Not-determined |
| KM_k141_99326 | Not-determined |
| KM_k141_21873 | Not-determined |
| KM_k141_266439 | Not-determined |
| KM_k141_54032 | Not-determined |
| KM_k141_54038 | Not-determined |
| KM_k141_99338 | Not-determined |
| KM_k141_54048 | Not-determined |
| KM_k141_266448 | Not-determined |
| KM_k141_21894 | Not-determined |
| KM_k141_21899 | Not-determined |
| KM_k141_266457 | Not-determined |
| KM_k141_99350 | Low-quality |
| KM_k141_99353 | Low-quality |
| KM_k141_254921 | Not-determined |
| KM_k141_54062 | Not-determined |
| KM_k141_266467 | Not-determined |
| KM_k141_266471 | Not-determined |
| KM_k141_21912 | Not-determined |
| KM_k141_99377 | Not-determined |
| KM_k141_266482 | Not-determined |
| KM_k141_266483 | Low-quality |
| KM_k141_99387 | Not-determined |
| KM_k141_99394 | Not-determined |
| KM_k141_254957 | Not-determined |
| KM_k141_266500 | Not-determined |
| KM_k141_99398 | Not-determined |
| KM_k141_254959 | Not-determined |
| KM_k141_266507 | Not-determined |
| KM_k141_54090 | Not-determined |
| KM_k141_54091 | Not-determined |
| KM_k141_266509 | Not-determined |
| KM_k141_99428 | Not-determined |
| KM_k141_254974 | Not-determined |
| KM_k141_254975 | Not-determined |
| KM_k141_99435 | Low-quality |
| KM_k141_21943 | Low-quality |
| KM_k141_54119 | Not-determined |
| KM_k141_255003 | Not-determined |
| 18-105-final-viral-combined_k141_175145\|\|full | High-quality |
| KM_k141_255009 | Not-determined |
| KM_k141_266512 | Not-determined |
| KM_k141_54142 | Not-determined |
| KM_k141_255015 | Not-determined |
| KM_k141_21957 | Low-quality |
| KM_k141_255017 | Not-determined |
| KM_k141_255026 | Not-determined |
| KM_k141_54155 | Not-determined |
| KM_k141_99483 | Not-determined |
| KM_k141_54162 | Not-determined |
| KM_k141_266523 | Not-determined |
| KM_k141_266524 | Not-determined |
| KM_k141_266533 | Not-determined |
| KM_k141_54186 | Not-determined |
| KM_k141_255037 | Not-determined |
| KM_k141_266542 | Not-determined |
| KM_k141_99512 | Not-determined |
| KM_k141_266543 | Low-quality |
| KM_k141_21976 | Not-determined |
| KM_k141_266544 | Not-determined |
| KM_k141_266549 | Not-determined |
| KM_k141_54198 | Not-determined |
| KM_k141_266555 | Not-determined |
| KM_k141_266559 | Not-determined |
| KM_k141_266568 | Not-determined |
| KM_k141_21985 | Not-determined |
| KM_k141_54216 | Not-determined |
| KM_k141_99541 | Not-determined |
| KM_k141_21988 | Not-determined |
| KM_k141_21991 | Not-determined |
| KM_k141_266592 | Not-determined |
| KM_k141_266595 | Not-determined |
| KM_k141_266600 | Not-determined |
| KM_k141_266604 | Not-determined |
| KM_k141_266614 | Not-determined |
| KM_k141_266619 | Not-determined |
| KM_k141_266620 | Not-determined |
| KM_k141_22004 | Low-quality |
| KM_k141_266623 | Not-determined |
| KM_k141_266624 | Not-determined |
| KM_k141_22008 | Not-determined |
| KM_k141_266634 | Not-determined |
| KM_k141_266636 | Not-determined |
| KM_k141_255083 | Not-determined |
| KM_k141_54248 | Not-determined |
| KM_k141_266642 | Not-determined |
| KM_k141_266648 | Not-determined |
| KM_k141_266650 | Not-determined |
| KM_k141_266655 | Not-determined |
| KM_k141_266664 | Not-determined |
| KM_k141_266666 | Not-determined |
| KM_k141_54265 | Not-determined |
| KM_k141_255100 | Not-determined |
| KM_k141_266669 | Not-determined |
| KM_k141_266671 | Not-determined |
| KM_k141_266685 | Not-determined |
| KM_k141_266687 | Not-determined |
| KM_k141_266698 | Not-determined |
| KM_k141_22028 | Not-determined |
| KM_k141_266703 | Not-determined |
| KM_k141_54272 | Not-determined |
| KM_k141_22033 | Not-determined |
| KM_k141_22040 | Not-determined |
| KM_k141_255128 | Not-determined |
| KM_k141_22043 | Not-determined |
| KM_k141_54291 | Not-determined |
| KM_k141_255158 | Not-determined |
| KM_k141_54297 | Not-determined |
| KM_k141_255163 | Not-determined |
| KM_k141_22085 | Not-determined |
| KM_k141_99571 | Not-determined |
| KM_k141_22102 | Not-determined |
| KM_k141_99579 | Not-determined |
| KM_k141_255176 | Not-determined |
| KM_k141_54347 | Not-determined |
| KM_k141_22115 | Not-determined |
| KM_k141_54349 | Not-determined |
| KM_k141_99585 | Not-determined |
| KM_k141_54352 | Low-quality |
| KM_k141_255203 | Not-determined |
| KM_k141_255212 | Not-determined |
| KM_k141_255216 | Not-determined |
| KM_k141_255232 | Not-determined |
| KM_k141_54383 | Not-determined |
| KM_k141_54391 | Not-determined |
| KM_k141_54406 | Not-determined |
| KM_k141_54407 | Not-determined |
| KM_k141_54413 | Not-determined |
| KM_k141_99622 | Not-determined |
| KM_k141_54414 | Not-determined |
| KM_k141_22173 | Not-determined |
| KM_k141_22180 | Not-determined |
| KM_k141_99635 | Not-determined |
| KM_k141_99636 | Not-determined |
| KM_k141_99637 | Not-determined |
| KM_k141_99638 | Not-determined |
| KM_k141_255271 | Low-quality |
| KM_k141_255274 | Not-determined |
| KM_k141_22195 | Not-determined |
| KM_k141_54446 | Not-determined |
| KM_k141_54448 | Low-quality |
| KM_k141_255301 | Not-determined |
| KM_k141_99670 | Not-determined |
| KM_k141_99672 | Not-determined |
| KM_k141_54474 | Not-determined |
| KM_k141_255332 | Not-determined |
| KM_k141_99709 | Not-determined |
| KM_k141_255344 | Not-determined |
| KM_k141_255351 | Not-determined |
| KM_k141_54503 | Not-determined |
| KM_k141_99719 | Not-determined |
| KM_k141_255369 | Low-quality |
| KM_k141_22215 | Not-determined |
| KM_k141_255375 | Not-determined |
| KM_k141_22220 | Not-determined |
| KM_k141_255377 | Not-determined |
| KM_k141_255383 | Not-determined |
| KM_k141_255391 | Low-quality |
| KM_k141_255396 | Not-determined |
| KM_k141_54547 | Not-determined |
| KM_k141_255424 | Not-determined |
| KM_k141_99746 | Not-determined |
| KM_k141_54562 | Low-quality |

| | |
|---|---|
| KM_k141_255432 | Not-determined |
| KM_k141_255433 | Not-determined |
| KM_k141_54567 | Not-determined |
| KM_k141_99754 | Not-determined |
| KM_k141_255435 | Low-quality |
| KM_k141_54575 | Not-determined |
| KM_k141_99756 | Not-determined |
| KM_k141_99759 | Low-quality |
| KM_k141_99760 | Not-determined |
| KM_k141_99762 | Not-determined |
| KM_k141_99769 | Not-determined |
| KM_k141_255467 | Not-determined |
| KM_k141_255474 | Not-determined |
| KM_k141_255483 | Not-determined |
| KM_k141_255489 | Not-determined |
| KM_k141_255494 | Not-determined |
| KM_k141_255510 | Not-determined |
| KM_k141_54682 | Not-determined |
| KM_k141_99785 | Not-determined |
| KM_k141_255545 | Not-determined |
| KM_k141_255547 | Not-determined |
| KM_k141_99808 | Not-determined |
| KM_k141_54726 | Not-determined |
| KM_k141_54730 | Not-determined |
| KM_k141_255576 | Not-determined |
| KM_k141_54739 | Not-determined |
| KM_k141_54750 | Low-quality |
| KM_k141_99836 | Low-quality |
| KM_k141_99837 | Not-determined |
| KM_k141_54752 | Not-determined |
| KM_k141_255593 | Not-determined |
| KM_k141_99847 | Low-quality |
| KM_k141_99849 | Not-determined |
| KM_k141_99850 | Not-determined |
| KM_k141_54766 | Not-determined |
| KM_k141_99856 | Not-determined |
| KM_k141_54779 | Low-quality |
| KM_k141_54784 | Not-determined |
| KM_k141_54802 | Not-determined |
| KM_k141_54809 | Not-determined |
| KM_k141_54812 | Not-determined |
| KM_k141_54816 | Not-determined |
| KM_k141_54829 | Not-determined |
| KM_k141_54845 | Not-determined |
| KM_k141_99901 | Not-determined |
| KM_k141_99907 | Not-determined |
| KM_k141_99914 | Not-determined |
| KM_k141_99917 | Not-determined |
| KM_k141_54916 | Not-determined |
| KM_k141_54919 | Not-determined |
| KM_k141_54928 | Not-determined |
| KM_k141_99933 | Not-determined |
| KM_k141_54949 | Not-determined |
| KM_k141_54950 | Not-determined |
| KM_k141_99949 | Low-quality |
| KM_k141_99958 | Not-determined |
| KM_k141_100007 | Low-quality |
| KM_k141_54962 | Not-determined |
| KM_k141_54963 | Not-determined |
| KM_k141_54971 | Low-quality |
| KM_k141_54979 | Not-determined |
| KM_k141_54980 | Not-determined |
| KM_k141_55005 | Not-determined |
| KM_k141_55032 | Not-determined |
| KM_k141_55075 | Not-determined |
| KM_k141_55111 | Not-determined |
| KM_k141_55118 | Not-determined |
| KM_k141_55147 | Low-quality |
| KM_k141_55157 | Low-quality |
| KM_k141_55165 | Not-determined |
| KM_k141_55179 | Not-determined |
| KM_k141_55210 | Not-determined |
| KM_k141_55224 | Not-determined |
| KM_k141_55250 | Not-determined |
| KM_k141_55279 | Not-determined |
| KM_k141_55288 | Not-determined |
| KM_k141_55311 | Not-determined |
| KM_k141_55324 | Not-determined |
| KM_k141_55348 | Not-determined |
| KM_k141_55351 | Not-determined |
| KM_k141_55367 | Not-determined |
| KM_k141_55389 | Not-determined |
| KM_k141_55390 | Not-determined |
| KM_k141_55448 | Not-determined |
| KM_k141_55450 | Not-determined |
| KM_k141_55451 | Not-determined |
| KM_k141_55453 | Not-determined |
| KM_k141_55474 | Not-determined |
| KM_k141_55493 | Not-determined |
| KM_k141_55495 | Not-determined |
| KM_k141_55504 | Not-determined |
| KM_k141_55515 | Not-determined |
| KM_k141_55524 | Not-determined |
| KM_k141_55547 | Not-determined |
| KM-final-viral-combined_k141_57987\|\|full | Medium-quality |
| 17-109_k141_135291 | Medium-quality |
| 17-109-final-viral-combined_k141_321355\|\|full | Not-determined |
| 17-109-final-viral-combined_k141_293627\|\|full | Not-determined |
| 17-109-final-viral-combined_k141_358807\|\|full | Low-quality |
| 17-109-final-viral-combined_k141_342900\|\|full | Not-determined |
| 17-109-final-viral-combined_k141_125982\|\|full | Not-determined |
| 17-109-final-viral-combined_k141_111981\|\|full | Not-determined |
| 17-109-final-viral-combined_k141_383488\|\|full | Not-determined |
| 17-109-final-viral-combined_k141_266071\|\|full | Not-determined |
| 17-109-final-viral-combined_k141_401364\|\|full | Not-determined |
| 17-109-final-viral-combined_k141_63056\|\|full | Medium-quality |
| 18-110_k141_57121 | High-quality |
| 17-109-final-viral-combined_k141_401374\|\|full | Not-determined |
| 17-109-final-viral-combined_k141_401375\|\|full | Not-determined |
| 18-110-final-viral-combined_k141_57121\|\|full | High-quality |
| 18-106_k141_54578 | Medium-quality |
| 17-109-final-viral-combined_k141_401396\|\|full | Not-determined |
| 18-106-final-viral-combined_k141_54578\|\|full | Medium-quality |
| 17-109-final-viral-combined_k141_401423\|\|full | Not-determined |
| 17-109-final-viral-combined_k141_401454\|\|full | Not-determined |
| 17-109-final-viral-combined_k141_217615\|\|full | Low-quality |
| 17-109-final-viral-combined_k141_200877\|\|full | Not-determined |
| 17-109-final-viral-combined_k141_83701\|\|full | Not-determined |
| 17-109-final-viral-combined_k141_385000\|\|full | Not-determined |
| 17-109-final-viral-combined_k141_16749\|\|full | Low-quality |
| 17-109-final-viral-combined_k141_83709\|\|full | Not-determined |
| 17-109-final-viral-combined_k141_50250\|\|full | Not-determined |
| 17-109-final-viral-combined_k141_21\|\|full | Not-determined |
| 17-109-final-viral-combined_k141_385009\|\|full | Not-determined |
| 17-109-final-viral-combined_k141_133922\|\|full | Not-determined |
| 17-109-final-viral-combined_k141_83728\|\|full | Not-determined |
| 17-109-final-viral-combined_k141_301330\|\|full | Low-quality |
| 17-109-final-viral-combined_k141_217634\|\|full | Not-determined |
| 17-109-final-viral-combined_k141_167405\|\|full | Low-quality |
| 17-109-final-viral-combined_k141_66958\|\|full | Low-quality |
| 17-109-final-viral-combined_k141_41\|\|full | Not-determined |
| 17-109-final-viral-combined_k141_234413\|\|full | Low-quality |
| 17-109-final-viral-combined_k141_385023\|\|full | Low-quality |
| 17-109-final-viral-combined_k141_200900\|\|full | Not-determined |
| 17-109-final-viral-combined_k141_351539\|\|full | Low-quality |
| 17-109-final-viral-combined_k141_117194\|\|full | Not-determined |
| 17-109-final-viral-combined_k141_368301\|\|full | Not-determined |
| 17-109-final-viral-combined_k141_184160\|\|full | Low-quality |
| 17-109-final-viral-combined_k141_100519\|\|full | Not-determined |
| 17-109-final-viral-combined_k141_16801\|\|full | Not-determined |
| 17-109-final-viral-combined_k141_351543\|\|full | Not-determined |
| 17-109-final-viral-combined_k141_251148\|\|full | Low-quality |
| 17-109-final-viral-combined_k141_33557\|\|full | Low-quality |
| 17-109-final-viral-combined_k141_117211\|\|full | Low-quality |
| 17-109-final-viral-combined_k141_368317\|\|full | Not-determined |
| 17-109-final-viral-combined_k141_67001\|\|full | Not-determined |
| 17-109-final-viral-combined_k141_16811\|\|full | Low-quality |
| 17-109-final-viral-combined_k141_67006\|\|full | Not-determined |
| 17-109-final-viral-combined_k141_217693\|\|full | Low-quality |
| 17-109-final-viral-combined_k141_117218\|\|full | Not-determined |
| 17-109-final-viral-combined_k141_67021\|\|full | Not-determined |
| 17-109-final-viral-combined_k141_150689\|\|full | Not-determined |
| 17-109-final-viral-combined_k141_217699\|\|full | Low-quality |
| 17-109-final-viral-combined_k141_33617\|\|full | Low-quality |
| 17-109-final-viral-combined_k141_385080\|\|full | Low-quality |
| 17-109-final-viral-combined_k141_251197\|\|full | Not-determined |
| 17-109-final-viral-combined_k141_251204\|\|full | Not-determined |
| 17-109-final-viral-combined_k141_125\|\|full | Low-quality |
| 17-109-final-viral-combined_k141_200985\|\|full | Not-determined |
| 17-109-final-viral-combined_k141_251216\|\|full | Low-quality |
| 17-109-final-viral-combined_k141_127\|\|full | Low-quality |
| 17-109-final-viral-combined_k141_217740\|\|full | Low-quality |
| 17-109-final-viral-combined_k141_167515\|\|full | Low-quality |
| 17-109_k141_44817 | Medium-quality |
| 17-109-final-viral-combined_k141_351571\|\|full | Low-quality |
| 17-109-final-viral-combined_k141_301396\|\|full | Low-quality |
| 17-109-final-viral-combined_k141_267898\|\|full | Not-determined |
| 17-109-final-viral-combined_k141_134023\|\|full | Not-determined |
| 17-109-final-viral-combined_k141_368375\|\|full | Low-quality |
| 17-109-final-viral-combined_k141_267908\|\|full | Low-quality |
| 17-109-final-viral-combined_k141_117276\|\|full | Not-determined |
| 17-109-final-viral-combined_k141_284745\|\|full | Not-determined |
| 17-109-final-viral-combined_k141_217780\|\|full | Low-quality |
| 17-109-final-viral-combined_k141_83766\|\|full | Not-determined |
| 17-109-final-viral-combined_k141_83769\|\|full | Not-determined |
| 17-109-final-viral-combined_k141_44817\|\|full | Medium-quality |
| 17-109-final-viral-combined_k141_301426\|\|full | Not-determined |
| 17-109-final-viral-combined_k141_117290\|\|full | Not-determined |
| 17-109-final-viral-combined_k141_334838\|\|full | Not-determined |
| 17-109-final-viral-combined_k141_351602\|\|full | Not-determined |
| 17-109-final-viral-combined_k141_301435\|\|full | Not-determined |
| 17-109-final-viral-combined_k141_83790\|\|full | Low-quality |
| 17-109-final-viral-combined_k141_267933\|\|full | Not-determined |
| 17-109-final-viral-combined_k141_385152\|\|full | Low-quality |
| 17-109-final-viral-combined_k141_201063\|\|full | Low-quality |
| 17-109-final-viral-combined_k141_217795\|\|full | Not-determined |
| 17-109-final-viral-combined_k141_284772\|\|full | Not-determined |
| 17-109-final-viral-combined_k141_83809\|\|full | Not-determined |
| 17-109-final-viral-combined_k141_50420\|\|full | Low-quality |
| 17-109-final-viral-combined_k141_50422\|\|full | Low-quality |
| 17-109-final-viral-combined_k141_201099\|\|full | Low-quality |
| 17-109-final-viral-combined_k141_150754\|\|full | Not-determined |
| 17-109-final-viral-combined_k141_83825\|\|full | Not-determined |
| 17-109-final-viral-combined_k141_301472\|\|full | Not-determined |
| 17-109-final-viral-combined_k141_251285\|\|full | Low-quality |
| 17-109-final-viral-combined_k141_150761\|\|full | Not-determined |
| 17-109-final-viral-combined_k141_67091\|\|full | Low-quality |
| 17-109-final-viral-combined_k141_100642\|\|full | Low-quality |
| 17-109-final-viral-combined_k141_117331\|\|full | Low-quality |
| 17-109-final-viral-combined_k141_301501\|\|full | Not-determined |
| 17-109-final-viral-combined_k141_385217\|\|full | Not-determined |
| 17-109-final-viral-combined_k141_50442\|\|full | Low-quality |
| 17-109-final-viral-combined_k141_318216\|\|full | Not-determined |
| 17-109-final-viral-combined_k141_217845\|\|full | Low-quality |
| 17-109-final-viral-combined_k141_117351\|\|full | Not-determined |
| 17-109-final-viral-combined_k141_33753\|\|full | Not-determined |
| 17-109-final-viral-combined_k141_368434\|\|full | Not-determined |
| 17-109-final-viral-combined_k141_318246\|\|full | Not-determined |
| 17-109-final-viral-combined_k141_284843\|\|full | Not-determined |
| 17-109-final-viral-combined_k141_368435\|\|full | Not-determined |
| 17-109-final-viral-combined_k141_83855\|\|full | Not-determined |
| 17-109-final-viral-combined_k141_16917\|\|full | Not-determined |
| 18-109-final-viral-combined_k141_189250\|\|full | High-quality |
| 17-109-final-viral-combined_k141_184339\|\|full | Low-quality |
| 17-109-final-viral-combined_k141_284847\|\|full | Not-determined |
| 17-109-final-viral-combined_k141_184348\|\|full | Not-determined |
| 17-109-final-viral-combined_k141_318265\|\|full | Low-quality |
| 17-109-final-viral-combined_k141_284880\|\|full | Low-quality |
| 17-109-final-viral-combined_k141_16923\|\|full | Low-quality |
| 17-109-final-viral-combined_k141_150834\|\|full | Low-quality |
| 17-109-final-viral-combined_k141_33771\|\|full | Not-determined |
| 17-109-final-viral-combined_k141_134144\|\|full | Not-determined |
| 17-109-final-viral-combined_k141_167650\|\|full | Low-quality |
| 17-109-final-viral-combined_k141_318268\|\|full | Not-determined |
| 17-109-final-viral-combined_k141_67153\|\|full | Low-quality |
| 17-109-final-viral-combined_k141_150847\|\|full | Low-quality |
| 17-109-final-viral-combined_k141_167657\|\|full | Not-determined |
| 17-109-final-viral-combined_k141_16943\|\|full | Low-quality |
| 17-109-final-viral-combined_k141_83904\|\|full | Low-quality |
| 17-109-final-viral-combined_k141_318281\|\|full | Low-quality |
| 17-109-final-viral-combined_k141_234462\|\|full | Not-determined |
| 17-109-final-viral-combined_k141_301591\|\|full | Not-determined |
| 17-109-final-viral-combined_k141_234470\|\|full | Not-determined |
| 17-109-final-viral-combined_k141_150862\|\|full | Low-quality |
| 17-109-final-viral-combined_k141_67168\|\|full | Not-determined |
| 17-109-final-viral-combined_k141_263\|\|full | Low-quality |
| 17-109-final-viral-combined_k141_33799\|\|full | Not-determined |
| 17-109-final-viral-combined_k141_100666\|\|full | Low-quality |
| 17-109-final-viral-combined_k141_16988\|\|full | Not-determined |
| 17-109-final-viral-combined_k141_100673\|\|full | Low-quality |
| 18-105-final-viral-combined_k141_18356\|\|full | Medium-quality |
| 17-109-final-viral-combined_k141_251374\|\|full | Low-quality |
| 17-109-final-viral-combined_k141_368465\|\|full | Not-determined |
| 17-109-final-viral-combined_k141_301618\|\|full | Low-quality |
| 17-109-final-viral-combined_k141_251377\|\|full | Not-determined |
| 18-104-final-viral-combined_k141_141607\|\|full | Medium-quality |
| 17-109-final-viral-combined_k141_368476\|\|full | Low-quality |
| 17-109-final-viral-combined_k141_184376\|\|full | Not-determined |
| 17-109-final-viral-combined_k141_83949\|\|full | Low-quality |
| 17-109-final-viral-combined_k141_351680\|\|full | Low-quality |
| 17-109-final-viral-combined_k141_301624\|\|full | Low-quality |
| 17-109-final-viral-combined_k141_67224\|\|full | Not-determined |
| 17-109-final-viral-combined_k141_16997\|\|full | Low-quality |
| 17-109-final-viral-combined_k141_318347\|\|full | Low-quality |
| 17-109-final-viral-combined_k141_117417\|\|full | Not-determined |
| 17-109-final-viral-combined_k141_67230\|\|full | Not-determined |
| 17-109-final-viral-combined_k141_67231\|\|full | Not-determined |
| 17-109-final-viral-combined_k141_334907\|\|full | Not-determined |
| 17-109-final-viral-combined_k141_251384\|\|full | Low-quality |
| 17-109-final-viral-combined_k141_301636\|\|full | Not-determined |
| 17-109-final-viral-combined_k141_251386\|\|full | Low-quality |
| 17-109-final-viral-combined_k141_284953\|\|full | Not-determined |
| 17-109-final-viral-combined_k141_251392\|\|full | Not-determined |
| 17-109-final-viral-combined_k141_284968\|\|full | Low-quality |
| 17-109-final-viral-combined_k141_297\|\|full | Low-quality |
| 17-109-final-viral-combined_k141_284970\|\|full | Not-determined |
| 17-109-final-viral-combined_k141_134216\|\|full | Not-determined |
| 17-109-final-viral-combined_k141_301668\|\|full | Not-determined |
| 17-109-final-viral-combined_k141_301677\|\|full | Not-determined |
| 17-109-final-viral-combined_k141_309\|\|full | Low-quality |
| 17-109-final-viral-combined_k141_67264\|\|full | Not-determined |
| 17-109-final-viral-combined_k141_268015\|\|full | Low-quality |
| 17-109-final-viral-combined_k141_251420\|\|full | Not-determined |
| 17-109-final-viral-combined_k141_385293\|\|full | Low-quality |
| 17-109-final-viral-combined_k141_313\|\|full | Not-determined |
| 17-109-final-viral-combined_k141_284987\|\|full | Not-determined |
| 17-109-final-viral-combined_k141_50556\|\|full | Not-determined |
| 17-109-final-viral-combined_k141_234583\|\|full | Not-determined |
| 17-109-final-viral-combined_k141_385313\|\|full | Low-quality |
| 17-109-final-viral-combined_k141_268025\|\|full | Not-determined |
| 17-109-final-viral-combined_k141_33881\|\|full | Not-determined |
| 17-109-final-viral-combined_k141_33888\|\|full | Not-determined |
| 17-109-final-viral-combined_k141_234584\|\|full | Not-determined |
| 17-109-final-viral-combined_k141_234588\|\|full | Not-determined |
| 17-109-final-viral-combined_k141_251436\|\|full | Not-determined |
| 17-109-final-viral-combined_k141_234598\|\|full | Not-determined |
| 17-109-final-viral-combined_k141_318401\|\|full | Not-determined |
| 17-109-final-viral-combined_k141_251449\|\|full | Not-determined |
| 17-109-final-viral-combined_k141_251464\|\|full | Low-quality |
| 17-109-final-viral-combined_k141_334947\|\|full | Low-quality |
| 18-110-final-viral-combined_k141_270359\|\|full | Medium-quality |
| 17-109-final-viral-combined_k141_380\|\|full | Low-quality |
| 17-109-final-viral-combined_k141_167761\|\|full | Not-determined |
| 17-109-final-viral-combined_k141_285092\|\|full | Low-quality |
| 17-109-final-viral-combined_k141_386\|\|full | Not-determined |
| 17-109-final-viral-combined_k141_167764\|\|full | Low-quality |
| 17-109-final-viral-combined_k141_318440\|\|full | Not-determined |
| 17-109-final-viral-combined_k141_334972\|\|full | Low-quality |
| 17-109-final-viral-combined_k141_334976\|\|full | Low-quality |
| 17-109-final-viral-combined_k141_385374\|\|full | Low-quality |
| 17-109-final-viral-combined_k141_100744\|\|full | Low-quality |
| 17-109-final-viral-combined_k141_385376\|\|full | Low-quality |
| 17-109-final-viral-combined_k141_67326\|\|full | Not-determined |
| 17-109-final-viral-combined_k141_100749\|\|full | Not-determined |
| 17-109-final-viral-combined_k141_334989\|\|full | Not-determined |
| 17-109-final-viral-combined_k141_50612\|\|full | Not-determined |
| 17-109-final-viral-combined_k141_201138\|\|full | Not-determined |
| 17-109-final-viral-combined_k141_67332\|\|full | Not-determined |
| 17-109-final-viral-combined_k141_201139\|\|full | Not-determined |
| 17-109-final-viral-combined_k141_234655\|\|full | Low-quality |
| 17-109-final-viral-combined_k141_268082\|\|full | Not-determined |
| 17-109-final-viral-combined_k141_325418\|\|full | Medium-quality |
| 17-109-final-viral-combined_k141_17160\|\|full | Not-determined |
| 17-109-final-viral-combined_k141_33984\|\|full | Not-determined |
| 17-109-final-viral-combined_k141_84100\|\|full | Low-quality |
| 17-109-final-viral-combined_k141_251564\|\|full | Low-quality |
| 17-109-final-viral-combined_k141_318478\|\|full | Not-determined |
| 17-109-final-viral-combined_k141_33996\|\|full | Not-determined |
| 17-109-final-viral-combined_k141_318484\|\|full | Low-quality |
| 17-109-final-viral-combined_k141_17178\|\|full | Low-quality |
| 17-109-final-viral-combined_k141_50653\|\|full | Not-determined |
| 17-109-final-viral-combined_k141_167831\|\|full | Not-determined |
| 17-109-final-viral-combined_k141_234699\|\|full | Low-quality |
| 17-109-final-viral-combined_k141_184510\|\|full | Low-quality |
| 17-109-final-viral-combined_k141_318505\|\|full | Not-determined |
| 17-109-final-viral-combined_k141_368581\|\|full | Not-determined |
| 17-109-final-viral-combined_k141_84118\|\|full | Not-determined |
| 17-109-final-viral-combined_k141_34029\|\|full | Not-determined |
| 17-109-final-viral-combined_k141_100825\|\|full | Not-determined |
| 17-109-final-viral-combined_k141_368611\|\|full | Low-quality |
| 17-109-final-viral-combined_k141_234728\|\|full | Low-quality |
| 17-109-final-viral-combined_k141_301769\|\|full | Low-quality |
| 17-109-final-viral-combined_k141_184526\|\|full | Low-quality |
| 17-109-final-viral-combined_k141_217996\|\|full | Not-determined |
| 17-109-final-viral-combined_k141_268128\|\|full | Not-determined |
| 17-109-final-viral-combined_k141_385450\|\|full | Not-determined |
| 17-109-final-viral-combined_k141_184536\|\|full | Low-quality |
| 17-109-final-viral-combined_k141_234755\|\|full | Not-determined |
| 17-109-final-viral-combined_k141_368652\|\|full | Low-quality |
| 17-109-final-viral-combined_k141_50693\|\|full | Not-determined |
| 17-109-final-viral-combined_k141_67384\|\|full | Low-quality |
| 17-109-final-viral-combined_k141_134382\|\|full | Low-quality |
| 17-109-final-viral-combined_k141_234767\|\|full | Not-determined |
| 17-109-final-viral-combined_k141_318546\|\|full | Not-determined |
| 17-109-final-viral-combined_k141_17243\|\|full | Low-quality |
| 17-109-final-viral-combined_k141_301791\|\|full | Low-quality |
| 17-109-final-viral-combined_k141_234773\|\|full | Low-quality |
| 17-109-final-viral-combined_k141_50712\|\|full | Not-determined |
| 17-109-final-viral-combined_k141_134415\|\|full | Not-determined |
| 17-109-final-viral-combined_k141_335054\|\|full | Not-determined |
| 17-109-final-viral-combined_k141_50742\|\|full | Low-quality |
| 17-109-final-viral-combined_k141_34097\|\|full | Low-quality |
| 17-109-final-viral-combined_k141_50745\|\|full | Not-determined |
| 17-109-final-viral-combined_k141_385472\|\|full | Not-determined |
| 17-109-final-viral-combined_k141_17272\|\|full | Low-quality |
| 17-109-final-viral-combined_k141_201220\|\|full | Not-determined |
| 17-109-final-viral-combined_k141_67394\|\|full | Not-determined |
| 17-109-final-viral-combined_k141_34115\|\|full | Not-determined |
| 17-109-final-viral-combined_k141_34117\|\|full | Low-quality |
| 17-109-final-viral-combined_k141_234796\|\|full | Not-determined |
| 17-109-final-viral-combined_k141_50756\|\|full | Low-quality |
| 17-109-final-viral-combined_k141_351747\|\|full | Not-determined |
| 17-109-final-viral-combined_k141_218077\|\|full | Low-quality |
| 17-109-final-viral-combined_k141_34125\|\|full | Low-quality |
| 17-109-final-viral-combined_k141_134453\|\|full | Low-quality |
| 17-109-final-viral-combined_k141_335082\|\|full | Not-determined |
| 17-109-final-viral-combined_k141_100913\|\|full | Low-quality |
| 17-109-final-viral-combined_k141_84196\|\|full | Not-determined |
| 17-109-final-viral-combined_k141_556\|\|full | Not-determined |
| 17-109-final-viral-combined_k141_218079\|\|full | Low-quality |
| 17-109-final-viral-combined_k141_335088\|\|full | Not-determined |
| 17-109-final-viral-combined_k141_335091\|\|full | Low-quality |
| 17-109-final-viral-combined_k141_100934\|\|full | Not-determined |
| 17-109-final-viral-combined_k141_134496\|\|full | Not-determined |
| 17-109-final-viral-combined_k141_17327\|\|full | Low-quality |
| 17-109-final-viral-combined_k141_368708\|\|full | Low-quality |
| 17-109-final-viral-combined_k141_201275\|\|full | Not-determined |
| 17-109-final-viral-combined_k141_134507\|\|full | Low-quality |
| 17-109-final-viral-combined_k141_50792\|\|full | Not-determined |
| 17-109-final-viral-combined_k141_218111\|\|full | Not-determined |
| 17-109-final-viral-combined_k141_368730\|\|full | Not-determined |
| 17-109-final-viral-combined_k141_218112\|\|full | Low-quality |
| 17-109-final-viral-combined_k141_301863\|\|full | Not-determined |
| 17-109-final-viral-combined_k141_335092\|\|full | Low-quality |
| 17-109-final-viral-combined_k141_134525\|\|full | Low-quality |
| 17-109-final-viral-combined_k141_285228\|\|full | Not-determined |
| 17-109-final-viral-combined_k141_335096\|\|full | Not-determined |
| 17-109-final-viral-combined_k141_301868\|\|full | Not-determined |
| 17-109-final-viral-combined_k141_617\|\|full | Not-determined |
| 17-109-final-viral-combined_k141_301874\|\|full | Not-determined |
| 17-109-final-viral-combined_k141_285237\|\|full | Not-determined |
| 17-109-final-viral-combined_k141_268220\|\|full | Not-determined |
| 17-109-final-viral-combined_k141_17351\|\|full | Low-quality |
| 17-109-final-viral-combined_k141_234840\|\|full | Not-determined |
| 17-109-final-viral-combined_k141_184608\|\|full | Not-determined |
| 17-109-final-viral-combined_k141_84238\|\|full | Not-determined |
| 17-109-final-viral-combined_k141_318712\|\|full | Low-quality |
| 17-109-final-viral-combined_k141_100983\|\|full | Not-determined |
| 17-109-final-viral-combined_k141_268232\|\|full | Not-determined |
| 17-109-final-viral-combined_k141_150997\|\|full | Not-determined |
| 17-109-final-viral-combined_k141_184621\|\|full | Not-determined |
| 17-109-final-viral-combined_k141_351767\|\|full | Not-determined |
| 17-109-final-viral-combined_k141_67462\|\|full | Low-quality |
| 17-109-final-viral-combined_k141_234862\|\|full | Not-determined |
| 17-109-final-viral-combined_k141_34179\|\|full | Not-determined |
| 17-109-final-viral-combined_k141_335103\|\|full | Not-determined |
| 17-109-final-viral-combined_k141_301905\|\|full | Low-quality |
| 17-109-final-viral-combined_k141_234871\|\|full | Low-quality |
| 17-109-final-viral-combined_k141_201347\|\|full | Not-determined |
| 17-109-final-viral-combined_k141_335106\|\|full | Low-quality |
| 17-109-final-viral-combined_k141_665\|\|full | Low-quality |
| 17-109-final-viral-combined_k141_34196\|\|full | Low-quality |
| 17-109-final-viral-combined_k141_385570\|\|full | Not-determined |
| 17-109-final-viral-combined_k141_67483\|\|full | Low-quality |
| 17-109-final-viral-combined_k141_351811\|\|full | Low-quality |
| 17-109-final-viral-combined_k141_134575\|\|full | Low-quality |
| 17-109-final-viral-combined_k141_67493\|\|full | Not-determined |
| 17-109-final-viral-combined_k141_234895\|\|full | Not-determined |
| 17-109-final-viral-combined_k141_50847\|\|full | Low-quality |
| 17-109-final-viral-combined_k141_134589\|\|full | Not-determined |
| 17-109-final-viral-combined_k141_84284\|\|full | Not-determined |
| 17-109-final-viral-combined_k141_34217\|\|full | Not-determined |
| 17-109-final-viral-combined_k141_268267\|\|full | Low-quality |
| 17-109-final-viral-combined_k141_50871\|\|full | Not-determined |
| 17-109-final-viral-combined_k141_67524\|\|full | Low-quality |
| 17-109-final-viral-combined_k141_67526\|\|full | Low-quality |
| 17-109-final-viral-combined_k141_368806\|\|full | Low-quality |
| 17-109-final-viral-combined_k141_50880\|\|full | Not-determined |
| 17-109-final-viral-combined_k141_385581\|\|full | Low-quality |
| 17-109-final-viral-combined_k141_17404\|\|full | Low-quality |
| 17-109-final-viral-combined_k141_268285\|\|full | Not-determined |
| 17-109-final-viral-combined_k141_201387\|\|full | Not-determined |
| 17-109-final-viral-combined_k141_34226\|\|full | Not-determined |
| 17-109-final-viral-combined_k141_368824\|\|full | Not-determined |
| 17-109-final-viral-combined_k141_335159\|\|full | Low-quality |
| 17-109-final-viral-combined_k141_285347\|\|full | Not-determined |
| 17-109-final-viral-combined_k141_34236\|\|full | Not-determined |
| 17-109-final-viral-combined_k141_368834\|\|full | Low-quality |
| 17-109-final-viral-combined_k141_117510\|\|full | Not-determined |
| 17-109-final-viral-combined_k141_218151\|\|full | Low-quality |
| 17-109-final-viral-combined_k141_268303\|\|full | Low-quality |
| 17-109-final-viral-combined_k141_151054\|\|full | Not-determined |
| 17-109-final-viral-combined_k141_84322\|\|full | Not-determined |
| 17-109-final-viral-combined_k141_151057\|\|full | Not-determined |
| 17-109-final-viral-combined_k141_335171\|\|full | Not-determined |
| 17-109-final-viral-combined_k141_268315\|\|full | Low-quality |
| 17-109-final-viral-combined_k141_101036\|\|full | Not-determined |
| 17-109-final-viral-combined_k141_50905\|\|full | Not-determined |
| 17-109-final-viral-combined_k141_117545\|\|full | Not-determined |
| 17-109-final-viral-combined_k141_50907\|\|full | Low-quality |
| 17-109-final-viral-combined_k141_134615\|\|full | Low-quality |
| 17-109-final-viral-combined_k141_50911\|\|full | Not-determined |
| 17-109-final-viral-combined_k141_50914\|\|full | Low-quality |
| 17-109-final-viral-combined_k141_201437\|\|full | Not-determined |
| 17-109-final-viral-combined_k141_134629\|\|full | Not-determined |
| 17-108-final-viral-combined_k141_112733\|\|full | High-quality |
| 17-109-final-viral-combined_k141_251702\|\|full | Low-quality |
| 17-109-final-viral-combined_k141_117573\|\|full | Not-determined |
| 17-109-final-viral-combined_k141_168074\|\|full | Low-quality |
| 17-109-final-viral-combined_k141_50919\|\|full | Low-quality |
| 17-109-final-viral-combined_k141_184716\|\|full | Not-determined |
| 17-109-final-viral-combined_k141_251716\|\|full | Low-quality |
| 17-109-final-viral-combined_k141_117595\|\|full | Low-quality |
| 17-109-final-viral-combined_k141_17531\|\|full | Low-quality |
| 17-109-final-viral-combined_k141_351889\|\|full | Low-quality |
| 17-109-final-viral-combined_k141_84358\|\|full | Not-determined |
| 17-109-final-viral-combined_k141_234980\|\|full | Low-quality |
| 17-109-final-viral-combined_k141_780\|\|full | Not-determined |
| 17-109-final-viral-combined_k141_184748\|\|full | Low-quality |
| 17-109-final-viral-combined_k141_285466\|\|full | Not-determined |
| 17-109-final-viral-combined_k141_184749\|\|full | Not-determined |
| 17-109-final-viral-combined_k141_268378\|\|full | Not-determined |
| 17-109-final-viral-combined_k141_335203\|\|full | Not-determined |
| 17-109-final-viral-combined_k141_34259\|\|full | Low-quality |
| 17-109-final-viral-combined_k141_368885\|\|full | Low-quality |
| 17-109-final-viral-combined_k141_351914\|\|full | Low-quality |
| 17-109-final-viral-combined_k141_84383\|\|full | Not-determined |
| 17-109-final-viral-combined_k141_201522\|\|full | Low-quality |
| 17-109-final-viral-combined_k141_385610\|\|full | Not-determined |
| 17-109-final-viral-combined_k141_335208\|\|full | Not-determined |
| 17-109-final-viral-combined_k141_218203\|\|full | Low-quality |
| 17-109-final-viral-combined_k141_17551\|\|full | Low-quality |
| 17-109-final-viral-combined_k141_335226\|\|full | Low-quality |
| 17-109-final-viral-combined_k141_34279\|\|full | Low-quality |
| 17-109-final-viral-combined_k141_301969\|\|full | Low-quality |
| 17-109-final-viral-combined_k141_34280\|\|full | Low-quality |
| 17-109-final-viral-combined_k141_797\|\|full | Low-quality |
| 17-109-final-viral-combined_k141_17565\|\|full | Not-determined |
| 17-109-final-viral-combined_k141_803\|\|full | Not-determined |
| 17-109-final-viral-combined_k141_201556\|\|full | Not-determined |
| 17-109-final-viral-combined_k141_318749\|\|full | Not-determined |
| 17-109-final-viral-combined_k141_117623\|\|full | Low-quality |
| 17-109-final-viral-combined_k141_301980\|\|full | Not-determined |
| 17-109-final-viral-combined_k141_351936\|\|full | Not-determined |
| 17-109-final-viral-combined_k141_285504\|\|full | Not-determined |
| 17-109-final-viral-combined_k141_385633\|\|full | Low-quality |
| 17-109-final-viral-combined_k141_168141\|\|full | Low-quality |
| 17-109-final-viral-combined_k141_67564\|\|full | Low-quality |
| 17-109-final-viral-combined_k141_351942\|\|full | Not-determined |
| 17-109-final-viral-combined_k141_84399\|\|full | Not-determined |
| 17-109-final-viral-combined_k141_318758\|\|full | Not-determined |
| 17-109-final-viral-combined_k141_184806\|\|full | Not-determined |
| 17-109-final-viral-combined_k141_67581\|\|full | Low-quality |
| 17-109-final-viral-combined_k141_285527\|\|full | Not-determined |
| 17-109-final-viral-combined_k141_351964\|\|full | Low-quality |
| 17-109-final-viral-combined_k141_168149\|\|full | Low-quality |
| 17-109-final-viral-combined_k141_151169\|\|full | Not-determined |
| 17-109-final-viral-combined_k141_318767\|\|full | Low-quality |
| 17-109-final-viral-combined_k141_201582\|\|full | Not-determined |
| 17-109-final-viral-combined_k141_235047\|\|full | Not-determined |
| 17-109-final-viral-combined_k141_67593\|\|full | Low-quality |
| 17-109-final-viral-combined_k141_151193\|\|full | Low-quality |
| 17-109-final-viral-combined_k141_385657\|\|full | Low-quality |
| 17-109-final-viral-combined_k141_168166\|\|full | Low-quality |
| 17-109-final-viral-combined_k141_268436\|\|full | Not-determined |
| 17-109-final-viral-combined_k141_318786\|\|full | Low-quality |
| 17-109-final-viral-combined_k141_84443\|\|full | Low-quality |
| 17-109-final-viral-combined_k141_201600\|\|full | Not-determined |
| 17-109-final-viral-combined_k141_836\|\|full | Low-quality |
| 17-109-final-viral-combined_k141_302024\|\|full | Not-determined |
| 17-109-final-viral-combined_k141_184847\|\|full | Low-quality |
| 17-109-final-viral-combined_k141_51000\|\|full | Low-quality |
| 17-109-final-viral-combined_k141_268450\|\|full | Low-quality |
| 17-109-final-viral-combined_k141_34330\|\|full | Low-quality |
| 17-109-final-viral-combined_k141_168176\|\|full | Low-quality |
| 17-109-final-viral-combined_k141_84475\|\|full | Not-determined |
| 17-109-final-viral-combined_k141_351969\|\|full | Low-quality |
| 17-109-final-viral-combined_k141_17598\|\|full | Not-determined |
| 17-109-final-viral-combined_k141_335268\|\|full | Low-quality |
| 17-109-final-viral-combined_k141_51012\|\|full | Not-determined |
| 17-109-final-viral-combined_k141_385697\|\|full | Not-determined |
| 17-109-final-viral-combined_k141_101110\|\|full | Not-determined |
| 17-109-final-viral-combined_k141_34361\|\|full | Not-determined |
| 17-109-final-viral-combined_k141_67661\|\|full | Low-quality |
| 17-109-final-viral-combined_k141_184870\|\|full | Not-determined |
| 17-109-final-viral-combined_k141_34369\|\|full | Not-determined |
| 17-109-final-viral-combined_k141_34370\|\|full | Not-determined |
| 17-109-final-viral-combined_k141_184872\|\|full | Not-determined |
| 17-109-final-viral-combined_k141_235099\|\|full | Low-quality |
| 17-109-final-viral-combined_k141_151256\|\|full | Low-quality |
| 17-109-final-viral-combined_k141_134775\|\|full | Not-determined |
| 18-101-final-viral-combined_k141_72167\|\|full | High-quality |
| 17-109-final-viral-combined_k141_235104\|\|full | Low-quality |
| 17-109-final-viral-combined_k141_51047\|\|full | Not-determined |
| 17-109-final-viral-combined_k141_134781\|\|full | Not-determined |
| 17-109-final-viral-combined_k141_318826\|\|full | Not-determined |
| 17-109-final-viral-combined_k141_385742\|\|full | Low-quality |
| 17-109-final-viral-combined_k141_117644\|\|full | Not-determined |
| 17-109-final-viral-combined_k141_318857\|\|full | Not-determined |
| 17-109-final-viral-combined_k141_51066\|\|full | Low-quality |
| 17-109-final-viral-combined_k141_168246\|\|full | Not-determined |
| 17-109-final-viral-combined_k141_318863\|\|full | Low-quality |
| 17-109-final-viral-combined_k141_251781\|\|full | Low-quality |
| 17-109-final-viral-combined_k141_34430\|\|full | Low-quality |
| 17-109-final-viral-combined_k141_318871\|\|full | Not-determined |
| 17-109-final-viral-combined_k141_34431\|\|full | Not-determined |
| 17-109-final-viral-combined_k141_168745\|\|full | Medium-quality |
| 17-109-final-viral-combined_k141_101135\|\|full | Low-quality |
| 17-109-final-viral-combined_k141_902\|\|full | Not-determined |
| 17-109-final-viral-combined_k141_201673\|\|full | Not-determined |
| 18-101_k141_185550 | Medium-quality |
| 17-109-final-viral-combined_k141_201683\|\|full | Not-determined |
| 17-109-final-viral-combined_k141_251810\|\|full | Low-quality |
| 17-109-final-viral-combined_k141_184945\|\|full | Not-determined |
| 17-109-final-viral-combined_k141_117667\|\|full | Low-quality |
| 17-109-final-viral-combined_k141_385757\|\|full | Not-determined |
| 17-109-final-viral-combined_k141_117670\|\|full | Not-determined |
| 17-109-final-viral-combined_k141_218282\|\|full | Low-quality |
| 17-109-final-viral-combined_k141_34457\|\|full | Not-determined |
| 17-109-final-viral-combined_k141_218287\|\|full | Low-quality |
| 17-109-final-viral-combined_k141_101189\|\|full | Not-determined |
| 17-109-final-viral-combined_k141_285644\|\|full | Not-determined |
| 17-109-final-viral-combined_k141_352031\|\|full | Low-quality |
| 17-109-final-viral-combined_k141_285652\|\|full | Not-determined |
| 17-109-final-viral-combined_k141_151368\|\|full | Not-determined |
| 17-109-final-viral-combined_k141_67765\|\|full | Low-quality |
| 17-109-final-viral-combined_k141_352051\|\|full | Low-quality |
| 17-109-final-viral-combined_k141_84515\|\|full | Low-quality |
| 17-109-final-viral-combined_k141_34491\|\|full | Low-quality |
| 17-109-final-viral-combined_k141_369073\|\|full | Low-quality |
| 17-109-final-viral-combined_k141_117704\|\|full | Not-determined |
| 17-109-final-viral-combined_k141_17687\|\|full | Not-determined |
| 17-109-final-viral-combined_k141_201691\|\|full | Low-quality |
| 17-109-final-viral-combined_k141_385802\|\|full | Not-determined |
| 17-109-final-viral-combined_k141_101223\|\|full | Low-quality |
| 17-109-final-viral-combined_k141_101227\|\|full | Not-determined |
| 17-109-final-viral-combined_k141_946\|\|full | Not-determined |
| 17-109-final-viral-combined_k141_302171\|\|full | Not-determined |
| 17-109-final-viral-combined_k141_34526\|\|full | Not-determined |

17-109-final-viral-combined_k141_84541||full Low-quality
17-109-final-viral-combined_k141_268597||full Low-quality
17-109-final-viral-combined_k141_302189||full Low-quality
17-109-final-viral-combined_k141_318947||full Not-determined
17-109-final-viral-combined_k141_34566||full Low-quality
17-109-final-viral-combined_k141_251840||full Low-quality
17-109-final-viral-combined_k141_218365||full Not-determined
17-109-final-viral-combined_k141_17744||full Low-quality
17-109-final-viral-combined_k141_976||full Not-determined
17-109-final-viral-combined_k141_285716||full Not-determined
17-109-final-viral-combined_k141_335362||full Not-determined
17-109-final-viral-combined_k141_101241||full Low-quality
17-109-final-viral-combined_k141_218383||full Not-determined
18-101-final-viral-combined_k141_185550||full Medium-quality
17-109-final-viral-combined_k141_268620||full Low-quality
17-109-final-viral-combined_k141_335366||full Not-determined
17-109-final-viral-combined_k141_251871||full Not-determined
17-109-final-viral-combined_k141_285729||full Not-determined
17-109-final-viral-combined_k141_385894||full Not-determined
17-109-final-viral-combined_k141_151420||full Low-quality
17-109-final-viral-combined_k141_67837||full Not-determined
17-109-final-viral-combined_k141_302234||full Not-determined
17-109-final-viral-combined_k141_17796||full Low-quality
17-109-final-viral-combined_k141_185034||full Low-quality
17-109-final-viral-combined_k141_17802||full Low-quality
17-109-final-viral-combined_k141_235232||full Not-determined
17-109-final-viral-combined_k141_51084||full Low-quality
17-109-final-viral-combined_k141_235236||full Not-determined
17-109-final-viral-combined_k141_268660||full Low-quality
17-109-final-viral-combined_k141_268669||full Not-determined
17-109-final-viral-combined_k141_335424||full Low-quality
17-109-final-viral-combined_k141_101274||full Not-determined
17-109-final-viral-combined_k141_117743||full Not-determined
17-109-final-viral-combined_k141_235250||full Low-quality
17-109-final-viral-combined_k141_101282||full Low-quality
17-109-final-viral-combined_k141_285753||full Not-determined
17-109-final-viral-combined_k141_34639||full Not-determined
17-109-final-viral-combined_k141_101305||full Not-determined
17-109-final-viral-combined_k141_1046||full Low-quality
17-109-final-viral-combined_k141_134817||full Low-quality
17-109-final-viral-combined_k141_285765||full Not-determined
17-109-final-viral-combined_k141_285769||full Not-determined
17-109-final-viral-combined_k141_335429||full Not-determined
17-109-final-viral-combined_k141_385927||full Low-quality
17-109-final-viral-combined_k141_369144||full Not-determined
17-109-final-viral-combined_k141_285778||full Not-determined
17-109-final-viral-combined_k141_117779||full Low-quality
17-109-final-viral-combined_k141_235276||full Low-quality
17-109-final-viral-combined_k141_285786||full Low-quality
17-109-final-viral-combined_k141_352164||full Low-quality
17-109-final-viral-combined_k141_101335||full Not-determined
17-109-final-viral-combined_k141_385958||full Not-determined
17-109-final-viral-combined_k141_17903||full Not-determined
17-109-final-viral-combined_k141_84630||full Low-quality
17-109-final-viral-combined_k141_285791||full Low-quality
17-109-final-viral-combined_k141_168303||full Low-quality
17-109-final-viral-combined_k141_134846||full Not-determined
17-109-final-viral-combined_k141_185086||full Not-determined
17-109-final-viral-combined_k141_51142||full Low-quality
17-109-final-viral-combined_k141_385975||full Low-quality
17-109-final-viral-combined_k141_151445||full Low-quality
17-109-final-viral-combined_k141_268752||full Low-quality
17-109-final-viral-combined_k141_84652||full Not-determined
17-109-final-viral-combined_k141_168328||full Low-quality
17-109-final-viral-combined_k141_385991||full Not-determined
17-109-final-viral-combined_k141_101364||full Low-quality
17-109-final-viral-combined_k141_302305||full Not-determined
17-109-final-viral-combined_k141_385995||full Not-determined
17-109-final-viral-combined_k141_117798||full Low-quality
17-109-final-viral-combined_k141_285800||full Low-quality
17-109-final-viral-combined_k141_134864||full Not-determined
17-109-final-viral-combined_k141_17940||full Not-determined
17-109-final-viral-combined_k141_84659||full Low-quality
17-109-final-viral-combined_k141_268774||full Not-determined
17-109-final-viral-combined_k141_268779||full Not-determined
17-109-final-viral-combined_k141_151482||full Low-quality
17-109-final-viral-combined_k141_218512||full Not-determined
18-105-final-viral-combined_k141_26061||full High-quality
17-109-final-viral-combined_k141_268782||full Not-determined
17-109-final-viral-combined_k141_352183||full Not-determined
17-109-final-viral-combined_k141_134875||full Not-determined
17-109-final-viral-combined_k141_134876||full Low-quality
17-109-final-viral-combined_k141_285815||full Low-quality
17-109-final-viral-combined_k141_34742||full Not-determined
17-109-final-viral-combined_k141_17955||full Not-determined
17-109-final-viral-combined_k141_268793||full Not-determined
17-109-final-viral-combined_k141_151529||full Low-quality
17-109-final-viral-combined_k141_84691||full Not-determined
17-109-final-viral-combined_k141_134897||full Not-determined
17-109-final-viral-combined_k141_268796||full Low-quality
17-109-final-viral-combined_k141_302330||full Not-determined
17-109-final-viral-combined_k141_285840||full Low-quality
17-109-final-viral-combined_k141_302331||full Low-quality
17-109-final-viral-combined_k141_386017||full Low-quality
17-109-final-viral-combined_k141_352224||full Low-quality
17-109-final-viral-combined_k141_117824||full Not-determined
17-109-final-viral-combined_k141_252018||full Low-quality
17-109-final-viral-combined_k141_319075||full Low-quality
17-109-final-viral-combined_k141_84710||full Low-quality
17-109-final-viral-combined_k141_335527||full Low-quality
17-109-final-viral-combined_k141_218539||full Low-quality
17-109-final-viral-combined_k141_117841||full Not-determined
17-109-final-viral-combined_k141_235405||full Not-determined
17-109-final-viral-combined_k141_185133||full Low-quality
17-109-final-viral-combined_k141_335545||full Low-quality
17-109-final-viral-combined_k141_34755||full Low-quality
17-109-final-viral-combined_k141_386023||full Low-quality
17-109-final-viral-combined_k141_134926||full Not-determined
17-109-final-viral-combined_k141_151587||full Low-quality
17-109-final-viral-combined_k141_151588||full Low-quality
17-109-final-viral-combined_k141_218586||full Low-quality
17-109-final-viral-combined_k141_319096||full Not-determined
17-109-final-viral-combined_k141_252070||full Not-determined
17-109-final-viral-combined_k141_352251||full Low-quality
17-109-final-viral-combined_k141_34773||full Not-determined
17-109-final-viral-combined_k141_34777||full Low-quality
17-109-final-viral-combined_k141_235432||full Low-quality
17-109-final-viral-combined_k141_117866||full Not-determined
17-109-final-viral-combined_k141_369183||full Low-quality
17-109-final-viral-combined_k141_252089||full Not-determined
17-109-final-viral-combined_k141_319106||full Low-quality
17-109-final-viral-combined_k141_1113||full Low-quality
17-109-final-viral-combined_k141_386064||full Not-determined
17-109-final-viral-combined_k141_101422||full Not-determined
17-109-final-viral-combined_k141_285887||full Not-determined
17-109-final-viral-combined_k141_352284||full Low-quality
17-109-final-viral-combined_k141_268849||full Not-determined
17-109-final-viral-combined_k141_67946||full Low-quality
17-109-final-viral-combined_k141_51258||full Low-quality
17-109-final-viral-combined_k141_386095||full Not-determined
17-109-final-viral-combined_k141_369257||full Low-quality
17-109-final-viral-combined_k141_117921||full Low-quality
17-109-final-viral-combined_k141_369261||full Not-determined
17-109-final-viral-combined_k141_1120||full Not-determined
17-109-final-viral-combined_k141_252154||full Not-determined
17-109-final-viral-combined_k141_185206||full Not-determined
17-109-final-viral-combined_k141_319170||full Low-quality
17-109-final-viral-combined_k141_1130||full Low-quality
17-109-final-viral-combined_k141_352313||full Not-determined
17-109-final-viral-combined_k141_352319||full Not-determined
17-109-final-viral-combined_k141_352322||full Not-determined
17-109-final-viral-combined_k141_369283||full Low-quality
17-109-final-viral-combined_k141_218648||full Low-quality
17-109-final-viral-combined_k141_268888||full Low-quality
17-109-final-viral-combined_k141_151655||full Low-quality
17-109-final-viral-combined_k141_84780||full Low-quality
17-109-final-viral-combined_k141_1154||full Not-determined
17-109-final-viral-combined_k141_151661||full Not-determined
17-109-final-viral-combined_k141_101456||full Low-quality
17-109-final-viral-combined_k141_352330||full Not-determined
17-109-final-viral-combined_k141_185251||full Low-quality
17-109-final-viral-combined_k141_101460||full Low-quality
17-109-final-viral-combined_k141_302388||full Not-determined
17-109-final-viral-combined_k141_302401||full Not-determined
17-109-final-viral-combined_k141_34862||full Low-quality
17-109-final-viral-combined_k141_185266||full Not-determined
17-109-final-viral-combined_k141_352366||full Not-determined
17-109-final-viral-combined_k141_369324||full Not-determined
17-109-final-viral-combined_k141_117947||full Not-determined
17-109-final-viral-combined_k141_369345||full Not-determined
17-109-final-viral-combined_k141_252173||full Not-determined
17-109-final-viral-combined_k141_101476||full Not-determined
17-109-final-viral-combined_k141_201923||full Low-quality
17-109-final-viral-combined_k141_252190||full Low-quality
17-109-final-viral-combined_k141_268939||full Low-quality
17-109-final-viral-combined_k141_252193||full Low-quality
17-109-final-viral-combined_k141_51338||full Low-quality
17-109-final-viral-combined_k141_218707||full Low-quality
17-109-final-viral-combined_k141_369377||full Low-quality
17-109-final-viral-combined_k141_319234||full Low-quality
17-109-final-viral-combined_k141_352399||full Low-quality
17-109-final-viral-combined_k141_319248||full Low-quality
17-109-final-viral-combined_k141_218731||full Low-quality
17-109-final-viral-combined_k141_1249||full Low-quality
17-109-final-viral-combined_k141_101494||full Low-quality
17-109-final-viral-combined_k141_84828||full Not-determined
17-109-final-viral-combined_k141_369412||full Not-determined
17-109-final-viral-combined_k141_84833||full Not-determined
17-109-final-viral-combined_k141_386199||full Low-quality
17-109-final-viral-combined_k141_319270||full Low-quality
17-109-final-viral-combined_k141_51374||full Not-determined
17-109-final-viral-combined_k141_118009||full Not-determined
17-109-final-viral-combined_k141_1265||full Not-determined
17-109-final-viral-combined_k141_18001||full Not-determined
17-109-final-viral-combined_k141_135001||full Low-quality
17-109-final-viral-combined_k141_386217||full Low-quality
17-109-final-viral-combined_k141_118020||full Not-determined
17-109-final-viral-combined_k141_201948||full Not-determined
17-109-final-viral-combined_k141_319292||full Low-quality
17-109-final-viral-combined_k141_185298||full Low-quality
17-109-final-viral-combined_k141_151690||full Not-determined
17-109-final-viral-combined_k141_369441||full Not-determined
17-109-final-viral-combined_k141_84867||full Low-quality
17-109-final-viral-combined_k141_268955||full Not-determined
17-109-final-viral-combined_k141_369451||full Low-quality
17-109-final-viral-combined_k141_185319||full Not-determined
17-109-final-viral-combined_k141_135013||full Low-quality
17-109-final-viral-combined_k141_252259||full Not-determined
17-109-final-viral-combined_k141_51427||full Low-quality
17-109-final-viral-combined_k141_18041||full Low-quality
17-109-final-viral-combined_k141_185334||full Not-determined
17-109-final-viral-combined_k141_68083||full Low-quality
17-109-final-viral-combined_k141_302502||full Low-quality
17-109-final-viral-combined_k141_352472||full Low-quality
17-109-final-viral-combined_k141_302509||full Not-determined
17-109-final-viral-combined_k141_386249||full Not-determined
17-109-final-viral-combined_k141_18068||full Low-quality
17-109-final-viral-combined_k141_252276||full Low-quality
17-109-final-viral-combined_k141_352480||full Not-determined
17-109-final-viral-combined_k141_252281||full Low-quality
17-109-final-viral-combined_k141_201969||full Low-quality
17-109-final-viral-combined_k141_302528||full Not-determined
17-109-final-viral-combined_k141_369506||full Low-quality
17-109-final-viral-combined_k141_151708||full Not-determined
17-109-final-viral-combined_k141_201977||full Not-determined
17-109-final-viral-combined_k141_168561||full Low-quality
17-109-final-viral-combined_k141_369514||full Not-determined
17-109-final-viral-combined_k141_252489||full Low-quality
17-109-final-viral-combined_k141_335661||full Not-determined
17-109-final-viral-combined_k141_252301||full Not-determined
17-109-final-viral-combined_k141_68119||full Low-quality
17-109-final-viral-combined_k141_151726||full Low-quality
17-109-final-viral-combined_k141_151727||full Not-determined
17-109-final-viral-combined_k141_84948||full Not-determined
17-109-final-viral-combined_k141_369530||full Low-quality
17-109-final-viral-combined_k141_84953||full Low-quality
17-109-final-viral-combined_k141_386284||full Not-determined
17-109-final-viral-combined_k141_202015||full Low-quality
17-109-final-viral-combined_k141_118091||full Not-determined
17-109-final-viral-combined_k141_285961||full Not-determined
17-109-final-viral-combined_k141_352503||full Not-determined
17-109-final-viral-combined_k141_118097||full Not-determined
17-109-final-viral-combined_k141_168601||full Low-quality
17-109-final-viral-combined_k141_84974||full Low-quality
17-109-final-viral-combined_k141_151733||full Not-determined
17-109-final-viral-combined_k141_18143||full Low-quality
17-109-final-viral-combined_k141_84980||full Low-quality
17-109-final-viral-combined_k141_202043||full Low-quality
17-109-final-viral-combined_k141_18163||full Low-quality
17-109-final-viral-combined_k141_151748||full Low-quality
17-109-final-viral-combined_k141_168615||full Low-quality
17-109-final-viral-combined_k141_101578||full Not-determined
17-109-final-viral-combined_k141_202063||full Not-determined
17-109-final-viral-combined_k141_352541||full Low-quality
17-109-final-viral-combined_k141_151775||full Low-quality
17-109-final-viral-combined_k141_235525||full Low-quality
17-109-final-viral-combined_k141_386364||full Low-quality
17-109-final-viral-combined_k141_218774||full Low-quality
17-109-final-viral-combined_k141_101591||full Not-determined
17-109-final-viral-combined_k141_51499||full Not-determined
17-109-final-viral-combined_k141_269081||full Low-quality
17-109-final-viral-combined_k141_386370||full Low-quality
17-109-final-viral-combined_k141_386371||full Not-determined
17-109-final-viral-combined_k141_235533||full Low-quality
17-109-final-viral-combined_k141_386381||full Low-quality
17-109-final-viral-combined_k141_118125||full Low-quality
17-109-final-viral-combined_k141_335753||full Not-determined
17-109-final-viral-combined_k141_218798||full Not-determined
17-109-final-viral-combined_k141_34919||full Low-quality
17-109-final-viral-combined_k141_335766||full Low-quality
17-109-final-viral-combined_k141_101615||full Not-determined
17-109-final-viral-combined_k141_51521||full Not-determined
17-109-final-viral-combined_k141_68224||full Low-quality
17-109-final-viral-combined_k141_185466||full Not-determined
17-109-final-viral-combined_k141_168692||full Not-determined
17-109-final-viral-combined_k141_202070||full Not-determined
17-109-final-viral-combined_k141_101622||full Low-quality
17-109-final-viral-combined_k141_135136||full Not-determined
17-109-final-viral-combined_k141_68229||full Not-determined
17-109-final-viral-combined_k141_218832||full Low-quality
17-109-final-viral-combined_k141_252396||full Not-determined
17-109-final-viral-combined_k141_68235||full Low-quality
17-109-final-viral-combined_k141_235587||full Low-quality
17-109-final-viral-combined_k141_252401||full Low-quality
17-109-final-viral-combined_k141_185488||full Low-quality
17-109-final-viral-combined_k141_34955||full Not-determined
17-109-final-viral-combined_k141_34959||full Low-quality
17-109-final-viral-combined_k141_369678||full Not-determined
17-109-final-viral-combined_k141_218837||full Low-quality
17-109-final-viral-combined_k141_135163||full Not-determined
17-109-final-viral-combined_k141_185508||full Low-quality
17-109-final-viral-combined_k141_286039||full Low-quality
17-109-final-viral-combined_k141_335802||full Not-determined
17-109-final-viral-combined_k141_252421||full Not-determined
17-109-final-viral-combined_k141_34972||full Not-determined
17-109-final-viral-combined_k141_235616||full Not-determined
17-109-final-viral-combined_k141_1486||full Not-determined
17-109-final-viral-combined_k141_335812||full Low-quality
17-109-final-viral-combined_k141_335813||full Low-quality
17-109-final-viral-combined_k141_319368||full Low-quality
17-109-final-viral-combined_k141_218872||full Not-determined
17-109-final-viral-combined_k141_85057||full Not-determined
17-109-final-viral-combined_k141_335824||full Not-determined
17-109-final-viral-combined_k141_185555||full Not-determined
17-109-final-viral-combined_k141_185557||full Low-quality
17-109-final-viral-combined_k141_369744||full Not-determined
17-109-final-viral-combined_k141_1529||full Low-quality
17-109-final-viral-combined_k141_202113||full Not-determined
17-109-final-viral-combined_k141_1537||full Not-determined
17-109-final-viral-combined_k141_68243||full Low-quality
17-109-final-viral-combined_k141_185580||full Not-determined
17-109-final-viral-combined_k141_1557||full Low-quality
17-109-final-viral-combined_k141_335862||full Not-determined
17-109-final-viral-combined_k141_135220||full Not-determined
17-108-final-viral-combined_k141_283220||full High-quality
17-109-final-viral-combined_k141_168746||full Low-quality
17-109-final-viral-combined_k141_51593||full Low-quality
17-109-final-viral-combined_k141_68258||full Low-quality
17-109-final-viral-combined_k141_118186||full Low-quality
17-109-final-viral-combined_k141_352580||full Not-determined
17-109-final-viral-combined_k141_135243||full Low-quality
17-109-final-viral-combined_k141_68273||full Low-quality
17-109-final-viral-combined_k141_386459||full Low-quality
17-109-final-viral-combined_k141_335881||full Low-quality
17-109-final-viral-combined_k141_352593||full Low-quality
17-109-final-viral-combined_k141_135265||full Not-determined
17-109-final-viral-combined_k141_335888||full Low-quality
17-109-final-viral-combined_k141_168768||full Not-determined
17-109-final-viral-combined_k141_335897||full Low-quality
17-109-final-viral-combined_k141_286126||full Low-quality
17-109-final-viral-combined_k141_319427||full Not-determined
17-109-final-viral-combined_k141_1598||full Low-quality
17-109-final-viral-combined_k141_118217||full Not-determined
17-109-final-viral-combined_k141_118218||full Not-determined
17-109-final-viral-combined_k141_1601||full Low-quality
17-109-final-viral-combined_k141_302776||full Not-determined
17-109-final-viral-combined_k141_286150||full Not-determined
17-109-final-viral-combined_k141_319448||full Not-determined
17-109-final-viral-combined_k141_369796||full Low-quality
17-109-final-viral-combined_k141_352629||full Low-quality
17-109-final-viral-combined_k141_235706||full Not-determined
17-109-final-viral-combined_k141_369802||full Low-quality
17-109-final-viral-combined_k141_369811||full Not-determined
17-109-final-viral-combined_k141_235718||full Not-determined
17-109-final-viral-combined_k141_85160||full Low-quality
18-109_k141_244866 Complete
17-109-final-viral-combined_k141_168808||full Low-quality
17-109-final-viral-combined_k141_352646||full Low-quality
17-109-final-viral-combined_k141_85169||full Not-determined
17-109-final-viral-combined_k141_352653||full Not-determined
17-109-final-viral-combined_k141_168810||full Low-quality
17-109-final-viral-combined_k141_151862||full Low-quality
17-109-final-viral-combined_k141_85178||full Not-determined
17-109-final-viral-combined_k141_369842||full Low-quality
17-109-final-viral-combined_k141_168823||full Low-quality
17-109-final-viral-combined_k141_1648||full Low-quality
17-109-final-viral-combined_k141_1651||full Not-determined
17-109-final-viral-combined_k141_286188||full Not-determined
17-109-final-viral-combined_k141_335946||full Not-determined
17-109-final-viral-combined_k141_168827||full Not-determined
17-109-final-viral-combined_k141_335956||full Not-determined
17-109-final-viral-combined_k141_352665||full Low-quality
17-109-final-viral-combined_k141_1667||full Not-determined
17-109-final-viral-combined_k141_335963||full Low-quality
17-109-final-viral-combined_k141_85197||full Low-quality
17-109-final-viral-combined_k141_51662||full Low-quality
17-109-final-viral-combined_k141_101820||full Not-determined
17-109-final-viral-combined_k141_269245||full Low-quality
17-109-final-viral-combined_k141_235769||full Low-quality
17-109-final-viral-combined_k141_135354||full Not-determined
17-109-final-viral-combined_k141_151925||full Low-quality
17-109-final-viral-combined_k141_35140||full Low-quality
17-109-final-viral-combined_k141_135366||full Not-determined
17-109-final-viral-combined_k141_151945||full Low-quality
17-109-final-viral-combined_k141_135369||full Not-determined
17-109-final-viral-combined_k141_168868||full Not-determined
17-109-final-viral-combined_k141_168874||full Not-determined
17-109-final-viral-combined_k141_269289||full Not-determined
17-109-final-viral-combined_k141_51696||full Low-quality
17-109-final-viral-combined_k141_252487||full Not-determined
17-109-final-viral-combined_k141_51699||full Not-determined
17-109-final-viral-combined_k141_185754||full Low-quality
17-109-final-viral-combined_k141_51704||full Low-quality
17-109-final-viral-combined_k141_51706||full Low-quality
17-109-final-viral-combined_k141_151985||full Low-quality
17-109-final-viral-combined_k141_386572||full Low-quality
17-109-final-viral-combined_k141_252526||full Low-quality
17-109-final-viral-combined_k141_219059||full Low-quality
17-109-final-viral-combined_k141_51730||full Low-quality
17-109-final-viral-combined_k141_168927||full Not-determined
17-109-final-viral-combined_k141_369896||full Low-quality
17-109-final-viral-combined_k141_235820||full Low-quality
17-109-final-viral-combined_k141_51746||full Low-quality
17-109-final-viral-combined_k141_85259||full Not-determined
17-109-final-viral-combined_k141_219078||full Low-quality
17-109-final-viral-combined_k141_1710||full Low-quality
18-106-final-viral-combined_k141_113986||full Medium-quality
17-109-final-viral-combined_k141_1715||full Low-quality
17-109-final-viral-combined_k141_369906||full Low-quality
17-109-final-viral-combined_k141_152043||full Not-determined
17-109-final-viral-combined_k141_51774||full Low-quality
17-109-final-viral-combined_k141_386595||full Not-determined
17-109-final-viral-combined_k141_319588||full Low-quality
17-109-final-viral-combined_k141_118311||full Not-determined
17-109-final-viral-combined_k141_235857||full Low-quality
17-109-final-viral-combined_k141_51779||full Not-determined
17-109-final-viral-combined_k141_68396||full Low-quality
17-109-final-viral-combined_k141_1735||full Low-quality
17-109-final-viral-combined_k141_68410||full Low-quality
17-109-final-viral-combined_k141_51789||full Low-quality
17-109-final-viral-combined_k141_302897||full Not-determined
17-109-final-viral-combined_k141_68423||full Not-determined
17-109-final-viral-combined_k141_252542||full Not-determined
17-109-final-viral-combined_k141_252544||full Not-determined
17-109-final-viral-combined_k141_35300||full Low-quality
17-109-final-viral-combined_k141_1769||full Low-quality
17-109-final-viral-combined_k141_118337||full Not-determined
17-109-final-viral-combined_k141_68434||full Not-determined
17-109-final-viral-combined_k141_202248||full Low-quality
17-109-final-viral-combined_k141_18256||full Low-quality
17-109-final-viral-combined_k141_18257||full Not-determined
17-109-final-viral-combined_k141_235880||full Low-quality
17-109-final-viral-combined_k141_352837||full Not-determined
17-109-final-viral-combined_k141_319660||full Not-determined
17-109-final-viral-combined_k141_18258||full Low-quality
17-109-final-viral-combined_k141_302937||full Not-determined
17-109-final-viral-combined_k141_152122||full Low-quality
17-109-final-viral-combined_k141_302939||full Low-quality
17-109-final-viral-combined_k141_235889||full Not-determined
17-109-final-viral-combined_k141_35323||full Low-quality
17-109-final-viral-combined_k141_118350||full Low-quality
17-109-final-viral-combined_k141_302942||full Low-quality
17-109-final-viral-combined_k141_202266||full Low-quality
17-109-final-viral-combined_k141_319671||full Not-determined
17-109-final-viral-combined_k141_101872||full Not-determined
17-109-final-viral-combined_k141_1792||full Not-determined
17-109-final-viral-combined_k141_336020||full Low-quality
17-109-final-viral-combined_k141_336022||full Not-determined
17-109-final-viral-combined_k141_302965||full Not-determined
17-109-final-viral-combined_k141_252596||full Not-determined
17-109-final-viral-combined_k141_252602||full Low-quality
17-109-final-viral-combined_k141_1802||full Not-determined
17-109-final-viral-combined_k141_202288||full Low-quality
17-109-final-viral-combined_k141_319692||full Low-quality
17-109-final-viral-combined_k141_118387||full Not-determined
17-109-final-viral-combined_k141_1813||full Low-quality
17-109-final-viral-combined_k141_319698||full Low-quality
17-109-final-viral-combined_k141_68477||full Not-determined
17-109-final-viral-combined_k141_135422||full Low-quality
17-109-final-viral-combined_k141_135426||full Low-quality
17-109-final-viral-combined_k141_1832||full Low-quality
17-109-final-viral-combined_k141_101905||full Not-determined
17-109-final-viral-combined_k141_135434||full Low-quality
17-109-final-viral-combined_k141_219156||full Low-quality
17-109-final-viral-combined_k141_1849||full Not-determined
17-109-final-viral-combined_k141_286243||full Low-quality
18-108-final-viral-combined_k141_23861||full Medium-quality
17-109-final-viral-combined_k141_303022||full Not-determined
17-109-final-viral-combined_k141_1863||full Low-quality
17-109-final-viral-combined_k141_235932||full Low-quality
17-109-final-viral-combined_k141_68509||full Low-quality
17-109-final-viral-combined_k141_319736||full Low-quality
17-109-final-viral-combined_k141_235945||full Not-determined
17-109-final-viral-combined_k141_152212||full Low-quality
17-109-final-viral-combined_k141_101942||full Not-determined
17-109-final-viral-combined_k141_101946||full Not-determined
17-109-final-viral-combined_k141_319746||full Not-determined
17-109-final-viral-combined_k141_219180||full Not-determined
17-109-final-viral-combined_k141_1875||full Low-quality
17-109-final-viral-combined_k141_1877||full Low-quality
17-109-final-viral-combined_k141_202330||full Low-quality
17-109-final-viral-combined_k141_51866||full Not-determined
17-109-final-viral-combined_k141_286285||full Not-determined
17-109-final-viral-combined_k141_85358||full Low-quality
17-109-final-viral-combined_k141_51867||full Low-quality
17-109-final-viral-combined_k141_235978||full Low-quality
17-109-final-viral-combined_k141_169057||full Not-determined
17-109-final-viral-combined_k141_370082||full Not-determined
17-109-final-viral-combined_k141_303071||full Low-quality
17-109-final-viral-combined_k141_352882||full Low-quality
17-109-final-viral-combined_k141_68518||full Not-determined
18-105-final-viral-combined_k141_205046||full High-quality
17-109-final-viral-combined_k141_303077||full Not-determined
17-109-final-viral-combined_k141_386726||full Not-determined
17-109-final-viral-combined_k141_85397||full Not-determined
17-109-final-viral-combined_k141_85398||full Low-quality
17-109-final-viral-combined_k141_135514||full Not-determined
17-109-final-viral-combined_k141_352897||full Low-quality
17-109-final-viral-combined_k141_18304||full Low-quality
17-109-final-viral-combined_k141_370100||full Low-quality
17-109-final-viral-combined_k141_1921||full Low-quality
17-109-final-viral-combined_k141_18318||full Not-determined
17-109-final-viral-combined_k141_352905||full Not-determined
17-109-final-viral-combined_k141_102029||full Not-determined
17-109-final-viral-combined_k141_370112||full Not-determined
17-109-final-viral-combined_k141_286386||full Low-quality
17-109-final-viral-combined_k141_370117||full Not-determined
17-109-final-viral-combined_k141_102036||full Low-quality
17-109-final-viral-combined_k141_118483||full Not-determined
17-109-final-viral-combined_k141_152275||full Not-determined
17-109-final-viral-combined_k141_135533||full Low-quality
17-109-final-viral-combined_k141_219219||full Not-determined
17-109-final-viral-combined_k141_35451||full Not-determined
17-109-final-viral-combined_k141_252735||full Low-quality
17-109-final-viral-combined_k141_202351||full Not-determined
17-109-final-viral-combined_k141_386785||full Low-quality
17-109-final-viral-combined_k141_252744||full Not-determined
17-109-final-viral-combined_k141_370127||full Not-determined
17-109-final-viral-combined_k141_269459||full Not-determined
17-109-final-viral-combined_k141_118510||full Not-determined
17-109-final-viral-combined_k141_152304||full Not-determined
17-109-final-viral-combined_k141_219245||full Not-determined
17-109-final-viral-combined_k141_202371||full Not-determined
17-109-final-viral-combined_k141_51930||full Low-quality
17-109-final-viral-combined_k141_236129||full Not-determined
17-109-final-viral-combined_k141_51936||full Low-quality
17-109-final-viral-combined_k141_352968||full Not-determined
17-109-final-viral-combined_k141_169137||full Low-quality
17-109-final-viral-combined_k141_85449||full Low-quality
17-109-final-viral-combined_k141_219280||full Low-quality
17-109-final-viral-combined_k141_252783||full Low-quality
17-109-final-viral-combined_k141_236180||full Low-quality
17-109-final-viral-combined_k141_1957||full Low-quality
17-109-final-viral-combined_k141_18357||full Low-quality
18-110-final-viral-combined_k141_77883||full Medium-quality
17-109-final-viral-combined_k141_152353||full Not-determined
17-109-final-viral-combined_k141_152366||full Low-quality
17-109-final-viral-combined_k141_35473||full Low-quality
17-109-final-viral-combined_k141_35488||full Low-quality
17-109-final-viral-combined_k141_236201||full Low-quality
17-109-final-viral-combined_k141_118546||full Low-quality
17-109-final-viral-combined_k141_118553||full Not-determined
17-109-final-viral-combined_k141_236215||full Low-quality
17-109-final-viral-combined_k141_252842||full Not-determined
17-109-final-viral-combined_k141_303251||full Not-determined
17-109-final-viral-combined_k141_219327||full Low-quality
17-109-final-viral-combined_k141_35517||full Not-determined
17-109-final-viral-combined_k141_51972||full Not-determined
17-109-final-viral-combined_k141_219335||full Not-determined
17-109-final-viral-combined_k141_236230||full Not-determined
17-109-final-viral-combined_k141_236231||full Not-determined
17-109-final-viral-combined_k141_135609||full Low-quality
17-109-final-viral-combined_k141_18387||full Not-determined
17-109-final-viral-combined_k141_135613||full Not-determined
17-109-final-viral-combined_k141_135614||full Low-quality
17-109-final-viral-combined_k141_370222||full Low-quality
17-109-final-viral-combined_k141_118575||full Not-determined
17-109-final-viral-combined_k141_202426||full Not-determined
17-109-final-viral-combined_k141_35556||full Not-determined
17-109-final-viral-combined_k141_1977||full Low-quality
17-109-final-viral-combined_k141_169162||full Not-determined
17-109-final-viral-combined_k141_135635||full Low-quality
17-109-final-viral-combined_k141_269487||full Low-quality
17-109-final-viral-combined_k141_135639||full Low-quality
17-109-final-viral-combined_k141_286577||full Not-determined
17-109-final-viral-combined_k141_303320||full Not-determined
17-109-final-viral-combined_k141_269492||full Not-determined
17-109-final-viral-combined_k141_353018||full Not-determined
17-109-final-viral-combined_k141_2015||full Low-quality
17-109-final-viral-combined_k141_152434||full Not-determined
17-109-final-viral-combined_k141_135660||full Low-quality
17-109-final-viral-combined_k141_386882||full Low-quality
17-109-final-viral-combined_k141_52052||full Not-determined
17-109-final-viral-combined_k141_52054||full Not-determined
17-109-final-viral-combined_k141_35585||full Not-determined
17-109-final-viral-combined_k141_52057||full Low-quality
17-109-final-viral-combined_k141_303341||full Not-determined
17-109-final-viral-combined_k141_186009||full Not-determined
17-109-final-viral-combined_k141_2024||full Not-determined
17-109-final-viral-combined_k141_386896||full Not-determined
17-109-final-viral-combined_k141_68665||full Low-quality
17-109-final-viral-combined_k141_303350||full Low-quality
17-109-final-viral-combined_k141_202482||full Not-determined
17-109-final-viral-combined_k141_353053||full Low-quality
17-109-final-viral-combined_k141_202483||full Not-determined
17-109-final-viral-combined_k141_336211||full Low-quality
17-109-final-viral-combined_k141_386907||full Low-quality
17-109-final-viral-combined_k141_353061||full Low-quality
17-109-final-viral-combined_k141_102092||full Low-quality
17-109-final-viral-combined_k141_85527||full Not-determined
17-109-final-viral-combined_k141_102098||full Not-determined
17-109-final-viral-combined_k141_2065||full Not-determined
17-109-final-viral-combined_k141_85540||full Not-determined
17-109-final-viral-combined_k141_269508||full Low-quality
17-109-final-viral-combined_k141_35622||full Low-quality
17-109-final-viral-combined_k141_18455||full Low-quality
17-109-final-viral-combined_k141_303370||full Not-determined
17-109-final-viral-combined_k141_102120||full Low-quality
17-109-final-viral-combined_k141_152507||full Not-determined
17-109-final-viral-combined_k141_353098||full Not-determined
17-109-final-viral-combined_k141_202497||full Low-quality
17-109-final-viral-combined_k141_353101||full Not-determined
17-109-final-viral-combined_k141_386943||full Not-determined
17-109-final-viral-combined_k141_152515||full Not-determined
18-103-final-viral-combined_k141_92053||full High-quality
17-109-final-viral-combined_k141_118674||full Low-quality
17-109-final-viral-combined_k141_186081||full Low-quality
17-109-final-viral-combined_k141_85582||full Low-quality
17-109-final-viral-combined_k141_118679||full Not-determined

| | |
|---|---|
| 17-109-final-viral-combined_k141_320025\|\|full | Not-determined |
| 17-109-final-viral-combined_k141_152524\|\|full | Low-quality |
| 17-109-final-viral-combined_k141_102131\|\|full | Not-determined |
| 17-109-final-viral-combined_k141_219431\|\|full | Not-determined |
| 17-109-final-viral-combined_k141_370330\|\|full | Low-quality |
| 17-109-final-viral-combined_k141_320037\|\|full | Not-determined |
| 17-109-final-viral-combined_k141_269542\|\|full | Not-determined |
| 17-109-final-viral-combined_k141_386973\|\|full | Low-quality |
| 17-109-final-viral-combined_k141_353150\|\|full | Not-determined |
| 17-109-final-viral-combined_k141_152557\|\|full | Low-quality |
| 17-109-final-viral-combined_k141_169185\|\|full | Low-quality |
| 17-109-final-viral-combined_k141_152564\|\|full | Low-quality |
| 17-109-final-viral-combined_k141_219461\|\|full | Not-determined |
| 17-109-final-viral-combined_k141_135765\|\|full | Low-quality |
| 17-109-final-viral-combined_k141_386990\|\|full | Not-determined |
| 17-109-final-viral-combined_k141_102151\|\|full | Low-quality |
| 17-109-final-viral-combined_k141_303431\|\|full | Low-quality |
| 17-109-final-viral-combined_k141_68794\|\|full | Not-determined |
| 17-109-final-viral-combined_k141_102157\|\|full | Low-quality |
| 17-109-final-viral-combined_k141_353178\|\|full | Low-quality |
| 17-109-final-viral-combined_k141_186138\|\|full | Not-determined |
| 17-109-final-viral-combined_k141_219482\|\|full | Not-determined |
| 17-109-final-viral-combined_k141_219488\|\|full | Low-quality |
| 17-109-final-viral-combined_k141_387022\|\|full | Low-quality |
| 17-109-final-viral-combined_k141_118726\|\|full | Low-quality |
| 17-109-final-viral-combined_k141_169212\|\|full | Low-quality |
| 17-109-final-viral-combined_k141_35696\|\|full | Not-determined |
| 17-109-final-viral-combined_k141_336320\|\|full | Low-quality |
| 17-109-final-viral-combined_k141_2104\|\|full | Not-determined |
| 17-109-final-viral-combined_k141_85666\|\|full | Not-determined |
| 17-109-final-viral-combined_k141_35707\|\|full | Not-determined |
| 17-109-final-viral-combined_k141_52191\|\|full | Low-quality |
| 17-109-final-viral-combined_k141_35709\|\|full | Not-determined |
| 17-109-final-viral-combined_k141_370416\|\|full | Not-determined |
| 17-109-final-viral-combined_k141_18494\|\|full | Not-determined |
| 17-109-final-viral-combined_k141_336341\|\|full | Low-quality |
| 17-109-final-viral-combined_k141_202566\|\|full | Not-determined |
| 17-109-final-viral-combined_k141_135793\|\|full | Not-determined |
| 17-109-final-viral-combined_k141_353216\|\|full | Low-quality |
| 17-109-final-viral-combined_k141_202568\|\|full | Low-quality |
| 17-109-final-viral-combined_k141_186194\|\|full | Low-quality |
| 17-109-final-viral-combined_k141_303482\|\|full | Not-determined |
| 17-109-final-viral-combined_k141_52226\|\|full | Not-determined |
| 17-109-final-viral-combined_k141_353217\|\|full | Not-determined |
| 17-109-final-viral-combined_k141_85692\|\|full | Not-determined |
| 17-109-final-viral-combined_k141_202574\|\|full | Low-quality |
| 17-109-final-viral-combined_k141_320112\|\|full | Not-determined |
| 17-109-final-viral-combined_k141_169242\|\|full | Low-quality |
| 17-109-final-viral-combined_k141_353239\|\|full | Low-quality |
| 17-109-final-viral-combined_k141_202595\|\|full | Low-quality |
| 17-109-final-viral-combined_k141_35754\|\|full | Low-quality |
| 17-109-final-viral-combined_k141_169257\|\|full | Not-determined |
| 17-109-final-viral-combined_k141_102257\|\|full | Not-determined |
| 17-109-final-viral-combined_k141_269612\|\|full | Low-quality |
| 17-109-final-viral-combined_k141_269622\|\|full | Not-determined |
| 17-109-final-viral-combined_k141_219548\|\|full | Not-determined |
| 17-109-final-viral-combined_k141_269626\|\|full | Low-quality |
| 17-109-final-viral-combined_k141_320126\|\|full | Not-determined |
| 17-109-final-viral-combined_k141_35770\|\|full | Low-quality |
| 17-109-final-viral-combined_k141_169281\|\|full | Low-quality |
| 17-109-final-viral-combined_k141_202633\|\|full | Not-determined |
| 17-109-final-viral-combined_k141_169282\|\|full | Low-quality |
| 17-109-final-viral-combined_k141_135817\|\|full | Not-determined |
| 17-109-final-viral-combined_k141_269636\|\|full | Not-determined |
| 17-109-final-viral-combined_k141_152683\|\|full | Not-determined |
| 17-109-final-viral-combined_k141_52277\|\|full | Not-determined |
| 17-109-final-viral-combined_k141_135833\|\|full | Low-quality |
| 17-109-final-viral-combined_k141_252964\|\|full | Not-determined |
| 17-109-final-viral-combined_k141_286743\|\|full | Not-determined |
| 17-109-final-viral-combined_k141_202667\|\|full | Low-quality |
| 17-109-final-viral-combined_k141_387076\|\|full | Not-determined |
| 17-109-final-viral-combined_k141_68898\|\|full | Low-quality |
| 17-109-final-viral-combined_k141_169312\|\|full | Low-quality |
| 17-109-final-viral-combined_k141_320155\|\|full | Low-quality |
| 17-109-final-viral-combined_k141_68899\|\|full | Not-determined |
| 17-109-final-viral-combined_k141_269663\|\|full | Not-determined |
| 17-109-final-viral-combined_k141_152709\|\|full | Not-determined |
| 17-109-final-viral-combined_k141_320157\|\|full | Low-quality |
| 17-109-final-viral-combined_k141_152710\|\|full | Low-quality |
| 17-109-final-viral-combined_k141_35796\|\|full | Low-quality |
| 17-109-final-viral-combined_k141_387081\|\|full | Low-quality |
| 17-109-final-viral-combined_k141_286761\|\|full | Not-determined |
| 17-109-final-viral-combined_k141_202707\|\|full | Not-determined |
| 17-109-final-viral-combined_k141_370486\|\|full | Low-quality |
| 17-109-final-viral-combined_k141_353300\|\|full | Not-determined |
| 17-109-final-viral-combined_k141_303550\|\|full | Low-quality |
| 17-109-final-viral-combined_k141_186315\|\|full | Not-determined |
| 17-109-final-viral-combined_k141_387111\|\|full | Low-quality |
| 17-109-final-viral-combined_k141_169352\|\|full | Low-quality |
| 17-109-final-viral-combined_k141_169358\|\|full | Low-quality |
| 17-109-final-viral-combined_k141_68938\|\|full | Low-quality |
| 17-109-final-viral-combined_k141_135856\|\|full | Low-quality |
| 17-109-final-viral-combined_k141_370512\|\|full | Not-determined |
| 17-109-final-viral-combined_k141_186337\|\|full | Low-quality |
| 17-109-final-viral-combined_k141_135859\|\|full | Not-determined |
| 17-109-final-viral-combined_k141_186343\|\|full | Not-determined |
| 17-109-final-viral-combined_k141_169382\|\|full | Not-determined |
| 17-109-final-viral-combined_k141_2232\|\|full | Low-quality |
| 17-109-final-viral-combined_k141_320223\|\|full | Low-quality |
| 17-109-final-viral-combined_k141_2235\|\|full | Not-determined |
| 17-109-final-viral-combined_k141_169399\|\|full | Low-quality |
| 17-109-final-viral-combined_k141_2237\|\|full | Low-quality |
| 17-109-final-viral-combined_k141_370543\|\|full | Not-determined |
| 17-109-final-viral-combined_k141_18570\|\|full | Low-quality |
| 17-109-final-viral-combined_k141_102344\|\|full | Low-quality |
| 17-109-final-viral-combined_k141_118930\|\|full | Low-quality |
| 17-109-final-viral-combined_k141_2254\|\|full | Low-quality |
| 17-109-final-viral-combined_k141_169418\|\|full | Low-quality |
| 17-109-final-viral-combined_k141_18587\|\|full | Not-determined |
| 17-109-final-viral-combined_k141_303617\|\|full | Low-quality |
| 17-109-final-viral-combined_k141_320255\|\|full | Not-determined |
| 17-109-final-viral-combined_k141_269763\|\|full | Not-determined |
| 17-109-final-viral-combined_k141_135932\|\|full | Not-determined |
| 17-109-final-viral-combined_k141_118957\|\|full | Not-determined |
| 17-109-final-viral-combined_k141_2274\|\|full | Low-quality |
| 17-109-final-viral-combined_k141_85785\|\|full | Low-quality |
| 17-109-final-viral-combined_k141_152789\|\|full | Not-determined |
| 17-109-final-viral-combined_k141_286843\|\|full | Low-quality |
| 17-109-final-viral-combined_k141_2282\|\|full | Not-determined |
| 17-109-final-viral-combined_k141_286851\|\|full | Not-determined |
| 17-109-final-viral-combined_k141_236277\|\|full | Not-determined |
| 17-109-final-viral-combined_k141_353427\|\|full | Not-determined |
| 17-109-final-viral-combined_k141_353428\|\|full | Low-quality |
| 17-109-final-viral-combined_k141_286858\|\|full | Low-quality |
| 17-109-final-viral-combined_k141_236279\|\|full | Not-determined |
| 17-109-final-viral-combined_k141_2302\|\|full | Not-determined |
| 17-109-final-viral-combined_k141_286860\|\|full | Low-quality |
| 17-109-final-viral-combined_k141_118981\|\|full | Not-determined |
| 17-109-final-viral-combined_k141_18632\|\|full | Not-determined |
| 17-109-final-viral-combined_k141_169430\|\|full | Not-determined |
| 17-109-final-viral-combined_k141_69022\|\|full | Not-determined |
| 17-109-final-viral-combined_k141_169431\|\|full | Low-quality |
| KM-final-viral-combined_k141_75892\|\|full | Medium-quality |
| 17-109-final-viral-combined_k141_2315\|\|full | Low-quality |
| 17-109-final-viral-combined_k141_186426\|\|full | Not-determined |
| 17-109-final-viral-combined_k141_102414\|\|full | Low-quality |
| 17-109-final-viral-combined_k141_336481\|\|full | Not-determined |
| 17-109-final-viral-combined_k141_303671\|\|full | Low-quality |
| 17-109-final-viral-combined_k141_102418\|\|full | Not-determined |
| 17-109-final-viral-combined_k141_186436\|\|full | Low-quality |
| 17-109-final-viral-combined_k141_152842\|\|full | Low-quality |
| 17-109-final-viral-combined_k141_119006\|\|full | Not-determined |
| 17-109-final-viral-combined_k141_135978\|\|full | Low-quality |
| 17-109-final-viral-combined_k141_152847\|\|full | Low-quality |
| 17-109-final-viral-combined_k141_135982\|\|full | Low-quality |
| 17-109-final-viral-combined_k141_269814\|\|full | Low-quality |
| 17-109-final-viral-combined_k141_186458\|\|full | Not-determined |
| 17-109-final-viral-combined_k141_102450\|\|full | Not-determined |
| 17-109-final-viral-combined_k141_119021\|\|full | Low-quality |
| 18-101-final-viral-combined_k141_100430\|\|full | Medium-quality |
| 17-109-final-viral-combined_k141_85827\|\|full | Not-determined |
| 17-109-final-viral-combined_k141_85828\|\|full | Not-determined |
| 17-109-final-viral-combined_k141_269833\|\|full | Not-determined |
| 17-109-final-viral-combined_k141_286910\|\|full | Not-determined |
| 17-109-final-viral-combined_k141_119039\|\|full | Not-determined |
| 17-109-final-viral-combined_k141_202816\|\|full | Not-determined |
| 17-109-final-viral-combined_k141_2349\|\|full | Low-quality |
| 17-109-final-viral-combined_k141_202825\|\|full | Not-determined |
| 17-109-final-viral-combined_k141_186497\|\|full | Low-quality |
| 17-109-final-viral-combined_k141_119057\|\|full | Not-determined |
| 17-109-final-viral-combined_k141_2362\|\|full | Not-determined |
| 17-109-final-viral-combined_k141_303723\|\|full | Low-quality |
| 17-109-final-viral-combined_k141_18685\|\|full | Low-quality |
| 17-109-final-viral-combined_k141_119078\|\|full | Not-determined |
| 17-109-final-viral-combined_k141_269877\|\|full | Not-determined |
| 17-109-final-viral-combined_k141_119082\|\|full | Low-quality |
| 17-109-final-viral-combined_k141_136023\|\|full | Not-determined |
| 17-109-final-viral-combined_k141_102469\|\|full | Low-quality |
| 17-109-final-viral-combined_k141_169455\|\|full | Low-quality |
| 17-109-final-viral-combined_k141_253084\|\|full | Not-determined |
| 17-109-final-viral-combined_k141_236365\|\|full | Low-quality |
| 17-109-final-viral-combined_k141_102475\|\|full | Not-determined |
| 17-109-final-viral-combined_k141_152912\|\|full | Not-determined |
| 17-109-final-viral-combined_k141_69155\|\|full | Not-determined |
| 17-109-final-viral-combined_k141_320344\|\|full | Low-quality |
| 17-109-final-viral-combined_k141_119106\|\|full | Not-determined |
| 17-109-final-viral-combined_k141_52488\|\|full | Low-quality |
| 17-109-final-viral-combined_k141_202865\|\|full | Not-determined |
| 17-109-final-viral-combined_k141_35950\|\|full | Low-quality |
| 17-109-final-viral-combined_k141_52491\|\|full | Not-determined |
| 17-109-final-viral-combined_k141_286977\|\|full | Not-determined |
| 17-109-final-viral-combined_k141_387284\|\|full | Low-quality |
| 17-109-final-viral-combined_k141_102505\|\|full | Not-determined |
| 17-109-final-viral-combined_k141_169520\|\|full | Low-quality |
| 17-109-final-viral-combined_k141_370664\|\|full | Low-quality |
| 17-109-final-viral-combined_k141_169524\|\|full | Low-quality |
| 17-109-final-viral-combined_k141_387291\|\|full | Low-quality |
| 17-109-final-viral-combined_k141_85940\|\|full | Not-determined |
| 17-109-final-viral-combined_k141_136118\|\|full | Low-quality |
| 17-109-final-viral-combined_k141_2381\|\|full | Not-determined |
| 17-109-final-viral-combined_k141_52523\|\|full | Low-quality |
| 17-109-final-viral-combined_k141_186643\|\|full | Not-determined |
| 17-109-final-viral-combined_k141_320387\|\|full | Not-determined |
| 17-109-final-viral-combined_k141_236456\|\|full | Not-determined |
| 17-109-final-viral-combined_k141_253154\|\|full | Low-quality |
| 17-109-final-viral-combined_k141_236462\|\|full | Low-quality |
| 17-109-final-viral-combined_k141_35982\|\|full | Not-determined |
| 17-109-final-viral-combined_k141_152996\|\|full | Not-determined |
| 17-109-final-viral-combined_k141_253156\|\|full | Low-quality |
| 17-109-final-viral-combined_k141_85999\|\|full | Not-determined |
| 17-109-final-viral-combined_k141_69245\|\|full | Not-determined |
| 17-109-final-viral-combined_k141_186677\|\|full | Not-determined |
| 17-109-final-viral-combined_k141_353558\|\|full | Low-quality |
| 17-109-final-viral-combined_k141_236477\|\|full | Low-quality |
| 17-109-final-viral-combined_k141_86007\|\|full | Low-quality |
| 17-109-final-viral-combined_k141_52571\|\|full | Low-quality |
| 17-109-final-viral-combined_k141_169553\|\|full | Not-determined |
| 17-109-final-viral-combined_k141_186696\|\|full | Not-determined |
| 17-109-final-viral-combined_k141_320424\|\|full | Not-determined |
| 17-109-final-viral-combined_k141_18702\|\|full | Not-determined |
| 17-109-final-viral-combined_k141_136153\|\|full | Low-quality |
| 17-109-final-viral-combined_k141_320431\|\|full | Not-determined |
| 17-109-final-viral-combined_k141_169558\|\|full | Low-quality |
| 17-109-final-viral-combined_k141_253180\|\|full | Low-quality |
| 17-109-final-viral-combined_k141_269979\|\|full | Low-quality |
| 17-109-final-viral-combined_k141_320448\|\|full | Not-determined |
| 17-109-final-viral-combined_k141_153027\|\|full | Low-quality |
| 17-109-final-viral-combined_k141_202936\|\|full | Not-determined |
| 17-109-final-viral-combined_k141_136169\|\|full | Not-determined |
| 17-109-final-viral-combined_k141_136172\|\|full | Not-determined |
| 17-109-final-viral-combined_k141_370743\|\|full | Not-determined |
| 17-109-final-viral-combined_k141_219655\|\|full | Low-quality |
| 17-109-final-viral-combined_k141_353595\|\|full | Not-determined |
| 17-109-final-viral-combined_k141_2440\|\|full | Not-determined |
| 17-109-final-viral-combined_k141_253199\|\|full | Not-determined |
| 17-109-final-viral-combined_k141_18733\|\|full | Low-quality |
| 17-109-final-viral-combined_k141_52623\|\|full | Not-determined |
| 17-109-final-viral-combined_k141_18738\|\|full | Not-determined |
| 17-109-final-viral-combined_k141_353630\|\|full | Low-quality |
| 17-109-final-viral-combined_k141_287089\|\|full | Low-quality |
| 17-109-final-viral-combined_k141_253233\|\|full | Low-quality |
| 17-109-final-viral-combined_k141_119202\|\|full | Low-quality |
| 17-109-final-viral-combined_k141_219676\|\|full | Not-determined |
| 17-109-final-viral-combined_k141_287093\|\|full | Not-determined |
| 17-109-final-viral-combined_k141_69358\|\|full | Not-determined |
| 17-109-final-viral-combined_k141_320495\|\|full | Low-quality |
| 17-109-final-viral-combined_k141_287096\|\|full | Not-determined |
| 17-109-final-viral-combined_k141_253250\|\|full | Low-quality |
| 17-109-final-viral-combined_k141_253254\|\|full | Low-quality |
| 17-109-final-viral-combined_k141_219699\|\|full | Not-determined |
| 17-109-final-viral-combined_k141_236592\|\|full | Low-quality |
| 17-109-final-viral-combined_k141_169609\|\|full | Low-quality |
| 17-109-final-viral-combined_k141_353665\|\|full | Low-quality |
| 17-109-final-viral-combined_k141_253268\|\|full | Low-quality |
| 17-109-final-viral-combined_k141_169633\|\|full | Low-quality |
| 17-109-final-viral-combined_k141_303944\|\|full | Not-determined |
| 17-109-final-viral-combined_k141_136219\|\|full | Low-quality |
| 17-109-final-viral-combined_k141_287115\|\|full | Not-determined |
| 17-109-final-viral-combined_k141_303957\|\|full | Low-quality |
| 17-109-final-viral-combined_k141_303959\|\|full | Low-quality |
| 17-109-final-viral-combined_k141_336640\|\|full | Low-quality |
| 17-109-final-viral-combined_k141_36038\|\|full | Not-determined |
| 17-109-final-viral-combined_k141_203020\|\|full | Not-determined |
| 17-109-final-viral-combined_k141_36040\|\|full | Low-quality |
| 17-109-final-viral-combined_k141_153101\|\|full | Not-determined |
| 18-103-final-viral-combined_k141_73262\|\|full | Medium-quality |
| 17-109-final-viral-combined_k141_136231\|\|full | Low-quality |
| 17-109-final-viral-combined_k141_18784\|\|full | Not-determined |
| 17-109-final-viral-combined_k141_36051\|\|full | Not-determined |
| 17-109-final-viral-combined_k141_287148\|\|full | Not-determined |
| 17-109-final-viral-combined_k141_18791\|\|full | Low-quality |
| 17-109-final-viral-combined_k141_52746\|\|full | Not-determined |
| 17-109-final-viral-combined_k141_236640\|\|full | Low-quality |
| 17-109-final-viral-combined_k141_169653\|\|full | Low-quality |
| 17-109-final-viral-combined_k141_18795\|\|full | Not-determined |
| 17-109-final-viral-combined_k141_236657\|\|full | Low-quality |
| 17-109-final-viral-combined_k141_2481\|\|full | Low-quality |
| 17-109-final-viral-combined_k141_69483\|\|full | Low-quality |
| 17-109-final-viral-combined_k141_18825\|\|full | Low-quality |
| 17-109-final-viral-combined_k141_304014\|\|full | Low-quality |
| 17-109-final-viral-combined_k141_203091\|\|full | Low-quality |
| 17-109-final-viral-combined_k141_304019\|\|full | Not-determined |
| 17-109-final-viral-combined_k141_353758\|\|full | Not-determined |
| 17-109-final-viral-combined_k141_236689\|\|full | Not-determined |
| 17-109-final-viral-combined_k141_236690\|\|full | Low-quality |
| 17-109-final-viral-combined_k141_2501\|\|full | Low-quality |
| 17-109-final-viral-combined_k141_169717\|\|full | Not-determined |
| 17-109-final-viral-combined_k141_353765\|\|full | Not-determined |
| 17-109-final-viral-combined_k141_169719\|\|full | Not-determined |
| 17-109-final-viral-combined_k141_304034\|\|full | Low-quality |
| 17-109-final-viral-combined_k141_136292\|\|full | Low-quality |
| 17-109-final-viral-combined_k141_370921\|\|full | Low-quality |
| 17-109-final-viral-combined_k141_253353\|\|full | Low-quality |
| 17-109-final-viral-combined_k141_36111\|\|full | Not-determined |
| 17-109-final-viral-combined_k141_236711\|\|full | Low-quality |
| 17-109-final-viral-combined_k141_304077\|\|full | Low-quality |
| 17-109-final-viral-combined_k141_136319\|\|full | Low-quality |
| 17-109-final-viral-combined_k141_169743\|\|full | Not-determined |
| 17-109-final-viral-combined_k141_2529\|\|full | Not-determined |
| 17-109-final-viral-combined_k141_86114\|\|full | Not-determined |
| 17-109-final-viral-combined_k141_86122\|\|full | Low-quality |
| 17-109-final-viral-combined_k141_270021\|\|full | Not-determined |
| 17-109-final-viral-combined_k141_270023\|\|full | Not-determined |
| 17-109-final-viral-combined_k141_370943\|\|full | Low-quality |
| 17-109-final-viral-combined_k141_320602\|\|full | Not-determined |
| 17-109-final-viral-combined_k141_136331\|\|full | Low-quality |
| 17-109-final-viral-combined_k141_169768\|\|full | Low-quality |
| 17-109-final-viral-combined_k141_304090\|\|full | Not-determined |
| 17-109-final-viral-combined_k141_18885\|\|full | Low-quality |
| 17-109-final-viral-combined_k141_119245\|\|full | Not-determined |
| 17-109-final-viral-combined_k141_320603\|\|full | Low-quality |
| 17-109-final-viral-combined_k141_36134\|\|full | Not-determined |
| 17-109-final-viral-combined_k141_219814\|\|full | Not-determined |
| 17-109-final-viral-combined_k141_2557\|\|full | Low-quality |
| 17-109-final-viral-combined_k141_169787\|\|full | Not-determined |
| 17-109-final-viral-combined_k141_219815\|\|full | Low-quality |
| 17-109-final-viral-combined_k141_153171\|\|full | Low-quality |
| 17-109-final-viral-combined_k141_136344\|\|full | Not-determined |
| 17-109-final-viral-combined_k141_219819\|\|full | Low-quality |
| 17-109-final-viral-combined_k141_353827\|\|full | Not-determined |
| 17-109-final-viral-combined_k141_36155\|\|full | Low-quality |
| 17-109-final-viral-combined_k141_353828\|\|full | Not-determined |
| 17-109-final-viral-combined_k141_304113\|\|full | Low-quality |
| 17-109-final-viral-combined_k141_169812\|\|full | Not-determined |
| 17-109-final-viral-combined_k141_69597\|\|full | Not-determined |
| 17-109-final-viral-combined_k141_69600\|\|full | Low-quality |
| 17-109-final-viral-combined_k141_203161\|\|full | Low-quality |
| 17-109-final-viral-combined_k141_387524\|\|full | Not-determined |
| 17-109-final-viral-combined_k141_371013\|\|full | Low-quality |
| 17-109-final-viral-combined_k141_186823\|\|full | Not-determined |
| 17-109-final-viral-combined_k141_69607\|\|full | Low-quality |
| 17-109-final-viral-combined_k141_371017\|\|full | Not-determined |
| 17-109-final-viral-combined_k141_236774\|\|full | Low-quality |
| 17-109-final-viral-combined_k141_320648\|\|full | Low-quality |
| 17-109-final-viral-combined_k141_387529\|\|full | Low-quality |
| 17-109-final-viral-combined_k141_2632\|\|full | Not-determined |
| 17-109-final-viral-combined_k141_236790\|\|full | Not-determined |
| 17-109-final-viral-combined_k141_320670\|\|full | Low-quality |
| 17-109-final-viral-combined_k141_287310\|\|full | Not-determined |
| 17-109-final-viral-combined_k141_371050\|\|full | Low-quality |
| 17-109-final-viral-combined_k141_69633\|\|full | Not-determined |
| 17-109-final-viral-combined_k141_304151\|\|full | Not-determined |
| 17-109-final-viral-combined_k141_387557\|\|full | Not-determined |
| 17-109-final-viral-combined_k141_320680\|\|full | Not-determined |
| 17-109-final-viral-combined_k141_304157\|\|full | Not-determined |
| 17-109-final-viral-combined_k141_86217\|\|full | Low-quality |
| 17-109-final-viral-combined_k141_320702\|\|full | Low-quality |
| 17-109-final-viral-combined_k141_86231\|\|full | Low-quality |
| 17-109-final-viral-combined_k141_169861\|\|full | Low-quality |
| 17-109-final-viral-combined_k141_86240\|\|full | Not-determined |
| 17-109-final-viral-combined_k141_102670\|\|full | Low-quality |
| 17-109-final-viral-combined_k141_69674\|\|full | Low-quality |
| 17-109-final-viral-combined_k141_102677\|\|full | Low-quality |
| 17-109-final-viral-combined_k141_153303\|\|full | Not-determined |
| 17-109-final-viral-combined_k141_320715\|\|full | Low-quality |
| 17-109-final-viral-combined_k141_253393\|\|full | Not-determined |
| 17-109-final-viral-combined_k141_304182\|\|full | Low-quality |
| 17-109-final-viral-combined_k141_186868\|\|full | Low-quality |
| 17-109-final-viral-combined_k141_287341\|\|full | Low-quality |
| 17-109-final-viral-combined_k141_153336\|\|full | Not-determined |
| 17-109-final-viral-combined_k141_102721\|\|full | Low-quality |
| 17-109-final-viral-combined_k141_102722\|\|full | Low-quality |
| 17-109-final-viral-combined_k141_69718\|\|full | Not-determined |
| 17-109-final-viral-combined_k141_102725\|\|full | Not-determined |
| 17-109-final-viral-combined_k141_236862\|\|full | Not-determined |
| 17-109-final-viral-combined_k141_203191\|\|full | Low-quality |
| 17-109-final-viral-combined_k141_253416\|\|full | Not-determined |
| 17-109-final-viral-combined_k141_186894\|\|full | Not-determined |
| 17-109-final-viral-combined_k141_169888\|\|full | Low-quality |
| 17-109-final-viral-combined_k141_186901\|\|full | Low-quality |
| 17-109-final-viral-combined_k141_253419\|\|full | Not-determined |
| 17-109-final-viral-combined_k141_336665\|\|full | Not-determined |
| 17-109-final-viral-combined_k141_153360\|\|full | Not-determined |
| 17-109-final-viral-combined_k141_136420\|\|full | Low-quality |
| 17-109-final-viral-combined_k141_287350\|\|full | Low-quality |
| 17-109-final-viral-combined_k141_69764\|\|full | Not-determined |
| 17-109-final-viral-combined_k141_253439\|\|full | Low-quality |
| 17-109-final-viral-combined_k141_236908\|\|full | Not-determined |
| 17-109-final-viral-combined_k141_136425\|\|full | Low-quality |
| 17-109-final-viral-combined_k141_2715\|\|full | Low-quality |
| 17-109-final-viral-combined_k141_136440\|\|full | Not-determined |
| 17-109-final-viral-combined_k141_219943\|\|full | Low-quality |
| 17-109-final-viral-combined_k141_336694\|\|full | Not-determined |
| 17-109-final-viral-combined_k141_52854\|\|full | Low-quality |
| 17-109-final-viral-combined_k141_169931\|\|full | Low-quality |
| 17-109-final-viral-combined_k141_69786\|\|full | Low-quality |
| 17-109-final-viral-combined_k141_270194\|\|full | Low-quality |
| 17-109-final-viral-combined_k141_219950\|\|full | Not-determined |
| 17-109-final-viral-combined_k141_153394\|\|full | Low-quality |
| 17-109-final-viral-combined_k141_52857\|\|full | Low-quality |
| 17-109-final-viral-combined_k141_186941\|\|full | Not-determined |
| 17-109-final-viral-combined_k141_304221\|\|full | Not-determined |
| 17-109-final-viral-combined_k141_336707\|\|full | Not-determined |
| 17-109-final-viral-combined_k141_119376\|\|full | Not-determined |
| 17-109-final-viral-combined_k141_304232\|\|full | Not-determined |
| 17-109-final-viral-combined_k141_203257\|\|full | Low-quality |
| 17-109-final-viral-combined_k141_236942\|\|full | Low-quality |
| 17-109-final-viral-combined_k141_253460\|\|full | Not-determined |
| 17-109-final-viral-combined_k141_287357\|\|full | Not-determined |
| 17-109-final-viral-combined_k141_36183\|\|full | Not-determined |
| 17-109-final-viral-combined_k141_186962\|\|full | Low-quality |
| 17-109-final-viral-combined_k141_86305\|\|full | Not-determined |
| 17-109-final-viral-combined_k141_153404\|\|full | Low-quality |
| 17-109-final-viral-combined_k141_186971\|\|full | Not-determined |
| 17-109-final-viral-combined_k141_371093\|\|full | Low-quality |
| 17-109-final-viral-combined_k141_203299\|\|full | Low-quality |
| 17-109-final-viral-combined_k141_186985\|\|full | Low-quality |
| 17-109-final-viral-combined_k141_52894\|\|full | Not-determined |
| 17-109-final-viral-combined_k141_253488\|\|full | Low-quality |
| 17-109-final-viral-combined_k141_336736\|\|full | Low-quality |
| 17-109-final-viral-combined_k141_52895\|\|full | Not-determined |
| 17-109-final-viral-combined_k141_187000\|\|full | Low-quality |
| 17-109-final-viral-combined_k141_336749\|\|full | Not-determined |
| 17-109-final-viral-combined_k141_119420\|\|full | Not-determined |
| 17-109-final-viral-combined_k141_52898\|\|full | Not-determined |
| 17-109-final-viral-combined_k141_2784\|\|full | Low-quality |
| 17-109-final-viral-combined_k141_219968\|\|full | Not-determined |
| 17-109-final-viral-combined_k141_169992\|\|full | Low-quality |
| 17-109-final-viral-combined_k141_187032\|\|full | Low-quality |
| 17-109-final-viral-combined_k141_287417\|\|full | Not-determined |
| 17-109-final-viral-combined_k141_203338\|\|full | Not-determined |
| 17-109-final-viral-combined_k141_69884\|\|full | Low-quality |
| 17-109-final-viral-combined_k141_270274\|\|full | Not-determined |
| 17-109-final-viral-combined_k141_371143\|\|full | Low-quality |
| 17-109-final-viral-combined_k141_203344\|\|full | Low-quality |
| 17-109-final-viral-combined_k141_219989\|\|full | Low-quality |
| 17-109-final-viral-combined_k141_336780\|\|full | Low-quality |
| 17-109-final-viral-combined_k141_36249\|\|full | Not-determined |
| 17-109-final-viral-combined_k141_287463\|\|full | Not-determined |
| 17-109-final-viral-combined_k141_18940\|\|full | Not-determined |
| 17-109-final-viral-combined_k141_102844\|\|full | Not-determined |
| 17-109-final-viral-combined_k141_203374\|\|full | Low-quality |
| 17-109-final-viral-combined_k141_86352\|\|full | Not-determined |
| 17-109-final-viral-combined_k141_270292\|\|full | Low-quality |
| 18-110_k141_134510 | Medium-quality |
| 17-109-final-viral-combined_k141_371194\|\|full | Low-quality |
| 17-109-final-viral-combined_k141_187110\|\|full | Low-quality |
| 17-109-final-viral-combined_k141_203407\|\|full | Low-quality |
| 17-109-final-viral-combined_k141_270322\|\|full | Not-determined |
| 17-109-final-viral-combined_k141_119499\|\|full | Low-quality |
| 17-109-final-viral-combined_k141_119504\|\|full | Low-quality |
| 17-109-final-viral-combined_k141_36300\|\|full | Not-determined |
| 17-109-final-viral-combined_k141_170058\|\|full | Low-quality |
| 17-109-final-viral-combined_k141_102890\|\|full | Not-determined |
| 17-109-final-viral-combined_k141_2829\|\|full | Not-determined |
| 17-109-final-viral-combined_k141_170067\|\|full | Not-determined |
| 17-109-final-viral-combined_k141_119531\|\|full | Not-determined |
| 17-109-final-viral-combined_k141_36320\|\|full | Low-quality |
| 17-109-final-viral-combined_k141_119538\|\|full | Low-quality |
| 17-109-final-viral-combined_k141_18970\|\|full | Low-quality |
| 17-109-final-viral-combined_k141_119540\|\|full | Low-quality |
| 17-109-final-viral-combined_k141_52961\|\|full | Low-quality |
| 17-109-final-viral-combined_k141_2844\|\|full | Low-quality |
| 17-109-final-viral-combined_k141_36322\|\|full | Not-determined |
| 17-109-final-viral-combined_k141_119547\|\|full | Low-quality |
| 17-109-final-viral-combined_k141_187160\|\|full | Low-quality |
| 17-109-final-viral-combined_k141_2849\|\|full | Not-determined |
| 17-109-final-viral-combined_k141_119555\|\|full | Not-determined |
| 18-110-final-viral-combined_k141_134510\|\|full | Medium-quality |
| 17-109-final-viral-combined_k141_119563\|\|full | Not-determined |
| 17-109-final-viral-combined_k141_136636\|\|full | Low-quality |
| 17-109-final-viral-combined_k141_136638\|\|full | Not-determined |
| 17-109-final-viral-combined_k141_86403\|\|full | Not-determined |
| 17-109-final-viral-combined_k141_2858\|\|full | Not-determined |
| 17-109-final-viral-combined_k141_287521\|\|full | Not-determined |
| 17-109-final-viral-combined_k141_52990\|\|full | Low-quality |
| 17-109-final-viral-combined_k141_320793\|\|full | Low-quality |
| 17-109-final-viral-combined_k141_52993\|\|full | Not-determined |
| 17-109-final-viral-combined_k141_170127\|\|full | Not-determined |
| 17-109-final-viral-combined_k141_187181\|\|full | Low-quality |
| 17-109-final-viral-combined_k141_371234\|\|full | Not-determined |
| 17-109-final-viral-combined_k141_86420\|\|full | Not-determined |
| 17-109-final-viral-combined_k141_253550\|\|full | Not-determined |
| 17-109-final-viral-combined_k141_170140\|\|full | Low-quality |
| 17-109-final-viral-combined_k141_170141\|\|full | Not-determined |
| 17-109-final-viral-combined_k141_320828\|\|full | Not-determined |
| 17-109-final-viral-combined_k141_86430\|\|full | Low-quality |
| 17-109-final-viral-combined_k141_170146\|\|full | Low-quality |
| 17-109-final-viral-combined_k141_187226\|\|full | Low-quality |
| 17-109-final-viral-combined_k141_102982\|\|full | Not-determined |
| 17-109-final-viral-combined_k141_119648\|\|full | Not-determined |
| 17-109-final-viral-combined_k141_253567\|\|full | Not-determined |
| 17-109-final-viral-combined_k141_102983\|\|full | Low-quality |
| 17-109-final-viral-combined_k141_237075\|\|full | Not-determined |
| 17-109-final-viral-combined_k141_36418\|\|full | Not-determined |
| 17-109-final-viral-combined_k141_36419\|\|full | Low-quality |
| 17-109-final-viral-combined_k141_220055\|\|full | Not-determined |
| 17-109-final-viral-combined_k141_320844\|\|full | Low-quality |
| 17-109-final-viral-combined_k141_304388\|\|full | Not-determined |
| 17-109-final-viral-combined_k141_86462\|\|full | Low-quality |
| 17-109-final-viral-combined_k141_270434\|\|full | Low-quality |
| 17-109-final-viral-combined_k141_237103\|\|full | Not-determined |
| 17-109-final-viral-combined_k141_2909\|\|full | Low-quality |
| 17-109-final-viral-combined_k141_203543\|\|full | Not-determined |
| 17-109-final-viral-combined_k141_203551\|\|full | Low-quality |
| 17-109-final-viral-combined_k141_187241\|\|full | Not-determined |
| 17-109-final-viral-combined_k141_2933\|\|full | Not-determined |
| 17-109-final-viral-combined_k141_304421\|\|full | Low-quality |
| 17-109-final-viral-combined_k141_2943\|\|full | Low-quality |
| 17-109-final-viral-combined_k141_270472\|\|full | Low-quality |
| 17-109-final-viral-combined_k141_371308\|\|full | Low-quality |
| 17-109-final-viral-combined_k141_304432\|\|full | Not-determined |
| 17-109-final-viral-combined_k141_287580\|\|full | Low-quality |
| 17-109-final-viral-combined_k141_170157\|\|full | Low-quality |
| 17-109-final-viral-combined_k141_371320\|\|full | Not-determined |
| 17-109-final-viral-combined_k141_53050\|\|full | Not-determined |
| 17-109-final-viral-combined_k141_304446\|\|full | Not-determined |
| 17-109-final-viral-combined_k141_336954\|\|full | Not-determined |
| 17-109-final-viral-combined_k141_170160\|\|full | Low-quality |
| 17-109-final-viral-combined_k141_270506\|\|full | Low-quality |
| 17-109-final-viral-combined_k141_336958\|\|full | Not-determined |
| 17-109-final-viral-combined_k141_70100\|\|full | Not-determined |
| 17-109-final-viral-combined_k141_153505\|\|full | Low-quality |
| 17-109-final-viral-combined_k141_36463\|\|full | Low-quality |
| 17-109-final-viral-combined_k141_371328\|\|full | Low-quality |
| 17-109-final-viral-combined_k141_170187\|\|full | Low-quality |
| 17-109-final-viral-combined_k141_2991\|\|full | Not-determined |
| 17-109-final-viral-combined_k141_304473\|\|full | Not-determined |
| 17-109-final-viral-combined_k141_220159\|\|full | Low-quality |
| 17-109-final-viral-combined_k141_103041\|\|full | Low-quality |
| 17-109-final-viral-combined_k141_371347\|\|full | Not-determined |
| 17-109-final-viral-combined_k141_387799\|\|full | Not-determined |
| 17-109-final-viral-combined_k141_187278\|\|full | Not-determined |
| 17-109-final-viral-combined_k141_53093\|\|full | Not-determined |
| 17-109-final-viral-combined_k141_270523\|\|full | Not-determined |
| 17-109-final-viral-combined_k141_320877\|\|full | Low-quality |
| 17-109-final-viral-combined_k141_320885\|\|full | Not-determined |
| 17-109-final-viral-combined_k141_119783\|\|full | Low-quality |
| 17-109-final-viral-combined_k141_187314\|\|full | Low-quality |
| 17-109-final-viral-combined_k141_337029\|\|full | Low-quality |
| 17-109-final-viral-combined_k141_320896\|\|full | Not-determined |
| 17-109-final-viral-combined_k141_136846\|\|full | Low-quality |
| 17-109-final-viral-combined_k141_387815\|\|full | Not-determined |
| 17-109-final-viral-combined_k141_354184\|\|full | Not-determined |
| 17-109-final-viral-combined_k141_3033\|\|full | Not-determined |
| 17-109-final-viral-combined_k141_354186\|\|full | Low-quality |
| 17-109-final-viral-combined_k141_19172\|\|full | Not-determined |
| 17-109-final-viral-combined_k141_119789\|\|full | Not-determined |
| 17-109-final-viral-combined_k141_203649\|\|full | Low-quality |
| 17-109-final-viral-combined_k141_387819\|\|full | Low-quality |
| 17-109-final-viral-combined_k141_270557\|\|full | Low-quality |
| 17-109-final-viral-combined_k141_203651\|\|full | Not-determined |
| 17-109-final-viral-combined_k141_19184\|\|full | Not-determined |
| 17-109-final-viral-combined_k141_270561\|\|full | Low-quality |
| 17-109-final-viral-combined_k141_320917\|\|full | Low-quality |
| 17-109-final-viral-combined_k141_86557\|\|full | Not-determined |
| 17-109-final-viral-combined_k141_337033\|\|full | Low-quality |
| 17-109-final-viral-combined_k141_371399\|\|full | Low-quality |
| 17-109-final-viral-combined_k141_270579\|\|full | Low-quality |
| 17-109-final-viral-combined_k141_337047\|\|full | Low-quality |
| 17-109-final-viral-combined_k141_253589\|\|full | Low-quality |
| 17-109-final-viral-combined_k141_70159\|\|full | Not-determined |
| 17-109-final-viral-combined_k141_287591\|\|full | Not-determined |
| 17-109-final-viral-combined_k141_153560\|\|full | Low-quality |
| 17-109-final-viral-combined_k141_170251\|\|full | Low-quality |
| 17-109-final-viral-combined_k141_304540\|\|full | Low-quality |
| 17-109-final-viral-combined_k141_220276\|\|full | Low-quality |
| 17-109-final-viral-combined_k141_36544\|\|full | Not-determined |
| 17-109-final-viral-combined_k141_70194\|\|full | Not-determined |
| 17-109-final-viral-combined_k141_237252\|\|full | Not-determined |
| 17-109-final-viral-combined_k141_136866\|\|full | Not-determined |
| 17-109-final-viral-combined_k141_287615\|\|full | Low-quality |
| 17-109-final-viral-combined_k141_220292\|\|full | Not-determined |
| 17-109-final-viral-combined_k141_36560\|\|full | Not-determined |
| 17-109-final-viral-combined_k141_170286\|\|full | Low-quality |
| 17-109-final-viral-combined_k141_203685\|\|full | Not-determined |
| 17-109-final-viral-combined_k141_220296\|\|full | Not-determined |
| 17-109-final-viral-combined_k141_70219\|\|full | Not-determined |
| 17-109-final-viral-combined_k141_270629\|\|full | Not-determined |
| 17-109-final-viral-combined_k141_136889\|\|full | Low-quality |
| 17-109-final-viral-combined_k141_220303\|\|full | Low-quality |
| 17-109-final-viral-combined_k141_19231\|\|full | Low-quality |
| 17-109-final-viral-combined_k141_220310\|\|full | Not-determined |
| 17-109-final-viral-combined_k141_3066\|\|full | Not-determined |
| 17-109-final-viral-combined_k141_304571\|\|full | Low-quality |
| 17-109-final-viral-combined_k141_337087\|\|full | Low-quality |
| 17-109-final-viral-combined_k141_70228\|\|full | Not-determined |
| 17-109-final-viral-combined_k141_220317\|\|full | Not-determined |
| 17-109-final-viral-combined_k141_153584\|\|full | Low-quality |
| 17-109-final-viral-combined_k141_220321\|\|full | Not-determined |
| 17-109-final-viral-combined_k141_371434\|\|full | Not-determined |
| 17-109-final-viral-combined_k141_86596\|\|full | Low-quality |
| 17-109-final-viral-combined_k141_270647\|\|full | Low-quality |
| 17-109-final-viral-combined_k141_220334\|\|full | Low-quality |
| 17-109-final-viral-combined_k141_86613\|\|full | Low-quality |
| 17-109-final-viral-combined_k141_187390\|\|full | Not-determined |
| 17-109-final-viral-combined_k141_387914\|\|full | Low-quality |
| 17-109-final-viral-combined_k141_253665\|\|full | Low-quality |
| 17-109-final-viral-combined_k141_253670\|\|full | Low-quality |
| 17-109-final-viral-combined_k141_203721\|\|full | Low-quality |
| 17-109-final-viral-combined_k141_86641\|\|full | Not-determined |
| 17-109-final-viral-combined_k141_153608\|\|full | Not-determined |
| 17-109-final-viral-combined_k141_203732\|\|full | Low-quality |
| 17-109-final-viral-combined_k141_3102\|\|full | Low-quality |
| 17-109-final-viral-combined_k141_3105\|\|full | Low-quality |
| 17-109-final-viral-combined_k141_253688\|\|full | Not-determined |
| 17-109-final-viral-combined_k141_170346\|\|full | Not-determined |
| 17-109-final-viral-combined_k141_119829\|\|full | Low-quality |
| 17-109-final-viral-combined_k141_220399\|\|full | Low-quality |

| | |
|---|---|
| 17-109-final-viral-combined_k141_203755\|\|full | Low-quality |
| 17-109-final-viral-combined_k141_387927\|\|full | Not-determined |
| 17-109-final-viral-combined_k141_3132\|\|full | Low-quality |
| 17-109-final-viral-combined_k141_53213\|\|full | Not-determined |
| 17-109-final-viral-combined_k141_203762\|\|full | Not-determined |
| 17-109-final-viral-combined_k141_220403\|\|full | Not-determined |
| 17-109-final-viral-combined_k141_203764\|\|full | Not-determined |
| 17-109-final-viral-combined_k141_103100\|\|full | Not-determined |
| 17-109-final-viral-combined_k141_153634\|\|full | Low-quality |
| 17-109-final-viral-combined_k141_220413\|\|full | Not-determined |
| 17-109-final-viral-combined_k141_136977\|\|full | Low-quality |
| 17-109-final-viral-combined_k141_53225\|\|full | Low-quality |
| 17-109-final-viral-combined_k141_136980\|\|full | Not-determined |
| 17-109-final-viral-combined_k141_3148\|\|full | Not-determined |
| 17-109-final-viral-combined_k141_119843\|\|full | Low-quality |
| 17-109-final-viral-combined_k141_119854\|\|full | Low-quality |
| 17-109-final-viral-combined_k141_86680\|\|full | Not-determined |
| 17-109-final-viral-combined_k141_136996\|\|full | Not-determined |
| 17-109-final-viral-combined_k141_304613\|\|full | Not-determined |
| 17-109-final-viral-combined_k141_187463\|\|full | Not-determined |
| 17-109-final-viral-combined_k141_187465\|\|full | Low-quality |
| 17-109-final-viral-combined_k141_287642\|\|full | Low-quality |
| 17-109-final-viral-combined_k141_304622\|\|full | Not-determined |
| 17-109-final-viral-combined_k141_270748\|\|full | Low-quality |
| 17-109-final-viral-combined_k141_170412\|\|full | Low-quality |
| 17-109-final-viral-combined_k141_53278\|\|full | Low-quality |
| 17-109-final-viral-combined_k141_270752\|\|full | Low-quality |
| 17-109-final-viral-combined_k141_187469\|\|full | Low-quality |
| 17-109-final-viral-combined_k141_304642\|\|full | Low-quality |
| 18-106-final-viral-combined_k141_140731\|\|full | Medium-quality |
| 17-109-final-viral-combined_k141_270763\|\|full | Low-quality |
| 17-109-final-viral-combined_k141_354280\|\|full | Low-quality |
| 17-109-final-viral-combined_k141_387994\|\|full | Not-determined |
| 17-109-final-viral-combined_k141_320996\|\|full | Not-determined |
| 17-109-final-viral-combined_k141_153689\|\|full | Low-quality |
| 17-109-final-viral-combined_k141_203838\|\|full | Low-quality |
| 17-109-final-viral-combined_k141_187512\|\|full | Not-determined |
| 17-109-final-viral-combined_k141_119868\|\|full | Not-determined |
| 17-109-final-viral-combined_k141_287692\|\|full | Low-quality |
| 17-109-final-viral-combined_k141_19362\|\|full | Low-quality |
| 17-109-final-viral-combined_k141_388007\|\|full | Low-quality |
| 17-109-final-viral-combined_k141_388008\|\|full | Not-determined |
| 17-109-final-viral-combined_k141_388009\|\|full | Not-determined |
| 17-109-final-viral-combined_k141_19368\|\|full | Not-determined |
| 17-109-final-viral-combined_k141_321017\|\|full | Low-quality |
| 17-109-final-viral-combined_k141_103152\|\|full | Not-determined |
| 17-109-final-viral-combined_k141_203861\|\|full | Low-quality |
| 17-109-final-viral-combined_k141_203864\|\|full | Not-determined |
| 17-109-final-viral-combined_k141_119887\|\|full | Not-determined |
| 17-109-final-viral-combined_k141_170468\|\|full | Not-determined |
| 17-109-final-viral-combined_k141_3213\|\|full | Not-determined |
| 17-109-final-viral-combined_k141_270790\|\|full | Low-quality |
| 17-109-final-viral-combined_k141_354315\|\|full | Not-determined |
| 17-109-final-viral-combined_k141_70335\|\|full | Low-quality |
| 17-109-final-viral-combined_k141_53346\|\|full | Low-quality |
| 17-109-final-viral-combined_k141_36753\|\|full | Low-quality |
| 17-109-final-viral-combined_k141_36754\|\|full | Not-determined |
| 17-109-final-viral-combined_k141_270799\|\|full | Low-quality |
| 17-109-final-viral-combined_k141_119927\|\|full | Low-quality |
| 17-109-final-viral-combined_k141_36760\|\|full | Not-determined |
| 17-109-final-viral-combined_k141_70366\|\|full | Not-determined |
| 17-109-final-viral-combined_k141_237274\|\|full | Low-quality |
| 17-109-final-viral-combined_k141_3232\|\|full | Low-quality |
| 17-109-final-viral-combined_k141_86776\|\|full | Low-quality |
| 17-109-final-viral-combined_k141_220491\|\|full | Not-determined |
| 17-109-final-viral-combined_k141_36771\|\|full | Not-determined |
| 17-109-final-viral-combined_k141_270821\|\|full | Not-determined |
| 17-109-final-viral-combined_k141_354337\|\|full | Not-determined |
| 17-109-final-viral-combined_k141_137097\|\|full | Not-determined |
| 17-109-final-viral-combined_k141_137098\|\|full | Low-quality |
| 17-109-final-viral-combined_k141_137109\|\|full | Not-determined |
| 17-109-final-viral-combined_k141_103216\|\|full | Not-determined |
| 17-109-final-viral-combined_k141_137111\|\|full | Low-quality |
| 17-109-final-viral-combined_k141_287733\|\|full | Not-determined |
| 17-109-final-viral-combined_k141_220510\|\|full | Low-quality |
| 17-109-final-viral-combined_k141_103224\|\|full | Low-quality |
| 17-109-final-viral-combined_k141_187576\|\|full | Low-quality |
| 17-109-final-viral-combined_k141_3252\|\|full | Not-determined |
| 17-109-final-viral-combined_k141_203935\|\|full | Not-determined |
| 17-109-final-viral-combined_k141_253753\|\|full | Not-determined |
| 17-109-final-viral-combined_k141_220534\|\|full | Not-determined |
| 17-109-final-viral-combined_k141_287740\|\|full | Not-determined |
| 17-109-final-viral-combined_k141_3269\|\|full | Low-quality |
| 17-109-final-viral-combined_k141_304715\|\|full | Low-quality |
| 17-109-final-viral-combined_k141_220556\|\|full | Low-quality |
| 17-109-final-viral-combined_k141_86826\|\|full | Not-determined |
| 17-109-final-viral-combined_k141_253773\|\|full | Not-determined |
| 17-109-final-viral-combined_k141_220573\|\|full | Not-determined |
| 17-109-final-viral-combined_k141_53406\|\|full | Not-determined |
| 17-109-final-viral-combined_k141_237333\|\|full | Not-determined |
| 17-109-final-viral-combined_k141_220575\|\|full | Low-quality |
| 17-109-final-viral-combined_k141_371601\|\|full | Not-determined |
| 17-109-final-viral-combined_k141_86842\|\|full | Not-determined |
| 17-109-final-viral-combined_k141_19464\|\|full | Not-determined |
| 17-109-final-viral-combined_k141_19468\|\|full | Low-quality |
| 17-109-final-viral-combined_k141_153792\|\|full | Low-quality |
| 17-109-final-viral-combined_k141_220598\|\|full | Not-determined |
| 17-109-final-viral-combined_k141_220602\|\|full | Not-determined |
| 17-109-final-viral-combined_k141_120002\|\|full | Not-determined |
| 17-109-final-viral-combined_k141_53417\|\|full | Not-determined |
| 17-109-final-viral-combined_k141_321116\|\|full | Low-quality |
| 17-109-final-viral-combined_k141_388107\|\|full | Not-determined |
| 17-109-final-viral-combined_k141_19479\|\|full | Low-quality |
| 17-109-final-viral-combined_k141_153804\|\|full | Low-quality |
| 17-109-final-viral-combined_k141_86871\|\|full | Not-determined |
| 17-109-final-viral-combined_k141_237382\|\|full | Not-determined |
| 17-109-final-viral-combined_k141_321142\|\|full | Not-determined |
| 17-109-final-viral-combined_k141_86876\|\|full | Low-quality |
| 17-109-final-viral-combined_k141_287800\|\|full | Not-determined |
| 17-109-final-viral-combined_k141_187595\|\|full | Low-quality |
| 17-109-final-viral-combined_k141_153815\|\|full | Low-quality |
| 17-109-final-viral-combined_k141_153821\|\|full | Not-determined |
| 17-109-final-viral-combined_k141_120047\|\|full | Not-determined |
| 17-109-final-viral-combined_k141_53455\|\|full | Low-quality |
| 17-109-final-viral-combined_k141_137190\|\|full | Low-quality |
| 17-109-final-viral-combined_k141_53474\|\|full | Low-quality |
| 17-109-final-viral-combined_k141_19548\|\|full | Low-quality |
| 17-109-final-viral-combined_k141_53485\|\|full | Low-quality |
| 17-109-final-viral-combined_k141_388137\|\|full | Not-determined |
| 17-109-final-viral-combined_k141_53494\|\|full | Not-determined |
| 17-109-final-viral-combined_k141_19556\|\|full | Low-quality |
| 17-109-final-viral-combined_k141_304787\|\|full | Not-determined |
| 17-109-final-viral-combined_k141_388144\|\|full | Low-quality |
| 17-109-final-viral-combined_k141_170610\|\|full | Low-quality |
| 17-109-final-viral-combined_k141_337466\|\|full | Not-determined |
| 17-109-final-viral-combined_k141_253824\|\|full | Low-quality |
| 17-109-final-viral-combined_k141_53511\|\|full | Not-determined |
| 17-109-final-viral-combined_k141_287860\|\|full | Low-quality |
| 17-109-final-viral-combined_k141_237442\|\|full | Low-quality |
| 17-109-final-viral-combined_k141_304804\|\|full | Not-determined |
| 17-109-final-viral-combined_k141_120072\|\|full | Not-determined |
| 17-109-final-viral-combined_k141_287867\|\|full | Low-quality |
| 17-109-final-viral-combined_k141_170642\|\|full | Low-quality |
| 17-109-final-viral-combined_k141_304828\|\|full | Low-quality |
| 17-109-final-viral-combined_k141_371686\|\|full | Low-quality |
| 17-109-final-viral-combined_k141_120099\|\|full | Low-quality |
| 17-109-final-viral-combined_k141_237485\|\|full | Low-quality |
| 17-109-final-viral-combined_k141_321193\|\|full | Low-quality |
| 17-109-final-viral-combined_k141_103343\|\|full | Low-quality |
| 17-109-final-viral-combined_k141_19598\|\|full | Low-quality |
| 17-109-final-viral-combined_k141_237492\|\|full | Low-quality |
| 17-109-final-viral-combined_k141_237502\|\|full | Not-determined |
| 17-109-final-viral-combined_k141_220648\|\|full | Not-determined |
| 17-109-final-viral-combined_k141_321216\|\|full | Not-determined |
| 17-109-final-viral-combined_k141_354505\|\|full | Not-determined |
| 17-109-final-viral-combined_k141_70539\|\|full | Not-determined |
| 17-109-final-viral-combined_k141_287910\|\|full | Low-quality |
| 17-109-final-viral-combined_k141_170672\|\|full | Low-quality |
| 17-109-final-viral-combined_k141_354513\|\|full | Low-quality |
| 17-109-final-viral-combined_k141_304873\|\|full | Low-quality |
| 17-109-final-viral-combined_k141_321232\|\|full | Not-determined |
| 17-109-final-viral-combined_k141_170685\|\|full | Not-determined |
| 17-109-final-viral-combined_k141_253887\|\|full | Low-quality |
| 17-109-final-viral-combined_k141_253888\|\|full | Low-quality |
| 17-109-final-viral-combined_k141_287937\|\|full | Not-determined |
| 17-109-final-viral-combined_k141_321245\|\|full | Low-quality |
| 17-109-final-viral-combined_k141_253896\|\|full | Not-determined |
| 17-109-final-viral-combined_k141_170701\|\|full | Low-quality |
| 17-109-final-viral-combined_k141_270893\|\|full | Low-quality |
| 17-109-final-viral-combined_k141_304895\|\|full | Low-quality |
| 17-109-final-viral-combined_k141_388206\|\|full | Not-determined |
| 17-109-final-viral-combined_k141_270898\|\|full | Not-determined |
| 17-109-final-viral-combined_k141_103405\|\|full | Low-quality |
| 17-109-final-viral-combined_k141_204097\|\|full | Low-quality |
| 17-109-final-viral-combined_k141_204100\|\|full | Low-quality |
| 17-109-final-viral-combined_k141_86934\|\|full | Not-determined |
| 17-109-final-viral-combined_k141_270909\|\|full | Not-determined |
| 17-109-final-viral-combined_k141_86937\|\|full | Not-determined |
| 17-109-final-viral-combined_k141_170717\|\|full | Not-determined |
| 17-109-final-viral-combined_k141_86940\|\|full | Not-determined |
| 17-109-final-viral-combined_k141_287973\|\|full | Not-determined |
| 17-109-final-viral-combined_k141_170722\|\|full | Low-quality |
| 17-109-final-viral-combined_k141_371792\|\|full | Low-quality |
| 17-109-final-viral-combined_k141_204119\|\|full | Not-determined |
| 17-109-final-viral-combined_k141_103444\|\|full | Not-determined |
| 17-109-final-viral-combined_k141_220692\|\|full | Not-determined |
| 17-109-final-viral-combined_k141_70575\|\|full | Not-determined |
| 17-109-final-viral-combined_k141_371798\|\|full | Low-quality |
| 17-109-final-viral-combined_k141_103457\|\|full | Not-determined |
| 17-109-final-viral-combined_k141_388257\|\|full | Not-determined |
| 17-109-final-viral-combined_k141_287999\|\|full | Not-determined |
| 17-109-final-viral-combined_k141_70599\|\|full | Low-quality |
| 17-109-final-viral-combined_k141_288008\|\|full | Low-quality |
| 17-109-final-viral-combined_k141_153939\|\|full | Not-determined |
| 17-109-final-viral-combined_k141_3428\|\|full | Low-quality |
| 17-109-final-viral-combined_k141_371809\|\|full | Low-quality |
| 17-109-final-viral-combined_k141_288014\|\|full | Not-determined |
| 17-109-final-viral-combined_k141_103484\|\|full | Low-quality |
| 17-109-final-viral-combined_k141_204160\|\|full | Not-determined |
| 17-109-final-viral-combined_k141_3435\|\|full | Not-determined |
| 17-109-final-viral-combined_k141_204184\|\|full | Not-determined |
| 17-109-final-viral-combined_k141_3440\|\|full | Not-determined |
| 17-109-final-viral-combined_k141_288026\|\|full | Not-determined |
| 17-109-final-viral-combined_k141_170778\|\|full | Not-determined |
| 17-109-final-viral-combined_k141_3454\|\|full | Low-quality |
| 17-109-final-viral-combined_k141_187868\|\|full | Low-quality |
| 17-109-final-viral-combined_k141_388296\|\|full | Not-determined |
| 17-109-final-viral-combined_k141_70643\|\|full | Low-quality |
| 17-109-final-viral-combined_k141_70648\|\|full | Not-determined |
| 17-109-final-viral-combined_k141_288048\|\|full | Low-quality |
| 18-109-final-viral-combined_k141_244866\|\|full | High-quality |
| 17-109-final-viral-combined_k141_204217\|\|full | Low-quality |
| 17-109-final-viral-combined_k141_19775\|\|full | Not-determined |
| 17-109-final-viral-combined_k141_237589\|\|full | Not-determined |
| 17-109-final-viral-combined_k141_19780\|\|full | Not-determined |
| 17-109-final-viral-combined_k141_288074\|\|full | Low-quality |
| 17-109-final-viral-combined_k141_337577\|\|full | Not-determined |
| 17-109-final-viral-combined_k141_187898\|\|full | Low-quality |
| 17-109-final-viral-combined_k141_86952\|\|full | Low-quality |
| 17-109-final-viral-combined_k141_137325\|\|full | Not-determined |
| 17-109-final-viral-combined_k141_204258\|\|full | Not-determined |
| 17-109-final-viral-combined_k141_3499\|\|full | Not-determined |
| 17-109-final-viral-combined_k141_86967\|\|full | Low-quality |
| 17-109-final-viral-combined_k141_220708\|\|full | Low-quality |
| 17-109-final-viral-combined_k141_304942\|\|full | Not-determined |
| 17-109-final-viral-combined_k141_354622\|\|full | Not-determined |
| 17-109-final-viral-combined_k141_321372\|\|full | Not-determined |
| 17-109-final-viral-combined_k141_70719\|\|full | Low-quality |
| 17-109-final-viral-combined_k141_388371\|\|full | Not-determined |
| 17-109-final-viral-combined_k141_86985\|\|full | Not-determined |
| 17-109-final-viral-combined_k141_103519\|\|full | Not-determined |
| 17-109-final-viral-combined_k141_288124\|\|full | Low-quality |
| 17-109-final-viral-combined_k141_304975\|\|full | Low-quality |
| 17-109-final-viral-combined_k141_321413\|\|full | Low-quality |
| 17-109-final-viral-combined_k141_170860\|\|full | Low-quality |
| 17-109-final-viral-combined_k141_53532\|\|full | Not-determined |
| 17-109-final-viral-combined_k141_170877\|\|full | Low-quality |
| 17-109-final-viral-combined_k141_187986\|\|full | Low-quality |
| 17-109-final-viral-combined_k141_220734\|\|full | Low-quality |
| 17-109-final-viral-combined_k141_220740\|\|full | Low-quality |
| 17-109-final-viral-combined_k141_103579\|\|full | Low-quality |
| 17-109-final-viral-combined_k141_254037\|\|full | Not-determined |
| 17-109-final-viral-combined_k141_305029\|\|full | Not-determined |
| 17-109-final-viral-combined_k141_254053\|\|full | Not-determined |
| 17-109-final-viral-combined_k141_371844\|\|full | Low-quality |
| 17-109-final-viral-combined_k141_371845\|\|full | Not-determined |
| 17-109-final-viral-combined_k141_270984\|\|full | Low-quality |
| 17-109-final-viral-combined_k141_103626\|\|full | Not-determined |
| 17-109-final-viral-combined_k141_53561\|\|full | Not-determined |
| 17-109-final-viral-combined_k141_170934\|\|full | Low-quality |
| 17-109-final-viral-combined_k141_70768\|\|full | Low-quality |
| 17-109-final-viral-combined_k141_3581\|\|full | Low-quality |
| 17-109-final-viral-combined_k141_70770\|\|full | Low-quality |
| 17-109-final-viral-combined_k141_321452\|\|full | Low-quality |
| 17-109-final-viral-combined_k141_220775\|\|full | Not-determined |
| 17-109-final-viral-combined_k141_271017\|\|full | Low-quality |
| 17-109-final-viral-combined_k141_170956\|\|full | Low-quality |
| 17-109-final-viral-combined_k141_337697\|\|full | Not-determined |
| 17-109-final-viral-combined_k141_137414\|\|full | Low-quality |
| 17-109-final-viral-combined_k141_237715\|\|full | Not-determined |
| 17-109-final-viral-combined_k141_3600\|\|full | Not-determined |
| 17-109-final-viral-combined_k141_288235\|\|full | Not-determined |
| 17-109-final-viral-combined_k141_19935\|\|full | Not-determined |
| 17-109-final-viral-combined_k141_154136\|\|full | Low-quality |
| 17-109-final-viral-combined_k141_37181\|\|full | Not-determined |
| 17-109-final-viral-combined_k141_204341\|\|full | Low-quality |
| 17-109-final-viral-combined_k141_220811\|\|full | Not-determined |
| 17-109-final-viral-combined_k141_154145\|\|full | Low-quality |
| 17-109-final-viral-combined_k141_3634\|\|full | Low-quality |
| 17-109-final-viral-combined_k141_53595\|\|full | Low-quality |
| 17-109-final-viral-combined_k141_305153\|\|full | Low-quality |
| 17-109-final-viral-combined_k141_170975\|\|full | Not-determined |
| 17-109-final-viral-combined_k141_103695\|\|full | Not-determined |
| 17-109-final-viral-combined_k141_288263\|\|full | Not-determined |
| 17-109-final-viral-combined_k141_288271\|\|full | Not-determined |
| 17-109-final-viral-combined_k141_103708\|\|full | Not-determined |
| 17-109-final-viral-combined_k141_188065\|\|full | Not-determined |
| 17-109-final-viral-combined_k141_321501\|\|full | Not-determined |
| 17-109-final-viral-combined_k141_288287\|\|full | Not-determined |
| 17-109-final-viral-combined_k141_321519\|\|full | Not-determined |
| 17-109-final-viral-combined_k141_3675\|\|full | Low-quality |
| 17-109-final-viral-combined_k141_371914\|\|full | Low-quality |
| 17-109-final-viral-combined_k141_388496\|\|full | Low-quality |
| 17-109-final-viral-combined_k141_271067\|\|full | Low-quality |
| 17-109-final-viral-combined_k141_371925\|\|full | Not-determined |
| 17-109-final-viral-combined_k141_103719\|\|full | Low-quality |
| 17-109-final-viral-combined_k141_305195\|\|full | Low-quality |
| 17-109-final-viral-combined_k141_120161\|\|full | Low-quality |
| 17-109-final-viral-combined_k141_337768\|\|full | Low-quality |
| 17-109-final-viral-combined_k141_87115\|\|full | Not-determined |
| 17-109-final-viral-combined_k141_254145\|\|full | Low-quality |
| 17-109-final-viral-combined_k141_204360\|\|full | Low-quality |
| 17-109-final-viral-combined_k141_154181\|\|full | Not-determined |
| 17-109-final-viral-combined_k141_254150\|\|full | Not-determined |
| 17-109-final-viral-combined_k141_271086\|\|full | Not-determined |
| 17-109-final-viral-combined_k141_321567\|\|full | Low-quality |
| 17-109-final-viral-combined_k141_3702\|\|full | Not-determined |
| 17-109-final-viral-combined_k141_204381\|\|full | Not-determined |
| 17-109-final-viral-combined_k141_137466\|\|full | Low-quality |
| 17-109-final-viral-combined_k141_154207\|\|full | Low-quality |
| 17-109-final-viral-combined_k141_53608\|\|full | Not-determined |
| 17-109-final-viral-combined_k141_354704\|\|full | Low-quality |
| 17-109-final-viral-combined_k141_337820\|\|full | Not-determined |
| 17-109-final-viral-combined_k141_271113\|\|full | Not-determined |
| 17-109-final-viral-combined_k141_137479\|\|full | Low-quality |
| 17-109-final-viral-combined_k141_337827\|\|full | Low-quality |
| 17-109-final-viral-combined_k141_271114\|\|full | Low-quality |
| 17-109-final-viral-combined_k141_337828\|\|full | Low-quality |
| 17-109-final-viral-combined_k141_120220\|\|full | Not-determined |
| 17-109-final-viral-combined_k141_271116\|\|full | Not-determined |
| 17-109-final-viral-combined_k141_371976\|\|full | Not-determined |
| 17-109-final-viral-combined_k141_137486\|\|full | Low-quality |
| 17-109-final-viral-combined_k141_237789\|\|full | Low-quality |
| 17-109-final-viral-combined_k141_337830\|\|full | Low-quality |
| 17-109-final-viral-combined_k141_53615\|\|full | Not-determined |
| 17-109-final-viral-combined_k141_354725\|\|full | Not-determined |
| 17-109-final-viral-combined_k141_271126\|\|full | Not-determined |
| 17-109-final-viral-combined_k141_20083\|\|full | Not-determined |
| 17-109-final-viral-combined_k141_220880\|\|full | Not-determined |
| 17-109-final-viral-combined_k141_321620\|\|full | Low-quality |
| 17-109-final-viral-combined_k141_371996\|\|full | Not-determined |
| 17-109-final-viral-combined_k141_188086\|\|full | Not-determined |
| 17-109-final-viral-combined_k141_354730\|\|full | Low-quality |
| 17-109-final-viral-combined_k141_305248\|\|full | Not-determined |
| 17-109-final-viral-combined_k141_288353\|\|full | Not-determined |
| 17-109-final-viral-combined_k141_20114\|\|full | Not-determined |
| 17-109-final-viral-combined_k141_220898\|\|full | Low-quality |
| 17-109-final-viral-combined_k141_345948\|\|full | Medium-quality |
| 17-109-final-viral-combined_k141_137512\|\|full | Low-quality |
| 17-109-final-viral-combined_k141_87171\|\|full | Low-quality |
| 17-109-final-viral-combined_k141_288373\|\|full | Not-determined |
| 17-109-final-viral-combined_k141_37271\|\|full | Not-determined |
| 17-109-final-viral-combined_k141_388574\|\|full | Not-determined |
| 17-109-final-viral-combined_k141_204468\|\|full | Not-determined |
| 17-109-final-viral-combined_k141_103806\|\|full | Not-determined |
| 17-109-final-viral-combined_k141_103812\|\|full | Low-quality |
| 17-109-final-viral-combined_k141_220930\|\|full | Low-quality |
| 17-109-final-viral-combined_k141_305284\|\|full | Not-determined |
| 17-109-final-viral-combined_k141_354785\|\|full | Not-determined |
| 17-109-final-viral-combined_k141_337911\|\|full | Low-quality |
| 17-109-final-viral-combined_k141_154278\|\|full | Low-quality |
| 17-109-final-viral-combined_k141_53701\|\|full | Low-quality |
| 17-109-final-viral-combined_k141_220936\|\|full | Low-quality |
| 17-109-final-viral-combined_k141_20173\|\|full | Low-quality |
| 17-109-final-viral-combined_k141_254232\|\|full | Not-determined |
| 17-109-final-viral-combined_k141_372071\|\|full | Not-determined |
| 17-109-final-viral-combined_k141_188137\|\|full | Not-determined |
| 17-109-final-viral-combined_k141_103826\|\|full | Low-quality |
| 17-109-final-viral-combined_k141_337935\|\|full | Not-determined |
| 17-109-final-viral-combined_k141_237865\|\|full | Not-determined |
| 17-109-final-viral-combined_k141_237871\|\|full | Low-quality |
| 17-109-final-viral-combined_k141_354794\|\|full | Not-determined |
| 17-109-final-viral-combined_k141_288487\|\|full | Not-determined |
| 17-109-final-viral-combined_k141_354796\|\|full | Not-determined |
| 17-109-final-viral-combined_k141_372110\|\|full | Not-determined |
| 17-109-final-viral-combined_k141_103856\|\|full | Not-determined |
| 17-109-final-viral-combined_k141_388665\|\|full | Low-quality |
| 17-109-final-viral-combined_k141_3822\|\|full | Not-determined |
| 17-109-final-viral-combined_k141_337974\|\|full | Low-quality |
| 17-109-final-viral-combined_k141_70956\|\|full | Low-quality |
| 17-109-final-viral-combined_k141_388678\|\|full | Not-determined |
| 17-109-final-viral-combined_k141_337979\|\|full | Not-determined |
| 17-109-final-viral-combined_k141_20224\|\|full | Not-determined |
| 17-109-final-viral-combined_k141_20230\|\|full | Low-quality |
| 17-109-final-viral-combined_k141_188215\|\|full | Low-quality |
| 17-109-final-viral-combined_k141_237922\|\|full | Not-determined |
| 17-109-final-viral-combined_k141_37350\|\|full | Low-quality |
| 17-109-final-viral-combined_k141_337999\|\|full | Low-quality |
| 17-109-final-viral-combined_k141_305373\|\|full | Low-quality |
| 17-109-final-viral-combined_k141_372138\|\|full | Not-determined |
| 17-109-final-viral-combined_k141_372146\|\|full | Low-quality |
| 17-109-final-viral-combined_k141_188232\|\|full | Not-determined |
| 17-109-final-viral-combined_k141_103866\|\|full | Not-determined |
| 17-109-final-viral-combined_k141_254321\|\|full | Not-determined |
| 17-109-final-viral-combined_k141_305384\|\|full | Low-quality |
| 17-109-final-viral-combined_k141_288525\|\|full | Low-quality |
| 17-109-final-viral-combined_k141_271250\|\|full | Low-quality |
| 17-109-final-viral-combined_k141_271260\|\|full | Not-determined |
| 17-109-final-viral-combined_k141_220976\|\|full | Low-quality |
| 17-109-final-viral-combined_k141_372155\|\|full | Not-determined |
| 17-109-final-viral-combined_k141_220979\|\|full | Not-determined |
| 17-109-final-viral-combined_k141_20279\|\|full | Not-determined |
| 17-109-final-viral-combined_k141_388713\|\|full | Not-determined |
| 17-109-final-viral-combined_k141_71004\|\|full | Not-determined |
| 17-109-final-viral-combined_k141_3824\|\|full | Low-quality |
| 17-109-final-viral-combined_k141_221016\|\|full | Low-quality |
| 17-109-final-viral-combined_k141_87271\|\|full | Not-determined |
| 17-109-final-viral-combined_k141_354878\|\|full | Low-quality |
| 17-109-final-viral-combined_k141_321767\|\|full | Not-determined |
| 17-109-final-viral-combined_k141_354884\|\|full | Not-determined |
| 17-109-final-viral-combined_k141_388740\|\|full | Not-determined |
| 17-109-final-viral-combined_k141_120319\|\|full | Low-quality |
| 17-109-final-viral-combined_k141_388751\|\|full | Not-determined |
| 17-109-final-viral-combined_k141_321788\|\|full | Not-determined |
| 17-109-final-viral-combined_k141_87289\|\|full | Low-quality |
| 17-109-final-viral-combined_k141_87292\|\|full | Not-determined |
| 17-109-final-viral-combined_k141_103933\|\|full | Not-determined |
| 17-109-final-viral-combined_k141_305461\|\|full | Low-quality |
| 17-109-final-viral-combined_k141_188344\|\|full | Low-quality |
| 17-109-final-viral-combined_k141_354900\|\|full | Low-quality |
| 17-109-final-viral-combined_k141_188346\|\|full | Low-quality |
| 17-109-final-viral-combined_k141_388777\|\|full | Low-quality |
| 17-109-final-viral-combined_k141_87303\|\|full | Low-quality |
| 17-109-final-viral-combined_k141_338091\|\|full | Not-determined |
| 17-109-final-viral-combined_k141_238042\|\|full | Not-determined |
| 17-109-final-viral-combined_k141_120350\|\|full | Low-quality |
| 17-109-final-viral-combined_k141_37379\|\|full | Low-quality |
| 17-109-final-viral-combined_k141_87308\|\|full | Low-quality |
| 17-109-final-viral-combined_k141_171076\|\|full | Low-quality |
| 17-109-final-viral-combined_k141_238049\|\|full | Not-determined |
| 17-109-final-viral-combined_k141_53850\|\|full | Not-determined |
| 17-109-final-viral-combined_k141_238052\|\|full | Not-determined |
| 17-109-final-viral-combined_k141_154428\|\|full | Low-quality |
| 17-109-final-viral-combined_k141_53857\|\|full | Not-determined |
| 17-109-final-viral-combined_k141_37386\|\|full | Not-determined |
| 17-109-final-viral-combined_k141_154431\|\|full | Not-determined |
| 17-109-final-viral-combined_k141_37389\|\|full | Low-quality |
| 17-109-final-viral-combined_k141_305468\|\|full | Low-quality |
| 17-109-final-viral-combined_k141_137731\|\|full | Low-quality |
| 17-109-final-viral-combined_k141_305472\|\|full | Not-determined |
| 17-109-final-viral-combined_k141_238069\|\|full | Not-determined |
| 17-109-final-viral-combined_k141_37407\|\|full | Not-determined |
| 17-109-final-viral-combined_k141_271398\|\|full | Low-quality |
| 17-109-final-viral-combined_k141_271400\|\|full | Low-quality |
| 17-109-final-viral-combined_k141_338127\|\|full | Not-determined |
| 17-109-final-viral-combined_k141_372234\|\|full | Not-determined |
| 17-109-final-viral-combined_k141_238083\|\|full | Low-quality |
| 17-109-final-viral-combined_k141_188354\|\|full | Low-quality |
| 17-109-final-viral-combined_k141_20393\|\|full | Low-quality |
| 17-109-final-viral-combined_k141_53886\|\|full | Low-quality |
| 17-109-final-viral-combined_k141_53888\|\|full | Low-quality |
| 17-109-final-viral-combined_k141_321842\|\|full | Low-quality |
| 17-109-final-viral-combined_k141_221051\|\|full | Low-quality |
| 17-109-final-viral-combined_k141_20412\|\|full | Not-determined |
| 17-109-final-viral-combined_k141_221052\|\|full | Not-determined |
| 17-109-final-viral-combined_k141_71093\|\|full | Low-quality |
| 17-109-final-viral-combined_k141_188381\|\|full | Not-determined |
| 17-109-final-viral-combined_k141_338156\|\|full | Not-determined |
| 17-109-final-viral-combined_k141_372250\|\|full | Low-quality |
| 17-109-final-viral-combined_k141_53903\|\|full | Not-determined |
| 17-109-final-viral-combined_k141_271436\|\|full | Low-quality |
| 17-109-final-viral-combined_k141_354968\|\|full | Not-determined |
| 17-109-final-viral-combined_k141_221066\|\|full | Not-determined |
| 17-109-final-viral-combined_k141_388860\|\|full | Not-determined |
| 17-109-final-viral-combined_k141_271448\|\|full | Low-quality |
| 17-109-final-viral-combined_k141_321863\|\|full | Not-determined |
| 17-109-final-viral-combined_k141_238144\|\|full | Low-quality |
| 17-109-final-viral-combined_k141_388869\|\|full | Low-quality |
| 17-109-final-viral-combined_k141_188407\|\|full | Low-quality |
| 17-109-final-viral-combined_k141_53919\|\|full | Not-determined |
| 17-109-final-viral-combined_k141_204538\|\|full | Not-determined |
| 17-109-final-viral-combined_k141_87367\|\|full | Not-determined |
| 17-109-final-viral-combined_k141_171190\|\|full | Not-determined |
| 17-109-final-viral-combined_k141_372279\|\|full | Low-quality |
| 17-109-final-viral-combined_k141_221105\|\|full | Not-determined |
| 17-109-final-viral-combined_k141_104011\|\|full | Low-quality |
| 17-109-final-viral-combined_k141_188429\|\|full | Low-quality |
| 17-109-final-viral-combined_k141_355014\|\|full | Low-quality |
| 17-109-final-viral-combined_k141_104014\|\|full | Not-determined |
| 17-109-final-viral-combined_k141_53950\|\|full | Not-determined |
| 17-109-final-viral-combined_k141_87392\|\|full | Not-determined |
| 17-109-final-viral-combined_k141_254407\|\|full | Not-determined |
| 17-109-final-viral-combined_k141_137819\|\|full | Low-quality |
| 17-109-final-viral-combined_k141_372283\|\|full | Low-quality |
| 17-109-final-viral-combined_k141_388909\|\|full | Not-determined |
| 17-109-final-viral-combined_k141_87408\|\|full | Low-quality |
| 17-109-final-viral-combined_k141_271497\|\|full | Not-determined |
| 17-109-final-viral-combined_k141_238215\|\|full | Low-quality |
| 17-109-final-viral-combined_k141_221148\|\|full | Not-determined |
| 17-109-final-viral-combined_k141_137842\|\|full | Not-determined |
| 17-109-final-viral-combined_k141_238224\|\|full | Not-determined |
| 17-109-final-viral-combined_k141_137853\|\|full | Low-quality |
| 17-109-final-viral-combined_k141_321951\|\|full | Not-determined |
| 17-109-final-viral-combined_k141_120476\|\|full | Not-determined |
| 17-109-final-viral-combined_k141_238242\|\|full | Low-quality |
| 17-109-final-viral-combined_k141_321953\|\|full | Not-determined |
| 17-109-final-viral-combined_k141_20486\|\|full | Not-determined |
| 17-109-final-viral-combined_k141_53993\|\|full | Low-quality |
| 17-109-final-viral-combined_k141_355025\|\|full | Not-determined |
| 17-109-final-viral-combined_k141_20491\|\|full | Low-quality |
| 17-109-final-viral-combined_k141_321967\|\|full | Low-quality |
| 17-109-final-viral-combined_k141_221173\|\|full | Not-determined |
| 17-109-final-viral-combined_k141_204583\|\|full | Not-determined |
| 17-109-final-viral-combined_k141_338233\|\|full | Not-determined |
| 17-109-final-viral-combined_k141_137877\|\|full | Low-quality |
| 17-109-final-viral-combined_k141_20507\|\|full | Not-determined |
| 17-109-final-viral-combined_k141_71200\|\|full | Low-quality |
| 17-109-final-viral-combined_k141_338243\|\|full | Not-determined |
| 17-109-final-viral-combined_k141_137896\|\|full | Low-quality |
| 17-109-final-viral-combined_k141_305576\|\|full | Not-determined |
| 17-109-final-viral-combined_k141_3963\|\|full | Low-quality |
| 17-109-final-viral-combined_k141_171266\|\|full | Low-quality |
| 17-109-final-viral-combined_k141_137901\|\|full | Not-determined |
| 17-109-final-viral-combined_k141_71228\|\|full | Not-determined |
| 17-109-final-viral-combined_k141_271562\|\|full | Not-determined |
| 17-109-final-viral-combined_k141_87459\|\|full | Not-determined |
| 17-109-final-viral-combined_k141_154508\|\|full | Not-determined |
| 17-109-final-viral-combined_k141_305598\|\|full | Not-determined |
| 17-109-final-viral-combined_k141_104098\|\|full | Not-determined |
| 17-109-final-viral-combined_k141_305606\|\|full | Low-quality |
| 17-109-final-viral-combined_k141_288608\|\|full | Not-determined |
| 17-109-final-viral-combined_k141_54021\|\|full | Not-determined |
| 17-109-final-viral-combined_k141_104108\|\|full | Not-determined |
| 17-109-final-viral-combined_k141_204644\|\|full | Low-quality |
| 18-106-final-viral-combined_k141_110950\|\|full | Medium-quality |
| 17-109-final-viral-combined_k141_154532\|\|full | Low-quality |
| 17-109-final-viral-combined_k141_137917\|\|full | Not-determined |
| 17-109-final-viral-combined_k141_221226\|\|full | Not-determined |
| 17-109-final-viral-combined_k141_288621\|\|full | Low-quality |
| 17-109-final-viral-combined_k141_338284\|\|full | Not-determined |
| 17-109-final-viral-combined_k141_154548\|\|full | Low-quality |
| 17-109-final-viral-combined_k141_3997\|\|full | Not-determined |
| 17-109-final-viral-combined_k141_355063\|\|full | Not-determined |
| 17-109-final-viral-combined_k141_372399\|\|full | Low-quality |
| 17-109-final-viral-combined_k141_204676\|\|full | Not-determined |
| 17-109-final-viral-combined_k141_322049\|\|full | Not-determined |
| 17-109-final-viral-combined_k141_305612\|\|full | Low-quality |
| 17-109-final-viral-combined_k141_71259\|\|full | Not-determined |
| 17-109-final-viral-combined_k141_305618\|\|full | Low-quality |
| 17-109-final-viral-combined_k141_305627\|\|full | Low-quality |
| 17-109-final-viral-combined_k141_87509\|\|full | Not-determined |
| 17-109-final-viral-combined_k141_188569\|\|full | Not-determined |
| 17-109-final-viral-combined_k141_254524\|\|full | Low-quality |
| 17-109-final-viral-combined_k141_355068\|\|full | Not-determined |
| 17-109-final-viral-combined_k141_87515\|\|full | Low-quality |
| 17-109-final-viral-combined_k141_104174\|\|full | Not-determined |
| 17-109-final-viral-combined_k141_355072\|\|full | Not-determined |
| 17-109-final-viral-combined_k141_305653\|\|full | Low-quality |
| 17-109-final-viral-combined_k141_254541\|\|full | Not-determined |
| 17-109-final-viral-combined_k141_87524\|\|full | Not-determined |
| 17-109-final-viral-combined_k141_171340\|\|full | Not-determined |
| 17-109-final-viral-combined_k141_305658\|\|full | Low-quality |
| 17-109-final-viral-combined_k141_104177\|\|full | Low-quality |
| 17-109-final-viral-combined_k141_54087\|\|full | Not-determined |
| 17-109-final-viral-combined_k141_154611\|\|full | Not-determined |
| 17-109-final-viral-combined_k141_137979\|\|full | Low-quality |
| 17-109-final-viral-combined_k141_104192\|\|full | Low-quality |
| 17-109-final-viral-combined_k141_4061\|\|full | Low-quality |
| 17-109-final-viral-combined_k141_372485\|\|full | Low-quality |
| 17-109-final-viral-combined_k141_71295\|\|full | Not-determined |
| 17-109-final-viral-combined_k141_171357\|\|full | Not-determined |
| 17-109-final-viral-combined_k141_288681\|\|full | Not-determined |
| 17-109-final-viral-combined_k141_322121\|\|full | Not-determined |
| 17-109-final-viral-combined_k141_4080\|\|full | Not-determined |
| 17-109-final-viral-combined_k141_372497\|\|full | Not-determined |
| 17-109-final-viral-combined_k141_188576\|\|full | Not-determined |
| 17-109-final-viral-combined_k141_372511\|\|full | Not-determined |
| 17-109-final-viral-combined_k141_154640\|\|full | Not-determined |
| 17-109-final-viral-combined_k141_4103\|\|full | Low-quality |
| 17-109-final-viral-combined_k141_271674\|\|full | Not-determined |
| 17-109-final-viral-combined_k141_372536\|\|full | Low-quality |
| 17-109-final-viral-combined_k141_171379\|\|full | Low-quality |
| 17-109-final-viral-combined_k141_388990\|\|full | Low-quality |
| 17-109-final-viral-combined_k141_388991\|\|full | Low-quality |
| 17-109-final-viral-combined_k141_271702\|\|full | Low-quality |
| 17-109-final-viral-combined_k141_171396\|\|full | Not-determined |
| 17-109-final-viral-combined_k141_171402\|\|full | Not-determined |
| 17-109-final-viral-combined_k141_271705\|\|full | Not-determined |
| 17-109-final-viral-combined_k141_305696\|\|full | Low-quality |
| 17-109-final-viral-combined_k141_104221\|\|full | Not-determined |
| 17-109-final-viral-combined_k141_221321\|\|full | Low-quality |
| 17-109-final-viral-combined_k141_221326\|\|full | Not-determined |
| 17-109-final-viral-combined_k141_54173\|\|full | Not-determined |
| 17-109-final-viral-combined_k141_171430\|\|full | Not-determined |
| 17-109-final-viral-combined_k141_372584\|\|full | Not-determined |
| 17-109-final-viral-combined_k141_188638\|\|full | Low-quality |
| 17-109-final-viral-combined_k141_389049\|\|full | Low-quality |
| 17-109-final-viral-combined_k141_322207\|\|full | Low-quality |
| 17-109-final-viral-combined_k141_305735\|\|full | Not-determined |
| 17-109-final-viral-combined_k141_188646\|\|full | Not-determined |
| 17-109-final-viral-combined_k141_37620\|\|full | Low-quality |
| 17-109-final-viral-combined_k141_355162\|\|full | Not-determined |
| 17-109-final-viral-combined_k141_71454\|\|full | Low-quality |
| 17-109-final-viral-combined_k141_20611\|\|full | Not-determined |
| 17-109-final-viral-combined_k141_37631\|\|full | Low-quality |
| 17-109-final-viral-combined_k141_20612\|\|full | Low-quality |
| 17-109-final-viral-combined_k141_138082\|\|full | Low-quality |
| 17-109-final-viral-combined_k141_54211\|\|full | Not-determined |
| 17-109-final-viral-combined_k141_389079\|\|full | Not-determined |
| 17-109-final-viral-combined_k141_188685\|\|full | Not-determined |
| 17-109-final-viral-combined_k141_221382\|\|full | Low-quality |
| 17-109-final-viral-combined_k141_71489\|\|full | Low-quality |
| 17-109-final-viral-combined_k141_221392\|\|full | Not-determined |
| 17-109-final-viral-combined_k141_138113\|\|full | Low-quality |
| 17-109-final-viral-combined_k141_54226\|\|full | Not-determined |
| 17-109-final-viral-combined_k141_20633\|\|full | Not-determined |
| 17-109-final-viral-combined_k141_154699\|\|full | Low-quality |
| 17-109-final-viral-combined_k141_204839\|\|full | Not-determined |
| 17-109-final-viral-combined_k141_204852\|\|full | Low-quality |
| 17-109-final-viral-combined_k141_221414\|\|full | Not-determined |
| 17-109-final-viral-combined_k141_204854\|\|full | Low-quality |
| 17-109-final-viral-combined_k141_254701\|\|full | Low-quality |
| 17-109-final-viral-combined_k141_71521\|\|full | Not-determined |
| 17-109-final-viral-combined_k141_238402\|\|full | Not-determined |
| 17-109-final-viral-combined_k141_338509\|\|full | Low-quality |
| 17-109-final-viral-combined_k141_372641\|\|full | Low-quality |
| 17-109-final-viral-combined_k141_37699\|\|full | Low-quality |
| 17-109-final-viral-combined_k141_154703\|\|full | Low-quality |
| 17-109-final-viral-combined_k141_171502\|\|full | Not-determined |

17-109-final-viral-combined_k141_4138||full Not-determined
17-109-final-viral-combined_k141_271832||full Not-determined
17-109-final-viral-combined_k141_4139||full Low-quality
17-109-final-viral-combined_k141_238428||full Low-quality
17-109-final-viral-combined_k141_120783||full Not-determined
18-102-final-viral-combined_k141_78670||full Medium-quality
17-109-final-viral-combined_k141_254731||full Not-determined
17-109-final-viral-combined_k141_271843||full Not-determined
17-109-final-viral-combined_k141_71568||full Not-determined
17-109-final-viral-combined_k141_305823||full Low-quality
17-109-final-viral-combined_k141_188758||full Not-determined
17-109-final-viral-combined_k141_54255||full Low-quality
17-109-final-viral-combined_k141_271855||full Low-quality
17-109-final-viral-combined_k141_171529||full Not-determined
17-109-final-viral-combined_k141_154726||full Low-quality
17-109-final-viral-combined_k141_254747||full Low-quality
17-109-final-viral-combined_k141_271871||full Low-quality
17-109-final-viral-combined_k141_389187||full Low-quality
17-109-final-viral-combined_k141_254754||full Low-quality
17-109-final-viral-combined_k141_221490||full Not-determined
17-109-final-viral-combined_k141_138238||full Low-quality
17-109-final-viral-combined_k141_271882||full Low-quality
17-109-final-viral-combined_k141_188778||full Not-determined
17-109-final-viral-combined_k141_254763||full Not-determined
17-109-final-viral-combined_k141_372654||full Low-quality
17-109-final-viral-combined_k141_305852||full Low-quality
17-109-final-viral-combined_k141_338566||full Low-quality
17-109-final-viral-combined_k141_104306||full Not-determined
17-109-final-viral-combined_k141_221515||full Low-quality
17-109-final-viral-combined_k141_322302||full Low-quality
17-109-final-viral-combined_k141_37768||full Low-quality
17-109-final-viral-combined_k141_271905||full Low-quality
17-109-final-viral-combined_k141_338589||full Not-determined
17-109-final-viral-combined_k141_120861||full Low-quality
17-109-final-viral-combined_k141_271907||full Low-quality
17-109-final-viral-combined_k141_389214||full Not-determined
17-109-final-viral-combined_k141_322310||full Low-quality
17-109-final-viral-combined_k141_238454||full Not-determined
17-109-final-viral-combined_k141_288871||full Not-determined
17-109-final-viral-combined_k141_54265||full Not-determined
17-109-final-viral-combined_k141_338634||full Not-determined
17-109-final-viral-combined_k141_322335||full Not-determined
17-109-final-viral-combined_k141_338635||full Not-determined
17-109-final-viral-combined_k141_154821||full Low-quality
17-109-final-viral-combined_k141_338639||full Not-determined
17-109-final-viral-combined_k141_238463||full Low-quality
17-109-final-viral-combined_k141_305903||full Low-quality
17-109-final-viral-combined_k141_104364||full Not-determined
17-109-final-viral-combined_k141_138294||full Not-determined
17-109-final-viral-combined_k141_271951||full Not-determined
17-109-final-viral-combined_k141_221589||full Not-determined
17-109-final-viral-combined_k141_238475||full Not-determined
17-109-final-viral-combined_k141_37818||full Low-quality
17-109-final-viral-combined_k141_54303||full Not-determined
17-109-final-viral-combined_k141_154861||full Not-determined
17-109-final-viral-combined_k141_221613||full Low-quality
17-109-final-viral-combined_k141_120925||full Low-quality
17-109-final-viral-combined_k141_154872||full Not-determined
17-109-final-viral-combined_k141_20857||full Low-quality
17-109-final-viral-combined_k141_355358||full Not-determined
17-109-final-viral-combined_k141_138323||full Low-quality
17-109-final-viral-combined_k141_20861||full Low-quality
17-109-final-viral-combined_k141_87694||full Low-quality
17-109-final-viral-combined_k141_54322||full Not-determined
17-109-final-viral-combined_k141_254831||full Low-quality
17-109-final-viral-combined_k141_154892||full Not-determined
17-109-final-viral-combined_k141_254836||full Not-determined
17-109-final-viral-combined_k141_204964||full Low-quality
17-109-final-viral-combined_k141_54331||full Low-quality
17-109-final-viral-combined_k141_254840||full Low-quality
17-109-final-viral-combined_k141_389252||full Low-quality
17-109-final-viral-combined_k141_4192||full Low-quality
17-109-final-viral-combined_k141_120940||full Low-quality
17-109-final-viral-combined_k141_87728||full Low-quality
17-109-final-viral-combined_k141_372760||full Low-quality
17-109-final-viral-combined_k141_154924||full Not-determined
17-109-final-viral-combined_k141_338704||full Not-determined
17-109-final-viral-combined_k141_221661||full Not-determined
17-109-final-viral-combined_k141_120950||full Low-quality
17-109-final-viral-combined_k141_4196||full Low-quality
17-109-final-viral-combined_k141_389268||full Low-quality
17-109-final-viral-combined_k141_154933||full Not-determined
17-109-final-viral-combined_k141_372788||full Low-quality
17-109-final-viral-combined_k141_322401||full Low-quality
17-109-final-viral-combined_k141_271964||full Low-quality
17-109-final-viral-combined_k141_87756||full Low-quality
17-109-final-viral-combined_k141_138363||full Low-quality
17-109-final-viral-combined_k141_205020||full Not-determined
17-109-final-viral-combined_k141_389288||full Low-quality
17-109-final-viral-combined_k141_221685||full Low-quality
17-109-final-viral-combined_k141_120962||full Not-determined
17-109-final-viral-combined_k141_4234||full Not-determined
17-109-final-viral-combined_k141_205028||full Not-determined
17-109-final-viral-combined_k141_138380||full Not-determined
17-109-final-viral-combined_k141_322432||full Not-determined
17-109-final-viral-combined_k141_306014||full Low-quality
17-109-final-viral-combined_k141_120970||full Low-quality
17-109-final-viral-combined_k141_4237||full Low-quality
17-109-final-viral-combined_k141_20923||full Not-determined
17-109-final-viral-combined_k141_205050||full Not-determined
17-109-final-viral-combined_k141_54364||full Low-quality
17-109-final-viral-combined_k141_254860||full Low-quality
17-109-final-viral-combined_k141_355434||full Low-quality
17-109-final-viral-combined_k141_154970||full Not-determined
17-109-final-viral-combined_k141_71784||full Not-determined
17-109-final-viral-combined_k141_4266||full Not-determined
17-109-final-viral-combined_k141_221728||full Not-determined
17-109-final-viral-combined_k141_389316||full Not-determined
17-109-final-viral-combined_k141_338773||full Not-determined
17-109-final-viral-combined_k141_306058||full Not-determined
17-109-final-viral-combined_k141_306059||full Not-determined
17-109-final-viral-combined_k141_238517||full Not-determined
17-109-final-viral-combined_k141_338777||full Not-determined
17-109-final-viral-combined_k141_120980||full Not-determined
17-109-final-viral-combined_k141_254870||full Not-determined
17-109-final-viral-combined_k141_306070||full Low-quality
17-109-final-viral-combined_k141_54403||full Not-determined
17-109-final-viral-combined_k141_389348||full Not-determined
17-109-final-viral-combined_k141_238531||full Not-determined
17-109-final-viral-combined_k141_120983||full Low-quality
17-109-final-viral-combined_k141_37927||full Not-determined
17-109-final-viral-combined_k141_289003||full Low-quality
17-109-final-viral-combined_k141_272080||full Low-quality
17-109-final-viral-combined_k141_104439||full Not-determined
17-109-final-viral-combined_k141_188934||full Low-quality
17-109-final-viral-combined_k141_138438||full Not-determined
17-109-final-viral-combined_k141_338824||full Low-quality
17-109-final-viral-combined_k141_4299||full Not-determined
17-109-final-viral-combined_k141_138441||full Not-determined
17-109-final-viral-combined_k141_221828||full Low-quality
17-109-final-viral-combined_k141_389390||full Not-determined
17-109-final-viral-combined_k141_4306||full Low-quality
17-109-final-viral-combined_k141_21049||full Not-determined
17-109-final-viral-combined_k141_205084||full Not-determined
17-109-final-viral-combined_k141_54451||full Not-determined
17-109-final-viral-combined_k141_104458||full Not-determined
17-109-final-viral-combined_k141_355500||full Not-determined
17-109-final-viral-combined_k141_322545||full Not-determined
17-109-final-viral-combined_k141_171544||full Not-determined
17-109-final-viral-combined_k141_289063||full Low-quality
17-109-final-viral-combined_k141_21071||full Not-determined
17-109-final-viral-combined_k141_372914||full Low-quality
17-109-final-viral-combined_k141_54478||full Not-determined
17-109-final-viral-combined_k141_104478||full Low-quality
17-109-final-viral-combined_k141_154984||full Not-determined
17-109-final-viral-combined_k141_4343||full Low-quality
17-109-final-viral-combined_k141_21094||full Not-determined
17-109-final-viral-combined_k141_171569||full Low-quality
17-109-final-viral-combined_k141_289096||full Not-determined
17-109-final-viral-combined_k141_138509||full Low-quality
17-109-final-viral-combined_k141_205123||full Not-determined
17-109-final-viral-combined_k141_189008||full Not-determined
17-109-final-viral-combined_k141_372930||full Low-quality
17-109-final-viral-combined_k141_71826||full Not-determined
17-109-final-viral-combined_k141_21117||full Not-determined
17-109-final-viral-combined_k141_21121||full Low-quality
17-109-final-viral-combined_k141_87881||full Low-quality
17-109-final-viral-combined_k141_87884||full Low-quality
17-109-final-viral-combined_k141_389414||full Not-determined
17-109-final-viral-combined_k141_54497||full Low-quality
17-109-final-viral-combined_k141_87890||full Not-determined
17-109-final-viral-combined_k141_189039||full Not-determined
17-109-final-viral-combined_k141_238663||full Not-determined
17-109-final-viral-combined_k141_21136||full Low-quality
17-109-final-viral-combined_k141_389428||full Not-determined
17-109-final-viral-combined_k141_306204||full Not-determined
17-109-final-viral-combined_k141_37998||full Not-determined
17-109-final-viral-combined_k141_171633||full Low-quality
17-109-final-viral-combined_k141_38003||full Low-quality
17-109-final-viral-combined_k141_171635||full Low-quality
17-109-final-viral-combined_k141_238685||full Low-quality
17-109-final-viral-combined_k141_4400||full Low-quality
17-109-final-viral-combined_k141_389469||full Not-determined
17-109-final-viral-combined_k141_289150||full Low-quality
17-109-final-viral-combined_k141_306213||full Not-determined
17-109-final-viral-combined_k141_389476||full Low-quality
17-109-final-viral-combined_k141_306220||full Not-determined
17-109-final-viral-combined_k141_104575||full Not-determined
17-109-final-viral-combined_k141_272171||full Not-determined
17-109-final-viral-combined_k141_238723||full Not-determined
17-109-final-viral-combined_k141_121072||full Not-determined
17-109-final-viral-combined_k141_38043||full Low-quality
17-109-final-viral-combined_k141_87967||full Low-quality
17-109-final-viral-combined_k141_87972||full Not-determined
17-109-final-viral-combined_k141_155020||full Low-quality
17-109-final-viral-combined_k141_38068||full Low-quality
17-109-final-viral-combined_k141_272225||full Not-determined
17-109-final-viral-combined_k141_104637||full Low-quality
17-109-final-viral-combined_k141_338946||full Not-determined
17-109-final-viral-combined_k141_121100||full Not-determined
17-109-final-viral-combined_k141_4486||full Low-quality
17-109-final-viral-combined_k141_338951||full Low-quality
17-109-final-viral-combined_k141_255048||full Low-quality
17-109-final-viral-combined_k141_373038||full Not-determined
17-109-final-viral-combined_k141_289231||full Not-determined
17-109-final-viral-combined_k141_4487||full Low-quality
17-109-final-viral-combined_k141_88012||full Not-determined
17-109-final-viral-combined_k141_71914||full Low-quality
17-109-final-viral-combined_k141_289260||full Low-quality
17-109-final-viral-combined_k141_389562||full Not-determined
17-109-final-viral-combined_k141_322640||full Not-determined
17-109-final-viral-combined_k141_373080||full Not-determined
17-109-final-viral-combined_k141_289275||full Not-determined
17-109-final-viral-combined_k141_238776||full Low-quality
17-109-final-viral-combined_k141_21234||full Low-quality
17-109-final-viral-combined_k141_322656||full Not-determined
17-109-final-viral-combined_k141_339025||full Not-determined
17-109-final-viral-combined_k141_272308||full Not-determined
17-109-final-viral-combined_k141_88033||full Not-determined
17-109-final-viral-combined_k141_339030||full Low-quality
17-109-final-viral-combined_k141_189237||full Low-quality
17-109-final-viral-combined_k141_189239||full Low-quality
17-109-final-viral-combined_k141_238821||full Not-determined
17-109-final-viral-combined_k141_171737||full Not-determined
17-109-final-viral-combined_k141_306306||full Not-determined
17-109-final-viral-combined_k141_21270||full Low-quality
17-109-final-viral-combined_k141_88066||full Not-determined
17-109-final-viral-combined_k141_171745||full Not-determined
17-109-final-viral-combined_k141_88070||full Not-determined
17-109-final-viral-combined_k141_121123||full Low-quality
17-109-final-viral-combined_k141_88074||full Not-determined
17-109-final-viral-combined_k141_289338||full Not-determined
17-109-final-viral-combined_k141_21278||full Low-quality
17-109-final-viral-combined_k141_339049||full Low-quality
17-109-final-viral-combined_k141_38141||full Low-quality
17-109-final-viral-combined_k141_71969||full Low-quality
17-109-final-viral-combined_k141_373131||full Low-quality
17-109-final-viral-combined_k141_104736||full Not-determined
17-109-final-viral-combined_k141_138772||full Not-determined
17-109-final-viral-combined_k141_54615||full Low-quality
17-109-final-viral-combined_k141_355626||full Low-quality
17-109-final-viral-combined_k141_322747||full Not-determined
17-109-final-viral-combined_k141_355641||full Low-quality
17-109-final-viral-combined_k141_255118||full Not-determined
17-109-final-viral-combined_k141_104793||full Low-quality
17-109-final-viral-combined_k141_4539||full Not-determined
17-109-final-viral-combined_k141_54656||full Low-quality
17-109-final-viral-combined_k141_4542||full Not-determined
17-109-final-viral-combined_k141_322764||full Low-quality
17-109-final-viral-combined_k141_54658||full Not-determined
17-109-final-viral-combined_k141_272355||full Low-quality
17-109-final-viral-combined_k141_155097||full Low-quality
17-109-final-viral-combined_k141_189248||full Not-determined
17-109-final-viral-combined_k141_4544||full Not-determined
17-109-final-viral-combined_k141_238870||full Not-determined
17-109-final-viral-combined_k141_306396||full Low-quality
17-109-final-viral-combined_k141_306397||full Low-quality
17-109-final-viral-combined_k141_21317||full Not-determined
17-109-final-viral-combined_k141_72032||full Low-quality
17-109-final-viral-combined_k141_289462||full Not-determined
17-109-final-viral-combined_k141_255164||full Low-quality
17-109-final-viral-combined_k141_72036||full Low-quality
17-109-final-viral-combined_k141_238890||full Not-determined
17-109-final-viral-combined_k141_373148||full Not-determined
17-109-final-viral-combined_k141_205350||full Low-quality
17-109-final-viral-combined_k141_322793||full Not-determined
17-109-final-viral-combined_k141_373179||full Not-determined
17-109-final-viral-combined_k141_339100||full Not-determined
17-109-final-viral-combined_k141_272413||full Low-quality
17-109-final-viral-combined_k141_306437||full Not-determined
17-109-final-viral-combined_k141_238919||full Low-quality
17-109-final-viral-combined_k141_322820||full Low-quality
17-109-final-viral-combined_k141_272428||full Not-determined
17-109-final-viral-combined_k141_289529||full Not-determined
17-109-final-viral-combined_k141_272441||full Not-determined
17-109-final-viral-combined_k141_255243||full Low-quality
17-109-final-viral-combined_k141_255245||full Low-quality
17-109-final-viral-combined_k141_289546||full Low-quality
17-109-final-viral-combined_k141_373204||full Low-quality
17-109-final-viral-combined_k141_88126||full Low-quality
17-109-final-viral-combined_k141_205402||full Low-quality
17-109-final-viral-combined_k141_155180||full Low-quality
17-109-final-viral-combined_k141_289567||full Low-quality
17-109-final-viral-combined_k141_121243||full Not-determined
17-109-final-viral-combined_k141_355701||full Not-determined
17-109-final-viral-combined_k141_272478||full Low-quality
17-109-final-viral-combined_k141_222049||full Not-determined
17-109-final-viral-combined_k141_155196||full Low-quality
17-109-final-viral-combined_k141_72132||full Not-determined
17-109-final-viral-combined_k141_155202||full Not-determined
17-109-final-viral-combined_k141_289576||full Low-quality
17-109-final-viral-combined_k141_205415||full Low-quality
17-109-final-viral-combined_k141_155210||full Not-determined
17-109-final-viral-combined_k141_155217||full Not-determined
17-109-final-viral-combined_k141_38315||full Low-quality
17-109-final-viral-combined_k141_322860||full Low-quality
17-109-final-viral-combined_k141_88161||full Low-quality
17-109-final-viral-combined_k141_339152||full Low-quality
17-109-final-viral-combined_k141_21405||full Low-quality
17-109-final-viral-combined_k141_255292||full Low-quality
17-109-final-viral-combined_k141_155236||full Not-determined
17-109-final-viral-combined_k141_289604||full Not-determined
17-109-final-viral-combined_k141_171915||full Not-determined
17-109-final-viral-combined_k141_38330||full Low-quality
17-109-final-viral-combined_k141_38335||full Low-quality
17-109-final-viral-combined_k141_272503||full Low-quality
17-109-final-viral-combined_k141_322886||full Not-determined
18-105-final-viral-combined_k141_3201||full Medium-quality
17-109-final-viral-combined_k141_171918||full Not-determined
17-109-final-viral-combined_k141_322892||full Not-determined
17-109-final-viral-combined_k141_72136||full Not-determined
17-109-final-viral-combined_k141_205465||full Low-quality
17-109-final-viral-combined_k141_155263||full Low-quality
17-109-final-viral-combined_k141_322906||full Low-quality
17-109-final-viral-combined_k141_389739||full Low-quality
17-109-final-viral-combined_k141_255313||full Not-determined
17-109-final-viral-combined_k141_38355||full Low-quality
17-109-final-viral-combined_k141_104953||full Not-determined
17-109-final-viral-combined_k141_155286||full Not-determined
17-109-final-viral-combined_k141_339179||full Not-determined
17-109-final-viral-combined_k141_155299||full Low-quality
17-109-final-viral-combined_k141_104972||full Low-quality
17-109-final-viral-combined_k141_189357||full Not-determined
17-109-final-viral-combined_k141_339185||full Not-determined
17-109-final-viral-combined_k141_389763||full Low-quality
17-109-final-viral-combined_k141_322930||full Low-quality
17-109-final-viral-combined_k141_38376||full Not-determined
17-109-final-viral-combined_k141_272528||full Not-determined
17-109-final-viral-combined_k141_389767||full Not-determined
17-109-final-viral-combined_k141_255327||full Not-determined
17-109-final-viral-combined_k141_389771||full Not-determined
17-109-final-viral-combined_k141_373375||full Not-determined
17-109-final-viral-combined_k141_155308||full Low-quality
17-109-final-viral-combined_k141_389774||full Not-determined
17-109-final-viral-combined_k141_105003||full Not-determined
17-109-final-viral-combined_k141_189367||full Not-determined
17-109-final-viral-combined_k141_105010||full Not-determined
17-109-final-viral-combined_k141_255343||full Low-quality
17-109-final-viral-combined_k141_138882||full Low-quality
17-109-final-viral-combined_k141_138893||full Low-quality
17-109-final-viral-combined_k141_373407||full Not-determined
17-109-final-viral-combined_k141_105041||full Not-determined
17-109-final-viral-combined_k141_339232||full Low-quality
17-109-final-viral-combined_k141_21483||full Not-determined
17-109-final-viral-combined_k141_38438||full Low-quality
17-109-final-viral-combined_k141_373418||full Not-determined
17-109-final-viral-combined_k141_171953||full Low-quality
17-109-final-viral-combined_k141_306484||full Low-quality
17-109-final-viral-combined_k141_355794||full Low-quality
17-109-final-viral-combined_k141_255397||full Low-quality
17-109-final-viral-combined_k141_155358||full Low-quality
17-109-final-viral-combined_k141_306496||full Low-quality
17-109-final-viral-combined_k141_306500||full Low-quality
17-109-final-viral-combined_k141_289662||full Low-quality
17-109-final-viral-combined_k141_138921||full Not-determined
17-109-final-viral-combined_k141_38465||full Not-determined
17-109-final-viral-combined_k141_323003||full Low-quality
17-109-final-viral-combined_k141_105090||full Low-quality
17-109-final-viral-combined_k141_289688||full Low-quality
17-109-final-viral-combined_k141_155378||full Low-quality
17-109-final-viral-combined_k141_323024||full Not-determined
17-109-final-viral-combined_k141_255417||full Low-quality
17-109-final-viral-combined_k141_272594||full Not-determined
17-109-final-viral-combined_k141_155385||full Not-determined
17-109-final-viral-combined_k141_54735||full Not-determined
17-109-final-viral-combined_k141_389813||full Not-determined
17-109-final-viral-combined_k141_88246||full Low-quality
17-109-final-viral-combined_k141_355841||full Not-determined
17-109-final-viral-combined_k141_105111||full Not-determined
17-109-final-viral-combined_k141_289716||full Low-quality
17-109-final-viral-combined_k141_155410||full Low-quality
17-109-final-viral-combined_k141_72256||full Not-determined
17-109-final-viral-combined_k141_105165||full Not-determined
17-109-final-viral-combined_k141_389848||full Low-quality
17-109-final-viral-combined_k141_255467||full Not-determined
17-109-final-viral-combined_k141_389856||full Not-determined
17-109-final-viral-combined_k141_155435||full Not-determined
18-110-final-viral-combined_k141_239791||full High-quality
17-109-final-viral-combined_k141_306574||full Low-quality
17-109-final-viral-combined_k141_21506||full Not-determined
17-109-final-viral-combined_k141_339346||full Low-quality
17-109-final-viral-combined_k141_21513||full Low-quality
17-109-final-viral-combined_k141_323100||full Not-determined
17-109-final-viral-combined_k141_88266||full Not-determined
17-109-final-viral-combined_k141_189535||full Low-quality
17-109-final-viral-combined_k141_355897||full Low-quality
17-109-final-viral-combined_k141_339379||full Not-determined
17-109-final-viral-combined_k141_171999||full Not-determined
17-109-final-viral-combined_k141_373554||full Low-quality
17-109-final-viral-combined_k141_373558||full Low-quality
17-109-final-viral-combined_k141_306622||full Not-determined
17-109-final-viral-combined_k141_88289||full Low-quality
17-109-final-viral-combined_k141_323121||full Not-determined
17-109-final-viral-combined_k141_88301||full Low-quality
17-109-final-viral-combined_k141_4670||full Low-quality
17-109-final-viral-combined_k141_323134||full Low-quality
17-109-final-viral-combined_k141_323135||full Low-quality
17-109-final-viral-combined_k141_4682||full Not-determined
17-109-final-viral-combined_k141_389889||full Low-quality
17-109-final-viral-combined_k141_4686||full Not-determined
17-109-final-viral-combined_k141_155504||full Not-determined
17-109-final-viral-combined_k141_54795||full Low-quality
17-109-final-viral-combined_k141_38581||full Low-quality
17-109-final-viral-combined_k141_222204||full Low-quality
17-109-final-viral-combined_k141_189593||full Not-determined
17-109-final-viral-combined_k141_323151||full Low-quality
17-109-final-viral-combined_k141_155524||full Low-quality
17-109-final-viral-combined_k141_88330||full Not-determined
17-109-final-viral-combined_k141_323156||full Low-quality
17-109-final-viral-combined_k141_38593||full Not-determined
17-109-final-viral-combined_k141_339434||full Not-determined
17-109-final-viral-combined_k141_373572||full Low-quality
17-109-final-viral-combined_k141_4723||full Not-determined
17-109-final-viral-combined_k141_222231||full Low-quality
17-109-final-viral-combined_k141_222236||full Low-quality
17-109-final-viral-combined_k141_355926||full Not-determined
17-109-final-viral-combined_k141_88358||full Low-quality
17-109-final-viral-combined_k141_373579||full Not-determined
17-109-final-viral-combined_k141_355944||full Not-determined
17-109-final-viral-combined_k141_54857||full Not-determined
17-109-final-viral-combined_k141_355952||full Low-quality
17-109-final-viral-combined_k141_355954||full Low-quality
17-109-final-viral-combined_k141_339461||full Not-determined
17-109-final-viral-combined_k141_389977||full Not-determined
17-109-final-viral-combined_k141_389988||full Not-determined
17-109-final-viral-combined_k141_38634||full Low-quality
17-109-final-viral-combined_k141_373608||full Low-quality
17-109-final-viral-combined_k141_355963||full Not-determined
17-109-final-viral-combined_k141_54869||full Low-quality
17-109-final-viral-combined_k141_105223||full Not-determined
17-109-final-viral-combined_k141_289764||full Not-determined
17-109-final-viral-combined_k141_88430||full Not-determined
17-109-final-viral-combined_k141_38653||full Not-determined
17-109-final-viral-combined_k141_105227||full Low-quality
17-109-final-viral-combined_k141_306697||full Low-quality
17-109-final-viral-combined_k141_390014||full Low-quality
17-109-final-viral-combined_k141_21615||full Not-determined
17-109-final-viral-combined_k141_306710||full Low-quality
17-109-final-viral-combined_k141_4758||full Low-quality
17-109-final-viral-combined_k141_339486||full Not-determined
17-109-final-viral-combined_k141_373631||full Low-quality
17-109-final-viral-combined_k141_21627||full Not-determined
17-109-final-viral-combined_k141_339204||full Low-quality
17-109-final-viral-combined_k141_72400||full Not-determined
17-109-final-viral-combined_k141_222257||full Not-determined
17-109-final-viral-combined_k141_189667||full Not-determined
17-109-final-viral-combined_k141_255691||full Not-determined
17-109-final-viral-combined_k141_390053||full Not-determined
17-109-final-viral-combined_k141_289803||full Low-quality
17-109-final-viral-combined_k141_189696||full Low-quality
17-109-final-viral-combined_k141_189698||full Low-quality
17-109-final-viral-combined_k141_21661||full Not-determined
17-109-final-viral-combined_k141_72428||full Low-quality
17-109-final-viral-combined_k141_339253||full Not-determined
17-109-final-viral-combined_k141_139072||full Not-determined
17-109-final-viral-combined_k141_289828||full Low-quality
17-109-final-viral-combined_k141_72442||full Low-quality
17-109-final-viral-combined_k141_21678||full Low-quality
17-109-final-viral-combined_k141_38722||full Not-determined
17-109-final-viral-combined_k141_21684||full Low-quality
17-109-final-viral-combined_k141_289834||full Low-quality
17-109-final-viral-combined_k141_189720||full Not-determined
17-109-final-viral-combined_k141_155582||full Low-quality
17-109-final-viral-combined_k141_38752||full Not-determined
17-109-final-viral-combined_k141_72463||full Not-determined
17-109-final-viral-combined_k141_289852||full Not-determined
17-109-final-viral-combined_k141_390112||full Low-quality
17-109-final-viral-combined_k141_339533||full Low-quality
17-109-final-viral-combined_k141_306798||full Not-determined
17-109-final-viral-combined_k141_255752||full Low-quality
17-109-final-viral-combined_k141_72493||full Low-quality
17-109-final-viral-combined_k141_139120||full Low-quality
17-109-final-viral-combined_k141_121419||full Low-quality
17-109-final-viral-combined_k141_72505||full Low-quality
17-109-final-viral-combined_k141_121424||full Not-determined
17-109-final-viral-combined_k141_272822||full Not-determined
17-109-final-viral-combined_k141_189767||full Low-quality
17-109-final-viral-combined_k141_272828||full Not-determined
17-109-final-viral-combined_k141_21723||full Not-determined
17-109-final-viral-combined_k141_121440||full Not-determined
17-109-final-viral-combined_k141_205586||full Not-determined
17-109-final-viral-combined_k141_54927||full Not-determined
17-109-final-viral-combined_k141_222362||full Not-determined
17-109-final-viral-combined_k141_222372||full Not-determined
17-109-final-viral-combined_k141_306854||full Not-determined
17-109-final-viral-combined_k141_72545||full Low-quality
17-109-final-viral-combined_k141_222388||full Low-quality
17-109-final-viral-combined_k141_356024||full Low-quality
17-109-final-viral-combined_k141_72549||full Low-quality
17-109-final-viral-combined_k141_339547||full Not-determined
17-109-final-viral-combined_k141_390174||full Not-determined
17-109-final-viral-combined_k141_339558||full Low-quality
17-109-final-viral-combined_k141_339566||full Not-determined
17-109-final-viral-combined_k141_205624||full Not-determined
17-109-final-viral-combined_k141_222405||full Not-determined
17-109-final-viral-combined_k141_239373||full Not-determined
17-109-final-viral-combined_k141_339584||full Low-quality
17-109-final-viral-combined_k141_323234||full Low-quality
17-109-final-viral-combined_k141_272881||full Not-determined
17-109-final-viral-combined_k141_155668||full Low-quality
17-109-final-viral-combined_k141_205638||full Low-quality
17-109-final-viral-combined_k141_339594||full Not-determined
17-109-final-viral-combined_k141_339601||full Not-determined
17-109-final-viral-combined_k141_239390||full Low-quality
17-109-final-viral-combined_k141_222408||full Low-quality
17-109-final-viral-combined_k141_72615||full Low-quality
17-109-final-viral-combined_k141_105292||full Not-determined
17-109-final-viral-combined_k141_88576||full Low-quality
17-109-final-viral-combined_k141_21768||full Low-quality
17-109-final-viral-combined_k141_289979||full Low-quality
17-109-final-viral-combined_k141_323243||full Not-determined
17-109-final-viral-combined_k141_172251||full Low-quality
17-109-final-viral-combined_k141_272916||full Low-quality
17-109-final-viral-combined_k141_21786||full Low-quality
17-109-final-viral-combined_k141_155702||full Not-determined
17-109-final-viral-combined_k141_88600||full Low-quality
17-109-final-viral-combined_k141_205675||full Not-determined
17-109-final-viral-combined_k141_121494||full Not-determined
17-109-final-viral-combined_k141_255858||full Low-quality
17-109-final-viral-combined_k141_21821||full Not-determined
17-109-final-viral-combined_k141_121513||full Low-quality
17-109-final-viral-combined_k141_72673||full Low-quality
17-109-final-viral-combined_k141_121521||full Not-determined
17-109-final-viral-combined_k141_172274||full Low-quality
17-109-final-viral-combined_k141_390269||full Not-determined
17-109-final-viral-combined_k141_307005||full Not-determined
17-109-final-viral-combined_k141_255883||full Low-quality
17-109-final-viral-combined_k141_189853||full Low-quality
17-109-final-viral-combined_k141_121534||full Not-determined
17-109-final-viral-combined_k141_272973||full Not-determined
17-109-final-viral-combined_k141_255913||full Low-quality
17-109-final-viral-combined_k141_5006||full Not-determined
17-109-final-viral-combined_k141_390299||full Low-quality
17-109-final-viral-combined_k141_5012||full Not-determined
17-109-final-viral-combined_k141_88673||full Not-determined
17-109-final-viral-combined_k141_121545||full Low-quality
17-109-final-viral-combined_k141_5016||full Low-quality
17-109-final-viral-combined_k141_121559||full Not-determined
17-109-final-viral-combined_k141_273000||full Not-determined
17-109-final-viral-combined_k141_21874||full Not-determined
17-109-final-viral-combined_k141_307092||full Low-quality
17-109-final-viral-combined_k141_121567||full Not-determined
17-109-final-viral-combined_k141_222556||full Not-determined
17-109-final-viral-combined_k141_172346||full Not-determined
17-109-final-viral-combined_k141_189931||full Not-determined
17-109-final-viral-combined_k141_38969||full Not-determined
17-109-final-viral-combined_k141_255951||full Low-quality
17-109-final-viral-combined_k141_290078||full Low-quality
17-109-final-viral-combined_k141_205758||full Low-quality
17-109-final-viral-combined_k141_205760||full Low-quality
17-109-final-viral-combined_k141_155781||full Low-quality
17-109-final-viral-combined_k141_290093||full Low-quality
17-109-final-viral-combined_k141_172359||full Low-quality
17-109-final-viral-combined_k141_290101||full Not-determined
17-109-final-viral-combined_k141_21897||full Low-quality
17-109-final-viral-combined_k141_222572||full Not-determined
17-109-final-viral-combined_k141_239426||full Low-quality
17-109-final-viral-combined_k141_273041||full Not-determined
17-109-final-viral-combined_k141_390363||full Low-quality
17-109-final-viral-combined_k141_290124||full Not-determined
17-109-final-viral-combined_k141_139162||full Not-determined
17-109-final-viral-combined_k141_323350||full Low-quality
17-109-final-viral-combined_k141_155808||full Low-quality
17-109-final-viral-combined_k141_255987||full Low-quality
17-109-final-viral-combined_k141_105456||full Low-quality
17-109-final-viral-combined_k141_239433||full Not-determined
17-109-final-viral-combined_k141_189988||full Low-quality
17-109-final-viral-combined_k141_21926||full Not-determined
17-109-final-viral-combined_k141_222614||full Not-determined
17-109-final-viral-combined_k141_189992||full Low-quality
17-109-final-viral-combined_k141_189996||full Not-determined
17-109-final-viral-combined_k141_373761||full Low-quality
17-109-final-viral-combined_k141_390399||full Not-determined
17-109-final-viral-combined_k141_190003||full Not-determined
17-109-final-viral-combined_k141_139209||full Not-determined
17-109-final-viral-combined_k141_339653||full Low-quality
17-109-final-viral-combined_k141_155853||full Low-quality
17-109-final-viral-combined_k141_39051||full Not-determined
17-109-final-viral-combined_k141_390420||full Not-determined
17-109-final-viral-combined_k141_105477||full Not-determined
17-109-final-viral-combined_k141_21957||full Not-determined
18-103-final-viral-combined_k141_20326||full Medium-quality
17-109-final-viral-combined_k141_21964||full Not-determined
17-109-final-viral-combined_k141_21967||full Not-determined
17-109-final-viral-combined_k141_339679||full Not-determined
17-109-final-viral-combined_k141_139237||full Low-quality
17-109-final-viral-combined_k141_339690||full Low-quality
17-109-final-viral-combined_k141_139249||full Not-determined
17-109-final-viral-combined_k141_155888||full Not-determined
17-109-final-viral-combined_k141_105523||full Not-determined
17-109-final-viral-combined_k141_88792||full Not-determined
17-109-final-viral-combined_k141_307204||full Not-determined
17-109-final-viral-combined_k141_155889||full Low-quality
17-109-final-viral-combined_k141_190066||full Not-determined
17-109-final-viral-combined_k141_5062||full Not-determined
17-109-final-viral-combined_k141_290193||full Not-determined
17-109-final-viral-combined_k141_390469||full Not-determined
17-109-final-viral-combined_k141_105532||full Not-determined
17-109-final-viral-combined_k141_373814||full Not-determined

| | |
|---|---|
| 17-109-final-viral-combined_k141_39083\|\|full | Not-determined |
| 17-109-final-viral-combined_k141_139268\|\|full | Low-quality |
| 17-109-final-viral-combined_k141_190090\|\|full | Low-quality |
| 17-109-final-viral-combined_k141_121644\|\|full | Low-quality |
| 17-109-final-viral-combined_k141_390490\|\|full | Low-quality |
| 17-109-final-viral-combined_k141_339725\|\|full | Low-quality |
| 17-109-final-viral-combined_k141_373835\|\|full | Not-determined |
| 17-109-final-viral-combined_k141_88840\|\|full | Not-determined |
| 17-109-final-viral-combined_k141_239546\|\|full | Low-quality |
| 17-109-final-viral-combined_k141_105568\|\|full | Not-determined |
| 17-109-final-viral-combined_k141_339732\|\|full | Not-determined |
| 17-109-final-viral-combined_k141_390500\|\|full | Not-determined |
| 17-109-final-viral-combined_k141_356066\|\|full | Not-determined |
| 17-109-final-viral-combined_k141_307223\|\|full | Not-determined |
| 17-109-final-viral-combined_k141_256091\|\|full | Low-quality |
| 17-109-final-viral-combined_k141_172534\|\|full | Low-quality |
| 17-109-final-viral-combined_k141_323473\|\|full | Low-quality |
| 17-109-final-viral-combined_k141_273191\|\|full | Low-quality |
| 17-109-final-viral-combined_k141_88866\|\|full | Low-quality |
| 17-109-final-viral-combined_k141_72960\|\|full | Not-determined |
| 17-109-final-viral-combined_k141_39098\|\|full | Not-determined |
| 17-109-final-viral-combined_k141_339761\|\|full | Not-determined |
| 17-109-final-viral-combined_k141_307256\|\|full | Low-quality |
| 17-109-final-viral-combined_k141_307257\|\|full | Low-quality |
| 17-109-final-viral-combined_k141_239601\|\|full | Low-quality |
| 17-108-final-viral-combined_k141_226165\|\|full | Medium-quality |
| 17-109-final-viral-combined_k141_22019\|\|full | Low-quality |
| 17-109-final-viral-combined_k141_222692\|\|full | Low-quality |
| 17-109-final-viral-combined_k141_356123\|\|full | Not-determined |
| 17-109-final-viral-combined_k141_256135\|\|full | Not-determined |
| 17-109-final-viral-combined_k141_22029\|\|full | Not-determined |
| 17-109-final-viral-combined_k141_172574\|\|full | Not-determined |
| 17-109-final-viral-combined_k141_290303\|\|full | Not-determined |
| 17-109-final-viral-combined_k141_105647\|\|full | Low-quality |
| 17-109-final-viral-combined_k141_323526\|\|full | Not-determined |
| 17-109-final-viral-combined_k141_390553\|\|full | Low-quality |
| 17-109-final-viral-combined_k141_290319\|\|full | Low-quality |
| 17-109-final-viral-combined_k141_290328\|\|full | Not-determined |
| 17-109-final-viral-combined_k141_307313\|\|full | Not-determined |
| 17-109-final-viral-combined_k141_239626\|\|full | Not-determined |
| 17-109-final-viral-combined_k141_307314\|\|full | Not-determined |
| 17-109-final-viral-combined_k141_307331\|\|full | Not-determined |
| 17-109-final-viral-combined_k141_22082\|\|full | Not-determined |
| 17-109-final-viral-combined_k141_390566\|\|full | Not-determined |
| 17-109-final-viral-combined_k141_72985\|\|full | Low-quality |
| 17-109-final-viral-combined_k141_256170\|\|full | Low-quality |
| 17-109-final-viral-combined_k141_156057\|\|full | Not-determined |
| 17-109-final-viral-combined_k141_139293\|\|full | Not-determined |
| 17-109-final-viral-combined_k141_121782\|\|full | Low-quality |
| 17-109-final-viral-combined_k141_156076\|\|full | Not-determined |
| 17-109-final-viral-combined_k141_72995\|\|full | Low-quality |
| 17-109-final-viral-combined_k141_290378\|\|full | Low-quality |
| 17-109-final-viral-combined_k141_139311\|\|full | Low-quality |
| 17-109-final-viral-combined_k141_121792\|\|full | Low-quality |
| 17-109-final-viral-combined_k141_139315\|\|full | Not-determined |
| 17-109-final-viral-combined_k141_323600\|\|full | Low-quality |
| 17-109-final-viral-combined_k141_273269\|\|full | Not-determined |
| 17-109-final-viral-combined_k141_323611\|\|full | Not-determined |
| 17-109-final-viral-combined_k141_22112\|\|full | Not-determined |
| 17-109-final-viral-combined_k141_22113\|\|full | Not-determined |
| 17-109-final-viral-combined_k141_222784\|\|full | Not-determined |
| 17-109-final-viral-combined_k141_323647\|\|full | Not-determined |
| 17-109-final-viral-combined_k141_239681\|\|full | Not-determined |
| 17-109-final-viral-combined_k141_323654\|\|full | Low-quality |
| 17-109-final-viral-combined_k141_22134\|\|full | Not-determined |
| 17-109-final-viral-combined_k141_88936\|\|full | Low-quality |
| 17-109-final-viral-combined_k141_73023\|\|full | Low-quality |
| 17-109-final-viral-combined_k141_172668\|\|full | Low-quality |
| 17-109-final-viral-combined_k141_88946\|\|full | Low-quality |
| 17-109-final-viral-combined_k141_390631\|\|full | Not-determined |
| [illegible] | [illegible] |
| 17-109-final-viral-combined_k141_256562\|\|full | Not-determined |
| 17-109-final-viral-combined_k141_206304\|\|full | Not-determined |
| 17-109-final-viral-combined_k141_256568\|\|full | Not-determined |
| 17-109-final-viral-combined_k141_73363\|\|full | Not-determined |
| 17-109-final-viral-combined_k141_39447\|\|full | Low-quality |
| 17-109-final-viral-combined_k141_139745\|\|full | Low-quality |
| 17-109-final-viral-combined_k141_106036\|\|full | Not-determined |
| 17-109-final-viral-combined_k141_139754\|\|full | Low-quality |
| 17-109-final-viral-combined_k141_106045\|\|full | Low-quality |
| 17-109-final-viral-combined_k141_206332\|\|full | Not-determined |
| 17-109-final-viral-combined_k141_290912\|\|full | Not-determined |
| 17-109-final-viral-combined_k141_39471\|\|full | Low-quality |
| 17-109-final-viral-combined_k141_290918\|\|full | Low-quality |
| 17-109-final-viral-combined_k141_223173\|\|full | Not-determined |
| 17-109-final-viral-combined_k141_356426\|\|full | Not-determined |
| 17-109-final-viral-combined_k141_122318\|\|full | Low-quality |
| 17-109-final-viral-combined_k141_240194\|\|full | Low-quality |
| 17-109-final-viral-combined_k141_223186\|\|full | Not-determined |
| 17-109-final-viral-combined_k141_89377\|\|full | Not-determined |
| 17-109-final-viral-combined_k141_39486\|\|full | Low-quality |
| 17-109-final-viral-combined_k141_307831\|\|full | Not-determined |
| 17-109-final-viral-combined_k141_73384\|\|full | Not-determined |
| 17-109-final-viral-combined_k141_240201\|\|full | Not-determined |
| 17-109-final-viral-combined_k141_256614\|\|full | Not-determined |
| 17-109-final-viral-combined_k141_223202\|\|full | Not-determined |
| 17-109-final-viral-combined_k141_73393\|\|full | Low-quality |
| 17-109-final-viral-combined_k141_391057\|\|full | Not-determined |
| 17-109-final-viral-combined_k141_39514\|\|full | Low-quality |
| 17-109-final-viral-combined_k141_5564\|\|full | Low-quality |
| 17-109-final-viral-combined_k141_5571\|\|full | Not-determined |
| 17-109-final-viral-combined_k141_89417\|\|full | Not-determined |
| 17-109-final-viral-combined_k141_324087\|\|full | Not-determined |
| 17-109-final-viral-combined_k141_173040\|\|full | Low-quality |
| 18-101-final-viral-combined_k141_50602\|\|full | Medium-quality |
| 17-109-final-viral-combined_k141_374272\|\|full | Not-determined |
| 17-109-final-viral-combined_k141_106080\|\|full | Low-quality |
| 17-109-final-viral-combined_k141_73434\|\|full | Not-determined |
| 17-109-final-viral-combined_k141_324113\|\|full | Low-quality |
| 17-109-final-viral-combined_k141_324119\|\|full | Low-quality |
| 17-109-final-viral-combined_k141_290988\|\|full | Not-determined |
| 17-109-final-viral-combined_k141_106113\|\|full | Low-quality |
| 17-109-final-viral-combined_k141_73483\|\|full | Low-quality |
| 17-109-final-viral-combined_k141_89447\|\|full | Low-quality |
| 17-109-final-viral-combined_k141_22697\|\|full | Not-determined |
| 17-109-final-viral-combined_k141_324172\|\|full | Low-quality |
| 17-109-final-viral-combined_k141_122373\|\|full | Low-quality |
| 17-109-final-viral-combined_k141_206464\|\|full | Low-quality |
| 18-101-final-viral-combined_k141_19031\|\|full | High-quality |
| 17-109-final-viral-combined_k141_356511\|\|full | Not-determined |
| 17-109-final-viral-combined_k141_206475\|\|full | Not-determined |
| 17-109-final-viral-combined_k141_139857\|\|full | Low-quality |
| 17-109-final-viral-combined_k141_190848\|\|full | Low-quality |
| 17-109-final-viral-combined_k141_73507\|\|full | Low-quality |
| 17-109-final-viral-combined_k141_39611\|\|full | Not-determined |
| 17-109-final-viral-combined_k141_391152\|\|full | Low-quality |
| 17-109-final-viral-combined_k141_89454\|\|full | Not-determined |
| 17-109-final-viral-combined_k141_223288\|\|full | Low-quality |
| 17-109-final-viral-combined_k141_307909\|\|full | Not-determined |
| 17-109-final-viral-combined_k141_223291\|\|full | Not-determined |
| 17-109-final-viral-combined_k141_374332\|\|full | Not-determined |
| 17-109-final-viral-combined_k141_190870\|\|full | Low-quality |
| 17-109-final-viral-combined_k141_39630\|\|full | Low-quality |
| 17-109-final-viral-combined_k141_240289\|\|full | Low-quality |
| 17-109-final-viral-combined_k141_240290\|\|full | Not-determined |
| 17-109-final-viral-combined_k141_89477\|\|full | Low-quality |
| 17-109-final-viral-combined_k141_156713\|\|full | Not-determined |
| 17-109-final-viral-combined_k141_256651\|\|full | Not-determined |
| 17-109-final-viral-combined_k141_307922\|\|full | Not-determined |
| 17-109-final-viral-combined_k141_73574\|\|full | Not-determined |
| 17-109-final-viral-combined_k141_206513\|\|full | Low-quality |
| 17-109-final-viral-combined_k141_39658\|\|full | Not-determined |
| 17-109-final-viral-combined_k141_307923\|\|full | Not-determined |
| 17-109-final-viral-combined_k141_240303\|\|full | Not-determined |
| 17-109-final-viral-combined_k141_5632\|\|full | Not-determined |
| 17-109-final-viral-combined_k141_206517\|\|full | Not-determined |
| 17-109-final-viral-combined_k141_356570\|\|full | Not-determined |
| [illegible] | [illegible] |

| | |
|---|---|
| 17-109-final-viral-combined_k141_224093\|\|full | Not-determined |
| 17-109-final-viral-combined_k141_6284\|\|full | Low-quality |
| 17-109-final-viral-combined_k141_140513\|\|full | Not-determined |
| 17-109-final-viral-combined_k141_173815\|\|full | Not-determined |
| 17-109-final-viral-combined_k141_207229\|\|full | Not-determined |
| 17-109-final-viral-combined_k141_274273\|\|full | Not-determined |
| 17-109-final-viral-combined_k141_74130\|\|full | Low-quality |
| 17-109-final-viral-combined_k141_106643\|\|full | Not-determined |
| 17-109-final-viral-combined_k141_391732\|\|full | Low-quality |
| 17-109-final-viral-combined_k141_324778\|\|full | Not-determined |
| 17-109-final-viral-combined_k141_224104\|\|full | Low-quality |
| 17-109-final-viral-combined_k141_173841\|\|full | Low-quality |
| 17-109-final-viral-combined_k141_173848\|\|full | Low-quality |
| 17-109-final-viral-combined_k141_274284\|\|full | Not-determined |
| 17-109-final-viral-combined_k141_40229\|\|full | Low-quality |
| 17-109-final-viral-combined_k141_157291\|\|full | Low-quality |
| 17-109-final-viral-combined_k141_123016\|\|full | Not-determined |
| 17-109-final-viral-combined_k141_106682\|\|full | Not-determined |
| 17-109-final-viral-combined_k141_157295\|\|full | Low-quality |
| 17-109-final-viral-combined_k141_224151\|\|full | Low-quality |
| 17-109-final-viral-combined_k141_324825\|\|full | Not-determined |
| 17-109-final-viral-combined_k141_90109\|\|full | Not-determined |
| 17-109-final-viral-combined_k141_90111\|\|full | Low-quality |
| 17-109-final-viral-combined_k141_90113\|\|full | Low-quality |
| 17-109-final-viral-combined_k141_308608\|\|full | Low-quality |
| 17-109-final-viral-combined_k141_6375\|\|full | Not-determined |
| 17-109-final-viral-combined_k141_308618\|\|full | Not-determined |
| 17-109-final-viral-combined_k141_375002\|\|full | Low-quality |
| 17-109-final-viral-combined_k141_123030\|\|full | Low-quality |
| 17-109-final-viral-combined_k141_40285\|\|full | Not-determined |
| 17-109-final-viral-combined_k141_140551\|\|full | Not-determined |
| 17-109-final-viral-combined_k141_340686\|\|full | Not-determined |
| 17-109-final-viral-combined_k141_257323\|\|full | Not-determined |
| 17-109-final-viral-combined_k141_391796\|\|full | Low-quality |
| 17-109-final-viral-combined_k141_123056\|\|full | Not-determined |
| 17-109-final-viral-combined_k141_6407\|\|full | Low-quality |
| 17-109-final-viral-combined_k141_106722\|\|full | Low-quality |
| 17-109-final-viral-combined_k141_123060\|\|full | Not-determined |
| 17-109-final-viral-combined_k141_56143\|\|full | Not-determined |
| 17-109-final-viral-combined_k141_207282\|\|full | Not-determined |
| 17-109-final-viral-combined_k141_56157\|\|full | Low-quality |
| 17-109-final-viral-combined_k141_308659\|\|full | Low-quality |
| 17-109-final-viral-combined_k141_56169\|\|full | Low-quality |
| 17-109-final-viral-combined_k141_23324\|\|full | Not-determined |
| 17-109-final-viral-combined_k141_40353\|\|full | Low-quality |
| 17-109-final-viral-combined_k141_106754\|\|full | Not-determined |
| 17-109-final-viral-combined_k141_257328\|\|full | Not-determined |
| 17-109-final-viral-combined_k141_157385\|\|full | Low-quality |
| 17-109-final-viral-combined_k141_375047\|\|full | Not-determined |
| 17-109-final-viral-combined_k141_340737\|\|full | Not-determined |
| 17-109-final-viral-combined_k141_257363\|\|full | Not-determined |
| 17-109-final-viral-combined_k141_224200\|\|full | Not-determined |
| 17-109-final-viral-combined_k141_274362\|\|full | Not-determined |
| 17-109-final-viral-combined_k141_308675\|\|full | Low-quality |
| 17-109-final-viral-combined_k141_173874\|\|full | Not-determined |
| 17-109-final-viral-combined_k141_173877\|\|full | Low-quality |
| 17-109-final-viral-combined_k141_224203\|\|full | Not-determined |
| 17-109-final-viral-combined_k141_257376\|\|full | Not-determined |
| 17-109-final-viral-combined_k141_340751\|\|full | Low-quality |
| 17-109-final-viral-combined_k141_56207\|\|full | Not-determined |
| 17-109-final-viral-combined_k141_291788\|\|full | Not-determined |
| 17-109-final-viral-combined_k141_40380\|\|full | Not-determined |
| 17-109-final-viral-combined_k141_123157\|\|full | Low-quality |
| 17-109-final-viral-combined_k141_357122\|\|full | Low-quality |
| 17-109-final-viral-combined_k141_257395\|\|full | Not-determined |
| 17-109-final-viral-combined_k141_90213\|\|full | Not-determined |
| 17-109-final-viral-combined_k141_375086\|\|full | Not-determined |
| 17-109-final-viral-combined_k141_40393\|\|full | Low-quality |
| 17-109-final-viral-combined_k141_257400\|\|full | Low-quality |
| 17-109-final-viral-combined_k141_274374\|\|full | Low-quality |
| 17-109-final-viral-combined_k141_6489\|\|full | Low-quality |
| 17-109-final-viral-combined_k141_123191\|\|full | Low-quality |
| 17-109-final-viral-combined_k141_140585\|\|full | Not-determined |
| 17-109-final-viral-combined_k141_140594\|\|full | Low-quality |
| 17-109-final-viral-combined_k141_6496\|\|full | Low-quality |
| 17-109-final-viral-combined_k141_391911\|\|full | Low-quality |
| 17-109-final-viral-combined_k141_224226\|\|full | Low-quality |
| 17-109-final-viral-combined_k141_340810\|\|full | Low-quality |
| 17-109-final-viral-combined_k141_391913\|\|full | Not-determined |
| 17-109-final-viral-combined_k141_106817\|\|full | Low-quality |
| 17-109-final-viral-combined_k141_56236\|\|full | Low-quality |
| 17-109-final-viral-combined_k141_257459\|\|full | Not-determined |
| 17-109-final-viral-combined_k141_56240\|\|full | Low-quality |
| 17-109-final-viral-combined_k141_207425\|\|full | Low-quality |
| 17-109-final-viral-combined_k141_157506\|\|full | Not-determined |
| 17-109-final-viral-combined_k141_157512\|\|full | Not-determined |
| 17-109-final-viral-combined_k141_257479\|\|full | Low-quality |
| 17-109-final-viral-combined_k141_274446\|\|full | Not-determined |
| 17-109-final-viral-combined_k141_257485\|\|full | Not-determined |
| 17-109-final-viral-combined_k141_357216\|\|full | Not-determined |
| 17-109-final-viral-combined_k141_6507\|\|full | Low-quality |
| 17-109-final-viral-combined_k141_157529\|\|full | Low-quality |
| 17-109-final-viral-combined_k141_375143\|\|full | Low-quality |
| 17-109-final-viral-combined_k141_308741\|\|full | Low-quality |
| 17-109-final-viral-combined_k141_23361\|\|full | Low-quality |
| 17-109-final-viral-combined_k141_140640\|\|full | Not-determined |
| 17-109-final-viral-combined_k141_90263\|\|full | Not-determined |
| 17-109-final-viral-combined_k141_257500\|\|full | Not-determined |
| 17-109-final-viral-combined_k141_308751\|\|full | Not-determined |
| 17-109-final-viral-combined_k141_291873\|\|full | Low-quality |
| 17-109-final-viral-combined_k141_274488\|\|full | Not-determined |
| 17-109-final-viral-combined_k141_191538\|\|full | Low-quality |
| 17-109-final-viral-combined_k141_56259\|\|full | Low-quality |
| 17-109-final-viral-combined_k141_23385\|\|full | Not-determined |
| 17-109-final-viral-combined_k141_74326\|\|full | Low-quality |
| 17-109-final-viral-combined_k141_207485\|\|full | Not-determined |
| 17-109-final-viral-combined_k141_240827\|\|full | Not-determined |
| 17-109-final-viral-combined_k141_257534\|\|full | Not-determined |
| 17-109-final-viral-combined_k141_40470\|\|full | Low-quality |
| 17-109-final-viral-combined_k141_357226\|\|full | Low-quality |
| 17-109-final-viral-combined_k141_106858\|\|full | Low-quality |
| 17-109-final-viral-combined_k141_191561\|\|full | Low-quality |
| 17-109-final-viral-combined_k141_340844\|\|full | Low-quality |
| 17-109-final-viral-combined_k141_173995\|\|full | Low-quality |
| 17-109-final-viral-combined_k141_357246\|\|full | Low-quality |
| 17-109-final-viral-combined_k141_106865\|\|full | Low-quality |
| 17-109-final-viral-combined_k141_274512\|\|full | Low-quality |
| 17-109-final-viral-combined_k141_157600\|\|full | Low-quality |
| 17-109-final-viral-combined_k141_357261\|\|full | Not-determined |
| 17-109-final-viral-combined_k141_291925\|\|full | Not-determined |
| 17-109-final-viral-combined_k141_240863\|\|full | Low-quality |
| 17-109-final-viral-combined_k141_140676\|\|full | Low-quality |
| 17-109-final-viral-combined_k141_257577\|\|full | Not-determined |
| 17-109-final-viral-combined_k141_6536\|\|full | Not-determined |
| 17-109-final-viral-combined_k141_140678\|\|full | Low-quality |
| 18-101-final-viral-combined_k141_169879\|\|full | Medium-quality |
| 17-109-final-viral-combined_k141_340871\|\|full | Not-determined |
| 17-109-final-viral-combined_k141_240870\|\|full | Low-quality |
| 17-109-final-viral-combined_k141_74394\|\|full | Low-quality |
| 17-109-final-viral-combined_k141_340882\|\|full | Not-determined |
| 17-109-final-viral-combined_k141_74405\|\|full | Not-determined |
| 17-109-final-viral-combined_k141_106888\|\|full | Low-quality |
| 17-109-final-viral-combined_k141_191584\|\|full | Not-determined |
| 17-109-final-viral-combined_k141_74414\|\|full | Low-quality |
| 17-109-final-viral-combined_k141_140715\|\|full | Not-determined |
| 17-109-final-viral-combined_k141_324970\|\|full | Low-quality |
| 17-109-final-viral-combined_k141_324978\|\|full | Not-determined |
| 17-109-final-viral-combined_k141_340915\|\|full | Low-quality |
| 17-109-final-viral-combined_k141_324993\|\|full | Low-quality |
| 17-109-final-viral-combined_k141_106901\|\|full | Low-quality |
| 17-109-final-viral-combined_k141_90354\|\|full | Not-determined |
| 17-109-final-viral-combined_k141_340935\|\|full | Not-determined |
| 17-109-final-viral-combined_k141_90363\|\|full | Low-quality |
| 17-109-final-viral-combined_k141_6547\|\|full | Not-determined |
| 17-109-final-viral-combined_k141_325007\|\|full | Not-determined |
| 17-109-final-viral-combined_k141_207599\|\|full | Not-determined |
| 17-109-final-viral-combined_k141_240932\|\|full | Not-determined |
| 17-109-final-viral-combined_k141_274577\|\|full | Low-quality |
| 17-109-final-viral-combined_k141_157645\|\|full | Not-determined |
| 17-109-final-viral-combined_k141_392063\|\|full | Not-determined |
| 17-109-final-viral-combined_k141_90382\|\|full | Not-determined |
| 17-109-final-viral-combined_k141_240948\|\|full | Low-quality |
| 17-109-final-viral-combined_k141_308866\|\|full | Not-determined |
| 17-109-final-viral-combined_k141_291998\|\|full | Not-determined |
| 17-109-final-viral-combined_k141_140766\|\|full | Not-determined |
| 17-109-final-viral-combined_k141_340956\|\|full | Low-quality |
| 17-109-final-viral-combined_k141_6591\|\|full | Not-determined |
| 17-109-final-viral-combined_k141_191626\|\|full | Low-quality |
| 17-109-final-viral-combined_k141_123310\|\|full | Not-determined |
| 17-109-final-viral-combined_k141_340959\|\|full | Not-determined |
| 17-109-final-viral-combined_k141_392085\|\|full | Low-quality |
| 17-109-final-viral-combined_k141_90401\|\|full | Low-quality |
| 17-109-final-viral-combined_k141_325046\|\|full | Low-quality |
| 17-109-final-viral-combined_k141_157704\|\|full | Low-quality |
| 17-109-final-viral-combined_k141_23453\|\|full | Low-quality |
| 17-109-final-viral-combined_k141_274608\|\|full | Not-determined |
| 17-109-final-viral-combined_k141_357360\|\|full | Low-quality |
| 17-109-final-viral-combined_k141_174106\|\|full | Low-quality |
| 17-109-final-viral-combined_k141_274610\|\|full | Low-quality |
| 17-109-final-viral-combined_k141_123322\|\|full | Not-determined |
| 17-109-final-viral-combined_k141_23464\|\|full | Not-determined |
| 17-109-final-viral-combined_k141_325061\|\|full | Not-determined |
| 17-109-final-viral-combined_k141_274620\|\|full | Low-quality |
| 17-109-final-viral-combined_k141_74476\|\|full | Low-quality |
| 17-109-final-viral-combined_k141_56356\|\|full | Not-determined |
| 17-109-final-viral-combined_k141_325070\|\|full | Low-quality |
| 17-109-final-viral-combined_k141_224383\|\|full | Not-determined |
| 17-109-final-viral-combined_k141_240997\|\|full | Low-quality |
| 17-109-final-viral-combined_k141_340996\|\|full | Low-quality |
| 17-109-final-viral-combined_k141_240998\|\|full | Low-quality |
| 17-109-final-viral-combined_k141_106984\|\|full | Not-determined |
| 17-109-final-viral-combined_k141_357369\|\|full | Low-quality |
| 17-109-final-viral-combined_k141_157741\|\|full | Low-quality |
| 17-109-final-viral-combined_k141_392111\|\|full | Low-quality |
| 17-109-final-viral-combined_k141_174133\|\|full | Not-determined |
| 17-109-final-viral-combined_k141_241022\|\|full | Low-quality |
| 17-109-final-viral-combined_k141_157773\|\|full | Not-determined |
| 17-109-final-viral-combined_k141_74507\|\|full | Low-quality |
| 17-109-final-viral-combined_k141_257661\|\|full | Not-determined |
| 17-109-final-viral-combined_k141_23492\|\|full | Low-quality |
| 17-109-final-viral-combined_k141_107016\|\|full | Low-quality |
| 17-109-final-viral-combined_k141_174158\|\|full | Low-quality |
| 17-109-final-viral-combined_k141_292024\|\|full | Not-determined |
| 17-109-final-viral-combined_k141_56376\|\|full | Low-quality |
| 17-109-final-viral-combined_k141_292028\|\|full | Low-quality |
| 17-109-final-viral-combined_k141_174159\|\|full | Low-quality |
| 17-109-final-viral-combined_k141_23502\|\|full | Not-determined |
| 17-109-final-viral-combined_k141_274711\|\|full | Not-determined |
| 17-109-final-viral-combined_k141_257685\|\|full | Not-determined |
| 17-109-final-viral-combined_k141_325132\|\|full | Low-quality |
| 17-109-final-viral-combined_k141_274723\|\|full | Not-determined |
| 17-109-final-viral-combined_k141_257693\|\|full | Low-quality |
| 17-109-final-viral-combined_k141_23509\|\|full | Not-determined |
| 17-109-final-viral-combined_k141_325137\|\|full | Low-quality |
| 17-109-final-viral-combined_k141_157796\|\|full | Not-determined |
| 17-109-final-viral-combined_k141_157802\|\|full | Low-quality |
| 17-109-final-viral-combined_k141_191708\|\|full | Not-determined |
| 17-109-final-viral-combined_k141_292049\|\|full | Low-quality |
| 17-109-final-viral-combined_k141_357446\|\|full | Not-determined |
| 17-109-final-viral-combined_k141_257705\|\|full | Not-determined |
| 17-109-final-viral-combined_k141_392151\|\|full | Low-quality |
| 17-109-final-viral-combined_k141_357458\|\|full | Not-determined |
| 17-109-final-viral-combined_k141_375276\|\|full | Not-determined |
| 17-109-final-viral-combined_k141_207721\|\|full | Not-determined |
| 17-109-final-viral-combined_k141_375290\|\|full | Low-quality |
| 17-109-final-viral-combined_k141_107039\|\|full | Low-quality |
| 17-109-final-viral-combined_k141_357464\|\|full | Not-determined |
| 17-109-final-viral-combined_k141_140830\|\|full | Not-determined |
| 17-109-final-viral-combined_k141_375292\|\|full | Not-determined |
| 17-109-final-viral-combined_k141_207743\|\|full | Not-determined |
| 17-109-final-viral-combined_k141_274754\|\|full | Low-quality |
| 17-109-final-viral-combined_k141_191753\|\|full | Not-determined |
| 17-109-final-viral-combined_k141_325164\|\|full | Not-determined |
| 17-109-final-viral-combined_k141_274766\|\|full | Not-determined |
| 17-109-final-viral-combined_k141_6705\|\|full | Low-quality |
| 17-109-final-viral-combined_k141_191766\|\|full | Low-quality |
| 17-109-final-viral-combined_k141_207771\|\|full | Low-quality |
| 17-109-final-viral-combined_k141_357490\|\|full | Not-determined |
| 17-109-final-viral-combined_k141_274774\|\|full | Not-determined |
| 17-109-final-viral-combined_k141_74553\|\|full | Low-quality |
| 17-109-final-viral-combined_k141_325185\|\|full | Not-determined |
| 17-109-final-viral-combined_k141_224480\|\|full | Not-determined |
| 17-109-final-viral-combined_k141_274786\|\|full | Low-quality |
| 17-109-final-viral-combined_k141_157843\|\|full | Not-determined |
| 17-109-final-viral-combined_k141_40614\|\|full | Low-quality |
| 17-109-final-viral-combined_k141_241092\|\|full | Low-quality |
| 18-105-final-viral-combined_k141_163165\|\|0_partial | High-quality |
| 17-109-final-viral-combined_k141_191790\|\|full | Not-determined |
| 17-109-final-viral-combined_k141_292167\|\|full | Low-quality |
| 17-109-final-viral-combined_k141_224500\|\|full | Low-quality |
| 17-109-final-viral-combined_k141_292178\|\|full | Not-determined |
| 17-109-final-viral-combined_k141_56477\|\|full | Not-determined |
| 17-109-final-viral-combined_k141_224509\|\|full | Not-determined |
| 17-109-final-viral-combined_k141_56484\|\|full | Not-determined |
| 17-109-final-viral-combined_k141_90523\|\|full | Not-determined |
| 17-109-final-viral-combined_k141_56495\|\|full | Not-determined |
| 17-109-final-viral-combined_k141_157915\|\|full | Not-determined |
| 17-109-final-viral-combined_k141_107121\|\|full | Low-quality |
| 18-109-final-viral-combined_k141_159432\|\|full | Medium-quality |
| 17-109-final-viral-combined_k141_257865\|\|full | Low-quality |
| 17-109-final-viral-combined_k141_40637\|\|full | Low-quality |
| 17-109-final-viral-combined_k141_107135\|\|full | Not-determined |
| 17-109-final-viral-combined_k141_292218\|\|full | Not-determined |
| 17-109-final-viral-combined_k141_40641\|\|full | Not-determined |
| 17-109-final-viral-combined_k141_357561\|\|full | Low-quality |
| 17-109-final-viral-combined_k141_325238\|\|full | Low-quality |
| 17-109-final-viral-combined_k141_107145\|\|full | Low-quality |
| 17-109-final-viral-combined_k141_107153\|\|full | Not-determined |
| 17-109-final-viral-combined_k141_375359\|\|full | Not-determined |
| 17-109-final-viral-combined_k141_90563\|\|full | Low-quality |
| 17-109-final-viral-combined_k141_23572\|\|full | Low-quality |
| 17-109-final-viral-combined_k141_375361\|\|full | Not-determined |
| 17-109-final-viral-combined_k141_224556\|\|full | Not-determined |
| 17-109-final-viral-combined_k141_241177\|\|full | Low-quality |
| 17-109-final-viral-combined_k141_107166\|\|full | Not-determined |
| 17-109-final-viral-combined_k141_241186\|\|full | Not-determined |
| 17-109-final-viral-combined_k141_23585\|\|full | Low-quality |
| 17-109-final-viral-combined_k141_341137\|\|full | Not-determined |
| 17-109-final-viral-combined_k141_107185\|\|full | Low-quality |
| 17-109-final-viral-combined_k141_90565\|\|full | Low-quality |
| 17-109-final-viral-combined_k141_23597\|\|full | Low-quality |
| 17-109-final-viral-combined_k141_392269\|\|full | Not-determined |
| 17-109-final-viral-combined_k141_375379\|\|full | Not-determined |
| 17-109-final-viral-combined_k141_40690\|\|full | Low-quality |
| 17-109-final-viral-combined_k141_309039\|\|full | Not-determined |
| 17-109-final-viral-combined_k141_23607\|\|full | Low-quality |
| 17-109-final-viral-combined_k141_140920\|\|full | Low-quality |
| 17-109-final-viral-combined_k141_123522\|\|full | Low-quality |
| 17-109-final-viral-combined_k141_23614\|\|full | Not-determined |
| 17-109-final-viral-combined_k141_56578\|\|full | Low-quality |
| 17-109-final-viral-combined_k141_375397\|\|full | Low-quality |
| 17-109-final-viral-combined_k141_123550\|\|full | Not-determined |
| 17-109-final-viral-combined_k141_23641\|\|full | Not-determined |
| 17-109-final-viral-combined_k141_375403\|\|full | Not-determined |
| 17-109-final-viral-combined_k141_357597\|\|full | Low-quality |
| 17-109-final-viral-combined_k141_157955\|\|full | Not-determined |
| 17-109-final-viral-combined_k141_257917\|\|full | Low-quality |
| 17-109-final-viral-combined_k141_157963\|\|full | Not-determined |
| 17-109-final-viral-combined_k141_157969\|\|full | Not-determined |
| 17-109-final-viral-combined_k141_74596\|\|full | Low-quality |
| 17-109-final-viral-combined_k141_6802\|\|full | Low-quality |
| 17-109-final-viral-combined_k141_123588\|\|full | Not-determined |
| 17-109-final-viral-combined_k141_309080\|\|full | Low-quality |
| 17-109-final-viral-combined_k141_140948\|\|full | Not-determined |
| 17-109-final-viral-combined_k141_56603\|\|full | Low-quality |
| 17-109-final-viral-combined_k141_224681\|\|full | Not-determined |
| 17-109-final-viral-combined_k141_392364\|\|full | Not-determined |
| 17-109-final-viral-combined_k141_191892\|\|full | Not-determined |
| 17-109-final-viral-combined_k141_224685\|\|full | Low-quality |
| 17-109-final-viral-combined_k141_23677\|\|full | Low-quality |
| 17-109-final-viral-combined_k141_325312\|\|full | Low-quality |
| 17-109-final-viral-combined_k141_40717\|\|full | Low-quality |
| 17-109-final-viral-combined_k141_23684\|\|full | Low-quality |
| 17-109-final-viral-combined_k141_6836\|\|full | Low-quality |
| 17-109-final-viral-combined_k141_123613\|\|full | Low-quality |
| 17-109-final-viral-combined_k141_56622\|\|full | Not-determined |
| 17-109-final-viral-combined_k141_241329\|\|full | Low-quality |
| 17-109-final-viral-combined_k141_392397\|\|full | Low-quality |
| 17-109-final-viral-combined_k141_158017\|\|full | Not-determined |
| 17-109-final-viral-combined_k141_123645\|\|full | Low-quality |
| 17-109-final-viral-combined_k141_191903\|\|full | Low-quality |
| 17-109-final-viral-combined_k141_257960\|\|full | Not-determined |
| 17-109-final-viral-combined_k141_325354\|\|full | Not-determined |
| 17-109-final-viral-combined_k141_23698\|\|full | Not-determined |
| 17-109-final-viral-combined_k141_357681\|\|full | Low-quality |
| 17-109-final-viral-combined_k141_292248\|\|full | Low-quality |
| 17-109-final-viral-combined_k141_375532\|\|full | Not-determined |
| 17-109-final-viral-combined_k141_357682\|\|full | Not-determined |
| 17-109-final-viral-combined_k141_56665\|\|full | Low-quality |
| 17-109-final-viral-combined_k141_309126\|\|full | Not-determined |
| 17-109-final-viral-combined_k141_141012\|\|full | Low-quality |
| 17-109-final-viral-combined_k141_23716\|\|full | Low-quality |
| 17-109-final-viral-combined_k141_309135\|\|full | Not-determined |
| 17-109-final-viral-combined_k141_23725\|\|full | Low-quality |
| 17-109-final-viral-combined_k141_224706\|\|full | Not-determined |
| 17-109-final-viral-combined_k141_241387\|\|full | Not-determined |
| 17-109-final-viral-combined_k141_6941\|\|full | Not-determined |
| 17-109-final-viral-combined_k141_241401\|\|full | Low-quality |
| 17-109-final-viral-combined_k141_158068\|\|full | Not-determined |
| 17-109-final-viral-combined_k141_357729\|\|full | Not-determined |
| 17-109-final-viral-combined_k141_123716\|\|full | Low-quality |
| 17-109-final-viral-combined_k141_275023\|\|full | Low-quality |
| 17-109-final-viral-combined_k141_174334\|\|full | Not-determined |
| 17-109-final-viral-combined_k141_191965\|\|full | Not-determined |
| 17-109-final-viral-combined_k141_224738\|\|full | Not-determined |
| 17-109-final-viral-combined_k141_56734\|\|full | Not-determined |
| 17-109-final-viral-combined_k141_325407\|\|full | Not-determined |
| 17-109-final-viral-combined_k141_224751\|\|full | Not-determined |
| 17-109-final-viral-combined_k141_6948\|\|full | Not-determined |
| 17-109-final-viral-combined_k141_141079\|\|full | Not-determined |
| 17-109-final-viral-combined_k141_192004\|\|full | Low-quality |
| 17-109-final-viral-combined_k141_241446\|\|full | Low-quality |
| 17-109-final-viral-combined_k141_207923\|\|full | Low-quality |
| 17-109-final-viral-combined_k141_375613\|\|full | Low-quality |
| 17-109-final-viral-combined_k141_174358\|\|full | Not-determined |
| 17-109-final-viral-combined_k141_141097\|\|full | Low-quality |
| 17-109-final-viral-combined_k141_275038\|\|full | Low-quality |
| 17-109-final-viral-combined_k141_141102\|\|full | Not-determined |
| 17-108_k141_192416 | High-quality |
| 17-109-final-viral-combined_k141_341324\|\|full | Not-determined |
| 17-109-final-viral-combined_k141_192017\|\|full | Not-determined |
| 17-109-final-viral-combined_k141_309184\|\|full | Not-determined |
| 17-109-final-viral-combined_k141_74742\|\|full | Low-quality |
| 17-109-final-viral-combined_k141_341325\|\|full | Low-quality |
| 17-109-final-viral-combined_k141_192021\|\|full | Not-determined |
| 17-109-final-viral-combined_k141_192023\|\|full | Not-determined |
| 17-109-final-viral-combined_k141_192024\|\|full | Not-determined |
| 17-109-final-viral-combined_k141_309188\|\|full | Not-determined |
| 17-109-final-viral-combined_k141_241513\|\|full | Not-determined |
| 17-109-final-viral-combined_k141_341336\|\|full | Not-determined |
| 17-109-final-viral-combined_k141_40848\|\|full | Not-determined |
| 17-109-final-viral-combined_k141_341347\|\|full | Not-determined |
| 17-109-final-viral-combined_k141_141143\|\|full | Not-determined |
| 17-109-final-viral-combined_k141_192038\|\|full | Not-determined |
| 17-109-final-viral-combined_k141_174408\|\|full | Low-quality |
| 17-109-final-viral-combined_k141_357806\|\|full | Low-quality |
| 17-109-final-viral-combined_k141_23861\|\|full | Not-determined |
| 17-109-final-viral-combined_k141_325445\|\|full | Not-determined |
| 17-109-final-viral-combined_k141_357813\|\|full | Low-quality |
| 17-109-final-viral-combined_k141_375700\|\|full | Not-determined |
| 17-109-final-viral-combined_k141_224849\|\|full | Low-quality |
| 17-109-final-viral-combined_k141_23875\|\|full | Not-determined |
| 17-109-final-viral-combined_k141_107315\|\|full | Not-determined |
| 17-109-final-viral-combined_k141_309214\|\|full | Low-quality |
| 17-109-final-viral-combined_k141_309217\|\|full | Not-determined |
| 17-109-final-viral-combined_k141_224854\|\|full | Not-determined |
| 17-109-final-viral-combined_k141_123803\|\|full | Low-quality |
| 17-109-final-viral-combined_k141_258040\|\|full | Low-quality |
| 17-109-final-viral-combined_k141_275119\|\|full | Low-quality |
| 17-109-final-viral-combined_k141_341380\|\|full | Not-determined |
| 17-109-final-viral-combined_k141_309237\|\|full | Not-determined |
| 17-109-final-viral-combined_k141_107354\|\|full | Not-determined |
| 17-109-final-viral-combined_k141_56856\|\|full | Low-quality |
| 17-109-final-viral-combined_k141_107356\|\|full | Not-determined |
| 17-109-final-viral-combined_k141_40915\|\|full | Low-quality |
| 17-109-final-viral-combined_k141_258059\|\|full | Low-quality |
| 17-109-final-viral-combined_k141_341408\|\|full | Not-determined |
| 17-109-final-viral-combined_k141_23919\|\|full | Low-quality |
| 17-109-final-viral-combined_k141_325470\|\|full | Low-quality |
| 17-109-final-viral-combined_k141_207952\|\|full | Not-determined |
| 17-109-final-viral-combined_k141_158198\|\|full | Low-quality |
| 17-109-final-viral-combined_k141_309275\|\|full | Low-quality |
| 17-109-final-viral-combined_k141_325487\|\|full | Low-quality |
| 17-109-final-viral-combined_k141_224893\|\|full | Low-quality |
| 17-109-final-viral-combined_k141_207977\|\|full | Not-determined |
| 17-109-final-viral-combined_k141_309293\|\|full | Not-determined |
| 17-109-final-viral-combined_k141_141172\|\|full | Low-quality |
| 17-109-final-viral-combined_k141_192125\|\|full | Not-determined |
| 17-109-final-viral-combined_k141_23928\|\|full | Low-quality |
| 17-109-final-viral-combined_k141_158246\|\|full | Not-determined |
| 17-109-final-viral-combined_k141_309328\|\|full | Low-quality |
| 17-109-final-viral-combined_k141_74838\|\|full | Not-determined |
| 17-109-final-viral-combined_k141_392513\|\|full | Low-quality |
| 17-109-final-viral-combined_k141_141200\|\|full | Not-determined |
| 17-109-final-viral-combined_k141_357932\|\|full | Not-determined |
| 17-109-final-viral-combined_k141_158265\|\|full | Low-quality |
| 17-109-final-viral-combined_k141_90628\|\|full | Low-quality |
| 17-109-final-viral-combined_k141_141218\|\|full | Not-determined |
| 17-109-final-viral-combined_k141_309365\|\|full | Not-determined |
| 17-109-final-viral-combined_k141_241559\|\|full | Not-determined |
| 17-109-final-viral-combined_k141_158276\|\|full | Low-quality |
| 17-109-final-viral-combined_k141_224911\|\|full | Not-determined |
| 17-109-final-viral-combined_k141_208007\|\|full | Not-determined |
| 17-109-final-viral-combined_k141_41015\|\|full | Not-determined |
| 17-109-final-viral-combined_k141_325547\|\|full | Not-determined |
| 17-109-final-viral-combined_k141_392558\|\|full | Low-quality |
| 17-109-final-viral-combined_k141_309394\|\|full | Low-quality |
| 17-109-final-viral-combined_k141_258130\|\|full | Not-determined |
| 17-109-final-viral-combined_k141_309396\|\|full | Low-quality |
| 17-109-final-viral-combined_k141_325559\|\|full | Low-quality |
| 17-109-final-viral-combined_k141_275244\|\|full | Low-quality |
| 17-109-final-viral-combined_k141_392568\|\|full | Low-quality |
| 17-109-final-viral-combined_k141_192144\|\|full | Not-determined |
| 17-109-final-viral-combined_k141_158329\|\|full | Low-quality |
| 17-109-final-viral-combined_k141_341469\|\|full | Not-determined |
| 17-109-final-viral-combined_k141_141246\|\|full | Low-quality |
| 17-109-final-viral-combined_k141_392591\|\|full | Not-determined |
| 17-109-final-viral-combined_k141_56943\|\|full | Not-determined |
| 17-109-final-viral-combined_k141_74906\|\|full | Not-determined |
| 17-109-final-viral-combined_k141_392597\|\|full | Not-determined |
| 17-109-final-viral-combined_k141_7035\|\|full | Not-determined |
| 17-109-final-viral-combined_k141_258151\|\|full | Low-quality |
| 17-109-final-viral-combined_k141_357980\|\|full | Not-determined |
| 17-109-final-viral-combined_k141_7038\|\|full | Not-determined |
| 17-109-final-viral-combined_k141_174546\|\|full | Not-determined |
| 17-109-final-viral-combined_k141_258156\|\|full | Low-quality |
| 17-109-final-viral-combined_k141_241612\|\|full | Not-determined |
| 17-109-final-viral-combined_k141_7045\|\|full | Not-determined |
| 17-109-final-viral-combined_k141_258167\|\|full | Not-determined |
| 17-109-final-viral-combined_k141_341509\|\|full | Low-quality |
| 17-109-final-viral-combined_k141_309457\|\|full | Low-quality |
| 17-109-final-viral-combined_k141_208073\|\|full | Low-quality |
| 17-109-final-viral-combined_k141_56950\|\|full | Low-quality |
| 17-109-final-viral-combined_k141_325590\|\|full | Low-quality |
| 17-109-final-viral-combined_k141_56953\|\|full | Not-determined |
| 17-109-final-viral-combined_k141_7053\|\|full | Not-determined |
| 17-109-final-viral-combined_k141_392609\|\|full | Low-quality |
| 17-109-final-viral-combined_k141_208077\|\|full | Low-quality |
| 17-109-final-viral-combined_k141_258199\|\|full | Not-determined |
| 17-109-final-viral-combined_k141_275301\|\|full | Not-determined |
| 17-109-final-viral-combined_k141_24060\|\|full | Not-determined |
| 17-109-final-viral-combined_k141_56986\|\|full | Not-determined |
| 17-109-final-viral-combined_k141_90731\|\|full | Not-determined |
| 17-109-final-viral-combined_k141_375876\|\|full | Not-determined |
| 17-109-final-viral-combined_k141_241656\|\|full | Low-quality |
| 17-109-final-viral-combined_k141_141301\|\|full | Low-quality |
| 17-109-final-viral-combined_k141_309525\|\|full | Not-determined |
| 17-109-final-viral-combined_k141_7061\|\|full | Low-quality |
| 17-109-final-viral-combined_k141_275355\|\|full | Low-quality |
| 17-109-final-viral-combined_k141_292370\|\|full | Not-determined |
| 17-109-final-viral-combined_k141_7068\|\|full | Not-determined |
| 17-109-final-viral-combined_k141_158445\|\|full | Not-determined |
| 17-109-final-viral-combined_k141_41150\|\|full | Low-quality |
| 17-109-final-viral-combined_k141_41152\|\|full | Low-quality |
| 17-109-final-viral-combined_k141_375894\|\|full | Low-quality |
| 17-109-final-viral-combined_k141_392657\|\|full | Not-determined |
| 17-109-final-viral-combined_k141_24103\|\|full | Not-determined |
| 17-109-final-viral-combined_k141_192233\|\|full | Low-quality |
| 17-109-final-viral-combined_k141_241695\|\|full | Low-quality |
| 17-109-final-viral-combined_k141_292404\|\|full | Low-quality |
| 17-109-final-viral-combined_k141_341576\|\|full | Not-determined |
| 17-109-final-viral-combined_k141_375911\|\|full | Not-determined |
| 17-109-final-viral-combined_k141_292405\|\|full | Not-determined |
| 17-109-final-viral-combined_k141_309582\|\|full | Low-quality |
| 17-109-final-viral-combined_k141_309583\|\|full | Low-quality |
| 17-109-final-viral-combined_k141_341578\|\|full | Not-determined |
| 17-109-final-viral-combined_k141_74957\|\|full | Low-quality |
| 17-109-final-viral-combined_k141_241720\|\|full | Low-quality |
| 17-109-final-viral-combined_k141_241721\|\|full | Low-quality |
| 17-109-final-viral-combined_k141_341582\|\|full | Low-quality |
| 18-105-final-viral-combined_k141_199135\|\|full | Medium-quality |
| 17-109-final-viral-combined_k141_123889\|\|full | Low-quality |
| 17-109-final-viral-combined_k141_208155\|\|full | Low-quality |
| 17-109-final-viral-combined_k141_241728\|\|full | Low-quality |
| 17-109-final-viral-combined_k141_57052\|\|full | Low-quality |
| 17-109-final-viral-combined_k141_325718\|\|full | Not-determined |
| 17-109-final-viral-combined_k141_341605\|\|full | Not-determined |
| 17-109-final-viral-combined_k141_24145\|\|full | Low-quality |
| 17-109-final-viral-combined_k141_275436\|\|full | Low-quality |
| 17-109-final-viral-combined_k141_141368\|\|full | Not-determined |
| 17-109-final-viral-combined_k141_341629\|\|full | Not-determined |
| 17-109-final-viral-combined_k141_192293\|\|full | Low-quality |
| 17-109-final-viral-combined_k141_57094\|\|full | Not-determined |
| 17-109-final-viral-combined_k141_90778\|\|full | Low-quality |
| 17-109-final-viral-combined_k141_57102\|\|full | Low-quality |
| 17-109-final-viral-combined_k141_225046\|\|full | Not-determined |
| 18-109-final-viral-combined_k141_139683\|\|full | High-quality |
| 17-109-final-viral-combined_k141_309621\|\|full | Not-determined |
| 17-109-final-viral-combined_k141_192314\|\|full | Not-determined |
| 17-109-final-viral-combined_k141_225052\|\|full | Not-determined |
| 17-109-final-viral-combined_k141_225055\|\|full | Low-quality |
| 17-109-final-viral-combined_k141_174703\|\|full | Low-quality |
| 17-109-final-viral-combined_k141_24243\|\|full | Low-quality |
| 17-109-final-viral-combined_k141_292496\|\|full | Not-determined |
| 17-109-final-viral-combined_k141_174714\|\|full | Not-determined |
| 17-109-final-viral-combined_k141_174716\|\|full | Not-determined |
| 17-109-final-viral-combined_k141_158555\|\|full | Low-quality |
| 17-109-final-viral-combined_k141_90819\|\|full | Not-determined |
| 17-109-final-viral-combined_k141_258339\|\|full | Low-quality |
| 17-109-final-viral-combined_k141_375992\|\|full | Not-determined |
| 17-109-final-viral-combined_k141_24247\|\|full | Low-quality |
| 17-109-final-viral-combined_k141_292510\|\|full | Low-quality |
| 17-109-final-viral-combined_k141_24254\|\|full | Not-determined |
| 17-109-final-viral-combined_k141_292767\|\|full | Low-quality |
| 17-109-final-viral-combined_k141_75029\|\|full | Low-quality |
| 17-109-final-viral-combined_k141_7226\|\|full | Low-quality |
| 17-109-final-viral-combined_k141_123953\|\|full | Not-determined |
| 17-109-final-viral-combined_k141_341716\|\|full | Not-determined |
| 17-109-final-viral-combined_k141_292523\|\|full | Not-determined |
| 17-109-final-viral-combined_k141_376009\|\|full | Not-determined |
| 17-109-final-viral-combined_k141_225092\|\|full | Not-determined |
| 17-109-final-viral-combined_k141_292531\|\|full | Not-determined |
| 17-109-final-viral-combined_k141_123973\|\|full | Low-quality |
| 17-109-final-viral-combined_k141_7251\|\|full | Low-quality |
| 17-109-final-viral-combined_k141_7257\|\|full | Low-quality |
| 17-109-final-viral-combined_k141_275568\|\|full | Not-determined |
| 18-106-final-viral-combined_k141_77065\|\|full | Medium-quality |
| 17-109-final-viral-combined_k141_7270\|\|full | Low-quality |
| 17-109-final-viral-combined_k141_341757\|\|full | Not-determined |
| 17-109-final-viral-combined_k141_57174\|\|full | Low-quality |
| 17-109-final-viral-combined_k141_7287\|\|full | Low-quality |
| 17-108-final-viral-combined_k141_192416\|\|full | High-quality |
| 17-109-final-viral-combined_k141_358036\|\|full | Low-quality |
| 17-109-final-viral-combined_k141_341762\|\|full | Low-quality |
| 17-109-final-viral-combined_k141_341763\|\|full | Low-quality |
| 17-109-final-viral-combined_k141_241872\|\|full | Low-quality |
| 17-109-final-viral-combined_k141_241877\|\|full | Not-determined |
| 17-109-final-viral-combined_k141_392791\|\|full | Not-determined |
| 17-109-final-viral-combined_k141_225176\|\|full | Not-determined |
| 17-109-final-viral-combined_k141_292629\|\|full | Low-quality |
| 17-109-final-viral-combined_k141_90895\|\|full | Not-determined |
| 17-109-final-viral-combined_k141_292630\|\|full | Not-determined |
| 17-109-final-viral-combined_k141_208323\|\|full | Not-determined |
| 17-109-final-viral-combined_k141_41219\|\|full | Low-quality |
| 17-109-final-viral-combined_k141_309749\|\|full | Low-quality |
| 17-109-final-viral-combined_k141_392829\|\|full | Not-determined |
| 17-109-final-viral-combined_k141_57237\|\|full | Not-determined |
| 17-109-final-viral-combined_k141_57238\|\|full | Low-quality |
| 17-109-final-viral-combined_k141_107623\|\|full | Low-quality |
| 17-109-final-viral-combined_k141_341822\|\|full | Low-quality |
| 17-109-final-viral-combined_k141_258462\|\|full | Not-determined |
| 17-109-final-viral-combined_k141_309769\|\|full | Not-determined |
| 17-109-final-viral-combined_k141_141590\|\|full | Not-determined |
| 17-109-final-viral-combined_k141_325839\|\|full | Low-quality |
| 17-109-final-viral-combined_k141_192416\|\|full | Not-determined |
| 17-109-final-viral-combined_k141_141602\|\|full | Not-determined |
| 17-109-final-viral-combined_k141_292659\|\|full | Not-determined |
| 17-109-final-viral-combined_k141_392867\|\|full | Not-determined |
| 17-109-final-viral-combined_k141_309794\|\|full | Not-determined |
| 17-109-final-viral-combined_k141_258512\|\|full | Low-quality |
| 17-109-final-viral-combined_k141_7337\|\|full | Not-determined |
| 17-109-final-viral-combined_k141_376088\|\|full | Low-quality |
| 17-109-final-viral-combined_k141_325856\|\|full | Low-quality |
| 17-109-final-viral-combined_k141_7342\|\|full | Low-quality |
| 17-109-final-viral-combined_k141_292685\|\|full | Low-quality |
| 17-109-final-viral-combined_k141_141624\|\|full | Not-determined |
| 17-109-final-viral-combined_k141_341867\|\|full | Low-quality |
| 17-109-final-viral-combined_k141_309841\|\|full | Low-quality |
| 17-109-final-viral-combined_k141_24322\|\|full | Low-quality |
| 17-109-final-viral-combined_k141_309851\|\|full | Not-determined |
| 17-109-final-viral-combined_k141_192482\|\|full | Low-quality |
| 17-109-final-viral-combined_k141_225264\|\|full | Not-determined |
| 17-109-final-viral-combined_k141_325902\|\|full | Not-determined |
| 17-109-final-viral-combined_k141_41296\|\|full | Not-determined |
| 17-109-final-viral-combined_k141_225280\|\|full | Low-quality |
| 17-109-final-viral-combined_k141_158660\|\|full | Low-quality |
| 17-109-final-viral-combined_k141_158667\|\|full | Low-quality |
| 17-109-final-viral-combined_k141_242030\|\|full | Low-quality |
| 17-109-final-viral-combined_k141_141678\|\|full | Not-determined |
| 17-109-final-viral-combined_k141_192506\|\|full | Not-determined |
| 17-109-final-viral-combined_k141_174797\|\|full | Not-determined |
| 17-109-final-viral-combined_k141_242052\|\|full | Not-determined |
| 17-109-final-viral-combined_k141_90998\|\|full | Not-determined |
| 17-109-final-viral-combined_k141_57334\|\|full | Not-determined |
| 17-109-final-viral-combined_k141_174808\|\|full | Low-quality |
| 17-109-final-viral-combined_k141_242069\|\|full | Low-quality |
| 17-109-final-viral-combined_k141_208372\|\|full | Low-quality |
| 17-109-final-viral-combined_k141_225333\|\|full | Not-determined |
| 17-109-final-viral-combined_k141_174831\|\|full | Not-determined |

| | |
|---|---|
| 17-109-final-viral-combined_k141_275699\|\|full | Low-quality |
| 17-109-final-viral-combined_k141_192574\|\|full | Not-determined |
| 17-109-final-viral-combined_k141_107716\|\|full | Low-quality |
| 17-109-final-viral-combined_k141_174846\|\|full | Not-determined |
| 17-109-final-viral-combined_k141_158711\|\|full | Low-quality |
| 17-109-final-viral-combined_k141_174865\|\|full | Not-determined |
| 17-109-final-viral-combined_k141_192589\|\|full | Low-quality |
| 17-109-final-viral-combined_k141_124016\|\|full | Low-quality |
| 17-109-final-viral-combined_k141_107738\|\|full | Not-determined |
| 17-109-final-viral-combined_k141_57394\|\|full | Low-quality |
| 17-109-final-viral-combined_k141_192601\|\|full | Low-quality |
| 17-109-final-viral-combined_k141_275732\|\|full | Low-quality |
| 17-109-final-viral-combined_k141_225395\|\|full | Not-determined |
| 17-109-final-viral-combined_k141_275743\|\|full | Not-determined |
| 17-109-final-viral-combined_k141_57413\|\|full | Low-quality |
| 17-109-final-viral-combined_k141_57435\|\|full | Low-quality |
| 17-109-final-viral-combined_k141_141754\|\|full | Low-quality |
| 17-109-final-viral-combined_k141_141755\|\|full | Low-quality |
| 17-109-final-viral-combined_k141_41423\|\|full | Low-quality |
| 17-109-final-viral-combined_k141_376177\|\|full | Not-determined |
| 17-109-final-viral-combined_k141_392939\|\|full | Low-quality |
| 17-109-final-viral-combined_k141_225454\|\|full | Low-quality |
| 17-109-final-viral-combined_k141_7541\|\|full | Not-determined |
| 17-109-final-viral-combined_k141_392950\|\|full | Low-quality |
| 17-109-final-viral-combined_k141_392955\|\|full | Low-quality |
| 17-109-final-viral-combined_k141_158722\|\|full | Not-determined |
| 17-109-final-viral-combined_k141_376191\|\|full | Not-determined |
| 17-109-final-viral-combined_k141_392957\|\|full | Low-quality |
| 17-109-final-viral-combined_k141_124093\|\|full | Not-determined |
| 17-109-final-viral-combined_k141_376196\|\|full | Low-quality |
| 17-109-final-viral-combined_k141_57449\|\|full | Not-determined |
| 17-109-final-viral-combined_k141_158735\|\|full | Not-determined |
| 17-109-final-viral-combined_k141_75113\|\|full | Low-quality |
| 17-109-final-viral-combined_k141_174907\|\|full | Low-quality |
| 17-109-final-viral-combined_k141_57468\|\|full | Not-determined |
| 17-109-final-viral-combined_k141_358127\|\|full | Low-quality |
| 17-109-final-viral-combined_k141_57471\|\|full | Not-determined |
| 17-109-final-viral-combined_k141_242164\|\|full | Not-determined |
| 17-109-final-viral-combined_k141_57475\|\|full | Low-quality |
| 17-109-final-viral-combined_k141_141857\|\|full | Low-quality |
| 17-109-final-viral-combined_k141_124131\|\|full | Not-determined |
| 17-109-final-viral-combined_k141_275828\|\|full | Low-quality |
| 17-109-final-viral-combined_k141_24433\|\|full | Not-determined |
| 17-109-final-viral-combined_k141_208504\|\|full | Not-determined |
| 17-109-final-viral-combined_k141_275836\|\|full | Not-determined |
| 17-109-final-viral-combined_k141_158772\|\|full | Not-determined |
| 17-109-final-viral-combined_k141_376237\|\|full | Not-determined |
| 17-109-final-viral-combined_k141_107863\|\|full | Low-quality |
| 17-109-final-viral-combined_k141_7620\|\|full | Not-determined |
| 17-109-final-viral-combined_k141_393018\|\|full | Low-quality |
| 17-109-final-viral-combined_k141_342029\|\|full | Low-quality |
| 17-109-final-viral-combined_k141_41488\|\|full | Low-quality |
| 17-109-final-viral-combined_k141_124165\|\|full | Low-quality |
| 17-109-final-viral-combined_k141_376272\|\|full | Not-determined |
| 17-109-final-viral-combined_k141_75176\|\|full | Not-determined |
| 17-109-final-viral-combined_k141_75185\|\|full | Not-determined |
| 17-109-final-viral-combined_k141_358180\|\|full | Low-quality |
| 17-109-final-viral-combined_k141_275894\|\|full | Not-determined |
| 17-109-final-viral-combined_k141_358183\|\|full | Low-quality |
| 17-109-final-viral-combined_k141_242218\|\|full | Not-determined |
| 17-109-final-viral-combined_k141_41530\|\|full | Not-determined |
| 17-109-final-viral-combined_k141_75191\|\|full | Not-determined |
| 17-109-final-viral-combined_k141_358188\|\|full | Not-determined |
| 17-109-final-viral-combined_k141_57507\|\|full | Low-quality |
| 17-109-final-viral-combined_k141_225537\|\|full | Not-determined |
| 17-109-final-viral-combined_k141_7666\|\|full | Not-determined |
| 17-109-final-viral-combined_k141_141881\|\|full | Low-quality |
| 17-109-final-viral-combined_k141_7668\|\|full | Not-determined |
| 17-109-final-viral-combined_k141_393074\|\|full | Low-quality |
| 17-109-final-viral-combined_k141_275929\|\|full | Low-quality |
| 17-109-final-viral-combined_k141_275930\|\|full | Not-determined |
| 17-109-final-viral-combined_k141_242259\|\|full | Not-determined |
| 17-109-final-viral-combined_k141_24519\|\|full | Low-quality |
| 17-109-final-viral-combined_k141_293044\|\|full | Low-quality |
| 17-109-final-viral-combined_k141_24521\|\|full | Not-determined |
| 17-109-final-viral-combined_k141_342095\|\|full | Not-determined |
| 17-109-final-viral-combined_k141_310135\|\|full | Not-determined |
| 17-109-final-viral-combined_k141_242274\|\|full | Not-determined |
| 17-109-final-viral-combined_k141_24526\|\|full | Low-quality |
| 17-109-final-viral-combined_k141_258759\|\|full | Not-determined |
| 17-109-final-viral-combined_k141_393098\|\|full | Low-quality |
| 17-109-final-viral-combined_k141_208523\|\|full | Low-quality |
| 17-109-final-viral-combined_k141_208524\|\|full | Low-quality |
| 17-109-final-viral-combined_k141_7687\|\|full | Not-determined |
| 17-109-final-viral-combined_k141_393109\|\|full | Low-quality |
| 17-109-final-viral-combined_k141_342133\|\|full | Low-quality |
| 17-109-final-viral-combined_k141_342134\|\|full | Not-determined |
| 17-109-final-viral-combined_k141_225589\|\|full | Low-quality |
| 17-109-final-viral-combined_k141_175032\|\|full | Low-quality |
| 17-109-final-viral-combined_k141_24560\|\|full | Not-determined |
| 17-109-final-viral-combined_k141_75237\|\|full | Low-quality |
| 17-109-final-viral-combined_k141_7722\|\|full | Not-determined |
| 17-109-final-viral-combined_k141_91123\|\|full | Low-quality |
| 17-109-final-viral-combined_k141_358269\|\|full | Not-determined |
| 17-109-final-viral-combined_k141_158878\|\|full | Not-determined |
| 17-109-final-viral-combined_k141_242298\|\|full | Low-quality |
| 17-109-final-viral-combined_k141_275994\|\|full | Low-quality |
| 17-109-final-viral-combined_k141_310156\|\|full | Low-quality |
| 17-109-final-viral-combined_k141_24590\|\|full | Not-determined |
| 17-109-final-viral-combined_k141_7739\|\|full | Low-quality |
| 17-109-final-viral-combined_k141_158887\|\|full | Low-quality |
| 17-109-final-viral-combined_k141_358303\|\|full | Not-determined |
| 17-109-final-viral-combined_k141_310172\|\|full | Not-determined |
| 17-109-final-viral-combined_k141_158892\|\|full | Not-determined |
| 17-109-final-viral-combined_k141_75304\|\|full | Low-quality |
| 17-109-final-viral-combined_k141_91175\|\|full | Not-determined |
| 17-109-final-viral-combined_k141_358314\|\|full | Low-quality |
| 17-109-final-viral-combined_k141_293130\|\|full | Low-quality |
| 17-109-final-viral-combined_k141_358315\|\|full | Not-determined |
| 17-109-final-viral-combined_k141_242329\|\|full | Low-quality |
| 17-109-final-viral-combined_k141_358321\|\|full | Low-quality |
| 17-109-final-viral-combined_k141_175083\|\|full | Not-determined |
| 17-109-final-viral-combined_k141_293143\|\|full | Low-quality |
| 17-109-final-viral-combined_k141_358326\|\|full | Low-quality |
| 17-109-final-viral-combined_k141_7773\|\|full | Not-determined |
| 17-109-final-viral-combined_k141_75351\|\|full | Not-determined |
| 17-109-final-viral-combined_k141_57665\|\|full | Low-quality |
| 17-109-final-viral-combined_k141_192844\|\|full | Low-quality |
| 17-109-final-viral-combined_k141_258818\|\|full | Not-determined |
| 17-109-final-viral-combined_k141_326159\|\|full | Low-quality |
| 17-109-final-viral-combined_k141_75367\|\|full | Not-determined |
| 17-109-final-viral-combined_k141_41679\|\|full | Not-determined |
| 17-109-final-viral-combined_k141_276054\|\|full | Not-determined |
| 17-109-final-viral-combined_k141_7800\|\|full | Low-quality |
| 17-109-final-viral-combined_k141_75379\|\|full | Not-determined |
| 17-109-final-viral-combined_k141_142017\|\|full | Low-quality |
| 17-109-final-viral-combined_k141_41697\|\|full | Not-determined |
| 17-109-final-viral-combined_k141_175144\|\|full | Low-quality |
| 17-109-final-viral-combined_k141_24666\|\|full | Not-determined |
| 18-105-final-viral-combined_k141_66017\|\|full | Medium-quality |
| 17-109-final-viral-combined_k141_142046\|\|full | Low-quality |
| 17-109-final-viral-combined_k141_57703\|\|full | Not-determined |
| 17-109-final-viral-combined_k141_142052\|\|full | Not-determined |
| 17-109-final-viral-combined_k141_293215\|\|full | Low-quality |
| 17-109-final-viral-combined_k141_57726\|\|full | Low-quality |
| 17-109-final-viral-combined_k141_376348\|\|full | Not-determined |
| 17-109-final-viral-combined_k141_192884\|\|full | Low-quality |
| 17-109-final-viral-combined_k141_124338\|\|full | Low-quality |
| 17-109-final-viral-combined_k141_142083\|\|full | Not-determined |
| 17-109-final-viral-combined_k141_158990\|\|full | Not-determined |
| 18-109-final-viral-combined_k141_79326\|\|0_partial | Medium-quality |
| 17-109-final-viral-combined_k141_258833\|\|full | Low-quality |
| 17-109-final-viral-combined_k141_75444\|\|full | Not-determined |
| 17-109-final-viral-combined_k141_326267\|\|full | Not-determined |
| 17-109-final-viral-combined_k141_242435\|\|full | Low-quality |
| 17-109-final-viral-combined_k141_342346\|\|full | Not-determined |
| 17-109-final-viral-combined_k141_192932\|\|full | Low-quality |
| 17-109-final-viral-combined_k141_75458\|\|full | Low-quality |
| 17-109-final-viral-combined_k141_41764\|\|full | Low-quality |
| 17-109-final-viral-combined_k141_24758\|\|full | Not-determined |
| 17-108-final-viral-combined_k141_221310\|\|full | High-quality |
| 17-109-final-viral-combined_k141_159015\|\|full | Not-determined |
| 17-109-final-viral-combined_k141_57793\|\|full | Not-determined |
| 17-109-final-viral-combined_k141_124356\|\|full | Low-quality |
| 17-109-final-viral-combined_k141_192967\|\|full | Low-quality |
| 17-109-final-viral-combined_k141_175163\|\|full | Low-quality |
| 17-109-final-viral-combined_k141_310274\|\|full | Low-quality |
| 17-109-final-viral-combined_k141_293286\|\|full | Not-determined |
| 17-109-final-viral-combined_k141_393277\|\|full | Not-determined |
| 17-109-final-viral-combined_k141_342386\|\|full | Not-determined |
| 17-109-final-viral-combined_k141_24792\|\|full | Low-quality |
| 17-109-final-viral-combined_k141_393282\|\|full | Not-determined |
| 17-109-final-viral-combined_k141_376377\|\|full | Not-determined |
| 17-109-final-viral-combined_k141_142123\|\|full | Low-quality |
| 17-109-final-viral-combined_k141_175188\|\|full | Low-quality |
| 17-109-final-viral-combined_k141_276187\|\|full | Not-determined |
| 17-109-final-viral-combined_k141_258925\|\|full | Not-determined |
| 17-109-final-viral-combined_k141_192997\|\|full | Not-determined |
| 17-109-final-viral-combined_k141_24813\|\|full | Low-quality |
| 17-109-final-viral-combined_k141_393318\|\|full | Low-quality |
| 17-109-final-viral-combined_k141_124385\|\|full | Not-determined |
| 17-109-final-viral-combined_k141_57828\|\|full | Low-quality |
| 17-109-final-viral-combined_k141_193023\|\|full | Not-determined |
| 17-109-final-viral-combined_k141_91284\|\|full | Not-determined |
| 17-109-final-viral-combined_k141_225744\|\|full | Low-quality |
| 17-109-final-viral-combined_k141_358429\|\|full | Not-determined |
| 17-109-final-viral-combined_k141_75489\|\|full | Low-quality |
| 17-109-final-viral-combined_k141_393326\|\|full | Low-quality |
| 17-109-final-viral-combined_k141_193043\|\|full | Not-determined |
| 17-109-final-viral-combined_k141_124392\|\|full | Not-determined |
| 17-109-final-viral-combined_k141_57851\|\|full | Low-quality |
| 17-109-final-viral-combined_k141_75502\|\|full | Not-determined |
| 17-109-final-viral-combined_k141_310313\|\|full | Low-quality |
| 17-109-final-viral-combined_k141_293323\|\|full | Low-quality |
| 17-109-final-viral-combined_k141_326302\|\|full | Not-determined |
| 17-109-final-viral-combined_k141_258963\|\|full | Not-determined |
| 17-109-final-viral-combined_k141_108031\|\|full | Low-quality |
| 17-109-final-viral-combined_k141_310327\|\|full | Not-determined |
| 17-109-final-viral-combined_k141_276249\|\|full | Not-determined |
| 17-109-final-viral-combined_k141_258975\|\|full | Low-quality |
| 17-109-final-viral-combined_k141_293338\|\|full | Low-quality |
| 17-109-final-viral-combined_k141_326325\|\|full | Low-quality |
| 17-109-final-viral-combined_k141_326326\|\|full | Low-quality |
| 17-109-final-viral-combined_k141_276263\|\|full | Not-determined |
| 17-109-final-viral-combined_k141_124409\|\|full | Not-determined |
| 17-109-final-viral-combined_k141_393348\|\|full | Low-quality |
| 17-109-final-viral-combined_k141_75534\|\|full | Low-quality |
| 17-109-final-viral-combined_k141_342430\|\|full | Not-determined |
| 17-109-final-viral-combined_k141_326327\|\|full | Low-quality |
| 17-109-final-viral-combined_k141_193111\|\|full | Low-quality |
| 17-109-final-viral-combined_k141_193113\|\|full | Not-determined |
| 17-109-final-viral-combined_k141_225818\|\|full | Not-determined |
| 17-109-final-viral-combined_k141_225824\|\|full | Low-quality |
| 17-109-final-viral-combined_k141_24896\|\|full | Not-determined |
| 17-109-final-viral-combined_k141_342449\|\|full | Low-quality |
| 17-109-final-viral-combined_k141_376483\|\|full | Not-determined |
| 17-109-final-viral-combined_k141_124442\|\|full | Not-determined |
| 17-109-final-viral-combined_k141_7995\|\|full | Not-determined |
| 17-109-final-viral-combined_k141_208770\|\|full | Not-determined |
| 17-109-final-viral-combined_k141_376490\|\|full | Low-quality |
| 17-109-final-viral-combined_k141_342471\|\|full | Not-determined |
| 17-109-final-viral-combined_k141_193168\|\|full | Low-quality |
| 17-109-final-viral-combined_k141_276295\|\|full | Not-determined |
| 17-109-final-viral-combined_k141_276296\|\|full | Low-quality |
| 17-106-final-viral-combined_k141_44402\|\|0_partial | Medium-quality |
| 17-109-final-viral-combined_k141_8011\|\|full | Low-quality |
| 17-109-final-viral-combined_k141_124466\|\|full | Not-determined |
| 17-109-final-viral-combined_k141_175331\|\|full | Not-determined |
| 17-109-final-viral-combined_k141_310414\|\|full | Not-determined |
| 17-109-final-viral-combined_k141_376497\|\|full | Not-determined |
| 17-109-final-viral-combined_k141_376500\|\|full | Low-quality |
| 17-109-final-viral-combined_k141_342481\|\|full | Low-quality |
| 17-109-final-viral-combined_k141_293370\|\|full | Low-quality |
| 17-109-final-viral-combined_k141_225855\|\|full | Low-quality |
| 17-109-final-viral-combined_k141_24956\|\|full | Not-determined |
| 17-109-final-viral-combined_k141_259057\|\|full | Not-determined |
| 17-109-final-viral-combined_k141_124487\|\|full | Low-quality |
| 17-109-final-viral-combined_k141_326390\|\|full | Not-determined |
| 17-109-final-viral-combined_k141_142256\|\|full | Not-determined |
| 17-109-final-viral-combined_k141_124501\|\|full | Low-quality |
| 17-109-final-viral-combined_k141_259079\|\|full | Low-quality |
| 17-109-final-viral-combined_k141_310446\|\|full | Not-determined |
| 17-109-final-viral-combined_k141_124517\|\|full | Low-quality |
| 17-109-final-viral-combined_k141_293415\|\|full | Low-quality |
| 17-109-final-viral-combined_k141_310455\|\|full | Low-quality |
| 17-109-final-viral-combined_k141_376537\|\|full | Low-quality |
| 17-109-final-viral-combined_k141_242557\|\|full | Not-determined |
| 17-109-final-viral-combined_k141_259094\|\|full | Not-determined |
| 17-109-final-viral-combined_k141_242558\|\|full | Not-determined |
| 17-109-final-viral-combined_k141_393454\|\|full | Not-determined |
| 17-109-final-viral-combined_k141_124531\|\|full | Low-quality |
| 17-109-final-viral-combined_k141_376546\|\|full | Not-determined |
| 17-109-final-viral-combined_k141_342521\|\|full | Not-determined |
| 17-109-final-viral-combined_k141_310471\|\|full | Low-quality |
| 17-109-final-viral-combined_k141_393475\|\|full | Low-quality |
| 17-109-final-viral-combined_k141_108149\|\|full | Not-determined |
| 17-109-final-viral-combined_k141_124549\|\|full | Not-determined |
| 17-109-final-viral-combined_k141_24999\|\|full | Low-quality |
| 17-109-final-viral-combined_k141_57958\|\|full | Not-determined |
| 17-109-final-viral-combined_k141_142273\|\|full | Not-determined |
| 17-109-final-viral-combined_k141_193271\|\|full | Not-determined |
| 17-109-final-viral-combined_k141_8134\|\|full | Low-quality |
| 17-109-final-viral-combined_k141_8137\|\|full | Low-quality |
| 17-109-final-viral-combined_k141_342557\|\|full | Not-determined |
| 17-109-final-viral-combined_k141_358560\|\|full | Not-determined |
| 17-109-final-viral-combined_k141_225962\|\|full | Low-quality |
| 17-109-final-viral-combined_k141_159162\|\|full | Low-quality |
| 17-109-final-viral-combined_k141_342570\|\|full | Not-determined |
| 17-109-final-viral-combined_k141_393497\|\|full | Low-quality |
| 17-109-final-viral-combined_k141_193312\|\|full | Low-quality |
| 17-109-final-viral-combined_k141_276398\|\|full | Not-determined |
| 17-109-final-viral-combined_k141_242616\|\|full | Not-determined |
| 17-109-final-viral-combined_k141_310517\|\|full | Not-determined |
| 17-109-final-viral-combined_k141_342578\|\|full | Low-quality |
| 17-109-final-viral-combined_k141_208881\|\|full | Not-determined |
| 17-109-final-viral-combined_k141_41999\|\|full | Not-determined |
| 17-109-final-viral-combined_k141_225972\|\|full | Low-quality |
| 17-109-final-viral-combined_k141_342581\|\|full | Not-determined |
| 17-109-final-viral-combined_k141_193317\|\|full | Low-quality |
| 17-109-final-viral-combined_k141_175404\|\|full | Low-quality |
| 17-109-final-viral-combined_k141_91433\|\|full | Not-determined |
| 17-109-final-viral-combined_k141_376584\|\|full | Low-quality |
| 17-109-final-viral-combined_k141_242652\|\|full | Not-determined |
| 17-109-final-viral-combined_k141_342616\|\|full | Not-determined |
| 17-109-final-viral-combined_k141_124560\|\|full | Low-quality |
| 17-109-final-viral-combined_k141_326505\|\|full | Not-determined |
| 17-109-final-viral-combined_k141_57989\|\|full | Low-quality |
| 17-109-final-viral-combined_k141_42015\|\|full | Not-determined |
| 17-109-final-viral-combined_k141_208921\|\|full | Low-quality |
| 17-109-final-viral-combined_k141_358572\|\|full | Low-quality |
| 17-109-final-viral-combined_k141_8166\|\|full | Low-quality |
| 17-109-final-viral-combined_k141_124575\|\|full | Low-quality |
| 17-109-final-viral-combined_k141_58000\|\|full | Not-determined |
| 17-109-final-viral-combined_k141_208922\|\|full | Low-quality |
| 17-109-final-viral-combined_k141_175444\|\|full | Low-quality |
| 17-109-final-viral-combined_k141_175446\|\|full | Not-determined |
| 17-109-final-viral-combined_k141_91452\|\|full | Low-quality |
| 17-109-final-viral-combined_k141_175447\|\|full | Low-quality |
| 17-109-final-viral-combined_k141_91457\|\|full | Low-quality |
| 17-109-final-viral-combined_k141_42038\|\|full | Not-determined |
| 17-109-final-viral-combined_k141_58023\|\|full | Not-determined |
| 17-109-final-viral-combined_k141_208950\|\|full | Not-determined |
| 17-109-final-viral-combined_k141_326543\|\|full | Not-determined |
| 17-109-final-viral-combined_k141_358604\|\|full | Not-determined |
| 17-109-final-viral-combined_k141_124594\|\|full | Not-determined |
| 17-109-final-viral-combined_k141_91470\|\|full | Not-determined |
| 17-109-final-viral-combined_k141_58028\|\|full | Low-quality |
| 17-109-final-viral-combined_k141_358613\|\|full | Low-quality |
| 17-109-final-viral-combined_k141_226056\|\|full | Not-determined |
| 17-109-final-viral-combined_k141_159187\|\|full | Low-quality |
| 17-109-final-viral-combined_k141_358624\|\|full | Not-determined |
| 17-109-final-viral-combined_k141_326559\|\|full | Not-determined |
| 17-109-final-viral-combined_k141_42050\|\|full | Low-quality |
| 17-109-final-viral-combined_k141_259192\|\|full | Not-determined |
| 17-109-final-viral-combined_k141_208983\|\|full | Low-quality |
| 17-109-final-viral-combined_k141_91479\|\|full | Low-quality |
| 17-109-final-viral-combined_k141_124618\|\|full | Low-quality |
| 17-109-final-viral-combined_k141_326567\|\|full | Low-quality |
| 17-109-final-viral-combined_k141_124630\|\|full | Low-quality |
| 17-109-final-viral-combined_k141_242717\|\|full | Not-determined |
| 17-109-final-viral-combined_k141_293547\|\|full | Not-determined |
| 17-109-final-viral-combined_k141_293553\|\|full | Low-quality |
| 17-109-final-viral-combined_k141_293557\|\|full | Low-quality |
| 17-109-final-viral-combined_k141_159232\|\|full | Not-determined |
| 17-109-final-viral-combined_k141_58097\|\|full | Not-determined |
| 17-109-final-viral-combined_k141_242718\|\|full | Low-quality |
| 17-109-final-viral-combined_k141_209030\|\|full | Not-determined |
| 17-109-final-viral-combined_k141_58102\|\|full | Not-determined |
| 17-109-final-viral-combined_k141_58105\|\|full | Low-quality |
| 17-109-final-viral-combined_k141_326623\|\|full | Not-determined |
| 17-109-final-viral-combined_k141_310615\|\|full | Not-determined |
| 17-109-final-viral-combined_k141_342710\|\|full | Not-determined |
| 17-109-final-viral-combined_k141_293570\|\|full | Low-quality |
| 17-109-final-viral-combined_k141_226129\|\|full | Not-determined |
| 17-109-final-viral-combined_k141_25191\|\|full | Low-quality |
| 17-109-final-viral-combined_k141_142480\|\|full | Low-quality |
| 17-109-final-viral-combined_k141_293587\|\|full | Low-quality |
| 17-109-final-viral-combined_k141_326650\|\|full | Low-quality |
| 17-109-final-viral-combined_k141_124655\|\|full | Not-determined |
| 17-109-final-viral-combined_k141_142527\|\|full | Low-quality |
| 17-109-final-viral-combined_k141_124676\|\|full | Not-determined |
| 17-109-final-viral-combined_k141_293637\|\|full | Not-determined |
| 17-109-final-viral-combined_k141_276519\|\|full | Low-quality |
| 17-109-final-viral-combined_k141_193460\|\|full | Not-determined |
| 17-109-final-viral-combined_k141_276527\|\|full | Not-determined |
| 17-109-final-viral-combined_k141_25243\|\|full | Not-determined |
| 17-109-final-viral-combined_k141_124706\|\|full | Low-quality |
| 17-109-final-viral-combined_k141_259195\|\|full | Low-quality |
| 17-109-final-viral-combined_k141_108346\|\|full | Low-quality |
| 17-109-final-viral-combined_k141_259206\|\|full | Low-quality |
| 17-109-final-viral-combined_k141_393604\|\|full | Not-determined |
| 17-109-final-viral-combined_k141_124726\|\|full | Low-quality |
| 17-109-final-viral-combined_k141_108361\|\|full | Not-determined |
| 17-109-final-viral-combined_k141_326707\|\|full | Low-quality |
| 17-109-final-viral-combined_k141_75682\|\|full | Low-quality |
| 17-109-final-viral-combined_k141_393616\|\|full | Low-quality |
| 17-109-final-viral-combined_k141_342789\|\|full | Not-determined |
| 17-109-final-viral-combined_k141_293682\|\|full | Not-determined |
| 17-109-final-viral-combined_k141_142570\|\|full | Not-determined |
| 17-109-final-viral-combined_k141_159271\|\|full | Low-quality |
| 17-109-final-viral-combined_k141_142586\|\|full | Low-quality |
| 17-109-final-viral-combined_k141_193531\|\|full | Low-quality |
| 17-109-final-viral-combined_k141_91569\|\|full | Not-determined |
| 17-109-final-viral-combined_k141_376660\|\|full | Low-quality |
| 17-109-final-viral-combined_k141_276582\|\|full | Not-determined |
| 17-109-final-viral-combined_k141_108375\|\|full | Not-determined |
| 17-109-final-viral-combined_k141_276585\|\|full | Low-quality |
| 17-109-final-viral-combined_k141_209121\|\|full | Low-quality |
| 17-109-final-viral-combined_k141_358752\|\|full | Not-determined |
| 18-106-final-viral-combined_k141_77276\|\|full | Medium-quality |
| 17-109-final-viral-combined_k141_326765\|\|full | Not-determined |
| 17-109-final-viral-combined_k141_175589\|\|full | Low-quality |
| 17-109-final-viral-combined_k141_358759\|\|full | Low-quality |
| 17-109-final-viral-combined_k141_276607\|\|full | Not-determined |
| 17-109-final-viral-combined_k141_124792\|\|full | Not-determined |
| 17-109-final-viral-combined_k141_326784\|\|full | Not-determined |
| 17-109-final-viral-combined_k141_276611\|\|full | Not-determined |
| 17-109-final-viral-combined_k141_376703\|\|full | Low-quality |
| 17-109-final-viral-combined_k141_259258\|\|full | Low-quality |
| 17-109-final-viral-combined_k141_276613\|\|full | Low-quality |
| 17-109-final-viral-combined_k141_124808\|\|full | Low-quality |
| 17-109-final-viral-combined_k141_91616\|\|full | Low-quality |
| 17-109-final-viral-combined_k141_175610\|\|full | Not-determined |
| 17-109-final-viral-combined_k141_58247\|\|full | Not-determined |
| 17-109-final-viral-combined_k141_293752\|\|full | Low-quality |
| 17-109-final-viral-combined_k141_75758\|\|full | Low-quality |
| 17-109-final-viral-combined_k141_159329\|\|full | Low-quality |
| 17-109-final-viral-combined_k141_276625\|\|full | Low-quality |
| 17-109-final-viral-combined_k141_276628\|\|full | Not-determined |
| 17-109-final-viral-combined_k141_42218\|\|full | Low-quality |
| 17-109-final-viral-combined_k141_42221\|\|full | Not-determined |
| 17-109-final-viral-combined_k141_142661\|\|full | Low-quality |
| 17-109-final-viral-combined_k141_159339\|\|full | Not-determined |
| 17-109-final-viral-combined_k141_376748\|\|full | Low-quality |
| 17-109-final-viral-combined_k141_259320\|\|full | Not-determined |
| 17-109-final-viral-combined_k141_58270\|\|full | Low-quality |
| 17-109-final-viral-combined_k141_293781\|\|full | Low-quality |
| 17-109-final-viral-combined_k141_58271\|\|full | Low-quality |
| 17-109-final-viral-combined_k141_108456\|\|full | Low-quality |
| 17-109-final-viral-combined_k141_25328\|\|full | Low-quality |
| 18-105-final-viral-combined_k141_197186\|\|full | Medium-quality |
| 17-109-final-viral-combined_k141_226239\|\|full | Not-determined |
| 17-109-final-viral-combined_k141_259346\|\|full | Not-determined |
| 17-109-final-viral-combined_k141_358784\|\|full | Low-quality |
| 17-109-final-viral-combined_k141_393712\|\|full | Low-quality |
| 17-109-final-viral-combined_k141_108480\|\|full | Not-determined |
| 17-109-final-viral-combined_k141_264077\|\|full | Medium-quality |
| 17-109-final-viral-combined_k141_242798\|\|full | Not-determined |
| 17-109_k141_264077 | Medium-quality |
| 17-109-final-viral-combined_k141_259368\|\|full | Not-determined |
| 17-109-final-viral-combined_k141_376786\|\|full | Not-determined |
| 17-109-final-viral-combined_k141_108486\|\|full | Not-determined |
| 17-109-final-viral-combined_k141_293815\|\|full | Low-quality |
| 17-109-final-viral-combined_k141_293816\|\|full | Low-quality |
| 17-109-final-viral-combined_k141_342855\|\|full | Not-determined |
| 17-109-final-viral-combined_k141_58319\|\|full | Not-determined |
| 17-109-final-viral-combined_k141_108509\|\|full | Not-determined |
| 17-109-final-viral-combined_k141_8456\|\|full | Low-quality |
| 17-109-final-viral-combined_k141_376807\|\|full | Not-determined |
| 17-109-final-viral-combined_k141_42344\|\|full | Low-quality |
| 17-109-final-viral-combined_k141_8462\|\|full | Low-quality |
| 17-109-final-viral-combined_k141_293840\|\|full | Low-quality |
| 17-109_k141_401401 | Complete |
| 17-109-final-viral-combined_k141_393752\|\|full | Not-determined |
| 17-109-final-viral-combined_k141_310803\|\|full | Not-determined |
| 17-109-final-viral-combined_k141_91698\|\|full | Not-determined |
| 17-109-final-viral-combined_k141_310807\|\|full | Low-quality |
| 17-109-final-viral-combined_k141_259427\|\|full | Low-quality |
| 17-109-final-viral-combined_k141_58337\|\|full | Low-quality |
| 17-109-final-viral-combined_k141_310817\|\|full | Low-quality |
| 17-109-final-viral-combined_k141_91714\|\|full | Low-quality |
| 17-109-final-viral-combined_k141_142756\|\|full | Low-quality |
| 17-109-final-viral-combined_k141_259446\|\|full | Low-quality |
| 17-109-final-viral-combined_k141_42376\|\|full | Not-determined |
| 17-109-final-viral-combined_k141_226356\|\|full | Not-determined |
| 17-109-final-viral-combined_k141_242906\|\|full | Low-quality |
| 17-109-final-viral-combined_k141_75896\|\|full | Low-quality |
| 17-109-final-viral-combined_k141_310849\|\|full | Low-quality |
| 17-109-final-viral-combined_k141_276693\|\|full | Not-determined |
| 17-109-final-viral-combined_k141_209202\|\|full | Low-quality |
| 17-109-final-viral-combined_k141_75907\|\|full | Low-quality |
| 17-109-final-viral-combined_k141_242912\|\|full | Not-determined |
| 17-109-final-viral-combined_k141_8495\|\|full | Not-determined |
| 17-109-final-viral-combined_k141_310864\|\|full | Low-quality |
| 17-109-final-viral-combined_k141_58370\|\|full | Low-quality |
| 17-109-final-viral-combined_k141_124966\|\|full | Not-determined |
| 17-109-final-viral-combined_k141_393791\|\|full | Not-determined |
| 17-109-final-viral-combined_k141_342921\|\|full | Low-quality |
| 17-109-final-viral-combined_k141_276734\|\|full | Not-determined |
| 17-109-final-viral-combined_k141_242952\|\|full | Not-determined |
| 17-109-final-viral-combined_k141_91762\|\|full | Low-quality |
| 17-109-final-viral-combined_k141_376861\|\|full | Low-quality |
| 17-109-final-viral-combined_k141_159457\|\|full | Low-quality |
| 17-109-final-viral-combined_k141_175769\|\|full | Low-quality |
| 17-109-final-viral-combined_k141_376865\|\|full | Not-determined |
| 17-109-final-viral-combined_k141_293892\|\|full | Not-determined |
| 17-109-final-viral-combined_k141_276744\|\|full | Low-quality |
| 17-109-final-viral-combined_k141_58403\|\|full | Low-quality |
| 17-109-final-viral-combined_k141_376867\|\|full | Low-quality |
| 17-109-final-viral-combined_k141_42402\|\|full | Not-determined |
| 17-109-final-viral-combined_k141_108569\|\|full | Low-quality |
| 17-109-final-viral-combined_k141_226444\|\|full | Low-quality |
| 17-109-final-viral-combined_k141_91800\|\|full | Low-quality |
| 17-109-final-viral-combined_k141_125009\|\|full | Not-determined |
| 17-109-final-viral-combined_k141_293913\|\|full | Low-quality |
| 17-109-final-viral-combined_k141_326976\|\|full | Low-quality |
| 17-109-final-viral-combined_k141_226457\|\|full | Not-determined |
| 17-109-final-viral-combined_k141_326980\|\|full | Low-quality |
| 17-109-final-viral-combined_k141_226464\|\|full | Not-determined |
| 17-109-final-viral-combined_k141_8548\|\|full | Low-quality |
| 17-109-final-viral-combined_k141_393853\|\|full | Low-quality |
| 17-109-final-viral-combined_k141_293924\|\|full | Low-quality |
| 17-109-final-viral-combined_k141_293929\|\|full | Not-determined |
| 17-109-final-viral-combined_k141_376910\|\|full | Low-quality |
| 17-109-final-viral-combined_k141_108610\|\|full | Not-determined |
| 17-109-final-viral-combined_k141_209293\|\|full | Not-determined |
| 17-109-final-viral-combined_k141_58436\|\|full | Not-determined |
| 17-109-final-viral-combined_k141_175852\|\|full | Not-determined |
| 17-109-final-viral-combined_k141_342952\|\|full | Low-quality |
| 17-109-final-viral-combined_k141_42431\|\|full | Not-determined |
| 17-109-final-viral-combined_k141_42434\|\|full | Low-quality |
| 17-109-final-viral-combined_k141_358941\|\|full | Not-determined |
| 17-109-final-viral-combined_k141_209320\|\|full | Low-quality |
| 17-109-final-viral-combined_k141_58459\|\|full | Not-determined |
| 17-109-final-viral-combined_k141_159568\|\|full | Not-determined |
| 17-109-final-viral-combined_k141_209325\|\|full | Not-determined |
| 17-109-final-viral-combined_k141_125071\|\|full | Low-quality |
| 17-109-final-viral-combined_k141_226505\|\|full | Low-quality |
| 17-109-final-viral-combined_k141_8570\|\|full | Not-determined |
| 17-109-final-viral-combined_k141_358965\|\|full | Low-quality |
| 17-109-final-viral-combined_k141_108651\|\|full | Not-determined |
| 17-109-final-viral-combined_k141_58476\|\|full | Low-quality |
| 17-109-final-viral-combined_k141_91840\|\|full | Not-determined |
| 17-109-final-viral-combined_k141_159583\|\|full | Low-quality |
| 17-109-final-viral-combined_k141_260036\|\|full | Medium-quality |
| 17-109-final-viral-combined_k141_159588\|\|full | Low-quality |
| 17-109-final-viral-combined_k141_8591\|\|full | Low-quality |
| 17-109-final-viral-combined_k141_8598\|\|full | Low-quality |
| 17-109-final-viral-combined_k141_175869\|\|full | Low-quality |
| 17-109-final-viral-combined_k141_25475\|\|full | Low-quality |
| 17-109-final-viral-combined_k141_76064\|\|full | Not-determined |
| 17-109-final-viral-combined_k141_293992\|\|full | Not-determined |
| 17-109-final-viral-combined_k141_343007\|\|full | Not-determined |
| 17-109-final-viral-combined_k141_125140\|\|full | Low-quality |
| 17-109-final-viral-combined_k141_175871\|\|full | Low-quality |
| 17-109-final-viral-combined_k141_25497\|\|full | Low-quality |
| 17-109-final-viral-combined_k141_294020\|\|full | Low-quality |
| 17-109-final-viral-combined_k141_58516\|\|full | Not-determined |
| 17-109-final-viral-combined_k141_209386\|\|full | Not-determined |
| 17-109-final-viral-combined_k141_294022\|\|full | Not-determined |
| 17-109-final-viral-combined_k141_226564\|\|full | Not-determined |
| 17-109-final-viral-combined_k141_393973\|\|full | Not-determined |
| 17-109-final-viral-combined_k141_193690\|\|full | Not-determined |
| 17-109-final-viral-combined_k141_393978\|\|full | Not-determined |
| 17-109-final-viral-combined_k141_125169\|\|full | Not-determined |
| 17-109-final-viral-combined_k141_193696\|\|full | Low-quality |
| 17-109-final-viral-combined_k141_327099\|\|full | Low-quality |
| 17-109-final-viral-combined_k141_294044\|\|full | Low-quality |
| 17-109-final-viral-combined_k141_343028\|\|full | Low-quality |
| 17-109-final-viral-combined_k141_359037\|\|full | Not-determined |
| 17-109-final-viral-combined_k141_108722\|\|full | Low-quality |
| 17-109-final-viral-combined_k141_226618\|\|full | Low-quality |
| 17-109-final-viral-combined_k141_8621\|\|full | Low-quality |
| 17-109-final-viral-combined_k141_377014\|\|full | Low-quality |
| 17-109-final-viral-combined_k141_142957\|\|full | Not-determined |
| 17-109-final-viral-combined_k141_359054\|\|full | Low-quality |
| 17-109-final-viral-combined_k141_343060\|\|full | Low-quality |
| 17-109-final-viral-combined_k141_175929\|\|full | Low-quality |
| 17-109-final-viral-combined_k141_343062\|\|full | Low-quality |
| 17-109-final-viral-combined_k141_25516\|\|full | Low-quality |
| 17-109-final-viral-combined_k141_226633\|\|full | Not-determined |
| 17-109-final-viral-combined_k141_108738\|\|full | Low-quality |
| 17-109-final-viral-combined_k141_76128\|\|full | Low-quality |
| 17-109-final-viral-combined_k141_359060\|\|full | Not-determined |
| 17-109-final-viral-combined_k141_175933\|\|full | Low-quality |
| 17-109-final-viral-combined_k141_142969\|\|full | Low-quality |
| 17-109-final-viral-combined_k141_175940\|\|full | Not-determined |
| 17-109-final-viral-combined_k141_209466\|\|full | Low-quality |
| 17-109-final-viral-combined_k141_91899\|\|full | Low-quality |
| 17-109-final-viral-combined_k141_359083\|\|full | Low-quality |
| 17-109-final-viral-combined_k141_343081\|\|full | Not-determined |
| 17-109-final-viral-combined_k141_159613\|\|full | Not-determined |
| 17-109-final-viral-combined_k141_108791\|\|full | Low-quality |
| 17-109-final-viral-combined_k141_76172\|\|full | Not-determined |
| 17-109-final-viral-combined_k141_125230\|\|full | Not-determined |
| 17-109-final-viral-combined_k141_175965\|\|full | Low-quality |
| 17-109-final-viral-combined_k141_294056\|\|full | Not-determined |
| 17-109-final-viral-combined_k141_226690\|\|full | Not-determined |
| 17-109-final-viral-combined_k141_8659\|\|full | Low-quality |
| 17-109-final-viral-combined_k141_327183\|\|full | Low-quality |
| 17-109-final-viral-combined_k141_159627\|\|full | Not-determined |
| 17-109-final-viral-combined_k141_25573\|\|full | Not-determined |
| 17-109-final-viral-combined_k141_176000\|\|full | Not-determined |
| 17-109-final-viral-combined_k141_209493\|\|full | Not-determined |
| 17-109-final-viral-combined_k141_42589\|\|full | Not-determined |
| 17-109-final-viral-combined_k141_25580\|\|full | Low-quality |
| 17-109-final-viral-combined_k141_176003\|\|full | Low-quality |
| 17-109-final-viral-combined_k141_226722\|\|full | Low-quality |
| 17-109-final-viral-combined_k141_108839\|\|full | Not-determined |
| 17-109-final-viral-combined_k141_76209\|\|full | Low-quality |
| 17-109-final-viral-combined_k141_327196\|\|full | Low-quality |
| 17-109-final-viral-combined_k141_209515\|\|full | Not-determined |
| 17-109-final-viral-combined_k141_276857\|\|full | Low-quality |
| 17-109-final-viral-combined_k141_76219\|\|full | Not-determined |
| 17-109-final-viral-combined_k141_327209\|\|full | Not-determined |
| 17-109-final-viral-combined_k141_42641\|\|full | Low-quality |
| 17-109-final-viral-combined_k141_91972\|\|full | Not-determined |
| 17-109-final-viral-combined_k141_311094\|\|full | Low-quality |
| 17-109-final-viral-combined_k141_243079\|\|full | Low-quality |
| 17-109-final-viral-combined_k141_394012\|\|full | Low-quality |
| 17-109-final-viral-combined_k141_209557\|\|full | Low-quality |
| 17-109-final-viral-combined_k141_343179\|\|full | Not-determined |
| 17-109-final-viral-combined_k141_91992\|\|full | Not-determined |
| 17-109-final-viral-combined_k141_343188\|\|full | Not-determined |
| 17-109-final-viral-combined_k141_143074\|\|full | Not-determined |
| 17-109-final-viral-combined_k141_343196\|\|full | Not-determined |
| 17-109-final-viral-combined_k141_377085\|\|full | Low-quality |
| 17-109-final-viral-combined_k141_259553\|\|full | Low-quality |
| 17-109-final-viral-combined_k141_25662\|\|full | Low-quality |
| 17-109-final-viral-combined_k141_343202\|\|full | Low-quality |
| 17-109-final-viral-combined_k141_276925\|\|full | Low-quality |
| 17-109-final-viral-combined_k141_394038\|\|full | Not-determined |
| 17-109-final-viral-combined_k141_243120\|\|full | Low-quality |
| 17-109-final-viral-combined_k141_209607\|\|full | Low-quality |
| 17-109-final-viral-combined_k141_377108\|\|full | Low-quality |
| 17-109-final-viral-combined_k141_193910\|\|full | Not-determined |
| 17-109-final-viral-combined_k141_226810\|\|full | Not-determined |
| 17-109-final-viral-combined_k141_343228\|\|full | Low-quality |
| 17-109-final-viral-combined_k141_25684\|\|full | Low-quality |
| 17-109-final-viral-combined_k141_394057\|\|full | Low-quality |
| 17-109-final-viral-combined_k141_343233\|\|full | Low-quality |
| 17-109-final-viral-combined_k141_394058\|\|full | Low-quality |
| 17-109-final-viral-combined_k141_343240\|\|full | Not-determined |
| 17-109-final-viral-combined_k141_108936\|\|full | Low-quality |
| 17-109-final-viral-combined_k141_377116\|\|full | Not-determined |
| 17-109-final-viral-combined_k141_209634\|\|full | Low-quality |
| 17-109-final-viral-combined_k141_176066\|\|full | Not-determined |
| 17-109-final-viral-combined_k141_209660\|\|full | Low-quality |
| 17-109-final-viral-combined_k141_108985\|\|full | Not-determined |

17-109-final-viral-combined_k141_125255||full Low-quality
17-109-final-viral-combined_k141_327346||full Low-quality
17-109-final-viral-combined_k141_176070||full Not-determined
17-109-final-viral-combined_k141_209671||full Low-quality
17-109-final-viral-combined_k141_226847||full Not-determined
17-109-final-viral-combined_k141_125256||full Not-determined
17-109-final-viral-combined_k141_394111||full Not-determined
17-109-final-viral-combined_k141_125275||full Low-quality
17-109-final-viral-combined_k141_209695||full Low-quality
17-109-final-viral-combined_k141_58715||full Not-determined
17-109-final-viral-combined_k141_327370||full Low-quality
17-109-final-viral-combined_k141_226861||full Low-quality
17-109-final-viral-combined_k141_8742||full Not-determined
17-109-final-viral-combined_k141_359111||full Not-determined
17-109-final-viral-combined_k141_277009||full Not-determined
17-109-final-viral-combined_k141_209716||full Low-quality
17-109-final-viral-combined_k141_209717||full Not-determined
17-109-final-viral-combined_k141_8762||full Not-determined
17-109-final-viral-combined_k141_176114||full Low-quality
17-109-final-viral-combined_k141_42716||full Not-determined
17-109-final-viral-combined_k141_294248||full Not-determined
17-109-final-viral-combined_k141_259629||full Low-quality
17-109-final-viral-combined_k141_394164||full Low-quality
17-109-final-viral-combined_k141_76440||full Not-determined
17-109-final-viral-combined_k141_176128||full Low-quality
17-109-final-viral-combined_k141_76444||full Not-determined
17-109-final-viral-combined_k141_92094||full Not-determined
17-109-final-viral-combined_k141_394185||full Low-quality
17-109-final-viral-combined_k141_194098||full Not-determined
17-109-final-viral-combined_k141_311290||full Not-determined
17-109-final-viral-combined_k141_42770||full Low-quality
17-109-final-viral-combined_k141_125327||full Not-determined
17-109-final-viral-combined_k141_277068||full Low-quality
17-109-final-viral-combined_k141_311293||full Low-quality
17-109-final-viral-combined_k141_176148||full Low-quality
17-109-final-viral-combined_k141_377254||full Low-quality
17-109-final-viral-combined_k141_377256||full Not-determined
17-109-final-viral-combined_k141_76493||full Not-determined
17-109-final-viral-combined_k141_226902||full Not-determined
17-109-final-viral-combined_k141_176156||full Not-determined
17-109-final-viral-combined_k141_327476||full Not-determined
17-109-final-viral-combined_k141_176166||full Low-quality
17-109-final-viral-combined_k141_159879||full Low-quality
17-109-final-viral-combined_k141_194155||full Low-quality
17-109-final-viral-combined_k141_25826||full Low-quality
17-109-final-viral-combined_k141_359154||full Low-quality
17-109-final-viral-combined_k141_394230||full Low-quality
17-109-final-viral-combined_k141_311328||full Not-determined
17-109-final-viral-combined_k141_394231||full Low-quality
17-109-final-viral-combined_k141_76526||full Low-quality
17-109-final-viral-combined_k141_277105||full Not-determined
17-109-final-viral-combined_k141_277106||full Not-determined
17-109-final-viral-combined_k141_343437||full Low-quality
17-109-final-viral-combined_k141_394234||full Not-determined
17-109-final-viral-combined_k141_243365||full Not-determined
17-109-final-viral-combined_k141_259713||full Not-determined
17-109-final-viral-combined_k141_277112||full Low-quality
17-109-final-viral-combined_k141_8870||full Low-quality
17-109-final-viral-combined_k141_343440||full Low-quality
17-109-final-viral-combined_k141_109126||full Not-determined
17-109-final-viral-combined_k141_159899||full Not-determined
17-109-final-viral-combined_k141_327518||full Not-determined
17-109-final-viral-combined_k141_42852||full Not-determined
17-109-final-viral-combined_k141_109130||full Not-determined
17-109-final-viral-combined_k141_359189||full Low-quality
17-109-final-viral-combined_k141_277148||full Not-determined
17-109-final-viral-combined_k141_8887||full Not-determined
17-109-final-viral-combined_k141_58849||full Low-quality
17-109-final-viral-combined_k141_226964||full Not-determined
17-109-final-viral-combined_k141_226969||full Low-quality
17-109-final-viral-combined_k141_243408||full Low-quality
17-109-final-viral-combined_k141_8902||full Low-quality
17-109-final-viral-combined_k141_327554||full Not-determined
17-109-final-viral-combined_k141_277168||full Not-determined
17-109-final-viral-combined_k141_311372||full Low-quality
17-109-final-viral-combined_k141_209813||full Low-quality
17-109-final-viral-combined_k141_42869||full Not-determined
17-109-final-viral-combined_k141_58867||full Not-determined
17-109-final-viral-combined_k141_311386||full Not-determined
17-109-final-viral-combined_k141_377323||full Low-quality
17-109-final-viral-combined_k141_42878||full Not-determined
17-109-final-viral-combined_k141_377339||full Not-determined
17-109-final-viral-combined_k141_227015||full Not-determined
17-109-final-viral-combined_k141_58879||full Low-quality
17-109-final-viral-combined_k141_92224||full Low-quality
17-109-final-viral-combined_k141_327586||full Low-quality
17-109-final-viral-combined_k141_343480||full Not-determined
17-109-final-viral-combined_k141_327593||full Low-quality
17-109-final-viral-combined_k141_343484||full Not-determined
17-109-final-viral-combined_k141_76613||full Not-determined
17-109-final-viral-combined_k141_42924||full Low-quality
17-109-final-viral-combined_k141_343487||full Not-determined
17-109-final-viral-combined_k141_8953||full Low-quality
17-109-final-viral-combined_k141_42925||full Not-determined
17-109-final-viral-combined_k141_176225||full Not-determined
17-109-final-viral-combined_k141_277198||full Low-quality
17-109-final-viral-combined_k141_311441||full Low-quality
17-108-final-viral-combined_k141_20271||full Medium-quality
17-109-final-viral-combined_k141_243436||full Not-determined
17-109-final-viral-combined_k141_294318||full Low-quality
17-109-final-viral-combined_k141_294327||full Low-quality
17-109-final-viral-combined_k141_311448||full Low-quality
17-109-final-viral-combined_k141_209881||full Not-determined
17-109-final-viral-combined_k141_311452||full Not-determined
17-109-final-viral-combined_k141_99704||full Medium-quality
17-109-final-viral-combined_k141_327635||full Low-quality
17-109-final-viral-combined_k141_25914||full Low-quality
17-109-final-viral-combined_k141_277235||full Low-quality
17-109-final-viral-combined_k141_359207||full Low-quality
17-109-final-viral-combined_k141_259797||full Not-determined
17-109-final-viral-combined_k141_109216||full Not-determined
17-109-final-viral-combined_k141_125337||full Not-determined
17-109-final-viral-combined_k141_277247||full Not-determined
17-109-final-viral-combined_k141_311467||full Low-quality
17-109-final-viral-combined_k141_109218||full Low-quality
17-109-final-viral-combined_k141_176279||full Not-determined
17-109-final-viral-combined_k141_311471||full Low-quality
17-109-final-viral-combined_k141_259806||full Not-determined
17-109-final-viral-combined_k141_377424||full Not-determined
17-109-final-viral-combined_k141_8965||full Not-determined
17-109-final-viral-combined_k141_243470||full Low-quality
17-109-final-viral-combined_k141_109223||full Not-determined
17-109-final-viral-combined_k141_160002||full Low-quality
17-109-final-viral-combined_k141_58922||full Not-determined
17-109-final-viral-combined_k141_343544||full Low-quality
17-109-final-viral-combined_k141_109227||full Low-quality
17-109-final-viral-combined_k141_259823||full Low-quality
17-109-final-viral-combined_k141_125364||full Not-determined
17-109-final-viral-combined_k141_327661||full Low-quality
17-109-final-viral-combined_k141_243477||full Low-quality
17-109-final-viral-combined_k141_343556||full Not-determined
17-109-final-viral-combined_k141_311480||full Not-determined
17-109-final-viral-combined_k141_294376||full Not-determined
17-109-final-viral-combined_k141_243482||full Not-determined
17-109-final-viral-combined_k141_359252||full Low-quality
17-109-final-viral-combined_k141_277272||full Low-quality
17-109-final-viral-combined_k141_359256||full Low-quality
17-109-final-viral-combined_k141_311519||full Not-determined
17-109-final-viral-combined_k141_209981||full Not-determined
17-109-final-viral-combined_k141_343602||full Low-quality
17-109-final-viral-combined_k141_76699||full Not-determined
17-109-final-viral-combined_k141_227139||full Low-quality
17-109-final-viral-combined_k141_343604||full Not-determined
17-109-final-viral-combined_k141_92341||full Low-quality
17-109-final-viral-combined_k141_209997||full Not-determined
17-109-final-viral-combined_k141_359270||full Low-quality
17-109-final-viral-combined_k141_210000||full Low-quality
17-109-final-viral-combined_k141_327700||full Low-quality
17-109-final-viral-combined_k141_176352||full Not-determined
17-109-final-viral-combined_k141_327702||full Low-quality
17-109-final-viral-combined_k141_176353||full Not-determined
17-109-final-viral-combined_k141_194161||full Low-quality
17-109-final-viral-combined_k141_343635||full Low-quality
17-109-final-viral-combined_k141_109291||full Not-determined
17-109-final-viral-combined_k141_9046||full Not-determined
17-109-final-viral-combined_k141_243542||full Low-quality
17-109-final-viral-combined_k141_343640||full Low-quality
17-109-final-viral-combined_k141_359283||full Low-quality
17-109-final-viral-combined_k141_377495||full Low-quality
17-109-final-viral-combined_k141_327726||full Not-determined
17-109-final-viral-combined_k141_125425||full Not-determined
17-109-final-viral-combined_k141_9056||full Low-quality
17-109-final-viral-combined_k141_9059||full Low-quality
17-109-final-viral-combined_k141_210039||full Not-determined
17-109-final-viral-combined_k141_109318||full Low-quality
17-109-final-viral-combined_k141_343654||full Not-determined
17-109-final-viral-combined_k141_210041||full Low-quality
17-109-final-viral-combined_k141_109330||full Low-quality
17-109-final-viral-combined_k141_210055||full Low-quality
17-109-final-viral-combined_k141_9065||full Not-determined
17-109-final-viral-combined_k141_25972||full Low-quality
17-109-final-viral-combined_k141_9071||full Not-determined
17-109-final-viral-combined_k141_243579||full Low-quality
17-109-final-viral-combined_k141_227205||full Not-determined
17-109-final-viral-combined_k141_277336||full Not-determined
17-109-final-viral-combined_k141_259850||full Not-determined
17-109-final-viral-combined_k141_377552||full Low-quality
17-109-final-viral-combined_k141_243593||full Low-quality
17-109-final-viral-combined_k141_359350||full Low-quality
17-109-final-viral-combined_k141_176426||full Low-quality
17-109-final-viral-combined_k141_243608||full Not-determined
17-109-final-viral-combined_k141_194180||full Low-quality
17-109-final-viral-combined_k141_43044||full Not-determined
17-109-final-viral-combined_k141_227238||full Low-quality
17-109-final-viral-combined_k141_176443||full Not-determined
17-109-final-viral-combined_k141_359373||full Low-quality
17-109-final-viral-combined_k141_227239||full Low-quality
17-109-final-viral-combined_k141_227240||full Low-quality
17-109-final-viral-combined_k141_359375||full Not-determined
17-109-final-viral-combined_k141_359386||full Not-determined
17-109-final-viral-combined_k141_210094||full Low-quality
17-109-final-viral-combined_k141_243632||full Low-quality
17-109-final-viral-combined_k141_359392||full Low-quality
17-109-final-viral-combined_k141_227275||full Low-quality
17-109-final-viral-combined_k141_58994||full Low-quality
17-109-final-viral-combined_k141_43110||full Low-quality
17-109-final-viral-combined_k141_277381||full Not-determined
17-109-final-viral-combined_k141_143132||full Low-quality
17-109-final-viral-combined_k141_59003||full Not-determined
17-109-final-viral-combined_k141_92475||full Low-quality
17-109-final-viral-combined_k141_160214||full Not-determined
17-109-final-viral-combined_k141_92477||full Not-determined
17-109-final-viral-combined_k141_227293||full Not-determined
17-109-final-viral-combined_k141_277394||full Not-determined
17-109-final-viral-combined_k141_311705||full Not-determined
17-109-final-viral-combined_k141_377587||full Not-determined
17-109-final-viral-combined_k141_294500||full Not-determined
17-109-final-viral-combined_k141_143173||full Not-determined
17-109-final-viral-combined_k141_176564||full Not-determined
17-109-final-viral-combined_k141_176569||full Low-quality
17-109-final-viral-combined_k141_311738||full Not-determined
17-109-final-viral-combined_k141_227334||full Low-quality
17-109-final-viral-combined_k141_243705||full Not-determined
17-109-final-viral-combined_k141_26101||full Not-determined
17-109-final-viral-combined_k141_59079||full Not-determined
17-109-final-viral-combined_k141_160255||full Not-determined
17-109-final-viral-combined_k141_92525||full Not-determined
17-109-final-viral-combined_k141_277420||full Not-determined
17-109-final-viral-combined_k141_26112||full Not-determined
17-109-final-viral-combined_k141_109501||full Low-quality
17-109-final-viral-combined_k141_143199||full Not-determined
17-109-final-viral-combined_k141_109514||full Not-determined
17-109-final-viral-combined_k141_109515||full Not-determined
17-109-final-viral-combined_k141_176618||full Not-determined
17-109-final-viral-combined_k141_59121||full Not-determined
17-109-final-viral-combined_k141_92575||full Low-quality
17-109-final-viral-combined_k141_343870||full Not-determined
17-109-final-viral-combined_k141_227395||full Low-quality
17-109-final-viral-combined_k141_294595||full Not-determined
17-109-final-viral-combined_k141_26156||full Not-determined
17-109-final-viral-combined_k141_343883||full Low-quality
17-109-final-viral-combined_k141_109530||full Not-determined
17-109-final-viral-combined_k141_59145||full Not-determined
17-109-final-viral-combined_k141_243750||full Low-quality
17-109-final-viral-combined_k141_311811||full Low-quality
17-109-final-viral-combined_k141_277454||full Not-determined
17-109-final-viral-combined_k141_227420||full Not-determined
17-109-final-viral-combined_k141_92612||full Low-quality
17-109-final-viral-combined_k141_294625||full Not-determined
17-109-final-viral-combined_k141_294629||full Low-quality
17-109-final-viral-combined_k141_59179||full Not-determined
17-109-final-viral-combined_k141_377656||full Low-quality
17-109-final-viral-combined_k141_194391||full Low-quality
17-109-final-viral-combined_k141_76911||full Not-determined
17-109-final-viral-combined_k141_143272||full Low-quality
17-109-final-viral-combined_k141_43241||full Low-quality
17-109-final-viral-combined_k141_260021||full Not-determined
17-109-final-viral-combined_k141_176644||full Not-determined
17-109-final-viral-combined_k141_194422||full Not-determined
17-109-final-viral-combined_k141_26209||full Not-determined
17-109-final-viral-combined_k141_160344||full Low-quality
17-109-final-viral-combined_k141_377699||full Low-quality
17-109-final-viral-combined_k141_359558||full Low-quality
17-109-final-viral-combined_k141_210270||full Not-determined
17-109-final-viral-combined_k141_277544||full Low-quality
17-109-final-viral-combined_k141_76936||full Low-quality
17-109-final-viral-combined_k141_176652||full Low-quality
17-109-final-viral-combined_k141_294666||full Not-determined
17-109-final-viral-combined_k141_359575||full Low-quality
17-109-final-viral-combined_k141_43283||full Low-quality
17-109-final-viral-combined_k141_194465||full Not-determined
17-109-final-viral-combined_k141_76975||full Low-quality
17-109-final-viral-combined_k141_43296||full Low-quality
17-109-final-viral-combined_k141_92666||full Low-quality
17-109-final-viral-combined_k141_43299||full Low-quality
17-109-final-viral-combined_k141_359610||full Not-determined
17-109-final-viral-combined_k141_76991||full Low-quality
17-109-final-viral-combined_k141_377776||full Low-quality
17-109-final-viral-combined_k141_321174||full High-quality
17-109-final-viral-combined_k141_9168||full Not-determined
17-109-final-viral-combined_k141_311914||full Not-determined
17-109-final-viral-combined_k141_327821||full Low-quality
17-109-final-viral-combined_k141_77027||full Low-quality
17-109-final-viral-combined_k141_311937||full Low-quality
17-109-final-viral-combined_k141_160387||full Low-quality
17-109-final-viral-combined_k141_43364||full Low-quality
17-109-final-viral-combined_k141_43365||full Not-determined
17-109-final-viral-combined_k141_59285||full Not-determined
17-109-final-viral-combined_k141_125581||full Not-determined
17-109-final-viral-combined_k141_394396||full Low-quality
17-109-final-viral-combined_k141_109590||full Low-quality
17-109-final-viral-combined_k141_109596||full Low-quality
17-109-final-viral-combined_k141_394404||full Not-determined
17-109-final-viral-combined_k141_59301||full Low-quality
17-109-final-viral-combined_k141_359625||full Low-quality
17-109-final-viral-combined_k141_311955||full Not-determined
17-109-final-viral-combined_k141_109607||full Not-determined
17-109-final-viral-combined_k141_377859||full Not-determined
17-109-final-viral-combined_k141_77085||full Low-quality
17-109-final-viral-combined_k141_311973||full Low-quality
17-109-final-viral-combined_k141_260119||full Not-determined
17-109-final-viral-combined_k141_277632||full Not-determined
17-109-final-viral-combined_k141_109628||full Low-quality
17-109-final-viral-combined_k141_344112||full Not-determined
17-109-final-viral-combined_k141_92672||full Not-determined
17-109-final-viral-combined_k141_210363||full Low-quality
17-109-final-viral-combined_k141_109639||full Not-determined
17-109-final-viral-combined_k141_176732||full Not-determined
17-109-final-viral-combined_k141_9224||full Low-quality
17-109-final-viral-combined_k141_176746||full Low-quality
17-109-final-viral-combined_k141_9231||full Low-quality
17-109-final-viral-combined_k141_359663||full Low-quality
17-109-final-viral-combined_k141_359670||full Low-quality
17-109-final-viral-combined_k141_176785||full Not-determined
17-109-final-viral-combined_k141_109667||full Low-quality
17-109-final-viral-combined_k141_176790||full Not-determined
17-109-final-viral-combined_k141_312026||full Low-quality
17-109-final-viral-combined_k141_194580||full Not-determined
17-109-final-viral-combined_k141_294804||full Not-determined
17-109-final-viral-combined_k141_327845||full Not-determined
17-109-final-viral-combined_k141_194581||full Not-determined
17-109-final-viral-combined_k141_160526||full Not-determined
17-109-final-viral-combined_k141_176804||full Not-determined
17-109-final-viral-combined_k141_344176||full Not-determined
17-109-final-viral-combined_k141_160530||full Low-quality
17-109-final-viral-combined_k141_394497||full Not-determined
17-109-final-viral-combined_k141_125693||full Low-quality
17-109-final-viral-combined_k141_243759||full Low-quality
17-109-final-viral-combined_k141_394500||full Not-determined
17-109-final-viral-combined_k141_194590||full Not-determined
17-109-final-viral-combined_k141_43496||full Low-quality
17-109-final-viral-combined_k141_109682||full Low-quality
17-109-final-viral-combined_k141_359687||full Not-determined
17-109-final-viral-combined_k141_377951||full Not-determined
17-109-final-viral-combined_k141_210447||full Low-quality
17-109-final-viral-combined_k141_77185||full Low-quality
17-109-final-viral-combined_k141_92715||full Not-determined
17-109-final-viral-combined_k141_143328||full Not-determined
17-109-final-viral-combined_k141_359697||full Not-determined
17-109-final-viral-combined_k141_377957||full Low-quality
17-109-final-viral-combined_k141_359698||full Not-determined
17-109-final-viral-combined_k141_77196||full Not-determined
17-109-final-viral-combined_k141_377962||full Low-quality
17-109-final-viral-combined_k141_394521||full Low-quality
17-109-final-viral-combined_k141_359699||full Not-determined
17-109-final-viral-combined_k141_59377||full Not-determined
17-109-final-viral-combined_k141_109701||full Not-determined
17-109-final-viral-combined_k141_277762||full Low-quality
17-109-final-viral-combined_k141_43513||full Not-determined
17-109-final-viral-combined_k141_312103||full Not-determined
17-109-final-viral-combined_k141_294861||full Low-quality
17-109-final-viral-combined_k141_344202||full Low-quality
17-109-final-viral-combined_k141_109744||full Not-determined
17-109-final-viral-combined_k141_176890||full Not-determined
17-109-final-viral-combined_k141_243825||full Low-quality
17-109-final-viral-combined_k141_143360||full Not-determined
17-109-final-viral-combined_k141_26460||full Low-quality
17-109-final-viral-combined_k141_77259||full Not-determined
17-109-final-viral-combined_k141_43568||full Not-determined
17-109-final-viral-combined_k141_92759||full Low-quality
17-109-final-viral-combined_k141_143363||full Low-quality
17-109-final-viral-combined_k141_312162||full Not-determined
17-109-final-viral-combined_k141_125760||full Low-quality
17-109-final-viral-combined_k141_77285||full Low-quality
17-109-final-viral-combined_k141_260206||full Low-quality
17-109-final-viral-combined_k141_378100||full Not-determined
17-109-final-viral-combined_k141_26505||full Not-determined
17-109-final-viral-combined_k141_160581||full Not-determined
17-109-final-viral-combined_k141_43623||full Not-determined
17-109-final-viral-combined_k141_227764||full Not-determined
17-109-final-viral-combined_k141_160604||full Low-quality
17-109-final-viral-combined_k141_227770||full Low-quality
17-109-final-viral-combined_k141_294961||full Not-determined
17-109-final-viral-combined_k141_344250||full Not-determined
17-109-final-viral-combined_k141_9366||full Low-quality
17-109-final-viral-combined_k141_260260||full Low-quality
17-109-final-viral-combined_k141_359814||full Low-quality
17-109-final-viral-combined_k141_312216||full Not-determined
17-109-final-viral-combined_k141_160620||full Low-quality
17-109-final-viral-combined_k141_328016||full Low-quality
17-109-final-viral-combined_k141_143387||full Low-quality
17-109-final-viral-combined_k141_92872||full Low-quality
17-109-final-viral-combined_k141_77333||full Not-determined
17-109-final-viral-combined_k141_194649||full Low-quality
17-109-final-viral-combined_k141_125795||full Not-determined
17-109-final-viral-combined_k141_312255||full Low-quality
17-109-final-viral-combined_k141_344303||full Not-determined
17-109-final-viral-combined_k141_312281||full Not-determined
17-109-final-viral-combined_k141_43696||full Low-quality
17-109-final-viral-combined_k141_160671||full Low-quality
17-109-final-viral-combined_k141_77365||full Low-quality
17-109-final-viral-combined_k141_160676||full Not-determined
17-109-final-viral-combined_k141_378178||full Low-quality
17-109-final-viral-combined_k141_59553||full Not-determined
17-109-final-viral-combined_k141_295047||full Not-determined
17-109-final-viral-combined_k141_77395||full Low-quality
17-109-final-viral-combined_k141_43727||full Not-determined
17-109-final-viral-combined_k141_394734||full Low-quality
17-109-final-viral-combined_k141_243844||full Not-determined
17-109-final-viral-combined_k141_328080||full Not-determined
17-109-final-viral-combined_k141_312310||full Not-determined
17-109-final-viral-combined_k141_92901||full Low-quality
17-109-final-viral-combined_k141_328086||full Not-determined
17-109-final-viral-combined_k141_260373||full Not-determined
17-109-final-viral-combined_k141_160705||full Low-quality
17-109-final-viral-combined_k141_92910||full Low-quality
17-109-final-viral-combined_k141_394747||full Low-quality
17-109-final-viral-combined_k141_295072||full Not-determined
17-109-final-viral-combined_k141_243876||full Low-quality
17-109-final-viral-combined_k141_394752||full Not-determined
17-109-final-viral-combined_k141_160730||full Not-determined
17-109-final-viral-combined_k141_227853||full Low-quality
17-109-final-viral-combined_k141_243886||full Not-determined
17-109-final-viral-combined_k141_277895||full Not-determined
17-109-final-viral-combined_k141_210535||full Low-quality
17-109-final-viral-combined_k141_312354||full Low-quality
17-109-final-viral-combined_k141_9434||full Not-determined
17-109-final-viral-combined_k141_109895||full Low-quality
17-109-final-viral-combined_k141_328103||full Not-determined
17-109-final-viral-combined_k141_378241||full Low-quality
17-109-final-viral-combined_k141_344364||full Not-determined
17-109-final-viral-combined_k141_92952||full Not-determined
17-109-final-viral-combined_k141_143456||full Low-quality
17-109-final-viral-combined_k141_43794||full Low-quality
17-109-final-viral-combined_k141_312392||full Not-determined
17-109-final-viral-combined_k141_43798||full Low-quality
17-109-final-viral-combined_k141_26691||full Not-determined
17-109-final-viral-combined_k141_109916||full Not-determined
17-109-final-viral-combined_k141_260417||full Low-quality
17-109-final-viral-combined_k141_312395||full Not-determined
17-109-final-viral-combined_k141_9462||full Not-determined
17-109-final-viral-combined_k141_277921||full Low-quality
17-109-final-viral-combined_k141_243926||full Not-determined
17-109-final-viral-combined_k141_59656||full Low-quality
17-109-final-viral-combined_k141_43819||full Low-quality
17-109-final-viral-combined_k141_227909||full Not-determined
17-109-final-viral-combined_k141_194721||full Low-quality
17-109-final-viral-combined_k141_243938||full Low-quality
17-109-final-viral-combined_k141_160755||full Low-quality
17-109-final-viral-combined_k141_394795||full Not-determined
17-109-final-viral-combined_k141_26748||full Not-determined
17-109-final-viral-combined_k141_160769||full Low-quality
17-109-final-viral-combined_k141_227931||full Low-quality
17-109-final-viral-combined_k141_143529||full Not-determined
17-109-final-viral-combined_k141_77521||full Low-quality
17-109-final-viral-combined_k141_210605||full Low-quality
17-109-final-viral-combined_k141_125940||full Not-determined
17-109-final-viral-combined_k141_260456||full Low-quality
17-109-final-viral-combined_k141_9529||full Low-quality
17-109-final-viral-combined_k141_125951||full Not-determined
17-109-final-viral-combined_k141_359982||full Not-determined
17-109-final-viral-combined_k141_295161||full Low-quality
17-109-final-viral-combined_k141_43858||full Not-determined
17-109-final-viral-combined_k141_59730||full Not-determined
17-109-final-viral-combined_k141_243972||full Not-determined
17-109-final-viral-combined_k141_243985||full Low-quality
17-109-final-viral-combined_k141_243986||full Not-determined
17-109-final-viral-combined_k141_59754||full Low-quality
17-109-final-viral-combined_k141_59763||full Not-determined
17-109-final-viral-combined_k141_77583||full Not-determined
17-109-final-viral-combined_k141_260478||full Not-determined
17-109-final-viral-combined_k141_43924||full Low-quality
17-109-final-viral-combined_k141_277988||full Not-determined
17-109-final-viral-combined_k141_277989||full Not-determined
17-109-final-viral-combined_k141_9603||full Not-determined
17-109-final-viral-combined_k141_328233||full Low-quality
17-109-final-viral-combined_k141_125960||full Low-quality
17-109-final-viral-combined_k141_77603||full Not-determined
17-109-final-viral-combined_k141_93100||full Low-quality
17-109-final-viral-combined_k141_228033||full Low-quality
17-109-final-viral-combined_k141_344481||full Low-quality
17-109-final-viral-combined_k141_260503||full Low-quality
17-109-final-viral-combined_k141_210663||full Not-determined
17-109-final-viral-combined_k141_26804||full Low-quality
17-109-final-viral-combined_k141_378278||full Not-determined
17-109-final-viral-combined_k141_378280||full Not-determined
17-109-final-viral-combined_k141_278014||full Not-determined
17-109-final-viral-combined_k141_312504||full Low-quality
17-109-final-viral-combined_k141_260513||full Low-quality
17-109-final-viral-combined_k141_295197||full Low-quality
17-109-final-viral-combined_k141_93114||full Not-determined
17-109-final-viral-combined_k141_59823||full Not-determined
17-109-final-viral-combined_k141_228046||full Low-quality
17-109-final-viral-combined_k141_26805||full Not-determined
17-109-final-viral-combined_k141_360070||full Low-quality
17-109-final-viral-combined_k141_394925||full Low-quality
17-109-final-viral-combined_k141_210669||full Not-determined
17-109-final-viral-combined_k141_378296||full Low-quality
17-109-final-viral-combined_k141_260518||full Not-determined
17-109-final-viral-combined_k141_160804||full Low-quality
17-109-final-viral-combined_k141_26813||full Not-determined
17-109-final-viral-combined_k141_177168||full Low-quality
17-109-final-viral-combined_k141_278028||full Not-determined
17-109-final-viral-combined_k141_328262||full Not-determined
17-109-final-viral-combined_k141_143565||full Not-determined
17-109-final-viral-combined_k141_228058||full Not-determined
17-109-final-viral-combined_k141_93153||full Not-determined
17-109-final-viral-combined_k141_59856||full Not-determined
17-109-final-viral-combined_k141_344499||full Not-determined
17-109-final-viral-combined_k141_77632||full Not-determined
17-109-final-viral-combined_k141_295230||full Low-quality
17-109-final-viral-combined_k141_394942||full Not-determined
17-109-final-viral-combined_k141_360111||full Not-determined
17-109-final-viral-combined_k141_328298||full Not-determined
17-109-final-viral-combined_k141_344528||full Low-quality
17-109-final-viral-combined_k141_244095||full Low-quality
17-109-final-viral-combined_k141_177197||full Low-quality
17-109-final-viral-combined_k141_44023||full Not-determined
17-109-final-viral-combined_k141_77675||full Low-quality
17-109-final-viral-combined_k141_160858||full Low-quality
17-109-final-viral-combined_k141_378328||full Not-determined
17-109-final-viral-combined_k141_295260||full Not-determined
17-109-final-viral-combined_k141_228087||full Low-quality
17-109-final-viral-combined_k141_344543||full Not-determined
17-109-final-viral-combined_k141_312573||full Not-determined
17-109-final-viral-combined_k141_295290||full Not-determined
17-109-final-viral-combined_k141_177230||full Not-determined
17-109-final-viral-combined_k141_378367||full Not-determined
17-109-final-viral-combined_k141_378369||full Not-determined
17-109-final-viral-combined_k141_77692||full Not-determined
17-109-final-viral-combined_k141_278067||full Low-quality
17-109-final-viral-combined_k141_312602||full Not-determined
17-109-final-viral-combined_k141_44083||full Not-determined
17-109-final-viral-combined_k141_344553||full Low-quality
17-109-final-viral-combined_k141_394981||full Not-determined
17-109-final-viral-combined_k141_160908||full Not-determined
17-109-final-viral-combined_k141_143636||full Low-quality
17-109-final-viral-combined_k141_26878||full Low-quality
17-109-final-viral-combined_k141_59953||full Not-determined
17-109-final-viral-combined_k141_210809||full Not-determined
17-109-final-viral-combined_k141_344563||full Low-quality
17-109-final-viral-combined_k141_160917||full Not-determined
17-109-final-viral-combined_k141_344572||full Not-determined
17-109-final-viral-combined_k141_378415||full Not-determined
17-109-final-viral-combined_k141_177253||full Low-quality
17-109-final-viral-combined_k141_160933||full Low-quality
17-109-final-viral-combined_k141_378423||full Not-determined
17-109-final-viral-combined_k141_328335||full Not-determined
17-109-final-viral-combined_k141_9666||full Low-quality
17-109-final-viral-combined_k141_109967||full Low-quality
17-109-final-viral-combined_k141_44136||full Not-determined
17-109-final-viral-combined_k141_401401||full High-quality
17-109-final-viral-combined_k141_328356||full Not-determined
17-109-final-viral-combined_k141_260673||full Low-quality
17-109-final-viral-combined_k141_44153||full Low-quality
17-109-final-viral-combined_k141_295361||full Low-quality
17-109-final-viral-combined_k141_143692||full Not-determined
17-109-final-viral-combined_k141_378477||full Low-quality
17-109-final-viral-combined_k141_228179||full Low-quality
17-109-final-viral-combined_k141_177313||full Not-determined
17-109-final-viral-combined_k141_59966||full Not-determined
17-109-final-viral-combined_k141_228184||full Low-quality
17-109-final-viral-combined_k141_378479||full Low-quality
17-109-final-viral-combined_k141_26968||full Low-quality
17-109-final-viral-combined_k141_77726||full Not-determined
17-109-final-viral-combined_k141_77729||full Not-determined
17-109-final-viral-combined_k141_109997||full Not-determined
17-109-final-viral-combined_k141_194848||full Low-quality
17-109-final-viral-combined_k141_378497||full Not-determined
17-109-final-viral-combined_k141_278131||full Low-quality
17-109-final-viral-combined_k141_295365||full Low-quality
17-109-final-viral-combined_k141_44191||full Not-determined
17-109-final-viral-combined_k141_194868||full Low-quality
17-109-final-viral-combined_k141_260738||full Low-quality
17-109-final-viral-combined_k141_360168||full Low-quality
17-109-final-viral-combined_k141_161015||full Low-quality
17-109-final-viral-combined_k141_77751||full Low-quality
17-109-final-viral-combined_k141_344653||full Low-quality
17-109-final-viral-combined_k141_295374||full Low-quality
17-109-final-viral-combined_k141_312704||full Not-determined
17-109-final-viral-combined_k141_228207||full Not-determined
17-109-final-viral-combined_k141_161018||full Not-determined
17-109-final-viral-combined_k141_93310||full Not-determined
17-109-final-viral-combined_k141_161022||full Low-quality
17-109-final-viral-combined_k141_260760||full Low-quality
17-109-final-viral-combined_k141_60031||full Not-determined
17-109-final-viral-combined_k141_210901||full Low-quality
17-109-final-viral-combined_k141_60037||full Not-determined
17-109-final-viral-combined_k141_278187||full Not-determined
17-109-final-viral-combined_k141_312737||full Low-quality
17-109-final-viral-combined_k141_177344||full Not-determined
17-109-final-viral-combined_k141_126106||full Low-quality
17-109-final-viral-combined_k141_9685||full Low-quality
17-109-final-viral-combined_k141_93346||full Low-quality
17-109-final-viral-combined_k141_26995||full Not-determined
17-109-final-viral-combined_k141_60057||full Low-quality
17-109-final-viral-combined_k141_44226||full Not-determined
17-109-final-viral-combined_k141_395157||full Not-determined
17-109-final-viral-combined_k141_60072||full Low-quality
17-109-final-viral-combined_k141_395159||full Not-determined
17-109-final-viral-combined_k141_328444||full Not-determined
17-109-final-viral-combined_k141_395167||full Not-determined
17-109-final-viral-combined_k141_378568||full Low-quality
17-109-final-viral-combined_k141_244239||full Low-quality
17-109-final-viral-combined_k141_60088||full Not-determined
17-109-final-viral-combined_k141_77770||full Not-determined
17-109-final-viral-combined_k141_60101||full Low-quality
17-109-final-viral-combined_k141_77777||full Not-determined
17-109-final-viral-combined_k141_244257||full Not-determined

| | |
|---|---|
| 17-109-final-viral-combined_k141_194969\|\|full | Low-quality |
| 17-109-final-viral-combined_k141_395204\|\|full | Low-quality |
| 17-109-final-viral-combined_k141_278298\|\|full | Not-determined |
| 17-109-final-viral-combined_k141_9733\|\|full | Low-quality |
| 17-109-final-viral-combined_k141_9735\|\|full | Low-quality |
| 17-109-final-viral-combined_k141_126165\|\|full | Low-quality |
| 17-109-final-viral-combined_k141_328480\|\|full | Not-determined |
| 17-109-final-viral-combined_k141_344787\|\|full | Low-quality |
| 18-101-final-viral-combined_k141_351889\|\|full | Medium-quality |
| 17-109-final-viral-combined_k141_395235\|\|full | Low-quality |
| 17-109-final-viral-combined_k141_260836\|\|full | Not-determined |
| 17-109-final-viral-combined_k141_378601\|\|full | Low-quality |
| 17-109-final-viral-combined_k141_328499\|\|full | Not-determined |
| 17-109-final-viral-combined_k141_344806\|\|full | Not-determined |
| 17-109-final-viral-combined_k141_143780\|\|full | Low-quality |
| 17-109-final-viral-combined_k141_260841\|\|full | Low-quality |
| 17-109-final-viral-combined_k141_126192\|\|full | Low-quality |
| 17-109-final-viral-combined_k141_60155\|\|full | Low-quality |
| 17-109-final-viral-combined_k141_195004\|\|full | Not-determined |
| 17-109-final-viral-combined_k141_110038\|\|full | Low-quality |
| 17-109-final-viral-combined_k141_260868\|\|full | Low-quality |
| 17-109-final-viral-combined_k141_9787\|\|full | Not-determined |
| 17-109-final-viral-combined_k141_210973\|\|full | Low-quality |
| 17-109-final-viral-combined_k141_395252\|\|full | Not-determined |
| 17-109-final-viral-combined_k141_210975\|\|full | Not-determined |
| 17-109-final-viral-combined_k141_278391\|\|full | Not-determined |
| 17-109-final-viral-combined_k141_278392\|\|full | Not-determined |
| 17-109-final-viral-combined_k141_260879\|\|full | Low-quality |
| 17-109-final-viral-combined_k141_126230\|\|full | Low-quality |
| 17-109-final-viral-combined_k141_60207\|\|full | Low-quality |
| 17-109-final-viral-combined_k141_312775\|\|full | Low-quality |
| 17-109-final-viral-combined_k141_9807\|\|full | Low-quality |
| 17-109-final-viral-combined_k141_27039\|\|full | Not-determined |
| 17-109-final-viral-combined_k141_60220\|\|full | Not-determined |
| 17-109-final-viral-combined_k141_228344\|\|full | Low-quality |
| 17-109-final-viral-combined_k141_278421\|\|full | Not-determined |
| 17-109-final-viral-combined_k141_27059\|\|full | Low-quality |
| 17-109-final-viral-combined_k141_211002\|\|full | Low-quality |
| 17-109-final-viral-combined_k141_143834\|\|full | Not-determined |
| 17-109-final-viral-combined_k141_395277\|\|full | Not-determined |
| 17-109-final-viral-combined_k141_211005\|\|full | Not-determined |
| 17-109-final-viral-combined_k141_60244\|\|full | Low-quality |
| 17-109-final-viral-combined_k141_195050\|\|full | Not-determined |
| 17-109-final-viral-combined_k141_378702\|\|full | Low-quality |
| 17-109-final-viral-combined_k141_126252\|\|full | Not-determined |
| 17-109-final-viral-combined_k141_143861\|\|full | Not-determined |
| 17-109-final-viral-combined_k141_211042\|\|full | Low-quality |
| 17-109-final-viral-combined_k141_395333\|\|full | Not-determined |
| 17-109-final-viral-combined_k141_195094\|\|full | Not-determined |
| 17-109-final-viral-combined_k141_44421\|\|full | Low-quality |
| 18-109-final-viral-combined_k141_235165\|\|full | High-quality |
| 17-109-final-viral-combined_k141_44435\|\|full | Low-quality |
| 17-109-final-viral-combined_k141_126272\|\|full | Not-determined |
| 17-109-final-viral-combined_k141_228435\|\|full | Low-quality |
| 17-109-final-viral-combined_k141_328566\|\|full | Not-determined |
| 17-109-final-viral-combined_k141_244376\|\|full | Low-quality |
| 17-109-final-viral-combined_k141_93479\|\|full | Not-determined |
| 17-109-final-viral-combined_k141_93480\|\|full | Not-determined |
| 17-109-final-viral-combined_k141_77913\|\|full | Low-quality |
| 17-109-final-viral-combined_k141_244385\|\|full | Low-quality |
| 17-109-final-viral-combined_k141_395370\|\|full | Not-determined |
| 17-109-final-viral-combined_k141_344875\|\|full | Low-quality |
| 17-109-final-viral-combined_k141_228452\|\|full | Low-quality |
| 17-109-final-viral-combined_k141_211063\|\|full | Not-determined |
| 17-109-final-viral-combined_k141_228454\|\|full | Not-determined |
| 17-109-final-viral-combined_k141_60302\|\|full | Low-quality |
| 17-109-final-viral-combined_k141_143889\|\|full | Low-quality |
| 17-109-final-viral-combined_k141_93500\|\|full | Low-quality |
| 17-109-final-viral-combined_k141_77922\|\|full | Not-determined |
| 17-109-final-viral-combined_k141_378746\|\|full | Not-determined |
| 17-109-final-viral-combined_k141_195133\|\|full | Not-determined |
| 17-109-final-viral-combined_k141_44453\|\|full | Not-determined |
| 17-109-final-viral-combined_k141_312831\|\|full | Low-quality |
| 17-109-final-viral-combined_k141_126282\|\|full | Low-quality |
| 17-109-final-viral-combined_k141_378760\|\|full | Low-quality |
| 17-109-final-viral-combined_k141_126289\|\|full | Not-determined |
| 17-109-final-viral-combined_k141_9911\|\|full | Not-determined |
| 17-109-final-viral-combined_k141_211088\|\|full | Low-quality |
| 17-109-final-viral-combined_k141_77942\|\|full | Not-determined |
| 17-109-final-viral-combined_k141_77945\|\|full | Low-quality |
| 17-109-final-viral-combined_k141_93521\|\|full | Low-quality |
| 17-109-final-viral-combined_k141_177525\|\|full | Not-determined |
| 17-109-final-viral-combined_k141_312860\|\|full | Not-determined |
| 17-109-final-viral-combined_k141_93534\|\|full | Not-determined |
| 17-109-final-viral-combined_k141_60358\|\|full | Not-determined |
| 17-109-final-viral-combined_k141_378779\|\|full | Low-quality |
| 17-109-final-viral-combined_k141_177548\|\|full | Low-quality |
| 17-109-final-viral-combined_k141_161180\|\|full | Not-determined |
| 17-109-final-viral-combined_k141_344903\|\|full | Low-quality |
| 17-109-final-viral-combined_k141_110188\|\|full | Low-quality |
| 17-109-final-viral-combined_k141_77990\|\|full | Not-determined |
| 17-109-final-viral-combined_k141_244433\|\|full | Low-quality |
| 17-109-final-viral-combined_k141_244439\|\|full | Low-quality |
| 17-109-final-viral-combined_k141_161194\|\|full | Not-determined |
| 17-109-final-viral-combined_k141_195200\|\|full | Not-determined |
| 17-109-final-viral-combined_k141_278569\|\|full | Low-quality |
| 17-109-final-viral-combined_k141_244454\|\|full | Low-quality |
| 17-109-final-viral-combined_k141_195209\|\|full | Not-determined |
| 17-109-final-viral-combined_k141_44549\|\|full | Low-quality |
| 17-109-final-viral-combined_k141_143990\|\|full | Not-determined |
| 17-109-final-viral-combined_k141_244463\|\|full | Not-determined |
| 17-109-final-viral-combined_k141_378825\|\|full | Not-determined |
| 17-109-final-viral-combined_k141_312915\|\|full | Low-quality |
| 17-109-final-viral-combined_k141_244480\|\|full | Low-quality |
| 17-109-final-viral-combined_k141_161229\|\|full | Low-quality |
| 17-109-final-viral-combined_k141_261009\|\|full | Not-determined |
| 17-109-final-viral-combined_k141_9971\|\|full | Not-determined |
| 17-109-final-viral-combined_k141_312937\|\|full | Low-quality |
| 17-109-final-viral-combined_k141_261013\|\|full | Not-determined |
| 17-109-final-viral-combined_k141_211176\|\|full | Not-determined |
| 17-109-final-viral-combined_k141_228601\|\|full | Low-quality |
| 17-109-final-viral-combined_k141_126375\|\|full | Low-quality |
| 17-109-final-viral-combined_k141_27161\|\|full | Not-determined |
| 17-109-final-viral-combined_k141_93588\|\|full | Low-quality |
| 17-109-final-viral-combined_k141_278630\|\|full | Not-determined |
| 17-109-final-viral-combined_k141_93591\|\|full | Low-quality |
| 17-109-final-viral-combined_k141_312952\|\|full | Low-quality |
| 18-102-final-viral-combined_k141_31084\|\|full | Medium-quality |
| 17-109-final-viral-combined_k141_110283\|\|full | Not-determined |
| 17-109-final-viral-combined_k141_360282\|\|full | Not-determined |
| 17-109-final-viral-combined_k141_261048\|\|full | Not-determined |
| 17-109-final-viral-combined_k141_211219\|\|full | Low-quality |
| 17-109-final-viral-combined_k141_345009\|\|full | Low-quality |
| 17-109-final-viral-combined_k141_328599\|\|full | Not-determined |
| 17-109-final-viral-combined_k141_195279\|\|full | Not-determined |
| 17-109-final-viral-combined_k141_244553\|\|full | Low-quality |
| 17-109-final-viral-combined_k141_78019\|\|full | Low-quality |
| 17-109-final-viral-combined_k141_328620\|\|full | Not-determined |
| 17-109-final-viral-combined_k141_177679\|\|full | Low-quality |
| 17-109-final-viral-combined_k141_195299\|\|full | Low-quality |
| 17-109-final-viral-combined_k141_261121\|\|full | Not-determined |
| 17-109-final-viral-combined_k141_295543\|\|full | Not-determined |
| 17-109-final-viral-combined_k141_78059\|\|full | Low-quality |
| 17-109-final-viral-combined_k141_228650\|\|full | Low-quality |
| 17-109-final-viral-combined_k141_44588\|\|full | Low-quality |
| 17-109-final-viral-combined_k141_10032\|\|full | Low-quality |
| 17-109-final-viral-combined_k141_261133\|\|full | Low-quality |
| 17-109-final-viral-combined_k141_93654\|\|full | Low-quality |
| 17-109-final-viral-combined_k141_328678\|\|full | Not-determined |
| 17-109-final-viral-combined_k141_328679\|\|full | Low-quality |
| 17-109-final-viral-combined_k141_27230\|\|full | Not-determined |
| 17-109-final-viral-combined_k141_295571\|\|full | Low-quality |
| 17-109-final-viral-combined_k141_378877\|\|full | Not-determined |
| 17-109-final-viral-combined_k141_278733\|\|full | Not-determined |
| 17-109-final-viral-combined_k141_261145\|\|full | Low-quality |
| 17-109-final-viral-combined_k141_78098\|\|full | Not-determined |
| 17-109-final-viral-combined_k141_295589\|\|full | Low-quality |
| 17-109-final-viral-combined_k141_345117\|\|full | Low-quality |
| 17-109-final-viral-combined_k141_211331\|\|full | Low-quality |
| 17-109-final-viral-combined_k141_60506\|\|full | Low-quality |
| 17-109-final-viral-combined_k141_345119\|\|full | Not-determined |
| 17-109-final-viral-combined_k141_60509\|\|full | Not-determined |
| 17-109-final-viral-combined_k141_93685\|\|full | Low-quality |
| 17-109-final-viral-combined_k141_161364\|\|full | Not-determined |
| 17-109-final-viral-combined_k141_144183\|\|full | Low-quality |
| 17-109-final-viral-combined_k141_177758\|\|full | Not-determined |
| 17-109-final-viral-combined_k141_228687\|\|full | Not-determined |
| 17-109-final-viral-combined_k141_228694\|\|full | Not-determined |
| 17-109-final-viral-combined_k141_44620\|\|full | Low-quality |
| 17-109-final-viral-combined_k141_261183\|\|full | Low-quality |
| 17-109-final-viral-combined_k141_44622\|\|full | Low-quality |
| 17-109-final-viral-combined_k141_60543\|\|full | Low-quality |
| 17-109-final-viral-combined_k141_360386\|\|full | Not-determined |
| 17-109-final-viral-combined_k141_78138\|\|full | Low-quality |
| 17-109-final-viral-combined_k141_44629\|\|full | Not-determined |
| 17-109-final-viral-combined_k141_144221\|\|full | Not-determined |
| 17-109-final-viral-combined_k141_360398\|\|full | Not-determined |
| 17-109-final-viral-combined_k141_44635\|\|full | Low-quality |
| 17-109-final-viral-combined_k141_93714\|\|full | Not-determined |
| 17-109-final-viral-combined_k141_261211\|\|full | Not-determined |
| 17-109-final-viral-combined_k141_312986\|\|full | Low-quality |
| 17-109-final-viral-combined_k141_345180\|\|full | Low-quality |
| 17-109-final-viral-combined_k141_78165\|\|full | Not-determined |
| 17-109-final-viral-combined_k141_244637\|\|full | Not-determined |
| 17-109-final-viral-combined_k141_10077\|\|full | Not-determined |
| 17-109-final-viral-combined_k141_161422\|\|full | Low-quality |
| 17-109-final-viral-combined_k141_360424\|\|full | Not-determined |
| 17-109-final-viral-combined_k141_261221\|\|full | Low-quality |
| 17-109-final-viral-combined_k141_27357\|\|full | Low-quality |
| 17-109-final-viral-combined_k141_126541\|\|full | Low-quality |
| 17-109-final-viral-combined_k141_295632\|\|full | Low-quality |
| 17-109-final-viral-combined_k141_93753\|\|full | Low-quality |
| 17-109-final-viral-combined_k141_278868\|\|full | Not-determined |
| 17-109-final-viral-combined_k141_313039\|\|full | Not-determined |
| 17-109-final-viral-combined_k141_211424\|\|full | Low-quality |
| 17-109-final-viral-combined_k141_360460\|\|full | Not-determined |
| 17-109-final-viral-combined_k141_44704\|\|full | Low-quality |
| 17-109-final-viral-combined_k141_195432\|\|full | Low-quality |
| 17-109-final-viral-combined_k141_211438\|\|full | Not-determined |
| 17-109-final-viral-combined_k141_27374\|\|full | Not-determined |
| 17-109-final-viral-combined_k141_177819\|\|full | Low-quality |
| 17-109-final-viral-combined_k141_395579\|\|full | Not-determined |
| 17-109-final-viral-combined_k141_27392\|\|full | Not-determined |
| 17-109-final-viral-combined_k141_328794\|\|full | Low-quality |
| 17-109-final-viral-combined_k141_360499\|\|full | Low-quality |
| 17-109-final-viral-combined_k141_211464\|\|full | Not-determined |
| 17-109-final-viral-combined_k141_379063\|\|full | Not-determined |
| 17-109-final-viral-combined_k141_110448\|\|full | Low-quality |
| 17-109-final-viral-combined_k141_161487\|\|full | Not-determined |
| 17-109-final-viral-combined_k141_60572\|\|full | Not-determined |
| 17-109-final-viral-combined_k141_27430\|\|full | Not-determined |
| 17-109-final-viral-combined_k141_126590\|\|full | Low-quality |
| 17-109-final-viral-combined_k141_328801\|\|full | Low-quality |
| 17-109-final-viral-combined_k141_10137\|\|full | Not-determined |
| 17-109-final-viral-combined_k141_44745\|\|full | Not-determined |
| 17-109-final-viral-combined_k141_360526\|\|full | Low-quality |
| 17-109-final-viral-combined_k141_78296\|\|full | Not-determined |
| 17-109-final-viral-combined_k141_379109\|\|full | Low-quality |
| 17-109-final-viral-combined_k141_295681\|\|full | Low-quality |
| 17-109-final-viral-combined_k141_244764\|\|full | Low-quality |
| 17-109-final-viral-combined_k141_328825\|\|full | Not-determined |
| 17-109-final-viral-combined_k141_228783\|\|full | Low-quality |
| 17-109-final-viral-combined_k141_144247\|\|full | Not-determined |
| 17-109-final-viral-combined_k141_295685\|\|full | Low-quality |
| 17-109-final-viral-combined_k141_44760\|\|full | Low-quality |
| 17-109-final-viral-combined_k141_228802\|\|full | Not-determined |
| 17-109-final-viral-combined_k141_244786\|\|full | Not-determined |
| 17-109-final-viral-combined_k141_44766\|\|full | Not-determined |
| 17-109-final-viral-combined_k141_295695\|\|full | Low-quality |
| 17-109-final-viral-combined_k141_328843\|\|full | Not-determined |
| 17-109-final-viral-combined_k141_44771\|\|full | Low-quality |
| 17-109-final-viral-combined_k141_261345\|\|full | Low-quality |
| 17-109-final-viral-combined_k141_144277\|\|full | Low-quality |
| 17-109-final-viral-combined_k141_27496\|\|full | Low-quality |
| 17-109-final-viral-combined_k141_328858\|\|full | Not-determined |
| 17-109-final-viral-combined_k141_177879\|\|full | Not-determined |
| 17-109-final-viral-combined_k141_144286\|\|full | Not-determined |
| 17-109-final-viral-combined_k141_395628\|\|full | Low-quality |
| 17-109-final-viral-combined_k141_177881\|\|full | Not-determined |
| 17-109-final-viral-combined_k141_295728\|\|full | Low-quality |
| 17-109-final-viral-combined_k141_328872\|\|full | Not-determined |
| 17-109-final-viral-combined_k141_144293\|\|full | Not-determined |
| 17-109-final-viral-combined_k141_78344\|\|full | Not-determined |
| 17-109-final-viral-combined_k141_328884\|\|full | Not-determined |
| 17-109-final-viral-combined_k141_379169\|\|full | Not-determined |
| 17-109-final-viral-combined_k141_228844\|\|full | Not-determined |
| 17-109-final-viral-combined_k141_161528\|\|full | Not-determined |
| 17-109-final-viral-combined_k141_10194\|\|full | Not-determined |
| 17-109-final-viral-combined_k141_244823\|\|full | Low-quality |
| 17-109-final-viral-combined_k141_345309\|\|full | Low-quality |
| 17-109-final-viral-combined_k141_93848\|\|full | Low-quality |
| 17-109-final-viral-combined_k141_244832\|\|full | Not-determined |
| 17-109-final-viral-combined_k141_228849\|\|full | Low-quality |
| 17-109-final-viral-combined_k141_244844\|\|full | Not-determined |
| 17-109-final-viral-combined_k141_244855\|\|full | Not-determined |
| 17-109-final-viral-combined_k141_345355\|\|full | Low-quality |
| 17-109-final-viral-combined_k141_93884\|\|full | Not-determined |
| 17-109-final-viral-combined_k141_228883\|\|full | Not-determined |
| 17-109-final-viral-combined_k141_261411\|\|full | Low-quality |
| 17-109-final-viral-combined_k141_313181\|\|full | Low-quality |
| 17-109-final-viral-combined_k141_27573\|\|full | Not-determined |
| 17-109-final-viral-combined_k141_211613\|\|full | Not-determined |
| 17-109-final-viral-combined_k141_228894\|\|full | Low-quality |
| 17-109-final-viral-combined_k141_60616\|\|full | Not-determined |
| 17-109-final-viral-combined_k141_379238\|\|full | Low-quality |
| 17-109-final-viral-combined_k141_228902\|\|full | Low-quality |
| 17-109-final-viral-combined_k141_360622\|\|full | Low-quality |
| 17-109-final-viral-combined_k141_78431\|\|full | Not-determined |
| 17-109-final-viral-combined_k141_60625\|\|full | Low-quality |
| 17-109-final-viral-combined_k141_10219\|\|full | Low-quality |
| 17-109-final-viral-combined_k141_60626\|\|full | Low-quality |
| 17-109-final-viral-combined_k141_379252\|\|full | Low-quality |
| 17-109-final-viral-combined_k141_261422\|\|full | Low-quality |
| 17-109-final-viral-combined_k141_93939\|\|full | Not-determined |
| 17-109-final-viral-combined_k141_110526\|\|full | Low-quality |
| 17-109-final-viral-combined_k141_279117\|\|full | Not-determined |
| 17-109-final-viral-combined_k141_395714\|\|full | Not-determined |
| 18-110-final-viral-combined_k141_140059\|\|full | High-quality |
| 17-109-final-viral-combined_k141_93953\|\|full | Low-quality |
| 17-109-final-viral-combined_k141_110563\|\|full | Low-quality |
| 17-109-final-viral-combined_k141_295794\|\|full | Low-quality |
| 17-109-final-viral-combined_k141_44844\|\|full | Not-determined |
| 17-109-final-viral-combined_k141_379319\|\|full | Low-quality |
| 17-109-final-viral-combined_k141_228954\|\|full | Low-quality |
| 17-109-final-viral-combined_k141_211679\|\|full | Not-determined |
| 17-109-final-viral-combined_k141_228957\|\|full | Low-quality |
| 17-109-final-viral-combined_k141_211685\|\|full | Not-determined |
| 17-109-final-viral-combined_k141_228962\|\|full | Low-quality |
| 17-109-final-viral-combined_k141_126730\|\|full | Not-determined |
| 17-109-final-viral-combined_k141_360657\|\|full | Low-quality |
| 17-109-final-viral-combined_k141_27668\|\|full | Low-quality |
| 17-109-final-viral-combined_k141_313233\|\|full | Low-quality |
| 17-109-final-viral-combined_k141_93996\|\|full | Low-quality |
| 17-109-final-viral-combined_k141_44869\|\|full | Not-determined |
| 17-109-final-viral-combined_k141_126751\|\|full | Not-determined |
| 17-109-final-viral-combined_k141_313254\|\|full | Low-quality |
| 17-109-final-viral-combined_k141_345467\|\|full | Not-determined |
| 17-109-final-viral-combined_k141_110579\|\|full | Low-quality |
| 17-109-final-viral-combined_k141_211723\|\|full | Not-determined |
| 17-109-final-viral-combined_k141_110594\|\|full | Low-quality |
| 17-109-final-viral-combined_k141_94018\|\|full | Low-quality |
| 17-109-final-viral-combined_k141_295857\|\|full | Not-determined |
| 17-109-final-viral-combined_k141_126794\|\|full | Not-determined |
| 17-109-final-viral-combined_k141_329084\|\|full | Low-quality |
| 17-109-final-viral-combined_k141_44914\|\|full | Low-quality |
| 17-109-final-viral-combined_k141_295863\|\|full | Not-determined |
| 17-109-final-viral-combined_k141_360683\|\|full | Low-quality |
| 17-109-final-viral-combined_k141_126808\|\|full | Low-quality |
| 17-109-final-viral-combined_k141_161610\|\|full | Low-quality |
| 17-109-final-viral-combined_k141_78539\|\|full | Not-determined |
| 17-109-final-viral-combined_k141_44934\|\|full | Low-quality |
| 17-109-final-viral-combined_k141_261501\|\|full | Low-quality |
| 17-109-final-viral-combined_k141_161630\|\|full | Low-quality |
| 17-109-final-viral-combined_k141_211761\|\|full | Not-determined |
| 17-109-final-viral-combined_k141_360705\|\|full | Low-quality |
| 17-109-final-viral-combined_k141_379406\|\|full | Not-determined |
| 17-109-final-viral-combined_k141_110634\|\|full | Not-determined |
| 17-109-final-viral-combined_k141_345529\|\|full | Low-quality |
| 17-109-final-viral-combined_k141_229081\|\|full | Low-quality |
| 17-109-final-viral-combined_k141_78559\|\|full | Not-determined |
| 17-109-final-viral-combined_k141_261531\|\|full | Low-quality |
| 17-109-final-viral-combined_k141_229089\|\|full | Low-quality |
| 17-109-final-viral-combined_k141_10341\|\|full | Low-quality |
| 17-109-final-viral-combined_k141_10346\|\|full | Low-quality |
| 17-109-final-viral-combined_k141_360730\|\|full | Low-quality |
| 17-109-final-viral-combined_k141_360733\|\|full | Not-determined |
| 17-109-final-viral-combined_k141_261546\|\|full | Low-quality |
| 17-109-final-viral-combined_k141_10359\|\|full | Not-determined |
| 17-109-final-viral-combined_k141_78588\|\|full | Not-determined |
| 17-109-final-viral-combined_k141_60785\|\|full | Not-determined |
| 17-109-final-viral-combined_k141_211788\|\|full | Not-determined |
| 17-109-final-viral-combined_k141_27708\|\|full | Not-determined |
| 17-109-final-viral-combined_k141_261553\|\|full | Low-quality |
| 17-109-final-viral-combined_k141_245064\|\|full | Not-determined |
| 17-109-final-viral-combined_k141_211794\|\|full | Low-quality |
| 17-109-final-viral-combined_k141_60801\|\|full | Low-quality |
| 17-109-final-viral-combined_k141_211800\|\|full | Low-quality |
| 17-109-final-viral-combined_k141_279240\|\|full | Low-quality |
| 17-109-final-viral-combined_k141_313344\|\|full | Not-determined |
| 17-109-final-viral-combined_k141_178007\|\|full | Not-determined |
| 17-109-final-viral-combined_k141_229152\|\|full | Not-determined |
| 17-109-final-viral-combined_k141_229154\|\|full | Low-quality |
| 17-109-final-viral-combined_k141_44991\|\|full | Low-quality |
| 17-109-final-viral-combined_k141_10395\|\|full | Not-determined |
| 17-109-final-viral-combined_k141_211825\|\|full | Low-quality |
| 17-109-final-viral-combined_k141_211827\|\|full | Low-quality |
| 17-109-final-viral-combined_k141_94115\|\|full | Low-quality |
| 17-109-final-viral-combined_k141_126912\|\|full | Not-determined |
| 17-109-final-viral-combined_k141_10406\|\|full | Low-quality |
| 17-109-final-viral-combined_k141_295914\|\|full | Not-determined |
| 17-109-final-viral-combined_k141_110683\|\|full | Low-quality |
| 17-109-final-viral-combined_k141_279262\|\|full | Not-determined |
| 17-109-final-viral-combined_k141_261587\|\|full | Not-determined |
| 17-109-final-viral-combined_k141_329184\|\|full | Low-quality |
| 17-109-final-viral-combined_k141_27762\|\|full | Low-quality |
| 17-109-final-viral-combined_k141_379515\|\|full | Low-quality |
| 17-109-final-viral-combined_k141_110708\|\|full | Low-quality |
| 17-109-final-viral-combined_k141_395892\|\|full | Not-determined |
| 17-109-final-viral-combined_k141_60839\|\|full | Low-quality |
| 17-109-final-viral-combined_k141_261620\|\|full | Not-determined |
| 17-109-final-viral-combined_k141_45020\|\|full | Not-determined |
| 17-109-final-viral-combined_k141_295965\|\|full | Not-determined |
| 17-109-final-viral-combined_k141_313383\|\|full | Not-determined |
| 17-109-final-viral-combined_k141_126943\|\|full | Low-quality |
| 17-109-final-viral-combined_k141_45042\|\|full | Not-determined |
| 17-109-final-viral-combined_k141_10431\|\|full | Low-quality |
| 17-109-final-viral-combined_k141_27834\|\|full | Not-determined |
| 17-109-final-viral-combined_k141_27844\|\|full | Not-determined |
| 17-109-final-viral-combined_k141_313398\|\|full | Low-quality |
| 17-109-final-viral-combined_k141_261677\|\|full | Low-quality |
| 17-109-final-viral-combined_k141_261688\|\|full | Not-determined |
| 17-109-final-viral-combined_k141_161678\|\|full | Low-quality |
| 17-109-final-viral-combined_k141_379591\|\|full | Low-quality |
| 17-109-final-viral-combined_k141_60846\|\|full | Not-determined |
| 17-109-final-viral-combined_k141_161688\|\|full | Not-determined |
| 17-109-final-viral-combined_k141_45086\|\|full | Low-quality |
| 17-109-final-viral-combined_k141_395992\|\|full | Not-determined |
| 17-109-final-viral-combined_k141_261719\|\|full | Low-quality |
| 17-109-final-viral-combined_k141_144400\|\|full | Not-determined |
| 17-109-final-viral-combined_k141_10495\|\|full | Not-determined |
| 17-109-final-viral-combined_k141_45098\|\|full | Not-determined |
| 17-109-final-viral-combined_k141_329217\|\|full | Not-determined |
| 17-109-final-viral-combined_k141_94212\|\|full | Not-determined |
| 17-109-final-viral-combined_k141_27879\|\|full | Not-determined |
| 17-109-final-viral-combined_k141_295978\|\|full | Low-quality |
| 17-109-final-viral-combined_k141_396011\|\|full | Low-quality |
| 17-109-final-viral-combined_k141_161723\|\|full | Not-determined |
| 17-109-final-viral-combined_k141_379629\|\|full | Low-quality |
| 17-109-final-viral-combined_k141_60891\|\|full | Low-quality |
| 17-109-final-viral-combined_k141_27883\|\|full | Low-quality |
| 17-109-final-viral-combined_k141_229310\|\|full | Not-determined |
| 17-109-final-viral-combined_k141_279368\|\|full | Not-determined |
| 17-109-final-viral-combined_k141_78676\|\|full | Not-determined |
| 17-109-final-viral-combined_k141_345597\|\|full | Not-determined |
| 17-109-final-viral-combined_k141_261783\|\|full | Low-quality |
| 17-109-final-viral-combined_k141_396034\|\|full | Not-determined |
| 17-109-final-viral-combined_k141_396046\|\|full | Not-determined |
| 17-109-final-viral-combined_k141_78710\|\|full | Not-determined |
| 17-109-final-viral-combined_k141_245206\|\|full | Not-determined |
| 17-109-final-viral-combined_k141_360922\|\|full | Not-determined |
| 17-109-final-viral-combined_k141_60927\|\|full | Not-determined |
| 17-109-final-viral-combined_k141_144496\|\|full | Low-quality |
| 17-109-final-viral-combined_k141_396073\|\|full | Not-determined |
| 17-109-final-viral-combined_k141_127091\|\|full | Low-quality |
| 17-109-final-viral-combined_k141_245216\|\|full | Not-determined |
| 17-109-final-viral-combined_k141_296040\|\|full | Low-quality |
| 18-101_k141_223620 | High-quality |
| 17-109-final-viral-combined_k141_110823\|\|full | Low-quality |
| 17-109-final-viral-combined_k141_60945\|\|full | Low-quality |
| 17-109-final-viral-combined_k141_144522\|\|full | Low-quality |
| 17-109-final-viral-combined_k141_195778\|\|full | Not-determined |
| 17-109-final-viral-combined_k141_178134\|\|full | Not-determined |
| 17-109-final-viral-combined_k141_195784\|\|full | Not-determined |
| 17-109-final-viral-combined_k141_229401\|\|full | Low-quality |
| 17-109-final-viral-combined_k141_345635\|\|full | Low-quality |
| 17-109-final-viral-combined_k141_345641\|\|full | Low-quality |
| 17-109-final-viral-combined_k141_94251\|\|full | Not-determined |
| 17-109-final-viral-combined_k141_329270\|\|full | Low-quality |
| 17-109-final-viral-combined_k141_60965\|\|full | Not-determined |
| 17-109-final-viral-combined_k141_110835\|\|full | Not-determined |
| 17-109-final-viral-combined_k141_195808\|\|full | Not-determined |
| 17-109-final-viral-combined_k141_313557\|\|full | Not-determined |
| 17-109-final-viral-combined_k141_195817\|\|full | Low-quality |
| 17-109-final-viral-combined_k141_161822\|\|full | Low-quality |
| 17-109-final-viral-combined_k141_345669\|\|full | Not-determined |
| 17-109-final-viral-combined_k141_45215\|\|full | Not-determined |
| 17-109-final-viral-combined_k141_379751\|\|full | Low-quality |
| 17-109-final-viral-combined_k141_60996\|\|full | Not-determined |
| 17-109-final-viral-combined_k141_127160\|\|full | Not-determined |
| 17-109-final-viral-combined_k141_261850\|\|full | Not-determined |
| 17-109-final-viral-combined_k141_345681\|\|full | Not-determined |
| 17-109-final-viral-combined_k141_61007\|\|full | Not-determined |
| 17-109-final-viral-combined_k141_61011\|\|full | Low-quality |
| 17-109-final-viral-combined_k141_178207\|\|full | Low-quality |
| 17-109-final-viral-combined_k141_379769\|\|full | Not-determined |
| 17-109-final-viral-combined_k141_127171\|\|full | Not-determined |
| 17-109-final-viral-combined_k141_379774\|\|full | Not-determined |
| 17-109-final-viral-combined_k141_212050\|\|full | Not-determined |
| 17-109-final-viral-combined_k141_127185\|\|full | Not-determined |
| 17-109-final-viral-combined_k141_110869\|\|full | Low-quality |
| 17-109-final-viral-combined_k141_94308\|\|full | Low-quality |
| 17-109-final-viral-combined_k141_45254\|\|full | Low-quality |
| 17-109-final-viral-combined_k141_361006\|\|full | Not-determined |
| 17-109-final-viral-combined_k141_127196\|\|full | Not-determined |
| 17-109-final-viral-combined_k141_379789\|\|full | Low-quality |
| 17-109-final-viral-combined_k141_195915\|\|full | Not-determined |
| 17-109-final-viral-combined_k141_78852\|\|full | Not-determined |
| 17-109-final-viral-combined_k141_127209\|\|full | Not-determined |
| 17-109-final-viral-combined_k141_195917\|\|full | Not-determined |
| 17-109-final-viral-combined_k141_229516\|\|full | Not-determined |
| 17-109-final-viral-combined_k141_279537\|\|full | Not-determined |
| 17-109-final-viral-combined_k141_78866\|\|full | Low-quality |
| 17-109-final-viral-combined_k141_212075\|\|full | Not-determined |
| 17-109-final-viral-combined_k141_61059\|\|full | Not-determined |
| 17-109-final-viral-combined_k141_379842\|\|full | Low-quality |
| 17-109-final-viral-combined_k141_229553\|\|full | Not-determined |
| 17-109-final-viral-combined_k141_27953\|\|full | Not-determined |
| 17-109-final-viral-combined_k141_229558\|\|full | Not-determined |
| 17-109-final-viral-combined_k141_229559\|\|full | Not-determined |
| 17-109-final-viral-combined_k141_345741\|\|full | Not-determined |
| 17-109-final-viral-combined_k141_78901\|\|full | Not-determined |
| 17-109-final-viral-combined_k141_161905\|\|full | Not-determined |
| 17-109-final-viral-combined_k141_379860\|\|full | Not-determined |
| 17-109-final-viral-combined_k141_78917\|\|full | Not-determined |
| 17-109-final-viral-combined_k141_94348\|\|full | Not-determined |
| 17-109-final-viral-combined_k141_144612\|\|full | Low-quality |
| 17-109-final-viral-combined_k141_178296\|\|full | Not-determined |
| 17-109-final-viral-combined_k141_178302\|\|full | Low-quality |
| 17-109-final-viral-combined_k141_45300\|\|full | Low-quality |
| 17-109-final-viral-combined_k141_94374\|\|full | Not-determined |
| 17-109-final-viral-combined_k141_396215\|\|full | Not-determined |
| 17-109-final-viral-combined_k141_379885\|\|full | Low-quality |
| 17-109-final-viral-combined_k141_94376\|\|full | Low-quality |
| 17-109-final-viral-combined_k141_45302\|\|full | Not-determined |
| 17-109-final-viral-combined_k141_379886\|\|full | Low-quality |
| 17-109-final-viral-combined_k141_229611\|\|full | Not-determined |
| 17-109-final-viral-combined_k141_127332\|\|full | Not-determined |
| 17-109-final-viral-combined_k141_161957\|\|full | Not-determined |
| 17-109-final-viral-combined_k141_94384\|\|full | Low-quality |
| 17-109-final-viral-combined_k141_61143\|\|full | Low-quality |
| 17-109-final-viral-combined_k141_178339\|\|full | Low-quality |
| 17-109-final-viral-combined_k141_396269\|\|full | Not-determined |
| 17-109-final-viral-combined_k141_379916\|\|full | Not-determined |
| 17-109-final-viral-combined_k141_329359\|\|full | Not-determined |
| 17-109-final-viral-combined_k141_61176\|\|full | Low-quality |
| 17-109-final-viral-combined_k141_61182\|\|full | Low-quality |
| 17-109-final-viral-combined_k141_313781\|\|full | Not-determined |
| 17-109-final-viral-combined_k141_162016\|\|full | Not-determined |
| 17-109-final-viral-combined_k141_127390\|\|full | Low-quality |
| 17-109-final-viral-combined_k141_279693\|\|full | Low-quality |
| 17-109-final-viral-combined_k141_245430\|\|full | Low-quality |
| 17-109-final-viral-combined_k141_396291\|\|full | Not-determined |
| 17-109-final-viral-combined_k141_78935\|\|full | Not-determined |
| 17-109-final-viral-combined_k141_78936\|\|full | Low-quality |
| 17-109-final-viral-combined_k141_178379\|\|full | Not-determined |
| 17-109-final-viral-combined_k141_45356\|\|full | Low-quality |
| 17-109-final-viral-combined_k141_94412\|\|full | Not-determined |
| 17-109-final-viral-combined_k141_162035\|\|full | Not-determined |
| 17-109-final-viral-combined_k141_212195\|\|full | Not-determined |
| 17-109-final-viral-combined_k141_78949\|\|full | Low-quality |
| 17-109-final-viral-combined_k141_127403\|\|full | Low-quality |
| 17-109-final-viral-combined_k141_94434\|\|full | Low-quality |
| 17-109-final-viral-combined_k141_162043\|\|full | Low-quality |
| 17-109-final-viral-combined_k141_127405\|\|full | Not-determined |
| 17-109-final-viral-combined_k141_61213\|\|full | Not-determined |
| 17-109-final-viral-combined_k141_127408\|\|full | Low-quality |
| 17-109-final-viral-combined_k141_144693\|\|full | Not-determined |
| 17-109-final-viral-combined_k141_229674\|\|full | Low-quality |
| 17-109-final-viral-combined_k141_178429\|\|full | Not-determined |
| 17-109-final-viral-combined_k141_111007\|\|full | Not-determined |
| 17-109-final-viral-combined_k141_245503\|\|full | Low-quality |
| 17-109-final-viral-combined_k141_10790\|\|full | Not-determined |
| 17-109-final-viral-combined_k141_94466\|\|full | Low-quality |
| 17-109-final-viral-combined_k141_178452\|\|full | Low-quality |
| 17-109-final-viral-combined_k141_313859\|\|full | Low-quality |
| 17-109-final-viral-combined_k141_144712\|\|full | Low-quality |
| 17-109-final-viral-combined_k141_61245\|\|full | Low-quality |
| 17-109-final-viral-combined_k141_162078\|\|full | Low-quality |
| 17-109-final-viral-combined_k141_61247\|\|full | Low-quality |
| 17-109-final-viral-combined_k141_212237\|\|full | Not-determined |
| 17-109-final-viral-combined_k141_111021\|\|full | Not-determined |
| 17-109-final-viral-combined_k141_178475\|\|full | Low-quality |
| 17-109-final-viral-combined_k141_361210\|\|full | Low-quality |
| 17-109-final-viral-combined_k141_162090\|\|full | Low-quality |
| 17-109-final-viral-combined_k141_61258\|\|full | Not-determined |
| 17-109-final-viral-combined_k141_229712\|\|full | Not-determined |
| 17-109-final-viral-combined_k141_229714\|\|full | Low-quality |
| 17-109-final-viral-combined_k141_261974\|\|full | Not-determined |
| 17-109-final-viral-combined_k141_396379\|\|full | Low-quality |
| 17-109-final-viral-combined_k141_396381\|\|full | Low-quality |
| 17-109-final-viral-combined_k141_245538\|\|full | Not-determined |
| 17-109-final-viral-combined_k141_144745\|\|full | Low-quality |
| 17-109-final-viral-combined_k141_61278\|\|full | Low-quality |
| 17-109-final-viral-combined_k141_329428\|\|full | Low-quality |
| 17-109-final-viral-combined_k141_162102\|\|full | Low-quality |
| 17-109-final-viral-combined_k141_144755\|\|full | Low-quality |
| 17-109-final-viral-combined_k141_127479\|\|full | Not-determined |
| 17-109-final-viral-combined_k141_379982\|\|full | Low-quality |
| 17-109-final-viral-combined_k141_396392\|\|full | Low-quality |
| 17-109-final-viral-combined_k141_196097\|\|full | Low-quality |
| 17-109-final-viral-combined_k141_144760\|\|full | Not-determined |
| 17-109-final-viral-combined_k141_229745\|\|full | Not-determined |
| 17-109-final-viral-combined_k141_396396\|\|full | Not-determined |
| 17-109-final-viral-combined_k141_144769\|\|full | Low-quality |
| 17-109-final-viral-combined_k141_229750\|\|full | Not-determined |
| 17-109-final-viral-combined_k141_329447\|\|full | Low-quality |
| 17-109-final-viral-combined_k141_262012\|\|full | Not-determined |
| 18-101-final-viral-combined_k141_92523\|\|full | Medium-quality |
| 17-109-final-viral-combined_k141_178542\|\|full | Low-quality |
| 17-109-final-viral-combined_k141_329451\|\|full | Low-quality |
| 17-109-final-viral-combined_k141_396429\|\|full | Not-determined |
| 17-109-final-viral-combined_k141_162155\|\|full | Not-determined |
| 17-109-final-viral-combined_k141_10831\|\|full | Not-determined |
| 17-109-final-viral-combined_k141_111100\|\|full | Low-quality |
| 17-109-final-viral-combined_k141_262051\|\|full | Low-quality |
| 17-109-final-viral-combined_k141_127543\|\|full | Low-quality |
| 17-109-final-viral-combined_k141_162170\|\|full | Low-quality |
| 17-109-final-viral-combined_k141_262062\|\|full | Not-determined |
| 17-109-final-viral-combined_k141_144821\|\|full | Not-determined |
| 17-109-final-viral-combined_k141_94590\|\|full | Not-determined |
| 17-109-final-viral-combined_k141_10854\|\|full | Not-determined |
| 17-109-final-viral-combined_k141_262075\|\|full | Low-quality |
| 17-109-final-viral-combined_k141_396456\|\|full | Low-quality |
| 17-109-final-viral-combined_k141_212317\|\|full | Not-determined |
| 17-109-final-viral-combined_k141_94597\|\|full | Low-quality |
| 17-109-final-viral-combined_k141_45547\|\|full | Low-quality |
| 17-109-final-viral-combined_k141_127577\|\|full | Not-determined |
| 17-109-final-viral-combined_k141_396478\|\|full | Low-quality |
| 17-109-final-viral-combined_k141_245666\|\|full | Not-determined |
| 17-109-final-viral-combined_k141_162211\|\|full | Low-quality |
| 17-109-final-viral-combined_k141_262125\|\|full | Low-quality |
| 17-109-final-viral-combined_k141_262126\|\|full | Not-determined |
| 17-109-final-viral-combined_k141_346067\|\|full | Low-quality |
| 17-109-final-viral-combined_k141_329501\|\|full | Low-quality |
| 17-109-final-viral-combined_k141_296290\|\|full | Low-quality |
| 17-109-final-viral-combined_k141_61331\|\|full | Not-determined |
| 17-109-final-viral-combined_k141_127601\|\|full | Not-determined |
| 17-109-final-viral-combined_k141_178584\|\|full | Not-determined |
| 17-109-final-viral-combined_k141_178589\|\|full | Low-quality |
| 17-109-final-viral-combined_k141_245693\|\|full | Low-quality |
| 17-109-final-viral-combined_k141_144865\|\|full | Low-quality |
| 17-109-final-viral-combined_k141_296313\|\|full | Not-determined |
| 17-109-final-viral-combined_k141_262144\|\|full | Not-determined |
| 17-109-final-viral-combined_k141_329529\|\|full | Low-quality |
| 17-109-final-viral-combined_k141_79062\|\|full | Low-quality |
| 17-109-final-viral-combined_k141_178608\|\|full | Not-determined |
| 17-109-final-viral-combined_k141_10881\|\|full | Low-quality |
| 17-109-final-viral-combined_k141_262155\|\|full | Not-determined |
| 17-109-final-viral-combined_k141_296345\|\|full | Not-determined |
| 17-109-final-viral-combined_k141_162235\|\|full | Low-quality |
| 17-109-final-viral-combined_k141_79080\|\|full | Low-quality |
| 17-109-final-viral-combined_k141_94636\|\|full | Not-determined |
| 18-109_k141_200936 | Medium-quality |
| 17-109-final-viral-combined_k141_127619\|\|full | Low-quality |
| 17-109-final-viral-combined_k141_262159\|\|full | Not-determined |
| 17-109-final-viral-combined_k141_178624\|\|full | Not-determined |
| 17-109-final-viral-combined_k141_61389\|\|full | Low-quality |
| 17-109-final-viral-combined_k141_313960\|\|full | Low-quality |
| 17-109-final-viral-combined_k141_212407\|\|full | Low-quality |

<table>
<tr><td>17-109-final-viral-combined_k141_346136||full</td><td>Low-quality</td></tr>
<tr><td>17-109-final-viral-combined_k141_111170||full</td><td>Low-quality</td></tr>
<tr><td>17-109-final-viral-combined_k141_127638||full</td><td>Not-determined</td></tr>
<tr><td>17-109-final-viral-combined_k141_196225||full</td><td>Low-quality</td></tr>
<tr><td>17-109-final-viral-combined_k141_61405||full</td><td>Low-quality</td></tr>
<tr><td>17-109-final-viral-combined_k141_346146||full</td><td>Not-determined</td></tr>
<tr><td>17-109-final-viral-combined_k141_61407||full</td><td>Not-determined</td></tr>
<tr><td>17-109-final-viral-combined_k141_396576||full</td><td>Low-quality</td></tr>
<tr><td>17-109-final-viral-combined_k141_329584||full</td><td>Not-determined</td></tr>
<tr><td>17-109-final-viral-combined_k141_45633||full</td><td>Not-determined</td></tr>
<tr><td>17-109-final-viral-combined_k141_127688||full</td><td>Not-determined</td></tr>
<tr><td>17-109-final-viral-combined_k141_396590||full</td><td>Not-determined</td></tr>
<tr><td>17-109-final-viral-combined_k141_61462||full</td><td>Low-quality</td></tr>
<tr><td>17-109-final-viral-combined_k141_296428||full</td><td>Not-determined</td></tr>
<tr><td>17-109-final-viral-combined_k141_396599||full</td><td>Not-determined</td></tr>
<tr><td>17-109-final-viral-combined_k141_361392||full</td><td>Low-quality</td></tr>
<tr><td>17-109-final-viral-combined_k141_178704||full</td><td>Not-determined</td></tr>
<tr><td>17-109-final-viral-combined_k141_61473||full</td><td>Not-determined</td></tr>
<tr><td>17-109-final-viral-combined_k141_144902||full</td><td>Low-quality</td></tr>
<tr><td>17-109-final-viral-combined_k141_11003||full</td><td>Low-quality</td></tr>
<tr><td>17-109-final-viral-combined_k141_212456||full</td><td>Low-quality</td></tr>
<tr><td>17-109-final-viral-combined_k141_279968||full</td><td>Not-determined</td></tr>
<tr><td>17-109-final-viral-combined_k141_262223||full</td><td>Low-quality</td></tr>
<tr><td>17-109-final-viral-combined_k141_279970||full</td><td>Low-quality</td></tr>
<tr><td>17-109-final-viral-combined_k141_396638||full</td><td>Not-determined</td></tr>
<tr><td>17-109-final-viral-combined_k141_212475||full</td><td>Low-quality</td></tr>
<tr><td>18-109-final-viral-combined_k141_200936||full</td><td>Medium-quality</td></tr>
<tr><td>17-109-final-viral-combined_k141_144938||full</td><td>Not-determined</td></tr>
<tr><td>17-109-final-viral-combined_k141_162358||full</td><td>Low-quality</td></tr>
<tr><td>17-109-final-viral-combined_k141_396648||full</td><td>Low-quality</td></tr>
<tr><td>17-109-final-viral-combined_k141_396656||full</td><td>Low-quality</td></tr>
<tr><td>17-109-final-viral-combined_k141_162365||full</td><td>Not-determined</td></tr>
<tr><td>17-109-final-viral-combined_k141_296497||full</td><td>Not-determined</td></tr>
<tr><td>17-109-final-viral-combined_k141_11031||full</td><td>Low-quality</td></tr>
<tr><td>17-109-final-viral-combined_k141_212506||full</td><td>Not-determined</td></tr>
<tr><td>17-109-final-viral-combined_k141_28076||full</td><td>Low-quality</td></tr>
<tr><td>17-109-final-viral-combined_k141_94755||full</td><td>Not-determined</td></tr>
<tr><td>17-109-final-viral-combined_k141_178743||full</td><td>Low-quality</td></tr>
<tr><td>17-109-final-viral-combined_k141_262263||full</td><td>Low-quality</td></tr>
<tr><td>17-109-final-viral-combined_k141_296512||full</td><td>Not-determined</td></tr>
<tr><td>17-109-final-viral-combined_k141_361448||full</td><td>Not-determined</td></tr>
<tr><td>17-109-final-viral-combined_k141_145007||full</td><td>Low-quality</td></tr>
<tr><td>17-109-final-viral-combined_k141_262291||full</td><td>Not-determined</td></tr>
<tr><td>17-109-final-viral-combined_k141_94786||full</td><td>Low-quality</td></tr>
<tr><td>17-109-final-viral-combined_k141_196294||full</td><td>Not-determined</td></tr>
<tr><td>17-109-final-viral-combined_k141_380203||full</td><td>Low-quality</td></tr>
<tr><td>17-109-final-viral-combined_k141_346211||full</td><td>Low-quality</td></tr>
<tr><td>17-109-final-viral-combined_k141_262316||full</td><td>Low-quality</td></tr>
<tr><td>17-109-final-viral-combined_k141_329718||full</td><td>Low-quality</td></tr>
<tr><td>17-109-final-viral-combined_k141_28135||full</td><td>Low-quality</td></tr>
<tr><td>17-109-final-viral-combined_k141_262323||full</td><td>Low-quality</td></tr>
<tr><td>17-109-final-viral-combined_k141_280014||full</td><td>Low-quality</td></tr>
<tr><td>17-109-final-viral-combined_k141_361500||full</td><td>Low-quality</td></tr>
<tr><td>17-109-final-viral-combined_k141_245863||full</td><td>Not-determined</td></tr>
<tr><td>17-109-final-viral-combined_k141_11089||full</td><td>Not-determined</td></tr>
<tr><td>17-109-final-viral-combined_k141_111370||full</td><td>Not-determined</td></tr>
<tr><td>17-109-final-viral-combined_k141_314092||full</td><td>Low-quality</td></tr>
<tr><td>17-109-final-viral-combined_k141_212597||full</td><td>Low-quality</td></tr>
<tr><td>17-109-final-viral-combined_k141_314102||full</td><td>Low-quality</td></tr>
<tr><td>17-109-final-viral-combined_k141_262365||full</td><td>Low-quality</td></tr>
<tr><td>17-109-final-viral-combined_k141_280051||full</td><td>Not-determined</td></tr>
<tr><td>17-109-final-viral-combined_k141_314112||full</td><td>Not-determined</td></tr>
<tr><td>17-109-final-viral-combined_k141_178836||full</td><td>Not-determined</td></tr>
<tr><td>17-109-final-viral-combined_k141_262380||full</td><td>Not-determined</td></tr>
<tr><td>17-109-final-viral-combined_k141_245906||full</td><td>Low-quality</td></tr>
<tr><td>17-109-final-viral-combined_k141_229938||full</td><td>Low-quality</td></tr>
<tr><td>17-109-final-viral-combined_k141_380265||full</td><td>Low-quality</td></tr>
<tr><td>17-109-final-viral-combined_k141_196360||full</td><td>Not-determined</td></tr>
<tr><td>17-109-final-viral-combined_k141_127844||full</td><td>Low-quality</td></tr>
<tr><td>17-109-final-viral-combined_k141_145127||full</td><td>Low-quality</td></tr>
<tr><td>17-109-final-viral-combined_k141_212647||full</td><td>Not-determined</td></tr>
<tr><td>17-109-final-viral-combined_k141_380294||full</td><td>Not-determined</td></tr>
<tr><td>17-109-final-viral-combined_k141_94846||full</td><td>Not-determined</td></tr>
<tr><td>17-109-final-viral-combined_k141_111442||full</td><td>Low-quality</td></tr>
<tr><td>17-109-final-viral-combined_k141_361595||full</td><td>Not-determined</td></tr>
<tr><td>17-109-final-viral-combined_k141_329763||full</td><td>Not-determined</td></tr>
<tr><td>17-109-final-viral-combined_k141_361604||full</td><td>Not-determined</td></tr>
<tr><td>17-109-final-viral-combined_k141_296597||full</td><td>Low-quality</td></tr>
<tr><td>17-109-final-viral-combined_k141_396844||full</td><td>Not-determined</td></tr>
<tr><td>17-109-final-viral-combined_k141_380319||full</td><td>Not-determined</td></tr>
<tr><td>17-109-final-viral-combined_k141_178893||full</td><td>Low-quality</td></tr>
<tr><td>17-109-final-viral-combined_k141_162535||full</td><td>Not-determined</td></tr>
<tr><td>17-109-final-viral-combined_k141_361623||full</td><td>Not-determined</td></tr>
<tr><td>17-109-final-viral-combined_k141_11226||full</td><td>Low-quality</td></tr>
<tr><td>17-109-final-viral-combined_k141_79288||full</td><td>Low-quality</td></tr>
<tr><td>17-109-final-viral-combined_k141_329791||full</td><td>Low-quality</td></tr>
<tr><td>17-109-final-viral-combined_k141_229981||full</td><td>Low-quality</td></tr>
<tr><td>17-109-final-viral-combined_k141_28185||full</td><td>Not-determined</td></tr>
<tr><td>17-109-final-viral-combined_k141_229984||full</td><td>Not-determined</td></tr>
<tr><td>17-109-final-viral-combined_k141_329804||full</td><td>Not-determined</td></tr>
<tr><td>17-109-final-viral-combined_k141_45789||full</td><td>Low-quality</td></tr>
<tr><td>17-109-final-viral-combined_k141_28205||full</td><td>Not-determined</td></tr>
<tr><td>17-109-final-viral-combined_k141_127916||full</td><td>Not-determined</td></tr>
<tr><td>17-109-final-viral-combined_k141_162553||full</td><td>Not-determined</td></tr>
<tr><td>17-109-final-viral-combined_k141_346376||full</td><td>Low-quality</td></tr>
<tr><td>17-109-final-viral-combined_k141_230008||full</td><td>Not-determined</td></tr>
<tr><td>17-109-final-viral-combined_k141_127919||full</td><td>Not-determined</td></tr>
<tr><td>17-109-final-viral-combined_k141_45811||full</td><td>Low-quality</td></tr>
<tr><td>17-109-final-viral-combined_k141_61595||full</td><td>Not-determined</td></tr>
<tr><td>17-109-final-viral-combined_k141_145209||full</td><td>Not-determined</td></tr>
<tr><td>17-109-final-viral-combined_k141_127925||full</td><td>Not-determined</td></tr>
<tr><td>17-109-final-viral-combined_k141_162562||full</td><td>Low-quality</td></tr>
<tr><td>17-109-final-viral-combined_k141_11258||full</td><td>Low-quality</td></tr>
<tr><td>17-109-final-viral-combined_k141_94933||full</td><td>Not-determined</td></tr>
<tr><td>17-109-final-viral-combined_k141_380381||full</td><td>Not-determined</td></tr>
<tr><td>17-109-final-viral-combined_k141_178995||full</td><td>Not-determined</td></tr>
<tr><td>17-109-final-viral-combined_k141_230048||full</td><td>Not-determined</td></tr>
<tr><td>17-109-final-viral-combined_k141_329862||full</td><td>Not-determined</td></tr>
<tr><td>17-109-final-viral-combined_k141_230055||full</td><td>Not-determined</td></tr>
<tr><td>17-109-final-viral-combined_k141_28252||full</td><td>Low-quality</td></tr>
<tr><td>17-109-final-viral-combined_k141_380417||full</td><td>Low-quality</td></tr>
<tr><td>17-109-final-viral-combined_k141_179008||full</td><td>Low-quality</td></tr>
<tr><td>17-109-final-viral-combined_k141_230066||full</td><td>Not-determined</td></tr>
<tr><td>17-109-final-viral-combined_k141_230071||full</td><td>Not-determined</td></tr>
<tr><td>17-109-final-viral-combined_k141_11293||full</td><td>Not-determined</td></tr>
<tr><td>17-109-final-viral-combined_k141_262572||full</td><td>Low-quality</td></tr>
<tr><td>17-109-final-viral-combined_k141_296706||full</td><td>Not-determined</td></tr>
<tr><td>17-109-final-viral-combined_k141_296729||full</td><td>Not-determined</td></tr>
<tr><td>17-109-final-viral-combined_k141_95037||full</td><td>Not-determined</td></tr>
<tr><td>17-109-final-viral-combined_k141_212781||full</td><td>Not-determined</td></tr>
<tr><td>17-109-final-viral-combined_k141_361728||full</td><td>Not-determined</td></tr>
<tr><td>17-109-final-viral-combined_k141_179070||full</td><td>Low-quality</td></tr>
<tr><td>17-109-final-viral-combined_k141_280272||full</td><td>Low-quality</td></tr>
<tr><td>17-109-final-viral-combined_k141_61684||full</td><td>Not-determined</td></tr>
<tr><td>17-109-final-viral-combined_k141_296760||full</td><td>Not-determined</td></tr>
<tr><td>17-109-final-viral-combined_k141_212804||full</td><td>Low-quality</td></tr>
<tr><td>17-109-final-viral-combined_k141_380463||full</td><td>Low-quality</td></tr>
<tr><td>17-109-final-viral-combined_k141_230147||full</td><td>Low-quality</td></tr>
<tr><td>17-109-final-viral-combined_k141_212809||full</td><td>Not-determined</td></tr>
<tr><td>17-109-final-viral-combined_k141_296771||full</td><td>Low-quality</td></tr>
<tr><td>17-109-final-viral-combined_k141_28314||full</td><td>Not-determined</td></tr>
<tr><td>17-109-final-viral-combined_k141_280286||full</td><td>Not-determined</td></tr>
<tr><td>17-109-final-viral-combined_k141_262615||full</td><td>Not-determined</td></tr>
<tr><td>17-109-final-viral-combined_k141_11300||full</td><td>Not-determined</td></tr>
<tr><td>17-109-final-viral-combined_k141_314230||full</td><td>Low-quality</td></tr>
<tr><td>17-109-final-viral-combined_k141_246000||full</td><td>Low-quality</td></tr>
<tr><td>17-109-final-viral-combined_k141_296790||full</td><td>Low-quality</td></tr>
<tr><td>17-109-final-viral-combined_k141_280303||full</td><td>Not-determined</td></tr>
<tr><td>17-109-final-viral-combined_k141_230171||full</td><td>Not-determined</td></tr>
<tr><td>17-109-final-viral-combined_k141_45968||full</td><td>Low-quality</td></tr>
<tr><td>17-109-final-viral-combined_k141_11320||full</td><td>Not-determined</td></tr>
<tr><td>17-109-final-viral-combined_k141_45973||full</td><td>Not-determined</td></tr>
<tr><td>17-109-final-viral-combined_k141_397001||full</td><td>Not-determined</td></tr>
<tr><td>17-109-final-viral-combined_k141_230189||full</td><td>Not-determined</td></tr>
<tr><td>17-109-final-viral-combined_k141_246021||full</td><td>Not-determined</td></tr>
<tr><td>17-109-final-viral-combined_k141_45990||full</td><td>Not-determined</td></tr>
<tr><td>18-101_k141_4277</td><td>Medium-quality</td></tr>
<tr><td>17-109-final-viral-combined_k141_79345||full</td><td>Not-determined</td></tr>
<tr><td>17-109-final-viral-combined_k141_95141||full</td><td>Not-determined</td></tr>
<tr><td>17-109-final-viral-combined_k141_46005||full</td><td>Low-quality</td></tr>
<tr><td>17-109-final-viral-combined_k141_397033||full</td><td>Low-quality</td></tr>
<tr><td>17-109-final-viral-combined_k141_212855||full</td><td>Not-determined</td></tr>
<tr><td>17-109-final-viral-combined_k141_397035||full</td><td>Not-determined</td></tr>
<tr><td>17-109-final-viral-combined_k141_212856||full</td><td>Low-quality</td></tr>
<tr><td>17-109-final-viral-combined_k141_230220||full</td><td>Low-quality</td></tr>
<tr><td>17-109-final-viral-combined_k141_397045||full</td><td>Low-quality</td></tr>
<tr><td>17-109-final-viral-combined_k141_79353||full</td><td>Low-quality</td></tr>
<tr><td>17-109-final-viral-combined_k141_262696||full</td><td>Not-determined</td></tr>
<tr><td>17-109-final-viral-combined_k141_262697||full</td><td>Low-quality</td></tr>
<tr><td>17-109-final-viral-combined_k141_380516||full</td><td>Not-determined</td></tr>
<tr><td>17-109-final-viral-combined_k141_111575||full</td><td>Not-determined</td></tr>
<tr><td>17-109-final-viral-combined_k141_128166||full</td><td>Not-determined</td></tr>
<tr><td>17-109-final-viral-combined_k141_28401||full</td><td>Low-quality</td></tr>
<tr><td>17-109-final-viral-combined_k141_314318||full</td><td>Not-determined</td></tr>
<tr><td>17-109-final-viral-combined_k141_329987||full</td><td>Low-quality</td></tr>
<tr><td>17-109-final-viral-combined_k141_397059||full</td><td>Not-determined</td></tr>
<tr><td>17-109-final-viral-combined_k141_179126||full</td><td>Not-determined</td></tr>
<tr><td>17-109-final-viral-combined_k141_95195||full</td><td>Low-quality</td></tr>
<tr><td>17-109-final-viral-combined_k141_212896||full</td><td>Low-quality</td></tr>
<tr><td>17-109-final-viral-combined_k141_380528||full</td><td>Not-determined</td></tr>
<tr><td>17-109-final-viral-combined_k141_179145||full</td><td>Not-determined</td></tr>
<tr><td>17-109-final-viral-combined_k141_95213||full</td><td>Low-quality</td></tr>
<tr><td>17-109-final-viral-combined_k141_95238||full</td><td>Low-quality</td></tr>
<tr><td>17-109-final-viral-combined_k141_280367||full</td><td>Not-determined</td></tr>
<tr><td>17-109-final-viral-combined_k141_46099||full</td><td>Not-determined</td></tr>
<tr><td>17-109-final-viral-combined_k141_111613||full</td><td>Not-determined</td></tr>
<tr><td>17-109-final-viral-combined_k141_262727||full</td><td>Not-determined</td></tr>
<tr><td>17-109-final-viral-combined_k141_128239||full</td><td>Not-determined</td></tr>
<tr><td>17-109-final-viral-combined_k141_330037||full</td><td>Not-determined</td></tr>
<tr><td>17-109-final-viral-combined_k141_212946||full</td><td>Low-quality</td></tr>
<tr><td>17-109-final-viral-combined_k141_162730||full</td><td>Not-determined</td></tr>
<tr><td>17-109-final-viral-combined_k141_111627||full</td><td>Low-quality</td></tr>
<tr><td>17-109-final-viral-combined_k141_262753||full</td><td>Not-determined</td></tr>
<tr><td>17-109-final-viral-combined_k141_179182||full</td><td>Low-quality</td></tr>
<tr><td>17-109-final-viral-combined_k141_397092||full</td><td>Not-determined</td></tr>
<tr><td>17-109-final-viral-combined_k141_246092||full</td><td>Not-determined</td></tr>
<tr><td>17-109-final-viral-combined_k141_145453||full</td><td>Low-quality</td></tr>
<tr><td>17-109-final-viral-combined_k141_28485||full</td><td>Not-determined</td></tr>
<tr><td>17-109-final-viral-combined_k141_128269||full</td><td>Low-quality</td></tr>
<tr><td>17-109-final-viral-combined_k141_330052||full</td><td>Low-quality</td></tr>
<tr><td>17-109-final-viral-combined_k141_314375||full</td><td>Not-determined</td></tr>
<tr><td>17-109-final-viral-combined_k141_380594||full</td><td>Not-determined</td></tr>
<tr><td>17-109-final-viral-combined_k141_111628||full</td><td>Low-quality</td></tr>
<tr><td>17-109-final-viral-combined_k141_262789||full</td><td>Not-determined</td></tr>
<tr><td>17-109-final-viral-combined_k141_95302||full</td><td>Low-quality</td></tr>
<tr><td>17-109-final-viral-combined_k141_246114||full</td><td>Low-quality</td></tr>
<tr><td>17-109-final-viral-combined_k141_61883||full</td><td>Not-determined</td></tr>
<tr><td>17-109-final-viral-combined_k141_346606||full</td><td>Not-determined</td></tr>
<tr><td>17-109-final-viral-combined_k141_361845||full</td><td>Low-quality</td></tr>
<tr><td>17-109-final-viral-combined_k141_79485||full</td><td>Low-quality</td></tr>
<tr><td>17-109-final-viral-combined_k141_162775||full</td><td>Not-determined</td></tr>
<tr><td>17-109-final-viral-combined_k141_95332||full</td><td>Low-quality</td></tr>
<tr><td>17-109-final-viral-combined_k141_346626||full</td><td>Low-quality</td></tr>
<tr><td>17-109-final-viral-combined_k141_111665||full</td><td>Low-quality</td></tr>
<tr><td>17-109-final-viral-combined_k141_196520||full</td><td>Low-quality</td></tr>
<tr><td>17-109-final-viral-combined_k141_314422||full</td><td>Low-quality</td></tr>
<tr><td>17-109-final-viral-combined_k141_145498||full</td><td>Not-determined</td></tr>
<tr><td>17-109-final-viral-combined_k141_280473||full</td><td>Low-quality</td></tr>
<tr><td>17-109-final-viral-combined_k141_280477||full</td><td>Not-determined</td></tr>
<tr><td>17-109-final-viral-combined_k141_111671||full</td><td>Not-determined</td></tr>
<tr><td>17-109-final-viral-combined_k141_95353||full</td><td>Not-determined</td></tr>
<tr><td>17-109-final-viral-combined_k141_162794||full</td><td>Low-quality</td></tr>
<tr><td>17-109-final-viral-combined_k141_145503||full</td><td>Not-determined</td></tr>
<tr><td>17-109-final-viral-combined_k141_246160||full</td><td>Low-quality</td></tr>
<tr><td>17-109-final-viral-combined_k141_346657||full</td><td>Low-quality</td></tr>
<tr><td>17-109-final-viral-combined_k141_213005||full</td><td>Not-determined</td></tr>
<tr><td>17-109-final-viral-combined_k141_79543||full</td><td>Not-determined</td></tr>
<tr><td>17-109-final-viral-combined_k141_361893||full</td><td>Not-determined</td></tr>
<tr><td>17-109-final-viral-combined_k141_262844||full</td><td>Not-determined</td></tr>
<tr><td>17-109-final-viral-combined_k141_179303||full</td><td>Not-determined</td></tr>
<tr><td>17-109-final-viral-combined_k141_128350||full</td><td>Not-determined</td></tr>
<tr><td>17-109-final-viral-combined_k141_111707||full</td><td>Low-quality</td></tr>
<tr><td>17-109-final-viral-combined_k141_397191||full</td><td>Not-determined</td></tr>
<tr><td>17-109-final-viral-combined_k141_46222||full</td><td>Not-determined</td></tr>
<tr><td>17-109-final-viral-combined_k141_46232||full</td><td>Low-quality</td></tr>
<tr><td>17-109-final-viral-combined_k141_297012||full</td><td>Not-determined</td></tr>
<tr><td>17-109-final-viral-combined_k141_314479||full</td><td>Low-quality</td></tr>
<tr><td>17-109-final-viral-combined_k141_128389||full</td><td>Low-quality</td></tr>
<tr><td>17-109-final-viral-combined_k141_297025||full</td><td>Low-quality</td></tr>
<tr><td>17-109-final-viral-combined_k141_145590||full</td><td>Not-determined</td></tr>
<tr><td>17-109-final-viral-combined_k141_246226||full</td><td>Not-determined</td></tr>
<tr><td>17-109-final-viral-combined_k141_346748||full</td><td>Not-determined</td></tr>
<tr><td>17-109-final-viral-combined_k141_246231||full</td><td>Low-quality</td></tr>
<tr><td>17-109-final-viral-combined_k141_346756||full</td><td>Low-quality</td></tr>
<tr><td>17-109-final-viral-combined_k141_62031||full</td><td>Not-determined</td></tr>
<tr><td>17-109-final-viral-combined_k141_397209||full</td><td>Not-determined</td></tr>
<tr><td>17-109-final-viral-combined_k141_128429||full</td><td>Not-determined</td></tr>
<tr><td>17-109-final-viral-combined_k141_46312||full</td><td>Not-determined</td></tr>
<tr><td>17-109-final-viral-combined_k141_314527||full</td><td>Low-quality</td></tr>
<tr><td>17-109-final-viral-combined_k141_297057||full</td><td>Low-quality</td></tr>
<tr><td>17-109-final-viral-combined_k141_380664||full</td><td>Not-determined</td></tr>
<tr><td>17-109-final-viral-combined_k141_262936||full</td><td>Low-quality</td></tr>
<tr><td>17-109-final-viral-combined_k141_46328||full</td><td>Low-quality</td></tr>
<tr><td>17-109-final-viral-combined_k141_346790||full</td><td>Not-determined</td></tr>
<tr><td>17-109-final-viral-combined_k141_162847||full</td><td>Low-quality</td></tr>
<tr><td>17-109-final-viral-combined_k141_62078||full</td><td>Low-quality</td></tr>
<tr><td>17-109-final-viral-combined_k141_330246||full</td><td>Not-determined</td></tr>
<tr><td>17-109-final-viral-combined_k141_346795||full</td><td>Not-determined</td></tr>
<tr><td>17-109-final-viral-combined_k141_397229||full</td><td>Low-quality</td></tr>
<tr><td>17-109-final-viral-combined_k141_79615||full</td><td>Not-determined</td></tr>
<tr><td>17-109-final-viral-combined_k141_213104||full</td><td>Not-determined</td></tr>
<tr><td>17-109-final-viral-combined_k141_397241||full</td><td>Not-determined</td></tr>
<tr><td>17-109-final-viral-combined_k141_262951||full</td><td>Not-determined</td></tr>
<tr><td>17-109-final-viral-combined_k141_213115||full</td><td>Low-quality</td></tr>
<tr><td>17-109-final-viral-combined_k141_246289||full</td><td>Not-determined</td></tr>
<tr><td>17-109-final-viral-combined_k141_262959||full</td><td>Low-quality</td></tr>
<tr><td>17-109-final-viral-combined_k141_280651||full</td><td>Not-determined</td></tr>
<tr><td>17-109-final-viral-combined_k141_213131||full</td><td>Low-quality</td></tr>
<tr><td>17-109-final-viral-combined_k141_380686||full</td><td>Not-determined</td></tr>
<tr><td>17-109-final-viral-combined_k141_314572||full</td><td>Low-quality</td></tr>
<tr><td>17-109-final-viral-combined_k141_280661||full</td><td>Low-quality</td></tr>
<tr><td>17-109-final-viral-combined_k141_330276||full</td><td>Not-determined</td></tr>
<tr><td>17-109-final-viral-combined_k141_280664||full</td><td>Not-determined</td></tr>
<tr><td>17-109-final-viral-combined_k141_162887||full</td><td>Not-determined</td></tr>
<tr><td>17-109-final-viral-combined_k141_361968||full</td><td>Low-quality</td></tr>
<tr><td>17-109-final-viral-combined_k141_297095||full</td><td>Not-determined</td></tr>
<tr><td>17-109-final-viral-combined_k141_361972||full</td><td>Not-determined</td></tr>
<tr><td>17-109-final-viral-combined_k141_46389||full</td><td>Low-quality</td></tr>
<tr><td>18-101-final-viral-combined_k141_4277||full</td><td>Medium-quality</td></tr>
<tr><td>17-109-final-viral-combined_k141_314578||full</td><td>Low-quality</td></tr>
<tr><td>17-109-final-viral-combined_k141_361979||full</td><td>Low-quality</td></tr>
<tr><td>17-109-final-viral-combined_k141_46401||full</td><td>Low-quality</td></tr>
<tr><td>17-109-final-viral-combined_k141_230330||full</td><td>Not-determined</td></tr>
<tr><td>17-109-final-viral-combined_k141_361985||full</td><td>Low-quality</td></tr>
<tr><td>17-109-final-viral-combined_k141_111834||full</td><td>Not-determined</td></tr>
<tr><td>17-109-final-viral-combined_k141_95463||full</td><td>Not-determined</td></tr>
<tr><td>17-109-final-viral-combined_k141_179401||full</td><td>Low-quality</td></tr>
<tr><td>17-109-final-viral-combined_k141_179403||full</td><td>Not-determined</td></tr>
<tr><td>17-109-final-viral-combined_k141_179408||full</td><td>Low-quality</td></tr>
<tr><td>17-109-final-viral-combined_k141_179411||full</td><td>Low-quality</td></tr>
<tr><td>17-109-final-viral-combined_k141_111855||full</td><td>Not-determined</td></tr>
<tr><td>17-109-final-viral-combined_k141_128544||full</td><td>Low-quality</td></tr>
<tr><td>17-109-final-viral-combined_k141_196598||full</td><td>Not-determined</td></tr>
<tr><td>17-109-final-viral-combined_k141_79694||full</td><td>Not-determined</td></tr>
<tr><td>17-109-final-viral-combined_k141_362016||full</td><td>Low-quality</td></tr>
<tr><td>17-109-final-viral-combined_k141_128573||full</td><td>Not-determined</td></tr>
<tr><td>17-109-final-viral-combined_k141_179434||full</td><td>Low-quality</td></tr>
<tr><td>17-109-final-viral-combined_k141_280691||full</td><td>Not-determined</td></tr>
<tr><td>17-109-final-viral-combined_k141_263061||full</td><td>Not-determined</td></tr>
<tr><td>17-109-final-viral-combined_k141_62181||full</td><td>Not-determined</td></tr>
<tr><td>17-109-final-viral-combined_k141_62191||full</td><td>Not-determined</td></tr>
<tr><td>17-109-final-viral-combined_k141_362034||full</td><td>Not-determined</td></tr>
<tr><td>17-109-final-viral-combined_k141_179450||full</td><td>Low-quality</td></tr>
<tr><td>17-109-final-viral-combined_k141_11658||full</td><td>Low-quality</td></tr>
<tr><td>17-109-final-viral-combined_k141_230358||full</td><td>Low-quality</td></tr>
<tr><td>17-109-final-viral-combined_k141_196623||full</td><td>Not-determined</td></tr>
<tr><td>17-109-final-viral-combined_k141_362042||full</td><td>Not-determined</td></tr>
<tr><td>17-109-final-viral-combined_k141_11666||full</td><td>Not-determined</td></tr>
<tr><td>17-109-final-viral-combined_k141_230364||full</td><td>Low-quality</td></tr>
<tr><td>17-109-final-viral-combined_k141_79734||full</td><td>Not-determined</td></tr>
<tr><td>17-109-final-viral-combined_k141_362051||full</td><td>Low-quality</td></tr>
<tr><td>17-109-final-viral-combined_k141_280726||full</td><td>Not-determined</td></tr>
<tr><td>17-109-final-viral-combined_k141_79737||full</td><td>Not-determined</td></tr>
<tr><td>17-109-final-viral-combined_k141_330342||full</td><td>Low-quality</td></tr>
<tr><td>17-109-final-viral-combined_k141_95521||full</td><td>Low-quality</td></tr>
<tr><td>17-109-final-viral-combined_k141_314659||full</td><td>Low-quality</td></tr>
<tr><td>17-109-final-viral-combined_k141_230384||full</td><td>Low-quality</td></tr>
<tr><td>17-109-final-viral-combined_k141_297185||full</td><td>Low-quality</td></tr>
<tr><td>17-109-final-viral-combined_k141_230389||full</td><td>Not-determined</td></tr>
<tr><td>17-109-final-viral-combined_k141_196651||full</td><td>Not-determined</td></tr>
<tr><td>17-109-final-viral-combined_k141_46496||full</td><td>Low-quality</td></tr>
<tr><td>17-109-final-viral-combined_k141_314679||full</td><td>Low-quality</td></tr>
<tr><td>17-109-final-viral-combined_k141_230416||full</td><td>Low-quality</td></tr>
<tr><td>17-109-final-viral-combined_k141_346984||full</td><td>Low-quality</td></tr>
<tr><td>17-109-final-viral-combined_k141_246376||full</td><td>Not-determined</td></tr>
<tr><td>17-109-final-viral-combined_k141_95538||full</td><td>Not-determined</td></tr>
<tr><td>17-109-final-viral-combined_k141_397280||full</td><td>Low-quality</td></tr>
<tr><td>17-109-final-viral-combined_k141_179533||full</td><td>Not-determined</td></tr>
<tr><td>17-109-final-viral-combined_k141_213271||full</td><td>Not-determined</td></tr>
<tr><td>17-109-final-viral-combined_k141_79795||full</td><td>Not-determined</td></tr>
<tr><td>17-109-final-viral-combined_k141_95554||full</td><td>Not-determined</td></tr>
<tr><td>17-109-final-viral-combined_k141_380787||full</td><td>Not-determined</td></tr>
<tr><td>17-109-final-viral-combined_k141_179570||full</td><td>Not-determined</td></tr>
<tr><td>17-109-final-viral-combined_k141_28763||full</td><td>Low-quality</td></tr>
<tr><td>17-109-final-viral-combined_k141_62254||full</td><td>Not-determined</td></tr>
<tr><td>17-109-final-viral-combined_k141_380790||full</td><td>Low-quality</td></tr>
<tr><td>17-109-final-viral-combined_k141_162981||full</td><td>Low-quality</td></tr>
<tr><td>17-109-final-viral-combined_k141_397315||full</td><td>Not-determined</td></tr>
<tr><td>17-109-final-viral-combined_k141_46526||full</td><td>Low-quality</td></tr>
<tr><td>17-109-final-viral-combined_k141_362088||full</td><td>Low-quality</td></tr>
<tr><td>17-109-final-viral-combined_k141_179597||full</td><td>Not-determined</td></tr>
<tr><td>17-109-final-viral-combined_k141_230453||full</td><td>Low-quality</td></tr>
<tr><td>17-109-final-viral-combined_k141_280820||full</td><td>Low-quality</td></tr>
<tr><td>17-109-final-viral-combined_k141_280821||full</td><td>Not-determined</td></tr>
<tr><td>17-109-final-viral-combined_k141_163001||full</td><td>Low-quality</td></tr>
<tr><td>17-109-final-viral-combined_k141_280824||full</td><td>Not-determined</td></tr>
<tr><td>17-109-final-viral-combined_k141_280833||full</td><td>Not-determined</td></tr>
<tr><td>17-109-final-viral-combined_k141_179626||full</td><td>Not-determined</td></tr>
<tr><td>17-109-final-viral-combined_k141_196766||full</td><td>Low-quality</td></tr>
<tr><td>17-109-final-viral-combined_k141_196770||full</td><td>Low-quality</td></tr>
<tr><td>17-109-final-viral-combined_k141_62305||full</td><td>Low-quality</td></tr>
<tr><td>17-109-final-viral-combined_k141_280850||full</td><td>Not-determined</td></tr>
<tr><td>17-109-final-viral-combined_k141_362131||full</td><td>Low-quality</td></tr>
<tr><td>17-109-final-viral-combined_k141_297276||full</td><td>Low-quality</td></tr>
<tr><td>17-109-final-viral-combined_k141_246431||full</td><td>Low-quality</td></tr>
<tr><td>17-109-final-viral-combined_k141_196785||full</td><td>Not-determined</td></tr>
<tr><td>17-109-final-viral-combined_k141_362132||full</td><td>Not-determined</td></tr>
<tr><td>17-109-final-viral-combined_k141_263164||full</td><td>Low-quality</td></tr>
<tr><td>17-109-final-viral-combined_k141_62323||full</td><td>Low-quality</td></tr>
<tr><td>17-109-final-viral-combined_k141_297278||full</td><td>Not-determined</td></tr>
<tr><td>17-109-final-viral-combined_k141_362133||full</td><td>Not-determined</td></tr>
<tr><td>17-109-final-viral-combined_k141_297284||full</td><td>Low-quality</td></tr>
<tr><td>17-109-final-viral-combined_k141_145817||full</td><td>Low-quality</td></tr>
<tr><td>17-109-final-viral-combined_k141_163046||full</td><td>Low-quality</td></tr>
<tr><td>17-109-final-viral-combined_k141_128731||full</td><td>Low-quality</td></tr>
<tr><td>17-109-final-viral-combined_k141_11787||full</td><td>Low-quality</td></tr>
<tr><td>17-109-final-viral-combined_k141_230479||full</td><td>Not-determined</td></tr>
<tr><td>17-109-final-viral-combined_k141_79899||full</td><td>Not-determined</td></tr>
<tr><td>17-109-final-viral-combined_k141_11801||full</td><td>Not-determined</td></tr>
<tr><td>17-109-final-viral-combined_k141_230493||full</td><td>Low-quality</td></tr>
<tr><td>17-109-final-viral-combined_k141_314686||full</td><td>Not-determined</td></tr>
<tr><td>17-109-final-viral-combined_k141_330539||full</td><td>Not-determined</td></tr>
<tr><td>17-109-final-viral-combined_k141_297311||full</td><td>Not-determined</td></tr>
<tr><td>17-109-final-viral-combined_k141_95562||full</td><td>Not-determined</td></tr>
<tr><td>17-109-final-viral-combined_k141_362142||full</td><td>Low-quality</td></tr>
<tr><td>17-109-final-viral-combined_k141_362143||full</td><td>Low-quality</td></tr>
<tr><td>17-109-final-viral-combined_k141_196843||full</td><td>Not-determined</td></tr>
<tr><td>17-109-final-viral-combined_k141_362164||full</td><td>Low-quality</td></tr>
<tr><td>17-109-final-viral-combined_k141_297325||full</td><td>Low-quality</td></tr>
<tr><td>17-109-final-viral-combined_k141_213427||full</td><td>Low-quality</td></tr>
<tr><td>17-109-final-viral-combined_k141_163130||full</td><td>Not-determined</td></tr>
<tr><td>17-109-final-viral-combined_k141_111991||full</td><td>Not-determined</td></tr>
<tr><td>17-109-final-viral-combined_k141_79965||full</td><td>Not-determined</td></tr>
<tr><td>17-109-final-viral-combined_k141_397406||full</td><td>Not-determined</td></tr>
<tr><td>17-109-final-viral-combined_k141_263219||full</td><td>Not-determined</td></tr>
<tr><td>17-109-final-viral-combined_k141_196869||full</td><td>Not-determined</td></tr>
<tr><td>17-109-final-viral-combined_k141_347163||full</td><td>Low-quality</td></tr>
<tr><td>17-109-final-viral-combined_k141_79981||full</td><td>Not-determined</td></tr>
<tr><td>17-109-final-viral-combined_k141_380868||full</td><td>Low-quality</td></tr>
<tr><td>17-109-final-viral-combined_k141_79985||full</td><td>Low-quality</td></tr>
<tr><td>17-109-final-viral-combined_k141_28832||full</td><td>Not-determined</td></tr>
<tr><td>17-109-final-viral-combined_k141_79986||full</td><td>Not-determined</td></tr>
<tr><td>17-109-final-viral-combined_k141_362205||full</td><td>Low-quality</td></tr>
<tr><td>17-109-final-viral-combined_k141_163170||full</td><td>Low-quality</td></tr>
<tr><td>17-109-final-viral-combined_k141_79993||full</td><td>Not-determined</td></tr>
<tr><td>17-109-final-viral-combined_k141_128796||full</td><td>Low-quality</td></tr>
<tr><td>17-109-final-viral-combined_k141_112023||full</td><td>Low-quality</td></tr>
<tr><td>17-109-final-viral-combined_k141_28842||full</td><td>Not-determined</td></tr>
<tr><td>17-109-final-viral-combined_k141_145908||full</td><td>Low-quality</td></tr>
<tr><td>17-109-final-viral-combined_k141_95616||full</td><td>Not-determined</td></tr>
<tr><td>17-109-final-viral-combined_k141_380902||full</td><td>Not-determined</td></tr>
<tr><td>17-109-final-viral-combined_k141_213484||full</td><td>Not-determined</td></tr>
<tr><td>17-109-final-viral-combined_k141_297352||full</td><td>Not-determined</td></tr>
<tr><td>17-109-final-viral-combined_k141_95642||full</td><td>Low-quality</td></tr>
<tr><td>17-109-final-viral-combined_k141_347221||full</td><td>Not-determined</td></tr>
<tr><td>17-109-final-viral-combined_k141_230592||full</td><td>Not-determined</td></tr>
<tr><td>17-109-final-viral-combined_k141_46690||full</td><td>Not-determined</td></tr>
<tr><td>17-109-final-viral-combined_k141_196936||full</td><td>Not-determined</td></tr>
<tr><td>17-109-final-viral-combined_k141_297376||full</td><td>Low-quality</td></tr>
<tr><td>17-109-final-viral-combined_k141_62406||full</td><td>Not-determined</td></tr>
<tr><td>17-109-final-viral-combined_k141_314771||full</td><td>Not-determined</td></tr>
<tr><td>17-109-final-viral-combined_k141_297379||full</td><td>Not-determined</td></tr>
<tr><td>17-109-final-viral-combined_k141_397440||full</td><td>Not-determined</td></tr>
<tr><td>17-109-final-viral-combined_k141_230595||full</td><td>Not-determined</td></tr>
<tr><td>17-109-final-viral-combined_k141_196946||full</td><td>Not-determined</td></tr>
<tr><td>17-109-final-viral-combined_k141_128830||full</td><td>Not-determined</td></tr>
<tr><td>17-109-final-viral-combined_k141_230599||full</td><td>Not-determined</td></tr>
<tr><td>17-109-final-viral-combined_k141_397452||full</td><td>Not-determined</td></tr>
<tr><td>17-109-final-viral-combined_k141_314785||full</td><td>Not-determined</td></tr>
<tr><td>17-109-final-viral-combined_k141_179845||full</td><td>Low-quality</td></tr>
<tr><td>17-109-final-viral-combined_k141_11930||full</td><td>Low-quality</td></tr>
<tr><td>17-109-final-viral-combined_k141_246532||full</td><td>Not-determined</td></tr>
<tr><td>17-109-final-viral-combined_k141_62433||full</td><td>Low-quality</td></tr>
<tr><td>17-109-final-viral-combined_k141_196978||full</td><td>Not-determined</td></tr>
<tr><td>17-109-final-viral-combined_k141_163239||full</td><td>Not-determined</td></tr>
<tr><td>17-109-final-viral-combined_k141_95672||full</td><td>Low-quality</td></tr>
<tr><td>17-109-final-viral-combined_k141_95679||full</td><td>Not-determined</td></tr>
<tr><td>17-109-final-viral-combined_k141_128853||full</td><td>Low-quality</td></tr>
<tr><td>17-109-final-viral-combined_k141_112127||full</td><td>Not-determined</td></tr>
<tr><td>17-109-final-viral-combined_k141_197012||full</td><td>Low-quality</td></tr>
<tr><td>17-109-final-viral-combined_k141_362312||full</td><td>Low-quality</td></tr>
<tr><td>17-109-final-viral-combined_k141_397477||full</td><td>Not-determined</td></tr>
<tr><td>17-109-final-viral-combined_k141_80064||full</td><td>Not-determined</td></tr>
<tr><td>17-109-final-viral-combined_k141_112142||full</td><td>Low-quality</td></tr>
<tr><td>17-109-final-viral-combined_k141_28905||full</td><td>Low-quality</td></tr>
<tr><td>17-109-final-viral-combined_k141_246585||full</td><td>Low-quality</td></tr>
<tr><td>17-109-final-viral-combined_k141_297465||full</td><td>Not-determined</td></tr>
<tr><td>17-109-final-viral-combined_k141_80078||full</td><td>Low-quality</td></tr>
<tr><td>17-109-final-viral-combined_k141_230665||full</td><td>Not-determined</td></tr>
<tr><td>17-109-final-viral-combined_k141_128883||full</td><td>Not-determined</td></tr>
<tr><td>17-109-final-viral-combined_k141_146012||full</td><td>Not-determined</td></tr>
<tr><td>17-109-final-viral-combined_k141_314838||full</td><td>Low-quality</td></tr>
<tr><td>17-109-final-viral-combined_k141_179916||full</td><td>Low-quality</td></tr>
<tr><td>17-109-final-viral-combined_k141_146017||full</td><td>Low-quality</td></tr>
<tr><td>17-109-final-viral-combined_k141_263319||full</td><td>Not-determined</td></tr>
<tr><td>17-109-final-viral-combined_k141_179926||full</td><td>Low-quality</td></tr>
<tr><td>17-109-final-viral-combined_k141_80084||full</td><td>Not-determined</td></tr>
<tr><td>17-109-final-viral-combined_k141_46785||full</td><td>Not-determined</td></tr>
<tr><td>17-109-final-viral-combined_k141_11988||full</td><td>Not-determined</td></tr>
<tr><td>17-109-final-viral-combined_k141_11998||full</td><td>Not-determined</td></tr>
<tr><td>17-109-final-viral-combined_k141_230686||full</td><td>Low-quality</td></tr>
<tr><td>17-109-final-viral-combined_k141_112191||full</td><td>Low-quality</td></tr>
<tr><td>17-109-final-viral-combined_k141_246645||full</td><td>Low-quality</td></tr>
<tr><td>17-109-final-viral-combined_k141_112203||full</td><td>Low-quality</td></tr>
<tr><td>17-109-final-viral-combined_k141_80099||full</td><td>Low-quality</td></tr>
<tr><td>17-109-final-viral-combined_k141_95732||full</td><td>Low-quality</td></tr>
<tr><td>17-109-final-viral-combined_k141_179970||full</td><td>Low-quality</td></tr>
<tr><td>17-109-final-viral-combined_k141_213592||full</td><td>Low-quality</td></tr>
<tr><td>17-109-final-viral-combined_k141_12049||full</td><td>Low-quality</td></tr>
<tr><td>17-109-final-viral-combined_k141_197039||full</td><td>Low-quality</td></tr>
<tr><td>17-109-final-viral-combined_k141_80132||full</td><td>Not-determined</td></tr>
<tr><td>17-109-final-viral-combined_k141_146059||full</td><td>Not-determined</td></tr>
<tr><td>17-109-final-viral-combined_k141_213597||full</td><td>Low-quality</td></tr>
<tr><td>17-109-final-viral-combined_k141_297529||full</td><td>Low-quality</td></tr>
<tr><td>17-109-final-viral-combined_k141_330636||full</td><td>Not-determined</td></tr>
<tr><td>17-109-final-viral-combined_k141_246687||full</td><td>Not-determined</td></tr>
<tr><td>17-109-final-viral-combined_k141_80160||full</td><td>Not-determined</td></tr>
<tr><td>17-109-final-viral-combined_k141_330641||full</td><td>Not-determined</td></tr>
<tr><td>17-109-final-viral-combined_k141_80163||full</td><td>Not-determined</td></tr>
<tr><td>17-109-final-viral-combined_k141_246697||full</td><td>Low-quality</td></tr>
<tr><td>17-109-final-viral-combined_k141_95793||full</td><td>Not-determined</td></tr>
<tr><td>17-109-final-viral-combined_k141_95796||full</td><td>Not-determined</td></tr>
<tr><td>17-109-final-viral-combined_k141_80174||full</td><td>Not-determined</td></tr>
<tr><td>17-109-final-viral-combined_k141_297571||full</td><td>Not-determined</td></tr>
<tr><td>17-109-final-viral-combined_k141_180019||full</td><td>Not-determined</td></tr>
<tr><td>17-109-final-viral-combined_k141_381028||full</td><td>Low-quality</td></tr>
<tr><td>17-109-final-viral-combined_k141_281152||full</td><td>Low-quality</td></tr>
<tr><td>17-109-final-viral-combined_k141_128932||full</td><td>Not-determined</td></tr>
<tr><td>17-109-final-viral-combined_k141_330680||full</td><td>Low-quality</td></tr>
<tr><td>17-109-final-viral-combined_k141_330684||full</td><td>Low-quality</td></tr>
<tr><td>17-109-final-viral-combined_k141_146118||full</td><td>Not-determined</td></tr>
<tr><td>17-109-final-viral-combined_k141_397590||full</td><td>Low-quality</td></tr>
<tr><td>17-109-final-viral-combined_k141_95834||full</td><td>Not-determined</td></tr>
<tr><td>17-109-final-viral-combined_k141_213644||full</td><td>Low-quality</td></tr>
<tr><td>17-109-final-viral-combined_k141_80211||full</td><td>Low-quality</td></tr>
<tr><td>17-109-final-viral-combined_k141_46887||full</td><td>Low-quality</td></tr>
<tr><td>17-109-final-viral-combined_k141_112250||full</td><td>Not-determined</td></tr>
<tr><td>17-109-final-viral-combined_k141_397624||full</td><td>Not-determined</td></tr>
<tr><td>17-109-final-viral-combined_k141_62638||full</td><td>Not-determined</td></tr>
<tr><td>17-109-final-viral-combined_k141_80236||full</td><td>Low-quality</td></tr>
<tr><td>17-109-final-viral-combined_k141_163413||full</td><td>Not-determined</td></tr>
<tr><td>17-109-final-viral-combined_k141_95850||full</td><td>Not-determined</td></tr>
<tr><td>17-109-final-viral-combined_k141_397635||full</td><td>Low-quality</td></tr>
<tr><td>17-109-final-viral-combined_k141_397641||full</td><td>Not-determined</td></tr>
<tr><td>17-109-final-viral-combined_k141_80251||full</td><td>Low-quality</td></tr>
<tr><td>17-109-final-viral-combined_k141_281225||full</td><td>Low-quality</td></tr>
<tr><td>17-109-final-viral-combined_k141_315015||full</td><td>Not-determined</td></tr>
<tr><td>17-109-final-viral-combined_k141_95853||full</td><td>Low-quality</td></tr>
<tr><td>17-109-final-viral-combined_k141_80261||full</td><td>Low-quality</td></tr>
<tr><td>17-109-final-viral-combined_k141_95854||full</td><td>Low-quality</td></tr>
<tr><td>17-109-final-viral-combined_k141_163428||full</td><td>Not-determined</td></tr>
<tr><td>17-109-final-viral-combined_k141_381084||full</td><td>Not-determined</td></tr>
<tr><td>17-109-final-viral-combined_k141_213666||full</td><td>Not-determined</td></tr>
<tr><td>17-109-final-viral-combined_k141_163430||full</td><td>Not-determined</td></tr>
<tr><td>18-109_k141_24084</td><td>High-quality</td></tr>
<tr><td>17-109-final-viral-combined_k141_146165||full</td><td>Not-determined</td></tr>
<tr><td>17-109-final-viral-combined_k141_129022||full</td><td>Low-quality</td></tr>
<tr><td>17-109-final-viral-combined_k141_197123||full</td><td>Not-determined</td></tr>
<tr><td>17-109-final-viral-combined_k141_263392||full</td><td>Low-quality</td></tr>
<tr><td>17-109-final-viral-combined_k141_29078||full</td><td>Not-determined</td></tr>
<tr><td>17-109-final-viral-combined_k141_163470||full</td><td>Low-quality</td></tr>
<tr><td>17-109-final-viral-combined_k141_397684||full</td><td>Low-quality</td></tr>
<tr><td>17-109-final-viral-combined_k141_12140||full</td><td>Low-quality</td></tr>
<tr><td>17-109-final-viral-combined_k141_112289||full</td><td>Low-quality</td></tr>
<tr><td>17-109-final-viral-combined_k141_163480||full</td><td>Low-quality</td></tr>
<tr><td>17-109-final-viral-combined_k141_80269||full</td><td>Low-quality</td></tr>
<tr><td>17-109-final-viral-combined_k141_62699||full</td><td>Not-determined</td></tr>
<tr><td>17-109-final-viral-combined_k141_197150||full</td><td>Low-quality</td></tr>
<tr><td>17-109-final-viral-combined_k141_12161||full</td><td>Not-determined</td></tr>
<tr><td>17-109-final-viral-combined_k141_330785||full</td><td>Low-quality</td></tr>
<tr><td>17-109-final-viral-combined_k141_381135||full</td><td>Low-quality</td></tr>
<tr><td>17-109-final-viral-combined_k141_62718||full</td><td>Not-determined</td></tr>
<tr><td>17-109-final-viral-combined_k141_197164||full</td><td>Not-determined</td></tr>
<tr><td>17-109-final-viral-combined_k141_263426||full</td><td>Low-quality</td></tr>
<tr><td>17-109-final-viral-combined_k141_347529||full</td><td>Not-determined</td></tr>
<tr><td>17-109-final-viral-combined_k141_213726||full</td><td>Not-determined</td></tr>
<tr><td>17-109-final-viral-combined_k141_12179||full</td><td>Low-quality</td></tr>
<tr><td>17-109-final-viral-combined_k141_246768||full</td><td>Not-determined</td></tr>
<tr><td>17-109-final-viral-combined_k141_29116||full</td><td>Low-quality</td></tr>
<tr><td>17-109-final-viral-combined_k141_146254||full</td><td>Low-quality</td></tr>
<tr><td>17-109-final-viral-combined_k141_213737||full</td><td>Low-quality</td></tr>
<tr><td>17-109-final-viral-combined_k141_95951||full</td><td>Not-determined</td></tr>
<tr><td>17-109-final-viral-combined_k141_263441||full</td><td>Not-determined</td></tr>
<tr><td>17-109-final-viral-combined_k141_263457||full</td><td>Not-determined</td></tr>
<tr><td>18-110_k141_163173</td><td>High-quality</td></tr>
<tr><td>17-109-final-viral-combined_k141_213759||full</td><td>Not-determined</td></tr>
<tr><td>17-109-final-viral-combined_k141_29147||full</td><td>Not-determined</td></tr>
<tr><td>17-109-final-viral-combined_k141_213761||full</td><td>Low-quality</td></tr>
<tr><td>17-109-final-viral-combined_k141_146283||full</td><td>Not-determined</td></tr>
<tr><td>17-109-final-viral-combined_k141_62761||full</td><td>Not-determined</td></tr>
<tr><td>17-109-final-viral-combined_k141_347575||full</td><td>Low-quality</td></tr>
<tr><td>17-109-final-viral-combined_k141_146289||full</td><td>Not-determined</td></tr>
<tr><td>17-109-final-viral-combined_k141_112345||full</td><td>Not-determined</td></tr>
<tr><td>17-109-final-viral-combined_k141_297578||full</td><td>Not-determined</td></tr>
<tr><td>17-109-final-viral-combined_k141_263487||full</td><td>Not-determined</td></tr>
<tr><td>17-109-final-viral-combined_k141_297589||full</td><td>Low-quality</td></tr>
<tr><td>17-109-final-viral-combined_k141_246838||full</td><td>Low-quality</td></tr>
<tr><td>17-109-final-viral-combined_k141_397759||full</td><td>Not-determined</td></tr>
<tr><td>17-109-final-viral-combined_k141_397766||full</td><td>Not-determined</td></tr>
<tr><td>17-109-final-viral-combined_k141_213782||full</td><td>Low-quality</td></tr>
<tr><td>17-109-final-viral-combined_k141_281370||full</td><td>Low-quality</td></tr>
<tr><td>17-109-final-viral-combined_k141_29196||full</td><td>Not-determined</td></tr>
<tr><td>17-109-final-viral-combined_k141_129156||full</td><td>Not-determined</td></tr>
<tr><td>17-109-final-viral-combined_k141_281375||full</td><td>Not-determined</td></tr>
<tr><td>17-109-final-viral-combined_k141_297635||full</td><td>Low-quality</td></tr>
<tr><td>17-109-final-viral-combined_k141_12228||full</td><td>Low-quality</td></tr>
<tr><td>17-109-final-viral-combined_k141_330889||full</td><td>Low-quality</td></tr>
<tr><td>17-109-final-viral-combined_k141_62780||full</td><td>Not-determined</td></tr>
<tr><td>17-109-final-viral-combined_k141_62784||full</td><td>Not-determined</td></tr>
<tr><td>17-109-final-viral-combined_k141_330895||full</td><td>Low-quality</td></tr>
<tr><td>17-109-final-viral-combined_k141_315230||full</td><td>Low-quality</td></tr>
<tr><td>17-109-final-viral-combined_k141_297646||full</td><td>Low-quality</td></tr>
<tr><td>17-109-final-viral-combined_k141_246855||full</td><td>Not-determined</td></tr>
<tr><td>17-109-final-viral-combined_k141_347627||full</td><td>Low-quality</td></tr>
<tr><td>17-109-final-viral-combined_k141_180214||full</td><td>Low-quality</td></tr>
<tr><td>17-109-final-viral-combined_k141_230907||full</td><td>Not-determined</td></tr>
<tr><td>17-109-final-viral-combined_k141_12254||full</td><td>Low-quality</td></tr>
<tr><td>17-109-final-viral-combined_k141_397817||full</td><td>Low-quality</td></tr>
<tr><td>17-109-final-viral-combined_k141_180229||full</td><td>Not-determined</td></tr>
<tr><td>17-109-final-viral-combined_k141_47125||full</td><td>Low-quality</td></tr>
<tr><td>17-109-final-viral-combined_k141_197274||full</td><td>Not-determined</td></tr>
<tr><td>17-109-final-viral-combined_k141_213835||full</td><td>Low-quality</td></tr>
<tr><td>17-109-final-viral-combined_k141_12276||full</td><td>Not-determined</td></tr>
<tr><td>17-109-final-viral-combined_k141_197296||full</td><td>Low-quality</td></tr>
<tr><td>17-109-final-viral-combined_k141_246893||full</td><td>Not-determined</td></tr>
<tr><td>17-109-final-viral-combined_k141_96093||full</td><td>Low-quality</td></tr>
<tr><td>17-109-final-viral-combined_k141_180255||full</td><td>Low-quality</td></tr>
<tr><td>17-109-final-viral-combined_k141_129194||full</td><td>Not-determined</td></tr>
<tr><td>17-109-final-viral-combined_k141_397845||full</td><td>Low-quality</td></tr>
<tr><td>17-109-final-viral-combined_k141_330953||full</td><td>Not-determined</td></tr>
<tr><td>17-109-final-viral-combined_k141_281458||full</td><td>Not-determined</td></tr>
<tr><td>17-109-final-viral-combined_k141_129210||full</td><td>Low-quality</td></tr>
<tr><td>17-109-final-viral-combined_k141_315281||full</td><td>Low-quality</td></tr>
<tr><td>17-109-final-viral-combined_k141_362562||full</td><td>Not-determined</td></tr>
<tr><td>17-109-final-viral-combined_k141_29271||full</td><td>Not-determined</td></tr>
<tr><td>17-109-final-viral-combined_k141_180263||full</td><td>Low-quality</td></tr>
<tr><td>17-109-final-viral-combined_k141_362570||full</td><td>Low-quality</td></tr>
<tr><td>17-109-final-viral-combined_k141_246928||full</td><td>Low-quality</td></tr>
<tr><td>17-109-final-viral-combined_k141_263564||full</td><td>Not-determined</td></tr>
<tr><td>17-109-final-viral-combined_k141_362576||full</td><td>Low-quality</td></tr>
<tr><td>17-109-final-viral-combined_k141_112405||full</td><td>Not-determined</td></tr>
<tr><td>17-109-final-viral-combined_k141_197314||full</td><td>Not-determined</td></tr>
<tr><td>17-109-final-viral-combined_k141_397879||full</td><td>Low-quality</td></tr>
<tr><td>17-109-final-viral-combined_k141_315309||full</td><td>Low-quality</td></tr>
<tr><td>17-109-final-viral-combined_k141_29295||full</td><td>Low-quality</td></tr>
<tr><td>17-109-final-viral-combined_k141_96148||full</td><td>Low-quality</td></tr>
<tr><td>17-109-final-viral-combined_k141_47180||full</td><td>Low-quality</td></tr>
<tr><td>17-109-final-viral-combined_k141_362616||full</td><td>Low-quality</td></tr>
<tr><td>17-109-final-viral-combined_k141_29313||full</td><td>Not-determined</td></tr>
</table>

| | |
|---|---|
| 17-109-final-viral-combined_k141_362626\|\|full | Not-determined |
| 17-109-final-viral-combined_k141_146410\|\|full | Not-determined |
| 17-109-final-viral-combined_k141_96174\|\|full | Low-quality |
| 17-109-final-viral-combined_k141_96177\|\|full | Low-quality |
| 17-109-final-viral-combined_k141_112435\|\|full | Low-quality |
| 17-109-final-viral-combined_k141_362627\|\|full | Low-quality |
| 17-109-final-viral-combined_k141_80380\|\|full | Not-determined |
| 18-110-final-viral-combined_k141_88439\|\|full | Medium-quality |
| 17-109-final-viral-combined_k141_12404\|\|full | Not-determined |
| 17-109-final-viral-combined_k141_163763\|\|full | Low-quality |
| 17-109-final-viral-combined_k141_281512\|\|full | Not-determined |
| 17-109-final-viral-combined_k141_197362\|\|full | Not-determined |
| 17-109-final-viral-combined_k141_347759\|\|full | Not-determined |
| 17-109-final-viral-combined_k141_331028\|\|full | Low-quality |
| 17-109-final-viral-combined_k141_12427\|\|full | Not-determined |
| 18-102-final-viral-combined_k141_248329\|\|full | High-quality |
| 17-109-final-viral-combined_k141_381330\|\|full | Low-quality |
| 17-109-final-viral-combined_k141_80405\|\|full | Low-quality |
| 17-109-final-viral-combined_k141_180358\|\|full | Not-determined |
| 17-109-final-viral-combined_k141_331039\|\|full | Low-quality |
| 17-109-final-viral-combined_k141_112480\|\|full | Not-determined |
| 17-109-final-viral-combined_k141_231085\|\|full | Not-determined |
| 17-109-final-viral-combined_k141_12446\|\|full | Not-determined |
| 17-109-final-viral-combined_k141_146482\|\|full | Low-quality |
| 17-109-final-viral-combined_k141_231102\|\|full | Not-determined |
| 17-109-final-viral-combined_k141_12449\|\|full | Low-quality |
| 17-109-final-viral-combined_k141_231109\|\|full | Low-quality |
| 17-109-final-viral-combined_k141_347814\|\|full | Not-determined |
| 17-109-final-viral-combined_k141_62886\|\|full | Not-determined |
| 17-109-final-viral-combined_k141_12462\|\|full | Low-quality |
| 17-109-final-viral-combined_k141_197428\|\|full | Not-determined |
| 17-109-final-viral-combined_k141_163821\|\|full | Not-determined |
| 17-109-final-viral-combined_k141_331090\|\|full | Low-quality |
| 17-109-final-viral-combined_k141_297735\|\|full | Low-quality |
| 17-109-final-viral-combined_k141_80457\|\|full | Not-determined |
| 17-109-final-viral-combined_k141_197436\|\|full | Not-determined |
| 17-109-final-viral-combined_k141_281615\|\|full | Not-determined |
| 17-109-final-viral-combined_k141_213911\|\|full | Low-quality |
| 17-109-final-viral-combined_k141_12468\|\|full | Not-determined |
| 17-109-final-viral-combined_k141_381385\|\|full | Not-determined |
| 17-109-final-viral-combined_k141_297753\|\|full | Not-determined |
| 17-109-final-viral-combined_k141_112535\|\|full | Not-determined |
| 17-109-final-viral-combined_k141_197442\|\|full | Low-quality |
| 17-109-final-viral-combined_k141_129352\|\|full | Not-determined |
| 17-109-final-viral-combined_k141_362736\|\|full | Low-quality |
| 17-109-final-viral-combined_k141_231154\|\|full | Not-determined |
| 17-109-final-viral-combined_k141_213960\|\|full | Low-quality |
| 17-109-final-viral-combined_k141_146573\|\|full | Low-quality |
| 17-109-final-viral-combined_k141_163847\|\|full | Not-determined |
| 17-109-final-viral-combined_k141_397951\|\|full | Not-determined |
| 17-109-final-viral-combined_k141_180386\|\|full | Low-quality |
| 17-109-final-viral-combined_k141_331143\|\|full | Not-determined |
| 17-109-final-viral-combined_k141_197490\|\|full | Not-determined |
| 17-109-final-viral-combined_k141_80526\|\|full | Low-quality |
| 17-109-final-viral-combined_k141_180396\|\|full | Low-quality |
| 17-109-final-viral-combined_k141_231179\|\|full | Low-quality |
| 17-109-final-viral-combined_k141_397960\|\|full | Not-determined |
| 17-109-final-viral-combined_k141_80527\|\|full | Low-quality |
| 17-109-final-viral-combined_k141_362773\|\|full | Not-determined |
| 17-109-final-viral-combined_k141_29358\|\|full | Low-quality |
| 17-109-final-viral-combined_k141_331154\|\|full | Not-determined |
| 17-109-final-viral-combined_k141_281688\|\|full | Not-determined |
| 17-109-final-viral-combined_k141_112627\|\|full | Low-quality |
| 17-109-final-viral-combined_k141_62948\|\|full | Not-determined |
| 17-109-final-viral-combined_k141_12538\|\|full | Low-quality |
| 17-109-final-viral-combined_k141_12539\|\|full | Not-determined |
| 17-109-final-viral-combined_k141_247109\|\|full | Not-determined |
| 17-109-final-viral-combined_k141_381479\|\|full | Low-quality |
| 17-109-final-viral-combined_k141_112654\|\|full | Not-determined |
| 17-109-final-viral-combined_k141_315408\|\|full | Not-determined |
| 17-109-final-viral-combined_k141_163920\|\|full | Low-quality |
| 17-109-final-viral-combined_k141_29418\|\|full | Low-quality |
| 17-109-final-viral-combined_k141_129420\|\|full | Not-determined |
| 17-109-final-viral-combined_k141_214051\|\|full | Not-determined |
| 17-109-final-viral-combined_k141_146614\|\|full | Low-quality |
| 17-109-final-viral-combined_k141_231244\|\|full | Low-quality |
| 17-109-final-viral-combined_k141_397990\|\|full | Low-quality |
| 17-109-final-viral-combined_k141_231250\|\|full | Not-determined |
| 17-109-final-viral-combined_k141_381513\|\|full | Low-quality |
| 17-109-final-viral-combined_k141_129430\|\|full | Low-quality |
| 17-109-final-viral-combined_k141_297800\|\|full | Low-quality |
| 17-109-final-viral-combined_k141_247165\|\|full | Low-quality |
| 17-109-final-viral-combined_k141_163946\|\|full | Not-determined |
| 17-109-final-viral-combined_k141_281744\|\|full | Not-determined |
| 17-109-final-viral-combined_k141_347939\|\|full | Low-quality |
| 17-109-final-viral-combined_k141_297804\|\|full | Not-determined |
| 17-109-final-viral-combined_k141_231263\|\|full | Not-determined |
| 17-109-final-viral-combined_k141_80562\|\|full | Low-quality |
| 17-109-final-viral-combined_k141_112674\|\|full | Not-determined |
| 17-109-final-viral-combined_k141_180451\|\|full | Low-quality |
| 17-109-final-viral-combined_k141_231271\|\|full | Not-determined |
| 17-109-final-viral-combined_k141_163962\|\|full | Not-determined |
| 17-109-final-viral-combined_k141_398009\|\|full | Not-determined |
| 17-109-final-viral-combined_k141_47378\|\|full | Low-quality |
| 17-109-final-viral-combined_k141_197519\|\|full | Not-determined |
| 17-109-final-viral-combined_k141_47381\|\|full | Low-quality |
| 17-109-final-viral-combined_k141_381545\|\|full | Low-quality |
| 17-109-final-viral-combined_k141_347964\|\|full | Not-determined |
| 17-109-final-viral-combined_k141_29482\|\|full | Low-quality |
| 17-109-final-viral-combined_k141_180490\|\|full | Not-determined |
| 17-109-final-viral-combined_k141_362844\|\|full | Not-determined |
| 17-109-final-viral-combined_k141_297814\|\|full | Low-quality |
| 17-109-final-viral-combined_k141_112722\|\|full | Not-determined |
| 17-109-final-viral-combined_k141_331259\|\|full | Low-quality |
| 17-109-final-viral-combined_k141_381561\|\|full | Low-quality |
| 17-109-final-viral-combined_k141_180499\|\|full | Not-determined |
| 17-109-final-viral-combined_k141_96432\|\|full | Low-quality |
| 17-109-final-viral-combined_k141_129498\|\|full | Not-determined |
| 17-109-final-viral-combined_k141_297839\|\|full | Not-determined |
| 17-109-final-viral-combined_k141_362863\|\|full | Not-determined |
| 17-109-final-viral-combined_k141_112750\|\|full | Low-quality |
| 17-109-final-viral-combined_k141_398044\|\|full | Low-quality |
| 17-109-final-viral-combined_k141_12610\|\|full | Not-determined |
| 17-109-final-viral-combined_k141_197561\|\|full | Low-quality |
| 17-109-final-viral-combined_k141_231325\|\|full | Low-quality |
| 17-109-final-viral-combined_k141_398055\|\|full | Not-determined |
| 17-109-final-viral-combined_k141_362887\|\|full | Low-quality |
| 17-109-final-viral-combined_k141_263735\|\|full | Not-determined |
| 17-109-final-viral-combined_k141_146671\|\|full | Low-quality |
| 17-109-final-viral-combined_k141_263740\|\|full | Low-quality |
| 17-109-final-viral-combined_k141_29581\|\|full | Not-determined |
| 17-109-final-viral-combined_k141_315485\|\|full | Low-quality |
| 17-109-final-viral-combined_k141_381630\|\|full | Low-quality |
| 17-109-final-viral-combined_k141_29582\|\|full | Not-determined |
| 17-109-final-viral-combined_k141_197599\|\|full | Not-determined |
| 17-109-final-viral-combined_k141_263753\|\|full | Low-quality |
| 17-109-final-viral-combined_k141_281846\|\|full | Not-determined |
| 17-109-final-viral-combined_k141_231348\|\|full | Low-quality |
| 17-109-final-viral-combined_k141_80630\|\|full | Not-determined |
| 17-109-final-viral-combined_k141_197614\|\|full | Not-determined |
| 17-109-final-viral-combined_k141_164112\|\|full | Not-determined |
| 17-109-final-viral-combined_k141_231355\|\|full | Low-quality |
| 17-109-final-viral-combined_k141_80641\|\|full | Low-quality |
| 17-109-final-viral-combined_k141_197625\|\|full | Not-determined |
| 17-109-final-viral-combined_k141_315532\|\|full | Low-quality |
| 17-109-final-viral-combined_k141_348047\|\|full | Low-quality |
| 17-109-final-viral-combined_k141_348049\|\|full | Low-quality |
| 17-109-final-viral-combined_k141_281870\|\|full | Not-determined |
| 17-109-final-viral-combined_k141_263798\|\|full | Not-determined |
| 18-110_k141_190781 | Medium-quality |
| 17-109-final-viral-combined_k141_180547\|\|full | Low-quality |
| 17-109-final-viral-combined_k141_263805\|\|full | Low-quality |
| 17-109-final-viral-combined_k141_348057\|\|full | Low-quality |
| 17-109-final-viral-combined_k141_80663\|\|full | Not-determined |
| 17-109-final-viral-combined_k141_331337\|\|full | Not-determined |
| 17-109-final-viral-combined_k141_381698\|\|full | Low-quality |
| 17-109-final-viral-combined_k141_297901\|\|full | Low-quality |
| 17-109-final-viral-combined_k141_62990\|\|full | Low-quality |
| 17-109-final-viral-combined_k141_96484\|\|full | Not-determined |
| 17-109-final-viral-combined_k141_129555\|\|full | Low-quality |
| 17-109-final-viral-combined_k141_281902\|\|full | Low-quality |
| 17-109-final-viral-combined_k141_63002\|\|full | Low-quality |
| 17-109-final-viral-combined_k141_164148\|\|full | Low-quality |
| 17-109-final-viral-combined_k141_231434\|\|full | Not-determined |
| 17-109-final-viral-combined_k141_197661\|\|full | Low-quality |
| 17-109-final-viral-combined_k141_331371\|\|full | Low-quality |
| 17-109-final-viral-combined_k141_247304\|\|full | Not-determined |
| 17-109-final-viral-combined_k141_112885\|\|full | Low-quality |
| 17-109-final-viral-combined_k141_96503\|\|full | Low-quality |
| 17-109-final-viral-combined_k141_281923\|\|full | Not-determined |
| 17-109-final-viral-combined_k141_63020\|\|full | Not-determined |
| 18-110-final-viral-combined_k141_190781\|\|full | Medium-quality |
| 17-109-final-viral-combined_k141_231447\|\|full | Not-determined |
| 17-109-final-viral-combined_k141_63023\|\|full | Not-determined |
| 17-109-final-viral-combined_k141_297943\|\|full | Not-determined |
| 17-109-final-viral-combined_k141_214160\|\|full | Not-determined |
| 17-109-final-viral-combined_k141_12668\|\|full | Not-determined |
| 17-109-final-viral-combined_k141_231457\|\|full | Not-determined |
| 17-109-final-viral-combined_k141_381741\|\|full | Not-determined |
| 17-109-final-viral-combined_k141_381754\|\|full | Low-quality |
| 17-109-final-viral-combined_k141_297960\|\|full | Not-determined |
| 17-109-final-viral-combined_k141_80718\|\|full | Low-quality |
| 17-109-final-viral-combined_k141_348107\|\|full | Not-determined |
| 17-109-final-viral-combined_k141_315563\|\|full | Low-quality |
| 17-109-final-viral-combined_k141_29699\|\|full | Not-determined |
| 17-109-final-viral-combined_k141_80722\|\|full | Low-quality |
| 17-109-final-viral-combined_k141_214198\|\|full | Not-determined |
| 17-109-final-viral-combined_k141_29704\|\|full | Not-determined |
| 17-109-final-viral-combined_k141_381776\|\|full | Low-quality |
| 17-109-final-viral-combined_k141_331421\|\|full | Low-quality |
| 17-109-final-viral-combined_k141_96566\|\|full | Not-determined |
| 17-109-final-viral-combined_k141_297984\|\|full | Low-quality |
| 17-109-final-viral-combined_k141_331426\|\|full | Not-determined |
| 17-109-final-viral-combined_k141_146782\|\|full | Low-quality |
| 17-109-final-viral-combined_k141_348123\|\|full | Not-determined |
| 17-109-final-viral-combined_k141_180641\|\|full | Not-determined |
| 17-109-final-viral-combined_k141_180643\|\|full | Not-determined |
| 17-109-final-viral-combined_k141_164221\|\|full | Low-quality |
| 17-109-final-viral-combined_k141_12710\|\|full | Low-quality |
| 17-109-final-viral-combined_k141_47635\|\|full | Not-determined |
| 17-109-final-viral-combined_k141_381786\|\|full | Not-determined |
| 17-109-final-viral-combined_k141_129632\|\|full | Low-quality |
| 17-109-final-viral-combined_k141_363056\|\|full | Low-quality |
| 17-109-final-viral-combined_k141_180671\|\|full | Low-quality |
| 17-109-final-viral-combined_k141_129636\|\|full | Low-quality |
| 17-109-final-viral-combined_k141_363065\|\|full | Not-determined |
| 18-105-final-viral-combined_k141_145067\|\|full | Medium-quality |
| 17-109-final-viral-combined_k141_363069\|\|full | Not-determined |
| 17-109-final-viral-combined_k141_298005\|\|full | Not-determined |
| 17-109-final-viral-combined_k141_164241\|\|full | Not-determined |
| 17-109-final-viral-combined_k141_231529\|\|full | Not-determined |
| 17-109-final-viral-combined_k141_263961\|\|full | Not-determined |
| 17-109-final-viral-combined_k141_331491\|\|full | Not-determined |
| 17-109-final-viral-combined_k141_381805\|\|full | Low-quality |
| 17-109-final-viral-combined_k141_180694\|\|full | Low-quality |
| 17-109-final-viral-combined_k141_96628\|\|full | Low-quality |
| 17-109-final-viral-combined_k141_63074\|\|full | Not-determined |
| 17-109-final-viral-combined_k141_47685\|\|full | Low-quality |
| 17-109-final-viral-combined_k141_164253\|\|full | Not-determined |
| 17-109-final-viral-combined_k141_146816\|\|full | Low-quality |
| 17-109-final-viral-combined_k141_29739\|\|full | Not-determined |
| 17-109-final-viral-combined_k141_231550\|\|full | Low-quality |
| 17-109-final-viral-combined_k141_197741\|\|full | Low-quality |
| 17-109-final-viral-combined_k141_381826\|\|full | Not-determined |
| 17-109-final-viral-combined_k141_231552\|\|full | Not-determined |
| 17-109-final-viral-combined_k141_146830\|\|full | Not-determined |
| 17-109-final-viral-combined_k141_231558\|\|full | Low-quality |
| 17-109-final-viral-combined_k141_214254\|\|full | Not-determined |
| 17-109-final-viral-combined_k141_315618\|\|full | Not-determined |
| 17-109-final-viral-combined_k141_47707\|\|full | Not-determined |
| 17-109-final-viral-combined_k141_363074\|\|full | Low-quality |
| 17-109-final-viral-combined_k141_348202\|\|full | Not-determined |
| 17-109-final-viral-combined_k141_47709\|\|full | Low-quality |
| 17-109-final-viral-combined_k141_214264\|\|full | Low-quality |
| 17-109-final-viral-combined_k141_263989\|\|full | Not-determined |
| 17-109-final-viral-combined_k141_315632\|\|full | Low-quality |
| 17-109-final-viral-combined_k141_29761\|\|full | Low-quality |
| 17-109-final-viral-combined_k141_96679\|\|full | Low-quality |
| 17-109-final-viral-combined_k141_12770\|\|full | Low-quality |
| 17-109-final-viral-combined_k141_315635\|\|full | Not-determined |
| 17-109-final-viral-combined_k141_282058\|\|full | Low-quality |
| 17-109-final-viral-combined_k141_264015\|\|full | Low-quality |
| 17-109-final-viral-combined_k141_381831\|\|full | Not-determined |
| 17-109-final-viral-combined_k141_12789\|\|full | Not-determined |
| 17-109-final-viral-combined_k141_80810\|\|full | Not-determined |
| 17-109-final-viral-combined_k141_247459\|\|full | Not-determined |
| 17-109-final-viral-combined_k141_282077\|\|full | Not-determined |
| 17-109-final-viral-combined_k141_264030\|\|full | Low-quality |
| 17-109-final-viral-combined_k141_129740\|\|full | Not-determined |
| 17-109-final-viral-combined_k141_112966\|\|full | Low-quality |
| 17-109-final-viral-combined_k141_146863\|\|full | Not-determined |
| 17-109-final-viral-combined_k141_112981\|\|full | Not-determined |
| 17-109-final-viral-combined_k141_164334\|\|full | Not-determined |
| 17-109-final-viral-combined_k141_197822\|\|full | Not-determined |
| 17-109-final-viral-combined_k141_381869\|\|full | Low-quality |
| 17-109-final-viral-combined_k141_164336\|\|full | Not-determined |
| 17-109-final-viral-combined_k141_298049\|\|full | Low-quality |
| 17-109-final-viral-combined_k141_247493\|\|full | Low-quality |
| 17-109-final-viral-combined_k141_282104\|\|full | Not-determined |
| 17-109-final-viral-combined_k141_197834\|\|full | Not-determined |
| 17-109-final-viral-combined_k141_398139\|\|full | Low-quality |
| 17-109-final-viral-combined_k141_12814\|\|full | Not-determined |
| 17-109-final-viral-combined_k141_315695\|\|full | Low-quality |
| 17-109-final-viral-combined_k141_80840\|\|full | Low-quality |
| 17-109-final-viral-combined_k141_214322\|\|full | Low-quality |
| 17-109-final-viral-combined_k141_180810\|\|full | Low-quality |
| 17-109-final-viral-combined_k141_381872\|\|full | Not-determined |
| 17-109-final-viral-combined_k141_298079\|\|full | Not-determined |
| 17-109-final-viral-combined_k141_247524\|\|full | Low-quality |
| 17-109-final-viral-combined_k141_197856\|\|full | Low-quality |
| 17-109-final-viral-combined_k141_180831\|\|full | Not-determined |
| 17-109-final-viral-combined_k141_282143\|\|full | Low-quality |
| 17-109-final-viral-combined_k141_363150\|\|full | Low-quality |
| 17-109-final-viral-combined_k141_214351\|\|full | Not-determined |
| 17-109-final-viral-combined_k141_348282\|\|full | Not-determined |
| 17-109-final-viral-combined_k141_113038\|\|full | Not-determined |
| 17-109-final-viral-combined_k141_298091\|\|full | Low-quality |
| 17-109-final-viral-combined_k141_113039\|\|full | Low-quality |
| 17-109-final-viral-combined_k141_96738\|\|full | Low-quality |
| 17-109-final-viral-combined_k141_180855\|\|full | Low-quality |
| 17-109-final-viral-combined_k141_363169\|\|full | Not-determined |
| 17-109-final-viral-combined_k141_315758\|\|full | Not-determined |
| 17-109-final-viral-combined_k141_247553\|\|full | Low-quality |
| 17-109-final-viral-combined_k141_113078\|\|full | Not-determined |
| 17-109-final-viral-combined_k141_282174\|\|full | Low-quality |
| 17-109-final-viral-combined_k141_47756\|\|full | Low-quality |
| 17-109-final-viral-combined_k141_197918\|\|full | Not-determined |
| 17-109-final-viral-combined_k141_164448\|\|full | Low-quality |
| 17-109-final-viral-combined_k141_231691\|\|full | Not-determined |
| 17-109-final-viral-combined_k141_247576\|\|full | Not-determined |
| 17-109-final-viral-combined_k141_264074\|\|full | Not-determined |
| 17-109-final-viral-combined_k141_63314\|\|full | Low-quality |
| 17-109-final-viral-combined_k141_180871\|\|full | Low-quality |
| 17-109-final-viral-combined_k141_146895\|\|full | Low-quality |
| 17-109-final-viral-combined_k141_63322\|\|full | Not-determined |
| 17-109-final-viral-combined_k141_113126\|\|full | Low-quality |
| 17-109-final-viral-combined_k141_315807\|\|full | Low-quality |
| 17-109-final-viral-combined_k141_282220\|\|full | Not-determined |
| 17-109-final-viral-combined_k141_363245\|\|full | Not-determined |
| 17-109-final-viral-combined_k141_214422\|\|full | Not-determined |
| 17-109-final-viral-combined_k141_146915\|\|full | Low-quality |
| 17-109-final-viral-combined_k141_146919\|\|full | Low-quality |
| 17-109-final-viral-combined_k141_180913\|\|full | Not-determined |
| 17-109-final-viral-combined_k141_315838\|\|full | Not-determined |
| 17-109-final-viral-combined_k141_96781\|\|full | Not-determined |
| 17-109-final-viral-combined_k141_197946\|\|full | Not-determined |
| 17-109-final-viral-combined_k141_96785\|\|full | Not-determined |
| 18-102-final-viral-combined_k141_13094\|\|0_partial | Medium-quality |
| 17-109-final-viral-combined_k141_197952\|\|full | Not-determined |
| 17-109-final-viral-combined_k141_247647\|\|full | Not-determined |
| 17-109-final-viral-combined_k141_146944\|\|full | Not-determined |
| 17-109-final-viral-combined_k141_315852\|\|full | Not-determined |
| 17-109-final-viral-combined_k141_180924\|\|full | Not-determined |
| 17-109-final-viral-combined_k141_282265\|\|full | Not-determined |
| 17-109-final-viral-combined_k141_197981\|\|full | Low-quality |
| 17-109-final-viral-combined_k141_47819\|\|full | Not-determined |
| 17-109-final-viral-combined_k141_398247\|\|full | Low-quality |
| 17-109-final-viral-combined_k141_197983\|\|full | Low-quality |
| 17-109-final-viral-combined_k141_363282\|\|full | Not-determined |
| 17-109-final-viral-combined_k141_80920\|\|full | Low-quality |
| 17-109-final-viral-combined_k141_80921\|\|full | Low-quality |
| 17-109-final-viral-combined_k141_331711\|\|full | Low-quality |
| 17-109-final-viral-combined_k141_231792\|\|full | Low-quality |
| 17-109-final-viral-combined_k141_180947\|\|full | Not-determined |
| 17-109-final-viral-combined_k141_381928\|\|full | Not-determined |
| 17-109-final-viral-combined_k141_348426\|\|full | Low-quality |
| 17-109-final-viral-combined_k141_231802\|\|full | Not-determined |
| 17-109-final-viral-combined_k141_198000\|\|full | Not-determined |
| 17-109-final-viral-combined_k141_113192\|\|full | Low-quality |
| 17-109-final-viral-combined_k141_164553\|\|full | Not-determined |
| 17-109-final-viral-combined_k141_164559\|\|full | Low-quality |
| 17-109-final-viral-combined_k141_63373\|\|full | Low-quality |
| 17-109-final-viral-combined_k141_282329\|\|full | Low-quality |
| 17-109-final-viral-combined_k141_282330\|\|full | Not-determined |
| 17-109-final-viral-combined_k141_282333\|\|full | Low-quality |
| 17-109-final-viral-combined_k141_96854\|\|full | Low-quality |
| 18-101_k141_244836 | Medium-quality |
| 17-109-final-viral-combined_k141_180978\|\|full | Not-determined |
| 17-109-final-viral-combined_k141_96857\|\|full | Not-determined |
| 17-109-final-viral-combined_k141_264133\|\|full | Low-quality |
| 17-109-final-viral-combined_k141_129801\|\|full | Low-quality |
| 17-109-final-viral-combined_k141_282348\|\|full | Not-determined |
| 17-109-final-viral-combined_k141_12991\|\|full | Low-quality |
| 17-109-final-viral-combined_k141_331766\|\|full | Low-quality |
| 17-109-final-viral-combined_k141_180988\|\|full | Low-quality |
| 17-109-final-viral-combined_k141_214563\|\|full | Not-determined |
| 17-109-final-viral-combined_k141_13003\|\|full | Not-determined |
| 17-109-final-viral-combined_k141_129808\|\|full | Low-quality |
| 17-109-final-viral-combined_k141_315917\|\|full | Not-determined |
| 17-109-final-viral-combined_k141_198032\|\|full | Low-quality |
| 17-109-final-viral-combined_k141_198034\|\|full | Low-quality |
| 17-109-final-viral-combined_k141_80945\|\|full | Low-quality |
| 17-109-final-viral-combined_k141_181030\|\|full | Not-determined |
| 17-109-final-viral-combined_k141_129833\|\|full | Low-quality |
| 17-109-final-viral-combined_k141_363300\|\|full | Low-quality |
| 17-109-final-viral-combined_k141_331792\|\|full | Not-determined |
| 17-109-final-viral-combined_k141_80969\|\|full | Low-quality |
| 17-109-final-viral-combined_k141_381987\|\|full | Not-determined |
| 17-109-final-viral-combined_k141_47887\|\|full | Not-determined |
| 17-109-final-viral-combined_k141_381999\|\|full | Not-determined |
| 17-109-final-viral-combined_k141_47896\|\|full | Not-determined |
| 17-109-final-viral-combined_k141_315951\|\|full | Low-quality |
| 17-109-final-viral-combined_k141_47899\|\|full | Low-quality |
| 17-109-final-viral-combined_k141_47900\|\|full | Low-quality |
| 17-109-final-viral-combined_k141_363325\|\|full | Not-determined |
| 17-109-final-viral-combined_k141_231880\|\|full | Low-quality |
| 17-109-final-viral-combined_k141_363328\|\|full | Not-determined |
| 17-109-final-viral-combined_k141_382004\|\|full | Low-quality |
| 17-109-final-viral-combined_k141_147028\|\|full | Low-quality |
| 17-109-final-viral-combined_k141_198096\|\|full | Not-determined |
| 17-109-final-viral-combined_k141_129860\|\|full | Low-quality |
| 17-109-final-viral-combined_k141_363344\|\|full | Not-determined |
| 17-109-final-viral-combined_k141_30086\|\|full | Low-quality |
| 17-109-final-viral-combined_k141_231904\|\|full | Low-quality |
| 17-109-final-viral-combined_k141_47935\|\|full | Low-quality |
| 17-109-final-viral-combined_k141_382032\|\|full | Low-quality |
| 17-109-final-viral-combined_k141_298258\|\|full | Not-determined |
| 17-109-final-viral-combined_k141_348553\|\|full | Not-determined |
| 17-109-final-viral-combined_k141_81020\|\|full | Not-determined |
| 17-109-final-viral-combined_k141_298269\|\|full | Low-quality |
| 17-109-final-viral-combined_k141_198127\|\|full | Not-determined |
| 17-109-final-viral-combined_k141_30120\|\|full | Not-determined |
| 17-109-final-viral-combined_k141_181120\|\|full | Not-determined |
| 17-109-final-viral-combined_k141_96959\|\|full | Not-determined |
| 17-109-final-viral-combined_k141_181127\|\|full | Not-determined |
| 17-109-final-viral-combined_k141_247836\|\|full | Not-determined |
| 17-109-final-viral-combined_k141_81054\|\|full | Low-quality |
| 17-109-final-viral-combined_k141_13125\|\|full | Not-determined |
| 17-109-final-viral-combined_k141_348571\|\|full | Low-quality |
| 17-109-final-viral-combined_k141_214712\|\|full | Not-determined |
| 17-109-final-viral-combined_k141_81073\|\|full | Not-determined |
| 17-109-final-viral-combined_k141_348575\|\|full | Low-quality |
| 17-109-final-viral-combined_k141_81081\|\|full | Low-quality |
| 17-109-final-viral-combined_k141_113352\|\|full | Not-determined |
| 17-109-final-viral-combined_k141_81082\|\|full | Not-determined |
| 17-109-final-viral-combined_k141_264241\|\|full | Low-quality |
| 17-109-final-viral-combined_k141_113358\|\|full | Low-quality |
| 17-109-final-viral-combined_k141_181144\|\|full | Not-determined |
| 17-109-final-viral-combined_k141_382063\|\|full | Not-determined |
| 17-109-final-viral-combined_k141_247869\|\|full | Low-quality |
| 17-109-final-viral-combined_k141_30146\|\|full | Not-determined |
| 17-109-final-viral-combined_k141_47992\|\|full | Not-determined |
| 17-109-final-viral-combined_k141_282458\|\|full | Not-determined |
| 17-109-final-viral-combined_k141_13157\|\|full | Not-determined |
| 17-109-final-viral-combined_k141_398335\|\|full | Not-determined |
| 17-109-final-viral-combined_k141_81110\|\|full | Low-quality |
| 17-109-final-viral-combined_k141_63508\|\|full | Low-quality |
| 17-109-final-viral-combined_k141_48000\|\|full | Low-quality |
| 17-109-final-viral-combined_k141_63514\|\|full | Not-determined |
| 17-109-final-viral-combined_k141_214749\|\|full | Low-quality |
| 17-109-final-viral-combined_k141_181179\|\|full | Low-quality |
| 17-109-final-viral-combined_k141_298329\|\|full | Not-determined |
| 17-109-final-viral-combined_k141_398365\|\|full | Low-quality |
| 17-109-final-viral-combined_k141_232021\|\|full | Low-quality |
| 17-109-final-viral-combined_k141_129958\|\|full | Low-quality |
| 17-109-final-viral-combined_k141_48022\|\|full | Not-determined |
| 17-109-final-viral-combined_k141_13181\|\|full | Not-determined |
| 17-109-final-viral-combined_k141_181185\|\|full | Low-quality |
| 17-109-final-viral-combined_k141_63546\|\|full | Low-quality |
| 17-109-final-viral-combined_k141_81142\|\|full | Not-determined |
| 17-109-final-viral-combined_k141_147105\|\|full | Not-determined |
| 17-109-final-viral-combined_k141_363392\|\|full | Not-determined |
| 17-109-final-viral-combined_k141_97051\|\|full | Low-quality |
| 17-109-final-viral-combined_k141_214783\|\|full | Not-determined |
| 17-109-final-viral-combined_k141_181207\|\|full | Not-determined |
| 17-109-final-viral-combined_k141_48031\|\|full | Not-determined |
| 17-109-final-viral-combined_k141_13200\|\|full | Low-quality |
| 17-109-final-viral-combined_k141_398392\|\|full | Low-quality |
| 17-109-final-viral-combined_k141_247922\|\|full | Not-determined |
| 17-109-final-viral-combined_k141_382105\|\|full | Not-determined |
| 17-109-final-viral-combined_k141_13212\|\|full | Not-determined |
| 17-109-final-viral-combined_k141_81159\|\|full | Not-determined |
| 17-109-final-viral-combined_k141_63577\|\|full | Not-determined |
| 17-109-final-viral-combined_k141_298393\|\|full | Not-determined |
| 17-109-final-viral-combined_k141_363426\|\|full | Not-determined |
| 17-109-final-viral-combined_k141_48068\|\|full | Low-quality |
| 17-109-final-viral-combined_k141_48073\|\|full | Low-quality |
| 17-109-final-viral-combined_k141_331983\|\|full | Not-determined |
| 17-109-final-viral-combined_k141_130002\|\|full | Not-determined |
| 17-109-final-viral-combined_k141_363436\|\|full | Not-determined |
| 17-109-final-viral-combined_k141_81198\|\|full | Low-quality |
| 17-109-final-viral-combined_k141_113423\|\|full | Not-determined |
| 17-109-final-viral-combined_k141_316122\|\|full | Low-quality |
| 17-109-final-viral-combined_k141_30163\|\|full | Low-quality |
| 17-109-final-viral-combined_k141_214850\|\|full | Low-quality |
| 17-109-final-viral-combined_k141_247953\|\|full | Not-determined |
| 17-109-final-viral-combined_k141_181244\|\|full | Not-determined |
| 17-109-final-viral-combined_k141_398446\|\|full | Low-quality |
| 17-109-final-viral-combined_k141_398447\|\|full | Not-determined |
| 17-109-final-viral-combined_k141_264309\|\|full | Not-determined |
| 17-109-final-viral-combined_k141_398449\|\|full | Low-quality |
| 17-109-final-viral-combined_k141_316154\|\|full | Low-quality |
| 17-109-final-viral-combined_k141_30184\|\|full | Not-determined |
| 17-109-final-viral-combined_k141_198224\|\|full | Not-determined |
| 17-109-final-viral-combined_k141_63599\|\|full | Not-determined |
| 17-109-final-viral-combined_k141_363474\|\|full | Low-quality |
| 17-109-final-viral-combined_k141_332026\|\|full | Not-determined |
| 17-109-final-viral-combined_k141_363475\|\|full | Not-determined |
| 17-109-final-viral-combined_k141_181294\|\|full | Not-determined |
| 17-109-final-viral-combined_k141_113466\|\|full | Low-quality |
| 17-109-final-viral-combined_k141_398476\|\|full | Not-determined |
| 17-109-final-viral-combined_k141_282590\|\|full | Low-quality |
| 17-109-final-viral-combined_k141_363514\|\|full | Low-quality |
| 17-109-final-viral-combined_k141_48142\|\|full | Not-determined |
| 17-109-final-viral-combined_k141_198239\|\|full | Low-quality |
| 17-109-final-viral-combined_k141_147163\|\|full | Not-determined |
| 17-109-final-viral-combined_k141_348657\|\|full | Not-determined |
| 17-109-final-viral-combined_k141_181326\|\|full | Low-quality |
| 17-109-final-viral-combined_k141_13307\|\|full | Low-quality |
| 17-109-final-viral-combined_k141_232106\|\|full | Not-determined |
| 17-109-final-viral-combined_k141_97120\|\|full | Low-quality |
| 17-109-final-viral-combined_k141_348660\|\|full | Not-determined |
| 17-109-final-viral-combined_k141_48145\|\|full | Not-determined |
| 17-109-final-viral-combined_k141_113504\|\|full | Low-quality |
| 17-109-final-viral-combined_k141_97127\|\|full | Low-quality |
| 17-109-final-viral-combined_k141_181359\|\|full | Low-quality |
| 17-109-final-viral-combined_k141_214894\|\|full | Low-quality |
| 18-102_k141_156439 | Medium-quality |
| 17-109-final-viral-combined_k141_298503\|\|full | Not-determined |
| 17-109-final-viral-combined_k141_13328\|\|full | Not-determined |
| 17-109-final-viral-combined_k141_164711\|\|full | Not-determined |
| 17-109-final-viral-combined_k141_97140\|\|full | Not-determined |
| 17-109-final-viral-combined_k141_316224\|\|full | Not-determined |
| 17-109-final-viral-combined_k141_382222\|\|full | Low-quality |
| 17-109-final-viral-combined_k141_316226\|\|full | Not-determined |
| 17-109-final-viral-combined_k141_164717\|\|full | Low-quality |
| 17-109-final-viral-combined_k141_214902\|\|full | Not-determined |
| 17-109-final-viral-combined_k141_13366\|\|full | Low-quality |
| 17-109-final-viral-combined_k141_348702\|\|full | Not-determined |
| 17-109-final-viral-combined_k141_130140\|\|full | Not-determined |
| 17-109-final-viral-combined_k141_214940\|\|full | Not-determined |
| 17-109-final-viral-combined_k141_332086\|\|full | Low-quality |
| 17-109-final-viral-combined_k141_113546\|\|full | Not-determined |
| 17-109-final-viral-combined_k141_214950\|\|full | Low-quality |
| 17-109-final-viral-combined_k141_63727\|\|full | Low-quality |
| 17-109-final-viral-combined_k141_248112\|\|full | Low-quality |
| 17-109-final-viral-combined_k141_164764\|\|full | Low-quality |
| 17-109-final-viral-combined_k141_81261\|\|full | Low-quality |
| 17-109-final-viral-combined_k141_198326\|\|full | Low-quality |
| 17-109-final-viral-combined_k141_81270\|\|full | Not-determined |
| 17-109-final-viral-combined_k141_332125\|\|full | Not-determined |
| 17-109-final-viral-combined_k141_181434\|\|full | Not-determined |
| 17-109-final-viral-combined_k141_164783\|\|full | Low-quality |
| 17-109-final-viral-combined_k141_147278\|\|full | Low-quality |
| 17-109-final-viral-combined_k141_147280\|\|full | Low-quality |
| 17-109-final-viral-combined_k141_316301\|\|full | Low-quality |
| 17-109-final-viral-combined_k141_363561\|\|full | Not-determined |
| 18-102-final-viral-combined_k141_156439\|\|full | Medium-quality |
| 17-109-final-viral-combined_k141_81290\|\|full | Not-determined |
| 17-109-final-viral-combined_k141_214996\|\|full | Not-determined |
| 17-109-final-viral-combined_k141_248146\|\|full | Not-determined |
| 17-109-final-viral-combined_k141_382280\|\|full | Not-determined |
| 17-109-final-viral-combined_k141_81306\|\|full | Low-quality |
| 17-109-final-viral-combined_k141_164819\|\|full | Not-determined |
| 17-109-final-viral-combined_k141_81324\|\|full | Low-quality |
| 17-109-final-viral-combined_k141_298580\|\|full | Low-quality |
| 17-109-final-viral-combined_k141_298583\|\|full | Low-quality |
| 17-109-final-viral-combined_k141_348840\|\|full | Low-quality |
| 17-109-final-viral-combined_k141_363579\|\|full | Low-quality |
| 17-109-final-viral-combined_k141_232268\|\|full | Not-determined |
| 17-109-final-viral-combined_k141_147318\|\|full | Not-determined |
| 17-109-final-viral-combined_k141_298595\|\|full | Not-determined |
| 17-109-final-viral-combined_k141_113647\|\|full | Not-determined |
| 17-109-final-viral-combined_k141_130241\|\|full | Not-determined |
| 17-109-final-viral-combined_k141_398536\|\|full | Low-quality |
| 17-109-final-viral-combined_k141_130244\|\|full | Low-quality |
| 17-109-final-viral-combined_k141_332192\|\|full | Not-determined |
| 17-109-final-viral-combined_k141_48259\|\|full | Not-determined |
| 17-109-final-viral-combined_k141_198426\|\|full | Low-quality |
| 17-109-final-viral-combined_k141_332195\|\|full | Not-determined |
| 17-109-final-viral-combined_k141_181486\|\|full | Low-quality |
| 17-109-final-viral-combined_k141_298635\|\|full | Low-quality |
| 17-109-final-viral-combined_k141_298637\|\|full | Low-quality |
| 17-109-final-viral-combined_k141_181497\|\|full | Low-quality |
| 17-109-final-viral-combined_k141_282664\|\|full | Not-determined |
| 17-109-final-viral-combined_k141_48289\|\|full | Not-determined |
| 17-109-final-viral-combined_k141_113680\|\|full | Low-quality |
| 17-109-final-viral-combined_k141_130280\|\|full | Not-determined |
| 17-109-final-viral-combined_k141_97265\|\|full | Not-determined |
| 17-109-final-viral-combined_k141_198456\|\|full | Low-quality |
| 17-109-final-viral-combined_k141_13530\|\|full | Not-determined |
| 17-109-final-viral-combined_k141_298686\|\|full | Not-determined |
| 17-109-final-viral-combined_k141_97283\|\|full | Low-quality |
| 18-109-final-viral-combined_k141_24084\|\|full | High-quality |
| 17-109-final-viral-combined_k141_232293\|\|full | Low-quality |
| 17-109-final-viral-combined_k141_30355\|\|full | Low-quality |
| 17-109-final-viral-combined_k141_316430\|\|full | Not-determined |
| 17-109-final-viral-combined_k141_248236\|\|full | Low-quality |
| 17-109-final-viral-combined_k141_232304\|\|full | Low-quality |
| 17-109-final-viral-combined_k141_264437\|\|full | Low-quality |
| 17-109-final-viral-combined_k141_30364\|\|full | Low-quality |
| 17-109-final-viral-combined_k141_264438\|\|full | Low-quality |
| 17-109-final-viral-combined_k141_164916\|\|full | Not-determined |
| 17-109-final-viral-combined_k141_164919\|\|full | Not-determined |
| 17-109-final-viral-combined_k141_30367\|\|full | Not-determined |
| 17-109-final-viral-combined_k141_264441\|\|full | Not-determined |
| 17-109-final-viral-combined_k141_181525\|\|full | Not-determined |
| 17-109-final-viral-combined_k141_282695\|\|full | Low-quality |
| 17-109-final-viral-combined_k141_30377\|\|full | Low-quality |
| 17-109-final-viral-combined_k141_63862\|\|full | Low-quality |
| 17-109-final-viral-combined_k141_81437\|\|full | Not-determined |
| 17-109-final-viral-combined_k141_348924\|\|full | Low-quality |
| 17-109-final-viral-combined_k141_316462\|\|full | Not-determined |
| 17-109-final-viral-combined_k141_13587\|\|full | Low-quality |
| 17-109-final-viral-combined_k141_363701\|\|full | Not-determined |
| 17-109-final-viral-combined_k141_215106\|\|full | Low-quality |
| 17-109-final-viral-combined_k141_298753\|\|full | Not-determined |
| 17-109-final-viral-combined_k141_164972\|\|full | Not-determined |
| 17-109-final-viral-combined_k141_147394\|\|full | Low-quality |
| 17-109-final-viral-combined_k141_382427\|\|full | Low-quality |
| 17-109-final-viral-combined_k141_398640\|\|full | Low-quality |
| 17-109-final-viral-combined_k141_48373\|\|full | Low-quality |
| 17-109-final-viral-combined_k141_348927\|\|full | Low-quality |
| 17-109-final-viral-combined_k141_13595\|\|full | Not-determined |
| 17-109-final-viral-combined_k141_113756\|\|full | Not-determined |
| 17-109-final-viral-combined_k141_181530\|\|full | Low-quality |
| 17-109-final-viral-combined_k141_332298\|\|full | Not-determined |
| 17-109-final-viral-combined_k141_363714\|\|full | Not-determined |
| 17-109-final-viral-combined_k141_316494\|\|full | Low-quality |
| 17-109-final-viral-combined_k141_232357\|\|full | Not-determined |
| 17-109-final-viral-combined_k141_97357\|\|full | Low-quality |
| 17-109-final-viral-combined_k141_113760\|\|full | Low-quality |
| 17-109-final-viral-combined_k141_97359\|\|full | Not-determined |
| 17-109-final-viral-combined_k141_332320\|\|full | Low-quality |
| 17-109-final-viral-combined_k141_382454\|\|full | Low-quality |
| 17-109-final-viral-combined_k141_81498\|\|full | Low-quality |
| 17-109-final-viral-combined_k141_232380\|\|full | Low-quality |
| 17-109-final-viral-combined_k141_181555\|\|full | Low-quality |
| 17-109-final-viral-combined_k141_198556\|\|full | Low-quality |
| 17-109-final-viral-combined_k141_232386\|\|full | Low-quality |
| 17-109-final-viral-combined_k141_316505\|\|full | Not-determined |
| 17-109-final-viral-combined_k141_348967\|\|full | Not-determined |
| 17-109-final-viral-combined_k141_332363\|\|full | Not-determined |
| 17-109-final-viral-combined_k141_48419\|\|full | Low-quality |
| 17-109-final-viral-combined_k141_398676\|\|full | Not-determined |
| 17-109-final-viral-combined_k141_398677\|\|full | Not-determined |
| 17-109-final-viral-combined_k141_48421\|\|full | Low-quality |
| 17-109-final-viral-combined_k141_181583\|\|full | Low-quality |
| 17-109-final-viral-combined_k141_48427\|\|full | Low-quality |

| | |
|---|---|
| 17-109-final-viral-combined_k141_282825\|\|full | Not-determined |
| 17-109-final-viral-combined_k141_130412\|\|full | Low-quality |
| 17-109-final-viral-combined_k141_147453\|\|full | Not-determined |
| 17-109-final-viral-combined_k141_63890\|\|full | Not-determined |
| 17-109-final-viral-combined_k141_232427\|\|full | Not-determined |
| 17-109-final-viral-combined_k141_181609\|\|full | Low-quality |
| 17-109-final-viral-combined_k141_63900\|\|full | Not-determined |
| 17-109-final-viral-combined_k141_147467\|\|full | Not-determined |
| 17-109-final-viral-combined_k141_63905\|\|full | Low-quality |
| 17-109-final-viral-combined_k141_48446\|\|full | Not-determined |
| 17-109-final-viral-combined_k141_198591\|\|full | Low-quality |
| 17-109-final-viral-combined_k141_81534\|\|full | Low-quality |
| 17-109-final-viral-combined_k141_63932\|\|full | Not-determined |
| 17-109-final-viral-combined_k141_232454\|\|full | Not-determined |
| 17-109-final-viral-combined_k141_198603\|\|full | Not-determined |
| 17-109-final-viral-combined_k141_215243\|\|full | Low-quality |
| 17-109-final-viral-combined_k141_97440\|\|full | Not-determined |
| 17-109-final-viral-combined_k141_282842\|\|full | Not-determined |
| 17-109-final-viral-combined_k141_48466\|\|full | Low-quality |
| 17-109-final-viral-combined_k141_232466\|\|full | Not-determined |
| 17-109-final-viral-combined_k141_113869\|\|full | Not-determined |
| 17-109-final-viral-combined_k141_181663\|\|full | Low-quality |
| 17-109-final-viral-combined_k141_316594\|\|full | Low-quality |
| 17-109-final-viral-combined_k141_398705\|\|full | Not-determined |
| 17-109-final-viral-combined_k141_282866\|\|full | Low-quality |
| 17-109-final-viral-combined_k141_97475\|\|full | Not-determined |
| 17-109-final-viral-combined_k141_113888\|\|full | Not-determined |
| 17-109-final-viral-combined_k141_248341\|\|full | Low-quality |
| 17-109-final-viral-combined_k141_198646\|\|full | Not-determined |
| 17-109-final-viral-combined_k141_248343\|\|full | Not-determined |
| 17-109-final-viral-combined_k141_97490\|\|full | Low-quality |
| 17-109-final-viral-combined_k141_130456\|\|full | Not-determined |
| 17-109-final-viral-combined_k141_332465\|\|full | Not-determined |
| 17-109-final-viral-combined_k141_215261\|\|full | Not-determined |
| 17-109-final-viral-combined_k141_30523\|\|full | Not-determined |
| 17-109-final-viral-combined_k141_349017\|\|full | Low-quality |
| 17-109-final-viral-combined_k141_332480\|\|full | Not-determined |
| 17-109-final-viral-combined_k141_30524\|\|full | Not-determined |
| 17-109-final-viral-combined_k141_316638\|\|full | Low-quality |
| 17-109-final-viral-combined_k141_282912\|\|full | Low-quality |
| 17-109-final-viral-combined_k141_113911\|\|full | Low-quality |
| 17-109-final-viral-combined_k141_248370\|\|full | Not-determined |
| 17-109-final-viral-combined_k141_215291\|\|full | Low-quality |
| 17-109-final-viral-combined_k141_215297\|\|full | Low-quality |
| 17-109-final-viral-combined_k141_282931\|\|full | Low-quality |
| 17-109-final-viral-combined_k141_113931\|\|full | Low-quality |
| 17-109-final-viral-combined_k141_130494\|\|full | Not-determined |
| 17-109-final-viral-combined_k141_30537\|\|full | Not-determined |
| 17-109-final-viral-combined_k141_113943\|\|full | Low-quality |
| 17-109-final-viral-combined_k141_298932\|\|full | Low-quality |
| 17-109-final-viral-combined_k141_282947\|\|full | Low-quality |
| 17-109-final-viral-combined_k141_13758\|\|full | Low-quality |
| 17-109-final-viral-combined_k141_63982\|\|full | Low-quality |
| 17-109-final-viral-combined_k141_13760\|\|full | Low-quality |
| 17-109-final-viral-combined_k141_130512\|\|full | Low-quality |
| 17-109-final-viral-combined_k141_97556\|\|full | Low-quality |
| 17-109-final-viral-combined_k141_215334\|\|full | Low-quality |
| 17-109-final-viral-combined_k141_316678\|\|full | Low-quality |
| 17-109-final-viral-combined_k141_30559\|\|full | Not-determined |
| 17-109-final-viral-combined_k141_165054\|\|full | Not-determined |
| 17-109-final-viral-combined_k141_282985\|\|full | Not-determined |
| 17-109-final-viral-combined_k141_113980\|\|full | Not-determined |
| 17-109-final-viral-combined_k141_298949\|\|full | Not-determined |
| 17-109-final-viral-combined_k141_130531\|\|full | Low-quality |
| 17-109-final-viral-combined_k141_215355\|\|full | Low-quality |
| 17-109-final-viral-combined_k141_363882\|\|full | Low-quality |
| 17-109-final-viral-combined_k141_181736\|\|full | Low-quality |
| 17-109-final-viral-combined_k141_97586\|\|full | Not-determined |
| 17-109-final-viral-combined_k141_363884\|\|full | Low-quality |
| 17-109-final-viral-combined_k141_13798\|\|full | Low-quality |
| 17-109-final-viral-combined_k141_97588\|\|full | Not-determined |
| 17-109-final-viral-combined_k141_181744\|\|full | Not-determined |
| 17-109-final-viral-combined_k141_232585\|\|full | Low-quality |
| 17-109-final-viral-combined_k141_64017\|\|full | Not-determined |
| 17-109-final-viral-combined_k141_113992\|\|full | Not-determined |
| 17-109-final-viral-combined_k141_30585\|\|full | Not-determined |
| 17-109-final-viral-combined_k141_81604\|\|full | Not-determined |
| 17-109-final-viral-combined_k141_64022\|\|full | Not-determined |
| 17-109-final-viral-combined_k141_398818\|\|full | Not-determined |
| 17-109-final-viral-combined_k141_13815\|\|full | Low-quality |
| 17-109-final-viral-combined_k141_147556\|\|full | Low-quality |
| 17-109-final-viral-combined_k141_298996\|\|full | Not-determined |
| 17-109-final-viral-combined_k141_165090\|\|full | Not-determined |
| 17-109-final-viral-combined_k141_114019\|\|full | Not-determined |
| 17-109-final-viral-combined_k141_147568\|\|full | Not-determined |
| 17-109-final-viral-combined_k141_97624\|\|full | Not-determined |
| 17-109-final-viral-combined_k141_147569\|\|full | Not-determined |
| 17-109-final-viral-combined_k141_147570\|\|full | Not-determined |
| 17-109-final-viral-combined_k141_332610\|\|full | Not-determined |
| 17-109-final-viral-combined_k141_13859\|\|full | Not-determined |
| 17-109-final-viral-combined_k141_48540\|\|full | Not-determined |
| 17-109-final-viral-combined_k141_283063\|\|full | Not-determined |
| 17-109-final-viral-combined_k141_30624\|\|full | Not-determined |
| 17-109-final-viral-combined_k141_363969\|\|full | Low-quality |
| 17-109-final-viral-combined_k141_215420\|\|full | Low-quality |
| 17-109-final-viral-combined_k141_165112\|\|full | Not-determined |
| 17-109-final-viral-combined_k141_349121\|\|full | Not-determined |
| 17-109-final-viral-combined_k141_316726\|\|full | Low-quality |
| 17-109-final-viral-combined_k141_198803\|\|full | Not-determined |
| 17-109-final-viral-combined_k141_81657\|\|full | Not-determined |
| 17-109-final-viral-combined_k141_81677\|\|full | Not-determined |
| 17-109-final-viral-combined_k141_198811\|\|full | Not-determined |
| 17-109-final-viral-combined_k141_398841\|\|full | Low-quality |
| 17-109-final-viral-combined_k141_382591\|\|full | Not-determined |
| 17-109-final-viral-combined_k141_81709\|\|full | Not-determined |
| 17-109-final-viral-combined_k141_97685\|\|full | Not-determined |
| 17-109-final-viral-combined_k141_332684\|\|full | Not-determined |
| 17-109-final-viral-combined_k141_165166\|\|full | Low-quality |
| 17-109-final-viral-combined_k141_299088\|\|full | Not-determined |
| 17-109-final-viral-combined_k141_364013\|\|full | Low-quality |
| 17-109-final-viral-combined_k141_30644\|\|full | Low-quality |
| 17-109-final-viral-combined_k141_130654\|\|full | Low-quality |
| 17-109-final-viral-combined_k141_398849\|\|full | Low-quality |
| 17-109-final-viral-combined_k141_398855\|\|full | Low-quality |
| 17-109-final-viral-combined_k141_264692\|\|full | Not-determined |
| 17-109-final-viral-combined_k141_316810\|\|full | Not-determined |
| 17-109-final-viral-combined_k141_398864\|\|full | Low-quality |
| 17-109-final-viral-combined_k141_13942\|\|full | Low-quality |
| 17-109-final-viral-combined_k141_316815\|\|full | Low-quality |
| 17-109-final-viral-combined_k141_264703\|\|full | Not-determined |
| 17-109-final-viral-combined_k141_232737\|\|full | Not-determined |
| 17-109-final-viral-combined_k141_215504\|\|full | Not-determined |
| 17-109-final-viral-combined_k141_215505\|\|full | Low-quality |
| 17-109-final-viral-combined_k141_382624\|\|full | Low-quality |
| 17-109-final-viral-combined_k141_13961\|\|full | Low-quality |
| 17-109-final-viral-combined_k141_114116\|\|full | Not-determined |
| 17-109-final-viral-combined_k141_181890\|\|full | Not-determined |
| 17-109-final-viral-combined_k141_283179\|\|full | Low-quality |
| 17-109-final-viral-combined_k141_181895\|\|full | Low-quality |
| 17-109-final-viral-combined_k141_130713\|\|full | Low-quality |
| 17-109-final-viral-combined_k141_382642\|\|full | Low-quality |
| 17-109-final-viral-combined_k141_81782\|\|full | Not-determined |
| 17-109-final-viral-combined_k141_13973\|\|full | Not-determined |
| 17-109-final-viral-combined_k141_398916\|\|full | Low-quality |
| 17-109-final-viral-combined_k141_81784\|\|full | Not-determined |
| 17-109-final-viral-combined_k141_114138\|\|full | Low-quality |
| 17-109-final-viral-combined_k141_198880\|\|full | Not-determined |
| 17-109-final-viral-combined_k141_198895\|\|full | Not-determined |
| 17-109-final-viral-combined_k141_114151\|\|full | Low-quality |
| 18-110-final-viral-combined_k141_163173\|\|full | High-quality |
| 17-109-final-viral-combined_k141_283211\|\|full | Low-quality |
| 17-109-final-viral-combined_k141_232777\|\|full | Not-determined |
| 17-109-final-viral-combined_k141_299184\|\|full | Low-quality |
| 17-109-final-viral-combined_k141_114168\|\|full | Low-quality |
| 17-109-final-viral-combined_k141_130746\|\|full | Not-determined |
| 17-109-final-viral-combined_k141_48623\|\|full | Not-determined |
| 17-109-final-viral-combined_k141_97758\|\|full | Not-determined |
| 17-109-final-viral-combined_k141_97759\|\|full | Not-determined |
| 17-109-final-viral-combined_k141_81813\|\|full | Not-determined |
| 17-109-final-viral-combined_k141_316864\|\|full | Low-quality |
| 17-109-final-viral-combined_k141_349295\|\|full | Not-determined |
| 17-109-final-viral-combined_k141_114184\|\|full | Low-quality |
| 17-109-final-viral-combined_k141_114189\|\|full | Not-determined |
| 17-109-final-viral-combined_k141_299234\|\|full | Not-determined |
| 17-109-final-viral-combined_k141_316891\|\|full | Low-quality |
| 17-109-final-viral-combined_k141_114191\|\|full | Not-determined |
| 17-109-final-viral-combined_k141_215585\|\|full | Not-determined |
| 17-109-final-viral-combined_k141_264792\|\|full | Low-quality |
| 17-109-final-viral-combined_k141_349316\|\|full | Not-determined |
| 17-109-final-viral-combined_k141_264795\|\|full | Not-determined |
| 17-109-final-viral-combined_k141_81843\|\|full | Low-quality |
| 17-109-final-viral-combined_k141_147699\|\|full | Not-determined |
| 17-109-final-viral-combined_k141_264816\|\|full | Not-determined |
| 18-101-final-viral-combined_k141_223620\|\|full | High-quality |
| 17-109-final-viral-combined_k141_48652\|\|full | Not-determined |
| 17-109-final-viral-combined_k141_30738\|\|full | Low-quality |
| 17-109-final-viral-combined_k141_215609\|\|full | Low-quality |
| 17-109-final-viral-combined_k141_48688\|\|full | Low-quality |
| 17-109-final-viral-combined_k141_398950\|\|full | Not-determined |
| 17-109-final-viral-combined_k141_182057\|\|full | Low-quality |
| 17-109-final-viral-combined_k141_97861\|\|full | Low-quality |
| 17-109-final-viral-combined_k141_130836\|\|full | Not-determined |
| 17-109-final-viral-combined_k141_382723\|\|full | Not-determined |
| 17-109-final-viral-combined_k141_283334\|\|full | Not-determined |
| 17-109-final-viral-combined_k141_165246\|\|full | Low-quality |
| 17-109-final-viral-combined_k141_81888\|\|full | Low-quality |
| 17-109-final-viral-combined_k141_64161\|\|full | Not-determined |
| 18-110_k141_30498 | Medium-quality |
| 17-109-final-viral-combined_k141_48715\|\|full | Not-determined |
| 17-109-final-viral-combined_k141_14090\|\|full | Low-quality |
| 17-109-final-viral-combined_k141_198962\|\|full | Not-determined |
| 17-109-final-viral-combined_k141_81903\|\|full | Low-quality |
| 17-109-final-viral-combined_k141_182084\|\|full | Not-determined |
| 17-109-final-viral-combined_k141_48726\|\|full | Not-determined |
| 17-109-final-viral-combined_k141_398963\|\|full | Not-determined |
| 17-109-final-viral-combined_k141_316914\|\|full | Low-quality |
| 17-109-final-viral-combined_k141_130851\|\|full | Low-quality |
| 17-109-final-viral-combined_k141_182113\|\|full | Low-quality |
| 17-109-final-viral-combined_k141_48738\|\|full | Low-quality |
| 17-109-final-viral-combined_k141_264890\|\|full | Not-determined |
| 17-109-final-viral-combined_k141_165279\|\|full | Low-quality |
| 17-109-final-viral-combined_k141_97929\|\|full | Not-determined |
| 17-109-final-viral-combined_k141_316926\|\|full | Low-quality |
| 17-109-final-viral-combined_k141_130876\|\|full | Low-quality |
| 17-109-final-viral-combined_k141_182141\|\|full | Low-quality |
| 17-109-final-viral-combined_k141_81942\|\|full | Not-determined |
| 17-109-final-viral-combined_k141_97935\|\|full | Not-determined |
| 17-109-final-viral-combined_k141_332890\|\|full | Low-quality |
| 17-109-final-viral-combined_k141_264927\|\|full | Low-quality |
| 17-109-final-viral-combined_k141_349406\|\|full | Not-determined |
| 17-109-final-viral-combined_k141_147773\|\|full | Low-quality |
| 17-109-final-viral-combined_k141_97949\|\|full | Not-determined |
| 17-109-final-viral-combined_k141_64240\|\|full | Low-quality |
| 18-110-final-viral-combined_k141_30498\|\|full | Medium-quality |
| 17-109-final-viral-combined_k141_30799\|\|full | Low-quality |
| 17-109-final-viral-combined_k141_264943\|\|full | Not-determined |
| 17-109-final-viral-combined_k141_349413\|\|full | Not-determined |
| 17-109-final-viral-combined_k141_299297\|\|full | Not-determined |
| 17-109-final-viral-combined_k141_81957\|\|full | Not-determined |
| 17-109-final-viral-combined_k141_316928\|\|full | Low-quality |
| 17-109-final-viral-combined_k141_316934\|\|full | Low-quality |
| 17-109-final-viral-combined_k141_81968\|\|full | Not-determined |
| 17-109-final-viral-combined_k141_382745\|\|full | Not-determined |
| 17-109-final-viral-combined_k141_398987\|\|full | Not-determined |
| 17-109-final-viral-combined_k141_382748\|\|full | Not-determined |
| 17-109-final-viral-combined_k141_48805\|\|full | Not-determined |
| 17-109-final-viral-combined_k141_97992\|\|full | Not-determined |
| 17-109-final-viral-combined_k141_264971\|\|full | Low-quality |
| 17-109-final-viral-combined_k141_364267\|\|full | Low-quality |
| 17-109-final-viral-combined_k141_64291\|\|full | Not-determined |
| 17-109-final-viral-combined_k141_130916\|\|full | Low-quality |
| 17-109-final-viral-combined_k141_182199\|\|full | Low-quality |
| 17-109-final-viral-combined_k141_332959\|\|full | Low-quality |
| 17-109-final-viral-combined_k141_264982\|\|full | Low-quality |
| 17-109-final-viral-combined_k141_199062\|\|full | Low-quality |
| 17-109-final-viral-combined_k141_199065\|\|full | Low-quality |
| 17-109-final-viral-combined_k141_30862\|\|full | Not-determined |
| 17-109-final-viral-combined_k141_264992\|\|full | Not-determined |
| 17-109-final-viral-combined_k141_382786\|\|full | Low-quality |
| 17-109-final-viral-combined_k141_14199\|\|full | Low-quality |
| 17-109-final-viral-combined_k141_265007\|\|full | Low-quality |
| 17-109-final-viral-combined_k141_332983\|\|full | Not-determined |
| 17-109-final-viral-combined_k141_48865\|\|full | Low-quality |
| 17-109-final-viral-combined_k141_332987\|\|full | Not-determined |
| 17-109-final-viral-combined_k141_232946\|\|full | Not-determined |
| 17-109-final-viral-combined_k141_283489\|\|full | Low-quality |
| 17-109-final-viral-combined_k141_182231\|\|full | Not-determined |
| 17-109-final-viral-combined_k141_265034\|\|full | Not-determined |
| 17-109-final-viral-combined_k141_333001\|\|full | Not-determined |
| 17-109-final-viral-combined_k141_182235\|\|full | Not-determined |
| 17-109-final-viral-combined_k141_30890\|\|full | Low-quality |
| 17-109-final-viral-combined_k141_130989\|\|full | Low-quality |
| 17-109-final-viral-combined_k141_130990\|\|full | Not-determined |
| 17-109-final-viral-combined_k141_283526\|\|full | Low-quality |
| 17-109-final-viral-combined_k141_14262\|\|full | Not-determined |
| 17-109-final-viral-combined_k141_248714\|\|full | Not-determined |
| 17-109-final-viral-combined_k141_30904\|\|full | Not-determined |
| 17-109-final-viral-combined_k141_48886\|\|full | Low-quality |
| 17-109-final-viral-combined_k141_265062\|\|full | Low-quality |
| 17-109-final-viral-combined_k141_48890\|\|full | Low-quality |
| 17-109-final-viral-combined_k141_382816\|\|full | Not-determined |
| 17-109-final-viral-combined_k141_30916\|\|full | Low-quality |
| 17-109-final-viral-combined_k141_199130\|\|full | Not-determined |
| 17-109-final-viral-combined_k141_215714\|\|full | Low-quality |
| 17-109-final-viral-combined_k141_82134\|\|full | Low-quality |
| 17-109-final-viral-combined_k141_349489\|\|full | Low-quality |
| 17-109-final-viral-combined_k141_131040\|\|full | Low-quality |
| 17-109-final-viral-combined_k141_131041\|\|full | Not-determined |
| 17-109-final-viral-combined_k141_30931\|\|full | Low-quality |
| 17-109-final-viral-combined_k141_233003\|\|full | Not-determined |
| 17-109-final-viral-combined_k141_399027\|\|full | Not-determined |
| 17-109-final-viral-combined_k141_131044\|\|full | Low-quality |
| 17-109-final-viral-combined_k141_299345\|\|full | Low-quality |
| 17-109-final-viral-combined_k141_215733\|\|full | Not-determined |
| 17-109-final-viral-combined_k141_114366\|\|full | Low-quality |
| 17-109-final-viral-combined_k141_14311\|\|full | Low-quality |
| 17-109-final-viral-combined_k141_317141\|\|full | Low-quality |
| 17-109-final-viral-combined_k141_165493\|\|full | Not-determined |
| 17-109-final-viral-combined_k141_399055\|\|full | Not-determined |
| 17-109-final-viral-combined_k141_64419\|\|full | Not-determined |
| 17-109-final-viral-combined_k141_364413\|\|full | Low-quality |
| 17-109-final-viral-combined_k141_349531\|\|full | Not-determined |
| 17-109-final-viral-combined_k141_165500\|\|full | Not-determined |
| 17-109-final-viral-combined_k141_233025\|\|full | Not-determined |
| 17-109-final-viral-combined_k141_283580\|\|full | Not-determined |
| 17-109-final-viral-combined_k141_182297\|\|full | Not-determined |
| 17-109-final-viral-combined_k141_131083\|\|full | Not-determined |
| 17-109-final-viral-combined_k141_349544\|\|full | Not-determined |
| 17-109-final-viral-combined_k141_64423\|\|full | Low-quality |
| 17-109-final-viral-combined_k141_333072\|\|full | Not-determined |
| 17-109-final-viral-combined_k141_30986\|\|full | Low-quality |
| 17-109-final-viral-combined_k141_114392\|\|full | Not-determined |
| 17-109-final-viral-combined_k141_299380\|\|full | Low-quality |
| 17-109-final-viral-combined_k141_165522\|\|full | Not-determined |
| 17-109-final-viral-combined_k141_317191\|\|full | Low-quality |
| 17-109-final-viral-combined_k141_382858\|\|full | Not-determined |
| 17-109-final-viral-combined_k141_215786\|\|full | Not-determined |
| 17-109-final-viral-combined_k141_265119\|\|full | Low-quality |
| 17-109-final-viral-combined_k141_299403\|\|full | Low-quality |
| 17-109-final-viral-combined_k141_349561\|\|full | Low-quality |
| 17-109-final-viral-combined_k141_317220\|\|full | Not-determined |
| 17-109-final-viral-combined_k141_333116\|\|full | Not-determined |
| 17-109-final-viral-combined_k141_48961\|\|full | Not-determined |
| 17-109-final-viral-combined_k141_215815\|\|full | Not-determined |
| 17-109-final-viral-combined_k141_233100\|\|full | Not-determined |
| 18-101-final-viral-combined_k141_244836\|\|full | Medium-quality |
| 17-109-final-viral-combined_k141_31033\|\|full | Not-determined |
| 17-109-final-viral-combined_k141_364489\|\|full | Low-quality |
| 17-109-final-viral-combined_k141_64499\|\|full | Not-determined |
| 17-109-final-viral-combined_k141_31044\|\|full | Not-determined |
| 17-109-final-viral-combined_k141_215843\|\|full | Not-determined |
| 17-109-final-viral-combined_k141_165604\|\|full | Low-quality |
| 17-109-final-viral-combined_k141_148018\|\|full | Low-quality |
| 17-109-final-viral-combined_k141_49000\|\|full | Not-determined |
| 17-109-final-viral-combined_k141_148021\|\|full | Not-determined |
| 17-109-final-viral-combined_k141_349617\|\|full | Low-quality |
| 17-109-final-viral-combined_k141_349621\|\|full | Low-quality |
| 17-109-final-viral-combined_k141_233139\|\|full | Low-quality |
| 17-109-final-viral-combined_k141_98086\|\|full | Not-determined |
| 17-109-final-viral-combined_k141_64546\|\|full | Not-determined |
| 17-109-final-viral-combined_k141_165617\|\|full | Not-determined |
| 17-109-final-viral-combined_k141_248808\|\|full | Low-quality |
| 17-109-final-viral-combined_k141_265127\|\|full | Not-determined |
| 17-109-final-viral-combined_k141_131185\|\|full | Low-quality |
| 17-109-final-viral-combined_k141_114503\|\|full | Low-quality |
| 17-109-final-viral-combined_k141_283639\|\|full | Low-quality |
| 17-109-final-viral-combined_k141_64574\|\|full | Low-quality |
| 17-109-final-viral-combined_k141_131210\|\|full | Low-quality |
| 17-109-final-viral-combined_k141_233212\|\|full | Not-determined |
| 17-109-final-viral-combined_k141_64596\|\|full | Not-determined |
| 17-109-final-viral-combined_k141_131226\|\|full | Not-determined |
| 17-109-final-viral-combined_k141_382953\|\|full | Not-determined |
| 17-109-final-viral-combined_k141_165644\|\|full | Low-quality |
| 17-109-final-viral-combined_k141_215927\|\|full | Not-determined |
| 17-109-final-viral-combined_k141_233219\|\|full | Not-determined |
| 17-109-final-viral-combined_k141_114567\|\|full | Low-quality |
| 17-109-final-viral-combined_k141_31140\|\|full | Low-quality |
| 17-109-final-viral-combined_k141_299467\|\|full | Not-determined |
| 17-109-final-viral-combined_k141_114579\|\|full | Not-determined |
| 17-109-final-viral-combined_k141_182456\|\|full | Not-determined |
| 17-109-final-viral-combined_k141_148114\|\|full | Not-determined |
| 17-109-final-viral-combined_k141_399191\|\|full | Not-determined |
| 17-109-final-viral-combined_k141_364582\|\|full | Low-quality |
| 17-109-final-viral-combined_k141_31158\|\|full | Low-quality |
| 17-109-final-viral-combined_k141_215975\|\|full | Low-quality |
| 17-109-final-viral-combined_k141_299519\|\|full | Not-determined |
| 17-109-final-viral-combined_k141_233287\|\|full | Low-quality |
| 17-109-final-viral-combined_k141_283671\|\|full | Not-determined |
| 17-109-final-viral-combined_k141_64632\|\|full | Low-quality |
| 17-109-final-viral-combined_k141_165702\|\|full | Low-quality |
| 17-109-final-viral-combined_k141_14504\|\|full | Low-quality |
| 17-109-final-viral-combined_k141_215980\|\|full | Low-quality |
| 17-109-final-viral-combined_k141_399208\|\|full | Not-determined |
| 17-109-final-viral-combined_k141_248891\|\|full | Low-quality |
| 17-109-final-viral-combined_k141_165717\|\|full | Not-determined |
| 17-109-final-viral-combined_k141_248892\|\|full | Low-quality |
| 17-109-final-viral-combined_k141_364617\|\|full | Not-determined |
| 17-109-final-viral-combined_k141_64654\|\|full | Not-determined |
| 17-109-final-viral-combined_k141_199260\|\|full | Not-determined |
| 17-109-final-viral-combined_k141_248918\|\|full | Low-quality |
| 17-109-final-viral-combined_k141_399238\|\|full | Low-quality |
| 17-109-final-viral-combined_k141_31201\|\|full | Low-quality |
| 17-109-final-viral-combined_k141_283709\|\|full | Not-determined |
| 17-109-final-viral-combined_k141_283716\|\|full | Not-determined |
| 17-109-final-viral-combined_k141_98189\|\|full | Not-determined |
| 17-109-final-viral-combined_k141_248941\|\|full | Not-determined |
| 17-109-final-viral-combined_k141_233335\|\|full | Low-quality |
| 17-109-final-viral-combined_k141_216026\|\|full | Not-determined |
| 17-109-final-viral-combined_k141_31241\|\|full | Not-determined |
| 17-109-final-viral-combined_k141_14524\|\|full | Low-quality |
| 17-109-final-viral-combined_k141_216056\|\|full | Not-determined |
| 17-109-final-viral-combined_k141_98228\|\|full | Low-quality |
| 17-109-final-viral-combined_k141_49070\|\|full | Not-determined |
| 17-109-final-viral-combined_k141_165742\|\|full | Not-determined |
| 17-109-final-viral-combined_k141_131394\|\|full | Low-quality |
| 17-109-final-viral-combined_k141_265168\|\|full | Low-quality |
| 17-109-final-viral-combined_k141_399299\|\|full | Not-determined |
| 17-109-final-viral-combined_k141_317442\|\|full | Low-quality |
| 17-109-final-viral-combined_k141_349808\|\|full | Low-quality |
| 17-109-final-viral-combined_k141_31270\|\|full | Not-determined |
| 17-109-final-viral-combined_k141_317446\|\|full | Low-quality |
| 17-109-final-viral-combined_k141_333309\|\|full | Low-quality |
| 17-109-final-viral-combined_k141_131446\|\|full | Not-determined |
| 17-109-final-viral-combined_k141_199321\|\|full | Low-quality |
| 17-109-final-viral-combined_k141_216092\|\|full | Low-quality |
| 17-109-final-viral-combined_k141_131472\|\|full | Not-determined |
| 17-109-final-viral-combined_k141_148304\|\|full | Not-determined |
| 17-109-final-viral-combined_k141_216116\|\|full | Not-determined |
| 17-109-final-viral-combined_k141_182565\|\|full | Not-determined |
| 17-109-final-viral-combined_k141_64828\|\|full | Not-determined |
| 17-109-final-viral-combined_k141_249051\|\|full | Low-quality |
| 17-109-final-viral-combined_k141_216123\|\|full | Not-determined |
| 17-109-final-viral-combined_k141_199360\|\|full | Low-quality |
| 17-109-final-viral-combined_k141_49121\|\|full | Low-quality |
| 17-109-final-viral-combined_k141_283803\|\|full | Low-quality |
| 17-109-final-viral-combined_k141_31356\|\|full | Not-determined |
| 17-109-final-viral-combined_k141_333382\|\|full | Low-quality |
| 17-109-final-viral-combined_k141_299628\|\|full | Not-determined |
| 17-109-final-viral-combined_k141_216165\|\|full | Not-determined |
| 17-109-final-viral-combined_k141_14638\|\|full | Not-determined |
| 17-109-final-viral-combined_k141_265263\|\|full | Low-quality |
| 17-109-final-viral-combined_k141_199387\|\|full | Not-determined |
| 17-109-final-viral-combined_k141_265278\|\|full | Not-determined |
| 17-109-final-viral-combined_k141_265283\|\|full | Low-quality |
| 17-109-final-viral-combined_k141_349871\|\|full | Low-quality |
| 17-109-final-viral-combined_k141_349875\|\|full | Not-determined |
| 17-109-final-viral-combined_k141_31428\|\|full | Low-quality |
| 17-109-final-viral-combined_k141_49167\|\|full | Not-determined |
| 17-109-final-viral-combined_k141_98378\|\|full | Not-determined |
| 17-109-final-viral-combined_k141_148383\|\|full | Not-determined |
| 17-109-final-viral-combined_k141_182631\|\|full | Low-quality |
| 17-109-final-viral-combined_k141_283848\|\|full | Low-quality |
| 17-109-final-viral-combined_k141_182641\|\|full | Low-quality |
| 17-109-final-viral-combined_k141_249144\|\|full | Low-quality |
| 17-109-final-viral-combined_k141_216224\|\|full | Low-quality |
| 17-109-final-viral-combined_k141_265320\|\|full | Low-quality |
| 17-109-final-viral-combined_k141_349928\|\|full | Not-determined |
| 17-109-final-viral-combined_k141_165836\|\|full | Low-quality |
| 17-109-final-viral-combined_k141_131498\|\|full | Low-quality |
| 17-109-final-viral-combined_k141_199439\|\|full | Low-quality |
| 17-109-final-viral-combined_k141_131500\|\|full | Low-quality |
| 17-109-final-viral-combined_k141_49219\|\|full | Not-determined |
| 17-109-final-viral-combined_k141_399487\|\|full | Not-determined |
| 17-109-final-viral-combined_k141_233510\|\|full | Not-determined |
| 17-109-final-viral-combined_k141_249172\|\|full | Not-determined |
| 17-109-final-viral-combined_k141_131511\|\|full | Low-quality |
| 17-109-final-viral-combined_k141_82530\|\|full | Not-determined |
| 17-109-final-viral-combined_k141_49238\|\|full | Low-quality |
| 17-109-final-viral-combined_k141_82537\|\|full | Low-quality |
| 17-109-final-viral-combined_k141_182671\|\|full | Not-determined |
| 17-109-final-viral-combined_k141_199460\|\|full | Not-determined |
| 17-109-final-viral-combined_k141_249198\|\|full | Not-determined |
| 17-109-final-viral-combined_k141_182683\|\|full | Low-quality |
| 17-109-final-viral-combined_k141_148440\|\|full | Not-determined |
| 17-109-final-viral-combined_k141_383076\|\|full | Low-quality |
| 17-109-final-viral-combined_k141_199476\|\|full | Not-determined |
| 17-109-final-viral-combined_k141_64988\|\|full | Not-determined |
| 17-109-final-viral-combined_k141_216287\|\|full | Low-quality |
| 17-109-final-viral-combined_k141_31466\|\|full | Low-quality |
| 17-109-final-viral-combined_k141_14735\|\|full | Not-determined |
| 17-109-final-viral-combined_k141_283899\|\|full | Not-determined |
| 17-109-final-viral-combined_k141_299740\|\|full | Low-quality |
| 17-109-final-viral-combined_k141_165909\|\|full | Not-determined |
| 17-109-final-viral-combined_k141_148470\|\|full | Not-determined |
| 17-109-final-viral-combined_k141_199485\|\|full | Not-determined |
| 17-109-final-viral-combined_k141_165925\|\|full | Not-determined |
| 17-109-final-viral-combined_k141_199504\|\|full | Low-quality |
| 17-109-final-viral-combined_k141_216293\|\|full | Low-quality |
| 17-109-final-viral-combined_k141_148494\|\|full | Low-quality |
| 17-109-final-viral-combined_k141_399513\|\|full | Low-quality |
| 17-109-final-viral-combined_k141_98457\|\|full | Low-quality |
| 17-109-final-viral-combined_k141_31488\|\|full | Low-quality |
| 17-109-final-viral-combined_k141_216329\|\|full | Low-quality |
| 17-109-final-viral-combined_k141_199549\|\|full | Low-quality |
| 17-109-final-viral-combined_k141_165948\|\|full | Not-determined |
| 17-109-final-viral-combined_k141_233617\|\|full | Not-determined |
| 17-109-final-viral-combined_k141_364927\|\|full | Low-quality |
| 17-109-final-viral-combined_k141_182774\|\|full | Not-determined |
| 17-109-final-viral-combined_k141_383163\|\|full | Low-quality |
| 17-109-final-viral-combined_k141_364929\|\|full | Low-quality |
| 17-109-final-viral-combined_k141_31534\|\|full | Not-determined |
| 17-109-final-viral-combined_k141_65070\|\|full | Low-quality |
| 17-109-final-viral-combined_k141_65072\|\|full | Not-determined |
| 17-109-final-viral-combined_k141_399533\|\|full | Not-determined |
| 17-109-final-viral-combined_k141_49332\|\|full | Low-quality |
| 17-109-final-viral-combined_k141_114797\|\|full | Low-quality |
| 17-109-final-viral-combined_k141_182784\|\|full | Not-determined |
| 17-109-final-viral-combined_k141_98495\|\|full | Low-quality |
| 17-109-final-viral-combined_k141_49343\|\|full | Not-determined |
| 17-109-final-viral-combined_k141_165975\|\|full | Not-determined |
| 17-109-final-viral-combined_k141_364955\|\|full | Not-determined |
| 17-109-final-viral-combined_k141_165991\|\|full | Not-determined |
| 17-109-final-viral-combined_k141_49354\|\|full | Low-quality |
| 17-109-final-viral-combined_k141_82644\|\|full | Not-determined |
| 17-109-final-viral-combined_k141_399567\|\|full | Not-determined |
| 17-109-final-viral-combined_k141_82658\|\|full | Low-quality |
| 17-109-final-viral-combined_k141_333544\|\|full | Low-quality |
| 17-109-final-viral-combined_k141_49379\|\|full | Not-determined |
| 17-109-final-viral-combined_k141_299812\|\|full | Not-determined |
| 17-109-final-viral-combined_k141_148583\|\|full | Not-determined |
| 17-109-final-viral-combined_k141_148585\|\|full | Low-quality |
| 17-109-final-viral-combined_k141_131616\|\|full | Not-determined |
| 17-109-final-viral-combined_k141_49408\|\|full | Not-determined |
| 17-109-final-viral-combined_k141_31593\|\|full | Low-quality |
| 17-109-final-viral-combined_k141_283230\|\|full | Low-quality |
| 17-109-final-viral-combined_k141_65170\|\|full | Low-quality |
| 17-109-final-viral-combined_k141_284006\|\|full | Not-determined |
| 17-109-final-viral-combined_k141_249291\|\|full | Not-determined |
| 17-109-final-viral-combined_k141_383236\|\|full | Not-determined |
| 17-109-final-viral-combined_k141_148608\|\|full | Low-quality |
| 17-109-final-viral-combined_k141_399616\|\|full | Low-quality |
| 17-109-final-viral-combined_k141_131652\|\|full | Low-quality |
| 17-109-final-viral-combined_k141_399617\|\|full | Low-quality |
| 17-109-final-viral-combined_k141_365054\|\|full | Not-determined |
| 17-109-final-viral-combined_k141_333615\|\|full | Not-determined |
| 17-109-final-viral-combined_k141_82720\|\|full | Low-quality |
| 17-109-final-viral-combined_k141_333620\|\|full | Low-quality |
| 17-109-final-viral-combined_k141_249310\|\|full | Low-quality |
| 17-109-final-viral-combined_k141_284051\|\|full | Not-determined |
| 17-109-final-viral-combined_k141_114901\|\|full | Low-quality |
| 17-109-final-viral-combined_k141_265465\|\|full | Low-quality |
| 18-109-final-viral-combined_k141_104872\|\|full | Medium-quality |
| 17-109-final-viral-combined_k141_265477\|\|full | Not-determined |
| 17-109-final-viral-combined_k141_216497\|\|full | Low-quality |
| 17-109-final-viral-combined_k141_350150\|\|full | Not-determined |
| 17-109-final-viral-combined_k141_31662\|\|full | Not-determined |
| 17-109-final-viral-combined_k141_333671\|\|full | Not-determined |
| 17-109-final-viral-combined_k141_82764\|\|full | Not-determined |
| 17-109-final-viral-combined_k141_65253\|\|full | Not-determined |
| 17-109-final-viral-combined_k141_299896\|\|full | Not-determined |
| 17-109-final-viral-combined_k141_148689\|\|full | Low-quality |
| 17-109-final-viral-combined_k141_399637\|\|full | Not-determined |
| 17-109-final-viral-combined_k141_148698\|\|full | Not-determined |
| 17-109-final-viral-combined_k141_65269\|\|full | Not-determined |
| 17-109-final-viral-combined_k141_65273\|\|full | Not-determined |
| 17-109-final-viral-combined_k141_199750\|\|full | Not-determined |
| 17-109-final-viral-combined_k141_114939\|\|full | Not-determined |
| 17-109-final-viral-combined_k141_249419\|\|full | Not-determined |
| 17-109-final-viral-combined_k141_98647\|\|full | Not-determined |
| 17-109-final-viral-combined_k141_131712\|\|full | Low-quality |
| 17-109-final-viral-combined_k141_49533\|\|full | Low-quality |
| 17-109-final-viral-combined_k141_265533\|\|full | Not-determined |
| 17-109-final-viral-combined_k141_65288\|\|full | Low-quality |
| 17-109-final-viral-combined_k141_249425\|\|full | Not-determined |
| 17-109-final-viral-combined_k141_216548\|\|full | Low-quality |
| 17-109-final-viral-combined_k141_148723\|\|full | Not-determined |
| 17-109-final-viral-combined_k141_98666\|\|full | Not-determined |
| 17-109-final-viral-combined_k141_365165\|\|full | Not-determined |
| 17-109-final-viral-combined_k141_49556\|\|full | Low-quality |
| 17-109-final-viral-combined_k141_82820\|\|full | Not-determined |
| 17-109-final-viral-combined_k141_216588\|\|full | Low-quality |
| 17-109-final-viral-combined_k141_299955\|\|full | Not-determined |
| 17-109-final-viral-combined_k141_65313\|\|full | Low-quality |
| 17-109-final-viral-combined_k141_14844\|\|full | Low-quality |
| 17-109-final-viral-combined_k141_299965\|\|full | Low-quality |
| 17-109-final-viral-combined_k141_114981\|\|full | Not-determined |
| 17-109-final-viral-combined_k141_350169\|\|full | Not-determined |
| 17-109-final-viral-combined_k141_98707\|\|full | Not-determined |
| 17-109-final-viral-combined_k141_31742\|\|full | Low-quality |
| 17-109-final-viral-combined_k141_31748\|\|full | Low-quality |
| 17-109-final-viral-combined_k141_399683\|\|full | Not-determined |
| 17-109-final-viral-combined_k141_365181\|\|full | Low-quality |
| 17-109-final-viral-combined_k141_399688\|\|full | Low-quality |
| 17-109-final-viral-combined_k141_115004\|\|full | Not-determined |
| 17-109-final-viral-combined_k141_199863\|\|full | Not-determined |
| 17-109-final-viral-combined_k141_131772\|\|full | Low-quality |
| 17-109-final-viral-combined_k141_333766\|\|full | Not-determined |
| 17-109-final-viral-combined_k141_115019\|\|full | Not-determined |
| 17-109-final-viral-combined_k141_49594\|\|full | Not-determined |
| 17-109-final-viral-combined_k141_14856\|\|full | Not-determined |
| 17-109-final-viral-combined_k141_82869\|\|full | Not-determined |
| 17-109-final-viral-combined_k141_98757\|\|full | Low-quality |
| 17-108_k141_320555 | Medium-quality |
| 17-108-final-viral-combined_k141_320555\|\|full | Medium-quality |
| 17-109-final-viral-combined_k141_365202\|\|full | Low-quality |
| 17-109-final-viral-combined_k141_183018\|\|full | Not-determined |
| 17-109-final-viral-combined_k141_299995\|\|full | Low-quality |
| 17-109-final-viral-combined_k141_98767\|\|full | Not-determined |
| 17-109-final-viral-combined_k141_317796\|\|full | Low-quality |
| 17-109-final-viral-combined_k141_284140\|\|full | Not-determined |
| 17-109-final-viral-combined_k141_284141\|\|full | Not-determined |
| 17-109-final-viral-combined_k141_233937\|\|full | Not-determined |
| 17-109-final-viral-combined_k141_333793\|\|full | Not-determined |
| 17-109-final-viral-combined_k141_365213\|\|full | Not-determined |
| 17-109-final-viral-combined_k141_216665\|\|full | Not-determined |
| 17-109-final-viral-combined_k141_115042\|\|full | Not-determined |
| 17-109-final-viral-combined_k141_333803\|\|full | Not-determined |
| 17-109-final-viral-combined_k141_333805\|\|full | Low-quality |
| 17-109-final-viral-combined_k141_216673\|\|full | Not-determined |
| 17-109-final-viral-combined_k141_98773\|\|full | Low-quality |
| 17-109-final-viral-combined_k141_115055\|\|full | Low-quality |
| 17-109-final-viral-combined_k141_299872\|\|full | Medium-quality |
| 17-109-final-viral-combined_k141_350226\|\|full | Not-determined |
| 17-109-final-viral-combined_k141_249514\|\|full | Not-determined |
| 17-109-final-viral-combined_k141_183036\|\|full | Low-quality |
| 17-109-final-viral-combined_k141_284177\|\|full | Low-quality |
| 17-109-final-viral-combined_k141_98787\|\|full | Not-determined |
| 17-109-final-viral-combined_k141_300033\|\|full | Low-quality |
| 17-109-final-viral-combined_k141_131825\|\|full | Low-quality |
| 17-109-final-viral-combined_k141_249516\|\|full | Not-determined |
| 17-109-final-viral-combined_k141_82942\|\|full | Not-determined |
| 17-109-final-viral-combined_k141_216685\|\|full | Low-quality |
| 17-109-final-viral-combined_k141_199898\|\|full | Not-determined |
| 17-109-final-viral-combined_k141_383404\|\|full | Not-determined |
| 17-109-final-viral-combined_k141_300047\|\|full | Not-determined |
| 17-109-final-viral-combined_k141_249522\|\|full | Low-quality |
| 17-109-final-viral-combined_k141_350257\|\|full | Not-determined |
| 17-109-final-viral-combined_k141_82962\|\|full | Low-quality |
| 17-109-final-viral-combined_k141_284194\|\|full | Not-determined |
| 17-109-final-viral-combined_k141_300052\|\|full | Low-quality |
| 17-109-final-viral-combined_k141_365258\|\|full | Not-determined |
| 17-109-final-viral-combined_k141_98793\|\|full | Low-quality |
| 17-109-final-viral-combined_k141_183068\|\|full | Low-quality |
| 17-109-final-viral-combined_k141_65376\|\|full | Low-quality |
| 17-109-final-viral-combined_k141_148792\|\|full | Not-determined |
| 17-109-final-viral-combined_k141_199921\|\|full | Low-quality |
| 17-109-final-viral-combined_k141_300074\|\|full | Low-quality |
| 17-109-final-viral-combined_k141_216731\|\|full | Not-determined |
| 17-109-final-viral-combined_k141_249540\|\|full | Low-quality |

| | |
|---|---|
| 17-109-final-viral-combined_k141_199937\|\|full | Low-quality |
| 17-109-final-viral-combined_k141_98818\|\|full | Not-determined |
| 17-109-final-viral-combined_k141_383463\|\|full | Not-determined |
| 17-109-final-viral-combined_k141_49723\|\|full | Not-determined |
| 17-109-final-viral-combined_k141_199948\|\|full | Not-determined |
| 17-109-final-viral-combined_k141_399770\|\|full | Low-quality |
| 17-109-final-viral-combined_k141_199955\|\|full | Low-quality |
| 17-109-final-viral-combined_k141_49740\|\|full | Not-determined |
| 17-109-final-viral-combined_k141_98847\|\|full | Low-quality |
| 17-109-final-viral-combined_k141_333867\|\|full | Not-determined |
| 17-109-final-viral-combined_k141_365317\|\|full | Not-determined |
| 17-109-final-viral-combined_k141_365323\|\|full | Not-determined |
| 17-109-final-viral-combined_k141_333887\|\|full | Low-quality |
| 17-109-final-viral-combined_k141_166253\|\|full | Low-quality |
| 17-109-final-viral-combined_k141_216781\|\|full | Low-quality |
| 17-109-final-viral-combined_k141_350343\|\|full | Not-determined |
| 17-109-final-viral-combined_k141_15016\|\|full | Not-determined |
| 17-109-final-viral-combined_k141_365342\|\|full | Not-determined |
| 17-109-final-viral-combined_k141_216787\|\|full | Low-quality |
| 17-109-final-viral-combined_k141_365346\|\|full | Not-determined |
| 17-109-final-viral-combined_k141_300159\|\|full | Not-determined |
| 17-109-final-viral-combined_k141_131937\|\|full | Not-determined |
| 17-109-final-viral-combined_k141_365360\|\|full | Low-quality |
| 17-109-final-viral-combined_k141_148867\|\|full | Low-quality |
| 17-109-final-viral-combined_k141_131947\|\|full | Not-determined |
| 17-109-final-viral-combined_k141_148869\|\|full | Low-quality |
| 17-109-final-viral-combined_k141_49803\|\|full | Low-quality |
| 17-109-final-viral-combined_k141_216804\|\|full | Low-quality |
| 17-109-final-viral-combined_k141_183146\|\|full | Low-quality |
| 17-109-final-viral-combined_k141_49813\|\|full | Low-quality |
| 17-109-final-viral-combined_k141_65457\|\|full | Low-quality |
| 17-109-final-viral-combined_k141_399807\|\|full | Not-determined |
| 17-109-final-viral-combined_k141_98904\|\|full | Not-determined |
| 17-109-final-viral-combined_k141_15082\|\|full | Low-quality |
| 17-109-final-viral-combined_k141_365380\|\|full | Not-determined |
| 17-109-final-viral-combined_k141_65468\|\|full | Not-determined |
| 17-109-final-viral-combined_k141_148897\|\|full | Not-determined |
| 17-109-final-viral-combined_k141_115143\|\|full | Not-determined |
| 17-109-final-viral-combined_k141_148908\|\|full | Low-quality |
| 17-109-final-viral-combined_k141_300215\|\|full | Low-quality |
| 17-109-final-viral-combined_k141_183173\|\|full | Low-quality |
| 17-109-final-viral-combined_k141_49841\|\|full | Low-quality |
| 17-109-final-viral-combined_k141_199962\|\|full | Not-determined |
| 17-109-final-viral-combined_k141_216850\|\|full | Not-determined |
| 17-109-final-viral-combined_k141_350420\|\|full | Low-quality |
| 17-109-final-viral-combined_k141_399835\|\|full | Low-quality |
| 17-109-final-viral-combined_k141_65512\|\|full | Low-quality |
| 17-109-final-viral-combined_k141_383579\|\|full | Low-quality |
| 17-109-final-viral-combined_k141_350430\|\|full | Not-determined |
| 17-109-final-viral-combined_k141_399843\|\|full | Low-quality |
| 17-109-final-viral-combined_k141_383583\|\|full | Low-quality |
| 17-109-final-viral-combined_k141_284320\|\|full | Not-determined |
| 17-109-final-viral-combined_k141_132031\|\|full | Low-quality |
| 17-109-final-viral-combined_k141_234094\|\|full | Low-quality |
| 17-109-final-viral-combined_k141_199989\|\|full | Not-determined |
| 17-109-final-viral-combined_k141_234097\|\|full | Not-determined |
| 17-109-final-viral-combined_k141_284325\|\|full | Not-determined |
| 17-109-final-viral-combined_k141_183212\|\|full | Low-quality |
| 17-109-final-viral-combined_k141_199996\|\|full | Not-determined |
| 17-109-final-viral-combined_k141_166359\|\|full | Not-determined |
| 17-109-final-viral-combined_k141_15178\|\|full | Low-quality |
| 17-109-final-viral-combined_k141_265696\|\|full | Not-determined |
| 17-109-final-viral-combined_k141_31975\|\|full | Not-determined |
| 17-109-final-viral-combined_k141_350458\|\|full | Not-determined |
| 17-109-final-viral-combined_k141_234116\|\|full | Low-quality |
| 17-109-final-viral-combined_k141_200013\|\|full | Not-determined |
| 17-109-final-viral-combined_k141_83129\|\|full | Not-determined |
| 17-109-final-viral-combined_k141_31986\|\|full | Not-determined |
| 17-109-final-viral-combined_k141_83150\|\|full | Not-determined |
| 17-109-final-viral-combined_k141_183236\|\|full | Low-quality |
| 17-109-final-viral-combined_k141_115233\|\|full | Not-determined |
| 17-109-final-viral-combined_k141_317966\|\|full | Not-determined |
| 17-109-final-viral-combined_k141_115236\|\|full | Not-determined |
| 17-109-final-viral-combined_k141_334019\|\|full | Low-quality |
| 17-109-final-viral-combined_k141_399919\|\|full | Low-quality |
| 17-109-final-viral-combined_k141_317976\|\|full | Low-quality |
| 17-109-final-viral-combined_k141_15225\|\|full | Low-quality |
| 17-109-final-viral-combined_k141_383614\|\|full | Not-determined |
| 17-109-final-viral-combined_k141_15233\|\|full | Not-determined |
| 17-109-final-viral-combined_k141_383624\|\|full | Not-determined |
| 17-109-final-viral-combined_k141_399944\|\|full | Not-determined |
| 17-109-final-viral-combined_k141_98966\|\|full | Not-determined |
| 17-109-final-viral-combined_k141_265779\|\|full | Not-determined |
| 17-109-final-viral-combined_k141_399952\|\|full | Low-quality |
| 17-109-final-viral-combined_k141_83188\|\|full | Not-determined |
| 17-109-final-viral-combined_k141_166417\|\|full | Not-determined |
| 17-109-final-viral-combined_k141_334057\|\|full | Low-quality |
| 17-109-final-viral-combined_k141_234200\|\|full | Not-determined |
| 17-109-final-viral-combined_k141_365442\|\|full | Low-quality |
| 17-109-final-viral-combined_k141_183326\|\|full | Low-quality |
| 17-109-final-viral-combined_k141_49969\|\|full | Low-quality |
| 17-109-final-viral-combined_k141_49973\|\|full | Low-quality |
| 17-109-final-viral-combined_k141_115302\|\|full | Low-quality |
| 17-109-final-viral-combined_k141_32034\|\|full | Low-quality |
| 17-109-final-viral-combined_k141_148930\|\|full | Low-quality |
| 17-109-final-viral-combined_k141_284468\|\|full | Not-determined |
| 17-109-final-viral-combined_k141_318034\|\|full | Low-quality |
| 17-109-final-viral-combined_k141_49989\|\|full | Low-quality |
| 17-109-final-viral-combined_k141_350537\|\|full | Low-quality |
| 17-109-final-viral-combined_k141_166464\|\|full | Not-determined |
| 17-109-final-viral-combined_k141_50002\|\|full | Low-quality |
| 17-109-final-viral-combined_k141_200118\|\|full | Low-quality |
| 17-109-final-viral-combined_k141_234239\|\|full | Not-determined |
| 17-109-final-viral-combined_k141_399981\|\|full | Not-determined |
| 17-109-final-viral-combined_k141_148954\|\|full | Low-quality |
| 17-109-final-viral-combined_k141_166477\|\|full | Not-determined |
| 17-109-final-viral-combined_k141_399989\|\|full | Low-quality |
| 17-109-final-viral-combined_k141_50012\|\|full | Low-quality |
| 17-109-final-viral-combined_k141_166486\|\|full | Low-quality |
| 17-109-final-viral-combined_k141_99057\|\|full | Low-quality |
| 17-109-final-viral-combined_k141_15318\|\|full | Not-determined |
| 17-109-final-viral-combined_k141_365480\|\|full | Low-quality |
| 17-109-final-viral-combined_k141_32106\|\|full | Low-quality |
| 17-109-final-viral-combined_k141_148981\|\|full | Low-quality |
| 17-109-final-viral-combined_k141_15357\|\|full | Not-determined |
| 17-109-final-viral-combined_k141_383671\|\|full | Low-quality |
| 17-109-final-viral-combined_k141_115384\|\|full | Low-quality |
| 17-109-final-viral-combined_k141_300361\|\|full | Not-determined |
| 17-109-final-viral-combined_k141_334152\|\|full | Not-determined |
| 17-109-final-viral-combined_k141_217002\|\|full | Low-quality |
| 17-109-final-viral-combined_k141_132177\|\|full | Not-determined |
| 17-109-final-viral-combined_k141_265901\|\|full | Not-determined |
| 17-109-final-viral-combined_k141_50028\|\|full | Low-quality |
| 17-109-final-viral-combined_k141_284524\|\|full | Low-quality |
| 17-109-final-viral-combined_k141_99126\|\|full | Low-quality |
| 17-109-final-viral-combined_k141_183461\|\|full | Not-determined |
| 17-109-final-viral-combined_k141_217031\|\|full | Low-quality |
| 17-109-final-viral-combined_k141_15430\|\|full | Not-determined |
| 17-109-final-viral-combined_k141_50052\|\|full | Low-quality |
| 17-109-final-viral-combined_k141_132238\|\|full | Low-quality |
| 17-109-final-viral-combined_k141_83366\|\|full | Not-determined |
| 17-109-final-viral-combined_k141_249787\|\|full | Not-determined |
| 17-109-final-viral-combined_k141_350620\|\|full | Not-determined |
| 17-109-final-viral-combined_k141_234325\|\|full | Not-determined |
| 17-109-final-viral-combined_k141_383756\|\|full | Low-quality |
| 17-109-final-viral-combined_k141_65755\|\|full | Not-determined |
| 17-109-final-viral-combined_k141_200189\|\|full | Low-quality |
| 17-109-final-viral-combined_k141_183494\|\|full | Not-determined |
| 17-109-final-viral-combined_k141_334165\|\|full | Not-determined |
| 17-109-final-viral-combined_k141_300415\|\|full | Low-quality |
| 17-109-final-viral-combined_k141_350630\|\|full | Low-quality |
| 17-109-final-viral-combined_k141_183519\|\|full | Low-quality |
| 17-109-final-viral-combined_k141_265966\|\|full | Not-determined |
| 17-109-final-viral-combined_k141_334186\|\|full | Not-determined |
| 17-109-final-viral-combined_k141_50100\|\|full | Not-determined |
| 17-109-final-viral-combined_k141_300429\|\|full | Low-quality |
| 17-109-final-viral-combined_k141_350639\|\|full | Low-quality |
| 17-109-final-viral-combined_k141_83419\|\|full | Not-determined |
| 17-109-final-viral-combined_k141_383784\|\|full | Low-quality |
| 17-109-final-viral-combined_k141_15480\|\|full | Not-determined |
| 17-109-final-viral-combined_k141_350656\|\|full | Low-quality |
| 17-109-final-viral-combined_k141_50133\|\|full | Low-quality |
| 17-109-final-viral-combined_k141_350666\|\|full | Low-quality |
| 17-109-final-viral-combined_k141_265983\|\|full | Not-determined |
| 17-109-final-viral-combined_k141_83456\|\|full | Low-quality |
| 17-109-final-viral-combined_k141_200245\|\|full | Not-determined |
| 17-109-final-viral-combined_k141_99239\|\|full | Not-determined |
| 17-109-final-viral-combined_k141_166656\|\|full | Not-determined |
| 17-109-final-viral-combined_k141_183584\|\|full | Not-determined |
| 17-109-final-viral-combined_k141_249861\|\|full | Low-quality |
| 17-109-final-viral-combined_k141_266011\|\|full | Not-determined |
| 17-109-final-viral-combined_k141_32216\|\|full | Low-quality |
| 17-109-final-viral-combined_k141_383822\|\|full | Not-determined |
| 17-109-final-viral-combined_k141_65807\|\|full | Low-quality |
| 17-109-final-viral-combined_k141_149037\|\|full | Not-determined |
| 17-109-final-viral-combined_k141_365515\|\|full | Low-quality |
| 17-109-final-viral-combined_k141_334282\|\|full | Low-quality |
| 17-109-final-viral-combined_k141_365527\|\|full | Low-quality |
| 17-109-final-viral-combined_k141_217152\|\|full | Not-determined |
| 17-109-final-viral-combined_k141_334290\|\|full | Not-determined |
| 17-109-final-viral-combined_k141_65828\|\|full | Not-determined |
| 17-109-final-viral-combined_k141_83483\|\|full | Low-quality |
| 17-109-final-viral-combined_k141_50197\|\|full | Not-determined |
| 17-109-final-viral-combined_k141_149051\|\|full | Not-determined |
| 17-109-final-viral-combined_k141_350735\|\|full | Not-determined |
| 17-109-final-viral-combined_k141_65836\|\|full | Low-quality |
| 17-109-final-viral-combined_k141_183652\|\|full | Not-determined |
| 17-109-final-viral-combined_k141_65845\|\|full | Low-quality |
| 17-109-final-viral-combined_k141_365531\|\|full | Low-quality |
| 17-109-final-viral-combined_k141_83510\|\|full | Low-quality |
| 18-109-final-viral-combined_k141_28138\|\|full | Medium-quality |
| 17-109-final-viral-combined_k141_65847\|\|full | Not-determined |
| 17-109-final-viral-combined_k141_83518\|\|full | Low-quality |
| 17-109-final-viral-combined_k141_365539\|\|full | Not-determined |
| 17-109-final-viral-combined_k141_132347\|\|full | Not-determined |
| 17-109-final-viral-combined_k141_300508\|\|full | Low-quality |
| 17-109-final-viral-combined_k141_166687\|\|full | Low-quality |
| 17-109-final-viral-combined_k141_132354\|\|full | Not-determined |
| 17-109-final-viral-combined_k141_99332\|\|full | Not-determined |
| 17-109-final-viral-combined_k141_166694\|\|full | Not-determined |
| 17-109-final-viral-combined_k141_83547\|\|full | Not-determined |
| 17-109-final-viral-combined_k141_115590\|\|full | Not-determined |
| 17-109-final-viral-combined_k141_365561\|\|full | Low-quality |
| 17-109-final-viral-combined_k141_15537\|\|full | Not-determined |
| 17-109-final-viral-combined_k141_99347\|\|full | Not-determined |
| 17-109-final-viral-combined_k141_115607\|\|full | Not-determined |
| 17-109-final-viral-combined_k141_15558\|\|full | Not-determined |
| 17-109-final-viral-combined_k141_149146\|\|full | Low-quality |
| 17-109-final-viral-combined_k141_200398\|\|full | Not-determined |
| 17-109-final-viral-combined_k141_83588\|\|full | Low-quality |
| 17-109-final-viral-combined_k141_350794\|\|full | Low-quality |
| 17-109-final-viral-combined_k141_32289\|\|full | Not-determined |
| 17-109-final-viral-combined_k141_166764\|\|full | Low-quality |
| 17-109-final-viral-combined_k141_83591\|\|full | Low-quality |
| 17-109-final-viral-combined_k141_166769\|\|full | Low-quality |
| 17-109-final-viral-combined_k141_266100\|\|full | Not-determined |
| 17-109-final-viral-combined_k141_217232\|\|full | Not-determined |
| 17-109-final-viral-combined_k141_115634\|\|full | Low-quality |
| 17-109-final-viral-combined_k141_166770\|\|full | Not-determined |
| 17-109-final-viral-combined_k141_115637\|\|full | Low-quality |
| 17-109-final-viral-combined_k141_217235\|\|full | Low-quality |
| 17-109-final-viral-combined_k141_350807\|\|full | Not-determined |
| 17-109-final-viral-combined_k141_132438\|\|full | Low-quality |
| 17-109-final-viral-combined_k141_300588\|\|full | Low-quality |
| 17-109-final-viral-combined_k141_83621\|\|full | Low-quality |
| 17-109-final-viral-combined_k141_99409\|\|full | Not-determined |
| 17-109-final-viral-combined_k141_83632\|\|full | Low-quality |
| 17-109-final-viral-combined_k141_115650\|\|full | Low-quality |
| 17-109-final-viral-combined_k141_334418\|\|full | Low-quality |
| 17-109-final-viral-combined_k141_99421\|\|full | Low-quality |
| 17-109-final-viral-combined_k141_32343\|\|full | Not-determined |
| 17-109-final-viral-combined_k141_99427\|\|full | Low-quality |
| 17-109-final-viral-combined_k141_266146\|\|full | Low-quality |
| 17-109-final-viral-combined_k141_249951\|\|full | Not-determined |
| 17-109-final-viral-combined_k141_200483\|\|full | Not-determined |
| 17-109-final-viral-combined_k141_32373\|\|full | Not-determined |
| 17-109-final-viral-combined_k141_149224\|\|full | Low-quality |
| 17-109-final-viral-combined_k141_149229\|\|full | Low-quality |
| 17-109-final-viral-combined_k141_365650\|\|full | Not-determined |
| 17-109-final-viral-combined_k141_15613\|\|full | Low-quality |
| 17-109-final-viral-combined_k141_32397\|\|full | Low-quality |
| 17-109-final-viral-combined_k141_166876\|\|full | Low-quality |
| 17-109-final-viral-combined_k141_15628\|\|full | Low-quality |
| 17-109-final-viral-combined_k141_250012\|\|full | Low-quality |
| 17-109-final-viral-combined_k141_183763\|\|full | Not-determined |
| 17-109-final-viral-combined_k141_266182\|\|full | Low-quality |
| 17-109-final-viral-combined_k141_300652\|\|full | Low-quality |
| 17-109-final-viral-combined_k141_132514\|\|full | Not-determined |
| 17-109-final-viral-combined_k141_300658\|\|full | Not-determined |
| 17-109-final-viral-combined_k141_149283\|\|full | Not-determined |
| 17-109-final-viral-combined_k141_365672\|\|full | Low-quality |
| 17-109-final-viral-combined_k141_115692\|\|full | Not-determined |
| 17-109-final-viral-combined_k141_66048\|\|full | Low-quality |
| 17-109-final-viral-combined_k141_365673\|\|full | Not-determined |
| 17-109-final-viral-combined_k141_200505\|\|full | Low-quality |
| 17-109-final-viral-combined_k141_217319\|\|full | Not-determined |
| 17-109-final-viral-combined_k141_66062\|\|full | Low-quality |
| 17-109-final-viral-combined_k141_300679\|\|full | Not-determined |
| 17-109-final-viral-combined_k141_384040\|\|full | Low-quality |
| 17-109-final-viral-combined_k141_166905\|\|full | Low-quality |
| 17-109-final-viral-combined_k141_183803\|\|full | Not-determined |
| 17-109-final-viral-combined_k141_365687\|\|full | Low-quality |
| 17-109-final-viral-combined_k141_250023\|\|full | Not-determined |
| 17-109-final-viral-combined_k141_32442\|\|full | Low-quality |
| 17-109-final-viral-combined_k141_15683\|\|full | Not-determined |
| 17-109-final-viral-combined_k141_183815\|\|full | Low-quality |
| 17-109-final-viral-combined_k141_66088\|\|full | Not-determined |
| 17-109-final-viral-combined_k141_266257\|\|full | Low-quality |
| 17-109-final-viral-combined_k141_250042\|\|full | Low-quality |
| 17-109-final-viral-combined_k141_334471\|\|full | Low-quality |
| 17-109-final-viral-combined_k141_99547\|\|full | Not-determined |
| 17-109-final-viral-combined_k141_350958\|\|full | Low-quality |
| 17-109-final-viral-combined_k141_99549\|\|full | Not-determined |
| 17-109-final-viral-combined_k141_250065\|\|full | Low-quality |
| 17-109-final-viral-combined_k141_365717\|\|full | Not-determined |
| 17-109-final-viral-combined_k141_132581\|\|full | Not-determined |
| 17-109-final-viral-combined_k141_32492\|\|full | Low-quality |
| 17-109-final-viral-combined_k141_115777\|\|full | Not-determined |
| 17-109-final-viral-combined_k141_200563\|\|full | Not-determined |
| 17-109-final-viral-combined_k141_300710\|\|full | Not-determined |
| 17-109-final-viral-combined_k141_115794\|\|full | Low-quality |
| 17-109-final-viral-combined_k141_334532\|\|full | Not-determined |
| 17-109-final-viral-combined_k141_300734\|\|full | Low-quality |
| 17-109-final-viral-combined_k141_132598\|\|full | Not-determined |
| 17-109-final-viral-combined_k141_132599\|\|full | Not-determined |
| 17-109-final-viral-combined_k141_166991\|\|full | Low-quality |
| 17-109-final-viral-combined_k141_66124\|\|full | Low-quality |
| 17-109-final-viral-combined_k141_365771\|\|full | Not-determined |
| 17-109-final-viral-combined_k141_115817\|\|full | Low-quality |
| 17-109-final-viral-combined_k141_107942\|\|full | Medium-quality |
| 17-109-final-viral-combined_k141_250105\|\|full | Not-determined |
| 17-109-final-viral-combined_k141_99590\|\|full | Low-quality |
| 17-109-final-viral-combined_k141_300778\|\|full | Low-quality |
| 17-109-final-viral-combined_k141_351016\|\|full | Not-determined |
| 17-109-final-viral-combined_k141_132648\|\|full | Not-determined |
| 17-109-final-viral-combined_k141_167034\|\|full | Not-determined |
| 17-109-final-viral-combined_k141_15722\|\|full | Not-determined |
| 17-109-final-viral-combined_k141_334592\|\|full | Low-quality |
| 17-109-final-viral-combined_k141_132660\|\|full | Low-quality |
| 17-109-final-viral-combined_k141_266372\|\|full | Low-quality |
| 17-109-final-viral-combined_k141_32594\|\|full | Low-quality |
| 17-109-final-viral-combined_k141_99626\|\|full | Not-determined |
| 17-109-final-viral-combined_k141_250147\|\|full | Low-quality |
| 17-109-final-viral-combined_k141_99631\|\|full | Low-quality |
| 17-109-final-viral-combined_k141_200616\|\|full | Low-quality |
| 17-109-final-viral-combined_k141_99639\|\|full | Low-quality |
| 17-109-final-viral-combined_k141_217428\|\|full | Low-quality |
| 17-109-final-viral-combined_k141_15737\|\|full | Low-quality |
| 17-109-final-viral-combined_k141_300828\|\|full | Low-quality |
| 17-109-final-viral-combined_k141_167073\|\|full | Low-quality |
| 17-109-final-viral-combined_k141_217436\|\|full | Low-quality |
| 17-109-final-viral-combined_k141_365849\|\|full | Low-quality |
| 17-109-final-viral-combined_k141_99666\|\|full | Not-determined |
| 17-109-final-viral-combined_k141_351094\|\|full | Low-quality |
| 17-109-final-viral-combined_k141_334642\|\|full | Low-quality |
| 17-109-final-viral-combined_k141_132699\|\|full | Low-quality |
| 17-109-final-viral-combined_k141_99678\|\|full | Low-quality |
| 17-109-final-viral-combined_k141_149427\|\|full | Not-determined |
| 17-109-final-viral-combined_k141_250192\|\|full | Not-determined |
| 17-109-final-viral-combined_k141_15777\|\|full | Not-determined |
| 17-109-final-viral-combined_k141_167109\|\|full | Not-determined |
| 17-109-final-viral-combined_k141_66290\|\|full | Not-determined |
| 17-109-final-viral-combined_k141_32644\|\|full | Not-determined |
| 17-109-final-viral-combined_k141_351122\|\|full | Not-determined |
| 17-109-final-viral-combined_k141_99697\|\|full | Not-determined |
| 17-109-final-viral-combined_k141_183957\|\|full | Not-determined |
| 17-109-final-viral-combined_k141_115942\|\|full | Low-quality |
| 17-109-final-viral-combined_k141_400125\|\|full | Not-determined |
| 17-109-final-viral-combined_k141_167146\|\|full | Not-determined |
| 17-109-final-viral-combined_k141_15808\|\|full | Low-quality |
| 17-109-final-viral-combined_k141_217506\|\|full | Low-quality |
| 17-109-final-viral-combined_k141_149491\|\|full | Low-quality |
| 17-109-final-viral-combined_k141_334664\|\|full | Low-quality |
| 17-109-final-viral-combined_k141_217509\|\|full | Not-determined |
| 17-109-final-viral-combined_k141_15837\|\|full | Not-determined |
| 17-109-final-viral-combined_k141_132751\|\|full | Not-determined |
| 17-109-final-viral-combined_k141_149511\|\|full | Not-determined |
| 17-106-final-viral-combined_k141_105462\|\|full | Medium-quality |
| 17-109-final-viral-combined_k141_351188\|\|full | Low-quality |
| 17-109-final-viral-combined_k141_200705\|\|full | Low-quality |
| 17-109-final-viral-combined_k141_132763\|\|full | Low-quality |
| 17-109-final-viral-combined_k141_183987\|\|full | Not-determined |
| 17-109-final-viral-combined_k141_400179\|\|full | Not-determined |
| 17-109-final-viral-combined_k141_365963\|\|full | Low-quality |
| 17-109-final-viral-combined_k141_15865\|\|full | Not-determined |
| 17-109-final-viral-combined_k141_32707\|\|full | Not-determined |
| 17-109-final-viral-combined_k141_384243\|\|full | Low-quality |
| 17-109-final-viral-combined_k141_15889\|\|full | Not-determined |
| 17-109-final-viral-combined_k141_384261\|\|full | Low-quality |
| 17-109-final-viral-combined_k141_116027\|\|full | Not-determined |
| 17-109-final-viral-combined_k141_66402\|\|full | Not-determined |
| 17-109-final-viral-combined_k141_66403\|\|full | Not-determined |
| 17-109-final-viral-combined_k141_351217\|\|full | Low-quality |
| 17-109-final-viral-combined_k141_334700\|\|full | Not-determined |
| 17-109-final-viral-combined_k141_167233\|\|full | Low-quality |
| 17-109-final-viral-combined_k141_116050\|\|full | Not-determined |
| 17-109-final-viral-combined_k141_32762\|\|full | Not-determined |
| 17-109-final-viral-combined_k141_116057\|\|full | Low-quality |
| 17-109-final-viral-combined_k141_149602\|\|full | Low-quality |
| 17-109-final-viral-combined_k141_32764\|\|full | Not-determined |
| 17-109-final-viral-combined_k141_266511\|\|full | Not-determined |
| 17-109-final-viral-combined_k141_384315\|\|full | Not-determined |
| 17-109-final-viral-combined_k141_132824\|\|full | Not-determined |
| 17-109-final-viral-combined_k141_366045\|\|full | Not-determined |
| 17-109-final-viral-combined_k141_334721\|\|full | Low-quality |
| 17-109-final-viral-combined_k141_384344\|\|full | Low-quality |
| 17-109-final-viral-combined_k141_366072\|\|full | Not-determined |
| 17-109-final-viral-combined_k141_99807\|\|full | Low-quality |
| 17-109-final-viral-combined_k141_149666\|\|full | Low-quality |
| 17-109-final-viral-combined_k141_334734\|\|full | Not-determined |
| 17-109-final-viral-combined_k141_149677\|\|full | Not-determined |
| 17-109-final-viral-combined_k141_16054\|\|full | Low-quality |
| 17-109-final-viral-combined_k141_16056\|\|full | Low-quality |
| 17-109-final-viral-combined_k141_200826\|\|full | Not-determined |
| 17-109-final-viral-combined_k141_366106\|\|full | Not-determined |
| 17-109-final-viral-combined_k141_116163\|\|full | Low-quality |
| 17-109-final-viral-combined_k141_366114\|\|full | Low-quality |
| 17-109-final-viral-combined_k141_149701\|\|full | Low-quality |
| 17-109-final-viral-combined_k141_334757\|\|full | Not-determined |
| 17-109-final-viral-combined_k141_99868\|\|full | Not-determined |
| 17-109-final-viral-combined_k141_384391\|\|full | Not-determined |
| 17-109-final-viral-combined_k141_149714\|\|full | Not-determined |
| 17-109-final-viral-combined_k141_116202\|\|full | Low-quality |
| 17-109-final-viral-combined_k141_167298\|\|full | Low-quality |
| 17-109-final-viral-combined_k141_99880\|\|full | Low-quality |
| 17-109-final-viral-combined_k141_366161\|\|full | Low-quality |
| 17-109-final-viral-combined_k141_66556\|\|full | Low-quality |
| 17-109-final-viral-combined_k141_116208\|\|full | Low-quality |
| 17-109-final-viral-combined_k141_351294\|\|full | Low-quality |
| 17-109-final-viral-combined_k141_32826\|\|full | Low-quality |
| 17-109-final-viral-combined_k141_200841\|\|full | Low-quality |
| 17-109-final-viral-combined_k141_167317\|\|full | Not-determined |
| 17-109-final-viral-combined_k141_66568\|\|full | Low-quality |
| 17-109-final-viral-combined_k141_366171\|\|full | Not-determined |
| 17-109_k141_184276 | High-quality |
| 17-109-final-viral-combined_k141_366173\|\|full | Not-determined |
| 17-109-final-viral-combined_k141_66581\|\|full | Not-determined |
| 17-109-final-viral-combined_k141_351312\|\|full | Not-determined |
| 17-109-final-viral-combined_k141_167325\|\|full | Low-quality |
| 17-109-final-viral-combined_k141_32847\|\|full | Low-quality |
| 17-109-final-viral-combined_k141_32850\|\|full | Low-quality |
| 17-109-final-viral-combined_k141_149756\|\|full | Not-determined |
| 17-109-final-viral-combined_k141_301096\|\|full | Low-quality |
| 17-109-final-viral-combined_k141_99925\|\|full | Not-determined |
| 17-109-final-viral-combined_k141_132952\|\|full | Low-quality |
| 17-109-final-viral-combined_k141_66622\|\|full | Not-determined |
| 17-109-final-viral-combined_k141_116246\|\|full | Not-determined |
| 17-109-final-viral-combined_k141_384460\|\|full | Not-determined |
| 17-109-final-viral-combined_k141_351322\|\|full | Low-quality |
| 17-109-final-viral-combined_k141_250493\|\|full | Low-quality |
| 17-109-final-viral-combined_k141_132974\|\|full | Not-determined |
| 17-109-final-viral-combined_k141_301117\|\|full | Low-quality |
| 17-109-final-viral-combined_k141_351334\|\|full | Not-determined |
| 17-109-final-viral-combined_k141_116268\|\|full | Low-quality |
| 17-109-final-viral-combined_k141_366225\|\|full | Not-determined |
| 17-109-final-viral-combined_k141_250505\|\|full | Low-quality |
| 17-109-final-viral-combined_k141_384492\|\|full | Not-determined |
| 17-109-final-viral-combined_k141_301129\|\|full | Low-quality |
| 17-109-final-viral-combined_k141_384509\|\|full | Low-quality |
| 17-109-final-viral-combined_k141_266567\|\|full | Not-determined |
| 17-109-final-viral-combined_k141_384520\|\|full | Not-determined |
| 17-109-final-viral-combined_k141_266585\|\|full | Low-quality |
| 17-109-final-viral-combined_k141_200867\|\|full | Low-quality |
| 17-109-final-viral-combined_k141_266599\|\|full | Not-determined |
| 17-109-final-viral-combined_k141_133044\|\|full | Low-quality |
| 17-109-final-viral-combined_k141_301165\|\|full | Low-quality |
| 17-109-final-viral-combined_k141_384563\|\|full | Low-quality |
| 17-109-final-viral-combined_k141_100021\|\|full | Low-quality |
| 17-109-final-viral-combined_k141_366298\|\|full | Low-quality |
| 17-109-final-viral-combined_k141_66708\|\|full | Low-quality |
| 17-109-final-viral-combined_k141_116336\|\|full | Low-quality |
| 17-109-final-viral-combined_k141_133065\|\|full | Not-determined |
| 17-109-final-viral-combined_k141_32939\|\|full | Low-quality |
| 17-108-final-viral-combined_k141_204522\|\|full | Medium-quality |
| 17-109-final-viral-combined_k141_351400\|\|full | Low-quality |
| 17-109-final-viral-combined_k141_66749\|\|full | Not-determined |
| 17-109-final-viral-combined_k141_149832\|\|full | Not-determined |
| 17-109-final-viral-combined_k141_351401\|\|full | Low-quality |
| 17-109-final-viral-combined_k141_184079\|\|full | Not-determined |
| 17-109-final-viral-combined_k141_100046\|\|full | Not-determined |
| 17-109-final-viral-combined_k141_149853\|\|full | Low-quality |
| 17-109-final-viral-combined_k141_66758\|\|full | Low-quality |
| 17-109-final-viral-combined_k141_184114\|\|full | Low-quality |
| 17-109-final-viral-combined_k141_366345\|\|full | Low-quality |
| 17-109-final-viral-combined_k141_351414\|\|full | Not-determined |
| 17-109-final-viral-combined_k141_184116\|\|full | Not-determined |
| 17-109-final-viral-combined_k141_16344\|\|full | Not-determined |
| 17-109-final-viral-combined_k141_32992\|\|full | Low-quality |
| 17-109-final-viral-combined_k141_266654\|\|full | Low-quality |
| 17-109-final-viral-combined_k141_250602\|\|full | Not-determined |
| 17-109-final-viral-combined_k141_184127\|\|full | Not-determined |
| 17-109-final-viral-combined_k141_16360\|\|full | Low-quality |
| 17-109-final-viral-combined_k141_149900\|\|full | Low-quality |
| 17-109-final-viral-combined_k141_266672\|\|full | Not-determined |
| 17-109-final-viral-combined_k141_66812\|\|full | Not-determined |
| 17-109-final-viral-combined_k141_250632\|\|full | Low-quality |
| 17-109-final-viral-combined_k141_133180\|\|full | Not-determined |
| 17-109-final-viral-combined_k141_133187\|\|full | Not-determined |
| 17-109-final-viral-combined_k141_266681\|\|full | Not-determined |
| 17-109-final-viral-combined_k141_149938\|\|full | Low-quality |
| 17-109-final-viral-combined_k141_133215\|\|full | Not-determined |
| 17-109-final-viral-combined_k141_400553\|\|full | Not-determined |
| 17-109-final-viral-combined_k141_100130\|\|full | Not-determined |
| 17-109-final-viral-combined_k141_250647\|\|full | Not-determined |
| 17-109-final-viral-combined_k141_149966\|\|full | Not-determined |
| 17-109-final-viral-combined_k141_149969\|\|full | Low-quality |
| 17-109-final-viral-combined_k141_250651\|\|full | Low-quality |
| 17-109-final-viral-combined_k141_116424\|\|full | Low-quality |
| 17-109-final-viral-combined_k141_366466\|\|full | Not-determined |
| 17-109-final-viral-combined_k141_116436\|\|full | Low-quality |
| 17-109-final-viral-combined_k141_366473\|\|full | Low-quality |
| 17-109-final-viral-combined_k141_366486\|\|full | Low-quality |
| 17-109-final-viral-combined_k141_100195\|\|full | Not-determined |
| 17-109-final-viral-combined_k141_100201\|\|full | Not-determined |
| 17-109-final-viral-combined_k141_66901\|\|full | Low-quality |
| 17-109-final-viral-combined_k141_301284\|\|full | Not-determined |
| 17-109-final-viral-combined_k141_400590\|\|full | Not-determined |
| 17-109-final-viral-combined_k141_33129\|\|full | Not-determined |
| 17-109-final-viral-combined_k141_301290\|\|full | Not-determined |
| 17-109-final-viral-combined_k141_400593\|\|full | Low-quality |
| 17-109-final-viral-combined_k141_384594\|\|full | Low-quality |
| 17-109-final-viral-combined_k141_116465\|\|full | Not-determined |
| 17-109-final-viral-combined_k141_100231\|\|full | Not-determined |
| 17-109-final-viral-combined_k141_16458\|\|full | Low-quality |
| 17-109-final-viral-combined_k141_16480\|\|full | Not-determined |
| 17-109-final-viral-combined_k141_400622\|\|full | Not-determined |
| 17-109-final-viral-combined_k141_133279\|\|full | Low-quality |
| 17-109-final-viral-combined_k141_33193\|\|full | Not-determined |
| 17-109-final-viral-combined_k141_250658\|\|full | Not-determined |
| 17-109-final-viral-combined_k141_250671\|\|full | Not-determined |
| 17-109-final-viral-combined_k141_400624\|\|full | Not-determined |
| 17-109-final-viral-combined_k141_400640\|\|full | Not-determined |
| 17-109-final-viral-combined_k141_16557\|\|full | Low-quality |
| 17-109-final-viral-combined_k141_133322\|\|full | Low-quality |
| 17-109-final-viral-combined_k141_116582\|\|full | Low-quality |
| 17-109-final-viral-combined_k141_150134\|\|full | Not-determined |
| 17-109-final-viral-combined_k141_366609\|\|full | Low-quality |
| 17-109-final-viral-combined_k141_384646\|\|full | Not-determined |
| 17-109-final-viral-combined_k141_250741\|\|full | Not-determined |
| 17-109-final-viral-combined_k141_100361\|\|full | Low-quality |
| 17-109-final-viral-combined_k141_133375\|\|full | Not-determined |
| 17-109-final-viral-combined_k141_250760\|\|full | Low-quality |
| 17-109-final-viral-combined_k141_250761\|\|full | Low-quality |
| 17-109-final-viral-combined_k141_400693\|\|full | Not-determined |
| 17-109-final-viral-combined_k141_366666\|\|full | Low-quality |
| 17-109-final-viral-combined_k141_400707\|\|full | Low-quality |
| 17-109-final-viral-combined_k141_384694\|\|full | Not-determined |
| 17-109-final-viral-combined_k141_116628\|\|full | Low-quality |
| 17-109-final-viral-combined_k141_266953\|\|full | Low-quality |
| 17-109-final-viral-combined_k141_150246\|\|full | Low-quality |
| 17-109-final-viral-combined_k141_150255\|\|full | Not-determined |
| 17-109-final-viral-combined_k141_116649\|\|full | Low-quality |
| 17-109-final-viral-combined_k141_16623\|\|full | Not-determined |
| 17-109-final-viral-combined_k141_116662\|\|full | Low-quality |
| 17-109-final-viral-combined_k141_33312\|\|full | Low-quality |
| 17-109-final-viral-combined_k141_33315\|\|full | Not-determined |
| 17-109-final-viral-combined_k141_266992\|\|full | Low-quality |
| 17-109-final-viral-combined_k141_133461\|\|full | Not-determined |
| 17-109-final-viral-combined_k141_16670\|\|full | Low-quality |
| 17-109-final-viral-combined_k141_366743\|\|full | Not-determined |
| 17-109-final-viral-combined_k141_16714\|\|full | Low-quality |
| 17-109-final-viral-combined_k141_250828\|\|full | Not-determined |
| 17-109-final-viral-combined_k141_250830\|\|full | Not-determined |
| 17-109-final-viral-combined_k141_400775\|\|full | Not-determined |
| 17-109-final-viral-combined_k141_384780\|\|full | Not-determined |
| 17-109-final-viral-combined_k141_400777\|\|full | Low-quality |
| 17-109-final-viral-combined_k141_400782\|\|full | Low-quality |
| 17-109-final-viral-combined_k141_267061\|\|full | Not-determined |
| 17-109-final-viral-combined_k141_16729\|\|full | Not-determined |
| 17-109-final-viral-combined_k141_384796\|\|full | Not-determined |
| 17-109-final-viral-combined_k141_150338\|\|full | Low-quality |
| 17-109-final-viral-combined_k141_366762\|\|full | Not-determined |
| 17-109-final-viral-combined_k141_250877\|\|full | Not-determined |
| 17-109-final-viral-combined_k141_150361\|\|full | Not-determined |
| 17-109-final-viral-combined_k141_150363\|\|full | Low-quality |
| 17-109-final-viral-combined_k141_116775\|\|full | Low-quality |
| 17-109-final-viral-combined_k141_150392\|\|full | Not-determined |
| 17-109-final-viral-combined_k141_400839\|\|full | Low-quality |
| 17-109-final-viral-combined_k141_366812\|\|full | Not-determined |
| 17-109-final-viral-combined_k141_250924\|\|full | Not-determined |
| 17-109-final-viral-combined_k141_400863\|\|full | Not-determined |
| 17-109-final-viral-combined_k141_133547\|\|full | Low-quality |
| 17-109-final-viral-combined_k141_116827\|\|full | Not-determined |
| 17-109-final-viral-combined_k141_150450\|\|full | Low-quality |
| 17-109-final-viral-combined_k141_366846\|\|full | Low-quality |
| 17-109-final-viral-combined_k141_400887\|\|full | Low-quality |
| 17-109-final-viral-combined_k141_133578\|\|full | Not-determined |
| 17-109-final-viral-combined_k141_267237\|\|full | Not-determined |
| 17-109-final-viral-combined_k141_116865\|\|full | Not-determined |
| 17-109-final-viral-combined_k141_150460\|\|full | Not-determined |
| 17-109-final-viral-combined_k141_251007\|\|full | Not-determined |
| 17-109-final-viral-combined_k141_267251\|\|full | Not-determined |
| 17-109-final-viral-combined_k141_267259\|\|full | Low-quality |
| 17-109-final-viral-combined_k141_150481\|\|full | Not-determined |
| 17-109-final-viral-combined_k141_366963\|\|full | Not-determined |
| 17-109-final-viral-combined_k141_33400\|\|full | Not-determined |
| 17-109-final-viral-combined_k141_133597\|\|full | Low-quality |
| 17-109-final-viral-combined_k141_251062\|\|full | Low-quality |
| 17-109-final-viral-combined_k141_267302\|\|full | Low-quality |
| 17-109-final-viral-combined_k141_150507\|\|full | Not-determined |
| 17-109-final-viral-combined_k141_367003\|\|full | Low-quality |
| 17-109-final-viral-combined_k141_367020\|\|full | Not-determined |
| 17-109-final-viral-combined_k141_33458\|\|full | Low-quality |
| 17-109-final-viral-combined_k141_116925\|\|full | Not-determined |
| 17-109-final-viral-combined_k141_150558\|\|full | Not-determined |
| 17-109-final-viral-combined_k141_400981\|\|full | Not-determined |
| 17-109-final-viral-combined_k141_400983\|\|full | Low-quality |
| 17-109-final-viral-combined_k141_400988\|\|full | Not-determined |
| 17-109-final-viral-combined_k141_267339\|\|full | Not-determined |
| 17-109-final-viral-combined_k141_133643\|\|full | Not-determined |
| 17-109-final-viral-combined_k141_267346\|\|full | Not-determined |
| 17-109-final-viral-combined_k141_267354\|\|full | Low-quality |
| 17-109-final-viral-combined_k141_117007\|\|full | Not-determined |
| 17-109-final-viral-combined_k141_150632\|\|full | Not-determined |
| 17-109-final-viral-combined_k141_150642\|\|full | Not-determined |
| 17-109-final-viral-combined_k141_267377\|\|full | Not-determined |
| 17-109-final-viral-combined_k141_117054\|\|full | Low-quality |
| 17-109-final-viral-combined_k141_117056\|\|full | Not-determined |
| 17-109-final-viral-combined_k141_117063\|\|full | Low-quality |
| 17-109-final-viral-combined_k141_367168\|\|full | Low-quality |
| 17-109-final-viral-combined_k141_267385\|\|full | Low-quality |
| 17-109-final-viral-combined_k141_401094\|\|full | Low-quality |
| 17-109-final-viral-combined_k141_367208\|\|full | Low-quality |
| 17-109-final-viral-combined_k141_117113\|\|full | Not-determined |
| 17-109-final-viral-combined_k141_267423\|\|full | Not-determined |
| 17-109-final-viral-combined_k141_401153\|\|full | Not-determined |
| 17-109-final-viral-combined_k141_401160\|\|full | Low-quality |
| 17-109-final-viral-combined_k141_133759\|\|full | Not-determined |
| 17-109-final-viral-combined_k141_133764\|\|full | Low-quality |
| 17-109-final-viral-combined_k141_367305\|\|full | Low-quality |
| 17-109-final-viral-combined_k141_133789\|\|full | Not-determined |
| 17-109-final-viral-combined_k141_401196\|\|full | Not-determined |
| 17-109-final-viral-combined_k141_401201\|\|full | Low-quality |
| 17-109-final-viral-combined_k141_401222\|\|full | Not-determined |
| 17-109-final-viral-combined_k141_401223\|\|full | Low-quality |
| 17-109-final-viral-combined_k141_367348\|\|full | Low-quality |
| 17-109-final-viral-combined_k141_367390\|\|full | Low-quality |
| 17-109-final-viral-combined_k141_367410\|\|full | Low-quality |
| 17-109-final-viral-combined_k141_401278\|\|full | Not-determined |

| | |
|---|---|
| 17-109-final-viral-combined_k141_267531\|\|full | Not-determined |
| 17-109-final-viral-combined_k141_133897\|\|full | Low-quality |
| 17-109-final-viral-combined_k141_267557\|\|full | Not-determined |
| 17-109-final-viral-combined_k141_401325\|\|full | Low-quality |
| 17-109-final-viral-combined_k141_401326\|\|full | Low-quality |
| 17-109-final-viral-combined_k141_401346\|\|full | Not-determined |
| 17-109-final-viral-combined_k141_401348\|\|full | Not-determined |
| 17-109-final-viral-combined_k141_367514\|\|full | Low-quality |
| 17-109-final-viral-combined_k141_367585\|\|full | Not-determined |
| 17-109-final-viral-combined_k141_367590\|\|full | Not-determined |
| 17-109-final-viral-combined_k141_267688\|\|full | Low-quality |
| 17-109-final-viral-combined_k141_367632\|\|full | Not-determined |
| 17-109-final-viral-combined_k141_367641\|\|full | Not-determined |
| 17-109-final-viral-combined_k141_267709\|\|full | Not-determined |
| 17-109-final-viral-combined_k141_367716\|\|full | Low-quality |
| 17-109-final-viral-combined_k141_367781\|\|full | Not-determined |
| 17-109-final-viral-combined_k141_367794\|\|full | Low-quality |
| 17-109-final-viral-combined_k141_367827\|\|full | Not-determined |
| 17-109-final-viral-combined_k141_267809\|\|full | Not-determined |
| 17-109-final-viral-combined_k141_368063\|\|full | Not-determined |
| 17-109-final-viral-combined_k141_368152\|\|full | Low-quality |
| 17-109-final-viral-combined_k141_368174\|\|full | Low-quality |
| 18-101-final-viral-combined_k141_94988\|\|1_partial | Medium-quality |
| 17-109-final-viral-combined_k141_368216\|\|full | Not-determined |
| 17-109-final-viral-combined_k141_368245\|\|full | Low-quality |
| 17-109-final-viral-combined_k141_16830\|\|0_partial | Low-quality |
| 17-109-final-viral-combined_k141_50426\|\|0_partial | Low-quality |
| 17-109-final-viral-combined_k141_234454\|\|0_partial | Not-determined |
| 17-109-final-viral-combined_k141_334917\|\|0_partial | Not-determined |
| 17-109-final-viral-combined_k141_335099\|\|0_partial | Low-quality |
| 17-109-final-viral-combined_k141_184276\|\|full | High-quality |
| 17-109-final-viral-combined_k141_34243\|\|0_partial | Low-quality |
| 17-109-final-viral-combined_k141_385599\|\|0_partial | Low-quality |
| 17-109-final-viral-combined_k141_218262\|\|0_partial | Low-quality |
| 18-101-final-viral-combined_k141_246923\|\|full | Medium-quality |
| 17-109-final-viral-combined_k141_84788\|\|0_partial | Not-determined |
| 17-109-final-viral-combined_k141_34886\|\|0_partial | Low-quality |
| 17-109-final-viral-combined_k141_35106\|\|0_partial | Low-quality |
| 17-109-final-viral-combined_k141_269316\|\|0_partial | Not-determined |
| 17-109-final-viral-combined_k141_235906\|\|0_partial | Not-determined |
| 17-109-final-viral-combined_k141_185931\|\|0_partial | Not-determined |
| 17-109-final-viral-combined_k141_303580\|\|0_partial | Low-quality |
| 17-109-final-viral-combined_k141_236407\|\|0_partial | Low-quality |
| 17-109-final-viral-combined_k141_36018\|\|0_partial | Not-determined |
| 17-109-final-viral-combined_k141_153141\|\|0_partial | Low-quality |
| 17-109-final-viral-combined_k141_236737\|\|0_partial | Low-quality |
| 17-109-final-viral-combined_k141_236986\|\|0_partial | Not-determined |
| 17-109-final-viral-combined_k141_220029\|\|0_partial | Not-determined |
| 18-102-final-viral-combined_k141_135014\|\|full | Medium-quality |
| 17-109-final-viral-combined_k141_120125\|\|0_partial | Not-determined |
| 17-109-final-viral-combined_k141_103489\|\|0_partial | Not-determined |
| 17-109-final-viral-combined_k141_304935\|\|0_partial | Low-quality |
| 17-109-final-viral-combined_k141_87166\|\|0_partial | Not-determined |
| 17-109-final-viral-combined_k141_338158\|\|0_partial | Low-quality |
| 17-109-final-viral-combined_k141_238433\|\|0_partial | Not-determined |
| 17-109-final-viral-combined_k141_138432\|\|0_partial | Not-determined |
| 17-109-final-viral-combined_k141_221903\|\|0_partial | Not-determined |
| 17-109-final-viral-combined_k141_339052\|\|0_partial | Not-determined |
| 17-109-final-viral-combined_k141_171971\|\|0_partial | Low-quality |
| 17-109-final-viral-combined_k141_155528\|\|0_partial | Low-quality |
| 17-109-final-viral-combined_k141_4752\|\|0_partial | Not-determined |
| 17-109-final-viral-combined_k141_373648\|\|0_partial | Not-determined |
| 17-109-final-viral-combined_k141_54992\|\|0_partial | Low-quality |
| 17-109-final-viral-combined_k141_323658\|\|0_partial | Not-determined |
| 17-109-final-viral-combined_k141_223051\|\|0_partial | Not-determined |
| 17-109-final-viral-combined_k141_273721\|\|0_partial | Low-quality |
| KM_k141_235536 | Medium-quality |
| 17-109-final-viral-combined_k141_357120\|\|0_partial | Low-quality |
| 17-109-final-viral-combined_k141_123195\|\|0_partial | Not-determined |
| 17-109-final-viral-combined_k141_324947\|\|0_partial | Low-quality |
| 17-109-final-viral-combined_k141_274850\|\|0_partial | Low-quality |
| 17-109-final-viral-combined_k141_74622\|\|0_partial | Low-quality |
| 17-109-final-viral-combined_k141_174292\|\|0_partial | Low-quality |
| 17-109-final-viral-combined_k141_141157\|\|0_partial | Not-determined |
| 17-109-final-viral-combined_k141_123991\|\|0_partial | Low-quality |
| 17-109-final-viral-combined_k141_124636\|\|0_partial | Not-determined |
| 17-109-final-viral-combined_k141_310910\|\|0_partial | Low-quality |
| 17-109-final-viral-combined_k141_259497\|\|0_partial | Low-quality |
| 17-109-final-viral-combined_k141_294260\|\|0_partial | Low-quality |
| 17-109-final-viral-combined_k141_277929\|\|0_partial | Low-quality |
| 17-109-final-viral-combined_k141_328463\|\|0_partial | Low-quality |
| 17-109-final-viral-combined_k141_378713\|\|0_partial | Low-quality |
| 17-109-final-viral-combined_k141_312983\|\|0_partial | Not-determined |
| 17-109-final-viral-combined_k141_27677\|\|0_partial | Low-quality |
| 17-109-final-viral-combined_k141_178228\|\|0_partial | Not-determined |
| 17-109-final-viral-combined_k141_361390\|\|0_partial | Low-quality |
| 17-109-final-viral-combined_k141_162415\|\|0_partial | Not-determined |
| 17-109-final-viral-combined_k141_230281\|\|0_partial | Low-quality |
| 17-109-final-viral-combined_k141_263260\|\|0_partial | Low-quality |
| 17-109-final-viral-combined_k141_330633\|\|0_partial | Low-quality |
| 17-109-final-viral-combined_k141_112207\|\|0_partial | Not-determined |
| 17-109-final-viral-combined_k141_112382\|\|0_partial | Not-determined |
| 17-109-final-viral-combined_k141_315391\|\|0_partial | Low-quality |
| 17-109-final-viral-combined_k141_331312\|\|0_partial | Low-quality |
| 17-109-final-viral-combined_k141_363126\|\|0_partial | Low-quality |
| 17-109-final-viral-combined_k141_181135\|\|0_partial | Low-quality |
| 18-102-final-viral-combined_k141_180640\|\|full | Medium-quality |
| 17-109-final-viral-combined_k141_364141\|\|0_partial | Low-quality |
| 17-109-final-viral-combined_k141_248567\|\|0_partial | Low-quality |
| 17-109-final-viral-combined_k141_114674\|\|0_partial | Low-quality |
| 17-109-final-viral-combined_k141_98930\|\|0_partial | Low-quality |
| 17-109-final-viral-combined_k141_148997\|\|0_partial | Not-determined |
| 17-109-final-viral-combined_k141_166684\|\|0_partial | Low-quality |
| 18-105-final-viral-combined_k141_180996\|\|full | Medium-quality |
| 17-109-final-viral-combined_k141_234346\|\|lt2gene | Low-quality |
| 17-109-final-viral-combined_k141_50222\|\|lt2gene | Low-quality |
| 17-109-final-viral-combined_k141_301311\|\|lt2gene | Low-quality |
| 17-109-final-viral-combined_k141_368290\|\|lt2gene | Not-determined |
| 17-109-final-viral-combined_k141_351523\|\|lt2gene | Low-quality |
| 17-109-final-viral-combined_k141_167435\|\|lt2gene | Not-determined |
| 17-109-final-viral-combined_k141_167441\|\|lt2gene | Low-quality |
| 17-109-final-viral-combined_k141_267846\|\|lt2gene | Not-determined |
| 17-109-final-viral-combined_k141_50347\|\|lt2gene | Low-quality |
| 17-109-final-viral-combined_k141_16813\|\|lt2gene | Low-quality |
| 17-109-final-viral-combined_k141_368328\|\|lt2gene | Low-quality |
| 17-109-final-viral-combined_k141_284655\|\|lt2gene | Low-quality |
| 17-109-final-viral-combined_k141_67015\|\|lt2gene | Low-quality |
| 17-109-final-viral-combined_k141_234451\|\|lt2gene | Low-quality |
| 17-109-final-viral-combined_k141_217705\|\|lt2gene | Not-determined |
| 17-109-final-viral-combined_k141_167499\|\|lt2gene | Low-quality |
| 17-109-final-viral-combined_k141_117237\|\|lt2gene | Low-quality |
| 17-109-final-viral-combined_k141_117241\|\|lt2gene | Not-determined |
| 17-109-final-viral-combined_k141_284732\|\|lt2gene | Not-determined |
| 17-109-final-viral-combined_k141_334817\|\|lt2gene | Low-quality |
| 17-109-final-viral-combined_k141_154\|\|lt2gene | Low-quality |
| 17-109-final-viral-combined_k141_385139\|\|lt2gene | Low-quality |
| 17-109-final-viral-combined_k141_251248\|\|lt2gene | Not-determined |
| 17-109-final-viral-combined_k141_301433\|\|lt2gene | Low-quality |
| 17-109-final-viral-combined_k141_267922\|\|lt2gene | Low-quality |
| 17-109-final-viral-combined_k141_201056\|\|lt2gene | Low-quality |
| 17-109-final-viral-combined_k141_134097\|\|lt2gene | Low-quality |
| 17-109-final-viral-combined_k141_184316\|\|lt2gene | Not-determined |
| 17-109-final-viral-combined_k141_150751\|\|lt2gene | Low-quality |
| 17-109-final-viral-combined_k141_117310\|\|lt2gene | Not-determined |
| 17-109-final-viral-combined_k141_150767\|\|lt2gene | Low-quality |
| 17-109-final-viral-combined_k141_368397\|\|lt2gene | Not-determined |
| 17-109-final-viral-combined_k141_117327\|\|lt2gene | Low-quality |
| 17-109-final-viral-combined_k141_67107\|\|lt2gene | Low-quality |
| 17-109-final-viral-combined_k141_217859\|\|lt2gene | Low-quality |
| 17-109-final-viral-combined_k141_385234\|\|lt2gene | Low-quality |
| 17-109-final-viral-combined_k141_318250\|\|lt2gene | Not-determined |
| 17-109-final-viral-combined_k141_83859\|\|lt2gene | Low-quality |
| 17-109-final-viral-combined_k141_167625\|\|lt2gene | Low-quality |
| 17-109-final-viral-combined_k141_251364\|\|lt2gene | Not-determined |
| 17-109-final-viral-combined_k141_234455\|\|lt2gene | Low-quality |
| 17-109-final-viral-combined_k141_83907\|\|lt2gene | Low-quality |
| 17-109-final-viral-combined_k141_217889\|\|lt2gene | Low-quality |
| 17-109-final-viral-combined_k141_334881\|\|lt2gene | Low-quality |
| 17-109-final-viral-combined_k141_318296\|\|lt2gene | Low-quality |
| 17-109-final-viral-combined_k141_334885\|\|lt2gene | Low-quality |
| 17-109-final-viral-combined_k141_254\|\|lt2gene | Low-quality |
| 17-109-final-viral-combined_k141_16963\|\|lt2gene | Low-quality |
| 17-109-final-viral-combined_k141_150875\|\|lt2gene | Low-quality |
| 17-109-final-viral-combined_k141_50512\|\|lt2gene | Not-determined |
| 17-109-final-viral-combined_k141_50518\|\|lt2gene | Low-quality |
| 17-109-final-viral-combined_k141_184375\|\|lt2gene | Not-determined |
| 17-109-final-viral-combined_k141_117418\|\|lt2gene | Not-determined |
| 17-109-final-viral-combined_k141_100707\|\|lt2gene | Low-quality |
| 17-109-final-viral-combined_k141_67261\|\|lt2gene | Low-quality |
| 17-109-final-viral-combined_k141_301698\|\|lt2gene | Not-determined |
| 17-109-final-viral-combined_k141_33873\|\|lt2gene | Low-quality |
| 17-109-final-viral-combined_k141_285014\|\|lt2gene | Low-quality |
| 17-109-final-viral-combined_k141_251465\|\|lt2gene | Low-quality |
| 17-109-final-viral-combined_k141_50574\|\|lt2gene | Low-quality |
| 17-109-final-viral-combined_k141_360\|\|lt2gene | Low-quality |
| 17-109-final-viral-combined_k141_167740\|\|lt2gene | Not-determined |
| 17-109-final-viral-combined_k141_334953\|\|lt2gene | Low-quality |
| 17-109-final-viral-combined_k141_84075\|\|lt2gene | Not-determined |
| 17-109-final-viral-combined_k141_50601\|\|lt2gene | Not-determined |
| 17-109-final-viral-combined_k141_50603\|\|lt2gene | Not-determined |
| 17-109-final-viral-combined_k141_84087\|\|lt2gene | Low-quality |
| 17-109-final-viral-combined_k141_100775\|\|lt2gene | Not-determined |
| 17-109-final-viral-combined_k141_201149\|\|lt2gene | Low-quality |
| 17-109-final-viral-combined_k141_84111\|\|lt2gene | Low-quality |
| 17-109-final-viral-combined_k141_385412\|\|lt2gene | Low-quality |
| 17-109-final-viral-combined_k141_318501\|\|lt2gene | Low-quality |
| 17-109-final-viral-combined_k141_34015\|\|lt2gene | Low-quality |
| 17-109-final-viral-combined_k141_34016\|\|lt2gene | Low-quality |
| 17-109-final-viral-combined_k141_318509\|\|lt2gene | Low-quality |
| 17-109-final-viral-combined_k141_335028\|\|lt2gene | Low-quality |
| 17-109-final-viral-combined_k141_84132\|\|lt2gene | Low-quality |
| 17-109-final-viral-combined_k141_385438\|\|lt2gene | Low-quality |
| 17-109-final-viral-combined_k141_184528\|\|lt2gene | Low-quality |
| 17-109-final-viral-combined_k141_485\|\|lt2gene | Low-quality |
| 17-109-final-viral-combined_k141_50690\|\|lt2gene | Low-quality |
| 17-109-final-viral-combined_k141_100851\|\|lt2gene | Not-determined |
| 17-109-final-viral-combined_k141_100859\|\|lt2gene | Low-quality |
| 17-109-final-viral-combined_k141_335053\|\|lt2gene | Low-quality |
| 17-109-final-viral-combined_k141_84170\|\|lt2gene | Low-quality |
| 17-109-final-viral-combined_k141_84189\|\|lt2gene | Not-determined |
| 17-109-final-viral-combined_k141_17283\|\|lt2gene | Low-quality |
| 17-109-final-viral-combined_k141_318609\|\|lt2gene | Low-quality |
| 17-109-final-viral-combined_k141_134465\|\|lt2gene | Low-quality |
| 17-109-final-viral-combined_k141_218098\|\|lt2gene | Not-determined |
| 17-109-final-viral-combined_k141_580\|\|lt2gene | Low-quality |
| 17-109-final-viral-combined_k141_50783\|\|lt2gene | Low-quality |
| 17-109-final-viral-combined_k141_201289\|\|lt2gene | Low-quality |
| 17-109-final-viral-combined_k141_134530\|\|lt2gene | Low-quality |
| 17-109-final-viral-combined_k141_285238\|\|lt2gene | Low-quality |
| 17-109-final-viral-combined_k141_268225\|\|lt2gene | Not-determined |
| 17-109-final-viral-combined_k141_67449\|\|lt2gene | Low-quality |
| 17-109-final-viral-combined_k141_351778\|\|lt2gene | Low-quality |
| 17-109-final-viral-combined_k141_151003\|\|lt2gene | Low-quality |
| 17-109-final-viral-combined_k141_201329\|\|lt2gene | Not-determined |
| 17-109-final-viral-combined_k141_234882\|\|lt2gene | Not-determined |
| 17-109-final-viral-combined_k141_335128\|\|lt2gene | Not-determined |
| 17-109-final-viral-combined_k141_351822\|\|lt2gene | Not-determined |
| 17-109-final-viral-combined_k141_101007\|\|lt2gene | Low-quality |
| 17-109-final-viral-combined_k141_151034\|\|lt2gene | Low-quality |
| 17-109-final-viral-combined_k141_84310\|\|lt2gene | Low-quality |
| 17-109-final-viral-combined_k141_84316\|\|lt2gene | Low-quality |
| 17-109-final-viral-combined_k141_17461\|\|lt2gene | Low-quality |
| 17-109-final-viral-combined_k141_151087\|\|lt2gene | Low-quality |
| 17-109-final-viral-combined_k141_745\|\|lt2gene | Low-quality |
| 17-109-final-viral-combined_k141_168064\|\|lt2gene | Not-determined |
| 17-109-final-viral-combined_k141_117583\|\|lt2gene | Not-determined |
| 17-109-final-viral-combined_k141_184737\|\|lt2gene | Low-quality |
| 17-109-final-viral-combined_k141_84361\|\|lt2gene | Low-quality |
| 17-109-final-viral-combined_k141_234979\|\|lt2gene | Low-quality |
| 17-109-final-viral-combined_k141_368879\|\|lt2gene | Low-quality |
| 17-109-final-viral-combined_k141_351931\|\|lt2gene | Not-determined |
| 17-109-final-viral-combined_k141_385636\|\|lt2gene | Low-quality |
| 17-109-final-viral-combined_k141_201584\|\|lt2gene | Low-quality |
| 17-109-final-viral-combined_k141_34333\|\|lt2gene | Low-quality |
| 17-109-final-viral-combined_k141_855\|\|lt2gene | Low-quality |
| 17-109-final-viral-combined_k141_862\|\|lt2gene | Low-quality |
| 17-109-final-viral-combined_k141_51019\|\|lt2gene | Low-quality |
| 17-109-final-viral-combined_k141_866\|\|lt2gene | Low-quality |
| 17-109-final-viral-combined_k141_51025\|\|lt2gene | Low-quality |
| 17-109-final-viral-combined_k141_871\|\|lt2gene | Low-quality |
| 17-109-final-viral-combined_k141_168203\|\|lt2gene | Low-quality |
| 17-109-final-viral-combined_k141_285603\|\|lt2gene | Low-quality |
| 17-109-final-viral-combined_k141_151287\|\|lt2gene | Low-quality |
| 17-109-final-viral-combined_k141_201654\|\|lt2gene | Low-quality |
| 17-109-final-viral-combined_k141_201656\|\|lt2gene | Low-quality |
| 17-109-final-viral-combined_k141_318878\|\|lt2gene | Low-quality |
| 17-109-final-viral-combined_k141_151330\|\|lt2gene | Not-determined |
| 17-109-final-viral-combined_k141_151347\|\|lt2gene | Low-quality |
| 17-109-final-viral-combined_k141_235145\|\|lt2gene | Low-quality |
| 17-109-final-viral-combined_k141_352060\|\|lt2gene | Low-quality |
| 17-109-final-viral-combined_k141_335332\|\|lt2gene | Low-quality |
| 17-109-final-viral-combined_k141_268586\|\|lt2gene | Not-determined |
| 17-109-final-viral-combined_k141_335335\|\|lt2gene | Not-determined |
| 17-109-final-viral-combined_k141_952\|\|lt2gene | Low-quality |
| 17-109-final-viral-combined_k141_34536\|\|lt2gene | Low-quality |
| 17-109-final-viral-combined_k141_285689\|\|lt2gene | Not-determined |
| 17-109-final-viral-combined_k141_34555\|\|lt2gene | Low-quality |
| 17-109-final-viral-combined_k141_335365\|\|lt2gene | Low-quality |
| 17-109-final-viral-combined_k141_235205\|\|lt2gene | Low-quality |
| 17-109-final-viral-combined_k141_17771\|\|lt2gene | Low-quality |
| 17-109-final-viral-combined_k141_251902\|\|lt2gene | Low-quality |
| 17-109-final-viral-combined_k141_101271\|\|lt2gene | Low-quality |
| 17-109-final-viral-combined_k141_335416\|\|lt2gene | Not-determined |
| 17-109-final-viral-combined_k141_17837\|\|lt2gene | Not-determined |
| 17-109-final-viral-combined_k141_218425\|\|lt2gene | Low-quality |
| 17-109-final-viral-combined_k141_101307\|\|lt2gene | Low-quality |
| 17-109-final-viral-combined_k141_385948\|\|lt2gene | Not-determined |
| 17-109-final-viral-combined_k141_235283\|\|lt2gene | Not-determined |
| 17-109-final-viral-combined_k141_235292\|\|lt2gene | Low-quality |
| 17-109-final-viral-combined_k141_185079\|\|lt2gene | Low-quality |
| 17-109-final-viral-combined_k141_185088\|\|lt2gene | Low-quality |
| 17-109-final-viral-combined_k141_302300\|\|lt2gene | Low-quality |
| 17-109-final-viral-combined_k141_101375\|\|lt2gene | Low-quality |
| 17-109-final-viral-combined_k141_151513\|\|lt2gene | Not-determined |
| 17-109-final-viral-combined_k141_335491\|\|lt2gene | Low-quality |
| 17-109-final-viral-combined_k141_352199\|\|lt2gene | Low-quality |
| 17-109-final-viral-combined_k141_117803\|\|lt2gene | Not-determined |
| 17-109-final-viral-combined_k141_185120\|\|lt2gene | Low-quality |
| 17-109-final-viral-combined_k141_51173\|\|lt2gene | Low-quality |
| 17-109-final-viral-combined_k141_285841\|\|lt2gene | Low-quality |
| 17-109-final-viral-combined_k141_352233\|\|lt2gene | Low-quality |
| 17-109-final-viral-combined_k141_235416\|\|lt2gene | Low-quality |
| 17-109-final-viral-combined_k141_51210\|\|lt2gene | Low-quality |
| 17-109-final-viral-combined_k141_285870\|\|lt2gene | Not-determined |
| 17-109-final-viral-combined_k141_268839\|\|lt2gene | Low-quality |
| 17-109-final-viral-combined_k141_185182\|\|lt2gene | Not-determined |
| 17-109-final-viral-combined_k141_218636\|\|lt2gene | Not-determined |
| 17-109-final-viral-combined_k141_1162\|\|lt2gene | Not-determined |
| 17-109-final-viral-combined_k141_369306\|\|lt2gene | Low-quality |
| 17-109-final-viral-combined_k141_302392\|\|lt2gene | Not-determined |
| 17-109-final-viral-combined_k141_185261\|\|lt2gene | Low-quality |
| 17-109-final-viral-combined_k141_34861\|\|lt2gene | Low-quality |
| 17-109-final-viral-combined_k141_51320\|\|lt2gene | Not-determined |
| 17-109-final-viral-combined_k141_51329\|\|lt2gene | Not-determined |
| 17-109-final-viral-combined_k141_335637\|\|lt2gene | Low-quality |
| 17-109-final-viral-combined_k141_268930\|\|lt2gene | Not-determined |
| 17-109-final-viral-combined_k141_302451\|\|lt2gene | Low-quality |
| 17-109-final-viral-combined_k141_51333\|\|lt2gene | Low-quality |
| 17-109-final-viral-combined_k141_68017\|\|lt2gene | Low-quality |
| 17-109-final-viral-combined_k141_84850\|\|lt2gene | Low-quality |
| 17-109-final-viral-combined_k141_51390\|\|lt2gene | Not-determined |
| 17-109-final-viral-combined_k141_118031\|\|lt2gene | Low-quality |
| 17-109-final-viral-combined_k141_84873\|\|lt2gene | Not-determined |
| 17-109-final-viral-combined_k141_369460\|\|lt2gene | Low-quality |
| 17-109-final-viral-combined_k141_135025\|\|lt2gene | Low-quality |
| 17-109-final-viral-combined_k141_369478\|\|lt2gene | Not-determined |
| 17-109-final-viral-combined_k141_84908\|\|lt2gene | Low-quality |
| 17-109-final-viral-combined_k141_335643\|\|lt2gene | Low-quality |
| 17-109-final-viral-combined_k141_151722\|\|lt2gene | Low-quality |
| 17-109-final-viral-combined_k141_352491\|\|lt2gene | Low-quality |
| 17-109-final-viral-combined_k141_252297\|\|lt2gene | Low-quality |
| 17-109-final-viral-combined_k141_269010\|\|lt2gene | Low-quality |
| 17-109-final-viral-combined_k141_18122\|\|lt2gene | Low-quality |
| 17-109-final-viral-combined_k141_18125\|\|lt2gene | Low-quality |
| 17-109-final-viral-combined_k141_168594\|\|lt2gene | Not-determined |
| 17-109-final-viral-combined_k141_269021\|\|lt2gene | Low-quality |
| 17-109-final-viral-combined_k141_135067\|\|lt2gene | Not-determined |
| 17-109-final-viral-combined_k141_386323\|\|lt2gene | Not-determined |
| 17-109-final-viral-combined_k141_302632\|\|lt2gene | Not-determined |
| 17-109-final-viral-combined_k141_101587\|\|lt2gene | Low-quality |
| 17-109-final-viral-combined_k141_101589\|\|lt2gene | Not-determined |
| 17-109-final-viral-combined_k141_101603\|\|lt2gene | Not-determined |
| 17-109-final-viral-combined_k141_18179\|\|lt2gene | Low-quality |
| 17-109-final-viral-combined_k141_101608\|\|lt2gene | Low-quality |
| 17-109-final-viral-combined_k141_335774\|\|lt2gene | Low-quality |
| 17-109-final-viral-combined_k141_51542\|\|lt2gene | Low-quality |
| 17-109-final-viral-combined_k141_185532\|\|lt2gene | Low-quality |
| 17-109-final-viral-combined_k141_18233\|\|lt2gene | Low-quality |
| 17-109-final-viral-combined_k141_1508\|\|lt2gene | Not-determined |
| 17-109-final-viral-combined_k141_335825\|\|lt2gene | Low-quality |
| 17-109-final-viral-combined_k141_369739\|\|lt2gene | Low-quality |
| 17-109-final-viral-combined_k141_35008\|\|lt2gene | Low-quality |
| 17-109-final-viral-combined_k141_286077\|\|lt2gene | Not-determined |
| 17-109-final-viral-combined_k141_185622\|\|lt2gene | Low-quality |
| 17-109-final-viral-combined_k141_168755\|\|lt2gene | Not-determined |
| 17-109-final-viral-combined_k141_168760\|\|lt2gene | Not-determined |
| 17-109-final-viral-combined_k141_235671\|\|lt2gene | Low-quality |
| 17-109-final-viral-combined_k141_286112\|\|lt2gene | Low-quality |
| 17-109-final-viral-combined_k141_369780\|\|lt2gene | Not-determined |
| 17-109-final-viral-combined_k141_185663\|\|lt2gene | Low-quality |
| 17-109-final-viral-combined_k141_352621\|\|lt2gene | Low-quality |
| 17-109-final-viral-combined_k141_235714\|\|lt2gene | Low-quality |
| 17-109-final-viral-combined_k141_1625\|\|lt2gene | Not-determined |
| 17-109-final-viral-combined_k141_218982\|\|lt2gene | Not-determined |
| 17-109-final-viral-combined_k141_151923\|\|lt2gene | Low-quality |
| 17-109-final-viral-combined_k141_35124\|\|lt2gene | Low-quality |
| 17-109-final-viral-combined_k141_35129\|\|lt2gene | Not-determined |
| 17-109-final-viral-combined_k141_219014\|\|lt2gene | Low-quality |
| 17-109-final-viral-combined_k141_269275\|\|lt2gene | Not-determined |
| 17-109-final-viral-combined_k141_168920\|\|lt2gene | Low-quality |
| 17-109-final-viral-combined_k141_352755\|\|lt2gene | Low-quality |
| 17-109-final-viral-combined_k141_85263\|\|lt2gene | Low-quality |
| 17-109-final-viral-combined_k141_352767\|\|lt2gene | Low-quality |
| 17-109-final-viral-combined_k141_118308\|\|lt2gene | Low-quality |
| 17-109-final-viral-combined_k141_35250\|\|lt2gene | Low-quality |
| 17-109-final-viral-combined_k141_51781\|\|lt2gene | Not-determined |
| 17-109-final-viral-combined_k141_35275\|\|lt2gene | Not-determined |
| 17-109-final-viral-combined_k141_252552\|\|lt2gene | Not-determined |
| 17-109-final-viral-combined_k141_18268\|\|lt2gene | Low-quality |
| 17-109-final-viral-combined_k141_35330\|\|lt2gene | Low-quality |
| 17-109-final-viral-combined_k141_252577\|\|lt2gene | Low-quality |
| 17-109-final-viral-combined_k141_302956\|\|lt2gene | Low-quality |
| 17-109-final-viral-combined_k141_369981\|\|lt2gene | Low-quality |
| 17-109-final-viral-combined_k141_286206\|\|lt2gene | Not-determined |
| 17-109-final-viral-combined_k141_336027\|\|lt2gene | Not-determined |
| 17-109-final-viral-combined_k141_302969\|\|lt2gene | Not-determined |
| 17-109-final-viral-combined_k141_1805\|\|lt2gene | Not-determined |
| 17-109-final-viral-combined_k141_336050\|\|lt2gene | Not-determined |
| 17-109-final-viral-combined_k141_336052\|\|lt2gene | Low-quality |
| 17-109-final-viral-combined_k141_252634\|\|lt2gene | Low-quality |
| 17-109-final-viral-combined_k141_152196\|\|lt2gene | Low-quality |
| 17-109-final-viral-combined_k141_152213\|\|lt2gene | Not-determined |
| 17-109-final-viral-combined_k141_185918\|\|lt2gene | Low-quality |
| 17-109-final-viral-combined_k141_236033\|\|lt2gene | Low-quality |
| 17-109-final-viral-combined_k141_219205\|\|lt2gene | Low-quality |
| 17-109-final-viral-combined_k141_386765\|\|lt2gene | Not-determined |
| 17-109-final-viral-combined_k141_102070\|\|lt2gene | Low-quality |
| 17-109-final-viral-combined_k141_336098\|\|lt2gene | Not-determined |
| 17-109-final-viral-combined_k141_102073\|\|lt2gene | Not-determined |
| 17-109-final-viral-combined_k141_202359\|\|lt2gene | Low-quality |
| 17-109-final-viral-combined_k141_135548\|\|lt2gene | Low-quality |
| 17-109-final-viral-combined_k141_68599\|\|lt2gene | Low-quality |
| 17-109-final-viral-combined_k141_185940\|\|lt2gene | Low-quality |
| 17-109-final-viral-combined_k141_185967\|\|lt2gene | Not-determined |
| 17-109-final-viral-combined_k141_18353\|\|lt2gene | Not-determined |
| 17-109-final-viral-combined_k141_236225\|\|lt2gene | Not-determined |
| 17-109-final-viral-combined_k141_286528\|\|lt2gene | Not-determined |
| 17-109-final-viral-combined_k141_370223\|\|lt2gene | Low-quality |
| 17-109-final-viral-combined_k141_202396\|\|lt2gene | Low-quality |
| 17-109-final-viral-combined_k141_370226\|\|lt2gene | Low-quality |
| 17-109-final-viral-combined_k141_152377\|\|lt2gene | Low-quality |
| 17-109-final-viral-combined_k141_219397\|\|lt2gene | Low-quality |
| 17-109-final-viral-combined_k141_370252\|\|lt2gene | Low-quality |
| 17-109-final-viral-combined_k141_202455\|\|lt2gene | Low-quality |
| 17-109-final-viral-combined_k141_286606\|\|lt2gene | Not-determined |
| 17-109-final-viral-combined_k141_68681\|\|lt2gene | Low-quality |
| 17-109-final-viral-combined_k141_152472\|\|lt2gene | Not-determined |
| 17-109-final-viral-combined_k141_152473\|\|lt2gene | Low-quality |
| 17-109-final-viral-combined_k141_35601\|\|lt2gene | Not-determined |
| 17-109-final-viral-combined_k141_186049\|\|lt2gene | Low-quality |
| 17-109-final-viral-combined_k141_319986\|\|lt2gene | Not-determined |
| 17-109-final-viral-combined_k141_336242\|\|lt2gene | Low-quality |
| 17-109-final-viral-combined_k141_386944\|\|lt2gene | Low-quality |
| 17-109-final-viral-combined_k141_370315\|\|lt2gene | Low-quality |
| 17-109-final-viral-combined_k141_68755\|\|lt2gene | Low-quality |
| 17-109-final-viral-combined_k141_219422\|\|lt2gene | Low-quality |
| 17-109-final-viral-combined_k141_353135\|\|lt2gene | Low-quality |
| 17-109-final-viral-combined_k141_68783\|\|lt2gene | Low-quality |
| 17-109-final-viral-combined_k141_85631\|\|lt2gene | Low-quality |
| 17-109-final-viral-combined_k141_102145\|\|lt2gene | Low-quality |
| 17-109-final-viral-combined_k141_386993\|\|lt2gene | Low-quality |
| 17-109-final-viral-combined_k141_386995\|\|lt2gene | Low-quality |
| 17-109-final-viral-combined_k141_370372\|\|lt2gene | Low-quality |
| 17-109-final-viral-combined_k141_336298\|\|lt2gene | Low-quality |
| 17-109-final-viral-combined_k141_85648\|\|lt2gene | Low-quality |
| 17-109-final-viral-combined_k141_370380\|\|lt2gene | Low-quality |
| 17-109-final-viral-combined_k141_186151\|\|lt2gene | Low-quality |
| 17-109-final-viral-combined_k141_320081\|\|lt2gene | Low-quality |
| 17-109-final-viral-combined_k141_169220\|\|lt2gene | Low-quality |
| 17-109-final-viral-combined_k141_219503\|\|lt2gene | Low-quality |
| 17-109-final-viral-combined_k141_35703\|\|lt2gene | Low-quality |
| 17-109-final-viral-combined_k141_202547\|\|lt2gene | Low-quality |
| 17-109-final-viral-combined_k141_135801\|\|lt2gene | Not-determined |
| 17-109-final-viral-combined_k141_186213\|\|lt2gene | Low-quality |
| 17-109-final-viral-combined_k141_269635\|\|lt2gene | Low-quality |
| 17-109-final-viral-combined_k141_303507\|\|lt2gene | Low-quality |
| 17-109-final-viral-combined_k141_320142\|\|lt2gene | Low-quality |
| 17-109-final-viral-combined_k141_202670\|\|lt2gene | Not-determined |
| 17-109-final-viral-combined_k141_219564\|\|lt2gene | Low-quality |
| 17-109-final-viral-combined_k141_219571\|\|lt2gene | Not-determined |
| 17-109-final-viral-combined_k141_52293\|\|lt2gene | Low-quality |
| 17-109-final-viral-combined_k141_370474\|\|lt2gene | Low-quality |
| 17-109-final-viral-combined_k141_152704\|\|lt2gene | Low-quality |
| 17-109-final-viral-combined_k141_118869\|\|lt2gene | Not-determined |
| 17-109-final-viral-combined_k141_353343\|\|lt2gene | Not-determined |
| 17-109-final-viral-combined_k141_118891\|\|lt2gene | Not-determined |
| 17-109-final-viral-combined_k141_286806\|\|lt2gene | Low-quality |
| 17-109-final-viral-combined_k141_387161\|\|lt2gene | Not-determined |
| 17-109-final-viral-combined_k141_336447\|\|lt2gene | Not-determined |
| 17-109-final-viral-combined_k141_18646\|\|lt2gene | Low-quality |
| 17-109-final-viral-combined_k141_35852\|\|lt2gene | Low-quality |
| 17-109-final-viral-combined_k141_387217\|\|lt2gene | Not-determined |
| 17-109-final-viral-combined_k141_387220\|\|lt2gene | Low-quality |
| 17-109-final-viral-combined_k141_35857\|\|lt2gene | Low-quality |
| 17-109-final-viral-combined_k141_253031\|\|lt2gene | Low-quality |
| 17-109-final-viral-combined_k141_269820\|\|lt2gene | Not-determined |
| 17-109-final-viral-combined_k141_69069\|\|lt2gene | Not-determined |
| 17-109-final-viral-combined_k141_370603\|\|lt2gene | Not-determined |
| 17-109-final-viral-combined_k141_35862\|\|lt2gene | Low-quality |
| 17-109-final-viral-combined_k141_186493\|\|lt2gene | Not-determined |
| 17-109-final-viral-combined_k141_69093\|\|lt2gene | Low-quality |
| 17-109-final-viral-combined_k141_253066\|\|lt2gene | Low-quality |
| 17-109-final-viral-combined_k141_119051\|\|lt2gene | Low-quality |
| 17-109-final-viral-combined_k141_236340\|\|lt2gene | Not-determined |
| 17-109-final-viral-combined_k141_35895\|\|lt2gene | Low-quality |
| 17-109-final-viral-combined_k141_236354\|\|lt2gene | Low-quality |
| 17-109-final-viral-combined_k141_303735\|\|lt2gene | Low-quality |
| 17-109-final-viral-combined_k141_253088\|\|lt2gene | Low-quality |
| 17-109-final-viral-combined_k141_102479\|\|lt2gene | Low-quality |
| 17-109-final-viral-combined_k141_35909\|\|lt2gene | Low-quality |
| 17-109-final-viral-combined_k141_387273\|\|lt2gene | Not-determined |
| 17-109-final-viral-combined_k141_35943\|\|lt2gene | Low-quality |
| 17-109-final-viral-combined_k141_169506\|\|lt2gene | Not-determined |
| 17-109-final-viral-combined_k141_119124\|\|lt2gene | Not-determined |
| 17-109-final-viral-combined_k141_102501\|\|lt2gene | Low-quality |
| 17-109-final-viral-combined_k141_336572\|\|lt2gene | Low-quality |
| 17-109-final-viral-combined_k141_236419\|\|lt2gene | Low-quality |
| 17-109-final-viral-combined_k141_186640\|\|lt2gene | Not-determined |
| 17-109-final-viral-combined_k141_69223\|\|lt2gene | Low-quality |
| 17-109-final-viral-combined_k141_152987\|\|lt2gene | Low-quality |
| 17-109-final-viral-combined_k141_202922\|\|lt2gene | Low-quality |
| 17-109-final-viral-combined_k141_52531\|\|lt2gene | Low-quality |
| 17-109-final-viral-combined_k141_353554\|\|lt2gene | Low-quality |
| 17-109-final-viral-combined_k141_186685\|\|lt2gene | Low-quality |
| 17-109-final-viral-combined_k141_186689\|\|lt2gene | Low-quality |
| 17-109-final-viral-combined_k141_69254\|\|lt2gene | Low-quality |
| 17-109-final-viral-combined_k141_236512\|\|lt2gene | Low-quality |
| 17-109-final-viral-combined_k141_303902\|\|lt2gene | Low-quality |
| 17-109-final-viral-combined_k141_202948\|\|lt2gene | Low-quality |
| 17-109-final-viral-combined_k141_287055\|\|lt2gene | Low-quality |
| 17-109-final-viral-combined_k141_387319\|\|lt2gene | Not-determined |
| 17-109-final-viral-combined_k141_236595\|\|lt2gene | Low-quality |
| 17-109-final-viral-combined_k141_153084\|\|lt2gene | Low-quality |
| 17-109-final-viral-combined_k141_136227\|\|lt2gene | Low-quality |
| 17-109-final-viral-combined_k141_203015\|\|lt2gene | Not-determined |
| 17-109-final-viral-combined_k141_69432\|\|lt2gene | Low-quality |
| 17-109-final-viral-combined_k141_18780\|\|lt2gene | Low-quality |
| 17-109-final-viral-combined_k141_36050\|\|lt2gene | Low-quality |
| 17-109-final-viral-combined_k141_203043\|\|lt2gene | Not-determined |
| 17-109-final-viral-combined_k141_304027\|\|lt2gene | Low-quality |
| 17-109-final-viral-combined_k141_69522\|\|lt2gene | Not-determined |
| 17-109-final-viral-combined_k141_136283\|\|lt2gene | Low-quality |
| 17-109-final-viral-combined_k141_2505\|\|lt2gene | Low-quality |
| 17-109-final-viral-combined_k141_236733\|\|lt2gene | Low-quality |
| 17-109-final-viral-combined_k141_153156\|\|lt2gene | Not-determined |
| 17-109-final-viral-combined_k141_102635\|\|lt2gene | Low-quality |
| 17-109-final-viral-combined_k141_304136\|\|lt2gene | Low-quality |
| 17-109-final-viral-combined_k141_86178\|\|lt2gene | Low-quality |
| 17-109-final-viral-combined_k141_119269\|\|lt2gene | Not-determined |
| 17-109-final-viral-combined_k141_253361\|\|lt2gene | Low-quality |
| 17-109-final-viral-combined_k141_387549\|\|lt2gene | Low-quality |
| 17-109-final-viral-combined_k141_69629\|\|lt2gene | Not-determined |
| 17-109-final-viral-combined_k141_219870\|\|lt2gene | Low-quality |
| 17-109-final-viral-combined_k141_287313\|\|lt2gene | Low-quality |
| 17-109-final-viral-combined_k141_169835\|\|lt2gene | Low-quality |
| 17-109-final-viral-combined_k141_353925\|\|lt2gene | Not-determined |
| 17-109-final-viral-combined_k141_353956\|\|lt2gene | Low-quality |
| 17-109-final-viral-combined_k141_136402\|\|lt2gene | Not-determined |
| 17-109-final-viral-combined_k141_219933\|\|lt2gene | Low-quality |
| 17-109-final-viral-combined_k141_52844\|\|lt2gene | Low-quality |
| 17-109-final-viral-combined_k141_153389\|\|lt2gene | Low-quality |
| 17-109-final-viral-combined_k141_153400\|\|lt2gene | Not-determined |
| 17-109-final-viral-combined_k141_203245\|\|lt2gene | Not-determined |
| 17-109-final-viral-combined_k141_169954\|\|lt2gene | Low-quality |
| 17-109-final-viral-combined_k141_287361\|\|lt2gene | Low-quality |
| 17-109-final-viral-combined_k141_253467\|\|lt2gene | Low-quality |
| 17-109-final-viral-combined_k141_287379\|\|lt2gene | Low-quality |
| 17-109-final-viral-combined_k141_371092\|\|lt2gene | Low-quality |
| 17-109-final-viral-combined_k141_119406\|\|lt2gene | Low-quality |
| 17-109-final-viral-combined_k141_219973\|\|lt2gene | Low-quality |
| 17-109-final-viral-combined_k141_270270\|\|lt2gene | Low-quality |
| 17-109-final-viral-combined_k141_287454\|\|lt2gene | Low-quality |
| 17-109-final-viral-combined_k141_336782\|\|lt2gene | Low-quality |
| 17-109-final-viral-combined_k141_336789\|\|lt2gene | Low-quality |
| 17-109-final-viral-combined_k141_18946\|\|lt2gene | Low-quality |
| 17-109-final-viral-combined_k141_287485\|\|lt2gene | Low-quality |
| 17-109-final-viral-combined_k141_270302\|\|lt2gene | Low-quality |
| 17-109-final-viral-combined_k141_18955\|\|lt2gene | Low-quality |
| 17-109-final-viral-combined_k141_18960\|\|lt2gene | Not-determined |
| 17-109-final-viral-combined_k141_170060\|\|lt2gene | Not-determined |
| 17-109-final-viral-combined_k141_203417\|\|lt2gene | Low-quality |
| 17-109-final-viral-combined_k141_287516\|\|lt2gene | Low-quality |
| 17-109-final-viral-combined_k141_52973\|\|lt2gene | Low-quality |
| 17-109-final-viral-combined_k141_336869\|\|lt2gene | Not-determined |
| 17-109-final-viral-combined_k141_203503\|\|lt2gene | Not-determined |
| 17-109-final-viral-combined_k141_119644\|\|lt2gene | Low-quality |
| 17-109-final-viral-combined_k141_253568\|\|lt2gene | Low-quality |
| 17-109-final-viral-combined_k141_220069\|\|lt2gene | Not-determined |
| 17-109-final-viral-combined_k141_119670\|\|lt2gene | Low-quality |
| 17-109-final-viral-combined_k141_70036\|\|lt2gene | Low-quality |
| 17-109-final-viral-combined_k141_187239\|\|lt2gene | Low-quality |
| 17-109-final-viral-combined_k141_70051\|\|lt2gene | Low-quality |
| 17-109-final-viral-combined_k141_237135\|\|lt2gene | Low-quality |
| 17-109-final-viral-combined_k141_304426\|\|lt2gene | Not-determined |
| 17-109-final-viral-combined_k141_2952\|\|lt2gene | Not-determined |
| 17-109-final-viral-combined_k141_19103\|\|lt2gene | Low-quality |
| 17-109-final-viral-combined_k141_153491\|\|lt2gene | Low-quality |
| 17-109-final-viral-combined_k141_387784\|\|lt2gene | Low-quality |
| 17-109-final-viral-combined_k141_371353\|\|lt2gene | Low-quality |
| 17-109-final-viral-combined_k141_36488\|\|lt2gene | Low-quality |
| 17-109-final-viral-combined_k141_19165\|\|lt2gene | Not-determined |
| 17-109-final-viral-combined_k141_86548\|\|lt2gene | Low-quality |
| 17-109-final-viral-combined_k141_270564\|\|lt2gene | Low-quality |
| 17-109-final-viral-combined_k141_36507\|\|lt2gene | Low-quality |
| 17-109-final-viral-combined_k141_187336\|\|lt2gene | Not-determined |
| 17-109-final-viral-combined_k141_270586\|\|lt2gene | Low-quality |
| 17-109-final-viral-combined_k141_53119\|\|lt2gene | Low-quality |
| 17-109-final-viral-combined_k141_304562\|\|lt2gene | Low-quality |
| 17-109-final-viral-combined_k141_220366\|\|lt2gene | Low-quality |
| 17-109-final-viral-combined_k141_53177\|\|lt2gene | Low-quality |
| 17-109-final-viral-combined_k141_270679\|\|lt2gene | Low-quality |
| 17-109-final-viral-combined_k141_136962\|\|lt2gene | Not-determined |
| 17-109-final-viral-combined_k141_53195\|\|lt2gene | Low-quality |
| 17-109-final-viral-combined_k141_337142\|\|lt2gene | Low-quality |
| 17-109-final-viral-combined_k141_371507\|\|lt2gene | Low-quality |
| 17-109-final-viral-combined_k141_53202\|\|lt2gene | Not-determined |
| 17-109-final-viral-combined_k141_270692\|\|lt2gene | Low-quality |
| 17-109-final-viral-combined_k141_337148\|\|lt2gene | Low-quality |
| 17-109-final-viral-combined_k141_371525\|\|lt2gene | Low-quality |
| 17-109-final-viral-combined_k141_3144\|\|lt2gene | Low-quality |
| 17-109-final-viral-combined_k141_387973\|\|lt2gene | Not-determined |
| 17-109-final-viral-combined_k141_170417\|\|lt2gene | Low-quality |
| 17-109-final-viral-combined_k141_86695\|\|lt2gene | Not-determined |
| 17-109-final-viral-combined_k141_137041\|\|lt2gene | Low-quality |
| 17-109-final-viral-combined_k141_53333\|\|lt2gene | Low-quality |
| 17-109-final-viral-combined_k141_19374\|\|lt2gene | Low-quality |
| 17-109-final-viral-combined_k141_103156\|\|lt2gene | Low-quality |
| 17-109-final-viral-combined_k141_70330\|\|lt2gene | Low-quality |
| 17-109-final-viral-combined_k141_388047\|\|lt2gene | Low-quality |
| 17-109-final-viral-combined_k141_137076\|\|lt2gene | Not-determined |
| 17-109-final-viral-combined_k141_70357\|\|lt2gene | Not-determined |
| 17-109-final-viral-combined_k141_270798\|\|lt2gene | Not-determined |
| 17-109-final-viral-combined_k141_86777\|\|lt2gene | Low-quality |
| 17-109-final-viral-combined_k141_253709\|\|lt2gene | Not-determined |
| 17-109-final-viral-combined_k141_103187\|\|lt2gene | Low-quality |
| 17-109-final-viral-combined_k141_220498\|\|lt2gene | Low-quality |
| 17-109-final-viral-combined_k141_220517\|\|lt2gene | Low-quality |
| 17-109-final-viral-combined_k141_153772\|\|lt2gene | Not-determined |
| 17-109-final-viral-combined_k141_53385\|\|lt2gene | Low-quality |
| 17-109-final-viral-combined_k141_36813\|\|lt2gene | Not-determined |
| 17-109-final-viral-combined_k141_220572\|\|lt2gene | Not-determined |
| 17-109-final-viral-combined_k141_53416\|\|lt2gene | Not-determined |
| 17-109-final-viral-combined_k141_120015\|\|lt2gene | Low-quality |
| 17-109-final-viral-combined_k141_103295\|\|lt2gene | Not-determined |
| 17-109-final-viral-combined_k141_53481\|\|lt2gene | Not-determined |
| 17-109-final-viral-combined_k141_337452\|\|lt2gene | Low-quality |
| 17-109-final-viral-combined_k141_220625\|\|lt2gene | Low-quality |
| 17-109-final-viral-combined_k141_170635\|\|lt2gene | Not-determined |
| 17-109-final-viral-combined_k141_371709\|\|lt2gene | Not-determined |

| | |
|---|---|
| 17-109-final-viral-combined_k141_253862\|\|lt2gene | Low-quality |
| 17-109-final-viral-combined_k141_153878\|\|lt2gene | Low-quality |
| 17-109-final-viral-combined_k141_220662\|\|lt2gene | Low-quality |
| 17-109-final-viral-combined_k141_103371\|\|lt2gene | Not-determined |
| 17-109-final-viral-combined_k141_3371\|\|lt2gene | Low-quality |
| 17-109-final-viral-combined_k141_371754\|\|lt2gene | Low-quality |
| 17-109-final-viral-combined_k141_204055\|\|lt2gene | Not-determined |
| 17-109-final-viral-combined_k141_103403\|\|lt2gene | Low-quality |
| 17-109-final-viral-combined_k141_304908\|\|lt2gene | Low-quality |
| 17-109-final-viral-combined_k141_253931\|\|lt2gene | Low-quality |
| 17-109-final-viral-combined_k141_287975\|\|lt2gene | Low-quality |
| 17-109-final-viral-combined_k141_187788\|\|lt2gene | Low-quality |
| 17-109-final-viral-combined_k141_287995\|\|lt2gene | Low-quality |
| 17-109-final-viral-combined_k141_170754\|\|lt2gene | Low-quality |
| 17-109-final-viral-combined_k141_137248\|\|lt2gene | Low-quality |
| 17-109-final-viral-combined_k141_153950\|\|lt2gene | Low-quality |
| 17-109-final-viral-combined_k141_3438\|\|lt2gene | Low-quality |
| 17-109-final-viral-combined_k141_288033\|\|lt2gene | Low-quality |
| 17-109-final-viral-combined_k141_70636\|\|lt2gene | Low-quality |
| 17-109-final-viral-combined_k141_3458\|\|lt2gene | Low-quality |
| 17-109-final-viral-combined_k141_204208\|\|lt2gene | Low-quality |
| 17-109-final-viral-combined_k141_321344\|\|lt2gene | Not-determined |
| 17-109-final-viral-combined_k141_388339\|\|lt2gene | Not-determined |
| 17-109-final-viral-combined_k141_204254\|\|lt2gene | Not-determined |
| 17-109-final-viral-combined_k141_137331\|\|lt2gene | Not-determined |
| 17-109-final-viral-combined_k141_19810\|\|lt2gene | Low-quality |
| 17-109-final-viral-combined_k141_270946\|\|lt2gene | Low-quality |
| 17-109-final-viral-combined_k141_103512\|\|lt2gene | Low-quality |
| 17-109-final-viral-combined_k141_237665\|\|lt2gene | Low-quality |
| 17-109-final-viral-combined_k141_321381\|\|lt2gene | Low-quality |
| 17-109-final-viral-combined_k141_304964\|\|lt2gene | Low-quality |
| 17-109-final-viral-combined_k141_304982\|\|lt2gene | Low-quality |
| 17-109-final-viral-combined_k141_354651\|\|lt2gene | Not-determined |
| 17-109-final-viral-combined_k141_154058\|\|lt2gene | Low-quality |
| 17-109-final-viral-combined_k141_270981\|\|lt2gene | Not-determined |
| 17-109-final-viral-combined_k141_137352\|\|lt2gene | Not-determined |
| 17-109-final-viral-combined_k141_337649\|\|lt2gene | Low-quality |
| 17-109-final-viral-combined_k141_388404\|\|lt2gene | Not-determined |
| 17-109-final-viral-combined_k141_19910\|\|lt2gene | Low-quality |
| 17-109-final-viral-combined_k141_337662\|\|lt2gene | Low-quality |
| 17-109-final-viral-combined_k141_154100\|\|lt2gene | Low-quality |
| 17-109-final-viral-combined_k141_305100\|\|lt2gene | Low-quality |
| 17-109-final-viral-combined_k141_254098\|\|lt2gene | Low-quality |
| 17-109-final-viral-combined_k141_154155\|\|lt2gene | Low-quality |
| 17-109-final-viral-combined_k141_305157\|\|lt2gene | Low-quality |
| 17-109-final-viral-combined_k141_237750\|\|lt2gene | Low-quality |
| 17-109-final-viral-combined_k141_371898\|\|lt2gene | Not-determined |
| 17-109-final-viral-combined_k141_70846\|\|lt2gene | Not-determined |
| 17-109-final-viral-combined_k141_37249\|\|lt2gene | Low-quality |
| 17-109-final-viral-combined_k141_20016\|\|lt2gene | Not-determined |
| 17-109-final-viral-combined_k141_3700\|\|lt2gene | Low-quality |
| 17-109-final-viral-combined_k141_87145\|\|lt2gene | Low-quality |
| 17-109-final-viral-combined_k141_371952\|\|lt2gene | Low-quality |
| 17-109-final-viral-combined_k141_254159\|\|lt2gene | Not-determined |
| 17-109-final-viral-combined_k141_220871\|\|lt2gene | Low-quality |
| 17-109-final-viral-combined_k141_354713\|\|lt2gene | Low-quality |
| 17-109-final-viral-combined_k141_53617\|\|lt2gene | Low-quality |
| 17-109-final-viral-combined_k141_137492\|\|lt2gene | Not-determined |
| 17-109-final-viral-combined_k141_237799\|\|lt2gene | Low-quality |
| 17-109-final-viral-combined_k141_120230\|\|lt2gene | Low-quality |
| 17-109-final-viral-combined_k141_288330\|\|lt2gene | Low-quality |
| 17-109-final-viral-combined_k141_372015\|\|lt2gene | Low-quality |
| 17-109-final-viral-combined_k141_337838\|\|lt2gene | Not-determined |
| 17-109-final-viral-combined_k141_3731\|\|lt2gene | Not-determined |
| 17-109-final-viral-combined_k141_220903\|\|lt2gene | Low-quality |
| 17-109-final-viral-combined_k141_220920\|\|lt2gene | Low-quality |
| 17-109-final-viral-combined_k141_288393\|\|lt2gene | Low-quality |
| 17-109-final-viral-combined_k141_20158\|\|lt2gene | Not-determined |
| 17-109-final-viral-combined_k141_337949\|\|lt2gene | Low-quality |
| [illegible] | [illegible] |

17-109-final-viral-combined_k141_174203||lt2gene Low-quality
17-109-final-viral-combined_k141_191767||lt2gene Low-quality
17-109-final-viral-combined_k141_123443||lt2gene Low-quality
17-109-final-viral-combined_k141_257765||lt2gene Low-quality
17-109-final-viral-combined_k141_257775||lt2gene Low-quality
17-109-final-viral-combined_k141_207804||lt2gene Not-determined
17-109-final-viral-combined_k141_292156||lt2gene Low-quality
17-109-final-viral-combined_k141_292166||lt2gene Low-quality
17-109-final-viral-combined_k141_325201||lt2gene Low-quality
17-109-final-viral-combined_k141_292201||lt2gene Not-determined
17-109-final-viral-combined_k141_56507||lt2gene Low-quality
17-109-final-viral-combined_k141_341091||lt2gene Low-quality
17-109-final-viral-combined_k141_241161||lt2gene Not-determined
17-109-final-viral-combined_k141_325287||lt2gene Low-quality
17-109-final-viral-combined_k141_224654||lt2gene Not-determined
17-109-final-viral-combined_k141_123595||lt2gene Low-quality
17-109-final-viral-combined_k141_6811||lt2gene Low-quality
17-109-final-viral-combined_k141_274912||lt2gene Not-determined
17-109-final-viral-combined_k141_40727||lt2gene Low-quality
17-109-final-viral-combined_k141_56653||lt2gene Not-determined
17-109-final-viral-combined_k141_107243||lt2gene Low-quality
17-109-final-viral-combined_k141_56655||lt2gene Low-quality
17-109-final-viral-combined_k141_6895||lt2gene Low-quality
17-109-final-viral-combined_k141_257993||lt2gene Low-quality
17-109-final-viral-combined_k141_274967||lt2gene Low-quality
17-109-final-viral-combined_k141_274980||lt2gene Not-determined
17-109-final-viral-combined_k141_74655||lt2gene Low-quality
17-109-final-viral-combined_k141_6942||lt2gene Low-quality
17-109-final-viral-combined_k141_191941||lt2gene Not-determined
17-109-final-viral-combined_k141_23766||lt2gene Not-determined
17-109-final-viral-combined_k141_191980||lt2gene Low-quality
17-109-final-viral-combined_k141_309166||lt2gene Low-quality
17-109-final-viral-combined_k141_341319||lt2gene Low-quality
17-109-final-viral-combined_k141_375598||lt2gene Not-determined
17-109-final-viral-combined_k141_375640||lt2gene Low-quality
17-109-final-viral-combined_k141_141101||lt2gene Low-quality
17-109-final-viral-combined_k141_192011||lt2gene Low-quality
17-109-final-viral-combined_k141_141107||lt2gene Not-determined
17-109-final-viral-combined_k141_74748||lt2gene Low-quality
17-109-final-viral-combined_k141_158142||lt2gene Not-determined
17-109-final-viral-combined_k141_375682||lt2gene Low-quality
17-109-final-viral-combined_k141_40869||lt2gene Low-quality
17-109-final-viral-combined_k141_192076||lt2gene Low-quality
17-109-final-viral-combined_k141_40891||lt2gene Low-quality
17-109-final-viral-combined_k141_309233||lt2gene Low-quality
17-109-final-viral-combined_k141_56851||lt2gene Low-quality
17-109-final-viral-combined_k141_275130||lt2gene Not-determined
17-109-final-viral-combined_k141_275137||lt2gene Low-quality
17-109-final-viral-combined_k141_258056||lt2gene Low-quality
17-109-final-viral-combined_k141_325484||lt2gene Low-quality
17-109-final-viral-combined_k141_192127||lt2gene Not-determined
17-109-final-viral-combined_k141_192132||lt2gene Low-quality
17-109-final-viral-combined_k141_325524||lt2gene Low-quality
17-109-final-viral-combined_k141_309331||lt2gene Low-quality
17-109-final-viral-combined_k141_74830||lt2gene Low-quality
17-109-final-viral-combined_k141_123874||lt2gene Low-quality
17-109-final-viral-combined_k141_74852||lt2gene Low-quality
17-109-final-viral-combined_k141_90632||lt2gene Low-quality
17-109-final-viral-combined_k141_375753||lt2gene Low-quality
17-109-final-viral-combined_k141_41001||lt2gene Low-quality
17-109-final-viral-combined_k141_141239||lt2gene Low-quality
17-109-final-viral-combined_k141_275243||lt2gene Low-quality
17-109-final-viral-combined_k141_74874||lt2gene Low-quality
17-109-final-viral-combined_k141_375796||lt2gene Low-quality
17-109-final-viral-combined_k141_375797||lt2gene Low-quality
17-109-final-viral-combined_k141_392590||lt2gene Low-quality
17-109-final-viral-combined_k141_341478||lt2gene Not-determined
17-109-final-viral-combined_k141_325575||lt2gene Not-determined
17-109-final-viral-combined_k141_174550||lt2gene Low-quality
17-109-final-viral-combined_k141_192189||lt2gene Low-quality
17-109-final-viral-combined_k141_309467||lt2gene Low-quality
17-109-final-viral-combined_k141_41071||lt2gene Low-quality
17-109-final-viral-combined_k141_309481||lt2gene Not-determined
17-109-final-viral-combined_k141_24050||lt2gene Low-quality
17-109-final-viral-combined_k141_258218||lt2gene Not-determined
17-109-final-viral-combined_k141_174603||lt2gene Not-determined
17-109-final-viral-combined_k141_90735||lt2gene Low-quality
17-109-final-viral-combined_k141_141305||lt2gene Not-determined
17-109-final-viral-combined_k141_158420||lt2gene Low-quality
17-109-final-viral-combined_k141_24097||lt2gene Not-determined
17-109-final-viral-combined_k141_241682||lt2gene Low-quality
17-109-final-viral-combined_k141_7101||lt2gene Low-quality
17-109-final-viral-combined_k141_158464||lt2gene Not-determined
17-109-final-viral-combined_k141_375935||lt2gene Not-determined
17-109-final-viral-combined_k141_275417||lt2gene Not-determined
17-109-final-viral-combined_k141_241730||lt2gene Low-quality
17-109-final-viral-combined_k141_123893||lt2gene Low-quality
17-109-final-viral-combined_k141_123894||lt2gene Low-quality
17-109-final-viral-combined_k141_24139||lt2gene Not-determined
17-109-final-viral-combined_k141_174669||lt2gene Low-quality
17-109-final-viral-combined_k141_90784||lt2gene Not-determined
17-109-final-viral-combined_k141_341670||lt2gene Low-quality
17-109-final-viral-combined_k141_225049||lt2gene Not-determined
17-109-final-viral-combined_k141_158550||lt2gene Not-determined
17-109-final-viral-combined_k141_208239||lt2gene Low-quality
17-109-final-viral-combined_k141_192340||lt2gene Low-quality
17-109-final-viral-combined_k141_225075||lt2gene Not-determined
17-109-final-viral-combined_k141_258337||lt2gene Low-quality
17-109-final-viral-combined_k141_24255||lt2gene Low-quality
17-109-final-viral-combined_k141_258353||lt2gene Not-determined
17-109-final-viral-combined_k141_57150||lt2gene Low-quality
17-109-final-viral-combined_k141_57167||lt2gene Low-quality
17-109-final-viral-combined_k141_7297||lt2gene Low-quality
17-109-final-viral-combined_k141_292626||lt2gene Low-quality
17-109-final-viral-combined_k141_225187||lt2gene Low-quality
17-109-final-viral-combined_k141_208326||lt2gene Low-quality
17-109-final-viral-combined_k141_141573||lt2gene Low-quality
17-109-final-viral-combined_k141_376050||lt2gene Not-determined
17-109-final-viral-combined_k141_392846||lt2gene Low-quality
17-109-final-viral-combined_k141_141646||lt2gene Low-quality
17-109-final-viral-combined_k141_241996||lt2gene Low-quality
17-109-final-viral-combined_k141_41281||lt2gene Low-quality
17-109-final-viral-combined_k141_90970||lt2gene Low-quality
17-109-final-viral-combined_k141_7377||lt2gene Not-determined
17-109-final-viral-combined_k141_158651||lt2gene Low-quality
17-109-final-viral-combined_k141_158657||lt2gene Low-quality
17-109-final-viral-combined_k141_41321||lt2gene Low-quality
17-109-final-viral-combined_k141_242038||lt2gene Low-quality
17-109-final-viral-combined_k141_124001||lt2gene Low-quality
17-109-final-viral-combined_k141_174841||lt2gene Low-quality
17-109-final-viral-combined_k141_91045||lt2gene Low-quality
17-109-final-viral-combined_k141_225349||lt2gene Not-determined
17-109-final-viral-combined_k141_124015||lt2gene Low-quality
17-109-final-viral-combined_k141_41416||lt2gene Not-determined
17-109-final-viral-combined_k141_225374||lt2gene Low-quality
17-109-final-viral-combined_k141_309926||lt2gene Low-quality
17-109-final-viral-combined_k141_107742||lt2gene Low-quality
17-109-final-viral-combined_k141_292879||lt2gene Low-quality
17-109-final-viral-combined_k141_7509||lt2gene Not-determined
17-109-final-viral-combined_k141_292884||lt2gene Not-determined
17-109-final-viral-combined_k141_141736||lt2gene Low-quality
17-109-final-viral-combined_k141_174890||lt2gene Low-quality
17-109-final-viral-combined_k141_124064||lt2gene Low-quality
17-109-final-viral-combined_k141_208452||lt2gene Low-quality
17-109-final-viral-combined_k141_309999||lt2gene Low-quality
17-109-final-viral-combined_k141_341960||lt2gene Low-quality
17-109-final-viral-combined_k141_275802||lt2gene Not-determined
17-109-final-viral-combined_k141_75103||lt2gene Low-quality
17-109-final-viral-combined_k141_7567||lt2gene Low-quality
17-109-final-viral-combined_k141_57464||lt2gene Low-quality
17-109-final-viral-combined_k141_174913||lt2gene Low-quality
17-109-final-viral-combined_k141_174924||lt2gene Low-quality
17-109-final-viral-combined_k141_75133||lt2gene Low-quality
17-109-final-viral-combined_k141_275829||lt2gene Low-quality
17-109-final-viral-combined_k141_292984||lt2gene Not-determined
17-109-final-viral-combined_k141_325978||lt2gene Low-quality
17-109-final-viral-combined_k141_310074||lt2gene Low-quality
17-109-final-viral-combined_k141_41484||lt2gene Not-determined
17-109-final-viral-combined_k141_376262||lt2gene Low-quality
17-109-final-viral-combined_k141_293003||lt2gene Low-quality
17-109-final-viral-combined_k141_124159||lt2gene Not-determined
17-109-final-viral-combined_k141_325998||lt2gene Low-quality
17-109-final-viral-combined_k141_258718||lt2gene Not-determined
17-109-final-viral-combined_k141_41529||lt2gene Low-quality
17-109-final-viral-combined_k141_358187||lt2gene Low-quality
17-109-final-viral-combined_k141_91097||lt2gene Low-quality
17-109-final-viral-combined_k141_7656||lt2gene Low-quality
17-109-final-viral-combined_k141_258741||lt2gene Not-determined
17-109-final-viral-combined_k141_24497||lt2gene Low-quality
17-109-final-viral-combined_k141_342076||lt2gene Low-quality
17-109-final-viral-combined_k141_158849||lt2gene Low-quality
17-109-final-viral-combined_k141_192731||lt2gene Not-determined
17-109-final-viral-combined_k141_57533||lt2gene Low-quality
17-109-final-viral-combined_k141_7684||lt2gene Low-quality
17-109-final-viral-combined_k141_141917||lt2gene Low-quality
17-109-final-viral-combined_k141_175024||lt2gene Low-quality
17-109-final-viral-combined_k141_175027||lt2gene Low-quality
17-109-final-viral-combined_k141_275972||lt2gene Low-quality
17-109-final-viral-combined_k141_393123||lt2gene Low-quality
17-109-final-viral-combined_k141_393125||lt2gene Low-quality
17-109-final-viral-combined_k141_326105||lt2gene Low-quality
17-109-final-viral-combined_k141_342190||lt2gene Not-determined
17-109-final-viral-combined_k141_175098||lt2gene Low-quality
17-109-final-viral-combined_k141_258811||lt2gene Low-quality
17-109-final-viral-combined_k141_175101||lt2gene Low-quality
17-109-final-viral-combined_k141_326188||lt2gene Not-determined
17-109-final-viral-combined_k141_41705||lt2gene Not-determined
17-109-final-viral-combined_k141_142035||lt2gene Low-quality
17-109-final-viral-combined_k141_358362||lt2gene Low-quality
17-109-final-viral-combined_k141_242387||lt2gene Low-quality
17-109-final-viral-combined_k141_24683||lt2gene Low-quality
17-109-final-viral-combined_k141_208660||lt2gene Low-quality
17-109-final-viral-combined_k141_41752||lt2gene Not-determined
17-109-final-viral-combined_k141_258827||lt2gene Low-quality
17-109-final-viral-combined_k141_242433||lt2gene Not-determined
17-109-final-viral-combined_k141_124369||lt2gene Low-quality
17-109-final-viral-combined_k141_242464||lt2gene Low-quality
17-109-final-viral-combined_k141_124384||lt2gene Low-quality
17-109-final-viral-combined_k141_193011||lt2gene Low-quality
17-109-final-viral-combined_k141_258938||lt2gene Low-quality
17-109-final-viral-combined_k141_242484||lt2gene Low-quality
17-109-final-viral-combined_k141_358423||lt2gene Not-determined
17-109-final-viral-combined_k141_91305||lt2gene Low-quality
17-109-final-viral-combined_k141_225759||lt2gene Low-quality
17-109-final-viral-combined_k141_326294||lt2gene Low-quality
17-109-final-viral-combined_k141_393338||lt2gene Not-determined
17-109-final-viral-combined_k141_225793||lt2gene Low-quality
17-109-final-viral-combined_k141_342425||lt2gene Low-quality
17-109-final-viral-combined_k141_24840||lt2gene Low-quality
17-109-final-viral-combined_k141_142190||lt2gene Not-determined
17-109-final-viral-combined_k141_310347||lt2gene Low-quality
17-109-final-viral-combined_k141_41820||lt2gene Not-determined
17-109-final-viral-combined_k141_376455||lt2gene Low-quality
17-109-final-viral-combined_k141_41832||lt2gene Low-quality
17-109-final-viral-combined_k141_393372||lt2gene Low-quality
17-109-final-viral-combined_k141_208775||lt2gene Not-determined
17-109-final-viral-combined_k141_310407||lt2gene Low-quality
17-109-final-viral-combined_k141_342474||lt2gene Not-determined
17-109-final-viral-combined_k141_342475||lt2gene Low-quality
17-109-final-viral-combined_k141_376498||lt2gene Not-determined
17-109-final-viral-combined_k141_108093||lt2gene Low-quality
17-109-final-viral-combined_k141_108095||lt2gene Low-quality
17-109-final-viral-combined_k141_326372||lt2gene Not-determined
17-109-final-viral-combined_k141_393412||lt2gene Low-quality
17-109-final-viral-combined_k141_310437||lt2gene Not-determined
17-109-final-viral-combined_k141_41892||lt2gene Not-determined
17-109-final-viral-combined_k141_342488||lt2gene Low-quality
17-109-final-viral-combined_k141_8063||lt2gene Not-determined
17-109-final-viral-combined_k141_159083||lt2gene Not-determined
17-109-final-viral-combined_k141_225901||lt2gene Low-quality
17-109-final-viral-combined_k141_326411||lt2gene Not-determined
17-109-final-viral-combined_k141_41941||lt2gene Not-determined
17-109-final-viral-combined_k141_208838||lt2gene Low-quality
17-109-final-viral-combined_k141_91390||lt2gene Not-determined
17-109-final-viral-combined_k141_208853||lt2gene Low-quality
17-109-final-viral-combined_k141_393488||lt2gene Low-quality
17-109-final-viral-combined_k141_142286||lt2gene Low-quality
17-109-final-viral-combined_k141_57970||lt2gene Low-quality
17-109-final-viral-combined_k141_393500||lt2gene Not-determined
17-109-final-viral-combined_k141_108186||lt2gene Low-quality
17-109-final-viral-combined_k141_310542||lt2gene Low-quality
17-109-final-viral-combined_k141_208917||lt2gene Low-quality
17-109-final-viral-combined_k141_57996||lt2gene Low-quality
17-109-final-viral-combined_k141_142365||lt2gene Not-determined
17-109-final-viral-combined_k141_42037||lt2gene Not-determined
17-109-final-viral-combined_k141_326533||lt2gene Not-determined
17-109-final-viral-combined_k141_242681||lt2gene Not-determined
17-109-final-viral-combined_k141_310597||lt2gene Not-determined
17-109-final-viral-combined_k141_310603||lt2gene Not-determined
17-109-final-viral-combined_k141_326582||lt2gene Low-quality
17-109-final-viral-combined_k141_342673||lt2gene Low-quality
17-109-final-viral-combined_k141_209031||lt2gene Low-quality
17-109-final-viral-combined_k141_42068||lt2gene Not-determined
17-109-final-viral-combined_k141_293636||lt2gene Low-quality
17-109-final-viral-combined_k141_58197||lt2gene Low-quality
17-109-final-viral-combined_k141_342752||lt2gene Low-quality
17-109-final-viral-combined_k141_108357||lt2gene Not-determined
17-109-final-viral-combined_k141_193500||lt2gene Low-quality
17-109-final-viral-combined_k141_108393||lt2gene Not-determined
17-109-final-viral-combined_k141_209167||lt2gene Low-quality
17-109-final-viral-combined_k141_159355||lt2gene Low-quality
17-109-final-viral-combined_k141_75798||lt2gene Low-quality
17-109-final-viral-combined_k141_159369||lt2gene Not-determined
17-109-final-viral-combined_k141_58276||lt2gene Low-quality
17-109-final-viral-combined_k141_242784||lt2gene Low-quality
17-109-final-viral-combined_k141_242819||lt2gene Low-quality
17-109-final-viral-combined_k141_242824||lt2gene Low-quality
17-109-final-viral-combined_k141_42319||lt2gene Low-quality
17-109-final-viral-combined_k141_310786||lt2gene Not-determined
17-109-final-viral-combined_k141_108522||lt2gene Not-determined
17-109-final-viral-combined_k141_8467||lt2gene Low-quality
17-109-final-viral-combined_k141_242853||lt2gene Low-quality
17-109-final-viral-combined_k141_142717||lt2gene Low-quality
17-109-final-viral-combined_k141_310809||lt2gene Low-quality
17-109-final-viral-combined_k141_342863||lt2gene Low-quality
17-109-final-viral-combined_k141_259443||lt2gene Not-determined
17-109-final-viral-combined_k141_193657||lt2gene Not-determined
17-109-final-viral-combined_k141_276708||lt2gene Not-determined
17-109-final-viral-combined_k141_226393||lt2gene Low-quality
17-109-final-viral-combined_k141_58388||lt2gene Low-quality
17-109-final-viral-combined_k141_242954||lt2gene Low-quality
17-109-final-viral-combined_k141_342926||lt2gene Low-quality
17-109-final-viral-combined_k141_175770||lt2gene Not-determined
17-109-final-viral-combined_k141_342937||lt2gene Low-quality
17-109-final-viral-combined_k141_159482||lt2gene Low-quality
17-109-final-viral-combined_k141_58414||lt2gene Low-quality
17-109-final-viral-combined_k141_75997||lt2gene Low-quality
17-109-final-viral-combined_k141_393874||lt2gene Not-determined
17-109-final-viral-combined_k141_159545||lt2gene Low-quality
17-109-final-viral-combined_k141_125048||lt2gene Not-determined
17-109-final-viral-combined_k141_209327||lt2gene Low-quality
17-109-final-viral-combined_k141_358969||lt2gene Low-quality
17-109-final-viral-combined_k141_91844||lt2gene Low-quality
17-109-final-viral-combined_k141_8588||lt2gene Not-determined
17-109-final-viral-combined_k141_376958||lt2gene Low-quality
17-109-final-viral-combined_k141_276791||lt2gene Not-determined
17-109-final-viral-combined_k141_359013||lt2gene Low-quality
17-109-final-viral-combined_k141_209388||lt2gene Low-quality
17-109-final-viral-combined_k141_125168||lt2gene Not-determined
17-109-final-viral-combined_k141_76086||lt2gene Low-quality
17-109-final-viral-combined_k141_376968||lt2gene Low-quality
17-109-final-viral-combined_k141_25520||lt2gene Low-quality
17-109-final-viral-combined_k141_175937||lt2gene Low-quality
17-109-final-viral-combined_k141_327151||lt2gene Not-determined
17-109-final-viral-combined_k141_226648||lt2gene Low-quality
17-109-final-viral-combined_k141_91889||lt2gene Low-quality
17-109-final-viral-combined_k141_143038||lt2gene Low-quality
17-109-final-viral-combined_k141_8683||lt2gene Low-quality
17-109-final-viral-combined_k141_343145||lt2gene Not-determined
17-109-final-viral-combined_k141_42623||lt2gene Not-determined
17-109-final-viral-combined_k141_25605||lt2gene Low-quality
17-109-final-viral-combined_k141_76280||lt2gene Low-quality
17-109-final-viral-combined_k141_327274||lt2gene Low-quality
17-109-final-viral-combined_k141_276929||lt2gene Not-determined
17-109-final-viral-combined_k141_394039||lt2gene Not-determined
17-109-final-viral-combined_k141_276938||lt2gene Low-quality
17-109-final-viral-combined_k141_58640||lt2gene Low-quality
17-109-final-viral-combined_k141_76331||lt2gene Low-quality
17-109-final-viral-combined_k141_343256||lt2gene Low-quality
17-109-final-viral-combined_k141_76365||lt2gene Low-quality
17-109-final-viral-combined_k141_58706||lt2gene Low-quality
17-109-final-viral-combined_k141_125260||lt2gene Low-quality
17-109-final-viral-combined_k141_259624||lt2gene Low-quality
17-109-final-viral-combined_k141_109002||lt2gene Low-quality
17-109-final-viral-combined_k141_243197||lt2gene Not-determined
17-109-final-viral-combined_k141_311249||lt2gene Low-quality
17-109-final-viral-combined_k141_92070||lt2gene Not-determined
17-109-final-viral-combined_k141_377209||lt2gene Low-quality
17-109-final-viral-combined_k141_25744||lt2gene Not-determined
17-109-final-viral-combined_k141_76412||lt2gene Low-quality
17-109-final-viral-combined_k141_359117||lt2gene Low-quality
17-109-final-viral-combined_k141_194060||lt2gene Low-quality
17-109-final-viral-combined_k141_359121||lt2gene Low-quality
17-109-final-viral-combined_k141_243249||lt2gene Low-quality
17-109-final-viral-combined_k141_194075||lt2gene Low-quality
17-109-final-viral-combined_k141_76443||lt2gene Low-quality
17-109-final-viral-combined_k141_42777||lt2gene Low-quality
17-109-final-viral-combined_k141_377257||lt2gene Low-quality
17-109-final-viral-combined_k141_109049||lt2gene Low-quality
17-109-final-viral-combined_k141_159847||lt2gene Not-determined
17-109-final-viral-combined_k141_42807||lt2gene Not-determined
17-109-final-viral-combined_k141_159860||lt2gene Not-determined
17-109-final-viral-combined_k141_243330||lt2gene Low-quality
17-109-final-viral-combined_k141_209761||lt2gene Low-quality
17-109-final-viral-combined_k141_25817||lt2gene Low-quality
17-109-final-viral-combined_k141_327482||lt2gene Low-quality
17-109-final-viral-combined_k141_243346||lt2gene Low-quality
17-109-final-viral-combined_k141_311331||lt2gene Not-determined
17-109-final-viral-combined_k141_176191||lt2gene Low-quality
17-109-final-viral-combined_k141_243423||lt2gene Not-determined
17-109-final-viral-combined_k141_277186||lt2gene Low-quality
17-109-final-viral-combined_k141_327597||lt2gene Low-quality
17-109-final-viral-combined_k141_327617||lt2gene Low-quality
17-109-final-viral-combined_k141_243440||lt2gene Low-quality
17-109-final-viral-combined_k141_277206||lt2gene Low-quality
17-109-final-viral-combined_k141_277215||lt2gene Low-quality
17-109-final-viral-combined_k141_343520||lt2gene Low-quality
17-109-final-viral-combined_k141_277230||lt2gene Low-quality
17-109-final-viral-combined_k141_277236||lt2gene Low-quality
17-109-final-viral-combined_k141_76657||lt2gene Not-determined
17-109-final-viral-combined_k141_160026||lt2gene Not-determined
17-109-final-viral-combined_k141_243500||lt2gene Low-quality
17-109-final-viral-combined_k141_343600||lt2gene Low-quality
17-109-final-viral-combined_k141_42972||lt2gene Not-determined
17-109-final-viral-combined_k141_343611||lt2gene Not-determined
17-109-final-viral-combined_k141_294418||lt2gene Not-determined
17-109-final-viral-combined_k141_343617||lt2gene Not-determined
17-109-final-viral-combined_k141_210003||lt2gene Low-quality
17-109-final-viral-combined_k141_125410||lt2gene Low-quality
17-109-final-viral-combined_k141_76724||lt2gene Low-quality
17-109-final-viral-combined_k141_359277||lt2gene Low-quality
17-109-final-viral-combined_k141_294427||lt2gene Not-determined
17-109-final-viral-combined_k141_210049||lt2gene Low-quality
17-109-final-viral-combined_k141_277326||lt2gene Low-quality
17-109-final-viral-combined_k141_343667||lt2gene Low-quality
17-109-final-viral-combined_k141_343690||lt2gene Low-quality
17-109-final-viral-combined_k141_243596||lt2gene Not-determined
17-109-final-viral-combined_k141_9131||lt2gene Low-quality
17-109-final-viral-combined_k141_43063||lt2gene Low-quality
17-109-final-viral-combined_k141_26026||lt2gene Low-quality
17-109-final-viral-combined_k141_109395||lt2gene Low-quality
17-109-final-viral-combined_k141_243642||lt2gene Low-quality
17-109-final-viral-combined_k141_277378||lt2gene Low-quality
17-109-final-viral-combined_k141_160204||lt2gene Low-quality
17-109-final-viral-combined_k141_26031||lt2gene Low-quality
17-109-final-viral-combined_k141_143157||lt2gene Not-determined
17-109-final-viral-combined_k141_343801||lt2gene Low-quality
17-109-final-viral-combined_k141_227320||lt2gene Low-quality
17-109-final-viral-combined_k141_194290||lt2gene Low-quality
17-109-final-viral-combined_k141_92531||lt2gene Not-determined
17-109-final-viral-combined_k141_294573||lt2gene Not-determined
17-109-final-viral-combined_k141_194307||lt2gene Low-quality
17-109-final-viral-combined_k141_210228||lt2gene Low-quality
17-109-final-viral-combined_k141_210232||lt2gene Low-quality
17-109-final-viral-combined_k141_294608||lt2gene Low-quality
17-109-final-viral-combined_k141_294614||lt2gene Low-quality
17-109-final-viral-combined_k141_26176||lt2gene Low-quality
17-109-final-viral-combined_k141_92609||lt2gene Low-quality
17-109-final-viral-combined_k141_76888||lt2gene Low-quality
17-109-final-viral-combined_k141_76926||lt2gene Not-determined
17-109-final-viral-combined_k141_176635||lt2gene Low-quality
17-109-final-viral-combined_k141_125568||lt2gene Not-determined
17-109-final-viral-combined_k141_343977||lt2gene Low-quality
17-109-final-viral-combined_k141_194453||lt2gene Low-quality
17-109-final-viral-combined_k141_344006||lt2gene Low-quality
17-109-final-viral-combined_k141_277576||lt2gene Not-determined
17-109-final-viral-combined_k141_277577||lt2gene Not-determined
17-109-final-viral-combined_k141_327826||lt2gene Low-quality
17-109-final-viral-combined_k141_9176||lt2gene Not-determined
17-109-final-viral-combined_k141_109572||lt2gene Low-quality
17-109-final-viral-combined_k141_260072||lt2gene Low-quality
17-109-final-viral-combined_k141_125597||lt2gene Low-quality
17-109-final-viral-combined_k141_277628||lt2gene Low-quality
17-109-final-viral-combined_k141_394438||lt2gene Low-quality
17-109-final-viral-combined_k141_260115||lt2gene Not-determined
17-109-final-viral-combined_k141_194514||lt2gene Low-quality
17-109-final-viral-combined_k141_277652||lt2gene Low-quality
17-109-final-viral-combined_k141_176770||lt2gene Not-determined
17-109-final-viral-combined_k141_160512||lt2gene Not-determined
17-109-final-viral-combined_k141_260174||lt2gene Low-quality
17-109-final-viral-combined_k141_43484||lt2gene Low-quality
17-109-final-viral-combined_k141_77189||lt2gene Low-quality
17-109-final-viral-combined_k141_277763||lt2gene Low-quality
17-109-final-viral-combined_k141_77237||lt2gene Not-determined
17-109-final-viral-combined_k141_92765||lt2gene Low-quality
17-109-final-viral-combined_k141_359752||lt2gene Low-quality
17-109-final-viral-combined_k141_92821||lt2gene Low-quality
17-109-final-viral-combined_k141_59447||lt2gene Not-determined
17-109-final-viral-combined_k141_294943||lt2gene Not-determined
17-109-final-viral-combined_k141_160600||lt2gene Low-quality
17-109-final-viral-combined_k141_92833||lt2gene Low-quality
17-109-final-viral-combined_k141_327995||lt2gene Low-quality
17-109-final-viral-combined_k141_312208||lt2gene Low-quality
17-109-final-viral-combined_k141_394689||lt2gene Not-determined
17-109-final-viral-combined_k141_77345||lt2gene Not-determined
17-109-final-viral-combined_k141_378143||lt2gene Low-quality
17-109-final-viral-combined_k141_59532||lt2gene Low-quality
17-109-final-viral-combined_k141_143423||lt2gene Low-quality
17-109-final-viral-combined_k141_125847||lt2gene Not-determined
17-109-final-viral-combined_k141_227834||lt2gene Low-quality
17-109-final-viral-combined_k141_43755||lt2gene Low-quality
17-109-final-viral-combined_k141_77424||lt2gene Low-quality
17-109-final-viral-combined_k141_227848||lt2gene Not-determined
17-109-final-viral-combined_k141_59608||lt2gene Low-quality
17-109-final-viral-combined_k141_177051||lt2gene Low-quality
17-109-final-viral-combined_k141_77462||lt2gene Low-quality
17-109-final-viral-combined_k141_92965||lt2gene Not-determined
17-109-final-viral-combined_k141_359894||lt2gene Low-quality
17-109-final-viral-combined_k141_227902||lt2gene Not-determined
17-109-final-viral-combined_k141_328151||lt2gene Low-quality
17-109-final-viral-combined_k141_177083||lt2gene Low-quality
17-109-final-viral-combined_k141_328157||lt2gene Low-quality
17-109-final-viral-combined_k141_125926||lt2gene Low-quality
17-109-final-viral-combined_k141_243976||lt2gene Not-determined
17-109-final-viral-combined_k141_9573||lt2gene Not-determined
17-109-final-viral-combined_k141_328196||lt2gene Low-quality
17-109-final-viral-combined_k141_260471||lt2gene Low-quality
17-109-final-viral-combined_k141_93078||lt2gene Low-quality
17-109-final-viral-combined_k141_228007||lt2gene Low-quality
17-109-final-viral-combined_k141_278006||lt2gene Low-quality
17-109-final-viral-combined_k141_93132||lt2gene Low-quality
17-109-final-viral-combined_k141_59843||lt2gene Low-quality
17-109-final-viral-combined_k141_328266||lt2gene Low-quality
17-109-final-viral-combined_k141_93146||lt2gene Low-quality
17-109-final-viral-combined_k141_59863||lt2gene Not-determined
17-109-final-viral-combined_k141_143585||lt2gene Not-determined
17-109-final-viral-combined_k141_77643||lt2gene Low-quality
17-109-final-viral-combined_k141_244089||lt2gene Low-quality
17-109-final-viral-combined_k141_228075||lt2gene Low-quality
17-109-final-viral-combined_k141_77666||lt2gene Not-determined
17-109-final-viral-combined_k141_378334||lt2gene Low-quality
17-109-final-viral-combined_k141_210729||lt2gene Low-quality
17-109-final-viral-combined_k141_394950||lt2gene Low-quality
17-109-final-viral-combined_k141_93187||lt2gene Low-quality
17-109-final-viral-combined_k141_295286||lt2gene Low-quality
17-109-final-viral-combined_k141_210776||lt2gene Low-quality
17-109-final-viral-combined_k141_44084||lt2gene Low-quality
17-109-final-viral-combined_k141_93216||lt2gene Not-determined
17-109-final-viral-combined_k141_244123||lt2gene Low-quality
17-109-final-viral-combined_k141_177264||lt2gene Not-determined
17-109-final-viral-combined_k141_93235||lt2gene Low-quality
17-109-final-viral-combined_k141_344587||lt2gene Not-determined
17-109-final-viral-combined_k141_143662||lt2gene Not-determined
17-109-final-viral-combined_k141_44174||lt2gene Low-quality
17-109-final-viral-combined_k141_26976||lt2gene Low-quality
17-109-final-viral-combined_k141_177334||lt2gene Low-quality
17-109-final-viral-combined_k141_360157||lt2gene Not-determined
17-109-final-viral-combined_k141_210878||lt2gene Low-quality
17-109-final-viral-combined_k141_344650||lt2gene Not-determined
17-109-final-viral-combined_k141_126075||lt2gene Low-quality
17-109-final-viral-combined_k141_44213||lt2gene Not-determined
17-109-final-viral-combined_k141_378527||lt2gene Not-determined
17-109-final-viral-combined_k141_278182||lt2gene Not-determined
17-109-final-viral-combined_k141_177393||lt2gene Low-quality
17-109-final-viral-combined_k141_177394||lt2gene Low-quality
17-109-final-viral-combined_k141_395165||lt2gene Not-determined
17-109-final-viral-combined_k141_9708||lt2gene Low-quality
17-109-final-viral-combined_k141_60091||lt2gene Low-quality
17-109-final-viral-combined_k141_60106||lt2gene Low-quality
17-109-final-viral-combined_k141_9744||lt2gene Low-quality
17-109-final-viral-combined_k141_328504||lt2gene Low-quality
17-109-final-viral-combined_k141_77857||lt2gene Not-determined
17-109-final-viral-combined_k141_278354||lt2gene Low-quality
17-109-final-viral-combined_k141_126228||lt2gene Low-quality
17-109-final-viral-combined_k141_161095||lt2gene Low-quality
17-109-final-viral-combined_k141_260893||lt2gene Not-determined
17-109-final-viral-combined_k141_27040||lt2gene Not-determined
17-109-final-viral-combined_k141_210987||lt2gene Low-quality
17-109-final-viral-combined_k141_93445||lt2gene Low-quality
17-109-final-viral-combined_k141_228360||lt2gene Low-quality
17-109-final-viral-combined_k141_211013||lt2gene Low-quality
17-109-final-viral-combined_k141_9839||lt2gene Low-quality
17-109-final-viral-combined_k141_110106||lt2gene Not-determined
17-109-final-viral-combined_k141_228439||lt2gene Low-quality
17-109-final-viral-combined_k141_44447||lt2gene Low-quality
17-109-final-viral-combined_k141_93495||lt2gene Not-determined
17-109-final-viral-combined_k141_295407||lt2gene Not-determined
17-109-final-viral-combined_k141_378751||lt2gene Low-quality
17-109-final-viral-combined_k141_177489||lt2gene Low-quality
17-109-final-viral-combined_k141_312846||lt2gene Low-quality
17-109-final-viral-combined_k141_126294||lt2gene Low-quality
17-109-final-viral-combined_k141_60343||lt2gene Low-quality
17-109-final-viral-combined_k141_126306||lt2gene Low-quality
17-109-final-viral-combined_k141_295432||lt2gene Low-quality
17-109-final-viral-combined_k141_228480||lt2gene Low-quality
17-109-final-viral-combined_k141_77980||lt2gene Low-quality
17-109-final-viral-combined_k141_44521||lt2gene Not-determined
17-109-final-viral-combined_k141_126360||lt2gene Not-determined
17-109-final-viral-combined_k141_9961||lt2gene Low-quality
17-109-final-viral-combined_k141_211134||lt2gene Not-determined
17-109-final-viral-combined_k141_344953||lt2gene Low-quality
17-109-final-viral-combined_k141_228564||lt2gene Low-quality
17-109-final-viral-combined_k141_261006||lt2gene Low-quality
17-109-final-viral-combined_k141_144035||lt2gene Low-quality
17-109-final-viral-combined_k141_344977||lt2gene Low-quality
17-109-final-viral-combined_k141_378843||lt2gene Not-determined
17-109-final-viral-combined_k141_228608||lt2gene Low-quality
17-109-final-viral-combined_k141_27187||lt2gene Not-determined
17-109-final-viral-combined_k141_295510||lt2gene Not-determined
17-109-final-viral-combined_k141_328606||lt2gene Low-quality
17-109-final-viral-combined_k141_211251||lt2gene Low-quality
17-109-final-viral-combined_k141_244590||lt2gene Low-quality
17-109-final-viral-combined_k141_345050||lt2gene Not-determined
17-109-final-viral-combined_k141_93634||lt2gene Low-quality
17-109-final-viral-combined_k141_177728||lt2gene Low-quality
17-109-final-viral-combined_k141_144194||lt2gene Low-quality
17-109-final-viral-combined_k141_360394||lt2gene Low-quality
17-109-final-viral-combined_k141_360397||lt2gene Low-quality
17-109-final-viral-combined_k141_328741||lt2gene Not-determined
17-109-final-viral-combined_k141_261225||lt2gene Not-determined
17-109-final-viral-combined_k141_278851||lt2gene Low-quality
17-109-final-viral-combined_k141_93735||lt2gene Low-quality
17-109-final-viral-combined_k141_345194||lt2gene Low-quality
17-109-final-viral-combined_k141_313029||lt2gene Low-quality
17-109-final-viral-combined_k141_244681||lt2gene Not-determined
17-109-final-viral-combined_k141_228718||lt2gene Low-quality
17-109-final-viral-combined_k141_78217||lt2gene Low-quality
17-109-final-viral-combined_k141_244692||lt2gene Low-quality
17-109-final-viral-combined_k141_295648||lt2gene Not-determined
17-109-final-viral-combined_k141_278878||lt2gene Low-quality
17-109-final-viral-combined_k141_395550||lt2gene Low-quality
17-109-final-viral-combined_k141_211435||lt2gene Low-quality
17-109-final-viral-combined_k141_211436||lt2gene Low-quality
17-109-final-viral-combined_k141_278897||lt2gene Low-quality
17-109-final-viral-combined_k141_93777||lt2gene Low-quality
17-109-final-viral-combined_k141_44729||lt2gene Low-quality
17-109-final-viral-combined_k141_360510||lt2gene Not-determined
17-109-final-viral-combined_k141_126594||lt2gene Low-quality
17-109-final-viral-combined_k141_395592||lt2gene Not-determined
17-109-final-viral-combined_k141_144249||lt2gene Low-quality
17-109-final-viral-combined_k141_228806||lt2gene Low-quality
17-109-final-viral-combined_k141_44768||lt2gene Low-quality
17-109-final-viral-combined_k141_328841||lt2gene Low-quality
17-109-final-viral-combined_k141_144270||lt2gene Low-quality
17-109-final-viral-combined_k141_78326||lt2gene Not-determined
17-109-final-viral-combined_k141_144281||lt2gene Low-quality
17-109-final-viral-combined_k141_144283||lt2gene Low-quality
17-109-final-viral-combined_k141_144294||lt2gene Low-quality
17-109-final-viral-combined_k141_278999||lt2gene Low-quality
17-109-final-viral-combined_k141_126648||lt2gene Low-quality
17-109-final-viral-combined_k141_27538||lt2gene Not-determined
17-109-final-viral-combined_k141_244824||lt2gene Low-quality
17-109-final-viral-combined_k141_10204||lt2gene Low-quality
17-109-final-viral-combined_k141_93853||lt2gene Low-quality
17-109-final-viral-combined_k141_126693||lt2gene Low-quality
17-109-final-viral-combined_k141_27562||lt2gene Low-quality
17-109-final-viral-combined_k141_93914||lt2gene Low-quality
17-109-final-viral-combined_k141_379249||lt2gene Low-quality
17-109-final-viral-combined_k141_328956||lt2gene Low-quality
17-109-final-viral-combined_k141_93934||lt2gene Low-quality
17-109-final-viral-combined_k141_27622||lt2gene Not-determined
17-109-final-viral-combined_k141_328969||lt2gene Low-quality
17-109-final-viral-combined_k141_44846||lt2gene Low-quality
17-109-final-viral-combined_k141_93978||lt2gene Low-quality
17-109-final-viral-combined_k141_27658||lt2gene Low-quality
17-109-final-viral-combined_k141_228966||lt2gene Low-quality
17-109-final-viral-combined_k141_379336||lt2gene Low-quality
17-109-final-viral-combined_k141_345465||lt2gene Not-determined
17-109-final-viral-combined_k141_211707||lt2gene Low-quality

17-109-final-viral-combined_k141_126756||lt2gene Low-quality
17-109-final-viral-combined_k141_44909||lt2gene Low-quality
17-109-final-viral-combined_k141_110611||lt2gene Low-quality
17-109-final-viral-combined_k141_360696||lt2gene Low-quality
17-109-final-viral-combined_k141_44944||lt2gene Not-determined
17-109-final-viral-combined_k141_126830||lt2gene Low-quality
17-109-final-viral-combined_k141_360711||lt2gene Low-quality
17-109-final-viral-combined_k141_27679||lt2gene Low-quality
17-109-final-viral-combined_k141_211781||lt2gene Low-quality
17-109-final-viral-combined_k141_110650||lt2gene Low-quality
17-109-final-viral-combined_k141_94102||lt2gene Low-quality
17-109-final-viral-combined_k141_110674||lt2gene Low-quality
17-109-final-viral-combined_k141_10388||lt2gene Low-quality
17-109-final-viral-combined_k141_261579||lt2gene Low-quality
17-109-final-viral-combined_k141_211837||lt2gene Low-quality
17-109-final-viral-combined_k141_261589||lt2gene Low-quality
17-109-final-viral-combined_k141_78606||lt2gene Low-quality
17-109-final-viral-combined_k141_229209||lt2gene Low-quality
17-109-final-viral-combined_k141_45041||lt2gene Low-quality
17-109-final-viral-combined_k141_395929||lt2gene Not-determined
17-109-final-viral-combined_k141_45053||lt2gene Low-quality
17-109-final-viral-combined_k141_379573||lt2gene Low-quality
17-109-final-viral-combined_k141_27847||lt2gene Not-determined
17-109-final-viral-combined_k141_229272||lt2gene Low-quality
17-109-final-viral-combined_k141_360863||lt2gene Not-determined
17-109-final-viral-combined_k141_211870||lt2gene Not-determined
17-109-final-viral-combined_k141_211871||lt2gene Not-determined
17-109-final-viral-combined_k141_229297||lt2gene Low-quality
17-109-final-viral-combined_k141_345580||lt2gene Low-quality
17-109-final-viral-combined_k141_360894||lt2gene Low-quality
17-109-final-viral-combined_k141_161764||lt2gene Low-quality
17-109-final-viral-combined_k141_60918||lt2gene Low-quality
17-109-final-viral-combined_k141_10541||lt2gene Low-quality
17-109-final-viral-combined_k141_144490||lt2gene Low-quality
17-109-final-viral-combined_k141_127082||lt2gene Low-quality
17-109-final-viral-combined_k141_345613||lt2gene Low-quality
17-109-final-viral-combined_k141_313514||lt2gene Low-quality
17-109-final-viral-combined_k141_161815||lt2gene Not-determined
17-109-final-viral-combined_k141_211985||lt2gene Low-quality
17-109-final-viral-combined_k141_313529||lt2gene Low-quality
17-109-final-viral-combined_k141_195791||lt2gene Low-quality
17-109-final-viral-combined_k141_178155||lt2gene Not-determined
17-109-final-viral-combined_k141_229416||lt2gene Low-quality
17-109-final-viral-combined_k141_94262||lt2gene Low-quality
17-109-final-viral-combined_k141_261851||lt2gene Low-quality
17-109-final-viral-combined_k141_178214||lt2gene Not-determined
17-109-final-viral-combined_k141_178216||lt2gene Not-determined
17-109-final-viral-combined_k141_360995||lt2gene Low-quality
17-109-final-viral-combined_k141_178224||lt2gene Not-determined
17-109-final-viral-combined_k141_45245||lt2gene Low-quality
17-109-final-viral-combined_k141_379799||lt2gene Not-determined
17-109-final-viral-combined_k141_10619||lt2gene Not-determined
17-109-final-viral-combined_k141_161865||lt2gene Not-determined
17-109-final-viral-combined_k141_144588||lt2gene Low-quality
17-109-final-viral-combined_k141_127235||lt2gene Low-quality
17-109-final-viral-combined_k141_229547||lt2gene Low-quality
17-109-final-viral-combined_k141_212086||lt2gene Not-determined
17-109-final-viral-combined_k141_45277||lt2gene Low-quality
17-109-final-viral-combined_k141_27958||lt2gene Low-quality
17-109-final-viral-combined_k141_345743||lt2gene Low-quality
17-109-final-viral-combined_k141_345751||lt2gene Low-quality
17-109-final-viral-combined_k141_178275||lt2gene Low-quality
17-109-final-viral-combined_k141_195978||lt2gene Low-quality
17-109-final-viral-combined_k141_396211||lt2gene Low-quality
17-109-final-viral-combined_k141_10662||lt2gene Low-quality
17-109-final-viral-combined_k141_229602||lt2gene Not-determined
17-109-final-viral-combined_k141_161972||lt2gene Low-quality
17-109-final-viral-combined_k141_94404||lt2gene Low-quality
17-109-final-viral-combined_k141_313751||lt2gene Low-quality
17-109-final-viral-combined_k141_329347||lt2gene Low-quality
17-109-final-viral-combined_k141_127363||lt2gene Low-quality
17-109-final-viral-combined_k141_10713||lt2gene Low-quality
17-109-final-viral-combined_k141_45350||lt2gene Low-quality
17-109-final-viral-combined_k141_379940||lt2gene Low-quality
17-109-final-viral-combined_k141_10737||lt2gene Low-quality
17-109-final-viral-combined_k141_94417||lt2gene Low-quality
17-109-final-viral-combined_k141_10751||lt2gene Low-quality
17-109-final-viral-combined_k141_245479||lt2gene Low-quality
17-109-final-viral-combined_k141_94449||lt2gene Not-determined
17-109-final-viral-combined_k141_178454||lt2gene Low-quality
17-109-final-viral-combined_k141_45427||lt2gene Not-determined
17-109-final-viral-combined_k141_94494||lt2gene Not-determined
17-109-final-viral-combined_k141_10798||lt2gene Low-quality
17-109-final-viral-combined_k141_361233||lt2gene Low-quality
17-109-final-viral-combined_k141_127476||lt2gene Low-quality
17-109-final-viral-combined_k141_144748||lt2gene Low-quality
17-109-final-viral-combined_k141_262010||lt2gene Not-determined
17-109-final-viral-combined_k141_229776||lt2gene Low-quality
17-109-final-viral-combined_k141_380010||lt2gene Not-determined
17-109-final-viral-combined_k141_380019||lt2gene Low-quality
17-109-final-viral-combined_k141_262046||lt2gene Low-quality
17-109-final-viral-combined_k141_45524||lt2gene Low-quality
17-109-final-viral-combined_k141_345991||lt2gene Low-quality
17-109-final-viral-combined_k141_61289||lt2gene Low-quality
17-109-final-viral-combined_k141_10856||lt2gene Low-quality
17-109-final-viral-combined_k141_196160||lt2gene Low-quality
17-109-final-viral-combined_k141_10861||lt2gene Not-determined
17-109-final-viral-combined_k141_10862||lt2gene Not-determined
17-109-final-viral-combined_k141_144830||lt2gene Low-quality
17-109-final-viral-combined_k141_10873||lt2gene Low-quality
17-109-final-viral-combined_k141_162207||lt2gene Not-determined
17-109-final-viral-combined_k141_127599||lt2gene Low-quality
17-109-final-viral-combined_k141_61354||lt2gene Low-quality
17-109-final-viral-combined_k141_212380||lt2gene Low-quality
17-109-final-viral-combined_k141_61366||lt2gene Low-quality
17-109-final-viral-combined_k141_396544||lt2gene Low-quality
17-109-final-viral-combined_k141_296401||lt2gene Low-quality
17-109-final-viral-combined_k141_127679||lt2gene Low-quality
17-109-final-viral-combined_k141_212428||lt2gene Not-determined
17-109-final-viral-combined_k141_10985||lt2gene Not-determined
17-109-final-viral-combined_k141_178697||lt2gene Not-determined
17-109-final-viral-combined_k141_94713||lt2gene Low-quality
17-109-final-viral-combined_k141_144907||lt2gene Low-quality
17-109-final-viral-combined_k141_11008||lt2gene Low-quality
17-109-final-viral-combined_k141_162332||lt2gene Low-quality
17-109-final-viral-combined_k141_11019||lt2gene Low-quality
17-109-final-viral-combined_k141_313997||lt2gene Low-quality
17-109-final-viral-combined_k141_45688||lt2gene Not-determined
17-109-final-viral-combined_k141_396661||lt2gene Low-quality
17-109-final-viral-combined_k141_94725||lt2gene Low-quality
17-109-final-viral-combined_k141_111287||lt2gene Low-quality
17-109-final-viral-combined_k141_28081||lt2gene Not-determined
17-109-final-viral-combined_k141_296509||lt2gene Not-determined
17-109-final-viral-combined_k141_79151||lt2gene Not-determined
17-109-final-viral-combined_k141_79157||lt2gene Not-determined
17-109-final-viral-combined_k141_329689||lt2gene Low-quality
17-109-final-viral-combined_k141_144996||lt2gene Low-quality
17-109-final-viral-combined_k141_329699||lt2gene Low-quality
17-109-final-viral-combined_k141_361468||lt2gene Low-quality
17-109-final-viral-combined_k141_245838||lt2gene Not-determined
17-109-final-viral-combined_k141_262322||lt2gene Not-determined
17-109-final-viral-combined_k141_380230||lt2gene Not-determined
17-109-final-viral-combined_k141_162429||lt2gene Low-quality
17-109-final-viral-combined_k141_361522||lt2gene Not-determined
17-109-final-viral-combined_k141_127817||lt2gene Not-determined
17-109-final-viral-combined_k141_296561||lt2gene Not-determined
17-109-final-viral-combined_k141_11161||lt2gene Low-quality
17-109-final-viral-combined_k141_229955||lt2gene Low-quality
17-109-final-viral-combined_k141_262409||lt2gene Low-quality
17-109-final-viral-combined_k141_145140||lt2gene Low-quality
17-109-final-viral-combined_k141_145148||lt2gene Low-quality
17-109-final-viral-combined_k141_79270||lt2gene Not-determined
17-109-final-viral-combined_k141_79272||lt2gene Not-determined
17-109-final-viral-combined_k141_212683||lt2gene Low-quality
17-109-final-viral-combined_k141_79291||lt2gene Low-quality
17-109-final-viral-combined_k141_280154||lt2gene Not-determined
17-109-final-viral-combined_k141_262475||lt2gene Low-quality
17-109-final-viral-combined_k141_45787||lt2gene Low-quality
17-109-final-viral-combined_k141_145206||lt2gene Low-quality
17-109-final-viral-combined_k141_94920||lt2gene Low-quality
17-109-final-viral-combined_k141_280165||lt2gene Low-quality
17-109-final-viral-combined_k141_230019||lt2gene Low-quality
17-109-final-viral-combined_k141_346409||lt2gene Low-quality
17-109-final-viral-combined_k141_127941||lt2gene Low-quality
17-109-final-viral-combined_k141_280179||lt2gene Low-quality
17-109-final-viral-combined_k141_45837||lt2gene Low-quality
17-109-final-viral-combined_k141_178984||lt2gene Not-determined
17-109-final-viral-combined_k141_28236||lt2gene Not-determined
17-109-final-viral-combined_k141_127953||lt2gene Low-quality
17-109-final-viral-combined_k141_162587||lt2gene Low-quality
17-109-final-viral-combined_k141_314143||lt2gene Low-quality
17-109-final-viral-combined_k141_380406||lt2gene Low-quality
17-109-final-viral-combined_k141_380421||lt2gene Low-quality
17-109-final-viral-combined_k141_145257||lt2gene Low-quality
17-109-final-viral-combined_k141_45875||lt2gene Low-quality
17-109-final-viral-combined_k141_128017||lt2gene Low-quality
17-109-final-viral-combined_k141_128023||lt2gene Low-quality
17-109-final-viral-combined_k141_230117||lt2gene Not-determined
17-109-final-viral-combined_k141_179048||lt2gene Low-quality
17-109-final-viral-combined_k141_361723||lt2gene Low-quality
17-109-final-viral-combined_k141_128054||lt2gene Low-quality
17-109-final-viral-combined_k141_245952||lt2gene Low-quality
17-109-final-viral-combined_k141_95057||lt2gene Low-quality
17-109-final-viral-combined_k141_95069||lt2gene Low-quality
17-109-final-viral-combined_k141_212807||lt2gene Not-determined
17-109-final-viral-combined_k141_45931||lt2gene Low-quality
17-109-final-viral-combined_k141_314225||lt2gene Low-quality
17-109-final-viral-combined_k141_245995||lt2gene Low-quality
17-109-final-viral-combined_k141_79310||lt2gene Low-quality
17-109-final-viral-combined_k141_280304||lt2gene Not-determined
17-109-final-viral-combined_k141_95109||lt2gene Low-quality
17-109-final-viral-combined_k141_11324||lt2gene Low-quality
17-109-final-viral-combined_k141_262674||lt2gene Low-quality
17-109-final-viral-combined_k141_230217||lt2gene Low-quality
17-109-final-viral-combined_k141_329936||lt2gene Low-quality
17-109-final-viral-combined_k141_380501||lt2gene Low-quality
17-109-final-viral-combined_k141_95164||lt2gene Low-quality
17-109-final-viral-combined_k141_179112||lt2gene Low-quality
17-109-final-viral-combined_k141_162666||lt2gene Not-determined
17-109-final-viral-combined_k141_196432||lt2gene Low-quality
17-109-final-viral-combined_k141_196443||lt2gene Low-quality
17-109-final-viral-combined_k141_128194||lt2gene Low-quality
17-109-final-viral-combined_k141_380543||lt2gene Not-determined
17-109-final-viral-combined_k141_346535||lt2gene Low-quality
17-109-final-viral-combined_k141_314359||lt2gene Low-quality
17-109-final-viral-combined_k141_162714||lt2gene Low-quality
17-109-final-viral-combined_k141_162717||lt2gene Low-quality
17-109-final-viral-combined_k141_361816||lt2gene Low-quality
17-109-final-viral-combined_k141_162736||lt2gene Low-quality
17-109-final-viral-combined_k141_280432||lt2gene Low-quality
17-109-final-viral-combined_k141_11477||lt2gene Not-determined
17-109-final-viral-combined_k141_246112||lt2gene Low-quality
17-109-final-viral-combined_k141_314381||lt2gene Low-quality
17-109-final-viral-combined_k141_46156||lt2gene Low-quality
17-109-final-viral-combined_k141_314404||lt2gene Low-quality
17-109-final-viral-combined_k141_79547||lt2gene Low-quality
17-109-final-viral-combined_k141_361902||lt2gene Low-quality
17-109-final-viral-combined_k141_361918||lt2gene Low-quality
17-109-final-viral-combined_k141_314459||lt2gene Low-quality
17-109-final-viral-combined_k141_246706||lt2gene Low-quality
17-109-final-viral-combined_k141_28577||lt2gene Low-quality
17-109-final-viral-combined_k141_162807||lt2gene Not-determined
17-109-final-viral-combined_k141_314493||lt2gene Low-quality
17-109-final-viral-combined_k141_128398||lt2gene Low-quality
17-109-final-viral-combined_k141_62025||lt2gene Low-quality
17-109-final-viral-combined_k141_11517||lt2gene Low-quality
17-109-final-viral-combined_k141_128420||lt2gene Not-determined
17-109-final-viral-combined_k141_11536||lt2gene Low-quality
17-109-final-viral-combined_k141_28630||lt2gene Low-quality
17-109-final-viral-combined_k141_28633||lt2gene Not-determined
17-109-final-viral-combined_k141_213095||lt2gene Low-quality
17-109-final-viral-combined_k141_297084||lt2gene Not-determined
17-109-final-viral-combined_k141_246296||lt2gene Not-determined
17-109-final-viral-combined_k141_46378||lt2gene Low-quality
17-109-final-viral-combined_k141_213156||lt2gene Low-quality
17-109-final-viral-combined_k141_179441||lt2gene Low-quality
17-109-final-viral-combined_k141_179444||lt2gene Low-quality
17-109-final-viral-combined_k141_128588||lt2gene Low-quality
17-109-final-viral-combined_k141_179460||lt2gene Not-determined
17-109-final-viral-combined_k141_230375||lt2gene Low-quality
17-109-final-viral-combined_k141_213226||lt2gene Low-quality
17-109-final-viral-combined_k141_314663||lt2gene Low-quality
17-109-final-viral-combined_k141_346948||lt2gene Not-determined
17-109-final-viral-combined_k141_79760||lt2gene Low-quality
17-109-final-viral-combined_k141_196650||lt2gene Not-determined
17-109-final-viral-combined_k141_196671||lt2gene Low-quality
17-109-final-viral-combined_k141_62237||lt2gene Low-quality
17-109-final-viral-combined_k141_79797||lt2gene Low-quality
17-109-final-viral-combined_k141_79808||lt2gene Not-determined
17-109-final-viral-combined_k141_179562||lt2gene Low-quality
17-109-final-viral-combined_k141_46524||lt2gene Not-determined
17-109-final-viral-combined_k141_347033||lt2gene Low-quality
17-109-final-viral-combined_k141_11745||lt2gene Not-determined
17-109-final-viral-combined_k141_397329||lt2gene Low-quality
17-109-final-viral-combined_k141_397334||lt2gene Low-quality
17-109-final-viral-combined_k141_263130||lt2gene Low-quality
17-109-final-viral-combined_k141_263131||lt2gene Low-quality
17-109-final-viral-combined_k141_196773||lt2gene Low-quality
17-109-final-viral-combined_k141_62308||lt2gene Low-quality
17-109-final-viral-combined_k141_213350||lt2gene Low-quality
17-109-final-viral-combined_k141_62314||lt2gene Low-quality
17-109-final-viral-combined_k141_163029||lt2gene Not-determined
17-109-final-viral-combined_k141_330521||lt2gene Low-quality
17-109-final-viral-combined_k141_11833||lt2gene Not-determined
17-109-final-viral-combined_k141_213437||lt2gene Low-quality
17-109-final-viral-combined_k141_145877||lt2gene Low-quality
17-109-final-viral-combined_k141_128793||lt2gene Low-quality
17-109-final-viral-combined_k141_145902||lt2gene Low-quality
17-109-final-viral-combined_k141_11860||lt2gene Low-quality
17-109-final-viral-combined_k141_280960||lt2gene Low-quality
17-109-final-viral-combined_k141_213501||lt2gene Low-quality
17-109-final-viral-combined_k141_314756||lt2gene Not-determined
17-109-final-viral-combined_k141_297386||lt2gene Low-quality
17-109-final-viral-combined_k141_112074||lt2gene Low-quality
17-109-final-viral-combined_k141_246525||lt2gene Low-quality
17-109-final-viral-combined_k141_179846||lt2gene Low-quality
17-109-final-viral-combined_k141_281008||lt2gene Low-quality
17-109-final-viral-combined_k141_197010||lt2gene Not-determined
17-109-final-viral-combined_k141_297454||lt2gene Low-quality
17-109-final-viral-combined_k141_46765||lt2gene Not-determined
17-109-final-viral-combined_k141_362368||lt2gene Not-determined
17-109-final-viral-combined_k141_314890||lt2gene Not-determined
17-109-final-viral-combined_k141_112205||lt2gene Low-quality
17-109-final-viral-combined_k141_46807||lt2gene Low-quality
17-109-final-viral-combined_k141_263339||lt2gene Low-quality
17-109-final-viral-combined_k141_28979||lt2gene Not-determined
17-109-final-viral-combined_k141_330635||lt2gene Low-quality
17-109-final-viral-combined_k141_146067||lt2gene Low-quality
17-109-final-viral-combined_k141_397559||lt2gene Not-determined
17-109-final-viral-combined_k141_179996||lt2gene Low-quality
17-109-final-viral-combined_k141_246705||lt2gene Not-determined
17-109-final-viral-combined_k141_281135||lt2gene Not-determined
17-109-final-viral-combined_k141_163348||lt2gene Low-quality
17-109-final-viral-combined_k141_347446||lt2gene Low-quality
17-109-final-viral-combined_k141_314966||lt2gene Low-quality
17-109-final-viral-combined_k141_281159||lt2gene Not-determined
17-109-final-viral-combined_k141_80223||lt2gene Low-quality
17-109-final-viral-combined_k141_315016||lt2gene Low-quality
17-109-final-viral-combined_k141_281232||lt2gene Low-quality
17-109-final-viral-combined_k141_381088||lt2gene Low-quality
17-109-final-viral-combined_k141_362483||lt2gene Not-determined
17-109-final-viral-combined_k141_46927||lt2gene Not-determined
17-109-final-viral-combined_k141_62678||lt2gene Low-quality
17-109-final-viral-combined_k141_381116||lt2gene Low-quality
17-109-final-viral-combined_k141_381117||lt2gene Not-determined
17-109-final-viral-combined_k141_95893||lt2gene Not-determined
17-109-final-viral-combined_k141_315046||lt2gene Low-quality
17-109-final-viral-combined_k141_213700||lt2gene Not-determined
17-109-final-viral-combined_k141_180131||lt2gene Not-determined
17-109-final-viral-combined_k141_12158||lt2gene Low-quality
17-109-final-viral-combined_k141_397715||lt2gene Low-quality
17-109-final-viral-combined_k141_281268||lt2gene Not-determined
17-109-final-viral-combined_k141_315083||lt2gene Low-quality
17-109-final-viral-combined_k141_281287||lt2gene Low-quality
17-109-final-viral-combined_k141_315105||lt2gene Low-quality
17-109-final-viral-combined_k141_95984||lt2gene Low-quality
17-109-final-viral-combined_k141_362528||lt2gene Low-quality
17-109-final-viral-combined_k141_246807||lt2gene Low-quality
17-109-final-viral-combined_k141_47032||lt2gene Low-quality
17-109-final-viral-combined_k141_163596||lt2gene Low-quality
17-109-final-viral-combined_k141_297631||lt2gene Low-quality
17-109-final-viral-combined_k141_213808||lt2gene Low-quality
17-109-final-viral-combined_k141_230875||lt2gene Low-quality
17-109-final-viral-combined_k141_315209||lt2gene Low-quality
17-109-final-viral-combined_k141_230886||lt2gene Low-quality
17-109-final-viral-combined_k141_297643||lt2gene Low-quality
17-109-final-viral-combined_k141_180219||lt2gene Low-quality
17-109-final-viral-combined_k141_315243||lt2gene Not-determined
17-109-final-viral-combined_k141_180247||lt2gene Low-quality
17-109-final-viral-combined_k141_397837||lt2gene Not-determined
17-109-final-viral-combined_k141_146330||lt2gene Low-quality
17-109-final-viral-combined_k141_12308||lt2gene Low-quality
17-109-final-viral-combined_k141_397862||lt2gene Low-quality
17-109-final-viral-combined_k141_246937||lt2gene Not-determined
17-109-final-viral-combined_k141_246940||lt2gene Low-quality
17-109-final-viral-combined_k141_231015||lt2gene Low-quality
17-109-final-viral-combined_k141_29299||lt2gene Low-quality
17-109-final-viral-combined_k141_12373||lt2gene Low-quality
17-109-final-viral-combined_k141_163739||lt2gene Low-quality
17-109-final-viral-combined_k141_381291||lt2gene Low-quality
17-109-final-viral-combined_k141_381294||lt2gene Not-determined
17-109-final-viral-combined_k141_146432||lt2gene Low-quality
17-109-final-viral-combined_k141_347742||lt2gene Not-determined
17-109-final-viral-combined_k141_29343||lt2gene Not-determined
17-109-final-viral-combined_k141_146440||lt2gene Low-quality
17-109-final-viral-combined_k141_362657||lt2gene Low-quality
17-109-final-viral-combined_k141_146465||lt2gene Low-quality
17-109-final-viral-combined_k141_381346||lt2gene Low-quality
17-109-final-viral-combined_k141_146478||lt2gene Low-quality
17-109-final-viral-combined_k141_146483||lt2gene Low-quality
17-109-final-viral-combined_k141_247011||lt2gene Low-quality
17-109-final-viral-combined_k141_62893||lt2gene Not-determined
17-109-final-viral-combined_k141_80497||lt2gene Low-quality
17-109-final-viral-combined_k141_247057||lt2gene Low-quality
17-109-final-viral-combined_k141_281644||lt2gene Low-quality
17-109-final-viral-combined_k141_381426||lt2gene Not-determined
17-109-final-viral-combined_k141_47271||lt2gene Not-determined
17-109-final-viral-combined_k141_62910||lt2gene Low-quality
17-109-final-viral-combined_k141_47296||lt2gene Not-determined
17-109-final-viral-combined_k141_47306||lt2gene Low-quality
17-109-final-viral-combined_k141_213976||lt2gene Not-determined
17-109-final-viral-combined_k141_129370||lt2gene Not-determined
17-109-final-viral-combined_k141_29367||lt2gene Low-quality
17-109-final-viral-combined_k141_112636||lt2gene Low-quality
17-109-final-viral-combined_k141_12556||lt2gene Low-quality
17-109-final-viral-combined_k141_214033||lt2gene Not-determined
17-109-final-viral-combined_k141_281726||lt2gene Low-quality
17-109-final-viral-combined_k141_381530||lt2gene Low-quality
17-109-final-viral-combined_k141_112726||lt2gene Not-determined
17-109-final-viral-combined_k141_362846||lt2gene Low-quality
17-109-final-viral-combined_k141_398031||lt2gene Not-determined
17-109-final-viral-combined_k141_398035||lt2gene Low-quality
17-109-final-viral-combined_k141_381569||lt2gene Low-quality
17-109-final-viral-combined_k141_331286||lt2gene Low-quality
17-109-final-viral-combined_k141_331307||lt2gene Not-determined
17-109-final-viral-combined_k141_297861||lt2gene Low-quality
17-109-final-viral-combined_k141_12609||lt2gene Low-quality
17-109-final-viral-combined_k141_281826||lt2gene Not-determined
17-109-final-viral-combined_k141_164104||lt2gene Not-determined
17-109-final-viral-combined_k141_112802||lt2gene Low-quality
17-109-final-viral-combined_k141_164128||lt2gene Low-quality
17-109-final-viral-combined_k141_112823||lt2gene Not-determined
17-109-final-viral-combined_k141_263810||lt2gene Low-quality
17-109-final-viral-combined_k141_231428||lt2gene Low-quality
17-109-final-viral-combined_k141_63005||lt2gene Low-quality
17-109-final-viral-combined_k141_47543||lt2gene Not-determined
17-109-final-viral-combined_k141_297935||lt2gene Low-quality
17-109-final-viral-combined_k141_12662||lt2gene Low-quality
17-109-final-viral-combined_k141_112909||lt2gene Low-quality
17-109-final-viral-combined_k141_80710||lt2gene Low-quality
17-109-final-viral-combined_k141_29703||lt2gene Low-quality
17-109-final-viral-combined_k141_214204||lt2gene Low-quality
17-109-final-viral-combined_k141_398091||lt2gene Low-quality
17-109-final-viral-combined_k141_197694||lt2gene Low-quality
17-109-final-viral-combined_k141_331459||lt2gene Low-quality
17-109-final-viral-combined_k141_197698||lt2gene Not-determined
17-109-final-viral-combined_k141_348137||lt2gene Low-quality
17-109-final-viral-combined_k141_363060||lt2gene Low-quality
17-109-final-viral-combined_k141_331478||lt2gene Low-quality
17-109-final-viral-combined_k141_146806||lt2gene Not-determined
17-109-final-viral-combined_k141_231530||lt2gene Low-quality
17-109-final-viral-combined_k141_231534||lt2gene Low-quality
17-109-final-viral-combined_k141_381814||lt2gene Low-quality
17-109-final-viral-combined_k141_146815||lt2gene Low-quality
17-109-final-viral-combined_k141_129655||lt2gene Low-quality
17-109-final-viral-combined_k141_398105||lt2gene Low-quality
17-109-final-viral-combined_k141_180707||lt2gene Low-quality
17-109-final-viral-combined_k141_214255||lt2gene Low-quality
17-109-final-viral-combined_k141_263987||lt2gene Low-quality
17-109-final-viral-combined_k141_214265||lt2gene Low-quality
17-109-final-viral-combined_k141_47720||lt2gene Low-quality
17-109-final-viral-combined_k141_247451||lt2gene Low-quality
17-109-final-viral-combined_k141_348232||lt2gene Low-quality
17-109-final-viral-combined_k141_363114||lt2gene Not-determined
17-109-final-viral-combined_k141_112953||lt2gene Low-quality
17-109-final-viral-combined_k141_381857||lt2gene Low-quality
17-109-final-viral-combined_k141_129767||lt2gene Low-quality
17-109-final-viral-combined_k141_197846||lt2gene Low-quality
17-109-final-viral-combined_k141_363153||lt2gene Not-determined
17-109-final-viral-combined_k141_231631||lt2gene Not-determined
17-109-final-viral-combined_k141_315741||lt2gene Not-determined
17-109-final-viral-combined_k141_47744||lt2gene Low-quality
17-109-final-viral-combined_k141_47760||lt2gene Low-quality
17-109-final-viral-combined_k141_331632||lt2gene Low-quality
17-109-final-viral-combined_k141_315779||lt2gene Not-determined
17-109-final-viral-combined_k141_29858||lt2gene Low-quality
17-109-final-viral-combined_k141_113114||lt2gene Low-quality
17-109-final-viral-combined_k141_29871||lt2gene Not-determined
17-109-final-viral-combined_k141_164455||lt2gene Low-quality
17-109-final-viral-combined_k141_315797||lt2gene Not-determined
17-109-final-viral-combined_k141_315809||lt2gene Not-determined
17-109-final-viral-combined_k141_146906||lt2gene Low-quality
17-109-final-viral-combined_k141_214428||lt2gene Low-quality
17-109-final-viral-combined_k141_96771||lt2gene Not-determined
17-109-final-viral-combined_k141_113146||lt2gene Not-determined
17-109-final-viral-combined_k141_146934||lt2gene Low-quality
17-109-final-viral-combined_k141_348403||lt2gene Low-quality
17-109-final-viral-combined_k141_63356||lt2gene Low-quality
17-109-final-viral-combined_k141_29974||lt2gene Low-quality
17-109-final-viral-combined_k141_331724||lt2gene Low-quality
17-109-final-viral-combined_k141_315890||lt2gene Low-quality
17-109-final-viral-combined_k141_12972||lt2gene Low-quality
17-109-final-viral-combined_k141_113201||lt2gene Low-quality
17-109-final-viral-combined_k141_12985||lt2gene Not-determined
17-109-final-viral-combined_k141_113214||lt2gene Not-determined
17-109-final-viral-combined_k141_298179||lt2gene Not-determined
17-109-final-viral-combined_k141_348460||lt2gene Low-quality
17-109-final-viral-combined_k141_30018||lt2gene Low-quality
17-109-final-viral-combined_k141_315919||lt2gene Low-quality
17-109-final-viral-combined_k141_63418||lt2gene Low-quality
17-109-final-viral-combined_k141_181027||lt2gene Low-quality
17-109-final-viral-combined_k141_331781||lt2gene Low-quality
17-109-final-viral-combined_k141_247756||lt2gene Not-determined
17-109-final-viral-combined_k141_113256||lt2gene Not-determined
17-109-final-viral-combined_k141_331812||lt2gene Low-quality
17-109-final-viral-combined_k141_129854||lt2gene Low-quality
17-109-final-viral-combined_k141_198085||lt2gene Low-quality
17-109-final-viral-combined_k141_81006||lt2gene Low-quality
17-109-final-viral-combined_k141_331875||lt2gene Low-quality
17-109-final-viral-combined_k141_129881||lt2gene Low-quality
17-109-final-viral-combined_k141_63457||lt2gene Low-quality
17-109-final-viral-combined_k141_47966||lt2gene Low-quality
17-109-final-viral-combined_k141_231949||lt2gene Low-quality
17-109-final-viral-combined_k141_214702||lt2gene Low-quality
17-109-final-viral-combined_k141_198144||lt2gene Low-quality
17-109-final-viral-combined_k141_63503||lt2gene Low-quality
17-109-final-viral-combined_k141_13161||lt2gene Not-determined
17-109-final-viral-combined_k141_231999||lt2gene Not-determined
17-109-final-viral-combined_k141_113373||lt2gene Low-quality
17-109-final-viral-combined_k141_316051||lt2gene Low-quality
17-109-final-viral-combined_k141_198162||lt2gene Low-quality
17-109-final-viral-combined_k141_282498||lt2gene Low-quality
17-109-final-viral-combined_k141_147084||lt2gene Low-quality
17-109-final-viral-combined_k141_113407||lt2gene Low-quality
17-109-final-viral-combined_k141_298366||lt2gene Low-quality
17-109-final-viral-combined_k141_48035||lt2gene Low-quality
17-109-final-viral-combined_k141_13210||lt2gene Not-determined
17-109-final-viral-combined_k141_97083||lt2gene Low-quality
17-109-final-viral-combined_k141_48101||lt2gene Not-determined
17-109-final-viral-combined_k141_382174||lt2gene Low-quality
17-109-final-viral-combined_k141_298435||lt2gene Low-quality
17-109-final-viral-combined_k141_316158||lt2gene Low-quality
17-109-final-viral-combined_k141_198234||lt2gene Low-quality
17-109-final-viral-combined_k141_198238||lt2gene Not-determined
17-109-final-viral-combined_k141_298456||lt2gene Not-determined
17-109-final-viral-combined_k141_147188||lt2gene Low-quality
17-109-final-viral-combined_k141_30224||lt2gene Low-quality
17-109-final-viral-combined_k141_181349||lt2gene Not-determined
17-109-final-viral-combined_k141_164686||lt2gene Low-quality
17-109-final-viral-combined_k141_13337||lt2gene Low-quality
17-109-final-viral-combined_k141_348678||lt2gene Low-quality
17-109-final-viral-combined_k141_348687||lt2gene Not-determined
17-109-final-viral-combined_k141_130126||lt2gene Low-quality
17-109-final-viral-combined_k141_97188||lt2gene Low-quality
17-109-final-viral-combined_k141_348720||lt2gene Low-quality
17-109-final-viral-combined_k141_248101||lt2gene Low-quality
17-109-final-viral-combined_k141_113551||lt2gene Not-determined
17-109-final-viral-combined_k141_348744||lt2gene Low-quality
17-109-final-viral-combined_k141_198333||lt2gene Low-quality
17-109-final-viral-combined_k141_248144||lt2gene Low-quality
17-109-final-viral-combined_k141_248145||lt2gene Low-quality
17-109-final-viral-combined_k141_30238||lt2gene Not-determined
17-109-final-viral-combined_k141_48201||lt2gene Low-quality
17-109-final-viral-combined_k141_232241||lt2gene Low-quality
17-109-final-viral-combined_k141_48214||lt2gene Low-quality
17-109-final-viral-combined_k141_48216||lt2gene Not-determined
17-109-final-viral-combined_k141_298608||lt2gene Low-quality
17-109-final-viral-combined_k141_181475||lt2gene Not-determined
17-109-final-viral-combined_k141_13469||lt2gene Low-quality
17-109-final-viral-combined_k141_164864||lt2gene Low-quality
17-109-final-viral-combined_k141_81394||lt2gene Not-determined
17-109-final-viral-combined_k141_298659||lt2gene Not-determined
17-109-final-viral-combined_k141_382355||lt2gene Low-quality
17-109-final-viral-combined_k141_63843||lt2gene Not-determined
17-109-final-viral-combined_k141_164899||lt2gene Not-determined
17-109-final-viral-combined_k141_298685||lt2gene Low-quality
17-109-final-viral-combined_k141_232298||lt2gene Not-determined
17-109-final-viral-combined_k141_97294||lt2gene Low-quality
17-109-final-viral-combined_k141_232301||lt2gene Low-quality
17-109-final-viral-combined_k141_316427||lt2gene Low-quality
17-109-final-viral-combined_k141_130331||lt2gene Low-quality
17-109-final-viral-combined_k141_332288||lt2gene Low-quality
17-109-final-viral-combined_k141_363703||lt2gene Low-quality
17-109-final-viral-combined_k141_282738||lt2gene Low-quality
17-109-final-viral-combined_k141_81454||lt2gene Not-determined
17-109-final-viral-combined_k141_113735||lt2gene Low-quality
17-109-final-viral-combined_k141_130370||lt2gene Low-quality
17-109-final-viral-combined_k141_316481||lt2gene Low-quality
17-109-final-viral-combined_k141_13613||lt2gene Low-quality
17-109-final-viral-combined_k141_248313||lt2gene Low-quality
17-109-final-viral-combined_k141_215144||lt2gene Not-determined
17-109-final-viral-combined_k141_198559||lt2gene Low-quality
17-109-final-viral-combined_k141_232388||lt2gene Low-quality
17-109-final-viral-combined_k141_232393||lt2gene Low-quality
17-109-final-viral-combined_k141_264523||lt2gene Low-quality
17-109-final-viral-combined_k141_198567||lt2gene Not-determined
17-109-final-viral-combined_k141_232423||lt2gene Low-quality
17-109-final-viral-combined_k141_13672||lt2gene Low-quality
17-109-final-viral-combined_k141_316560||lt2gene Low-quality
17-109-final-viral-combined_k141_398697||lt2gene Low-quality
17-109-final-viral-combined_k141_113841||lt2gene Not-determined
17-109-final-viral-combined_k141_363815||lt2gene Low-quality
17-109-final-viral-combined_k141_147499||lt2gene Not-determined
17-109-final-viral-combined_k141_248339||lt2gene Not-determined
17-109-final-viral-combined_k141_232512||lt2gene Not-determined
17-109-final-viral-combined_k141_13748||lt2gene Low-quality
17-109-final-viral-combined_k141_130504||lt2gene Not-determined
17-109-final-viral-combined_k141_316673||lt2gene Not-determined
17-109-final-viral-combined_k141_130516||lt2gene Low-quality
17-109-final-viral-combined_k141_248422||lt2gene Not-determined
17-109-final-viral-combined_k141_181723||lt2gene Low-quality
17-109-final-viral-combined_k141_282989||lt2gene Low-quality
17-109-final-viral-combined_k141_181733||lt2gene Low-quality
17-109-final-viral-combined_k141_232582||lt2gene Low-quality
17-109-final-viral-combined_k141_30580||lt2gene Low-quality
17-109-final-viral-combined_k141_349064||lt2gene Low-quality
17-109-final-viral-combined_k141_298965||lt2gene Not-determined
17-109-final-viral-combined_k141_332569||lt2gene Low-quality
17-109-final-viral-combined_k141_97616||lt2gene Not-determined
17-109-final-viral-combined_k141_64039||lt2gene Not-determined
17-109-final-viral-combined_k141_13844||lt2gene Low-quality
17-109-final-viral-combined_k141_114040||lt2gene Not-determined
17-109-final-viral-combined_k141_283082||lt2gene Not-determined
17-109-final-viral-combined_k141_81653||lt2gene Low-quality
17-109-final-viral-combined_k141_147601||lt2gene Low-quality
17-109-final-viral-combined_k141_264647||lt2gene Not-determined
17-109-final-viral-combined_k141_349133||lt2gene Not-determined
17-109-final-viral-combined_k141_181816||lt2gene Not-determined
17-109-final-viral-combined_k141_299044||lt2gene Not-determined
17-109-final-viral-combined_k141_264669||lt2gene Not-determined
17-109-final-viral-combined_k141_165140||lt2gene Not-determined
17-109-final-viral-combined_k141_81695||lt2gene Low-quality
17-109-final-viral-combined_k141_332683||lt2gene Low-quality
17-109-final-viral-combined_k141_215473||lt2gene Not-determined
17-109-final-viral-combined_k141_264707||lt2gene Not-determined
17-109-final-viral-combined_k141_81764||lt2gene Low-quality
17-109-final-viral-combined_k141_181887||lt2gene Not-determined
17-109-final-viral-combined_k141_398902||lt2gene Low-quality
17-109-final-viral-combined_k141_332767||lt2gene Low-quality
17-109-final-viral-combined_k141_114133||lt2gene Low-quality
17-109-final-viral-combined_k141_382666||lt2gene Not-determined
17-109-final-viral-combined_k141_364036||lt2gene Low-quality
17-109-final-viral-combined_k141_382670||lt2gene Not-determined
17-109-final-viral-combined_k141_349253||lt2gene Not-determined
17-109-final-viral-combined_k141_264768||lt2gene Low-quality
17-109-final-viral-combined_k141_130747||lt2gene Low-quality
17-109-final-viral-combined_k141_349272||lt2gene Not-determined
17-109-final-viral-combined_k141_316862||lt2gene Not-determined
17-109-final-viral-combined_k141_48627||lt2gene Low-quality
17-109-final-viral-combined_k141_215561||lt2gene Low-quality
17-109-final-viral-combined_k141_364082||lt2gene Low-quality
17-109-final-viral-combined_k141_299228||lt2gene Low-quality
17-109-final-viral-combined_k141_264796||lt2gene Low-quality
17-109-final-viral-combined_k141_48659||lt2gene Low-quality
17-109-final-viral-combined_k141_364135||lt2gene Low-quality
17-109-final-viral-combined_k141_332821||lt2gene Low-quality
17-109-final-viral-combined_k141_14071||lt2gene Low-quality
17-109-final-viral-combined_k141_165253||lt2gene Low-quality
17-109-final-viral-combined_k141_283340||lt2gene Low-quality

| | |
|---|---|
| 17-109-final-viral-combined_k141_332847\|\|lt2gene | Low-quality |
| 17-109-final-viral-combined_k141_14101\|\|lt2gene | Low-quality |
| 17-109-final-viral-combined_k141_382740\|\|lt2gene | Low-quality |
| 17-109-final-viral-combined_k141_364164\|\|lt2gene | Low-quality |
| 17-109-final-viral-combined_k141_248594\|\|lt2gene | Not-determined |
| 17-109-final-viral-combined_k141_248600\|\|lt2gene | Low-quality |
| 17-109-final-viral-combined_k141_130882\|\|lt2gene | Not-determined |
| 17-109-final-viral-combined_k141_232895\|\|lt2gene | Low-quality |
| 17-109-final-viral-combined_k141_264952\|\|lt2gene | Not-determined |
| 17-109-final-viral-combined_k141_199039\|\|lt2gene | Low-quality |
| 17-109-final-viral-combined_k141_199040\|\|lt2gene | Low-quality |
| 17-109-final-viral-combined_k141_382746\|\|lt2gene | Low-quality |
| 17-109-final-viral-combined_k141_364262\|\|lt2gene | Not-determined |
| 17-109-final-viral-combined_k141_64277\|\|lt2gene | Not-determined |
| 17-109-final-viral-combined_k141_364270\|\|lt2gene | Low-quality |
| 17-109-final-viral-combined_k141_316964\|\|lt2gene | Low-quality |
| 17-109-final-viral-combined_k141_364295\|\|lt2gene | Low-quality |
| 17-109-final-viral-combined_k141_30874\|\|lt2gene | Low-quality |
| 17-109-final-viral-combined_k141_64344\|\|lt2gene | Low-quality |
| 17-109-final-viral-combined_k141_317068\|\|lt2gene | Low-quality |
| 17-109-final-viral-combined_k141_82104\|\|lt2gene | Low-quality |
| 17-109-final-viral-combined_k141_48922\|\|lt2gene | Not-determined |
| 17-109-final-viral-combined_k141_182315\|\|lt2gene | Low-quality |
| 17-109-final-viral-combined_k141_114403\|\|lt2gene | Not-determined |
| 17-109-final-viral-combined_k141_299412\|\|lt2gene | Low-quality |
| 17-109-final-viral-combined_k141_165580\|\|lt2gene | Not-determined |
| 17-109-final-viral-combined_k141_165597\|\|lt2gene | Low-quality |
| 17-109-final-viral-combined_k141_349611\|\|lt2gene | Not-determined |
| 17-109-final-viral-combined_k141_182403\|\|lt2gene | Low-quality |
| 17-109-final-viral-combined_k141_233140\|\|lt2gene | Low-quality |
| 17-109-final-viral-combined_k141_31088\|\|lt2gene | Low-quality |
| 17-109-final-viral-combined_k141_14432\|\|lt2gene | Low-quality |
| 17-109-final-viral-combined_k141_114507\|\|lt2gene | Low-quality |
| 17-109-final-viral-combined_k141_114510\|\|lt2gene | Low-quality |
| 17-109-final-viral-combined_k141_31143\|\|lt2gene | Low-quality |
| 17-109-final-viral-combined_k141_233262\|\|lt2gene | Low-quality |
| 17-109-final-viral-combined_k141_31156\|\|lt2gene | Low-quality |
| 17-109-final-viral-combined_k141_248888\|\|lt2gene | Not-determined |
| 17-109-final-viral-combined_k141_399227\|\|lt2gene | Low-quality |
| 17-109-final-viral-combined_k141_98182\|\|lt2gene | Low-quality |
| 17-109-final-viral-combined_k141_31198\|\|lt2gene | Low-quality |
| 17-109-final-viral-combined_k141_364631\|\|lt2gene | Low-quality |
| 17-109-final-viral-combined_k141_233329\|\|lt2gene | Low-quality |
| 17-109-final-viral-combined_k141_31215\|\|lt2gene | Not-determined |
| 17-109-final-viral-combined_k141_349749\|\|lt2gene | Low-quality |
| 17-109-final-viral-combined_k141_233353\|\|lt2gene | Low-quality |
| 17-109-final-viral-combined_k141_98207\|\|lt2gene | Low-quality |
| 17-109-final-viral-combined_k141_82343\|\|lt2gene | Not-determined |
| 17-109-final-viral-combined_k141_233360\|\|lt2gene | Low-quality |
| 17-109-final-viral-combined_k141_148209\|\|lt2gene | Low-quality |
| 17-109-final-viral-combined_k141_265154\|\|lt2gene | Low-quality |
| 17-109-final-viral-combined_k141_333306\|\|lt2gene | Low-quality |
| 17-109-final-viral-combined_k141_199331\|\|lt2gene | Not-determined |
| 17-109-final-viral-combined_k141_31300\|\|lt2gene | Low-quality |
| 17-109-final-viral-combined_k141_399342\|\|lt2gene | Low-quality |
| 17-109-final-viral-combined_k141_31316\|\|lt2gene | Low-quality |
| 17-109-final-viral-combined_k141_249036\|\|lt2gene | Not-determined |
| 17-109-final-viral-combined_k141_364737\|\|lt2gene | Low-quality |
| 17-109-final-viral-combined_k141_364749\|\|lt2gene | Not-determined |
| 17-109-final-viral-combined_k141_14624\|\|lt2gene | Low-quality |
| 17-109-final-viral-combined_k141_364779\|\|lt2gene | Low-quality |
| 17-109-final-viral-combined_k141_182594\|\|lt2gene | Not-determined |
| 17-109-final-viral-combined_k141_199378\|\|lt2gene | Low-quality |
| 17-109-final-viral-combined_k141_349858\|\|lt2gene | Not-determined |
| 17-109-final-viral-combined_k141_249105\|\|lt2gene | Low-quality |
| 17-109-final-viral-combined_k141_383007\|\|lt2gene | Not-determined |
| 17-109-final-viral-combined_k141_114691\|\|lt2gene | Not-determined |
| 17-109-final-viral-combined_k141_349893\|\|lt2gene | Not-determined |
| 17-109-final-viral-combined_k141_199434\|\|lt2gene | Low-quality |
| 17-109-final-viral-combined_k141_349930\|\|lt2gene | Low-quality |
| 17-109-final-viral-combined_k141_64949\|\|lt2gene | Not-determined |
| 17-109-final-viral-combined_k141_82531\|\|lt2gene | Not-determined |
| 17-109-final-viral-combined_k141_299711\|\|lt2gene | Low-quality |
| 17-109-final-viral-combined_k141_131539\|\|lt2gene | Low-quality |
| 17-109-final-viral-combined_k141_317540\|\|lt2gene | Low-quality |
| 17-109-final-viral-combined_k141_82555\|\|lt2gene | Low-quality |
| 17-109-final-viral-combined_k141_114729\|\|lt2gene | Not-determined |
| 17-109-final-viral-combined_k141_383082\|\|lt2gene | Low-quality |
| 17-109-final-viral-combined_k141_148457\|\|lt2gene | Low-quality |
| 17-109-final-viral-combined_k141_333415\|\|lt2gene | Not-determined |
| 17-109-final-viral-combined_k141_364881\|\|lt2gene | Low-quality |
| 17-109-final-viral-combined_k141_249231\|\|lt2gene | Low-quality |
| 17-109-final-viral-combined_k141_98430\|\|lt2gene | Low-quality |
| 17-109-final-viral-combined_k141_383132\|\|lt2gene | Low-quality |
| 17-109-final-viral-combined_k141_399520\|\|lt2gene | Low-quality |
| 17-109-final-viral-combined_k141_65058\|\|lt2gene | Not-determined |
| 17-109-final-viral-combined_k141_333474\|\|lt2gene | Low-quality |
| 17-109-final-viral-combined_k141_333486\|\|lt2gene | Low-quality |
| 17-109-final-viral-combined_k141_299807\|\|lt2gene | Low-quality |
| 17-109-final-viral-combined_k141_199596\|\|lt2gene | Low-quality |
| 17-109-final-viral-combined_k141_233648\|\|lt2gene | Low-quality |
| 17-109-final-viral-combined_k141_31554\|\|lt2gene | Low-quality |
| 17-109-final-viral-combined_k141_283924\|\|lt2gene | Not-determined |
| 17-109-final-viral-combined_k141_65123\|\|lt2gene | Not-determined |
| 17-109-final-viral-combined_k141_216401\|\|lt2gene | Low-quality |
| 17-109-final-viral-combined_k141_249272\|\|lt2gene | Low-quality |
| 17-109-final-viral-combined_k141_199653\|\|lt2gene | Low-quality |
| 17-109-final-viral-combined_k141_233689\|\|lt2gene | Low-quality |
| 17-109-final-viral-combined_k141_98541\|\|lt2gene | Low-quality |
| 17-109-final-viral-combined_k141_199680\|\|lt2gene | Low-quality |
| 17-109-final-viral-combined_k141_249296\|\|lt2gene | Low-quality |
| 17-109-final-viral-combined_k141_131651\|\|lt2gene | Low-quality |
| 17-109-final-viral-combined_k141_82712\|\|lt2gene | Not-determined |
| 17-109-final-viral-combined_k141_249305\|\|lt2gene | Low-quality |
| 17-109-final-viral-combined_k141_350119\|\|lt2gene | Low-quality |
| 17-109-final-viral-combined_k141_333618\|\|lt2gene | Low-quality |
| 17-109-final-viral-combined_k141_31608\|\|lt2gene | Low-quality |
| 17-109-final-viral-combined_k141_265453\|\|lt2gene | Low-quality |
| 17-109-final-viral-combined_k141_49486\|\|lt2gene | Low-quality |
| 17-109-final-viral-combined_k141_131673\|\|lt2gene | Not-determined |
| 17-109-final-viral-combined_k141_166082\|\|lt2gene | Low-quality |
| 17-109-final-viral-combined_k141_199728\|\|lt2gene | Low-quality |
| 17-109-final-viral-combined_k141_166093\|\|lt2gene | Low-quality |
| 17-109-final-viral-combined_k141_82785\|\|lt2gene | Not-determined |
| 17-109-final-viral-combined_k141_317715\|\|lt2gene | Not-determined |
| 17-109-final-viral-combined_k141_383312\|\|lt2gene | Not-determined |
| 17-109-final-viral-combined_k141_98690\|\|lt2gene | Low-quality |
| 17-109-final-viral-combined_k141_399670\|\|lt2gene | Not-determined |
| 17-109-final-viral-combined_k141_299971\|\|lt2gene | Low-quality |
| 17-109-final-viral-combined_k141_333762\|\|lt2gene | Low-quality |
| 17-109-final-viral-combined_k141_131787\|\|lt2gene | Not-determined |
| 17-109-final-viral-combined_k141_14880\|\|lt2gene | Low-quality |
| 17-109-final-viral-combined_k141_98768\|\|lt2gene | Low-quality |
| 17-109-final-viral-combined_k141_365206\|\|lt2gene | Low-quality |
| 17-109-final-viral-combined_k141_216660\|\|lt2gene | Not-determined |
| 17-109-final-viral-combined_k141_284138\|\|lt2gene | Low-quality |
| 17-109-final-viral-combined_k141_399738\|\|lt2gene | Not-determined |
| 17-109-final-viral-combined_k141_333800\|\|lt2gene | Low-quality |
| 17-109-final-viral-combined_k141_383392\|\|lt2gene | Low-quality |
| 17-109-final-viral-combined_k141_333806\|\|lt2gene | Low-quality |
| 17-109-final-viral-combined_k141_98777\|\|lt2gene | Low-quality |
| 17-109-final-viral-combined_k141_300032\|\|lt2gene | Not-determined |
| 17-109-final-viral-combined_k141_82950\|\|lt2gene | Low-quality |
| 17-109-final-viral-combined_k141_350248\|\|lt2gene | Not-determined |
| 17-109-final-viral-combined_k141_82974\|\|lt2gene | Low-quality |
| 17-109-final-viral-combined_k141_49684\|\|lt2gene | Low-quality |
| 17-109-final-viral-combined_k141_234017\|\|lt2gene | Low-quality |
| 17-109-final-viral-combined_k141_383474\|\|lt2gene | Low-quality |
| 17-109-final-viral-combined_k141_115093\|\|lt2gene | Low-quality |
| 17-109-final-viral-combined_k141_98859\|\|lt2gene | Not-determined |
| 17-109-final-viral-combined_k141_65426\|\|lt2gene | Not-determined |
| 17-109-final-viral-combined_k141_131911\|\|lt2gene | Low-quality |
| 17-109-final-viral-combined_k141_31854\|\|lt2gene | Low-quality |
| 17-109-final-viral-combined_k141_14998\|\|lt2gene | Not-determined |
| 17-109-final-viral-combined_k141_83029\|\|lt2gene | Low-quality |
| 17-109-final-viral-combined_k141_365338\|\|lt2gene | Not-determined |
| 17-109-final-viral-combined_k141_83035\|\|lt2gene | Not-determined |
| 17-109-final-viral-combined_k141_300145\|\|lt2gene | Low-quality |
| 17-109-final-viral-combined_k141_399781\|\|lt2gene | Low-quality |
| 17-109-final-viral-combined_k141_234044\|\|lt2gene | Not-determined |
| 17-109-final-viral-combined_k141_365351\|\|lt2gene | Low-quality |
| 17-109-final-viral-combined_k141_300174\|\|lt2gene | Not-determined |
| 17-109-final-viral-combined_k141_317876\|\|lt2gene | Low-quality |
| 17-109-final-viral-combined_k141_284257\|\|lt2gene | Not-determined |
| 17-109-final-viral-combined_k141_317884\|\|lt2gene | Not-determined |
| 17-109-final-viral-combined_k141_65481\|\|lt2gene | Low-quality |
| 17-109-final-viral-combined_k141_365411\|\|lt2gene | Low-quality |
| 17-109-final-viral-combined_k141_234072\|\|lt2gene | Not-determined |
| 17-109-final-viral-combined_k141_284297\|\|lt2gene | Not-determined |
| 17-109-final-viral-combined_k141_15129\|\|lt2gene | Low-quality |
| 17-109-final-viral-combined_k141_350441\|\|lt2gene | Not-determined |
| 17-109-final-viral-combined_k141_317917\|\|lt2gene | Not-determined |
| 17-109-final-viral-combined_k141_399867\|\|lt2gene | Low-quality |
| 17-109-final-viral-combined_k141_265700\|\|lt2gene | Low-quality |
| 17-109-final-viral-combined_k141_399875\|\|lt2gene | Low-quality |
| 17-109-final-viral-combined_k141_83128\|\|lt2gene | Low-quality |
| 17-109-final-viral-combined_k141_83160\|\|lt2gene | Low-quality |
| 17-109-final-viral-combined_k141_65577\|\|lt2gene | Low-quality |
| 17-109-final-viral-combined_k141_49908\|\|lt2gene | Not-determined |
| 17-109-final-viral-combined_k141_383605\|\|lt2gene | Low-quality |
| 17-109-final-viral-combined_k141_249618\|\|lt2gene | Low-quality |
| 17-109-final-viral-combined_k141_98934\|\|lt2gene | Low-quality |
| 17-109-final-viral-combined_k141_200037\|\|lt2gene | Low-quality |
| 17-109-final-viral-combined_k141_83177\|\|lt2gene | Low-quality |
| 17-109-final-viral-combined_k141_98960\|\|lt2gene | Not-determined |
| 17-109-final-viral-combined_k141_166412\|\|lt2gene | Not-determined |
| 17-109-final-viral-combined_k141_115262\|\|lt2gene | Low-quality |
| 17-109-final-viral-combined_k141_249639\|\|lt2gene | Not-determined |
| 17-109-final-viral-combined_k141_132105\|\|lt2gene | Not-determined |
| 17-109-final-viral-combined_k141_200085\|\|lt2gene | Not-determined |
| 17-109-final-viral-combined_k141_399957\|\|lt2gene | Not-determined |
| 17-109-final-viral-combined_k141_166466\|\|lt2gene | Not-determined |
| 17-109-final-viral-combined_k141_99053\|\|lt2gene | Low-quality |
| 17-109-final-viral-combined_k141_32091\|\|lt2gene | Low-quality |
| 17-109-final-viral-combined_k141_300326\|\|lt2gene | Low-quality |
| 17-109-final-viral-combined_k141_183390\|\|lt2gene | Not-determined |
| 17-109-final-viral-combined_k141_32109\|\|lt2gene | Low-quality |
| 17-109-final-viral-combined_k141_99091\|\|lt2gene | Low-quality |
| 17-109-final-viral-combined_k141_132142\|\|lt2gene | Low-quality |
| 17-109-final-viral-combined_k141_166557\|\|lt2gene | Not-determined |
| 17-109-final-viral-combined_k141_183422\|\|lt2gene | Not-determined |
| 17-109-final-viral-combined_k141_265913\|\|lt2gene | Low-quality |
| 17-109-final-viral-combined_k141_265934\|\|lt2gene | Low-quality |
| 17-109-final-viral-combined_k141_350594\|\|lt2gene | Not-determined |
| 17-109-final-viral-combined_k141_383707\|\|lt2gene | Not-determined |
| 17-109-final-viral-combined_k141_183470\|\|lt2gene | Not-determined |
| 17-109-final-viral-combined_k141_65764\|\|lt2gene | Not-determined |
| 17-109-final-viral-combined_k141_234332\|\|lt2gene | Low-quality |
| 17-109-final-viral-combined_k141_65771\|\|lt2gene | Low-quality |
| 17-109-final-viral-combined_k141_50110\|\|lt2gene | Low-quality |
| 17-109-final-viral-combined_k141_300443\|\|lt2gene | Not-determined |
| 17-109-final-viral-combined_k141_99193\|\|lt2gene | Low-quality |
| 17-109-final-viral-combined_k141_50124\|\|lt2gene | Low-quality |
| 17-109-final-viral-combined_k141_334225\|\|lt2gene | Low-quality |
| 17-109-final-viral-combined_k141_183567\|\|lt2gene | Low-quality |
| 17-109-final-viral-combined_k141_265986\|\|lt2gene | Not-determined |
| 17-109-final-viral-combined_k141_200239\|\|lt2gene | Not-determined |
| 17-109-final-viral-combined_k141_99226\|\|lt2gene | Low-quality |
| 17-109-final-viral-combined_k141_149005\|\|lt2gene | Low-quality |
| 17-109-final-viral-combined_k141_183600\|\|lt2gene | Not-determined |
| 17-109-final-viral-combined_k141_266020\|\|lt2gene | Low-quality |
| 17-109-final-viral-combined_k141_132310\|\|lt2gene | Low-quality |
| 17-109-final-viral-combined_k141_149016\|\|lt2gene | Not-determined |
| 17-109-final-viral-combined_k141_132315\|\|lt2gene | Low-quality |
| 17-109-final-viral-combined_k141_149018\|\|lt2gene | Not-determined |
| 17-109-final-viral-combined_k141_200281\|\|lt2gene | Low-quality |
| 17-109-final-viral-combined_k141_115484\|\|lt2gene | Low-quality |
| 17-109-final-viral-combined_k141_383842\|\|lt2gene | Low-quality |
| 17-109-final-viral-combined_k141_65832\|\|lt2gene | Low-quality |
| 17-109-final-viral-combined_k141_183671\|\|lt2gene | Low-quality |
| 17-109-final-viral-combined_k141_200360\|\|lt2gene | Low-quality |
| 17-109-final-viral-combined_k141_115559\|\|lt2gene | Low-quality |
| 17-109-final-viral-combined_k141_99331\|\|lt2gene | Low-quality |
| 17-109-final-viral-combined_k141_334334\|\|lt2gene | Low-quality |
| 17-109-final-viral-combined_k141_383872\|\|lt2gene | Low-quality |
| 17-109-final-viral-combined_k141_166721\|\|lt2gene | Low-quality |
| 17-109-final-viral-combined_k141_266072\|\|lt2gene | Low-quality |
| 17-109-final-viral-combined_k141_266085\|\|lt2gene | Low-quality |
| 17-109-final-viral-combined_k141_65900\|\|lt2gene | Not-determined |
| 17-109-final-viral-combined_k141_32293\|\|lt2gene | Low-quality |
| 17-109-final-viral-combined_k141_300573\|\|lt2gene | Low-quality |
| 17-109-final-viral-combined_k141_83600\|\|lt2gene | Not-determined |
| 17-109-final-viral-combined_k141_266127\|\|lt2gene | Low-quality |
| 17-109-final-viral-combined_k141_32341\|\|lt2gene | Not-determined |
| 17-109-final-viral-combined_k141_65995\|\|lt2gene | Not-determined |
| 17-109-final-viral-combined_k141_350894\|\|lt2gene | Low-quality |
| 17-109-final-viral-combined_k141_149262\|\|lt2gene | Not-determined |
| 17-109-final-viral-combined_k141_384026\|\|lt2gene | Low-quality |
| 17-109-final-viral-combined_k141_183797\|\|lt2gene | Low-quality |
| 17-109-final-viral-combined_k141_266247\|\|lt2gene | Low-quality |
| 17-109-final-viral-combined_k141_32441\|\|lt2gene | Low-quality |
| 17-109-final-viral-combined_k141_166930\|\|lt2gene | Not-determined |
| 17-109-final-viral-combined_k141_32474\|\|lt2gene | Low-quality |
| 17-109-final-viral-combined_k141_99560\|\|lt2gene | Low-quality |
| 17-109-final-viral-combined_k141_115782\|\|lt2gene | Low-quality |
| 17-109-final-viral-combined_k141_334528\|\|lt2gene | Low-quality |
| 17-109-final-viral-combined_k141_217349\|\|lt2gene | Low-quality |
| 17-109-final-viral-combined_k141_300776\|\|lt2gene | Low-quality |
| 17-109-final-viral-combined_k141_351021\|\|lt2gene | Low-quality |
| 17-109-final-viral-combined_k141_217400\|\|lt2gene | Low-quality |
| 17-109-final-viral-combined_k141_334590\|\|lt2gene | Low-quality |
| 17-109-final-viral-combined_k141_334602\|\|lt2gene | Low-quality |
| 17-109-final-viral-combined_k141_351055\|\|lt2gene | Not-determined |
| 17-109-final-viral-combined_k141_351061\|\|lt2gene | Low-quality |
| 17-109-final-viral-combined_k141_99641\|\|lt2gene | Not-determined |
| 17-109-final-viral-combined_k141_99650\|\|lt2gene | Low-quality |
| 17-109-final-viral-combined_k141_66219\|\|lt2gene | Low-quality |
| 17-109-final-viral-combined_k141_200636\|\|lt2gene | Low-quality |
| 17-109-final-viral-combined_k141_167088\|\|lt2gene | Low-quality |
| 17-109-final-viral-combined_k141_200672\|\|lt2gene | Low-quality |
| 17-109-final-viral-combined_k141_250194\|\|lt2gene | Low-quality |
| 17-109-final-viral-combined_k141_99695\|\|lt2gene | Low-quality |
| 17-109-final-viral-combined_k141_250235\|\|lt2gene | Low-quality |
| 17-109-final-viral-combined_k141_334666\|\|lt2gene | Low-quality |
| 17-109-final-viral-combined_k141_66356\|\|lt2gene | Low-quality |
| 17-109-final-viral-combined_k141_115998\|\|lt2gene | Not-determined |
| 17-109-final-viral-combined_k141_116020\|\|lt2gene | Low-quality |
| 17-109-final-viral-combined_k141_66382\|\|lt2gene | Low-quality |
| 17-109-final-viral-combined_k141_99707\|\|lt2gene | Low-quality |
| 17-109-final-viral-combined_k141_384250\|\|lt2gene | Low-quality |
| 17-109-final-viral-combined_k141_66398\|\|lt2gene | Not-determined |
| 17-109-final-viral-combined_k141_66411\|\|lt2gene | Low-quality |
| 17-109-final-viral-combined_k141_266487\|\|lt2gene | Low-quality |
| 17-109-final-viral-combined_k141_200773\|\|lt2gene | Low-quality |
| 17-109-final-viral-combined_k141_384317\|\|lt2gene | Low-quality |
| 17-109-final-viral-combined_k141_351241\|\|lt2gene | Low-quality |
| 17-109-final-viral-combined_k141_149637\|\|lt2gene | Low-quality |
| 17-109-final-viral-combined_k141_384339\|\|lt2gene | Low-quality |
| 17-109-final-viral-combined_k141_116116\|\|lt2gene | Low-quality |
| 17-109-final-viral-combined_k141_384369\|\|lt2gene | Low-quality |
| 17-109-final-viral-combined_k141_351254\|\|lt2gene | Not-determined |
| 17-109-final-viral-combined_k141_266550\|\|lt2gene | Low-quality |
| 17-109-final-viral-combined_k141_334753\|\|lt2gene | Low-quality |
| 17-109-final-viral-combined_k141_366110\|\|lt2gene | Low-quality |
| 17-109-final-viral-combined_k141_149725\|\|lt2gene | Low-quality |
| 17-109-final-viral-combined_k141_32797\|\|lt2gene | Low-quality |
| 17-109-final-viral-combined_k141_66562\|\|lt2gene | Low-quality |
| 17-109-final-viral-combined_k141_99899\|\|lt2gene | Not-determined |
| 17-109-final-viral-combined_k141_384421\|\|lt2gene | Not-determined |
| 17-109-final-viral-combined_k141_16145\|\|lt2gene | Low-quality |
| 17-109-final-viral-combined_k141_366183\|\|lt2gene | Not-determined |
| 17-109-final-viral-combined_k141_301079\|\|lt2gene | Low-quality |
| 17-109-final-viral-combined_k141_167331\|\|lt2gene | Not-determined |
| 17-109-final-viral-combined_k141_301100\|\|lt2gene | Low-quality |
| 17-109-final-viral-combined_k141_250511\|\|lt2gene | Low-quality |
| 17-109-final-viral-combined_k141_250512\|\|lt2gene | Low-quality |
| 17-109-final-viral-combined_k141_116305\|\|lt2gene | Low-quality |
| 17-109-final-viral-combined_k141_32897\|\|lt2gene | Low-quality |
| 17-109-final-viral-combined_k141_400378\|\|lt2gene | Not-determined |
| 17-109-final-viral-combined_k141_167387\|\|lt2gene | Low-quality |
| 17-109-final-viral-combined_k141_366294\|\|lt2gene | Low-quality |
| 17-109-final-viral-combined_k141_366295\|\|lt2gene | Low-quality |
| 17-109-final-viral-combined_k141_100032\|\|lt2gene | Not-determined |
| 17-109-final-viral-combined_k141_116351\|\|lt2gene | Low-quality |
| 17-109-final-viral-combined_k141_32955\|\|lt2gene | Low-quality |
| 17-109-final-viral-combined_k141_149829\|\|lt2gene | Low-quality |
| 17-109-final-viral-combined_k141_32966\|\|lt2gene | Low-quality |
| 17-109-final-viral-combined_k141_149885\|\|lt2gene | Low-quality |
| 17-109-final-viral-combined_k141_16364\|\|lt2gene | Low-quality |
| 17-109-final-viral-combined_k141_66805\|\|lt2gene | Low-quality |
| 17-109-final-viral-combined_k141_301241\|\|lt2gene | Low-quality |
| 17-109-final-viral-combined_k141_400521\|\|lt2gene | Low-quality |
| 17-109-final-viral-combined_k141_149928\|\|lt2gene | Low-quality |
| 17-109-final-viral-combined_k141_33099\|\|lt2gene | Low-quality |
| 17-109-final-viral-combined_k141_66914\|\|lt2gene | Low-quality |
| 17-109-final-viral-combined_k141_116452\|\|lt2gene | Low-quality |
| 17-109-final-viral-combined_k141_133242\|\|lt2gene | Low-quality |
| 17-109-final-viral-combined_k141_351475\|\|lt2gene | Not-determined |
| 17-109-final-viral-combined_k141_33146\|\|lt2gene | Low-quality |
| 17-109-final-viral-combined_k141_266765\|\|lt2gene | Low-quality |
| 17-109-final-viral-combined_k141_33182\|\|lt2gene | Low-quality |
| 17-109-final-viral-combined_k141_351505\|\|lt2gene | Not-determined |
| 17-109-final-viral-combined_k141_366610\|\|lt2gene | Not-determined |
| 17-109-final-viral-combined_k141_400683\|\|lt2gene | Low-quality |
| 17-109-final-viral-combined_k141_100383\|\|lt2gene | Low-quality |
| 17-109-final-viral-combined_k141_400691\|\|lt2gene | Low-quality |
| 17-109-final-viral-combined_k141_16595\|\|lt2gene | Low-quality |
| 17-109-final-viral-combined_k141_100412\|\|lt2gene | Low-quality |
| 17-109-final-viral-combined_k141_16617\|\|lt2gene | Low-quality |
| 17-109-final-viral-combined_k141_16631\|\|lt2gene | Low-quality |
| 17-109-final-viral-combined_k141_384719\|\|lt2gene | Low-quality |
| 17-109-final-viral-combined_k141_267027\|\|lt2gene | Low-quality |
| 17-109-final-viral-combined_k141_400843\|\|lt2gene | Low-quality |
| 17-109-final-viral-combined_k141_400852\|\|lt2gene | Low-quality |
| 17-109-final-viral-combined_k141_251016\|\|lt2gene | Not-determined |
| 17-109-final-viral-combined_k141_33382\|\|lt2gene | Low-quality |
| 17-109-final-viral-combined_k141_116900\|\|lt2gene | Not-determined |
| 17-109-final-viral-combined_k141_150531\|\|lt2gene | Low-quality |
| 17-109-final-viral-combined_k141_367032\|\|lt2gene | Low-quality |
| 17-109-final-viral-combined_k141_400975\|\|lt2gene | Low-quality |
| 17-109-final-viral-combined_k141_116966\|\|lt2gene | Low-quality |
| 17-109-final-viral-combined_k141_133655\|\|lt2gene | Low-quality |
| 17-109-final-viral-combined_k141_401045\|\|lt2gene | Low-quality |
| 17-109-final-viral-combined_k141_117019\|\|lt2gene | Low-quality |
| 17-109-final-viral-combined_k141_117066\|\|lt2gene | Low-quality |
| 17-109-final-viral-combined_k141_367240\|\|lt2gene | Low-quality |
| 17-109-final-viral-combined_k141_133702\|\|lt2gene | Low-quality |
| 17-109-final-viral-combined_k141_117149\|\|lt2gene | Low-quality |
| 17-109-final-viral-combined_k141_367277\|\|lt2gene | Low-quality |
| 17-109-final-viral-combined_k141_133810\|\|lt2gene | Low-quality |
| 17-109-final-viral-combined_k141_367331\|\|lt2gene | Low-quality |
| 17-109-final-viral-combined_k141_367351\|\|lt2gene | Not-determined |
| 17-109-final-viral-combined_k141_133856\|\|lt2gene | Not-determined |
| 17-109-final-viral-combined_k141_133894\|\|lt2gene | Low-quality |
| 17-109-final-viral-combined_k141_401317\|\|lt2gene | Low-quality |
| 17-109-final-viral-combined_k141_367452\|\|lt2gene | Low-quality |
| 17-109-final-viral-combined_k141_367471\|\|lt2gene | Not-determined |
| 17-109-final-viral-combined_k141_401338\|\|lt2gene | Low-quality |
| 17-109-final-viral-combined_k141_401354\|\|lt2gene | Low-quality |
| 17-109-final-viral-combined_k141_367516\|\|lt2gene | Not-determined |
| 17-109-final-viral-combined_k141_267727\|\|lt2gene | Low-quality |
| 17-109-final-viral-combined_k141_267738\|\|lt2gene | Low-quality |
| 17-109-final-viral-combined_k141_367832\|\|lt2gene | Low-quality |
| 17-109-final-viral-combined_k141_367961\|\|lt2gene | Low-quality |
| 17-109-final-viral-combined_k141_368071\|\|lt2gene | Not-determined |
| 17-109_k141_334780 | Not-determined |
| 17-109_k141_2 | Not-determined |
| 17-109_k141_284564 | Not-determined |
| 17-109_k141_217609 | Not-determined |
| 17-109_k141_167392 | Not-determined |
| 17-109_k141_167393 | Not-determined |
| 17-109_k141_234349 | Not-determined |
| 17-109_k141_167394 | Not-determined |
| 17-109_k141_100442 | Not-determined |
| 17-109_k141_133915 | Not-determined |
| 17-109_k141_33488 | Not-determined |
| 17-109_k141_50227 | Not-determined |
| 17-109_k141_9 | Not-determined |
| 17-109_k141_50228 | Not-determined |
| 17-109_k141_16744 | Not-determined |
| 17-109_k141_217618 | Not-determined |
| 17-109_k141_100450 | Not-determined |
| 17-109_k141_16745 | Not-determined |
| 17-109_k141_200879 | Not-determined |
| 17-109_k141_385002 | Not-determined |
| 17-109_k141_16746 | Not-determined |
| 17-109_k141_385004 | Not-determined |
| 17-109_k141_251101 | Not-determined |
| 17-109_k141_33493 | Not-determined |
| 17-109_k141_16749 | Low-quality |
| 17-109_k141_284576 | Low-quality |
| 17-109_k141_83709 | Not-determined |
| 17-109_k141_234364 | Not-determined |
| 17-109_k141_50249 | Not-determined |
| 17-109_k141_217626 | Not-determined |
| 17-109_k141_100465 | Not-determined |
| 17-109_k141_234366 | Not-determined |
| 17-109_k141_83713 | Not-determined |
| 17-109_k141_100468 | Low-quality |
| 17-109_k141_385008 | Not-determined |
| 17-109_k141_83722 | Not-determined |
| 17-109_k141_50261 | Not-determined |
| 17-109_k141_133925 | Not-determined |
| 17-109_k141_301325 | Not-determined |
| 17-109_k141_50272 | Not-determined |
| 17-109_k141_133941 | Not-determined |
| 17-109_k141_133944 | Not-determined |
| 17-109_k141_301334 | Not-determined |
| 17-109_k141_334802 | Not-determined |
| 17-109_k141_267832 | Not-determined |
| 17-109_k141_368286 | Not-determined |
| 17-109_k141_368290 | Not-determined |
| 17-109_k141_368292 | Not-determined |
| 17-109_k141_66958 | Low-quality |
| 17-109_k141_167406 | Not-determined |
| 17-109_k141_100496 | Not-determined |
| 17-109_k141_66964 | Not-determined |
| 17-109_k141_234397 | Not-determined |
| 17-109_k141_16791 | Not-determined |
| 17-109_k141_234400 | Not-determined |
| 17-109_k141_100503 | Not-determined |
| 17-109_k141_301350 | Not-determined |
| 17-109_k141_117184 | Not-determined |
| 17-109_k141_117189 | Not-determined |
| 17-109_k141_167423 | Not-determined |
| 17-109_k141_66973 | Not-determined |
| 17-109_k141_301356 | Not-determined |
| 17-109_k141_234416 | Not-determined |
| 17-109_k141_351535 | Not-determined |
| 17-109_k141_83755 | Not-determined |
| 17-109_k141_234420 | Not-determined |
| 17-109_k141_167432 | Not-determined |
| 17-109_k141_385029 | Not-determined |
| 17-109_k141_167438 | Not-determined |
| 17-109_k141_167439 | Not-determined |
| 17-109_k141_251140 | Not-determined |
| 17-109_k141_33549 | Not-determined |
| 17-109_k141_117196 | Not-determined |
| 17-109_k141_33550 | Not-determined |
| 17-109_k141_16805 | Not-determined |
| 17-109_k141_16806 | Not-determined |
| 17-109_k141_200916 | Not-determined |
| 17-109_k141_251143 | Not-determined |
| 17-109_k141_184173 | Not-determined |
| 17-109_k141_66983 | Not-determined |
| 17-109_k141_16808 | Not-determined |
| 17-109_k141_200920 | Not-determined |
| 17-109_k141_301366 | Not-determined |
| 17-109_k141_251147 | Not-determined |
| 17-109_k141_167446 | Not-determined |
| 17-109_k141_368309 | Not-determined |
| 17-109_k141_301367 | Not-determined |
| 17-109_k141_267846 | Not-determined |
| 17-109_k141_117202 | Not-determined |
| 17-109_k141_33559 | Low-quality |
| 17-109_k141_368313 | Not-determined |
| 17-109_k141_184180 | Not-determined |
| 17-109_k141_33561 | Not-determined |
| 17-109_k141_184182 | Not-determined |
| 17-109_k141_385044 | Not-determined |
| 17-109_k141_66996 | Not-determined |
| 17-109_k141_66998 | Low-quality |
| 17-109_k141_251155 | Not-determined |
| 17-109_k141_100532 | Not-determined |
| 17-109_k141_385050 | Not-determined |
| 17-109_k141_184190 | Not-determined |
| 17-109_k141_368319 | Not-determined |
| 17-109_k141_368321 | Not-determined |
| 17-109_k141_251160 | Not-determined |
| 17-109_k141_385057 | Not-determined |
| 17-109_k141_33575 | Not-determined |
| 17-109_k141_184196 | Low-quality |
| 17-109_k141_33576 | Not-determined |
| 17-109_k141_33577 | Not-determined |
| 17-109_k141_167464 | Not-determined |
| 17-109_k141_33578 | Not-determined |
| 17-109_k141_33580 | Not-determined |
| 17-109_k141_385062 | Not-determined |
| 17-109_k141_251174 | Not-determined |
| 17-109_k141_76 | Not-determined |
| 17-109_k141_234437 | Not-determined |
| 17-109_k141_67008 | Not-determined |
| 17-109_k141_184209 | Not-determined |
| 17-109_k141_33591 | Not-determined |
| 17-109_k141_50360 | Low-quality |
| 17-109_k141_385073 | Not-determined |
| 17-109_k141_284659 | Not-determined |
| 17-109_k141_284660 | Not-determined |
| 17-109_k141_133980 | Not-determined |
| 17-109_k141_284663 | Not-determined |
| 17-109_k141_301383 | Not-determined |
| 17-109_k141_87 | Not-determined |
| 17-109_k141_50368 | Not-determined |
| 17-109_k141_33604 | Not-determined |
| 17-109_k141_301386 | Not-determined |
| 17-109_k141_93 | Not-determined |
| 17-109_k141_217691 | Not-determined |
| 17-109_k141_133987 | Not-determined |
| 17-109_k141_133990 | Not-determined |
| 17-109_k141_167486 | Not-determined |
| 17-109_k141_318084 | Not-determined |
| 17-109_k141_150692 | Not-determined |
| 17-109_k141_117233 | Not-determined |
| 17-109_k141_33612 | Not-determined |
| 17-109_k141_167503 | Not-determined |
| 17-109_k141_217713 | Not-determined |
| 17-109_k141_301393 | Not-determined |
| 17-109_k141_117237 | Low-quality |
| 17-109_k141_301394 | Not-determined |
| 17-109_k141_67025 | Not-determined |
| 17-109_k141_67026 | Not-determined |
| 17-109_k141_67027 | Not-determined |
| 17-109_k141_251201 | Not-determined |
| 17-109_k141_33623 | Not-determined |
| 17-109_k141_217724 | Low-quality |
| 17-109_k141_318102 | Not-determined |
| 17-109_k141_318107 | Not-determined |
| 17-109_k141_284690 | Not-determined |
| 17-109_k141_100571 | Not-determined |
| 17-109_k141_200982 | Not-determined |
| 17-109_k141_184262 | Not-determined |
| 17-109_k141_100579 | Not-determined |
| 17-109_k141_368344 | Low-quality |
| 17-109_k141_217740 | Low-quality |
| 17-109_k141_134004 | Not-determined |
| 17-109_k141_267888 | Not-determined |
| 17-109_k141_117247 | Not-determined |
| 17-109_k141_368352 | Not-determined |
| 17-109_k141_251226 | Not-determined |
| 17-109_k141_284707 | Not-determined |
| 17-109_k141_167529 | Not-determined |
| 17-109_k141_184271 | Not-determined |
| 17-109_k141_217745 | Low-quality |
| 17-109_k141_184272 | Not-determined |
| 17-109_k141_284723 | Not-determined |
| 17-109_k141_351570 | Low-quality |
| 17-109_k141_385094 | Not-determined |
| 17-109_k141_117260 | Not-determined |
| 17-109_k141_368362 | Not-determined |
| 17-109_k141_334807 | Not-determined |
| 17-109_k141_134016 | Not-determined |
| 17-109_k141_134017 | Not-determined |
| 17-109_k141_351574 | Not-determined |
| 17-109_k141_217748 | Not-determined |
| 17-109_k141_284735 | Not-determined |
| 17-109_k141_16833 | Not-determined |
| 17-109_k141_385115 | Not-determined |
| 17-109_k141_150 | Not-determined |
| 17-109_k141_151 | Not-determined |
| 17-109_k141_134030 | Not-determined |
| 17-109_k141_155 | Not-determined |
| 17-109_k141_301406 | Not-determined |
| 17-109_k141_217758 | Not-determined |
| 17-109_k141_217760 | Not-determined |
| 17-109_k141_217764 | Not-determined |
| 17-109_k141_368383 | Not-determined |
| 17-109_k141_217768 | Not-determined |
| 17-109_k141_201020 | Not-determined |
| 17-109_k141_167 | Not-determined |
| 17-109_k141_385125 | Not-determined |
| 17-109_k141_117282 | Not-determined |
| 17-109_k141_201032 | Not-determined |
| 17-109_k141_201033 | Not-determined |
| 17-109_k141_33660 | Not-determined |
| 17-109_k141_33664 | Not-determined |
| 17-109_k141_134057 | Not-determined |
| 17-109_k141_134059 | Not-determined |
| 17-109_k141_134062 | Not-determined |
| 17-109_k141_385145 | Low-quality |
| 17-109_k141_134063 | Not-determined |
| 17-109_k141_117289 | Not-determined |
| 17-109_k141_251249 | Not-determined |
| 18-102-final-viral-combined_k141_246480\|\|full | Medium-quality |
| 17-109_k141_334833 | Not-determined |
| 17-109_k141_334834 | Not-determined |
| 17-109_k141_251256 | Not-determined |
| 17-109_k141_251264 | Not-determined |
| 17-109_k141_201055 | Not-determined |
| 17-109_k141_267922 | Low-quality |
| 17-109_k141_267924 | Not-determined |
| 17-109_k141_195 | Low-quality |
| 17-109_k141_197 | Not-determined |
| 17-109_k141_351607 | Not-determined |
| 17-109_k141_33680 | Not-determined |
| 17-109_k141_167543 | Not-determined |
| 17-109_k141_201062 | Low-quality |
| 17-109_k141_301449 | Not-determined |
| 17-109_k141_267933 | Not-determined |
| 17-109_k141_100598 | Not-determined |

| | |
|---|---|
| 17-109_k141_301451 | Not-determined |
| 17-109_k141_167549 | Not-determined |
| 17-109_k141_134086 | Not-determined |
| 17-109_k141_167553 | Not-determined |
| 17-109_k141_301458 | Not-determined |
| 17-109_k141_217794 | Not-determined |
| 17-109_k141_201075 | Not-determined |
| 17-109_k141_184300 | Not-determined |
| 17-109_k141_201081 | Low-quality |
| 17-109_k141_351619 | Not-determined |
| 17-109_k141_167559 | Not-determined |
| 17-109_k141_184308 | Not-determined |
| 17-109_k141_385174 | Not-determined |
| 17-109_k141_351622 | Not-determined |
| 17-109_k141_134109 | Not-determined |
| 17-109_k141_351627 | Low-quality |
| 17-109_k141_201091 | Not-determined |
| 17-109_k141_318167 | Not-determined |
| 17-109_k141_150742 | Not-determined |
| 17-109_k141_217810 | Not-determined |
| 17-109_k141_201105 | Not-determined |
| 17-109_k141_251283 | Not-determined |
| 17-109_k141_117310 | Not-determined |
| 17-109_k141_83826 | Not-determined |
| 17-109_k141_67080 | Not-determined |
| 17-109_k141_201107 | Not-determined |
| 17-109_k141_301477 | Not-determined |
| 17-109_k141_301478 | Not-determined |
| 17-109_k141_16869 | Not-determined |
| 17-109_k141_301480 | Not-determined |
| 17-109_k141_301481 | Not-determined |
| 17-109_k141_301484 | Low-quality |
| 17-109_k141_150765 | Not-determined |
| 17-109_k141_301487 | Not-determined |
| 17-109_k141_67094 | Low-quality |
| 17-109_k141_201121 | Not-determined |
| 17-109_k141_67095 | Not-determined |
| 17-109_k141_100630 | Not-determined |
| 17-109_k141_67096 | Not-determined |
| 17-109_k141_100631 | Not-determined |
| 17-109_k141_167600 | Not-determined |
| 17-109_k141_167601 | Not-determined |
| 17-109_k141_33699 | Not-determined |
| 17-109_k141_100638 | Not-determined |
| 17-109_k141_67105 | Not-determined |
| 17-109_k141_33708 | Not-determined |
| 17-109_k141_67107 | Low-quality |
| 17-109_k141_351637 | Not-determined |
| 17-109_k141_167603 | Not-determined |
| 17-109_k141_100643 | Not-determined |
| 17-109_k141_33720 | Not-determined |
| 17-109_k141_33721 | Low-quality |
| 17-109_k141_50435 | Not-determined |
| 17-109_k141_33724 | Low-quality |
| 17-109_k141_16894 | Not-determined |
| 17-109_k141_251311 | Not-determined |
| 17-109_k141_33729 | Low-quality |
| 17-109_k141_16897 | Not-determined |
| 17-109_k141_16899 | Not-determined |
| 17-109_k141_134138 | Not-determined |
| 17-109_k141_167618 | Not-determined |
| 17-109_k141_385220 | Not-determined |
| 17-109_k141_284816 | Not-determined |
| 17-109_k141_83851 | Not-determined |
| 17-109_k141_50444 | Not-determined |
| 17-109_k141_368417 | Not-determined |
| 17-109_k141_368418 | Not-determined |
| 17-109_k141_33737 | Not-determined |
| 17-109_k141_50445 | Not-determined |
| 17-109_k141_117345 | Not-determined |
| 17-109_k141_167621 | Not-determined |
| 17-109_k141_284828 | Not-determined |
| 17-109_k141_33745 | Not-determined |
| 17-109_k141_385228 | Not-determined |
| 17-109_k141_33748 | Not-determined |
| 17-109_k141_318244 | Not-determined |
| 17-109_k141_251335 | Not-determined |
| 17-109_k141_318246 | Not-determined |
| 17-109_k141_318247 | Not-determined |
| 17-109_k141_385238 | Low-quality |
| 17-109_k141_318253 | Not-determined |
| 17-109_k141_83859 | Low-quality |
| 17-109_k141_184340 | Not-determined |
| 17-109_k141_50471 | Not-determined |
| 17-109_k141_368451 | Not-determined |
| 17-109_k141_351653 | Not-determined |
| 17-109_k141_368455 | Not-determined |
| 17-109_k141_368458 | Not-determined |
| 17-109_k141_50478 | Not-determined |
| 17-109_k141_150813 | Not-determined |
| 17-109_k141_167643 | Not-determined |
| 17-109_k141_351667 | Not-determined |
| 17-109_k141_150819 | Not-determined |
| 17-109_k141_67136 | Not-determined |
| 17-109_k141_318259 | Not-determined |
| 17-109_k141_284867 | Not-determined |
| 17-109_k141_150825 | Not-determined |
| 17-109_k141_385250 | Not-determined |
| 17-109_k141_284875 | Not-determined |
| 17-109_k141_385252 | Not-determined |
| 17-109_k141_184365 | Not-determined |
| 17-109_k141_385253 | Not-determined |
| 17-109_k141_385255 | Not-determined |
| 17-109_k141_334849 | Not-determined |
| 17-109_k141_385257 | Not-determined |
| 17-109_k141_217869 | Not-determined |
| 17-109_k141_217871 | Not-determined |
| 17-109_k141_301576 | Not-determined |
| 17-109_k141_217876 | Not-determined |
| 17-109_k141_150844 | Low-quality |
| 17-109_k141_134147 | Not-determined |
| 17-109_k141_16936 | Low-quality |
| 17-109_k141_301583 | Low-quality |
| 17-109_k141_267950 | Not-determined |
| 17-109_k141_301584 | Not-determined |
| 17-109_k141_67154 | Low-quality |
| 17-109_k141_234458 | Not-determined |
| 17-109_k141_67156 | Not-determined |
| 17-109_k141_16942 | Not-determined |
| 17-109_k141_67160 | Not-determined |
| 17-109_k141_267959 | Not-determined |
| 17-109_k141_16944 | Not-determined |
| 17-109_k141_117369 | Not-determined |
| 17-109_k141_117372 | Low-quality |
| 17-109_k141_334876 | Not-determined |
| 17-109_k141_217887 | Low-quality |
| 17-109_k141_117377 | Not-determined |
| 17-109_k141_217889 | Low-quality |
| 17-109_k141_33776 | Not-determined |
| 17-109_k141_301594 | Not-determined |
| 17-109_k141_284901 | Not-determined |
| 17-109_k141_50499 | Not-determined |
| 17-109_k141_257 | Not-determined |
| 17-109_k141_33789 | Not-determined |
| 17-109_k141_16964 | Not-determined |
| 17-109_k141_217898 | Not-determined |
| 17-109_k141_217899 | Not-determined |
| 17-109_k141_83915 | Low-quality |
| 17-109_k141_117382 | Not-determined |
| 17-109_k141_100652 | Not-determined |
| 17-109_k141_217902 | Not-determined |
| 17-109_k141_16967 | Not-determined |
| 17-109_k141_83928 | Not-determined |
| 17-109_k141_318304 | Not-determined |
| 17-109_k141_100659 | Not-determined |
| 17-109_k141_279 | Not-determined |
| 17-109_k141_83935 | Not-determined |
| 17-109_k141_16975 | Not-determined |
| 17-109_k141_67183 | Not-determined |
| 17-109_k141_16986 | Not-determined |
| 17-109_k141_16987 | Not-determined |
| 17-109_k141_117385 | Not-determined |
| 17-109_k141_67190 | Low-quality |
| 17-109_k141_117386 | Not-determined |
| 17-109_k141_67196 | Not-determined |
| 17-109_k141_50523 | Not-determined |
| 17-109_k141_234496 | Not-determined |
| 17-109_k141_16993 | Not-determined |
| 17-109_k141_100675 | Not-determined |
| 17-109_k141_67202 | Not-determined |
| 17-109_k141_301615 | Not-determined |
| 17-109_k141_301617 | Not-determined |
| 17-109_k141_294 | Not-determined |
| 17-109_k141_334895 | Not-determined |
| 17-109_k141_284910 | Not-determined |
| 17-109_k141_267972 | Not-determined |
| 17-109_k141_334897 | Not-determined |
| 17-109_k141_368473 | Not-determined |
| 17-109_k141_83949 | Low-quality |
| 17-109_k141_67216 | Not-determined |
| 17-109_k141_318343 | Low-quality |
| 17-109_k141_134155 | Low-quality |
| 17-109_k141_334902 | Not-determined |
| 17-109_k141_234528 | Not-determined |
| 17-109_k141_184385 | Not-determined |
| 17-109_k141_83958 | Not-determined |
| 17-109_k141_117406 | Not-determined |
| 17-109_k141_234531 | Not-determined |
| 17-109_k141_234538 | Not-determined |
| 17-109_k141_267976 | Not-determined |
| 17-109_k141_134168 | Not-determined |
| 17-109_k141_67228 | Low-quality |
| 17-109_k141_67229 | Not-determined |
| 17-109_k141_385271 | Not-determined |
| 17-109_k141_117425 | Not-determined |
| 17-109_k141_100688 | Not-determined |
| 17-109_k141_33815 | Not-determined |
| 17-109_k141_385275 | Not-determined |
| 17-109_k141_351695 | Not-determined |
| 17-109_k141_67243 | Not-determined |
| 17-109_k141_33825 | Low-quality |
| 17-109_k141_301638 | Not-determined |
| 17-109_k141_217959 | Not-determined |
| 17-109_k141_267999 | Not-determined |
| 17-109_k141_368496 | Not-determined |
| 17-109_k141_284951 | Not-determined |
| 17-109_k141_217961 | Not-determined |
| 17-109_k141_17017 | Not-determined |
| 17-109_k141_100705 | Not-determined |
| 17-109_k141_234557 | Not-determined |
| 17-109_k141_368503 | Not-determined |
| 17-109_k141_251395 | Not-determined |
| 17-109_k141_100711 | Not-determined |
| 17-109_k141_33840 | Not-determined |
| 17-109_k141_368506 | Low-quality |
| 17-109_k141_100714 | Not-determined |
| 17-109_k141_318367 | Not-determined |
| 17-109_k141_351710 | Not-determined |
| 17-109_k141_234564 | Not-determined |
| 17-109_k141_351713 | Not-determined |
| 17-109_k141_351717 | Not-determined |
| 17-109_k141_297 | Low-quality |
| 17-109_k141_368515 | Not-determined |
| 17-109_k141_134216 | Not-determined |
| 17-109_k141_67255 | Not-determined |
| 17-109_k141_304 | Not-determined |
| 17-109_k141_83970 | Not-determined |
| 17-109_k141_83972 | Not-determined |
| 17-109_k141_351719 | Low-quality |
| 17-109_k141_134226 | Not-determined |
| 17-109_k141_368517 | Low-quality |
| 17-109_k141_310 | Not-determined |
| 17-109_k141_234576 | Not-determined |
| 17-109_k141_301684 | Not-determined |
| 17-109_k141_385296 | Low-quality |
| 17-109_k141_385298 | Not-determined |
| 17-109_k141_100727 | Not-determined |
| 17-109_k141_284987 | Not-determined |
| 17-109_k141_317 | Not-determined |
| 17-109_k141_301703 | Not-determined |
| 17-109_k141_84004 | Not-determined |
| 17-109_k141_319 | Not-determined |
| 17-109_k141_167696 | Not-determined |
| 17-109_k141_50560 | Not-determined |
| 17-109_k141_323 | Not-determined |
| 17-109_k141_134243 | Not-determined |
| 17-109_k141_100730 | Not-determined |
| 17-109_k141_84007 | Low-quality |
| 17-109_k141_385313 | Low-quality |
| 17-109_k141_285005 | Not-determined |
| 17-109_k141_285007 | Not-determined |
| 17-109_k141_167700 | Not-determined |
| 17-109_k141_84018 | Not-determined |
| 17-109_k141_84020 | Not-determined |
| 17-109_k141_33883 | Not-determined |
| 17-109_k141_301712 | Not-determined |
| 17-109_k141_84021 | Not-determined |
| 17-109_k141_301713 | Not-determined |
| 17-109_k141_33885 | Not-determined |
| 17-109_k141_67294 | Not-determined |
| 17-109_k141_341 | Low-quality |
| 17-109_k141_84036 | Not-determined |
| 17-109_k141_301734 | Low-quality |
| 17-109_k141_301736 | Not-determined |
| 17-109_k141_234598 | Not-determined |
| 17-109_k141_84044 | Not-determined |
| 17-109_k141_84047 | Not-determined |
| 17-109_k141_84049 | Not-determined |
| 17-109_k141_33915 | Not-determined |
| 17-109_k141_117436 | Not-determined |
| 17-109_k141_318401 | Not-determined |
| 17-109_k141_268053 | Not-determined |
| 17-109_k141_334932 | Low-quality |
| 17-109_k141_184425 | Low-quality |
| 17-109_k141_285061 | Not-determined |
| 17-109_k141_84060 | Not-determined |
| 17-109_k141_84061 | Not-determined |
| 17-109_k141_355 | Not-determined |
| 17-109_k141_251467 | Low-quality |
| 17-109_k141_184433 | Not-determined |
| 17-109_k141_67308 | Not-determined |
| 17-109_k141_318408 | Not-determined |
| 17-109_k141_334943 | Not-determined |
| 17-109_k141_33933 | Low-quality |
| 17-109_k141_167738 | Not-determined |
| 17-109_k141_234618 | Not-determined |
| 17-109_k141_184445 | Not-determined |
| 17-109_k141_17071 | Not-determined |
| 17-109_k141_17077 | Not-determined |
| 17-109_k141_17080 | Not-determined |
| 17-109_k141_285069 | Not-determined |
| 17-109_k141_334951 | Not-determined |
| 17-109_k141_334955 | Not-determined |
| 17-109_k141_368532 | Not-determined |
| 17-109_k141_167753 | Not-determined |
| 17-109_k141_167756 | Not-determined |
| 17-109_k141_334957 | Not-determined |
| 17-109_k141_372 | Not-determined |
| 17-109_k141_17092 | Low-quality |
| 17-109_k141_375 | Not-determined |
| 17-109_k141_50582 | Not-determined |
| 17-109_k141_17096 | Not-determined |
| 17-109_k141_268071 | Not-determined |
| 17-109_k141_17098 | Not-determined |
| 17-109_k141_368536 | Not-determined |
| 17-109_k141_17099 | Not-determined |
| 17-109_k141_318426 | Not-determined |
| 17-109_k141_234642 | Not-determined |
| 17-109_k141_318428 | Not-determined |
| 17-109_k141_251513 | Not-determined |
| 17-109_k141_251514 | Not-determined |
| 17-109_k141_184456 | Not-determined |
| 17-109_k141_368549 | Not-determined |
| 17-109_k141_134277 | Not-determined |
| 17-109_k141_368554 | Not-determined |
| 17-109_k141_33945 | Not-determined |
| 17-109_k141_50606 | Not-determined |
| 17-109_k141_251525 | Not-determined |
| 17-109_k141_368558 | Not-determined |
| 17-109_k141_318450 | Low-quality |
| 17-109_k141_334972 | Low-quality |
| 17-109_k141_184471 | Not-determined |
| 17-109_k141_334974 | Not-determined |
| 17-109_k141_67317 | Not-determined |
| 17-109_k141_385376 | Low-quality |
| 17-109_k141_334983 | Not-determined |
| 17-109_k141_167770 | Not-determined |
| 17-109_k141_134294 | Not-determined |
| 17-109_k141_167779 | Not-determined |
| 17-109_k141_234654 | Not-determined |
| 17-109_k141_167780 | Not-determined |
| 17-109_k141_17151 | Not-determined |
| 17-109_k141_50619 | Not-determined |
| 17-109_k141_184482 | Not-determined |
| 17-109_k141_184483 | Not-determined |
| 17-109_k141_50622 | Not-determined |
| 17-109_k141_50624 | Not-determined |
| 17-109_k141_100766 | Not-determined |
| 17-109_k141_418 | Low-quality |
| 17-109_k141_318456 | Not-determined |
| 17-109_k141_318463 | Not-determined |
| 17-109_k141_201146 | Not-determined |
| 17-109_k141_385394 | Not-determined |
| 17-109_k141_268091 | Not-determined |
| 17-109_k141_17160 | Not-determined |
| 17-109_k141_301754 | Not-determined |
| 17-109_k141_17166 | Not-determined |
| 17-109_k141_67339 | Not-determined |
| 17-109_k141_301761 | Not-determined |
| 17-109_k141_50633 | Not-determined |
| 17-109_k141_84100 | Low-quality |
| 17-109_k141_184493 | Not-determined |
| 17-109_k141_385407 | Not-determined |
| 17-109_k141_318473 | Not-determined |
| 17-109_k141_268106 | Not-determined |
| 17-109_k141_234682 | Not-determined |
| 17-109_k141_368569 | Not-determined |
| 17-109_k141_50642 | Not-determined |
| 17-109_k141_100784 | Not-determined |
| 17-109_k141_268110 | Not-determined |
| 17-109_k141_318480 | Not-determined |
| 17-109_k141_33995 | Not-determined |
| 17-109_k141_318481 | Not-determined |
| 17-109_k141_84108 | Not-determined |
| 17-109_k141_50650 | Not-determined |
| 17-109_k141_84111 | Low-quality |
| 17-109_k141_251575 | Not-determined |
| 17-109_k141_33998 | Not-determined |
| 17-109_k141_34001 | Not-determined |
| 17-109_k141_426 | Not-determined |
| 17-109_k141_84114 | Not-determined |
| 17-109_k141_84117 | Not-determined |
| 17-109_k141_318501 | Low-quality |
| 17-109_k141_201158 | Not-determined |
| 17-109_k141_100801 | Not-determined |
| 17-109_k141_433 | Not-determined |
| 17-109_k141_84118 | Not-determined |
| 17-109_k141_17193 | Not-determined |
| 17-109_k141_50657 | Not-determined |
| 17-109_k141_444 | Low-quality |
| 17-109_k141_335028 | Low-quality |
| 17-109_k141_234715 | Not-determined |
| 17-109_k141_134337 | Not-determined |
| 17-109_k141_318513 | Not-determined |
| 17-109_k141_34023 | Not-determined |
| 17-109_k141_67360 | Not-determined |
| 17-109_k141_67361 | Not-determined |
| 17-109_k141_447 | Not-determined |
| 17-109_k141_448 | Not-determined |
| 17-109_k141_67364 | Not-determined |
| 17-109_k141_217990 | Not-determined |
| 17-109_k141_201161 | Not-determined |
| 17-109_k141_368602 | Not-determined |
| 17-109_k141_268124 | Not-determined |
| 17-109_k141_84132 | Low-quality |
| 17-109_k141_84133 | Not-determined |
| 17-109_k141_251596 | Not-determined |
| 17-109_k141_456 | Not-determined |
| 17-109_k141_234725 | Not-determined |
| 17-109_k141_368617 | Not-determined |
| 17-109_k141_17209 | Not-determined |
| 17-109_k141_301769 | Low-quality |
| 17-109_k141_301770 | Not-determined |
| 17-109_k141_184526 | Low-quality |
| 17-109_k141_368624 | Not-determined |
| 17-109_k141_251601 | Not-determined |
| 17-109_k141_285133 | Not-determined |
| 17-109_k141_67373 | Not-determined |
| 17-109_k141_201176 | Not-determined |
| 17-109_k141_471 | Not-determined |
| 17-109_k141_217997 | Not-determined |
| 17-109_k141_368632 | Not-determined |
| 17-109_k141_478 | Not-determined |
| 17-109_k141_84160 | Not-determined |
| 17-109_k141_268131 | Not-determined |
| 17-109_k141_268134 | Not-determined |
| 17-109_k141_234749 | Not-determined |
| 17-109_k141_218009 | Not-determined |
| 17-109_k141_285149 | Not-determined |
| 17-109_k141_184532 | Not-determined |
| 17-109_k141_184538 | Not-determined |
| 17-109_k141_184540 | Not-determined |
| 17-109_k141_487 | Not-determined |
| 17-109_k141_234757 | Low-quality |
| 17-109_k141_134376 | Not-determined |
| 17-109_k141_100839 | Not-determined |
| 17-109_k141_67384 | Low-quality |
| 17-109_k141_50696 | Not-determined |
| 17-109_k141_67385 | Not-determined |
| 17-109_k141_497 | Not-determined |
| 17-109_k141_335044 | Not-determined |
| 17-109_k141_218025 | Not-determined |
| 17-109_k141_100849 | Not-determined |
| 17-109_k141_17241 | Low-quality |
| 17-109_k141_285163 | Not-determined |
| 17-109_k141_285165 | Not-determined |
| 17-109_k141_100851 | Not-determined |
| 17-109_k141_318546 | Not-determined |
| 17-109_k141_17242 | Not-determined |
| 17-109_k141_167869 | Not-determined |
| 17-109_k141_34067 | Not-determined |
| 17-109_k141_318552 | Not-determined |
| 17-109_k141_201184 | Low-quality |
| 17-109_k141_318556 | Not-determined |
| 17-109_k141_201189 | Not-determined |
| 17-109_k141_201192 | Not-determined |
| 17-109_k141_234773 | Low-quality |
| 17-109_k141_234774 | Not-determined |
| 17-109_k141_234776 | Not-determined |
| 17-109_k141_34073 | Not-determined |
| 17-109_k141_150924 | Not-determined |
| 17-109_k141_134406 | Not-determined |
| 17-109_k141_50712 | Not-determined |
| 17-109_k141_285183 | Not-determined |
| 17-109_k141_167892 | Low-quality |
| 17-109_k141_167895 | Not-determined |
| 17-109_k141_167897 | Not-determined |
| 17-109_k141_50714 | Not-determined |
| 17-109_k141_335057 | Not-determined |
| 17-109_k141_167904 | Not-determined |
| 17-109_k141_268176 | Not-determined |
| 17-109_k141_318576 | Not-determined |
| 17-109_k141_508 | Not-determined |
| 17-109_k141_335059 | Not-determined |
| 17-109_k141_34093 | Not-determined |
| 17-109_k141_301810 | Not-determined |
| 17-109_k141_201208 | Not-determined |
| 17-109_k141_167917 | Not-determined |
| 17-109_k141_218042 | Not-determined |
| 17-109_k141_218047 | Not-determined |
| 17-109_k141_518 | Not-determined |
| 17-109_k141_218048 | Low-quality |
| 17-109_k141_167921 | Low-quality |
| 17-109_k141_84170 | Low-quality |
| 17-109_k141_134424 | Not-determined |
| 17-109_k141_525 | Not-determined |
| 17-109_k141_201212 | Not-determined |
| 17-109_k141_150961 | Not-determined |
| 17-109_k141_134429 | Not-determined |
| 17-109_k141_218055 | Not-determined |
| 17-109_k141_67390 | Not-determined |
| 17-109_k141_167925 | Low-quality |
| 17-109_k141_34107 | Not-determined |
| 17-109_k141_34115 | Not-determined |
| 17-109_k141_34116 | Not-determined |
| 17-109_k141_84185 | Not-determined |
| 17-109_k141_201222 | Not-determined |
| 17-109_k141_268196 | Not-determined |
| 17-109_k141_368675 | Not-determined |
| 17-109_k141_268197 | Not-determined |
| 17-109_k141_100897 | Not-determined |
| 17-109_k141_201225 | Not-determined |
| 17-109_k141_100905 | Not-determined |
| 17-109_k141_201227 | Not-determined |
| 17-109_k141_234806 | Not-determined |
| 17-109_k141_351747 | Not-determined |
| 17-109_k141_318600 | Not-determined |
| 17-109_k141_34126 | Not-determined |
| 17-109_k141_134453 | Low-quality |
| 17-109_k141_318609 | Low-quality |
| 17-109_k141_540 | Not-determined |
| 17-109_k141_368682 | Not-determined |
| 17-109_k141_545 | Not-determined |
| 17-109_k141_34140 | Not-determined |
| 17-109_k141_318618 | Not-determined |
| 17-109_k141_554 | Low-quality |
| 17-109_k141_50768 | Not-determined |
| 17-109_k141_318625 | Not-determined |
| 17-109_k141_50772 | Not-determined |
| 17-109_k141_50776 | Not-determined |
| 17-109_k141_84208 | Not-determined |
| 17-109_k141_218080 | Low-quality |
| 17-109_k141_218086 | Not-determined |
| 17-109_k141_201262 | Not-determined |
| 17-109_k141_100923 | Not-determined |
| 17-109_k141_201266 | Not-determined |
| 17-109_k141_318645 | Not-determined |
| 17-109_k141_167948 | Not-determined |
| 17-109_k141_318646 | Not-determined |
| 17-109_k141_201269 | Not-determined |
| 17-109_k141_167950 | Not-determined |
| 17-109_k141_167951 | Not-determined |
| 17-109_k141_318649 | Not-determined |
| 17-109_k141_351760 | Not-determined |
| 17-109_k141_218096 | Not-determined |
| 17-109_k141_17317 | Not-determined |
| 17-109_k141_100936 | Not-determined |
| 17-109_k141_17322 | Not-determined |
| 17-109_k141_134497 | Low-quality |
| 17-109_k141_34151 | Not-determined |
| 17-109_k141_134500 | Low-quality |
| 17-109_k141_318651 | Not-determined |
| 17-109_k141_50785 | Not-determined |
| 17-109_k141_318659 | Not-determined |
| 17-109_k141_67431 | Low-quality |
| 17-109_k141_368713 | Not-determined |
| 17-109_k141_318668 | Not-determined |
| 17-109_k141_368722 | Not-determined |
| 17-109_k141_318672 | Not-determined |
| 17-109_k141_201281 | Not-determined |
| 17-109_k141_368725 | Not-determined |
| 17-109_k141_601 | Not-determined |
| 17-109_k141_17342 | Not-determined |
| 17-109_k141_385487 | Not-determined |
| 17-109_k141_368731 | Not-determined |
| 17-109_k141_268212 | Not-determined |
| 17-109_k141_134517 | Not-determined |
| 17-109_k141_167981 | Not-determined |
| 17-109_k141_218112 | Low-quality |
| 17-109_k141_50797 | Not-determined |
| 17-109_k141_184575 | Not-determined |
| 17-109_k141_84221 | Not-determined |
| 17-109_k141_201291 | Not-determined |
| 17-109_k141_612 | Not-determined |
| 17-109_k141_335092 | Low-quality |
| 17-109_k141_84225 | Not-determined |
| 17-109_k141_50812 | Not-determined |
| 17-109_k141_385508 | Not-determined |
| 17-109_k141_134529 | Not-determined |
| 17-109_k141_385510 | Not-determined |
| 17-109_k141_385511 | Low-quality |
| 17-109_k141_318693 | Not-determined |
| 17-109_k141_385517 | Not-determined |
| 17-109_k141_50818 | Not-determined |
| 17-109_k141_318701 | Not-determined |
| 17-109_k141_167988 | Not-determined |
| 17-109_k141_201304 | Not-determined |
| 17-109_k141_385522 | Not-determined |
| 17-109_k141_234838 | Not-determined |
| 17-109_k141_100974 | Not-determined |
| 17-109_k141_34161 | Not-determined |
| 17-109_k141_636 | Not-determined |
| 17-109_k141_385539 | Not-determined |
| 17-109_k141_285256 | Not-determined |
| 17-109_k141_318709 | Not-determined |
| 17-109_k141_285260 | Not-determined |
| 17-109_k141_100979 | Not-determined |
| 17-109_k141_150995 | Not-determined |
| 17-109_k141_84237 | Not-determined |
| 17-109_k141_285264 | Not-determined |
| 17-109_k141_84245 | Not-determined |
| 17-109_k141_285270 | Not-determined |
| 17-109_k141_84253 | Not-determined |
| 17-109_k141_201321 | Not-determined |
| 17-109_k141_318727 | Not-determined |
| 17-109_k141_268244 | Not-determined |
| 17-109_k141_351776 | Not-determined |
| 17-109_k141_285281 | Not-determined |
| 17-109_k141_234860 | Low-quality |
| 17-109_k141_184646 | Not-determined |
| 17-109_k141_151007 | Not-determined |
| 17-109_k141_84261 | Not-determined |
| 17-109_k141_184649 | Not-determined |
| 17-109_k141_201329 | Not-determined |
| 17-109_k141_661 | Not-determined |
| 17-109_k141_385562 | Not-determined |
| 17-109_k141_351799 | Not-determined |
| 17-109_k141_34180 | Not-determined |
| 17-109_k141_201339 | Not-determined |

| | |
|---|---|
| 17-109_k141_201343 | Not-determined |
| 17-109_k141_50836 | Not-determined |
| 17-109_k141_17388 | Not-determined |
| 17-109_k141_67468 | Not-determined |
| 17-109_k141_67470 | Not-determined |
| 17-109_k141_234871 | Low-quality |
| 17-109_k141_234875 | Not-determined |
| 17-109_k141_335110 | Not-determined |
| 17-109_k141_234879 | Not-determined |
| 17-109_k141_670 | Not-determined |
| 17-109_k141_184659 | Not-determined |
| 17-109_k141_234885 | Not-determined |
| 17-109_k141_117462 | Not-determined |
| 17-109_k141_234887 | Not-determined |
| 17-109_k141_680 | Not-determined |
| 17-109_k141_385576 | Not-determined |
| 17-109_k141_134574 | Not-determined |
| 17-109_k141_285298 | Not-determined |
| 17-109_k141_285299 | Not-determined |
| 17-109_k141_682 | Not-determined |
| 17-109_k141_684 | Low-quality |
| 17-109_k141_686 | Not-determined |
| 17-109_k141_100999 | Not-determined |
| 17-109_k141_368778 | Not-determined |
| 17-109_k141_368780 | Not-determined |
| 17-109_k141_67502 | Not-determined |
| 17-109_k141_50846 | Not-determined |
| 17-109_k141_234901 | Not-determined |
| 17-109_k141_201363 | Not-determined |
| 17-109_k141_268261 | Not-determined |
| 17-109_k141_34213 | Not-determined |
| 17-109_k141_134594 | Not-determined |
| 17-109_k141_117480 | Not-determined |
| 17-109_k141_84295 | Not-determined |
| 17-109_k141_84299 | Not-determined |
| 17-109_k141_67519 | Not-determined |
| 17-109_k141_50866 | Not-determined |
| 17-109_k141_50869 | Low-quality |
| 17-109_k141_218117 | Not-determined |
| 17-109_k141_151019 | Not-determined |
| 17-109_k141_50871 | Not-determined |
| 17-109_k141_703 | Not-determined |
| 17-109_k141_218122 | Not-determined |
| 17-109_k141_335137 | Not-determined |
| 17-109_k141_67524 | Low-quality |
| 17-109_k141_17395 | Not-determined |
| 17-109_k141_17396 | Not-determined |
| 17-109_k141_151030 | Not-determined |
| 17-109_k141_67528 | Not-determined |
| 17-109_k141_151031 | Not-determined |
| 17-109_k141_50874 | Not-determined |
| 17-109_k141_335150 | Not-determined |
| 17-109_k141_67532 | Not-determined |
| 17-109_k141_151035 | Not-determined |
| 17-109_k141_50879 | Not-determined |
| 17-109_k141_268283 | Not-determined |
| 17-109_k141_17404 | Low-quality |
| 17-109_k141_268287 | Not-determined |
| 17-109_k141_268290 | Not-determined |
| 17-109_k141_67541 | Low-quality |
| 17-109_k141_218132 | Not-determined |
| 17-109_k141_201395 | Low-quality |
| 17-109_k141_201396 | Not-determined |
| 17-109_k141_335159 | Low-quality |
| 17-109_k141_251668 | Low-quality |
| 17-109_k141_251669 | Not-determined |
| 17-109_k141_285346 | Not-determined |
| 17-109_k141_117504 | Not-determined |
| 17-109_k141_17412 | Not-determined |
| 17-109_k141_151046 | Low-quality |
| 17-109_k141_17417 | Not-determined |
| 17-109_k141_218144 | Not-determined |
| 17-109_k141_34238 | Not-determined |
| 17-109_k141_368835 | Not-determined |
| 17-109_k141_218147 | Not-determined |
| 17-109_k141_17429 | Not-determined |
| 17-109_k141_151053 | Not-determined |
| 17-109_k141_84316 | Low-quality |
| 17-109_k141_285355 | Not-determined |
| 17-109_k141_117514 | Not-determined |
| 17-109_k141_117515 | Not-determined |
| 17-109_k141_17435 | Not-determined |
| 17-109_k141_151055 | Not-determined |
| 17-109_k141_17438 | Not-determined |
| 17-109_k141_268309 | Not-determined |
| 17-109_k141_117525 | Not-determined |
| 17-109_k141_251676 | Not-determined |
| 17-109_k141_251678 | Not-determined |
| 17-109_k141_84334 | Low-quality |
| 17-109_k141_50897 | Not-determined |
| 17-109_k141_17442 | Not-determined |
| 17-109_k141_201434 | Not-determined |
| 17-109_k141_168019 | Not-determined |
| 17-109_k141_101015 | Not-determined |
| 17-109_k141_234960 | Not-determined |
| 17-109_k141_234961 | Not-determined |
| 17-109_k141_101018 | Not-determined |
| 17-109_k141_117537 | Not-determined |
| 17-109_k141_101026 | Not-determined |
| 17-109_k141_285378 | Not-determined |
| 17-109_k141_117541 | Not-determined |
| 17-109_k141_17463 | Not-determined |
| 17-109_k141_117542 | Not-determined |
| 17-109_k141_101035 | Not-determined |
| 17-109_k141_101039 | Not-determined |
| 17-109_k141_134613 | Not-determined |
| 17-109_k141_134619 | Not-determined |
| 17-109_k141_351845 | Not-determined |
| 17-109_k141_134621 | Not-determined |
| 17-109_k141_728 | Not-determined |
| 17-109_k141_151081 | Low-quality |
| 17-109_k141_168028 | Not-determined |
| 17-109_k141_168029 | Not-determined |
| 17-109_k141_285394 | Low-quality |
| 17-109_k141_134634 | Not-determined |
| 17-109_k141_168042 | Not-determined |
| 17-109_k141_117567 | Not-determined |
| 17-109_k141_285412 | Not-determined |
| 17-109_k141_168054 | Not-determined |
| 17-109_k141_134642 | Not-determined |
| 17-109_k141_17504 | Not-determined |
| 17-109_k141_285422 | Low-quality |
| 17-109_k141_753 | Not-determined |
| 17-109_k141_134649 | Not-determined |
| 17-109_k141_117577 | Not-determined |
| 17-109_k141_268333 | Not-determined |
| 17-109_k141_134652 | Low-quality |
| 17-109_k141_168072 | Not-determined |
| 17-109_k141_201446 | Not-determined |
| 17-109_k141_50919 | Low-quality |
| 17-109_k141_184714 | Not-determined |
| 17-109_k141_50925 | Not-determined |
| 17-109_k141_184719 | Not-determined |
| 17-109_k141_184722 | Not-determined |
| 17-109_k141_766 | Not-determined |
| 17-109_k141_335191 | Not-determined |
| 17-109_k141_134671 | Not-determined |
| 17-109_k141_351878 | Not-determined |
| 17-109_k141_17528 | Low-quality |
| 17-109_k141_351886 | Not-determined |
| 17-109_k141_776 | Not-determined |
| 17-109_k141_184733 | Not-determined |
| 17-109_k141_268342 | Not-determined |
| 17-109_k141_17531 | Low-quality |
| 17-109_k141_201482 | Not-determined |
| 17-109_k141_351889 | Low-quality |
| 17-109_k141_84363 | Not-determined |
| 17-109_k141_368854 | Not-determined |
| 17-109_k141_368855 | Not-determined |
| 17-109_k141_268359 | Not-determined |
| 17-109_k141_234967 | Not-determined |
| 17-109_k141_117610 | Not-determined |
| 17-109_k141_201501 | Not-determined |
| 17-109_k141_234977 | Not-determined |
| 17-109_k141_184747 | Not-determined |
| 17-109_k141_151118 | Not-determined |
| 17-109_k141_168123 | Not-determined |
| 17-109_k141_151119 | Not-determined |
| 17-109_k141_184748 | Low-quality |
| 17-109_k141_368877 | Not-determined |
| 17-109_k141_301955 | Not-determined |
| 17-109_k141_17543 | Low-quality |
| 17-109_k141_335203 | Not-determined |
| 17-109_k141_84380 | Not-determined |
| 17-109_k141_84381 | Not-determined |
| 17-109_k141_134676 | Low-quality |
| 17-109_k141_268392 | Not-determined |
| 17-109_k141_218203 | Low-quality |
| 17-109_k141_34267 | Low-quality |
| 17-109_k141_34271 | Not-determined |
| 17-109_k141_335220 | Not-determined |
| 17-109_k141_792 | Not-determined |
| 17-109_k141_184763 | Not-determined |
| 17-109_k141_184765 | Not-determined |
| 17-109_k141_285486 | Not-determined |
| 17-109_k141_184769 | Not-determined |
| 17-109_k141_184770 | Not-determined |
| 17-109_k141_235012 | Not-determined |
| 17-109_k141_34279 | Low-quality |
| 17-109_k141_385613 | Not-determined |
| 17-109_k141_335229 | Not-determined |
| 17-109_k141_17564 | Low-quality |
| 17-109_k141_351927 | Not-determined |
| 17-109_k141_798 | Not-determined |
| 17-109_k141_34286 | Not-determined |
| 17-109_k141_151132 | Not-determined |
| 17-109_k141_17571 | Not-determined |
| 17-109_k141_34297 | Not-determined |
| 17-109_k141_134694 | Not-determined |
| 17-109_k141_67549 | Not-determined |
| 17-109_k141_101076 | Not-determined |
| 17-109_k141_318751 | Not-determined |
| 17-109_k141_67554 | Not-determined |
| 17-109_k141_385619 | Low-quality |
| 17-109_k141_235021 | Not-determined |
| 17-109_k141_101078 | Not-determined |
| 17-109_k141_134697 | Not-determined |
| 17-109_k141_285501 | Not-determined |
| 17-109_k141_385625 | Not-determined |
| 17-109_k141_67558 | Not-determined |
| 17-109_k141_201563 | Not-determined |
| 17-109_k141_184793 | Low-quality |
| 17-109_k141_235027 | Not-determined |
| 17-109_k141_285505 | Low-quality |
| 17-109_k141_201565 | Not-determined |
| 17-109_k141_184798 | Not-determined |
| 17-109_k141_318757 | Low-quality |
| 17-109_k141_101082 | Not-determined |
| 17-109_k141_67566 | Not-determined |
| 17-109_k141_351943 | Low-quality |
| 17-109_k141_351945 | Not-determined |
| 17-109_k141_268407 | Not-determined |
| 17-109_k141_17589 | Not-determined |
| 17-109_k141_808 | Not-determined |
| 17-109_k141_201575 | Not-determined |
| 17-109_k141_67576 | Not-determined |
| 17-109_k141_302004 | Low-quality |
| 17-109_k141_184817 | Not-determined |
| 17-109_k141_151164 | Not-determined |
| 17-109_k141_235035 | Not-determined |
| 17-109_k141_302007 | Not-determined |
| 17-109_k141_50963 | Not-determined |
| 17-109_k141_235042 | Not-determined |
| 17-109_k141_318767 | Low-quality |
| 17-109_k141_184824 | Not-determined |
| 17-109_k141_84418 | Not-determined |
| 17-109_k141_168158 | Low-quality |
| 17-109_k141_151174 | Not-determined |
| 17-109_k141_201584 | Low-quality |
| 17-109_k141_67586 | Not-determined |
| 17-109_k141_813 | Not-determined |
| 17-109_k141_816 | Not-determined |
| 17-109_k141_368928 | Not-determined |
| 17-109_k141_50973 | Not-determined |
| 17-109_k141_285535 | Not-determined |
| 17-109_k141_50974 | Low-quality |
| 17-109_k141_368931 | Low-quality |
| 17-109_k141_134710 | Low-quality |
| 17-109_k141_151188 | Not-determined |
| 17-109_k141_67598 | Not-determined |
| 17-109_k141_218233 | Not-determined |
| 17-109_k141_134715 | Not-determined |
| 17-109_k141_151196 | Not-determined |
| 17-109_k141_84432 | Not-determined |
| 17-109_k141_235055 | Low-quality |
| 17-109_k141_134720 | Not-determined |
| 17-109_k141_84433 | Not-determined |
| 17-109_k141_34306 | Not-determined |
| 17-109_k141_67607 | Not-determined |
| 17-109_k141_134723 | Not-determined |
| 17-109_k141_368938 | Not-determined |
| 17-109_k141_385660 | Low-quality |
| 17-109_k141_218247 | Not-determined |
| 17-109_k141_368940 | Not-determined |
| 17-109_k141_235064 | Not-determined |
| 17-109_k141_34311 | Not-determined |
| 17-109_k141_218255 | Not-determined |
| 17-109_k141_151209 | Not-determined |
| 17-109_k141_235076 | Not-determined |
| 17-109_k141_335240 | Not-determined |
| 17-109_k141_67632 | Low-quality |
| 17-109_k141_335241 | Not-determined |
| 17-109_k141_318792 | Not-determined |
| 17-109_k141_302017 | Not-determined |
| 17-109_k141_335248 | Not-determined |
| 17-109_k141_67638 | Not-determined |
| 17-109_k141_84452 | Not-determined |
| 17-109_k141_385681 | Not-determined |
| 17-109_k141_268444 | Low-quality |
| 17-109_k141_385683 | Not-determined |
| 17-109_k141_842 | Not-determined |
| 17-109_k141_849 | Not-determined |
| 17-109_k141_235083 | Not-determined |
| 17-109_k141_285563 | Not-determined |
| 17-109_k141_151225 | Not-determined |
| 17-109_k141_17593 | Not-determined |
| 17-109_k141_17594 | Not-determined |
| 17-109_k141_235089 | Low-quality |
| 17-109_k141_168184 | Not-determined |
| 17-109_k141_854 | Not-determined |
| 17-109_k141_235091 | Not-determined |
| 17-109_k141_134740 | Not-determined |
| 17-109_k141_134741 | Not-determined |
| 17-109_k141_84479 | Low-quality |
| 17-109_k141_268456 | Not-determined |
| 17-109_k141_285564 | Low-quality |
| 17-109_k141_17602 | Not-determined |
| 17-109_k141_67646 | Not-determined |
| 17-109_k141_17604 | Not-determined |
| 17-109_k141_251729 | Not-determined |
| 17-109_k141_67649 | Not-determined |
| 17-109_k141_51013 | Not-determined |
| 17-109_k141_67650 | Not-determined |
| 17-109_k141_67653 | Not-determined |
| 17-109_k141_101111 | Not-determined |
| 17-109_k141_385699 | Not-determined |
| 17-109_k141_318804 | Not-determined |
| 17-109_k141_151239 | Not-determined |
| 17-109_k141_385702 | Not-determined |
| 17-109_k141_34366 | Not-determined |
| 17-109_k141_268475 | Not-determined |
| 17-109_k141_201619 | Not-determined |
| 17-109_k141_51023 | Not-determined |
| 17-109_k141_235094 | Not-determined |
| 17-109_k141_67669 | Not-determined |
| 17-109_k141_67671 | Not-determined |
| 17-109_k141_876 | Not-determined |
| 17-109_k141_351985 | Low-quality |
| 17-109_k141_34369 | Not-determined |
| 17-109_k141_285586 | Not-determined |
| 17-109_k141_251748 | Not-determined |
| 17-109_k141_168197 | Not-determined |
| 17-109_k141_184877 | Not-determined |
| 17-109_k141_268484 | Not-determined |
| 17-109_k141_302033 | Low-quality |
| 17-109_k141_302035 | Low-quality |
| 17-109_k141_151262 | Not-determined |
| 17-109_k141_251757 | Not-determined |
| 17-109_k141_168201 | Not-determined |
| 17-109_k141_385727 | Not-determined |
| 17-109_k141_201629 | Not-determined |
| 17-109_k141_385729 | Not-determined |
| 17-109_k141_51044 | Not-determined |
| 17-109_k141_168206 | Not-determined |
| 17-109_k141_318827 | Not-determined |
| 17-109_k141_168213 | Not-determined |
| 17-109_k141_302049 | Not-determined |
| 17-109_k141_168215 | Not-determined |
| 17-109_k141_101123 | Not-determined |
| 17-109_k141_368977 | Not-determined |
| 17-109_k141_184889 | Low-quality |
| 17-109_k141_318839 | Not-determined |
| 17-109_k141_34398 | Not-determined |
| 17-109_k141_168223 | Not-determined |
| 17-109_k141_168224 | Not-determined |
| 17-109_k141_168225 | Not-determined |
| 17-109_k141_151279 | Low-quality |
| 17-109_k141_151281 | Not-determined |
| 17-109_k141_34402 | Not-determined |
| 17-109_k141_368983 | Not-determined |
| 17-109_k141_318847 | Not-determined |
| 17-109_k141_168233 | Not-determined |
| 17-109_k141_285618 | Not-determined |
| 17-109_k141_201657 | Not-determined |
| 17-109_k141_201658 | Not-determined |
| 17-109_k141_201659 | Not-determined |
| 17-109_k141_201662 | Not-determined |
| 17-109_k141_368990 | Not-determined |
| 17-109_k141_251780 | Not-determined |
| 17-109_k141_368991 | Not-determined |
| 17-109_k141_368992 | Not-determined |
| 17-109_k141_352004 | Not-determined |
| 17-109_k141_134801 | Not-determined |
| 17-109_k141_285626 | Not-determined |
| 17-109_k141_51065 | Not-determined |
| 17-109_k141_352005 | Low-quality |
| 17-109_k141_151299 | Low-quality |
| 17-109_k141_268512 | Not-determined |
| 17-109_k141_51068 | Not-determined |
| 17-109_k141_151305 | Not-determined |
| 17-109_k141_251781 | Low-quality |
| 17-109_k141_34427 | Not-determined |
| 17-109_k141_34429 | Not-determined |
| 17-109_k141_67703 | Not-determined |
| 17-109_k141_67706 | Not-determined |
| 17-109_k141_302103 | Not-determined |
| 17-109_k141_251791 | Not-determined |
| 17-109_k141_117650 | Not-determined |
| 17-108-final-viral-combined_k141_17029\|\|full | Medium-quality |
| 17-109_k141_894 | Not-determined |
| 17-109_k141_285632 | Not-determined |
| 17-109_k141_117658 | Low-quality |
| 17-109_k141_318876 | Not-determined |
| 17-109_k141_302106 | Not-determined |
| 17-109_k141_151316 | Not-determined |
| 17-109_k141_302107 | Not-determined |
| 17-109_k141_34435 | Not-determined |
| 17-109_k141_34436 | Not-determined |
| 17-109_k141_201670 | Not-determined |
| 17-109_k141_302109 | Not-determined |
| 17-109_k141_285636 | Low-quality |
| 17-109_k141_235130 | Not-determined |
| 17-109_k141_251802 | Not-determined |
| 17-109_k141_302117 | Not-determined |
| 17-109_k141_302120 | Not-determined |
| 17-109_k141_201673 | Not-determined |
| 17-109_k141_235136 | Not-determined |
| 17-109_k141_101153 | Not-determined |
| 17-109_k141_369014 | Not-determined |
| 17-109_k141_151324 | Not-determined |
| 17-109_k141_67715 | Not-determined |
| 17-109_k141_151330 | Not-determined |
| 17-109_k141_251821 | Not-determined |
| 17-109_k141_369029 | Not-determined |
| 17-109_k141_218267 | Not-determined |
| 17-109_k141_17628 | Not-determined |
| 17-109_k141_151347 | Low-quality |
| 17-109_k141_268553 | Not-determined |
| 17-109_k141_218273 | Low-quality |
| 17-109_k141_218274 | Not-determined |
| 17-109_k141_385769 | Not-determined |
| 17-109_k141_385770 | Not-determined |
| 17-109_k141_268561 | Not-determined |
| 17-109_k141_218282 | Low-quality |
| 17-109_k141_17658 | Not-determined |
| 17-109_k141_101188 | Low-quality |
| 17-109_k141_101191 | Not-determined |
| 17-109_k141_318888 | Not-determined |
| 17-109_k141_318889 | Not-determined |
| 17-109_k141_34469 | Not-determined |
| 17-109_k141_101198 | Not-determined |
| 17-109_k141_369054 | Not-determined |
| 17-109_k141_84504 | Not-determined |
| 17-109_k141_369055 | Not-determined |
| 17-109_k141_34472 | Low-quality |
| 17-109_k141_34473 | Not-determined |
| 17-109_k141_385795 | Not-determined |
| 17-109_k141_34475 | Not-determined |
| 17-109_k141_285657 | Low-quality |
| 17-109_k141_67755 | Not-determined |
| 17-109_k141_67757 | Not-determined |
| 17-109_k141_101207 | Not-determined |
| 17-109_k141_318915 | Not-determined |
| 17-109_k141_34479 | Not-determined |
| 17-109_k141_302153 | Not-determined |
| 17-109_k141_218309 | Not-determined |
| 17-109_k141_352051 | Low-quality |
| 17-109_k141_235180 | Not-determined |
| 17-109_k141_84517 | Not-determined |
| 17-109_k141_101216 | Not-determined |
| 17-109_k141_17681 | Not-determined |
| 17-109_k141_369073 | Low-quality |
| 17-109_k141_318936 | Low-quality |
| 17-109_k141_67773 | Low-quality |
| 17-109_k141_352060 | Low-quality |
| 17-109_k141_201691 | Low-quality |
| 17-109_k141_268582 | Not-determined |
| 17-109_k141_352062 | Not-determined |
| 17-109_k141_285662 | Not-determined |
| 17-109_k141_117709 | Not-determined |
| 17-109_k141_335332 | Low-quality |
| 17-109_k141_935 | Not-determined |
| 17-109_k141_335334 | Not-determined |
| 17-109_k141_101223 | Low-quality |
| 17-109_k141_117711 | Not-determined |
| 17-109_k141_352070 | Not-determined |
| 17-109_k141_34513 | Not-determined |
| 17-109_k141_941 | Not-determined |
| 17-109_k141_385810 | Not-determined |
| 17-109_k141_17706 | Not-determined |
| 17-109_k141_201706 | Not-determined |
| 17-109_k141_84537 | Not-determined |
| 17-109_k141_201709 | Not-determined |
| 17-109_k141_385819 | Not-determined |
| 17-109_k141_151391 | Not-determined |
| 17-109_k141_117720 | Not-determined |
| 17-109_k141_117723 | Not-determined |
| 17-109_k141_335348 | Not-determined |
| 17-109_k141_235185 | Not-determined |
| 17-109_k141_235186 | Not-determined |
| 17-109_k141_201721 | Not-determined |
| 17-109_k141_285682 | Not-determined |
| 17-109_k141_385836 | Not-determined |
| 17-109_k141_84544 | Not-determined |
| 17-109_k141_34543 | Not-determined |
| 17-109_k141_285688 | Not-determined |
| 17-109_k141_385839 | Not-determined |
| 17-109_k141_151394 | Low-quality |
| 17-109_k141_385840 | Low-quality |
| 17-109_k141_201732 | Not-determined |
| 17-109_k141_218349 | Not-determined |
| 17-109_k141_17717 | Not-determined |
| 17-109_k141_201741 | Not-determined |
| 17-109_k141_285699 | Not-determined |
| 17-109_k141_268602 | Not-determined |
| 17-109_k141_151403 | Not-determined |
| 17-109_k141_302198 | Not-determined |
| 17-109_k141_17732 | Not-determined |
| 17-109_k141_335352 | Not-determined |
| 17-109_k141_201744 | Not-determined |
| 17-109_k141_218360 | Not-determined |
| 17-109_k141_385874 | Not-determined |
| 17-109_k141_385875 | Not-determined |
| 17-109_k141_184998 | Not-determined |
| 17-109_k141_218366 | Not-determined |
| 17-109_k141_976 | Not-determined |
| 17-109_k141_251845 | Not-determined |
| 17-109_k141_369090 | Not-determined |
| 17-109_k141_285715 | Not-determined |
| 17-109_k141_218368 | Not-determined |
| 17-109_k141_235200 | Not-determined |
| 17-109_k141_84557 | Not-determined |
| 17-109_k141_218378 | Not-determined |
| 17-109_k141_302206 | Not-determined |
| 17-109_k141_251853 | Not-determined |
| 17-109_k141_34581 | Not-determined |
| 17-109_k141_34585 | Not-determined |
| 17-109_k141_218383 | Not-determined |
| 17-109_k141_268620 | Low-quality |
| 17-109_k141_995 | Not-determined |
| 17-109_k141_101252 | Not-determined |
| 17-109_k141_218388 | Not-determined |
| 17-109_k141_285725 | Not-determined |
| 17-109_k141_34588 | Not-determined |
| 17-109_k141_268625 | Not-determined |
| 17-109_k141_84568 | Not-determined |
| 17-109_k141_251866 | Not-determined |
| 17-109_k141_268626 | Not-determined |
| 17-109_k141_251869 | Not-determined |
| 17-109_k141_285727 | Not-determined |
| 17-109_k141_352080 | Not-determined |
| 17-109_k141_218393 | Not-determined |
| 17-109_k141_302215 | Low-quality |
| 17-109_k141_34599 | Low-quality |
| 17-109_k141_34600 | Not-determined |
| 17-109_k141_335384 | Not-determined |
| 17-109_k141_352087 | Not-determined |
| 17-109_k141_151420 | Low-quality |
| 17-109_k141_67823 | Not-determined |
| 17-109_k141_251881 | Low-quality |
| 17-109_k141_385900 | Low-quality |
| 17-109_k141_302224 | Not-determined |
| 17-109_k141_385905 | Not-determined |
| 17-109_k141_17789 | Not-determined |
| 17-109_k141_251889 | Low-quality |
| 17-109_k141_302230 | Not-determined |
| 17-109_k141_352101 | Not-determined |
| 17-109_k141_235221 | Not-determined |
| 17-109_k141_268651 | Not-determined |
| 17-109_k141_268653 | Not-determined |
| 17-109_k141_268656 | Not-determined |
| 17-109_k141_17798 | Not-determined |
| 17-109_k141_251907 | Not-determined |
| 17-109_k141_34618 | Not-determined |
| 17-109_k141_34619 | Low-quality |
| 17-109_k141_34621 | Not-determined |
| 17-109_k141_251909 | Not-determined |
| 17-109_k141_34624 | Not-determined |
| 17-109_k141_51085 | Not-determined |
| 17-109_k141_335408 | Not-determined |
| 17-109_k141_335410 | Not-determined |
| 17-109_k141_51097 | Not-determined |
| 17-109_k141_302241 | Not-determined |
| 17-109_k141_335416 | Not-determined |
| 17-109_k141_335422 | Not-determined |
| 17-109_k141_51105 | Not-determined |
| 17-109_k141_369105 | Not-determined |
| 17-109_k141_1031 | Not-determined |
| 17-109_k141_1032 | Not-determined |
| 17-109_k141_67864 | Not-determined |
| 17-109_k141_1033 | Not-determined |
| 17-109_k141_302246 | Not-determined |
| 17-109_k141_335424 | Low-quality |
| 17-109_k141_335425 | Not-determined |
| 17-109_k141_168273 | Not-determined |
| 17-109_k141_185059 | Not-determined |
| 17-109_k141_101279 | Not-determined |
| 17-109_k141_101280 | Not-determined |
| 17-109_k141_34633 | Not-determined |
| 17-109_k141_1039 | Not-determined |
| 17-109_k141_268675 | Not-determined |
| 17-109_k141_101287 | Not-determined |
| 17-109_k141_218425 | Low-quality |
| 17-109_k141_117752 | Low-quality |
| 17-109_k141_101294 | Not-determined |
| 17-109_k141_369120 | Not-determined |
| 17-109_k141_352125 | Not-determined |
| 17-109_k141_34647 | Not-determined |
| 17-109_k141_235270 | Not-determined |
| 17-109_k141_369123 | Low-quality |
| 17-109_k141_168288 | Not-determined |
| 17-109_k141_302261 | Not-determined |
| 17-109_k141_117764 | Not-determined |
| 17-109_k141_168294 | Low-quality |
| 17-109_k141_268700 | Not-determined |
| 17-109_k141_352135 | Not-determined |
| 17-109_k141_352144 | Not-determined |
| 17-109_k141_352145 | Not-determined |
| 17-109_k141_352146 | Not-determined |
| 17-109_k141_17871 | Not-determined |
| 17-109_k141_134817 | Low-quality |
| 17-109_k141_134818 | Not-determined |
| 17-109_k141_168301 | Not-determined |
| 17-109_k141_117775 | Not-determined |
| 17-109_k141_101309 | Not-determined |
| 17-109_k141_285775 | Not-determined |
| 17-109_k141_17888 | Not-determined |
| 17-109_k141_335433 | Not-determined |
| 17-109_k141_335434 | Not-determined |

| | |
|---|---|
| 17-109_k141_335435 | Not-determined |
| 17-109_k141_235276 | Low-quality |
| 17-109_k141_369154 | Not-determined |
| 17-109_k141_101330 | Not-determined |
| 17-109_k141_385947 | Not-determined |
| 17-109_k141_302274 | Not-determined |
| 17-109_k141_235283 | Not-determined |
| 17-109_k141_235286 | Not-determined |
| 17-109_k141_318995 | Not-determined |
| 17-109_k141_101335 | Not-determined |
| 17-109_k141_352166 | Not-determined |
| 17-109_k141_17904 | Low-quality |
| 17-109_k141_134833 | Not-determined |
| 17-109_k141_134838 | Not-determined |
| 17-109_k141_385965 | Not-determined |
| 17-109_k141_34695 | Not-determined |
| 17-109_k141_67871 | Low-quality |
| 17-109_k141_352169 | Not-determined |
| 17-109_k141_134842 | Not-determined |
| 17-109_k141_385971 | Not-determined |
| 17-109_k141_168303 | Low-quality |
| 17-109_k141_235313 | Not-determined |
| 17-109_k141_201756 | Not-determined |
| 17-109_k141_51142 | Low-quality |
| 17-109_k141_84643 | Not-determined |
| 17-109_k141_268745 | Not-determined |
| 17-109_k141_84648 | Not-determined |
| 17-109_k141_34707 | Not-determined |
| 17-109_k141_235316 | Low-quality |
| 17-109_k141_268750 | Low-quality |
| 17-109_k141_385987 | Not-determined |
| 17-109_k141_235319 | Not-determined |
| 17-109_k141_319028 | Not-determined |
| 17-109_k141_168325 | Not-determined |
| 17-109_k141_218491 | Not-determined |
| 17-109_k141_67883 | Not-determined |
| 17-109_k141_268753 | Not-determined |
| 17-109_k141_335466 | Not-determined |
| 17-109_k141_17920 | Not-determined |
| 17-109_k141_268756 | Low-quality |
| 17-109_k141_117795 | Not-determined |
| 17-109_k141_168331 | Not-determined |
| 17-109_k141_101364 | Low-quality |
| 17-109_k141_268764 | Not-determined |
| 17-109_k141_67892 | Not-determined |
| 17-109_k141_117798 | Low-quality |
| 17-109_k141_218503 | Not-determined |
| 17-109_k141_285800 | Low-quality |
| 17-109_k141_17934 | Not-determined |
| 17-109_k141_235335 | Not-determined |
| 17-109_k141_386000 | Not-determined |
| 17-109_k141_134864 | Not-determined |
| 17-109_k141_268771 | Not-determined |
| 17-109_k141_268773 | Not-determined |
| 17-109_k141_168336 | Not-determined |
| 17-109_k141_34724 | Not-determined |
| 17-109_k141_168338 | Not-determined |
| 17-109_k141_151479 | Not-determined |
| 17-109_k141_151487 | Not-determined |
| 17-109_k141_319043 | Not-determined |
| 17-109_k141_218507 | Not-determined |
| 17-109_k141_218508 | Not-determined |
| 17-109_k141_218509 | Not-determined |
| 17-109_k141_101370 | Not-determined |
| 17-109_k141_252006 | Not-determined |
| 17-109_k141_151498 | Not-determined |
| KM-final-viral-combined_k141_130215\|\|full | Medium-quality |
| 17-109_k141_235370 | Not-determined |
| 17-109_k141_235371 | Not-determined |
| 17-109_k141_352185 | Not-determined |
| 17-109_k141_151514 | Not-determined |
| 17-109_k141_34736 | Not-determined |
| 17-109_k141_285821 | Low-quality |
| 17-109_k141_352202 | Not-determined |
| 17-109_k141_134890 | Not-determined |
| 17-109_k141_352205 | Not-determined |
| 17-109_k141_201796 | Not-determined |
| 17-109_k141_319059 | Not-determined |
| 17-109_k141_1064 | Low-quality |
| 17-109_k141_218526 | Not-determined |
| 17-109_k141_185123 | Not-determined |
| 17-109_k141_218532 | Not-determined |
| 17-109_k141_386008 | Not-determined |
| 17-109_k141_117815 | Not-determined |
| 17-109_k141_201813 | Not-determined |
| 17-109_k141_67906 | Not-determined |
| 17-109_k141_302331 | Low-quality |
| 17-109_k141_17979 | Not-determined |
| 17-109_k141_151544 | Not-determined |
| 17-109_k141_319070 | Not-determined |
| 17-109_k141_201818 | Not-determined |
| 17-109_k141_319071 | Not-determined |
| 17-109_k141_335526 | Not-determined |
| 17-109_k141_302337 | Not-determined |
| 17-109_k141_201825 | Not-determined |
| 17-109_k141_117831 | Not-determined |
| 17-109_k141_352230 | Not-determined |
| 17-109_k141_319075 | Low-quality |
| 17-109_k141_17983 | Not-determined |
| 17-109_k141_67913 | Low-quality |
| 17-109_k141_252023 | Not-determined |
| 17-109_k141_201832 | Not-determined |
| 17-109_k141_67919 | Not-determined |
| 17-109_k141_218543 | Not-determined |
| 17-109_k141_151557 | Not-determined |
| 17-109_k141_67922 | Not-determined |
| 17-109_k141_335533 | Not-determined |
| 17-109_k141_335539 | Not-determined |
| 17-109_k141_201837 | Not-determined |
| 17-109_k141_134907 | Not-determined |
| 17-109_k141_235414 | Not-determined |
| 17-109_k141_51193 | Not-determined |
| 17-109_k141_235416 | Low-quality |
| 17-109_k141_252041 | Low-quality |
| 17-109_k141_34758 | Not-determined |
| 17-109_k141_51198 | Not-determined |
| 17-109_k141_34759 | Not-determined |
| 17-109_k141_335549 | Not-determined |
| 17-109_k141_34762 | Not-determined |
| 17-109_k141_51201 | Not-determined |
| 17-109_k141_285853 | Not-determined |
| 17-109_k141_51207 | Not-determined |
| 17-109_k141_201851 | Low-quality |
| 17-109_k141_134927 | Not-determined |
| 17-109_k141_218577 | Not-determined |
| 17-109_k141_218578 | Not-determined |
| 17-109_k141_335556 | Not-determined |
| 17-109_k141_319097 | Not-determined |
| 17-109_k141_268820 | Not-determined |
| 17-109_k141_34767 | Not-determined |
| 17-109_k141_252067 | Not-determined |
| 17-109_k141_252068 | Not-determined |
| 17-109_k141_268824 | Not-determined |
| 17-109_k141_218592 | Not-determined |
| 17-109_k141_369162 | Not-determined |
| 17-109_k141_218594 | Not-determined |
| 17-109_k141_168361 | Not-determined |
| 17-109_k141_352255 | Not-determined |
| 17-109_k141_34774 | Not-determined |
| 17-109_k141_369172 | Not-determined |
| 17-109_k141_168366 | Not-determined |
| 17-109_k141_134932 | Not-determined |
| 17-109_k141_335573 | Not-determined |
| 17-109_k141_352260 | Not-determined |
| 17-109_k141_386049 | Not-determined |
| 17-109_k141_352261 | Not-determined |
| 17-109_k141_218606 | Not-determined |
| 17-109_k141_335578 | Not-determined |
| 17-109_k141_369183 | Low-quality |
| 17-109_k141_1108 | Not-determined |
| 17-109_k141_268844 | Not-determined |
| 17-109_k141_252091 | Not-determined |
| 17-109_k141_134937 | Not-determined |
| 17-109_k141_235441 | Not-determined |
| 17-109_k141_34786 | Not-determined |
| 17-109_k141_235456 | Not-determined |
| 17-109_k141_1113 | Low-quality |
| 17-109_k141_218619 | Not-determined |
| 17-109_k141_185171 | Not-determined |
| 17-109_k141_185172 | Not-determined |
| 17-109_k141_201878 | Not-determined |
| 17-109_k141_185175 | Not-determined |
| 17-109_k141_252117 | Not-determined |
| 17-109_k141_252118 | Not-determined |
| 17-109_k141_302369 | Low-quality |
| 17-109_k141_302371 | Not-determined |
| 17-109_k141_134955 | Not-determined |
| 17-109_k141_117897 | Not-determined |
| 17-109_k141_134956 | Not-determined |
| 17-109_k141_151638 | Not-determined |
| 17-109_k141_369215 | Not-determined |
| 17-109_k141_151644 | Not-determined |
| 17-109_k141_352285 | Not-determined |
| 17-109_k141_151650 | Not-determined |
| 17-109_k141_185197 | Not-determined |
| 17-109_k141_369224 | Not-determined |
| 17-109_k141_117909 | Not-determined |
| 17-109_k141_369233 | Not-determined |
| 17-109_k141_369235 | Not-determined |
| 17-109_k141_352301 | Not-determined |
| 17-109_k141_369239 | Not-determined |
| 17-109_k141_335599 | Not-determined |
| 17-109_k141_268863 | Not-determined |
| 17-109_k141_218637 | Not-determined |
| 17-109_k141_101427 | Not-determined |
| 17-109_k141_369248 | Not-determined |
| 17-109_k141_218640 | Not-determined |
| 17-109_k141_386097 | Not-determined |
| 17-109_k141_101438 | Not-determined |
| 17-109_k141_1120 | Not-determined |
| 17-109_k141_369264 | Not-determined |
| 17-109_k141_1128 | Not-determined |
| 17-109_k141_51269 | Low-quality |
| 17-109_k141_335612 | Not-determined |
| 17-109_k141_51278 | Not-determined |
| 17-109_k141_168417 | Not-determined |
| 17-109_k141_34822 | Not-determined |
| 17-109_k141_386112 | Not-determined |
| 17-109_k141_319176 | Not-determined |
| 17-109_k141_319177 | Not-determined |
| 17-109_k141_1139 | Not-determined |
| 17-109_k141_51285 | Not-determined |
| 17-109_k141_84776 | Not-determined |
| 17-109_k141_369283 | Low-quality |
| 17-109_k141_218645 | Not-determined |
| 17-109_k141_386122 | Not-determined |
| 17-109_k141_369288 | Not-determined |
| 17-109_k141_101446 | Not-determined |
| 17-109_k141_386128 | Not-determined |
| 17-109_k141_185233 | Not-determined |
| 17-109_k141_101447 | Not-determined |
| 17-109_k141_1153 | Not-determined |
| 17-109_k141_151656 | Not-determined |
| 17-109_k141_151658 | Not-determined |
| 17-109_k141_185244 | Not-determined |
| 17-109_k141_185246 | Not-determined |
| 17-109_k141_369297 | Not-determined |
| 17-109_k141_1158 | Not-determined |
| 17-109_k141_101456 | Low-quality |
| 17-109_k141_168427 | Not-determined |
| 17-109_k141_1163 | Not-determined |
| 17-109_k141_34835 | Not-determined |
| 17-109_k141_352336 | Not-determined |
| 17-109_k141_319186 | Not-determined |
| 17-109_k141_352337 | Low-quality |
| 17-109_k141_302382 | Not-determined |
| 17-109_k141_302383 | Not-determined |
| 17-109_k141_352342 | Not-determined |
| 17-109_k141_352344 | Not-determined |
| 17-109_k141_34846 | Not-determined |
| 17-109_k141_34847 | Not-determined |
| 17-109_k141_319192 | Not-determined |
| 17-109_k141_268906 | Not-determined |
| 17-109_k141_34861 | Low-quality |
| 17-109_k141_51308 | Not-determined |
| 17-109_k141_302402 | Not-determined |
| 17-109_k141_51314 | Not-determined |
| 17-109_k141_185269 | Not-determined |
| 17-109_k141_319207 | Low-quality |
| 17-109_k141_1187 | Not-determined |
| 17-109_k141_218678 | Not-determined |
| 17-109_k141_51321 | Not-determined |
| 17-109_k141_201900 | Not-determined |
| 17-109_k141_335631 | Not-determined |
| 17-109_k141_369337 | Not-determined |
| 17-109_k141_1193 | Not-determined |
| 17-109_k141_302419 | Not-determined |
| 17-109_k141_335632 | Not-determined |
| 17-109_k141_168463 | Not-determined |
| 17-109_k141_369346 | Not-determined |
| 17-109_k141_168466 | Not-determined |
| 17-109_k141_117951 | Not-determined |
| 17-109_k141_101474 | Not-determined |
| 17-109_k141_218683 | Low-quality |
| 17-109_k141_168473 | Not-determined |
| 17-109_k141_252178 | Not-determined |
| 17-109_k141_319215 | Not-determined |
| 17-109_k141_319216 | Not-determined |
| 17-109_k141_34885 | Not-determined |
| 17-109_k141_117965 | Not-determined |
| 17-109_k141_218699 | Not-determined |
| 17-109_k141_268929 | Not-determined |
| 17-109_k141_369362 | Not-determined |
| 17-109_k141_369363 | Not-determined |
| 17-109_k141_319219 | Not-determined |
| 17-109_k141_1208 | Not-determined |
| 17-109_k141_51332 | Not-determined |
| 17-109_k141_218707 | Low-quality |
| 17-109_k141_319228 | Not-determined |
| 17-109_k141_151671 | Not-determined |
| 17-109_k141_84794 | Not-determined |
| 17-109_k141_252196 | Not-determined |
| 17-109_k141_218709 | Not-determined |
| 17-109_k141_84795 | Not-determined |
| 17-109_k141_1223 | Not-determined |
| 17-109_k141_369377 | Low-quality |
| 17-109_k141_1228 | Not-determined |
| 17-109_k141_201938 | Low-quality |
| 17-109_k141_168489 | Not-determined |
| 17-109_k141_352397 | Not-determined |
| 17-109_k141_319242 | Not-determined |
| 17-109_k141_369380 | Not-determined |
| 17-109_k141_51346 | Not-determined |
| 17-109_k141_369382 | Not-determined |
| 17-109_k141_369383 | Not-determined |
| 17-109_k141_51347 | Not-determined |
| 17-109_k141_218728 | Not-determined |
| 17-109_k141_319253 | Not-determined |
| 17-109_k141_218729 | Not-determined |
| 17-109_k141_84815 | Not-determined |
| 17-109_k141_1247 | Not-determined |
| 17-109_k141_168511 | Not-determined |
| 17-109_k141_352411 | Not-determined |
| 17-109_k141_352413 | Not-determined |
| 17-109_k141_185274 | Not-determined |
| 17-109_k141_101495 | Not-determined |
| 17-109_k141_369404 | Not-determined |
| 17-109_k141_117981 | Not-determined |
| 17-109_k141_68041 | Not-determined |
| 17-109_k141_352416 | Not-determined |
| 17-109_k141_369411 | Not-determined |
| 17-109_k141_51371 | Not-determined |
| 17-109_k141_84833 | Not-determined |
| 17-109_k141_185283 | Not-determined |
| 17-109_k141_68052 | Not-determined |
| 17-109_k141_68053 | Not-determined |
| 17-109_k141_185288 | Not-determined |
| 17-109_k141_302466 | Not-determined |
| 17-109_k141_252238 | Not-determined |
| 17-109_k141_17992 | Low-quality |
| 17-109_k141_386205 | Not-determined |
| 17-109_k141_51374 | Not-determined |
| 17-109_k141_51376 | Not-determined |
| 17-109_k141_252244 | Not-determined |
| 17-109_k141_252245 | Not-determined |
| 17-109_k141_151679 | Not-determined |
| 17-109_k141_17998 | Not-determined |
| 17-109_k141_302483 | Not-determined |
| 17-109_k141_151687 | Not-determined |
| 17-109_k141_386217 | Low-quality |
| 17-109_k141_168532 | Not-determined |
| 17-109_k141_84857 | Not-determined |
| 17-109_k141_319291 | Not-determined |
| 17-109_k141_51390 | Not-determined |
| 17-109_k141_185295 | Not-determined |
| 17-109_k141_118030 | Not-determined |
| 17-109_k141_185304 | Low-quality |
| 17-109_k141_101525 | Not-determined |
| 17-109_k141_84867 | Low-quality |
| 17-109_k141_51406 | Not-determined |
| 17-109_k141_185313 | Not-determined |
| 17-109_k141_185318 | Not-determined |
| 17-109_k141_51414 | Not-determined |
| 17-109_k141_18031 | Not-determined |
| 17-109_k141_18039 | Not-determined |
| 17-109_k141_51423 | Not-determined |
| 17-109_k141_51424 | Not-determined |
| 17-109_k141_101530 | Not-determined |
| 17-109_k141_68068 | Not-determined |
| 17-109_k141_51427 | Low-quality |
| 17-109_k141_268977 | Low-quality |
| 17-109_k141_18042 | Not-determined |
| 17-109_k141_1300 | Not-determined |
| 17-109_k141_302500 | Not-determined |
| 17-109_k141_168535 | Not-determined |
| 17-109_k141_18054 | Not-determined |
| 17-109_k141_302502 | Low-quality |
| 17-109_k141_18055 | Not-determined |
| 17-109_k141_185346 | Not-determined |
| 17-109_k141_352472 | Low-quality |
| 17-109_k141_369479 | Low-quality |
| 17-109_k141_51437 | Not-determined |
| 17-109_k141_352477 | Not-determined |
| 17-109_k141_386254 | Not-determined |
| 17-109_k141_101565 | Not-determined |
| 17-109_k141_268982 | Low-quality |
| 17-109_k141_369490 | Not-determined |
| 17-109_k141_168542 | Not-determined |
| 17-109_k141_185367 | Not-determined |
| 17-109_k141_118053 | Not-determined |
| 17-109_k141_84901 | Not-determined |
| 17-109_k141_168543 | Not-determined |
| 17-109_k141_386259 | Not-determined |
| 17-109_k141_235490 | Not-determined |
| 17-109_k141_369496 | Low-quality |
| 17-109_k141_335641 | Not-determined |
| 17-109_k141_84914 | Not-determined |
| 17-109_k141_51442 | Not-determined |
| 17-109_k141_84918 | Not-determined |
| 17-109_k141_168551 | Not-determined |
| 17-109_k141_235498 | Not-determined |
| 17-109_k141_168553 | Not-determined |
| 17-109_k141_201974 | Not-determined |
| 17-109_k141_369507 | Not-determined |
| 17-109_k141_201975 | Not-determined |
| 17-109_k141_302531 | Low-quality |
| 17-109_k141_201979 | Not-determined |
| 17-109_k141_84926 | Not-determined |
| 17-109_k141_185387 | Not-determined |
| 17-109_k141_302540 | Not-determined |
| 17-109_k141_168564 | Not-determined |
| 17-109_k141_352489 | Low-quality |
| 17-109_k141_269001 | Not-determined |
| 17-109_k141_18111 | Not-determined |
| 17-109_k141_369523 | Not-determined |
| 17-109_k141_185405 | Not-determined |
| 17-109_k141_68113 | Low-quality |
| 17-109_k141_68117 | Not-determined |
| 17-109_k141_151725 | Not-determined |
| 17-109_k141_252308 | Not-determined |
| 17-109_k141_252310 | Not-determined |
| 17-109_k141_168577 | Not-determined |
| 17-109_k141_352496 | Low-quality |
| 17-109_k141_335667 | Not-determined |
| 17-109_k141_352497 | Not-determined |
| 17-109_k141_168581 | Not-determined |
| 17-109_k141_18127 | Low-quality |
| 17-109_k141_335677 | Not-determined |
| 17-109_k141_118079 | Not-determined |
| 17-109_k141_335681 | Not-determined |
| 17-109_k141_18132 | Not-determined |
| 17-109_k141_302577 | Low-quality |
| 17-109_k141_135054 | Not-determined |
| 17-109_k141_369537 | Not-determined |
| 17-109_k141_252332 | Not-determined |
| 17-109_k141_302579 | Low-quality |
| 17-109_k141_202017 | Not-determined |
| 17-109_k141_285964 | Not-determined |
| 17-109_k141_202019 | Not-determined |
| 17-109_k141_51478 | Not-determined |
| 17-109_k141_352511 | Not-determined |
| 17-109_k141_18135 | Low-quality |
| 17-109_k141_352514 | Not-determined |
| 17-109_k141_151731 | Not-determined |
| 17-109_k141_135066 | Not-determined |
| 17-109_k141_84974 | Low-quality |
| 17-109_k141_135067 | Not-determined |
| 17-109_k141_285969 | Not-determined |
| 17-109_k141_285970 | Not-determined |
| 17-109_k141_135069 | Not-determined |
| 17-109_k141_1339 | Not-determined |
| 17-109_k141_1341 | Low-quality |
| 17-109_k141_302588 | Not-determined |
| 17-109_k141_1343 | Not-determined |
| 17-109_k141_202028 | Not-determined |
| 17-109_k141_202029 | Not-determined |
| 17-109_k141_319300 | Not-determined |
| 17-109_k141_135073 | Not-determined |
| 17-109_k141_202036 | Not-determined |
| 17-109_k141_302600 | Not-determined |
| 17-109_k141_168608 | Not-determined |
| 17-109_k141_302602 | Not-determined |
| 17-109_k141_302611 | Low-quality |
| 17-109_k141_269053 | Not-determined |
| 17-109_k141_335700 | Not-determined |
| 17-109_k141_1349 | Not-determined |
| 17-109_k141_352538 | Not-determined |
| 17-109_k141_218742 | Not-determined |
| 17-109_k141_202047 | Not-determined |
| 17-109_k141_269055 | Not-determined |
| 17-109_k141_84985 | Not-determined |
| 17-109_k141_101568 | Not-determined |
| 17-109_k141_319325 | Not-determined |
| 17-109_k141_68160 | Not-determined |
| 17-109_k141_151752 | Not-determined |
| 17-109_k141_68162 | Low-quality |
| 17-109_k141_218754 | Not-determined |
| 17-109_k141_218757 | Low-quality |
| 17-109_k141_202052 | Not-determined |
| 17-109_k141_386335 | Not-determined |
| 17-109_k141_84991 | Not-determined |
| 17-109_k141_285979 | Not-determined |
| 17-109_k141_202057 | Not-determined |
| 17-109_k141_168624 | Low-quality |
| 17-109_k141_386340 | Not-determined |
| 17-109_k141_319332 | Not-determined |
| 17-109_k141_386341 | Not-determined |
| 17-109_k141_386345 | Not-determined |
| 17-109_k141_68176 | Not-determined |
| 17-109_k141_51489 | Not-determined |
| 17-109_k141_386350 | Not-determined |
| 17-109_k141_185417 | Not-determined |
| 17-109_k141_269060 | Not-determined |
| 17-109_k141_185419 | Not-determined |
| 17-109_k141_319344 | Not-determined |
| 17-109_k141_369595 | Not-determined |
| 17-109_k141_218773 | Not-determined |
| 17-109_k141_269062 | Not-determined |
| 17-109_k141_51494 | Not-determined |
| 17-109_k141_269067 | Low-quality |
| 17-109_k141_168636 | Not-determined |
| 17-109_k141_335723 | Not-determined |
| 17-109_k141_252346 | Not-determined |
| 17-109_k141_335726 | Low-quality |
| 17-109_k141_168640 | Not-determined |
| 17-109_k141_1361 | Not-determined |
| 17-109_k141_168649 | Not-determined |
| 17-109_k141_118115 | Low-quality |
| 17-109_k141_1370 | Not-determined |
| 17-109_k141_369614 | Not-determined |
| 17-109_k141_151793 | Not-determined |
| 17-109_k141_185434 | Not-determined |
| 17-109_k141_1378 | Not-determined |
| 17-109_k141_85008 | Not-determined |
| 17-109_k141_101593 | Not-determined |
| 17-109_k141_168656 | Not-determined |
| 17-109_k141_335739 | Not-determined |
| 17-109_k141_151796 | Not-determined |
| 17-109_k141_51506 | Not-determined |
| 17-109_k141_286002 | Not-determined |
| 17-109_k141_286005 | Not-determined |
| 17-109_k141_51512 | Not-determined |
| 17-109_k141_34897 | Low-quality |
| 17-109_k141_235536 | Not-determined |
| 17-109_k141_34898 | Not-determined |
| 17-109_k141_101602 | Not-determined |
| 17-109_k141_101603 | Not-determined |
| 17-109_k141_34905 | Not-determined |
| 17-109_k141_218787 | Not-determined |
| 17-109_k141_185452 | Not-determined |
| 17-109_k141_218790 | Not-determined |
| 17-109_k141_151813 | Not-determined |
| 17-109_k141_85009 | Not-determined |
| 17-109_k141_168681 | Not-determined |
| 17-109_k141_168684 | Not-determined |
| 17-109_k141_286023 | Not-determined |
| 17-109_k141_369629 | Not-determined |
| 17-109_k141_1406 | Not-determined |
| 17-109_k141_269101 | Not-determined |
| 17-109_k141_85017 | Not-determined |
| 17-109_k141_1408 | Not-determined |
| 17-109_k141_269103 | Not-determined |
| 17-109_k141_269105 | Not-determined |
| 17-109_k141_85020 | Not-determined |
| 17-109_k141_302658 | Not-determined |
| 17-109_k141_18195 | Not-determined |
| 17-109_k141_235560 | Not-determined |
| 17-109_k141_386401 | Not-determined |
| 17-109_k141_68216 | Low-quality |
| 17-109_k141_302662 | Not-determined |
| 17-109_k141_235562 | Not-determined |
| 17-109_k141_235563 | Not-determined |
| 17-109_k141_168689 | Not-determined |
| 17-109_k141_218806 | Not-determined |
| 17-109_k141_335773 | Not-determined |
| 17-109_k141_218808 | Not-determined |
| 17-109_k141_269111 | Not-determined |
| 17-109_k141_218813 | Not-determined |
| 17-109_k141_335776 | Not-determined |
| 17-109_k141_151826 | Not-determined |
| 17-109_k141_118130 | Not-determined |
| 17-109_k141_302672 | Not-determined |
| 17-109_k141_34929 | Not-determined |
| 17-109_k141_252381 | Not-determined |
| 17-109_k141_386405 | Not-determined |
| 17-109_k141_135126 | Not-determined |
| 17-109_k141_335781 | Not-determined |
| 17-109_k141_235572 | Not-determined |
| 17-109_k141_68224 | Low-quality |
| 17-109_k141_185467 | Not-determined |
| 17-109_k141_1424 | Low-quality |
| 17-109_k141_101622 | Low-quality |
| 17-109_k141_168693 | Not-determined |
| 17-109_k141_18216 | Not-determined |
| 17-109_k141_68229 | Not-determined |
| 17-109_k141_34945 | Not-determined |
| 17-109_k141_168702 | Not-determined |
| 17-109_k141_269130 | Not-determined |
| 17-109_k141_202078 | Not-determined |
| 17-109_k141_34954 | Not-determined |
| 17-109_k141_1442 | Not-determined |
| 17-109_k141_1448 | Not-determined |
| 17-109_k141_252405 | Not-determined |
| 17-109_k141_168708 | Low-quality |
| 17-109_k141_335797 | Not-determined |
| 17-109_k141_18223 | Not-determined |
| 17-109_k141_1459 | Not-determined |
| 17-109_k141_185507 | Not-determined |
| 17-109_k141_202091 | Not-determined |
| 17-109_k141_1461 | Not-determined |
| 17-109_k141_369681 | Not-determined |
| 17-109_k141_202094 | Not-determined |
| 17-109_k141_1464 | Not-determined |
| 17-109_k141_286039 | Low-quality |
| 17-109_k141_369684 | Not-determined |
| 17-109_k141_101645 | Not-determined |
| 17-109_k141_185509 | Low-quality |
| 17-109_k141_135167 | Not-determined |
| 17-109_k141_101646 | Not-determined |
| 17-109_k141_235601 | Not-determined |
| 17-109_k141_51558 | Not-determined |
| 17-109_k141_235612 | Not-determined |
| 17-109_k141_286045 | Not-determined |
| 17-109_k141_51568 | Not-determined |
| 17-109_k141_352557 | Not-determined |
| 17-109_k141_369711 | Not-determined |
| 17-109_k141_118138 | Not-determined |
| 17-109_k141_335810 | Not-determined |
| 17-109_k141_118141 | Not-determined |
| 17-109_k141_135169 | Low-quality |
| 17-109_k141_302691 | Low-quality |
| 17-109_k141_185543 | Not-determined |
| 17-109_k141_18227 | Not-determined |
| 17-109_k141_319362 | Not-determined |
| 17-109_k141_286054 | Not-determined |
| 17-109_k141_369720 | Not-determined |
| 17-109_k141_252438 | Not-determined |
| 17-109_k141_18234 | Not-determined |
| 17-109_k141_185545 | Not-determined |

| | |
|---|---|
| 17-109_k141_335823 | Not-determined |
| 17-109_k141_185547 | Not-determined |
| 17-109_k141_235633 | Not-determined |
| 17-109_k141_1504 | Not-determined |
| 17-109_k141_235635 | Not-determined |
| 17-109_k141_85060 | Not-determined |
| 17-109_k141_218877 | Not-determined |
| 17-109_k141_85062 | Not-determined |
| 17-109_k141_302711 | Not-determined |
| 17-109_k141_135184 | Not-determined |
| 17-109_k141_101676 | Not-determined |
| 17-109_k141_302714 | Not-determined |
| 17-109_k141_1510 | Not-determined |
| 17-109_k141_218883 | Not-determined |
| 17-109_k141_335825 | Low-quality |
| 17-109_k141_218884 | Not-determined |
| 17-109_k141_85068 | Not-determined |
| 17-109_k141_202104 | Not-determined |
| 17-109_k141_101684 | Not-determined |
| 17-109_k141_185560 | Low-quality |
| 17-109_k141_369745 | Not-determined |
| 17-109_k141_135193 | Not-determined |
| 17-109_k141_202106 | Low-quality |
| 17-109_k141_302726 | Low-quality |
| 17-109_k141_335833 | Not-determined |
| 17-109_k141_135202 | Not-determined |
| 17-109_k141_218891 | Not-determined |
| 17-109_k141_1536 | Not-determined |
| 17-109_k141_34998 | Not-determined |
| 17-109_k141_101694 | Not-determined |
| 17-109_k141_218899 | Not-determined |
| 17-109_k141_386424 | Not-determined |
| 17-109_k141_386425 | Not-determined |
| 17-109_k141_369761 | Not-determined |
| 17-109_k141_369766 | Not-determined |
| 17-109_k141_185576 | Not-determined |
| 17-109_k141_51575 | Not-determined |
| 17-109_k141_335849 | Not-determined |
| 17-109_k141_269148 | Low-quality |
| 17-109_k141_135209 | Not-determined |
| 17-109_k141_269155 | Not-determined |
| 17-109_k141_319398 | Not-determined |
| 17-109_k141_68253 | Not-determined |
| 17-109_k141_135217 | Not-determined |
| 17-109_k141_135219 | Not-determined |
| 17-109_k141_302762 | Low-quality |
| 17-109_k141_185590 | Not-determined |
| 17-109_k141_135221 | Not-determined |
| 17-109_k141_302764 | Not-determined |
| 17-109_k141_51590 | Not-determined |
| 17-109_k141_302767 | Not-determined |
| 17-109_k141_118174 | Not-determined |
| 17-109_k141_51596 | Not-determined |
| 17-109_k141_135228 | Not-determined |
| 17-109_k141_101736 | Not-determined |
| 17-109_k141_185601 | Not-determined |
| 17-109_k141_101739 | Not-determined |
| 17-109_k141_352570 | Not-determined |
| 17-109_k141_185603 | Not-determined |
| 17-109_k141_68260 | Not-determined |
| 17-109_k141_35045 | Not-determined |
| 17-109_k141_68265 | Not-determined |
| 17-109_k141_319410 | Not-determined |
| 17-109_k141_68270 | Not-determined |
| 17-109_k141_135243 | Low-quality |
| 17-109_k141_386453 | Low-quality |
| 17-109_k141_386457 | Not-determined |
| 17-109_k141_168751 | Not-determined |
| 17-109_k141_185621 | Not-determined |
| 17-109_k141_168754 | Not-determined |
| 17-109_k141_135252 | Not-determined |
| 17-109_k141_269174 | Not-determined |
| 17-109_k141_118195 | Not-determined |
| 17-109_k141_386464 | Not-determined |
| 17-109_k141_286092 | Not-determined |
| 17-109_k141_218952 | Not-determined |
| 17-109_k141_135263 | Not-determined |
| 17-109_k141_286097 | Not-determined |
| 17-109_k141_101745 | Not-determined |
| 17-109_k141_235677 | Not-determined |
| 17-109_k141_185643 | Not-determined |
| 17-109_k141_352603 | Not-determined |
| 17-109_k141_135268 | Not-determined |
| 17-109_k141_185648 | Not-determined |
| 17-109_k141_101754 | Not-determined |
| 17-109_k141_319419 | Not-determined |
| 17-109_k141_1591 | Low-quality |
| 17-109_k141_135284 | Not-determined |
| 17-109_k141_35052 | Not-determined |
| 17-109_k141_369779 | Not-determined |
| 17-109_k141_235695 | Not-determined |
| 17-109_k141_168775 | Not-determined |
| 17-109_k141_335897 | Low-quality |
| 17-109_k141_235700 | Not-determined |
| 17-109_k141_369786 | Not-determined |
| 17-109_k141_185665 | Not-determined |
| 17-109_k141_386490 | Not-determined |
| 17-109_k141_85121 | Low-quality |
| 17-109_k141_319435 | Low-quality |
| 17-109_k141_51617 | Not-determined |
| 17-109_k141_118217 | Not-determined |
| 17-109_k141_185668 | Not-determined |
| 17-109_k141_51622 | Not-determined |
| 17-109_k141_386506 | Not-determined |
| 17-109_k141_1601 | Low-quality |
| 17-109_k141_118223 | Not-determined |
| 17-109_k141_85143 | Not-determined |
| 17-109_k141_386509 | Not-determined |
| 17-109_k141_35089 | Not-determined |
| 17-109_k141_35091 | Not-determined |
| 17-109_k141_302780 | Not-determined |
| 17-109_k141_185673 | Not-determined |
| 17-109_k141_218956 | Not-determined |
| 17-109_k141_286154 | Not-determined |
| 17-109_k141_352629 | Low-quality |
| 17-109_k141_386518 | Not-determined |
| 17-109_k141_352630 | Not-determined |
| 17-109_k141_185678 | Low-quality |
| 17-109_k141_235708 | Not-determined |
| 17-109_k141_369805 | Not-determined |
| 17-109_k141_335915 | Not-determined |
| 17-109_k141_218959 | Not-determined |
| 17-109_k141_302789 | Not-determined |
| 17-109_k141_101766 | Not-determined |
| 17-109_k141_302791 | Not-determined |
| 17-109_k141_101768 | Not-determined |
| 17-109_k141_85159 | Not-determined |
| 17-109_k141_352644 | Not-determined |
| 17-109_k141_235719 | Not-determined |
| 17-109_k141_235720 | Not-determined |
| 17-109_k141_286165 | Not-determined |
| 17-109_k141_302794 | Not-determined |
| 17-109_k141_352646 | Low-quality |
| 17-109_k141_335924 | Not-determined |
| 17-109_k141_151848 | Not-determined |
| 17-109_k141_286171 | Not-determined |
| 17-109_k141_101781 | Not-determined |
| 17-109_k141_352654 | Not-determined |
| 17-109_k141_335931 | Not-determined |
| 17-109_k141_386531 | Not-determined |
| 18-102-final-viral-combined_k141_165833\|\|full | Medium-quality |
| 17-109_k141_185696 | Not-determined |
| 17-109_k141_286173 | Not-determined |
| 17-109_k141_185707 | Not-determined |
| 17-109_k141_151863 | Not-determined |
| 17-109_k141_386536 | Not-determined |
| 17-109_k141_1638 | Not-determined |
| 17-109_k141_386538 | Not-determined |
| 17-109_k141_286176 | Not-determined |
| 17-109_k141_85181 | Not-determined |
| 17-109_k141_185711 | Not-determined |
| 17-109_k141_369844 | Not-determined |
| 17-109_k141_135306 | Not-determined |
| 17-109_k141_269218 | Not-determined |
| 17-109_k141_135307 | Not-determined |
| 17-109_k141_302811 | Not-determined |
| 17-109_k141_185715 | Low-quality |
| 17-109_k141_286188 | Not-determined |
| 17-109_k141_286189 | Not-determined |
| 17-109_k141_302822 | Not-determined |
| 17-109_k141_135321 | Not-determined |
| 17-109_k141_202135 | Not-determined |
| 17-109_k141_202139 | Not-determined |
| 17-109_k141_202141 | Not-determined |
| 17-109_k141_1660 | Not-determined |
| 17-109_k141_335954 | Not-determined |
| 17-109_k141_101808 | Not-determined |
| 17-109_k141_269235 | Not-determined |
| 17-109_k141_202145 | Not-determined |
| 17-109_k141_202147 | Low-quality |
| 17-109_k141_151898 | Not-determined |
| 17-109_k141_168846 | Not-determined |
| 17-109_k141_168847 | Not-determined |
| 17-109_k141_369866 | Not-determined |
| 17-109_k141_85195 | Not-determined |
| 17-109_k141_352667 | Not-determined |
| 17-109_k141_151901 | Not-determined |
| 17-109_k141_51663 | Not-determined |
| 17-109_k141_118248 | Not-determined |
| 17-109_k141_51664 | Not-determined |
| 17-109_k141_135344 | Not-determined |
| 17-109_k141_1670 | Not-determined |
| 17-109_k141_168853 | Not-determined |
| 17-109_k141_35117 | Not-determined |
| 17-109_k141_85206 | Not-determined |
| 17-109_k141_151913 | Not-determined |
| 17-109_k141_1675 | Not-determined |
| 17-109_k141_235765 | Not-determined |
| 17-109_k141_135349 | Not-determined |
| 17-109_k141_269252 | Not-determined |
| 17-109_k141_135352 | Not-determined |
| 17-109_k141_269255 | Not-determined |
| 17-109_k141_319467 | Not-determined |
| 17-109_k141_151924 | Not-determined |
| 17-109_k141_35124 | Low-quality |
| 17-109_k141_219001 | Low-quality |
| 17-109_k141_235772 | Not-determined |
| 17-109_k141_219004 | Not-determined |
| 17-109_k141_51684 | Not-determined |
| 17-109_k141_68304 | Low-quality |
| 17-109_k141_35128 | Low-quality |
| 17-109_k141_85215 | Not-determined |
| 17-109_k141_319474 | Not-determined |
| 17-109_k141_319477 | Not-determined |
| 17-109_k141_269271 | Not-determined |
| 17-109_k141_85218 | Not-determined |
| 17-109_k141_386558 | Not-determined |
| 17-109_k141_68316 | Not-determined |
| 17-109_k141_335968 | Not-determined |
| 17-109_k141_202179 | Not-determined |
| 17-109_k141_219013 | Not-determined |
| 17-109_k141_85225 | Not-determined |
| 17-109_k141_335975 | Not-determined |
| 17-109_k141_319486 | Not-determined |
| 17-109_k141_35147 | Not-determined |
| 17-109_k141_335984 | Not-determined |
| 17-109_k141_319492 | Not-determined |
| 17-109_k141_319493 | Not-determined |
| 17-109_k141_302835 | Not-determined |
| 17-109_k141_269283 | Not-determined |
| 17-109_k141_68321 | Not-determined |
| 17-109_k141_185747 | Low-quality |
| 17-109_k141_68325 | Not-determined |
| 17-109_k141_219031 | Not-determined |
| 17-109_k141_151960 | Not-determined |
| 17-109_k141_352676 | Not-determined |
| 17-109_k141_151962 | Not-determined |
| 17-109_k141_302848 | Not-determined |
| 17-109_k141_101852 | Not-determined |
| 17-109_k141_302849 | Low-quality |
| 17-109_k141_352680 | Not-determined |
| 17-109_k141_302853 | Not-determined |
| 17-109_k141_35170 | Not-determined |
| 17-109_k141_202193 | Not-determined |
| 17-109_k141_185757 | Not-determined |
| 17-109_k141_168891 | Not-determined |
| 17-109_k141_168892 | Not-determined |
| 17-109_k141_352689 | Not-determined |
| 17-109_k141_151976 | Low-quality |
| 17-109_k141_135392 | Not-determined |
| 17-109_k141_51713 | Not-determined |
| 17-109_k141_151984 | Not-determined |
| 17-109_k141_168904 | Not-determined |
| 17-109_k141_269305 | Not-determined |
| 17-109_k141_118253 | Not-determined |
| 17-109_k141_269307 | Not-determined |
| 17-109_k141_269308 | Not-determined |
| 17-109_k141_369883 | Not-determined |
| 17-109_k141_168913 | Not-determined |
| 17-109_k141_302869 | Not-determined |
| 17-109_k141_319533 | Not-determined |
| 17-109_k141_151998 | Low-quality |
| 17-109_k141_302873 | Not-determined |
| 17-109_k141_152008 | Not-determined |
| 17-109_k141_302874 | Not-determined |
| 17-109_k141_352706 | Not-determined |
| 17-109_k141_152013 | Not-determined |
| 17-109_k141_68337 | Not-determined |
| 17-109_k141_118270 | Not-determined |
| 17-109_k141_68345 | Not-determined |
| 17-109_k141_219064 | Not-determined |
| 17-109_k141_269316 | Not-determined |
| 17-109_k141_85244 | Not-determined |
| 17-109_k141_319555 | Not-determined |
| 17-109_k141_85246 | Not-determined |
| 17-109_k141_269321 | Not-determined |
| 17-109_k141_269323 | Not-determined |
| 17-109_k141_269325 | Not-determined |
| 17-109_k141_369896 | Low-quality |
| 17-109_k141_369897 | Not-determined |
| 17-109_k141_235820 | Low-quality |
| 17-109_k141_118277 | Not-determined |
| 17-109_k141_369899 | Not-determined |
| 17-109_k141_1701 | Not-determined |
| 17-109_k141_352743 | Not-determined |
| 17-109_k141_319576 | Not-determined |
| 17-109_k141_269329 | Not-determined |
| 17-109_k141_235830 | Not-determined |
| 17-109_k141_202201 | Not-determined |
| 17-109_k141_386576 | Not-determined |
| 17-109_k141_118287 | Not-determined |
| 17-109_k141_202204 | Not-determined |
| 17-109_k141_1713 | Not-determined |
| 17-109_k141_235838 | Not-determined |
| 17-109_k141_369906 | Low-quality |
| 17-109_k141_352766 | Not-determined |
| 17-109_k141_386586 | Not-determined |
| 17-109_k141_118288 | Not-determined |
| 17-109_k141_386587 | Not-determined |
| 17-109_k141_51763 | Not-determined |
| 17-109_k141_68373 | Not-determined |
| 17-109_k141_35238 | Not-determined |
| 17-109_k141_235851 | Not-determined |
| 17-109_k141_185812 | Not-determined |
| 17-109_k141_18251 | Not-determined |
| 17-109_k141_51772 | Not-determined |
| 17-109_k141_1720 | Not-determined |
| 17-109_k141_118305 | Low-quality |
| 17-109_k141_68379 | Not-determined |
| 17-109_k141_235857 | Low-quality |
| 17-109_k141_168942 | Not-determined |
| 17-109_k141_168943 | Not-determined |
| 17-109_k141_235859 | Not-determined |
| 17-109_k141_185818 | Not-determined |
| 17-109_k141_152055 | Not-determined |
| 17-109_k141_319595 | Not-determined |
| 17-109_k141_35246 | Not-determined |
| 17-109_k141_68389 | Not-determined |
| 17-109_k141_68390 | Not-determined |
| 17-109_k141_319598 | Not-determined |
| 17-109_k141_269335 | Not-determined |
| 17-109_k141_35251 | Not-determined |
| 17-109_k141_386608 | Not-determined |
| 17-109_k141_386611 | Not-determined |
| 17-109_k141_386613 | Not-determined |
| 17-109_k141_269337 | Not-determined |
| 17-109_k141_152058 | Not-determined |
| 17-109_k141_68400 | Not-determined |
| 17-109_k141_51781 | Not-determined |
| 17-109_k141_152064 | Not-determined |
| 17-109_k141_85283 | Not-determined |
| 17-109_k141_202230 | Not-determined |
| 17-109_k141_85290 | Not-determined |
| 17-109_k141_185823 | Low-quality |
| 17-109_k141_51789 | Low-quality |
| 17-109_k141_51792 | Not-determined |
| 17-109_k141_302891 | Not-determined |
| 17-109_k141_319630 | Not-determined |
| 17-109_k141_319631 | Not-determined |
| 17-109_k141_369932 | Not-determined |
| 17-109_k141_18253 | Low-quality |
| 17-109_k141_68422 | Not-determined |
| 17-109_k141_302899 | Low-quality |
| 17-109_k141_85301 | Not-determined |
| 17-109_k141_319637 | Not-determined |
| 17-109_k141_152085 | Not-determined |
| 17-109_k141_118318 | Not-determined |
| 17-109_k141_35292 | Low-quality |
| 17-109_k141_118323 | Not-determined |
| 17-109_k141_1757 | Not-determined |
| 17-109_k141_68429 | Not-determined |
| 17-109_k141_118326 | Not-determined |
| 17-109_k141_352812 | Not-determined |
| 17-109_k141_386628 | Not-determined |
| 17-109_k141_302913 | Not-determined |
| 17-109_k141_51803 | Not-determined |
| 17-109_k141_219109 | Not-determined |
| 17-109_k141_152104 | Not-determined |
| 17-109_k141_185844 | Not-determined |
| 17-109_k141_369943 | Not-determined |
| 17-109_k141_185846 | Not-determined |
| 17-109_k141_118334 | Not-determined |
| 17-109_k141_85314 | Low-quality |
| 17-109_k141_319656 | Not-determined |
| 17-109_k141_85316 | Not-determined |
| 17-109_k141_168965 | Not-determined |
| 17-109_k141_152109 | Not-determined |
| 17-109_k141_369945 | Not-determined |
| 17-109_k141_35306 | Not-determined |
| 17-109_k141_235874 | Not-determined |
| 17-109_k141_369947 | Not-determined |
| 17-109_k141_369951 | Low-quality |
| 17-109_k141_319657 | Low-quality |
| 17-109_k141_118341 | Not-determined |
| 17-109_k141_302931 | Not-determined |
| 17-109_k141_152117 | Not-determined |
| 17-109_k141_118346 | Not-determined |
| 17-109_k141_168975 | Low-quality |
| 17-109_k141_352842 | Not-determined |
| 17-109_k141_352844 | Not-determined |
| 17-109_k141_68444 | Not-determined |
| 17-109_k141_319667 | Not-determined |
| 17-109_k141_35322 | Not-determined |
| 17-109_k141_302940 | Not-determined |
| 17-109_k141_269380 | Not-determined |
| 17-109_k141_152133 | Not-determined |
| 17-109_k141_18274 | Not-determined |
| 17-109_k141_152136 | Not-determined |
| 17-109_k141_35329 | Not-determined |
| 17-109_k141_35330 | Low-quality |
| 17-109_k141_18277 | Not-determined |
| 17-109_k141_202264 | Not-determined |
| 17-109_k141_18281 | Not-determined |
| 17-109_k141_35337 | Not-determined |
| 17-109_k141_18285 | Not-determined |
| 17-109_k141_269396 | Not-determined |
| 17-109_k141_35346 | Not-determined |
| 17-109_k141_51821 | Not-determined |
| 17-109_k141_1786 | Not-determined |
| 17-109_k141_252581 | Not-determined |
| 17-109_k141_202276 | Not-determined |
| 17-109_k141_252584 | Not-determined |
| 17-109_k141_202278 | Not-determined |
| 17-109_k141_202284 | Not-determined |
| 17-109_k141_85341 | Not-determined |
| 17-109_k141_302961 | Not-determined |
| 17-109_k141_336016 | Not-determined |
| 17-109_k141_286210 | Not-determined |
| 17-109_k141_336019 | Not-determined |
| 17-109_k141_168986 | Not-determined |
| 17-109_k141_369990 | Not-determined |
| 17-109_k141_336022 | Not-determined |
| 17-109_k141_286215 | Not-determined |
| 17-109_k141_185860 | Not-determined |
| 17-109_k141_369991 | Not-determined |
| 17-109_k141_286217 | Not-determined |
| 17-109_k141_336025 | Low-quality |
| 17-109_k141_118377 | Not-determined |
| 17-109_k141_101879 | Low-quality |
| 17-109_k141_302969 | Not-determined |
| 17-109_k141_118380 | Not-determined |
| 17-109_k141_152170 | Not-determined |
| 17-109_k141_302972 | Not-determined |
| 17-109_k141_269417 | Not-determined |
| 17-109_k141_152172 | Not-determined |
| 17-109_k141_302979 | Not-determined |
| 17-109_k141_202289 | Not-determined |
| 17-109_k141_269420 | Not-determined |
| 17-109_k141_269423 | Not-determined |
| 17-109_k141_370006 | Not-determined |
| 17-109_k141_235907 | Not-determined |
| 17-109_k141_286226 | Not-determined |
| 17-109_k141_219138 | Not-determined |
| 17-109_k141_286230 | Not-determined |
| 17-109_k141_302992 | Not-determined |
| 17-109_k141_302993 | Not-determined |
| 17-109_k141_336040 | Not-determined |
| 17-109_k141_219145 | Not-determined |
| 17-109_k141_336042 | Not-determined |
| 17-109_k141_202307 | Not-determined |
| 17-109_k141_135431 | Not-determined |
| 17-109_k141_302997 | Not-determined |
| 17-109_k141_169006 | Not-determined |
| 17-109_k141_336052 | Low-quality |
| 17-109_k141_51826 | Not-determined |
| 17-109_k141_51828 | Not-determined |
| 17-109_k141_319710 | Not-determined |
| 17-109_k141_101910 | Not-determined |
| 17-109_k141_1842 | Not-determined |
| 17-109_k141_1843 | Not-determined |
| 17-109_k141_51841 | Not-determined |
| 17-109_k141_202323 | Not-determined |
| 17-109_k141_235922 | Not-determined |
| 17-109_k141_68492 | Low-quality |
| 17-109_k141_370045 | Not-determined |
| 17-109_k141_35397 | Not-determined |
| 17-109_k141_51845 | Not-determined |
| 17-109_k141_185880 | Not-determined |
| 17-109_k141_1849 | Not-determined |
| 17-109_k141_303018 | Not-determined |
| 17-109_k141_319728 | Not-determined |
| 17-109_k141_286243 | Low-quality |
| 17-109_k141_219165 | Not-determined |
| 17-109_k141_1856 | Not-determined |
| 17-109_k141_219167 | Not-determined |
| 17-106-final-viral-combined_k141_25007\|\|full | Medium-quality |
| 17-109_k141_319735 | Low-quality |
| 17-109_k141_219175 | Not-determined |
| 17-109_k141_1868 | Not-determined |
| 17-109_k141_101930 | Low-quality |
| 17-109_k141_286254 | Low-quality |
| 17-109_k141_101937 | Low-quality |
| 17-109_k141_235944 | Not-determined |
| 17-109_k141_303027 | Not-determined |
| 17-109_k141_152209 | Not-determined |
| 17-109_k141_1871 | Not-determined |
| 17-109_k141_118406 | Not-determined |
| 17-109_k141_68512 | Not-determined |
| 18-105-final-viral-combined_k141_173345\|\|full | Medium-quality |
| 17-109_k141_235953 | Not-determined |
| 17-109_k141_235956 | Not-determined |
| 17-109_k141_51854 | Not-determined |
| 17-109_k141_101947 | Not-determined |
| 17-109_k141_286274 | Not-determined |
| 17-109_k141_152222 | Not-determined |
| 17-109_k141_336075 | Not-determined |
| 17-109_k141_152228 | Low-quality |
| 17-109_k141_370050 | Not-determined |
| 17-109_k141_370051 | Not-determined |
| 17-109_k141_252647 | Not-determined |
| 17-109_k141_303039 | Not-determined |
| 17-109_k141_152233 | Not-determined |
| 17-109_k141_336081 | Not-determined |
| 17-109_k141_85355 | Not-determined |
| 17-109_k141_319751 | Not-determined |
| 17-109_k141_169042 | Not-determined |
| 17-109_k141_1876 | Not-determined |
| 17-109_k141_235975 | Not-determined |
| 17-109_k141_202330 | Low-quality |
| 17-109_k141_252658 | Low-quality |
| 17-109_k141_219185 | Not-determined |
| 17-109_k141_202335 | Not-determined |
| 17-109_k141_370062 | Not-determined |
| 17-109_k141_386702 | Not-determined |
| 17-109_k141_352862 | Not-determined |
| 17-109_k141_135460 | Low-quality |
| 17-109_k141_386704 | Not-determined |
| 17-109_k141_1886 | Not-determined |
| 17-109_k141_85359 | Low-quality |
| 17-109_k141_51868 | Not-determined |
| 17-109_k141_386710 | Low-quality |
| 17-109_k141_185928 | Not-determined |
| 17-109_k141_303059 | Not-determined |
| 17-109_k141_352872 | Not-determined |
| 17-109_k141_85367 | Low-quality |
| 17-109_k141_1894 | Not-determined |
| 17-109_k141_101967 | Low-quality |
| 17-109_k141_303067 | Not-determined |
| 17-109_k141_286297 | Not-determined |
| 17-109_k141_352880 | Not-determined |
| 17-109_k141_101974 | Not-determined |
| 17-109_k141_101976 | Not-determined |
| 17-109_k141_135478 | Not-determined |
| 17-109_k141_1899 | Not-determined |
| 17-109_k141_135481 | Not-determined |
| 17-109_k141_169065 | Not-determined |
| 17-109_k141_68520 | Not-determined |
| 17-109_k141_68524 | Not-determined |
| 17-109_k141_51878 | Not-determined |
| 17-109_k141_169075 | Not-determined |
| 17-109_k141_169078 | Not-determined |
| 17-109_k141_286314 | Not-determined |
| 17-109_k141_303077 | Not-determined |
| 17-109_k141_303078 | Not-determined |
| 17-109_k141_85394 | Not-determined |
| 17-109_k141_386725 | Not-determined |
| 17-109_k141_252679 | Not-determined |
| 17-109_k141_286325 | Not-determined |
| 17-109_k141_252686 | Not-determined |
| 17-109_k141_303095 | Low-quality |
| 17-109_k141_85401 | Not-determined |
| 17-109_k141_303099 | Not-determined |
| 17-109_k141_352897 | Low-quality |
| 17-109_k141_286341 | Not-determined |
| 17-109_k141_386746 | Not-determined |
| 17-109_k141_236020 | Not-determined |
| 17-109_k141_286348 | Not-determined |
| 17-109_k141_386750 | Not-determined |
| 17-109_k141_169087 | Not-determined |
| 17-109_k141_135519 | Low-quality |
| 17-109_k141_236029 | Not-determined |
| 17-109_k141_286355 | Not-determined |
| 17-109_k141_85409 | Not-determined |
| 17-109_k141_236035 | Not-determined |
| 17-109_k141_152261 | Not-determined |
| 17-109_k141_236042 | Not-determined |
| 17-109_k141_51890 | Not-determined |
| 17-109_k141_152268 | Not-determined |
| 17-109_k141_319806 | Not-determined |
| 17-109_k141_85425 | Not-determined |
| 17-109_k141_169093 | Not-determined |
| 17-109_k141_319808 | Not-determined |
| 17-109_k141_51900 | Not-determined |
| 17-109_k141_35426 | Not-determined |
| 17-109_k141_370115 | Not-determined |
| 17-109_k141_286386 | Low-quality |
| 17-109_k141_219210 | Not-determined |
| 17-109_k141_68575 | Low-quality |
| 17-109_k141_102048 | Not-determined |
| 17-109_k141_68580 | Not-determined |
| 17-109_k141_352918 | Not-determined |
| 17-109_k141_152282 | Not-determined |
| 17-109_k141_219220 | Not-determined |
| 17-109_k141_386774 | Not-determined |
| 17-109_k141_352922 | Not-determined |
| 17-109_k141_35450 | Not-determined |
| 17-109_k141_269439 | Not-determined |
| 17-109_k141_51911 | Not-determined |
| 17-109_k141_68582 | Not-determined |
| 17-109_k141_269446 | Not-determined |
| 17-109_k141_219227 | Not-determined |
| 17-109_k141_202348 | Not-determined |
| 17-109_k141_303133 | Not-determined |
| 17-109_k141_252735 | Low-quality |
| 17-109_k141_35453 | Not-determined |
| 17-109_k141_102071 | Not-determined |
| 17-109_k141_370126 | Not-determined |
| 17-109_k141_202354 | Not-determined |
| 17-109_k141_202355 | Not-determined |
| 17-109_k141_202358 | Not-determined |
| 17-109_k141_202359 | Low-quality |
| 17-109_k141_35460 | Not-determined |
| 17-109_k141_236100 | Not-determined |
| 17-109_k141_252746 | Not-determined |
| 17-109_k141_370128 | Not-determined |
| 17-109_k141_370129 | Not-determined |
| 17-109_k141_252751 | Not-determined |
| 17-109_k141_152304 | Not-determined |
| 17-109_k141_236107 | Not-determined |

| | |
|---|---|
| 17-109_k141_152308 | Not-determined |
| 17-109_k141_303152 | Not-determined |
| 17-109_k141_386795 | Not-determined |
| 17-109_k141_236113 | Not-determined |
| 17-109_k141_336120 | Not-determined |
| 17-109_k141_386798 | Not-determined |
| 17-109_k141_386801 | Not-determined |
| 17-109_k141_336128 | Not-determined |
| 17-109_k141_336130 | Not-determined |
| 17-109_k141_336133 | Not-determined |
| 17-109_k141_370144 | Not-determined |
| 17-109_k141_336134 | Not-determined |
| 17-109_k141_252761 | Not-determined |
| 17-109_k141_303162 | Not-determined |
| 17-109_k141_152320 | Not-determined |
| 17-109_k141_202378 | Not-determined |
| 17-109_k141_135566 | Not-determined |
| 17-109_k141_169123 | Not-determined |
| 17-109_k141_202382 | Not-determined |
| 17-109_k141_236130 | Not-determined |
| 17-109_k141_236132 | Not-determined |
| 17-109_k141_336142 | Not-determined |
| 17-109_k141_236138 | Not-determined |
| 17-109_k141_236141 | Not-determined |
| 17-109_k141_252779 | Not-determined |
| 17-109_k141_336150 | Not-determined |
| 17-109_k141_286432 | Not-determined |
| 17-109_k141_85446 | Not-determined |
| 17-109_k141_252781 | Low-quality |
| 17-109_k141_85449 | Low-quality |
| 17-109_k141_185950 | Not-determined |
| 17-109_k141_319825 | Not-determined |
| 17-109_k141_303194 | Not-determined |
| 17-109_k141_85461 | Not-determined |
| 17-109_k141_185967 | Not-determined |
| 17-109_k141_303207 | Not-determined |
| 17-109_k141_269481 | Not-determined |
| 17-109_k141_236171 | Not-determined |
| 17-109_k141_68615 | Not-determined |
| 17-109_k141_319844 | Not-determined |
| 17-109_k141_252783 | Low-quality |
| 17-109_k141_252787 | Not-determined |
| 17-109_k141_252788 | Not-determined |
| 17-109_k141_236180 | Low-quality |
| 17-109_k141_319848 | Not-determined |
| 17-109_k141_219304 | Not-determined |
| 17-109_k141_18359 | Not-determined |
| 17-109_k141_219306 | Not-determined |
| 17-109_k141_336176 | Not-determined |
| 17-109_k141_303221 | Not-determined |
| 18-102-final-viral-combined_k141_201205\|\|full | High-quality |
| 17-109_k141_386817 | Not-determined |
| 17-109_k141_236192 | Not-determined |
| 17-109_k141_252804 | Not-determined |
| 17-109_k141_386823 | Not-determined |
| 17-109_k141_35473 | Low-quality |
| 17-109_k141_370173 | Not-determined |
| 17-109_k141_336184 | Not-determined |
| 17-109_k141_118532 | Not-determined |
| 17-109_k141_118534 | Not-determined |
| 17-109_k141_386838 | Not-determined |
| 17-109_k141_252832 | Not-determined |
| 17-109_k141_319873 | Low-quality |
| 17-109_k141_35489 | Not-determined |
| 17-109_k141_370188 | Not-determined |
| 17-109_k141_118545 | Not-determined |
| 17-109_k141_370193 | Not-determined |
| 17-109_k141_286486 | Not-determined |
| 17-109_k141_319882 | Not-determined |
| 17-109_k141_118555 | Not-determined |
| 17-109_k141_286501 | Not-determined |
| 17-109_k141_319895 | Not-determined |
| 17-109_k141_286506 | Not-determined |
| 17-109_k141_51959 | Not-determined |
| 17-109_k141_252855 | Not-determined |
| 17-109_k141_319904 | Not-determined |
| 17-109_k141_219325 | Not-determined |
| 17-109_k141_118565 | Not-determined |
| 17-109_k141_303254 | Not-determined |
| 17-109_k141_303259 | Not-determined |
| 17-109_k141_135603 | Not-determined |
| 17-109_k141_35523 | Not-determined |
| 17-109_k141_35525 | Not-determined |
| 17-109_k141_303262 | Not-determined |
| 17-109_k141_252872 | Not-determined |
| 17-109_k141_1964 | Not-determined |
| 17-109_k141_51984 | Not-determined |
| 17-109_k141_370219 | Not-determined |
| 17-109_k141_18389 | Not-determined |
| 17-109_k141_135614 | Low-quality |
| 17-109_k141_35531 | Not-determined |
| 17-109_k141_118577 | Not-determined |
| 17-109_k141_219345 | Not-determined |
| 17-109_k141_135621 | Not-determined |
| 17-109_k141_68628 | Not-determined |
| 17-109_k141_202405 | Not-determined |
| 17-109_k141_286547 | Not-determined |
| 17-109_k141_52003 | Not-determined |
| 17-109_k141_303274 | Not-determined |
| 17-109_k141_386852 | Not-determined |
| 17-109_k141_303280 | Not-determined |
| 17-109_k141_303284 | Not-determined |
| 17-109_k141_386860 | Not-determined |
| 17-109_k141_52006 | Not-determined |
| 17-109_k141_18404 | Not-determined |
| 17-109_k141_169149 | Not-determined |
| 17-109_k141_52008 | Not-determined |
| 17-109_k141_18406 | Not-determined |
| 17-109_k141_286555 | Not-determined |
| 17-109_k141_18408 | Not-determined |
| 17-109_k141_219370 | Not-determined |
| 17-109_k141_68639 | Not-determined |
| 17-109_k141_169151 | Not-determined |
| 17-109_k141_35555 | Not-determined |
| 17-109_k141_152390 | Not-determined |
| 17-109_k141_202440 | Low-quality |
| 17-109_k141_303306 | Not-determined |
| 17-109_k141_352989 | Not-determined |
| 17-109_k141_68644 | Not-determined |
| 17-109_k141_352990 | Not-determined |
| 17-109_k141_1977 | Low-quality |
| 17-109_k141_303312 | Not-determined |
| 17-109_k141_352996 | Not-determined |
| 17-109_k141_118611 | Not-determined |
| 17-109_k141_353000 | Not-determined |
| 17-109_k141_219390 | Not-determined |
| 17-109_k141_252894 | Not-determined |
| 17-109_k141_1981 | Not-determined |
| 17-109_k141_52028 | Not-determined |
| 17-109_k141_169167 | Not-determined |
| 17-109_k141_152407 | Low-quality |
| 17-109_k141_319945 | Not-determined |
| 17-109_k141_2002 | Not-determined |
| 17-109_k141_319946 | Not-determined |
| 17-109_k141_219400 | Not-determined |
| 17-109_k141_52033 | Not-determined |
| 17-109_k141_269488 | Not-determined |
| 17-109_k141_286577 | Not-determined |
| 17-109_k141_35561 | Not-determined |
| 17-109_k141_35562 | Not-determined |
| 17-109_k141_18435 | Not-determined |
| 17-109_k141_85491 | Not-determined |
| 17-109_k141_353014 | Not-determined |
| 17-109_k141_353015 | Not-determined |
| 17-109_k141_370235 | Low-quality |
| 17-109_k141_269493 | Not-determined |
| 17-109_k141_370240 | Not-determined |
| 17-109_k141_35572 | Not-determined |
| 17-109_k141_68657 | Not-determined |
| 17-109_k141_219417 | Not-determined |
| 17-109_k141_303333 | Not-determined |
| 17-109_k141_370262 | Not-determined |
| 17-109_k141_202449 | Not-determined |
| 17-109_k141_202453 | Not-determined |
| 17-109_k141_202455 | Low-quality |
| 17-109_k141_52057 | Low-quality |
| 17-109_k141_303341 | Not-determined |
| 17-109_k141_52058 | Not-determined |
| 17-109_k141_370275 | Not-determined |
| 17-109_k141_202461 | Not-determined |
| 17-109_k141_202464 | Not-determined |
| 17-109_k141_252926 | Not-determined |
| 17-109_k141_202466 | Not-determined |
| 17-109_k141_386895 | Not-determined |
| 17-109_k141_135686 | Not-determined |
| 17-109_k141_2029 | Not-determined |
| 17-109_k141_68665 | Low-quality |
| 17-109_k141_353032 | Not-determined |
| 17-109_k141_336199 | Not-determined |
| 17-109_k141_353033 | Not-determined |
| 17-109_k141_2033 | Not-determined |
| 17-109_k141_52074 | Not-determined |
| 17-109_k141_118646 | Low-quality |
| 17-109_k141_186026 | Not-determined |
| 17-109_k141_135695 | Not-determined |
| 17-109_k141_286599 | Not-determined |
| 17-109_k141_68668 | Not-determined |
| 17-109_k141_152455 | Not-determined |
| 17-109_k141_68670 | Not-determined |
| 17-109_k141_286604 | Low-quality |
| 17-109_k141_353049 | Not-determined |
| 17-109_k141_370292 | Not-determined |
| 17-109_k141_152465 | Not-determined |
| 17-109_k141_152466 | Not-determined |
| 17-109_k141_2044 | Not-determined |
| 17-109_k141_186044 | Not-determined |
| 17-109_k141_286626 | Not-determined |
| 17-109_k141_152472 | Not-determined |
| 17-109_k141_35601 | Not-determined |
| 17-109_k141_35602 | Not-determined |
| 17-109_k141_118654 | Not-determined |
| 17-109_k141_269501 | Not-determined |
| 17-109_k141_2057 | Not-determined |
| 17-109_k141_85533 | Not-determined |
| 17-109_k141_102101 | Not-determined |
| 17-109_k141_319984 | Not-determined |
| 17-109_k141_353071 | Not-determined |
| 17-109_k141_386915 | Not-determined |
| 17-109_k141_18448 | Not-determined |
| 17-109_k141_386917 | Not-determined |
| 17-109_k141_386923 | Not-determined |
| 17-109_k141_102105 | Not-determined |
| 17-109_k141_386925 | Not-determined |
| 17-109_k141_102107 | Not-determined |
| 17-109_k141_35626 | Low-quality |
| 17-109_k141_336229 | Not-determined |
| 17-109_k141_85549 | Not-determined |
| 17-109_k141_386928 | Not-determined |
| 17-109_k141_303370 | Not-determined |
| 17-109_k141_152492 | Not-determined |
| 17-109_k141_319993 | Not-determined |
| 17-109_k141_35645 | Not-determined |
| 17-109_k141_303376 | Not-determined |
| 17-109_k141_303378 | Not-determined |
| 17-109_k141_152498 | Not-determined |
| 17-109_k141_303382 | Not-determined |
| 17-109_k141_320002 | Not-determined |
| 17-109_k141_152504 | Not-determined |
| 17-109_k141_202487 | Not-determined |
| 17-109_k141_152506 | Not-determined |
| 17-109_k141_35658 | Not-determined |
| 17-109_k141_68738 | Not-determined |
| 17-109_k141_353102 | Not-determined |
| 17-109_k141_303389 | Not-determined |
| 17-109_k141_286662 | Not-determined |
| 17-109_k141_269525 | Not-determined |
| 17-109_k141_303391 | Not-determined |
| 17-109_k141_303392 | Not-determined |
| 17-109_k141_320017 | Not-determined |
| 17-109_k141_353106 | Not-determined |
| 17-109_k141_68746 | Not-determined |
| 17-109_k141_386949 | Not-determined |
| 17-109_k141_102128 | Not-determined |
| 17-109_k141_386953 | Not-determined |
| 17-109_k141_118674 | Low-quality |
| 17-109_k141_353111 | Not-determined |
| 17-109_k141_286678 | Not-determined |
| 17-109_k141_353113 | Low-quality |
| 17-109_k141_269537 | Not-determined |
| 17-109_k141_370317 | Not-determined |
| 17-109_k141_52113 | Not-determined |
| 17-109_k141_85585 | Not-determined |
| 17-109_k141_303394 | Not-determined |
| 17-109_k141_336264 | Not-determined |
| 17-109_k141_68758 | Not-determined |
| 17-109_k141_320027 | Not-determined |
| 17-109_k141_219422 | Low-quality |
| 17-109_k141_303397 | Not-determined |
| 17-109_k141_186086 | Not-determined |
| 17-109_k141_152524 | Low-quality |
| 17-109_k141_320031 | Not-determined |
| 17-109_k141_186089 | Not-determined |
| 17-109_k141_353127 | Not-determined |
| 17-109_k141_186092 | Not-determined |
| 17-109_k141_303406 | Not-determined |
| 17-109_k141_370328 | Low-quality |
| 17-109_k141_52122 | Not-determined |
| 17-109_k141_303417 | Not-determined |
| 17-109_k141_85599 | Not-determined |
| 17-109_k141_152535 | Not-determined |
| 17-109_k141_85602 | Not-determined |
| 17-109_k141_135744 | Not-determined |
| 17-109_k141_85604 | Not-determined |
| 17-109_k141_85609 | Not-determined |
| 17-109_k141_320047 | Low-quality |
| 17-109_k141_152546 | Not-determined |
| 17-109_k141_286702 | Not-determined |
| 17-109_k141_219441 | Not-determined |
| 17-109_k141_135758 | Not-determined |
| 17-109_k141_219447 | Not-determined |
| 17-109_k141_269555 | Not-determined |
| 17-109_k141_68783 | Low-quality |
| 17-109_k141_386987 | Not-determined |
| 17-109_k141_219452 | Low-quality |
| 17-109_k141_152561 | Not-determined |
| 17-109_k141_85627 | Low-quality |
| 17-109_k141_152563 | Not-determined |
| 17-109_k141_85631 | Low-quality |
| 17-109_k141_52141 | Not-determined |
| 17-109_k141_118714 | Not-determined |
| 17-109_k141_152570 | Not-determined |
| 17-109_k141_118719 | Not-determined |
| 17-109_k141_202507 | Not-determined |
| 17-109_k141_102152 | Not-determined |
| 17-109_k141_336288 | Not-determined |
| 17-109_k141_152577 | Not-determined |
| 17-109_k141_320059 | Not-determined |
| 17-109_k141_353169 | Not-determined |
| 17-109_k141_320061 | Not-determined |
| 17-109_k141_186124 | Not-determined |
| 17-109_k141_252939 | Not-determined |
| 17-109_k141_252940 | Not-determined |
| 17-109_k141_303436 | Not-determined |
| 17-109_k141_152582 | Not-determined |
| 17-109_k141_370371 | Not-determined |
| 17-109_k141_85641 | Not-determined |
| 17-109_k141_52165 | Not-determined |
| 17-109_k141_152590 | Not-determined |
| 17-109_k141_269567 | Not-determined |
| 17-109_k141_68804 | Not-determined |
| 17-109_k141_387008 | Low-quality |
| 17-109_k141_219480 | Not-determined |
| 17-109_k141_186142 | Not-determined |
| 17-109_k141_85652 | Not-determined |
| 17-109_k141_202521 | Low-quality |
| 17-109_k141_186145 | Not-determined |
| 17-109_k141_118726 | Low-quality |
| 17-109_k141_387027 | Not-determined |
| 17-109_k141_135781 | Not-determined |
| 17-109_k141_18464 | Not-determined |
| 17-109_k141_370389 | Not-determined |
| 17-109_k141_186173 | Not-determined |
| 17-109_k141_35705 | Not-determined |
| 17-109_k141_68822 | Not-determined |
| 17-109_k141_102202 | Low-quality |
| 17-109_k141_68825 | Not-determined |
| 17-109_k141_370407 | Not-determined |
| 17-109_k141_85672 | Not-determined |
| 17-109_k141_370413 | Not-determined |
| 17-109_k141_35709 | Not-determined |
| 17-109_k141_152625 | Not-determined |
| 17-109_k141_18482 | Low-quality |
| 17-109_k141_2107 | Not-determined |
| 17-109_k141_269571 | Not-determined |
| 17-109_k141_336328 | Not-determined |
| 17-109_k141_336329 | Not-determined |
| 17-109_k141_152629 | Not-determined |
| 17-109_k141_269572 | Not-determined |
| 17-109_k141_152630 | Not-determined |
| 17-109_k141_353215 | Not-determined |
| 17-109_k141_336335 | Not-determined |
| 17-109_k141_85685 | Not-determined |
| 17-109_k141_219518 | Not-determined |
| 17-109_k141_68832 | Not-determined |
| 17-109_k141_85688 | Low-quality |
| 17-109_k141_68834 | Not-determined |
| 17-109_k141_52211 | Not-determined |
| 17-109_k141_370423 | Not-determined |
| 17-109_k141_186183 | Not-determined |
| 17-109_k141_186184 | Not-determined |
| 17-109_k141_336345 | Not-determined |
| 17-109_k141_169227 | Not-determined |
| 17-109_k141_68843 | Not-determined |
| 17-109_k141_303475 | Not-determined |
| 17-109_k141_35726 | Not-determined |
| 17-109_k141_303481 | Not-determined |
| 17-109_k141_135803 | Not-determined |
| 17-109_k141_118751 | Low-quality |
| 17-109_k141_52229 | Not-determined |
| 17-109_k141_353220 | Not-determined |
| 17-109_k141_102217 | Not-determined |
| 17-109_k141_85698 | Low-quality |
| 17-109_k141_269589 | Not-determined |
| 17-109_k141_52237 | Low-quality |
| 17-109_k141_152665 | Not-determined |
| 17-109_k141_102239 | Not-determined |
| 17-109_k141_186216 | Not-determined |
| 17-109_k141_68882 | Not-determined |
| 17-109_k141_202591 | Not-determined |
| 17-109_k141_202593 | Not-determined |
| 17-109_k141_35744 | Not-determined |
| 17-109_k141_269601 | Not-determined |
| 17-109_k141_2149 | Not-determined |
| 17-109_k141_169247 | Not-determined |
| 17-109_k141_35751 | Not-determined |
| 17-109_k141_269607 | Not-determined |
| 17-109_k141_336354 | Not-determined |
| 17-109_k141_269609 | Not-determined |
| 17-109_k141_202596 | Not-determined |
| 17-109_k141_2154 | Not-determined |
| 17-109_k141_2155 | Not-determined |
| 17-109_k141_35754 | Low-quality |
| 17-109_k141_2159 | Not-determined |
| 17-109_k141_2166 | Low-quality |
| 17-109_k141_202615 | Not-determined |
| 17-109_k141_202616 | Not-determined |
| 17-109_k141_353256 | Not-determined |
| 17-109_k141_186236 | Not-determined |
| 17-109_k141_102264 | Not-determined |
| 17-109_k141_353258 | Not-determined |
| 17-109_k141_336364 | Not-determined |
| 17-109_k141_2173 | Not-determined |
| 17-109_k141_202618 | Not-determined |
| 17-109_k141_336367 | Not-determined |
| 17-109_k141_169266 | Not-determined |
| 17-109_k141_202620 | Not-determined |
| 17-109_k141_102270 | Not-determined |
| 17-109_k141_269620 | Not-determined |
| 17-109_k141_118787 | Not-determined |
| 17-109_k141_118791 | Not-determined |
| 17-109_k141_186249 | Not-determined |
| 17-109_k141_336376 | Not-determined |
| 17-109_k141_186251 | Not-determined |
| 17-109_k141_2181 | Not-determined |
| 17-109_k141_320134 | Not-determined |
| 17-109_k141_202630 | Not-determined |
| 17-109_k141_52264 | Not-determined |
| 17-109_k141_118804 | Not-determined |
| 17-109_k141_202635 | Not-determined |
| 17-109_k141_387037 | Not-determined |
| 17-109_k141_303507 | Low-quality |
| 17-109_k141_320142 | Low-quality |
| 17-109_k141_387040 | Not-determined |
| 17-109_k141_135816 | Not-determined |
| 17-109_k141_370461 | Not-determined |
| 17-109_k141_186269 | Not-determined |
| 17-109_k141_152677 | Not-determined |
| 17-109_k141_353278 | Not-determined |
| 17-109_k141_286734 | Low-quality |
| 17-109_k141_135822 | Not-determined |
| 17-109_k141_35784 | Not-determined |
| 17-109_k141_35787 | Not-determined |
| 17-109_k141_353282 | Low-quality |
| 17-109_k141_152687 | Low-quality |
| 17-109_k141_387059 | Not-determined |
| 17-109_k141_152689 | Not-determined |
| 17-109_k141_202654 | Not-determined |
| 17-109_k141_169298 | Not-determined |
| 17-109_k141_370471 | Not-determined |
| 17-109_k141_336391 | Not-determined |
| 17-109_k141_202661 | Not-determined |
| 17-109_k141_202664 | Not-determined |
| 17-109_k141_286743 | Not-determined |
| 17-109_k141_186279 | Not-determined |
| 17-109_k141_118826 | Not-determined |
| 17-109_k141_202671 | Not-determined |
| 17-109_k141_202674 | Not-determined |
| 17-109_k141_252971 | Not-determined |
| 17-109_k141_152694 | Not-determined |
| 17-109_k141_102300 | Not-determined |
| 17-109_k141_353288 | Low-quality |
| 17-109_k141_202684 | Not-determined |
| 17-109_k141_186286 | Not-determined |
| 17-109_k141_202695 | Not-determined |
| 17-109_k141_152700 | Not-determined |
| 17-109_k141_370478 | Not-determined |
| 17-109_k141_320159 | Not-determined |
| 17-109_k141_202704 | Not-determined |
| 17-109_k141_35794 | Not-determined |
| 17-109_k141_387080 | Not-determined |
| 17-109_k141_269674 | Not-determined |
| 17-109_k141_269678 | Low-quality |
| 17-109_k141_286766 | Not-determined |
| 17-109_k141_118851 | Not-determined |
| 17-109_k141_202710 | Not-determined |
| 17-109_k141_286769 | Not-determined |
| 17-109_k141_202718 | Not-determined |
| 17-109_k141_68917 | Not-determined |
| 17-109_k141_320167 | Low-quality |
| 17-109_k141_85723 | Not-determined |
| 17-109_k141_135849 | Not-determined |
| 17-109_k141_320169 | Not-determined |
| 17-109_k141_169335 | Not-determined |
| 17-109_k141_186302 | Not-determined |
| 17-109_k141_370489 | Not-determined |
| 17-109_k141_186305 | Low-quality |
| 17-109_k141_387095 | Not-determined |
| 17-109_k141_118858 | Not-determined |
| 17-109_k141_85738 | Not-determined |
| 17-109_k141_387103 | Not-determined |
| 17-109_k141_118862 | Not-determined |
| 17-109_k141_303553 | Not-determined |
| 17-109_k141_85743 | Not-determined |
| 17-109_k141_320185 | Not-determined |
| 17-109_k141_286780 | Not-determined |
| 17-109_k141_202734 | Not-determined |
| 17-109_k141_202738 | Not-determined |
| 17-109_k141_370502 | Low-quality |
| 17-109_k141_186328 | Not-determined |
| 17-109_k141_303569 | Not-determined |
| 17-109_k141_269707 | Not-determined |
| 17-109_k141_169358 | Low-quality |
| 17-109_k141_202751 | Not-determined |
| 17-109_k141_269723 | Not-determined |
| 17-109_k141_353341 | Not-determined |
| 17-109_k141_169361 | Not-determined |
| 17-109_k141_169364 | Not-determined |
| 17-109_k141_118876 | Not-determined |
| 17-109_k141_118878 | Not-determined |
| 17-109_k141_118882 | Not-determined |
| 17-109_k141_118885 | Not-determined |
| 17-109_k141_2228 | Not-determined |
| 17-109_k141_135871 | Not-determined |
| 17-109_k141_135877 | Not-determined |
| 17-109_k141_169383 | Not-determined |
| 17-109_k141_169384 | Not-determined |
| 17-109_k141_186356 | Not-determined |
| 17-109_k141_135886 | Not-determined |
| 17-109_k141_186357 | Not-determined |
| 17-109_k141_135887 | Not-determined |
| 17-109_k141_387147 | Not-determined |
| 17-109_k141_387148 | Not-determined |
| 17-109_k141_186360 | Not-determined |
| 17-109_k141_135890 | Not-determined |
| 17-109_k141_135893 | Low-quality |
| 17-109_k141_169391 | Not-determined |
| 17-109_k141_18556 | Not-determined |
| 17-109_k141_135899 | Not-determined |
| 17-109_k141_186364 | Not-determined |
| 17-109_k141_102330 | Not-determined |
| 17-109_k141_135903 | Not-determined |
| 17-109_k141_286798 | Not-determined |
| 17-109_k141_102332 | Not-determined |
| 17-109_k141_286800 | Not-determined |
| 17-109_k141_102336 | Not-determined |
| 17-109_k141_169402 | Not-determined |
| 17-109_k141_286806 | Low-quality |
| 17-109_k141_269742 | Not-determined |
| 17-109_k141_286807 | Not-determined |
| 17-109_k141_118903 | Not-determined |
| 17-109_k141_286813 | Not-determined |
| 17-109_k141_2247 | Not-determined |
| 17-109_k141_320240 | Low-quality |
| 17-109_k141_370553 | Not-determined |
| 17-109_k141_269751 | Not-determined |
| 17-109_k141_152744 | Low-quality |
| 17-109_k141_169416 | Not-determined |
| 17-109_k141_269757 | Not-determined |
| 17-109_k141_253012 | Not-determined |
| 17-109_k141_269758 | Not-determined |
| 17-109_k141_118925 | Not-determined |
| 17-109_k141_353354 | Not-determined |
| 17-109_k141_253013 | Not-determined |
| 17-109_k141_102346 | Not-determined |
| 17-109_k141_320250 | Not-determined |
| 17-109_k141_353360 | Low-quality |
| 17-109_k141_52361 | Not-determined |
| 17-109_k141_336413 | Not-determined |
| 17-109_k141_169423 | Not-determined |
| 17-109_k141_102358 | Not-determined |
| 17-109_k141_68977 | Not-determined |
| 17-109_k141_169426 | Low-quality |
| 17-109_k141_152774 | Not-determined |
| 17-109_k141_68978 | Not-determined |
| 17-109_k141_353383 | Not-determined |
| 17-109_k141_269762 | Not-determined |
| 17-109_k141_85775 | Not-determined |
| 17-109_k141_286832 | Not-determined |
| 17-109_k141_85778 | Not-determined |
| 17-109_k141_303632 | Not-determined |
| 17-109_k141_353396 | Not-determined |
| 17-109_k141_269772 | Not-determined |
| 17-109_k141_102365 | Not-determined |
| 17-109_k141_336436 | Not-determined |
| 17-109_k141_118958 | Not-determined |
| 17-109_k141_68999 | Not-determined |
| 17-109_k141_370564 | Not-determined |
| 17-109_k141_152784 | Not-determined |
| 17-109_k141_69004 | Not-determined |
| 17-109_k141_336448 | Not-determined |
| 17-109_k141_102371 | Not-determined |
| 17-109_k141_35834 | Not-determined |
| 17-109_k141_336450 | Not-determined |
| 17-109_k141_186382 | Not-determined |
| 17-109_k141_135942 | Not-determined |
| 17-109_k141_353422 | Low-quality |
| 17-109_k141_303635 | Not-determined |
| 17-109_k141_18609 | Not-determined |
| 17-109_k141_336462 | Not-determined |
| 17-109_k141_186392 | Low-quality |
| 17-109_k141_303648 | Not-determined |
| 17-109_k141_336470 | Not-determined |
| 17-109_k141_387194 | Not-determined |
| 17-109_k141_152807 | Not-determined |
| 17-109_k141_52368 | Not-determined |
| 17-109_k141_303656 | Not-determined |
| 17-109_k141_2304 | Not-determined |
| 17-109_k141_2308 | Not-determined |
| 17-109_k141_186409 | Not-determined |
| 17-109_k141_18640 | Not-determined |
| 17-109_k141_186413 | Not-determined |
| 17-109_k141_353445 | Not-determined |
| 17-109_k141_152822 | Not-determined |
| 17-109_k141_353447 | Not-determined |
| 17-109_k141_169431 | Low-quality |
| 17-109_k141_186419 | Not-determined |
| 17-109_k141_152826 | Not-determined |
| 17-109_k141_69032 | Not-determined |
| 17-109_k141_336474 | Not-determined |
| 17-109_k141_336475 | Not-determined |
| 17-109_k141_186425 | Not-determined |
| 17-109_k141_152831 | Not-determined |
| 17-109_k141_336477 | Not-determined |
| 17-109_k141_387209 | Not-determined |
| 17-109_k141_236282 | Low-quality |
| 17-109_k141_353453 | Not-determined |
| 17-109_k141_118997 | Not-determined |
| 17-109_k141_2318 | Not-determined |
| 17-109_k141_336482 | Low-quality |

| | |
|---|---|
| 17-109_k141_236290 | Not-determined |
| 17-109_k141_320279 | Not-determined |
| 17-109_k141_286882 | Not-determined |
| 17-109_k141_2329 | Not-determined |
| 17-109_k141_269802 | Low-quality |
| 17-109_k141_85806 | Not-determined |
| 17-109_k141_52412 | Low-quality |
| 17-109_k141_2334 | Not-determined |
| 17-109_k141_253029 | Not-determined |
| 17-109_k141_253030 | Low-quality |
| 17-109_k141_269806 | Not-determined |
| 17-109_k141_353473 | Not-determined |
| 17-109_k141_119012 | Not-determined |
| 17-109_k141_85810 | Not-determined |
| 17-109_k141_18664 | Not-determined |
| 17-109_k141_102429 | Not-determined |
| 17-109_k141_102431 | Not-determined |
| 17-109_k141_353476 | Not-determined |
| 17-109_k141_336495 | Not-determined |
| 17-109_k141_119014 | Not-determined |
| 17-109_k141_85813 | Not-determined |
| 17-109_k141_69059 | Not-determined |
| 17-109_k141_69061 | Not-determined |
| 17-109_k141_186450 | Not-determined |
| 17-109_k141_102437 | Not-determined |
| 17-109_k141_119019 | Not-determined |
| 17-109_k141_52423 | Not-determined |
| 17-109_k141_269814 | Low-quality |
| 17-109_k141_2338 | Not-determined |
| 17-109_k141_320293 | Not-determined |
| 17-109_k141_186468 | Not-determined |
| 17-109_k141_236304 | Not-determined |
| 17-109_k141_186471 | Not-determined |
| 17-109_k141_102452 | Not-determined |
| 17-109_k141_135987 | Not-determined |
| 17-109_k141_102455 | Not-determined |
| 17-109_k141_69079 | Not-determined |
| 17-109_k141_119026 | Not-determined |
| 17-109_k141_320300 | Not-determined |
| 18-101-final-viral-combined_k141_332236\|\|full | Medium-quality |
| 17-109_k141_135997 | Not-determined |
| 17-109_k141_202809 | Not-determined |
| 17-109_k141_286909 | Not-determined |
| 17-109_k141_303690 | Not-determined |
| 17-109_k141_2347 | Not-determined |
| 17-109_k141_320304 | Not-determined |
| 17-109_k141_202816 | Not-determined |
| 17-109_k141_85833 | Not-determined |
| 17-109_k141_286915 | Not-determined |
| 17-109_k141_286920 | Not-determined |
| 17-109_k141_186493 | Not-determined |
| 17-109_k141_85845 | Not-determined |
| 17-109_k141_286930 | Not-determined |
| 17-109_k141_152884 | Not-determined |
| 17-109_k141_286935 | Not-determined |
| 17-109_k141_269862 | Low-quality |
| 17-109_k141_136018 | Not-determined |
| 17-109_k141_136019 | Not-determined |
| 17-109_k141_152889 | Not-determined |
| 17-109_k141_269868 | Not-determined |
| 17-109_k141_186509 | Not-determined |
| 17-109_k141_236321 | Not-determined |
| 17-109_k141_236322 | Not-determined |
| 17-109_k141_269872 | Not-determined |
| 17-109_k141_320320 | Not-determined |
| 17-109_k141_303722 | Not-determined |
| 17-109_k141_303723 | Low-quality |
| 17-109_k141_85860 | Not-determined |
| 17-109_k141_336527 | Not-determined |
| 17-109_k141_186517 | Not-determined |
| 17-109_k141_303731 | Not-determined |
| 17-109_k141_119081 | Not-determined |
| 17-109_k141_186526 | Not-determined |
| 17-109_k141_35895 | Low-quality |
| 17-109_k141_269880 | Not-determined |
| 17-109_k141_152893 | Not-determined |
| 17-109_k141_136023 | Not-determined |
| 17-109_k141_336539 | Not-determined |
| 17-109_k141_236348 | Not-determined |
| 17-109_k141_286958 | Not-determined |
| 17-109_k141_152896 | Not-determined |
| 17-109_k141_186537 | Not-determined |
| 17-109_k141_303736 | Not-determined |
| 17-109_k141_269895 | Not-determined |
| 17-109_k141_136039 | Not-determined |
| 17-109_k141_69127 | Not-determined |
| 17-109_k141_69128 | Not-determined |
| 17-109_k141_236364 | Not-determined |
| 17-109_k141_169460 | Not-determined |
| 17-109_k141_353482 | Not-determined |
| 17-109_k141_186546 | Not-determined |
| 17-109_k141_253088 | Low-quality |
| 17-109_k141_85885 | Not-determined |
| 17-109_k141_35912 | Not-determined |
| 17-109_k141_35914 | Not-determined |
| 17-109_k141_69139 | Not-determined |
| 17-109_k141_186552 | Not-determined |
| 17-109_k141_136056 | Not-determined |
| 17-109_k141_353491 | Not-determined |
| 17-109_k141_69142 | Not-determined |
| 17-109_k141_152922 | Not-determined |
| 17-109_k141_102491 | Not-determined |
| 17-109_k141_387256 | Not-determined |
| 17-109_k141_186560 | Not-determined |
| 17-109_k141_236380 | Not-determined |
| 17-109_k141_102495 | Not-determined |
| 17-109_k141_69145 | Not-determined |
| 17-109_k141_303746 | Not-determined |
| 17-109_k141_303749 | Not-determined |
| 17-109_k141_387265 | Not-determined |
| 17-109_k141_353495 | Not-determined |
| 17-109_k141_320340 | Not-determined |
| 17-109_k141_35918 | Not-determined |
| 17-109_k141_336550 | Not-determined |
| 17-109_k141_69158 | Not-determined |
| 17-109_k141_69163 | Not-determined |
| 17-109_k141_336552 | Not-determined |
| 17-109_k141_202846 | Not-determined |
| 17-109_k141_336560 | Not-determined |
| 17-109_k141_52467 | Not-determined |
| 17-109_k141_336562 | Not-determined |
| 17-109_k141_336563 | Not-determined |
| 17-109_k141_303771 | Not-determined |
| 17-109_k141_286960 | Not-determined |
| 17-109_k141_186589 | Not-determined |
| 17-109_k141_303775 | Not-determined |
| 17-109_k141_336564 | Not-determined |
| 17-109_k141_136077 | Not-determined |
| 17-109_k141_136078 | Low-quality |
| 17-109_k141_136081 | Low-quality |
| 17-109_k141_136083 | Low-quality |
| 17-109_k141_52486 | Not-determined |
| 17-109_k141_370656 | Not-determined |
| 17-109_k141_286966 | Not-determined |
| 17-109_k141_169496 | Not-determined |
| 17-109_k141_286969 | Not-determined |
| 17-109_k141_119120 | Not-determined |
| 17-109_k141_169501 | Not-determined |
| 17-109_k141_320351 | Low-quality |
| 17-109_k141_186617 | Not-determined |
| 17-109_k141_69175 | Not-determined |
| 17-109_k141_35956 | Not-determined |
| 17-109_k141_35957 | Not-determined |
| 17-109_k141_303794 | Not-determined |
| 17-109_k141_119123 | Not-determined |
| 17-109_k141_136097 | Not-determined |
| 17-109_k141_286984 | Not-determined |
| 17-109_k141_169515 | Not-determined |
| 17-109_k141_102503 | Not-determined |
| 17-109_k141_52502 | Not-determined |
| 17-109_k141_85929 | Not-determined |
| 17-109_k141_202881 | Not-determined |
| 17-109_k141_269908 | Not-determined |
| 17-109_k141_336566 | Not-determined |
| 17-109_k141_353518 | Not-determined |
| 17-109_k141_136106 | Low-quality |
| 17-109_k141_52513 | Not-determined |
| 17-109_k141_320369 | Not-determined |
| 17-109_k141_136111 | Not-determined |
| 17-109_k141_169524 | Low-quality |
| 17-109_k141_387291 | Low-quality |
| 17-109_k141_202895 | Not-determined |
| 17-109_k141_52516 | Not-determined |
| 17-109_k141_85943 | Not-determined |
| 17-109_k141_85946 | Low-quality |
| 17-109_k141_236437 | Not-determined |
| 17-109_k141_85947 | Not-determined |
| 17-109_k141_52520 | Not-determined |
| 17-109_k141_52521 | Not-determined |
| 17-109_k141_236441 | Not-determined |
| 17-109_k141_353524 | Not-determined |
| 17-109_k141_320377 | Not-determined |
| 17-109_k141_152960 | Not-determined |
| 17-109_k141_85957 | Not-determined |
| 17-109_k141_219608 | Not-determined |
| 17-109_k141_320383 | Not-determined |
| 17-109_k141_69210 | Not-determined |
| 17-109_k141_320385 | Not-determined |
| 17-109_k141_236451 | Not-determined |
| 17-109_k141_186649 | Not-determined |
| 17-109_k141_236452 | Low-quality |
| 17-109_k141_85963 | Not-determined |
| 17-109_k141_219612 | Not-determined |
| 17-109_k141_102510 | Not-determined |
| 17-109_k141_102512 | Not-determined |
| 17-109_k141_102516 | Not-determined |
| 17-109_k141_269911 | Not-determined |
| 17-109_k141_152979 | Not-determined |
| 17-109_k141_320392 | Not-determined |
| 17-109_k141_85986 | Not-determined |
| 17-109_k141_69229 | Not-determined |
| 17-109_k141_152987 | Low-quality |
| 17-109_k141_35980 | Not-determined |
| 17-109_k141_152988 | Not-determined |
| 17-109_k141_152991 | Not-determined |
| 17-109_k141_202918 | Not-determined |
| 17-109_k141_35989 | Not-determined |
| 17-109_k141_35990 | Not-determined |
| 17-109_k141_269934 | Not-determined |
| 17-109_k141_236474 | Not-determined |
| 17-109_k141_303824 | Not-determined |
| 17-109_k141_85992 | Not-determined |
| 17-109_k141_85995 | Not-determined |
| 17-109_k141_253156 | Low-quality |
| 17-109_k141_52540 | Not-determined |
| 17-109_k141_269942 | Not-determined |
| 17-109_k141_186663 | Not-determined |
| 17-109_k141_52544 | Low-quality |
| 17-109_k141_269952 | Not-determined |
| 17-109_k141_186673 | Not-determined |
| 17-109_k141_320395 | Not-determined |
| 17-109_k141_219635 | Not-determined |
| 17-109_k141_269957 | Not-determined |
| 17-109_k141_320399 | Not-determined |
| 17-109_k141_52554 | Not-determined |
| 17-109_k141_353564 | Not-determined |
| 17-109_k141_186685 | Low-quality |
| 17-109_k141_303848 | Not-determined |
| 17-109_k141_35998 | Not-determined |
| 17-109_k141_303851 | Not-determined |
| 17-109_k141_86010 | Not-determined |
| 17-109_k141_236486 | Not-determined |
| 17-109_k141_320404 | Not-determined |
| 17-109_k141_219648 | Not-determined |
| 17-109_k141_36005 | Not-determined |
| 17-109_k141_236492 | Not-determined |
| 17-109_k141_102529 | Not-determined |
| 17-109_k141_119163 | Not-determined |
| 17-109_k141_236494 | Not-determined |
| 17-109_k141_320413 | Low-quality |
| 17-109_k141_36011 | Not-determined |
| 17-109_k141_18697 | Not-determined |
| 17-109_k141_69269 | Not-determined |
| 17-109_k141_186698 | Not-determined |
| 17-109_k141_353576 | Not-determined |
| 17-109_k141_353580 | Not-determined |
| 17-109_k141_269977 | Not-determined |
| 17-109_k141_69284 | Not-determined |
| 17-109_k141_153003 | Not-determined |
| 17-109_k141_236503 | Not-determined |
| 17-109_k141_269978 | Not-determined |
| 17-109_k141_18715 | Not-determined |
| 17-109_k141_387307 | Not-determined |
| 17-109_k141_153014 | Not-determined |
| 17-109_k141_153015 | Not-determined |
| 17-109_k141_69296 | Not-determined |
| 17-109_k141_236511 | Not-determined |
| 17-109_k141_52589 | Not-determined |
| 17-109_k141_287041 | Not-determined |
| 17-109_k141_52593 | Not-determined |
| 17-109_k141_52594 | Not-determined |
| 17-109_k141_236517 | Not-determined |
| 17-109_k141_370736 | Not-determined |
| 17-109_k141_253185 | Not-determined |
| 17-109_k141_253187 | Not-determined |
| 17-109_k141_353589 | Low-quality |
| 17-109_k141_136169 | Not-determined |
| 17-109_k141_236523 | Not-determined |
| 17-109_k141_69309 | Low-quality |
| 17-109_k141_86031 | Not-determined |
| 17-109_k141_219657 | Not-determined |
| 17-109_k141_52607 | Not-determined |
| 17-109_k141_320461 | Not-determined |
| 17-109_k141_69325 | Low-quality |
| 17-109_k141_320463 | Not-determined |
| 17-109_k141_119170 | Not-determined |
| 17-109_k141_119171 | Not-determined |
| 17-109_k141_69329 | Not-determined |
| 17-109_k141_136181 | Not-determined |
| 17-109_k141_52616 | Not-determined |
| 17-109_k141_287051 | Not-determined |
| 17-109_k141_202948 | Low-quality |
| 17-109_k141_119182 | Not-determined |
| 17-109_k141_169583 | Not-determined |
| 17-109_k141_119183 | Not-determined |
| 17-109_k141_153048 | Not-determined |
| 17-109_k141_18736 | Not-determined |
| 17-109_k141_18737 | Not-determined |
| 17-109_k141_287060 | Not-determined |
| 17-109_k141_320473 | Not-determined |
| 17-109_k141_253206 | Not-determined |
| 17-109_k141_52625 | Not-determined |
| 17-109_k141_153062 | Not-determined |
| 17-109_k141_236537 | Not-determined |
| 17-109_k141_387311 | Not-determined |
| 17-109_k141_320475 | Not-determined |
| 17-109_k141_353631 | Not-determined |
| 17-109_k141_353636 | Not-determined |
| 17-109_k141_236550 | Not-determined |
| 17-109_k141_370773 | Low-quality |
| 17-109_k141_303919 | Not-determined |
| 17-109_k141_169599 | Not-determined |
| 17-109_k141_69344 | Not-determined |
| 17-109_k141_236562 | Not-determined |
| 17-109_k141_202980 | Low-quality |
| 17-109_k141_236565 | Not-determined |
| 17-109_k141_253233 | Low-quality |
| 17-109_k141_69357 | Not-determined |
| 17-109_k141_320495 | Low-quality |
| 17-109_k141_236572 | Not-determined |
| 17-109_k141_320503 | Not-determined |
| 17-109_k141_387338 | Not-determined |
| 17-109_k141_69371 | Not-determined |
| 17-109_k141_136212 | Not-determined |
| 17-109_k141_219689 | Not-determined |
| 17-109_k141_387342 | Not-determined |
| 17-109_k141_387343 | Not-determined |
| 17-109_k141_336605 | Not-determined |
| 17-109_k141_320516 | Not-determined |
| 17-109_k141_69376 | Not-determined |
| 17-109_k141_219693 | Not-determined |
| 17-109_k141_253251 | Not-determined |
| 17-109_k141_320521 | Not-determined |
| 17-109_k141_320523 | Not-determined |
| 17-109_k141_119224 | Not-determined |
| 17-109_k141_69391 | Not-determined |
| 17-109_k141_253268 | Low-quality |
| 17-109_k141_253273 | Not-determined |
| 17-109_k141_69402 | Not-determined |
| 17-109_k141_18749 | Not-determined |
| 17-109_k141_303939 | Not-determined |
| 17-109_k141_153084 | Low-quality |
| 17-109_k141_336618 | Not-determined |
| 17-109_k141_353682 | Not-determined |
| 17-109_k141_287110 | Low-quality |
| 17-109_k141_353685 | Not-determined |
| 17-109_k141_236606 | Not-determined |
| 17-109_k141_69419 | Not-determined |
| 17-109_k141_169629 | Not-determined |
| 17-109_k141_353692 | Not-determined |
| 17-109_k141_303945 | Not-determined |
| 17-109_k141_353696 | Not-determined |
| 17-109_k141_202999 | Not-determined |
| 17-109_k141_253276 | Low-quality |
| 17-109_k141_353702 | Not-determined |
| 17-109_k141_336634 | Not-determined |
| 17-109_k141_52712 | Not-determined |
| 17-109_k141_303957 | Low-quality |
| 17-109_k141_336638 | Not-determined |
| 17-109_k141_287119 | Not-determined |
| 17-109_k141_253282 | Not-determined |
| 17-109_k141_336640 | Low-quality |
| 17-109_k141_353715 | Not-determined |
| 17-109_k141_253290 | Not-determined |
| 17-109_k141_370819 | Not-determined |
| 17-109_k141_36046 | Not-determined |
| 17-109_k141_203029 | Not-determined |
| 17-109_k141_18781 | Not-determined |
| 17-109_k141_136235 | Not-determined |
| 17-109_k141_2460 | Not-determined |
| 17-109_k141_69437 | Not-determined |
| 17-109_k141_86041 | Not-determined |
| 17-109_k141_203044 | Not-determined |
| 17-109_k141_253300 | Not-determined |
| 17-109_k141_253301 | Not-determined |
| 17-109_k141_253302 | Not-determined |
| 17-109_k141_136245 | Not-determined |
| 17-109_k141_86051 | Not-determined |
| 17-109_k141_253305 | Not-determined |
| 17-109_k141_203056 | Not-determined |
| 17-109_k141_203060 | Not-determined |
| 17-109_k141_219746 | Not-determined |
| 17-109_k141_219747 | Not-determined |
| 17-109_k141_219748 | Not-determined |
| 17-109_k141_203063 | Not-determined |
| 17-109_k141_36057 | Not-determined |
| 17-109_k141_36059 | Not-determined |
| 17-109_k141_2478 | Not-determined |
| 17-109_k141_169663 | Not-determined |
| 17-109_k141_236651 | Not-determined |
| 17-109_k141_236652 | Not-determined |
| 17-109_k141_52758 | Not-determined |
| 17-109_k141_236655 | Not-determined |
| 17-109_k141_18805 | Not-determined |
| 17-109_k141_52763 | Not-determined |
| 17-109_k141_387385 | Not-determined |
| 17-109_k141_370853 | Not-determined |
| 17-109_k141_153135 | Not-determined |
| 17-109_k141_18808 | Not-determined |
| 17-109_k141_52769 | Not-determined |
| 17-109_k141_18813 | Not-determined |
| 17-109_k141_387397 | Not-determined |
| 17-109_k141_236671 | Not-determined |
| 17-109_k141_169693 | Low-quality |
| 17-109_k141_18821 | Not-determined |
| 17-109_k141_52778 | Not-determined |
| 17-109_k141_387404 | Not-determined |
| 17-109_k141_253324 | Not-determined |
| 17-109_k141_219760 | Not-determined |
| 17-109_k141_320534 | Not-determined |
| 17-109_k141_253325 | Not-determined |
| 17-109_k141_304010 | Not-determined |
| 17-109_k141_370876 | Not-determined |
| 17-109_k141_169699 | Not-determined |
| 17-109_k141_353744 | Not-determined |
| 17-109_k141_69487 | Not-determined |
| 17-109_k141_69491 | Not-determined |
| 17-109_k141_169704 | Not-determined |
| 17-109_k141_18834 | Not-determined |
| 17-109_k141_2496 | Not-determined |
| 17-109_k141_320540 | Not-determined |
| 17-109_k141_304017 | Not-determined |
| 17-109_k141_353754 | Not-determined |
| 17-109_k141_52779 | Not-determined |
| 17-109_k141_219777 | Not-determined |
| 17-109_k141_18841 | Not-determined |
| 17-109_k141_304024 | Not-determined |
| 17-109_k141_36089 | Not-determined |
| 17-109_k141_52785 | Low-quality |
| 17-109_k141_69510 | Not-determined |
| 17-109_k141_18846 | Not-determined |
| 17-109_k141_203103 | Not-determined |
| 17-109_k141_353761 | Not-determined |
| 17-109_k141_287181 | Not-determined |
| 17-109_k141_18850 | Not-determined |
| 17-109_k141_320558 | Not-determined |
| 17-109_k141_287185 | Not-determined |
| 17-109_k141_136275 | Not-determined |
| 17-109_k141_169720 | Not-determined |
| 17-109_k141_304034 | Low-quality |
| 17-109_k141_304035 | Not-determined |
| 17-109_k141_69529 | Not-determined |
| 17-109_k141_320563 | Not-determined |
| 17-109_k141_203121 | Not-determined |
| 17-109_k141_86088 | Not-determined |
| 17-109_k141_2503 | Not-determined |
| 17-109_k141_169724 | Not-determined |
| 17-109_k141_304039 | Low-quality |
| 17-109_k141_86093 | Not-determined |
| 17-109_k141_219803 | Not-determined |
| 17-109_k141_219804 | Low-quality |
| 17-109_k141_136290 | Not-determined |
| 17-109_k141_304048 | Not-determined |
| 17-109_k141_304052 | Not-determined |
| 17-109_k141_52798 | Low-quality |
| 17-109_k141_186717 | Low-quality |
| 17-109_k141_370921 | Low-quality |
| 17-109_k141_18865 | Not-determined |
| 17-109_k141_253353 | Low-quality |
| 17-109_k141_18867 | Not-determined |
| 17-109_k141_304062 | Not-determined |
| 17-109_k141_387427 | Not-determined |
| 17-109_k141_169732 | Not-determined |
| 17-109_k141_52806 | Not-determined |
| 17-109_k141_370926 | Not-determined |
| 17-109_k141_304072 | Not-determined |
| 17-109_k141_36119 | Low-quality |
| 17-109_k141_186738 | Not-determined |
| 17-109_k141_287211 | Not-determined |
| 17-109_k141_136319 | Low-quality |
| 17-109_k141_102616 | Not-determined |
| 17-109_k141_287216 | Not-determined |
| 17-109_k141_304084 | Not-determined |
| 17-109_k141_320590 | Not-determined |
| 17-109_k141_270011 | Not-determined |
| 17-109_k141_169754 | Not-determined |
| 17-109_k141_18877 | Not-determined |
| 17-109_k141_270017 | Not-determined |
| 17-109_k141_387454 | Not-determined |
| 17-109_k141_2539 | Low-quality |
| 17-109_k141_2540 | Not-determined |
| 17-109_k141_304089 | Not-determined |
| 17-109_k141_219808 | Not-determined |
| 17-109_k141_136326 | Not-determined |
| 17-109_k141_287227 | Not-determined |
| 17-109_k141_370940 | Not-determined |
| 17-109_k141_2551 | Not-determined |
| 17-109_k141_370941 | Not-determined |
| 17-109_k141_136330 | Not-determined |
| 17-109_k141_304093 | Not-determined |
| 17-109_k141_119244 | Not-determined |
| 17-109_k141_270037 | Not-determined |
| 17-109_k141_287238 | Not-determined |
| 17-109_k141_304098 | Not-determined |
| 17-109_k141_18893 | Not-determined |
| 17-109_k141_119245 | Not-determined |
| 17-109_k141_186768 | Not-determined |
| 17-109_k141_136337 | Not-determined |
| 17-109_k141_186773 | Not-determined |
| 17-109_k141_186776 | Not-determined |
| 17-109_k141_353801 | Not-determined |
| 17-109_k141_136340 | Not-determined |
| 17-109_k141_353804 | Low-quality |
| 17-109_k141_153161 | Low-quality |
| 17-109_k141_370960 | Not-determined |
| 17-109_k141_153163 | Not-determined |
| 17-109_k141_219815 | Low-quality |
| 17-109_k141_102635 | Low-quality |
| 17-109_k141_2566 | Not-determined |
| 17-109_k141_2568 | Not-determined |
| 17-109_k141_119258 | Not-determined |
| 17-109_k141_36150 | Not-determined |
| 17-109_k141_270044 | Not-determined |
| 17-109_k141_219820 | Not-determined |
| 17-109_k141_219824 | Not-determined |
| 17-109_k141_119262 | Not-determined |
| 17-109_k141_219826 | Low-quality |
| 17-109_k141_387478 | Not-determined |
| 17-109_k141_119264 | Not-determined |
| 17-109_k141_387481 | Low-quality |
| 17-109_k141_270053 | Not-determined |
| 17-109_k141_36155 | Low-quality |
| 17-109_k141_36156 | Not-determined |
| 17-109_k141_119266 | Not-determined |
| 17-109_k141_153180 | Not-determined |
| 17-109_k141_236742 | Not-determined |
| 17-109_k141_186792 | Not-determined |
| 17-109_k141_2579 | Not-determined |
| 17-109_k141_169811 | Not-determined |
| 17-109_k141_270069 | Not-determined |
| 17-109_k141_353839 | Not-determined |
| 17-109_k141_353841 | Low-quality |
| 17-109_k141_86147 | Not-determined |
| 17-109_k141_69583 | Low-quality |
| 17-109_k141_304118 | Not-determined |
| 17-109_k141_353851 | Not-determined |
| 17-109_k141_153205 | Not-determined |
| 17-109_k141_186815 | Not-determined |
| 17-109_k141_186817 | Not-determined |
| 17-109_k141_387519 | Not-determined |
| 17-109_k141_153208 | Not-determined |
| 17-109_k141_287268 | Not-determined |
| 17-109_k141_69600 | Low-quality |
| 17-109_k141_320634 | Not-determined |
| 17-109_k141_371013 | Low-quality |
| 17-109_k141_2613 | Not-determined |
| 17-109_k141_203166 | Not-determined |
| 17-109_k141_2617 | Not-determined |
| 17-109_k141_186829 | Not-determined |
| 17-109_k141_203168 | Not-determined |
| 17-109_k141_2623 | Not-determined |
| 17-109_k141_371019 | Not-determined |
| 17-109_k141_86180 | Not-determined |
| 17-109_k141_236774 | Low-quality |
| 17-109_k141_287293 | Not-determined |
| 17-109_k141_371033 | Low-quality |
| 17-109_k141_353879 | Not-determined |
| 17-109_k141_387537 | Not-determined |
| 17-109_k141_153231 | Not-determined |
| 17-109_k141_387538 | Not-determined |
| 17-109_k141_371040 | Not-determined |
| 17-109_k141_253363 | Not-determined |
| 17-109_k141_320665 | Not-determined |
| 17-109_k141_2641 | Not-determined |
| 17-109_k141_219864 | Not-determined |
| 17-109_k141_287309 | Not-determined |
| 17-109_k141_304143 | Not-determined |
| 17-109_k141_387549 | Low-quality |
| 17-109_k141_69631 | Not-determined |
| 17-109_k141_219870 | Low-quality |
| 17-109_k141_236795 | Not-determined |
| 17-109_k141_287317 | Not-determined |
| 17-109_k141_86190 | Not-determined |
| 17-109_k141_353904 | Not-determined |
| 17-109_k141_270104 | Not-determined |
| 17-109_k141_270105 | Not-determined |
| 17-109_k141_353908 | Not-determined |
| 17-109_k141_86194 | Low-quality |
| 17-109_k141_387558 | Not-determined |
| 17-109_k141_86196 | Not-determined |
| 17-109_k141_36160 | Not-determined |
| 17-109_k141_387561 | Not-determined |
| 17-109_k141_387562 | Not-determined |
| 17-109_k141_387565 | Not-determined |
| 17-109_k141_219874 | Low-quality |
| 17-109_k141_119293 | Not-determined |
| 17-109_k141_86206 | Not-determined |
| 17-109_k141_2674 | Not-determined |
| 17-109_k141_86208 | Not-determined |
| 17-109_k141_86210 | Not-determined |
| 17-109_k141_387572 | Not-determined |
| 17-109_k141_119302 | Not-determined |
| 17-109_k141_2678 | Not-determined |
| 17-109_k141_102645 | Not-determined |
| 17-109_k141_169836 | Not-determined |
| 17-109_k141_119309 | Not-determined |
| 17-109_k141_86222 | Not-determined |
| 17-109_k141_236826 | Not-determined |
| 17-109_k141_153271 | Not-determined |
| 17-109_k141_169843 | Not-determined |
| 17-109_k141_219888 | Not-determined |
| 17-109_k141_236832 | Not-determined |
| 17-109_k141_169853 | Not-determined |
| 17-109_k141_353934 | Not-determined |
| 17-109_k141_169858 | Not-determined |
| 17-109_k141_387592 | Not-determined |
| 17-109_k141_102658 | Not-determined |
| 17-109_k141_353941 | Not-determined |
| 17-109_k141_353944 | Not-determined |

| | |
|---|---|
| 17-109_k141_69662 | Not-determined |
| 17-109_k141_169861 | Low-quality |
| 17-109_k141_86234 | Not-determined |
| 17-109_k141_86235 | Not-determined |
| 17-109_k141_69666 | Not-determined |
| 17-109_k141_69667 | Low-quality |
| 17-109_k141_69669 | Low-quality |
| 17-109_k141_353967 | Not-determined |
| 17-109_k141_169876 | Not-determined |
| 17-109_k141_353971 | Not-determined |
| 17-109_k141_136354 | Not-determined |
| 17-109_k141_236849 | Not-determined |
| 17-109_k141_153302 | Not-determined |
| 17-109_k141_270154 | Not-determined |
| 17-109_k141_102680 | Not-determined |
| 17-109_k141_86253 | Not-determined |
| 17-109_k141_2711 | Not-determined |
| 17-109_k141_353982 | Not-determined |
| 17-109_k141_320713 | Not-determined |
| 17-109_k141_69694 | Not-determined |
| 17-109_k141_320716 | Not-determined |
| 17-109_k141_387603 | Not-determined |
| 17-109_k141_102696 | Not-determined |
| 17-109_k141_320721 | Not-determined |
| 17-109_k141_320723 | Not-determined |
| 17-109_k141_119327 | Not-determined |
| 17-109_k141_387609 | Not-determined |
| 17-109_k141_69708 | Not-determined |
| 17-109_k141_387614 | Not-determined |
| 17-109_k141_153320 | Not-determined |
| 17-109_k141_136375 | Low-quality |
| 17-109_k141_69714 | Not-determined |
| 17-109_k141_102710 | Not-determined |
| 17-109_k141_102712 | Not-determined |
| 17-109_k141_136382 | Not-determined |
| 17-109_k141_153325 | Not-determined |
| 17-109_k141_136384 | Not-determined |
| 17-109_k141_219891 | Not-determined |
| 17-109_k141_153328 | Not-determined |
| 17-109_k141_136389 | Not-determined |
| 17-109_k141_354025 | Not-determined |
| 17-109_k141_354026 | Low-quality |
| 17-109_k141_253412 | Not-determined |
| 17-109_k141_253413 | Not-determined |
| 17-109_k141_304186 | Not-determined |
| 17-109_k141_253414 | Not-determined |
| 17-109_k141_136401 | Not-determined |
| 17-109_k141_102726 | Not-determined |
| 17-109_k141_336646 | Not-determined |
| 17-109_k141_236864 | Not-determined |
| 17-109_k141_186880 | Not-determined |
| 17-109_k141_270173 | Not-determined |
| 17-109_k141_371063 | Not-determined |
| 17-109_k141_186883 | Not-determined |
| 17-109_k141_203195 | Not-determined |
| 17-109_k141_236872 | Not-determined |
| 17-109_k141_270180 | Not-determined |
| 17-109_k141_354044 | Not-determined |
| 17-109_k141_153353 | Not-determined |
| 17-109_k141_136415 | Not-determined |
| 17-109_k141_336664 | Not-determined |
| 17-109_k141_236887 | Not-determined |
| 17-109_k141_186901 | Low-quality |
| 17-109_k141_371072 | Not-determined |
| 17-109_k141_304213 | Not-determined |
| 17-109_k141_186904 | Not-determined |
| 17-109_k141_169892 | Low-quality |
| 17-109_k141_186907 | Not-determined |
| 17-109_k141_169894 | Not-determined |
| 17-109_k141_119344 | Low-quality |
| 17-109_k141_203211 | Not-determined |
| 17-109_k141_219933 | Low-quality |
| 17-109_k141_169897 | Not-determined |
| 17-109_k141_186916 | Not-determined |
| 17-109_k141_119353 | Not-determined |
| 17-109_k141_219938 | Not-determined |
| 17-109_k141_153368 | Not-determined |
| 17-109_k141_69756 | Not-determined |
| 17-109_k141_69758 | Not-determined |
| 17-109_k141_119356 | Not-determined |
| 17-109_k141_336682 | Not-determined |
| 17-109_k141_136433 | Not-determined |
| 17-109_k141_2717 | Not-determined |
| 17-109_k141_169923 | Not-determined |
| 17-109_k141_102745 | Not-determined |
| 17-109_k141_102746 | Not-determined |
| 17-109_k141_219944 | Not-determined |
| 17-109_k141_236921 | Not-determined |
| 17-109_k141_219948 | Not-determined |
| 17-109_k141_186939 | Not-determined |
| 17-109_k141_169940 | Not-determined |
| 17-109_k141_153395 | Not-determined |
| 17-109_k141_169941 | Not-determined |
| 17-109_k141_52857 | Low-quality |
| 17-109_k141_354096 | Not-determined |
| 17-109_k141_387662 | Not-determined |
| 17-109_k141_69803 | Not-determined |
| 17-109_k141_153400 | Not-determined |
| 17-109_k141_336703 | Not-determined |
| 17-109_k141_69808 | Not-determined |
| 17-109_k141_304229 | Not-determined |
| 17-109_k141_270198 | Not-determined |
| 17-109_k141_52868 | Not-determined |
| 17-109_k141_236938 | Not-determined |
| 17-109_k141_86284 | Not-determined |
| 17-109_k141_203252 | Low-quality |
| 17-109_k141_270202 | Low-quality |
| 17-109_k141_52875 | Not-determined |
| 17-109_k141_119378 | Not-determined |
| 17-109_k141_336716 | Not-determined |
| 17-109_k141_86291 | Low-quality |
| 17-109_k141_336718 | Not-determined |
| 17-109_k141_270203 | Not-determined |
| 17-109_k141_52884 | Not-determined |
| 17-109_k141_169958 | Not-determined |
| 17-109_k141_2730 | Not-determined |
| 17-109_k141_203266 | Not-determined |
| 17-109_k141_270207 | Not-determined |
| 17-109_k141_2734 | Low-quality |
| 17-109_k141_203268 | Low-quality |
| 17-109_k141_169960 | Not-determined |
| 17-109_k141_36176 | Not-determined |
| 17-109_k141_119386 | Not-determined |
| 17-109_k141_287358 | Not-determined |
| 17-109_k141_2741 | Not-determined |
| 17-109_k141_287361 | Low-quality |
| 17-109_k141_270215 | Not-determined |
| 17-109_k141_387670 | Not-determined |
| 17-109_k141_36186 | Not-determined |
| 17-109_k141_169968 | Low-quality |
| 17-109_k141_136467 | Not-determined |
| 17-109_k141_2749 | Not-determined |
| 17-109_k141_387680 | Not-determined |
| 17-109_k141_387682 | Not-determined |
| 17-109_k141_270226 | Not-determined |
| 17-109_k141_287372 | Not-determined |
| 17-109_k141_354100 | Not-determined |
| 17-109_k141_36197 | Not-determined |
| 17-109_k141_287376 | Not-determined |
| 17-109_k141_186972 | Not-determined |
| 17-109_k141_86312 | Not-determined |
| 17-109_k141_36199 | Not-determined |
| 17-109_k141_287379 | Low-quality |
| 17-109_k141_236981 | Not-determined |
| 17-109_k141_287382 | Not-determined |
| 17-109_k141_86318 | Not-determined |
| 17-109_k141_287385 | Not-determined |
| 17-109_k141_86322 | Not-determined |
| 17-109_k141_287387 | Not-determined |
| 17-109_k141_371096 | Not-determined |
| 17-109_k141_2763 | Not-determined |
| 17-109_k141_253488 | Low-quality |
| 17-109_k141_86329 | Not-determined |
| 17-109_k141_270234 | Not-determined |
| 17-109_k141_371108 | Not-determined |
| 17-109_k141_187005 | Not-determined |
| 17-109_k141_119407 | Not-determined |
| 17-109_k141_187011 | Not-determined |
| 17-109_k141_169984 | Low-quality |
| 17-109_k141_119414 | Not-determined |
| 17-109_k141_187018 | Not-determined |
| 17-109_k141_102806 | Not-determined |
| 17-109_k141_270254 | Not-determined |
| 17-109_k141_69855 | Not-determined |
| 17-109_k141_270255 | Not-determined |
| 17-109_k141_387702 | Not-determined |
| 17-109_k141_336755 | Not-determined |
| 17-109_k141_270258 | Low-quality |
| 17-109_k141_169990 | Not-determined |
| 17-109_k141_219969 | Not-determined |
| 17-109_k141_119427 | Not-determined |
| 17-109_k141_2789 | Not-determined |
| 17-109_k141_169997 | Not-determined |
| 17-109_k141_153441 | Not-determined |
| 17-109_k141_69867 | Not-determined |
| 17-109_k141_2793 | Not-determined |
| 17-109_k141_219975 | Not-determined |
| 17-109_k141_69870 | Not-determined |
| 17-109_k141_304316 | Low-quality |
| 17-109_k141_270264 | Not-determined |
| 17-109_k141_219983 | Not-determined |
| 17-109_k141_102828 | Not-determined |
| 17-109_k141_136514 | Not-determined |
| 17-109_k141_336770 | Not-determined |
| 17-109_k141_69885 | Not-determined |
| 17-109_k141_287433 | Not-determined |
| 17-109_k141_371135 | Not-determined |
| 17-109_k141_270280 | Low-quality |
| 17-109_k141_187044 | Not-determined |
| 17-109_k141_203348 | Not-determined |
| 17-109_k141_170010 | Not-determined |
| 17-109_k141_153472 | Not-determined |
| 17-109_k141_203357 | Low-quality |
| 18-105_k141_62679 | High-quality |
| 17-109_k141_336782 | Low-quality |
| 17-109_k141_203363 | Not-determined |
| 17-109_k141_371154 | Not-determined |
| 17-109_k141_18935 | Not-determined |
| 17-109_k141_387712 | Low-quality |
| 17-109_k141_187068 | Not-determined |
| 17-109_k141_220021 | Not-determined |
| 17-109_k141_287481 | Not-determined |
| 17-109_k141_203379 | Not-determined |
| 17-109_k141_336795 | Not-determined |
| 17-109_k141_119452 | Not-determined |
| 17-109_k141_86339 | Low-quality |
| 17-109_k141_69948 | Not-determined |
| 17-109_k141_371183 | Not-determined |
| 17-109_k141_371187 | Not-determined |
| 17-109_k141_86352 | Not-determined |
| 17-109_k141_270301 | Not-determined |
| 17-109_k141_270302 | Low-quality |
| 17-109_k141_36278 | Not-determined |
| 17-109_k141_371192 | Not-determined |
| 17-109_k141_203402 | Not-determined |
| 17-109_k141_270313 | Not-determined |
| 17-109_k141_371198 | Not-determined |
| 17-109_k141_187115 | Not-determined |
| 17-109_k141_119488 | Not-determined |
| 17-109_k141_119490 | Low-quality |
| 17-109_k141_253494 | Not-determined |
| 17-109_k141_2824 | Not-determined |
| 17-109_k141_320743 | Not-determined |
| 17-109_k141_253497 | Not-determined |
| 17-109_k141_253499 | Not-determined |
| 17-109_k141_270334 | Not-determined |
| 17-109_k141_36288 | Not-determined |
| 17-109_k141_270340 | Not-determined |
| 17-109_k141_304347 | Not-determined |
| 17-109_k141_86368 | Not-determined |
| 17-109_k141_69987 | Not-determined |
| 17-109_k141_36295 | Low-quality |
| 17-109_k141_336813 | Not-determined |
| 17-109_k141_36296 | Not-determined |
| 17-109_k141_253517 | Not-determined |
| 17-109_k141_102882 | Not-determined |
| 17-109_k141_320766 | Not-determined |
| 17-109_k141_119510 | Low-quality |
| 17-109_k141_102892 | Not-determined |
| 17-109_k141_52959 | Not-determined |
| 17-109_k141_18966 | Not-determined |
| 17-109_k141_2835 | Not-determined |
| 17-109_k141_119526 | Low-quality |
| 17-109_k141_119529 | Not-determined |
| 17-109_k141_102902 | Not-determined |
| 17-109_k141_203415 | Not-determined |
| 17-109_k141_336829 | Not-determined |
| 17-109_k141_36315 | Not-determined |
| 17-109_k141_270357 | Not-determined |
| 17-109_k141_136602 | Not-determined |
| 17-109_k141_253536 | Not-determined |
| 17-109_k141_187137 | Not-determined |
| 17-109_k141_18970 | Low-quality |
| 17-109_k141_18978 | Not-determined |
| 17-109_k141_270370 | Not-determined |
| 17-109_k141_187153 | Not-determined |
| 17-109_k141_336838 | Not-determined |
| 17-109_k141_170080 | Not-determined |
| 17-109_k141_52966 | Not-determined |
| 17-109_k141_136614 | Not-determined |
| 17-109_k141_36337 | Not-determined |
| 17-109_k141_270394 | Not-determined |
| 17-109_k141_119559 | Not-determined |
| 17-109_k141_136627 | Not-determined |
| 17-109_k141_36348 | Not-determined |
| 17-109_k141_336847 | Not-determined |
| 17-109_k141_136630 | Not-determined |
| 17-109_k141_52973 | Low-quality |
| 17-109_k141_203446 | Not-determined |
| 17-109_k141_119563 | Not-determined |
| 17-109_k141_203449 | Not-determined |
| 17-109_k141_203450 | Not-determined |
| 17-109_k141_203451 | Not-determined |
| 17-109_k141_36358 | Not-determined |
| 17-109_k141_170093 | Not-determined |
| 17-109_k141_19013 | Not-determined |
| 17-109_k141_320791 | Not-determined |
| 17-109_k141_102913 | Not-determined |
| 17-109_k141_86392 | Not-determined |
| 17-109_k141_170109 | Not-determined |
| 17-109_k141_86396 | Not-determined |
| 17-109_k141_237013 | Not-determined |
| 17-109_k141_52989 | Not-determined |
| 17-109_k141_336867 | Not-determined |
| 17-109_k141_136649 | Not-determined |
| 17-109_k141_36377 | Not-determined |
| 17-109_k141_119592 | Not-determined |
| 17-109_k141_102918 | Not-determined |
| 17-109_k141_36382 | Not-determined |
| 17-109_k141_320801 | Not-determined |
| 17-109_k141_2868 | Not-determined |
| 17-109_k141_237032 | Not-determined |
| 17-109_k141_336871 | Not-determined |
| 17-109_k141_336873 | Not-determined |
| 17-109_k141_287536 | Low-quality |
| 17-109_k141_387737 | Low-quality |
| 17-109_k141_2874 | Not-determined |
| 17-109_k141_36393 | Not-determined |
| 17-109_k141_119599 | Low-quality |
| 17-109_k141_102938 | Low-quality |
| 17-109_k141_336875 | Low-quality |
| 17-109_k141_237046 | Not-determined |
| 17-109_k141_19038 | Not-determined |
| 17-109_k141_237049 | Not-determined |
| 17-109_k141_336881 | Not-determined |
| 17-109_k141_336883 | Not-determined |
| 17-109_k141_36399 | Not-determined |
| 17-109_k141_53001 | Not-determined |
| 17-109_k141_19045 | Not-determined |
| 17-109_k141_371246 | Not-determined |
| 17-109_k141_237060 | Not-determined |
| 17-109_k141_371247 | Not-determined |
| 17-109_k141_119612 | Not-determined |
| 17-109_k141_53011 | Not-determined |
| 17-109_k141_102958 | Not-determined |
| 17-109_k141_287563 | Not-determined |
| 17-109_k141_371252 | Not-determined |
| 17-109_k141_287565 | Not-determined |
| 17-109_k141_270424 | Not-determined |
| 17-109_k141_119622 | Not-determined |
| 17-109_k141_304367 | Low-quality |
| 17-109_k141_220030 | Not-determined |
| 17-109_k141_53029 | Not-determined |
| 17-109_k141_320820 | Not-determined |
| 17-109_k141_187214 | Not-determined |
| 17-109_k141_170141 | Low-quality |
| 17-109_k141_102971 | Not-determined |
| 17-109_k141_19049 | Not-determined |
| 17-109_k141_19054 | Not-determined |
| 17-109_k141_320835 | Not-determined |
| 17-109_k141_304379 | Not-determined |
| 17-109_k141_320836 | Not-determined |
| 17-109_k141_237067 | Not-determined |
| 17-109_k141_86445 | Not-determined |
| 17-109_k141_102983 | Low-quality |
| 17-109_k141_86447 | Not-determined |
| 17-109_k141_102986 | Not-determined |
| 17-109_k141_70005 | Not-determined |
| 17-109_k141_136675 | Not-determined |
| 17-109_k141_253570 | Not-determined |
| 17-109_k141_70007 | Low-quality |
| 17-109_k141_220055 | Not-determined |
| 17-109_k141_320844 | Low-quality |
| 17-109_k141_320845 | Not-determined |
| 17-109_k141_371264 | Not-determined |
| 17-109_k141_36431 | Not-determined |
| 17-109_k141_387766 | Not-determined |
| 17-109_k141_387769 | Not-determined |
| 17-109_k141_220070 | Not-determined |
| 17-109_k141_237087 | Not-determined |
| 17-109_k141_320851 | Low-quality |
| 17-109_k141_70024 | Low-quality |
| 17-109_k141_70027 | Not-determined |
| 17-109_k141_304386 | Not-determined |
| 17-109_k141_103007 | Not-determined |
| 17-109_k141_86462 | Low-quality |
| 17-109_k141_119679 | Not-determined |
| 17-109_k141_304396 | Not-determined |
| 17-109_k141_119686 | Not-determined |
| 17-109_k141_70036 | Low-quality |
| 17-109_k141_270437 | Not-determined |
| 17-109_k141_270440 | Low-quality |
| 17-109_k141_70039 | Not-determined |
| 17-109_k141_220087 | Not-determined |
| 17-109_k141_2909 | Low-quality |
| 17-109_k141_203541 | Low-quality |
| 17-109_k141_304403 | Not-determined |
| 17-109_k141_304405 | Not-determined |
| 17-109_k141_86471 | Not-determined |
| 17-109_k141_103031 | Not-determined |
| 17-109_k141_270452 | Low-quality |
| 17-109_k141_203544 | Not-determined |
| 17-109_k141_70051 | Low-quality |
| 17-109_k141_2913 | Not-determined |
| 17-109_k141_237134 | Not-determined |
| 17-109_k141_203551 | Low-quality |
| 17-109_k141_237135 | Low-quality |
| 17-109_k141_119717 | Not-determined |
| 17-109_k141_136729 | Not-determined |
| 17-109_k141_354157 | Not-determined |
| 17-109_k141_270459 | Not-determined |
| 17-109_k141_203568 | Not-determined |
| 17-109_k141_304421 | Low-quality |
| 17-109_k141_220123 | Not-determined |
| 17-109_k141_187253 | Not-determined |
| 17-109_k141_304422 | Not-determined |
| 17-109_k141_220124 | Not-determined |
| 17-109_k141_2941 | Not-determined |
| 17-109_k141_136746 | Not-determined |
| 17-109_k141_2943 | Low-quality |
| 17-109_k141_371295 | Not-determined |
| 17-109_k141_304431 | Not-determined |
| 17-109_k141_371298 | Not-determined |
| 17-109_k141_86496 | Not-determined |
| 17-109_k141_136747 | Not-determined |
| 17-109_k141_203581 | Low-quality |
| 17-109_k141_203582 | Not-determined |
| 17-109_k141_70079 | Not-determined |
| 17-109_k141_270486 | Not-determined |
| 17-109_k141_2957 | Not-determined |
| 17-109_k141_19086 | Low-quality |
| 17-109_k141_136760 | Not-determined |
| 17-109_k141_287574 | Not-determined |
| 17-109_k141_70083 | Low-quality |
| 17-109_k141_70088 | Low-quality |
| 17-109_k141_19099 | Not-determined |
| 17-109_k141_170157 | Low-quality |
| 17-109_k141_86512 | Not-determined |
| 17-109_k141_2960 | Not-determined |
| 17-109_k141_153481 | Not-determined |
| 17-109_k141_304446 | Not-determined |
| 17-109_k141_170160 | Low-quality |
| 17-109_k141_119755 | Not-determined |
| 17-109_k141_153491 | Low-quality |
| 17-109_k141_220139 | Not-determined |
| 17-109_k141_336960 | Not-determined |
| 17-109_k141_220142 | Not-determined |
| 17-109_k141_270509 | Not-determined |
| 17-109_k141_136790 | Not-determined |
| 17-109_k141_36456 | Not-determined |
| 17-109_k141_203601 | Not-determined |
| 17-109_k141_36461 | Not-determined |
| 17-109_k141_2985 | Not-determined |
| 17-109_k141_203604 | Low-quality |
| 17-109_k141_36463 | Low-quality |
| 17-109_k141_371328 | Low-quality |
| 17-109_k141_220147 | Not-determined |
| 17-109_k141_220152 | Not-determined |
| 17-109_k141_86533 | Not-determined |
| 17-109_k141_136799 | Not-determined |
| 17-109_k141_220157 | Not-determined |
| 17-109_k141_86535 | Not-determined |
| 17-109_k141_304477 | Not-determined |
| 17-109_k141_336977 | Not-determined |
| 17-109_k141_86537 | Not-determined |
| 17-109_k141_170203 | Not-determined |
| 17-109_k141_86539 | Low-quality |
| 17-109_k141_103041 | Low-quality |
| 17-109_k141_203623 | Not-determined |
| 17-109_k141_220162 | Low-quality |
| 17-109_k141_3004 | Not-determined |
| 17-109_k141_136811 | Not-determined |
| 17-109_k141_336986 | Not-determined |
| 17-109_k141_103049 | Not-determined |
| 17-109_k141_304488 | Not-determined |
| 17-109_k141_19143 | Not-determined |
| 17-109_k141_53092 | Not-determined |
| 17-109_k141_36479 | Not-determined |
| 17-109_k141_19145 | Not-determined |
| 17-109_k141_220177 | Not-determined |
| 17-109_k141_336999 | Not-determined |
| 17-109_k141_3017 | Not-determined |
| 17-109_k141_237200 | Not-determined |
| 17-109_k141_119779 | Not-determined |
| 17-109_k141_170210 | Not-determined |
| 17-109_k141_187301 | Low-quality |
| 17-109_k141_220185 | Not-determined |
| 17-109_k141_53106 | Not-determined |
| 17-109_k141_220198 | Not-determined |
| 17-109_k141_119786 | Not-determined |
| 17-109_k141_220202 | Low-quality |
| 17-109_k141_220205 | Not-determined |
| 17-109_k141_19159 | Not-determined |
| 17-109_k141_136840 | Not-determined |
| 17-109_k141_337024 | Not-determined |
| 17-109_k141_187315 | Not-determined |
| 17-109_k141_270550 | Not-determined |
| 17-109_k141_320896 | Not-determined |
| 17-109_k141_337031 | Not-determined |
| 17-109_k141_354185 | Low-quality |
| 17-109_k141_119790 | Not-determined |
| 17-109_k141_354189 | Not-determined |
| 17-109_k141_86553 | Not-determined |
| 17-109_k141_86554 | Not-determined |
| 17-109_k141_320898 | Not-determined |
| 17-109_k141_320900 | Not-determined |
| 17-109_k141_19182 | Not-determined |
| 17-109_k141_19183 | Not-determined |
| 17-109_k141_371394 | Not-determined |
| 17-109_k141_153531 | Not-determined |
| 17-109_k141_320908 | Not-determined |
| 17-109_k141_153537 | Not-determined |
| 17-109_k141_170231 | Not-determined |
| 17-109_k141_187329 | Not-determined |
| 17-109_k141_187330 | Not-determined |
| 17-109_k141_387825 | Low-quality |
| 17-109_k141_19191 | Not-determined |
| 17-109_k141_220223 | Not-determined |
| 17-109_k141_270568 | Not-determined |
| 17-109_k141_270573 | Not-determined |
| 17-109_k141_220234 | Not-determined |
| 17-109_k141_36513 | Not-determined |
| 17-109_k141_36515 | Not-determined |
| 17-109_k141_220237 | Low-quality |
| 17-109_k141_70141 | Low-quality |
| 17-109_k141_36520 | Not-determined |
| 17-109_k141_371399 | Low-quality |
| 17-109_k141_70143 | Not-determined |
| 17-109_k141_70148 | Not-determined |
| 17-109_k141_70152 | Low-quality |
| 17-109_k141_304527 | Low-quality |
| 17-109_k141_371405 | Not-determined |
| 17-109_k141_237225 | Not-determined |
| 17-109_k141_187347 | Not-determined |
| 17-109_k141_70160 | Not-determined |
| 17-109_k141_187348 | Not-determined |
| 17-109_k141_387837 | Not-determined |
| 17-109_k141_220248 | Not-determined |
| 17-109_k141_187352 | Not-determined |
| 17-109_k141_220249 | Not-determined |
| 17-109_k141_70165 | Not-determined |
| 17-109_k141_36527 | Not-determined |
| 17-109_k141_153565 | Not-determined |
| 17-109_k141_70177 | Not-determined |
| 17-109_k141_287598 | Not-determined |
| 17-109_k141_237239 | Not-determined |
| 17-109_k141_220260 | Not-determined |
| 17-109_k141_387857 | Not-determined |
| 17-109_k141_53113 | Not-determined |
| 17-109_k141_220264 | Not-determined |
| 17-109_k141_170258 | Not-determined |
| 17-109_k141_36537 | Not-determined |
| 17-109_k141_287606 | Not-determined |
| 17-109_k141_53123 | Not-determined |
| 17-109_k141_237246 | Not-determined |
| 17-109_k141_53124 | Not-determined |
| 17-109_k141_70192 | Not-determined |
| 17-109_k141_287611 | Not-determined |
| 17-109_k141_86573 | Not-determined |
| 17-109_k141_253619 | Not-determined |
| 17-109_k141_119816 | Not-determined |
| 17-109_k141_70203 | Not-determined |
| 17-109_k141_253622 | Low-quality |
| 17-109_k141_3060 | Not-determined |
| 17-109_k141_304550 | Not-determined |
| 17-109_k141_70205 | Not-determined |
| 17-109_k141_170284 | Not-determined |
| 17-109_k141_387869 | Not-determined |
| 17-109_k141_371422 | Not-determined |
| 17-109_k141_387875 | Not-determined |
| 17-109_k141_36565 | Not-determined |
| 17-109_k141_170286 | Low-quality |
| 17-109_k141_36568 | Not-determined |
| 17-109_k141_337075 | Not-determined |
| 17-109_k141_136885 | Not-determined |
| 17-109_k141_36570 | Not-determined |
| 17-109_k141_119826 | Not-determined |
| 17-109_k141_86588 | Not-determined |
| 17-109_k141_337082 | Not-determined |
| 17-109_k141_136889 | Low-quality |
| 17-109_k141_36576 | Not-determined |
| 17-109_k141_337084 | Not-determined |
| 17-109_k141_86593 | Not-determined |
| 17-109_k141_220304 | Not-determined |
| 17-109_k141_36578 | Low-quality |
| 17-109_k141_170297 | Low-quality |
| 17-109_k141_237262 | Not-determined |
| 17-109_k141_387881 | Low-quality |
| 17-109_k141_270631 | Not-determined |
| 17-109_k141_70228 | Not-determined |
| 17-109_k141_70230 | Not-determined |
| 17-109_k141_270636 | Not-determined |
| 17-109_k141_19251 | Not-determined |
| 17-109_k141_187374 | Low-quality |
| 17-109_k141_387888 | Not-determined |
| 17-109_k141_70243 | Not-determined |
| 17-109_k141_253652 | Not-determined |
| 17-109_k141_253655 | Not-determined |
| 17-109_k141_19262 | Not-determined |
| 17-109_k141_153599 | Not-determined |
| 17-109_k141_70257 | Not-determined |
| 17-109_k141_270665 | Not-determined |
| 17-109_k141_187393 | Not-determined |
| 17-109_k141_3077 | Not-determined |
| 17-109_k141_387906 | Not-determined |
| 17-109_k141_387907 | Not-determined |
| 17-109_k141_203706 | Low-quality |
| 17-109_k141_3079 | Not-determined |
| 17-109_k141_36589 | Not-determined |
| 17-109_k141_270670 | Not-determined |
| 17-109_k141_203708 | Not-determined |
| 17-109_k141_270671 | Not-determined |
| 17-109_k141_371448 | Low-quality |
| 17-109_k141_253656 | Low-quality |
| 17-109_k141_3082 | Not-determined |
| 17-109_k141_371453 | Not-determined |
| 17-109_k141_3086 | Not-determined |
| 17-109_k141_220349 | Not-determined |
| 17-109_k141_371454 | Not-determined |

| | |
|---|---|
| 17-109_k141_337107 | Low-quality |
| 17-109_k141_220351 | Not-determined |
| 17-109_k141_19280 | Not-determined |
| 17-109_k141_203713 | Low-quality |
| 17-109_k141_371463 | Not-determined |
| 17-109_k141_36593 | Low-quality |
| 17-109_k141_53158 | Not-determined |
| 17-109_k141_203717 | Not-determined |
| 17-109_k141_136928 | Not-determined |
| 17-109_k141_187415 | Not-determined |
| 17-109_k141_3093 | Not-determined |
| 17-109_k141_3095 | Not-determined |
| 17-109_k141_371470 | Not-determined |
| 17-109_k141_253675 | Not-determined |
| 17-109_k141_287631 | Not-determined |
| 17-109_k141_337125 | Not-determined |
| 17-109_k141_170323 | Not-determined |
| 17-109_k141_220378 | Not-determined |
| 17-109_k141_103073 | Not-determined |
| 17-109_k141_170329 | Not-determined |
| 17-109_k141_103077 | Not-determined |
| 17-109_k141_170342 | Not-determined |
| 17-109_k141_220389 | Low-quality |
| 17-109_k141_36620 | Not-determined |
| 17-109_k141_136967 | Not-determined |
| 17-109_k141_203746 | Not-determined |
| 17-109_k141_136968 | Not-determined |
| 17-109_k141_270687 | Not-determined |
| 17-109_k141_86661 | Not-determined |
| 17-109_k141_337135 | Not-determined |
| 17-109_k141_371499 | Not-determined |
| 17-109_k141_371501 | Not-determined |
| 17-109_k141_220399 | Low-quality |
| 17-109_k141_337142 | Low-quality |
| 17-109_k141_371503 | Not-determined |
| 17-109_k141_387918 | Not-determined |
| 17-109_k141_3122 | Not-determined |
| 17-109_k141_153631 | Not-determined |
| 17-109_k141_53207 | Not-determined |
| 17-109_k141_36642 | Not-determined |
| 17-109_k141_187447 | Not-determined |
| 17-109_k141_253700 | Not-determined |
| 17-109_k141_337152 | Not-determined |
| 17-109_k141_253703 | Not-determined |
| 17-109_k141_3136 | Not-determined |
| 17-109_k141_371522 | Not-determined |
| 17-109_k141_387936 | Not-determined |
| 17-109_k141_220406 | Not-determined |
| 17-109_k141_387939 | Not-determined |
| 17-109_k141_387940 | Not-determined |
| 17-109_k141_203773 | Not-determined |
| 17-109_k141_170382 | Not-determined |
| 17-109_k141_136979 | Not-determined |
| 17-109_k141_136982 | Not-determined |
| 17-109_k141_136985 | Not-determined |
| 17-109_k141_220424 | Not-determined |
| 17-109_k141_270710 | Not-determined |
| 17-109_k141_387950 | Not-determined |
| 17-109_k141_304603 | Not-determined |
| 17-109_k141_203780 | Not-determined |
| 17-109_k141_119848 | Not-determined |
| 17-109_k141_203782 | Not-determined |
| 17-109_k141_3160 | Not-determined |
| 17-109_k141_270716 | Not-determined |
| 17-109_k141_320982 | Not-determined |
| 17-109_k141_86681 | Not-determined |
| 17-109_k141_387958 | Not-determined |
| 17-109_k141_387959 | Not-determined |
| 17-109_k141_19311 | Not-determined |
| 17-109_k141_187460 | Not-determined |
| 17-109_k141_304618 | Not-determined |
| 17-109_k141_203799 | Not-determined |
| 17-109_k141_270729 | Not-determined |
| 17-109_k141_53258 | Not-determined |
| 17-109_k141_170395 | Not-determined |
| 17-109_k141_287644 | Not-determined |
| 17-109_k141_287646 | Not-determined |
| 17-109_k141_354238 | Not-determined |
| 17-109_k141_287648 | Not-determined |
| 17-109_k141_3174 | Not-determined |
| 17-109_k141_220440 | Not-determined |
| 17-109_k141_354246 | Low-quality |
| 17-109_k141_3179 | Not-determined |
| 17-109_k141_137017 | Not-determined |
| 17-109_k141_337199 | Not-determined |
| 17-109_k141_3185 | Not-determined |
| 17-109_k141_19331 | Not-determined |
| 17-109_k141_137024 | Not-determined |
| 17-109_k141_70272 | Not-determined |
| 17-109_k141_153665 | Not-determined |
| 17-109_k141_36670 | Not-determined |
| 17-109_k141_220446 | Not-determined |
| 17-109_k141_137030 | Not-determined |
| 17-109_k141_270754 | Not-determined |
| 17-109_k141_354267 | Low-quality |
| 17-109_k141_3207 | Not-determined |
| 17-109_k141_287660 | Not-determined |
| 17-109_k141_70277 | Not-determined |
| 17-109_k141_70280 | Not-determined |
| 17-109_k141_287664 | Not-determined |
| 17-109_k141_354271 | Not-determined |
| 17-109_k141_287668 | Not-determined |
| 17-109_k141_70282 | Not-determined |
| 17-109_k141_354274 | Not-determined |
| 17-109_k141_354277 | Not-determined |
| 17-109_k141_304640 | Low-quality |
| 17-109_k141_103114 | Not-determined |
| 17-109_k141_103115 | Not-determined |
| 17-109_k141_304641 | Not-determined |
| 17-109_k141_86701 | Not-determined |
| 17-109_k141_337214 | Not-determined |
| 17-109_k141_86706 | Not-determined |
| 17-109_k141_86708 | Not-determined |
| 17-109_k141_137041 | Low-quality |
| 17-109_k141_337218 | Not-determined |
| 17-109_k141_19337 | Not-determined |
| 17-109_k141_170442 | Not-determined |
| 17-109_k141_320995 | Not-determined |
| 17-109_k141_270768 | Not-determined |
| 17-109_k141_19341 | Not-determined |
| 17-109_k141_337225 | Not-determined |
| 17-109_k141_86714 | Not-determined |
| 17-109_k141_304658 | Not-determined |
| 17-109_k141_354289 | Not-determined |
| 17-109_k141_103127 | Not-determined |
| 17-109_k141_187496 | Not-determined |
| 17-109_k141_304660 | Not-determined |
| 17-109_k141_354292 | Not-determined |
| 17-109_k141_354293 | Not-determined |
| 17-109_k141_137048 | Low-quality |
| 17-109_k141_387996 | Low-quality |
| 17-109_k141_304666 | Not-determined |
| 17-109_k141_103130 | Not-determined |
| 17-109_k141_304667 | Not-determined |
| 17-109_k141_203839 | Low-quality |
| 17-109_k141_287676 | Not-determined |
| 17-109_k141_388001 | Not-determined |
| 17-109_k141_153696 | Not-determined |
| 17-109_k141_287687 | Not-determined |
| 17-109_k141_187517 | Not-determined |
| 17-109_k141_137056 | Not-determined |
| 17-109_k141_287692 | Low-quality |
| 17-109_k141_19363 | Not-determined |
| 17-109_k141_19364 | Not-determined |
| 17-109_k141_103137 | Not-determined |
| 17-109_k141_103138 | Not-determined |
| 17-109_k141_153705 | Not-determined |
| 17-109_k141_388008 | Not-determined |
| 17-109_k141_287702 | Not-determined |
| 17-109_k141_53322 | Not-determined |
| 17-109_k141_321018 | Not-determined |
| 17-109_k141_170459 | Not-determined |
| 17-109_k141_270772 | Not-determined |
| 17-109_k141_36742 | Not-determined |
| 17-109_k141_321035 | Not-determined |
| 17-109_k141_388025 | Not-determined |
| 17-109_k141_170465 | Low-quality |
| 17-109_k141_103158 | Not-determined |
| 17-109_k141_170469 | Not-determined |
| 17-109_k141_337279 | Not-determined |
| 17-109_k141_70331 | Not-determined |
| 17-109_k141_187546 | Not-determined |
| 17-109_k141_70333 | Low-quality |
| 17-109_k141_270791 | Not-determined |
| 17-109_k141_187547 | Not-determined |
| 17-109_k141_70336 | Not-determined |
| 17-109_k141_170478 | Not-determined |
| 17-109_k141_388041 | Not-determined |
| 17-109_k141_86738 | Not-determined |
| 17-109_k141_337286 | Not-determined |
| 17-109_k141_170488 | Not-determined |
| 17-109_k141_170491 | Low-quality |
| 17-109_k141_388050 | Not-determined |
| 17-109_k141_170494 | Not-determined |
| 17-109_k141_86746 | Not-determined |
| 17-109_k141_354333 | Not-determined |
| 17-109_k141_137078 | Not-determined |
| 17-109_k141_119917 | Not-determined |
| 17-109_k141_119918 | Not-determined |
| 17-109_k141_119921 | Not-determined |
| 17-109_k141_119922 | Not-determined |
| 17-109_k141_137080 | Not-determined |
| 17-109_k141_270806 | Not-determined |
| 17-109_k141_86764 | Not-determined |
| 17-109_k141_119932 | Not-determined |
| 17-109_k141_86768 | Not-determined |
| 17-109_k141_103181 | Not-determined |
| 17-109_k141_270809 | Not-determined |
| 17-109_k141_388053 | Low-quality |
| 17-109_k141_170504 | Not-determined |
| 17-109_k141_253705 | Low-quality |
| 17-109_k141_203884 | Not-determined |
| 17-109_k141_270817 | Not-determined |
| 17-109_k141_170510 | Not-determined |
| 17-109_k141_337294 | Not-determined |
| 17-109_k141_137094 | Not-determined |
| 17-109_k141_103194 | Not-determined |
| 17-109_k141_337300 | Not-determined |
| 17-109_k141_170518 | Not-determined |
| 17-109_k141_220495 | Not-determined |
| 17-109_k141_36777 | Low-quality |
| 17-109_k141_270823 | Not-determined |
| 17-109_k141_388055 | Not-determined |
| 17-109_k141_220503 | Not-determined |
| 17-109_k141_388057 | Not-determined |
| 17-109_k141_153750 | Not-determined |
| 17-109_k141_19392 | Low-quality |
| 17-109_k141_119947 | Not-determined |
| 17-109_k141_53371 | Not-determined |
| 17-109_k141_19394 | Not-determined |
| 17-109_k141_388064 | Not-determined |
| 17-109_k141_388066 | Not-determined |
| 17-109_k141_203906 | Not-determined |
| 17-109_k141_70376 | Not-determined |
| 17-109_k141_86788 | Not-determined |
| 17-109_k141_86790 | Not-determined |
| 17-109_k141_388068 | Low-quality |
| 17-109_k141_203918 | Not-determined |
| 17-109_k141_36793 | Not-determined |
| 17-109_k141_237292 | Not-determined |
| 17-109_k141_321049 | Not-determined |
| 17-109_k141_354355 | Low-quality |
| 17-109_k141_53381 | Not-determined |
| 17-109_k141_19415 | Not-determined |
| 17-109_k141_203925 | Not-determined |
| 17-109_k141_153770 | Low-quality |
| 17-109_k141_203929 | Not-determined |
| 17-109_k141_70401 | Not-determined |
| 17-109_k141_203932 | Not-determined |
| 17-109_k141_220524 | Not-determined |
| 17-109_k141_203933 | Not-determined |
| 17-109_k141_187576 | Low-quality |
| 17-109_k141_253744 | Not-determined |
| 17-109_k141_3253 | Not-determined |
| 17-109_k141_170557 | Not-determined |
| 17-109_k141_187583 | Not-determined |
| 17-109_k141_187584 | Not-determined |
| 17-109_k141_170558 | Not-determined |
| 17-109_k141_119982 | Not-determined |
| 17-109_k141_237310 | Not-determined |
| 17-109_k141_371570 | Not-determined |
| 17-109_k141_237312 | Not-determined |
| 17-109_k141_304707 | Not-determined |
| 17-109_k141_354376 | Low-quality |
| 17-109_k141_253754 | Not-determined |
| 17-109_k141_119988 | Not-determined |
| 17-109_k141_53389 | Not-determined |
| 17-109_k141_220535 | Low-quality |
| 17-109_k141_203942 | Not-determined |
| 17-109_k141_304715 | Low-quality |
| 17-109_k141_253762 | Not-determined |
| 17-109_k141_36817 | Not-determined |
| 17-109_k141_371586 | Not-determined |
| 17-109_k141_220549 | Not-determined |
| 17-109_k141_3277 | Not-determined |
| 17-109_k141_70422 | Not-determined |
| 17-109_k141_354392 | Not-determined |
| 17-109_k141_354393 | Not-determined |
| 17-109_k141_19441 | Not-determined |
| 17-109_k141_253768 | Not-determined |
| 17-109_k141_354398 | Not-determined |
| 17-109_k141_253769 | Not-determined |
| 17-109_k141_354400 | Not-determined |
| 17-109_k141_371594 | Not-determined |
| 17-109_k141_237332 | Not-determined |
| 17-109_k141_203960 | Not-determined |
| 17-109_k141_321084 | Not-determined |
| 17-109_k141_321087 | Not-determined |
| 17-109_k141_253777 | Not-determined |
| 17-109_k141_220575 | Low-quality |
| 17-109_k141_237341 | Not-determined |
| 17-109_k141_321095 | Not-determined |
| 17-109_k141_321096 | Not-determined |
| 17-109_k141_86843 | Not-determined |
| 17-109_k141_371607 | Not-determined |
| 17-109_k141_371612 | Not-determined |
| 17-109_k141_287768 | Not-determined |
| 17-109_k141_321105 | Not-determined |
| 17-109_k141_103269 | Low-quality |
| 17-109_k141_337380 | Not-determined |
| 17-109_k141_371614 | Not-determined |
| 17-109_k141_153788 | Not-determined |
| 17-109_k141_337384 | Low-quality |
| 17-109_k141_371616 | Not-determined |
| 17-109_k141_203964 | Not-determined |
| 17-109_k141_203965 | Not-determined |
| 17-109_k141_203966 | Not-determined |
| 17-109_k141_253801 | Not-determined |
| 17-109_k141_137140 | Not-determined |
| 17-109_k141_36861 | Not-determined |
| 17-109_k141_304738 | Not-determined |
| 17-109_k141_237363 | Not-determined |
| 17-109_k141_137147 | Not-determined |
| 17-109_k141_304740 | Not-determined |
| 17-109_k141_237371 | Not-determined |
| 17-109_k141_203977 | Not-determined |
| 17-109_k141_137151 | Low-quality |
| 17-109_k141_287786 | Not-determined |
| 17-109_k141_287787 | Not-determined |
| 17-109_k141_170608 | Not-determined |
| 17-109_k141_137156 | Not-determined |
| 17-109_k141_137158 | Low-quality |
| 17-109_k141_321120 | Not-determined |
| 17-109_k141_287796 | Not-determined |
| 17-109_k141_120018 | Not-determined |
| 17-109_k141_304765 | Low-quality |
| 17-109_k141_237375 | Not-determined |
| 17-109_k141_388113 | Not-determined |
| 17-109_k141_237378 | Not-determined |
| 17-109_k141_70454 | Not-determined |
| 17-109_k141_86873 | Low-quality |
| 17-109_k141_19487 | Not-determined |
| 17-109_k141_86874 | Not-determined |
| 17-109_k141_19492 | Low-quality |
| 17-109_k141_70465 | Not-determined |
| 17-109_k141_103279 | Not-determined |
| 17-109_k141_103280 | Not-determined |
| 17-109_k141_3328 | Not-determined |
| 17-109_k141_337417 | Not-determined |
| 17-109_k141_86893 | Not-determined |
| 17-109_k141_203999 | Not-determined |
| 17-109_k141_3329 | Not-determined |
| 17-109_k141_103293 | Not-determined |
| 17-109_k141_287818 | Not-determined |
| 17-109_k141_204003 | Not-determined |
| 17-109_k141_253810 | Not-determined |
| 17-109_k141_253811 | Not-determined |
| 17-109_k141_153828 | Not-determined |
| 17-109_k141_371648 | Not-determined |
| 17-109_k141_354438 | Not-determined |
| 17-109_k141_53455 | Low-quality |
| 17-109_k141_19523 | Not-determined |
| 17-109_k141_237412 | Not-determined |
| 17-109_k141_388119 | Not-determined |
| 17-109_k141_36888 | Not-determined |
| 17-109_k141_19538 | Not-determined |
| 17-109_k141_187632 | Low-quality |
| 17-109_k141_53474 | Low-quality |
| 17-109_k141_53478 | Not-determined |
| 17-109_k141_354447 | Not-determined |
| 17-109_k141_187639 | Low-quality |
| 17-109_k141_354449 | Not-determined |
| 17-109_k141_337439 | Not-determined |
| 17-109_k141_287836 | Not-determined |
| 17-109_k141_388135 | Not-determined |
| 17-109_k141_237422 | Not-determined |
| 17-109_k141_337443 | Not-determined |
| 17-109_k141_120063 | Not-determined |
| 17-109_k141_36902 | Not-determined |
| 17-109_k141_120068 | Not-determined |
| 17-109_k141_53492 | Not-determined |
| 17-109_k141_187647 | Not-determined |
| 17-109_k141_53493 | Not-determined |
| 17-109_k141_3357 | Not-determined |
| 17-109_k141_371668 | Not-determined |
| 17-109_k141_388139 | Not-determined |
| 17-109_k141_337452 | Low-quality |
| 17-109_k141_220616 | Not-determined |
| 17-109_k141_287842 | Not-determined |
| 17-109_k141_388142 | Not-determined |
| 17-109_k141_388144 | Low-quality |
| 17-109_k141_103307 | Not-determined |
| 17-109_k141_70491 | Not-determined |
| 17-109_k141_103318 | Not-determined |
| 17-109_k141_304803 | Not-determined |
| 17-109_k141_103324 | Not-determined |
| 17-109_k141_304809 | Not-determined |
| 17-109_k141_220623 | Not-determined |
| 17-109_k141_70512 | Not-determined |
| 17-109_k141_187673 | Not-determined |
| 17-109_k141_388168 | Not-determined |
| 17-109_k141_237461 | Not-determined |
| 17-109_k141_237467 | Not-determined |
| 17-109_k141_237470 | Not-determined |
| 17-109_k141_103335 | Not-determined |
| 17-109_k141_287867 | Low-quality |
| 17-109_k141_253841 | Not-determined |
| 17-109_k141_304823 | Not-determined |
| 17-109_k141_120088 | Not-determined |
| 17-109_k141_253844 | Not-determined |
| 17-109_k141_287872 | Not-determined |
| 17-109_k141_120097 | Low-quality |
| 17-109_k141_103342 | Not-determined |
| 17-109_k141_270844 | Not-determined |
| 17-109_k141_36958 | Not-determined |
| 17-109_k141_371686 | Low-quality |
| 17-109_k141_187690 | Not-determined |
| 17-109_k141_187692 | Not-determined |
| 17-109_k141_120099 | Low-quality |
| 17-109_k141_287883 | Not-determined |
| 17-109_k141_354476 | Low-quality |
| 17-109_k141_371692 | Not-determined |
| 17-109_k141_237485 | Low-quality |
| 17-109_k141_287888 | Not-determined |
| 17-109_k141_187695 | Not-determined |
| 17-109_k141_270851 | Not-determined |
| 17-109_k141_321194 | Not-determined |
| 17-109_k141_120105 | Not-determined |
| 17-109_k141_187699 | Not-determined |
| 17-109_k141_19595 | Not-determined |
| 17-109_k141_187702 | Not-determined |
| 17-109_k141_19597 | Not-determined |
| 17-109_k141_237492 | Low-quality |
| 17-109_k141_36967 | Not-determined |
| 17-109_k141_237500 | Not-determined |
| 17-109_k141_19605 | Not-determined |
| 17-109_k141_19608 | Not-determined |
| 17-109_k141_304853 | Not-determined |
| 17-109_k141_19613 | Not-determined |
| 17-109_k141_103357 | Not-determined |
| 17-109_k141_253859 | Not-determined |
| 17-109_k141_321216 | Not-determined |
| 17-109_k141_220654 | Not-determined |
| 17-109_k141_354510 | Not-determined |
| 17-109_k141_304863 | Not-determined |
| 17-109_k141_170670 | Not-determined |
| 17-109_k141_287914 | Not-determined |
| 17-109_k141_220661 | Not-determined |
| 17-109_k141_371728 | Not-determined |
| 17-109_k141_253873 | Not-determined |
| 17-109_k141_70551 | Not-determined |
| 17-109_k141_153892 | Not-determined |
| 17-109_k141_371733 | Not-determined |
| 17-109_k141_304874 | Not-determined |
| 17-109_k141_321225 | Not-determined |
| 17-109_k141_287924 | Not-determined |
| 17-109_k141_103376 | Not-determined |
| 17-109_k141_70563 | Not-determined |
| 17-109_k141_287929 | Not-determined |
| 17-109_k141_371744 | Not-determined |
| 17-109_k141_253885 | Not-determined |
| 17-109_k141_103382 | Not-determined |
| 17-109_k141_204037 | Low-quality |
| 17-109_k141_103386 | Not-determined |
| 17-109_k141_220690 | Not-determined |
| 17-109_k141_37014 | Not-determined |
| 17-109_k141_270887 | Not-determined |
| 17-109_k141_170691 | Not-determined |
| 17-109_k141_19638 | Not-determined |
| 17-109_k141_103393 | Not-determined |
| 17-109_k141_204055 | Not-determined |
| 17-109_k141_388198 | Not-determined |
| 17-109_k141_19644 | Not-determined |
| 17-109_k141_304882 | Not-determined |
| 17-109_k141_187751 | Not-determined |
| 17-109_k141_3386 | Not-determined |
| 17-109_k141_321245 | Low-quality |
| 17-109_k141_253894 | Not-determined |
| 17-109_k141_388200 | Not-determined |
| 17-109_k141_170699 | Low-quality |
| 17-109_k141_304889 | Not-determined |
| 17-109_k141_253903 | Not-determined |
| 17-109_k141_270896 | Not-determined |
| 17-109_k141_304895 | Low-quality |
| 17-109_k141_321247 | Not-determined |
| 17-109_k141_388209 | Not-determined |
| 17-109_k141_86922 | Not-determined |
| 17-109_k141_204096 | Not-determined |
| 17-109_k141_270903 | Not-determined |
| 17-109_k141_253920 | Not-determined |
| 17-109_k141_3399 | Not-determined |
| 17-109_k141_19668 | Not-determined |
| 17-109_k141_287951 | Low-quality |
| 17-109_k141_103422 | Not-determined |
| 17-109_k141_19673 | Not-determined |
| 17-109_k141_103428 | Not-determined |
| 17-109_k141_287957 | Not-determined |
| 17-109_k141_287958 | Not-determined |
| 17-109_k141_103436 | Not-determined |
| 17-109_k141_204109 | Not-determined |
| 17-109_k141_388242 | Not-determined |
| 17-109_k141_304908 | Low-quality |
| 17-109_k141_337479 | Not-determined |
| 17-109_k141_253931 | Low-quality |
| 17-109_k141_19691 | Not-determined |
| 17-109_k141_170721 | Low-quality |
| 17-109_k141_187783 | Not-determined |
| 17-109_k141_287977 | Not-determined |
| 17-109_k141_204119 | Not-determined |
| 17-109_k141_187788 | Low-quality |
| 17-109_k141_304931 | Not-determined |
| 17-109_k141_321264 | Not-determined |
| 17-109_k141_103448 | Not-determined |
| 17-109_k141_103449 | Not-determined |
| 17-109_k141_103454 | Not-determined |
| 17-109_k141_19705 | Not-determined |
| 17-109_k141_70575 | Not-determined |
| 17-109_k141_337499 | Not-determined |
| 17-109_k141_204130 | Not-determined |
| 17-109_k141_371798 | Low-quality |
| 17-109_k141_19708 | Not-determined |
| 17-109_k141_371799 | Not-determined |
| 17-109_k141_388251 | Not-determined |
| 17-109_k141_388254 | Not-determined |
| 17-109_k141_103460 | Not-determined |
| 17-109_k141_70588 | Not-determined |
| 17-109_k141_287994 | Not-determined |
| 17-109_k141_170744 | Not-determined |
| 17-109_k141_204144 | Not-determined |
| 17-109_k141_187815 | Not-determined |
| 17-109_k141_170749 | Not-determined |
| 17-109_k141_153936 | Not-determined |
| 17-109_k141_103475 | Not-determined |
| 17-109_k141_237532 | Not-determined |
| 17-109_k141_153938 | Not-determined |
| 17-109_k141_170751 | Not-determined |
| 17-109_k141_103482 | Not-determined |
| 17-109_k141_70612 | Not-determined |
| 17-109_k141_19723 | Low-quality |
| 17-109_k141_153941 | Not-determined |
| 17-109_k141_371808 | Low-quality |
| 17-109_k141_170753 | Not-determined |
| 17-109_k141_3428 | Low-quality |
| 17-109_k141_153947 | Not-determined |
| 17-109_k141_19725 | Not-determined |
| 17-109_k141_337519 | Not-determined |
| 17-109_k141_170756 | Not-determined |
| 17-109_k141_288016 | Not-determined |
| 17-109_k141_153955 | Not-determined |
| 17-109_k141_137257 | Not-determined |
| 17-109_k141_204164 | Not-determined |
| 17-109_k141_371812 | Low-quality |
| 17-109_k141_237543 | Not-determined |
| 17-109_k141_187837 | Not-determined |
| 17-109_k141_237545 | Not-determined |
| 17-109_k141_237550 | Not-determined |
| 17-109_k141_388273 | Not-determined |
| 17-109_k141_204182 | Not-determined |
| 17-109_k141_321272 | Not-determined |
| 17-109_k141_103488 | Low-quality |
| 17-109_k141_288033 | Low-quality |
| 17-109_k141_204189 | Not-determined |
| 17-109_k141_204190 | Not-determined |
| 17-109_k141_354549 | Not-determined |
| 17-109_k141_70636 | Low-quality |
| 17-109_k141_3452 | Not-determined |
| 17-109_k141_321291 | Not-determined |
| 17-109_k141_321292 | Not-determined |
| 17-109_k141_170781 | Not-determined |
| 17-109_k141_187868 | Low-quality |
| 17-109_k141_237571 | Not-determined |
| 17-109_k141_237578 | Not-determined |
| 17-109_k141_337540 | Not-determined |
| 17-109_k141_19761 | Not-determined |
| 17-109_k141_70645 | Not-determined |
| 17-109_k141_3467 | Not-determined |
| 17-109_k141_204211 | Not-determined |
| 17-109_k141_337547 | Not-determined |
| 17-109_k141_388308 | Not-determined |
| 17-109_k141_70655 | Not-determined |
| 17-109_k141_19770 | Not-determined |
| 17-109_k141_253958 | Not-determined |
| 17-109_k141_187884 | Not-determined |
| 17-109_k141_354565 | Not-determined |
| 17-109_k141_3481 | Not-determined |
| 17-109_k141_137311 | Not-determined |
| 17-109_k141_321336 | Not-determined |
| 17-109_k141_337562 | Not-determined |
| 17-109_k141_253963 | Not-determined |
| 17-109_k141_388313 | Not-determined |
| 17-109_k141_37037 | Not-determined |
| 17-109_k141_354572 | Not-determined |
| 17-109_k141_388314 | Not-determined |
| 17-109_k141_19783 | Not-determined |
| 17-109_k141_288078 | Not-determined |
| 17-109_k141_253971 | Not-determined |
| 17-109_k141_321352 | Not-determined |
| 17-109_k141_103492 | Not-determined |
| 17-109_k141_70695 | Not-determined |
| 17-109_k141_19798 | Not-determined |
| 17-109_k141_253983 | Not-determined |
| 17-109_k141_237616 | Not-determined |
| 17-109_k141_153992 | Not-determined |
| 17-109_k141_86955 | Not-determined |
| 17-109_k141_253985 | Not-determined |
| 17-109_k141_321355 | Not-determined |
| 17-109_k141_204255 | Not-determined |
| 17-109_k141_70699 | Not-determined |
| 17-109_k141_187906 | Not-determined |
| 17-109_k141_3494 | Not-determined |
| 17-109_k141_253989 | Not-determined |
| 17-109_k141_354610 | Not-determined |
| 17-109_k141_86960 | Not-determined |
| 17-109_k141_270922 | Not-determined |
| 17-109_k141_86962 | Not-determined |
| 17-109_k141_154004 | Not-determined |
| 17-109_k141_220707 | Not-determined |
| 17-109_k141_304939 | Not-determined |
| 17-109_k141_270930 | Not-determined |

| | |
|---|---|
| 17-109_k141_354618 | Not-determined |
| 17-109_k141_70710 | Not-determined |
| 17-109_k141_70711 | Not-determined |
| 17-109_k141_388354 | Not-determined |
| 17-109_k141_86972 | Not-determined |
| 17-109_k141_204264 | Low-quality |
| 17-109_k141_270939 | Not-determined |
| 17-109_k141_237649 | Not-determined |
| 17-109_k141_86976 | Not-determined |
| 17-109_k141_237653 | Not-determined |
| 17-109_k141_70717 | Not-determined |
| 17-109_k141_288097 | Not-determined |
| 17-109_k141_354624 | Low-quality |
| 17-109_k141_237661 | Not-determined |
| 17-109_k141_137339 | Not-determined |
| 17-109_k141_170823 | Not-determined |
| 17-109_k141_154022 | Not-determined |
| 17-109_k141_170826 | Not-determined |
| 17-109_k141_270953 | Not-determined |
| 17-109_k141_304959 | Not-determined |
| 17-109_k141_270954 | Low-quality |
| 17-109_k141_220716 | Not-determined |
| 17-109_k141_304961 | Not-determined |
| 17-109_k141_170830 | Not-determined |
| 17-109_k141_19821 | Not-determined |
| 17-109_k141_187928 | Not-determined |
| 17-109_k141_86986 | Not-determined |
| 17-109_k141_304968 | Not-determined |
| 17-109_k141_86988 | Not-determined |
| 17-109_k141_103523 | Not-determined |
| 17-109_k141_86990 | Not-determined |
| 17-109_k141_37071 | Not-determined |
| 17-109_k141_321388 | Not-determined |
| 17-109_k141_37072 | Not-determined |
| 17-109_k141_103525 | Not-determined |
| 17-109_k141_321400 | Not-determined |
| 17-109_k141_388382 | Low-quality |
| 17-109_k141_388384 | Not-determined |
| 17-109_k141_288124 | Low-quality |
| 17-109_k141_337595 | Not-determined |
| 17-109_k141_304975 | Low-quality |
| 17-109_k141_187943 | Not-determined |
| 17-109_k141_321405 | Not-determined |
| 17-109_k141_70738 | Not-determined |
| 17-109_k141_304977 | Not-determined |
| 17-109_k141_154030 | Not-determined |
| 17-109_k141_304979 | Low-quality |
| 17-109_k141_70740 | Not-determined |
| 17-109_k141_321411 | Not-determined |
| 17-109_k141_337601 | Not-determined |
| 17-109_k141_170849 | Not-determined |
| 17-109_k141_19835 | Not-determined |
| 17-109_k141_37082 | Not-determined |
| 17-109_k141_337603 | Not-determined |
| 17-109_k141_388388 | Not-determined |
| 17-109_k141_204300 | Not-determined |
| 17-109_k141_86998 | Low-quality |
| 17-109_k141_204306 | Not-determined |
| 17-109_k141_388395 | Not-determined |
| 17-109_k141_321417 | Not-determined |
| 17-109_k141_187962 | Not-determined |
| 17-109_k141_304997 | Not-determined |
| 17-109_k141_187973 | Not-determined |
| 17-109_k141_187974 | Not-determined |
| 17-109_k141_170885 | Not-determined |
| 17-109_k141_170887 | Not-determined |
| 17-109_k141_187990 | Not-determined |
| 17-109_k141_187996 | Not-determined |
| 17-109_k141_170894 | Not-determined |
| 17-109_k141_371834 | Low-quality |
| 17-109_k141_305013 | Not-determined |
| 17-109_k141_19872 | Not-determined |
| 17-109_k141_154044 | Not-determined |
| 17-109_k141_188009 | Not-determined |
| 17-109_k141_220752 | Not-determined |
| 17-109_k141_254032 | Not-determined |
| 17-109_k141_19881 | Not-determined |
| 17-109_k141_103588 | Not-determined |
| 17-109_k141_154058 | Low-quality |
| 17-109_k141_270979 | Not-determined |
| 17-109_k141_3531 | Not-determined |
| 17-109_k141_3535 | Not-determined |
| 17-109_k141_137351 | Not-determined |
| 17-109_k141_137352 | Not-determined |
| 17-109_k141_337627 | Not-determined |
| 17-109_k141_53541 | Not-determined |
| 17-109_k141_103600 | Not-determined |
| 17-109_k141_337637 | Not-determined |
| 17-109_k141_188026 | Not-determined |
| 17-109_k141_371839 | Not-determined |
| 17-109_k141_154070 | Not-determined |
| 17-109_k141_170902 | Not-determined |
| 17-109_k141_3548 | Not-determined |
| 17-109_k141_305046 | Not-determined |
| 17-109_k141_305047 | Not-determined |
| 17-109_k141_3552 | Low-quality |
| 17-109_k141_37095 | Not-determined |
| 17-109_k141_103612 | Low-quality |
| 17-109_k141_19884 | Not-determined |
| 17-109_k141_53547 | Not-determined |
| 17-109_k141_53548 | Not-determined |
| 17-109_k141_305057 | Not-determined |
| 17-109_k141_37103 | Not-determined |
| 17-109_k141_288188 | Not-determined |
| 17-109_k141_103616 | Not-determined |
| 17-109_k141_103620 | Not-determined |
| 17-109_k141_254068 | Not-determined |
| 17-109_k141_53549 | Low-quality |
| 17-109_k141_305066 | Not-determined |
| 17-109_k141_170910 | Not-determined |
| 17-109_k141_288200 | Low-quality |
| 17-109_k141_170916 | Not-determined |
| 17-109_k141_37111 | Not-determined |
| 17-109_k141_170919 | Not-determined |
| 17-109_k141_137389 | Low-quality |
| 17-109_k141_170920 | Not-determined |
| 17-109_k141_371852 | Not-determined |
| 17-109_k141_254076 | Not-determined |
| 17-109_k141_305073 | Not-determined |
| 17-109_k141_137391 | Not-determined |
| 17-109_k141_37115 | Not-determined |
| 17-109_k141_270991 | Not-determined |
| 17-109_k141_19897 | Not-determined |
| 17-109_k141_53556 | Not-determined |
| 17-109_k141_70762 | Not-determined |
| 17-109_k141_19903 | Not-determined |
| 17-109_k141_53559 | Not-determined |
| 17-109_k141_170933 | Not-determined |
| 17-109_k141_305085 | Not-determined |
| 17-109_k141_19906 | Not-determined |
| 17-109_k141_37126 | Not-determined |
| 17-109_k141_53561 | Not-determined |
| 17-109_k141_19908 | Not-determined |
| 17-109_k141_37127 | Not-determined |
| 17-109_k141_19909 | Not-determined |
| 17-109_k141_53563 | Not-determined |
| 17-109_k141_19911 | Not-determined |
| 17-109_k141_321444 | Low-quality |
| 17-109_k141_37131 | Not-determined |
| 17-109_k141_37133 | Not-determined |
| 17-109_k141_137394 | Not-determined |
| 17-109_k141_37138 | Not-determined |
| 17-109_k141_37141 | Not-determined |
| 17-109_k141_288209 | Not-determined |
| 17-109_k141_53572 | Not-determined |
| 17-109_k141_53574 | Not-determined |
| 17-109_k141_137396 | Not-determined |
| 17-109_k141_305093 | Low-quality |
| 17-109_k141_220769 | Low-quality |
| 17-109_k141_70778 | Not-determined |
| 17-109_k141_154103 | Not-determined |
| 17-109_k141_337677 | Not-determined |
| 17-109_k141_371872 | Not-determined |
| 17-109_k141_371877 | Not-determined |
| 17-109_k141_37151 | Not-determined |
| 17-109_k141_220781 | Not-determined |
| 17-109_k141_3594 | Not-determined |
| 17-109_k141_37154 | Not-determined |
| 17-109_k141_37155 | Not-determined |
| 17-109_k141_220784 | Not-determined |
| 17-109_k141_103634 | Not-determined |
| 17-109_k141_3599 | Not-determined |
| 17-109_k141_103636 | Not-determined |
| 17-109_k141_204309 | Not-determined |
| 17-109_k141_154122 | Not-determined |
| 17-109_k141_103644 | Not-determined |
| 17-109_k141_170962 | Not-determined |
| 17-109_k141_37172 | Not-determined |
| 17-109_k141_103652 | Not-determined |
| 17-109_k141_288231 | Not-determined |
| 17-109_k141_254101 | Not-determined |
| 17-109_k141_254102 | Not-determined |
| 17-109_k141_103661 | Not-determined |
| 17-109_k141_154133 | Not-determined |
| 17-109_k141_237722 | Not-determined |
| 17-109_k141_288235 | Not-determined |
| 17-109_k141_237723 | Not-determined |
| 17-109_k141_70796 | Not-determined |
| 17-109_k141_305136 | Not-determined |
| 17-109_k141_19937 | Not-determined |
| 17-109_k141_154137 | Not-determined |
| 17-109_k141_220798 | Not-determined |
| 17-109_k141_103674 | Not-determined |
| 17-109_k141_3613 | Not-determined |
| 17-109_k141_37190 | Not-determined |
| 17-109_k141_37194 | Not-determined |
| 17-109_k141_337710 | Not-determined |
| 17-109_k141_388449 | Low-quality |
| 17-109_k141_70814 | Not-determined |
| 17-109_k141_154147 | Not-determined |
| 17-109_k141_220817 | Not-determined |
| 17-109_k141_70820 | Not-determined |
| 17-109_k141_53588 | Not-determined |
| 17-109_k141_288257 | Not-determined |
| 17-109_k141_154151 | Not-determined |
| 17-109_k141_170971 | Not-determined |
| 17-109_k141_337717 | Not-determined |
| 17-109_k141_19962 | Not-determined |
| 17-109_k141_337720 | Not-determined |
| 17-109_k141_53593 | Not-determined |
| 17-109_k141_337721 | Not-determined |
| 17-109_k141_337724 | Not-determined |
| 17-109_k141_103691 | Not-determined |
| 17-109_k141_321484 | Low-quality |
| 17-109_k141_103694 | Not-determined |
| 17-109_k141_337726 | Low-quality |
| 17-109_k141_337727 | Not-determined |
| 17-109_k141_70833 | Not-determined |
| 17-109_k141_19967 | Low-quality |
| 17-109_k141_154158 | Not-determined |
| 17-109_k141_19969 | Not-determined |
| 17-109_k141_321493 | Not-determined |
| 17-109_k141_3638 | Not-determined |
| 17-109_k141_103708 | Not-determined |
| 17-109_k141_321496 | Low-quality |
| 17-109_k141_87054 | Not-determined |
| 17-109_k141_288275 | Not-determined |
| 17-109_k141_237751 | Not-determined |
| 17-109_k141_220831 | Not-determined |
| 17-109_k141_305170 | Not-determined |
| 17-109_k141_220832 | Not-determined |
| 17-109_k141_288281 | Not-determined |
| 17-109_k141_271034 | Not-determined |
| 17-109_k141_271035 | Not-determined |
| 17-109_k141_371898 | Not-determined |
| 17-109_k141_137436 | Not-determined |
| 17-109_k141_3653 | Not-determined |
| 17-109_k141_137437 | Not-determined |
| 17-109_k141_87062 | Not-determined |
| 17-109_k141_19997 | Not-determined |
| 17-109_k141_237753 | Not-determined |
| 17-109_k141_137441 | Not-determined |
| 17-109_k141_87082 | Not-determined |
| 17-109_k141_321511 | Not-determined |
| 17-109_k141_87084 | Not-determined |
| 17-109_k141_37222 | Not-determined |
| 17-109_k141_154169 | Not-determined |
| 17-109_k141_288295 | Not-determined |
| 17-109_k141_137445 | Not-determined |
| 17-109_k141_288299 | Not-determined |
| 17-109_k141_103714 | Not-determined |
| 17-109_k141_388491 | Not-determined |
| 17-109_k141_87096 | Not-determined |
| 17-109_k141_204353 | Not-determined |
| 17-109_k141_70854 | Not-determined |
| 17-109_k141_337760 | Not-determined |
| 17-109_k141_271068 | Not-determined |
| 17-109_k141_3682 | Not-determined |
| 17-109_k141_321528 | Not-determined |
| 17-109_k141_388505 | Not-determined |
| 17-109_k141_3687 | Not-determined |
| 17-109_k141_120162 | Not-determined |
| 17-109_k141_321536 | Not-determined |
| 17-109_k141_70868 | Not-determined |
| 17-109_k141_271078 | Not-determined |
| 17-109_k141_87112 | Not-determined |
| 17-109_k141_37249 | Low-quality |
| 17-109_k141_70873 | Not-determined |
| 17-109_k141_371941 | Not-determined |
| 17-109_k141_204359 | Not-determined |
| 17-109_k141_120170 | Not-determined |
| 17-109_k141_337773 | Not-determined |
| 17-109_k141_321553 | Not-determined |
| 17-109_k141_204369 | Not-determined |
| 17-109_k141_321567 | Low-quality |
| 17-109_k141_288324 | Low-quality |
| 17-109_k141_204379 | Not-determined |
| 17-109_k141_321572 | Not-determined |
| 17-109_k141_20039 | Not-determined |
| 17-109_k141_87142 | Not-determined |
| 17-109_k141_337809 | Low-quality |
| 17-109_k141_271098 | Not-determined |
| 17-109_k141_321581 | Not-determined |
| 17-109_k141_271102 | Not-determined |
| 17-109_k141_154205 | Not-determined |
| 17-109_k141_3703 | Not-determined |
| 17-109_k141_321588 | Not-determined |
| 17-109_k141_354696 | Not-determined |
| 17-109_k141_337811 | Not-determined |
| 17-109_k141_20050 | Not-determined |
| 17-109_k141_337816 | Not-determined |
| 17-109_k141_220858 | Not-determined |
| 17-109_k141_20056 | Not-determined |
| 17-109_k141_321596 | Low-quality |
| 17-109_k141_220864 | Not-determined |
| 17-109_k141_20061 | Not-determined |
| 17-109_k141_87150 | Not-determined |
| 17-109_k141_137468 | Not-determined |
| 17-109_k141_103755 | Low-quality |
| 17-109_k141_237777 | Not-determined |
| 17-109_k141_337818 | Not-determined |
| 17-109_k141_103765 | Not-determined |
| 17-109_k141_103768 | Not-determined |
| 17-109_k141_371967 | Not-determined |
| 17-109_k141_371968 | Not-determined |
| 17-109_k141_305218 | Not-determined |
| 17-109_k141_337828 | Low-quality |
| 17-109_k141_305221 | Not-determined |
| 17-109_k141_354721 | Low-quality |
| 17-109_k141_305222 | Not-determined |
| 17-109_k141_237789 | Low-quality |
| 17-109_k141_70875 | Not-determined |
| 17-109_k141_120223 | Not-determined |
| 17-109_k141_53616 | Not-determined |
| 17-109_k141_53617 | Low-quality |
| 17-109_k141_337832 | Not-determined |
| 17-109_k141_53618 | Not-determined |
| 17-109_k141_120226 | Not-determined |
| 17-109_k141_354722 | Low-quality |
| 17-109_k141_70878 | Not-determined |
| 17-109_k141_137489 | Not-determined |
| 17-109_k141_321614 | Not-determined |
| 17-109_k141_70885 | Not-determined |
| 17-109_k141_137499 | Not-determined |
| 17-109_k141_20089 | Not-determined |
| 17-109_k141_321623 | Low-quality |
| 17-109_k141_70895 | Not-determined |
| 17-109_k141_321625 | Low-quality |
| 17-109_k141_371999 | Not-determined |
| 17-109_k141_204417 | Not-determined |
| 17-109_k141_321628 | Not-determined |
| 17-109_k141_321631 | Low-quality |
| 17-109_k141_321633 | Not-determined |
| 17-109_k141_354730 | Low-quality |
| 17-109_k141_220893 | Not-determined |
| 17-109_k141_288346 | Not-determined |
| 17-109_k141_103772 | Not-determined |
| 17-109_k141_53640 | Not-determined |
| 17-109_k141_188095 | Not-determined |
| 17-109_k141_305254 | Not-determined |
| 17-109_k141_20099 | Not-determined |
| 17-109_k141_237809 | Not-determined |
| 17-109_k141_87169 | Not-determined |
| 17-109_k141_254168 | Not-determined |
| 17-109_k141_288357 | Not-determined |
| 17-109_k141_288358 | Not-determined |
| 17-109_k141_20107 | Not-determined |
| 17-109_k141_372014 | Not-determined |
| 17-109_k141_254174 | Not-determined |
| 17-109_k141_254175 | Not-determined |
| 17-109_k141_204434 | Not-determined |
| 17-109_k141_337838 | Not-determined |
| 17-109_k141_288362 | Not-determined |
| 17-109_k141_70912 | Not-determined |
| 17-109_k141_204437 | Not-determined |
| 17-109_k141_70915 | Not-determined |
| 17-109_k141_288366 | Not-determined |
| 17-109_k141_372023 | Not-determined |
| 17-109_k141_337844 | Not-determined |
| 17-109_k141_337845 | Not-determined |
| 17-109_k141_137512 | Low-quality |
| 17-109_k141_271144 | Not-determined |
| 17-109_k141_254190 | Not-determined |
| 17-109_k141_87173 | Not-determined |
| 17-109_k141_271148 | Not-determined |
| 17-109_k141_254194 | Not-determined |
| 17-109_k141_20121 | Not-determined |
| 17-109_k141_220907 | Not-determined |
| 17-109_k141_354745 | Not-determined |
| 17-109_k141_3746 | Not-determined |
| 17-109_k141_87177 | Low-quality |
| 17-109_k141_103784 | Not-determined |
| 17-109_k141_137521 | Low-quality |
| 17-109_k141_372034 | Not-determined |
| 17-109_k141_204451 | Not-determined |
| 17-109_k141_137528 | Not-determined |
| 17-109_k141_220917 | Not-determined |
| 17-109_k141_70929 | Not-determined |
| 17-109_k141_288389 | Not-determined |
| 17-109_k141_288393 | Low-quality |
| 17-109_k141_137532 | Not-determined |
| 17-109_k141_288401 | Low-quality |
| 17-109_k141_354771 | Not-determined |
| 17-109_k141_103809 | Not-determined |
| 17-109_k141_354777 | Not-determined |
| 17-109_k141_103813 | Low-quality |
| 17-109_k141_37278 | Not-determined |
| 17-109_k141_372044 | Not-determined |
| 17-109_k141_305276 | Not-determined |
| 17-109_k141_3764 | Not-determined |
| 17-109_k141_271166 | Low-quality |
| 17-109_k141_3766 | Not-determined |
| 17-109_k141_388585 | Not-determined |
| 17-109_k141_3767 | Not-determined |
| 17-109_k141_372048 | Not-determined |
| 17-109_k141_337889 | Not-determined |
| 17-109_k141_354784 | Low-quality |
| 17-109_k141_37281 | Not-determined |
| 17-109_k141_137549 | Low-quality |
| 17-109_k141_337898 | Not-determined |
| 17-109_k141_188134 | Not-determined |
| 17-109_k141_305295 | Not-determined |
| 17-109_k141_337907 | Not-determined |
| 17-109_k141_372066 | Not-determined |
| 17-109_k141_271173 | Not-determined |
| 17-109_k141_137559 | Low-quality |
| 17-109_k141_388604 | Not-determined |
| 17-109_k141_271176 | Not-determined |
| 17-109_k141_37291 | Not-determined |
| 17-109_k141_388605 | Not-determined |
| 17-109_k141_53701 | Low-quality |
| 17-109_k141_337917 | Not-determined |
| 17-109_k141_20173 | Low-quality |
| 17-109_k141_254232 | Not-determined |
| 17-109_k141_103817 | Low-quality |
| 17-109_k141_271183 | Not-determined |
| 17-109_k141_372076 | Not-determined |
| 17-109_k141_271184 | Not-determined |
| 17-109_k141_372079 | Not-determined |
| 17-109_k141_388615 | Not-determined |
| 17-109_k141_372081 | Not-determined |
| 17-109_k141_372082 | Not-determined |
| 17-109_k141_271189 | Not-determined |
| 17-109_k141_288449 | Not-determined |
| 17-109_k141_20186 | Not-determined |
| 17-109_k141_337922 | Not-determined |
| 17-109_k141_254241 | Not-determined |
| 17-109_k141_103829 | Low-quality |
| 17-109_k141_53719 | Not-determined |
| 17-109_k141_337924 | Not-determined |
| 17-109_k141_321657 | Not-determined |
| 17-109_k141_337925 | Not-determined |
| 17-109_k141_237859 | Not-determined |
| 17-109_k141_188160 | Low-quality |
| 17-109_k141_37309 | Not-determined |
| 17-109_k141_372101 | Not-determined |
| 17-109_k141_288468 | Not-determined |
| 17-109_k141_37311 | Not-determined |
| 17-109_k141_271203 | Not-determined |
| 17-109_k141_37313 | Not-determined |
| 17-109_k141_3792 | Not-determined |
| 17-109_k141_237869 | Not-determined |
| 17-109_k141_254251 | Not-determined |
| 17-109_k141_237874 | Not-determined |
| 17-109_k141_3800 | Not-determined |
| 17-109_k141_254259 | Not-determined |
| 17-109_k141_103844 | Not-determined |
| 17-109_k141_3801 | Not-determined |
| 17-109_k141_103845 | Not-determined |
| 17-109_k141_288489 | Not-determined |
| 17-109_k141_3805 | Not-determined |
| 17-109_k141_70938 | Not-determined |
| 17-109_k141_354802 | Not-determined |
| 17-109_k141_70939 | Not-determined |
| 17-109_k141_337956 | Not-determined |
| 17-109_k141_154336 | Not-determined |
| 17-109_k141_137573 | Not-determined |
| 17-109_k141_354810 | Not-determined |
| 17-109_k141_137577 | Not-determined |
| 17-109_k141_254268 | Not-determined |
| 17-109_k141_53735 | Not-determined |
| 17-109_k141_254270 | Not-determined |
| 17-109_k141_372126 | Not-determined |
| 17-109_k141_337968 | Not-determined |
| 17-109_k141_372129 | Not-determined |
| 17-109_k141_337971 | Not-determined |
| 17-109_k141_288497 | Not-determined |
| 17-109_k141_137592 | Not-determined |
| 17-109_k141_170997 | Not-determined |
| 17-109_k141_170998 | Not-determined |
| 17-109_k141_188199 | Not-determined |
| 17-109_k141_37338 | Not-determined |
| 17-109_k141_120239 | Not-determined |
| 17-109_k141_188201 | Not-determined |
| 17-109_k141_20214 | Not-determined |
| 17-109_k141_70956 | Low-quality |
| 17-109_k141_120244 | Not-determined |
| 17-109_k141_354835 | Not-determined |
| 17-109_k141_237904 | Not-determined |
| 17-109_k141_354842 | Not-determined |
| 17-109_k141_354843 | Not-determined |
| 17-109_k141_388685 | Not-determined |
| 17-109_k141_388686 | Not-determined |
| 17-109_k141_87217 | Not-determined |
| 17-109_k141_337990 | Not-determined |
| 17-109_k141_305361 | Not-determined |
| 17-109_k141_171018 | Not-determined |
| 17-109_k141_288509 | Not-determined |
| 17-109_k141_254306 | Not-determined |
| 17-109_k141_103859 | Not-determined |
| 17-109_k141_338000 | Low-quality |
| 17-109_k141_254315 | Not-determined |
| 17-109_k141_171034 | Not-determined |
| 17-109_k141_20242 | Low-quality |
| 17-109_k141_271233 | Not-determined |
| 17-109_k141_305383 | Not-determined |
| 17-109_k141_321705 | Not-determined |
| 17-109_k141_171041 | Not-determined |
| 17-109_k141_372145 | Not-determined |
| 17-109_k141_20249 | Not-determined |
| 17-109_k141_103864 | Not-determined |
| 17-109_k141_171044 | Not-determined |
| 17-109_k141_87230 | Not-determined |
| 17-109_k141_87231 | Not-determined |
| 17-109_k141_120255 | Not-determined |
| 17-109_k141_171048 | Not-determined |
| 17-109_k141_220960 | Not-determined |
| 17-109_k141_237939 | Not-determined |
| 17-109_k141_204505 | Not-determined |
| 17-109_k141_171056 | Not-determined |
| 17-109_k141_171059 | Not-determined |
| 17-109_k141_271242 | Not-determined |
| 17-109_k141_137608 | Not-determined |
| 17-109_k141_120259 | Not-determined |
| 17-109_k141_237945 | Not-determined |
| 17-109_k141_271243 | Low-quality |
| 17-109_k141_388698 | Not-determined |
| 17-109_k141_271246 | Not-determined |
| 17-109_k141_137617 | Not-determined |
| 17-109_k141_271250 | Low-quality |
| 17-109_k141_237952 | Not-determined |
| 17-109_k141_137618 | Not-determined |
| 17-109_k141_220969 | Not-determined |
| 17-109_k141_87251 | Not-determined |
| 17-109_k141_288533 | Not-determined |
| 17-109_k141_70973 | Not-determined |
| 17-109_k141_154364 | Low-quality |
| 17-109_k141_20266 | Not-determined |
| 17-109_k141_305401 | Not-determined |
| 17-109_k141_271264 | Not-determined |
| 17-109_k141_137642 | Not-determined |
| 17-109_k141_354851 | Not-determined |
| 17-109_k141_305408 | Not-determined |
| 17-109_k141_220989 | Not-determined |
| 17-109_k141_103902 | Not-determined |
| 17-109_k141_154381 | Low-quality |
| 17-109_k141_120284 | Not-determined |
| 17-109_k141_70991 | Not-determined |
| 17-109_k141_305411 | Not-determined |
| 17-109_k141_137648 | Not-determined |
| 17-109_k141_220996 | Not-determined |
| 17-109_k141_271282 | Not-determined |
| 17-109_k141_71002 | Not-determined |
| 17-109_k141_71005 | Not-determined |
| 17-109_k141_221009 | Not-determined |
| 17-109_k141_120296 | Not-determined |
| 17-109_k141_305422 | Not-determined |
| 17-109_k141_71008 | Not-determined |
| 17-109_k141_120297 | Not-determined |
| 17-109_k141_388723 | Low-quality |
| 17-109_k141_71010 | Not-determined |
| 17-109_k141_53776 | Not-determined |
| 17-109_k141_338048 | Not-determined |
| 17-109_k141_388727 | Not-determined |
| 17-109_k141_237999 | Not-determined |
| 17-109_k141_53783 | Not-determined |
| 17-109_k141_221016 | Low-quality |
| 17-109_k141_3834 | Not-determined |
| 17-109_k141_321758 | Not-determined |
| 17-109_k141_3835 | Low-quality |
| 17-109_k141_238005 | Not-determined |
| 17-109_k141_254336 | Not-determined |
| 17-109_k141_120311 | Not-determined |
| 17-109_k141_87271 | Not-determined |
| 17-109_k141_338059 | Not-determined |
| 17-109_k141_321765 | Not-determined |
| 17-109_k141_188302 | Not-determined |
| 17-109_k141_3841 | Not-determined |
| 17-109_k141_188304 | Not-determined |
| 17-109_k141_71013 | Not-determined |
| 17-109_k141_271310 | Not-determined |
| 17-109_k141_338062 | Not-determined |
| 17-109_k141_254343 | Not-determined |
| 17-109_k141_87282 | Not-determined |
| 17-109_k141_354886 | Not-determined |
| 17-109_k141_53796 | Not-determined |
| 17-109_k141_120315 | Not-determined |
| 17-109_k141_3851 | Not-determined |
| 17-109_k141_3852 | Low-quality |
| 17-109_k141_372172 | Not-determined |
| 17-109_k141_120317 | Not-determined |
| 17-109_k141_120318 | Not-determined |
| 17-109_k141_338073 | Not-determined |
| 17-109_k141_305434 | Not-determined |
| 17-109_k141_120323 | Not-determined |
| 17-109_k141_321784 | Not-determined |
| 17-109_k141_137680 | Not-determined |
| 17-109_k141_271316 | Not-determined |
| 17-109_k141_372181 | Not-determined |
| 17-109_k141_120325 | Not-determined |
| 17-109_k141_372183 | Low-quality |
| 17-109_k141_305444 | Not-determined |
| 17-109_k141_238025 | Not-determined |
| 17-109_k141_305447 | Not-determined |
| 17-109_k141_53819 | Not-determined |
| 17-109_k141_120334 | Not-determined |
| 17-109_k141_20311 | Not-determined |
| 17-109_k141_120337 | Not-determined |
| 17-109_k141_388762 | Not-determined |

| | |
|---|---|
| 17-109_k141_188341 | Not-determined |
| 17-109_k141_20320 | Not-determined |
| 17-109_k141_271333 | Not-determined |
| 17-109_k141_154407 | Low-quality |
| 17-109_k141_53839 | Not-determined |
| 17-109_k141_20327 | Not-determined |
| 17-109_k141_271341 | Not-determined |
| 17-109_k141_338087 | Low-quality |
| 17-109_k141_3864 | Not-determined |
| 17-109_k141_271346 | Low-quality |
| 17-109_k141_271347 | Not-determined |
| 17-109_k141_20337 | Not-determined |
| 17-109_k141_103943 | Not-determined |
| 17-109_k141_103944 | Not-determined |
| 17-109_k141_20341 | Not-determined |
| 17-109_k141_71035 | Not-determined |
| 17-109_k141_338096 | Not-determined |
| 17-109_k141_238047 | Not-determined |
| 17-109_k141_120357 | Not-determined |
| 17-109_k141_71041 | Not-determined |
| 17-109_k141_120359 | Not-determined |
| 17-109_k141_71044 | Low-quality |
| 17-109_k141_171084 | Not-determined |
| 17-109_k141_372205 | Not-determined |
| 17-109_k141_171087 | Not-determined |
| 17-109_k141_238053 | Not-determined |
| 17-109_k141_372209 | Not-determined |
| 17-109_k141_238054 | Low-quality |
| 17-109_k141_388790 | Not-determined |
| 17-109_k141_271370 | Not-determined |
| 17-109_k141_388795 | Not-determined |
| 17-109_k141_87319 | Not-determined |
| 17-109_k141_137729 | Not-determined |
| 17-109_k141_137730 | Not-determined |
| 17-109_k141_137731 | Low-quality |
| 17-109_k141_120373 | Low-quality |
| 17-109_k141_20374 | Not-determined |
| 17-109_k141_354935 | Not-determined |
| 17-109_k141_271386 | Not-determined |
| 17-109_k141_271388 | Not-determined |
| 17-109_k141_37402 | Not-determined |
| 17-109_k141_53870 | Not-determined |
| 17-109_k141_372221 | Not-determined |
| 17-109_k141_53872 | Not-determined |
| 17-109_k141_305481 | Low-quality |
| [illegible] | [illegible] |

| | |
|---|---|
| 17-109_k141_254696 | Not-determined |
| 17-109_k141_188718 | Not-determined |
| 17-109_k141_71519 | Not-determined |
| 17-109_k141_338508 | Not-determined |
| 17-109_k141_221431 | Not-determined |
| 17-109_k141_71525 | Not-determined |
| 17-109_k141_188724 | Not-determined |
| 17-109_k141_254702 | Not-determined |
| 17-109_k141_389125 | Not-determined |
| 17-109_k141_288747 | Not-determined |
| 17-109_k141_389129 | Not-determined |
| 17-109_k141_138170 | Not-determined |
| 17-109_k141_138173 | Not-determined |
| 17-109_k141_71535 | Not-determined |
| 17-109_k141_389134 | Not-determined |
| 17-109_k141_71538 | Not-determined |
| 17-109_k141_305758 | Not-determined |
| 17-109_k141_188741 | Not-determined |
| 17-109_k141_355216 | Not-determined |
| 17-109_k141_305762 | Not-determined |
| 17-109_k141_37686 | Not-determined |
| 17-109_k141_20676 | Not-determined |
| 17-109_k141_254707 | Not-determined |
| 17-109_k141_238411 | Not-determined |
| 17-109_k141_238413 | Not-determined |
| 17-109_k141_338516 | Not-determined |
| 17-109_k141_238418 | Not-determined |
| 17-109_k141_4128 | Not-determined |
| 17-109_k141_254711 | Not-determined |
| 17-109_k141_4131 | Not-determined |
| 17-109_k141_37698 | Not-determined |
| 17-109_k141_389155 | Not-determined |
| 17-109_k141_338523 | Not-determined |
| 17-109_k141_305784 | Low-quality |
| 17-109_k141_54244 | Low-quality |
| 17-109_k141_37715 | Not-determined |
| 17-109_k141_71560 | Not-determined |
| 17-109_k141_120777 | Not-determined |
| 17-109_k141_4143 | Not-determined |
| 17-109_k141_4146 | Not-determined |
| 17-109_k141_221457 | Not-determined |
| 17-109_k141_305806 | Not-determined |
| 17-109_k141_221461 | Not-determined |
| 17-109_k141_288785 | Not-determined |
| 18-102-final-viral-combined_k141_72725\|\|full | Medium-quality |
| 17-109_k141_221463 | Low-quality |
| 17-109_k141_355263 | Not-determined |
| 17-109_k141_188747 | Not-determined |
| 17-109_k141_188750 | Not-determined |
| 17-109_k141_221469 | Low-quality |
| 17-109_k141_71567 | Not-determined |
| 17-109_k141_322267 | Not-determined |
| 17-109_k141_154709 | Not-determined |
| 17-109_k141_254740 | Not-determined |
| 17-109_k141_37732 | Not-determined |
| 17-109_k141_271846 | Not-determined |
| 17-109_k141_271847 | Not-determined |
| 17-109_k141_120796 | Not-determined |
| 17-109_k141_389164 | Not-determined |
| 17-109_k141_271856 | Low-quality |
| 17-109_k141_54260 | Not-determined |
| 17-109_k141_71577 | Not-determined |
| 17-109_k141_71579 | Not-determined |
| 17-109_k141_305833 | Not-determined |
| 17-109_k141_120822 | Not-determined |
| 17-109_k141_254753 | Not-determined |
| 17-109_k141_389184 | Not-determined |
| 17-109_k141_221474 | Not-determined |
| 17-109_k141_271877 | Not-determined |
| 17-109_k141_71598 | Not-determined |
| 17-109_k141_154748 | Not-determined |
| 17-109_k141_221484 | Not-determined |
| 17-109_k141_305846 | Not-determined |
| 17-109_k141_204881 | Low-quality |
| 17-109_k141_271882 | Low-quality |
| 17-109_k141_71609 | Not-determined |
| 17-109_k141_20726 | Not-determined |
| 17-109_k141_120844 | Low-quality |
| 17-109_k141_20727 | Not-determined |
| 17-109_k141_138250 | Not-determined |
| 17-109_k141_338566 | Low-quality |
| 17-109_k141_254771 | Not-determined |
| 17-109_k141_138256 | Not-determined |
| 17-109_k141_4166 | Not-determined |
| 17-109_k141_154773 | Low-quality |
| 17-109_k141_188783 | Not-determined |
| 17-109_k141_20745 | Not-determined |
| 17-109_k141_271901 | Low-quality |
| 17-109_k141_204889 | Not-determined |
| 17-109_k141_120849 | Not-determined |
| 17-109_k141_204892 | Not-determined |
| 17-109_k141_221505 | Not-determined |
| 17-109_k141_221506 | Not-determined |
| 17-109_k141_322293 | Not-determined |
| 17-109_k141_221512 | Not-determined |
| 17-109_k141_20751 | Not-determined |
| 17-109_k141_221513 | Not-determined |
| 17-109_k141_238449 | Low-quality |
| 17-109_k141_322300 | Low-quality |
| 17-109_k141_322301 | Not-determined |
| 17-109_k141_322303 | Not-determined |
| 17-109_k141_355285 | Not-determined |
| 17-109_k141_221518 | Not-determined |
| 17-109_k141_271905 | Low-quality |
| 17-109_k141_338589 | Not-determined |
| 17-109_k141_271909 | Low-quality |
| 17-109_k141_154794 | Low-quality |
| 17-109_k141_372682 | Not-determined |
| 17-109_k141_154797 | Not-determined |
| 17-109_k141_271911 | Not-determined |
| 17-109_k141_104329 | Not-determined |
| 17-109_k141_389219 | Not-determined |
| 17-109_k141_138278 | Not-determined |
| 17-109_k141_20763 | Not-determined |
| 17-109_k141_20767 | Not-determined |
| 17-109_k141_271916 | Not-determined |
| 17-109_k141_204901 | Not-determined |
| 17-109_k141_305873 | Not-determined |
| 17-109_k141_372692 | Not-determined |
| 17-109_k141_372693 | Not-determined |
| 17-109_k141_188795 | Low-quality |
| 17-109_k141_71660 | Not-determined |
| 17-109_k141_271924 | Not-determined |
| 17-109_k141_221545 | Not-determined |
| 17-109_k141_288845 | Not-determined |
| 17-109_k141_271926 | Not-determined |
| 17-109_k141_322318 | Not-determined |
| 17-109_k141_138285 | Not-determined |
| 17-109_k141_288851 | Not-determined |
| 17-109_k141_71667 | Not-determined |
| 17-109_k141_104347 | Not-determined |
| 17-109_k141_188811 | Not-determined |
| 17-109_k141_188812 | Low-quality |
| 17-109_k141_305879 | Not-determined |
| 17-109_k141_254789 | Not-determined |
| 17-109_k141_305881 | Not-determined |
| 17-109_k141_254791 | Not-determined |
| 17-109_k141_372703 | Not-determined |
| 17-109_k141_254794 | Not-determined |
| 17-109_k141_20788 | Not-determined |
| 17-109_k141_271939 | Not-determined |
| 17-109_k141_271940 | Not-determined |
| 17-109_k141_20790 | Not-determined |
| 17-109_k141_305887 | Not-determined |
| 17-109_k141_288874 | Not-determined |
| 17-109_k141_154817 | Not-determined |
| 17-109_k141_54268 | Not-determined |
| 17-109_k141_54270 | Not-determined |
| 17-109_k141_288882 | Not-determined |
| 17-109_k141_20798 | Low-quality |
| 17-109_k141_204910 | Not-determined |
| 17-109_k141_338636 | Not-determined |
| 17-109_k141_20806 | Not-determined |
| 17-109_k141_288892 | Not-determined |
| 17-109_k141_20808 | Not-determined |
| 17-109_k141_254810 | Not-determined |
| 17-109_k141_20812 | Not-determined |
| 17-109_k141_271947 | Low-quality |
| 17-109_k141_20821 | Not-determined |
| 17-109_k141_254815 | Not-determined |
| 17-109_k141_54282 | Low-quality |
| 17-109_k141_254818 | Not-determined |
| 17-109_k141_138295 | Not-determined |
| 17-109_k141_254819 | Not-determined |
| 17-109_k141_322353 | Not-determined |
| 17-109_k141_305906 | Not-determined |
| 17-109_k141_355324 | Not-determined |
| 17-109_k141_288903 | Not-determined |
| 17-109_k141_288904 | Not-determined |
| 17-109_k141_20824 | Not-determined |
| 17-109_k141_338655 | Not-determined |
| 17-109_k141_154842 | Not-determined |
| 17-109_k141_188855 | Not-determined |
| 17-109_k141_188856 | Low-quality |
| 17-109_k141_87674 | Not-determined |
| 17-109_k141_54291 | Low-quality |
| 17-109_k141_204924 | Low-quality |
| 17-109_k141_54294 | Not-determined |
| 17-109_k141_322361 | Not-determined |
| 17-109_k141_20839 | Not-determined |
| 17-109_k141_87678 | Not-determined |
| 17-109_k141_322366 | Not-determined |
| 17-109_k141_322367 | Not-determined |
| 17-109_k141_204930 | Not-determined |
| 17-109_k141_87679 | Not-determined |
| 17-109_k141_54300 | Not-determined |
| 17-109_k141_372718 | Not-determined |
| 17-109_k141_221603 | Not-determined |
| 17-109_k141_221606 | Not-determined |
| 17-109_k141_87684 | Not-determined |
| 17-109_k141_322371 | Not-determined |
| 17-109_k141_338676 | Low-quality |
| 17-109_k141_120915 | Not-determined |
| 17-109_k141_120918 | Not-determined |
| 17-109_k141_355356 | Not-determined |
| 17-109_k141_355357 | Not-determined |
| 17-109_k141_288921 | Low-quality |
| 17-109_k141_154866 | Not-determined |
| 17-109_k141_188874 | Low-quality |
| 17-109_k141_20855 | Not-determined |
| 17-109_k141_20856 | Not-determined |
| 17-109_k141_138320 | Not-determined |
| 17-109_k141_221621 | Not-determined |
| 17-109_k141_20861 | Low-quality |
| 17-109_k141_54311 | Not-determined |
| 17-109_k141_20862 | Not-determined |
| 17-109_k141_4178 | Not-determined |
| 17-109_k141_54314 | Not-determined |
| 17-109_k141_20865 | Not-determined |
| 17-109_k141_154882 | Not-determined |
| 17-109_k141_389236 | Not-determined |
| 17-109_k141_204954 | Not-determined |
| 17-109_k141_288947 | Not-determined |
| 17-109_k141_372744 | Not-determined |
| 17-109_k141_288952 | Not-determined |
| 17-109_k141_221642 | Not-determined |
| 17-109_k141_54327 | Not-determined |
| 17-109_k141_288959 | Not-determined |
| 17-109_k141_71693 | Not-determined |
| 17-109_k141_204970 | Not-determined |
| 17-109_k141_154899 | Not-determined |
| 17-109_k141_54332 | Not-determined |
| 17-109_k141_238488 | Not-determined |
| 17-109_k141_4192 | Low-quality |
| 17-109_k141_238493 | Not-determined |
| 17-109_k141_355361 | Not-determined |
| 17-109_k141_54341 | Not-determined |
| 17-109_k141_238497 | Low-quality |
| 17-109_k141_87728 | Low-quality |
| 17-109_k141_372760 | Low-quality |
| 17-109_k141_20886 | Not-determined |
| 17-109_k141_204999 | Low-quality |
| 17-109_k141_389265 | Not-determined |
| 17-109_k141_355372 | Not-determined |
| 17-109_k141_87735 | Not-determined |
| 17-109_k141_20893 | Not-determined |
| 17-109_k141_221668 | Not-determined |
| 17-109_k141_372770 | Not-determined |
| 17-109_k141_87739 | Not-determined |
| 17-109_k141_37862 | Not-determined |
| 17-109_k141_37866 | Not-determined |
| 17-109_k141_372778 | Not-determined |
| 17-109_k141_221670 | Low-quality |
| 17-109_k141_205017 | Not-determined |
| 17-109_k141_87745 | Not-determined |
| 17-109_k141_372783 | Not-determined |
| 17-109_k141_305970 | Not-determined |
| 17-109_k141_20898 | Not-determined |
| 17-109_k141_372784 | Not-determined |
| 17-109_k141_138346 | Not-determined |
| 17-109_k141_71730 | Not-determined |
| 17-109_k141_338715 | Not-determined |
| 17-109_k141_305974 | Not-determined |
| 17-109_k141_138355 | Not-determined |
| 17-109_k141_37874 | Not-determined |
| 17-109_k141_154945 | Not-determined |
| 17-109_k141_372801 | Not-determined |
| 17-109_k141_305984 | Not-determined |
| 17-109_k141_37879 | Not-determined |
| 17-109_k141_305987 | Not-determined |
| 17-109_k141_4221 | Low-quality |
| 17-109_k141_221683 | Not-determined |
| 17-109_k141_87764 | Not-determined |
| 17-109_k141_154951 | Not-determined |
| 17-109_k141_355390 | Not-determined |
| 17-109_k141_138375 | Not-determined |
| 17-109_k141_322421 | Not-determined |
| 17-109_k141_271988 | Low-quality |
| 17-109_k141_322424 | Not-determined |
| 17-109_k141_154960 | Not-determined |
| 17-109_k141_322431 | Not-determined |
| 17-109_k141_71748 | Not-determined |
| 17-109_k141_306009 | Not-determined |
| 17-109_k141_221693 | Not-determined |
| 17-109_k141_306014 | Low-quality |
| 17-109_k141_54348 | Not-determined |
| 17-109_k141_205033 | Not-determined |
| 17-109_k141_138386 | Not-determined |
| 17-109_k141_71759 | Not-determined |
| 17-109_k141_355414 | Not-determined |
| 17-109_k141_355415 | Not-determined |
| 17-109_k141_271995 | Not-determined |
| 17-109_k141_54354 | Not-determined |
| 17-109_k141_306031 | Not-determined |
| 17-109_k141_138395 | Not-determined |
| 17-109_k141_338736 | Not-determined |
| 17-109_k141_205049 | Not-determined |
| 17-109_k141_272006 | Not-determined |
| 17-109_k141_355421 | Not-determined |
| 17-109_k141_272007 | Not-determined |
| 17-109_k141_355425 | Not-determined |
| 17-109_k141_54364 | Low-quality |
| 17-109_k141_355430 | Not-determined |
| 17-109_k141_54365 | Not-determined |
| 17-109_k141_4255 | Low-quality |
| 17-109_k141_154970 | Not-determined |
| 17-109_k141_71776 | Not-determined |
| 17-109_k141_221716 | Not-determined |
| 17-109_k141_288966 | Not-determined |
| 17-109_k141_272011 | Not-determined |
| 17-109_k141_71782 | Not-determined |
| 17-109_k141_254864 | Not-determined |
| 17-109_k141_221722 | Not-determined |
| 17-109_k141_221723 | Not-determined |
| 17-109_k141_254867 | Low-quality |
| 17-109_k141_322469 | Not-determined |
| 17-109_k141_338759 | Not-determined |
| 17-109_k141_87770 | Not-determined |
| 17-109_k141_322471 | Not-determined |
| 17-109_k141_338761 | Not-determined |
| 17-109_k141_338763 | Not-determined |
| 17-109_k141_272019 | Not-determined |
| 17-109_k141_272020 | Not-determined |
| 17-109_k141_338765 | Not-determined |
| 17-109_k141_338766 | Not-determined |
| 17-109_k141_205053 | Low-quality |
| 17-109_k141_288976 | Not-determined |
| 17-109_k141_338771 | Not-determined |
| 17-109_k141_54394 | Not-determined |
| 17-109_k141_20959 | Not-determined |
| 17-109_k141_221742 | Low-quality |
| 17-109_k141_322487 | Not-determined |
| 17-109_k141_355443 | Not-determined |
| 17-109_k141_4284 | Not-determined |
| 17-109_k141_288978 | Not-determined |
| 17-109_k141_4287 | Not-determined |
| 17-109_k141_221748 | Not-determined |
| 17-109_k141_20977 | Not-determined |
| 17-109_k141_338781 | Not-determined |
| 17-109_k141_389336 | Not-determined |
| 17-109_k141_254873 | Not-determined |
| 17-109_k141_37911 | Not-determined |
| 17-109_k141_288983 | Not-determined |
| 17-109_k141_54398 | Not-determined |
| 17-109_k141_288984 | Low-quality |
| 17-109_k141_37913 | Low-quality |
| 17-109_k141_37916 | Not-determined |
| 17-109_k141_254879 | Not-determined |
| 17-109_k141_138424 | Not-determined |
| 17-109_k141_306070 | Low-quality |
| 17-109_k141_20982 | Not-determined |
| 17-109_k141_104400 | Low-quality |
| 17-109_k141_188912 | Not-determined |
| 17-109_k141_389351 | Not-determined |
| 17-109_k141_188914 | Not-determined |
| 17-109_k141_272060 | Not-determined |
| 17-109_k141_54410 | Not-determined |
| 17-109_k141_389357 | Not-determined |
| 17-109_k141_120982 | Not-determined |
| 17-109_k141_104409 | Not-determined |
| 17-109_k141_355451 | Not-determined |
| 17-109_k141_87786 | Not-determined |
| 17-109_k141_221773 | Not-determined |
| 17-109_k141_20994 | Not-determined |
| 17-109_k141_254888 | Not-determined |
| 17-109_k141_306084 | Not-determined |
| 17-109_k141_20996 | Not-determined |
| 17-109_k141_20997 | Not-determined |
| 17-109_k141_87799 | Not-determined |
| 17-109_k141_272068 | Not-determined |
| 17-109_k141_372851 | Not-determined |
| 17-109_k141_372854 | Not-determined |
| 17-109_k141_87806 | Not-determined |
| 17-109_k141_104419 | Low-quality |
| 17-109_k141_289003 | Low-quality |
| 17-109_k141_221794 | Not-determined |
| 17-109_k141_289006 | Not-determined |
| 17-109_k141_289007 | Not-determined |
| 17-109_k141_21023 | Not-determined |
| 17-109_k141_355469 | Not-determined |
| 17-109_k141_289012 | Not-determined |
| 17-109_k141_21026 | Not-determined |
| 17-109_k141_254927 | Not-determined |
| 17-109_k141_205058 | Not-determined |
| 17-109_k141_355476 | Not-determined |
| 17-109_k141_338812 | Not-determined |
| 17-109_k141_338814 | Not-determined |
| 17-109_k141_37935 | Not-determined |
| 17-109_k141_289025 | Not-determined |
| 17-109_k141_104432 | Not-determined |
| 17-109_k141_87822 | Not-determined |
| 17-109_k141_121015 | Not-determined |
| 17-109_k141_54422 | Not-determined |
| 17-109_k141_272087 | Not-determined |
| 17-109_k141_389379 | Not-determined |
| 17-109_k141_238555 | Not-determined |
| 17-109_k141_87829 | Not-determined |
| 17-109_k141_87830 | Not-determined |
| 17-109_k141_188934 | Low-quality |
| 17-109_k141_37950 | Not-determined |
| 17-109_k141_4299 | Not-determined |
| 17-109_k141_221821 | Not-determined |
| 17-109_k141_221822 | Not-determined |
| 17-109_k141_254954 | Not-determined |
| 17-109_k141_254958 | Not-determined |
| 17-109_k141_322532 | Not-determined |
| 17-109_k141_87845 | Not-determined |
| 17-109_k141_188954 | Not-determined |
| 17-109_k141_188956 | Not-determined |
| 17-109_k141_4306 | Low-quality |
| 17-109_k141_138453 | Not-determined |
| 17-109_k141_221834 | Not-determined |
| 17-109_k141_254962 | Not-determined |
| 17-109_k141_4313 | Not-determined |
| 17-109_k141_171531 | Not-determined |
| 17-109_k141_87855 | Not-determined |
| 17-109_k141_21057 | Not-determined |
| 17-109_k141_87856 | Not-determined |
| 17-109_k141_188970 | Not-determined |
| 17-109_k141_104459 | Not-determined |
| 17-109_k141_87865 | Not-determined |
| 17-109_k141_188974 | Not-determined |
| 17-109_k141_188977 | Not-determined |
| 17-109_k141_171541 | Not-determined |
| 17-109_k141_289060 | Not-determined |
| 17-109_k141_372902 | Not-determined |
| 17-109_k141_138470 | Not-determined |
| 17-109_k141_138471 | Low-quality |
| 17-109_k141_322549 | Not-determined |
| 17-109_k141_221848 | Not-determined |
| 17-109_k141_322550 | Not-determined |
| 17-109_k141_188987 | Not-determined |
| 17-109_k141_138476 | Not-determined |
| 17-109_k141_171550 | Not-determined |
| 17-109_k141_171552 | Not-determined |
| 17-109_k141_221856 | Not-determined |
| 17-109_k141_372908 | Not-determined |
| 17-109_k141_322554 | Not-determined |
| 17-109_k141_221860 | Not-determined |
| 17-109_k141_21073 | Low-quality |
| 17-109_k141_154979 | Low-quality |
| 17-109_k141_355510 | Not-determined |
| 17-109_k141_37964 | Not-determined |
| 17-109_k141_289074 | Not-determined |
| 17-109_k141_272096 | Not-determined |
| 17-109_k141_289080 | Not-determined |
| 17-109_k141_171556 | Not-determined |
| 17-109_k141_372914 | Low-quality |
| 17-109_k141_238594 | Not-determined |
| 17-109_k141_4333 | Low-quality |
| 17-109_k141_37970 | Not-determined |
| 17-109_k141_4342 | Not-determined |
| 17-109_k141_171560 | Not-determined |
| 17-109_k141_104480 | Low-quality |
| 17-109_k141_4343 | Low-quality |
| 17-109_k141_289088 | Not-determined |
| 17-109_k141_205103 | Low-quality |
| 17-109_k141_171563 | Not-determined |
| 17-109_k141_205114 | Not-determined |
| 17-109_k141_21096 | Not-determined |
| 17-109_k141_254970 | Not-determined |
| 17-109_k141_289095 | Not-determined |
| 17-109_k141_37984 | Not-determined |
| 17-109_k141_306143 | Not-determined |
| 17-109_k141_138511 | Not-determined |
| 17-109_k141_189004 | Not-determined |
| 17-109_k141_238613 | Not-determined |
| 17-109_k141_171577 | Not-determined |
| 17-109_k141_289103 | Not-determined |
| 17-109_k141_238614 | Not-determined |
| 17-109_k141_138516 | Not-determined |
| 17-109_k141_289105 | Not-determined |
| 17-109_k141_272106 | Not-determined |
| 17-109_k141_171579 | Not-determined |
| 17-109_k141_104505 | Not-determined |
| 17-109_k141_21101 | Not-determined |
| 17-109_k141_21106 | Not-determined |
| 17-109_k141_238629 | Not-determined |
| 17-109_k141_54494 | Not-determined |
| 17-109_k141_171596 | Not-determined |
| 17-109_k141_71829 | Not-determined |
| 17-109_k141_306165 | Not-determined |
| 17-109_k141_372949 | Not-determined |
| 17-109_k141_238638 | Not-determined |
| 17-109_k141_338829 | Not-determined |
| 17-109_k141_238641 | Not-determined |
| 17-109_k141_221865 | Not-determined |
| 17-109_k141_221868 | Low-quality |
| 17-109_k141_189030 | Not-determined |
| 17-109_k141_238654 | Not-determined |
| 17-109_k141_221873 | Not-determined |
| 17-109_k141_238657 | Not-determined |
| 17-109_k141_54501 | Not-determined |
| 17-109_k141_255003 | Low-quality |
| 17-109_k141_238659 | Not-determined |
| 17-109_k141_4362 | Not-determined |
| 17-109_k141_87891 | Not-determined |
| 17-109_k141_4363 | Not-determined |
| 17-109_k141_171616 | Low-quality |
| 17-109_k141_389420 | Not-determined |
| 17-109_k141_4368 | Not-determined |
| 17-109_k141_389423 | Low-quality |
| 17-109_k141_389424 | Not-determined |
| 17-109_k141_104524 | Low-quality |
| 17-109_k141_272137 | Not-determined |
| 17-109_k141_189045 | Not-determined |
| 17-109_k141_255011 | Not-determined |
| 17-109_k141_255012 | Not-determined |
| 17-109_k141_104528 | Not-determined |
| 17-109_k141_289118 | Not-determined |
| 17-109_k141_189050 | Not-determined |
| 17-109_k141_171624 | Not-determined |
| 17-109_k141_221892 | Not-determined |
| 17-109_k141_138543 | Not-determined |
| 17-109_k141_306204 | Not-determined |
| 17-109_k141_389431 | Not-determined |
| 17-109_k141_189054 | Not-determined |
| 17-109_k141_221895 | Not-determined |
| 17-109_k141_372961 | Low-quality |
| 17-109_k141_37999 | Not-determined |
| 17-109_k141_338853 | Not-determined |
| 17-109_k141_289128 | Not-determined |
| 17-109_k141_138549 | Not-determined |
| 17-109_k141_21153 | Not-determined |
| 17-109_k141_4389 | Not-determined |
| 17-109_k141_189064 | Low-quality |
| 17-109_k141_38005 | Not-determined |
| 17-109_k141_54538 | Not-determined |
| 17-109_k141_255023 | Not-determined |
| 17-109_k141_389454 | Not-determined |
| 17-109_k141_205149 | Not-determined |
| 17-109_k141_87908 | Not-determined |
| 17-109_k141_87909 | Not-determined |
| 17-109_k141_38011 | Not-determined |
| 17-109_k141_4394 | Not-determined |
| 17-109_k141_272158 | Not-determined |
| 17-109_k141_54546 | Not-determined |
| 17-109_k141_205150 | Not-determined |
| 17-109_k141_104558 | Not-determined |
| 17-109_k141_238689 | Not-determined |
| 17-109_k141_87926 | Not-determined |
| 17-109_k141_189078 | Not-determined |
| 17-109_k141_338891 | Low-quality |
| 17-109_k141_87929 | Not-determined |
| 17-109_k141_138566 | Not-determined |
| 17-109_k141_38019 | Not-determined |
| 17-109_k141_238709 | Not-determined |
| 17-109_k141_338902 | Not-determined |
| 17-109_k141_138572 | Not-determined |
| 17-109_k141_138574 | Not-determined |
| 17-109_k141_289167 | Not-determined |
| 17-109_k141_389489 | Not-determined |
| 17-109_k141_289173 | Low-quality |
| 17-109_k141_372987 | Not-determined |
| 17-109_k141_205189 | Not-determined |
| 17-109_k141_87942 | Low-quality |
| 17-109_k141_171667 | Not-determined |
| 17-109_k141_104581 | Not-determined |
| 17-109_k141_54573 | Not-determined |
| 17-109_k141_71849 | Not-determined |
| 17-109_k141_272178 | Not-determined |
| 17-109_k141_205204 | Not-determined |
| 17-109_k141_322573 | Not-determined |
| 17-109_k141_322575 | Not-determined |
| 17-109_k141_4436 | Not-determined |
| 17-109_k141_389508 | Not-determined |
| 17-109_k141_4441 | Not-determined |
| 17-109_k141_272188 | Not-determined |
| 17-109_k141_272189 | Not-determined |
| 17-109_k141_121089 | Not-determined |
| 17-109_k141_255041 | Not-determined |
| 17-109_k141_289197 | Not-determined |
| 17-109_k141_155010 | Not-determined |
| 17-109_k141_322588 | Not-determined |
| 17-109_k141_138583 | Not-determined |
| 17-109_k141_238750 | Not-determined |
| 17-109_k141_272197 | Not-determined |
| 17-109_k141_338926 | Not-determined |
| 17-109_k141_322591 | Not-determined |
| 17-109_k141_87968 | Not-determined |
| 17-109_k141_289210 | Not-determined |
| 17-109_k141_272205 | Not-determined |
| 17-109_k141_338933 | Not-determined |
| 17-109_k141_21184 | Not-determined |
| 17-109_k141_338936 | Not-determined |
| 17-109_k141_21185 | Not-determined |
| 17-109_k141_138598 | Not-determined |
| 17-109_k141_87978 | Not-determined |
| 17-109_k141_138599 | Not-determined |
| 17-109_k141_21189 | Not-determined |
| 17-109_k141_155025 | Low-quality |
| 17-109_k141_87982 | Not-determined |
| 17-109_k141_138605 | Not-determined |
| 17-109_k141_138606 | Not-determined |
| 17-109_k141_4460 | Not-determined |
| 17-109_k141_138611 | Not-determined |
| 17-109_k141_104626 | Not-determined |
| 17-109_k141_205230 | Not-determined |
| 17-109_k141_338939 | Not-determined |

| | |
|---|---|
| 17-109_k141_171704 | Not-determined |
| 17-109_k141_138616 | Not-determined |
| 17-109_k141_289228 | Not-determined |
| 17-109_k141_338944 | Not-determined |
| 17-109_k141_138618 | Not-determined |
| 17-109_k141_71867 | Not-determined |
| 17-109_k141_4472 | Low-quality |
| 17-109_k141_71869 | Not-determined |
| 17-109_k141_87989 | Low-quality |
| 17-109_k141_189136 | Not-determined |
| 17-109_k141_189140 | Not-determined |
| 17-109_k141_104645 | Not-determined |
| 17-109_k141_104649 | Low-quality |
| 17-109_k141_71878 | Not-determined |
| 17-109_k141_272234 | Not-determined |
| 17-109_k141_4486 | Low-quality |
| 17-109_k141_189156 | Low-quality |
| 17-109_k141_272246 | Not-determined |
| 17-109_k141_189161 | Low-quality |
| 17-109_k141_88005 | Not-determined |
| 17-109_k141_322600 | Not-determined |
| 17-109_k141_4487 | Low-quality |
| 17-109_k141_38090 | Not-determined |
| 17-109_k141_71901 | Not-determined |
| 17-109_k141_373051 | Not-determined |
| 17-109_k141_71907 | Not-determined |
| 17-109_k141_71908 | Not-determined |
| 17-109_k141_338966 | Not-determined |
| 17-109_k141_322616 | Not-determined |
| 17-109_k141_373059 | Low-quality |
| 17-109_k141_189181 | Not-determined |
| 17-109_k141_255059 | Not-determined |
| 17-109_k141_255061 | Low-quality |
| 17-109_k141_373066 | Not-determined |
| 17-109_k141_4500 | Not-determined |
| 17-109_k141_4503 | Not-determined |
| 17-109_k141_71935 | Not-determined |
| 17-109_k141_289261 | Not-determined |
| 17-109_k141_322635 | Not-determined |
| 17-109_k141_255070 | Not-determined |
| 17-109_k141_373075 | Not-determined |
| 17-109_k141_389564 | Not-determined |
| 17-109_k141_389565 | Not-determined |
| 17-109_k141_189201 | Not-determined |
| 17-109_k141_138674 | Not-determined |
| 17-109_k141_306248 | Not-determined |
| 17-109_k141_389572 | Not-determined |
| 17-109_k141_322645 | Low-quality |
| 17-109_k141_338991 | Not-determined |
| 17-109_k141_238774 | Not-determined |
| 17-109_k141_272290 | Not-determined |
| 17-109_k141_238785 | Not-determined |
| 17-109_k141_238786 | Not-determined |
| 17-109_k141_205267 | Not-determined |
| 17-109_k141_189219 | Not-determined |
| 17-109_k141_21239 | Not-determined |
| 17-109_k141_205273 | Not-determined |
| 17-109_k141_289286 | Not-determined |
| 17-109_k141_104697 | Not-determined |
| 17-109_k141_322661 | Not-determined |
| 17-109_k141_322668 | Not-determined |
| 17-109_k141_355529 | Not-determined |
| 17-109_k141_272308 | Not-determined |
| 17-109_k141_238797 | Not-determined |
| 17-109_k141_355535 | Not-determined |
| 17-109_k141_289306 | Not-determined |
| 17-109_k141_373110 | Not-determined |
| 17-109_k141_238802 | Not-determined |
| 17-109_k141_21257 | Low-quality |
| 17-109_k141_138701 | Not-determined |
| 17-109_k141_339039 | Not-determined |
| 17-109_k141_189239 | Low-quality |
| 17-109_k141_389605 | Not-determined |
| 17-109_k141_306293 | Low-quality |
| 17-109_k141_71942 | Not-determined |
| 17-109_k141_138703 | Low-quality |
| 17-109_k141_138705 | Not-determined |
| 17-109_k141_21261 | Not-determined |
| 17-109_k141_4530 | Low-quality |
| 17-109_k141_71951 | Not-determined |
| 17-109_k141_171734 | Not-determined |
| 17-109_k141_289315 | Not-determined |
| 17-109_k141_355547 | Not-determined |
| 17-109_k141_389618 | Not-determined |
| 17-109_k141_355550 | Not-determined |
| 17-109_k141_272328 | Not-determined |
| 17-109_k141_88063 | Not-determined |
| 17-109_k141_38102 | Not-determined |
| 17-109_k141_138719 | Not-determined |
| 17-109_k141_138720 | Not-determined |
| 17-109_k141_255084 | Low-quality |
| 17-109_k141_171743 | Not-determined |
| 17-109_k141_355556 | Not-determined |
| 17-109_k141_38121 | Not-determined |
| 17-109_k141_322703 | Not-determined |
| 17-109_k141_322705 | Low-quality |
| 17-109_k141_88088 | Not-determined |
| 17-109_k141_355577 | Not-determined |
| 17-109_k141_322708 | Not-determined |
| 17-109_k141_289340 | Not-determined |
| 17-109_k141_88091 | Not-determined |
| 17-109_k141_306321 | Not-determined |
| 17-109_k141_138744 | Not-determined |
| 17-109_k141_355589 | Not-determined |
| 17-109_k141_306326 | Low-quality |
| 17-109_k141_306327 | Not-determined |
| 17-109_k141_289349 | Not-determined |
| 17-109_k141_221899 | Not-determined |
| 17-109_k141_38145 | Low-quality |
| 17-109_k141_289356 | Not-determined |
| 17-109_k141_171769 | Not-determined |
| 17-109_k141_71974 | Not-determined |
| 17-109_k141_54585 | Not-determined |
| 17-109_k141_373136 | Low-quality |
| 17-109_k141_322721 | Not-determined |
| 17-109_k141_289365 | Not-determined |
| 17-109_k141_138752 | Not-determined |
| 17-109_k141_171785 | Not-determined |
| 17-109_k141_171786 | Not-determined |
| 17-109_k141_71977 | Low-quality |
| 17-109_k141_104744 | Not-determined |
| 17-109_k141_389637 | Not-determined |
| 17-109_k141_355610 | Not-determined |
| 17-109_k141_54595 | Not-determined |
| 17-109_k141_389639 | Not-determined |
| 17-109_k141_355616 | Not-determined |
| 17-109_k141_38157 | Not-determined |
| 17-109_k141_389642 | Low-quality |
| 17-109_k141_104753 | Not-determined |
| 17-109_k141_38162 | Not-determined |
| 17-109_k141_322739 | Not-determined |
| 17-109_k141_322743 | Low-quality |
| 17-109_k141_71981 | Not-determined |
| 17-109_k141_104762 | Not-determined |
| 17-109_k141_289388 | Not-determined |
| 17-109_k141_389651 | Not-determined |
| 17-109_k141_38171 | Not-determined |
| 17-109_k141_104768 | Not-determined |
| 17-109_k141_21282 | Not-determined |
| 17-109_k141_21283 | Not-determined |
| 17-109_k141_255103 | Low-quality |
| 17-109_k141_355628 | Not-determined |
| 17-109_k141_255105 | Not-determined |
| 17-109_k141_21287 | Not-determined |
| 17-109_k141_238838 | Not-determined |
| 17-109_k141_389661 | Not-determined |
| 17-109_k141_104771 | Not-determined |
| 17-109_k141_72000 | Not-determined |
| 17-109_k141_255107 | Not-determined |
| 17-109_k141_389666 | Not-determined |
| 17-109_k141_138796 | Not-determined |
| 17-109_k141_306360 | Not-determined |
| 17-109_k141_72011 | Low-quality |
| 17-109_k141_289400 | Not-determined |
| 17-109_k141_205289 | Not-determined |
| 17-109_k141_54635 | Not-determined |
| 17-109_k141_255118 | Not-determined |
| 17-109_k141_355649 | Not-determined |
| 17-109_k141_255124 | Not-determined |
| 17-109_k141_205293 | Not-determined |
| 17-109_k141_255129 | Not-determined |
| 17-109_k141_221917 | Not-determined |
| 17-109_k141_255132 | Not-determined |
| 17-109_k141_54654 | Not-determined |
| 17-109_k141_155091 | Not-determined |
| 17-109_k141_289411 | Not-determined |
| 17-109_k141_121142 | Not-determined |
| 17-109_k141_238850 | Not-determined |
| 17-109_k141_306378 | Not-determined |
| 17-109_k141_289420 | Not-determined |
| 17-109_k141_306383 | Not-determined |
| 17-109_k141_389681 | Not-determined |
| 17-109_k141_289423 | Not-determined |
| 17-109_k141_4543 | Not-determined |
| 17-109_k141_255146 | Not-determined |
| 17-109_k141_205309 | Not-determined |
| 17-109_k141_138820 | Not-determined |
| 17-109_k141_138821 | Not-determined |
| 17-109_k141_104826 | Not-determined |
| 17-109_k141_38187 | Low-quality |
| 17-109_k141_189246 | Not-determined |
| 17-109_k141_306393 | Not-determined |
| 17-109_k141_155099 | Not-determined |
| 17-109_k141_289432 | Not-determined |
| 17-109_k141_389691 | Not-determined |
| 17-109_k141_38193 | Not-determined |
| 17-109_k141_189254 | Not-determined |
| 17-109_k141_38197 | Not-determined |
| 17-109_k141_389696 | Not-determined |
| 17-109_k141_38200 | Not-determined |
| 17-109_k141_189257 | Not-determined |
| 17-109_k141_121151 | Not-determined |
| 17-109_k141_306399 | Not-determined |
| 17-109_k141_155111 | Not-determined |
| 17-109_k141_21314 | Not-determined |
| 17-109_k141_189259 | Low-quality |
| 17-109_k141_221930 | Not-determined |
| 17-109_k141_4557 | Not-determined |
| 17-109_k141_38208 | Not-determined |
| 17-109_k141_389700 | Not-determined |
| 17-109_k141_373141 | Not-determined |
| 17-109_k141_189266 | Not-determined |
| 17-109_k141_238884 | Not-determined |
| 17-109_k141_205338 | Not-determined |
| 17-109_k141_155121 | Not-determined |
| 17-109_k141_121164 | Not-determined |
| 17-109_k141_221946 | Not-determined |
| 17-109_k141_289464 | Not-determined |
| 17-109_k141_72036 | Low-quality |
| 17-109_k141_72037 | Not-determined |
| 17-109_k141_289466 | Not-determined |
| 17-109_k141_238890 | Not-determined |
| 17-109_k141_339076 | Not-determined |
| 17-109_k141_121178 | Not-determined |
| 17-109_k141_289468 | Not-determined |
| 17-109_k141_38220 | Not-determined |
| 17-109_k141_38223 | Not-determined |
| 17-109_k141_88124 | Not-determined |
| 17-109_k141_205346 | Not-determined |
| 17-109_k141_373164 | Not-determined |
| 17-109_k141_72069 | Not-determined |
| 17-109_k141_373166 | Low-quality |
| 17-109_k141_373168 | Not-determined |
| 17-109_k141_339092 | Not-determined |
| 17-109_k141_221970 | Not-determined |
| 17-109_k141_289501 | Not-determined |
| 17-109_k141_72081 | Low-quality |
| 17-109_k141_221975 | Not-determined |
| 17-109_k141_373186 | Not-determined |
| 17-109_k141_121213 | Not-determined |
| 17-109_k141_221978 | Not-determined |
| 17-109_k141_339104 | Not-determined |
| 17-109_k141_373190 | Not-determined |
| 17-109_k141_21347 | Not-determined |
| 17-109_k141_339111 | Not-determined |
| 17-109_k141_373196 | Not-determined |
| 17-109_k141_373197 | Not-determined |
| 17-109_k141_373198 | Not-determined |
| 17-109_k141_21351 | Not-determined |
| 17-109_k141_306436 | Not-determined |
| 17-109_k141_189314 | Not-determined |
| 17-109_k141_255211 | Not-determined |
| 17-109_k141_272417 | Not-determined |
| 17-109_k141_138841 | Not-determined |
| 17-109_k141_138842 | Not-determined |
| 17-109_k141_255218 | Not-determined |
| 17-109_k141_272426 | Not-determined |
| 17-109_k141_255224 | Not-determined |
| 17-109_k141_339131 | Not-determined |
| 17-109_k141_355660 | Not-determined |
| 17-109_k141_38245 | Not-determined |
| 17-109_k141_289524 | Not-determined |
| 17-109_k141_221990 | Not-determined |
| 17-109_k141_238923 | Not-determined |
| 17-109_k141_104851 | Not-determined |
| 17-109_k141_322829 | Not-determined |
| 17-109_k141_322833 | Not-determined |
| 17-109_k141_171847 | Not-determined |
| 17-109_k141_322840 | Not-determined |
| 17-109_k141_21358 | Not-determined |
| 17-109_k141_289540 | Not-determined |
| 17-109_k141_72115 | Not-determined |
| 17-109_k141_272451 | Low-quality |
| 17-109_k141_389716 | Not-determined |
| 17-109_k141_373200 | Not-determined |
| 17-109_k141_21362 | Not-determined |
| 17-109_k141_155168 | Not-determined |
| 17-109_k141_4582 | Not-determined |
| 17-109_k141_155171 | Not-determined |
| 17-109_k141_322844 | Not-determined |
| 17-109_k141_373206 | Not-determined |
| 17-109_k141_289548 | Not-determined |
| 17-109_k141_155178 | Low-quality |
| 17-109_k141_238949 | Not-determined |
| 17-109_k141_373213 | Not-determined |
| 17-109_k141_355683 | Low-quality |
| 17-109_k141_38278 | Not-determined |
| 17-109_k141_121234 | Not-determined |
| 17-109_k141_38280 | Not-determined |
| 17-109_k141_38282 | Not-determined |
| 17-109_k141_171870 | Not-determined |
| 17-109_k141_222031 | Not-determined |
| 17-109_k141_355697 | Not-determined |
| 17-109_k141_205404 | Not-determined |
| 17-109_k141_205406 | Not-determined |
| 17-109_k141_355700 | Not-determined |
| 17-109_k141_272470 | Not-determined |
| 17-109_k141_272472 | Not-determined |
| 17-109_k141_171880 | Low-quality |
| 17-109_k141_155188 | Not-determined |
| 17-109_k141_339148 | Not-determined |
| 17-109_k141_238965 | Not-determined |
| 17-109_k141_4597 | Not-determined |
| 17-109_k141_289575 | Not-determined |
| 17-109_k141_272480 | Not-determined |
| 17-109_k141_155201 | Not-determined |
| 17-109_k141_222053 | Not-determined |
| 17-109_k141_289576 | Low-quality |
| 17-109_k141_238970 | Not-determined |
| 17-109_k141_171883 | Low-quality |
| 17-109_k141_255272 | Not-determined |
| 17-109_k141_205414 | Not-determined |
| 17-109_k141_38303 | Not-determined |
| 17-109_k141_88142 | Not-determined |
| 17-109_k141_355717 | Low-quality |
| 17-109_k141_21377 | Not-determined |
| 17-109_k141_38311 | Low-quality |
| 17-109_k141_222064 | Not-determined |
| 17-109_k141_205418 | Not-determined |
| 17-109_k141_272489 | Not-determined |
| 17-109_k141_21382 | Not-determined |
| 17-109_k141_205422 | Not-determined |
| 17-109_k141_272494 | Not-determined |
| 17-109_k141_205427 | Not-determined |
| 17-109_k141_289588 | Not-determined |
| 17-109_k141_322856 | Not-determined |
| 17-109_k141_21396 | Not-determined |
| 17-109_k141_121250 | Not-determined |
| 17-109_k141_104892 | Not-determined |
| 17-109_k141_205431 | Low-quality |
| 17-109_k141_4614 | Not-determined |
| 17-109_k141_104895 | Not-determined |
| 17-109_k141_38328 | Not-determined |
| 17-109_k141_238989 | Not-determined |
| 17-109_k141_238990 | Not-determined |
| 17-109_k141_322863 | Not-determined |
| 17-109_k141_255287 | Not-determined |
| 17-109_k141_104904 | Not-determined |
| 17-109_k141_289597 | Low-quality |
| 17-109_k141_339152 | Low-quality |
| 17-109_k141_104907 | Not-determined |
| 17-109_k141_339153 | Not-determined |
| 17-109_k141_289602 | Not-determined |
| 17-109_k141_21405 | Low-quality |
| 17-109_k141_322874 | Not-determined |
| 17-109_k141_88172 | Not-determined |
| 17-109_k141_104917 | Not-determined |
| 17-109_k141_373280 | Not-determined |
| 17-109_k141_88179 | Not-determined |
| 17-109_k141_104927 | Not-determined |
| 17-109_k141_38335 | Low-quality |
| 17-109_k141_355741 | Not-determined |
| 17-109_k141_104946 | Not-determined |
| 17-109_k141_355749 | Not-determined |
| 17-109_k141_272508 | Not-determined |
| 17-109_k141_189334 | Not-determined |
| 17-109_k141_72136 | Not-determined |
| 17-109_k141_205465 | Low-quality |
| 17-109_k141_239009 | Not-determined |
| 17-109_k141_272513 | Not-determined |
| 17-109_k141_373315 | Not-determined |
| 17-109_k141_239012 | Not-determined |
| 17-109_k141_155277 | Not-determined |
| 17-109_k141_21436 | Not-determined |
| 17-109_k141_239019 | Not-determined |
| 17-109_k141_189349 | Not-determined |
| 17-109_k141_104954 | Not-determined |
| 17-109_k141_272521 | Not-determined |
| 17-109_k141_239023 | Not-determined |
| 17-109_k141_239026 | Not-determined |
| 17-109_k141_104960 | Low-quality |
| 17-109_k141_339169 | Not-determined |
| 17-109_k141_54661 | Not-determined |
| 17-109_k141_171947 | Not-determined |
| 17-109_k141_373336 | Not-determined |
| 17-109_k141_104971 | Not-determined |
| 17-109_k141_21452 | Not-determined |
| 17-109_k141_138858 | Not-determined |
| 17-109_k141_54666 | Not-determined |
| 17-109_k141_38374 | Not-determined |
| 17-109_k141_373345 | Not-determined |
| 17-109_k141_339185 | Not-determined |
| 17-109_k141_389763 | Low-quality |
| 17-109_k141_272525 | Not-determined |
| 17-109_k141_322933 | Not-determined |
| 17-109_k141_322934 | Not-determined |
| 17-109_k141_38379 | Not-determined |
| 17-109_k141_373363 | Not-determined |
| 17-109_k141_38381 | Not-determined |
| 17-109_k141_373364 | Not-determined |
| 17-109_k141_322939 | Not-determined |
| 17-109_k141_339201 | Not-determined |
| 17-109_k141_373369 | Not-determined |
| 17-109_k141_389767 | Not-determined |
| 17-109_k141_155308 | Low-quality |
| 17-109_k141_255331 | Not-determined |
| 17-109_k141_21467 | Not-determined |
| 17-109_k141_88189 | Not-determined |
| 17-109_k141_255337 | Not-determined |
| 17-109_k141_155312 | Not-determined |
| 17-109_k141_105004 | Not-determined |
| 17-109_k141_189367 | Not-determined |
| 17-109_k141_121267 | Not-determined |
| 17-109_k141_355772 | Not-determined |
| 17-109_k141_121271 | Not-determined |
| 17-109_k141_355780 | Not-determined |
| 17-109_k141_355783 | Not-determined |
| 17-109_k141_105024 | Not-determined |
| 17-109_k141_138887 | Not-determined |
| 17-109_k141_373399 | Not-determined |
| 17-109_k141_155320 | Not-determined |
| 17-109_k141_38412 | Not-determined |
| 17-109_k141_339210 | Not-determined |
| 17-109_k141_38413 | Not-determined |
| 17-109_k141_38415 | Not-determined |
| 17-109_k141_339212 | Not-determined |
| 17-109_k141_155328 | Not-determined |
| 17-109_k141_38422 | Not-determined |
| 17-109_k141_222086 | Not-determined |
| 17-109_k141_255370 | Low-quality |
| 17-109_k141_339222 | Not-determined |
| 17-109_k141_105036 | Not-determined |
| 17-109_k141_38426 | Not-determined |
| 17-109_k141_38427 | Not-determined |
| 17-109_k141_105037 | Not-determined |
| 17-109_k141_339226 | Low-quality |
| 17-109_k141_105038 | Not-determined |
| 17-109_k141_222090 | Not-determined |
| 17-109_k141_339228 | Not-determined |
| 17-109_k141_189393 | Not-determined |
| 17-109_k141_189395 | Not-determined |
| 17-109_k141_255383 | Not-determined |
| 17-109_k141_272562 | Not-determined |
| 17-109_k141_21482 | Not-determined |
| 17-109_k141_138903 | Not-determined |
| 17-109_k141_155341 | Not-determined |
| 17-109_k141_54700 | Not-determined |
| 17-109_k141_373421 | Not-determined |
| 17-109_k141_155350 | Not-determined |
| 17-109_k141_373426 | Not-determined |
| 17-109_k141_171954 | Low-quality |
| 17-109_k141_339253 | Not-determined |
| 17-109_k141_306487 | Not-determined |
| 17-109_k141_88211 | Low-quality |
| 17-109_k141_54714 | Not-determined |
| 17-109_k141_189402 | Not-determined |
| 17-109_k141_54715 | Not-determined |
| 17-109_k141_88220 | Not-determined |
| 17-109_k141_155358 | Low-quality |
| 17-109_k141_155359 | Not-determined |
| 17-109_k141_88223 | Not-determined |
| 17-109_k141_105059 | Not-determined |
| 17-109_k141_138921 | Not-determined |
| 17-109_k141_339275 | Not-determined |
| 17-109_k141_138927 | Not-determined |
| 17-109_k141_323003 | Low-quality |
| 17-109_k141_373448 | Not-determined |
| 17-109_k141_189420 | Not-determined |
| 17-109_k141_255409 | Not-determined |
| 17-109_k141_306515 | Not-determined |
| 17-109_k141_306516 | Low-quality |
| 17-109_k141_88227 | Not-determined |
| 17-109_k141_189428 | Not-determined |
| 17-109_k141_323022 | Not-determined |
| 17-109_k141_88230 | Not-determined |
| 17-109_k141_121284 | Not-determined |
| 17-109_k141_323024 | Not-determined |
| 17-109_k141_373464 | Not-determined |
| 17-109_k141_255417 | Low-quality |
| 17-109_k141_121295 | Not-determined |
| 17-109_k141_54729 | Not-determined |
| 17-109_k141_4637 | Low-quality |
| 17-109_k141_373469 | Not-determined |
| 17-109_k141_121301 | Not-determined |
| 17-109_k141_155392 | Not-determined |
| 17-109_k141_105110 | Not-determined |
| 17-109_k141_289696 | Not-determined |
| 17-109_k141_21498 | Not-determined |
| 17-109_k141_255437 | Not-determined |
| 17-109_k141_54741 | Not-determined |
| 17-109_k141_189452 | Not-determined |
| 17-109_k141_255441 | Low-quality |
| 17-109_k141_155402 | Not-determined |
| 17-109_k141_21503 | Not-determined |
| 17-109_k141_272612 | Not-determined |
| 17-109_k141_306531 | Not-determined |
| 17-109_k141_272620 | Not-determined |
| 17-109_k141_72238 | Low-quality |
| 17-109_k141_306539 | Not-determined |
| 17-109_k141_272621 | Not-determined |
| 17-109_k141_323043 | Not-determined |
| 17-109_k141_38501 | Not-determined |
| 17-109_k141_272631 | Not-determined |
| 17-109_k141_4643 | Not-determined |
| 17-109_k141_339292 | Not-determined |
| 17-109_k141_289716 | Low-quality |
| 17-109_k141_105140 | Not-determined |
| 17-109_k141_121318 | Not-determined |
| 17-109_k141_54744 | Not-determined |
| 17-109_k141_389840 | Not-determined |
| 17-109_k141_88251 | Not-determined |
| 17-109_k141_373500 | Not-determined |
| 17-109_k141_105148 | Not-determined |
| 17-109_k141_155415 | Not-determined |
| 17-109_k141_306552 | Not-determined |
| 17-109_k141_272637 | Not-determined |
| 17-109_k141_289726 | Not-determined |
| 17-109_k141_355868 | Not-determined |
| 17-109_k141_72247 | Not-determined |
| 17-109_k141_339319 | Not-determined |
| 17-109_k141_339320 | Not-determined |
| 17-109_k141_72261 | Not-determined |
| 17-109_k141_255472 | Not-determined |
| 17-109_k141_38528 | Not-determined |
| 17-109_k141_222119 | Not-determined |
| 17-109_k141_121327 | Not-determined |
| 17-109_k141_4645 | Not-determined |
| 17-109_k141_339336 | Not-determined |
| 17-109_k141_38537 | Not-determined |
| 17-109_k141_272665 | Not-determined |
| 17-109_k141_189503 | Not-determined |
| 17-109_k141_306581 | Not-determined |
| 17-109_k141_21509 | Not-determined |
| 17-109_k141_88257 | Not-determined |
| 17-109_k141_272672 | Not-determined |
| 17-109_k141_189513 | Not-determined |
| 17-109_k141_4653 | Not-determined |
| 17-109_k141_54752 | Not-determined |
| 17-109_k141_54754 | Not-determined |
| 17-109_k141_171978 | Not-determined |
| 17-109_k141_239116 | Not-determined |
| 17-109_k141_171982 | Low-quality |
| 17-109_k141_54760 | Not-determined |
| 17-109_k141_339352 | Not-determined |
| 17-109_k141_339353 | Not-determined |
| 17-109_k141_355889 | Not-determined |
| 17-109_k141_339354 | Not-determined |
| 17-109_k141_339355 | Not-determined |
| 17-109_k141_155466 | Not-determined |
| 17-109_k141_272692 | Not-determined |
| 17-109_k141_272693 | Not-determined |
| 17-109_k141_38546 | Not-determined |
| 17-109_k141_239129 | Not-determined |
| 17-109_k141_189535 | Low-quality |
| 17-109_k141_54771 | Low-quality |
| 17-109_k141_339371 | Not-determined |
| 17-109_k141_171989 | Not-determined |
| 17-109_k141_339373 | Not-determined |
| 17-109_k141_88274 | Not-determined |
| 17-109_k141_38550 | Low-quality |
| 17-109_k141_306609 | Low-quality |
| 17-109_k141_171995 | Not-determined |
| 17-109_k141_72284 | Not-determined |
| 17-109_k141_355898 | Not-determined |
| 17-109_k141_72288 | Not-determined |
| 17-109_k141_306617 | Not-determined |
| 17-109_k141_339384 | Not-determined |
| 17-109_k141_255524 | Not-determined |
| 17-109_k141_373555 | Not-determined |
| 17-109_k141_389877 | Not-determined |
| 17-109_k141_339393 | Not-determined |
| 17-109_k141_54778 | Not-determined |
| 17-109_k141_306625 | Not-determined |
| 17-109_k141_323120 | Not-determined |
| 17-109_k141_88290 | Low-quality |
| 17-109_k141_138989 | Not-determined |
| 17-109_k141_355902 | Not-determined |
| 17-109_k141_138998 | Low-quality |
| 17-109_k141_172005 | Not-determined |
| 17-109_k141_189559 | Not-determined |
| 17-109_k141_155490 | Not-determined |
| 17-109_k141_323132 | Not-determined |
| 17-109_k141_323135 | Low-quality |
| 17-109_k141_72313 | Not-determined |
| 17-109_k141_38571 | Low-quality |
| 17-109_k141_38573 | Not-determined |
| 17-109_k141_189571 | Not-determined |
| 17-109_k141_38576 | Not-determined |
| 17-109_k141_139002 | Not-determined |
| 17-109_k141_139003 | Not-determined |
| 17-109_k141_189575 | Low-quality |
| 17-109_k141_4686 | Not-determined |
| 17-109_k141_172024 | Not-determined |
| 17-109_k141_339413 | Not-determined |
| 17-109_k141_339415 | Not-determined |
| 17-109_k141_189581 | Not-determined |
| 17-109_k141_255549 | Not-determined |
| 17-109_k141_72323 | Not-determined |
| 17-109_k141_54799 | Not-determined |
| 17-109_k141_155523 | Not-determined |
| 17-109_k141_189592 | Low-quality |
| 17-109_k141_306651 | Not-determined |
| 17-109_k141_389912 | Not-determined |
| 17-109_k141_4703 | Not-determined |
| 17-109_k141_88326 | Not-determined |
| 17-109_k141_189595 | Not-determined |
| 17-109_k141_189596 | Not-determined |
| 17-109_k141_38587 | Not-determined |
| 17-109_k141_306658 | Not-determined |

| | |
|---|---|
| 17-109_k141_4706 | Not-determined |
| 17-109_k141_389918 | Not-determined |
| 17-109_k141_323156 | Low-quality |
| 17-109_k141_72332 | Not-determined |
| 17-109_k141_306665 | Not-determined |
| 17-109_k141_189603 | Not-determined |
| 17-109_k141_373564 | Not-determined |
| 17-109_k141_373570 | Not-determined |
| 17-109_k141_88337 | Not-determined |
| 17-109_k141_389922 | Not-determined |
| 17-109_k141_373572 | Low-quality |
| 17-109_k141_4719 | Not-determined |
| 17-109_k141_54824 | Not-determined |
| 17-109_k141_389928 | Not-determined |
| 17-109_k141_339441 | Not-determined |
| 17-109_k141_4725 | Low-quality |
| 17-109_k141_389938 | Not-determined |
| 17-109_k141_389939 | Not-determined |
| 17-109_k141_389943 | Not-determined |
| 17-109_k141_4737 | Not-determined |
| 17-109_k141_355935 | Not-determined |
| 17-109_k141_289752 | Not-determined |
| 17-109_k141_139008 | Not-determined |
| 17-109_k141_255585 | Not-determined |
| 17-109_k141_54838 | Not-determined |
| 17-109_k141_189632 | Not-determined |
| 17-109_k141_389947 | Not-determined |
| 17-109_k141_21572 | Not-determined |
| 17-109_k141_38613 | Low-quality |
| 17-109_k141_54855 | Not-determined |
| 17-109_k141_38614 | Not-determined |
| 17-109_k141_306675 | Not-determined |
| 17-109_k141_21578 | Not-determined |
| 17-109_k141_306684 | Not-determined |
| 17-109_k141_72347 | Not-determined |
| 17-109_k141_339457 | Not-determined |
| 17-109_k141_72350 | Not-determined |
| 17-109_k141_306690 | Low-quality |
| 17-109_k141_88381 | Not-determined |
| 17-109_k141_389972 | Not-determined |
| 17-109_k141_255622 | Not-determined |
| 17-109_k141_389975 | Not-determined |
| 17-109_k141_121343 | Not-determined |
| 17-109_k141_389984 | Not-determined |
| 17-109_k141_189649 | Not-determined |
| 17-109_k141_389986 | Not-determined |
| 17-109_k141_121350 | Not-determined |
| 17-109_k141_88406 | Not-determined |
| 17-109_k141_172070 | Not-determined |
| 17-109_k141_373608 | Low-quality |
| 17-109_k141_373609 | Not-determined |
| 17-109_k141_121359 | Not-determined |
| 17-109_k141_38642 | Not-determined |
| 17-109_k141_189660 | Not-determined |
| 17-109_k141_189664 | Not-determined |
| 17-109_k141_88419 | Not-determined |
| 17-109_k141_38649 | Not-determined |
| 17-109_k141_54869 | Low-quality |
| 17-109_k141_105222 | Low-quality |
| 17-109_k141_339471 | Not-determined |
| 17-109_k141_172079 | Not-determined |
| 17-109_k141_239177 | Low-quality |
| 17-109_k141_105227 | Low-quality |
| 17-109_k141_38658 | Not-determined |
| 17-109_k141_255659 | Not-determined |
| 17-109_k141_255662 | Not-determined |
| 17-109_k141_54879 | Not-determined |
| 17-109_k141_38660 | Not-determined |
| 17-109_k141_306697 | Low-quality |
| 17-109_k141_306700 | Low-quality |
| 17-109_k141_339476 | Not-determined |
| 17-109_k141_390020 | Low-quality |
| 17-109_k141_255676 | Not-determined |
| 17-109_k141_390023 | Not-determined |
| 17-109_k141_38674 | Not-determined |
| 17-109_k141_289784 | Not-determined |
| 17-109_k141_38675 | Not-determined |
| 17-109_k141_205508 | Not-determined |
| 17-109_k141_54893 | Not-determined |
| 17-109_k141_4758 | Low-quality |
| 17-109_k141_72399 | Not-determined |
| 17-109_k141_4759 | Not-determined |
| 17-109_k141_339486 | Not-determined |
| 17-109_k141_88443 | Low-quality |
| 17-109_k141_205518 | Not-determined |
| 17-109_k141_54904 | Not-determined |
| 17-109_k141_390035 | Not-determined |
| 17-109_k141_38691 | Not-determined |
| 17-109_k141_4766 | Not-determined |
| 17-109_k141_21633 | Not-determined |
| 17-109_k141_72401 | Not-determined |
| 17-109_k141_139053 | Not-determined |
| 17-109_k141_72403 | Not-determined |
| 17-109_k141_4777 | Not-determined |
| 17-109_k141_38702 | Low-quality |
| 17-109_k141_38704 | Not-determined |
| 17-109_k141_38705 | Not-determined |
| 17-109_k141_222264 | Not-determined |
| 17-109_k141_21642 | Not-determined |
| 17-109_k141_289796 | Not-determined |
| 17-109_k141_4786 | Not-determined |
| 17-109_k141_4788 | Not-determined |
| 17-109_k141_21649 | Not-determined |
| 17-109_k141_72415 | Not-determined |
| 17-109_k141_222279 | Not-determined |
| 17-109_k141_4792 | Not-determined |
| 17-109_k141_390054 | Not-determined |
| 17-109_k141_339499 | Not-determined |
| 17-109_k141_139070 | Not-determined |
| 17-109_k141_339501 | Not-determined |
| 17-109_k141_4797 | Not-determined |
| 17-109_k141_88468 | Not-determined |
| 17-109_k141_222287 | Not-determined |
| 17-109_k141_239242 | Not-determined |
| 17-109_k141_189699 | Not-determined |
| 17-109_k141_88472 | Not-determined |
| 17-109_k141_239249 | Not-determined |
| 17-109_k141_239250 | Not-determined |
| 17-109_k141_272749 | Not-determined |
| 17-109_k141_272751 | Not-determined |
| 17-109_k141_172097 | Not-determined |
| 17-109_k141_289818 | Not-determined |
| 17-109_k141_139072 | Not-determined |
| 17-109_k141_255699 | Low-quality |
| 17-109_k141_189708 | Not-determined |
| 17-109_k141_172101 | Not-determined |
| 17-109_k141_205526 | Not-determined |
| 17-109_k141_239264 | Not-determined |
| 17-109_k141_38717 | Not-determined |
| 17-109_k141_21676 | Not-determined |
| 17-109_k141_289828 | Low-quality |
| 17-109_k141_172114 | Not-determined |
| 17-109_k141_355976 | Not-determined |
| 17-109_k141_355978 | Low-quality |
| 17-109_k141_88495 | Not-determined |
| 17-109_k141_239270 | Not-determined |
| 17-109_k141_72448 | Not-determined |
| 17-109_k141_390081 | Low-quality |
| 17-109_k141_4810 | Not-determined |
| 17-109_k141_222311 | Not-determined |
| 17-109_k141_355985 | Not-determined |
| 17-109_k141_289832 | Low-quality |
| 17-109_k141_255707 | Not-determined |
| 17-109_k141_255713 | Not-determined |
| 17-109_k141_155582 | Low-quality |
| 17-109_k141_355995 | Not-determined |
| 17-109_k141_390093 | Not-determined |
| 17-109_k141_255715 | Not-determined |
| 17-109_k141_390094 | Not-determined |
| 17-109_k141_373652 | Not-determined |
| 17-109_k141_21701 | Not-determined |
| 17-109_k141_38736 | Not-determined |
| 17-109_k141_272784 | Not-determined |
| 17-109_k141_21703 | Not-determined |
| 17-109_k141_272785 | Not-determined |
| 17-109_k141_21706 | Not-determined |
| 17-109_k141_289842 | Not-determined |
| 17-109_k141_172135 | Not-determined |
| 17-109_k141_38749 | Not-determined |
| 17-109_k141_139101 | Low-quality |
| 17-109_k141_38752 | Not-determined |
| 17-109_k141_255730 | Low-quality |
| 17-109_k141_205560 | Not-determined |
| 17-109_k141_373659 | Not-determined |
| 17-109_k141_205562 | Not-determined |
| 17-109_k141_255736 | Not-determined |
| 17-109_k141_306781 | Not-determined |
| 17-109_k141_289858 | Not-determined |
| 17-109_k141_72478 | Not-determined |
| 17-109_k141_72479 | Not-determined |
| 17-109_k141_390117 | Not-determined |
| 17-109_k141_373671 | Not-determined |
| 17-109_k141_4814 | Not-determined |
| 17-109_k141_289869 | Not-determined |
| 17-109_k141_239281 | Not-determined |
| 17-109_k141_121407 | Not-determined |
| 17-109_k141_72486 | Not-determined |
| 17-109_k141_306801 | Not-determined |
| 17-109_k141_72490 | Not-determined |
| 17-109_k141_189746 | Not-determined |
| 17-109_k141_289881 | Not-determined |
| 17-109_k141_373679 | Not-determined |
| 17-109_k141_189754 | Not-determined |
| 17-109_k141_390134 | Not-determined |
| 17-109_k141_72496 | Low-quality |
| 17-109_k141_139120 | Low-quality |
| 17-109_k141_255760 | Not-determined |
| 17-109_k141_390137 | Not-determined |
| 17-109_k141_121419 | Low-quality |
| 17-109_k141_390138 | Not-determined |
| 17-109_k141_72501 | Not-determined |
| 17-109_k141_72503 | Not-determined |
| 17-109_k141_272817 | Not-determined |
| 17-109_k141_390142 | Not-determined |
| 17-109_k141_222344 | Not-determined |
| 17-109_k141_373686 | Not-determined |
| 17-109_k141_239293 | Not-determined |
| 17-109_k141_390149 | Low-quality |
| 17-109_k141_306819 | Not-determined |
| 17-109_k141_306821 | Not-determined |
| 17-109_k141_289894 | Not-determined |
| 17-109_k141_306824 | Low-quality |
| 17-109_k141_189779 | Not-determined |
| 17-109_k141_272831 | Not-determined |
| 17-109_k141_54913 | Not-determined |
| 17-109_k141_189780 | Low-quality |
| 17-109_k141_189781 | Low-quality |
| 17-109_k141_373695 | Not-determined |
| 17-109_k141_272837 | Not-determined |
| 17-109_k141_239314 | Not-determined |
| 17-109_k141_72516 | Not-determined |
| 17-109_k141_323171 | Not-determined |
| 17-109_k141_373704 | Not-determined |
| 17-109_k141_54929 | Not-determined |
| 17-109_k141_189791 | Not-determined |
| 17-109_k141_356012 | Not-determined |
| 17-109_k141_323180 | Not-determined |
| 17-109_k141_21737 | Not-determined |
| 17-109_k141_306844 | Not-determined |
| 17-109_k141_222372 | Not-determined |
| 17-109_k141_88517 | Not-determined |
| 17-109_k141_172206 | Not-determined |
| 17-109_k141_121452 | Not-determined |
| 17-109_k141_306854 | Not-determined |
| 17-109_k141_373725 | Not-determined |
| 17-109_k141_390169 | Not-determined |
| 17-109_k141_289921 | Not-determined |
| 17-109_k141_172207 | Low-quality |
| 17-109_k141_4875 | Low-quality |
| 17-109_k141_356024 | Low-quality |
| 17-109_k141_339542 | Not-determined |
| 17-109_k141_239352 | Not-determined |
| 17-109_k141_356033 | Not-determined |
| 17-109_k141_4882 | Not-determined |
| 17-109_k141_323216 | Not-determined |
| 17-109_k141_38793 | Not-determined |
| 17-109_k141_390178 | Not-determined |
| 17-109_k141_38795 | Not-determined |
| 17-109_k141_255793 | Not-determined |
| 17-109_k141_323220 | Not-determined |
| 17-109_k141_255794 | Not-determined |
| 17-109_k141_323221 | Not-determined |
| 17-109_k141_306886 | Not-determined |
| 17-109_k141_222402 | Low-quality |
| 17-109_k141_155656 | Not-determined |
| 17-109_k141_339577 | Not-determined |
| 17-109_k141_222407 | Not-determined |
| 17-109_k141_172225 | Not-determined |
| 17-109_k141_272868 | Not-determined |
| 17-109_k141_323229 | Not-determined |
| 17-109_k141_72586 | Low-quality |
| 17-109_k141_272871 | Not-determined |
| 17-109_k141_255811 | Not-determined |
| 17-109_k141_88543 | Not-determined |
| 17-109_k141_205631 | Not-determined |
| 17-109_k141_21741 | Not-determined |
| 17-109_k141_205633 | Low-quality |
| 17-109_k141_155665 | Not-determined |
| 17-109_k141_21743 | Not-determined |
| 17-109_k141_289958 | Not-determined |
| 17-109_k141_72593 | Not-determined |
| 17-109_k141_239379 | Not-determined |
| 17-109_k141_323234 | Low-quality |
| 17-109_k141_21749 | Not-determined |
| 17-109_k141_323235 | Not-determined |
| 17-109_k141_390186 | Not-determined |
| 17-109_k141_306914 | Not-determined |
| 17-109_k141_155668 | Low-quality |
| 17-109_k141_88553 | Not-determined |
| 17-109_k141_356042 | Not-determined |
| 17-109_k141_390188 | Not-determined |
| 17-109_k141_339594 | Low-quality |
| 17-109_k141_72599 | Not-determined |
| 17-109_k141_88557 | Not-determined |
| 17-109_k141_4909 | Low-quality |
| 17-109_k141_72606 | Not-determined |
| 17-109_k141_189801 | Low-quality |
| 17-109_k141_189802 | Not-determined |
| 17-109_k141_189804 | Not-determined |
| 17-109_k141_356051 | Low-quality |
| 17-109_k141_306920 | Not-determined |
| 17-109_k141_239390 | Low-quality |
| 17-109_k141_390203 | Not-determined |
| 17-109_k141_356053 | Not-determined |
| 17-109_k141_21758 | Not-determined |
| 17-109_k141_4921 | Not-determined |
| 17-109_k141_189808 | Not-determined |
| 17-109_k141_189809 | Not-determined |
| 17-109_k141_4923 | Not-determined |
| 17-109_k141_4924 | Low-quality |
| 17-109_k141_88573 | Not-determined |
| 17-109_k141_38843 | Not-determined |
| 17-109_k141_21760 | Low-quality |
| 17-109_k141_4928 | Not-determined |
| 17-109_k141_72621 | Not-determined |
| 17-109_k141_105293 | Not-determined |
| 17-109_k141_172232 | Not-determined |
| 17-109_k141_272901 | Not-determined |
| 17-109_k141_255825 | Not-determined |
| 17-109_k141_390219 | Not-determined |
| 17-109_k141_189820 | Not-determined |
| 17-109_k141_189822 | Not-determined |
| 17-109_k141_339606 | Not-determined |
| 17-109_k141_339607 | Not-determined |
| 17-109_k141_289978 | Not-determined |
| 17-109_k141_255832 | Not-determined |
| 17-109_k141_172248 | Not-determined |
| 17-109_k141_272908 | Not-determined |
| 17-109_k141_105304 | Not-determined |
| 17-109_k141_88592 | Not-determined |
| 17-109_k141_289987 | Not-determined |
| 17-109_k141_72644 | Not-determined |
| 17-109_k141_272914 | Not-determined |
| 17-109_k141_4937 | Not-determined |
| 17-109_k141_4939 | Not-determined |
| 17-109_k141_255839 | Not-determined |
| 17-109_k141_272916 | Low-quality |
| 17-109_k141_255842 | Not-determined |
| 17-109_k141_155685 | Not-determined |
| 17-109_k141_72655 | Not-determined |
| 17-109_k141_105315 | Not-determined |
| 17-109_k141_72658 | Not-determined |
| 17-109_k141_390228 | Not-determined |
| 17-109_k141_272921 | Not-determined |
| 17-109_k141_339611 | Not-determined |
| 17-109_k141_72661 | Not-determined |
| 17-109_k141_222423 | Not-determined |
| 17-109_k141_390240 | Not-determined |
| 17-109_k141_21798 | Not-determined |
| 17-109_k141_323265 | Not-determined |
| 17-109_k141_323269 | Not-determined |
| 17-109_k141_339628 | Not-determined |
| 17-109_k141_323270 | Not-determined |
| 17-109_k141_290025 | Not-determined |
| 17-109_k141_205685 | Not-determined |
| 17-109_k141_121499 | Not-determined |
| 17-109_k141_139135 | Not-determined |
| 17-109_k141_306987 | Not-determined |
| 17-109_k141_306988 | Not-determined |
| 17-109_k141_139139 | Not-determined |
| 17-109_k141_222442 | Low-quality |
| 17-109_k141_172267 | Not-determined |
| 17-109_k141_306995 | Not-determined |
| 17-109_k141_172270 | Not-determined |
| 17-109_k141_222445 | Not-determined |
| 17-109_k141_323279 | Not-determined |
| 17-109_k141_255873 | Not-determined |
| 17-109_k141_222454 | Not-determined |
| 17-109_k141_222455 | Not-determined |
| 17-109_k141_4972 | Not-determined |
| 17-109_k141_21823 | Not-determined |
| 17-109_k141_222470 | Not-determined |
| 17-109_k141_205702 | Low-quality |
| 17-109_k141_172276 | Not-determined |
| 17-109_k141_222472 | Not-determined |
| 17-109_k141_323299 | Not-determined |
| 17-109_k141_121525 | Not-determined |
| 17-109_k141_105355 | Not-determined |
| 17-109_k141_72685 | Not-determined |
| 17-109_k141_88621 | Not-determined |
| 17-109_k141_172288 | Not-determined |
| 17-109_k141_390274 | Not-determined |
| 17-109_k141_88623 | Not-determined |
| 17-109_k141_323311 | Not-determined |
| 17-109_k141_88625 | Not-determined |
| 17-109_k141_72687 | Not-determined |
| 17-109_k141_4980 | Not-determined |
| 17-109_k141_307010 | Not-determined |
| 17-109_k141_4985 | Not-determined |
| 17-109_k141_4987 | Not-determined |
| 17-109_k141_4992 | Low-quality |
| 17-109_k141_222487 | Not-determined |
| 17-109_k141_255892 | Not-determined |
| 17-109_k141_72701 | Not-determined |
| 17-109_k141_172293 | Not-determined |
| 17-109_k141_88640 | Not-determined |
| 17-109_k141_205717 | Low-quality |
| 17-109_k141_290041 | Not-determined |
| 17-109_k141_255901 | Not-determined |
| 17-109_k141_4999 | Not-determined |
| 17-109_k141_255904 | Low-quality |
| 17-109_k141_222498 | Not-determined |
| 17-109_k141_72707 | Not-determined |
| 17-109_k141_272969 | Not-determined |
| 17-109_k141_390287 | Not-determined |
| 17-109_k141_222501 | Not-determined |
| 17-109_k141_307032 | Not-determined |
| 17-109_k141_307034 | Not-determined |
| 17-109_k141_272973 | Not-determined |
| 17-109_k141_155750 | Not-determined |
| 17-109_k141_155751 | Not-determined |
| 17-109_k141_272979 | Not-determined |
| 17-109_k141_88651 | Not-determined |
| 17-109_k141_307049 | Not-determined |
| 17-109_k141_307050 | Not-determined |
| 17-109_k141_189883 | Not-determined |
| 17-109_k141_390299 | Low-quality |
| 17-109_k141_390304 | Not-determined |
| 17-109_k141_105384 | Not-determined |
| 17-109_k141_72729 | Not-determined |
| 17-109_k141_21832 | Not-determined |
| 17-109_k141_21833 | Not-determined |
| 17-109_k141_307062 | Not-determined |
| 17-109_k141_222527 | Low-quality |
| 17-109_k141_72739 | Not-determined |
| 17-109_k141_390313 | Not-determined |
| 17-109_k141_21846 | Not-determined |
| 17-109_k141_390314 | Not-determined |
| 17-109_k141_222531 | Not-determined |
| 17-109_k141_121544 | Not-determined |
| 17-109_k141_21853 | Not-determined |
| 17-109_k141_21860 | Not-determined |
| 17-109_k141_88676 | Low-quality |
| 17-109_k141_105408 | Not-determined |
| 17-109_k141_189914 | Not-determined |
| 17-109_k141_72754 | Not-determined |
| 17-109_k141_38946 | Not-determined |
| 17-109_k141_390329 | Not-determined |
| 17-109_k141_390330 | Not-determined |
| 17-109_k141_390331 | Not-determined |
| 17-109_k141_390332 | Not-determined |
| 17-109_k141_5019 | Not-determined |
| 17-109_k141_189915 | Not-determined |
| 17-109_k141_21868 | Not-determined |
| 17-109_k141_21873 | Not-determined |
| 17-109_k141_155760 | Not-determined |
| 17-109_k141_155761 | Not-determined |
| 17-109_k141_273005 | Not-determined |
| 17-109_k141_121564 | Low-quality |
| 17-109_k141_38965 | Low-quality |
| 17-109_k141_189930 | Not-determined |
| 17-109_k141_255924 | Not-determined |
| 17-109_k141_390342 | Not-determined |
| 17-109_k141_307104 | Not-determined |
| 17-109_k141_121575 | Not-determined |
| 17-109_k141_121576 | Low-quality |
| 17-109_k141_307110 | Not-determined |
| 17-109_k141_307113 | Not-determined |
| 17-109_k141_390351 | Not-determined |
| 17-109_k141_273017 | Not-determined |
| 17-109_k141_273021 | Not-determined |
| 17-109_k141_390355 | Not-determined |
| 17-109_k141_155774 | Not-determined |
| 17-109_k141_290080 | Not-determined |
| 17-109_k141_222561 | Not-determined |
| 17-109_k141_390359 | Not-determined |
| 17-109_k141_290089 | Not-determined |
| 17-109_k141_105427 | Not-determined |
| 17-109_k141_255962 | Not-determined |
| 17-109_k141_273035 | Not-determined |
| 17-109_k141_205766 | Not-determined |
| 17-109_k141_205769 | Not-determined |
| 17-109_k141_88719 | Not-determined |
| 17-109_k141_323327 | Not-determined |
| 17-109_k141_222577 | Not-determined |
| 17-109_k141_323330 | Not-determined |
| 17-109_k141_222582 | Not-determined |
| 17-109_k141_72783 | Not-determined |
| 17-109_k141_189975 | Not-determined |
| 17-109_k141_323338 | Not-determined |
| 17-109_k141_307155 | Not-determined |
| 17-109_k141_390375 | Not-determined |
| 17-109_k141_189977 | Not-determined |
| 17-109_k141_373733 | Not-determined |
| 17-109_k141_121617 | Not-determined |
| 17-109_k141_222594 | Not-determined |
| 17-109_k141_38996 | Not-determined |
| 17-109_k141_54955 | Not-determined |
| 17-109_k141_373739 | Not-determined |
| 17-109_k141_155808 | Low-quality |
| 17-109_k141_390386 | Not-determined |
| 17-109_k141_105456 | Low-quality |
| 17-109_k141_189983 | Not-determined |
| 17-109_k141_189984 | Not-determined |
| 17-109_k141_172404 | Not-determined |
| 17-109_k141_239436 | Not-determined |
| 17-109_k141_189990 | Not-determined |
| 17-109_k141_239437 | Not-determined |
| 17-109_k141_72812 | Not-determined |
| 17-109_k141_155817 | Not-determined |
| 17-109_k141_155819 | Not-determined |
| 17-109_k141_21929 | Not-determined |
| 17-109_k141_155822 | Not-determined |
| 17-109_k141_273075 | Not-determined |
| 17-109_k141_172418 | Not-determined |
| 17-109_k141_256004 | Low-quality |
| 17-109_k141_307174 | Not-determined |
| 17-109_k141_256005 | Not-determined |
| 17-109_k141_373761 | Low-quality |
| 17-109_k141_373762 | Not-determined |
| 17-109_k141_307176 | Low-quality |
| 17-109_k141_54974 | Not-determined |
| 17-109_k141_39015 | Low-quality |
| 17-109_k141_390400 | Low-quality |
| 17-109_k141_155835 | Not-determined |
| 17-109_k141_256015 | Not-determined |
| 17-109_k141_390407 | Not-determined |
| 17-109_k141_256017 | Not-determined |
| 17-109_k141_290159 | Not-determined |
| 17-109_k141_373772 | Not-determined |
| 17-109_k141_155841 | Not-determined |
| 17-109_k141_290169 | Not-determined |
| 17-109_k141_190014 | Not-determined |
| 17-109_k141_190017 | Not-determined |
| 17-109_k141_39033 | Not-determined |
| 17-109_k141_290180 | Not-determined |
| 17-109_k141_39040 | Not-determined |
| 17-109_k141_339661 | Not-determined |
| 17-109_k141_88757 | Not-determined |
| 17-109_k141_21952 | Not-determined |
| 17-109_k141_105468 | Not-determined |
| 17-109_k141_88761 | Not-determined |
| 17-109_k141_88763 | Not-determined |
| 17-109_k141_39052 | Not-determined |
| 17-109_k141_256029 | Not-determined |
| 17-109_k141_323385 | Not-determined |
| 17-109_k141_190031 | Not-determined |
| 17-109_k141_172429 | Not-determined |
| 17-109_k141_172432 | Not-determined |
| 18-105-final-viral-combined_k141_62679\|\|full | High-quality |
| 17-109_k141_239473 | Not-determined |
| 17-109_k141_105481 | Not-determined |
| 17-109_k141_105486 | Not-determined |
| 17-109_k141_139219 | Not-determined |
| 17-109_k141_172439 | Low-quality |
| 17-109_k141_105488 | Not-determined |
| 17-109_k141_139222 | Not-determined |
| 17-109_k141_323394 | Not-determined |
| 17-109_k141_139225 | Not-determined |
| 17-109_k141_155866 | Not-determined |
| 17-109_k141_21973 | Not-determined |
| 17-109_k141_155869 | Low-quality |
| 17-109_k141_323407 | Not-determined |
| 17-109_k141_373797 | Not-determined |
| 17-109_k141_323411 | Not-determined |
| 17-109_k141_373799 | Not-determined |
| 17-109_k141_190048 | Not-determined |
| 17-109_k141_88780 | Not-determined |
| 17-109_k141_5050 | Not-determined |
| 17-109_k141_339683 | Low-quality |
| 17-109_k141_139240 | Not-determined |
| 17-109_k141_239502 | Not-determined |
| 17-109_k141_190057 | Not-determined |
| 17-109_k141_190058 | Low-quality |
| 17-109_k141_105509 | Not-determined |
| 17-109_k141_190062 | Not-determined |
| 17-109_k141_105510 | Not-determined |
| 17-109_k141_190065 | Not-determined |
| 17-109_k141_256043 | Not-determined |
| 17-109_k141_172466 | Not-determined |
| 17-109_k141_390454 | Not-determined |
| 17-109_k141_105517 | Not-determined |
| 17-109_k141_105520 | Not-determined |
| 17-109_k141_155888 | Not-determined |
| 17-109_k141_273131 | Not-determined |
| 17-109_k141_373810 | Low-quality |
| 17-109_k141_273133 | Not-determined |
| 17-109_k141_339702 | Low-quality |
| 17-109_k141_155889 | Low-quality |
| 17-109_k141_390462 | Not-determined |
| 17-109_k141_139254 | Not-determined |
| 17-109_k141_39071 | Not-determined |
| 17-109_k141_273141 | Low-quality |
| 17-109_k141_239511 | Not-determined |
| 17-109_k141_21976 | Not-determined |
| 17-109_k141_273148 | Not-determined |
| 17-109_k141_239518 | Not-determined |
| 17-109_k141_105532 | Not-determined |
| 17-109_k141_121634 | Low-quality |
| 17-109_k141_390477 | Not-determined |
| 17-109_k141_139272 | Not-determined |
| 17-109_k141_290205 | Not-determined |
| 17-109_k141_172484 | Not-determined |
| 17-109_k141_72872 | Not-determined |
| 17-109_k141_72873 | Not-determined |
| 17-109_k141_290208 | Not-determined |
| 17-109_k141_105536 | Not-determined |
| 17-109_k141_105538 | Low-quality |
| 17-109_k141_290215 | Not-determined |
| 17-109_k141_273163 | Not-determined |
| 17-109_k141_273164 | Not-determined |
| 17-109_k141_190093 | Low-quality |
| 17-109_k141_88820 | Not-determined |
| 17-109_k141_190098 | Not-determined |
| 17-109_k141_5088 | Not-determined |
| 17-109_k141_273168 | Not-determined |
| 17-109_k141_121654 | Not-determined |
| 17-109_k141_290225 | Not-determined |
| 17-109_k141_339724 | Not-determined |

| | |
|---|---|
| 17-109_k141_339725 | Low-quality |
| 17-109_k141_72901 | Not-determined |
| 17-109_k141_290230 | Low-quality |
| 17-109_k141_323461 | Low-quality |
| 17-109_k141_339730 | Low-quality |
| 17-109_k141_190102 | Not-determined |
| 17-109_k141_256070 | Not-determined |
| 17-109_k141_256071 | Not-determined |
| 17-109_k141_105560 | Not-determined |
| 17-109_k141_5104 | Not-determined |
| 17-109_k141_373836 | Not-determined |
| 17-109_k141_105566 | Not-determined |
| 17-109_k141_356061 | Not-determined |
| 17-109_k141_72925 | Not-determined |
| 17-109_k141_172519 | Not-determined |
| 17-109_k141_323467 | Not-determined |
| 17-109_k141_323469 | Not-determined |
| 17-109_k141_88842 | Not-determined |
| 17-109_k141_290246 | Not-determined |
| 17-109_k141_172522 | Not-determined |
| 17-109_k141_121665 | Not-determined |
| 17-109_k141_373847 | Not-determined |
| 17-109_k141_390503 | Not-determined |
| 17-109_k141_155925 | Not-determined |
| 17-109_k141_72937 | Not-determined |
| 17-109_k141_105579 | Not-determined |
| 17-109_k141_72942 | Not-determined |
| 17-109_k141_105586 | Not-determined |
| 17-109_k141_88853 | Not-determined |
| 17-109_k141_88854 | Low-quality |
| 17-109_k141_222651 | Not-determined |
| 17-109_k141_105589 | Not-determined |
| 17-109_k141_356079 | Not-determined |
| 17-109_k141_239560 | Not-determined |
| 17-109_k141_88860 | Not-determined |
| 17-109_k141_105590 | Not-determined |
| 17-109_k141_356084 | Not-determined |
| 17-109_k141_323475 | Not-determined |
| 17-109_k141_121685 | Not-determined |
| 17-109_k141_121686 | Not-determined |
| 17-109_k141_273191 | Low-quality |
| 17-109_k141_339749 | Low-quality |
| 17-109_k141_239565 | Not-determined |
| 17-109_k141_88871 | Not-determined |
| 17-109_k141_121692 | Low-quality |
| 17-109_k141_290271 | Not-determined |
| 17-109_k141_373862 | Low-quality |
| 17-109_k141_356095 | Not-determined |
| 17-109_k141_239572 | Not-determined |
| 17-109_k141_205776 | Not-determined |
| 17-109_k141_273205 | Not-determined |
| 17-109_k141_307252 | Not-determined |
| 17-109_k141_88876 | Not-determined |
| 17-109_k141_273208 | Not-determined |
| 17-109_k141_88877 | Not-determined |
| 17-109_k141_205779 | Not-determined |
| 17-109_k141_373874 | Not-determined |
| 17-109_k141_323481 | Not-determined |
| 17-109_k141_222669 | Not-determined |
| 17-109_k141_239590 | Not-determined |
| 17-109_k141_307254 | Not-determined |
| 17-109_k141_222671 | Not-determined |
| 17-109_k141_323487 | Not-determined |
| 17-109_k141_172553 | Not-determined |
| 17-109_k141_356101 | Not-determined |
| 17-109_k141_239598 | Not-determined |
| 17-109_k141_356102 | Not-determined |
| 17-109_k141_356103 | Not-determined |
| 17-109_k141_105609 | Not-determined |
| 17-109_k141_307257 | Low-quality |
| 17-109_k141_222678 | Not-determined |
| 17-109_k141_222679 | Not-determined |
| 17-109_k141_356106 | Not-determined |
| 17-109_k141_105616 | Not-determined |
| 17-109_k141_339775 | Not-determined |
| 17-109_k141_105621 | Not-determined |
| 17-109_k141_205800 | Not-determined |
| 17-109_k141_205801 | Not-determined |
| 17-109_k141_121728 | Not-determined |
| 17-109_k141_373898 | Not-determined |
| 17-109_k141_22022 | Not-determined |
| 17-109_k141_390535 | Not-determined |
| 17-109_k141_172568 | Not-determined |
| 17-109_k141_155977 | Not-determined |
| 17-109_k141_290284 | Not-determined |
| 17-109_k141_290290 | Not-determined |
| 17-109_k141_22035 | Not-determined |
| 17-109_k141_307285 | Not-determined |
| 17-109_k141_105642 | Not-determined |
| 17-109_k141_205814 | Not-determined |
| 17-109_k141_205815 | Not-determined |
| 17-109_k141_256146 | Not-determined |
| 17-109_k141_139283 | Not-determined |
| 17-109_k141_105647 | Low-quality |
| 17-109_k141_323513 | Not-determined |
| 17-109_k141_323514 | Low-quality |
| 17-109_k141_339794 | Not-determined |
| 17-109_k141_105655 | Not-determined |
| 17-109_k141_390540 | Not-determined |
| 17-109_k141_139284 | Not-determined |
| 17-109_k141_205816 | Not-determined |
| 17-109_k141_155996 | Not-determined |
| 17-109_k141_22045 | Not-determined |
| 17-109_k141_323531 | Not-determined |
| 17-109_k141_139292 | Not-determined |
| 17-109_k141_121744 | Not-determined |
| 17-109_k141_156012 | Low-quality |
| 17-109_k141_307292 | Not-determined |
| 17-109_k141_390553 | Low-quality |
| 17-109_k141_156016 | Not-determined |
| 17-109_k141_72972 | Not-determined |
| 17-109_k141_307301 | Not-determined |
| 17-109_k141_323537 | Not-determined |
| 17-109_k141_121753 | Not-determined |
| 17-109_k141_121757 | Low-quality |
| 17-109_k141_121758 | Not-determined |
| 17-109_k141_307308 | Not-determined |
| 17-109_k141_323547 | Not-determined |
| 17-109_k141_323548 | Not-determined |
| 17-109_k141_323550 | Low-quality |
| 17-109_k141_172606 | Not-determined |
| 17-109_k141_239623 | Not-determined |
| 17-109_k141_273212 | Not-determined |
| 17-109_k141_323563 | Low-quality |
| 17-109_k141_273220 | Not-determined |
| 17-109_k141_22074 | Not-determined |
| 17-109_k141_323568 | Not-determined |
| 17-109_k141_307330 | Low-quality |
| 17-109_k141_222740 | Not-determined |
| 17-109_k141_222741 | Not-determined |
| 17-109_k141_273227 | Not-determined |
| 17-109_k141_156057 | Not-determined |
| 17-109_k141_273232 | Not-determined |
| 17-109_k141_290353 | Not-determined |
| 17-109_k141_172624 | Not-determined |
| 17-109_k141_156063 | Not-determined |
| 17-109_k141_390576 | Not-determined |
| 17-109_k141_156065 | Not-determined |
| 17-109_k141_121782 | Low-quality |
| 17-109_k141_290357 | Not-determined |
| 17-109_k141_239648 | Not-determined |
| 17-109_k141_239650 | Not-determined |
| 17-109_k141_239651 | Not-determined |
| 17-109_k141_273243 | Not-determined |
| 17-109_k141_88896 | Not-determined |
| 17-109_k141_273247 | Not-determined |
| 17-109_k141_323588 | Not-determined |
| 17-109_k141_323589 | Not-determined |
| 17-109_k141_156073 | Not-determined |
| 17-109_k141_88902 | Not-determined |
| 17-109_k141_290372 | Not-determined |
| 17-109_k141_273264 | Not-determined |
| 17-109_k141_121787 | Not-determined |
| 17-109_k141_307336 | Not-determined |
| 17-109_k141_72996 | Not-determined |
| 17-109_k141_290378 | Low-quality |
| 17-109_k141_307338 | Not-determined |
| 17-109_k141_190166 | Not-determined |
| 17-109_k141_22093 | Not-determined |
| 17-109_k141_22100 | Not-determined |
| 17-109_k141_73004 | Not-determined |
| 17-109_k141_105682 | Not-determined |
| 17-109_k141_256204 | Not-determined |
| 17-109_k141_22104 | Not-determined |
| 17-109_k141_307350 | Not-determined |
| 17-109_k141_22108 | Not-determined |
| 17-109_k141_22111 | Not-determined |
| 17-109_k141_205842 | Not-determined |
| 17-109_k141_205843 | Not-determined |
| 17-109_k141_390589 | Not-determined |
| 17-109_k141_88920 | Not-determined |
| 17-109_k141_273280 | Not-determined |
| 17-109_k141_273281 | Not-determined |
| 17-109_k141_172640 | Not-determined |
| 17-109_k141_390597 | Low-quality |
| 17-109_k141_22125 | Not-determined |
| 17-109_k141_88927 | Not-determined |
| 17-109_k141_121825 | Not-determined |
| 17-109_k141_205863 | Not-determined |
| 17-109_k141_121827 | Not-determined |
| 17-109_k141_205867 | Not-determined |
| 17-109_k141_256212 | Not-determined |
| 17-109_k141_172648 | Not-determined |
| 17-109_k141_256220 | Not-determined |
| 17-109_k141_172656 | Not-determined |
| 17-109_k141_205871 | Not-determined |
| 17-109_k141_172660 | Not-determined |
| 17-109_k141_172663 | Not-determined |
| 17-109_k141_88933 | Not-determined |
| 17-109_k141_390619 | Not-determined |
| 17-109_k141_390625 | Not-determined |
| 17-109_k141_73020 | Not-determined |
| 17-109_k141_156098 | Not-determined |
| 17-109_k141_256239 | Not-determined |
| 17-109_k141_88941 | Not-determined |
| 17-109_k141_307387 | Not-determined |
| 17-109_k141_172669 | Not-determined |
| 17-109_k141_222810 | Not-determined |
| 17-109_k141_121845 | Not-determined |
| 17-109_k141_307388 | Not-determined |
| 17-109_k141_121850 | Not-determined |
| 17-109_k141_88947 | Low-quality |
| 17-109_k141_172677 | Not-determined |
| 17-109_k141_172678 | Not-determined |
| 17-109_k141_172680 | Not-determined |
| 17-109_k141_390635 | Not-determined |
| 17-109_k141_55017 | Not-determined |
| 17-109_k141_222832 | Not-determined |
| 17-109_k141_55020 | Not-determined |
| 17-109_k141_273314 | Not-determined |
| 17-109_k141_222836 | Not-determined |
| 17-109_k141_55027 | Not-determined |
| 17-109_k141_273322 | Not-determined |
| 17-109_k141_390649 | Not-determined |
| 17-109_k141_390651 | Not-determined |
| 17-109_k141_55034 | Not-determined |
| 17-109_k141_190203 | Not-determined |
| 17-109_k141_190207 | Not-determined |
| 17-109_k141_390661 | Not-determined |
| 17-109_k141_55044 | Not-determined |
| 17-109_k141_205907 | Not-determined |
| 17-109_k141_190215 | Not-determined |
| 17-109_k141_390666 | Not-determined |
| 17-109_k141_55054 | Not-determined |
| 17-109_k141_139359 | Not-determined |
| 17-109_k141_88983 | Not-determined |
| 17-109_k141_156153 | Not-determined |
| 17-109_k141_339804 | Not-determined |
| 17-109_k141_339809 | Not-determined |
| 17-109_k141_172691 | Not-determined |
| 17-109_k141_172693 | Not-determined |
| 17-109_k141_39122 | Not-determined |
| 17-109_k141_307410 | Not-determined |
| 17-109_k141_156166 | Not-determined |
| 17-109_k141_39128 | Not-determined |
| 17-109_k141_307419 | Not-determined |
| 17-109_k141_139378 | Not-determined |
| 17-109_k141_39131 | Low-quality |
| 17-109_k141_139381 | Not-determined |
| 17-109_k141_290440 | Not-determined |
| 17-109_k141_307434 | Not-determined |
| 17-109_k141_390686 | Not-determined |
| 17-109_k141_139387 | Not-determined |
| 17-109_k141_390687 | Not-determined |
| 17-109_k141_307437 | Not-determined |
| 17-109_k141_55079 | Not-determined |
| 17-109_k141_39147 | Not-determined |
| 17-109_k141_55081 | Not-determined |
| 17-109_k141_323690 | Not-determined |
| 17-109_k141_55082 | Not-determined |
| 17-109_k141_89003 | Not-determined |
| 17-109_k141_290448 | Not-determined |
| 17-109_k141_390704 | Not-determined |
| 17-109_k141_323697 | Not-determined |
| 17-109_k141_55097 | Not-determined |
| 17-109_k141_290454 | Not-determined |
| 17-109_k141_5115 | Not-determined |
| 17-109_k141_121885 | Not-determined |
| 17-109_k141_39162 | Not-determined |
| 17-109_k141_273357 | Not-determined |
| 17-109_k141_256291 | Not-determined |
| 17-109_k141_139420 | Not-determined |
| 17-109_k141_73045 | Not-determined |
| 17-109_k141_339822 | Not-determined |
| 17-109_k141_156198 | Not-determined |
| 17-109_k141_273365 | Not-determined |
| 17-109_k141_5134 | Not-determined |
| 17-109_k141_55122 | Not-determined |
| 17-109_k141_273366 | Not-determined |
| 17-109_k141_222930 | Low-quality |
| 17-109_k141_390715 | Not-determined |
| 17-109_k141_307450 | Not-determined |
| 17-109_k141_390720 | Not-determined |
| 17-109_k141_390724 | Not-determined |
| 17-109_k141_73057 | Not-determined |
| 17-109_k141_205957 | Not-determined |
| 17-109_k141_156211 | Not-determined |
| 17-109_k141_55138 | Not-determined |
| 17-109_k141_390731 | Not-determined |
| 17-109_k141_273376 | Not-determined |
| 17-109_k141_39177 | Not-determined |
| 17-109_k141_339839 | Not-determined |
| 17-109_k141_356187 | Not-determined |
| 17-109_k141_323741 | Not-determined |
| 17-109_k141_39182 | Not-determined |
| 17-109_k141_323745 | Not-determined |
| 17-109_k141_290460 | Not-determined |
| 17-109_k141_323751 | Not-determined |
| 17-109_k141_273385 | Not-determined |
| 17-109_k141_290464 | Low-quality |
| 17-109_k141_339857 | Not-determined |
| 17-109_k141_323754 | Not-determined |
| 17-109_k141_5151 | Not-determined |
| 17-109_k141_239774 | Not-determined |
| 17-109_k141_39200 | Not-determined |
| 17-109_k141_139456 | Not-determined |
| 17-109_k141_156228 | Not-determined |
| 17-109_k141_121921 | Not-determined |
| 17-109_k141_222949 | Not-determined |
| 17-109_k141_39214 | Not-determined |
| 17-109_k141_222951 | Not-determined |
| 17-109_k141_39217 | Not-determined |
| 17-109_k141_290474 | Low-quality |
| 17-109_k141_222958 | Not-determined |
| 17-109_k141_356216 | Not-determined |
| 17-109_k141_307471 | Not-determined |
| 17-109_k141_390745 | Not-determined |
| 17-109_k141_172736 | Low-quality |
| 17-109_k141_5180 | Low-quality |
| 17-109_k141_5181 | Low-quality |
| 17-109_k141_222969 | Not-determined |
| 17-109_k141_307482 | Low-quality |
| 17-109_k141_323771 | Not-determined |
| 17-109_k141_290504 | Not-determined |
| 17-109_k141_323773 | Not-determined |
| 17-109_k141_89035 | Not-determined |
| 17-109_k141_172754 | Not-determined |
| 17-109_k141_290516 | Not-determined |
| 17-109_k141_89037 | Not-determined |
| 17-109_k141_89039 | Not-determined |
| 17-109_k141_323785 | Not-determined |
| 17-109_k141_222981 | Not-determined |
| 17-109_k141_323787 | Not-determined |
| 17-109_k141_139474 | Not-determined |
| 17-109_k141_273387 | Not-determined |
| 17-109_k141_273388 | Not-determined |
| 17-109_k141_5190 | Not-determined |
| 17-109_k141_323791 | Not-determined |
| 17-109_k141_373970 | Low-quality |
| 17-109_k141_239818 | Not-determined |
| 17-109_k141_5198 | Not-determined |
| 17-109_k141_5200 | Not-determined |
| 17-109_k141_323802 | Not-determined |
| 17-109_k141_190307 | Not-determined |
| 17-109_k141_222996 | Low-quality |
| 17-109_k141_190308 | Not-determined |
| 17-109_k141_121959 | Low-quality |
| 17-109_k141_222999 | Low-quality |
| 17-109_k141_273404 | Not-determined |
| 17-109_k141_22179 | Not-determined |
| 17-109_k141_172770 | Not-determined |
| 17-109_k141_390754 | Not-determined |
| 17-109_k141_390757 | Not-determined |
| 17-109_k141_273407 | Not-determined |
| 17-109_k141_273412 | Not-determined |
| 17-109_k141_5206 | Not-determined |
| 17-109_k141_55184 | Not-determined |
| 17-109_k141_55185 | Not-determined |
| 17-109_k141_390767 | Not-determined |
| 17-109_k141_323819 | Not-determined |
| 17-109_k141_121977 | Low-quality |
| 17-109_k141_223017 | Not-determined |
| 17-109_k141_39259 | Not-determined |
| 17-109_k141_339891 | Not-determined |
| 17-109_k141_156241 | Not-determined |
| 17-109_k141_307488 | Not-determined |
| 17-109_k141_156242 | Not-determined |
| 17-109_k141_39262 | Not-determined |
| 17-109_k141_256306 | Not-determined |
| 17-109_k141_223022 | Not-determined |
| 17-109_k141_223025 | Not-determined |
| 17-109_k141_156248 | Not-determined |
| 17-109_k141_139483 | Not-determined |
| 17-109_k141_22183 | Not-determined |
| 17-109_k141_89077 | Not-determined |
| 17-109_k141_22185 | Not-determined |
| 17-109_k141_22187 | Not-determined |
| 17-109_k141_373984 | Low-quality |
| 17-109_k141_55204 | Not-determined |
| 17-109_k141_172793 | Not-determined |
| 17-109_k141_55206 | Not-determined |
| 17-109_k141_323836 | Not-determined |
| 17-109_k141_39282 | Low-quality |
| 17-109_k141_339910 | Not-determined |
| 17-109_k141_390778 | Not-determined |
| 17-109_k141_323838 | Low-quality |
| 17-109_k141_139490 | Low-quality |
| 17-109_k141_22200 | Not-determined |
| 17-109_k141_39288 | Not-determined |
| 17-109_k141_89092 | Not-determined |
| 17-109_k141_223032 | Not-determined |
| 17-109_k141_390779 | Not-determined |
| 17-109_k141_256324 | Not-determined |
| 17-109_k141_172810 | Not-determined |
| 17-109_k141_290542 | Not-determined |
| 17-109_k141_256326 | Not-determined |
| 17-109_k141_172815 | Not-determined |
| 17-109_k141_55215 | Not-determined |
| 17-109_k141_190354 | Not-determined |
| 17-109_k141_55217 | Not-determined |
| 17-109_k141_390789 | Not-determined |
| 17-109_k141_190360 | Not-determined |
| 17-109_k141_323871 | Not-determined |
| 17-109_k141_22216 | Not-determined |
| 17-109_k141_390790 | Not-determined |
| 17-109_k141_172821 | Not-determined |
| 17-109_k141_73062 | Not-determined |
| 17-109_k141_273437 | Not-determined |
| 17-109_k141_373990 | Not-determined |
| 17-109_k141_89112 | Not-determined |
| 17-109_k141_5231 | Not-determined |
| 17-109_k141_105716 | Not-determined |
| 17-109_k141_122032 | Not-determined |
| 17-109_k141_172831 | Not-determined |
| 17-109_k141_73072 | Not-determined |
| 17-109_k141_5237 | Not-determined |
| 17-109_k141_339933 | Not-determined |
| 17-109_k141_339936 | Not-determined |
| 17-109_k141_105723 | Not-determined |
| 17-109_k141_323886 | Not-determined |
| 17-109_k141_55246 | Not-determined |
| 17-109_k141_307545 | Low-quality |
| 17-109_k141_390813 | Not-determined |
| 17-109_k141_390816 | Low-quality |
| 17-109_k141_374000 | Low-quality |
| 17-109_k141_172842 | Not-determined |
| 17-109_k141_374004 | Not-determined |
| 17-109_k141_374005 | Not-determined |
| 17-109_k141_273458 | Not-determined |
| 17-109_k141_374007 | Not-determined |
| 17-109_k141_307555 | Not-determined |
| 17-109_k141_290556 | Not-determined |
| 17-109_k141_156294 | Not-determined |
| 17-109_k141_307556 | Not-determined |
| 17-109_k141_5243 | Not-determined |
| 17-109_k141_307568 | Not-determined |
| 17-109_k141_256347 | Low-quality |
| 17-109_k141_256351 | Not-determined |
| 17-109_k141_5253 | Not-determined |
| 17-109_k141_5258 | Not-determined |
| 17-109_k141_390843 | Not-determined |
| 17-109_k141_390846 | Not-determined |
| 17-109_k141_290563 | Not-determined |
| 17-109_k141_5268 | Not-determined |
| 17-109_k141_5269 | Not-determined |
| 17-109_k141_55253 | Not-determined |
| 17-109_k141_190428 | Not-determined |
| 17-109_k141_206021 | Low-quality |
| 17-109_k141_307591 | Not-determined |
| 17-109_k141_55257 | Not-determined |
| 17-109_k141_307592 | Not-determined |
| 17-109_k141_256369 | Not-determined |
| 17-109_k141_390851 | Low-quality |
| 17-109_k141_190434 | Not-determined |
| 17-109_k141_390856 | Not-determined |
| 17-109_k141_89131 | Not-determined |
| 17-109_k141_55259 | Not-determined |
| 17-109_k141_339988 | Not-determined |
| 17-109_k141_139524 | Not-determined |
| 17-109_k141_223053 | Not-determined |
| 17-109_k141_239890 | Not-determined |
| 17-109_k141_206028 | Not-determined |
| 17-109_k141_190440 | Not-determined |
| 17-109_k141_390869 | Not-determined |
| 17-109_k141_290583 | Not-determined |
| 17-109_k141_156311 | Low-quality |
| 17-109_k141_273481 | Not-determined |
| 17-109_k141_105771 | Not-determined |
| 17-109_k141_239898 | Not-determined |
| 17-109_k141_89146 | Not-determined |
| 17-109_k141_206033 | Not-determined |
| 17-109_k141_156319 | Not-determined |
| 17-109_k141_139537 | Not-determined |
| 17-109_k141_239902 | Not-determined |
| 17-109_k141_156323 | Not-determined |
| 17-109_k141_156326 | Not-determined |
| 17-109_k141_256380 | Not-determined |
| 17-109_k141_156330 | Not-determined |
| 17-109_k141_339995 | Not-determined |
| 17-109_k141_139545 | Not-determined |
| 17-109_k141_190462 | Not-determined |
| 17-109_k141_139548 | Not-determined |
| 17-109_k141_73102 | Not-determined |
| 17-109_k141_190463 | Not-determined |
| 17-109_k141_374019 | Not-determined |
| 17-109_k141_139552 | Low-quality |
| 17-109_k141_139553 | Not-determined |
| 17-109_k141_139555 | Not-determined |
| 17-109_k141_190473 | Not-determined |
| 17-109_k141_374027 | Not-determined |
| 17-109_k141_73108 | Not-determined |
| 17-109_k141_307621 | Low-quality |
| 17-109_k141_256402 | Not-determined |
| 17-109_k141_105778 | Not-determined |
| 17-109_k141_290607 | Not-determined |
| 17-109_k141_190478 | Not-determined |
| 17-109_k141_5295 | Not-determined |
| 17-109_k141_339999 | Low-quality |
| 17-109_k141_256407 | Not-determined |
| 17-109_k141_156355 | Not-determined |
| 17-109_k141_256408 | Not-determined |
| 17-109_k141_190484 | Not-determined |
| 17-109_k141_374037 | Not-determined |
| 17-109_k141_73117 | Not-determined |
| 17-109_k141_273504 | Not-determined |
| 17-109_k141_273505 | Not-determined |
| 17-109_k141_5303 | Not-determined |
| 17-109_k141_55288 | Not-determined |
| 17-109_k141_206074 | Not-determined |
| 17-109_k141_190497 | Not-determined |
| 17-109_k141_273515 | Not-determined |
| 17-109_k141_55292 | Not-determined |
| 17-109_k141_374049 | Low-quality |
| 17-109_k141_290622 | Not-determined |
| 17-109_k141_55294 | Not-determined |
| 17-109_k141_273524 | Not-determined |
| 17-109_k141_55295 | Not-determined |
| 17-109_k141_239926 | Not-determined |
| 17-109_k141_172861 | Not-determined |
| 17-109_k141_239931 | Not-determined |
| 17-109_k141_89168 | Not-determined |
| 17-109_k141_55300 | Not-determined |
| 17-109_k141_356253 | Low-quality |
| 17-109_k141_105794 | Not-determined |
| 17-109_k141_273536 | Not-determined |
| 17-109_k141_273539 | Not-determined |
| 17-109_k141_190512 | Not-determined |
| 17-109_k141_89172 | Not-determined |
| 17-109_k141_22288 | Not-determined |
| 17-109_k141_273545 | Low-quality |
| 17-109_k141_273547 | Not-determined |
| 17-109_k141_89174 | Low-quality |
| 17-109_k141_307638 | Not-determined |
| 17-109_k141_273550 | Not-determined |
| 17-109_k141_5322 | Not-determined |
| 17-109_k141_206097 | Not-determined |
| 17-109_k141_374053 | Not-determined |
| 17-109_k141_273555 | Not-determined |
| 17-109_k141_239944 | Not-determined |
| 17-109_k141_273556 | Not-determined |
| 17-109_k141_206100 | Low-quality |
| 17-109_k141_190529 | Not-determined |
| 17-109_k141_356276 | Not-determined |
| 17-109_k141_139578 | Not-determined |
| 17-109_k141_5337 | Not-determined |
| 17-109_k141_22304 | Not-determined |
| 17-109_k141_356281 | Not-determined |
| 17-109_k141_356282 | Not-determined |
| 17-109_k141_290647 | Not-determined |
| 17-109_k141_172875 | Not-determined |
| 17-109_k141_307655 | Not-determined |
| 17-109_k141_73152 | Not-determined |
| 17-109_k141_206114 | Not-determined |
| 17-109_k141_156374 | Not-determined |
| 17-109_k141_139593 | Not-determined |
| 17-109_k141_290656 | Low-quality |
| 17-109_k141_356293 | Not-determined |
| 17-109_k141_55306 | Not-determined |
| 17-109_k141_139599 | Low-quality |
| 17-109_k141_356295 | Low-quality |
| 17-109_k141_307665 | Not-determined |
| 17-109_k141_206120 | Not-determined |
| 17-109_k141_307668 | Not-determined |
| 17-109_k141_206122 | Not-determined |
| 17-109_k141_206123 | Not-determined |
| 17-109_k141_55315 | Not-determined |
| 17-109_k141_156385 | Not-determined |
| 17-109_k141_139604 | Not-determined |
| 17-109_k141_390908 | Not-determined |
| 17-109_k141_156391 | Not-determined |
| 17-109_k141_55327 | Not-determined |
| 17-109_k141_290667 | Not-determined |
| 17-109_k141_55331 | Not-determined |
| 17-109_k141_307679 | Low-quality |
| 17-109_k141_156399 | Not-determined |
| 17-109_k141_89209 | Low-quality |
| 17-109_k141_122081 | Low-quality |
| 17-109_k141_22308 | Not-determined |
| 17-109_k141_290674 | Not-determined |
| 17-109_k141_374076 | Not-determined |
| 17-109_k141_172887 | Low-quality |
| 17-109_k141_156402 | Not-determined |
| 17-109_k141_290677 | Not-determined |
| 17-109_k141_105838 | Not-determined |
| 17-109_k141_105839 | Not-determined |
| 17-109_k141_374082 | Not-determined |
| 17-109_k141_307688 | Not-determined |
| 17-109_k141_139623 | Low-quality |
| 17-109_k141_105844 | Not-determined |
| 17-109_k141_356329 | Not-determined |
| 17-109_k141_356331 | Not-determined |
| 17-109_k141_256466 | Not-determined |
| 17-109_k141_73169 | Not-determined |
| 17-109_k141_122099 | Not-determined |
| 17-109_k141_5375 | Not-determined |
| 17-109_k141_356338 | Not-determined |
| 17-109_k141_307696 | Not-determined |
| 17-109_k141_307697 | Not-determined |
| 17-109_k141_156414 | Not-determined |

| | |
|---|---|
| 17-109_k141_139636 | Not-determined |
| 17-109_k141_105860 | Not-determined |
| 17-109_k141_256480 | Not-determined |
| 17-109_k141_356344 | Not-determined |
| 17-109_k141_374101 | Not-determined |
| 17-109_k141_273573 | Not-determined |
| 17-109_k141_139641 | Not-determined |
| 17-109_k141_172905 | Not-determined |
| 17-109_k141_172906 | Not-determined |
| 17-109_k141_139646 | Not-determined |
| 17-109_k141_73193 | Not-determined |
| 17-109_k141_156427 | Not-determined |
| 17-109_k141_139650 | Not-determined |
| 17-109_k141_122119 | Not-determined |
| 17-109_k141_22330 | Low-quality |
| 17-109_k141_156435 | Not-determined |
| 17-109_k141_374114 | Not-determined |
| 17-109_k141_73197 | Not-determined |
| 17-109_k141_273593 | Not-determined |
| 17-109_k141_206184 | Not-determined |
| 17-109_k141_374118 | Low-quality |
| 17-109_k141_22340 | Not-determined |
| 17-109_k141_22345 | Not-determined |
| 17-109_k141_22347 | Not-determined |
| 17-109_k141_190561 | Low-quality |
| 17-109_k141_139659 | Not-determined |
| 17-109_k141_307710 | Not-determined |
| 17-109_k141_374124 | Not-determined |
| 17-109_k141_22350 | Not-determined |
| 17-109_k141_172937 | Not-determined |
| 17-109_k141_307715 | Not-determined |
| 17-109_k141_240024 | Not-determined |
| 17-109_k141_374133 | Not-determined |
| 17-109_k141_374134 | Not-determined |
| 17-109_k141_172948 | Not-determined |
| 17-109_k141_390916 | Not-determined |
| 17-109_k141_156461 | Not-determined |
| 17-109_k141_240033 | Not-determined |
| 17-109_k141_307728 | Not-determined |
| 17-109_k141_156466 | Not-determined |
| 17-109_k141_5397 | Not-determined |
| 17-109_k141_55355 | Low-quality |
| 17-109_k141_206216 | Low-quality |
| 17-109_k141_273613 | Low-quality |
| 17-109_k141_156471 | Not-determined |
| 17-109_k141_323990 | Not-determined |
| 17-109_k141_156478 | Not-determined |
| 17-109_k141_273615 | Not-determined |
| 17-109_k141_73221 | Not-determined |
| 17-109_k141_73222 | Not-determined |
| 17-109_k141_105904 | Not-determined |
| 17-109_k141_307740 | Low-quality |
| 17-109_k141_307741 | Low-quality |
| 17-109_k141_190595 | Not-determined |
| 17-109_k141_206232 | Not-determined |
| 17-109_k141_273628 | Not-determined |
| 17-109_k141_5412 | Not-determined |
| 17-109_k141_55369 | Low-quality |
| 17-109_k141_374153 | Not-determined |
| 17-109_k141_139675 | Not-determined |
| 17-109_k141_22385 | Not-determined |
| 17-109_k141_206235 | Low-quality |
| 17-109_k141_105919 | Not-determined |
| 17-109_k141_73236 | Not-determined |
| 17-109_k141_223056 | Not-determined |
| 17-109_k141_105924 | Not-determined |
| 17-109_k141_374154 | Low-quality |
| 17-109_k141_139684 | Not-determined |
| 17-109_k141_105934 | Not-determined |
| 17-109_k141_290714 | Low-quality |
| 17-109_k141_256502 | Not-determined |
| 17-109_k141_290720 | Not-determined |
| 17-109_k141_5436 | Low-quality |
| 17-109_k141_256505 | Not-determined |
| 17-109_k141_5440 | Not-determined |
| 17-109_k141_5441 | Not-determined |
| 17-109_k141_5442 | Not-determined |
| 17-109_k141_390941 | Not-determined |
| 17-109_k141_390943 | Not-determined |
| 17-109_k141_5446 | Low-quality |
| 17-109_k141_290729 | Not-determined |
| 17-109_k141_190632 | Not-determined |
| 17-109_k141_22408 | Not-determined |
| 17-109_k141_105955 | Not-determined |
| 17-109_k141_22411 | Not-determined |
| 17-109_k141_374189 | Not-determined |
| 17-109_k141_256513 | Not-determined |
| 17-109_k141_22415 | Low-quality |
| 17-109_k141_223071 | Not-determined |
| 17-109_k141_22421 | Not-determined |
| 17-109_k141_240082 | Low-quality |
| 17-109_k141_89256 | Not-determined |
| 17-109_k141_190648 | Not-determined |
| 17-109_k141_89257 | Not-determined |
| 17-109_k141_206251 | Not-determined |
| 17-109_k141_89259 | Not-determined |
| 17-109_k141_5449 | Low-quality |
| 17-109_k141_172963 | Low-quality |
| 17-109_k141_39311 | Not-determined |
| 17-109_k141_172964 | Not-determined |
| 17-109_k141_324014 | Not-determined |
| 17-109_k141_256528 | Not-determined |
| 17-109_k141_240088 | Not-determined |
| 17-109_k141_206255 | Not-determined |
| 17-109_k141_290745 | Not-determined |
| 17-109_k141_240089 | Not-determined |
| 17-109_k141_256533 | Not-determined |
| 17-109_k141_340090 | Not-determined |
| 17-109_k141_122152 | Low-quality |
| 17-109_k141_256536 | Not-determined |
| 17-109_k141_223082 | Not-determined |
| 17-109_k141_22440 | Not-determined |
| 17-109_k141_356380 | Not-determined |
| 17-109_k141_5455 | Not-determined |
| 17-109_k141_256546 | Not-determined |
| 17-109_k141_89276 | Not-determined |
| 17-109_k141_190675 | Not-determined |
| 17-109_k141_190677 | Not-determined |
| 17-109_k141_340097 | Not-determined |
| 17-109_k141_340098 | Not-determined |
| 17-109_k141_89285 | Not-determined |
| 17-109_k141_39322 | Low-quality |
| 17-109_k141_290759 | Low-quality |
| 17-109_k141_55390 | Not-determined |
| 17-109_k141_206269 | Not-determined |
| 17-109_k141_156498 | Low-quality |
| 17-109_k141_290764 | Not-determined |
| 17-109_k141_273642 | Not-determined |
| 17-109_k141_89289 | Not-determined |
| 17-109_k141_73289 | Not-determined |
| 17-109_k141_340109 | Not-determined |
| 17-109_k141_55401 | Not-determined |
| 17-109_k141_273645 | Low-quality |
| 17-109_k141_206278 | Not-determined |
| 17-109_k141_273649 | Not-determined |
| 17-109_k141_223098 | Not-determined |
| 17-109_k141_356391 | Not-determined |
| 17-109_k141_240122 | Not-determined |
| 17-109_k141_223102 | Not-determined |
| 17-109_k141_39337 | Not-determined |
| 17-109_k141_39339 | Not-determined |
| 17-109_k141_223106 | Not-determined |
| 17-109_k141_273663 | Not-determined |
| 17-109_k141_73304 | Not-determined |
| 17-109_k141_290791 | Not-determined |
| 17-109_k141_156500 | Low-quality |
| 17-109_k141_105981 | Not-determined |
| 17-109_k141_290793 | Not-determined |
| 17-109_k141_172980 | Not-determined |
| 17-109_k141_240140 | Not-determined |
| 17-109_k141_172984 | Not-determined |
| 17-109_k141_89308 | Not-determined |
| 17-109_k141_290808 | Not-determined |
| 17-109_k141_290809 | Not-determined |
| 17-109_k141_290810 | Not-determined |
| 17-109_k141_122206 | Low-quality |
| 17-109_k141_390955 | Not-determined |
| 17-109_k141_39349 | Not-determined |
| 17-109_k141_390958 | Low-quality |
| 17-109_k141_390964 | Not-determined |
| 17-109_k141_73335 | Not-determined |
| 17-109_k141_172996 | Not-determined |
| 17-109_k141_390968 | Not-determined |
| 17-109_k141_22506 | Not-determined |
| 17-109_k141_156509 | Not-determined |
| 17-109_k141_55413 | Not-determined |
| 17-109_k141_89324 | Not-determined |
| 17-109_k141_22512 | Not-determined |
| 17-109_k141_39363 | Not-determined |
| 17-109_k141_190711 | Not-determined |
| 17-109_k141_55416 | Not-determined |
| 17-109_k141_190712 | Not-determined |
| 17-109_k141_240161 | Not-determined |
| 17-109_k141_39367 | Not-determined |
| 17-109_k141_39368 | Not-determined |
| 17-109_k141_390978 | Low-quality |
| 17-109_k141_240165 | Not-determined |
| 17-109_k141_173000 | Not-determined |
| 17-109_k141_39369 | Not-determined |
| 17-109_k141_273683 | Not-determined |
| 17-109_k141_223123 | Not-determined |
| 17-109_k141_55422 | Not-determined |
| 17-109_k141_5468 | Not-determined |
| 17-109_k141_55429 | Not-determined |
| 17-109_k141_122246 | Not-determined |
| 17-109_k141_156533 | Not-determined |
| 17-109_k141_39394 | Not-determined |
| 17-109_k141_273692 | Not-determined |
| 17-109_k141_173020 | Not-determined |
| 17-109_k141_39398 | Not-determined |
| 17-109_k141_190740 | Low-quality |
| 17-109_k141_273694 | Not-determined |
| 17-109_k141_39400 | Not-determined |
| 17-109_k141_122249 | Not-determined |
| 17-109_k141_156541 | Not-determined |
| 17-109_k141_156542 | Low-quality |
| 17-109_k141_273700 | Not-determined |
| 17-109_k141_290853 | Not-determined |
| 17-109_k141_324035 | Not-determined |
| 17-109_k141_206280 | Not-determined |
| 17-109_k141_206281 | Not-determined |
| 17-109_k141_324040 | Not-determined |
| 17-109_k141_139715 | Low-quality |
| 17-109_k141_391001 | Not-determined |
| 17-109_k141_324044 | Not-determined |
| 17-109_k141_290862 | Not-determined |
| 17-109_k141_307809 | Not-determined |
| 17-109_k141_256551 | Not-determined |
| 17-109_k141_156548 | Not-determined |
| 17-109_k141_290865 | Not-determined |
| 17-109_k141_206290 | Not-determined |
| 17-109_k141_273713 | Not-determined |
| 17-109_k141_340163 | Not-determined |
| 17-109_k141_290867 | Not-determined |
| 17-109_k141_5500 | Not-determined |
| 17-109_k141_156555 | Not-determined |
| 17-109_k141_223134 | Low-quality |
| 17-109_k141_156556 | Not-determined |
| 17-109_k141_39415 | Not-determined |
| 17-109_k141_206299 | Not-determined |
| 17-109_k141_290869 | Low-quality |
| 17-109_k141_73348 | Not-determined |
| 17-109_k141_307814 | Not-determined |
| 17-109_k141_240172 | Not-determined |
| 17-109_k141_256563 | Not-determined |
| 17-109_k141_73349 | Not-determined |
| 17-109_k141_324053 | Not-determined |
| 17-109_k141_324054 | Not-determined |
| 17-109_k141_206306 | Not-determined |
| 17-109_k141_223143 | Not-determined |
| 17-109_k141_290875 | Not-determined |
| 17-109_k141_256578 | Not-determined |
| 17-109_k141_324075 | Not-determined |
| 17-109_k141_340187 | Not-determined |
| 17-109_k141_122271 | Not-determined |
| 17-109_k141_256583 | Low-quality |
| 17-109_k141_139741 | Not-determined |
| 17-109_k141_89355 | Low-quality |
| 17-109_k141_190757 | Not-determined |
| 17-109_k141_139745 | Low-quality |
| 17-109_k141_122277 | Not-determined |
| 17-109_k141_206326 | Not-determined |
| 17-109_k141_22574 | Not-determined |
| 17-109_k141_39455 | Not-determined |
| 17-109_k141_156579 | Not-determined |
| 17-109_k141_156581 | Not-determined |
| 17-109_k141_122288 | Not-determined |
| 17-109_k141_5514 | Not-determined |
| 17-109_k141_39460 | Not-determined |
| 17-109_k141_122297 | Not-determined |
| 17-109_k141_391011 | Not-determined |
| 17-109_k141_139762 | Not-determined |
| 17-109_k141_340215 | Not-determined |
| 17-109_k141_5516 | Not-determined |
| 17-109_k141_240185 | Not-determined |
| 17-109_k141_190768 | Not-determined |
| 17-109_k141_5521 | Not-determined |
| 17-109_k141_156598 | Not-determined |
| 17-109_k141_55446 | Low-quality |
| 17-109_k141_139779 | Not-determined |
| 17-109_k141_5527 | Not-determined |
| 17-109_k141_391023 | Not-determined |
| 17-109_k141_290927 | Not-determined |
| 17-109_k141_290929 | Not-determined |
| 17-109_k141_55453 | Not-determined |
| 17-109_k141_5533 | Low-quality |
| 17-109_k141_89376 | Low-quality |
| 17-109_k141_89377 | Not-determined |
| 17-109_k141_223195 | Not-determined |
| 17-109_k141_307833 | Low-quality |
| 17-109_k141_391052 | Not-determined |
| 17-109_k141_156611 | Not-determined |
| 17-109_k141_73389 | Not-determined |
| 17-109_k141_290948 | Not-determined |
| 17-109_k141_5553 | Not-determined |
| 17-109_k141_374231 | Not-determined |
| 17-109_k141_307838 | Not-determined |
| 17-109_k141_206359 | Not-determined |
| 17-109_k141_5559 | Not-determined |
| 17-109_k141_206366 | Not-determined |
| 17-109_k141_156620 | Not-determined |
| 17-109_k141_5562 | Not-determined |
| 17-109_k141_223210 | Not-determined |
| 17-109_k141_356455 | Not-determined |
| 17-109_k141_89405 | Not-determined |
| 17-109_k141_22618 | Not-determined |
| 17-109_k141_340262 | Not-determined |
| 17-109_k141_290975 | Not-determined |
| 17-109_k141_374250 | Not-determined |
| 17-109_k141_89411 | Not-determined |
| 17-109_k141_5566 | Not-determined |
| 17-109_k141_374258 | Low-quality |
| 17-109_k141_223227 | Not-determined |
| 17-109_k141_39522 | Not-determined |
| 17-109_k141_324086 | Not-determined |
| 17-109_k141_356467 | Not-determined |
| 17-109_k141_139795 | Not-determined |
| 17-109_k141_374266 | Not-determined |
| 17-109_k141_89423 | Not-determined |
| 17-109_k141_22649 | Not-determined |
| 17-109_k141_391071 | Not-determined |
| 17-108-final-viral-combined_k141_46816\|\|full | Medium-quality |
| 17-109_k141_5582 | Not-determined |
| 17-109_k141_73417 | Not-determined |
| 17-109_k141_22653 | Not-determined |
| 17-109_k141_139802 | Not-determined |
| 17-109_k141_223242 | Not-determined |
| 17-109_k141_223243 | Low-quality |
| 17-109_k141_139804 | Not-determined |
| 17-109_k141_22659 | Not-determined |
| 17-109_k141_22660 | Not-determined |
| 17-109_k141_39539 | Not-determined |
| 17-109_k141_324105 | Not-determined |
| 17-109_k141_73428 | Not-determined |
| 17-109_k141_391077 | Not-determined |
| 17-109_k141_122352 | Not-determined |
| 17-109_k141_39541 | Not-determined |
| 17-109_k141_206405 | Not-determined |
| 17-109_k141_190814 | Not-determined |
| 17-109_k141_5587 | Not-determined |
| 17-109_k141_223249 | Not-determined |
| 17-109_k141_206407 | Not-determined |
| 17-109_k141_173054 | Not-determined |
| 17-109_k141_73436 | Not-determined |
| 17-109_k141_223253 | Not-determined |
| 17-109_k141_223254 | Not-determined |
| 17-109_k141_391086 | Not-determined |
| 17-109_k141_206417 | Not-determined |
| 17-109_k141_223261 | Not-determined |
| 17-109_k141_240246 | Not-determined |
| 17-109_k141_39560 | Not-determined |
| 17-109_k141_156653 | Not-determined |
| 17-109_k141_307848 | Not-determined |
| 17-109_k141_73454 | Not-determined |
| 17-109_k141_156655 | Not-determined |
| 17-109_k141_324144 | Not-determined |
| 17-109_k141_240263 | Not-determined |
| 17-109_k141_206439 | Not-determined |
| 17-109_k141_240266 | Not-determined |
| 17-109_k141_73467 | Not-determined |
| 17-109_k141_206443 | Low-quality |
| 17-109_k141_391099 | Low-quality |
| 17-109_k141_22679 | Not-determined |
| 17-109_k141_173085 | Low-quality |
| 17-109_k141_206448 | Not-determined |
| 17-109_k141_324159 | Not-determined |
| 17-109_k141_73478 | Not-determined |
| 17-109_k141_391102 | Not-determined |
| 17-109_k141_89439 | Not-determined |
| 17-109_k141_5594 | Not-determined |
| 17-109_k141_106120 | Not-determined |
| 17-109_k141_291014 | Not-determined |
| 17-109_k141_139834 | Not-determined |
| 17-109_k141_89452 | Not-determined |
| 17-109_k141_73488 | Not-determined |
| 17-109_k141_291019 | Not-determined |
| 17-109_k141_73489 | Not-determined |
| 17-109_k141_39590 | Not-determined |
| 17-109_k141_22704 | Not-determined |
| 17-109_k141_206461 | Low-quality |
| 17-109_k141_307864 | Low-quality |
| 17-109_k141_106125 | Not-determined |
| 17-109_k141_106126 | Not-determined |
| 17-109_k141_106127 | Not-determined |
| 17-109_k141_291031 | Not-determined |
| 17-109_k141_206470 | Not-determined |
| 17-109_k141_324189 | Not-determined |
| 17-109_k141_22716 | Not-determined |
| 17-109_k141_374303 | Not-determined |
| 17-109_k141_106139 | Not-determined |
| 17-109_k141_206475 | Not-determined |
| 17-109_k141_55480 | Not-determined |
| 17-109_k141_374309 | Not-determined |
| 17-109_k141_139855 | Not-determined |
| 17-109_k141_356514 | Not-determined |
| 17-109_k141_307877 | Not-determined |
| 17-109_k141_291047 | Not-determined |
| 17-109_k141_307879 | Not-determined |
| 17-109_k141_106150 | Not-determined |
| 17-109_k141_139864 | Not-determined |
| 17-109_k141_324198 | Not-determined |
| 17-109_k141_356521 | Not-determined |
| 17-109_k141_106153 | Not-determined |
| 17-109_k141_139866 | Not-determined |
| 17-109_k141_22737 | Not-determined |
| 17-109_k141_39610 | Not-determined |
| 17-109_k141_273733 | Not-determined |
| 17-109_k141_173134 | Not-determined |
| 17-109_k141_55489 | Not-determined |
| 17-109_k141_156690 | Not-determined |
| 17-109_k141_156693 | Low-quality |
| 17-109_k141_356535 | Not-determined |
| 17-109_k141_223284 | Not-determined |
| 17-109_k141_106168 | Not-determined |
| 17-109_k141_73530 | Not-determined |
| 17-109_k141_324220 | Not-determined |
| 17-109_k141_356547 | Not-determined |
| 17-109_k141_291068 | Low-quality |
| 17-109_k141_22746 | Not-determined |
| 17-109_k141_89471 | Not-determined |
| 17-109_k141_89472 | Not-determined |
| 17-109_k141_106186 | Not-determined |
| 17-109_k141_139898 | Low-quality |
| 17-109_k141_240284 | Not-determined |
| 17-109_k141_139907 | Not-determined |
| 17-109_k141_106189 | Not-determined |
| 17-109_k141_391178 | Not-determined |
| 17-109_k141_39631 | Not-determined |
| 17-109_k141_173142 | Not-determined |
| 17-109_k141_356563 | Not-determined |
| 17-109_k141_391185 | Not-determined |
| 17-109_k141_156708 | Not-determined |
| 17-109_k141_206504 | Not-determined |
| 17-109_k141_223305 | Not-determined |
| 17-109_k141_256641 | Not-determined |
| 17-109_k141_291093 | Not-determined |
| 17-109_k141_89495 | Not-determined |
| 17-109_k141_256644 | Not-determined |
| 17-109_k141_55528 | Not-determined |
| 17-109_k141_374362 | Not-determined |
| 17-109_k141_39655 | Not-determined |
| 17-109_k141_89498 | Low-quality |
| 17-109_k141_73570 | Not-determined |
| 17-109_k141_22765 | Not-determined |
| 17-109_k141_223313 | Not-determined |
| 17-109_k141_190895 | Not-determined |
| 17-109_k141_156726 | Not-determined |
| 17-109_k141_256653 | Not-determined |
| 17-109_k141_190902 | Not-determined |
| 17-109_k141_89509 | Not-determined |
| 17-109_k141_39662 | Low-quality |
| 17-109_k141_156728 | Not-determined |
| 17-109_k141_89511 | Not-determined |
| 17-109_k141_122425 | Not-determined |
| 17-109_k141_391210 | Not-determined |
| 17-109_k141_240303 | Not-determined |
| 17-109_k141_122428 | Not-determined |
| 17-109_k141_122429 | Not-determined |
| 17-109_k141_256659 | Low-quality |
| 17-109_k141_256661 | Not-determined |
| 17-109_k141_307933 | Not-determined |
| 17-109_k141_206520 | Not-determined |
| 17-109_k141_240310 | Not-determined |
| 17-109_k141_39675 | Low-quality |
| 17-109_k141_206523 | Low-quality |
| 17-109_k141_240312 | Not-determined |
| 17-109_k141_391227 | Not-determined |
| 17-109_k141_122432 | Not-determined |
| 17-109_k141_256670 | Not-determined |
| 17-109_k141_190919 | Not-determined |
| 17-109_k141_122435 | Not-determined |
| 17-109_k141_223336 | Not-determined |
| 17-109_k141_391230 | Not-determined |
| 17-109_k141_324259 | Not-determined |
| 17-109_k141_122436 | Low-quality |
| 17-109_k141_89535 | Not-determined |
| 17-109_k141_190922 | Not-determined |
| 17-109_k141_307936 | Not-determined |
| 17-109_k141_190924 | Not-determined |
| 17-109_k141_256677 | Not-determined |
| 17-109_k141_122441 | Not-determined |
| 17-109_k141_391234 | Not-determined |
| 17-109_k141_256681 | Not-determined |
| 17-109_k141_223342 | Not-determined |
| 17-109_k141_291105 | Not-determined |
| 17-109_k141_324269 | Not-determined |
| 17-109_k141_89541 | Not-determined |
| 17-109_k141_206532 | Not-determined |
| 17-109_k141_173203 | Not-determined |
| 17-109_k141_391236 | Low-quality |
| 17-109_k141_206533 | Not-determined |
| 17-109_k141_223346 | Not-determined |
| 17-109_k141_391239 | Not-determined |
| 17-109_k141_256689 | Not-determined |
| 17-109_k141_122445 | Not-determined |
| 17-109_k141_55540 | Low-quality |
| 17-109_k141_173209 | Not-determined |
| 17-109_k141_190938 | Not-determined |
| 17-109_k141_190940 | Not-determined |
| 17-109_k141_206543 | Not-determined |
| 17-109_k141_39696 | Not-determined |
| 17-109_k141_223353 | Not-determined |
| 17-109_k141_55549 | Not-determined |
| 17-109_k141_122458 | Not-determined |
| 17-109_k141_223356 | Not-determined |
| 17-109_k141_307942 | Not-determined |
| 17-109_k141_324287 | Not-determined |
| 17-109_k141_324288 | Not-determined |
| 17-109_k141_89564 | Not-determined |
| 17-109_k141_55556 | Not-determined |
| 17-109_k141_156771 | Not-determined |
| 17-109_k141_156775 | Not-determined |
| 17-109_k141_391251 | Not-determined |
| 17-109_k141_89569 | Not-determined |
| 17-109_k141_307953 | Not-determined |
| 17-109_k141_139932 | Not-determined |
| 17-109_k141_122469 | Not-determined |
| 17-109_k141_324293 | Not-determined |
| 17-109_k141_256701 | Not-determined |
| 17-109_k141_256705 | Not-determined |
| 17-109_k141_39704 | Not-determined |
| 17-109_k141_173225 | Not-determined |
| 17-109_k141_307961 | Not-determined |
| 17-109_k141_223377 | Not-determined |
| 17-109_k141_324297 | Not-determined |
| 17-109_k141_206564 | Not-determined |
| 17-109_k141_256712 | Not-determined |
| 17-109_k141_190958 | Not-determined |
| 17-109_k141_55563 | Low-quality |
| 17-109_k141_356582 | Not-determined |
| 17-109_k141_39715 | Not-determined |
| 17-109_k141_122483 | Not-determined |
| 17-109_k141_89593 | Not-determined |
| 17-109_k141_55572 | Not-determined |
| 17-109_k141_5655 | Low-quality |
| 17-109_k141_173237 | Not-determined |
| 17-109_k141_173238 | Not-determined |
| 17-109_k141_256723 | Low-quality |
| 17-109_k141_122490 | Not-determined |
| 17-109_k141_55587 | Not-determined |
| 17-109_k141_156797 | Not-determined |
| 17-109_k141_173248 | Not-determined |
| 17-109_k141_223405 | Not-determined |
| 17-109_k141_223406 | Not-determined |
| 17-109_k141_256732 | Not-determined |
| 17-109_k141_223411 | Not-determined |
| 17-109_k141_256734 | Not-determined |
| 17-109_k141_291124 | Not-determined |
| 17-109_k141_391298 | Low-quality |
| 17-109_k141_73637 | Not-determined |
| 17-109_k141_55595 | Not-determined |
| 17-109_k141_173258 | Not-determined |
| 17-109_k141_173260 | Not-determined |
| 17-109_k141_223420 | Not-determined |
| 17-109_k141_256746 | Not-determined |
| 17-109_k141_5687 | Not-determined |
| 17-109_k141_106219 | Not-determined |
| 17-109_k141_256749 | Not-determined |
| 17-109_k141_22788 | Not-determined |
| 17-109_k141_324314 | Not-determined |
| 17-109_k141_273759 | Not-determined |
| 17-109_k141_5695 | Not-determined |
| 17-109_k141_324321 | Not-determined |
| 17-109_k141_106233 | Not-determined |
| 17-109_k141_307997 | Low-quality |
| 17-109_k141_307999 | Not-determined |
| 17-109_k141_173273 | Not-determined |
| 17-109_k141_291145 | Not-determined |
| 17-109_k141_273772 | Not-determined |
| 17-109_k141_291146 | Low-quality |
| 17-109_k141_122504 | Low-quality |
| 17-109_k141_291147 | Not-determined |
| 17-109_k141_273774 | Not-determined |
| 17-109_k141_308012 | Not-determined |
| 17-109_k141_55607 | Low-quality |
| 17-109_k141_22807 | Not-determined |
| 17-109_k141_240384 | Low-quality |
| 17-109_k141_122517 | Not-determined |
| 17-109_k141_240390 | Not-determined |
| 17-109_k141_308019 | Not-determined |
| 17-109_k141_324340 | Not-determined |
| 17-109_k141_22817 | Not-determined |
| 17-109_k141_374383 | Not-determined |
| 17-109_k141_55611 | Not-determined |
| 17-109_k141_324345 | Not-determined |
| 17-109_k141_256780 | Not-determined |
| 17-109_k141_223447 | Not-determined |
| 17-109_k141_256782 | Not-determined |
| 17-109_k141_273788 | Not-determined |
| 17-109_k141_324349 | Not-determined |
| 17-109_k141_173306 | Low-quality |
| 17-109_k141_291169 | Not-determined |
| 17-109_k141_5740 | Not-determined |
| 17-109_k141_5741 | Not-determined |
| 17-109_k141_206582 | Not-determined |
| 17-109_k141_223456 | Not-determined |
| 17-109_k141_240407 | Not-determined |
| 17-109_k141_173317 | Not-determined |
| 17-109_k141_223458 | Not-determined |
| 17-109_k141_173319 | Not-determined |
| 17-109_k141_273808 | Not-determined |
| 17-109_k141_106252 | Not-determined |
| 17-109_k141_240419 | Not-determined |
| 17-109_k141_273819 | Not-determined |
| 17-109_k141_156849 | Not-determined |
| 17-109_k141_122532 | Not-determined |
| 17-109_k141_156850 | Not-determined |
| 17-109_k141_273827 | Not-determined |
| 17-109_k141_73661 | Low-quality |

| | |
|---|---|
| 17-109_k141_139982 | Not-determined |
| 17-109_k141_240428 | Not-determined |
| 17-109_k141_73665 | Not-determined |
| 17-109_k141_139989 | Not-determined |
| 17-109_k141_374411 | Not-determined |
| 17-109_k141_139991 | Not-determined |
| 17-109_k141_223464 | Not-determined |
| 17-109_k141_273829 | Not-determined |
| 17-109_k141_374417 | Not-determined |
| 17-109_k141_156861 | Not-determined |
| 17-109_k141_140000 | Not-determined |
| 17-109_k141_273837 | Not-determined |
| 17-109_k141_291185 | Not-determined |
| 17-109_k141_22833 | Not-determined |
| 17-109_k141_73676 | Low-quality |
| 17-109_k141_22834 | Not-determined |
| 17-109_k141_140008 | Not-determined |
| 17-109_k141_291196 | Not-determined |
| 17-109_k141_106271 | Not-determined |
| 17-109_k141_391311 | Not-determined |
| 17-109_k141_140012 | Low-quality |
| 17-109_k141_340267 | Not-determined |
| 17-109_k141_73682 | Low-quality |
| 17-109_k141_156885 | Not-determined |
| 17-109_k141_140015 | Not-determined |
| 17-109_k141_240447 | Not-determined |
| 17-109_k141_356611 | Not-determined |
| 17-109_k141_391317 | Low-quality |
| 17-109_k141_5797 | Not-determined |
| 17-109_k141_273859 | Low-quality |
| 17-109_k141_140017 | Low-quality |
| 17-109_k141_340271 | Not-determined |
| 17-109_k141_5799 | Not-determined |
| 17-109_k141_340273 | Not-determined |
| 17-109_k141_291211 | Not-determined |
| 17-109_k141_5802 | Low-quality |
| 17-109_k141_89612 | Not-determined |
| 17-109_k141_240457 | Not-determined |
| 17-109_k141_356613 | Not-determined |
| 17-109_k141_273875 | Low-quality |
| 17-109_k141_5813 | Not-determined |
| 17-109_k141_340290 | Not-determined |
| 17-109_k141_223504 | Not-determined |
| 17-109_k141_140027 | Not-determined |
| 17-109_k141_156895 | Not-determined |
| 17-109_k141_22868 | Low-quality |
| 17-109_k141_223508 | Not-determined |
| 17-109_k141_308056 | Not-determined |
| 17-109_k141_356626 | Low-quality |
| 17-109_k141_106293 | Not-determined |
| 17-109_k141_122581 | Not-determined |
| 17-109_k141_156898 | Not-determined |
| 17-109_k141_5821 | Not-determined |
| 17-109_k141_240476 | Low-quality |
| 17-109_k141_240477 | Not-determined |
| 17-109_k141_324388 | Not-determined |
| 17-109_k141_191003 | Not-determined |
| 17-109_k141_291227 | Not-determined |
| 17-109_k141_206614 | Not-determined |
| 17-109_k141_340301 | Low-quality |
| 17-109_k141_240487 | Not-determined |
| 17-109_k141_356635 | Not-determined |
| 17-109_k141_89634 | Not-determined |
| 17-109_k141_39869 | Not-determined |
| 17-109_k141_223525 | Low-quality |
| 17-109_k141_55672 | Not-determined |
| 17-109_k141_374457 | Not-determined |
| 17-109_k141_173371 | Low-quality |
| 17-109_k141_55675 | Not-determined |
| 17-109_k141_340317 | Not-determined |
| 17-109_k141_191032 | Not-determined |
| 17-109_k141_340322 | Not-determined |
| 17-109_k141_291247 | Not-determined |
| 17-109_k141_173383 | Not-determined |
| 17-109_k141_55685 | Not-determined |
| 17-109_k141_223533 | Not-determined |
| 17-109_k141_55693 | Not-determined |
| 17-109_k141_55694 | Not-determined |
| 17-109_k141_73720 | Not-determined |
| 17-109_k141_55698 | Not-determined |
| 17-109_k141_55702 | Not-determined |
| 17-109_k141_122603 | Not-determined |
| 17-109_k141_256812 | Not-determined |
| 17-109_k141_173394 | Not-determined |
| 17-109_k141_5832 | Low-quality |
| 17-109_k141_55708 | Not-determined |
| 17-109_k141_324410 | Not-determined |
| 17-109_k141_55710 | Not-determined |
| 17-109_k141_173401 | Not-determined |
| 17-109_k141_324414 | Not-determined |
| 17-109_k141_340329 | Not-determined |
| 17-109_k141_291258 | Not-determined |
| 17-109_k141_206658 | Low-quality |
| 17-109_k141_22891 | Not-determined |
| 17-109_k141_324422 | Low-quality |
| 17-109_k141_191062 | Low-quality |
| 17-109_k141_256823 | Not-determined |
| 17-109_k141_308087 | Low-quality |
| 17-109_k141_308089 | Low-quality |
| 17-109_k141_106299 | Low-quality |
| 17-109_k141_122617 | Low-quality |
| 17-109_k141_324429 | Not-determined |
| 17-109_k141_308092 | Not-determined |
| 17-109_k141_256829 | Not-determined |
| 17-109_k141_374493 | Not-determined |
| 17-109_k141_106307 | Not-determined |
| 17-109_k141_291281 | Not-determined |
| 17-109_k141_324440 | Not-determined |
| 17-109_k141_206682 | Not-determined |
| 17-109_k141_39897 | Not-determined |
| 17-109_k141_256833 | Not-determined |
| 17-109_k141_308097 | Not-determined |
| 17-109_k141_106327 | Not-determined |
| 17-109_k141_191071 | Not-determined |
| 17-109_k141_308101 | Low-quality |
| 17-109_k141_106328 | Not-determined |
| 17-109_k141_206697 | Not-determined |
| 17-109_k141_223595 | Not-determined |
| 17-109_k141_206700 | Not-determined |
| 17-109_k141_122642 | Not-determined |
| 17-109_k141_173420 | Not-determined |
| 17-109_k141_39903 | Not-determined |
| 17-109_k141_356674 | Not-determined |
| 17-109_k141_39906 | Not-determined |
| 17-109_k141_308104 | Not-determined |
| 17-109_k141_191074 | Not-determined |
| 17-109_k141_5863 | Not-determined |
| 17-109_k141_223601 | Not-determined |
| 17-109_k141_308106 | Not-determined |
| 17-109_k141_374507 | Not-determined |
| 17-109_k141_73745 | Low-quality |
| 17-109_k141_308109 | Not-determined |
| 17-109_k141_122655 | Not-determined |
| 17-109_k141_89685 | Not-determined |
| 17-109_k141_122661 | Low-quality |
| 17-109_k141_55728 | Not-determined |
| 17-109_k141_391336 | Not-determined |
| 17-109_k141_308123 | Not-determined |
| 17-109_k141_308124 | Not-determined |
| 17-109_k141_391341 | Not-determined |
| 17-109_k141_191090 | Not-determined |
| 17-109_k141_374519 | Not-determined |
| 17-109_k141_308130 | Not-determined |
| 17-109_k141_55738 | Not-determined |
| 17-109_k141_122669 | Not-determined |
| 17-109_k141_291323 | Not-determined |
| 17-109_k141_5876 | Not-determined |
| 17-109_k141_291325 | Not-determined |
| 17-109_k141_122672 | Not-determined |
| 17-109_k141_374523 | Not-determined |
| 17-109_k141_22926 | Not-determined |
| 17-109_k141_22927 | Not-determined |
| 17-109_k141_5883 | Not-determined |
| 17-109_k141_308142 | Not-determined |
| 17-109_k141_73769 | Not-determined |
| 17-109_k141_140095 | Not-determined |
| 17-109_k141_22937 | Not-determined |
| 17-109_k141_374533 | Not-determined |
| 17-109_k141_122674 | Not-determined |
| 17-109_k141_223629 | Not-determined |
| 17-109_k141_308151 | Not-determined |
| 17-109_k141_140104 | Low-quality |
| 17-109_k141_374542 | Low-quality |
| 17-109_k141_374544 | Not-determined |
| 17-109_k141_391361 | Not-determined |
| 17-109_k141_308154 | Low-quality |
| 17-109_k141_374550 | Low-quality |
| 17-109_k141_273899 | Not-determined |
| 17-109_k141_191121 | Not-determined |
| 17-109_k141_22953 | Not-determined |
| 17-109_k141_5903 | Not-determined |
| 17-109_k141_73786 | Not-determined |
| 17-109_k141_391373 | Not-determined |
| 17-109_k141_191126 | Not-determined |
| 17-109_k141_173477 | Not-determined |
| 17-109_k141_191129 | Not-determined |
| 17-109_k141_273902 | Not-determined |
| 17-109_k141_374563 | Not-determined |
| 17-109_k141_191136 | Not-determined |
| 17-109_k141_374567 | Low-quality |
| 17-109_k141_374570 | Not-determined |
| 17-109_k141_206735 | Not-determined |
| 17-109_k141_308161 | Not-determined |
| 17-109_k141_140130 | Not-determined |
| 17-109_k141_223639 | Not-determined |
| 17-109_k141_308163 | Not-determined |
| 17-109_k141_256899 | Low-quality |
| 17-109_k141_356699 | Not-determined |
| 17-109_k141_223647 | Low-quality |
| 17-109_k141_391384 | Low-quality |
| 17-109_k141_356705 | Not-determined |
| 17-109_k141_356711 | Not-determined |
| 17-109_k141_122698 | Not-determined |
| 17-109_k141_308182 | Not-determined |
| 17-109_k141_240545 | Not-determined |
| 17-109_k141_140139 | Not-determined |
| 17-109_k141_89739 | Not-determined |
| 17-109_k141_140143 | Not-determined |
| 17-109_k141_73803 | Not-determined |
| 17-109_k141_206749 | Not-determined |
| 17-109_k141_73805 | Not-determined |
| 17-109_k141_73806 | Not-determined |
| 17-109_k141_5938 | Not-determined |
| 17-109_k141_356724 | Not-determined |
| 17-109_k141_356728 | Not-determined |
| 17-109_k141_22982 | Not-determined |
| 17-109_k141_5946 | Not-determined |
| 17-109_k141_73823 | Not-determined |
| 17-109_k141_206758 | Not-determined |
| 17-109_k141_391402 | Not-determined |
| 17-109_k141_106353 | Not-determined |
| 17-109_k141_391405 | Not-determined |
| 17-109_k141_140162 | Not-determined |
| 17-109_k141_106361 | Not-determined |
| 17-109_k141_206764 | Low-quality |
| 17-109_k141_308201 | Not-determined |
| 17-109_k141_374594 | Not-determined |
| 17-109_k141_140167 | Not-determined |
| 17-109_k141_173511 | Low-quality |
| 17-109_k141_73836 | Not-determined |
| 17-109_k141_173516 | Not-determined |
| 17-109_k141_173517 | Not-determined |
| 17-109_k141_273935 | Not-determined |
| 17-109_k141_308211 | Not-determined |
| 17-109_k141_55789 | Not-determined |
| 17-109_k141_308212 | Not-determined |
| 17-109_k141_89771 | Not-determined |
| 17-109_k141_256913 | Not-determined |
| 17-109_k141_374609 | Not-determined |
| 17-109_k141_256915 | Not-determined |
| 17-109_k141_173527 | Low-quality |
| 17-109_k141_55796 | Not-determined |
| 17-109_k141_156960 | Not-determined |
| 17-109_k141_140183 | Low-quality |
| 17-109_k141_256920 | Not-determined |
| 17-109_k141_256922 | Not-determined |
| 17-109_k141_308219 | Low-quality |
| 17-109_k141_206787 | Not-determined |
| 17-109_k141_374615 | Not-determined |
| 17-109_k141_191146 | Not-determined |
| 17-109_k141_206792 | Not-determined |
| 17-109_k141_73846 | Low-quality |
| 17-109_k141_391426 | Not-determined |
| 17-109_k141_256930 | Not-determined |
| 17-109_k141_374629 | Not-determined |
| 17-109_k141_5966 | Not-determined |
| 17-109_k141_5968 | Not-determined |
| 17-109_k141_39955 | Not-determined |
| 17-109_k141_106375 | Low-quality |
| 17-109_k141_374640 | Not-determined |
| 17-109_k141_374641 | Not-determined |
| 17-109_k141_308234 | Low-quality |
| 17-109_k141_173556 | Not-determined |
| 17-109_k141_5981 | Not-determined |
| 17-109_k141_106385 | Not-determined |
| 17-109_k141_206816 | Low-quality |
| 17-109_k141_273951 | Not-determined |
| 17-109_k141_273953 | Low-quality |
| 17-109_k141_273955 | Not-determined |
| 17-109_k141_223697 | Not-determined |
| 17-109_k141_256941 | Low-quality |
| 17-109_k141_89804 | Not-determined |
| 17-109_k141_240614 | Not-determined |
| 17-109_k141_55812 | Not-determined |
| 17-109_k141_273964 | Not-determined |
| 17-109_k141_240616 | Low-quality |
| 17-109_k141_173565 | Not-determined |
| 17-109_k141_256947 | Low-quality |
| 17-109_k141_39992 | Not-determined |
| 17-109_k141_291374 | Not-determined |
| 17-109_k141_206835 | Not-determined |
| 17-109_k141_223713 | Not-determined |
| 17-109_k141_256952 | Not-determined |
| 17-109_k141_240626 | Low-quality |
| 17-109_k141_223718 | Not-determined |
| 17-109_k141_340344 | Not-determined |
| 17-109_k141_6001 | Not-determined |
| 17-109_k141_173575 | Not-determined |
| 17-109_k141_55820 | Not-determined |
| 17-109_k141_324487 | Not-determined |
| 17-109_k141_340348 | Not-determined |
| 17-109_k141_340352 | Not-determined |
| 17-109_k141_391441 | Low-quality |
| 17-109_k141_391442 | Not-determined |
| 17-109_k141_55826 | Not-determined |
| 17-109_k141_6012 | Low-quality |
| 17-109_k141_273987 | Not-determined |
| 17-109_k141_206848 | Low-quality |
| 17-109_k141_73868 | Not-determined |
| 17-109_k141_340361 | Not-determined |
| 17-109_k141_106401 | Not-determined |
| 17-109_k141_240650 | Not-determined |
| 17-109_k141_140247 | Not-determined |
| 17-109_k141_340363 | Not-determined |
| 17-109_k141_240653 | Not-determined |
| 17-109_k141_223727 | Not-determined |
| 17-109_k141_240655 | Not-determined |
| 17-109_k141_391451 | Not-determined |
| 17-109_k141_206856 | Not-determined |
| 17-109_k141_391455 | Not-determined |
| 17-109_k141_106412 | Not-determined |
| 17-109_k141_356798 | Not-determined |
| 17-109_k141_223734 | Not-determined |
| 17-109_k141_191167 | Low-quality |
| 17-109_k141_223735 | Not-determined |
| 17-109_k141_374679 | Not-determined |
| 17-109_k141_206864 | Not-determined |
| 17-109_k141_191170 | Not-determined |
| 17-109_k141_356805 | Not-determined |
| 17-109_k141_291396 | Not-determined |
| 17-109_k141_291397 | Not-determined |
| 17-109_k141_391460 | Not-determined |
| 17-109_k141_73881 | Not-determined |
| 17-109_k141_356809 | Not-determined |
| 17-109_k141_356811 | Not-determined |
| 17-109_k141_356812 | Not-determined |
| 17-109_k141_191175 | Not-determined |
| 17-109_k141_191176 | Not-determined |
| 17-109_k141_73884 | Not-determined |
| 17-109_k141_391466 | Not-determined |
| 17-109_k141_356816 | Not-determined |
| 17-109_k141_223754 | Not-determined |
| 17-109_k141_340379 | Not-determined |
| 17-109_k141_391471 | Not-determined |
| 17-109_k141_223759 | Not-determined |
| 17-109_k141_391475 | Low-quality |
| 17-109_k141_356818 | Not-determined |
| 17-109_k141_223760 | Not-determined |
| 17-109_k141_291413 | Not-determined |
| 17-109_k141_73890 | Not-determined |
| 17-109_k141_140254 | Low-quality |
| 17-109_k141_106438 | Not-determined |
| 17-109_k141_223763 | Not-determined |
| 17-109_k141_89842 | Not-determined |
| 17-109_k141_223765 | Not-determined |
| 17-109_k141_391482 | Low-quality |
| 17-109_k141_324508 | Not-determined |
| 17-109_k141_55863 | Not-determined |
| 17-109_k141_206888 | Not-determined |
| 17-109_k141_240672 | Not-determined |
| 17-109_k141_340397 | Not-determined |
| 17-109_k141_240676 | Not-determined |
| 17-109_k141_324515 | Not-determined |
| 17-109_k141_206894 | Not-determined |
| 17-109_k141_122728 | Not-determined |
| 17-109_k141_140263 | Not-determined |
| 17-109_k141_140264 | Not-determined |
| 17-109_k141_122731 | Not-determined |
| 17-109_k141_291437 | Not-determined |
| 17-109_k141_374725 | Not-determined |
| 17-109_k141_324521 | Not-determined |
| 17-109_k141_122738 | Not-determined |
| 17-109_k141_256968 | Low-quality |
| 17-109_k141_223774 | Low-quality |
| 17-109_k141_240686 | Not-determined |
| 17-109_k141_206900 | Not-determined |
| 17-109_k141_340409 | Not-determined |
| 17-109_k141_274049 | Not-determined |
| 17-109_k141_308304 | Not-determined |
| 17-109_k141_240695 | Not-determined |
| 17-109_k141_324536 | Not-determined |
| 17-109_k141_23033 | Not-determined |
| 17-109_k141_324538 | Not-determined |
| 17-109_k141_206908 | Not-determined |
| 17-109_k141_191195 | Not-determined |
| 17-109_k141_206912 | Low-quality |
| 17-109_k141_206914 | Not-determined |
| 17-109_k141_291458 | Low-quality |
| 17-109_k141_308320 | Not-determined |
| 17-109_k141_6062 | Not-determined |
| 17-109_k141_191199 | Not-determined |
| 17-109_k141_257001 | Not-determined |
| 17-109_k141_173608 | Not-determined |
| 17-109_k141_291471 | Low-quality |
| 17-109_k141_156981 | Not-determined |
| 17-109_k141_257004 | Not-determined |
| 17-109_k141_156989 | Not-determined |
| 17-109_k141_156993 | Not-determined |
| 17-109_k141_257006 | Not-determined |
| 17-109_k141_324563 | Not-determined |
| 17-109_k141_274064 | Low-quality |
| 17-109_k141_6077 | Not-determined |
| 17-109_k141_122775 | Low-quality |
| 17-109_k141_206931 | Not-determined |
| 17-109_k141_340437 | Not-determined |
| 17-109_k141_374752 | Not-determined |
| 17-109_k141_73913 | Not-determined |
| 17-109_k141_89859 | Not-determined |
| 17-109_k141_55902 | Not-determined |
| 17-109_k141_157010 | Not-determined |
| 17-109_k141_191217 | Not-determined |
| 17-109_k141_356854 | Not-determined |
| 17-109_k141_324569 | Not-determined |
| 17-109_k141_308328 | Not-determined |
| 17-109_k141_40049 | Not-determined |
| 17-109_k141_274089 | Not-determined |
| 17-109_k141_191239 | Not-determined |
| 17-109_k141_340443 | Not-determined |
| 17-109_k141_340444 | Not-determined |
| 17-109_k141_191243 | Not-determined |
| 17-109_k141_308339 | Not-determined |
| 17-109_k141_122796 | Not-determined |
| 17-109_k141_157028 | Not-determined |
| 17-109_k141_157032 | Not-determined |
| 17-109_k141_191251 | Low-quality |
| 17-109_k141_374763 | Low-quality |
| 17-109_k141_291491 | Not-determined |
| 17-109_k141_122799 | Not-determined |
| 17-109_k141_191259 | Low-quality |
| 17-109_k141_308350 | Not-determined |
| 17-109_k141_340464 | Not-determined |
| 17-109_k141_206962 | Not-determined |
| 17-109_k141_23106 | Low-quality |
| 17-109_k141_140307 | Not-determined |
| 17-109_k141_73923 | Not-determined |
| 17-109_k141_340468 | Not-determined |
| 17-109_k141_391526 | Not-determined |
| 17-109_k141_140310 | Not-determined |
| 17-109_k141_191264 | Not-determined |
| 17-109_k141_40071 | Not-determined |
| 17-109_k141_240715 | Not-determined |
| 17-109_k141_240717 | Not-determined |
| 17-109_k141_106482 | Not-determined |
| 17-109_k141_106484 | Not-determined |
| 17-109_k141_257039 | Not-determined |
| 17-109_k141_173614 | Not-determined |
| 17-109_k141_257043 | Not-determined |
| 17-109_k141_140337 | Not-determined |
| 17-109_k141_157063 | Not-determined |
| 17-109_k141_122806 | Not-determined |
| 17-109_k141_257050 | Not-determined |
| 17-109_k141_257052 | Not-determined |
| 17-109_k141_122809 | Not-determined |
| 17-109_k141_274101 | Not-determined |
| 17-109_k141_374789 | Not-determined |
| 17-109_k141_206981 | Low-quality |
| 17-109_k141_291516 | Not-determined |
| 17-109_k141_157074 | Low-quality |
| 17-109_k141_173617 | Not-determined |
| 17-109_k141_223821 | Not-determined |
| 17-109_k141_356906 | Not-determined |
| 17-109_k141_23138 | Not-determined |
| 17-109_k141_391555 | Not-determined |
| 17-109_k141_122825 | Low-quality |
| 17-109_k141_257078 | Not-determined |
| 17-109_k141_206997 | Not-determined |
| 17-109_k141_291521 | Not-determined |
| 17-109_k141_291527 | Not-determined |
| 17-109_k141_40088 | Not-determined |
| 17-109_k141_6100 | Not-determined |
| 17-109_k141_122839 | Not-determined |
| 17-109_k141_23142 | Not-determined |
| 17-109_k141_6101 | Not-determined |
| 17-109_k141_40094 | Not-determined |
| 17-109_k141_291534 | Not-determined |
| 17-109_k141_173638 | Not-determined |
| 17-109_k141_122846 | Not-determined |
| 17-109_k141_191305 | Not-determined |
| 17-109_k141_356920 | Low-quality |
| 17-109_k141_173641 | Low-quality |
| 17-109_k141_191312 | Not-determined |
| 17-109_k141_73955 | Not-determined |
| 17-109_k141_207006 | Not-determined |
| 17-109_k141_140363 | Not-determined |
| 17-109_k141_324607 | Not-determined |
| 17-109_k141_324609 | Not-determined |
| 17-109_k141_374816 | Not-determined |
| 17-109_k141_191313 | Low-quality |
| 17-109_k141_291551 | Not-determined |
| 17-109_k141_157120 | Not-determined |
| 17-109_k141_207025 | Not-determined |
| 17-109_k141_89886 | Not-determined |
| 17-109_k141_207027 | Not-determined |
| 17-109_k141_308398 | Not-determined |
| 17-109_k141_191326 | Not-determined |
| 17-109_k141_6117 | Not-determined |
| 17-109_k141_89893 | Not-determined |
| 17-109_k141_207037 | Not-determined |
| 17-109_k141_140375 | Not-determined |
| 17-109_k141_223861 | Not-determined |
| 17-109_k141_240736 | Not-determined |
| 17-109_k141_6123 | Not-determined |
| 17-109_k141_106522 | Not-determined |
| 17-109_k141_89909 | Not-determined |
| 17-109_k141_257118 | Not-determined |
| 17-109_k141_291567 | Not-determined |
| 17-109_k141_122875 | Not-determined |
| 17-109_k141_257126 | Not-determined |
| 17-109_k141_6132 | Not-determined |
| 17-109_k141_340494 | Not-determined |
| 17-109_k141_240750 | Not-determined |
| 17-109_k141_291576 | Low-quality |
| 17-109_k141_106527 | Not-determined |
| 17-109_k141_240753 | Not-determined |
| 17-109_k141_374829 | Not-determined |
| 17-109_k141_308437 | Not-determined |
| 17-109_k141_6136 | Not-determined |
| 17-109_k141_140386 | Not-determined |
| 17-109_k141_374832 | Not-determined |
| 17-109_k141_374835 | Not-determined |
| 17-109_k141_6144 | Not-determined |
| 17-109_k141_374840 | Not-determined |
| 17-109_k141_240766 | Not-determined |
| 17-109_k141_240768 | Not-determined |
| 17-109_k141_240769 | Not-determined |
| 17-109_k141_157140 | Not-determined |
| 17-109_k141_89929 | Low-quality |
| 17-109_k141_23190 | Not-determined |
| 17-109_k141_223901 | Not-determined |
| 17-109_k141_6152 | Not-determined |
| 17-109_k141_157144 | Low-quality |
| 17-109_k141_140397 | Not-determined |
| 17-109_k141_257135 | Not-determined |
| 17-109_k141_89931 | Not-determined |
| 17-109_k141_56000 | Not-determined |
| 17-109_k141_223907 | Not-determined |
| 17-109_k141_356973 | Not-determined |
| 17-109_k141_56004 | Not-determined |
| 17-109_k141_6158 | Not-determined |
| 17-109_k141_140409 | Not-determined |
| 17-109_k141_223913 | Not-determined |
| 17-109_k141_191363 | Not-determined |
| 17-109_k141_73995 | Low-quality |
| 17-109_k141_340518 | Not-determined |
| 17-109_k141_106550 | Not-determined |
| 17-109_k141_274156 | Not-determined |
| 17-109_k141_356986 | Not-determined |
| 17-109_k141_122885 | Not-determined |
| 17-109_k141_122887 | Not-determined |
| 17-109_k141_274159 | Not-determined |
| 17-109_k141_40107 | Low-quality |
| 17-109_k141_173671 | Not-determined |
| 17-109_k141_240789 | Not-determined |
| 17-109_k141_257144 | Not-determined |
| 17-109_k141_191382 | Low-quality |
| 17-109_k141_74009 | Not-determined |
| 17-109_k141_122897 | Not-determined |
| 17-109_k141_74013 | Not-determined |
| 17-109_k141_308477 | Not-determined |
| 17-109_k141_6167 | Low-quality |
| 17-109_k141_308478 | Not-determined |
| 17-109_k141_324638 | Not-determined |
| 17-109_k141_122904 | Not-determined |
| 17-109_k141_291626 | Not-determined |
| 17-109_k141_308483 | Not-determined |
| 17-109_k141_291628 | Not-determined |
| 17-109_k141_340538 | Not-determined |
| 17-109_k141_74021 | Not-determined |
| 17-109_k141_122908 | Not-determined |
| 17-109_k141_140438 | Not-determined |
| 17-109_k141_74024 | Not-determined |
| 17-109_k141_340545 | Not-determined |
| 17-109_k141_223943 | Not-determined |
| 17-109_k141_223944 | Not-determined |
| 17-109_k141_340547 | Not-determined |
| 17-109_k141_89945 | Not-determined |
| 17-109_k141_291647 | Not-determined |
| 17-109_k141_106572 | Not-determined |
| 17-109_k141_122920 | Not-determined |
| 17-109_k141_291658 | Not-determined |
| 17-109_k141_74040 | Not-determined |
| 17-109_k141_173680 | Not-determined |
| 17-109_k141_173683 | Not-determined |
| 17-109_k141_6185 | Not-determined |
| 17-109_k141_308493 | Not-determined |
| 17-109_k141_106591 | Not-determined |
| 17-109_k141_357035 | Not-determined |
| 17-109_k141_340570 | Not-determined |
| 17-109_k141_223965 | Not-determined |
| 17-109_k141_340571 | Not-determined |
| 17-109_k141_106597 | Not-determined |
| 17-109_k141_173689 | Not-determined |
| 17-109_k141_74043 | Not-determined |
| 17-109_k141_74046 | Not-determined |
| 17-109_k141_74047 | Not-determined |
| 17-109_k141_74049 | Not-determined |
| 17-109_k141_391618 | Not-determined |
| 17-109_k141_340578 | Not-determined |
| 17-109_k141_207122 | Not-determined |
| 17-109_k141_324653 | Not-determined |
| 17-109_k141_122934 | Not-determined |
| 17-109_k141_74060 | Not-determined |
| 17-109_k141_56029 | Low-quality |
| 17-109_k141_274202 | Not-determined |
| 17-109_k141_74069 | Not-determined |
| 17-109_k141_74070 | Not-determined |
| 17-109_k141_6195 | Not-determined |
| 17-109_k141_257177 | Not-determined |
| 17-109_k141_257181 | Not-determined |

| | |
|---|---|
| 17-109_k141_106603 | Not-determined |
| 17-109_k141_89996 | Not-determined |
| 17-109_k141_324672 | Not-determined |
| 17-109_k141_340599 | Not-determined |
| 17-109_k141_74079 | Not-determined |
| 17-109_k141_274213 | Not-determined |
| 17-109_k141_391650 | Low-quality |
| 17-109_k141_391651 | Not-determined |
| 17-109_k141_173730 | Not-determined |
| 17-109_k141_257186 | Not-determined |
| 17-109_k141_374882 | Not-determined |
| 17-109_k141_191443 | Low-quality |
| 17-109_k141_340606 | Not-determined |
| 17-109_k141_40156 | Not-determined |
| 17-109_k141_6205 | Low-quality |
| 17-109_k141_308512 | Low-quality |
| 17-109_k141_40160 | Not-determined |
| 17-109_k141_374889 | Low-quality |
| 17-109_k141_173738 | Not-determined |
| 17-109_k141_324684 | Not-determined |
| 17-109_k141_257197 | Not-determined |
| 17-109_k141_324687 | Not-determined |
| 17-109_k141_90006 | Not-determined |
| 17-109_k141_40176 | Not-determined |
| 17-109_k141_157174 | Not-determined |
| 17-109_k141_191454 | Low-quality |
| 17-109_k141_90016 | Not-determined |
| 17-109_k141_340619 | Not-determined |
| 17-109_k141_74100 | Not-determined |
| 17-109_k141_90020 | Not-determined |
| 17-109_k141_391671 | Not-determined |
| 17-109_k141_90021 | Not-determined |
| 17-109_k141_308532 | Not-determined |
| 17-109_k141_340628 | Not-determined |
| 17-109_k141_308534 | Not-determined |
| 17-109_k141_157185 | Not-determined |
| 17-109_k141_391677 | Low-quality |
| 17-109_k141_106622 | Not-determined |
| 17-109_k141_340631 | Not-determined |
| 17-109_k141_40196 | Not-determined |
| 17-109_k141_122990 | Low-quality |
| 17-109_k141_23292 | Not-determined |
| 17-109_k141_391682 | Not-determined |
| 17-109_k141_224005 | Not-determined |
| 17-109_k141_224008 | Not-determined |
| 17-109_k141_140468 | Not-determined |
| 17-109_k141_224010 | Not-determined |
| 17-109_k141_106635 | Not-determined |
| 17-109_k141_157200 | Not-determined |
| 17-109_k141_157202 | Not-determined |
| 17-109_k141_357105 | Not-determined |
| 17-109_k141_157203 | Not-determined |
| 17-109_k141_308547 | Not-determined |
| 17-109_k141_224022 | Not-determined |
| 17-109_k141_340633 | Not-determined |
| 17-109_k141_257209 | Low-quality |
| 17-109_k141_324706 | Not-determined |
| 17-109_k141_257213 | Not-determined |
| 17-109_k141_391699 | Not-determined |
| 17-109_k141_324708 | Low-quality |
| 17-109_k141_374913 | Low-quality |
| 17-109_k141_173763 | Not-determined |
| 17-109_k141_224036 | Not-determined |
| 17-109_k141_207152 | Not-determined |
| 17-109_k141_123006 | Low-quality |
| 17-109_k141_140484 | Low-quality |
| 17-109_k141_23312 | Not-determined |
| 17-109_k141_207154 | Not-determined |
| 17-109_k141_224040 | Not-determined |
| 17-109_k141_340649 | Not-determined |
| 17-109_k141_157213 | Not-determined |
| 17-109_k141_90038 | Not-determined |
| 17-109_k141_90039 | Not-determined |
| 17-109_k141_207159 | Not-determined |
| 17-109_k141_374921 | Not-determined |
| 17-109_k141_123011 | Not-determined |
| 17-109_k141_207163 | Not-determined |
| 17-109_k141_207164 | Low-quality |
| 17-109_k141_191464 | Not-determined |
| 17-109_k141_324725 | Not-determined |
| 17-109_k141_173767 | Low-quality |
| 17-109_k141_6241 | Not-determined |
| 17-109_k141_324734 | Not-determined |
| 17-109_k141_173775 | Not-determined |
| 17-109_k141_224059 | Not-determined |
| 17-109_k141_207180 | Not-determined |
| 17-109_k141_207184 | Not-determined |
| 17-109_k141_324738 | Not-determined |
| 17-109_k141_140502 | Not-determined |
| 17-109_k141_74112 | Low-quality |
| 17-109_k141_157231 | Low-quality |
| 17-109_k141_157233 | Not-determined |
| 17-109_k141_374949 | Not-determined |
| 17-109_k141_257247 | Not-determined |
| 17-109_k141_140507 | Not-determined |
| 17-109_k141_257253 | Not-determined |
| 17-109_k141_6268 | Not-determined |
| 17-109_k141_257265 | Not-determined |
| 17-109_k141_224088 | Not-determined |
| 17-109_k141_6277 | Not-determined |
| 17-109_k141_291700 | Not-determined |
| 17-109_k141_324750 | Not-determined |
| 17-109_k141_391716 | Not-determined |
| 17-109_k141_324752 | Not-determined |
| 17-109_k141_391723 | Not-determined |
| 17-109_k141_140516 | Not-determined |
| 17-109_k141_157266 | Low-quality |
| 17-109_k141_324758 | Not-determined |
| 17-109_k141_224095 | Not-determined |
| 17-109_k141_40207 | Not-determined |
| 17-109_k141_308566 | Low-quality |
| 17-109_k141_324771 | Not-determined |
| 17-109_k141_140530 | Not-determined |
| 17-109_k141_40212 | Not-determined |
| 17-109_k141_324781 | Low-quality |
| 17-109_k141_157270 | Not-determined |
| 17-109_k141_308574 | Not-determined |
| 17-109_k141_274281 | Not-determined |
| 17-109_k141_173834 | Not-determined |
| 17-109_k141_224111 | Not-determined |
| 17-109_k141_374953 | Not-determined |
| 17-109_k141_90094 | Not-determined |
| 17-109_k141_157278 | Not-determined |
| 17-109_k141_224126 | Not-determined |
| 17-109_k141_224128 | Not-determined |
| 17-109_k141_40223 | Not-determined |
| 17-109_k141_340665 | Not-determined |
| 17-109_k141_308583 | Not-determined |
| 17-109_k141_308584 | Not-determined |
| 17-109_k141_6333 | Not-determined |
| 17-109_k141_308587 | Not-determined |
| 17-109_k141_40229 | Low-quality |
| 17-109_k141_224139 | Not-determined |
| 17-109_k141_74144 | Not-determined |
| 17-109_k141_308592 | Not-determined |
| 17-109_k141_391762 | Not-determined |
| 17-109_k141_40233 | Not-determined |
| 17-109_k141_90102 | Not-determined |
| 17-109_k141_173857 | Not-determined |
| 17-109_k141_74151 | Not-determined |
| 17-109_k141_40240 | Not-determined |
| 17-109_k141_308598 | Not-determined |
| 17-109_k141_74153 | Not-determined |
| 17-109_k141_74154 | Not-determined |
| 17-109_k141_324813 | Not-determined |
| 17-109_k141_324815 | Not-determined |
| 17-109_k141_257296 | Not-determined |
| 17-109_k141_6353 | Not-determined |
| 17-109_k141_324825 | Not-determined |
| 17-109_k141_90111 | Low-quality |
| 17-109_k141_291708 | Not-determined |
| 17-109_k141_207240 | Not-determined |
| 17-109_k141_291709 | Low-quality |
| 17-109_k141_90115 | Low-quality |
| 17-109_k141_308606 | Not-determined |
| 17-109_k141_157317 | Not-determined |
| 17-109_k141_257306 | Not-determined |
| 17-109_k141_257307 | Not-determined |
| 17-109_k141_374990 | Low-quality |
| 17-109_k141_6377 | Not-determined |
| 17-109_k141_374992 | Not-determined |
| 17-109_k141_40274 | Low-quality |
| 17-109_k141_40277 | Not-determined |
| 17-109_k141_6390 | Not-determined |
| 17-109_k141_123023 | Not-determined |
| 17-109_k141_375002 | Low-quality |
| 17-109_k141_340677 | Not-determined |
| 17-109_k141_140549 | Not-determined |
| 17-109_k141_40285 | Not-determined |
| 17-109_k141_207243 | Not-determined |
| 17-109_k141_40291 | Not-determined |
| 17-109_k141_123040 | Not-determined |
| 17-109_k141_340689 | Not-determined |
| 17-109_k141_106711 | Not-determined |
| 17-109_k141_207253 | Low-quality |
| 17-109_k141_74187 | Not-determined |
| 17-109_k141_207254 | Low-quality |
| 17-109_k141_90134 | Not-determined |
| 17-109_k141_291729 | Not-determined |
| 17-109_k141_391804 | Low-quality |
| 17-109_k141_191473 | Low-quality |
| 17-109_k141_274335 | Not-determined |
| 17-109_k141_40318 | Not-determined |
| 17-109_k141_274336 | Not-determined |
| 17-109_k141_40323 | Not-determined |
| 17-109_k141_40326 | Not-determined |
| 17-109_k141_106728 | Not-determined |
| 17-109_k141_157346 | Low-quality |
| 17-109_k141_375021 | Not-determined |
| 17-109_k141_375023 | Not-determined |
| 17-109_k141_56156 | Not-determined |
| 17-109_k141_308651 | Not-determined |
| 17-109_k141_106735 | Not-determined |
| 17-109_k141_340704 | Not-determined |
| 17-109_k141_207294 | Not-determined |
| 17-109_k141_40348 | Not-determined |
| 17-109_k141_6420 | Not-determined |
| 17-109_k141_375032 | Low-quality |
| 17-109_k141_106748 | Low-quality |
| 17-109_k141_207300 | Not-determined |
| 17-109_k141_224176 | Not-determined |
| 17-109_k141_324862 | Not-determined |
| 17-109_k141_375035 | Not-determined |
| 17-109_k141_157369 | Not-determined |
| 17-109_k141_140564 | Not-determined |
| 17-109_k141_40354 | Not-determined |
| 17-109_k141_340718 | Low-quality |
| 17-109_k141_324867 | Not-determined |
| 17-109_k141_106759 | Not-determined |
| 17-109_k141_391811 | Low-quality |
| 17-109_k141_6445 | Not-determined |
| 17-109_k141_391820 | Not-determined |
| 17-109_k141_257336 | Not-determined |
| 17-109_k141_324887 | Not-determined |
| 17-109_k141_391827 | Not-determined |
| 17-109_k141_207323 | Not-determined |
| 17-109_k141_74201 | Not-determined |
| 17-109_k141_391831 | Not-determined |
| 17-109_k141_90159 | Not-determined |
| 17-109_k141_340734 | Not-determined |
| 17-109_k141_90162 | Not-determined |
| 17-109_k141_391839 | Not-determined |
| 17-109_k141_324900 | Not-determined |
| 17-109_k141_375057 | Not-determined |
| 17-109_k141_391841 | Not-determined |
| 17-109_k141_340741 | Not-determined |
| 17-109_k141_6452 | Not-determined |
| 17-109_k141_375063 | Not-determined |
| 17-109_k141_207339 | Not-determined |
| 17-109_k141_123137 | Low-quality |
| 17-109_k141_375066 | Low-quality |
| 17-109_k141_375067 | Not-determined |
| 17-109_k141_274359 | Not-determined |
| 17-109_k141_6459 | Low-quality |
| 17-109_k141_90190 | Low-quality |
| 17-109_k141_157416 | Not-determined |
| 17-109_k141_90193 | Not-determined |
| 17-109_k141_173882 | Not-determined |
| 17-109_k141_324917 | Not-determined |
| 17-109_k141_207349 | Not-determined |
| 17-109_k141_106782 | Not-determined |
| 17-109_k141_308694 | Not-determined |
| 17-109_k141_391870 | Not-determined |
| 17-109_k141_391872 | Not-determined |
| 17-109_k141_308697 | Not-determined |
| 17-109_k141_257374 | Not-determined |
| 17-109_k141_207353 | Not-determined |
| 17-109_k141_56206 | Low-quality |
| 17-109_k141_340751 | Low-quality |
| 17-109_k141_308700 | Not-determined |
| 17-109_k141_40384 | Not-determined |
| 17-109_k141_40385 | Not-determined |
| 17-109_k141_340760 | Not-determined |
| 17-109_k141_123163 | Not-determined |
| 17-109_k141_391879 | Not-determined |
| 17-109_k141_123168 | Low-quality |
| 17-109_k141_375080 | Not-determined |
| 17-109_k141_391883 | Not-determined |
| 17-109_k141_324935 | Not-determined |
| 17-109_k141_357136 | Not-determined |
| 17-109_k141_23329 | Low-quality |
| 17-109_k141_140578 | Not-determined |
| 17-109_k141_157443 | Not-determined |
| 17-109_k141_40391 | Not-determined |
| 17-109_k141_340777 | Not-determined |
| 17-109_k141_340778 | Not-determined |
| 17-109_k141_140579 | Not-determined |
| 17-109_k141_257401 | Not-determined |
| 17-109_k141_207393 | Not-determined |
| 17-109_k141_40394 | Not-determined |
| 17-109_k141_274374 | Low-quality |
| 17-109_k141_340784 | Not-determined |
| 17-109_k141_123188 | Not-determined |
| 17-109_k141_224210 | Not-determined |
| 17-109_k141_6489 | Low-quality |
| 17-109_k141_274384 | Not-determined |
| 17-109_k141_157460 | Not-determined |
| 17-109_k141_240813 | Not-determined |
| 17-109_k141_340791 | Not-determined |
| 17-109_k141_257421 | Not-determined |
| 17-109_k141_257427 | Low-quality |
| 17-109_k141_375093 | Low-quality |
| 17-109_k141_224224 | Not-determined |
| 17-109_k141_357151 | Not-determined |
| 17-109_k141_274401 | Not-determined |
| 17-109_k141_40411 | Not-determined |
| 17-109_k141_157473 | Not-determined |
| 17-109_k141_357155 | Not-determined |
| 17-109_k141_6501 | Not-determined |
| 17-109_k141_140610 | Not-determined |
| 17-109_k141_90247 | Not-determined |
| 17-109_k141_291815 | Not-determined |
| 17-109_k141_157477 | Not-determined |
| 17-109_k141_140617 | Not-determined |
| 17-109_k141_340814 | Not-determined |
| 17-109_k141_357170 | Not-determined |
| 17-109_k141_90251 | Not-determined |
| 17-109_k141_140619 | Not-determined |
| 17-109_k141_191485 | Not-determined |
| 17-109_k141_74258 | Not-determined |
| 17-109_k141_274412 | Not-determined |
| 17-109_k141_257448 | Not-determined |
| 17-109_k141_224244 | Low-quality |
| 17-109_k141_257455 | Not-determined |
| 17-109_k141_207417 | Not-determined |
| 17-109_k141_140624 | Low-quality |
| 17-109_k141_224248 | Not-determined |
| 17-109_k141_74260 | Not-determined |
| 17-109_k141_56236 | Low-quality |
| 17-109_k141_274423 | Not-determined |
| 17-109_k141_291830 | Not-determined |
| 17-109_k141_340827 | Not-determined |
| 17-109_k141_274425 | Low-quality |
| 17-109_k141_106827 | Not-determined |
| 17-109_k141_106828 | Not-determined |
| 17-109_k141_340833 | Not-determined |
| 17-109_k141_157492 | Not-determined |
| 17-109_k141_191495 | Not-determined |
| 17-109_k141_291843 | Not-determined |
| 17-109_k141_106834 | Not-determined |
| 17-109_k141_40422 | Not-determined |
| 17-109_k141_157499 | Not-determined |
| 17-109_k141_257469 | Low-quality |
| 17-109_k141_207428 | Not-determined |
| 17-109_k141_74272 | Not-determined |
| 17-109_k141_106838 | Not-determined |
| 17-109_k141_191510 | Not-determined |
| 17-109_k141_375113 | Not-determined |
| 17-109_k141_274444 | Not-determined |
| 17-109_k141_90260 | Not-determined |
| 17-109_k141_291850 | Low-quality |
| 17-109_k141_173914 | Not-determined |
| 17-109_k141_74285 | Not-determined |
| 17-109_k141_291858 | Not-determined |
| 17-109_k141_357205 | Not-determined |
| 17-109_k141_157519 | Not-determined |
| 17-109_k141_357207 | Not-determined |
| 17-109_k141_308734 | Not-determined |
| 17-109_k141_357208 | Not-determined |
| 17-109_k141_207448 | Not-determined |
| 17-109_k141_157523 | Not-determined |
| 17-109_k141_257483 | Not-determined |
| 17-109_k141_157526 | Not-determined |
| 17-109_k141_207458 | Not-determined |
| 17-109_k141_224265 | Low-quality |
| 17-109_k141_173928 | Low-quality |
| 17-109_k141_375140 | Not-determined |
| 17-109_k141_23351 | Not-determined |
| 17-109_k141_74298 | Not-determined |
| 17-109_k141_173934 | Not-determined |
| 17-109_k141_74301 | Not-determined |
| 17-109_k141_274461 | Not-determined |
| 17-109_k141_224271 | Not-determined |
| 17-109_k141_274465 | Not-determined |
| 17-109_k141_74308 | Not-determined |
| 17-109_k141_173941 | Not-determined |
| 17-109_k141_375151 | Low-quality |
| 17-109_k141_274469 | Not-determined |
| 17-109_k141_308746 | Not-determined |
| 17-109_k141_191518 | Not-determined |
| 17-109_k141_23365 | Not-determined |
| 17-109_k141_40443 | Not-determined |
| 17-109_k141_140647 | Not-determined |
| 17-109_k141_191529 | Not-determined |
| 17-109_k141_207478 | Not-determined |
| 17-109_k141_308753 | Not-determined |
| 17-109_k141_207481 | Not-determined |
| 17-109_k141_375159 | Not-determined |
| 17-109_k141_157543 | Not-determined |
| 17-109_k141_375162 | Not-determined |
| 17-109_k141_140653 | Not-determined |
| 17-109_k141_23375 | Low-quality |
| 17-109_k141_274491 | Not-determined |
| 17-109_k141_90278 | Not-determined |
| 17-109_k141_257514 | Not-determined |
| 17-109_k141_56259 | Low-quality |
| 17-109_k141_257519 | Not-determined |
| 17-109_k141_140667 | Not-determined |
| 17-109_k141_157564 | Not-determined |
| 17-109_k141_257528 | Not-determined |
| 17-109_k141_207492 | Not-determined |
| 17-109_k141_74333 | Not-determined |
| 17-109_k141_40468 | Not-determined |
| 17-109_k141_257532 | Not-determined |
| 17-109_k141_90296 | Not-determined |
| 17-109_k141_308776 | Not-determined |
| 17-109_k141_291885 | Not-determined |
| 17-109_k141_240829 | Not-determined |
| 17-109_k141_207499 | Not-determined |
| 17-109_k141_40471 | Not-determined |
| 17-109_k141_173985 | Low-quality |
| 17-109_k141_291887 | Not-determined |
| 17-109_k141_157585 | Not-determined |
| 17-109_k141_90314 | Not-determined |
| 17-109_k141_74340 | Not-determined |
| 17-109_k141_106858 | Low-quality |
| 17-109_k141_257543 | Not-determined |
| 17-109_k141_56274 | Not-determined |
| 17-109_k141_191561 | Low-quality |
| 17-109_k141_257549 | Not-determined |
| 17-109_k141_257552 | Not-determined |
| 17-109_k141_340846 | Not-determined |
| 17-109_k141_391954 | Not-determined |
| 17-109_k141_357247 | Not-determined |
| 17-109_k141_391956 | Low-quality |
| 17-109_k141_291912 | Not-determined |
| 17-109_k141_357252 | Not-determined |
| 17-109_k141_274512 | Low-quality |
| 17-109_k141_257558 | Low-quality |
| 17-109_k141_90330 | Not-determined |
| 17-109_k141_340855 | Low-quality |
| 17-109_k141_174002 | Not-determined |
| 17-109_k141_90333 | Not-determined |
| 17-109_k141_340860 | Not-determined |
| 17-109_k141_257568 | Not-determined |
| 17-109_k141_56280 | Not-determined |
| 17-109_k141_240849 | Not-determined |
| 17-109_k141_174005 | Low-quality |
| 17-109_k141_240857 | Not-determined |
| 17-109_k141_174011 | Not-determined |
| 17-109_k141_357275 | Not-determined |
| 17-109_k141_6525 | Not-determined |
| 17-109_k141_6528 | Not-determined |
| 17-109_k141_240869 | Not-determined |
| 17-109_k141_391989 | Not-determined |
| 17-109_k141_357280 | Not-determined |
| 17-109_k141_257580 | Not-determined |
| 17-109_k141_74380 | Not-determined |
| 17-109_k141_391999 | Not-determined |
| 17-109_k141_140678 | Low-quality |
| 17-109_k141_140683 | Not-determined |
| 17-109_k141_274528 | Not-determined |
| 17-109_k141_357294 | Low-quality |
| 17-109_k141_240870 | Low-quality |
| 17-109_k141_340875 | Not-determined |
| 17-109_k141_274533 | Low-quality |
| 17-109_k141_56315 | Not-determined |
| 17-109_k141_340882 | Not-determined |
| 17-109_k141_274542 | Not-determined |
| 17-109_k141_74403 | Not-determined |
| 17-109_k141_207544 | Not-determined |
| 17-109_k141_191577 | Not-determined |
| 17-109_k141_56323 | Not-determined |
| 17-109_k141_106888 | Low-quality |
| 17-109_k141_23414 | Not-determined |
| 17-109_k141_106890 | Not-determined |
| 17-109_k141_274552 | Not-determined |
| 17-109_k141_74410 | Not-determined |
| 17-109_k141_324960 | Not-determined |
| 17-109_k141_56329 | Not-determined |
| 17-109_k141_40511 | Not-determined |
| 17-109_k141_56334 | Not-determined |
| 17-109_k141_308823 | Not-determined |
| 17-109_k141_56337 | Not-determined |
| 17-109_k141_274561 | Not-determined |
| 17-109_k141_56342 | Not-determined |
| 17-109_k141_240890 | Not-determined |
| 17-109_k141_191590 | Not-determined |
| 17-109_k141_324978 | Not-determined |
| 17-109_k141_324982 | Not-determined |
| 17-109_k141_324983 | Not-determined |
| 17-109_k141_123232 | Low-quality |
| 17-109_k141_74423 | Low-quality |
| 17-109_k141_207560 | Not-determined |
| 17-109_k141_106894 | Not-determined |
| 17-109_k141_106899 | Not-determined |
| 17-109_k141_340913 | Not-determined |
| 17-109_k141_191603 | Not-determined |
| 17-109_k141_191604 | Not-determined |
| 17-109_k141_123241 | Not-determined |
| 17-109_k141_357330 | Not-determined |
| 17-109_k141_191608 | Not-determined |
| 17-109_k141_191610 | Not-determined |
| 17-109_k141_191611 | Not-determined |
| 17-109_k141_392011 | Low-quality |
| 17-109_k141_90353 | Not-determined |
| 17-109_k141_106905 | Not-determined |
| 17-109_k141_324994 | Not-determined |
| 17-109_k141_123245 | Not-determined |
| 17-109_k141_90355 | Not-determined |
| 17-109_k141_240910 | Low-quality |
| 17-109_k141_90357 | Not-determined |
| 17-109_k141_140735 | Not-determined |
| 17-109_k141_40544 | Not-determined |
| 17-109_k141_207571 | Not-determined |
| 17-109_k141_240915 | Low-quality |
| 17-109_k141_291949 | Not-determined |
| 17-109_k141_74446 | Not-determined |
| 17-109_k141_340932 | Not-determined |
| 17-109_k141_392033 | Not-determined |
| 17-109_k141_157623 | Not-determined |
| 17-109_k141_140739 | Not-determined |
| 17-109_k141_291954 | Not-determined |
| 17-109_k141_106915 | Not-determined |
| 17-109_k141_392038 | Not-determined |
| 17-109_k141_174061 | Not-determined |
| 17-109_k141_106916 | Not-determined |
| 17-109_k141_207578 | Not-determined |
| 17-109_k141_90366 | Low-quality |
| 17-109_k141_6544 | Not-determined |
| 17-109_k141_40552 | Not-determined |
| 17-109_k141_23423 | Not-determined |
| 17-109_k141_325004 | Not-determined |
| 17-109_k141_274565 | Not-determined |
| 17-109_k141_106928 | Not-determined |
| 17-109_k141_240927 | Not-determined |
| 17-109_k141_240931 | Not-determined |
| 17-109_k141_191615 | Not-determined |
| 17-109_k141_392047 | Not-determined |
| 17-109_k141_106932 | Not-determined |
| 17-109_k141_140751 | Not-determined |
| 17-109_k141_123273 | Not-determined |
| 17-109_k141_325017 | Not-determined |
| 17-109_k141_6559 | Low-quality |
| 17-109_k141_74461 | Not-determined |
| 17-109_k141_174068 | Not-determined |
| 17-109_k141_392062 | Not-determined |
| 17-109_k141_274579 | Not-determined |
| 17-109_k141_106935 | Low-quality |
| 17-109_k141_106937 | Not-determined |
| 17-109_k141_123275 | Not-determined |
| 17-109_k141_325023 | Not-determined |
| 17-109_k141_123279 | Not-determined |
| 17-109_k141_325025 | Not-determined |
| 17-109_k141_308865 | Low-quality |
| 17-109_k141_257608 | Not-determined |
| 17-109_k141_207617 | Not-determined |
| 17-109_k141_40566 | Not-determined |
| 17-109_k141_174083 | Low-quality |
| 17-109_k141_207620 | Not-determined |
| 17-109_k141_106947 | Not-determined |
| 17-109_k141_40568 | Not-determined |
| 17-109_k141_106950 | Not-determined |
| 17-109_k141_340947 | Not-determined |
| 17-109_k141_90396 | Not-determined |
| 17-109_k141_207628 | Not-determined |
| 17-109_k141_308873 | Low-quality |
| 17-109_k141_240956 | Not-determined |
| 17-109_k141_240958 | Not-determined |
| 17-109_k141_157667 | Not-determined |
| 17-109_k141_240961 | Not-determined |
| 17-109_k141_174092 | Not-determined |
| 17-109_k141_224334 | Not-determined |
| 17-109_k141_308884 | Low-quality |
| 17-109_k141_157676 | Not-determined |
| 17-109_k141_174095 | Not-determined |
| 17-109_k141_157679 | Not-determined |
| 17-109_k141_392079 | Not-determined |
| 17-109_k141_157681 | Not-determined |
| 17-109_k141_191626 | Low-quality |
| 17-109_k141_157683 | Not-determined |
| 17-109_k141_157684 | Not-determined |
| 17-109_k141_357353 | Not-determined |
| 17-109_k141_274598 | Not-determined |
| 17-109_k141_274604 | Not-determined |
| 17-109_k141_90400 | Not-determined |
| 17-109_k141_340968 | Not-determined |
| 17-109_k141_123316 | Not-determined |
| 17-109_k141_325046 | Low-quality |
| 17-109_k141_174106 | Low-quality |
| 17-109_k141_224363 | Not-determined |
| 17-109_k141_106959 | Low-quality |
| 17-109_k141_224365 | Not-determined |
| 17-109_k141_340977 | Not-determined |
| 17-109_k141_340979 | Not-determined |
| 17-109_k141_123328 | Not-determined |
| 17-109_k141_123329 | Not-determined |
| 17-109_k141_274616 | Not-determined |
| 17-109_k141_23464 | Not-determined |
| 17-109_k141_224374 | Not-determined |
| 17-109_k141_340984 | Low-quality |
| 17-109_k141_392090 | Not-determined |
| 17-109_k141_56354 | Not-determined |
| 17-109_k141_392094 | Not-determined |
| 17-109_k141_392100 | Not-determined |
| 17-109_k141_174115 | Not-determined |
| 17-109_k141_274636 | Not-determined |
| 17-109_k141_392103 | Low-quality |
| 17-109_k141_157735 | Not-determined |
| 17-109_k141_274637 | Low-quality |
| 17-109_k141_340996 | Low-quality |
| 17-109_k141_106983 | Not-determined |
| 17-109_k141_241000 | Not-determined |
| 17-109_k141_106988 | Not-determined |
| 17-109_k141_90417 | Not-determined |
| 17-109_k141_357369 | Low-quality |
| 17-109_k141_308920 | Not-determined |
| 17-109_k141_392111 | Low-quality |
| 17-109_k141_274649 | Not-determined |

| | |
|---|---|
| 17-109_k141_241011 | Not-determined |
| 17-109_k141_90420 | Not-determined |
| 17-109_k141_274652 | Not-determined |
| 17-109_k141_174134 | Not-determined |
| 17-109_k141_6623 | Not-determined |
| 17-109_k141_6624 | Not-determined |
| 17-109_k141_274654 | Not-determined |
| 17-109_k141_90424 | Not-determined |
| 17-109_k141_274657 | Not-determined |
| 17-109_k141_341001 | Not-determined |
| 17-109_k141_90431 | Not-determined |
| 17-109_k141_274662 | Not-determined |
| 17-109_k141_357381 | Not-determined |
| 17-109_k141_257637 | Not-determined |
| 17-109_k141_40582 | Not-determined |
| 17-109_k141_74496 | Not-determined |
| 17-109_k141_207661 | Not-determined |
| 17-109_k141_357386 | Low-quality |
| 17-109_k141_74502 | Not-determined |
| 17-109_k141_140789 | Not-determined |
| 17-109_k141_341014 | Not-determined |
| 17-109_k141_123343 | Not-determined |
| 17-109_k141_123345 | Not-determined |
| 17-109_k141_274679 | Not-determined |
| 17-109_k141_207671 | Not-determined |
| 17-109_k141_308946 | Not-determined |
| 17-109_k141_157772 | Not-determined |
| 17-109_k141_207676 | Not-determined |
| 17-109_k141_207680 | Not-determined |
| 17-109_k141_23485 | Not-determined |
| 17-109_k141_191646 | Not-determined |
| 17-109_k141_140797 | Not-determined |
| 17-109_k141_375221 | Not-determined |
| 17-109_k141_74507 | Low-quality |
| 17-109_k141_341026 | Not-determined |
| 17-109_k141_357407 | Not-determined |
| 17-109_k141_292015 | Not-determined |
| 17-109_k141_191658 | Low-quality |
| 17-109_k141_292019 | Not-determined |
| 17-109_k141_257672 | Not-determined |
| 17-109_k141_292021 | Not-determined |
| 17-109_k141_274703 | Not-determined |
| 17-109_k141_325115 | Not-determined |
| 17-109_k141_174158 | Low-quality |
| 17-109_k141_257676 | Not-determined |
| 17-109_k141_6645 | Not-determined |
| 17-109_k141_292026 | Not-determined |
| 17-109_k141_325122 | Low-quality |
| 17-109_k141_224418 | Low-quality |
| 17-109_k141_325124 | Low-quality |
| 17-109_k141_308968 | Not-determined |
| 17-109_k141_292029 | Not-determined |
| 17-109_k141_191686 | Not-determined |
| 17-109_k141_341039 | Not-determined |
| 17-109_k141_90468 | Low-quality |
| 17-109_k141_375235 | Not-determined |
| 17-109_k141_341043 | Not-determined |
| 17-109_k141_123350 | Not-determined |
| 17-109_k141_6662 | Not-determined |
| 17-109_k141_308974 | Low-quality |
| 17-109_k141_308976 | Not-determined |
| 17-109_k141_241039 | Not-determined |
| 17-109_k141_224430 | Not-determined |
| 17-109_k141_392127 | Not-determined |
| 17-109_k141_74534 | Not-determined |
| 17-109_k141_257693 | Low-quality |
| 17-109_k141_392133 | Not-determined |
| 17-109_k141_392135 | Not-determined |
| 17-109_k141_207702 | Not-determined |
| 17-109_k141_107035 | Not-determined |
| 17-109_k141_23515 | Not-determined |
| 17-109_k141_207707 | Low-quality |
| 17-109_k141_56400 | Low-quality |
| 17-109_k141_207711 | Not-determined |
| 17-109_k141_357448 | Not-determined |
| 17-109_k141_308990 | Low-quality |
| 17-109_k141_140801 | Not-determined |
| 17-109_k141_357456 | Not-determined |
| 17-109_k141_157807 | Not-determined |
| 17-109_k141_274735 | Not-determined |
| 17-109_k141_257715 | Not-determined |
| 17-109_k141_308999 | Not-determined |
| 17-109_k141_257716 | Not-determined |
| 17-109_k141_309000 | Not-determined |
| 17-109_k141_123381 | Not-determined |
| 17-109_k141_357459 | Not-determined |
| 17-109_k141_309003 | Not-determined |
| 17-109_k141_292066 | Not-determined |
| 17-109_k141_224442 | Not-determined |
| 17-109_k141_375284 | Not-determined |
| 17-109_k141_207723 | Low-quality |
| 17-109_k141_292074 | Low-quality |
| 17-109_k141_174196 | Not-determined |
| 17-109_k141_191735 | Not-determined |
| 17-109_k141_357464 | Not-determined |
| 17-109_k141_6695 | Low-quality |
| 17-109_k141_357467 | Not-determined |
| 17-109_k141_107049 | Not-determined |
| 17-109_k141_325156 | Not-determined |
| 17-109_k141_392177 | Not-determined |
| 17-109_k141_309024 | Not-determined |
| 17-109_k141_392180 | Not-determined |
| 17-109_k141_56413 | Not-determined |
| 17-109_k141_107063 | Not-determined |
| 17-109_k141_274762 | Not-determined |
| 17-109_k141_274764 | Not-determined |
| 17-109_k141_140849 | Not-determined |
| 17-109_k141_207758 | Not-determined |
| 17-109_k141_325165 | Not-determined |
| 17-109_k141_74548 | Not-determined |
| 17-109_k141_157818 | Not-determined |
| 17-109_k141_6703 | Not-determined |
| 17-109_k141_207763 | Not-determined |
| 17-109_k141_224474 | Not-determined |
| 17-109_k141_392192 | Not-determined |
| 17-109_k141_56430 | Not-determined |
| 17-109_k141_292101 | Not-determined |
| 17-109_k141_140861 | Not-determined |
| 17-109_k141_392198 | Low-quality |
| 17-109_k141_123436 | Not-determined |
| 17-109_k141_207771 | Low-quality |
| 17-109_k141_241079 | Not-determined |
| 17-109_k141_207773 | Low-quality |
| 17-109_k141_207775 | Not-determined |
| 17-109_k141_123439 | Not-determined |
| 17-109_k141_123440 | Not-determined |
| 17-109_k141_292105 | Not-determined |
| 17-109_k141_107100 | Not-determined |
| 17-109_k141_257754 | Not-determined |
| 17-109_k141_241088 | Not-determined |
| 17-109_k141_107105 | Not-determined |
| 17-109_k141_207786 | Not-determined |
| 17-109_k141_392206 | Not-determined |
| 17-109_k141_357500 | Not-determined |
| 17-109_k141_174228 | Not-determined |
| 17-109_k141_325186 | Not-determined |
| 17-109_k141_224482 | Not-determined |
| 17-109_k141_292122 | Low-quality |
| 17-109_k141_224485 | Not-determined |
| 17-109_k141_174238 | Low-quality |
| 17-109_k141_224486 | Not-determined |
| 17-109_k141_392211 | Not-determined |
| 17-109_k141_375338 | Not-determined |
| 17-109_k141_40614 | Low-quality |
| 17-109_k141_292128 | Not-determined |
| 17-109_k141_274793 | Low-quality |
| 17-109_k141_257777 | Not-determined |
| 17-109_k141_40621 | Not-determined |
| 17-109_k141_40622 | Low-quality |
| 17-109_k141_174244 | Not-determined |
| 17-109_k141_241102 | Not-determined |
| 17-109_k141_56448 | Not-determined |
| 17-109_k141_341064 | Not-determined |
| 17-109_k141_392223 | Not-determined |
| 17-109_k141_274801 | Not-determined |
| 17-109_k141_174250 | Low-quality |
| 17-109_k141_392230 | Low-quality |
| 17-109_k141_157853 | Not-determined |
| 17-109_k141_274809 | Not-determined |
| 17-109_k141_123459 | Not-determined |
| 17-109_k141_274812 | Not-determined |
| 17-109_k141_174261 | Not-determined |
| 17-109_k141_292144 | Not-determined |
| 17-109_k141_157869 | Low-quality |
| 17-109_k141_274820 | Not-determined |
| 17-109_k141_392233 | Not-determined |
| 17-109_k141_292155 | Not-determined |
| 17-109_k141_292156 | Low-quality |
| 17-109_k141_56466 | Not-determined |
| 17-109_k141_6735 | Not-determined |
| 17-109_k141_241131 | Not-determined |
| 17-109_k141_56470 | Not-determined |
| 17-109_k141_140882 | Not-determined |
| 17-109_k141_56475 | Not-determined |
| 17-109_k141_224497 | Not-determined |
| 17-109_k141_157876 | Not-determined |
| 17-109_k141_224498 | Low-quality |
| 17-109_k141_174272 | Not-determined |
| 17-109_k141_292167 | Low-quality |
| 17-109_k141_257816 | Not-determined |
| 17-109_k141_23533 | Not-determined |
| 17-109_k141_257818 | Not-determined |
| 17-109_k141_174276 | Not-determined |
| 17-109_k141_257830 | Not-determined |
| 17-109_k141_174279 | Not-determined |
| 17-109_k141_292178 | Not-determined |
| 17-109_k141_325196 | Not-determined |
| 17-109_k141_56481 | Not-determined |
| 17-109_k141_257837 | Low-quality |
| 17-109_k141_392243 | Not-determined |
| 17-109_k141_392244 | Not-determined |
| 17-109_k141_90509 | Not-determined |
| 17-109_k141_140907 | Low-quality |
| 17-109_k141_23550 | Not-determined |
| 17-109_k141_23552 | Not-determined |
| 17-109_k141_224517 | Not-determined |
| 17-109_k141_292184 | Not-determined |
| 17-109_k141_257847 | Not-determined |
| 17-109_k141_257849 | Not-determined |
| 17-109_k141_257850 | Not-determined |
| 17-109_k141_123468 | Not-determined |
| 17-109_k141_23556 | Not-determined |
| 17-109_k141_23558 | Not-determined |
| 17-109_k141_56492 | Not-determined |
| 17-109_k141_174288 | Low-quality |
| 17-109_k141_224521 | Not-determined |
| 17-109_k141_157917 | Not-determined |
| 17-109_k141_224528 | Not-determined |
| 17-109_k141_224529 | Not-determined |
| 17-109_k141_90533 | Not-determined |
| 17-109_k141_90534 | Not-determined |
| 17-109_k141_56507 | Low-quality |
| 17-109_k141_357542 | Not-determined |
| 17-109_k141_6746 | Not-determined |
| 17-109_k141_257861 | Not-determined |
| 17-109_k141_224538 | Not-determined |
| 17-109_k141_6750 | Not-determined |
| 17-109_k141_40636 | Low-quality |
| 17-109_k141_224545 | Not-determined |
| 17-109_k141_375352 | Not-determined |
| 17-109_k141_375353 | Not-determined |
| 17-109_k141_157923 | Low-quality |
| 17-109_k141_241147 | Not-determined |
| 17-109_k141_107139 | Not-determined |
| 17-109_k141_341091 | Low-quality |
| 17-109_k141_107142 | Not-determined |
| 17-109_k141_341094 | Not-determined |
| 17-109_k141_123482 | Not-determined |
| 17-109_k141_123487 | Not-determined |
| 17-109_k141_224555 | Not-determined |
| 17-109_k141_241163 | Not-determined |
| 17-109_k141_90560 | Not-determined |
| 17-109_k141_157935 | Not-determined |
| 17-109_k141_341119 | Not-determined |
| 17-109_k141_241166 | Low-quality |
| 17-109_k141_241167 | Not-determined |
| 17-109_k141_107155 | Not-determined |
| 17-109_k141_23567 | Not-determined |
| 17-109_k141_341128 | Not-determined |
| 17-109_k141_241174 | Not-determined |
| 17-109_k141_191830 | Not-determined |
| 17-109_k141_357573 | Not-determined |
| 17-109_k141_224570 | Not-determined |
| 17-109_k141_107180 | Not-determined |
| 17-109_k141_40660 | Not-determined |
| 17-109_k141_241188 | Not-determined |
| 17-109_k141_140912 | Low-quality |
| 17-109_k141_157939 | Not-determined |
| 17-109_k141_123499 | Not-determined |
| 17-109_k141_157953 | Not-determined |
| 17-109_k141_325254 | Not-determined |
| 17-109_k141_241204 | Not-determined |
| 17-109_k141_375376 | Not-determined |
| 17-109_k141_241205 | Not-determined |
| 17-109_k141_341147 | Not-determined |
| 17-109_k141_123516 | Not-determined |
| 17-109_k141_241209 | Low-quality |
| 17-109_k141_40682 | Not-determined |
| 17-109_k141_325265 | Not-determined |
| 17-109_k141_392273 | Not-determined |
| 17-109_k141_357578 | Not-determined |
| 17-109_k141_357579 | Not-determined |
| 17-109_k141_6768 | Not-determined |
| 17-109_k141_56563 | Not-determined |
| 17-109_k141_40689 | Not-determined |
| 17-109_k141_107194 | Not-determined |
| 17-109_k141_325270 | Not-determined |
| 17-109_k141_23611 | Low-quality |
| 17-109_k141_375382 | Not-determined |
| 17-109_k141_274856 | Low-quality |
| 17-109_k141_325273 | Not-determined |
| 17-109_k141_274858 | Not-determined |
| 17-109_k141_123529 | Not-determined |
| 17-109_k141_224587 | Not-determined |
| 17-109_k141_392292 | Not-determined |
| 17-109_k141_274864 | Not-determined |
| 17-109_k141_123533 | Not-determined |
| 17-109_k141_257880 | Not-determined |
| 17-109_k141_325278 | Not-determined |
| 17-109_k141_392295 | Not-determined |
| 17-109_k141_257883 | Not-determined |
| 17-109_k141_241225 | Not-determined |
| 17-109_k141_257884 | Not-determined |
| 17-109_k141_140933 | Low-quality |
| 17-109_k141_241229 | Not-determined |
| 17-109_k141_257887 | Not-determined |
| 17-109_k141_392296 | Not-determined |
| 17-109_k141_257895 | Low-quality |
| 17-109_k141_375400 | Not-determined |
| 17-109_k141_341163 | Not-determined |
| 17-109_k141_23638 | Not-determined |
| 17-109_k141_341167 | Not-determined |
| 17-109_k141_341170 | Not-determined |
| 17-109_k141_309047 | Not-determined |
| 17-109_k141_274871 | Not-determined |
| 17-109_k141_123557 | Not-determined |
| 17-109_k141_56593 | Not-determined |
| 17-109_k141_224617 | Not-determined |
| 17-109_k141_392303 | Low-quality |
| 17-109_k141_392304 | Not-determined |
| 17-109_k141_23648 | Not-determined |
| 17-109_k141_224622 | Not-determined |
| 17-109_k141_241256 | Not-determined |
| 17-109_k141_224632 | Not-determined |
| 17-109_k141_375420 | Not-determined |
| 17-109_k141_375421 | Not-determined |
| 17-109_k141_392321 | Not-determined |
| 17-109_k141_6792 | Not-determined |
| 17-109_k141_174291 | Not-determined |
| 17-109_k141_157978 | Not-determined |
| 17-109_k141_375429 | Not-determined |
| 17-109_k141_375435 | Low-quality |
| 17-109_k141_241268 | Not-determined |
| 17-109_k141_123579 | Not-determined |
| 17-109_k141_274878 | Not-determined |
| 17-109_k141_107219 | Not-determined |
| 17-109_k141_274882 | Not-determined |
| 17-109_k141_274883 | Not-determined |
| 17-109_k141_107221 | Not-determined |
| 17-109_k141_392329 | Not-determined |
| 17-109_k141_274885 | Not-determined |
| 17-109_k141_191851 | Not-determined |
| 17-109_k141_157991 | Not-determined |
| 17-109_k141_392330 | Not-determined |
| 17-109_k141_325296 | Not-determined |
| 17-109_k141_375443 | Not-determined |
| 17-109_k141_274890 | Not-determined |
| 17-109_k141_325298 | Not-determined |
| 17-109_k141_309077 | Not-determined |
| 17-109_k141_274902 | Not-determined |
| 17-109_k141_23670 | Not-determined |
| 17-109_k141_6811 | Low-quality |
| 17-109_k141_23672 | Not-determined |
| 17-109_k141_123598 | Low-quality |
| 17-109_k141_23675 | Low-quality |
| 17-109_k141_6816 | Not-determined |
| 17-109_k141_309086 | Not-determined |
| 17-109_k141_392355 | Not-determined |
| 17-109_k141_274922 | Not-determined |
| 17-109_k141_392358 | Not-determined |
| 17-109_k141_140950 | Not-determined |
| 17-109_k141_241289 | Low-quality |
| 17-109_k141_274926 | Not-determined |
| 17-109_k141_224680 | Not-determined |
| 17-109_k141_309094 | Not-determined |
| 17-109_k141_191885 | Not-determined |
| 17-109_k141_309098 | Not-determined |
| 17-109_k141_74612 | Not-determined |
| 17-109_k141_158006 | Not-determined |
| 17-109_k141_140960 | Not-determined |
| 17-109_k141_191889 | Not-determined |
| 17-109_k141_241303 | Not-determined |
| 17-109_k141_392366 | Not-determined |
| 17-109_k141_241307 | Low-quality |
| 17-109_k141_309105 | Not-determined |
| 17-109_k141_325313 | Not-determined |
| 17-109_k141_191895 | Not-determined |
| 17-109_k141_6827 | Not-determined |
| 17-109_k141_207816 | Not-determined |
| 17-109_k141_6834 | Not-determined |
| 17-109_k141_375478 | Not-determined |
| 17-109_k141_123609 | Not-determined |
| 17-109_k141_375480 | Not-determined |
| 17-109_k141_6837 | Not-determined |
| 17-109_k141_309120 | Not-determined |
| 17-109_k141_257950 | Not-determined |
| 17-109_k141_392390 | Not-determined |
| 17-109_k141_257953 | Low-quality |
| 17-109_k141_292222 | Not-determined |
| 17-109_k141_392395 | Not-determined |
| 17-109_k141_56628 | Not-determined |
| 17-109_k141_325341 | Not-determined |
| 17-109_k141_341253 | Not-determined |
| 17-109_k141_56629 | Not-determined |
| 17-109_k141_123639 | Not-determined |
| 17-109_k141_292229 | Not-determined |
| 17-109_k141_292233 | Not-determined |
| 17-109_k141_325343 | Not-determined |
| 17-109_k141_191900 | Not-determined |
| 17-109_k141_375510 | Not-determined |
| 17-109_k141_123649 | Not-determined |
| 17-109_k141_357662 | Not-determined |
| 17-109_k141_191904 | Not-determined |
| 17-109_k141_123655 | Not-determined |
| 17-109_k141_140997 | Not-determined |
| 17-109_k141_123659 | Not-determined |
| 17-109_k141_56652 | Not-determined |
| 17-109_k141_23691 | Not-determined |
| 17-109_k141_341271 | Not-determined |
| 17-109_k141_107244 | Not-determined |
| 17-109_k141_56661 | Not-determined |
| 17-109_k141_274931 | Not-determined |
| 17-109_k141_107250 | Not-determined |
| 17-109_k141_274935 | Not-determined |
| 17-109_k141_341277 | Not-determined |
| 17-109_k141_23703 | Not-determined |
| 17-109_k141_158026 | Low-quality |
| 17-109_k141_158027 | Not-determined |
| 17-109_k141_375520 | Not-determined |
| 17-109_k141_325365 | Not-determined |
| 17-109_k141_325368 | Not-determined |
| 17-109_k141_107261 | Not-determined |
| 17-109_k141_375526 | Not-determined |
| 17-109_k141_257982 | Low-quality |
| 17-109_k141_375530 | Not-determined |
| 17-109_k141_23712 | Not-determined |
| 17-109_k141_292250 | Not-determined |
| 17-109_k141_107273 | Not-determined |
| 17-109_k141_241359 | Low-quality |
| 17-109_k141_375537 | Not-determined |
| 17-109_k141_40741 | Low-quality |
| 17-109_k141_40742 | Not-determined |
| 17-109_k141_141014 | Not-determined |
| 17-109_k141_107277 | Not-determined |
| 17-109_k141_6902 | Not-determined |
| 17-109_k141_257995 | Not-determined |
| 17-109_k141_241374 | Not-determined |
| 17-109_k141_158054 | Not-determined |
| 17-109_k141_74627 | Low-quality |
| 17-109_k141_74628 | Not-determined |
| 17-109_k141_207849 | Not-determined |
| 17-109_k141_258003 | Not-determined |
| 17-109_k141_274969 | Not-determined |
| 17-109_k141_341289 | Not-determined |
| 17-109_k141_258006 | Not-determined |
| 17-109_k141_274980 | Not-determined |
| 17-109_k141_191922 | Not-determined |
| 17-109_k141_274982 | Not-determined |
| 17-109_k141_309149 | Not-determined |
| 17-109_k141_341303 | Not-determined |
| 17-109_k141_357706 | Low-quality |
| 17-109_k141_392414 | Not-determined |
| 17-109_k141_224706 | Not-determined |
| 17-109_k141_141035 | Not-determined |
| 17-109_k141_375562 | Not-determined |
| 17-109_k141_141039 | Not-determined |
| 17-109_k141_56694 | Not-determined |
| 17-109_k141_6928 | Not-determined |
| 17-109_k141_6929 | Not-determined |
| 17-109_k141_241388 | Not-determined |
| 17-109_k141_241390 | Not-determined |
| 17-109_k141_191928 | Not-determined |
| 17-109_k141_6935 | Not-determined |
| 17-109_k141_241395 | Not-determined |
| 17-109_k141_275013 | Not-determined |
| 17-109_k141_375569 | Low-quality |
| 17-109_k141_6940 | Not-determined |
| 17-109_k141_174296 | Not-determined |
| 17-109_k141_375572 | Not-determined |
| 17-109_k141_74660 | Not-determined |
| 17-109_k141_224717 | Not-determined |
| 17-109_k141_241404 | Not-determined |
| 17-109_k141_207860 | Not-determined |
| 17-109_k141_141051 | Not-determined |
| 17-109_k141_325396 | Not-determined |
| 17-109_k141_241411 | Not-determined |
| 17-109_k141_191941 | Not-determined |
| 17-109_k141_158073 | Not-determined |
| 17-109_k141_141057 | Low-quality |
| 17-109_k141_392438 | Not-determined |
| 17-109_k141_191948 | Not-determined |
| 17-109_k141_191950 | Not-determined |
| 17-109_k141_123724 | Not-determined |
| 17-109_k141_191953 | Not-determined |
| 17-109_k141_392443 | Not-determined |
| 17-109_k141_392448 | Not-determined |
| 17-109_k141_158080 | Not-determined |
| 17-109_k141_392451 | Not-determined |
| 17-109_k141_141063 | Low-quality |
| 17-109_k141_191964 | Not-determined |
| 17-109_k141_74668 | Not-determined |
| 17-109_k141_275035 | Not-determined |
| 17-109_k141_357748 | Not-determined |
| 17-109_k141_224736 | Low-quality |
| 17-109_k141_224737 | Not-determined |
| 17-109_k141_207881 | Not-determined |
| 17-109_k141_224739 | Not-determined |
| 17-109_k141_191976 | Not-determined |
| 17-109_k141_224745 | Not-determined |
| 17-109_k141_23775 | Not-determined |
| 17-109_k141_74687 | Not-determined |
| 17-109_k141_123739 | Not-determined |
| 17-109_k141_325411 | Not-determined |
| 17-109_k141_23785 | Not-determined |
| 17-109_k141_74688 | Not-determined |
| 17-109_k141_23790 | Not-determined |
| 17-109_k141_6956 | Low-quality |
| 17-109_k141_192004 | Low-quality |
| 17-109_k141_375599 | Not-determined |
| 17-109_k141_174342 | Low-quality |
| 17-109_k141_241447 | Not-determined |
| 17-109_k141_74698 | Not-determined |
| 17-109_k141_207928 | Not-determined |
| 17-109_k141_6957 | Not-determined |
| 17-109_k141_174355 | Low-quality |
| 17-109_k141_375613 | Low-quality |
| 17-109_k141_224792 | Not-determined |
| 17-109_k141_158112 | Low-quality |
| 17-109_k141_158115 | Not-determined |
| 17-109_k141_357774 | Not-determined |
| 17-109_k141_357775 | Not-determined |
| 17-109_k141_375625 | Not-determined |
| 17-109_k141_158118 | Not-determined |
| 17-109_k141_357777 | Not-determined |
| 17-109_k141_357778 | Not-determined |
| 17-109_k141_241464 | Not-determined |
| 17-109_k141_141098 | Not-determined |
| 17-109_k141_292283 | Not-determined |
| 17-109_k141_292285 | Not-determined |
| 17-109_k141_90571 | Not-determined |
| 17-109_k141_141106 | Not-determined |
| 17-109_k141_74737 | Low-quality |
| 17-109_k141_241495 | Low-quality |
| 17-109_k141_224805 | Not-determined |
| 17-109_k141_40808 | Not-determined |
| 17-109_k141_224807 | Not-determined |
| 17-109_k141_224808 | Not-determined |
| 17-109_k141_325421 | Not-determined |
| 17-109_k141_23845 | Not-determined |
| 17-109_k141_74741 | Not-determined |
| 17-109_k141_241499 | Not-determined |
| 17-109_k141_174382 | Not-determined |
| 17-109_k141_325423 | Not-determined |
| 17-109_k141_141109 | Not-determined |
| 17-109_k141_174384 | Not-determined |
| 17-109_k141_292292 | Not-determined |
| 17-109_k141_192020 | Not-determined |
| 17-109_k141_241504 | Not-determined |
| 17-109_k141_74745 | Not-determined |
| 17-109_k141_141114 | Not-determined |
| 17-109_k141_74748 | Low-quality |
| 17-109_k141_141116 | Not-determined |
| 17-109_k141_325434 | Not-determined |
| 17-109_k141_74752 | Not-determined |
| 17-109_k141_174392 | Not-determined |
| 17-109_k141_275047 | Not-determined |
| 17-109_k141_325442 | Not-determined |
| 17-109_k141_309189 | Not-determined |
| 17-109_k141_74763 | Not-determined |
| 17-109_k141_275049 | Not-determined |
| 17-109_k141_309193 | Not-determined |
| 17-109_k141_141132 | Not-determined |
| 17-109_k141_309197 | Not-determined |
| 17-109_k141_158133 | Not-determined |
| 17-109_k141_40843 | Not-determined |
| 17-109_k141_192029 | Not-determined |
| 17-109_k141_23859 | Not-determined |
| 17-109_k141_40850 | Not-determined |
| 17-109_k141_275059 | Not-determined |
| 17-109_k141_275065 | Not-determined |
| 17-109_k141_90598 | Not-determined |
| 17-109_k141_275069 | Not-determined |
| 17-109_k141_40862 | Low-quality |
| 17-109_k141_357790 | Low-quality |
| 17-109_k141_275074 | Not-determined |
| 17-109_k141_375683 | Not-determined |
| 17-109_k141_357794 | Not-determined |
| 17-109_k141_375693 | Not-determined |
| 17-109_k141_107295 | Not-determined |
| 17-109_k141_158163 | Not-determined |
| 17-109_k141_174414 | Not-determined |
| 17-109_k141_174418 | Not-determined |
| 17-109_k141_174420 | Not-determined |
| 17-109_k141_192048 | Not-determined |
| 17-109_k141_325448 | Low-quality |
| 17-109_k141_158172 | Not-determined |
| 17-109_k141_224847 | Not-determined |
| 17-109_k141_192052 | Not-determined |
| 17-109_k141_192053 | Not-determined |
| 17-109_k141_341361 | Not-determined |
| 17-109_k141_224849 | Low-quality |
| 17-109_k141_375701 | Low-quality |
| 17-109_k141_158179 | Not-determined |
| 17-109_k141_258025 | Not-determined |
| 17-109_k141_90610 | Not-determined |
| 17-109_k141_90612 | Not-determined |
| 17-109_k141_40875 | Not-determined |
| 17-109_k141_40877 | Not-determined |
| 17-109_k141_107318 | Low-quality |
| 17-109_k141_107323 | Not-determined |
| 17-109_k141_56825 | Not-determined |
| 17-109_k141_224854 | Not-determined |
| 17-109_k141_224858 | Not-determined |
| 17-109_k141_158194 | Not-determined |
| 17-109_k141_40879 | Not-determined |
| 17-109_k141_357834 | Not-determined |
| 17-109_k141_23895 | Not-determined |
| 17-109_k141_40893 | Not-determined |
| 17-109_k141_56838 | Not-determined |

| | |
|---|---|
| 17-109_k141_107339 | Not-determined |
| 17-109_k141_56840 | Not-determined |
| 17-109_k141_325464 | Not-determined |
| 17-109_k141_6963 | Not-determined |
| 17-109_k141_275120 | Not-determined |
| 17-109_k141_123814 | Not-determined |
| 17-109_k141_6969 | Not-determined |
| 17-109_k141_275123 | Not-determined |
| 17-109_k141_56851 | Low-quality |
| 17-109_k141_258048 | Not-determined |
| 17-109_k141_275125 | Not-determined |
| 17-109_k141_341386 | Not-determined |
| 17-109_k141_275128 | Not-determined |
| 17-109_k141_357866 | Not-determined |
| 17-109_k141_241533 | Not-determined |
| 17-109_k141_123823 | Not-determined |
| 17-109_k141_192095 | Low-quality |
| 17-109_k141_309249 | Not-determined |
| 17-109_k141_123828 | Not-determined |
| 17-109_k141_40908 | Low-quality |
| 17-109_k141_275137 | Low-quality |
| 17-109_k141_325468 | Not-determined |
| 17-109_k141_56864 | Not-determined |
| 17-109_k141_341404 | Not-determined |
| 17-109_k141_56871 | Not-determined |
| 17-109_k141_341408 | Not-determined |
| 17-109_k141_23919 | Low-quality |
| 17-109_k141_309267 | Not-determined |
| 17-109_k141_309268 | Not-determined |
| 17-109_k141_40927 | Not-determined |
| 17-109_k141_275161 | Not-determined |
| 17-109_k141_275164 | Not-determined |
| 17-109_k141_392475 | Not-determined |
| 17-109_k141_107371 | Not-determined |
| 17-109_k141_158199 | Not-determined |
| 17-109_k141_107377 | Not-determined |
| 17-109_k141_158203 | Not-determined |
| 17-109_k141_174456 | Not-determined |
| 17-109_k141_23926 | Not-determined |
| 17-109_k141_341420 | Low-quality |
| 17-109_k141_309279 | Not-determined |
| 17-109_k141_325487 | Low-quality |
| 17-109_k141_392489 | Not-determined |
| 17-109_k141_309280 | Not-determined |
| 17-109_k141_158216 | Not-determined |
| 17-109_k141_325490 | Not-determined |
| 17-109_k141_325494 | Not-determined |
| 17-109_k141_192121 | Not-determined |
| 17-109_k141_141163 | Not-determined |
| 17-109_k141_258077 | Not-determined |
| 17-109_k141_56880 | Not-determined |
| 17-109_k141_357897 | Not-determined |
| 17-109_k141_141169 | Low-quality |
| 17-109_k141_325506 | Not-determined |
| 17-109_k141_56892 | Not-determined |
| 17-109_k141_56893 | Not-determined |
| 17-109_k141_357914 | Not-determined |
| 17-109_k141_192135 | Not-determined |
| 17-109_k141_275202 | Low-quality |
| 17-109_k141_23928 | Low-quality |
| 17-109_k141_158241 | Not-determined |
| 17-109_k141_23935 | Not-determined |
| 17-109_k141_309324 | Not-determined |
| 17-109_k141_325520 | Not-determined |
| 17-109_k141_158250 | Not-determined |
| 17-109_k141_23943 | Not-determined |
| 17-109_k141_56903 | Not-determined |
| 17-109_k141_325527 | Not-determined |
| 17-109_k141_341447 | Not-determined |
| 17-109_k141_141187 | Not-determined |
| 17-109_k141_74829 | Not-determined |
| 17-109_k141_141190 | Low-quality |
| 17-109_k141_56915 | Not-determined |
| 17-109_k141_341457 | Not-determined |
| 17-109_k141_357922 | Not-determined |
| 17-109_k141_357923 | Not-determined |
| 17-109_k141_141199 | Not-determined |
| 17-109_k141_275221 | Not-determined |
| 17-109_k141_275222 | Not-determined |
| 17-109_k141_309348 | Not-determined |
| 17-109_k141_357927 | Not-determined |
| 17-109_k141_123868 | Not-determined |
| 17-109_k141_357930 | Not-determined |
| 17-109_k141_107418 | Not-determined |
| 17-109_k141_357935 | Not-determined |
| 17-109_k141_107420 | Not-determined |
| 17-109_k141_7021 | Not-determined |
| 17-109_k141_7022 | Not-determined |
| 17-109_k141_7023 | Not-determined |
| 17-109_k141_174477 | Low-quality |
| 17-109_k141_90622 | Not-determined |
| 17-109_k141_174481 | Not-determined |
| 17-109_k141_174484 | Not-determined |
| 17-109_k141_141219 | Not-determined |
| 17-109_k141_74852 | Low-quality |
| 17-109_k141_141223 | Not-determined |
| 17-109_k141_158270 | Not-determined |
| 17-109_k141_207989 | Not-determined |
| 17-109_k141_241558 | Low-quality |
| 17-109_k141_107443 | Not-determined |
| 17-109_k141_357959 | Not-determined |
| 17-109_k141_325542 | Not-determined |
| 17-109_k141_375756 | Not-determined |
| 17-109_k141_107450 | Not-determined |
| 17-109_k141_275227 | Not-determined |
| 17-109_k141_325546 | Not-determined |
| 17-109_k141_107451 | Not-determined |
| 17-109_k141_158279 | Not-determined |
| 17-109_k141_174496 | Not-determined |
| 17-109_k141_224901 | Not-determined |
| 17-109_k141_41005 | Low-quality |
| 17-109_k141_41010 | Low-quality |
| 17-109_k141_158285 | Not-determined |
| 17-109_k141_258116 | Not-determined |
| 17-109_k141_208005 | Not-determined |
| 17-109_k141_23980 | Not-determined |
| 17-109_k141_224910 | Not-determined |
| 17-109_k141_258120 | Not-determined |
| 17-109_k141_392550 | Not-determined |
| 17-109_k141_258123 | Not-determined |
| 17-109_k141_141242 | Not-determined |
| 17-109_k141_41018 | Not-determined |
| 17-109_k141_158293 | Low-quality |
| 17-109_k141_392555 | Not-determined |
| 17-109_k141_309386 | Not-determined |
| 17-109_k141_309387 | Not-determined |
| 17-109_k141_224924 | Not-determined |
| 17-109_k141_74865 | Not-determined |
| 17-109_k141_174515 | Not-determined |
| 17-109_k141_158305 | Not-determined |
| 17-109_k141_357966 | Not-determined |
| 17-109_k141_357967 | Not-determined |
| 17-109_k141_392565 | Not-determined |
| 17-109_k141_392566 | Not-determined |
| 17-109_k141_41040 | Not-determined |
| 17-109_k141_275244 | Low-quality |
| 17-109_k141_158321 | Not-determined |
| 17-109_k141_174521 | Not-determined |
| 17-109_k141_375794 | Not-determined |
| 17-109_k141_174524 | Not-determined |
| 17-109_k141_375797 | Low-quality |
| 17-109_k141_208022 | Not-determined |
| 17-109_k141_174535 | Not-determined |
| 17-109_k141_141247 | Not-determined |
| 17-109_k141_56932 | Not-determined |
| 17-109_k141_208027 | Not-determined |
| 17-109_k141_74897 | Not-determined |
| 17-109_k141_56934 | Not-determined |
| 17-109_k141_241586 | Not-determined |
| 17-109_k141_158339 | Not-determined |
| 17-109_k141_309427 | Not-determined |
| 17-109_k141_23987 | Low-quality |
| 17-109_k141_375812 | Low-quality |
| 17-109_k141_56944 | Not-determined |
| 17-109_k141_241596 | Not-determined |
| 17-109_k141_192163 | Not-determined |
| 17-109_k141_392597 | Not-determined |
| 17-109_k141_208047 | Not-determined |
| 17-109_k141_275257 | Not-determined |
| 17-109_k141_309432 | Not-determined |
| 17-109_k141_74924 | Not-determined |
| 17-109_k141_158357 | Not-determined |
| 17-109_k141_341494 | Not-determined |
| 17-109_k141_309442 | Not-determined |
| 17-109_k141_258154 | Not-determined |
| 17-109_k141_241604 | Low-quality |
| 17-109_k141_7041 | Not-determined |
| 17-109_k141_41054 | Not-determined |
| 17-109_k141_90666 | Low-quality |
| 17-109_k141_41057 | Low-quality |
| 17-109_k141_174550 | Low-quality |
| 17-109_k141_258160 | Not-determined |
| 17-109_k141_258161 | Not-determined |
| 17-109_k141_208070 | Not-determined |
| 17-109_k141_309453 | Not-determined |
| 17-109_k141_41060 | Not-determined |
| 17-109_k141_325577 | Not-determined |
| 17-109_k141_341510 | Not-determined |
| 17-109_k141_107510 | Not-determined |
| 17-109_k141_325583 | Not-determined |
| 17-109_k141_107511 | Not-determined |
| 17-109_k141_141260 | Not-determined |
| 17-109_k141_241619 | Low-quality |
| 17-109_k141_341514 | Not-determined |
| 17-109_k141_241622 | Not-determined |
| 17-109_k141_375833 | Not-determined |
| 17-109_k141_325588 | Not-determined |
| 17-109_k141_174570 | Not-determined |
| 17-109_k141_258177 | Not-determined |
| 17-109_k141_158395 | Not-determined |
| 17-109_k141_258180 | Not-determined |
| 17-109_k141_56955 | Low-quality |
| 17-109_k141_141273 | Not-determined |
| 17-109_k141_174575 | Not-determined |
| 17-109_k141_90684 | Not-determined |
| 17-109_k141_208079 | Not-determined |
| 17-109_k141_392614 | Not-determined |
| 17-109_k141_241630 | Low-quality |
| 17-109_k141_241631 | Not-determined |
| 17-109_k141_292325 | Not-determined |
| 17-109_k141_375849 | Not-determined |
| 17-109_k141_341516 | Not-determined |
| 17-109_k141_275311 | Not-determined |
| 17-109_k141_341523 | Not-determined |
| 17-109_k141_309501 | Not-determined |
| 17-109_k141_325611 | Not-determined |
| 17-109_k141_341525 | Not-determined |
| 17-109_k141_158407 | Not-determined |
| 17-109_k141_275316 | Not-determined |
| 17-109_k141_341529 | Low-quality |
| 17-109_k141_24056 | Not-determined |
| 17-109_k141_375859 | Not-determined |
| 17-109_k141_375862 | Not-determined |
| 17-109_k141_224932 | Not-determined |
| 17-109_k141_325622 | Not-determined |
| 17-109_k141_258219 | Not-determined |
| 17-109_k141_392625 | Not-determined |
| 17-109_k141_208081 | Not-determined |
| 17-109_k141_90707 | Low-quality |
| 17-109_k141_309520 | Not-determined |
| 17-109_k141_392637 | Not-determined |
| 17-109_k141_56980 | Not-determined |
| 17-109_k141_174603 | Not-determined |
| 17-109_k141_141297 | Not-determined |
| 17-109_k141_275337 | Not-determined |
| 17-109_k141_90720 | Not-determined |
| 17-109_k141_292347 | Not-determined |
| 17-109_k141_141299 | Not-determined |
| 17-109_k141_292348 | Not-determined |
| 17-109_k141_90725 | Low-quality |
| 17-109_k141_375875 | Not-determined |
| 17-109_k141_292352 | Not-determined |
| 17-109_k141_224953 | Not-determined |
| 17-109_k141_341552 | Not-determined |
| 17-109_k141_325642 | Not-determined |
| 17-109_k141_375880 | Not-determined |
| 17-109_k141_357998 | Not-determined |
| 17-109_k141_309526 | Low-quality |
| 17-109_k141_258249 | Not-determined |
| 17-109_k141_208111 | Not-determined |
| 17-109_k141_208114 | Not-determined |
| 17-109_k141_292363 | Low-quality |
| 17-109_k141_309531 | Not-determined |
| 17-109_k141_56993 | Not-determined |
| 17-109_k141_275355 | Low-quality |
| 17-109_k141_158426 | Not-determined |
| 17-109_k141_174618 | Not-determined |
| 17-109_k141_309542 | Low-quality |
| 17-109_k141_325649 | Low-quality |
| 17-109_k141_41132 | Not-determined |
| 17-109_k141_57001 | Not-determined |
| 17-109_k141_325652 | Not-determined |
| 17-109_k141_292375 | Not-determined |
| 17-109_k141_7076 | Not-determined |
| 17-109_k141_375884 | Not-determined |
| 17-109_k141_375886 | Not-determined |
| 17-109_k141_7084 | Not-determined |
| 17-109_k141_275371 | Not-determined |
| 17-109_k141_275375 | Not-determined |
| 17-109_k141_141333 | Not-determined |
| 17-109_k141_41152 | Low-quality |
| 17-109_k141_141334 | Not-determined |
| 17-109_k141_375892 | Not-determined |
| 17-109_k141_7092 | Not-determined |
| 17-109_k141_275382 | Not-determined |
| 17-109_k141_7094 | Not-determined |
| 17-109_k141_192229 | Low-quality |
| 17-109_k141_325677 | Not-determined |
| 17-109_k141_292396 | Not-determined |
| 17-109_k141_224974 | Not-determined |
| 17-109_k141_192233 | Low-quality |
| 17-109_k141_375903 | Not-determined |
| 17-109_k141_174634 | Not-determined |
| 17-109_k141_275388 | Not-determined |
| 17-109_k141_309577 | Low-quality |
| 17-109_k141_192241 | Not-determined |
| 17-109_k141_74944 | Not-determined |
| 17-109_k141_174645 | Not-determined |
| 17-109_k141_325695 | Not-determined |
| 17-109_k141_325696 | Not-determined |
| 17-109_k141_74951 | Not-determined |
| 17-109_k141_375919 | Not-determined |
| 17-109_k141_174652 | Not-determined |
| 17-109_k141_375923 | Low-quality |
| 17-109_k141_241714 | Low-quality |
| 17-109_k141_7122 | Not-determined |
| 17-109_k141_174660 | Not-determined |
| 17-109_k141_57042 | Not-determined |
| 17-109_k141_241720 | Low-quality |
| 17-109_k141_158464 | Not-determined |
| 17-109_k141_241722 | Not-determined |
| 17-109_k141_57044 | Not-determined |
| 17-109_k141_241723 | Not-determined |
| 17-109_k141_141348 | Not-determined |
| 17-109_k141_341586 | Not-determined |
| 17-109_k141_341587 | Not-determined |
| 17-109_k141_375939 | Not-determined |
| 17-109_k141_341592 | Not-determined |
| 17-109_k141_208155 | Low-quality |
| 17-109_k141_241728 | Low-quality |
| 17-109_k141_275417 | Not-determined |
| 17-109_k141_275418 | Not-determined |
| 17-109_k141_241732 | Not-determined |
| 17-109_k141_192265 | Low-quality |
| 17-109_k141_41167 | Not-determined |
| 17-109_k141_158476 | Not-determined |
| 17-109_k141_7137 | Not-determined |
| 17-109_k141_192273 | Not-determined |
| 17-109_k141_158483 | Not-determined |
| 17-109_k141_158484 | Not-determined |
| 17-109_k141_275427 | Not-determined |
| 17-109_k141_192277 | Not-determined |
| 17-109_k141_392700 | Not-determined |
| 17-109_k141_158488 | Not-determined |
| 17-109_k141_123897 | Not-determined |
| 17-109_k141_123900 | Not-determined |
| 17-109_k141_141358 | Not-determined |
| 17-109_k141_7155 | Not-determined |
| 17-109_k141_24153 | Not-determined |
| 17-109_k141_241747 | Not-determined |
| 17-109_k141_224991 | Not-determined |
| 17-109_k141_24156 | Not-determined |
| 17-109_k141_392712 | Low-quality |
| 17-109_k141_392714 | Not-determined |
| 17-109_k141_24161 | Not-determined |
| 17-109_k141_107526 | Not-determined |
| 17-109_k141_275444 | Not-determined |
| 17-109_k141_325742 | Not-determined |
| 17-109_k141_309606 | Not-determined |
| 17-109_k141_74987 | Not-determined |
| 17-109_k141_7169 | Not-determined |
| 17-109_k141_325743 | Not-determined |
| 17-109_k141_141377 | Not-determined |
| 17-109_k141_107537 | Not-determined |
| 17-109_k141_292453 | Not-determined |
| 17-109_k141_309616 | Not-determined |
| 17-109_k141_141385 | Not-determined |
| 17-109_k141_275452 | Low-quality |
| 17-109_k141_141389 | Not-determined |
| 17-109_k141_341640 | Not-determined |
| 17-109_k141_141392 | Not-determined |
| 17-109_k141_292463 | Not-determined |
| 17-109_k141_341642 | Not-determined |
| 17-109_k141_57089 | Not-determined |
| 17-109_k141_158520 | Not-determined |
| 17-109_k141_192306 | Not-determined |
| 17-109_k141_158521 | Not-determined |
| 17-109_k141_208204 | Low-quality |
| 17-109_k141_57093 | Not-determined |
| 17-109_k141_392719 | Not-determined |
| 17-109_k141_341648 | Not-determined |
| 17-109_k141_141396 | Not-determined |
| 17-109_k141_292470 | Not-determined |
| 17-109_k141_392721 | Not-determined |
| 17-109_k141_158528 | Not-determined |
| 17-109_k141_24200 | Not-determined |
| 17-109_k141_158529 | Not-determined |
| 17-109_k141_158530 | Low-quality |
| 17-109_k141_90778 | Low-quality |
| 17-109_k141_90781 | Not-determined |
| 17-109_k141_174684 | Not-determined |
| 17-109_k141_341663 | Not-determined |
| 17-109_k141_24219 | Not-determined |
| 17-109_k141_341667 | Not-determined |
| 17-109_k141_24223 | Not-determined |
| 17-109_k141_341668 | Not-determined |
| 17-109_k141_275470 | Not-determined |
| 17-109_k141_107563 | Not-determined |
| 17-109_k141_225041 | Not-determined |
| 17-109_k141_90788 | Not-determined |
| 17-109_k141_174693 | Low-quality |
| 17-109_k141_75009 | Not-determined |
| 17-109_k141_7177 | Low-quality |
| 17-109_k141_241777 | Not-determined |
| 17-109_k141_375955 | Not-determined |
| 17-109_k141_174699 | Not-determined |
| 17-109_k141_158540 | Not-determined |
| 17-109_k141_309630 | Not-determined |
| 17-109_k141_75022 | Not-determined |
| 17-109_k141_24239 | Not-determined |
| 17-109_k141_275504 | Not-determined |
| 17-109_k141_292486 | Not-determined |
| 17-109_k141_7191 | Not-determined |
| 17-109_k141_375959 | Not-determined |
| 17-109_k141_57109 | Not-determined |
| 17-109_k141_275509 | Not-determined |
| 17-109_k141_158551 | Not-determined |
| 17-109_k141_174703 | Low-quality |
| 17-109_k141_192325 | Not-determined |
| 17-109_k141_7201 | Not-determined |
| 17-109_k141_141409 | Not-determined |
| 17-109_k141_24243 | Low-quality |
| 17-109_k141_275514 | Not-determined |
| 17-109_k141_174713 | Not-determined |
| 17-109_k141_241779 | Not-determined |
| 17-109_k141_192335 | Not-determined |
| 17-109_k141_225066 | Low-quality |
| 17-109_k141_192339 | Low-quality |
| 17-109_k141_392750 | Not-determined |
| 17-109_k141_275517 | Not-determined |
| 17-109_k141_192341 | Not-determined |
| 17-109_k141_392752 | Not-determined |
| 17-109_k141_90817 | Not-determined |
| 17-109_k141_292500 | Not-determined |
| 17-109_k141_90818 | Not-determined |
| 17-109_k141_275523 | Not-determined |
| 17-109_k141_7212 | Not-determined |
| 17-109_k141_123936 | Not-determined |
| 17-109_k141_292507 | Not-determined |
| 17-109_k141_107579 | Not-determined |
| 17-109_k141_123938 | Not-determined |
| 17-109_k141_57120 | Not-determined |
| 17-109_k141_57122 | Not-determined |
| 17-109_k141_57128 | Not-determined |
| 17-109_k141_141421 | Not-determined |
| 17-109_k141_7218 | Not-determined |
| 17-109_k141_225076 | Low-quality |
| 17-109_k141_225078 | Not-determined |
| 17-109_k141_225080 | Not-determined |
| 17-109_k141_309643 | Not-determined |
| 17-109_k141_392761 | Not-determined |
| 17-109_k141_141425 | Not-determined |
| 17-109_k141_24248 | Not-determined |
| 17-109_k141_7223 | Not-determined |
| 17-109_k141_24251 | Not-determined |
| 17-109_k141_24252 | Not-determined |
| 17-109_k141_258348 | Not-determined |
| 17-109_k141_341710 | Not-determined |
| 17-109_k141_292521 | Not-determined |
| 17-109_k141_275533 | Not-determined |
| 17-109_k141_392772 | Not-determined |
| 17-109_k141_107587 | Not-determined |
| 17-109_k141_225090 | Not-determined |
| 17-109_k141_341715 | Not-determined |
| 17-109_k141_57145 | Not-determined |
| 17-109_k141_275537 | Not-determined |
| 17-109_k141_241816 | Not-determined |
| 17-109_k141_90836 | Not-determined |
| 17-109_k141_241817 | Not-determined |
| 17-109_k141_57152 | Not-determined |
| 17-109_k141_90838 | Not-determined |
| 17-109_k141_107591 | Not-determined |
| 17-109_k141_208274 | Low-quality |
| 17-109_k141_123963 | Not-determined |
| 17-109_k141_225095 | Low-quality |
| 17-109_k141_208277 | Not-determined |
| 17-109_k141_107601 | Not-determined |
| 17-109_k141_292531 | Not-determined |
| 17-109_k141_258360 | Not-determined |
| 17-109_k141_57162 | Not-determined |
| 17-109_k141_292539 | Not-determined |
| 17-109_k141_7241 | Not-determined |
| 17-109_k141_292546 | Low-quality |
| 17-109_k141_258377 | Not-determined |
| 17-109_k141_7253 | Not-determined |
| 17-109_k141_174743 | Not-determined |
| 17-109_k141_7255 | Not-determined |
| 17-109_k141_141460 | Not-determined |
| 17-109_k141_292556 | Not-determined |
| 17-109_k141_174752 | Not-determined |
| 17-109_k141_292558 | Not-determined |
| 17-109_k141_258384 | Not-determined |
| 17-109_k141_241845 | Not-determined |
| 17-109_k141_341736 | Low-quality |
| 17-109_k141_141466 | Not-determined |
| 17-109_k141_275570 | Not-determined |
| 17-109_k141_158579 | Not-determined |
| 17-109_k141_158580 | Not-determined |
| 17-109_k141_341742 | Not-determined |
| 17-109_k141_208288 | Not-determined |
| 17-109_k141_341745 | Not-determined |
| 17-109_k141_225116 | Not-determined |
| 17-109_k141_275581 | Not-determined |
| 17-109_k141_275583 | Not-determined |
| 17-109_k141_275584 | Not-determined |
| 17-109_k141_275587 | Not-determined |
| 17-109_k141_192350 | Not-determined |
| 17-109_k141_309664 | Not-determined |
| 17-109_k141_275591 | Not-determined |
| 17-109_k141_309668 | Not-determined |
| 17-109_k141_309672 | Not-determined |
| 17-109_k141_174766 | Low-quality |
| 17-109_k141_341753 | Not-determined |
| 17-109_k141_292588 | Not-determined |
| 17-109_k141_292589 | Not-determined |
| 17-109_k141_275595 | Not-determined |
| 17-109_k141_7282 | Not-determined |
| 17-109_k141_325765 | Not-determined |
| 17-109_k141_392785 | Not-determined |
| 17-109_k141_392786 | Not-determined |
| 17-109_k141_275600 | Not-determined |
| 17-109_k141_292594 | Not-determined |
| 17-109_k141_225153 | Not-determined |
| 17-109_k141_309693 | Not-determined |
| 17-109_k141_57181 | Not-determined |
| 17-109_k141_392789 | Not-determined |
| 17-109_k141_192377 | Not-determined |
| 17-109_k141_107603 | Not-determined |
| 17-109_k141_392790 | Not-determined |
| 17-109_k141_141505 | Not-determined |
| 17-109_k141_309700 | Not-determined |
| 17-109_k141_241875 | Not-determined |
| 17-109_k141_241877 | Not-determined |
| 17-109_k141_90890 | Not-determined |
| 17-109_k141_141511 | Not-determined |
| 17-109_k141_309711 | Not-determined |
| 17-109_k141_90892 | Not-determined |
| 17-109_k141_309714 | Not-determined |
| 17-109_k141_325792 | Not-determined |
| 17-109_k141_258416 | Not-determined |
| 17-109_k141_258419 | Not-determined |
| 17-109_k141_309719 | Not-determined |
| 17-109_k141_57197 | Not-determined |
| 17-109_k141_225168 | Not-determined |
| 17-109_k141_141519 | Not-determined |
| 17-109_k141_325796 | Not-determined |
| 17-109_k141_258424 | Not-determined |
| 17-109_k141_292617 | Not-determined |
| 17-109_k141_392795 | Not-determined |
| 17-109_k141_158609 | Not-determined |
| 17-109_k141_141522 | Not-determined |
| 17-109_k141_392797 | Not-determined |
| 17-109_k141_341775 | Not-determined |
| 17-109_k141_41212 | Low-quality |
| 17-109_k141_392803 | Not-determined |
| 17-109_k141_123983 | Not-determined |
| 17-109_k141_341784 | Not-determined |
| 17-109_k141_241912 | Not-determined |
| 17-109_k141_7312 | Not-determined |
| 17-109_k141_57224 | Not-determined |
| 17-109_k141_107615 | Low-quality |
| 17-109_k141_225188 | Not-determined |
| 17-109_k141_107616 | Not-determined |
| 17-109_k141_309744 | Not-determined |
| 17-109_k141_325819 | Not-determined |
| 17-109_k141_325820 | Not-determined |
| 17-109_k141_225193 | Not-determined |
| 17-109_k141_292637 | Not-determined |
| 17-109_k141_341800 | Not-determined |
| 17-109_k141_341810 | Not-determined |
| 17-109_k141_90918 | Not-determined |
| 17-109_k141_141569 | Not-determined |
| 17-109_k141_258450 | Not-determined |
| 17-109_k141_141574 | Not-determined |
| 17-109_k141_57249 | Not-determined |
| 17-109_k141_57252 | Not-determined |
| 17-109_k141_158637 | Not-determined |
| 17-109_k141_107632 | Not-determined |
| 17-109_k141_258471 | Not-determined |
| 17-109_k141_309773 | Not-determined |
| 17-109_k141_325839 | Low-quality |
| 17-109_k141_376055 | Not-determined |
| 17-109_k141_309777 | Low-quality |
| 17-109_k141_208341 | Not-determined |
| 17-109_k141_376058 | Not-determined |
| 17-109_k141_325844 | Not-determined |
| 17-109_k141_258479 | Not-determined |
| 17-109_k141_7321 | Not-determined |
| 17-109_k141_258482 | Not-determined |
| 17-109_k141_258483 | Not-determined |
| 17-109_k141_376067 | Not-determined |
| 17-109_k141_325852 | Not-determined |
| 17-109_k141_341836 | Not-determined |
| 17-109_k141_392854 | Not-determined |
| 17-109_k141_392855 | Not-determined |
| 17-109_k141_41246 | Not-determined |
| 17-109_k141_41248 | Not-determined |
| 17-109_k141_90930 | Not-determined |
| 17-109_k141_192431 | Not-determined |
| 17-109_k141_392864 | Not-determined |
| 17-109_k141_258501 | Not-determined |
| 17-109_k141_376075 | Not-determined |
| 17-109_k141_292670 | Low-quality |
| 17-109_k141_41255 | Not-determined |
| 17-109_k141_292671 | Not-determined |
| 17-109_k141_41261 | Not-determined |
| 17-109_k141_392867 | Not-determined |
| 17-109_k141_392870 | Not-determined |
| 17-109_k141_90949 | Not-determined |
| 17-109_k141_309800 | Not-determined |
| 17-109_k141_57275 | Not-determined |
| 17-109_k141_292693 | Low-quality |
| 17-109_k141_325866 | Not-determined |
| 17-109_k141_241979 | Not-determined |
| 17-109_k141_309810 | Low-quality |
| 17-109_k141_376099 | Not-determined |
| 17-109_k141_225214 | Not-determined |
| 17-109_k141_292698 | Not-determined |
| 17-109_k141_341854 | Not-determined |

| | |
|---|---|
| 17-109_k141_241982 | Not-determined |
| 17-109_k141_57285 | Not-determined |
| 17-109_k141_241983 | Not-determined |
| 17-109_k141_309817 | Not-determined |
| 17-109_k141_292704 | Not-determined |
| 17-109_k141_392884 | Not-determined |
| 17-109_k141_325869 | Not-determined |
| 17-109_k141_341861 | Not-determined |
| 17-109_k141_141629 | Not-determined |
| 17-109_k141_57288 | Not-determined |
| 17-109_k141_309831 | Not-determined |
| 17-109_k141_225231 | Not-determined |
| 17-109_k141_24322 | Low-quality |
| 17-109_k141_141643 | Not-determined |
| 17-109_k141_141644 | Low-quality |
| 17-109_k141_192469 | Not-determined |
| 17-109_k141_174783 | Not-determined |
| 17-109_k141_24335 | Not-determined |
| 17-109_k141_174785 | Not-determined |
| 17-109_k141_225239 | Not-determined |
| 17-109_k141_141649 | Not-determined |
| 17-109_k141_141650 | Low-quality |
| 17-109_k141_309853 | Not-determined |
| 17-109_k141_225247 | Not-determined |
| 17-109_k141_309855 | Not-determined |
| 17-109_k141_57303 | Not-determined |
| 17-109_k141_24337 | Not-determined |
| 17-109_k141_225249 | Not-determined |
| 17-109_k141_24341 | Not-determined |
| 17-109_k141_90984 | Low-quality |
| 17-109_k141_275612 | Not-determined |
| 17-109_k141_90987 | Not-determined |
| 17-109_k141_275614 | Not-determined |
| 17-109_k141_258533 | Not-determined |
| 17-109_k141_275617 | Not-determined |
| 17-109_k141_325891 | Not-determined |
| 17-109_k141_275619 | Not-determined |
| 17-109_k141_7371 | Not-determined |
| 17-109_k141_107636 | Not-determined |
| 17-109_k141_192489 | Not-determined |
| 17-109_k141_376116 | Not-determined |
| 17-109_k141_325898 | Not-determined |
| 17-109_k141_376121 | Not-determined |
| 17-109_k141_325901 | Not-determined |
| 17-109_k141_275624 | Not-determined |
| 17-109_k141_158652 | Not-determined |
| 17-109_k141_325902 | Not-determined |
| 17-109_k141_7380 | Not-determined |
| 17-109_k141_24358 | Not-determined |
| 17-109_k141_242026 | Not-determined |
| 17-109_k141_292728 | Not-determined |
| 17-109_k141_225273 | Not-determined |
| 17-109_k141_192503 | Not-determined |
| 17-109_k141_325912 | Not-determined |
| 17-109_k141_275628 | Not-determined |
| 17-109_k141_225280 | Low-quality |
| 17-109_k141_275631 | Not-determined |
| 17-109_k141_325920 | Not-determined |
| 17-109_k141_208364 | Not-determined |
| 17-109_k141_7392 | Low-quality |
| 17-109_k141_341889 | Not-determined |
| 17-109_k141_275637 | Not-determined |
| 17-109_k141_107649 | Not-determined |
| 17-109_k141_292747 | Not-determined |
| 17-109_k141_225293 | Not-determined |
| 17-109_k141_107653 | Low-quality |
| 17-109_k141_158674 | Not-determined |
| 17-109_k141_41318 | Not-determined |
| 17-109_k141_57313 | Not-determined |
| 17-109_k141_158687 | Not-determined |
| 17-109_k141_275649 | Not-determined |
| 17-109_k141_41330 | Not-determined |
| 17-109_k141_292759 | Not-determined |
| 17-109_k141_158693 | Not-determined |
| 17-109_k141_192516 | Not-determined |
| 17-109_k141_174797 | Not-determined |
| 17-109_k141_309872 | Not-determined |
| 17-109_k141_192526 | Not-determined |
| 17-109_k141_292767 | Not-determined |
| 17-109_k141_107677 | Not-determined |
| 17-109_k141_57327 | Not-determined |
| 17-109_k141_174799 | Low-quality |
| 17-109_k141_242058 | Not-determined |
| 17-109_k141_192528 | Not-determined |
| 17-109_k141_242061 | Low-quality |
| 17-109_k141_41361 | Not-determined |
| 17-109_k141_24386 | Not-determined |
| 17-109_k141_41362 | Not-determined |
| 17-109_k141_225311 | Not-determined |
| 17-109_k141_41363 | Not-determined |
| 17-109_k141_225313 | Not-determined |
| 17-109_k141_41367 | Not-determined |
| 17-109_k141_208370 | Not-determined |
| 17-109_k141_174814 | Not-determined |
| 17-109_k141_242069 | Low-quality |
| 17-109_k141_41371 | Not-determined |
| 17-109_k141_275672 | Not-determined |
| 17-109_k141_242072 | Not-determined |
| 17-109_k141_41373 | Not-determined |
| 17-109_k141_225324 | Not-determined |
| 17-109_k141_292791 | Not-determined |
| 17-109_k141_7435 | Not-determined |
| 17-109_k141_107697 | Not-determined |
| 17-109_k141_7437 | Not-determined |
| 17-109_k141_292795 | Low-quality |
| 17-109_k141_275688 | Not-determined |
| 17-109_k141_7445 | Not-determined |
| 17-109_k141_242084 | Low-quality |
| 17-109_k141_292802 | Not-determined |
| 17-109_k141_107700 | Not-determined |
| 17-109_k141_225329 | Not-determined |
| 17-109_k141_57344 | Not-determined |
| 17-109_k141_192560 | Not-determined |
| 17-109_k141_309902 | Not-determined |
| 17-109_k141_57351 | Not-determined |
| 17-109_k141_174833 | Not-determined |
| 17-109_k141_292816 | Not-determined |
| 17-109_k141_41406 | Not-determined |
| 17-109_k141_275707 | Not-determined |
| 17-109_k141_309920 | Not-determined |
| 17-109_k141_174842 | Not-determined |
| 17-109_k141_275709 | Not-determined |
| 17-109_k141_91046 | Not-determined |
| 17-109_k141_275712 | Not-determined |
| 17-109_k141_225346 | Not-determined |
| 17-109_k141_7456 | Not-determined |
| 17-109_k141_107723 | Not-determined |
| 17-109_k141_358064 | Not-determined |
| 17-109_k141_174853 | Not-determined |
| 17-109_k141_158711 | Low-quality |
| 17-109_k141_91053 | Low-quality |
| 17-109_k141_225358 | Not-determined |
| 17-109_k141_24416 | Not-determined |
| 17-109_k141_124012 | Low-quality |
| 17-109_k141_91059 | Not-determined |
| 17-109_k141_392906 | Not-determined |
| 17-109_k141_41413 | Not-determined |
| 17-109_k141_392907 | Not-determined |
| 17-109_k141_57373 | Not-determined |
| 17-109_k141_41416 | Not-determined |
| 17-109_k141_91065 | Not-determined |
| 17-109_k141_107735 | Not-determined |
| 17-109_k141_124020 | Low-quality |
| 17-109_k141_192592 | Not-determined |
| 17-109_k141_358082 | Not-determined |
| 17-109_k141_174869 | Low-quality |
| 17-109_k141_225375 | Not-determined |
| 17-109_k141_124028 | Not-determined |
| 17-109_k141_392916 | Not-determined |
| 17-109_k141_107741 | Not-determined |
| 17-109_k141_309932 | Not-determined |
| 17-109_k141_309933 | Not-determined |
| 17-109_k141_392931 | Not-determined |
| 17-109_k141_258553 | Not-determined |
| 17-109_k141_292843 | Not-determined |
| 17-109_k141_225386 | Not-determined |
| 17-109_k141_7474 | Not-determined |
| 17-109_k141_258556 | Not-determined |
| 17-109_k141_7475 | Low-quality |
| 17-109_k141_107757 | Not-determined |
| 17-109_k141_292855 | Not-determined |
| 17-109_k141_242117 | Not-determined |
| 17-109_k141_225398 | Not-determined |
| 17-109_k141_57407 | Not-determined |
| 17-109_k141_124051 | Not-determined |
| 17-109_k141_107770 | Not-determined |
| 17-109_k141_258569 | Not-determined |
| 17-109_k141_341910 | Low-quality |
| 17-109_k141_292874 | Not-determined |
| 17-109_k141_292876 | Not-determined |
| 17-109_k141_275753 | Not-determined |
| 17-109_k141_174877 | Not-determined |
| 17-109_k141_57421 | Not-determined |
| 17-109_k141_7509 | Not-determined |
| 17-109_k141_309944 | Not-determined |
| 17-109_k141_258589 | Not-determined |
| 17-109_k141_107793 | Not-determined |
| 17-109_k141_341923 | Not-determined |
| 17-109_k141_57436 | Not-determined |
| 17-109_k141_208412 | Not-determined |
| 17-109_k141_242120 | Not-determined |
| 17-109_k141_174891 | Not-determined |
| 17-109_k141_174894 | Not-determined |
| 17-109_k141_258601 | Not-determined |
| 17-109_k141_7515 | Not-determined |
| 17-109_k141_309953 | Not-determined |
| 17-109_k141_309955 | Not-determined |
| 17-109_k141_208425 | Low-quality |
| 17-109_k141_208426 | Not-determined |
| 17-109_k141_107817 | Not-determined |
| 17-109_k141_292906 | Not-determined |
| 17-109_k141_107831 | Low-quality |
| 17-109_k141_192677 | Not-determined |
| 17-109_k141_225442 | Not-determined |
| 17-109_k141_141774 | Not-determined |
| 17-109_k141_225444 | Not-determined |
| 17-109_k141_275770 | Not-determined |
| 17-109_k141_41433 | Not-determined |
| 17-109_k141_192685 | Not-determined |
| 17-109_k141_309982 | Not-determined |
| 17-109_k141_124062 | Not-determined |
| 17-109_k141_392941 | Not-determined |
| 17-109_k141_7530 | Low-quality |
| 17-109_k141_41444 | Not-determined |
| 17-109_k141_225456 | Not-determined |
| 17-109_k141_158716 | Not-determined |
| 17-109_k141_75086 | Not-determined |
| 17-109_k141_192694 | Not-determined |
| 17-109_k141_392949 | Not-determined |
| 17-109_k141_376186 | Not-determined |
| 17-109_k141_208464 | Not-determined |
| 17-109_k141_192697 | Not-determined |
| 17-109_k141_376187 | Not-determined |
| 17-109_k141_275788 | Not-determined |
| 17-109_k141_258637 | Low-quality |
| 17-109_k141_208467 | Not-determined |
| 17-109_k141_124090 | Not-determined |
| 17-109_k141_192704 | Not-determined |
| 17-109_k141_392957 | Low-quality |
| 17-109_k141_392959 | Not-determined |
| 17-109_k141_57443 | Low-quality |
| 17-109_k141_392961 | Not-determined |
| 17-109_k141_341958 | Low-quality |
| 17-109_k141_376200 | Not-determined |
| 17-109_k141_376202 | Not-determined |
| 17-109_k141_7560 | Not-determined |
| 17-109_k141_7563 | Not-determined |
| 17-109_k141_158736 | Not-determined |
| 17-109_k141_275809 | Not-determined |
| 17-109_k141_158738 | Not-determined |
| 17-109_k141_158739 | Not-determined |
| 17-109_k141_242160 | Not-determined |
| 17-109_k141_57457 | Not-determined |
| 17-109_k141_141831 | Not-determined |
| 17-109_k141_57458 | Not-determined |
| 17-109_k141_341976 | Not-determined |
| 17-109_k141_57463 | Not-determined |
| 17-109_k141_341978 | Not-determined |
| 17-109_k141_292939 | Not-determined |
| 17-109_k141_7574 | Not-determined |
| 17-109_k141_292942 | Not-determined |
| 17-109_k141_292943 | Not-determined |
| 17-109_k141_174907 | Low-quality |
| 17-109_k141_7579 | Not-determined |
| 17-109_k141_341983 | Not-determined |
| 17-109_k141_341984 | Not-determined |
| 17-109_k141_376214 | Not-determined |
| 17-109_k141_75117 | Not-determined |
| 17-109_k141_358125 | Not-determined |
| 17-109_k141_225490 | Not-determined |
| 17-109_k141_124114 | Not-determined |
| 17-109_k141_310040 | Not-determined |
| 17-109_k141_392987 | Not-determined |
| 17-109_k141_124116 | Not-determined |
| 17-109_k141_358127 | Low-quality |
| 17-109_k141_376219 | Not-determined |
| 17-109_k141_141847 | Not-determined |
| 17-109_k141_376224 | Not-determined |
| 17-109_k141_258679 | Not-determined |
| 17-109_k141_376227 | Not-determined |
| 17-109_k141_7591 | Not-determined |
| 17-109_k141_158751 | Not-determined |
| 17-109_k141_358144 | Not-determined |
| 17-109_k141_258686 | Not-determined |
| 17-109_k141_292965 | Not-determined |
| 17-109_k141_342006 | Not-determined |
| 17-109_k141_7604 | Not-determined |
| 17-109_k141_208485 | Not-determined |
| 17-109_k141_141871 | Not-determined |
| 17-109_k141_208488 | Not-determined |
| 17-109_k141_292978 | Not-determined |
| 17-109_k141_107845 | Not-determined |
| 17-109_k141_107846 | Not-determined |
| 17-109_k141_174941 | Low-quality |
| 17-109_k141_208504 | Not-determined |
| 17-109_k141_275842 | Not-determined |
| 17-109_k141_393009 | Not-determined |
| 17-109_k141_342023 | Not-determined |
| 17-109_k141_275847 | Not-determined |
| 17-109_k141_24451 | Not-determined |
| 17-109_k141_242187 | Low-quality |
| 17-109_k141_342025 | Not-determined |
| 17-109_k141_158787 | Not-determined |
| 17-109_k141_107866 | Low-quality |
| 17-109_k141_242190 | Not-determined |
| 17-109_k141_24456 | Not-determined |
| 17-109_k141_376245 | Not-determined |
| 17-109_k141_7623 | Not-determined |
| 17-109_k141_393015 | Not-determined |
| 17-109_k141_158790 | Not-determined |
| 17-109_k141_393016 | Not-determined |
| 17-109_k141_24461 | Low-quality |
| 17-109_k141_325978 | Low-quality |
| 17-109_k141_41479 | Not-determined |
| 17-109_k141_325982 | Not-determined |
| 17-109_k141_376256 | Not-determined |
| 17-109_k141_41487 | Not-determined |
| 17-109_k141_107888 | Not-determined |
| 17-109_k141_158793 | Not-determined |
| 17-109_k141_342035 | Not-determined |
| 17-109_k141_376268 | Not-determined |
| 17-109_k141_174969 | Not-determined |
| 17-109_k141_107891 | Not-determined |
| 17-109_k141_325994 | Not-determined |
| 17-109_k141_325995 | Not-determined |
| 17-109_k141_325998 | Low-quality |
| 17-109_k141_41503 | Not-determined |
| 17-109_k141_342043 | Not-determined |
| 17-109_k141_358172 | Low-quality |
| 17-109_k141_326013 | Not-determined |
| 17-109_k141_258712 | Not-determined |
| 17-109_k141_310088 | Not-determined |
| 17-109_k141_242199 | Not-determined |
| 17-109_k141_75183 | Not-determined |
| 17-109_k141_107904 | Not-determined |
| 17-109_k141_310092 | Not-determined |
| 17-109_k141_75185 | Not-determined |
| 17-109_k141_242205 | Not-determined |
| 17-109_k141_242206 | Not-determined |
| 17-109_k141_41514 | Not-determined |
| 17-109_k141_275891 | Not-determined |
| 17-109_k141_41516 | Not-determined |
| 17-109_k141_258722 | Not-determined |
| 17-109_k141_41517 | Not-determined |
| 17-109_k141_358180 | Low-quality |
| 17-109_k141_24471 | Not-determined |
| 17-109_k141_24474 | Not-determined |
| 17-109_k141_258726 | Not-determined |
| 17-109_k141_91091 | Not-determined |
| 17-109_k141_41525 | Not-determined |
| 17-109_k141_91092 | Not-determined |
| 17-109_k141_41526 | Not-determined |
| 17-109_k141_41529 | Low-quality |
| 17-109_k141_242219 | Not-determined |
| 17-109_k141_310103 | Not-determined |
| 17-109_k141_326032 | Not-determined |
| 17-109_k141_342054 | Low-quality |
| 17-109_k141_225526 | Not-determined |
| 17-109_k141_342055 | Low-quality |
| 17-109_k141_275906 | Not-determined |
| 17-109_k141_393047 | Not-determined |
| 17-109_k141_225531 | Not-determined |
| 17-109_k141_7656 | Low-quality |
| 17-109_k141_342060 | Not-determined |
| 17-109_k141_258741 | Not-determined |
| 17-109_k141_124200 | Not-determined |
| 17-109_k141_24493 | Not-determined |
| 17-109_k141_141879 | Not-determined |
| 17-109_k141_158830 | Low-quality |
| 17-109_k141_7664 | Not-determined |
| 17-109_k141_158833 | Not-determined |
| 17-109_k141_393050 | Not-determined |
| 17-109_k141_275917 | Not-determined |
| 17-109_k141_310105 | Low-quality |
| 17-109_k141_124204 | Not-determined |
| 17-109_k141_242242 | Not-determined |
| 17-109_k141_225544 | Not-determined |
| 17-109_k141_7669 | Not-determined |
| 17-109_k141_7670 | Not-determined |
| 17-109_k141_393061 | Not-determined |
| 17-109_k141_57514 | Not-determined |
| 17-109_k141_57516 | Not-determined |
| 17-109_k141_41557 | Not-determined |
| 17-109_k141_24510 | Not-determined |
| 17-109_k141_41565 | Not-determined |
| 17-109_k141_310112 | Not-determined |
| 17-109_k141_326060 | Not-determined |
| 17-109_k141_393074 | Low-quality |
| 17-109_k141_358224 | Not-determined |
| 17-109_k141_342085 | Not-determined |
| 17-109_k141_310114 | Not-determined |
| 17-109_k141_41575 | Not-determined |
| 17-109_k141_124220 | Not-determined |
| 17-109_k141_41582 | Not-determined |
| 17-109_k141_358237 | Not-determined |
| 17-109_k141_124223 | Not-determined |
| 17-109_k141_358240 | Not-determined |
| 17-109_k141_124225 | Not-determined |
| 17-109_k141_141891 | Not-determined |
| 17-109_k141_174991 | Not-determined |
| 17-109_k141_192746 | Not-determined |
| 17-109_k141_342105 | Not-determined |
| 17-109_k141_24529 | Not-determined |
| 17-109_k141_358254 | Not-determined |
| 17-109_k141_293055 | Not-determined |
| 17-109_k141_192758 | Not-determined |
| 17-109_k141_293064 | Not-determined |
| 17-109_k141_41601 | Not-determined |
| 17-109_k141_141902 | Not-determined |
| 17-109_k141_208523 | Low-quality |
| 17-109_k141_293068 | Not-determined |
| 17-109_k141_124250 | Not-determined |
| 17-109_k141_175008 | Not-determined |
| 17-109_k141_342126 | Not-determined |
| 17-109_k141_175011 | Not-determined |
| 17-109_k141_293081 | Not-determined |
| 17-109_k141_141911 | Not-determined |
| 17-109_k141_293084 | Not-determined |
| 17-109_k141_208534 | Low-quality |
| 17-109_k141_208537 | Not-determined |
| 17-109_k141_208539 | Not-determined |
| 17-109_k141_57562 | Not-determined |
| 17-109_k141_7709 | Not-determined |
| 17-109_k141_208544 | Not-determined |
| 17-109_k141_24555 | Not-determined |
| 17-109_k141_7712 | Not-determined |
| 17-109_k141_57568 | Not-determined |
| 17-109_k141_393113 | Not-determined |
| 17-109_k141_342135 | Not-determined |
| 17-109_k141_326083 | Not-determined |
| 17-109_k141_293104 | Not-determined |
| 17-109_k141_192800 | Not-determined |
| 17-109_k141_393117 | Not-determined |
| 17-109_k141_208547 | Not-determined |
| 17-109_k141_7718 | Not-determined |
| 17-109_k141_158859 | Not-determined |
| 17-109_k141_192805 | Not-determined |
| 17-109_k141_158860 | Not-determined |
| 17-109_k141_24563 | Not-determined |
| 17-109_k141_57574 | Not-determined |
| 17-109_k141_57577 | Not-determined |
| 17-109_k141_57578 | Not-determined |
| 17-109_k141_57580 | Not-determined |
| 17-109_k141_91121 | Not-determined |
| 17-109_k141_192811 | Not-determined |
| 17-109_k141_41613 | Not-determined |
| 17-109_k141_192818 | Not-determined |
| 17-109_k141_192820 | Not-determined |
| 17-109_k141_293112 | Not-determined |
| 17-109_k141_192823 | Not-determined |
| 17-109_k141_175046 | Low-quality |
| 17-109_k141_393134 | Not-determined |
| 17-109_k141_175047 | Not-determined |
| 17-109_k141_393135 | Not-determined |
| 17-109_k141_275995 | Not-determined |
| 17-109_k141_242301 | Low-quality |
| 17-109_k141_75262 | Low-quality |
| 17-109_k141_275998 | Not-determined |
| 17-109_k141_342158 | Not-determined |
| 17-109_k141_57591 | Not-determined |
| 17-109_k141_124253 | Not-determined |
| 17-109_k141_91150 | Not-determined |
| 17-109_k141_124256 | Not-determined |
| 17-109_k141_7744 | Not-determined |
| 17-109_k141_75272 | Not-determined |
| 17-109_k141_24596 | Not-determined |
| 17-109_k141_276012 | Not-determined |
| 17-109_k141_91153 | Not-determined |
| 17-109_k141_24599 | Not-determined |
| 17-109_k141_124269 | Not-determined |
| 17-109_k141_57611 | Not-determined |
| 17-109_k141_376275 | Not-determined |
| 17-109_k141_75282 | Not-determined |
| 17-109_k141_258767 | Not-determined |
| 17-109_k141_258768 | Not-determined |
| 17-109_k141_376278 | Not-determined |
| 17-109_k141_276019 | Not-determined |
| 17-109_k141_158887 | Low-quality |
| 17-109_k141_393143 | Not-determined |
| 17-109_k141_75291 | Not-determined |
| 17-109_k141_310170 | Not-determined |
| 17-109_k141_393151 | Not-determined |
| 17-109_k141_358306 | Not-determined |
| 17-109_k141_91173 | Not-determined |
| 17-109_k141_75300 | Not-determined |
| 17-109_k141_376285 | Not-determined |
| 17-109_k141_41636 | Not-determined |
| 17-109_k141_158893 | Not-determined |
| 17-109_k141_225596 | Not-determined |
| 17-109_k141_41637 | Not-determined |
| 17-109_k141_225597 | Not-determined |
| 17-109_k141_57631 | Not-determined |
| 17-109_k141_158894 | Not-determined |
| 17-109_k141_7747 | Not-determined |
| 17-109_k141_91181 | Not-determined |
| 17-109_k141_258773 | Not-determined |
| 17-109_k141_393168 | Not-determined |
| 17-109_k141_91186 | Not-determined |
| 17-109_k141_310190 | Not-determined |
| 17-109_k141_293128 | Not-determined |
| 17-109_k141_258779 | Not-determined |
| 17-109_k141_326132 | Not-determined |
| 17-109_k141_258786 | Not-determined |
| 17-109_k141_7751 | Not-determined |
| 17-109_k141_342208 | Not-determined |
| 17-109_k141_376301 | Not-determined |
| 17-109_k141_57651 | Not-determined |
| 17-109_k141_342210 | Not-determined |
| 17-109_k141_57652 | Not-determined |
| 17-109_k141_57653 | Not-determined |
| 17-109_k141_342214 | Not-determined |
| 17-109_k141_41656 | Not-determined |
| 17-109_k141_7760 | Low-quality |
| 17-109_k141_258802 | Not-determined |
| 17-109_k141_342220 | Not-determined |
| 17-109_k141_342222 | Not-determined |
| 17-109_k141_376319 | Not-determined |
| 17-109_k141_310205 | Not-determined |
| 17-109_k141_141961 | Low-quality |
| 17-109_k141_175094 | Not-determined |
| 17-109_k141_175095 | Not-determined |
| 17-109_k141_175098 | Low-quality |
| 17-109_k141_141966 | Low-quality |
| 17-109_k141_258806 | Not-determined |
| 17-109_k141_326152 | Not-determined |
| 17-109_k141_258811 | Low-quality |
| 17-109_k141_225623 | Not-determined |
| 17-109_k141_175100 | Not-determined |
| 17-109_k141_175108 | Not-determined |
| 17-109_k141_258820 | Not-determined |
| 17-109_k141_141981 | Not-determined |
| 17-109_k141_393195 | Not-determined |
| 17-109_k141_175111 | Not-determined |
| 17-109_k141_141987 | Not-determined |
| 17-109_k141_158925 | Not-determined |
| 17-109_k141_124273 | Not-determined |
| 17-109_k141_175121 | Not-determined |
| 17-109_k141_124276 | Not-determined |
| 17-109_k141_276033 | Not-determined |
| 17-109_k141_124279 | Low-quality |
| 17-109_k141_24635 | Low-quality |
| 17-109_k141_7782 | Low-quality |
| 17-109_k141_158934 | Not-determined |
| 17-109_k141_225633 | Not-determined |
| 17-109_k141_326161 | Not-determined |
| 17-109_k141_342225 | Not-determined |
| 17-109_k141_342227 | Not-determined |
| 17-109_k141_342229 | Not-determined |
| 17-109_k141_7792 | Not-determined |
| 17-109_k141_124286 | Not-determined |
| 17-109_k141_7794 | Not-determined |
| 17-109_k141_326170 | Not-determined |
| 17-109_k141_326173 | Not-determined |
| 17-109_k141_7796 | Not-determined |
| 17-109_k141_276055 | Not-determined |
| 17-109_k141_326176 | Not-determined |
| 17-109_k141_393202 | Not-determined |
| 17-109_k141_124293 | Not-determined |
| 17-109_k141_75378 | Not-determined |
| 17-109_k141_225646 | Not-determined |
| 17-109_k141_326184 | Not-determined |
| 17-109_k141_326187 | Not-determined |
| 17-109_k141_7806 | Not-determined |
| 17-109_k141_7807 | Not-determined |
| 17-109_k141_124295 | Not-determined |
| 17-109_k141_326188 | Not-determined |
| 17-109_k141_393206 | Not-determined |
| 17-109_k141_225653 | Not-determined |
| 17-109_k141_142015 | Not-determined |
| 17-109_k141_225657 | Not-determined |
| 17-109_k141_242355 | Not-determined |
| 17-109_k141_124308 | Not-determined |
| 17-109_k141_326195 | Not-determined |
| 17-109_k141_293173 | Low-quality |
| 17-109_k141_326196 | Not-determined |
| 17-109_k141_326200 | Low-quality |
| 17-109_k141_175144 | Low-quality |
| 17-109_k141_175149 | Not-determined |
| 17-109_k141_175151 | Not-determined |
| 17-109_k141_326204 | Not-determined |
| 17-109_k141_208632 | Not-determined |
| 17-109_k141_293182 | Not-determined |
| 17-109_k141_276078 | Not-determined |
| 17-109_k141_24670 | Not-determined |
| 17-109_k141_326211 | Not-determined |
| 17-109_k141_7821 | Not-determined |
| 17-109_k141_142038 | Low-quality |
| 17-109_k141_208639 | Not-determined |
| 17-109_k141_342269 | Not-determined |
| 17-109_k141_376331 | Not-determined |
| 17-109_k141_326221 | Not-determined |
| 17-109_k141_124312 | Not-determined |
| 17-109_k141_342274 | Not-determined |
| 17-109_k141_24676 | Low-quality |
| 17-109_k141_376336 | Not-determined |
| 17-109_k141_326225 | Not-determined |
| 17-109_k141_358354 | Not-determined |
| 17-109_k141_124313 | Not-determined |
| 17-109_k141_124314 | Not-determined |
| 17-109_k141_124315 | Not-determined |
| 17-109_k141_142049 | Not-determined |
| 17-109_k141_158957 | Not-determined |
| 17-109_k141_192871 | Not-determined |
| 17-109_k141_358365 | Not-determined |
| 17-109_k141_57705 | Not-determined |
| 17-109_k141_75424 | Not-determined |
| 17-109_k141_242389 | Not-determined |
| 17-109_k141_342280 | Not-determined |
| 17-109_k141_358369 | Not-determined |

| | |
|---|---|
| 17-109_k141_7834 | Not-determined |
| 17-109_k141_310217 | Not-determined |
| 17-109_k141_242398 | Not-determined |
| 17-109_k141_208654 | Not-determined |
| 17-109_k141_57717 | Not-determined |
| 17-109_k141_7843 | Not-determined |
| 17-109_k141_7844 | Not-determined |
| 17-109_k141_158980 | Not-determined |
| 17-109_k141_24692 | Not-determined |
| 17-109_k141_342292 | Not-determined |
| 17-109_k141_158981 | Not-determined |
| 17-109_k141_24694 | Not-determined |
| 17-109_k141_342297 | Not-determined |
| 17-109_k141_192884 | Low-quality |
| 17-109_k141_326238 | Not-determined |
| 17-109_k141_124338 | Low-quality |
| 17-109_k141_242413 | Not-determined |
| 17-109_k141_326241 | Not-determined |
| 17-109_k141_142077 | Not-determined |
| 17-109_k141_326243 | Not-determined |
| 17-109_k141_293220 | Not-determined |
| 17-109_k141_293231 | Not-determined |
| 17-109_k141_293233 | Not-determined |
| 17-109_k141_107932 | Not-determined |
| 17-109_k141_326250 | Not-determined |
| 17-109_k141_158993 | Not-determined |
| 17-109_k141_358393 | Not-determined |
| 17-109_k141_242431 | Not-determined |
| 17-109_k141_276109 | Low-quality |
| 17-109_k141_276110 | Not-determined |
| 17-109_k141_342319 | Not-determined |
| 17-109_k141_326253 | Not-determined |
| 17-109_k141_342326 | Not-determined |
| 17-109_k141_342329 | Low-quality |
| 17-109_k141_142100 | Not-determined |
| 17-109_k141_57751 | Not-determined |
| 17-109_k141_393250 | Not-determined |
| 17-109_k141_310239 | Not-determined |
| 17-109_k141_24726 | Not-determined |
| 17-109_k141_293250 | Not-determined |
| 17-109_k141_342341 | Not-determined |
| 17-109_k141_293252 | Not-determined |
| 17-109_k141_192928 | Not-determined |
| 17-109_k141_258850 | Not-determined |
| 17-109_k141_276118 | Low-quality |
| 17-109_k141_342343 | Not-determined |
| 17-109_k141_7867 | Not-determined |
| 17-109_k141_24731 | Not-determined |
| 17-109_k141_342348 | Not-determined |
| 17-109_k141_57771 | Not-determined |
| 17-109_k141_192935 | Not-determined |
| 17-109_k141_225701 | Not-determined |
| 17-109_k141_225703 | Not-determined |
| 17-109_k141_225704 | Not-determined |
| 17-109_k141_192942 | Not-determined |
| 17-109_k141_192945 | Not-determined |
| 17-109_k141_57783 | Not-determined |
| 17-109_k141_75459 | Not-determined |
| 17-109_k141_342356 | Not-determined |
| 17-109_k141_342358 | Not-determined |
| 17-109_k141_342360 | Not-determined |
| 17-109_k141_258880 | Not-determined |
| 17-109_k141_57794 | Not-determined |
| 17-109_k141_376353 | Not-determined |
| 17-109_k141_107948 | Not-determined |
| 17-109_k141_276156 | Not-determined |
| 17-109_k141_376361 | Not-determined |
| 17-109_k141_192967 | Low-quality |
| 17-109_k141_225714 | Not-determined |
| 17-109_k141_124363 | Not-determined |
| 17-109_k141_293276 | Not-determined |
| 17-109_k141_293277 | Not-determined |
| 17-109_k141_75474 | Low-quality |
| 17-109_k141_376366 | Not-determined |
| 17-109_k141_124366 | Not-determined |
| 17-109_k141_293282 | Not-determined |
| 17-109_k141_124367 | Not-determined |
| 17-109_k141_258897 | Not-determined |
| 17-109_k141_24780 | Not-determined |
| 17-109_k141_192980 | Not-determined |
| 17-109_k141_276172 | Low-quality |
| 17-109_k141_276173 | Not-determined |
| 17-109_k141_159044 | Low-quality |
| 17-109_k141_242464 | Low-quality |
| 17-109_k141_342386 | Low-quality |
| 17-109_k141_175175 | Not-determined |
| 17-109_k141_310282 | Not-determined |
| 17-109_k141_57806 | Not-determined |
| 17-109_k141_258913 | Low-quality |
| 17-109_k141_393284 | Low-quality |
| 17-109_k141_107980 | Not-determined |
| 17-109_k141_258918 | Not-determined |
| 17-109_k141_24803 | Not-determined |
| 17-109_k141_107983 | Not-determined |
| 17-109_k141_310293 | Not-determined |
| 17-109_k141_142132 | Not-determined |
| 17-109_k141_358419 | Not-determined |
| 17-109_k141_41801 | Not-determined |
| 17-109_k141_192992 | Not-determined |
| 17-109_k141_208695 | Not-determined |
| 17-109_k141_393299 | Not-determined |
| 17-109_k141_393300 | Not-determined |
| 17-109_k141_24807 | Low-quality |
| 17-109_k141_393303 | Not-determined |
| 17-109_k141_91254 | Not-determined |
| 17-109_k141_276190 | Not-determined |
| 17-109_k141_107996 | Not-determined |
| 17-109_k141_242471 | Not-determined |
| 17-109_k141_7911 | Low-quality |
| 17-109_k141_91257 | Not-determined |
| 17-109_k141_310304 | Not-determined |
| 17-109_k141_393312 | Not-determined |
| 17-109_k141_310306 | Not-determined |
| 17-109_k141_124378 | Not-determined |
| 17-109_k141_393317 | Not-determined |
| 17-109_k141_175193 | Not-determined |
| 17-109_k141_7916 | Not-determined |
| 17-109_k141_193004 | Not-determined |
| 17-109_k141_376388 | Low-quality |
| 17-109_k141_57830 | Not-determined |
| 17-109_k141_376390 | Not-determined |
| 17-109_k141_376393 | Not-determined |
| 17-109_k141_258939 | Not-determined |
| 17-109_k141_24827 | Not-determined |
| 17-109_k141_142157 | Not-determined |
| 17-109_k141_142161 | Not-determined |
| 17-109_k141_24830 | Low-quality |
| 17-109_k141_242487 | Not-determined |
| 17-109_k141_258943 | Not-determined |
| 17-109_k141_342397 | Not-determined |
| 17-109_k141_91280 | Not-determined |
| 17-109_k141_175205 | Not-determined |
| 17-109_k141_358428 | Not-determined |
| 17-109_k141_57838 | Not-determined |
| 17-109_k141_393325 | Not-determined |
| 17-109_k141_393327 | Not-determined |
| 17-109_k141_175213 | Not-determined |
| 17-109_k141_91292 | Not-determined |
| 17-109_k141_276211 | Not-determined |
| 17-109_k141_358435 | Not-determined |
| 17-109_k141_142169 | Not-determined |
| 17-109_k141_242500 | Not-determined |
| 17-109_k141_193047 | Not-determined |
| 17-109_k141_193050 | Not-determined |
| 17-109_k141_124396 | Not-determined |
| 17-109_k141_91307 | Not-determined |
| 17-109_k141_193052 | Not-determined |
| 17-109_k141_91310 | Not-determined |
| 17-109_k141_75499 | Not-determined |
| 17-109_k141_358451 | Not-determined |
| 17-109_k141_225765 | Not-determined |
| 17-109_k141_91312 | Not-determined |
| 17-109_k141_310312 | Not-determined |
| 17-109_k141_376405 | Not-determined |
| 17-109_k141_142177 | Not-determined |
| 17-109_k141_175231 | Not-determined |
| 17-109_k141_57860 | Not-determined |
| 17-109_k141_7948 | Not-determined |
| 17-109_k141_91319 | Low-quality |
| 17-109_k141_193078 | Low-quality |
| 17-109_k141_208708 | Not-determined |
| 17-109_k141_142181 | Not-determined |
| 17-109_k141_342414 | Not-determined |
| 17-109_k141_208710 | Not-determined |
| 17-109_k141_276232 | Not-determined |
| 17-109_k141_258957 | Not-determined |
| 17-109_k141_7954 | Not-determined |
| 17-109_k141_293326 | Not-determined |
| 17-109_k141_142184 | Not-determined |
| 17-109_k141_242511 | Not-determined |
| 17-109_k141_258961 | Not-determined |
| 17-109_k141_376421 | Not-determined |
| 17-109_k141_310319 | Not-determined |
| 17-109_k141_208721 | Low-quality |
| 17-109_k141_75519 | Not-determined |
| 17-109_k141_208724 | Not-determined |
| 17-109_k141_393345 | Not-determined |
| 17-109_k141_242512 | Not-determined |
| 17-109_k141_208727 | Not-determined |
| 17-109_k141_108036 | Not-determined |
| 17-109_k141_91325 | Not-determined |
| 17-109_k141_310331 | Low-quality |
| 17-109_k141_208732 | Not-determined |
| 17-109_k141_326322 | Not-determined |
| 17-109_k141_91326 | Not-determined |
| 17-109_k141_208735 | Not-determined |
| 17-109_k141_108044 | Not-determined |
| 17-109_k141_258980 | Not-determined |
| 17-109_k141_276257 | Not-determined |
| 17-109_k141_24843 | Not-determined |
| 17-109_k141_75532 | Not-determined |
| 17-109_k141_276262 | Not-determined |
| 17-109_k141_326326 | Low-quality |
| 17-109_k141_393348 | Low-quality |
| 17-109_k141_293352 | Not-determined |
| 17-109_k141_310334 | Not-determined |
| 17-109_k141_175270 | Not-determined |
| 17-109_k141_276266 | Not-determined |
| 17-109_k141_258992 | Not-determined |
| 17-109_k141_208739 | Not-determined |
| 17-109_k141_75546 | Not-determined |
| 17-109_k141_108060 | Not-determined |
| 17-109_k141_276276 | Not-determined |
| 17-109_k141_342431 | Not-determined |
| 17-109_k141_342432 | Not-determined |
| 17-109_k141_41817 | Not-determined |
| 17-109_k141_376453 | Not-determined |
| 17-109_k141_24870 | Not-determined |
| 17-109_k141_175284 | Not-determined |
| 17-109_k141_376455 | Low-quality |
| 17-109_k141_24872 | Not-determined |
| 17-109_k141_175288 | Not-determined |
| 17-109_k141_193113 | Not-determined |
| 17-109_k141_259003 | Not-determined |
| 17-109_k141_41824 | Not-determined |
| 17-109_k141_108073 | Not-determined |
| 17-109_k141_108074 | Not-determined |
| 17-109_k141_193122 | Not-determined |
| 17-109_k141_376471 | Not-determined |
| 17-109_k141_108077 | Not-determined |
| 17-109_k141_175308 | Not-determined |
| 17-109_k141_358474 | Not-determined |
| 17-109_k141_175309 | Not-determined |
| 17-109_k141_225828 | Not-determined |
| 17-109_k141_208755 | Not-determined |
| 17-109_k141_193131 | Not-determined |
| 17-109_k141_193132 | Not-determined |
| 17-109_k141_41833 | Not-determined |
| 17-109_k141_342453 | Not-determined |
| 17-109_k141_193133 | Not-determined |
| 17-109_k141_393359 | Not-determined |
| 17-109_k141_7991 | Not-determined |
| 17-109_k141_310371 | Not-determined |
| 17-109_k141_358481 | Not-determined |
| 17-109_k141_175310 | Not-determined |
| 17-109_k141_193141 | Not-determined |
| 17-109_k141_326346 | Not-determined |
| 17-109_k141_124439 | Not-determined |
| 17-109_k141_175311 | Not-determined |
| 17-109_k141_24898 | Low-quality |
| 17-109_k141_193143 | Low-quality |
| 17-109_k141_41847 | Not-determined |
| 17-109_k141_193144 | Not-determined |
| 17-109_k141_310379 | Not-determined |
| 17-109_k141_24914 | Not-determined |
| 17-109_k141_175317 | Not-determined |
| 17-109_k141_193153 | Not-determined |
| 17-109_k141_8005 | Not-determined |
| 17-109_k141_208772 | Not-determined |
| 17-109_k141_124454 | Not-determined |
| 17-109_k141_310393 | Not-determined |
| 17-109_k141_310396 | Not-determined |
| 17-109_k141_24920 | Not-determined |
| 17-109_k141_193167 | Not-determined |
| 17-109_k141_376495 | Not-determined |
| 17-109_k141_142239 | Not-determined |
| 17-109_k141_259021 | Not-determined |
| 17-109_k141_225844 | Not-determined |
| 17-109_k141_276297 | Not-determined |
| 17-109_k141_225847 | Low-quality |
| 17-109_k141_193179 | Not-determined |
| 17-109_k141_259033 | Low-quality |
| 17-109_k141_124467 | Not-determined |
| 17-109_k141_57885 | Not-determined |
| 17-109_k141_208786 | Not-determined |
| 17-109_k141_342476 | Not-determined |
| 17-109_k141_57888 | Not-determined |
| 17-109_k141_293360 | Not-determined |
| 17-109_k141_108087 | Not-determined |
| 17-109_k141_276309 | Not-determined |
| 17-109_k141_24945 | Not-determined |
| 17-109_k141_310419 | Not-determined |
| 17-109_k141_342486 | Not-determined |
| 17-109_k141_108097 | Not-determined |
| 17-109_k141_8025 | Not-determined |
| 17-109_k141_393410 | Not-determined |
| 17-109_k141_310430 | Not-determined |
| 17-109_k141_124480 | Not-determined |
| 17-109_k141_293378 | Not-determined |
| 17-109_k141_326381 | Not-determined |
| 17-109_k141_310432 | Not-determined |
| 17-109_k141_24954 | Not-determined |
| 17-109_k141_41888 | Not-determined |
| 17-109_k141_376517 | Not-determined |
| 17-109_k141_242525 | Not-determined |
| 17-109_k141_193191 | Low-quality |
| 17-109_k141_259062 | Not-determined |
| 17-109_k141_208807 | Not-determined |
| 17-109_k141_57910 | Not-determined |
| 17-109_k141_159061 | Not-determined |
| 17-109_k141_242529 | Not-determined |
| 17-109_k141_175342 | Not-determined |
| 17-109_k141_175343 | Not-determined |
| 17-109_k141_225876 | Not-determined |
| 17-109_k141_8042 | Not-determined |
| 17-109_k141_276335 | Not-determined |
| 17-109_k141_24966 | Low-quality |
| 17-109_k141_293400 | Not-determined |
| 17-109_k141_242536 | Not-determined |
| 17-109_k141_276340 | Not-determined |
| 17-109_k141_276342 | Not-determined |
| 17-109_k141_393429 | Low-quality |
| 17-109_k141_208817 | Not-determined |
| 17-109_k141_41914 | Not-determined |
| 17-109_k141_276344 | Not-determined |
| 17-109_k141_259079 | Low-quality |
| 17-109_k141_124503 | Low-quality |
| 17-109_k141_124504 | Not-determined |
| 17-109_k141_8063 | Not-determined |
| 17-109_k141_225885 | Not-determined |
| 17-109_k141_225888 | Not-determined |
| 17-109_k141_225894 | Not-determined |
| 17-109_k141_310451 | Not-determined |
| 17-109_k141_225897 | Not-determined |
| 17-109_k141_293414 | Not-determined |
| 17-109_k141_225900 | Not-determined |
| 17-109_k141_159088 | Not-determined |
| 17-109_k141_8077 | Not-determined |
| 17-109_k141_225901 | Low-quality |
| 17-109_k141_358512 | Not-determined |
| 17-109_k141_193226 | Not-determined |
| 17-109_k141_41930 | Not-determined |
| 17-109_k141_310457 | Not-determined |
| 17-109_k141_326400 | Not-determined |
| 17-109_k141_225908 | Not-determined |
| 17-109_k141_376529 | Low-quality |
| 17-109_k141_24977 | Not-determined |
| 17-109_k141_310458 | Not-determined |
| 17-109_k141_193235 | Not-determined |
| 17-109_k141_376535 | Not-determined |
| 17-109_k141_326406 | Not-determined |
| 17-109_k141_225914 | Not-determined |
| 17-109_k141_310463 | Not-determined |
| 17-109_k141_91367 | Not-determined |
| 17-109_k141_124530 | Not-determined |
| 17-109_k141_393451 | Not-determined |
| 17-109_k141_393453 | Not-determined |
| 17-109_k141_242559 | Not-determined |
| 17-109_k141_342515 | Not-determined |
| 17-109_k141_393457 | Not-determined |
| 17-109_k141_24978 | Not-determined |
| 17-109_k141_24979 | Not-determined |
| 17-109_k141_208830 | Not-determined |
| 17-109_k141_342519 | Not-determined |
| 17-109_k141_242565 | Not-determined |
| 17-109_k141_108128 | Low-quality |
| 17-109_k141_376546 | Not-determined |
| 17-109_k141_259098 | Not-determined |
| 17-109_k141_57937 | Not-determined |
| 17-109_k141_24990 | Not-determined |
| 17-109_k141_91376 | Not-determined |
| 17-109_k141_57945 | Not-determined |
| 17-109_k141_193255 | Not-determined |
| 17-109_k141_259101 | Not-determined |
| 17-109_k141_242577 | Not-determined |
| 17-109_k141_259107 | Not-determined |
| 17-109_k141_225933 | Not-determined |
| 17-109_k141_175382 | Not-determined |
| 17-109_k141_358529 | Not-determined |
| 17-109_k141_41955 | Not-determined |
| 17-109_k141_358530 | Not-determined |
| 17-109_k141_358531 | Not-determined |
| 17-109_k141_41958 | Not-determined |
| 17-109_k141_142270 | Not-determined |
| 17-109_k141_124542 | Not-determined |
| 17-109_k141_326434 | Not-determined |
| 17-109_k141_159115 | Not-determined |
| 17-109_k141_310485 | Not-determined |
| 17-109_k141_393479 | Not-determined |
| 17-109_k141_393480 | Low-quality |
| 17-109_k141_25003 | Not-determined |
| 17-109_k141_25004 | Not-determined |
| 17-109_k141_358541 | Not-determined |
| 17-109_k141_108160 | Not-determined |
| 17-109_k141_91404 | Low-quality |
| 17-109_k141_159127 | Not-determined |
| 17-109_k141_393488 | Low-quality |
| 17-109_k141_25016 | Not-determined |
| 17-109_k141_159134 | Not-determined |
| 17-109_k141_225954 | Not-determined |
| 17-109_k141_108168 | Not-determined |
| 17-109_k141_208857 | Low-quality |
| 17-109_k141_8137 | Low-quality |
| 17-109_k141_276373 | Not-determined |
| 17-109_k141_342559 | Not-determined |
| 17-109_k141_208865 | Low-quality |
| 17-109_k141_142299 | Not-determined |
| 17-109_k141_259152 | Not-determined |
| 17-109_k141_276385 | Not-determined |
| 17-109_k141_276386 | Low-quality |
| 17-109_k141_326464 | Not-determined |
| 17-109_k141_57973 | Not-determined |
| 17-109_k141_242603 | Not-determined |
| 17-109_k141_342571 | Low-quality |
| 17-109_k141_293430 | Not-determined |
| 17-109_k141_310507 | Not-determined |
| 17-109_k141_159168 | Not-determined |
| 17-109_k141_293434 | Not-determined |
| 17-109_k141_259157 | Not-determined |
| 17-109_k141_310514 | Not-determined |
| 17-109_k141_175388 | Not-determined |
| 17-109_k141_310516 | Not-determined |
| 17-109_k141_259162 | Not-determined |
| 17-109_k141_193313 | Not-determined |
| 17-109_k141_242615 | Not-determined |
| 17-109_k141_108183 | Not-determined |
| 17-109_k141_310518 | Not-determined |
| 17-109_k141_310521 | Not-determined |
| 17-109_k141_259171 | Not-determined |
| 17-109_k141_225972 | Low-quality |
| 17-109_k141_108188 | Not-determined |
| 17-109_k141_57984 | Not-determined |
| 17-109_k141_75578 | Not-determined |
| 17-109_k141_159173 | Not-determined |
| 17-109_k141_159174 | Not-determined |
| 17-109_k141_326477 | Not-determined |
| 17-109_k141_159176 | Not-determined |
| 17-109_k141_159178 | Not-determined |
| 17-109_k141_326481 | Not-determined |
| 17-109_k141_208892 | Not-determined |
| 17-109_k141_142314 | Not-determined |
| 17-109_k141_242627 | Not-determined |
| 17-109_k141_142320 | Not-determined |
| 17-109_k141_175407 | Not-determined |
| 17-109_k141_91434 | Not-determined |
| 17-109_k141_124558 | Not-determined |
| 17-109_k141_8145 | Not-determined |
| 17-109_k141_225991 | Not-determined |
| 17-109_k141_310535 | Not-determined |
| 17-109_k141_25056 | Not-determined |
| 17-109_k141_293443 | Not-determined |
| 17-109_k141_142332 | Not-determined |
| 17-109_k141_208899 | Not-determined |
| 17-109_k141_242653 | Not-determined |
| 17-109_k141_326498 | Not-determined |
| 17-109_k141_208906 | Not-determined |
| 17-109_k141_25063 | Not-determined |
| 17-109_k141_310542 | Low-quality |
| 17-109_k141_242657 | Not-determined |
| 17-109_k141_8159 | Not-determined |
| 17-109_k141_226006 | Not-determined |
| 17-109_k141_25066 | Not-determined |
| 17-109_k141_208911 | Not-determined |
| 17-109_k141_310543 | Not-determined |
| 17-109_k141_310544 | Low-quality |
| 17-109_k141_226010 | Not-determined |
| 17-109_k141_142344 | Not-determined |
| 17-109_k141_25068 | Not-determined |
| 17-109_k141_310550 | Not-determined |
| 17-109_k141_226015 | Not-determined |
| 17-109_k141_57989 | Low-quality |
| 17-109_k141_242666 | Low-quality |
| 17-109_k141_208918 | Not-determined |
| 17-109_k141_124563 | Not-determined |
| 17-109_k141_175433 | Not-determined |
| 17-109_k141_310560 | Not-determined |
| 17-109_k141_124570 | Not-determined |
| 17-109_k141_326519 | Not-determined |
| 17-109_k141_175440 | Not-determined |
| 17-109_k141_124575 | Low-quality |
| 17-109_k141_208922 | Low-quality |
| 17-109_k141_208925 | Not-determined |
| 17-109_k141_108210 | Not-determined |
| 17-109_k141_8170 | Not-determined |
| 17-109_k141_358580 | Not-determined |
| 17-109_k141_358582 | Not-determined |
| 17-109_k141_342641 | Not-determined |
| 17-109_k141_91454 | Not-determined |
| 17-109_k141_326528 | Not-determined |
| 17-109_k141_259183 | Not-determined |
| 17-109_k141_326530 | Low-quality |
| 17-109_k141_8177 | Not-determined |
| 17-109_k141_58019 | Not-determined |
| 17-109_k141_259184 | Not-determined |
| 17-109_k141_58023 | Not-determined |
| 17-109_k141_226034 | Not-determined |
| 17-109_k141_25099 | Not-determined |
| 17-109_k141_208950 | Not-determined |
| 17-109_k141_193362 | Not-determined |
| 17-109_k141_142388 | Not-determined |
| 17-109_k141_25103 | Not-determined |
| 17-109_k141_358602 | Not-determined |
| 17-109_k141_226046 | Not-determined |
| 17-109_k141_8189 | Not-determined |
| 17-109_k141_358606 | Not-determined |
| 17-109_k141_25109 | Low-quality |
| 17-109_k141_25110 | Not-determined |
| 17-109_k141_124602 | Not-determined |
| 17-109_k141_159185 | Not-determined |
| 17-109_k141_25121 | Not-determined |
| 17-109_k141_293502 | Not-determined |
| 17-109_k141_208961 | Not-determined |
| 17-109_k141_358628 | Low-quality |
| 17-109_k141_293507 | Not-determined |
| 17-109_k141_293508 | Not-determined |
| 17-109_k141_293511 | Low-quality |
| 17-109_k141_208970 | Not-determined |
| 17-109_k141_293515 | Not-determined |
| 17-109_k141_358640 | Not-determined |
| 17-109_k141_358641 | Not-determined |
| 17-109_k141_8206 | Not-determined |
| 17-109_k141_310579 | Not-determined |
| 17-109_k141_310580 | Not-determined |
| 17-109_k141_293528 | Not-determined |
| 17-109_k141_58059 | Not-determined |
| 17-109_k141_8220 | Not-determined |
| 17-109_k141_326565 | Not-determined |
| 17-109_k141_242709 | Not-determined |
| 17-109_k141_326566 | Not-determined |
| 17-109_k141_193373 | Not-determined |
| 17-109_k141_193374 | Not-determined |
| 17-109_k141_124620 | Not-determined |
| 17-109_k141_226086 | Not-determined |
| 17-109_k141_58064 | Not-determined |
| 17-109_k141_226089 | Not-determined |
| 17-109_k141_310590 | Not-determined |
| 17-109_k141_8227 | Not-determined |
| 17-109_k141_193380 | Not-determined |
| 17-109_k141_310593 | Not-determined |
| 17-109_k141_142428 | Not-determined |
| 17-109_k141_124631 | Not-determined |
| 17-109_k141_276414 | Low-quality |
| 17-109_k141_209007 | Not-determined |
| 17-109_k141_25168 | Not-determined |
| 17-109_k141_58082 | Not-determined |
| 17-109_k141_342669 | Low-quality |
| 17-109_k141_8240 | Not-determined |
| 17-109_k141_142443 | Not-determined |
| 17-109_k141_276426 | Not-determined |
| 17-109_k141_326585 | Not-determined |
| 17-109_k141_58089 | Not-determined |
| 17-109_k141_226100 | Low-quality |
| 17-109_k141_326589 | Not-determined |
| 17-109_k141_25181 | Not-determined |
| 17-109_k141_226103 | Not-determined |
| 17-109_k141_75589 | Low-quality |
| 17-109_k141_342685 | Not-determined |
| 17-109_k141_108239 | Not-determined |
| 17-109_k141_91482 | Not-determined |
| 17-109_k141_58094 | Not-determined |
| 17-109_k141_193392 | Not-determined |
| 17-109_k141_342691 | Not-determined |
| 17-109_k141_193395 | Not-determined |
| 17-109_k141_326605 | Not-determined |
| 17-109_k141_193396 | Not-determined |
| 17-109_k141_58102 | Not-determined |
| 17-109_k141_326610 | Not-determined |
| 17-109_k141_175472 | Not-determined |
| 17-109_k141_242723 | Not-determined |
| 17-109_k141_209037 | Not-determined |
| 17-109_k141_142459 | Not-determined |
| 17-109_k141_276445 | Not-determined |
| 17-109_k141_193409 | Not-determined |
| 17-109_k141_326620 | Not-determined |
| 17-109_k141_326623 | Not-determined |
| 17-109_k141_58108 | Not-determined |
| 17-109_k141_175481 | Not-determined |
| 17-109_k141_342707 | Not-determined |
| 17-109_k141_175486 | Not-determined |
| 17-109_k141_310620 | Not-determined |
| 17-109_k141_326629 | Low-quality |
| 17-109_k141_326630 | Not-determined |
| 17-109_k141_193418 | Not-determined |
| 17-109_k141_42059 | Not-determined |
| 17-109_k141_342712 | Not-determined |
| 17-109_k141_310624 | Low-quality |
| 17-109_k141_242728 | Not-determined |
| 17-109_k141_276458 | Not-determined |
| 17-109_k141_293575 | Not-determined |
| 17-109_k141_25192 | Not-determined |
| 17-109_k141_42072 | Not-determined |
| 17-109_k141_393511 | Not-determined |
| 17-109_k141_108256 | Not-determined |
| 17-109_k141_25195 | Low-quality |
| 17-109_k141_276463 | Not-determined |
| 17-109_k141_193432 | Not-determined |
| 17-109_k141_226135 | Not-determined |
| 17-109_k141_326637 | Not-determined |
| 17-109_k141_142496 | Not-determined |
| 17-109_k141_42079 | Not-determined |
| 17-109_k141_393522 | Not-determined |
| 17-109_k141_326641 | Not-determined |
| 17-109_k141_393523 | Not-determined |
| 17-109_k141_142502 | Not-determined |
| 17-109_k141_209050 | Not-determined |
| 17-109_k141_209052 | Not-determined |
| 17-109_k141_142508 | Not-determined |
| 17-109_k141_42087 | Not-determined |
| 17-109_k141_276472 | Not-determined |

| | |
|---|---|
| 17-109_k141_25211 | Not-determined |
| 17-109_k141_91485 | Not-determined |
| 17-109_k141_242735 | Not-determined |
| 17-109_k141_293601 | Not-determined |
| 17-109_k141_293604 | Not-determined |
| 17-109_k141_242738 | Not-determined |
| 17-109_k141_310646 | Low-quality |
| 17-109_k141_326653 | Not-determined |
| 17-109_k141_142520 | Not-determined |
| 17-109_k141_124650 | Not-determined |
| 17-109_k141_393538 | Low-quality |
| 17-109_k141_75629 | Not-determined |
| 17-109_k141_393540 | Not-determined |
| 17-109_k141_142525 | Not-determined |
| 17-109_k141_226153 | Not-determined |
| 17-109_k141_8250 | Not-determined |
| 17-109_k141_276489 | Not-determined |
| 17-109_k141_393544 | Not-determined |
| 17-109_k141_393546 | Not-determined |
| 17-109_k141_58159 | Not-determined |
| 17-109_k141_8259 | Low-quality |
| 17-109_k141_226165 | Not-determined |
| 17-109_k141_226166 | Not-determined |
| 17-109_k141_108301 | Not-determined |
| 17-109_k141_142528 | Not-determined |
| 17-109_k141_393553 | Not-determined |
| 17-109_k141_393555 | Low-quality |
| 17-109_k141_58171 | Low-quality |
| 17-109_k141_159246 | Not-determined |
| 17-109_k141_293635 | Not-determined |
| 17-109_k141_58175 | Not-determined |
| 17-109_k141_393567 | Not-determined |
| 17-109_k141_209059 | Not-determined |
| 17-109_k141_108321 | Not-determined |
| 17-109_k141_8279 | Not-determined |
| 17-109_k141_91504 | Not-determined |
| 17-109_k141_25230 | Not-determined |
| 17-109_k141_393576 | Not-determined |
| 17-109_k141_108326 | Not-determined |
| 17-109_k141_108327 | Not-determined |
| 17-109_k141_58181 | Not-determined |
| 17-109_k141_326677 | Not-determined |
| 17-109_k141_175536 | Not-determined |
| 17-109_k141_108331 | Not-determined |
| 17-109_k141_175537 | Not-determined |
| 17-109_k141_124692 | Not-determined |
| 17-109_k141_91510 | Not-determined |
| 17-109_k141_58185 | Not-determined |
| 17-109_k141_276524 | Not-determined |
| 17-109_k141_91514 | Not-determined |
| 17-109_k141_310669 | Not-determined |
| 17-109_k141_393590 | Not-determined |
| 17-109_k141_124705 | Not-determined |
| 17-109_k141_276536 | Not-determined |
| 17-109_k141_159262 | Not-determined |
| 17-109_k141_276537 | Not-determined |
| 17-109_k141_393593 | Not-determined |
| 17-109_k141_393595 | Not-determined |
| 17-109_k141_58198 | Not-determined |
| 17-109_k141_259198 | Not-determined |
| 17-109_k141_75666 | Low-quality |
| 17-109_k141_58202 | Not-determined |
| 17-109_k141_142534 | Not-determined |
| 17-109_k141_58204 | Not-determined |
| 17-109_k141_293650 | Not-determined |
| 17-109_k141_175552 | Not-determined |
| 17-109_k141_293654 | Low-quality |
| 17-109_k141_276546 | Not-determined |
| 17-109_k141_124718 | Not-determined |
| 17-109_k141_342747 | Not-determined |
| 17-109_k141_326695 | Not-determined |
| 17-109_k141_342748 | Not-determined |
| 17-109_k141_25246 | Not-determined |
| 17-109_k141_25247 | Not-determined |
| 17-109_k141_342752 | Low-quality |
| 17-109_k141_8319 | Not-determined |
| 17-109_k141_342755 | Low-quality |
| 17-109_k141_108350 | Not-determined |
| 17-109_k141_293661 | Low-quality |
| 17-109_k141_393604 | Not-determined |
| 17-109_k141_293663 | Low-quality |
| 17-109_k141_142547 | Not-determined |
| 17-109_k141_58218 | Not-determined |
| 17-109_k141_58220 | Not-determined |
| 17-109_k141_358717 | Not-determined |
| 17-109_k141_326700 | Not-determined |
| 17-109_k141_358718 | Not-determined |
| 17-109_k141_91534 | Not-determined |
| 17-109_k141_393608 | Not-determined |
| 17-109_k141_259209 | Not-determined |
| 17-109_k141_342766 | Not-determined |
| 17-109_k141_91541 | Not-determined |
| 17-109_k141_175567 | Not-determined |
| 17-109_k141_358725 | Not-determined |
| 17-109_k141_226169 | Not-determined |
| 17-109_k141_42139 | Not-determined |
| 17-109_k141_259214 | Not-determined |
| 17-109_k141_108358 | Not-determined |
| 17-109_k141_193498 | Not-determined |
| 17-109_k141_226178 | Not-determined |
| 17-109_k141_310688 | Not-determined |
| 17-109_k141_376622 | Not-determined |
| 17-109_k141_142561 | Not-determined |
| 17-109_k141_142562 | Not-determined |
| 17-109_k141_108367 | Not-determined |
| 17-109_k141_142563 | Not-determined |
| 17-109_k141_358732 | Low-quality |
| 17-109_k141_310692 | Not-determined |
| 17-109_k141_358738 | Not-determined |
| 17-109_k141_342798 | Not-determined |
| 17-109_k141_209091 | Low-quality |
| 17-109_k141_376635 | Not-determined |
| 17-109_k141_226197 | Not-determined |
| 17-109_k141_142574 | Low-quality |
| 17-109_k141_293689 | Not-determined |
| 17-109_k141_376637 | Not-determined |
| 17-109_k141_42166 | Not-determined |
| 17-109_k141_42167 | Not-determined |
| 17-109_k141_393639 | Not-determined |
| 17-109_k141_75706 | Not-determined |
| 17-109_k141_376649 | Not-determined |
| 17-109_k141_342806 | Not-determined |
| 17-109_k141_75711 | Not-determined |
| 17-109_k141_259252 | Not-determined |
| 17-109_k141_42186 | Not-determined |
| 17-109_k141_310725 | Low-quality |
| 17-109_k141_310729 | Not-determined |
| 17-109_k141_142587 | Not-determined |
| 17-109_k141_75723 | Not-determined |
| 17-109_k141_42202 | Not-determined |
| 17-109_k141_342813 | Not-determined |
| 17-109_k141_75725 | Not-determined |
| 17-109_k141_75726 | Not-determined |
| 17-109_k141_25276 | Low-quality |
| 17-109_k141_376668 | Not-determined |
| 17-109_k141_276584 | Not-determined |
| 17-109_k141_326759 | Not-determined |
| 17-109_k141_209121 | Low-quality |
| 17-109_k141_209122 | Not-determined |
| 17-109_k141_142594 | Not-determined |
| 17-109_k141_326763 | Not-determined |
| 17-109_k141_358755 | Not-determined |
| 17-109_k141_193552 | Low-quality |
| 17-109_k141_124781 | Not-determined |
| 17-109_k141_108386 | Not-determined |
| 17-109_k141_75734 | Low-quality |
| 17-109_k141_193554 | Not-determined |
| 17-109_k141_293723 | Not-determined |
| 17-109_k141_193555 | Not-determined |
| 17-109_k141_393663 | Not-determined |
| 17-109_k141_42211 | Not-determined |
| 17-109_k141_91594 | Not-determined |
| 17-109_k141_42215 | Not-determined |
| 17-109_k141_342818 | Not-determined |
| 17-109_k141_293730 | Not-determined |
| 17-109_k141_108398 | Not-determined |
| 17-109_k141_108403 | Not-determined |
| 17-109_k141_108405 | Not-determined |
| 17-109_k141_25289 | Not-determined |
| 17-109_k141_124790 | Low-quality |
| 17-109_k141_159295 | Low-quality |
| 17-109_k141_25293 | Not-determined |
| 17-109_k141_326779 | Not-determined |
| 17-109_k141_159297 | Not-determined |
| 17-109_k141_209138 | Not-determined |
| 17-109_k141_310738 | Not-determined |
| 17-109_k141_358762 | Low-quality |
| 17-109_k141_108408 | Not-determined |
| 17-109_k141_175603 | Not-determined |
| 17-109_k141_310743 | Low-quality |
| 17-109_k141_358768 | Not-determined |
| 17-109_k141_108412 | Not-determined |
| 17-109_k141_376703 | Low-quality |
| 17-109_k141_142635 | Not-determined |
| 17-109_k141_124802 | Not-determined |
| 17-109_k141_259260 | Not-determined |
| 17-109_k141_159308 | Not-determined |
| 17-109_k141_91609 | Not-determined |
| 17-109_k141_142638 | Not-determined |
| 17-109_k141_8333 | Not-determined |
| 17-109_k141_293746 | Low-quality |
| 17-109_k141_276613 | Low-quality |
| 17-109_k141_209154 | Not-determined |
| 17-109_k141_276614 | Not-determined |
| 17-109_k141_376708 | Not-determined |
| 17-109_k141_276615 | Not-determined |
| 17-109_k141_376715 | Not-determined |
| 17-109_k141_159322 | Not-determined |
| 17-109_k141_209167 | Low-quality |
| 17-109_k141_124818 | Not-determined |
| 17-109_k141_175618 | Not-determined |
| 17-109_k141_91629 | Not-determined |
| 17-109_k141_175621 | Not-determined |
| 17-109_k141_259280 | Not-determined |
| 17-109_k141_159329 | Low-quality |
| 17-109_k141_259287 | Low-quality |
| 17-109_k141_376740 | Not-determined |
| 17-109_k141_393686 | Not-determined |
| 17-109_k141_293760 | Not-determined |
| 17-109_k141_259292 | Not-determined |
| 17-109_k141_326799 | Not-determined |
| 17-109_k141_124836 | Not-determined |
| 17-109_k141_159338 | Not-determined |
| 17-109_k141_108442 | Not-determined |
| 17-109_k141_124849 | Not-determined |
| 17-109_k141_393698 | Not-determined |
| 17-109_k141_108454 | Not-determined |
| 17-109_k141_175646 | Not-determined |
| 17-109_k141_159348 | Not-determined |
| 17-109_k141_259312 | Not-determined |
| 17-109_k141_75789 | Not-determined |
| 17-109_k141_276645 | Not-determined |
| 17-109_k141_159354 | Not-determined |
| 17-109_k141_376761 | Not-determined |
| 17-109_k141_159355 | Low-quality |
| 17-109_k141_376762 | Not-determined |
| 17-109_k141_8374 | Not-determined |
| 17-109_k141_209196 | Not-determined |
| 17-109_k141_242758 | Not-determined |
| 17-109_k141_25312 | Not-determined |
| 17-109_k141_8381 | Not-determined |
| 17-109_k141_124865 | Not-determined |
| 17-109_k141_124867 | Not-determined |
| 17-109_k141_259323 | Not-determined |
| 17-109_k141_175663 | Not-determined |
| 17-109_k141_75798 | Low-quality |
| 17-109_k141_242762 | Not-determined |
| 17-109_k141_124869 | Not-determined |
| 17-109_k141_376770 | Not-determined |
| 17-109_k141_25316 | Not-determined |
| 17-109_k141_8388 | Not-determined |
| 17-109_k141_75804 | Not-determined |
| 17-109_k141_25320 | Low-quality |
| 17-109_k141_42250 | Not-determined |
| 17-109_k141_159373 | Low-quality |
| 17-109_k141_376774 | Not-determined |
| 17-109_k141_75810 | Not-determined |
| 17-109_k141_25322 | Not-determined |
| 17-109_k141_326806 | Low-quality |
| 17-109_k141_326808 | Not-determined |
| 17-109_k141_91643 | Not-determined |
| 17-109_k141_259336 | Low-quality |
| 17-109_k141_393704 | Not-determined |
| 17-109_k141_108466 | Not-determined |
| 17-109_k141_58292 | Not-determined |
| 17-109_k141_358786 | Not-determined |
| 17-109_k141_293794 | Not-determined |
| 17-109_k141_226245 | Not-determined |
| 17-109_k141_358795 | Not-determined |
| 17-109_k141_58295 | Not-determined |
| 17-109_k141_25332 | Not-determined |
| 17-109_k141_242794 | Not-determined |
| 17-109_k141_342838 | Not-determined |
| 17-109_k141_91659 | Not-determined |
| 18-104-final-viral-combined_k141_113721\|\|full | Medium-quality |
| 17-109_k141_193566 | Not-determined |
| 17-109_k141_58307 | Not-determined |
| 17-109_k141_58310 | Not-determined |
| 17-109_k141_42285 | Low-quality |
| 17-109_k141_193579 | Not-determined |
| 17-109_k141_91677 | Not-determined |
| 17-109_k141_226259 | Not-determined |
| 17-109_k141_376789 | Not-determined |
| 17-109_k141_25349 | Not-determined |
| 17-109_k141_124904 | Not-determined |
| 17-109_k141_124907 | Not-determined |
| 17-109_k141_326845 | Not-determined |
| 17-109_k141_91682 | Not-determined |
| 17-109_k141_193585 | Not-determined |
| 17-109_k141_293816 | Low-quality |
| 17-109_k141_8439 | Not-determined |
| 17-109_k141_293820 | Not-determined |
| 17-109_k141_25356 | Not-determined |
| 17-109_k141_8447 | Not-determined |
| 17-109_k141_108507 | Not-determined |
| 17-109_k141_58323 | Not-determined |
| 17-109_k141_293827 | Not-determined |
| 17-109_k141_42317 | Not-determined |
| 17-109_k141_276653 | Not-determined |
| 17-109_k141_193604 | Not-determined |
| 17-109_k141_276660 | Low-quality |
| 17-109_k141_259389 | Not-determined |
| 17-109_k141_42334 | Not-determined |
| 17-109_k141_310789 | Not-determined |
| 17-109_k141_226288 | Not-determined |
| 17-109_k141_242847 | Not-determined |
| 17-109_k141_75821 | Not-determined |
| 17-109_k141_259396 | Not-determined |
| 17-109_k141_226289 | Not-determined |
| 17-109_k141_310794 | Not-determined |
| 17-109_k141_310795 | Not-determined |
| 17-109_k141_293839 | Not-determined |
| 17-109_k141_75829 | Not-determined |
| 17-109_k141_8467 | Low-quality |
| 17-109_k141_259404 | Not-determined |
| 17-109_k141_276670 | Not-determined |
| 17-109_k141_193619 | Not-determined |
| 17-109_k141_108535 | Not-determined |
| 17-109_k141_276680 | Not-determined |
| 17-109_k141_226310 | Not-determined |
| 17-109_k141_142722 | Not-determined |
| 17-109_k141_326869 | Not-determined |
| 17-109_k141_326870 | Not-determined |
| 17-109_k141_293858 | Not-determined |
| 17-109_k141_310805 | Low-quality |
| 17-109_k141_142731 | Not-determined |
| 17-109_k141_259425 | Not-determined |
| 17-109_k141_193637 | Not-determined |
| 17-109_k141_75866 | Not-determined |
| 17-109_k141_175724 | Not-determined |
| 17-109_k141_175725 | Not-determined |
| 17-109_k141_108557 | Not-determined |
| 17-109_k141_242868 | Not-determined |
| 17-109_k141_393756 | Not-determined |
| 17-109_k141_393758 | Not-determined |
| 17-109_k141_242872 | Not-determined |
| 17-109_k141_259439 | Not-determined |
| 17-109_k141_242873 | Low-quality |
| 17-109_k141_175729 | Not-determined |
| 17-109_k141_25391 | Not-determined |
| 17-109_k141_310821 | Low-quality |
| 17-109_k141_58352 | Not-determined |
| 17-109_k141_91714 | Low-quality |
| 17-109_k141_242883 | Not-determined |
| 17-109_k141_159402 | Not-determined |
| 17-109_k141_226336 | Low-quality |
| 17-109_k141_42368 | Not-determined |
| 17-109_k141_91720 | Low-quality |
| 17-109_k141_259447 | Not-determined |
| 17-109_k141_91721 | Not-determined |
| 17-109_k141_108561 | Not-determined |
| 17-109_k141_75869 | Not-determined |
| 17-109_k141_91724 | Not-determined |
| 17-109_k141_358824 | Not-determined |
| 17-109_k141_108565 | Not-determined |
| 17-109_k141_259451 | Not-determined |
| 17-109_k141_259452 | Not-determined |
| 17-109_k141_259454 | Not-determined |
| 17-109_k141_159409 | Not-determined |
| 17-109_k141_42372 | Not-determined |
| 17-109_k141_142766 | Not-determined |
| 17-109_k141_193650 | Not-determined |
| 17-109_k141_226350 | Not-determined |
| 17-109_k141_75885 | Not-determined |
| 17-109_k141_393777 | Low-quality |
| 17-109_k141_310841 | Not-determined |
| 17-109_k141_342886 | Not-determined |
| 17-109_k141_393778 | Not-determined |
| 17-109_k141_8488 | Not-determined |
| 17-109_k141_342891 | Not-determined |
| 17-109_k141_142781 | Not-determined |
| 17-109_k141_75899 | Not-determined |
| 17-109_k141_209202 | Low-quality |
| 17-109_k141_259465 | Not-determined |
| 17-109_k141_25411 | Not-determined |
| 17-109_k141_376832 | Not-determined |
| 17-109_k141_25417 | Not-determined |
| 17-109_k141_326902 | Not-determined |
| 17-109_k141_276708 | Not-determined |
| 17-109_k141_376839 | Not-determined |
| 17-109_k141_242921 | Not-determined |
| 17-109_k141_259475 | Not-determined |
| 17-109_k141_276716 | Not-determined |
| 17-109_k141_175753 | Low-quality |
| 17-109_k141_226384 | Not-determined |
| 17-109_k141_8504 | Not-determined |
| 17-109_k141_75922 | Not-determined |
| 17-109_k141_91729 | Not-determined |
| 17-109_k141_75927 | Not-determined |
| 17-109_k141_8507 | Not-determined |
| 17-109_k141_393791 | Not-determined |
| 17-109_k141_358831 | Not-determined |
| 17-109_k141_209209 | Not-determined |
| 17-109_k141_242929 | Not-determined |
| 17-109_k141_276723 | Not-determined |
| 17-109_k141_342915 | Low-quality |
| 17-109_k141_342919 | Not-determined |
| 17-109_k141_8519 | Not-determined |
| 17-109_k141_58385 | Low-quality |
| 17-109_k141_91747 | Not-determined |
| 17-109_k141_124980 | Not-determined |
| 17-109_k141_124982 | Low-quality |
| 17-109_k141_124984 | Low-quality |
| 17-109_k141_358843 | Not-determined |
| 17-109_k141_376856 | Not-determined |
| 17-109_k141_226408 | Not-determined |
| 17-109_k141_159448 | Not-determined |
| 17-109_k141_124989 | Not-determined |
| 17-109_k141_91764 | Not-determined |
| 17-109_k141_242954 | Low-quality |
| 17-109_k141_159451 | Not-determined |
| 17-109_k141_326923 | Not-determined |
| 17-109_k141_175759 | Not-determined |
| 17-109_k141_342926 | Low-quality |
| 17-109_k141_175764 | Not-determined |
| 17-109_k141_358862 | Not-determined |
| 17-109_k141_242966 | Not-determined |
| 17-109_k141_326931 | Not-determined |
| 17-109_k141_276741 | Not-determined |
| 17-109_k141_326932 | Not-determined |
| 17-109_k141_310901 | Not-determined |
| 17-109_k141_209234 | Not-determined |
| 17-109_k141_242971 | Not-determined |
| 17-109_k141_8531 | Not-determined |
| 17-109_k141_310905 | Not-determined |
| 17-109_k141_310909 | Not-determined |
| 17-109_k141_393819 | Not-determined |
| 17-109_k141_142813 | Not-determined |
| 17-109_k141_8533 | Low-quality |
| 17-109_k141_376867 | Low-quality |
| 17-109_k141_142817 | Not-determined |
| 17-109_k141_42403 | Not-determined |
| 17-109_k141_75975 | Low-quality |
| 17-109_k141_91788 | Not-determined |
| 17-109_k141_91789 | Not-determined |
| 17-109_k141_175781 | Not-determined |
| 17-109_k141_393828 | Not-determined |
| 17-109_k141_8538 | Not-determined |
| 17-109_k141_25443 | Not-determined |
| 17-109_k141_209253 | Low-quality |
| 17-109_k141_42404 | Low-quality |
| 17-109_k141_8543 | Not-determined |
| 17-109_k141_209254 | Not-determined |
| 17-109_k141_25447 | Not-determined |
| 17-109_k141_242986 | Not-determined |
| 17-109_k141_242987 | Not-determined |
| 17-109_k141_393833 | Not-determined |
| 17-109_k141_108569 | Low-quality |
| 17-109_k141_175784 | Not-determined |
| 17-109_k141_226444 | Low-quality |
| 17-109_k141_75990 | Not-determined |
| 17-109_k141_175791 | Not-determined |
| 17-109_k141_58416 | Low-quality |
| 17-109_k141_326971 | Not-determined |
| 17-109_k141_175793 | Not-determined |
| 17-109_k141_358905 | Not-determined |
| 17-109_k141_108577 | Not-determined |
| 17-109_k141_393838 | Not-determined |
| 17-109_k141_76002 | Not-determined |
| 17-109_k141_393843 | Not-determined |
| 17-109_k141_25452 | Not-determined |
| 17-109_k141_226458 | Not-determined |
| 17-109_k141_159502 | Not-determined |
| 17-109_k141_326983 | Not-determined |
| 17-109_k141_25457 | Not-determined |
| 17-109_k141_125022 | Not-determined |
| 17-109_k141_175805 | Not-determined |
| 17-109_k141_125027 | Not-determined |
| 17-109_k141_8548 | Low-quality |
| 17-109_k141_8550 | Not-determined |
| 17-109_k141_76017 | Not-determined |
| 17-109_k141_393853 | Low-quality |
| 17-109_k141_209275 | Not-determined |
| 17-109_k141_175809 | Not-determined |
| 17-109_k141_108587 | Not-determined |
| 17-109_k141_276756 | Not-determined |
| 17-109_k141_125040 | Not-determined |
| 17-109_k141_393862 | Low-quality |
| 17-109_k141_276761 | Not-determined |
| 17-109_k141_175821 | Not-determined |
| 17-109_k141_226472 | Not-determined |
| 17-109_k141_393869 | Not-determined |
| 17-109_k141_209287 | Not-determined |
| 17-109_k141_393874 | Not-determined |
| 17-109_k141_393875 | Not-determined |
| 17-109_k141_293933 | Not-determined |
| 17-109_k141_376911 | Not-determined |
| 17-109_k141_376914 | Not-determined |
| 17-109_k141_376917 | Not-determined |
| 17-109_k141_108613 | Low-quality |
| 17-109_k141_342947 | Not-determined |
| 17-109_k141_108614 | Not-determined |
| 17-109_k141_175842 | Not-determined |
| 17-109_k141_108618 | Not-determined |
| 17-109_k141_293946 | Low-quality |
| 17-109_k141_108619 | Not-determined |
| 17-109_k141_175845 | Low-quality |
| 17-109_k141_276770 | Not-determined |
| 17-109_k141_209301 | Not-determined |
| 17-109_k141_159554 | Low-quality |
| 17-109_k141_209304 | Not-determined |
| 17-109_k141_58451 | Not-determined |
| 17-109_k141_58455 | Not-determined |
| 17-109_k141_358947 | Not-determined |
| 17-109_k141_108633 | Not-determined |
| 17-109_k141_108634 | Not-determined |
| 17-109_k141_108635 | Not-determined |
| 17-109_k141_108636 | Not-determined |
| 17-109_k141_142853 | Not-determined |
| 17-109_k141_108638 | Not-determined |
| 17-109_k141_159571 | Not-determined |
| 17-109_k141_42444 | Not-determined |
| 17-109_k141_327033 | Not-determined |
| 17-109_k141_58462 | Not-determined |
| 17-109_k141_108645 | Not-determined |
| 17-109_k141_209329 | Not-determined |
| 17-109_k141_293966 | Not-determined |
| 17-109_k141_327038 | Not-determined |
| 17-109_k141_58470 | Not-determined |
| 17-109_k141_91830 | Not-determined |
| 17-109_k141_91832 | Not-determined |
| 17-109_k141_125080 | Not-determined |
| 17-109_k141_108649 | Not-determined |
| 17-109_k141_376931 | Not-determined |
| 17-109_k141_376933 | Not-determined |
| 17-109_k141_91841 | Not-determined |
| 17-109_k141_159583 | Low-quality |
| 17-109_k141_358973 | Not-determined |
| 17-109_k141_25460 | Not-determined |
| 17-109_k141_25461 | Not-determined |
| 17-109_k141_226531 | Not-determined |
| 17-109_k141_58491 | Not-determined |
| 17-109_k141_25464 | Not-determined |
| 17-109_k141_376946 | Not-determined |
| 17-109_k141_108660 | Not-determined |
| 17-109_k141_25466 | Not-determined |
| 17-109_k141_142891 | Low-quality |
| 17-109_k141_159588 | Low-quality |
| 17-109_k141_142892 | Not-determined |
| 17-109_k141_8589 | Not-determined |
| 17-109_k141_276786 | Not-determined |
| 17-109_k141_25474 | Low-quality |
| 17-109_k141_142902 | Not-determined |
| 17-109_k141_25475 | Low-quality |
| 17-109_k141_393929 | Not-determined |
| 17-109_k141_276795 | Not-determined |
| 17-109_k141_175870 | Not-determined |
| 17-109_k141_342993 | Not-determined |
| 17-109_k141_76065 | Not-determined |
| 17-109_k141_25478 | Not-determined |
| 17-109_k141_91857 | Low-quality |
| 17-109_k141_293996 | Not-determined |
| 17-109_k141_393934 | Not-determined |
| 17-109_k141_209362 | Not-determined |
| 17-109_k141_142920 | Not-determined |
| 17-109_k141_142922 | Not-determined |
| 17-109_k141_276799 | Not-determined |
| 17-109_k141_142927 | Not-determined |
| 17-109_k141_393944 | Not-determined |
| 17-109_k141_393945 | Not-determined |
| 17-109_k141_125134 | Not-determined |
| 17-109_k141_58512 | Not-determined |
| 17-109_k141_393953 | Not-determined |
| 17-109_k141_209374 | Low-quality |
| 17-109_k141_276816 | Not-determined |
| 17-109_k141_209379 | Not-determined |
| 17-109_k141_294013 | Not-determined |
| 17-109_k141_276819 | Not-determined |
| 17-109_k141_343010 | Not-determined |
| 17-109_k141_327071 | Not-determined |
| 17-109_k141_108687 | Low-quality |
| 17-109_k141_276824 | Not-determined |
| 17-109_k141_393963 | Not-determined |
| 17-109_k141_42494 | Not-determined |
| 17-109_k141_175887 | Not-determined |
| 17-109_k141_193685 | Not-determined |
| 17-109_k141_276833 | Not-determined |
| 17-109_k141_193686 | Not-determined |
| 17-109_k141_125164 | Not-determined |
| 17-109_k141_76079 | Not-determined |
| 17-109_k141_310935 | Not-determined |
| 17-109_k141_343026 | Not-determined |
| 17-109_k141_108709 | Not-determined |
| 17-109_k141_294037 | Not-determined |
| 17-109_k141_393974 | Not-determined |
| 17-109_k141_175896 | Not-determined |
| 17-109_k141_76083 | Low-quality |
| 17-109_k141_393978 | Not-determined |
| 17-109_k141_226570 | Not-determined |
| 17-109_k141_193694 | Not-determined |
| 17-109_k141_125169 | Not-determined |
| 17-109_k141_209405 | Not-determined |
| 17-109_k141_42516 | Not-determined |
| 17-109_k141_76091 | Not-determined |
| 17-109_k141_76095 | Not-determined |
| 17-109_k141_76097 | Not-determined |
| 17-109_k141_327094 | Not-determined |
| 17-109_k141_76101 | Not-determined |
| 17-109_k141_376971 | Not-determined |
| 17-109_k141_42525 | Low-quality |
| 17-109_k141_310951 | Not-determined |
| 17-109_k141_310957 | Not-determined |
| 17-109_k141_327102 | Not-determined |
| 17-109_k141_376982 | Not-determined |
| 17-109_k141_343028 | Low-quality |
| 17-109_k141_359034 | Not-determined |
| 17-109_k141_226600 | Not-determined |
| 17-109_k141_376986 | Not-determined |
| 17-109_k141_343032 | Low-quality |

| | |
|---|---|
| 17-109_k141_193712 | Not-determined |
| 17-109_k141_175905 | Not-determined |
| 17-109_k141_8611 | Not-determined |
| 17-109_k141_142936 | Not-determined |
| 17-109_k141_310966 | Not-determined |
| 17-109_k141_175908 | Not-determined |
| 17-109_k141_226605 | Not-determined |
| 17-109_k141_343036 | Not-determined |
| 17-109_k141_209421 | Not-determined |
| 17-109_k141_142942 | Not-determined |
| 17-109_k141_243016 | Not-determined |
| 17-109_k141_76109 | Not-determined |
| 17-109_k141_359040 | Not-determined |
| 17-109_k141_193724 | Not-determined |
| 17-109_k141_359042 | Not-determined |
| 17-109_k141_377000 | Not-determined |
| 17-109_k141_226619 | Not-determined |
| 17-109_k141_243024 | Not-determined |
| 17-109_k141_359048 | Not-determined |
| 17-109_k141_209434 | Not-determined |
| 17-109_k141_193732 | Not-determined |
| 17-109_k141_310986 | Not-determined |
| 17-109_k141_108728 | Not-determined |
| 17-109_k141_175921 | Not-determined |
| 17-109_k141_91867 | Not-determined |
| 17-109_k141_8631 | Not-determined |
| 17-109_k141_359057 | Not-determined |
| 17-109_k141_310989 | Not-determined |
| 17-109_k141_108732 | Not-determined |
| 17-109_k141_209449 | Low-quality |
| 17-109_k141_76127 | Not-determined |
| 17-109_k141_175929 | Low-quality |
| 17-109_k141_25516 | Low-quality |
| 17-109_k141_377019 | Not-determined |
| 17-109_k141_209457 | Not-determined |
| 17-109_k141_25520 | Low-quality |
| 17-109_k141_327149 | Not-determined |
| 17-109_k141_327150 | Not-determined |
| 17-109_k141_125214 | Not-determined |
| 17-109_k141_76134 | Not-determined |
| 17-109_k141_91884 | Not-determined |
| 17-109_k141_377024 | Not-determined |
| 17-109_k141_91886 | Not-determined |
| 17-109_k141_142977 | Not-determined |
| 17-109_k141_359072 | Not-determined |
| 17-109_k141_243037 | Not-determined |
| 17-109_k141_327157 | Not-determined |
| 17-109_k141_209464 | Not-determined |
| 17-109_k141_108758 | Not-determined |
| 17-109_k141_8653 | Not-determined |
| 17-109_k141_359075 | Not-determined |
| 17-109_k141_91892 | Not-determined |
| 17-109_k141_76145 | Not-determined |
| 17-109_k141_108765 | Not-determined |
| 17-109_k141_25534 | Not-determined |
| 17-109_k141_25537 | Not-determined |
| 17-109_k141_226662 | Not-determined |
| 17-109_k141_377040 | Not-determined |
| 17-109_k141_359081 | Not-determined |
| 17-109_k141_193752 | Not-determined |
| 17-109_k141_142998 | Not-determined |
| 17-109_k141_311026 | Low-quality |
| 17-109_k141_91905 | Not-determined |
| 17-109_k141_226665 | Not-determined |
| 17-109_k141_91907 | Not-determined |
| 17-109_k141_76163 | Low-quality |
| 17-109_k141_91910 | Not-determined |
| 17-109_k141_91913 | Not-determined |
| 17-109_k141_175953 | Low-quality |
| 17-109_k141_159612 | Not-determined |
| 17-109_k141_108787 | Not-determined |
| 17-109_k141_108789 | Not-determined |
| 17-109_k141_226673 | Not-determined |
| 17-109_k141_108791 | Low-quality |
| 17-109_k141_175956 | Not-determined |
| 17-109_k141_76172 | Not-determined |
| 17-109_k141_42563 | Not-determined |
| 17-109_k141_327170 | Not-determined |
| 17-109_k141_76185 | Not-determined |
| 17-109_k141_193770 | Not-determined |
| 17-109_k141_25563 | Not-determined |
| 17-109_k141_327183 | Low-quality |
| 17-109_k141_108826 | Not-determined |
| 17-109_k141_159636 | Not-determined |
| 17-109_k141_159638 | Not-determined |
| 17-109_k141_175998 | Low-quality |
| 17-109_k141_175999 | Not-determined |
| 17-109_k141_25580 | Low-quality |
| 17-109_k141_311061 | Not-determined |
| 17-109_k141_8667 | Not-determined |
| 17-109_k141_311064 | Not-determined |
| 17-109_k141_294072 | Not-determined |
| 17-109_k141_311066 | Low-quality |
| 17-109_k141_42597 | Not-determined |
| 17-109_k141_159647 | Not-determined |
| 17-109_k141_209505 | Not-determined |
| 17-109_k141_8676 | Not-determined |
| 17-109_k141_209507 | Not-determined |
| 17-109_k141_294079 | Not-determined |
| 17-109_k141_76209 | Low-quality |
| 17-109_k141_159655 | Not-determined |
| 17-109_k141_8684 | Not-determined |
| 17-109_k141_42610 | Not-determined |
| 17-109_k141_42611 | Not-determined |
| 17-109_k141_226729 | Not-determined |
| 17-109_k141_327200 | Low-quality |
| 17-109_k141_209523 | Not-determined |
| 17-109_k141_108848 | Not-determined |
| 17-109_k141_226731 | Not-determined |
| 17-109_k141_343149 | Not-determined |
| 17-109_k141_76220 | Not-determined |
| 17-109_k141_343157 | Not-determined |
| 17-109_k141_193816 | Not-determined |
| 17-109_k141_76230 | Not-determined |
| 17-109_k141_176008 | Not-determined |
| 17-109_k141_91960 | Not-determined |
| 17-109_k141_276868 | Not-determined |
| 17-109_k141_25601 | Not-determined |
| 17-109_k141_76236 | Not-determined |
| 17-109_k141_108858 | Not-determined |
| 17-109_k141_311088 | Not-determined |
| 17-109_k141_25607 | Not-determined |
| 17-109_k141_91969 | Not-determined |
| 17-109_k141_243059 | Not-determined |
| 17-109_k141_25612 | Not-determined |
| 17-109_k141_327212 | Not-determined |
| 17-109_k141_209542 | Not-determined |
| 17-109_k141_226765 | Not-determined |
| 17-109_k141_327216 | Not-determined |
| 17-109_k141_58561 | Not-determined |
| 17-109_k141_25620 | Not-determined |
| 17-109_k141_259512 | Not-determined |
| 17-109_k141_25627 | Not-determined |
| 17-109_k141_108868 | Not-determined |
| 17-109_k141_76257 | Not-determined |
| 17-109_k141_259515 | Not-determined |
| 17-109_k141_226775 | Not-determined |
| 17-109_k141_243074 | Not-determined |
| 17-109_k141_91982 | Not-determined |
| 17-109_k141_25629 | Low-quality |
| 17-109_k141_243079 | Low-quality |
| 17-109_k141_25631 | Not-determined |
| 17-109_k141_394012 | Low-quality |
| 17-109_k141_343177 | Not-determined |
| 17-109_k141_259525 | Not-determined |
| 17-109_k141_25640 | Not-determined |
| 17-109_k141_91990 | Not-determined |
| 17-109_k141_143059 | Low-quality |
| 17-109_k141_243095 | Not-determined |
| 17-109_k141_42654 | Not-determined |
| 17-109_k141_58575 | Low-quality |
| 17-109_k141_91997 | Low-quality |
| 17-109_k141_193857 | Not-determined |
| 17-109_k141_209568 | Not-determined |
| 17-109_k141_143067 | Not-determined |
| 17-109_k141_209575 | Not-determined |
| 17-109_k141_76273 | Low-quality |
| 17-109_k141_294123 | Not-determined |
| 17-109_k141_92010 | Not-determined |
| 17-109_k141_92012 | Not-determined |
| 17-109_k141_58584 | Not-determined |
| 17-109_k141_193865 | Not-determined |
| 17-109_k141_377076 | Not-determined |
| 17-109_k141_394021 | Not-determined |
| 17-109_k141_276913 | Not-determined |
| 17-109_k141_42671 | Not-determined |
| 17-109_k141_159701 | Not-determined |
| 17-109_k141_209584 | Not-determined |
| 17-109_k141_159704 | Not-determined |
| 17-109_k141_209586 | Not-determined |
| 17-109_k141_209590 | Not-determined |
| 17-109_k141_193881 | Not-determined |
| 17-109_k141_259555 | Not-determined |
| 17-109_k141_108901 | Not-determined |
| 17-109_k141_209593 | Not-determined |
| 17-109_k141_377091 | Not-determined |
| 17-109_k141_143089 | Not-determined |
| 17-109_k141_327268 | Not-determined |
| 17-109_k141_143092 | Not-determined |
| 17-109_k141_327272 | Not-determined |
| 17-109_k141_193891 | Not-determined |
| 17-109_k141_76292 | Not-determined |
| 17-109_k141_311120 | Not-determined |
| 17-109_k141_209603 | Not-determined |
| 17-109_k141_343206 | Not-determined |
| 17-109_k141_394040 | Not-determined |
| 17-109_k141_276932 | Not-determined |
| 17-109_k141_294140 | Not-determined |
| 17-109_k141_377103 | Not-determined |
| 17-109_k141_143104 | Not-determined |
| 17-109_k141_159731 | Not-determined |
| 17-109_k141_25671 | Not-determined |
| 17-109_k141_294148 | Low-quality |
| 17-109_k141_108923 | Not-determined |
| 17-109_k141_226808 | Not-determined |
| 17-109_k141_259567 | Low-quality |
| 17-109_k141_259569 | Not-determined |
| 17-109_k141_377113 | Not-determined |
| 17-109_k141_58626 | Not-determined |
| 17-109_k141_394053 | Not-determined |
| 17-109_k141_25684 | Low-quality |
| 17-109_k141_311145 | Not-determined |
| 17-109_k141_193920 | Not-determined |
| 17-109_k141_108931 | Not-determined |
| 17-109_k141_25688 | Not-determined |
| 17-109_k141_311149 | Not-determined |
| 17-109_k141_394057 | Low-quality |
| 17-109_k141_193924 | Not-determined |
| 17-109_k141_159756 | Not-determined |
| 17-109_k141_58642 | Not-determined |
| 17-109_k141_58643 | Not-determined |
| 17-109_k141_294163 | Not-determined |
| 17-109_k141_311160 | Not-determined |
| 17-109_k141_327309 | Not-determined |
| 17-109_k141_311164 | Not-determined |
| 17-109_k141_327311 | Not-determined |
| 17-109_k141_311166 | Not-determined |
| 17-109_k141_377118 | Not-determined |
| 17-109_k141_311172 | Not-determined |
| 17-109_k141_76332 | Not-determined |
| 17-109_k141_176048 | Not-determined |
| 17-109_k141_327320 | Not-determined |
| 17-109_k141_343258 | Not-determined |
| 17-109_k141_377123 | Not-determined |
| 17-109_k141_394078 | Not-determined |
| 17-109_k141_226827 | Not-determined |
| 17-109_k141_377129 | Not-determined |
| 17-109_k141_394082 | Not-determined |
| 17-109_k141_327326 | Not-determined |
| 17-109_k141_226831 | Not-determined |
| 17-109_k141_327327 | Low-quality |
| 17-109_k141_327328 | Not-determined |
| 17-109_k141_209642 | Not-determined |
| 17-109_k141_159775 | Not-determined |
| 17-109_k141_343269 | Not-determined |
| 17-109_k141_343274 | Not-determined |
| 17-109_k141_327336 | Not-determined |
| 17-109_k141_343277 | Not-determined |
| 17-109_k141_193955 | Not-determined |
| 17-109_k141_243140 | Not-determined |
| 17-109_k141_125247 | Not-determined |
| 17-109_k141_311196 | Not-determined |
| 17-109_k141_108964 | Low-quality |
| 17-109_k141_176060 | Not-determined |
| 17-109_k141_276953 | Not-determined |
| 17-109_k141_125250 | Not-determined |
| 17-109_k141_343283 | Not-determined |
| 17-109_k141_276956 | Not-determined |
| 17-109_k141_311200 | Not-determined |
| 17-109_k141_209654 | Not-determined |
| 17-109_k141_377148 | Not-determined |
| 17-109_k141_311201 | Not-determined |
| 17-109_k141_108974 | Not-determined |
| 17-109_k141_193964 | Not-determined |
| 17-109_k141_108977 | Not-determined |
| 17-109_k141_343291 | Low-quality |
| 17-109_k141_259577 | Not-determined |
| 17-109_k141_176066 | Not-determined |
| 17-109_k141_343293 | Not-determined |
| 17-109_k141_76364 | Not-determined |
| 17-109_k141_108979 | Not-determined |
| 17-109_k141_25720 | Not-determined |
| 17-109_k141_327346 | Low-quality |
| 17-109_k141_108987 | Not-determined |
| 17-109_k141_58698 | Not-determined |
| 17-109_k141_209671 | Low-quality |
| 17-109_k141_209675 | Not-determined |
| 17-109_k141_243149 | Not-determined |
| 17-109_k141_8709 | Not-determined |
| 17-109_k141_243151 | Not-determined |
| 17-109_k141_226851 | Not-determined |
| 17-109_k141_209681 | Not-determined |
| 17-109_k141_226853 | Not-determined |
| 17-109_k141_343309 | Not-determined |
| 17-109_k141_8716 | Not-determined |
| 17-109_k141_8717 | Not-determined |
| 17-109_k141_176078 | Not-determined |
| 17-109_k141_159781 | Not-determined |
| 17-109_k141_276984 | Not-determined |
| 17-109_k141_76378 | Not-determined |
| 17-109_k141_259623 | Not-determined |
| 17-109_k141_359101 | Not-determined |
| 17-109_k141_359103 | Not-determined |
| 17-109_k141_108990 | Not-determined |
| 17-109_k141_194013 | Not-determined |
| 17-109_k141_8729 | Not-determined |
| 17-109_k141_377185 | Not-determined |
| 17-109_k141_209700 | Not-determined |
| 17-109_k141_311238 | Not-determined |
| 17-109_k141_243185 | Not-determined |
| 17-109_k141_109001 | Not-determined |
| 17-109_k141_243192 | Not-determined |
| 17-109_k141_125292 | Not-determined |
| 17-109_k141_226861 | Low-quality |
| 17-109_k141_159807 | Not-determined |
| 17-109_k141_294218 | Not-determined |
| 17-109_k141_92060 | Not-determined |
| 17-109_k141_159809 | Not-determined |
| 17-109_k141_343338 | Not-determined |
| 17-109_k141_159810 | Not-determined |
| 17-109_k141_394129 | Not-determined |
| 17-109_k141_58731 | Not-determined |
| 17-109_k141_327376 | Not-determined |
| 17-109_k141_25733 | Not-determined |
| 17-109_k141_42688 | Not-determined |
| 17-109_k141_209711 | Not-determined |
| 17-109_k141_277012 | Not-determined |
| 17-109_k141_194031 | Not-determined |
| 17-109_k141_159823 | Not-determined |
| 17-109_k141_243216 | Not-determined |
| 17-109_k141_42700 | Not-determined |
| 17-109_k141_25738 | Not-determined |
| 17-109_k141_343361 | Not-determined |
| 17-109_k141_377205 | Not-determined |
| 17-109_k141_243235 | Not-determined |
| 17-109_k141_209718 | Not-determined |
| 17-109_k141_194046 | Low-quality |
| 17-109_k141_377209 | Low-quality |
| 17-109_k141_243237 | Low-quality |
| 17-109_k141_194048 | Not-determined |
| 17-109_k141_394150 | Not-determined |
| 17-109_k141_209721 | Low-quality |
| 17-109_k141_209722 | Not-determined |
| 17-109_k141_76409 | Not-determined |
| 17-109_k141_377210 | Not-determined |
| 17-109_k141_194052 | Not-determined |
| 17-109_k141_42721 | Not-determined |
| 17-109_k141_8772 | Not-determined |
| 17-109_k141_377212 | Not-determined |
| 17-109_k141_125316 | Not-determined |
| 17-109_k141_311275 | Not-determined |
| 17-109_k141_76421 | Not-determined |
| 17-109_k141_42730 | Not-determined |
| 17-109_k141_76424 | Not-determined |
| 17-109_k141_359125 | Not-determined |
| 17-109_k141_194068 | Not-determined |
| 17-109_k141_243253 | Not-determined |
| 17-109_k141_259629 | Low-quality |
| 17-109_k141_259635 | Not-determined |
| 17-109_k141_327402 | Not-determined |
| 17-109_k141_277048 | Not-determined |
| 17-109_k141_311283 | Not-determined |
| 17-109_k141_259647 | Not-determined |
| 17-109_k141_25763 | Not-determined |
| 17-109_k141_109010 | Not-determined |
| 17-109_k141_109011 | Not-determined |
| 17-109_k141_243270 | Not-determined |
| 17-109_k141_194091 | Not-determined |
| 17-109_k141_76447 | Low-quality |
| 17-109_k141_109022 | Not-determined |
| 17-109_k141_25772 | Not-determined |
| 17-109_k141_109028 | Not-determined |
| 17-109_k141_394185 | Low-quality |
| 17-109_k141_394186 | Not-determined |
| 17-109_k141_8783 | Not-determined |
| 17-109_k141_311292 | Not-determined |
| 17-109_k141_8788 | Not-determined |
| 17-109_k141_42776 | Not-determined |
| 17-109_k141_394192 | Not-determined |
| 17-109_k141_25785 | Low-quality |
| 17-109_k141_394193 | Not-determined |
| 17-109_k141_92102 | Not-determined |
| 17-109_k141_25791 | Low-quality |
| 17-109_k141_377246 | Not-determined |
| 17-109_k141_327438 | Low-quality |
| 17-109_k141_76460 | Not-determined |
| 17-109_k141_243293 | Not-determined |
| 17-109_k141_92106 | Not-determined |
| 17-109_k141_377252 | Not-determined |
| 17-109_k141_25797 | Low-quality |
| 17-109_k141_327446 | Not-determined |
| 17-109_k141_243302 | Not-determined |
| 17-109_k141_377253 | Not-determined |
| 17-109_k141_92109 | Not-determined |
| 17-109_k141_25804 | Low-quality |
| 17-109_k141_109041 | Not-determined |
| 17-109_k141_76485 | Not-determined |
| 17-109_k141_42792 | Not-determined |
| 17-109_k141_243310 | Not-determined |
| 17-109_k141_377257 | Low-quality |
| 17-109_k141_109045 | Not-determined |
| 17-109_k141_42796 | Not-determined |
| 17-109_k141_327459 | Not-determined |
| 17-109_k141_76490 | Low-quality |
| 17-109_k141_243318 | Not-determined |
| 17-109_k141_159844 | Not-determined |
| 17-109_k141_311303 | Not-determined |
| 17-109_k141_377264 | Not-determined |
| 17-109_k141_327464 | Not-determined |
| 17-109_k141_76493 | Not-determined |
| 17-109_k141_194125 | Not-determined |
| 17-109_k141_394205 | Not-determined |
| 17-109_k141_42802 | Not-determined |
| 17-109_k141_25809 | Not-determined |
| 17-109_k141_109057 | Not-determined |
| 17-109_k141_42807 | Not-determined |
| 17-109_k141_159858 | Not-determined |
| 17-109_k141_76503 | Low-quality |
| 17-109_k141_327474 | Not-determined |
| 17-109_k141_243326 | Not-determined |
| 17-109_k141_76507 | Not-determined |
| 17-109_k141_109072 | Not-determined |
| 17-109_k141_76508 | Not-determined |
| 17-109_k141_311318 | Not-determined |
| 17-109_k141_194138 | Not-determined |
| 17-109_k141_377280 | Not-determined |
| 17-109_k141_8821 | Not-determined |
| 17-109_k141_109082 | Not-determined |
| 17-109_k141_25821 | Not-determined |
| 17-109_k141_259691 | Not-determined |
| 17-109_k141_159879 | Low-quality |
| 17-109_k141_92127 | Not-determined |
| 17-109_k141_259693 | Not-determined |
| 17-109_k141_176169 | Not-determined |
| 17-109_k141_209775 | Not-determined |
| 17-109_k141_8842 | Low-quality |
| 17-109_k141_8843 | Not-determined |
| 17-109_k141_243347 | Low-quality |
| 17-109_k141_92135 | Low-quality |
| 17-109_k141_209782 | Not-determined |
| 17-109_k141_109103 | Not-determined |
| 17-109_k141_8847 | Not-determined |
| 17-109_k141_25833 | Not-determined |
| 17-109_k141_8850 | Low-quality |
| 17-109_k141_327497 | Not-determined |
| 17-109_k141_327506 | Not-determined |
| 17-109_k141_42828 | Not-determined |
| 17-109_k141_8863 | Not-determined |
| 17-109_k141_42830 | Low-quality |
| 17-109_k141_259715 | Not-determined |
| 17-109_k141_243371 | Not-determined |
| 17-109_k141_243372 | Not-determined |
| 17-109_k141_109124 | Not-determined |
| 17-109_k141_92160 | Not-determined |
| 17-109_k141_343442 | Not-determined |
| 17-109_k141_394247 | Not-determined |
| 17-109_k141_226934 | Not-determined |
| 17-109_k141_42850 | Not-determined |
| 17-109_k141_277133 | Not-determined |
| 17-109_k141_243389 | Low-quality |
| 17-109_k141_243390 | Not-determined |
| 17-109_k141_58827 | Not-determined |
| 17-109_k141_226948 | Not-determined |
| 17-109_k141_25864 | Not-determined |
| 17-109_k141_359188 | Not-determined |
| 17-109_k141_394258 | Not-determined |
| 17-109_k141_394259 | Not-determined |
| 17-109_k141_58838 | Low-quality |
| 17-109_k141_92167 | Not-determined |
| 17-109_k141_159909 | Not-determined |
| 17-109_k141_92168 | Not-determined |
| 17-109_k141_109139 | Low-quality |
| 17-109_k141_109141 | Not-determined |
| 17-109_k141_343459 | Not-determined |
| 17-109_k141_226961 | Not-determined |
| 17-109_k141_311351 | Not-determined |
| 17-109_k141_76549 | Not-determined |
| 17-109_k141_176199 | Not-determined |
| 17-109_k141_327535 | Low-quality |
| 17-109_k141_92179 | Not-determined |
| 17-109_k141_277156 | Not-determined |
| 17-109_k141_176203 | Not-determined |
| 17-109_k141_343464 | Not-determined |
| 17-109_k141_76556 | Not-determined |
| 17-109_k141_159922 | Not-determined |
| 17-109_k141_259735 | Not-determined |
| 17-109_k141_25872 | Not-determined |
| 17-109_k141_8900 | Not-determined |
| 17-109_k141_259738 | Not-determined |
| 17-109_k141_159927 | Not-determined |
| 17-109_k141_92187 | Not-determined |
| 17-109_k141_25881 | Not-determined |
| 17-109_k141_327549 | Not-determined |
| 17-109_k141_259750 | Not-determined |
| 17-109_k141_92195 | Not-determined |
| 17-109_k141_8902 | Low-quality |
| 17-109_k141_176212 | Not-determined |
| 17-109_k141_8903 | Not-determined |
| 17-109_k141_277166 | Not-determined |
| 17-109_k141_277167 | Not-determined |
| 17-109_k141_327556 | Not-determined |
| 17-109_k141_209824 | Not-determined |
| 17-109_k141_8923 | Not-determined |
| 17-109_k141_377319 | Not-determined |
| 17-109_k141_311389 | Not-determined |
| 17-109_k141_42877 | Not-determined |
| 17-109_k141_311392 | Not-determined |
| 17-109_k141_109147 | Not-determined |
| 17-109_k141_109149 | Not-determined |
| 17-109_k141_226997 | Not-determined |
| 17-109_k141_394266 | Not-determined |
| 17-109_k141_311400 | Not-determined |
| 17-109_k141_259768 | Low-quality |
| 17-109_k141_311401 | Not-determined |
| 17-109_k141_159949 | Not-determined |
| 17-109_k141_394273 | Not-determined |
| 17-109_k141_259772 | Not-determined |
| 17-109_k141_227006 | Not-determined |
| 17-109_k141_377338 | Not-determined |
| 17-109_k141_42884 | Not-determined |
| 17-109_k141_243425 | Not-determined |
| 17-109_k141_227012 | Not-determined |
| 17-109_k141_227014 | Not-determined |
| 17-109_k141_92214 | Not-determined |
| 17-109_k141_377342 | Not-determined |
| 17-109_k141_8933 | Not-determined |
| 17-109_k141_227019 | Not-determined |
| 17-109_k141_109162 | Not-determined |
| 17-109_k141_209846 | Not-determined |
| 17-109_k141_76587 | Not-determined |
| 17-109_k141_8939 | Not-determined |
| 17-109_k141_227036 | Not-determined |
| 17-109_k141_209856 | Not-determined |
| 17-109_k141_76601 | Not-determined |
| 17-109_k141_377360 | Not-determined |
| 17-109_k141_92229 | Not-determined |
| 17-109_k141_92231 | Not-determined |
| 17-109_k141_159971 | Not-determined |
| 17-109_k141_58912 | Not-determined |
| 17-109_k141_311428 | Not-determined |
| 17-109_k141_227055 | Not-determined |
| 17-109_k141_377376 | Not-determined |
| 17-109_k141_176225 | Not-determined |
| 17-109_k141_277198 | Low-quality |
| 17-109_k141_311433 | Not-determined |
| 17-109_k141_176229 | Not-determined |
| 17-109_k141_209867 | Not-determined |
| 17-109_k141_42933 | Not-determined |
| 17-109_k141_227068 | Not-determined |
| 17-109_k141_176237 | Not-determined |
| 17-109_k141_209875 | Not-determined |
| 17-109_k141_311442 | Not-determined |
| 17-109_k141_311445 | Not-determined |
| 17-109_k141_294314 | Not-determined |
| 17-109_k141_243435 | Not-determined |
| 17-109_k141_25904 | Not-determined |
| 17-109_k141_25906 | Not-determined |
| 17-109_k141_109180 | Not-determined |
| 17-109_k141_92243 | Not-determined |
| 17-109_k141_359201 | Not-determined |
| 17-109_k141_327614 | Not-determined |
| 17-109_k141_327615 | Not-determined |
| 17-109_k141_109184 | Not-determined |
| 17-109_k141_327620 | Not-determined |
| 17-109_k141_76636 | Low-quality |
| 17-109_k141_76637 | Not-determined |
| 17-109_k141_294328 | Not-determined |
| 17-109_k141_377387 | Not-determined |
| 17-109_k141_176259 | Not-determined |
| 17-109_k141_243439 | Not-determined |
| 17-109_k141_243445 | Not-determined |
| 17-109_k141_311457 | Not-determined |
| 17-109_k141_76640 | Not-determined |
| 17-109_k141_277220 | Not-determined |
| 17-109_k141_377395 | Not-determined |
| 17-109_k141_159979 | Not-determined |
| 17-109_k141_243456 | Not-determined |
| 17-109_k141_209894 | Not-determined |
| 17-109_k141_109208 | Not-determined |
| 17-109_k141_159985 | Not-determined |
| 17-109_k141_259793 | Not-determined |
| 17-109_k141_76650 | Not-determined |
| 17-109_k141_277230 | Low-quality |
| 17-109_k141_159993 | Low-quality |
| 17-109_k141_327646 | Not-determined |
| 17-109_k141_259799 | Not-determined |
| 17-109_k141_109215 | Not-determined |
| 17-109_k141_227079 | Not-determined |
| 17-109_k141_277243 | Not-determined |
| 17-109_k141_76663 | Not-determined |
| 17-109_k141_8959 | Not-determined |
| 17-109_k141_227082 | Not-determined |
| 17-109_k141_359224 | Not-determined |
| 17-109_k141_243463 | Not-determined |
| 17-109_k141_209916 | Low-quality |
| 17-109_k141_359225 | Not-determined |
| 17-109_k141_294355 | Not-determined |
| 17-109_k141_327655 | Not-determined |
| 17-109_k141_294362 | Not-determined |
| 17-109_k141_8966 | Not-determined |
| 17-109_k141_8968 | Not-determined |
| 17-109_k141_259815 | Not-determined |
| 17-109_k141_243471 | Not-determined |
| 17-109_k141_160007 | Not-determined |
| 17-109_k141_294367 | Not-determined |
| 17-109_k141_227102 | Not-determined |
| 17-109_k141_58928 | Not-determined |
| 17-109_k141_227105 | Not-determined |
| 17-109_k141_160013 | Not-determined |

| | |
|---|---|
| 17-109_k141_311478 | Not-determined |
| 17-109_k141_227109 | Not-determined |
| 17-109_k141_125368 | Not-determined |
| 17-109_k141_227114 | Not-determined |
| 17-109_k141_327675 | Not-determined |
| 17-109_k141_277268 | Not-determined |
| 17-109_k141_294384 | Not-determined |
| 17-109_k141_359249 | Not-determined |
| 17-109_k141_343564 | Not-determined |
| 17-109_k141_343570 | Not-determined |
| 17-109_k141_160035 | Not-determined |
| 17-109_k141_343572 | Not-determined |
| 17-109_k141_343573 | Not-determined |
| 17-109_k141_327687 | Not-determined |
| 17-109_k141_359256 | Low-quality |
| 17-109_k141_92316 | Not-determined |
| 17-109_k141_311499 | Not-determined |
| 17-109_k141_92320 | Low-quality |
| 17-109_k141_76689 | Not-determined |
| 17-109_k141_76691 | Low-quality |
| 17-109_k141_76692 | Not-determined |
| 17-109_k141_92325 | Not-determined |
| 17-109_k141_311509 | Not-determined |
| 17-109_k141_76697 | Not-determined |
| 17-109_k141_42966 | Not-determined |
| 17-109_k141_343599 | Not-determined |
| 17-109_k141_160056 | Not-determined |
| 17-109_k141_125397 | Not-determined |
| 17-109_k141_9003 | Not-determined |
| 17-109_k141_311518 | Not-determined |
| 17-109_k141_227133 | Low-quality |
| 17-109_k141_343601 | Not-determined |
| 17-109_k141_125405 | Not-determined |
| 17-109_k141_160065 | Not-determined |
| 17-109_k141_377442 | Not-determined |
| 17-109_k141_109257 | Not-determined |
| 17-109_k141_343605 | Not-determined |
| 17-109_k141_109259 | Not-determined |
| 17-109_k141_294412 | Not-determined |
| 17-109_k141_209995 | Not-determined |
| 17-109_k141_109263 | Not-determined |
| 17-109_k141_58937 | Not-determined |
| 17-109_k141_377455 | Not-determined |
| 17-109_k141_9027 | Not-determined |
| 17-109_k141_109267 | Not-determined |
| 17-109_k141_343620 | Not-determined |
| 17-109_k141_210000 | Low-quality |
| 17-109_k141_58942 | Not-determined |
| 17-109_k141_210002 | Not-determined |
| 17-109_k141_160082 | Not-determined |
| 17-109_k141_327700 | Low-quality |
| 17-109_k141_377461 | Not-determined |
| 17-109_k141_227150 | Not-determined |
| 17-109_k141_109280 | Not-determined |
| 17-109_k141_43003 | Not-determined |
| 17-109_k141_76718 | Low-quality |
| 17-109_k141_210008 | Not-determined |
| 17-109_k141_125415 | Not-determined |
| 17-109_k141_76726 | Not-determined |
| 17-109_k141_25925 | Not-determined |
| 17-109_k141_176362 | Not-determined |
| 17-109_k141_160091 | Not-determined |
| 17-109_k141_25929 | Low-quality |
| 17-109_k141_76738 | Not-determined |
| 17-109_k141_176366 | Not-determined |
| 17-109_k141_311548 | Not-determined |
| 17-109_k141_259839 | Not-determined |
| 17-109_k141_210033 | Not-determined |
| 17-109_k141_76750 | Not-determined |
| 17-109_k141_243542 | Low-quality |
| 17-109_k141_210035 | Not-determined |
| 17-109_k141_343640 | Low-quality |
| 17-109_k141_76754 | Not-determined |
| 17-109_k141_210036 | Not-determined |
| 17-109_k141_9050 | Not-determined |
| 17-109_k141_227165 | Not-determined |
| 17-109_k141_109303 | Low-quality |
| 17-109_k141_343644 | Not-determined |
| 17-109_k141_9051 | Low-quality |
| 17-109_k141_76760 | Not-determined |
| 17-109_k141_160107 | Not-determined |
| 17-109_k141_125424 | Not-determined |
| 17-109_k141_160115 | Not-determined |
| 17-109_k141_109312 | Not-determined |
| 17-109_k141_311579 | Not-determined |
| 17-109_k141_243558 | Not-determined |
| 17-109_k141_359292 | Not-determined |
| 17-109_k141_9059 | Low-quality |
| 17-109_k141_277304 | Low-quality |
| 17-109_k141_277306 | Not-determined |
| 17-109_k141_294424 | Not-determined |
| 17-109_k141_277310 | Not-determined |
| 17-109_k141_294425 | Not-determined |
| 17-109_k141_243567 | Not-determined |
| 17-109_k141_377509 | Not-determined |
| 17-109_k141_25961 | Not-determined |
| 17-109_k141_294430 | Not-determined |
| 17-109_k141_109324 | Not-determined |
| 17-109_k141_294433 | Not-determined |
| 17-109_k141_43020 | Not-determined |
| 17-109_k141_125442 | Low-quality |
| 17-109_k141_277317 | Not-determined |
| 17-109_k141_125444 | Not-determined |
| 17-109_k141_43022 | Not-determined |
| 17-109_k141_109332 | Not-determined |
| 17-109_k141_9069 | Not-determined |
| 17-109_k141_377532 | Not-determined |
| 17-109_k141_243577 | Not-determined |
| 17-109_k141_25975 | Not-determined |
| 17-109_k141_92406 | Not-determined |
| 17-109_k141_160141 | Not-determined |
| 17-109_k141_311603 | Not-determined |
| 17-109_k141_160142 | Not-determined |
| 17-109_k141_377542 | Not-determined |
| 17-109_k141_343676 | Not-determined |
| 17-109_k141_343678 | Not-determined |
| 17-109_k141_125466 | Not-determined |
| 17-109_k141_160147 | Not-determined |
| 17-109_k141_160148 | Not-determined |
| 17-109_k141_327771 | Not-determined |
| 17-109_k141_277336 | Not-determined |
| 17-109_k141_243585 | Not-determined |
| 17-109_k141_327774 | Not-determined |
| 17-109_k141_9083 | Not-determined |
| 17-109_k141_227211 | Not-determined |
| 17-109_k141_259851 | Not-determined |
| 17-109_k141_343689 | Not-determined |
| 17-109_k141_243587 | Not-determined |
| 17-109_k141_9086 | Not-determined |
| 17-109_k141_259854 | Not-determined |
| 17-109_k141_294450 | Not-determined |
| 17-109_k141_125473 | Not-determined |
| 17-109_k141_294451 | Not-determined |
| 17-109_k141_343692 | Not-determined |
| 17-109_k141_259862 | Not-determined |
| 17-109_k141_343695 | Not-determined |
| 17-109_k141_311627 | Not-determined |
| 17-109_k141_9104 | Not-determined |
| 17-109_k141_259867 | Not-determined |
| 17-109_k141_125483 | Not-determined |
| 17-109_k141_311632 | Not-determined |
| 17-109_k141_160167 | Not-determined |
| 17-109_k141_125489 | Not-determined |
| 17-109_k141_76792 | Low-quality |
| 17-109_k141_343710 | Not-determined |
| 17-109_k141_25991 | Not-determined |
| 17-109_k141_43043 | Not-determined |
| 17-109_k141_243613 | Not-determined |
| 17-109_k141_194182 | Not-determined |
| 17-109_k141_160173 | Not-determined |
| 17-109_k141_109376 | Not-determined |
| 17-109_k141_259882 | Not-determined |
| 17-109_k141_227237 | Not-determined |
| 17-109_k141_9124 | Not-determined |
| 17-109_k141_43060 | Not-determined |
| 17-109_k141_176449 | Not-determined |
| 17-109_k141_176450 | Not-determined |
| 17-109_k141_9130 | Not-determined |
| 17-109_k141_76806 | Not-determined |
| 17-109_k141_227243 | Not-determined |
| 17-109_k141_227244 | Not-determined |
| 17-109_k141_43063 | Low-quality |
| 17-109_k141_259893 | Not-determined |
| 17-109_k141_176454 | Not-determined |
| 17-109_k141_311673 | Not-determined |
| 17-109_k141_92423 | Not-determined |
| 17-109_k141_109392 | Not-determined |
| 17-109_k141_26015 | Not-determined |
| 17-109_k141_76817 | Not-determined |
| 17-109_k141_92429 | Not-determined |
| 17-109_k141_194197 | Not-determined |
| 17-109_k141_311677 | Not-determined |
| 17-109_k141_176467 | Not-determined |
| 17-109_k141_359383 | Not-determined |
| 17-109_k141_43075 | Not-determined |
| 17-109_k141_26028 | Not-determined |
| 17-109_k141_43079 | Not-determined |
| 17-109_k141_109394 | Not-determined |
| 17-109_k141_109395 | Low-quality |
| 17-109_k141_359389 | Not-determined |
| 17-109_k141_43083 | Not-determined |
| 17-109_k141_210097 | Not-determined |
| 17-109_k141_243632 | Low-quality |
| 17-109_k141_294481 | Not-determined |
| 17-109_k141_210103 | Not-determined |
| 17-109_k141_76828 | Not-determined |
| 17-109_k141_227250 | Low-quality |
| 17-109_k141_176490 | Not-determined |
| 17-109_k141_76829 | Not-determined |
| 17-109_k141_377559 | Not-determined |
| 17-109_k141_109397 | Low-quality |
| 17-109_k141_343770 | Not-determined |
| 17-109_k141_243645 | Not-determined |
| 17-109_k141_194223 | Not-determined |
| 17-109_k141_176497 | Not-determined |
| 17-109_k141_377568 | Not-determined |
| 17-109_k141_210116 | Not-determined |
| 17-109_k141_311687 | Not-determined |
| 17-109_k141_259927 | Not-determined |
| 17-109_k141_58983 | Not-determined |
| 17-109_k141_210121 | Not-determined |
| 17-109_k141_43097 | Not-determined |
| 17-109_k141_194246 | Not-determined |
| 17-109_k141_43108 | Not-determined |
| 17-109_k141_143127 | Not-determined |
| 17-109_k141_58994 | Low-quality |
| 17-109_k141_58995 | Not-determined |
| 17-109_k141_59000 | Not-determined |
| 17-109_k141_92474 | Not-determined |
| 17-109_k141_377569 | Not-determined |
| 17-109_k141_377570 | Not-determined |
| 17-109_k141_26038 | Not-determined |
| 17-109_k141_243670 | Not-determined |
| 17-109_k141_26040 | Not-determined |
| 17-109_k141_176522 | Not-determined |
| 17-109_k141_243675 | Low-quality |
| 17-109_k141_143146 | Not-determined |
| 17-109_k141_227288 | Not-determined |
| 17-109_k141_143157 | Not-determined |
| 17-109_k141_143158 | Not-determined |
| 17-109_k141_176540 | Not-determined |
| 17-109_k141_259973 | Low-quality |
| 17-109_k141_311706 | Not-determined |
| 17-109_k141_176543 | Low-quality |
| 17-109_k141_259974 | Not-determined |
| 17-109_k141_294499 | Not-determined |
| 17-109_k141_176544 | Not-determined |
| 17-109_k141_359428 | Not-determined |
| 17-109_k141_311707 | Not-determined |
| 17-109_k141_259979 | Not-determined |
| 17-109_k141_59027 | Not-determined |
| 17-109_k141_176551 | Low-quality |
| 17-109_k141_143173 | Not-determined |
| 17-109_k141_359439 | Not-determined |
| 17-109_k141_343805 | Not-determined |
| 17-109_k141_227299 | Not-determined |
| 17-109_k141_176553 | Not-determined |
| 17-109_k141_359442 | Not-determined |
| 17-109_k141_294505 | Not-determined |
| 17-109_k141_377593 | Not-determined |
| 17-109_k141_210145 | Not-determined |
| 17-109_k141_59038 | Not-determined |
| 17-109_k141_26067 | Not-determined |
| 17-109_k141_109457 | Not-determined |
| 17-109_k141_227317 | Not-determined |
| 17-109_k141_311729 | Not-determined |
| 17-109_k141_59055 | Not-determined |
| 17-109_k141_26080 | Not-determined |
| 17-109_k141_210155 | Not-determined |
| 17-109_k141_92507 | Not-determined |
| 17-109_k141_294520 | Not-determined |
| 17-109_k141_109459 | Not-determined |
| 17-109_k141_294525 | Not-determined |
| 17-109_k141_176575 | Low-quality |
| 17-109_k141_59064 | Not-determined |
| 17-109_k141_194280 | Not-determined |
| 17-109_k141_227332 | Low-quality |
| 17-109_k141_210160 | Not-determined |
| 17-109_k141_194283 | Not-determined |
| 17-109_k141_26092 | Not-determined |
| 17-109_k141_243697 | Not-determined |
| 17-109_k141_92520 | Not-determined |
| 17-109_k141_109469 | Not-determined |
| 17-109_k141_227337 | Not-determined |
| 17-109_k141_59069 | Not-determined |
| 17-109_k141_59070 | Not-determined |
| 17-109_k141_43149 | Not-determined |
| 17-109_k141_294542 | Not-determined |
| 17-109_k141_227338 | Not-determined |
| 17-109_k141_210167 | Not-determined |
| 17-109_k141_227340 | Not-determined |
| 17-109_k141_243706 | Not-determined |
| 17-109_k141_109479 | Not-determined |
| 17-109_k141_176587 | Not-determined |
| 17-109_k141_26100 | Not-determined |
| 17-109_k141_311751 | Not-determined |
| 17-109_k141_109481 | Low-quality |
| 17-109_k141_227344 | Not-determined |
| 17-109_k141_311753 | Low-quality |
| 17-109_k141_59081 | Not-determined |
| 17-109_k141_109488 | Not-determined |
| 17-109_k141_210171 | Not-determined |
| 17-109_k141_109494 | Not-determined |
| 17-109_k141_311761 | Not-determined |
| 17-109_k141_160260 | Not-determined |
| 17-109_k141_92528 | Not-determined |
| 17-109_k141_227359 | Not-determined |
| 17-109_k141_227361 | Not-determined |
| 17-109_k141_294554 | Not-determined |
| 17-109_k141_160271 | Not-determined |
| 17-109_k141_92534 | Not-determined |
| 17-109_k141_210185 | Not-determined |
| 17-109_k141_210187 | Not-determined |
| 17-109_k141_277428 | Not-determined |
| 17-109_k141_294559 | Not-determined |
| 17-109_k141_92542 | Not-determined |
| 17-109_k141_160278 | Not-determined |
| 17-109_k141_26123 | Not-determined |
| 17-109_k141_377607 | Not-determined |
| 17-109_k141_294569 | Not-determined |
| 17-109_k141_227368 | Not-determined |
| 17-109_k141_160280 | Not-determined |
| 17-109_k141_26127 | Not-determined |
| 17-109_k141_109511 | Low-quality |
| 17-109_k141_59115 | Not-determined |
| 17-109_k141_176613 | Not-determined |
| 17-109_k141_26133 | Not-determined |
| 17-109_k141_160286 | Not-determined |
| 17-109_k141_143209 | Not-determined |
| 17-109_k141_26138 | Not-determined |
| 17-109_k141_160290 | Not-determined |
| 17-109_k141_43190 | Not-determined |
| 17-109_k141_160294 | Not-determined |
| 17-109_k141_243741 | Not-determined |
| 17-109_k141_311790 | Not-determined |
| 17-109_k141_43193 | Not-determined |
| 17-109_k141_227378 | Not-determined |
| 17-109_k141_277438 | Not-determined |
| 17-109_k141_243747 | Not-determined |
| 17-109_k141_343857 | Low-quality |
| 17-109_k141_377628 | Not-determined |
| 17-109_k141_194330 | Not-determined |
| 17-109_k141_109518 | Not-determined |
| 17-109_k141_294592 | Not-determined |
| 17-109_k141_194332 | Not-determined |
| 17-109_k141_277441 | Not-determined |
| 17-109_k141_210215 | Not-determined |
| 17-109_k141_109525 | Not-determined |
| 17-109_k141_343874 | Not-determined |
| 17-109_k141_76864 | Not-determined |
| 17-109_k141_194344 | Not-determined |
| 17-109_k141_76865 | Not-determined |
| 17-109_k141_210222 | Not-determined |
| 17-109_k141_294597 | Low-quality |
| 17-109_k141_59147 | Not-determined |
| 17-109_k141_343888 | Not-determined |
| 17-109_k141_277451 | Not-determined |
| 17-109_k141_294603 | Not-determined |
| 17-109_k141_26161 | Not-determined |
| 17-109_k141_311815 | Not-determined |
| 17-109_k141_76879 | Not-determined |
| 17-109_k141_377645 | Not-determined |
| 17-109_k141_377646 | Not-determined |
| 17-109_k141_59153 | Not-determined |
| 17-109_k141_26170 | Not-determined |
| 17-109_k141_59158 | Low-quality |
| 17-109_k141_59162 | Not-determined |
| 17-109_k141_59163 | Not-determined |
| 17-109_k141_160311 | Not-determined |
| 17-109_k141_343910 | Not-determined |
| 17-109_k141_311838 | Not-determined |
| 17-109_k141_125525 | Low-quality |
| 17-109_k141_194371 | Not-determined |
| 17-109_k141_227422 | Not-determined |
| 17-109_k141_176630 | Not-determined |
| 17-109_k141_227426 | Not-determined |
| 17-109_k141_76890 | Not-determined |
| 17-109_k141_227427 | Not-determined |
| 17-109_k141_294630 | Not-determined |
| 17-109_k141_125532 | Not-determined |
| 17-109_k141_160315 | Not-determined |
| 17-109_k141_294632 | Not-determined |
| 17-109_k141_377659 | Not-determined |
| 17-109_k141_76900 | Not-determined |
| 17-109_k141_377660 | Not-determined |
| 17-109_k141_294634 | Not-determined |
| 17-109_k141_377661 | Not-determined |
| 17-109_k141_76902 | Low-quality |
| 17-109_k141_359532 | Not-determined |
| 17-109_k141_160328 | Not-determined |
| 17-109_k141_359535 | Not-determined |
| 17-109_k141_359537 | Not-determined |
| 17-109_k141_343934 | Not-determined |
| 17-109_k141_160336 | Not-determined |
| 17-109_k141_43229 | Not-determined |
| 17-109_k141_260004 | Not-determined |
| 17-109_k141_76912 | Not-determined |
| 17-109_k141_343945 | Low-quality |
| 17-109_k141_26198 | Low-quality |
| 17-109_k141_59190 | Not-determined |
| 17-109_k141_76924 | Not-determined |
| 17-109_k141_343951 | Not-determined |
| 17-109_k141_143288 | Not-determined |
| 17-109_k141_311881 | Not-determined |
| 17-109_k141_377693 | Not-determined |
| 17-109_k141_359555 | Low-quality |
| 17-109_k141_311884 | Low-quality |
| 17-109_k141_194418 | Not-determined |
| 17-109_k141_176636 | Not-determined |
| 17-109_k141_311888 | Not-determined |
| 17-109_k141_125561 | Low-quality |
| 17-109_k141_176641 | Not-determined |
| 17-109_k141_59207 | Not-determined |
| 17-109_k141_210256 | Not-determined |
| 17-109_k141_143300 | Not-determined |
| 17-109_k141_76933 | Not-determined |
| 17-109_k141_194425 | Not-determined |
| 17-109_k141_343969 | Not-determined |
| 17-109_k141_277532 | Not-determined |
| 17-109_k141_277535 | Not-determined |
| 17-109_k141_26219 | Not-determined |
| 17-109_k141_359558 | Low-quality |
| 17-109_k141_311895 | Low-quality |
| 17-109_k141_59224 | Not-determined |
| 17-109_k141_59225 | Not-determined |
| 17-109_k141_59228 | Not-determined |
| 17-109_k141_311902 | Not-determined |
| 17-109_k141_160351 | Not-determined |
| 17-109_k141_210274 | Not-determined |
| 17-109_k141_394307 | Not-determined |
| 17-109_k141_359565 | Not-determined |
| 17-109_k141_43256 | Not-determined |
| 17-109_k141_59235 | Not-determined |
| 17-109_k141_210280 | Not-determined |
| 17-109_k141_311907 | Not-determined |
| 17-109_k141_76940 | Not-determined |
| 17-109_k141_76948 | Not-determined |
| 17-109_k141_377735 | Not-determined |
| 17-109_k141_76950 | Not-determined |
| 17-109_k141_277551 | Not-determined |
| 17-109_k141_76952 | Not-determined |
| 17-109_k141_176666 | Not-determined |
| 17-109_k141_359575 | Low-quality |
| 17-109_k141_359576 | Not-determined |
| 17-109_k141_394329 | Not-determined |
| 17-109_k141_109545 | Not-determined |
| 17-109_k141_76965 | Low-quality |
| 17-109_k141_343999 | Not-determined |
| 17-109_k141_9138 | Not-determined |
| 17-109_k141_359603 | Not-determined |
| 17-109_k141_210295 | Not-determined |
| 17-109_k141_344007 | Not-determined |
| 17-109_k141_394347 | Not-determined |
| 17-109_k141_76985 | Low-quality |
| 17-109_k141_227462 | Not-determined |
| 17-109_k141_227464 | Not-determined |
| 17-109_k141_9146 | Not-determined |
| 17-109_k141_210303 | Not-determined |
| 17-109_k141_277582 | Not-determined |
| 17-109_k141_9156 | Not-determined |
| 17-109_k141_26243 | Low-quality |
| 17-109_k141_227473 | Not-determined |
| 17-109_k141_76998 | Not-determined |
| 17-109_k141_77006 | Not-determined |
| 17-109_k141_344016 | Not-determined |
| 17-109_k141_9170 | Not-determined |
| 17-109_k141_227486 | Not-determined |
| 17-109_k141_77015 | Not-determined |
| 17-109_k141_344022 | Not-determined |
| 17-109_k141_311917 | Not-determined |
| 17-109_k141_77021 | Not-determined |
| 17-109_k141_311922 | Not-determined |
| 17-109_k141_176705 | Not-determined |
| 17-109_k141_43338 | Not-determined |
| 17-109_k141_59252 | Not-determined |
| 17-109_k141_160383 | Not-determined |
| 17-109_k141_9178 | Not-determined |
| 17-109_k141_26267 | Low-quality |
| 17-109_k141_26271 | Not-determined |
| 17-109_k141_311936 | Not-determined |
| 17-109_k141_26273 | Not-determined |
| 17-109_k141_327837 | Not-determined |
| 17-109_k141_9182 | Not-determined |
| 17-109_k141_109570 | Not-determined |
| 17-109_k141_210317 | Not-determined |
| 17-109_k141_344037 | Not-determined |
| 17-109_k141_394368 | Not-determined |
| 17-109_k141_109579 | Not-determined |
| 17-109_k141_394375 | Not-determined |
| 17-109_k141_26292 | Not-determined |
| 17-109_k141_210326 | Not-determined |
| 17-109_k141_377797 | Low-quality |
| 17-109_k141_43380 | Low-quality |
| 17-109_k141_277597 | Not-determined |
| 17-109_k141_377799 | Not-determined |
| 17-109_k141_277600 | Not-determined |
| 17-109_k141_260067 | Low-quality |
| 17-109_k141_377803 | Not-determined |
| 17-109_k141_277601 | Not-determined |
| 17-109_k141_394395 | Not-determined |
| 17-109_k141_125585 | Not-determined |
| 17-109_k141_394396 | Low-quality |
| 17-109_k141_210330 | Not-determined |
| 17-109_k141_26304 | Not-determined |
| 17-109_k141_109594 | Not-determined |
| 17-109_k141_394400 | Not-determined |
| 17-109_k141_59292 | Not-determined |
| 17-109_k141_160419 | Not-determined |
| 17-109_k141_77050 | Not-determined |
| 17-109_k141_344066 | Not-determined |
| 17-109_k141_59301 | Low-quality |
| 17-109_k141_43402 | Not-determined |
| 17-109_k141_359625 | Low-quality |
| 17-109_k141_277620 | Low-quality |
| 17-109_k141_109608 | Not-determined |
| 17-109_k141_109609 | Not-determined |
| 17-109_k141_377835 | Not-determined |
| 17-109_k141_377837 | Not-determined |
| 17-109_k141_377841 | Not-determined |
| 17-109_k141_227544 | Not-determined |
| 17-109_k141_9211 | Not-determined |
| 17-109_k141_125599 | Not-determined |
| 17-109_k141_377844 | Not-determined |
| 17-109_k141_125602 | Not-determined |
| 17-109_k141_394421 | Not-determined |
| 17-109_k141_260088 | Not-determined |
| 17-109_k141_160433 | Not-determined |
| 17-109_k141_176714 | Not-determined |
| 17-109_k141_125606 | Not-determined |
| 17-109_k141_311969 | Not-determined |
| 17-109_k141_260093 | Not-determined |
| 17-109_k141_43410 | Not-determined |
| 17-109_k141_260099 | Not-determined |
| 17-109_k141_77084 | Not-determined |
| 17-109_k141_260110 | Not-determined |
| 17-109_k141_109613 | Not-determined |
| 17-109_k141_109614 | Not-determined |
| 17-109_k141_176721 | Not-determined |
| 17-109_k141_109621 | Not-determined |
| 17-109_k141_26344 | Not-determined |
| 17-109_k141_210346 | Not-determined |
| 17-109_k141_77098 | Not-determined |
| 17-109_k141_311981 | Not-determined |
| 17-109_k141_311986 | Not-determined |
| 17-109_k141_377867 | Low-quality |
| 17-109_k141_160457 | Low-quality |
| 17-109_k141_377873 | Not-determined |
| 17-109_k141_92669 | Not-determined |
| 17-109_k141_77111 | Not-determined |
| 17-109_k141_92671 | Not-determined |
| 17-109_k141_377880 | Not-determined |
| 17-109_k141_77113 | Not-determined |
| 17-109_k141_26365 | Not-determined |
| 17-109_k141_194542 | Not-determined |
| 17-109_k141_176731 | Not-determined |
| 17-109_k141_377882 | Not-determined |
| 17-109_k141_377884 | Not-determined |
| 17-109_k141_109643 | Not-determined |
| 17-109_k141_176738 | Not-determined |
| 17-109_k141_9224 | Low-quality |
| 17-109_k141_77121 | Not-determined |
| 17-109_k141_109647 | Not-determined |
| 17-109_k141_311992 | Not-determined |
| 17-109_k141_77124 | Not-determined |
| 17-109_k141_26374 | Low-quality |
| 17-109_k141_377890 | Not-determined |
| 17-109_k141_176746 | Low-quality |
| 17-109_k141_359654 | Not-determined |
| 17-109_k141_311994 | Low-quality |
| 17-109_k141_210391 | Not-determined |
| 17-109_k141_109659 | Not-determined |
| 17-109_k141_26383 | Not-determined |
| 17-109_k141_294783 | Not-determined |
| 17-109_k141_125640 | Not-determined |
| 17-109_k141_194566 | Not-determined |
| 17-109_k141_194567 | Not-determined |
| 17-109_k141_160467 | Not-determined |
| 17-109_k141_160472 | Not-determined |
| 17-109_k141_125648 | Not-determined |
| 17-109_k141_377905 | Not-determined |
| 17-109_k141_160476 | Not-determined |
| 17-109_k141_176767 | Not-determined |
| 17-109_k141_359663 | Low-quality |
| 17-109_k141_344155 | Not-determined |
| 17-109_k141_394465 | Not-determined |
| 17-109_k141_327841 | Not-determined |
| 17-109_k141_227594 | Not-determined |
| 17-109_k141_160491 | Not-determined |
| 17-109_k141_359675 | Not-determined |
| 17-109_k141_277682 | Not-determined |
| 17-109_k141_394473 | Not-determined |
| 17-109_k141_277683 | Not-determined |
| 17-109_k141_176787 | Not-determined |
| 17-109_k141_394482 | Not-determined |
| 17-109_k141_312024 | Not-determined |
| 17-109_k141_312025 | Not-determined |
| 17-109_k141_176791 | Not-determined |
| 17-109_k141_43457 | Not-determined |
| 17-109_k141_194578 | Not-determined |
| 17-109_k141_9261 | Not-determined |
| 17-109_k141_312031 | Not-determined |
| 17-109_k141_43461 | Not-determined |
| 17-109_k141_260153 | Low-quality |
| 17-109_k141_344164 | Not-determined |
| 17-109_k141_344165 | Not-determined |
| 17-109_k141_344166 | Not-determined |
| 17-109_k141_9265 | Not-determined |
| 17-109_k141_312037 | Not-determined |
| 17-109_k141_160521 | Not-determined |

| | |
|---|---|
| 17-109_k141_260167 | Not-determined |
| 17-109_k141_327862 | Not-determined |
| 17-109_k141_125682 | Not-determined |
| 17-109_k141_277694 | Not-determined |
| 17-109_k141_59324 | Not-determined |
| 17-109_k141_294822 | Not-determined |
| 17-109_k141_160530 | Low-quality |
| 17-109_k141_260181 | Not-determined |
| 17-109_k141_312056 | Not-determined |
| 17-109_k141_312058 | Not-determined |
| 17-109_k141_9290 | Not-determined |
| 17-109_k141_77145 | Not-determined |
| 17-109_k141_59332 | Low-quality |
| 17-109_k141_210422 | Not-determined |
| 17-109_k141_59333 | Not-determined |
| 17-109_k141_77148 | Not-determined |
| 17-109_k141_377925 | Low-quality |
| 17-109_k141_125693 | Low-quality |
| 17-109_k141_277707 | Not-determined |
| 17-109_k141_77153 | Not-determined |
| 17-109_k141_176824 | Not-determined |
| 17-109_k141_210429 | Not-determined |
| 17-109_k141_26396 | Not-determined |
| 17-109_k141_160544 | Not-determined |
| 17-109_k141_26397 | Not-determined |
| 17-109_k141_77159 | Low-quality |
| 17-109_k141_277716 | Not-determined |
| 17-109_k141_312067 | Not-determined |
| 17-109_k141_26406 | Not-determined |
| 17-109_k141_243764 | Not-determined |
| 17-109_k141_59352 | Not-determined |
| 17-109_k141_227658 | Not-determined |
| 17-109_k141_227664 | Low-quality |
| 17-109_k141_77172 | Not-determined |
| 17-109_k141_92710 | Not-determined |
| 17-109_k141_26413 | Not-determined |
| 17-109_k141_77183 | Not-determined |
| 17-109_k141_277743 | Not-determined |
| 17-109_k141_312075 | Not-determined |
| 17-109_k141_359692 | Not-determined |
| 17-109_k141_210447 | Low-quality |
| 17-109_k141_359696 | Not-determined |
| 17-109_k141_77187 | Not-determined |
| 17-109_k141_359697 | Not-determined |
| 17-109_k141_160551 | Not-determined |
| 17-109_k141_160554 | Not-determined |
| 17-109_k141_312088 | Not-determined |
| 17-109_k141_210461 | Not-determined |
| 17-109_k141_294856 | Not-determined |
| 17-109_k141_294857 | Low-quality |
| 17-109_k141_210463 | Not-determined |
| 17-109_k141_394519 | Not-determined |
| 17-109_k141_160558 | Not-determined |
| 17-109_k141_294858 | Not-determined |
| 17-109_k141_312091 | Not-determined |
| 17-109_k141_394520 | Not-determined |
| 17-109_k141_312094 | Not-determined |
| 17-109_k141_125715 | Low-quality |
| 17-109_k141_312096 | Not-determined |
| 17-109_k141_394522 | Low-quality |
| 17-109_k141_210469 | Not-determined |
| 17-109_k141_194600 | Not-determined |
| 17-109_k141_59380 | Not-determined |
| 17-109_k141_160571 | Low-quality |
| 17-109_k141_210474 | Low-quality |
| 17-109_k141_92738 | Not-determined |
| 17-109_k141_243792 | Not-determined |
| 17-109_k141_277772 | Not-determined |
| 17-109_k141_43519 | Not-determined |
| 17-109_k141_59398 | Low-quality |
| 17-109_k141_9311 | Not-determined |
| 17-109_k141_294859 | Not-determined |
| 17-109_k141_227667 | Not-determined |
| 17-109_k141_377985 | Not-determined |
| 17-109_k141_143342 | Not-determined |
| 17-109_k141_210490 | Not-determined |
| 17-109_k141_109731 | Not-determined |
| 17-109_k141_77229 | Not-determined |
| 17-109_k141_243801 | Not-determined |
| 17-109_k141_176867 | Not-determined |
| 17-109_k141_43540 | Not-determined |
| 17-109_k141_312124 | Not-determined |
| 17-109_k141_394537 | Not-determined |
| 17-109_k141_277810 | Not-determined |
| 17-109_k141_344206 | Not-determined |
| 17-109_k141_143352 | Not-determined |
| 17-109_k141_378003 | Not-determined |
| 17-109_k141_176875 | Not-determined |
| 17-109_k141_344210 | Not-determined |
| 17-109_k141_176877 | Not-determined |
| 17-109_k141_344214 | Not-determined |
| 17-109_k141_26449 | Not-determined |
| 17-109_k141_109748 | Not-determined |
| 17-109_k141_344215 | Not-determined |
| 17-109_k141_227687 | Not-determined |
| 17-109_k141_43546 | Not-determined |
| 17-109_k141_109753 | Not-determined |
| 17-109_k141_109756 | Not-determined |
| 17-109_k141_394548 | Not-determined |
| 17-109_k141_210524 | Not-determined |
| 17-109_k141_109761 | Not-determined |
| 17-109_k141_109764 | Not-determined |
| 17-109_k141_327906 | Not-determined |
| 17-109_k141_243825 | Low-quality |
| 17-109_k141_344226 | Not-determined |
| 17-109_k141_227697 | Not-determined |
| 17-109_k141_26459 | Not-determined |
| 17-109_k141_312145 | Not-determined |
| 17-109_k141_312146 | Not-determined |
| 17-109_k141_194639 | Not-determined |
| 17-109_k141_294878 | Not-determined |
| 17-109_k141_378025 | Not-determined |
| 17-109_k141_394557 | Not-determined |
| 17-109_k141_176898 | Not-determined |
| 17-109_k141_176901 | Not-determined |
| 17-109_k141_176905 | Not-determined |
| 17-109_k141_125754 | Not-determined |
| 17-109_k141_394562 | Low-quality |
| 17-109_k141_312148 | Not-determined |
| 17-109_k141_327922 | Not-determined |
| 17-109_k141_359744 | Not-determined |
| 17-109_k141_125757 | Not-determined |
| 17-109_k141_77264 | Low-quality |
| 17-109_k141_227714 | Not-determined |
| 17-109_k141_394574 | Not-determined |
| 17-109_k141_59415 | Not-determined |
| 17-109_k141_92764 | Not-determined |
| 17-109_k141_394584 | Not-determined |
| 17-109_k141_143367 | Not-determined |
| 17-109_k141_227720 | Low-quality |
| 17-109_k141_394589 | Not-determined |
| 17-109_k141_294896 | Not-determined |
| 17-109_k141_394591 | Not-determined |
| 17-109_k141_77272 | Not-determined |
| 17-109_k141_109801 | Not-determined |
| 17-109_k141_327939 | Not-determined |
| 17-109_k141_378058 | Not-determined |
| 17-109_k141_109803 | Not-determined |
| 17-109_k141_378060 | Not-determined |
| 17-109_k141_143374 | Not-determined |
| 17-109_k141_59430 | Not-determined |
| 17-109_k141_260200 | Not-determined |
| 17-109_k141_359760 | Not-determined |
| 17-109_k141_77288 | Not-determined |
| 17-109_k141_227737 | Low-quality |
| 17-109_k141_312175 | Low-quality |
| 17-109_k141_359766 | Not-determined |
| 17-109_k141_359767 | Not-determined |
| 17-109_k141_394611 | Not-determined |
| 17-109_k141_394612 | Not-determined |
| 17-109_k141_43587 | Low-quality |
| 17-109_k141_26487 | Not-determined |
| 17-109_k141_26488 | Not-determined |
| 17-109_k141_77294 | Not-determined |
| 17-109_k141_312180 | Not-determined |
| 17-109_k141_92798 | Not-determined |
| 17-109_k141_77297 | Not-determined |
| 17-109_k141_312183 | Low-quality |
| 17-109_k141_312185 | Not-determined |
| 17-109_k141_43596 | Not-determined |
| 17-109_k141_26494 | Not-determined |
| 17-109_k141_26495 | Not-determined |
| 17-109_k141_312192 | Not-determined |
| 17-109_k141_77306 | Not-determined |
| 17-109_k141_277837 | Not-determined |
| 17-109_k141_327958 | Not-determined |
| 17-109_k141_294909 | Low-quality |
| 17-109_k141_109832 | Not-determined |
| 17-109_k141_394627 | Not-determined |
| 17-109_k141_294918 | Not-determined |
| 17-109_k141_92821 | Low-quality |
| 17-109_k141_26505 | Not-determined |
| 17-109_k141_26506 | Not-determined |
| 17-109_k141_394632 | Low-quality |
| 17-109_k141_59449 | Not-determined |
| 17-109_k141_394634 | Not-determined |
| 17-109_k141_9337 | Not-determined |
| 17-109_k141_43620 | Not-determined |
| 17-109_k141_176938 | Not-determined |
| 17-109_k141_160583 | Low-quality |
| 17-109_k141_26513 | Not-determined |
| 17-109_k141_26516 | Not-determined |
| 17-109_k141_26518 | Not-determined |
| 17-109_k141_176943 | Not-determined |
| 17-109_k141_312195 | Low-quality |
| 17-109_k141_43628 | Not-determined |
| 17-109_k141_26525 | Not-determined |
| 17-109_k141_294940 | Not-determined |
| 17-109_k141_59457 | Not-determined |
| 17-109_k141_294948 | Not-determined |
| 17-109_k141_394661 | Not-determined |
| 17-109_k141_160604 | Low-quality |
| 17-109_k141_43632 | Not-determined |
| 17-109_k141_59468 | Not-determined |
| 17-109_k141_294953 | Not-determined |
| 17-109_k141_359802 | Not-determined |
| 17-109_k141_92834 | Not-determined |
| 17-109_k141_394668 | Not-determined |
| 17-109_k141_294958 | Not-determined |
| 17-109_k141_26537 | Not-determined |
| 17-109_k141_160611 | Not-determined |
| 17-109_k141_9358 | Not-determined |
| 17-109_k141_160613 | Not-determined |
| 17-109_k141_59478 | Not-determined |
| 17-109_k141_92837 | Not-determined |
| 17-109_k141_125777 | Not-determined |
| 17-109_k141_294964 | Not-determined |
| 17-109_k141_227782 | Low-quality |
| 17-109_k141_43646 | Not-determined |
| 17-109_k141_26552 | Not-determined |
| 17-109_k141_92850 | Not-determined |
| 17-109_k141_260264 | Not-determined |
| 17-109_k141_394685 | Not-determined |
| 17-109_k141_312215 | Not-determined |
| 17-109_k141_359814 | Low-quality |
| 17-109_k141_312217 | Not-determined |
| 17-109_k141_312221 | Not-determined |
| 17-109_k141_394694 | Not-determined |
| 17-109_k141_9373 | Not-determined |
| 17-109_k141_43658 | Not-determined |
| 17-109_k141_143380 | Not-determined |
| 17-109_k141_26562 | Not-determined |
| 17-109_k141_328014 | Not-determined |
| 17-109_k141_143385 | Not-determined |
| 17-109_k141_312236 | Not-determined |
| 17-109_k141_59486 | Not-determined |
| 17-109_k141_92872 | Low-quality |
| 17-109_k141_59487 | Low-quality |
| 17-109_k141_160640 | Not-determined |
| 17-109_k141_59491 | Low-quality |
| 17-109_k141_344272 | Not-determined |
| 17-109_k141_160641 | Not-determined |
| 17-109_k141_359825 | Not-determined |
| 17-109_k141_359826 | Not-determined |
| 17-109_k141_125792 | Not-determined |
| 17-109_k141_43672 | Not-determined |
| 17-109_k141_176978 | Not-determined |
| 17-109_k141_176979 | Not-determined |
| 17-109_k141_143395 | Not-determined |
| 17-109_k141_59504 | Not-determined |
| 17-109_k141_160651 | Not-determined |
| 17-109_k141_160652 | Not-determined |
| 17-109_k141_43682 | Not-determined |
| 17-109_k141_176994 | Not-determined |
| 17-109_k141_26583 | Not-determined |
| 17-109_k141_394713 | Not-determined |
| 17-109_k141_344296 | Low-quality |
| 17-109_k141_277849 | Not-determined |
| 17-109_k141_359853 | Not-determined |
| 17-109_k141_177001 | Low-quality |
| 17-109_k141_43693 | Not-determined |
| 17-109_k141_125806 | Low-quality |
| 17-109_k141_59523 | Not-determined |
| 17-109_k141_312271 | Not-determined |
| 17-109_k141_59524 | Not-determined |
| 17-109_k141_344309 | Not-determined |
| 17-109_k141_312272 | Not-determined |
| 17-109_k141_394721 | Not-determined |
| 17-109_k141_59528 | Not-determined |
| 17-109_k141_295018 | Not-determined |
| 17-109_k141_77345 | Not-determined |
| 17-109_k141_125813 | Not-determined |
| 17-109_k141_160659 | Not-determined |
| 17-109_k141_77349 | Not-determined |
| 17-109_k141_378143 | Low-quality |
| 17-109_k141_295024 | Not-determined |
| 17-109_k141_77351 | Low-quality |
| 17-109_k141_160665 | Not-determined |
| 17-109_k141_260333 | Not-determined |
| 17-109_k141_143414 | Low-quality |
| 17-109_k141_26602 | Not-determined |
| 17-109_k141_143415 | Not-determined |
| 17-109_k141_312300 | Not-determined |
| 17-109_k141_160676 | Not-determined |
| 17-109_k141_312304 | Not-determined |
| 17-109_k141_26607 | Not-determined |
| 17-109_k141_160678 | Not-determined |
| 17-109_k141_77377 | Not-determined |
| 17-109_k141_9394 | Not-determined |
| 17-109_k141_77378 | Not-determined |
| 17-109_k141_9395 | Low-quality |
| 17-109_k141_9400 | Not-determined |
| 17-109_k141_227796 | Not-determined |
| 17-109_k141_125842 | Not-determined |
| 17-109_k141_227797 | Not-determined |
| 17-109_k141_9402 | Not-determined |
| 17-109_k141_227798 | Not-determined |
| 17-109_k141_227799 | Not-determined |
| 17-109_k141_26623 | Not-determined |
| 17-109_k141_9404 | Not-determined |
| 17-109_k141_26624 | Not-determined |
| 17-109_k141_378174 | Not-determined |
| 17-109_k141_59550 | Not-determined |
| 17-109_k141_260353 | Not-determined |
| 17-109_k141_260354 | Not-determined |
| 17-109_k141_378180 | Not-determined |
| 17-109_k141_227808 | Not-determined |
| 17-109_k141_77392 | Not-determined |
| 17-109_k141_227809 | Not-determined |
| 17-109_k141_43722 | Not-determined |
| 17-109_k141_43723 | Not-determined |
| 17-109_k141_295047 | Not-determined |
| 17-109_k141_295048 | Not-determined |
| 17-109_k141_260359 | Not-determined |
| 17-109_k141_109866 | Not-determined |
| 17-109_k141_378197 | Not-determined |
| 17-109_k141_394735 | Not-determined |
| 17-109_k141_177020 | Not-determined |
| 17-109_k141_177022 | Not-determined |
| 17-109_k141_77399 | Not-determined |
| 17-109_k141_243850 | Not-determined |
| 17-109_k141_109879 | Not-determined |
| 17-109_k141_77405 | Not-determined |
| 17-109_k141_277863 | Not-determined |
| 17-109_k141_160700 | Not-determined |
| 17-109_k141_328086 | Not-determined |
| 17-109_k141_26631 | Not-determined |
| 17-109_k141_295054 | Not-determined |
| 17-109_k141_295055 | Not-determined |
| 17-109_k141_143425 | Not-determined |
| 17-109_k141_125860 | Not-determined |
| 17-109_k141_59580 | Not-determined |
| 17-109_k141_243856 | Not-determined |
| 17-109_k141_125869 | Low-quality |
| 17-109_k141_125870 | Not-determined |
| 17-109_k141_77409 | Not-determined |
| 17-109_k141_26641 | Low-quality |
| 17-109_k141_394741 | Not-determined |
| 17-109_k141_59583 | Not-determined |
| 17-109_k141_227833 | Low-quality |
| 17-109_k141_59585 | Not-determined |
| 17-109_k141_77416 | Not-determined |
| 17-109_k141_243873 | Not-determined |
| 17-109_k141_77419 | Not-determined |
| 17-109_k141_26649 | Not-determined |
| 17-109_k141_312334 | Not-determined |
| 17-109_k141_227839 | Not-determined |
| 17-109_k141_227841 | Not-determined |
| 17-109_k141_312339 | Not-determined |
| 17-109_k141_312341 | Low-quality |
| 17-109_k141_295076 | Not-determined |
| 17-109_k141_59598 | Not-determined |
| 17-109_k141_344355 | Low-quality |
| 17-109_k141_92934 | Not-determined |
| 17-109_k141_394758 | Not-determined |
| 17-109_k141_59603 | Not-determined |
| 17-109_k141_210528 | Not-determined |
| 17-109_k141_328098 | Not-determined |
| 17-109_k141_394761 | Not-determined |
| 17-109_k141_295085 | Not-determined |
| 17-109_k141_394763 | Not-determined |
| 17-109_k141_277893 | Not-determined |
| 17-109_k141_9432 | Low-quality |
| 17-109_k141_243884 | Not-determined |
| 17-109_k141_210532 | Low-quality |
| 17-109_k141_59610 | Not-determined |
| 17-109_k141_277895 | Not-determined |
| 17-109_k141_77441 | Not-determined |
| 17-109_k141_177048 | Not-determined |
| 17-109_k141_59617 | Not-determined |
| 17-109_k141_92950 | Not-determined |
| 17-109_k141_59619 | Not-determined |
| 17-109_k141_9439 | Not-determined |
| 17-109_k141_243894 | Not-determined |
| 17-109_k141_26669 | Not-determined |
| 17-109_k141_227868 | Not-determined |
| 17-109_k141_210549 | Not-determined |
| 17-109_k141_77447 | Not-determined |
| 17-109_k141_295097 | Not-determined |
| 17-109_k141_9450 | Low-quality |
| 17-109_k141_378242 | Not-determined |
| 17-109_k141_177053 | Not-determined |
| 17-109_k141_59626 | Not-determined |
| 17-109_k141_43771 | Not-determined |
| 17-109_k141_378243 | Not-determined |
| 17-109_k141_344376 | Not-determined |
| 17-109_k141_43776 | Not-determined |
| 17-109_k141_92957 | Not-determined |
| 17-109_k141_177060 | Not-determined |
| 17-109_k141_125890 | Not-determined |
| 17-109_k141_344380 | Not-determined |
| 17-109_k141_312375 | Not-determined |
| 17-109_k141_77460 | Not-determined |
| 17-109_k141_77465 | Not-determined |
| 17-109_k141_312379 | Not-determined |
| 17-109_k141_143456 | Low-quality |
| 17-109_k141_344386 | Not-determined |
| 17-109_k141_77469 | Not-determined |
| 17-109_k141_243907 | Not-determined |
| 17-109_k141_277904 | Not-determined |
| 17-109_k141_312387 | Not-determined |
| 17-109_k141_194701 | Low-quality |
| 17-109_k141_43794 | Low-quality |
| 17-109_k141_59637 | Not-determined |
| 17-109_k141_43795 | Not-determined |
| 17-109_k141_9453 | Not-determined |
| 17-109_k141_243915 | Not-determined |
| 17-109_k141_177066 | Not-determined |
| 17-109_k141_26692 | Not-determined |
| 17-109_k141_109916 | Not-determined |
| 17-109_k141_9457 | Not-determined |
| 17-109_k141_194709 | Not-determined |
| 17-109_k141_359894 | Low-quality |
| 17-109_k141_243921 | Not-determined |
| 17-109_k141_77480 | Not-determined |
| 17-109_k141_43807 | Not-determined |
| 17-109_k141_9467 | Not-determined |
| 17-109_k141_277921 | Low-quality |
| 17-109_k141_277922 | Not-determined |
| 17-109_k141_344398 | Not-determined |
| 17-109_k141_260427 | Not-determined |
| 17-109_k141_227899 | Low-quality |
| 17-109_k141_260436 | Not-determined |
| 17-109_k141_260439 | Not-determined |
| 17-109_k141_26711 | Not-determined |
| 17-109_k141_92975 | Not-determined |
| 17-109_k141_295121 | Not-determined |
| 17-109_k141_43817 | Not-determined |
| 17-109_k141_295122 | Not-determined |
| 17-109_k141_394788 | Not-determined |
| 17-109_k141_177075 | Not-determined |
| 17-109_k141_77497 | Not-determined |
| 17-109_k141_26718 | Not-determined |
| 17-109_k141_194721 | Low-quality |
| 17-109_k141_160753 | Not-determined |
| 17-109_k141_177083 | Low-quality |
| 17-109_k141_160754 | Not-determined |
| 17-109_k141_92985 | Not-determined |
| 17-109_k141_243943 | Not-determined |
| 17-109_k141_26731 | Not-determined |
| 17-109_k141_210584 | Low-quality |
| 17-109_k141_109950 | Not-determined |
| 17-109_k141_59666 | Not-determined |
| 17-109_k141_328158 | Not-determined |
| 17-109_k141_9498 | Not-determined |
| 17-109_k141_227923 | Not-determined |
| 17-109_k141_328161 | Not-determined |
| 17-109_k141_143512 | Not-determined |
| 17-109_k141_9505 | Not-determined |
| 17-109_k141_125927 | Not-determined |
| 17-109_k141_77518 | Not-determined |
| 17-109_k141_125931 | Low-quality |
| 17-109_k141_359944 | Not-determined |
| 17-109_k141_93005 | Not-determined |
| 17-109_k141_177106 | Not-determined |
| 17-109_k141_344423 | Not-determined |
| 17-109_k141_243947 | Not-determined |
| 17-109_k141_9511 | Not-determined |
| 17-109_k141_160772 | Not-determined |
| 17-109_k141_26755 | Not-determined |
| 17-109_k141_394808 | Not-determined |
| 17-109_k141_344428 | Not-determined |
| 17-109_k141_59690 | Low-quality |
| 17-109_k141_143530 | Not-determined |
| 17-109_k141_177115 | Not-determined |
| 17-109_k141_227942 | Not-determined |
| 17-109_k141_227943 | Not-determined |
| 17-109_k141_359960 | Not-determined |
| 17-109_k141_93021 | Not-determined |
| 17-109_k141_394814 | Not-determined |
| 17-109_k141_227949 | Not-determined |
| 17-109_k141_125938 | Not-determined |
| 17-109_k141_177127 | Not-determined |
| 17-109_k141_394822 | Not-determined |
| 17-109_k141_260451 | Not-determined |
| 17-109_k141_143541 | Not-determined |
| 17-109_k141_26765 | Not-determined |
| 17-109_k141_359968 | Low-quality |
| 17-109_k141_43847 | Not-determined |
| 17-109_k141_260454 | Not-determined |
| 17-109_k141_260455 | Not-determined |
| 17-109_k141_59708 | Not-determined |
| 17-109_k141_77536 | Not-determined |
| 17-109_k141_277940 | Not-determined |
| 17-109_k141_243960 | Not-determined |
| 17-109_k141_210610 | Low-quality |
| 17-109_k141_359977 | Low-quality |
| 17-109_k141_210616 | Not-determined |
| 17-109_k141_125950 | Low-quality |
| 17-109_k141_43852 | Not-determined |
| 17-109_k141_77542 | Not-determined |
| 17-109_k141_359978 | Not-determined |
| 17-109_k141_26779 | Not-determined |
| 17-109_k141_312443 | Not-determined |
| 17-109_k141_227958 | Not-determined |
| 17-109_k141_394835 | Not-determined |
| 17-109_k141_77549 | Not-determined |
| 17-109_k141_210622 | Not-determined |
| 17-109_k141_344442 | Not-determined |
| 17-109_k141_227964 | Not-determined |
| 17-109_k141_43861 | Not-determined |
| 17-109_k141_295163 | Not-determined |
| 17-109_k141_125955 | Not-determined |
| 17-109_k141_243969 | Not-determined |
| 17-109_k141_394843 | Not-determined |
| 17-109_k141_328176 | Not-determined |
| 17-109_k141_77559 | Not-determined |
| 17-109_k141_328178 | Not-determined |
| 17-109_k141_243973 | Not-determined |
| 17-109_k141_227967 | Not-determined |
| 17-109_k141_295170 | Not-determined |
| 17-109_k141_394847 | Not-determined |
| 17-109_k141_93048 | Not-determined |
| 17-109_k141_227976 | Not-determined |
| 17-109_k141_210642 | Not-determined |
| 17-109_k141_227980 | Low-quality |
| 17-109_k141_210646 | Not-determined |
| 17-109_k141_394863 | Not-determined |
| 17-109_k141_394868 | Not-determined |
| 17-109_k141_59751 | Not-determined |
| 17-109_k141_277956 | Not-determined |
| 17-109_k141_43887 | Not-determined |
| 17-109_k141_328198 | Not-determined |
| 17-109_k141_93077 | Not-determined |
| 17-109_k141_228006 | Not-determined |
| 17-109_k141_228007 | Low-quality |
| 17-109_k141_177152 | Not-determined |
| 17-109_k141_344463 | Not-determined |
| 17-109_k141_344467 | Not-determined |
| 17-109_k141_59766 | Not-determined |
| 17-109_k141_43909 | Not-determined |
| 17-109_k141_43912 | Not-determined |
| 17-109_k141_210653 | Not-determined |
| 17-109_k141_59779 | Not-determined |
| 17-109_k141_77588 | Not-determined |
| 17-109_k141_228025 | Not-determined |
| 17-109_k141_328222 | Not-determined |
| 17-109_k141_59787 | Not-determined |
| 17-109_k141_277989 | Not-determined |
| 17-109_k141_59789 | Not-determined |
| 17-109_k141_26798 | Not-determined |
| 17-109_k141_228030 | Low-quality |
| 17-109_k141_344479 | Not-determined |
| 17-109_k141_93091 | Not-determined |
| 17-109_k141_77600 | Not-determined |
| 17-109_k141_244011 | Not-determined |
| 17-109_k141_244012 | Not-determined |
| 17-109_k141_328233 | Low-quality |
| 17-109_k141_260493 | Not-determined |
| 17-109_k141_394898 | Not-determined |
| 17-109_k141_277994 | Not-determined |
| 17-109_k141_378268 | Not-determined |
| 17-109_k141_394903 | Low-quality |
| 17-109_k141_394904 | Not-determined |
| 17-109_k141_312496 | Not-determined |
| 17-109_k141_278002 | Low-quality |
| 17-109_k141_394906 | Low-quality |
| 17-109_k141_278004 | Not-determined |
| 17-109_k141_360064 | Not-determined |
| 17-109_k141_278008 | Not-determined |
| 17-109_k141_9626 | Not-determined |
| 17-109_k141_260507 | Not-determined |
| 17-109_k141_9629 | Not-determined |
| 17-109_k141_378279 | Not-determined |
| 17-109_k141_344486 | Not-determined |
| 17-109_k141_312503 | Not-determined |
| 17-109_k141_244029 | Not-determined |
| 17-109_k141_295192 | Low-quality |
| 17-109_k141_394921 | Not-determined |
| 17-109_k141_260513 | Low-quality |
| 17-109_k141_295197 | Low-quality |
| 17-109_k141_378291 | Not-determined |
| 17-109_k141_378292 | Not-determined |
| 17-109_k141_160796 | Low-quality |
| 17-109_k141_278020 | Not-determined |
| 17-109_k141_93121 | Not-determined |
| 17-109_k141_160798 | Not-determined |
| 17-109_k141_360070 | Low-quality |
| 17-109_k141_278022 | Not-determined |
| 17-109_k141_43952 | Low-quality |
| 17-109_k141_177167 | Not-determined |
| 17-109_k141_394934 | Not-determined |
| 17-109_k141_244039 | Not-determined |
| 17-109_k141_328261 | Not-determined |
| 17-109_k141_244045 | Not-determined |
| 17-109_k141_378309 | Not-determined |
| 17-109_k141_312515 | Not-determined |
| 17-109_k141_26816 | Not-determined |
| 17-109_k141_244051 | Not-determined |
| 17-109_k141_59846 | Not-determined |
| 17-109_k141_59847 | Not-determined |
| 17-109_k141_160817 | Not-determined |
| 17-109_k141_43974 | Not-determined |
| 17-109_k141_312522 | Not-determined |
| 17-109_k141_59853 | Not-determined |
| 17-109_k141_328270 | Low-quality |
| 17-109_k141_295227 | Not-determined |
| 17-109_k141_394941 | Not-determined |
| 17-109_k141_143566 | Not-determined |

| | |
|---|---|
| 17-109_k141_210699 | Not-determined |
| 17-109_k141_125992 | Not-determined |
| 17-109_k141_125993 | Not-determined |
| 17-109_k141_312542 | Not-determined |
| 17-109_k141_328288 | Not-determined |
| 17-109_k141_278042 | Not-determined |
| 17-109_k141_344502 | Not-determined |
| 17-109_k141_143583 | Not-determined |
| 17-109_k141_77642 | Not-determined |
| 17-109_k141_295230 | Low-quality |
| 17-109_k141_59870 | Not-determined |
| 17-109_k141_43996 | Not-determined |
| 17-109_k141_93164 | Not-determined |
| 17-109_k141_394943 | Not-determined |
| 17-109_k141_360108 | Not-determined |
| 17-109_k141_77648 | Not-determined |
| 17-109_k141_244082 | Not-determined |
| 17-109_k141_77651 | Not-determined |
| 17-109_k141_295235 | Not-determined |
| 17-109_k141_9636 | Not-determined |
| 17-109_k141_344514 | Not-determined |
| 17-109_k141_360112 | Not-determined |
| 17-109_k141_360113 | Not-determined |
| 17-109_k141_328297 | Not-determined |
| 17-109_k141_9642 | Not-determined |
| 17-109_k141_260561 | Low-quality |
| 17-109_k141_228070 | Not-determined |
| 17-109_k141_194758 | Not-determined |
| 17-109_k141_228071 | Not-determined |
| 17-109_k141_378317 | Not-determined |
| 17-109_k141_244093 | Not-determined |
| 17-109_k141_378322 | Not-determined |
| 17-109_k141_59882 | Low-quality |
| 17-109_k141_143607 | Not-determined |
| 17-109_k141_228077 | Not-determined |
| 17-109_k141_160855 | Not-determined |
| 17-109_k141_143609 | Not-determined |
| 17-109_k141_295254 | Not-determined |
| 17-109_k141_44024 | Not-determined |
| 17-109_k141_328313 | Not-determined |
| 17-109_k141_210719 | Not-determined |
| 17-109_k141_378331 | Low-quality |
| 17-109_k141_260565 | Not-determined |
| 17-109_k141_44031 | Not-determined |
| 17-109_k141_328321 | Not-determined |
| 17-109_k141_210730 | Not-determined |
| 17-109_k141_26832 | Not-determined |
| 17-109_k141_378338 | Not-determined |
| 17-109_k141_26835 | Not-determined |
| 17-109_k141_378340 | Not-determined |
| 17-109_k141_360146 | Not-determined |
| 17-109_k141_378342 | Not-determined |
| 17-109_k141_378344 | Not-determined |
| 17-109_k141_210738 | Not-determined |
| 17-109_k141_378349 | Not-determined |
| 17-109_k141_177208 | Not-determined |
| 17-109_k141_26850 | Not-determined |
| 17-109_k141_177213 | Not-determined |
| 17-109_k141_210745 | Not-determined |
| 17-109_k141_44049 | Not-determined |
| 17-109_k141_295272 | Not-determined |
| 17-109_k141_194784 | Not-determined |
| 17-109_k141_194787 | Not-determined |
| 17-109_k141_194789 | Not-determined |
| 17-109_k141_278053 | Low-quality |
| 17-109_k141_177218 | Not-determined |
| 17-109_k141_295282 | Not-determined |
| 17-109_k141_394969 | Not-determined |
| 17-109_k141_278057 | Not-determined |
| 17-109_k141_194795 | Not-determined |
| 17-109_k141_93184 | Not-determined |
| 17-109_k141_44055 | Not-determined |
| 17-109_k141_44057 | Not-determined |
| 17-109_k141_312583 | Not-determined |
| 17-109_k141_44064 | Not-determined |
| 17-109_k141_59923 | Not-determined |
| 17-109_k141_160883 | Not-determined |
| 17-109_k141_295297 | Not-determined |
| 17-109_k141_77689 | Not-determined |
| 17-109_k141_210773 | Not-determined |
| 17-109_k141_59926 | Not-determined |
| 17-109_k141_59927 | Not-determined |
| 17-109_k141_295304 | Not-determined |
| 17-109_k141_210783 | Not-determined |
| 17-109_k141_160894 | Not-determined |
| 17-109_k141_160901 | Not-determined |
| 17-109_k141_93216 | Not-determined |
| 17-109_k141_160904 | Not-determined |
| 17-109_k141_210800 | Not-determined |
| 17-109_k141_394981 | Not-determined |
| 17-109_k141_210802 | Not-determined |
| 17-109_k141_378388 | Not-determined |
| 17-109_k141_295313 | Not-determined |
| 17-109_k141_260623 | Not-determined |
| 17-109_k141_344557 | Not-determined |
| 17-109_k141_378390 | Not-determined |
| 17-109_k141_394987 | Not-determined |
| 17-109_k141_59948 | Not-determined |
| 17-109_k141_278072 | Not-determined |
| 17-109_k141_210808 | Not-determined |
| 17-109_k141_394997 | Not-determined |
| 17-109_k141_177242 | Not-determined |
| 17-109_k141_26878 | Low-quality |
| 17-109_k141_210810 | Not-determined |
| 17-109_k141_26887 | Not-determined |
| 17-109_k141_210813 | Low-quality |
| 17-109_k141_295325 | Not-determined |
| 17-109_k141_26890 | Not-determined |
| 17-109_k141_260640 | Not-determined |
| 17-109_k141_44113 | Not-determined |
| 17-109_k141_260646 | Not-determined |
| 17-109_k141_378407 | Not-determined |
| 17-109_k141_210820 | Low-quality |
| 17-109_k141_210821 | Not-determined |
| 17-109_k141_210822 | Not-determined |
| 17-109_k141_344571 | Not-determined |
| 17-109_k141_260654 | Low-quality |
| 17-109_k141_244113 | Not-determined |
| 17-109_k141_395004 | Not-determined |
| 17-109_k141_210826 | Not-determined |
| 17-109_k141_210829 | Not-determined |
| 17-109_k141_26911 | Not-determined |
| 17-109_k141_228104 | Not-determined |
| 17-109_k141_228108 | Not-determined |
| 17-109_k141_312644 | Not-determined |
| 17-109_k141_295334 | Not-determined |
| 17-109_k141_177259 | Not-determined |
| 17-109_k141_9655 | Not-determined |
| 17-109_k141_395026 | Not-determined |
| 17-109_k141_244140 | Low-quality |
| 17-109_k141_278087 | Not-determined |
| 17-109_k141_312649 | Not-determined |
| 17-109_k141_228123 | Not-determined |
| 17-109_k141_210836 | Not-determined |
| 17-109_k141_160943 | Not-determined |
| 17-109_k141_378430 | Low-quality |
| 17-109_k141_278093 | Not-determined |
| 17-109_k141_328345 | Not-determined |
| 17-109_k141_109967 | Low-quality |
| 17-109_k141_160951 | Not-determined |
| 17-109_k141_93243 | Not-determined |
| 17-109_k141_143660 | Not-determined |
| 17-109_k141_109968 | Not-determined |
| 17-109_k141_228142 | Not-determined |
| 17-109_k141_160954 | Not-determined |
| 17-109_k141_143663 | Not-determined |
| 17-109_k141_295344 | Not-determined |
| 17-109_k141_228145 | Not-determined |
| 17-109_k141_295348 | Not-determined |
| 17-109_k141_244145 | Not-determined |
| 17-109_k141_77706 | Not-determined |
| 17-109_k141_143668 | Not-determined |
| 17-109_k141_344604 | Not-determined |
| 17-109_k141_378446 | Not-determined |
| 17-109_k141_328350 | Not-determined |
| 17-109_k141_378448 | Not-determined |
| 17-109_k141_378450 | Not-determined |
| 17-109_k141_160967 | Low-quality |
| 17-109_k141_278112 | Not-determined |
| 17-109_k141_44139 | Not-determined |
| 17-109_k141_295355 | Not-determined |
| 17-109_k141_126032 | Not-determined |
| 17-109_k141_126034 | Not-determined |
| 17-109_k141_93261 | Not-determined |
| 17-109_k141_228156 | Not-determined |
| 17-109_k141_109985 | Not-determined |
| 17-109_k141_93264 | Not-determined |
| 17-109_k141_378466 | Not-determined |
| 17-109_k141_44156 | Low-quality |
| 17-109_k141_143688 | Not-determined |
| 17-109_k141_93271 | Not-determined |
| 17-109_k141_143690 | Not-determined |
| 17-109_k141_260683 | Not-determined |
| 17-109_k141_210857 | Not-determined |
| 17-109_k141_126040 | Not-determined |
| 17-109_k141_295361 | Low-quality |
| 17-109_k141_328362 | Not-determined |
| 17-109_k141_44165 | Not-determined |
| 17-109_k141_328368 | Not-determined |
| 17-109_k141_378477 | Low-quality |
| 17-109_k141_160982 | Not-determined |
| 17-109_k141_260696 | Not-determined |
| 17-109_k141_278122 | Not-determined |
| 17-109_k141_160987 | Not-determined |
| 17-109_k141_26965 | Not-determined |
| 17-109_k141_278124 | Not-determined |
| 17-109_k141_260702 | Not-determined |
| 17-109_k141_59967 | Not-determined |
| 17-109_k141_344631 | Not-determined |
| 17-109_k141_59970 | Not-determined |
| 17-109_k141_395071 | Not-determined |
| 17-109_k141_194842 | Not-determined |
| 17-109_k141_260707 | Not-determined |
| 17-109_k141_109995 | Not-determined |
| 17-109_k141_395074 | Not-determined |
| 17-109_k141_378486 | Not-determined |
| 17-109_k141_177330 | Not-determined |
| 17-109_k141_378487 | Not-determined |
| 17-109_k141_260711 | Not-determined |
| 17-109_k141_143705 | Not-determined |
| 17-109_k141_395076 | Not-determined |
| 17-109_k141_210867 | Not-determined |
| 17-109_k141_93279 | Low-quality |
| 17-109_k141_77734 | Not-determined |
| 17-109_k141_126068 | Not-determined |
| 17-109_k141_228193 | Not-determined |
| 17-109_k141_395082 | Not-determined |
| 17-109_k141_395083 | Not-determined |
| 17-109_k141_395085 | Low-quality |
| 17-109_k141_210878 | Low-quality |
| 17-109_k141_295366 | Not-determined |
| 17-109_k141_228202 | Not-determined |
| 17-109_k141_395094 | Not-determined |
| 17-109_k141_395096 | Low-quality |
| 17-109_k141_260737 | Not-determined |
| 17-109_k141_194870 | Not-determined |
| 17-109_k141_328394 | Not-determined |
| 17-109_k141_194872 | Not-determined |
| 17-109_k141_44198 | Not-determined |
| 17-109_k141_260741 | Not-determined |
| 17-109_k141_161016 | Not-determined |
| 17-109_k141_360171 | Not-determined |
| 17-109_k141_344655 | Not-determined |
| 17-109_k141_228208 | Not-determined |
| 17-109_k141_228216 | Not-determined |
| 17-109_k141_228218 | Not-determined |
| 17-109_k141_194887 | Not-determined |
| 17-109_k141_93313 | Not-determined |
| 17-109_k141_143718 | Not-determined |
| 17-109_k141_278169 | Not-determined |
| 17-109_k141_161020 | Low-quality |
| 17-109_k141_312726 | Not-determined |
| 17-109_k141_210900 | Not-determined |
| 17-109_k141_395114 | Not-determined |
| 17-109_k141_360214 | Not-determined |
| 17-109_k141_278179 | Not-determined |
| 17-109_k141_228241 | Low-quality |
| 17-109_k141_344689 | Not-determined |
| 17-109_k141_60038 | Not-determined |
| 17-109_k141_93330 | Not-determined |
| 17-109_k141_395132 | Not-determined |
| 17-109_k141_244201 | Not-determined |
| 17-109_k141_278196 | Not-determined |
| 17-109_k141_161036 | Not-determined |
| 17-109_k141_312741 | Not-determined |
| 17-109_k141_395137 | Not-determined |
| 17-109_k141_194908 | Not-determined |
| 17-109_k141_395138 | Not-determined |
| 17-109_k141_312745 | Not-determined |
| 17-109_k141_244207 | Not-determined |
| 17-109_k141_395141 | Not-determined |
| 17-109_k141_60050 | Not-determined |
| 17-109_k141_278200 | Low-quality |
| 17-109_k141_395147 | Not-determined |
| 17-109_k141_244212 | Not-determined |
| 17-109_k141_177359 | Low-quality |
| 17-109_k141_328416 | Not-determined |
| 17-109_k141_126115 | Not-determined |
| 17-109_k141_177361 | Not-determined |
| 17-109_k141_312750 | Not-determined |
| 17-109_k141_93351 | Not-determined |
| 17-109_k141_194918 | Not-determined |
| 17-109_k141_177367 | Low-quality |
| 17-109_k141_244218 | Not-determined |
| 17-109_k141_328424 | Not-determined |
| 17-109_k141_177379 | Not-determined |
| 17-109_k141_260780 | Not-determined |
| 17-109_k141_228252 | Not-determined |
| 17-109_k141_44232 | Not-determined |
| 17-109_k141_194931 | Not-determined |
| 17-109_k141_60072 | Low-quality |
| 17-109_k141_260787 | Not-determined |
| 17-109_k141_278215 | Not-determined |
| 17-109_k141_244229 | Not-determined |
| 17-109_k141_328436 | Not-determined |
| 17-109_k141_260790 | Not-determined |
| 17-109_k141_278220 | Not-determined |
| 17-109_k141_328438 | Not-determined |
| 17-109_k141_260795 | Not-determined |
| 17-109_k141_194950 | Not-determined |
| 17-109_k141_344712 | Not-determined |
| 17-109_k141_60082 | Not-determined |
| 17-109_k141_126151 | Not-determined |
| 17-109_k141_9705 | Not-determined |
| 17-109_k141_126155 | Not-determined |
| 17-109_k141_93377 | Not-determined |
| 17-109_k141_60092 | Not-determined |
| 17-109_k141_60094 | Not-determined |
| 17-109_k141_60097 | Not-determined |
| 17-109_k141_60098 | Not-determined |
| 17-109_k141_278257 | Not-determined |
| 17-109_k141_210927 | Not-determined |
| 17-109_k141_44254 | Not-determined |
| 17-109_k141_77786 | Not-determined |
| 17-109_k141_60116 | Not-determined |
| 17-109_k141_44263 | Not-determined |
| 17-109_k141_161068 | Not-determined |
| 17-109_k141_395204 | Low-quality |
| 17-109_k141_44274 | Not-determined |
| 17-109_k141_244277 | Low-quality |
| 17-109_k141_278287 | Not-determined |
| 17-109_k141_278292 | Not-determined |
| 17-109_k141_378581 | Not-determined |
| 17-109_k141_77802 | Not-determined |
| 17-109_k141_395211 | Not-determined |
| 17-109_k141_328469 | Not-determined |
| 17-109_k141_9733 | Low-quality |
| 17-109_k141_395221 | Not-determined |
| 17-109_k141_244285 | Not-determined |
| 17-109_k141_9735 | Low-quality |
| 17-109_k141_378592 | Not-determined |
| 17-109_k141_44291 | Not-determined |
| 17-109_k141_260828 | Not-determined |
| 17-109_k141_44298 | Not-determined |
| 17-109_k141_161078 | Not-determined |
| 17-109_k141_44299 | Not-determined |
| 17-109_k141_328482 | Not-determined |
| 17-109_k141_60124 | Not-determined |
| 17-109_k141_161083 | Not-determined |
| 17-109_k141_328488 | Not-determined |
| 17-109_k141_244295 | Not-determined |
| 17-109_k141_344794 | Not-determined |
| 17-109_k141_161085 | Not-determined |
| 17-109_k141_77829 | Not-determined |
| 17-109_k141_278330 | Not-determined |
| 17-109_k141_9752 | Not-determined |
| 17-109_k141_344804 | Not-determined |
| 17-109_k141_44319 | Not-determined |
| 17-109_k141_278336 | Not-determined |
| 17-109_k141_344805 | Not-determined |
| 17-109_k141_60143 | Not-determined |
| 17-109_k141_328504 | Low-quality |
| 17-109_k141_9762 | Not-determined |
| 17-109_k141_378613 | Not-determined |
| 17-109_k141_77837 | Not-determined |
| 17-109_k141_44327 | Not-determined |
| 17-109_k141_77843 | Not-determined |
| 17-109_k141_328507 | Not-determined |
| 17-109_k141_60154 | Not-determined |
| 17-109_k141_378621 | Not-determined |
| 17-109_k141_260841 | Low-quality |
| 17-109_k141_378622 | Not-determined |
| 17-109_k141_143789 | Not-determined |
| 17-109_k141_210953 | Not-determined |
| 17-109_k141_143792 | Low-quality |
| 17-109_k141_210954 | Not-determined |
| 17-109_k141_60159 | Not-determined |
| 17-109_k141_126196 | Low-quality |
| 17-109_k141_344813 | Not-determined |
| 17-109_k141_143800 | Not-determined |
| 17-109_k141_44337 | Not-determined |
| 17-109_k141_378635 | Low-quality |
| 17-109_k141_344821 | Not-determined |
| 17-109_k141_9769 | Not-determined |
| 17-109_k141_194989 | Not-determined |
| 17-109_k141_110025 | Not-determined |
| 17-109_k141_395244 | Not-determined |
| 17-109_k141_93399 | Not-determined |
| 17-109_k141_77863 | Not-determined |
| 17-109_k141_60169 | Not-determined |
| 17-109_k141_194994 | Not-determined |
| 17-109_k141_328520 | Not-determined |
| 17-109_k141_194997 | Not-determined |
| 17-109_k141_93405 | Not-determined |
| 17-109_k141_27015 | Not-determined |
| 17-109_k141_195002 | Not-determined |
| 17-109_k141_344828 | Not-determined |
| 17-109_k141_93414 | Not-determined |
| 17-109_k141_195007 | Not-determined |
| 17-109_k141_60185 | Not-determined |
| 17-109_k141_110044 | Not-determined |
| 17-109_k141_143804 | Not-determined |
| 17-109_k141_260873 | Not-determined |
| 17-109_k141_60198 | Not-determined |
| 17-109_k141_228317 | Not-determined |
| 17-109_k141_210974 | Not-determined |
| 17-109_k141_228321 | Not-determined |
| 17-109_k141_395254 | Not-determined |
| 17-109_k141_9791 | Not-determined |
| 17-109_k141_44362 | Not-determined |
| 17-109_k141_260877 | Not-determined |
| 17-109_k141_195025 | Not-determined |
| 17-109_k141_378650 | Not-determined |
| 17-109_k141_228325 | Not-determined |
| 17-109_k141_126227 | Not-determined |
| 17-109_k141_44374 | Not-determined |
| 17-109_k141_228330 | Not-determined |
| 17-109_k141_312774 | Low-quality |
| 17-109_k141_378655 | Not-determined |
| 17-109_k141_9799 | Not-determined |
| 17-109_k141_93443 | Not-determined |
| 17-109_k141_195038 | Not-determined |
| 17-109_k141_378666 | Low-quality |
| 17-109_k141_143821 | Not-determined |
| 17-109_k141_60207 | Low-quality |
| 17-109_k141_278398 | Not-determined |
| 17-109_k141_244310 | Low-quality |
| 17-109_k141_228334 | Not-determined |
| 17-109_k141_9815 | Not-determined |
| 17-109_k141_126242 | Not-determined |
| 17-109_k141_244321 | Not-determined |
| 17-109_k141_27042 | Not-determined |
| 17-109_k141_60219 | Not-determined |
| 17-109_k141_27049 | Not-determined |
| 17-109_k141_27055 | Not-determined |
| 17-109_k141_77892 | Not-determined |
| 17-109_k141_278421 | Not-determined |
| 17-109_k141_27058 | Not-determined |
| 17-109_k141_211002 | Low-quality |
| 17-109_k141_27062 | Not-determined |
| 17-109_k141_395273 | Not-determined |
| 17-109_k141_260905 | Not-determined |
| 17-109_k141_244336 | Not-determined |
| 17-109_k141_77901 | Not-determined |
| 17-109_k141_278432 | Not-determined |
| 17-109_k141_143838 | Not-determined |
| 17-109_k141_77906 | Not-determined |
| 17-109_k141_60242 | Not-determined |
| 17-109_k141_260909 | Not-determined |
| 17-109_k141_395278 | Not-determined |
| 17-109_k141_395279 | Not-determined |
| 17-109_k141_395283 | Low-quality |
| 17-109_k141_244348 | Not-determined |
| 17-109_k141_211010 | Not-determined |
| 17-109_k141_395287 | Not-determined |
| 17-109_k141_110083 | Not-determined |
| 17-109_k141_60250 | Not-determined |
| 17-109_k141_143848 | Not-determined |
| 17-109_k141_110090 | Not-determined |
| 17-109_k141_378703 | Not-determined |
| 17-109_k141_161119 | Not-determined |
| 17-109_k141_195057 | Not-determined |
| 17-109_k141_260942 | Not-determined |
| 17-109_k141_161132 | Low-quality |
| 17-109_k141_9847 | Not-determined |
| 17-109_k141_126250 | Not-determined |
| 17-109_k141_195072 | Not-determined |
| 17-109_k141_228394 | Not-determined |
| 17-109_k141_395314 | Not-determined |
| 17-109_k141_228400 | Not-determined |
| 17-109_k141_161143 | Not-determined |
| 17-109_k141_228403 | Not-determined |
| 17-109_k141_9852 | Not-determined |
| 17-109_k141_211035 | Not-determined |
| 17-109_k141_228406 | Not-determined |
| 17-109_k141_195084 | Not-determined |
| 17-109_k141_44418 | Not-determined |
| 17-109_k141_278459 | Not-determined |
| 17-109_k141_110101 | Not-determined |
| 17-109_k141_161159 | Not-determined |
| 17-109_k141_278462 | Not-determined |
| 17-109_k141_244366 | Not-determined |
| 17-109_k141_177442 | Not-determined |
| 17-109_k141_228421 | Not-determined |
| 17-109_k141_9865 | Not-determined |
| 17-109_k141_195097 | Not-determined |
| 17-109_k141_126261 | Not-determined |
| 17-109_k141_110108 | Low-quality |
| 17-109_k141_395350 | Not-determined |
| 17-109_k141_126263 | Low-quality |
| 17-109_k141_9870 | Not-determined |
| 17-109_k141_177456 | Not-determined |
| 17-109_k141_195109 | Not-determined |
| 17-109_k141_60288 | Not-determined |
| 17-109_k141_228434 | Not-determined |
| 17-109_k141_126271 | Not-determined |
| 17-109_k141_195112 | Not-determined |
| 17-109_k141_395353 | Not-determined |
| 17-109_k141_378714 | Low-quality |
| 17-109_k141_395356 | Low-quality |
| 17-109_k141_278486 | Not-determined |
| 17-109_k141_143870 | Not-determined |
| 17-109_k141_395359 | Not-determined |
| 17-109_k141_344864 | Not-determined |
| 17-109_k141_93481 | Not-determined |
| 17-109_k141_211052 | Not-determined |
| 17-109_k141_143874 | Not-determined |
| 17-109_k141_278491 | Low-quality |
| 17-109_k141_244385 | Low-quality |
| 17-109_k141_77914 | Not-determined |
| 17-109_k141_143877 | Not-determined |
| 17-109_k141_344872 | Not-determined |
| 17-109_k141_60295 | Low-quality |
| 17-109_k141_110131 | Not-determined |
| 17-109_k141_344875 | Low-quality |
| 17-109_k141_295393 | Not-determined |
| 17-109_k141_378732 | Not-determined |
| 17-109_k141_44446 | Not-determined |
| 17-109_k141_328573 | Not-determined |
| 17-109_k141_44447 | Low-quality |
| 17-109_k141_344878 | Not-determined |
| 17-109_k141_211063 | Not-determined |
| 17-109_k141_278497 | Not-determined |
| 17-109_k141_228455 | Not-determined |
| 17-109_k141_93497 | Not-determined |
| 17-109_k141_93500 | Low-quality |
| 17-109_k141_77923 | Not-determined |
| 17-109_k141_93505 | Not-determined |
| 17-109_k141_378750 | Not-determined |
| 17-109_k141_278514 | Not-determined |
| 17-109_k141_378755 | Not-determined |
| 17-109_k141_312834 | Not-determined |
| 17-109_k141_60321 | Not-determined |
| 17-109_k141_143909 | Low-quality |
| 17-109_k141_143910 | Not-determined |
| 17-109_k141_211080 | Not-determined |
| 17-109_k141_60326 | Not-determined |
| 17-109_k141_312841 | Not-determined |
| 17-109_k141_60329 | Not-determined |
| 17-109_k141_177492 | Not-determined |
| 17-109_k141_195153 | Not-determined |
| 17-109_k141_295418 | Not-determined |
| 17-109_k141_228460 | Not-determined |
| 17-109_k141_44480 | Not-determined |
| 17-109_k141_143923 | Low-quality |
| 17-109_k141_126300 | Not-determined |
| 17-109_k141_177505 | Not-determined |
| 17-109_k141_395390 | Not-determined |
| 17-109_k141_77937 | Not-determined |
| 17-109_k141_77938 | Not-determined |
| 17-109_k141_126305 | Not-determined |
| 17-109_k141_110138 | Not-determined |
| 17-109_k141_211096 | Not-determined |
| 17-109_k141_77945 | Low-quality |
| 17-109_k141_143929 | Not-determined |
| 17-109_k141_228476 | Not-determined |
| 17-109_k141_77952 | Low-quality |
| 17-109_k141_312861 | Not-determined |
| 17-109_k141_312864 | Not-determined |
| 17-109_k141_295444 | Not-determined |
| 17-109_k141_177533 | Not-determined |
| 17-109_k141_295445 | Not-determined |
| 17-109_k141_93528 | Not-determined |
| 17-109_k141_27079 | Not-determined |
| 17-109_k141_44502 | Not-determined |
| 17-109_k141_110168 | Not-determined |
| 17-109_k141_143953 | Not-determined |
| 17-109_k141_195178 | Not-determined |
| 17-109_k141_244419 | Not-determined |
| 17-109_k141_143961 | Low-quality |
| 17-109_k141_9927 | Not-determined |
| 17-109_k141_93543 | Not-determined |
| 17-109_k141_312884 | Not-determined |
| 17-109_k141_312885 | Not-determined |
| 17-109_k141_77983 | Not-determined |
| 17-109_k141_177552 | Not-determined |
| 17-109_k141_312892 | Not-determined |
| 17-109_k141_195190 | Not-determined |
| 17-109_k141_126319 | Not-determined |
| 17-109_k141_44528 | Not-determined |
| 17-109_k141_278551 | Not-determined |
| 17-109_k141_344900 | Not-determined |
| 17-109_k141_260964 | Not-determined |
| 17-109_k141_177559 | Not-determined |
| 17-109_k141_244428 | Not-determined |
| 17-109_k141_177560 | Not-determined |
| 17-109_k141_260966 | Not-determined |
| 17-109_k141_228510 | Not-determined |
| 17-109_k141_244431 | Not-determined |
| 17-109_k141_295466 | Low-quality |
| 17-109_k141_278554 | Not-determined |
| 17-109_k141_228516 | Not-determined |
| 17-109_k141_344911 | Not-determined |
| 17-109_k141_211108 | Not-determined |
| 17-109_k141_177567 | Not-determined |
| 17-109_k141_126333 | Not-determined |
| 17-109_k141_244439 | Low-quality |
| 17-109_k141_244441 | Not-determined |
| 17-109_k141_60377 | Not-determined |
| 17-109_k141_378795 | Not-determined |
| 17-109_k141_44529 | Not-determined |
| 17-109_k141_44531 | Not-determined |
| 17-109_k141_27103 | Not-determined |
| 17-109_k141_27106 | Low-quality |
| 17-109_k141_260981 | Not-determined |
| 17-109_k141_60382 | Not-determined |
| 17-109_k141_44536 | Not-determined |
| 17-109_k141_177581 | Low-quality |
| 17-109_k141_378804 | Not-determined |
| 17-109_k141_278572 | Not-determined |
| 17-109_k141_126352 | Not-determined |
| 17-109_k141_344930 | Not-determined |
| 17-109_k141_228539 | Not-determined |
| 17-109_k141_110214 | Not-determined |
| 17-109_k141_177590 | Not-determined |
| 17-109_k141_244455 | Low-quality |
| 17-109_k141_110221 | Not-determined |
| 17-109_k141_260990 | Not-determined |
| 17-109_k141_211123 | Not-determined |

| | |
|---|---|
| 17-109_k141_143984 | Not-determined |
| 17-109_k141_126367 | Not-determined |
| 17-109_k141_110226 | Not-determined |
| 17-109_k141_143990 | Not-determined |
| 17-109_k141_228549 | Not-determined |
| 17-109_k141_211128 | Not-determined |
| 17-109_k141_344945 | Not-determined |
| 17-109_k141_161213 | Not-determined |
| 17-109_k141_195220 | Not-determined |
| 17-109_k141_344957 | Not-determined |
| 17-109_k141_60419 | Not-determined |
| 17-109_k141_195225 | Not-determined |
| 17-109_k141_278594 | Not-determined |
| 17-109_k141_211140 | Not-determined |
| 17-109_k141_27135 | Low-quality |
| 17-109_k141_195228 | Not-determined |
| 17-109_k141_211144 | Not-determined |
| 17-109_k141_360242 | Not-determined |
| 17-109_k141_244476 | Not-determined |
| 17-109_k141_278598 | Not-determined |
| 17-109_k141_60425 | Not-determined |
| 17-109_k141_360249 | Not-determined |
| 17-109_k141_60429 | Not-determined |
| 17-109_k141_27150 | Not-determined |
| 17-109_k141_144018 | Not-determined |
| 17-109_k141_228576 | Not-determined |
| 17-109_k141_378832 | Not-determined |
| 17-109_k141_177604 | Not-determined |
| 17-109_k141_211164 | Not-determined |
| 17-109_k141_244483 | Not-determined |
| 17-109_k141_211166 | Not-determined |
| 17-109_k141_228589 | Not-determined |
| 17-109_k141_261006 | Low-quality |
| 17-109_k141_244491 | Not-determined |
| 17-109_k141_211172 | Not-determined |
| 17-109_k141_278606 | Not-determined |
| 17-109_k141_144035 | Low-quality |
| 17-109_k141_211174 | Not-determined |
| 17-109_k141_228594 | Not-determined |
| 17-109_k141_60446 | Low-quality |
| 17-109_k141_211175 | Not-determined |
| 17-109_k141_395426 | Not-determined |
| 17-109_k141_144044 | Not-determined |
| 17-109_k141_312944 | Not-determined |
| 17-109_k141_378837 | Not-determined |
| 17-109_k141_211181 | Not-determined |
| 17-109_k141_144052 | Not-determined |
| 17-109_k141_278625 | Not-determined |
| 17-109_k141_395434 | Not-determined |
| 17-109_k141_60462 | Not-determined |
| 17-109_k141_110267 | Not-determined |
| 17-109_k141_126380 | Low-quality |
| 17-109_k141_244507 | Not-determined |
| 17-109_k141_244508 | Not-determined |
| 17-109_k141_177637 | Not-determined |
| 17-109_k141_195244 | Not-determined |
| 17-109_k141_93591 | Low-quality |
| 17-109_k141_144058 | Not-determined |
| 17-109_k141_244521 | Not-determined |
| 17-109_k141_295484 | Low-quality |
| 17-109_k141_144060 | Not-determined |
| 17-109_k141_195252 | Not-determined |
| 17-109_k141_244527 | Not-determined |
| 17-109_k141_60467 | Not-determined |
| 17-109_k141_378851 | Not-determined |
| 17-109_k141_110272 | Not-determined |
| 17-109_k141_378853 | Not-determined |
| 17-109_k141_295491 | Not-determined |
| 17-109_k141_211195 | Not-determined |
| 17-109_k141_27180 | Not-determined |
| 17-109_k141_295497 | Not-determined |
| 17-109_k141_228612 | Not-determined |
| 18-101-final-viral-combined_k141_263562\|\|full | Medium-quality |
| 17-109_k141_312967 | Not-determined |
| 17-109_k141_27187 | Not-determined |
| 17-109_k141_328585 | Not-determined |
| 17-109_k141_345005 | Not-determined |
| 17-109_k141_328586 | Not-determined |
| 17-109_k141_211205 | Not-determined |
| 17-109_k141_27190 | Not-determined |
| 17-109_k141_261041 | Not-determined |
| 17-109_k141_27192 | Not-determined |
| 17-109_k141_161246 | Not-determined |
| 17-109_k141_395459 | Not-determined |
| 17-109_k141_295514 | Not-determined |
| 17-109_k141_161248 | Not-determined |
| 17-109_k141_144078 | Not-determined |
| 17-109_k141_161251 | Not-determined |
| 17-109_k141_395467 | Not-determined |
| 17-109_k141_278655 | Low-quality |
| 17-109_k141_9981 | Not-determined |
| 17-109_k141_161258 | Not-determined |
| 17-109_k141_345013 | Not-determined |
| 17-109_k141_9989 | Not-determined |
| 17-109_k141_278665 | Not-determined |
| 17-109_k141_78015 | Not-determined |
| 17-109_k141_261075 | Not-determined |
| 17-109_k141_211236 | Not-determined |
| 17-109_k141_177668 | Not-determined |
| 17-109_k141_395479 | Not-determined |
| 17-109_k141_211237 | Low-quality |
| 17-109_k141_211241 | Not-determined |
| 17-109_k141_144103 | Not-determined |
| 17-109_k141_110320 | Not-determined |
| 17-109_k141_110322 | Not-determined |
| 17-109_k141_261088 | Not-determined |
| 17-109_k141_177676 | Not-determined |
| 17-109_k141_328621 | Not-determined |
| 17-109_k141_78031 | Not-determined |
| 17-109_k141_211252 | Not-determined |
| 17-109_k141_261098 | Not-determined |
| 17-109_k141_328623 | Not-determined |
| 17-109_k141_195292 | Not-determined |
| 17-109_k141_93605 | Not-determined |
| 17-109_k141_195296 | Not-determined |
| 17-109_k141_161287 | Not-determined |
| 17-109_k141_244579 | Not-determined |
| 17-109_k141_161293 | Not-determined |
| 17-109_k141_93613 | Not-determined |
| 17-109_k141_244581 | Not-determined |
| 17-109_k141_211258 | Not-determined |
| 17-109_k141_195307 | Not-determined |
| 17-109_k141_244590 | Low-quality |
| 17-109_k141_93629 | Not-determined |
| 17-109_k141_27209 | Not-determined |
| 17-109_k141_345053 | Low-quality |
| 17-109_k141_93634 | Low-quality |
| 17-109_k141_10018 | Not-determined |
| 17-109_k141_345057 | Not-determined |
| 17-109_k141_345058 | Not-determined |
| 17-109_k141_144122 | Not-determined |
| 17-109_k141_60472 | Not-determined |
| 17-109_k141_93637 | Not-determined |
| 17-109_k141_78057 | Not-determined |
| 17-109_k141_378866 | Low-quality |
| 17-109_k141_93641 | Not-determined |
| 17-109_k141_110339 | Not-determined |
| 17-109_k141_144128 | Not-determined |
| 17-109_k141_360315 | Not-determined |
| 17-109_k141_177704 | Not-determined |
| 17-109_k141_44587 | Not-determined |
| 17-109_k141_261128 | Not-determined |
| 17-109_k141_161316 | Not-determined |
| 17-109_k141_195325 | Not-determined |
| 17-109_k141_244620 | Not-determined |
| 17-109_k141_10033 | Low-quality |
| 17-109_k141_93646 | Not-determined |
| 17-109_k141_161321 | Not-determined |
| 17-109_k141_295556 | Not-determined |
| 17-109_k141_278713 | Not-determined |
| 17-109_k141_110344 | Low-quality |
| 17-109_k141_228657 | Not-determined |
| 17-109_k141_177723 | Not-determined |
| 17-109_k141_360323 | Not-determined |
| 17-109_k141_27222 | Low-quality |
| 17-109_k141_211295 | Not-determined |
| 17-109_k141_126410 | Low-quality |
| 17-109_k141_126411 | Not-determined |
| 17-109_k141_144144 | Not-determined |
| 17-109_k141_211297 | Not-determined |
| 17-109_k141_161330 | Not-determined |
| 17-109_k141_345085 | Low-quality |
| 17-109_k141_144148 | Not-determined |
| 17-109_k141_328679 | Low-quality |
| 17-109_k141_27230 | Not-determined |
| 17-109_k141_126416 | Not-determined |
| 17-109_k141_211308 | Not-determined |
| 17-109_k141_211310 | Not-determined |
| 17-109_k141_78096 | Not-determined |
| 17-109_k141_60495 | Not-determined |
| 17-109_k141_126420 | Not-determined |
| 17-109_k141_161345 | Not-determined |
| 17-109_k141_345097 | Not-determined |
| 17-109_k141_211315 | Not-determined |
| 17-109_k141_345101 | Not-determined |
| 17-109_k141_378879 | Not-determined |
| 17-109_k141_228668 | Not-determined |
| 17-109_k141_161353 | Not-determined |
| 17-109_k141_161355 | Not-determined |
| 17-109_k141_211322 | Not-determined |
| 17-109_k141_360346 | Not-determined |
| 17-109_k141_295582 | Not-determined |
| 17-109_k141_345109 | Not-determined |
| 17-109_k141_228673 | Not-determined |
| 17-109_k141_44598 | Not-determined |
| 17-109_k141_195336 | Not-determined |
| 17-109_k141_27247 | Not-determined |
| 17-109_k141_27253 | Not-determined |
| 17-109_k141_27254 | Not-determined |
| 17-109_k141_278741 | Not-determined |
| 17-109_k141_278743 | Not-determined |
| 17-109_k141_295590 | Not-determined |
| 17-109_k141_378890 | Not-determined |
| 17-109_k141_295591 | Not-determined |
| 17-109_k141_78106 | Not-determined |
| 17-109_k141_60510 | Not-determined |
| 17-109_k141_93684 | Not-determined |
| 17-109_k141_278748 | Not-determined |
| 17-109_k141_378896 | Not-determined |
| 17-109_k141_93685 | Low-quality |
| 17-109_k141_161364 | Not-determined |
| 17-109_k141_10049 | Not-determined |
| 17-109_k141_93692 | Not-determined |
| 17-109_k141_261169 | Not-determined |
| 17-109_k141_93694 | Not-determined |
| 17-109_k141_27271 | Not-determined |
| 17-109_k141_228687 | Not-determined |
| 17-109_k141_161368 | Not-determined |
| 17-109_k141_312975 | Not-determined |
| 17-109_k141_44603 | Not-determined |
| 17-109_k141_161370 | Not-determined |
| 17-109_k141_312977 | Not-determined |
| 17-109_k141_44605 | Not-determined |
| 17-109_k141_60534 | Not-determined |
| 17-109_k141_161374 | Not-determined |
| 17-109_k141_360368 | Not-determined |
| 17-109_k141_360369 | Not-determined |
| 17-109_k141_144198 | Low-quality |
| 17-109_k141_228689 | Low-quality |
| 17-109_k141_27285 | Not-determined |
| 17-109_k141_211356 | Not-determined |
| 17-109_k141_345129 | Not-determined |
| 17-109_k141_161380 | Not-determined |
| 17-109_k141_177775 | Not-determined |
| 17-109_k141_378926 | Not-determined |
| 17-109_k141_27290 | Not-determined |
| 17-109_k141_278771 | Not-determined |
| 17-109_k141_228694 | Not-determined |
| 17-109_k141_177779 | Not-determined |
| 17-109_k141_261183 | Low-quality |
| 17-109_k141_345137 | Not-determined |
| 17-109_k141_161385 | Not-determined |
| 17-109_k141_44622 | Low-quality |
| 17-109_k141_27295 | Not-determined |
| 17-109_k141_27297 | Not-determined |
| 17-109_k141_161389 | Not-determined |
| 17-109_k141_144211 | Not-determined |
| 17-109_k141_228701 | Not-determined |
| 17-109_k141_261192 | Not-determined |
| 17-109_k141_126479 | Not-determined |
| 17-109_k141_126482 | Not-determined |
| 17-109_k141_78139 | Low-quality |
| 17-109_k141_261201 | Not-determined |
| 17-109_k141_211360 | Not-determined |
| 17-109_k141_360391 | Not-determined |
| 17-109_k141_44633 | Not-determined |
| 17-109_k141_126495 | Low-quality |
| 17-109_k141_110367 | Not-determined |
| 17-109_k141_345152 | Not-determined |
| 17-109_k141_360398 | Not-determined |
| 17-109_k141_360404 | Not-determined |
| 17-109_k141_110373 | Low-quality |
| 17-109_k141_27309 | Not-determined |
| 17-109_k141_27314 | Not-determined |
| 17-109_k141_93709 | Not-determined |
| 17-109_k141_278805 | Not-determined |
| 17-109_k141_345160 | Not-determined |
| 17-109_k141_345162 | Low-quality |
| 17-109_k141_378957 | Not-determined |
| 17-109_k141_328733 | Not-determined |
| 17-109_k141_93716 | Not-determined |
| 17-109_k141_10074 | Low-quality |
| 17-109_k141_261208 | Not-determined |
| 17-109_k141_378965 | Not-determined |
| 17-109_k141_345173 | Not-determined |
| 17-109_k141_295614 | Not-determined |
| 17-109_k141_378969 | Not-determined |
| 17-109_k141_110385 | Not-determined |
| 17-109_k141_126509 | Not-determined |
| 17-109_k141_126511 | Not-determined |
| 17-109_k141_126513 | Not-determined |
| 17-109_k141_126515 | Not-determined |
| 17-109_k141_395503 | Not-determined |
| 17-109_k141_27339 | Not-determined |
| 17-109_k141_378977 | Not-determined |
| 17-109_k141_312990 | Not-determined |
| 17-109_k141_44664 | Not-determined |
| 17-109_k141_161413 | Not-determined |
| 17-109_k141_211365 | Not-determined |
| 17-109_k141_27345 | Not-determined |
| 17-109_k141_195352 | Not-determined |
| 17-109_k141_27349 | Not-determined |
| 17-109_k141_195359 | Low-quality |
| 17-109_k141_195363 | Not-determined |
| 17-109_k141_244651 | Not-determined |
| 17-109_k141_195368 | Not-determined |
| 17-109_k141_78177 | Not-determined |
| 17-109_k141_211375 | Not-determined |
| 17-109_k141_195372 | Not-determined |
| 17-109_k141_395516 | Low-quality |
| 17-109_k141_177793 | Not-determined |
| 17-109_k141_261229 | Not-determined |
| 17-109_k141_78181 | Not-determined |
| 17-109_k141_44682 | Not-determined |
| 17-109_k141_328751 | Not-determined |
| 17-109_k141_295625 | Not-determined |
| 17-109_k141_93732 | Not-determined |
| 17-109_k141_295626 | Not-determined |
| 17-109_k141_78187 | Not-determined |
| 17-109_k141_177798 | Not-determined |
| 17-109_k141_379006 | Not-determined |
| 17-109_k141_278844 | Not-determined |
| 17-109_k141_195384 | Not-determined |
| 17-109_k141_345186 | Not-determined |
| 17-109_k141_126542 | Not-determined |
| 17-109_k141_228756 | Not-determined |
| 17-109_k141_395526 | Low-quality |
| 17-109_k141_211391 | Not-determined |
| 17-109_k141_228711 | Not-determined |
| 17-109_k141_93735 | Low-quality |
| 17-109_k141_278854 | Not-determined |
| 17-109_k141_44692 | Not-determined |
| 17-109_k141_244682 | Not-determined |
| 17-109_k141_395533 | Not-determined |
| 17-109_k141_211403 | Low-quality |
| 17-109_k141_93743 | Not-determined |
| 17-109_k141_278856 | Not-determined |
| 17-109_k141_195402 | Not-determined |
| 17-109_k141_261250 | Not-determined |
| 17-109_k141_126557 | Not-determined |
| 17-109_k141_261252 | Not-determined |
| 17-109_k141_295640 | Not-determined |
| 17-109_k141_261254 | Not-determined |
| 17-109_k141_78212 | Low-quality |
| 17-109_k141_395536 | Not-determined |
| 17-109_k141_228728 | Not-determined |
| 17-109_k141_195407 | Not-determined |
| 17-109_k141_93753 | Low-quality |
| 17-109_k141_78220 | Not-determined |
| 17-109_k141_78221 | Not-determined |
| 17-109_k141_328773 | Not-determined |
| 17-109_k141_278868 | Not-determined |
| 17-109_k141_295645 | Not-determined |
| 17-109_k141_93756 | Not-determined |
| 17-109_k141_244695 | Not-determined |
| 17-109_k141_244696 | Not-determined |
| 17-109_k141_10096 | Not-determined |
| 17-109_k141_360456 | Not-determined |
| 17-109_k141_211420 | Not-determined |
| 17-109_k141_126571 | Not-determined |
| 17-109_k141_360459 | Not-determined |
| 17-109_k141_244702 | Low-quality |
| 17-109_k141_278877 | Not-determined |
| 17-109_k141_278880 | Not-determined |
| 17-109_k141_27367 | Not-determined |
| 17-109_k141_395548 | Not-determined |
| 17-109_k141_195421 | Not-determined |
| 17-109_k141_195422 | Not-determined |
| 17-109_k141_27371 | Not-determined |
| 17-109_k141_78246 | Not-determined |
| 17-109_k141_244712 | Low-quality |
| 17-109_k141_395562 | Not-determined |
| 17-109_k141_395565 | Not-determined |
| 17-109_k141_211437 | Not-determined |
| 17-109_k141_10117 | Not-determined |
| 17-109_k141_395569 | Not-determined |
| 17-109_k141_395571 | Not-determined |
| 17-109_k141_44707 | Not-determined |
| 17-109_k141_161466 | Not-determined |
| 17-109_k141_261285 | Not-determined |
| 17-109_k141_278900 | Not-determined |
| 17-109_k141_78259 | Not-determined |
| 17-109_k141_211445 | Not-determined |
| 17-109_k141_93776 | Not-determined |
| 17-109_k141_278902 | Not-determined |
| 17-109_k141_360489 | Not-determined |
| 17-109_k141_379035 | Not-determined |
| 17-109_k141_211450 | Not-determined |
| 17-109_k141_360493 | Not-determined |
| 17-109_k141_211454 | Not-determined |
| 17-109_k141_177819 | Low-quality |
| 17-109_k141_278911 | Not-determined |
| 17-109_k141_328792 | Not-determined |
| 17-109_k141_328793 | Not-determined |
| 17-109_k141_211459 | Not-determined |
| 17-109_k141_211462 | Not-determined |
| 17-109_k141_93786 | Not-determined |
| 17-109_k141_379046 | Not-determined |
| 17-109_k141_110441 | Not-determined |
| 17-109_k141_244731 | Not-determined |
| 17-109_k141_313044 | Not-determined |
| 17-109_k141_278918 | Not-determined |
| 17-109_k141_110445 | Low-quality |
| 17-109_k141_10125 | Not-determined |
| 17-109_k141_379059 | Not-determined |
| 17-109_k141_244739 | Not-determined |
| 17-109_k141_244742 | Not-determined |
| 17-109_k141_211466 | Not-determined |
| 17-109_k141_295671 | Not-determined |
| 17-109_k141_360510 | Not-determined |
| 17-109_k141_228747 | Not-determined |
| 17-109_k141_78288 | Low-quality |
| 17-109_k141_211475 | Not-determined |
| 17-109_k141_110452 | Not-determined |
| 17-109_k141_228749 | Not-determined |
| 17-109_k141_278921 | Low-quality |
| 17-109_k141_278928 | Not-determined |
| 17-109_k141_27422 | Not-determined |
| 17-109_k141_110460 | Not-determined |
| 17-109_k141_27425 | Not-determined |
| 17-109_k141_278931 | Not-determined |
| 17-109_k141_379078 | Not-determined |
| 17-109_k141_10135 | Not-determined |
| 17-109_k141_278936 | Not-determined |
| 17-109_k141_60573 | Not-determined |
| 17-109_k141_360523 | Not-determined |
| 17-109_k141_126591 | Not-determined |
| 17-109_k141_27433 | Not-determined |
| 17-109_k141_379088 | Not-determined |
| 17-109_k141_379098 | Not-determined |
| 17-109_k141_278943 | Not-determined |
| 17-109_k141_379104 | Not-determined |
| 17-109_k141_78296 | Not-determined |
| 17-109_k141_379109 | Low-quality |
| 17-109_k141_228768 | Not-determined |
| 17-109_k141_78299 | Not-determined |
| 17-109_k141_228770 | Low-quality |
| 17-109_k141_395586 | Not-determined |
| 17-109_k141_228772 | Not-determined |
| 17-109_k141_60588 | Not-determined |
| 17-109_k141_78304 | Not-determined |
| 17-109_k141_60590 | Not-determined |
| 17-109_k141_144231 | Not-determined |
| 17-109_k141_360534 | Not-determined |
| 17-109_k141_27456 | Not-determined |
| 17-109_k141_126600 | Not-determined |
| 17-109_k141_126602 | Not-determined |
| 17-109_k141_27459 | Not-determined |
| 17-109_k141_27463 | Low-quality |
| 17-109_k141_278952 | Low-quality |
| 17-109_k141_244770 | Not-determined |
| 17-109_k141_78308 | Not-determined |
| 17-109_k141_78310 | Not-determined |
| 17-109_k141_244773 | Not-determined |
| 17-109_k141_27470 | Not-determined |
| 17-109_k141_10146 | Not-determined |
| 17-109_k141_10153 | Not-determined |
| 17-109_k141_228798 | Not-determined |
| 17-109_k141_228800 | Not-determined |
| 17-109_k141_211509 | Low-quality |
| 17-109_k141_313093 | Not-determined |
| 17-109_k141_211510 | Low-quality |
| 17-109_k141_177855 | Not-determined |
| 17-109_k141_211513 | Not-determined |
| 17-109_k141_44765 | Not-determined |
| 17-109_k141_278957 | Not-determined |
| 17-109_k141_144257 | Low-quality |
| 17-109_k141_27485 | Not-determined |
| 17-109_k141_313096 | Not-determined |
| 17-109_k141_313098 | Not-determined |
| 17-109_k141_44768 | Low-quality |
| 17-109_k141_27487 | Not-determined |
| 17-109_k141_395604 | Not-determined |
| 17-109_k141_328841 | Low-quality |
| 17-109_k141_27490 | Not-determined |
| 17-109_k141_144262 | Not-determined |
| 17-109_k141_261335 | Not-determined |
| 17-109_k141_10167 | Not-determined |
| 17-109_k141_360549 | Not-determined |
| 17-109_k141_328849 | Not-determined |
| 17-109_k141_360550 | Not-determined |
| 17-109_k141_93806 | Not-determined |
| 17-109_k141_228818 | Not-determined |
| 17-109_k141_144277 | Low-quality |
| 17-109_k141_395617 | Not-determined |
| 17-109_k141_379141 | Not-determined |
| 17-109_k141_261353 | Not-determined |
| 17-109_k141_395621 | Not-determined |
| 17-109_k141_360556 | Not-determined |
| 17-109_k141_360558 | Not-determined |
| 17-109_k141_345284 | Not-determined |
| 17-109_k141_10178 | Not-determined |
| 17-109_k141_110472 | Not-determined |
| 17-109_k141_360561 | Not-determined |
| 17-109_k141_328867 | Not-determined |
| 17-109_k141_10181 | Not-determined |
| 17-109_k141_360564 | Not-determined |
| 17-109_k141_313123 | Not-determined |
| 17-109_k141_211540 | Low-quality |
| 17-109_k141_126629 | Not-determined |
| 17-109_k141_313124 | Not-determined |
| 17-109_k141_110487 | Not-determined |
| 17-109_k141_228843 | Not-determined |
| 17-109_k141_379161 | Not-determined |
| 17-109_k141_211546 | Not-determined |
| 17-109_k141_328881 | Not-determined |
| 17-109_k141_360566 | Not-determined |
| 17-109_k141_345292 | Not-determined |
| 17-109_k141_110495 | Not-determined |
| 17-109_k141_395638 | Not-determined |
| 17-109_k141_395640 | Not-determined |
| 17-109_k141_328895 | Not-determined |
| 17-109_k141_279002 | Not-determined |
| 17-109_k141_144305 | Not-determined |
| 17-109_k141_78352 | Not-determined |
| 17-109_k141_144308 | Not-determined |
| 17-109_k141_395653 | Not-determined |
| 17-109_k141_93825 | Not-determined |
| 17-109_k141_295749 | Not-determined |
| 17-109_k141_177914 | Not-determined |
| 17-109_k141_177915 | Not-determined |
| 17-109_k141_126654 | Not-determined |
| 17-109_k141_10192 | Not-determined |
| 17-109_k141_93833 | Low-quality |
| 17-109_k141_27523 | Not-determined |
| 17-109_k141_261359 | Low-quality |
| 17-109_k141_10200 | Low-quality |
| 17-109_k141_78361 | Not-determined |
| 17-109_k141_27534 | Not-determined |
| 17-109_k141_244818 | Not-determined |
| 17-109_k141_211558 | Not-determined |
| 17-109_k141_211561 | Not-determined |
| 17-109_k141_379192 | Low-quality |
| 17-109_k141_379194 | Not-determined |
| 17-109_k141_177927 | Not-determined |
| 17-109_k141_177932 | Not-determined |
| 17-109_k141_261369 | Not-determined |
| 17-109_k141_360579 | Low-quality |
| 17-109_k141_78383 | Not-determined |
| 17-109_k141_144341 | Not-determined |
| 17-109_k141_161552 | Not-determined |
| 17-109_k141_10205 | Not-determined |
| 17-109_k141_360585 | Low-quality |
| 17-109_k141_295789 | Not-determined |
| 17-109_k141_228849 | Low-quality |
| 17-109_k141_228850 | Not-determined |
| 17-109_k141_126667 | Not-determined |
| 17-109_k141_244838 | Not-determined |
| 17-109_k141_328900 | Not-determined |
| 17-109_k141_211588 | Not-determined |
| 17-109_k141_244844 | Not-determined |
| 17-109_k141_126674 | Not-determined |
| 17-109_k141_345327 | Not-determined |
| 17-109_k141_126679 | Not-determined |
| 17-109_k141_211596 | Not-determined |
| 17-109_k141_244853 | Not-determined |
| 17-109_k141_328910 | Not-determined |
| 17-109_k141_161565 | Not-determined |
| 17-109_k141_78410 | Not-determined |
| 17-109_k141_126688 | Not-determined |
| 17-109_k141_44793 | Not-determined |
| 17-109_k141_328915 | Not-determined |
| 17-109_k141_395686 | Not-determined |
| 17-109_k141_328916 | Not-determined |
| 17-109_k141_279043 | Low-quality |
| 17-109_k141_44797 | Not-determined |
| 17-109_k141_27562 | Low-quality |
| 17-109_k141_279046 | Not-determined |
| 17-109_k141_328923 | Not-determined |
| 17-109_k141_313175 | Not-determined |
| 17-109_k141_78415 | Not-determined |
| 17-109_k141_345359 | Not-determined |
| 17-109_k141_244873 | Not-determined |
| 17-109_k141_244874 | Low-quality |
| 17-109_k141_10210 | Not-determined |
| 17-109_k141_379220 | Not-determined |
| 17-109_k141_279067 | Not-determined |
| 17-109_k141_379225 | Not-determined |
| 17-109_k141_244883 | Not-determined |
| 17-109_k141_10216 | Not-determined |
| 17-109_k141_379227 | Not-determined |
| 17-109_k141_360605 | Not-determined |
| 17-109_k141_345374 | Not-determined |
| 17-109_k141_360610 | Not-determined |
| 17-109_k141_78422 | Not-determined |
| 17-109_k141_360614 | Not-determined |
| 17-109_k141_144366 | Not-determined |
| 17-109_k141_78426 | Not-determined |
| 17-109_k141_228901 | Not-determined |
| 17-109_k141_93898 | Not-determined |
| 17-109_k141_228905 | Not-determined |
| 17-109_k141_328943 | Not-determined |
| 17-109_k141_93908 | Not-determined |
| 17-109_k141_78435 | Not-determined |
| 17-109_k141_10217 | Not-determined |
| 17-109_k141_211636 | Not-determined |
| 17-109_k141_10220 | Not-determined |
| 17-109_k141_27610 | Not-determined |
| 17-109_k141_78446 | Low-quality |
| 17-109_k141_10223 | Not-determined |
| 17-109_k141_244911 | Not-determined |
| 17-109_k141_279092 | Not-determined |
| 17-109_k141_345378 | Not-determined |
| 17-109_k141_228925 | Not-determined |
| 17-109_k141_279096 | Not-determined |
| 17-109_k141_244917 | Not-determined |
| 17-109_k141_345382 | Not-determined |
| 17-109_k141_78448 | Not-determined |
| 17-109_k141_279100 | Low-quality |

| | |
|---|---|
| 17-109_k141_211640 | Not-determined |
| 17-109_k141_395698 | Not-determined |
| 17-109_k141_161577 | Not-determined |
| 17-109_k141_211647 | Not-determined |
| 17-109_k141_395701 | Not-determined |
| 17-109_k141_228938 | Not-determined |
| 17-109_k141_379257 | Not-determined |
| 17-109_k141_228939 | Not-determined |
| 17-109_k141_379261 | Not-determined |
| 17-109_k141_110526 | Low-quality |
| 17-109_k141_360647 | Not-determined |
| 17-109_k141_261429 | Not-determined |
| 17-109_k141_360649 | Not-determined |
| 17-109_k141_10236 | Not-determined |
| 17-109_k141_60637 | Not-determined |
| 17-109_k141_10241 | Not-determined |
| 17-109_k141_110537 | Not-determined |
| 17-109_k141_328979 | Not-determined |
| 18-101_k141_185046 | Medium-quality |
| 17-109_k141_78457 | Not-determined |
| 17-109_k141_211665 | Not-determined |
| 17-109_k141_27639 | Low-quality |
| 17-109_k141_110543 | Not-determined |
| 17-109_k141_379283 | Not-determined |
| 17-109_k141_211667 | Not-determined |
| 17-109_k141_44822 | Low-quality |
| 17-109_k141_27641 | Not-determined |
| 17-109_k141_44823 | Not-determined |
| 17-109_k141_110547 | Low-quality |
| 17-109_k141_379287 | Not-determined |
| 17-109_k141_60648 | Not-determined |
| 17-109_k141_93954 | Not-determined |
| 17-109_k141_328989 | Not-determined |
| 17-109_k141_60650 | Not-determined |
| 17-109_k141_345413 | Not-determined |
| 17-109_k141_110558 | Not-determined |
| 17-109_k141_110560 | Not-determined |
| 17-109_k141_10250 | Not-determined |
| 17-109_k141_244952 | Not-determined |
| 17-109_k141_395732 | Not-determined |
| 17-109_k141_379305 | Not-determined |
| 17-109_k141_44837 | Not-determined |
| 17-109_k141_93959 | Not-determined |
| 17-109_k141_379310 | Not-determined |
| 17-109_k141_60669 | Not-determined |
| 17-109_k141_345427 | Not-determined |
| 17-109_k141_60675 | Not-determined |
| 17-109_k141_93966 | Not-determined |
| 17-109_k141_60678 | Not-determined |
| 17-109_k141_228957 | Low-quality |
| 17-109_k141_211683 | Not-determined |
| 17-109_k141_345435 | Not-determined |
| 17-109_k141_345439 | Not-determined |
| 17-109_k141_93978 | Low-quality |
| 17-109_k141_379331 | Not-determined |
| 17-109_k141_211686 | Not-determined |
| 17-109_k141_78492 | Not-determined |
| 17-109_k141_345442 | Low-quality |
| 17-109_k141_60683 | Not-determined |
| 17-109_k141_279122 | Not-determined |
| 17-109_k141_228965 | Not-determined |
| 17-109_k141_329026 | Not-determined |
| 17-109_k141_60684 | Low-quality |
| 17-109_k141_60685 | Not-determined |
| 17-109_k141_78500 | Not-determined |
| 17-109_k141_228969 | Not-determined |
| 17-109_k141_10274 | Not-determined |
| 17-109_k141_379337 | Not-determined |
| 17-109_k141_244983 | Not-determined |
| 17-109_k141_360659 | Not-determined |
| 17-109_k141_345465 | Not-determined |
| 17-109_k141_110574 | Not-determined |
| 17-109_k141_110575 | Not-determined |
| 17-109_k141_211697 | Low-quality |
| 17-109_k141_279136 | Not-determined |
| 17-109_k141_211699 | Not-determined |
| 17-109_k141_228978 | Not-determined |
| 17-109_k141_94004 | Not-determined |
| 17-109_k141_295810 | Not-determined |
| 17-109_k141_60705 | Low-quality |
| 17-109_k141_228980 | Not-determined |
| 17-109_k141_27670 | Not-determined |
| 17-109_k141_228982 | Not-determined |
| 17-109_k141_228986 | Not-determined |
| 17-109_k141_161584 | Not-determined |
| 17-109_k141_10285 | Not-determined |
| 17-109_k141_313254 | Low-quality |
| 17-109_k141_279146 | Not-determined |
| 17-109_k141_161587 | Not-determined |
| 17-109_k141_244997 | Low-quality |
| 17-109_k141_228993 | Not-determined |
| 17-109_k141_345469 | Not-determined |
| 17-109_k141_295817 | Not-determined |
| 17-109_k141_245007 | Not-determined |
| 17-109_k141_245008 | Not-determined |
| 17-109_k141_313257 | Not-determined |
| 17-109_k141_60723 | Not-determined |
| 17-109_k141_295827 | Not-determined |
| 17-109_k141_211724 | Not-determined |
| 17-109_k141_313266 | Not-determined |
| 17-109_k141_126764 | Not-determined |
| 17-109_k141_395767 | Not-determined |
| 17-109_k141_126766 | Not-determined |
| 17-109_k141_245015 | Not-determined |
| 17-109_k141_10301 | Not-determined |
| 17-109_k141_161601 | Not-determined |
| 17-109_k141_245018 | Not-determined |
| 17-109_k141_261445 | Not-determined |
| 17-109_k141_379353 | Not-determined |
| 17-109_k141_126776 | Low-quality |
| 17-109_k141_229021 | Not-determined |
| 17-109_k141_345496 | Not-determined |
| 17-109_k141_126788 | Not-determined |
| 17-109_k141_395776 | Not-determined |
| 17-109_k141_110607 | Not-determined |
| 17-109_k141_345505 | Not-determined |
| 17-109_k141_78526 | Not-determined |
| 17-109_k141_44906 | Not-determined |
| 17-109_k141_94044 | Not-determined |
| 17-109_k141_44909 | Low-quality |
| 17-109_k141_44910 | Not-determined |
| 17-109_k141_360682 | Not-determined |
| 17-109_k141_78530 | Not-determined |
| 17-109_k141_261471 | Not-determined |
| 17-109_k141_94056 | Not-determined |
| 17-109_k141_313276 | Not-determined |
| 17-109_k141_44916 | Not-determined |
| 17-109_k141_144383 | Not-determined |
| 17-109_k141_379382 | Not-determined |
| 17-109_k141_78533 | Not-determined |
| 17-109_k141_10318 | Low-quality |
| 17-109_k141_379385 | Not-determined |
| 17-109_k141_313283 | Low-quality |
| 17-109_k141_94063 | Low-quality |
| 17-109_k141_126811 | Not-determined |
| 17-109_k141_211730 | Not-determined |
| 17-109_k141_360685 | Not-determined |
| 17-109_k141_211733 | Not-determined |
| 17-109_k141_279201 | Not-determined |
| 17-109_k141_313292 | Not-determined |
| 17-109_k141_313293 | Not-determined |
| 17-109_k141_211735 | Not-determined |
| 17-109_k141_161625 | Not-determined |
| 17-109_k141_313300 | Not-determined |
| 17-109_k141_360695 | Not-determined |
| 17-109_k141_261492 | Not-determined |
| 17-109_k141_211746 | Not-determined |
| 17-109_k141_313308 | Not-determined |
| 17-109_k141_110614 | Not-determined |
| 17-109_k141_161629 | Not-determined |
| 17-109_k141_161630 | Low-quality |
| 17-109_k141_161634 | Not-determined |
| 17-109_k141_360702 | Not-determined |
| 17-109_k141_60735 | Not-determined |
| 17-109_k141_295879 | Low-quality |
| 17-109_k141_60736 | Not-determined |
| 17-109_k141_211761 | Not-determined |
| 17-109_k141_161638 | Not-determined |
| 17-109_k141_345516 | Not-determined |
| 17-109_k141_78556 | Not-determined |
| 17-109_k141_329113 | Not-determined |
| 17-109_k141_261517 | Not-determined |
| 17-109_k141_60755 | Not-determined |
| 17-109_k141_261520 | Not-determined |
| 17-109_k141_60758 | Not-determined |
| 17-109_k141_245052 | Not-determined |
| 17-109_k141_60763 | Not-determined |
| 17-109_k141_177969 | Low-quality |
| 17-109_k141_211771 | Not-determined |
| 17-109_k141_360723 | Not-determined |
| 17-109_k141_177972 | Not-determined |
| 17-109_k141_211775 | Not-determined |
| 17-109_k141_177980 | Not-determined |
| 17-109_k141_229101 | Not-determined |
| 17-109_k141_360727 | Not-determined |
| 17-109_k141_211781 | Low-quality |
| 17-109_k141_329134 | Not-determined |
| 17-109_k141_211782 | Not-determined |
| 17-109_k141_195480 | Not-determined |
| 17-109_k141_229105 | Not-determined |
| 17-109_k141_177988 | Not-determined |
| 17-109_k141_60770 | Not-determined |
| 17-109_k141_229110 | Not-determined |
| 17-109_k141_195490 | Not-determined |
| 17-109_k141_329140 | Not-determined |
| 17-109_k141_229116 | Not-determined |
| 17-109_k141_126862 | Not-determined |
| 17-109_k141_126863 | Not-determined |
| 17-109_k141_78580 | Not-determined |
| 17-109_k141_44966 | Not-determined |
| 17-109_k141_313324 | Not-determined |
| 17-109_k141_10361 | Not-determined |
| 17-109_k141_60779 | Not-determined |
| 17-109_k141_161646 | Not-determined |
| 17-109_k141_126872 | Not-determined |
| 17-109_k141_195501 | Not-determined |
| 17-109_k141_379445 | Not-determined |
| 17-109_k141_379447 | Not-determined |
| 17-109_k141_395827 | Not-determined |
| 17-109_k141_94087 | Not-determined |
| 17-109_k141_360752 | Not-determined |
| 17-109_k141_10367 | Not-determined |
| 17-109_k141_60799 | Not-determined |
| 17-109_k141_10372 | Not-determined |
| 17-109_k141_379454 | Not-determined |
| 17-109_k141_177993 | Not-determined |
| 17-109_k141_10379 | Not-determined |
| 17-109_k141_10381 | Not-determined |
| 17-109_k141_345548 | Not-determined |
| 17-109_k141_44982 | Not-determined |
| 17-109_k141_126898 | Not-determined |
| 17-109_k141_126899 | Not-determined |
| 17-109_k141_44983 | Not-determined |
| 17-109_k141_395840 | Not-determined |
| 17-109_k141_245067 | Low-quality |
| 17-109_k141_178009 | Not-determined |
| 17-109_k141_211808 | Not-determined |
| 17-109_k141_44988 | Not-determined |
| 17-109_k141_360758 | Not-determined |
| 17-109_k141_195519 | Not-determined |
| 17-109_k141_379469 | Not-determined |
| 17-109_k141_379471 | Not-determined |
| 17-109_k141_261579 | Low-quality |
| 17-109_k141_245072 | Not-determined |
| 17-109_k141_195526 | Low-quality |
| 17-109_k141_395854 | Not-determined |
| 17-109_k141_195530 | Not-determined |
| 17-109_k141_229159 | Not-determined |
| 17-109_k141_279254 | Not-determined |
| 17-109_k141_279256 | Not-determined |
| 17-109_k141_229164 | Not-determined |
| 17-109_k141_27740 | Not-determined |
| 17-109_k141_295906 | Not-determined |
| 17-109_k141_27742 | Not-determined |
| 17-109_k141_313348 | Not-determined |
| 17-109_k141_195537 | Not-determined |
| 17-109_k141_379487 | Not-determined |
| 17-109_k141_94115 | Low-quality |
| 17-109_k141_279257 | Low-quality |
| 17-109_k141_379489 | Low-quality |
| 17-109_k141_313354 | Not-determined |
| 17-109_k141_211833 | Not-determined |
| 17-109_k141_295915 | Not-determined |
| 17-109_k141_126920 | Not-determined |
| 17-109_k141_110685 | Not-determined |
| 17-109_k141_379501 | Not-determined |
| 17-109_k141_395863 | Not-determined |
| 17-109_k141_379504 | Not-determined |
| 17-109_k141_229185 | Not-determined |
| 17-109_k141_395873 | Not-determined |
| 17-109_k141_110692 | Not-determined |
| 17-109_k141_395874 | Not-determined |
| 17-109_k141_94138 | Not-determined |
| 17-109_k141_94139 | Not-determined |
| 17-109_k141_295931 | Not-determined |
| 17-109_k141_379506 | Not-determined |
| 17-109_k141_94141 | Not-determined |
| 17-109_k141_195552 | Not-determined |
| 17-109_k141_261600 | Not-determined |
| 17-109_k141_229192 | Not-determined |
| 17-109_k141_195556 | Not-determined |
| 17-109_k141_94147 | Not-determined |
| 17-109_k141_261604 | Not-determined |
| 17-109_k141_78605 | Not-determined |
| 17-109_k141_195562 | Not-determined |
| 17-109_k141_110704 | Not-determined |
| 17-109_k141_245080 | Not-determined |
| 17-109_k141_195563 | Not-determined |
| 17-109_k141_78609 | Not-determined |
| 17-109_k141_60831 | Not-determined |
| 17-109_k141_78611 | Not-determined |
| 17-109_k141_195568 | Not-determined |
| 17-109_k141_245086 | Not-determined |
| 17-109_k141_78618 | Not-determined |
| 17-109_k141_178047 | Not-determined |
| 17-109_k141_295938 | Not-determined |
| 17-109_k141_60836 | Not-determined |
| 17-109_k141_78621 | Not-determined |
| 17-109_k141_45015 | Not-determined |
| 17-109_k141_45018 | Not-determined |
| 17-109_k141_110725 | Not-determined |
| 17-109_k141_279281 | Not-determined |
| 17-109_k141_27792 | Low-quality |
| 17-109_k141_110727 | Not-determined |
| 17-109_k141_261623 | Not-determined |
| 17-109_k141_395905 | Not-determined |
| 17-109_k141_10416 | Not-determined |
| 17-109_k141_94167 | Not-determined |
| 17-109_k141_110732 | Not-determined |
| 17-109_k141_45025 | Not-determined |
| 17-109_k141_229215 | Not-determined |
| 17-109_k141_295959 | Not-determined |
| 17-109_k141_379532 | Not-determined |
| 17-109_k141_261630 | Not-determined |
| 17-109_k141_360785 | Not-determined |
| 17-109_k141_360786 | Not-determined |
| 17-109_k141_45033 | Not-determined |
| 17-109_k141_379545 | Low-quality |
| 17-109_k141_195586 | Not-determined |
| 17-109_k141_10426 | Not-determined |
| 17-109_k141_313384 | Not-determined |
| 17-109_k141_245102 | Low-quality |
| 17-109_k141_313386 | Not-determined |
| 17-109_k141_195589 | Not-determined |
| 17-109_k141_45043 | Low-quality |
| 17-109_k141_261650 | Not-determined |
| 17-109_k141_279300 | Not-determined |
| 17-109_k141_110754 | Not-determined |
| 17-109_k141_126947 | Not-determined |
| 17-109_k141_329197 | Not-determined |
| 17-109_k141_45047 | Not-determined |
| 17-109_k141_94186 | Low-quality |
| 17-109_k141_360794 | Not-determined |
| 17-109_k141_261657 | Not-determined |
| 17-109_k141_27835 | Not-determined |
| 17-109_k141_229251 | Not-determined |
| 17-109_k141_178060 | Not-determined |
| 17-109_k141_110760 | Not-determined |
| 17-109_k141_379568 | Not-determined |
| 17-109_k141_211849 | Not-determined |
| 17-109_k141_395940 | Not-determined |
| 17-109_k141_27844 | Not-determined |
| 17-109_k141_229272 | Low-quality |
| 17-109_k141_360805 | Not-determined |
| 17-109_k141_360806 | Not-determined |
| 17-109_k141_360808 | Not-determined |
| 17-109_k141_229277 | Not-determined |
| 17-109_k141_10469 | Not-determined |
| 17-109_k141_45064 | Not-determined |
| 17-109_k141_245120 | Not-determined |
| 17-109_k141_211857 | Low-quality |
| 17-109_k141_161674 | Not-determined |
| 17-109_k141_10471 | Not-determined |
| 17-109_k141_10475 | Not-determined |
| 17-109_k141_161676 | Not-determined |
| 17-109_k141_27856 | Not-determined |
| 17-109_k141_395968 | Not-determined |
| 17-109_k141_229287 | Not-determined |
| 17-109_k141_229289 | Not-determined |
| 17-109_k141_379583 | Not-determined |
| 17-109_k141_245125 | Not-determined |
| 17-109_k141_161678 | Low-quality |
| 17-109_k141_395983 | Not-determined |
| 17-109_k141_45078 | Not-determined |
| 17-109_k141_161687 | Not-determined |
| 17-109_k141_360848 | Not-determined |
| 17-109_k141_261707 | Not-determined |
| 17-109_k141_379594 | Low-quality |
| 17-109_k141_45085 | Not-determined |
| 17-109_k141_195653 | Not-determined |
| 17-109_k141_144393 | Not-determined |
| 17-109_k141_261711 | Not-determined |
| 17-109_k141_144394 | Not-determined |
| 17-109_k141_379599 | Not-determined |
| 17-109_k141_10489 | Not-determined |
| 17-109_k141_261718 | Not-determined |
| 17-109_k141_45088 | Not-determined |
| 17-109_k141_195654 | Low-quality |
| 17-109_k141_45090 | Not-determined |
| 17-109_k141_360863 | Not-determined |
| 17-109_k141_261723 | Not-determined |
| 17-109_k141_261725 | Not-determined |
| 17-109_k141_144403 | Not-determined |
| 17-109_k141_60851 | Not-determined |
| 17-109_k141_45094 | Not-determined |
| 17-109_k141_161702 | Not-determined |
| 17-109_k141_94208 | Not-determined |
| 17-109_k141_45095 | Not-determined |
| 17-109_k141_279335 | Not-determined |
| 17-109_k141_211875 | Not-determined |
| 17-109_k141_10497 | Not-determined |
| 17-109_k141_27878 | Low-quality |
| 17-109_k141_94214 | Not-determined |
| 17-109_k141_161710 | Not-determined |
| 17-109_k141_60868 | Not-determined |
| 17-109_k141_329227 | Not-determined |
| 17-109_k141_229294 | Not-determined |
| 17-109_k141_78654 | Not-determined |
| 17-109_k141_279343 | Low-quality |
| 17-109_k141_245166 | Not-determined |
| 17-109_k141_379628 | Not-determined |
| 17-109_k141_195667 | Not-determined |
| 17-109_k141_279350 | Not-determined |
| 17-109_k141_60882 | Not-determined |
| 17-109_k141_313414 | Low-quality |
| 17-109_k141_261739 | Not-determined |
| 17-109_k141_295983 | Not-determined |
| 17-109_k141_313418 | Not-determined |
| 17-109_k141_261742 | Not-determined |
| 17-109_k141_45125 | Not-determined |
| 17-109_k141_295989 | Not-determined |
| 17-109_k141_245175 | Not-determined |
| 17-109_k141_10504 | Not-determined |
| 17-109_k141_144425 | Low-quality |
| 17-109_k141_245177 | Not-determined |
| 17-109_k141_195679 | Not-determined |
| 17-109_k141_245178 | Not-determined |
| 17-109_k141_78661 | Not-determined |
| 17-109_k141_60887 | Not-determined |
| 17-109_k141_144427 | Not-determined |
| 17-109_k141_396016 | Not-determined |
| 17-109_k141_10509 | Not-determined |
| 17-109_k141_261750 | Not-determined |
| 17-109_k141_295992 | Low-quality |
| 17-109_k141_360886 | Not-determined |
| 17-109_k141_261752 | Not-determined |
| 17-109_k141_60891 | Low-quality |
| 17-109_k141_127028 | Not-determined |
| 17-109_k141_229304 | Low-quality |
| 17-109_k141_313432 | Not-determined |
| 17-109_k141_313434 | Not-determined |
| 17-109_k141_127031 | Not-determined |
| 17-109_k141_229308 | Not-determined |
| 17-109_k141_127035 | Not-determined |
| 17-109_k141_94218 | Not-determined |
| 17-109_k141_94219 | Not-determined |
| 17-109_k141_45144 | Not-determined |
| 17-109_k141_279364 | Low-quality |
| 17-109_k141_144443 | Low-quality |
| 17-109_k141_27889 | Not-determined |
| 17-109_k141_60894 | Not-determined |
| 17-109_k141_279371 | Low-quality |
| 17-109_k141_144451 | Not-determined |
| 17-109_k141_60897 | Not-determined |
| 17-109_k141_279373 | Not-determined |
| 17-109_k141_345584 | Low-quality |
| 17-109_k141_313445 | Not-determined |
| 17-109_k141_379650 | Not-determined |
| 17-109_k141_195702 | Not-determined |
| 17-109_k141_144460 | Not-determined |
| 17-109_k141_296019 | Not-determined |
| 17-109_k141_229328 | Not-determined |
| 17-109_k141_379658 | Not-determined |
| 17-109_k141_396032 | Low-quality |
| 17-109_k141_45149 | Not-determined |
| 17-109_k141_110777 | Low-quality |
| 17-109_k141_195706 | Not-determined |
| 17-109_k141_60905 | Not-determined |
| 17-109_k141_10521 | Not-determined |
| 17-109_k141_379669 | Not-determined |
| 17-109_k141_60907 | Not-determined |
| 17-109_k141_127038 | Not-determined |
| 17-109_k141_178084 | Not-determined |
| 17-109_k141_60910 | Not-determined |
| 17-109_k141_45158 | Not-determined |
| 17-109_k141_110786 | Not-determined |
| 17-109_k141_261783 | Low-quality |
| 17-109_k141_313453 | Not-determined |
| 17-109_k141_161756 | Not-determined |
| 17-109_k141_60913 | Not-determined |
| 17-109_k141_313457 | Not-determined |
| 17-109_k141_127049 | Not-determined |
| 17-109_k141_27907 | Not-determined |
| 17-109_k141_60916 | Not-determined |
| 17-109_k141_10533 | Not-determined |
| 17-109_k141_211912 | Low-quality |
| 17-109_k141_296027 | Not-determined |
| 17-109_k141_211919 | Not-determined |
| 17-109_k141_313468 | Not-determined |
| 17-109_k141_296032 | Not-determined |
| 17-109_k141_360902 | Not-determined |
| 17-109_k141_279404 | Not-determined |
| 17-109_k141_10544 | Not-determined |
| 17-109_k141_27921 | Not-determined |
| 17-109_k141_178097 | Not-determined |
| 17-109_k141_127062 | Not-determined |
| 17-109_k141_161772 | Not-determined |
| 17-109_k141_396055 | Not-determined |
| 17-109_k141_396060 | Not-determined |
| 17-109_k141_313472 | Not-determined |
| 17-109_k141_229359 | Not-determined |
| 17-109_k141_78713 | Not-determined |
| 17-109_k141_279419 | Not-determined |
| 17-109_k141_211941 | Not-determined |
| 17-109_k141_313486 | Not-determined |
| 17-109_k141_229375 | Not-determined |
| 17-109_k141_195754 | Not-determined |
| 17-109_k141_127092 | Not-determined |
| 17-109_k141_144499 | Not-determined |
| 17-109_k141_396077 | Not-determined |
| 17-109_k141_211953 | Not-determined |
| 17-109_k141_110819 | Not-determined |
| 17-109_k141_178112 | Not-determined |
| 17-109_k141_45196 | Not-determined |
| 17-109_k141_245221 | Not-determined |
| 17-109_k141_360938 | Not-determined |
| 17-109_k141_94244 | Not-determined |
| 17-109_k141_94246 | Not-determined |
| 17-109_k141_94247 | Not-determined |
| 17-109_k141_10570 | Low-quality |
| 17-109_k141_161797 | Not-determined |
| 17-109_k141_229392 | Not-determined |
| 17-109_k141_396093 | Low-quality |
| 17-109_k141_27926 | Not-determined |
| 17-109_k141_345627 | Not-determined |
| 17-109_k141_195776 | Not-determined |
| 17-109_k141_127111 | Not-determined |
| 17-109_k141_195777 | Not-determined |
| 17-109_k141_329245 | Not-determined |
| 17-109_k141_127114 | Not-determined |
| 17-109_k141_110827 | Not-determined |
| 17-109_k141_296042 | Not-determined |
| 17-109_k141_296043 | Not-determined |
| 17-109_k141_144521 | Not-determined |
| 17-109_k141_178127 | Not-determined |
| 17-109_k141_279438 | Not-determined |
| 17-109_k141_360949 | Not-determined |
| 17-109_k141_296047 | Not-determined |
| 17-109_k141_94249 | Low-quality |
| 17-109_k141_144536 | Not-determined |
| 17-109_k141_345637 | Not-determined |
| 17-109_k141_195786 | Not-determined |
| 17-109_k141_60960 | Not-determined |
| 17-109_k141_279453 | Not-determined |
| 17-109_k141_60964 | Not-determined |
| 17-109_k141_279457 | Not-determined |
| 17-109_k141_279458 | Not-determined |
| 17-109_k141_261829 | Low-quality |
| 17-109_k141_127134 | Not-determined |
| 17-109_k141_195797 | Not-determined |
| 17-109_k141_329270 | Low-quality |
| 17-109_k141_78770 | Not-determined |
| 17-109_k141_279466 | Not-determined |
| 17-109_k141_212011 | Not-determined |
| 17-109_k141_229413 | Not-determined |
| 17-109_k141_178154 | Not-determined |
| 17-109_k141_345654 | Not-determined |
| 17-109_k141_313551 | Not-determined |
| 17-109_k141_212018 | Not-determined |
| 17-109_k141_195812 | Not-determined |
| 17-109_k141_178163 | Not-determined |
| 17-109_k141_360957 | Not-determined |
| 17-109_k141_178168 | Not-determined |
| 17-109_k141_195817 | Low-quality |
| 17-109_k141_360959 | Not-determined |
| 17-109_k141_261836 | Low-quality |
| 17-109_k141_313565 | Not-determined |
| 17-109_k141_212027 | Not-determined |
| 17-109_k141_161823 | Not-determined |
| 17-109_k141_261839 | Not-determined |
| 17-109_k141_212032 | Not-determined |
| 17-109_k141_60976 | Not-determined |
| 17-109_k141_229438 | Not-determined |
| 17-109_k141_60978 | Not-determined |
| 17-109_k141_396129 | Not-determined |
| 17-109_k141_178173 | Low-quality |
| 17-109_k141_60982 | Not-determined |
| 17-109_k141_212038 | Not-determined |
| 17-109_k141_245261 | Not-determined |
| 17-109_k141_212040 | Not-determined |
| 17-109_k141_245265 | Not-determined |
| 17-109_k141_360977 | Not-determined |
| 17-109_k141_45212 | Not-determined |
| 17-109_k141_195840 | Not-determined |
| 17-109_k141_195842 | Not-determined |
| 17-109_k141_396139 | Not-determined |
| 17-109_k141_229451 | Not-determined |
| 17-109_k141_229453 | Not-determined |
| 17-109_k141_345678 | Not-determined |
| 17-109_k141_261854 | Not-determined |
| 17-109_k141_229457 | Not-determined |
| 17-109_k141_329309 | Not-determined |
| 17-109_k141_178197 | Not-determined |
| 17-109_k141_279496 | Not-determined |
| 17-109_k141_178200 | Not-determined |
| 17-109_k141_78821 | Not-determined |
| 17-109_k141_178204 | Not-determined |
| 17-109_k141_61011 | Low-quality |
| 17-109_k141_329316 | Not-determined |
| 17-109_k141_195868 | Not-determined |
| 17-109_k141_229472 | Not-determined |
| 17-109_k141_78829 | Not-determined |
| 17-109_k141_195871 | Not-determined |
| 17-109_k141_178213 | Not-determined |
| 17-109_k141_345694 | Not-determined |
| 17-109_k141_61017 | Not-determined |
| 17-109_k141_379773 | Not-determined |
| 17-109_k141_61019 | Not-determined |
| 17-109_k141_396162 | Not-determined |
| 17-109_k141_396163 | Not-determined |
| 17-109_k141_10588 | Not-determined |
| 17-109_k141_127180 | Not-determined |
| 17-109_k141_61027 | Not-determined |
| 17-109_k141_229482 | Low-quality |
| 17-109_k141_195884 | Not-determined |
| 17-109_k141_360995 | Low-quality |
| 17-109_k141_195890 | Not-determined |
| 17-109_k141_396184 | Not-determined |
| 17-109_k141_94290 | Not-determined |

| | |
|---|---|
| 17-109_k141_379785 | Not-determined |
| 17-109_k141_195894 | Not-determined |
| 17-109_k141_329325 | Not-determined |
| 17-109_k141_144578 | Not-determined |
| 17-109_k141_45245 | Low-quality |
| 17-109_k141_94297 | Not-determined |
| 17-109_k141_229494 | Not-determined |
| 17-109_k141_110878 | Not-determined |
| 17-109_k141_45254 | Low-quality |
| 17-109_k141_195906 | Low-quality |
| 17-109_k141_361006 | Not-determined |
| 17-109_k141_313625 | Not-determined |
| 17-109_k141_78848 | Low-quality |
| 17-109_k141_361012 | Not-determined |
| 17-109_k141_229511 | Not-determined |
| 17-109_k141_61039 | Not-determined |
| 17-109_k141_345707 | Not-determined |
| 17-109_k141_127207 | Not-determined |
| 17-109_k141_379792 | Not-determined |
| 17-109_k141_279536 | Not-determined |
| 17-109_k141_379793 | Not-determined |
| 17-109_k141_10608 | Not-determined |
| 17-109_k141_212060 | Not-determined |
| 17-109_k141_78853 | Not-determined |
| 17-109_k141_45270 | Not-determined |
| 17-109_k141_313638 | Not-determined |
| 17-109_k141_361018 | Not-determined |
| 17-109_k141_379806 | Not-determined |
| 17-109_k141_195924 | Not-determined |
| 17-109_k141_78859 | Not-determined |
| 17-109_k141_195927 | Not-determined |
| 17-109_k141_61048 | Not-determined |
| 17-109_k141_178237 | Not-determined |
| 17-109_k141_110899 | Not-determined |
| 17-109_k141_161851 | Not-determined |
| 17-109_k141_229522 | Not-determined |
| 17-109_k141_61053 | Not-determined |
| 17-109_k141_212073 | Not-determined |
| 17-109_k141_144586 | Not-determined |
| 17-109_k141_229525 | Not-determined |
| 17-109_k141_178247 | Not-determined |
| 17-109_k141_110912 | Not-determined |
| 17-109_k141_279557 | Not-determined |
| 17-109_k141_78872 | Not-determined |
| 17-109_k141_212075 | Not-determined |
| 17-109_k141_212077 | Not-determined |
| 17-109_k141_78878 | Not-determined |
| 17-109_k141_212079 | Not-determined |
| 17-109_k141_78881 | Not-determined |
| 17-109_k141_195942 | Not-determined |
| 17-109_k141_10630 | Low-quality |
| 17-109_k141_161875 | Not-determined |
| 17-109_k141_144596 | Not-determined |
| 17-109_k141_27947 | Not-determined |
| 17-109_k141_61071 | Not-determined |
| 17-109_k141_10637 | Not-determined |
| 17-109_k141_229551 | Low-quality |
| 17-109_k141_178255 | Not-determined |
| 17-109_k141_127256 | Not-determined |
| 17-109_k141_127261 | Not-determined |
| 17-109_k141_212088 | Not-determined |
| 17-109_k141_379848 | Not-determined |
| 17-109_k141_279577 | Not-determined |
| 17-109_k141_195960 | Not-determined |
| 17-109_k141_313664 | Low-quality |
| 17-109_k141_94335 | Not-determined |
| 17-109_k141_161890 | Not-determined |
| 17-109_k141_10649 | Low-quality |
| 17-109_k141_161895 | Not-determined |
| 17-109_k141_27966 | Not-determined |
| 17-109_k141_178266 | Low-quality |
| 17-109_k141_110942 | Not-determined |
| 17-109_k141_78904 | Not-determined |
| 17-109_k141_27976 | Not-determined |
| 17-109_k141_361068 | Not-determined |
| 17-109_k141_178275 | Low-quality |
| 17-109_k141_178280 | Not-determined |
| 17-109_k141_161910 | Low-quality |
| 17-109_k141_78917 | Not-determined |
| 17-109_k141_396202 | Not-determined |
| 17-109_k141_94348 | Not-determined |
| 17-109_k141_229570 | Low-quality |
| 17-109_k141_144608 | Not-determined |
| 17-109_k141_313689 | Not-determined |
| 17-109_k141_144611 | Not-determined |
| 17-109_k141_279607 | Not-determined |
| 17-109_k141_144612 | Low-quality |
| 17-109_k141_144615 | Not-determined |
| 17-109_k141_161931 | Not-determined |
| 17-109_k141_161933 | Not-determined |
| 17-109_k141_212116 | Low-quality |
| 17-109_k141_229581 | Not-determined |
| 17-109_k141_144622 | Not-determined |
| 17-109_k141_144626 | Not-determined |
| 17-109_k141_229590 | Not-determined |
| 17-109_k141_229592 | Not-determined |
| 17-109_k141_212124 | Not-determined |
| 17-109_k141_10661 | Not-determined |
| 17-109_k141_396216 | Not-determined |
| 17-109_k141_10664 | Not-determined |
| 17-109_k141_144630 | Not-determined |
| 17-109_k141_396220 | Not-determined |
| 17-109_k141_94377 | Not-determined |
| 17-109_k141_10665 | Not-determined |
| 17-109_k141_212134 | Low-quality |
| 17-109_k141_94381 | Not-determined |
| 17-109_k141_329341 | Not-determined |
| 17-109_k141_10670 | Not-determined |
| 17-109_k141_313703 | Not-determined |
| 17-109_k141_345781 | Not-determined |
| 17-109_k141_178312 | Not-determined |
| 17-109_k141_313712 | Not-determined |
| 17-109_k141_361088 | Not-determined |
| 17-109_k141_161951 | Not-determined |
| 17-109_k141_229607 | Not-determined |
| 17-109_k141_361090 | Not-determined |
| 17-109_k141_279630 | Not-determined |
| 17-109_k141_195983 | Not-determined |
| 17-109_k141_195985 | Not-determined |
| 17-109_k141_296125 | Not-determined |
| 17-109_k141_345802 | Not-determined |
| 17-109_k141_313728 | Low-quality |
| 17-109_k141_10677 | Not-determined |
| 17-109_k141_379893 | Not-determined |
| 17-109_k141_45313 | Not-determined |
| 17-109_k141_61136 | Not-determined |
| 17-109_k141_379897 | Not-determined |
| 17-109_k141_279636 | Not-determined |
| 17-109_k141_345809 | Not-determined |
| 17-109_k141_361105 | Not-determined |
| 17-109_k141_229614 | Not-determined |
| 17-109_k141_361108 | Not-determined |
| 17-109_k141_94384 | Low-quality |
| 17-109_k141_195996 | Not-determined |
| 17-109_k141_229618 | Low-quality |
| 17-109_k141_144642 | Not-determined |
| 17-109_k141_245399 | Not-determined |
| 17-109_k141_94395 | Low-quality |
| 17-109_k141_61144 | Not-determined |
| 17-109_k141_10684 | Not-determined |
| 17-109_k141_212159 | Not-determined |
| 17-109_k141_279647 | Not-determined |
| 17-109_k141_111005 | Not-determined |
| 17-109_k141_345818 | Not-determined |
| 17-109_k141_229622 | Not-determined |
| 17-109_k141_212170 | Not-determined |
| 17-109_k141_45322 | Not-determined |
| 17-109_k141_196005 | Not-determined |
| 17-109_k141_229627 | Not-determined |
| 17-109_k141_161982 | Not-determined |
| 17-109_k141_94404 | Low-quality |
| 17-109_k141_396250 | Not-determined |
| 17-109_k141_379906 | Not-determined |
| 17-109_k141_245405 | Not-determined |
| 17-109_k141_279661 | Not-determined |
| 17-109_k141_313751 | Low-quality |
| 17-109_k141_245406 | Not-determined |
| 17-109_k141_229636 | Not-determined |
| 17-109_k141_196010 | Not-determined |
| 17-109_k141_396260 | Low-quality |
| 17-109_k141_178348 | Not-determined |
| 17-109_k141_345833 | Not-determined |
| 17-109_k141_313770 | Not-determined |
| 17-109_k141_313772 | Not-determined |
| 17-109_k141_245425 | Low-quality |
| 17-109_k141_279681 | Not-determined |
| 17-109_k141_396274 | Not-determined |
| 17-109_k141_261892 | Not-determined |
| 17-109_k141_178358 | Not-determined |
| 17-109_k141_296161 | Not-determined |
| 17-109_k141_329371 | Not-determined |
| 17-109_k141_379926 | Not-determined |
| 17-109_k141_61190 | Not-determined |
| 17-109_k141_361118 | Not-determined |
| 17-109_k141_144678 | Not-determined |
| 17-109_k141_162024 | Not-determined |
| 17-109_k141_361120 | Not-determined |
| 17-109_k141_162026 | Not-determined |
| 17-109_k141_329391 | Not-determined |
| 17-109_k141_396292 | Not-determined |
| 17-109_k141_162029 | Not-determined |
| 17-109_k141_379935 | Not-determined |
| 17-109_k141_10730 | Not-determined |
| 17-109_k141_162034 | Not-determined |
| 17-109_k141_379939 | Not-determined |
| 17-109_k141_245440 | Not-determined |
| 17-109_k141_396296 | Not-determined |
| 17-109_k141_396297 | Not-determined |
| 17-109_k141_212179 | Not-determined |
| 17-109_k141_212181 | Not-determined |
| 17-109_k141_329407 | Not-determined |
| 17-109_k141_10749 | Not-determined |
| 17-109_k141_162035 | Not-determined |
| 17-109_k141_245449 | Not-determined |
| 17-109_k141_94418 | Not-determined |
| 17-109_k141_329413 | Not-determined |
| 17-109_k141_61204 | Not-determined |
| 17-109_k141_212186 | Not-determined |
| 17-109_k141_396305 | Not-determined |
| 17-109_k141_94426 | Not-determined |
| 17-109_k141_178402 | Not-determined |
| 17-109_k141_245462 | Low-quality |
| 17-109_k141_127403 | Low-quality |
| 17-109_k141_245466 | Not-determined |
| 17-109_k141_396314 | Not-determined |
| 17-109_k141_261916 | Not-determined |
| 17-109_k141_313817 | Not-determined |
| 17-109_k141_61211 | Low-quality |
| 17-109_k141_178412 | Not-determined |
| 17-109_k141_261923 | Not-determined |
| 17-109_k141_212208 | Not-determined |
| 17-109_k141_361164 | Not-determined |
| 17-109_k141_10772 | Not-determined |
| 17-109_k141_245480 | Not-determined |
| 17-109_k141_61221 | Not-determined |
| 17-109_k141_127412 | Not-determined |
| 17-109_k141_162053 | Not-determined |
| 17-109_k141_61230 | Not-determined |
| 17-109_k141_212214 | Not-determined |
| 17-109_k141_78961 | Not-determined |
| 17-109_k141_144697 | Not-determined |
| 17-109_k141_279734 | Not-determined |
| 17-109_k141_144701 | Not-determined |
| 17-109_k141_61236 | Not-determined |
| 17-109_k141_261942 | Not-determined |
| 17-109_k141_245497 | Not-determined |
| 17-109_k141_45403 | Not-determined |
| 17-109_k141_296222 | Low-quality |
| 17-109_k141_296223 | Not-determined |
| 17-109_k141_229687 | Low-quality |
| 17-109_k141_144706 | Not-determined |
| 17-109_k141_396336 | Not-determined |
| 17-109_k141_10780 | Not-determined |
| 17-109_k141_313840 | Not-determined |
| 17-109_k141_127423 | Not-determined |
| 17-109_k141_279749 | Not-determined |
| 17-109_k141_61240 | Low-quality |
| 17-109_k141_361178 | Not-determined |
| 17-109_k141_61241 | Not-determined |
| 17-109_k141_313846 | Not-determined |
| 17-109_k141_361180 | Not-determined |
| 17-109_k141_379963 | Not-determined |
| 17-109_k141_127431 | Not-determined |
| 17-109_k141_127433 | Not-determined |
| 17-109_k141_313852 | Not-determined |
| 17-109_k141_45415 | Low-quality |
| 17-109_k141_345915 | Not-determined |
| 17-109_k141_345917 | Not-determined |
| 17-109_k141_396347 | Not-determined |
| 17-109_k141_379970 | Not-determined |
| 17-109_k141_345920 | Not-determined |
| 17-109_k141_10792 | Not-determined |
| 17-109_k141_94473 | Not-determined |
| 17-109_k141_361194 | Not-determined |
| 17-109_k141_144713 | Not-determined |
| 17-109_k141_279752 | Not-determined |
| 17-109_k141_279755 | Not-determined |
| 17-109_k141_361196 | Not-determined |
| 17-109_k141_279758 | Not-determined |
| 17-109_k141_212235 | Not-determined |
| 17-109_k141_345929 | Low-quality |
| 17-109_k141_127441 | Not-determined |
| 17-109_k141_396351 | Low-quality |
| 17-109_k141_94477 | Not-determined |
| 17-109_k141_313871 | Low-quality |
| 17-109_k141_345932 | Not-determined |
| 17-109_k141_144722 | Not-determined |
| 17-109_k141_313874 | Not-determined |
| 17-109_k141_361204 | Not-determined |
| 17-109_k141_127446 | Not-determined |
| 17-109_k141_61254 | Not-determined |
| 17-109_k141_111032 | Not-determined |
| 17-109_k141_396357 | Not-determined |
| 17-109_k141_111034 | Not-determined |
| 17-109_k141_111035 | Not-determined |
| 17-109_k141_178478 | Not-determined |
| 17-109_k141_361211 | Not-determined |
| 17-109_k141_178480 | Not-determined |
| 17-109_k141_178481 | Not-determined |
| 17-109_k141_45434 | Not-determined |
| 17-109_k141_127456 | Not-determined |
| 17-109_k141_229713 | Not-determined |
| 17-109_k141_45437 | Not-determined |
| 17-109_k141_396370 | Not-determined |
| 17-109_k141_279781 | Not-determined |
| 17-109_k141_279782 | Not-determined |
| 17-109_k141_127463 | Not-determined |
| 17-109_k141_94503 | Not-determined |
| 17-109_k141_396374 | Not-determined |
| 17-109_k141_313893 | Not-determined |
| 17-109_k141_245531 | Low-quality |
| 17-109_k141_313898 | Not-determined |
| 17-109_k141_396378 | Not-determined |
| 17-109_k141_127469 | Not-determined |
| 17-109_k141_61269 | Not-determined |
| 17-109_k141_229724 | Not-determined |
| 17-109_k141_196072 | Not-determined |
| 17-109_k141_45454 | Not-determined |
| 17-109_k141_279791 | Not-determined |
| 17-109_k141_45456 | Not-determined |
| 17-109_k141_178499 | Not-determined |
| 17-109_k141_61274 | Not-determined |
| 17-109_k141_144748 | Low-quality |
| 17-109_k141_144752 | Not-determined |
| 17-109_k141_379981 | Not-determined |
| 17-109_k141_144757 | Not-determined |
| 17-109_k141_144758 | Not-determined |
| 17-109_k141_245554 | Not-determined |
| 17-109_k141_94527 | Not-determined |
| 17-109_k141_45461 | Not-determined |
| 17-109_k141_162113 | Not-determined |
| 17-109_k141_45464 | Not-determined |
| 17-109_k141_396392 | Low-quality |
| 17-109_k141_313917 | Not-determined |
| 17-109_k141_45468 | Not-determined |
| 17-109_k141_45470 | Not-determined |
| 17-109_k141_144760 | Not-determined |
| 17-109_k141_162122 | Not-determined |
| 17-109_k141_379993 | Not-determined |
| 17-109_k141_361247 | Not-determined |
| 17-109_k141_127494 | Not-determined |
| 17-109_k141_111070 | Not-determined |
| 17-109_k141_45481 | Low-quality |
| 17-109_k141_196106 | Not-determined |
| 17-109_k141_162131 | Not-determined |
| 17-109_k141_396410 | Not-determined |
| 17-109_k141_78969 | Not-determined |
| 17-109_k141_10812 | Not-determined |
| 17-109_k141_380001 | Not-determined |
| 17-109_k141_127506 | Not-determined |
| 17-109_k141_45493 | Not-determined |
| 17-109_k141_380007 | Not-determined |
| 17-109_k141_45501 | Not-determined |
| 17-109_k141_78978 | Not-determined |
| 17-109_k141_162145 | Not-determined |
| 17-109_k141_313946 | Not-determined |
| 17-109_k141_313947 | Not-determined |
| 17-109_k141_396420 | Not-determined |
| 17-109_k141_111084 | Not-determined |
| 17-109_k141_245580 | Not-determined |
| 17-109_k141_262019 | Low-quality |
| 17-109_k141_245583 | Not-determined |
| 17-109_k141_279804 | Not-determined |
| 17-109_k141_345954 | Not-determined |
| 17-109_k141_245587 | Not-determined |
| 17-109_k141_229770 | Low-quality |
| 17-109_k141_345960 | Not-determined |
| 17-109_k141_279816 | Not-determined |
| 17-109_k141_94575 | Not-determined |
| 17-109_k141_262038 | Not-determined |
| 17-109_k141_162159 | Low-quality |
| 17-109_k141_380015 | Not-determined |
| 17-109_k141_396435 | Not-determined |
| 17-109_k141_262042 | Not-determined |
| 17-109_k141_45507 | Low-quality |
| 17-109_k141_262043 | Not-determined |
| 17-109_k141_127538 | Not-determined |
| 17-109_k141_262045 | Not-determined |
| 17-109_k141_245599 | Not-determined |
| 17-109_k141_262046 | Low-quality |
| 17-109_k141_45513 | Not-determined |
| 17-109_k141_111098 | Not-determined |
| 17-109_k141_196118 | Low-quality |
| 17-109_k141_396441 | Not-determined |
| 17-109_k141_396446 | Not-determined |
| 17-109_k141_79007 | Not-determined |
| 17-109_k141_162170 | Low-quality |
| 17-109_k141_79008 | Not-determined |
| 17-109_k141_162177 | Not-determined |
| 17-109_k141_79014 | Not-determined |
| 17-109_k141_196131 | Not-determined |
| 17-109_k141_162180 | Not-determined |
| 17-109_k141_196135 | Not-determined |
| 17-109_k141_279838 | Not-determined |
| 17-109_k141_196137 | Not-determined |
| 17-109_k141_162184 | Low-quality |
| 17-109_k141_144817 | Not-determined |
| 17-109_k141_196141 | Not-determined |
| 17-109_k141_144820 | Not-determined |
| 17-109_k141_396455 | Not-determined |
| 17-109_k141_229792 | Not-determined |
| 17-109_k141_10850 | Not-determined |
| 17-109_k141_380037 | Not-determined |
| 17-109_k141_10852 | Not-determined |
| 17-109_k141_162189 | Not-determined |
| 17-109_k141_196151 | Not-determined |
| 17-109_k141_262076 | Not-determined |
| 17-109_k141_396456 | Low-quality |
| 17-109_k141_162190 | Not-determined |
| 17-109_k141_94594 | Low-quality |
| 17-109_k141_196154 | Not-determined |
| 17-109_k141_262080 | Not-determined |
| 17-109_k141_345999 | Not-determined |
| 17-109_k141_196156 | Not-determined |
| 17-109_k141_262083 | Not-determined |
| 17-109_k141_396459 | Not-determined |
| 17-109_k141_396460 | Not-determined |
| 17-109_k141_245635 | Low-quality |
| 17-109_k141_262085 | Not-determined |
| 17-109_k141_196160 | Low-quality |
| 17-109_k141_127557 | Not-determined |
| 17-109_k141_111113 | Not-determined |
| 17-109_k141_144826 | Not-determined |
| 17-109_k141_10860 | Not-determined |
| 17-109_k141_212319 | Not-determined |
| 17-109_k141_262089 | Not-determined |
| 17-109_k141_111117 | Low-quality |
| 17-109_k141_396468 | Not-determined |
| 17-109_k141_10866 | Not-determined |
| 17-109_k141_144833 | Not-determined |
| 17-109_k141_127571 | Not-determined |
| 17-109_k141_279860 | Not-determined |
| 17-109_k141_245649 | Not-determined |
| 17-109_k141_79033 | Not-determined |
| 17-109_k141_296277 | Not-determined |
| 17-109_k141_396478 | Low-quality |
| 17-109_k141_45559 | Not-determined |
| 17-109_k141_61316 | Not-determined |
| 17-109_k141_245660 | Not-determined |
| 17-109_k141_45561 | Low-quality |
| 17-109_k141_262103 | Not-determined |
| 17-109_k141_346035 | Not-determined |
| 17-109_k141_196168 | Not-determined |
| 17-109_k141_212339 | Not-determined |
| 17-109_k141_79046 | Low-quality |
| 17-109_k141_346041 | Not-determined |
| 17-109_k141_94610 | Not-determined |
| 17-109_k141_262113 | Not-determined |
| 17-109_k141_346045 | Not-determined |
| 17-109_k141_162206 | Not-determined |
| 17-109_k141_262115 | Low-quality |
| 17-109_k141_162210 | Not-determined |
| 17-109_k141_245678 | Not-determined |
| 17-109_k141_162211 | Low-quality |
| 17-109_k141_279881 | Not-determined |
| 17-109_k141_212348 | Not-determined |
| 17-109_k141_279886 | Not-determined |
| 17-109_k141_94621 | Not-determined |
| 17-109_k141_245685 | Not-determined |
| 17-109_k141_245689 | Not-determined |
| 17-109_k141_229838 | Not-determined |
| 17-109_k141_229840 | Not-determined |
| 17-109_k141_329499 | Not-determined |
| 17-109_k141_27983 | Not-determined |
| 17-109_k141_196186 | Not-determined |
| 17-109_k141_127599 | Low-quality |
| 17-109_k141_178576 | Not-determined |
| 17-109_k141_178583 | Not-determined |
| 17-109_k141_196191 | Not-determined |
| 17-109_k141_329516 | Not-determined |
| 17-109_k141_329518 | Not-determined |
| 17-109_k141_279895 | Not-determined |
| 17-109_k141_178589 | Low-quality |
| 17-109_k141_346078 | Not-determined |
| 17-109_k141_61344 | Not-determined |
| 17-109_k141_144862 | Not-determined |
| 17-109_k141_178592 | Not-determined |
| 17-109_k141_144864 | Not-determined |
| 17-109_k141_245694 | Not-determined |
| 17-109_k141_144866 | Not-determined |
| 17-109_k141_144871 | Not-determined |
| 17-109_k141_279906 | Not-determined |
| 17-109_k141_144875 | Not-determined |
| 17-109_k141_212380 | Low-quality |
| 17-109_k141_396511 | Not-determined |
| 17-109_k141_346092 | Not-determined |
| 17-109_k141_127605 | Not-determined |
| 17-109_k141_178611 | Not-determined |
| 17-109_k141_127608 | Not-determined |
| 17-109_k141_380076 | Not-determined |
| 17-109_k141_380077 | Not-determined |
| 17-109_k141_10889 | Not-determined |
| 17-109_k141_396522 | Not-determined |
| 17-109_k141_79077 | Not-determined |
| 17-109_k141_94631 | Not-determined |
| 17-109_k141_380080 | Not-determined |
| 17-109_k141_396531 | Not-determined |
| 17-109_k141_10898 | Low-quality |
| 17-109_k141_162235 | Low-quality |
| 17-109_k141_296352 | Not-determined |
| 17-109_k141_79082 | Not-determined |
| 17-109_k141_61382 | Not-determined |
| 17-109_k141_45574 | Not-determined |
| 17-109_k141_396541 | Not-determined |
| 17-109_k141_79085 | Not-determined |
| 17-109_k141_28012 | Not-determined |
| 17-109_k141_245723 | Not-determined |
| 17-109_k141_10908 | Not-determined |
| 17-109_k141_296366 | Not-determined |
| 17-109_k141_28015 | Not-determined |
| 17-109_k141_245724 | Not-determined |
| 17-109_k141_346124 | Not-determined |
| 17-109_k141_296371 | Not-determined |
| 17-109_k141_380098 | Not-determined |
| 17-109_k141_79094 | Not-determined |
| 17-109_k141_296373 | Not-determined |
| 17-109_k141_313960 | Low-quality |
| 17-109_k141_79098 | Not-determined |
| 17-109_k141_10925 | Not-determined |
| 17-109_k141_10927 | Not-determined |
| 17-109_k141_45589 | Not-determined |
| 17-109_k141_127626 | Low-quality |
| 17-109_k141_61398 | Not-determined |
| 17-109_k141_212407 | Low-quality |
| 17-109_k141_79107 | Not-determined |
| 17-109_k141_28038 | Not-determined |
| 17-109_k141_127635 | Not-determined |
| 17-109_k141_162251 | Not-determined |
| 17-109_k141_162253 | Not-determined |
| 17-109_k141_262178 | Not-determined |
| 17-109_k141_196224 | Not-determined |
| 17-109_k141_61401 | Not-determined |
| 17-109_k141_10938 | Not-determined |
| 17-109_k141_380115 | Not-determined |
| 17-109_k141_380116 | Not-determined |
| 17-109_k141_94667 | Not-determined |
| 17-109_k141_346144 | Not-determined |
| 17-109_k141_262180 | Low-quality |
| 17-109_k141_380120 | Not-determined |
| 17-109_k141_162265 | Not-determined |
| 17-109_k141_127653 | Not-determined |
| 17-109_k141_127654 | Not-determined |
| 17-109_k141_262190 | Not-determined |
| 17-109_k141_10951 | Not-determined |
| 17-109_k141_111188 | Not-determined |
| 17-109_k141_127657 | Low-quality |
| 17-109_k141_61419 | Not-determined |
| 17-109_k141_178656 | Not-determined |
| 17-109_k141_61420 | Not-determined |
| 17-109_k141_127660 | Not-determined |
| 17-109_k141_45623 | Not-determined |
| 17-109_k141_45630 | Not-determined |
| 17-109_k141_178663 | Not-determined |
| 17-109_k141_296401 | Low-quality |
| 17-109_k141_178666 | Not-determined |
| 17-109_k141_329590 | Not-determined |
| 17-109_k141_245742 | Not-determined |
| 17-109_k141_45637 | Not-determined |
| 17-109_k141_45639 | Not-determined |
| 17-109_k141_45644 | Not-determined |
| 17-109_k141_61441 | Not-determined |
| 17-109_k141_178677 | Not-determined |
| 17-109_k141_61444 | Not-determined |
| 17-109_k141_296416 | Not-determined |
| 17-109_k141_212424 | Not-determined |
| 17-109_k141_178685 | Not-determined |
| 17-109_k141_45654 | Not-determined |
| 17-109_k141_127694 | Not-determined |
| 17-109_k141_61455 | Not-determined |
| 17-109_k141_296419 | Not-determined |
| 17-109_k141_94694 | Not-determined |
| 17-109_k141_61460 | Not-determined |
| 17-109_k141_396597 | Not-determined |
| 17-109_k141_313973 | Low-quality |
| 17-109_k141_396598 | Not-determined |
| 17-109_k141_61462 | Low-quality |
| 17-109_k141_178692 | Not-determined |
| 17-109_k141_313983 | Not-determined |
| 17-109_k141_313984 | Not-determined |
| 17-109_k141_111218 | Not-determined |
| 17-109_k141_61468 | Not-determined |
| 17-109_k141_10992 | Not-determined |
| 17-109_k141_127716 | Not-determined |
| 17-109_k141_380145 | Not-determined |
| 17-109_k141_313991 | Low-quality |
| 17-109_k141_162310 | Not-determined |
| 17-109_k141_396602 | Low-quality |
| 17-109_k141_212452 | Not-determined |
| 17-109_k141_11000 | Not-determined |
| 17-109_k141_212453 | Not-determined |
| 17-109_k141_329619 | Low-quality |
| 17-109_k141_396606 | Not-determined |
| 17-109_k141_329624 | Not-determined |
| 17-109_k141_61474 | Not-determined |
| 17-109_k141_144904 | Not-determined |
| 17-109_k141_144905 | Not-determined |
| 17-109_k141_212456 | Low-quality |
| 17-109_k141_144909 | Not-determined |
| 17-109_k141_262219 | Low-quality |
| 17-109_k141_45679 | Not-determined |
| 17-109_k141_162331 | Not-determined |
| 17-109_k141_45681 | Low-quality |
| 17-109_k141_162332 | Low-quality |
| 17-109_k141_262223 | Low-quality |
| 17-109_k141_11014 | Not-determined |
| 17-109_k141_162334 | Not-determined |
| 17-109_k141_296451 | Not-determined |
| 17-109_k141_162338 | Not-determined |
| 17-109_k141_11020 | Not-determined |

| | |
|---|---|
| 17-109_k141_162341 | Not-determined |
| 17-109_k141_144918 | Not-determined |
| 17-109_k141_279972 | Not-determined |
| 17-109_k141_162344 | Not-determined |
| 17-109_k141_11021 | Not-determined |
| 17-109_k141_144922 | Not-determined |
| 17-109_k141_279976 | Not-determined |
| 17-109_k141_396635 | Not-determined |
| 17-109_k141_262235 | Not-determined |
| 17-109_k141_380166 | Not-determined |
| 17-109_k141_162356 | Not-determined |
| 17-109_k141_212481 | Not-determined |
| 17-109_k141_296467 | Not-determined |
| 17-109_k141_144934 | Low-quality |
| 17-109_k141_296469 | Not-determined |
| 17-109_k141_396644 | Not-determined |
| 17-109_k141_279989 | Low-quality |
| 17-109_k141_144941 | Not-determined |
| 17-109_k141_162358 | Low-quality |
| 17-109_k141_45685 | Not-determined |
| 17-109_k141_212490 | Not-determined |
| 17-109_k141_296477 | Not-determined |
| 17-109_k141_380179 | Not-determined |
| 17-109_k141_361401 | Not-determined |
| 17-109_k141_162363 | Not-determined |
| 17-109_k141_111250 | Not-determined |
| 17-109_k141_212496 | Low-quality |
| 17-109_k141_178720 | Not-determined |
| 17-109_k141_262253 | Low-quality |
| 17-109_k141_329650 | Not-determined |
| 17-109_k141_229864 | Not-determined |
| 17-109_k141_346165 | Not-determined |
| 17-109_k141_361415 | Not-determined |
| 17-109_k141_245786 | Not-determined |
| 17-109_k141_144950 | Not-determined |
| 17-109_k141_245787 | Not-determined |
| 17-109_k141_11028 | Not-determined |
| 17-109_k141_111269 | Not-determined |
| 17-109_k141_361418 | Not-determined |
| 17-109_k141_111275 | Not-determined |
| 17-109_k141_396676 | Not-determined |
| 17-109_k141_79141 | Not-determined |
| 17-109_k141_178737 | Not-determined |
| 17-109_k141_396679 | Not-determined |
| 17-109_k141_94740 | Not-determined |
| 17-109_k141_28072 | Not-determined |
| 17-109_k141_296501 | Not-determined |
| 17-109_k141_94743 | Not-determined |
| 17-109_k141_396686 | Not-determined |
| 17-109_k141_346180 | Not-determined |
| 17-109_k141_11047 | Not-determined |
| 17-109_k141_346181 | Not-determined |
| 17-109_k141_162403 | Not-determined |
| 17-109_k141_329676 | Not-determined |
| 17-109_k141_396694 | Not-determined |
| 17-109_k141_245814 | Not-determined |
| 17-109_k141_245815 | Not-determined |
| 17-109_k141_245819 | Not-determined |
| 17-109_k141_212530 | Not-determined |
| 17-109_k141_144983 | Not-determined |
| 17-109_k141_361448 | Not-determined |
| 17-109_k141_346201 | Not-determined |
| 17-109_k141_127725 | Not-determined |
| 17-109_k141_127733 | Not-determined |
| 17-109_k141_127739 | Not-determined |
| 17-109_k141_361460 | Not-determined |
| 17-109_k141_144996 | Low-quality |
| 17-109_k141_11058 | Not-determined |
| 17-109_k141_314055 | Not-determined |
| 17-109_k141_28115 | Not-determined |
| 17-109_k141_94775 | Low-quality |
| 17-109_k141_329695 | Not-determined |
| 17-109_k141_396701 | Low-quality |
| 17-109_k141_178760 | Not-determined |
| 17-109_k141_262290 | Not-determined |
| 17-109_k141_361461 | Low-quality |
| 17-109_k141_178762 | Not-determined |
| 17-109_k141_361463 | Not-determined |
| 17-109_k141_396705 | Not-determined |
| 17-109_k141_361467 | Not-determined |
| 17-109_k141_361468 | Low-quality |
| 17-109_k141_245832 | Not-determined |
| 17-109_k141_361473 | Not-determined |
| 17-109_k141_145011 | Not-determined |
| 17-109_k141_262293 | Not-determined |
| 17-109_k141_79184 | Not-determined |
| 17-109_k141_28124 | Low-quality |
| 17-109_k141_11070 | Not-determined |
| 17-109_k141_212556 | Not-determined |
| 17-109_k141_145014 | Not-determined |
| 17-109_k141_361477 | Not-determined |
| 17-109_k141_212562 | Not-determined |
| 17-109_k141_196291 | Not-determined |
| 17-109_k141_346211 | Low-quality |
| 17-109_k141_79199 | Not-determined |
| 17-109_k141_145027 | Not-determined |
| 17-109_k141_329718 | Low-quality |
| 17-109_k141_262321 | Not-determined |
| 17-109_k141_11082 | Not-determined |
| 17-109_k141_28141 | Not-determined |
| 17-109_k141_212572 | Not-determined |
| 17-109_k141_127766 | Not-determined |
| 17-109_k141_145030 | Low-quality |
| 17-109_k141_28144 | Not-determined |
| 17-109_k141_127768 | Not-determined |
| 17-109_k141_127774 | Not-determined |
| 17-109_k141_346245 | Not-determined |
| 17-109_k141_11088 | Not-determined |
| 17-109_k141_280021 | Not-determined |
| 17-109_k141_314088 | Not-determined |
| 17-109_k141_212586 | Not-determined |
| 17-109_k141_145038 | Not-determined |
| 17-109_k141_245871 | Not-determined |
| 17-109_k141_11092 | Not-determined |
| 17-109_k141_396738 | Not-determined |
| 17-109_k141_145043 | Not-determined |
| 17-109_k141_296529 | Not-determined |
| 17-109_k141_314089 | Not-determined |
| 17-109_k141_212590 | Not-determined |
| 17-109_k141_145046 | Not-determined |
| 17-109_k141_296535 | Not-determined |
| 17-109_k141_178814 | Not-determined |
| 17-109_k141_111354 | Not-determined |
| 17-109_k141_361515 | Not-determined |
| 17-109_k141_380227 | Not-determined |
| 17-109_k141_361516 | Not-determined |
| 17-109_k141_94795 | Not-determined |
| 17-109_k141_280034 | Low-quality |
| 17-109_k141_280035 | Not-determined |
| 17-109_k141_11100 | Not-determined |
| 17-109_k141_145054 | Not-determined |
| 17-109_k141_396760 | Not-determined |
| 17-109_k141_127801 | Not-determined |
| 17-109_k141_396764 | Not-determined |
| 17-109_k141_111370 | Not-determined |
| 17-109_k141_111371 | Not-determined |
| 17-109_k141_314094 | Not-determined |
| 17-109_k141_296551 | Not-determined |
| 17-109_k141_314101 | Not-determined |
| 17-109_k141_280040 | Not-determined |
| 17-109_k141_196327 | Not-determined |
| 17-109_k141_314102 | Low-quality |
| 17-109_k141_45717 | Not-determined |
| 17-109_k141_127813 | Not-determined |
| 17-109_k141_45723 | Not-determined |
| 17-109_k141_145065 | Not-determined |
| 17-109_k141_262366 | Low-quality |
| 17-109_k141_145069 | Not-determined |
| 17-109_k141_145073 | Not-determined |
| 17-109_k141_229923 | Low-quality |
| 17-109_k141_245899 | Not-determined |
| 17-109_k141_145078 | Not-determined |
| 17-109_k141_162460 | Not-determined |
| 17-109_k141_329745 | Not-determined |
| 17-109_k141_229927 | Not-determined |
| 17-109_k141_280055 | Not-determined |
| 17-109_k141_229929 | Not-determined |
| 17-109_k141_45727 | Not-determined |
| 17-109_k141_329748 | Not-determined |
| 17-109_k141_396792 | Low-quality |
| 17-109_k141_346258 | Not-determined |
| 17-109_k141_346259 | Not-determined |
| 17-109_k141_245906 | Low-quality |
| 17-109_k141_196342 | Not-determined |
| 17-109_k141_346261 | Not-determined |
| 17-109_k141_380260 | Not-determined |
| 17-109_k141_245911 | Not-determined |
| 17-109_k141_145097 | Not-determined |
| 17-109_k141_162464 | Not-determined |
| 17-109_k141_280067 | Low-quality |
| 17-109_k141_61493 | Not-determined |
| 17-109_k141_45746 | Not-determined |
| 17-109_k141_45747 | Not-determined |
| 17-109_k141_280072 | Not-determined |
| 17-109_k141_127840 | Not-determined |
| 17-109_k141_196354 | Not-determined |
| 17-109_k141_45750 | Not-determined |
| 17-109_k141_196357 | Not-determined |
| 17-109_k141_346278 | Not-determined |
| 17-109_k141_11163 | Not-determined |
| 17-109_k141_127844 | Low-quality |
| 17-109_k141_314133 | Not-determined |
| 17-109_k141_145109 | Not-determined |
| 17-109_k141_79210 | Not-determined |
| 17-109_k141_127848 | Not-determined |
| 17-109_k141_145111 | Not-determined |
| 17-109_k141_296573 | Not-determined |
| 17-109_k141_11167 | Low-quality |
| 17-109_k141_396820 | Low-quality |
| 17-109_k141_396822 | Not-determined |
| 17-109_k141_280094 | Not-determined |
| 17-109_k141_280095 | Not-determined |
| 17-109_k141_127852 | Low-quality |
| 17-109_k141_262399 | Not-determined |
| 17-109_k141_11175 | Not-determined |
| 17-109_k141_127854 | Not-determined |
| 17-109_k141_79226 | Low-quality |
| 17-109_k141_11177 | Not-determined |
| 17-109_k141_79227 | Not-determined |
| 17-109_k141_162492 | Not-determined |
| 17-109_k141_229955 | Low-quality |
| 17-109_k141_111415 | Not-determined |
| 17-109_k141_296583 | Not-determined |
| 17-109_k141_111419 | Not-determined |
| 17-109_k141_11187 | Not-determined |
| 17-109_k141_145126 | Low-quality |
| 17-109_k141_94827 | Not-determined |
| 17-109_k141_94828 | Not-determined |
| 17-109_k141_94829 | Low-quality |
| 17-109_k141_79241 | Not-determined |
| 17-109_k141_111435 | Not-determined |
| 17-109_k141_361590 | Not-determined |
| 17-109_k141_280110 | Not-determined |
| 17-109_k141_11194 | Not-determined |
| 17-109_k141_111438 | Not-determined |
| 17-109_k141_262418 | Not-determined |
| 17-109_k141_212658 | Not-determined |
| 17-109_k141_212659 | Not-determined |
| 17-109_k141_111441 | Not-determined |
| 17-109_k141_94845 | Not-determined |
| 17-109_k141_45755 | Not-determined |
| 17-109_k141_79248 | Not-determined |
| 17-109_k141_79250 | Not-determined |
| 17-109_k141_361600 | Not-determined |
| 17-109_k141_94851 | Low-quality |
| 17-109_k141_45763 | Not-determined |
| 17-109_k141_296588 | Not-determined |
| 17-109_k141_329767 | Low-quality |
| 17-109_k141_79264 | Not-determined |
| 17-109_k141_162519 | Not-determined |
| 17-109_k141_111452 | Low-quality |
| 17-109_k141_380309 | Not-determined |
| 17-109_k141_111455 | Not-determined |
| 17-109_k141_178880 | Not-determined |
| 17-109_k141_329772 | Not-determined |
| 17-109_k141_111457 | Not-determined |
| 17-109_k141_346300 | Not-determined |
| 17-109_k141_61524 | Not-determined |
| 17-109_k141_262438 | Not-determined |
| 17-109_k141_346308 | Not-determined |
| 17-109_k141_361606 | Low-quality |
| 17-109_k141_127873 | Not-determined |
| 17-109_k141_94852 | Low-quality |
| 17-109_k141_127874 | Not-determined |
| 17-109_k141_162531 | Not-determined |
| 17-109_k141_111461 | Not-determined |
| 17-109_k141_396854 | Not-determined |
| 17-109_k141_162534 | Not-determined |
| 17-109_k141_61533 | Not-determined |
| 17-109_k141_396856 | Not-determined |
| 17-109_k141_296603 | Not-determined |
| 17-109_k141_361614 | Not-determined |
| 17-109_k141_380321 | Low-quality |
| 17-109_k141_61535 | Low-quality |
| 17-109_k141_346324 | Not-determined |
| 17-109_k141_178897 | Not-determined |
| 17-109_k141_361621 | Not-determined |
| 17-109_k141_396859 | Not-determined |
| 17-109_k141_380323 | Low-quality |
| 17-109_k141_178902 | Not-determined |
| 17-109_k141_396864 | Not-determined |
| 17-109_k141_61537 | Not-determined |
| 17-109_k141_212669 | Not-determined |
| 17-109_k141_61538 | Not-determined |
| 17-109_k141_396866 | Not-determined |
| 17-109_k141_346333 | Not-determined |
| 17-109_k141_178905 | Not-determined |
| 17-109_k141_329784 | Not-determined |
| 17-109_k141_45772 | Not-determined |
| 17-109_k141_145154 | Not-determined |
| 17-109_k141_94871 | Not-determined |
| 17-109_k141_11225 | Not-determined |
| 17-109_k141_79289 | Not-determined |
| 17-109_k141_61551 | Not-determined |
| 17-109_k141_396874 | Not-determined |
| 17-109_k141_280156 | Not-determined |
| 17-109_k141_229979 | Low-quality |
| 17-109_k141_11230 | Not-determined |
| 17-109_k141_79295 | Not-determined |
| 17-109_k141_178923 | Not-determined |
| 17-109_k141_396875 | Not-determined |
| 17-109_k141_145161 | Not-determined |
| 17-109_k141_11235 | Not-determined |
| 17-109_k141_79300 | Not-determined |
| 17-109_k141_145163 | Not-determined |
| 17-109_k141_11238 | Not-determined |
| 17-109_k141_11239 | Not-determined |
| 17-109_k141_111492 | Not-determined |
| 17-109_k141_28194 | Not-determined |
| 17-109_k141_145177 | Not-determined |
| 17-109_k141_396889 | Not-determined |
| 17-109_k141_45780 | Not-determined |
| 17-109_k141_262490 | Not-determined |
| 17-109_k141_380346 | Not-determined |
| 17-109_k141_361646 | Not-determined |
| 17-109_k141_127899 | Not-determined |
| 17-109_k141_61571 | Not-determined |
| 17-109_k141_94889 | Not-determined |
| 17-109_k141_61574 | Not-determined |
| 17-109_k141_94892 | Not-determined |
| 17-109_k141_61577 | Not-determined |
| 17-109_k141_127902 | Not-determined |
| 17-109_k141_229994 | Not-determined |
| 17-109_k141_28206 | Not-determined |
| 17-109_k141_28207 | Not-determined |
| 17-109_k141_262501 | Not-determined |
| 17-109_k141_346363 | Not-determined |
| 17-109_k141_94901 | Not-determined |
| 17-109_k141_162553 | Not-determined |
| 17-109_k141_61587 | Not-determined |
| 17-109_k141_61589 | Not-determined |
| 17-109_k141_94908 | Not-determined |
| 17-109_k141_329823 | Not-determined |
| 17-109_k141_296628 | Not-determined |
| 17-109_k141_296630 | Not-determined |
| 17-109_k141_145204 | Not-determined |
| 17-109_k141_11248 | Not-determined |
| 17-109_k141_28220 | Low-quality |
| 17-109_k141_94916 | Not-determined |
| 17-109_k141_230010 | Not-determined |
| 17-109_k141_280163 | Not-determined |
| 17-109_k141_346384 | Not-determined |
| 17-109_k141_178952 | Not-determined |
| 17-109_k141_230019 | Low-quality |
| 17-109_k141_94926 | Not-determined |
| 17-109_k141_329833 | Not-determined |
| 17-109_k141_61599 | Not-determined |
| 17-109_k141_11258 | Low-quality |
| 17-109_k141_178958 | Low-quality |
| 17-109_k141_178963 | Not-determined |
| 17-109_k141_45824 | Low-quality |
| 17-109_k141_145216 | Not-determined |
| 17-109_k141_178969 | Not-determined |
| 17-109_k141_346404 | Not-determined |
| 17-109_k141_329849 | Not-determined |
| 17-109_k141_280176 | Not-determined |
| 17-109_k141_396912 | Not-determined |
| 17-109_k141_94941 | Not-determined |
| 17-109_k141_230040 | Not-determined |
| 17-109_k141_329851 | Not-determined |
| 17-109_k141_329852 | Not-determined |
| 17-109_k141_230043 | Not-determined |
| 17-109_k141_361681 | Not-determined |
| 17-109_k141_178980 | Not-determined |
| 17-109_k141_162577 | Not-determined |
| 17-109_k141_212710 | Not-determined |
| 17-109_k141_361683 | Not-determined |
| 17-109_k141_94943 | Low-quality |
| 17-109_k141_329854 | Not-determined |
| 17-109_k141_45837 | Low-quality |
| 17-109_k141_245916 | Not-determined |
| 17-109_k141_212720 | Not-determined |
| 17-109_k141_262532 | Not-determined |
| 17-109_k141_346428 | Low-quality |
| 17-109_k141_45845 | Not-determined |
| 17-109_k141_127957 | Low-quality |
| 17-109_k141_380386 | Not-determined |
| 17-109_k141_262540 | Not-determined |
| 17-109_k141_245926 | Low-quality |
| 17-109_k141_212728 | Not-determined |
| 17-109_k141_314140 | Not-determined |
| 17-109_k141_212730 | Not-determined |
| 17-109_k141_245929 | Not-determined |
| 17-109_k141_262550 | Not-determined |
| 17-109_k141_212746 | Not-determined |
| 17-109_k141_296669 | Not-determined |
| 17-109_k141_329863 | Low-quality |
| 17-109_k141_111522 | Low-quality |
| 17-109_k141_296675 | Not-determined |
| 17-109_k141_94970 | Not-determined |
| 17-109_k141_314154 | Not-determined |
| 17-109_k141_314155 | Not-determined |
| 17-109_k141_314158 | Not-determined |
| 17-109_k141_296682 | Low-quality |
| 17-109_k141_380421 | Low-quality |
| 17-109_k141_329871 | Not-determined |
| 17-109_k141_94981 | Not-determined |
| 17-109_k141_280230 | Not-determined |
| 17-109_k141_94983 | Not-determined |
| 17-109_k141_230069 | Low-quality |
| 17-109_k141_145233 | Not-determined |
| 17-109_k141_280233 | Not-determined |
| 17-109_k141_127982 | Not-determined |
| 17-109_k141_329881 | Not-determined |
| 17-109_k141_127984 | Not-determined |
| 17-109_k141_329882 | Not-determined |
| 17-109_k141_11285 | Not-determined |
| 17-109_k141_280238 | Low-quality |
| 17-109_k141_61633 | Not-determined |
| 17-109_k141_127988 | Not-determined |
| 17-109_k141_28274 | Not-determined |
| 17-109_k141_296701 | Not-determined |
| 17-109_k141_127996 | Not-determined |
| 17-109_k141_11294 | Low-quality |
| 17-109_k141_230087 | Not-determined |
| 17-109_k141_262572 | Low-quality |
| 17-109_k141_280250 | Low-quality |
| 17-109_k141_380430 | Not-determined |
| 17-109_k141_95002 | Not-determined |
| 17-109_k141_296714 | Not-determined |
| 17-109_k141_262578 | Not-determined |
| 17-109_k141_230094 | Not-determined |
| 17-109_k141_61644 | Low-quality |
| 17-109_k141_212774 | Not-determined |
| 17-109_k141_145271 | Low-quality |
| 17-109_k141_45876 | Not-determined |
| 17-109_k141_196412 | Not-determined |
| 17-109_k141_296736 | Not-determined |
| 17-109_k141_95026 | Not-determined |
| 17-109_k141_280262 | Not-determined |
| 17-109_k141_61664 | Not-determined |
| 17-109_k141_128023 | Low-quality |
| 17-109_k141_230114 | Not-determined |
| 17-109_k141_95036 | Not-determined |
| 17-109_k141_314207 | Not-determined |
| 17-109_k141_361716 | Not-determined |
| 17-109_k141_262602 | Not-determined |
| 17-109_k141_196416 | Not-determined |
| 17-109_k141_380457 | Not-determined |
| 17-109_k141_28289 | Not-determined |
| 17-109_k141_61672 | Not-determined |
| 17-109_k141_28290 | Not-determined |
| 17-109_k141_128039 | Not-determined |
| 17-109_k141_145291 | Not-determined |
| 17-109_k141_179062 | Not-determined |
| 17-109_k141_296745 | Low-quality |
| 17-109_k141_314211 | Not-determined |
| 17-109_k141_179065 | Not-determined |
| 17-109_k141_95046 | Not-determined |
| 17-109_k141_314214 | Not-determined |
| 17-109_k141_95048 | Not-determined |
| 17-109_k141_179067 | Not-determined |
| 17-109_k141_361729 | Not-determined |
| 17-109_k141_145302 | Low-quality |
| 17-109_k141_230128 | Not-determined |
| 17-109_k141_95057 | Low-quality |
| 17-109_k141_28301 | Not-determined |
| 17-109_k141_280272 | Low-quality |
| 17-109_k141_230131 | Not-determined |
| 17-109_k141_95060 | Not-determined |
| 17-109_k141_95066 | Not-determined |
| 17-109_k141_296757 | Not-determined |
| 17-109_k141_61685 | Not-determined |
| 17-109_k141_45907 | Not-determined |
| 17-109_k141_245962 | Not-determined |
| 17-109_k141_28307 | Not-determined |
| 17-109_k141_61690 | Not-determined |
| 17-109_k141_45913 | Not-determined |
| 17-109_k141_230145 | Not-determined |
| 17-109_k141_296770 | Not-determined |
| 17-109_k141_230147 | Low-quality |
| 17-109_k141_212809 | Not-determined |
| 17-109_k141_212811 | Not-determined |
| 17-109_k141_212813 | Not-determined |
| 17-109_k141_212815 | Low-quality |
| 17-109_k141_361767 | Not-determined |
| 17-109_k141_396949 | Not-determined |
| 17-109_k141_361777 | Not-determined |
| 17-109_k141_280285 | Not-determined |
| 17-109_k141_396965 | Not-determined |
| 17-109_k141_280291 | Not-determined |
| 17-109_k141_11300 | Not-determined |
| 17-109_k141_262616 | Not-determined |
| 17-109_k141_245992 | Not-determined |
| 17-109_k141_396969 | Not-determined |
| 17-109_k141_262620 | Not-determined |
| 17-109_k141_212830 | Not-determined |
| 17-109_k141_280297 | Not-determined |
| 17-109_k141_280299 | Not-determined |
| 17-109_k141_262624 | Not-determined |
| 17-109_k141_45951 | Not-determined |
| 17-109_k141_145335 | Not-determined |
| 17-109_k141_28330 | Low-quality |
| 17-109_k141_28331 | Not-determined |
| 17-109_k141_28332 | Not-determined |
| 17-109_k141_230163 | Not-determined |
| 17-109_k141_162606 | Not-determined |
| 17-109_k141_314239 | Not-determined |
| 17-109_k141_314240 | Not-determined |
| 17-109_k141_61713 | Not-determined |
| 17-109_k141_45955 | Not-determined |
| 17-109_k141_314241 | Not-determined |
| 17-109_k141_95111 | Not-determined |
| 17-109_k141_61718 | Not-determined |
| 17-109_k141_61719 | Not-determined |
| 17-109_k141_61720 | Not-determined |
| 17-109_k141_230173 | Not-determined |
| 17-109_k141_262642 | Not-determined |
| 17-109_k141_128081 | Not-determined |
| 17-109_k141_396991 | Not-determined |
| 17-109_k141_95116 | Not-determined |
| 17-109_k141_28340 | Not-determined |
| 17-109_k141_11319 | Not-determined |
| 17-109_k141_28346 | Not-determined |
| 17-109_k141_296813 | Not-determined |
| 17-109_k141_262652 | Not-determined |
| 17-109_k141_95126 | Not-determined |
| 17-109_k141_45975 | Not-determined |
| 17-109_k141_230187 | Not-determined |
| 17-109_k141_79333 | Not-determined |
| 17-109_k141_11334 | Low-quality |
| 17-109_k141_230192 | Not-determined |
| 17-109_k141_28362 | Not-determined |
| 17-109_k141_61748 | Not-determined |
| 17-109_k141_314262 | Not-determined |
| 17-109_k141_314263 | Low-quality |
| 17-109_k141_329925 | Not-determined |
| 17-109_k141_11340 | Not-determined |
| 17-109_k141_45988 | Not-determined |
| 17-109_k141_230195 | Not-determined |
| 17-109_k141_329927 | Not-determined |
| 17-109_k141_179084 | Not-determined |
| 17-109_k141_212853 | Not-determined |
| 17-109_k141_262669 | Not-determined |
| 17-109_k141_346477 | Not-determined |
| 17-109_k141_380486 | Not-determined |
| 17-109_k141_262675 | Not-determined |
| 17-109_k141_296828 | Not-determined |
| 17-109_k141_28373 | Not-determined |
| 17-109_k141_280353 | Not-determined |
| 17-109_k141_179099 | Not-determined |
| 17-109_k141_262678 | Not-determined |
| 17-109_k141_28376 | Not-determined |
| 17-109_k141_329933 | Not-determined |
| 17-109_k141_61760 | Not-determined |
| 17-109_k141_28379 | Not-determined |
| 17-109_k141_46005 | Low-quality |
| 17-109_k141_314282 | Not-determined |
| 17-109_k141_111541 | Not-determined |
| 17-109_k141_230218 | Not-determined |
| 17-109_k141_11350 | Low-quality |
| 17-109_k141_296834 | Not-determined |
| 17-109_k141_329938 | Not-determined |
| 17-109_k141_61766 | Low-quality |
| 17-109_k141_262689 | Not-determined |
| 17-109_k141_11358 | Not-determined |
| 17-109_k141_380502 | Not-determined |
| 17-109_k141_329939 | Not-determined |
| 17-109_k141_397043 | Not-determined |
| 17-109_k141_262691 | Not-determined |
| 17-109_k141_262694 | Low-quality |
| 17-109_k141_11363 | Not-determined |
| 17-109_k141_296846 | Not-determined |
| 17-109_k141_11365 | Not-determined |
| 17-109_k141_128138 | Not-determined |
| 17-109_k141_296852 | Not-determined |
| 17-109_k141_314297 | Not-determined |
| 17-109_k141_346505 | Not-determined |
| 17-109_k141_380513 | Not-determined |
| 17-109_k141_79366 | Not-determined |
| 17-109_k141_46030 | Low-quality |
| 17-109_k141_380515 | Not-determined |
| 17-109_k141_61771 | Low-quality |
| 17-109_k141_11383 | Not-determined |
| 17-109_k141_11384 | Not-determined |
| 17-109_k141_95173 | Not-determined |
| 17-109_k141_246031 | Not-determined |
| 17-109_k141_11386 | Not-determined |
| 17-109_k141_128162 | Low-quality |
| 17-109_k141_95177 | Not-determined |
| 17-109_k141_46045 | Not-determined |
| 17-109_k141_11393 | Not-determined |
| 17-109_k141_380517 | Low-quality |
| 17-109_k141_212873 | Not-determined |
| 17-109_k141_28402 | Not-determined |
| 17-109_k141_380521 | Not-determined |
| 17-109_k141_212877 | Not-determined |
| 17-109_k141_128170 | Not-determined |
| 17-109_k141_262700 | Not-determined |
| 17-109_k141_11397 | Not-determined |
| 17-109_k141_11399 | Low-quality |
| 17-109_k141_397065 | Not-determined |
| 17-109_k141_196427 | Not-determined |
| 17-109_k141_11401 | Not-determined |
| 17-109_k141_145406 | Low-quality |
| 17-109_k141_314323 | Not-determined |
| 17-109_k141_162675 | Not-determined |
| 17-109_k141_128180 | Not-determined |
| 17-109_k141_46060 | Not-determined |
| 17-109_k141_111594 | Not-determined |
| 17-109_k141_95199 | Not-determined |
| 17-109_k141_314327 | Not-determined |
| 17-109_k141_196439 | Not-determined |
| 17-109_k141_61800 | Not-determined |
| 17-109_k141_380529 | Not-determined |
| 17-109_k141_128185 | Not-determined |
| 17-109_k141_61801 | Not-determined |
| 17-109_k141_11412 | Not-determined |
| 17-109_k141_380533 | Not-determined |

| | |
|---|---|
| 17-109_k141_346516 | Not-determined |
| 17-109_k141_61802 | Not-determined |
| 17-109_k141_330009 | Not-determined |
| 17-109_k141_11416 | Not-determined |
| 17-109_k141_230253 | Not-determined |
| 17-109_k141_128191 | Not-determined |
| 17-109_k141_11417 | Not-determined |
| 17-109_k141_128193 | Not-determined |
| 17-109_k141_11418 | Not-determined |
| 17-109_k141_330012 | Low-quality |
| 17-109_k141_46074 | Not-determined |
| 17-109_k141_162680 | Not-determined |
| 17-109_k141_11428 | Not-determined |
| 17-109_k141_212912 | Not-determined |
| 17-109_k141_296887 | Not-determined |
| 17-109_k141_46084 | Not-determined |
| 17-109_k141_95218 | Not-determined |
| 17-109_k141_196457 | Not-determined |
| 17-109_k141_296888 | Low-quality |
| 17-109_k141_61823 | Not-determined |
| 17-109_k141_95219 | Not-determined |
| 17-109_k141_380547 | Not-determined |
| 17-109_k141_296892 | Not-determined |
| 17-109_k141_95225 | Not-determined |
| 17-109_k141_330021 | Not-determined |
| 17-109_k141_212916 | Not-determined |
| 17-109_k141_296896 | Not-determined |
| 17-109_k141_380554 | Not-determined |
| 17-109_k141_246066 | Not-determined |
| 17-109_k141_95231 | Low-quality |
| 17-109_k141_11444 | Not-determined |
| 17-109_k141_128209 | Not-determined |
| 17-109_k141_128210 | Not-determined |
| 17-109_k141_95234 | Not-determined |
| 17-109_k141_79410 | Not-determined |
| 17-109_k141_346535 | Low-quality |
| 17-109_k141_95240 | Not-determined |
| 17-109_k141_397077 | Not-determined |
| 17-109_k141_280371 | Not-determined |
| 17-109_k141_296904 | Not-determined |
| 17-109_k141_95248 | Low-quality |
| 17-109_k141_280374 | Not-determined |
| 17-109_k141_46100 | Not-determined |
| 17-109_k141_330030 | Not-determined |
| 17-109_k141_145424 | Not-determined |
| 17-109_k141_380569 | Not-determined |
| 17-109_k141_61831 | Not-determined |
| 17-109_k141_79425 | Not-determined |
| 17-109_k141_230277 | Not-determined |
| 17-109_k141_380571 | Low-quality |
| 17-109_k141_230280 | Not-determined |
| 17-109_k141_46110 | Not-determined |
| 17-109_k141_361812 | Not-determined |
| 17-109_k141_262732 | Not-determined |
| 17-109_k141_46118 | Not-determined |
| 17-109_k141_346552 | Not-determined |
| 17-109_k141_162721 | Not-determined |
| 17-109_k141_95272 | Not-determined |
| 17-109_k141_162723 | Not-determined |
| 17-109_k141_280395 | Not-determined |
| 17-109_k141_162727 | Not-determined |
| 17-109_k141_346558 | Not-determined |
| 17-109_k141_280400 | Low-quality |
| 17-109_k141_346561 | Low-quality |
| 17-109_k141_79444 | Not-determined |
| 17-109_k141_196503 | Not-determined |
| 17-109_k141_346566 | Not-determined |
| 17-109_k141_46142 | Not-determined |
| 17-109_k141_79450 | Not-determined |
| 17-109_k141_162739 | Not-determined |
| 17-109_k141_128256 | Not-determined |
| 17-109_k141_380585 | Not-determined |
| 17-109_k141_397091 | Not-determined |
| 17-109_k141_28478 | Low-quality |
| 17-109_k141_280425 | Not-determined |
| 17-109_k141_397096 | Not-determined |
| 17-109_k141_246093 | Not-determined |
| 17-109_k141_95296 | Not-determined |
| 17-109_k141_79456 | Not-determined |
| 17-109_k141_162744 | Not-determined |
| 17-109_k141_162748 | Not-determined |
| 17-109_k141_162749 | Not-determined |
| 17-109_k141_262779 | Not-determined |
| 17-109_k141_361820 | Not-determined |
| 17-109_k141_296933 | Not-determined |
| 17-109_k141_212967 | Not-determined |
| 17-109_k141_212968 | Not-determined |
| 17-109_k141_280432 | Low-quality |
| 17-109_k141_11480 | Not-determined |
| 17-109_k141_179200 | Not-determined |
| 17-109_k141_196506 | Not-determined |
| 17-109_k141_330055 | Not-determined |
| 17-109_k141_162763 | Not-determined |
| 17-109_k141_145463 | Not-determined |
| 17-109_k141_162764 | Not-determined |
| 17-109_k141_380597 | Not-determined |
| 17-109_k141_145464 | Not-determined |
| 17-109_k141_330057 | Low-quality |
| 17-109_k141_314380 | Not-determined |
| 17-109_k141_145468 | Not-determined |
| 17-109_k141_314383 | Not-determined |
| 17-109_k141_111635 | Not-determined |
| 17-109_k141_330061 | Not-determined |
| 17-109_k141_95304 | Not-determined |
| 17-109_k141_262798 | Not-determined |
| 17-109_k141_95307 | Not-determined |
| 17-109_k141_330065 | Not-determined |
| 17-109_k141_296951 | Not-determined |
| 17-109_k141_111638 | Not-determined |
| 17-109_k141_162772 | Not-determined |
| 17-109_k141_212977 | Not-determined |
| 17-109_k141_95310 | Not-determined |
| 17-109_k141_128284 | Not-determined |
| 17-109_k141_330072 | Not-determined |
| 17-109_k141_145479 | Not-determined |
| 17-109_k141_28506 | Not-determined |
| 17-109_k141_246117 | Not-determined |
| 17-109_k141_28508 | Not-determined |
| 17-109_k141_280446 | Not-determined |
| 17-109_k141_330079 | Not-determined |
| 17-109_k141_79485 | Low-quality |
| 17-109_k141_246122 | Not-determined |
| 17-109_k141_212992 | Not-determined |
| 17-109_k141_95316 | Not-determined |
| 17-109_k141_179235 | Not-determined |
| 17-109_k141_330087 | Not-determined |
| 17-109_k141_46159 | Not-determined |
| 17-109_k141_397126 | Not-determined |
| 17-109_k141_330095 | Not-determined |
| 17-109_k141_46162 | Not-determined |
| 17-109_k141_162787 | Not-determined |
| 17-109_k141_79500 | Not-determined |
| 17-109_k141_46163 | Not-determined |
| 17-109_k141_46165 | Not-determined |
| 17-109_k141_314404 | Low-quality |
| 17-109_k141_314405 | Not-determined |
| 17-109_k141_314406 | Not-determined |
| 17-109_k141_46168 | Not-determined |
| 17-109_k141_61916 | Not-determined |
| 17-109_k141_314409 | Not-determined |
| 17-109_k141_61917 | Not-determined |
| 17-109_k141_61920 | Not-determined |
| 17-109_k141_246143 | Not-determined |
| 17-109_k141_330109 | Not-determined |
| 17-109_k141_61925 | Not-determined |
| 17-109_k141_179244 | Not-determined |
| 17-109_k141_61927 | Not-determined |
| 17-109_k141_246626 | Low-quality |
| 17-109_k141_95346 | Not-determined |
| 17-109_k141_179247 | Not-determined |
| 17-109_k141_330120 | Not-determined |
| 17-109_k141_61932 | Not-determined |
| 17-109_k141_179254 | Not-determined |
| 17-109_k141_280473 | Low-quality |
| 17-109_k141_397147 | Not-determined |
| 17-109_k141_262812 | Not-determined |
| 17-109_k141_280476 | Not-determined |
| 17-109_k141_296976 | Not-determined |
| 17-109_k141_111675 | Not-determined |
| 17-109_k141_61944 | Not-determined |
| 17-109_k141_361882 | Not-determined |
| 17-109_k141_79521 | Not-determined |
| 17-109_k141_162798 | Not-determined |
| 17-109_k141_346655 | Not-determined |
| 17-109_k141_79523 | Not-determined |
| 17-109_k141_346656 | Not-determined |
| 17-109_k141_280486 | Not-determined |
| 17-109_k141_111682 | Not-determined |
| 17-109_k141_262827 | Not-determined |
| 17-109_k141_79527 | Low-quality |
| 17-109_k141_128330 | Not-determined |
| 17-109_k141_397156 | Not-determined |
| 17-109_k141_361884 | Not-determined |
| 17-109_k141_346659 | Not-determined |
| 17-109_k141_179271 | Not-determined |
| 17-109_k141_61960 | Not-determined |
| 17-109_k141_330146 | Not-determined |
| 17-109_k141_280493 | Not-determined |
| 17-109_k141_196539 | Not-determined |
| 17-109_k141_145509 | Not-determined |
| 17-109_k141_330153 | Not-determined |
| 17-109_k141_179283 | Not-determined |
| 17-109_k141_196543 | Not-determined |
| 17-109_k141_196547 | Not-determined |
| 17-109_k141_79543 | Not-determined |
| 17-109_k141_145515 | Not-determined |
| 17-109_k141_280508 | Not-determined |
| 17-109_k141_46195 | Not-determined |
| 17-109_k141_46198 | Not-determined |
| 17-109_k141_61963 | Not-determined |
| 17-109_k141_280513 | Not-determined |
| 17-109_k141_280514 | Not-determined |
| 17-109_k141_79552 | Not-determined |
| 17-109_k141_46204 | Low-quality |
| 17-109_k141_397174 | Not-determined |
| 17-109_k141_61966 | Low-quality |
| 17-109_k141_95383 | Not-determined |
| 17-109_k141_46207 | Not-determined |
| 17-109_k141_145529 | Not-determined |
| 17-109_k141_111706 | Not-determined |
| 17-109_k141_213017 | Not-determined |
| 17-109_k141_128351 | Not-determined |
| 17-109_k141_61973 | Not-determined |
| 17-109_k141_61975 | Not-determined |
| 17-109_k141_79567 | Not-determined |
| 17-109_k141_95392 | Not-determined |
| 17-109_k141_111707 | Low-quality |
| 17-109_k141_145535 | Low-quality |
| 17-109_k141_213027 | Not-determined |
| 17-109_k141_213028 | Low-quality |
| 17-109_k141_361917 | Not-determined |
| 17-109_k141_314444 | Not-determined |
| 17-109_k141_361919 | Not-determined |
| 17-109_k141_46222 | Not-determined |
| 17-109_k141_79576 | Low-quality |
| 17-109_k141_361921 | Not-determined |
| 17-109_k141_145543 | Not-determined |
| 17-109_k141_179312 | Not-determined |
| 17-109_k141_179313 | Not-determined |
| 17-109_k141_95399 | Not-determined |
| 17-109_k141_61981 | Not-determined |
| 17-109_k141_314450 | Not-determined |
| 17-109_k141_145549 | Not-determined |
| 17-109_k141_314454 | Not-determined |
| 17-109_k141_128358 | Not-determined |
| 17-109_k141_297009 | Not-determined |
| 17-109_k141_262863 | Not-determined |
| 17-109_k141_314459 | Low-quality |
| 17-109_k141_145550 | Not-determined |
| 17-109_k141_330177 | Not-determined |
| 17-109_k141_28546 | Not-determined |
| 17-109_k141_213041 | Not-determined |
| 17-109_k141_128367 | Not-determined |
| 17-109_k141_213043 | Not-determined |
| 17-109_k141_262865 | Not-determined |
| 17-109_k141_213044 | Low-quality |
| 17-109_k141_346710 | Not-determined |
| 17-109_k141_280550 | Not-determined |
| 17-109_k141_179320 | Not-determined |
| 17-109_k141_314460 | Low-quality |
| 17-109_k141_262875 | Not-determined |
| 17-109_k141_346714 | Not-determined |
| 17-109_k141_330183 | Not-determined |
| 17-109_k141_213053 | Not-determined |
| 17-109_k141_111731 | Not-determined |
| 17-109_k141_314468 | Not-determined |
| 17-109_k141_297017 | Not-determined |
| 17-109_k141_314469 | Low-quality |
| 17-109_k141_145564 | Not-determined |
| 17-109_k141_361948 | Not-determined |
| 17-109_k141_330190 | Not-determined |
| 17-109_k141_46249 | Not-determined |
| 17-109_k141_361950 | Not-determined |
| 17-109_k141_179337 | Not-determined |
| 17-109_k141_297021 | Not-determined |
| 17-109_k141_179339 | Low-quality |
| 17-109_k141_280556 | Not-determined |
| 17-109_k141_380620 | Not-determined |
| 17-109_k141_162801 | Not-determined |
| 17-109_k141_380621 | Not-determined |
| 17-109_k141_314482 | Not-determined |
| 17-109_k141_262891 | Not-determined |
| 17-109_k141_111745 | Not-determined |
| 17-109_k141_111746 | Not-determined |
| 17-109_k141_28571 | Not-determined |
| 17-109_k141_28572 | Not-determined |
| 17-109_k141_330205 | Not-determined |
| 17-109_k141_128391 | Not-determined |
| 17-109_k141_246210 | Not-determined |
| 17-109_k141_46267 | Not-determined |
| 17-109_k141_28579 | Not-determined |
| 17-109_k141_162805 | Not-determined |
| 17-109_k141_297026 | Not-determined |
| 17-109_k141_111758 | Not-determined |
| 17-109_k141_262894 | Not-determined |
| 17-109_k141_314494 | Not-determined |
| 17-109_k141_28591 | Not-determined |
| 17-109_k141_314497 | Not-determined |
| 17-109_k141_46284 | Not-determined |
| 17-109_k141_246229 | Low-quality |
| 17-109_k141_179360 | Not-determined |
| 17-109_k141_46291 | Not-determined |
| 17-109_k141_380643 | Low-quality |
| 17-109_k141_179362 | Not-determined |
| 17-109_k141_262910 | Not-determined |
| 17-109_k141_314514 | Low-quality |
| 17-109_k141_46302 | Not-determined |
| 17-109_k141_46304 | Not-determined |
| 17-109_k141_128423 | Not-determined |
| 17-109_k141_280604 | Not-determined |
| 17-109_k141_128425 | Not-determined |
| 17-109_k141_162833 | Not-determined |
| 17-109_k141_380656 | Not-determined |
| 17-109_k141_145620 | Not-determined |
| 17-109_k141_262919 | Not-determined |
| 17-109_k141_79606 | Not-determined |
| 17-109_k141_145626 | Not-determined |
| 17-109_k141_297050 | Not-determined |
| 17-109_k141_346778 | Not-determined |
| 17-109_k141_46314 | Not-determined |
| 17-109_k141_145631 | Not-determined |
| 17-109_k141_213071 | Not-determined |
| 17-109_k141_280610 | Not-determined |
| 17-109_k141_346780 | Not-determined |
| 17-109_k141_246255 | Not-determined |
| 17-109_k141_397214 | Not-determined |
| 17-109_k141_179383 | Not-determined |
| 17-109_k141_314529 | Not-determined |
| 17-109_k141_346782 | Not-determined |
| 17-109_k141_46319 | Not-determined |
| 17-109_k141_330224 | Not-determined |
| 17-109_k141_314531 | Not-determined |
| 17-109_k141_297058 | Not-determined |
| 17-109_k141_397218 | Not-determined |
| 17-109_k141_62068 | Not-determined |
| 17-109_k141_11548 | Low-quality |
| 17-109_k141_297063 | Not-determined |
| 17-109_k141_330235 | Not-determined |
| 17-109_k141_62073 | Not-determined |
| 17-109_k141_213086 | Not-determined |
| 17-109_k141_11556 | Not-determined |
| 17-109_k141_11559 | Not-determined |
| 17-109_k141_330239 | Not-determined |
| 17-109_k141_397225 | Not-determined |
| 17-109_k141_28650 | Not-determined |
| 17-109_k141_297078 | Not-determined |
| 17-109_k141_230285 | Not-determined |
| 17-109_k141_346793 | Not-determined |
| 17-109_k141_128453 | Not-determined |
| 17-109_k141_62078 | Low-quality |
| 17-109_k141_145657 | Low-quality |
| 17-109_k141_95416 | Not-determined |
| 17-109_k141_145661 | Not-determined |
| 17-109_k141_11583 | Not-determined |
| 17-109_k141_95423 | Not-determined |
| 17-109_k141_213103 | Not-determined |
| 17-109_k141_346811 | Not-determined |
| 17-109_k141_346812 | Not-determined |
| 17-109_k141_346814 | Not-determined |
| 17-109_k141_62087 | Not-determined |
| 17-109_k141_230292 | Not-determined |
| 17-109_k141_346815 | Not-determined |
| 17-109_k141_95427 | Not-determined |
| 17-109_k141_213107 | Not-determined |
| 17-109_k141_213108 | Not-determined |
| 17-109_k141_330259 | Not-determined |
| 17-109_k141_213109 | Not-determined |
| 17-109_k141_330260 | Not-determined |
| 17-109_k141_280646 | Not-determined |
| 17-109_k141_28662 | Not-determined |
| 17-109_k141_62099 | Not-determined |
| 17-109_k141_128472 | Not-determined |
| 17-109_k141_28667 | Not-determined |
| 17-109_k141_95442 | Not-determined |
| 17-109_k141_162872 | Low-quality |
| 17-109_k141_380678 | Low-quality |
| 17-109_k141_46366 | Low-quality |
| 17-109_k141_262965 | Not-determined |
| 17-109_k141_280657 | Low-quality |
| 17-109_k141_128485 | Not-determined |
| 17-109_k141_162877 | Not-determined |
| 17-109_k141_280659 | Not-determined |
| 17-109_k141_128489 | Not-determined |
| 17-109_k141_128490 | Not-determined |
| 17-109_k141_361959 | Not-determined |
| 17-109_k141_62103 | Not-determined |
| 17-109_k141_128494 | Not-determined |
| 17-109_k141_246313 | Not-determined |
| 17-109_k141_28684 | Low-quality |
| 17-109_k141_361963 | Not-determined |
| 17-109_k141_330278 | Low-quality |
| 17-109_k141_79647 | Not-determined |
| 17-109_k141_280665 | Not-determined |
| 17-109_k141_62108 | Not-determined |
| 17-109_k141_196567 | Not-determined |
| 17-109_k141_62113 | Not-determined |
| 17-109_k141_380696 | Low-quality |
| 17-109_k141_28690 | Not-determined |
| 17-109_k141_346840 | Not-determined |
| 17-109_k141_196571 | Not-determined |
| 17-109_k141_380698 | Not-determined |
| 17-109_k141_79662 | Not-determined |
| 17-109_k141_62121 | Low-quality |
| 17-109_k141_111828 | Not-determined |
| 17-109_k141_162908 | Not-determined |
| 17-109_k141_230322 | Not-determined |
| 18-102-final-viral-combined_k141_158821\|\|full | Medium-quality |
| 17-109_k141_361977 | Not-determined |
| 17-109_k141_196576 | Not-determined |
| 17-109_k141_314576 | Not-determined |
| 17-109_k141_361978 | Not-determined |
| 17-109_k141_380704 | Not-determined |
| 17-109_k141_111832 | Not-determined |
| 17-109_k141_361979 | Low-quality |
| 17-109_k141_46398 | Not-determined |
| 17-109_k141_361982 | Not-determined |
| 17-109_k141_79671 | Not-determined |
| 17-109_k141_28701 | Not-determined |
| 17-109_k141_128522 | Not-determined |
| 17-109_k141_230336 | Not-determined |
| 17-109_k141_79676 | Not-determined |
| 17-109_k141_297105 | Low-quality |
| 17-109_k141_230337 | Not-determined |
| 17-109_k141_11622 | Not-determined |
| 17-109_k141_361988 | Not-determined |
| 17-109_k141_46409 | Not-determined |
| 17-109_k141_314580 | Not-determined |
| 17-109_k141_11628 | Not-determined |
| 17-109_k141_314585 | Not-determined |
| 17-109_k141_361995 | Not-determined |
| 17-109_k141_162930 | Not-determined |
| 17-109_k141_263021 | Not-determined |
| 17-109_k141_346865 | Not-determined |
| 17-109_k141_314598 | Not-determined |
| 17-109_k141_213180 | Not-determined |
| 17-109_k141_346869 | Not-determined |
| 17-109_k141_380721 | Low-quality |
| 17-109_k141_62148 | Not-determined |
| 17-109_k141_196595 | Not-determined |
| 17-109_k141_346879 | Not-determined |
| 17-109_k141_62155 | Low-quality |
| 17-109_k141_263043 | Not-determined |
| 17-109_k141_179415 | Not-determined |
| 17-109_k141_95476 | Not-determined |
| 17-109_k141_362011 | Not-determined |
| 17-109_k141_380729 | Low-quality |
| 17-109_k141_362016 | Low-quality |
| 17-109_k141_111869 | Not-determined |
| 17-109_k141_380732 | Not-determined |
| 17-109_k141_145713 | Not-determined |
| 17-109_k141_263052 | Not-determined |
| 17-109_k141_297128 | Not-determined |
| 17-109_k141_145716 | Not-determined |
| 17-109_k141_314625 | Low-quality |
| 17-109_k141_314627 | Not-determined |
| 17-109_k141_28718 | Not-determined |
| 17-109_k141_314630 | Not-determined |
| 17-109_k141_230350 | Not-determined |
| 17-109_k141_145724 | Not-determined |
| 17-109_k141_297134 | Not-determined |
| 17-109_k141_79705 | Low-quality |
| 17-109_k141_230357 | Not-determined |
| 17-109_k141_95489 | Not-determined |
| 17-109_k141_11643 | Not-determined |
| 17-109_k141_11645 | Not-determined |
| 17-109_k141_46447 | Not-determined |
| 17-109_k141_11648 | Not-determined |
| 17-109_k141_79714 | Low-quality |
| 17-109_k141_297152 | Not-determined |
| 17-109_k141_62188 | Not-determined |
| 17-109_k141_297155 | Not-determined |
| 17-109_k141_11655 | Not-determined |
| 17-109_k141_330315 | Not-determined |
| 17-109_k141_128598 | Not-determined |
| 17-109_k141_145742 | Not-determined |
| 17-109_k141_362037 | Low-quality |
| 17-109_k141_397261 | Not-determined |
| 17-109_k141_145748 | Not-determined |
| 17-109_k141_28724 | Not-determined |
| 17-109_k141_346934 | Not-determined |
| 17-109_k141_95509 | Not-determined |
| 17-109_k141_330323 | Not-determined |
| 17-109_k141_79726 | Not-determined |
| 17-109_k141_11662 | Not-determined |
| 17-109_k141_128610 | Low-quality |
| 17-109_k141_280710 | Not-determined |
| 17-109_k141_145753 | Not-determined |
| 17-109_k141_362043 | Not-determined |
| 17-109_k141_179460 | Not-determined |
| 17-109_k141_95514 | Not-determined |
| 17-109_k141_280712 | Not-determined |
| 17-109_k141_362048 | Not-determined |
| 17-109_k141_246339 | Not-determined |
| 17-109_k141_11669 | Not-determined |
| 17-109_k141_62203 | Not-determined |
| 17-109_k141_314655 | Low-quality |
| 17-109_k141_62209 | Not-determined |
| 17-109_k141_128623 | Not-determined |
| 17-109_k141_246346 | Not-determined |
| 17-109_k141_79739 | Not-determined |
| 17-109_k141_280730 | Not-determined |
| 17-109_k141_213227 | Low-quality |
| 17-109_k141_330341 | Not-determined |
| 17-109_k141_213231 | Not-determined |
| 17-109_k141_79746 | Not-determined |
| 17-109_k141_362057 | Not-determined |
| 17-109_k141_213234 | Not-determined |
| 17-109_k141_362058 | Not-determined |
| 17-109_k141_179481 | Not-determined |
| 17-109_k141_346945 | Not-determined |
| 17-109_k141_196642 | Not-determined |
| 17-109_k141_330351 | Not-determined |
| 17-109_k141_230390 | Not-determined |
| 17-109_k141_46486 | Not-determined |
| 17-109_k141_46487 | Not-determined |
| 17-109_k141_330353 | Not-determined |
| 17-109_k141_346952 | Not-determined |
| 17-109_k141_62216 | Not-determined |
| 17-109_k141_11682 | Not-determined |
| 17-109_k141_330361 | Not-determined |
| 17-109_k141_179500 | Not-determined |
| 17-109_k141_330368 | Not-determined |
| 17-109_k141_230402 | Not-determined |
| 17-109_k141_46500 | Not-determined |
| 17-109_k141_297212 | Not-determined |
| 17-109_k141_230412 | Not-determined |
| 17-109_k141_46508 | Low-quality |
| 17-109_k141_196671 | Low-quality |
| 17-109_k141_346979 | Not-determined |
| 17-109_k141_196679 | Not-determined |
| 17-109_k141_380753 | Not-determined |
| 17-109_k141_346984 | Low-quality |
| 17-109_k141_380754 | Not-determined |
| 17-109_k141_179524 | Not-determined |
| 17-109_k141_297222 | Not-determined |
| 17-109_k141_330401 | Not-determined |
| 17-109_k141_230431 | Not-determined |
| 17-109_k141_280762 | Not-determined |
| 17-109_k141_28743 | Low-quality |
| 17-109_k141_62232 | Not-determined |
| 17-109_k141_280764 | Not-determined |
| 17-109_k141_28744 | Not-determined |
| 17-109_k141_79783 | Not-determined |
| 17-109_k141_62233 | Not-determined |
| 17-109_k141_28747 | Not-determined |
| 17-109_k141_263102 | Not-determined |
| 17-109_k141_79785 | Not-determined |
| 17-109_k141_263103 | Not-determined |
| 17-109_k141_263104 | Not-determined |
| 17-109_k141_397278 | Not-determined |
| 17-109_k141_79788 | Not-determined |
| 17-109_k141_362062 | Not-determined |
| 17-109_k141_346996 | Not-determined |
| 17-109_k141_397282 | Not-determined |
| 17-109_k141_28754 | Not-determined |
| 17-109_k141_46514 | Not-determined |
| 17-109_k141_179545 | Not-determined |
| 17-109_k141_196706 | Not-determined |
| 17-109_k141_246386 | Not-determined |
| 17-109_k141_380779 | Not-determined |
| 17-109_k141_128639 | Not-determined |
| 17-109_k141_79805 | Not-determined |
| 17-109_k141_246394 | Not-determined |
| 17-109_k141_179558 | Not-determined |
| 17-109_k141_179559 | Not-determined |
| 17-109_k141_95550 | Not-determined |
| 17-109_k141_46524 | Not-determined |
| 17-109_k141_230451 | Not-determined |
| 17-109_k141_230452 | Low-quality |
| 17-109_k141_280787 | Not-determined |
| 17-109_k141_79814 | Not-determined |
| 17-109_k141_246401 | Not-determined |
| 17-109_k141_213296 | Not-determined |
| 17-109_k141_95559 | Not-determined |
| 17-109_k141_79821 | Not-determined |
| 17-109_k141_280794 | Not-determined |
| 17-109_k141_246402 | Not-determined |
| 17-109_k141_162973 | Not-determined |
| 17-109_k141_145770 | Not-determined |
| 17-109_k141_330449 | Not-determined |
| 17-109_k141_145772 | Not-determined |
| 17-109_k141_330450 | Not-determined |
| 17-109_k141_111902 | Not-determined |
| 17-109_k141_128657 | Not-determined |
| 17-109_k141_347022 | Not-determined |
| 17-109_k141_62256 | Not-determined |
| 17-109_k141_179579 | Not-determined |
| 17-109_k141_280803 | Not-determined |
| 17-109_k141_246405 | Not-determined |
| 17-109_k141_162985 | Low-quality |
| 17-109_k141_362088 | Low-quality |
| 17-109_k141_46527 | Not-determined |
| 17-109_k141_46530 | Not-determined |
| 17-109_k141_62264 | Not-determined |
| 17-109_k141_179591 | Not-determined |
| 17-109_k141_330461 | Not-determined |
| 17-109_k141_11738 | Not-determined |
| 17-109_k141_62268 | Not-determined |
| 17-109_k141_11739 | Not-determined |
| 17-109_k141_397321 | Not-determined |
| 17-109_k141_246414 | Not-determined |
| 17-109_k141_62274 | Not-determined |
| 17-109_k141_111919 | Not-determined |
| 17-109_k141_330470 | Not-determined |
| 17-109_k141_111921 | Not-determined |
| 17-109_k141_79839 | Not-determined |
| 17-109_k141_179607 | Not-determined |
| 17-109_k141_330477 | Not-determined |
| 17-109_k141_347048 | Not-determined |

| | |
|---|---|
| 17-109_k141_62289 | Not-determined |
| 17-109_k141_111932 | Not-determined |
| 17-109_k141_111933 | Not-determined |
| 17-109_k141_196751 | Not-determined |
| 17-109_k141_362115 | Not-determined |
| 17-109_k141_330486 | Not-determined |
| 17-109_k141_213320 | Low-quality |
| 17-109_k141_280832 | Not-determined |
| 17-109_k141_163011 | Not-determined |
| 17-109_k141_196762 | Not-determined |
| 17-109_k141_380820 | Not-determined |
| 17-109_k141_163014 | Low-quality |
| 17-109_k141_347066 | Not-determined |
| 17-109_k141_11748 | Low-quality |
| 17-109_k141_79865 | Not-determined |
| 17-109_k141_79866 | Not-determined |
| 17-109_k141_213337 | Not-determined |
| 17-109_k141_79867 | Not-determined |
| 17-109_k141_380832 | Not-determined |
| 17-109_k141_297264 | Not-determined |
| 17-109_k141_263154 | Not-determined |
| 17-109_k141_280846 | Low-quality |
| 17-109_k141_145805 | Not-determined |
| 17-109_k141_347071 | Not-determined |
| 17-109_k141_11756 | Not-determined |
| 17-109_k141_246430 | Not-determined |
| 17-109_k141_11759 | Not-determined |
| 17-109_k141_62305 | Low-quality |
| 17-109_k141_46570 | Not-determined |
| 17-109_k141_11763 | Not-determined |
| 17-109_k141_163022 | Not-determined |
| 17-109_k141_213350 | Low-quality |
| 17-109_k141_46574 | Not-determined |
| 17-109_k141_297275 | Not-determined |
| 17-109_k141_79886 | Not-determined |
| 17-109_k141_163027 | Not-determined |
| 17-109_k141_179635 | Not-determined |
| 17-109_k141_163031 | Not-determined |
| 17-109_k141_362131 | Low-quality |
| 17-109_k141_196784 | Not-determined |
| 17-109_k141_246432 | Not-determined |
| 17-109_k141_179642 | Not-determined |
| 17-109_k141_246434 | Not-determined |
| 17-109_k141_179645 | Not-determined |
| 17-109_k141_263165 | Not-determined |
| 17-109_k141_111955 | Not-determined |
| 17-109_k141_230463 | Not-determined |
| 17-109_k141_196794 | Not-determined |
| 17-109_k141_263169 | Not-determined |
| 17-109_k141_196795 | Not-determined |
| 17-109_k141_79894 | Not-determined |
| 17-109_k141_145817 | Low-quality |
| 17-109_k141_263174 | Not-determined |
| 17-109_k141_230472 | Not-determined |
| 17-109_k141_230473 | Not-determined |
| 17-109_k141_179655 | Not-determined |
| 17-109_k141_62337 | Not-determined |
| 17-109_k141_297289 | Not-determined |
| 17-109_k141_163053 | Not-determined |
| 17-109_k141_196799 | Not-determined |
| 17-109_k141_263182 | Not-determined |
| 17-109_k141_263186 | Not-determined |
| 17-109_k141_62351 | Not-determined |
| 17-109_k141_196808 | Not-determined |
| 17-109_k141_163064 | Not-determined |
| 17-109_k141_280874 | Not-determined |
| 17-109_k141_163069 | Not-determined |
| 17-109_k141_230491 | Not-determined |
| 17-109_k141_263195 | Not-determined |
| 17-109_k141_11801 | Not-determined |
| 17-109_k141_46620 | Not-determined |
| 17-109_k141_213382 | Not-determined |
| 17-109_k141_179686 | Not-determined |
| 17-109_k141_128742 | Not-determined |
| 17-109_k141_314691 | Not-determined |
| 17-109_k141_128744 | Not-determined |
| 17-109_k141_397352 | Not-determined |
| 17-109_k141_79913 | Not-determined |
| 17-109_k141_62366 | Not-determined |
| 17-109_k141_213387 | Not-determined |
| 17-109_k141_95562 | Not-determined |
| 17-109_k141_213390 | Not-determined |
| 17-109_k141_213391 | Not-determined |
| 17-109_k141_95564 | Not-determined |
| 17-109_k141_95566 | Not-determined |
| 17-109_k141_362139 | Not-determined |
| 17-109_k141_297318 | Not-determined |
| 17-109_k141_297319 | Not-determined |
| 17-109_k141_145834 | Not-determined |
| 17-109_k141_196834 | Not-determined |
| 17-109_k141_79925 | Not-determined |
| 17-109_k141_62376 | Low-quality |
| 17-109_k141_163090 | Not-determined |
| 17-109_k141_163096 | Not-determined |
| 17-109_k141_314707 | Not-determined |
| 17-109_k141_196840 | Not-determined |
| 17-109_k141_314712 | Not-determined |
| 17-109_k141_79939 | Not-determined |
| 17-109_k141_246436 | Not-determined |
| 17-109_k141_79941 | Not-determined |
| 17-109_k141_397369 | Not-determined |
| 17-109_k141_362162 | Not-determined |
| 17-109_k141_163114 | Not-determined |
| 17-109_k141_213422 | Not-determined |
| 17-109_k141_95568 | Not-determined |
| 17-109_k141_11845 | Not-determined |
| 17-109_k141_362164 | Low-quality |
| 17-109_k141_179725 | Low-quality |
| 17-109_k141_347124 | Not-determined |
| 17-109_k141_95577 | Not-determined |
| 17-109_k141_230509 | Not-determined |
| 17-109_k141_297325 | Low-quality |
| 17-109_k141_163119 | Not-determined |
| 17-109_k141_95580 | Not-determined |
| 17-109_k141_362170 | Not-determined |
| 17-109_k141_163122 | Not-determined |
| 17-109_k141_362172 | Not-determined |
| 17-109_k141_179732 | Not-determined |
| 17-109_k141_145864 | Not-determined |
| 17-109_k141_330561 | Not-determined |
| 17-109_k141_145866 | Not-determined |
| 17-109_k141_145868 | Not-determined |
| 17-109_k141_347134 | Not-determined |
| 17-109_k141_163129 | Not-determined |
| 17-109_k141_347136 | Not-determined |
| 17-109_k141_163130 | Not-determined |
| 17-109_k141_397386 | Not-determined |
| 17-109_k141_347140 | Not-determined |
| 17-109_k141_397390 | Not-determined |
| 17-109_k141_213436 | Not-determined |
| 17-109_k141_46639 | Not-determined |
| 17-109_k141_213437 | Low-quality |
| 17-109_k141_179741 | Low-quality |
| 17-109_k141_145876 | Not-determined |
| 17-109_k141_179742 | Not-determined |
| 17-109_k141_128757 | Low-quality |
| 17-109_k141_79964 | Not-determined |
| 17-109_k141_163141 | Not-determined |
| 17-109_k141_179748 | Not-determined |
| 17-109_k141_28803 | Not-determined |
| 17-109_k141_347158 | Not-determined |
| 17-109_k141_347160 | Not-determined |
| 17-109_k141_347161 | Not-determined |
| 17-109_k141_179756 | Not-determined |
| 17-109_k141_28806 | Not-determined |
| 17-109_k141_179759 | Low-quality |
| 17-109_k141_163154 | Not-determined |
| 17-109_k141_380855 | Not-determined |
| 17-109_k141_397413 | Not-determined |
| 17-109_k141_128778 | Not-determined |
| 17-109_k141_28818 | Not-determined |
| 17-109_k141_362191 | Not-determined |
| 17-109_k141_196880 | Not-determined |
| 17-109_k141_280909 | Not-determined |
| 17-109_k141_213448 | Not-determined |
| 17-109_k141_213452 | Not-determined |
| 17-109_k141_380874 | Not-determined |
| 17-109_k141_179770 | Not-determined |
| 17-109_k141_79988 | Not-determined |
| 17-109_k141_380877 | Not-determined |
| 17-109_k141_263237 | Not-determined |
| 17-109_k141_280920 | Not-determined |
| 17-109_k141_145892 | Not-determined |
| 17-109_k141_347181 | Not-determined |
| 17-109_k141_79993 | Not-determined |
| 17-109_k141_330603 | Not-determined |
| 17-109_k141_95615 | Not-determined |
| 17-109_k141_297339 | Not-determined |
| 17-109_k141_397431 | Not-determined |
| 17-109_k141_145905 | Not-determined |
| 17-109_k141_145908 | Low-quality |
| 17-109_k141_246467 | Not-determined |
| 17-109_k141_362216 | Not-determined |
| 17-109_k141_213469 | Not-determined |
| 17-109_k141_179792 | Not-determined |
| 17-109_k141_362221 | Not-determined |
| 17-109_k141_28859 | Not-determined |
| 17-109_k141_163191 | Not-determined |
| 17-109_k141_380907 | Not-determined |
| 17-109_k141_213477 | Not-determined |
| 17-109_k141_179801 | Not-determined |
| 17-109_k141_95631 | Not-determined |
| 17-109_k141_330625 | Not-determined |
| 17-109_k141_80001 | Not-determined |
| 17-109_k141_330626 | Not-determined |
| 17-109_k141_380916 | Not-determined |
| 17-109_k141_80003 | Not-determined |
| 17-109_k141_297347 | Not-determined |
| 17-109_k141_362235 | Not-determined |
| 17-109_k141_213485 | Not-determined |
| 17-109_k141_380918 | Low-quality |
| 17-109_k141_213489 | Not-determined |
| 17-109_k141_179815 | Not-determined |
| 17-109_k141_95635 | Not-determined |
| 17-109_k141_112053 | Not-determined |
| 17-109_k141_196920 | Not-determined |
| 17-109_k141_11874 | Not-determined |
| 17-109_k141_179819 | Low-quality |
| 17-109_k141_362238 | Not-determined |
| 17-109_k141_179822 | Not-determined |
| 17-109_k141_280956 | Not-determined |
| 17-109_k141_196924 | Not-determined |
| 17-109_k141_280959 | Not-determined |
| 17-109_k141_362240 | Low-quality |
| 17-109_k141_80013 | Not-determined |
| 17-109_k141_230579 | Not-determined |
| 17-109_k141_280963 | Not-determined |
| 17-109_k141_230587 | Not-determined |
| 17-109_k141_280964 | Not-determined |
| 17-109_k141_380924 | Not-determined |
| 17-109_k141_80021 | Not-determined |
| 17-109_k141_297365 | Not-determined |
| 17-109_k141_80026 | Low-quality |
| 17-109_k141_380930 | Not-determined |
| 17-109_k141_80027 | Not-determined |
| 17-109_k141_263258 | Low-quality |
| 17-109_k141_128806 | Not-determined |
| 17-109_k141_362253 | Not-determined |
| 17-109_k141_347222 | Not-determined |
| 17-109_k141_280973 | Not-determined |
| 17-109_k141_80032 | Low-quality |
| 17-109_k141_297373 | Not-determined |
| 17-109_k141_246492 | Not-determined |
| 17-109_k141_246493 | Not-determined |
| 17-109_k141_314765 | Not-determined |
| 17-109_k141_347232 | Not-determined |
| 17-109_k141_163218 | Not-determined |
| 17-109_k141_297376 | Low-quality |
| 17-109_k141_297378 | Not-determined |
| 17-109_k141_95653 | Not-determined |
| 17-109_k141_314772 | Not-determined |
| 17-109_k141_128816 | Not-determined |
| 17-109_k141_347237 | Not-determined |
| 17-109_k141_314773 | Low-quality |
| 17-109_k141_314776 | Not-determined |
| 17-109_k141_128825 | Not-determined |
| 17-109_k141_280987 | Not-determined |
| 17-109_k141_62415 | Not-determined |
| 17-109_k141_11916 | Not-determined |
| 17-109_k141_314780 | Not-determined |
| 17-109_k141_213517 | Not-determined |
| 17-109_k141_213519 | Not-determined |
| 17-109_k141_179837 | Not-determined |
| 17-109_k141_128833 | Not-determined |
| 17-109_k141_95658 | Not-determined |
| 17-109_k141_62425 | Not-determined |
| 17-109_k141_347251 | Low-quality |
| 17-109_k141_62428 | Not-determined |
| 17-109_k141_213523 | Not-determined |
| 17-109_k141_246524 | Not-determined |
| 17-109_k141_196959 | Low-quality |
| 17-109_k141_230600 | Not-determined |
| 17-109_k141_397452 | Low-quality |
| 17-109_k141_297399 | Not-determined |
| 17-109_k141_179845 | Low-quality |
| 17-109_k141_112086 | Not-determined |
| 17-109_k141_362272 | Not-determined |
| 17-109_k141_196982 | Not-determined |
| 17-109_k141_297420 | Not-determined |
| 17-109_k141_246534 | Low-quality |
| 17-109_k141_347281 | Not-determined |
| 17-109_k141_263262 | Not-determined |
| 17-109_k141_362286 | Not-determined |
| 17-109_k141_297427 | Not-determined |
| 17-109_k141_297430 | Low-quality |
| 17-109_k141_281013 | Not-determined |
| 17-109_k141_263270 | Not-determined |
| 17-109_k141_314806 | Not-determined |
| 17-109_k141_112115 | Not-determined |
| 17-109_k141_62450 | Not-determined |
| 17-109_k141_62452 | Not-determined |
| 17-109_k141_246547 | Not-determined |
| 17-109_k141_362302 | Not-determined |
| 17-109_k141_179889 | Not-determined |
| 17-109_k141_11942 | Not-determined |
| 17-109_k141_213566 | Not-determined |
| 17-109_k141_297446 | Not-determined |
| 17-109_k141_263287 | Not-determined |
| 17-109_k141_163267 | Not-determined |
| 17-109_k141_80058 | Not-determined |
| 17-109_k141_197008 | Not-determined |
| 17-109_k141_362311 | Not-determined |
| 17-109_k141_112132 | Not-determined |
| 17-109_k141_297449 | Not-determined |
| 17-109_k141_112134 | Not-determined |
| 17-109_k141_263290 | Not-determined |
| 17-109_k141_297450 | Not-determined |
| 17-109_k141_281045 | Not-determined |
| 17-109_k141_246570 | Not-determined |
| 17-109_k141_246572 | Not-determined |
| 17-109_k141_246573 | Low-quality |
| 17-109_k141_281048 | Not-determined |
| 17-109_k141_397477 | Not-determined |
| 17-109_k141_297455 | Low-quality |
| 17-109_k141_28901 | Not-determined |
| 17-109_k141_163272 | Not-determined |
| 17-109_k141_112141 | Not-determined |
| 17-109_k141_28904 | Not-determined |
| 17-109_k141_297461 | Not-determined |
| 17-109_k141_397483 | Not-determined |
| 17-109_k141_263297 | Not-determined |
| 17-109_k141_80068 | Not-determined |
| 17-109_k141_145995 | Not-determined |
| 17-109_k141_347313 | Not-determined |
| 17-109_k141_246585 | Low-quality |
| 17-109_k141_112153 | Not-determined |
| 17-109_k141_112154 | Not-determined |
| 17-109_k141_263306 | Not-determined |
| 17-109_k141_80075 | Low-quality |
| 17-109_k141_246588 | Not-determined |
| 17-109_k141_230664 | Not-determined |
| 17-109_k141_112157 | Not-determined |
| 17-109_k141_362329 | Not-determined |
| 17-109_k141_62478 | Not-determined |
| 17-109_k141_297467 | Not-determined |
| 17-109_k141_281063 | Not-determined |
| 17-109_k141_380935 | Not-determined |
| 17-109_k141_347319 | Not-determined |
| 17-109_k141_246596 | Not-determined |
| 17-109_k141_297474 | Not-determined |
| 17-109_k141_281073 | Not-determined |
| 17-109_k141_11962 | Not-determined |
| 17-109_k141_112170 | Not-determined |
| 17-109_k141_179921 | Not-determined |
| 17-109_k141_112172 | Low-quality |
| 17-109_k141_297485 | Not-determined |
| 17-109_k141_28933 | Not-determined |
| 17-109_k141_314847 | Not-determined |
| 17-109_k141_11972 | Not-determined |
| 17-109_k141_62498 | Not-determined |
| 17-109_k141_46778 | Not-determined |
| 17-109_k141_163279 | Not-determined |
| 17-109_k141_62499 | Not-determined |
| 17-109_k141_112182 | Not-determined |
| 17-109_k141_179926 | Low-quality |
| 17-109_k141_163285 | Not-determined |
| 17-109_k141_213573 | Not-determined |
| 17-109_k141_297506 | Not-determined |
| 17-109_k141_163291 | Not-determined |
| 17-109_k141_46786 | Not-determined |
| 17-109_k141_347337 | Not-determined |
| 17-109_k141_179936 | Not-determined |
| 17-109_k141_46790 | Not-determined |
| 17-109_k141_362376 | Not-determined |
| 17-109_k141_213585 | Not-determined |
| 17-109_k141_11994 | Not-determined |
| 17-109_k141_362380 | Not-determined |
| 17-109_k141_179943 | Not-determined |
| 17-109_k141_362381 | Not-determined |
| 17-109_k141_95700 | Not-determined |
| 17-109_k141_347347 | Not-determined |
| 17-109_k141_179946 | Not-determined |
| 17-109_k141_95702 | Not-determined |
| 17-109_k141_362385 | Not-determined |
| 17-109_k141_95704 | Not-determined |
| 17-109_k141_380976 | Not-determined |
| 17-109_k141_12002 | Not-determined |
| 17-109_k141_95710 | Not-determined |
| 17-109_k141_230681 | Not-determined |
| 17-109_k141_179953 | Not-determined |
| 17-109_k141_95712 | Not-determined |
| 17-109_k141_230683 | Not-determined |
| 17-109_k141_314880 | Not-determined |
| 17-109_k141_179956 | Not-determined |
| 17-109_k141_397504 | Not-determined |
| 17-109_k141_179958 | Not-determined |
| 17-109_k141_314882 | Not-determined |
| 17-109_k141_112191 | Low-quality |
| 17-109_k141_163305 | Not-determined |
| 17-109_k141_362394 | Not-determined |
| 17-109_k141_347360 | Not-determined |
| 17-109_k141_146031 | Not-determined |
| 17-109_k141_146036 | Not-determined |
| 17-109_k141_95729 | Not-determined |
| 17-109_k141_347366 | Low-quality |
| 17-109_k141_347367 | Not-determined |
| 17-109_k141_397516 | Not-determined |
| 17-109_k141_347370 | Not-determined |
| 17-109_k141_28952 | Not-determined |
| 17-109_k141_80099 | Low-quality |
| 17-109_k141_397520 | Not-determined |
| 17-109_k141_146041 | Not-determined |
| 17-109_k141_28955 | Low-quality |
| 17-109_k141_12032 | Not-determined |
| 17-109_k141_28962 | Not-determined |
| 17-109_k141_28965 | Not-determined |
| 17-109_k141_80110 | Not-determined |
| 17-109_k141_314903 | Not-determined |
| 17-109_k141_28970 | Not-determined |
| 17-109_k141_380980 | Not-determined |
| 17-109_k141_12039 | Not-determined |
| 17-109_k141_362404 | Not-determined |
| 17-109_k141_380982 | Not-determined |
| 17-109_k141_197027 | Not-determined |
| 17-109_k141_146052 | Not-determined |
| 17-109_k141_314908 | Not-determined |
| 17-109_k141_128911 | Not-determined |
| 17-109_k141_314910 | Not-determined |
| 17-109_k141_95747 | Not-determined |
| 17-109_k141_128913 | Not-determined |
| 17-109_k141_263343 | Not-determined |
| 17-109_k141_146057 | Not-determined |
| 17-109_k141_213592 | Low-quality |
| 17-109_k141_179975 | Not-determined |
| 17-109_k141_380991 | Not-determined |
| 17-109_k141_197032 | Not-determined |
| 17-109_k141_46823 | Not-determined |
| 17-109_k141_213596 | Not-determined |
| 17-109_k141_197033 | Not-determined |
| 17-109_k141_246667 | Not-determined |
| 17-109_k141_80130 | Not-determined |
| 17-109_k141_397534 | Not-determined |
| 17-109_k141_380999 | Not-determined |
| 17-109_k141_230715 | Not-determined |
| 17-109_k141_80133 | Not-determined |
| 17-109_k141_397538 | Not-determined |
| 17-109_k141_28999 | Not-determined |
| 17-109_k141_281100 | Not-determined |
| 17-109_k141_179991 | Not-determined |
| 17-109_k141_12052 | Not-determined |
| 17-109_k141_230724 | Not-determined |
| 17-109_k141_281101 | Low-quality |
| 17-109_k141_347398 | Low-quality |
| 17-109_k141_163328 | Not-determined |
| 17-109_k141_281105 | Not-determined |
| 17-109_k141_347400 | Not-determined |
| 17-109_k141_397543 | Not-determined |
| 17-109_k141_281110 | Not-determined |
| 17-109_k141_281111 | Not-determined |
| 17-109_k141_397548 | Not-determined |
| 17-109_k141_62544 | Low-quality |
| 17-109_k141_95778 | Not-determined |
| 17-109_k141_347406 | Not-determined |
| 17-109_k141_281116 | Not-determined |
| 17-109_k141_80150 | Low-quality |
| 17-109_k141_314921 | Not-determined |
| 17-109_k141_347412 | Not-determined |
| 17-109_k141_80160 | Low-quality |
| 17-109_k141_146085 | Not-determined |
| 17-109_k141_213619 | Not-determined |
| 17-109_k141_281124 | Not-determined |
| 17-109_k141_362416 | Not-determined |
| 17-109_k141_95787 | Not-determined |
| 17-109_k141_397564 | Not-determined |
| 17-109_k141_263374 | Not-determined |
| 17-109_k141_246705 | Not-determined |
| 17-109_k141_146097 | Not-determined |
| 17-109_k141_362423 | Not-determined |
| 17-109_k141_347429 | Not-determined |
| 17-109_k141_281135 | Not-determined |
| 17-109_k141_397569 | Not-determined |
| 17-109_k141_281139 | Not-determined |
| 17-109_k141_80177 | Not-determined |
| 17-109_k141_362432 | Not-determined |
| 17-109_k141_347437 | Not-determined |
| 17-109_k141_95812 | Not-determined |
| 17-109_k141_281147 | Not-determined |
| 17-109_k141_180031 | Not-determined |
| 17-109_k141_146115 | Not-determined |
| 17-109_k141_180040 | Not-determined |
| 17-109_k141_128926 | Not-determined |
| 17-109_k141_95823 | Not-determined |
| 17-109_k141_112220 | Not-determined |
| 17-109_k141_163362 | Not-determined |
| 17-109_k141_95827 | Not-determined |
| 17-109_k141_330678 | Not-determined |
| 17-109_k141_230740 | Not-determined |
| 17-109_k141_213635 | Not-determined |
| 17-109_k141_362439 | Not-determined |
| 17-109_k141_281158 | Not-determined |
| 17-109_k141_213637 | Not-determined |
| 17-109_k141_213639 | Low-quality |
| 17-109_k141_95833 | Not-determined |
| 17-109_k141_314971 | Not-determined |
| 17-109_k141_180047 | Not-determined |
| 17-109_k141_112222 | Not-determined |
| 17-109_k141_330690 | Not-determined |
| 17-109_k141_12078 | Low-quality |
| 17-109_k141_314979 | Not-determined |
| 17-109_k141_347469 | Not-determined |
| 17-109_k141_62608 | Not-determined |
| 17-109_k141_330693 | Not-determined |
| 17-109_k141_397600 | Not-determined |
| 17-109_k141_128943 | Not-determined |
| 17-109_k141_12087 | Not-determined |
| 17-109_k141_163376 | Not-determined |
| 17-109_k141_330698 | Not-determined |
| 17-109_k141_397604 | Not-determined |
| 17-109_k141_95839 | Not-determined |
| 17-109_k141_46888 | Not-determined |
| 17-109_k141_347479 | Not-determined |
| 17-109_k141_80219 | Not-determined |
| 17-109_k141_281189 | Not-determined |
| 17-109_k141_180075 | Not-determined |
| 17-109_k141_163401 | Not-determined |
| 17-109_k141_381057 | Not-determined |
| 17-109_k141_163408 | Not-determined |
| 17-109_k141_397628 | Not-determined |
| 17-109_k141_397629 | Not-determined |
| 17-109_k141_397630 | Not-determined |
| 17-109_k141_112259 | Not-determined |
| 17-109_k141_230765 | Not-determined |
| 17-109_k141_62643 | Not-determined |
| 17-109_k141_381066 | Not-determined |
| 17-109_k141_347505 | Not-determined |
| 17-109_k141_46905 | Not-determined |
| 17-109_k141_112264 | Not-determined |
| 17-109_k141_46912 | Not-determined |
| 17-109_k141_146147 | Not-determined |
| 17-109_k141_12110 | Not-determined |
| 17-109_k141_381078 | Not-determined |
| 17-109_k141_80255 | Not-determined |
| 17-109_k141_397648 | Not-determined |
| 17-109_k141_381079 | Not-determined |
| 17-109_k141_397649 | Not-determined |
| 17-109_k141_128983 | Not-determined |
| 17-109_k141_330733 | Not-determined |
| 17-109_k141_330734 | Not-determined |
| 17-109_k141_62658 | Not-determined |
| 17-109_k141_197095 | Low-quality |
| 17-109_k141_230779 | Low-quality |
| 17-109_k141_281235 | Not-determined |
| 17-109_k141_381087 | Not-determined |
| 17-109_k141_197100 | Not-determined |
| 17-109_k141_381090 | Not-determined |
| 17-109_k141_397659 | Not-determined |
| 17-109_k141_315023 | Not-determined |
| 17-109_k141_29066 | Not-determined |
| 17-109_k141_330742 | Not-determined |
| 17-109_k141_46922 | Not-determined |
| 17-109_k141_197106 | Not-determined |
| 17-109_k141_95865 | Not-determined |
| 17-109_k141_197107 | Not-determined |
| 17-109_k141_180099 | Not-determined |
| 17-109_k141_129001 | Not-determined |
| 17-109_k141_397669 | Not-determined |
| 17-109_k141_397671 | Not-determined |
| 17-109_k141_397672 | Not-determined |
| 17-109_k141_230795 | Not-determined |
| 17-109_k141_397676 | Not-determined |
| 17-109_k141_129009 | Not-determined |
| 17-109_k141_381097 | Not-determined |
| 17-109_k141_163444 | Not-determined |
| 17-109_k141_197116 | Not-determined |
| 17-109_k141_95874 | Not-determined |
| 17-109_k141_330752 | Not-determined |
| 17-109_k141_95881 | Not-determined |
| 17-109_k141_381111 | Not-determined |
| 17-109_k141_163459 | Not-determined |
| 17-109_k141_146181 | Not-determined |
| 17-109_k141_263392 | Low-quality |
| 17-109_k141_230804 | Not-determined |
| 17-109_k141_146183 | Not-determined |
| 17-109_k141_46943 | Not-determined |
| 17-109_k141_315039 | Not-determined |
| 17-109_k141_146185 | Not-determined |
| 17-109_k141_95890 | Not-determined |
| 17-109_k141_129033 | Not-determined |
| 17-109_k141_129034 | Low-quality |
| 17-109_k141_397684 | Low-quality |
| 17-109_k141_95895 | Not-determined |
| 17-109_k141_29086 | Not-determined |
| 17-109_k141_29087 | Not-determined |
| 17-109_k141_80267 | Not-determined |
| 17-109_k141_397688 | Not-determined |
| 17-109_k141_246743 | Not-determined |
| 17-109_k141_112289 | Low-quality |
| 17-109_k141_180128 | Not-determined |
| 17-109_k141_330768 | Not-determined |
| 17-109_k141_397698 | Not-determined |
| 17-109_k141_263397 | Not-determined |
| 17-109_k141_12148 | Low-quality |
| 17-109_k141_213698 | Not-determined |
| 17-109_k141_246751 | Not-determined |
| 17-109_k141_163487 | Not-determined |
| 17-109_k141_213701 | Not-determined |
| 17-109_k141_46956 | Low-quality |
| 17-109_k141_246754 | Not-determined |
| 17-109_k141_180130 | Not-determined |
| 17-109_k141_246756 | Not-determined |
| 17-109_k141_362503 | Not-determined |
| 17-109_k141_330781 | Not-determined |
| 17-109_k141_397704 | Not-determined |
| 17-109_k141_330783 | Not-determined |
| 17-109_k141_362509 | Not-determined |
| 17-109_k141_315071 | Not-determined |
| 17-109_k141_397709 | Not-determined |
| 17-109_k141_197150 | Low-quality |
| 17-109_k141_197151 | Not-determined |

| | |
|---|---|
| 17-109_k141_146220 | Not-determined |
| 17-109_k141_330785 | Low-quality |
| 17-109_k141_213706 | Not-determined |
| 17-109_k141_129061 | Not-determined |
| 17-109_k141_163502 | Not-determined |
| 17-109_k141_12168 | Not-determined |
| 17-109_k141_62719 | Not-determined |
| 17-109_k141_112313 | Not-determined |
| 17-109_k141_397719 | Not-determined |
| 17-109_k141_397720 | Not-determined |
| 17-109_k141_347526 | Not-determined |
| 17-109_k141_95930 | Not-determined |
| 17-109_k141_330800 | Not-determined |
| 17-109_k141_315081 | Not-determined |
| 17-109_k141_163520 | Not-determined |
| 17-109_k141_29105 | Not-determined |
| 17-109_k141_62725 | Not-determined |
| 17-109_k141_263426 | Low-quality |
| 17-109_k141_397724 | Not-determined |
| 17-109_k141_163523 | Not-determined |
| 17-109_k141_281270 | Not-determined |
| 17-109_k141_315086 | Not-determined |
| 17-109_k141_146249 | Not-determined |
| 17-109_k141_315089 | Not-determined |
| 17-109_k141_146251 | Not-determined |
| 17-109_k141_62733 | Not-determined |
| 17-109_k141_12182 | Not-determined |
| 17-109_k141_146253 | Not-determined |
| 17-109_k141_163534 | Low-quality |
| 17-109_k141_315092 | Not-determined |
| 17-109_k141_281287 | Low-quality |
| 17-109_k141_362517 | Low-quality |
| 17-109_k141_95952 | Not-determined |
| 17-109_k141_263441 | Not-determined |
| 17-109_k141_95953 | Not-determined |
| 17-109_k141_263444 | Not-determined |
| 17-109_k141_180149 | Not-determined |
| 17-109_k141_263445 | Low-quality |
| 17-109_k141_213747 | Not-determined |
| 17-109_k141_95957 | Not-determined |
| 17-109_k141_330837 | Not-determined |
| 17-109_k141_281305 | Not-determined |
| 17-109_k141_95965 | Not-determined |
| 17-109_k141_347549 | Not-determined |
| 17-109_k141_315117 | Not-determined |
| 17-109_k141_330841 | Not-determined |
| 17-109_k141_146274 | Not-determined |
| 17-109_k141_330846 | Not-determined |
| 17-109_k141_297547 | Not-determined |
| 17-109_k141_95978 | Not-determined |
| 17-109_k141_29147 | Not-determined |
| 17-109_k141_297558 | Not-determined |
| 17-109_k141_29148 | Not-determined |
| 17-109_k141_80279 | Low-quality |
| 17-109_k141_362527 | Not-determined |
| 17-109_k141_315140 | Not-determined |
| 17-109_k141_297559 | Low-quality |
| 17-109_k141_347569 | Not-determined |
| 17-109_k141_95987 | Not-determined |
| 17-109_k141_315144 | Not-determined |
| 17-109_k141_129111 | Not-determined |
| 17-109_k141_129112 | Not-determined |
| 17-109_k141_246810 | Not-determined |
| 17-109_k141_197205 | Not-determined |
| 17-109_k141_29160 | Not-determined |
| 17-109_k141_230827 | Not-determined |
| 17-109_k141_263476 | Not-determined |
| 17-109_k141_29167 | Not-determined |
| 17-109_k141_146291 | Not-determined |
| 17-109_k141_230836 | Not-determined |
| 17-109_k141_163559 | Not-determined |
| 17-109_k141_297581 | Low-quality |
| 17-109_k141_347588 | Not-determined |
| 17-109_k141_381164 | Not-determined |
| 17-109_k141_362541 | Not-determined |
| 17-109_k141_281327 | Not-determined |
| 17-109_k141_29170 | Low-quality |
| 17-109_k141_281328 | Not-determined |
| 17-109_k141_315165 | Not-determined |
| 17-109_k141_29173 | Not-determined |
| 17-109_k141_281330 | Not-determined |
| 17-109_k141_163572 | Not-determined |
| 17-109_k141_315168 | Not-determined |
| 17-109_k141_347596 | Not-determined |
| 17-109_k141_246830 | Not-determined |
| 17-109_k141_197215 | Not-determined |
| 17-109_k141_330855 | Not-determined |
| 17-109_k141_263487 | Not-determined |
| 17-109_k141_297585 | Not-determined |
| 17-109_k141_281336 | Not-determined |
| 17-109_k141_146310 | Not-determined |
| 17-109_k141_129120 | Not-determined |
| 17-109_k141_397763 | Not-determined |
| 17-109_k141_330867 | Not-determined |
| 17-109_k141_163590 | Not-determined |
| 17-109_k141_80304 | Not-determined |
| 17-109_k141_163596 | Low-quality |
| 17-109_k141_397766 | Not-determined |
| 17-109_k141_163598 | Not-determined |
| 17-109_k141_230844 | Not-determined |
| 17-109_k141_29180 | Not-determined |
| 17-109_k141_129129 | Not-determined |
| 17-109_k141_47056 | Not-determined |
| 17-109_k141_297616 | Not-determined |
| 17-109_k141_112358 | Not-determined |
| 17-109_k141_213785 | Not-determined |
| 17-109_k141_129136 | Low-quality |
| 17-109_k141_12204 | Not-determined |
| 17-109_k141_230861 | Not-determined |
| 17-109_k141_197231 | Not-determined |
| 17-109_k141_47064 | Not-determined |
| 17-109_k141_230865 | Not-determined |
| 17-109_k141_230866 | Not-determined |
| 17-109_k141_96047 | Not-determined |
| 17-109_k141_347619 | Not-determined |
| 17-109_k141_163615 | Not-determined |
| 17-109_k141_397793 | Not-determined |
| 17-109_k141_281372 | Not-determined |
| 17-109_k141_112373 | Not-determined |
| 17-109_k141_129155 | Not-determined |
| 17-109_k141_281373 | Low-quality |
| 17-109_k141_213803 | Not-determined |
| 17-109_k141_297631 | Low-quality |
| 17-109_k141_297632 | Not-determined |
| 17-109_k141_315203 | Not-determined |
| 17-109_k141_381194 | Not-determined |
| 17-109_k141_163630 | Not-determined |
| 17-109_k141_230877 | Not-determined |
| 17-109_k141_297638 | Not-determined |
| 17-109_k141_12229 | Not-determined |
| 17-109_k141_230881 | Not-determined |
| 17-109_k141_197240 | Not-determined |
| 17-109_k141_330887 | Not-determined |
| 17-109_k141_397806 | Not-determined |
| 17-109_k141_263532 | Not-determined |
| 17-109_k141_397808 | Not-determined |
| 17-109_k141_197253 | Not-determined |
| 17-109_k141_47096 | Low-quality |
| 17-109_k141_315224 | Not-determined |
| 17-109_k141_12239 | Not-determined |
| 17-109_k141_96060 | Not-determined |
| 17-109_k141_12243 | Not-determined |
| 17-109_k141_315230 | Low-quality |
| 17-109_k141_230899 | Not-determined |
| 17-109_k141_29216 | Not-determined |
| 17-109_k141_29220 | Not-determined |
| 17-109_k141_330910 | Not-determined |
| 17-109_k141_47112 | Not-determined |
| 17-109_k141_297648 | Low-quality |
| 17-109_k141_47115 | Not-determined |
| 17-109_k141_246857 | Low-quality |
| 17-109_k141_197269 | Not-determined |
| 17-109_k141_197271 | Not-determined |
| 17-109_k141_180219 | Low-quality |
| 17-109_k141_47117 | Not-determined |
| 17-109_k141_62798 | Not-determined |
| 17-109_k141_297652 | Not-determined |
| 17-109_k141_163643 | Not-determined |
| 17-109_k141_47120 | Low-quality |
| 17-109_k141_281415 | Not-determined |
| 17-109_k141_163645 | Not-determined |
| 17-109_k141_397819 | Not-determined |
| 17-109_k141_246865 | Not-determined |
| 17-109_k141_129177 | Not-determined |
| 17-109_k141_62806 | Not-determined |
| 17-109_k141_246872 | Not-determined |
| 17-109_k141_246873 | Not-determined |
| 17-109_k141_12258 | Not-determined |
| 17-109_k141_397827 | Not-determined |
| 17-109_k141_397828 | Not-determined |
| 17-109_k141_47131 | Not-determined |
| 17-109_k141_180235 | Not-determined |
| 17-109_k141_180239 | Not-determined |
| 17-109_k141_213830 | Not-determined |
| 17-109_k141_281426 | Not-determined |
| 17-109_k141_213832 | Not-determined |
| 17-109_k141_29246 | Not-determined |
| 17-109_k141_347661 | Not-determined |
| 17-109_k141_29247 | Not-determined |
| 17-109_k141_129188 | Not-determined |
| 17-109_k141_12277 | Not-determined |
| 17-109_k141_397835 | Not-determined |
| 17-109_k141_397836 | Not-determined |
| 17-109_k141_180249 | Not-determined |
| 17-109_k141_197296 | Low-quality |
| 17-109_k141_230933 | Not-determined |
| 17-109_k141_96095 | Not-determined |
| 17-109_k141_281446 | Not-determined |
| 17-109_k141_330943 | Not-determined |
| 17-109_k141_246897 | Not-determined |
| 17-109_k141_246900 | Not-determined |
| 17-109_k141_96097 | Not-determined |
| 17-109_k141_397845 | Low-quality |
| 17-109_k141_397849 | Not-determined |
| 17-109_k141_47151 | Not-determined |
| 17-109_k141_12301 | Not-determined |
| 17-109_k141_281454 | Not-determined |
| 17-109_k141_129204 | Low-quality |
| 17-109_k141_12305 | Not-determined |
| 17-109_k141_112388 | Not-determined |
| 17-109_k141_129208 | Not-determined |
| 17-109_k141_12309 | Not-determined |
| 17-109_k141_315279 | Not-determined |
| 17-109_k141_12311 | Not-determined |
| 17-109_k141_315282 | Not-determined |
| 17-109_k141_62839 | Not-determined |
| 17-109_k141_146338 | Not-determined |
| 17-109_k141_180258 | Not-determined |
| 17-109_k141_230966 | Not-determined |
| 17-109_k141_62843 | Not-determined |
| 17-109_k141_96110 | Not-determined |
| 17-109_k141_281463 | Not-determined |
| 17-109_k141_381228 | Low-quality |
| 17-109_k141_297688 | Not-determined |
| 17-109_k141_347683 | Not-determined |
| 17-109_k141_246919 | Low-quality |
| 17-109_k141_246923 | Not-determined |
| 17-109_k141_96118 | Not-determined |
| 17-109_k141_96120 | Not-determined |
| 17-109_k141_230982 | Low-quality |
| 17-109_k141_29280 | Not-determined |
| 17-109_k141_397863 | Not-determined |
| 17-109_k141_29283 | Not-determined |
| 17-109_k141_96126 | Not-determined |
| 17-109_k141_29286 | Not-determined |
| 17-109_k141_146367 | Low-quality |
| 17-109_k141_96129 | Not-determined |
| 17-109_k141_12322 | Not-determined |
| 17-109_k141_347694 | Not-determined |
| 17-109_k141_397871 | Low-quality |
| 17-109_k141_129237 | Not-determined |
| 17-109_k141_112392 | Not-determined |
| 17-109_k141_281479 | Not-determined |
| 17-109_k141_347702 | Not-determined |
| 17-109_k141_12335 | Not-determined |
| 17-109_k141_381249 | Not-determined |
| 17-109_k141_163696 | Not-determined |
| 17-109_k141_230997 | Not-determined |
| 17-109_k141_362592 | Not-determined |
| 17-109_k141_231007 | Not-determined |
| 17-109_k141_330987 | Not-determined |
| 17-109_k141_362595 | Not-determined |
| 17-109_k141_362596 | Not-determined |
| 17-109_k141_163704 | Not-determined |
| 17-109_k141_12351 | Not-determined |
| 17-109_k141_330989 | Not-determined |
| 17-109_k141_80346 | Not-determined |
| 17-109_k141_12356 | Not-determined |
| 17-109_k141_231022 | Not-determined |
| 17-109_k141_347714 | Not-determined |
| 17-109_k141_80351 | Not-determined |
| 17-109_k141_163715 | Not-determined |
| 17-109_k141_47170 | Not-determined |
| 17-109_k141_197330 | Not-determined |
| 17-109_k141_96156 | Not-determined |
| 17-109_k141_362603 | Low-quality |
| 17-109_k141_12367 | Not-determined |
| 17-109_k141_180306 | Not-determined |
| 17-109_k141_362606 | Low-quality |
| 17-109_k141_180308 | Not-determined |
| 17-109_k141_362608 | Not-determined |
| 17-109_k141_96160 | Not-determined |
| 17-109_k141_246966 | Not-determined |
| 17-109_k141_263600 | Not-determined |
| 17-109_k141_80356 | Not-determined |
| 17-109_k141_47181 | Not-determined |
| 17-109_k141_381269 | Not-determined |
| 17-109_k141_80359 | Not-determined |
| 17-109_k141_381275 | Not-determined |
| 17-109_k141_80362 | Not-determined |
| 17-109_k141_129264 | Not-determined |
| 17-109_k141_47187 | Not-determined |
| 17-109_k141_246981 | Not-determined |
| 17-109_k141_163734 | Not-determined |
| 17-109_k141_281488 | Not-determined |
| 17-109_k141_163736 | Not-determined |
| 17-109_k141_231032 | Not-determined |
| 17-109_k141_315338 | Not-determined |
| 17-109_k141_381287 | Not-determined |
| 17-109_k141_12386 | Not-determined |
| 17-109_k141_163741 | Not-determined |
| 17-109_k141_129273 | Not-determined |
| 17-109_k141_12389 | Not-determined |
| 17-109_k141_231034 | Not-determined |
| 17-109_k141_12393 | Not-determined |
| 17-109_k141_12396 | Not-determined |
| 17-109_k141_47199 | Not-determined |
| 17-109_k141_29332 | Not-determined |
| 17-109_k141_381295 | Not-determined |
| 17-109_k141_96186 | Not-determined |
| 17-109_k141_80385 | Not-determined |
| 17-109_k141_129279 | Low-quality |
| 17-106-final-viral-combined_k141_36459\|\|full | Medium-quality |
| 17-109_k141_163755 | Not-determined |
| 17-109_k141_231048 | Not-determined |
| 17-109_k141_12405 | Low-quality |
| 17-109_k141_112450 | Not-determined |
| 17-109_k141_347739 | Not-determined |
| 17-109_k141_12406 | Not-determined |
| 17-109_k141_197356 | Not-determined |
| 17-109_k141_347740 | Not-determined |
| 17-109_k141_62875 | Not-determined |
| 17-109_k141_129284 | Not-determined |
| 17-109_k141_381308 | Low-quality |
| 17-109_k141_163763 | Low-quality |
| 17-109_k141_80388 | Not-determined |
| 17-109_k141_12410 | Not-determined |
| 17-109_k141_315360 | Not-determined |
| 17-109_k141_146439 | Not-determined |
| 17-109_k141_146440 | Low-quality |
| 17-109_k141_281511 | Not-determined |
| 17-109_k141_197361 | Not-determined |
| 17-109_k141_281514 | Not-determined |
| 17-109_k141_197362 | Not-determined |
| 17-109_k141_231065 | Not-determined |
| 17-109_k141_96190 | Not-determined |
| 17-109_k141_315368 | Not-determined |
| 17-109_k141_197371 | Not-determined |
| 17-109_k141_281520 | Not-determined |
| 17-109_k141_146455 | Not-determined |
| 17-109_k141_146456 | Not-determined |
| 17-109_k141_231028 | Low-quality |
| 17-109_k141_96196 | Not-determined |
| 17-109_k141_80400 | Not-determined |
| 17-109_k141_197380 | Not-determined |
| 17-109_k141_231070 | Not-determined |
| 17-109_k141_180360 | Not-determined |
| 17-109_k141_315384 | Not-determined |
| 17-109_k141_129298 | Not-determined |
| 17-109_k141_96204 | Not-determined |
| 17-109_k141_146465 | Low-quality |
| 17-109_k141_146466 | Not-determined |
| 17-109_k141_96206 | Not-determined |
| 17-109_k141_163780 | Not-determined |
| 17-109_k141_197384 | Not-determined |
| 17-109_k141_96212 | Not-determined |
| 17-109_k141_197386 | Not-determined |
| 17-109_k141_80418 | Not-determined |
| 17-109_k141_362662 | Not-determined |
| 17-109_k141_281549 | Not-determined |
| 17-109_k141_80421 | Not-determined |
| 17-109_k141_347783 | Low-quality |
| 17-109_k141_12442 | Not-determined |
| 17-109_k141_231096 | Not-determined |
| 17-109_k141_80433 | Not-determined |
| 17-109_k141_163801 | Not-determined |
| 17-109_k141_96225 | Not-determined |
| 17-109_k141_213884 | Not-determined |
| 17-109_k141_347800 | Low-quality |
| 17-109_k141_96229 | Not-determined |
| 17-109_k141_96230 | Not-determined |
| 17-109_k141_163807 | Not-determined |
| 17-109_k141_197410 | Not-determined |
| 17-109_k141_197412 | Not-determined |
| 17-109_k141_96232 | Not-determined |
| 17-109_k141_80439 | Not-determined |
| 17-109_k141_12454 | Not-determined |
| 17-109_k141_247016 | Not-determined |
| 17-109_k141_281582 | Not-determined |
| 17-109_k141_12456 | Not-determined |
| 17-109_k141_347811 | Not-determined |
| 17-109_k141_96238 | Not-determined |
| 17-109_k141_281585 | Not-determined |
| 17-109_k141_146500 | Not-determined |
| 17-109_k141_362699 | Not-determined |
| 17-109_k141_146501 | Not-determined |
| 17-109_k141_247022 | Not-determined |
| 17-109_k141_281593 | Not-determined |
| 17-109_k141_347816 | Not-determined |
| 17-109_k141_297721 | Not-determined |
| 17-109_k141_129316 | Not-determined |
| 17-109_k141_331083 | Not-determined |
| 17-109_k141_347820 | Not-determined |
| 17-109_k141_347821 | Not-determined |
| 17-109_k141_146519 | Not-determined |
| 17-109_k141_297731 | Not-determined |
| 17-109_k141_281606 | Not-determined |
| 17-109_k141_112515 | Not-determined |
| 17-109_k141_96250 | Not-determined |
| 17-109_k141_62893 | Not-determined |
| 17-109_k141_381379 | Not-determined |
| 17-109_k141_331092 | Not-determined |
| 17-109_k141_12464 | Not-determined |
| 17-109_k141_213904 | Not-determined |
| 17-109_k141_96259 | Low-quality |
| 17-109_k141_180375 | Not-determined |
| 17-109_k141_281614 | Not-determined |
| 17-109_k141_12466 | Not-determined |
| 17-109_k141_347832 | Not-determined |
| 17-109_k141_347834 | Not-determined |
| 17-109_k141_347836 | Not-determined |
| 17-109_k141_80473 | Not-determined |
| 17-109_k141_331106 | Not-determined |
| 17-109_k141_96271 | Not-determined |
| 17-109_k141_197441 | Not-determined |
| 17-109_k141_331107 | Not-determined |
| 17-109_k141_112538 | Not-determined |
| 17-109_k141_381395 | Not-determined |
| 17-109_k141_297757 | Not-determined |
| 17-109_k141_96277 | Not-determined |
| 17-109_k141_347839 | Not-determined |
| 17-109_k141_129347 | Low-quality |
| 17-109_k141_96279 | Not-determined |
| 17-109_k141_281623 | Low-quality |
| 17-109_k141_381402 | Low-quality |
| 17-109_k141_297761 | Not-determined |
| 17-109_k141_96281 | Low-quality |
| 17-109_k141_96283 | Not-determined |
| 17-109_k141_231128 | Not-determined |
| 17-109_k141_80488 | Not-determined |
| 17-109_k141_213927 | Not-determined |
| 17-109_k141_381405 | Not-determined |
| 17-109_k141_231132 | Not-determined |
| 17-109_k141_62902 | Not-determined |
| 17-109_k141_381412 | Not-determined |
| 17-109_k141_62904 | Not-determined |
| 17-109_k141_247048 | Not-determined |
| 17-109_k141_247049 | Not-determined |
| 17-109_k141_47260 | Not-determined |
| 17-109_k141_397920 | Not-determined |
| 17-109_k141_12497 | Not-determined |
| 17-109_k141_281644 | Low-quality |
| 17-109_k141_112573 | Not-determined |
| 17-109_k141_96302 | Not-determined |
| 17-109_k141_231150 | Not-determined |
| 17-109_k141_146557 | Low-quality |
| 17-109_k141_197460 | Not-determined |
| 17-109_k141_397935 | Not-determined |
| 17-109_k141_62909 | Not-determined |
| 17-109_k141_62910 | Low-quality |
| 17-109_k141_129357 | Not-determined |
| 17-109_k141_397937 | Not-determined |
| 17-109_k141_362750 | Not-determined |
| 17-109_k141_96315 | Not-determined |
| 17-109_k141_362752 | Not-determined |
| 17-109_k141_146572 | Not-determined |
| 17-109_k141_12515 | Not-determined |
| 17-109_k141_213969 | Not-determined |
| 17-109_k141_231169 | Not-determined |
| 17-109_k141_362757 | Low-quality |
| 17-109_k141_80515 | Not-determined |
| 17-109_k141_281661 | Not-determined |
| 17-109_k141_146579 | Not-determined |
| 17-109_k141_381437 | Low-quality |
| 17-109_k141_47296 | Not-determined |
| 17-109_k141_96333 | Not-determined |
| 17-109_k141_347881 | Not-determined |
| 17-109_k141_281668 | Not-determined |
| 17-109_k141_281675 | Not-determined |
| 17-109_k141_331145 | Not-determined |
| 17-109_k141_96336 | Not-determined |
| 17-109_k141_381450 | Not-determined |
| 17-109_k141_213980 | Not-determined |
| 17-109_k141_381453 | Not-determined |
| 17-109_k141_62941 | Not-determined |
| 17-109_k141_397958 | Not-determined |
| 17-109_k141_213983 | Not-determined |
| 17-109_k141_47317 | Not-determined |
| 17-109_k141_180398 | Not-determined |
| 17-109_k141_47318 | Not-determined |
| 17-109_k141_247072 | Not-determined |
| 17-109_k141_213989 | Not-determined |
| 17-109_k141_112606 | Not-determined |
| 17-109_k141_247078 | Not-determined |
| 17-109_k141_47322 | Not-determined |
| 17-109_k141_331157 | Not-determined |
| 17-109_k141_47330 | Not-determined |
| 17-109_k141_112619 | Not-determined |
| 17-109_k141_47332 | Not-determined |
| 17-109_k141_281688 | Not-determined |
| 17-109_k141_231191 | Not-determined |
| 17-109_k141_180407 | Not-determined |
| 17-109_k141_297769 | Not-determined |
| 17-109_k141_231194 | Not-determined |
| 17-109_k141_80542 | Not-determined |
| 17-109_k141_297771 | Not-determined |
| 17-109_k141_29375 | Not-determined |
| 17-109_k141_29376 | Low-quality |
| 17-109_k141_163883 | Low-quality |
| 17-109_k141_62950 | Not-determined |
| 17-109_k141_112638 | Not-determined |
| 17-109_k141_362796 | Not-determined |
| 17-109_k141_163891 | Not-determined |
| 17-109_k141_12543 | Not-determined |
| 17-109_k141_231213 | Not-determined |
| 17-109_k141_12547 | Not-determined |
| 17-109_k141_96360 | Not-determined |
| 17-109_k141_12550 | Not-determined |
| 17-109_k141_96365 | Not-determined |
| 17-109_k141_112650 | Not-determined |
| 17-109_k141_381482 | Not-determined |
| 17-109_k141_214023 | Not-determined |
| 17-109_k141_214024 | Not-determined |
| 17-109_k141_381485 | Low-quality |
| 17-109_k141_214025 | Not-determined |
| 17-109_k141_214027 | Not-determined |
| 17-109_k141_12556 | Low-quality |
| 17-109_k141_96368 | Not-determined |
| 17-109_k141_163908 | Low-quality |
| 17-109_k141_112655 | Not-determined |
| 17-109_k141_231230 | Not-determined |
| 17-109_k141_397981 | Not-determined |
| 17-109_k141_214034 | Not-determined |
| 17-109_k141_347898 | Not-determined |
| 17-109_k141_129408 | Not-determined |
| 17-109_k141_180428 | Not-determined |
| 17-109_k141_12561 | Not-determined |
| 17-109_k141_331180 | Not-determined |
| 17-109_k141_331186 | Not-determined |
| 17-109_k141_180430 | Not-determined |
| 17-109_k141_80549 | Not-determined |
| 17-109_k141_214044 | Not-determined |
| 17-109_k141_247132 | Not-determined |
| 17-109_k141_129419 | Not-determined |
| 17-109_k141_315412 | Low-quality |
| 17-109_k141_381496 | Low-quality |
| 17-109_k141_146606 | Low-quality |
| 17-109_k141_163923 | Not-determined |
| 17-109_k141_247141 | Not-determined |
| 17-109_k141_29422 | Not-determined |
| 17-109_k141_347902 | Not-determined |
| 17-109_k141_315421 | Not-determined |
| 17-109_k141_347906 | Not-determined |
| 17-109_k141_331196 | Not-determined |
| 17-109_k141_381504 | Not-determined |
| 17-109_k141_331198 | Not-determined |
| 17-109_k141_281728 | Not-determined |
| 17-109_k141_281732 | Not-determined |
| 17-109_k141_247155 | Not-determined |
| 17-109_k141_281733 | Not-determined |
| 17-109_k141_129424 | Not-determined |
| 17-109_k141_347913 | Not-determined |
| 17-109_k141_315424 | Not-determined |
| 17-109_k141_263631 | Low-quality |
| 17-109_k141_263633 | Not-determined |
| 17-109_k141_29427 | Not-determined |
| 17-109_k141_315436 | Not-determined |
| 17-109_k141_381518 | Not-determined |
| 17-109_k141_281745 | Not-determined |
| 17-109_k141_47359 | Low-quality |
| 17-109_k141_247168 | Low-quality |
| 17-109_k141_331220 | Not-determined |
| 17-109_k141_129442 | Not-determined |
| 17-109_k141_80560 | Not-determined |
| 17-109_k141_362827 | Not-determined |
| 17-109_k141_146621 | Not-determined |
| 17-109_k141_247178 | Not-determined |
| 17-109_k141_29437 | Not-determined |
| 17-109_k141_347950 | Not-determined |
| 17-109_k141_231270 | Not-determined |
| 17-109_k141_381533 | Not-determined |
| 17-109_k141_231272 | Not-determined |
| 17-109_k141_129447 | Not-determined |
| 17-109_k141_146633 | Not-determined |
| 17-109_k141_129450 | Not-determined |
| 17-109_k141_129452 | Not-determined |
| 17-109_k141_180468 | Not-determined |
| 17-109_k141_47375 | Not-determined |
| 17-109_k141_263654 | Low-quality |
| 17-109_k141_231283 | Not-determined |
| 17-109_k141_231286 | Not-determined |
| 17-109_k141_398016 | Not-determined |
| 17-109_k141_331248 | Not-determined |
| 17-109_k141_231295 | Not-determined |
| 17-109_k141_347968 | Not-determined |
| 17-109_k141_112710 | Not-determined |
| 17-109_k141_347972 | Not-determined |
| 17-109_k141_381549 | Not-determined |
| 17-109_k141_180489 | Not-determined |
| 17-109_k141_47392 | Not-determined |
| 17-109_k141_29483 | Not-determined |
| 17-109_k141_263687 | Not-determined |
| 17-109_k141_347981 | Not-determined |
| 17-109_k141_163975 | Low-quality |
| 17-109_k141_231311 | Not-determined |
| 17-109_k141_163976 | Not-determined |
| 17-109_k141_231312 | Not-determined |
| 17-109_k141_398027 | Not-determined |
| 17-109_k141_247232 | Not-determined |
| 17-109_k141_247233 | Not-determined |
| 17-109_k141_47407 | Not-determined |
| 17-109_k141_163979 | Not-determined |
| 17-109_k141_297816 | Not-determined |
| 17-109_k141_281782 | Low-quality |
| 17-109_k141_347985 | Not-determined |
| 17-109_k141_331272 | Not-determined |
| 17-109_k141_129489 | Not-determined |
| 17-109_k141_129490 | Not-determined |
| 17-109_k141_163983 | Not-determined |

| | |
|---|---|
| 17-109_k141_381570 | Not-determined |
| 17-109_k141_180502 | Not-determined |
| 17-109_k141_29503 | Not-determined |
| 17-109_k141_129492 | Not-determined |
| 17-109_k141_281789 | Not-determined |
| 17-109_k141_80597 | Not-determined |
| 17-109_k141_96428 | Low-quality |
| 17-109_k141_381574 | Low-quality |
| 17-109_k141_80602 | Not-determined |
| 17-109_k141_197539 | Not-determined |
| 17-109_k141_381580 | Not-determined |
| 17-109_k141_381581 | Not-determined |
| 17-109_k141_80605 | Not-determined |
| 17-109_k141_163998 | Not-determined |
| 17-109_k141_381584 | Not-determined |
| 17-109_k141_381585 | Not-determined |
| 17-109_k141_80609 | Not-determined |
| 17-109_k141_96438 | Not-determined |
| 17-109_k141_47429 | Not-determined |
| 17-109_k141_164001 | Not-determined |
| 17-109_k141_129506 | Not-determined |
| 17-109_k141_331292 | Not-determined |
| 17-109_k141_348003 | Not-determined |
| 17-109_k141_164006 | Not-determined |
| 17-109_k141_348005 | Not-determined |
| 17-109_k141_331298 | Not-determined |
| 17-109_k141_29523 | Not-determined |
| 17-109_k141_164011 | Not-determined |
| 17-109_k141_331301 | Not-determined |
| 17-109_k141_297844 | Not-determined |
| 17-109_k141_164014 | Not-determined |
| 17-109_k141_164017 | Not-determined |
| 17-109_k141_348015 | Not-determined |
| 17-109_k141_297851 | Not-determined |
| 17-109_k141_263714 | Not-determined |
| 17-109_k141_281820 | Not-determined |
| 17-109_k141_29539 | Not-determined |
| 17-109_k141_146644 | Not-determined |
| 17-109_k141_231316 | Not-determined |
| 17-109_k141_146649 | Not-determined |
| 17-109_k141_297862 | Not-determined |
| 17-109_k141_164022 | Not-determined |
| 17-109_k141_381592 | Not-determined |
| 17-109_k141_146651 | Not-determined |
| 17-109_k141_297865 | Not-determined |
| 17-109_k141_146654 | Not-determined |
| 17-109_k141_263723 | Not-determined |
| 17-109_k141_398048 | Not-determined |
| 17-109_k141_164027 | Not-determined |
| 17-109_k141_29543 | Not-determined |
| 17-109_k141_281828 | Low-quality |
| 17-109_k141_112753 | Low-quality |
| 17-109_k141_231331 | Not-determined |
| 17-109_k141_197568 | Not-determined |
| 17-109_k141_381605 | Not-determined |
| 17-109_k141_29556 | Not-determined |
| 17-109_k141_164049 | Not-determined |
| 17-109_k141_362903 | Not-determined |
| 17-109_k141_47441 | Not-determined |
| 17-109_k141_315476 | Not-determined |
| 17-109_k141_47442 | Not-determined |
| 17-109_k141_315477 | Not-determined |
| 17-109_k141_129529 | Not-determined |
| 17-109_k141_29575 | Not-determined |
| 17-109_k141_164068 | Low-quality |
| 17-109_k141_362917 | Not-determined |
| 17-109_k141_362918 | Not-determined |
| 17-109_k141_146673 | Not-determined |
| 17-109_k141_129540 | Not-determined |
| 17-109_k141_381630 | Low-quality |
| 17-109_k141_263746 | Not-determined |
| 17-109_k141_315489 | Not-determined |
| 17-109_k141_112780 | Not-determined |
| 17-109_k141_281843 | Not-determined |
| 17-109_k141_315495 | Not-determined |
| 17-109_k141_263753 | Low-quality |
| 17-109_k141_129549 | Not-determined |
| 17-109_k141_281846 | Not-determined |
| 17-109_k141_129552 | Not-determined |
| 17-109_k141_146681 | Not-determined |
| 17-109_k141_281847 | Not-determined |
| 17-109_k141_12637 | Not-determined |
| 17-109_k141_214069 | Not-determined |
| 17-109_k141_263758 | Not-determined |
| 17-109_k141_29591 | Not-determined |
| 17-109_k141_315501 | Low-quality |
| 17-109_k141_47469 | Not-determined |
| 17-109_k141_29595 | Not-determined |
| 17-109_k141_112793 | Not-determined |
| 17-109_k141_164090 | Not-determined |
| 17-109_k141_381644 | Not-determined |
| 17-109_k141_112799 | Not-determined |
| 17-109_k141_47473 | Not-determined |
| 17-109_k141_29598 | Not-determined |
| 17-109_k141_231348 | Low-quality |
| 17-109_k141_96451 | Not-determined |
| 17-109_k141_281860 | Not-determined |
| 17-109_k141_80631 | Not-determined |
| 17-109_k141_281862 | Not-determined |
| 17-109_k141_315511 | Not-determined |
| 17-109_k141_164105 | Not-determined |
| 17-109_k141_348032 | Not-determined |
| 17-109_k141_348033 | Not-determined |
| 17-109_k141_231357 | Not-determined |
| 17-109_k141_315520 | Not-determined |
| 17-109_k141_331324 | Not-determined |
| 17-109_k141_214084 | Not-determined |
| 17-109_k141_381667 | Not-determined |
| 17-109_k141_29626 | Not-determined |
| 17-109_k141_80637 | Not-determined |
| 17-109_k141_315528 | Not-determined |
| 17-109_k141_381682 | Not-determined |
| 17-109_k141_80645 | Not-determined |
| 17-109_k141_381685 | Not-determined |
| 17-109_k141_263791 | Not-determined |
| 17-109_k141_180531 | Not-determined |
| 17-109_k141_164128 | Low-quality |
| 17-109_k141_381691 | Not-determined |
| 17-109_k141_231375 | Not-determined |
| 17-109_k141_80653 | Not-determined |
| 17-109_k141_80654 | Not-determined |
| 17-109_k141_231378 | Not-determined |
| 17-109_k141_263798 | Not-determined |
| 17-109_k141_112831 | Not-determined |
| 17-109_k141_96466 | Not-determined |
| 17-109_k141_231380 | Not-determined |
| 17-109_k141_263802 | Not-determined |
| 17-109_k141_96470 | Not-determined |
| 17-109_k141_47508 | Not-determined |
| 17-109_k141_62969 | Not-determined |
| 17-109_k141_180548 | Not-determined |
| 17-109_k141_297879 | Not-determined |
| 17-109_k141_112840 | Not-determined |
| 17-109_k141_231388 | Not-determined |
| 17-109_k141_96476 | Not-determined |
| 17-109_k141_47514 | Not-determined |
| 17-109_k141_331338 | Not-determined |
| 17-109_k141_112845 | Not-determined |
| 17-109_k141_62974 | Not-determined |
| 17-109_k141_297883 | Not-determined |
| 17-109_k141_146711 | Not-determined |
| 17-109_k141_197636 | Not-determined |
| 17-109_k141_62978 | Not-determined |
| 17-109_k141_47518 | Low-quality |
| 17-109_k141_381705 | Low-quality |
| 17-109_k141_263816 | Not-determined |
| 17-109_k141_263819 | Not-determined |
| 17-109_k141_247271 | Not-determined |
| 17-109_k141_297901 | Low-quality |
| 17-109_k141_146717 | Low-quality |
| 17-109_k141_247274 | Not-determined |
| 17-109_k141_197650 | Not-determined |
| 17-109_k141_112855 | Not-determined |
| 17-109_k141_231413 | Not-determined |
| 17-109_k141_348075 | Not-determined |
| 17-109_k141_214116 | Not-determined |
| 17-109_k141_231417 | Not-determined |
| 17-109_k141_297909 | Not-determined |
| 17-109_k141_62994 | Not-determined |
| 17-109_k141_297911 | Not-determined |
| 17-109_k141_62996 | Not-determined |
| 17-109_k141_247286 | Low-quality |
| 17-109_k141_331363 | Not-determined |
| 17-109_k141_231421 | Not-determined |
| 17-109_k141_362970 | Not-determined |
| 17-109_k141_112870 | Not-determined |
| 17-109_k141_112871 | Not-determined |
| 17-109_k141_180572 | Low-quality |
| 17-109_k141_331367 | Not-determined |
| 17-109_k141_331368 | Not-determined |
| 17-109_k141_63001 | Not-determined |
| 17-109_k141_263840 | Not-determined |
| 17-109_k141_247290 | Not-determined |
| 17-109_k141_348084 | Not-determined |
| 17-109_k141_47535 | Not-determined |
| 17-109_k141_214121 | Not-determined |
| 17-109_k141_112872 | Not-determined |
| 17-109_k141_197660 | Not-determined |
| 17-109_k141_297923 | Not-determined |
| 17-109_k141_297924 | Low-quality |
| 17-109_k141_297925 | Not-determined |
| 17-109_k141_297926 | Low-quality |
| 17-109_k141_197664 | Not-determined |
| 17-109_k141_47541 | Low-quality |
| 17-109_k141_348090 | Low-quality |
| 17-109_k141_180596 | Not-determined |
| 17-109_k141_263855 | Not-determined |
| 17-109_k141_180604 | Not-determined |
| 17-109_k141_146744 | Not-determined |
| 17-109_k141_146746 | Not-determined |
| 17-109_k141_331376 | Not-determined |
| 17-109_k141_47566 | Not-determined |
| 17-109_k141_164170 | Not-determined |
| 17-109_k141_164172 | Not-determined |
| 17-109_k141_231441 | Not-determined |
| 17-109_k141_146754 | Not-determined |
| 17-109_k141_231443 | Not-determined |
| 18-101-final-viral-combined_k141_164894\|\|full | Medium-quality |
| 17-109_k141_197678 | Not-determined |
| 17-109_k141_80682 | Not-determined |
| 17-109_k141_112903 | Not-determined |
| 17-109_k141_80684 | Not-determined |
| 17-109_k141_12662 | Low-quality |
| 17-109_k141_214153 | Not-determined |
| 17-109_k141_214155 | Not-determined |
| 17-109_k141_381732 | Not-determined |
| 17-109_k141_12667 | Not-determined |
| 17-109_k141_29673 | Not-determined |
| 17-109_k141_47583 | Not-determined |
| 17-109_k141_214165 | Not-determined |
| 17-109_k141_129584 | Not-determined |
| 17-109_k141_197682 | Low-quality |
| 17-109_k141_231459 | Not-determined |
| 17-109_k141_281938 | Not-determined |
| 17-109_k141_281940 | Not-determined |
| 17-109_k141_247321 | Not-determined |
| 17-109_k141_214172 | Not-determined |
| 17-109_k141_297949 | Not-determined |
| 17-109_k141_331396 | Not-determined |
| 17-109_k141_112907 | Not-determined |
| 17-109_k141_281947 | Not-determined |
| 17-109_k141_80702 | Not-determined |
| 17-109_k141_297954 | Not-determined |
| 17-109_k141_381748 | Not-determined |
| 17-109_k141_297955 | Not-determined |
| 17-109_k141_112911 | Not-determined |
| 17-109_k141_80706 | Not-determined |
| 17-109_k141_263889 | Not-determined |
| 17-109_k141_80712 | Not-determined |
| 17-109_k141_112913 | Not-determined |
| 17-109_k141_129600 | Not-determined |
| 17-109_k141_47595 | Not-determined |
| 17-109_k141_381756 | Not-determined |
| 17-109_k141_96538 | Low-quality |
| 17-109_k141_231472 | Not-determined |
| 17-109_k141_96541 | Low-quality |
| 17-109_k141_381759 | Not-determined |
| 17-109_k141_12681 | Not-determined |
| 17-109_k141_297962 | Not-determined |
| 17-109_k141_281953 | Not-determined |
| 17-109_k141_398073 | Not-determined |
| 17-109_k141_96545 | Not-determined |
| 17-109_k141_231478 | Not-determined |
| 17-109_k141_129612 | Not-determined |
| 17-109_k141_231480 | Not-determined |
| 17-109_k141_231481 | Not-determined |
| 17-109_k141_63051 | Not-determined |
| 17-109_k141_214188 | Not-determined |
| 17-109_k141_281957 | Not-determined |
| 17-109_k141_80720 | Not-determined |
| 17-109_k141_297975 | Not-determined |
| 17-109_k141_381767 | Not-determined |
| 17-109_k141_263913 | Not-determined |
| 17-109_k141_29697 | Not-determined |
| 17-109_k141_315563 | Low-quality |
| 17-109_k141_315565 | Not-determined |
| 17-109_k141_281966 | Not-determined |
| 17-109_k141_348114 | Not-determined |
| 17-109_k141_29701 | Not-determined |
| 17-109_k141_348116 | Not-determined |
| 17-109_k141_214195 | Not-determined |
| 17-109_k141_29703 | Low-quality |
| 17-109_k141_348120 | Not-determined |
| 17-109_k141_331421 | Low-quality |
| 17-109_k141_363049 | Not-determined |
| 17-109_k141_214201 | Not-determined |
| 17-109_k141_315577 | Not-determined |
| 17-109_k141_12709 | Not-determined |
| 17-109_k141_214209 | Not-determined |
| 17-109_k141_297990 | Not-determined |
| 17-109_k141_96574 | Not-determined |
| 17-109_k141_180643 | Not-determined |
| 17-109_k141_214210 | Not-determined |
| 17-109_k141_281984 | Low-quality |
| 17-109_k141_80732 | Low-quality |
| 17-109_k141_348128 | Not-determined |
| 17-109_k141_263932 | Not-determined |
| 17-109_k141_281989 | Not-determined |
| 17-109_k141_263934 | Not-determined |
| 17-109_k141_214215 | Not-determined |
| 17-109_k141_12714 | Not-determined |
| 17-109_k141_231507 | Not-determined |
| 17-109_k141_263935 | Not-determined |
| 17-109_k141_331446 | Not-determined |
| 17-109_k141_12716 | Not-determined |
| 17-109_k141_180662 | Not-determined |
| 17-109_k141_96585 | Low-quality |
| 17-109_k141_164229 | Not-determined |
| 17-109_k141_214223 | Not-determined |
| 17-109_k141_331451 | Not-determined |
| 17-109_k141_12721 | Not-determined |
| 17-109_k141_247346 | Not-determined |
| 17-109_k141_263940 | Not-determined |
| 17-109_k141_12725 | Not-determined |
| 17-109_k141_363052 | Not-determined |
| 17-109_k141_146802 | Not-determined |
| 17-109_k141_214225 | Low-quality |
| 17-109_k141_231512 | Not-determined |
| 17-109_k141_146804 | Not-determined |
| 17-109_k141_12729 | Not-determined |
| 17-109_k141_281999 | Not-determined |
| 17-109_k141_129632 | Low-quality |
| 17-109_k141_197699 | Not-determined |
| 17-109_k141_197702 | Low-quality |
| 17-109_k141_331463 | Not-determined |
| 17-109_k141_263945 | Low-quality |
| 17-109_k141_263948 | Not-determined |
| 17-109_k141_282005 | Not-determined |
| 17-109_k141_348145 | Not-determined |
| 17-109_k141_214235 | Not-determined |
| 17-109_k141_129641 | Not-determined |
| 17-109_k141_180681 | Not-determined |
| 17-109_k141_363071 | Not-determined |
| 17-109_k141_129644 | Not-determined |
| 17-109_k141_363072 | Not-determined |
| 17-109_k141_298005 | Not-determined |
| 17-109_k141_298011 | Not-determined |
| 17-109_k141_247366 | Not-determined |
| 17-109_k141_348164 | Not-determined |
| 17-109_k141_298016 | Not-determined |
| 17-109_k141_164247 | Not-determined |
| 17-109_k141_47669 | Not-determined |
| 17-109_k141_96628 | Low-quality |
| 17-109_k141_381807 | Not-determined |
| 17-109_k141_180695 | Not-determined |
| 17-109_k141_29716 | Not-determined |
| 17-109_k141_12754 | Not-determined |
| 17-109_k141_214244 | Not-determined |
| 17-109_k141_331496 | Not-determined |
| 17-109_k141_331498 | Not-determined |
| 17-109_k141_164248 | Not-determined |
| 17-109_k141_96632 | Not-determined |
| 17-109_k141_47678 | Not-determined |
| 17-109_k141_381820 | Not-determined |
| 17-109_k141_381821 | Not-determined |
| 17-109_k141_47680 | Not-determined |
| 17-109_k141_29727 | Not-determined |
| 17-109_k141_47683 | Not-determined |
| 17-109_k141_129655 | Low-quality |
| 17-109_k141_282033 | Not-determined |
| 17-109_k141_146816 | Low-quality |
| 17-109_k141_164255 | Not-determined |
| 17-109_k141_214249 | Not-determined |
| 17-109_k141_197742 | Not-determined |
| 17-109_k141_146825 | Not-determined |
| 17-109_k141_47692 | Not-determined |
| 17-109_k141_214252 | Not-determined |
| 17-109_k141_263974 | Not-determined |
| 17-109_k141_214253 | Not-determined |
| 17-109_k141_96659 | Not-determined |
| 17-109_k141_96660 | Not-determined |
| 17-109_k141_63087 | Not-determined |
| 17-109_k141_63089 | Not-determined |
| 17-109_k141_263978 | Not-determined |
| 17-109_k141_129682 | Not-determined |
| 17-109_k141_214256 | Not-determined |
| 17-109_k141_348189 | Not-determined |
| 17-109_k141_315620 | Not-determined |
| 17-109_k141_164274 | Not-determined |
| 17-109_k141_96670 | Not-determined |
| 17-109_k141_247403 | Not-determined |
| 17-109_k141_63100 | Not-determined |
| 17-109_k141_47708 | Not-determined |
| 17-109_k141_363080 | Not-determined |
| 17-109_k141_331522 | Not-determined |
| 17-109_k141_12764 | Not-determined |
| 17-109_k141_348204 | Not-determined |
| 17-109_k141_282045 | Not-determined |
| 17-109_k141_247417 | Not-determined |
| 17-109_k141_129695 | Not-determined |
| 17-109_k141_348207 | Not-determined |
| 17-109_k141_348209 | Not-determined |
| 17-109_k141_247423 | Not-determined |
| 17-109_k141_197764 | Not-determined |
| 17-109_k141_231582 | Not-determined |
| 17-109_k141_129697 | Not-determined |
| 17-109_k141_112929 | Not-determined |
| 17-109_k141_63110 | Not-determined |
| 17-109_k141_63111 | Not-determined |
| 17-109_k141_63112 | Not-determined |
| 17-109_k141_348218 | Not-determined |
| 17-109_k141_96681 | Not-determined |
| 17-109_k141_247433 | Not-determined |
| 17-109_k141_264005 | Not-determined |
| 17-109_k141_47722 | Not-determined |
| 17-109_k141_12775 | Not-determined |
| 17-109_k141_214277 | Not-determined |
| 17-109_k141_264012 | Not-determined |
| 17-109_k141_129714 | Not-determined |
| 17-109_k141_282065 | Not-determined |
| 17-109_k141_47732 | Not-determined |
| 17-109_k141_12784 | Not-determined |
| 17-109_k141_214286 | Not-determined |
| 17-109_k141_247451 | Low-quality |
| 17-109_k141_381831 | Not-determined |
| 17-109_k141_214294 | Low-quality |
| 17-109_k141_12790 | Low-quality |
| 17-109_k141_63129 | Not-determined |
| 17-109_k141_363107 | Not-determined |
| 17-109_k141_264025 | Low-quality |
| 17-109_k141_63135 | Not-determined |
| 17-109_k141_363111 | Not-determined |
| 17-109_k141_12799 | Low-quality |
| 17-109_k141_247467 | Low-quality |
| 17-109_k141_331539 | Not-determined |
| 17-109_k141_180763 | Not-determined |
| 17-109_k141_331543 | Not-determined |
| 17-109_k141_331544 | Not-determined |
| 17-109_k141_197800 | Not-determined |
| 17-109_k141_381846 | Not-determined |
| 17-109_k141_180770 | Not-determined |
| 17-109_k141_247475 | Not-determined |
| 17-109_k141_282090 | Not-determined |
| 17-109_k141_282099 | Not-determined |
| 17-109_k141_197819 | Not-determined |
| 17-109_k141_331558 | Not-determined |
| 17-109_k141_164326 | Not-determined |
| 17-109_k141_348250 | Not-determined |
| 17-109_k141_247486 | Not-determined |
| 17-109_k141_180790 | Not-determined |
| 17-109_k141_247490 | Not-determined |
| 17-109_k141_348258 | Not-determined |
| 17-109_k141_381869 | Low-quality |
| 17-109_k141_112987 | Not-determined |
| 17-109_k141_180796 | Not-determined |
| 17-109_k141_129764 | Not-determined |
| 17-109_k141_80836 | Not-determined |
| 17-109_k141_63153 | Not-determined |
| 17-109_k141_348261 | Not-determined |
| 17-109_k141_129765 | Not-determined |
| 17-109_k141_197833 | Not-determined |
| 17-109_k141_180799 | Not-determined |
| 17-109_k141_398136 | Not-determined |
| 17-109_k141_331585 | Not-determined |
| 17-109_k141_129773 | Not-determined |
| 17-109_k141_398139 | Low-quality |
| 17-109_k141_164353 | Not-determined |
| 17-109_k141_214317 | Not-determined |
| 17-109_k141_298066 | Not-determined |
| 17-109_k141_315695 | Low-quality |
| 17-109_k141_113002 | Not-determined |
| 17-109_k141_247508 | Not-determined |
| 17-109_k141_129784 | Not-determined |
| 17-109_k141_29777 | Not-determined |
| 17-109_k141_282117 | Not-determined |
| 17-109_k141_298075 | Not-determined |
| 17-109_k141_282122 | Not-determined |
| 17-109_k141_180810 | Low-quality |
| 17-109_k141_282126 | Not-determined |
| 17-109_k141_282127 | Not-determined |
| 17-109_k141_247513 | Not-determined |
| 17-109_k141_197850 | Not-determined |
| 17-109_k141_282130 | Not-determined |
| 17-109_k141_12840 | Not-determined |
| 17-109_k141_398154 | Not-determined |
| 17-109_k141_164379 | Not-determined |
| 17-109_k141_164380 | Not-determined |
| 17-109_k141_214339 | Not-determined |
| 17-109_k141_282138 | Not-determined |
| 17-109_k141_381884 | Not-determined |
| 17-109_k141_298080 | Not-determined |
| 17-109_k141_398159 | Not-determined |
| 17-109_k141_214343 | Not-determined |
| 17-109_k141_231624 | Not-determined |
| 17-109_k141_315722 | Not-determined |
| 17-109_k141_214349 | Not-determined |
| 17-109_k141_63214 | Not-determined |
| 17-109_k141_315729 | Not-determined |
| 17-109_k141_180837 | Not-determined |
| 17-109_k141_96717 | Not-determined |
| 17-109_k141_348286 | Not-determined |
| 17-109_k141_247530 | Not-determined |
| 17-109_k141_164395 | Not-determined |
| 17-109_k141_298091 | Low-quality |
| 17-109_k141_96724 | Not-determined |
| 17-109_k141_96725 | Not-determined |
| 17-109_k141_12861 | Not-determined |
| 17-109_k141_298094 | Not-determined |
| 17-109_k141_197868 | Not-determined |
| 17-109_k141_197870 | Not-determined |
| 17-109_k141_298097 | Not-determined |
| 17-109_k141_29802 | Not-determined |
| 17-109_k141_282158 | Not-determined |
| 17-109_k141_180849 | Not-determined |
| 17-109_k141_247539 | Not-determined |
| 17-109_k141_381911 | Low-quality |
| 17-109_k141_63229 | Low-quality |
| 17-109_k141_247544 | Not-determined |
| 17-109_k141_29810 | Not-determined |
| 17-109_k141_398175 | Not-determined |
| 17-109_k141_315750 | Low-quality |
| 17-109_k141_96736 | Not-determined |
| 17-109_k141_298104 | Not-determined |
| 17-109_k141_113061 | Not-determined |
| 17-109_k141_247546 | Low-quality |
| 17-109_k141_197883 | Not-determined |
| 17-109_k141_298107 | Not-determined |
| 17-109_k141_113069 | Not-determined |
| 17-109_k141_348304 | Low-quality |
| 17-109_k141_96745 | Not-determined |
| 17-109_k141_363170 | Not-determined |
| 17-109_k141_348312 | Not-determined |
| 17-109_k141_398189 | Not-determined |
| 17-109_k141_247554 | Not-determined |
| 17-109_k141_63261 | Not-determined |
| 17-109_k141_63264 | Not-determined |
| 17-109_k141_63265 | Not-determined |
| 17-109_k141_231663 | Low-quality |
| 17-109_k141_247562 | Not-determined |
| 17-109_k141_113089 | Not-determined |
| 17-109_k141_80871 | Not-determined |
| 17-109_k141_47756 | Low-quality |
| 17-109_k141_363196 | Not-determined |
| 17-109_k141_164434 | Not-determined |
| 17-109_k141_398199 | Not-determined |
| 17-109_k141_96763 | Not-determined |
| 17-109_k141_47760 | Low-quality |
| 17-109_k141_331622 | Not-determined |
| 17-109_k141_164439 | Not-determined |
| 17-109_k141_113098 | Not-determined |
| 17-109_k141_96764 | Not-determined |
| 17-109_k141_315772 | Not-determined |
| 17-109_k141_63290 | Not-determined |
| 17-109_k141_331629 | Not-determined |
| 17-109_k141_29849 | Not-determined |
| 17-109_k141_47770 | Low-quality |
| 17-109_k141_47773 | Not-determined |
| 17-109_k141_214395 | Not-determined |
| 17-109_k141_398208 | Not-determined |
| 17-109_k141_214396 | Not-determined |
| 17-109_k141_214397 | Not-determined |
| 17-109_k141_113107 | Not-determined |
| 17-109_k141_398210 | Not-determined |
| 17-109_k141_247571 | Not-determined |
| 17-109_k141_47780 | Not-determined |
| 17-109_k141_129788 | Not-determined |
| 17-109_k141_214407 | Not-determined |
| 17-109_k141_164448 | Low-quality |
| 17-109_k141_214409 | Not-determined |
| 17-109_k141_398215 | Low-quality |
| 17-109_k141_264071 | Not-determined |
| 17-109_k141_331644 | Not-determined |
| 17-109_k141_164455 | Low-quality |
| 17-109_k141_264075 | Not-determined |
| 17-109_k141_29879 | Not-determined |
| 17-109_k141_29880 | Not-determined |
| 17-109_k141_331650 | Not-determined |
| 17-109_k141_331651 | Not-determined |
| 17-109_k141_231696 | Not-determined |
| 17-109_k141_282204 | Not-determined |
| 17-109_k141_146893 | Not-determined |
| 17-109_k141_231699 | Not-determined |
| 17-109_k141_29887 | Not-determined |
| 17-109_k141_164464 | Not-determined |
| 17-109_k141_164465 | Not-determined |
| 17-109_k141_331655 | Not-determined |
| 17-109_k141_180873 | Not-determined |
| 17-109_k141_29891 | Not-determined |
| 17-109_k141_315801 | Not-determined |
| 17-109_k141_247599 | Not-determined |
| 17-109_k141_331661 | Not-determined |
| 17-109_k141_214414 | Low-quality |
| 17-109_k141_363238 | Not-determined |
| 17-109_k141_214417 | Low-quality |
| 17-109_k141_315811 | Not-determined |
| 17-109_k141_164479 | Not-determined |
| 17-109_k141_164481 | Low-quality |
| 17-109_k141_282222 | Not-determined |
| 17-109_k141_315817 | Not-determined |
| 17-109_k141_47800 | Not-determined |
| 17-109_k141_363254 | Not-determined |
| 17-109_k141_197945 | Not-determined |
| 17-109_k141_231726 | Not-determined |
| 17-109_k141_96767 | Low-quality |
| 17-109_k141_331665 | Not-determined |
| 17-109_k141_146917 | Not-determined |
| 17-109_k141_214439 | Not-determined |
| 17-109_k141_96777 | Not-determined |
| 17-109_k141_315831 | Not-determined |
| 17-109_k141_315832 | Not-determined |
| 17-109_k141_247630 | Not-determined |
| 17-109_k141_331671 | Not-determined |
| 17-109_k141_282248 | Not-determined |
| 17-109_k141_247635 | Not-determined |
| 17-109_k141_247637 | Not-determined |
| 17-109_k141_113159 | Not-determined |
| 17-109_k141_164516 | Not-determined |
| 17-109_k141_348370 | Not-determined |
| 17-109_k141_164519 | Not-determined |

| | |
|---|---|
| 17-109_k141_282255 | Low-quality |
| 17-109_k141_214446 | Not-determined |
| 18-101-final-viral-combined_k141_185046\|\|full | Medium-quality |
| 17-109_k141_247646 | Not-determined |
| 17-109_k141_146940 | Not-determined |
| 17-109_k141_315850 | Not-determined |
| 17-109_k141_96792 | Not-determined |
| 17-109_k141_197952 | Not-determined |
| 17-109_k141_214452 | Not-determined |
| 17-109_k141_29931 | Not-determined |
| 17-109_k141_164530 | Low-quality |
| 17-109_k141_146945 | Not-determined |
| 17-109_k141_247652 | Low-quality |
| 17-109_k141_231746 | Not-determined |
| 17-109_k141_63333 | Low-quality |
| 17-109_k141_29945 | Not-determined |
| 17-109_k141_264089 | Not-determined |
| 17-109_k141_348384 | Not-determined |
| 17-109_k141_197965 | Not-determined |
| 17-109_k141_47809 | Not-determined |
| 17-109_k141_180926 | Not-determined |
| 17-109_k141_315861 | Not-determined |
| 17-109_k141_146956 | Not-determined |
| 17-109_k141_298147 | Not-determined |
| 17-109_k141_146958 | Not-determined |
| 17-109_k141_231766 | Not-determined |
| 17-109_k141_96795 | Not-determined |
| 17-109_k141_231768 | Not-determined |
| 17-109_k141_315864 | Not-determined |
| 17-109_k141_80907 | Not-determined |
| 17-109_k141_315867 | Not-determined |
| 17-109_k141_363277 | Not-determined |
| 17-109_k141_214480 | Not-determined |
| 17-109_k141_282279 | Not-determined |
| 17-109_k141_113173 | Not-determined |
| 17-109_k141_63350 | Not-determined |
| 17-109_k141_363281 | Low-quality |
| 17-109_k141_214490 | Low-quality |
| 17-109_k141_282284 | Not-determined |
| 17-109_k141_197984 | Not-determined |
| 17-109_k141_96805 | Not-determined |
| 17-109_k141_214497 | Not-determined |
| 17-109_k141_214498 | Not-determined |
| 17-109_k141_282290 | Not-determined |
| 17-109_k141_12945 | Not-determined |
| 17-109_k141_214501 | Not-determined |
| 17-109_k141_348417 | Not-determined |
| 17-109_k141_231797 | Not-determined |
| 17-109_k141_298159 | Low-quality |
| 17-109_k141_231801 | Not-determined |
| 17-109_k141_363287 | Not-determined |
| 17-109_k141_12956 | Not-determined |
| 17-109_k141_12965 | Not-determined |
| 17-109_k141_180953 | Not-determined |
| 17-109_k141_348432 | Not-determined |
| 17-109_k141_180956 | Not-determined |
| 17-109_k141_96829 | Not-determined |
| 17-109_k141_315890 | Low-quality |
| 17-109_k141_381931 | Not-determined |
| 17-109_k141_315891 | Not-determined |
| 17-109_k141_47849 | Not-determined |
| 17-109_k141_298173 | Not-determined |
| 17-109_k141_47851 | Not-determined |
| 17-109_k141_264123 | Not-determined |
| 17-109_k141_47853 | Not-determined |
| 17-109_k141_282324 | Not-determined |
| 17-109_k141_12972 | Low-quality |
| 17-109_k141_29996 | Not-determined |
| 17-109_k141_30001 | Not-determined |
| 17-109_k141_331749 | Not-determined |
| 17-109_k141_180972 | Not-determined |
| 17-109_k141_198018 | Not-determined |
| 17-109_k141_315900 | Not-determined |
| 17-109_k141_30007 | Not-determined |
| 17-109_k141_30008 | Low-quality |
| 17-109_k141_12982 | Not-determined |
| 17-109_k141_12984 | Not-determined |
| 17-109_k141_348457 | Not-determined |
| 17-109_k141_315904 | Not-determined |
| 17-109_k141_113211 | Not-determined |
| 17-109_k141_282335 | Not-determined |
| 17-109_k141_348461 | Not-determined |
| 17-109_k141_63393 | Low-quality |
| 17-109_k141_331760 | Not-determined |
| 17-109_k141_96864 | Not-determined |
| 17-109_k141_247722 | Not-determined |
| 17-109_k141_30026 | Low-quality |
| 17-109_k141_12991 | Low-quality |
| 17-109_k141_247725 | Not-determined |
| 17-109_k141_12996 | Not-determined |
| 17-109_k141_381936 | Not-determined |
| 17-109_k141_348467 | Not-determined |
| 17-109_k141_13002 | Not-determined |
| 17-109_k141_63413 | Not-determined |
| 17-109_k141_331772 | Not-determined |
| 17-109_k141_381957 | Not-determined |
| 17-109_k141_264141 | Not-determined |
| 17-109_k141_80933 | Not-determined |
| 17-109_k141_264142 | Not-determined |
| 17-109_k141_13013 | Not-determined |
| 17-109_k141_264145 | Not-determined |
| 17-109_k141_381961 | Not-determined |
| 17-109_k141_13017 | Not-determined |
| 17-109_k141_381962 | Low-quality |
| 17-109_k141_381963 | Not-determined |
| 17-109_k141_13018 | Not-determined |
| 17-109_k141_63421 | Not-determined |
| 17-109_k141_348474 | Not-determined |
| 17-109_k141_298185 | Not-determined |
| 17-109_k141_80951 | Not-determined |
| 17-109_k141_113239 | Not-determined |
| 17-109_k141_231837 | Not-determined |
| 17-109_k141_247755 | Not-determined |
| 17-109_k141_231840 | Not-determined |
| 17-109_k141_198051 | Low-quality |
| 17-109_k141_331786 | Not-determined |
| 17-109_k141_231845 | Not-determined |
| 17-109_k141_164586 | Not-determined |
| 17-109_k141_315939 | Not-determined |
| 17-109_k141_231850 | Not-determined |
| 17-109_k141_331790 | Low-quality |
| 17-109_k141_214597 | Low-quality |
| 17-109_k141_214598 | Not-determined |
| 17-109_k141_129830 | Not-determined |
| 17-109_k141_146996 | Not-determined |
| 17-109_k141_47866 | Not-determined |
| 17-109_k141_214600 | Not-determined |
| 17-109_k141_47868 | Not-determined |
| 17-109_k141_181038 | Not-determined |
| 17-109_k141_47872 | Not-determined |
| 17-109_k141_247771 | Not-determined |
| 17-109_k141_198070 | Not-determined |
| 17-109_k141_47876 | Not-determined |
| 17-109_k141_231861 | Not-determined |
| 17-109_k141_348498 | Not-determined |
| 17-109_k141_47878 | Not-determined |
| 17-109_k141_331800 | Not-determined |
| 17-109_k141_398256 | Not-determined |
| 17-109_k141_181048 | Not-determined |
| 17-109_k141_398258 | Not-determined |
| 17-109_k141_247775 | Not-determined |
| 17-109_k141_129838 | Not-determined |
| 17-109_k141_398259 | Not-determined |
| 17-109_k141_129839 | Not-determined |
| 17-109_k141_181051 | Low-quality |
| 17-109_k141_30042 | Not-determined |
| 17-109_k141_113271 | Not-determined |
| 17-109_k141_247781 | Not-determined |
| 17-109_k141_398263 | Low-quality |
| 17-109_k141_30047 | Not-determined |
| 17-109_k141_214615 | Not-determined |
| 17-109_k141_181057 | Not-determined |
| 17-109_k141_113274 | Not-determined |
| 17-109_k141_113275 | Not-determined |
| 17-109_k141_231875 | Low-quality |
| 17-109_k141_231876 | Low-quality |
| 17-109_k141_381987 | Not-determined |
| 17-109_k141_214622 | Not-determined |
| 17-109_k141_381989 | Not-determined |
| 17-109_k141_181064 | Not-determined |
| 17-109_k141_113282 | Not-determined |
| 17-109_k141_80983 | Low-quality |
| 17-109_k141_264169 | Not-determined |
| 17-109_k141_181066 | Not-determined |
| 17-109_k141_214628 | Not-determined |
| 17-109_k141_331815 | Not-determined |
| 17-109_k141_264174 | Not-determined |
| 17-109_k141_398276 | Not-determined |
| 17-109_k141_315952 | Not-determined |
| 17-109_k141_47899 | Low-quality |
| 17-109_k141_129857 | Not-determined |
| 17-109_k141_164618 | Not-determined |
| 17-109_k141_47900 | Low-quality |
| 17-109_k141_164620 | Not-determined |
| 17-109_k141_113292 | Not-determined |
| 17-109_k141_113293 | Not-determined |
| 17-109_k141_63436 | Not-determined |
| 17-109_k141_282382 | Not-determined |
| 17-109_k141_13046 | Not-determined |
| 17-109_k141_264182 | Not-determined |
| 17-109_k141_264183 | Not-determined |
| 17-109_k141_214643 | Not-determined |
| 17-109_k141_363333 | Not-determined |
| 17-109_k141_363334 | Not-determined |
| 17-109_k141_282390 | Not-determined |
| 17-109_k141_80996 | Low-quality |
| 17-109_k141_147025 | Not-determined |
| 17-109_k141_282392 | Low-quality |
| 17-109_k141_13055 | Not-determined |
| 17-109_k141_348531 | Not-determined |
| 17-109_k141_331843 | Not-determined |
| 17-109_k141_264188 | Not-determined |
| 17-109_k141_315968 | Not-determined |
| 17-109_k141_147030 | Low-quality |
| 17-109_k141_13065 | Not-determined |
| 17-109_k141_63440 | Not-determined |
| 17-109_k141_382015 | Not-determined |
| 17-109_k141_147034 | Not-determined |
| 17-109_k141_264198 | Not-determined |
| 17-109_k141_264199 | Not-determined |
| 17-109_k141_382023 | Not-determined |
| 17-109_k141_298248 | Not-determined |
| 17-109_k141_47930 | Not-determined |
| 17-109_k141_147037 | Not-determined |
| 17-109_k141_298250 | Not-determined |
| 17-109_k141_264202 | Not-determined |
| 17-109_k141_63444 | Low-quality |
| 17-109_k141_315984 | Not-determined |
| 17-109_k141_63445 | Not-determined |
| 17-109_k141_47933 | Not-determined |
| 17-109_k141_214666 | Not-determined |
| 17-109_k141_298254 | Not-determined |
| 17-109_k141_315986 | Not-determined |
| 17-109_k141_164637 | Not-determined |
| 17-109_k141_315987 | Not-determined |
| 17-109_k141_198099 | Not-determined |
| 17-109_k141_315989 | Not-determined |
| 17-109_k141_231904 | Low-quality |
| 17-109_k141_348547 | Low-quality |
| 17-109_k141_282405 | Not-determined |
| 17-109_k141_47935 | Low-quality |
| 17-109_k141_382036 | Not-determined |
| 17-109_k141_231914 | Not-determined |
| 17-109_k141_214675 | Not-determined |
| 17-109_k141_348550 | Low-quality |
| 17-109_k141_231916 | Not-determined |
| 17-109_k141_81009 | Not-determined |
| 17-109_k141_129881 | Low-quality |
| 17-109_k141_47939 | Not-determined |
| 17-109_k141_198113 | Not-determined |
| 17-109_k141_81011 | Not-determined |
| 17-109_k141_398287 | Not-determined |
| 17-109_k141_214682 | Not-determined |
| 17-109_k141_81015 | Not-determined |
| 17-109_k141_96937 | Not-determined |
| 17-109_k141_181106 | Not-determined |
| 17-109_k141_96938 | Not-determined |
| 17-109_k141_147060 | Low-quality |
| 17-109_k141_81019 | Not-determined |
| 17-109_k141_30116 | Not-determined |
| 17-109_k141_181111 | Not-determined |
| 17-109_k141_81021 | Not-determined |
| 17-109_k141_298271 | Not-determined |
| 17-109_k141_247823 | Not-determined |
| 17-109_k141_81022 | Not-determined |
| 17-109_k141_282434 | Not-determined |
| 17-109_k141_129895 | Not-determined |
| 17-109_k141_63460 | Not-determined |
| 17-109_k141_113317 | Not-determined |
| 17-109_k141_231933 | Not-determined |
| 17-109_k141_231936 | Not-determined |
| 17-109_k141_198125 | Low-quality |
| 17-109_k141_13115 | Not-determined |
| 17-109_k141_282444 | Not-determined |
| 17-109_k141_231940 | Not-determined |
| 17-109_k141_282446 | Not-determined |
| 17-109_k141_382046 | Not-determined |
| 17-109_k141_214695 | Not-determined |
| 17-109_k141_331882 | Not-determined |
| 17-109_k141_247831 | Not-determined |
| 17-109_k141_247833 | Not-determined |
| 17-109_k141_129906 | Not-determined |
| 17-109_k141_81045 | Not-determined |
| 17-109_k141_113336 | Not-determined |
| 17-109_k141_214697 | Not-determined |
| 17-109_k141_231952 | Not-determined |
| 17-109_k141_198131 | Not-determined |
| 17-109_k141_113338 | Not-determined |
| 17-109_k141_113340 | Not-determined |
| 17-109_k141_63469 | Low-quality |
| 17-109_k141_214700 | Not-determined |
| 17-109_k141_13122 | Not-determined |
| 17-109_k141_214703 | Not-determined |
| 17-109_k141_247843 | Not-determined |
| 17-109_k141_398293 | Not-determined |
| 17-109_k141_129910 | Low-quality |
| 17-109_k141_348578 | Not-determined |
| 17-109_k141_231959 | Not-determined |
| 17-109_k141_81080 | Not-determined |
| 17-109_k141_198142 | Not-determined |
| 17-109_k141_13135 | Not-determined |
| 17-109_k141_63489 | Not-determined |
| 17-109_k141_214726 | Not-determined |
| 17-109_k141_198149 | Not-determined |
| 17-109_k141_298304 | Not-determined |
| 17-109_k141_398301 | Not-determined |
| 17-109_k141_81082 | Not-determined |
| 17-109_k141_129924 | Not-determined |
| 17-109_k141_129925 | Not-determined |
| 17-109_k141_214734 | Not-determined |
| 17-109_k141_81088 | Not-determined |
| 17-109_k141_47979 | Not-determined |
| 17-109_k141_316019 | Not-determined |
| 17-109_k141_13155 | Low-quality |
| 17-109_k141_231974 | Low-quality |
| 17-109_k141_398321 | Not-determined |
| 17-109_k141_47990 | Not-determined |
| 17-109_k141_231979 | Not-determined |
| 17-109_k141_264253 | Not-determined |
| 17-109_k141_96995 | Not-determined |
| 17-109_k141_181152 | Not-determined |
| 17-109_k141_398326 | Not-determined |
| 17-109_k141_316024 | Not-determined |
| 17-109_k141_63500 | Not-determined |
| 17-109_k141_398329 | Not-determined |
| 17-109_k141_63503 | Low-quality |
| 17-109_k141_382071 | Not-determined |
| 17-109_k141_316029 | Not-determined |
| 17-109_k141_13158 | Not-determined |
| 17-109_k141_13159 | Not-determined |
| 17-109_k141_129942 | Not-determined |
| 17-109_k141_231989 | Low-quality |
| 17-109_k141_181164 | Not-determined |
| 17-109_k141_48000 | Low-quality |
| 17-109_k141_181166 | Not-determined |
| 17-109_k141_48001 | Not-determined |
| 17-109_k141_231999 | Not-determined |
| 17-109_k141_282475 | Not-determined |
| 17-109_k141_113370 | Not-determined |
| 17-109_k141_63513 | Not-determined |
| 17-109_k141_282479 | Not-determined |
| 17-109_k141_232005 | Not-determined |
| 17-109_k141_398351 | Not-determined |
| 17-109_k141_282482 | Not-determined |
| 17-109_k141_81123 | Not-determined |
| 17-109_k141_48006 | Not-determined |
| 17-109_k141_214752 | Not-determined |
| 17-109_k141_214753 | Not-determined |
| 17-109_k141_214755 | Not-determined |
| 17-109_k141_398353 | Not-determined |
| 17-109_k141_316053 | Not-determined |
| 17-109_k141_331919 | Not-determined |
| 17-109_k141_48010 | Not-determined |
| 17-109_k141_63524 | Not-determined |
| 17-109_k141_316056 | Not-determined |
| 17-109_k141_232017 | Not-determined |
| 17-109_k141_398359 | Not-determined |
| 17-109_k141_198164 | Not-determined |
| 17-109_k141_331927 | Not-determined |
| 17-109_k141_198167 | Not-determined |
| 17-109_k141_181183 | Low-quality |
| 17-109_k141_97022 | Not-determined |
| 17-109_k141_282500 | Low-quality |
| 17-109_k141_63529 | Not-determined |
| 17-109_k141_147077 | Not-determined |
| 17-109_k141_298334 | Not-determined |
| 17-109_k141_97026 | Not-determined |
| 17-109_k141_97027 | Not-determined |
| 17-109_k141_63533 | Not-determined |
| 17-109_k141_382095 | Not-determined |
| 17-109_k141_81126 | Not-determined |
| 17-109_k141_147089 | Not-determined |
| 17-109_k141_232029 | Not-determined |
| 17-109_k141_331936 | Not-determined |
| 17-109_k141_48020 | Not-determined |
| 17-109_k141_147097 | Not-determined |
| 17-109_k141_331939 | Not-determined |
| 17-109_k141_113382 | Not-determined |
| 17-109_k141_63542 | Not-determined |
| 17-109_k141_129964 | Not-determined |
| 17-109_k141_81138 | Not-determined |
| 17-109_k141_13183 | Not-determined |
| 17-109_k141_63549 | Not-determined |
| 17-109_k141_181193 | Low-quality |
| 17-109_k141_147106 | Not-determined |
| 17-109_k141_13190 | Not-determined |
| 17-109_k141_298356 | Not-determined |
| 17-109_k141_81144 | Not-determined |
| 17-109_k141_97051 | Low-quality |
| 17-109_k141_363396 | Not-determined |
| 17-109_k141_331962 | Not-determined |
| 17-109_k141_181201 | Not-determined |
| 17-109_k141_13194 | Not-determined |
| 17-109_k141_398380 | Not-determined |
| 17-109_k141_181205 | Not-determined |
| 17-109_k141_63560 | Not-determined |
| 17-109_k141_81150 | Not-determined |
| 17-109_k141_147121 | Not-determined |
| 17-109_k141_63568 | Not-determined |
| 17-109_k141_48032 | Not-determined |
| 17-109_k141_147126 | Not-determined |
| 17-109_k141_398392 | Low-quality |
| 17-109_k141_147135 | Not-determined |
| 17-109_k141_214795 | Not-determined |
| 17-109_k141_316090 | Low-quality |
| 17-109_k141_247926 | Not-determined |
| 17-109_k141_214796 | Not-determined |
| 17-109_k141_316095 | Not-determined |
| 17-109_k141_348635 | Not-determined |
| 17-109_k141_48049 | Low-quality |
| 17-109_k141_398415 | Not-determined |
| 17-109_k141_363417 | Not-determined |
| 17-109_k141_363420 | Not-determined |
| 17-109_k141_63582 | Low-quality |
| 17-109_k141_382118 | Not-determined |
| 17-109_k141_97062 | Not-determined |
| 17-109_k141_382121 | Not-determined |
| 17-109_k141_232050 | Not-determined |
| 17-109_k141_181218 | Not-determined |
| 17-109_k141_363426 | Not-determined |
| 17-109_k141_214820 | Not-determined |
| 17-109_k141_48066 | Not-determined |
| 17-109_k141_232060 | Not-determined |
| 17-109_k141_81188 | Not-determined |
| 17-109_k141_232061 | Not-determined |
| 17-109_k141_48068 | Low-quality |
| 17-109_k141_398426 | Not-determined |
| 17-109_k141_214835 | Not-determined |
| 17-109_k141_81194 | Low-quality |
| 17-109_k141_298405 | Not-determined |
| 17-109_k141_130007 | Not-determined |
| 17-109_k141_13254 | Not-determined |
| 17-109_k141_13255 | Not-determined |
| 17-109_k141_398438 | Not-determined |
| 17-109_k141_316122 | Low-quality |
| 17-109_k141_331990 | Not-determined |
| 17-109_k141_113431 | Not-determined |
| 17-109_k141_30161 | Not-determined |
| 17-109_k141_398442 | Not-determined |
| 17-109_k141_214847 | Not-determined |
| 17-109_k141_214848 | Not-determined |
| 17-109_k141_363444 | Not-determined |
| 17-109_k141_363445 | Not-determined |
| 17-109_k141_264296 | Not-determined |
| 17-109_k141_232064 | Not-determined |
| 17-109_k141_48083 | Not-determined |
| 17-109_k141_130016 | Not-determined |
| 17-109_k141_331997 | Not-determined |
| 17-109_k141_382133 | Not-determined |
| 17-109_k141_48087 | Not-determined |
| 17-109_k141_398446 | Low-quality |
| 17-109_k141_316135 | Not-determined |
| 17-109_k141_181246 | Not-determined |
| 17-109_k141_264303 | Low-quality |
| 17-109_k141_232075 | Not-determined |
| 17-109_k141_382141 | Not-determined |
| 17-109_k141_382145 | Not-determined |
| 17-109_k141_332008 | Not-determined |
| 17-109_k141_13271 | Not-determined |
| 17-109_k141_63591 | Not-determined |
| 17-109_k141_48091 | Not-determined |
| 17-109_k141_382151 | Not-determined |
| 17-109_k141_332013 | Low-quality |
| 17-109_k141_181268 | Not-determined |
| 17-109_k141_113450 | Not-determined |
| 17-109_k141_113451 | Not-determined |
| 17-109_k141_398463 | Not-determined |
| 17-109_k141_181272 | Not-determined |
| 17-109_k141_13287 | Not-determined |
| 17-109_k141_81201 | Not-determined |
| 17-109_k141_181274 | Not-determined |
| 17-109_k141_332024 | Low-quality |
| 17-109_k141_282577 | Not-determined |
| 17-109_k141_97087 | Not-determined |
| 17-109_k141_81208 | Not-determined |
| 17-109_k141_48103 | Not-determined |
| 17-109_k141_63608 | Not-determined |
| 17-109_k141_81212 | Not-determined |
| 17-109_k141_264336 | Not-determined |
| 17-109_k141_63612 | Not-determined |
| 17-109_k141_97096 | Not-determined |
| 17-109_k141_264341 | Not-determined |
| 17-109_k141_97099 | Not-determined |
| 17-109_k141_81215 | Not-determined |
| 17-109_k141_282586 | Not-determined |
| 17-109_k141_48113 | Not-determined |
| 17-109_k141_13294 | Not-determined |
| 17-109_k141_48115 | Not-determined |
| 17-109_k141_282589 | Not-determined |
| 17-109_k141_48116 | Not-determined |
| 17-109_k141_113473 | Not-determined |
| 17-109_k141_13295 | Not-determined |
| 17-109_k141_97104 | Not-determined |
| 17-109_k141_363502 | Not-determined |
| 17-109_k141_298440 | Not-determined |
| 17-109_k141_363504 | Not-determined |
| 17-109_k141_298449 | Not-determined |
| 17-109_k141_363513 | Not-determined |
| 17-109_k141_214869 | Not-determined |
| 17-109_k141_147157 | Not-determined |
| 17-109_k141_214873 | Not-determined |
| 17-109_k141_147158 | Not-determined |
| 17-109_k141_232093 | Not-determined |
| 17-109_k141_63632 | Not-determined |
| 17-109_k141_81238 | Not-determined |
| 17-109_k141_382181 | Not-determined |
| 17-109_k141_298466 | Low-quality |
| 17-109_k141_316172 | Not-determined |
| 17-109_k141_348655 | Not-determined |
| 17-109_k141_214880 | Not-determined |
| 17-109_k141_298468 | Not-determined |
| 17-109_k141_63638 | Not-determined |
| 17-109_k141_198242 | Low-quality |
| 17-109_k141_198243 | Low-quality |
| 17-109_k141_181329 | Not-determined |
| 17-109_k141_363525 | Not-determined |
| 17-109_k141_382189 | Not-determined |
| 17-109_k141_63644 | Not-determined |
| 17-109_k141_316181 | Low-quality |
| 17-109_k141_181336 | Not-determined |
| 17-109_k141_316188 | Not-determined |
| 17-109_k141_97121 | Not-determined |
| 17-109_k141_164671 | Not-determined |
| 17-109_k141_30212 | Not-determined |
| 17-109_k141_48146 | Not-determined |
| 17-109_k141_298486 | Not-determined |
| 17-109_k141_382202 | Low-quality |
| 17-109_k141_48149 | Not-determined |
| 17-109_k141_298488 | Not-determined |
| 17-109_k141_248016 | Not-determined |
| 17-109_k141_30215 | Not-determined |
| 17-109_k141_316198 | Not-determined |
| 17-109_k141_147186 | Low-quality |
| 17-109_k141_248021 | Not-determined |
| 17-109_k141_164678 | Not-determined |
| 17-109_k141_198259 | Not-determined |
| 17-109_k141_30223 | Not-determined |
| 17-109_k141_30224 | Low-quality |
| 17-109_k141_363540 | Low-quality |
| 17-109_k141_232123 | Not-determined |
| 17-109_k141_30228 | Not-determined |
| 17-109_k141_181353 | Not-determined |
| 17-109_k141_264372 | Not-determined |
| 17-109_k141_214894 | Low-quality |
| 17-109_k141_164698 | Not-determined |
| 17-109_k141_130096 | Not-determined |
| 17-109_k141_316218 | Low-quality |
| 17-109_k141_198269 | Not-determined |
| 17-109_k141_232138 | Not-determined |
| 17-109_k141_198270 | Not-determined |
| 17-109_k141_113514 | Not-determined |
| 17-109_k141_198273 | Not-determined |
| 17-109_k141_298504 | Not-determined |
| 17-109_k141_298505 | Not-determined |
| 17-109_k141_232142 | Not-determined |
| 17-109_k141_113520 | Low-quality |
| 17-109_k141_13328 | Not-determined |
| 17-109_k141_232148 | Not-determined |
| 17-109_k141_248039 | Not-determined |
| 17-109_k141_348665 | Not-determined |
| 17-109_k141_13337 | Low-quality |
| 17-109_k141_97142 | Not-determined |
| 17-109_k141_332053 | Not-determined |
| 17-109_k141_97146 | Not-determined |
| 17-109_k141_232163 | Not-determined |
| 17-109_k141_147227 | Not-determined |
| 17-109_k141_232167 | Not-determined |
| 17-109_k141_13350 | Not-determined |
| 17-109_k141_348678 | Low-quality |
| 17-109_k141_63671 | Not-determined |
| 17-109_k141_13352 | Not-determined |
| 17-109_k141_248061 | Not-determined |
| 17-109_k141_147232 | Not-determined |
| 17-109_k141_147235 | Not-determined |
| 17-109_k141_248064 | Low-quality |
| 17-109_k141_97158 | Not-determined |
| 17-109_k141_181375 | Not-determined |
| 17-109_k141_198288 | Not-determined |
| 17-109_k141_130124 | Not-determined |
| 17-109_k141_382236 | Not-determined |
| 17-109_k141_316245 | Not-determined |
| 17-109_k141_113541 | Not-determined |
| 17-109_k141_130131 | Low-quality |
| 17-109_k141_13369 | Not-determined |
| 17-109_k141_164725 | Not-determined |
| 17-109_k141_316251 | Not-determined |
| 17-109_k141_332075 | Not-determined |
| 17-109_k141_198296 | Not-determined |
| 17-109_k141_97179 | Not-determined |
| 17-109_k141_130140 | Not-determined |
| 17-109_k141_232171 | Not-determined |
| 17-109_k141_164730 | Not-determined |
| 17-109_k141_298535 | Not-determined |
| 17-109_k141_214935 | Not-determined |
| 17-109_k141_214941 | Not-determined |
| 17-109_k141_316264 | Not-determined |
| 17-109_k141_63691 | Not-determined |
| 17-109_k141_316271 | Not-determined |
| 17-109_k141_13381 | Not-determined |
| 17-109_k141_248097 | Not-determined |
| 17-109_k141_248098 | Not-determined |
| 17-109_k141_382242 | Not-determined |
| 17-109_k141_214950 | Low-quality |
| 17-109_k141_214951 | Not-determined |
| 17-109_k141_382247 | Not-determined |
| 17-109_k141_348729 | Not-determined |
| 17-109_k141_63703 | Not-determined |
| 17-109_k141_164750 | Not-determined |
| 17-109_k141_63707 | Not-determined |

| | |
|---|---|
| 17-109_k141_382251 | Not-determined |
| 17-109_k141_63711 | Not-determined |
| 17-109_k141_63714 | Not-determined |
| 17-109_k141_332105 | Not-determined |
| 17-109_k141_113551 | Not-determined |
| 17-109_k141_63724 | Not-determined |
| 17-109_k141_248106 | Not-determined |
| 17-109_k141_113557 | Not-determined |
| 17-109_k141_181413 | Not-determined |
| 17-109_k141_332115 | Low-quality |
| 17-109_k141_214964 | Not-determined |
| 17-109_k141_13406 | Not-determined |
| 17-109_k141_214969 | Not-determined |
| 17-109_k141_348755 | Not-determined |
| 17-109_k141_164767 | Not-determined |
| 17-109_k141_214973 | Not-determined |
| 17-109_k141_147263 | Low-quality |
| 17-109_k141_147264 | Not-determined |
| 17-109_k141_97192 | Not-determined |
| 17-109_k141_348769 | Not-determined |
| 17-109_k141_248128 | Not-determined |
| 17-109_k141_214977 | Not-determined |
| 17-109_k141_363557 | Not-determined |
| 17-109_k141_113585 | Not-determined |
| 17-109_k141_48162 | Not-determined |
| 17-109_k141_348785 | Not-determined |
| 17-109_k141_348786 | Not-determined |
| 17-109_k141_81286 | Not-determined |
| 17-109_k141_248138 | Not-determined |
| 17-109_k141_13414 | Not-determined |
| 17-109_k141_214980 | Not-determined |
| 17-109_k141_214983 | Not-determined |
| 17-109_k141_248142 | Not-determined |
| 17-109_k141_63733 | Not-determined |
| 17-109_k141_214987 | Not-determined |
| 17-109_k141_248144 | Low-quality |
| 17-109_k141_13425 | Not-determined |
| 17-109_k141_81293 | Not-determined |
| 17-109_k141_232227 | Not-determined |
| 17-109_k141_63744 | Not-determined |
| 17-109_k141_332162 | Not-determined |
| 17-109_k141_30242 | Not-determined |
| 17-109_k141_348809 | Not-determined |
| 17-109_k141_382280 | Not-determined |
| 17-109_k141_13432 | Not-determined |
| 17-109_k141_348815 | Not-determined |
| 17-109_k141_316323 | Not-determined |
| 17-109_k141_147285 | Not-determined |
| 17-109_k141_81308 | Not-determined |
| 17-109_k141_398498 | Not-determined |
| 17-109_k141_48195 | Low-quality |
| 17-109_k141_181458 | Not-determined |
| 17-109_k141_30258 | Not-determined |
| 17-109_k141_382297 | Not-determined |
| 17-109_k141_147294 | Not-determined |
| 17-109_k141_198353 | Not-determined |
| 17-109_k141_30261 | Not-determined |
| 17-109_k141_398506 | Not-determined |
| 17-109_k141_348834 | Not-determined |
| 17-109_k141_348837 | Not-determined |
| 17-109_k141_363567 | Not-determined |
| 17-109_k141_298580 | Low-quality |
| 17-109_k141_264397 | Not-determined |
| 17-109_k141_264399 | Not-determined |
| 17-109_k141_198374 | Not-determined |
| 17-109_k141_316329 | Not-determined |
| 17-109_k141_164822 | Not-determined |
| 17-109_k141_348840 | Low-quality |
| 17-109_k141_130217 | Not-determined |
| 17-109_k141_48219 | Not-determined |
| 17-109_k141_48222 | Not-determined |
| 17-109_k141_348844 | Low-quality |
| 17-109_k141_63767 | Not-determined |
| 17-109_k141_198385 | Not-determined |
| 17-109_k141_130221 | Not-determined |
| 17-109_k141_363581 | Not-determined |
| 17-109_k141_198389 | Not-determined |
| 17-109_k141_113629 | Not-determined |
| 17-109_k141_382328 | Not-determined |
| 17-109_k141_348853 | Not-determined |
| 17-109_k141_30293 | Not-determined |
| 17-109_k141_382329 | Not-determined |
| 17-109_k141_164830 | Not-determined |
| 17-109_k141_248158 | Not-determined |
| 17-109_k141_264417 | Not-determined |
| 17-109_k141_164835 | Not-determined |
| 17-109_k141_248160 | Low-quality |
| 17-109_k141_248161 | Not-determined |
| 17-109_k141_48240 | Not-determined |
| 17-109_k141_248165 | Not-determined |
| 17-109_k141_398530 | Not-determined |
| 17-109_k141_264422 | Not-determined |
| 17-109_k141_147325 | Not-determined |
| 17-109_k141_81344 | Not-determined |
| 17-109_k141_30304 | Not-determined |
| 17-109_k141_147326 | Not-determined |
| 17-109_k141_316355 | Not-determined |
| 17-109_k141_363595 | Not-determined |
| 17-109_k141_316357 | Not-determined |
| 17-109_k141_48256 | Not-determined |
| 17-109_k141_298608 | Low-quality |
| 17-109_k141_164849 | Not-determined |
| 17-109_k141_13457 | Not-determined |
| 17-109_k141_332187 | Not-determined |
| 17-109_k141_298614 | Not-determined |
| 17-109_k141_130244 | Low-quality |
| 17-109_k141_164856 | Not-determined |
| 17-109_k141_13464 | Not-determined |
| 17-109_k141_181474 | Not-determined |
| 17-109_k141_30317 | Not-determined |
| 17-109_k141_298626 | Not-determined |
| 17-109_k141_30320 | Not-determined |
| 17-109_k141_298627 | Not-determined |
| 17-109_k141_147342 | Not-determined |
| 17-109_k141_13469 | Low-quality |
| 17-109_k141_30321 | Low-quality |
| 17-109_k141_164864 | Low-quality |
| 17-109_k141_97236 | Not-determined |
| 17-109_k141_198432 | Not-determined |
| 17-109_k141_282647 | Not-determined |
| 17-109_k141_181492 | Not-determined |
| 17-109_k141_97241 | Not-determined |
| 17-109_k141_48268 | Not-determined |
| 17-109_k141_181493 | Not-determined |
| 17-109_k141_181495 | Not-determined |
| 17-109_k141_130263 | Not-determined |
| 17-109_k141_97248 | Not-determined |
| 17-109_k141_13480 | Not-determined |
| 17-109_k141_48271 | Not-determined |
| 17-109_k141_48273 | Not-determined |
| 17-109_k141_282662 | Not-determined |
| 17-109_k141_48276 | Not-determined |
| 17-109_k141_48278 | Not-determined |
| 17-109_k141_215022 | Not-determined |
| 17-109_k141_332213 | Not-determined |
| 17-109_k141_282664 | Not-determined |
| 17-109_k141_81383 | Not-determined |
| 17-109_k141_363617 | Not-determined |
| 17-109_k141_282666 | Not-determined |
| 17-109_k141_81389 | Not-determined |
| 17-109_k141_282673 | Not-determined |
| 17-109_k141_316401 | Low-quality |
| 17-109_k141_248192 | Not-determined |
| 17-109_k141_164888 | Not-determined |
| 17-109_k141_113679 | Not-determined |
| 17-109_k141_382355 | Low-quality |
| 17-109_k141_298663 | Not-determined |
| 17-109_k141_398566 | Not-determined |
| 17-109_k141_48294 | Low-quality |
| 17-109_k141_181512 | Not-determined |
| 17-109_k141_282677 | Not-determined |
| 17-109_k141_398568 | Not-determined |
| 17-109_k141_248199 | Not-determined |
| 17-109_k141_97266 | Not-determined |
| 17-109_k141_215046 | Not-determined |
| 17-109_k141_198456 | Low-quality |
| 17-109_k141_215047 | Not-determined |
| 17-109_k141_298671 | Not-determined |
| 17-109_k141_363634 | Not-determined |
| 17-109_k141_81401 | Not-determined |
| 17-109_k141_382362 | Not-determined |
| 17-109_k141_248200 | Not-determined |
| 17-109_k141_232282 | Not-determined |
| 17-109_k141_382364 | Not-determined |
| 17-109_k141_398578 | Not-determined |
| 17-109_k141_13518 | Not-determined |
| 17-109_k141_248204 | Not-determined |
| 17-109_k141_13522 | Not-determined |
| 17-109_k141_282683 | Not-determined |
| 17-109_k141_48303 | Not-determined |
| 17-109_k141_130296 | Not-determined |
| 17-109_k141_215057 | Not-determined |
| 17-109_k141_48304 | Not-determined |
| 17-109_k141_398585 | Not-determined |
| 17-109_k141_48305 | Not-determined |
| 17-109_k141_298678 | Not-determined |
| 17-109_k141_81408 | Not-determined |
| 17-109_k141_81409 | Low-quality |
| 17-109_k141_147362 | Not-determined |
| 17-109_k141_130301 | Not-determined |
| 17-109_k141_382374 | Low-quality |
| 17-109_k141_13532 | Not-determined |
| 17-109_k141_363645 | Not-determined |
| 17-109_k141_164902 | Not-determined |
| 17-109_k141_164904 | Not-determined |
| 17-109_k141_97280 | Not-determined |
| 17-109_k141_382378 | Not-determined |
| 17-109_k141_332228 | Not-determined |
| 17-109_k141_30346 | Not-determined |
| 17-109_k141_198475 | Not-determined |
| 17-109_k141_248224 | Not-determined |
| 18-105-final-viral-combined_k141_83412\|\|full | Medium-quality |
| 17-109_k141_48320 | Not-determined |
| 17-109_k141_97291 | Not-determined |
| 17-109_k141_298693 | Not-determined |
| 17-109_k141_130323 | Not-determined |
| 17-109_k141_48324 | Not-determined |
| 17-109_k141_13542 | Low-quality |
| 17-109_k141_48330 | Not-determined |
| 17-109_k141_264428 | Low-quality |
| 17-109_k141_48332 | Not-determined |
| 17-109_k141_298701 | Not-determined |
| 17-109_k141_81415 | Not-determined |
| 17-109_k141_298702 | Not-determined |
| 17-109_k141_232305 | Low-quality |
| 17-109_k141_332250 | Not-determined |
| 17-109_k141_264437 | Low-quality |
| 17-109_k141_30363 | Not-determined |
| 17-109_k141_398598 | Not-determined |
| 17-109_k141_30364 | Low-quality |
| 17-109_k141_30365 | Not-determined |
| 17-109_k141_398600 | Not-determined |
| 17-109_k141_264438 | Low-quality |
| 17-109_k141_348902 | Not-determined |
| 17-109_k141_130345 | Not-determined |
| 17-109_k141_97312 | Not-determined |
| 17-109_k141_97313 | Not-determined |
| 17-109_k141_348908 | Low-quality |
| 17-109_k141_316447 | Low-quality |
| 17-109_k141_264443 | Not-determined |
| 17-109_k141_81420 | Not-determined |
| 17-109_k141_332265 | Not-determined |
| 17-109_k141_13562 | Not-determined |
| 17-109_k141_398613 | Not-determined |
| 17-109_k141_164925 | Not-determined |
| 17-109_k141_363677 | Low-quality |
| 17-109_k141_348915 | Not-determined |
| 17-109_k141_13568 | Not-determined |
| 17-109_k141_298727 | Not-determined |
| 17-109_k141_81428 | Not-determined |
| 17-109_k141_164928 | Not-determined |
| 17-109_k141_282710 | Not-determined |
| 17-109_k141_63858 | Low-quality |
| 17-109_k141_63859 | Not-determined |
| 17-109_k141_264457 | Not-determined |
| 17-109_k141_198489 | Not-determined |
| 17-109_k141_264459 | Not-determined |
| 17-109_k141_382401 | Not-determined |
| 17-109_k141_164940 | Not-determined |
| 17-109_k141_348922 | Not-determined |
| 17-109_k141_13577 | Not-determined |
| 17-109_k141_48348 | Not-determined |
| 17-109_k141_164944 | Not-determined |
| 17-109_k141_113726 | Not-determined |
| 17-109_k141_282725 | Not-determined |
| 17-109_k141_282728 | Not-determined |
| 17-109_k141_264464 | Not-determined |
| 17-109_k141_363699 | Not-determined |
| 17-109_k141_48364 | Not-determined |
| 17-109_k141_363703 | Low-quality |
| 17-109_k141_282736 | Not-determined |
| 17-109_k141_198505 | Not-determined |
| 17-109_k141_264471 | Not-determined |
| 17-109_k141_215106 | Low-quality |
| 17-109_k141_232322 | Not-determined |
| 17-109_k141_30402 | Not-determined |
| 17-109_k141_198506 | Not-determined |
| 17-109_k141_232323 | Not-determined |
| 17-109_k141_147385 | Not-determined |
| 17-109_k141_30403 | Not-determined |
| 17-109_k141_113737 | Not-determined |
| 17-109_k141_113738 | Not-determined |
| 17-109_k141_282743 | Not-determined |
| 17-109_k141_97326 | Not-determined |
| 17-109_k141_215108 | Not-determined |
| 17-109_k141_232333 | Not-determined |
| 17-109_k141_316471 | Not-determined |
| 17-109_k141_198517 | Not-determined |
| 17-109_k141_248273 | Not-determined |
| 17-109_k141_48373 | Low-quality |
| 17-109_k141_298766 | Not-determined |
| 17-109_k141_147408 | Not-determined |
| 17-109_k141_232345 | Not-determined |
| 17-109_k141_316479 | Not-determined |
| 17-109_k141_164977 | Not-determined |
| 17-109_k141_298769 | Not-determined |
| 17-109_k141_298771 | Not-determined |
| 17-109_k141_298773 | Not-determined |
| 17-109_k141_232347 | Not-determined |
| 17-109_k141_147410 | Not-determined |
| 17-109_k141_298774 | Not-determined |
| 17-109_k141_13595 | Not-determined |
| 17-109_k141_13596 | Not-determined |
| 17-109_k141_164982 | Not-determined |
| 17-109_k141_147414 | Not-determined |
| 17-109_k141_348935 | Not-determined |
| 17-109_k141_13599 | Low-quality |
| 17-109_k141_264493 | Not-determined |
| 17-109_k141_398643 | Not-determined |
| 17-109_k141_147419 | Not-determined |
| 17-109_k141_398646 | Not-determined |
| 17-109_k141_348938 | Not-determined |
| 17-109_k141_48397 | Not-determined |
| 17-109_k141_316493 | Not-determined |
| 17-109_k141_81484 | Not-determined |
| 17-109_k141_348944 | Not-determined |
| 17-109_k141_165000 | Low-quality |
| 17-109_k141_282775 | Not-determined |
| 17-109_k141_181542 | Not-determined |
| 17-109_k141_232358 | Low-quality |
| 17-109_k141_97354 | Not-determined |
| 17-109_k141_48405 | Not-determined |
| 17-109_k141_248295 | Not-determined |
| 17-109_k141_48406 | Not-determined |
| 17-109_k141_215135 | Not-determined |
| 17-109_k141_248300 | Not-determined |
| 17-109_k141_198548 | Not-determined |
| 17-109_k141_147428 | Not-determined |
| 17-109_k141_363741 | Not-determined |
| 17-109_k141_382449 | Not-determined |
| 17-109_k141_215139 | Not-determined |
| 17-109_k141_215142 | Not-determined |
| 17-109_k141_332322 | Not-determined |
| 17-109_k141_215143 | Not-determined |
| 17-109_k141_348959 | Not-determined |
| 17-109_k141_298779 | Not-determined |
| 17-109_k141_198557 | Not-determined |
| 17-109_k141_81507 | Low-quality |
| 17-109_k141_81509 | Low-quality |
| 17-109_k141_97371 | Not-determined |
| 17-109_k141_147435 | Not-determined |
| 17-109_k141_215153 | Not-determined |
| 17-109_k141_382461 | Not-determined |
| 17-109_k141_13633 | Not-determined |
| 17-109_k141_63867 | Low-quality |
| 17-109_k141_382468 | Not-determined |
| 17-109_k141_13638 | Not-determined |
| 17-109_k141_282789 | Not-determined |
| 17-109_k141_63869 | Not-determined |
| 17-109_k141_48410 | Not-determined |
| 17-109_k141_282796 | Not-determined |
| 17-109_k141_332347 | Not-determined |
| 17-109_k141_113788 | Not-determined |
| 17-109_k141_264525 | Not-determined |
| 17-109_k141_382480 | Not-determined |
| 17-109_k141_181568 | Not-determined |
| 17-109_k141_48415 | Not-determined |
| 17-109_k141_282801 | Not-determined |
| 17-109_k141_215175 | Not-determined |
| 17-109_k141_63875 | Not-determined |
| 17-109_k141_63876 | Not-determined |
| 17-109_k141_398671 | Not-determined |
| 17-109_k141_130395 | Not-determined |
| 17-109_k141_13653 | Not-determined |
| 17-109_k141_147440 | Not-determined |
| 17-109_k141_30448 | Not-determined |
| 17-109_k141_30455 | Not-determined |
| 17-109_k141_97400 | Low-quality |
| 17-109_k141_130412 | Low-quality |
| 17-109_k141_398687 | Not-determined |
| 17-109_k141_97402 | Not-determined |
| 17-109_k141_363774 | Not-determined |
| 17-109_k141_113826 | Not-determined |
| 17-109_k141_48437 | Not-determined |
| 17-109_k141_63891 | Not-determined |
| 17-109_k141_215211 | Not-determined |
| 17-109_k141_298809 | Not-determined |
| 17-109_k141_298810 | Not-determined |
| 17-109_k141_363780 | Not-determined |
| 17-109_k141_97408 | Not-determined |
| 17-109_k141_97410 | Not-determined |
| 17-109_k141_130420 | Not-determined |
| 17-109_k141_113833 | Not-determined |
| 17-109_k141_298820 | Not-determined |
| 17-109_k141_316559 | Not-determined |
| 17-109_k141_181629 | Not-determined |
| 17-109_k141_332402 | Not-determined |
| 17-109_k141_48448 | Not-determined |
| 17-109_k141_63910 | Not-determined |
| 17-109_k141_232446 | Not-determined |
| 17-109_k141_316568 | Not-determined |
| 17-109_k141_332413 | Not-determined |
| 17-109_k141_332417 | Low-quality |
| 17-109_k141_316577 | Not-determined |
| 17-109_k141_232452 | Not-determined |
| 17-109_k141_398698 | Low-quality |
| 17-109_k141_363802 | Not-determined |
| 17-109_k141_113847 | Low-quality |
| 17-109_k141_232456 | Low-quality |
| 17-109_k141_363805 | Not-determined |
| 17-109_k141_113849 | Not-determined |
| 17-109_k141_48461 | Not-determined |
| 17-109_k141_113852 | Not-determined |
| 17-109_k141_298839 | Not-determined |
| 17-109_k141_298840 | Not-determined |
| 17-109_k141_97441 | Not-determined |
| 17-109_k141_215252 | Not-determined |
| 17-109_k141_147495 | Not-determined |
| 17-109_k141_30496 | Not-determined |
| 17-109_k141_13695 | Not-determined |
| 17-109_k141_198614 | Not-determined |
| 17-109_k141_30497 | Not-determined |
| 17-109_k141_48473 | Not-determined |
| 17-109_k141_363819 | Not-determined |
| 17-109_k141_348982 | Not-determined |
| 17-109_k141_282853 | Not-determined |
| 17-109_k141_282855 | Not-determined |
| 17-109_k141_316593 | Not-determined |
| 17-109_k141_81544 | Not-determined |
| 17-109_k141_248332 | Not-determined |
| 17-109_k141_130439 | Not-determined |
| 17-109_k141_130440 | Not-determined |
| 17-109_k141_316595 | Not-determined |
| 17-109_k141_198709 | Low-quality |
| 17-109_k141_248339 | Not-determined |
| 17-109_k141_332454 | Not-determined |
| 17-109_k141_248340 | Not-determined |
| 17-109_k141_232478 | Not-determined |
| 17-109_k141_81553 | Not-determined |
| 17-109_k141_264539 | Not-determined |
| 17-109_k141_264540 | Not-determined |
| 17-109_k141_348988 | Not-determined |
| 17-109_k141_130450 | Low-quality |
| 17-109_k141_97480 | Not-determined |
| 17-109_k141_30510 | Not-determined |
| 17-109_k141_198644 | Not-determined |
| 17-109_k141_316603 | Not-determined |
| 17-109_k141_382545 | Not-determined |
| 17-109_k141_130452 | Not-determined |
| 17-109_k141_198646 | Not-determined |
| 17-109_k141_316610 | Not-determined |
| 17-109_k141_348999 | Low-quality |
| 17-109_k141_248347 | Not-determined |
| 17-109_k141_298880 | Not-determined |
| 17-109_k141_113899 | Not-determined |
| 17-109_k141_398732 | Low-quality |
| 17-109_k141_130462 | Not-determined |
| 17-109_k141_181673 | Low-quality |
| 17-109_k141_398734 | Not-determined |
| 17-109_k141_316620 | Not-determined |
| 17-109_k141_198658 | Not-determined |
| 17-109_k141_332472 | Not-determined |
| 17-109_k141_398738 | Not-determined |
| 17-109_k141_248351 | Low-quality |
| 17-109_k141_298892 | Not-determined |
| 17-109_k141_181680 | Not-determined |
| 17-109_k141_298894 | Not-determined |
| 17-109_k141_382568 | Not-determined |
| 17-109_k141_248362 | Not-determined |
| 17-109_k141_97505 | Not-determined |
| 17-109_k141_13718 | Not-determined |
| 17-109_k141_215280 | Not-determined |
| 17-109_k141_332486 | Not-determined |
| 17-109_k141_198677 | Not-determined |
| 17-109_k141_13726 | Not-determined |
| 17-109_k141_48492 | Not-determined |
| 17-109_k141_97515 | Not-determined |
| 17-109_k141_282914 | Not-determined |
| 17-109_k141_97520 | Not-determined |
| 17-109_k141_113914 | Not-determined |
| 17-109_k141_232505 | Not-determined |
| 17-109_k141_232507 | Not-determined |
| 17-109_k141_232508 | Not-determined |
| 17-109_k141_264591 | Not-determined |
| 17-109_k141_97538 | Not-determined |
| 17-109_k141_48501 | Low-quality |
| 17-109_k141_232515 | Low-quality |
| 17-109_k141_113930 | Not-determined |
| 17-109_k141_264597 | Not-determined |
| 17-109_k141_48506 | Not-determined |
| 17-109_k141_215304 | Not-determined |
| 17-109_k141_215305 | Not-determined |
| 17-109_k141_316648 | Not-determined |
| 17-109_k141_63971 | Not-determined |
| 17-109_k141_349044 | Low-quality |
| 17-109_k141_130494 | Not-determined |
| 17-109_k141_398766 | Not-determined |
| 17-109_k141_13749 | Not-determined |
| 17-109_k141_30536 | Not-determined |
| 17-109_k141_48510 | Not-determined |
| 17-109_k141_215308 | Not-determined |
| 17-109_k141_232531 | Not-determined |
| 17-109_k141_97549 | Not-determined |
| 17-109_k141_215314 | Not-determined |
| 17-109_k141_215316 | Not-determined |
| 17-109_k141_363856 | Not-determined |
| 17-109_k141_165029 | Not-determined |
| 17-109_k141_113957 | Not-determined |
| 17-109_k141_198702 | Not-determined |
| 17-109_k141_97551 | Not-determined |
| 17-109_k141_48522 | Not-determined |
| 17-109_k141_215331 | Not-determined |
| 17-109_k141_248405 | Not-determined |
| 17-109_k141_48526 | Not-determined |
| 17-109_k141_215333 | Not-determined |
| 17-109_k141_198708 | Not-determined |
| 17-109_k141_165034 | Not-determined |
| 17-109_k141_232547 | Not-determined |
| 17-109_k141_48530 | Not-determined |
| 17-109_k141_97554 | Low-quality |
| 17-109_k141_48533 | Not-determined |
| 17-109_k141_97556 | Low-quality |
| 17-109_k141_81580 | Not-determined |
| 17-109_k141_13777 | Not-determined |
| 17-109_k141_13779 | Not-determined |
| 17-109_k141_81584 | Low-quality |
| 17-109_k141_147544 | Not-determined |
| 17-109_k141_363875 | Not-determined |
| 17-109_k141_298942 | Not-determined |
| 17-109_k141_165057 | Not-determined |
| 17-109_k141_97573 | Not-determined |
| 17-109_k141_165063 | Not-determined |
| 17-109_k141_349055 | Not-determined |
| 17-109_k141_64011 | Low-quality |
| 17-109_k141_13789 | Not-determined |
| 17-109_k141_13791 | Not-determined |
| 17-109_k141_398793 | Not-determined |
| 17-109_k141_113987 | Low-quality |
| 17-109_k141_13794 | Not-determined |
| 17-109_k141_363882 | Low-quality |
| 17-109_k141_398795 | Not-determined |
| 17-109_k141_232581 | Not-determined |
| 17-109_k141_332555 | Not-determined |
| 17-109_k141_283002 | Not-determined |
| 17-109_k141_232582 | Low-quality |
| 17-109_k141_349063 | Not-determined |
| 17-109_k141_316692 | Not-determined |
| 17-109_k141_130543 | Not-determined |
| 17-109_k141_97593 | Not-determined |
| 17-109_k141_30583 | Low-quality |
| 17-109_k141_363893 | Not-determined |
| 17-109_k141_13805 | Not-determined |
| 17-109_k141_363897 | Not-determined |
| 17-109_k141_198733 | Not-determined |
| 17-109_k141_232595 | Not-determined |
| 17-109_k141_13812 | Not-determined |
| 17-109_k141_30588 | Not-determined |
| 17-109_k141_97603 | Not-determined |
| 17-109_k141_165077 | Not-determined |
| 17-109_k141_298979 | Not-determined |
| 17-109_k141_13818 | Not-determined |
| 17-109_k141_298981 | Not-determined |
| 17-109_k141_316699 | Not-determined |
| 17-109_k141_332582 | Not-determined |
| 17-109_k141_283031 | Not-determined |
| 17-109_k141_64026 | Not-determined |
| 17-109_k141_349085 | Not-determined |
| 17-109_k141_30595 | Not-determined |
| 17-109_k141_30597 | Low-quality |
| 17-109_k141_398827 | Not-determined |
| 17-109_k141_165088 | Not-determined |
| 17-109_k141_130573 | Not-determined |
| 17-109_k141_363923 | Not-determined |
| 17-109_k141_165090 | Not-determined |
| 17-109_k141_147564 | Not-determined |
| 17-109_k141_332592 | Not-determined |
| 17-109_k141_130583 | Not-determined |
| 17-109_k141_363933 | Not-determined |
| 17-109_k141_349098 | Not-determined |
| 17-109_k141_232633 | Not-determined |
| 17-109_k141_349099 | Not-determined |
| 17-109_k141_97625 | Not-determined |
| 17-109_k141_283052 | Not-determined |
| 17-109_k141_181780 | Not-determined |
| 17-109_k141_215396 | Not-determined |
| 17-109_k141_363942 | Not-determined |
| 17-109_k141_363944 | Not-determined |
| 17-109_k141_114030 | Not-determined |
| 17-109_k141_147578 | Not-determined |
| 17-109_k141_363950 | Not-determined |
| 17-109_k141_147579 | Not-determined |
| 17-109_k141_81629 | Not-determined |
| 17-109_k141_81630 | Low-quality |
| 17-109_k141_81631 | Not-determined |
| 17-109_k141_299000 | Not-determined |
| 17-109_k141_13861 | Not-determined |
| 17-109_k141_363961 | Not-determined |
| 17-109_k141_215406 | Not-determined |
| 17-109_k141_283063 | Not-determined |
| 17-109_k141_232654 | Low-quality |
| 17-109_k141_283067 | Not-determined |
| 17-109_k141_363967 | Not-determined |
| 17-109_k141_316713 | Not-determined |
| 17-109_k141_81639 | Low-quality |
| 17-109_k141_363970 | Low-quality |
| 17-109_k141_232662 | Not-determined |
| 17-109_k141_114040 | Not-determined |
| 17-109_k141_363974 | Not-determined |
| 17-109_k141_332639 | Not-determined |
| 17-109_k141_349120 | Not-determined |
| 17-109_k141_232672 | Not-determined |
| 17-109_k141_147586 | Not-determined |
| 17-109_k141_232677 | Not-determined |
| 17-109_k141_232679 | Not-determined |
| 17-109_k141_215431 | Not-determined |
| 17-109_k141_181799 | Low-quality |
| 17-109_k141_13881 | Not-determined |
| 17-109_k141_13883 | Not-determined |
| 17-109_k141_349128 | Not-determined |

| | |
|---|---|
| 17-109_k141_332650 | Not-determined |
| 17-109_k141_264649 | Low-quality |
| 17-109_k141_332655 | Not-determined |
| 17-109_k141_264654 | Not-determined |
| 17-109_k141_13896 | Not-determined |
| 17-109_k141_332658 | Not-determined |
| 17-109_k141_13898 | Not-determined |
| 17-109_k141_264658 | Not-determined |
| 17-109_k141_130594 | Not-determined |
| 17-109_k141_81665 | Not-determined |
| 17-109_k141_349135 | Low-quality |
| 17-109_k141_181816 | Not-determined |
| 17-109_k141_97659 | Not-determined |
| 17-109_k141_81682 | Not-determined |
| 17-109_k141_332673 | Not-determined |
| 17-109_k141_382585 | Not-determined |
| 17-109_k141_198816 | Low-quality |
| 17-109_k141_165145 | Not-determined |
| 17-109_k141_264675 | Not-determined |
| 17-109_k141_97660 | Not-determined |
| 17-109_k141_382587 | Not-determined |
| 17-109_k141_64060 | Low-quality |
| 17-109_k141_114061 | Not-determined |
| 17-109_k141_97663 | Not-determined |
| 17-109_k141_299058 | Not-determined |
| 17-109_k141_398841 | Low-quality |
| 17-109_k141_64067 | Not-determined |
| 17-109_k141_97668 | Not-determined |
| 17-109_k141_316775 | Not-determined |
| 17-109_k141_114067 | Not-determined |
| 17-109_k141_114068 | Not-determined |
| 17-109_k141_97672 | Not-determined |
| 17-109_k141_349153 | Not-determined |
| 17-109_k141_13919 | Not-determined |
| 17-109_k141_114075 | Not-determined |
| 17-109_k141_299064 | Not-determined |
| 17-109_k141_349158 | Not-determined |
| 17-109_k141_283115 | Not-determined |
| 17-109_k141_299074 | Not-determined |
| 17-109_k141_198830 | Not-determined |
| 17-109_k141_316791 | Not-determined |
| 17-109_k141_97682 | Not-determined |
| 17-109_k141_13923 | Not-determined |
| 17-109_k141_283123 | Not-determined |
| 17-109_k141_130644 | Not-determined |
| 17-109_k141_299083 | Not-determined |
| 17-109_k141_364011 | Low-quality |
| 17-109_k141_332686 | Not-determined |
| 17-109_k141_364013 | Low-quality |
| 17-109_k141_248467 | Not-determined |
| 17-109_k141_283131 | Not-determined |
| 17-109_k141_283132 | Not-determined |
| 17-109_k141_30646 | Not-determined |
| 17-109_k141_198833 | Low-quality |
| 17-109_k141_283133 | Not-determined |
| 17-109_k141_97693 | Not-determined |
| 17-109_k141_248473 | Not-determined |
| 17-109_k141_299099 | Not-determined |
| 17-109_k141_349178 | Not-determined |
| 17-109_k141_349179 | Not-determined |
| 17-109_k141_48553 | Not-determined |
| 17-109_k141_97699 | Not-determined |
| 17-109_k141_165183 | Not-determined |
| 17-109_k141_130663 | Not-determined |
| 17-109_k141_181863 | Not-determined |
| 17-109_k141_283143 | Not-determined |
| 17-109_k141_181866 | Not-determined |
| 17-109_k141_64100 | Not-determined |
| 17-109_k141_64102 | Not-determined |
| 17-109_k141_215475 | Not-determined |
| 17-109_k141_64106 | Not-determined |
| 17-109_k141_398855 | Low-quality |
| 17-109_k141_97706 | Not-determined |
| 17-109_k141_81733 | Not-determined |
| 17-109_k141_299118 | Not-determined |
| 17-109_k141_299121 | Not-determined |
| 17-109_k141_398863 | Not-determined |
| 17-109_k141_215490 | Not-determined |
| 17-109_k141_198837 | Not-determined |
| 17-109_k141_332715 | Low-quality |
| 17-109_k141_264698 | Not-determined |
| 17-109_k141_349193 | Not-determined |
| 17-109_k141_130686 | Not-determined |
| 17-109_k141_264700 | Not-determined |
| 17-109_k141_198846 | Not-determined |
| 17-109_k141_13942 | Low-quality |
| 17-109_k141_130688 | Not-determined |
| 17-109_k141_198847 | Not-determined |
| 17-109_k141_81752 | Not-determined |
| 17-109_k141_215501 | Not-determined |
| 17-109_k141_13949 | Low-quality |
| 17-109_k141_264705 | Not-determined |
| 17-109_k141_114106 | Not-determined |
| 17-109_k141_332733 | Not-determined |
| 17-109_k141_349206 | Not-determined |
| 17-109_k141_30659 | Not-determined |
| 17-109_k141_264711 | Not-determined |
| 17-109_k141_382622 | Not-determined |
| 17-109_k141_81759 | Not-determined |
| 17-109_k141_382623 | Not-determined |
| 17-109_k141_283171 | Not-determined |
| 17-109_k141_332737 | Not-determined |
| 17-109_k141_181883 | Not-determined |
| 17-109_k141_215508 | Not-determined |
| 17-109_k141_332739 | Low-quality |
| 17-109_k141_283173 | Not-determined |
| 17-109_k141_349211 | Not-determined |
| 17-109_k141_232743 | Not-determined |
| 17-109_k141_215512 | Not-determined |
| 17-109_k141_30667 | Not-determined |
| 17-109_k141_316820 | Not-determined |
| 17-109_k141_232745 | Not-determined |
| 17-109_k141_13956 | Not-determined |
| 17-109_k141_382624 | Low-quality |
| 17-109_k141_181889 | Low-quality |
| 17-109_k141_398884 | Not-determined |
| 17-109_k141_232747 | Not-determined |
| 17-109_k141_232748 | Not-determined |
| 17-109_k141_382632 | Not-determined |
| 17-109_k141_398889 | Not-determined |
| 17-109_k141_299141 | Not-determined |
| 17-109_k141_198861 | Not-determined |
| 17-109_k141_382635 | Not-determined |
| 17-109_k141_382636 | Not-determined |
| 17-109_k141_232752 | Not-determined |
| 17-109_k141_299147 | Not-determined |
| 17-109_k141_13967 | Not-determined |
| 17-109_k141_30683 | Not-determined |
| 17-109_k141_81778 | Not-determined |
| 17-109_k141_48587 | Not-determined |
| 17-109_k141_332758 | Not-determined |
| 17-109_k141_232758 | Low-quality |
| 17-109_k141_232759 | Not-determined |
| 17-109_k141_13973 | Not-determined |
| 17-109_k141_215534 | Not-determined |
| 17-109_k141_264733 | Not-determined |
| 17-109_k141_248514 | Not-determined |
| 17-109_k141_30694 | Not-determined |
| 17-109_k141_232762 | Not-determined |
| 17-109_k141_30696 | Not-determined |
| 17-109_k141_264737 | Not-determined |
| 17-109_k141_332769 | Not-determined |
| 17-109_k141_264739 | Not-determined |
| 17-109_k141_283193 | Not-determined |
| 17-109_k141_30700 | Not-determined |
| 17-109_k141_81785 | Not-determined |
| 17-109_k141_97729 | Not-determined |
| 17-109_k141_248533 | Not-determined |
| 17-109_k141_30709 | Low-quality |
| 17-109_k141_198887 | Not-determined |
| 17-109_k141_198888 | Not-determined |
| 17-109_k141_382661 | Low-quality |
| 17-109_k141_364029 | Not-determined |
| 17-109_k141_283199 | Not-determined |
| 17-109_k141_97734 | Not-determined |
| 17-109_k141_382666 | Not-determined |
| 17-109_k141_13983 | Not-determined |
| 17-109_k141_349239 | Low-quality |
| 17-109_k141_198892 | Not-determined |
| 17-109_k141_264761 | Not-determined |
| 17-109_k141_299171 | Not-determined |
| 17-109_k141_30721 | Low-quality |
| 17-109_k141_30722 | Not-determined |
| 17-109_k141_181937 | Not-determined |
| 17-109_k141_316858 | Not-determined |
| 17-109_k141_364041 | Not-determined |
| 17-109_k141_398930 | Not-determined |
| 17-109_k141_114155 | Low-quality |
| 17-109_k141_364046 | Not-determined |
| 17-109_k141_114160 | Not-determined |
| 17-109_k141_283215 | Not-determined |
| 17-109_k141_364056 | Not-determined |
| 17-109_k141_114168 | Low-quality |
| 17-109_k141_299187 | Not-determined |
| 17-109_k141_48618 | Not-determined |
| 17-109_k141_181956 | Not-determined |
| 17-109_k141_299189 | Not-determined |
| 17-109_k141_97749 | Not-determined |
| 17-109_k141_283226 | Not-determined |
| 17-109_k141_81801 | Not-determined |
| 17-109_k141_264772 | Not-determined |
| 17-109_k141_283229 | Not-determined |
| 17-109_k141_283230 | Not-determined |
| 17-109_k141_349277 | Not-determined |
| 17-109_k141_232786 | Not-determined |
| 17-109_k141_198915 | Not-determined |
| 17-109_k141_198917 | Not-determined |
| 17-109_k141_299213 | Not-determined |
| 17-109_k141_364078 | Not-determined |
| 17-109_k141_349292 | Not-determined |
| 17-109_k141_316871 | Not-determined |
| 17-109_k141_232799 | Low-quality |
| 17-109_k141_215561 | Low-quality |
| 17-109_k141_248551 | Not-determined |
| 17-109_k141_81820 | Not-determined |
| 17-109_k141_364087 | Not-determined |
| 17-109_k141_299233 | Not-determined |
| 17-109_k141_349299 | Not-determined |
| 17-109_k141_147660 | Low-quality |
| 17-109_k141_364094 | Not-determined |
| 17-109_k141_81831 | Not-determined |
| 17-109_k141_316894 | Not-determined |
| 17-109_k141_283256 | Not-determined |
| 17-109_k141_349304 | Not-determined |
| 17-109_k141_130770 | Not-determined |
| 17-109_k141_283260 | Not-determined |
| 17-109_k141_349307 | Not-determined |
| 17-109_k141_299244 | Not-determined |
| 17-109_k141_215577 | Not-determined |
| 17-109_k141_283262 | Not-determined |
| 17-109_k141_130776 | Not-determined |
| 17-109_k141_130778 | Not-determined |
| 17-109_k141_283274 | Not-determined |
| 17-109_k141_264792 | Low-quality |
| 17-109_k141_130782 | Not-determined |
| 17-109_k141_382695 | Not-determined |
| 17-109_k141_14032 | Not-determined |
| 17-109_k141_232831 | Low-quality |
| 17-109_k141_147690 | Low-quality |
| 17-109_k141_232832 | Low-quality |
| 17-109_k141_349322 | Not-determined |
| 17-109_k141_364112 | Not-determined |
| 17-109_k141_232835 | Not-determined |
| 17-109_k141_364114 | Not-determined |
| 17-109_k141_232839 | Low-quality |
| 17-109_k141_97807 | Not-determined |
| 17-109_k141_182004 | Low-quality |
| 17-109_k141_283290 | Not-determined |
| 17-109_k141_283292 | Not-determined |
| 17-109_k141_130792 | Not-determined |
| 17-109_k141_332794 | Not-determined |
| 17-109_k141_299267 | Not-determined |
| 17-109_k141_30736 | Not-determined |
| 17-109_k141_165219 | Not-determined |
| 17-109_k141_264823 | Not-determined |
| 17-109_k141_349325 | Not-determined |
| 17-109_k141_130805 | Not-determined |
| 17-109_k141_264831 | Not-determined |
| 17-109_k141_283310 | Not-determined |
| 17-109_k141_232858 | Not-determined |
| 17-109_k141_364126 | Not-determined |
| 17-109_k141_182034 | Not-determined |
| 17-109_k141_264836 | Not-determined |
| 17-109_k141_283317 | Not-determined |
| 17-109_k141_299291 | Not-determined |
| 17-109_k141_130819 | Not-determined |
| 17-109_k141_182041 | Not-determined |
| 17-109_k141_81864 | Not-determined |
| 17-109_k141_48675 | Not-determined |
| 17-109_k141_48676 | Not-determined |
| 17-109_k141_48678 | Not-determined |
| 17-109_k141_398948 | Not-determined |
| 17-109_k141_364137 | Not-determined |
| 17-109_k141_97848 | Not-determined |
| 17-109_k141_97849 | Not-determined |
| 17-109_k141_364140 | Not-determined |
| 17-109_k141_147715 | Not-determined |
| 17-109_k141_14068 | Not-determined |
| 17-109_k141_198933 | Not-determined |
| 17-109_k141_130834 | Not-determined |
| 17-109_k141_198936 | Low-quality |
| 17-109_k141_165237 | Low-quality |
| 17-109_k141_198940 | Not-determined |
| 17-109_k141_81880 | Not-determined |
| 17-109_k141_147731 | Not-determined |
| 17-109_k141_398960 | Not-determined |
| 17-109_k141_130837 | Not-determined |
| 17-109_k141_64147 | Not-determined |
| 17-109_k141_198947 | Not-determined |
| 17-109_k141_165246 | Low-quality |
| 17-109_k141_64157 | Not-determined |
| 17-109_k141_283340 | Low-quality |
| 17-109_k141_165255 | Not-determined |
| 17-109_k141_382729 | Not-determined |
| 17-109_k141_64163 | Not-determined |
| 17-109_k141_14093 | Not-determined |
| 17-109_k141_332850 | Low-quality |
| 17-109_k141_14095 | Not-determined |
| 17-109_k141_198964 | Not-determined |
| 17-109_k141_97872 | Not-determined |
| 17-109_k141_48719 | Not-determined |
| 17-109_k141_97873 | Not-determined |
| 17-109_k141_81899 | Not-determined |
| 17-109_k141_264851 | Not-determined |
| 17-109_k141_264855 | Not-determined |
| 17-109_k141_364152 | Low-quality |
| 17-109_k141_382738 | Not-determined |
| 17-109_k141_283352 | Not-determined |
| 17-109_k141_198972 | Not-determined |
| 17-109_k141_182086 | Not-determined |
| 17-109_k141_398963 | Not-determined |
| 17-109_k141_48734 | Not-determined |
| 17-109_k141_364171 | Not-determined |
| 17-109_k141_364172 | Not-determined |
| 17-109_k141_316914 | Low-quality |
| 17-109_k141_398970 | Low-quality |
| 17-109_k141_264878 | Low-quality |
| 17-109_k141_215648 | Not-determined |
| 17-109_k141_398974 | Not-determined |
| 17-109_k141_64180 | Not-determined |
| 17-109_k141_182112 | Not-determined |
| 17-109_k141_283374 | Not-determined |
| 17-109_k141_130853 | Not-determined |
| 17-109_k141_182117 | Not-determined |
| 17-109_k141_248600 | Low-quality |
| 17-109_k141_130857 | Not-determined |
| 17-109_k141_64182 | Not-determined |
| 17-109_k141_198982 | Not-determined |
| 17-109_k141_248604 | Not-determined |
| 17-109_k141_81927 | Not-determined |
| 17-109_k141_97927 | Not-determined |
| 17-109_k141_316918 | Not-determined |
| 17-109_k141_198986 | Low-quality |
| 17-109_k141_198987 | Not-determined |
| 17-109_k141_48746 | Not-determined |
| 17-109_k141_48748 | Not-determined |
| 17-109_k141_64190 | Not-determined |
| 17-109_k141_165285 | Not-determined |
| 17-109_k141_130876 | Low-quality |
| 17-109_k141_349391 | Not-determined |
| 17-109_k141_64204 | Not-determined |
| 17-109_k141_48756 | Not-determined |
| 17-109_k141_165296 | Not-determined |
| 17-109_k141_147763 | Not-determined |
| 17-109_k141_64209 | Not-determined |
| 17-109_k141_64212 | Not-determined |
| 17-109_k141_30779 | Not-determined |
| 17-109_k141_165304 | Not-determined |
| 17-109_k141_283396 | Not-determined |
| 17-109_k141_165306 | Not-determined |
| 17-109_k141_64220 | Low-quality |
| 17-109_k141_64221 | Not-determined |
| 17-109_k141_165309 | Low-quality |
| 17-109_k141_97940 | Low-quality |
| 17-109_k141_182152 | Not-determined |
| 17-109_k141_48762 | Low-quality |
| 17-109_k141_165315 | Not-determined |
| 17-109_k141_264927 | Low-quality |
| 17-109_k141_48769 | Not-determined |
| 17-109_k141_165318 | Not-determined |
| 17-109_k141_165319 | Not-determined |
| 17-109_k141_349409 | Low-quality |
| 17-109_k141_64239 | Not-determined |
| 17-109_k141_97951 | Low-quality |
| 17-109_k141_332895 | Not-determined |
| 17-109_k141_147780 | Not-determined |
| 17-109_k141_332900 | Not-determined |
| 17-109_k141_30797 | Not-determined |
| 17-109_k141_114242 | Not-determined |
| 17-109_k141_364230 | Not-determined |
| 17-109_k141_364231 | Not-determined |
| 17-109_k141_165328 | Not-determined |
| 17-109_k141_165330 | Not-determined |
| 17-109_k141_147788 | Not-determined |
| 17-109_k141_165332 | Not-determined |
| 17-109_k141_264950 | Not-determined |
| 17-109_k141_264951 | Not-determined |
| 17-109_k141_97979 | Not-determined |
| 17-109_k141_349421 | Not-determined |
| 17-109_k141_147798 | Not-determined |
| 17-109_k141_165341 | Not-determined |
| 17-109_k141_165342 | Not-determined |
| 17-109_k141_48793 | Not-determined |
| 17-109_k141_349426 | Not-determined |
| 17-109_k141_316943 | Not-determined |
| 17-109_k141_283426 | Not-determined |
| 17-109_k141_48794 | Not-determined |
| 17-109_k141_48795 | Not-determined |
| 17-109_k141_97984 | Not-determined |
| 17-109_k141_48798 | Not-determined |
| 17-109_k141_199037 | Not-determined |
| 17-109_k141_283432 | Not-determined |
| 17-109_k141_48799 | Not-determined |
| 17-109_k141_199039 | Low-quality |
| 17-109_k141_364248 | Not-determined |
| 17-109_k141_30823 | Not-determined |
| 17-109_k141_349434 | Not-determined |
| 17-109_k141_199042 | Low-quality |
| 17-109_k141_398986 | Low-quality |
| 17-109_k141_382746 | Low-quality |
| 17-109_k141_199044 | Not-determined |
| 17-109_k141_364251 | Low-quality |
| 17-109_k141_364252 | Not-determined |
| 17-109_k141_299308 | Not-determined |
| 17-109_k141_48805 | Not-determined |
| 17-109_k141_64266 | Not-determined |
| 17-109_k141_130897 | Not-determined |
| 17-109_k141_316956 | Not-determined |
| 17-109_k141_316957 | Not-determined |
| 17-109_k141_349440 | Low-quality |
| 17-109_k141_64271 | Not-determined |
| 17-109_k141_64272 | Not-determined |
| 17-109_k141_14159 | Not-determined |
| 17-109_k141_147820 | Not-determined |
| 17-109_k141_64277 | Not-determined |
| 17-109_k141_64282 | Not-determined |
| 17-109_k141_248632 | Not-determined |
| 17-109_k141_199050 | Not-determined |
| 17-109_k141_14169 | Not-determined |
| 17-109_k141_364276 | Not-determined |
| 17-109_k141_199055 | Not-determined |
| 17-109_k141_364278 | Low-quality |
| 17-109_k141_248640 | Low-quality |
| 17-109_k141_399008 | Not-determined |
| 17-109_k141_81988 | Not-determined |
| 17-109_k141_349469 | Not-determined |
| 17-109_k141_316974 | Not-determined |
| 17-109_k141_332949 | Not-determined |
| 17-109_k141_64296 | Not-determined |
| 17-109_k141_82000 | Not-determined |
| 17-109_k141_48833 | Not-determined |
| 17-109_k141_248654 | Low-quality |
| 17-109_k141_82001 | Not-determined |
| 17-109_k141_64300 | Not-determined |
| 17-109_k141_64301 | Not-determined |
| 17-109_k141_316983 | Not-determined |
| 17-109_k141_165386 | Not-determined |
| 17-109_k141_48837 | Not-determined |
| 17-109_k141_332958 | Not-determined |
| 17-109_k141_248663 | Not-determined |
| 17-109_k141_364292 | Not-determined |
| 17-109_k141_64309 | Not-determined |
| 17-109_k141_283461 | Not-determined |
| 17-109_k141_130926 | Not-determined |
| 17-109_k141_98019 | Low-quality |
| 17-109_k141_332960 | Low-quality |
| 17-109_k141_264983 | Not-determined |
| 17-109_k141_364298 | Not-determined |
| 17-109_k141_364300 | Not-determined |
| 17-109_k141_232936 | Low-quality |
| 17-109_k141_147856 | Not-determined |
| 17-109_k141_14189 | Not-determined |
| 17-109_k141_332977 | Low-quality |
| 17-109_k141_264988 | Not-determined |
| 17-109_k141_264991 | Not-determined |
| 17-109_k141_14191 | Not-determined |
| 17-109_k141_248685 | Not-determined |
| 17-109_k141_248686 | Not-determined |
| 17-109_k141_232938 | Not-determined |
| 17-109_k141_232940 | Not-determined |
| 17-109_k141_264992 | Not-determined |
| 17-109_k141_82032 | Not-determined |
| 17-109_k141_48850 | Not-determined |
| 17-109_k141_199074 | Not-determined |
| 17-109_k141_48853 | Not-determined |
| 17-109_k141_317017 | Low-quality |
| 17-109_k141_82039 | Not-determined |
| 17-109_k141_48855 | Not-determined |
| 17-109_k141_165402 | Not-determined |
| 17-109_k141_114294 | Not-determined |
| 17-109_k141_265004 | Not-determined |
| 17-109_k141_265005 | Not-determined |
| 17-109_k141_114296 | Not-determined |
| 17-109_k141_82041 | Not-determined |
| 17-109_k141_317022 | Not-determined |
| 17-109_k141_30874 | Low-quality |
| 17-109_k141_130942 | Not-determined |
| 17-109_k141_14207 | Not-determined |
| 17-109_k141_82046 | Not-determined |
| 17-109_k141_14208 | Not-determined |
| 17-109_k141_64342 | Not-determined |
| 17-109_k141_364304 | Not-determined |
| 17-109_k141_114302 | Low-quality |
| 17-109_k141_382794 | Not-determined |
| 17-109_k141_382795 | Not-determined |
| 17-109_k141_248689 | Not-determined |
| 17-109_k141_199086 | Not-determined |
| 17-109_k141_248690 | Low-quality |
| 17-109_k141_199087 | Not-determined |
| 17-109_k141_114304 | Not-determined |
| 17-109_k141_265011 | Not-determined |
| 17-109_k141_14212 | Not-determined |
| 17-109_k141_265012 | Not-determined |
| 17-109_k141_48867 | Not-determined |
| 17-109_k141_248692 | Not-determined |
| 17-109_k141_114308 | Not-determined |
| 17-109_k141_165415 | Not-determined |
| 17-109_k141_82063 | Not-determined |
| 17-109_k141_82064 | Not-determined |
| 17-109_k141_114311 | Not-determined |
| 17-109_k141_265022 | Not-determined |
| 17-109_k141_64353 | Not-determined |
| 17-109_k141_114313 | Not-determined |
| 17-109_k141_265024 | Not-determined |
| 17-109_k141_64358 | Not-determined |
| 17-109_k141_199108 | Not-determined |
| 17-109_k141_215688 | Not-determined |
| 17-109_k141_82069 | Not-determined |
| 17-109_k141_283491 | Not-determined |
| 17-109_k141_232960 | Not-determined |
| 17-109_k141_14244 | Not-determined |
| 17-109_k141_30880 | Not-determined |
| 17-109_k141_232969 | Not-determined |
| 17-109_k141_364337 | Not-determined |
| 17-109_k141_232973 | Not-determined |
| 17-109_k141_364338 | Not-determined |
| 17-109_k141_30884 | Not-determined |
| 17-109_k141_48879 | Not-determined |
| 17-109_k141_283504 | Not-determined |
| 17-109_k141_317057 | Not-determined |
| 17-109_k141_399010 | Not-determined |
| 17-109_k141_130978 | Not-determined |
| 17-109_k141_364348 | Not-determined |
| 17-109_k141_130981 | Not-determined |
| 17-109_k141_114321 | Not-determined |
| 17-109_k141_248709 | Not-determined |
| 17-109_k141_248710 | Not-determined |
| 17-109_k141_82093 | Not-determined |
| 17-109_k141_283517 | Not-determined |
| 17-109_k141_14256 | Not-determined |
| 17-109_k141_165436 | Not-determined |
| 17-109_k141_165437 | Not-determined |
| 17-109_k141_130989 | Low-quality |
| 17-109_k141_399015 | Not-determined |
| 17-109_k141_283524 | Not-determined |
| 17-109_k141_82101 | Not-determined |
| 17-109_k141_14262 | Not-determined |
| 17-109_k141_130994 | Not-determined |
| 17-109_k141_64370 | Not-determined |
| 17-109_k141_199125 | Not-determined |
| 17-109_k141_283535 | Not-determined |
| 17-109_k141_333019 | Not-determined |
| 17-109_k141_165446 | Not-determined |
| 17-109_k141_82113 | Not-determined |
| 17-109_k141_248720 | Not-determined |
| 17-109_k141_215700 | Not-determined |
| 17-109_k141_114338 | Not-determined |
| 17-109_k141_317093 | Not-determined |
| 17-109_k141_165448 | Not-determined |
| 17-109_k141_147901 | Not-determined |
| 17-109_k141_165449 | Not-determined |
| 17-109_k141_283541 | Not-determined |
| 17-109_k141_82122 | Not-determined |
| 17-109_k141_215706 | Not-determined |
| 17-109_k141_283544 | Not-determined |
| 17-109_k141_382816 | Not-determined |
| 17-109_k141_165453 | Not-determined |
| 17-109_k141_82125 | Not-determined |
| 17-109_k141_30916 | Low-quality |
| 17-109_k141_14267 | Not-determined |
| 17-109_k141_82131 | Not-determined |
| 17-109_k141_382818 | Not-determined |
| 17-109_k141_165462 | Not-determined |
| 17-109_k141_382821 | Not-determined |
| 17-109_k141_232988 | Not-determined |
| 17-109_k141_147917 | Not-determined |
| 17-109_k141_165464 | Not-determined |
| 17-109_k141_232993 | Not-determined |
| 17-109_k141_64402 | Not-determined |
| 17-109_k141_48899 | Not-determined |
| 17-109_k141_131032 | Not-determined |
| 17-109_k141_131036 | Not-determined |
| 17-109_k141_182275 | Not-determined |
| 17-109_k141_131037 | Not-determined |
| 17-109_k141_131038 | Not-determined |
| 17-109_k141_382834 | Not-determined |
| 17-109_k141_147934 | Not-determined |
| 17-109_k141_232998 | Not-determined |
| 17-109_k141_364384 | Not-determined |
| 17-109_k141_382838 | Not-determined |
| 17-109_k141_131043 | Not-determined |
| 17-109_k141_82152 | Not-determined |
| 17-109_k141_147943 | Not-determined |
| 17-109_k141_199142 | Not-determined |
| 17-109_k141_199143 | Not-determined |
| 17-109_k141_233006 | Not-determined |
| 17-109_k141_299345 | Low-quality |
| 17-109_k141_364394 | Not-determined |
| 17-109_k141_399033 | Not-determined |
| 17-109_k141_30942 | Not-determined |
| 17-109_k141_364396 | Not-determined |
| 17-109_k141_364397 | Not-determined |
| 17-109_k141_147955 | Not-determined |
| 17-109_k141_114361 | Not-determined |
| 17-109_k141_399044 | Not-determined |
| 17-109_k141_349519 | Not-determined |
| 17-109_k141_283566 | Not-determined |
| 17-109_k141_48909 | Not-determined |
| 17-109_k141_14306 | Not-determined |
| 17-109_k141_215748 | Not-determined |
| 17-109_k141_399049 | Not-determined |
| 17-109_k141_147965 | Not-determined |
| 17-109_k141_233022 | Not-determined |
| 17-109_k141_182280 | Not-determined |
| 17-109_k141_248763 | Not-determined |
| 17-109_k141_364410 | Not-determined |
| 17-109_k141_182285 | Not-determined |
| 17-109_k141_14321 | Not-determined |
| 17-109_k141_48918 | Not-determined |

| | |
|---|---|
| 17-109_k141_399056 | Not-determined |
| 17-109_k141_131071 | Not-determined |
| 17-109_k141_333053 | Not-determined |
| 17-109_k141_82159 | Not-determined |
| 17-109_k141_299363 | Not-determined |
| 17-109_k141_14323 | Not-determined |
| 17-109_k141_114379 | Low-quality |
| 17-109_k141_233028 | Not-determined |
| 17-109_k141_30972 | Not-determined |
| 17-109_k141_233031 | Not-determined |
| 17-109_k141_82169 | Not-determined |
| 17-109_k141_333064 | Not-determined |
| 17-109_k141_30977 | Not-determined |
| 17-109_k141_333068 | Not-determined |
| 17-109_k141_165505 | Not-determined |
| 17-109_k141_233036 | Low-quality |
| 17-109_k141_233037 | Not-determined |
| 17-109_k141_30981 | Not-determined |
| 17-109_k141_64428 | Not-determined |
| 17-109_k141_233043 | Not-determined |
| 17-109_k141_30988 | Not-determined |
| 17-109_k141_114392 | Not-determined |
| 17-109_k141_233048 | Not-determined |
| 17-109_k141_114395 | Not-determined |
| 17-109_k141_199148 | Not-determined |
| 17-109_k141_317187 | Not-determined |
| 17-109_k141_333083 | Not-determined |
| 17-109_k141_64438 | Not-determined |
| 17-109_k141_82185 | Not-determined |
| 17-109_k141_165524 | Not-determined |
| 17-109_k141_82186 | Not-determined |
| 17-109_k141_233054 | Not-determined |
| 17-109_k141_349551 | Not-determined |
| 17-109_k141_215776 | Not-determined |
| 17-109_k141_382860 | Not-determined |
| 17-109_k141_317197 | Not-determined |
| 17-109_k141_382861 | Not-determined |
| 17-109_k141_364446 | Not-determined |
| 17-109_k141_333087 | Not-determined |
| 17-109_k141_382864 | Not-determined |
| 17-109_k141_14334 | Low-quality |
| 17-109_k141_317199 | Not-determined |
| 17-109_k141_165531 | Not-determined |
| 17-109_k141_349555 | Not-determined |
| 17-109_k141_14337 | Not-determined |
| 17-109_k141_299388 | Not-determined |
| 17-109_k141_199169 | Not-determined |
| 17-109_k141_283611 | Not-determined |
| 17-109_k141_31007 | Not-determined |
| 17-109_k141_82200 | Not-determined |
| 17-109_k141_364464 | Not-determined |
| 17-109_k141_131108 | Not-determined |
| 17-109_k141_364468 | Not-determined |
| 17-109_k141_333096 | Not-determined |
| 17-109_k141_114411 | Not-determined |
| 17-109_k141_199172 | Not-determined |
| 17-109_k141_48941 | Not-determined |
| 17-109_k141_199174 | Low-quality |
| 17-109_k141_165549 | Not-determined |
| 17-109_k141_182334 | Low-quality |
| 17-109_k141_199179 | Not-determined |
| 17-109_k141_215793 | Not-determined |
| 17-109_k141_299406 | Not-determined |
| 17-109_k141_364475 | Not-determined |
| 17-109_k141_233081 | Not-determined |
| 17-109_k141_182340 | Not-determined |
| 17-109_k141_48954 | Not-determined |
| 17-109_k141_399087 | Not-determined |
| 17-109_k141_299411 | Not-determined |
| 17-109_k141_299412 | Low-quality |
| 17-109_k141_399090 | Not-determined |
| 17-109_k141_114429 | Not-determined |
| 17-109_k141_364484 | Not-determined |
| 17-109_k141_399093 | Not-determined |
| 17-109_k141_31020 | Not-determined |
| 17-109_k141_64477 | Not-determined |
| 17-109_k141_233087 | Not-determined |
| 17-109_k141_64481 | Not-determined |
| 17-109_k141_14365 | Low-quality |
| 17-109_k141_233092 | Not-determined |
| 17-109_k141_31026 | Not-determined |
| 17-109_k141_147979 | Not-determined |
| 17-109_k141_233096 | Not-determined |
| 17-109_k141_233099 | Not-determined |
| 17-109_k141_48963 | Not-determined |
| 17-109_k141_199186 | Not-determined |
| 17-109_k141_114448 | Not-determined |
| 17-109_k141_98050 | Not-determined |
| 17-109_k141_64493 | Not-determined |
| 17-109_k141_147992 | Not-determined |
| 17-109_k141_317241 | Low-quality |
| 17-109_k141_233107 | Not-determined |
| 17-109_k141_333129 | Not-determined |
| 17-109_k141_147996 | Not-determined |
| 17-109_k141_64499 | Not-determined |
| 17-109_k141_233111 | Not-determined |
| 17-109_k141_182366 | Not-determined |
| 17-109_k141_147999 | Not-determined |
| 17-109_k141_131126 | Low-quality |
| 17-109_k141_48973 | Not-determined |
| 17-109_k141_364499 | Not-determined |
| 17-109_k141_182372 | Low-quality |
| 17-109_k141_182374 | Not-determined |
| 17-109_k141_31049 | Not-determined |
| 17-109_k141_382896 | Not-determined |
| 17-109_k141_64505 | Not-determined |
| 17-109_k141_31051 | Low-quality |
| 17-109_k141_98070 | Not-determined |
| 17-109_k141_317253 | Not-determined |
| 17-109_k141_382899 | Not-determined |
| 17-109_k141_233120 | Low-quality |
| 17-109_k141_114462 | Not-determined |
| 17-109_k141_114463 | Not-determined |
| 17-109_k141_382904 | Not-determined |
| 17-109_k141_14377 | Not-determined |
| 17-109_k141_98078 | Low-quality |
| 17-109_k141_233128 | Not-determined |
| 17-109_k141_349604 | Not-determined |
| 17-109_k141_182389 | Not-determined |
| 17-109_k141_349606 | Not-determined |
| 17-109_k141_364510 | Not-determined |
| 17-109_k141_182395 | Not-determined |
| 17-109_k141_349611 | Not-determined |
| 17-109_k141_317264 | Not-determined |
| 17-109_k141_131142 | Not-determined |
| 17-109_k141_182396 | Not-determined |
| 17-109_k141_114477 | Not-determined |
| 17-109_k141_49001 | Low-quality |
| 17-109_k141_14402 | Not-determined |
| 17-109_k141_333137 | Not-determined |
| 17-109_k141_148022 | Not-determined |
| 17-109_k141_49003 | Not-determined |
| 17-109_k141_49005 | Not-determined |
| 17-109_k141_14407 | Not-determined |
| 17-109_k141_14410 | Not-determined |
| 17-109_k141_114490 | Low-quality |
| 17-109_k141_131158 | Not-determined |
| 17-109_k141_131160 | Not-determined |
| 17-109_k141_182416 | Not-determined |
| 17-109_k141_131161 | Not-determined |
| 17-109_k141_283625 | Not-determined |
| 17-109_k141_14420 | Not-determined |
| 17-109_k141_317269 | Not-determined |
| 17-109_k141_98086 | Not-determined |
| 17-109_k141_248795 | Not-determined |
| 17-109_k141_248798 | Not-determined |
| 17-109_k141_215868 | Low-quality |
| 17-109_k141_333146 | Not-determined |
| 17-109_k141_31071 | Not-determined |
| 17-109_k141_283635 | Not-determined |
| 17-109_k141_283636 | Not-determined |
| 17-109_k141_283637 | Not-determined |
| 17-109_k141_182430 | Low-quality |
| 17-109_k141_98092 | Not-determined |
| 17-109_k141_31084 | Not-determined |
| 17-109_k141_182438 | Not-determined |
| 17-109_k141_14429 | Not-determined |
| 17-109_k141_14435 | Not-determined |
| 17-109_k141_131183 | Not-determined |
| 17-109_k141_131184 | Not-determined |
| 17-109_k141_233161 | Not-determined |
| 17-109_k141_148060 | Low-quality |
| 17-109_k141_131186 | Not-determined |
| 17-109_k141_114504 | Not-determined |
| 17-109_k141_233167 | Not-determined |
| 17-109_k141_31097 | Not-determined |
| 17-109_k141_131190 | Not-determined |
| 17-109_k141_215893 | Not-determined |
| 17-109_k141_98104 | Not-determined |
| 17-109_k141_131194 | Not-determined |
| 17-109_k141_349645 | Not-determined |
| 17-109_k141_215901 | Low-quality |
| 17-109_k141_317305 | Not-determined |
| 17-109_k141_248837 | Not-determined |
| 17-109_k141_64576 | Not-determined |
| 17-109_k141_31106 | Not-determined |
| 17-109_k141_64579 | Not-determined |
| 17-109_k141_317313 | Low-quality |
| 17-109_k141_248848 | Not-determined |
| 17-109_k141_14463 | Not-determined |
| 17-109_k141_233188 | Not-determined |
| 17-109_k141_31111 | Not-determined |
| 17-109_k141_233192 | Not-determined |
| 17-109_k141_248852 | Not-determined |
| 17-109_k141_248853 | Not-determined |
| 17-109_k141_82274 | Low-quality |
| 17-109_k141_382948 | Not-determined |
| 17-109_k141_317324 | Not-determined |
| 17-109_k141_114531 | Not-determined |
| 17-109_k141_233202 | Not-determined |
| 17-109_k141_317331 | Not-determined |
| 17-109_k141_233204 | Not-determined |
| 17-109_k141_333193 | Not-determined |
| 17-109_k141_299453 | Not-determined |
| 17-109_k141_317334 | Not-determined |
| 17-109_k141_98115 | Not-determined |
| 17-109_k141_399154 | Not-determined |
| 17-109_k141_333197 | Not-determined |
| 17-109_k141_399156 | Not-determined |
| 17-109_k141_399158 | Not-determined |
| 17-109_k141_333200 | Not-determined |
| 17-109_k141_233216 | Not-determined |
| 17-109_k141_14479 | Not-determined |
| 17-109_k141_114554 | Not-determined |
| 17-109_k141_333204 | Not-determined |
| 17-109_k141_399166 | Not-determined |
| 17-109_k141_333207 | Not-determined |
| 17-109_k141_14481 | Low-quality |
| 17-109_k141_399171 | Not-determined |
| 17-109_k141_333212 | Not-determined |
| 17-109_k141_349670 | Not-determined |
| 17-109_k141_215931 | Not-determined |
| 17-109_k141_399174 | Not-determined |
| 17-109_k141_114566 | Not-determined |
| 17-109_k141_233229 | Not-determined |
| 17-109_k141_182444 | Not-determined |
| 17-109_k141_114568 | Not-determined |
| 17-109_k141_82295 | Not-determined |
| 17-109_k141_349672 | Not-determined |
| 17-109_k141_14487 | Low-quality |
| 17-109_k141_364546 | Low-quality |
| 17-109_k141_14488 | Low-quality |
| 17-109_k141_364549 | Not-determined |
| 17-109_k141_31133 | Not-determined |
| 17-109_k141_31135 | Not-determined |
| 17-109_k141_31136 | Not-determined |
| 17-109_k141_248870 | Not-determined |
| 17-109_k141_148099 | Not-determined |
| 17-109_k141_148102 | Not-determined |
| 17-109_k141_98126 | Not-determined |
| 17-109_k141_182458 | Not-determined |
| 17-109_k141_233245 | Not-determined |
| 17-109_k141_148119 | Not-determined |
| 17-109_k141_114594 | Low-quality |
| 17-109_k141_98138 | Not-determined |
| 17-109_k141_399202 | Not-determined |
| 17-109_k141_399203 | Not-determined |
| 17-109_k141_64623 | Not-determined |
| 17-109_k141_299492 | Not-determined |
| 17-109_k141_148133 | Not-determined |
| 17-109_k141_299496 | Not-determined |
| 17-109_k141_165673 | Not-determined |
| 17-109_k141_114608 | Not-determined |
| 17-109_k141_364588 | Not-determined |
| 17-109_k141_233262 | Low-quality |
| 17-109_k141_114610 | Not-determined |
| 17-109_k141_233264 | Low-quality |
| 17-109_k141_283655 | Not-determined |
| 17-109_k141_317371 | Not-determined |
| 17-109_k141_148140 | Not-determined |
| 17-109_k141_283657 | Not-determined |
| 17-109_k141_148141 | Not-determined |
| 17-109_k141_64629 | Not-determined |
| 17-109_k141_283661 | Not-determined |
| 17-109_k141_165688 | Not-determined |
| 17-109_k141_349690 | Not-determined |
| 17-109_k141_349692 | Not-determined |
| 17-109_k141_364594 | Low-quality |
| 17-109_k141_299509 | Not-determined |
| 17-109_k141_317381 | Low-quality |
| 17-109_k141_14498 | Not-determined |
| 17-109_k141_364601 | Not-determined |
| 17-109_k141_98152 | Not-determined |
| 17-109_k141_283672 | Not-determined |
| 17-109_k141_49015 | Not-determined |
| 17-109_k141_14503 | Not-determined |
| 17-109_k141_148162 | Low-quality |
| 17-109_k141_148165 | Low-quality |
| 17-109_k141_64639 | Not-determined |
| 17-109_k141_165712 | Not-determined |
| 17-109_k141_131291 | Not-determined |
| 17-109_k141_349712 | Not-determined |
| 17-109_k141_399218 | Not-determined |
| 17-109_k141_364613 | Not-determined |
| 17-109_k141_31176 | Not-determined |
| 17-109_k141_248893 | Not-determined |
| 17-109_k141_399221 | Not-determined |
| 17-109_k141_399223 | Not-determined |
| 17-109_k141_248899 | Not-determined |
| 17-109_k141_349735 | Not-determined |
| 17-109_k141_82309 | Low-quality |
| 17-109_k141_399231 | Low-quality |
| 17-109_k141_248904 | Low-quality |
| 17-109_k141_349740 | Not-determined |
| 17-109_k141_399232 | Not-determined |
| 17-109_k141_215992 | Not-determined |
| 17-109_k141_364625 | Not-determined |
| 17-109_k141_283693 | Not-determined |
| 17-109_k141_49040 | Not-determined |
| 17-109_k141_248909 | Not-determined |
| 17-109_k141_283697 | Not-determined |
| 17-109_k141_248911 | Not-determined |
| 17-109_k141_364627 | Not-determined |
| 17-109_k141_364628 | Not-determined |
| 17-109_k141_148187 | Low-quality |
| 17-109_k141_31195 | Not-determined |
| 17-109_k141_233318 | Not-determined |
| 17-109_k141_148190 | Not-determined |
| 17-109_k141_31198 | Low-quality |
| 17-109_k141_399236 | Not-determined |
| 17-109_k141_82326 | Not-determined |
| 17-109_k141_31199 | Not-determined |
| 17-109_k141_49049 | Not-determined |
| 17-109_k141_31201 | Low-quality |
| 17-109_k141_31203 | Not-determined |
| 17-109_k141_399241 | Not-determined |
| 17-109_k141_233326 | Not-determined |
| 17-109_k141_399243 | Low-quality |
| 17-109_k141_131330 | Not-determined |
| 17-109_k141_233327 | Not-determined |
| 17-109_k141_64679 | Not-determined |
| 17-109_k141_148192 | Not-determined |
| 17-109_k141_248938 | Not-determined |
| 17-109_k141_364636 | Not-determined |
| 17-109_k141_248940 | Not-determined |
| 17-109_k141_364637 | Not-determined |
| 17-109_k141_299558 | Not-determined |
| 17-109_k141_233335 | Low-quality |
| 17-109_k141_364639 | Not-determined |
| 17-109_k141_349748 | Not-determined |
| 17-109_k141_399252 | Low-quality |
| 17-109_k141_233337 | Not-determined |
| 17-109_k141_31225 | Not-determined |
| 17-109_k141_349750 | Low-quality |
| 17-109_k141_283720 | Not-determined |
| 17-109_k141_31233 | Not-determined |
| 17-109_k141_31234 | Not-determined |
| 17-109_k141_64684 | Not-determined |
| 17-109_k141_349753 | Not-determined |
| 17-109_k141_31236 | Not-determined |
| 17-109_k141_98206 | Not-determined |
| 17-109_k141_199269 | Low-quality |
| 17-109_k141_114665 | Not-determined |
| 17-109_k141_199270 | Not-determined |
| 17-109_k141_82343 | Not-determined |
| 17-109_k141_216041 | Low-quality |
| 17-109_k141_82347 | Not-determined |
| 17-109_k141_131362 | Not-determined |
| 17-109_k141_216043 | Not-determined |
| 17-109_k141_364650 | Not-determined |
| 17-109_k141_317401 | Not-determined |
| 17-109_k141_317402 | Not-determined |
| 17-109_k141_98212 | Not-determined |
| 17-109_k141_98213 | Not-determined |
| 17-109_k141_31247 | Low-quality |
| 17-109_k141_98214 | Not-determined |
| 17-109_k141_98215 | Not-determined |
| 17-109_k141_233367 | Not-determined |
| 17-109_k141_199285 | Not-determined |
| 17-109_k141_199287 | Not-determined |
| 17-109_k141_98218 | Not-determined |
| 17-109_k141_199290 | Not-determined |
| 17-109_k141_165736 | Not-determined |
| 17-109_k141_14527 | Low-quality |
| 17-109_k141_82368 | Not-determined |
| 17-109_k141_14531 | Not-determined |
| 17-109_k141_349787 | Not-determined |
| 17-109_k141_64715 | Not-determined |
| 17-109_k141_14535 | Not-determined |
| 17-109_k141_399276 | Not-determined |
| 17-109_k141_49070 | Not-determined |
| 17-109_k141_265150 | Not-determined |
| 17-109_k141_317415 | Not-determined |
| 17-109_k141_317416 | Not-determined |
| 17-109_k141_64721 | Not-determined |
| 17-109_k141_216062 | Not-determined |
| 17-109_k141_399280 | Not-determined |
| 17-109_k141_399281 | Not-determined |
| 17-109_k141_248979 | Not-determined |
| 17-109_k141_349792 | Not-determined |
| 17-109_k141_265156 | Not-determined |
| 17-109_k141_399286 | Not-determined |
| 17-109_k141_265158 | Not-determined |
| 17-109_k141_148216 | Not-determined |
| 17-109_k141_98236 | Not-determined |
| 17-109_k141_82374 | Not-determined |
| 17-109_k141_82377 | Not-determined |
| 17-109_k141_399293 | Not-determined |
| 17-109_k141_64733 | Not-determined |
| 17-109_k141_248990 | Not-determined |
| 17-109_k141_265166 | Not-determined |
| 17-109_k141_182489 | Not-determined |
| 17-109_k141_199292 | Not-determined |
| 17-109_k141_349795 | Not-determined |
| 17-109_k141_216070 | Not-determined |
| 17-109_k141_333261 | Not-determined |
| 17-109_k141_64741 | Not-determined |
| 17-109_k141_199297 | Not-determined |
| 17-109_k141_98243 | Not-determined |
| 17-109_k141_64748 | Low-quality |
| 17-109_k141_283757 | Not-determined |
| 17-109_k141_199306 | Not-determined |
| 17-109_k141_131413 | Not-determined |
| 17-109_k141_98254 | Not-determined |
| 17-109_k141_131416 | Not-determined |
| 17-109_k141_283765 | Not-determined |
| 17-109_k141_82401 | Not-determined |
| 17-109_k141_249010 | Not-determined |
| 17-109_k141_349809 | Not-determined |
| 17-109_k141_148245 | Not-determined |
| 17-109_k141_364691 | Not-determined |
| 17-109_k141_148248 | Low-quality |
| 17-109_k141_333289 | Not-determined |
| 17-109_k141_148252 | Not-determined |
| 17-109_k141_182503 | Not-determined |
| 17-109_k141_182504 | Not-determined |
| 17-109_k141_199311 | Not-determined |
| 17-109_k141_283768 | Not-determined |
| 17-109_k141_333294 | Not-determined |
| 17-109_k141_148257 | Not-determined |
| 17-109_k141_131440 | Not-determined |
| 17-109_k141_14561 | Not-determined |
| 17-109_k141_131444 | Not-determined |
| 17-109_k141_199314 | Not-determined |
| 17-109_k141_216073 | Not-determined |
| 17-109_k141_182517 | Not-determined |
| 17-109_k141_199320 | Not-determined |
| 17-109_k141_98277 | Not-determined |
| 17-109_k141_82411 | Not-determined |
| 17-109_k141_399334 | Not-determined |
| 17-109_k141_216077 | Not-determined |
| 17-109_k141_216080 | Not-determined |
| 17-109_k141_14564 | Not-determined |
| 17-109_k141_31280 | Not-determined |
| 17-109_k141_98279 | Not-determined |
| 17-109_k141_199321 | Low-quality |
| 17-109_k141_216084 | Not-determined |
| 17-109_k141_364710 | Not-determined |
| 17-109_k141_131447 | Not-determined |
| 17-109_k141_148279 | Not-determined |
| 17-109_k141_265193 | Not-determined |
| 17-109_k141_31291 | Not-determined |
| 17-109_k141_31294 | Not-determined |
| 17-109_k141_64778 | Not-determined |
| 17-109_k141_31295 | Not-determined |
| 17-109_k141_333324 | Not-determined |
| 17-109_k141_216091 | Not-determined |
| 17-109_k141_31299 | Not-determined |
| 17-109_k141_148286 | Not-determined |
| 17-109_k141_317469 | Not-determined |
| 17-109_k141_399345 | Not-determined |
| 17-109_k141_333331 | Not-determined |
| 17-109_k141_216092 | Low-quality |
| 17-109_k141_216093 | Low-quality |
| 17-109_k141_399348 | Not-determined |
| 17-109_k141_233395 | Not-determined |
| 17-109_k141_82424 | Not-determined |
| 17-109_k141_31307 | Not-determined |
| 17-109_k141_199343 | Not-determined |
| 17-109_k141_317479 | Not-determined |
| 17-109_k141_14583 | Not-determined |
| 17-109_k141_131468 | Not-determined |
| 17-109_k141_216100 | Not-determined |
| 17-109_k141_14587 | Not-determined |
| 17-109_k141_333339 | Not-determined |
| 17-109_k141_333341 | Not-determined |
| 17-109_k141_31314 | Not-determined |
| 17-109_k141_14593 | Not-determined |
| 17-109_k141_333346 | Not-determined |
| 17-109_k141_148304 | Not-determined |
| 17-109_k141_265206 | Not-determined |
| 17-109_k141_216112 | Not-determined |
| 17-109_k141_31325 | Not-determined |
| 17-109_k141_64816 | Not-determined |
| 17-109_k141_399380 | Not-determined |
| 17-109_k141_49108 | Not-determined |
| 17-109_k141_265223 | Not-determined |
| 17-109_k141_364748 | Not-determined |
| 17-109_k141_399398 | Not-determined |
| 17-109_k141_31343 | Not-determined |
| 17-109_k141_165765 | Not-determined |
| 17-109_k141_199359 | Not-determined |
| 17-109_k141_364756 | Not-determined |
| 17-109_k141_165769 | Not-determined |
| 17-109_k141_64834 | Not-determined |
| 17-109_k141_182585 | Not-determined |
| 17-109_k141_148344 | Not-determined |
| 17-109_k141_216132 | Not-determined |
| 17-109_k141_216133 | Not-determined |
| 17-109_k141_148350 | Not-determined |
| 17-109_k141_299611 | Not-determined |
| 17-109_k141_98313 | Not-determined |
| 17-109_k141_31367 | Not-determined |
| 17-109_k141_299615 | Low-quality |
| 17-109_k141_216145 | Not-determined |
| 17-109_k141_399412 | Not-determined |
| 17-109_k141_148353 | Not-determined |
| 17-109_k141_31372 | Not-determined |
| 17-109_k141_283813 | Not-determined |
| 17-109_k141_233408 | Not-determined |
| 17-109_k141_399416 | Not-determined |
| 17-109_k141_399417 | Not-determined |
| 17-109_k141_199374 | Not-determined |
| 17-109_k141_64853 | Not-determined |
| 17-109_k141_31375 | Not-determined |
| 17-109_k141_216148 | Not-determined |
| 17-109_k141_182590 | Low-quality |
| 17-109_k141_364776 | Not-determined |
| 17-109_k141_216150 | Not-determined |
| 17-109_k141_265239 | Not-determined |
| 17-109_k141_182593 | Not-determined |
| 17-109_k141_31379 | Not-determined |
| 17-109_k141_349839 | Not-determined |
| 17-109_k141_216151 | Not-determined |
| 17-109_k141_165776 | Not-determined |
| 17-109_k141_249087 | Not-determined |
| 17-109_k141_98327 | Not-determined |
| 17-109_k141_216159 | Not-determined |
| 17-109_k141_233418 | Not-determined |
| 17-109_k141_165782 | Not-determined |
| 17-109_k141_31387 | Not-determined |
| 17-109_k141_49142 | Not-determined |
| 17-109_k141_299638 | Not-determined |
| 17-109_k141_216163 | Not-determined |
| 17-109_k141_265253 | Not-determined |
| 17-109_k141_349858 | Not-determined |
| 17-109_k141_199379 | Not-determined |
| 17-109_k141_14638 | Not-determined |
| 17-109_k141_265263 | Low-quality |
| 17-109_k141_64874 | Not-determined |
| 17-109_k141_349863 | Not-determined |
| 17-109_k141_82458 | Not-determined |
| 17-109_k141_265268 | Not-determined |
| 17-109_k141_14642 | Not-determined |
| 17-109_k141_14643 | Not-determined |
| 17-109_k141_249092 | Not-determined |
| 17-109_k141_31410 | Not-determined |
| 17-109_k141_182614 | Low-quality |
| 17-109_k141_265276 | Not-determined |
| 17-109_k141_249097 | Not-determined |
| 17-109_k141_364825 | Not-determined |
| 17-109_k141_249101 | Not-determined |
| 17-109_k141_249102 | Not-determined |
| 17-109_k141_49157 | Not-determined |
| 17-109_k141_98360 | Low-quality |
| 17-109_k141_382997 | Not-determined |
| 17-109_k141_283830 | Not-determined |
| 17-109_k141_364833 | Not-determined |
| 17-109_k141_283831 | Not-determined |
| 17-109_k141_333394 | Not-determined |
| 17-109_k141_283833 | Not-determined |
| 17-109_k141_98367 | Not-determined |
| 17-109_k141_165811 | Not-determined |
| 17-109_k141_283835 | Low-quality |
| 17-109_k141_98369 | Not-determined |
| 17-109_k141_265288 | Not-determined |
| 17-109_k141_249108 | Not-determined |
| 17-109_k141_249109 | Not-determined |
| 17-109_k141_233451 | Not-determined |
| 17-109_k141_31427 | Not-determined |
| 17-109_k141_114682 | Not-determined |
| 17-109_k141_165816 | Not-determined |
| 17-109_k141_14653 | Not-determined |
| 17-109_k141_299676 | Low-quality |
| 17-109_k141_199390 | Not-determined |
| 17-109_k141_31433 | Not-determined |
| 17-109_k141_31435 | Not-determined |
| 17-109_k141_399459 | Not-determined |
| 17-109_k141_148378 | Not-determined |
| 17-109_k141_199396 | Not-determined |
| 17-109_k141_114691 | Not-determined |
| 17-109_k141_182631 | Low-quality |
| 17-109_k141_182632 | Not-determined |
| 17-109_k141_299685 | Not-determined |
| 17-109_k141_364841 | Not-determined |
| 17-109_k141_265302 | Not-determined |
| 17-109_k141_364842 | Not-determined |
| 17-109_k141_114696 | Low-quality |
| 17-109_k141_364846 | Not-determined |
| 17-109_k141_283848 | Low-quality |
| 17-109_k141_364847 | Not-determined |
| 17-109_k141_182639 | Not-determined |
| 17-109_k141_216215 | Not-determined |
| 17-109_k141_283853 | Not-determined |
| 17-109_k141_283854 | Not-determined |
| 17-109_k141_349907 | Not-determined |
| 17-109_k141_249140 | Not-determined |
| 17-109_k141_349913 | Not-determined |
| 17-109_k141_349915 | Not-determined |
| 17-109_k141_64920 | Not-determined |
| 17-109_k141_283864 | Not-determined |
| 17-109_k141_148399 | Not-determined |
| 17-109_k141_14676 | Not-determined |
| 17-109_k141_216224 | Low-quality |
| 17-109_k141_265310 | Not-determined |
| 17-109_k141_265311 | Not-determined |
| 17-109_k141_148403 | Not-determined |
| 17-109_k141_199425 | Not-determined |
| 17-109_k141_148409 | Not-determined |

| | |
|---|---|
| 17-109_k141_249921 | Not-determined |
| 17-109_k141_64932 | Not-determined |
| 17-109_k141_49208 | Not-determined |
| 17-109_k141_199433 | Low-quality |
| 17-109_k141_216232 | Not-determined |
| 17-109_k141_249153 | Not-determined |
| 17-109_k141_399473 | Not-determined |
| 17-109_k141_148418 | Not-determined |
| 17-109_k141_131498 | Low-quality |
| 17-109_k141_64940 | Low-quality |
| 17-109_k141_131499 | Not-determined |
| 17-109_k141_249162 | Not-determined |
| 17-109_k141_399480 | Not-determined |
| 17-109_k141_82518 | Not-determined |
| 17-109_k141_199439 | Low-quality |
| 17-109_k141_131500 | Low-quality |
| 17-109_k141_364854 | Not-determined |
| 17-109_k141_182650 | Not-determined |
| 17-109_k141_64952 | Not-determined |
| 17-109_k141_383041 | Not-determined |
| 17-109_k141_182658 | Not-determined |
| 17-109_k141_383042 | Not-determined |
| 17-109_k141_383043 | Not-determined |
| 17-109_k141_165852 | Not-determined |
| 17-109_k141_49226 | Low-quality |
| 17-109_k141_383045 | Not-determined |
| 17-109_k141_165859 | Not-determined |
| 17-109_k141_114716 | Not-determined |
| 17-109_k141_114718 | Not-determined |
| 17-109_k141_249177 | Not-determined |
| 17-109_k141_399493 | Not-determined |
| 17-109_k141_49233 | Not-determined |
| 17-109_k141_216251 | Not-determined |
| 17-109_k141_82532 | Not-determined |
| 17-109_k141_349950 | Not-determined |
| 17-109_k141_131518 | Not-determined |
| 17-109_k141_131521 | Not-determined |
| 17-109_k141_216255 | Not-determined |
| 17-109_k141_299711 | Low-quality |
| 17-109_k141_199449 | Not-determined |
| 17-109_k141_82540 | Not-determined |
| 17-109_k141_199452 | Not-determined |
| 17-109_k141_131528 | Not-determined |
| 17-109_k141_383065 | Not-determined |
| 17-109_k141_165872 | Not-determined |
| 17-109_k141_82544 | Not-determined |
| 17-109_k141_64975 | Low-quality |
| 17-109_k141_299715 | Not-determined |
| 17-109_k141_265338 | Low-quality |
| 17-109_k141_148436 | Not-determined |
| 17-109_k141_317530 | Not-determined |
| 17-109_k141_49253 | Not-determined |
| 17-109_k141_265340 | Not-determined |
| 17-109_k141_14717 | Not-determined |
| 17-109_k141_265341 | Not-determined |
| 17-109_k141_283888 | Not-determined |
| 17-109_k141_182682 | Not-determined |
| 17-109_k141_49260 | Not-determined |
| 17-109_k141_148440 | Not-determined |
| 17-109_k141_182685 | Not-determined |
| 17-109_k141_317545 | Not-determined |
| 17-109_k141_14729 | Not-determined |
| 17-109_k141_148445 | Not-determined |
| 17-109_k141_31463 | Not-determined |
| 17-109_k141_64987 | Not-determined |
| 17-109_k141_199477 | Not-determined |
| 17-109_k141_233554 | Not-determined |
| 17-109_k141_265350 | Not-determined |
| 17-109_k141_383082 | Low-quality |
| 17-109_k141_233558 | Not-determined |
| 17-109_k141_49278 | Not-determined |
| 17-109_k141_199481 | Not-determined |
| 17-109_k141_31468 | Not-determined |
| 17-109_k141_49282 | Not-determined |
| 17-109_k141_82567 | Not-determined |
| 17-109_k141_317560 | Not-determined |
| 17-109_k141_216289 | Not-determined |
| 17-109_k141_317562 | Not-determined |
| 17-109_k141_233563 | Not-determined |
| 17-109_k141_31478 | Not-determined |
| 17-109_k141_317566 | Not-determined |
| 17-109_k141_383089 | Not-determined |
| 17-109_k141_49295 | Not-determined |
| 17-109_k141_249218 | Not-determined |
| 17-109_k141_82573 | Not-determined |
| 17-109_k141_399506 | Not-determined |
| 17-109_k141_283902 | Not-determined |
| 17-109_k141_383092 | Not-determined |
| 17-109_k141_283903 | Not-determined |
| 17-109_k141_98423 | Not-determined |
| 17-109_k141_148475 | Not-determined |
| 17-109_k141_349970 | Not-determined |
| 17-109_k141_349971 | Not-determined |
| 17-109_k141_98428 | Not-determined |
| 17-109_k141_148479 | Not-determined |
| 17-109_k141_65009 | Not-determined |
| 17-109_k141_333435 | Not-determined |
| 17-109_k141_383107 | Not-determined |
| 17-109_k141_383110 | Low-quality |
| 17-109_k141_14760 | Not-determined |
| 17-109_k141_383113 | Not-determined |
| 17-109_k141_364896 | Not-determined |
| 17-109_k141_148493 | Not-determined |
| 17-109_k141_364904 | Not-determined |
| 17-109_k141_165933 | Not-determined |
| 17-109_k141_216298 | Not-determined |
| 17-109_k141_399512 | Not-determined |
| 17-109_k141_299766 | Not-determined |
| 17-109_k141_364908 | Not-determined |
| 17-109_k141_399513 | Low-quality |
| 17-109_k141_299768 | Not-determined |
| 17-109_k141_182730 | Not-determined |
| 17-109_k141_216307 | Not-determined |
| 17-109_k141_233593 | Not-determined |
| 17-109_k141_49315 | Not-determined |
| 17-109_k141_233597 | Not-determined |
| 17-109_k141_98457 | Low-quality |
| 17-109_k141_199523 | Not-determined |
| 17-109_k141_148508 | Not-determined |
| 17-109_k141_65036 | Not-determined |
| 17-109_k141_31493 | Not-determined |
| 17-109_k141_216319 | Not-determined |
| 17-109_k141_114753 | Not-determined |
| 17-109_k141_31499 | Not-determined |
| 17-109_k141_383145 | Not-determined |
| 17-109_k141_333462 | Not-determined |
| 17-109_k141_98468 | Not-determined |
| 17-109_k141_14782 | Not-determined |
| 17-109_k141_199535 | Not-determined |
| 17-109_k141_14783 | Not-determined |
| 17-109_k141_165944 | Not-determined |
| 17-109_k141_14785 | Not-determined |
| 17-109_k141_383147 | Not-determined |
| 17-109_k141_165947 | Not-determined |
| 17-109_k141_31513 | Not-determined |
| 17-109_k141_82598 | Not-determined |
| 17-109_k141_350000 | Low-quality |
| 17-109_k141_114762 | Not-determined |
| 17-109_k141_114763 | Not-determined |
| 17-109_k141_216343 | Not-determined |
| 17-109_k141_199554 | Not-determined |
| 17-109_k141_31523 | Not-determined |
| 17-109_k141_49322 | Not-determined |
| 17-109_k141_233619 | Not-determined |
| 17-109_k141_98490 | Not-determined |
| 17-109_k141_364927 | Low-quality |
| 17-109_k141_98491 | Not-determined |
| 17-109_k141_364928 | Not-determined |
| 17-109_k141_399532 | Not-determined |
| 17-109_k141_383166 | Not-determined |
| 17-109_k141_31530 | Not-determined |
| 17-109_k141_182779 | Low-quality |
| 17-109_k141_216357 | Not-determined |
| 17-109_k141_265373 | Not-determined |
| 17-109_k141_114778 | Not-determined |
| 17-109_k141_216363 | Not-determined |
| 17-109_k141_333484 | Not-determined |
| 17-109_k141_216365 | Not-determined |
| 17-109_k141_333489 | Not-determined |
| 17-109_k141_317627 | Not-determined |
| 17-109_k141_350015 | Not-determined |
| 17-109_k141_333490 | Not-determined |
| 17-109_k141_399534 | Not-determined |
| 17-109_k141_317630 | Not-determined |
| 17-109_k141_350021 | Not-determined |
| 17-109_k141_350024 | Not-determined |
| 17-109_k141_65079 | Not-determined |
| 17-109_k141_199587 | Not-determined |
| 17-109_k141_82612 | Not-determined |
| 17-109_k141_364947 | Not-determined |
| 17-109_k141_399536 | Not-determined |
| 17-109_k141_283913 | Not-determined |
| 17-109_k141_350028 | Not-determined |
| 17-109_k141_65087 | Low-quality |
| 17-109_k141_333496 | Not-determined |
| 17-109_k141_399545 | Not-determined |
| 17-109_k141_65092 | Not-determined |
| 17-109_k141_265384 | Not-determined |
| 17-109_k141_82618 | Not-determined |
| 17-109_k141_82619 | Not-determined |
| 17-109_k141_317638 | Not-determined |
| 17-109_k141_31554 | Low-quality |
| 17-109_k141_82622 | Low-quality |
| 17-109_k141_216375 | Not-determined |
| 17-109_k141_233654 | Not-determined |
| 17-109_k141_216379 | Not-determined |
| 17-109_k141_31557 | Not-determined |
| 17-109_k141_31559 | Not-determined |
| 17-109_k141_283926 | Not-determined |
| 17-109_k141_317645 | Not-determined |
| 17-109_k141_165974 | Not-determined |
| 17-109_k141_364951 | Not-determined |
| 17-109_k141_114797 | Low-quality |
| 17-109_k141_49339 | Not-determined |
| 17-109_k141_65105 | Not-determined |
| 17-109_k141_65107 | Not-determined |
| 17-109_k141_216385 | Not-determined |
| 17-109_k141_82625 | Not-determined |
| 17-109_k141_216388 | Not-determined |
| 17-109_k141_98495 | Low-quality |
| 17-109_k141_65111 | Not-determined |
| 17-109_k141_165979 | Not-determined |
| 17-109_k141_82630 | Not-determined |
| 17-109_k141_265396 | Not-determined |
| 17-109_k141_199612 | Not-determined |
| 17-109_k141_165987 | Not-determined |
| 17-109_k141_31579 | Low-quality |
| 17-109_k141_265402 | Not-determined |
| 17-109_k141_333522 | Not-determined |
| 17-109_k141_364965 | Not-determined |
| 17-109_k141_49354 | Low-quality |
| 17-109_k141_98511 | Not-determined |
| 17-109_k141_199620 | Not-determined |
| 17-109_k141_333529 | Not-determined |
| 17-109_k141_333531 | Low-quality |
| 17-109_k141_399561 | Not-determined |
| 17-109_k141_114805 | Not-determined |
| 17-109_k141_283947 | Not-determined |
| 17-109_k141_399564 | Not-determined |
| 17-109_k141_49365 | Not-determined |
| 17-109_k141_364970 | Not-determined |
| 17-109_k141_114809 | Not-determined |
| 17-109_k141_283950 | Not-determined |
| 17-109_k141_317671 | Not-determined |
| 17-109_k141_216401 | Low-quality |
| 17-109_k141_148575 | Not-determined |
| 17-109_k141_131596 | Low-quality |
| 17-109_k141_199644 | Not-determined |
| 17-109_k141_350053 | Not-determined |
| 17-109_k141_399573 | Not-determined |
| 17-109_k141_383196 | Not-determined |
| 17-109_k141_364989 | Not-determined |
| 17-109_k141_148580 | Not-determined |
| 17-109_k141_49384 | Not-determined |
| 17-109_k141_283972 | Not-determined |
| 17-109_k141_333549 | Not-determined |
| 17-109_k141_333551 | Not-determined |
| 17-109_k141_49387 | Not-determined |
| 17-109_k141_65144 | Not-determined |
| 17-109_k141_333553 | Not-determined |
| 17-109_k141_148587 | Not-determined |
| 17-109_k141_148591 | Not-determined |
| 17-109_k141_399577 | Not-determined |
| 17-109_k141_182807 | Low-quality |
| 17-109_k141_283982 | Not-determined |
| 17-109_k141_49398 | Not-determined |
| 17-109_k141_216428 | Not-determined |
| 17-109_k141_350075 | Not-determined |
| 17-109_k141_383225 | Not-determined |
| 17-109_k141_365021 | Not-determined |
| 17-109_k141_65157 | Not-determined |
| 17-109_k141_383229 | Not-determined |
| 17-109_k141_165997 | Not-determined |
| 17-109_k141_283993 | Not-determined |
| 17-109_k141_233695 | Not-determined |
| 17-109_k141_333583 | Not-determined |
| 17-109_k141_265432 | Not-determined |
| 17-109_k141_166002 | Not-determined |
| 17-109_k141_350086 | Low-quality |
| 17-109_k141_350087 | Not-determined |
| 17-109_k141_350089 | Not-determined |
| 17-109_k141_399596 | Not-determined |
| 17-109_k141_31593 | Low-quality |
| 17-109_k141_199663 | Not-determined |
| 17-109_k141_284000 | Not-determined |
| 17-109_k141_114849 | Not-determined |
| 17-109_k141_114850 | Not-determined |
| 17-109_k141_82696 | Not-determined |
| 17-109_k141_199669 | Not-determined |
| 17-109_k141_299847 | Not-determined |
| 17-109_k141_82698 | Not-determined |
| 17-109_k141_284003 | Not-determined |
| 17-109_k141_65166 | Not-determined |
| 17-109_k141_399601 | Not-determined |
| 17-109_k141_65168 | Not-determined |
| 17-109_k141_233713 | Not-determined |
| 17-109_k141_114859 | Low-quality |
| 17-109_k141_399604 | Not-determined |
| 17-109_k141_82699 | Not-determined |
| 17-109_k141_399605 | Not-determined |
| 17-109_k141_249288 | Not-determined |
| 17-109_k141_199680 | Low-quality |
| 17-109_k141_399608 | Not-determined |
| 17-109_k141_249290 | Low-quality |
| 17-109_k141_350102 | Not-determined |
| 17-109_k141_233729 | Not-determined |
| 17-109_k141_249295 | Not-determined |
| 17-109_k141_216434 | Not-determined |
| 17-109_k141_265450 | Not-determined |
| 17-109_k141_131646 | Not-determined |
| 17-109_k141_199694 | Not-determined |
| 17-109_k141_65181 | Not-determined |
| 17-109_k141_383244 | Not-determined |
| 17-109_k141_65184 | Not-determined |
| 17-109_k141_182837 | Not-determined |
| 17-109_k141_333612 | Not-determined |
| 17-109_k141_216443 | Not-determined |
| 17-109_k141_284022 | Not-determined |
| 17-109_k141_82719 | Not-determined |
| 17-109_k141_114877 | Not-determined |
| 17-109_k141_166030 | Not-determined |
| 17-109_k141_233735 | Not-determined |
| 17-109_k141_199703 | Low-quality |
| 17-109_k141_148613 | Not-determined |
| 17-109_k141_114880 | Not-determined |
| 17-109_k141_333618 | Low-quality |
| 17-109_k141_249308 | Not-determined |
| 17-109_k141_166033 | Low-quality |
| 17-109_k141_383260 | Not-determined |
| 17-109_k141_399628 | Not-determined |
| 17-109_k141_284044 | Not-determined |
| 17-109_k141_31608 | Low-quality |
| 17-109_k141_166037 | Not-determined |
| 17-109_k141_98561 | Not-determined |
| 17-109_k141_233738 | Not-determined |
| 17-109_k141_65217 | Not-determined |
| 17-109_k141_82737 | Low-quality |
| 17-109_k141_31612 | Not-determined |
| 17-109_k141_333630 | Not-determined |
| 17-109_k141_365079 | Not-determined |
| 17-109_k141_98567 | Not-determined |
| 17-109_k141_49450 | Not-determined |
| 17-109_k141_333639 | Not-determined |
| 17-109_k141_365085 | Not-determined |
| 17-109_k141_216464 | Not-determined |
| 17-109_k141_49456 | Not-determined |
| 17-109_k141_216467 | Not-determined |
| 17-109_k141_49458 | Not-determined |
| 17-109_k141_365090 | Low-quality |
| 17-109_k141_216472 | Not-determined |
| 17-109_k141_365095 | Not-determined |
| 17-109_k141_365098 | Low-quality |
| 17-109_k141_82747 | Not-determined |
| 17-109_k141_82748 | Not-determined |
| 17-109_k141_365103 | Not-determined |
| 17-109_k141_31621 | Not-determined |
| 17-109_k141_166051 | Not-determined |
| 17-109_k141_31627 | Not-determined |
| 17-109_k141_265468 | Not-determined |
| 17-109_k141_98581 | Not-determined |
| 17-109_k141_148649 | Not-determined |
| 17-109_k141_148651 | Not-determined |
| 17-109_k141_383278 | Not-determined |
| 17-109_k141_265475 | Not-determined |
| 17-109_k141_182886 | Not-determined |
| 17-109_k141_166060 | Not-determined |
| 17-109_k141_233778 | Not-determined |
| 17-109_k141_182890 | Not-determined |
| 17-109_k141_148659 | Not-determined |
| 17-109_k141_383283 | Low-quality |
| 17-109_k141_383284 | Not-determined |
| 17-109_k141_333663 | Not-determined |
| 17-109_k141_249346 | Not-determined |
| 17-109_k141_216486 | Not-determined |
| 17-109_k141_249351 | Not-determined |
| 17-109_k141_216492 | Not-determined |
| 17-109_k141_249356 | Not-determined |
| 17-109_k141_98587 | Not-determined |
| 17-109_k141_216494 | Not-determined |
| 17-109_k141_49502 | Not-determined |
| 17-109_k141_166081 | Not-determined |
| 17-109_k141_166083 | Not-determined |
| 17-109_k141_65245 | Not-determined |
| 17-109_k141_98596 | Not-determined |
| 17-109_k141_98598 | Not-determined |
| 17-109_k141_82762 | Not-determined |
| 17-109_k141_350155 | Not-determined |
| 17-109_k141_98602 | Not-determined |
| 17-109_k141_265495 | Not-determined |
| 17-109_k141_49509 | Not-determined |
| 17-109_k141_148685 | Not-determined |
| 17-109_k141_265497 | Not-determined |
| 17-109_k141_82769 | Not-determined |
| 17-109_k141_199719 | Not-determined |
| 17-109_k141_284085 | Not-determined |
| 17-109_k141_148693 | Not-determined |
| 17-109_k141_299900 | Not-determined |
| 17-109_k141_299902 | Not-determined |
| 17-109_k141_233793 | Not-determined |
| 17-109_k141_31675 | Not-determined |
| 17-109_k141_265504 | Not-determined |
| 17-109_k141_284088 | Not-determined |
| 17-109_k141_333680 | Not-determined |
| 17-109_k141_216514 | Not-determined |
| 17-109_k141_49524 | Not-determined |
| 17-109_k141_216517 | Not-determined |
| 17-109_k141_49530 | Not-determined |
| 17-109_k141_199734 | Not-determined |
| 17-109_k141_265517 | Not-determined |
| 17-109_k141_65272 | Not-determined |
| 17-109_k141_148702 | Not-determined |
| 17-109_k141_249408 | Not-determined |
| 17-109_k141_182920 | Not-determined |
| 17-109_k141_284095 | Not-determined |
| 17-109_k141_98635 | Low-quality |
| 17-109_k141_98640 | Low-quality |
| 17-109_k141_333692 | Not-determined |
| 17-109_k141_333693 | Not-determined |
| 17-109_k141_114938 | Low-quality |
| 17-109_k141_233817 | Not-determined |
| 17-109_k141_365124 | Not-determined |
| 17-109_k141_365125 | Not-determined |
| 17-109_k141_299906 | Low-quality |
| 17-109_k141_98646 | Not-determined |
| 17-109_k141_284101 | Not-determined |
| 17-109_k141_114947 | Not-determined |
| 17-109_k141_49533 | Low-quality |
| 17-109_k141_82783 | Not-determined |
| 17-109_k141_216543 | Not-determined |
| 17-109_k141_14823 | Not-determined |
| 17-109_k141_65287 | Not-determined |
| 17-109_k141_148718 | Not-determined |
| 17-109_k141_299917 | Not-determined |
| 17-109_k141_98654 | Not-determined |
| 17-109_k141_199773 | Not-determined |
| 17-109_k141_317702 | Not-determined |
| 17-109_k141_299925 | Not-determined |
| 17-109_k141_284116 | Not-determined |
| 17-109_k141_284119 | Not-determined |
| 17-109_k141_82797 | Not-determined |
| 17-109_k141_31725 | Not-determined |
| 17-109_k141_299929 | Not-determined |
| 17-109_k141_199779 | Not-determined |
| 17-109_k141_299931 | Low-quality |
| 17-109_k141_299934 | Not-determined |
| 17-109_k141_82806 | Not-determined |
| 17-109_k141_49553 | Not-determined |
| 17-109_k141_148733 | Not-determined |
| 17-109_k141_299939 | Not-determined |
| 17-109_k141_14831 | Not-determined |
| 17-109_k141_82813 | Not-determined |
| 17-109_k141_98675 | Not-determined |
| 17-109_k141_98676 | Not-determined |
| 17-109_k141_14834 | Not-determined |
| 17-109_k141_216584 | Not-determined |
| 17-109_k141_49556 | Low-quality |
| 17-109_k141_199802 | Low-quality |
| 17-109_k141_199803 | Not-determined |
| 17-109_k141_399657 | Not-determined |
| 17-109_k141_199804 | Not-determined |
| 17-109_k141_131743 | Not-determined |
| 17-109_k141_131744 | Not-determined |
| 17-109_k141_233853 | Not-determined |
| 17-109_k141_98682 | Not-determined |
| 17-109_k141_365169 | Not-determined |
| 17-109_k141_333712 | Not-determined |
| 17-109_k141_233858 | Not-determined |
| 17-109_k141_49562 | Not-determined |
| 17-109_k141_383318 | Not-determined |
| 17-109_k141_65311 | Not-determined |
| 17-109_k141_199816 | Not-determined |
| 17-109_k141_98685 | Not-determined |
| 17-109_k141_233861 | Not-determined |
| 17-109_k141_383321 | Not-determined |
| 17-109_k141_65314 | Not-determined |
| 17-109_k141_166101 | Not-determined |
| 17-109_k141_350160 | Not-determined |
| 17-109_k141_131750 | Not-determined |
| 17-109_k141_216597 | Not-determined |
| 17-109_k141_131751 | Not-determined |
| 17-109_k141_199823 | Not-determined |
| 17-109_k141_216599 | Not-determined |
| 17-109_k141_249452 | Not-determined |
| 17-109_k141_14844 | Low-quality |
| 17-109_k141_98698 | Not-determined |
| 17-109_k141_216604 | Not-determined |
| 17-109_k141_216605 | Not-determined |
| 17-109_k141_333729 | Not-determined |
| 17-109_k141_131759 | Not-determined |
| 17-109_k141_350169 | Not-determined |
| 17-109_k141_399669 | Low-quality |
| 17-109_k141_299970 | Not-determined |
| 17-109_k141_383334 | Not-determined |
| 17-109_k141_233879 | Not-determined |
| 17-109_k141_31744 | Not-determined |
| 17-109_k141_166105 | Not-determined |
| 17-109_k141_299976 | Not-determined |
| 17-109_k141_166106 | Not-determined |
| 17-109_k141_233882 | Not-determined |
| 17-109_k141_317755 | Not-determined |
| 17-109_k141_233886 | Not-determined |
| 17-109_k141_383341 | Not-determined |
| 17-109_k141_299982 | Not-determined |
| 17-109_k141_199845 | Not-determined |
| 17-109_k141_383347 | Not-determined |
| 17-109_k141_166109 | Not-determined |
| 17-109_k141_333741 | Not-determined |
| 17-109_k141_299988 | Not-determined |
| 17-109_k141_333742 | Not-determined |
| 17-109_k141_216626 | Not-determined |
| 17-109_k141_65316 | Not-determined |
| 17-109_k141_49580 | Not-determined |
| 17-109_k141_166116 | Not-determined |
| 17-109_k141_31757 | Not-determined |
| 17-109_k141_182986 | Not-determined |
| 17-109_k141_182987 | Not-determined |
| 17-109_k141_216632 | Not-determined |
| 17-109_k141_233894 | Not-determined |
| 17-109_k141_199858 | Not-determined |
| 17-109_k141_131767 | Not-determined |
| 17-109_k141_199860 | Not-determined |
| 17-109_k141_233896 | Low-quality |
| 17-109_k141_131771 | Not-determined |
| 17-109_k141_317774 | Not-determined |
| 17-109_k141_115012 | Not-determined |
| 17-109_k141_98742 | Not-determined |
| 17-109_k141_333757 | Not-determined |
| 17-109_k141_166129 | Not-determined |
| 17-109_k141_98751 | Not-determined |
| 17-109_k141_333769 | Not-determined |
| 17-109_k141_233910 | Low-quality |
| 17-109_k141_233911 | Not-determined |
| 17-109_k141_399704 | Not-determined |
| 17-109_k141_399705 | Not-determined |
| 17-109_k141_333774 | Not-determined |
| 17-109_k141_333775 | Not-determined |
| 17-109_k141_317788 | Not-determined |
| 17-109_k141_98757 | Low-quality |
| 17-109_k141_14862 | Not-determined |
| 17-109_k141_317790 | Not-determined |
| 17-109_k141_399712 | Not-determined |
| 17-109_k141_333778 | Not-determined |
| 17-109_k141_14869 | Not-determined |
| 17-109_k141_115024 | Not-determined |
| 17-109_k141_399717 | Not-determined |
| 17-109_k141_199867 | Not-determined |
| 17-109_k141_399723 | Not-determined |
| 17-109_k141_233923 | Not-determined |
| 17-109_k141_233925 | Not-determined |
| 17-109_k141_65329 | Not-determined |
| 17-109_k141_233927 | Not-determined |
| 17-109_k141_265555 | Not-determined |
| 17-109_k141_233932 | Not-determined |
| 17-109_k141_14884 | Not-determined |
| 17-109_k141_365206 | Low-quality |
| 17-109_k141_249486 | Not-determined |
| 17-109_k141_284140 | Not-determined |
| 17-109_k141_333792 | Not-determined |
| 17-109_k141_284142 | Not-determined |
| 17-109_k141_383381 | Not-determined |
| 17-109_k141_399737 | Not-determined |
| 17-109_k141_399738 | Not-determined |
| 17-109_k141_383383 | Not-determined |
| 17-109_k141_399740 | Not-determined |
| 17-109_k141_14896 | Not-determined |
| 17-109_k141_131798 | Not-determined |
| 17-109_k141_300007 | Not-determined |
| 17-109_k141_14900 | Not-determined |
| 17-109_k141_166161 | Not-determined |
| 17-109_k141_115038 | Not-determined |
| 17-109_k141_333801 | Not-determined |
| 17-109_k141_383387 | Not-determined |
| 17-109_k141_82910 | Not-determined |
| 17-109_k141_333802 | Not-determined |
| 17-109_k141_300008 | Not-determined |
| 17-109_k141_199888 | Not-determined |
| 17-109_k141_383392 | Low-quality |
| 17-109_k141_383394 | Not-determined |
| 17-109_k141_82919 | Not-determined |
| 17-109_k141_317809 | Not-determined |
| 17-109_k141_166173 | Not-determined |
| 17-109_k141_383396 | Low-quality |
| 17-109_k141_265575 | Not-determined |
| 17-109_k141_216672 | Not-determined |
| 17-109_k141_115052 | Not-determined |
| 17-109_k141_284157 | Not-determined |
| 17-109_k141_333811 | Not-determined |
| 17-109_k141_350225 | Low-quality |
| 17-109_k141_98776 | Not-determined |
| 17-109_k141_265581 | Not-determined |
| 17-109_k141_284166 | Not-determined |
| 17-109_k141_249509 | Not-determined |
| 17-109_k141_365228 | Low-quality |
| 17-109_k141_350227 | Not-determined |
| 17-109_k141_265586 | Not-determined |
| 17-109_k141_82931 | Not-determined |
| 17-109_k141_265587 | Not-determined |
| 17-109_k141_98784 | Not-determined |
| 17-109_k141_300029 | Not-determined |
| 17-109_k141_183036 | Low-quality |
| 17-109_k141_131821 | Not-determined |
| 17-109_k141_265598 | Not-determined |
| 17-109_k141_65352 | Not-determined |
| 17-109_k141_216685 | Low-quality |
| 17-109_k141_365246 | Not-determined |
| 17-109_k141_216686 | Not-determined |
| 17-109_k141_199898 | Not-determined |
| 17-109_k141_65358 | Not-determined |
| 17-109_k141_365249 | Not-determined |

| | |
|---|---|
| 17-109_k141_199902 | Not-determined |
| 17-109_k141_317835 | Not-determined |
| 17-109_k141_199905 | Not-determined |
| 17-109_k141_49667 | Not-determined |
| 17-109_k141_399744 | Low-quality |
| 17-109_k141_317839 | Not-determined |
| 17-109_k141_365251 | Not-determined |
| 17-109_k141_399746 | Low-quality |
| 17-109_k141_199912 | Not-determined |
| 17-109_k141_216700 | Not-determined |
| 17-109_k141_216701 | Not-determined |
| 17-109_k141_115080 | Not-determined |
| 17-109_k141_49673 | Not-determined |
| 17-109_k141_49674 | Not-determined |
| 17-109_k141_350259 | Not-determined |
| 17-109_k141_131846 | Not-determined |
| 17-109_k141_148782 | Not-determined |
| 17-109_k141_148783 | Not-determined |
| 17-109_k141_216703 | Not-determined |
| 17-109_k141_284199 | Not-determined |
| 17-109_k141_31783 | Not-determined |
| 17-109_k141_383423 | Not-determined |
| 17-109_k141_333839 | Not-determined |
| 17-109_k141_333840 | Not-determined |
| 17-109_k141_98799 | Not-determined |
| 17-109_k141_65375 | Not-determined |
| 17-109_k141_300064 | Not-determined |
| 17-109_k141_233988 | Not-determined |
| 17-109_k141_233991 | Not-determined |
| 17-109_k141_183072 | Not-determined |
| 17-109_k141_333848 | Not-determined |
| 17-109_k141_365270 | Not-determined |
| 17-109_k141_49690 | Not-determined |
| 17-109_k141_233998 | Not-determined |
| 17-109_k141_31801 | Low-quality |
| 17-109_k141_350270 | Not-determined |
| 17-109_k141_317852 | Low-quality |
| 17-109_k141_234002 | Not-determined |
| 17-109_k141_216726 | Not-determined |
| 17-109_k141_234005 | Not-determined |
| 17-109_k141_82985 | Not-determined |
| 17-109_k141_49695 | Not-determined |
| 17-109_k141_365277 | Low-quality |
| 17-109_k141_14927 | Not-determined |
| 17-109_k141_365281 | Not-determined |
| 17-109_k141_166214 | Not-determined |
| 17-109_k141_383444 | Low-quality |
| 17-109_k141_49703 | Not-determined |
| 17-109_k141_216740 | Not-determined |
| 17-109_k141_49707 | Not-determined |
| 17-109_k141_131881 | Not-determined |
| 17-109_k141_49712 | Not-determined |
| 17-109_k141_383462 | Not-determined |
| 17-109_k141_183101 | Not-determined |
| 17-109_k141_98818 | Not-determined |
| 17-109_k141_300091 | Not-determined |
| 17-109_k141_49716 | Not-determined |
| 17-109_k141_383465 | Not-determined |
| 17-109_k141_49718 | Not-determined |
| 17-109_k141_265642 | Not-determined |
| 17-109_k141_183115 | Not-determined |
| 17-109_k141_399767 | Not-determined |
| 17-109_k141_365296 | Not-determined |
| 17-109_k141_98831 | Not-determined |
| 17-109_k141_98833 | Not-determined |
| 17-109_k141_65398 | Not-determined |
| 17-109_k141_365304 | Low-quality |
| 17-109_k141_166240 | Not-determined |
| 17-109_k141_365305 | Not-determined |
| 17-109_k141_65402 | Not-determined |
| 17-109_k141_49725 | Low-quality |
| 17-109_k141_300113 | Not-determined |
| 17-109_k141_265650 | Low-quality |
| 17-109_k141_98837 | Not-determined |
| 17-109_k141_183126 | Not-determined |
| 17-109_k141_333860 | Low-quality |
| 17-109_k141_31836 | Not-determined |
| 17-109_k141_115084 | Not-determined |
| 17-109_k141_350318 | Not-determined |
| 17-109_k141_131897 | Not-determined |
| 17-109_k141_49744 | Not-determined |
| 17-109_k141_365313 | Not-determined |
| 17-109_k141_115091 | Not-determined |
| 17-109_k141_14973 | Low-quality |
| 17-109_k141_115093 | Low-quality |
| 17-109_k141_265660 | Not-determined |
| 17-109_k141_216772 | Not-determined |
| 17-109_k141_284229 | Not-determined |
| 17-109_k141_148829 | Not-determined |
| 17-109_k141_284233 | Not-determined |
| 17-109_k141_14982 | Not-determined |
| 17-109_k141_115096 | Not-determined |
| 17-109_k141_333870 | Not-determined |
| 17-109_k141_98853 | Not-determined |
| 17-109_k141_14983 | Not-determined |
| 17-109_k141_131903 | Not-determined |
| 17-109_k141_65424 | Not-determined |
| 17-109_k141_183144 | Not-determined |
| 17-109_k141_350330 | Not-determined |
| 17-109_k141_49761 | Not-determined |
| 17-109_k141_216777 | Not-determined |
| 17-109_k141_284243 | Not-determined |
| 17-109_k141_383499 | Not-determined |
| 17-109_k141_31856 | Not-determined |
| 17-109_k141_350337 | Not-determined |
| 17-109_k141_49765 | Not-determined |
| 17-109_k141_14995 | Not-determined |
| 17-109_k141_333886 | Not-determined |
| 17-109_k141_131917 | Not-determined |
| 17-109_k141_31861 | Not-determined |
| 17-109_k141_131919 | Not-determined |
| 17-109_k141_31863 | Not-determined |
| 17-109_k141_166254 | Not-determined |
| 17-109_k141_166256 | Not-determined |
| 17-109_k141_300140 | Not-determined |
| 17-109_k141_216781 | Low-quality |
| 17-109_k141_234036 | Not-determined |
| 17-109_k141_365342 | Not-determined |
| 17-109_k141_300145 | Low-quality |
| 17-109_k141_49785 | Not-determined |
| 17-109_k141_31871 | Not-determined |
| 17-109_k141_333896 | Not-determined |
| 17-109_k141_300147 | Not-determined |
| 17-109_k141_383513 | Not-determined |
| 17-109_k141_216787 | Low-quality |
| 17-109_k141_350347 | Not-determined |
| 17-109_k141_216789 | Not-determined |
| 17-109_k141_216790 | Not-determined |
| 17-109_k141_49794 | Not-determined |
| 17-109_k141_15025 | Not-determined |
| 17-109_k141_350350 | Not-determined |
| 17-109_k141_383519 | Not-determined |
| 17-109_k141_365346 | Not-determined |
| 17-109_k141_115116 | Not-determined |
| 17-109_k141_49797 | Not-determined |
| 17-109_k141_115118 | Not-determined |
| 17-109_k141_65431 | Not-determined |
| 17-109_k141_383534 | Not-determined |
| 17-109_k141_98877 | Not-determined |
| 17-109_k141_300168 | Not-determined |
| 17-109_k141_383538 | Not-determined |
| 17-109_k141_365359 | Not-determined |
| 17-109_k141_15039 | Not-determined |
| 17-109_k141_383541 | Not-determined |
| 17-109_k141_15043 | Low-quality |
| 17-109_k141_166289 | Not-determined |
| 17-109_k141_148864 | Not-determined |
| 17-109_k141_98889 | Not-determined |
| 17-109_k141_148866 | Not-determined |
| 17-109_k141_83038 | Not-determined |
| 17-109_k141_65447 | Not-determined |
| 17-109_k141_166296 | Not-determined |
| 17-109_k141_49803 | Low-quality |
| 17-109_k141_131952 | Not-determined |
| 17-109_k141_131953 | Not-determined |
| 17-109_k141_216804 | Low-quality |
| 17-109_k141_131956 | Not-determined |
| 17-109_k141_65456 | Not-determined |
| 17-109_k141_333930 | Not-determined |
| 17-109_k141_148876 | Not-determined |
| 17-109_k141_183154 | Not-determined |
| 17-109_k141_98906 | Not-determined |
| 17-109_k141_131969 | Not-determined |
| 17-109_k141_300203 | Not-determined |
| 17-109_k141_98916 | Not-determined |
| 17-109_k141_183161 | Not-determined |
| 17-109_k141_166306 | Low-quality |
| 17-109_k141_216810 | Not-determined |
| 17-109_k141_317872 | Not-determined |
| 17-109_k141_216814 | Not-determined |
| 17-109_k141_234056 | Not-determined |
| 17-109_k141_166309 | Not-determined |
| 17-109_k141_148894 | Not-determined |
| 17-109_k141_131986 | Not-determined |
| 17-109_k141_317876 | Low-quality |
| 17-109_k141_31898 | Not-determined |
| 17-109_k141_148898 | Not-determined |
| 17-109_k141_216819 | Not-determined |
| 17-109_k141_31900 | Not-determined |
| 17-109_k141_284258 | Not-determined |
| 17-109_k141_115146 | Not-determined |
| 17-109_k141_317884 | Not-determined |
| 17-109_k141_365394 | Not-determined |
| 17-109_k141_65478 | Not-determined |
| 17-109_k141_131993 | Not-determined |
| 17-109_k141_31908 | Not-determined |
| 17-109_k141_183171 | Not-determined |
| 17-109_k141_115148 | Low-quality |
| 17-109_k141_148914 | Not-determined |
| 17-109_k141_115151 | Not-determined |
| 17-109_k141_49822 | Not-determined |
| 17-109_k141_166318 | Not-determined |
| 17-109_k141_31912 | Not-determined |
| 17-109_k141_365402 | Not-determined |
| 17-109_k141_115157 | Not-determined |
| 17-109_k141_31917 | Not-determined |
| 17-109_k141_284273 | Not-determined |
| 17-109_k141_166325 | Not-determined |
| 17-109_k141_115162 | Not-determined |
| 17-109_k141_132008 | Not-determined |
| 17-109_k141_115164 | Not-determined |
| 17-109_k141_115168 | Not-determined |
| 17-109_k141_166329 | Not-determined |
| 17-109_k141_350404 | Not-determined |
| 17-109_k141_383553 | Not-determined |
| 17-109_k141_383554 | Not-determined |
| 17-109_k141_183186 | Not-determined |
| 17-109_k141_216838 | Not-determined |
| 17-109_k141_115181 | Not-determined |
| 17-109_k141_216839 | Not-determined |
| 17-109_k141_216840 | Not-determined |
| 17-109_k141_300231 | Not-determined |
| 17-109_k141_216844 | Not-determined |
| 17-109_k141_166338 | Not-determined |
| 17-109_k141_65502 | Not-determined |
| 17-109_k141_183190 | Not-determined |
| 17-109_k141_284293 | Not-determined |
| 17-109_k141_15119 | Not-determined |
| 17-109_k141_15120 | Not-determined |
| 17-109_k141_350408 | Not-determined |
| 17-109_k141_132023 | Not-determined |
| 17-109_k141_31924 | Not-determined |
| 17-109_k141_383569 | Not-determined |
| 17-109_k141_383570 | Not-determined |
| 17-109_k141_31928 | Not-determined |
| 17-109_k141_300237 | Not-determined |
| 17-109_k141_249563 | Not-determined |
| 17-109_k141_31934 | Not-determined |
| 17-109_k141_15136 | Not-determined |
| 17-109_k141_234085 | Not-determined |
| 17-109_k141_83077 | Not-determined |
| 17-109_k141_333941 | Not-determined |
| 17-109_k141_350424 | Not-determined |
| 17-109_k141_115198 | Not-determined |
| 17-109_k141_183198 | Not-determined |
| 17-109_k141_317910 | Not-determined |
| 17-109_k141_383579 | Low-quality |
| 17-109_k141_350431 | Not-determined |
| 17-109_k141_216865 | Not-determined |
| 17-109_k141_98927 | Not-determined |
| 17-109_k141_98928 | Not-determined |
| 17-109_k141_132031 | Low-quality |
| 17-109_k141_132032 | Not-determined |
| 17-109_k141_31950 | Not-determined |
| 17-109_k141_333948 | Not-determined |
| 17-109_k141_15155 | Not-determined |
| 17-109_k141_199989 | Not-determined |
| 17-109_k141_83098 | Not-determined |
| 17-109_k141_317917 | Not-determined |
| 17-109_k141_15161 | Not-determined |
| 17-109_k141_65532 | Not-determined |
| 17-109_k141_216875 | Not-determined |
| 17-109_k141_115215 | Not-determined |
| 17-109_k141_333954 | Not-determined |
| 17-109_k141_15165 | Not-determined |
| 17-109_k141_15168 | Not-determined |
| 17-109_k141_83101 | Low-quality |
| 17-109_k141_300271 | Not-determined |
| 17-109_k141_15177 | Low-quality |
| 17-109_k141_83106 | Not-determined |
| 17-109_k141_265695 | Not-determined |
| 17-109_k141_115218 | Not-determined |
| 17-109_k141_132043 | Not-determined |
| 17-109_k141_132046 | Not-determined |
| 17-109_k141_333969 | Not-determined |
| 17-109_k141_83120 | Not-determined |
| 17-109_k141_300284 | Not-determined |
| 17-109_k141_31977 | Not-determined |
| 17-109_k141_399873 | Not-determined |
| 17-109_k141_65544 | Not-determined |
| 17-109_k141_300290 | Low-quality |
| 17-109_k141_234122 | Not-determined |
| 17-109_k141_166364 | Not-determined |
| 17-109_k141_317944 | Not-determined |
| 17-109_k141_300294 | Not-determined |
| 17-109_k141_300296 | Not-determined |
| 17-109_k141_166372 | Low-quality |
| 17-109_k141_216890 | Not-determined |
| 17-109_k141_234129 | Not-determined |
| 17-109_k141_317952 | Not-determined |
| 17-109_k141_31988 | Not-determined |
| 17-109_k141_317956 | Not-determined |
| 17-109_k141_317957 | Not-determined |
| 17-109_k141_350475 | Not-determined |
| 17-109_k141_333999 | Not-determined |
| 17-109_k141_317964 | Not-determined |
| 17-109_k141_83147 | Not-determined |
| 17-109_k141_334003 | Not-determined |
| 17-109_k141_284353 | Not-determined |
| 17-109_k141_234141 | Not-determined |
| 17-109_k141_284357 | Not-determined |
| 17-109_k141_15197 | Not-determined |
| 17-109_k141_383591 | Not-determined |
| 17-109_k141_183236 | Low-quality |
| 17-109_k141_284360 | Not-determined |
| 17-109_k141_334011 | Not-determined |
| 17-109_k141_83157 | Not-determined |
| 17-109_k141_15200 | Not-determined |
| 17-109_k141_216907 | Not-determined |
| 17-109_k141_65573 | Not-determined |
| 17-109_k141_383594 | Not-determined |
| 17-109_k141_383595 | Not-determined |
| 17-109_k141_83162 | Not-determined |
| 17-109_k141_265731 | Not-determined |
| 17-109_k141_350489 | Low-quality |
| 17-109_k141_399912 | Not-determined |
| 17-109_k141_65582 | Not-determined |
| 17-109_k141_399914 | Not-determined |
| 17-109_k141_399919 | Low-quality |
| 17-109_k141_15220 | Not-determined |
| 17-109_k141_32019 | Not-determined |
| 17-109_k141_183255 | Not-determined |
| 17-109_k141_249616 | Not-determined |
| 17-109_k141_399925 | Not-determined |
| 17-109_k141_399927 | Not-determined |
| 17-109_k141_216933 | Low-quality |
| 17-109_k141_284395 | Low-quality |
| 17-109_k141_216935 | Not-determined |
| 17-109_k141_49917 | Low-quality |
| 17-109_k141_65592 | Not-determined |
| 17-109_k141_284399 | Not-determined |
| 17-109_k141_399932 | Not-determined |
| 17-109_k141_49922 | Not-determined |
| 17-109_k141_284402 | Not-determined |
| 17-109_k141_183267 | Not-determined |
| 17-109_k141_15232 | Not-determined |
| 17-109_k141_383613 | Not-determined |
| 17-109_k141_65600 | Not-determined |
| 17-109_k141_32026 | Not-determined |
| 17-109_k141_383618 | Not-determined |
| 17-109_k141_317991 | Not-determined |
| 17-109_k141_350494 | Not-determined |
| 17-109_k141_383620 | Not-determined |
| 17-109_k141_383623 | Not-determined |
| 17-109_k141_249635 | Not-determined |
| 17-109_k141_249637 | Not-determined |
| 17-109_k141_334030 | Low-quality |
| 17-109_k141_98957 | Not-determined |
| 17-109_k141_15239 | Not-determined |
| 17-109_k141_83177 | Low-quality |
| 17-109_k141_284417 | Not-determined |
| 17-109_k141_98963 | Not-determined |
| 17-109_k141_49937 | Not-determined |
| 17-109_k141_318004 | Low-quality |
| 17-109_k141_284420 | Not-determined |
| 17-109_k141_83183 | Not-determined |
| 17-109_k141_132090 | Not-determined |
| 17-109_k141_132097 | Not-determined |
| 17-109_k141_383633 | Not-determined |
| 17-109_k141_318010 | Not-determined |
| 17-109_k141_200045 | Not-determined |
| 17-109_k141_49942 | Low-quality |
| 17-109_k141_350513 | Low-quality |
| 17-109_k141_98978 | Not-determined |
| 17-109_k141_183299 | Not-determined |
| 17-109_k141_115266 | Not-determined |
| 17-109_k141_132106 | Not-determined |
| 17-109_k141_284441 | Not-determined |
| 17-109_k141_234191 | Not-determined |
| 17-109_k141_49953 | Not-determined |
| 17-109_k141_365429 | Not-determined |
| 17-109_k141_350527 | Not-determined |
| 17-109_k141_334057 | Low-quality |
| 17-109_k141_365436 | Not-determined |
| 17-109_k141_200063 | Not-determined |
| 17-109_k141_334062 | Not-determined |
| 17-109_k141_98982 | Not-determined |
| 17-109_k141_200066 | Not-determined |
| 17-109_k141_132125 | Not-determined |
| 17-109_k141_234202 | Not-determined |
| 17-109_k141_166441 | Not-determined |
| 17-109_k141_365443 | Not-determined |
| 17-109_k141_15267 | Not-determined |
| 17-109_k141_98994 | Not-determined |
| 17-109_k141_98995 | Not-determined |
| 17-109_k141_166447 | Not-determined |
| 17-109_k141_166448 | Not-determined |
| 17-109_k141_183335 | Not-determined |
| 17-109_k141_399957 | Not-determined |
| 17-109_k141_115300 | Not-determined |
| 17-109_k141_115302 | Low-quality |
| 17-109_k141_49976 | Not-determined |
| 17-109_k141_32033 | Not-determined |
| 17-109_k141_265792 | Not-determined |
| 17-109_k141_49977 | Low-quality |
| 17-109_k141_284464 | Low-quality |
| 17-109_k141_365449 | Not-determined |
| 17-109_k141_99008 | Not-determined |
| 17-109_k141_32038 | Not-determined |
| 17-109_k141_115313 | Not-determined |
| 17-109_k141_49986 | Not-determined |
| 17-109_k141_265799 | Not-determined |
| 17-109_k141_83220 | Low-quality |
| 17-109_k141_115319 | Not-determined |
| 17-109_k141_200103 | Not-determined |
| 17-109_k141_83221 | Not-determined |
| 17-109_k141_200108 | Not-determined |
| 17-109_k141_32052 | Not-determined |
| 17-109_k141_234235 | Not-determined |
| 17-109_k141_399970 | Not-determined |
| 17-109_k141_83223 | Not-determined |
| 17-109_k141_284483 | Not-determined |
| 17-109_k141_216953 | Not-determined |
| 17-109_k141_49999 | Not-determined |
| 17-109_k141_350537 | Low-quality |
| 17-109_k141_265812 | Not-determined |
| 17-109_k141_166465 | Not-determined |
| 17-109_k141_265813 | Not-determined |
| 17-109_k141_83231 | Not-determined |
| 17-109_k141_265814 | Not-determined |
| 17-109_k141_300304 | Not-determined |
| 17-109_k141_350539 | Not-determined |
| 17-109_k141_50004 | Not-determined |
| 17-109_k141_334097 | Not-determined |
| 17-109_k141_50005 | Not-determined |
| 17-109_k141_216965 | Not-determined |
| 17-109_k141_249663 | Not-determined |
| 17-109_k141_234241 | Not-determined |
| 17-109_k141_99036 | Not-determined |
| 17-109_k141_249667 | Low-quality |
| 17-109_k141_99040 | Not-determined |
| 17-109_k141_83245 | Not-determined |
| 17-109_k141_166474 | Not-determined |
| 17-109_k141_399984 | Not-determined |
| 17-109_k141_166476 | Not-determined |
| 17-109_k141_148954 | Low-quality |
| 17-109_k141_350545 | Not-determined |
| 17-109_k141_83251 | Not-determined |
| 17-109_k141_99052 | Not-determined |
| 17-109_k141_99053 | Low-quality |
| 17-109_k141_249683 | Not-determined |
| 17-109_k141_365474 | Not-determined |
| 17-109_k141_383645 | Not-determined |
| 17-109_k141_265824 | Not-determined |
| 17-109_k141_399996 | Not-determined |
| 17-109_k141_15311 | Not-determined |
| 17-109_k141_166496 | Not-determined |
| 17-109_k141_265829 | Not-determined |
| 17-109_k141_400000 | Not-determined |
| 17-109_k141_400004 | Not-determined |
| 17-109_k141_249694 | Not-determined |
| 17-109_k141_265840 | Not-determined |
| 17-109_k141_334112 | Not-determined |
| 17-109_k141_32079 | Not-determined |
| 17-109_k141_83271 | Not-determined |
| 17-109_k141_249704 | Not-determined |
| 17-109_k141_99079 | Not-determined |
| 17-109_k141_15330 | Not-determined |
| 17-109_k141_166507 | Not-determined |
| 17-109_k141_15333 | Not-determined |
| 17-109_k141_300324 | Not-determined |
| 17-109_k141_166510 | Not-determined |
| 17-109_k141_183390 | Not-determined |
| 17-109_k141_284497 | Not-determined |
| 17-109_k141_32103 | Not-determined |
| 17-109_k141_65621 | Not-determined |
| 17-109_k141_265851 | Low-quality |
| 17-109_k141_65624 | Not-determined |
| 17-109_k141_32109 | Low-quality |
| 17-109_k141_99081 | Not-determined |
| 17-109_k141_183402 | Not-determined |
| 17-109_k141_148977 | Not-determined |
| 17-109_k141_15349 | Not-determined |
| 17-109_k141_32113 | Not-determined |
| 17-109_k141_65631 | Not-determined |
| 17-109_k141_334120 | Not-determined |
| 17-109_k141_15356 | Not-determined |
| 17-109_k141_115378 | Not-determined |
| 17-109_k141_166533 | Not-determined |
| 17-109_k141_99091 | Low-quality |
| 17-109_k141_334124 | Not-determined |
| 17-109_k141_300330 | Not-determined |
| 17-109_k141_15361 | Low-quality |
| 17-109_k141_334126 | Not-determined |
| 17-109_k141_300334 | Not-determined |
| 17-109_k141_166547 | Not-determined |
| 17-109_k141_132141 | Not-determined |
| 17-109_k141_265868 | Not-determined |
| 17-109_k141_334129 | Not-determined |
| 17-109_k141_32137 | Not-determined |
| 17-109_k141_166557 | Not-determined |
| 17-109_k141_265875 | Not-determined |
| 17-109_k141_166561 | Not-determined |
| 17-109_k141_234283 | Not-determined |
| 17-109_k141_166562 | Not-determined |
| 17-109_k141_300353 | Not-determined |
| 17-109_k141_115392 | Not-determined |
| 17-109_k141_65644 | Not-determined |
| 17-109_k141_166566 | Not-determined |
| 17-109_k141_265880 | Not-determined |
| 17-109_k141_300357 | Not-determined |
| 17-109_k141_334145 | Not-determined |
| 17-109_k141_249761 | Not-determined |
| 17-109_k141_265885 | Not-determined |
| 17-109_k141_65653 | Not-determined |
| 17-109_k141_132171 | Not-determined |
| 17-109_k141_334150 | Not-determined |
| 17-109_k141_65655 | Not-determined |
| 17-109_k141_166575 | Not-determined |
| 17-109_k141_265893 | Not-determined |
| 17-109_k141_32142 | Not-determined |
| 17-109_k141_65661 | Not-determined |
| 17-109_k141_334153 | Not-determined |
| 17-109_k141_32147 | Not-determined |
| 17-109_k141_65668 | Not-determined |
| 17-109_k141_183431 | Not-determined |
| 17-109_k141_65672 | Not-determined |
| 17-109_k141_83343 | Not-determined |
| 17-109_k141_265902 | Not-determined |
| 17-109_k141_15395 | Not-determined |
| 17-109_k141_65676 | Not-determined |
| 17-109_k141_32157 | Not-determined |
| 17-109_k141_50028 | Low-quality |
| 17-109_k141_200139 | Not-determined |
| 17-109_k141_300377 | Low-quality |
| 17-109_k141_15408 | Not-determined |
| 17-109_k141_132193 | Not-determined |
| 17-109_k141_65686 | Not-determined |
| 17-109_k141_32161 | Not-determined |
| 17-109_k141_32165 | Not-determined |
| 17-109_k141_65696 | Low-quality |
| 17-109_k141_183455 | Not-determined |
| 17-109_k141_383703 | Not-determined |
| 17-109_k141_300393 | Not-determined |
| 17-109_k141_166604 | Not-determined |
| 17-109_k141_166605 | Not-determined |
| 17-109_k141_166608 | Not-determined |
| 17-109_k141_217024 | Not-determined |
| 17-109_k141_300400 | Not-determined |
| 17-109_k141_249785 | Not-determined |
| 17-109_k141_115400 | Not-determined |
| 17-109_k141_234315 | Not-determined |
| 17-109_k141_350610 | Not-determined |
| 17-109_k141_132226 | Not-determined |
| 17-109_k141_83346 | Not-determined |
| 17-109_k141_383730 | Not-determined |
| 17-109_k141_99162 | Not-determined |
| 17-109_k141_83354 | Not-determined |
| 17-109_k141_15449 | Not-determined |
| 17-109_k141_83359 | Not-determined |
| 17-109_k141_132249 | Not-determined |
| 17-109_k141_200179 | Not-determined |
| 17-109_k141_15452 | Not-determined |
| 17-109_k141_50067 | Not-determined |
| 17-109_k141_15458 | Not-determined |
| 17-109_k141_15459 | Not-determined |
| 17-109_k141_249796 | Not-determined |
| 17-109_k141_249797 | Not-determined |
| 17-109_k141_50072 | Not-determined |
| 17-109_k141_183496 | Not-determined |
| 17-109_k141_83388 | Not-determined |
| 17-109_k141_50083 | Not-determined |
| 17-109_k141_200198 | Not-determined |
| 17-109_k141_334178 | Not-determined |
| 17-109_k141_115427 | Not-determined |
| 17-109_k141_284537 | Not-determined |
| 17-109_k141_132262 | Not-determined |
| 17-109_k141_50098 | Not-determined |
| 17-109_k141_166620 | Not-determined |
| 17-109_k141_83408 | Not-determined |
| 17-109_k141_83410 | Not-determined |
| 17-109_k141_166626 | Not-determined |
| 17-109_k141_200214 | Low-quality |
| 17-109_k141_284553 | Low-quality |
| 17-109_k141_265968 | Not-determined |
| 17-109_k141_15472 | Not-determined |
| 17-109_k141_50105 | Not-determined |
| 17-109_k141_265970 | Low-quality |
| 17-109_k141_249834 | Not-determined |
| 17-109_k141_284561 | Not-determined |
| 17-109_k141_132278 | Not-determined |
| 17-109_k141_217077 | Not-determined |
| 17-109_k141_249841 | Not-determined |
| 17-109_k141_50108 | Not-determined |
| 17-109_k141_334198 | Not-determined |
| 17-109_k141_50110 | Low-quality |
| 17-109_k141_99193 | Low-quality |
| 17-109_k141_32176 | Not-determined |
| 17-109_k141_15483 | Not-determined |
| 17-109_k141_265975 | Not-determined |
| 17-109_k141_115448 | Not-determined |
| 17-109_k141_99203 | Not-determined |
| 17-109_k141_32179 | Not-determined |
| 17-109_k141_65780 | Not-determined |
| 17-109_k141_350656 | Low-quality |
| 17-109_k141_83436 | Not-determined |
| 17-109_k141_334210 | Not-determined |
| 17-109_k141_249848 | Not-determined |
| 17-109_k141_50129 | Not-determined |

| | |
|---|---|
| 17-109_k141_99205 | Not-determined |
| 17-109_k141_350664 | Not-determined |
| 17-109_k141_334222 | Not-determined |
| 17-109_k141_334223 | Not-determined |
| 17-109_k141_65794 | Not-determined |
| 17-109_k141_383814 | Not-determined |
| 17-109_k141_166639 | Not-determined |
| 17-109_k141_200236 | Not-determined |
| 17-109_k141_83451 | Not-determined |
| 17-109_k141_83452 | Not-determined |
| 17-109_k141_166647 | Not-determined |
| 17-109_k141_265987 | Not-determined |
| 17-109_k141_200242 | Not-determined |
| 17-109_k141_32195 | Not-determined |
| 17-109_k141_99226 | Low-quality |
| 17-109_k141_83456 | Low-quality |
| 17-109_k141_265997 | Not-determined |
| 17-109_k141_350680 | Not-determined |
| 17-109_k141_200251 | Not-determined |
| 17-109_k141_183584 | Not-determined |
| 17-109_k141_115466 | Not-determined |
| 17-109_k141_249858 | Not-determined |
| 17-109_k141_149007 | Not-determined |
| 17-109_k141_166666 | Not-determined |
| 17-109_k141_149010 | Not-determined |
| 17-109_k141_350692 | Not-determined |
| 17-109_k141_65796 | Not-determined |
| 17-109_k141_200266 | Not-determined |
| 17-109_k141_334250 | Not-determined |
| 17-109_k141_365493 | Not-determined |
| 17-109_k141_83472 | Not-determined |
| 17-109_k141_83473 | Not-determined |
| 17-109_k141_99259 | Not-determined |
| 17-109_k141_266020 | Low-quality |
| 17-109_k141_149015 | Not-determined |
| 17-109_k141_149016 | Not-determined |
| 17-109_k141_149017 | Not-determined |
| 17-109_k141_132315 | Low-quality |
| 17-109_k141_200280 | Not-determined |
| 17-109_k141_115482 | Not-determined |
| 17-109_k141_149020 | Not-determined |
| 17-109_k141_50159 | Not-determined |
| 17-109_k141_115484 | Low-quality |
| 17-109_k141_266032 | Not-determined |
| 17-109_k141_200283 | Not-determined |
| 17-109_k141_266035 | Not-determined |
| 17-109_k141_132321 | Not-determined |
| 17-109_k141_132322 | Not-determined |
| 17-109_k141_365506 | Not-determined |
| 17-109_k141_365509 | Not-determined |
| 17-109_k141_350708 | Not-determined |
| 17-109_k141_300456 | Not-determined |
| 17-109_k141_334274 | Not-determined |
| 17-109_k141_115492 | Not-determined |
| 17-109_k141_99279 | Not-determined |
| 17-109_k141_249876 | Low-quality |
| 17-109_k141_300461 | Not-determined |
| 17-109_k141_200307 | Low-quality |
| 17-109_k141_365515 | Low-quality |
| 17-109_k141_183624 | Not-determined |
| 17-109_k141_334286 | Not-determined |
| 17-109_k141_217149 | Not-determined |
| 17-109_k141_149041 | Not-determined |
| 17-109_k141_65821 | Not-determined |
| 17-109_k141_300468 | Not-determined |
| 17-109_k141_365527 | Low-quality |
| 17-109_k141_334289 | Not-determined |
| 17-109_k141_83491 | Not-determined |
| 17-109_k141_115519 | Not-determined |
| 17-109_k141_200325 | Not-determined |
| 17-109_k141_149060 | Not-determined |
| 17-109_k141_300476 | Not-determined |
| 17-109_k141_83497 | Not-determined |
| 17-109_k141_217162 | Not-determined |
| 17-109_k141_350743 | Not-determined |
| 17-109_k141_365530 | Not-determined |
| 17-109_k141_217172 | Not-determined |
| 17-109_k141_115536 | Not-determined |
| 17-109_k141_300494 | Not-determined |
| 17-109_k141_99302 | Not-determined |
| 17-109_k141_83511 | Not-determined |
| 17-109_k141_99307 | Not-determined |
| 17-109_k141_32238 | Not-determined |
| 17-109_k141_217183 | Low-quality |
| 17-109_k141_132338 | Not-determined |
| 17-109_k141_83517 | Not-determined |
| 17-109_k141_115547 | Not-determined |
| 17-109_k141_249893 | Not-determined |
| 18-102-final-viral-combined_k141_75144\|\|0_partial | Medium-quality |
| 17-109_k141_32244 | Not-determined |
| 17-109_k141_99315 | Not-determined |
| 17-109_k141_65850 | Not-determined |
| 17-109_k141_365538 | Not-determined |
| 17-109_k141_132342 | Not-determined |
| 17-109_k141_300503 | Not-determined |
| 17-109_k141_350755 | Low-quality |
| 17-109_k141_149097 | Not-determined |
| 17-109_k141_183670 | Not-determined |
| 17-109_k141_300504 | Not-determined |
| 17-109_k141_183671 | Low-quality |
| 17-109_k141_115557 | Not-determined |
| 17-109_k141_99324 | Not-determined |
| 17-109_k141_115562 | Not-determined |
| 17-109_k141_115563 | Not-determined |
| 17-109_k141_132352 | Not-determined |
| 17-109_k141_200366 | Not-determined |
| 17-109_k141_115569 | Not-determined |
| 17-109_k141_83537 | Not-determined |
| 17-109_k141_15517 | Not-determined |
| 17-109_k141_334320 | Not-determined |
| 17-109_k141_149110 | Not-determined |
| 17-109_k141_365550 | Not-determined |
| 17-109_k141_115582 | Not-determined |
| 17-109_k141_365553 | Not-determined |
| 17-109_k141_300522 | Not-determined |
| 17-109_k141_15529 | Not-determined |
| 17-109_k141_365560 | Not-determined |
| 17-109_k141_149116 | Not-determined |
| 17-109_k141_132374 | Not-determined |
| 17-109_k141_383868 | Not-determined |
| 17-109_k141_149118 | Not-determined |
| 17-109_k141_15535 | Not-determined |
| 17-109_k141_149121 | Not-determined |
| 17-109_k141_266047 | Not-determined |
| 17-109_k141_99340 | Not-determined |
| 17-109_k141_149131 | Not-determined |
| 17-109_k141_83562 | Not-determined |
| 17-109_k141_115596 | Not-determined |
| 17-109_k141_32259 | Not-determined |
| 17-109_k141_300525 | Not-determined |
| 17-109_k141_183702 | Not-determined |
| 17-109_k141_217190 | Not-determined |
| 17-109_k141_334346 | Not-determined |
| 17-109_k141_15548 | Not-determined |
| 17-109_k141_249906 | Not-determined |
| 17-109_k141_249907 | Low-quality |
| 17-109_k141_249908 | Not-determined |
| 17-109_k141_83574 | Not-determined |
| 17-109_k141_65889 | Not-determined |
| 17-109_k141_249916 | Not-determined |
| 17-109_k141_183710 | Not-determined |
| 17-109_k141_200386 | Not-determined |
| 17-109_k141_132411 | Not-determined |
| 17-109_k141_15560 | Not-determined |
| 17-109_k141_149138 | Not-determined |
| 17-109_k141_32268 | Not-determined |
| 17-109_k141_166734 | Not-determined |
| 17-109_k141_217205 | Not-determined |
| 17-109_k141_334367 | Not-determined |
| 17-109_k141_149144 | Not-determined |
| 17-109_k141_15568 | Not-determined |
| 17-109_k141_217213 | Low-quality |
| 17-109_k141_266075 | Not-determined |
| 17-109_k141_149146 | Low-quality |
| 17-109_k141_115614 | Not-determined |
| 17-109_k141_334376 | Not-determined |
| 17-109_k141_149153 | Not-determined |
| 17-109_k141_32282 | Not-determined |
| 17-109_k141_149154 | Not-determined |
| 17-109_k141_249925 | Not-determined |
| 17-109_k141_166757 | Not-determined |
| 17-109_k141_300546 | Low-quality |
| 17-109_k141_266088 | Not-determined |
| 17-109_k141_383900 | Not-determined |
| 17-109_k141_217223 | Not-determined |
| 17-109_k141_149161 | Not-determined |
| 17-109_k141_99364 | Not-determined |
| 17-109_k141_300553 | Not-determined |
| 17-109_k141_200413 | Low-quality |
| 17-109_k141_99374 | Not-determined |
| 17-109_k141_32293 | Low-quality |
| 17-109_k141_300562 | Not-determined |
| 17-109_k141_83593 | Not-determined |
| 17-109_k141_200415 | Not-determined |
| 17-109_k141_300572 | Not-determined |
| 17-109_k141_266108 | Not-determined |
| 17-109_k141_99378 | Not-determined |
| 17-109_k141_65914 | Not-determined |
| 17-109_k141_65916 | Not-determined |
| 17-109_k141_99382 | Not-determined |
| 17-109_k141_132431 | Not-determined |
| 17-109_k141_217242 | Not-determined |
| 17-109_k141_217244 | Not-determined |
| 17-109_k141_83609 | Not-determined |
| 17-109_k141_132440 | Not-determined |
| 17-109_k141_132441 | Not-determined |
| 17-109_k141_32310 | Not-determined |
| 17-109_k141_132452 | Low-quality |
| 17-109_k141_183720 | Not-determined |
| 17-109_k141_183721 | Not-determined |
| 17-109_k141_350824 | Not-determined |
| 17-109_k141_300598 | Not-determined |
| 17-109_k141_350827 | Not-determined |
| 17-109_k141_334404 | Not-determined |
| 17-109_k141_99398 | Not-determined |
| 17-109_k141_149189 | Not-determined |
| 17-109_k141_132468 | Not-determined |
| 17-109_k141_266118 | Not-determined |
| 17-109_k141_217257 | Not-determined |
| 17-109_k141_334407 | Not-determined |
| 17-109_k141_365590 | Low-quality |
| 17-109_k141_32322 | Not-determined |
| 17-109_k141_99409 | Not-determined |
| 17-109_k141_334413 | Not-determined |
| 17-109_k141_383936 | Low-quality |
| 17-109_k141_65962 | Not-determined |
| 17-109_k141_266127 | Low-quality |
| 17-109_k141_300619 | Not-determined |
| 17-109_k141_350847 | Not-determined |
| 17-109_k141_15601 | Not-determined |
| 17-109_k141_115650 | Low-quality |
| 17-109_k141_115653 | Not-determined |
| 17-109_k141_200444 | Not-determined |
| 17-109_k141_32334 | Not-determined |
| 17-109_k141_115658 | Not-determined |
| 17-109_k141_200445 | Not-determined |
| 17-109_k141_183732 | Not-determined |
| 17-109_k141_266132 | Not-determined |
| 17-109_k141_65982 | Not-determined |
| 17-109_k141_166813 | Low-quality |
| 17-109_k141_383958 | Not-determined |
| 17-109_k141_99423 | Not-determined |
| 17-109_k141_149203 | Not-determined |
| 17-109_k141_115666 | Not-determined |
| 17-109_k141_166816 | Not-determined |
| 17-109_k141_383962 | Not-determined |
| 17-109_k141_183736 | Not-determined |
| 17-109_k141_149207 | Not-determined |
| 17-109_k141_334436 | Not-determined |
| 17-109_k141_183741 | Not-determined |
| 17-109_k141_166829 | Not-determined |
| 17-109_k141_32356 | Not-determined |
| 17-109_k141_383967 | Not-determined |
| 17-109_k141_183748 | Not-determined |
| 17-109_k141_249948 | Low-quality |
| 17-109_k141_383970 | Not-determined |
| 17-109_k141_183754 | Not-determined |
| 17-109_k141_132477 | Not-determined |
| 17-109_k141_183755 | Not-determined |
| 17-109_k141_200480 | Not-determined |
| 17-109_k141_249964 | Not-determined |
| 17-109_k141_266155 | Not-determined |
| 17-109_k141_383987 | Not-determined |
| 17-109_k141_66006 | Not-determined |
| 17-109_k141_132490 | Not-determined |
| 17-109_k141_149218 | Not-determined |
| 17-109_k141_149221 | Not-determined |
| 17-109_k141_249972 | Low-quality |
| 17-109_k141_15606 | Not-determined |
| 17-109_k141_15608 | Not-determined |
| 17-109_k141_32376 | Not-determined |
| 17-109_k141_149228 | Not-determined |
| 17-109_k141_350881 | Not-determined |
| 17-109_k141_365633 | Not-determined |
| 17-109_k141_32377 | Not-determined |
| 17-109_k141_32378 | Not-determined |
| 17-109_k141_365638 | Not-determined |
| 17-109_k141_350886 | Not-determined |
| 17-109_k141_32388 | Not-determined |
| 17-109_k141_99448 | Not-determined |
| 17-109_k141_15613 | Low-quality |
| 17-109_k141_32392 | Not-determined |
| 17-109_k141_200501 | Not-determined |
| 17-109_k141_383996 | Not-determined |
| 17-109_k141_99449 | Not-determined |
| 17-109_k141_99450 | Not-determined |
| 17-109_k141_266163 | Not-determined |
| 17-109_k141_249993 | Not-determined |
| 17-109_k141_149239 | Not-determined |
| 17-109_k141_365663 | Not-determined |
| 17-109_k141_266167 | Not-determined |
| 17-109_k141_83660 | Not-determined |
| 17-109_k141_217286 | Not-determined |
| 17-109_k141_149242 | Not-determined |
| 17-109_k141_266169 | Not-determined |
| 17-109_k141_166868 | Not-determined |
| 17-109_k141_149244 | Not-determined |
| 17-109_k141_83662 | Not-determined |
| 17-109_k141_32407 | Not-determined |
| 17-109_k141_149248 | Not-determined |
| 17-109_k141_132501 | Not-determined |
| 17-109_k141_99458 | Not-determined |
| 17-109_k141_149252 | Not-determined |
| 17-109_k141_166875 | Not-determined |
| 17-109_k141_132504 | Not-determined |
| 17-109_k141_32411 | Not-determined |
| 17-109_k141_149255 | Not-determined |
| 17-109_k141_32412 | Not-determined |
| 17-109_k141_217298 | Not-determined |
| 17-109_k141_166881 | Not-determined |
| 17-109_k141_300652 | Not-determined |
| 17-109_k141_66036 | Not-determined |
| 17-109_k141_149265 | Not-determined |
| 17-109_k141_300653 | Not-determined |
| 17-109_k141_15644 | Low-quality |
| 17-109_k141_83667 | Not-determined |
| 17-109_k141_250015 | Low-quality |
| 17-109_k141_266196 | Not-determined |
| 17-109_k141_300660 | Not-determined |
| 17-109_k141_32418 | Not-determined |
| 17-109_k141_66046 | Not-determined |
| 17-109_k141_384023 | Not-determined |
| 17-109_k141_365672 | Low-quality |
| 17-109_k141_99485 | Not-determined |
| 17-109_k141_365673 | Not-determined |
| 17-109_k141_166904 | Low-quality |
| 17-109_k141_99492 | Not-determined |
| 17-109_k141_200509 | Not-determined |
| 17-109_k141_266212 | Not-determined |
| 17-109_k141_384030 | Not-determined |
| 17-109_k141_217320 | Not-determined |
| 17-109_k141_115698 | Not-determined |
| 17-109_k141_217326 | Not-determined |
| 17-109_k141_15657 | Not-determined |
| 17-109_k141_217327 | Not-determined |
| 17-109_k141_15658 | Not-determined |
| 17-109_k141_217328 | Low-quality |
| 17-109_k141_15661 | Not-determined |
| 17-109_k141_384040 | Low-quality |
| 17-109_k141_400031 | Not-determined |
| 17-109_k141_300680 | Not-determined |
| 17-109_k141_266219 | Not-determined |
| 17-109_k141_166905 | Low-quality |
| 17-109_k141_384051 | Not-determined |
| 17-109_k141_99499 | Low-quality |
| 17-109_k141_365680 | Not-determined |
| 17-109_k141_15671 | Not-determined |
| 17-109_k141_350929 | Not-determined |
| 17-109_k141_217338 | Not-determined |
| 17-109_k141_149303 | Not-determined |
| 17-109_k141_132533 | Not-determined |
| 17-109_k141_132536 | Not-determined |
| 17-109_k141_99509 | Not-determined |
| 17-109_k141_149307 | Not-determined |
| 17-109_k141_15676 | Not-determined |
| 17-109_k141_99515 | Not-determined |
| 17-109_k141_149309 | Not-determined |
| 17-109_k141_250036 | Not-determined |
| 17-109_k141_132548 | Not-determined |
| 17-109_k141_149316 | Not-determined |
| 17-109_k141_115727 | Not-determined |
| 17-109_k141_32442 | Low-quality |
| 17-109_k141_266253 | Low-quality |
| 17-109_k141_15685 | Not-determined |
| 17-109_k141_166934 | Not-determined |
| 17-109_k141_266257 | Low-quality |
| 17-109_k141_183818 | Not-determined |
| 17-109_k141_334474 | Not-determined |
| 17-109_k141_166936 | Not-determined |
| 17-109_k141_32447 | Not-determined |
| 17-109_k141_99524 | Not-determined |
| 17-109_k141_166937 | Not-determined |
| 17-109_k141_300700 | Not-determined |
| 17-109_k141_266267 | Not-determined |
| 17-109_k141_15690 | Not-determined |
| 17-109_k141_334483 | Not-determined |
| 17-109_k141_166945 | Not-determined |
| 17-109_k141_183841 | Not-determined |
| 17-109_k141_15692 | Not-determined |
| 17-109_k141_365704 | Low-quality |
| 17-109_k141_132559 | Not-determined |
| 17-109_k141_250048 | Not-determined |
| 17-109_k141_115763 | Not-determined |
| 17-109_k141_32466 | Not-determined |
| 17-109_k141_266284 | Not-determined |
| 17-109_k141_115768 | Not-determined |
| 17-109_k141_183857 | Not-determined |
| 17-109_k141_32467 | Not-determined |
| 17-109_k141_250057 | Not-determined |
| 17-109_k141_183862 | Not-determined |
| 17-109_k141_66091 | Low-quality |
| 17-109_k141_334504 | Not-determined |
| 17-109_k141_266297 | Not-determined |
| 17-109_k141_266298 | Not-determined |
| 17-109_k141_200561 | Not-determined |
| 17-109_k141_350970 | Not-determined |
| 17-109_k141_266305 | Not-determined |
| 17-109_k141_334511 | Not-determined |
| 17-109_k141_334513 | Not-determined |
| 17-109_k141_365735 | Not-determined |
| 17-109_k141_99562 | Not-determined |
| 17-109_k141_166970 | Not-determined |
| 17-109_k141_334531 | Not-determined |
| 17-109_k141_32506 | Not-determined |
| 17-109_k141_200570 | Not-determined |
| 17-109_k141_365761 | Not-determined |
| 17-109_k141_300723 | Not-determined |
| 17-109_k141_334541 | Not-determined |
| 17-109_k141_384079 | Not-determined |
| 17-109_k141_132601 | Not-determined |
| 17-109_k141_149368 | Not-determined |
| 17-109_k141_384087 | Not-determined |
| 17-109_k141_334560 | Not-determined |
| 17-109_k141_183889 | Not-determined |
| 17-109_k141_183891 | Not-determined |
| 17-109_k141_115818 | Low-quality |
| 17-109_k141_384096 | Not-determined |
| 17-109_k141_115820 | Not-determined |
| 17-109_k141_300757 | Not-determined |
| 17-109_k141_350995 | Not-determined |
| 18-105-final-viral-combined_k141_183113\|\|0_partial | Medium-quality |
| 17-109_k141_300758 | Low-quality |
| 17-109_k141_217358 | Not-determined |
| 17-109_k141_250098 | Not-determined |
| 17-109_k141_266328 | Not-determined |
| 17-109_k141_266332 | Not-determined |
| 17-109_k141_351002 | Not-determined |
| 17-109_k141_266335 | Not-determined |
| 17-109_k141_32548 | Not-determined |
| 17-109_k141_217368 | Not-determined |
| 17-109_k141_183904 | Not-determined |
| 17-109_k141_32555 | Not-determined |
| 17-109_k141_167018 | Low-quality |
| 17-109_k141_99592 | Not-determined |
| 17-109_k141_32565 | Low-quality |
| 17-109_k141_132636 | Not-determined |
| 17-109_k141_266349 | Not-determined |
| 17-109_k141_351023 | Not-determined |
| 17-109_k141_351026 | Not-determined |
| 17-109_k141_32571 | Not-determined |
| 17-109_k141_300792 | Not-determined |
| 17-109_k141_32576 | Low-quality |
| 17-109_k141_66159 | Not-determined |
| 17-109_k141_183922 | Not-determined |
| 17-109_k141_300798 | Not-determined |
| 17-109_k141_132652 | Not-determined |
| 17-109_k141_300800 | Not-determined |
| 17-109_k141_250113 | Not-determined |
| 17-109_k141_250114 | Not-determined |
| 17-109_k141_167032 | Not-determined |
| 17-109_k141_15714 | Not-determined |
| 17-109_k141_300811 | Not-determined |
| 17-109_k141_200606 | Not-determined |
| 17-109_k141_250125 | Not-determined |
| 17-109_k141_99607 | Not-determined |
| 17-109_k141_115865 | Not-determined |
| 17-109_k141_266366 | Not-determined |
| 17-109_k141_15721 | Not-determined |
| 17-109_k141_250138 | Not-determined |
| 17-109_k141_250139 | Not-determined |
| 17-109_k141_66182 | Not-determined |
| 17-109_k141_266372 | Low-quality |
| 17-109_k141_183938 | Not-determined |
| 17-109_k141_266374 | Not-determined |
| 17-109_k141_99624 | Not-determined |
| 17-109_k141_66188 | Not-determined |
| 17-109_k141_99625 | Not-determined |
| 17-109_k141_266378 | Not-determined |
| 17-109_k141_266379 | Not-determined |
| 17-109_k141_99633 | Not-determined |
| 17-109_k141_351058 | Not-determined |
| 17-109_k141_400049 | Not-determined |
| 17-109_k141_66196 | Not-determined |
| 17-109_k141_32606 | Not-determined |
| 17-109_k141_384137 | Not-determined |
| 17-109_k141_66200 | Not-determined |
| 17-109_k141_217427 | Not-determined |
| 17-109_k141_250156 | Not-determined |
| 17-109_k141_365804 | Not-determined |
| 17-109_k141_32612 | Not-determined |
| 17-109_k141_66204 | Not-determined |
| 17-109_k141_266392 | Not-determined |
| 17-109_k141_66207 | Not-determined |
| 17-109_k141_250160 | Not-determined |
| 17-109_k141_351071 | Not-determined |
| 17-109_k141_400061 | Not-determined |
| 17-109_k141_149394 | Not-determined |
| 17-109_k141_200634 | Not-determined |
| 17-109_k141_250166 | Low-quality |
| 17-109_k141_266404 | Not-determined |
| 17-109_k141_334615 | Not-determined |
| 17-109_k141_200638 | Not-determined |
| 17-109_k141_149402 | Not-determined |
| 17-109_k141_132667 | Not-determined |
| 17-109_k141_115893 | Not-determined |
| 17-109_k141_167088 | Low-quality |
| 17-109_k141_132672 | Not-determined |
| 17-109_k141_66232 | Not-determined |
| 17-109_k141_200646 | Not-determined |
| 17-109_k141_200647 | Not-determined |
| 17-109_k141_99656 | Not-determined |
| 17-109_k141_167094 | Not-determined |
| 17-109_k141_200655 | Not-determined |
| 17-109_k141_351077 | Not-determined |
| 17-109_k141_400080 | Not-determined |
| 17-109_k141_400081 | Not-determined |
| 17-109_k141_351080 | Not-determined |
| 17-109_k141_200661 | Not-determined |
| 17-109_k141_66244 | Not-determined |
| 17-109_k141_300843 | Not-determined |
| 17-109_k141_66246 | Not-determined |
| 17-109_k141_200665 | Not-determined |
| 17-109_k141_217456 | Not-determined |
| 17-109_k141_250185 | Not-determined |
| 17-109_k141_400086 | Not-determined |
| 17-109_k141_115905 | Not-determined |
| 17-109_k141_66258 | Not-determined |
| 17-109_k141_149415 | Not-determined |
| 17-109_k141_351089 | Not-determined |
| 17-109_k141_132685 | Not-determined |
| 17-109_k141_351090 | Not-determined |
| 17-109_k141_32642 | Not-determined |
| 17-109_k141_132690 | Not-determined |
| 17-109_k141_15770 | Not-determined |
| 17-109_k141_217463 | Not-determined |
| 17-109_k141_200674 | Not-determined |
| 17-109_k141_400096 | Not-determined |
| 17-109_k141_300857 | Not-determined |
| 17-109_k141_115916 | Low-quality |
| 17-109_k141_351102 | Not-determined |
| 17-109_k141_300859 | Low-quality |
| 17-109_k141_334646 | Not-determined |
| 17-109_k141_149426 | Not-determined |
| 17-109_k141_15773 | Not-determined |
| 17-109_k141_115923 | Not-determined |
| 17-109_k141_66274 | Not-determined |
| 17-109_k141_365862 | Not-determined |
| 17-109_k141_365870 | Low-quality |
| 17-109_k141_200685 | Not-determined |
| 17-109_k141_217475 | Not-determined |
| 17-109_k141_217476 | Not-determined |
| 17-109_k141_66282 | Not-determined |
| 17-109_k141_132713 | Not-determined |
| 17-109_k141_115929 | Not-determined |
| 17-109_k141_132716 | Not-determined |
| 17-109_k141_365872 | Not-determined |
| 17-109_k141_400109 | Not-determined |
| 17-109_k141_351119 | Not-determined |
| 17-109_k141_149439 | Not-determined |
| 17-109_k141_351123 | Not-determined |
| 17-109_k141_15787 | Not-determined |
| 17-109_k141_400117 | Not-determined |
| 17-109_k141_149442 | Not-determined |
| 17-109_k141_400118 | Not-determined |
| 17-109_k141_115940 | Not-determined |
| 17-109_k141_183956 | Not-determined |
| 17-109_k141_149450 | Not-determined |
| 17-109_k141_351129 | Not-determined |
| 17-109_k141_167135 | Not-determined |
| 17-109_k141_351133 | Not-determined |
| 17-109_k141_149461 | Not-determined |
| 17-109_k141_250236 | Not-determined |
| 17-109_k141_167148 | Low-quality |
| 17-109_k141_115948 | Low-quality |
| 17-109_k141_400130 | Not-determined |
| 17-109_k141_15811 | Not-determined |
| 17-109_k141_149471 | Not-determined |
| 17-109_k141_400131 | Not-determined |
| 17-109_k141_384189 | Not-determined |
| 17-109_k141_365912 | Not-determined |
| 17-109_k141_15823 | Not-determined |
| 17-109_k141_384198 | Not-determined |
| 17-109_k141_66332 | Not-determined |
| 17-109_k141_351169 | Low-quality |
| 17-109_k141_250276 | Not-determined |
| 17-109_k141_400139 | Not-determined |
| 17-109_k141_334668 | Not-determined |
| 17-109_k141_66344 | Not-determined |
| 17-109_k141_250289 | Not-determined |
| 17-109_k141_334672 | Not-determined |
| 17-109_k141_132752 | Not-determined |
| 17-109_k141_66349 | Low-quality |
| 17-109_k141_351183 | Not-determined |
| 17-109_k141_149510 | Not-determined |
| 17-109_k141_365942 | Not-determined |
| 17-109_k141_149515 | Not-determined |
| 17-109_k141_132758 | Not-determined |
| 17-109_k141_200705 | Low-quality |
| 17-109_k141_250300 | Not-determined |
| 17-109_k141_365948 | Not-determined |
| 17-109_k141_183984 | Not-determined |
| 17-109_k141_217527 | Not-determined |
| 17-109_k141_250306 | Low-quality |
| 17-109_k141_149523 | Not-determined |
| 17-109_k141_15852 | Not-determined |
| 17-109_k141_400160 | Not-determined |
| 17-109_k141_266426 | Not-determined |
| 17-109_k141_400162 | Not-determined |
| 17-109_k141_400164 | Not-determined |
| 17-109_k141_250315 | Not-determined |
| 17-109_k141_200712 | Not-determined |
| 17-109_k141_217531 | Not-determined |
| 17-109_k141_149529 | Not-determined |
| 17-109_k141_116012 | Low-quality |
| 17-109_k141_200716 | Not-determined |
| 17-109_k141_32697 | Not-determined |
| 17-109_k141_266439 | Not-determined |
| 17-109_k141_266440 | Not-determined |
| 17-109_k141_66382 | Low-quality |

| | |
|---|---|
| 17-109_k141_66383 | Not-determined |
| 17-109_k141_167188 | Not-determined |
| 17-109_k141_32709 | Not-determined |
| 17-109_k141_400186 | Not-determined |
| 17-109_k141_15876 | Not-determined |
| 17-109_k141_365974 | Not-determined |
| 17-109_k141_351202 | Not-determined |
| 17-109_k141_149552 | Not-determined |
| 17-109_k141_365979 | Not-determined |
| 17-109_k141_200729 | Not-determined |
| 17-109_k141_200732 | Not-determined |
| 17-109_k141_217556 | Low-quality |
| 17-109_k141_149561 | Not-determined |
| 17-109_k141_15891 | Not-determined |
| 17-109_k141_200744 | Not-determined |
| 17-109_k141_217559 | Not-determined |
| 17-109_k141_384261 | Low-quality |
| 17-109_k141_167205 | Not-determined |
| 17-109_k141_99722 | Not-determined |
| 17-109_k141_32732 | Not-determined |
| 17-109_k141_200750 | Not-determined |
| 17-109_k141_167209 | Not-determined |
| 17-109_k141_250331 | Not-determined |
| 17-109_k141_15895 | Not-determined |
| 17-109_k141_250333 | Low-quality |
| 17-109_k141_15898 | Not-determined |
| 17-109_k141_15899 | Not-determined |
| 17-109_k141_351214 | Not-determined |
| 17-109_k141_250334 | Not-determined |
| 17-109_k141_365998 | Not-determined |
| 17-109_k141_167218 | Not-determined |
| 17-109_k141_384270 | Not-determined |
| 17-109_k141_250336 | Not-determined |
| 17-109_k141_400194 | Not-determined |
| 17-109_k141_250337 | Not-determined |
| 17-109_k141_15902 | Not-determined |
| 17-109_k141_15904 | Not-determined |
| 17-109_k141_99737 | Not-determined |
| 17-109_k141_99739 | Not-determined |
| 17-109_k141_15908 | Not-determined |
| 17-109_k141_366006 | Not-determined |
| 17-109_k141_366007 | Not-determined |
| 17-109_k141_15910 | Not-determined |
| 17-109_k141_400202 | Not-determined |
| 17-109_k141_217576 | Low-quality |
| 17-109_k141_132779 | Not-determined |
| 17-109_k141_116039 | Not-determined |
| 17-109_k141_149575 | Not-determined |
| 17-109_k141_116041 | Not-determined |
| 17-109_k141_116042 | Not-determined |
| 17-109_k141_99745 | Not-determined |
| 17-109_k141_132784 | Not-determined |
| 17-109_k141_132785 | Not-determined |
| 17-109_k141_384283 | Low-quality |
| 17-109_k141_66423 | Not-determined |
| 17-109_k141_184014 | Not-determined |
| 17-109_k141_266489 | Not-determined |
| 17-109_k141_132787 | Not-determined |
| 17-109_k141_149587 | Not-determined |
| 17-109_k141_132790 | Not-determined |
| 17-109_k141_132791 | Not-determined |
| 17-109_k141_351226 | Not-determined |
| 17-109_k141_167233 | Low-quality |
| 17-109_k141_32760 | Not-determined |
| 17-109_k141_15937 | Not-determined |
| 17-109_k141_66445 | Not-determined |
| 17-109_k141_400211 | Not-determined |
| 17-109_k141_167236 | Not-determined |
| 17-109_k141_99762 | Not-determined |
| 17-109_k141_400213 | Not-determined |
| 17-109_k141_149599 | Not-determined |
| 17-109_k141_149600 | Not-determined |
| 17-109_k141_66449 | Not-determined |
| 17-109_k141_66450 | Not-determined |
| 17-109_k141_167245 | Low-quality |
| 17-109_k141_15948 | Not-determined |
| 17-109_k141_149602 | Low-quality |
| 17-109_k141_132804 | Not-determined |
| 17-109_k141_351230 | Not-determined |
| 17-109_k141_300983 | Not-determined |
| 17-109_k141_266514 | Not-determined |
| 17-109_k141_99772 | Not-determined |
| 17-109_k141_200780 | Low-quality |
| 17-109_k141_15959 | Not-determined |
| 17-109_k141_66468 | Not-determined |
| 17-109_k141_384317 | Low-quality |
| 17-109_k141_200785 | Not-determined |
| 17-109_k141_149615 | Low-quality |
| 17-109_k141_149618 | Not-determined |
| 17-109_k141_149619 | Not-determined |
| 17-109_k141_250379 | Not-determined |
| 17-109_k141_250383 | Not-determined |
| 17-109_k141_384329 | Not-determined |
| 17-109_k141_99787 | Not-determined |
| 17-109_k141_300998 | Not-determined |
| 17-109_k141_15979 | Not-determined |
| 17-109_k141_116089 | Not-determined |
| 17-109_k141_132827 | Not-determined |
| 17-109_k141_351245 | Not-determined |
| 17-109_k141_266534 | Low-quality |
| 17-109_k141_132833 | Not-determined |
| 17-109_k141_400242 | Not-determined |
| 17-109_k141_200793 | Not-determined |
| 17-109_k141_184036 | Not-determined |
| 17-109_k141_132836 | Low-quality |
| 17-109_k141_116112 | Not-determined |
| 17-109_k141_366048 | Not-determined |
| 17-109_k141_99799 | Not-determined |
| 17-109_k141_149644 | Not-determined |
| 17-109_k141_366054 | Not-determined |
| 17-109_k141_400260 | Not-determined |
| 17-109_k141_334728 | Low-quality |
| 17-109_k141_400268 | Not-determined |
| 17-109_k141_149651 | Not-determined |
| 17-109_k141_15999 | Not-determined |
| 17-109_k141_366077 | Not-determined |
| 17-109_k141_400280 | Not-determined |
| 17-109_k141_384359 | Not-determined |
| 17-109_k141_116129 | Not-determined |
| 17-109_k141_351254 | Not-determined |
| 17-109_k141_384372 | Not-determined |
| 17-109_k141_16034 | Not-determined |
| 17-109_k141_167256 | Not-determined |
| 17-109_k141_99833 | Not-determined |
| 17-109_k141_116142 | Not-determined |
| 17-109_k141_334739 | Not-determined |
| 17-109_k141_366084 | Not-determined |
| 17-109_k141_149679 | Low-quality |
| 17-109_k141_200818 | Not-determined |
| 17-109_k141_266550 | Low-quality |
| 17-109_k141_116150 | Not-determined |
| 17-109_k141_366087 | Not-determined |
| 17-109_k141_200822 | Not-determined |
| 17-109_k141_66496 | Not-determined |
| 17-109_k141_301044 | Not-determined |
| 17-109_k141_116158 | Not-determined |
| 17-109_k141_149694 | Not-determined |
| 17-109_k141_200826 | Not-determined |
| 17-109_k141_116162 | Not-determined |
| 17-109_k141_200827 | Not-determined |
| 17-109_k141_301053 | Not-determined |
| 17-109_k141_200829 | Not-determined |
| 17-109_k141_301054 | Not-determined |
| 17-109_k141_66514 | Not-determined |
| 17-109_k141_116170 | Not-determined |
| 17-109_k141_149707 | Not-determined |
| 17-109_k141_116174 | Not-determined |
| 17-109_k141_149711 | Not-determined |
| 17-109_k141_149712 | Not-determined |
| 17-109_k141_66533 | Not-determined |
| 17-109_k141_116182 | Not-determined |
| 17-109_k141_66535 | Not-determined |
| 17-109_k141_334771 | Not-determined |
| 17-109_k141_384392 | Not-determined |
| 17-109_k141_250430 | Not-determined |
| 17-109_k141_351262 | Not-determined |
| 17-109_k141_66545 | Not-determined |
| 17-109_k141_116205 | Not-determined |
| 17-109_k141_32802 | Not-determined |
| 17-109_k141_384396 | Not-determined |
| 17-109_k141_351279 | Not-determined |
| 17-109_k141_200835 | Low-quality |
| 17-109_k141_250433 | Not-determined |
| 17-109_k141_32817 | Not-determined |
| 17-109_k141_66557 | Not-determined |
| 17-109_k141_384402 | Not-determined |
| 17-109_k141_32818 | Not-determined |
| 17-109_k141_32819 | Not-determined |
| 17-109_k141_66559 | Not-determined |
| 17-109_k141_99894 | Not-determined |
| 17-109_k141_351287 | Not-determined |
| 17-109_k141_99896 | Not-determined |
| 17-109_k141_384408 | Not-determined |
| 17-109_k141_99897 | Not-determined |
| 17-109_k141_16117 | Not-determined |
| 17-109_k141_384409 | Low-quality |
| 17-109_k141_32822 | Not-determined |
| 17-109_k141_116209 | Low-quality |
| 17-109_k141_116210 | Not-determined |
| 17-109_k141_16121 | Low-quality |
| 17-109_k141_116211 | Not-determined |
| 17-109_k141_366168 | Not-determined |
| 17-109_k141_99910 | Not-determined |
| 17-109_k141_16133 | Not-determined |
| 17-109_k141_250449 | Not-determined |
| 17-109_k141_32841 | Not-determined |
| 17-109_k141_351305 | Not-determined |
| 17-109_k141_66576 | Not-determined |
| 17-109_k141_351309 | Not-determined |
| 17-109_k141_132908 | Not-determined |
| 17-109_k141_366174 | Not-determined |
| 17-109_k141_301068 | Not-determined |
| 17-109_k141_301069 | Not-determined |
| 17-109_k141_301070 | Not-determined |
| 17-109_k141_400293 | Not-determined |
| 17-109_k141_167326 | Not-determined |
| 17-109_k141_16152 | Not-determined |
| 17-109_k141_132918 | Not-determined |
| 17-109_k141_250462 | Not-determined |
| 17-109_k141_250464 | Not-determined |
| 17-109_k141_66592 | Not-determined |
| 17-109_k141_132924 | Not-determined |
| 17-109_k141_366197 | Not-determined |
| 17-109_k141_301091 | Not-determined |
| 17-109_k141_167330 | Not-determined |
| 17-109_k141_132932 | Not-determined |
| 17-109_k141_99923 | Not-determined |
| 17-109_k141_66603 | Not-determined |
| 17-109_k141_301097 | Not-determined |
| 17-109_k141_116231 | Not-determined |
| 17-109_k141_301101 | Not-determined |
| 17-109_k141_116236 | Not-determined |
| 17-109_k141_66608 | Not-determined |
| 17-109_k141_301105 | Not-determined |
| 17-109_k141_66613 | Not-determined |
| 17-109_k141_250472 | Not-determined |
| 17-109_k141_132947 | Low-quality |
| 17-109_k141_149774 | Low-quality |
| 17-109_k141_301115 | Not-determined |
| 17-109_k141_99933 | Not-determined |
| 17-109_k141_99938 | Not-determined |
| 17-109_k141_384452 | Not-determined |
| 17-109_k141_250487 | Not-determined |
| 17-109_k141_132959 | Not-determined |
| 17-109_k141_99947 | Not-determined |
| 17-109_k141_250493 | Low-quality |
| 17-109_k141_66646 | Not-determined |
| 17-109_k141_66649 | Not-determined |
| 17-109_k141_366210 | Not-determined |
| 17-109_k141_116272 | Not-determined |
| 17-109_k141_400328 | Not-determined |
| 17-109_k141_200860 | Not-determined |
| 17-109_k141_200862 | Not-determined |
| 17-109_k141_384492 | Not-determined |
| 17-109_k141_366232 | Not-determined |
| 17-109_k141_149820 | Not-determined |
| 17-109_k141_16189 | Not-determined |
| 17-109_k141_66684 | Not-determined |
| 17-109_k141_167357 | Not-determined |
| 17-109_k141_301137 | Not-determined |
| 17-109_k141_16196 | Not-determined |
| 17-109_k141_266566 | Not-determined |
| 17-109_k141_366245 | Not-determined |
| 17-109_k141_116302 | Not-determined |
| 17-109_k141_133021 | Not-determined |
| 17-109_k141_384509 | Low-quality |
| 17-109_k141_384513 | Not-determined |
| 17-109_k141_32889 | Not-determined |
| 17-109_k141_250528 | Not-determined |
| 17-109_k141_250536 | Not-determined |
| 17-109_k141_400368 | Not-determined |
| 17-109_k141_266583 | Not-determined |
| 17-109_k141_16216 | Not-determined |
| 17-109_k141_266588 | Not-determined |
| 17-109_k141_400379 | Not-determined |
| 17-109_k141_266595 | Not-determined |
| 17-109_k141_266597 | Low-quality |
| 17-109_k141_32908 | Not-determined |
| 17-109_k141_384541 | Not-determined |
| 17-109_k141_32914 | Not-determined |
| 17-109_k141_99999 | Not-determined |
| 17-109_k141_133042 | Not-determined |
| 17-109_k141_301156 | Not-determined |
| 17-109_k141_116323 | Not-determined |
| 17-109_k141_301158 | Not-determined |
| 17-109_k141_133046 | Not-determined |
| 17-109_k141_400391 | Not-determined |
| 17-109_k141_133047 | Not-determined |
| 17-109_k141_301162 | Not-determined |
| 17-109_k141_400393 | Not-determined |
| 17-109_k141_400397 | Not-determined |
| 17-109_k141_366295 | Low-quality |
| 17-109_k141_266617 | Not-determined |
| 17-109_k141_32928 | Low-quality |
| 17-109_k141_266623 | Not-determined |
| 17-109_k141_384570 | Not-determined |
| 17-109_k141_133061 | Not-determined |
| 17-109_k141_366298 | Low-quality |
| 17-109_k141_66708 | Low-quality |
| 17-109_k141_116336 | Low-quality |
| 17-109_k141_66711 | Not-determined |
| 17-109_k141_116338 | Not-determined |
| 17-109_k141_133068 | Not-determined |
| 17-109_k141_66720 | Not-determined |
| 17-109_k141_366306 | Not-determined |
| 17-109_k141_66724 | Not-determined |
| 17-109_k141_133078 | Not-determined |
| 17-109_k141_32943 | Not-determined |
| 17-109_k141_32945 | Not-determined |
| 17-109_k141_366317 | Not-determined |
| 17-109_k141_351387 | Not-determined |
| 17-109_k141_400422 | Not-determined |
| 17-109_k141_366319 | Not-determined |
| 17-109_k141_16249 | Not-determined |
| 17-109_k141_66733 | Not-determined |
| 17-109_k141_66735 | Not-determined |
| 17-109_k141_116374 | Not-determined |
| 17-109_k141_301171 | Not-determined |
| 17-109_k141_351399 | Not-determined |
| 17-109_k141_301176 | Not-determined |
| 17-109_k141_301177 | Not-determined |
| 17-109_k141_32963 | Not-determined |
| 17-109_k141_250561 | Not-determined |
| 17-109_k141_32967 | Not-determined |
| 17-109_k141_301179 | Not-determined |
| 17-109_k141_133113 | Not-determined |
| 17-109_k141_32972 | Not-determined |
| 17-109_k141_250572 | Not-determined |
| 17-109_k141_400453 | Not-determined |
| 17-109_k141_66754 | Not-determined |
| 17-109_k141_149839 | Not-determined |
| 17-109_k141_149842 | Not-determined |
| 17-109_k141_366323 | Not-determined |
| 17-109_k141_16289 | Not-determined |
| 17-109_k141_184092 | Not-determined |
| 17-109_k141_184094 | Not-determined |
| 17-109_k141_366337 | Not-determined |
| 17-109_k141_301203 | Not-determined |
| 17-109_k141_149858 | Not-determined |
| 17-109_k141_16302 | Not-determined |
| 17-109_k141_184102 | Not-determined |
| 17-109_k141_149868 | Not-determined |
| 17-109_k141_100054 | Not-determined |
| 17-109_k141_184109 | Not-determined |
| 17-109_k141_301213 | Not-determined |
| 17-109_k141_149874 | Not-determined |
| 17-109_k141_184114 | Low-quality |
| 17-109_k141_32987 | Not-determined |
| 17-109_k141_66778 | Not-determined |
| 17-109_k141_250582 | Not-determined |
| 17-109_k141_16339 | Low-quality |
| 17-109_k141_266640 | Not-determined |
| 17-109_k141_16343 | Not-determined |
| 17-109_k141_133131 | Not-determined |
| 17-109_k141_133134 | Not-determined |
| 17-109_k141_250594 | Not-determined |
| 17-109_k141_400495 | Not-determined |
| 17-109_k141_250598 | Not-determined |
| 17-109_k141_133140 | Low-quality |
| 17-109_k141_66796 | Not-determined |
| 17-109_k141_400500 | Not-determined |
| 17-109_k141_250602 | Not-determined |
| 17-109_k141_266656 | Not-determined |
| 17-109_k141_33001 | Not-determined |
| 17-109_k141_400506 | Not-determined |
| 17-109_k141_16364 | Low-quality |
| 17-109_k141_351428 | Not-determined |
| 17-109_k141_66800 | Not-determined |
| 17-109_k141_33009 | Low-quality |
| 17-109_k141_351437 | Not-determined |
| 17-109_k141_100099 | Not-determined |
| 17-109_k141_33022 | Low-quality |
| 17-109_k141_366387 | Not-determined |
| 17-109_k141_133174 | Not-determined |
| 17-109_k141_66816 | Not-determined |
| 17-109_k141_366391 | Not-determined |
| 17-109_k141_33034 | Not-determined |
| 17-109_k141_133181 | Not-determined |
| 17-109_k141_250641 | Not-determined |
| 17-109_k141_366399 | Not-determined |
| 17-109_k141_149919 | Low-quality |
| 17-109_k141_366410 | Not-determined |
| 17-109_k141_400529 | Not-determined |
| 17-109_k141_149929 | Not-determined |
| 17-109_k141_149938 | Low-quality |
| 17-109_k141_133201 | Not-determined |
| 17-109_k141_266694 | Not-determined |
| 17-109_k141_133206 | Not-determined |
| 17-109_k141_100121 | Not-determined |
| 17-109_k141_116396 | Not-determined |
| 17-109_k141_116403 | Not-determined |
| 17-109_k141_66825 | Not-determined |
| 17-109_k141_66826 | Not-determined |
| 17-109_k141_116406 | Low-quality |
| 17-109_k141_133229 | Not-determined |
| 17-109_k141_149957 | Not-determined |
| 17-109_k141_33069 | Low-quality |
| 17-109_k141_149962 | Not-determined |
| 17-109_k141_149963 | Not-determined |
| 17-109_k141_266699 | Not-determined |
| 17-109_k141_66836 | Not-determined |
| 17-109_k141_100136 | Not-determined |
| 17-109_k141_66842 | Not-determined |
| 17-109_k141_149964 | Not-determined |
| 17-109_k141_100142 | Not-determined |
| 17-109_k141_66850 | Not-determined |
| 17-109_k141_100147 | Not-determined |
| 17-109_k141_366452 | Not-determined |
| 17-109_k141_366456 | Not-determined |
| 17-109_k141_100155 | Not-determined |
| 17-109_k141_149977 | Not-determined |
| 17-109_k141_266715 | Low-quality |
| 17-109_k141_149987 | Not-determined |
| 17-109_k141_366475 | Not-determined |
| 17-109_k141_400570 | Not-determined |
| 17-109_k141_400576 | Not-determined |
| 17-109_k141_400578 | Not-determined |
| 17-109_k141_400581 | Not-determined |
| 17-109_k141_66865 | Not-determined |
| 17-109_k141_351458 | Not-determined |
| 17-109_k141_66869 | Not-determined |
| 17-109_k141_351461 | Not-determined |
| 17-109_k141_351465 | Not-determined |
| 17-109_k141_351466 | Not-determined |
| 17-109_k141_301266 | Not-determined |
| 17-109_k141_351469 | Not-determined |
| 17-109_k141_351473 | Not-determined |
| 17-109_k141_266739 | Not-determined |
| 17-109_k141_266741 | Not-determined |
| 17-109_k141_366495 | Not-determined |
| 17-109_k141_66895 | Not-determined |
| 17-109_k141_33115 | Not-determined |
| 17-109_k141_301273 | Not-determined |
| 17-109_k141_100201 | Not-determined |
| 17-109_k141_100207 | Not-determined |
| 17-109_k141_116450 | Low-quality |
| 17-109_k141_16436 | Not-determined |
| 17-109_k141_366528 | Low-quality |
| 17-109_k141_16439 | Not-determined |
| 17-109_k141_133242 | Low-quality |
| 17-109_k141_16444 | Not-determined |
| 17-109_k141_266756 | Not-determined |
| 17-109_k141_266759 | Not-determined |
| 17-109_k141_100224 | Not-determined |
| 17-109_k141_366539 | Not-determined |
| 17-109_k141_16454 | Not-determined |
| 17-109_k141_384599 | Not-determined |
| 17-109_k141_400605 | Not-determined |
| 17-109_k141_400606 | Not-determined |
| 17-109_k141_150015 | Not-determined |
| 17-109_k141_384605 | Low-quality |
| 17-109_k141_266768 | Not-determined |
| 17-109_k141_66947 | Not-determined |
| 17-109_k141_366551 | Not-determined |
| 17-109_k141_351484 | Not-determined |
| 17-109_k141_33156 | Not-determined |
| 17-109_k141_150026 | Not-determined |
| 17-109_k141_366554 | Not-determined |
| 17-109_k141_100254 | Not-determined |
| 17-109_k141_33165 | Not-determined |
| 17-109_k141_100256 | Not-determined |
| 17-109_k141_266792 | Not-determined |
| 17-109_k141_100258 | Not-determined |
| 17-109_k141_33172 | Not-determined |
| 17-109_k141_116476 | Not-determined |
| 17-109_k141_266801 | Not-determined |
| 17-109_k141_400619 | Not-determined |
| 17-109_k141_266808 | Not-determined |
| 17-109_k141_150046 | Not-determined |
| 17-109_k141_100275 | Not-determined |
| 17-109_k141_16483 | Not-determined |
| 17-109_k141_266825 | Not-determined |
| 17-109_k141_133285 | Not-determined |
| 17-109_k141_116497 | Not-determined |
| 17-109_k141_16496 | Not-determined |
| 17-109_k141_100282 | Not-determined |
| 17-109_k141_133296 | Not-determined |
| 17-109_k141_116516 | Not-determined |
| 17-109_k141_16519 | Not-determined |
| 17-109_k141_250675 | Not-determined |
| 17-109_k141_116519 | Not-determined |
| 17-109_k141_266846 | Not-determined |
| 17-109_k141_150086 | Low-quality |
| 17-109_k141_116527 | Not-determined |
| 17-109_k141_250690 | Not-determined |
| 17-109_k141_250696 | Low-quality |
| 17-109_k141_116537 | Not-determined |
| 17-109_k141_250700 | Not-determined |
| 17-109_k141_16541 | Not-determined |
| 17-109_k141_133311 | Not-determined |
| 17-109_k141_100303 | Not-determined |
| 17-109_k141_266871 | Not-determined |
| 17-109_k141_266885 | Not-determined |
| 17-109_k141_400639 | Not-determined |
| 17-109_k141_100317 | Not-determined |
| 17-109_k141_16557 | Low-quality |
| 17-109_k141_366587 | Not-determined |
| 17-109_k141_366589 | Not-determined |
| 17-109_k141_16571 | Not-determined |
| 17-109_k141_100330 | Not-determined |
| 17-109_k141_250736 | Low-quality |
| 17-109_k141_100340 | Not-determined |
| 17-109_k141_133329 | Not-determined |
| 17-109_k141_150126 | Not-determined |
| 17-109_k141_116599 | Not-determined |
| 17-109_k141_133331 | Not-determined |
| 17-109_k141_116600 | Not-determined |
| 17-109_k141_150131 | Not-determined |
| 17-109_k141_100348 | Low-quality |
| 17-109_k141_100350 | Not-determined |
| 17-109_k141_266896 | Not-determined |
| 17-109_k141_266897 | Not-determined |
| 17-109_k141_400667 | Not-determined |
| 17-109_k141_366606 | Not-determined |
| 17-109_k141_384645 | Not-determined |
| 17-109_k141_150141 | Not-determined |
| 17-109_k141_400676 | Not-determined |
| 17-109_k141_33209 | Not-determined |
| 17-109_k141_133365 | Low-quality |
| 17-109_k141_384656 | Low-quality |
| 17-109_k141_384658 | Not-determined |
| 17-109_k141_384663 | Not-determined |
| 17-109_k141_366630 | Not-determined |
| 17-109_k141_100366 | Not-determined |
| 17-109_k141_100367 | Not-determined |
| 17-109_k141_266923 | Not-determined |
| 17-109_k141_250759 | Not-determined |
| 17-109_k141_266928 | Not-determined |
| 17-109_k141_150178 | Not-determined |
| 17-109_k141_100380 | Not-determined |
| 17-109_k141_100381 | Not-determined |
| 17-109_k141_33227 | Low-quality |
| 17-109_k141_384683 | Not-determined |
| 17-109_k141_400690 | Not-determined |
| 17-109_k141_400691 | Low-quality |
| 17-109_k141_100388 | Not-determined |
| 17-109_k141_400700 | Not-determined |
| 17-109_k141_400704 | Not-determined |
| 17-109_k141_366652 | Not-determined |
| 17-109_k141_133385 | Not-determined |
| 17-109_k141_33254 | Not-determined |
| 17-109_k141_366672 | Not-determined |
| 17-109_k141_250789 | Not-determined |
| 17-109_k141_400708 | Not-determined |
| 17-109_k141_366674 | Low-quality |
| 17-109_k141_133399 | Not-determined |
| 17-109_k141_116617 | Not-determined |
| 17-109_k141_400714 | Not-determined |
| 17-109_k141_366679 | Not-determined |
| 17-109_k141_116619 | Not-determined |
| 17-109_k141_133405 | Not-determined |
| 17-109_k141_16594 | Not-determined |
| 17-109_k141_33271 | Not-determined |
| 17-109_k141_250804 | Not-determined |
| 17-109_k141_116628 | Low-quality |
| 17-109_k141_266953 | Low-quality |
| 17-109_k141_116633 | Low-quality |
| 17-109_k141_384699 | Not-determined |
| 17-109_k141_33284 | Not-determined |
| 17-109_k141_100411 | Not-determined |
| 17-109_k141_100412 | Low-quality |
| 17-109_k141_400736 | Not-determined |
| 17-109_k141_33292 | Not-determined |
| 17-109_k141_116644 | Not-determined |
| 17-109_k141_33299 | Not-determined |
| 17-109_k141_33300 | Not-determined |
| 17-109_k141_33303 | Low-quality |
| 17-109_k141_366687 | Not-determined |
| 17-109_k141_100421 | Not-determined |
| 17-109_k141_133433 | Not-determined |
| 17-109_k141_266978 | Low-quality |
| 17-109_k141_16636 | Not-determined |
| 17-109_k141_116660 | Not-determined |
| 17-109_k141_116666 | Not-determined |
| 17-109_k141_384719 | Low-quality |
| 17-109_k141_116683 | Not-determined |
| 17-109_k141_150290 | Not-determined |
| 17-109_k141_266993 | Not-determined |
| 17-109_k141_384729 | Not-determined |
| 17-109_k141_266998 | Not-determined |
| 17-109_k141_16670 | Low-quality |
| 17-109_k141_16671 | Not-determined |
| 17-109_k141_267002 | Not-determined |
| 17-109_k141_267007 | Not-determined |
| 17-109_k141_267008 | Not-determined |
| 17-109_k141_267009 | Not-determined |
| 17-109_k141_384743 | Not-determined |
| 17-109_k141_267017 | Not-determined |
| 17-109_k141_384749 | Low-quality |
| 17-109_k141_133489 | Not-determined |
| 17-109_k141_116713 | Not-determined |
| 17-109_k141_267034 | Not-determined |
| 17-109_k141_384770 | Not-determined |
| 17-109_k141_400756 | Not-determined |
| 17-109_k141_400760 | Not-determined |
| 17-109_k141_400761 | Not-determined |
| 17-109_k141_366747 | Not-determined |
| 17-109_k141_16716 | Not-determined |
| 17-109_k141_250836 | Not-determined |
| 17-109_k141_400775 | Not-determined |
| 17-109_k141_400777 | Low-quality |
| 17-109_k141_400779 | Not-determined |
| 17-109_k141_116751 | Not-determined |
| 17-109_k141_384789 | Not-determined |
| 17-109_k141_250855 | Not-determined |
| 17-109_k141_150330 | Not-determined |
| 17-109_k141_250867 | Not-determined |
| 17-109_k141_366757 | Not-determined |
| 17-109_k141_150341 | Not-determined |

| | |
|---|---|
| 17-109_k141_250876 | Not-determined |
| 17-109_k141_384805 | Not-determined |
| 17-109_k141_250877 | Not-determined |
| 17-109_k141_384807 | Not-determined |
| 17-109_k141_267086 | Not-determined |
| 17-109_k141_133508 | Not-determined |
| 17-109_k141_400806 | Not-determined |
| 17-109_k141_267091 | Not-determined |
| 17-109_k141_150360 | Not-determined |
| 17-109_k141_250884 | Not-determined |
| 17-109_k141_400818 | Not-determined |
| 17-109_k141_133509 | Not-determined |
| 17-109_k141_150362 | Not-determined |
| 17-109_k141_267097 | Not-determined |
| 17-109_k141_250900 | Not-determined |
| 17-109_k141_150373 | Not-determined |
| 17-109_k141_267102 | Not-determined |
| 17-109_k141_116785 | Not-determined |
| 17-109_k141_384842 | Not-determined |
| 17-109_k141_267117 | Not-determined |
| 17-109_k141_366810 | Not-determined |
| 17-109_k141_267131 | Not-determined |
| 17-109_k141_150395 | Not-determined |
| 17-109_k141_267133 | Not-determined |
| 17-109_k141_116798 | Low-quality |
| 17-109_k141_116799 | Not-determined |
| 17-109_k141_267140 | Not-determined |
| 17-109_k141_33357 | Not-determined |
| 17-109_k141_250911 | Low-quality |
| 17-109_k141_384863 | Not-determined |
| 17-109_k141_384865 | Not-determined |
| 17-109_k141_400849 | Not-determined |
| 17-109_k141_267153 | Not-determined |
| 17-109_k141_116812 | Not-determined |
| 17-109_k141_384871 | Not-determined |
| 17-109_k141_400856 | Not-determined |
| 17-109_k141_267158 | Not-determined |
| 17-109_k141_400858 | Not-determined |
| 17-109_k141_400861 | Not-determined |
| 17-109_k141_116824 | Low-quality |
| 17-109_k141_267174 | Not-determined |
| 17-109_k141_150437 | Not-determined |
| 17-109_k141_267178 | Not-determined |
| 17-109_k141_400870 | Not-determined |
| 17-109_k141_150441 | Not-determined |
| 17-109_k141_366839 | Not-determined |
| 17-109_k141_116839 | Not-determined |
| 17-109_k141_384898 | Not-determined |
| 17-109_k141_150458 | Not-determined |
| 17-109_k141_366859 | Not-determined |
| 17-109_k141_366872 | Not-determined |
| 17-109_k141_400885 | Not-determined |
| 17-109_k141_384908 | Not-determined |
| 17-109_k141_384910 | Not-determined |
| 17-109_k141_384913 | Not-determined |
| 17-109_k141_133583 | Not-determined |
| 17-109_k141_250982 | Not-determined |
| 17-109_k141_267238 | Not-determined |
| 17-109_k141_366887 | Not-determined |
| 17-109_k141_366897 | Not-determined |
| 17-109_k141_267246 | Low-quality |
| 17-109_k141_366908 | Not-determined |
| 17-109_k141_384921 | Low-quality |
| 17-109_k141_116869 | Low-quality |
| 17-109_k141_366914 | Not-determined |
| 17-109_k141_366926 | Not-determined |
| 17-109_k141_150476 | Low-quality |
| 17-109_k141_400926 | Not-determined |
| 17-109_k141_267270 | Not-determined |
| 17-109_k141_267271 | Not-determined |
| 17-109_k141_400934 | Not-determined |
| 17-109_k141_33375 | Not-determined |
| 17-109_k141_267275 | Not-determined |
| 17-109_k141_150484 | Not-determined |
| 17-109_k141_133586 | Not-determined |
| 17-109_k141_133587 | Not-determined |
| 17-109_k141_133589 | Not-determined |
| 17-109_k141_33382 | Low-quality |
| 17-109_k141_366960 | Not-determined |
| 17-109_k141_366963 | Not-determined |
| 17-109_k141_133592 | Not-determined |
| 17-109_k141_400953 | Not-determined |
| 17-109_k141_33401 | Not-determined |
| 17-109_k141_366974 | Not-determined |
| 17-109_k141_150496 | Not-determined |
| 17-109_k141_150498 | Not-determined |
| 17-109_k141_366995 | Not-determined |
| 17-109_k141_366996 | Not-determined |
| 17-109_k141_150513 | Not-determined |
| 17-109_k141_150517 | Not-determined |
| 17-109_k141_133609 | Not-determined |
| 17-109_k141_367008 | Not-determined |
| 17-109_k141_367014 | Not-determined |
| 17-109_k141_33448 | Not-determined |
| 17-109_k141_116916 | Low-quality |
| 17-109_k141_367041 | Not-determined |
| 17-109_k141_367042 | Not-determined |
| 17-109_k141_150541 | Not-determined |
| 17-109_k141_33458 | Low-quality |
| 17-109_k141_367044 | Not-determined |
| 17-109_k141_133626 | Not-determined |
| 17-109_k141_367060 | Not-determined |
| 17-109_k141_367062 | Not-determined |
| 17-109_k141_33465 | Not-determined |
| 17-109_k141_150548 | Not-determined |
| 17-109_k141_150551 | Not-determined |
| 17-109_k141_150552 | Not-determined |
| 17-109_k141_33471 | Not-determined |
| 17-109_k141_116937 | Not-determined |
| 17-109_k141_150569 | Not-determined |
| 17-109_k141_367082 | Not-determined |
| 17-109_k141_150572 | Not-determined |
| 17-109_k141_367088 | Not-determined |
| 17-109_k141_367090 | Not-determined |
| 17-109_k141_150576 | Not-determined |
| 17-109_k141_384963 | Not-determined |
| 17-109_k141_150580 | Not-determined |
| 17-109_k141_367102 | Not-determined |
| 17-109_k141_400966 | Not-determined |
| 17-109_k141_400980 | Not-determined |
| 17-109_k141_116958 | Not-determined |
| 17-109_k141_400997 | Not-determined |
| 17-109_k141_401004 | Not-determined |
| 17-109_k141_133643 | Not-determined |
| 17-109_k141_267345 | Not-determined |
| 17-109_k141_133653 | Not-determined |
| 17-109_k141_401016 | Not-determined |
| 17-109_k141_116981 | Not-determined |
| 17-109_k141_267351 | Not-determined |
| 17-109_k141_401029 | Not-determined |
| 17-109_k141_367137 | Not-determined |
| 17-109_k141_401040 | Not-determined |
| 17-109_k141_401056 | Not-determined |
| 17-109_k141_267373 | Not-determined |
| 17-109_k141_117018 | Not-determined |
| 17-109_k141_133689 | Not-determined |
| 17-109_k141_117021 | Not-determined |
| 17-109_k141_401066 | Not-determined |
| 17-109_k141_117028 | Not-determined |
| 17-109_k141_150646 | Not-determined |
| 17-109_k141_117043 | Not-determined |
| 17-109_k141_401080 | Not-determined |
| 17-109_k141_401087 | Not-determined |
| 17-109_k141_367168 | Low-quality |
| 17-109_k141_401090 | Not-determined |
| 17-109_k141_117068 | Not-determined |
| 17-109_k141_367174 | Not-determined |
| 17-109_k141_267383 | Not-determined |
| 17-109_k141_117075 | Not-determined |
| 17-109_k141_367192 | Not-determined |
| 17-109_k141_117082 | Low-quality |
| 17-109_k141_367203 | Not-determined |
| 17-109_k141_117086 | Not-determined |
| 17-109_k141_267395 | Not-determined |
| 17-109_k141_267406 | Not-determined |
| 17-109_k141_133699 | Not-determined |
| 17-109_k141_367217 | Not-determined |
| 17-109_k141_367219 | Low-quality |
| 17-109_k141_117111 | Not-determined |
| 17-109_k141_117115 | Not-determined |
| 17-109_k141_401132 | Low-quality |
| 17-109_k141_367234 | Not-determined |
| 17-109_k141_117135 | Not-determined |
| 17-109_k141_401151 | Not-determined |
| 17-109_k141_367247 | Not-determined |
| 17-109_k141_367250 | Not-determined |
| 17-109_k141_267436 | Not-determined |
| 17-109_k141_367252 | Not-determined |
| 17-109_k141_267438 | Not-determined |
| 17-109_k141_367254 | Not-determined |
| 17-109_k141_117149 | Low-quality |
| 17-109_k141_367259 | Not-determined |
| 17-109_k141_367262 | Not-determined |
| 17-109_k141_367265 | Not-determined |
| 17-109_k141_267450 | Not-determined |
| 17-109_k141_401163 | Not-determined |
| 17-109_k141_133742 | Not-determined |
| 17-109_k141_367276 | Not-determined |
| 17-109_k141_117154 | Not-determined |
| 17-109_k141_401171 | Not-determined |
| 17-109_k141_367291 | Not-determined |
| 17-109_k141_133762 | Low-quality |
| 17-109_k141_367306 | Not-determined |
| 17-109_k141_133772 | Not-determined |
| 17-109_k141_133781 | Not-determined |
| 17-109_k141_267482 | Not-determined |
| 17-109_k141_267484 | Not-determined |
| 17-109_k141_267486 | Not-determined |
| 17-109_k141_267492 | Not-determined |
| 17-109_k141_401175 | Not-determined |
| 17-109_k141_133790 | Low-quality |
| 17-109_k141_133806 | Low-quality |
| 17-109_k141_133821 | Not-determined |
| 17-109_k141_133824 | Not-determined |
| 17-109_k141_133825 | Not-determined |
| 17-109_k141_401197 | Not-determined |
| 17-109_k141_367321 | Not-determined |
| 17-109_k141_133827 | Not-determined |
| 17-109_k141_401199 | Not-determined |
| 17-109_k141_401202 | Not-determined |
| 17-109_k141_367329 | Low-quality |
| 17-109_k141_367335 | Not-determined |
| 17-109_k141_401219 | Not-determined |
| 17-109_k141_401223 | Low-quality |
| 17-109_k141_401226 | Not-determined |
| 17-109_k141_401233 | Low-quality |
| 17-109_k141_401236 | Not-determined |
| 17-109_k141_401243 | Not-determined |
| 17-109_k141_401247 | Not-determined |
| 17-109_k141_367350 | Not-determined |
| 17-109_k141_401260 | Not-determined |
| 17-109_k141_133833 | Not-determined |
| 17-109_k141_133838 | Not-determined |
| 17-109_k141_133841 | Not-determined |
| 17-109_k141_133847 | Not-determined |
| 17-109_k141_367381 | Not-determined |
| 17-109_k141_133856 | Not-determined |
| 17-109_k141_367407 | Not-determined |
| 17-109_k141_133866 | Not-determined |
| 17-109_k141_267512 | Not-determined |
| 17-109_k141_367426 | Not-determined |
| 17-109_k141_401281 | Not-determined |
| 17-109_k141_267515 | Not-determined |
| 17-109_k141_267526 | Not-determined |
| 17-109_k141_401283 | Not-determined |
| 17-109_k141_367442 | Not-determined |
| 17-109_k141_133890 | Not-determined |
| 17-109_k141_267538 | Not-determined |
| 17-109_k141_133899 | Not-determined |
| 17-109_k141_267550 | Not-determined |
| 17-109_k141_401317 | Low-quality |
| 17-109_k141_267568 | Low-quality |
| 17-109_k141_367472 | Not-determined |
| 17-109_k141_367490 | Not-determined |
| 17-109_k141_367497 | Not-determined |
| 17-109_k141_267587 | Not-determined |
| 17-109_k141_267591 | Not-determined |
| 17-109_k141_267592 | Not-determined |
| 17-109_k141_401364 | Not-determined |
| 17-109_k141_267599 | Not-determined |
| 17-109_k141_367521 | Not-determined |
| 17-109_k141_267603 | Not-determined |
| 17-109_k141_367538 | Not-determined |
| 17-109_k141_267620 | Not-determined |
| 17-109_k141_367556 | Not-determined |
| 17-109_k141_367558 | Not-determined |
| 17-109_k141_267642 | Not-determined |
| 17-109_k141_267644 | Not-determined |
| 17-108-final-viral-combined_k141_172889\|\|full | Medium-quality |
| 17-109_k141_367603 | Not-determined |
| 17-109_k141_367616 | Not-determined |
| 17-109_k141_367617 | Not-determined |
| 17-109_k141_367620 | Not-determined |
| 17-109_k141_267680 | Not-determined |
| 17-109_k141_267683 | Not-determined |
| 17-109_k141_367640 | Not-determined |
| 17-109_k141_367649 | Not-determined |
| 17-109_k141_267703 | Not-determined |
| 17-109_k141_267717 | Not-determined |
| 17-109_k141_267721 | Not-determined |
| 17-109_k141_267727 | Low-quality |
| 17-109_k141_367663 | Not-determined |
| 17-109_k141_367665 | Not-determined |
| 17-109_k141_267739 | Not-determined |
| 17-109_k141_267749 | Not-determined |
| 17-109_k141_367689 | Not-determined |
| 17-109_k141_367718 | Not-determined |
| 17-109_k141_367740 | Not-determined |
| 17-108-final-viral-combined_k141_195425\|\|full | Medium-quality |
| 17-109_k141_367750 | Not-determined |
| 17-109_k141_367760 | Low-quality |
| 17-109_k141_367786 | Not-determined |
| 17-109_k141_367796 | Not-determined |
| 17-109_k141_367808 | Not-determined |
| 17-109_k141_401381 | Not-determined |
| 17-109_k141_367818 | Not-determined |
| 17-109_k141_401387 | Not-determined |
| 17-109_k141_267807 | Not-determined |
| 17-109_k141_401391 | Not-determined |
| 17-109_k141_401392 | Not-determined |
| 17-109_k141_367840 | Not-determined |
| 17-109_k141_267817 | Not-determined |
| 17-109_k141_367850 | Not-determined |
| 17-109_k141_367886 | Low-quality |
| 17-109_k141_367891 | Not-determined |
| 17-109_k141_367908 | Not-determined |
| 18-101-final-viral-combined_k141_279492\|\|full | Medium-quality |
| 17-109_k141_367919 | Not-determined |
| 17-109_k141_401413 | Not-determined |
| 17-109_k141_367926 | Not-determined |
| 17-109_k141_367928 | Not-determined |
| 17-109_k141_401422 | Not-determined |
| 17-109_k141_401423 | Not-determined |
| 17-109_k141_401426 | Not-determined |
| 17-109_k141_367950 | Not-determined |
| 17-109_k141_401437 | Not-determined |
| 17-109_k141_401445 | Not-determined |
| 17-109_k141_367958 | Not-determined |
| 17-109_k141_367960 | Not-determined |
| 17-109_k141_401453 | Not-determined |
| 17-109_k141_401460 | Not-determined |
| 17-109_k141_401477 | Not-determined |
| 17-109_k141_401478 | Not-determined |
| 17-109_k141_401479 | Not-determined |
| 17-109_k141_401480 | Not-determined |
| 17-109_k141_401484 | Not-determined |
| 17-109_k141_367973 | Not-determined |
| 17-109_k141_401503 | Not-determined |
| 17-109_k141_401506 | Not-determined |
| 17-109_k141_401510 | Not-determined |
| 17-109_k141_367983 | Not-determined |
| 17-109_k141_401521 | Not-determined |
| 17-109_k141_401524 | Not-determined |
| 17-109_k141_401525 | Not-determined |
| 17-109_k141_401529 | Not-determined |
| 17-109_k141_401532 | Not-determined |
| 17-109_k141_401536 | Not-determined |
| 17-109_k141_401537 | Not-determined |
| 17-109_k141_368003 | Not-determined |
| 17-109_k141_401561 | Not-determined |
| 17-109_k141_401565 | Not-determined |
| 17-109_k141_368024 | Not-determined |
| 17-109_k141_401572 | Not-determined |
| 17-109_k141_401586 | Not-determined |
| 17-109_k141_401587 | Not-determined |
| 17-109_k141_401608 | Not-determined |
| 17-109_k141_401611 | Not-determined |
| 17-109_k141_368037 | Low-quality |
| 17-109_k141_401630 | Not-determined |
| 17-109_k141_401631 | Not-determined |
| 17-109_k141_401632 | Not-determined |
| 17-109_k141_401644 | Not-determined |
| 17-109_k141_401646 | Not-determined |
| 17-109_k141_401653 | Not-determined |
| 17-109_k141_368058 | Not-determined |
| 17-109_k141_401661 | Not-determined |
| 17-109_k141_401666 | Not-determined |
| 17-109_k141_401671 | Not-determined |
| 17-109_k141_401673 | Not-determined |
| 17-109_k141_368071 | Not-determined |
| 17-109_k141_401686 | Not-determined |
| 17-109_k141_368084 | Not-determined |
| 17-109_k141_401702 | Low-quality |
| 17-109_k141_401703 | Not-determined |
| 17-109_k141_368087 | Not-determined |
| 17-109_k141_368088 | Low-quality |
| 17-109_k141_401713 | Not-determined |
| 17-109_k141_368089 | Not-determined |
| 17-109_k141_401715 | Not-determined |
| 17-109_k141_368136 | Not-determined |
| 17-109_k141_368145 | Low-quality |
| 17-109_k141_368169 | Not-determined |
| 17-109_k141_368197 | Not-determined |
| 17-109_k141_368209 | Not-determined |
| 17-109_k141_368215 | Not-determined |
| 17-109_k141_368217 | Not-determined |
| 17-109_k141_368231 | Not-determined |
| 17-109_k141_368232 | Not-determined |
| 17-109_k141_368249 | Not-determined |
| 17-109_k141_368253 | Not-determined |
| 18-102-final-viral-combined_k141_135289\|\|full | Not-determined |
| 17-109-final-viral-combined_k141_245755\|\|full | Medium-quality |
| 18-102-final-viral-combined_k141_125246\|\|full | Not-determined |
| 18-102-final-viral-combined_k141_177256\|\|full | Not-determined |
| 18-102-final-viral-combined_k141_128045\|\|full | Low-quality |
| 18-102-final-viral-combined_k141_15783\|\|full | Not-determined |
| 18-110_k141_167678 | Medium-quality |
| 18-110-final-viral-combined_k141_167678\|\|full | Medium-quality |
| 18-102-final-viral-combined_k141_50086\|\|full | Not-determined |
| 18-102-final-viral-combined_k141_235094\|\|full | Low-quality |
| 18-109-final-viral-combined_k141_121062\|\|full | Medium-quality |
| 18-102-final-viral-combined_k141_244863\|\|full | Not-determined |
| 18-110-final-viral-combined_k141_126492\|\|full | Medium-quality |
| 18-103-final-viral-combined_k141_135801\|\|full | Medium-quality |
| 18-101-final-viral-combined_k141_102801\|\|full | Medium-quality |
| 18-110_k141_152831 | Medium-quality |
| 18-102-final-viral-combined_k141_248325\|\|full | Not-determined |
| 18-104-final-viral-combined_k141_12422\|\|full | Medium-quality |
| 18-109-final-viral-combined_k141_239784\|\|full | Medium-quality |
| 18-102-final-viral-combined_k141_248339\|\|full | Not-determined |
| 18-102-final-viral-combined_k141_113920\|\|full | Not-determined |
| 18-102-final-viral-combined_k141_82854\|\|full | Low-quality |
| 18-102-final-viral-combined_k141_62146\|\|full | Not-determined |
| 18-102-final-viral-combined_k141_31082\|\|full | Not-determined |
| 18-102-final-viral-combined_k141_31083\|\|full | Not-determined |
| 18-102-final-viral-combined_k141_5\|\|full | Not-determined |
| 18-102-final-viral-combined_k141_196775\|\|full | Not-determined |
| 18-102-final-viral-combined_k141_72505\|\|full | Not-determined |
| 18-102-final-viral-combined_k141_134641\|\|full | Not-determined |
| 18-102-final-viral-combined_k141_186421\|\|full | Low-quality |
| 18-102-final-viral-combined_k141_72507\|\|full | Not-determined |
| 18-102-final-viral-combined_k141_62159\|\|full | Low-quality |
| 18-102-final-viral-combined_k141_196781\|\|full | Low-quality |
| 18-102-final-viral-combined_k141_155361\|\|full | Low-quality |
| 18-102-final-viral-combined_k141_134646\|\|full | Not-determined |
| 18-102-final-viral-combined_k141_72515\|\|full | Not-determined |
| 18-102-final-viral-combined_k141_217502\|\|full | Not-determined |
| 18-102-final-viral-combined_k141_207148\|\|full | Not-determined |
| 18-102-final-viral-combined_k141_51801\|\|full | Low-quality |
| 18-109_k141_151112 | Medium-quality |
| 18-102-final-viral-combined_k141_155386\|\|full | Low-quality |
| 18-102-final-viral-combined_k141_155387\|\|full | Not-determined |
| 18-102-final-viral-combined_k141_207156\|\|full | Not-determined |
| 18-102-final-viral-combined_k141_103640\|\|full | Not-determined |
| 18-102-final-viral-combined_k141_41482\|\|full | Low-quality |
| 18-102-final-viral-combined_k141_62183\|\|full | Low-quality |
| 18-102-final-viral-combined_k141_176098\|\|full | Not-determined |
| 18-102-final-viral-combined_k141_20743\|\|full | Low-quality |
| 18-102-final-viral-combined_k141_113951\|\|full | Not-determined |
| 18-102-final-viral-combined_k141_113952\|\|full | Low-quality |
| 18-102-final-viral-combined_k141_82879\|\|full | Not-determined |
| 18-102-final-viral-combined_k141_51811\|\|full | Low-quality |
| 18-102-final-viral-combined_k141_227855\|\|full | Not-determined |
| 18-102-final-viral-combined_k141_72535\|\|full | Low-quality |
| 18-102-final-viral-combined_k141_94\|\|full | Low-quality |
| 18-102-final-viral-combined_k141_176111\|\|full | Not-determined |
| 18-102-final-viral-combined_k141_134682\|\|full | Low-quality |
| 18-103-final-viral-combined_k141_135428\|\|full | Medium-quality |
| 18-102-final-viral-combined_k141_134693\|\|full | Low-quality |
| 18-102-final-viral-combined_k141_134695\|\|full | Not-determined |
| 18-102-final-viral-combined_k141_82924\|\|full | Low-quality |
| 18-102-final-viral-combined_k141_155402\|\|full | Low-quality |
| 18-102-final-viral-combined_k141_176138\|\|full | Not-determined |
| 18-102-final-viral-combined_k141_155413\|\|full | Low-quality |
| 18-102-final-viral-combined_k141_145007\|\|full | Low-quality |
| 18-102-final-viral-combined_k141_165747\|\|full | Not-determined |
| 18-102-final-viral-combined_k141_186481\|\|full | Not-determined |
| 18-102-final-viral-combined_k141_41557\|\|full | Not-determined |
| 18-102-final-viral-combined_k141_72542\|\|full | Not-determined |
| 18-102-final-viral-combined_k141_41559\|\|full | Not-determined |
| 18-102-final-viral-combined_k141_51857\|\|full | Not-determined |
| 18-102-final-viral-combined_k141_62199\|\|full | Low-quality |
| 18-102-final-viral-combined_k141_134747\|\|full | Not-determined |
| 18-102-final-viral-combined_k141_196818\|\|full | Not-determined |
| 18-102-final-viral-combined_k141_20766\|\|full | Low-quality |
| 18-102-final-viral-combined_k141_176176\|\|full | Not-determined |
| 18-102-final-viral-combined_k141_113980\|\|full | Not-determined |
| 18-102-final-viral-combined_k141_41586\|\|full | Low-quality |
| 18-102-final-viral-combined_k141_134758\|\|full | Low-quality |
| 18-102-final-viral-combined_k141_10444\|\|full | Not-determined |
| 18-102-final-viral-combined_k141_134759\|\|full | Low-quality |
| 18-102-final-viral-combined_k141_227900\|\|full | Low-quality |
| 18-102-final-viral-combined_k141_41598\|\|full | Not-determined |
| 18-102-final-viral-combined_k141_176190\|\|full | Low-quality |
| 18-102-final-viral-combined_k141_134776\|\|full | Not-determined |
| 18-102-final-viral-combined_k141_103785\|\|full | Low-quality |
| 18-102-final-viral-combined_k141_93257\|\|full | Not-determined |
| 18-102-final-viral-combined_k141_124\|\|full | Not-determined |
| 18-102-final-viral-combined_k141_227911\|\|full | Low-quality |
| 18-102-final-viral-combined_k141_20776\|\|full | Low-quality |
| 18-102-final-viral-combined_k141_51931\|\|full | Not-determined |
| 18-102-final-viral-combined_k141_20783\|\|full | Not-determined |
| 18-102-final-viral-combined_k141_134788\|\|full | Low-quality |
| 18-102-final-viral-combined_k141_227938\|\|full | Low-quality |
| 18-102-final-viral-combined_k141_31161\|\|full | Low-quality |
| 18-102-final-viral-combined_k141_165806\|\|full | Low-quality |
| 18-102-final-viral-combined_k141_238254\|\|full | Not-determined |
| 18-102-final-viral-combined_k141_196869\|\|full | Low-quality |
| 18-102-final-viral-combined_k141_31180\|\|full | Low-quality |
| 18-102-final-viral-combined_k141_51952\|\|full | Low-quality |
| 18-102-final-viral-combined_k141_130\|\|full | Not-determined |
| 18-102-final-viral-combined_k141_176263\|\|full | Low-quality |
| 18-102-final-viral-combined_k141_51954\|\|full | Low-quality |
| 18-102-final-viral-combined_k141_196876\|\|full | Low-quality |
| 18-102-final-viral-combined_k141_10499\|\|full | Not-determined |
| 18-102-final-viral-combined_k141_186567\|\|full | Low-quality |
| 18-102-final-viral-combined_k141_114024\|\|full | Low-quality |
| 18-102-final-viral-combined_k141_82995\|\|full | Low-quality |
| 18-102-final-viral-combined_k141_165832\|\|full | Not-determined |
| 18-102-final-viral-combined_k141_41626\|\|full | Not-determined |
| 18-102-final-viral-combined_k141_150\|\|full | Not-determined |
| 18-102-final-viral-combined_k141_134805\|\|full | Low-quality |
| 18-102-final-viral-combined_k141_51984\|\|full | Low-quality |
| 18-102-final-viral-combined_k141_176300\|\|full | Not-determined |
| 18-102-final-viral-combined_k141_196901\|\|full | Not-determined |
| 18-102-final-viral-combined_k141_31196\|\|full | Not-determined |
| 18-102-final-viral-combined_k141_155495\|\|full | Not-determined |
| 18-102-final-viral-combined_k141_31212\|\|full | Not-determined |
| 18-109-final-viral-combined_k141_151112\|\|full | Medium-quality |
| 18-102-final-viral-combined_k141_196924\|\|full | Low-quality |
| 18-102-final-viral-combined_k141_155497\|\|full | Not-determined |
| 18-102-final-viral-combined_k141_238298\|\|full | Low-quality |
| 18-102-final-viral-combined_k141_83017\|\|full | Low-quality |
| 18-102-final-viral-combined_k141_93339\|\|full | Not-determined |
| 18-102-final-viral-combined_k141_72568\|\|full | Not-determined |
| 18-102-final-viral-combined_k141_62281\|\|full | Not-determined |
| 18-102-final-viral-combined_k141_83023\|\|full | Not-determined |
| 18-102-final-viral-combined_k141_228003\|\|full | Low-quality |
| 18-102-final-viral-combined_k141_238301\|\|full | Low-quality |
| 18-102-final-viral-combined_k141_124410\|\|full | Low-quality |
| 18-102-final-viral-combined_k141_83036\|\|full | Low-quality |
| 18-102-final-viral-combined_k141_62294\|\|full | Low-quality |
| 18-102-final-viral-combined_k141_52005\|\|full | Low-quality |
| 18-102-final-viral-combined_k141_93344\|\|full | Low-quality |
| 18-102-final-viral-combined_k141_103853\|\|full | Low-quality |
| 18-102-final-viral-combined_k141_114045\|\|full | Low-quality |
| 18-102-final-viral-combined_k141_207191\|\|full | Low-quality |
| 18-102-final-viral-combined_k141_103858\|\|full | Low-quality |
| 18-102-final-viral-combined_k141_196964\|\|full | Not-determined |
| 18-102-final-viral-combined_k141_41682\|\|full | Not-determined |
| 18-102-final-viral-combined_k141_209\|\|full | Low-quality |
| 18-102-final-viral-combined_k141_114055\|\|full | Not-determined |
| 18-102-final-viral-combined_k141_155523\|\|full | Not-determined |
| 18-102-final-viral-combined_k141_238345\|\|full | Not-determined |
| 18-102-final-viral-combined_k141_228013\|\|full | Not-determined |
| 18-102-final-viral-combined_k141_72603\|\|full | Not-determined |
| 18-102-final-viral-combined_k141_41720\|\|full | Low-quality |
| 18-102-final-viral-combined_k141_228022\|\|full | Low-quality |
| 18-102-final-viral-combined_k141_217726\|\|full | Low-quality |
| 18-102-final-viral-combined_k141_93396\|\|full | Low-quality |
| 18-102-final-viral-combined_k141_134821\|\|full | Low-quality |
| 18-106-final-viral-combined_k141_49909\|\|full | Medium-quality |
| 18-102-final-viral-combined_k141_124426\|\|full | Low-quality |
| 18-102-final-viral-combined_k141_41729\|\|full | Not-determined |
| 18-110-final-viral-combined_k141_152831\|\|full | Medium-quality |
| 18-102-final-viral-combined_k141_207221\|\|full | Not-determined |
| 18-102-final-viral-combined_k141_176384\|\|full | Low-quality |
| 18-102-final-viral-combined_k141_228051\|\|full | Not-determined |
| 18-102-final-viral-combined_k141_228073\|\|full | Low-quality |
| 18-102-final-viral-combined_k141_93424\|\|full | Low-quality |
| 18-102-final-viral-combined_k141_176397\|\|full | Low-quality |
| 18-102-final-viral-combined_k141_228081\|\|full | Low-quality |
| 18-102-final-viral-combined_k141_238357\|\|full | Low-quality |
| 18-102-final-viral-combined_k141_242\|\|full | Not-determined |
| 18-102-final-viral-combined_k141_243\|\|full | Not-determined |
| 18-102-final-viral-combined_k141_83063\|\|full | Low-quality |
| 18-102-final-viral-combined_k141_114071\|\|full | Not-determined |
| 18-102-final-viral-combined_k141_134874\|\|full | Low-quality |
| 18-102-final-viral-combined_k141_114077\|\|full | Low-quality |
| 18-102-final-viral-combined_k141_83067\|\|full | Not-determined |
| 18-102-final-viral-combined_k141_103902\|\|full | Not-determined |
| 18-102-final-viral-combined_k141_31312\|\|full | Not-determined |
| 18-102-final-viral-combined_k141_134893\|\|full | Not-determined |
| 18-102-final-viral-combined_k141_41774\|\|full | Not-determined |
| 18-102-final-viral-combined_k141_134900\|\|full | Low-quality |
| 18-102-final-viral-combined_k141_20879\|\|full | Not-determined |
| 18-102-final-viral-combined_k141_103913\|\|full | Low-quality |
| 18-102-final-viral-combined_k141_134912\|\|full | Low-quality |
| 18-102-final-viral-combined_k141_62366\|\|full | Low-quality |
| 18-102-final-viral-combined_k141_124460\|\|full | Not-determined |
| 18-102-final-viral-combined_k141_103924\|\|full | Low-quality |
| 18-102-final-viral-combined_k141_62374\|\|full | Low-quality |
| 18-102-final-viral-combined_k141_31352\|\|full | Not-determined |
| 18-102-final-viral-combined_k141_176415\|\|full | Low-quality |
| 18-102-final-viral-combined_k141_176422\|\|full | Not-determined |
| 18-102-final-viral-combined_k141_217740\|\|full | Not-determined |
| 18-102-final-viral-combined_k141_31370\|\|full | Low-quality |
| 18-102-final-viral-combined_k141_217742\|\|full | Not-determined |
| 18-102-final-viral-combined_k141_186703\|\|full | Low-quality |
| 18-102-final-viral-combined_k141_124488\|\|full | Not-determined |
| 18-102-final-viral-combined_k141_228148\|\|full | Not-determined |
| 18-102-final-viral-combined_k141_298\|\|full | Not-determined |
| 18-102-final-viral-combined_k141_41818\|\|full | Low-quality |
| 18-102-final-viral-combined_k141_145164\|\|full | Low-quality |
| 18-102-final-viral-combined_k141_124498\|\|full | Low-quality |
| 18-102-final-viral-combined_k141_72674\|\|full | Not-determined |
| 18-102-final-viral-combined_k141_103941\|\|full | Low-quality |
| 18-102-final-viral-combined_k141_31398\|\|full | Not-determined |
| 18-102-final-viral-combined_k141_114092\|\|full | Low-quality |
| 18-102-final-viral-combined_k141_207251\|\|full | Low-quality |
| 18-102-final-viral-combined_k141_186718\|\|full | Low-quality |
| 18-102-final-viral-combined_k141_145178\|\|full | Not-determined |
| 18-102-final-viral-combined_k141_155612\|\|full | Low-quality |
| 18-102-final-viral-combined_k141_83111\|\|full | Not-determined |
| 18-102-final-viral-combined_k141_238421\|\|full | Low-quality |
| 18-102-final-viral-combined_k141_134964\|\|full | Low-quality |
| 18-102-final-viral-combined_k141_238425\|\|full | Low-quality |
| 18-102-final-viral-combined_k141_217769\|\|full | Not-determined |
| 18-102-final-viral-combined_k141_134971\|\|full | Low-quality |
| 18-102-final-viral-combined_k141_155615\|\|full | Low-quality |
| 18-102-final-viral-combined_k141_103956\|\|full | Not-determined |
| 18-102-final-viral-combined_k141_41861\|\|full | Not-determined |
| 18-102-final-viral-combined_k141_93497\|\|full | Not-determined |
| 18-102-final-viral-combined_k141_145207\|\|full | Low-quality |
| 18-102-final-viral-combined_k141_134976\|\|full | Low-quality |
| 18-102-final-viral-combined_k141_238439\|\|full | Not-determined |
| 18-102-final-viral-combined_k141_155626\|\|full | Not-determined |
| 18-102-final-viral-combined_k141_155632\|\|full | Not-determined |
| 18-102-final-viral-combined_k141_134979\|\|full | Not-determined |
| 18-102-final-viral-combined_k141_186736\|\|full | Low-quality |
| 18-102-final-viral-combined_k141_238444\|\|full | Low-quality |
| 18-102-final-viral-combined_k141_186737\|\|full | Not-determined |
| 18-102-final-viral-combined_k141_176455\|\|full | Low-quality |
| 18-102-final-viral-combined_k141_124554\|\|full | Not-determined |
| 18-102-final-viral-combined_k141_31426\|\|full | Low-quality |
| 17-108-final-viral-combined_k141_282227\|\|full | Medium-quality |
| 18-102-final-viral-combined_k141_93509\|\|full | Not-determined |
| 18-102-final-viral-combined_k141_228178\|\|full | Not-determined |
| 18-102-final-viral-combined_k141_228180\|\|full | Not-determined |

| | |
|---|---|
| 18-102-final-viral-combined_k141_320\|\|full | Not-determined |
| 18-102-final-viral-combined_k141_124575\|\|full | Not-determined |
| 18-102-final-viral-combined_k141_10559\|\|full | Low-quality |
| 18-102-final-viral-combined_k141_10564\|\|full | Not-determined |
| 18-102-final-viral-combined_k141_326\|\|full | Low-quality |
| 18-102-final-viral-combined_k141_62416\|\|full | Not-determined |
| 18-102-final-viral-combined_k141_145250\|\|full | Low-quality |
| 18-102-final-viral-combined_k141_10576\|\|full | Not-determined |
| 18-102-final-viral-combined_k141_41914\|\|full | Not-determined |
| 18-102-final-viral-combined_k141_335\|\|full | Low-quality |
| 18-102-final-viral-combined_k141_41924\|\|full | Not-determined |
| 18-102-final-viral-combined_k141_207276\|\|full | Low-quality |
| 18-102-final-viral-combined_k141_41929\|\|full | Low-quality |
| KM-final-viral-combined_k141_70983\|\|full | Medium-quality |
| 18-102-final-viral-combined_k141_207286\|\|full | Low-quality |
| 18-102-final-viral-combined_k141_207287\|\|full | Low-quality |
| 18-102-final-viral-combined_k141_217831\|\|full | Low-quality |
| 18-102-final-viral-combined_k141_186785\|\|full | Low-quality |
| 18-102-final-viral-combined_k141_354\|\|full | Not-determined |
| 18-109-final-viral-combined_k141_31068\|\|full | Medium-quality |
| 18-102-final-viral-combined_k141_135024\|\|full | Not-determined |
| 18-102-final-viral-combined_k141_10608\|\|full | Not-determined |
| 18-102-final-viral-combined_k141_62480\|\|full | Low-quality |
| 18-102-final-viral-combined_k141_72774\|\|full | Not-determined |
| 18-102-final-viral-combined_k141_217849\|\|full | Low-quality |
| 18-102-final-viral-combined_k141_114138\|\|full | Not-determined |
| 18-102-final-viral-combined_k141_41972\|\|full | Low-quality |
| 18-102-final-viral-combined_k141_228233\|\|full | Low-quality |
| 18-102-final-viral-combined_k141_145329\|\|full | Low-quality |
| 18-102-final-viral-combined_k141_145331\|\|full | Not-determined |
| 18-102-final-viral-combined_k141_124630\|\|full | Low-quality |
| 18-102-final-viral-combined_k141_83135\|\|full | Low-quality |
| 18-102-final-viral-combined_k141_207345\|\|full | Low-quality |
| 18-102-final-viral-combined_k141_145339\|\|full | Low-quality |
| 18-102-final-viral-combined_k141_135056\|\|full | Not-determined |
| 18-102-final-viral-combined_k141_83144\|\|full | Low-quality |
| 18-102-final-viral-combined_k141_145341\|\|full | Not-determined |
| 18-102-final-viral-combined_k141_145344\|\|full | Low-quality |
| 18-102-final-viral-combined_k141_135062\|\|full | Not-determined |
| 18-102-final-viral-combined_k141_72810\|\|full | Low-quality |
| 18-102-final-viral-combined_k141_41989\|\|full | Not-determined |
| 18-102-final-viral-combined_k141_155690\|\|full | Low-quality |
| 18-102-final-viral-combined_k141_93575\|\|full | Not-determined |
| 18-102-final-viral-combined_k141_238519\|\|full | Not-determined |
| 18-102-final-viral-combined_k141_72828\|\|full | Low-quality |
| 18-102-final-viral-combined_k141_114150\|\|full | Not-determined |
| 18-102-final-viral-combined_k141_238538\|\|full | Not-determined |
| 18-102-final-viral-combined_k141_176516\|\|full | Not-determined |
| 18-102-final-viral-combined_k141_135100\|\|full | Low-quality |
| 18-102-final-viral-combined_k141_371\|\|full | Not-determined |
| 18-102-final-viral-combined_k141_165960\|\|full | Low-quality |
| 18-102-final-viral-combined_k141_124700\|\|full | Not-determined |
| 18-102-final-viral-combined_k141_228280\|\|full | Low-quality |
| 18-102-final-viral-combined_k141_124701\|\|full | Not-determined |
| 18-102-final-viral-combined_k141_114182\|\|full | Low-quality |
| 18-102-final-viral-combined_k141_83184\|\|full | Not-determined |
| 18-102-final-viral-combined_k141_93614\|\|full | Not-determined |
| 18-102-final-viral-combined_k141_155737\|\|full | Low-quality |
| 18-102-final-viral-combined_k141_72847\|\|full | Not-determined |
| 18-102-final-viral-combined_k141_145361\|\|full | Low-quality |
| 18-102-final-viral-combined_k141_42043\|\|full | Low-quality |
| 18-102-final-viral-combined_k141_166006\|\|full | Not-determined |
| 18-102-final-viral-combined_k141_155748\|\|full | Low-quality |
| 18-102-final-viral-combined_k141_186809\|\|full | Low-quality |
| 18-102-final-viral-combined_k141_20971\|\|full | Not-determined |
| 18-102-final-viral-combined_k141_135112\|\|full | Low-quality |
| 18-102-final-viral-combined_k141_42053\|\|full | Low-quality |
| 18-102-final-viral-combined_k141_155751\|\|full | Low-quality |
| 18-102-final-viral-combined_k141_83203\|\|full | Not-determined |
| 18-102-final-viral-combined_k141_135121\|\|full | Not-determined |
| 18-102-final-viral-combined_k141_135125\|\|full | Low-quality |
| 18-102-final-viral-combined_k141_10631\|\|full | Not-determined |
| 18-102-final-viral-combined_k141_10633\|\|full | Low-quality |
| 18-102-final-viral-combined_k141_10634\|\|full | Not-determined |
| 18-102-final-viral-combined_k141_83225\|\|full | Not-determined |
| 18-102-final-viral-combined_k141_207420\|\|full | Low-quality |
| 18-102-final-viral-combined_k141_155769\|\|full | Not-determined |
| 18-102-final-viral-combined_k141_42096\|\|full | Low-quality |
| 18-102-final-viral-combined_k141_83242\|\|full | Low-quality |
| 18-102-final-viral-combined_k141_83247\|\|full | Low-quality |
| 18-102-final-viral-combined_k141_10651\|\|full | Low-quality |
| 18-102-final-viral-combined_k141_83254\|\|full | Not-determined |
| 18-102-final-viral-combined_k141_145428\|\|full | Not-determined |
| 18-102-final-viral-combined_k141_186842\|\|full | Low-quality |
| 18-102-final-viral-combined_k141_228309\|\|full | Low-quality |
| 18-110-final-viral-combined_k141_146562\|\|full | Medium-quality |
| 18-102-final-viral-combined_k141_10659\|\|full | Low-quality |
| 18-102-final-viral-combined_k141_207441\|\|full | Not-determined |
| 18-102-final-viral-combined_k141_31489\|\|full | Not-determined |
| 18-102-final-viral-combined_k141_420\|\|full | Low-quality |
| 18-102-final-viral-combined_k141_42126\|\|full | Not-determined |
| 18-102-final-viral-combined_k141_155788\|\|full | Low-quality |
| 18-102-final-viral-combined_k141_114239\|\|full | Not-determined |
| 18-102-final-viral-combined_k141_72911\|\|full | Low-quality |
| 18-102-final-viral-combined_k141_421\|\|full | Low-quality |
| 18-102-final-viral-combined_k141_196986\|\|full | Low-quality |
| 18-102-final-viral-combined_k141_155790\|\|full | Not-determined |
| 18-102-final-viral-combined_k141_228325\|\|full | Low-quality |
| 18-102-final-viral-combined_k141_10680\|\|full | Low-quality |
| 18-102-final-viral-combined_k141_453\|\|full | Low-quality |
| 18-102-final-viral-combined_k141_124746\|\|full | Low-quality |
| 18-102-final-viral-combined_k141_93682\|\|full | Not-determined |
| 18-102-final-viral-combined_k141_10698\|\|full | Not-determined |
| 18-102-final-viral-combined_k141_93690\|\|full | Low-quality |
| 18-102-final-viral-combined_k141_238687\|\|full | Not-determined |
| 18-102-final-viral-combined_k141_238689\|\|full | Low-quality |
| 18-102-final-viral-combined_k141_21055\|\|full | Low-quality |
| 18-102-final-viral-combined_k141_470\|\|full | Not-determined |
| 18-102-final-viral-combined_k141_21060\|\|full | Low-quality |
| 18-102-final-viral-combined_k141_124758\|\|full | Low-quality |
| 18-102-final-viral-combined_k141_72949\|\|full | Not-determined |
| 18-101-final-viral-combined_k141_186878\|\|full | Medium-quality |
| 18-102-final-viral-combined_k141_186930\|\|full | Not-determined |
| 18-102-final-viral-combined_k141_72950\|\|full | Not-determined |
| 18-102-final-viral-combined_k141_114260\|\|full | Not-determined |
| 18-102-final-viral-combined_k141_482\|\|full | Low-quality |
| 18-102-final-viral-combined_k141_10736\|\|full | Not-determined |
| 18-102-final-viral-combined_k141_186956\|\|full | Low-quality |
| 18-102-final-viral-combined_k141_10740\|\|full | Low-quality |
| 18-102-final-viral-combined_k141_197015\|\|full | Not-determined |
| 18-102-final-viral-combined_k141_166098\|\|full | Not-determined |
| 18-102-final-viral-combined_k141_42138\|\|full | Not-determined |
| 18-102-final-viral-combined_k141_135149\|\|full | Low-quality |
| 18-102-final-viral-combined_k141_155834\|\|full | Not-determined |
| 18-102-final-viral-combined_k141_238724\|\|full | Low-quality |
| 18-102-final-viral-combined_k141_124775\|\|full | Not-determined |
| 18-102-final-viral-combined_k141_42156\|\|full | Not-determined |
| 17-108-final-viral-combined_k141_119215\|\|full | Medium-quality |
| 18-102-final-viral-combined_k141_207488\|\|full | Low-quality |
| 18-102-final-viral-combined_k141_72991\|\|full | Not-determined |
| 18-102-final-viral-combined_k141_52213\|\|full | Not-determined |
| 18-102-final-viral-combined_k141_124813\|\|full | Low-quality |
| 18-102-final-viral-combined_k141_10778\|\|full | Not-determined |
| 18-102-final-viral-combined_k141_207526\|\|full | Low-quality |
| 18-102-final-viral-combined_k141_10791\|\|full | Low-quality |
| 18-102-final-viral-combined_k141_155924\|\|full | Low-quality |
| KM-final-viral-combined_k141_199440\|\|full | Medium-quality |
| 18-102-final-viral-combined_k141_197030\|\|full | Not-determined |
| 18-102-final-viral-combined_k141_42204\|\|full | Not-determined |
| 18-102-final-viral-combined_k141_21150\|\|full | Not-determined |
| 18-102-final-viral-combined_k141_197043\|\|full | Not-determined |
| 18-102-final-viral-combined_k141_145530\|\|full | Not-determined |
| 18-102-final-viral-combined_k141_124864\|\|full | Low-quality |
| 18-102-final-viral-combined_k141_207584\|\|full | Not-determined |
| 18-102-final-viral-combined_k141_145564\|\|full | Low-quality |
| 18-102-final-viral-combined_k141_145565\|\|full | Not-determined |
| 18-102-final-viral-combined_k141_124874\|\|full | Low-quality |
| 18-102-final-viral-combined_k141_83324\|\|full | Not-determined |
| 18-102-final-viral-combined_k141_52265\|\|full | Not-determined |
| 18-102-final-viral-combined_k141_73061\|\|full | Not-determined |
| 18-102-final-viral-combined_k141_83335\|\|full | Low-quality |
| 18-102-final-viral-combined_k141_52266\|\|full | Not-determined |
| 18-102-final-viral-combined_k141_73063\|\|full | Low-quality |
| 18-102-final-viral-combined_k141_145579\|\|full | Low-quality |
| 18-102-final-viral-combined_k141_83340\|\|full | Low-quality |
| 18-102-final-viral-combined_k141_104126\|\|full | Low-quality |
| 18-102-final-viral-combined_k141_31654\|\|full | Low-quality |
| 18-102-final-viral-combined_k141_83352\|\|full | Not-determined |
| 18-102-final-viral-combined_k141_52284\|\|full | Not-determined |
| 18-102-final-viral-combined_k141_238834\|\|full | Not-determined |
| 18-102-final-viral-combined_k141_62615\|\|full | Not-determined |
| 18-102-final-viral-combined_k141_42287\|\|full | Low-quality |
| 18-102-final-viral-combined_k141_155952\|\|full | Low-quality |
| 18-102-final-viral-combined_k141_31664\|\|full | Not-determined |
| 18-102-final-viral-combined_k141_124921\|\|full | Not-determined |
| 18-102-final-viral-combined_k141_42307\|\|full | Not-determined |
| 18-102-final-viral-combined_k141_52304\|\|full | Not-determined |
| 18-102-final-viral-combined_k141_31677\|\|full | Not-determined |
| 18-102-final-viral-combined_k141_572\|\|full | Not-determined |
| 18-102-final-viral-combined_k141_42313\|\|full | Low-quality |
| 18-102-final-viral-combined_k141_42317\|\|full | Low-quality |
| 18-102-final-viral-combined_k141_31693\|\|full | Not-determined |
| 18-102-final-viral-combined_k141_21267\|\|full | Not-determined |
| 18-102-final-viral-combined_k141_42342\|\|full | Not-determined |
| 18-102-final-viral-combined_k141_31707\|\|full | Not-determined |
| 18-102-final-viral-combined_k141_155997\|\|full | Not-determined |
| 18-102-final-viral-combined_k141_73132\|\|full | Not-determined |
| 18-102-final-viral-combined_k141_176676\|\|full | Not-determined |
| 18-102-final-viral-combined_k141_166175\|\|full | Not-determined |
| 18-102-final-viral-combined_k141_42359\|\|full | Not-determined |
| 18-102-final-viral-combined_k141_156015\|\|full | Not-determined |
| 18-102-final-viral-combined_k141_145631\|\|full | Low-quality |
| 18-102-final-viral-combined_k141_228435\|\|full | Low-quality |
| 18-102-final-viral-combined_k141_156017\|\|full | Not-determined |
| 18-102-final-viral-combined_k141_238912\|\|full | Low-quality |
| 18-102-final-viral-combined_k141_218100\|\|full | Low-quality |
| 18-102-final-viral-combined_k141_207659\|\|full | Not-determined |
| 18-102-final-viral-combined_k141_197066\|\|full | Low-quality |
| 18-102-final-viral-combined_k141_187086\|\|full | Not-determined |
| 18-102-final-viral-combined_k141_166193\|\|full | Not-determined |
| 18-102-final-viral-combined_k141_135228\|\|full | Not-determined |
| 18-102-final-viral-combined_k141_52343\|\|full | Low-quality |
| 18-102-final-viral-combined_k141_187089\|\|full | Low-quality |
| 18-102-final-viral-combined_k141_42371\|\|full | Low-quality |
| 18-102-final-viral-combined_k141_156052\|\|full | Not-determined |
| 18-102-final-viral-combined_k141_104192\|\|full | Not-determined |
| 18-102-final-viral-combined_k141_207696\|\|full | Not-determined |
| 18-102-final-viral-combined_k141_42391\|\|full | Low-quality |
| 18-102-final-viral-combined_k141_104200\|\|full | Low-quality |
| 18-102-final-viral-combined_k141_104201\|\|full | Low-quality |
| 18-102-final-viral-combined_k141_93753\|\|full | Low-quality |
| 18-102-final-viral-combined_k141_228451\|\|full | Not-determined |
| 18-102-final-viral-combined_k141_207720\|\|full | Low-quality |
| 18-102-final-viral-combined_k141_31743\|\|full | Low-quality |
| 18-102-final-viral-combined_k141_228464\|\|full | Low-quality |
| 18-102-final-viral-combined_k141_93762\|\|full | Low-quality |
| 18-102-final-viral-combined_k141_166234\|\|full | Not-determined |
| 18-102-final-viral-combined_k141_135284\|\|full | Not-determined |
| 18-102-final-viral-combined_k141_31753\|\|full | Low-quality |
| 18-102-final-viral-combined_k141_104208\|\|full | Not-determined |
| 18-102-final-viral-combined_k141_10849\|\|full | Not-determined |
| 18-102-final-viral-combined_k141_176727\|\|full | Low-quality |
| 18-102-final-viral-combined_k141_207735\|\|full | Low-quality |
| 18-102-final-viral-combined_k141_156069\|\|full | Low-quality |
| 18-101-final-viral-combined_k141_340558\|\|full | Medium-quality |
| 18-102-final-viral-combined_k141_228478\|\|full | Low-quality |
| 18-102-final-viral-combined_k141_176760\|\|full | Not-determined |
| 18-102-final-viral-combined_k141_52347\|\|full | Not-determined |
| 18-102-final-viral-combined_k141_73186\|\|full | Not-determined |
| 18-102-final-viral-combined_k141_93800\|\|full | Not-determined |
| 18-102-final-viral-combined_k141_197149\|\|full | Not-determined |
| 18-102-final-viral-combined_k141_207761\|\|full | Not-determined |
| 18-102-final-viral-combined_k141_52356\|\|full | Low-quality |
| 18-102-final-viral-combined_k141_197154\|\|full | Low-quality |
| 18-102-final-viral-combined_k141_21370\|\|full | Low-quality |
| 18-102-final-viral-combined_k141_21377\|\|full | Low-quality |
| 18-102-final-viral-combined_k141_207765\|\|full | Not-determined |
| 18-102-final-viral-combined_k141_93814\|\|full | Low-quality |
| 18-102-final-viral-combined_k141_166270\|\|full | Low-quality |
| 18-102-final-viral-combined_k141_21384\|\|full | Low-quality |
| 18-102-final-viral-combined_k141_10932\|\|full | Low-quality |
| 18-102-final-viral-combined_k141_228507\|\|full | Low-quality |
| 18-102-final-viral-combined_k141_705\|\|full | Not-determined |
| 18-102-final-viral-combined_k141_197167\|\|full | Low-quality |
| 18-102-final-viral-combined_k141_21394\|\|full | Not-determined |
| 18-102-final-viral-combined_k141_145713\|\|full | Low-quality |
| 17-109-final-viral-combined_k141_129554\|\|full | Medium-quality |
| 18-102-final-viral-combined_k141_83423\|\|full | Not-determined |
| 18-102-final-viral-combined_k141_114313\|\|full | Low-quality |
| 18-102-final-viral-combined_k141_228531\|\|full | Low-quality |
| 18-102-final-viral-combined_k141_125003\|\|full | Not-determined |
| 18-102-final-viral-combined_k141_83433\|\|full | Not-determined |
| 18-102-final-viral-combined_k141_10972\|\|full | Low-quality |
| 18-102-final-viral-combined_k141_724\|\|full | Low-quality |
| 18-102-final-viral-combined_k141_176790\|\|full | Not-determined |
| 18-102-final-viral-combined_k141_207784\|\|full | Not-determined |
| 18-102-final-viral-combined_k141_156102\|\|full | Not-determined |
| 18-102-final-viral-combined_k141_736\|\|full | Low-quality |
| 18-102-final-viral-combined_k141_31803\|\|full | Not-determined |
| 18-102-final-viral-combined_k141_42498\|\|full | Low-quality |
| 18-102-final-viral-combined_k141_166358\|\|full | Not-determined |
| 18-102-final-viral-combined_k141_228587\|\|full | Not-determined |
| 18-102-final-viral-combined_k141_21440\|\|full | Not-determined |
| 18-102-final-viral-combined_k141_62655\|\|full | Low-quality |
| 18-102-final-viral-combined_k141_228612\|\|full | Low-quality |
| 18-102-final-viral-combined_k141_104277\|\|full | Not-determined |
| 18-102-final-viral-combined_k141_11046\|\|full | Low-quality |
| 18-102-final-viral-combined_k141_114351\|\|full | Not-determined |
| 18-102-final-viral-combined_k141_228645\|\|full | Not-determined |
| 18-102-final-viral-combined_k141_21497\|\|full | Not-determined |
| 18-102-final-viral-combined_k141_187140\|\|full | Low-quality |
| 18-102-final-viral-combined_k141_42586\|\|full | Not-determined |
| 18-102-final-viral-combined_k141_114361\|\|full | Low-quality |
| 18-102-final-viral-combined_k141_114372\|\|full | Not-determined |
| 18-102-final-viral-combined_k141_187162\|\|full | Low-quality |
| 18-102-final-viral-combined_k141_21529\|\|full | Not-determined |
| 18-102-final-viral-combined_k141_11070\|\|full | Not-determined |
| 18-102-final-viral-combined_k141_187180\|\|full | Not-determined |
| 18-102-final-viral-combined_k141_176886\|\|full | Not-determined |
| 18-102-final-viral-combined_k141_176896\|\|full | Low-quality |
| 18-102-final-viral-combined_k141_93834\|\|full | Low-quality |
| 18-102-final-viral-combined_k141_145788\|\|full | Low-quality |
| 18-102-final-viral-combined_k141_166415\|\|full | Not-determined |
| 18-102-final-viral-combined_k141_145792\|\|full | Low-quality |
| 18-102-final-viral-combined_k141_176905\|\|full | Low-quality |
| 18-102-final-viral-combined_k141_187210\|\|full | Low-quality |
| 18-102-final-viral-combined_k141_104308\|\|full | Not-determined |
| 18-102-final-viral-combined_k141_187211\|\|full | Not-determined |
| 18-102-final-viral-combined_k141_93841\|\|full | Low-quality |
| 18-102-final-viral-combined_k141_228710\|\|full | Not-determined |
| 18-102-final-viral-combined_k141_813\|\|full | Low-quality |
| 18-102-final-viral-combined_k141_73231\|\|full | Not-determined |
| 18-102-final-viral-combined_k141_145813\|\|full | Not-determined |
| 18-102-final-viral-combined_k141_114418\|\|full | Low-quality |
| 18-102-final-viral-combined_k141_207840\|\|full | Low-quality |
| 18-102-final-viral-combined_k141_145816\|\|full | Low-quality |
| 18-102-final-viral-combined_k141_197236\|\|full | Not-determined |
| 18-102-final-viral-combined_k141_207852\|\|full | Low-quality |
| 18-102-final-viral-combined_k141_42655\|\|full | Low-quality |
| 18-102-final-viral-combined_k141_125071\|\|full | Low-quality |
| 18-102-final-viral-combined_k141_156116\|\|full | Low-quality |
| 18-102-final-viral-combined_k141_218125\|\|full | Not-determined |
| 18-102-final-viral-combined_k141_42662\|\|full | Low-quality |
| 18-102-final-viral-combined_k141_218132\|\|full | Low-quality |
| 18-102-final-viral-combined_k141_114448\|\|full | Not-determined |
| 18-102-final-viral-combined_k141_156134\|\|full | Not-determined |
| 18-102-final-viral-combined_k141_166457\|\|full | Low-quality |
| 18-102-final-viral-combined_k141_52440\|\|full | Low-quality |
| 18-102-final-viral-combined_k141_21591\|\|full | Low-quality |
| 18-102-final-viral-combined_k141_114465\|\|full | Not-determined |
| 18-102-final-viral-combined_k141_228742\|\|full | Low-quality |
| 18-102-final-viral-combined_k141_62712\|\|full | Low-quality |
| 18-102-final-viral-combined_k141_52462\|\|full | Not-determined |
| 18-102-final-viral-combined_k141_73280\|\|full | Not-determined |
| 18-102-final-viral-combined_k141_207876\|\|full | Low-quality |
| 18-110-final-viral-combined_k141_117281\|\|full | Medium-quality |
| 18-102-final-viral-combined_k141_52472\|\|full | Not-determined |
| 18-102-final-viral-combined_k141_156165\|\|full | Low-quality |
| 18-102-final-viral-combined_k141_52473\|\|full | Not-determined |
| 18-102-final-viral-combined_k141_42687\|\|full | Low-quality |
| 18-102-final-viral-combined_k141_218169\|\|full | Not-determined |
| 18-102-final-viral-combined_k141_125081\|\|full | Not-determined |
| 18-102-final-viral-combined_k141_104355\|\|full | Not-determined |
| 18-102-final-viral-combined_k141_73300\|\|full | Not-determined |
| 18-102-final-viral-combined_k141_62727\|\|full | Not-determined |
| 18-102-final-viral-combined_k141_176935\|\|full | Not-determined |
| 18-102-final-viral-combined_k141_11099\|\|full | Low-quality |
| 18-102-final-viral-combined_k141_125094\|\|full | Not-determined |
| 18-102-final-viral-combined_k141_62736\|\|full | Low-quality |
| 18-102-final-viral-combined_k141_228760\|\|full | Low-quality |
| 18-102-final-viral-combined_k141_187219\|\|full | Low-quality |
| 18-102-final-viral-combined_k141_83556\|\|full | Low-quality |
| 18-102-final-viral-combined_k141_885\|\|full | Low-quality |
| 18-102-final-viral-combined_k141_228768\|\|full | Low-quality |
| 18-102-final-viral-combined_k141_895\|\|full | Not-determined |
| 18-102-final-viral-combined_k141_218200\|\|full | Not-determined |
| 18-102-final-viral-combined_k141_176970\|\|full | Not-determined |
| 18-102-final-viral-combined_k141_52499\|\|full | Low-quality |
| 18-102-final-viral-combined_k141_145913\|\|full | Not-determined |
| 18-102-final-viral-combined_k141_114525\|\|full | Not-determined |
| 18-102-final-viral-combined_k141_83568\|\|full | Not-determined |
| 18-102-final-viral-combined_k141_898\|\|full | Low-quality |
| 18-102-final-viral-combined_k141_176994\|\|full | Low-quality |
| 18-102-final-viral-combined_k141_83595\|\|full | Not-determined |
| 18-102-final-viral-combined_k141_166507\|\|full | Not-determined |
| 18-102-final-viral-combined_k141_218221\|\|full | Not-determined |
| 18-102-final-viral-combined_k141_104387\|\|full | Not-determined |
| 18-102-final-viral-combined_k141_923\|\|full | Low-quality |
| 18-102-final-viral-combined_k141_31918\|\|full | Low-quality |
| 18-102-final-viral-combined_k141_11138\|\|full | Low-quality |
| 18-102-final-viral-combined_k141_125138\|\|full | Not-determined |
| 18-102-final-viral-combined_k141_83614\|\|full | Low-quality |
| 18-102-final-viral-combined_k141_228839\|\|full | Low-quality |
| 18-102-final-viral-combined_k141_135364\|\|full | Not-determined |
| 18-102-final-viral-combined_k141_11146\|\|full | Not-determined |
| 18-102-final-viral-combined_k141_177032\|\|full | Low-quality |
| 18-102-final-viral-combined_k141_104402\|\|full | Low-quality |
| 18-102-final-viral-combined_k141_228847\|\|full | Not-determined |
| 18-102-final-viral-combined_k141_62776\|\|full | Low-quality |
| 18-102-final-viral-combined_k141_207914\|\|full | Not-determined |
| 18-102-final-viral-combined_k141_125151\|\|full | Low-quality |
| 18-102-final-viral-combined_k141_197284\|\|full | Low-quality |
| 18-102-final-viral-combined_k141_228871\|\|full | Not-determined |
| 18-102-final-viral-combined_k141_125155\|\|full | Not-determined |
| 18-102-final-viral-combined_k141_218287\|\|full | Not-determined |
| 18-102-final-viral-combined_k141_11168\|\|full | Not-determined |
| 18-102-final-viral-combined_k141_125184\|\|full | Not-determined |
| 18-102-final-viral-combined_k141_135386\|\|full | Not-determined |
| 18-102-final-viral-combined_k141_177097\|\|full | Not-determined |
| 18-102-final-viral-combined_k141_166548\|\|full | Low-quality |
| 18-102-final-viral-combined_k141_104439\|\|full | Low-quality |
| 18-102-final-viral-combined_k141_135396\|\|full | Low-quality |
| 18-102-final-viral-combined_k141_62782\|\|full | Low-quality |
| 17-109-final-viral-combined_k141_260671\|\|full | Medium-quality |
| 18-102-final-viral-combined_k141_941\|\|full | Not-determined |
| 18-102-final-viral-combined_k141_62783\|\|full | Not-determined |
| 18-102-final-viral-combined_k141_114586\|\|full | Low-quality |
| 18-102-final-viral-combined_k141_166564\|\|full | Not-determined |
| 18-102-final-viral-combined_k141_145936\|\|full | Not-determined |
| 18-102-final-viral-combined_k141_156218\|\|full | Not-determined |
| 18-102-final-viral-combined_k141_104459\|\|full | Not-determined |
| 18-102-final-viral-combined_k141_73345\|\|full | Not-determined |
| 18-102-final-viral-combined_k141_83679\|\|full | Low-quality |
| 18-102-final-viral-combined_k141_11204\|\|full | Not-determined |
| 18-102-final-viral-combined_k141_228916\|\|full | Not-determined |
| 18-102-final-viral-combined_k141_177130\|\|full | Low-quality |
| 18-102-final-viral-combined_k141_228917\|\|full | Not-determined |
| 18-102-final-viral-combined_k141_197296\|\|full | Not-determined |
| 18-102-final-viral-combined_k141_104467\|\|full | Low-quality |
| 18-102-final-viral-combined_k141_93974\|\|full | Low-quality |
| 18-102-final-viral-combined_k141_42769\|\|full | Not-determined |
| 18-102-final-viral-combined_k141_238945\|\|full | Not-determined |
| 18-102-final-viral-combined_k141_83696\|\|full | Not-determined |
| 18-102-final-viral-combined_k141_207941\|\|full | Not-determined |
| 18-102-final-viral-combined_k141_83698\|\|full | Not-determined |
| 18-102-final-viral-combined_k141_145952\|\|full | Not-determined |
| 18-102-final-viral-combined_k141_238956\|\|full | Not-determined |
| 18-102-final-viral-combined_k141_145960\|\|full | Not-determined |
| 18-102-final-viral-combined_k141_83705\|\|full | Not-determined |
| 18-102-final-viral-combined_k141_125210\|\|full | Low-quality |
| 18-101-final-viral-combined_k141_106746\|\|full | Medium-quality |
| 18-102-final-viral-combined_k141_197313\|\|full | Low-quality |
| 18-102-final-viral-combined_k141_114609\|\|full | Not-determined |
| 18-102-final-viral-combined_k141_93988\|\|full | Low-quality |
| 18-102-final-viral-combined_k141_207961\|\|full | Low-quality |
| 18-102-final-viral-combined_k141_156272\|\|full | Low-quality |
| 18-102-final-viral-combined_k141_156273\|\|full | Low-quality |
| 18-102-final-viral-combined_k141_135455\|\|full | Not-determined |
| 18-102-final-viral-combined_k141_125228\|\|full | Not-determined |
| 18-102-final-viral-combined_k141_238981\|\|full | Not-determined |
| 18-102-final-viral-combined_k141_125229\|\|full | Low-quality |
| 18-102-final-viral-combined_k141_11237\|\|full | Not-determined |
| 18-102-final-viral-combined_k141_21734\|\|full | Low-quality |
| 18-102-final-viral-combined_k141_177155\|\|full | Low-quality |
| 18-102-final-viral-combined_k141_207975\|\|full | Low-quality |
| 18-102-final-viral-combined_k141_228940\|\|full | Not-determined |
| 18-102-final-viral-combined_k141_21748\|\|full | Not-determined |
| 18-102-final-viral-combined_k141_11256\|\|full | Low-quality |
| 18-102-final-viral-combined_k141_156287\|\|full | Low-quality |
| 18-102-final-viral-combined_k141_239010\|\|full | Not-determined |
| 18-102-final-viral-combined_k141_166595\|\|full | Not-determined |
| 18-102-final-viral-combined_k141_135478\|\|full | Low-quality |
| 18-102-final-viral-combined_k141_62824\|\|full | Not-determined |
| 18-102-final-viral-combined_k141_239023\|\|full | Not-determined |
| 18-102-final-viral-combined_k141_52608\|\|full | Low-quality |
| 18-102-final-viral-combined_k141_197350\|\|full | Not-determined |
| 18-102-final-viral-combined_k141_32010\|\|full | Low-quality |
| 18-102-final-viral-combined_k141_94000\|\|full | Low-quality |
| 18-102-final-viral-combined_k141_228974\|\|full | Low-quality |
| 18-102-final-viral-combined_k141_73372\|\|full | Not-determined |
| 18-102-final-viral-combined_k141_32021\|\|full | Not-determined |
| 18-102-final-viral-combined_k141_125274\|\|full | Not-determined |
| 18-102-final-viral-combined_k141_218351\|\|full | Low-quality |
| 18-102-final-viral-combined_k141_125294\|\|full | Not-determined |
| 18-102-final-viral-combined_k141_218355\|\|full | Not-determined |
| 18-102-final-viral-combined_k141_21767\|\|full | Low-quality |
| 18-102-final-viral-combined_k141_52646\|\|full | Not-determined |
| 18-102-final-viral-combined_k141_32027\|\|full | Not-determined |
| 18-102-final-viral-combined_k141_208008\|\|full | Not-determined |
| 18-102-final-viral-combined_k141_42835\|\|full | Low-quality |
| 18-102-final-viral-combined_k141_188780\|\|full | Medium-quality |
| 18-102-final-viral-combined_k141_125322\|\|full | Low-quality |
| 18-102-final-viral-combined_k141_156320\|\|full | Low-quality |
| 18-102-final-viral-combined_k141_73395\|\|full | Not-determined |
| 18-102-final-viral-combined_k141_42847\|\|full | Low-quality |
| 18-102-final-viral-combined_k141_32050\|\|full | Low-quality |
| 18-102-final-viral-combined_k141_156323\|\|full | Low-quality |
| 18-102-final-viral-combined_k141_11336\|\|full | Not-determined |
| 18-102-final-viral-combined_k141_83755\|\|full | Not-determined |
| 18-102-final-viral-combined_k141_73404\|\|full | Not-determined |
| 18-102-final-viral-combined_k141_125365\|\|full | Not-determined |
| 18-102-final-viral-combined_k141_73410\|\|full | Not-determined |
| 18-102-final-viral-combined_k141_156334\|\|full | Not-determined |
| 18-102-final-viral-combined_k141_208078\|\|full | Low-quality |
| 18-102-final-viral-combined_k141_114669\|\|full | Low-quality |
| 18-102-final-viral-combined_k141_21791\|\|full | Low-quality |
| 18-102-final-viral-combined_k141_58121\|\|full | Medium-quality |
| 18-102-final-viral-combined_k141_104574\|\|full | Low-quality |
| 18-102-final-viral-combined_k141_114705\|\|full | Low-quality |
| 18-102-final-viral-combined_k141_114710\|\|full | Low-quality |
| 18-102-final-viral-combined_k141_146095\|\|full | Low-quality |
| 18-102-final-viral-combined_k141_125370\|\|full | Low-quality |
| 18-102-final-viral-combined_k141_11372\|\|full | Not-determined |
| 18-102-final-viral-combined_k141_996\|\|full | Not-determined |
| 18-102-final-viral-combined_k141_166684\|\|full | Not-determined |
| 18-102-final-viral-combined_k141_239032\|\|full | Low-quality |
| 18-102-final-viral-combined_k141_104611\|\|full | Low-quality |
| 18-102-final-viral-combined_k141_1005\|\|full | Not-determined |
| 18-102-final-viral-combined_k141_42934\|\|full | Not-determined |
| 18-102-final-viral-combined_k141_177333\|\|full | Low-quality |
| 18-102-final-viral-combined_k141_83872\|\|full | Low-quality |
| 18-104-final-viral-combined_k141_22012\|\|full | Medium-quality |
| 18-102-final-viral-combined_k141_1018\|\|full | Not-determined |
| 18-102-final-viral-combined_k141_62841\|\|full | Low-quality |
| 18-102-final-viral-combined_k141_166703\|\|full | Not-determined |
| 18-102-final-viral-combined_k141_73504\|\|full | Not-determined |
| 18-102-final-viral-combined_k141_83885\|\|full | Not-determined |
| 18-102-final-viral-combined_k141_156417\|\|full | Low-quality |
| 18-102-final-viral-combined_k141_114733\|\|full | Low-quality |
| 18-102-final-viral-combined_k141_73512\|\|full | Low-quality |
| 18-102-final-viral-combined_k141_11436\|\|full | Low-quality |
| 18-102-final-viral-combined_k141_218432\|\|full | Low-quality |
| 18-102-final-viral-combined_k141_229062\|\|full | Low-quality |
| 18-102-final-viral-combined_k141_83896\|\|full | Low-quality |
| 18-102-final-viral-combined_k141_156420\|\|full | Not-determined |
| 18-102-final-viral-combined_k141_229077\|\|full | Low-quality |
| 18-102-final-viral-combined_k141_73542\|\|full | Not-determined |
| 18-102-final-viral-combined_k141_146104\|\|full | Not-determined |
| 18-102-final-viral-combined_k141_229085\|\|full | Not-determined |
| 18-102-final-viral-combined_k141_104673\|\|full | Low-quality |
| 18-102-final-viral-combined_k141_104674\|\|full | Not-determined |
| 18-102-final-viral-combined_k141_11465\|\|full | Not-determined |
| 18-102-final-viral-combined_k141_166730\|\|full | Low-quality |
| 18-102-final-viral-combined_k141_146120\|\|full | Low-quality |
| 18-102-final-viral-combined_k141_239083\|\|full | Not-determined |
| 18-102-final-viral-combined_k141_114768\|\|full | Low-quality |
| 18-102-final-viral-combined_k141_83918\|\|full | Not-determined |
| 18-102-final-viral-combined_k141_21832\|\|full | Not-determined |
| 18-102-final-viral-combined_k141_146139\|\|full | Not-determined |
| 18-102-final-viral-combined_k141_52691\|\|full | Not-determined |
| 17-109_k141_188744 | Medium-quality |
| 18-102-final-viral-combined_k141_43000\|\|full | Not-determined |
| 18-102-final-viral-combined_k141_1049\|\|full | Low-quality |
| 18-102-final-viral-combined_k141_104682\|\|full | Low-quality |
| 18-102-final-viral-combined_k141_229099\|\|full | Not-determined |
| 18-102-final-viral-combined_k141_239122\|\|full | Not-determined |
| 18-102-final-viral-combined_k141_229101\|\|full | Not-determined |
| 18-102-final-viral-combined_k141_114796\|\|full | Not-determined |
| 18-102-final-viral-combined_k141_114807\|\|full | Not-determined |
| 18-102-final-viral-combined_k141_32122\|\|full | Low-quality |
| 18-102-final-viral-combined_k141_187344\|\|full | Low-quality |
| 18-102-final-viral-combined_k141_114846\|\|full | Not-determined |
| 18-102-final-viral-combined_k141_146167\|\|full | Not-determined |
| 18-102-final-viral-combined_k141_187356\|\|full | Low-quality |
| 18-102-final-viral-combined_k141_62908\|\|full | Not-determined |
| 18-102-final-viral-combined_k141_187375\|\|full | Low-quality |
| 18-102-final-viral-combined_k141_73629\|\|full | Not-determined |
| 18-102-final-viral-combined_k141_135613\|\|full | Not-determined |
| 18-102-final-viral-combined_k141_114862\|\|full | Not-determined |
| 18-102-final-viral-combined_k141_187376\|\|full | Not-determined |
| 18-102-final-viral-combined_k141_135615\|\|full | Not-determined |
| 18-102-final-viral-combined_k141_177379\|\|full | Not-determined |
| 18-102-final-viral-combined_k141_177382\|\|full | Not-determined |
| 18-102-final-viral-combined_k141_177393\|\|full | Not-determined |
| 18-102-final-viral-combined_k141_166817\|\|full | Not-determined |
| 18-102-final-viral-combined_k141_229120\|\|full | Not-determined |
| 18-102-final-viral-combined_k141_21918\|\|full | Low-quality |
| 18-102-final-viral-combined_k141_187414\|\|full | Low-quality |
| 18-102-final-viral-combined_k141_146195\|\|full | Low-quality |
| 18-102-final-viral-combined_k141_166825\|\|full | Not-determined |
| 18-102-final-viral-combined_k141_52764\|\|full | Low-quality |
| 18-102-final-viral-combined_k141_125414\|\|full | Low-quality |
| 18-102-final-viral-combined_k141_166835\|\|full | Not-determined |
| 18-102-final-viral-combined_k141_146212\|\|full | Low-quality |
| 18-102-final-viral-combined_k141_166845\|\|full | Low-quality |
| 18-102-final-viral-combined_k141_146215\|\|full | Not-determined |
| 18-102-final-viral-combined_k141_197534\|\|full | Low-quality |
| 18-102-final-viral-combined_k141_135640\|\|full | Not-determined |
| 18-102-final-viral-combined_k141_104711\|\|full | Not-determined |
| 18-102-final-viral-combined_k141_52777\|\|full | Low-quality |
| 18-102-final-viral-combined_k141_32127\|\|full | Not-determined |
| 18-102-final-viral-combined_k141_104716\|\|full | Not-determined |
| 18-102-final-viral-combined_k141_43083\|\|full | Low-quality |
| 18-102-final-viral-combined_k141_125430\|\|full | Not-determined |
| 18-102-final-viral-combined_k141_32132\|\|full | Not-determined |
| 18-102-final-viral-combined_k141_32135\|\|full | Not-determined |
| 18-102-final-viral-combined_k141_156480\|\|full | Not-determined |
| 18-102-final-viral-combined_k141_166869\|\|full | Low-quality |
| 18-102-final-viral-combined_k141_208192\|\|full | Not-determined |
| 18-102-final-viral-combined_k141_166875\|\|full | Low-quality |
| 18-102-final-viral-combined_k141_84063\|\|full | Not-determined |
| 18-102-final-viral-combined_k141_208204\|\|full | Not-determined |
| 18-102-final-viral-combined_k141_73720\|\|full | Low-quality |
| 18-102-final-viral-combined_k141_84076\|\|full | Low-quality |
| 18-102-final-viral-combined_k141_125449\|\|full | Low-quality |
| 18-102-final-viral-combined_k141_94221\|\|full | Not-determined |
| 18-102-final-viral-combined_k141_146262\|\|full | Low-quality |
| 18-102-final-viral-combined_k141_125463\|\|full | Low-quality |
| 18-102-final-viral-combined_k141_146270\|\|full | Not-determined |
| 18-102-final-viral-combined_k141_43127\|\|full | Not-determined |
| 18-102-final-viral-combined_k141_146275\|\|full | Not-determined |
| 18-102-final-viral-combined_k141_52813\|\|full | Not-determined |
| 18-102-final-viral-combined_k141_177407\|\|full | Not-determined |
| 18-102-final-viral-combined_k141_73739\|\|full | Low-quality |
| 18-102-final-viral-combined_k141_43132\|\|full | Low-quality |
| 18-102-final-viral-combined_k141_125489\|\|full | Not-determined |
| 18-102-final-viral-combined_k141_125492\|\|full | Low-quality |
| 18-102-final-viral-combined_k141_62941\|\|full | Not-determined |
| 18-102-final-viral-combined_k141_84096\|\|full | Not-determined |
| 18-102-final-viral-combined_k141_43140\|\|full | Not-determined |
| 18-102-final-viral-combined_k141_146301\|\|full | Low-quality |
| 18-102-final-viral-combined_k141_43142\|\|full | Low-quality |
| 18-102-final-viral-combined_k141_125493\|\|full | Low-quality |
| 18-102-final-viral-combined_k141_187473\|\|full | Not-determined |
| 18-102-final-viral-combined_k141_73770\|\|full | Not-determined |
| 18-102-final-viral-combined_k141_239246\|\|full | Low-quality |
| 18-102-final-viral-combined_k141_1184\|\|full | Low-quality |
| 18-102-final-viral-combined_k141_177448\|\|full | Low-quality |
| 18-102-final-viral-combined_k141_73780\|\|full | Not-determined |
| 18-102-final-viral-combined_k141_52837\|\|full | Low-quality |
| 18-102-final-viral-combined_k141_229172\|\|full | Low-quality |
| 18-102-final-viral-combined_k141_94265\|\|full | Low-quality |
| 18-102-final-viral-combined_k141_177471\|\|full | Not-determined |
| 18-102-final-viral-combined_k141_187497\|\|full | Low-quality |
| 18-102-final-viral-combined_k141_146330\|\|full | Low-quality |
| 18-102-final-viral-combined_k141_43181\|\|full | Not-determined |
| 18-102-final-viral-combined_k141_1205\|\|full | Not-determined |
| 18-102-final-viral-combined_k141_11636\|\|full | Low-quality |
| 18-102-final-viral-combined_k141_22016\|\|full | Low-quality |
| 18-102-final-viral-combined_k141_177475\|\|full | Low-quality |
| 18-102-final-viral-combined_k141_187505\|\|full | Low-quality |
| 17-109-final-viral-combined_k141_188744\|\|full | Medium-quality |
| 18-102-final-viral-combined_k141_177484\|\|full | Low-quality |
| 18-102-final-viral-combined_k141_166968\|\|full | Not-determined |
| 18-102-final-viral-combined_k141_52866\|\|full | Low-quality |
| 18-102-final-viral-combined_k141_1222\|\|full | Not-determined |
| 18-102-final-viral-combined_k141_1226\|\|full | Not-determined |
| 18-102-final-viral-combined_k141_197589\|\|full | Low-quality |
| 18-102-final-viral-combined_k141_22042\|\|full | Not-determined |

| | |
|---|---|
| 18-102-final-viral-combined_k141_177509\|\|full | Not-determined |
| 18-102-final-viral-combined_k141_22054\|\|full | Not-determined |
| 18-102-final-viral-combined_k141_84133\|\|full | Not-determined |
| 18-102-final-viral-combined_k141_229220\|\|full | Low-quality |
| 18-102-final-viral-combined_k141_1252\|\|full | Not-determined |
| 18-102-final-viral-combined_k141_135722\|\|full | Not-determined |
| 18-102-final-viral-combined_k141_239308\|\|full | Low-quality |
| 18-102-final-viral-combined_k141_146394\|\|full | Not-determined |
| 18-102-final-viral-combined_k141_43265\|\|full | Not-determined |
| 18-102-final-viral-combined_k141_239314\|\|full | Low-quality |
| 18-102-final-viral-combined_k141_167041\|\|full | Low-quality |
| 18-102-final-viral-combined_k141_135750\|\|full | Not-determined |
| 18-102-final-viral-combined_k141_104771\|\|full | Low-quality |
| 18-102-final-viral-combined_k141_125523\|\|full | Not-determined |
| 18-102-final-viral-combined_k141_135755\|\|full | Not-determined |
| 18-102-final-viral-combined_k141_135760\|\|full | Low-quality |
| 18-102-final-viral-combined_k141_229242\|\|full | Not-determined |
| 18-102-final-viral-combined_k141_63005\|\|full | Low-quality |
| 18-102-final-viral-combined_k141_73894\|\|full | Low-quality |
| 18-102-final-viral-combined_k141_63013\|\|full | Not-determined |
| 18-102-final-viral-combined_k141_11701\|\|full | Low-quality |
| 18-102-final-viral-combined_k141_229248\|\|full | Not-determined |
| 18-102-final-viral-combined_k141_167083\|\|full | Not-determined |
| 18-102-final-viral-combined_k141_63018\|\|full | Low-quality |
| 18-102-final-viral-combined_k141_208294\|\|full | Not-determined |
| 18-102-final-viral-combined_k141_197650\|\|full | Low-quality |
| 18-102-final-viral-combined_k141_146419\|\|full | Low-quality |
| 18-102-final-viral-combined_k141_104824\|\|full | Low-quality |
| 18-102-final-viral-combined_k141_104828\|\|full | Not-determined |
| 18-102-final-viral-combined_k141_167099\|\|full | Not-determined |
| 18-102-final-viral-combined_k141_208298\|\|full | Not-determined |
| 18-102-final-viral-combined_k141_197665\|\|full | Low-quality |
| 18-102-final-viral-combined_k141_208318\|\|full | Not-determined |
| 18-102-final-viral-combined_k141_197668\|\|full | Not-determined |
| 18-102-final-viral-combined_k141_229294\|\|full | Not-determined |
| 18-102-final-viral-combined_k141_135804\|\|full | Not-determined |
| 18-102-final-viral-combined_k141_104842\|\|full | Low-quality |
| 18-102-final-viral-combined_k141_239384\|\|full | Low-quality |
| 18-102-final-viral-combined_k141_167119\|\|full | Not-determined |
| 18-102-final-viral-combined_k141_239400\|\|full | Low-quality |
| 18-102-final-viral-combined_k141_43341\|\|full | Not-determined |
| 18-102-final-viral-combined_k141_197679\|\|full | Low-quality |
| 18-102-final-viral-combined_k141_11741\|\|full | Not-determined |
| 18-102-final-viral-combined_k141_229308\|\|full | Low-quality |
| 18-102-final-viral-combined_k141_114947\|\|full | Not-determined |
| 18-102-final-viral-combined_k141_11745\|\|full | Not-determined |
| 18-102-final-viral-combined_k141_63035\|\|full | Not-determined |
| 18-102-final-viral-combined_k141_1282\|\|full | Low-quality |
| 18-102-final-viral-combined_k141_94424\|\|full | Not-determined |
| 18-102-final-viral-combined_k141_229315\|\|full | Not-determined |
| 18-102-final-viral-combined_k141_11746\|\|full | Not-determined |
| 18-102-final-viral-combined_k141_167133\|\|full | Low-quality |
| 18-102-final-viral-combined_k141_104871\|\|full | Not-determined |
| 18-102-final-viral-combined_k141_94435\|\|full | Not-determined |
| 18-102-final-viral-combined_k141_114961\|\|full | Not-determined |
| 18-102-final-viral-combined_k141_197689\|\|full | Not-determined |
| 18-102-final-viral-combined_k141_43375\|\|full | Low-quality |
| 18-102-final-viral-combined_k141_114964\|\|full | Not-determined |
| 18-102-final-viral-combined_k141_125573\|\|full | Not-determined |
| 18-102-final-viral-combined_k141_239411\|\|full | Not-determined |
| 18-102-final-viral-combined_k141_208360\|\|full | Not-determined |
| 18-102-final-viral-combined_k141_229361\|\|full | Low-quality |
| 18-102-final-viral-combined_k141_84292\|\|full | Not-determined |
| 18-102-final-viral-combined_k141_146447\|\|full | Not-determined |
| 18-102-final-viral-combined_k141_94477\|\|full | Not-determined |
| 18-102-final-viral-combined_k141_94478\|\|full | Not-determined |
| 18-102-final-viral-combined_k141_94479\|\|full | Low-quality |
| 18-102-final-viral-combined_k141_208374\|\|full | Low-quality |
| 18-102-final-viral-combined_k141_11828\|\|full | Not-determined |
| 18-102-final-viral-combined_k141_167195\|\|full | Low-quality |
| 18-102-final-viral-combined_k141_1309\|\|full | Low-quality |
| 18-102-final-viral-combined_k141_63118\|\|full | Not-determined |
| 18-102-final-viral-combined_k141_218586\|\|full | Low-quality |
| 18-102-final-viral-combined_k141_52921\|\|full | Low-quality |
| 18-102-final-viral-combined_k141_63137\|\|full | Not-determined |
| 18-102-final-viral-combined_k141_187621\|\|full | Low-quality |
| 18-102-final-viral-combined_k141_104892\|\|full | Not-determined |
| 18-102-final-viral-combined_k141_197702\|\|full | Low-quality |
| 18-102-final-viral-combined_k141_208405\|\|full | Low-quality |
| 18-102-final-viral-combined_k141_146492\|\|full | Not-determined |
| 18-102-final-viral-combined_k141_94510\|\|full | Not-determined |
| 18-102-final-viral-combined_k141_239433\|\|full | Not-determined |
| 18-102-final-viral-combined_k141_1351\|\|full | Not-determined |
| 18-102-final-viral-combined_k141_146496\|\|full | Not-determined |
| 18-102-final-viral-combined_k141_63150\|\|full | Not-determined |
| 18-102-final-viral-combined_k141_167271\|\|full | Not-determined |
| 18-102-final-viral-combined_k141_208428\|\|full | Low-quality |
| 18-102-final-viral-combined_k141_84315\|\|full | Not-determined |
| 18-102-final-viral-combined_k141_115001\|\|full | Not-determined |
| 18-102-final-viral-combined_k141_84325\|\|full | Not-determined |
| 18-102-final-viral-combined_k141_73944\|\|full | Not-determined |
| 18-102-final-viral-combined_k141_94533\|\|full | Not-determined |
| 18-102-final-viral-combined_k141_187687\|\|full | Not-determined |
| 18-102-final-viral-combined_k141_218671\|\|full | Not-determined |
| 18-102-final-viral-combined_k141_135925\|\|full | Not-determined |
| 18-102-final-viral-combined_k141_197733\|\|full | Low-quality |
| 18-102-final-viral-combined_k141_239450\|\|full | Not-determined |
| 18-102-final-viral-combined_k141_218675\|\|full | Not-determined |
| 18-102-final-viral-combined_k141_43459\|\|full | Not-determined |
| 18-102-final-viral-combined_k141_218682\|\|full | Not-determined |
| 18-102-final-viral-combined_k141_73982\|\|full | Not-determined |
| 18-102-final-viral-combined_k141_115036\|\|full | Low-quality |
| 18-102-final-viral-combined_k141_197753\|\|full | Not-determined |
| 18-102-final-viral-combined_k141_146543\|\|full | Low-quality |
| 18-102-final-viral-combined_k141_63228\|\|full | Low-quality |
| 18-102-final-viral-combined_k141_156635\|\|full | Not-determined |
| 18-102-final-viral-combined_k141_167341\|\|full | Not-determined |
| 18-102-final-viral-combined_k141_84370\|\|full | Not-determined |
| 18-102-final-viral-combined_k141_156647\|\|full | Not-determined |
| 18-102-final-viral-combined_k141_43516\|\|full | Not-determined |
| 18-102-final-viral-combined_k141_156648\|\|full | Low-quality |
| 18-102-final-viral-combined_k141_63266\|\|full | Not-determined |
| 18-102-final-viral-combined_k141_197773\|\|full | Not-determined |
| 18-102-final-viral-combined_k141_229408\|\|full | Not-determined |
| 18-102-final-viral-combined_k141_208516\|\|full | Low-quality |
| 18-102-final-viral-combined_k141_94623\|\|full | Low-quality |
| 18-102-final-viral-combined_k141_43533\|\|full | Not-determined |
| 18-102-final-viral-combined_k141_239481\|\|full | Low-quality |
| 18-102-final-viral-combined_k141_115057\|\|full | Not-determined |
| 18-102-final-viral-combined_k141_167373\|\|full | Not-determined |
| 18-102-final-viral-combined_k141_167374\|\|full | Not-determined |
| 18-102-final-viral-combined_k141_63293\|\|full | Low-quality |
| 18-102-final-viral-combined_k141_22278\|\|full | Not-determined |
| 18-102-final-viral-combined_k141_104975\|\|full | Low-quality |
| 18-102-final-viral-combined_k141_115072\|\|full | Not-determined |
| 18-102-final-viral-combined_k141_197804\|\|full | Low-quality |
| 18-102-final-viral-combined_k141_43548\|\|full | Not-determined |
| 18-102-final-viral-combined_k141_43554\|\|full | Not-determined |
| 18-102-final-viral-combined_k141_167397\|\|full | Low-quality |
| 18-102-final-viral-combined_k141_146573\|\|full | Not-determined |
| 18-102-final-viral-combined_k141_43564\|\|full | Not-determined |
| 18-102-final-viral-combined_k141_167401\|\|full | Not-determined |
| 18-102-final-viral-combined_k141_43568\|\|full | Not-determined |
| 18-102-final-viral-combined_k141_1393\|\|full | Not-determined |
| 18-102-final-viral-combined_k141_84390\|\|full | Not-determined |
| 18-102-final-viral-combined_k141_84393\|\|full | Not-determined |
| 18-102-final-viral-combined_k141_167408\|\|full | Not-determined |
| 18-102-final-viral-combined_k141_1397\|\|full | Low-quality |
| 18-102-final-viral-combined_k141_105003\|\|full | Not-determined |
| 18-102-final-viral-combined_k141_22311\|\|full | Not-determined |
| 18-102-final-viral-combined_k141_115092\|\|full | Not-determined |
| 18-102-final-viral-combined_k141_1406\|\|full | Low-quality |
| 18-102-final-viral-combined_k141_43591\|\|full | Not-determined |
| 18-102-final-viral-combined_k141_115095\|\|full | Low-quality |
| 18-102-final-viral-combined_k141_197827\|\|full | Not-determined |
| 18-102-final-viral-combined_k141_1410\|\|full | Not-determined |
| 18-102-final-viral-combined_k141_43600\|\|full | Low-quality |
| 18-102-final-viral-combined_k141_94664\|\|full | Not-determined |
| 18-102-final-viral-combined_k141_115107\|\|full | Low-quality |
| 18-102-final-viral-combined_k141_167427\|\|full | Low-quality |
| 18-102-final-viral-combined_k141_229446\|\|full | Low-quality |
| 18-102-final-viral-combined_k141_1430\|\|full | Low-quality |
| 18-102-final-viral-combined_k141_187770\|\|full | Not-determined |
| 18-102-final-viral-combined_k141_53022\|\|full | Low-quality |
| 18-102-final-viral-combined_k141_187775\|\|full | Not-determined |
| 18-102-final-viral-combined_k141_197833\|\|full | Low-quality |
| 18-102-final-viral-combined_k141_167434\|\|full | Low-quality |
| 18-102-final-viral-combined_k141_218773\|\|full | Not-determined |
| 18-102-final-viral-combined_k141_187778\|\|full | Not-determined |
| 18-102-final-viral-combined_k141_135983\|\|full | Not-determined |
| 18-102-final-viral-combined_k141_135985\|\|full | Low-quality |
| 18-102-final-viral-combined_k141_135986\|\|full | Not-determined |
| 18-102-final-viral-combined_k141_53044\|\|full | Not-determined |
| 18-102-final-viral-combined_k141_22364\|\|full | Low-quality |
| 18-102-final-viral-combined_k141_187786\|\|full | Low-quality |
| 18-102-final-viral-combined_k141_125678\|\|full | Not-determined |
| 18-102-final-viral-combined_k141_115164\|\|full | Low-quality |
| 18-102-final-viral-combined_k141_22374\|\|full | Not-determined |
| 18-102-final-viral-combined_k141_63389\|\|full | Not-determined |
| 18-102-final-viral-combined_k141_63390\|\|full | Not-determined |
| 18-102-final-viral-combined_k141_146632\|\|full | Not-determined |
| 18-102-final-viral-combined_k141_146634\|\|full | Low-quality |
| 18-102-final-viral-combined_k141_167488\|\|full | Low-quality |
| 18-102-final-viral-combined_k141_125707\|\|full | Low-quality |
| 18-102-final-viral-combined_k141_43641\|\|full | Low-quality |
| 18-102-final-viral-combined_k141_94685\|\|full | Not-determined |
| 18-102-final-viral-combined_k141_125719\|\|full | Low-quality |
| 18-102-final-viral-combined_k141_43644\|\|full | Low-quality |
| 18-102-final-viral-combined_k141_229496\|\|full | Low-quality |
| 18-102-final-viral-combined_k141_136002\|\|full | Not-determined |
| 18-102-final-viral-combined_k141_1449\|\|full | Not-determined |
| 18-102-final-viral-combined_k141_167514\|\|full | Not-determined |
| 18-102-final-viral-combined_k141_105012\|\|full | Low-quality |
| 18-102-final-viral-combined_k141_94696\|\|full | Not-determined |
| 18-102-final-viral-combined_k141_218829\|\|full | Low-quality |
| 18-102-final-viral-combined_k141_115194\|\|full | Low-quality |
| 18-102-final-viral-combined_k141_84498\|\|full | Not-determined |
| 18-102-final-viral-combined_k141_1459\|\|full | Low-quality |
| 18-102-final-viral-combined_k141_84508\|\|full | Not-determined |
| 18-102-final-viral-combined_k141_156727\|\|full | Not-determined |
| 18-102-final-viral-combined_k141_167532\|\|full | Not-determined |
| 18-102-final-viral-combined_k141_84514\|\|full | Low-quality |
| 18-102-final-viral-combined_k141_105038\|\|full | Low-quality |
| 18-102-final-viral-combined_k141_84526\|\|full | Low-quality |
| 18-104-final-viral-combined_k141_47565\|\|full | Medium-quality |
| 18-102-final-viral-combined_k141_11945\|\|full | Not-determined |
| 18-102-final-viral-combined_k141_105039\|\|full | Low-quality |
| 18-102-final-viral-combined_k141_187881\|\|full | Not-determined |
| 18-102-final-viral-combined_k141_115207\|\|full | Not-determined |
| 18-102-final-viral-combined_k141_208583\|\|full | Not-determined |
| 18-102-final-viral-combined_k141_167563\|\|full | Low-quality |
| 18-102-final-viral-combined_k141_239502\|\|full | Not-determined |
| 18-102-final-viral-combined_k141_125749\|\|full | Low-quality |
| 18-102-final-viral-combined_k141_115240\|\|full | Low-quality |
| 18-102-final-viral-combined_k141_239510\|\|full | Low-quality |
| 18-102-final-viral-combined_k141_115247\|\|full | Not-determined |
| 18-102-final-viral-combined_k141_11979\|\|full | Not-determined |
| 18-102-final-viral-combined_k141_105049\|\|full | Low-quality |
| 18-102-final-viral-combined_k141_53171\|\|full | Not-determined |
| 18-102-final-viral-combined_k141_11991\|\|full | Not-determined |
| 18-102-final-viral-combined_k141_187909\|\|full | Not-determined |
| 18-102-final-viral-combined_k141_53185\|\|full | Low-quality |
| 18-102-final-viral-combined_k141_146713\|\|full | Not-determined |
| 18-102-final-viral-combined_k141_208625\|\|full | Low-quality |
| 18-102-final-viral-combined_k141_136090\|\|full | Low-quality |
| 18-102-final-viral-combined_k141_12003\|\|full | Not-determined |
| 18-102-final-viral-combined_k141_53198\|\|full | Not-determined |
| 18-102-final-viral-combined_k141_1465\|\|full | Not-determined |
| 18-102-final-viral-combined_k141_156762\|\|full | Low-quality |
| 18-102-final-viral-combined_k141_105064\|\|full | Not-determined |
| 18-102-final-viral-combined_k141_197878\|\|full | Not-determined |
| 18-102-final-viral-combined_k141_105070\|\|full | Low-quality |
| 18-102-final-viral-combined_k141_53216\|\|full | Not-determined |
| 18-102-final-viral-combined_k141_136097\|\|full | Low-quality |
| 18-102-final-viral-combined_k141_43707\|\|full | Not-determined |
| 18-102-final-viral-combined_k141_177885\|\|full | Not-determined |
| 18-102-final-viral-combined_k141_53225\|\|full | Low-quality |
| 18-102-final-viral-combined_k141_105093\|\|full | Not-determined |
| 18-102-final-viral-combined_k141_197894\|\|full | Not-determined |
| 18-102-final-viral-combined_k141_43740\|\|full | Low-quality |
| 18-102-final-viral-combined_k141_115255\|\|full | Not-determined |
| 18-102-final-viral-combined_k141_167617\|\|full | Low-quality |
| 18-102-final-viral-combined_k141_63487\|\|full | Not-determined |
| 18-102-final-viral-combined_k141_1505\|\|full | Low-quality |
| 18-102-final-viral-combined_k141_115273\|\|full | Not-determined |
| 18-109-final-viral-combined_k141_130902\|\|full | Medium-quality |
| 18-102-final-viral-combined_k141_94788\|\|full | Not-determined |
| 18-102-final-viral-combined_k141_84607\|\|full | Low-quality |
| 18-102-final-viral-combined_k141_43767\|\|full | Not-determined |
| 18-102-final-viral-combined_k141_1512\|\|full | Low-quality |
| 18-102-final-viral-combined_k141_146761\|\|full | Not-determined |
| 18-102-final-viral-combined_k141_1514\|\|full | Not-determined |
| 18-102-final-viral-combined_k141_177911\|\|full | Not-determined |
| 18-102-final-viral-combined_k141_136122\|\|full | Low-quality |
| 18-102-final-viral-combined_k141_239597\|\|full | Low-quality |
| 18-102-final-viral-combined_k141_125782\|\|full | Not-determined |
| 18-102-final-viral-combined_k141_125784\|\|full | Not-determined |
| 18-102-final-viral-combined_k141_187928\|\|full | Low-quality |
| 18-102-final-viral-combined_k141_177940\|\|full | Not-determined |
| 18-102-final-viral-combined_k141_43799\|\|full | Low-quality |
| 18-102-final-viral-combined_k141_239621\|\|full | Not-determined |
| 18-102-final-viral-combined_k141_105115\|\|full | Not-determined |
| 18-102-final-viral-combined_k141_32453\|\|full | Low-quality |
| 18-102-final-viral-combined_k141_136150\|\|full | Low-quality |
| 18-102-final-viral-combined_k141_12029\|\|full | Not-determined |
| 18-102-final-viral-combined_k141_43822\|\|full | Not-determined |
| 18-102-final-viral-combined_k141_208664\|\|full | Low-quality |
| 18-102-final-viral-combined_k141_32469\|\|full | Not-determined |
| 18-102-final-viral-combined_k141_218894\|\|full | Low-quality |
| 18-102-final-viral-combined_k141_156851\|\|full | Not-determined |
| 18-102-final-viral-combined_k141_156853\|\|full | Not-determined |
| 18-102-final-viral-combined_k141_63552\|\|full | Low-quality |
| 18-102-final-viral-combined_k141_125828\|\|full | Not-determined |
| 18-102-final-viral-combined_k141_63555\|\|full | Not-determined |
| 18-102-final-viral-combined_k141_208682\|\|full | Low-quality |
| 18-102-final-viral-combined_k141_239653\|\|full | Low-quality |
| 18-102-final-viral-combined_k141_177980\|\|full | Low-quality |
| 18-102-final-viral-combined_k141_177981\|\|full | Not-determined |
| 18-110-final-viral-combined_k141_134284\|\|full | Medium-quality |
| 18-102-final-viral-combined_k141_156864\|\|full | Low-quality |
| 18-102-final-viral-combined_k141_177997\|\|full | Low-quality |
| 18-102-final-viral-combined_k141_22422\|\|full | Low-quality |
| 18-102-final-viral-combined_k141_32509\|\|full | Not-determined |
| 18-102-final-viral-combined_k141_187995\|\|full | Low-quality |
| 18-102-final-viral-combined_k141_22437\|\|full | Not-determined |
| 18-102-final-viral-combined_k141_63599\|\|full | Low-quality |
| 18-102-final-viral-combined_k141_32547\|\|full | Low-quality |
| 18-102-final-viral-combined_k141_218932\|\|full | Low-quality |
| 18-102-final-viral-combined_k141_125886\|\|full | Not-determined |
| 18-102-final-viral-combined_k141_218935\|\|full | Low-quality |
| 18-102-final-viral-combined_k141_43913\|\|full | Not-determined |
| 18-102-final-viral-combined_k141_136231\|\|full | Low-quality |
| 18-102-final-viral-combined_k141_1555\|\|full | Not-determined |
| 18-102-final-viral-combined_k141_136246\|\|full | Not-determined |
| 18-102-final-viral-combined_k141_125901\|\|full | Not-determined |
| 18-102-final-viral-combined_k141_43922\|\|full | Not-determined |
| 18-102-final-viral-combined_k141_188028\|\|full | Low-quality |
| 18-102-final-viral-combined_k141_105170\|\|full | Not-determined |
| 18-102-final-viral-combined_k141_105177\|\|full | Low-quality |
| 18-101_k141_106115 | Medium-quality |
| 18-102-final-viral-combined_k141_43943\|\|full | Low-quality |
| 18-102-final-viral-combined_k141_1574\|\|full | Not-determined |
| 18-102-final-viral-combined_k141_32597\|\|full | Not-determined |
| 18-102-final-viral-combined_k141_94877\|\|full | Not-determined |
| 18-102-final-viral-combined_k141_229630\|\|full | Low-quality |
| 18-102-final-viral-combined_k141_43953\|\|full | Not-determined |
| 18-102-final-viral-combined_k141_53299\|\|full | Low-quality |
| 18-102-final-viral-combined_k141_167708\|\|full | Low-quality |
| 18-102-final-viral-combined_k141_239696\|\|full | Low-quality |
| 18-102-final-viral-combined_k141_156920\|\|full | Low-quality |
| 18-102-final-viral-combined_k141_63635\|\|full | Low-quality |
| 18-102-final-viral-combined_k141_125920\|\|full | Low-quality |
| 18-102-final-viral-combined_k141_94885\|\|full | Not-determined |
| 18-102-final-viral-combined_k141_136273\|\|full | Not-determined |
| 18-102-final-viral-combined_k141_32620\|\|full | Not-determined |
| 18-102-final-viral-combined_k141_136275\|\|full | Low-quality |
| 18-102-final-viral-combined_k141_125947\|\|full | Not-determined |
| 18-102-final-viral-combined_k141_53322\|\|full | Low-quality |
| 18-102-final-viral-combined_k141_218969\|\|full | Not-determined |
| 18-102-final-viral-combined_k141_197947\|\|full | Low-quality |
| 18-102-final-viral-combined_k141_125948\|\|full | Not-determined |
| 18-102-final-viral-combined_k141_167739\|\|full | Low-quality |
| 18-102-final-viral-combined_k141_136283\|\|full | Not-determined |
| 18-102-final-viral-combined_k141_167745\|\|full | Not-determined |
| 18-102-final-viral-combined_k141_105215\|\|full | Not-determined |
| 18-102-final-viral-combined_k141_188090\|\|full | Low-quality |
| 18-102-final-viral-combined_k141_105222\|\|full | Low-quality |
| 18-102-final-viral-combined_k141_1661\|\|full | Not-determined |
| 18-102-final-viral-combined_k141_1668\|\|full | Low-quality |
| 18-102-final-viral-combined_k141_53345\|\|full | Low-quality |
| 18-102-final-viral-combined_k141_167768\|\|full | Not-determined |
| 18-102-final-viral-combined_k141_94965\|\|full | Not-determined |
| 18-102-final-viral-combined_k141_94967\|\|full | Low-quality |
| 18-102-final-viral-combined_k141_178079\|\|full | Not-determined |
| 18-102-final-viral-combined_k141_125971\|\|full | Low-quality |
| 18-102-final-viral-combined_k141_44026\|\|full | Low-quality |
| 18-102-final-viral-combined_k141_208723\|\|full | Low-quality |
| 18-102-final-viral-combined_k141_208725\|\|full | Low-quality |
| 18-102-final-viral-combined_k141_219000\|\|full | Not-determined |
| 18-102-final-viral-combined_k141_22579\|\|full | Not-determined |
| 18-102-final-viral-combined_k141_63678\|\|full | Low-quality |
| 18-102-final-viral-combined_k141_198037\|\|full | Low-quality |
| 18-102-final-viral-combined_k141_239774\|\|full | Not-determined |
| 18-102-final-viral-combined_k141_178106\|\|full | Low-quality |
| 18-102-final-viral-combined_k141_188142\|\|full | Not-determined |
| 18-102-final-viral-combined_k141_44053\|\|full | Not-determined |
| 18-102-final-viral-combined_k141_198048\|\|full | Low-quality |
| 18-102-final-viral-combined_k141_63690\|\|full | Low-quality |
| 18-102-final-viral-combined_k141_198050\|\|full | Not-determined |
| 18-102-final-viral-combined_k141_188155\|\|full | Low-quality |
| 18-102-final-viral-combined_k141_22589\|\|full | Not-determined |
| 18-102-final-viral-combined_k141_136365\|\|full | Low-quality |
| 18-102-final-viral-combined_k141_136373\|\|full | Not-determined |
| 18-102-final-viral-combined_k141_115451\|\|full | Low-quality |
| 18-102-final-viral-combined_k141_63708\|\|full | Low-quality |
| 18-102-final-viral-combined_k141_229683\|\|full | Not-determined |
| 18-102-final-viral-combined_k141_74070\|\|full | Low-quality |
| 18-102-final-viral-combined_k141_44106\|\|full | Low-quality |
| 18-102-final-viral-combined_k141_22614\|\|full | Not-determined |
| 18-102-final-viral-combined_k141_84640\|\|full | Low-quality |
| 18-102-final-viral-combined_k141_1715\|\|full | Low-quality |
| 18-102-final-viral-combined_k141_1716\|\|full | Low-quality |
| 18-102-final-viral-combined_k141_1718\|\|full | Low-quality |
| 18-102-final-viral-combined_k141_53414\|\|full | Not-determined |
| 18-102-final-viral-combined_k141_229724\|\|full | Not-determined |
| 18-102-final-viral-combined_k141_105323\|\|full | Low-quality |
| 18-102-final-viral-combined_k141_22625\|\|full | Not-determined |
| 18-102-final-viral-combined_k141_1741\|\|full | Low-quality |
| 18-102-final-viral-combined_k141_95005\|\|full | Low-quality |
| 18-102-final-viral-combined_k141_239811\|\|full | Low-quality |
| 18-102-final-viral-combined_k141_95012\|\|full | Low-quality |
| 18-102-final-viral-combined_k141_1745\|\|full | Low-quality |
| 18-102-final-viral-combined_k141_178214\|\|full | Low-quality |
| 18-102-final-viral-combined_k141_74108\|\|full | Low-quality |
| 18-102-final-viral-combined_k141_178216\|\|full | Not-determined |
| 18-102-final-viral-combined_k141_12128\|\|full | Low-quality |
| 18-102-final-viral-combined_k141_208743\|\|full | Not-determined |
| 18-102-final-viral-combined_k141_74119\|\|full | Low-quality |
| 18-102-final-viral-combined_k141_115492\|\|full | Not-determined |
| 18-102-final-viral-combined_k141_84659\|\|full | Low-quality |
| 18-102-final-viral-combined_k141_1768\|\|full | Low-quality |
| 18-102-final-viral-combined_k141_239835\|\|full | Low-quality |
| 18-102-final-viral-combined_k141_178223\|\|full | Not-determined |
| 18-102-final-viral-combined_k141_125981\|\|full | Low-quality |
| 18-102-final-viral-combined_k141_44128\|\|full | Not-determined |
| 18-102-final-viral-combined_k141_53444\|\|full | Low-quality |
| 18-102-final-viral-combined_k141_74134\|\|full | Not-determined |
| 18-102-final-viral-combined_k141_63786\|\|full | Low-quality |
| 18-102-final-viral-combined_k141_63787\|\|full | Low-quality |
| 18-102-final-viral-combined_k141_239859\|\|full | Not-determined |
| 18-102-final-viral-combined_k141_63791\|\|full | Not-determined |
| 18-102-final-viral-combined_k141_1784\|\|full | Low-quality |
| 18-102-final-viral-combined_k141_208778\|\|full | Low-quality |
| 18-102-final-viral-combined_k141_208779\|\|full | Not-determined |
| 18-102-final-viral-combined_k141_63797\|\|full | Low-quality |
| 18-102-final-viral-combined_k141_126003\|\|full | Low-quality |
| 18-102-final-viral-combined_k141_84683\|\|full | Low-quality |
| 18-102-final-viral-combined_k141_239862\|\|full | Low-quality |
| 18-102-final-viral-combined_k141_115553\|\|full | Low-quality |
| 18-102-final-viral-combined_k141_198099\|\|full | Not-determined |
| 18-102-final-viral-combined_k141_178260\|\|full | Low-quality |
| 18-102-final-viral-combined_k141_126026\|\|full | Low-quality |
| 18-102-final-viral-combined_k141_84716\|\|full | Not-determined |
| 18-102-final-viral-combined_k141_84723\|\|full | Not-determined |
| 18-102-final-viral-combined_k141_84725\|\|full | Not-determined |
| 18-102-final-viral-combined_k141_167841\|\|full | Low-quality |
| 18-102-final-viral-combined_k141_12191\|\|full | Low-quality |
| 18-102-final-viral-combined_k141_115577\|\|full | Not-determined |
| 18-102-final-viral-combined_k141_126044\|\|full | Not-determined |
| 18-102-final-viral-combined_k141_229820\|\|full | Low-quality |
| 18-102-final-viral-combined_k141_198109\|\|full | Not-determined |
| 18-102-final-viral-combined_k141_115597\|\|full | Not-determined |
| 18-102-final-viral-combined_k141_178297\|\|full | Low-quality |
| 18-102-final-viral-combined_k141_178298\|\|full | Low-quality |
| 18-102-final-viral-combined_k141_136491\|\|full | Low-quality |
| 18-102-final-viral-combined_k141_178300\|\|full | Low-quality |
| 18-102-final-viral-combined_k141_136495\|\|full | Low-quality |
| 18-102-final-viral-combined_k141_115606\|\|full | Not-determined |
| 18-102-final-viral-combined_k141_239885\|\|full | Low-quality |
| 18-102-final-viral-combined_k141_126071\|\|full | Not-determined |
| 18-102-final-viral-combined_k141_74183\|\|full | Not-determined |
| 18-102-final-viral-combined_k141_115627\|\|full | Not-determined |
| 18-102-final-viral-combined_k141_74189\|\|full | Not-determined |
| 18-102-final-viral-combined_k141_95065\|\|full | Low-quality |
| 18-102-final-viral-combined_k141_53511\|\|full | Not-determined |
| 18-102-final-viral-combined_k141_22643\|\|full | Not-determined |
| 18-102-final-viral-combined_k141_44194\|\|full | Low-quality |
| 18-102-final-viral-combined_k141_74206\|\|full | Low-quality |
| 18-102-final-viral-combined_k141_136542\|\|full | Low-quality |
| 18-102-final-viral-combined_k141_229853\|\|full | Not-determined |
| 18-102-final-viral-combined_k141_198142\|\|full | Not-determined |
| 18-102-final-viral-combined_k141_178335\|\|full | Low-quality |
| 18-102-final-viral-combined_k141_146848\|\|full | Low-quality |
| 18-102-final-viral-combined_k141_178339\|\|full | Not-determined |
| 18-102-final-viral-combined_k141_178341\|\|full | Low-quality |
| 18-102-final-viral-combined_k141_22662\|\|full | Low-quality |
| 18-102-final-viral-combined_k141_229864\|\|full | Not-determined |
| 18-102-final-viral-combined_k141_167879\|\|full | Not-determined |
| 18-102-final-viral-combined_k141_105472\|\|full | Not-determined |
| 18-102-final-viral-combined_k141_22674\|\|full | Low-quality |
| 18-102-final-viral-combined_k141_105479\|\|full | Low-quality |
| 18-102-final-viral-combined_k141_44243\|\|full | Low-quality |
| 18-102-final-viral-combined_k141_208814\|\|full | Not-determined |
| 18-102-final-viral-combined_k141_22684\|\|full | Low-quality |
| 18-102-final-viral-combined_k141_22693\|\|full | Not-determined |
| 18-102-final-viral-combined_k141_208826\|\|full | Not-determined |
| 18-102-final-viral-combined_k141_208837\|\|full | Low-quality |
| 18-102-final-viral-combined_k141_208842\|\|full | Low-quality |
| 18-102-final-viral-combined_k141_198179\|\|full | Not-determined |
| 18-102-final-viral-combined_k141_44268\|\|full | Low-quality |
| 18-102-final-viral-combined_k141_44276\|\|full | Low-quality |
| 18-102-final-viral-combined_k141_74248\|\|full | Low-quality |
| 18-102-final-viral-combined_k141_208852\|\|full | Low-quality |
| 18-102-final-viral-combined_k141_84771\|\|full | Low-quality |
| 18-102-final-viral-combined_k141_84787\|\|full | Low-quality |
| 18-102-final-viral-combined_k141_63865\|\|full | Not-determined |
| 18-102-final-viral-combined_k141_74276\|\|full | Not-determined |
| 18-102-final-viral-combined_k141_32731\|\|full | Low-quality |
| 18-102-final-viral-combined_k141_105551\|\|full | Low-quality |
| 18-102-final-viral-combined_k141_198209\|\|full | Low-quality |
| 18-102-final-viral-combined_k141_178381\|\|full | Not-determined |
| 18-102-final-viral-combined_k141_219094\|\|full | Not-determined |
| 18-102-final-viral-combined_k141_219098\|\|full | Low-quality |
| 18-102-final-viral-combined_k141_188216\|\|full | Low-quality |
| 18-102-final-viral-combined_k141_136578\|\|full | Low-quality |
| 18-102-final-viral-combined_k141_219102\|\|full | Not-determined |
| 18-102_k141_192667 | Medium-quality |
| 18-102-final-viral-combined_k141_53597\|\|full | Low-quality |
| 18-102-final-viral-combined_k141_126136\|\|full | Low-quality |
| 18-102-final-viral-combined_k141_53599\|\|full | Low-quality |
| 18-102-final-viral-combined_k141_146888\|\|full | Low-quality |
| 18-102-final-viral-combined_k141_105555\|\|full | Low-quality |
| 18-102-final-viral-combined_k141_84840\|\|full | Low-quality |
| 18-102-final-viral-combined_k141_157116\|\|full | Low-quality |
| 18-102-final-viral-combined_k141_53612\|\|full | Not-determined |
| 18-102-final-viral-combined_k141_63879\|\|full | Not-determined |
| 18-102-final-viral-combined_k141_1929\|\|full | Not-determined |
| 18-102-final-viral-combined_k141_229916\|\|full | Low-quality |
| 18-102-final-viral-combined_k141_126147\|\|full | Low-quality |
| 18-102-final-viral-combined_k141_63894\|\|full | Not-determined |
| 18-102-final-viral-combined_k141_84863\|\|full | Not-determined |
| 18-102-final-viral-combined_k141_53648\|\|full | Not-determined |
| 18-102-final-viral-combined_k141_146901\|\|full | Not-determined |
| 18-102-final-viral-combined_k141_105590\|\|full | Low-quality |
| 18-102-final-viral-combined_k141_239953\|\|full | Not-determined |
| 18-102-final-viral-combined_k141_74307\|\|full | Low-quality |
| 18-102-final-viral-combined_k141_229950\|\|full | Not-determined |
| 18-102-final-viral-combined_k141_157159\|\|full | Not-determined |
| 18-102-final-viral-combined_k141_22758\|\|full | Not-determined |
| 18-102-final-viral-combined_k141_53671\|\|full | Low-quality |
| 18-102-final-viral-combined_k141_53675\|\|full | Not-determined |
| 18-102-final-viral-combined_k141_1970\|\|full | Low-quality |
| 18-102-final-viral-combined_k141_229959\|\|full | Not-determined |
| 18-102-final-viral-combined_k141_136617\|\|full | Low-quality |
| 18-102-final-viral-combined_k141_229961\|\|full | Not-determined |
| 18-102-final-viral-combined_k141_229970\|\|full | Low-quality |
| 18-102-final-viral-combined_k141_22763\|\|full | Not-determined |
| 18-102-final-viral-combined_k141_229971\|\|full | Low-quality |
| 18-102-final-viral-combined_k141_74334\|\|full | Not-determined |
| 18-102-final-viral-combined_k141_84885\|\|full | Low-quality |
| 18-102-final-viral-combined_k141_178405\|\|full | Low-quality |
| 18-102-final-viral-combined_k141_178421\|\|full | Low-quality |
| 18-102-final-viral-combined_k141_95111\|\|full | Low-quality |
| 18-102-final-viral-combined_k141_219117\|\|full | Low-quality |
| 18-102-final-viral-combined_k141_74353\|\|full | Low-quality |
| 18-102-final-viral-combined_k141_74355\|\|full | Low-quality |
| 18-102-final-viral-combined_k141_126210\|\|full | Not-determined |
| 18-102-final-viral-combined_k141_44385\|\|full | Not-determined |
| 18-102-final-viral-combined_k141_126217\|\|full | Low-quality |
| 18-102-final-viral-combined_k141_53736\|\|full | Not-determined |
| 18-102-final-viral-combined_k141_44402\|\|full | Low-quality |
| 18-102-final-viral-combined_k141_178447\|\|full | Not-determined |
| 18-102-final-viral-combined_k141_178448\|\|full | Low-quality |
| 18-102-final-viral-combined_k141_240006\|\|full | Low-quality |
| 18-102-final-viral-combined_k141_126249\|\|full | Low-quality |
| 18-102-final-viral-combined_k141_115757\|\|full | Low-quality |
| 18-102-final-viral-combined_k141_188317\|\|full | Not-determined |
| 18-102-final-viral-combined_k141_2029\|\|full | Not-determined |
| 18-102-final-viral-combined_k141_209027\|\|full | Not-determined |
| 18-102-final-viral-combined_k141_105666\|\|full | Low-quality |
| 18-102-final-viral-combined_k141_115764\|\|full | Not-determined |
| 18-102-final-viral-combined_k141_64021\|\|full | Not-determined |
| 18-102-final-viral-combined_k141_240034\|\|full | Low-quality |
| 18-102-final-viral-combined_k141_146942\|\|full | Not-determined |
| 18-102-final-viral-combined_k141_53778\|\|full | Low-quality |
| 18-102-final-viral-combined_k141_44488\|\|full | Not-determined |
| 18-102-final-viral-combined_k141_188342\|\|full | Low-quality |
| 18-102-final-viral-combined_k141_53791\|\|full | Low-quality |
| 18-102-final-viral-combined_k141_53792\|\|full | Not-determined |
| 18-102-final-viral-combined_k141_230035\|\|full | Low-quality |
| 18-102-final-viral-combined_k141_84969\|\|full | Not-determined |
| 18-102-final-viral-combined_k141_188360\|\|full | Not-determined |
| 18-102-final-viral-combined_k141_64044\|\|full | Not-determined |
| 18-102-final-viral-combined_k141_53804\|\|full | Not-determined |
| KM-final-viral-combined_k141_225356\|\|full | Medium-quality |
| 18-102-final-viral-combined_k141_32768\|\|full | Low-quality |
| 18-102-final-viral-combined_k141_198251\|\|full | Not-determined |
| 18-102-final-viral-combined_k141_105689\|\|full | Not-determined |
| 18-102-final-viral-combined_k141_74392\|\|full | Not-determined |
| 18-102-final-viral-combined_k141_230063\|\|full | Low-quality |
| 18-102-final-viral-combined_k141_64062\|\|full | Low-quality |
| 18-102-final-viral-combined_k141_230074\|\|full | Not-determined |
| 18-102-final-viral-combined_k141_32777\|\|full | Not-determined |
| 18-102-final-viral-combined_k141_95249\|\|full | Not-determined |
| 18-102-final-viral-combined_k141_188393\|\|full | Low-quality |
| 18-102-final-viral-combined_k141_219159\|\|full | Low-quality |
| 18-102-final-viral-combined_k141_167926\|\|full | Not-determined |
| 18-102-final-viral-combined_k141_198264\|\|full | Not-determined |
| 18-102-final-viral-combined_k141_178485\|\|full | Low-quality |
| 18-102-final-viral-combined_k141_219177\|\|full | Low-quality |
| 18-102-final-viral-combined_k141_136758\|\|full | Low-quality |
| 18-102-final-viral-combined_k141_126259\|\|full | Not-determined |
| 18-102-final-viral-combined_k141_157293\|\|full | Low-quality |
| 18-102-final-viral-combined_k141_115821\|\|full | Low-quality |
| 18-102-final-viral-combined_k141_209040\|\|full | Not-determined |
| 18-102-final-viral-combined_k141_22839\|\|full | Not-determined |
| 18-102-final-viral-combined_k141_12265\|\|full | Not-determined |
| 18-102-final-viral-combined_k141_85047\|\|full | Low-quality |
| 18-102-final-viral-combined_k141_157319\|\|full | Not-determined |
| 18-102-final-viral-combined_k141_136795\|\|full | Not-determined |
| 18-102-final-viral-combined_k141_157322\|\|full | Low-quality |
| 18-102-final-viral-combined_k141_85059\|\|full | Not-determined |
| 18-102-final-viral-combined_k141_240063\|\|full | Low-quality |
| 18-102-final-viral-combined_k141_126276\|\|full | Low-quality |
| 18-102-final-viral-combined_k141_44523\|\|full | Low-quality |
| 18-102-final-viral-combined_k141_22869\|\|full | Not-determined |
| 18-102-final-viral-combined_k141_178529\|\|full | Low-quality |
| 18-102-final-viral-combined_k141_44529\|\|full | Low-quality |
| 18-102-final-viral-combined_k141_105771\|\|full | Not-determined |
| 18-102-final-viral-combined_k141_219245\|\|full | Not-determined |
| 18-102-final-viral-combined_k141_230131\|\|full | Low-quality |
| 18-102-final-viral-combined_k141_178546\|\|full | Not-determined |
| 18-102-final-viral-combined_k141_105775\|\|full | Not-determined |
| 18-102-final-viral-combined_k141_230135\|\|full | Not-determined |
| 18-102-final-viral-combined_k141_12277\|\|full | Low-quality |
| 18-102-final-viral-combined_k141_230138\|\|full | Low-quality |
| 18-102-final-viral-combined_k141_64152\|\|full | Not-determined |
| 18-102-final-viral-combined_k141_95330\|\|full | Not-determined |
| 18-102-final-viral-combined_k141_146948\|\|full | Not-determined |
| 18-102-final-viral-combined_k141_157359\|\|full | Not-determined |
| 18-102-final-viral-combined_k141_85104\|\|full | Low-quality |
| 18-102-final-viral-combined_k141_240117\|\|full | Low-quality |
| 18-102-final-viral-combined_k141_168014\|\|full | Low-quality |
| 18-102-final-viral-combined_k141_209057\|\|full | Low-quality |
| 18-102-final-viral-combined_k141_178591\|\|full | Low-quality |
| 18-102-final-viral-combined_k141_22908\|\|full | Low-quality |
| 18-102-final-viral-combined_k141_95357\|\|full | Not-determined |
| 18-102-final-viral-combined_k141_230197\|\|full | Not-determined |
| 18-102-final-viral-combined_k141_2078\|\|full | Low-quality |
| 18-101-final-viral-combined_k141_106115\|\|full | Medium-quality |
| 18-102-final-viral-combined_k141_136860\|\|full | Not-determined |
| 18-102-final-viral-combined_k141_12309\|\|full | Low-quality |
| 18-102-final-viral-combined_k141_95378\|\|full | Not-determined |
| 18-102-final-viral-combined_k141_240150\|\|full | Not-determined |
| 18-102-final-viral-combined_k141_105794\|\|full | Low-quality |
| 18-102-final-viral-combined_k141_240154\|\|full | Not-determined |
| 18-102-final-viral-combined_k141_44570\|\|full | Not-determined |
| 18-102-final-viral-combined_k141_85129\|\|full | Low-quality |
| 18-102-final-viral-combined_k141_240161\|\|full | Not-determined |

| | |
|---|---|
| 18-102-final-viral-combined_k141_22946||full | Not-determined |
| 18-102-final-viral-combined_k141_44602||full | Not-determined |
| 18-102-final-viral-combined_k141_240169||full | Not-determined |
| 18-102-final-viral-combined_k141_146981||full | Low-quality |
| 18-102-final-viral-combined_k141_230261||full | Low-quality |
| 18-102-final-viral-combined_k141_230263||full | Not-determined |
| 18-102-final-viral-combined_k141_188458||full | Not-determined |
| 18-102-final-viral-combined_k141_157411||full | Not-determined |
| 18-102-final-viral-combined_k141_85159||full | Low-quality |
| 18-102-final-viral-combined_k141_12331||full | Not-determined |
| 18-102-final-viral-combined_k141_230270||full | Not-determined |
| 18-102-final-viral-combined_k141_157422||full | Not-determined |
| 18-102-final-viral-combined_k141_64241||full | Not-determined |
| 18-102-final-viral-combined_k141_168080||full | Low-quality |
| 18-102-final-viral-combined_k141_168083||full | Low-quality |
| 18-102-final-viral-combined_k141_230278||full | Low-quality |
| 18-102-final-viral-combined_k141_53968||full | Low-quality |
| 18-102-final-viral-combined_k141_95474||full | Not-determined |
| 18-109-final-viral-combined_k141_163980||0_partial | Medium-quality |
| 18-102-final-viral-combined_k141_219310||full | Low-quality |
| 18-102-final-viral-combined_k141_240189||full | Low-quality |
| 18-102-final-viral-combined_k141_188514||full | Low-quality |
| 18-102-final-viral-combined_k141_219315||full | Low-quality |
| 18-102-final-viral-combined_k141_44627||full | Not-determined |
| 18-102-final-viral-combined_k141_32844||full | Low-quality |
| 18-102-final-viral-combined_k141_240214||full | Not-determined |
| 18-102-final-viral-combined_k141_198338||full | Low-quality |
| 18-102-final-viral-combined_k141_85209||full | Low-quality |
| 18-102-final-viral-combined_k141_74465||full | Not-determined |
| 18-102-final-viral-combined_k141_32869||full | Not-determined |
| 18-102-final-viral-combined_k141_147003||full | Not-determined |
| 18-102-final-viral-combined_k141_219325||full | Low-quality |
| 18-102-final-viral-combined_k141_157471||full | Low-quality |
| 18-102-final-viral-combined_k141_74469||full | Not-determined |
| 18-102-final-viral-combined_k141_53986||full | Not-determined |
| 18-102-final-viral-combined_k141_137000||full | Not-determined |
| 18-102-final-viral-combined_k141_105844||full | Low-quality |
| 18-102-final-viral-combined_k141_230341||full | Not-determined |
| 18-102-final-viral-combined_k141_188530||full | Low-quality |
| 18-102-final-viral-combined_k141_137024||full | Not-determined |
| 18-102-final-viral-combined_k141_168175||full | Low-quality |
| 18-102-final-viral-combined_k141_12401||full | Low-quality |
| 18-102-final-viral-combined_k141_168180||full | Low-quality |
| 18-102-final-viral-combined_k141_198383||full | Low-quality |
| 18-102-final-viral-combined_k141_168182||full | Not-determined |
| 18-101-final-viral-combined_k141_231234||full | Medium-quality |
| 18-102-final-viral-combined_k141_230359||full | Not-determined |
| 18-102-final-viral-combined_k141_126447||full | Not-determined |
| 18-102-final-viral-combined_k141_230369||full | Not-determined |
| 18-102-final-viral-combined_k141_85214||full | Not-determined |
| 18-102-final-viral-combined_k141_2125||full | Not-determined |
| 18-102-final-viral-combined_k141_230370||full | Not-determined |
| 18-102-final-viral-combined_k141_188570||full | Not-determined |
| 18-102-final-viral-combined_k141_85222||full | Not-determined |
| 18-102-final-viral-combined_k141_2137||full | Not-determined |
| 18-102-final-viral-combined_k141_126452||full | Not-determined |
| 18-102-final-viral-combined_k141_32950||full | Not-determined |
| 18-102-final-viral-combined_k141_230378||full | Not-determined |
| 18-102-final-viral-combined_k141_105906||full | Not-determined |
| 18-102-final-viral-combined_k141_44670||full | Not-determined |
| 18-102-final-viral-combined_k141_85229||full | Low-quality |
| 18-102-final-viral-combined_k141_137055||full | Low-quality |
| 18-102-final-viral-combined_k141_126482||full | Low-quality |
| 18-102-final-viral-combined_k141_115905||full | Low-quality |
| 18-102-final-viral-combined_k141_157508||full | Low-quality |
| 18-102-final-viral-combined_k141_188592||full | Low-quality |
| 18-102-final-viral-combined_k141_157509||full | Not-determined |
| 18-102-final-viral-combined_k141_95554||full | Low-quality |
| 18-102-final-viral-combined_k141_188602||full | Not-determined |
| 18-102-final-viral-combined_k141_2159||full | Not-determined |
| 18-102-final-viral-combined_k141_44692||full | Not-determined |
| 18-102-final-viral-combined_k141_219340||full | Not-determined |
| 18-102-final-viral-combined_k141_2162||full | Low-quality |
| 18-102-final-viral-combined_k141_105951||full | Not-determined |
| 18-102-final-viral-combined_k141_198413||full | Not-determined |
| 18-102-final-viral-combined_k141_95568||full | Low-quality |
| 18-102-final-viral-combined_k141_105956||full | Not-determined |
| 18-102-final-viral-combined_k141_64312||full | Not-determined |
| 18-102-final-viral-combined_k141_137082||full | Low-quality |
| 18-102-final-viral-combined_k141_157537||full | Not-determined |
| 18-102-final-viral-combined_k141_168221||full | Not-determined |
| 18-102-final-viral-combined_k141_64327||full | Low-quality |
| 18-102-final-viral-combined_k141_157549||full | Low-quality |
| 18-102-final-viral-combined_k141_198429||full | Low-quality |
| 18-102-final-viral-combined_k141_157552||full | Not-determined |
| 18-102-final-viral-combined_k141_240261||full | Not-determined |
| 18-102-final-viral-combined_k141_44731||full | Not-determined |
| 18-102-final-viral-combined_k141_115974||full | Low-quality |
| 18-102-final-viral-combined_k141_64342||full | Not-determined |
| 18-102-final-viral-combined_k141_126542||full | Low-quality |
| 18-102-final-viral-combined_k141_95603||full | Low-quality |
| 18-102-final-viral-combined_k141_33039||full | Not-determined |
| 18-102-final-viral-combined_k141_2179||full | Not-determined |
| 18-102-final-viral-combined_k141_54088||full | Not-determined |
| 18-102-final-viral-combined_k141_198450||full | Not-determined |
| 18-102-final-viral-combined_k141_240262||full | Not-determined |
| 18-102-final-viral-combined_k141_126564||full | Not-determined |
| 18-102-final-viral-combined_k141_95614||full | Not-determined |
| 18-102-final-viral-combined_k141_12490||full | Not-determined |
| 18-102-final-viral-combined_k141_168275||full | Not-determined |
| 18-102-final-viral-combined_k141_230449||full | Low-quality |
| 18-102-final-viral-combined_k141_230454||full | Not-determined |
| 18-102-final-viral-combined_k141_126577||full | Low-quality |
| 18-102-final-viral-combined_k141_23097||full | Low-quality |
| 18-102-final-viral-combined_k141_230460||full | Not-determined |
| 18-102-final-viral-combined_k141_240304||full | Not-determined |
| 18-102-final-viral-combined_k141_23111||full | Not-determined |
| 18-102-final-viral-combined_k141_157614||full | Not-determined |
| 18-102-final-viral-combined_k141_54125||full | Not-determined |
| 18-102-final-viral-combined_k141_209184||full | Low-quality |
| 18-102-final-viral-combined_k141_74650||full | Low-quality |
| 18-102-final-viral-combined_k141_74654||full | Not-determined |
| 18-102-final-viral-combined_k141_54153||full | Not-determined |
| 18-102-final-viral-combined_k141_116047||full | Not-determined |
| 18-102-final-viral-combined_k141_64366||full | Low-quality |
| 18-102-final-viral-combined_k141_116049||full | Not-determined |
| 18-102-final-viral-combined_k141_44829||full | Not-determined |
| 18-102-final-viral-combined_k141_219434||full | Low-quality |
| 18-102-final-viral-combined_k141_33073||full | Low-quality |
| 18-102-final-viral-combined_k141_188699||full | Low-quality |
| 18-109-final-viral-combined_k141_40779||full | Medium-quality |
| 18-102-final-viral-combined_k141_12552||full | Low-quality |
| 18-102-final-viral-combined_k141_116057||full | Not-determined |
| 18-102-final-viral-combined_k141_198481||full | Low-quality |
| 18-102-final-viral-combined_k141_116060||full | Low-quality |
| 18-102-final-viral-combined_k141_190624||full | Medium-quality |
| 18-102-final-viral-combined_k141_137121||full | Not-determined |
| 18-102-final-viral-combined_k141_188718||full | Not-determined |
| 18-102-final-viral-combined_k141_74703||full | Low-quality |
| 18-102-final-viral-combined_k141_137132||full | Not-determined |
| 18-102-final-viral-combined_k141_137135||full | Low-quality |
| 18-102-final-viral-combined_k141_168351||full | Not-determined |
| 18-102-final-viral-combined_k141_44869||full | Not-determined |
| 18-102-final-viral-combined_k141_168357||full | Low-quality |
| 18-102-final-viral-combined_k141_209211||full | Not-determined |
| 18-102-final-viral-combined_k141_33093||full | Low-quality |
| 18-102-final-viral-combined_k141_126611||full | Not-determined |
| 18-102-final-viral-combined_k141_33105||full | Low-quality |
| 18-102-final-viral-combined_k141_12583||full | Low-quality |
| 18-102-final-viral-combined_k141_2276||full | Low-quality |
| 18-102-final-viral-combined_k141_126628||full | Low-quality |
| 18-102-final-viral-combined_k141_168373||full | Low-quality |
| 18-102-final-viral-combined_k141_192667||full | Medium-quality |
| 18-102-final-viral-combined_k141_2285||full | Low-quality |
| 18-102-final-viral-combined_k141_157626||full | Low-quality |
| 18-102-final-viral-combined_k141_126651||full | Not-determined |
| 18-102-final-viral-combined_k141_95726||full | Not-determined |
| 18-102-final-viral-combined_k141_147124||full | Not-determined |
| 18-102-final-viral-combined_k141_116090||full | Not-determined |
| 18-102-final-viral-combined_k141_209241||full | Not-determined |
| 18-102-final-viral-combined_k141_126664||full | Not-determined |
| 18-102-final-viral-combined_k141_12604||full | Not-determined |
| 18-105-final-viral-combined_k141_117095||full | Medium-quality |
| 18-102-final-viral-combined_k141_147138||full | Low-quality |
| 18-102-final-viral-combined_k141_95760||full | Not-determined |
| 18-102-final-viral-combined_k141_157648||full | Not-determined |
| 18-102-final-viral-combined_k141_147154||full | Not-determined |
| 18-102-final-viral-combined_k141_12612||full | Low-quality |
| 18-102-final-viral-combined_k141_188794||full | Not-determined |
| 18-102-final-viral-combined_k141_147156||full | Not-determined |
| 18-102-final-viral-combined_k141_12619||full | Not-determined |
| 18-102-final-viral-combined_k141_147159||full | Not-determined |
| 18-102-final-viral-combined_k141_219527||full | Not-determined |
| 18-102-final-viral-combined_k141_198548||full | Low-quality |
| 18-102-final-viral-combined_k141_209294||full | Not-determined |
| 18-102-final-viral-combined_k141_178657||full | Not-determined |
| 18-102-final-viral-combined_k141_126674||full | Low-quality |
| 18-102-final-viral-combined_k141_230561||full | Low-quality |
| 18-102-final-viral-combined_k141_230566||full | Not-determined |
| 18-102-final-viral-combined_k141_23162||full | Not-determined |
| 18-102-final-viral-combined_k141_44945||full | Not-determined |
| 18-102-final-viral-combined_k141_230575||full | Low-quality |
| 18-102-final-viral-combined_k141_54246||full | Not-determined |
| 18-102-final-viral-combined_k141_44955||full | Not-determined |
| 18-102-final-viral-combined_k141_12658||full | Not-determined |
| 18-102-final-viral-combined_k141_54252||full | Not-determined |
| 18-102-final-viral-combined_k141_23178||full | Low-quality |
| 18-102-final-viral-combined_k141_219559||full | Not-determined |
| 18-102-final-viral-combined_k141_44964||full | Low-quality |
| 18-102-final-viral-combined_k141_209338||full | Low-quality |
| 18-102-final-viral-combined_k141_23200||full | Low-quality |
| 18-102-final-viral-combined_k141_230588||full | Low-quality |
| 18-102-final-viral-combined_k141_240370||full | Not-determined |
| 18-102-final-viral-combined_k141_54297||full | Not-determined |
| 18-102-final-viral-combined_k141_95834||full | Low-quality |
| 18-102-final-viral-combined_k141_168441||full | Low-quality |
| 18-102-final-viral-combined_k141_44984||full | Not-determined |
| 18-102-final-viral-combined_k141_106025||full | Low-quality |
| 18-102-final-viral-combined_k141_230601||full | Low-quality |
| 18-102-final-viral-combined_k141_198595||full | Low-quality |
| 18-102-final-viral-combined_k141_157703||full | Low-quality |
| 18-102-final-viral-combined_k141_209353||full | Low-quality |
| 18-102-final-viral-combined_k141_23219||full | Low-quality |
| 18-102-final-viral-combined_k141_95863||full | Low-quality |
| 18-102-final-viral-combined_k141_54301||full | Not-determined |
| 18-102-final-viral-combined_k141_209367||full | Not-determined |
| 18-102-final-viral-combined_k141_2347||full | Low-quality |
| 18-102-final-viral-combined_k141_198596||full | Low-quality |
| 18-102-final-viral-combined_k141_64388||full | Not-determined |
| 18-102-final-viral-combined_k141_147186||full | Low-quality |
| 18-102-final-viral-combined_k141_106041||full | Low-quality |
| 18-102-final-viral-combined_k141_106043||full | Not-determined |
| 18-102-final-viral-combined_k141_147200||full | Not-determined |
| 18-102-final-viral-combined_k141_178727||full | Not-determined |
| 18-102-final-viral-combined_k141_54331||full | Not-determined |
| 18-102-final-viral-combined_k141_116114||full | Low-quality |
| 18-102-final-viral-combined_k141_106078||full | Not-determined |
| 18-102-final-viral-combined_k141_240385||full | Not-determined |
| 18-102-final-viral-combined_k141_54348||full | Not-determined |
| 18-102-final-viral-combined_k141_116129||full | Low-quality |
| 18-102-final-viral-combined_k141_147228||full | Not-determined |
| 18-102-final-viral-combined_k141_230652||full | Not-determined |
| 18-102-final-viral-combined_k141_178742||full | Not-determined |
| 18-102-final-viral-combined_k141_157787||full | Low-quality |
| 18-102-final-viral-combined_k141_240394||full | Not-determined |
| 18-102-final-viral-combined_k141_219656||full | Low-quality |
| 18-102-final-viral-combined_k141_230661||full | Low-quality |
| 18-102-final-viral-combined_k141_178753||full | Low-quality |
| 18-102-final-viral-combined_k141_209441||full | Not-determined |
| 18-102-final-viral-combined_k141_178756||full | Not-determined |
| 18-102-final-viral-combined_k141_157794||full | Not-determined |
| 18-102-final-viral-combined_k141_178765||full | Low-quality |
| 18-102-final-viral-combined_k141_230685||full | Not-determined |
| 18-102-final-viral-combined_k141_85474||full | Low-quality |
| 18-102-final-viral-combined_k141_23254||full | Not-determined |
| 18-102-final-viral-combined_k141_230688||full | Low-quality |
| 18-102-final-viral-combined_k141_168524||full | Not-determined |
| 18-102-final-viral-combined_k141_219676||full | Low-quality |
| 18-102-final-viral-combined_k141_157818||full | Low-quality |
| 18-102-final-viral-combined_k141_85490||full | Not-determined |
| 18-102-final-viral-combined_k141_23267||full | Low-quality |
| 18-102-final-viral-combined_k141_12754||full | Not-determined |
| 18-102-final-viral-combined_k141_188890||full | Low-quality |
| 18-102-final-viral-combined_k141_23273||full | Low-quality |
| 18-102-final-viral-combined_k141_74830||full | Not-determined |
| 18-102-final-viral-combined_k141_23281||full | Not-determined |
| 18-102-final-viral-combined_k141_219715||full | Low-quality |
| 18-102-final-viral-combined_k141_33220||full | Not-determined |
| 18-102-final-viral-combined_k141_230731||full | Low-quality |
| 18-102-final-viral-combined_k141_147277||full | Not-determined |
| 18-102-final-viral-combined_k141_230746||full | Not-determined |
| 18-102-final-viral-combined_k141_45082||full | Low-quality |
| 18-102-final-viral-combined_k141_64430||full | Low-quality |
| 18-102-final-viral-combined_k141_147294||full | Not-determined |
| 18-102-final-viral-combined_k141_147296||full | Low-quality |
| 18-102-final-viral-combined_k141_198663||full | Not-determined |
| 18-102-final-viral-combined_k141_33248||full | Low-quality |
| 18-102-final-viral-combined_k141_116220||full | Low-quality |
| 18-102-final-viral-combined_k141_33253||full | Not-determined |
| 18-102-final-viral-combined_k141_23321||full | Not-determined |
| 18-102-final-viral-combined_k141_157886||full | Not-determined |
| 18-102-final-viral-combined_k141_178830||full | Low-quality |
| 18-102-final-viral-combined_k141_157887||full | Low-quality |
| 18-102-final-viral-combined_k141_85528||full | Not-determined |
| 18-102-final-viral-combined_k141_198680||full | Not-determined |
| 18-102-final-viral-combined_k141_178844||full | Not-determined |
| 18-102-final-viral-combined_k141_126699||full | Not-determined |
| 18-102-final-viral-combined_k141_198686||full | Low-quality |
| 18-102-final-viral-combined_k141_54429||full | Low-quality |
| 18-102-final-viral-combined_k141_2387||full | Low-quality |
| 18-102-final-viral-combined_k141_45090||full | Not-determined |
| 18-102-final-viral-combined_k141_126715||full | Not-determined |
| 18-102-final-viral-combined_k141_2391||full | Not-determined |
| 18-102-final-viral-combined_k141_33282||full | Low-quality |
| 18-102-final-viral-combined_k141_85560||full | Low-quality |
| 18-102-final-viral-combined_k141_147301||full | Low-quality |
| 18-102-final-viral-combined_k141_178886||full | Not-determined |
| 18-102-final-viral-combined_k141_12850||full | Not-determined |
| 18-102-final-viral-combined_k141_209501||full | Low-quality |
| 18-102-final-viral-combined_k141_54457||full | Not-determined |
| 18-102-final-viral-combined_k141_85585||full | Low-quality |
| 18-102-final-viral-combined_k141_240512||full | Low-quality |
| 18-102-final-viral-combined_k141_33298||full | Low-quality |
| 18-102-final-viral-combined_k141_188974||full | Not-determined |
| 18-102-final-viral-combined_k141_2402||full | Not-determined |
| 18-102-final-viral-combined_k141_147333||full | Not-determined |
| 18-102-final-viral-combined_k141_33318||full | Low-quality |
| 18-102-final-viral-combined_k141_137280||full | Not-determined |
| 18-102-final-viral-combined_k141_74891||full | Low-quality |
| 18-102-final-viral-combined_k141_137289||full | Not-determined |
| 18-102-final-viral-combined_k141_64481||full | Not-determined |
| 18-102-final-viral-combined_k141_54488||full | Low-quality |
| 18-102-final-viral-combined_k141_64492||full | Not-determined |
| KM-final-viral-combined_k141_222715||full | Medium-quality |
| 18-102-final-viral-combined_k141_189009||full | Low-quality |
| 18-102-final-viral-combined_k141_23369||full | Low-quality |
| 18-102-final-viral-combined_k141_54503||full | Low-quality |
| 18-102-final-viral-combined_k141_64503||full | Low-quality |
| 18-102-final-viral-combined_k141_137303||full | Low-quality |
| 18-102-final-viral-combined_k141_85617||full | Not-determined |
| 18-102-final-viral-combined_k141_64521||full | Not-determined |
| 18-102-final-viral-combined_k141_137315||full | Low-quality |
| 18-102-final-viral-combined_k141_240562||full | Not-determined |
| 18-102-final-viral-combined_k141_74941||full | Low-quality |
| 18-102-final-viral-combined_k141_198713||full | Not-determined |
| 18-102-final-viral-combined_k141_12877||full | Low-quality |
| 18-102-final-viral-combined_k141_74946||full | Low-quality |
| 18-102-final-viral-combined_k141_85654||full | Not-determined |
| 18-102-final-viral-combined_k141_147347||full | Not-determined |
| 18-102-final-viral-combined_k141_168612||full | Not-determined |
| 18-110-final-viral-combined_k141_219796||full | Medium-quality |
| 18-102-final-viral-combined_k141_137370||full | Low-quality |
| 18-102-final-viral-combined_k141_54571||full | Not-determined |
| 18-102-final-viral-combined_k141_33419||full | Not-determined |
| 18-102-final-viral-combined_k141_95991||full | Not-determined |
| 18-102-final-viral-combined_k141_33422||full | Low-quality |
| 18-102-final-viral-combined_k141_23395||full | Not-determined |
| 18-102-final-viral-combined_k141_45243||full | Low-quality |
| 18-102-final-viral-combined_k141_23404||full | Not-determined |
| 18-102-final-viral-combined_k141_219799||full | Low-quality |
| 18-102-final-viral-combined_k141_178960||full | Not-determined |
| 18-102-final-viral-combined_k141_2465||full | Low-quality |
| 18-102-final-viral-combined_k141_137385||full | Low-quality |
| 18-102-final-viral-combined_k141_2467||full | Low-quality |
| 18-102-final-viral-combined_k141_189053||full | Not-determined |
| 18-102-final-viral-combined_k141_33448||full | Low-quality |
| 18-102-final-viral-combined_k141_2481||full | Not-determined |
| 18-102-final-viral-combined_k141_33454||full | Not-determined |
| 18-102-final-viral-combined_k141_137400||full | Not-determined |
| 18-102-final-viral-combined_k141_2484||full | Not-determined |
| 18-102-final-viral-combined_k141_168654||full | Not-determined |
| 18-102-final-viral-combined_k141_126823||full | Not-determined |
| 18-102-final-viral-combined_k141_168665||full | Low-quality |
| 18-102-final-viral-combined_k141_147393||full | Low-quality |
| 18-102-final-viral-combined_k141_179009||full | Not-determined |
| 18-102-final-viral-combined_k141_189073||full | Not-determined |
| 18-102-final-viral-combined_k141_126840||full | Not-determined |
| 18-102-final-viral-combined_k141_54633||full | Low-quality |
| 18-102-final-viral-combined_k141_33489||full | Not-determined |
| 18-102-final-viral-combined_k141_147415||full | Not-determined |
| 18-102-final-viral-combined_k141_179018||full | Low-quality |
| 18-102-final-viral-combined_k141_240606||full | Not-determined |
| 18-102-final-viral-combined_k141_12944||full | Not-determined |
| 18-102-final-viral-combined_k141_2508||full | Low-quality |
| 18-102-final-viral-combined_k141_179020||full | Not-determined |
| 18-102-final-viral-combined_k141_64630||full | Not-determined |
| 18-102-final-viral-combined_k141_137432||full | Low-quality |
| 18-102-final-viral-combined_k141_147436||full | Low-quality |
| 18-102-final-viral-combined_k141_75006||full | Not-determined |
| 18-102-final-viral-combined_k141_179022||full | Not-determined |
| KM-final-viral-combined_k141_136622||full | Medium-quality |
| 18-102-final-viral-combined_k141_147438||full | Not-determined |
| 18-102-final-viral-combined_k141_33520||full | Low-quality |
| 18-102-final-viral-combined_k141_198826||full | Not-determined |
| 18-102-final-viral-combined_k141_12955||full | Low-quality |
| 18-102-final-viral-combined_k141_33534||full | Not-determined |
| 18-102-final-viral-combined_k141_2533||full | Low-quality |
| 18-102-final-viral-combined_k141_85743||full | Not-determined |
| 18-102-final-viral-combined_k141_106223||full | Not-determined |
| 18-102-final-viral-combined_k141_64651||full | Not-determined |
| 18-102-final-viral-combined_k141_189141||full | Low-quality |
| 18-102-final-viral-combined_k141_85745||full | Low-quality |
| 18-102-final-viral-combined_k141_12966||full | Not-determined |
| 18-102-final-viral-combined_k141_126898||full | Low-quality |
| 18-102-final-viral-combined_k141_2540||full | Not-determined |
| 18-102-final-viral-combined_k141_137449||full | Low-quality |
| 18-102-final-viral-combined_k141_85751||full | Low-quality |
| 18-102-final-viral-combined_k141_168708||full | Not-determined |
| 18-102-final-viral-combined_k141_85755||full | Not-determined |
| 18-102-final-viral-combined_k141_209596||full | Low-quality |
| 18-102-final-viral-combined_k141_2546||full | Low-quality |
| 18-102-final-viral-combined_k141_106238||full | Not-determined |
| 18-102-final-viral-combined_k141_230865||full | Low-quality |
| 18-102-final-viral-combined_k141_230869||full | Not-determined |
| 18-102-final-viral-combined_k141_209604||full | Not-determined |
| 18-102-final-viral-combined_k141_85772||full | Low-quality |
| 18-102-final-viral-combined_k141_106249||full | Low-quality |
| 18-102-final-viral-combined_k141_168733||full | Not-determined |
| 18-102-final-viral-combined_k141_2548||full | Low-quality |
| 18-102-final-viral-combined_k141_64668||full | Not-determined |
| 18-102-final-viral-combined_k141_45357||full | Low-quality |
| 18-102-final-viral-combined_k141_168749||full | Low-quality |
| 18-102-final-viral-combined_k141_126931||full | Low-quality |
| 18-102-final-viral-combined_k141_116332||full | Not-determined |
| 18-102-final-viral-combined_k141_106259||full | Not-determined |
| 18-102-final-viral-combined_k141_209630||full | Not-determined |
| 18-102-final-viral-combined_k141_157992||full | Not-determined |
| 18-102-final-viral-combined_k141_157996||full | Not-determined |
| 18-102-final-viral-combined_k141_189206||full | Low-quality |
| 18-102-final-viral-combined_k141_147523||full | Not-determined |
| 18-102-final-viral-combined_k141_168773||full | Not-determined |
| 18-102-final-viral-combined_k141_126948||full | Not-determined |
| 18-102-final-viral-combined_k141_75075||full | Low-quality |
| 18-102-final-viral-combined_k141_33582||full | Low-quality |
| 18-102-final-viral-combined_k141_137489||full | Low-quality |
| 18-102-final-viral-combined_k141_2569||full | Low-quality |
| 18-102-final-viral-combined_k141_147531||full | Low-quality |
| 18-102-final-viral-combined_k141_23480||full | Low-quality |
| 18-102-final-viral-combined_k141_33587||full | Not-determined |
| 18-102-final-viral-combined_k141_240629||full | Low-quality |
| 18-102-final-viral-combined_k141_126952||full | Low-quality |
| 18-102-final-viral-combined_k141_106275||full | Not-determined |
| 18-102-final-viral-combined_k141_96169||full | Not-determined |
| 18-102-final-viral-combined_k141_106282||full | Low-quality |
| 18-102-final-viral-combined_k141_23515||full | Low-quality |
| 18-102-final-viral-combined_k141_240641||full | Not-determined |
| 18-102-final-viral-combined_k141_54682||full | Not-determined |
| 18-102-final-viral-combined_k141_168785||full | Low-quality |
| 18-102-final-viral-combined_k141_33631||full | Not-determined |
| 18-102-final-viral-combined_k141_179069||full | Not-determined |
| 17-106_k141_114971 | Medium-quality |
| 18-102-final-viral-combined_k141_179072||full | Not-determined |
| 18-102-final-viral-combined_k141_85820||full | Not-determined |
| 18-102-final-viral-combined_k141_179079||full | Not-determined |
| 18-102-final-viral-combined_k141_158036||full | Low-quality |
| 18-102-final-viral-combined_k141_85826||full | Low-quality |
| 18-102-final-viral-combined_k141_33655||full | Not-determined |
| 18-102-final-viral-combined_k141_158052||full | Not-determined |
| 18-102-final-viral-combined_k141_64729||full | Not-determined |
| 18-102-final-viral-combined_k141_168799||full | Low-quality |
| 18-102-final-viral-combined_k141_85845||full | Low-quality |
| 18-102-final-viral-combined_k141_85851||full | Low-quality |
| 18-102-final-viral-combined_k141_209654||full | Not-determined |
| 18-102-final-viral-combined_k141_168820||full | Not-determined |
| 18-102-final-viral-combined_k141_64739||full | Low-quality |
| 18-102-final-viral-combined_k141_75089||full | Not-determined |
| 18-102-final-viral-combined_k141_147578||full | Not-determined |
| 18-102-final-viral-combined_k141_209667||full | Low-quality |
| 18-102-final-viral-combined_k141_85855||full | Low-quality |
| 18-102-final-viral-combined_k141_240670||full | Not-determined |
| 18-102-final-viral-combined_k141_33687||full | Not-determined |
| 18-102-final-viral-combined_k141_75094||full | Not-determined |
| 18-102-final-viral-combined_k141_85859||full | Not-determined |
| 18-102-final-viral-combined_k141_179134||full | Not-determined |
| 18-102-final-viral-combined_k141_179141||full | Not-determined |
| 18-102-final-viral-combined_k141_23550||full | Not-determined |
| 18-102-final-viral-combined_k141_230913||full | Not-determined |
| 18-102-final-viral-combined_k141_189327||full | Not-determined |
| 18-102-final-viral-combined_k141_75123||full | Low-quality |
| 18-102-final-viral-combined_k141_96287||full | Low-quality |
| 18-102-final-viral-combined_k141_96299||full | Not-determined |
| 18-102-final-viral-combined_k141_23559||full | Not-determined |
| 18-102-final-viral-combined_k141_116364||full | Not-determined |
| 18-102-final-viral-combined_k141_209697||full | Low-quality |
| 18-102-final-viral-combined_k141_85905||full | Not-determined |
| 18-102-final-viral-combined_k141_116376||full | Low-quality |
| 17-106-final-viral-combined_k141_114971||full | Medium-quality |
| 18-102-final-viral-combined_k141_106352||full | Low-quality |
| 18-102-final-viral-combined_k141_168888||full | Low-quality |
| 18-102-final-viral-combined_k141_179184||full | Not-determined |
| 18-102-final-viral-combined_k141_2624||full | Not-determined |
| 18-102-final-viral-combined_k141_168890||full | Not-determined |
| 18-102-final-viral-combined_k141_54768||full | Not-determined |
| 18-102-final-viral-combined_k141_147651||full | Low-quality |
| 18-102-final-viral-combined_k141_45479||full | Low-quality |
| 18-102-final-viral-combined_k141_230925||full | Not-determined |
| 18-102-final-viral-combined_k141_54775||full | Low-quality |
| 18-110_k141_118779 | Medium-quality |
| 18-102-final-viral-combined_k141_198877||full | Low-quality |
| 18-102-final-viral-combined_k141_45489||full | Not-determined |
| 18-102-final-viral-combined_k141_33758||full | Not-determined |
| 18-102-final-viral-combined_k141_158119||full | Low-quality |
| 18-102-final-viral-combined_k141_198897||full | Low-quality |
| 18-102-final-viral-combined_k141_158124||full | Not-determined |
| 18-102-final-viral-combined_k141_33782||full | Low-quality |
| 18-102-final-viral-combined_k141_2653||full | Not-determined |
| 18-102-final-viral-combined_k141_168949||full | Low-quality |
| 18-102-final-viral-combined_k141_147694||full | Low-quality |
| 18-102-final-viral-combined_k141_158148||full | Low-quality |
| 18-102-final-viral-combined_k141_106388||full | Not-determined |
| 18-102-final-viral-combined_k141_85981||full | Low-quality |
| 18-102-final-viral-combined_k141_198899||full | Low-quality |
| 18-102-final-viral-combined_k141_147708||full | Not-determined |
| 18-102-final-viral-combined_k141_96323||full | Low-quality |
| 18-102-final-viral-combined_k141_198911||full | Not-determined |
| 18-102-final-viral-combined_k141_189390||full | Low-quality |
| 18-102-final-viral-combined_k141_23623||full | Low-quality |
| 18-102-final-viral-combined_k141_179263||full | Low-quality |
| 18-102-final-viral-combined_k141_45544||full | Low-quality |
| 18-102-final-viral-combined_k141_127089||full | Low-quality |
| 18-102-final-viral-combined_k141_230967||full | Not-determined |
| 18-102-final-viral-combined_k141_127090||full | Not-determined |
| 18-102-final-viral-combined_k141_45550||full | Not-determined |
| 18-102-final-viral-combined_k141_179272||full | Not-determined |
| 18-102-final-viral-combined_k141_13137||full | Not-determined |
| 18-102-final-viral-combined_k141_127096||full | Not-determined |
| 18-102-final-viral-combined_k141_33824||full | Not-determined |
| 18-102-final-viral-combined_k141_86013||full | Not-determined |
| 18-102-final-viral-combined_k141_189412||full | Not-determined |
| 18-102-final-viral-combined_k141_13147||full | Not-determined |
| 18-102-final-viral-combined_k141_179285||full | Low-quality |
| 18-102-final-viral-combined_k141_127112||full | Low-quality |
| 18-102-final-viral-combined_k141_116424||full | Low-quality |
| 18-102-final-viral-combined_k141_96348||full | Low-quality |
| 18-102-final-viral-combined_k141_96359||full | Low-quality |
| 18-102-final-viral-combined_k141_45594||full | Low-quality |
| 18-102-final-viral-combined_k141_2718||full | Low-quality |
| 18-102-final-viral-combined_k141_219847||full | Low-quality |
| 18-102-final-viral-combined_k141_54850||full | Not-determined |
| 18-102-final-viral-combined_k141_179326||full | Low-quality |
| 18-102-final-viral-combined_k141_230976||full | Low-quality |
| 18-102-final-viral-combined_k141_33834||full | Low-quality |
| 18-102-final-viral-combined_k141_158151||full | Low-quality |
| 18-102-final-viral-combined_k141_54858||full | Low-quality |
| 18-102-final-viral-combined_k141_198922||full | Not-determined |
| 18-102-final-viral-combined_k141_179334||full | Low-quality |
| 18-102-final-viral-combined_k141_158155||full | Not-determined |
| 18-102-final-viral-combined_k141_179335||full | Low-quality |
| 18-102-final-viral-combined_k141_158174||full | Not-determined |
| 18-102-final-viral-combined_k141_116441||full | Low-quality |
| 18-102-final-viral-combined_k141_179354||full | Not-determined |
| 18-102-final-viral-combined_k141_209819||full | Low-quality |
| 18-102-final-viral-combined_k141_13193||full | Not-determined |
| 18-102-final-viral-combined_k141_33884||full | Low-quality |
| 18-102-final-viral-combined_k141_13200||full | Low-quality |
| 18-102-final-viral-combined_k141_127208||full | Not-determined |
| 18-102-final-viral-combined_k141_209835||full | Low-quality |
| 18-102-final-viral-combined_k141_137665||full | Not-determined |
| 18-102-final-viral-combined_k141_13225||full | Not-determined |
| 18-102-final-viral-combined_k141_116459||full | Low-quality |
| 18-102-final-viral-combined_k141_106420||full | Low-quality |
| 18-102-final-viral-combined_k141_106423||full | Not-determined |
| 18-102-final-viral-combined_k141_198975||full | Not-determined |
| 18-102-final-viral-combined_k141_198978||full | Low-quality |
| 18-102-final-viral-combined_k141_106430||full | Low-quality |
| 18-102-final-viral-combined_k141_240746||full | Low-quality |
| 18-102-final-viral-combined_k141_2737||full | Not-determined |
| 18-102-final-viral-combined_k141_231027||full | Not-determined |
| 18-102-final-viral-combined_k141_198987||full | Not-determined |
| 18-102-final-viral-combined_k141_2738||full | Not-determined |
| 18-102-final-viral-combined_k141_33953||full | Not-determined |
| 18-102-final-viral-combined_k141_106435||full | Not-determined |
| 18-102-final-viral-combined_k141_33957||full | Low-quality |
| 18-102-final-viral-combined_k141_198995||full | Low-quality |
| 18-102-final-viral-combined_k141_158224||full | Not-determined |
| 18-102-final-viral-combined_k141_179404||full | Low-quality |
| 18-102-final-viral-combined_k141_240765||full | Not-determined |
| 18-102-final-viral-combined_k141_137697||full | Not-determined |
| 18-102-final-viral-combined_k141_2756||full | Not-determined |
| 18-102-final-viral-combined_k141_106457||full | Low-quality |
| 18-102-final-viral-combined_k141_240771||full | Not-determined |
| 18-102-final-viral-combined_k141_147800||full | Not-determined |
| 18-102-final-viral-combined_k141_240791||full | Not-determined |
| 18-102-final-viral-combined_k141_116462||full | Not-determined |
| 18-102-final-viral-combined_k141_54886||full | Low-quality |
| 18-102-final-viral-combined_k141_106473||full | Low-quality |
| 18-102-final-viral-combined_k141_189557||full | Not-determined |
| 18-102-final-viral-combined_k141_116477||full | Low-quality |
| 18-102-final-viral-combined_k141_179459||full | Low-quality |
| 18-102-final-viral-combined_k141_179463||full | Low-quality |
| 18-102-final-viral-combined_k141_106475||full | Not-determined |
| 18-102-final-viral-combined_k141_158269||full | Not-determined |
| 18-102-final-viral-combined_k141_45680||full | Not-determined |
| 18-102-final-viral-combined_k141_106491||full | Low-quality |
| 18-102-final-viral-combined_k141_137781||full | Not-determined |
| 18-102-final-viral-combined_k141_231096||full | Low-quality |
| 18-102-final-viral-combined_k141_137784||full | Not-determined |
| 18-102-final-viral-combined_k141_137787||full | Not-determined |
| 18-102-final-viral-combined_k141_96530||full | Low-quality |
| 18-102-final-viral-combined_k141_147842||full | Low-quality |
| 18-102-final-viral-combined_k141_219932||full | Low-quality |
| 18-102-final-viral-combined_k141_2813||full | Not-determined |
| 18-102-final-viral-combined_k141_158292||full | Not-determined |
| 18-102-final-viral-combined_k141_231102||full | Low-quality |
| 18-102-final-viral-combined_k141_34016||full | Low-quality |
| 18-102-final-viral-combined_k141_231103||full | Not-determined |
| 18-102-final-viral-combined_k141_219944||full | Not-determined |
| 18-102-final-viral-combined_k141_127304||full | Not-determined |
| 18-102-final-viral-combined_k141_96545||full | Not-determined |
| 18-102-final-viral-combined_k141_116506||full | Low-quality |
| 18-102-final-viral-combined_k141_86090||full | Not-determined |
| 18-102-final-viral-combined_k141_34022||full | Not-determined |
| 18-102-final-viral-combined_k141_2842||full | Not-determined |
| 18-102-final-viral-combined_k141_137840||full | Low-quality |
| 18-102-final-viral-combined_k141_2848||full | Not-determined |
| 18-102-final-viral-combined_k141_34033||full | Not-determined |
| 18-102-final-viral-combined_k141_189634||full | Not-determined |
| 18-102-final-viral-combined_k141_96554||full | Not-determined |
| 18-102-final-viral-combined_k141_231133||full | Low-quality |
| 18-102-final-viral-combined_k141_34047||full | Low-quality |
| 18-102-final-viral-combined_k141_64904||full | Not-determined |
| 18-102-final-viral-combined_k141_137868||full | Low-quality |
| 18-102-final-viral-combined_k141_116543||full | Low-quality |
| 18-102-final-viral-combined_k141_116545||full | Not-determined |
| 18-102-final-viral-combined_k141_240907||full | Not-determined |
| 18-102-final-viral-combined_k141_106609||full | Low-quality |
| 18-102-final-viral-combined_k141_64909||full | Low-quality |
| 18-102-final-viral-combined_k141_240941||full | Low-quality |
| 18-102-final-viral-combined_k141_127345||full | Low-quality |
| 18-102-final-viral-combined_k141_219956||full | Low-quality |
| 18-102-final-viral-combined_k141_34081||full | Low-quality |
| 18-102-final-viral-combined_k141_2881||full | Not-determined |
| 18-102-final-viral-combined_k141_127382||full | Not-determined |
| 18-102-final-viral-combined_k141_169219||full | Not-determined |
| 18-102-final-viral-combined_k141_106636||full | Not-determined |
| 18-102-final-viral-combined_k141_127392||full | Not-determined |
| 18-102-final-viral-combined_k141_179490||full | Not-determined |
| 18-102-final-viral-combined_k141_64924||full | Low-quality |
| 18-102-final-viral-combined_k141_127402||full | Low-quality |

| | |
|---|---|
| 18-102-final-viral-combined_k141_137945\|\|full | Not-determined |
| 18-102-final-viral-combined_k141_64936\|\|full | Low-quality |
| 18-102-final-viral-combined_k141_137951\|\|full | Low-quality |
| 18-102-final-viral-combined_k141_75160\|\|full | Low-quality |
| 18-102-final-viral-combined_k141_137952\|\|full | Low-quality |
| 18-102-final-viral-combined_k141_219987\|\|full | Not-determined |
| 18-102-final-viral-combined_k141_2910\|\|full | Not-determined |
| 18-102-final-viral-combined_k141_23706\|\|full | Low-quality |
| 18-102-final-viral-combined_k141_179506\|\|full | Not-determined |
| 18-102-final-viral-combined_k141_219995\|\|full | Not-determined |
| 18-102-final-viral-combined_k141_2913\|\|full | Not-determined |
| 18-102-final-viral-combined_k141_209941\|\|full | Not-determined |
| 18-102-final-viral-combined_k141_23712\|\|full | Low-quality |
| 18-102-final-viral-combined_k141_75177\|\|full | Not-determined |
| 18-102-final-viral-combined_k141_209943\|\|full | Not-determined |
| 18-102-final-viral-combined_k141_220001\|\|full | Not-determined |
| 18-102-final-viral-combined_k141_199123\|\|full | Not-determined |
| 18-102-final-viral-combined_k141_127414\|\|full | Not-determined |
| 18-102-final-viral-combined_k141_64970\|\|full | Not-determined |
| 18-102-final-viral-combined_k141_220008\|\|full | Low-quality |
| 18-102-final-viral-combined_k141_147885\|\|full | Not-determined |
| 18-102-final-viral-combined_k141_199126\|\|full | Low-quality |
| 18-102-final-viral-combined_k141_116628\|\|full | Not-determined |
| 18-102-final-viral-combined_k141_220011\|\|full | Low-quality |
| 18-102-final-viral-combined_k141_127438\|\|full | Not-determined |
| 18-102-final-viral-combined_k141_86151\|\|full | Not-determined |
| 18-102-final-viral-combined_k141_106670\|\|full | Not-determined |
| 18-102-final-viral-combined_k141_23739\|\|full | Not-determined |
| 18-102-final-viral-combined_k141_127443\|\|full | Not-determined |
| 18-102-final-viral-combined_k141_96579\|\|full | Not-determined |
| 18-102-final-viral-combined_k141_199142\|\|full | Low-quality |
| 18-102-final-viral-combined_k141_116642\|\|full | Not-determined |
| 18-102-final-viral-combined_k141_75203\|\|full | Not-determined |
| 18-102-final-viral-combined_k141_54926\|\|full | Not-determined |
| 18-102-final-viral-combined_k141_231177\|\|full | Low-quality |
| 18-102-final-viral-combined_k141_189775\|\|full | Low-quality |
| 18-102-final-viral-combined_k141_86178\|\|full | Low-quality |
| 17-108-final-viral-combined_k141_299954\|\|full | Medium-quality |
| 18-102-final-viral-combined_k141_106702\|\|full | Low-quality |
| 18-102-final-viral-combined_k141_189798\|\|full | Not-determined |
| 18-102-final-viral-combined_k141_65024\|\|full | Not-determined |
| 18-102-final-viral-combined_k141_231194\|\|full | Low-quality |
| 18-102-final-viral-combined_k141_127472\|\|full | Not-determined |
| 18-102-final-viral-combined_k141_86189\|\|full | Low-quality |
| 18-102-final-viral-combined_k141_96601\|\|full | Low-quality |
| 18-102-final-viral-combined_k141_127476\|\|full | Low-quality |
| 18-102-final-viral-combined_k141_116666\|\|full | Not-determined |
| 18-102-final-viral-combined_k141_96616\|\|full | Low-quality |
| 18-102-final-viral-combined_k141_138006\|\|full | Low-quality |
| 18-102-final-viral-combined_k141_2972\|\|full | Low-quality |
| 18-102-final-viral-combined_k141_210006\|\|full | Low-quality |
| 18-102-final-viral-combined_k141_169338\|\|full | Low-quality |
| KM-final-viral-combined_k141_140517\|\|full | Medium-quality |
| 18-102-final-viral-combined_k141_75241\|\|full | Not-determined |
| 18-102-final-viral-combined_k141_34126\|\|full | Low-quality |
| 18-102-final-viral-combined_k141_210010\|\|full | Low-quality |
| 18-102-final-viral-combined_k141_231221\|\|full | Low-quality |
| 18-102-final-viral-combined_k141_138022\|\|full | Not-determined |
| 18-102-final-viral-combined_k141_45747\|\|full | Not-determined |
| 18-102-final-viral-combined_k141_147934\|\|full | Not-determined |
| 18-102-final-viral-combined_k141_34133\|\|full | Low-quality |
| 18-102-final-viral-combined_k141_179539\|\|full | Not-determined |
| 18-102-final-viral-combined_k141_75262\|\|full | Not-determined |
| 18-102-final-viral-combined_k141_65083\|\|full | Low-quality |
| 18-102-final-viral-combined_k141_147965\|\|full | Not-determined |
| 18-102-final-viral-combined_k141_65091\|\|full | Low-quality |
| 18-102-final-viral-combined_k141_210054\|\|full | Low-quality |
| 18-102-final-viral-combined_k141_210062\|\|full | Low-quality |
| 18-102-final-viral-combined_k141_45767\|\|full | Low-quality |
| 18-102-final-viral-combined_k141_179583\|\|full | Low-quality |
| 18-102-final-viral-combined_k141_45769\|\|full | Not-determined |
| 18-102-final-viral-combined_k141_169367\|\|full | Not-determined |
| 18-102-final-viral-combined_k141_179586\|\|full | Not-determined |
| 18-102-final-viral-combined_k141_199156\|\|full | Not-determined |
| 18-102-final-viral-combined_k141_65109\|\|full | Low-quality |
| 18-102-final-viral-combined_k141_189858\|\|full | Not-determined |
| 18-102-final-viral-combined_k141_148016\|\|full | Not-determined |
| 18-102-final-viral-combined_k141_169385\|\|full | Not-determined |
| 18-102-final-viral-combined_k141_169395\|\|full | Low-quality |
| 18-102-final-viral-combined_k141_106721\|\|full | Not-determined |
| 18-102-final-viral-combined_k141_13295\|\|full | Not-determined |
| 18-102-final-viral-combined_k141_86263\|\|full | Not-determined |
| 18-102-final-viral-combined_k141_75313\|\|full | Low-quality |
| 18-102-final-viral-combined_k141_138055\|\|full | Low-quality |
| 18-102-final-viral-combined_k141_116752\|\|full | Low-quality |
| 18-102-final-viral-combined_k141_106740\|\|full | Low-quality |
| 18-102-final-viral-combined_k141_75321\|\|full | Low-quality |
| 18-102-final-viral-combined_k141_116763\|\|full | Low-quality |
| 18-102-final-viral-combined_k141_54998\|\|full | Low-quality |
| 18-102-final-viral-combined_k141_231313\|\|full | Not-determined |
| 18-102-final-viral-combined_k141_210115\|\|full | Not-determined |
| 18-102-final-viral-combined_k141_75331\|\|full | Not-determined |
| 18-102-final-viral-combined_k141_55010\|\|full | Not-determined |
| 18-102-final-viral-combined_k141_231320\|\|full | Not-determined |
| 18-102-final-viral-combined_k141_65162\|\|full | Low-quality |
| 18-102-final-viral-combined_k141_158408\|\|full | Low-quality |
| 18-102-final-viral-combined_k141_169424\|\|full | Not-determined |
| 18-102-final-viral-combined_k141_210133\|\|full | Not-determined |
| 18-102-final-viral-combined_k141_127582\|\|full | Low-quality |
| 18-102-final-viral-combined_k141_86291\|\|full | Low-quality |
| 18-102-final-viral-combined_k141_45858\|\|full | Low-quality |
| 18-102-final-viral-combined_k141_158422\|\|full | Low-quality |
| 18-102-final-viral-combined_k141_45862\|\|full | Low-quality |
| 18-102-final-viral-combined_k141_96679\|\|full | Low-quality |
| 18-102-final-viral-combined_k141_231346\|\|full | Low-quality |
| 18-102-final-viral-combined_k141_179675\|\|full | Not-determined |
| 18-102-final-viral-combined_k141_231353\|\|full | Not-determined |
| 18-102-final-viral-combined_k141_231356\|\|full | Not-determined |
| 18-102-final-viral-combined_k141_210184\|\|full | Low-quality |
| 18-102-final-viral-combined_k141_34249\|\|full | Not-determined |
| 18-102-final-viral-combined_k141_96717\|\|full | Not-determined |
| 18-102-final-viral-combined_k141_231364\|\|full | Low-quality |
| 18-102-final-viral-combined_k141_65228\|\|full | Not-determined |
| 18-102-final-viral-combined_k141_96727\|\|full | Low-quality |
| 18-102-final-viral-combined_k141_116825\|\|full | Low-quality |
| 18-102-final-viral-combined_k141_45927\|\|full | Not-determined |
| 18-102-final-viral-combined_k141_241066\|\|full | Not-determined |
| 18-102-final-viral-combined_k141_179726\|\|full | Low-quality |
| 18-102-final-viral-combined_k141_169465\|\|full | Low-quality |
| 18-102-final-viral-combined_k141_148103\|\|full | Not-determined |
| 18-102-final-viral-combined_k141_189864\|\|full | Low-quality |
| 18-102-final-viral-combined_k141_3083\|\|full | Low-quality |
| 18-102-final-viral-combined_k141_231375\|\|full | Low-quality |
| 18-102-final-viral-combined_k141_199244\|\|full | Low-quality |
| 18-102-final-viral-combined_k141_148107\|\|full | Low-quality |
| 18-102-final-viral-combined_k141_34277\|\|full | Low-quality |
| 18-102-final-viral-combined_k141_148123\|\|full | Not-determined |
| 18-102-final-viral-combined_k141_65281\|\|full | Not-determined |
| 18-102-final-viral-combined_k141_189883\|\|full | Low-quality |
| 18-102-final-viral-combined_k141_106843\|\|full | Not-determined |
| 18-102-final-viral-combined_k141_75353\|\|full | Low-quality |
| 18-102-final-viral-combined_k141_34301\|\|full | Low-quality |
| 18-102-final-viral-combined_k141_210253\|\|full | Not-determined |
| 18-102-final-viral-combined_k141_138110\|\|full | Low-quality |
| 18-102-final-viral-combined_k141_210255\|\|full | Low-quality |
| 18-102-final-viral-combined_k141_210257\|\|full | Not-determined |
| 18-102-final-viral-combined_k141_138121\|\|full | Not-determined |
| 18-102-final-viral-combined_k141_138130\|\|full | Not-determined |
| 18-102-final-viral-combined_k141_45992\|\|full | Not-determined |
| 18-102-final-viral-combined_k141_3137\|\|full | Low-quality |
| 18-102-final-viral-combined_k141_231416\|\|full | Not-determined |
| 18-102-final-viral-combined_k141_169515\|\|full | Low-quality |
| 18-102-final-viral-combined_k141_179825\|\|full | Low-quality |
| 18-102-final-viral-combined_k141_75390\|\|full | Low-quality |
| 18-102-final-viral-combined_k141_138159\|\|full | Not-determined |
| 18-102-final-viral-combined_k141_116853\|\|full | Low-quality |
| 18-102-final-viral-combined_k141_241100\|\|full | Not-determined |
| 18-102-final-viral-combined_k141_158518\|\|full | Not-determined |
| 18-102-final-viral-combined_k141_241105\|\|full | Low-quality |
| 18-102-final-viral-combined_k141_189922\|\|full | Not-determined |
| 18-102-final-viral-combined_k141_138171\|\|full | Not-determined |
| 18-102-final-viral-combined_k141_231459\|\|full | Low-quality |
| 18-102-final-viral-combined_k141_116858\|\|full | Not-determined |
| 18-102-final-viral-combined_k141_158524\|\|full | Not-determined |
| 18-102-final-viral-combined_k141_65373\|\|full | Low-quality |
| 18-102-final-viral-combined_k141_75415\|\|full | Low-quality |
| 18-102-final-viral-combined_k141_13432\|\|full | Not-determined |
| 18-102-final-viral-combined_k141_210302\|\|full | Not-determined |
| 18-102-final-viral-combined_k141_116869\|\|full | Not-determined |
| 18-102-final-viral-combined_k141_231468\|\|full | Not-determined |
| 18-102-final-viral-combined_k141_189938\|\|full | Low-quality |
| 18-102-final-viral-combined_k141_13438\|\|full | Low-quality |
| 18-102-final-viral-combined_k141_75436\|\|full | Low-quality |
| 18-102-final-viral-combined_k141_210319\|\|full | Low-quality |
| 18-102-final-viral-combined_k141_116885\|\|full | Not-determined |
| 18-102-final-viral-combined_k141_34377\|\|full | Not-determined |
| 18-102-final-viral-combined_k141_231484\|\|full | Not-determined |
| 18-102-final-viral-combined_k141_46031\|\|full | Not-determined |
| 18-102-final-viral-combined_k141_86410\|\|full | Low-quality |
| 18-102-final-viral-combined_k141_127633\|\|full | Low-quality |
| 18-102-final-viral-combined_k141_241108\|\|full | Low-quality |
| 18-102-final-viral-combined_k141_46035\|\|full | Not-determined |
| 18-102-final-viral-combined_k141_241110\|\|full | Low-quality |
| 18-102-final-viral-combined_k141_127635\|\|full | Not-determined |
| 18-102-final-viral-combined_k141_23941\|\|full | Low-quality |
| 18-102-final-viral-combined_k141_86421\|\|full | Not-determined |
| 18-102-final-viral-combined_k141_65399\|\|full | Low-quality |
| 18-102-final-viral-combined_k141_169563\|\|full | Low-quality |
| 18-102-final-viral-combined_k141_86427\|\|full | Not-determined |
| 18-102-final-viral-combined_k141_34411\|\|full | Low-quality |
| 18-102-final-viral-combined_k141_96748\|\|full | Not-determined |
| 18-102-final-viral-combined_k141_158552\|\|full | Low-quality |
| 18-102-final-viral-combined_k141_75470\|\|full | Not-determined |
| 18-102-final-viral-combined_k141_3222\|\|full | Low-quality |
| 18-102-final-viral-combined_k141_189955\|\|full | Low-quality |
| 18-102-final-viral-combined_k141_189957\|\|full | Not-determined |
| 18-102-final-viral-combined_k141_86447\|\|full | Low-quality |
| 18-102-final-viral-combined_k141_3228\|\|full | Low-quality |
| 18-102-final-viral-combined_k141_158560\|\|full | Not-determined |
| 18-102-final-viral-combined_k141_169596\|\|full | Low-quality |
| 18-102-final-viral-combined_k141_231517\|\|full | Low-quality |
| 18-102-final-viral-combined_k141_241134\|\|full | Not-determined |
| 18-102-final-viral-combined_k141_106933\|\|full | Low-quality |
| 18-102-final-viral-combined_k141_148151\|\|full | Not-determined |
| 18-102-final-viral-combined_k141_75495\|\|full | Low-quality |
| 18-102-final-viral-combined_k141_148154\|\|full | Not-determined |
| 18-102-final-viral-combined_k141_179868\|\|full | Low-quality |
| 18-102-final-viral-combined_k141_75508\|\|full | Not-determined |
| 18-102-final-viral-combined_k141_46079\|\|full | Not-determined |
| 18-102-final-viral-combined_k141_75509\|\|full | Low-quality |
| 18-102-final-viral-combined_k141_220022\|\|full | Low-quality |
| 18-102-final-viral-combined_k141_189984\|\|full | Not-determined |
| 18-102-final-viral-combined_k141_148174\|\|full | Low-quality |
| 18-102-final-viral-combined_k141_13466\|\|full | Not-determined |
| 18-102-final-viral-combined_k141_241167\|\|full | Low-quality |
| 18-102-final-viral-combined_k141_46113\|\|full | Not-determined |
| 18-102-final-viral-combined_k141_13500\|\|full | Low-quality |
| 18-102-final-viral-combined_k141_96789\|\|full | Low-quality |
| 18-102-final-viral-combined_k141_46120\|\|full | Low-quality |
| 18-102-final-viral-combined_k141_220051\|\|full | Not-determined |
| 18-102-final-viral-combined_k141_158598\|\|full | Not-determined |
| 18-102-final-viral-combined_k141_65444\|\|full | Low-quality |
| 18-102-final-viral-combined_k141_231581\|\|full | Not-determined |
| 18-102-final-viral-combined_k141_96806\|\|full | Not-determined |
| 18-102-final-viral-combined_k141_65450\|\|full | Low-quality |
| 18-102-final-viral-combined_k141_127654\|\|full | Not-determined |
| 18-102-final-viral-combined_k141_241187\|\|full | Low-quality |
| 18-102-final-viral-combined_k141_158619\|\|full | Not-determined |
| 18-102-final-viral-combined_k141_231589\|\|full | Low-quality |
| 18-102-final-viral-combined_k141_3284\|\|full | Not-determined |
| 18-102-final-viral-combined_k141_220063\|\|full | Low-quality |
| 18-102-final-viral-combined_k141_23974\|\|full | Low-quality |
| 18-102-final-viral-combined_k141_3286\|\|full | Not-determined |
| 18-102-final-viral-combined_k141_96816\|\|full | Low-quality |
| 18-102-final-viral-combined_k141_65478\|\|full | Low-quality |
| 18-102-final-viral-combined_k141_190037\|\|full | Not-determined |
| 18-102-final-viral-combined_k141_96823\|\|full | Low-quality |
| 18-102-final-viral-combined_k141_210420\|\|full | Not-determined |
| 18-102-final-viral-combined_k141_107004\|\|full | Not-determined |
| 18-102-final-viral-combined_k141_210423\|\|full | Not-determined |
| 18-102-final-viral-combined_k141_148236\|\|full | Low-quality |
| 18-102-final-viral-combined_k141_169693\|\|full | Low-quality |
| 18-102-final-viral-combined_k141_65502\|\|full | Low-quality |
| 18-102-final-viral-combined_k141_169698\|\|full | Low-quality |
| 18-102-final-viral-combined_k141_96832\|\|full | Not-determined |
| 18-102-final-viral-combined_k141_96834\|\|full | Not-determined |
| 18-102-final-viral-combined_k141_75589\|\|full | Not-determined |
| 18-102-final-viral-combined_k141_231626\|\|full | Low-quality |
| 18-102-final-viral-combined_k141_127678\|\|full | Not-determined |
| 18-102-final-viral-combined_k141_107010\|\|full | Low-quality |
| 18-102-final-viral-combined_k141_148244\|\|full | Low-quality |
| 18-102-final-viral-combined_k141_220097\|\|full | Low-quality |
| 18-102-final-viral-combined_k141_86532\|\|full | Not-determined |
| 18-102-final-viral-combined_k141_179898\|\|full | Not-determined |
| 18-102-final-viral-combined_k141_210450\|\|full | Low-quality |
| 18-102-final-viral-combined_k141_148255\|\|full | Low-quality |
| 18-102-final-viral-combined_k141_34471\|\|full | Low-quality |
| 18-102-final-viral-combined_k141_231651\|\|full | Low-quality |
| 18-102-final-viral-combined_k141_55168\|\|full | Low-quality |
| 18-102-final-viral-combined_k141_13591\|\|full | Not-determined |
| 18-102-final-viral-combined_k141_55174\|\|full | Not-determined |
| 18-102-final-viral-combined_k141_117132\|\|full | Not-determined |
| 18-102-final-viral-combined_k141_107023\|\|full | Low-quality |
| 18-102-final-viral-combined_k141_107024\|\|full | Low-quality |
| 18-102-final-viral-combined_k141_96846\|\|full | Not-determined |
| 18-102-final-viral-combined_k141_231657\|\|full | Not-determined |
| 18-102-final-viral-combined_k141_148273\|\|full | Low-quality |
| 18-102-final-viral-combined_k141_107034\|\|full | Not-determined |
| 18-102-final-viral-combined_k141_3324\|\|full | Low-quality |
| 18-102-final-viral-combined_k141_148279\|\|full | Not-determined |
| 18-102-final-viral-combined_k141_96861\|\|full | Low-quality |
| 18-102-final-viral-combined_k141_75606\|\|full | Low-quality |
| 18-102-final-viral-combined_k141_24037\|\|full | Low-quality |
| 18-102-final-viral-combined_k141_96867\|\|full | Low-quality |
| 18-102-final-viral-combined_k141_86561\|\|full | Low-quality |
| 18-102-final-viral-combined_k141_138218\|\|full | Not-determined |
| 18-105-final-viral-combined_k141_167746\|\|full | Medium-quality |
| 18-102-final-viral-combined_k141_231668\|\|full | Low-quality |
| 18-102-final-viral-combined_k141_46201\|\|full | Not-determined |
| 18-102-final-viral-combined_k141_65541\|\|full | Low-quality |
| 18-102-final-viral-combined_k141_96874\|\|full | Not-determined |
| 18-102-final-viral-combined_k141_199406\|\|full | Low-quality |
| 18-102-final-viral-combined_k141_199412\|\|full | Low-quality |
| 18-102-final-viral-combined_k141_96892\|\|full | Not-determined |
| 18-102-final-viral-combined_k141_241238\|\|full | Not-determined |
| 18-102-final-viral-combined_k141_220135\|\|full | Not-determined |
| 18-102-final-viral-combined_k141_34516\|\|full | Low-quality |
| 18-102-final-viral-combined_k141_169746\|\|full | Not-determined |
| 18-102-final-viral-combined_k141_148339\|\|full | Not-determined |
| 18-102-final-viral-combined_k141_65565\|\|full | Not-determined |
| 18-102-final-viral-combined_k141_220152\|\|full | Low-quality |
| 18-102-final-viral-combined_k141_65567\|\|full | Low-quality |
| 18-102-final-viral-combined_k141_107106\|\|full | Low-quality |
| 18-102-final-viral-combined_k141_158645\|\|full | Low-quality |
| 18-102-final-viral-combined_k141_55213\|\|full | Not-determined |
| 18-102-final-viral-combined_k141_117194\|\|full | Not-determined |
| 18-102-final-viral-combined_k141_231716\|\|full | Not-determined |
| 18-102-final-viral-combined_k141_220160\|\|full | Not-determined |
| 18-102-final-viral-combined_k141_75648\|\|full | Not-determined |
| 18-102-final-viral-combined_k141_86610\|\|full | Low-quality |
| 18-102-final-viral-combined_k141_190130\|\|full | Not-determined |
| 18-102-final-viral-combined_k141_75686\|\|full | Low-quality |
| 18-102-final-viral-combined_k141_107128\|\|full | Low-quality |
| 18-102-final-viral-combined_k141_210509\|\|full | Low-quality |
| 18-102-final-viral-combined_k141_138267\|\|full | Low-quality |
| 18-102-final-viral-combined_k141_65611\|\|full | Low-quality |
| 18-102-final-viral-combined_k141_55238\|\|full | Low-quality |
| 18-102-final-viral-combined_k141_138271\|\|full | Low-quality |
| 18-102-final-viral-combined_k141_55241\|\|full | Low-quality |
| 18-102-final-viral-combined_k141_199446\|\|full | Not-determined |
| 18-102-final-viral-combined_k141_13696\|\|full | Low-quality |
| 18-102-final-viral-combined_k141_210517\|\|full | Low-quality |
| 18-102-final-viral-combined_k141_13704\|\|full | Not-determined |
| 18-102-final-viral-combined_k141_117225\|\|full | Low-quality |
| 18-102-final-viral-combined_k141_107140\|\|full | Low-quality |
| 18-102-final-viral-combined_k141_220191\|\|full | Not-determined |
| 18-102-final-viral-combined_k141_86635\|\|full | Not-determined |
| 18-102-final-viral-combined_k141_190170\|\|full | Not-determined |
| 18-102-final-viral-combined_k141_148424\|\|full | Not-determined |
| 18-102-final-viral-combined_k141_46269\|\|full | Not-determined |
| 18-102-final-viral-combined_k141_148431\|\|full | Not-determined |
| 18-102-final-viral-combined_k141_158709\|\|full | Not-determined |
| 18-102-final-viral-combined_k141_86640\|\|full | Not-determined |
| 18-102-final-viral-combined_k141_199451\|\|full | Low-quality |
| 18-102-final-viral-combined_k141_97011\|\|full | Not-determined |
| 18-102-final-viral-combined_k141_3403\|\|full | Not-determined |
| 18-102-final-viral-combined_k141_117253\|\|full | Not-determined |
| 18-102-final-viral-combined_k141_231778\|\|full | Low-quality |
| 18-102-final-viral-combined_k141_97024\|\|full | Not-determined |
| 18-102-final-viral-combined_k141_138318\|\|full | Low-quality |
| 18-102-final-viral-combined_k141_75724\|\|full | Not-determined |
| 18-102-final-viral-combined_k141_199467\|\|full | Low-quality |
| 18-102-final-viral-combined_k141_231788\|\|full | Not-determined |
| 18-102-final-viral-combined_k141_180095\|\|full | Not-determined |
| 18-102-final-viral-combined_k141_3418\|\|full | Not-determined |
| 18-102-final-viral-combined_k141_241271\|\|full | Low-quality |
| 18-102-final-viral-combined_k141_138331\|\|full | Not-determined |
| 18-102-final-viral-combined_k141_148481\|\|full | Not-determined |
| 18-102-final-viral-combined_k141_55307\|\|full | Not-determined |
| 18-102-final-viral-combined_k141_65621\|\|full | Low-quality |
| 18-102-final-viral-combined_k141_55317\|\|full | Not-determined |
| 18-102-final-viral-combined_k141_148494\|\|full | Not-determined |
| 18-102-final-viral-combined_k141_46354\|\|full | Not-determined |
| 18-102-final-viral-combined_k141_65640\|\|full | Not-determined |
| 18-102-final-viral-combined_k141_199489\|\|full | Not-determined |
| 18-102-final-viral-combined_k141_86674\|\|full | Not-determined |
| 18-102-final-viral-combined_k141_13779\|\|full | Low-quality |
| 18-102-final-viral-combined_k141_180136\|\|full | Low-quality |
| 18-102-final-viral-combined_k141_241310\|\|full | Low-quality |
| 18-102-final-viral-combined_k141_169849\|\|full | Low-quality |
| 18-102-final-viral-combined_k141_75766\|\|full | Not-determined |
| 18-102-final-viral-combined_k141_138359\|\|full | Not-determined |
| 18-102-final-viral-combined_k141_24119\|\|full | Low-quality |
| 18-102-final-viral-combined_k141_169863\|\|full | Not-determined |
| 18-102-final-viral-combined_k141_3459\|\|full | Not-determined |
| 18-102-final-viral-combined_k141_97051\|\|full | Low-quality |
| 18-102-final-viral-combined_k141_46378\|\|full | Not-determined |
| 18-102-final-viral-combined_k141_241319\|\|full | Low-quality |
| 18-102-final-viral-combined_k141_117271\|\|full | Not-determined |
| 18-102-final-viral-combined_k141_117277\|\|full | Not-determined |
| 18-102-final-viral-combined_k141_13817\|\|full | Low-quality |
| 18-102-final-viral-combined_k141_231862\|\|full | Not-determined |
| 18-102-final-viral-combined_k141_190257\|\|full | Not-determined |
| 18-102-final-viral-combined_k141_3475\|\|full | Not-determined |
| 18-102-final-viral-combined_k141_241334\|\|full | Not-determined |
| 18-102-final-viral-combined_k141_86682\|\|full | Not-determined |
| 18-102-final-viral-combined_k141_210576\|\|full | Not-determined |
| 18-102-final-viral-combined_k141_180175\|\|full | Low-quality |
| 18-102-final-viral-combined_k141_3476\|\|full | Not-determined |
| 18-102-final-viral-combined_k141_13826\|\|full | Low-quality |
| 18-102-final-viral-combined_k141_138389\|\|full | Low-quality |
| 18-102-final-viral-combined_k141_138390\|\|full | Low-quality |
| 18-102-final-viral-combined_k141_138394\|\|full | Not-determined |
| 18-102-final-viral-combined_k141_86684\|\|full | Low-quality |
| 18-102-final-viral-combined_k141_65692\|\|full | Not-determined |
| 18-102-final-viral-combined_k141_86686\|\|full | Low-quality |
| 18-102-final-viral-combined_k141_34659\|\|full | Low-quality |
| 18-102-final-viral-combined_k141_138398\|\|full | Low-quality |
| 18-102-final-viral-combined_k141_210602\|\|full | Low-quality |
| 18-102-final-viral-combined_k141_190273\|\|full | Not-determined |
| 18-102-final-viral-combined_k141_138401\|\|full | Low-quality |
| 18-102-final-viral-combined_k141_75829\|\|full | Low-quality |
| 18-102-final-viral-combined_k141_34675\|\|full | Not-determined |
| 18-102-final-viral-combined_k141_55337\|\|full | Not-determined |
| 18-102-final-viral-combined_k141_75836\|\|full | Not-determined |
| 18-102-final-viral-combined_k141_148574\|\|full | Not-determined |
| 18-102-final-viral-combined_k141_107229\|\|full | Not-determined |
| 18-102-final-viral-combined_k141_55342\|\|full | Low-quality |
| 18-102-final-viral-combined_k141_190297\|\|full | Low-quality |
| 18-102-final-viral-combined_k141_55360\|\|full | Low-quality |
| 18-102-final-viral-combined_k141_210627\|\|full | Not-determined |
| 18-102-final-viral-combined_k141_3541\|\|full | Not-determined |
| 18-102-final-viral-combined_k141_86723\|\|full | Not-determined |
| 18-102-final-viral-combined_k141_180209\|\|full | Low-quality |
| 18-102-final-viral-combined_k141_210640\|\|full | Not-determined |
| 18-102-final-viral-combined_k141_241344\|\|full | Low-quality |
| 18-102-final-viral-combined_k141_190324\|\|full | Not-determined |
| 18-102-final-viral-combined_k141_210642\|\|full | Not-determined |
| 18-102-final-viral-combined_k141_210643\|\|full | Not-determined |
| 18-102-final-viral-combined_k141_169957\|\|full | Not-determined |
| 18-102-final-viral-combined_k141_241349\|\|full | Not-determined |
| 18-102-final-viral-combined_k141_117386\|\|full | Not-determined |
| 18-102-final-viral-combined_k141_107276\|\|full | Low-quality |
| 18-102-final-viral-combined_k141_55397\|\|full | Not-determined |
| 18-102-final-viral-combined_k141_24217\|\|full | Low-quality |
| 18-102-final-viral-combined_k141_180228\|\|full | Low-quality |
| 18-102-final-viral-combined_k141_55413\|\|full | Low-quality |
| 18-102-final-viral-combined_k141_107281\|\|full | Not-determined |
| 18-102-final-viral-combined_k141_190359\|\|full | Low-quality |
| 18-102-final-viral-combined_k141_46456\|\|full | Low-quality |
| 18-102-final-viral-combined_k141_169991\|\|full | Low-quality |
| 18-102-final-viral-combined_k141_169996\|\|full | Low-quality |
| 18-102-final-viral-combined_k141_117431\|\|full | Not-determined |
| 18-102-final-viral-combined_k141_190390\|\|full | Not-determined |
| 18-102-final-viral-combined_k141_55458\|\|full | Not-determined |
| 18-102-final-viral-combined_k141_158803\|\|full | Low-quality |
| 18-102-final-viral-combined_k141_107291\|\|full | Low-quality |
| 18-102-final-viral-combined_k141_180257\|\|full | Not-determined |
| 18-102-final-viral-combined_k141_241384\|\|full | Not-determined |
| 18-102-final-viral-combined_k141_138460\|\|full | Not-determined |
| 18-102-final-viral-combined_k141_3564\|\|full | Not-determined |
| 18-102-final-viral-combined_k141_3569\|\|full | Low-quality |
| 18-102-final-viral-combined_k141_65800\|\|full | Low-quality |
| 18-102-final-viral-combined_k141_117450\|\|full | Not-determined |
| 18-102-final-viral-combined_k141_3572\|\|full | Low-quality |
| 18-102-final-viral-combined_k141_231962\|\|full | Low-quality |
| 18-102-final-viral-combined_k141_231965\|\|full | Not-determined |
| 18-102-final-viral-combined_k141_13895\|\|full | Low-quality |
| 18-102-final-viral-combined_k141_180302\|\|full | Not-determined |
| 18-102-final-viral-combined_k141_13899\|\|full | Low-quality |
| 18-102-final-viral-combined_k141_13912\|\|full | Low-quality |
| 18-102-final-viral-combined_k141_180321\|\|full | Not-determined |
| 18-101-final-viral-combined_k141_331544\|\|0_partial | Complete |
| 18-102-final-viral-combined_k141_13921\|\|full | Not-determined |
| 18-102-final-viral-combined_k141_97178\|\|full | Not-determined |
| 18-102-final-viral-combined_k141_180322\|\|full | Not-determined |
| 18-102-final-viral-combined_k141_13935\|\|full | Low-quality |
| 18-102-final-viral-combined_k141_170018\|\|full | Not-determined |
| 18-102-final-viral-combined_k141_117490\|\|full | Not-determined |
| 18-102-final-viral-combined_k141_13947\|\|full | Low-quality |
| 18-110-final-viral-combined_k141_118779\|\|full | Medium-quality |
| 17-109-final-viral-combined_k141_191570\|\|full | Medium-quality |
| 18-102-final-viral-combined_k141_107332\|\|full | Not-determined |
| 18-102-final-viral-combined_k141_210736\|\|full | Not-determined |
| 18-102-final-viral-combined_k141_107335\|\|full | Low-quality |
| 18-102-final-viral-combined_k141_3658\|\|full | Low-quality |
| 18-102-final-viral-combined_k141_138542\|\|full | Not-determined |
| 18-102-final-viral-combined_k141_190451\|\|full | Not-determined |
| 18-102-final-viral-combined_k141_148743\|\|full | Not-determined |
| 18-102-final-viral-combined_k141_86823\|\|full | Low-quality |
| 18-102-final-viral-combined_k141_220326\|\|full | Not-determined |
| 18-102-final-viral-combined_k141_190463\|\|full | Low-quality |
| 18-102-final-viral-combined_k141_138568\|\|full | Not-determined |
| 18-102-final-viral-combined_k141_75930\|\|full | Low-quality |
| 18-102-final-viral-combined_k141_107369\|\|full | Not-determined |
| 18-102-final-viral-combined_k141_210781\|\|full | Not-determined |
| 18-102-final-viral-combined_k141_180380\|\|full | Low-quality |
| 18-102-final-viral-combined_k141_138570\|\|full | Not-determined |
| 18-102-final-viral-combined_k141_107373\|\|full | Low-quality |
| 18-102-final-viral-combined_k141_34846\|\|full | Not-determined |
| 18-102-final-viral-combined_k141_46544\|\|full | Not-determined |
| 18-102-final-viral-combined_k141_14008\|\|full | Not-determined |
| 18-102-final-viral-combined_k141_107396\|\|full | Not-determined |
| 18-102-final-viral-combined_k141_46557\|\|full | Low-quality |
| 18-102-final-viral-combined_k141_55631\|\|full | Low-quality |
| 18-102-final-viral-combined_k141_34875\|\|full | Not-determined |
| 18-102-final-viral-combined_k141_86883\|\|full | Not-determined |
| 18-102-final-viral-combined_k141_117526\|\|full | Not-determined |
| 18-102-final-viral-combined_k141_148779\|\|full | Low-quality |
| 18-102-final-viral-combined_k141_117532\|\|full | Not-determined |
| 18-102-final-viral-combined_k141_199511\|\|full | Low-quality |
| 18-102-final-viral-combined_k141_180391\|\|full | Low-quality |
| 18-102-final-viral-combined_k141_158849\|\|full | Not-determined |
| 18-102-final-viral-combined_k141_107444\|\|full | Low-quality |
| 18-102-final-viral-combined_k141_117545\|\|full | Low-quality |
| 18-102-final-viral-combined_k141_34894\|\|full | Not-determined |
| 18-102-final-viral-combined_k141_55659\|\|full | Not-determined |
| 18-102-final-viral-combined_k141_158853\|\|full | Not-determined |
| 18-102-final-viral-combined_k141_107467\|\|full | Low-quality |
| 18-102-final-viral-combined_k141_55686\|\|full | Low-quality |
| 18-102-final-viral-combined_k141_107470\|\|full | Low-quality |
| 18-102-final-viral-combined_k141_86909\|\|full | Low-quality |
| 18-102-final-viral-combined_k141_107474\|\|full | Not-determined |
| 18-102-final-viral-combined_k141_107477\|\|full | Not-determined |
| 18-102-final-viral-combined_k141_190500\|\|full | Low-quality |
| 18-102-final-viral-combined_k141_24432\|\|full | Low-quality |
| 18-102-final-viral-combined_k141_158882\|\|full | Not-determined |
| 18-102-final-viral-combined_k141_127729\|\|full | Not-determined |
| 18-102-final-viral-combined_k141_158887\|\|full | Low-quality |
| 18-102-final-viral-combined_k141_46614\|\|full | Not-determined |
| 18-102-final-viral-combined_k141_86930\|\|full | Not-determined |
| 18-102-final-viral-combined_k141_148812\|\|full | Not-determined |
| 18-102-final-viral-combined_k141_76027\|\|full | Low-quality |
| 18-102-final-viral-combined_k141_220407\|\|full | Not-determined |
| 18-102-final-viral-combined_k141_107496\|\|full | Not-determined |
| 18-102-final-viral-combined_k141_158897\|\|full | Not-determined |
| 18-102-final-viral-combined_k141_55720\|\|full | Low-quality |
| 18-102-final-viral-combined_k141_107498\|\|full | Not-determined |
| 18-102-final-viral-combined_k141_24448\|\|full | Not-determined |
| 18-102-final-viral-combined_k141_158907\|\|full | Not-determined |
| 18-102-final-viral-combined_k141_117584\|\|full | Low-quality |
| 18-102-final-viral-combined_k141_76036\|\|full | Not-determined |
| 18-102-final-viral-combined_k141_127758\|\|full | Low-quality |
| 18-102-final-viral-combined_k141_24455\|\|full | Low-quality |
| 18-102-final-viral-combined_k141_86952\|\|full | Low-quality |
| 18-102-final-viral-combined_k141_117594\|\|full | Low-quality |
| 18-102-final-viral-combined_k141_148847\|\|full | Not-determined |
| 18-102-final-viral-combined_k141_117614\|\|full | Not-determined |
| 18-102-final-viral-combined_k141_190568\|\|full | Low-quality |
| 18-102-final-viral-combined_k141_34956\|\|full | Not-determined |
| 18-102-final-viral-combined_k141_158916\|\|full | Low-quality |
| 18-102-final-viral-combined_k141_76071\|\|full | Not-determined |
| 18-102-final-viral-combined_k141_86970\|\|full | Not-determined |
| 18-106-final-viral-combined_k141_119017\|\|full | Medium-quality |
| 18-102-final-viral-combined_k141_148856\|\|full | Low-quality |
| 18-102-final-viral-combined_k141_138711\|\|full | Not-determined |
| 18-102-final-viral-combined_k141_117652\|\|full | Low-quality |
| 18-102-final-viral-combined_k141_158941\|\|full | Not-determined |
| 18-102-final-viral-combined_k141_24494\|\|full | Low-quality |
| 18-102-final-viral-combined_k141_24495\|\|full | Not-determined |
| 18-102-final-viral-combined_k141_199552\|\|full | Low-quality |
| 18-102-final-viral-combined_k141_34994\|\|full | Not-determined |
| 18-102-final-viral-combined_k141_24506\|\|full | Not-determined |
| 18-102-final-viral-combined_k141_220494\|\|full | Not-determined |
| 18-102-final-viral-combined_k141_158960\|\|full | Not-determined |
| 18-102-final-viral-combined_k141_46695\|\|full | Low-quality |
| 18-102-final-viral-combined_k141_87041\|\|full | Low-quality |
| 18-102-final-viral-combined_k141_232081\|\|full | Not-determined |
| 18-102-final-viral-combined_k141_24522\|\|full | Not-determined |
| 18-102-final-viral-combined_k141_232090\|\|full | Low-quality |
| 18-102-final-viral-combined_k141_24530\|\|full | Not-determined |
| 18-102-final-viral-combined_k141_148882\|\|full | Low-quality |
| 18-102-final-viral-combined_k141_87089\|\|full | Not-determined |
| 18-102-final-viral-combined_k141_180514\|\|full | Not-determined |
| 18-102-final-viral-combined_k141_180515\|\|full | Low-quality |
| 18-102-final-viral-combined_k141_65897\|\|full | Low-quality |
| 18-102-final-viral-combined_k141_65910\|\|full | Not-determined |
| 18-104-final-viral-combined_k141_112093\|\|full | Medium-quality |
| 18-102-final-viral-combined_k141_232128\|\|full | Low-quality |
| 18-102-final-viral-combined_k141_159006\|\|full | Not-determined |
| 18-102-final-viral-combined_k141_138761\|\|full | Low-quality |
| 18-102-final-viral-combined_k141_87111\|\|full | Not-determined |
| 18-102-final-viral-combined_k141_76165\|\|full | Not-determined |
| 18-102-final-viral-combined_k141_97320\|\|full | Not-determined |
| 18-102-final-viral-combined_k141_190645\|\|full | Not-determined |
| 18-102-final-viral-combined_k141_199567\|\|full | Not-determined |
| 18-102-final-viral-combined_k141_76205\|\|full | Not-determined |
| 18-102-final-viral-combined_k141_138776\|\|full | Not-determined |
| 18-102-final-viral-combined_k141_35054\|\|full | Not-determined |
| 18-102-final-viral-combined_k141_46761\|\|full | Not-determined |
| 18-102-final-viral-combined_k141_65917\|\|full | Not-determined |
| 18-102-final-viral-combined_k141_14210\|\|full | Low-quality |
| 18-102-final-viral-combined_k141_138808\|\|full | Not-determined |
| 18-102-final-viral-combined_k141_65919\|\|full | Not-determined |
| 18-102-final-viral-combined_k141_159057\|\|full | Not-determined |
| 18-102-final-viral-combined_k141_97340\|\|full | Low-quality |
| 18-102-final-viral-combined_k141_76250\|\|full | Not-determined |
| 18-102-final-viral-combined_k141_199582\|\|full | Not-determined |
| 18-102-final-viral-combined_k141_180558\|\|full | Low-quality |
| 18-102-final-viral-combined_k141_180560\|\|full | Not-determined |
| 18-102-final-viral-combined_k141_76266\|\|full | Low-quality |
| 18-102-final-viral-combined_k141_14237\|\|full | Low-quality |
| 18-102-final-viral-combined_k141_159079\|\|full | Low-quality |
| 18-102-final-viral-combined_k141_220527\|\|full | Low-quality |
| 18-102-final-viral-combined_k141_220533\|\|full | Not-determined |
| 18-106-final-viral-combined_k141_38764\|\|full | Medium-quality |
| 18-102-final-viral-combined_k141_65945\|\|full | Not-determined |
| 18-102-final-viral-combined_k141_148944\|\|full | Not-determined |
| 18-102-final-viral-combined_k141_3745\|\|full | Not-determined |
| 18-102-final-viral-combined_k141_220539\|\|full | Not-determined |
| 18-102-final-viral-combined_k141_97380\|\|full | Low-quality |
| 18-102-final-viral-combined_k141_138869\|\|full | Low-quality |
| 18-102-final-viral-combined_k141_35153\|\|full | Not-determined |
| 18-102-final-viral-combined_k141_127900\|\|full | Not-determined |
| 18-102-final-viral-combined_k141_138870\|\|full | Low-quality |
| 18-102-final-viral-combined_k141_55860\|\|full | Not-determined |
| 18-102-final-viral-combined_k141_241494\|\|full | Not-determined |
| 18-102-final-viral-combined_k141_138875\|\|full | Not-determined |
| 18-102-final-viral-combined_k141_87140\|\|full | Low-quality |
| 18-102-final-viral-combined_k141_138886\|\|full | Not-determined |
| 18-102-final-viral-combined_k141_180610\|\|full | Not-determined |
| 18-102-final-viral-combined_k141_117791\|\|full | Low-quality |
| 18-102-final-viral-combined_k141_76323\|\|full | Low-quality |
| 18-102-final-viral-combined_k141_14260\|\|full | Low-quality |
| 18-102-final-viral-combined_k141_138888\|\|full | Not-determined |
| 18-102-final-viral-combined_k141_65980\|\|full | Low-quality |
| 18-102-final-viral-combined_k141_14270\|\|full | Low-quality |
| 18-102-final-viral-combined_k141_3789\|\|full | Low-quality |
| 18-102-final-viral-combined_k141_87150\|\|full | Not-determined |
| 18-102-final-viral-combined_k141_87153\|\|full | Low-quality |
| 18-102-final-viral-combined_k141_107668\|\|full | Not-determined |
| 18-102-final-viral-combined_k141_97408\|\|full | Not-determined |
| 18-102-final-viral-combined_k141_14278\|\|full | Not-determined |
| 18-102-final-viral-combined_k141_232212\|\|full | Not-determined |
| 18-102-final-viral-combined_k141_97411\|\|full | Low-quality |
| 18-102-final-viral-combined_k141_46858\|\|full | Not-determined |
| 18-102-final-viral-combined_k141_220567\|\|full | Not-determined |
| 18-102-final-viral-combined_k141_232220\|\|full | Not-determined |
| 18-102-final-viral-combined_k141_190714\|\|full | Low-quality |

| | |
|---|---|
| 18-102-final-viral-combined_k141_190717\|\|full | Not-determined |
| 18-102-final-viral-combined_k141_35196\|\|full | Low-quality |
| 18-102-final-viral-combined_k141_14310\|\|full | Low-quality |
| 18-102-final-viral-combined_k141_24619\|\|full | Low-quality |
| 18-102-final-viral-combined_k141_46880\|\|full | Not-determined |
| 18-102-final-viral-combined_k141_199638\|\|full | Low-quality |
| 18-102-final-viral-combined_k141_117840\|\|full | Low-quality |
| 18-102-final-viral-combined_k141_149022\|\|full | Not-determined |
| 18-102-final-viral-combined_k141_46897\|\|full | Low-quality |
| 18-102-final-viral-combined_k141_220599\|\|full | Low-quality |
| 18-102-final-viral-combined_k141_107738\|\|full | Not-determined |
| 17-108_k141_303398 | Medium-quality |
| 18-102-final-viral-combined_k141_180648\|\|full | Low-quality |
| 18-102-final-viral-combined_k141_232280\|\|full | Low-quality |
| 18-102-final-viral-combined_k141_232282\|\|full | Not-determined |
| 18-102-final-viral-combined_k141_210869\|\|full | Low-quality |
| 18-102-final-viral-combined_k141_14320\|\|full | Not-determined |
| 18-102-final-viral-combined_k141_210874\|\|full | Not-determined |
| 18-102-final-viral-combined_k141_241532\|\|full | Not-determined |
| 18-102-final-viral-combined_k141_199666\|\|full | Low-quality |
| 18-102-final-viral-combined_k141_97494\|\|full | Low-quality |
| 18-102-final-viral-combined_k141_87176\|\|full | Not-determined |
| 18-102-final-viral-combined_k141_55984\|\|full | Not-determined |
| 18-102-final-viral-combined_k141_180688\|\|full | Low-quality |
| 18-102-final-viral-combined_k141_232295\|\|full | Low-quality |
| 18-102-final-viral-combined_k141_35243\|\|full | Low-quality |
| 18-102-final-viral-combined_k141_46955\|\|full | Not-determined |
| 18-102-final-viral-combined_k141_159159\|\|full | Low-quality |
| 18-102-final-viral-combined_k141_24636\|\|full | Low-quality |
| 18-102-final-viral-combined_k141_14360\|\|full | Not-determined |
| 18-102-final-viral-combined_k141_87191\|\|full | Not-determined |
| 18-102-final-viral-combined_k141_220664\|\|full | Not-determined |
| 18-102-final-viral-combined_k141_66044\|\|full | Low-quality |
| 18-102-final-viral-combined_k141_35262\|\|full | Low-quality |
| 18-102-final-viral-combined_k141_138944\|\|full | Not-determined |
| 18-102-final-viral-combined_k141_117891\|\|full | Low-quality |
| 18-102-final-viral-combined_k141_241558\|\|full | Low-quality |
| 18-102-final-viral-combined_k141_159183\|\|full | Low-quality |
| 18-102-final-viral-combined_k141_87233\|\|full | Low-quality |
| 18-102-final-viral-combined_k141_66069\|\|full | Low-quality |
| 18-102-final-viral-combined_k141_199744\|\|full | Not-determined |
| 18-102-final-viral-combined_k141_47025\|\|full | Not-determined |
| 18-102-final-viral-combined_k141_241588\|\|full | Low-quality |
| 18-102-final-viral-combined_k141_66073\|\|full | Not-determined |
| 18-102-final-viral-combined_k141_180733\|\|full | Low-quality |
| 18-102-final-viral-combined_k141_117904\|\|full | Not-determined |
| 18-102-final-viral-combined_k141_87266\|\|full | Not-determined |
| 18-102-final-viral-combined_k141_232360\|\|full | Low-quality |
| 18-102-final-viral-combined_k141_138991\|\|full | Not-determined |
| 18-102-final-viral-combined_k141_241591\|\|full | Low-quality |
| 18-102-final-viral-combined_k141_117905\|\|full | Low-quality |
| 18-102-final-viral-combined_k141_24691\|\|full | Not-determined |
| 18-102-final-viral-combined_k141_97553\|\|full | Low-quality |
| 18-102-final-viral-combined_k141_47045\|\|full | Not-determined |
| 18-102-final-viral-combined_k141_97556\|\|full | Low-quality |
| 18-102-final-viral-combined_k141_138999\|\|full | Low-quality |
| 18-102-final-viral-combined_k141_199763\|\|full | Not-determined |
| 18-102-final-viral-combined_k141_97558\|\|full | Not-determined |
| 18-102-final-viral-combined_k141_159229\|\|full | Not-determined |
| 18-102-final-viral-combined_k141_180755\|\|full | Not-determined |
| 18-102-final-viral-combined_k141_107762\|\|full | Not-determined |
| 18-102-final-viral-combined_k141_56075\|\|full | Low-quality |
| 18-102-final-viral-combined_k141_170154\|\|full | Low-quality |
| 18-102-final-viral-combined_k141_127951\|\|full | Not-determined |
| 18-102-final-viral-combined_k141_24708\|\|full | Not-determined |
| 18-102-final-viral-combined_k141_24712\|\|full | Low-quality |
| 18-102-final-viral-combined_k141_127956\|\|full | Not-determined |
| 18-102-final-viral-combined_k141_117916\|\|full | Low-quality |
| 18-102-final-viral-combined_k141_232429\|\|full | Low-quality |
| 18-102-final-viral-combined_k141_170180\|\|full | Not-determined |
| 18-102-final-viral-combined_k141_3848\|\|full | Low-quality |
| 18-102-final-viral-combined_k141_127985\|\|full | Low-quality |
| 18-102-final-viral-combined_k141_87316\|\|full | Not-determined |
| 18-102-final-viral-combined_k141_24734\|\|full | Low-quality |
| 18-102-final-viral-combined_k141_180762\|\|full | Not-determined |
| 18-102-final-viral-combined_k141_24736\|\|full | Low-quality |
| 18-102-final-viral-combined_k141_199781\|\|full | Low-quality |
| 18-102-final-viral-combined_k141_66124\|\|full | Not-determined |
| 18-102-final-viral-combined_k141_117953\|\|full | Low-quality |
| 18-102-final-viral-combined_k141_199793\|\|full | Low-quality |
| 18-102-final-viral-combined_k141_117964\|\|full | Not-determined |
| 18-102-final-viral-combined_k141_139036\|\|full | Not-determined |
| 18-102-final-viral-combined_k141_180785\|\|full | Not-determined |
| 18-102-final-viral-combined_k141_24760\|\|full | Low-quality |
| 18-102-final-viral-combined_k141_180791\|\|full | Not-determined |
| 18-102-final-viral-combined_k141_24764\|\|full | Low-quality |
| 18-102-final-viral-combined_k141_87363\|\|full | Low-quality |
| 18-102-final-viral-combined_k141_118024\|\|full | Not-determined |
| 18-102-final-viral-combined_k141_76584\|\|full | Low-quality |
| 18-102-final-viral-combined_k141_87390\|\|full | Not-determined |
| 18-102-final-viral-combined_k141_149073\|\|full | Low-quality |
| 18-102-final-viral-combined_k141_107826\|\|full | Not-determined |
| 18-102-final-viral-combined_k141_210966\|\|full | Not-determined |
| 18-102-final-viral-combined_k141_35389\|\|full | Not-determined |
| 18-102-final-viral-combined_k141_35397\|\|full | Low-quality |
| 18-102-final-viral-combined_k141_35400\|\|full | Not-determined |
| 18-102-final-viral-combined_k141_24812\|\|full | Not-determined |
| 18-102-final-viral-combined_k141_210973\|\|full | Not-determined |
| 18-102-final-viral-combined_k141_35409\|\|full | Not-determined |
| 18-102-final-viral-combined_k141_241696\|\|full | Not-determined |
| 18-102-final-viral-combined_k141_170300\|\|full | Low-quality |
| 18-102-final-viral-combined_k141_66224\|\|full | Low-quality |
| 18-102-final-viral-combined_k141_199903\|\|full | Not-determined |
| 18-102-final-viral-combined_k141_3888\|\|full | Low-quality |
| 18-102-final-viral-combined_k141_118078\|\|full | Not-determined |
| 18-102-final-viral-combined_k141_66238\|\|full | Low-quality |
| 18-102-final-viral-combined_k141_220768\|\|full | Low-quality |
| 18-102-final-viral-combined_k141_47102\|\|full | Low-quality |
| 18-102-final-viral-combined_k141_35435\|\|full | Low-quality |
| 18-102-final-viral-combined_k141_66250\|\|full | Low-quality |
| 18-102-final-viral-combined_k141_139148\|\|full | Low-quality |
| 18-102-final-viral-combined_k141_47115\|\|full | Low-quality |
| 18-102-final-viral-combined_k141_87445\|\|full | Not-determined |
| 18-102-final-viral-combined_k141_232509\|\|full | Low-quality |
| 18-102-final-viral-combined_k141_180873\|\|full | Low-quality |
| 18-102-final-viral-combined_k141_97654\|\|full | Low-quality |
| 18-102-final-viral-combined_k141_87457\|\|full | Low-quality |
| 18-102-final-viral-combined_k141_211018\|\|full | Not-determined |
| 18-102-final-viral-combined_k141_35474\|\|full | Not-determined |
| 18-102-final-viral-combined_k141_14455\|\|full | Not-determined |
| 18-102-final-viral-combined_k141_3899\|\|full | Low-quality |
| 18-102-final-viral-combined_k141_47154\|\|full | Not-determined |
| 18-102-final-viral-combined_k141_170351\|\|full | Not-determined |
| 18-102-final-viral-combined_k141_170357\|\|full | Not-determined |
| 18-102-final-viral-combined_k141_211040\|\|full | Not-determined |
| 18-102-final-viral-combined_k141_199927\|\|full | Low-quality |
| 18-102-final-viral-combined_k141_56250\|\|full | Low-quality |
| 18-102-final-viral-combined_k141_118114\|\|full | Not-determined |
| 18-102-final-viral-combined_k141_35486\|\|full | Low-quality |
| 18-102-final-viral-combined_k141_107872\|\|full | Low-quality |
| 18-102-final-viral-combined_k141_97678\|\|full | Not-determined |
| 18-102-final-viral-combined_k141_170387\|\|full | Not-determined |
| 18-102-final-viral-combined_k141_107883\|\|full | Not-determined |
| 18-102-final-viral-combined_k141_87482\|\|full | Low-quality |
| 18-102-final-viral-combined_k141_35498\|\|full | Not-determined |
| 18-102-final-viral-combined_k141_35499\|\|full | Low-quality |
| 18-102-final-viral-combined_k141_170404\|\|full | Not-determined |
| 18-102-final-viral-combined_k141_56276\|\|full | Not-determined |
| 18-102-final-viral-combined_k141_76645\|\|full | Low-quality |
| 18-102-final-viral-combined_k141_97696\|\|full | Not-determined |
| 18-102-final-viral-combined_k141_35511\|\|full | Low-quality |
| 18-102-final-viral-combined_k141_97702\|\|full | Low-quality |
| 18-102-final-viral-combined_k141_14516\|\|full | Low-quality |
| 18-102-final-viral-combined_k141_76653\|\|full | Low-quality |
| 18-102-final-viral-combined_k141_97708\|\|full | Not-determined |
| 18-102-final-viral-combined_k141_35519\|\|full | Low-quality |
| 18-102-final-viral-combined_k141_87527\|\|full | Not-determined |
| 18-102-final-viral-combined_k141_170479\|\|full | Low-quality |
| 18-102-final-viral-combined_k141_76684\|\|full | Low-quality |
| 18-102-final-viral-combined_k141_190817\|\|full | Low-quality |
| 18-102-final-viral-combined_k141_76689\|\|full | Low-quality |
| 18-102-final-viral-combined_k141_35532\|\|full | Low-quality |
| 18-102-final-viral-combined_k141_107895\|\|full | Not-determined |
| 18-102-final-viral-combined_k141_3915\|\|full | Low-quality |
| 18-102-final-viral-combined_k141_180993\|\|full | Not-determined |
| 18-102-final-viral-combined_k141_170486\|\|full | Low-quality |
| 18-102-final-viral-combined_k141_56322\|\|full | Not-determined |
| 18-102-final-viral-combined_k141_199973\|\|full | Low-quality |
| 18-102-final-viral-combined_k141_35554\|\|full | Not-determined |
| 18-102-final-viral-combined_k141_56333\|\|full | Low-quality |
| 18-102-final-viral-combined_k141_181005\|\|full | Not-determined |
| 18-102-final-viral-combined_k141_76699\|\|full | Low-quality |
| 18-102-final-viral-combined_k141_35567\|\|full | Not-determined |
| 18-102-final-viral-combined_k141_190856\|\|full | Low-quality |
| 18-102-final-viral-combined_k141_56363\|\|full | Low-quality |
| 18-102-final-viral-combined_k141_190890\|\|full | Low-quality |
| 18-102-final-viral-combined_k141_190897\|\|full | Not-determined |
| 18-102-final-viral-combined_k141_35619\|\|full | Low-quality |
| 18-102-final-viral-combined_k141_87581\|\|full | Not-determined |
| 18-102-final-viral-combined_k141_190904\|\|full | Low-quality |
| 18-102-final-viral-combined_k141_220926\|\|full | Not-determined |
| 18-102-final-viral-combined_k141_181066\|\|full | Not-determined |
| 18-102-final-viral-combined_k141_87607\|\|full | Not-determined |
| 18-102-final-viral-combined_k141_3948\|\|full | Not-determined |
| 18-102-final-viral-combined_k141_181075\|\|full | Not-determined |
| 18-102-final-viral-combined_k141_47288\|\|full | Low-quality |
| 18-102-final-viral-combined_k141_211108\|\|full | Low-quality |
| 18-102-final-viral-combined_k141_149183\|\|full | Not-determined |
| 18-102-final-viral-combined_k141_211114\|\|full | Low-quality |
| 18-102-final-viral-combined_k141_159352\|\|full | Low-quality |
| 18-102-final-viral-combined_k141_149185\|\|full | Low-quality |
| 18-102-final-viral-combined_k141_211118\|\|full | Low-quality |
| 18-102-final-viral-combined_k141_159359\|\|full | Not-determined |
| 18-102-final-viral-combined_k141_56447\|\|full | Not-determined |
| 18-102-final-viral-combined_k141_47312\|\|full | Not-determined |
| 18-102-final-viral-combined_k141_87630\|\|full | Low-quality |
| 18-102-final-viral-combined_k141_190938\|\|full | Not-determined |
| 18-102-final-viral-combined_k141_107975\|\|full | Not-determined |
| 18-102-final-viral-combined_k141_190946\|\|full | Low-quality |
| 18-102-final-viral-combined_k141_149219\|\|full | Low-quality |
| 18-102-final-viral-combined_k141_107976\|\|full | Low-quality |
| 18-102-final-viral-combined_k141_107981\|\|full | Low-quality |
| 18-102-final-viral-combined_k141_47327\|\|full | Low-quality |
| 18-102-final-viral-combined_k141_87662\|\|full | Not-determined |
| 18-102-final-viral-combined_k141_159408\|\|full | Not-determined |
| 18-102-final-viral-combined_k141_128063\|\|full | Low-quality |
| 18-102-final-viral-combined_k141_159410\|\|full | Not-determined |
| 18-102-final-viral-combined_k141_66483\|\|full | Low-quality |
| 18-102-final-viral-combined_k141_128072\|\|full | Not-determined |
| 18-102-final-viral-combined_k141_87681\|\|full | Not-determined |
| 18-102-final-viral-combined_k141_47352\|\|full | Low-quality |
| 18-102-final-viral-combined_k141_190982\|\|full | Low-quality |
| 18-102-final-viral-combined_k141_128082\|\|full | Not-determined |
| 18-102-final-viral-combined_k141_87688\|\|full | Low-quality |
| 18-102-final-viral-combined_k141_200091\|\|full | Low-quality |
| 18-102-final-viral-combined_k141_87696\|\|full | Not-determined |
| 18-102-final-viral-combined_k141_149262\|\|full | Not-determined |
| 18-102-final-viral-combined_k141_159460\|\|full | Not-determined |
| 18-102-final-viral-combined_k141_200107\|\|full | Not-determined |
| 18-102-final-viral-combined_k141_170669\|\|full | Low-quality |
| 18-102-final-viral-combined_k141_4045\|\|full | Low-quality |
| 18-102-final-viral-combined_k141_139331\|\|full | Not-determined |
| 18-102-final-viral-combined_k141_56532\|\|full | Not-determined |
| 18-102-final-viral-combined_k141_149280\|\|full | Not-determined |
| 18-102-final-viral-combined_k141_4050\|\|full | Not-determined |
| 18-102-final-viral-combined_k141_97892\|\|full | Low-quality |
| 18-102-final-viral-combined_k141_159488\|\|full | Not-determined |
| 18-102-final-viral-combined_k141_4052\|\|full | Low-quality |
| 18-102-final-viral-combined_k141_181154\|\|full | Not-determined |
| 18-102-final-viral-combined_k141_232546\|\|full | Low-quality |
| 18-102-final-viral-combined_k141_211188\|\|full | Not-determined |
| 18-102-final-viral-combined_k141_181162\|\|full | Not-determined |
| 18-102-final-viral-combined_k141_191011\|\|full | Not-determined |
| 18-102-final-viral-combined_k141_170698\|\|full | Low-quality |
| 18-102-final-viral-combined_k141_97899\|\|full | Low-quality |
| 18-102-final-viral-combined_k141_14614\|\|full | Low-quality |
| 18-102-final-viral-combined_k141_170700\|\|full | Low-quality |
| 18-102-final-viral-combined_k141_181171\|\|full | Low-quality |
| 18-102-final-viral-combined_k141_97908\|\|full | Not-determined |
| 18-102-final-viral-combined_k141_14626\|\|full | Not-determined |
| 18-102-final-viral-combined_k141_108065\|\|full | Low-quality |
| 18-102-final-viral-combined_k141_4086\|\|full | Not-determined |
| 18-102-final-viral-combined_k141_97927\|\|full | Low-quality |
| 18-102-final-viral-combined_k141_220960\|\|full | Not-determined |
| 18-102-final-viral-combined_k141_139377\|\|full | Not-determined |
| 18-102-final-viral-combined_k141_47398\|\|full | Not-determined |
| 18-102-final-viral-combined_k141_4087\|\|full | Low-quality |
| 18-102-final-viral-combined_k141_170737\|\|full | Not-determined |
| 18-102-final-viral-combined_k141_14662\|\|full | Not-determined |
| 18-102-final-viral-combined_k141_108079\|\|full | Low-quality |
| 18-102-final-viral-combined_k141_128151\|\|full | Low-quality |
| 18-102-final-viral-combined_k141_76821\|\|full | Not-determined |
| 18-102-final-viral-combined_k141_159535\|\|full | Low-quality |
| 18-102-final-viral-combined_k141_76827\|\|full | Low-quality |
| 18-102-final-viral-combined_k141_24948\|\|full | Low-quality |
| 18-102-final-viral-combined_k141_128171\|\|full | Not-determined |
| 18-102-final-viral-combined_k141_76837\|\|full | Not-determined |
| 18-102-final-viral-combined_k141_200155\|\|full | Low-quality |
| 18-102-final-viral-combined_k141_191035\|\|full | Low-quality |
| 18-102-final-viral-combined_k141_76838\|\|full | Low-quality |
| 18-102-final-viral-combined_k141_220983\|\|full | Not-determined |
| 18-102-final-viral-combined_k141_128175\|\|full | Low-quality |
| 18-102-final-viral-combined_k141_191046\|\|full | Not-determined |
| 18-102-final-viral-combined_k141_24976\|\|full | Not-determined |
| 18-102-final-viral-combined_k141_211250\|\|full | Low-quality |
| 18-102-final-viral-combined_k141_108114\|\|full | Low-quality |
| 18-102-final-viral-combined_k141_97989\|\|full | Not-determined |
| 18-102-final-viral-combined_k141_66535\|\|full | Not-determined |
| 18-102-final-viral-combined_k141_47481\|\|full | Not-determined |
| 18-102-final-viral-combined_k141_211253\|\|full | Not-determined |
| 18-102-final-viral-combined_k141_211258\|\|full | Not-determined |
| 18-102-final-viral-combined_k141_76866\|\|full | Not-determined |
| 18-102-final-viral-combined_k141_14706\|\|full | Low-quality |
| 18-102-final-viral-combined_k141_47505\|\|full | Low-quality |
| 18-102-final-viral-combined_k141_221006\|\|full | Not-determined |
| 18-102-final-viral-combined_k141_35755\|\|full | Not-determined |
| 18-102-final-viral-combined_k141_181264\|\|full | Low-quality |
| 18-102-final-viral-combined_k141_221012\|\|full | Not-determined |
| 18-102-final-viral-combined_k141_170777\|\|full | Low-quality |
| 18-102-final-viral-combined_k141_66557\|\|full | Low-quality |
| 18-102-final-viral-combined_k141_211280\|\|full | Not-determined |
| 18-102-final-viral-combined_k141_108144\|\|full | Not-determined |
| 18-102-final-viral-combined_k141_4166\|\|full | Low-quality |
| 18-102-final-viral-combined_k141_170786\|\|full | Not-determined |
| 18-102-final-viral-combined_k141_241825\|\|full | Low-quality |
| 18-102-final-viral-combined_k141_35763\|\|full | Low-quality |
| 18-102-final-viral-combined_k141_149354\|\|full | Low-quality |
| 18-102-final-viral-combined_k141_24995\|\|full | Low-quality |
| 18-102-final-viral-combined_k141_76881\|\|full | Not-determined |
| 18-102-final-viral-combined_k141_241834\|\|full | Low-quality |
| 18-102-final-viral-combined_k141_76883\|\|full | Not-determined |
| 18-102-final-viral-combined_k141_232608\|\|full | Low-quality |
| 18-102-final-viral-combined_k141_221040\|\|full | Not-determined |
| 18-102-final-viral-combined_k141_108166\|\|full | Low-quality |
| 18-102-final-viral-combined_k141_211299\|\|full | Not-determined |
| 18-102-final-viral-combined_k141_76888\|\|full | Low-quality |
| 18-102-final-viral-combined_k141_56566\|\|full | Not-determined |
| 18-102-final-viral-combined_k141_56569\|\|full | Not-determined |
| 18-102-final-viral-combined_k141_118488\|\|full | Low-quality |
| 18-102-final-viral-combined_k141_108174\|\|full | Low-quality |
| 18-102-final-viral-combined_k141_66596\|\|full | Not-determined |
| 18-102-final-viral-combined_k141_221044\|\|full | Not-determined |
| 18-102-final-viral-combined_k141_108193\|\|full | Low-quality |
| 18-102-final-viral-combined_k141_181344\|\|full | Not-determined |
| 18-102-final-viral-combined_k141_98067\|\|full | Not-determined |
| 18-102-final-viral-combined_k141_118502\|\|full | Low-quality |
| 18-102-final-viral-combined_k141_221062\|\|full | Low-quality |
| 18-102-final-viral-combined_k141_87794\|\|full | Low-quality |
| 18-102-final-viral-combined_k141_241874\|\|full | Low-quality |
| 18-102-final-viral-combined_k141_66631\|\|full | Not-determined |
| 18-102-final-viral-combined_k141_66640\|\|full | Low-quality |
| 18-102-final-viral-combined_k141_35834\|\|full | Low-quality |
| 18-102-final-viral-combined_k141_241885\|\|full | Not-determined |
| 18-102-final-viral-combined_k141_139409\|\|full | Low-quality |
| 18-102-final-viral-combined_k141_221083\|\|full | Low-quality |
| 18-102-final-viral-combined_k141_4224\|\|full | Low-quality |
| 18-102-final-viral-combined_k141_87812\|\|full | Low-quality |
| 18-102-final-viral-combined_k141_98107\|\|full | Low-quality |
| 18-102-final-viral-combined_k141_25014\|\|full | Not-determined |
| 18-102-final-viral-combined_k141_211310\|\|full | Low-quality |
| 18-102-final-viral-combined_k141_25020\|\|full | Low-quality |
| 18-102-final-viral-combined_k141_35871\|\|full | Low-quality |
| 18-102-final-viral-combined_k141_66656\|\|full | Low-quality |
| 18-102-final-viral-combined_k141_211349\|\|full | Low-quality |
| 18-102-final-viral-combined_k141_200190\|\|full | Not-determined |
| 18-102-final-viral-combined_k141_25030\|\|full | Not-determined |
| 18-102-final-viral-combined_k141_76991\|\|full | Not-determined |
| 18-102-final-viral-combined_k141_191148\|\|full | Low-quality |
| 18-102-final-viral-combined_k141_56646\|\|full | Not-determined |
| 18-102-final-viral-combined_k141_77008\|\|full | Not-determined |
| 18-102-final-viral-combined_k141_159625\|\|full | Low-quality |
| 18-102-final-viral-combined_k141_211355\|\|full | Low-quality |
| 18-102-final-viral-combined_k141_35909\|\|full | Not-determined |
| 18-102-final-viral-combined_k141_35913\|\|full | Low-quality |
| 18-102-final-viral-combined_k141_181388\|\|full | Not-determined |
| 18-102-final-viral-combined_k141_139486\|\|full | Not-determined |
| 18-102-final-viral-combined_k141_87857\|\|full | Low-quality |
| 17-108-final-viral-combined_k141_117756\|\|full | Medium-quality |
| 18-102-final-viral-combined_k141_232690\|\|full | Not-determined |
| 18-102-final-viral-combined_k141_98163\|\|full | Not-determined |
| 18-102-final-viral-combined_k141_232693\|\|full | Low-quality |
| 18-102-final-viral-combined_k141_221109\|\|full | Not-determined |
| 18-102-final-viral-combined_k141_56673\|\|full | Low-quality |
| 18-102-final-viral-combined_k141_66704\|\|full | Not-determined |
| 18-102-final-viral-combined_k141_25059\|\|full | Not-determined |
| 18-102-final-viral-combined_k141_170875\|\|full | Not-determined |
| 18-102-final-viral-combined_k141_47618\|\|full | Not-determined |
| 18-102-final-viral-combined_k141_98168\|\|full | Low-quality |
| 18-102-final-viral-combined_k141_159664\|\|full | Not-determined |
| 18-102-final-viral-combined_k141_170886\|\|full | Low-quality |
| 18-102-final-viral-combined_k141_170898\|\|full | Not-determined |
| 18-102-final-viral-combined_k141_149527\|\|full | Not-determined |
| 18-102-final-viral-combined_k141_118594\|\|full | Low-quality |
| 18-102-final-viral-combined_k141_56710\|\|full | Not-determined |
| 18-102-final-viral-combined_k141_118595\|\|full | Not-determined |
| 18-102-final-viral-combined_k141_170901\|\|full | Not-determined |
| 18-102-final-viral-combined_k141_14783\|\|full | Low-quality |
| 18-102-final-viral-combined_k141_66709\|\|full | Not-determined |
| 18-102-final-viral-combined_k141_98198\|\|full | Low-quality |
| 18-102-final-viral-combined_k141_66729\|\|full | Not-determined |
| 18-102-final-viral-combined_k141_118606\|\|full | Not-determined |
| 18-102-final-viral-combined_k141_77096\|\|full | Not-determined |
| 18-102-final-viral-combined_k141_4259\|\|full | Not-determined |
| 18-102-final-viral-combined_k141_139580\|\|full | Low-quality |
| 17-108-final-viral-combined_k141_303398\|\|full | Medium-quality |
| 18-102-final-viral-combined_k141_56750\|\|full | Not-determined |
| 18-102-final-viral-combined_k141_149542\|\|full | Not-determined |
| 18-102-final-viral-combined_k141_98219\|\|full | Not-determined |
| 18-102-final-viral-combined_k141_87905\|\|full | Not-determined |
| 18-102-final-viral-combined_k141_77122\|\|full | Low-quality |
| 18-102-final-viral-combined_k141_159683\|\|full | Not-determined |
| 18-102-final-viral-combined_k141_98229\|\|full | Low-quality |
| 18-102-final-viral-combined_k141_36021\|\|full | Not-determined |
| 18-102-final-viral-combined_k141_159684\|\|full | Low-quality |
| 18-102-final-viral-combined_k141_118625\|\|full | Not-determined |
| 18-102-final-viral-combined_k141_170934\|\|full | Low-quality |
| 18-102-final-viral-combined_k141_14859\|\|full | Low-quality |
| 18-102-final-viral-combined_k141_149576\|\|full | Low-quality |
| 18-102-final-viral-combined_k141_159707\|\|full | Low-quality |
| 18-102-final-viral-combined_k141_200273\|\|full | Not-determined |
| 18-102-final-viral-combined_k141_191238\|\|full | Not-determined |
| 18-102-final-viral-combined_k141_211366\|\|full | Not-determined |
| 18-102-final-viral-combined_k141_14878\|\|full | Not-determined |
| 18-102-final-viral-combined_k141_47640\|\|full | Low-quality |
| 18-102-final-viral-combined_k141_47641\|\|full | Low-quality |
| 18-102-final-viral-combined_k141_128367\|\|full | Low-quality |
| 18-102-final-viral-combined_k141_56786\|\|full | Low-quality |
| 18-102-final-viral-combined_k141_232748\|\|full | Low-quality |
| 18-102-final-viral-combined_k141_232750\|\|full | Low-quality |
| 18-102-final-viral-combined_k141_159714\|\|full | Low-quality |
| 18-102-final-viral-combined_k141_200308\|\|full | Not-determined |
| 18-102-final-viral-combined_k141_232760\|\|full | Not-determined |
| 18-102-final-viral-combined_k141_191255\|\|full | Not-determined |
| 18-102-final-viral-combined_k141_139615\|\|full | Not-determined |
| 18-102-final-viral-combined_k141_98266\|\|full | Not-determined |
| 18-102-final-viral-combined_k141_170995\|\|full | Low-quality |
| 18-101-final-viral-combined_k141_224578\|\|full | Medium-quality |
| 18-102-final-viral-combined_k141_171011\|\|full | Low-quality |
| 18-102-final-viral-combined_k141_149618\|\|full | Not-determined |
| 18-102-final-viral-combined_k141_191265\|\|full | Low-quality |
| 18-102-final-viral-combined_k141_159729\|\|full | Low-quality |
| 18-102-final-viral-combined_k141_128381\|\|full | Not-determined |
| 18-102-final-viral-combined_k141_25168\|\|full | Low-quality |
| 18-102-final-viral-combined_k141_159752\|\|full | Not-determined |
| 18-102-final-viral-combined_k141_128395\|\|full | Not-determined |
| 18-102-final-viral-combined_k141_36066\|\|full | Low-quality |
| 18-102-final-viral-combined_k141_159774\|\|full | Not-determined |
| 18-102-final-viral-combined_k141_241948\|\|full | Not-determined |
| 18-102-final-viral-combined_k141_139667\|\|full | Not-determined |
| 18-102-final-viral-combined_k141_221256\|\|full | Not-determined |
| 18-102-final-viral-combined_k141_241952\|\|full | Low-quality |
| 18-102-final-viral-combined_k141_149648\|\|full | Not-determined |
| 18-102-final-viral-combined_k141_241957\|\|full | Low-quality |
| 18-102-final-viral-combined_k141_56830\|\|full | Low-quality |
| 18-102-final-viral-combined_k141_4315\|\|full | Low-quality |
| 18-102-final-viral-combined_k141_36107\|\|full | Not-determined |
| 18-102-final-viral-combined_k141_139721\|\|full | Not-determined |
| 18-102-final-viral-combined_k141_25223\|\|full | Low-quality |
| 18-102-final-viral-combined_k141_171045\|\|full | Not-determined |
| 18-102-final-viral-combined_k141_77198\|\|full | Low-quality |
| 18-102-final-viral-combined_k141_4318\|\|full | Low-quality |
| 18-102-final-viral-combined_k141_232833\|\|full | Low-quality |
| 18-102-final-viral-combined_k141_77207\|\|full | Low-quality |
| 18-102-final-viral-combined_k141_200366\|\|full | Low-quality |
| 18-102-final-viral-combined_k141_118703\|\|full | Not-determined |
| 18-102-final-viral-combined_k141_211498\|\|full | Low-quality |
| 18-102-final-viral-combined_k141_66769\|\|full | Not-determined |
| 18-102-final-viral-combined_k141_128426\|\|full | Low-quality |
| 18-102-final-viral-combined_k141_25249\|\|full | Low-quality |
| 18-102-final-viral-combined_k141_171058\|\|full | Low-quality |
| 18-102-final-viral-combined_k141_128440\|\|full | Low-quality |
| 18-102-final-viral-combined_k141_36151\|\|full | Low-quality |
| 18-102-final-viral-combined_k141_128443\|\|full | Low-quality |
| 18-102-final-viral-combined_k141_98367\|\|full | Not-determined |
| 18-102-final-viral-combined_k141_14941\|\|full | Not-determined |
| 18-102-final-viral-combined_k141_139766\|\|full | Low-quality |
| 18-102-final-viral-combined_k141_139768\|\|full | Not-determined |
| 18-102-final-viral-combined_k141_221367\|\|full | Not-determined |
| 18-102-final-viral-combined_k141_47775\|\|full | Not-determined |
| 18-102-final-viral-combined_k141_159835\|\|full | Not-determined |
| 18-102-final-viral-combined_k141_181464\|\|full | Not-determined |
| 18-102-final-viral-combined_k141_47781\|\|full | Low-quality |
| 18-102-final-viral-combined_k141_118723\|\|full | Not-determined |
| 18-102-final-viral-combined_k141_87977\|\|full | Low-quality |
| 18-102-final-viral-combined_k141_47790\|\|full | Low-quality |
| 18-102-final-viral-combined_k141_77255\|\|full | Low-quality |
| 18-102-final-viral-combined_k141_87990\|\|full | Low-quality |
| 18-102-final-viral-combined_k141_14989\|\|full | Low-quality |
| 18-102-final-viral-combined_k141_4361\|\|full | Low-quality |
| 18-102-final-viral-combined_k141_118728\|\|full | Low-quality |
| 18-102-final-viral-combined_k141_149708\|\|full | Not-determined |
| 18-102-final-viral-combined_k141_4366\|\|full | Not-determined |
| 18-102-final-viral-combined_k141_211561\|\|full | Not-determined |
| 18-102-final-viral-combined_k141_181512\|\|full | Not-determined |
| 18-102-final-viral-combined_k141_66785\|\|full | Not-determined |
| 18-102-final-viral-combined_k141_171111\|\|full | Low-quality |
| 18-102-final-viral-combined_k141_66794\|\|full | Low-quality |
| 18-102-final-viral-combined_k141_66800\|\|full | Not-determined |
| 18-102-final-viral-combined_k141_98436\|\|full | Low-quality |
| 18-102-final-viral-combined_k141_77288\|\|full | Not-determined |
| 18-102-final-viral-combined_k141_15017\|\|full | Not-determined |
| 18-102-final-viral-combined_k141_98456\|\|full | Not-determined |
| 18-102-final-viral-combined_k141_25332\|\|full | Not-determined |
| 18-102-final-viral-combined_k141_232953\|\|full | Low-quality |
| 18-102-final-viral-combined_k141_139835\|\|full | Not-determined |
| 18-102-final-viral-combined_k141_77324\|\|full | Low-quality |
| 18-102-final-viral-combined_k141_242080\|\|full | Not-determined |
| 18-102-final-viral-combined_k141_77330\|\|full | Low-quality |
| 18-102-final-viral-combined_k141_25347\|\|full | Low-quality |
| 18-102-final-viral-combined_k141_4376\|\|full | Low-quality |
| 18-102-final-viral-combined_k141_56972\|\|full | Not-determined |
| 18-102-final-viral-combined_k141_242087\|\|full | Low-quality |
| 18-102-final-viral-combined_k141_149769\|\|full | Not-determined |
| 18-102-final-viral-combined_k141_159874\|\|full | Low-quality |
| 18-102-final-viral-combined_k141_139857\|\|full | Not-determined |
| 18-102-final-viral-combined_k141_211577\|\|full | Not-determined |
| 18-102-final-viral-combined_k141_242102\|\|full | Low-quality |
| 18-102-final-viral-combined_k141_25353\|\|full | Low-quality |
| 18-110-final-viral-combined_k141_211119\|\|full | Medium-quality |
| 18-102-final-viral-combined_k141_242108\|\|full | Low-quality |
| 18-102-final-viral-combined_k141_25356\|\|full | Not-determined |
| 18-102-final-viral-combined_k141_15070\|\|full | Low-quality |
| 18-102-final-viral-combined_k141_191300\|\|full | Low-quality |
| 18-102-final-viral-combined_k141_77355\|\|full | Not-determined |
| 18-102-final-viral-combined_k141_139881\|\|full | Not-determined |
| 18-102-final-viral-combined_k141_139882\|\|full | Low-quality |
| 18-102-final-viral-combined_k141_221491\|\|full | Not-determined |
| 18-102-final-viral-combined_k141_191313\|\|full | Low-quality |
| 18-102-final-viral-combined_k141_77370\|\|full | Not-determined |
| 18-102-final-viral-combined_k141_36288\|\|full | Low-quality |
| 18-102-final-viral-combined_k141_233032\|\|full | Low-quality |
| 18-102-final-viral-combined_k141_77376\|\|full | Not-determined |
| 18-102-final-viral-combined_k141_233034\|\|full | Not-determined |
| 18-102-final-viral-combined_k141_108280\|\|full | Low-quality |
| 18-102-final-viral-combined_k141_98523\|\|full | Not-determined |
| 18-102-final-viral-combined_k141_211612\|\|full | Low-quality |
| 18-102-final-viral-combined_k141_15122\|\|full | Low-quality |
| 18-102-final-viral-combined_k141_47947\|\|full | Not-determined |
| 18-102-final-viral-combined_k141_98531\|\|full | Not-determined |
| 18-102-final-viral-combined_k141_242133\|\|full | Low-quality |
| 18-102-final-viral-combined_k141_139931\|\|full | Low-quality |
| 18-102-final-viral-combined_k141_98548\|\|full | Not-determined |
| 18-102-final-viral-combined_k141_15140\|\|full | Not-determined |
| 18-102-final-viral-combined_k141_200449\|\|full | Low-quality |
| 18-102-final-viral-combined_k141_88013\|\|full | Low-quality |
| 18-102-final-viral-combined_k141_98562\|\|full | Low-quality |
| 18-102-final-viral-combined_k141_118817\|\|full | Not-determined |
| 18-102-final-viral-combined_k141_88019\|\|full | Not-determined |
| 18-102-final-viral-combined_k141_139949\|\|full | Not-determined |
| 18-102-final-viral-combined_k141_149846\|\|full | Not-determined |
| 18-102-final-viral-combined_k141_200464\|\|full | Not-determined |
| 18-102-final-viral-combined_k141_36327\|\|full | Not-determined |
| 18-102-final-viral-combined_k141_233085\|\|full | Low-quality |
| 18-102-final-viral-combined_k141_47978\|\|full | Not-determined |
| 18-102-final-viral-combined_k141_15148\|\|full | Low-quality |
| 18-102-final-viral-combined_k141_88034\|\|full | Not-determined |
| 18-102-final-viral-combined_k141_36343\|\|full | Not-determined |
| 18-102-final-viral-combined_k141_191337\|\|full | Not-determined |
| 18-102-final-viral-combined_k141_200475\|\|full | Low-quality |
| 18-102-final-viral-combined_k141_211668\|\|full | Low-quality |
| 18-102-final-viral-combined_k141_66891\|\|full | Not-determined |
| 18-102-final-viral-combined_k141_47984\|\|full | Not-determined |
| 18-102-final-viral-combined_k141_211682\|\|full | Low-quality |
| 18-102-final-viral-combined_k141_15218\|\|full | Not-determined |
| 18-102-final-viral-combined_k141_149857\|\|full | Not-determined |
| 18-102-final-viral-combined_k141_98623\|\|full | Not-determined |
| 18-102-final-viral-combined_k141_128555\|\|full | Not-determined |
| 18-102-final-viral-combined_k141_66962\|\|full | Low-quality |
| 18-102-final-viral-combined_k141_159968\|\|full | Low-quality |
| 18-102-final-viral-combined_k141_211717\|\|full | Not-determined |
| 18-102-final-viral-combined_k141_48029\|\|full | Not-determined |
| 18-102-final-viral-combined_k141_36379\|\|full | Not-determined |
| 18-102-final-viral-combined_k141_36383\|\|full | Not-determined |
| 18-102-final-viral-combined_k141_57001\|\|full | Not-determined |
| 18-102-final-viral-combined_k141_191355\|\|full | Low-quality |
| 18-102-final-viral-combined_k141_15251\|\|full | Not-determined |
| 18-102-final-viral-combined_k141_48037\|\|full | Low-quality |
| 18-102-final-viral-combined_k141_159991\|\|full | Not-determined |
| 18-102-final-viral-combined_k141_200504\|\|full | Not-determined |
| 18-102-final-viral-combined_k141_36395\|\|full | Low-quality |
| 18-102-final-viral-combined_k141_57036\|\|full | Not-determined |
| 18-102-final-viral-combined_k141_200508\|\|full | Not-determined |
| 17-108-final-viral-combined_k141_116589\|\|full | Medium-quality |
| 18-102-final-viral-combined_k141_57049\|\|full | Low-quality |
| 18-102-final-viral-combined_k141_98670\|\|full | Not-determined |
| 18-102-final-viral-combined_k141_233111\|\|full | Not-determined |
| 18-102-final-viral-combined_k141_242247\|\|full | Low-quality |
| 18-102-final-viral-combined_k141_242248\|\|full | Not-determined |
| 18-102-final-viral-combined_k141_200524\|\|full | Not-determined |
| 18-102-final-viral-combined_k141_67014\|\|full | Not-determined |
| 18-102-final-viral-combined_k141_57065\|\|full | Not-determined |
| 18-102-final-viral-combined_k141_118912\|\|full | Not-determined |
| 18-102-final-viral-combined_k141_57068\|\|full | Low-quality |
| 18-102-final-viral-combined_k141_98683\|\|full | Not-determined |
| 18-102-final-viral-combined_k141_191396\|\|full | Not-determined |
| 18-102-final-viral-combined_k141_200527\|\|full | Not-determined |
| 18-102-final-viral-combined_k141_160025\|\|full | Low-quality |
| 18-102-final-viral-combined_k141_149937\|\|full | Not-determined |
| 18-102-final-viral-combined_k141_233131\|\|full | Not-determined |
| 18-102-final-viral-combined_k141_36430\|\|full | Low-quality |
| 18-102-final-viral-combined_k141_118935\|\|full | Low-quality |
| 18-102-final-viral-combined_k141_181598\|\|full | Not-determined |
| 18-102-final-viral-combined_k141_88097\|\|full | Low-quality |
| 18-102-final-viral-combined_k141_191428\|\|full | Not-determined |
| 18-102-final-viral-combined_k141_171237\|\|full | Low-quality |
| 18-102-final-viral-combined_k141_171239\|\|full | Low-quality |
| 18-102-final-viral-combined_k141_57090\|\|full | Low-quality |
| 18-102-final-viral-combined_k141_233161\|\|full | Not-determined |
| 18-102-final-viral-combined_k141_15364\|\|full | Not-determined |
| 18-102-final-viral-combined_k141_118963\|\|full | Low-quality |
| 18-102-final-viral-combined_k141_77534\|\|full | Low-quality |
| 18-102-final-viral-combined_k141_36449\|\|full | Low-quality |
| 18-102-final-viral-combined_k141_221554\|\|full | Not-determined |
| 18-102-final-viral-combined_k141_221555\|\|full | Not-determined |
| 18-102-final-viral-combined_k141_242259\|\|full | Low-quality |
| 18-102-final-viral-combined_k141_77557\|\|full | Low-quality |
| 18-102-final-viral-combined_k141_88132\|\|full | Not-determined |
| 18-102-final-viral-combined_k141_25482\|\|full | Not-determined |
| 18-102-final-viral-combined_k141_200575\|\|full | Low-quality |
| 18-102-final-viral-combined_k141_171248\|\|full | Low-quality |
| 18-102-final-viral-combined_k141_233219\|\|full | Not-determined |
| 18-102-final-viral-combined_k141_221584\|\|full | Not-determined |
| 18-102-final-viral-combined_k141_200595\|\|full | Low-quality |
| 18-102-final-viral-combined_k141_67085\|\|full | Low-quality |
| 18-102-final-viral-combined_k141_4414\|\|full | Low-quality |
| 18-102-final-viral-combined_k141_150024\|\|full | Low-quality |
| 18-102-final-viral-combined_k141_25487\|\|full | Not-determined |
| 18-102-final-viral-combined_k141_57188\|\|full | Low-quality |
| 18-102-final-viral-combined_k141_160094\|\|full | Low-quality |
| 18-102-final-viral-combined_k141_57212\|\|full | Not-determined |
| 18-102-final-viral-combined_k141_171299\|\|full | Low-quality |
| 18-102-final-viral-combined_k141_88180\|\|full | Not-determined |
| 18-102-final-viral-combined_k141_57222\|\|full | Low-quality |
| 18-102-final-viral-combined_k141_88194\|\|full | Not-determined |
| 18-102-final-viral-combined_k141_242363\|\|full | Low-quality |
| 18-102-final-viral-combined_k141_171337\|\|full | Low-quality |
| 18-102-final-viral-combined_k141_171352\|\|full | Low-quality |
| 18-102-final-viral-combined_k141_242371\|\|full | Low-quality |
| 18-102-final-viral-combined_k141_108366\|\|full | Not-determined |

18-102-final-viral-combined_k141_88223||full Not-determined
18-102-final-viral-combined_k141_128592||full Low-quality
18-102-final-viral-combined_k141_108383||full Not-determined
18-102-final-viral-combined_k141_233268||full Low-quality
18-102-final-viral-combined_k141_88241||full Not-determined
18-102-final-viral-combined_k141_221602||full Low-quality
18-102-final-viral-combined_k141_67141||full Low-quality
18-102-final-viral-combined_k141_139965||full Not-determined
18-102-final-viral-combined_k141_233304||full Not-determined
18-102-final-viral-combined_k141_191507||full Not-determined
18-102-final-viral-combined_k141_242402||full Not-determined
18-102-final-viral-combined_k141_139984||full Low-quality
18-102-final-viral-combined_k141_181711||full Not-determined
18-102-final-viral-combined_k141_36615||full Low-quality
18-102-final-viral-combined_k141_242420||full Low-quality
18-102-final-viral-combined_k141_67200||full Low-quality
18-102-final-viral-combined_k141_211898||full Low-quality
18-102-final-viral-combined_k141_36626||full Not-determined
18-102-final-viral-combined_k141_108417||full Not-determined
18-102-final-viral-combined_k141_140001||full Low-quality
18-102-final-viral-combined_k141_88255||full Not-determined
18-102-final-viral-combined_k141_140013||full Low-quality
18-102-final-viral-combined_k141_211925||full Low-quality
18-102-final-viral-combined_k141_119063||full Low-quality
18-102-final-viral-combined_k141_211936||full Low-quality
18-102-final-viral-combined_k141_171449||full Low-quality
18-102-final-viral-combined_k141_233342||full Low-quality
18-102-final-viral-combined_k141_221676||full Low-quality
18-102-final-viral-combined_k141_150057||full Low-quality
18-102-final-viral-combined_k141_221684||full Low-quality
18-102-final-viral-combined_k141_181770||full Low-quality
18-102-final-viral-combined_k141_171494||full Low-quality
18-102-final-viral-combined_k141_48227||full Not-determined
18-102-final-viral-combined_k141_181780||full Not-determined
18-102-final-viral-combined_k141_119120||full Not-determined
18-102-final-viral-combined_k141_57290||full Low-quality
18-102-final-viral-combined_k141_242461||full Not-determined
18-102-final-viral-combined_k141_4466||full Not-determined
18-102-final-viral-combined_k141_67221||full Not-determined
18-102-final-viral-combined_k141_233384||full Not-determined
18-102-final-viral-combined_k141_160149||full Low-quality
18-102-final-viral-combined_k141_67223||full Not-determined
18-102-final-viral-combined_k141_211975||full Low-quality
18-102-final-viral-combined_k141_233403||full Low-quality
18-102-final-viral-combined_k141_15522||full Not-determined
18-102-final-viral-combined_k141_98810||full Low-quality
18-102-final-viral-combined_k141_119146||full Not-determined
18-102-final-viral-combined_k141_119152||full Low-quality
18-102-final-viral-combined_k141_77700||full Not-determined
18-102-final-viral-combined_k141_119156||full Not-determined
18-102-final-viral-combined_k141_119160||full Low-quality
18-102-final-viral-combined_k141_171527||full Low-quality
18-102-final-viral-combined_k141_67256||full Not-determined
18-109-final-viral-combined_k141_234716||full Medium-quality
18-102-final-viral-combined_k141_242489||full Low-quality
18-102-final-viral-combined_k141_160172||full Not-determined
18-102-final-viral-combined_k141_119169||full Low-quality
18-102-final-viral-combined_k141_150157||full Not-determined
18-102-final-viral-combined_k141_150161||full Low-quality
18-102-final-viral-combined_k141_98849||full Not-determined
18-102-final-viral-combined_k141_36711||full Low-quality
18-102-final-viral-combined_k141_221713||full Not-determined
18-102-final-viral-combined_k141_242504||full Not-determined
18-102-final-viral-combined_k141_171550||full Low-quality
18-102-final-viral-combined_k141_77765||full Not-determined
18-102-final-viral-combined_k141_150176||full Low-quality
18-102-final-viral-combined_k141_77766||full Low-quality
18-102-final-viral-combined_k141_200797||full Low-quality
18-102-final-viral-combined_k141_150185||full Low-quality
18-102-final-viral-combined_k141_57298||full Low-quality
KM-final-viral-combined_k141_258087||full Medium-quality
18-102-final-viral-combined_k141_48303||full Low-quality
18-102-final-viral-combined_k141_48305||full Low-quality
18-102-final-viral-combined_k141_212024||full Low-quality
18-102-final-viral-combined_k141_150199||full Low-quality
18-102-final-viral-combined_k141_200823||full Not-determined
18-102-final-viral-combined_k141_67277||full Low-quality
18-102-final-viral-combined_k141_242535||full Not-determined
18-102-final-viral-combined_k141_57319||full Low-quality
18-102-final-viral-combined_k141_150221||full Not-determined
18-102-final-viral-combined_k141_67295||full Not-determined
18-102-final-viral-combined_k141_221758||full Not-determined
18-102-final-viral-combined_k141_67303||full Low-quality
18-102-final-viral-combined_k141_212041||full Not-determined
18-102-final-viral-combined_k141_4517||full Low-quality
18-102-final-viral-combined_k141_191674||full Low-quality
18-102-final-viral-combined_k141_15613||full Low-quality
18-102-final-viral-combined_k141_57371||full Low-quality
18-102-final-viral-combined_k141_128679||full Not-determined
18-102-final-viral-combined_k141_119256||full Not-determined
18-102-final-viral-combined_k141_233528||full Not-determined
18-102-final-viral-combined_k141_191697||full Not-determined
18-102-final-viral-combined_k141_200859||full Low-quality
18-102-final-viral-combined_k141_119266||full Low-quality
18-102-final-viral-combined_k141_191703||full Not-determined
18-102-final-viral-combined_k141_200862||full Not-determined
18-102-final-viral-combined_k141_128702||full Not-determined
18-102-final-viral-combined_k141_128706||full Low-quality
18-102-final-viral-combined_k141_140183||full Not-determined
18-102-final-viral-combined_k141_57400||full Low-quality
18-102-final-viral-combined_k141_200867||full Not-determined
18-102-final-viral-combined_k141_200871||full Not-determined
18-102-final-viral-combined_k141_150279||full Low-quality
18-102-final-viral-combined_k141_140195||full Not-determined
18-102-final-viral-combined_k141_119286||full Not-determined
18-102-final-viral-combined_k141_160257||full Not-determined
18-102-final-viral-combined_k141_57424||full Not-determined
18-102-final-viral-combined_k141_108483||full Not-determined
18-102-final-viral-combined_k141_36866||full Not-determined
17-109-final-viral-combined_k141_7174||full Medium-quality
18-102-final-viral-combined_k141_88466||full Not-determined
18-102-final-viral-combined_k141_127080||full Medium-quality
18-102-final-viral-combined_k141_140213||full Low-quality
18-102-final-viral-combined_k141_200890||full Not-determined
18-102-final-viral-combined_k141_119331||full Low-quality
18-102-final-viral-combined_k141_200892||full Low-quality
18-102-final-viral-combined_k141_57453||full Low-quality
18-102-final-viral-combined_k141_140218||full Not-determined
18-102-final-viral-combined_k141_212116||full Not-determined
18-102-final-viral-combined_k141_181852||full Low-quality
18-102-final-viral-combined_k141_36882||full Low-quality
18-102-final-viral-combined_k141_98921||full Not-determined
18-102-final-viral-combined_k141_108518||full Not-determined
18-102-final-viral-combined_k141_36889||full Low-quality
18-102-final-viral-combined_k141_77921||full Not-determined
18-102-final-viral-combined_k141_150293||full Low-quality
18-102-final-viral-combined_k141_171651||full Not-determined
18-102-final-viral-combined_k141_233590||full Not-determined
18-102-final-viral-combined_k141_233591||full Not-determined
18-102-final-viral-combined_k141_128739||full Low-quality
18-102-final-viral-combined_k141_15697||full Low-quality
18-102-final-viral-combined_k141_98947||full Low-quality
18-102-final-viral-combined_k141_98952||full Not-determined
18-102-final-viral-combined_k141_57504||full Not-determined
18-102-final-viral-combined_k141_98961||full Low-quality
18-102-final-viral-combined_k141_233610||full Not-determined
18-102-final-viral-combined_k141_77969||full Not-determined
18-102-final-viral-combined_k141_57511||full Low-quality
18-102-final-viral-combined_k141_212124||full Not-determined
18-102-final-viral-combined_k141_171685||full Not-determined
18-102-final-viral-combined_k141_57518||full Not-determined
18-102-final-viral-combined_k141_77979||full Not-determined
18-102-final-viral-combined_k141_119408||full Not-determined
18-102-final-viral-combined_k141_108558||full Low-quality
18-102-final-viral-combined_k141_171691||full Low-quality
18-102-final-viral-combined_k141_181875||full Low-quality
18-102-final-viral-combined_k141_57529||full Low-quality
18-102-final-viral-combined_k141_242699||full Not-determined
18-102-final-viral-combined_k141_200952||full Low-quality
18-102-final-viral-combined_k141_140306||full Not-determined
18-102-final-viral-combined_k141_119423||full Not-determined
18-102-final-viral-combined_k141_77988||full Not-determined
18-102-final-viral-combined_k141_108573||full Low-quality
18-102-final-viral-combined_k141_108574||full Low-quality
18-102-final-viral-combined_k141_171731||full Not-determined
18-102-final-viral-combined_k141_140312||full Not-determined
18-102-final-viral-combined_k141_77998||full Not-determined
18-102-final-viral-combined_k141_15708||full Not-determined
18-102-final-viral-combined_k141_212138||full Low-quality
18-102-final-viral-combined_k141_108594||full Low-quality
18-102-final-viral-combined_k141_48440||full Low-quality
18-102-final-viral-combined_k141_150355||full Low-quality
18-102-final-viral-combined_k141_108597||full Not-determined
18-102-final-viral-combined_k141_171746||full Not-determined
18-102-final-viral-combined_k141_191753||full Not-determined
18-102-final-viral-combined_k141_4614||full Low-quality
18-102-final-viral-combined_k141_88649||full Not-determined
18-102-final-viral-combined_k141_191761||full Not-determined
18-102-final-viral-combined_k141_171772||full Low-quality
18-102-final-viral-combined_k141_119479||full Not-determined
18-102-final-viral-combined_k141_171789||full Not-determined
18-102-final-viral-combined_k141_128756||full Not-determined
18-102-final-viral-combined_k141_221803||full Low-quality
18-102-final-viral-combined_k141_36931||full Low-quality
18-102-final-viral-combined_k141_160276||full Not-determined
18-102-final-viral-combined_k141_201017||full Not-determined
18-102-final-viral-combined_k141_160282||full Low-quality
18-102-final-viral-combined_k141_150394||full Not-determined
18-102-final-viral-combined_k141_78068||full Low-quality
18-102-final-viral-combined_k141_150396||full Low-quality
18-102-final-viral-combined_k141_150414||full Not-determined
18-102-final-viral-combined_k141_88675||full Not-determined
18-102-final-viral-combined_k141_67341||full Low-quality
18-102-final-viral-combined_k141_233689||full Not-determined
18-102-final-viral-combined_k141_4638||full Not-determined
18-102-final-viral-combined_k141_191778||full Not-determined
18-102-final-viral-combined_k141_221846||full Not-determined
18-102-final-viral-combined_k141_119517||full Low-quality
18-102-final-viral-combined_k141_88690||full Low-quality
18-102-final-viral-combined_k141_233722||full Low-quality
18-102-final-viral-combined_k141_88716||full Low-quality
18-102-final-viral-combined_k141_99010||full Low-quality
18-102-final-viral-combined_k141_36976||full Low-quality
18-102-final-viral-combined_k141_67354||full Not-determined
18-102-final-viral-combined_k141_36981||full Low-quality
18-102-final-viral-combined_k141_67360||full Low-quality
18-102-final-viral-combined_k141_78153||full Not-determined
18-102-final-viral-combined_k141_48531||full Not-determined
18-102-final-viral-combined_k141_15836||full Low-quality
18-102-final-viral-combined_k141_108667||full Not-determined
18-102-final-viral-combined_k141_128817||full Low-quality
18-102-final-viral-combined_k141_128823||full Low-quality
18-102-final-viral-combined_k141_171856||full Low-quality
18-102-final-viral-combined_k141_128825||full Low-quality
18-102-final-viral-combined_k141_140430||full Not-determined
18-102-final-viral-combined_k141_233773||full Low-quality
18-102-final-viral-combined_k141_99034||full Low-quality
18-102-final-viral-combined_k141_191852||full Low-quality
18-102-final-viral-combined_k141_99036||full Not-determined
17-109_k141_161239 Medium-quality
18-102-final-viral-combined_k141_201110||full Not-determined
18-102-final-viral-combined_k141_221948||full Not-determined
18-102-final-viral-combined_k141_140443||full Low-quality
18-102-final-viral-combined_k141_78204||full Low-quality
18-102-final-viral-combined_k141_181962||full Not-determined
18-102-final-viral-combined_k141_99059||full Not-determined
18-102-final-viral-combined_k141_181974||full Low-quality
18-102-final-viral-combined_k141_233791||full Low-quality
18-102-final-viral-combined_k141_171889||full Not-determined
18-102-final-viral-combined_k141_108714||full Not-determined
18-102-final-viral-combined_k141_221963||full Not-determined
18-102-final-viral-combined_k141_140471||full Not-determined
18-102-final-viral-combined_k141_171908||full Low-quality
18-102-final-viral-combined_k141_48617||full Not-determined
18-102-final-viral-combined_k141_119623||full Not-determined
18-102-final-viral-combined_k141_140475||full Not-determined
18-102-final-viral-combined_k141_233822||full Not-determined
18-102-final-viral-combined_k141_181995||full Low-quality
18-102-final-viral-combined_k141_181996||full Not-determined
18-102-final-viral-combined_k141_4688||full Not-determined
18-102-final-viral-combined_k141_233832||full Not-determined
18-102-final-viral-combined_k141_242767||full Not-determined
18-102-final-viral-combined_k141_119651||full Low-quality
18-102-final-viral-combined_k141_150532||full Low-quality
18-102-final-viral-combined_k141_119655||full Low-quality
18-102-final-viral-combined_k141_212182||full Not-determined
18-102-final-viral-combined_k141_150538||full Not-determined
18-102-final-viral-combined_k141_212198||full Low-quality
18-102-final-viral-combined_k141_108774||full Not-determined
18-102-final-viral-combined_k141_212204||full Low-quality
18-102-final-viral-combined_k141_171965||full Low-quality
18-102-final-viral-combined_k141_182029||full Not-determined
18-102-final-viral-combined_k141_4724||full Low-quality
18-102-final-viral-combined_k141_57611||full Not-determined
18-102-final-viral-combined_k141_57615||full Low-quality
18-102-final-viral-combined_k141_15924||full Low-quality
17-109-final-viral-combined_k141_161239||full Medium-quality
18-102-final-viral-combined_k141_67419||full Not-determined
18-102-final-viral-combined_k141_67426||full Not-determined
18-102-final-viral-combined_k141_78253||full Not-determined
18-102-final-viral-combined_k141_171994||full Low-quality
18-102-final-viral-combined_k141_182050||full Not-determined
18-102-final-viral-combined_k141_48682||full Not-determined
18-102-final-viral-combined_k141_78262||full Not-determined
18-102-final-viral-combined_k141_57643||full Not-determined
18-102-final-viral-combined_k141_119706||full Low-quality
18-102-final-viral-combined_k141_67445||full Low-quality
18-102-final-viral-combined_k141_78276||full Not-determined
18-102-final-viral-combined_k141_191961||full Not-determined
18-102-final-viral-combined_k141_212262||full Not-determined
18-102-final-viral-combined_k141_242836||full Not-determined
18-102-final-viral-combined_k141_78289||full Not-determined
18-102-final-viral-combined_k141_15948||full Not-determined
18-102-final-viral-combined_k141_78296||full Not-determined
18-102-final-viral-combined_k141_150582||full Low-quality
18-102-final-viral-combined_k141_15956||full Not-determined
18-102-final-viral-combined_k141_57661||full Not-determined
18-102-final-viral-combined_k141_242863||full Not-determined
18-102-final-viral-combined_k141_99137||full Not-determined
18-102-final-viral-combined_k141_88832||full Not-determined
18-102-final-viral-combined_k141_25514||full Low-quality
17-108-final-viral-combined_k141_53907||full Medium-quality
18-102-final-viral-combined_k141_119739||full Low-quality
18-102-final-viral-combined_k141_88844||full Low-quality
18-102-final-viral-combined_k141_191972||full Not-determined
18-102-final-viral-combined_k141_67467||full Low-quality
18-102-final-viral-combined_k141_172078||full Not-determined
18-102-final-viral-combined_k141_242876||full Not-determined
18-102-final-viral-combined_k141_233876||full Low-quality
18-102-final-viral-combined_k141_67473||full Low-quality
18-102-final-viral-combined_k141_172083||full Low-quality
18-102-final-viral-combined_k141_15995||full Low-quality
18-102-final-viral-combined_k141_201227||full Not-determined
18-102-final-viral-combined_k141_191979||full Low-quality
18-102-final-viral-combined_k141_25547||full Not-determined
18-102-final-viral-combined_k141_48786||full Not-determined
18-102-final-viral-combined_k141_150644||full Not-determined
18-102-final-viral-combined_k141_88875||full Not-determined
18-102-final-viral-combined_k141_201236||full Not-determined
18-102-final-viral-combined_k141_78359||full Not-determined
18-102-final-viral-combined_k141_119759||full Not-determined
18-102-final-viral-combined_k141_150658||full Not-determined
18-102-final-viral-combined_k141_140603||full Low-quality
18-102-final-viral-combined_k141_182157||full Low-quality
18-102-final-viral-combined_k141_172092||full Not-determined
18-102-final-viral-combined_k141_172103||full Not-determined
18-102-final-viral-combined_k141_172104||full Not-determined
18-102-final-viral-combined_k141_16036||full Not-determined
18-102-final-viral-combined_k141_67502||full Low-quality
18-102-final-viral-combined_k141_140621||full Low-quality
18-102-final-viral-combined_k141_48831||full Low-quality
18-102-final-viral-combined_k141_182182||full Low-quality
18-102-final-viral-combined_k141_172108||full Not-determined
18-102-final-viral-combined_k141_78386||full Not-determined
18-102-final-viral-combined_k141_78391||full Not-determined
18-102-final-viral-combined_k141_182206||full Low-quality
18-102-final-viral-combined_k141_4741||full Low-quality
18-102-final-viral-combined_k141_140641||full Not-determined
18-102-final-viral-combined_k141_99175||full Not-determined
18-102-final-viral-combined_k141_119807||full Not-determined
18-102-final-viral-combined_k141_67528||full Not-determined
18-102-final-viral-combined_k141_182221||full Not-determined
18-102-final-viral-combined_k141_108870||full Low-quality
18-102-final-viral-combined_k141_108876||full Low-quality
18-102-final-viral-combined_k141_233930||full Low-quality
18-102-final-viral-combined_k141_201258||full Not-determined
18-102-final-viral-combined_k141_233936||full Not-determined
18-102-final-viral-combined_k141_48871||full Low-quality
18-102-final-viral-combined_k141_172130||full Low-quality
18-102-final-viral-combined_k141_160431||full Low-quality
18-102-final-viral-combined_k141_140658||full Low-quality
18-102-final-viral-combined_k141_48890||full Low-quality
18-102-final-viral-combined_k141_4772||full Not-determined
18-102-final-viral-combined_k141_119820||full Low-quality
18-102-final-viral-combined_k141_140664||full Low-quality
18-102-final-viral-combined_k141_57705||full Low-quality
18-102-final-viral-combined_k141_119833||full Low-quality
18-102-final-viral-combined_k141_37171||full Not-determined
18-102-final-viral-combined_k141_48914||full Not-determined
18-102-final-viral-combined_k141_119843||full Low-quality
18-102-final-viral-combined_k141_48929||full Not-determined
18-102-final-viral-combined_k141_140700||full Not-determined
18-102-final-viral-combined_k141_108908||full Not-determined
18-102-final-viral-combined_k141_182246||full Low-quality
18-102-final-viral-combined_k141_182250||full Low-quality
18-102-final-viral-combined_k141_48942||full Low-quality
18-102-final-viral-combined_k141_88973||full Not-determined
18-102-final-viral-combined_k141_128980||full Low-quality
18-102-final-viral-combined_k141_57745||full Low-quality
18-102-final-viral-combined_k141_48950||full Low-quality
18-102-final-viral-combined_k141_99252||full Low-quality
18-102-final-viral-combined_k141_150760||full Not-determined
18-102-final-viral-combined_k141_67563||full Low-quality
18-102-final-viral-combined_k141_99265||full Not-determined
18-102-final-viral-combined_k141_192021||full Not-determined
18-102-final-viral-combined_k141_160507||full Not-determined
18-102-final-viral-combined_k141_78449||full Not-determined
18-102-final-viral-combined_k141_172182||full Not-determined
18-102-final-viral-combined_k141_88994||full Low-quality
18-102-final-viral-combined_k141_129016||full Low-quality
18-102-final-viral-combined_k141_160535||full Not-determined
18-102-final-viral-combined_k141_99311||full Not-determined
18-102-final-viral-combined_k141_172207||full Not-determined
18-102-final-viral-combined_k141_160544||full Not-determined
18-102-final-viral-combined_k141_129038||full Not-determined
18-102-final-viral-combined_k141_108971||full Not-determined
18-102-final-viral-combined_k141_37218||full Low-quality
18-102-final-viral-combined_k141_212307||full Not-determined
18-102-final-viral-combined_k141_49020||full Not-determined
18-102-final-viral-combined_k141_37228||full Not-determined
18-102-final-viral-combined_k141_108980||full Not-determined
18-102-final-viral-combined_k141_99321||full Low-quality
18-102-final-viral-combined_k141_37241||full Low-quality
18-102-final-viral-combined_k141_234016||full Not-determined
18-102-final-viral-combined_k141_182326||full Not-determined
18-102-final-viral-combined_k141_37244||full Not-determined
18-102-final-viral-combined_k141_37246||full Not-determined
18-102-final-viral-combined_k141_37247||full Not-determined
18-102-final-viral-combined_k141_108998||full Low-quality
18-102-final-viral-combined_k141_37249||full Low-quality
18-102-final-viral-combined_k141_109011||full Not-determined
18-102-final-viral-combined_k141_192072||full Low-quality
18-102-final-viral-combined_k141_212352||full Not-determined
18-102-final-viral-combined_k141_109014||full Not-determined
18-102-final-viral-combined_k141_192083||full Low-quality
18-102-final-viral-combined_k141_150874||full Not-determined
18-102-final-viral-combined_k141_89082||full Low-quality
18-102-final-viral-combined_k141_37304||full Not-determined
18-102-final-viral-combined_k141_201344||full Not-determined
18-102-final-viral-combined_k141_201346||full Not-determined
18-102-final-viral-combined_k141_109036||full Low-quality
18-102-final-viral-combined_k141_16106||full Not-determined
18-102-final-viral-combined_k141_150901||full Low-quality
18-102-final-viral-combined_k141_201349||full Not-determined
18-102-final-viral-combined_k141_57825||full Low-quality
18-102-final-viral-combined_k141_109045||full Low-quality
18-102-final-viral-combined_k141_172264||full Low-quality
18-102-final-viral-combined_k141_140864||full Not-determined
18-102-final-viral-combined_k141_119951||full Low-quality
18-102-final-viral-combined_k141_99371||full Not-determined
18-102-final-viral-combined_k141_16127||full Not-determined
18-102-final-viral-combined_k141_49049||full Not-determined
18-102-final-viral-combined_k141_89127||full Not-determined
18-102-final-viral-combined_k141_99378||full Low-quality
18-102-final-viral-combined_k141_89131||full Low-quality
18-102-final-viral-combined_k141_78542||full Low-quality
18-102-final-viral-combined_k141_99389||full Low-quality
18-102-final-viral-combined_k141_57858||full Not-determined
18-102-final-viral-combined_k141_16152||full Not-determined
18-102-final-viral-combined_k141_212378||full Low-quality
18-102-final-viral-combined_k141_49073||full Not-determined
18-102-final-viral-combined_k141_172301||full Not-determined
18-102-final-viral-combined_k141_192100||full Not-determined
18-102-final-viral-combined_k141_172306||full Not-determined
18-102-final-viral-combined_k141_16173||full Not-determined
18-102-final-viral-combined_k141_67625||full Low-quality
18-102-final-viral-combined_k141_67629||full Low-quality
18-102-final-viral-combined_k141_172316||full Low-quality
18-102-final-viral-combined_k141_160618||full Not-determined
18-102-final-viral-combined_k141_78549||full Not-determined
18-102-final-viral-combined_k141_192137||full Not-determined
18-102-final-viral-combined_k141_109123||full Not-determined
18-102-final-viral-combined_k141_16182||full Not-determined
18-102-final-viral-combined_k141_25606||full Not-determined
18-102-final-viral-combined_k141_150934||full Not-determined
18-102-final-viral-combined_k141_16192||full Low-quality
18-102-final-viral-combined_k141_120005||full Low-quality
18-102-final-viral-combined_k141_109143||full Low-quality
18-102-final-viral-combined_k141_140885||full Not-determined
18-102-final-viral-combined_k141_192167||full Not-determined
18-102-final-viral-combined_k141_37328||full Low-quality
18-102-final-viral-combined_k141_37335||full Low-quality
18-102-final-viral-combined_k141_160671||full Low-quality
18-102-final-viral-combined_k141_120028||full Not-determined
18-102-final-viral-combined_k141_67681||full Not-determined
18-102-final-viral-combined_k141_4822||full Low-quality
18-102-final-viral-combined_k141_109161||full Low-quality
18-102-final-viral-combined_k141_78578||full Not-determined
18-102-final-viral-combined_k141_49136||full Low-quality
18-102-final-viral-combined_k141_172400||full Not-determined
18-102-final-viral-combined_k141_49152||full Low-quality
18-102-final-viral-combined_k141_25646||full Not-determined
18-102-final-viral-combined_k141_49153||full Low-quality
18-102-final-viral-combined_k141_172422||full Not-determined
18-102-final-viral-combined_k141_37389||full Low-quality
18-102-final-viral-combined_k141_160697||full Low-quality
18-102-final-viral-combined_k141_222013||full Not-determined
18-102-final-viral-combined_k141_78594||full Low-quality
18-106_k141_106109 Medium-quality
18-102-final-viral-combined_k141_120068||full Low-quality
18-102-final-viral-combined_k141_57914||full Not-determined
18-102-final-viral-combined_k141_109207||full Not-determined
18-102-final-viral-combined_k141_25678||full Low-quality
18-102-final-viral-combined_k141_182420||full Low-quality
18-102-final-viral-combined_k141_25681||full Low-quality
18-102-final-viral-combined_k141_57925||full Not-determined
18-102-final-viral-combined_k141_172463||full Not-determined
18-102-final-viral-combined_k141_160731||full Low-quality
18-102-final-viral-combined_k141_192212||full Not-determined
18-102-final-viral-combined_k141_120085||full Low-quality
18-102-final-viral-combined_k141_120091||full Not-determined
18-102-final-viral-combined_k141_49196||full Low-quality
18-102-final-viral-combined_k141_234088||full Low-quality
18-102-final-viral-combined_k141_172481||full Not-determined
18-102-final-viral-combined_k141_37461||full Low-quality
18-102-final-viral-combined_k141_201467||full Not-determined
18-102-final-viral-combined_k141_57941||full Low-quality
18-102-final-viral-combined_k141_25688||full Low-quality
18-102-final-viral-combined_k141_25694||full Not-determined
18-102-final-viral-combined_k141_243050||full Not-determined
18-102-final-viral-combined_k141_67798||full Low-quality
18-102-final-viral-combined_k141_49225||full Low-quality
18-102-final-viral-combined_k141_222054||full Not-determined
18-102-final-viral-combined_k141_89224||full Low-quality
18-102-final-viral-combined_k141_99461||full Not-determined
18-102-final-viral-combined_k141_222073||full Low-quality
17-106-final-viral-combined_k141_89249||full Medium-quality
18-102-final-viral-combined_k141_192263||full Low-quality
18-102-final-viral-combined_k141_16239||full Low-quality
18-102-final-viral-combined_k141_243091||full Not-determined
18-102-final-viral-combined_k141_89273||full Low-quality
18-102-final-viral-combined_k141_243093||full Low-quality
18-102-final-viral-combined_k141_120171||full Not-determined
18-102-final-viral-combined_k141_212430||full Not-determined
18-102-final-viral-combined_k141_129085||full Not-determined
18-102-final-viral-combined_k141_120181||full Not-determined
18-102-final-viral-combined_k141_151032||full Low-quality
18-102-final-viral-combined_k141_192275||full Low-quality
18-102-final-viral-combined_k141_78648||full Low-quality
18-102-final-viral-combined_k141_49291||full Not-determined
18-102-final-viral-combined_k141_192277||full Low-quality
18-102-final-viral-combined_k141_172584||full Not-determined
18-102-final-viral-combined_k141_16268||full Low-quality
18-102-final-viral-combined_k141_89284||full Low-quality
18-102-final-viral-combined_k141_120197||full Not-determined
18-101-final-viral-combined_k141_227003||full Medium-quality
18-102-final-viral-combined_k141_129097||full Low-quality
18-102-final-viral-combined_k141_67869||full Not-determined
18-102-final-viral-combined_k141_160777||full Not-determined
18-102-final-viral-combined_k141_212466||full Low-quality
18-102-final-viral-combined_k141_99534||full Not-determined
18-102-final-viral-combined_k141_16288||full Not-determined
18-102-final-viral-combined_k141_99539||full Not-determined
18-102-final-viral-combined_k141_99542||full Not-determined
18-102-final-viral-combined_k141_89340||full Not-determined
18-102-final-viral-combined_k141_109217||full Not-determined
18-102-final-viral-combined_k141_49352||full Low-quality
18-102-final-viral-combined_k141_234256||full Low-quality
18-102-final-viral-combined_k141_109238||full Not-determined
18-102-final-viral-combined_k141_58013||full Not-determined
18-102-final-viral-combined_k141_109245||full Not-determined
18-102-final-viral-combined_k141_16312||full Not-determined
18-102-final-viral-combined_k141_129161||full Not-determined
18-102-final-viral-combined_k141_89351||full Low-quality
18-102-final-viral-combined_k141_109249||full Not-determined
18-102-final-viral-combined_k141_120238||full Low-quality
18-102-final-viral-combined_k141_109257||full Low-quality
18-102-final-viral-combined_k141_120242||full Low-quality
18-102-final-viral-combined_k141_67947||full Not-determined
18-102-final-viral-combined_k141_222170||full Not-determined
18-102-final-viral-combined_k141_78668||full Not-determined
18-102-final-viral-combined_k141_89387||full Low-quality
18-102-final-viral-combined_k141_89393||full Not-determined
18-102-final-viral-combined_k141_58074||full Low-quality
18-102-final-viral-combined_k141_234348||full Not-determined
18-102-final-viral-combined_k141_109321||full Low-quality
18-102-final-viral-combined_k141_234349||full Low-quality
18-105-final-viral-combined_k141_106597||full Medium-quality
18-102-final-viral-combined_k141_25819||full Low-quality
18-102-final-viral-combined_k141_201632||full Not-determined
18-102-final-viral-combined_k141_182618||full Low-quality
18-102-final-viral-combined_k141_89419||full Not-determined
18-102-final-viral-combined_k141_109336||full Low-quality
18-102-final-viral-combined_k141_182620||full Low-quality
18-102-final-viral-combined_k141_4884||full Not-determined
18-102-final-viral-combined_k141_67984||full Not-determined
18-102-final-viral-combined_k141_172729||full Not-determined
18-102-final-viral-combined_k141_129198||full Low-quality
18-102-final-viral-combined_k141_151204||full Low-quality
18-102-final-viral-combined_k141_99668||full Low-quality
18-102-final-viral-combined_k141_243234||full Not-determined
18-102-final-viral-combined_k141_192331||full Low-quality
18-102-final-viral-combined_k141_4894||full Not-determined
18-102-final-viral-combined_k141_192332||full Low-quality
18-102-final-viral-combined_k141_234377||full Not-determined
18-102-final-viral-combined_k141_49408||full Not-determined
18-102-final-viral-combined_k141_68003||full Not-determined
18-102-final-viral-combined_k141_89452||full Low-quality
18-102-final-viral-combined_k141_160904||full Low-quality
18-102-final-viral-combined_k141_222185||full Not-determined
18-102-final-viral-combined_k141_192346||full Low-quality
18-102-final-viral-combined_k141_222193||full Low-quality
18-102-final-viral-combined_k141_99695||full Low-quality
18-102-final-viral-combined_k141_16389||full Not-determined
18-102-final-viral-combined_k141_37467||full Low-quality
18-102-final-viral-combined_k141_151227||full Low-quality
18-102-final-viral-combined_k141_192361||full Low-quality
18-102-final-viral-combined_k141_68033||full Not-determined
18-102-final-viral-combined_k141_201691||full Not-determined
18-102-final-viral-combined_k141_234407||full Not-determined
18-102-final-viral-combined_k141_120272||full Not-determined
18-102-final-viral-combined_k141_222222||full Low-quality
18-102-final-viral-combined_k141_151263||full Low-quality
18-102-final-viral-combined_k141_37487||full Low-quality
18-102-final-viral-combined_k141_120286||full Low-quality
18-102-final-viral-combined_k141_172827||full Low-quality
18-102-final-viral-combined_k141_234420||full Low-quality
18-102-final-viral-combined_k141_222234||full Low-quality
18-102-final-viral-combined_k141_89472||full Low-quality
18-102-final-viral-combined_k141_68058||full Not-determined
17-108-final-viral-combined_k141_154845||full Medium-quality
18-102-final-viral-combined_k141_129218||full Low-quality
18-102-final-viral-combined_k141_172851||full Not-determined
18-102-final-viral-combined_k141_160918||full Low-quality
18-102-final-viral-combined_k141_192398||full Not-determined
18-102-final-viral-combined_k141_49506||full Not-determined
18-102-final-viral-combined_k141_151321||full Not-determined
18-102-final-viral-combined_k141_201710||full Low-quality
18-102-final-viral-combined_k141_201716||full Low-quality
18-102-final-viral-combined_k141_129231||full Low-quality
18-102-final-viral-combined_k141_89495||full Low-quality
18-102-final-viral-combined_k141_78773||full Not-determined
18-102-final-viral-combined_k141_49535||full Low-quality
18-102-final-viral-combined_k141_49536||full Low-quality
18-102-final-viral-combined_k141_89508||full Low-quality
18-102-final-viral-combined_k141_192418||full Not-determined
18-102-final-viral-combined_k141_25914||full Not-determined
18-102-final-viral-combined_k141_172892||full Not-determined
18-102-final-viral-combined_k141_89509||full Not-determined
18-102-final-viral-combined_k141_49553||full Not-determined
18-102-final-viral-combined_k141_37539||full Low-quality
18-106-final-viral-combined_k141_106109||full Medium-quality
18-102-final-viral-combined_k141_4925||full Not-determined
18-102-final-viral-combined_k141_151373||full Not-determined
18-102-final-viral-combined_k141_172914||full Not-determined
18-102-final-viral-combined_k141_160960||full Low-quality
18-102-final-viral-combined_k141_25941||full Not-determined
18-102-final-viral-combined_k141_192434||full Low-quality
18-102-final-viral-combined_k141_89525||full Low-quality

| | |
|---|---|
| 18-102-final-viral-combined_k141_172945\|\|full | Low-quality |
| 18-102-final-viral-combined_k141_49613\|\|full | Low-quality |
| 18-102-final-viral-combined_k141_160969\|\|full | Not-determined |
| 18-102-final-viral-combined_k141_172956\|\|full | Low-quality |
| 18-102-final-viral-combined_k141_99775\|\|full | Not-determined |
| 18-102-final-viral-combined_k141_160979\|\|full | Not-determined |
| 18-102-final-viral-combined_k141_172963\|\|full | Not-determined |
| 18-102-final-viral-combined_k141_120333\|\|full | Not-determined |
| 18-102-final-viral-combined_k141_120338\|\|full | Low-quality |
| 18-102-final-viral-combined_k141_192452\|\|full | Not-determined |
| 18-102-final-viral-combined_k141_58210\|\|full | Not-determined |
| 18-102-final-viral-combined_k141_160999\|\|full | Low-quality |
| 18-102-final-viral-combined_k141_89572\|\|full | Not-determined |
| 18-102-final-viral-combined_k141_120359\|\|full | Not-determined |
| 18-102-final-viral-combined_k141_212617\|\|full | Low-quality |
| 18-102-final-viral-combined_k141_16577\|\|full | Low-quality |
| 18-102-final-viral-combined_k141_234546\|\|full | Not-determined |
| 18-102-final-viral-combined_k141_58218\|\|full | Not-determined |
| 18-102-final-viral-combined_k141_129275\|\|full | Not-determined |
| 18-102-final-viral-combined_k141_58227\|\|full | Low-quality |
| 18-102-final-viral-combined_k141_141077\|\|full | Not-determined |
| 18-102-final-viral-combined_k141_243322\|\|full | Not-determined |
| 18-102-final-viral-combined_k141_234563\|\|full | Not-determined |
| 18-102-final-viral-combined_k141_234568\|\|full | Not-determined |
| 18-102-final-viral-combined_k141_58232\|\|full | Not-determined |
| 18-102-final-viral-combined_k141_234570\|\|full | Not-determined |
| 18-102-final-viral-combined_k141_68165\|\|full | Low-quality |
| 18-102-final-viral-combined_k141_109462\|\|full | Not-determined |
| 18-102-final-viral-combined_k141_68175\|\|full | Low-quality |
| 18-102-final-viral-combined_k141_243335\|\|full | Low-quality |
| 18-102-final-viral-combined_k141_68178\|\|full | Not-determined |
| 18-102-final-viral-combined_k141_192509\|\|full | Not-determined |
| 18-102-final-viral-combined_k141_173070\|\|full | Not-determined |
| 18-102-final-viral-combined_k141_129309\|\|full | Not-determined |
| 18-102-final-viral-combined_k141_173086\|\|full | Not-determined |
| 18-102-final-viral-combined_k141_234608\|\|full | Not-determined |
| 18-102-final-viral-combined_k141_5014\|\|full | Low-quality |
| 18-102-final-viral-combined_k141_129330\|\|full | Low-quality |
| 18-102-final-viral-combined_k141_151471\|\|full | Low-quality |
| 18-102-final-viral-combined_k141_5015\|\|full | Not-determined |
| 18-102-final-viral-combined_k141_25989\|\|full | Low-quality |
| 18-102-final-viral-combined_k141_129338\|\|full | Low-quality |
| 18-102-final-viral-combined_k141_78806\|\|full | Not-determined |
| 18-102-final-viral-combined_k141_5024\|\|full | Not-determined |
| 18-102-final-viral-combined_k141_89697\|\|full | Not-determined |
| 18-102-final-viral-combined_k141_234641\|\|full | Low-quality |
| 18-102-final-viral-combined_k141_68198\|\|full | Not-determined |
| 18-102-final-viral-combined_k141_49625\|\|full | Not-determined |
| 18-102-final-viral-combined_k141_109485\|\|full | Low-quality |
| 18-102-final-viral-combined_k141_234655\|\|full | Low-quality |
| 18-102-final-viral-combined_k141_129371\|\|full | Not-determined |
| 18-102-final-viral-combined_k141_16642\|\|full | Low-quality |
| 18-102-final-viral-combined_k141_173108\|\|full | Low-quality |
| 18-102-final-viral-combined_k141_212690\|\|full | Low-quality |
| 18-102-final-viral-combined_k141_212699\|\|full | Low-quality |
| 18-102-final-viral-combined_k141_68212\|\|full | Low-quality |
| 18-102-final-viral-combined_k141_161060\|\|full | Not-determined |
| 18-102-final-viral-combined_k141_182771\|\|full | Low-quality |
| 18-102-final-viral-combined_k141_5039\|\|full | Low-quality |
| 18-102-final-viral-combined_k141_120416\|\|full | Low-quality |
| 18-102-final-viral-combined_k141_151483\|\|full | Low-quality |
| 18-102-final-viral-combined_k141_49652\|\|full | Not-determined |
| 18-102-final-viral-combined_k141_129425\|\|full | Not-determined |
| 18-102-final-viral-combined_k141_222401\|\|full | Low-quality |
| 18-102-final-viral-combined_k141_99890\|\|full | Not-determined |
| 18-102-final-viral-combined_k141_151514\|\|full | Low-quality |
| 18-102-final-viral-combined_k141_120437\|\|full | Not-determined |
| 18-102-final-viral-combined_k141_99910\|\|full | Not-determined |
| 18-102-final-viral-combined_k141_182801\|\|full | Not-determined |
| 18-102-final-viral-combined_k141_161131\|\|full | Low-quality |
| 18-102-final-viral-combined_k141_151529\|\|full | Not-determined |
| 18-102-final-viral-combined_k141_120457\|\|full | Not-determined |
| 18-102-final-viral-combined_k141_151538\|\|full | Low-quality |
| 18-102-final-viral-combined_k141_182820\|\|full | Low-quality |
| 18-102-final-viral-combined_k141_234725\|\|full | Not-determined |
| 18-102-final-viral-combined_k141_37686\|\|full | Low-quality |
| 18-102-final-viral-combined_k141_212775\|\|full | Low-quality |
| 18-102-final-viral-combined_k141_49703\|\|full | Not-determined |
| 18-102-final-viral-combined_k141_222419\|\|full | Not-determined |
| 18-102-final-viral-combined_k141_161154\|\|full | Low-quality |
| 18-102-final-viral-combined_k141_78873\|\|full | Low-quality |
| 18-102-final-viral-combined_k141_234743\|\|full | Low-quality |
| 18-102-final-viral-combined_k141_49731\|\|full | Low-quality |
| 18-102-final-viral-combined_k141_99955\|\|full | Not-determined |
| 18-102-final-viral-combined_k141_49732\|\|full | Not-determined |
| 18-102-final-viral-combined_k141_5097\|\|full | Not-determined |
| 18-102-final-viral-combined_k141_129485\|\|full | Low-quality |
| 18-102-final-viral-combined_k141_99957\|\|full | Not-determined |
| 18-102-final-viral-combined_k141_234766\|\|full | Not-determined |
| 18-102-final-viral-combined_k141_99960\|\|full | Low-quality |
| 18-102-final-viral-combined_k141_5106\|\|full | Not-determined |
| 18-102-final-viral-combined_k141_222476\|\|full | Not-determined |
| 18-102-final-viral-combined_k141_99969\|\|full | Low-quality |
| 18-102-final-viral-combined_k141_129488\|\|full | Not-determined |
| 18-102-final-viral-combined_k141_243366\|\|full | Not-determined |
| 18-102-final-viral-combined_k141_182856\|\|full | Not-determined |
| 18-102-final-viral-combined_k141_234780\|\|full | Not-determined |
| 18-102-final-viral-combined_k141_173195\|\|full | Not-determined |
| 18-102-final-viral-combined_k141_16746\|\|full | Low-quality |
| 18-102-final-viral-combined_k141_201973\|\|full | Low-quality |
| 18-102-final-viral-combined_k141_120547\|\|full | Low-quality |
| 18-102-final-viral-combined_k141_234788\|\|full | Not-determined |
| 18-102-final-viral-combined_k141_89714\|\|full | Not-determined |
| 18-102-final-viral-combined_k141_120569\|\|full | Not-determined |
| 18-102-final-viral-combined_k141_68245\|\|full | Not-determined |
| 18-102-final-viral-combined_k141_89734\|\|full | Not-determined |
| 18-102-final-viral-combined_k141_129511\|\|full | Low-quality |
| 18-102-final-viral-combined_k141_49806\|\|full | Low-quality |
| 18-102-final-viral-combined_k141_182908\|\|full | Low-quality |
| 18-102-final-viral-combined_k141_161269\|\|full | Low-quality |
| 18-102-final-viral-combined_k141_49817\|\|full | Low-quality |
| 18-102-final-viral-combined_k141_5167\|\|full | Not-determined |
| 18-102-final-viral-combined_k141_161273\|\|full | Not-determined |
| 18-102-final-viral-combined_k141_120603\|\|full | Not-determined |
| 18-102-final-viral-combined_k141_151646\|\|full | Not-determined |
| 18-102-final-viral-combined_k141_5176\|\|full | Low-quality |
| 18-102-final-viral-combined_k141_248318\|\|full | High-quality |
| 18-102-final-viral-combined_k141_212873\|\|full | Low-quality |
| 18-102-final-viral-combined_k141_151664\|\|full | Low-quality |
| 18-102-final-viral-combined_k141_212882\|\|full | Low-quality |
| 18-102-final-viral-combined_k141_49828\|\|full | Not-determined |
| 18-102-final-viral-combined_k141_182935\|\|full | Not-determined |
| 18-102-final-viral-combined_k141_68266\|\|full | Low-quality |
| 18-102-final-viral-combined_k141_89769\|\|full | Low-quality |
| 18-102-final-viral-combined_k141_68269\|\|full | Low-quality |
| 18-102-final-viral-combined_k141_151690\|\|full | Low-quality |
| 18-102-final-viral-combined_k141_78900\|\|full | Low-quality |
| 18-102-final-viral-combined_k141_16826\|\|full | Low-quality |
| 18-102-final-viral-combined_k141_49862\|\|full | Low-quality |
| 18-102-final-viral-combined_k141_222526\|\|full | Low-quality |
| 18-102-final-viral-combined_k141_182956\|\|full | Low-quality |
| 18-102-final-viral-combined_k141_16835\|\|full | Low-quality |
| 18-102-final-viral-combined_k141_182958\|\|full | Not-determined |
| 18-102-final-viral-combined_k141_202014\|\|full | Low-quality |
| 18-102-final-viral-combined_k141_161324\|\|full | Not-determined |
| 18-102-final-viral-combined_k141_212944\|\|full | Low-quality |
| 18-102-final-viral-combined_k141_202025\|\|full | Not-determined |
| 18-102-final-viral-combined_k141_58353\|\|full | Low-quality |
| 18-102-final-viral-combined_k141_49891\|\|full | Low-quality |
| 18-102-final-viral-combined_k141_58366\|\|full | Not-determined |
| 18-102-final-viral-combined_k141_89806\|\|full | Not-determined |
| 18-102-final-viral-combined_k141_173283\|\|full | Not-determined |
| 18-102-final-viral-combined_k141_120670\|\|full | Not-determined |
| 18-102-final-viral-combined_k141_173300\|\|full | Low-quality |
| 18-102-final-viral-combined_k141_89814\|\|full | Not-determined |
| 18-102-final-viral-combined_k141_109623\|\|full | Low-quality |
| 17-109-final-viral-combined_k141_320777\|\|full | Medium-quality |
| 18-102-final-viral-combined_k141_89825\|\|full | Not-determined |
| 18-102-final-viral-combined_k141_151735\|\|full | Not-determined |
| 18-101-final-viral-combined_k141_57370\|\|full | Medium-quality |
| 18-102-final-viral-combined_k141_141200\|\|full | Low-quality |
| 18-102-final-viral-combined_k141_109655\|\|full | Not-determined |
| 18-102-final-viral-combined_k141_141230\|\|full | Low-quality |
| 18-102-final-viral-combined_k141_68331\|\|full | Not-determined |
| 18-102-final-viral-combined_k141_222556\|\|full | Not-determined |
| 18-102-final-viral-combined_k141_68337\|\|full | Low-quality |
| 18-102-final-viral-combined_k141_109666\|\|full | Not-determined |
| 18-102-final-viral-combined_k141_100064\|\|full | Low-quality |
| 18-102-final-viral-combined_k141_192715\|\|full | Low-quality |
| 18-102-final-viral-combined_k141_109685\|\|full | Low-quality |
| 18-102-final-viral-combined_k141_37816\|\|full | Low-quality |
| 18-102-final-viral-combined_k141_173375\|\|full | Low-quality |
| 18-102-final-viral-combined_k141_213017\|\|full | Low-quality |
| 18-102-final-viral-combined_k141_213021\|\|full | Not-determined |
| 18-102-final-viral-combined_k141_26106\|\|full | Low-quality |
| 18-102-final-viral-combined_k141_141268\|\|full | Not-determined |
| 18-102-final-viral-combined_k141_58482\|\|full | Not-determined |
| 18-102-final-viral-combined_k141_151818\|\|full | Not-determined |
| 18-102-final-viral-combined_k141_16974\|\|full | Low-quality |
| 18-102-final-viral-combined_k141_151827\|\|full | Low-quality |
| 18-102-final-viral-combined_k141_78964\|\|full | Not-determined |
| 18-102-final-viral-combined_k141_173426\|\|full | Not-determined |
| 18-102-final-viral-combined_k141_141292\|\|full | Not-determined |
| 18-102-final-viral-combined_k141_89921\|\|full | Low-quality |
| 18-102-final-viral-combined_k141_89922\|\|full | Low-quality |
| 18-102-final-viral-combined_k141_151847\|\|full | Not-determined |
| 18-102-final-viral-combined_k141_173439\|\|full | Low-quality |
| 18-102-final-viral-combined_k141_192737\|\|full | Low-quality |
| 18-102-final-viral-combined_k141_109730\|\|full | Low-quality |
| 18-102-final-viral-combined_k141_173446\|\|full | Not-determined |
| 18-102-final-viral-combined_k141_17005\|\|full | Not-determined |
| 18-102-final-viral-combined_k141_109751\|\|full | Low-quality |
| 18-102-final-viral-combined_k141_5247\|\|full | Low-quality |
| 18-102-final-viral-combined_k141_120721\|\|full | Low-quality |
| 18-102-final-viral-combined_k141_173451\|\|full | Not-determined |
| 18-102-final-viral-combined_k141_50023\|\|full | Low-quality |
| 18-102-final-viral-combined_k141_161472\|\|full | Not-determined |
| 18-102-final-viral-combined_k141_222648\|\|full | Low-quality |
| 18-102-final-viral-combined_k141_109771\|\|full | Low-quality |
| 18-102-final-viral-combined_k141_120741\|\|full | Low-quality |
| 18-102-final-viral-combined_k141_173480\|\|full | Low-quality |
| 18-102-final-viral-combined_k141_68385\|\|full | Not-determined |
| 18-102-final-viral-combined_k141_234897\|\|full | Not-determined |
| 18-102-final-viral-combined_k141_222676\|\|full | Low-quality |
| 18-102-final-viral-combined_k141_173490\|\|full | Not-determined |
| 18-102-final-viral-combined_k141_79020\|\|full | Low-quality |
| 18-102-final-viral-combined_k141_120761\|\|full | Not-determined |
| 18-102-final-viral-combined_k141_26136\|\|full | Not-determined |
| 18-102-final-viral-combined_k141_192755\|\|full | Low-quality |
| 18-102-final-viral-combined_k141_243479\|\|full | Not-determined |
| 18-102-final-viral-combined_k141_202117\|\|full | Not-determined |
| 18-102-final-viral-combined_k141_243481\|\|full | Low-quality |
| 18-102-final-viral-combined_k141_17027\|\|full | Not-determined |
| 18-102-final-viral-combined_k141_68413\|\|full | Not-determined |
| 18-102-final-viral-combined_k141_161534\|\|full | Low-quality |
| 18-102-final-viral-combined_k141_68414\|\|full | Not-determined |
| 18-102-final-viral-combined_k141_202134\|\|full | Not-determined |
| 18-102-final-viral-combined_k141_17040\|\|full | Low-quality |
| 18-102-final-viral-combined_k141_68422\|\|full | Low-quality |
| 18-102-final-viral-combined_k141_202136\|\|full | Not-determined |
| 18-102-final-viral-combined_k141_17046\|\|full | Not-determined |
| 18-102-final-viral-combined_k141_100189\|\|full | Not-determined |
| 18-102-final-viral-combined_k141_17056\|\|full | Not-determined |
| 18-102-final-viral-combined_k141_37916\|\|full | Not-determined |
| 18-102-final-viral-combined_k141_234916\|\|full | Low-quality |
| 18-102-final-viral-combined_k141_213082\|\|full | Not-determined |
| 18-102-final-viral-combined_k141_58527\|\|full | Low-quality |
| 18-102-final-viral-combined_k141_100192\|\|full | Not-determined |
| 18-102-final-viral-combined_k141_58540\|\|full | Low-quality |
| 18-102-final-viral-combined_k141_161553\|\|full | Not-determined |
| 18-102-final-viral-combined_k141_192816\|\|full | Low-quality |
| 18-102-final-viral-combined_k141_202192\|\|full | Not-determined |
| 18-102-final-viral-combined_k141_234921\|\|full | Low-quality |
| 18-102-final-viral-combined_k141_151917\|\|full | Not-determined |
| 18-102-final-viral-combined_k141_213111\|\|full | Not-determined |
| 18-102-final-viral-combined_k141_17089\|\|full | Low-quality |
| 18-102-final-viral-combined_k141_213118\|\|full | Not-determined |
| 18-102-final-viral-combined_k141_151921\|\|full | Low-quality |
| 18-102-final-viral-combined_k141_17102\|\|full | Not-determined |
| 18-102-final-viral-combined_k141_192841\|\|full | Low-quality |
| 18-102-final-viral-combined_k141_17106\|\|full | Not-determined |
| 18-102-final-viral-combined_k141_50146\|\|full | Low-quality |
| 18-102-final-viral-combined_k141_192856\|\|full | Low-quality |
| 18-102-final-viral-combined_k141_173601\|\|full | Not-determined |
| 18-102-final-viral-combined_k141_243583\|\|full | Low-quality |
| 18-102-final-viral-combined_k141_141431\|\|full | Not-determined |
| 18-102-final-viral-combined_k141_222744\|\|full | Not-determined |
| 18-102-final-viral-combined_k141_120850\|\|full | Low-quality |
| 18-102-final-viral-combined_k141_243594\|\|full | Not-determined |
| 18-102-final-viral-combined_k141_202244\|\|full | Not-determined |
| 18-109-final-viral-combined_k141_136642\|\|0_partial | High-quality |
| 18-102-final-viral-combined_k141_17154\|\|full | Not-determined |
| 18-102-final-viral-combined_k141_141457\|\|full | Not-determined |
| 18-102-final-viral-combined_k141_151957\|\|full | Low-quality |
| 18-102-final-viral-combined_k141_141464\|\|full | Low-quality |
| 18-102-final-viral-combined_k141_141468\|\|full | Not-determined |
| 18-102-final-viral-combined_k141_202253\|\|full | Low-quality |
| 18-102-final-viral-combined_k141_120870\|\|full | Low-quality |
| 18-102-final-viral-combined_k141_141474\|\|full | Not-determined |
| 18-102-final-viral-combined_k141_90076\|\|full | Low-quality |
| 18-102-final-viral-combined_k141_38046\|\|full | Low-quality |
| 18-102-final-viral-combined_k141_213186\|\|full | Low-quality |
| 18-102-final-viral-combined_k141_120873\|\|full | Not-determined |
| 18-102-final-viral-combined_k141_100234\|\|full | Low-quality |
| 18-102-final-viral-combined_k141_202265\|\|full | Not-determined |
| 18-102-final-viral-combined_k141_243638\|\|full | Low-quality |
| 18-102-final-viral-combined_k141_5327\|\|full | Low-quality |
| 18-102-final-viral-combined_k141_79120\|\|full | Not-determined |
| 18-102-final-viral-combined_k141_68542\|\|full | Not-determined |
| 18-102-final-viral-combined_k141_152012\|\|full | Not-determined |
| 18-102-final-viral-combined_k141_90141\|\|full | Not-determined |
| 18-102-final-viral-combined_k141_68560\|\|full | Not-determined |
| 18-104-final-viral-combined_k141_51079\|\|0_partial | High-quality |
| 18-102-final-viral-combined_k141_243647\|\|full | Not-determined |
| 18-102-final-viral-combined_k141_129814\|\|full | Low-quality |
| 18-102-final-viral-combined_k141_235071\|\|full | Not-determined |
| 18-102-final-viral-combined_k141_5351\|\|full | Not-determined |
| 18-102-final-viral-combined_k141_192934\|\|full | Low-quality |
| 18-102-final-viral-combined_k141_50216\|\|full | Not-determined |
| 18-102-final-viral-combined_k141_129815\|\|full | Low-quality |
| 18-102-final-viral-combined_k141_222802\|\|full | Not-determined |
| 18-102-final-viral-combined_k141_243669\|\|full | Not-determined |
| 18-102-final-viral-combined_k141_202342\|\|full | Low-quality |
| 18-102-final-viral-combined_k141_141503\|\|full | Low-quality |
| 18-102-final-viral-combined_k141_100294\|\|full | Low-quality |
| 18-102-final-viral-combined_k141_222826\|\|full | Low-quality |
| 18-102-final-viral-combined_k141_79193\|\|full | Low-quality |
| 18-102-final-viral-combined_k141_202360\|\|full | Low-quality |
| 18-102-final-viral-combined_k141_68607\|\|full | Not-determined |
| 18-102-final-viral-combined_k141_38194\|\|full | Low-quality |
| 18-102-final-viral-combined_k141_173656\|\|full | Low-quality |
| 18-102-final-viral-combined_k141_100335\|\|full | Low-quality |
| 18-102-final-viral-combined_k141_90200\|\|full | Not-determined |
| 18-102-final-viral-combined_k141_235110\|\|full | Low-quality |
| 18-102-final-viral-combined_k141_68620\|\|full | Not-determined |
| 18-102-final-viral-combined_k141_100339\|\|full | Not-determined |
| 18-102-final-viral-combined_k141_173665\|\|full | Low-quality |
| 18-102-final-viral-combined_k141_183128\|\|full | Low-quality |
| 18-102-final-viral-combined_k141_68625\|\|full | Not-determined |
| 18-102-final-viral-combined_k141_152044\|\|full | Low-quality |
| 18-102-final-viral-combined_k141_79224\|\|full | Low-quality |
| 18-102-final-viral-combined_k141_213214\|\|full | Low-quality |
| 18-102-final-viral-combined_k141_100348\|\|full | Not-determined |
| 18-102-final-viral-combined_k141_141569\|\|full | Low-quality |
| 18-102-final-viral-combined_k141_213219\|\|full | Not-determined |
| 18-102-final-viral-combined_k141_38201\|\|full | Not-determined |
| 18-102-final-viral-combined_k141_129867\|\|full | Low-quality |
| 18-102-final-viral-combined_k141_100369\|\|full | Not-determined |
| 18-102-final-viral-combined_k141_141584\|\|full | Low-quality |
| 18-102-final-viral-combined_k141_183158\|\|full | Low-quality |
| 18-102-final-viral-combined_k141_90248\|\|full | Low-quality |
| 18-102-final-viral-combined_k141_26245\|\|full | Low-quality |
| 18-102-final-viral-combined_k141_141598\|\|full | Low-quality |
| 18-102-final-viral-combined_k141_79272\|\|full | Low-quality |
| 18-102-final-viral-combined_k141_100397\|\|full | Not-determined |
| 18-102-final-viral-combined_k141_141602\|\|full | Low-quality |
| 18-102-final-viral-combined_k141_173690\|\|full | Low-quality |
| 18-102-final-viral-combined_k141_183185\|\|full | Not-determined |
| 18-102-final-viral-combined_k141_202429\|\|full | Not-determined |
| 18-102-final-viral-combined_k141_183196\|\|full | Not-determined |
| 18-102-final-viral-combined_k141_50366\|\|full | Not-determined |
| 18-102-final-viral-combined_k141_202436\|\|full | Low-quality |
| 18-102-final-viral-combined_k141_121064\|\|full | Low-quality |
| 18-102-final-viral-combined_k141_90297\|\|full | Not-determined |
| 18-102-final-viral-combined_k141_50378\|\|full | Not-determined |
| 18-102-final-viral-combined_k141_68718\|\|full | Not-determined |
| 18-102-final-viral-combined_k141_129885\|\|full | Not-determined |
| 18-102-final-viral-combined_k141_68735\|\|full | Low-quality |
| 18-110-final-viral-combined_k141_41861\|\|full | Medium-quality |
| 18-102-final-viral-combined_k141_193048\|\|full | Not-determined |
| 18-102-final-viral-combined_k141_213315\|\|full | Low-quality |
| 18-102-final-viral-combined_k141_50419\|\|full | Not-determined |
| 18-102-final-viral-combined_k141_129915\|\|full | Low-quality |
| 18-102-final-viral-combined_k141_50424\|\|full | Not-determined |
| 18-102-final-viral-combined_k141_26313\|\|full | Low-quality |
| 18-102-final-viral-combined_k141_173770\|\|full | Not-determined |
| 18-102-final-viral-combined_k141_193074\|\|full | Not-determined |
| 18-102-final-viral-combined_k141_213345\|\|full | Low-quality |
| 18-102-final-viral-combined_k141_193076\|\|full | Low-quality |
| 18-102-final-viral-combined_k141_152126\|\|full | Low-quality |
| 18-102-final-viral-combined_k141_38298\|\|full | Low-quality |
| 18-102-final-viral-combined_k141_90346\|\|full | Not-determined |
| 18-102-final-viral-combined_k141_100489\|\|full | Low-quality |
| 18-102-final-viral-combined_k141_129954\|\|full | Not-determined |
| 18-102-final-viral-combined_k141_235239\|\|full | Not-determined |
| 18-102-final-viral-combined_k141_90358\|\|full | Low-quality |
| 18-102-final-viral-combined_k141_152145\|\|full | Low-quality |
| 18-102-final-viral-combined_k141_152148\|\|full | Not-determined |
| 18-102-final-viral-combined_k141_235248\|\|full | Not-determined |
| 18-102-final-viral-combined_k141_129963\|\|full | Low-quality |
| 18-102-final-viral-combined_k141_50492\|\|full | Not-determined |
| 18-102-final-viral-combined_k141_161610\|\|full | Not-determined |
| 18-102-final-viral-combined_k141_68836\|\|full | Low-quality |
| 18-102-final-viral-combined_k141_129978\|\|full | Low-quality |
| 18-102-final-viral-combined_k141_109824\|\|full | Not-determined |
| 18-102-final-viral-combined_k141_79360\|\|full | Low-quality |
| 18-102-final-viral-combined_k141_68847\|\|full | Not-determined |
| 18-102-final-viral-combined_k141_79365\|\|full | Low-quality |
| 18-102-final-viral-combined_k141_152163\|\|full | Low-quality |
| 18-102-final-viral-combined_k141_202474\|\|full | Low-quality |
| 18-102-final-viral-combined_k141_213400\|\|full | Low-quality |
| 18-102-final-viral-combined_k141_68862\|\|full | Low-quality |
| 18-102-final-viral-combined_k141_38356\|\|full | Low-quality |
| 18-102-final-viral-combined_k141_17292\|\|full | Not-determined |
| 18-102-final-viral-combined_k141_130015\|\|full | Low-quality |
| 18-102-final-viral-combined_k141_161651\|\|full | Not-determined |
| 18-102-final-viral-combined_k141_68880\|\|full | Low-quality |
| 18-102-final-viral-combined_k141_141666\|\|full | Not-determined |
| 18-102-final-viral-combined_k141_79429\|\|full | Low-quality |
| 18-102-final-viral-combined_k141_100507\|\|full | Not-determined |
| 18-102-final-viral-combined_k141_90429\|\|full | Not-determined |
| 18-102-final-viral-combined_k141_109866\|\|full | Low-quality |
| 18-102-final-viral-combined_k141_141675\|\|full | Low-quality |
| 18-102-final-viral-combined_k141_79441\|\|full | Not-determined |
| 18-102-final-viral-combined_k141_109886\|\|full | Low-quality |
| 18-102-final-viral-combined_k141_26346\|\|full | Low-quality |
| 18-102-final-viral-combined_k141_68918\|\|full | Not-determined |
| 18-102-final-viral-combined_k141_58749\|\|full | Low-quality |
| 18-102-final-viral-combined_k141_26355\|\|full | Low-quality |
| 18-102-final-viral-combined_k141_38404\|\|full | Not-determined |
| 18-102-final-viral-combined_k141_50541\|\|full | Not-determined |
| 18-102-final-viral-combined_k141_38414\|\|full | Low-quality |
| 18-102-final-viral-combined_k141_58765\|\|full | Low-quality |
| 18-102-final-viral-combined_k141_173897\|\|full | Not-determined |
| 18-102-final-viral-combined_k141_213451\|\|full | Not-determined |
| 18-102-final-viral-combined_k141_173907\|\|full | Low-quality |
| 18-102-final-viral-combined_k141_193189\|\|full | Low-quality |
| 18-102-final-viral-combined_k141_50568\|\|full | Not-determined |
| 18-102-final-viral-combined_k141_213466\|\|full | Low-quality |
| 18-102-final-viral-combined_k141_26369\|\|full | Low-quality |
| 18-102-final-viral-combined_k141_100543\|\|full | Not-determined |
| 18-102-final-viral-combined_k141_130027\|\|full | Low-quality |
| 18-102-final-viral-combined_k141_38449\|\|full | Not-determined |
| 18-102-final-viral-combined_k141_121206\|\|full | Not-determined |
| 18-102-final-viral-combined_k141_130040\|\|full | Low-quality |
| 18-102-final-viral-combined_k141_161745\|\|full | Low-quality |
| 18-102-final-viral-combined_k141_130049\|\|full | Not-determined |
| 18-102-final-viral-combined_k141_202552\|\|full | Low-quality |
| 18-102-final-viral-combined_k141_152318\|\|full | Not-determined |
| 18-102-final-viral-combined_k141_223006\|\|full | Low-quality |
| 18-102-final-viral-combined_k141_235354\|\|full | Low-quality |
| 18-102-final-viral-combined_k141_26417\|\|full | Not-determined |
| 18-102-final-viral-combined_k141_235355\|\|full | Not-determined |
| 18-102-final-viral-combined_k141_152329\|\|full | Low-quality |
| 18-102-final-viral-combined_k141_130071\|\|full | Not-determined |
| 18-102-final-viral-combined_k141_58810\|\|full | Not-determined |
| 18-102-final-viral-combined_k141_5520\|\|full | Not-determined |
| 18-102-final-viral-combined_k141_141756\|\|full | Low-quality |
| 18-102-final-viral-combined_k141_235375\|\|full | Low-quality |
| 18-102-final-viral-combined_k141_121234\|\|full | Low-quality |
| 18-102-final-viral-combined_k141_121237\|\|full | Not-determined |
| 18-102-final-viral-combined_k141_173945\|\|full | Not-determined |
| 18-102-final-viral-combined_k141_243719\|\|full | Low-quality |
| 18-102-final-viral-combined_k141_100608\|\|full | Low-quality |
| 18-102-final-viral-combined_k141_58822\|\|full | Not-determined |
| 18-102-final-viral-combined_k141_110011\|\|full | Not-determined |
| 18-102-final-viral-combined_k141_58823\|\|full | Not-determined |
| 18-102-final-viral-combined_k141_26468\|\|full | Not-determined |
| 18-102-final-viral-combined_k141_38538\|\|full | Low-quality |
| 18-102-final-viral-combined_k141_17401\|\|full | Low-quality |
| 18-102-final-viral-combined_k141_121276\|\|full | Not-determined |
| 18-102-final-viral-combined_k141_5568\|\|full | Low-quality |
| 18-102-final-viral-combined_k141_38557\|\|full | Low-quality |
| 18-102-final-viral-combined_k141_243742\|\|full | Low-quality |
| 18-102-final-viral-combined_k141_235434\|\|full | Not-determined |
| 18-102-final-viral-combined_k141_130098\|\|full | Low-quality |
| 18-102-final-viral-combined_k141_243777\|\|full | Low-quality |
| 18-102-final-viral-combined_k141_50701\|\|full | Low-quality |
| 18-102-final-viral-combined_k141_69054\|\|full | Low-quality |
| 18-102-final-viral-combined_k141_243791\|\|full | Not-determined |
| 18-102-final-viral-combined_k141_38612\|\|full | Low-quality |
| 18-102-final-viral-combined_k141_193268\|\|full | Not-determined |
| 18-102-final-viral-combined_k141_90463\|\|full | Low-quality |
| 18-102-final-viral-combined_k141_79556\|\|full | Not-determined |
| 18-102-final-viral-combined_k141_26489\|\|full | Low-quality |
| 18-102-final-viral-combined_k141_90472\|\|full | Not-determined |
| 18-102-final-viral-combined_k141_79562\|\|full | Low-quality |
| 18-102-final-viral-combined_k141_50722\|\|full | Not-determined |
| 18-102-final-viral-combined_k141_213543\|\|full | Not-determined |
| 18-102-final-viral-combined_k141_213549\|\|full | Low-quality |
| 18-102-final-viral-combined_k141_5621\|\|full | Not-determined |
| 18-102-final-viral-combined_k141_183295\|\|full | Low-quality |
| 18-102-final-viral-combined_k141_235468\|\|full | Not-determined |
| 18-102-final-viral-combined_k141_141865\|\|full | Not-determined |
| 18-102-final-viral-combined_k141_79567\|\|full | Not-determined |
| 18-102-final-viral-combined_k141_17488\|\|full | Not-determined |
| 18-102-final-viral-combined_k141_130159\|\|full | Low-quality |
| 18-102-final-viral-combined_k141_50746\|\|full | Low-quality |
| 18-102-final-viral-combined_k141_50748\|\|full | Not-determined |
| 18-102-final-viral-combined_k141_17496\|\|full | Not-determined |
| 18-102-final-viral-combined_k141_174080\|\|full | Not-determined |
| 18-102-final-viral-combined_k141_17499\|\|full | Low-quality |
| 18-102-final-viral-combined_k141_202578\|\|full | Low-quality |
| 18-102-final-viral-combined_k141_223120\|\|full | Low-quality |
| 18-102-final-viral-combined_k141_243874\|\|full | Low-quality |
| 18-102-final-viral-combined_k141_235486\|\|full | Not-determined |
| 18-102-final-viral-combined_k141_90501\|\|full | Not-determined |
| 18-102-final-viral-combined_k141_243886\|\|full | Not-determined |
| 18-102-final-viral-combined_k141_193350\|\|full | Not-determined |
| 18-102-final-viral-combined_k141_235500\|\|full | Not-determined |
| 18-102-final-viral-combined_k141_235503\|\|full | Not-determined |
| 18-102-final-viral-combined_k141_58920\|\|full | Not-determined |
| 18-102-final-viral-combined_k141_161799\|\|full | Not-determined |
| 18-102-final-viral-combined_k141_174123\|\|full | Not-determined |
| 18-102-final-viral-combined_k141_174128\|\|full | Low-quality |
| 18-102-final-viral-combined_k141_69079\|\|full | Not-determined |
| 18-102-final-viral-combined_k141_58925\|\|full | Low-quality |
| 18-102-final-viral-combined_k141_202627\|\|full | Not-determined |
| 18-102-final-viral-combined_k141_213573\|\|full | Not-determined |
| 18-102-final-viral-combined_k141_121391\|\|full | Not-determined |
| 18-102-final-viral-combined_k141_50799\|\|full | Low-quality |
| 18-102-final-viral-combined_k141_235536\|\|full | Low-quality |
| 18-102-final-viral-combined_k141_100827\|\|full | Low-quality |
| 18-102-final-viral-combined_k141_110030\|\|full | Low-quality |
| 18-102-final-viral-combined_k141_223159\|\|full | Not-determined |
| 18-102-final-viral-combined_k141_79579\|\|full | Not-determined |
| 18-102-final-viral-combined_k141_202639\|\|full | Not-determined |
| 18-102-final-viral-combined_k141_130215\|\|full | Low-quality |
| 18-102-final-viral-combined_k141_110035\|\|full | Low-quality |
| 18-102-final-viral-combined_k141_141936\|\|full | Low-quality |
| 18-102-final-viral-combined_k141_213603\|\|full | Not-determined |
| 18-102-final-viral-combined_k141_235562\|\|full | Low-quality |
| 18-102-final-viral-combined_k141_243930\|\|full | Not-determined |
| 18-102-final-viral-combined_k141_17532\|\|full | Low-quality |
| 18-102-final-viral-combined_k141_38694\|\|full | Not-determined |
| 18-102-final-viral-combined_k141_130220\|\|full | Low-quality |
| 18-102-final-viral-combined_k141_141971\|\|full | Not-determined |
| 18-102-final-viral-combined_k141_79588\|\|full | Not-determined |
| 18-102-final-viral-combined_k141_100845\|\|full | Not-determined |
| 18-102-final-viral-combined_k141_90541\|\|full | Low-quality |
| 18-102-final-viral-combined_k141_110072\|\|full | Low-quality |
| 18-102-final-viral-combined_k141_152491\|\|full | Low-quality |
| 18-102-final-viral-combined_k141_223204\|\|full | Low-quality |
| 18-102-final-viral-combined_k141_243965\|\|full | Low-quality |
| 18-102-final-viral-combined_k141_50882\|\|full | Not-determined |
| 18-102-final-viral-combined_k141_79599\|\|full | Low-quality |
| 18-102-final-viral-combined_k141_50884\|\|full | Low-quality |
| 18-102-final-viral-combined_k141_130241\|\|full | Low-quality |
| 18-102-final-viral-combined_k141_5702\|\|full | Low-quality |
| 18-102-final-viral-combined_k141_110080\|\|full | Not-determined |
| 18-102-final-viral-combined_k141_90557\|\|full | Not-determined |
| 18-102-final-viral-combined_k141_152508\|\|full | Not-determined |
| 18-102-final-viral-combined_k141_174208\|\|full | Not-determined |
| 18-102-final-viral-combined_k141_5721\|\|full | Not-determined |
| 18-102-final-viral-combined_k141_193425\|\|full | Not-determined |
| 18-102-final-viral-combined_k141_90574\|\|full | Not-determined |
| 18-102-final-viral-combined_k141_90582\|\|full | Low-quality |
| 18-102-final-viral-combined_k141_58991\|\|full | Low-quality |
| 18-102-final-viral-combined_k141_243971\|\|full | Low-quality |
| 18-102-final-viral-combined_k141_223240\|\|full | Not-determined |
| 18-102-final-viral-combined_k141_193460\|\|full | Not-determined |
| 18-102-final-viral-combined_k141_142047\|\|full | Not-determined |
| 18-102-final-viral-combined_k141_183428\|\|full | Not-determined |
| 18-102-final-viral-combined_k141_17628\|\|full | Not-determined |
| 18-102-final-viral-combined_k141_121434\|\|full | Low-quality |
| 18-102-final-viral-combined_k141_235633\|\|full | Not-determined |
| 18-102-final-viral-combined_k141_38796\|\|full | Low-quality |
| 18-102-final-viral-combined_k141_38802\|\|full | Not-determined |
| 18-102-final-viral-combined_k141_183459\|\|full | Not-determined |
| 18-102-final-viral-combined_k141_17670\|\|full | Low-quality |
| 18-102-final-viral-combined_k141_17679\|\|full | Low-quality |
| 18-102-final-viral-combined_k141_202792\|\|full | Not-determined |
| 18-102-final-viral-combined_k141_235644\|\|full | Not-determined |
| 18-102-final-viral-combined_k141_17687\|\|full | Low-quality |
| 18-102-final-viral-combined_k141_202794\|\|full | Low-quality |
| 18-102-final-viral-combined_k141_202799\|\|full | Low-quality |
| 18-102-final-viral-combined_k141_243989\|\|full | Not-determined |
| 18-102-final-viral-combined_k141_89742\|\|full | Medium-quality |
| 18-102-final-viral-combined_k141_59042\|\|full | Not-determined |
| 18-102-final-viral-combined_k141_235663\|\|full | Low-quality |
| 18-102-final-viral-combined_k141_213715\|\|full | Not-determined |
| 18-102-final-viral-combined_k141_121463\|\|full | Not-determined |
| 18-102-final-viral-combined_k141_121465\|\|full | Not-determined |
| 18-102-final-viral-combined_k141_79680\|\|full | Not-determined |
| 18-102-final-viral-combined_k141_79681\|\|full | Low-quality |
| 18-102-final-viral-combined_k141_121476\|\|full | Not-determined |
| 18-102-final-viral-combined_k141_152585\|\|full | Low-quality |
| 18-102-final-viral-combined_k141_130272\|\|full | Low-quality |
| 18-102-final-viral-combined_k141_244004\|\|full | Not-determined |
| 18-102-final-viral-combined_k141_121480\|\|full | Not-determined |
| 18-102-final-viral-combined_k141_5772\|\|full | Low-quality |
| 18-102-final-viral-combined_k141_174272\|\|full | Not-determined |
| 18-102-final-viral-combined_k141_130287\|\|full | Low-quality |
| 18-102-final-viral-combined_k141_38811\|\|full | Not-determined |
| 18-102-final-viral-combined_k141_152605\|\|full | Not-determined |
| 18-102-final-viral-combined_k141_183504\|\|full | Not-determined |
| 18-102-final-viral-combined_k141_213750\|\|full | Low-quality |
| 18-102-final-viral-combined_k141_100951\|\|full | Not-determined |
| 18-102-final-viral-combined_k141_235701\|\|full | Low-quality |
| 18-102-final-viral-combined_k141_244030\|\|full | Low-quality |
| 18-102-final-viral-combined_k141_59071\|\|full | Not-determined |
| 18-102-final-viral-combined_k141_100954\|\|full | Not-determined |
| 18-102-final-viral-combined_k141_121503\|\|full | Not-determined |
| 18-102-final-viral-combined_k141_244037\|\|full | Not-determined |
| 18-102-final-viral-combined_k141_183511\|\|full | Not-determined |
| 18-102-final-viral-combined_k141_152630\|\|full | Low-quality |
| 18-102-final-viral-combined_k141_100964\|\|full | Low-quality |
| 18-102-final-viral-combined_k141_213776\|\|full | Not-determined |
| 18-102-final-viral-combined_k141_79707\|\|full | Not-determined |
| 18-102-final-viral-combined_k141_69242\|\|full | Low-quality |
| 18-102-final-viral-combined_k141_79715\|\|full | Low-quality |
| 18-102-final-viral-combined_k141_235762\|\|full | Not-determined |
| 18-102-final-viral-combined_k141_174302\|\|full | Low-quality |
| 18-102-final-viral-combined_k141_193557\|\|full | Not-determined |
| 18-102-final-viral-combined_k141_79732\|\|full | Not-determined |
| 18-102-final-viral-combined_k141_59127\|\|full | Not-determined |
| 18-102-final-viral-combined_k141_152651\|\|full | Not-determined |
| 18-102-final-viral-combined_k141_152660\|\|full | Low-quality |
| 18-102-final-viral-combined_k141_142089\|\|full | Low-quality |
| 18-102-final-viral-combined_k141_152666\|\|full | Not-determined |
| 18-102-final-viral-combined_k141_223353\|\|full | Low-quality |
| 18-102-final-viral-combined_k141_223362\|\|full | Not-determined |
| 18-102-final-viral-combined_k141_142120\|\|full | Low-quality |
| 18-102-final-viral-combined_k141_26530\|\|full | Low-quality |
| 18-102-final-viral-combined_k141_142132\|\|full | Low-quality |
| 18-102-final-viral-combined_k141_202871\|\|full | Low-quality |
| 18-102-final-viral-combined_k141_235809\|\|full | Not-determined |
| 18-102-final-viral-combined_k141_69280\|\|full | Not-determined |
| 18-102-final-viral-combined_k141_69282\|\|full | Not-determined |
| 18-102-final-viral-combined_k141_51011\|\|full | Not-determined |
| 18-102-final-viral-combined_k141_244126\|\|full | Low-quality |
| 18-102-final-viral-combined_k141_17703\|\|full | Not-determined |
| 18-102-final-viral-combined_k141_69286\|\|full | Not-determined |
| 18-102-final-viral-combined_k141_235828\|\|full | Not-determined |
| 18-102-final-viral-combined_k141_244131\|\|full | Not-determined |
| 18-102-final-viral-combined_k141_202899\|\|full | Not-determined |
| 18-102-final-viral-combined_k141_100992\|\|full | Not-determined |
| 18-102-final-viral-combined_k141_79788\|\|full | Not-determined |
| 18-102-final-viral-combined_k141_59174\|\|full | Low-quality |
| 18-102-final-viral-combined_k141_235836\|\|full | Low-quality |
| 18-102-final-viral-combined_k141_51048\|\|full | Not-determined |
| 18-102-final-viral-combined_k141_101010\|\|full | Low-quality |
| 18-102-final-viral-combined_k141_51054\|\|full | Low-quality |
| 18-102-final-viral-combined_k141_213809\|\|full | Low-quality |
| 18-102-final-viral-combined_k141_223445\|\|full | Not-determined |
| 17-109-final-viral-combined_k141_164656\|\|0_partial | Medium-quality |
| 18-102-final-viral-combined_k141_38880\|\|full | Not-determined |
| 18-102-final-viral-combined_k141_174407\|\|full | Low-quality |
| 18-102-final-viral-combined_k141_17746\|\|full | Not-determined |

| | |
|---|---|
| 18-102-final-viral-combined_k141_79831\|\|full | Not-determined |
| 18-102-final-viral-combined_k141_213824\|\|full | Not-determined |
| 18-102-final-viral-combined_k141_235874\|\|full | Not-determined |
| 18-102-final-viral-combined_k141_183598\|\|full | Low-quality |
| 18-102-final-viral-combined_k141_202956\|\|full | Not-determined |
| 18-102-final-viral-combined_k141_79838\|\|full | Not-determined |
| 18-102-final-viral-combined_k141_69362\|\|full | Not-determined |
| 18-102-final-viral-combined_k141_38922\|\|full | Not-determined |
| 18-102-final-viral-combined_k141_101096\|\|full | Low-quality |
| 18-102-final-viral-combined_k141_235897\|\|full | Low-quality |
| 18-102-final-viral-combined_k141_235901\|\|full | Not-determined |
| 18-102-final-viral-combined_k141_69390\|\|full | Not-determined |
| 18-102-final-viral-combined_k141_183633\|\|full | Not-determined |
| 18-102-final-viral-combined_k141_183634\|\|full | Low-quality |
| 18-102-final-viral-combined_k141_69396\|\|full | Not-determined |
| 18-102-final-viral-combined_k141_235916\|\|full | Low-quality |
| 18-102-final-viral-combined_k141_161895\|\|full | Low-quality |
| 18-102-final-viral-combined_k141_59241\|\|full | Not-determined |
| 18-102-final-viral-combined_k141_59245\|\|full | Low-quality |
| 17-106-final-viral-combined_k141_122742\|\|0_partial | Medium-quality |
| 18-102-final-viral-combined_k141_17772\|\|full | Low-quality |
| 18-102-final-viral-combined_k141_193717\|\|full | Not-determined |
| 18-102-final-viral-combined_k141_213881\|\|full | Not-determined |
| 18-102-final-viral-combined_k141_235927\|\|full | Not-determined |
| 18-102-final-viral-combined_k141_244177\|\|full | Not-determined |
| 18-102-final-viral-combined_k141_110261\|\|full | Low-quality |
| 18-102-final-viral-combined_k141_5804\|\|full | Low-quality |
| 18-102-final-viral-combined_k141_38933\|\|full | Low-quality |
| 18-102-final-viral-combined_k141_213893\|\|full | Not-determined |
| 18-102-final-viral-combined_k141_130362\|\|full | Low-quality |
| 18-102-final-viral-combined_k141_193719\|\|full | Not-determined |
| 18-102-final-viral-combined_k141_130373\|\|full | Low-quality |
| 18-102-final-viral-combined_k141_161940\|\|full | Not-determined |
| 18-102-final-viral-combined_k141_130374\|\|full | Not-determined |
| 18-102-final-viral-combined_k141_235953\|\|full | Not-determined |
| 18-102-final-viral-combined_k141_69470\|\|full | Low-quality |
| 18-102-final-viral-combined_k141_121663\|\|full | Low-quality |
| 18-102-final-viral-combined_k141_69474\|\|full | Not-determined |
| 18-102-final-viral-combined_k141_193748\|\|full | Not-determined |
| 18-102-final-viral-combined_k141_223527\|\|full | Not-determined |
| 18-102-final-viral-combined_k141_174465\|\|full | Not-determined |
| 18-102-final-viral-combined_k141_213916\|\|full | Low-quality |
| 18-102-final-viral-combined_k141_203031\|\|full | Not-determined |
| 18-102-final-viral-combined_k141_90670\|\|full | Not-determined |
| 18-102-final-viral-combined_k141_26642\|\|full | Not-determined |
| 18-102-final-viral-combined_k141_203034\|\|full | Low-quality |
| 18-102-final-viral-combined_k141_79863\|\|full | Not-determined |
| 18-102-final-viral-combined_k141_59341\|\|full | Low-quality |
| 18-102-final-viral-combined_k141_90687\|\|full | Low-quality |
| 18-102-final-viral-combined_k141_152757\|\|full | Not-determined |
| 18-102-final-viral-combined_k141_152760\|\|full | Not-determined |
| 18-102-final-viral-combined_k141_193804\|\|full | Low-quality |
| 18-102-final-viral-combined_k141_223576\|\|full | Not-determined |
| 18-102-final-viral-combined_k141_69542\|\|full | Not-determined |
| 17-109-final-viral-combined_k141_302876\|\|full | Medium-quality |
| 18-102-final-viral-combined_k141_236015\|\|full | Low-quality |
| 18-102-final-viral-combined_k141_121736\|\|full | Low-quality |
| 18-102-final-viral-combined_k141_236030\|\|full | Not-determined |
| 18-102-final-viral-combined_k141_236033\|\|full | Not-determined |
| 18-102-final-viral-combined_k141_39032\|\|full | Low-quality |
| 18-102-final-viral-combined_k141_203058\|\|full | Low-quality |
| 18-102-final-viral-combined_k141_203061\|\|full | Not-determined |
| 18-102-final-viral-combined_k141_69567\|\|full | Low-quality |
| 18-102-final-viral-combined_k141_183765\|\|full | Low-quality |
| 18-102-final-viral-combined_k141_152804\|\|full | Low-quality |
| 18-102-final-viral-combined_k141_193830\|\|full | Not-determined |
| 18-102-final-viral-combined_k141_51199\|\|full | Not-determined |
| 18-102-final-viral-combined_k141_39058\|\|full | Low-quality |
| 18-102-final-viral-combined_k141_110310\|\|full | Low-quality |
| 18-102-final-viral-combined_k141_183777\|\|full | Low-quality |
| 18-102-final-viral-combined_k141_142260\|\|full | Not-determined |
| 18-102-final-viral-combined_k141_152835\|\|full | Not-determined |
| 18-102-final-viral-combined_k141_236061\|\|full | Not-determined |
| 18-102-final-viral-combined_k141_203095\|\|full | Not-determined |
| 18-102-final-viral-combined_k141_79886\|\|full | Low-quality |
| 18-102-final-viral-combined_k141_130454\|\|full | Not-determined |
| [illegible] | [illegible] |

| | |
|---|---|
| 17-106-final-viral-combined_k141_33229\|\|0_partial | Medium-quality |
| 18-102-final-viral-combined_k141_143245\|\|full | Low-quality |
| 18-102-final-viral-combined_k141_214950\|\|full | Not-determined |
| 18-102-final-viral-combined_k141_224440\|\|full | Low-quality |
| 18-102-final-viral-combined_k141_102340\|\|full | Low-quality |
| 18-102-final-viral-combined_k141_214961\|\|full | Low-quality |
| 18-102-final-viral-combined_k141_244663\|\|full | Low-quality |
| 18-102-final-viral-combined_k141_237024\|\|full | Not-determined |
| 18-102-final-viral-combined_k141_91979\|\|full | Not-determined |
| 18-102-final-viral-combined_k141_244676\|\|full | Not-determined |
| 18-102-final-viral-combined_k141_162826\|\|full | Not-determined |
| 18-102-final-viral-combined_k141_91982\|\|full | Not-determined |
| 18-102-final-viral-combined_k141_6629\|\|full | Not-determined |
| 18-102-final-viral-combined_k141_60558\|\|full | Low-quality |
| 18-102-final-viral-combined_k141_27469\|\|full | Not-determined |
| 18-102-final-viral-combined_k141_70726\|\|full | Not-determined |
| 18-102-final-viral-combined_k141_70749\|\|full | Not-determined |
| 18-102-final-viral-combined_k141_214970\|\|full | Not-determined |
| 18-102-final-viral-combined_k141_122674\|\|full | Not-determined |
| 18-102-final-viral-combined_k141_244710\|\|full | Low-quality |
| 18-102-final-viral-combined_k141_70754\|\|full | Not-determined |
| 18-102-final-viral-combined_k141_131353\|\|full | Low-quality |
| 18-102-final-viral-combined_k141_237086\|\|full | Low-quality |
| 18-102-final-viral-combined_k141_131356\|\|full | Low-quality |
| 18-102-final-viral-combined_k141_122677\|\|full | Not-determined |
| 18-102-final-viral-combined_k141_70762\|\|full | Low-quality |
| 18-102-final-viral-combined_k141_40386\|\|full | Not-determined |
| 18-102-final-viral-combined_k141_162854\|\|full | Not-determined |
| 18-102-final-viral-combined_k141_60600\|\|full | Not-determined |
| 18-102-final-viral-combined_k141_40394\|\|full | Low-quality |
| 18-102-final-viral-combined_k141_153527\|\|full | Low-quality |
| 18-102-final-viral-combined_k141_92064\|\|full | Low-quality |
| 18-102-final-viral-combined_k141_18895\|\|full | Low-quality |
| 18-102-final-viral-combined_k141_143326\|\|full | Low-quality |
| 18-102-final-viral-combined_k141_153532\|\|full | Not-determined |
| 18-102-final-viral-combined_k141_215015\|\|full | Not-determined |
| 18-102-final-viral-combined_k141_237107\|\|full | Low-quality |
| 18-102-final-viral-combined_k141_40418\|\|full | Not-determined |
| 18-102-final-viral-combined_k141_111267\|\|full | Low-quality |
| 18-102-final-viral-combined_k141_162871\|\|full | Not-determined |
| 18-102-final-viral-combined_k141_237129\|\|full | Not-determined |
| 18-102-final-viral-combined_k141_224525\|\|full | Not-determined |
| 18-102-final-viral-combined_k141_60621\|\|full | Low-quality |
| 18-102-final-viral-combined_k141_244728\|\|full | Low-quality |
| 18-102-final-viral-combined_k141_122719\|\|full | Not-determined |
| 18-102-final-viral-combined_k141_224531\|\|full | Low-quality |
| 18-102-final-viral-combined_k141_122722\|\|full | Low-quality |
| 18-102-final-viral-combined_k141_237147\|\|full | Not-determined |
| 18-102-final-viral-combined_k141_153569\|\|full | Low-quality |
| 18-102-final-viral-combined_k141_111285\|\|full | Low-quality |
| 18-102-final-viral-combined_k141_60641\|\|full | Not-determined |
| 18-102-final-viral-combined_k141_70819\|\|full | Low-quality |
| 18-102-final-viral-combined_k141_6685\|\|full | Not-determined |
| 18-102-final-viral-combined_k141_122750\|\|full | Low-quality |
| 18-102-final-viral-combined_k141_143412\|\|full | Not-determined |
| 18-102-final-viral-combined_k141_153601\|\|full | Not-determined |
| 18-102-final-viral-combined_k141_131405\|\|full | Not-determined |
| 18-102-final-viral-combined_k141_122755\|\|full | Low-quality |
| 18-102-final-viral-combined_k141_80784\|\|full | Low-quality |
| 18-102-final-viral-combined_k141_215058\|\|full | Not-determined |
| 18-102-final-viral-combined_k141_244754\|\|full | Not-determined |
| 18-102-final-viral-combined_k141_92129\|\|full | Not-determined |
| 18-102-final-viral-combined_k141_60674\|\|full | Low-quality |
| 18-102-final-viral-combined_k141_244765\|\|full | Not-determined |
| 18-102-final-viral-combined_k141_215071\|\|full | Low-quality |
| 18-102-final-viral-combined_k141_131442\|\|full | Not-determined |
| 18-102-final-viral-combined_k141_92145\|\|full | Low-quality |
| 18-102-final-viral-combined_k141_237242\|\|full | Low-quality |
| 18-102-final-viral-combined_k141_122798\|\|full | Not-determined |
| 18-102-final-viral-combined_k141_92149\|\|full | Low-quality |
| 18-102-final-viral-combined_k141_80803\|\|full | Low-quality |
| 18-102-final-viral-combined_k141_203716\|\|full | Not-determined |
| 18-102-final-viral-combined_k141_27524\|\|full | Not-determined |
| 18-102-final-viral-combined_k141_102403\|\|full | Not-determined |
| 18-102-final-viral-combined_k141_153649\|\|full | Not-determined |
| 18-102-final-viral-combined_k141_162889\|\|full | Low-quality |
| 18-102-final-viral-combined_k141_122813\|\|full | Low-quality |
| 18-102-final-viral-combined_k141_60715\|\|full | Not-determined |
| 18-102-final-viral-combined_k141_80826\|\|full | Not-determined |
| 18-102-final-viral-combined_k141_80840\|\|full | Not-determined |
| 18-102-final-viral-combined_k141_162923\|\|full | Not-determined |
| 18-102-final-viral-combined_k141_111343\|\|full | Not-determined |
| 18-102-final-viral-combined_k141_80859\|\|full | Not-determined |
| 18-102-final-viral-combined_k141_237260\|\|full | Not-determined |
| 18-102-final-viral-combined_k141_92216\|\|full | Low-quality |
| 18-102-final-viral-combined_k141_80876\|\|full | Low-quality |
| 18-102-final-viral-combined_k141_70868\|\|full | Not-determined |
| 18-102-final-viral-combined_k141_175769\|\|full | Low-quality |
| 18-102-final-viral-combined_k141_122837\|\|full | Low-quality |
| 18-102-final-viral-combined_k141_80880\|\|full | Not-determined |
| 18-102-final-viral-combined_k141_175778\|\|full | Not-determined |
| 18-102-final-viral-combined_k141_18998\|\|full | Not-determined |
| 18-102-final-viral-combined_k141_215185\|\|full | Not-determined |
| 18-102-final-viral-combined_k141_122847\|\|full | Not-determined |
| 18-102-final-viral-combined_k141_215192\|\|full | Low-quality |
| 18-102-final-viral-combined_k141_60767\|\|full | Not-determined |
| 18-102-final-viral-combined_k141_122858\|\|full | Not-determined |
| 18-102-final-viral-combined_k141_203789\|\|full | Not-determined |
| 18-102-final-viral-combined_k141_224554\|\|full | Low-quality |
| 18-102-final-viral-combined_k141_237298\|\|full | Low-quality |
| 18-102-final-viral-combined_k141_244831\|\|full | Low-quality |
| 18-102-final-viral-combined_k141_60773\|\|full | Low-quality |
| 18-102-final-viral-combined_k141_111384\|\|full | Low-quality |
| 18-102-final-viral-combined_k141_80903\|\|full | Low-quality |
| 18-102-final-viral-combined_k141_175796\|\|full | Low-quality |
| 18-102-final-viral-combined_k141_215206\|\|full | Not-determined |
| 18-102-final-viral-combined_k141_19018\|\|full | Not-determined |
| 18-102-final-viral-combined_k141_175806\|\|full | Low-quality |
| 18-102-final-viral-combined_k141_80910\|\|full | Not-determined |
| 18-102-final-viral-combined_k141_195046\|\|full | Not-determined |
| 18-102-final-viral-combined_k141_122869\|\|full | Not-determined |
| 18-102-final-viral-combined_k141_215216\|\|full | Low-quality |
| 18-102-final-viral-combined_k141_111400\|\|full | Low-quality |
| 18-102-final-viral-combined_k141_111402\|\|full | Low-quality |
| 18-102-final-viral-combined_k141_19036\|\|full | Not-determined |
| 18-102-final-viral-combined_k141_175830\|\|full | Not-determined |
| 18-102-final-viral-combined_k141_111407\|\|full | Not-determined |
| 18-102-final-viral-combined_k141_131480\|\|full | Not-determined |
| 18-102-final-viral-combined_k141_237319\|\|full | Low-quality |
| 18-102-final-viral-combined_k141_184987\|\|full | Not-determined |
| 18-102-final-viral-combined_k141_143546\|\|full | Not-determined |
| 18-102-final-viral-combined_k141_102501\|\|full | Not-determined |
| 18-102-final-viral-combined_k141_80965\|\|full | Low-quality |
| 18-102-final-viral-combined_k141_131484\|\|full | Low-quality |
| 18-102-final-viral-combined_k141_215267\|\|full | Not-determined |
| 18-102-final-viral-combined_k141_102520\|\|full | Low-quality |
| 18-102-final-viral-combined_k141_70929\|\|full | Not-determined |
| 18-102-final-viral-combined_k141_80987\|\|full | Not-determined |
| 18-102-final-viral-combined_k141_215270\|\|full | Low-quality |
| 18-102-final-viral-combined_k141_92277\|\|full | Low-quality |
| 18-102-final-viral-combined_k141_19077\|\|full | Low-quality |
| 18-102-final-viral-combined_k141_27604\|\|full | Low-quality |
| 18-102-final-viral-combined_k141_40579\|\|full | Not-determined |
| 18-102-final-viral-combined_k141_111437\|\|full | Not-determined |
| 18-102-final-viral-combined_k141_237374\|\|full | Low-quality |
| 18-102-final-viral-combined_k141_40593\|\|full | Not-determined |
| 18-102-final-viral-combined_k141_143576\|\|full | Low-quality |
| 18-102-final-viral-combined_k141_81016\|\|full | Low-quality |
| 18-102-final-viral-combined_k141_185016\|\|full | Not-determined |
| 18-102-final-viral-combined_k141_237386\|\|full | Not-determined |
| 18-102-final-viral-combined_k141_195143\|\|full | Not-determined |
| 18-102-final-viral-combined_k141_40604\|\|full | Low-quality |
| 18-102-final-viral-combined_k141_131519\|\|full | Not-determined |
| 18-102-final-viral-combined_k141_237403\|\|full | Low-quality |
| 18-102-final-viral-combined_k141_27607\|\|full | Low-quality |
| 18-102-final-viral-combined_k141_102604\|\|full | Not-determined |
| 18-102-final-viral-combined_k141_153812\|\|full | Low-quality |
| 18-102-final-viral-combined_k141_224591\|\|full | Not-determined |
| 18-102-final-viral-combined_k141_237414\|\|full | Low-quality |
| 18-102-final-viral-combined_k141_203849\|\|full | Low-quality |
| 18-102-final-viral-combined_k141_143591\|\|full | Not-determined |
| 18-102-final-viral-combined_k141_60879\|\|full | Low-quality |
| 18-102-final-viral-combined_k141_102609\|\|full | Low-quality |
| 18-102-final-viral-combined_k141_244858\|\|full | Not-determined |
| 18-102-final-viral-combined_k141_19155\|\|full | Not-determined |
| 18-102-final-viral-combined_k141_203864\|\|full | Not-determined |
| 18-102-final-viral-combined_k141_131561\|\|full | Not-determined |
| 18-102-final-viral-combined_k141_185038\|\|full | Not-determined |
| 18-102-final-viral-combined_k141_143597\|\|full | Not-determined |
| 18-102-final-viral-combined_k141_40628\|\|full | Low-quality |
| 18-102-final-viral-combined_k141_185042\|\|full | Not-determined |
| 18-102-final-viral-combined_k141_224593\|\|full | Low-quality |
| 18-102-final-viral-combined_k141_102643\|\|full | Low-quality |
| 18-102-final-viral-combined_k141_175888\|\|full | Not-determined |
| 18-102-final-viral-combined_k141_237454\|\|full | Not-determined |
| 18-102-final-viral-combined_k141_237456\|\|full | Low-quality |
| 18-102-final-viral-combined_k141_224603\|\|full | Low-quality |
| 18-102-final-viral-combined_k141_102666\|\|full | Not-determined |
| 18-102-final-viral-combined_k141_175906\|\|full | Low-quality |
| 18-102-final-viral-combined_k141_203935\|\|full | Not-determined |
| 18-102-final-viral-combined_k141_175921\|\|full | Low-quality |
| 18-102-final-viral-combined_k141_175922\|\|full | Not-determined |
| 18-102-final-viral-combined_k141_215377\|\|full | Not-determined |
| 18-102-final-viral-combined_k141_175924\|\|full | Low-quality |
| 18-102-final-viral-combined_k141_111492\|\|full | Not-determined |
| 18-102-final-viral-combined_k141_102700\|\|full | Low-quality |
| 18-102-final-viral-combined_k141_81074\|\|full | Low-quality |
| 18-102-final-viral-combined_k141_40697\|\|full | Not-determined |
| 18-102-final-viral-combined_k141_185082\|\|full | Low-quality |
| 18-102-final-viral-combined_k141_143638\|\|full | Not-determined |
| 18-102-final-viral-combined_k141_215393\|\|full | Not-determined |
| 18-102-final-viral-combined_k141_215394\|\|full | Not-determined |
| 18-102-final-viral-combined_k141_162987\|\|full | Not-determined |
| 18-102-final-viral-combined_k141_19212\|\|full | Low-quality |
| 18-102-final-viral-combined_k141_215396\|\|full | Not-determined |
| 18-102-final-viral-combined_k141_71024\|\|full | Not-determined |
| 18-102-final-viral-combined_k141_19215\|\|full | Not-determined |
| 18-102-final-viral-combined_k141_71035\|\|full | Not-determined |
| 18-102-final-viral-combined_k141_40729\|\|full | Low-quality |
| 18-102-final-viral-combined_k141_163004\|\|full | Low-quality |
| 18-102-final-viral-combined_k141_195237\|\|full | Not-determined |
| 18-102-final-viral-combined_k141_19230\|\|full | Not-determined |
| 18-102-final-viral-combined_k141_153862\|\|full | Low-quality |
| 18-102-final-viral-combined_k141_244902\|\|full | Low-quality |
| 18-102-final-viral-combined_k141_81124\|\|full | Not-determined |
| 18-102-final-viral-combined_k141_6860\|\|full | Not-determined |
| 18-102-final-viral-combined_k141_153878\|\|full | Not-determined |
| 18-102-final-viral-combined_k141_185122\|\|full | Not-determined |
| 18-102-final-viral-combined_k141_6864\|\|full | Low-quality |
| 18-102-final-viral-combined_k141_143686\|\|full | Not-determined |
| 18-102-final-viral-combined_k141_195278\|\|full | Not-determined |
| 18-102-final-viral-combined_k141_81136\|\|full | Low-quality |
| 18-102-final-viral-combined_k141_215443\|\|full | Low-quality |
| 18-102-final-viral-combined_k141_215446\|\|full | Low-quality |
| 18-102-final-viral-combined_k141_195284\|\|full | Low-quality |
| 18-102-final-viral-combined_k141_19266\|\|full | Not-determined |
| 18-102-final-viral-combined_k141_237530\|\|full | Low-quality |
| 18-102-final-viral-combined_k141_224704\|\|full | Not-determined |
| 18-102-final-viral-combined_k141_19277\|\|full | Low-quality |
| 18-102-final-viral-combined_k141_102750\|\|full | Not-determined |
| 18-102-final-viral-combined_k141_237556\|\|full | Low-quality |
| 18-102-final-viral-combined_k141_60999\|\|full | Low-quality |
| 18-102-final-viral-combined_k141_92333\|\|full | Low-quality |
| 18-102-final-viral-combined_k141_19294\|\|full | Low-quality |
| 18-102-final-viral-combined_k141_40809\|\|full | Low-quality |
| 18-102-final-viral-combined_k141_153938\|\|full | Not-determined |
| 18-102-final-viral-combined_k141_81196\|\|full | Low-quality |
| 18-102-final-viral-combined_k141_204011\|\|full | Not-determined |
| 18-102-final-viral-combined_k141_71124\|\|full | Not-determined |
| 18-102-final-viral-combined_k141_40814\|\|full | Low-quality |
| 18-102-final-viral-combined_k141_185186\|\|full | Low-quality |
| 18-102-final-viral-combined_k141_244911\|\|full | Low-quality |
| 18-102-final-viral-combined_k141_195330\|\|full | Not-determined |
| 18-102-final-viral-combined_k141_102779\|\|full | Not-determined |
| 18-102-final-viral-combined_k141_195339\|\|full | Low-quality |
| 18-102-final-viral-combined_k141_244920\|\|full | Low-quality |
| 18-102-final-viral-combined_k141_6924\|\|full | Low-quality |
| 18-102-final-viral-combined_k141_244922\|\|full | Low-quality |
| 18-102-final-viral-combined_k141_163086\|\|full | Not-determined |
| 18-102-final-viral-combined_k141_131584\|\|full | Not-determined |
| 18-102-final-viral-combined_k141_215492\|\|full | Not-determined |
| 18-102-final-viral-combined_k141_143729\|\|full | Low-quality |
| 18-102-final-viral-combined_k141_131605\|\|full | Not-determined |
| 18-102-final-viral-combined_k141_163106\|\|full | Not-determined |
| 18-102-final-viral-combined_k141_71160\|\|full | Not-determined |
| 18-102-final-viral-combined_k141_153997\|\|full | Not-determined |
| 18-102-final-viral-combined_k141_224787\|\|full | Not-determined |
| 18-102-final-viral-combined_k141_195361\|\|full | Low-quality |
| 18-102-final-viral-combined_k141_71170\|\|full | Low-quality |
| 18-102-final-viral-combined_k141_224795\|\|full | Not-determined |
| 18-102-final-viral-combined_k141_195364\|\|full | Not-determined |
| 18-102-final-viral-combined_k141_122952\|\|full | Low-quality |
| 18-102-final-viral-combined_k141_131633\|\|full | Low-quality |
| 18-102-final-viral-combined_k141_102832\|\|full | Not-determined |
| 18-102-final-viral-combined_k141_244956\|\|full | Not-determined |
| 18-102-final-viral-combined_k141_131640\|\|full | Low-quality |
| 18-102-final-viral-combined_k141_185270\|\|full | Low-quality |
| 18-102-final-viral-combined_k141_61021\|\|full | Low-quality |
| 18-102-final-viral-combined_k141_92394\|\|full | Not-determined |
| 18-102-final-viral-combined_k141_27725\|\|full | Low-quality |
| 18-102-final-viral-combined_k141_195417\|\|full | Low-quality |
| 18-102-final-viral-combined_k141_92416\|\|full | Not-determined |
| 18-102-final-viral-combined_k141_215551\|\|full | Not-determined |
| 18-102-final-viral-combined_k141_111605\|\|full | Low-quality |
| 18-102-final-viral-combined_k141_27737\|\|full | Not-determined |
| 18-102-final-viral-combined_k141_123023\|\|full | Low-quality |
| 18-102-final-viral-combined_k141_176001\|\|full | Not-determined |
| 18-102-final-viral-combined_k141_131656\|\|full | Low-quality |
| 18-102-final-viral-combined_k141_224824\|\|full | Not-determined |
| 18-102-final-viral-combined_k141_81327\|\|full | Low-quality |
| 18-102-final-viral-combined_k141_6993\|\|full | Low-quality |
| 18-102-final-viral-combined_k141_204120\|\|full | Low-quality |
| 18-102-final-viral-combined_k141_81341\|\|full | Not-determined |
| 18-102-final-viral-combined_k141_71266\|\|full | Low-quality |
| 18-102-final-viral-combined_k141_81349\|\|full | Not-determined |
| 18-102-final-viral-combined_k141_71269\|\|full | Not-determined |
| 18-102-final-viral-combined_k141_61049\|\|full | Not-determined |
| 18-102-final-viral-combined_k141_176033\|\|full | Not-determined |
| 18-102-final-viral-combined_k141_163222\|\|full | Not-determined |
| 18-102-final-viral-combined_k141_61070\|\|full | Low-quality |
| 18-102-final-viral-combined_k141_215607\|\|full | Low-quality |
| 18-102-final-viral-combined_k141_102873\|\|full | Low-quality |
| 18-102-final-viral-combined_k141_131680\|\|full | Low-quality |
| 18-102-final-viral-combined_k141_224854\|\|full | Low-quality |
| 18-102-final-viral-combined_k141_71295\|\|full | Low-quality |
| 18-102-final-viral-combined_k141_224856\|\|full | Low-quality |
| 18-102-final-viral-combined_k141_111660\|\|full | Not-determined |
| 18-102-final-viral-combined_k141_123052\|\|full | Not-determined |
| 18-102-final-viral-combined_k141_102894\|\|full | Low-quality |
| 18-102-final-viral-combined_k141_195510\|\|full | Not-determined |
| 18-102-final-viral-combined_k141_204159\|\|full | Not-determined |
| 18-102-final-viral-combined_k141_245018\|\|full | Low-quality |
| 18-102-final-viral-combined_k141_111661\|\|full | Low-quality |
| 18-102-final-viral-combined_k141_27840\|\|full | Low-quality |
| 18-102-final-viral-combined_k141_71337\|\|full | Not-determined |
| 18-102-final-viral-combined_k141_154025\|\|full | Not-determined |
| 18-102-final-viral-combined_k141_245027\|\|full | Not-determined |
| 18-102-final-viral-combined_k141_185336\|\|full | Not-determined |
| 18-102-final-viral-combined_k141_111684\|\|full | Low-quality |
| 18-102-final-viral-combined_k141_40851\|\|full | Low-quality |
| 18-102-final-viral-combined_k141_40852\|\|full | Low-quality |
| 18-102-final-viral-combined_k141_61142\|\|full | Not-determined |
| 18-102-final-viral-combined_k141_71362\|\|full | Low-quality |
| 18-102-final-viral-combined_k141_111713\|\|full | Low-quality |
| 18-102-final-viral-combined_k141_131746\|\|full | Low-quality |
| 18-102-final-viral-combined_k141_61183\|\|full | Low-quality |
| 18-102-final-viral-combined_k141_224950\|\|full | Not-determined |
| 18-102-final-viral-combined_k141_111722\|\|full | Low-quality |
| 18-102-final-viral-combined_k141_131753\|\|full | Not-determined |
| 18-102-final-viral-combined_k141_40905\|\|full | Not-determined |
| 18-102-final-viral-combined_k141_61207\|\|full | Not-determined |
| 18-102-final-viral-combined_k141_163282\|\|full | Not-determined |
| 18-102-final-viral-combined_k141_185389\|\|full | Not-determined |
| 18-102-final-viral-combined_k141_7028\|\|full | Not-determined |
| 18-102-final-viral-combined_k141_185403\|\|full | Low-quality |
| 18-102-final-viral-combined_k141_71405\|\|full | Low-quality |
| 18-102-final-viral-combined_k141_237658\|\|full | Not-determined |
| 18-102-final-viral-combined_k141_71409\|\|full | Not-determined |
| 18-102-final-viral-combined_k141_154102\|\|full | Not-determined |
| 18-102-final-viral-combined_k141_245134\|\|full | Low-quality |
| 18-102-final-viral-combined_k141_195655\|\|full | Not-determined |
| 18-102-final-viral-combined_k141_92460\|\|full | Not-determined |
| 18-102-final-viral-combined_k141_111776\|\|full | Low-quality |
| 18-102-final-viral-combined_k141_131794\|\|full | Low-quality |
| 18-102-final-viral-combined_k141_245151\|\|full | Low-quality |
| 18-102-final-viral-combined_k141_185452\|\|full | Not-determined |
| 18-102-final-viral-combined_k141_245156\|\|full | Not-determined |
| 18-102-final-viral-combined_k141_131801\|\|full | Low-quality |
| 18-102-final-viral-combined_k141_245157\|\|full | Not-determined |
| 18-102-final-viral-combined_k141_215686\|\|full | Not-determined |
| 18-102-final-viral-combined_k141_195670\|\|full | Not-determined |
| 18-102-final-viral-combined_k141_61268\|\|full | Not-determined |
| 18-102-final-viral-combined_k141_92481\|\|full | Low-quality |
| 18-102-final-viral-combined_k141_215696\|\|full | Not-determined |
| 18-102-final-viral-combined_k141_111810\|\|full | Not-determined |
| 18-102-final-viral-combined_k141_237698\|\|full | Low-quality |
| 18-102-final-viral-combined_k141_103018\|\|full | Not-determined |
| 18-102-final-viral-combined_k141_27854\|\|full | Low-quality |
| 18-102-final-viral-combined_k141_7057\|\|full | Low-quality |
| 18-102-final-viral-combined_k141_111814\|\|full | Low-quality |
| 18-102-final-viral-combined_k141_19429\|\|full | Not-determined |
| 18-102-final-viral-combined_k141_111820\|\|full | Low-quality |
| 18-102-final-viral-combined_k141_195714\|\|full | Low-quality |
| 18-102-final-viral-combined_k141_185470\|\|full | Not-determined |
| 18-102-final-viral-combined_k141_143853\|\|full | Low-quality |
| 18-102-final-viral-combined_k141_154174\|\|full | Not-determined |
| 18-102-final-viral-combined_k141_185482\|\|full | Low-quality |
| 18-102-final-viral-combined_k141_154182\|\|full | Not-determined |
| 18-102-final-viral-combined_k141_27882\|\|full | Low-quality |
| 18-102-final-viral-combined_k141_143880\|\|full | Low-quality |
| 18-102-final-viral-combined_k141_237737\|\|full | Not-determined |
| 18-102-final-viral-combined_k141_163296\|\|full | Low-quality |
| 18-102-final-viral-combined_k141_195740\|\|full | Not-determined |
| 18-102-final-viral-combined_k141_123198\|\|full | Not-determined |
| 18-102-final-viral-combined_k141_215759\|\|full | Low-quality |
| 18-102-final-viral-combined_k141_27903\|\|full | Low-quality |
| 18-102-final-viral-combined_k141_40943\|\|full | Low-quality |
| 18-102-final-viral-combined_k141_19485\|\|full | Not-determined |
| 18-102-final-viral-combined_k141_195756\|\|full | Not-determined |
| 18-102-final-viral-combined_k141_163328\|\|full | Not-determined |
| 18-102-final-viral-combined_k141_163331\|\|full | Low-quality |
| 18-102-final-viral-combined_k141_19501\|\|full | Not-determined |
| 18-102-final-viral-combined_k141_92578\|\|full | Low-quality |
| 18-102-final-viral-combined_k141_195781\|\|full | Not-determined |
| 18-102-final-viral-combined_k141_19531\|\|full | Not-determined |
| 18-102-final-viral-combined_k141_81542\|\|full | Low-quality |
| 18-102-final-viral-combined_k141_237800\|\|full | Not-determined |
| 18-102-final-viral-combined_k141_103063\|\|full | Low-quality |
| 18-102-final-viral-combined_k141_215808\|\|full | Not-determined |
| 18-102-final-viral-combined_k141_19544\|\|full | Not-determined |
| 18-102-final-viral-combined_k141_195818\|\|full | Not-determined |
| 18-102-final-viral-combined_k141_215814\|\|full | Low-quality |
| 18-102-final-viral-combined_k141_143956\|\|full | Not-determined |
| 18-102-final-viral-combined_k141_195832\|\|full | Not-determined |
| 18-102-final-viral-combined_k141_19571\|\|full | Not-determined |
| 18-102-final-viral-combined_k141_143977\|\|full | Low-quality |
| 18-102-final-viral-combined_k141_27968\|\|full | Low-quality |
| 18-102-final-viral-combined_k141_19573\|\|full | Not-determined |
| 18-102-final-viral-combined_k141_81580\|\|full | Low-quality |
| 18-102-final-viral-combined_k141_131901\|\|full | Not-determined |
| 18-102-final-viral-combined_k141_19579\|\|full | Not-determined |
| 18-102-final-viral-combined_k141_131909\|\|full | Not-determined |
| 18-102-final-viral-combined_k141_154318\|\|full | Low-quality |
| 18-102-final-viral-combined_k141_7155\|\|full | Not-determined |
| 18-102-final-viral-combined_k141_71522\|\|full | Low-quality |
| 18-102-final-viral-combined_k141_144027\|\|full | Low-quality |
| 18-102-final-viral-combined_k141_225108\|\|full | Low-quality |
| 18-102-final-viral-combined_k141_71527\|\|full | Low-quality |
| 18-102-final-viral-combined_k141_103120\|\|full | Not-determined |
| 18-102-final-viral-combined_k141_61390\|\|full | Low-quality |
| 18-102-final-viral-combined_k141_131927\|\|full | Low-quality |
| 18-102-final-viral-combined_k141_71532\|\|full | Low-quality |
| 18-102-final-viral-combined_k141_7178\|\|full | Not-determined |
| 18-102-final-viral-combined_k141_81654\|\|full | Low-quality |
| 18-102-final-viral-combined_k141_81662\|\|full | Low-quality |
| 18-102-final-viral-combined_k141_131953\|\|full | Not-determined |
| 18-102-final-viral-combined_k141_131954\|\|full | Not-determined |
| 18-102-final-viral-combined_k141_144077\|\|full | Low-quality |
| 18-102-final-viral-combined_k141_225147\|\|full | Not-determined |
| 18-102-final-viral-combined_k141_71556\|\|full | Low-quality |
| 18-102-final-viral-combined_k141_154367\|\|full | Low-quality |
| 18-102-final-viral-combined_k141_7205\|\|full | Low-quality |
| 18-102-final-viral-combined_k141_27994\|\|full | Not-determined |
| 18-102-final-viral-combined_k141_204291\|\|full | Not-determined |
| 18-102-final-viral-combined_k141_81697\|\|full | Low-quality |
| 18-102-final-viral-combined_k141_123253\|\|full | Not-determined |
| 18-102-final-viral-combined_k141_19628\|\|full | Not-determined |
| 18-102-final-viral-combined_k141_7232\|\|full | Low-quality |
| 18-102-final-viral-combined_k141_41115\|\|full | Low-quality |
| 18-102-final-viral-combined_k141_28022\|\|full | Not-determined |
| 18-102-final-viral-combined_k141_163450\|\|full | Low-quality |
| 18-102-final-viral-combined_k141_7235\|\|full | Low-quality |
| 18-102-final-viral-combined_k141_154395\|\|full | Low-quality |
| 18-102-final-viral-combined_k141_245224\|\|full | Not-determined |
| 18-102-final-viral-combined_k141_41125\|\|full | Not-determined |
| 18-102-final-viral-combined_k141_81744\|\|full | Not-determined |
| 18-102-final-viral-combined_k141_131984\|\|full | Not-determined |
| 18-102-final-viral-combined_k141_81757\|\|full | Low-quality |
| 18-102-final-viral-combined_k141_154423\|\|full | Low-quality |
| 18-102-final-viral-combined_k141_28043\|\|full | Low-quality |
| 18-102-final-viral-combined_k141_204328\|\|full | Not-determined |
| 18-102-final-viral-combined_k141_81771\|\|full | Low-quality |
| 18-102-final-viral-combined_k141_144140\|\|full | Low-quality |
| 18-102-final-viral-combined_k141_144143\|\|full | Low-quality |
| 18-102-final-viral-combined_k141_19692\|\|full | Not-determined |
| 18-102-final-viral-combined_k141_195936\|\|full | Low-quality |
| 18-102-final-viral-combined_k141_195938\|\|full | Not-determined |
| 18-102-final-viral-combined_k141_92670\|\|full | Not-determined |
| KM_k141_116605 | Medium-quality |
| 18-102-final-viral-combined_k141_195944\|\|full | Not-determined |
| 18-102-final-viral-combined_k141_225239\|\|full | Low-quality |
| 18-102-final-viral-combined_k141_41161\|\|full | Low-quality |
| 18-102-final-viral-combined_k141_195950\|\|full | Not-determined |
| 18-102-final-viral-combined_k141_61484\|\|full | Not-determined |
| KM-final-viral-combined_k141_116605\|\|full | Medium-quality |
| 18-102-final-viral-combined_k141_144188\|\|full | Not-determined |
| 18-102-final-viral-combined_k141_28066\|\|full | Low-quality |
| 18-102-final-viral-combined_k141_185527\|\|full | Not-determined |
| 18-102-final-viral-combined_k141_225281\|\|full | Low-quality |
| 18-102-final-viral-combined_k141_185529\|\|full | Not-determined |
| 18-102-final-viral-combined_k141_111888\|\|full | Not-determined |
| 18-102-final-viral-combined_k141_225300\|\|full | Not-determined |
| 18-102-final-viral-combined_k141_237892\|\|full | Low-quality |
| 18-102-final-viral-combined_k141_28092\|\|full | Low-quality |
| 18-102-final-viral-combined_k141_92744\|\|full | Low-quality |
| 18-102-final-viral-combined_k141_163550\|\|full | Low-quality |
| 18-102-final-viral-combined_k141_196024\|\|full | Low-quality |
| 18-102-final-viral-combined_k141_123328\|\|full | Low-quality |
| 18-102-final-viral-combined_k141_185563\|\|full | Not-determined |
| 18-102-final-viral-combined_k141_196041\|\|full | Low-quality |
| 18-102-final-viral-combined_k141_163569\|\|full | Not-determined |
| 18-102-final-viral-combined_k141_163573\|\|full | Low-quality |
| 18-102-final-viral-combined_k141_215871\|\|full | Low-quality |
| 18-102-final-viral-combined_k141_215877\|\|full | Low-quality |
| 18-102-final-viral-combined_k141_163580\|\|full | Low-quality |
| 18-102-final-viral-combined_k141_41215\|\|full | Low-quality |
| 18-102-final-viral-combined_k141_111915\|\|full | Low-quality |
| 18-102-final-viral-combined_k141_111917\|\|full | Low-quality |
| 18-102-final-viral-combined_k141_144235\|\|full | Low-quality |
| 18-102-final-viral-combined_k141_237909\|\|full | Not-determined |
| 18-102-final-viral-combined_k141_196082\|\|full | Low-quality |
| 18-102-final-viral-combined_k141_7293\|\|full | Not-determined |
| 18-102-final-viral-combined_k141_215886\|\|full | Not-determined |
| 18-102-final-viral-combined_k141_196097\|\|full | Not-determined |
| 18-102-final-viral-combined_k141_111946\|\|full | Low-quality |
| 18-102-final-viral-combined_k141_7311\|\|full | Low-quality |
| 18-102-final-viral-combined_k141_41235\|\|full | Low-quality |
| 18-102-final-viral-combined_k141_111951\|\|full | Not-determined |
| 18-102-final-viral-combined_k141_215921\|\|full | Not-determined |
| 17-108-final-viral-combined_k141_84326\|\|full | Medium-quality |
| 18-102-final-viral-combined_k141_154544\|\|full | Not-determined |
| 18-102-final-viral-combined_k141_123390\|\|full | Low-quality |
| 18-102-final-viral-combined_k141_111986\|\|full | Low-quality |
| 18-102-final-viral-combined_k141_154561\|\|full | Low-quality |
| 18-102-final-viral-combined_k141_196152\|\|full | Low-quality |
| 18-102-final-viral-combined_k141_154582\|\|full | Low-quality |
| 18-102-final-viral-combined_k141_81880\|\|full | Low-quality |
| 18-102-final-viral-combined_k141_154585\|\|full | Low-quality |
| 18-102-final-viral-combined_k141_19876\|\|full | Not-determined |
| 18-102-final-viral-combined_k141_112005\|\|full | Not-determined |
| 18-102-final-viral-combined_k141_71695\|\|full | Low-quality |
| 18-102-final-viral-combined_k141_28213\|\|full | Low-quality |
| 18-102-final-viral-combined_k141_71707\|\|full | Not-determined |
| 18-102-final-viral-combined_k141_163637\|\|full | Not-determined |
| 18-102-final-viral-combined_k141_185639\|\|full | Not-determined |
| 18-102-final-viral-combined_k141_71727\|\|full | Low-quality |
| 18-102-final-viral-combined_k141_132099\|\|full | Not-determined |
| 18-102-final-viral-combined_k141_185654\|\|full | Not-determined |
| 18-102-final-viral-combined_k141_92796\|\|full | Not-determined |
| 18-102-final-viral-combined_k141_245271\|\|full | Low-quality |
| 18-102-final-viral-combined_k141_132115\|\|full | Low-quality |
| 18-102-final-viral-combined_k141_204382\|\|full | Not-determined |
| 18-102-final-viral-combined_k141_71784\|\|full | Low-quality |
| 18-102-final-viral-combined_k141_144334\|\|full | Not-determined |
| 18-102-final-viral-combined_k141_7392\|\|full | Low-quality |
| 18-102-final-viral-combined_k141_41291\|\|full | Low-quality |
| 18-102-final-viral-combined_k141_7401\|\|full | Not-determined |
| 18-102-final-viral-combined_k141_92823\|\|full | Low-quality |
| 18-102-final-viral-combined_k141_185668\|\|full | Not-determined |
| 18-102-final-viral-combined_k141_92832\|\|full | Low-quality |
| 18-102-final-viral-combined_k141_41310\|\|full | Low-quality |
| 18-102-final-viral-combined_k141_185685\|\|full | Low-quality |
| 18-102-final-viral-combined_k141_28309\|\|full | Low-quality |
| 18-102-final-viral-combined_k141_204412\|\|full | Low-quality |
| 18-102-final-viral-combined_k141_28319\|\|full | Not-determined |
| 18-102-final-viral-combined_k141_245322\|\|full | Low-quality |
| 18-102-final-viral-combined_k141_216070\|\|full | Not-determined |
| 18-102-final-viral-combined_k141_225456\|\|full | Low-quality |
| 18-102-final-viral-combined_k141_238015\|\|full | Low-quality |
| 18-102-final-viral-combined_k141_154673\|\|full | Low-quality |
| 18-102-final-viral-combined_k141_185718\|\|full | Low-quality |
| 18-102-final-viral-combined_k141_71861\|\|full | Not-determined |
| 18-102-final-viral-combined_k141_7444\|\|full | Low-quality |
| 18-102-final-viral-combined_k141_112065\|\|full | Low-quality |
| 18-102-final-viral-combined_k141_154675\|\|full | Not-determined |
| 18-102-final-viral-combined_k141_28343\|\|full | Low-quality |
| 18-102-final-viral-combined_k141_112071\|\|full | Not-determined |
| 18-102-final-viral-combined_k141_154687\|\|full | Not-determined |
| 18-102-final-viral-combined_k141_28345\|\|full | Not-determined |
| 18-102-final-viral-combined_k141_81965\|\|full | Not-determined |
| 18-102-final-viral-combined_k141_81973\|\|full | Low-quality |
| 18-102-final-viral-combined_k141_132212\|\|full | Low-quality |
| 18-102-final-viral-combined_k141_204431\|\|full | Not-determined |
| 18-102-final-viral-combined_k141_71926\|\|full | Not-determined |
| 18-102-final-viral-combined_k141_112101\|\|full | Low-quality |
| 18-102-final-viral-combined_k141_204432\|\|full | Low-quality |
| 18-102-final-viral-combined_k141_132225\|\|full | Not-determined |
| 18-102-final-viral-combined_k141_61668\|\|full | Not-determined |
| 18-102-final-viral-combined_k141_61670\|\|full | Not-determined |
| 18-102-final-viral-combined_k141_92915\|\|full | Low-quality |
| 18-102-final-viral-combined_k141_61690\|\|full | Low-quality |
| 18-102-final-viral-combined_k141_123516\|\|full | Low-quality |
| 18-102-final-viral-combined_k141_123517\|\|full | Not-determined |
| 18-102-final-viral-combined_k141_41354\|\|full | Not-determined |
| 18-102-final-viral-combined_k141_92929\|\|full | Not-determined |
| 18-102-final-viral-combined_k141_71970\|\|full | Low-quality |
| 18-102-final-viral-combined_k141_28395\|\|full | Low-quality |
| 18-102-final-viral-combined_k141_238038\|\|full | Low-quality |
| 18-102-final-viral-combined_k141_19982\|\|full | Not-determined |
| 18-102-final-viral-combined_k141_123531\|\|full | Not-determined |
| 18-102-final-viral-combined_k141_163750\|\|full | Not-determined |
| 18-102-final-viral-combined_k141_41381\|\|full | Low-quality |
| 18-102-final-viral-combined_k141_103284\|\|full | Low-quality |
| 18-102-final-viral-combined_k141_92936\|\|full | Not-determined |
| 18-102-final-viral-combined_k141_238076\|\|full | Low-quality |
| 18-102-final-viral-combined_k141_144474\|\|full | Low-quality |
| 18-102-final-viral-combined_k141_20009\|\|full | Low-quality |
| 18-102-final-viral-combined_k141_238082\|\|full | Not-determined |
| 18-102-final-viral-combined_k141_163780\|\|full | Not-determined |
| 18-102-final-viral-combined_k141_112134\|\|full | Not-determined |
| 18-102-final-viral-combined_k141_28422\|\|full | Not-determined |
| 18-102-final-viral-combined_k141_7507\|\|full | Not-determined |
| 18-102-final-viral-combined_k141_185838\|\|full | Not-determined |
| 18-102-final-viral-combined_k141_123589\|\|full | Not-determined |
| 18-102-final-viral-combined_k141_245360\|\|full | Not-determined |
| 18-102-final-viral-combined_k141_82053\|\|full | Low-quality |
| 18-102-final-viral-combined_k141_72010\|\|full | Not-determined |
| 18-102-final-viral-combined_k141_82057\|\|full | Not-determined |
| 18-102-final-viral-combined_k141_92983\|\|full | Low-quality |
| 18-102-final-viral-combined_k141_185867\|\|full | Not-determined |
| 18-102-final-viral-combined_k141_154822\|\|full | Low-quality |
| 18-102-final-viral-combined_k141_82059\|\|full | Not-determined |
| 18-102-final-viral-combined_k141_61782\|\|full | Low-quality |
| 18-102-final-viral-combined_k141_103337\|\|full | Low-quality |
| 18-102-final-viral-combined_k141_82062\|\|full | Low-quality |
| 18-102-final-viral-combined_k141_7552\|\|full | Low-quality |
| 18-102-final-viral-combined_k141_41427\|\|full | Not-determined |
| 18-102-final-viral-combined_k141_154839\|\|full | Low-quality |
| 18-102-final-viral-combined_k141_163854\|\|full | Not-determined |
| 18-102-final-viral-combined_k141_216197\|\|full | Not-determined |
| 18-102-final-viral-combined_k141_82115\|\|full | Low-quality |
| 18-102-final-viral-combined_k141_61792\|\|full | Not-determined |
| 18-102-final-viral-combined_k141_7570\|\|full | Not-determined |
| 18-102-final-viral-combined_k141_123613\|\|full | Not-determined |
| 18-102-final-viral-combined_k141_61802\|\|full | Not-determined |
| 18-102-final-viral-combined_k141_216224\|\|full | Not-determined |
| 18-102-final-viral-combined_k141_163855\|\|full | Low-quality |
| 18-102-final-viral-combined_k141_238105\|\|full | Not-determined |
| 18-102-final-viral-combined_k141_7586\|\|full | Not-determined |
| 18-102-final-viral-combined_k141_103367\|\|full | Not-determined |
| 18-102-final-viral-combined_k141_238112\|\|full | Low-quality |
| 18-102-final-viral-combined_k141_132311\|\|full | Not-determined |
| 18-102-final-viral-combined_k141_245410\|\|full | Low-quality |
| 18-102-final-viral-combined_k141_216235\|\|full | Low-quality |
| 18-102-final-viral-combined_k141_7598\|\|full | Not-determined |
| 18-102-final-viral-combined_k141_238125\|\|full | Low-quality |
| 18-102-final-viral-combined_k141_216248\|\|full | Not-determined |
| 18-102-final-viral-combined_k141_112201\|\|full | Low-quality |
| 18-102-final-viral-combined_k141_163891\|\|full | Low-quality |
| 18-102-final-viral-combined_k141_82138\|\|full | Low-quality |
| 18-102-final-viral-combined_k141_72087\|\|full | Not-determined |
| 18-102-final-viral-combined_k141_93035\|\|full | Low-quality |
| 18-102-final-viral-combined_k141_82146\|\|full | Low-quality |
| 18-102-final-viral-combined_k141_112222\|\|full | Low-quality |
| 18-102-final-viral-combined_k141_72103\|\|full | Not-determined |
| 18-102-final-viral-combined_k141_112229\|\|full | Not-determined |
| 18-102-final-viral-combined_k141_112239\|\|full | Not-determined |
| 18-102-final-viral-combined_k141_112240\|\|full | Not-determined |
| 18-102-final-viral-combined_k141_7633\|\|full | Low-quality |

18-102-final-viral-combined_k141_238190||full Low-quality
18-102-final-viral-combined_k141_144534||full Not-determined
18-102-final-viral-combined_k141_196382||full Low-quality
18-102-final-viral-combined_k141_163927||full Low-quality
18-102-final-viral-combined_k141_144542||full Low-quality
18-102-final-viral-combined_k141_82217||full Low-quality
18-102-final-viral-combined_k141_163935||full Low-quality
18-102-final-viral-combined_k141_112284||full Not-determined
18-102-final-viral-combined_k141_132428||full Low-quality
18-102-final-viral-combined_k141_225668||full Not-determined
18-102-final-viral-combined_k141_123728||full Not-determined
18-102-final-viral-combined_k141_112289||full Not-determined
18-102-final-viral-combined_k141_7669||full Not-determined
18-102-final-viral-combined_k141_82252||full Not-determined
18-102-final-viral-combined_k141_82256||full Low-quality
18-102-final-viral-combined_k141_112298||full Low-quality
18-102-final-viral-combined_k141_7683||full Not-determined
18-102-final-viral-combined_k141_144609||full Low-quality
18-102-final-viral-combined_k141_112301||full Not-determined
18-102-final-viral-combined_k141_204611||full Not-determined
18-102-final-viral-combined_k141_144616||full Not-determined
18-102-final-viral-combined_k141_204622||full Not-determined
18-102-final-viral-combined_k141_225691||full Low-quality
18-102-final-viral-combined_k141_61891||full Low-quality
18-102-final-viral-combined_k141_103443||full Not-determined
18-102-final-viral-combined_k141_163986||full Low-quality
18-102-final-viral-combined_k141_112333||full Low-quality
18-102-final-viral-combined_k141_82315||full Low-quality
18-102-final-viral-combined_k141_82329||full Low-quality
18-102-final-viral-combined_k141_7714||full Not-determined
18-102-final-viral-combined_k141_132494||full Low-quality
18-102-final-viral-combined_k141_204654||full Low-quality
18-102-final-viral-combined_k141_132507||full Low-quality
18-102-final-viral-combined_k141_144655||full Low-quality
18-102-final-viral-combined_k141_216304||full Not-determined
18-102-final-viral-combined_k141_245445||full Not-determined
18-102-final-viral-combined_k141_103450||full Low-quality
18-102-final-viral-combined_k141_7737||full Low-quality
18-102-final-viral-combined_k141_103460||full Not-determined
18-102-final-viral-combined_k141_28491||full Low-quality
18-102-final-viral-combined_k141_186051||full Low-quality
18-102-final-viral-combined_k141_216316||full Low-quality
18-102-final-viral-combined_k141_112393||full Not-determined
18-102-final-viral-combined_k141_245454||full Low-quality
18-102-final-viral-combined_k141_196468||full Low-quality
18-102-final-viral-combined_k141_196473||full Low-quality
18-102-final-viral-combined_k141_28511||full Low-quality
18-102-final-viral-combined_k141_132565||full Not-determined
18-102-final-viral-combined_k141_103504||full Not-determined
18-102-final-viral-combined_k141_196488||full Low-quality
18-102-final-viral-combined_k141_132570||full Not-determined
18-102-final-viral-combined_k141_216331||full Not-determined
18-102-final-viral-combined_k141_186092||full Not-determined
18-102-final-viral-combined_k141_196500||full Not-determined
18-102-final-viral-combined_k141_28537||full Not-determined
18-102-final-viral-combined_k141_204702||full Not-determined
18-102-final-viral-combined_k141_123785||full Low-quality
18-102-final-viral-combined_k141_123788||full Low-quality
18-102-final-viral-combined_k141_20106||full Not-determined
18-102-final-viral-combined_k141_186112||full Not-determined
18-102-final-viral-combined_k141_93115||full Not-determined
18-102-final-viral-combined_k141_196549||full Not-determined
18-102-final-viral-combined_k141_155050||full Low-quality
18-102-final-viral-combined_k141_204764||full Low-quality
18-102-final-viral-combined_k141_132646||full Not-determined
18-102-final-viral-combined_k141_204772||full Low-quality
18-102-final-viral-combined_k141_196574||full Not-determined
18-102-final-viral-combined_k141_20163||full Low-quality
18-102-final-viral-combined_k141_245516||full Low-quality
18-102-final-viral-combined_k141_225864||full Low-quality
18-102-final-viral-combined_k141_20176||full Not-determined
18-102-final-viral-combined_k141_72297||full Low-quality
18-102-final-viral-combined_k141_82408||full Not-determined
18-102-final-viral-combined_k141_196599||full Not-determined
18-102-final-viral-combined_k141_82409||full Low-quality
18-102-final-viral-combined_k141_20179||full Low-quality
18-102-final-viral-combined_k141_123836||full Low-quality
18-102-final-viral-combined_k141_245534||full Not-determined
18-102-final-viral-combined_k141_62002||full Not-determined
18-102-final-viral-combined_k141_72323||full Low-quality
18-102-final-viral-combined_k141_72327||full Not-determined
18-102-final-viral-combined_k141_245542||full Not-determined
18-102-final-viral-combined_k141_132681||full Not-determined
18-102-final-viral-combined_k141_132687||full Low-quality
18-102-final-viral-combined_k141_62014||full Not-determined
18-102-final-viral-combined_k141_204808||full Not-determined
18-102-final-viral-combined_k141_245564||full Not-determined
18-102-final-viral-combined_k141_225887||full Low-quality
18-102-final-viral-combined_k141_20217||full Low-quality
18-102-final-viral-combined_k141_144742||full Not-determined
18-102-final-viral-combined_k141_20221||full Not-determined
18-102-final-viral-combined_k141_196628||full Not-determined
18-102-final-viral-combined_k141_216390||full Low-quality
18-102-final-viral-combined_k141_20228||full Not-determined
18-102-final-viral-combined_k141_204829||full Not-determined
18-102-final-viral-combined_k141_204834||full Low-quality
18-102-final-viral-combined_k141_62060||full Low-quality
18-102-final-viral-combined_k141_164110||full Not-determined
18-102-final-viral-combined_k141_28678||full Low-quality
18-102-final-viral-combined_k141_72393||full Not-determined
18-102-final-viral-combined_k141_20260||full Not-determined
18-102-final-viral-combined_k141_72401||full Low-quality
18-102-final-viral-combined_k141_123881||full Low-quality
18-102-final-viral-combined_k141_196687||full Low-quality
18-102-final-viral-combined_k141_72413||full Not-determined
18-102-final-viral-combined_k141_103553||full Not-determined
18-102-final-viral-combined_k141_164123||full Low-quality
18-102-final-viral-combined_k141_186205||full Low-quality
18-102-final-viral-combined_k141_93139||full Low-quality
18-102-final-viral-combined_k141_216466||full Not-determined
18-102-final-viral-combined_k141_245586||full Low-quality
18-102-final-viral-combined_k141_28721||full Not-determined
18-102-final-viral-combined_k141_216475||full Not-determined
18-102-final-viral-combined_k141_93143||full Not-determined
18-102-final-viral-combined_k141_132838||full Low-quality
18-102-final-viral-combined_k141_132842||full Not-determined
18-102-final-viral-combined_k141_93148||full Not-determined
18-102-final-viral-combined_k141_132846||full Low-quality
18-102-final-viral-combined_k141_155152||full Low-quality
18-102-final-viral-combined_k141_245594||full Low-quality
18-102-final-viral-combined_k141_155173||full Not-determined
18-102-final-viral-combined_k141_112474||full Low-quality
18-102-final-viral-combined_k141_132864||full Low-quality
18-102-final-viral-combined_k141_216521||full Low-quality
18-102-final-viral-combined_k141_164163||full Low-quality
18-102-final-viral-combined_k141_196708||full Not-determined
18-102-final-viral-combined_k141_204870||full Not-determined
18-102-final-viral-combined_k141_132867||full Low-quality
18-102-final-viral-combined_k141_164179||full Low-quality
18-102-final-viral-combined_k141_204894||full Not-determined
18-102-final-viral-combined_k141_216555||full Low-quality
18-102-final-viral-combined_k141_132901||full Not-determined
18-102-final-viral-combined_k141_20375||full Low-quality
18-102-final-viral-combined_k141_155210||full Low-quality
18-102-final-viral-combined_k141_186226||full Low-quality
18-102-final-viral-combined_k141_245680||full Not-determined
18-102-final-viral-combined_k141_20388||full Low-quality
18-102-final-viral-combined_k141_132917||full Low-quality
18-102-final-viral-combined_k141_82520||full Low-quality
18-102-final-viral-combined_k141_216584||full Not-determined
18-102-final-viral-combined_k141_245682||full Low-quality
18-102-final-viral-combined_k141_186248||full Not-determined
18-102-final-viral-combined_k141_186262||full Not-determined
18-102-final-viral-combined_k141_132948||full Not-determined
18-102-final-viral-combined_k141_164213||full Not-determined
18-102-final-viral-combined_k141_196729||full Not-determined
18-102-final-viral-combined_k141_155302||full Low-quality
17-109_k141_235125 Medium-quality
18-102-final-viral-combined_k141_155303||full Low-quality
18-102-final-viral-combined_k141_144798||full Not-determined
18-102-final-viral-combined_k141_20422||full Low-quality
18-102-final-viral-combined_k141_112556||full Low-quality
18-102-final-viral-combined_k141_245706||full Not-determined
18-102-final-viral-combined_k141_123972||full Not-determined
18-102-final-viral-combined_k141_112572||full Low-quality
18-102-final-viral-combined_k141_186293||full Low-quality
18-102-final-viral-combined_k141_196754||full Not-determined
18-102-final-viral-combined_k141_112577||full Not-determined
18-102-final-viral-combined_k141_123987||full Low-quality
18-102-final-viral-combined_k141_123990||full Not-determined
18-102-final-viral-combined_k141_245718||full Not-determined
18-102-final-viral-combined_k141_164216||full Low-quality
18-102-final-viral-combined_k141_123993||full Not-determined
18-102-final-viral-combined_k141_164221||full Not-determined
18-102-final-viral-combined_k141_20439||full Not-determined
18-102-final-viral-combined_k141_186346||full Low-quality
18-102-final-viral-combined_k141_186349||full Not-determined
18-102-final-viral-combined_k141_205027||full Low-quality
KM_k141_106671 Medium-quality
18-102-final-viral-combined_k141_82535||full Not-determined
18-102-final-viral-combined_k141_225923||full Low-quality
18-102-final-viral-combined_k141_225925||full Not-determined
18-102-final-viral-combined_k141_112634||full Not-determined
18-102-final-viral-combined_k141_245756||full Low-quality
18-102-final-viral-combined_k141_225929||full Not-determined
18-102-final-viral-combined_k141_82554||full Not-determined
18-102-final-viral-combined_k141_216747||full Not-determined
18-102-final-viral-combined_k141_216751||full Not-determined
18-102-final-viral-combined_k141_124048||full Low-quality
18-102-final-viral-combined_k141_112677||full Not-determined
18-102-final-viral-combined_k141_112684||full Not-determined
18-102-final-viral-combined_k141_20498||full Not-determined
18-102-final-viral-combined_k141_124059||full Low-quality
18-102-final-viral-combined_k141_225950||full Low-quality
18-102-final-viral-combined_k141_216773||full Not-determined
18-102-final-viral-combined_k141_216787||full Not-determined
18-102-final-viral-combined_k141_82606||full Low-quality
18-102-final-viral-combined_k141_112713||full Not-determined
18-102-final-viral-combined_k141_216841||full Not-determined
18-102-final-viral-combined_k141_20530||full Low-quality
18-102-final-viral-combined_k141_112723||full Low-quality
18-102-final-viral-combined_k141_82620||full Low-quality
18-102-final-viral-combined_k141_20537||full Low-quality
18-102-final-viral-combined_k141_164260||full Low-quality
18-102-final-viral-combined_k141_164275||full Not-determined
18-102-final-viral-combined_k141_216889||full Not-determined
18-102-final-viral-combined_k141_245789||full Low-quality
18-102-final-viral-combined_k141_112759||full Not-determined
18-102-final-viral-combined_k141_82646||full Not-determined
18-102-final-viral-combined_k141_205090||full Low-quality
18-102-final-viral-combined_k141_164310||full Low-quality
18-102-final-viral-combined_k141_20614||full Not-determined
18-102-final-viral-combined_k141_124075||full Low-quality
18-102-final-viral-combined_k141_28958||full Not-determined
18-102-final-viral-combined_k141_144945||full Low-quality
18-102-final-viral-combined_k141_226010||full Low-quality
18-102-final-viral-combined_k141_226022||full Not-determined
18-102-final-viral-combined_k141_20663||full Low-quality
18-102-final-viral-combined_k141_82703||full Low-quality
18-102-final-viral-combined_k141_164366||full Low-quality
18-102-final-viral-combined_k141_164379||full Not-determined
18-102-final-viral-combined_k141_82735||full Low-quality
18-102-final-viral-combined_k141_216934||full Not-determined
18-102-final-viral-combined_k141_112795||full Not-determined
18-102-final-viral-combined_k141_112809||full Not-determined
18-102-final-viral-combined_k141_226117||full Not-determined
18-102-final-viral-combined_k141_205246||full Not-determined
18-102-final-viral-combined_k141_226129||full Low-quality
18-102-final-viral-combined_k141_205268||full Low-quality
18-102-final-viral-combined_k141_245876||full Low-quality
18-102-final-viral-combined_k141_132996||full Not-determined
18-102-final-viral-combined_k141_164434||full Not-determined
18-102-final-viral-combined_k141_7770||full Not-determined
18-102-final-viral-combined_k141_164448||full Low-quality
18-102-final-viral-combined_k141_29076||full Low-quality
18-102-final-viral-combined_k141_133021||full Not-determined
18-102-final-viral-combined_k141_216983||full Not-determined
18-102-final-viral-combined_k141_205319||full Low-quality
18-102-final-viral-combined_k141_7787||full Low-quality
18-102-final-viral-combined_k141_124270||full Not-determined
18-102-final-viral-combined_k141_82804||full Low-quality
18-102-final-viral-combined_k141_226222||full Low-quality
18-102-final-viral-combined_k141_82826||full Not-determined
18-102-final-viral-combined_k141_217037||full Low-quality
18-102-final-viral-combined_k141_133124||full Not-determined
18-102-final-viral-combined_k141_133130||full Low-quality
18-102-final-viral-combined_k141_29138||full Not-determined
18-102-final-viral-combined_k141_217053||full Low-quality
18-102-final-viral-combined_k141_164479||full Low-quality
18-102-final-viral-combined_k141_7837||full Low-quality
18-102-final-viral-combined_k141_226306||full Not-determined
18-102-final-viral-combined_k141_7839||full Not-determined
18-102-final-viral-combined_k141_7842||full Not-determined
18-102-final-viral-combined_k141_29160||full Not-determined
18-102-final-viral-combined_k141_226332||full Not-determined
18-102-final-viral-combined_k141_164502||full Low-quality
18-102-final-viral-combined_k141_245989||full Low-quality
18-102-final-viral-combined_k141_164528||full Not-determined
18-102-final-viral-combined_k141_133211||full Not-determined
18-102-final-viral-combined_k141_164539||full Low-quality
18-102-final-viral-combined_k141_246024||full Low-quality
18-102-final-viral-combined_k141_226399||full Low-quality
18-102-final-viral-combined_k141_217157||full Not-determined
18-102-final-viral-combined_k141_217159||full Low-quality
18-102-final-viral-combined_k141_205387||full Not-determined
18-102-final-viral-combined_k141_217162||full Not-determined
18-102-final-viral-combined_k141_217174||full Not-determined
18-102-final-viral-combined_k141_112956||full Low-quality
18-102-final-viral-combined_k141_164596||full Low-quality
18-102-final-viral-combined_k141_112986||full Low-quality
18-102-final-viral-combined_k141_246059||full Not-determined
18-102-final-viral-combined_k141_217189||full Not-determined
18-102-final-viral-combined_k141_7910||full Not-determined
18-102-final-viral-combined_k141_7919||full Not-determined
18-102-final-viral-combined_k141_226455||full Not-determined
18-102-final-viral-combined_k141_217206||full Low-quality
18-102-final-viral-combined_k141_217207||full Low-quality
18-102-final-viral-combined_k141_164659||full Low-quality
18-102-final-viral-combined_k141_217213||full Not-determined
18-102-final-viral-combined_k141_217215||full Not-determined
18-102-final-viral-combined_k141_29221||full Not-determined
18-102-final-viral-combined_k141_29225||full Low-quality
18-102-final-viral-combined_k141_217219||full Not-determined
18-102-final-viral-combined_k141_164677||full Not-determined
18-102-final-viral-combined_k141_133353||full Low-quality
18-102-final-viral-combined_k141_29229||full Not-determined
18-102-final-viral-combined_k141_113070||full Not-determined
18-102-final-viral-combined_k141_29241||full Not-determined
18-102-final-viral-combined_k141_113088||full Low-quality
18-102-final-viral-combined_k141_113103||full Low-quality
18-102-final-viral-combined_k141_164738||full Not-determined
18-102-final-viral-combined_k141_217239||full Low-quality
18-102-final-viral-combined_k141_164749||full Low-quality
18-102-final-viral-combined_k141_226508||full Not-determined
18-102-final-viral-combined_k141_113129||full Not-determined
18-102-final-viral-combined_k141_164757||full Low-quality
18-102-final-viral-combined_k141_226522||full Low-quality
18-102-final-viral-combined_k141_113146||full Not-determined
18-102-final-viral-combined_k141_113150||full Not-determined
18-102-final-viral-combined_k141_205507||full Not-determined
18-102-final-viral-combined_k141_217259||full Not-determined
18-102-final-viral-combined_k141_113184||full Low-quality
18-102-final-viral-combined_k141_217286||full Low-quality
18-102-final-viral-combined_k141_246158||full Low-quality
18-102-final-viral-combined_k141_246159||full Low-quality
18-102-final-viral-combined_k141_205538||full Not-determined
18-102-final-viral-combined_k141_226585||full Low-quality
18-102-final-viral-combined_k141_246181||full Low-quality
18-102-final-viral-combined_k141_246190||full Not-determined
18-102-final-viral-combined_k141_7979||full Low-quality
18-102-final-viral-combined_k141_205576||full Low-quality
18-102-final-viral-combined_k141_217338||full Not-determined
18-102-final-viral-combined_k141_164859||full Not-determined
18-102-final-viral-combined_k141_7987||full Not-determined
18-102-final-viral-combined_k141_133374||full Low-quality
18-102-final-viral-combined_k141_226616||full Low-quality
18-102-final-viral-combined_k141_217364||full Not-determined
18-102-final-viral-combined_k141_113275||full Not-determined
18-102-final-viral-combined_k141_113279||full Not-determined
18-102-final-viral-combined_k141_8022||full Low-quality
18-102-final-viral-combined_k141_205616||full Not-determined
18-102-final-viral-combined_k141_205617||full Low-quality
18-102-final-viral-combined_k141_29367||full Low-quality
18-102-final-viral-combined_k141_217371||full Low-quality
18-102-final-viral-combined_k141_226688||full Low-quality
18-102-final-viral-combined_k141_133459||full Not-determined
18-102-final-viral-combined_k141_8038||full Low-quality
18-102-final-viral-combined_k141_29383||full Low-quality
18-102-final-viral-combined_k141_246243||full Not-determined
18-102-final-viral-combined_k141_164961||full Low-quality
18-102-final-viral-combined_k141_29399||full Not-determined
18-102-final-viral-combined_k141_205672||full Not-determined
18-102-final-viral-combined_k141_29416||full Low-quality
18-102-final-viral-combined_k141_226762||full Low-quality
18-102-final-viral-combined_k141_226780||full Not-determined
18-102-final-viral-combined_k141_29464||full Low-quality
18-102-final-viral-combined_k141_165041||full Not-determined
18-102-final-viral-combined_k141_205717||full Not-determined
18-102-final-viral-combined_k141_205720||full Not-determined
18-102-final-viral-combined_k141_205721||full Low-quality
18-102-final-viral-combined_k141_8088||full Low-quality
18-102-final-viral-combined_k141_29476||full Not-determined
18-102-final-viral-combined_k141_165059||full Low-quality
18-102-final-viral-combined_k141_8097||full Not-determined
18-102-final-viral-combined_k141_205725||full Not-determined
18-102-final-viral-combined_k141_205727||full Not-determined
18-102-final-viral-combined_k141_8112||full Not-determined
18-102-final-viral-combined_k141_29508||full Low-quality
18-102-final-viral-combined_k141_8129||full Not-determined
18-102-final-viral-combined_k141_217413||full Low-quality
18-102-final-viral-combined_k141_29534||full Low-quality
KM-final-viral-combined_k141_106671||full Medium-quality
18-102-final-viral-combined_k141_29550||full Low-quality
18-102-final-viral-combined_k141_226862||full Low-quality
18-102-final-viral-combined_k141_29561||full Not-determined
18-102-final-viral-combined_k141_246322||full Not-determined
18-102-final-viral-combined_k141_8216||full Low-quality
18-102-final-viral-combined_k141_226896||full Not-determined
18-102-final-viral-combined_k141_165169||full Not-determined
18-102-final-viral-combined_k141_165171||full Not-determined
18-102-final-viral-combined_k141_8232||full Not-determined
18-102-final-viral-combined_k141_113401||full Low-quality
18-102-final-viral-combined_k141_226912||full Low-quality
18-102-final-viral-combined_k141_205807||full Not-determined
18-102-final-viral-combined_k141_217465||full Low-quality
18-102-final-viral-combined_k141_113432||full Not-determined
18-102-final-viral-combined_k141_217466||full Low-quality
18-102-final-viral-combined_k141_165218||full Low-quality
18-102-final-viral-combined_k141_226950||full Low-quality
18-102-final-viral-combined_k141_113438||full Low-quality
18-102-final-viral-combined_k141_165262||full Low-quality
18-102-final-viral-combined_k141_226978||full Not-determined
18-102-final-viral-combined_k141_113463||full Not-determined
18-102-final-viral-combined_k141_113474||full Low-quality
18-102-final-viral-combined_k141_113475||full Not-determined
18-102-final-viral-combined_k141_227005||full Low-quality
18-102-final-viral-combined_k141_246367||full Not-determined
18-102-final-viral-combined_k141_165283||full Low-quality
18-102-final-viral-combined_k141_246421||full Low-quality
18-102-final-viral-combined_k141_113486||full Not-determined
18-102-final-viral-combined_k141_227056||full Not-determined
18-102-final-viral-combined_k141_246423||full Not-determined
18-102-final-viral-combined_k141_227064||full Not-determined
18-102-final-viral-combined_k141_8273||full Low-quality
18-102-final-viral-combined_k141_165353||full Low-quality
18-102-final-viral-combined_k141_133608||full Low-quality
18-102-final-viral-combined_k141_205951||full Low-quality
18-102-final-viral-combined_k141_113521||full Not-determined
18-102-final-viral-combined_k141_227099||full Not-determined
18-102-final-viral-combined_k141_165391||full Low-quality
18-102-final-viral-combined_k141_165394||full Not-determined
18-102-final-viral-combined_k141_113535||full Not-determined
18-102-final-viral-combined_k141_29690||full Not-determined
18-102-final-viral-combined_k141_133634||full Low-quality
18-102-final-viral-combined_k141_133643||full Not-determined
18-102-final-viral-combined_k141_205997||full Not-determined
18-102-final-viral-combined_k141_165412||full Low-quality
18-102-final-viral-combined_k141_29714||full Not-determined
18-102-final-viral-combined_k141_227127||full Not-determined
18-102-final-viral-combined_k141_8312||full Not-determined
18-102-final-viral-combined_k141_133661||full Low-quality
18-109-final-viral-combined_k141_90728||full Medium-quality
18-102-final-viral-combined_k141_29753||full Low-quality
18-102-final-viral-combined_k141_8340||full Not-determined
18-102-final-viral-combined_k141_113610||full Low-quality
18-102-final-viral-combined_k141_133692||full Low-quality
18-102-final-viral-combined_k141_29775||full Low-quality
18-102-final-viral-combined_k141_165483||full Not-determined
18-102-final-viral-combined_k141_133709||full Low-quality
18-102-final-viral-combined_k141_165492||full Low-quality
18-102-final-viral-combined_k141_29790||full Not-determined
18-102-final-viral-combined_k141_165503||full Not-determined
18-101-final-viral-combined_k141_52643||full Medium-quality
18-102-final-viral-combined_k141_227216||full Low-quality
18-102-final-viral-combined_k141_8388||full Not-determined
18-102-final-viral-combined_k141_246569||full Low-quality
18-102-final-viral-combined_k141_246575||full Low-quality
18-102-final-viral-combined_k141_165526||full Low-quality
18-102-final-viral-combined_k141_246591||full Low-quality
18-102-final-viral-combined_k141_227244||full Not-determined
18-102-final-viral-combined_k141_165546||full Low-quality
18-102-final-viral-combined_k141_133746||full Not-determined
18-102-final-viral-combined_k141_227271||full Low-quality
18-102-final-viral-combined_k141_133750||full Not-determined
18-102-final-viral-combined_k141_246632||full Low-quality
18-102-final-viral-combined_k141_227313||full Not-determined
18-102-final-viral-combined_k141_227317||full Not-determined
18-102-final-viral-combined_k141_8426||full Not-determined
18-102-final-viral-combined_k141_29859||full Low-quality
18-102-final-viral-combined_k141_29860||full Low-quality
18-102-final-viral-combined_k141_246671||full Not-determined
18-102-final-viral-combined_k141_29862||full Not-determined
18-102-final-viral-combined_k141_227326||full Low-quality
18-102-final-viral-combined_k141_206168||full Not-determined
18-102-final-viral-combined_k141_133810||full Not-determined
18-102-final-viral-combined_k141_133813||full Low-quality
18-102-final-viral-combined_k141_227333||full Not-determined
18-102-final-viral-combined_k141_8451||full Not-determined
18-102-final-viral-combined_k141_113695||full Low-quality
18-102-final-viral-combined_k141_133823||full Low-quality
18-102-final-viral-combined_k141_133834||full Not-determined
18-102-final-viral-combined_k141_206191||full Not-determined
18-102-final-viral-combined_k141_133861||full Not-determined
18-102-final-viral-combined_k141_29923||full Low-quality
18-102-final-viral-combined_k141_133879||full Low-quality
18-102-final-viral-combined_k141_113741||full Not-determined
18-102-final-viral-combined_k141_206249||full Low-quality
18-102-final-viral-combined_k141_133909||full Not-determined
18-102-final-viral-combined_k141_113768||full Low-quality
18-102-final-viral-combined_k141_133918||full Not-determined
18-102-final-viral-combined_k141_133924||full Not-determined
18-102-final-viral-combined_k141_206256||full Low-quality
18-102-final-viral-combined_k141_8504||full Not-determined
18-102-final-viral-combined_k141_206259||full Low-quality
18-102-final-viral-combined_k141_133959||full Not-determined
18-102-final-viral-combined_k141_246776||full Low-quality
18-102-final-viral-combined_k141_134000||full Low-quality
18-102-final-viral-combined_k141_246796||full Low-quality
18-102-final-viral-combined_k141_8548||full Not-determined
18-102-final-viral-combined_k141_134048||full Not-determined
18-102-final-viral-combined_k141_165642||full Not-determined
18-102-final-viral-combined_k141_134060||full Not-determined
18-102-final-viral-combined_k141_134064||full Low-quality
18-102-final-viral-combined_k141_134069||full Low-quality
18-102-final-viral-combined_k141_113830||full Not-determined
18-102-final-viral-combined_k141_134076||full Low-quality
18-102-final-viral-combined_k141_30102||full Not-determined
18-102-final-viral-combined_k141_206329||full Low-quality
18-102-final-viral-combined_k141_8582||full Not-determined
18-102-final-viral-combined_k141_227428||full Low-quality
18-102-final-viral-combined_k141_165691||full Low-quality
18-102-final-viral-combined_k141_246909||full Low-quality
18-102-final-viral-combined_k141_165699||full Low-quality
18-102-final-viral-combined_k141_113848||full Low-quality
18-102-final-viral-combined_k141_206395||full Low-quality
18-102-final-viral-combined_k141_30164||full Not-determined
18-102-final-viral-combined_k141_30189||full Low-quality
18-102-final-viral-combined_k141_227490||full Low-quality
18-102-final-viral-combined_k141_246942||full Low-quality
18-102-final-viral-combined_k141_30245||full Not-determined
18-102-final-viral-combined_k141_113900||full Low-quality
18-102-final-viral-combined_k141_134213||full Low-quality
18-102-final-viral-combined_k141_206479||full Not-determined
18-102-final-viral-combined_k141_134229||full Low-quality
18-102-final-viral-combined_k141_30299||full Not-determined
18-102-final-viral-combined_k141_134240||full Low-quality
18-102-final-viral-combined_k141_30319||full Not-determined
18-102-final-viral-combined_k141_134269||full Low-quality
18-102-final-viral-combined_k141_247035||full Low-quality
18-102-final-viral-combined_k141_134319||full Not-determined
18-102-final-viral-combined_k141_134335||full Low-quality
18-102-final-viral-combined_k141_227625||full Low-quality
18-102-final-viral-combined_k141_227631||full Low-quality
18-102-final-viral-combined_k141_227635||full Low-quality
18-102-final-viral-combined_k141_30391||full Not-determined
18-102-final-viral-combined_k141_206540||full Not-determined
18-102-final-viral-combined_k141_134370||full Not-determined
18-102-final-viral-combined_k141_30395||full Low-quality
18-102-final-viral-combined_k141_206571||full Not-determined
18-102-final-viral-combined_k141_247105||full Not-determined
18-102-final-viral-combined_k141_30443||full Low-quality
18-102-final-viral-combined_k141_30449||full Low-quality
18-102-final-viral-combined_k141_30454||full Not-determined
18-102-final-viral-combined_k141_134426||full Low-quality
18-102-final-viral-combined_k141_227741||full Not-determined
18-102-final-viral-combined_k141_206609||full Low-quality
18-102-final-viral-combined_k141_8625||full Not-determined
18-102-final-viral-combined_k141_8653||full Not-determined
18-102-final-viral-combined_k141_8661||full Low-quality
18-102-final-viral-combined_k141_134475||full Low-quality
18-102-final-viral-combined_k141_134484||full Low-quality
18-102-final-viral-combined_k141_30547||full Low-quality
18-102-final-viral-combined_k141_30558||full Not-determined
18-102-final-viral-combined_k141_134488||full Not-determined
18-102-final-viral-combined_k141_30577||full Not-determined
18-102-final-viral-combined_k141_227784||full Not-determined
18-102-final-viral-combined_k141_206698||full Not-determined
18-102-final-viral-combined_k141_8687||full Not-determined
18-102-final-viral-combined_k141_134512||full Low-quality
18-102-final-viral-combined_k141_134524||full Low-quality
17-109-final-viral-combined_k141_235125||full Medium-quality
18-102-final-viral-combined_k141_227809||full Low-quality
18-102-final-viral-combined_k141_227820||full Low-quality
18-102-final-viral-combined_k141_30628||full Low-quality
18-102-final-viral-combined_k141_30630||full Low-quality
18-102-final-viral-combined_k141_30654||full Not-determined
18-102-final-viral-combined_k141_8780||full Not-determined
18-102-final-viral-combined_k141_134607||full Low-quality
18-102-final-viral-combined_k141_30672||full Not-determined
18-102-final-viral-combined_k141_134610||full Not-determined
18-102-final-viral-combined_k141_30700||full Low-quality
18-102-final-viral-combined_k141_30730||full Low-quality
18-102-final-viral-combined_k141_8865||full Low-quality
18-102-final-viral-combined_k141_247157||full Low-quality
18-102-final-viral-combined_k141_206725||full Low-quality
18-102-final-viral-combined_k141_8893||full Low-quality
18-110-final-viral-combined_k141_134098||full Medium-quality
18-102-final-viral-combined_k141_206749||full Low-quality
18-102-final-viral-combined_k141_206788||full Low-quality
18-102-final-viral-combined_k141_247242||full Low-quality
18-102-final-viral-combined_k141_8962||full Not-determined
18-102-final-viral-combined_k141_247268||full Not-determined
18-102-final-viral-combined_k141_30796||full Not-determined
18-102-final-viral-combined_k141_8991||full Not-determined
18-102-final-viral-combined_k141_30838||full Not-determined
18-102-final-viral-combined_k141_206808||full Low-quality
18-102-final-viral-combined_k141_247305||full Low-quality
18-102-final-viral-combined_k141_9012||full Not-determined
18-102-final-viral-combined_k141_247318||full Not-determined
18-102-final-viral-combined_k141_30873||full Low-quality
18-102-final-viral-combined_k141_247325||full Not-determined
18-102-final-viral-combined_k141_9037||full Not-determined
18-102-final-viral-combined_k141_247369||full Low-quality
18-102-final-viral-combined_k141_247383||full Not-determined
18-102-final-viral-combined_k141_247385||full Not-determined
18-102-final-viral-combined_k141_9078||full Low-quality
18-102-final-viral-combined_k141_247404||full Not-determined
18-102-final-viral-combined_k141_247413||full Low-quality
18-102-final-viral-combined_k141_30962||full Low-quality
18-102-final-viral-combined_k141_9144||full Low-quality
18-102-final-viral-combined_k141_247494||full Not-determined
18-102-final-viral-combined_k141_9160||full Not-determined
18-102-final-viral-combined_k141_206839||full Not-determined
18-102-final-viral-combined_k141_206841||full Not-determined
18-102-final-viral-combined_k141_206843||full Not-determined
18-102-final-viral-combined_k141_206864||full Not-determined
18-102-final-viral-combined_k141_247589||full Not-determined
18-102-final-viral-combined_k141_247601||full Not-determined
18-102-final-viral-combined_k141_247637||full Low-quality
18-102-final-viral-combined_k141_9320||full Not-determined
18-102-final-viral-combined_k141_206961||full Not-determined
18-102-final-viral-combined_k141_9340||full Not-determined
18-102-final-viral-combined_k141_247697||full Not-determined
18-102-final-viral-combined_k141_247699||full Not-determined
18-102-final-viral-combined_k141_247781||full Low-quality
18-102-final-viral-combined_k141_247791||full Not-determined
18-102-final-viral-combined_k141_9365||full Not-determined
18-102-final-viral-combined_k141_247803||full Not-determined
18-102-final-viral-combined_k141_247807||full Low-quality
18-102-final-viral-combined_k141_247826||full Not-determined
18-102-final-viral-combined_k141_9462||full Not-determined
18-102-final-viral-combined_k141_9487||full Low-quality
18-102-final-viral-combined_k141_247915||full Not-determined
18-102-final-viral-combined_k141_9499||full Not-determined
18-102-final-viral-combined_k141_207036||full Not-determined
18-102-final-viral-combined_k141_247998||full Not-determined
18-102-final-viral-combined_k141_207046||full Not-determined
18-102-final-viral-combined_k141_207078||full Not-determined
18-102-final-viral-combined_k141_248064||full Not-determined
18-102-final-viral-combined_k141_248067||full Not-determined
18-102-final-viral-combined_k141_248074||full Not-determined
18-102-final-viral-combined_k141_9563||full Not-determined
18-102-final-viral-combined_k141_207090||full Not-determined
18-102-final-viral-combined_k141_207108||full Not-determined
18-102-final-viral-combined_k141_9599||full Low-quality
18-102-final-viral-combined_k141_9612||full Low-quality
18-102-final-viral-combined_k141_248200||full Low-quality
18-102-final-viral-combined_k141_248228||full Low-quality
18-102-final-viral-combined_k141_9644||full Not-determined

| | |
|---|---|
| 18-102-final-viral-combined_k141_248250\|\|full | Not-determined |
| 18-102-final-viral-combined_k141_9665\|\|full | Not-determined |
| 18-102-final-viral-combined_k141_248278\|\|full | Low-quality |
| 18-102-final-viral-combined_k141_9694\|\|full | Not-determined |
| 18-102-final-viral-combined_k141_9695\|\|full | Low-quality |
| 18-102-final-viral-combined_k141_248295\|\|full | Not-determined |
| 18-102-final-viral-combined_k141_9708\|\|full | Low-quality |
| 18-102-final-viral-combined_k141_9726\|\|full | Not-determined |
| 18-102-final-viral-combined_k141_9760\|\|full | Low-quality |
| 18-102-final-viral-combined_k141_9888\|\|full | Not-determined |
| 18-102-final-viral-combined_k141_9974\|\|full | Not-determined |
| 18-102-final-viral-combined_k141_9987\|\|full | Not-determined |
| 18-102-final-viral-combined_k141_10000\|\|full | Not-determined |
| 18-102-final-viral-combined_k141_10005\|\|full | Not-determined |
| 18-102-final-viral-combined_k141_10017\|\|full | Not-determined |
| 18-102-final-viral-combined_k141_10020\|\|full | Not-determined |
| 18-102-final-viral-combined_k141_10041\|\|full | Not-determined |
| 18-102-final-viral-combined_k141_10064\|\|full | Not-determined |
| 18-102-final-viral-combined_k141_10156\|\|full | Low-quality |
| 18-102-final-viral-combined_k141_10194\|\|full | Low-quality |
| 18-102-final-viral-combined_k141_10207\|\|full | Low-quality |
| 18-102-final-viral-combined_k141_10277\|\|full | Low-quality |
| 18-102-final-viral-combined_k141_10324\|\|full | Not-determined |
| 18-102-final-viral-combined_k141_239431\|\|0_partial | Low-quality |
| 18-102-final-viral-combined_k141_88248\|\|0_partial | Not-determined |
| 18-102-final-viral-combined_k141_113943\|\|0_partial | Not-determined |
| 18-102-final-viral-combined_k141_72534\|\|0_partial | Low-quality |
| 18-102-final-viral-combined_k141_126440\|\|full | Medium-quality |
| 18-102-final-viral-combined_k141_72636\|\|0_partial | Low-quality |
| 18-102-final-viral-combined_k141_368\|\|0_partial | Not-determined |
| 18-102-final-viral-combined_k141_104019\|\|0_partial | Low-quality |
| 18-102-final-viral-combined_k141_196980\|\|0_partial | Not-determined |
| 18-102-final-viral-combined_k141_187023\|\|0_partial | Low-quality |
| 18-102-final-viral-combined_k141_104174\|\|0_partial | Not-determined |
| 18-102-final-viral-combined_k141_218102\|\|0_partial | Not-determined |
| 17-109-final-viral-combined_k141_368562\|\|full | Medium-quality |
| 18-102-final-viral-combined_k141_692\|\|0_partial | Low-quality |
| 18-102-final-viral-combined_k141_83469\|\|0_partial | Low-quality |
| 18-102-final-viral-combined_k141_166678\|\|0_partial | Low-quality |
| 18-102-final-viral-combined_k141_125615\|\|0_partial | Low-quality |
| 18-102-final-viral-combined_k141_32320\|\|0_partial | Not-determined |
| 18-102-final-viral-combined_k141_43699\|\|0_partial | Low-quality |
| 18-102-final-viral-combined_k141_157082\|\|0_partial | Not-determined |
| 18-102-final-viral-combined_k141_208918\|\|0_partial | Low-quality |
| 18-102-final-viral-combined_k141_115711\|\|0_partial | Not-determined |
| 18-102-final-viral-combined_k141_219136\|\|0_partial | Not-determined |
| 18-106-final-viral-combined_k141_65225\|\|full | Medium-quality |
| 18-102-final-viral-combined_k141_147022\|\|0_partial | Low-quality |
| 18-102-final-viral-combined_k141_198409\|\|0_partial | Not-determined |
| 18-102-final-viral-combined_k141_95583\|\|0_partial | Not-determined |
| 18-102-final-viral-combined_k141_209106\|\|0_partial | Not-determined |
| 18-102-final-viral-combined_k141_23092\|\|0_partial | Not-determined |
| 18-102-final-viral-combined_k141_33092\|\|0_partial | Not-determined |
| 18-102-final-viral-combined_k141_74730\|\|0_partial | Not-determined |
| 18-102-final-viral-combined_k141_188796\|\|0_partial | Low-quality |
| 18-105-final-viral-combined_k141_52741\|\|0_partial | Medium-quality |
| 18-102-final-viral-combined_k141_116405\|\|0_partial | Not-determined |
| 18-105-final-viral-combined_k141_156329\|\|full | Medium-quality |
| 18-102-final-viral-combined_k141_75144\|\|1_partial | Low-quality |
| 18-102-final-viral-combined_k141_138182\|\|0_partial | Not-determined |
| 18-102-final-viral-combined_k141_55179\|\|0_partial | Not-determined |
| 18-102-final-viral-combined_k141_210534\|\|0_partial | Not-determined |
| 18-102-final-viral-combined_k141_86680\|\|0_partial | Low-quality |
| 18-102-final-viral-combined_k141_86805\|\|0_partial | Not-determined |
| 18-102-final-viral-combined_k141_210794\|\|0_partial | Not-determined |
| 18-102-final-viral-combined_k141_170048\|\|0_partial | Not-determined |
| 18-102-final-viral-combined_k141_159290\|\|0_partial | Not-determined |
| KM-final-viral-combined_k141_66123\|\|full | Medium-quality |
| 18-102-final-viral-combined_k141_241768\|\|0_partial | Low-quality |
| 18-102-final-viral-combined_k141_108203\|\|0_partial | Low-quality |
| 18-102-final-viral-combined_k141_200530\|\|0_partial | Not-determined |
| 18-102-final-viral-combined_k141_221524\|\|0_partial | Not-determined |
| 18-102-final-viral-combined_k141_181670\|\|0_partial | Not-determined |
| 18-102-final-viral-combined_k141_128644\|\|0_partial | Not-determined |
| 18-102-final-viral-combined_k141_160216\|\|0_partial | Low-quality |
| 18-102-final-viral-combined_k141_171832\|\|0_partial | Low-quality |
| 17-109-final-viral-combined_k141_71235\|\|full | Medium-quality |
| 18-102-final-viral-combined_k141_78223\|\|0_partial | Low-quality |
| 18-102-final-viral-combined_k141_16054\|\|0_partial | Low-quality |
| 18-102-final-viral-combined_k141_150929\|\|0_partial | Low-quality |
| 18-102-final-viral-combined_k141_4805\|\|0_partial | Not-determined |
| 18-102-final-viral-combined_k141_99753\|\|0_partial | Low-quality |
| 18-102-final-viral-combined_k141_26158\|\|0_partial | Low-quality |
| 18-102-final-viral-combined_k141_161578\|\|0_partial | Not-determined |
| 18-102-final-viral-combined_k141_121213\|\|0_partial | Not-determined |
| 18-102-final-viral-combined_k141_202570\|\|0_partial | Low-quality |
| 18-102-final-viral-combined_k141_213786\|\|0_partial | Low-quality |
| 18-102-final-viral-combined_k141_142224\|\|0_partial | Low-quality |
| 18-102-final-viral-combined_k141_193979\|\|0_partial | Low-quality |
| 18-102-final-viral-combined_k141_79982\|\|0_partial | Not-determined |
| 18-102-final-viral-combined_k141_223850\|\|0_partial | Low-quality |
| 18-102-final-viral-combined_k141_51566\|\|0_partial | Not-determined |
| 18-102-final-viral-combined_k141_131116\|\|0_partial | Low-quality |
| 18-102-final-viral-combined_k141_203488\|\|0_partial | Not-determined |
| 18-102-final-viral-combined_k141_18817\|\|0_partial | Low-quality |
| 18-102-final-viral-combined_k141_175971\|\|0_partial | Not-determined |
| KM_k141_124308 | Medium-quality |
| 17-109_k141_383851 | Medium-quality |
| 18-102-final-viral-combined_k141_204345\|\|0_partial | Low-quality |
| 18-102-final-viral-combined_k141_225412\|\|0_partial | Not-determined |
| 18-102-final-viral-combined_k141_154612\|\|0_partial | Low-quality |
| 18-102-final-viral-combined_k141_112045\|\|0_partial | Low-quality |
| 18-102-final-viral-combined_k141_196261\|\|0_partial | Low-quality |
| 18-102-final-viral-combined_k141_61928\|\|0_partial | Not-determined |
| 18-102-final-viral-combined_k141_216377\|\|0_partial | Not-determined |
| 18-102-final-viral-combined_k141_103506\|\|0_partial | Low-quality |
| 18-102-final-viral-combined_k141_103506\|\|1_partial | Not-determined |
| 18-102-final-viral-combined_k141_132972\|\|0_partial | Low-quality |
| 17-109-final-viral-combined_k141_41160\|\|full | Medium-quality |
| 18-102-final-viral-combined_k141_133600\|\|0_partial | Low-quality |
| 18-102-final-viral-combined_k141_133733\|\|0_partial | Low-quality |
| KM-final-viral-combined_k141_124308\|\|full | Medium-quality |
| 18-102-final-viral-combined_k141_9600\|\|0_partial | Low-quality |
| 18-102-final-viral-combined_k141_82853\|\|lt2gene | Not-determined |
| 18-102-final-viral-combined_k141_72501\|\|lt2gene | Low-quality |
| 18-102-final-viral-combined_k141_124286\|\|lt2gene | Low-quality |
| 18-102-final-viral-combined_k141_20740\|\|lt2gene | Low-quality |
| 18-102-final-viral-combined_k141_86\|\|lt2gene | Low-quality |
| 18-102-final-viral-combined_k141_113962\|\|lt2gene | Not-determined |
| 18-102-final-viral-combined_k141_176117\|\|lt2gene | Low-quality |
| 18-102-final-viral-combined_k141_186460\|\|lt2gene | Low-quality |
| 18-102-final-viral-combined_k141_217580\|\|lt2gene | Not-determined |
| 18-102-final-viral-combined_k141_41571\|\|lt2gene | Low-quality |
| 18-102-final-viral-combined_k141_134752\|\|lt2gene | Low-quality |
| 18-102-final-viral-combined_k141_51882\|\|lt2gene | Low-quality |
| 18-102-final-viral-combined_k141_134766\|\|lt2gene | Low-quality |
| 18-102-final-viral-combined_k141_106\|\|lt2gene | Low-quality |
| 18-102-final-viral-combined_k141_51906\|\|lt2gene | Low-quality |
| 18-102-final-viral-combined_k141_176211\|\|lt2gene | Low-quality |
| 18-102-final-viral-combined_k141_145072\|\|lt2gene | Low-quality |
| 18-102-final-viral-combined_k141_155476\|\|lt2gene | Not-determined |
| 18-102-final-viral-combined_k141_113999\|\|lt2gene | Low-quality |
| 18-102-final-viral-combined_k141_196851\|\|lt2gene | Low-quality |
| 18-102-final-viral-combined_k141_196857\|\|lt2gene | Low-quality |
| 18-102-final-viral-combined_k141_176243\|\|lt2gene | Low-quality |
| 18-102-final-viral-combined_k141_93268\|\|lt2gene | Not-determined |
| 18-102-final-viral-combined_k141_176258\|\|lt2gene | Not-determined |
| 18-102-final-viral-combined_k141_10500\|\|lt2gene | Low-quality |
| 18-102-final-viral-combined_k141_186574\|\|lt2gene | Low-quality |
| 18-102-final-viral-combined_k141_176279\|\|lt2gene | Low-quality |
| 18-102-final-viral-combined_k141_51965\|\|lt2gene | Low-quality |
| 18-102-final-viral-combined_k141_186580\|\|lt2gene | Low-quality |
| 18-102-final-viral-combined_k141_238295\|\|lt2gene | Low-quality |
| 18-102-final-viral-combined_k141_31207\|\|lt2gene | Low-quality |
| 18-102-final-viral-combined_k141_176312\|\|lt2gene | Low-quality |
| 18-102-final-viral-combined_k141_176317\|\|lt2gene | Low-quality |
| 18-102-final-viral-combined_k141_20815\|\|lt2gene | Low-quality |
| 18-102-final-viral-combined_k141_196928\|\|lt2gene | Low-quality |
| 18-102-final-viral-combined_k141_155507\|\|lt2gene | Low-quality |
| 18-102-final-viral-combined_k141_62311\|\|lt2gene | Low-quality |
| 18-102-final-viral-combined_k141_62334\|\|lt2gene | Low-quality |
| 18-102-final-viral-combined_k141_186652\|\|lt2gene | Not-determined |
| 18-102-final-viral-combined_k141_134814\|\|lt2gene | Low-quality |
| 18-102-final-viral-combined_k141_134815\|\|lt2gene | Low-quality |
| 18-102-final-viral-combined_k141_103882\|\|lt2gene | Low-quality |
| 18-102-final-viral-combined_k141_228070\|\|lt2gene | Not-determined |
| 18-102-final-viral-combined_k141_41767\|\|lt2gene | Low-quality |
| 18-102-final-viral-combined_k141_41792\|\|lt2gene | Not-determined |
| 18-102-final-viral-combined_k141_103925\|\|lt2gene | Low-quality |
| 18-102-final-viral-combined_k141_134935\|\|lt2gene | Low-quality |
| 18-102-final-viral-combined_k141_186705\|\|lt2gene | Low-quality |
| 18-102-final-viral-combined_k141_217753\|\|lt2gene | Low-quality |
| 18-102-final-viral-combined_k141_72691\|\|lt2gene | Low-quality |
| 18-102-final-viral-combined_k141_228168\|\|lt2gene | Not-determined |
| 18-102-final-viral-combined_k141_31414\|\|lt2gene | Low-quality |
| 18-102-final-viral-combined_k141_83129\|\|lt2gene | Low-quality |
| 18-102-final-viral-combined_k141_186734\|\|lt2gene | Not-determined |
| 18-102-final-viral-combined_k141_176467\|\|lt2gene | Low-quality |
| 18-102-final-viral-combined_k141_93528\|\|lt2gene | Low-quality |
| 18-102-final-viral-combined_k141_334\|\|lt2gene | Not-determined |
| 18-102-final-viral-combined_k141_339\|\|lt2gene | Low-quality |
| 18-102-final-viral-combined_k141_207277\|\|lt2gene | Not-determined |
| 18-102-final-viral-combined_k141_41933\|\|lt2gene | Low-quality |
| 18-102-final-viral-combined_k141_124628\|\|lt2gene | Low-quality |
| 18-102-final-viral-combined_k141_155672\|\|lt2gene | Low-quality |
| 18-102-final-viral-combined_k141_20942\|\|lt2gene | Low-quality |
| 18-102-final-viral-combined_k141_42049\|\|lt2gene | Low-quality |
| 18-102-final-viral-combined_k141_72881\|\|lt2gene | Low-quality |
| 18-102-final-viral-combined_k141_207423\|\|lt2gene | Low-quality |
| 18-102-final-viral-combined_k141_104041\|\|lt2gene | Low-quality |
| 18-102-final-viral-combined_k141_145399\|\|lt2gene | Low-quality |
| 18-102-final-viral-combined_k141_42105\|\|lt2gene | Not-determined |
| 18-102-final-viral-combined_k141_238594\|\|lt2gene | Low-quality |
| 18-102-final-viral-combined_k141_217996\|\|lt2gene | Low-quality |
| 18-102-final-viral-combined_k141_186895\|\|lt2gene | Not-determined |
| 18-102-final-viral-combined_k141_62546\|\|lt2gene | Not-determined |
| 18-102-final-viral-combined_k141_62558\|\|lt2gene | Low-quality |
| 18-102-final-viral-combined_k141_62561\|\|lt2gene | Low-quality |
| 18-102-final-viral-combined_k141_466\|\|lt2gene | Low-quality |
| 18-102-final-viral-combined_k141_197000\|\|lt2gene | Low-quality |
| 18-102-final-viral-combined_k141_21070\|\|lt2gene | Not-determined |
| 18-102-final-viral-combined_k141_491\|\|lt2gene | Low-quality |
| 18-102-final-viral-combined_k141_10747\|\|lt2gene | Not-determined |
| 18-102-final-viral-combined_k141_207467\|\|lt2gene | Not-determined |
| 18-102-final-viral-combined_k141_238754\|\|lt2gene | Low-quality |
| 18-102-final-viral-combined_k141_218051\|\|lt2gene | Low-quality |
| 18-102-final-viral-combined_k141_31605\|\|lt2gene | Low-quality |
| 18-102-final-viral-combined_k141_176619\|\|lt2gene | Low-quality |
| 18-102-final-viral-combined_k141_145534\|\|lt2gene | Low-quality |
| 18-102-final-viral-combined_k141_124875\|\|lt2gene | Low-quality |
| 18-102-final-viral-combined_k141_52268\|\|lt2gene | Low-quality |
| 18-102-final-viral-combined_k141_104138\|\|lt2gene | Low-quality |
| 18-102-final-viral-combined_k141_52295\|\|lt2gene | Low-quality |
| 18-102-final-viral-combined_k141_10801\|\|lt2gene | Low-quality |
| 18-102-final-viral-combined_k141_31673\|\|lt2gene | Not-determined |
| 18-102-final-viral-combined_k141_187028\|\|lt2gene | Low-quality |
| 18-102-final-viral-combined_k141_93749\|\|lt2gene | Not-determined |
| 18-102-final-viral-combined_k141_145651\|\|lt2gene | Low-quality |
| 18-102-final-viral-combined_k141_228455\|\|lt2gene | Low-quality |
| 18-102-final-viral-combined_k141_42424\|\|lt2gene | Not-determined |
| 18-102-final-viral-combined_k141_10908\|\|lt2gene | Low-quality |
| 18-102-final-viral-combined_k141_207774\|\|lt2gene | Low-quality |
| 18-102-final-viral-combined_k141_21397\|\|lt2gene | Low-quality |
| 18-102-final-viral-combined_k141_124993\|\|lt2gene | Low-quality |
| 18-102-final-viral-combined_k141_228533\|\|lt2gene | Low-quality |
| 18-102-final-viral-combined_k141_42477\|\|lt2gene | Low-quality |
| 18-102-final-viral-combined_k141_104247\|\|lt2gene | Low-quality |
| 18-102-final-viral-combined_k141_42488\|\|lt2gene | Low-quality |
| 18-102-final-viral-combined_k141_42505\|\|lt2gene | Not-determined |
| 18-102-final-viral-combined_k141_145721\|\|lt2gene | Low-quality |
| 18-102-final-viral-combined_k141_145745\|\|lt2gene | Low-quality |
| 18-102-final-viral-combined_k141_93830\|\|lt2gene | Not-determined |
| 18-102-final-viral-combined_k141_11091\|\|lt2gene | Low-quality |
| 18-102-final-viral-combined_k141_197220\|\|lt2gene | Low-quality |
| 18-102-final-viral-combined_k141_21556\|\|lt2gene | Low-quality |
| 18-102-final-viral-combined_k141_823\|\|lt2gene | Low-quality |
| 18-102-final-viral-combined_k141_156176\|\|lt2gene | Not-determined |
| 18-102-final-viral-combined_k141_145892\|\|lt2gene | Low-quality |
| 18-102-final-viral-combined_k141_228793\|\|lt2gene | Not-determined |
| 18-102-final-viral-combined_k141_93912\|\|lt2gene | Low-quality |
| 18-102-final-viral-combined_k141_228845\|\|lt2gene | Low-quality |
| 18-102-final-viral-combined_k141_93920\|\|lt2gene | Low-quality |
| 18-102-final-viral-combined_k141_83653\|\|lt2gene | Not-determined |
| 18-102-final-viral-combined_k141_73320\|\|lt2gene | Not-determined |
| 18-102-final-viral-combined_k141_166587\|\|lt2gene | Not-determined |
| 18-102-final-viral-combined_k141_31978\|\|lt2gene | Not-determined |
| 18-102-final-viral-combined_k141_114602\|\|lt2gene | Low-quality |
| 18-102-final-viral-combined_k141_42791\|\|lt2gene | Low-quality |
| 18-102-final-viral-combined_k141_177158\|\|lt2gene | Low-quality |
| 18-102-final-viral-combined_k141_21749\|\|lt2gene | Not-determined |
| 18-102-final-viral-combined_k141_21758\|\|lt2gene | Low-quality |
| 18-102-final-viral-combined_k141_197342\|\|lt2gene | Low-quality |
| 18-102-final-viral-combined_k141_166603\|\|lt2gene | Low-quality |
| 18-102-final-viral-combined_k141_166606\|\|lt2gene | Low-quality |
| 18-102-final-viral-combined_k141_62829\|\|lt2gene | Low-quality |
| 18-102-final-viral-combined_k141_62836\|\|lt2gene | Low-quality |
| 18-102-final-viral-combined_k141_208047\|\|lt2gene | Not-determined |
| 18-102-final-viral-combined_k141_114661\|\|lt2gene | Not-determined |
| 18-102-final-viral-combined_k141_208089\|\|lt2gene | Low-quality |
| 18-102-final-viral-combined_k141_73455\|\|lt2gene | Low-quality |
| 18-102-final-viral-combined_k141_997\|\|lt2gene | Low-quality |
| 18-102-final-viral-combined_k141_1004\|\|lt2gene | Not-determined |
| 18-102-final-viral-combined_k141_83871\|\|lt2gene | Not-determined |
| 18-102-final-viral-combined_k141_62857\|\|lt2gene | Low-quality |
| 18-102-final-viral-combined_k141_73510\|\|lt2gene | Low-quality |
| 18-102-final-viral-combined_k141_11456\|\|lt2gene | Not-determined |
| 18-102-final-viral-combined_k141_239096\|\|lt2gene | Low-quality |
| 18-102-final-viral-combined_k141_32109\|\|lt2gene | Low-quality |
| 18-102-final-viral-combined_k141_1056\|\|lt2gene | Low-quality |
| 18-102-final-viral-combined_k141_11505\|\|lt2gene | Not-determined |
| 18-102-final-viral-combined_k141_166797\|\|lt2gene | Low-quality |
| 18-102-final-viral-combined_k141_43047\|\|lt2gene | Low-quality |
| 18-102-final-viral-combined_k141_52740\|\|lt2gene | Low-quality |
| 18-102-final-viral-combined_k141_94153\|\|lt2gene | Low-quality |
| 18-102-final-viral-combined_k141_177392\|\|lt2gene | Low-quality |
| 18-102-final-viral-combined_k141_187400\|\|lt2gene | Not-determined |
| 18-102-final-viral-combined_k141_11579\|\|lt2gene | Low-quality |
| 18-102-final-viral-combined_k141_104705\|\|lt2gene | Low-quality |
| 18-102-final-viral-combined_k141_52768\|\|lt2gene | Low-quality |
| 18-102-final-viral-combined_k141_1125\|\|lt2gene | Low-quality |
| 18-102-final-viral-combined_k141_84042\|\|lt2gene | Low-quality |
| 18-102-final-viral-combined_k141_104748\|\|lt2gene | Low-quality |
| 18-102-final-viral-combined_k141_239179\|\|lt2gene | Low-quality |
| 18-102-final-viral-combined_k141_52815\|\|lt2gene | Low-quality |
| 18-102-final-viral-combined_k141_84092\|\|lt2gene | Low-quality |
| 18-102-final-viral-combined_k141_43176\|\|lt2gene | Low-quality |
| 18-102-final-viral-combined_k141_22009\|\|lt2gene | Low-quality |
| 18-102-final-viral-combined_k141_166962\|\|lt2gene | Not-determined |
| 18-102-final-viral-combined_k141_1219\|\|lt2gene | Low-quality |
| 18-102-final-viral-combined_k141_177521\|\|lt2gene | Not-determined |
| 18-102-final-viral-combined_k141_94294\|\|lt2gene | Low-quality |
| 18-102-final-viral-combined_k141_208282\|\|lt2gene | Not-determined |
| 18-102-final-viral-combined_k141_125521\|\|lt2gene | Not-determined |
| 18-102-final-viral-combined_k141_43283\|\|lt2gene | Not-determined |
| 18-102-final-viral-combined_k141_187542\|\|lt2gene | Not-determined |
| 18-102-final-viral-combined_k141_197638\|\|lt2gene | Low-quality |
| 18-102-final-viral-combined_k141_135787\|\|lt2gene | Low-quality |
| 18-102-final-viral-combined_k141_146428\|\|lt2gene | Not-determined |
| 18-102-final-viral-combined_k141_239377\|\|lt2gene | Not-determined |
| 18-102-final-viral-combined_k141_239381\|\|lt2gene | Low-quality |
| 18-102-final-viral-combined_k141_229297\|\|lt2gene | Low-quality |
| 18-102-final-viral-combined_k141_104867\|\|lt2gene | Low-quality |
| 18-102-final-viral-combined_k141_94422\|\|lt2gene | Low-quality |
| 18-102-final-viral-combined_k141_135821\|\|lt2gene | Low-quality |
| 18-102-final-viral-combined_k141_11770\|\|lt2gene | Low-quality |
| 18-102-final-viral-combined_k141_11777\|\|lt2gene | Low-quality |
| 18-102-final-viral-combined_k141_63076\|\|lt2gene | Low-quality |
| 18-102-final-viral-combined_k141_177659\|\|lt2gene | Low-quality |
| 18-102-final-viral-combined_k141_11849\|\|lt2gene | Low-quality |
| 18-102-final-viral-combined_k141_187606\|\|lt2gene | Low-quality |
| 18-102-final-viral-combined_k141_177677\|\|lt2gene | Low-quality |
| 18-102-final-viral-combined_k141_208406\|\|lt2gene | Low-quality |
| 18-102-final-viral-combined_k141_197723\|\|lt2gene | Not-determined |
| 18-102-final-viral-combined_k141_22153\|\|lt2gene | Not-determined |
| 18-102-final-viral-combined_k141_167286\|\|lt2gene | Low-quality |
| 18-102-final-viral-combined_k141_94531\|\|lt2gene | Low-quality |
| 18-102-final-viral-combined_k141_63174\|\|lt2gene | Not-determined |
| 18-102-final-viral-combined_k141_187689\|\|lt2gene | Low-quality |
| 18-102-final-viral-combined_k141_73959\|\|lt2gene | Not-determined |
| 18-102-final-viral-combined_k141_22194\|\|lt2gene | Low-quality |
| 18-102-final-viral-combined_k141_94562\|\|lt2gene | Low-quality |
| 18-102-final-viral-combined_k141_104938\|\|lt2gene | Low-quality |
| 18-102-final-viral-combined_k141_94579\|\|lt2gene | Low-quality |
| 18-102-final-viral-combined_k141_32286\|\|lt2gene | Low-quality |
| 18-102-final-viral-combined_k141_94583\|\|lt2gene | Low-quality |
| 18-102-final-viral-combined_k141_104954\|\|lt2gene | Not-determined |
| 18-102-final-viral-combined_k141_218704\|\|lt2gene | Low-quality |
| 18-102-final-viral-combined_k141_229406\|\|lt2gene | Low-quality |
| 18-102-final-viral-combined_k141_94624\|\|lt2gene | Low-quality |
| 18-102-final-viral-combined_k141_94634\|\|lt2gene | Low-quality |
| 18-102-final-viral-combined_k141_43598\|\|lt2gene | Not-determined |
| 18-102-final-viral-combined_k141_135976\|\|lt2gene | Not-determined |
| 18-102-final-viral-combined_k141_53041\|\|lt2gene | Low-quality |
| 18-102-final-viral-combined_k141_43639\|\|lt2gene | Low-quality |
| 18-102-final-viral-combined_k141_94689\|\|lt2gene | Not-determined |
| 18-102-final-viral-combined_k141_229503\|\|lt2gene | Low-quality |
| 18-102-final-viral-combined_k141_53100\|\|lt2gene | Low-quality |
| 18-102-final-viral-combined_k141_197863\|\|lt2gene | Not-determined |
| 18-102-final-viral-combined_k141_53167\|\|lt2gene | Low-quality |
| 18-102-final-viral-combined_k141_146690\|\|lt2gene | Not-determined |
| 18-102-final-viral-combined_k141_156768\|\|lt2gene | Low-quality |
| 18-102-final-viral-combined_k141_1497\|\|lt2gene | Low-quality |
| 18-102-final-viral-combined_k141_94769\|\|lt2gene | Low-quality |
| 18-102-final-viral-combined_k141_105106\|\|lt2gene | Not-determined |
| 18-102-final-viral-combined_k141_115259\|\|lt2gene | Low-quality |
| 18-102-final-viral-combined_k141_105108\|\|lt2gene | Not-determined |
| 18-102-final-viral-combined_k141_197925\|\|lt2gene | Low-quality |
| 18-102-final-viral-combined_k141_43788\|\|lt2gene | Not-determined |
| 18-102-final-viral-combined_k141_105130\|\|lt2gene | Low-quality |
| 18-102-final-viral-combined_k141_105141\|\|lt2gene | Low-quality |
| 18-102-final-viral-combined_k141_22460\|\|lt2gene | Low-quality |
| 18-102-final-viral-combined_k141_136223\|\|lt2gene | Low-quality |
| 18-102-final-viral-combined_k141_218941\|\|lt2gene | Low-quality |
| 18-102-final-viral-combined_k141_188029\|\|lt2gene | Low-quality |
| 18-102-final-viral-combined_k141_84613\|\|lt2gene | Low-quality |
| 18-102-final-viral-combined_k141_43972\|\|lt2gene | Low-quality |
| 18-102-final-viral-combined_k141_136270\|\|lt2gene | Low-quality |
| 18-102-final-viral-combined_k141_136276\|\|lt2gene | Not-determined |
| 18-102-final-viral-combined_k141_239729\|\|lt2gene | Low-quality |
| 18-102-final-viral-combined_k141_105211\|\|lt2gene | Low-quality |
| 18-102-final-viral-combined_k141_53326\|\|lt2gene | Low-quality |
| 18-102-final-viral-combined_k141_197978\|\|lt2gene | Not-determined |
| 18-102-final-viral-combined_k141_178066\|\|lt2gene | Low-quality |
| 18-102-final-viral-combined_k141_115427\|\|lt2gene | Low-quality |
| 18-102-final-viral-combined_k141_178133\|\|lt2gene | Low-quality |
| 18-102-final-viral-combined_k141_178153\|\|lt2gene | Low-quality |
| 18-102-final-viral-combined_k141_12118\|\|lt2gene | Low-quality |
| 18-102-final-viral-combined_k141_105373\|\|lt2gene | Low-quality |
| 18-102-final-viral-combined_k141_198067\|\|lt2gene | Low-quality |
| 18-102-final-viral-combined_k141_115525\|\|lt2gene | Low-quality |
| 18-102-final-viral-combined_k141_115541\|\|lt2gene | Low-quality |
| 18-102-final-viral-combined_k141_136463\|\|lt2gene | Low-quality |
| 18-102-final-viral-combined_k141_198096\|\|lt2gene | Low-quality |
| 18-102-final-viral-combined_k141_105431\|\|lt2gene | Low-quality |
| 18-102-final-viral-combined_k141_178296\|\|lt2gene | Not-determined |
| 18-102-final-viral-combined_k141_126055\|\|lt2gene | Not-determined |
| 18-102-final-viral-combined_k141_115607\|\|lt2gene | Low-quality |
| 18-102-final-viral-combined_k141_126075\|\|lt2gene | Not-determined |
| 18-102-final-viral-combined_k141_95056\|\|lt2gene | Low-quality |
| 18-102-final-viral-combined_k141_239892\|\|lt2gene | Low-quality |
| 18-102-final-viral-combined_k141_22639\|\|lt2gene | Low-quality |
| 18-102-final-viral-combined_k141_115653\|\|lt2gene | Low-quality |
| 18-102-final-viral-combined_k141_115664\|\|lt2gene | Not-determined |
| 18-102-final-viral-combined_k141_188177\|\|lt2gene | Low-quality |
| 18-102-final-viral-combined_k141_198145\|\|lt2gene | Not-determined |
| 18-102-final-viral-combined_k141_105462\|\|lt2gene | Low-quality |
| 18-102-final-viral-combined_k141_105476\|\|lt2gene | Low-quality |
| 18-102-final-viral-combined_k141_95080\|\|lt2gene | Low-quality |
| 18-102-final-viral-combined_k141_208832\|\|lt2gene | Low-quality |
| 18-102-final-viral-combined_k141_32691\|\|lt2gene | Low-quality |
| 18-102-final-viral-combined_k141_22731\|\|lt2gene | Low-quality |
| 18-102-final-viral-combined_k141_53634\|\|lt2gene | Low-quality |
| 18-102-final-viral-combined_k141_53652\|\|lt2gene | Low-quality |
| 18-102-final-viral-combined_k141_84876\|\|lt2gene | Low-quality |
| 18-102-final-viral-combined_k141_136613\|\|lt2gene | Not-determined |
| 18-102-final-viral-combined_k141_146906\|\|lt2gene | Low-quality |
| 18-102-final-viral-combined_k141_44342\|\|lt2gene | Low-quality |
| 18-102-final-viral-combined_k141_84891\|\|lt2gene | Low-quality |
| 18-102-final-viral-combined_k141_105621\|\|lt2gene | Low-quality |
| 18-102-final-viral-combined_k141_2014\|\|lt2gene | Low-quality |
| 18-102-final-viral-combined_k141_136673\|\|lt2gene | Low-quality |
| 18-102-final-viral-combined_k141_64002\|\|lt2gene | Low-quality |
| 18-102-final-viral-combined_k141_209010\|\|lt2gene | Low-quality |
| 18-102-final-viral-combined_k141_136713\|\|lt2gene | Low-quality |
| 18-102-final-viral-combined_k141_188338\|\|lt2gene | Not-determined |
| 18-102-final-viral-combined_k141_188378\|\|lt2gene | Low-quality |
| 18-102-final-viral-combined_k141_188379\|\|lt2gene | Not-determined |
| 18-102-final-viral-combined_k141_64059\|\|lt2gene | Low-quality |
| 18-102-final-viral-combined_k141_230068\|\|lt2gene | Low-quality |
| 18-102-final-viral-combined_k141_64072\|\|lt2gene | Low-quality |
| 18-102-final-viral-combined_k141_74408\|\|lt2gene | Not-determined |
| 18-102-final-viral-combined_k141_167950\|\|lt2gene | Not-determined |
| 18-102-final-viral-combined_k141_167952\|\|lt2gene | Low-quality |
| 18-102-final-viral-combined_k141_115830\|\|lt2gene | Low-quality |
| 18-102-final-viral-combined_k141_105724\|\|lt2gene | Low-quality |
| 18-102-final-viral-combined_k141_115833\|\|lt2gene | Low-quality |
| 18-102-final-viral-combined_k141_167991\|\|lt2gene | Not-determined |
| 18-102-final-viral-combined_k141_230122\|\|lt2gene | Low-quality |
| 18-102-final-viral-combined_k141_105778\|\|lt2gene | Low-quality |
| 18-102-final-viral-combined_k141_146951\|\|lt2gene | Not-determined |
| 18-102-final-viral-combined_k141_230169\|\|lt2gene | Low-quality |
| 18-102-final-viral-combined_k141_64204\|\|lt2gene | Not-determined |
| 18-102-final-viral-combined_k141_115860\|\|lt2gene | Low-quality |
| 18-102-final-viral-combined_k141_44584\|\|lt2gene | Not-determined |
| 18-102-final-viral-combined_k141_95443\|\|lt2gene | Low-quality |
| 18-102-final-viral-combined_k141_126334\|\|lt2gene | Low-quality |
| 18-102-final-viral-combined_k141_157430\|\|lt2gene | Not-determined |
| 18-102-final-viral-combined_k141_64262\|\|lt2gene | Low-quality |
| 18-102-final-viral-combined_k141_240182\|\|lt2gene | Low-quality |
| 18-102-final-viral-combined_k141_126401\|\|lt2gene | Low-quality |
| 18-102-final-viral-combined_k141_32841\|\|lt2gene | Low-quality |
| 18-102-final-viral-combined_k141_105846\|\|lt2gene | Low-quality |
| 18-102-final-viral-combined_k141_74516\|\|lt2gene | Low-quality |
| 18-102-final-viral-combined_k141_95515\|\|lt2gene | Not-determined |
| 18-102-final-viral-combined_k141_95530\|\|lt2gene | Low-quality |
| 18-102-final-viral-combined_k141_147055\|\|lt2gene | Low-quality |
| 18-102-final-viral-combined_k141_64281\|\|lt2gene | Low-quality |
| 18-102-final-viral-combined_k141_115921\|\|lt2gene | Not-determined |
| 18-102-final-viral-combined_k141_105942\|\|lt2gene | Low-quality |
| 18-102-final-viral-combined_k141_64319\|\|lt2gene | Low-quality |
| 18-102-final-viral-combined_k141_95573\|\|lt2gene | Not-determined |
| 18-102-final-viral-combined_k141_115957\|\|lt2gene | Low-quality |
| 18-102-final-viral-combined_k141_137094\|\|lt2gene | Low-quality |
| 18-102-final-viral-combined_k141_198435\|\|lt2gene | Not-determined |
| 18-102-final-viral-combined_k141_33045\|\|lt2gene | Low-quality |
| 18-102-final-viral-combined_k141_74582\|\|lt2gene | Low-quality |
| 18-102-final-viral-combined_k141_230432\|\|lt2gene | Low-quality |
| 18-102-final-viral-combined_k141_95623\|\|lt2gene | Not-determined |
| 18-102-final-viral-combined_k141_85307\|\|lt2gene | Not-determined |
| 18-102-final-viral-combined_k141_240311\|\|lt2gene | Not-determined |
| 18-102-final-viral-combined_k141_219416\|\|lt2gene | Low-quality |
| 18-102-final-viral-combined_k141_168320\|\|lt2gene | Low-quality |
| 18-102-final-viral-combined_k141_219435\|\|lt2gene | Low-quality |
| 18-102-final-viral-combined_k141_188704\|\|lt2gene | Low-quality |
| 18-102-final-viral-combined_k141_198487\|\|lt2gene | Not-determined |
| 18-102-final-viral-combined_k141_44846\|\|lt2gene | Low-quality |
| 18-102-final-viral-combined_k141_230539\|\|lt2gene | Low-quality |
| 18-102-final-viral-combined_k141_2263\|\|lt2gene | Not-determined |
| 18-102-final-viral-combined_k141_230543\|\|lt2gene | Low-quality |
| 18-102-final-viral-combined_k141_168369\|\|lt2gene | Not-determined |
| 18-102-final-viral-combined_k141_12601\|\|lt2gene | Not-determined |
| 18-102-final-viral-combined_k141_12618\|\|lt2gene | Low-quality |
| 18-102-final-viral-combined_k141_209302\|\|lt2gene | Low-quality |
| 18-102-final-viral-combined_k141_44938\|\|lt2gene | Not-determined |
| 18-102-final-viral-combined_k141_178671\|\|lt2gene | Low-quality |
| 18-102-final-viral-combined_k141_209305\|\|lt2gene | Not-determined |
| 18-102-final-viral-combined_k141_23169\|\|lt2gene | Low-quality |
| 18-102-final-viral-combined_k141_23202\|\|lt2gene | Low-quality |
| 18-102-final-viral-combined_k141_45039\|\|lt2gene | Low-quality |
| 18-102-final-viral-combined_k141_116164\|\|lt2gene | Low-quality |
| 18-102-final-viral-combined_k141_168530\|\|lt2gene | Low-quality |
| 18-102-final-viral-combined_k141_45051\|\|lt2gene | Low-quality |
| 18-102-final-viral-combined_k141_45068\|\|lt2gene | Low-quality |
| 18-102-final-viral-combined_k141_33223\|\|lt2gene | Low-quality |
| 18-102-final-viral-combined_k141_106134\|\|lt2gene | Not-determined |
| 18-102-final-viral-combined_k141_147290\|\|lt2gene | Not-determined |
| 18-102-final-viral-combined_k141_137239\|\|lt2gene | Low-quality |
| 18-102-final-viral-combined_k141_85534\|\|lt2gene | Low-quality |
| 18-102-final-viral-combined_k141_126714\|\|lt2gene | Low-quality |
| 18-102-final-viral-combined_k141_126718\|\|lt2gene | Low-quality |
| 18-102-final-viral-combined_k141_33288\|\|lt2gene | Not-determined |
| 18-102-final-viral-combined_k141_54459\|\|lt2gene | Low-quality |
| 18-102-final-viral-combined_k141_126752\|\|lt2gene | Low-quality |
| 18-102-final-viral-combined_k141_137279\|\|lt2gene | Not-determined |
| 18-102-final-viral-combined_k141_219786\|\|lt2gene | Low-quality |
| 18-102-final-viral-combined_k141_54510\|\|lt2gene | Not-determined |
| 18-102-final-viral-combined_k141_2433\|\|lt2gene | Low-quality |
| 18-102-final-viral-combined_k141_54558\|\|lt2gene | Low-quality |
| 18-102-final-viral-combined_k141_189029\|\|lt2gene | Low-quality |
| 18-102-final-viral-combined_k141_168624\|\|lt2gene | Not-determined |
| 18-102-final-viral-combined_k141_64601\|\|lt2gene | Low-quality |
| 18-102-final-viral-combined_k141_189064\|\|lt2gene | Low-quality |
| 18-102-final-viral-combined_k141_23455\|\|lt2gene | Not-determined |
| 18-102-final-viral-combined_k141_126857\|\|lt2gene | Low-quality |
| 18-102-final-viral-combined_k141_64629\|\|lt2gene | Low-quality |
| 18-102-final-viral-combined_k141_96046\|\|lt2gene | Low-quality |
| 18-102-final-viral-combined_k141_96053\|\|lt2gene | Low-quality |
| 18-102-final-viral-combined_k141_189136\|\|lt2gene | Low-quality |
| 18-102-final-viral-combined_k141_106240\|\|lt2gene | Low-quality |
| 18-102-final-viral-combined_k141_64667\|\|lt2gene | Low-quality |
| 18-102-final-viral-combined_k141_2562\|\|lt2gene | Low-quality |
| 18-102-final-viral-combined_k141_147513\|\|lt2gene | Not-determined |
| 18-102-final-viral-combined_k141_116350\|\|lt2gene | Low-quality |
| 18-102-final-viral-combined_k141_126985\|\|lt2gene | Low-quality |
| 18-102-final-viral-combined_k141_127001\|\|lt2gene | Low-quality |
| 18-102-final-viral-combined_k141_168794\|\|lt2gene | Low-quality |
| 18-102-final-viral-combined_k141_168795\|\|lt2gene | Low-quality |
| 18-102-final-viral-combined_k141_158050\|\|lt2gene | Not-determined |
| 18-102-final-viral-combined_k141_33682\|\|lt2gene | Low-quality |
| 18-102-final-viral-combined_k141_240674\|\|lt2gene | Low-quality |
| 18-102-final-viral-combined_k141_13067\|\|lt2gene | Not-determined |
| 18-102-final-viral-combined_k141_147601\|\|lt2gene | Low-quality |
| 18-102-final-viral-combined_k141_147608\|\|lt2gene | Low-quality |
| 18-102-final-viral-combined_k141_240715\|\|lt2gene | Low-quality |
| 18-102-final-viral-combined_k141_116391\|\|lt2gene | Not-determined |
| 18-102-final-viral-combined_k141_179198\|\|lt2gene | Not-determined |
| 18-102-final-viral-combined_k141_85951\|\|lt2gene | Not-determined |
| 18-102-final-viral-combined_k141_147689\|\|lt2gene | Not-determined |
| 18-102-final-viral-combined_k141_158143\|\|lt2gene | Not-determined |
| 18-102-final-viral-combined_k141_2658\|\|lt2gene | Not-determined |
| 18-102-final-viral-combined_k141_168978\|\|lt2gene | Low-quality |
| 18-102-final-viral-combined_k141_230969\|\|lt2gene | Low-quality |
| 18-102-final-viral-combined_k141_198916\|\|lt2gene | Low-quality |
| 18-102-final-viral-combined_k141_127093\|\|lt2gene | Low-quality |
| 18-102-final-viral-combined_k141_45580\|\|lt2gene | Low-quality |
| 18-102-final-viral-combined_k141_86040\|\|lt2gene | Low-quality |
| 18-102-final-viral-combined_k141_96408\|\|lt2gene | Low-quality |
| 18-102-final-viral-combined_k141_96414\|\|lt2gene | Low-quality |
| 18-102-final-viral-combined_k141_96420\|\|lt2gene | Low-quality |
| 18-102-final-viral-combined_k141_158162\|\|lt2gene | Low-quality |
| 18-102-final-viral-combined_k141_127172\|\|lt2gene | Low-quality |
| 18-102-final-viral-combined_k141_158212\|\|lt2gene | Not-determined |
| 18-102-final-viral-combined_k141_198979\|\|lt2gene | Low-quality |
| 18-102-final-viral-combined_k141_147776\|\|lt2gene | Not-determined |
| 18-102-final-viral-combined_k141_169024\|\|lt2gene | Low-quality |
| 18-102-final-viral-combined_k141_189510\|\|lt2gene | Not-determined |
| 18-102-final-viral-combined_k141_106470\|\|lt2gene | Low-quality |
| 18-102-final-viral-combined_k141_179439\|\|lt2gene | Low-quality |
| 18-102-final-viral-combined_k141_169057\|\|lt2gene | Not-determined |
| 18-102-final-viral-combined_k141_169072\|\|lt2gene | Low-quality |
| 18-102-final-viral-combined_k141_231091\|\|lt2gene | Low-quality |
| 18-102-final-viral-combined_k141_169088\|\|lt2gene | Not-determined |
| 18-102-final-viral-combined_k141_45705\|\|lt2gene | Low-quality |
| 18-102-final-viral-combined_k141_34017\|\|lt2gene | Low-quality |
| 18-102-final-viral-combined_k141_127322\|\|lt2gene | Low-quality |
| 18-102-final-viral-combined_k141_106555\|\|lt2gene | Low-quality |
| 18-102-final-viral-combined_k141_137836\|\|lt2gene | Low-quality |
| 18-102-final-viral-combined_k141_231124\|\|lt2gene | Low-quality |
| 18-102-final-viral-combined_k141_158310\|\|lt2gene | Low-quality |
| 18-102-final-viral-combined_k141_158346\|\|lt2gene | Low-quality |
| 18-102-final-viral-combined_k141_219962\|\|lt2gene | Low-quality |
| 18-102-final-viral-combined_k141_34100\|\|lt2gene | Not-determined |
| 18-102-final-viral-combined_k141_2900\|\|lt2gene | Low-quality |
| 18-102-final-viral-combined_k141_64925\|\|lt2gene | Low-quality |
| 18-102-final-viral-combined_k141_23720\|\|lt2gene | Low-quality |
| 18-102-final-viral-combined_k141_189765\|\|lt2gene | Low-quality |
| 18-102-final-viral-combined_k141_199144\|\|lt2gene | Not-determined |
| 18-102-final-viral-combined_k141_209991\|\|lt2gene | Not-determined |
| 18-102-final-viral-combined_k141_179525\|\|lt2gene | Low-quality |
| 18-102-final-viral-combined_k141_147917\|\|lt2gene | Low-quality |
| 18-102-final-viral-combined_k141_34130\|\|lt2gene | Not-determined |
| 18-102-final-viral-combined_k141_179545\|\|lt2gene | Not-determined |
| 18-102-final-viral-combined_k141_179547\|\|lt2gene | Low-quality |
| 18-102-final-viral-combined_k141_231245\|\|lt2gene | Low-quality |
| 18-102-final-viral-combined_k141_34137\|\|lt2gene | Low-quality |
| 18-102-final-viral-combined_k141_189853\|\|lt2gene | Not-determined |
| 18-102-final-viral-combined_k141_23779\|\|lt2gene | Not-determined |
| 18-102-final-viral-combined_k141_179597\|\|lt2gene | Low-quality |
| 18-102-final-viral-combined_k141_148013\|\|lt2gene | Low-quality |
| 18-102-final-viral-combined_k141_34154\|\|lt2gene | Low-quality |
| 18-102-final-viral-combined_k141_148029\|\|lt2gene | Low-quality |
| 18-102-final-viral-combined_k141_75297\|\|lt2gene | Low-quality |
| 18-102-final-viral-combined_k141_127550\|\|lt2gene | Low-quality |
| 18-102-final-viral-combined_k141_169396\|\|lt2gene | Low-quality |
| 18-102-final-viral-combined_k141_169399\|\|lt2gene | Not-determined |
| 18-102-final-viral-combined_k141_86264\|\|lt2gene | Low-quality |
| 18-102-final-viral-combined_k141_34166\|\|lt2gene | Low-quality |
| 18-102-final-viral-combined_k141_34177\|\|lt2gene | Low-quality |
| 18-102-final-viral-combined_k141_179638\|\|lt2gene | Low-quality |
| 18-102-final-viral-combined_k141_106783\|\|lt2gene | Low-quality |
| 18-102-final-viral-combined_k141_106809\|\|lt2gene | Low-quality |
| 18-102-final-viral-combined_k141_34233\|\|lt2gene | Low-quality |
| 18-102-final-viral-combined_k141_158442\|\|lt2gene | Not-determined |
| 18-102-final-viral-combined_k141_169441\|\|lt2gene | Not-determined |
| 18-102-final-viral-combined_k141_179720\|\|lt2gene | Not-determined |
| 18-102-final-viral-combined_k141_55034\|\|lt2gene | Low-quality |
| 18-102-final-viral-combined_k141_210202\|\|lt2gene | Not-determined |
| 18-102-final-viral-combined_k141_55056\|\|lt2gene | Low-quality |
| 18-102-final-viral-combined_k141_23840\|\|lt2gene | Low-quality |
| 18-102-final-viral-combined_k141_13367\|\|lt2gene | Not-determined |
| 18-102-final-viral-combined_k141_65290\|\|lt2gene | Not-determined |
| 18-102-final-viral-combined_k141_158465\|\|lt2gene | Not-determined |
| 18-102-final-viral-combined_k141_231399\|\|lt2gene | Not-determined |
| 18-102-final-viral-combined_k141_158475\|\|lt2gene | Not-determined |
| 18-102-final-viral-combined_k141_65310\|\|lt2gene | Not-determined |
| 18-102-final-viral-combined_k141_3124\|\|lt2gene | Low-quality |
| 18-102-final-viral-combined_k141_65318\|\|lt2gene | Low-quality |
| 18-102-final-viral-combined_k141_158494\|\|lt2gene | Not-determined |
| 18-102-final-viral-combined_k141_199291\|\|lt2gene | Low-quality |

| | |
|---|---|
| 18-102-final-viral-combined_k141_86365\|\|lt2gene | Low-quality |
| 18-102-final-viral-combined_k141_138158\|\|lt2gene | Low-quality |
| 18-102-final-viral-combined_k141_65363\|\|lt2gene | Low-quality |
| 18-102-final-viral-combined_k141_116857\|\|lt2gene | Low-quality |
| 18-102-final-viral-combined_k141_23930\|\|lt2gene | Not-determined |
| 18-102-final-viral-combined_k141_34368\|\|lt2gene | Low-quality |
| 18-102-final-viral-combined_k141_210323\|\|lt2gene | Low-quality |
| 18-102-final-viral-combined_k141_75445\|\|lt2gene | Low-quality |
| 18-102-final-viral-combined_k141_46036\|\|lt2gene | Not-determined |
| 18-102-final-viral-combined_k141_199323\|\|lt2gene | Low-quality |
| 18-102-final-viral-combined_k141_34420\|\|lt2gene | Low-quality |
| 18-102-final-viral-combined_k141_23963\|\|lt2gene | Low-quality |
| 18-102-final-viral-combined_k141_158567\|\|lt2gene | Low-quality |
| 18-102-final-viral-combined_k141_96767\|\|lt2gene | Low-quality |
| 18-102-final-viral-combined_k141_86454\|\|lt2gene | Low-quality |
| 18-102-final-viral-combined_k141_231597\|\|lt2gene | Low-quality |
| 18-102-final-viral-combined_k141_86485\|\|lt2gene | Low-quality |
| 18-102-final-viral-combined_k141_65498\|\|lt2gene | Not-determined |
| 18-102-final-viral-combined_k141_127679\|\|lt2gene | Low-quality |
| 18-102-final-viral-combined_k141_231628\|\|lt2gene | Low-quality |
| 18-102-final-viral-combined_k141_117106\|\|lt2gene | Low-quality |
| 18-102-final-viral-combined_k141_231637\|\|lt2gene | Low-quality |
| 18-102-final-viral-combined_k141_3322\|\|lt2gene | Low-quality |
| 18-102-final-viral-combined_k141_24011\|\|lt2gene | Not-determined |
| 18-102-final-viral-combined_k141_13601\|\|lt2gene | Low-quality |
| 18-102-final-viral-combined_k141_96876\|\|lt2gene | Low-quality |
| 18-102-final-viral-combined_k141_190095\|\|lt2gene | Low-quality |
| 18-102-final-viral-combined_k141_190115\|\|lt2gene | Low-quality |
| 18-102-final-viral-combined_k141_220193\|\|lt2gene | Low-quality |
| 18-102-final-viral-combined_k141_158708\|\|lt2gene | Low-quality |
| 18-102-final-viral-combined_k141_241259\|\|lt2gene | Low-quality |
| 18-102-final-viral-combined_k141_241261\|\|lt2gene | Low-quality |
| 18-102-final-viral-combined_k141_46307\|\|lt2gene | Not-determined |
| 18-102-final-viral-combined_k141_46321\|\|lt2gene | Low-quality |
| 18-102-final-viral-combined_k141_13764\|\|lt2gene | Not-determined |
| 18-102-final-viral-combined_k141_97045\|\|lt2gene | Low-quality |
| 18-102-final-viral-combined_k141_180126\|\|lt2gene | Not-determined |
| 18-102-final-viral-combined_k141_46361\|\|lt2gene | Low-quality |
| 18-102-final-viral-combined_k141_231831\|\|lt2gene | Low-quality |
| 18-102-final-viral-combined_k141_138365\|\|lt2gene | Low-quality |
| 18-102-final-viral-combined_k141_13802\|\|lt2gene | Low-quality |
| 18-102-final-viral-combined_k141_13819\|\|lt2gene | Low-quality |
| 18-102-final-viral-combined_k141_180167\|\|lt2gene | Low-quality |
| 18-102-final-viral-combined_k141_107193\|\|lt2gene | Not-determined |
| 18-102-final-viral-combined_k141_210587\|\|lt2gene | Low-quality |
| 18-102-final-viral-combined_k141_75818\|\|lt2gene | Low-quality |
| 18-102-final-viral-combined_k141_34691\|\|lt2gene | Not-determined |
| 18-102-final-viral-combined_k141_210631\|\|lt2gene | Not-determined |
| 18-102-final-viral-combined_k141_231917\|\|lt2gene | Low-quality |
| 18-102-final-viral-combined_k141_107282\|\|lt2gene | Low-quality |
| 18-102-final-viral-combined_k141_34744\|\|lt2gene | Low-quality |
| 18-102-final-viral-combined_k141_148660\|\|lt2gene | Low-quality |
| 18-102-final-viral-combined_k141_97164\|\|lt2gene | Low-quality |
| 18-102-final-viral-combined_k141_158809\|\|lt2gene | Low-quality |
| 18-102-final-viral-combined_k141_55480\|\|lt2gene | Not-determined |
| 18-102-final-viral-combined_k141_220274\|\|lt2gene | Low-quality |
| 18-102-final-viral-combined_k141_24273\|\|lt2gene | Low-quality |
| 18-102-final-viral-combined_k141_180316\|\|lt2gene | Low-quality |
| 18-102-final-viral-combined_k141_231983\|\|lt2gene | Low-quality |
| 18-102-final-viral-combined_k141_180333\|\|lt2gene | Low-quality |
| 18-102-final-viral-combined_k141_190411\|\|lt2gene | Low-quality |
| 18-102-final-viral-combined_k141_46532\|\|lt2gene | Low-quality |
| 18-102-final-viral-combined_k141_232004\|\|lt2gene | Low-quality |
| 18-102-final-viral-combined_k141_24291\|\|lt2gene | Low-quality |
| 18-102-final-viral-combined_k141_97233\|\|lt2gene | Not-determined |
| 18-102-final-viral-combined_k141_97236\|\|lt2gene | Low-quality |
| 18-102-final-viral-combined_k141_3663\|\|lt2gene | Low-quality |
| 18-102-final-viral-combined_k141_138548\|\|lt2gene | Not-determined |
| 18-102-final-viral-combined_k141_13999\|\|lt2gene | Low-quality |
| 18-102-final-viral-combined_k141_180386\|\|lt2gene | Low-quality |
| 18-102-final-viral-combined_k141_55605\|\|lt2gene | Low-quality |
| 18-102-final-viral-combined_k141_75940\|\|lt2gene | Low-quality |
| 18-102-final-viral-combined_k141_3684\|\|lt2gene | Not-determined |
| 18-102-final-viral-combined_k141_138608\|\|lt2gene | Low-quality |
| 18-102-final-viral-combined_k141_34908\|\|lt2gene | Low-quality |
| 18-102-final-viral-combined_k141_86926\|\|lt2gene | Low-quality |
| 18-102-final-viral-combined_k141_127735\|\|lt2gene | Not-determined |
| 18-102-final-viral-combined_k141_180461\|\|lt2gene | Low-quality |
| 18-102-final-viral-combined_k141_55757\|\|lt2gene | Low-quality |
| 18-102-final-viral-combined_k141_199539\|\|lt2gene | Low-quality |
| 18-102-final-viral-combined_k141_46673\|\|lt2gene | Low-quality |
| 18-102-final-viral-combined_k141_55770\|\|lt2gene | Low-quality |
| 18-102-final-viral-combined_k141_34986\|\|lt2gene | Low-quality |
| 18-102-final-viral-combined_k141_232070\|\|lt2gene | Not-determined |
| 18-102-final-viral-combined_k141_46703\|\|lt2gene | Low-quality |
| 18-102-final-viral-combined_k141_232083\|\|lt2gene | Not-determined |
| 18-102-final-viral-combined_k141_14090\|\|lt2gene | Low-quality |
| 18-102-final-viral-combined_k141_24537\|\|lt2gene | Not-determined |
| 18-102-final-viral-combined_k141_180507\|\|lt2gene | Low-quality |
| 18-102-final-viral-combined_k141_14155\|\|lt2gene | Low-quality |
| 18-102-final-viral-combined_k141_87135\|\|lt2gene | Low-quality |
| 18-102-final-viral-combined_k141_14199\|\|lt2gene | Low-quality |
| 18-102-final-viral-combined_k141_14200\|\|lt2gene | Not-determined |
| 18-102-final-viral-combined_k141_127848\|\|lt2gene | Low-quality |
| 18-102-final-viral-combined_k141_14214\|\|lt2gene | Low-quality |
| 18-102-final-viral-combined_k141_117719\|\|lt2gene | Low-quality |
| 18-102-final-viral-combined_k141_220513\|\|lt2gene | Low-quality |
| 18-102-final-viral-combined_k141_180572\|\|lt2gene | Not-determined |
| 18-102-final-viral-combined_k141_76275\|\|lt2gene | Low-quality |
| 18-102-final-viral-combined_k141_97382\|\|lt2gene | Low-quality |
| 18-102-final-viral-combined_k141_14256\|\|lt2gene | Low-quality |
| 18-102-final-viral-combined_k141_76319\|\|lt2gene | Low-quality |
| 18-102-final-viral-combined_k141_46850\|\|lt2gene | Not-determined |
| 18-102-final-viral-combined_k141_107702\|\|lt2gene | Not-determined |
| 18-102-final-viral-combined_k141_3820\|\|lt2gene | Not-determined |
| 18-102-final-viral-combined_k141_35191\|\|lt2gene | Low-quality |
| 18-102-final-viral-combined_k141_76383\|\|lt2gene | Low-quality |
| 18-102-final-viral-combined_k141_138921\|\|lt2gene | Not-determined |
| 18-102-final-viral-combined_k141_97480\|\|lt2gene | Low-quality |
| 18-102-final-viral-combined_k141_87163\|\|lt2gene | Low-quality |
| 18-102-final-viral-combined_k141_117880\|\|lt2gene | Low-quality |
| 18-102-final-viral-combined_k141_220657\|\|lt2gene | Low-quality |
| 18-102-final-viral-combined_k141_76407\|\|lt2gene | Low-quality |
| 18-102-final-viral-combined_k141_76409\|\|lt2gene | Low-quality |
| 18-102-final-viral-combined_k141_76419\|\|lt2gene | Low-quality |
| 18-102-final-viral-combined_k141_220667\|\|lt2gene | Low-quality |
| 18-102-final-viral-combined_k141_66043\|\|lt2gene | Low-quality |
| 18-102-final-viral-combined_k141_14394\|\|lt2gene | Not-determined |
| 18-102-final-viral-combined_k141_138964\|\|lt2gene | Low-quality |
| 18-102-final-viral-combined_k141_14413\|\|lt2gene | Low-quality |
| 18-102-final-viral-combined_k141_180728\|\|lt2gene | Not-determined |
| 18-102-final-viral-combined_k141_24683\|\|lt2gene | Not-determined |
| 18-102-final-viral-combined_k141_24693\|\|lt2gene | Low-quality |
| 18-102-final-viral-combined_k141_56058\|\|lt2gene | Low-quality |
| 18-102-final-viral-combined_k141_56068\|\|lt2gene | Low-quality |
| 18-102-final-viral-combined_k141_3844\|\|lt2gene | Not-determined |
| 18-102-final-viral-combined_k141_232432\|\|lt2gene | Low-quality |
| 18-102-final-viral-combined_k141_117929\|\|lt2gene | Low-quality |
| 18-102-final-viral-combined_k141_170197\|\|lt2gene | Low-quality |
| 18-102-final-viral-combined_k141_66150\|\|lt2gene | Not-determined |
| 18-102-final-viral-combined_k141_128021\|\|lt2gene | Not-determined |
| 18-102-final-viral-combined_k141_87370\|\|lt2gene | Low-quality |
| 18-102-final-viral-combined_k141_66164\|\|lt2gene | Low-quality |
| 18-102-final-viral-combined_k141_170281\|\|lt2gene | Low-quality |
| 18-102-final-viral-combined_k141_180810\|\|lt2gene | Low-quality |
| 18-102-final-viral-combined_k141_87396\|\|lt2gene | Low-quality |
| 18-102-final-viral-combined_k141_220720\|\|lt2gene | Low-quality |
| 18-102-final-viral-combined_k141_139091\|\|lt2gene | Low-quality |
| 18-102-final-viral-combined_k141_56197\|\|lt2gene | Not-determined |
| 18-102-final-viral-combined_k141_97630\|\|lt2gene | Low-quality |
| 18-102-final-viral-combined_k141_232508\|\|lt2gene | Low-quality |
| 18-102-final-viral-combined_k141_118103\|\|lt2gene | Not-determined |
| 18-102-final-viral-combined_k141_56232\|\|lt2gene | Not-determined |
| 18-102-final-viral-combined_k141_211037\|\|lt2gene | Not-determined |
| 18-102-final-viral-combined_k141_232517\|\|lt2gene | Not-determined |
| 18-102-final-viral-combined_k141_118118\|\|lt2gene | Low-quality |
| 18-102-final-viral-combined_k141_180927\|\|lt2gene | Low-quality |
| 18-102-final-viral-combined_k141_220869\|\|lt2gene | Not-determined |
| 18-102-final-viral-combined_k141_76675\|\|lt2gene | Low-quality |
| 18-102-final-viral-combined_k141_56305\|\|lt2gene | Low-quality |
| 18-102-final-viral-combined_k141_76683\|\|lt2gene | Not-determined |
| 18-102-final-viral-combined_k141_56354\|\|lt2gene | Not-determined |
| 18-102-final-viral-combined_k141_56373\|\|lt2gene | Low-quality |
| 18-102-final-viral-combined_k141_118275\|\|lt2gene | Low-quality |
| 18-102-final-viral-combined_k141_56411\|\|lt2gene | Low-quality |
| 18-102-final-viral-combined_k141_118300\|\|lt2gene | Low-quality |
| 18-102-final-viral-combined_k141_56444\|\|lt2gene | Not-determined |
| 18-102-final-viral-combined_k141_159370\|\|lt2gene | Low-quality |
| 18-102-final-viral-combined_k141_35660\|\|lt2gene | Not-determined |
| 18-102-final-viral-combined_k141_97834\|\|lt2gene | Low-quality |
| 18-102-final-viral-combined_k141_4013\|\|lt2gene | Low-quality |
| 18-102-final-viral-combined_k141_97849\|\|lt2gene | Low-quality |
| 18-102-final-viral-combined_k141_128095\|\|lt2gene | Not-determined |
| 18-102-final-viral-combined_k141_170677\|\|lt2gene | Not-determined |
| 18-102-final-viral-combined_k141_108043\|\|lt2gene | Low-quality |
| 18-102-final-viral-combined_k141_149306\|\|lt2gene | Low-quality |
| 18-102-final-viral-combined_k141_118408\|\|lt2gene | Low-quality |
| 18-102-final-viral-combined_k141_170713\|\|lt2gene | Low-quality |
| 18-102-final-viral-combined_k141_14633\|\|lt2gene | Low-quality |
| 18-102-final-viral-combined_k141_211242\|\|lt2gene | Low-quality |
| 18-102-final-viral-combined_k141_108095\|\|lt2gene | Low-quality |
| 18-102-final-viral-combined_k141_108128\|\|lt2gene | Low-quality |
| 18-102-final-viral-combined_k141_211272\|\|lt2gene | Not-determined |
| 18-102-final-viral-combined_k141_170772\|\|lt2gene | Low-quality |
| 18-102-final-viral-combined_k141_170780\|\|lt2gene | Not-determined |
| 18-102-final-viral-combined_k141_181282\|\|lt2gene | Low-quality |
| 18-102-final-viral-combined_k141_66574\|\|lt2gene | Low-quality |
| 18-102-final-viral-combined_k141_76884\|\|lt2gene | Low-quality |
| 18-102-final-viral-combined_k141_221041\|\|lt2gene | Low-quality |
| 18-102-final-viral-combined_k141_128269\|\|lt2gene | Low-quality |
| 18-102-final-viral-combined_k141_76905\|\|lt2gene | Low-quality |
| 18-102-final-viral-combined_k141_108190\|\|lt2gene | Low-quality |
| 18-102-final-viral-combined_k141_232617\|\|lt2gene | Low-quality |
| 18-102-final-viral-combined_k141_4203\|\|lt2gene | Not-determined |
| 18-102-final-viral-combined_k141_118506\|\|lt2gene | Not-determined |
| 18-102-final-viral-combined_k141_35826\|\|lt2gene | Low-quality |
| 18-102-final-viral-combined_k141_4234\|\|lt2gene | Not-determined |
| 18-102-final-viral-combined_k141_76965\|\|lt2gene | Low-quality |
| 18-102-final-viral-combined_k141_76999\|\|lt2gene | Not-determined |
| 18-102-final-viral-combined_k141_35905\|\|lt2gene | Not-determined |
| 18-102-final-viral-combined_k141_139479\|\|lt2gene | Not-determined |
| 18-102-final-viral-combined_k141_25039\|\|lt2gene | Low-quality |
| 18-102-final-viral-combined_k141_77044\|\|lt2gene | Low-quality |
| 18-102-final-viral-combined_k141_66692\|\|lt2gene | Low-quality |
| 18-102-final-viral-combined_k141_170854\|\|lt2gene | Low-quality |
| 18-102-final-viral-combined_k141_170869\|\|lt2gene | Low-quality |
| 18-102-final-viral-combined_k141_87869\|\|lt2gene | Low-quality |
| 18-102-final-viral-combined_k141_98190\|\|lt2gene | Low-quality |
| 18-102-final-viral-combined_k141_56742\|\|lt2gene | Not-determined |
| 18-102-final-viral-combined_k141_14843\|\|lt2gene | Low-quality |
| 18-102-final-viral-combined_k141_4279\|\|lt2gene | Low-quality |
| 18-102-final-viral-combined_k141_4283\|\|lt2gene | Low-quality |
| 18-102-final-viral-combined_k141_4300\|\|lt2gene | Low-quality |
| 18-102-final-viral-combined_k141_159725\|\|lt2gene | Low-quality |
| 18-102-final-viral-combined_k141_98288\|\|lt2gene | Not-determined |
| 18-102-final-viral-combined_k141_159744\|\|lt2gene | Low-quality |
| 18-102-final-viral-combined_k141_241946\|\|lt2gene | Low-quality |
| 18-102-final-viral-combined_k141_221253\|\|lt2gene | Low-quality |
| 18-102-final-viral-combined_k141_139665\|\|lt2gene | Low-quality |
| 18-102-final-viral-combined_k141_139673\|\|lt2gene | Low-quality |
| 18-102-final-viral-combined_k141_77179\|\|lt2gene | Low-quality |
| 18-102-final-viral-combined_k141_139715\|\|lt2gene | Not-determined |
| 18-102-final-viral-combined_k141_211466\|\|lt2gene | Low-quality |
| 18-102-final-viral-combined_k141_241986\|\|lt2gene | Low-quality |
| 18-102-final-viral-combined_k141_25225\|\|lt2gene | Not-determined |
| 18-102-final-viral-combined_k141_4327\|\|lt2gene | Low-quality |
| 18-102-final-viral-combined_k141_242004\|\|lt2gene | Low-quality |
| 18-102-final-viral-combined_k141_221346\|\|lt2gene | Low-quality |
| 18-102-final-viral-combined_k141_77230\|\|lt2gene | Not-determined |
| 18-102-final-viral-combined_k141_181475\|\|lt2gene | Not-determined |
| 18-102-final-viral-combined_k141_242034\|\|lt2gene | Not-determined |
| 18-102-final-viral-combined_k141_4350\|\|lt2gene | Low-quality |
| 18-102-final-viral-combined_k141_232924\|\|lt2gene | Not-determined |
| 18-102-final-viral-combined_k141_221396\|\|lt2gene | Not-determined |
| 18-102-final-viral-combined_k141_56916\|\|lt2gene | Low-quality |
| 18-102-final-viral-combined_k141_221413\|\|lt2gene | Low-quality |
| 18-102-final-viral-combined_k141_159844\|\|lt2gene | Low-quality |
| 18-102-final-viral-combined_k141_181539\|\|lt2gene | Low-quality |
| 18-102-final-viral-combined_k141_139842\|\|lt2gene | Low-quality |
| 18-102-final-viral-combined_k141_108219\|\|lt2gene | Low-quality |
| 18-102-final-viral-combined_k141_232982\|\|lt2gene | Low-quality |
| 18-102-final-viral-combined_k141_139859\|\|lt2gene | Low-quality |
| 18-102-final-viral-combined_k141_98493\|\|lt2gene | Low-quality |
| 18-102-final-viral-combined_k141_108271\|\|lt2gene | Low-quality |
| 18-102-final-viral-combined_k141_108302\|\|lt2gene | Low-quality |
| 18-102-final-viral-combined_k141_36334\|\|lt2gene | Low-quality |
| 18-102-final-viral-combined_k141_88030\|\|lt2gene | Low-quality |
| 18-102-final-viral-combined_k141_118840\|\|lt2gene | Low-quality |
| 18-102-final-viral-combined_k141_66907\|\|lt2gene | Low-quality |
| 18-102-final-viral-combined_k141_159939\|\|lt2gene | Not-determined |
| 18-102-final-viral-combined_k141_15239\|\|lt2gene | Not-determined |
| 18-102-final-viral-combined_k141_171206\|\|lt2gene | Low-quality |
| 18-102-final-viral-combined_k141_159969\|\|lt2gene | Low-quality |
| 18-102-final-viral-combined_k141_242218\|\|lt2gene | Low-quality |
| 18-102-final-viral-combined_k141_48048\|\|lt2gene | Low-quality |
| 18-102-final-viral-combined_k141_48051\|\|lt2gene | Low-quality |
| 18-102-final-viral-combined_k141_57053\|\|lt2gene | Not-determined |
| 18-102-final-viral-combined_k141_15330\|\|lt2gene | Low-quality |
| 18-102-final-viral-combined_k141_171229\|\|lt2gene | Low-quality |
| 18-102-final-viral-combined_k141_77514\|\|lt2gene | Low-quality |
| 18-102-final-viral-combined_k141_15351\|\|lt2gene | Low-quality |
| 18-102-final-viral-combined_k141_57104\|\|lt2gene | Low-quality |
| 18-102-final-viral-combined_k141_48141\|\|lt2gene | Low-quality |
| 18-102-final-viral-combined_k141_25485\|\|lt2gene | Low-quality |
| 18-102-final-viral-combined_k141_221599\|\|lt2gene | Low-quality |
| 18-102-final-viral-combined_k141_200629\|\|lt2gene | Low-quality |
| 18-102-final-viral-combined_k141_15437\|\|lt2gene | Not-determined |
| 18-102-final-viral-combined_k141_36545\|\|lt2gene | Not-determined |
| 18-102-final-viral-combined_k141_36550\|\|lt2gene | Low-quality |
| 18-102-final-viral-combined_k141_67203\|\|lt2gene | Low-quality |
| 18-102-final-viral-combined_k141_211929\|\|lt2gene | Low-quality |
| 18-102-final-viral-combined_k141_171435\|\|lt2gene | Low-quality |
| 18-102-final-viral-combined_k141_211951\|\|lt2gene | Low-quality |
| 18-102-final-viral-combined_k141_233339\|\|lt2gene | Not-determined |
| 18-102-final-viral-combined_k141_119080\|\|lt2gene | Low-quality |
| 18-102-final-viral-combined_k141_119103\|\|lt2gene | Low-quality |
| 18-102-final-viral-combined_k141_200740\|\|lt2gene | Low-quality |
| 18-102-final-viral-combined_k141_181794\|\|lt2gene | Not-determined |
| 18-102-final-viral-combined_k141_88312\|\|lt2gene | Low-quality |
| 18-102-final-viral-combined_k141_15553\|\|lt2gene | Low-quality |
| 18-102-final-viral-combined_k141_98836\|\|lt2gene | Not-determined |
| 18-102-final-viral-combined_k141_4500\|\|lt2gene | Low-quality |
| 18-102-final-viral-combined_k141_150142\|\|lt2gene | Low-quality |
| 18-102-final-viral-combined_k141_242495\|\|lt2gene | Low-quality |
| 18-102-final-viral-combined_k141_98861\|\|lt2gene | Low-quality |
| 18-102-final-viral-combined_k141_36729\|\|lt2gene | Not-determined |
| 18-102-final-viral-combined_k141_200801\|\|lt2gene | Low-quality |
| 18-102-final-viral-combined_k141_160208\|\|lt2gene | Not-determined |
| 18-102-final-viral-combined_k141_36771\|\|lt2gene | Low-quality |
| 18-102-final-viral-combined_k141_119204\|\|lt2gene | Low-quality |
| 18-102-final-viral-combined_k141_57365\|\|lt2gene | Not-determined |
| 18-102-final-viral-combined_k141_200854\|\|lt2gene | Not-determined |
| 18-102-final-viral-combined_k141_36815\|\|lt2gene | Low-quality |
| 18-102-final-viral-combined_k141_160226\|\|lt2gene | Not-determined |
| 18-102-final-viral-combined_k141_242588\|\|lt2gene | Low-quality |
| 18-102-final-viral-combined_k141_171592\|\|lt2gene | Low-quality |
| 18-102-final-viral-combined_k141_128728\|\|lt2gene | Low-quality |
| 18-102-final-viral-combined_k141_119301\|\|lt2gene | Low-quality |
| 18-102-final-viral-combined_k141_88453\|\|lt2gene | Not-determined |
| 18-102-final-viral-combined_k141_212095\|\|lt2gene | Low-quality |
| 18-102-final-viral-combined_k141_57445\|\|lt2gene | Low-quality |
| 18-102-final-viral-combined_k141_108514\|\|lt2gene | Low-quality |
| 18-102-final-viral-combined_k141_181867\|\|lt2gene | Not-determined |
| 18-102-final-viral-combined_k141_88529\|\|lt2gene | Low-quality |
| 18-102-final-viral-combined_k141_150326\|\|lt2gene | Low-quality |
| 18-102-final-viral-combined_k141_88578\|\|lt2gene | Low-quality |
| 18-102-final-viral-combined_k141_57534\|\|lt2gene | Low-quality |
| 18-102-final-viral-combined_k141_119425\|\|lt2gene | Low-quality |
| 18-102-final-viral-combined_k141_57555\|\|lt2gene | Low-quality |
| 18-102-final-viral-combined_k141_4596\|\|lt2gene | Low-quality |
| 18-102-final-viral-combined_k141_171736\|\|lt2gene | Low-quality |
| 18-102-final-viral-combined_k141_171741\|\|lt2gene | Low-quality |
| 18-102-final-viral-combined_k141_191750\|\|lt2gene | Not-determined |
| 18-102-final-viral-combined_k141_171754\|\|lt2gene | Low-quality |
| 18-102-final-viral-combined_k141_171785\|\|lt2gene | Low-quality |
| 18-102-final-viral-combined_k141_221796\|\|lt2gene | Low-quality |
| 18-102-final-viral-combined_k141_119486\|\|lt2gene | Low-quality |
| 18-102-final-viral-combined_k141_221808\|\|lt2gene | Low-quality |
| 18-102-final-viral-combined_k141_67332\|\|lt2gene | Low-quality |
| 18-102-final-viral-combined_k141_48505\|\|lt2gene | Low-quality |
| 18-102-final-viral-combined_k141_150403\|\|lt2gene | Not-determined |
| 18-102-final-viral-combined_k141_242723\|\|lt2gene | Low-quality |
| 18-102-final-viral-combined_k141_233695\|\|lt2gene | Low-quality |
| 18-102-final-viral-combined_k141_88699\|\|lt2gene | Low-quality |
| 18-102-final-viral-combined_k141_98989\|\|lt2gene | Low-quality |
| 18-102-final-viral-combined_k141_201054\|\|lt2gene | Low-quality |
| 18-102-final-viral-combined_k141_150448\|\|lt2gene | Not-determined |
| 18-102-final-viral-combined_k141_171837\|\|lt2gene | Low-quality |
| 18-102-final-viral-combined_k141_191833\|\|lt2gene | Low-quality |
| 18-102-final-viral-combined_k141_48550\|\|lt2gene | Not-determined |
| 18-102-final-viral-combined_k141_108671\|\|lt2gene | Not-determined |
| 18-102-final-viral-combined_k141_108677\|\|lt2gene | Low-quality |
| 18-102-final-viral-combined_k141_108683\|\|lt2gene | Low-quality |
| 18-102-final-viral-combined_k141_150497\|\|lt2gene | Not-determined |
| 18-102-final-viral-combined_k141_140451\|\|lt2gene | Not-determined |
| 18-102-final-viral-combined_k141_108706\|\|lt2gene | Low-quality |
| 18-102-final-viral-combined_k141_171883\|\|lt2gene | Not-determined |
| 18-102-final-viral-combined_k141_171888\|\|lt2gene | Low-quality |
| 18-102-final-viral-combined_k141_99072\|\|lt2gene | Low-quality |
| 18-102-final-viral-combined_k141_212158\|\|lt2gene | Low-quality |
| 18-102-final-viral-combined_k141_171918\|\|lt2gene | Not-determined |
| 18-102-final-viral-combined_k141_99107\|\|lt2gene | Low-quality |
| 18-102-final-viral-combined_k141_212187\|\|lt2gene | Low-quality |
| 18-102-final-viral-combined_k141_212225\|\|lt2gene | Low-quality |
| 18-102-final-viral-combined_k141_212239\|\|lt2gene | Low-quality |
| 18-102-final-viral-combined_k141_37040\|\|lt2gene | Not-determined |
| 18-102-final-viral-combined_k141_119710\|\|lt2gene | Low-quality |
| 18-102-final-viral-combined_k141_128912\|\|lt2gene | Low-quality |
| 18-102-final-viral-combined_k141_128916\|\|lt2gene | Low-quality |
| 18-102-final-viral-combined_k141_119716\|\|lt2gene | Low-quality |
| 18-102-final-viral-combined_k141_25509\|\|lt2gene | Not-determined |
| 18-102-final-viral-combined_k141_119733\|\|lt2gene | Low-quality |
| 18-102-final-viral-combined_k141_25540\|\|lt2gene | Not-determined |
| 18-102-final-viral-combined_k141_182163\|\|lt2gene | Not-determined |
| 18-102-final-viral-combined_k141_191999\|\|lt2gene | Not-determined |
| 18-102-final-viral-combined_k141_242899\|\|lt2gene | Not-determined |
| 18-102-final-viral-combined_k141_160391\|\|lt2gene | Low-quality |
| 18-102-final-viral-combined_k141_140636\|\|lt2gene | Not-determined |
| 18-102-final-viral-combined_k141_172113\|\|lt2gene | Not-determined |
| 18-102-final-viral-combined_k141_48877\|\|lt2gene | Low-quality |
| 18-102-final-viral-combined_k141_99193\|\|lt2gene | Not-determined |
| 18-102-final-viral-combined_k141_172157\|\|lt2gene | Low-quality |
| 18-102-final-viral-combined_k141_37172\|\|lt2gene | Low-quality |
| 18-102-final-viral-combined_k141_128969\|\|lt2gene | Not-determined |
| 18-102-final-viral-combined_k141_160473\|\|lt2gene | Low-quality |
| 18-102-final-viral-combined_k141_48948\|\|lt2gene | Not-determined |
| 18-102-final-viral-combined_k141_233953\|\|lt2gene | Low-quality |
| 18-102-final-viral-combined_k141_48965\|\|lt2gene | Low-quality |
| 18-102-final-viral-combined_k141_201266\|\|lt2gene | Low-quality |
| 18-102-final-viral-combined_k141_99277\|\|lt2gene | Low-quality |
| 18-102-final-viral-combined_k141_48994\|\|lt2gene | Low-quality |
| 18-102-final-viral-combined_k141_37211\|\|lt2gene | Not-determined |
| 18-102-final-viral-combined_k141_150799\|\|lt2gene | Low-quality |
| 18-102-final-viral-combined_k141_242929\|\|lt2gene | Not-determined |
| 18-102-final-viral-combined_k141_234002\|\|lt2gene | Not-determined |
| 18-102-final-viral-combined_k141_57798\|\|lt2gene | Low-quality |
| 18-102-final-viral-combined_k141_57801\|\|lt2gene | Low-quality |
| 18-102-final-viral-combined_k141_99336\|\|lt2gene | Low-quality |
| 18-102-final-viral-combined_k141_16091\|\|lt2gene | Not-determined |
| 18-102-final-viral-combined_k141_192073\|\|lt2gene | Not-determined |
| 18-102-final-viral-combined_k141_242936\|\|lt2gene | Low-quality |
| 18-102-final-viral-combined_k141_109041\|\|lt2gene | Low-quality |
| 18-102-final-viral-combined_k141_49029\|\|lt2gene | Not-determined |
| 18-102-final-viral-combined_k141_172297\|\|lt2gene | Not-determined |
| 18-102-final-viral-combined_k141_57879\|\|lt2gene | Not-determined |
| 18-102-final-viral-combined_k141_109141\|\|lt2gene | Not-determined |
| 18-102-final-viral-combined_k141_120015\|\|lt2gene | Not-determined |
| 18-102-final-viral-combined_k141_172383\|\|lt2gene | Low-quality |
| 18-102-final-viral-combined_k141_37359\|\|lt2gene | Low-quality |
| 18-102-final-viral-combined_k141_4825\|\|lt2gene | Low-quality |
| 18-102-final-viral-combined_k141_182404\|\|lt2gene | Low-quality |
| 18-102-final-viral-combined_k141_140923\|\|lt2gene | Low-quality |
| 18-102-final-viral-combined_k141_120074\|\|lt2gene | Low-quality |
| 18-102-final-viral-combined_k141_37450\|\|lt2gene | Low-quality |
| 18-102-final-viral-combined_k141_37453\|\|lt2gene | Low-quality |
| 18-102-final-viral-combined_k141_120096\|\|lt2gene | Low-quality |
| 18-102-final-viral-combined_k141_78634\|\|lt2gene | Low-quality |
| 18-102-final-viral-combined_k141_182502\|\|lt2gene | Low-quality |
| 18-102-final-viral-combined_k141_120142\|\|lt2gene | Low-quality |
| 18-102-final-viral-combined_k141_49270\|\|lt2gene | Low-quality |
| 18-102-final-viral-combined_k141_212425\|\|lt2gene | Not-determined |
| 18-102-final-viral-combined_k141_129082\|\|lt2gene | Low-quality |
| 18-102-final-viral-combined_k141_140957\|\|lt2gene | Low-quality |
| 18-102-final-viral-combined_k141_192282\|\|lt2gene | Low-quality |
| 18-102-final-viral-combined_k141_222099\|\|lt2gene | Low-quality |
| 18-102-final-viral-combined_k141_16284\|\|lt2gene | Low-quality |
| 18-102-final-viral-combined_k141_172619\|\|lt2gene | Low-quality |
| 18-102-final-viral-combined_k141_172684\|\|lt2gene | Low-quality |
| 18-102-final-viral-combined_k141_234290\|\|lt2gene | Low-quality |
| 18-102-final-viral-combined_k141_16348\|\|lt2gene | Not-determined |
| 18-102-final-viral-combined_k141_192309\|\|lt2gene | Low-quality |
| 18-102-final-viral-combined_k141_160804\|\|lt2gene | Not-determined |
| 18-102-final-viral-combined_k141_109318\|\|lt2gene | Low-quality |
| 18-102-final-viral-combined_k141_67978\|\|lt2gene | Low-quality |
| 18-102-final-viral-combined_k141_49377\|\|lt2gene | Low-quality |
| 18-102-final-viral-combined_k141_89432\|\|lt2gene | Not-determined |
| 18-102-final-viral-combined_k141_234382\|\|lt2gene | Low-quality |
| 18-102-final-viral-combined_k141_192370\|\|lt2gene | Low-quality |
| 18-102-final-viral-combined_k141_222229\|\|lt2gene | Low-quality |
| 18-102-final-viral-combined_k141_234424\|\|lt2gene | Low-quality |
| 18-102-final-viral-combined_k141_234475\|\|lt2gene | Low-quality |
| 18-102-final-viral-combined_k141_234494\|\|lt2gene | Low-quality |
| 18-102-final-viral-combined_k141_192417\|\|lt2gene | Low-quality |
| 18-102-final-viral-combined_k141_172901\|\|lt2gene | Not-determined |
| 18-102-final-viral-combined_k141_37547\|\|lt2gene | Not-determined |
| 18-102-final-viral-combined_k141_160959\|\|lt2gene | Not-determined |
| 18-102-final-viral-combined_k141_16514\|\|lt2gene | Low-quality |
| 18-102-final-viral-combined_k141_4950\|\|lt2gene | Low-quality |
| 18-102-final-viral-combined_k141_89584\|\|lt2gene | Low-quality |
| 18-102-final-viral-combined_k141_173024\|\|lt2gene | Low-quality |
| 18-102-final-viral-combined_k141_109449\|\|lt2gene | Low-quality |
| 18-102-final-viral-combined_k141_89624\|\|lt2gene | Low-quality |
| 18-102-final-viral-combined_k141_141096\|\|lt2gene | Low-quality |
| 18-102-final-viral-combined_k141_201802\|\|lt2gene | Not-determined |
| 18-102-final-viral-combined_k141_37654\|\|lt2gene | Low-quality |
| 18-102-final-viral-combined_k141_243337\|\|lt2gene | Low-quality |
| 18-102-final-viral-combined_k141_222353\|\|lt2gene | Low-quality |
| 18-102-final-viral-combined_k141_129306\|\|lt2gene | Low-quality |
| 18-102-final-viral-combined_k141_25981\|\|lt2gene | Low-quality |
| 18-102-final-viral-combined_k141_151466\|\|lt2gene | Not-determined |
| 18-102-final-viral-combined_k141_201838\|\|lt2gene | Not-determined |
| 18-102-final-viral-combined_k141_5023\|\|lt2gene | Not-determined |
| 18-102-final-viral-combined_k141_5026\|\|lt2gene | Low-quality |
| 18-102-final-viral-combined_k141_173104\|\|lt2gene | Low-quality |
| 18-102-final-viral-combined_k141_16640\|\|lt2gene | Low-quality |
| 18-102-final-viral-combined_k141_182763\|\|lt2gene | Low-quality |
| 18-102-final-viral-combined_k141_120422\|\|lt2gene | Low-quality |
| 18-102-final-viral-combined_k141_173167\|\|lt2gene | Not-determined |
| 18-102-final-viral-combined_k141_234740\|\|lt2gene | Low-quality |
| 18-102-final-viral-combined_k141_120499\|\|lt2gene | Low-quality |
| 18-102-final-viral-combined_k141_222467\|\|lt2gene | Low-quality |
| 18-102-final-viral-combined_k141_49735\|\|lt2gene | Low-quality |
| 18-102-final-viral-combined_k141_49748\|\|lt2gene | Not-determined |
| 18-102-final-viral-combined_k141_212828\|\|lt2gene | Low-quality |
| 18-102-final-viral-combined_k141_58279\|\|lt2gene | Low-quality |
| 18-102-final-viral-combined_k141_120551\|\|lt2gene | Low-quality |
| 18-102-final-viral-combined_k141_120565\|\|lt2gene | Low-quality |
| 18-102-final-viral-combined_k141_5148\|\|lt2gene | Low-quality |
| 18-102-final-viral-combined_k141_151612\|\|lt2gene | Low-quality |
| 18-102-final-viral-combined_k141_58306\|\|lt2gene | Low-quality |
| 18-102-final-viral-combined_k141_201984\|\|lt2gene | Low-quality |
| 18-102-final-viral-combined_k141_16771\|\|lt2gene | Low-quality |
| 18-102-final-viral-combined_k141_49830\|\|lt2gene | Low-quality |
| 18-102-final-viral-combined_k141_202012\|\|lt2gene | Low-quality |
| 18-102-final-viral-combined_k141_37777\|\|lt2gene | Low-quality |
| 18-102-final-viral-combined_k141_49870\|\|lt2gene | Low-quality |
| 18-102-final-viral-combined_k141_78916\|\|lt2gene | Low-quality |
| 18-102-final-viral-combined_k141_78925\|\|lt2gene | Not-determined |
| 18-102-final-viral-combined_k141_100020\|\|lt2gene | Low-quality |
| 18-102-final-viral-combined_k141_58375\|\|lt2gene | Low-quality |
| 18-102-final-viral-combined_k141_120655\|\|lt2gene | Low-quality |
| 18-102-final-viral-combined_k141_89812\|\|lt2gene | Low-quality |
| 18-102-final-viral-combined_k141_89815\|\|lt2gene | Low-quality |
| 18-102-final-viral-combined_k141_109631\|\|lt2gene | Low-quality |
| 18-102-final-viral-combined_k141_173329\|\|lt2gene | Low-quality |
| 18-102-final-viral-combined_k141_161424\|\|lt2gene | Not-determined |
| 18-102-final-viral-combined_k141_89858\|\|lt2gene | Low-quality |
| 18-102-final-viral-combined_k141_213014\|\|lt2gene | Not-determined |
| 18-102-final-viral-combined_k141_222589\|\|lt2gene | Low-quality |
| 18-102-final-viral-combined_k141_49985\|\|lt2gene | Low-quality |
| 18-102-final-viral-combined_k141_37860\|\|lt2gene | Low-quality |
| 18-102-final-viral-combined_k141_5225\|\|lt2gene | Low-quality |
| 18-102-final-viral-combined_k141_213051\|\|lt2gene | Low-quality |
| 18-102-final-viral-combined_k141_234873\|\|lt2gene | Not-determined |
| 18-102-final-viral-combined_k141_109775\|\|lt2gene | Low-quality |
| 18-102-final-viral-combined_k141_79051\|\|lt2gene | Not-determined |
| 18-102-final-viral-combined_k141_68487\|\|lt2gene | Not-determined |
| 18-102-final-viral-combined_k141_129678\|\|lt2gene | Low-quality |
| 18-102-final-viral-combined_k141_90030\|\|lt2gene | Low-quality |
| 18-102-final-viral-combined_k141_192827\|\|lt2gene | Low-quality |
| 18-102-final-viral-combined_k141_129721\|\|lt2gene | Low-quality |
| 18-102-final-viral-combined_k141_222736\|\|lt2gene | Low-quality |
| 18-102-final-viral-combined_k141_213153\|\|lt2gene | Low-quality |
| 18-102-final-viral-combined_k141_129748\|\|lt2gene | Not-determined |
| 18-102-final-viral-combined_k141_151958\|\|lt2gene | Low-quality |
| 18-102-final-viral-combined_k141_38050\|\|lt2gene | Not-determined |
| 18-102-final-viral-combined_k141_5323\|\|lt2gene | Low-quality |
| 18-102-final-viral-combined_k141_243642\|\|lt2gene | Low-quality |
| 18-102-final-viral-combined_k141_152016\|\|lt2gene | Low-quality |
| 18-102-final-viral-combined_k141_50214\|\|lt2gene | Low-quality |
| 18-102-final-viral-combined_k141_50236\|\|lt2gene | Low-quality |
| 18-102-final-viral-combined_k141_79165\|\|lt2gene | Not-determined |
| 18-102-final-viral-combined_k141_100289\|\|lt2gene | Low-quality |
| 18-102-final-viral-combined_k141_141514\|\|lt2gene | Low-quality |
| 18-102-final-viral-combined_k141_235090\|\|lt2gene | Low-quality |
| 18-102-final-viral-combined_k141_38161\|\|lt2gene | Low-quality |
| 18-102-final-viral-combined_k141_90169\|\|lt2gene | Low-quality |
| 18-102-final-viral-combined_k141_68600\|\|lt2gene | Low-quality |
| 18-102-final-viral-combined_k141_202361\|\|lt2gene | Not-determined |
| 18-102-final-viral-combined_k141_235119\|\|lt2gene | Low-quality |
| 18-102-final-viral-combined_k141_141572\|\|lt2gene | Not-determined |
| 18-102-final-viral-combined_k141_100377\|\|lt2gene | Low-quality |
| 18-102-final-viral-combined_k141_121038\|\|lt2gene | Low-quality |
| 18-102-final-viral-combined_k141_26262\|\|lt2gene | Not-determined |
| 18-102-final-viral-combined_k141_50368\|\|lt2gene | Low-quality |
| 18-102-final-viral-combined_k141_90303\|\|lt2gene | Low-quality |
| 18-102-final-viral-combined_k141_58707\|\|lt2gene | Low-quality |
| 18-102-final-viral-combined_k141_100470\|\|lt2gene | Low-quality |
| 18-102-final-viral-combined_k141_129937\|\|lt2gene | Low-quality |
| 18-102-final-viral-combined_k141_121073\|\|lt2gene | Not-determined |
| 18-102-final-viral-combined_k141_38326\|\|lt2gene | Low-quality |
| 18-102-final-viral-combined_k141_68870\|\|lt2gene | Low-quality |
| 18-102-final-viral-combined_k141_141649\|\|lt2gene | Not-determined |
| 18-102-final-viral-combined_k141_58760\|\|lt2gene | Low-quality |
| 18-102-final-viral-combined_k141_109911\|\|lt2gene | Not-determined |
| 18-102-final-viral-combined_k141_161731\|\|lt2gene | Low-quality |
| 18-102-final-viral-combined_k141_100549\|\|lt2gene | Low-quality |
| 18-102-final-viral-combined_k141_38483\|\|lt2gene | Low-quality |
| 18-102-final-viral-combined_k141_69023\|\|lt2gene | Low-quality |
| 18-102-final-viral-combined_k141_26424\|\|lt2gene | Low-quality |
| 18-102-final-viral-combined_k141_26431\|\|lt2gene | Not-determined |
| 18-102-final-viral-combined_k141_5526\|\|lt2gene | Low-quality |
| 18-102-final-viral-combined_k141_38497\|\|lt2gene | Not-determined |
| 18-102-final-viral-combined_k141_38518\|\|lt2gene | Low-quality |
| 18-102-final-viral-combined_k141_235425\|\|lt2gene | Low-quality |
| 18-102-final-viral-combined_k141_17436\|\|lt2gene | Low-quality |
| 18-102-final-viral-combined_k141_174038\|\|lt2gene | Low-quality |
| 18-102-final-viral-combined_k141_121330\|\|lt2gene | Low-quality |
| 18-102-final-viral-combined_k141_130156\|\|lt2gene | Low-quality |
| 18-102-final-viral-combined_k141_100732\|\|lt2gene | Not-determined |
| 18-102-final-viral-combined_k141_235491\|\|lt2gene | Low-quality |
| 18-102-final-viral-combined_k141_193352\|\|lt2gene | Not-determined |
| 18-102-final-viral-combined_k141_183319\|\|lt2gene | Not-determined |
| 18-102-final-viral-combined_k141_69068\|\|lt2gene | Low-quality |
| 18-102-final-viral-combined_k141_174122\|\|lt2gene | Low-quality |
| 18-102-final-viral-combined_k141_121381\|\|lt2gene | Low-quality |
| 18-102-final-viral-combined_k141_223152\|\|lt2gene | Low-quality |
| 18-102-final-viral-combined_k141_58960\|\|lt2gene | Low-quality |
| 18-102-final-viral-combined_k141_183361\|\|lt2gene | Low-quality |
| 18-102-final-viral-combined_k141_202644\|\|lt2gene | Low-quality |
| 18-102-final-viral-combined_k141_174164\|\|lt2gene | Low-quality |
| 18-102-final-viral-combined_k141_235582\|\|lt2gene | Not-determined |
| 18-102-final-viral-combined_k141_202696\|\|lt2gene | Not-determined |
| 18-102-final-viral-combined_k141_38742\|\|lt2gene | Not-determined |
| 18-102-final-viral-combined_k141_243961\|\|lt2gene | Low-quality |
| 18-102-final-viral-combined_k141_100872\|\|lt2gene | Low-quality |
| 18-102-final-viral-combined_k141_100877\|\|lt2gene | Not-determined |
| 18-102-final-viral-combined_k141_100885\|\|lt2gene | Low-quality |
| 18-102-final-viral-combined_k141_142014\|\|lt2gene | Low-quality |
| 18-102-final-viral-combined_k141_100891\|\|lt2gene | Low-quality |
| 18-102-final-viral-combined_k141_174231\|\|lt2gene | Low-quality |
| 18-102-final-viral-combined_k141_90613\|\|lt2gene | Low-quality |
| 18-102-final-viral-combined_k141_17602\|\|lt2gene | Low-quality |
| 18-102-final-viral-combined_k141_59025\|\|lt2gene | Low-quality |
| 18-102-final-viral-combined_k141_69125\|\|lt2gene | Low-quality |
| 18-102-final-viral-combined_k141_213708\|\|lt2gene | Low-quality |
| 18-102-final-viral-combined_k141_130262\|\|lt2gene | Low-quality |
| 18-102-final-viral-combined_k141_79669\|\|lt2gene | Not-determined |
| 18-102-final-viral-combined_k141_59041\|\|lt2gene | Low-quality |
| 18-102-final-viral-combined_k141_5743\|\|lt2gene | Low-quality |
| 18-102-final-viral-combined_k141_223319\|\|lt2gene | Not-determined |
| 18-102-final-viral-combined_k141_100926\|\|lt2gene | Not-determined |
| 18-102-final-viral-combined_k141_130312\|\|lt2gene | Low-quality |
| 18-102-final-viral-combined_k141_193543\|\|lt2gene | Low-quality |
| 18-102-final-viral-combined_k141_235753\|\|lt2gene | Low-quality |
| 18-102-final-viral-combined_k141_50951\|\|lt2gene | Low-quality |
| 18-102-final-viral-combined_k141_50952\|\|lt2gene | Low-quality |
| 18-102-final-viral-combined_k141_223384\|\|lt2gene | Low-quality |
| 18-102-final-viral-combined_k141_223399\|\|lt2gene | Low-quality |
| 18-102-final-viral-combined_k141_235812\|\|lt2gene | Not-determined |
| 18-102-final-viral-combined_k141_142159\|\|lt2gene | Not-determined |
| 18-102-final-viral-combined_k141_101039\|\|lt2gene | Not-determined |
| 18-102-final-viral-combined_k141_26548\|\|lt2gene | Not-determined |
| 18-102-final-viral-combined_k141_223471\|\|lt2gene | Low-quality |
| 18-102-final-viral-combined_k141_121615\|\|lt2gene | Low-quality |
| 18-102-final-viral-combined_k141_130336\|\|lt2gene | Low-quality |
| 18-102-final-viral-combined_k141_213854\|\|lt2gene | Low-quality |
| 18-102-final-viral-combined_k141_69412\|\|lt2gene | Not-determined |
| 18-102-final-viral-combined_k141_223491\|\|lt2gene | Low-quality |
| 18-102-final-viral-combined_k141_26608\|\|lt2gene | Low-quality |
| 18-102-final-viral-combined_k141_121658\|\|lt2gene | Low-quality |
| 18-102-final-viral-combined_k141_223513\|\|lt2gene | Low-quality |
| 18-102-final-viral-combined_k141_203003\|\|lt2gene | Not-determined |
| 18-102-final-viral-combined_k141_59310\|\|lt2gene | Low-quality |
| 18-102-final-viral-combined_k141_101145\|\|lt2gene | Low-quality |
| 18-102-final-viral-combined_k141_244187\|\|lt2gene | Not-determined |
| 18-102-final-viral-combined_k141_90660\|\|lt2gene | Low-quality |
| 18-102-final-viral-combined_k141_152723\|\|lt2gene | Low-quality |
| 18-102-final-viral-combined_k141_193771\|\|lt2gene | Low-quality |

| | |
|---|---|
| 18-102-final-viral-combined_k141_59346||lt2gene | Low-quality |
| 18-102-final-viral-combined_k141_121696||lt2gene | Low-quality |
| 18-102-final-viral-combined_k141_69566||lt2gene | Low-quality |
| 18-102-final-viral-combined_k141_26678||lt2gene | Low-quality |
| 18-102-final-viral-combined_k141_5847||lt2gene | Not-determined |
| 18-102-final-viral-combined_k141_51231||lt2gene | Low-quality |
| 18-102-final-viral-combined_k141_142275||lt2gene | Not-determined |
| 18-102-final-viral-combined_k141_130478||lt2gene | Low-quality |
| 18-102-final-viral-combined_k141_90751||lt2gene | Not-determined |
| 18-102-final-viral-combined_k141_152872||lt2gene | Low-quality |
| 18-102-final-viral-combined_k141_69664||lt2gene | Low-quality |
| 18-102-final-viral-combined_k141_193930||lt2gene | Low-quality |
| 18-102-final-viral-combined_k141_90773||lt2gene | Not-determined |
| 18-102-final-viral-combined_k141_79950||lt2gene | Low-quality |
| 18-102-final-viral-combined_k141_5915||lt2gene | Low-quality |
| 18-102-final-viral-combined_k141_69741||lt2gene | Low-quality |
| 18-102-final-viral-combined_k141_203173||lt2gene | Low-quality |
| 18-102-final-viral-combined_k141_223747||lt2gene | Low-quality |
| 18-102-final-viral-combined_k141_214119||lt2gene | Low-quality |
| 18-102-final-viral-combined_k141_51381||lt2gene | Not-determined |
| 18-102-final-viral-combined_k141_51393||lt2gene | Low-quality |
| 18-102-final-viral-combined_k141_203232||lt2gene | Low-quality |
| 18-102-final-viral-combined_k141_5996||lt2gene | Low-quality |
| 18-102-final-viral-combined_k141_69800||lt2gene | Low-quality |
| 18-102-final-viral-combined_k141_162109||lt2gene | Low-quality |
| 18-102-final-viral-combined_k141_18053||lt2gene | Low-quality |
| 18-102-final-viral-combined_k141_153084||lt2gene | Low-quality |
| 18-102-final-viral-combined_k141_26742||lt2gene | Not-determined |
| 18-102-final-viral-combined_k141_69862||lt2gene | Low-quality |
| 18-102-final-viral-combined_k141_236217||lt2gene | Low-quality |
| 18-102-final-viral-combined_k141_184110||lt2gene | Low-quality |
| 18-102-final-viral-combined_k141_153122||lt2gene | Not-determined |
| 18-102-final-viral-combined_k141_39340||lt2gene | Low-quality |
| 18-102-final-viral-combined_k141_69883||lt2gene | Not-determined |
| 18-102-final-viral-combined_k141_184169||lt2gene | Low-quality |
| 18-102-final-viral-combined_k141_214238||lt2gene | Low-quality |
| 18-102-final-viral-combined_k141_162188||lt2gene | Low-quality |
| 18-102-final-viral-combined_k141_214295||lt2gene | Low-quality |
| 18-102-final-viral-combined_k141_18160||lt2gene | Low-quality |
| 18-102-final-viral-combined_k141_51509||lt2gene | Not-determined |
| 18-102-final-viral-combined_k141_194163||lt2gene | Not-determined |
| 18-102-final-viral-combined_k141_110706||lt2gene | Not-determined |
| 18-102-final-viral-combined_k141_101572||lt2gene | Low-quality |
| 18-102-final-viral-combined_k141_18227||lt2gene | Low-quality |
| 18-102-final-viral-combined_k141_59696||lt2gene | Low-quality |
| 18-102-final-viral-combined_k141_101588||lt2gene | Low-quality |
| 18-102-final-viral-combined_k141_101589||lt2gene | Not-determined |
| 18-102-final-viral-combined_k141_122056||lt2gene | Not-determined |
| 18-102-final-viral-combined_k141_153222||lt2gene | Low-quality |
| 18-102-final-viral-combined_k141_6235||lt2gene | Not-determined |
| 18-102-final-viral-combined_k141_223912||lt2gene | Low-quality |
| 18-102-final-viral-combined_k141_184391||lt2gene | Not-determined |
| 18-102-final-viral-combined_k141_26939||lt2gene | Low-quality |
| 18-102-final-viral-combined_k141_214330||lt2gene | Not-determined |
| 18-102-final-viral-combined_k141_214336||lt2gene | Low-quality |
| 18-102-final-viral-combined_k141_194282||lt2gene | Low-quality |
| 18-102-final-viral-combined_k141_101667||lt2gene | Low-quality |
| 18-102-final-viral-combined_k141_39628||lt2gene | Low-quality |
| 18-102-final-viral-combined_k141_91191||lt2gene | Low-quality |
| 18-102-final-viral-combined_k141_162342||lt2gene | Low-quality |
| 18-102-final-viral-combined_k141_69992||lt2gene | Not-determined |
| 18-102-final-viral-combined_k141_91246||lt2gene | Low-quality |
| 18-102-final-viral-combined_k141_142653||lt2gene | Not-determined |
| 18-102-final-viral-combined_k141_110771||lt2gene | Low-quality |
| 18-102-final-viral-combined_k141_51645||lt2gene | Low-quality |
| 18-102-final-viral-combined_k141_194326||lt2gene | Not-determined |
| 18-102-final-viral-combined_k141_162403||lt2gene | Low-quality |
| 18-102-final-viral-combined_k141_51666||lt2gene | Low-quality |
| 18-102-final-viral-combined_k141_130964||lt2gene | Not-determined |
| 18-102-final-viral-combined_k141_162421||lt2gene | Low-quality |
| 18-102-final-viral-combined_k141_59805||lt2gene | Not-determined |
| 18-102-final-viral-combined_k141_101780||lt2gene | Low-quality |
| 18-102-final-viral-combined_k141_236507||lt2gene | Low-quality |
| 18-102-final-viral-combined_k141_224020||lt2gene | Low-quality |
| 18-102-final-viral-combined_k141_80291||lt2gene | Low-quality |
| 18-102-final-viral-combined_k141_80321||lt2gene | Low-quality |
| 18-102-final-viral-combined_k141_27110||lt2gene | Low-quality |
| 18-102-final-viral-combined_k141_236541||lt2gene | Low-quality |
| 18-102-final-viral-combined_k141_80343||lt2gene | Low-quality |
| 18-102-final-viral-combined_k141_110870||lt2gene | Low-quality |
| 18-102-final-viral-combined_k141_244440||lt2gene | Low-quality |
| 18-102-final-viral-combined_k141_59874||lt2gene | Low-quality |
| 18-102-final-viral-combined_k141_194472||lt2gene | Low-quality |
| 18-102-final-viral-combined_k141_91352||lt2gene | Not-determined |
| 18-102-final-viral-combined_k141_214471||lt2gene | Not-determined |
| 18-102-final-viral-combined_k141_214479||lt2gene | Low-quality |
| 18-102-final-viral-combined_k141_236624||lt2gene | Low-quality |
| 18-102-final-viral-combined_k141_91403||lt2gene | Low-quality |
| 18-102-final-viral-combined_k141_194546||lt2gene | Low-quality |
| 18-102-final-viral-combined_k141_203309||lt2gene | Not-determined |
| 18-102-final-viral-combined_k141_194584||lt2gene | Not-determined |
| 18-102-final-viral-combined_k141_194585||lt2gene | Low-quality |
| 18-102-final-viral-combined_k141_18542||lt2gene | Low-quality |
| 18-102-final-viral-combined_k141_214527||lt2gene | Low-quality |
| 18-102-final-viral-combined_k141_91452||lt2gene | Low-quality |
| 18-102-final-viral-combined_k141_194613||lt2gene | Low-quality |
| 18-102-final-viral-combined_k141_39972||lt2gene | Low-quality |
| 18-102-final-viral-combined_k141_39974||lt2gene | Not-determined |
| 18-102-final-viral-combined_k141_39987||lt2gene | Not-determined |
| 18-102-final-viral-combined_k141_203381||lt2gene | Low-quality |
| 18-102-final-viral-combined_k141_91516||lt2gene | Not-determined |
| 18-102-final-viral-combined_k141_91529||lt2gene | Not-determined |
| 18-102-final-viral-combined_k141_142929||lt2gene | Low-quality |
| 18-102-final-viral-combined_k141_101933||lt2gene | Low-quality |
| 18-102-final-viral-combined_k141_91551||lt2gene | Low-quality |
| 18-102-final-viral-combined_k141_40039||lt2gene | Low-quality |
| 18-102-final-viral-combined_k141_175321||lt2gene | Low-quality |
| 18-102-final-viral-combined_k141_27289||lt2gene | Low-quality |
| 18-102-final-viral-combined_k141_122473||lt2gene | Not-determined |
| 18-102-final-viral-combined_k141_142994||lt2gene | Low-quality |
| 18-102-final-viral-combined_k141_175366||lt2gene | Low-quality |
| 18-102-final-viral-combined_k141_194762||lt2gene | Low-quality |
| 18-102-final-viral-combined_k141_27342||lt2gene | Low-quality |
| 18-102-final-viral-combined_k141_70430||lt2gene | Not-determined |
| 18-102-final-viral-combined_k141_40165||lt2gene | Low-quality |
| 18-102-final-viral-combined_k141_40186||lt2gene | Not-determined |
| 18-102-final-viral-combined_k141_175408||lt2gene | Low-quality |
| 18-102-final-viral-combined_k141_40228||lt2gene | Low-quality |
| 18-102-final-viral-combined_k141_40236||lt2gene | Low-quality |
| 18-102-final-viral-combined_k141_40263||lt2gene | Low-quality |
| 18-102-final-viral-combined_k141_175506||lt2gene | Low-quality |
| 18-102-final-viral-combined_k141_236859||lt2gene | Low-quality |
| 18-102-final-viral-combined_k141_184785||lt2gene | Low-quality |
| 18-102-final-viral-combined_k141_224326||lt2gene | Low-quality |
| 18-102-final-viral-combined_k141_224335||lt2gene | Low-quality |
| 18-102-final-viral-combined_k141_224362||lt2gene | Low-quality |
| 18-102-final-viral-combined_k141_91881||lt2gene | Not-determined |
| 18-102-final-viral-combined_k141_184830||lt2gene | Low-quality |
| 18-102-final-viral-combined_k141_91897||lt2gene | Low-quality |
| 18-102-final-viral-combined_k141_153474||lt2gene | Low-quality |
| 18-102-final-viral-combined_k141_143177||lt2gene | Low-quality |
| 18-102-final-viral-combined_k141_111162||lt2gene | Not-determined |
| 18-102-final-viral-combined_k141_153488||lt2gene | Not-determined |
| 18-102-final-viral-combined_k141_60495||lt2gene | Not-determined |
| 18-102-final-viral-combined_k141_91936||lt2gene | Low-quality |
| 18-102-final-viral-combined_k141_70657||lt2gene | Low-quality |
| 18-102-final-viral-combined_k141_236992||lt2gene | Not-determined |
| 18-102-final-viral-combined_k141_70678||lt2gene | Low-quality |
| 18-102-final-viral-combined_k141_40363||lt2gene | Low-quality |
| 18-102-final-viral-combined_k141_27454||lt2gene | Low-quality |
| 18-102-final-viral-combined_k141_102359||lt2gene | Not-determined |
| 18-102-final-viral-combined_k141_237039||lt2gene | Low-quality |
| 18-102-final-viral-combined_k141_237054||lt2gene | Low-quality |
| 18-102-final-viral-combined_k141_60586||lt2gene | Low-quality |
| 18-102-final-viral-combined_k141_102389||lt2gene | Not-determined |
| 18-102-final-viral-combined_k141_194951||lt2gene | Not-determined |
| 18-102-final-viral-combined_k141_111268||lt2gene | Low-quality |
| 18-102-final-viral-combined_k141_60619||lt2gene | Low-quality |
| 18-102-final-viral-combined_k141_153566||lt2gene | Low-quality |
| 18-102-final-viral-combined_k141_60666||lt2gene | Low-quality |
| 18-102-final-viral-combined_k141_60677||lt2gene | Not-determined |
| 18-102-final-viral-combined_k141_175710||lt2gene | Low-quality |
| 18-102-final-viral-combined_k141_203734||lt2gene | Low-quality |
| 18-102-final-viral-combined_k141_195006||lt2gene | Low-quality |
| 18-102-final-viral-combined_k141_184895||lt2gene | Low-quality |
| 18-102-final-viral-combined_k141_244782||lt2gene | Low-quality |
| 18-102-final-viral-combined_k141_195014||lt2gene | Low-quality |
| 18-102-final-viral-combined_k141_6744||lt2gene | Low-quality |
| 18-102-final-viral-combined_k141_244811||lt2gene | Low-quality |
| 18-102-final-viral-combined_k141_162959||lt2gene | Low-quality |
| 18-102-final-viral-combined_k141_184962||lt2gene | Not-determined |
| 18-102-final-viral-combined_k141_215233||lt2gene | Low-quality |
| 18-102-final-viral-combined_k141_143535||lt2gene | Not-determined |
| 18-102-final-viral-combined_k141_80979||lt2gene | Low-quality |
| 18-102-final-viral-combined_k141_195101||lt2gene | Low-quality |
| 18-102-final-viral-combined_k141_195103||lt2gene | Low-quality |
| 18-102-final-viral-combined_k141_70930||lt2gene | Not-determined |
| 18-102-final-viral-combined_k141_60842||lt2gene | Not-determined |
| 18-102-final-viral-combined_k141_102539||lt2gene | Low-quality |
| 18-102-final-viral-combined_k141_102567||lt2gene | Low-quality |
| 18-102-final-viral-combined_k141_81015||lt2gene | Low-quality |
| 18-102-final-viral-combined_k141_143578||lt2gene | Low-quality |
| 18-102-final-viral-combined_k141_19131||lt2gene | Low-quality |
| 18-102-final-viral-combined_k141_175858||lt2gene | Low-quality |
| 18-102-final-viral-combined_k141_203874||lt2gene | Not-determined |
| 18-102-final-viral-combined_k141_224600||lt2gene | Low-quality |
| 18-102-final-viral-combined_k141_81064||lt2gene | Low-quality |
| 18-102-final-viral-combined_k141_175915||lt2gene | Low-quality |
| 18-102-final-viral-combined_k141_40678||lt2gene | Low-quality |
| 18-102-final-viral-combined_k141_185065||lt2gene | Low-quality |
| 18-102-final-viral-combined_k141_40699||lt2gene | Low-quality |
| 18-102-final-viral-combined_k141_70998||lt2gene | Low-quality |
| 18-102-final-viral-combined_k141_185078||lt2gene | Low-quality |
| 18-102-final-viral-combined_k141_60956||lt2gene | Not-determined |
| 18-102-final-viral-combined_k141_195227||lt2gene | Low-quality |
| 18-102-final-viral-combined_k141_102725||lt2gene | Not-determined |
| 18-102-final-viral-combined_k141_71077||lt2gene | Low-quality |
| 18-102-final-viral-combined_k141_71092||lt2gene | Not-determined |
| 18-102-final-viral-combined_k141_102741||lt2gene | Low-quality |
| 18-102-final-viral-combined_k141_81189||lt2gene | Low-quality |
| 18-102-final-viral-combined_k141_102774||lt2gene | Low-quality |
| 18-102-final-viral-combined_k141_195333||lt2gene | Not-determined |
| 18-102-final-viral-combined_k141_131575||lt2gene | Not-determined |
| 18-102-final-viral-combined_k141_244931||lt2gene | Low-quality |
| 18-102-final-viral-combined_k141_153978||lt2gene | Not-determined |
| 18-102-final-viral-combined_k141_215502||lt2gene | Low-quality |
| 18-102-final-viral-combined_k141_111553||lt2gene | Not-determined |
| 18-102-final-viral-combined_k141_204067||lt2gene | Low-quality |
| 18-102-final-viral-combined_k141_122990||lt2gene | Low-quality |
| 18-102-final-viral-combined_k141_102861||lt2gene | Not-determined |
| 18-102-final-viral-combined_k141_195413||lt2gene | Low-quality |
| 18-102-final-viral-combined_k141_27746||lt2gene | Low-quality |
| 18-102-final-viral-combined_k141_61036||lt2gene | Low-quality |
| 18-102-final-viral-combined_k141_163218||lt2gene | Low-quality |
| 18-102-final-viral-combined_k141_224836||lt2gene | Low-quality |
| 18-102-final-viral-combined_k141_61087||lt2gene | Not-determined |
| 18-102-final-viral-combined_k141_215625||lt2gene | Not-determined |
| 18-102-final-viral-combined_k141_143745||lt2gene | Not-determined |
| 18-102-final-viral-combined_k141_224899||lt2gene | Low-quality |
| 18-102-final-viral-combined_k141_40849||lt2gene | Not-determined |
| 18-102-final-viral-combined_k141_143786||lt2gene | Low-quality |
| 18-102-final-viral-combined_k141_245068||lt2gene | Low-quality |
| 18-102-final-viral-combined_k141_143793||lt2gene | Not-determined |
| 18-102-final-viral-combined_k141_40883||lt2gene | Low-quality |
| 18-102-final-viral-combined_k141_102921||lt2gene | Not-determined |
| 18-102-final-viral-combined_k141_61180||lt2gene | Low-quality |
| 18-102-final-viral-combined_k141_111719||lt2gene | Not-determined |
| 18-102-final-viral-combined_k141_245092||lt2gene | Not-determined |
| 18-102-final-viral-combined_k141_143807||lt2gene | Low-quality |
| 18-102-final-viral-combined_k141_185420||lt2gene | Low-quality |
| 18-102-final-viral-combined_k141_111765||lt2gene | Not-determined |
| 18-102-final-viral-combined_k141_143826||lt2gene | Low-quality |
| 18-102-final-viral-combined_k141_71435||lt2gene | Low-quality |
| 18-102-final-viral-combined_k141_102982||lt2gene | Low-quality |
| 18-102-final-viral-combined_k141_195707||lt2gene | Low-quality |
| 18-102-final-viral-combined_k141_7082||lt2gene | Not-determined |
| 18-102-final-viral-combined_k141_143867||lt2gene | Not-determined |
| 18-102-final-viral-combined_k141_81537||lt2gene | Not-determined |
| 18-102-final-viral-combined_k141_154254||lt2gene | Low-quality |
| 18-102-final-viral-combined_k141_225052||lt2gene | Low-quality |
| 18-102-final-viral-combined_k141_131896||lt2gene | Low-quality |
| 18-102-final-viral-combined_k141_61374||lt2gene | Low-quality |
| 18-102-final-viral-combined_k141_81660||lt2gene | Low-quality |
| 18-102-final-viral-combined_k141_144078||lt2gene | Not-determined |
| 18-102-final-viral-combined_k141_204279||lt2gene | Low-quality |
| 18-102-final-viral-combined_k141_154369||lt2gene | Not-determined |
| 18-102-final-viral-combined_k141_19624||lt2gene | Low-quality |
| 18-102-final-viral-combined_k141_41101||lt2gene | Low-quality |
| 18-102-final-viral-combined_k141_7236||lt2gene | Low-quality |
| 18-102-final-viral-combined_k141_154464||lt2gene | Low-quality |
| 18-102-final-viral-combined_k141_195971||lt2gene | Low-quality |
| 18-102-final-viral-combined_k141_19725||lt2gene | Not-determined |
| 18-102-final-viral-combined_k141_71630||lt2gene | Low-quality |
| 18-102-final-viral-combined_k141_185539||lt2gene | Not-determined |
| 18-102-final-viral-combined_k141_144196||lt2gene | Low-quality |
| 18-102-final-viral-combined_k141_28095||lt2gene | Low-quality |
| 18-102-final-viral-combined_k141_196031||lt2gene | Low-quality |
| 18-102-final-viral-combined_k141_111906||lt2gene | Low-quality |
| 18-102-final-viral-combined_k141_154491||lt2gene | Low-quality |
| 18-102-final-viral-combined_k141_19780||lt2gene | Low-quality |
| 18-102-final-viral-combined_k141_196084||lt2gene | Not-determined |
| 18-102-final-viral-combined_k141_144248||lt2gene | Not-determined |
| 18-102-final-viral-combined_k141_144258||lt2gene | Low-quality |
| 18-102-final-viral-combined_k141_196131||lt2gene | Not-determined |
| 18-102-final-viral-combined_k141_71668||lt2gene | Low-quality |
| 18-102-final-viral-combined_k141_196138||lt2gene | Not-determined |
| 18-102-final-viral-combined_k141_215940||lt2gene | Low-quality |
| 18-102-final-viral-combined_k141_19849||lt2gene | Low-quality |
| 18-102-final-viral-combined_k141_196147||lt2gene | Not-determined |
| 18-102-final-viral-combined_k141_196159||lt2gene | Low-quality |
| 18-102-final-viral-combined_k141_154580||lt2gene | Low-quality |
| 18-102-final-viral-combined_k141_92811||lt2gene | Low-quality |
| 18-102-final-viral-combined_k141_7382||lt2gene | Low-quality |
| 18-102-final-viral-combined_k141_7405||lt2gene | Low-quality |
| 18-102-final-viral-combined_k141_132180||lt2gene | Low-quality |
| 18-102-final-viral-combined_k141_132187||lt2gene | Not-determined |
| 18-102-final-viral-combined_k141_225474||lt2gene | Low-quality |
| 18-102-final-viral-combined_k141_185726||lt2gene | Not-determined |
| 18-102-final-viral-combined_k141_216092||lt2gene | Not-determined |
| 18-102-final-viral-combined_k141_216094||lt2gene | Not-determined |
| 18-102-final-viral-combined_k141_216099||lt2gene | Low-quality |
| 18-102-final-viral-combined_k141_28353||lt2gene | Not-determined |
| 18-102-final-viral-combined_k141_71919||lt2gene | Low-quality |
| 18-102-final-viral-combined_k141_7476||lt2gene | Low-quality |
| 18-102-final-viral-combined_k141_154723||lt2gene | Not-determined |
| 18-102-final-viral-combined_k141_144466||lt2gene | Low-quality |
| 18-102-final-viral-combined_k141_19988||lt2gene | Low-quality |
| 18-102-final-viral-combined_k141_132287||lt2gene | Low-quality |
| 18-102-final-viral-combined_k141_112104||lt2gene | Low-quality |
| 18-102-final-viral-combined_k141_154779||lt2gene | Low-quality |
| 18-102-final-viral-combined_k141_185837||lt2gene | Not-determined |
| 18-102-final-viral-combined_k141_154798||lt2gene | Not-determined |
| 18-102-final-viral-combined_k141_92985||lt2gene | Not-determined |
| 18-102-final-viral-combined_k141_185872||lt2gene | Low-quality |
| 18-102-final-viral-combined_k141_20044||lt2gene | Not-determined |
| 18-102-final-viral-combined_k141_245382||lt2gene | Low-quality |
| 18-102-final-viral-combined_k141_204552||lt2gene | Low-quality |
| 18-102-final-viral-combined_k141_154868||lt2gene | Low-quality |
| 18-102-final-viral-combined_k141_7580||lt2gene | Low-quality |
| 18-102-final-viral-combined_k141_103364||lt2gene | Not-determined |
| 18-102-final-viral-combined_k141_20070||lt2gene | Low-quality |
| 18-102-final-viral-combined_k141_112207||lt2gene | Low-quality |
| 18-102-final-viral-combined_k141_238165||lt2gene | Low-quality |
| 18-102-final-viral-combined_k141_72106||lt2gene | Not-determined |
| 18-102-final-viral-combined_k141_72118||lt2gene | Low-quality |
| 18-102-final-viral-combined_k141_186013||lt2gene | Low-quality |
| 18-102-final-viral-combined_k141_225649||lt2gene | Low-quality |
| 18-102-final-viral-combined_k141_132414||lt2gene | Not-determined |
| 18-102-final-viral-combined_k141_225664||lt2gene | Low-quality |
| 18-102-final-viral-combined_k141_112288||lt2gene | Low-quality |
| 18-102-final-viral-combined_k141_144596||lt2gene | Low-quality |
| 18-102-final-viral-combined_k141_123741||lt2gene | Low-quality |
| 18-102-final-viral-combined_k141_154933||lt2gene | Low-quality |
| 18-102-final-viral-combined_k141_112358||lt2gene | Not-determined |
| 18-102-final-viral-combined_k141_216287||lt2gene | Low-quality |
| 18-102-final-viral-combined_k141_132505||lt2gene | Low-quality |
| 18-102-final-viral-combined_k141_93094||lt2gene | Low-quality |
| 18-102-final-viral-combined_k141_132564||lt2gene | Low-quality |
| 18-102-final-viral-combined_k141_82342||lt2gene | Low-quality |
| 18-102-final-viral-combined_k141_123775||lt2gene | Low-quality |
| 18-102-final-viral-combined_k141_132599||lt2gene | Low-quality |
| 18-102-final-viral-combined_k141_132630||lt2gene | Low-quality |
| 18-102-final-viral-combined_k141_204760||lt2gene | Low-quality |
| 18-102-final-viral-combined_k141_82389||lt2gene | Low-quality |
| 18-102-final-viral-combined_k141_196577||lt2gene | Low-quality |
| 18-102-final-viral-combined_k141_132669||lt2gene | Low-quality |
| 18-102-final-viral-combined_k141_72302||lt2gene | Not-determined |
| 18-102-final-viral-combined_k141_62003||lt2gene | Low-quality |
| 18-102-final-viral-combined_k141_62020||lt2gene | Not-determined |
| 18-102-final-viral-combined_k141_72370||lt2gene | Low-quality |
| 18-102-final-viral-combined_k141_216406||lt2gene | Not-determined |
| 18-102-final-viral-combined_k141_196657||lt2gene | Low-quality |
| 18-102-final-viral-combined_k141_20263||lt2gene | Low-quality |
| 18-102-final-viral-combined_k141_216477||lt2gene | Low-quality |
| 18-102-final-viral-combined_k141_216486||lt2gene | Low-quality |
| 18-102-final-viral-combined_k141_93197||lt2gene | Low-quality |
| 18-102-final-viral-combined_k141_216558||lt2gene | Not-determined |
| 18-102-final-viral-combined_k141_82473||lt2gene | Low-quality |
| 18-102-final-viral-combined_k141_112520||lt2gene | Low-quality |
| 18-102-final-viral-combined_k141_245681||lt2gene | Low-quality |
| 18-102-final-viral-combined_k141_20391||lt2gene | Not-determined |
| 18-102-final-viral-combined_k141_82526||lt2gene | Low-quality |
| 18-102-final-viral-combined_k141_186269||lt2gene | Low-quality |
| 18-102-final-viral-combined_k141_20399||lt2gene | Low-quality |
| 18-102-final-viral-combined_k141_144819||lt2gene | Not-determined |
| 18-102-final-viral-combined_k141_164222||lt2gene | Low-quality |
| 18-102-final-viral-combined_k141_124030||lt2gene | Not-determined |
| 18-102-final-viral-combined_k141_82536||lt2gene | Low-quality |
| 18-102-final-viral-combined_k141_186373||lt2gene | Not-determined |
| 18-102-final-viral-combined_k141_216748||lt2gene | Low-quality |
| 18-102-final-viral-combined_k141_112697||lt2gene | Low-quality |
| 18-102-final-viral-combined_k141_216827||lt2gene | Low-quality |
| 18-102-final-viral-combined_k141_164319||lt2gene | Not-determined |
| 18-102-final-viral-combined_k141_205122||lt2gene | Low-quality |
| 18-102-final-viral-combined_k141_124094||lt2gene | Low-quality |
| 18-102-final-viral-combined_k141_205174||lt2gene | Not-determined |
| 18-102-final-viral-combined_k141_82713||lt2gene | Not-determined |
| 18-102-final-viral-combined_k141_28991||lt2gene | Low-quality |
| 18-102-final-viral-combined_k141_226098||lt2gene | Not-determined |
| 18-102-final-viral-combined_k141_20708||lt2gene | Low-quality |
| 18-102-final-viral-combined_k141_112814||lt2gene | Not-determined |
| 18-102-final-viral-combined_k141_245836||lt2gene | Low-quality |
| 18-102-final-viral-combined_k141_164393||lt2gene | Not-determined |
| 18-102-final-viral-combined_k141_164427||lt2gene | Low-quality |
| 18-102-final-viral-combined_k141_205304||lt2gene | Low-quality |
| 18-102-final-viral-combined_k141_205317||lt2gene | Not-determined |
| 18-102-final-viral-combined_k141_133047||lt2gene | Not-determined |
| 18-102-final-viral-combined_k141_226214||lt2gene | Low-quality |
| 18-102-final-viral-combined_k141_205344||lt2gene | Not-determined |
| 18-102-final-viral-combined_k141_133104||lt2gene | Low-quality |
| 18-102-final-viral-combined_k141_217036||lt2gene | Low-quality |
| 18-102-final-viral-combined_k141_226294||lt2gene | Low-quality |
| 18-102-final-viral-combined_k141_246000||lt2gene | Not-determined |
| 18-102-final-viral-combined_k141_133249||lt2gene | Low-quality |
| 18-102-final-viral-combined_k141_164616||lt2gene | Low-quality |
| 18-102-final-viral-combined_k141_246226||lt2gene | Low-quality |
| 18-102-final-viral-combined_k141_113331||lt2gene | Low-quality |
| 18-102-final-viral-combined_k141_217396||lt2gene | Not-determined |
| 18-102-final-viral-combined_k141_217397||lt2gene | Low-quality |
| 18-102-final-viral-combined_k141_165016||lt2gene | Not-determined |
| 18-102-final-viral-combined_k141_165035||lt2gene | Not-determined |
| 18-102-final-viral-combined_k141_29480||lt2gene | Low-quality |
| 18-102-final-viral-combined_k141_29483||lt2gene | Not-determined |
| 18-102-final-viral-combined_k141_8134||lt2gene | Not-determined |
| 18-102-final-viral-combined_k141_29522||lt2gene | Low-quality |
| 18-102-final-viral-combined_k141_205750||lt2gene | Low-quality |
| 18-102-final-viral-combined_k141_226847||lt2gene | Low-quality |
| 18-102-final-viral-combined_k141_8214||lt2gene | Low-quality |
| 18-102-final-viral-combined_k141_165159||lt2gene | Low-quality |
| 18-102-final-viral-combined_k141_113411||lt2gene | Low-quality |
| 18-102-final-viral-combined_k141_165198||lt2gene | Not-determined |
| 18-102-final-viral-combined_k141_226936||lt2gene | Low-quality |
| 18-102-final-viral-combined_k141_246357||lt2gene | Not-determined |
| 18-102-final-viral-combined_k141_8304||lt2gene | Low-quality |
| 18-102-final-viral-combined_k141_29699||lt2gene | Not-determined |
| 18-102-final-viral-combined_k141_246470||lt2gene | Low-quality |
| 18-102-final-viral-combined_k141_113565||lt2gene | Low-quality |
| 18-102-final-viral-combined_k141_206051||lt2gene | Low-quality |
| 18-102-final-viral-combined_k141_8336||lt2gene | Not-determined |
| 18-102-final-viral-combined_k141_246492||lt2gene | Low-quality |
| 18-102-final-viral-combined_k141_29762||lt2gene | Low-quality |
| 18-102-final-viral-combined_k141_29793||lt2gene | Low-quality |
| 18-102-final-viral-combined_k141_246532||lt2gene | Low-quality |
| 18-102-final-viral-combined_k141_133835||lt2gene | Low-quality |
| 18-102-final-viral-combined_k141_113712||lt2gene | Not-determined |
| 18-102-final-viral-combined_k141_206207||lt2gene | Low-quality |
| 18-102-final-viral-combined_k141_133890||lt2gene | Low-quality |
| 18-102-final-viral-combined_k141_227378||lt2gene | Low-quality |
| 18-102-final-viral-combined_k141_133937||lt2gene | Low-quality |
| 18-102-final-viral-combined_k141_29996||lt2gene | Not-determined |
| 18-102-final-viral-combined_k141_134012||lt2gene | Low-quality |
| 18-102-final-viral-combined_k141_30048||lt2gene | Not-determined |
| 18-102-final-viral-combined_k141_246840||lt2gene | Low-quality |
| 18-102-final-viral-combined_k141_246910||lt2gene | Low-quality |
| 18-102-final-viral-combined_k141_134187||lt2gene | Not-determined |
| 18-102-final-viral-combined_k141_113893||lt2gene | Low-quality |
| 18-102-final-viral-combined_k141_227551||lt2gene | Low-quality |
| 18-102-final-viral-combined_k141_30288||lt2gene | Low-quality |
| 18-102-final-viral-combined_k141_30289||lt2gene | Low-quality |
| 18-102-final-viral-combined_k141_246990||lt2gene | Not-determined |
| 18-102-final-viral-combined_k141_134278||lt2gene | Low-quality |
| 18-102-final-viral-combined_k141_134357||lt2gene | Low-quality |
| 18-102-final-viral-combined_k141_227638||lt2gene | Low-quality |
| 18-102-final-viral-combined_k141_134375||lt2gene | Low-quality |
| 18-102-final-viral-combined_k141_30462||lt2gene | Low-quality |
| 18-102-final-viral-combined_k141_30471||lt2gene | Low-quality |
| 18-102-final-viral-combined_k141_30473||lt2gene | Low-quality |
| 18-102-final-viral-combined_k141_134445||lt2gene | Not-determined |
| 18-102-final-viral-combined_k141_30499||lt2gene | Low-quality |
| 18-102-final-viral-combined_k141_8657||lt2gene | Not-determined |
| 18-102-final-viral-combined_k141_134492||lt2gene | Low-quality |
| 18-102-final-viral-combined_k141_134507||lt2gene | Low-quality |
| 18-102-final-viral-combined_k141_134533||lt2gene | Not-determined |
| 18-102-final-viral-combined_k141_134623||lt2gene | Low-quality |
| 18-102-final-viral-combined_k141_8884||lt2gene | Low-quality |
| 18-102-final-viral-combined_k141_247193||lt2gene | Low-quality |
| 18-102-final-viral-combined_k141_206738||lt2gene | Low-quality |
| 18-102-final-viral-combined_k141_8907||lt2gene | Low-quality |
| 18-102-final-viral-combined_k141_8924||lt2gene | Low-quality |
| 18-102-final-viral-combined_k141_30781||lt2gene | Not-determined |
| 18-102-final-viral-combined_k141_30812||lt2gene | Low-quality |
| 18-102-final-viral-combined_k141_30861||lt2gene | Low-quality |
| 18-102-final-viral-combined_k141_247340||lt2gene | Low-quality |
| 18-102-final-viral-combined_k141_247493||lt2gene | Not-determined |
| 18-102-final-viral-combined_k141_31059||lt2gene | Not-determined |
| 18-102-final-viral-combined_k141_9215||lt2gene | Low-quality |
| 18-102-final-viral-combined_k141_9321||lt2gene | Low-quality |
| 18-102-final-viral-combined_k141_247684||lt2gene | Not-determined |
| 18-102-final-viral-combined_k141_9478||lt2gene | Low-quality |
| 18-102-final-viral-combined_k141_248076||lt2gene | Not-determined |
| 18-102-final-viral-combined_k141_9622||lt2gene | Low-quality |
| 18-102-final-viral-combined_k141_9650||lt2gene | Low-quality |
| 18-102-final-viral-combined_k141_248256||lt2gene | Low-quality |
| 18-102-final-viral-combined_k141_9877||lt2gene | Low-quality |
| 18-102-final-viral-combined_k141_9882||lt2gene | Low-quality |
| 18-102-final-viral-combined_k141_10025||lt2gene | Low-quality |
| 18-102-final-viral-combined_k141_10113||lt2gene | Low-quality |
| 18-102-final-viral-combined_k141_10133||lt2gene | Low-quality |
| 18-102-final-viral-combined_k141_10189||lt2gene | Low-quality |
| 18-102-final-viral-combined_k141_10340||lt2gene | Low-quality |
| 18-102_k141_31071 | Not-determined |
| 18-102_k141_82853 | Not-determined |
| 18-102_k141_207128 | Not-determined |
| 18-102_k141_41430 | Low-quality |
| 18-102_k141_31077 | Not-determined |
| 18-102_k141_207131 | Low-quality |
| 18-102_k141_31080 | Not-determined |
| 18-102_k141_176064 | Not-determined |
| 18-102_k141_124281 | Not-determined |
| 18-102_k141_113929 | Not-determined |
| 18-102_k141_103579 | Low-quality |
| 18-102_k141_72512 | Not-determined |
| 18-102_k141_41445 | Not-determined |
| 18-102_k141_93225 | Not-determined |
| 18-102_k141_217500 | Not-determined |
| 18-102_k141_10380 | Not-determined |
| 18-102_k141_10382 | Not-determined |
| 18-102_k141_186435 | Not-determined |
| 18-102_k141_72521 | Not-determined |
| 18-102_k141_93238 | Not-determined |
| 18-102_k141_41463 | Not-determined |
| 18-102_k141_217502 | Not-determined |
| 18-102_k141_72526 | Not-determined |
| 18-102_k141_20721 | Low-quality |
| 18-102_k141_155375 | Not-determined |
| 18-102_k141_207138 | Not-determined |
| 18-102_k141_37 | Not-determined |
| 18-102_k141_93247 | Not-determined |
| 18-102_k141_217510 | Not-determined |
| 18-102_k141_217512 | Not-determined |
| 18-102_k141_134664 | Not-determined |
| 18-102_k141_62163 | Not-determined |
| 18-102_k141_82870 | Not-determined |
| 18-102_k141_155384 | Low-quality |
| 18-102_k141_155386 | Low-quality |
| 18-102_k141_50 | Not-determined |
| 18-102_k141_103640 | Not-determined |
| 18-102_k141_61 | Not-determined |
| 18-102_k141_155391 | Low-quality |
| 18-102_k141_103643 | Not-determined |
| 18-102_k141_20738 | Not-determined |
| 18-102_k141_63 | Not-determined |
| 18-102_k141_155396 | Not-determined |
| 18-102_k141_176089 | Not-determined |
| 18-102_k141_124316 | Not-determined |
| 18-102_k141_20743 | Low-quality |
| 18-102_k141_186440 | Not-determined |
| 18-102_k141_186442 | Not-determined |
| 18-102_k141_82875 | Low-quality |
| 18-102_k141_80 | Not-determined |
| 18-102_k141_51811 | Low-quality |
| 18-102_k141_51816 | Not-determined |
| 18-102_k141_85 | Not-determined |
| 18-102_k141_41493 | Not-determined |
| 18-102_k141_217540 | Not-determined |
| 18-102_k141_51821 | Not-determined |
| 18-102_k141_41495 | Not-determined |
| 18-102_k141_62195 | Not-determined |
| 18-102_k141_103666 | Not-determined |
| 18-102_k141_113961 | Not-determined |
| 18-102_k141_82912 | Not-determined |
| 18-102_k141_165734 | Not-determined |
| 18-102_k141_176123 | Not-determined |
| 18-102_k141_103679 | Not-determined |
| 18-102_k141_51836 | Not-determined |
| 18-102_k141_134693 | Low-quality |
| 18-102_k141_103690 | Not-determined |
| 18-102_k141_238222 | Not-determined |
| 18-102_k141_186459 | Not-determined |
| 18-102_k141_186460 | Low-quality |
| 18-102_k141_186463 | Not-determined |
| 18-102_k141_186466 | Not-determined |
| 18-102_k141_238226 | Low-quality |
| 18-102_k141_41517 | Not-determined |
| 18-102_k141_134704 | Not-determined |
| 18-102_k141_176139 | Not-determined |
| 18-102_k141_155408 | Low-quality |
| 18-102_k141_217571 | Not-determined |
| 18-102_k141_134723 | Not-determined |
| 18-102_k141_217573 | Not-determined |
| 18-102_k141_41536 | Not-determined |
| 18-102_k141_165743 | Not-determined |
| 18-102_k141_41545 | Not-determined |
| 18-102_k141_176150 | Not-determined |
| 18-102_k141_176154 | Not-determined |
| 18-102_k141_51847 | Not-determined |
| 18-102_k141_51849 | Not-determined |
| 18-102_k141_20752 | Not-determined |
| 18-102_k141_72547 | Not-determined |
| 18-102_k141_155427 | Not-determined |
| 18-102_k141_51862 | Not-determined |
| 18-102_k141_41564 | Not-determined |
| 18-102_k141_165768 | Not-determined |
| 18-102_k141_103739 | Not-determined |
| 18-102_k141_41573 | Not-determined |
| 18-102_k141_155435 | Not-determined |
| 18-102_k141_41579 | Not-determined |
| 18-102_k141_82948 | Not-determined |
| 18-102_k141_113978 | Not-determined |
| 18-102_k141_217602 | Not-determined |
| 18-102_k141_165778 | Not-determined |
| 18-102_k141_196819 | Not-determined |
| 18-102_k141_51878 | Low-quality |
| 18-102_k141_145042 | Not-determined |
| 18-102_k141_217605 | Not-determined |
| 18-102_k141_176178 | Not-determined |
| 18-102_k141_113981 | Not-determined |
| 18-102_k141_227881 | Not-determined |
| 18-102_k141_51881 | Not-determined |
| 18-102_k141_165786 | Not-determined |
| 18-102_k141_227887 | Not-determined |
| 18-102_k141_134758 | Low-quality |
| 18-102_k141_227890 | Not-determined |
| 18-102_k141_103758 | Not-determined |
| 18-102_k141_10445 | Not-determined |
| 18-102_k141_145045 | Not-determined |
| 18-102_k141_31124 | Not-determined |
| 18-102_k141_103762 | Not-determined |
| 18-102_k141_62216 | Not-determined |
| 18-102_k141_227896 | Not-determined |
| 18-102_k141_103764 | Not-determined |
| 18-102_k141_62219 | Low-quality |
| 18-102_k141_155453 | Not-determined |
| 18-102_k141_31132 | Not-determined |
| 18-102_k141_186508 | Not-determined |
| 18-102_k141_41596 | Not-determined |
| 18-102_k141_51892 | Not-determined |
| 18-102_k141_31134 | Not-determined |
| 18-102_k141_31135 | Not-determined |
| 18-102_k141_41600 | Not-determined |
| 18-102_k141_10459 | Not-determined |
| 18-102_k141_186519 | Not-determined |
| 18-102_k141_186520 | Not-determined |
| 18-102_k141_10464 | Not-determined |
| 18-102_k141_10465 | Not-determined |
| 18-102_k141_186523 | Not-determined |
| 18-102_k141_51902 | Not-determined |
| 18-102_k141_103775 | Not-determined |
| 18-102_k141_134774 | Not-determined |

| | |
|---|---|
| 18-102_k141_176190 | Low-quality |
| 18-102_k141_145065 | Not-determined |
| 18-102_k141_176191 | Low-quality |
| 18-102_k141_51909 | Not-determined |
| 18-102_k141_217622 | Low-quality |
| 18-102_k141_118 | Not-determined |
| 18-102_k141_134779 | Not-determined |
| 18-102_k141_31147 | Not-determined |
| 18-102_k141_176201 | Not-determined |
| 18-102_k141_20770 | Not-determined |
| 18-102_k141_124374 | Not-determined |
| 18-102_k141_227912 | Not-determined |
| 18-102_k141_20774 | Not-determined |
| 18-102_k141_103794 | Low-quality |
| 18-102_k141_227917 | Not-determined |
| 18-102_k141_227921 | Not-determined |
| 18-102_k141_134783 | Not-determined |
| 18-102_k141_196842 | Not-determined |
| 18-102_k141_103797 | Not-determined |
| 18-102_k141_31157 | Not-determined |
| 18-102_k141_155468 | Not-determined |
| 18-102_k141_176226 | Not-determined |
| 18-102_k141_31160 | Low-quality |
| 18-102_k141_103800 | Not-determined |
| 18-102_k141_145072 | Low-quality |
| 18-102_k141_196846 | Not-determined |
| 18-102_k141_165798 | Not-determined |
| 18-102_k141_165799 | Not-determined |
| 18-102_k141_114003 | Not-determined |
| 18-102_k141_186545 | Not-determined |
| 18-102_k141_176236 | Not-determined |
| 18-102_k141_186546 | Not-determined |
| 18-102_k141_20789 | Not-determined |
| 18-102_k141_196858 | Not-determined |
| 18-102_k141_155486 | Not-determined |
| 18-102_k141_186553 | Not-determined |
| 18-102_k141_165814 | Not-determined |
| 18-102_k141_196864 | Not-determined |
| 18-102_k141_176243 | Low-quality |
| 18-102_k141_31172 | Not-determined |
| 18-102_k141_31174 | Not-determined |
| 18-102_k141_165817 | Not-determined |
| 18-102_k141_127 | Not-determined |
| 18-102_k141_51943 | Low-quality |
| 18-102_k141_217647 | Not-determined |
| 18-102_k141_176250 | Not-determined |
| 18-102_k141_62231 | Not-determined |
| 18-102_k141_176254 | Low-quality |
| 18-102_k141_31181 | Not-determined |
| 18-102_k141_20804 | Not-determined |
| 18-102_k141_165821 | Not-determined |
| 18-102_k141_51953 | Not-determined |
| 18-102_k141_196876 | Low-quality |
| 18-102_k141_176267 | Not-determined |
| 18-102_k141_41618 | Not-determined |
| 18-102_k141_186566 | Not-determined |
| 18-102_k141_82985 | Not-determined |
| 18-102_k141_227973 | Not-determined |
| 18-102_k141_124400 | Not-determined |
| 18-102_k141_103814 | Not-determined |
| 18-102_k141_124403 | Not-determined |
| 18-102_k141_103816 | Not-determined |
| 18-102_k141_51955 | Not-determined |
| 18-102_k141_72562 | Not-determined |
| 18-102_k141_72563 | Not-determined |
| 18-102_k141_176280 | Not-determined |
| 18-102_k141_83000 | Not-determined |
| 18-102_k141_72564 | Not-determined |
| 18-102_k141_72565 | Not-determined |
| 18-102_k141_62245 | Not-determined |
| 18-102_k141_51959 | Not-determined |
| 18-102_k141_227982 | Not-determined |
| 18-102_k141_62252 | Not-determined |
| 18-102_k141_186581 | Not-determined |
| 18-102_k141_51971 | Not-determined |
| 18-102_k141_227991 | Not-determined |
| 18-102_k141_196896 | Not-determined |
| 18-102_k141_20812 | Not-determined |
| 18-102_k141_227995 | Not-determined |
| 18-102_k141_51978 | Not-determined |
| 18-102_k141_10521 | Not-determined |
| 18-102_k141_196899 | Not-determined |
| 18-102_k141_134806 | Not-determined |
| 18-102_k141_93304 | Not-determined |
| 18-102_k141_161 | Not-determined |
| 18-102_k141_176307 | Not-determined |
| 18-102_k141_62263 | Not-determined |
| 18-102_k141_93312 | Not-determined |
| 18-102_k141_31197 | Not-determined |
| 18-102_k141_62267 | Not-determined |
| 18-102_k141_186596 | Not-determined |
| 18-102_k141_62274 | Not-determined |
| 18-102_k141_176313 | Not-determined |
| 18-102_k141_83009 | Not-determined |
| 18-102_k141_93329 | Not-determined |
| 18-102_k141_93334 | Not-determined |
| 18-102_k141_207177 | Low-quality |
| 18-102_k141_196928 | Low-quality |
| 18-102_k141_207182 | Not-determined |
| 18-102_k141_181 | Not-determined |
| 18-102_k141_93342 | Not-determined |
| 18-102_k141_51990 | Not-determined |
| 18-102_k141_217701 | Not-determined |
| 18-102_k141_51992 | Not-determined |
| 18-102_k141_41652 | Not-determined |
| 18-102_k141_72571 | Not-determined |
| 18-102_k141_20819 | Not-determined |
| 18-102_k141_103830 | Not-determined |
| 18-102_k141_155514 | Not-determined |
| 18-102_k141_72578 | Not-determined |
| 18-102_k141_176341 | Not-determined |
| 18-102_k141_31227 | Not-determined |
| 18-102_k141_238318 | Not-determined |
| 18-102_k141_196947 | Low-quality |
| 18-102_k141_103840 | Not-determined |
| 18-102_k141_103845 | Not-determined |
| 18-102_k141_103851 | Not-determined |
| 18-102_k141_93348 | Not-determined |
| 18-102_k141_31243 | Not-determined |
| 18-102_k141_41672 | Not-determined |
| 18-102_k141_62308 | Not-determined |
| 18-102_k141_83047 | Not-determined |
| 18-102_k141_83050 | Not-determined |
| 18-102_k141_238330 | Low-quality |
| 18-102_k141_145078 | Not-determined |
| 18-102_k141_72586 | Low-quality |
| 18-102_k141_31257 | Low-quality |
| 18-102_k141_201 | Not-determined |
| 18-102_k141_202 | Not-determined |
| 18-102_k141_217715 | Not-determined |
| 18-102_k141_217719 | Not-determined |
| 18-102_k141_41696 | Not-determined |
| 18-102_k141_155516 | Not-determined |
| 18-102_k141_186633 | Not-determined |
| 18-102_k141_206 | Not-determined |
| 18-102_k141_208 | Not-determined |
| 18-102_k141_93373 | Not-determined |
| 18-102_k141_72594 | Not-determined |
| 18-102_k141_114057 | Not-determined |
| 18-102_k141_186642 | Not-determined |
| 18-102_k141_62327 | Not-determined |
| 18-102_k141_155525 | Not-determined |
| 18-102_k141_155526 | Not-determined |
| 18-102_k141_62334 | Low-quality |
| 18-102_k141_93383 | Low-quality |
| 18-102_k141_93389 | Not-determined |
| 18-102_k141_217726 | Low-quality |
| 18-102_k141_228024 | Not-determined |
| 18-102_k141_155530 | Not-determined |
| 18-102_k141_217735 | Not-determined |
| 18-110_k141_58515 | Medium-quality |
| 18-102_k141_52017 | Not-determined |
| 18-102_k141_155547 | Not-determined |
| 18-102_k141_134829 | Not-determined |
| 18-102_k141_41728 | Not-determined |
| 18-102_k141_31278 | Not-determined |
| 18-102_k141_52021 | Not-determined |
| 18-102_k141_103888 | Not-determined |
| 18-102_k141_52023 | Not-determined |
| 18-102_k141_72610 | Not-determined |
| 18-102_k141_20850 | Not-determined |
| 18-102_k141_165854 | Not-determined |
| 18-102_k141_41734 | Not-determined |
| 18-102_k141_93415 | Not-determined |
| 18-102_k141_93418 | Not-determined |
| 18-102_k141_228061 | Not-determined |
| 18-102_k141_228065 | Not-determined |
| 18-102_k141_228075 | Not-determined |
| 18-102_k141_186672 | Not-determined |
| 18-102_k141_186674 | Not-determined |
| 18-102_k141_176397 | Low-quality |
| 18-102_k141_228078 | Not-determined |
| 18-102_k141_186683 | Not-determined |
| 18-102_k141_52049 | Not-determined |
| 18-102_k141_239 | Low-quality |
| 18-102_k141_241 | Not-determined |
| 18-102_k141_238357 | Low-quality |
| 18-102_k141_176402 | Not-determined |
| 18-102_k141_52056 | Not-determined |
| 18-102_k141_228087 | Low-quality |
| 18-102_k141_83064 | Not-determined |
| 18-102_k141_134869 | Not-determined |
| 18-102_k141_228100 | Not-determined |
| 18-102_k141_52078 | Not-determined |
| 18-102_k141_83080 | Not-determined |
| 18-102_k141_83081 | Not-determined |
| 18-102_k141_238378 | Not-determined |
| 18-102_k141_83086 | Not-determined |
| 18-102_k141_20874 | Not-determined |
| 18-102_k141_83090 | Not-determined |
| 18-102_k141_186689 | Not-determined |
| 18-102_k141_263 | Not-determined |
| 18-102_k141_228112 | Not-determined |
| 18-102_k141_238395 | Not-determined |
| 18-102_k141_228114 | Not-determined |
| 18-102_k141_134909 | Not-determined |
| 18-102_k141_238403 | Not-determined |
| 18-102_k141_93443 | Not-determined |
| 18-102_k141_52093 | Not-determined |
| 18-102_k141_93446 | Not-determined |
| 18-102_k141_238406 | Low-quality |
| 18-102_k141_93448 | Not-determined |
| 18-102_k141_238407 | Not-determined |
| 18-102_k141_41787 | Not-determined |
| 18-102_k141_134915 | Not-determined |
| 18-102_k141_134916 | Not-determined |
| 18-102_k141_31346 | Not-determined |
| 18-102_k141_134917 | Not-determined |
| 18-102_k141_103917 | Not-determined |
| 18-102_k141_124458 | Not-determined |
| 18-102_k141_282 | Not-determined |
| 18-102_k141_72642 | Not-determined |
| 18-102_k141_62366 | Low-quality |
| 18-102_k141_103920 | Not-determined |
| 18-102_k141_93454 | Not-determined |
| 18-102_k141_176412 | Not-determined |
| 18-102_k141_289 | Not-determined |
| 18-102_k141_103924 | Low-quality |
| 18-102_k141_103925 | Low-quality |
| 18-102_k141_93467 | Not-determined |
| 18-102_k141_103927 | Not-determined |
| 18-102_k141_134927 | Not-determined |
| 18-102_k141_72662 | Not-determined |
| 18-102_k141_176423 | Not-determined |
| 18-102_k141_145153 | Not-determined |
| 18-102_k141_103937 | Not-determined |
| 18-102_k141_217742 | Not-determined |
| 18-102_k141_145156 | Not-determined |
| 18-102_k141_134942 | Not-determined |
| 18-102_k141_52114 | Not-determined |
| 18-102_k141_155589 | Not-determined |
| 18-102_k141_52120 | Not-determined |
| 18-102_k141_207248 | Not-determined |
| 18-102_k141_52129 | Not-determined |
| 18-102_k141_155596 | Not-determined |
| 18-102_k141_124494 | Not-determined |
| 18-102_k141_31377 | Not-determined |
| 18-102_k141_114085 | Low-quality |
| 18-102_k141_145165 | Not-determined |
| 18-102_k141_41821 | Low-quality |
| 18-102_k141_228159 | Not-determined |
| 18-102_k141_41822 | Not-determined |
| 18-102_k141_31394 | Not-determined |
| 18-102_k141_124505 | Not-determined |
| 18-102_k141_228162 | Not-determined |
| 18-102_k141_155604 | Not-determined |
| 18-102_k141_72681 | Not-determined |
| 18-102_k141_238410 | Not-determined |
| 18-102_k141_155607 | Not-determined |
| 18-102_k141_72684 | Not-determined |
| 18-102_k141_72686 | Not-determined |
| 18-102_k141_41831 | Not-determined |
| 18-102_k141_186718 | Low-quality |
| 18-102_k141_52154 | Not-determined |
| 18-102_k141_124512 | Not-determined |
| 18-102_k141_155613 | Not-determined |
| 18-102_k141_186720 | Not-determined |
| 18-102_k141_238418 | Not-determined |
| 18-102_k141_31406 | Not-determined |
| 18-102_k141_238420 | Not-determined |
| 18-102_k141_72699 | Not-determined |
| 18-102_k141_72704 | Not-determined |
| 18-102_k141_134971 | Low-quality |
| 18-102_k141_31413 | Not-determined |
| 18-102_k141_31414 | Low-quality |
| 18-102_k141_83129 | Low-quality |
| 18-102_k141_176437 | Not-determined |
| 18-102_k141_114120 | Not-determined |
| 18-102_k141_145221 | Not-determined |
| 18-102_k141_145223 | Not-determined |
| 18-102_k141_155638 | Not-determined |
| 18-102_k141_41863 | Not-determined |
| 18-102_k141_186743 | Not-determined |
| 18-102_k141_41864 | Not-determined |
| 18-102_k141_41867 | Not-determined |
| 18-102_k141_186744 | Not-determined |
| 18-102_k141_103969 | Not-determined |
| 18-102_k141_41873 | Not-determined |
| 18-102_k141_228177 | Not-determined |
| 18-102_k141_217774 | Not-determined |
| 18-102_k141_103974 | Not-determined |
| 18-102_k141_31426 | Low-quality |
| 18-102_k141_41878 | Not-determined |
| 18-102_k141_41882 | Not-determined |
| 18-102_k141_155659 | Low-quality |
| 18-102_k141_124564 | Not-determined |
| 18-102_k141_315 | Not-determined |
| 18-102_k141_103977 | Not-determined |
| 18-102_k141_41894 | Not-determined |
| 18-102_k141_319 | Not-determined |
| 18-102_k141_135002 | Not-determined |
| 18-102_k141_93524 | Not-determined |
| 18-102_k141_217802 | Not-determined |
| 18-102_k141_176467 | Low-quality |
| 18-102_k141_217806 | Not-determined |
| 18-102_k141_62394 | Not-determined |
| 18-102_k141_31462 | Not-determined |
| 18-102_k141_93536 | Not-determined |
| 18-102_k141_329 | Not-determined |
| 18-102_k141_10566 | Low-quality |
| 18-102_k141_331 | Not-determined |
| 18-102_k141_10569 | Not-determined |
| 18-102_k141_186773 | Not-determined |
| 18-102_k141_165883 | Not-determined |
| 18-102_k141_217818 | Not-determined |
| 18-102_k141_135013 | Not-determined |
| 18-102_k141_165897 | Not-determined |
| 18-102_k141_207277 | Not-determined |
| 18-102_k141_217824 | Not-determined |
| 18-102_k141_10584 | Not-determined |
| 18-102_k141_186776 | Not-determined |
| 18-102_k141_93554 | Not-determined |
| 18-102_k141_41935 | Not-determined |
| 18-102_k141_72730 | Not-determined |
| 18-102_k141_93563 | Not-determined |
| 18-102_k141_72732 | Not-determined |
| 18-102_k141_165916 | Not-determined |
| 18-102_k141_93565 | Not-determined |
| 18-102_k141_93566 | Not-determined |
| 18-102_k141_165921 | Not-determined |
| 18-102_k141_124587 | Not-determined |
| 18-102_k141_62446 | Not-determined |
| 18-102_k141_351 | Not-determined |
| 18-102_k141_62447 | Not-determined |
| 18-102_k141_217834 | Not-determined |
| 18-102_k141_217836 | Low-quality |
| 18-102_k141_62452 | Not-determined |
| 18-102_k141_72749 | Not-determined |
| 18-102_k141_72750 | Not-determined |
| 18-102_k141_228202 | Not-determined |
| 18-102_k141_72755 | Not-determined |
| 18-102_k141_165929 | Not-determined |
| 18-102_k141_228214 | Not-determined |
| 18-102_k141_124610 | Not-determined |
| 18-102_k141_207316 | Not-determined |
| 18-102_k141_10594 | Not-determined |
| 18-102_k141_72761 | Not-determined |
| 18-102_k141_145306 | Low-quality |
| 18-102_k141_10604 | Not-determined |
| 18-102_k141_165943 | Not-determined |
| 18-102_k141_41967 | Not-determined |
| 18-102_k141_124623 | Not-determined |
| 18-102_k141_207340 | Not-determined |
| 18-102_k141_145321 | Not-determined |
| 18-102_k141_217849 | Low-quality |
| 18-102_k141_238475 | Low-quality |
| 18-102_k141_186793 | Not-determined |
| 18-102_k141_228236 | Low-quality |
| 18-102_k141_238483 | Not-determined |
| 18-102_k141_124628 | Low-quality |
| 18-102_k141_41984 | Not-determined |
| 18-102_k141_238487 | Not-determined |
| 18-102_k141_124630 | Low-quality |
| 18-102_k141_62495 | Not-determined |
| 18-102_k141_124634 | Low-quality |
| 18-102_k141_176475 | Not-determined |
| 18-102_k141_155672 | Low-quality |
| 18-102_k141_83144 | Low-quality |
| 18-102_k141_31475 | Not-determined |
| 18-102_k141_72792 | Not-determined |
| 18-102_k141_31476 | Not-determined |
| 18-102_k141_31477 | Not-determined |
| 18-102_k141_31480 | Not-determined |
| 18-102_k141_83158 | Not-determined |
| 18-102_k141_145344 | Low-quality |
| 18-102_k141_124656 | Not-determined |
| 18-102_k141_135063 | Not-determined |
| 18-102_k141_155679 | Not-determined |
| 18-102_k141_72809 | Not-determined |
| 18-102_k141_83164 | Not-determined |
| 18-102_k141_217858 | Not-determined |
| 18-102_k141_176481 | Not-determined |
| 18-102_k141_207357 | Not-determined |
| 18-102_k141_207359 | Not-determined |
| 18-102_k141_217870 | Not-determined |
| 18-102_k141_176496 | Not-determined |
| 18-102_k141_135095 | Not-determined |
| 18-102_k141_135098 | Not-determined |
| 18-102_k141_124678 | Not-determined |
| 18-102_k141_124680 | Not-determined |
| 18-102_k141_93581 | Not-determined |
| 18-102_k141_238532 | Not-determined |
| 18-102_k141_207378 | Not-determined |
| 18-102_k141_238533 | Low-quality |
| 18-102_k141_114146 | Not-determined |
| 18-102_k141_228273 | Not-determined |
| 18-102_k141_114154 | Not-determined |
| 18-102_k141_155721 | Not-determined |
| 18-102_k141_124702 | Not-determined |
| 18-102_k141_124706 | Not-determined |
| 18-102_k141_217891 | Not-determined |
| 18-102_k141_207388 | Not-determined |
| 18-102_k141_20943 | Not-determined |
| 18-102_k141_155726 | Not-determined |
| 18-102_k141_155728 | Not-determined |
| 18-102_k141_114175 | Not-determined |
| 18-102_k141_72838 | Not-determined |
| 18-102_k141_135102 | Not-determined |
| 18-102_k141_135103 | Not-determined |
| 18-102_k141_155730 | Not-determined |
| 18-102_k141_155732 | Not-determined |
| 18-102_k141_166002 | Not-determined |
| 18-102_k141_207406 | Not-determined |
| 18-102_k141_155741 | Not-determined |
| 18-102_k141_155742 | Not-determined |
| 18-102_k141_166004 | Not-determined |
| 18-102_k141_186801 | Not-determined |
| 18-102_k141_42045 | Not-determined |
| 18-102_k141_166005 | Not-determined |
| 18-102_k141_217914 | Not-determined |
| 18-102_k141_42050 | Low-quality |
| 18-102_k141_83195 | Not-determined |
| 18-102_k141_217917 | Not-determined |
| 18-102_k141_114190 | Not-determined |
| 18-102_k141_186807 | Not-determined |
| 18-102_k141_207413 | Not-determined |
| 18-102_k141_114193 | Not-determined |
| 18-102_k141_217924 | Not-determined |
| 18-102_k141_135106 | Not-determined |
| 18-102_k141_166010 | Not-determined |
| 18-102_k141_186812 | Not-determined |
| 18-102_k141_217927 | Not-determined |
| 18-102_k141_166013 | Not-determined |
| 18-102_k141_42058 | Not-determined |
| 18-102_k141_83205 | Not-determined |
| 18-102_k141_186818 | Not-determined |
| 18-102_k141_72872 | Not-determined |
| 18-102_k141_93646 | Not-determined |
| 18-102_k141_93648 | Not-determined |
| 18-102_k141_207423 | Low-quality |
| 18-102_k141_238558 | Not-determined |
| 18-102_k141_238559 | Not-determined |
| 18-102_k141_104040 | Not-determined |
| 18-102_k141_217945 | Low-quality |
| 18-102_k141_72887 | Low-quality |
| 18-102_k141_42081 | Not-determined |
| 18-102_k141_238572 | Not-determined |
| 18-102_k141_145401 | Not-determined |
| 18-102_k141_21001 | Not-determined |
| 18-102_k141_42095 | Not-determined |
| 18-102_k141_238576 | Not-determined |
| 18-102_k141_145410 | Not-determined |
| 18-102_k141_238586 | Not-determined |
| 18-102_k141_72898 | Not-determined |
| 18-102_k141_114210 | Not-determined |
| 18-102_k141_217962 | Not-determined |
| 18-102_k141_145427 | Not-determined |
| 18-102_k141_42105 | Not-determined |
| 18-102_k141_114217 | Not-determined |
| 18-102_k141_408 | Not-determined |
| 18-102_k141_42112 | Not-determined |
| 18-102_k141_83253 | Not-determined |
| 18-102_k141_10651 | Low-quality |
| 18-102_k141_155782 | Not-determined |
| 18-102_k141_42120 | Not-determined |
| 18-102_k141_72902 | Not-determined |
| 18-102_k141_72907 | Not-determined |
| 18-102_k141_114224 | Low-quality |
| 18-102_k141_114225 | Not-determined |
| 18-102_k141_238617 | Not-determined |
| 18-102_k141_10661 | Not-determined |
| 18-102_k141_186882 | Not-determined |
| 18-102_k141_420 | Low-quality |
| 18-102_k141_145441 | Not-determined |
| 18-102_k141_42131 | Not-determined |
| 18-102_k141_10668 | Not-determined |
| 18-102_k141_228314 | Not-determined |
| 18-102_k141_228321 | Not-determined |
| 18-102_k141_217985 | Not-determined |
| 18-102_k141_196989 | Not-determined |
| 18-102_k141_10675 | Not-determined |
| 18-102_k141_207455 | Not-determined |
| 18-102_k141_217990 | Low-quality |
| 18-102_k141_429 | Not-determined |
| 18-102_k141_238640 | Not-determined |
| 18-102_k141_145444 | Low-quality |
| 18-102_k141_176558 | Not-determined |
| 18-102_k141_238642 | Not-determined |
| 18-102_k141_238643 | Not-determined |
| 18-102_k141_238644 | Not-determined |
| 18-102_k141_238645 | Not-determined |
| 18-102_k141_238646 | Not-determined |
| 18-102_k141_433 | Not-determined |
| 18-102_k141_93666 | Not-determined |
| 18-102_k141_186885 | Not-determined |
| 18-102_k141_238649 | Not-determined |
| 18-102_k141_176568 | Not-determined |
| 18-102_k141_444 | Not-determined |
| 18-102_k141_176571 | Not-determined |
| 18-102_k141_124733 | Not-determined |
| 18-102_k141_186892 | Not-determined |
| 18-102_k141_446 | Low-quality |
| 18-102_k141_238656 | Not-determined |
| 18-102_k141_72933 | Not-determined |
| 18-102_k141_238658 | Not-determined |
| 18-102_k141_166046 | Not-determined |
| 18-102_k141_228333 | Not-determined |
| 18-102_k141_176578 | Not-determined |
| 18-102_k141_10682 | Not-determined |
| 18-102_k141_10684 | Not-determined |
| 18-102_k141_155806 | Not-determined |
| 18-102_k141_10688 | Not-determined |
| 18-102_k141_31522 | Not-determined |
| 18-102_k141_124747 | Not-determined |
| 18-102_k141_31524 | Low-quality |
| 18-102_k141_186912 | Not-determined |
| 18-102_k141_238683 | Low-quality |
| 18-102_k141_10705 | Not-determined |
| 18-102_k141_72942 | Not-determined |
| 18-102_k141_72948 | Not-determined |
| 18-102_k141_124754 | Not-determined |
| 18-102_k141_10715 | Not-determined |
| 18-102_k141_238696 | Not-determined |
| 18-102_k141_10723 | Not-determined |
| 18-102_k141_31529 | Not-determined |
| 18-102_k141_31531 | Not-determined |
| 18-102_k141_114257 | Not-determined |
| 18-102_k141_72952 | Not-determined |
| 18-102_k141_114266 | Not-determined |
| 18-102_k141_114269 | Low-quality |
| 18-102_k141_479 | Not-determined |
| 18-102_k141_166082 | Not-determined |
| 18-102_k141_21076 | Not-determined |
| 18-102_k141_10738 | Not-determined |
| 18-102_k141_21077 | Not-determined |
| 18-102_k141_166086 | Not-determined |
| 18-102_k141_186953 | Not-determined |
| 18-102_k141_186956 | Low-quality |
| 18-102_k141_238720 | Not-determined |
| 18-102_k141_10741 | Not-determined |
| 18-102_k141_496 | Not-determined |
| 18-102_k141_21085 | Not-determined |
| 18-102_k141_21089 | Not-determined |
| 18-102_k141_10748 | Low-quality |
| 18-102_k141_21091 | Not-determined |
| 18-102_k141_31574 | Low-quality |
| 18-102_k141_31577 | Not-determined |
| 18-102_k141_10754 | Low-quality |
| 18-102_k141_52187 | Not-determined |
| 18-102_k141_52189 | Not-determined |
| 18-102_k141_207473 | Not-determined |
| 18-102_k141_52196 | Not-determined |
| 18-102_k141_21102 | Not-determined |
| 18-102_k141_124785 | Not-determined |
| 18-102_k141_207481 | Low-quality |
| 18-102_k141_238731 | Not-determined |
| 18-102_k141_52200 | Not-determined |
| 18-102_k141_124793 | Not-determined |
| 18-102_k141_52211 | Not-determined |
| 18-102_k141_186963 | Not-determined |
| 18-102_k141_21107 | Low-quality |
| 18-102_k141_238735 | Not-determined |
| 18-102_k141_218016 | Not-determined |
| 18-102_k141_218018 | Not-determined |
| 18-102_k141_10766 | Not-determined |
| 18-102_k141_124808 | Not-determined |
| 18-102_k141_124810 | Not-determined |
| 18-102_k141_124813 | Low-quality |
| 18-102_k141_42163 | Not-determined |
| 18-102_k141_124818 | Not-determined |
| 18-102_k141_218026 | Low-quality |
| 18-102_k141_73002 | Not-determined |
| 18-102_k141_62577 | Not-determined |
| 18-102_k141_207506 | Not-determined |
| 18-102_k141_155873 | Not-determined |
| 18-102_k141_238758 | Not-determined |
| 18-102_k141_186981 | Not-determined |
| 18-102_k141_207513 | Not-determined |
| 18-102_k141_238764 | Not-determined |
| 18-102_k141_207517 | Not-determined |
| 18-102_k141_21125 | Not-determined |
| 18-102_k141_155889 | Not-determined |
| 18-102_k141_187001 | Not-determined |
| 18-102_k141_218047 | Not-determined |
| 18-102_k141_207527 | Not-determined |
| 18-102_k141_187005 | Not-determined |
| 18-102_k141_73008 | Not-determined |
| 18-102_k141_21136 | Not-determined |
| 18-102_k141_187008 | Not-determined |
| 18-102_k141_124828 | Not-determined |
| 18-102_k141_187009 | Not-determined |
| 18-102_k141_124833 | Not-determined |
| 18-102_k141_145468 | Not-determined |
| 18-102_k141_124837 | Not-determined |
| 18-102_k141_83273 | Not-determined |
| 18-102_k141_155914 | Not-determined |
| 18-102_k141_207536 | Not-determined |
| 18-102_k141_228344 | Low-quality |
| 18-102_k141_155925 | Not-determined |

| | |
|---|---|
| 18-102_k141_145488 | Not-determined |
| 18-102_k141_42191 | Not-determined |
| 18-102_k141_145492 | Not-determined |
| 18-102_k141_531 | Not-determined |
| 18-102_k141_207550 | Not-determined |
| 18-102_k141_31621 | Not-determined |
| 18-102_k141_73013 | Not-determined |
| 18-102_k141_155931 | Not-determined |
| 18-102_k141_73016 | Not-determined |
| 18-102_k141_197038 | Not-determined |
| 18-102_k141_145512 | Not-determined |
| 18-102_k141_228376 | Not-determined |
| 18-102_k141_21155 | Not-determined |
| 18-102_k141_21157 | Low-quality |
| 18-102_k141_124860 | Low-quality |
| 18-102_k141_207569 | Low-quality |
| 18-102_k141_176616 | Not-determined |
| 18-102_k141_145536 | Not-determined |
| 18-102_k141_52225 | Not-determined |
| 18-102_k141_52226 | Not-determined |
| 18-102_k141_21170 | Not-determined |
| 18-102_k141_52235 | Not-determined |
| 18-102_k141_135165 | Low-quality |
| 18-102_k141_228407 | Not-determined |
| 18-102_k141_83302 | Low-quality |
| 18-102_k141_73036 | Not-determined |
| 18-102_k141_83306 | Not-determined |
| 18-102_k141_145559 | Low-quality |
| 18-102_k141_207603 | Not-determined |
| 18-102_k141_31640 | Low-quality |
| 18-102_k141_73046 | Not-determined |
| 18-102_k141_31642 | Low-quality |
| 18-102_k141_145565 | Not-determined |
| 18-102_k141_42271 | Not-determined |
| 18-102_k141_83326 | Low-quality |
| 18-102_k141_42276 | Not-determined |
| 18-102_k141_238814 | Not-determined |
| 18-102_k141_52267 | Not-determined |
| 18-102_k141_52268 | Low-quality |
| 18-102_k141_145577 | Not-determined |
| 18-102_k141_207622 | Not-determined |
| 18-102_k141_83352 | Not-determined |
| 18-102_k141_104131 | Not-determined |
| 18-102_k141_166109 | Not-determined |
| 18-102_k141_21197 | Not-determined |
| 18-102_k141_62613 | Not-determined |
| 18-102_k141_124897 | Not-determined |
| 18-102_k141_176627 | Not-determined |
| 18-102_k141_124901 | Not-determined |
| 18-102_k141_21204 | Not-determined |
| 18-102_k141_238842 | Not-determined |
| 18-102_k141_238852 | Low-quality |
| 18-102_k141_238857 | Not-determined |
| 18-102_k141_155948 | Not-determined |
| 18-102_k141_42286 | Not-determined |
| 18-102_k141_21214 | Not-determined |
| 18-102_k141_73098 | Low-quality |
| 18-102_k141_238867 | Not-determined |
| 18-102_k141_104138 | Low-quality |
| 18-102_k141_238870 | Not-determined |
| 18-102_k141_104139 | Not-determined |
| 18-102_k141_10801 | Low-quality |
| 18-102_k141_73106 | Not-determined |
| 18-102_k141_42300 | Not-determined |
| 18-102_k141_83371 | Not-determined |
| 18-102_k141_31673 | Not-determined |
| 18-102_k141_31674 | Not-determined |
| 18-102_k141_21228 | Not-determined |
| 18-102_k141_238884 | Not-determined |
| 18-102_k141_104159 | Not-determined |
| 18-102_k141_31680 | Not-determined |
| 18-102_k141_21238 | Low-quality |
| 18-102_k141_155968 | Not-determined |
| 18-102_k141_52313 | Not-determined |
| 18-102_k141_124931 | Low-quality |
| 18-102_k141_124932 | Not-determined |
| 18-102_k141_104169 | Not-determined |
| 18-102_k141_21243 | Not-determined |
| 18-102_k141_21244 | Not-determined |
| 18-102_k141_155975 | Not-determined |
| 18-102_k141_155976 | Not-determined |
| 18-102_k141_21251 | Not-determined |
| 18-102_k141_73123 | Not-determined |
| 18-102_k141_135199 | Not-determined |
| 18-102_k141_135202 | Not-determined |
| 18-102_k141_42313 | Low-quality |
| 18-102_k141_21258 | Low-quality |
| 18-102_k141_21260 | Not-determined |
| 18-102_k141_575 | Not-determined |
| 18-102_k141_187030 | Not-determined |
| 18-102_k141_145606 | Not-determined |
| 18-102_k141_166133 | Not-determined |
| 18-102_k141_187036 | Not-determined |
| 18-102_k141_124960 | Not-determined |
| 18-102_k141_187044 | Not-determined |
| 18-102_k141_21274 | Not-determined |
| 18-102_k141_155993 | Not-determined |
| 18-102_k141_166150 | Low-quality |
| 18-102_k141_166152 | Not-determined |
| 18-102_k141_238888 | Not-determined |
| 18-102_k141_166154 | Low-quality |
| 18-102_k141_155999 | Not-determined |
| 18-102_k141_93703 | Not-determined |
| 18-102_k141_176674 | Not-determined |
| 18-102_k141_145620 | Not-determined |
| 18-102_k141_73133 | Low-quality |
| 18-102_k141_176681 | Not-determined |
| 18-102_k141_156009 | Low-quality |
| 18-102_k141_156012 | Not-determined |
| 18-102_k141_93711 | Not-determined |
| 18-102_k141_93712 | Not-determined |
| 18-102_k141_218061 | Not-determined |
| 18-102_k141_218064 | Not-determined |
| 18-102_k141_601 | Not-determined |
| 18-102_k141_228424 | Not-determined |
| 18-102_k141_238902 | Not-determined |
| 18-102_k141_228426 | Not-determined |
| 18-102_k141_228428 | Not-determined |
| 18-102_k141_228440 | Not-determined |
| 18-102_k141_156023 | Not-determined |
| 18-102_k141_620 | Not-determined |
| 18-102_k141_21297 | Not-determined |
| 18-102_k141_176698 | Not-determined |
| 18-102_k141_218098 | Not-determined |
| 18-102_k141_218099 | Not-determined |
| 18-102_k141_197064 | Not-determined |
| 18-102_k141_135224 | Not-determined |
| 18-102_k141_135225 | Not-determined |
| 18-102_k141_187080 | Not-determined |
| 18-102_k141_207659 | Not-determined |
| 18-102_k141_176704 | Not-determined |
| 18-102_k141_207661 | Not-determined |
| 18-102_k141_197068 | Not-determined |
| 18-102_k141_156032 | Not-determined |
| 18-102_k141_52340 | Not-determined |
| 18-102_k141_21322 | Not-determined |
| 18-102_k141_135233 | Not-determined |
| 18-102_k141_104181 | Not-determined |
| 18-102_k141_197085 | Not-determined |
| 18-102_k141_21329 | Not-determined |
| 18-102_k141_135241 | Not-determined |
| 18-102_k141_156046 | Not-determined |
| 18-102_k141_187092 | Not-determined |
| 18-102_k141_238917 | Not-determined |
| 18-102_k141_124988 | Not-determined |
| 18-102_k141_207688 | Not-determined |
| 18-102_k141_218105 | Not-determined |
| 18-102_k141_176719 | Not-determined |
| 18-102_k141_207691 | Not-determined |
| 18-102_k141_176723 | Not-determined |
| 18-102_k141_156053 | Not-determined |
| 18-102_k141_93744 | Not-determined |
| 18-102_k141_42391 | Low-quality |
| 18-102_k141_228445 | Not-determined |
| 18-102_k141_228448 | Not-determined |
| 18-102_k141_660 | Not-determined |
| 18-102_k141_135257 | Not-determined |
| 18-102_k141_31735 | Not-determined |
| 18-102_k141_207710 | Not-determined |
| 18-102_k141_197111 | Not-determined |
| 18-102_k141_156059 | Not-determined |
| 18-102_k141_135265 | Not-determined |
| 18-102_k141_21336 | Not-determined |
| 18-102_k141_228452 | Not-determined |
| 18-102_k141_135277 | Not-determined |
| 18-102_k141_166230 | Not-determined |
| 18-102_k141_145656 | Not-determined |
| 18-102_k141_145657 | Not-determined |
| 18-102_k141_93759 | Not-determined |
| 18-102_k141_31749 | Not-determined |
| 18-102_k141_135285 | Low-quality |
| 18-102_k141_166235 | Not-determined |
| 18-102_k141_197131 | Not-determined |
| 18-102_k141_197133 | Not-determined |
| 18-102_k141_187098 | Not-determined |
| 18-102_k141_684 | Not-determined |
| 18-102_k141_21342 | Not-determined |
| 18-102_k141_42410 | Not-determined |
| 18-102_k141_73155 | Not-determined |
| 18-102_k141_10869 | Not-determined |
| 18-102_k141_10871 | Not-determined |
| 18-102_k141_207743 | Not-determined |
| 18-102_k141_21348 | Not-determined |
| 18-102_k141_176743 | Not-determined |
| 18-102_k141_73162 | Not-determined |
| 18-102_k141_156080 | Not-determined |
| 18-102_k141_73174 | Not-determined |
| 18-102_k141_21354 | Low-quality |
| 18-102_k141_10886 | Not-determined |
| 18-102_k141_228487 | Not-determined |
| 18-102_k141_10888 | Not-determined |
| 18-102_k141_93792 | Not-determined |
| 18-102_k141_21361 | Not-determined |
| 18-102_k141_93798 | Not-determined |
| 18-102_k141_10901 | Not-determined |
| 18-102_k141_10906 | Not-determined |
| 18-102_k141_114280 | Not-determined |
| 18-102_k141_228490 | Not-determined |
| 18-102_k141_42430 | Not-determined |
| 18-102_k141_698 | Low-quality |
| 18-102_k141_228493 | Not-determined |
| 18-102_k141_207764 | Not-determined |
| 18-102_k141_114290 | Not-determined |
| 18-102_k141_114291 | Not-determined |
| 18-102_k141_104226 | Low-quality |
| 18-102_k141_114295 | Not-determined |
| 18-102_k141_207765 | Not-determined |
| 18-102_k141_145687 | Not-determined |
| 18-102_k141_145693 | Not-determined |
| 18-102_k141_228501 | Not-determined |
| 18-102_k141_21384 | Low-quality |
| 18-102_k141_42442 | Not-determined |
| 18-102_k141_42444 | Not-determined |
| 18-102_k141_93816 | Not-determined |
| 18-102_k141_93818 | Low-quality |
| 18-102_k141_708 | Low-quality |
| 18-102_k141_42450 | Not-determined |
| 18-102_k141_166284 | Low-quality |
| 18-102_k141_21395 | Not-determined |
| 18-102_k141_42458 | Not-determined |
| 18-102_k141_197176 | Not-determined |
| 18-102_k141_21402 | Not-determined |
| 18-102_k141_62630 | Not-determined |
| 18-102_k141_42464 | Not-determined |
| 18-110-final-viral-combined_k141_58515\|\|full | Medium-quality |
| 18-102_k141_10946 | Not-determined |
| 18-102_k141_124993 | Low-quality |
| 18-102_k141_166305 | Not-determined |
| 18-102_k141_10957 | Not-determined |
| 18-102_k141_125000 | Not-determined |
| 18-102_k141_83432 | Not-determined |
| 18-102_k141_228536 | Not-determined |
| 18-102_k141_135312 | Not-determined |
| 18-102_k141_104242 | Low-quality |
| 18-102_k141_31786 | Not-determined |
| 18-102_k141_176779 | Not-determined |
| 18-102_k141_31788 | Not-determined |
| 18-102_k141_31789 | Not-determined |
| 18-102_k141_31793 | Not-determined |
| 18-102_k141_31799 | Not-determined |
| 18-102_k141_83445 | Not-determined |
| 18-102_k141_10979 | Not-determined |
| 18-102_k141_176790 | Not-determined |
| 18-102_k141_156099 | Not-determined |
| 18-102_k141_176792 | Not-determined |
| 18-102_k141_228559 | Not-determined |
| 18-102_k141_176797 | Not-determined |
| 18-102_k141_228563 | Not-determined |
| 18-102_k141_83450 | Not-determined |
| 18-102_k141_62643 | Not-determined |
| 18-102_k141_62647 | Not-determined |
| 18-102_k141_83457 | Low-quality |
| 18-102_k141_62651 | Not-determined |
| 18-102_k141_197203 | Not-determined |
| 18-102_k141_42488 | Low-quality |
| 18-102_k141_156112 | Low-quality |
| 18-102_k141_10993 | Not-determined |
| 18-102_k141_228576 | Not-determined |
| 18-102_k141_104264 | Not-determined |
| 18-102_k141_166362 | Not-determined |
| 18-102_k141_31815 | Not-determined |
| 18-102_k141_207800 | Not-determined |
| 18-102_k141_42514 | Not-determined |
| 18-102_k141_207804 | Not-determined |
| 18-102_k141_31826 | Not-determined |
| 18-102_k141_31827 | Not-determined |
| 18-102_k141_21446 | Low-quality |
| 18-102_k141_11017 | Not-determined |
| 18-102_k141_31841 | Low-quality |
| 18-102_k141_228600 | Not-determined |
| 18-102_k141_11025 | Not-determined |
| 18-102_k141_42534 | Not-determined |
| 18-102_k141_145732 | Not-determined |
| 18-102_k141_42538 | Not-determined |
| 18-102_k141_228614 | Not-determined |
| 18-102_k141_21460 | Low-quality |
| 18-102_k141_21470 | Not-determined |
| 18-102_k141_228629 | Not-determined |
| 18-102_k141_176818 | Not-determined |
| 18-102_k141_145747 | Not-determined |
| 18-102_k141_145748 | Low-quality |
| 18-102_k141_93825 | Not-determined |
| 18-102_k141_42562 | Not-determined |
| 18-102_k141_93827 | Not-determined |
| 18-102_k141_145751 | Not-determined |
| 18-102_k141_228639 | Not-determined |
| 18-102_k141_228640 | Not-determined |
| 18-102_k141_42568 | Not-determined |
| 18-102_k141_228642 | Not-determined |
| 18-102_k141_93831 | Not-determined |
| 18-102_k141_176822 | Not-determined |
| 18-102_k141_218111 | Not-determined |
| 18-102_k141_145757 | Not-determined |
| 18-102_k141_176827 | Not-determined |
| 18-102_k141_31846 | Not-determined |
| 18-102_k141_176829 | Not-determined |
| 18-102_k141_187138 | Not-determined |
| 18-102_k141_104283 | Not-determined |
| 18-102_k141_104287 | Not-determined |
| 18-102_k141_187144 | Not-determined |
| 18-102_k141_11056 | Not-determined |
| 18-102_k141_11057 | Not-determined |
| 18-102_k141_42588 | Not-determined |
| 18-102_k141_52368 | Not-determined |
| 18-102_k141_176841 | Not-determined |
| 18-102_k141_31853 | Not-determined |
| 18-102_k141_176844 | Not-determined |
| 18-102_k141_176851 | Not-determined |
| 18-102_k141_176855 | Not-determined |
| 18-102_k141_176857 | Not-determined |
| 18-102_k141_176858 | Not-determined |
| 18-102_k141_42597 | Low-quality |
| 18-102_k141_42598 | Not-determined |
| 18-102_k141_187159 | Not-determined |
| 18-102_k141_228675 | Not-determined |
| 18-102_k141_114373 | Not-determined |
| 18-102_k141_145782 | Not-determined |
| 18-102_k141_52374 | Not-determined |
| 18-102_k141_187176 | Not-determined |
| 18-102_k141_788 | Not-determined |
| 18-102_k141_21529 | Not-determined |
| 18-102_k141_207823 | Low-quality |
| 18-102_k141_207827 | Low-quality |
| 18-102_k141_21534 | Not-determined |
| 18-102_k141_52382 | Not-determined |
| 18-102_k141_166394 | Not-determined |
| 18-102_k141_83484 | Not-determined |
| 18-102_k141_166395 | Not-determined |
| 18-102_k141_11083 | Low-quality |
| 18-102_k141_187199 | Not-determined |
| 18-102_k141_114398 | Not-determined |
| 18-102_k141_83493 | Not-determined |
| 18-102_k141_114404 | Not-determined |
| 18-102_k141_176894 | Not-determined |
| 18-102_k141_125032 | Not-determined |
| 18-102_k141_228689 | Not-determined |
| 18-102_k141_166408 | Not-determined |
| 18-102_k141_799 | Not-determined |
| 18-102_k141_52404 | Not-determined |
| 18-102_k141_93834 | Low-quality |
| 18-102_k141_187203 | Not-determined |
| 18-102_k141_62673 | Not-determined |
| 18-102_k141_166418 | Not-determined |
| 18-102_k141_62675 | Not-determined |
| 18-102_k141_166422 | Not-determined |
| 18-102_k141_187209 | Not-determined |
| 18-102_k141_62685 | Not-determined |
| 18-102_k141_52413 | Not-determined |
| 18-102_k141_52419 | Not-determined |
| 18-102_k141_83523 | Not-determined |
| 18-102_k141_83527 | Not-determined |
| 18-102_k141_21556 | Low-quality |
| 18-102_k141_228728 | Not-determined |
| 18-102_k141_21558 | Not-determined |
| 18-102_k141_207838 | Not-determined |
| 18-102_k141_197227 | Not-determined |
| 18-102_k141_197228 | Not-determined |
| 18-102_k141_114424 | Not-determined |
| 18-102_k141_104314 | Not-determined |
| 18-102_k141_125056 | Not-determined |
| 18-102_k141_125066 | Not-determined |
| 18-102_k141_823 | Low-quality |
| 18-102_k141_104317 | Not-determined |
| 18-102_k141_207849 | Not-determined |
| 18-102_k141_207851 | Low-quality |
| 18-102_k141_207852 | Low-quality |
| 18-102_k141_52426 | Not-determined |
| 18-102_k141_52427 | Not-determined |
| 18-102_k141_826 | Not-determined |
| 18-102_k141_218121 | Not-determined |
| 18-102_k141_73239 | Not-determined |
| 18-102_k141_218123 | Low-quality |
| 18-102_k141_218124 | Not-determined |
| 18-102_k141_114435 | Not-determined |
| 18-102_k141_114438 | Not-determined |
| 18-102_k141_73246 | Not-determined |
| 18-102_k141_156124 | Not-determined |
| 18-102_k141_73249 | Not-determined |
| 18-102_k141_145833 | Low-quality |
| 18-102_k141_836 | Low-quality |
| 18-102_k141_145834 | Not-determined |
| 18-102_k141_218131 | Not-determined |
| 18-102_k141_207854 | Low-quality |
| 18-102_k141_42669 | Not-determined |
| 18-102_k141_114454 | Low-quality |
| 18-102_k141_145843 | Not-determined |
| 18-102_k141_218149 | Not-determined |
| 18-102_k141_156143 | Not-determined |
| 18-102_k141_52450 | Not-determined |
| 18-102_k141_114462 | Not-determined |
| 18-102_k141_207868 | Not-determined |
| 18-102_k141_21593 | Not-determined |
| 18-102_k141_156155 | Not-determined |
| 18-102_k141_52458 | Not-determined |
| 18-102_k141_207871 | Not-determined |
| 18-102_k141_21596 | Not-determined |
| 18-102_k141_73276 | Not-determined |
| 18-102_k141_52463 | Not-determined |
| 18-102_k141_156160 | Not-determined |
| 18-102_k141_238934 | Not-determined |
| 18-102_k141_21599 | Not-determined |
| 18-102_k141_238937 | Not-determined |
| 18-102_k141_21604 | Not-determined |
| 18-102_k141_52472 | Not-determined |
| 18-102_k141_42686 | Not-determined |
| 18-102_k141_52474 | Not-determined |
| 18-102_k141_145860 | Not-determined |
| 18-102_k141_93849 | Not-determined |
| 18-102_k141_145869 | Low-quality |
| 18-102_k141_176919 | Not-determined |
| 18-102_k141_207883 | Not-determined |
| 18-102_k141_207884 | Not-determined |
| 18-102_k141_156177 | Not-determined |
| 18-102_k141_104347 | Not-determined |
| 18-102_k141_104349 | Not-determined |
| 18-102_k141_135323 | Not-determined |
| 18-102_k141_218180 | Not-determined |
| 18-102_k141_125088 | Not-determined |
| 18-102_k141_125091 | Not-determined |
| 18-102_k141_228757 | Not-determined |
| 18-102_k141_114493 | Not-determined |
| 18-102_k141_176930 | Low-quality |
| 18-102_k141_176931 | Not-determined |
| 18-102_k141_21630 | Low-quality |
| 18-102_k141_884 | Not-determined |
| 18-102_k141_62737 | Not-determined |
| 18-102_k141_228760 | Low-quality |
| 18-102_k141_218187 | Not-determined |
| 18-102_k141_166481 | Not-determined |
| 18-102_k141_218190 | Not-determined |
| 18-102_k141_218193 | Not-determined |
| 18-102_k141_83556 | Low-quality |
| 18-102_k141_93858 | Not-determined |
| 18-102_k141_228768 | Low-quality |
| 18-102_k141_114514 | Low-quality |
| 18-102_k141_228773 | Not-determined |
| 18-102_k141_31882 | Not-determined |
| 18-102_k141_228774 | Not-determined |
| 18-102_k141_228777 | Not-determined |
| 18-102_k141_125104 | Not-determined |
| 18-102_k141_42710 | Low-quality |
| 18-102_k141_42712 | Not-determined |
| 18-102_k141_104369 | Not-determined |
| 18-102_k141_114521 | Not-determined |
| 18-102_k141_93873 | Not-determined |
| 18-102_k141_228791 | Not-determined |
| 18-102_k141_228794 | Not-determined |
| 18-102_k141_114531 | Not-determined |
| 18-102_k141_83568 | Not-determined |
| 18-102_k141_52507 | Not-determined |
| 18-102_k141_62758 | Not-determined |
| 18-102_k141_176987 | Not-determined |
| 18-102_k141_218211 | Not-determined |
| 18-102_k141_218213 | Not-determined |
| 18-102_k141_218214 | Not-determined |
| 18-102_k141_218215 | Not-determined |
| 18-102_k141_228811 | Low-quality |
| 18-102_k141_228815 | Not-determined |
| 18-102_k141_104376 | Not-determined |
| 18-102_k141_93890 | Not-determined |
| 18-102_k141_166510 | Not-determined |
| 18-102_k141_83602 | Not-determined |
| 18-102_k141_177018 | Not-determined |
| 18-102_k141_93893 | Not-determined |
| 18-102_k141_166516 | Not-determined |
| 18-102_k141_166518 | Not-determined |
| 18-102_k141_21635 | Low-quality |
| 18-102_k141_207892 | Not-determined |
| 18-102_k141_104390 | Not-determined |
| 18-102_k141_135352 | Not-determined |
| 18-102_k141_21643 | Not-determined |
| 18-102_k141_21646 | Not-determined |
| 18-102_k141_177027 | Not-determined |
| 18-102_k141_207899 | Not-determined |
| 18-102_k141_21648 | Not-determined |
| 18-102_k141_923 | Low-quality |
| 18-102_k141_93904 | Not-determined |
| 18-102_k141_11138 | Low-quality |
| 18-102_k141_11142 | Not-determined |
| 18-102_k141_31924 | Low-quality |
| 18-102_k141_125139 | Not-determined |
| 18-102_k141_125146 | Not-determined |
| 18-102_k141_83620 | Not-determined |
| 18-102_k141_207907 | Not-determined |
| 18-102_k141_83624 | Not-determined |
| 18-102_k141_11149 | Not-determined |
| 18-102_k141_62776 | Low-quality |
| 18-102_k141_21672 | Low-quality |
| 18-102_k141_218258 | Low-quality |
| 18-102_k141_177054 | Not-determined |
| 18-102_k141_927 | Low-quality |
| 18-102_k141_177056 | Not-determined |
| 18-102_k141_930 | Not-determined |
| 18-102_k141_932 | Not-determined |
| 18-102_k141_93917 | Low-quality |
| 18-102_k141_93920 | Low-quality |
| 18-102_k141_177061 | Not-determined |
| 18-102_k141_93929 | Not-determined |
| 18-102_k141_93931 | Not-determined |
| 18-102_k141_197291 | Not-determined |
| 18-102_k141_114569 | Not-determined |
| 18-102_k141_114570 | Not-determined |
| 18-102_k141_228872 | Not-determined |
| 18-102_k141_218287 | Not-determined |
| 18-102_k141_114578 | Not-determined |
| 18-102_k141_125166 | Not-determined |
| 18-102_k141_177072 | Not-determined |
| 18-102_k141_135383 | Not-determined |
| 18-102_k141_177081 | Not-determined |
| 18-102_k141_207919 | Not-determined |
| 18-102_k141_52521 | Not-determined |
| 18-102_k141_73318 | Not-determined |
| 18-102_k141_177087 | Not-determined |
| 18-102_k141_73325 | Not-determined |
| 18-102_k141_93940 | Not-determined |
| 18-102_k141_93941 | Not-determined |
| 18-102_k141_93944 | Not-determined |
| 18-102_k141_228891 | Not-determined |
| 18-102_k141_228892 | Not-determined |
| 18-102_k141_228895 | Not-determined |
| 18-102_k141_177097 | Not-determined |
| 18-102_k141_52544 | Not-determined |
| 18-102_k141_52547 | Not-determined |
| 18-102_k141_177103 | Not-determined |
| 18-102_k141_187253 | Not-determined |
| 18-102_k141_145932 | Not-determined |
| 18-102_k141_145933 | Not-determined |
| 18-102_k141_228901 | Not-determined |
| 18-102_k141_156214 | Not-determined |
| 18-102_k141_62785 | Not-determined |
| 18-102_k141_166565 | Not-determined |
| 18-102_k141_145936 | Not-determined |
| 18-102_k141_21695 | Not-determined |
| 18-102_k141_177111 | Not-determined |
| 18-102_k141_31948 | Not-determined |
| 18-102_k141_156226 | Not-determined |
| 18-102_k141_166579 | Not-determined |
| 18-102_k141_166585 | Not-determined |
| 18-102_k141_135405 | Not-determined |
| 18-102_k141_187263 | Low-quality |
| 18-102_k141_21703 | Not-determined |
| 18-102_k141_31957 | Not-determined |
| 18-102_k141_177119 | Not-determined |
| 18-102_k141_156239 | Low-quality |
| 18-102_k141_83676 | Not-determined |
| 18-102_k141_83680 | Not-determined |
| 18-102_k141_228915 | Not-determined |
| 18-102_k141_31968 | Not-determined |
| 18-102_k141_207931 | Not-determined |
| 18-102_k141_73354 | Not-determined |
| 18-102_k141_31970 | Not-determined |
| 18-102_k141_218299 | Not-determined |
| 18-102_k141_177130 | Low-quality |
| 18-102_k141_145943 | Not-determined |
| 18-102_k141_145946 | Not-determined |
| 18-102_k141_228925 | Not-determined |
| 18-102_k141_207937 | Not-determined |
| 18-102_k141_135417 | Not-determined |
| 18-102_k141_135418 | Not-determined |
| 18-102_k141_83693 | Not-determined |
| 18-102_k141_135420 | Not-determined |
| 18-102_k141_104475 | Not-determined |
| 18-102_k141_207945 | Not-determined |
| 18-102_k141_187265 | Not-determined |
| 18-102_k141_135426 | Not-determined |
| 18-102_k141_156257 | Not-determined |
| 18-102_k141_145954 | Not-determined |
| 18-102_k141_156261 | Not-determined |
| 18-102_k141_228934 | Not-determined |
| 18-102_k141_21721 | Not-determined |
| 18-102_k141_114605 | Not-determined |
| 18-102_k141_114606 | Not-determined |
| 18-102_k141_135438 | Not-determined |
| 18-102_k141_197306 | Not-determined |
| 18-102_k141_228936 | Not-determined |
| 18-102_k141_197307 | Not-determined |
| 18-102_k141_52569 | Not-determined |
| 18-102_k141_238965 | Not-determined |
| 18-102_k141_197309 | Not-determined |
| 18-102_k141_238968 | Not-determined |
| 18-102_k141_145973 | Not-determined |
| 18-102_k141_21729 | Not-determined |
| 18-102_k141_31997 | Not-determined |
| 18-102_k141_145980 | Not-determined |
| 18-102_k141_125225 | Not-determined |
| 18-102_k141_145982 | Not-determined |
| 18-102_k141_156274 | Not-determined |
| 18-102_k141_145991 | Not-determined |
| 18-102_k141_135458 | Not-determined |

| | |
|---|---|
| 18-102_k141_125229 | Low-quality |
| 18-102_k141_104487 | Not-determined |
| 18-102_k141_145993 | Not-determined |
| 18-102_k141_11241 | Not-determined |
| 18-102_k141_52586 | Not-determined |
| 18-102_k141_238987 | Not-determined |
| 18-102_k141_146000 | Not-determined |
| 18-102_k141_238990 | Not-determined |
| 18-102_k141_146002 | Low-quality |
| 18-102_k141_197321 | Low-quality |
| 18-102_k141_42793 | Not-determined |
| 18-102_k141_146010 | Not-determined |
| 18-102_k141_228938 | Low-quality |
| 18-102_k141_197330 | Not-determined |
| 18-102_k141_104499 | Low-quality |
| 18-102_k141_135468 | Not-determined |
| 18-102_k141_62820 | Not-determined |
| 18-102_k141_11264 | Not-determined |
| 18-102_k141_239012 | Not-determined |
| 18-102_k141_146027 | Not-determined |
| 18-102_k141_239016 | Not-determined |
| 18-102_k141_21762 | Low-quality |
| 18-102_k141_207985 | Not-determined |
| 18-102_k141_207986 | Not-determined |
| 18-102_k141_166602 | Not-determined |
| 18-102_k141_104506 | Not-determined |
| 18-102_k141_125249 | Not-determined |
| 18-102_k141_166604 | Not-determined |
| 18-102_k141_83721 | Not-determined |
| 18-102_k141_218330 | Not-determined |
| 18-102_k141_207992 | Not-determined |
| 18-102_k141_197352 | Not-determined |
| 18-102_k141_11289 | Not-determined |
| 18-102_k141_197355 | Not-determined |
| 18-102_k141_156301 | Not-determined |
| 18-102_k141_52622 | Not-determined |
| 18-102_k141_166613 | Low-quality |
| 18-102_k141_177188 | Not-determined |
| 18-102_k141_32021 | Not-determined |
| 18-102_k141_73379 | Not-determined |
| 18-102_k141_187278 | Not-determined |
| 18-102_k141_177202 | Not-determined |
| 18-102_k141_135496 | Not-determined |
| 18-102_k141_146049 | Not-determined |
| 18-102_k141_187284 | Not-determined |
| 18-102_k141_32025 | Not-determined |
| 18-102_k141_125302 | Not-determined |
| 18-102_k141_52640 | Not-determined |
| 18-102_k141_208003 | Not-determined |
| 18-102_k141_114628 | Not-determined |
| 18-102_k141_177208 | Not-determined |
| 18-102_k141_83729 | Not-determined |
| 18-102_k141_208005 | Not-determined |
| 18-102_k141_208006 | Low-quality |
| 18-102_k141_104532 | Not-determined |
| 18-102_k141_197372 | Not-determined |
| 18-102_k141_125315 | Not-determined |
| 18-102_k141_52649 | Low-quality |
| 18-102_k141_52652 | Not-determined |
| 18-102_k141_11297 | Not-determined |
| 18-102_k141_52654 | Not-determined |
| 18-102_k141_52655 | Not-determined |
| 18-102_k141_104537 | Not-determined |
| 18-102_k141_187306 | Not-determined |
| 18-102_k141_208018 | Not-determined |
| 18-102_k141_32044 | Not-determined |
| 18-102_k141_125322 | Low-quality |
| 18-102_k141_11319 | Low-quality |
| 18-102_k141_21769 | Not-determined |
| 18-102_k141_83741 | Not-determined |
| 18-102_k141_52667 | Not-determined |
| 18-102_k141_11326 | Not-determined |
| 18-102_k141_228991 | Not-determined |
| 18-102_k141_208026 | Not-determined |
| 18-102_k141_228993 | Not-determined |
| 18-102_k141_177225 | Not-determined |
| 18-102_k141_208032 | Not-determined |
| 18-102_k141_83747 | Low-quality |
| 18-102_k141_83749 | Not-determined |
| 18-102_k141_166659 | Not-determined |
| 18-102_k141_32057 | Not-determined |
| 18-102_k141_104546 | Not-determined |
| 18-102_k141_125346 | Not-determined |
| 18-102_k141_125348 | Not-determined |
| 18-102_k141_11335 | Not-determined |
| 18-102_k141_208044 | Not-determined |
| 18-102_k141_208047 | Not-determined |
| 18-102_k141_83753 | Not-determined |
| 18-102_k141_104547 | Not-determined |
| 18-102_k141_42875 | Not-determined |
| 18-102_k141_146075 | Not-determined |
| 18-102_k141_177254 | Low-quality |
| 18-102_k141_42884 | Not-determined |
| 18-102_k141_94002 | Low-quality |
| 18-102_k141_73413 | Not-determined |
| 18-102_k141_73414 | Not-determined |
| 18-102_k141_208066 | Not-determined |
| 18-102_k141_104569 | Not-determined |
| 18-102_k141_83777 | Not-determined |
| 18-102_k141_94011 | Not-determined |
| 18-102_k141_83780 | Not-determined |
| 18-102_k141_104572 | Not-determined |
| 18-102_k141_104573 | Low-quality |
| 18-102_k141_11345 | Not-determined |
| 18-102_k141_114671 | Not-determined |
| 18-102_k141_177266 | Not-determined |
| 18-102_k141_114676 | Not-determined |
| 18-102_k141_177272 | Not-determined |
| 18-102_k141_197400 | Not-determined |
| 18-102_k141_177278 | Not-determined |
| 18-102_k141_229027 | Not-determined |
| 18-102_k141_21796 | Not-determined |
| 18-102_k141_177286 | Not-determined |
| 18-102_k141_229035 | Not-determined |
| 18-102_k141_229037 | Not-determined |
| 18-102_k141_218395 | Not-determined |
| 18-102_k141_197409 | Not-determined |
| 18-102_k141_94028 | Not-determined |
| 18-102_k141_229043 | Not-determined |
| 18-102_k141_986 | Not-determined |
| 18-102_k141_197410 | Low-quality |
| 18-102_k141_208096 | Not-determined |
| 18-102_k141_229053 | Not-determined |
| 18-102_k141_83800 | Not-determined |
| 18-102_k141_218407 | Not-determined |
| 18-102_k141_156376 | Not-determined |
| 18-102_k141_135504 | Not-determined |
| 18-102_k141_94037 | Not-determined |
| 18-102_k141_177300 | Not-determined |
| 18-102_k141_135517 | Not-determined |
| 18-102_k141_42906 | Not-determined |
| 18-102_k141_156387 | Not-determined |
| 18-102_k141_42908 | Not-determined |
| 18-102_k141_146098 | Not-determined |
| 18-102_k141_114711 | Low-quality |
| 18-102_k141_83828 | Not-determined |
| 18-102_k141_156396 | Not-determined |
| 18-102_k141_991 | Not-determined |
| 18-102_k141_992 | Not-determined |
| 18-102_k141_104599 | Not-determined |
| 18-102_k141_104600 | Not-determined |
| 18-102_k141_104602 | Not-determined |
| 18-102_k141_229058 | Not-determined |
| 18-102_k141_11374 | Not-determined |
| 18-102_k141_177311 | Not-determined |
| 18-102_k141_73469 | Not-determined |
| 18-102_k141_114718 | Not-determined |
| 18-102_k141_177316 | Not-determined |
| 18-102_k141_83846 | Not-determined |
| 18-102_k141_83847 | Not-determined |
| 18-102_k141_73478 | Not-determined |
| 18-102_k141_997 | Low-quality |
| 18-102_k141_135529 | Not-determined |
| 18-102_k141_42925 | Not-determined |
| 18-102_k141_83855 | Not-determined |
| 18-102_k141_135533 | Low-quality |
| 18-102_k141_125372 | Low-quality |
| 18-102_k141_42930 | Not-determined |
| 18-102_k141_1004 | Not-determined |
| 18-102_k141_166690 | Not-determined |
| 18-102_k141_166696 | Not-determined |
| 18-102_k141_83872 | Low-quality |
| 18-102_k141_239040 | Not-determined |
| 18-102_k141_125377 | Not-determined |
| 18-102_k141_73497 | Not-determined |
| 18-102_k141_73498 | Not-determined |
| 18-102_k141_125379 | Not-determined |
| 18-102_k141_104629 | Not-determined |
| 18-102_k141_42948 | Not-determined |
| 18-102_k141_135544 | Not-determined |
| 18-102_k141_135545 | Not-determined |
| 18-102_k141_114727 | Not-determined |
| 18-102_k141_11425 | Not-determined |
| 18-102_k141_156419 | Not-determined |
| 18-102_k141_239053 | Not-determined |
| 18-102_k141_114732 | Not-determined |
| 18-102_k141_218429 | Not-determined |
| 18-102_k141_239057 | Not-determined |
| 18-102_k141_125387 | Not-determined |
| 18-102_k141_11432 | Not-determined |
| 18-102_k141_125391 | Not-determined |
| 18-102_k141_94064 | Not-determined |
| 18-102_k141_62868 | Not-determined |
| 18-102_k141_125394 | Not-determined |
| 18-102_k141_73518 | Not-determined |
| 18-102_k141_187317 | Not-determined |
| 18-102_k141_94067 | Not-determined |
| 18-102_k141_73527 | Not-determined |
| 18-102_k141_42965 | Not-determined |
| 18-102_k141_229071 | Not-determined |
| 18-102_k141_73531 | Not-determined |
| 18-102_k141_73533 | Not-determined |
| 18-102_k141_104650 | Low-quality |
| 18-102_k141_104655 | Not-determined |
| 18-102_k141_42980 | Not-determined |
| 18-102_k141_229078 | Not-determined |
| 18-102_k141_104660 | Not-determined |
| 18-102_k141_94080 | Not-determined |
| 18-102_k141_229084 | Not-determined |
| 18-102_k141_197434 | Not-determined |
| 18-102_k141_11456 | Not-determined |
| 18-102_k141_114755 | Not-determined |
| 18-102_k141_166729 | Not-determined |
| 18-102_k141_21825 | Not-determined |
| 18-102_k141_104672 | Not-determined |
| 18-102_k141_229097 | Not-determined |
| 18-102_k141_197442 | Not-determined |
| 18-102_k141_166730 | Low-quality |
| 18-102_k141_83911 | Not-determined |
| 18-102_k141_146120 | Low-quality |
| 18-102_k141_83915 | Not-determined |
| 18-102_k141_208132 | Not-determined |
| 18-102_k141_94106 | Not-determined |
| 18-102_k141_11482 | Low-quality |
| 18-102_k141_73562 | Not-determined |
| 18-102_k141_208138 | Low-quality |
| 18-102_k141_146128 | Not-determined |
| 18-102_k141_21832 | Not-determined |
| 18-102_k141_73566 | Low-quality |
| 18-102_k141_32112 | Not-determined |
| 18-102_k141_114778 | Not-determined |
| 18-102_k141_83923 | Not-determined |
| 18-102_k141_197455 | Not-determined |
| 18-102_k141_239106 | Not-determined |
| 18-102_k141_239107 | Low-quality |
| 18-102_k141_197462 | Not-determined |
| 18-101-final-viral-combined_k141_248622\|\|full | Medium-quality |
| 18-102_k141_42998 | Not-determined |
| 18-102_k141_146154 | Not-determined |
| 18-102_k141_73585 | Not-determined |
| 18-102_k141_83946 | Not-determined |
| 18-102_k141_21859 | Not-determined |
| 18-102_k141_21860 | Not-determined |
| 18-102_k141_166756 | Not-determined |
| 18-102_k141_156459 | Not-determined |
| 18-102_k141_83959 | Not-determined |
| 18-102_k141_11507 | Not-determined |
| 18-102_k141_11508 | Low-quality |
| 18-102_k141_11510 | Not-determined |
| 18-102_k141_52701 | Not-determined |
| 18-102_k141_135583 | Not-determined |
| 18-102_k141_1070 | Not-determined |
| 18-102_k141_114826 | Not-determined |
| 18-102_k141_114828 | Not-determined |
| 18-102_k141_11522 | Not-determined |
| 18-102_k141_11525 | Not-determined |
| 18-102_k141_94110 | Not-determined |
| 18-102_k141_135597 | Not-determined |
| 18-102_k141_94115 | Not-determined |
| 18-102_k141_94116 | Not-determined |
| 18-102_k141_73602 | Not-determined |
| 18-102_k141_94119 | Not-determined |
| 18-102_k141_11536 | Not-determined |
| 18-102_k141_73603 | Not-determined |
| 18-102_k141_166783 | Not-determined |
| 18-102_k141_83974 | Low-quality |
| 18-102_k141_114845 | Not-determined |
| 18-102_k141_166790 | Not-determined |
| 18-102_k141_197473 | Not-determined |
| 18-102_k141_83980 | Not-determined |
| 18-102_k141_83981 | Not-determined |
| 18-102_k141_1088 | Not-determined |
| 18-102_k141_1090 | Not-determined |
| 18-102_k141_197480 | Not-determined |
| 18-102_k141_1092 | Not-determined |
| 18-102_k141_83982 | Not-determined |
| 18-102_k141_177349 | Low-quality |
| 18-102_k141_177355 | Low-quality |
| 18-102_k141_21889 | Not-determined |
| 18-102_k141_43041 | Not-determined |
| 18-102_k141_177366 | Not-determined |
| 18-102_k141_197483 | Not-determined |
| 18-102_k141_52740 | Low-quality |
| 18-102_k141_135614 | Not-determined |
| 18-102_k141_114862 | Not-determined |
| 18-102_k141_177378 | Not-determined |
| 18-102_k141_73639 | Low-quality |
| 18-102_k141_73643 | Not-determined |
| 18-102_k141_73645 | Not-determined |
| 18-102_k141_125409 | Not-determined |
| 18-102_k141_146170 | Not-determined |
| 18-102_k141_187386 | Not-determined |
| 18-102_k141_94153 | Low-quality |
| 18-102_k141_21912 | Not-determined |
| 18-102_k141_146178 | Not-determined |
| 18-102_k141_197509 | Not-determined |
| 18-102_k141_52749 | Not-determined |
| 18-102_k141_11573 | Not-determined |
| 18-102_k141_94168 | Not-determined |
| 18-102_k141_84005 | Not-determined |
| 18-102_k141_84006 | Not-determined |
| 18-102_k141_229127 | Not-determined |
| 18-102_k141_21922 | Not-determined |
| 18-102_k141_84009 | Not-determined |
| 18-102_k141_21933 | Low-quality |
| 18-102_k141_114881 | Not-determined |
| 18-102_k141_125415 | Not-determined |
| 18-102_k141_146210 | Low-quality |
| 18-102_k141_208160 | Not-determined |
| 18-102_k141_166836 | Not-determined |
| 18-102_k141_135635 | Not-determined |
| 18-102_k141_135636 | Not-determined |
| 18-102_k141_197525 | Not-determined |
| 18-102_k141_197529 | Not-determined |
| 18-102_k141_166846 | Not-determined |
| 18-102_k141_197533 | Not-determined |
| 18-102_k141_43056 | Not-determined |
| 18-102_k141_73678 | Not-determined |
| 18-102_k141_104702 | Not-determined |
| 18-102_k141_73686 | Not-determined |
| 18-102_k141_43071 | Not-determined |
| 18-102_k141_52773 | Not-determined |
| 18-102_k141_43075 | Not-determined |
| 18-102_k141_21936 | Not-determined |
| 18-102_k141_125431 | Not-determined |
| 18-102_k141_208181 | Not-determined |
| 18-102_k141_1130 | Not-determined |
| 18-102_k141_239150 | Not-determined |
| 18-102_k141_239151 | Not-determined |
| 18-102_k141_208183 | Low-quality |
| 18-102_k141_84051 | Not-determined |
| 18-102_k141_32139 | Not-determined |
| 18-102_k141_94199 | Not-determined |
| 18-102_k141_94201 | Not-determined |
| 18-102_k141_84056 | Not-determined |
| 18-102_k141_104735 | Not-determined |
| 18-102_k141_197547 | Not-determined |
| 18-102_k141_104738 | Not-determined |
| 18-102_k141_135668 | Not-determined |
| 18-102_k141_156484 | Low-quality |
| 18-102_k141_166870 | Not-determined |
| 18-102_k141_135669 | Not-determined |
| 18-102_k141_156486 | Low-quality |
| 18-102_k141_156487 | Not-determined |
| 18-102_k141_32153 | Not-determined |
| 18-102_k141_135673 | Not-determined |
| 18-102_k141_135675 | Not-determined |
| 18-102_k141_146250 | Not-determined |
| 18-102_k141_32157 | Not-determined |
| 18-102_k141_104742 | Low-quality |
| 18-102_k141_135685 | Not-determined |
| 18-102_k141_135686 | Not-determined |
| 18-102_k141_43093 | Low-quality |
| 18-102_k141_187424 | Not-determined |
| 18-102_k141_73702 | Not-determined |
| 18-102_k141_73709 | Not-determined |
| 18-102_k141_166875 | Low-quality |
| 18-102_k141_197562 | Not-determined |
| 18-102_k141_43115 | Not-determined |
| 18-102_k141_21961 | Not-determined |
| 18-102_k141_11602 | Not-determined |
| 18-102_k141_239180 | Not-determined |
| 18-102_k141_62919 | Not-determined |
| 18-102_k141_187441 | Not-determined |
| 18-102_k141_52802 | Not-determined |
| 18-102_k141_62921 | Not-determined |
| 18-102_k141_84076 | Low-quality |
| 18-102_k141_197571 | Not-determined |
| 18-102_k141_166881 | Low-quality |
| 18-102_k141_104768 | Not-determined |
| 18-102_k141_146260 | Not-determined |
| 18-102_k141_166885 | Not-determined |
| 18-102_k141_62930 | Not-determined |
| 18-102_k141_208213 | Not-determined |
| 18-102_k141_208217 | Not-determined |
| 18-102_k141_156504 | Not-determined |
| 18-102_k141_239196 | Low-quality |
| 18-102_k141_1159 | Not-determined |
| 18-102_k141_94227 | Not-determined |
| 18-102_k141_239200 | Not-determined |
| 18-102_k141_166892 | Low-quality |
| 18-102_k141_218455 | Not-determined |
| 18-102_k141_125467 | Not-determined |
| 18-102_k141_166900 | Not-determined |
| 18-102_k141_125473 | Not-determined |
| 18-102_k141_43131 | Not-determined |
| 18-102_k141_125485 | Low-quality |
| 18-102_k141_43132 | Low-quality |
| 18-102_k141_125490 | Low-quality |
| 18-102_k141_32188 | Not-determined |
| 18-102_k141_32192 | Not-determined |
| 18-102_k141_187460 | Not-determined |
| 18-102_k141_166907 | Not-determined |
| 18-102_k141_166909 | Not-determined |
| 18-102_k141_218467 | Not-determined |
| 18-102_k141_43138 | Not-determined |
| 18-102_k141_177416 | Not-determined |
| 18-102_k141_146298 | Not-determined |
| 18-102_k141_43142 | Low-quality |
| 18-102_k141_43146 | Not-determined |
| 18-102_k141_43149 | Not-determined |
| 18-102_k141_21976 | Not-determined |
| 18-102_k141_73766 | Not-determined |
| 18-102_k141_177428 | Not-determined |
| 18-102_k141_187467 | Not-determined |
| 18-102_k141_84112 | Not-determined |
| 18-102_k141_43153 | Not-determined |
| 18-102_k141_239233 | Not-determined |
| 18-102_k141_229148 | Not-determined |
| 18-102_k141_229150 | Not-determined |
| 18-102_k141_239242 | Not-determined |
| 18-102_k141_229159 | Not-determined |
| 18-102_k141_11606 | Not-determined |
| 18-102_k141_187477 | Not-determined |
| 18-102_k141_32200 | Not-determined |
| 18-102_k141_125500 | Not-determined |
| 18-102_k141_32206 | Not-determined |
| 18-102_k141_218496 | Not-determined |
| 18-102_k141_218497 | Not-determined |
| 18-102_k141_177459 | Not-determined |
| 18-102_k141_166937 | Not-determined |
| 18-102_k141_32213 | Not-determined |
| 18-102_k141_166940 | Not-determined |
| 18-102_k141_43174 | Not-determined |
| 18-102_k141_11634 | Not-determined |
| 18-102_k141_22006 | Not-determined |
| 18-102_k141_187497 | Low-quality |
| 18-102_k141_43179 | Not-determined |
| 18-102_k141_94274 | Not-determined |
| 18-102_k141_239269 | Not-determined |
| 18-102_k141_43184 | Not-determined |
| 18-102_k141_94277 | Not-determined |
| 18-102_k141_52842 | Low-quality |
| 18-102_k141_22018 | Not-determined |
| 18-102_k141_43188 | Low-quality |
| 18-102_k141_218529 | Not-determined |
| 18-102_k141_187505 | Low-quality |
| 18-102_k141_187506 | Not-determined |
| 18-102_k141_43196 | Not-determined |
| 18-102_k141_208234 | Not-determined |
| 18-102_k141_177484 | Low-quality |
| 18-102_k141_73809 | Not-determined |
| 18-102_k141_52862 | Not-determined |
| 18-102_k141_43208 | Not-determined |
| 18-102_k141_218545 | Not-determined |
| 18-102_k141_62951 | Not-determined |
| 18-102_k141_62954 | Not-determined |
| 18-102_k141_62956 | Not-determined |
| 18-102_k141_177488 | Not-determined |
| 18-102_k141_62964 | Not-determined |
| 18-102_k141_43222 | Not-determined |
| 18-102_k141_43228 | Not-determined |
| 18-102_k141_239300 | Not-determined |
| 18-102_k141_43230 | Not-determined |
| 18-102_k141_197590 | Low-quality |
| 18-102_k141_156507 | Not-determined |
| 18-102_k141_156509 | Not-determined |
| 18-102_k141_43234 | Low-quality |
| 18-102_k141_1228 | Not-determined |
| 18-102_k141_197599 | Not-determined |
| 18-102_k141_43242 | Not-determined |
| 18-102_k141_22045 | Not-determined |
| 18-102_k141_146345 | Not-determined |
| 18-102_k141_52884 | Not-determined |
| 18-102_k141_208254 | Not-determined |
| 18-102_k141_1238 | Not-determined |
| 18-102_k141_177518 | Not-determined |
| 18-102_k141_177520 | Not-determined |
| 18-102_k141_208258 | Not-determined |
| 18-102_k141_43257 | Not-determined |
| 18-102_k141_84139 | Low-quality |
| 18-102_k141_73832 | Not-determined |
| 18-102_k141_146382 | Not-determined |
| 18-102_k141_73841 | Not-determined |
| 18-102_k141_11688 | Not-determined |
| 18-102_k141_187518 | Not-determined |
| 18-102_k141_197615 | Not-determined |
| 18-102_k141_135725 | Not-determined |
| 18-102_k141_239311 | Not-determined |
| 18-102_k141_229236 | Not-determined |
| 18-102_k141_135728 | Not-determined |
| 18-102_k141_197619 | Not-determined |
| 18-102_k141_239314 | Low-quality |
| 18-102_k141_239316 | Not-determined |
| 18-102_k141_62994 | Not-determined |
| 18-102_k141_73866 | Low-quality |
| 18-102_k141_135739 | Not-determined |
| 18-102_k141_167038 | Not-determined |
| 18-102_k141_177548 | Not-determined |
| 18-102_k141_73870 | Not-determined |
| 18-102_k141_22062 | Low-quality |
| 18-102_k141_146401 | Not-determined |
| 18-102_k141_197626 | Not-determined |
| 18-102_k141_135745 | Low-quality |
| 18-102_k141_208275 | Not-determined |
| 18-102_k141_73876 | Not-determined |
| 18-102_k141_208283 | Not-determined |
| 18-102_k141_135752 | Not-determined |
| 18-102_k141_197632 | Not-determined |
| 18-102_k141_177565 | Not-determined |
| 18-102_k141_104772 | Not-determined |
| 18-102_k141_104774 | Not-determined |
| 18-102_k141_22071 | Not-determined |
| 18-102_k141_43284 | Not-determined |
| 18-102_k141_22073 | Not-determined |
| 18-102_k141_22075 | Not-determined |
| 18-102_k141_114925 | Not-determined |
| 18-102_k141_135760 | Low-quality |
| 18-102_k141_94324 | Not-determined |
| 18-102_k141_114927 | Not-determined |
| 18-102_k141_135763 | Low-quality |
| 18-102_k141_22083 | Not-determined |
| 18-102_k141_73903 | Not-determined |
| 18-102_k141_135779 | Not-determined |
| 18-102_k141_104804 | Not-determined |
| 18-102_k141_43308 | Not-determined |
| 18-102_k141_156570 | Not-determined |
| 18-102_k141_43310 | Not-determined |
| 18-102_k141_22091 | Not-determined |
| 18-102_k141_73911 | Not-determined |
| 18-102_k141_84191 | Not-determined |
| 18-102_k141_94347 | Not-determined |
| 18-102_k141_43316 | Not-determined |
| 18-102_k141_104814 | Low-quality |
| 18-102_k141_239346 | Not-determined |
| 18-102_k141_94354 | Not-determined |
| 18-102_k141_43324 | Not-determined |
| 18-102_k141_146409 | Not-determined |
| 18-102_k141_187571 | Not-determined |
| 18-102_k141_197641 | Low-quality |
| 18-102_k141_11718 | Not-determined |
| 18-102_k141_94362 | Not-determined |
| 18-102_k141_125534 | Not-determined |
| 18-102_k141_94364 | Not-determined |
| 18-102_k141_43331 | Not-determined |
| 18-102_k141_43332 | Not-determined |
| 18-102_k141_125539 | Not-determined |
| 18-102_k141_11725 | Low-quality |
| 18-102_k141_125546 | Not-determined |
| 18-102_k141_239352 | Low-quality |
| 18-102_k141_125547 | Not-determined |
| 18-102_k141_146424 | Not-determined |
| 18-102_k141_125550 | Not-determined |
| 18-102_k141_125552 | Not-determined |
| 18-102_k141_11731 | Not-determined |
| 18-102_k141_73931 | Not-determined |
| 18-102_k141_197656 | Not-determined |
| 18-102_k141_167106 | Not-determined |
| 18-102_k141_167107 | Not-determined |
| 18-102_k141_239358 | Not-determined |
| 18-102_k141_135789 | Not-determined |
| 18-102_k141_197664 | Not-determined |
| 18-102_k141_229287 | Not-determined |
| 18-102_k141_94383 | Not-determined |
| 18-102_k141_156590 | Low-quality |
| 18-102_k141_239370 | Not-determined |
| 18-102_k141_229291 | Low-quality |
| 18-102_k141_94387 | Not-determined |
| 18-102_k141_94391 | Not-determined |
| 18-102_k141_208321 | Not-determined |
| 18-102_k141_156594 | Not-determined |
| 18-102_k141_229297 | Low-quality |
| 18-102_k141_239388 | Not-determined |
| 18-102_k141_239391 | Not-determined |
| 18-102_k141_94404 | Not-determined |
| 18-102_k141_104847 | Not-determined |
| 18-102_k141_239392 | Not-determined |
| 18-102_k141_135814 | Not-determined |
| 18-102_k141_167121 | Not-determined |
| 18-102_k141_114939 | Not-determined |
| 18-102_k141_167129 | Not-determined |
| 18-102_k141_239407 | Not-determined |
| 18-102_k141_229307 | Not-determined |
| 18-102_k141_1279 | Not-determined |
| 18-102_k141_63032 | Not-determined |
| 18-102_k141_94422 | Low-quality |
| 18-102_k141_135819 | Low-quality |
| 18-102_k141_84250 | Not-determined |
| 18-102_k141_84251 | Not-determined |
| 18-102_k141_114951 | Not-determined |
| 18-102_k141_114952 | Not-determined |
| 18-102_k141_84255 | Not-determined |
| 18-102_k141_94432 | Not-determined |
| 18-102_k141_208338 | Not-determined |
| 18-102_k141_229323 | Not-determined |
| 18-102_k141_94438 | Not-determined |
| 18-102_k141_94446 | Not-determined |
| 18-102_k141_218570 | Not-determined |
| 18-102_k141_218574 | Not-determined |
| 18-102_k141_218578 | Not-determined |
| 18-102_k141_11770 | Low-quality |
| 18-102_k141_229335 | Not-determined |
| 18-102_k141_167156 | Not-determined |
| 18-102_k141_43367 | Not-determined |
| 18-102_k141_135832 | Not-determined |
| 18-102_k141_167159 | Not-determined |
| 18-102_k141_94456 | Not-determined |
| 18-102_k141_135844 | Low-quality |
| 18-102_k141_167178 | Not-determined |

| | |
|---|---|
| 18-102_k141_11777 | Low-quality |
| 18-102_k141_167179 | Not-determined |
| 18-102_k141_208359 | Not-determined |
| 18-102_k141_104876 | Not-determined |
| 18-102_k141_167187 | Not-determined |
| 18-102_k141_104877 | Not-determined |
| 18-102_k141_229412 | Not-determined |
| 18-102_k141_135849 | Not-determined |
| 18-102_k141_229354 | Not-determined |
| 18-102_k141_229355 | Not-determined |
| 18-102_k141_229416 | Not-determined |
| 18-102_k141_43399 | Not-determined |
| 18-102_k141_177627 | Not-determined |
| 18-102_k141_208362 | Not-determined |
| 18-102_k141_229363 | Not-determined |
| 18-102_k141_52890 | Not-determined |
| 18-102_k141_63082 | Not-determined |
| 18-102_k141_43409 | Low-quality |
| 18-102_k141_11804 | Not-determined |
| 18-102_k141_94476 | Not-determined |
| 18-102_k141_43418 | Not-determined |
| 18-102_k141_43419 | Not-determined |
| 18-102_k141_187585 | Not-determined |
| 18-102_k141_177638 | Not-determined |
| 18-102_k141_63100 | Not-determined |
| 18-102_k141_1307 | Not-determined |
| 18-102_k141_1308 | Not-determined |
| 18-102_k141_229367 | Not-determined |
| 18-102_k141_135856 | Not-determined |
| 18-102_k141_177663 | Not-determined |
| 18-102_k141_11842 | Not-determined |
| 18-102_k141_187605 | Not-determined |
| 18-102_k141_187606 | Low-quality |
| 18-102_k141_94498 | Not-determined |
| 18-102_k141_135875 | Not-determined |
| 18-102_k141_177680 | Not-determined |
| 18-102_k141_63127 | Not-determined |
| 18-102_k141_114994 | Not-determined |
| 18-102_k141_22141 | Not-determined |
| 18-102_k141_63136 | Not-determined |
| 18-102_k141_167230 | Low-quality |
| 18-102_k141_208391 | Not-determined |
| 18-102_k141_52920 | Low-quality |
| 18-102_k141_1327 | Not-determined |
| 18-102_k141_229384 | Not-determined |
| 18-102_k141_167234 | Low-quality |
| 18-102_k141_229391 | Not-determined |
| 18-102_k141_229396 | Not-determined |
| 18-102_k141_135888 | Not-determined |
| 18-102_k141_197707 | Not-determined |
| 18-102_k141_197708 | Not-determined |
| 18-102_k141_94503 | Not-determined |
| 18-102_k141_197720 | Low-quality |
| 18-102_k141_208415 | Not-determined |
| 18-102_k141_197725 | Not-determined |
| 18-102_k141_208417 | Not-determined |
| 18-102_k141_94514 | Not-determined |
| 18-102_k141_52951 | Not-determined |
| 18-102_k141_218618 | Not-determined |
| 18-102_k141_167269 | Not-determined |
| 18-102_k141_218624 | Not-determined |
| 18-102_k141_218629 | Not-determined |
| 18-102_k141_177713 | Not-determined |
| 18-102_k141_197730 | Not-determined |
| 18-102_k141_104901 | Not-determined |
| 18-102_k141_104902 | Not-determined |
| 18-102_k141_208433 | Not-determined |
| 18-102_k141_84305 | Not-determined |
| 18-102_k141_177718 | Not-determined |
| 18-102_k141_104904 | Not-determined |
| 18-102_k141_146501 | Not-determined |
| 18-102_k141_208441 | Not-determined |
| 18-102_k141_167286 | Low-quality |
| 18-102_k141_208444 | Not-determined |
| 18-102_k141_22166 | Not-determined |
| 18-102_k141_32258 | Not-determined |
| 18-102_k141_187662 | Not-determined |
| 18-102_k141_52964 | Not-determined |
| 18-102_k141_43440 | Not-determined |
| 18-102_k141_135909 | Not-determined |
| 18-102_k141_32261 | Not-determined |
| 18-102_k141_22170 | Not-determined |
| 18-102_k141_22176 | Not-determined |
| 18-102_k141_218655 | Not-determined |
| 18-102_k141_208457 | Not-determined |
| 18-102_k141_177737 | Not-determined |
| 18-102_k141_218659 | Not-determined |
| 18-102_k141_84326 | Not-determined |
| 18-102_k141_218660 | Not-determined |
| 18-102_k141_146518 | Not-determined |
| 18-102_k141_208467 | Not-determined |
| 18-102_k141_84333 | Not-determined |
| 18-102_k141_22190 | Not-determined |
| 18-102_k141_187699 | Not-determined |
| 18-102_k141_94545 | Low-quality |
| 18-102_k141_63200 | Not-determined |
| 18-102_k141_239448 | Not-determined |
| 18-102_k141_239449 | Not-determined |
| 18-102_k141_177752 | Not-determined |
| 18-102_k141_22196 | Not-determined |
| 18-102_k141_73961 | Not-determined |
| 18-102_k141_43456 | Not-determined |
| 18-102_k141_187715 | Not-determined |
| 18-102_k141_32273 | Not-determined |
| 18-102_k141_73970 | Not-determined |
| 18-102_k141_73975 | Not-determined |
| 18-102_k141_94560 | Not-determined |
| 18-102_k141_94561 | Not-determined |
| 18-102_k141_115005 | Not-determined |
| 18-102_k141_115008 | Low-quality |
| 18-102_k141_115009 | Not-determined |
| 18-102_k141_208494 | Not-determined |
| 18-102_k141_43472 | Not-determined |
| 18-102_k141_115020 | Not-determined |
| 18-102_k141_177776 | Low-quality |
| 18-102_k141_43477 | Not-determined |
| 18-102_k141_43481 | Not-determined |
| 18-102_k141_125620 | Not-determined |
| 18-102_k141_73986 | Not-determined |
| 18-102_k141_146527 | Not-determined |
| 18-102_k141_32283 | Low-quality |
| 18-102_k141_146530 | Not-determined |
| 18-102_k141_177788 | Not-determined |
| 18-102_k141_22225 | Not-determined |
| 18-102_k141_73994 | Not-determined |
| 18-102_k141_177790 | Not-determined |
| 18-102_k141_22226 | Not-determined |
| 18-102_k141_177791 | Not-determined |
| 18-102_k141_177793 | Not-determined |
| 18-102_k141_63227 | Not-determined |
| 18-102_k141_104954 | Not-determined |
| 18-102_k141_104955 | Not-determined |
| 18-102_k141_115042 | Not-determined |
| 18-102_k141_146541 | Not-determined |
| 18-102_k141_115048 | Not-determined |
| 18-102_k141_32295 | Not-determined |
| 18-102_k141_32297 | Not-determined |
| 18-102_k141_177800 | Not-determined |
| 18-102_k141_32298 | Not-determined |
| 18-102_k141_167340 | Not-determined |
| 18-102_k141_22233 | Not-determined |
| 18-102_k141_177812 | Not-determined |
| 18-102_k141_177813 | Not-determined |
| 18-102_k141_218695 | Not-determined |
| 18-102_k141_187736 | Not-determined |
| 18-102_k141_52976 | Low-quality |
| 18-102_k141_167342 | Not-determined |
| 18-102_k141_63241 | Not-determined |
| 18-102_k141_84370 | Not-determined |
| 18-102_k141_52982 | Not-determined |
| 18-102_k141_22248 | Not-determined |
| 18-102_k141_94610 | Not-determined |
| 18-102_k141_177822 | Not-determined |
| 18-102_k141_229399 | Not-determined |
| 18-102_k141_43527 | Not-determined |
| 18-102_k141_146553 | Not-determined |
| 18-102_k141_239474 | Not-determined |
| 18-102_k141_239475 | Not-determined |
| 18-102_k141_229406 | Low-quality |
| 18-102_k141_94616 | Not-determined |
| 18-102_k141_229408 | Not-determined |
| 18-102_k141_229412 | Not-determined |
| 18-102_k141_1355 | Not-determined |
| 18-102_k141_94620 | Not-determined |
| 18-102_k141_22256 | Not-determined |
| 18-102_k141_167359 | Not-determined |
| 18-102_k141_63277 | Not-determined |
| 18-102_k141_43535 | Low-quality |
| 18-102_k141_135941 | Not-determined |
| 18-102_k141_135943 | Not-determined |
| 18-102_k141_208526 | Not-determined |
| 18-102_k141_135949 | Not-determined |
| 18-102_k141_1372 | Not-determined |
| 18-102_k141_1375 | Not-determined |
| 18-102_k141_63293 | Low-quality |
| 18-102_k141_22280 | Not-determined |
| 18-102_k141_22281 | Not-determined |
| 18-102_k141_229433 | Low-quality |
| 18-102_k141_104975 | Low-quality |
| 18-102_k141_197802 | Not-determined |
| 18-102_k141_135959 | Not-determined |
| 18-102_k141_197804 | Low-quality |
| 18-102_k141_1391 | Not-determined |
| 18-102_k141_135965 | Not-determined |
| 18-102_k141_53010 | Not-determined |
| 18-102_k141_53013 | Not-determined |
| 18-102_k141_53015 | Not-determined |
| 18-102_k141_167402 | Not-determined |
| 18-102_k141_94645 | Not-determined |
| 18-102_k141_22299 | Low-quality |
| 18-102_k141_218728 | Not-determined |
| 18-102_k141_197814 | Not-determined |
| 18-102_k141_115083 | Not-determined |
| 18-102_k141_84395 | Not-determined |
| 18-102_k141_167410 | Not-determined |
| 18-102_k141_84397 | Not-determined |
| 18-102_k141_218740 | Low-quality |
| 18-102_k141_105002 | Not-determined |
| 18-102_k141_115086 | Not-determined |
| 18-102_k141_1397 | Low-quality |
| 18-102_k141_146593 | Not-determined |
| 18-102_k141_63320 | Not-determined |
| 18-102_k141_63327 | Not-determined |
| 18-102_k141_218749 | Not-determined |
| 18-102_k141_63337 | Not-determined |
| 18-102_k141_84411 | Not-determined |
| 18-102_k141_22318 | Not-determined |
| 18-102_k141_1417 | Not-determined |
| 18-102_k141_115113 | Not-determined |
| 18-102_k141_1425 | Not-determined |
| 18-102_k141_94673 | Not-determined |
| 18-102_k141_218760 | Not-determined |
| 18-102_k141_1428 | Not-determined |
| 18-102_k141_84434 | Not-determined |
| 18-102_k141_1430 | Low-quality |
| 18-102_k141_1432 | Not-determined |
| 18-102_k141_115118 | Not-determined |
| 18-102_k141_22336 | Not-determined |
| 18-102_k141_84441 | Low-quality |
| 18-102_k141_115128 | Not-determined |
| 18-102_k141_22342 | Not-determined |
| 18-102_k141_22345 | Not-determined |
| 18-102_k141_135981 | Low-quality |
| 18-102_k141_187772 | Not-determined |
| 18-102_k141_63368 | Not-determined |
| 18-102_k141_167430 | Not-determined |
| 18-102_k141_197833 | Low-quality |
| 18-102_k141_43618 | Not-determined |
| 18-102_k141_53032 | Low-quality |
| 18-102_k141_229461 | Not-determined |
| 18-102_k141_218779 | Not-determined |
| 18-102_k141_218781 | Not-determined |
| 18-102_k141_218782 | Not-determined |
| 18-102_k141_187785 | Not-determined |
| 18-102_k141_218785 | Not-determined |
| 18-102_k141_22358 | Not-determined |
| 18-102_k141_197840 | Not-determined |
| 18-102_k141_167453 | Not-determined |
| 18-102_k141_115157 | Not-determined |
| 18-102_k141_229471 | Not-determined |
| 18-102_k141_187786 | Low-quality |
| 18-102_k141_125676 | Not-determined |
| 18-102_k141_53049 | Not-determined |
| 18-102_k141_84460 | Not-determined |
| 18-102_k141_11867 | Not-determined |
| 18-102_k141_84465 | Not-determined |
| 18-102_k141_53055 | Not-determined |
| 18-102_k141_218802 | Not-determined |
| 18-102_k141_115174 | Not-determined |
| 18-102_k141_84478 | Not-determined |
| 18-102_k141_146623 | Not-determined |
| 18-102_k141_146631 | Not-determined |
| 18-102_k141_22385 | Not-determined |
| 18-102_k141_125695 | Low-quality |
| 18-102_k141_167488 | Low-quality |
| 18-102_k141_146640 | Not-determined |
| 18-102_k141_146642 | Not-determined |
| 18-102_k141_167494 | Not-determined |
| 18-102_k141_229487 | Not-determined |
| 18-102_k141_84482 | Not-determined |
| 18-102_k141_125707 | Low-quality |
| 18-102_k141_167503 | Low-quality |
| 18-102_k141_156704 | Low-quality |
| 18-102_k141_94686 | Low-quality |
| 18-102_k141_187824 | Not-determined |
| 18-102_k141_53076 | Not-determined |
| 18-102_k141_43651 | Not-determined |
| 18-102_k141_32326 | Not-determined |
| 18-102_k141_218820 | Not-determined |
| 18-102_k141_167512 | Not-determined |
| 18-102_k141_229504 | Not-determined |
| 18-102_k141_115190 | Not-determined |
| 18-102_k141_218829 | Low-quality |
| 18-102_k141_218831 | Not-determined |
| 18-102_k141_218838 | Not-determined |
| 18-102_k141_105028 | Not-determined |
| 18-102_k141_11921 | Not-determined |
| 18-102_k141_229511 | Not-determined |
| 18-102_k141_1459 | Low-quality |
| 18-102_k141_11929 | Not-determined |
| 18-102_k141_156726 | Not-determined |
| 18-102_k141_167539 | Low-quality |
| 18-102_k141_84519 | Not-determined |
| 18-102_k141_167541 | Not-determined |
| 18-102_k141_32345 | Not-determined |
| 18-102_k141_32359 | Not-determined |
| 18-102_k141_229523 | Not-determined |
| 18-102_k141_115197 | Not-determined |
| 18-102_k141_11944 | Not-determined |
| 18-102_k141_84533 | Not-determined |
| 18-102_k141_177851 | Not-determined |
| 18-102_k141_53146 | Not-determined |
| 18-102_k141_105039 | Low-quality |
| 18-102_k141_136036 | Not-determined |
| 18-102_k141_115207 | Not-determined |
| 18-102_k141_32370 | Not-determined |
| 18-102_k141_136044 | Not-determined |
| 18-102_k141_115212 | Not-determined |
| 18-102_k141_167551 | Not-determined |
| 18-102_k141_84536 | Not-determined |
| 18-102_k141_94706 | Not-determined |
| 18-102_k141_94707 | Not-determined |
| 18-102_k141_94708 | Not-determined |
| 18-102_k141_94709 | Not-determined |
| 18-102_k141_115216 | Not-determined |
| 18-102_k141_115221 | Not-determined |
| 18-102_k141_167563 | Low-quality |
| 18-102_k141_167564 | Not-determined |
| 18-102_k141_125749 | Low-quality |
| 18-102_k141_136056 | Not-determined |
| 18-102_k141_146668 | Not-determined |
| 18-102_k141_146672 | Not-determined |
| 18-102_k141_156738 | Not-determined |
| 18-102_k141_53162 | Not-determined |
| 18-102_k141_115244 | Not-determined |
| 18-102_k141_115245 | Not-determined |
| 18-102_k141_136071 | Not-determined |
| 18-102_k141_146680 | Not-determined |
| 18-102_k141_11979 | Not-determined |
| 18-102_k141_239516 | Not-determined |
| 18-102_k141_115252 | Not-determined |
| 18-102_k141_11985 | Not-determined |
| 18-102_k141_239520 | Not-determined |
| 18-102_k141_94716 | Low-quality |
| 18-102_k141_53180 | Not-determined |
| 18-102_k141_105056 | Not-determined |
| 18-102_k141_53191 | Not-determined |
| 18-102_k141_146722 | Not-determined |
| 18-102_k141_32388 | Not-determined |
| 18-102_k141_208641 | Not-determined |
| 18-102_k141_208643 | Not-determined |
| 18-102_k141_94742 | Not-determined |
| 18-102_k141_53200 | Not-determined |
| 18-102_k141_94749 | Not-determined |
| 18-102_k141_94757 | Not-determined |
| 18-102_k141_156760 | Not-determined |
| 18-102_k141_146731 | Not-determined |
| 18-102_k141_146732 | Not-determined |
| 18-102_k141_218841 | Not-determined |
| 18-102_k141_218843 | Not-determined |
| 18-102_k141_156762 | Low-quality |
| 18-102_k141_53214 | Not-determined |
| 18-102_k141_146736 | Low-quality |
| 18-102_k141_167576 | Not-determined |
| 18-102_k141_167577 | Not-determined |
| 18-102_k141_197876 | Low-quality |
| 18-102_k141_177874 | Not-determined |
| 18-102_k141_177880 | Not-determined |
| 18-102_k141_136096 | Not-determined |
| 18-102_k141_177882 | Not-determined |
| 18-102_k141_84567 | Not-determined |
| 18-102_k141_1484 | Not-determined |
| 18-102_k141_156767 | Not-determined |
| 18-102_k141_63447 | Not-determined |
| 18-102_k141_177886 | Not-determined |
| 18-102_k141_1488 | Not-determined |
| 18-102_k141_136099 | Low-quality |
| 18-102_k141_156782 | Not-determined |
| 18-102_k141_32425 | Not-determined |
| 18-102_k141_84580 | Not-determined |
| 18-102_k141_105092 | Not-determined |
| 18-102_k141_43729 | Not-determined |
| 18-102_k141_63465 | Low-quality |
| 18-102_k141_63469 | Not-determined |
| 18-102_k141_84585 | Not-determined |
| 18-102_k141_63470 | Not-determined |
| 18-102_k141_94771 | Not-determined |
| 18-102_k141_146749 | Not-determined |
| 18-102_k141_136111 | Not-determined |
| 18-102_k141_156792 | Not-determined |
| 18-102_k141_197901 | Not-determined |
| 18-102_k141_136114 | Not-determined |
| 18-102_k141_63483 | Not-determined |
| 18-102_k141_84599 | Not-determined |
| 18-102_k141_115266 | Not-determined |
| 18-102_k141_156808 | Not-determined |
| 18-102_k141_197915 | Not-determined |
| 18-102_k141_197916 | Low-quality |
| 18-102_k141_146755 | Not-determined |
| 18-102_k141_146758 | Not-determined |
| 18-102_k141_229537 | Not-determined |
| 18-102_k141_125776 | Not-determined |
| 18-102_k141_177909 | Not-determined |
| 18-102_k141_115285 | Not-determined |
| 18-102_k141_197930 | Not-determined |
| 18-102_k141_187913 | Not-determined |
| 18-102_k141_94792 | Not-determined |
| 18-102_k141_1515 | Not-determined |
| 18-102_k141_197932 | Low-quality |
| 18-102_k141_156812 | Not-determined |
| 18-102_k141_187916 | Low-quality |
| 18-102_k141_187917 | Not-determined |
| 18-102_k141_115289 | Low-quality |
| 18-102_k141_63499 | Not-determined |
| 18-102_k141_32438 | Low-quality |
| 18-102_k141_229552 | Not-determined |
| 18-102_k141_229553 | Not-determined |
| 18-102_k141_125789 | Not-determined |
| 18-102_k141_43781 | Not-determined |
| 18-102_k141_156828 | Low-quality |
| 18-102_k141_229562 | Not-determined |
| 18-102_k141_136132 | Not-determined |
| 18-102_k141_239610 | Not-determined |
| 18-102_k141_177938 | Not-determined |
| 18-102_k141_43797 | Not-determined |
| 18-102_k141_12016 | Low-quality |
| 18-102_k141_63518 | Not-determined |
| 18-102_k141_239617 | Low-quality |
| 18-102_k141_218858 | Not-determined |
| 18-102_k141_115311 | Not-determined |
| 18-102_k141_94824 | Not-determined |
| 18-102_k141_239622 | Not-determined |
| 18-102_k141_187937 | Not-determined |
| 18-102_k141_177951 | Not-determined |
| 18-102_k141_43807 | Not-determined |
| 18-102_k141_208657 | Not-determined |
| 18-102_k141_218872 | Not-determined |
| 18-102_k141_218873 | Not-determined |
| 18-102_k141_63532 | Not-determined |
| 18-102_k141_43820 | Not-determined |
| 18-102_k141_105131 | Low-quality |
| 18-102_k141_177961 | Not-determined |
| 18-102_k141_105133 | Not-determined |
| 18-102_k141_177962 | Not-determined |
| 18-102_k141_218880 | Not-determined |
| 18-102_k141_32457 | Low-quality |
| 18-102_k141_32458 | Low-quality |
| 18-102_k141_136157 | Not-determined |
| 18-102_k141_208665 | Low-quality |
| 18-102_k141_156841 | Low-quality |
| 18-102_k141_115327 | Not-determined |
| 18-102_k141_43828 | Not-determined |
| 18-102_k141_43831 | Not-determined |
| 18-102_k141_32465 | Not-determined |
| 18-102_k141_63546 | Low-quality |
| 18-102_k141_32472 | Not-determined |
| 18-102_k141_43839 | Low-quality |
| 18-102_k141_136171 | Not-determined |
| 18-102_k141_218894 | Low-quality |
| 18-102_k141_136174 | Not-determined |
| 18-102_k141_156852 | Not-determined |
| 18-102_k141_125815 | Not-determined |
| 18-102_k141_187956 | Not-determined |
| 18-102_k141_12050 | Not-determined |
| 18-102_k141_94832 | Not-determined |
| 18-102_k141_105141 | Low-quality |
| 18-102_k141_12051 | Not-determined |
| 18-102_k141_125827 | Not-determined |
| 18-102_k141_12052 | Not-determined |
| 18-102_k141_12053 | Not-determined |
| 18-102_k141_208676 | Not-determined |
| 18-102_k141_63552 | Low-quality |
| 18-102_k141_105146 | Not-determined |
| 18-102_k141_32477 | Not-determined |
| 18-102_k141_63556 | Not-determined |
| 18-102_k141_105147 | Low-quality |
| 18-102_k141_177972 | Not-determined |
| 18-102_k141_43863 | Not-determined |
| 18-102_k141_125835 | Not-determined |
| 18-102_k141_239653 | Low-quality |
| 18-102_k141_115351 | Not-determined |
| 18-102_k141_239655 | Not-determined |
| 18-102_k141_43867 | Not-determined |
| 18-102_k141_43869 | Not-determined |
| 18-102_k141_177986 | Not-determined |
| 18-102_k141_115353 | Not-determined |
| 18-102_k141_177987 | Low-quality |
| 18-102_k141_156864 | Low-quality |
| 18-102_k141_239664 | Not-determined |
| 18-102_k141_156866 | Not-determined |
| 18-102_k141_12074 | Not-determined |
| 18-102_k141_12077 | Not-determined |
| 18-102_k141_125843 | Not-determined |
| 18-102_k141_115357 | Low-quality |
| 18-102_k141_115360 | Not-determined |
| 18-102_k141_22426 | Not-determined |
| 18-102_k141_32515 | Not-determined |
| 18-102_k141_187991 | Not-determined |
| 18-102_k141_178012 | Not-determined |
| 18-102_k141_218911 | Not-determined |
| 18-102_k141_229574 | Not-determined |
| 18-102_k141_63579 | Low-quality |
| 18-102_k141_22439 | Not-determined |
| 18-102_k141_22442 | Not-determined |
| 18-102_k141_22445 | Not-determined |
| 18-102_k141_32534 | Low-quality |
| 18-102_k141_22449 | Not-determined |
| 18-102_k141_43897 | Not-determined |
| 18-102_k141_1536 | Not-determined |
| 18-102_k141_12085 | Not-determined |
| 18-102_k141_136222 | Not-determined |
| 18-102_k141_22469 | Low-quality |
| 18-102_k141_43907 | Not-determined |
| 18-102_k141_218933 | Not-determined |
| 18-102_k141_43913 | Not-determined |
| 18-102_k141_188021 | Not-determined |
| 18-102_k141_239668 | Low-quality |
| 18-102_k141_43919 | Not-determined |
| 18-102_k141_32560 | Low-quality |
| 18-102_k141_136247 | Low-quality |
| 18-102_k141_218947 | Not-determined |
| 18-102_k141_22498 | Not-determined |
| 18-102_k141_1567 | Not-determined |
| 18-102_k141_218949 | Not-determined |
| 18-102_k141_218950 | Not-determined |
| 18-102_k141_94851 | Not-determined |
| 18-102_k141_188028 | Low-quality |
| 18-102_k141_105164 | Not-determined |
| 18-102_k141_43929 | Low-quality |
| 18-102_k141_22509 | Not-determined |
| 18-102_k141_22511 | Not-determined |
| 18-102_k141_105172 | Not-determined |
| 18-102_k141_32577 | Not-determined |
| 18-102_k141_32580 | Not-determined |
| 18-102_k141_63607 | Not-determined |
| 18-102_k141_22516 | Not-determined |
| 18-102_k141_188048 | Not-determined |
| 18-102_k141_43942 | Not-determined |
| 18-102_k141_84610 | Not-determined |
| 18-102_k141_32591 | Not-determined |
| 18-102_k141_84611 | Not-determined |
| 18-102_k141_53287 | Not-determined |
| 18-102_k141_136256 | Not-determined |
| 18-102_k141_188055 | Not-determined |
| 18-102_k141_136257 | Not-determined |
| 18-102_k141_156912 | Not-determined |
| 18-102_k141_84622 | Not-determined |
| 18-102_k141_84625 | Not-determined |
| 18-102_k141_188065 | Not-determined |
| 18-102_k141_22530 | Not-determined |
| 18-102_k141_229628 | Not-determined |
| 18-102_k141_32604 | Not-determined |
| 18-102_k141_156916 | Low-quality |
| 18-102_k141_63630 | Not-determined |
| 18-102_k141_43964 | Not-determined |
| 18-102_k141_63635 | Low-quality |
| 18-102_k141_156925 | Not-determined |
| 18-102_k141_32612 | Not-determined |
| 18-102_k141_32613 | Not-determined |
| 18-102_k141_167715 | Not-determined |
| 18-102_k141_156933 | Not-determined |
| 18-102_k141_74007 | Not-determined |
| 18-102_k141_32619 | Not-determined |
| 18-102_k141_43975 | Not-determined |
| 18-102_k141_43977 | Not-determined |
| 18-102_k141_94885 | Not-determined |
| 18-102_k141_156942 | Not-determined |
| 18-102_k141_167721 | Not-determined |
| 18-102_k141_94894 | Not-determined |
| 18-102_k141_1612 | Not-determined |
| 18-102_k141_229654 | Not-determined |
| 18-102_k141_94902 | Not-determined |
| 18-102_k141_125945 | Low-quality |
| 18-102_k141_136278 | Not-determined |
| 18-102_k141_94914 | Not-determined |
| 18-102_k141_167737 | Low-quality |
| 18-102_k141_125948 | Not-determined |
| 18-102_k141_197949 | Not-determined |
| 18-102_k141_239730 | Low-quality |
| 18-102_k141_197950 | Low-quality |
| 18-102_k141_125952 | Not-determined |
| 18-102_k141_167740 | Not-determined |
| 18-102_k141_167745 | Not-determined |
| 18-102_k141_197957 | Not-determined |
| 18-102_k141_156950 | Not-determined |
| 18-102_k141_94925 | Not-determined |
| 18-102_k141_197961 | Not-determined |
| 18-102_k141_178041 | Not-determined |
| 18-102_k141_197962 | Not-determined |
| 18-102_k141_178042 | Not-determined |
| 18-102_k141_136298 | Not-determined |
| 18-102_k141_178050 | Not-determined |
| 18-102_k141_188084 | Not-determined |
| 18-102_k141_188085 | Not-determined |
| 18-102_k141_218986 | Not-determined |
| 18-102_k141_188087 | Not-determined |
| 18-102_k141_239734 | Not-determined |
| 18-102_k141_239738 | Not-determined |
| 18-102_k141_197968 | Not-determined |
| 18-102_k141_1649 | Not-determined |
| 18-102_k141_1650 | Not-determined |
| 18-102_k141_197971 | Low-quality |
| 18-102_k141_178054 | Not-determined |
| 18-102_k141_1657 | Not-determined |
| 18-102_k141_94951 | Not-determined |
| 18-102_k141_188101 | Not-determined |
| 18-102_k141_178061 | Not-determined |
| 18-102_k141_167757 | Not-determined |
| 18-102_k141_197984 | Not-determined |

| | |
|---|---|
| 18-102_k141_53341 | Not-determined |
| 18-102_k141_22550 | Not-determined |
| 18-102_k141_1662 | Not-determined |
| 18-102_k141_188108 | Not-determined |
| 18-102_k141_136309 | Low-quality |
| 18-102_k141_105232 | Not-determined |
| 18-102_k141_74010 | Not-determined |
| 18-102_k141_136317 | Not-determined |
| 18-102_k141_44009 | Not-determined |
| 18-102_k141_167777 | Not-determined |
| 18-102_k141_44016 | Not-determined |
| 18-102_k141_198011 | Not-determined |
| 18-102_k141_239760 | Not-determined |
| 18-102_k141_167780 | Not-determined |
| 18-102_k141_74026 | Not-determined |
| 18-102_k141_22560 | Not-determined |
| 18-102_k141_22561 | Low-quality |
| 18-102_k141_53374 | Not-determined |
| 18-102_k141_178094 | Not-determined |
| 18-102_k141_44030 | Not-determined |
| 18-102_k141_44032 | Not-determined |
| 18-102_k141_167795 | Not-determined |
| 18-102_k141_22569 | Not-determined |
| 18-102_k141_136325 | Not-determined |
| 18-102_k141_208710 | Not-determined |
| 18-102_k141_218991 | Not-determined |
| 18-102_k141_198027 | Not-determined |
| 18-102_k141_44033 | Low-quality |
| 18-102_k141_63665 | Not-determined |
| 18-102_k141_74042 | Not-determined |
| 18-102_k141_44037 | Not-determined |
| 18-102_k141_44038 | Not-determined |
| 18-102_k141_218998 | Not-determined |
| 18-102_k141_1687 | Not-determined |
| 18-102_k141_219000 | Not-determined |
| 18-102_k141_105274 | Not-determined |
| 18-102_k141_136341 | Not-determined |
| 18-102_k141_105275 | Not-determined |
| 18-102_k141_22582 | Not-determined |
| 18-102_k141_74055 | Not-determined |
| 18-102_k141_44043 | Low-quality |
| 18-102_k141_219001 | Not-determined |
| 18-102_k141_115421 | Not-determined |
| 18-102_k141_22584 | Not-determined |
| 18-102_k141_136345 | Low-quality |
| 18-102_k141_44046 | Not-determined |
| 18-102_k141_74066 | Not-determined |
| 18-102_k141_239780 | Not-determined |
| 18-102_k141_198043 | Not-determined |
| 18-102_k141_198044 | Not-determined |
| 18-102_k141_53393 | Not-determined |
| 18-102_k141_53397 | Not-determined |
| 18-102_k141_178121 | Not-determined |
| 18-102_k141_188152 | Not-determined |
| 18-102_k141_115443 | Not-determined |
| 18-102_k141_136355 | Not-determined |
| 18-102_k141_44060 | Not-determined |
| 18-102_k141_105282 | Not-determined |
| 18-102_k141_44062 | Not-determined |
| 18-102_k141_63691 | Not-determined |
| 18-102_k141_105283 | Not-determined |
| 18-102_k141_136372 | Low-quality |
| 18-102_k141_156958 | Not-determined |
| 18-102_k141_136376 | Not-determined |
| 18-102_k141_136381 | Not-determined |
| 18-102_k141_136387 | Not-determined |
| 18-102_k141_44087 | Not-determined |
| 18-102_k141_156963 | Low-quality |
| 18-102_k141_84632 | Not-determined |
| 18-102_k141_74071 | Not-determined |
| 18-102_k141_105301 | Low-quality |
| 18-102_k141_44100 | Not-determined |
| 18-102_k141_105304 | Low-quality |
| 18-102_k141_178133 | Low-quality |
| 18-102_k141_229693 | Not-determined |
| 18-102_k141_1706 | Not-determined |
| 18-102_k141_178135 | Not-determined |
| 18-102_k141_239794 | Low-quality |
| 18-102_k141_22617 | Not-determined |
| 18-102_k141_94993 | Not-determined |
| 18-102_k141_136398 | Not-determined |
| 18-102_k141_74083 | Not-determined |
| 18-102_k141_229699 | Not-determined |
| 18-102_k141_74092 | Not-determined |
| 18-102_k141_63717 | Not-determined |
| 18-102_k141_53416 | Not-determined |
| 18-102_k141_229716 | Not-determined |
| 18-102_k141_219015 | Not-determined |
| 18-102_k141_63729 | Not-determined |
| 18-102_k141_63731 | Not-determined |
| 18-102_k141_63734 | Not-determined |
| 18-102_k141_115476 | Low-quality |
| 18-102_k141_156996 | Not-determined |
| 18-102_k141_157000 | Low-quality |
| 18-102_k141_219018 | Not-determined |
| 18-102_k141_1741 | Low-quality |
| 18-102_k141_105330 | Not-determined |
| 18-102_k141_178186 | Not-determined |
| 18-102_k141_239812 | Not-determined |
| 18-102_k141_239813 | Not-determined |
| 18-102_k141_178193 | Not-determined |
| 18-102_k141_219023 | Not-determined |
| 18-102_k141_239815 | Not-determined |
| 18-102_k141_157021 | Not-determined |
| 18-102_k141_239819 | Not-determined |
| 18-102_k141_178210 | Not-determined |
| 18-102_k141_63752 | Low-quality |
| 18-102_k141_63764 | Not-determined |
| 18-102_k141_136437 | Not-determined |
| 18-102_k141_136443 | Not-determined |
| 18-102_k141_12140 | Not-determined |
| 18-102_k141_157027 | Not-determined |
| 18-102_k141_157028 | Not-determined |
| 18-102_k141_105342 | Not-determined |
| 18-102_k141_84661 | Not-determined |
| 18-102_k141_115494 | Not-determined |
| 18-102_k141_178224 | Not-determined |
| 18-102_k141_105365 | Not-determined |
| 18-102_k141_178227 | Not-determined |
| 18-102_k141_198064 | Low-quality |
| 18-102_k141_157047 | Not-determined |
| 18-102_k141_44139 | Not-determined |
| 18-102_k141_219036 | Not-determined |
| 18-102_k141_198070 | Not-determined |
| 18-102_k141_105386 | Not-determined |
| 18-102_k141_63786 | Low-quality |
| 18-102_k141_105388 | Not-determined |
| 18-102_k141_115530 | Not-determined |
| 18-102_k141_208766 | Not-determined |
| 18-102_k141_208767 | Not-determined |
| 18-102_k141_208769 | Low-quality |
| 18-102_k141_208770 | Not-determined |
| 18-102_k141_115536 | Not-determined |
| 18-102_k141_105392 | Not-determined |
| 18-102_k141_12152 | Not-determined |
| 18-102_k141_63792 | Not-determined |
| 18-102_k141_115541 | Low-quality |
| 18-102_k141_105395 | Not-determined |
| 18-102_k141_125999 | Not-determined |
| 18-102_k141_178235 | Not-determined |
| 18-102_k141_208786 | Not-determined |
| 18-102_k141_115544 | Not-determined |
| 18-102_k141_115547 | Not-determined |
| 18-102_k141_126003 | Low-quality |
| 18-102_k141_53477 | Not-determined |
| 18-102_k141_198078 | Not-determined |
| 18-102_k141_126011 | Not-determined |
| 18-102_k141_84686 | Low-quality |
| 18-102_k141_198082 | Not-determined |
| 18-102_k141_74151 | Not-determined |
| 18-102_k141_126016 | Not-determined |
| 18-102_k141_1818 | Not-determined |
| 18-102_k141_32642 | Not-determined |
| 18-102_k141_32646 | Not-determined |
| 18-102_k141_178244 | Not-determined |
| 18-102_k141_1828 | Not-determined |
| 18-102_k141_208802 | Low-quality |
| 18-102_k141_198096 | Low-quality |
| 18-102_k141_126026 | Low-quality |
| 18-102_k141_157071 | Not-determined |
| 18-102_k141_12170 | Not-determined |
| 18-102_k141_178265 | Not-determined |
| 18-102_k141_178266 | Not-determined |
| 18-102_k141_157075 | Not-determined |
| 18-102_k141_229797 | Not-determined |
| 18-102_k141_126043 | Not-determined |
| 18-102_k141_53488 | Not-determined |
| 18-102_k141_229802 | Not-determined |
| 18-102_k141_167841 | Low-quality |
| 18-102_k141_1859 | Low-quality |
| 18-102_k141_167855 | Not-determined |
| 18-102_k141_115575 | Low-quality |
| 18-102_k141_12199 | Not-determined |
| 18-102_k141_1870 | Not-determined |
| 18-102_k141_115578 | Not-determined |
| 18-102_k141_84732 | Not-determined |
| 18-102_k141_229813 | Low-quality |
| 18-102_k141_136474 | Low-quality |
| 18-102_k141_115581 | Low-quality |
| 18-102_k141_178281 | Low-quality |
| 18-102_k141_136479 | Not-determined |
| 18-102_k141_229821 | Not-determined |
| 18-102_k141_239880 | Not-determined |
| 18-102_k141_198107 | Not-determined |
| 18-102_k141_229829 | Not-determined |
| 18-102_k141_105432 | Not-determined |
| 18-102_k141_1877 | Not-determined |
| 18-102_k141_44153 | Not-determined |
| 18-102_k141_115602 | Not-determined |
| 18-102_k141_115603 | Not-determined |
| 18-102_k141_1881 | Not-determined |
| 18-102_k141_115605 | Not-determined |
| 18-102_k141_44160 | Not-determined |
| 18-102_k141_95023 | Not-determined |
| 18-102_k141_178298 | Low-quality |
| 18-102_k141_126059 | Low-quality |
| 18-102_k141_219064 | Not-determined |
| 18-102_k141_95030 | Low-quality |
| 18-102_k141_136497 | Not-determined |
| 18-102_k141_167868 | Not-determined |
| 18-102_k141_44173 | Not-determined |
| 18-102_k141_95039 | Not-determined |
| 18-102_k141_219073 | Not-determined |
| 18-102_k141_74178 | Low-quality |
| 18-102_k141_105448 | Not-determined |
| 18-102_k141_136504 | Not-determined |
| 18-102_k141_126075 | Not-determined |
| 18-102_k141_32669 | Not-determined |
| 18-102_k141_219085 | Not-determined |
| 18-102_k141_95059 | Not-determined |
| 18-102_k141_32672 | Not-determined |
| 18-102_k141_146807 | Low-quality |
| 18-102_k141_115635 | Not-determined |
| 18-102_k141_239892 | Low-quality |
| 18-102_k141_198120 | Not-determined |
| 18-102_k141_115641 | Not-determined |
| 18-102_k141_44185 | Not-determined |
| 18-102_k141_53512 | Not-determined |
| 18-102_k141_136527 | Not-determined |
| 18-102_k141_146823 | Not-determined |
| 18-102_k141_239904 | Not-determined |
| 18-102_k141_53518 | Not-determined |
| 18-102_k141_105456 | Not-determined |
| 18-102_k141_229851 | Not-determined |
| 18-102_k141_136542 | Low-quality |
| 18-102_k141_53531 | Not-determined |
| 18-102_k141_136547 | Not-determined |
| 18-102_k141_126089 | Not-determined |
| 18-102_k141_136554 | Not-determined |
| 18-102_k141_146842 | Not-determined |
| 18-102_k141_178337 | Low-quality |
| 18-102_k141_126093 | Not-determined |
| 18-102_k141_53550 | Not-determined |
| 18-102_k141_1890 | Not-determined |
| 18-102_k141_74212 | Not-determined |
| 18-102_k141_22654 | Not-determined |
| 18-102_k141_22658 | Not-determined |
| 18-102_k141_198148 | Not-determined |
| 18-102_k141_198149 | Not-determined |
| 18-102_k141_74217 | Not-determined |
| 18-102_k141_178344 | Not-determined |
| 18-102_k141_229863 | Not-determined |
| 18-102_k141_74221 | Not-determined |
| 18-102_k141_1896 | Not-determined |
| 18-102_k141_198154 | Not-determined |
| 18-102_k141_22670 | Not-determined |
| 18-102_k141_44231 | Not-determined |
| 18-102_k141_167884 | Not-determined |
| 18-102_k141_44236 | Not-determined |
| 18-102_k141_44238 | Not-determined |
| 18-102_k141_208810 | Not-determined |
| 18-102_k141_1902 | Not-determined |
| 18-102_k141_22678 | Not-determined |
| 18-102_k141_84741 | Not-determined |
| 18-102_k141_208813 | Not-determined |
| 18-102_k141_126105 | Not-determined |
| 18-102_k141_84745 | Not-determined |
| 18-102_k141_126107 | Not-determined |
| 18-102_k141_167898 | Not-determined |
| 18-102_k141_95084 | Not-determined |
| 18-102_k141_105487 | Not-determined |
| 18-102_k141_105491 | Not-determined |
| 18-102_k141_63840 | Not-determined |
| 18-102_k141_146877 | Not-determined |
| 18-102_k141_146878 | Not-determined |
| 18-102_k141_105504 | Not-determined |
| 18-102_k141_22713 | Not-determined |
| 18-102_k141_32688 | Low-quality |
| 18-102_k141_198188 | Not-determined |
| 18-102_k141_105517 | Low-quality |
| 18-102_k141_22717 | Not-determined |
| 18-102_k141_22724 | Not-determined |
| 18-102_k141_208856 | Not-determined |
| 18-102_k141_84775 | Not-determined |
| 18-102_k141_105532 | Not-determined |
| 18-102_k141_63861 | Not-determined |
| 18-102_k141_105534 | Not-determined |
| 18-102_k141_53566 | Not-determined |
| 18-102_k141_84781 | Not-determined |
| 18-102_k141_63864 | Not-determined |
| 18-102_k141_84782 | Not-determined |
| 18-102_k141_84784 | Not-determined |
| 18-102_k141_198197 | Not-determined |
| 18-102_k141_188209 | Not-determined |
| 18-102_k141_188210 | Low-quality |
| 18-102_k141_126126 | Not-determined |
| 18-102_k141_63866 | Not-determined |
| 18-102_k141_126133 | Not-determined |
| 18-102_k141_198203 | Low-quality |
| 18-102_k141_105547 | Not-determined |
| 18-102_k141_84808 | Not-determined |
| 18-102_k141_219100 | Not-determined |
| 18-102_k141_84817 | Not-determined |
| 18-102_k141_136576 | Not-determined |
| 18-102_k141_188218 | Not-determined |
| 18-102_k141_208885 | Not-determined |
| 18-102_k141_84826 | Not-determined |
| 18-102_k141_84827 | Not-determined |
| 18-102_k141_1910 | Not-determined |
| 18-102_k141_84829 | Not-determined |
| 18-102_k141_157104 | Not-determined |
| 18-102_k141_1914 | Not-determined |
| 18-102_k141_208893 | Not-determined |
| 18-102_k141_146888 | Low-quality |
| 18-102_k141_208900 | Not-determined |
| 18-102_k141_157112 | Not-determined |
| 18-102_k141_157116 | Low-quality |
| 18-102_k141_126144 | Not-determined |
| 18-102_k141_188248 | Not-determined |
| 18-102_k141_157124 | Not-determined |
| 18-102_k141_157126 | Not-determined |
| 18-102_k141_229911 | Not-determined |
| 18-102_k141_105569 | Not-determined |
| 18-102_k141_126147 | Low-quality |
| 18-102_k141_126149 | Not-determined |
| 18-102_k141_126150 | Low-quality |
| 18-102_k141_44307 | Low-quality |
| 18-102_k141_44309 | Low-quality |
| 18-102_k141_229924 | Not-determined |
| 18-102_k141_126156 | Not-determined |
| 18-102_k141_229931 | Not-determined |
| 18-102_k141_178401 | Not-determined |
| 18-102_k141_229934 | Not-determined |
| 18-102_k141_126159 | Not-determined |
| 18-102_k141_178403 | Not-determined |
| 18-102_k141_84862 | Not-determined |
| 18-102_k141_126163 | Not-determined |
| 18-102_k141_53636 | Not-determined |
| 18-102_k141_178404 | Not-determined |
| 18-102_k141_22754 | Not-determined |
| 18-102_k141_239926 | Not-determined |
| 18-102_k141_239932 | Not-determined |
| 18-102_k141_53652 | Low-quality |
| 18-102_k141_74291 | Not-determined |
| 18-102_k141_229942 | Not-determined |
| 18-102_k141_105588 | Not-determined |
| 18-102_k141_84877 | Not-determined |
| 18-102_k141_157150 | Not-determined |
| 18-102_k141_53664 | Low-quality |
| 18-102_k141_188274 | Not-determined |
| 18-102_k141_239954 | Not-determined |
| 18-102_k141_1956 | Not-determined |
| 18-102_k141_1957 | Not-determined |
| 18-102_k141_126175 | Not-determined |
| 18-102_k141_44320 | Low-quality |
| 18-102_k141_44321 | Not-determined |
| 18-102_k141_53674 | Not-determined |
| 18-102_k141_239976 | Not-determined |
| 18-102_k141_239978 | Not-determined |
| 18-102_k141_53685 | Low-quality |
| 18-102_k141_74333 | Not-determined |
| 18-102_k141_53696 | Not-determined |
| 18-102_k141_74334 | Not-determined |
| 18-102_k141_146914 | Not-determined |
| 18-102_k141_229974 | Not-determined |
| 18-102_k141_239991 | Not-determined |
| 18-102_k141_229975 | Low-quality |
| 18-102_k141_229976 | Not-determined |
| 18-102_k141_178405 | Low-quality |
| 18-102_k141_229978 | Not-determined |
| 18-102_k141_84887 | Low-quality |
| 18-102_k141_63957 | Not-determined |
| 18-102_k141_178412 | Not-determined |
| 18-102_k141_157166 | Not-determined |
| 18-102_k141_157168 | Not-determined |
| 18-102_k141_136642 | Not-determined |
| 18-102_k141_136645 | Not-determined |
| 18-102_k141_178420 | Not-determined |
| 18-102_k141_219105 | Not-determined |
| 18-102_k141_229989 | Low-quality |
| 18-102_k141_229991 | Not-determined |
| 18-102_k141_157175 | Not-determined |
| 18-102_k141_240001 | Not-determined |
| 18-102_k141_146922 | Not-determined |
| 18-102_k141_146923 | Not-determined |
| 18-102_k141_53717 | Not-determined |
| 18-102_k141_74346 | Not-determined |
| 18-102_k141_53722 | Not-determined |
| 18-102_k141_53726 | Not-determined |
| 18-102_k141_44379 | Not-determined |
| 18-102_k141_188276 | Not-determined |
| 18-102_k141_63972 | Not-determined |
| 18-102_k141_74351 | Not-determined |
| 18-102_k141_44380 | Not-determined |
| 18-102_k141_44383 | Not-determined |
| 18-102_k141_84923 | Not-determined |
| 18-102_k141_126211 | Not-determined |
| 18-102_k141_105634 | Not-determined |
| 18-102_k141_105635 | Not-determined |
| 18-102_k141_95126 | Not-determined |
| 18-102_k141_95127 | Not-determined |
| 18-102_k141_136666 | Not-determined |
| 18-102_k141_2014 | Low-quality |
| 18-102_k141_188288 | Not-determined |
| 18-102_k141_105645 | Not-determined |
| 18-102_k141_188290 | Not-determined |
| 18-102_k141_84946 | Not-determined |
| 18-102_k141_105646 | Not-determined |
| 18-102_k141_136680 | Not-determined |
| 18-102_k141_105650 | Not-determined |
| 18-102_k141_53747 | Not-determined |
| 18-102_k141_84949 | Not-determined |
| 18-102_k141_208958 | Not-determined |
| 18-102_k141_84952 | Not-determined |
| 18-102_k141_208959 | Not-determined |
| 18-102_k141_115721 | Not-determined |
| 18-102_k141_95140 | Not-determined |
| 18-102_k141_126222 | Not-determined |
| 18-102_k141_208973 | Not-determined |
| 18-102_k141_22794 | Not-determined |
| 18-102_k141_157215 | Not-determined |
| 18-102_k141_22796 | Not-determined |
| 18-102_k141_115732 | Not-determined |
| 18-102_k141_115734 | Not-determined |
| 18-102_k141_230001 | Not-determined |
| 18-102_k141_115736 | Not-determined |
| 18-102_k141_230002 | Not-determined |
| 18-102_k141_136696 | Not-determined |
| 18-102_k141_136697 | Not-determined |
| 18-102_k141_115740 | Not-determined |
| 18-102_k141_44410 | Not-determined |
| 18-102_k141_208984 | Not-determined |
| 18-102_k141_22803 | Low-quality |
| 18-102_k141_188299 | Not-determined |
| 18-102_k141_95159 | Not-determined |
| 18-102_k141_188310 | Low-quality |
| 18-102_k141_208999 | Not-determined |
| 18-102_k141_240014 | Not-determined |
| 18-102_k141_240017 | Not-determined |
| 18-102_k141_115749 | Not-determined |
| 18-102_k141_44445 | Not-determined |
| 18-102_k141_146932 | Not-determined |
| 18-102_k141_44448 | Not-determined |
| 18-102_k141_105665 | Not-determined |
| 18-102_k141_136713 | Low-quality |
| 18-102_k141_136716 | Not-determined |
| 18-102_k141_157232 | Not-determined |
| 18-102_k141_2041 | Not-determined |
| 18-102_k141_64019 | Not-determined |
| 18-102_k141_44454 | Not-determined |
| 18-102_k141_2046 | Not-determined |
| 18-102_k141_2050 | Low-quality |
| 18-102_k141_84966 | Not-determined |
| 18-102_k141_230009 | Not-determined |
| 18-102_k141_95179 | Not-determined |
| 18-102_k141_240027 | Not-determined |
| 18-102_k141_230012 | Low-quality |
| 18-102_k141_240039 | Not-determined |
| 18-102_k141_157246 | Not-determined |
| 18-102_k141_32753 | Not-determined |
| 18-102_k141_157248 | Not-determined |
| 18-102_k141_53780 | Not-determined |
| 18-102_k141_44487 | Low-quality |
| 18-102_k141_2062 | Not-determined |
| 18-102_k141_230031 | Not-determined |
| 18-102_k141_157254 | Not-determined |
| 18-102_k141_53791 | Low-quality |
| 18-102_k141_32757 | Not-determined |
| 18-102_k141_53792 | Not-determined |
| 18-102_k141_2065 | Not-determined |
| 18-102_k141_105683 | Not-determined |
| 18-102_k141_198239 | Not-determined |
| 18-102_k141_95214 | Not-determined |
| 18-102_k141_178480 | Not-determined |
| 18-102_k141_64043 | Not-determined |
| 18-102_k141_198243 | Not-determined |
| 18-102_k141_198245 | Not-determined |
| 18-102_k141_22818 | Not-determined |
| 18-102_k141_188368 | Not-determined |
| 18-102_k141_22820 | Not-determined |
| 18-102_k141_84992 | Not-determined |
| 18-102_k141_105690 | Low-quality |
| 18-102_k141_64056 | Not-determined |
| 18-102_k141_74392 | Not-determined |
| 18-102_k141_53813 | Not-determined |
| 18-102_k141_74397 | Not-determined |
| 18-102_k141_95245 | Not-determined |
| 18-102_k141_188387 | Not-determined |
| 18-102_k141_157269 | Low-quality |
| 18-102_k141_64071 | Not-determined |
| 18-102_k141_188390 | Not-determined |
| 18-102_k141_157272 | Not-determined |
| 18-102_k141_219168 | Not-determined |
| 18-102_k141_44510 | Not-determined |
| 18-102_k141_188406 | Not-determined |
| 18-102_k141_198269 | Not-determined |
| 18-102_k141_95259 | Not-determined |
| 18-102_k141_115805 | Not-determined |
| 18-102_k141_198273 | Low-quality |
| 18-102_k141_188415 | Not-determined |
| 18-102_k141_115807 | Low-quality |
| 18-102_k141_95270 | Not-determined |
| 18-102_k141_74420 | Not-determined |
| 18-102_k141_12250 | Low-quality |
| 18-102_k141_178491 | Low-quality |
| 18-102_k141_178492 | Not-determined |
| 18-102_k141_178494 | Not-determined |
| 18-102_k141_219182 | Not-determined |
| 18-102_k141_12257 | Not-determined |
| 18-102_k141_95276 | Not-determined |
| 18-102_k141_198281 | Not-determined |
| 18-102_k141_219186 | Not-determined |
| 18-102_k141_167950 | Not-determined |
| 18-102_k141_167954 | Not-determined |
| 18-102_k141_115812 | Not-determined |
| 18-102_k141_178504 | Low-quality |
| 18-102_k141_95287 | Not-determined |
| 18-102_k141_178509 | Not-determined |
| 18-102_k141_219201 | Not-determined |
| 18-102_k141_12262 | Low-quality |
| 18-102_k141_167965 | Not-determined |
| 18-102_k141_53816 | Not-determined |
| 18-102_k141_53819 | Not-determined |
| 18-102_k141_12265 | Not-determined |
| 18-102_k141_53822 | Not-determined |
| 18-102_k141_85032 | Not-determined |
| 18-102_k141_219213 | Not-determined |
| 18-102_k141_85035 | Not-determined |
| 18-102_k141_85036 | Not-determined |
| 18-102_k141_85038 | Low-quality |
| 18-102_k141_219216 | Not-determined |
| 18-102_k141_157306 | Not-determined |
| 18-102_k141_105722 | Not-determined |
| 18-102_k141_105723 | Not-determined |
| 18-102_k141_136784 | Not-determined |
| 18-102_k141_105729 | Not-determined |
| 18-102_k141_240051 | Not-determined |
| 18-102_k141_230095 | Not-determined |
| 18-102_k141_53837 | Low-quality |
| 18-102_k141_240053 | Not-determined |
| 18-102_k141_22856 | Not-determined |
| 18-102_k141_53843 | Not-determined |
| 18-102_k141_105739 | Not-determined |
| 18-102_k141_22858 | Low-quality |
| 18-102_k141_178514 | Not-determined |
| 18-102_k141_85055 | Not-determined |
| 18-102_k141_22860 | Not-determined |
| 18-102_k141_157324 | Not-determined |
| 18-102_k141_64119 | Low-quality |
| 18-102_k141_240063 | Low-quality |
| 18-102_k141_64120 | Low-quality |
| 18-102_k141_105760 | Not-determined |
| 18-102_k141_178536 | Not-determined |
| 18-102_k141_157340 | Not-determined |
| 18-102_k141_105764 | Not-determined |
| 18-102_k141_126279 | Not-determined |
| 18-102_k141_64136 | Low-quality |
| 18-102_k141_44540 | Low-quality |
| 18-102_k141_167997 | Not-determined |
| 18-102_k141_105772 | Not-determined |
| 18-102_k141_167998 | Not-determined |
| 18-102_k141_44544 | Not-determined |
| 18-102_k141_178549 | Not-determined |
| 18-102_k141_95311 | Not-determined |
| 18-102_k141_64144 | Not-determined |
| 18-102_k141_230137 | Not-determined |
| 18-102_k141_95318 | Not-determined |
| 18-102_k141_240084 | Not-determined |
| 18-102_k141_230144 | Not-determined |
| 18-102_k141_240087 | Not-determined |
| 18-102_k141_64157 | Not-determined |
| 18-102_k141_85093 | Not-determined |
| 18-102_k141_64164 | Not-determined |
| 18-102_k141_240094 | Not-determined |
| 18-102_k141_146953 | Not-determined |
| 18-102_k141_168007 | Not-determined |
| 18-102_k141_157369 | Not-determined |
| 18-102_k141_64188 | Low-quality |
| 18-102_k141_240119 | Low-quality |
| 18-102_k141_157378 | Not-determined |
| 18-102_k141_168023 | Not-determined |
| 18-102_k141_209062 | Not-determined |
| 18-102_k141_168028 | Not-determined |
| 18-102_k141_12300 | Not-determined |
| 18-102_k141_168033 | Not-determined |
| 18-102_k141_168036 | Not-determined |
| 18-102_k141_178593 | Not-determined |
| 18-102_k141_178594 | Not-determined |
| 18-102_k141_22909 | Not-determined |
| 18-102_k141_209072 | Not-determined |
| 18-102_k141_64204 | Not-determined |
| 18-102_k141_95360 | Not-determined |
| 18-102_k141_178599 | Not-determined |
| 18-102_k141_64211 | Not-determined |
| 18-102_k141_64212 | Not-determined |
| 18-102_k141_178608 | Not-determined |
| 17-106-final-viral-combined_k141_71647\|\|full | Medium-quality |
| 18-102_k141_115850 | Not-determined |
| 18-102_k141_12316 | Not-determined |

| | |
|---|---|
| 18-102_k141_157403 | Not-determined |
| 18-102_k141_136873 | Not-determined |
| 18-102_k141_178618 | Not-determined |
| 18-102_k141_230215 | Not-determined |
| 18-102_k141_136895 | Not-determined |
| 18-102_k141_95397 | Not-determined |
| 18-102_k141_209092 | Not-determined |
| 18-102_k141_105799 | Not-determined |
| 18-102_k141_95404 | Not-determined |
| 18-102_k141_136903 | Not-determined |
| 18-102_k141_115871 | Low-quality |
| 18-102_k141_126317 | Not-determined |
| 18-102_k141_219290 | Not-determined |
| 18-102_k141_146962 | Not-determined |
| 18-102_k141_85123 | Not-determined |
| 18-102_k141_95421 | Not-determined |
| 18-102_k141_219294 | Not-determined |
| 18-102_k141_85125 | Not-determined |
| 18-102_k141_44569 | Not-determined |
| 18-102_k141_240157 | Not-determined |
| 18-102_k141_44575 | Not-determined |
| 18-102_k141_95434 | Not-determined |
| 18-102_k141_85129 | Low-quality |
| 18-102_k141_44582 | Not-determined |
| 18-102_k141_32803 | Not-determined |
| 18-102_k141_32805 | Not-determined |
| 18-102_k141_126334 | Low-quality |
| 18-102_k141_230241 | Low-quality |
| 18-102_k141_230242 | Low-quality |
| 18-102_k141_198283 | Not-determined |
| 18-102_k141_230243 | Not-determined |
| 18-102_k141_146973 | Low-quality |
| 18-102_k141_146975 | Not-determined |
| 18-102_k141_64223 | Not-determined |
| 18-102_k141_44601 | Low-quality |
| 18-102_k141_126342 | Not-determined |
| 18-102_k141_32812 | Not-determined |
| 18-102_k141_32818 | Low-quality |
| 18-102_k141_198293 | Not-determined |
| 18-102_k141_240169 | Not-determined |
| 18-102_k141_53940 | Not-determined |
| 18-102_k141_230257 | Not-determined |
| 18-102_k141_198300 | Not-determined |
| 18-102_k141_188454 | Not-determined |
| 18-102_k141_12320 | Not-determined |
| 18-102_k141_32820 | Not-determined |
| 18-102_k141_240174 | Low-quality |
| 18-102_k141_168061 | Not-determined |
| 18-102_k141_95459 | Not-determined |
| 18-102_k141_95461 | Not-determined |
| 18-102_k141_85165 | Low-quality |
| 18-102_k141_157420 | Not-determined |
| 18-102_k141_168069 | Not-determined |
| 18-102_k141_188477 | Not-determined |
| 18-102_k141_146990 | Not-determined |
| 18-102_k141_188486 | Not-determined |
| 18-102_k141_136921 | Not-determined |
| 18-102_k141_64244 | Not-determined |
| 18-102_k141_64246 | Not-determined |
| 18-102_k141_74430 | Not-determined |
| 18-102_k141_157430 | Not-determined |
| 18-102_k141_188496 | Not-determined |
| 18-102_k141_126380 | Not-determined |
| 18-102_k141_64263 | Not-determined |
| 18-102_k141_22981 | Not-determined |
| 18-102_k141_22983 | Not-determined |
| 18-102_k141_157442 | Not-determined |
| 18-102_k141_219303 | Not-determined |
| 18-102_k141_44613 | Not-determined |
| 18-102_k141_219307 | Not-determined |
| 18-102_k141_126386 | Not-determined |
| 18-102_k141_168105 | Not-determined |
| 18-102_k141_126388 | Not-determined |
| 18-102_k141_188499 | Not-determined |
| 18-102_k141_95471 | Not-determined |
| 18-102_k141_53968 | Low-quality |
| 18-102_k141_219310 | Low-quality |
| 18-102_k141_168110 | Not-determined |
| 18-102_k141_85194 | Not-determined |
| 18-102_k141_240186 | Low-quality |
| 18-102_k141_126399 | Not-determined |
| 18-102_k141_136948 | Not-determined |
| 18-102_k141_44624 | Not-determined |
| 18-102_k141_219313 | Not-determined |
| 18-102_k141_126407 | Not-determined |
| 18-102_k141_198325 | Not-determined |
| 18-102_k141_168122 | Not-determined |
| 18-102_k141_53977 | Not-determined |
| 18-102_k141_136957 | Not-determined |
| 18-102_k141_32833 | Not-determined |
| 18-102_k141_188522 | Not-determined |
| 18-102_k141_32838 | Not-determined |
| 18-102_k141_219319 | Low-quality |
| 18-102_k141_136965 | Not-determined |
| 18-102_k141_95496 | Not-determined |
| 18-102_k141_157458 | Not-determined |
| 18-102_k141_2097 | Not-determined |
| 18-102_k141_23003 | Not-determined |
| 18-102_k141_136982 | Not-determined |
| 18-102_k141_23008 | Not-determined |
| 18-102_k141_219321 | Not-determined |
| 18-102_k141_2101 | Not-determined |
| 18-102_k141_219323 | Not-determined |
| 18-102_k141_230324 | Not-determined |
| 18-102_k141_2104 | Not-determined |
| 18-102_k141_74465 | Not-determined |
| 18-102_k141_230326 | Not-determined |
| 18-102_k141_12385 | Not-determined |
| 18-102_k141_2108 | Not-determined |
| 18-102_k141_230330 | Not-determined |
| 18-102_k141_147006 | Not-determined |
| 18-102_k141_230332 | Not-determined |
| 18-102_k141_230334 | Not-determined |
| 18-102_k141_126428 | Not-determined |
| 18-102_k141_198348 | Not-determined |
| 18-102_k141_74476 | Not-determined |
| 18-102_k141_137008 | Low-quality |
| 18-102_k141_230342 | Not-determined |
| 18-102_k141_168159 | Not-determined |
| 18-102_k141_168160 | Not-determined |
| 18-102_k141_32878 | Not-determined |
| 18-102_k141_74496 | Not-determined |
| 18-102_k141_188532 | Not-determined |
| 18-102_k141_2118 | Not-determined |
| 18-102_k141_32887 | Not-determined |
| 18-102_k141_12400 | Not-determined |
| 18-102_k141_168176 | Not-determined |
| 18-102_k141_32892 | Not-determined |
| 18-102_k141_188540 | Not-determined |
| 18-102_k141_147025 | Not-determined |
| 18-102_k141_105854 | Not-determined |
| 18-102_k141_198378 | Not-determined |
| 18-102_k141_2121 | Not-determined |
| 18-102_k141_32896 | Not-determined |
| 18-102_k141_126444 | Not-determined |
| 18-102_k141_32910 | Not-determined |
| 18-102_k141_32911 | Not-determined |
| 18-102_k141_230360 | Not-determined |
| 18-102_k141_105867 | Not-determined |
| 18-102_k141_32915 | Not-determined |
| 18-102_k141_12416 | Not-determined |
| 18-102_k141_64271 | Not-determined |
| 18-102_k141_32930 | Not-determined |
| 18-102_k141_147035 | Not-determined |
| 18-102_k141_147039 | Not-determined |
| 18-102_k141_2129 | Not-determined |
| 18-102_k141_198397 | Not-determined |
| 18-102_k141_12420 | Not-determined |
| 18-102_k141_147046 | Not-determined |
| 18-102_k141_105887 | Not-determined |
| 18-102_k141_95516 | Not-determined |
| 18-102_k141_168203 | Not-determined |
| 18-102_k141_2137 | Not-determined |
| 18-102_k141_95519 | Not-determined |
| 18-102_k141_2138 | Not-determined |
| 18-102_k141_95520 | Not-determined |
| 18-102_k141_2142 | Not-determined |
| 18-102_k141_95524 | Not-determined |
| 18-102_k141_105897 | Not-determined |
| 18-102_k141_95525 | Not-determined |
| 18-102_k141_95529 | Not-determined |
| 18-102_k141_178641 | Not-determined |
| 18-102_k141_147055 | Low-quality |
| 18-102_k141_32955 | Not-determined |
| 18-102_k141_44665 | Not-determined |
| 18-102_k141_230386 | Not-determined |
| 18-102_k141_32958 | Not-determined |
| 18-102_k141_44666 | Not-determined |
| 18-102_k141_230390 | Not-determined |
| 18-102_k141_137054 | Not-determined |
| 18-102_k141_147062 | Not-determined |
| 18-102_k141_85229 | Low-quality |
| 18-102_k141_2155 | Not-determined |
| 18-102_k141_64280 | Low-quality |
| 18-102_k141_2156 | Not-determined |
| 18-102_k141_137055 | Low-quality |
| 18-102_k141_32982 | Not-determined |
| 18-102_k141_115909 | Not-determined |
| 18-102_k141_32985 | Not-determined |
| 18-102_k141_105929 | Not-determined |
| 18-102_k141_32988 | Not-determined |
| 18-102_k141_105934 | Not-determined |
| 18-102_k141_105936 | Not-determined |
| 18-102_k141_95554 | Low-quality |
| 18-102_k141_115916 | Low-quality |
| 18-102_k141_115920 | Not-determined |
| 18-102_k141_74540 | Not-determined |
| 18-102_k141_74542 | Not-determined |
| 18-102_k141_137070 | Not-determined |
| 18-102_k141_44694 | Not-determined |
| 18-102_k141_115934 | Not-determined |
| 18-102_k141_115935 | Not-determined |
| 18-102_k141_23054 | Not-determined |
| 18-102_k141_85244 | Not-determined |
| 18-102_k141_115940 | Not-determined |
| 18-102_k141_240245 | Not-determined |
| 18-102_k141_64306 | Low-quality |
| 18-102_k141_219351 | Not-determined |
| 18-102_k141_157534 | Not-determined |
| 18-102_k141_105956 | Not-determined |
| 18-102_k141_219356 | Not-determined |
| 18-102_k141_219360 | Not-determined |
| 18-102_k141_85256 | Low-quality |
| 18-102_k141_54016 | Not-determined |
| 18-102_k141_23059 | Not-determined |
| 18-102_k141_137086 | Not-determined |
| 18-102_k141_85263 | Not-determined |
| 18-102_k141_105964 | Not-determined |
| 18-102_k141_137089 | Not-determined |
| 18-102_k141_219371 | Not-determined |
| 18-102_k141_115952 | Not-determined |
| 18-102_k141_64325 | Low-quality |
| 18-102_k141_115957 | Low-quality |
| 18-102_k141_23069 | Not-determined |
| 18-102_k141_219387 | Not-determined |
| 18-102_k141_12456 | Not-determined |
| 18-102_k141_54033 | Low-quality |
| 18-102_k141_64334 | Not-determined |
| 18-102_k141_105975 | Low-quality |
| 18-102_k141_157550 | Not-determined |
| 18-102_k141_240257 | Low-quality |
| 18-102_k141_240259 | Not-determined |
| 18-102_k141_85275 | Low-quality |
| 18-102_k141_230404 | Not-determined |
| 18-102_k141_44732 | Not-determined |
| 18-102_k141_115974 | Low-quality |
| 18-102_k141_33023 | Not-determined |
| 18-102_k141_105983 | Low-quality |
| 18-102_k141_44734 | Not-determined |
| 18-102_k141_12469 | Not-determined |
| 18-102_k141_95594 | Not-determined |
| 18-102_k141_157570 | Not-determined |
| 18-102_k141_209111 | Not-determined |
| 18-102_k141_209112 | Not-determined |
| 18-102_k141_74567 | Not-determined |
| 18-102_k141_168252 | Low-quality |
| 18-102_k141_44762 | Not-determined |
| 18-102_k141_33044 | Not-determined |
| 18-102_k141_33045 | Low-quality |
| 18-102_k141_198441 | Not-determined |
| 18-102_k141_188633 | Not-determined |
| 18-102_k141_54076 | Not-determined |
| 18-102_k141_198445 | Not-determined |
| 18-102_k141_198447 | Not-determined |
| 18-102_k141_198449 | Not-determined |
| 18-102_k141_168264 | Not-determined |
| 18-102_k141_12482 | Not-determined |
| 18-102_k141_198453 | Not-determined |
| 18-102_k141_188658 | Not-determined |
| 18-102_k141_240270 | Low-quality |
| 18-102_k141_240273 | Low-quality |
| 18-102_k141_209138 | Not-determined |
| 18-102_k141_188664 | Not-determined |
| 18-102_k141_209144 | Not-determined |
| 18-102_k141_157599 | Not-determined |
| 18-102_k141_44781 | Not-determined |
| 18-102_k141_230449 | Low-quality |
| 18-102_k141_116012 | Not-determined |
| 18-102_k141_85286 | Not-determined |
| 18-102_k141_54109 | Not-determined |
| 18-102_k141_95621 | Not-determined |
| 18-102_k141_126577 | Low-quality |
| 18-102_k141_209154 | Not-determined |
| 18-102_k141_23097 | Low-quality |
| 18-102_k141_240285 | Low-quality |
| 18-102_k141_219409 | Not-determined |
| 18-102_k141_12514 | Not-determined |
| 18-102_k141_12516 | Not-determined |
| 18-102_k141_12520 | Low-quality |
| 18-102_k141_240294 | Not-determined |
| 18-102_k141_240295 | Not-determined |
| 18-102_k141_188671 | Not-determined |
| 18-102_k141_230462 | Not-determined |
| 18-102_k141_44800 | Low-quality |
| 18-102_k141_240301 | Not-determined |
| 18-102_k141_240307 | Not-determined |
| 18-102_k141_74613 | Not-determined |
| 18-102_k141_74621 | Not-determined |
| 18-102_k141_198457 | Not-determined |
| 18-102_k141_2216 | Not-determined |
| 18-102_k141_23113 | Not-determined |
| 18-102_k141_74632 | Not-determined |
| 18-102_k141_12544 | Not-determined |
| 18-102_k141_54129 | Not-determined |
| 18-102_k141_95643 | Not-determined |
| 18-102_k141_95645 | Not-determined |
| 18-102_k141_230485 | Not-determined |
| 18-102_k141_209187 | Not-determined |
| 18-102_k141_44808 | Not-determined |
| 18-102_k141_209189 | Not-determined |
| 18-102_k141_74653 | Not-determined |
| 18-102_k141_219416 | Low-quality |
| 18-102_k141_209195 | Not-determined |
| 18-102_k141_198470 | Not-determined |
| 18-102_k141_95647 | Not-determined |
| 18-102_k141_209200 | Low-quality |
| 18-102_k141_85339 | Not-determined |
| 18-102_k141_188691 | Not-determined |
| 18-102_k141_85354 | Not-determined |
| 18-102_k141_219433 | Not-determined |
| 18-102_k141_2228 | Not-determined |
| 18-102_k141_12549 | Not-determined |
| 18-102_k141_116055 | Not-determined |
| 18-102_k141_74678 | Not-determined |
| 18-102_k141_116060 | Low-quality |
| 18-102_k141_116061 | Not-determined |
| 18-102_k141_12556 | Not-determined |
| 18-102_k141_33082 | Not-determined |
| 18-102_k141_116067 | Not-determined |
| 18-102_k141_12562 | Not-determined |
| 18-102_k141_198487 | Not-determined |
| 18-102_k141_188709 | Not-determined |
| 18-102_k141_198489 | Not-determined |
| 18-102_k141_219456 | Not-determined |
| 18-102_k141_2235 | Not-determined |
| 18-102_k141_230505 | Not-determined |
| 18-102_k141_2236 | Not-determined |
| 18-102_k141_188713 | Not-determined |
| 18-102_k141_105999 | Not-determined |
| 18-102_k141_74698 | Not-determined |
| 18-102_k141_230509 | Not-determined |
| 18-102_k141_188716 | Not-determined |
| 18-102_k141_219466 | Low-quality |
| 18-102_k141_147080 | Not-determined |
| 18-102_k141_219471 | Not-determined |
| 18-102_k141_106012 | Not-determined |
| 18-102_k141_44853 | Not-determined |
| 18-102_k141_147083 | Not-determined |
| 18-102_k141_230529 | Not-determined |
| 18-102_k141_95659 | Not-determined |
| 18-102_k141_44868 | Not-determined |
| 18-102_k141_126608 | Not-determined |
| 18-102_k141_219483 | Not-determined |
| 18-102_k141_209208 | Not-determined |
| 18-102_k141_44878 | Not-determined |
| 18-102_k141_2247 | Low-quality |
| 18-102_k141_2248 | Not-determined |
| 18-102_k141_44881 | Not-determined |
| 18-102_k141_230539 | Low-quality |
| 18-102_k141_2253 | Not-determined |
| 18-102_k141_74723 | Not-determined |
| 18-102_k141_33093 | Low-quality |
| 18-102_k141_209213 | Not-determined |
| 18-102_k141_44883 | Not-determined |
| 18-102_k141_2257 | Low-quality |
| 18-102_k141_74727 | Not-determined |
| 18-102_k141_219486 | Not-determined |
| 18-102_k141_219487 | Not-determined |
| 18-102_k141_126613 | Not-determined |
| 18-102_k141_12577 | Not-determined |
| 18-102_k141_95686 | Not-determined |
| 18-102_k141_95691 | Low-quality |
| 18-102_k141_95692 | Not-determined |
| 18-102_k141_126619 | Not-determined |
| 18-102_k141_33113 | Not-determined |
| 18-102_k141_12584 | Low-quality |
| 18-102_k141_219492 | Not-determined |
| 18-102_k141_198513 | Not-determined |
| 18-102_k141_168368 | Not-determined |
| 18-102_k141_188764 | Not-determined |
| 18-102_k141_188766 | Not-determined |
| 18-102_k141_126632 | Not-determined |
| 18-102_k141_54181 | Not-determined |
| 18-102_k141_2287 | Not-determined |
| 18-102_k141_168381 | Low-quality |
| 18-102_k141_168384 | Not-determined |
| 18-102_k141_188775 | Not-determined |
| 18-102_k141_116074 | Not-determined |
| 18-102_k141_188779 | Not-determined |
| 18-102_k141_168387 | Not-determined |
| 18-102_k141_54186 | Not-determined |
| 18-102_k141_230550 | Not-determined |
| 18-102_k141_116083 | Low-quality |
| 18-102_k141_147113 | Not-determined |
| 18-102_k141_198534 | Low-quality |
| 18-102_k141_198539 | Not-determined |
| 18-102_k141_95727 | Not-determined |
| 18-102_k141_95734 | Not-determined |
| 18-102_k141_23128 | Not-determined |
| 18-102_k141_12600 | Not-determined |
| 18-102_k141_219505 | Not-determined |
| 18-102_k141_209242 | Not-determined |
| 18-102_k141_54216 | Low-quality |
| 18-102_k141_137153 | Not-determined |
| 18-102_k141_209246 | Not-determined |
| 18-102_k141_219518 | Not-determined |
| 18-102_k141_240321 | Not-determined |
| 18-102_k141_12606 | Not-determined |
| 18-102_k141_209257 | Not-determined |
| 18-102_k141_95761 | Not-determined |
| 18-102_k141_157652 | Not-determined |
| 18-102_k141_12612 | Low-quality |
| 18-102_k141_219523 | Not-determined |
| 18-102_k141_157662 | Not-determined |
| 18-102_k141_219526 | Not-determined |
| 18-102_k141_95773 | Not-determined |
| 18-102_k141_209277 | Not-determined |
| 18-102_k141_147157 | Low-quality |
| 18-102_k141_2300 | Not-determined |
| 18-102_k141_157679 | Not-determined |
| 18-102_k141_2302 | Not-determined |
| 18-102_k141_240328 | Not-determined |
| 18-102_k141_95789 | Not-determined |
| 18-102_k141_219529 | Not-determined |
| 18-102_k141_209295 | Not-determined |
| 18-102_k141_126675 | Not-determined |
| 18-102_k141_198553 | Not-determined |
| 18-102_k141_219534 | Not-determined |
| 18-102_k141_209301 | Not-determined |
| 18-102_k141_209302 | Low-quality |
| 18-102_k141_137160 | Not-determined |
| 18-102_k141_198554 | Not-determined |
| 18-102_k141_178664 | Not-determined |
| 18-102_k141_44937 | Not-determined |
| 18-102_k141_178666 | Not-determined |
| 18-102_k141_178669 | Not-determined |
| 18-102_k141_178670 | Not-determined |
| 18-102_k141_126682 | Not-determined |
| 18-102_k141_74752 | Not-determined |
| 18-102_k141_209305 | Not-determined |
| 18-102_k141_209306 | Low-quality |
| 18-102_k141_74754 | Not-determined |
| 18-102_k141_230567 | Not-determined |
| 18-102_k141_74755 | Not-determined |
| 18-102_k141_137174 | Not-determined |
| 18-102_k141_95792 | Not-determined |
| 18-102_k141_54244 | Not-determined |
| 18-102_k141_12645 | Not-determined |
| 18-102_k141_95797 | Not-determined |
| 18-102_k141_178683 | Not-determined |
| 18-102_k141_178687 | Not-determined |
| 18-102_k141_240359 | Not-determined |
| 18-102_k141_74764 | Not-determined |
| 18-102_k141_33141 | Not-determined |
| 18-102_k141_219554 | Not-determined |
| 18-102_k141_178697 | Not-determined |
| 18-102_k141_178698 | Not-determined |
| 18-102_k141_54256 | Not-determined |
| 18-102_k141_54257 | Not-determined |
| 18-102_k141_12664 | Low-quality |
| 18-102_k141_23178 | Low-quality |
| 18-102_k141_12667 | Not-determined |
| 18-102_k141_12668 | Not-determined |
| 18-102_k141_54265 | Not-determined |
| 18-102_k141_33153 | Not-determined |
| 18-102_k141_54268 | Not-determined |
| 18-102_k141_147167 | Not-determined |
| 18-102_k141_95810 | Not-determined |
| 18-102_k141_230583 | Not-determined |
| 18-102_k141_95813 | Low-quality |
| 18-102_k141_2334 | Not-determined |
| 18-102_k141_188801 | Not-determined |
| 18-102_k141_54279 | Not-determined |
| 18-102_k141_198567 | Not-determined |
| 18-102_k141_168426 | Low-quality |
| 18-102_k141_23201 | Not-determined |
| 18-102_k141_44971 | Low-quality |
| 18-102_k141_198579 | Not-determined |
| 18-102_k141_74794 | Not-determined |
| 18-102_k141_137195 | Not-determined |
| 18-102_k141_198581 | Not-determined |
| 18-102_k141_12682 | Not-determined |
| 18-102_k141_168434 | Not-determined |
| 18-102_k141_95824 | Not-determined |
| 18-102_k141_44978 | Not-determined |
| 18-102_k141_12686 | Not-determined |
| 18-102_k141_95833 | Not-determined |
| 18-102_k141_12694 | Not-determined |
| 18-102_k141_12696 | Not-determined |
| 18-102_k141_168441 | Low-quality |
| 18-102_k141_44987 | Not-determined |
| 18-102_k141_23212 | Not-determined |
| 18-102_k141_168445 | Not-determined |
| 18-102_k141_240373 | Not-determined |
| 18-102_k141_33177 | Not-determined |
| 18-102_k141_209351 | Not-determined |
| 18-102_k141_106031 | Not-determined |
| 18-102_k141_33186 | Low-quality |
| 18-102_k141_157708 | Not-determined |
| 18-102_k141_209363 | Not-determined |
| 18-102_k141_230612 | Not-determined |
| 18-102_k141_23225 | Not-determined |
| 18-102_k141_23226 | Not-determined |
| 18-102_k141_116102 | Not-determined |
| 18-102_k141_219607 | Not-determined |
| 18-102_k141_33190 | Not-determined |
| 18-102_k141_85425 | Not-determined |
| 18-102_k141_188829 | Not-determined |
| 18-102_k141_116103 | Not-determined |
| 18-102_k141_85427 | Not-determined |
| 18-102_k141_54304 | Not-determined |
| 18-102_k141_157721 | Not-determined |
| 18-102_k141_188832 | Not-determined |
| 18-102_k141_85432 | Not-determined |
| 18-102_k141_188834 | Not-determined |
| 18-102_k141_178704 | Not-determined |
| 18-102_k141_64393 | Not-determined |
| 18-102_k141_106041 | Low-quality |
| 18-102_k141_198602 | Not-determined |
| 18-102_k141_188841 | Not-determined |
| 18-102_k141_137229 | Not-determined |
| 18-102_k141_198604 | Not-determined |
| 18-102_k141_178711 | Low-quality |
| 18-102_k141_33194 | Not-determined |
| 18-102_k141_209380 | Not-determined |
| 18-102_k141_157728 | Low-quality |
| 18-102_k141_147192 | Not-determined |
| 18-102_k141_106043 | Not-determined |
| 18-102_k141_95877 | Not-determined |
| 18-102_k141_106048 | Not-determined |
| 18-102_k141_178720 | Not-determined |
| 18-102_k141_157731 | Not-determined |
| 18-102_k141_33201 | Not-determined |
| 18-102_k141_157732 | Not-determined |
| 18-102_k141_157733 | Not-determined |
| 18-102_k141_230631 | Not-determined |
| 18-102_k141_106053 | Not-determined |
| 18-102_k141_106060 | Not-determined |
| 18-102_k141_230635 | Not-determined |
| 18-102_k141_219622 | Not-determined |
| 18-102_k141_209387 | Not-determined |
| 18-102_k141_168457 | Not-determined |
| 18-102_k141_198616 | Not-determined |
| 18-102_k141_44999 | Not-determined |
| 18-102_k141_178730 | Not-determined |
| 18-102_k141_147208 | Not-determined |
| 18-102_k141_168466 | Not-determined |
| 18-102_k141_188855 | Not-determined |
| 18-102_k141_33214 | Not-determined |
| 18-102_k141_230649 | Not-determined |
| 18-102_k141_147220 | Not-determined |
| 18-102_k141_45003 | Not-determined |
| 18-102_k141_54347 | Not-determined |
| 18-102_k141_230650 | Not-determined |
| 18-102_k141_209410 | Not-determined |
| 18-102_k141_240391 | Not-determined |
| 18-102_k141_188866 | Not-determined |
| 18-102_k141_230652 | Not-determined |
| 18-102_k141_178737 | Not-determined |
| 18-102_k141_209424 | Not-determined |
| 18-102_k141_219651 | Not-determined |
| 18-102_k141_219654 | Not-determined |
| 18-102_k141_178745 | Not-determined |
| 18-102_k141_168477 | Not-determined |
| 18-102_k141_147230 | Not-determined |
| 18-102_k141_157791 | Not-determined |
| 18-102_k141_95883 | Not-determined |
| 18-102_k141_178753 | Low-quality |
| 18-102_k141_188874 | Not-determined |
| 18-102_k141_12707 | Low-quality |
| 18-102_k141_230667 | Not-determined |
| 18-102_k141_54379 | Not-determined |
| 18-102_k141_219663 | Not-determined |
| 18-102_k141_12712 | Not-determined |
| 18-102_k141_116151 | Not-determined |
| 18-102_k141_219666 | Not-determined |
| 18-102_k141_54383 | Not-determined |
| 18-102_k141_168482 | Low-quality |
| 18-102_k141_168486 | Not-determined |
| 18-102_k141_116154 | Low-quality |
| 18-102_k141_240397 | Not-determined |
| 18-102_k141_85453 | Not-determined |
| 18-102_k141_168491 | Not-determined |
| 18-102_k141_230674 | Not-determined |
| 18-102_k141_230675 | Not-determined |
| 18-102_k141_188879 | Low-quality |
| 18-102_k141_54390 | Not-determined |
| 18-102_k141_188882 | Not-determined |
| 18-102_k141_230677 | Not-determined |
| 18-102_k141_168498 | Not-determined |
| 18-102_k141_85461 | Not-determined |
| 18-102_k141_178765 | Low-quality |
| 18-102_k141_116158 | Not-determined |
| 18-102_k141_188889 | Not-determined |
| 18-102_k141_240402 | Not-determined |
| 18-102_k141_23238 | Not-determined |
| 18-102_k141_168507 | Not-determined |
| 18-102_k141_168511 | Not-determined |
| 18-102_k141_178775 | Low-quality |
| 18-102_k141_116169 | Low-quality |
| 18-102_k141_23250 | Not-determined |
| 18-102_k141_157807 | Not-determined |
| 18-102_k141_240414 | Not-determined |
| 18-102_k141_12739 | Not-determined |
| 18-102_k141_147249 | Not-determined |
| 18-102_k141_240417 | Not-determined |
| 18-102_k141_85479 | Low-quality |
| 18-102_k141_178790 | Not-determined |
| 18-102_k141_178791 | Not-determined |

| | |
|---|---|
| 18-102_k141_74815 | Not-determined |
| 18-102_k141_219679 | Not-determined |
| 18-102_k141_168533 | Not-determined |
| 18-102_k141_230707 | Not-determined |
| 18-102_k141_147261 | Not-determined |
| 18-102_k141_230710 | Not-determined |
| 18-102_k141_178803 | Not-determined |
| 18-102_k141_240441 | Not-determined |
| 18-102_k141_85496 | Not-determined |
| 18-102_k141_178807 | Not-determined |
| 18-102_k141_219695 | Not-determined |
| 18-102_k141_85499 | Not-determined |
| 18-102_k141_85500 | Not-determined |
| 18-102_k141_12761 | Not-determined |
| 18-102_k141_12765 | Not-determined |
| 18-102_k141_219709 | Not-determined |
| 18-102_k141_45059 | Not-determined |
| 18-102_k141_219712 | Not-determined |
| 18-102_k141_209462 | Not-determined |
| 18-102_k141_85510 | Not-determined |
| 18-102_k141_85511 | Not-determined |
| 18-102_k141_45071 | Low-quality |
| 18-102_k141_198654 | Not-determined |
| 18-102_k141_240451 | Not-determined |
| 18-102_k141_209471 | Not-determined |
| 18-102_k141_230726 | Not-determined |
| 18-102_k141_230730 | Not-determined |
| 18-102_k141_45079 | Not-determined |
| 18-102_k141_33223 | Low-quality |
| 18-102_k141_33224 | Not-determined |
| 18-102_k141_198658 | Not-determined |
| 18-102_k141_33227 | Not-determined |
| 18-102_k141_230741 | Not-determined |
| 18-102_k141_64423 | Not-determined |
| 18-102_k141_74839 | Not-determined |
| 18-102_k141_64425 | Not-determined |
| 18-102_k141_157859 | Not-determined |
| 18-102_k141_106134 | Not-determined |
| 18-102_k141_230750 | Not-determined |
| 18-102_k141_230751 | Not-determined |
| 18-102_k141_147288 | Not-determined |
| 18-102_k141_45084 | Not-determined |
| 18-102_k141_188904 | Not-determined |
| 18-102_k141_64435 | Not-determined |
| 18-102_k141_106142 | Not-determined |
| 18-102_k141_137240 | Not-determined |
| 18-102_k141_137241 | Not-determined |
| 18-102_k141_106146 | Not-determined |
| 18-102_k141_23307 | Not-determined |
| 18-102_k141_137249 | Not-determined |
| 18-102_k141_157873 | Not-determined |
| 18-102_k141_116212 | Not-determined |
| 18-102_k141_219735 | Not-determined |
| 18-102_k141_85517 | Low-quality |
| 18-102_k141_116214 | Not-determined |
| 18-102_k141_240474 | Not-determined |
| 18-102_k141_116220 | Low-quality |
| 18-102_k141_230770 | Low-quality |
| 18-102_k141_64440 | Not-determined |
| 18-102_k141_106158 | Not-determined |
| 18-102_k141_168562 | Not-determined |
| 18-102_k141_33263 | Not-determined |
| 18-102_k141_64451 | Not-determined |
| 18-102_k141_168576 | Not-determined |
| 18-102_k141_240493 | Low-quality |
| 18-102_k141_64457 | Not-determined |
| 18-102_k141_33269 | Not-determined |
| 18-102_k141_64459 | Not-determined |
| 18-102_k141_198677 | Not-determined |
| 18-102_k141_64460 | Not-determined |
| 18-102_k141_12810 | Not-determined |
| 18-102_k141_168591 | Not-determined |
| 18-102_k141_85529 | Not-determined |
| 18-102_k141_219761 | Not-determined |
| 18-102_k141_106166 | Not-determined |
| 18-102_k141_85533 | Not-determined |
| 18-102_k141_168601 | Not-determined |
| 18-102_k141_23336 | Low-quality |
| 18-102_k141_209476 | Not-determined |
| 18-102_k141_106173 | Not-determined |
| 18-102_k141_209477 | Not-determined |
| 18-102_k141_157899 | Not-determined |
| 18-102_k141_198690 | Not-determined |
| 18-102_k141_54428 | Not-determined |
| 18-102_k141_126710 | Low-quality |
| 18-102_k141_157903 | Not-determined |
| 18-102_k141_209483 | Not-determined |
| 18-102_k141_219768 | Low-quality |
| 18-102_k141_126716 | Not-determined |
| 18-102_k141_33280 | Not-determined |
| 18-102_k141_45099 | Not-determined |
| 18-102_k141_85561 | Not-determined |
| 18-102_k141_2393 | Not-determined |
| 18-102_k141_12846 | Not-determined |
| 18-102_k141_33286 | Not-determined |
| 18-102_k141_178888 | Not-determined |
| 18-102_k141_54443 | Not-determined |
| 18-102_k141_178892 | Not-determined |
| 18-102_k141_33295 | Not-determined |
| 18-102_k141_188967 | Not-determined |
| 18-102_k141_240503 | Not-determined |
| 18-102_k141_54451 | Not-determined |
| 18-102_k141_209503 | Not-determined |
| 18-102_k141_2397 | Not-determined |
| 18-102_k141_126740 | Not-determined |
| 18-102_k141_85570 | Not-determined |
| 18-102_k141_85571 | Not-determined |
| 18-102_k141_45122 | Not-determined |
| 18-102_k141_54457 | Not-determined |
| 18-102_k141_126744 | Not-determined |
| 18-102_k141_54458 | Not-determined |
| 18-102_k141_85577 | Not-determined |
| 18-102_k141_85581 | Not-determined |
| 18-102_k141_54465 | Not-determined |
| 18-102_k141_54469 | Not-determined |
| 18-102_k141_54472 | Not-determined |
| 18-102_k141_147319 | Not-determined |
| 18-102_k141_33304 | Not-determined |
| 18-102_k141_74859 | Not-determined |
| 18-102_k141_137274 | Not-determined |
| 18-102_k141_74860 | Not-determined |
| 18-102_k141_137275 | Not-determined |
| 18-102_k141_240515 | Low-quality |
| 18-102_k141_33312 | Not-determined |
| 18-102_k141_45138 | Not-determined |
| 18-102_k141_74867 | Not-determined |
| 18-102_k141_147334 | Not-determined |
| 18-102_k141_74875 | Not-determined |
| 18-102_k141_85593 | Not-determined |
| 18-102_k141_85600 | Not-determined |
| 18-102_k141_137285 | Not-determined |
| 18-102_k141_45154 | Not-determined |
| 18-102_k141_178920 | Not-determined |
| 18-102_k141_147337 | Not-determined |
| 18-102_k141_188992 | Not-determined |
| 18-102_k141_23364 | Not-determined |
| 18-102_k141_12859 | Not-determined |
| 18-102_k141_33326 | Not-determined |
| 18-102_k141_12863 | Not-determined |
| 18-102_k141_219783 | Not-determined |
| 18-102_k141_189001 | Not-determined |
| 18-102_k141_54492 | Not-determined |
| 18-102_k141_240532 | Not-determined |
| 18-102_k141_198703 | Not-determined |
| 18-102_k141_74904 | Not-determined |
| 18-102_k141_240539 | Not-determined |
| 18-102_k141_189012 | Not-determined |
| 18-102_k141_116225 | Not-determined |
| 18-102_k141_45167 | Low-quality |
| 18-102_k141_95918 | Not-determined |
| 18-102_k141_64506 | Not-determined |
| 18-102_k141_85610 | Not-determined |
| 18-102_k141_85611 | Not-determined |
| 18-102_k141_106189 | Not-determined |
| 18-102_k141_45179 | Not-determined |
| 18-102_k141_33361 | Not-determined |
| 18-102_k141_116241 | Not-determined |
| 18-102_k141_95923 | Not-determined |
| 18-102_k141_2428 | Low-quality |
| 18-102_k141_116246 | Not-determined |
| 18-102_k141_137308 | Not-determined |
| 18-102_k141_85621 | Not-determined |
| 18-102_k141_64518 | Not-determined |
| 18-102_k141_33368 | Not-determined |
| 18-102_k141_33370 | Not-determined |
| 18-102_k141_85629 | Not-determined |
| 18-102_k141_54522 | Low-quality |
| 18-102_k141_85632 | Not-determined |
| 18-102_k141_240561 | Not-determined |
| 18-102_k141_2432 | Not-determined |
| 18-102_k141_95939 | Not-determined |
| 18-102_k141_116263 | Not-determined |
| 18-102_k141_126758 | Not-determined |
| 18-102_k141_116269 | Not-determined |
| 18-102_k141_137322 | Low-quality |
| 18-102_k141_95953 | Low-quality |
| 18-102_k141_33380 | Not-determined |
| 18-102_k141_54539 | Not-determined |
| 18-102_k141_116278 | Low-quality |
| 18-102_k141_116281 | Not-determined |
| 18-102_k141_116282 | Not-determined |
| 18-102_k141_54554 | Not-determined |
| 18-102_k141_54556 | Not-determined |
| 18-102_k141_54558 | Low-quality |
| 18-102_k141_54559 | Not-determined |
| 18-102_k141_33392 | Not-determined |
| 18-102_k141_137339 | Not-determined |
| 18-102_k141_95979 | Not-determined |
| 18-102_k141_23378 | Not-determined |
| 18-102_k141_168611 | Not-determined |
| 18-102_k141_74952 | Not-determined |
| 18-102_k141_147353 | Not-determined |
| 18-102_k141_74961 | Not-determined |
| 18-102_k141_147356 | Not-determined |
| 18-102_k141_74963 | Not-determined |
| 18-102_k141_23394 | Not-determined |
| 18-102_k141_45241 | Not-determined |
| 18-102_k141_198722 | Not-determined |
| 18-102_k141_147365 | Not-determined |
| 18-102_k141_137370 | Low-quality |
| 18-102_k141_74969 | Not-determined |
| 18-102_k141_54572 | Not-determined |
| 18-102_k141_95992 | Not-determined |
| 18-102_k141_74972 | Not-determined |
| 18-102_k141_2463 | Not-determined |
| 18-102_k141_33423 | Not-determined |
| 18-102_k141_64574 | Not-determined |
| 18-102_k141_2464 | Not-determined |
| 18-102_k141_189032 | Not-determined |
| 18-102_k141_147375 | Not-determined |
| 18-102_k141_137382 | Not-determined |
| 18-102_k141_168617 | Not-determined |
| 18-102_k141_96009 | Low-quality |
| 18-102_k141_198732 | Not-determined |
| 18-102_k141_33442 | Not-determined |
| 18-102_k141_147388 | Not-determined |
| 18-102_k141_178974 | Not-determined |
| 18-102_k141_198734 | Not-determined |
| 18-102_k141_209518 | Not-determined |
| 18-102_k141_85673 | Not-determined |
| 18-102_k141_137390 | Low-quality |
| 18-102_k141_2478 | Not-determined |
| 18-102_k141_64580 | Not-determined |
| 18-102_k141_12900 | Not-determined |
| 18-102_k141_33449 | Not-determined |
| 18-102_k141_168646 | Low-quality |
| 18-102_k141_178980 | Not-determined |
| 18-102_k141_178983 | Low-quality |
| 18-102_k141_126819 | Not-determined |
| 18-102_k141_12916 | Not-determined |
| 18-102_k141_178992 | Not-determined |
| 18-102_k141_240593 | Not-determined |
| 18-102_k141_23436 | Not-determined |
| 18-102_k141_23439 | Not-determined |
| 18-102_k141_189060 | Not-determined |
| 18-102_k141_96018 | Not-determined |
| 18-102_k141_230793 | Not-determined |
| 18-102_k141_230794 | Not-determined |
| 18-102_k141_179004 | Low-quality |
| 18-102_k141_189070 | Not-determined |
| 18-102_k141_198769 | Not-determined |
| 18-102_k141_64607 | Not-determined |
| 18-102_k141_179009 | Not-determined |
| 18-102_k141_23449 | Not-determined |
| 18-102_k141_54623 | Not-determined |
| 18-102_k141_64614 | Not-determined |
| 18-102_k141_240599 | Not-determined |
| 18-102_k141_168672 | Not-determined |
| 18-102_k141_147401 | Not-determined |
| 18-102_k141_240603 | Not-determined |
| 18-102_k141_189076 | Not-determined |
| 18-102_k141_219825 | Not-determined |
| 18-102_k141_12924 | Not-determined |
| 18-102_k141_189078 | Low-quality |
| 18-102_k141_33485 | Not-determined |
| 18-102_k141_85709 | Not-determined |
| 18-102_k141_23464 | Low-quality |
| 18-102_k141_219830 | Not-determined |
| 18-102_k141_96025 | Not-determined |
| 18-102_k141_2492 | Not-determined |
| 18-102_k141_23466 | Not-determined |
| 18-102_k141_198786 | Low-quality |
| 18-102_k141_85713 | Not-determined |
| 18-102_k141_179016 | Not-determined |
| 18-102_k141_198800 | Low-quality |
| 18-102_k141_126859 | Not-determined |
| 18-102_k141_230828 | Not-determined |
| 18-102_k141_45304 | Not-determined |
| 18-102_k141_209542 | Low-quality |
| 18-102_k141_45306 | Not-determined |
| 18-102_k141_209548 | Low-quality |
| 18-102_k141_96039 | Not-determined |
| 18-102_k141_209551 | Not-determined |
| 18-102_k141_137436 | Not-determined |
| 18-102_k141_2513 | Not-determined |
| 18-102_k141_240613 | Low-quality |
| 18-102_k141_75010 | Not-determined |
| 18-102_k141_75012 | Not-determined |
| 18-102_k141_230841 | Not-determined |
| 18-102_k141_2515 | Not-determined |
| 18-102_k141_230842 | Not-determined |
| 18-102_k141_198819 | Not-determined |
| 18-102_k141_179028 | Not-determined |
| 18-102_k141_209554 | Not-determined |
| 18-102_k141_189110 | Not-determined |
| 18-102_k141_189112 | Not-determined |
| 18-102_k141_12952 | Not-determined |
| 18-102_k141_2522 | Not-determined |
| 18-102_k141_126871 | Not-determined |
| 18-102_k141_96053 | Low-quality |
| 18-102_k141_209568 | Not-determined |
| 18-102_k141_2528 | Not-determined |
| 18-102_k141_85733 | Not-determined |
| 18-102_k141_85734 | Not-determined |
| 18-102_k141_157942 | Not-determined |
| 18-102_k141_189129 | Not-determined |
| 18-102_k141_85737 | Not-determined |
| 18-102_k141_209571 | Not-determined |
| 18-102_k141_96064 | Not-determined |
| 18-102_k141_147445 | Not-determined |
| 18-102_k141_64650 | Not-determined |
| 18-102_k141_12960 | Not-determined |
| 18-102_k141_126890 | Not-determined |
| 18-102_k141_85743 | Low-quality |
| 18-102_k141_64651 | Not-determined |
| 18-102_k141_147458 | Not-determined |
| 18-102_k141_157951 | Not-determined |
| 18-102_k141_209578 | Not-determined |
| 18-102_k141_126897 | Not-determined |
| 18-102_k141_126898 | Low-quality |
| 18-102_k141_45320 | Not-determined |
| 18-102_k141_33542 | Not-determined |
| 18-102_k141_209584 | Not-determined |
| 18-102_k141_106236 | Not-determined |
| 18-102_k141_230852 | Not-determined |
| 18-102_k141_96086 | Not-determined |
| 18-102_k141_157967 | Not-determined |
| 18-102_k141_85751 | Low-quality |
| 18-102_k141_157969 | Not-determined |
| 18-102_k141_230855 | Low-quality |
| 18-102_k141_33553 | Not-determined |
| 18-102_k141_189158 | Not-determined |
| 18-102_k141_126905 | Not-determined |
| 18-102_k141_168711 | Not-determined |
| 18-102_k141_33561 | Not-determined |
| 18-102_k141_230861 | Not-determined |
| 18-102_k141_2546 | Low-quality |
| 18-102_k141_157975 | Not-determined |
| 18-102_k141_33569 | Not-determined |
| 18-102_k141_33571 | Not-determined |
| 18-102_k141_45340 | Not-determined |
| 18-102_k141_33578 | Not-determined |
| 18-102_k141_189182 | Not-determined |
| 18-102_k141_45347 | Not-determined |
| 18-102_k141_106251 | Low-quality |
| 18-102_k141_45353 | Not-determined |
| 18-102_k141_189194 | Not-determined |
| 18-102_k141_168739 | Not-determined |
| 18-102_k141_64667 | Low-quality |
| 18-102_k141_85786 | Not-determined |
| 18-102_k141_168740 | Not-determined |
| 18-102_k141_230878 | Not-determined |
| 18-102_k141_209614 | Not-determined |
| 18-102_k141_116324 | Not-determined |
| 18-102_k141_2557 | Low-quality |
| 18-102_k141_12980 | Not-determined |
| 18-102_k141_168751 | Not-determined |
| 18-102_k141_230882 | Not-determined |
| 18-102_k141_45363 | Not-determined |
| 18-102_k141_230884 | Not-determined |
| 18-102_k141_209624 | Low-quality |
| 18-102_k141_126930 | Not-determined |
| 18-102_k141_126931 | Low-quality |
| 18-102_k141_157981 | Not-determined |
| 18-102_k141_126934 | Not-determined |
| 18-102_k141_96099 | Not-determined |
| 18-102_k141_64681 | Not-determined |
| 18-102_k141_147500 | Not-determined |
| 18-102_k141_45375 | Not-determined |
| 18-102_k141_13001 | Not-determined |
| 18-102_k141_126940 | Not-determined |
| 18-102_k141_147503 | Not-determined |
| 18-102_k141_96102 | Not-determined |
| 18-102_k141_96103 | Not-determined |
| 18-102_k141_96110 | Not-determined |
| 18-102_k141_64689 | Not-determined |
| 18-102_k141_13011 | Not-determined |
| 18-102_k141_96113 | Not-determined |
| 18-102_k141_137479 | Not-determined |
| 18-102_k141_189206 | Low-quality |
| 18-102_k141_189207 | Not-determined |
| 18-102_k141_147518 | Not-determined |
| 18-102_k141_147519 | Not-determined |
| 18-102_k141_116349 | Not-determined |
| 18-102_k141_147520 | Not-determined |
| 18-102_k141_116350 | Low-quality |
| 18-102_k141_189209 | Low-quality |
| 18-102_k141_126944 | Not-determined |
| 18-102_k141_158001 | Not-determined |
| 18-102_k141_23471 | Not-determined |
| 18-102_k141_13023 | Not-determined |
| 18-102_k141_179043 | Not-determined |
| 18-102_k141_85813 | Low-quality |
| 18-102_k141_33585 | Not-determined |
| 18-102_k141_85816 | Not-determined |
| 18-102_k141_2569 | Low-quality |
| 18-102_k141_13030 | Not-determined |
| 18-102_k141_137493 | Not-determined |
| 18-102_k141_106272 | Not-determined |
| 18-102_k141_168779 | Not-determined |
| 18-102_k141_33589 | Not-determined |
| 18-102_k141_137497 | Not-determined |
| 18-102_k141_96140 | Not-determined |
| 18-102_k141_147535 | Not-determined |
| 18-102_k141_23493 | Not-determined |
| 18-102_k141_147540 | Not-determined |
| 18-102_k141_54670 | Not-determined |
| 18-102_k141_33604 | Not-determined |
| 18-102_k141_96149 | Not-determined |
| 18-102_k141_189238 | Not-determined |
| 18-102_k141_126959 | Not-determined |
| 18-102_k141_189240 | Not-determined |
| 18-102_k141_147551 | Not-determined |
| 18-102_k141_198852 | Not-determined |
| 18-102_k141_33611 | Not-determined |
| 18-102_k141_158013 | Low-quality |
| 18-102_k141_33623 | Not-determined |
| 18-102_k141_158018 | Not-determined |
| 18-102_k141_137509 | Not-determined |
| 18-102_k141_126972 | Not-determined |
| 18-102_k141_189253 | Not-determined |
| 18-102_k141_23516 | Not-determined |
| 18-102_k141_137512 | Low-quality |
| 18-102_k141_179071 | Not-determined |
| 18-102_k141_158030 | Not-determined |
| 18-102_k141_64696 | Not-determined |
| 18-102_k141_240646 | Not-determined |
| 18-102_k141_240647 | Not-determined |
| 18-102_k141_96205 | Not-determined |
| 18-102_k141_96211 | Not-determined |
| 18-102_k141_189273 | Not-determined |
| 18-102_k141_45401 | Not-determined |
| 18-102_k141_189276 | Not-determined |
| 18-102_k141_96214 | Low-quality |
| 18-102_k141_168792 | Not-determined |
| 18-102_k141_189279 | Not-determined |
| 18-102_k141_85835 | Not-determined |
| 18-102_k141_168795 | Low-quality |
| 18-102_k141_45409 | Not-determined |
| 18-102_k141_33661 | Not-determined |
| 18-102_k141_33662 | Low-quality |
| 18-102_k141_96222 | Low-quality |
| 18-102_k141_127011 | Not-determined |
| 18-102_k141_127014 | Not-determined |
| 18-102_k141_106322 | Not-determined |
| 18-102_k141_137528 | Not-determined |
| 18-102_k141_127018 | Not-determined |
| 18-102_k141_127020 | Not-determined |
| 18-102_k141_96234 | Not-determined |
| 18-102_k141_168805 | Not-determined |
| 18-102_k141_209652 | Not-determined |
| 18-102_k141_168814 | Not-determined |
| 18-102_k141_96243 | Not-determined |
| 18-102_k141_33667 | Not-determined |
| 18-102_k141_147570 | Not-determined |
| 18-102_k141_54706 | Not-determined |
| 18-102_k141_13043 | Not-determined |
| 18-102_k141_179114 | Not-determined |
| 18-102_k141_33673 | Not-determined |
| 18-102_k141_23530 | Not-determined |
| 18-102_k141_96258 | Not-determined |
| 18-102_k141_13047 | Not-determined |
| 18-102_k141_54716 | Low-quality |
| 18-102_k141_189308 | Low-quality |
| 18-102_k141_45424 | Not-determined |
| 18-102_k141_64751 | Not-determined |
| 18-102_k141_33688 | Not-determined |
| 18-102_k141_96269 | Not-determined |
| 18-102_k141_45432 | Not-determined |
| 18-102_k141_75101 | Not-determined |
| 18-102_k141_45433 | Not-determined |
| 18-102_k141_45434 | Not-determined |
| 18-102_k141_45435 | Not-determined |
| 18-102_k141_33694 | Not-determined |
| 18-102_k141_127034 | Low-quality |
| 18-102_k141_127035 | Not-determined |
| 18-102_k141_13066 | Not-determined |
| 18-102_k141_240679 | Not-determined |
| 18-102_k141_2573 | Not-determined |
| 18-102_k141_13069 | Not-determined |
| 18-102_k141_13070 | Not-determined |
| 18-102_k141_45442 | Not-determined |
| 18-102_k141_13072 | Not-determined |
| 18-102_k141_147592 | Not-determined |
| 18-102_k141_189318 | Low-quality |
| 18-102_k141_33705 | Not-determined |
| 18-102_k141_127036 | Not-determined |
| 18-102_k141_2580 | Not-determined |
| 18-102_k141_240692 | Not-determined |
| 18-102_k141_127038 | Not-determined |
| 18-102_k141_75108 | Not-determined |
| 18-102_k141_96275 | Not-determined |
| 18-102_k141_209680 | Not-determined |
| 18-102_k141_209682 | Not-determined |
| 18-102_k141_2590 | Not-determined |
| 18-102_k141_168839 | Not-determined |
| 18-102_k141_75120 | Not-determined |
| 18-102_k141_2596 | Not-determined |
| 18-102_k141_209687 | Low-quality |
| 18-102_k141_147611 | Not-determined |
| 18-102_k141_75128 | Not-determined |
| 18-102_k141_54727 | Not-determined |
| 18-102_k141_147612 | Not-determined |
| 18-102_k141_45448 | Low-quality |
| 18-102_k141_147616 | Not-determined |
| 18-102_k141_147617 | Not-determined |
| 18-102_k141_189340 | Not-determined |
| 18-102_k141_127066 | Not-determined |
| 18-102_k141_96302 | Low-quality |
| 18-102_k141_54742 | Not-determined |
| 18-102_k141_116354 | Not-determined |
| 18-102_k141_240713 | Not-determined |
| 18-102_k141_189349 | Not-determined |
| 18-102_k141_137591 | Not-determined |
| 18-102_k141_189350 | Low-quality |
| 18-102_k141_179158 | Not-determined |
| 18-102_k141_85901 | Not-determined |
| 18-102_k141_147632 | Not-determined |
| 18-102_k141_168878 | Not-determined |
| 18-102_k141_85906 | Not-determined |
| 18-102_k141_147644 | Not-determined |
| 18-102_k141_168885 | Not-determined |
| 18-102_k141_45474 | Not-determined |
| 18-102_k141_116377 | Not-determined |
| 18-102_k141_85908 | Not-determined |
| 18-102_k141_116379 | Not-determined |
| KM-final-viral-combined_k141_157468\|\|full | High-quality |
| 18-102_k141_116380 | Not-determined |
| 18-102_k141_158104 | Not-determined |
| 18-102_k141_158105 | Not-determined |
| 18-102_k141_158108 | Not-determined |
| 18-102_k141_23566 | Not-determined |
| 18-102_k141_23573 | Not-determined |
| 18-102_k141_179183 | Not-determined |
| 18-102_k141_179185 | Not-determined |
| 18-102_k141_116395 | Not-determined |
| 18-102_k141_106364 | Not-determined |
| 18-102_k141_116400 | Not-determined |
| 18-102_k141_23580 | Not-determined |
| 18-102_k141_179193 | Not-determined |
| 18-102_k141_209728 | Not-determined |
| 18-102_k141_147653 | Not-determined |
| 18-102_k141_147655 | Low-quality |
| 18-102_k141_106366 | Not-determined |
| 18-102_k141_198875 | Low-quality |
| 18-102_k141_230925 | Not-determined |
| 18-102_k141_45483 | Not-determined |
| 18-102_k141_147663 | Not-determined |
| 18-102_k141_85926 | Not-determined |
| 18-102_k141_33753 | Low-quality |
| 18-102_k141_168912 | Not-determined |
| 18-102_k141_85932 | Not-determined |
| 18-102_k141_45489 | Not-determined |
| 18-102_k141_45490 | Not-determined |
| 18-102_k141_2640 | Not-determined |
| 18-102_k141_85941 | Not-determined |
| 18-102_k141_209743 | Not-determined |
| 18-102_k141_209746 | Low-quality |
| 18-102_k141_85951 | Not-determined |
| 18-102_k141_198897 | Low-quality |
| 18-102_k141_209748 | Not-determined |
| 18-102_k141_85967 | Not-determined |
| 18-102_k141_158138 | Not-determined |
| 18-102_k141_23608 | Not-determined |
| 18-102_k141_13103 | Low-quality |
| 18-102_k141_2656 | Not-determined |
| 18-102_k141_158140 | Low-quality |
| 18-102_k141_179232 | Not-determined |
| 18-102_k141_33793 | Not-determined |
| 18-102_k141_106375 | Not-determined |
| 18-102_k141_179236 | Not-determined |
| 18-102_k141_33796 | Not-determined |
| 18-102_k141_106379 | Not-determined |
| 18-102_k141_106380 | Low-quality |
| 18-102_k141_85974 | Not-determined |
| 18-102_k141_45509 | Not-determined |
| 18-102_k141_168955 | Not-determined |
| 18-102_k141_85978 | Low-quality |
| 18-102_k141_2666 | Not-determined |
| 18-102_k141_168957 | Not-determined |
| 18-102_k141_96315 | Not-determined |
| 18-102_k141_2668 | Not-determined |
| 18-102_k141_2669 | Not-determined |
| 18-102_k141_54796 | Not-determined |
| 18-102_k141_106402 | Not-determined |
| 18-102_k141_168965 | Not-determined |
| 18-102_k141_54805 | Low-quality |
| 18-102_k141_45524 | Not-determined |
| 18-102_k141_45527 | Not-determined |
| 18-102_k141_45532 | Low-quality |
| 18-102_k141_23623 | Low-quality |
| 18-102_k141_23624 | Not-determined |
| 18-102_k141_209762 | Not-determined |
| 18-102_k141_2694 | Not-determined |
| 18-102_k141_2697 | Not-determined |
| 18-102_k141_230974 | Not-determined |
| 18-102_k141_198917 | Not-determined |
| 18-102_k141_127093 | Low-quality |
| 18-102_k141_54826 | Not-determined |

| | |
|---|---|
| 18-102_k141_54828 | Not-determined |
| 18-102_k141_179277 | Not-determined |
| 18-102_k141_45559 | Not-determined |
| 18-102_k141_86015 | Not-determined |
| 18-102_k141_45563 | Not-determined |
| 18-102_k141_127099 | Not-determined |
| 18-102_k141_13144 | Not-determined |
| 18-102_k141_2710 | Not-determined |
| 18-102_k141_179285 | Low-quality |
| 18-102_k141_54836 | Not-determined |
| 18-102_k141_127114 | Not-determined |
| 18-102_k141_127116 | Not-determined |
| 18-102_k141_116424 | Low-quality |
| 18-102_k141_179297 | Not-determined |
| 18-102_k141_209771 | Not-determined |
| 18-102_k141_209776 | Not-determined |
| 18-102_k141_86033 | Not-determined |
| 18-102_k141_86034 | Not-determined |
| 18-102_k141_209794 | Not-determined |
| 18-102_k141_2720 | Not-determined |
| 18-102_k141_86056 | Not-determined |
| 18-102_k141_96375 | Not-determined |
| 18-102_k141_179315 | Not-determined |
| 18-102_k141_54847 | Not-determined |
| 18-102_k141_179324 | Not-determined |
| 18-102_k141_230976 | Low-quality |
| 18-102_k141_179329 | Not-determined |
| 18-102_k141_127156 | Not-determined |
| 18-102_k141_96388 | Not-determined |
| 18-102_k141_230989 | Not-determined |
| 18-102_k141_158152 | Not-determined |
| 18-102_k141_116432 | Low-quality |
| 18-102_k141_116433 | Low-quality |
| 18-102_k141_96408 | Low-quality |
| 18-102_k141_96411 | Not-determined |
| 18-102_k141_13153 | Not-determined |
| 18-102_k141_96413 | Not-determined |
| 18-102_k141_189428 | Not-determined |
| 18-102_k141_116440 | Not-determined |
| 18-102_k141_189430 | Not-determined |
| 18-102_k141_96417 | Not-determined |
| 18-102_k141_179335 | Low-quality |
| 18-102_k141_158164 | Not-determined |
| 18-102_k141_33847 | Not-determined |
| 18-102_k141_23670 | Not-determined |
| 18-102_k141_127176 | Not-determined |
| 18-102_k141_23679 | Not-determined |
| 18-102_k141_158175 | Not-determined |
| 18-102_k141_127182 | Not-determined |
| 18-102_k141_13178 | Not-determined |
| 18-102_k141_116443 | Not-determined |
| 18-102_k141_33873 | Not-determined |
| 18-102_k141_116446 | Not-determined |
| 18-102_k141_198940 | Not-determined |
| 18-102_k141_209821 | Not-determined |
| 18-102_k141_209823 | Low-quality |
| 18-102_k141_13194 | Not-determined |
| 18-102_k141_147738 | Not-determined |
| 18-102_k141_189443 | Low-quality |
| 18-102_k141_64760 | Not-determined |
| 18-102_k141_64767 | Not-determined |
| 18-102_k141_231006 | Low-quality |
| 18-102_k141_96438 | Not-determined |
| 18-102_k141_33895 | Low-quality |
| 18-102_k141_147742 | Not-determined |
| 18-102_k141_33898 | Not-determined |
| 18-102_k141_96449 | Not-determined |
| 18-102_k141_127212 | Not-determined |
| 18-102_k141_96452 | Not-determined |
| 18-102_k141_231016 | Not-determined |
| 18-102_k141_2722 | Not-determined |
| 18-102_k141_13206 | Not-determined |
| 18-102_k141_158199 | Not-determined |
| 18-102_k141_189461 | Not-determined |
| 18-102_k141_33915 | Not-determined |
| 18-102_k141_147751 | Not-determined |
| 18-102_k141_116454 | Not-determined |
| 18-102_k141_64790 | Not-determined |
| 18-102_k141_158203 | Not-determined |
| 18-102_k141_209835 | Low-quality |
| 18-102_k141_33919 | Not-determined |
| 18-102_k141_231021 | Not-determined |
| 18-102_k141_137665 | Not-determined |
| 18-102_k141_231022 | Not-determined |
| 18-102_k141_209842 | Not-determined |
| 18-102_k141_96462 | Not-determined |
| 18-102_k141_127228 | Low-quality |
| 18-102_k141_127234 | Not-determined |
| 18-102_k141_33927 | Low-quality |
| 18-102_k141_147768 | Not-determined |
| 18-102_k141_179391 | Not-determined |
| 18-102_k141_179394 | Not-determined |
| 18-102_k141_179396 | Not-determined |
| 18-102_k141_127248 | Not-determined |
| 18-102_k141_13237 | Not-determined |
| 18-102_k141_189475 | Not-determined |
| 18-102_k141_64819 | Not-determined |
| 18-102_k141_127256 | Not-determined |
| 18-102_k141_147780 | Not-determined |
| 18-102_k141_189479 | Not-determined |
| 18-102_k141_169009 | Not-determined |
| 18-102_k141_198989 | Not-determined |
| 18-102_k141_106434 | Not-determined |
| 18-102_k141_45617 | Not-determined |
| 18-102_k141_137687 | Not-determined |
| 18-102_k141_179401 | Not-determined |
| 18-102_k141_169024 | Low-quality |
| 18-102_k141_2751 | Not-determined |
| 18-102_k141_33967 | Not-determined |
| 18-102_k141_33968 | Not-determined |
| 18-102_k141_179407 | Not-determined |
| 18-102_k141_240766 | Not-determined |
| 18-102_k141_158226 | Low-quality |
| 18-102_k141_33971 | Not-determined |
| 18-102_k141_33972 | Not-determined |
| 18-102_k141_158232 | Not-determined |
| 18-102_k141_209878 | Not-determined |
| 18-102_k141_179412 | Not-determined |
| 18-102_k141_2762 | Not-determined |
| 18-102_k141_209879 | Not-determined |
| 18-102_k141_147788 | Not-determined |
| 18-102_k141_209880 | Not-determined |
| 18-102_k141_189506 | Not-determined |
| 18-102_k141_169037 | Not-determined |
| 18-102_k141_54871 | Not-determined |
| 18-102_k141_147791 | Not-determined |
| 18-102_k141_189509 | Not-determined |
| 18-102_k141_209884 | Not-determined |
| 18-102_k141_45627 | Not-determined |
| 18-102_k141_147798 | Low-quality |
| 18-102_k141_240781 | Not-determined |
| 18-102_k141_179420 | Not-determined |
| 18-102_k141_179422 | Not-determined |
| 18-102_k141_2770 | Not-determined |
| 18-102_k141_189523 | Low-quality |
| 18-102_k141_2771 | Not-determined |
| 18-102_k141_189526 | Not-determined |
| 18-102_k141_240791 | Not-determined |
| 18-102_k141_231058 | Not-determined |
| 18-102_k141_2778 | Not-determined |
| 18-102_k141_2782 | Not-determined |
| 18-102_k141_219875 | Not-determined |
| 18-102_k141_106469 | Not-determined |
| 18-102_k141_158249 | Not-determined |
| 18-102_k141_137732 | Not-determined |
| 18-102_k141_219887 | Not-determined |
| 18-102_k141_54886 | Low-quality |
| 18-102_k141_106474 | Not-determined |
| 18-102_k141_54889 | Low-quality |
| 18-102_k141_45653 | Not-determined |
| 18-102_k141_240809 | Not-determined |
| 18-102_k141_116477 | Low-quality |
| 18-102_k141_96487 | Not-determined |
| 18-102_k141_209921 | Not-determined |
| 18-102_k141_169052 | Not-determined |
| 18-102_k141_96491 | Not-determined |
| 18-102_k141_96492 | Not-determined |
| 18-102_k141_64847 | Low-quality |
| 18-102_k141_169057 | Not-determined |
| 18-102_k141_116487 | Not-determined |
| 18-102_k141_64854 | Not-determined |
| 18-102_k141_179467 | Not-determined |
| 18-102_k141_179468 | Not-determined |
| 18-102_k141_169067 | Not-determined |
| 18-102_k141_64855 | Not-determined |
| 18-102_k141_169072 | Low-quality |
| 18-102_k141_179475 | Not-determined |
| 18-102_k141_147807 | Not-determined |
| 18-102_k141_231080 | Not-determined |
| 18-102_k141_240817 | Not-determined |
| 18-102_k141_147809 | Not-determined |
| 18-102_k141_189576 | Not-determined |
| 18-102_k141_189577 | Not-determined |
| 18-102_k141_96510 | Not-determined |
| 18-102_k141_96512 | Not-determined |
| 18-102_k141_2809 | Not-determined |
| 18-102_k141_64864 | Not-determined |
| 18-102_k141_137781 | Not-determined |
| 18-102_k141_231095 | Not-determined |
| 18-102_k141_169096 | Not-determined |
| 18-102_k141_169102 | Not-determined |
| 18-102_k141_45695 | Low-quality |
| 18-102_k141_45697 | Not-determined |
| 18-102_k141_127279 | Not-determined |
| 18-102_k141_147835 | Not-determined |
| 18-102_k141_137797 | Not-determined |
| 18-102_k141_240831 | Not-determined |
| 18-102_k141_45707 | Not-determined |
| 18-102_k141_96533 | Not-determined |
| 18-102_k141_219928 | Not-determined |
| 18-102_k141_240837 | Not-determined |
| 18-102_k141_219933 | Not-determined |
| 18-102_k141_137807 | Not-determined |
| 18-102_k141_34011 | Not-determined |
| 18-102_k141_45711 | Not-determined |
| 18-102_k141_169117 | Not-determined |
| 18-102_k141_96540 | Not-determined |
| 18-102_k141_106518 | Not-determined |
| 18-102_k141_86077 | Not-determined |
| 18-102_k141_158292 | Not-determined |
| 18-102_k141_96543 | Not-determined |
| 18-102_k141_96544 | Not-determined |
| 18-102_k141_199050 | Not-determined |
| 18-102_k141_86085 | Not-determined |
| 18-102_k141_240850 | Not-determined |
| 18-102_k141_240854 | Low-quality |
| 18-102_k141_127308 | Not-determined |
| 18-102_k141_169131 | Not-determined |
| 18-102_k141_147854 | Not-determined |
| 18-102_k141_231115 | Not-determined |
| 18-102_k141_106536 | Not-determined |
| 18-102_k141_96547 | Not-determined |
| 18-102_k141_86095 | Not-determined |
| 18-102_k141_64900 | Not-determined |
| 18-102_k141_127323 | Not-determined |
| 18-102_k141_189622 | Not-determined |
| 18-102_k141_169142 | Low-quality |
| 18-102_k141_96550 | Not-determined |
| 18-102_k141_116518 | Not-determined |
| 18-102_k141_106556 | Low-quality |
| 18-102_k141_231123 | Not-determined |
| 18-102_k141_137841 | Not-determined |
| 18-102_k141_169147 | Low-quality |
| 18-102_k141_189630 | Not-determined |
| 18-102_k141_2848 | Not-determined |
| 18-102_k141_127332 | Not-determined |
| 18-102_k141_231129 | Not-determined |
| 18-102_k141_158295 | Not-determined |
| 18-102_k141_34039 | Not-determined |
| 18-102_k141_96560 | Not-determined |
| 18-102_k141_96564 | Not-determined |
| 18-102_k141_240892 | Not-determined |
| 18-102_k141_34052 | Not-determined |
| 18-102_k141_34055 | Not-determined |
| 18-102_k141_106576 | Not-determined |
| 18-102_k141_189649 | Not-determined |
| 18-102_k141_106583 | Not-determined |
| 18-102_k141_158309 | Not-determined |
| 18-102_k141_34058 | Not-determined |
| 18-102_k141_189665 | Not-determined |
| 18-102_k141_116544 | Not-determined |
| 18-102_k141_240907 | Not-determined |
| 18-102_k141_106597 | Not-determined |
| 18-102_k141_189674 | Not-determined |
| 18-102_k141_189676 | Not-determined |
| 18-102_k141_2859 | Not-determined |
| 18-102_k141_2860 | Not-determined |
| 18-102_k141_106614 | Low-quality |
| 18-102_k141_106618 | Not-determined |
| 18-102_k141_169175 | Not-determined |
| 18-102_k141_231138 | Not-determined |
| 18-102_k141_137885 | Not-determined |
| 18-102_k141_116578 | Not-determined |
| 18-102_k141_240945 | Not-determined |
| 18-102_k141_240946 | Not-determined |
| 18-102_k141_169194 | Not-determined |
| 18-102_k141_219955 | Not-determined |
| 18-102_k141_158347 | Not-determined |
| 18-102_k141_137889 | Not-determined |
| 18-102_k141_116596 | Not-determined |
| 18-102_k141_219959 | Not-determined |
| 18-102_k141_116601 | Not-determined |
| 18-102_k141_86124 | Not-determined |
| 18-102_k141_158353 | Not-determined |
| 18-102_k141_34084 | Not-determined |
| 18-102_k141_169214 | Not-determined |
| 18-102_k141_64914 | Not-determined |
| 18-102_k141_64918 | Not-determined |
| 18-102_k141_34100 | Not-determined |
| 18-102_k141_199090 | Not-determined |
| 18-102_k141_169220 | Low-quality |
| 18-102_k141_106636 | Not-determined |
| 18-102_k141_137919 | Not-determined |
| 18-102_k141_86131 | Not-determined |
| 18-102_k141_2889 | Not-determined |
| 18-102_k141_169224 | Not-determined |
| 18-102_k141_2893 | Low-quality |
| 18-102_k141_189707 | Low-quality |
| 18-102_k141_189709 | Not-determined |
| 18-102_k141_2900 | Low-quality |
| 18-102_k141_127399 | Not-determined |
| 18-102_k141_219968 | Not-determined |
| 18-102_k141_137940 | Not-determined |
| 18-102_k141_169236 | Not-determined |
| 18-102_k141_127404 | Not-determined |
| 18-102_k141_219982 | Not-determined |
| 18-102_k141_137952 | Low-quality |
| 18-102_k141_231146 | Not-determined |
| 18-102_k141_64953 | Not-determined |
| 18-102_k141_219997 | Not-determined |
| 18-102_k141_23709 | Low-quality |
| 18-102_k141_75175 | Not-determined |
| 18-102_k141_179512 | Not-determined |
| 18-102_k141_231151 | Not-determined |
| 18-102_k141_23716 | Not-determined |
| 18-102_k141_75181 | Not-determined |
| 18-102_k141_189738 | Not-determined |
| 18-102_k141_189739 | Not-determined |
| 18-102_k141_209951 | Not-determined |
| 18-102_k141_209952 | Not-determined |
| 18-102_k141_23728 | Not-determined |
| 18-102_k141_220005 | Not-determined |
| 18-102_k141_189744 | Not-determined |
| 18-102_k141_23730 | Not-determined |
| 18-102_k141_64973 | Not-determined |
| 18-102_k141_127429 | Not-determined |
| 18-102_k141_189758 | Not-determined |
| 18-102_k141_75196 | Not-determined |
| 18-102_k141_106660 | Not-determined |
| 18-102_k141_147894 | Not-determined |
| 18-102_k141_158383 | Not-determined |
| 18-102_k141_86152 | Not-determined |
| 18-102_k141_189767 | Low-quality |
| 18-102_k141_199134 | Not-determined |
| 18-102_k141_34113 | Not-determined |
| 18-102_k141_106669 | Not-determined |
| 18-102_k141_34115 | Not-determined |
| 18-102_k141_209969 | Not-determined |
| 18-102_k141_96577 | Not-determined |
| 18-102_k141_158395 | Not-determined |
| 18-102_k141_96579 | Not-determined |
| 18-102_k141_199142 | Low-quality |
| 18-102_k141_54909 | Not-determined |
| 18-102_k141_169265 | Not-determined |
| 18-102_k141_127447 | Low-quality |
| 18-102_k141_116643 | Not-determined |
| 18-102_k141_106683 | Not-determined |
| 18-102_k141_116654 | Low-quality |
| 18-102_k141_189772 | Not-determined |
| 18-102_k141_54930 | Not-determined |
| 18-102_k141_96594 | Not-determined |
| 18-102_k141_2939 | Not-determined |
| 18-102_k141_169284 | Not-determined |
| 18-102_k141_231177 | Low-quality |
| 18-102_k141_96597 | Not-determined |
| 18-102_k141_96598 | Not-determined |
| 18-102_k141_189778 | Not-determined |
| 18-102_k141_231184 | Not-determined |
| 18-102_k141_189791 | Not-determined |
| 18-102_k141_75210 | Not-determined |
| 18-102_k141_86182 | Not-determined |
| 18-102_k141_65021 | Not-determined |
| 18-102_k141_106704 | Not-determined |
| 18-102_k141_137994 | Not-determined |
| 18-102_k141_189804 | Not-determined |
| 18-102_k141_65029 | Not-determined |
| 18-102_k141_65030 | Not-determined |
| 18-102_k141_45725 | Not-determined |
| 18-102_k141_231198 | Not-determined |
| 18-102_k141_138000 | Not-determined |
| 18-102_k141_2958 | Not-determined |
| 18-102_k141_2959 | Not-determined |
| 18-102_k141_65042 | Not-determined |
| 18-102_k141_169323 | Not-determined |
| 18-102_k141_169325 | Not-determined |
| 18-102_k141_127488 | Not-determined |
| 18-102_k141_169326 | Not-determined |
| 18-102_k141_96613 | Not-determined |
| 18-102_k141_96614 | Not-determined |
| 18-102_k141_65050 | Not-determined |
| 18-102_k141_147903 | Not-determined |
| 18-102_k141_75228 | Not-determined |
| 18-102_k141_2972 | Low-quality |
| 18-102_k141_127497 | Not-determined |
| 18-102_k141_34123 | Not-determined |
| 18-102_k141_34124 | Not-determined |
| 18-102_k141_127501 | Not-determined |
| 18-102_k141_45737 | Not-determined |
| 18-102_k141_231208 | Not-determined |
| 18-102_k141_169338 | Low-quality |
| 18-102_k141_179534 | Not-determined |
| 18-102_k141_127509 | Not-determined |
| 18-102_k141_65060 | Not-determined |
| 18-102_k141_45741 | Not-determined |
| 18-101_k141_236105 | Medium-quality |
| 18-102_k141_138016 | Not-determined |
| 18-102_k141_96640 | Not-determined |
| 18-102_k141_231218 | Not-determined |
| 18-102_k141_96642 | Not-determined |
| 18-102_k141_138018 | Not-determined |
| 18-102_k141_240968 | Not-determined |
| 18-102_k141_210013 | Low-quality |
| 18-102_k141_179537 | Not-determined |
| 18-102_k141_189839 | Not-determined |
| 18-102_k141_189841 | Not-determined |
| 18-102_k141_169342 | Not-determined |
| 18-102_k141_138033 | Not-determined |
| 18-102_k141_189849 | Not-determined |
| 18-102_k141_231232 | Not-determined |
| 18-102_k141_169349 | Not-determined |
| 18-102_k141_147943 | Not-determined |
| 18-102_k141_147950 | Not-determined |
| 18-102_k141_116691 | Not-determined |
| 18-102_k141_54941 | Not-determined |
| 18-102_k141_147957 | Not-determined |
| 18-102_k141_65081 | Not-determined |
| 18-102_k141_210046 | Not-determined |
| 18-102_k141_240982 | Not-determined |
| 18-102_k141_210049 | Not-determined |
| 18-102_k141_210051 | Not-determined |
| 18-102_k141_179568 | Not-determined |
| 18-102_k141_2984 | Not-determined |
| 18-102_k141_2986 | Not-determined |
| 18-102_k141_2987 | Not-determined |
| 18-102_k141_179581 | Not-determined |
| 18-102_k141_147986 | Not-determined |
| 18-102_k141_86225 | Low-quality |
| 18-102_k141_23779 | Not-determined |
| 18-102_k141_65107 | Low-quality |
| 18-102_k141_86227 | Not-determined |
| 18-102_k141_86229 | Not-determined |
| 18-102_k141_45779 | Not-determined |
| 18-102_k141_147998 | Not-determined |
| 18-102_k141_210072 | Not-determined |
| 18-102_k141_116697 | Not-determined |
| 18-102_k141_148004 | Low-quality |
| 18-102_k141_116699 | Not-determined |
| 18-102_k141_75280 | Not-determined |
| 18-102_k141_179595 | Not-determined |
| 18-102_k141_34152 | Not-determined |
| 18-102_k141_231269 | Not-determined |
| 18-102_k141_34155 | Not-determined |
| 18-102_k141_34159 | Not-determined |
| 18-102_k141_169385 | Not-determined |
| 18-102_k141_86250 | Not-determined |
| 18-102_k141_210083 | Not-determined |
| 18-102_k141_127543 | Not-determined |
| 18-102_k141_75295 | Not-determined |
| 18-102_k141_179614 | Not-determined |
| 18-102_k141_148029 | Low-quality |
| 18-102_k141_54980 | Not-determined |
| 18-102_k141_106719 | Not-determined |
| 18-102_k141_13291 | Low-quality |
| 18-102_k141_13292 | Not-determined |
| 18-102_k141_13293 | Not-determined |
| 18-102_k141_116729 | Not-determined |
| 18-102_k141_169397 | Not-determined |
| 18-102_k141_86255 | Not-determined |
| 18-102_k141_210092 | Not-determined |
| 18-102_k141_86257 | Not-determined |
| 18-102_k141_116733 | Low-quality |
| 18-102_k141_106724 | Not-determined |
| 18-102_k141_45786 | Not-determined |
| 18-102_k141_127552 | Not-determined |
| 18-102_k141_86264 | Low-quality |
| 18-102_k141_13297 | Not-determined |
| 18-102_k141_116741 | Not-determined |
| 18-102_k141_65142 | Not-determined |
| 18-102_k141_116745 | Low-quality |
| 18-102_k141_116748 | Not-determined |
| 18-102_k141_116750 | Not-determined |
| 18-102_k141_169411 | Not-determined |
| 18-102_k141_106739 | Not-determined |
| 18-102_k141_127556 | Not-determined |
| 18-102_k141_169417 | Not-determined |
| 18-102_k141_241008 | Low-quality |
| 18-102_k141_199166 | Low-quality |
| 18-102_k141_45810 | Not-determined |
| 18-102_k141_34163 | Not-determined |
| 18-102_k141_210109 | Not-determined |
| 18-102_k141_3027 | Not-determined |
| 18-102_k141_116770 | Not-determined |
| 18-102_k141_231310 | Not-determined |
| 18-102_k141_148066 | Not-determined |
| 18-102_k141_45825 | Not-determined |
| 18-102_k141_106752 | Not-determined |
| 18-102_k141_65158 | Not-determined |
| 18-102_k141_75332 | Not-determined |
| 18-102_k141_210120 | Low-quality |
| 18-102_k141_34177 | Low-quality |
| 18-102_k141_55017 | Not-determined |
| 18-102_k141_210123 | Not-determined |
| 18-102_k141_65163 | Not-determined |
| 18-102_k141_34186 | Not-determined |
| 18-102_k141_34187 | Not-determined |
| 18-102_k141_158409 | Not-determined |
| 18-102_k141_169424 | Not-determined |
| 18-102_k141_65176 | Not-determined |
| 18-102_k141_169433 | Not-determined |
| 18-102_k141_210132 | Not-determined |
| 18-102_k141_65178 | Not-determined |
| 18-102_k141_106787 | Not-determined |
| 18-102_k141_231331 | Not-determined |
| 18-102_k141_241041 | Not-determined |
| 18-102_k141_179650 | Not-determined |
| 18-102_k141_127582 | Low-quality |
| 18-102_k141_106794 | Not-determined |
| 18-102_k141_106799 | Not-determined |
| 18-102_k141_116796 | Not-determined |
| 18-102_k141_106803 | Not-determined |
| 18-102_k141_199193 | Not-determined |
| 18-102_k141_116805 | Not-determined |
| 18-102_k141_199195 | Not-determined |
| 18-102_k141_158421 | Not-determined |
| 18-102_k141_199197 | Not-determined |
| 18-102_k141_34216 | Not-determined |
| 18-102_k141_96676 | Low-quality |
| 18-102_k141_86296 | Not-determined |
| 18-102_k141_45865 | Not-determined |
| 18-102_k141_210158 | Not-determined |
| 18-102_k141_116809 | Not-determined |
| 18-102_k141_127587 | Not-determined |
| 18-102_k141_3054 | Not-determined |
| 18-102_k141_34222 | Not-determined |
| 18-102_k141_96679 | Low-quality |
| 18-102_k141_96682 | Low-quality |
| 18-102_k141_86303 | Not-determined |
| 18-102_k141_106808 | Not-determined |
| 18-102_k141_199210 | Not-determined |
| 18-102_k141_34230 | Not-determined |
| 18-102_k141_158431 | Not-determined |
| 18-102_k141_199224 | Not-determined |
| 18-102_k141_45899 | Not-determined |
| 18-102_k141_199229 | Not-determined |
| 18-102_k141_127603 | Not-determined |
| 18-102_k141_179687 | Not-determined |
| 18-102_k141_13319 | Not-determined |
| 18-102_k141_45907 | Not-determined |
| 18-102_k141_23813 | Not-determined |
| 18-102_k141_13324 | Low-quality |
| 18-102_k141_13333 | Not-determined |
| 18-102_k141_148084 | Not-determined |
| 18-102_k141_210188 | Not-determined |
| 18-102_k141_45910 | Not-determined |
| 18-102_k141_96719 | Not-determined |
| 18-102_k141_96722 | Not-determined |
| 18-102_k141_45914 | Not-determined |
| 18-102_k141_241063 | Not-determined |
| 18-102_k141_116823 | Not-determined |
| 18-102_k141_55030 | Not-determined |
| 18-102_k141_45922 | Not-determined |
| 18-102_k141_86313 | Low-quality |
| 18-102_k141_138087 | Not-determined |
| 18-102_k141_199235 | Not-determined |
| 18-102_k141_45926 | Not-determined |
| 18-102_k141_199236 | Not-determined |
| 18-102_k141_199238 | Not-determined |
| 18-102_k141_138093 | Not-determined |
| 18-102_k141_116826 | Not-determined |
| 18-102_k141_55038 | Not-determined |
| 18-102_k141_55039 | Not-determined |
| 18-102_k141_148090 | Not-determined |
| 18-102_k141_210208 | Not-determined |
| 18-102_k141_241066 | Not-determined |
| 18-102_k141_55046 | Not-determined |
| 18-102_k141_65252 | Not-determined |
| 18-102_k141_169459 | Not-determined |
| 18-102_k141_148099 | Not-determined |
| 18-102_k141_210221 | Not-determined |
| 18-102_k141_23836 | Not-determined |
| 18-102_k141_34258 | Low-quality |
| 18-102_k141_179732 | Not-determined |
| 18-102_k141_34266 | Not-determined |
| 18-102_k141_34272 | Not-determined |
| 18-102_k141_199244 | Low-quality |
| 18-102_k141_179739 | Not-determined |
| 18-102_k141_148109 | Not-determined |
| 18-102_k141_148113 | Low-quality |
| 18-102_k141_3094 | Low-quality |
| 18-102_k141_231386 | Not-determined |
| 18-102_k141_231387 | Not-determined |
| 18-102_k141_231388 | Not-determined |
| 18-102_k141_148121 | Not-determined |
| 18-102_k141_34288 | Not-determined |
| 18-102_k141_210229 | Not-determined |
| 18-102_k141_199254 | Low-quality |
| 18-102_k141_210233 | Not-determined |
| 18-102_k141_65287 | Not-determined |
| 18-102_k141_45966 | Not-determined |
| 18-102_k141_189885 | Not-determined |
| 18-102_k141_55085 | Low-quality |
| 18-102_k141_55090 | Not-determined |
| 18-102_k141_158458 | Not-determined |
| 18-102_k141_3114 | Not-determined |
| 18-102_k141_55093 | Not-determined |
| 18-102_k141_138107 | Not-determined |
| 18-102_k141_55094 | Not-determined |
| 18-102_k141_189889 | Not-determined |
| 18-102_k141_23862 | Low-quality |
| 18-102_k141_199265 | Not-determined |
| 18-102_k141_189891 | Low-quality |
| 18-102_k141_231398 | Not-determined |
| 18-102_k141_210244 | Not-determined |
| 18-102_k141_210245 | Not-determined |
| 18-102_k141_23869 | Not-determined |
| 18-102_k141_106852 | Not-determined |
| 18-102_k141_34300 | Not-determined |
| 18-102_k141_158473 | Not-determined |
| 18-102_k141_3127 | Not-determined |

| | |
|---|---|
| 18-102_k141_241094 | Not-determined |
| 18-102_k141_34303 | Not-determined |
| 18-102_k141_158477 | Not-determined |
| 18-102_k141_65310 | Not-determined |
| 18-102_k141_116842 | Not-determined |
| 18-102_k141_65313 | Not-determined |
| 18-102_k141_23884 | Not-determined |
| 18-102_k141_169486 | Not-determined |
| 18-102_k141_199277 | Not-determined |
| 18-102_k141_34308 | Not-determined |
| 18-102_k141_65321 | Not-determined |
| 18-102_k141_158489 | Not-determined |
| 18-102_k141_158490 | Not-determined |
| 18-102_k141_158491 | Not-determined |
| 18-102_k141_169500 | Not-determined |
| 18-102_k141_231414 | Not-determined |
| 18-102_k141_13386 | Not-determined |
| 18-102_k141_23898 | Not-determined |
| 18-102_k141_3137 | Low-quality |
| 18-102_k141_13392 | Not-determined |
| 18-102_k141_199295 | Not-determined |
| 18-102_k141_231419 | Low-quality |
| 18-102_k141_231420 | Not-determined |
| 18-102_k141_75380 | Not-determined |
| 18-102_k141_199300 | Not-determined |
| 18-102_k141_75384 | Not-determined |
| 18-102_k141_86365 | Low-quality |
| 18-102_k141_23900 | Not-determined |
| 18-102_k141_86366 | Not-determined |
| 18-102_k141_86369 | Not-determined |
| 18-102_k141_45997 | Not-determined |
| 18-102_k141_23907 | Not-determined |
| 18-102_k141_45998 | Not-determined |
| 18-102_k141_189911 | Not-determined |
| 18-102_k141_199306 | Low-quality |
| 18-102_k141_86372 | Not-determined |
| 18-102_k141_46007 | Low-quality |
| 18-102_k141_116853 | Low-quality |
| 18-102_k141_65359 | Not-determined |
| 18-102_k141_158519 | Not-determined |
| 18-102_k141_3174 | Not-determined |
| 18-102_k141_34356 | Not-determined |
| 18-102_k141_46020 | Not-determined |
| 18-102_k141_179837 | Not-determined |
| 18-102_k141_46023 | Not-determined |
| 18-102_k141_231458 | Low-quality |
| 18-102_k141_75402 | Not-determined |
| 18-102_k141_116857 | Low-quality |
| 18-102_k141_23938 | Not-determined |
| 18-102_k141_138176 | Not-determined |
| 18-102_k141_116861 | Low-quality |
| 18-102_k141_116864 | Not-determined |
| 18-102_k141_189934 | Not-determined |
| 18-102_k141_158532 | Not-determined |
| 18-102_k141_127620 | Not-determined |
| 18-102_k141_106856 | Not-determined |
| 18-102_k141_75433 | Not-determined |
| 18-102_k141_127627 | Not-determined |
| 18-102_k141_3192 | Not-determined |
| 18-102_k141_86394 | Not-determined |
| 18-102_k141_75443 | Not-determined |
| 18-102_k141_106874 | Not-determined |
| 18-102_k141_169545 | Not-determined |
| 18-102_k141_86401 | Not-determined |
| 18-102_k141_86404 | Not-determined |
| 18-102_k141_210330 | Not-determined |
| 18-102_k141_231482 | Not-determined |
| 18-102_k141_55127 | Not-determined |
| 18-102_k141_46031 | Not-determined |
| 18-102_k141_241108 | Low-quality |
| 18-102_k141_75457 | Not-determined |
| 18-102_k141_106884 | Low-quality |
| 18-102_k141_106887 | Not-determined |
| 18-102_k141_241117 | Not-determined |
| 18-102_k141_241118 | Not-determined |
| 18-102_k141_23943 | Not-determined |
| 18-102_k141_34385 | Not-determined |
| 18-102_k141_116899 | Not-determined |
| 18-102_k141_106893 | Not-determined |
| 18-102_k141_55147 | Not-determined |
| 18-102_k141_55149 | Not-determined |
| 18-102_k141_116904 | Not-determined |
| 18-102_k141_199312 | Not-determined |
| 18-102_k141_55151 | Not-determined |
| 18-102_k141_3206 | Not-determined |
| 18-102_k141_169569 | Not-determined |
| 18-102_k141_241125 | Not-determined |
| 18-102_k141_106904 | Not-determined |
| 18-102_k141_241131 | Not-determined |
| 18-102_k141_169573 | Not-determined |
| 18-102_k141_96739 | Not-determined |
| 18-102_k141_106908 | Low-quality |
| 18-102_k141_158548 | Not-determined |
| 18-102_k141_231506 | Low-quality |
| 18-102_k141_116931 | Not-determined |
| 18-102_k141_96746 | Low-quality |
| 18-102_k141_96747 | Not-determined |
| 18-102_k141_75471 | Not-determined |
| 18-102_k141_158553 | Not-determined |
| 18-102_k141_116938 | Not-determined |
| 18-102_k141_106917 | Not-determined |
| 18-102_k141_75477 | Not-determined |
| 18-102_k141_148146 | Not-determined |
| 18-102_k141_86443 | Not-determined |
| 18-102_k141_34419 | Not-determined |
| 18-102_k141_3226 | Not-determined |
| 18-102_k141_158555 | Not-determined |
| 18-102_k141_179854 | Not-determined |
| 18-102_k141_169592 | Not-determined |
| 18-102_k141_96753 | Not-determined |
| 18-102_k141_158559 | Not-determined |
| 18-102_k141_199329 | Not-determined |
| 18-102_k141_3230 | Not-determined |
| 18-102_k141_75490 | Low-quality |
| 18-102_k141_106930 | Not-determined |
| 18-102_k141_158569 | Not-determined |
| 18-102_k141_46066 | Not-determined |
| 18-102_k141_169601 | Not-determined |
| 18-102_k141_189961 | Not-determined |
| 18-102_k141_116959 | Low-quality |
| 18-102_k141_241136 | Not-determined |
| 18-102_k141_241137 | Not-determined |
| 18-102_k141_116969 | Not-determined |
| 18-102_k141_199343 | Not-determined |
| 18-102_k141_148156 | Not-determined |
| 18-102_k141_116973 | Not-determined |
| 18-102_k141_3246 | Not-determined |
| 18-102_k141_148158 | Not-determined |
| 18-102_k141_116974 | Not-determined |
| 18-102_k141_13450 | Not-determined |
| 18-102_k141_106939 | Not-determined |
| 18-102_k141_231535 | Not-determined |
| 18-102_k141_199349 | Low-quality |
| 18-102_k141_169621 | Not-determined |
| 18-102_k141_189980 | Not-determined |
| 18-102_k141_210372 | Not-determined |
| 18-102_k141_199352 | Not-determined |
| 18-102_k141_106946 | Not-determined |
| 18-102_k141_231539 | Not-determined |
| 18-102_k141_106950 | Not-determined |
| 18-102_k141_179874 | Not-determined |
| 18-102_k141_241147 | Not-determined |
| 18-102_k141_241148 | Not-determined |
| 18-102_k141_199357 | Low-quality |
| 18-102_k141_75516 | Not-determined |
| 18-102_k141_3258 | Not-determined |
| 18-102_k141_13464 | Low-quality |
| 18-102_k141_148177 | Not-determined |
| 18-102_k141_231556 | Not-determined |
| 18-102_k141_231561 | Not-determined |
| 18-102_k141_220027 | Not-determined |
| 18-102_k141_231564 | Not-determined |
| 18-102_k141_75543 | Not-determined |
| 18-102_k141_220035 | Low-quality |
| 18-102_k141_46108 | Not-determined |
| 18-102_k141_46111 | Not-determined |
| 18-102_k141_127643 | Not-determined |
| 18-102_k141_148197 | Not-determined |
| 18-102_k141_117008 | Not-determined |
| 18-102_k141_210403 | Not-determined |
| 18-102_k141_158588 | Not-determined |
| 18-102_k141_117016 | Not-determined |
| 18-102_k141_169661 | Not-determined |
| 18-102_k141_46115 | Not-determined |
| 18-102_k141_190010 | Not-determined |
| 18-102_k141_210407 | Not-determined |
| 18-102_k141_158592 | Not-determined |
| 18-102_k141_210410 | Not-determined |
| 18-102_k141_117022 | Not-determined |
| 18-102_k141_169672 | Not-determined |
| 18-102_k141_158612 | Not-determined |
| 18-102_k141_158613 | Not-determined |
| 18-102_k141_190030 | Low-quality |
| 18-102_k141_241181 | Low-quality |
| 18-102_k141_148200 | Not-determined |
| 18-102_k141_65456 | Not-determined |
| 18-102_k141_220062 | Not-determined |
| 18-102_k141_241185 | Low-quality |
| 18-102_k141_117038 | Not-determined |
| 18-102_k141_75566 | Not-determined |
| 18-102_k141_106981 | Not-determined |
| 18-102_k141_75569 | Not-determined |
| 18-102_k141_3281 | Not-determined |
| 18-102_k141_158632 | Not-determined |
| 18-102_k141_241190 | Low-quality |
| 18-102_k141_241195 | Not-determined |
| 18-102_k141_241201 | Not-determined |
| 18-102_k141_13521 | Not-determined |
| 18-102_k141_117060 | Not-determined |
| 18-102_k141_231606 | Not-determined |
| 18-102_k141_23975 | Not-determined |
| 18-102_k141_241208 | Not-determined |
| 18-102_k141_86471 | Not-determined |
| 18-102_k141_148231 | Not-determined |
| 18-102_k141_148232 | Low-quality |
| 18-102_k141_179883 | Not-determined |
| 18-102_k141_13534 | Not-determined |
| 18-102_k141_65487 | Not-determined |
| 18-102_k141_210422 | Not-determined |
| 18-102_k141_169692 | Not-determined |
| 18-102_k141_65495 | Not-determined |
| 18-102_k141_220076 | Not-determined |
| 18-102_k141_86502 | Not-determined |
| 18-102_k141_23983 | Not-determined |
| 18-102_k141_169693 | Low-quality |
| 18-102_k141_23988 | Not-determined |
| 18-102_k141_231613 | Not-determined |
| 18-102_k141_210440 | Not-determined |
| 18-102_k141_13553 | Not-determined |
| 18-102_k141_96832 | Not-determined |
| 18-102_k141_75589 | Not-determined |
| 18-102_k141_75590 | Not-determined |
| 18-102_k141_117098 | Not-determined |
| 18-102_k141_127678 | Not-determined |
| 18-102_k141_117103 | Not-determined |
| 18-102_k141_86523 | Not-determined |
| 18-102_k141_220091 | Not-determined |
| 18-102_k141_220094 | Not-determined |
| 18-102_k141_231629 | Low-quality |
| 18-102_k141_24005 | Not-determined |
| 18-102_k141_13570 | Not-determined |
| 18-102_k141_107015 | Not-determined |
| 18-102_k141_231637 | Low-quality |
| 18-102_k141_117120 | Low-quality |
| 18-102_k141_55165 | Not-determined |
| 18-102_k141_220110 | Not-determined |
| 18-102_k141_13583 | Not-determined |
| 18-102_k141_117126 | Not-determined |
| 18-102_k141_96842 | Not-determined |
| 18-102_k141_138183 | Not-determined |
| 18-102_k141_24016 | Not-determined |
| 18-102_k141_210465 | Not-determined |
| 18-102_k141_107027 | Not-determined |
| 18-102_k141_34484 | Not-determined |
| 18-102_k141_24024 | Not-determined |
| 18-102_k141_34488 | Not-determined |
| 18-102_k141_138192 | Not-determined |
| 18-102_k141_179916 | Not-determined |
| 18-102_k141_96854 | Low-quality |
| 18-102_k141_13614 | Not-determined |
| 18-102_k141_75596 | Not-determined |
| 18-102_k141_210475 | Not-determined |
| 18-102_k141_96858 | Not-determined |
| 18-102_k141_148276 | Not-determined |
| 18-102_k141_34494 | Not-determined |
| 18-102_k141_13625 | Not-determined |
| 18-102_k141_138200 | Low-quality |
| 18-102_k141_220121 | Not-determined |
| 18-102_k141_138206 | Not-determined |
| 18-102_k141_96866 | Not-determined |
| 18-102_k141_138208 | Not-determined |
| 18-102_k141_148284 | Not-determined |
| 18-102_k141_13639 | Not-determined |
| 18-102_k141_96868 | Not-determined |
| 18-102_k141_138215 | Not-determined |
| 18-102_k141_46196 | Not-determined |
| 18-102_k141_231667 | Not-determined |
| 18-102_k141_179922 | Low-quality |
| 18-102_k141_65534 | Not-determined |
| 18-102_k141_199385 | Not-determined |
| 18-102_k141_107049 | Not-determined |
| 18-102_k141_107059 | Not-determined |
| 18-102_k141_231668 | Low-quality |
| 18-102_k141_107065 | Not-determined |
| 18-102_k141_46206 | Not-determined |
| 18-102_k141_231673 | Low-quality |
| 18-102_k141_117141 | Not-determined |
| 18-102_k141_179942 | Not-determined |
| 18-102_k141_138224 | Not-determined |
| 18-102_k141_231682 | Low-quality |
| 18-102_k141_169703 | Not-determined |
| 18-102_k141_117146 | Not-determined |
| 18-102_k141_241222 | Not-determined |
| 18-102_k141_179952 | Not-determined |
| 18-102_k141_13663 | Not-determined |
| 18-102_k141_231690 | Not-determined |
| 18-102_k141_13667 | Not-determined |
| 18-102_k141_107087 | Not-determined |
| 18-102_k141_241226 | Not-determined |
| 18-102_k141_34505 | Not-determined |
| 18-102_k141_179964 | Not-determined |
| 18-102_k141_169719 | Not-determined |
| 18-102_k141_107095 | Not-determined |
| 18-102_k141_241235 | Not-determined |
| 18-102_k141_169725 | Not-determined |
| 18-102_k141_96901 | Not-determined |
| 18-102_k141_96903 | Not-determined |
| 18-102_k141_65547 | Not-determined |
| 18-102_k141_190095 | Low-quality |
| 18-102_k141_241245 | Not-determined |
| 18-102_k141_179979 | Not-determined |
| 18-102_k141_220140 | Not-determined |
| 18-102_k141_96917 | Not-determined |
| 18-102_k141_148336 | Not-determined |
| 18-102_k141_179989 | Not-determined |
| 18-102_k141_117174 | Not-determined |
| 18-102_k141_179990 | Not-determined |
| 18-102_k141_86571 | Not-determined |
| 18-102_k141_179996 | Not-determined |
| 18-102_k141_117185 | Not-determined |
| 18-102_k141_169753 | Not-determined |
| 18-102_k141_180003 | Not-determined |
| 18-102_k141_220153 | Not-determined |
| 18-102_k141_86582 | Not-determined |
| 18-102_k141_180009 | Not-determined |
| 18-102_k141_180011 | Not-determined |
| 18-102_k141_148346 | Not-determined |
| 18-102_k141_180012 | Not-determined |
| 18-102_k141_96939 | Not-determined |
| 18-102_k141_96944 | Low-quality |
| 18-102_k141_55208 | Not-determined |
| 18-102_k141_86595 | Not-determined |
| 18-102_k141_13678 | Low-quality |
| 18-102_k141_75640 | Not-determined |
| 18-102_k141_148354 | Not-determined |
| 18-102_k141_3358 | Not-determined |
| 18-102_k141_158657 | Not-determined |
| 18-102_k141_158658 | Not-determined |
| 18-102_k141_148369 | Not-determined |
| 18-102_k141_117207 | Not-determined |
| 18-102_k141_55216 | Not-determined |
| 18-102_k141_65591 | Not-determined |
| 18-102_k141_3365 | Not-determined |
| 18-102_k141_86602 | Not-determined |
| 18-102_k141_148382 | Not-determined |
| 18-102_k141_210486 | Not-determined |
| 18-102_k141_148385 | Not-determined |
| 18-102_k141_148388 | Not-determined |
| 18-102_k141_180034 | Not-determined |
| 18-102_k141_220176 | Low-quality |
| 18-102_k141_86611 | Not-determined |
| 18-102_k141_148392 | Not-determined |
| 18-102_k141_169786 | Not-determined |
| 18-102_k141_86618 | Not-determined |
| 18-102_k141_199439 | Not-determined |
| 18-102_k141_180048 | Not-determined |
| 18-102_k141_180052 | Not-determined |
| 18-102_k141_210499 | Not-determined |
| 18-102_k141_158682 | Not-determined |
| 18-102_k141_107130 | Not-determined |
| 18-102_k141_180063 | Not-determined |
| 18-102_k141_46238 | Not-determined |
| 18-102_k141_46239 | Not-determined |
| 18-102_k141_46240 | Not-determined |
| 18-102_k141_46242 | Not-determined |
| 18-102_k141_65613 | Not-determined |
| 18-102_k141_46245 | Not-determined |
| 18-102_k141_169802 | Not-determined |
| 18-102_k141_46246 | Not-determined |
| 18-102_k141_46247 | Not-determined |
| 18-102_k141_138271 | Low-quality |
| 18-102_k141_190154 | Not-determined |
| 18-102_k141_55242 | Not-determined |
| 18-102_k141_13694 | Not-determined |
| 18-102_k141_180074 | Not-determined |
| 18-102_k141_13696 | Low-quality |
| 18-102_k141_190160 | Not-determined |
| 18-102_k141_55254 | Not-determined |
| 18-102_k141_34525 | Not-determined |
| 18-102_k141_24065 | Not-determined |
| 18-102_k141_158693 | Not-determined |
| 18-102_k141_210517 | Low-quality |
| 18-102_k141_96994 | Not-determined |
| 18-102_k141_138279 | Not-determined |
| 18-102_k141_55258 | Not-determined |
| 18-102_k141_220187 | Not-determined |
| 18-102_k141_46261 | Not-determined |
| 18-102_k141_86632 | Not-determined |
| 18-102_k141_148405 | Not-determined |
| 18-102_k141_231735 | Not-determined |
| 18-102_k141_210525 | Not-determined |
| 18-102_k141_117229 | Low-quality |
| 18-102_k141_75700 | Not-determined |
| 18-102_k141_107143 | Not-determined |
| 18-102_k141_75701 | Not-determined |
| 18-102_k141_55266 | Not-determined |
| 18-102_k141_138288 | Not-determined |
| 18-102_k141_210529 | Not-determined |
| 18-102_k141_55267 | Low-quality |
| 18-102_k141_148412 | Not-determined |
| 18-102_k141_220194 | Not-determined |
| 18-102_k141_13713 | Not-determined |
| 18-102_k141_190174 | Not-determined |
| 18-102_k141_107150 | Not-determined |
| 18-102_k141_24073 | Not-determined |
| 18-102_k141_148430 | Not-determined |
| 18-102_k141_220199 | Low-quality |
| 18-102_k141_231755 | Not-determined |
| 18-102_k141_158707 | Not-determined |
| 18-102_k141_220202 | Not-determined |
| 18-102_k141_46274 | Not-determined |
| 18-102_k141_190191 | Not-determined |
| 18-102_k141_24082 | Not-determined |
| 18-102_k141_220206 | Low-quality |
| 18-102_k141_199451 | Low-quality |
| 18-102_k141_3402 | Not-determined |
| 18-102_k141_97006 | Not-determined |
| 18-102_k141_46280 | Not-determined |
| 18-102_k141_148443 | Not-determined |
| 18-102_k141_34549 | Not-determined |
| 18-102_k141_97015 | Not-determined |
| 18-102_k141_107175 | Not-determined |
| 18-102_k141_46293 | Not-determined |
| 18-102_k141_55294 | Not-determined |
| 18-102_k141_75712 | Low-quality |
| 18-102_k141_46299 | Not-determined |
| 18-102_k141_138311 | Not-determined |
| 18-102_k141_34560 | Not-determined |
| 18-102_k141_138316 | Not-determined |
| 18-102_k141_34570 | Not-determined |
| 18-102_k141_46308 | Not-determined |
| 18-102_k141_241265 | Not-determined |
| 18-102_k141_117269 | Not-determined |
| 18-102_k141_97028 | Not-determined |
| 18-102_k141_34575 | Not-determined |
| 18-102_k141_97032 | Not-determined |
| 18-102_k141_34577 | Not-determined |
| 18-102_k141_3414 | Not-determined |
| 18-102_k141_13744 | Low-quality |
| 18-102_k141_180089 | Not-determined |
| 18-102_k141_46316 | Not-determined |
| 18-102_k141_34586 | Not-determined |
| 18-102_k141_13753 | Not-determined |
| 18-102_k141_180103 | Not-determined |
| 18-102_k141_190216 | Not-determined |
| 18-102_k141_241279 | Not-determined |
| 18-102_k141_241282 | Not-determined |
| 18-102_k141_148485 | Not-determined |
| 18-102_k141_241283 | Not-determined |
| 18-102_k141_241285 | Not-determined |
| 18-102_k141_180108 | Not-determined |
| 18-102_k141_180112 | Not-determined |
| 18-102_k141_75746 | Not-determined |
| 18-102_k141_55313 | Not-determined |
| 18-102_k141_180114 | Not-determined |
| 18-102_k141_199477 | Not-determined |
| 18-102_k141_180116 | Not-determined |
| 18-102_k141_180117 | Not-determined |
| 18-102_k141_46337 | Low-quality |
| 18-102_k141_46340 | Not-determined |
| 18-102_k141_231813 | Not-determined |
| 18-102_k141_231815 | Not-determined |
| 18-102_k141_46350 | Not-determined |
| 18-102_k141_97047 | Low-quality |
| 18-102_k141_199482 | Low-quality |
| 18-102_k141_65639 | Not-determined |
| 18-102_k141_13775 | Not-determined |
| 18-102_k141_46361 | Low-quality |
| 18-102_k141_46364 | Not-determined |
| 18-102_k141_241299 | Not-determined |
| 18-102_k141_138352 | Not-determined |
| 18-102_k141_180130 | Not-determined |
| 18-102_k141_65644 | Not-determined |
| 18-102_k141_148504 | Not-determined |
| 18-102_k141_138356 | Low-quality |
| 18-102_k141_169847 | Not-determined |
| 18-102_k141_158718 | Not-determined |
| 18-102_k141_3451 | Not-determined |
| 18-102_k141_180141 | Not-determined |
| 18-102_k141_3452 | Not-determined |
| 18-102_k141_190232 | Not-determined |
| 18-102_k141_138359 | Not-determined |
| 18-102_k141_34620 | Not-determined |
| 18-102_k141_34621 | Not-determined |
| 18-102_k141_138363 | Not-determined |
| 18-102_k141_46373 | Low-quality |
| 18-102_k141_231842 | Not-determined |
| 18-102_k141_24125 | Not-determined |
| 18-102_k141_46376 | Not-determined |
| 18-102_k141_158735 | Not-determined |
| 18-102_k141_75782 | Not-determined |
| 18-102_k141_107183 | Not-determined |
| 18-102_k141_138365 | Low-quality |
| 18-102_k141_138366 | Not-determined |
| 18-102_k141_13791 | Not-determined |
| 18-102_k141_148522 | Not-determined |
| 18-102_k141_210544 | Not-determined |
| 18-102_k141_97055 | Not-determined |
| 18-102_k141_148527 | Not-determined |
| 18-102_k141_97058 | Not-determined |
| 18-102_k141_3467 | Low-quality |
| 18-102_k141_148529 | Not-determined |
| 18-102_k141_24138 | Not-determined |
| 18-102_k141_117271 | Not-determined |
| 18-102_k141_97063 | Not-determined |
| 18-102_k141_210556 | Not-determined |
| 18-102_k141_117275 | Not-determined |
| 18-102_k141_210559 | Not-determined |
| 18-102_k141_180152 | Not-determined |
| 18-102_k141_231860 | Not-determined |
| 18-102_k141_241324 | Not-determined |
| 18-102_k141_65665 | Low-quality |
| 18-102_k141_65666 | Not-determined |
| 18-102_k141_75808 | Not-determined |
| 18-102_k141_13812 | Not-determined |
| 18-102_k141_180162 | Not-determined |
| 18-102_k141_241327 | Not-determined |
| 18-102_k141_169878 | Not-determined |
| 18-102_k141_138373 | Not-determined |
| 18-102_k141_180168 | Not-determined |
| 18-102_k141_97083 | Not-determined |
| 18-102_k141_34652 | Not-determined |
| 18-102_k141_65671 | Not-determined |
| 18-102_k141_138379 | Not-determined |
| 18-102_k141_117290 | Not-determined |
| 18-102_k141_180174 | Not-determined |
| 18-102_k141_169892 | Not-determined |
| 18-102_k141_24156 | Not-determined |
| 18-102_k141_138384 | Not-determined |
| 18-102_k141_148543 | Not-determined |
| 18-102_k141_210577 | Not-determined |
| 18-102_k141_148545 | Not-determined |
| 18-102_k141_65684 | Not-determined |
| 18-102_k141_169895 | Not-determined |
| 18-102_k141_210587 | Low-quality |
| 18-102_k141_210588 | Not-determined |
| 18-102_k141_86684 | Low-quality |
| 18-102_k141_97094 | Not-determined |
| 18-102_k141_231874 | Not-determined |
| 18-102_k141_97096 | Low-quality |
| 18-102_k141_210598 | Not-determined |
| 18-102_k141_3488 | Not-determined |
| 18-102_k141_117305 | Low-quality |
| 18-102_k141_138396 | Low-quality |
| 18-102_k141_34659 | Low-quality |
| 18-102_k141_231881 | Not-determined |
| 18-102_k141_148556 | Not-determined |
| 18-102_k141_24175 | Not-determined |
| 18-102_k141_55324 | Not-determined |
| 18-102_k141_3497 | Low-quality |
| 18-102_k141_97105 | Not-determined |
| 18-102_k141_158763 | Not-determined |
| 18-102_k141_138411 | Not-determined |
| 18-102_k141_65696 | Not-determined |
| 18-102_k141_117323 | Not-determined |
| 18-102_k141_65699 | Not-determined |
| 18-102_k141_3510 | Not-determined |
| 18-102_k141_138417 | Low-quality |
| 18-102_k141_138424 | Low-quality |
| 18-102_k141_180178 | Not-determined |
| 18-102_k141_169917 | Not-determined |
| 18-102_k141_24183 | Not-determined |
| 18-102_k141_169921 | Not-determined |
| 18-102_k141_75847 | Not-determined |
| 18-102_k141_148575 | Not-determined |
| 18-102_k141_169926 | Not-determined |
| 18-102_k141_107232 | Not-determined |
| 18-102_k141_117343 | Not-determined |
| 18-102_k141_138437 | Not-determined |
| 18-102_k141_107239 | Not-determined |
| 18-102_k141_3527 | Not-determined |
| 18-102_k141_169936 | Not-determined |
| 18-102_k141_117355 | Not-determined |
| 18-102_k141_220227 | Not-determined |
| 18-102_k141_210621 | Not-determined |
| 18-102_k141_220230 | Not-determined |
| 18-102_k141_46394 | Not-determined |
| 18-102_k141_13838 | Not-determined |
| 18-102_k141_46397 | Not-determined |
| 18-102_k141_46398 | Not-determined |
| 18-102_k141_34699 | Not-determined |
| 18-102_k141_46401 | Not-determined |
| 18-102_k141_190313 | Not-determined |
| 18-102_k141_180200 | Not-determined |
| 18-102_k141_107243 | Not-determined |
| 18-102_k141_148590 | Low-quality |
| 18-102_k141_180204 | Low-quality |
| 18-102_k141_169945 | Not-determined |
| 18-102_k141_117376 | Not-determined |
| 18-102_k141_86738 | Not-determined |
| 18-102_k141_148596 | Not-determined |
| 18-102_k141_190326 | Not-determined |
| 18-102_k141_210643 | Not-determined |
| 18-102_k141_241346 | Not-determined |
| 18-102_k141_13858 | Not-determined |
| 18-102_k141_107259 | Not-determined |
| 18-102_k141_210648 | Not-determined |
| 18-102_k141_46409 | Not-determined |
| 18-102_k141_107272 | Not-determined |
| 18-102_k141_107275 | Not-determined |
| 18-102_k141_180224 | Not-determined |
| 18-102_k141_117397 | Not-determined |
| 18-102_k141_24217 | Low-quality |
| 18-102_k141_55405 | Not-determined |
| 18-102_k141_190344 | Not-determined |
| 18-102_k141_210654 | Not-determined |
| 18-102_k141_190347 | Low-quality |
| 18-102_k141_190348 | Not-determined |
| 18-102_k141_180228 | Low-quality |

| | |
|---|---|
| 18-102_k141_169971 | Not-determined |
| 18-102_k141_169972 | Not-determined |
| 18-102_k141_46435 | Not-determined |
| 18-102_k141_46436 | Not-determined |
| 18-102_k141_97114 | Not-determined |
| 18-102_k141_231918 | Not-determined |
| 18-102_k141_190358 | Not-determined |
| 18-102_k141_46444 | Not-determined |
| 18-102_k141_46449 | Not-determined |
| 18-102_k141_55420 | Not-determined |
| 18-102_k141_24226 | Not-determined |
| 18-102_k141_190363 | Not-determined |
| 18-102_k141_190365 | Low-quality |
| 18-102_k141_241356 | Not-determined |
| 18-102_k141_3545 | Not-determined |
| 18-102_k141_97128 | Not-determined |
| 18-102_k141_117423 | Not-determined |
| 18-102_k141_75869 | Not-determined |
| 18-102_k141_86759 | Not-determined |
| 18-102_k141_117427 | Not-determined |
| 18-102_k141_138450 | Not-determined |
| 18-102_k141_55434 | Not-determined |
| 18-102_k141_3553 | Not-determined |
| 18-102_k141_24234 | Not-determined |
| 18-102_k141_158796 | Not-determined |
| 18-102_k141_170005 | Not-determined |
| 18-102_k141_55440 | Not-determined |
| 18-102_k141_65773 | Not-determined |
| 18-102_k141_170007 | Not-determined |
| 18-102_k141_158797 | Not-determined |
| 18-102_k141_34750 | Not-determined |
| 18-102_k141_170009 | Low-quality |
| 18-102_k141_55443 | Low-quality |
| 18-102_k141_158801 | Not-determined |
| 18-102_k141_190388 | Not-determined |
| 18-102_k141_107290 | Not-determined |
| 18-102_k141_158803 | Low-quality |
| 18-102_k141_46484 | Low-quality |
| 18-102_k141_65788 | Not-determined |
| 18-102_k141_180270 | Not-determined |
| 18-102_k141_148681 | Not-determined |
| 18-102_k141_180281 | Low-quality |
| 18-102_k141_75897 | Low-quality |
| 18-102_k141_34787 | Not-determined |
| 18-102_k141_34790 | Not-determined |
| 18-102_k141_138468 | Not-determined |
| 18-102_k141_241398 | Not-determined |
| 18-102_k141_241399 | Not-determined |
| 18-102_k141_34796 | Not-determined |
| 18-102_k141_65800 | Low-quality |
| 18-102_k141_34804 | Not-determined |
| 18-102_k141_231962 | Low-quality |
| 18-102_k141_241410 | Not-determined |
| 18-102_k141_241411 | Not-determined |
| 18-102_k141_180293 | Not-determined |
| 18-102_k141_107313 | Not-determined |
| 18-102_k141_148688 | Not-determined |
| 18-102_k141_55476 | Not-determined |
| 18-102_k141_210704 | Not-determined |
| 18-102_k141_34820 | Not-determined |
| 18-102_k141_34823 | Not-determined |
| 18-102_k141_158806 | Not-determined |
| 18-102_k141_34824 | Not-determined |
| 18-102_k141_148692 | Not-determined |
| 18-102_k141_24253 | Low-quality |
| 18-102_k141_241421 | Not-determined |
| 18-102_k141_241422 | Not-determined |
| 18-102_k141_3576 | Not-determined |
| 18-102_k141_158816 | Not-determined |
| 18-102_k141_75906 | Not-determined |
| 18-102_k141_3582 | Not-determined |
| 18-102_k141_3587 | Low-quality |
| 18-102_k141_13907 | Not-determined |
| 18-102_k141_13909 | Not-determined |
| 18-102_k141_138485 | Not-determined |
| 18-102_k141_138486 | Not-determined |
| 18-102_k141_75915 | Not-determined |
| 18-102_k141_13912 | Low-quality |
| 18-102_k141_180310 | Not-determined |
| 18-102_k141_3598 | Not-determined |
| 18-102_k141_180316 | Low-quality |
| 18-102_k141_65819 | Not-determined |
| 18-102_k141_231987 | Not-determined |
| 18-102_k141_220299 | Not-determined |
| 18-102_k141_148713 | Low-quality |
| 18-102_k141_97180 | Not-determined |
| 18-102_k141_231993 | Not-determined |
| 18-102_k141_97184 | Not-determined |
| 18-102_k141_220306 | Not-determined |
| 18-102_k141_117472 | Not-determined |
| 18-102_k141_117473 | Not-determined |
| 18-102_k141_13924 | Not-determined |
| 18-102_k141_220307 | Not-determined |
| 18-102_k141_180325 | Not-determined |
| 18-102_k141_210710 | Not-determined |
| 18-102_k141_180332 | Not-determined |
| 18-102_k141_220315 | Not-determined |
| 18-102_k141_13931 | Not-determined |
| 18-102_k141_3618 | Not-determined |
| 18-102_k141_190412 | Not-determined |
| 18-102_k141_3620 | Not-determined |
| 18-102_k141_180338 | Not-determined |
| 18-102_k141_232006 | Not-determined |
| 18-102_k141_86801 | Not-determined |
| 18-102_k141_220322 | Not-determined |
| 18-102_k141_117490 | Not-determined |
| 18-102_k141_3631 | Not-determined |
| 18-102_k141_190421 | Not-determined |
| 18-102_k141_232018 | Not-determined |
| 18-102_k141_13947 | Low-quality |
| 18-102_k141_232022 | Not-determined |
| 18-102_k141_55525 | Not-determined |
| 18-102_k141_170033 | Not-determined |
| 18-102_k141_97207 | Not-determined |
| 18-102_k141_210729 | Not-determined |
| 18-102_k141_232030 | Not-determined |
| 18-102_k141_190426 | Not-determined |
| 18-102_k141_190427 | Not-determined |
| 18-102_k141_55535 | Not-determined |
| 18-102_k141_55536 | Not-determined |
| 18-102_k141_24284 | Not-determined |
| 18-102_k141_24285 | Low-quality |
| 18-102_k141_190430 | Not-determined |
| 18-101-final-viral-combined_k141_236105\|\|full | Medium-quality |
| 18-102_k141_180359 | Not-determined |
| 18-102_k141_190437 | Not-determined |
| 18-102_k141_210736 | Not-determined |
| 18-102_k141_24298 | Not-determined |
| 18-102_k141_210744 | Not-determined |
| 18-102_k141_97228 | Not-determined |
| 18-102_k141_3658 | Low-quality |
| 18-102_k141_190448 | Not-determined |
| 18-102_k141_210747 | Not-determined |
| 18-102_k141_3663 | Low-quality |
| 18-102_k141_148747 | Not-determined |
| 18-102_k141_210759 | Not-determined |
| 18-102_k141_180374 | Not-determined |
| 18-102_k141_138548 | Not-determined |
| 18-102_k141_190455 | Not-determined |
| 18-102_k141_210764 | Not-determined |
| 18-102_k141_138551 | Not-determined |
| 18-102_k141_55555 | Not-determined |
| 18-102_k141_107361 | Not-determined |
| 18-102_k141_13967 | Not-determined |
| 18-102_k141_190463 | Low-quality |
| 18-102_k141_3673 | Not-determined |
| 18-102_k141_220327 | Not-determined |
| 18-102_k141_210774 | Not-determined |
| 18-102_k141_75926 | Low-quality |
| 18-102_k141_75929 | Low-quality |
| 18-102_k141_220339 | Not-determined |
| 18-102_k141_34834 | Not-determined |
| 18-102_k141_190473 | Not-determined |
| 18-102_k141_24331 | Not-determined |
| 18-102_k141_55584 | Not-determined |
| 18-102_k141_34844 | Not-determined |
| 18-102_k141_55587 | Not-determined |
| 18-102_k141_210792 | Not-determined |
| 18-102_k141_34856 | Not-determined |
| 18-102_k141_46544 | Not-determined |
| 18-102_k141_107388 | Not-determined |
| 18-102_k141_55597 | Not-determined |
| 18-102_k141_180385 | Not-determined |
| 18-102_k141_75938 | Not-determined |
| 18-102_k141_107399 | Not-determined |
| 18-102_k141_138578 | Not-determined |
| 18-102_k141_55615 | Not-determined |
| 18-102_k141_24336 | Not-determined |
| 18-102_k141_75947 | Not-determined |
| 18-102_k141_3679 | Not-determined |
| 18-102_k141_46566 | Not-determined |
| 18-102_k141_107415 | Not-determined |
| 18-102_k141_3686 | Not-determined |
| 18-102_k141_46571 | Not-determined |
| 18-102_k141_138591 | Not-determined |
| 18-102_k141_158836 | Not-determined |
| 18-102_k141_117522 | Not-determined |
| 18-102_k141_158838 | Not-determined |
| 18-102_k141_24359 | Not-determined |
| 18-102_k141_14015 | Low-quality |
| 18-102_k141_14017 | Not-determined |
| 18-102_k141_14018 | Not-determined |
| 18-102_k141_199501 | Not-determined |
| 18-102_k141_148777 | Low-quality |
| 18-102_k141_117529 | Not-determined |
| 18-102_k141_117530 | Not-determined |
| 18-102_k141_148779 | Low-quality |
| 18-102_k141_107435 | Not-determined |
| 18-102_k141_148784 | Not-determined |
| 18-102_k141_75972 | Not-determined |
| 18-102_k141_148787 | Not-determined |
| 18-102_k141_24371 | Not-determined |
| 18-102_k141_24376 | Not-determined |
| 18-102_k141_127705 | Low-quality |
| 18-102_k141_34884 | Not-determined |
| 18-102_k141_107444 | Low-quality |
| 18-102_k141_55652 | Not-determined |
| 18-102_k141_199515 | Not-determined |
| 18-102_k141_75991 | Low-quality |
| 18-102_k141_14034 | Not-determined |
| 18-102_k141_3711 | Not-determined |
| 18-102_k141_127727 | Not-determined |
| 18-102_k141_199522 | Not-determined |
| 18-102_k141_3714 | Not-determined |
| 18-102_k141_148800 | Not-determined |
| 18-102_k141_75999 | Low-quality |
| 18-102_k141_107451 | Low-quality |
| 18-102_k141_24408 | Not-determined |
| 18-102_k141_86899 | Not-determined |
| 18-102_k141_76003 | Not-determined |
| 18-102_k141_199531 | Low-quality |
| 18-102_k141_199533 | Not-determined |
| 18-102_k141_55678 | Not-determined |
| 18-102_k141_55681 | Not-determined |
| 18-102_k141_117576 | Not-determined |
| 18-102_k141_14045 | Not-determined |
| 18-102_k141_190491 | Not-determined |
| 18-102_k141_14046 | Not-determined |
| 18-102_k141_220388 | Not-determined |
| 18-102_k141_180412 | Not-determined |
| 18-102_k141_220391 | Not-determined |
| 18-102_k141_158863 | Not-determined |
| 18-102_k141_158865 | Low-quality |
| 18-102_k141_46604 | Not-determined |
| 18-102_k141_14057 | Not-determined |
| 18-102_k141_220402 | Not-determined |
| 18-102_k141_180424 | Not-determined |
| 18-102_k141_46605 | Not-determined |
| 18-102_k141_24436 | Not-determined |
| 18-102_k141_24439 | Not-determined |
| 18-102_k141_46607 | Not-determined |
| 18-102_k141_34903 | Not-determined |
| 18-102_k141_34908 | Low-quality |
| 18-102_k141_34909 | Low-quality |
| 18-102_k141_46609 | Not-determined |
| 18-102_k141_138637 | Not-determined |
| 18-102_k141_190515 | Not-determined |
| 18-102_k141_158887 | Low-quality |
| 18-102_k141_86929 | Not-determined |
| 18-102_k141_127735 | Not-determined |
| 18-102_k141_86935 | Not-determined |
| 18-102_k141_86936 | Not-determined |
| 18-102_k141_190527 | Not-determined |
| 18-102_k141_127736 | Not-determined |
| 18-102_k141_127737 | Not-determined |
| 18-102_k141_148811 | Not-determined |
| 18-102_k141_86940 | Not-determined |
| 18-102_k141_76027 | Low-quality |
| 18-102_k141_148815 | Not-determined |
| 18-102_k141_220411 | Not-determined |
| 18-102_k141_76030 | Not-determined |
| 18-102_k141_46628 | Not-determined |
| 18-102_k141_180455 | Low-quality |
| 18-102_k141_55719 | Not-determined |
| 18-102_k141_220421 | Not-determined |
| 18-102_k141_180461 | Low-quality |
| 18-102_k141_148822 | Not-determined |
| 18-102_k141_180468 | Not-determined |
| 18-102_k141_55724 | Not-determined |
| 18-102_k141_148825 | Low-quality |
| 18-102_k141_127750 | Not-determined |
| 18-102_k141_107506 | Not-determined |
| 18-102_k141_127757 | Not-determined |
| 18-102_k141_190562 | Low-quality |
| 18-102_k141_55732 | Not-determined |
| 18-102_k141_138678 | Not-determined |
| 18-102_k141_138681 | Not-determined |
| 18-102_k141_220437 | Not-determined |
| 18-102_k141_180476 | Not-determined |
| 18-102_k141_117595 | Not-determined |
| 18-102_k141_55743 | Not-determined |
| 18-102_k141_220444 | Low-quality |
| 18-102_k141_180478 | Not-determined |
| 18-102_k141_117602 | Not-determined |
| 18-102_k141_86954 | Low-quality |
| 18-102_k141_127775 | Low-quality |
| 18-102_k141_148838 | Not-determined |
| 18-102_k141_138692 | Not-determined |
| 18-102_k141_55747 | Not-determined |
| 18-102_k141_220446 | Not-determined |
| 18-102_k141_220448 | Not-determined |
| 18-102_k141_107520 | Not-determined |
| 18-102_k141_138695 | Low-quality |
| 18-102_k141_76063 | Not-determined |
| 18-102_k141_65848 | Not-determined |
| 18-102_k141_86969 | Not-determined |
| 18-102_k141_117618 | Not-determined |
| 18-102_k141_117619 | Low-quality |
| 18-102_k141_220452 | Not-determined |
| 18-102_k141_190574 | Low-quality |
| 18-102_k141_220455 | Not-determined |
| 18-102_k141_158921 | Not-determined |
| 18-102_k141_127787 | Not-determined |
| 18-102_k141_220460 | Not-determined |
| 18-102_k141_210824 | Not-determined |
| 18-102_k141_232059 | Not-determined |
| 18-102_k141_232062 | Not-determined |
| 18-102_k141_210832 | Not-determined |
| 18-102_k141_117639 | Not-determined |
| 18-102_k141_55760 | Low-quality |
| 18-102_k141_220464 | Not-determined |
| 18-102_k141_107542 | Not-determined |
| 18-102_k141_158939 | Not-determined |
| 18-102_k141_76088 | Low-quality |
| 18-102_k141_76091 | Not-determined |
| 18-102_k141_220472 | Low-quality |
| 18-102_k141_46669 | Not-determined |
| 18-102_k141_220473 | Not-determined |
| 18-102_k141_220480 | Low-quality |
| 18-102_k141_87008 | Not-determined |
| 18-102_k141_220485 | Not-determined |
| 18-102_k141_55773 | Not-determined |
| 18-102_k141_199547 | Not-determined |
| 18-102_k141_127821 | Not-determined |
| 18-102_k141_199549 | Not-determined |
| 18-102_k141_24495 | Not-determined |
| 18-102_k141_138735 | Not-determined |
| 18-102_k141_232068 | Not-determined |
| 18-102_k141_65866 | Not-determined |
| 18-102_k141_127824 | Not-determined |
| 18-102_k141_190591 | Low-quality |
| 18-102_k141_55780 | Not-determined |
| 18-102_k141_76107 | Low-quality |
| 18-102_k141_232072 | Not-determined |
| 18-102_k141_190595 | Not-determined |
| 18-102_k141_55784 | Not-determined |
| 18-102_k141_65874 | Not-determined |
| 18-102_k141_76113 | Not-determined |
| 18-102_k141_158956 | Not-determined |
| 18-102_k141_65876 | Not-determined |
| 18-102_k141_87032 | Not-determined |
| 18-102_k141_190604 | Not-determined |
| 18-102_k141_87034 | Low-quality |
| 18-102_k141_190605 | Not-determined |
| 18-102_k141_87036 | Not-determined |
| 18-102_k141_232079 | Not-determined |
| 18-102_k141_158961 | Not-determined |
| 18-102_k141_55795 | Not-determined |
| 18-102_k141_107569 | Not-determined |
| 18-102_k141_97265 | Not-determined |
| 18-102_k141_107571 | Not-determined |
| 18-102_k141_107577 | Not-determined |
| 18-102_k141_24524 | Not-determined |
| 18-102_k141_87054 | Not-determined |
| 18-102_k141_232088 | Not-determined |
| 18-102_k141_232089 | Not-determined |
| 18-102_k141_14083 | Not-determined |
| 18-102_k141_46710 | Not-determined |
| 18-102_k141_232091 | Low-quality |
| 18-102_k141_107584 | Not-determined |
| 18-102_k141_232101 | Not-determined |
| 18-102_k141_158977 | Low-quality |
| 18-102_k141_232106 | Low-quality |
| 18-102_k141_76122 | Not-determined |
| 18-102_k141_65884 | Not-determined |
| 18-102_k141_24543 | Not-determined |
| 18-102_k141_87081 | Not-determined |
| 18-102_k141_97274 | Not-determined |
| 18-102_k141_158991 | Not-determined |
| 18-102_k141_55800 | Not-determined |
| 18-102_k141_117668 | Not-determined |
| 18-102_k141_180505 | Not-determined |
| 18-102_k141_14107 | Not-determined |
| 18-102_k141_232109 | Not-determined |
| 18-102_k141_180511 | Not-determined |
| 18-102_k141_107595 | Low-quality |
| 18-102_k141_180513 | Low-quality |
| 18-102_k141_107596 | Not-determined |
| 18-102_k141_46734 | Not-determined |
| 18-102_k141_46741 | Not-determined |
| 18-102_k141_180527 | Not-determined |
| 18-102_k141_55811 | Not-determined |
| 18-102_k141_190631 | Not-determined |
| 18-102_k141_76155 | Not-determined |
| 18-102_k141_117683 | Not-determined |
| 18-102_k141_138761 | Low-quality |
| 18-102_k141_76159 | Low-quality |
| 18-102_k141_117696 | Not-determined |
| 18-102_k141_107611 | Not-determined |
| 18-102_k141_107613 | Not-determined |
| 18-102_k141_76169 | Not-determined |
| 18-102_k141_97305 | Not-determined |
| 18-102_k141_87123 | Low-quality |
| 18-102_k141_190639 | Not-determined |
| 18-102_k141_87130 | Not-determined |
| 18-102_k141_159025 | Not-determined |
| 18-102_k141_159028 | Not-determined |
| 18-102_k141_190643 | Not-determined |
| 18-102_k141_14189 | Not-determined |
| 18-102_k141_199564 | Not-determined |
| 18-102_k141_14199 | Low-quality |
| 18-102_k141_127844 | Not-determined |
| 18-102_k141_14200 | Not-determined |
| 18-102_k141_138774 | Not-determined |
| 18-102_k141_127846 | Not-determined |
| 18-102_k141_35052 | Not-determined |
| 18-102_k141_138782 | Not-determined |
| 18-102_k141_138783 | Not-determined |
| 18-102_k141_127853 | Not-determined |
| 18-102_k141_138784 | Not-determined |
| 18-102_k141_55834 | Not-determined |
| 18-102_k141_199572 | Low-quality |
| 18-102_k141_35058 | Not-determined |
| 18-102_k141_55836 | Not-determined |
| 18-102_k141_55840 | Not-determined |
| 18-102_k141_138796 | Not-determined |
| 18-102_k141_148902 | Not-determined |
| 18-102_k141_138797 | Not-determined |
| 18-102_k141_46768 | Not-determined |
| 18-102_k141_241444 | Not-determined |
| 18-102_k141_232140 | Not-determined |
| 18-102_k141_35075 | Not-determined |
| 18-102_k141_97335 | Not-determined |
| 18-102_k141_76233 | Low-quality |
| 18-102_k141_46783 | Not-determined |
| 18-102_k141_138819 | Not-determined |
| 18-102_k141_35076 | Low-quality |
| 18-102_k141_159053 | Not-determined |
| 18-102_k141_46790 | Low-quality |
| 18-102_k141_65920 | Not-determined |
| 18-102_k141_117716 | Not-determined |
| 18-102_k141_46795 | Not-determined |
| 18-102_k141_46798 | Not-determined |
| 18-102_k141_76250 | Not-determined |
| 18-102_k141_76254 | Not-determined |
| 18-102_k141_35098 | Not-determined |
| 18-102_k141_199589 | Not-determined |
| 18-102_k141_199590 | Low-quality |
| 18-102_k141_35110 | Not-determined |
| 18-102_k141_14240 | Not-determined |
| 18-102_k141_241450 | Not-determined |
| 18-102_k141_117746 | Not-determined |
| 18-102_k141_117747 | Not-determined |
| 18-102_k141_138839 | Not-determined |
| 18-102_k141_241452 | Not-determined |
| 18-102_k141_117751 | Not-determined |
| 18-102_k141_241454 | Not-determined |
| 18-102_k141_117752 | Not-determined |
| 18-102_k141_76270 | Not-determined |
| 18-102_k141_138841 | Not-determined |
| 18-102_k141_65935 | Not-determined |
| 18-102_k141_232159 | Not-determined |
| 18-102_k141_117759 | Low-quality |
| 18-102_k141_138846 | Not-determined |
| 18-102_k141_241458 | Not-determined |
| 18-102_k141_232167 | Not-determined |
| 18-102_k141_127862 | Not-determined |
| 18-102_k141_35121 | Not-determined |
| 18-102_k141_76282 | Not-determined |
| 18-102_k141_159090 | Not-determined |
| 18-102_k141_138854 | Not-determined |
| 18-102_k141_148944 | Not-determined |
| 18-102_k141_117767 | Not-determined |
| 18-102_k141_127884 | Not-determined |
| 18-102_k141_97373 | Not-determined |
| 18-102_k141_117770 | Not-determined |
| 18-102_k141_127886 | Not-determined |
| 18-102_k141_180593 | Not-determined |
| 18-102_k141_3752 | Not-determined |
| 18-102_k141_241476 | Not-determined |
| 18-102_k141_180599 | Not-determined |
| 18-102_k141_55848 | Not-determined |
| 18-102_k141_220542 | Not-determined |
| 18-102_k141_159101 | Not-determined |
| 18-102_k141_35138 | Not-determined |
| 18-102_k141_35142 | Not-determined |
| 18-102_k141_199596 | Not-determined |
| 18-102_k141_97384 | Not-determined |
| 18-102_k141_241484 | Not-determined |
| 18-102_k141_127901 | Not-determined |
| 18-102_k141_3762 | Not-determined |
| 18-102_k141_97389 | Not-determined |
| 18-102_k141_55863 | Not-determined |
| 18-102_k141_117785 | Not-determined |
| 18-102_k141_35167 | Not-determined |
| 18-102_k141_117787 | Not-determined |
| 18-102_k141_107648 | Not-determined |
| 18-102_k141_14255 | Not-determined |
| 18-102_k141_117788 | Not-determined |
| 18-102_k141_14256 | Low-quality |
| 18-102_k141_107651 | Not-determined |
| 18-102_k141_87145 | Not-determined |
| 18-102_k141_117795 | Not-determined |
| 18-102_k141_55883 | Not-determined |
| 18-102_k141_55884 | Not-determined |
| 18-102_k141_3780 | Not-determined |
| 18-102_k141_97395 | Not-determined |
| 18-102_k141_3783 | Not-determined |
| 18-102_k141_3785 | Not-determined |
| 18-102_k141_138890 | Not-determined |
| 18-102_k141_55892 | Low-quality |
| 18-102_k141_97402 | Not-determined |
| 18-102_k141_55897 | Not-determined |
| 18-102_k141_46844 | Not-determined |
| 18-102_k141_14271 | Not-determined |
| 18-102_k141_232199 | Not-determined |
| 18-102_k141_76345 | Not-determined |
| 18-102_k141_148952 | Not-determined |
| 18-102_k141_159106 | Not-determined |
| 18-102_k141_76351 | Not-determined |
| 18-102_k141_232206 | Low-quality |
| 18-102_k141_65986 | Not-determined |
| 18-102_k141_117803 | Not-determined |
| 18-102_k141_107673 | Not-determined |
| 18-102_k141_107678 | Not-determined |
| 18-102_k141_55909 | Low-quality |
| 18-102_k141_14283 | Not-determined |
| 18-102_k141_241515 | Not-determined |
| 18-102_k141_24589 | Not-determined |
| 18-102_k141_148961 | Not-determined |
| 18-102_k141_107688 | Not-determined |
| 18-102_k141_107693 | Low-quality |
| 18-102_k141_107694 | Not-determined |
| 18-102_k141_76371 | Not-determined |
| 18-102_k141_232217 | Not-determined |
| 18-102_k141_159118 | Not-determined |
| 18-102_k141_65995 | Not-determined |
| 18-102_k141_46865 | Not-determined |
| 18-102_k141_76374 | Not-determined |
| 18-102_k141_148970 | Not-determined |
| 18-102_k141_220571 | Not-determined |
| 18-102_k141_46867 | Not-determined |
| 18-102_k141_97421 | Low-quality |
| 18-102_k141_107702 | Not-determined |
| 18-102_k141_76379 | Not-determined |
| 18-102_k141_232226 | Not-determined |
| 18-102_k141_3820 | Not-determined |
| 18-102_k141_190718 | Not-determined |
| 18-102_k141_148988 | Not-determined |
| 18-102_k141_107707 | Not-determined |
| 18-102_k141_148991 | Not-determined |
| 18-102_k141_24609 | Not-determined |
| 18-102_k141_148995 | Not-determined |
| 18-102_k141_24611 | Not-determined |
| 18-102_k141_220577 | Not-determined |
| 18-102_k141_14306 | Not-determined |
| 18-102_k141_97435 | Not-determined |
| 18-102_k141_24613 | Low-quality |
| 18-102_k141_14307 | Not-determined |
| 18-102_k141_35202 | Not-determined |
| 18-102_k141_55944 | Not-determined |
| 18-102_k141_220583 | Not-determined |
| 18-102_k141_199633 | Not-determined |
| 18-102_k141_149006 | Not-determined |
| 18-102_k141_232247 | Not-determined |
| 18-102_k141_66010 | Not-determined |
| 18-102_k141_220587 | Not-determined |
| 18-102_k141_159144 | Not-determined |
| 18-102_k141_3833 | Not-determined |
| 18-102_k141_97449 | Not-determined |
| 18-102_k141_232258 | Not-determined |
| 18-102_k141_138916 | Not-determined |
| 18-102_k141_46893 | Not-determined |
| 18-102_k141_97468 | Not-determined |
| 18-102_k141_46897 | Low-quality |
| 18-102_k141_46907 | Not-determined |
| 18-102_k141_210856 | Not-determined |
| 18-102_k141_117847 | Not-determined |
| 18-102_k141_210857 | Not-determined |
| 18-102_k141_199646 | Not-determined |
| 18-102_k141_55954 | Not-determined |
| 18-102_k141_149043 | Not-determined |
| 18-102_k141_232281 | Not-determined |
| 18-102_k141_117865 | Not-determined |
| 18-102_k141_55966 | Low-quality |
| 18-102_k141_220630 | Not-determined |
| 18-102_k141_190742 | Not-determined |
| 18-102_k141_180677 | Not-determined |
| 18-102_k141_220638 | Not-determined |
| 18-102_k141_149057 | Low-quality |
| 18-102_k141_190761 | Not-determined |
| 18-102_k141_46930 | Not-determined |
| 18-102_k141_149061 | Not-determined |
| 18-102_k141_14331 | Not-determined |
| 18-102_k141_210878 | Low-quality |
| 18-102_k141_14333 | Not-determined |
| 18-102_k141_210885 | Not-determined |
| 18-102_k141_210886 | Low-quality |
| 18-102_k141_199670 | Not-determined |
| 18-102_k141_97498 | Not-determined |
| 18-102_k141_55985 | Not-determined |
| 18-102_k141_220654 | Low-quality |
| 18-102_k141_55989 | Not-determined |
| 18-102_k141_24629 | Low-quality |
| 18-102_k141_220657 | Low-quality |
| 18-102_k141_241543 | Not-determined |
| 18-102_k141_35242 | Low-quality |
| 18-102_k141_14350 | Not-determined |
| 18-102_k141_199673 | Not-determined |

| | |
|---|---|
| 18-102_k141_14357 | Not-determined |
| 18-102_k141_232299 | Not-determined |
| 18-102_k141_46953 | Not-determined |
| 18-102_k141_46954 | Not-determined |
| 18-102_k141_76412 | Not-determined |
| 18-102_k141_66030 | Not-determined |
| 18-102_k141_190772 | Not-determined |
| 18-102_k141_97508 | Not-determined |
| 18-102_k141_46962 | Not-determined |
| 18-102_k141_56006 | Not-determined |
| 18-102_k141_56008 | Low-quality |
| 18-102_k141_76420 | Not-determined |
| 18-102_k141_159165 | Not-determined |
| 18-102_k141_24637 | Not-determined |
| 18-102_k141_35249 | Not-determined |
| 18-102_k141_232304 | Not-determined |
| 18-102_k141_14371 | Not-determined |
| 18-102_k141_138936 | Not-determined |
| 18-102_k141_97526 | Not-determined |
| 18-102_k141_138938 | Not-determined |
| 18-102_k141_159176 | Not-determined |
| 18-102_k141_35261 | Not-determined |
| 18-102_k141_66059 | Not-determined |
| 18-102_k141_76448 | Not-determined |
| 18-102_k141_220680 | Not-determined |
| 18-102_k141_24651 | Not-determined |
| 18-102_k141_87219 | Not-determined |
| 18-102_k141_14387 | Not-determined |
| 18-102_k141_14389 | Not-determined |
| 18-102_k141_35268 | Low-quality |
| 18-102_k141_138950 | Not-determined |
| 18-102_k141_199715 | Not-determined |
| 18-102_k141_14394 | Not-determined |
| 18-102_k141_138953 | Not-determined |
| 18-102_k141_14395 | Not-determined |
| 18-102_k141_138957 | Not-determined |
| 18-102_k141_127919 | Not-determined |
| 18-102_k141_87237 | Not-determined |
| 18-102_k141_180707 | Not-determined |
| 18-102_k141_87240 | Not-determined |
| 18-102_k141_56032 | Not-determined |
| 18-102_k141_138964 | Low-quality |
| 18-102_k141_97546 | Not-determined |
| 18-102_k141_56035 | Not-determined |
| 18-102_k141_199726 | Not-determined |
| 18-102_k141_24659 | Not-determined |
| 18-102_k141_76458 | Not-determined |
| 18-102_k141_47005 | Not-determined |
| 18-102_k141_76461 | Not-determined |
| 18-102_k141_138972 | Not-determined |
| 18-102_k141_159205 | Low-quality |
| 18-102_k141_138975 | Not-determined |
| 18-102_k141_35286 | Low-quality |
| 18-102_k141_232354 | Not-determined |
| 18-102_k141_47025 | Not-determined |
| 18-102_k141_87260 | Not-determined |
| 18-102_k141_47027 | Not-determined |
| 18-102_k141_180733 | Low-quality |
| 18-102_k141_180738 | Not-determined |
| 18-102_k141_24683 | Not-determined |
| 18-102_k141_199750 | Not-determined |
| 18-102_k141_56047 | Not-determined |
| 18-102_k141_24690 | Not-determined |
| 18-102_k141_117905 | Low-quality |
| 18-102_k141_199758 | Not-determined |
| 18-102_k141_24693 | Low-quality |
| 18-102_k141_199762 | Not-determined |
| 18-102_k141_220686 | Not-determined |
| 18-102_k141_170142 | Not-determined |
| 18-102_k141_232379 | Not-determined |
| 18-102_k141_170146 | Not-determined |
| 18-102_k141_97562 | Not-determined |
| 18-102_k141_66099 | Not-determined |
| 18-102_k141_107754 | Low-quality |
| 18-102_k141_35302 | Not-determined |
| 18-102_k141_66105 | Low-quality |
| 18-102_k141_107759 | Not-determined |
| 18-102_k141_66111 | Not-determined |
| 18-102_k141_97575 | Not-determined |
| 18-102_k141_76487 | Low-quality |
| 18-102_k141_220694 | Low-quality |
| 18-102_k141_24705 | Not-determined |
| 18-102_k141_220696 | Not-determined |
| 18-102_k141_56086 | Not-determined |
| 18-102_k141_3837 | Not-determined |
| 18-102_k141_3841 | Not-determined |
| 18-102_k141_159249 | Not-determined |
| 18-102_k141_170164 | Not-determined |
| 18-102_k141_35321 | Not-determined |
| 18-102_k141_241619 | Not-determined |
| 18-102_k141_170170 | Not-determined |
| 18-102_k141_241629 | Not-determined |
| 18-102_k141_241633 | Not-determined |
| 18-102_k141_66116 | Not-determined |
| 18-102_k141_159263 | Not-determined |
| 18-102_k141_127968 | Low-quality |
| 18-102_k141_117914 | Not-determined |
| 18-102_k141_232430 | Not-determined |
| 18-102_k141_232432 | Low-quality |
| 18-102_k141_159274 | Not-determined |
| 18-102_k141_87301 | Not-determined |
| 18-102_k141_127978 | Low-quality |
| 18-102_k141_170196 | Not-determined |
| 18-102_k141_117933 | Not-determined |
| 18-102_k141_117940 | Not-determined |
| 18-102_k141_66126 | Not-determined |
| 18-102_k141_232445 | Not-determined |
| 18-102_k141_97586 | Not-determined |
| 18-102_k141_117944 | Not-determined |
| 18-102_k141_97589 | Not-determined |
| 18-102_k141_117948 | Not-determined |
| 18-102_k141_199786 | Not-determined |
| 18-102_k141_66134 | Not-determined |
| 18-102_k141_76520 | Not-determined |
| 18-102_k141_56106 | Not-determined |
| 18-102_k141_170229 | Not-determined |
| 18-102_k141_87334 | Not-determined |
| 18-102_k141_87335 | Not-determined |
| 18-102_k141_66137 | Not-determined |
| 18-102_k141_66138 | Not-determined |
| 18-102_k141_97598 | Not-determined |
| 18-102_k141_170231 | Not-determined |
| 18-102_k141_117970 | Not-determined |
| 18-102_k141_117975 | Low-quality |
| 18-102_k141_180782 | Low-quality |
| 18-102_k141_139040 | Not-determined |
| 18-102_k141_76540 | Not-determined |
| 18-102_k141_127999 | Low-quality |
| 18-102_k141_128000 | Not-determined |
| 18-102_k141_76545 | Not-determined |
| 18-102_k141_76547 | Not-determined |
| 18-102_k141_170252 | Not-determined |
| 18-102_k141_47058 | Low-quality |
| 18-102_k141_56129 | Low-quality |
| 18-102_k141_128008 | Not-determined |
| 18-102_k141_14431 | Not-determined |
| 18-102_k141_128012 | Low-quality |
| 18-102_k141_139050 | Low-quality |
| 18-102_k141_56133 | Not-determined |
| 18-102_k141_180795 | Not-determined |
| 18-102_k141_139055 | Not-determined |
| 18-102_k141_97619 | Not-determined |
| 18-102_k141_3869 | Not-determined |
| 18-102_k141_170265 | Not-determined |
| 18-102_k141_170269 | Not-determined |
| 18-102_k141_87360 | Not-determined |
| 18-102_k141_210924 | Not-determined |
| 18-102_k141_180806 | Not-determined |
| 18-102_k141_199826 | Not-determined |
| 18-102_k141_24771 | Not-determined |
| 18-102_k141_128021 | Not-determined |
| 18-102_k141_199830 | Not-determined |
| 18-102_k141_139067 | Not-determined |
| 18-102_k141_87370 | Low-quality |
| 18-102_k141_76576 | Not-determined |
| 18-102_k141_118028 | Not-determined |
| 18-102_k141_128036 | Not-determined |
| 18-102_k141_76582 | Not-determined |
| 18-102_k141_66176 | Not-determined |
| 18-102_k141_24786 | Low-quality |
| 18-102_k141_128042 | Not-determined |
| 18-102_k141_118034 | Not-determined |
| 18-102_k141_180810 | Low-quality |
| 18-102_k141_180814 | Not-determined |
| 18-102_k141_199856 | Not-determined |
| 18-102_k141_149077 | Not-determined |
| 18-102_k141_87402 | Not-determined |
| 18-102_k141_66187 | Not-determined |
| 18-102_k141_180827 | Not-determined |
| 18-102_k141_210943 | Low-quality |
| 18-102_k141_199863 | Not-determined |
| 18-102_k141_220729 | Not-determined |
| 18-102_k141_139089 | Not-determined |
| 18-102_k141_66202 | Not-determined |
| 18-102_k141_66205 | Not-determined |
| 18-102_k141_139093 | Not-determined |
| 18-102_k141_149091 | Not-determined |
| 18-102_k141_118052 | Not-determined |
| 18-102_k141_66208 | Not-determined |
| 18-102_k141_56154 | Not-determined |
| 18-102_k141_180832 | Not-determined |
| 18-102_k141_56156 | Not-determined |
| 18-102_k141_149102 | Not-determined |
| 18-102_k141_220747 | Not-determined |
| 18-102_k141_210964 | Not-determined |
| 18-102_k141_210967 | Not-determined |
| 18-102_k141_66219 | Not-determined |
| 18-102_k141_149113 | Low-quality |
| 18-102_k141_149114 | Not-determined |
| 18-102_k141_220753 | Low-quality |
| 18-102_k141_47075 | Not-determined |
| 18-102_k141_232459 | Not-determined |
| 18-102_k141_180852 | Not-determined |
| 18-102_k141_56171 | Not-determined |
| 18-102_k141_35399 | Not-determined |
| 18-102_k141_56175 | Not-determined |
| 18-102_k141_241694 | Low-quality |
| 18-102_k141_139116 | Not-determined |
| 18-102_k141_210976 | Not-determined |
| 18-102_k141_56183 | Low-quality |
| 18-102_k141_56185 | Not-determined |
| 18-102_k141_35412 | Not-determined |
| 18-102_k141_118067 | Not-determined |
| 18-102_k141_118070 | Not-determined |
| 18-102_k141_232476 | Not-determined |
| 18-102_k141_232477 | Not-determined |
| 18-102_k141_139127 | Not-determined |
| 18-102_k141_66224 | Low-quality |
| 18-102_k141_66228 | Not-determined |
| 18-102_k141_118072 | Not-determined |
| 18-102_k141_118076 | Not-determined |
| 18-102_k141_35432 | Not-determined |
| 18-102_k141_199907 | Not-determined |
| 18-102_k141_210988 | Not-determined |
| 18-102_k141_232495 | Not-determined |
| 18-102_k141_118084 | Not-determined |
| 18-102_k141_170311 | Not-determined |
| 18-102_k141_35440 | Not-determined |
| 18-102_k141_87427 | Not-determined |
| 18-102_k141_35448 | Not-determined |
| 18-102_k141_87431 | Not-determined |
| 18-102_k141_66257 | Not-determined |
| 18-102_k141_66264 | Not-determined |
| 18-102_k141_170318 | Not-determined |
| 18-102_k141_97637 | Low-quality |
| 18-102_k141_35460 | Not-determined |
| 18-102_k141_47120 | Not-determined |
| 18-102_k141_211007 | Not-determined |
| 18-102_k141_97651 | Not-determined |
| 18-102_k141_35467 | Not-determined |
| 18-102_k141_180873 | Low-quality |
| 18-102_k141_211012 | Not-determined |
| 18-102_k141_47133 | Not-determined |
| 18-102_k141_87457 | Low-quality |
| 18-102_k141_180876 | Not-determined |
| 18-102_k141_56231 | Not-determined |
| 18-102_k141_180884 | Not-determined |
| 18-102_k141_211017 | Not-determined |
| 18-102_k141_76612 | Not-determined |
| 18-102_k141_139182 | Low-quality |
| 18-102_k141_66287 | Not-determined |
| 18-102_k141_211022 | Not-determined |
| 18-102_k141_170348 | Not-determined |
| 18-102_k141_66291 | Not-determined |
| 18-102_k141_220791 | Not-determined |
| 18-102_k141_211032 | Not-determined |
| 18-102_k141_220800 | Not-determined |
| 18-102_k141_220802 | Not-determined |
| 18-102_k141_76616 | Not-determined |
| 18-102_k141_76617 | Not-determined |
| 18-102_k141_47159 | Not-determined |
| 18-102_k141_220803 | Not-determined |
| 18-102_k141_56250 | Low-quality |
| 18-102_k141_47165 | Not-determined |
| 18-102_k141_220814 | Not-determined |
| 18-102_k141_107859 | Not-determined |
| 18-102_k141_107860 | Low-quality |
| 18-102_k141_107861 | Not-determined |
| 18-102_k141_241702 | Not-determined |
| 18-102_k141_76632 | Not-determined |
| 18-102_k141_56261 | Not-determined |
| 18-102_k141_211055 | Not-determined |
| 18-102_k141_118122 | Not-determined |
| 18-102_k141_97672 | Not-determined |
| 18-102_k141_211058 | Low-quality |
| 18-102_k141_97674 | Not-determined |
| 18-102_k141_76636 | Not-determined |
| 18-102_k141_118124 | Not-determined |
| 18-102_k141_211061 | Not-determined |
| 18-102_k141_87476 | Low-quality |
| 18-102_k141_170380 | Low-quality |
| 18-102_k141_170384 | Not-determined |
| 18-102_k141_47180 | Low-quality |
| 18-102_k141_139193 | Not-determined |
| 18-102_k141_118145 | Low-quality |
| 18-102_k141_14495 | Not-determined |
| 18-102_k141_14496 | Not-determined |
| 18-102_k141_170393 | Not-determined |
| 18-102_k141_47188 | Not-determined |
| 18-102_k141_97683 | Not-determined |
| 18-102_k141_241715 | Not-determined |
| 18-102_k141_87481 | Not-determined |
| 18-102_k141_87482 | Low-quality |
| 18-102_k141_66304 | Not-determined |
| 18-102_k141_118165 | Not-determined |
| 18-102_k141_87492 | Not-determined |
| 18-102_k141_180942 | Not-determined |
| 18-102_k141_220850 | Not-determined |
| 18-102_k141_97692 | Not-determined |
| 18-102_k141_180944 | Not-determined |
| 18-102_k141_76647 | Low-quality |
| 18-102_k141_170435 | Not-determined |
| 18-102_k141_159294 | Not-determined |
| 18-102_k141_190806 | Not-determined |
| 18-102_k141_190810 | Not-determined |
| 18-102_k141_220857 | Not-determined |
| 18-102_k141_66323 | Not-determined |
| 18-102_k141_35515 | Low-quality |
| 18-102_k141_180950 | Not-determined |
| 18-102_k141_220862 | Not-determined |
| 18-102_k141_180954 | Low-quality |
| 18-102_k141_220865 | Not-determined |
| 18-102_k141_66334 | Not-determined |
| 18-102_k141_159299 | Not-determined |
| 18-102_k141_159303 | Not-determined |
| 18-102_k141_97706 | Low-quality |
| 18-102_k141_87525 | Not-determined |
| 18-102_k141_199943 | Not-determined |
| 18-102_k141_35517 | Not-determined |
| 18-102_k141_190815 | Not-determined |
| 18-102_k141_159305 | Not-determined |
| 18-102_k141_87529 | Not-determined |
| 18-102_k141_220876 | Not-determined |
| 18-102_k141_66348 | Not-determined |
| 18-102_k141_170482 | Not-determined |
| 18-102_k141_199958 | Not-determined |
| 18-102_k141_76690 | Not-determined |
| 18-102_k141_24828 | Not-determined |
| 18-102_k141_76694 | Not-determined |
| 18-102_k141_66363 | Not-determined |
| 18-102_k141_35532 | Low-quality |
| 18-102_k141_220880 | Not-determined |
| 18-102_k141_66370 | Not-determined |
| 18-102_k141_107901 | Not-determined |
| 18-102_k141_66374 | Not-determined |
| 18-102_k141_170489 | Not-determined |
| 18-102_k141_24832 | Not-determined |
| 18-102_k141_97728 | Not-determined |
| 18-102_k141_97733 | Not-determined |
| 18-102_k141_241764 | Not-determined |
| 18-102_k141_66382 | Not-determined |
| 18-102_k141_220895 | Not-determined |
| 18-102_k141_3928 | Not-determined |
| 18-102_k141_56336 | Not-determined |
| 18-102_k141_76700 | Not-determined |
| 18-102_k141_97742 | Not-determined |
| 18-102_k141_220896 | Not-determined |
| 18-102_k141_47213 | Not-determined |
| 18-102_k141_118203 | Not-determined |
| 18-102_k141_3934 | Not-determined |
| 18-102_k141_190837 | Not-determined |
| 18-102_k141_190839 | Not-determined |
| 18-102_k141_170491 | Not-determined |
| 18-102_k141_199986 | Not-determined |
| 18-102_k141_97748 | Not-determined |
| 18-102_k141_181013 | Low-quality |
| 18-102_k141_35569 | Not-determined |
| 18-102_k141_220904 | Not-determined |
| 18-102_k141_24847 | Not-determined |
| 18-102_k141_190842 | Not-determined |
| 18-102_k141_220906 | Not-determined |
| 18-102_k141_181030 | Not-determined |
| 18-102_k141_118227 | Not-determined |
| 18-102_k141_170495 | Not-determined |
| 18-102_k141_47241 | Not-determined |
| 18-102_k141_118239 | Low-quality |
| 18-102_k141_97764 | Not-determined |
| 18-102_k141_35594 | Not-determined |
| 18-102_k141_97766 | Low-quality |
| 18-102_k141_220925 | Not-determined |
| 18-102_k141_200000 | Not-determined |
| 18-102_k141_200005 | Not-determined |
| 18-102_k141_35607 | Not-determined |
| 18-102_k141_76725 | Not-determined |
| 18-102_k141_56369 | Not-determined |
| 18-102_k141_200010 | Not-determined |
| 18-102_k141_190882 | Not-determined |
| 18-102_k141_56371 | Not-determined |
| 18-102_k141_87563 | Not-determined |
| 18-102_k141_200013 | Not-determined |
| 18-102_k141_107916 | Not-determined |
| 18-102_k141_87576 | Not-determined |
| 18-102_k141_107919 | Not-determined |
| 18-102_k141_76737 | Not-determined |
| 18-102_k141_97772 | Not-determined |
| 18-102_k141_56386 | Low-quality |
| 18-102_k141_181052 | Not-determined |
| 18-102_k141_76745 | Not-determined |
| 18-102_k141_170538 | Not-determined |
| 18-102_k141_76754 | Not-determined |
| 18-102_k141_139236 | Not-determined |
| 18-102_k141_118259 | Not-determined |
| 18-102_k141_87594 | Not-determined |
| 18-102_k141_170552 | Not-determined |
| 18-102_k141_149147 | Not-determined |
| 18-102_k141_107925 | Not-determined |
| 18-102_k141_190907 | Not-determined |
| 18-102_k141_149148 | Not-determined |
| 18-102_k141_211091 | Not-determined |
| 18-102_k141_87597 | Not-determined |
| 18-102_k141_118268 | Not-determined |
| 18-102_k141_76763 | Not-determined |
| 18-102_k141_200036 | Not-determined |
| 18-102_k141_56406 | Not-determined |
| 18-102_k141_159324 | Not-determined |
| 18-102_k141_149156 | Low-quality |
| 18-102_k141_118279 | Not-determined |
| 18-102_k141_76777 | Not-determined |
| 18-102_k141_107943 | Not-determined |
| 18-102_k141_200039 | Not-determined |
| 18-102_k141_149160 | Not-determined |
| 18-102_k141_149162 | Not-determined |
| 18-102_k141_107948 | Not-determined |
| 18-102_k141_200041 | Not-determined |
| 18-102_k141_87616 | Low-quality |
| 18-102_k141_3954 | Not-determined |
| 18-102_k141_56415 | Not-determined |
| 18-102_k141_190933 | Not-determined |
| 18-102_k141_118292 | Not-determined |
| 18-102_k141_170578 | Not-determined |
| 18-102_k141_47282 | Not-determined |
| 18-102_k141_47289 | Not-determined |
| 18-102_k141_56427 | Not-determined |
| 18-102_k141_159349 | Not-determined |
| 18-102_k141_56429 | Not-determined |
| 18-102_k141_35642 | Not-determined |
| 18-102_k141_181087 | Not-determined |
| 18-102_k141_97794 | Not-determined |
| 18-102_k141_159357 | Not-determined |
| 18-102_k141_118329 | Not-determined |
| 18-102_k141_211121 | Not-determined |
| 18-102_k141_56443 | Not-determined |
| 18-102_k141_211126 | Not-determined |
| 18-102_k141_56448 | Not-determined |
| 18-102_k141_97801 | Not-determined |
| 18-102_k141_14536 | Not-determined |
| 18-102_k141_24878 | Not-determined |
| 18-102_k141_107963 | Low-quality |
| 18-102_k141_87627 | Not-determined |
| 18-102_k141_66437 | Not-determined |
| 18-102_k141_56466 | Not-determined |
| 18-102_k141_47314 | Not-determined |
| 18-102_k141_3979 | Not-determined |
| 18-102_k141_3986 | Not-determined |
| 18-102_k141_47323 | Low-quality |
| 18-102_k141_56475 | Not-determined |
| 18-102_k141_56478 | Not-determined |
| 18-102_k141_190950 | Not-determined |
| 18-102_k141_211138 | Not-determined |
| 18-102_k141_159374 | Not-determined |
| 18-102_k141_3987 | Not-determined |
| 18-102_k141_97828 | Not-determined |
| 18-102_k141_190962 | Not-determined |
| 18-102_k141_14561 | Low-quality |
| 18-102_k141_3998 | Low-quality |
| 18-102_k141_3999 | Not-determined |
| 18-102_k141_56501 | Not-determined |
| 18-102_k141_149236 | Not-determined |
| 18-102_k141_35666 | Not-determined |
| 18-102_k141_170624 | Not-determined |
| 18-102_k141_118356 | Not-determined |
| 18-102_k141_4009 | Not-determined |
| 18-102_k141_87659 | Not-determined |
| 18-102_k141_128056 | Not-determined |
| 18-102_k141_4021 | Not-determined |
| 18-102_k141_4022 | Not-determined |
| 18-102_k141_118371 | Not-determined |
| 18-102_k141_66482 | Not-determined |
| 18-102_k141_97843 | Low-quality |
| 18-102_k141_190978 | Not-determined |
| 18-102_k141_118378 | Not-determined |
| 18-102_k141_128073 | Not-determined |
| 18-102_k141_97850 | Not-determined |
| 18-102_k141_170635 | Not-determined |
| 18-102_k141_47353 | Not-determined |
| 18-102_k141_159423 | Low-quality |
| 18-102_k141_97860 | Not-determined |
| 18-102_k141_149250 | Not-determined |
| 18-102_k141_97870 | Not-determined |
| 18-102_k141_200092 | Not-determined |
| 18-102_k141_159445 | Not-determined |
| 18-102_k141_159447 | Not-determined |
| 18-102_k141_170656 | Not-determined |
| 18-102_k141_170658 | Not-determined |
| 18-102_k141_128095 | Not-determined |
| 18-102_k141_139312 | Low-quality |
| 18-102_k141_190998 | Not-determined |
| 18-102_k141_128097 | Low-quality |
| 18-102_k141_200102 | Not-determined |
| 18-102_k141_159462 | Not-determined |
| 18-102_k141_35726 | Not-determined |
| 18-102_k141_139323 | Not-determined |
| 18-102_k141_128107 | Low-quality |
| 18-102_k141_14587 | Not-determined |
| 18-102_k141_149270 | Not-determined |
| 18-102_k141_139330 | Not-determined |
| 18-102_k141_159470 | Not-determined |
| 18-102_k141_170672 | Low-quality |
| 18-102_k141_139334 | Not-determined |
| 18-102_k141_211171 | Not-determined |
| 18-102_k141_128118 | Low-quality |
| 18-102_k141_97886 | Not-determined |
| 18-102_k141_14601 | Not-determined |
| 18-102_k141_139335 | Not-determined |
| 18-102_k141_211177 | Not-determined |
| 18-102_k141_14606 | Not-determined |
| 18-102_k141_200113 | Not-determined |
| 18-102_k141_149284 | Not-determined |
| 18-102_k141_118396 | Not-determined |
| 18-102_k141_87706 | Not-determined |
| 18-102_k141_139340 | Not-determined |
| 18-102_k141_159483 | Not-determined |
| 18-102_k141_128135 | Not-determined |
| 18-102_k141_170687 | Low-quality |
| 18-102_k141_97891 | Not-determined |
| 18-102_k141_149287 | Not-determined |
| 18-102_k141_97895 | Not-determined |
| 18-102_k141_232541 | Not-determined |
| 18-102_k141_149297 | Not-determined |
| 18-102_k141_159493 | Not-determined |
| 18-102_k141_139347 | Not-determined |
| 18-102_k141_232549 | Not-determined |
| 18-102_k141_118407 | Not-determined |
| 18-102_k141_200139 | Not-determined |
| 18-102_k141_14608 | Not-determined |
| 18-102_k141_181165 | Not-determined |
| 18-102_k141_191012 | Not-determined |
| 18-102_k141_76797 | Not-determined |
| 18-102_k141_191015 | Not-determined |
| 18-102_k141_191016 | Not-determined |
| 18-102_k141_191017 | Not-determined |
| 18-102_k141_211195 | Not-determined |
| 18-102_k141_220941 | Low-quality |
| 18-102_k141_97899 | Low-quality |
| 18-102_k141_211198 | Not-determined |
| 18-102_k141_108050 | Not-determined |
| 18-102_k141_220944 | Low-quality |
| 18-102_k141_220946 | Not-determined |
| 18-102_k141_232556 | Not-determined |
| 18-102_k141_139363 | Not-determined |
| 18-102_k141_14620 | Not-determined |
| 18-102_k141_108057 | Not-determined |
| 18-102_k141_47382 | Not-determined |
| 18-102_k141_108061 | Not-determined |
| 18-102_k141_97912 | Not-determined |
| 18-102_k141_211211 | Not-determined |
| 18-102_k141_232571 | Not-determined |
| 18-102_k141_108072 | Not-determined |
| 18-102_k141_200143 | Not-determined |
| 18-102_k141_170723 | Not-determined |
| 18-102_k141_170725 | Not-determined |
| 18-102_k141_139376 | Not-determined |
| 18-102_k141_14644 | Not-determined |
| 18-102_k141_220958 | Not-determined |
| 18-102_k141_97927 | Low-quality |
| 18-102_k141_14648 | Not-determined |
| 18-102_k141_97929 | Not-determined |
| 18-102_k141_211228 | Not-determined |
| 18-102_k141_170732 | Not-determined |
| 18-102_k141_159518 | Not-determined |
| 18-102_k141_97936 | Not-determined |
| 18-102_k141_14655 | Not-determined |
| 18-102_k141_159525 | Not-determined |
| 18-102_k141_108079 | Low-quality |
| 18-102_k141_128151 | Low-quality |
| 18-102_k141_14667 | Not-determined |
| 18-102_k141_47415 | Not-determined |
| 18-102_k141_170747 | Not-determined |
| 18-102_k141_181205 | Not-determined |
| 18-102_k141_159534 | Not-determined |
| 18-102_k141_47431 | Not-determined |
| 18-102_k141_14678 | Not-determined |
| 18-102_k141_47436 | Not-determined |
| 18-102_k141_118447 | Not-determined |
| 18-102_k141_4109 | Not-determined |
| 18-102_k141_47443 | Not-determined |
| 18-102_k141_241783 | Not-determined |
| 18-102_k141_241785 | Not-determined |
| 18-102_k141_4113 | Not-determined |
| 18-102_k141_159549 | Not-determined |
| 18-102_k141_97957 | Not-determined |
| 18-102_k141_118455 | Not-determined |
| 18-102_k141_128170 | Not-determined |
| 18-102_k141_241794 | Not-determined |
| 18-102_k141_87718 | Not-determined |
| 18-102_k141_181235 | Not-determined |
| 18-102_k141_97965 | Not-determined |
| 18-102_k141_220991 | Low-quality |
| 18-102_k141_76848 | Not-determined |
| 18-102_k141_128177 | Not-determined |
| 18-102_k141_14688 | Not-determined |
| 18-102_k141_108111 | Not-determined |
| 18-102_k141_4129 | Not-determined |
| 18-102_k141_66529 | Not-determined |
| 18-102_k141_97983 | Not-determined |
| 18-102_k141_47472 | Not-determined |
| 18-102_k141_4133 | Not-determined |
| 18-102_k141_97985 | Not-determined |
| 18-102_k141_128190 | Not-determined |

| | |
|---|---|
| 18-102_k141_128193 | Not-determined |
| 18-102_k141_118461 | Not-determined |
| 18-102_k141_97994 | Not-determined |
| 18-102_k141_128196 | Not-determined |
| 18-102_k141_47482 | Not-determined |
| 18-102_k141_4146 | Not-determined |
| 18-102_k141_76864 | Not-determined |
| 18-102_k141_220999 | Not-determined |
| 18-102_k141_128209 | Not-determined |
| 18-102_k141_76870 | Not-determined |
| 18-102_k141_98008 | Not-determined |
| 18-102_k141_76873 | Not-determined |
| 18-102_k141_66546 | Not-determined |
| 18-102_k141_191069 | Not-determined |
| 18-102_k141_221009 | Not-determined |
| 18-102_k141_24985 | Not-determined |
| 18-102_k141_118474 | Not-determined |
| 18-102_k141_221013 | Not-determined |
| 18-102_k141_191075 | Not-determined |
| 18-102_k141_108142 | Not-determined |
| 18-102_k141_221024 | Not-determined |
| 18-102_k141_191082 | Not-determined |
| 18-102_k141_170792 | Not-determined |
| 18-102_k141_66568 | Not-determined |
| 18-102_k141_149354 | Low-quality |
| 18-102_k141_35767 | Not-determined |
| 18-102_k141_66574 | Low-quality |
| 18-102_k141_47513 | Not-determined |
| 18-102_k141_181294 | Not-determined |
| 18-102_k141_149357 | Not-determined |
| 18-102_k141_181298 | Not-determined |
| 18-102_k141_149360 | Not-determined |
| 18-102_k141_47523 | Not-determined |
| 18-102_k141_232602 | Not-determined |
| 18-102_k141_128237 | Not-determined |
| 18-102_k141_98018 | Not-determined |
| 18-102_k141_35776 | Not-determined |
| 18-102_k141_170799 | Not-determined |
| 18-102_k141_241832 | Not-determined |
| 18-102_k141_47527 | Not-determined |
| 18-102_k141_98024 | Low-quality |
| 18-102_k141_47529 | Not-determined |
| 18-102_k141_241837 | Low-quality |
| 18-102_k141_47534 | Not-determined |
| 18-102_k141_108155 | Not-determined |
| 18-102_k141_47540 | Not-determined |
| 18-102_k141_87772 | Not-determined |
| 18-102_k141_241846 | Not-determined |
| 18-102_k141_241847 | Not-determined |
| 18-102_k141_149366 | Not-determined |
| 18-102_k141_98039 | Not-determined |
| 18-102_k141_170808 | Low-quality |
| 18-102_k141_149375 | Low-quality |
| 18-102_k141_35789 | Not-determined |
| 18-102_k141_241856 | Not-determined |
| 18-102_k141_56572 | Not-determined |
| 18-102_k141_139390 | Not-determined |
| 18-102_k141_149382 | Not-determined |
| 18-102_k141_181312 | Not-determined |
| 18-102_k141_128269 | Low-quality |
| 18-102_k141_108175 | Not-determined |
| 18-102_k141_76904 | Not-determined |
| 18-102_k141_56579 | Not-determined |
| 18-102_k141_98044 | Not-determined |
| 18-102_k141_56581 | Not-determined |
| 18-102_k141_128276 | Not-determined |
| 18-102_k141_128280 | Not-determined |
| 18-102_k141_128282 | Not-determined |
| 18-102_k141_221050 | Not-determined |
| 18-102_k141_108185 | Not-determined |
| 18-102_k141_108187 | Not-determined |
| 18-102_k141_221052 | Not-determined |
| 18-102_k141_128288 | Not-determined |
| 18-102_k141_181331 | Not-determined |
| 18-102_k141_66612 | Not-determined |
| 18-102_k141_108193 | Low-quality |
| 18-102_k141_241866 | Not-determined |
| 18-102_k141_221057 | Not-determined |
| 18-102_k141_181337 | Not-determined |
| 18-102_k141_191116 | Not-determined |
| 18-102_k141_232621 | Not-determined |
| 18-102_k141_170824 | Not-determined |
| 18-102_k141_170825 | Not-determined |
| 18-102_k141_191120 | Not-determined |
| 18-102_k141_98060 | Not-determined |
| 18-102_k141_98061 | Not-determined |
| 18-102_k141_98064 | Not-determined |
| 18-102_k141_149398 | Not-determined |
| 18-102_k141_4208 | Not-determined |
| 18-102_k141_4210 | Not-determined |
| 18-102_k141_149402 | Low-quality |
| 18-102_k141_87796 | Not-determined |
| 18-102_k141_76915 | Not-determined |
| 18-102_k141_149404 | Not-determined |
| 18-102_k141_232637 | Not-determined |
| 18-102_k141_181356 | Not-determined |
| 18-102_k141_221074 | Not-determined |
| 18-102_k141_35837 | Not-determined |
| 18-102_k141_87806 | Not-determined |
| 18-102_k141_181363 | Not-determined |
| 18-102_k141_14735 | Not-determined |
| 18-102_k141_181364 | Not-determined |
| 18-102_k141_221078 | Not-determined |
| 18-102_k141_139412 | Not-determined |
| 18-102_k141_221089 | Not-determined |
| 18-102_k141_25007 | Not-determined |
| 18-102_k141_35843 | Not-determined |
| 18-102_k141_35844 | Not-determined |
| 18-102_k141_35845 | Not-determined |
| 18-102_k141_35848 | Not-determined |
| 18-102_k141_98100 | Not-determined |
| 18-102_k141_35851 | Not-determined |
| 18-102_k141_35853 | Not-determined |
| 18-102_k141_118536 | Not-determined |
| 18-102_k141_118537 | Not-determined |
| 18-102_k141_149433 | Not-determined |
| 18-102_k141_118539 | Not-determined |
| 18-102_k141_76947 | Not-determined |
| 18-102_k141_4236 | Not-determined |
| 18-102_k141_4237 | Not-determined |
| 18-102_k141_118544 | Not-determined |
| 18-102_k141_139434 | Low-quality |
| 18-102_k141_87826 | Not-determined |
| 18-102_k141_139437 | Not-determined |
| 18-102_k141_221098 | Not-determined |
| 18-102_k141_221099 | Not-determined |
| 18-102_k141_200182 | Low-quality |
| 18-102_k141_76956 | Not-determined |
| 18-102_k141_87833 | Not-determined |
| 18-102_k141_159607 | Low-quality |
| 18-102_k141_211318 | Not-determined |
| 18-102_k141_159610 | Not-determined |
| 18-102_k141_98121 | Not-determined |
| 18-102_k141_159613 | Not-determined |
| 18-102_k141_98124 | Not-determined |
| 18-102_k141_149452 | Not-determined |
| 18-102_k141_76979 | Not-determined |
| 18-102_k141_211341 | Not-determined |
| 18-102_k141_35883 | Not-determined |
| 18-102_k141_56625 | Not-determined |
| 18-102_k141_191140 | Not-determined |
| 18-102_k141_76982 | Not-determined |
| 18-102_k141_211349 | Low-quality |
| 18-102_k141_35887 | Not-determined |
| 18-102_k141_56631 | Not-determined |
| 18-102_k141_47568 | Not-determined |
| 18-102_k141_76990 | Not-determined |
| 18-102_k141_56638 | Not-determined |
| 18-102_k141_47577 | Not-determined |
| 18-102_k141_56640 | Not-determined |
| 18-102_k141_4245 | Not-determined |
| 18-102_k141_200197 | Not-determined |
| 18-102_k141_149468 | Not-determined |
| 18-102_k141_35899 | Not-determined |
| 18-102_k141_139469 | Low-quality |
| 18-102_k141_200206 | Not-determined |
| 18-102_k141_98130 | Not-determined |
| 18-102_k141_98131 | Not-determined |
| 18-102_k141_200210 | Not-determined |
| 18-102_k141_159624 | Not-determined |
| 18-102_k141_200212 | Low-quality |
| 18-102_k141_191162 | Not-determined |
| 18-102_k141_35912 | Not-determined |
| 18-102_k141_159632 | Not-determined |
| 18-102_k141_35914 | Not-determined |
| 18-102_k141_35915 | Not-determined |
| 18-102_k141_98147 | Not-determined |
| 18-102_k141_98151 | Not-determined |
| 18-102_k141_139485 | Not-determined |
| 18-102_k141_35922 | Not-determined |
| 18-102_k141_77025 | Not-determined |
| 18-102_k141_66680 | Not-determined |
| 18-102_k141_181389 | Not-determined |
| 18-102_k141_149487 | Not-determined |
| 18-102_k141_56654 | Not-determined |
| 18-102_k141_87857 | Low-quality |
| 18-102_k141_66688 | Not-determined |
| 18-102_k141_191179 | Not-determined |
| 18-102_k141_77040 | Not-determined |
| 18-102_k141_159651 | Not-determined |
| 18-102_k141_98163 | Not-determined |
| 18-102_k141_77049 | Not-determined |
| 18-102_k141_35940 | Not-determined |
| 18-102_k141_170858 | Not-determined |
| 18-102_k141_159657 | Not-determined |
| 18-102_k141_149503 | Not-determined |
| 18-102_k141_118554 | Not-determined |
| 18-102_k141_221112 | Not-determined |
| 18-102_k141_170860 | Not-determined |
| 18-102_k141_56669 | Not-determined |
| 18-102_k141_47605 | Not-determined |
| 18-102_k141_35944 | Not-determined |
| 18-102_k141_232704 | Not-determined |
| 18-102_k141_170866 | Low-quality |
| 18-102_k141_35953 | Not-determined |
| 18-102_k141_118568 | Not-determined |
| 18-102_k141_170876 | Not-determined |
| 18-102_k141_87860 | Not-determined |
| 18-102_k141_139523 | Not-determined |
| 18-102_k141_200231 | Not-determined |
| 18-102_k141_98168 | Low-quality |
| 18-102_k141_139532 | Low-quality |
| 18-102_k141_25070 | Not-determined |
| 18-102_k141_191207 | Not-determined |
| 18-102_k141_87874 | Not-determined |
| 18-102_k141_170898 | Not-determined |
| 18-102_k141_56706 | Low-quality |
| 18-102_k141_191220 | Not-determined |
| 18-102_k141_14778 | Not-determined |
| 18-102_k141_87888 | Not-determined |
| 18-102_k141_170901 | Not-determined |
| 18-102_k141_98180 | Not-determined |
| 18-102_k141_118597 | Not-determined |
| 18-102_k141_170908 | Not-determined |
| 18-102_k141_98196 | Not-determined |
| 18-102_k141_200248 | Not-determined |
| 18-102_k141_77090 | Not-determined |
| 18-102_k141_25084 | Not-determined |
| 18-102_k141_56728 | Low-quality |
| 18-102_k141_14800 | Not-determined |
| 18-102_k141_14808 | Not-determined |
| 18-102_k141_36002 | Not-determined |
| 18-102_k141_36003 | Not-determined |
| 18-102_k141_98208 | Not-determined |
| 18-102_k141_98212 | Not-determined |
| 18-102_k141_139574 | Not-determined |
| 18-102_k141_14819 | Not-determined |
| 18-102_k141_14820 | Not-determined |
| 18-102_k141_4260 | Not-determined |
| 18-102_k141_77097 | Low-quality |
| 18-102_k141_77099 | Not-determined |
| 18-102_k141_77100 | Not-determined |
| 18-102_k141_4263 | Not-determined |
| 18-102_k141_77109 | Not-determined |
| 18-102_k141_149545 | Not-determined |
| 18-102_k141_87904 | Not-determined |
| 18-102_k141_98220 | Not-determined |
| 18-102_k141_118617 | Not-determined |
| 18-102_k141_14831 | Not-determined |
| 18-102_k141_241890 | Not-determined |
| 18-102_k141_241893 | Not-determined |
| 18-102_k141_25091 | Not-determined |
| 18-102_k141_66738 | Not-determined |
| 18-102_k141_98224 | Not-determined |
| 18-102_k141_200258 | Low-quality |
| 18-102_k141_36019 | Not-determined |
| 18-102_k141_241900 | Not-determined |
| 18-102_k141_77130 | Not-determined |
| 18-102_k141_118624 | Not-determined |
| 18-102_k141_77132 | Not-determined |
| 18-102_k141_77135 | Not-determined |
| 18-102_k141_36028 | Not-determined |
| 18-102_k141_159687 | Not-determined |
| 18-102_k141_14859 | Low-quality |
| 18-102_k141_159696 | Not-determined |
| 18-102_k141_149576 | Low-quality |
| 18-102_k141_221191 | Not-determined |
| 18-102_k141_159698 | Not-determined |
| 18-102_k141_159703 | Not-determined |
| 18-102_k141_232719 | Not-determined |
| 18-102_k141_66760 | Not-determined |
| 18-102_k141_118633 | Not-determined |
| 18-102_k141_159707 | Low-quality |
| 18-102_k141_221198 | Not-determined |
| 18-102_k141_4279 | Low-quality |
| 18-102_k141_77155 | Low-quality |
| 18-102_k141_128351 | Not-determined |
| 18-102_k141_4283 | Low-quality |
| 18-102_k141_98239 | Not-determined |
| 18-102_k141_170973 | Low-quality |
| 18-102_k141_4294 | Not-determined |
| 18-102_k141_170974 | Not-determined |
| 18-102_k141_118646 | Low-quality |
| 18-102_k141_200291 | Not-determined |
| 18-102_k141_4300 | Low-quality |
| 18-102_k141_56789 | Not-determined |
| 18-102_k141_56794 | Not-determined |
| 18-102_k141_87934 | Not-determined |
| 18-102_k141_232751 | Not-determined |
| 18-102_k141_87936 | Not-determined |
| 18-102_k141_128375 | Not-determined |
| 18-102_k141_232756 | Not-determined |
| 18-102_k141_159715 | Not-determined |
| 18-102_k141_56805 | Not-determined |
| 18-102_k141_211380 | Not-determined |
| 18-102_k141_14905 | Not-determined |
| 18-102_k141_14908 | Not-determined |
| 18-102_k141_200314 | Not-determined |
| 18-102_k141_200318 | Not-determined |
| 18-102_k141_232773 | Low-quality |
| 18-102_k141_232776 | Not-determined |
| 18-102_k141_139620 | Not-determined |
| 18-102_k141_25156 | Not-determined |
| 18-102_k141_159720 | Not-determined |
| 18-102_k141_139626 | Not-determined |
| 18-102_k141_149609 | Low-quality |
| 18-102_k141_159727 | Not-determined |
| 18-102_k141_149614 | Not-determined |
| 18-102_k141_149616 | Not-determined |
| 18-102_k141_98279 | Not-determined |
| 18-102_k141_149618 | Not-determined |
| 18-102_k141_191265 | Low-quality |
| 18-102_k141_98282 | Not-determined |
| 18-102_k141_98283 | Not-determined |
| 18-102_k141_98286 | Not-determined |
| 18-102_k141_159732 | Not-determined |
| 18-102_k141_191271 | Not-determined |
| 18-102_k141_98289 | Not-determined |
| 18-102_k141_36060 | Not-determined |
| 18-102_k141_118666 | Not-determined |
| 18-102_k141_47676 | Not-determined |
| 18-102_k141_149634 | Not-determined |
| 18-102_k141_159750 | Not-determined |
| 18-102_k141_221237 | Not-determined |
| 18-102_k141_211411 | Not-determined |
| 18-102_k141_47685 | Not-determined |
| 18-102_k141_118674 | Not-determined |
| 18-102_k141_118675 | Not-determined |
| 18-102_k141_211414 | Not-determined |
| 18-102_k141_25173 | Not-determined |
| 18-102_k141_232784 | Not-determined |
| 18-102_k141_98301 | Not-determined |
| 18-102_k141_139658 | Low-quality |
| 18-102_k141_128404 | Not-determined |
| 18-102_k141_118679 | Low-quality |
| 18-102_k141_181398 | Not-determined |
| 18-102_k141_232796 | Not-determined |
| 18-102_k141_159773 | Not-determined |
| 18-102_k141_118687 | Not-determined |
| 18-102_k141_241948 | Not-determined |
| 18-102_k141_211433 | Not-determined |
| 18-102_k141_159777 | Not-determined |
| 18-102_k141_139669 | Not-determined |
| 18-102_k141_181406 | Not-determined |
| 18-102_k141_211438 | Not-determined |
| 18-102_k141_56818 | Not-determined |
| 18-102_k141_221264 | Not-determined |
| 18-102_k141_98325 | Not-determined |
| 18-102_k141_221267 | Not-determined |
| 18-102_k141_36070 | Not-determined |
| 18-102_k141_56821 | Not-determined |
| 18-102_k141_221271 | Not-determined |
| 18-102_k141_36074 | Not-determined |
| 18-102_k141_241961 | Not-determined |
| 18-102_k141_241962 | Not-determined |
| 18-102_k141_241963 | Low-quality |
| 18-102_k141_98327 | Not-determined |
| 18-102_k141_221274 | Not-determined |
| 18-102_k141_241965 | Not-determined |
| 18-102_k141_128413 | Not-determined |
| 18-102_k141_98337 | Not-determined |
| 18-102_k141_232817 | Not-determined |
| 18-102_k141_77161 | Not-determined |
| 18-102_k141_25202 | Low-quality |
| 18-102_k141_98340 | Not-determined |
| 18-102_k141_36095 | Not-determined |
| 18-102_k141_77171 | Not-determined |
| 18-102_k141_139701 | Not-determined |
| 18-102_k141_149660 | Not-determined |
| 18-102_k141_36099 | Not-determined |
| 18-102_k141_36101 | Not-determined |
| 18-102_k141_77177 | Not-determined |
| 18-102_k141_221289 | Low-quality |
| 18-102_k141_77181 | Low-quality |
| 18-102_k141_232827 | Not-determined |
| 18-102_k141_241975 | Not-determined |
| 18-102_k141_159800 | Not-determined |
| 18-102_k141_98359 | Not-determined |
| 18-102_k141_241979 | Not-determined |
| 18-102_k141_171037 | Not-determined |
| 18-102_k141_25223 | Low-quality |
| 18-102_k141_77193 | Not-determined |
| 18-102_k141_211475 | Not-determined |
| 18-102_k141_14929 | Not-determined |
| 18-102_k141_241989 | Not-determined |
| 18-102_k141_47719 | Not-determined |
| 18-102_k141_4318 | Low-quality |
| 18-102_k141_241992 | Not-determined |
| 18-102_k141_211481 | Not-determined |
| 18-102_k141_77200 | Not-determined |
| 18-102_k141_56861 | Low-quality |
| 18-102_k141_241997 | Not-determined |
| 18-102_k141_221321 | Low-quality |
| 18-102_k141_232833 | Low-quality |
| 18-102_k141_171050 | Not-determined |
| 18-102_k141_56867 | Not-determined |
| 18-102_k141_171051 | Not-determined |
| 18-102_k141_232837 | Not-determined |
| 18-102_k141_159816 | Not-determined |
| 18-102_k141_56872 | Not-determined |
| 18-102_k141_159818 | Not-determined |
| 18-102_k141_159821 | Not-determined |
| 18-102_k141_66778 | Not-determined |
| 18-102_k141_118710 | Not-determined |
| 18-102_k141_25252 | Not-determined |
| 18-102_k141_56876 | Not-determined |
| 18-102_k141_56877 | Not-determined |
| 18-102_k141_56879 | Low-quality |
| 18-102_k141_211510 | Not-determined |
| 18-102_k141_87965 | Not-determined |
| 18-102_k141_36145 | Low-quality |
| 18-102_k141_221337 | Not-determined |
| 18-102_k141_200383 | Not-determined |
| 18-102_k141_242007 | Not-determined |
| 18-102_k141_128440 | Low-quality |
| 18-102_k141_139745 | Low-quality |
| 18-102_k141_242009 | Not-determined |
| 18-102_k141_200387 | Not-determined |
| 18-102_k141_128449 | Not-determined |
| 18-102_k141_128452 | Not-determined |
| 18-102_k141_98371 | Not-determined |
| 18-102_k141_232863 | Not-determined |
| 18-102_k141_36161 | Not-determined |
| 18-102_k141_139761 | Not-determined |
| 18-102_k141_36167 | Not-determined |
| 18-102_k141_149670 | Not-determined |
| 18-102_k141_191273 | Not-determined |
| 18-102_k141_139762 | Not-determined |
| 18-102_k141_221351 | Not-determined |
| 18-102_k141_221353 | Not-determined |
| 18-102_k141_56889 | Not-determined |
| 18-102_k141_56895 | Not-determined |
| 18-102_k141_221359 | Not-determined |
| 18-102_k141_149672 | Low-quality |
| 18-102_k141_159828 | Not-determined |
| 18-102_k141_159830 | Not-determined |
| 18-102_k141_181454 | Not-determined |
| 18-102_k141_139770 | Not-determined |
| 18-102_k141_242028 | Not-determined |
| 18-102_k141_139774 | Not-determined |
| 18-102_k141_171091 | Not-determined |
| 18-102_k141_181462 | Not-determined |
| 18-102_k141_47777 | Not-determined |
| 18-102_k141_211521 | Not-determined |
| 18-102_k141_171093 | Not-determined |
| 18-102_k141_171094 | Not-determined |
| 18-102_k141_47782 | Not-determined |
| 18-102_k141_232893 | Low-quality |
| 18-102_k141_36190 | Not-determined |
| 18-102_k141_14971 | Not-determined |
| 18-102_k141_211527 | Not-determined |
| 18-102_k141_181475 | Not-determined |
| 18-102_k141_87979 | Not-determined |
| 18-102_k141_36193 | Low-quality |
| 18-102_k141_149697 | Not-determined |
| 18-102_k141_211529 | Not-determined |
| 18-102_k141_98379 | Not-determined |
| 18-102_k141_47801 | Not-determined |
| 18-102_k141_77255 | Low-quality |
| 18-102_k141_36196 | Not-determined |
| 18-102_k141_77256 | Not-determined |
| 18-102_k141_25280 | Not-determined |
| 18-102_k141_36208 | Not-determined |
| 18-102_k141_36210 | Not-determined |
| 18-102_k141_36213 | Not-determined |
| 18-102_k141_4358 | Not-determined |
| 18-102_k141_181489 | Not-determined |
| 18-102_k141_56902 | Not-determined |
| 18-102_k141_4361 | Low-quality |
| 18-102_k141_98391 | Not-determined |
| 18-102_k141_232927 | Not-determined |
| 18-102_k141_181497 | Low-quality |
| 18-102_k141_56907 | Not-determined |
| 18-102_k141_56908 | Not-determined |
| 18-102_k141_77270 | Not-determined |
| 18-102_k141_118731 | Not-determined |
| 18-102_k141_181504 | Not-determined |
| 18-102_k141_47828 | Not-determined |
| 18-102_k141_77277 | Not-determined |
| 18-102_k141_149716 | Not-determined |
| 18-102_k141_181517 | Not-determined |
| 18-102_k141_4368 | Not-determined |
| 18-102_k141_4369 | Not-determined |
| 18-102_k141_232934 | Not-determined |
| 18-102_k141_221417 | Not-determined |
| 18-102_k141_15002 | Not-determined |
| 18-102_k141_98430 | Not-determined |
| 18-102_k141_171120 | Not-determined |
| 18-102_k141_25303 | Low-quality |
| 18-102_k141_56934 | Not-determined |
| 18-102_k141_25305 | Not-determined |
| 18-102_k141_98437 | Not-determined |
| 18-102_k141_242048 | Not-determined |
| 18-102_k141_221434 | Not-determined |
| 18-102_k141_159843 | Not-determined |
| 18-102_k141_181539 | Low-quality |
| 18-102_k141_15015 | Not-determined |
| 18-102_k141_171128 | Low-quality |
| 18-102_k141_77291 | Not-determined |
| 18-102_k141_159850 | Not-determined |
| 18-102_k141_159855 | Not-determined |
| 18-102_k141_221449 | Not-determined |
| 18-102_k141_221452 | Not-determined |
| 18-102_k141_47863 | Low-quality |
| 18-102_k141_47864 | Not-determined |
| 18-102_k141_242059 | Not-determined |
| 18-102_k141_56949 | Not-determined |
| 18-102_k141_149738 | Not-determined |
| 18-102_k141_77311 | Not-determined |
| 18-102_k141_98460 | Not-determined |
| 18-102_k141_77314 | Not-determined |
| 18-102_k141_139834 | Low-quality |
| 18-102_k141_36235 | Not-determined |
| 18-102_k141_98467 | Not-determined |
| 18-102_k141_221466 | Not-determined |
| 18-102_k141_181546 | Not-determined |
| 18-102_k141_181549 | Not-determined |
| 18-102_k141_47875 | Not-determined |
| 18-102_k141_108205 | Not-determined |
| 18-102_k141_108208 | Not-determined |
| 18-102_k141_98477 | Not-determined |
| 18-102_k141_66859 | Not-determined |
| 18-102_k141_108212 | Not-determined |
| 18-102_k141_15037 | Not-determined |
| 18-102_k141_25344 | Not-determined |
| 18-102_k141_149761 | Not-determined |
| 18-102_k141_149763 | Not-determined |
| 18-102_k141_159873 | Not-determined |
| 18-102_k141_149765 | Low-quality |
| 18-102_k141_15045 | Not-determined |
| 18-102_k141_211566 | Not-determined |
| 18-102_k141_56972 | Not-determined |
| 18-102_k141_221477 | Not-determined |
| 18-102_k141_139852 | Not-determined |
| 18-102_k141_108224 | Low-quality |
| 18-102_k141_221482 | Not-determined |
| 18-102_k141_211574 | Not-determined |
| 18-102_k141_47886 | Not-determined |
| 18-102_k141_191294 | Not-determined |
| 18-102_k141_15052 | Not-determined |
| 18-102_k141_15059 | Not-determined |
| 18-102_k141_25353 | Low-quality |
| 18-102_k141_36244 | Not-determined |
| 18-102_k141_233005 | Not-determined |
| 18-102_k141_149791 | Not-determined |
| 18-102_k141_36255 | Not-determined |
| 18-102_k141_98500 | Not-determined |
| 18-102_k141_98501 | Not-determined |
| 18-102_k141_242111 | Not-determined |
| 18-102_k141_108235 | Not-determined |
| 18-102_k141_77345 | Not-determined |
| 18-102_k141_77347 | Not-determined |
| 18-102_k141_221488 | Not-determined |
| 18-102_k141_47896 | Not-determined |
| 18-102_k141_149795 | Not-determined |
| 18-102_k141_108251 | Not-determined |
| 18-102_k141_98514 | Not-determined |
| 18-102_k141_139888 | Not-determined |
| 18-102_k141_36270 | Not-determined |
| 18-102_k141_77369 | Not-determined |
| 18-102_k141_149808 | Not-determined |
| 18-102_k141_25365 | Not-determined |
| 18-102_k141_98521 | Low-quality |
| 18-102_k141_233028 | Not-determined |
| 18-102_k141_47913 | Not-determined |
| 18-102_k141_159882 | Not-determined |
| 18-102_k141_47923 | Not-determined |
| 18-102_k141_36293 | Not-determined |
| 18-102_k141_211605 | Not-determined |
| 18-102_k141_200420 | Not-determined |
| 18-102_k141_211606 | Low-quality |
| 18-102_k141_233034 | Not-determined |
| 18-102_k141_77379 | Low-quality |
| 18-102_k141_200424 | Not-determined |
| 18-102_k141_47939 | Not-determined |
| 18-102_k141_108284 | Not-determined |
| 18-102_k141_211612 | Low-quality |
| 18-102_k141_108289 | Not-determined |
| 18-102_k141_139919 | Not-determined |
| 18-102_k141_108295 | Not-determined |
| 18-102_k141_15129 | Not-determined |
| 18-102_k141_211625 | Not-determined |
| 18-102_k141_221511 | Not-determined |
| 18-102_k141_15133 | Not-determined |
| 18-102_k141_47953 | Not-determined |
| 18-102_k141_221521 | Not-determined |
| 18-102_k141_200436 | Not-determined |
| 18-102_k141_139936 | Not-determined |
| 18-102_k141_242136 | Not-determined |
| 18-102_k141_36307 | Low-quality |
| 18-102_k141_159915 | Not-determined |
| 18-102_k141_36309 | Not-determined |
| 18-102_k141_36310 | Not-determined |
| 18-102_k141_200445 | Not-determined |
| 18-102_k141_77404 | Not-determined |
| 18-102_k141_242144 | Not-determined |
| 18-102_k141_118802 | Not-determined |
| 18-102_k141_118803 | Not-determined |
| 18-102_k141_87998 | Not-determined |

| | |
|---|---|
| 18-102_k141_77408 | Not-determined |
| 18-102_k141_88002 | Not-determined |
| 18-102_k141_15143 | Not-determined |
| 18-102_k141_200451 | Not-determined |
| 18-102_k141_118805 | Low-quality |
| 18-102_k141_36318 | Not-determined |
| 18-102_k141_149840 | Not-determined |
| 18-102_k141_211649 | Not-determined |
| 18-102_k141_47962 | Not-determined |
| 18-102_k141_118812 | Not-determined |
| 18-102_k141_77433 | Not-determined |
| 18-102_k141_149845 | Not-determined |
| 18-102_k141_77437 | Not-determined |
| 18-102_k141_233079 | Not-determined |
| 18-102_k141_77439 | Not-determined |
| 18-102_k141_118828 | Not-determined |
| 18-102_k141_149850 | Not-determined |
| 18-102_k141_77441 | Not-determined |
| 18-102_k141_149851 | Not-determined |
| 18-102_k141_118831 | Not-determined |
| 18-102_k141_200465 | Not-determined |
| 18-102_k141_233085 | Low-quality |
| 18-102_k141_233087 | Low-quality |
| 18-102_k141_128500 | Not-determined |
| 18-102_k141_233089 | Not-determined |
| 18-102_k141_15147 | Not-determined |
| 18-102_k141_88033 | Not-determined |
| 18-102_k141_171144 | Not-determined |
| 18-102_k141_15153 | Not-determined |
| 18-102_k141_200471 | Not-determined |
| 18-102_k141_15168 | Not-determined |
| 18-102_k141_66884 | Not-determined |
| 18-102_k141_4390 | Not-determined |
| 18-102_k141_128531 | Low-quality |
| 18-102_k141_36355 | Not-determined |
| 18-102_k141_15178 | Not-determined |
| 18-102_k141_66899 | Not-determined |
| 18-102_k141_47981 | Not-determined |
| 18-102_k141_128547 | Not-determined |
| 18-102_k141_171167 | Not-determined |
| 18-102_k141_159938 | Not-determined |
| 18-102_k141_242170 | Not-determined |
| 18-102_k141_159946 | Not-determined |
| 18-102_k141_159951 | Not-determined |
| 18-102_k141_171186 | Not-determined |
| 18-102_k141_98611 | Not-determined |
| 18-102_k141_191343 | Not-determined |
| 18-102_k141_66943 | Not-determined |
| 18-102_k141_242180 | Not-determined |
| 18-102_k141_47997 | Not-determined |
| 18-102_k141_66952 | Not-determined |
| 18-102_k141_171199 | Not-determined |
| 18-102_k141_191349 | Not-determined |
| 18-102_k141_242187 | Not-determined |
| 18-102_k141_242188 | Not-determined |
| 18-102_k141_242191 | Not-determined |
| 18-102_k141_36361 | Not-determined |
| 18-102_k141_211697 | Not-determined |
| 18-102_k141_56984 | Not-determined |
| 18-102_k141_149873 | Not-determined |
| 18-102_k141_36367 | Not-determined |
| 18-102_k141_36368 | Not-determined |
| 18-102_k141_98631 | Not-determined |
| 18-102_k141_15238 | Not-determined |
| 18-102_k141_48023 | Not-determined |
| 18-102_k141_36371 | Not-determined |
| 18-102_k141_15240 | Not-determined |
| 18-102_k141_25394 | Not-determined |
| 18-102_k141_56990 | Not-determined |
| 18-102_k141_171206 | Low-quality |
| 18-102_k141_171207 | Not-determined |
| 18-102_k141_25402 | Not-determined |
| 18-102_k141_118866 | Not-determined |
| 18-102_k141_211719 | Not-determined |
| 18-102_k141_25410 | Not-determined |
| 18-102_k141_181559 | Not-determined |
| 18-102_k141_149890 | Not-determined |
| 18-102_k141_98651 | Not-determined |
| 18-102_k141_181570 | Not-determined |
| 18-102_k141_233102 | Not-determined |
| 18-102_k141_181573 | Not-determined |
| 18-102_k141_15251 | Not-determined |
| 18-102_k141_57013 | Not-determined |
| 18-102_k141_181576 | Not-determined |
| 18-102_k141_57016 | Low-quality |
| 18-102_k141_159985 | Not-determined |
| 18-102_k141_48039 | Not-determined |
| 18-102_k141_159987 | Not-determined |
| 18-102_k141_159988 | Not-determined |
| 18-102_k141_181581 | Not-determined |
| 18-102_k141_159992 | Not-determined |
| 18-102_k141_181583 | Not-determined |
| 18-102_k141_48047 | Not-determined |
| 18-102_k141_25421 | Not-determined |
| 18-102_k141_181585 | Not-determined |
| 18-102_k141_48053 | Not-determined |
| 18-102_k141_48054 | Not-determined |
| 18-102_k141_98657 | Not-determined |
| 18-102_k141_118869 | Not-determined |
| 18-102_k141_118872 | Not-determined |
| 18-102_k141_15269 | Not-determined |
| 18-102_k141_211761 | Not-determined |
| 18-102_k141_25436 | Not-determined |
| 18-102_k141_15274 | Not-determined |
| 18-102_k141_118881 | Not-determined |
| 18-102_k141_118884 | Not-determined |
| 18-102_k141_66993 | Not-determined |
| 18-102_k141_57044 | Not-determined |
| 18-102_k141_211768 | Not-determined |
| 18-102_k141_191378 | Low-quality |
| 18-102_k141_242237 | Not-determined |
| 18-102_k141_15285 | Not-determined |
| 18-102_k141_171219 | Not-determined |
| 18-102_k141_200520 | Not-determined |
| 18-102_k141_191384 | Not-determined |
| 18-102_k141_242246 | Not-determined |
| 18-102_k141_118906 | Not-determined |
| 18-102_k141_200523 | Not-determined |
| 18-102_k141_98671 | Not-determined |
| 18-102_k141_191390 | Not-determined |
| 18-102_k141_118908 | Not-determined |
| 18-102_k141_118909 | Not-determined |
| 18-102_k141_242251 | Not-determined |
| 18-102_k141_242252 | Not-determined |
| 18-102_k141_57067 | Not-determined |
| 18-102_k141_15301 | Not-determined |
| 18-102_k141_118912 | Not-determined |
| 18-102_k141_160025 | Low-quality |
| 18-102_k141_160027 | Not-determined |
| 18-102_k141_149938 | Not-determined |
| 18-102_k141_57077 | Not-determined |
| 18-102_k141_88078 | Not-determined |
| 18-102_k141_98697 | Not-determined |
| 18-102_k141_36430 | Low-quality |
| 18-102_k141_57084 | Not-determined |
| 18-102_k141_15322 | Not-determined |
| 18-102_k141_171226 | Not-determined |
| 18-102_k141_160037 | Not-determined |
| 18-102_k141_15328 | Not-determined |
| 18-102_k141_15329 | Not-determined |
| 18-102_k141_36433 | Not-determined |
| 18-102_k141_36434 | Not-determined |
| 18-102_k141_171228 | Not-determined |
| 18-102_k141_171229 | Low-quality |
| 18-102_k141_88090 | Not-determined |
| 18-102_k141_88092 | Not-determined |
| 18-102_k141_149951 | Not-determined |
| 18-102_k141_25451 | Not-determined |
| 18-102_k141_77497 | Not-determined |
| 18-102_k141_25452 | Not-determined |
| 18-102_k141_36444 | Not-determined |
| 18-102_k141_181602 | Not-determined |
| 18-102_k141_4395 | Not-determined |
| 18-102_k141_15333 | Not-determined |
| 18-102_k141_4399 | Not-determined |
| 18-102_k141_15334 | Not-determined |
| 18-102_k141_233153 | Not-determined |
| 18-102_k141_15335 | Not-determined |
| 18-102_k141_118947 | Not-determined |
| 18-102_k141_221529 | Not-determined |
| 18-102_k141_4406 | Not-determined |
| 18-102_k141_4409 | Not-determined |
| 18-102_k141_77509 | Not-determined |
| 18-102_k141_77511 | Not-determined |
| 18-102_k141_57092 | Not-determined |
| 18-102_k141_233161 | Not-determined |
| 18-102_k141_221537 | Not-determined |
| 18-102_k141_57098 | Not-determined |
| 18-102_k141_181615 | Not-determined |
| 18-102_k141_57099 | Not-determined |
| 18-102_k141_200545 | Not-determined |
| 18-102_k141_57102 | Not-determined |
| 18-102_k141_57103 | Not-determined |
| 18-102_k141_160061 | Not-determined |
| 18-102_k141_15361 | Not-determined |
| 18-102_k141_57105 | Not-determined |
| 18-102_k141_200548 | Not-determined |
| 18-102_k141_200549 | Low-quality |
| 18-102_k141_160067 | Not-determined |
| 18-102_k141_200552 | Not-determined |
| 18-102_k141_57110 | Not-determined |
| 18-102_k141_242258 | Not-determined |
| 18-102_k141_211798 | Low-quality |
| 18-102_k141_233182 | Not-determined |
| 18-102_k141_77541 | Not-determined |
| 18-102_k141_57112 | Not-determined |
| 18-102_k141_200558 | Low-quality |
| 18-102_k141_200559 | Not-determined |
| 18-102_k141_25471 | Not-determined |
| 18-102_k141_200567 | Not-determined |
| 18-102_k141_233201 | Not-determined |
| 18-102_k141_57145 | Not-determined |
| 18-102_k141_67046 | Low-quality |
| 18-102_k141_221565 | Low-quality |
| 18-102_k141_36472 | Not-determined |
| 18-102_k141_67051 | Not-determined |
| 18-102_k141_57149 | Not-determined |
| 18-102_k141_221571 | Not-determined |
| 18-102_k141_181650 | Not-determined |
| 18-102_k141_15391 | Not-determined |
| 18-102_k141_88136 | Not-determined |
| 18-102_k141_149985 | Not-determined |
| 18-102_k141_57165 | Not-determined |
| 18-102_k141_15401 | Low-quality |
| 18-102_k141_36483 | Not-determined |
| 18-102_k141_88142 | Not-determined |
| 18-102_k141_181654 | Low-quality |
| 18-102_k141_200583 | Not-determined |
| 18-102_k141_200585 | Not-determined |
| 18-102_k141_77580 | Not-determined |
| 18-102_k141_242288 | Not-determined |
| 18-102_k141_242290 | Not-determined |
| 18-102_k141_171248 | Low-quality |
| 18-102_k141_242296 | Not-determined |
| 18-102_k141_15412 | Not-determined |
| 18-102_k141_15414 | Not-determined |
| 18-102_k141_171255 | Not-determined |
| 18-102_k141_48155 | Not-determined |
| 18-102_k141_48157 | Not-determined |
| 18-102_k141_15416 | Not-determined |
| 18-102_k141_67086 | Not-determined |
| 18-102_k141_4416 | Not-determined |
| 18-102_k141_15423 | Not-determined |
| 18-102_k141_25485 | Low-quality |
| 18-102_k141_4420 | Low-quality |
| 18-102_k141_242314 | Not-determined |
| 18-102_k141_211833 | Not-determined |
| 18-102_k141_221593 | Not-determined |
| 18-102_k141_128568 | Not-determined |
| 18-102_k141_128571 | Low-quality |
| 18-102_k141_242318 | Not-determined |
| 18-102_k141_200608 | Not-determined |
| 18-102_k141_242319 | Not-determined |
| 18-102_k141_57190 | Not-determined |
| 18-102_k141_200610 | Not-determined |
| 18-102_k141_57195 | Not-determined |
| 18-102_k141_128573 | Not-determined |
| 18-102_k141_36507 | Not-determined |
| 18-102_k141_57209 | Not-determined |
| 18-102_k141_200618 | Not-determined |
| 18-102_k141_200619 | Not-determined |
| 18-102_k141_200622 | Not-determined |
| 18-102_k141_242338 | Not-determined |
| 18-102_k141_200631 | Not-determined |
| 18-102_k141_15433 | Not-determined |
| 18-102_k141_15435 | Not-determined |
| 18-102_k141_128581 | Not-determined |
| 18-102_k141_36521 | Not-determined |
| 18-102_k141_200639 | Not-determined |
| 18-102_k141_36523 | Not-determined |
| 18-102_k141_200640 | Not-determined |
| 18-102_k141_171312 | Not-determined |
| 18-102_k141_150048 | Not-determined |
| 18-102_k141_57237 | Low-quality |
| 18-102_k141_242353 | Not-determined |
| 18-102_k141_88196 | Not-determined |
| 18-102_k141_36536 | Not-determined |
| 18-102_k141_88208 | Not-determined |
| 18-102_k141_36539 | Not-determined |
| 18-102_k141_67095 | Not-determined |
| 18-102_k141_88216 | Not-determined |
| 18-102_k141_171338 | Not-determined |
| 18-102_k141_57246 | Not-determined |
| 18-102_k141_36564 | Not-determined |
| 18-102_k141_200657 | Not-determined |
| 18-102_k141_108366 | Not-determined |
| 18-102_k141_211856 | Not-determined |
| 18-102_k141_191455 | Not-determined |
| 18-102_k141_4439 | Not-determined |
| 18-102_k141_88226 | Not-determined |
| 18-102_k141_88227 | Not-determined |
| 18-102_k141_108376 | Low-quality |
| 18-102_k141_88228 | Not-determined |
| 18-102_k141_242380 | Low-quality |
| 18-102_k141_67109 | Not-determined |
| 18-102_k141_108378 | Not-determined |
| 18-102_k141_171359 | Not-determined |
| 18-102_k141_108386 | Not-determined |
| 18-102_k141_36581 | Not-determined |
| 18-102_k141_118982 | Not-determined |
| 18-102_k141_211879 | Not-determined |
| 18-102_k141_211883 | Not-determined |
| 18-102_k141_211886 | Not-determined |
| 18-102_k141_211887 | Not-determined |
| 18-102_k141_118992 | Not-determined |
| 18-102_k141_118996 | Not-determined |
| 18-102_k141_88243 | Not-determined |
| 18-102_k141_67140 | Not-determined |
| 18-102_k141_128628 | Not-determined |
| 18-102_k141_171376 | Not-determined |
| 18-102_k141_181671 | Not-determined |
| 18-102_k141_67142 | Not-determined |
| 18-102_k141_128630 | Not-determined |
| 18-102_k141_67146 | Not-determined |
| 18-102_k141_98711 | Not-determined |
| 18-102_k141_128637 | Not-determined |
| 18-102_k141_160125 | Not-determined |
| 18-102_k141_242392 | Not-determined |
| 18-102_k141_67156 | Not-determined |
| 18-102_k141_139957 | Not-determined |
| 18-102_k141_119013 | Not-determined |
| 18-102_k141_191493 | Not-determined |
| 18-102_k141_67166 | Not-determined |
| 18-102_k141_181692 | Low-quality |
| 18-102_k141_139965 | Not-determined |
| 18-102_k141_119023 | Not-determined |
| 18-102_k141_242395 | Not-determined |
| 18-102_k141_242398 | Not-determined |
| 18-102_k141_181702 | Not-determined |
| 18-102_k141_67176 | Not-determined |
| 18-102_k141_181706 | Not-determined |
| 18-102_k141_139977 | Not-determined |
| 18-102_k141_67183 | Not-determined |
| 18-102_k141_233308 | Not-determined |
| 18-102_k141_242404 | Not-determined |
| 18-102_k141_160136 | Not-determined |
| 18-102_k141_181716 | Not-determined |
| 18-102_k141_171412 | Not-determined |
| 18-102_k141_181717 | Not-determined |
| 18-102_k141_36612 | Not-determined |
| 18-102_k141_181720 | Not-determined |
| 18-102_k141_139990 | Not-determined |
| 18-102_k141_36614 | Not-determined |
| 18-102_k141_160137 | Not-determined |
| 18-102_k141_221625 | Low-quality |
| 18-102_k141_36619 | Not-determined |
| 18-102_k141_181734 | Not-determined |
| 18-102_k141_119048 | Not-determined |
| 18-102_k141_48164 | Not-determined |
| 18-102_k141_181736 | Not-determined |
| 18-102_k141_211897 | Not-determined |
| 18-102_k141_181739 | Not-determined |
| 18-102_k141_211901 | Not-determined |
| 18-102_k141_200678 | Not-determined |
| 18-102_k141_200680 | Not-determined |
| 18-102_k141_48168 | Not-determined |
| 18-102_k141_48169 | Not-determined |
| 18-102_k141_211919 | Not-determined |
| 18-102_k141_67205 | Not-determined |
| 18-102_k141_211921 | Not-determined |
| 18-102_k141_211923 | Not-determined |
| 18-102_k141_108425 | Not-determined |
| 18-102_k141_221647 | Not-determined |
| 18-102_k141_108427 | Not-determined |
| 18-102_k141_108430 | Not-determined |
| 18-102_k141_108431 | Not-determined |
| 18-102_k141_15444 | Not-determined |
| 18-102_k141_191553 | Not-determined |
| 18-102_k141_233326 | Not-determined |
| 18-102_k141_221658 | Not-determined |
| 18-102_k141_211929 | Low-quality |
| 18-102_k141_171433 | Not-determined |
| 18-102_k141_221661 | Not-determined |
| 18-102_k141_191564 | Not-determined |
| 18-102_k141_191566 | Not-determined |
| 18-102_k141_57268 | Not-determined |
| 18-102_k141_48197 | Not-determined |
| 18-102_k141_48199 | Not-determined |
| 18-102_k141_171445 | Not-determined |
| 18-102_k141_211943 | Not-determined |
| 18-102_k141_171447 | Not-determined |
| 18-102_k141_48204 | Not-determined |
| 18-102_k141_171449 | Low-quality |
| 18-102_k141_119079 | Not-determined |
| 18-102_k141_119084 | Not-determined |
| 18-102_k141_4464 | Not-determined |
| 18-102_k141_191581 | Not-determined |
| 18-102_k141_77655 | Not-determined |
| 18-102_k141_15461 | Not-determined |
| 18-102_k141_77656 | Not-determined |
| 18-102_k141_171468 | Not-determined |
| 18-102_k141_200719 | Not-determined |
| 18-102_k141_77663 | Not-determined |
| 18-102_k141_119099 | Not-determined |
| 18-102_k141_119100 | Not-determined |
| 18-102_k141_181764 | Not-determined |
| 18-102_k141_119103 | Low-quality |
| 18-102_k141_171481 | Not-determined |
| 18-102_k141_98782 | Not-determined |
| 18-102_k141_48214 | Not-determined |
| 18-102_k141_119110 | Not-determined |
| 18-102_k141_57272 | Not-determined |
| 18-102_k141_119111 | Not-determined |
| 18-102_k141_15473 | Not-determined |
| 18-102_k141_181775 | Not-determined |
| 18-102_k141_36650 | Not-determined |
| 18-102_k141_171494 | Low-quality |
| 18-102_k141_119114 | Not-determined |
| 18-102_k141_200740 | Low-quality |
| 18-102_k141_57282 | Low-quality |
| 18-102_k141_57283 | Not-determined |
| 18-102_k141_36655 | Not-determined |
| 18-102_k141_171498 | Low-quality |
| 18-102_k141_15485 | Not-determined |
| 18-102_k141_191610 | Low-quality |
| 18-102_k141_119123 | Not-determined |
| 18-102_k141_171505 | Not-determined |
| 18-102_k141_77669 | Not-determined |
| 18-102_k141_67223 | Not-determined |
| 18-102_k141_15497 | Not-determined |
| 18-102_k141_98803 | Low-quality |
| 18-102_k141_119135 | Not-determined |
| 18-102_k141_200754 | Not-determined |
| 18-102_k141_233394 | Not-determined |
| 18-102_k141_160161 | Not-determined |
| 18-102_k141_67228 | Not-determined |
| 18-102_k141_15505 | Not-determined |
| 18-102_k141_15506 | Not-determined |
| 18-102_k141_77681 | Not-determined |
| 18-102_k141_119140 | Not-determined |
| 18-102_k141_140033 | Not-determined |
| 18-102_k141_242466 | Not-determined |
| 18-102_k141_77689 | Not-determined |
| 18-102_k141_88303 | Not-determined |
| 18-102_k141_15520 | Not-determined |
| 18-102_k141_181786 | Not-determined |
| 18-102_k141_98810 | Low-quality |
| 18-102_k141_200766 | Not-determined |
| 18-102_k141_200767 | Not-determined |
| 18-102_k141_67241 | Not-determined |
| 18-102_k141_67246 | Not-determined |
| 18-102_k141_98816 | Not-determined |
| 18-102_k141_88310 | Not-determined |
| 18-102_k141_181801 | Not-determined |
| 18-102_k141_150114 | Not-determined |
| 18-102_k141_88316 | Low-quality |
| 18-102_k141_88319 | Not-determined |
| 18-102_k141_4488 | Not-determined |
| 18-102_k141_15538 | Not-determined |
| 18-102_k141_98834 | Not-determined |
| 18-102_k141_48265 | Not-determined |
| 18-102_k141_15555 | Low-quality |
| 18-102_k141_233419 | Not-determined |
| 18-102_k141_242484 | Not-determined |
| 18-102_k141_4502 | Not-determined |
| 18-102_k141_140059 | Not-determined |
| 18-102_k141_181806 | Not-determined |
| 18-102_k141_150134 | Not-determined |
| 18-102_k141_200788 | Not-determined |
| 18-102_k141_200791 | Not-determined |
| 18-102_k141_150159 | Not-determined |
| 18-102_k141_150168 | Not-determined |
| 18-102_k141_98856 | Not-determined |
| 18-102_k141_77758 | Not-determined |
| 18-102_k141_88334 | Not-determined |
| 18-102_k141_242513 | Low-quality |
| 18-102_k141_15582 | Not-determined |
| 18-102_k141_242515 | Not-determined |
| 18-102_k141_98861 | Low-quality |
| 18-102_k141_15584 | Not-determined |
| 18-102_k141_36730 | Low-quality |
| 18-102_k141_171557 | Low-quality |
| 18-102_k141_98865 | Not-determined |
| 18-102_k141_15588 | Not-determined |
| 18-102_k141_181818 | Not-determined |
| 18-102_k141_233439 | Not-determined |
| 18-102_k141_77769 | Not-determined |
| 18-102_k141_4507 | Not-determined |
| 18-102_k141_233445 | Not-determined |
| 18-102_k141_36738 | Not-determined |
| 18-102_k141_98887 | Not-determined |
| 18-102_k141_77782 | Not-determined |
| 18-102_k141_150196 | Low-quality |
| 18-102_k141_77795 | Not-determined |
| 18-102_k141_57302 | Not-determined |
| 18-102_k141_181833 | Not-determined |
| 18-102_k141_200811 | Not-determined |
| 18-102_k141_36747 | Not-determined |
| 18-102_k141_212024 | Low-quality |
| 18-102_k141_88368 | Not-determined |
| 18-102_k141_191631 | Not-determined |
| 18-102_k141_212025 | Not-determined |
| 18-102_k141_88371 | Not-determined |
| 18-102_k141_191632 | Not-determined |
| 18-102_k141_150206 | Not-determined |
| 18-102_k141_119186 | Not-determined |
| 18-102_k141_67277 | Low-quality |
| 18-102_k141_67279 | Low-quality |
| 18-102_k141_242534 | Not-determined |
| 18-102_k141_77800 | Not-determined |
| 18-102_k141_160214 | Not-determined |
| 18-102_k141_242535 | Not-determined |
| 18-102_k141_140094 | Not-determined |
| 18-102_k141_88391 | Not-determined |
| 18-102_k141_233477 | Not-determined |
| 18-102_k141_233479 | Low-quality |
| 18-102_k141_48324 | Low-quality |
| 18-102_k141_88395 | Not-determined |
| 18-102_k141_36771 | Low-quality |
| 18-102_k141_119208 | Not-determined |
| 18-102_k141_221752 | Not-determined |
| 18-102_k141_57330 | Not-determined |
| 18-102_k141_150232 | Not-determined |
| 18-102_k141_221755 | Not-determined |
| 18-102_k141_191664 | Not-determined |
| 18-102_k141_77814 | Not-determined |
| 18-102_k141_36778 | Not-determined |
| 18-102_k141_233493 | Not-determined |
| 18-102_k141_36781 | Not-determined |
| 18-102_k141_48343 | Not-determined |
| 18-102_k141_67303 | Low-quality |
| 18-102_k141_140117 | Not-determined |
| 18-102_k141_128646 | Not-determined |
| 18-102_k141_128649 | Low-quality |
| 18-102_k141_140122 | Not-determined |
| 18-102_k141_15598 | Not-determined |
| 18-102_k141_212044 | Not-determined |
| 18-102_k141_242552 | Not-determined |
| 18-102_k141_233508 | Low-quality |
| 18-102_k141_57351 | Not-determined |
| 18-102_k141_233513 | Low-quality |
| 18-102_k141_191681 | Not-determined |
| 18-102_k141_119238 | Low-quality |
| 18-102_k141_212060 | Not-determined |
| 18-102_k141_119239 | Not-determined |
| 18-102_k141_242561 | Low-quality |
| 18-102_k141_36797 | Low-quality |
| 18-102_k141_15609 | Low-quality |
| 18-102_k141_15612 | Not-determined |
| 18-102_k141_48350 | Not-determined |
| 18-102_k141_57369 | Not-determined |
| 18-102_k141_119253 | Not-determined |
| 18-102_k141_48351 | Not-determined |
| 18-102_k141_48352 | Not-determined |
| 18-102_k141_140148 | Not-determined |
| 18-102_k141_48355 | Low-quality |
| 18-102_k141_36804 | Not-determined |
| 18-102_k141_77834 | Not-determined |
| 18-102_k141_77836 | Not-determined |
| 18-102_k141_119259 | Not-determined |
| 18-102_k141_15620 | Not-determined |
| 18-102_k141_242572 | Not-determined |
| 18-102_k141_191695 | Not-determined |
| 18-102_k141_119263 | Not-determined |
| 18-102_k141_160220 | Not-determined |
| 18-102_k141_200858 | Not-determined |
| 18-102_k141_108470 | Not-determined |
| 18-102_k141_150263 | Not-determined |
| 18-102_k141_48376 | Not-determined |
| 18-102_k141_150268 | Not-determined |
| 18-102_k141_160226 | Not-determined |
| 18-102_k141_140179 | Not-determined |
| 18-102_k141_191706 | Not-determined |
| 18-102_k141_150273 | Not-determined |
| 18-102_k141_191708 | Not-determined |
| 18-102_k141_140182 | Not-determined |
| 18-102_k141_150276 | Not-determined |
| 18-102_k141_57399 | Not-determined |
| 18-102_k141_57400 | Low-quality |
| 18-102_k141_242593 | Not-determined |
| 18-102_k141_150277 | Not-determined |
| 18-102_k141_36837 | Not-determined |
| 18-102_k141_171586 | Not-determined |
| 18-102_k141_36839 | Not-determined |
| 18-102_k141_77846 | Not-determined |
| 18-102_k141_150279 | Low-quality |
| 18-102_k141_200872 | Not-determined |
| 18-102_k141_77851 | Not-determined |
| 18-102_k141_77853 | Not-determined |
| 18-102_k141_119282 | Not-determined |
| 18-102_k141_128717 | Not-determined |
| 18-102_k141_119285 | Not-determined |
| 18-102_k141_48414 | Not-determined |
| 18-102_k141_77867 | Not-determined |
| 18-102_k141_77869 | Not-determined |
| 18-102_k141_57408 | Not-determined |
| 18-102_k141_88430 | Not-determined |
| 18-102_k141_36848 | Not-determined |
| 18-102_k141_36851 | Not-determined |
| 18-102_k141_77874 | Not-determined |
| 18-102_k141_88438 | Not-determined |
| 18-102_k141_140200 | Low-quality |
| 18-102_k141_88440 | Not-determined |
| 18-102_k141_128728 | Low-quality |
| 18-102_k141_212087 | Not-determined |
| 18-102_k141_242612 | Not-determined |
| 18-102_k141_57422 | Not-determined |
| 18-102_k141_88450 | Not-determined |
| 18-102_k141_57424 | Not-determined |
| 18-102_k141_36861 | Not-determined |
| 18-102_k141_242619 | Not-determined |
| 18-102_k141_140207 | Not-determined |
| 18-102_k141_242620 | Not-determined |
| 18-102_k141_36866 | Not-determined |
| 18-102_k141_57429 | Not-determined |
| 18-102_k141_57431 | Not-determined |
| 18-102_k141_119321 | Not-determined |
| 18-102_k141_15638 | Not-determined |
| 18-102_k141_88464 | Not-determined |

| | |
|---|---|
| 18-102_k141_108495 | Not-determined |
| 18-102_k141_119324 | Not-determined |
| 18-102_k141_108497 | Not-determined |
| 18-102_k141_140212 | Not-determined |
| 18-102_k141_98889 | Low-quality |
| 18-102_k141_98890 | Not-determined |
| 18-102_k141_212095 | Low-quality |
| 18-102_k141_108501 | Not-determined |
| 18-102_k141_88475 | Not-determined |
| 18-102_k141_77892 | Not-determined |
| 18-102_k141_119330 | Not-determined |
| 18-102_k141_88479 | Not-determined |
| 18-102_k141_15654 | Not-determined |
| 18-102_k141_98896 | Not-determined |
| 18-102_k141_233545 | Not-determined |
| 18-102_k141_15658 | Not-determined |
| 18-102_k141_108507 | Not-determined |
| 18-102_k141_57453 | Low-quality |
| 18-102_k141_242642 | Not-determined |
| 18-102_k141_108511 | Not-determined |
| 18-102_k141_88484 | Not-determined |
| 18-102_k141_200898 | Not-determined |
| 18-102_k141_88488 | Not-determined |
| 18-102_k141_119339 | Not-determined |
| 18-102_k141_212115 | Not-determined |
| 18-102_k141_200902 | Low-quality |
| 18-102_k141_200903 | Not-determined |
| 18-102_k141_119343 | Not-determined |
| 18-102_k141_88493 | Not-determined |
| 18-102_k141_171641 | Not-determined |
| 18-102_k141_233555 | Not-determined |
| 18-102_k141_140227 | Not-determined |
| 18-102_k141_150290 | Low-quality |
| 18-102_k141_57462 | Low-quality |
| 18-102_k141_233563 | Not-determined |
| 18-102_k141_140231 | Not-determined |
| 18-102_k141_191718 | Not-determined |
| 18-102_k141_77912 | Not-determined |
| 18-102_k141_119356 | Not-determined |
| 18-102_k141_200913 | Not-determined |
| 18-102_k141_233574 | Not-determined |
| 18-102_k141_77922 | Not-determined |
| 18-102_k141_140245 | Not-determined |
| 18-102_k141_119362 | Low-quality |
| 18-102_k141_88510 | Not-determined |
| 18-102_k141_140249 | Not-determined |
| 18-102_k141_98926 | Low-quality |
| 18-102_k141_77928 | Not-determined |
| 18-102_k141_200921 | Not-determined |
| 18-102_k141_88518 | Low-quality |
| 18-102_k141_88521 | Low-quality |
| 18-102_k141_88523 | Not-determined |
| 18-102_k141_200925 | Not-determined |
| 18-102_k141_191732 | Not-determined |
| 18-102_k141_242646 | Not-determined |
| 18-102_k141_36899 | Not-determined |
| 18-102_k141_242649 | Not-determined |
| 18-102_k141_233589 | Not-determined |
| 18-102_k141_57484 | Not-determined |
| 18-102_k141_88532 | Not-determined |
| 18-102_k141_57487 | Not-determined |
| 18-102_k141_242655 | Not-determined |
| 18-102_k141_108526 | Not-determined |
| 18-102_k141_242658 | Not-determined |
| 18-102_k141_88543 | Not-determined |
| 18-102_k141_242661 | Not-determined |
| 18-102_k141_57493 | Not-determined |
| 18-102_k141_77940 | Not-determined |
| 18-102_k141_242668 | Not-determined |
| 18-102_k141_119390 | Not-determined |
| 18-102_k141_98949 | Not-determined |
| 18-102_k141_150332 | Low-quality |
| 18-102_k141_171671 | Not-determined |
| 18-102_k141_98959 | Not-determined |
| 18-102_k141_98960 | Not-determined |
| 18-102_k141_150337 | Low-quality |
| 18-102_k141_88560 | Not-determined |
| 18-102_k141_119402 | Not-determined |
| 18-102_k141_88567 | Not-determined |
| 18-102_k141_57511 | Low-quality |
| 18-102_k141_57513 | Not-determined |
| 18-102_k141_77977 | Low-quality |
| 18-102_k141_140285 | Not-determined |
| 18-102_k141_212122 | Not-determined |
| 18-102_k141_15703 | Not-determined |
| 18-102_k141_88580 | Not-determined |
| 18-102_k141_88581 | Not-determined |
| 18-102_k141_140294 | Not-determined |
| 18-102_k141_108563 | Not-determined |
| 18-102_k141_171693 | Not-determined |
| 18-102_k141_171694 | Not-determined |
| 18-102_k141_57528 | Not-determined |
| 18-102_k141_108564 | Not-determined |
| 18-102_k141_242693 | Not-determined |
| 18-102_k141_108565 | Not-determined |
| 18-102_k141_140295 | Low-quality |
| 18-102_k141_88587 | Not-determined |
| 18-102_k141_57531 | Not-determined |
| 18-102_k141_57532 | Not-determined |
| 18-102_k141_181879 | Not-determined |
| 18-102_k141_57533 | Not-determined |
| 18-102_k141_88590 | Not-determined |
| 18-102_k141_57534 | Low-quality |
| 18-102_k141_88595 | Not-determined |
| 18-102_k141_233644 | Not-determined |
| 18-102_k141_171711 | Not-determined |
| 18-102_k141_200957 | Not-determined |
| 18-102_k141_233649 | Not-determined |
| 18-102_k141_108570 | Not-determined |
| 18-102_k141_233654 | Not-determined |
| 18-102_k141_233655 | Not-determined |
| 18-102_k141_88606 | Not-determined |
| 18-102_k141_171719 | Not-determined |
| 18-102_k141_108575 | Not-determined |
| 18-102_k141_233662 | Not-determined |
| 18-102_k141_200968 | Low-quality |
| 18-102_k141_57552 | Not-determined |
| 18-102_k141_57554 | Not-determined |
| 18-102_k141_88620 | Not-determined |
| 18-102_k141_171735 | Not-determined |
| 18-102_k141_181897 | Not-determined |
| 18-102_k141_77998 | Not-determined |
| 18-102_k141_171737 | Low-quality |
| 18-102_k141_88626 | Low-quality |
| 18-102_k141_181899 | Not-determined |
| 18-102_k141_4606 | Not-determined |
| 18-102_k141_150348 | Not-determined |
| 18-102_k141_15710 | Not-determined |
| 18-102_k141_242706 | Not-determined |
| 18-102_k141_191747 | Not-determined |
| 18-102_k141_150353 | Not-determined |
| 18-102_k141_140318 | Not-determined |
| 18-102_k141_191750 | Not-determined |
| 18-102_k141_212143 | Not-determined |
| 18-102_k141_221774 | Not-determined |
| 18-102_k141_88633 | Not-determined |
| 18-102_k141_212150 | Not-determined |
| 18-102_k141_140326 | Not-determined |
| 18-102_k141_119463 | Low-quality |
| 18-102_k141_78005 | Not-determined |
| 18-102_k141_88639 | Not-determined |
| 18-102_k141_221776 | Not-determined |
| 18-102_k141_78007 | Not-determined |
| 18-102_k141_171756 | Not-determined |
| 18-102_k141_108610 | Not-determined |
| 18-102_k141_108611 | Not-determined |
| 18-102_k141_221782 | Not-determined |
| 18-102_k141_108613 | Not-determined |
| 18-102_k141_88650 | Not-determined |
| 18-102_k141_88652 | Not-determined |
| 18-102_k141_78026 | Not-determined |
| 18-102_k141_221783 | Not-determined |
| 18-102_k141_119474 | Low-quality |
| 18-102_k141_78029 | Low-quality |
| 18-102_k141_108625 | Not-determined |
| 18-102_k141_78033 | Not-determined |
| 18-102_k141_119482 | Not-determined |
| 18-102_k141_221797 | Not-determined |
| 18-102_k141_171788 | Not-determined |
| 18-102_k141_88662 | Not-determined |
| 18-102_k141_119486 | Low-quality |
| 18-102_k141_191769 | Not-determined |
| 18-102_k141_48482 | Not-determined |
| 18-102_k141_78041 | Not-determined |
| 18-102_k141_78042 | Low-quality |
| 18-102_k141_78049 | Not-determined |
| 18-102_k141_36932 | Not-determined |
| 18-102_k141_150385 | Not-determined |
| 18-102_k141_108642 | Not-determined |
| 18-102_k141_67333 | Not-determined |
| 18-102_k141_160282 | Low-quality |
| 18-102_k141_15762 | Not-determined |
| 18-102_k141_171799 | Not-determined |
| 18-102_k141_78069 | Not-determined |
| 18-102_k141_150396 | Low-quality |
| 18-102_k141_78072 | Not-determined |
| 18-102_k141_15773 | Not-determined |
| 18-102_k141_15774 | Not-determined |
| 18-102_k141_171809 | Not-determined |
| 18-102_k141_119514 | Not-determined |
| 18-102_k141_78080 | Not-determined |
| 18-102_k141_108646 | Not-determined |
| 18-102_k141_221843 | Not-determined |
| 18-102_k141_160302 | Not-determined |
| 18-102_k141_221845 | Low-quality |
| 18-102_k141_201026 | Not-determined |
| 18-102_k141_191778 | Not-determined |
| 18-102_k141_191785 | Not-determined |
| 18-102_k141_150417 | Not-determined |
| 18-102_k141_233704 | Not-determined |
| 18-102_k141_201034 | Not-determined |
| 18-102_k141_119518 | Not-determined |
| 18-102_k141_233711 | Not-determined |
| 18-102_k141_78103 | Not-determined |
| 18-102_k141_150428 | Not-determined |
| 18-102_k141_150429 | Not-determined |
| 18-102_k141_88690 | Low-quality |
| 18-102_k141_140362 | Not-determined |
| 18-102_k141_98978 | Not-determined |
| 18-102_k141_233718 | Not-determined |
| 18-102_k141_140367 | Not-determined |
| 18-102_k141_242733 | Not-determined |
| 18-102_k141_201042 | Not-determined |
| 18-102_k141_233723 | Low-quality |
| 18-102_k141_150437 | Not-determined |
| 18-102_k141_88701 | Not-determined |
| 18-102_k141_88703 | Not-determined |
| 18-102_k141_78116 | Not-determined |
| 18-102_k141_242736 | Not-determined |
| 18-102_k141_242737 | Not-determined |
| 18-102_k141_242741 | Not-determined |
| 18-102_k141_98988 | Not-determined |
| 18-102_k141_88710 | Not-determined |
| 18-102_k141_221863 | Not-determined |
| 18-102_k141_221872 | Not-determined |
| 18-102_k141_36953 | Not-determined |
| 18-102_k141_221880 | Not-determined |
| 18-102_k141_150456 | Not-determined |
| 18-102_k141_36961 | Not-determined |
| 18-102_k141_36962 | Not-determined |
| 18-102_k141_78127 | Not-determined |
| 18-102_k141_201070 | Not-determined |
| 18-102_k141_88733 | Not-determined |
| 18-102_k141_150470 | Low-quality |
| 18-102_k141_201077 | Not-determined |
| 18-102_k141_150475 | Not-determined |
| 18-102_k141_150476 | Not-determined |
| 18-102_k141_88743 | Not-determined |
| 18-102_k141_15821 | Not-determined |
| 18-102_k141_78150 | Not-determined |
| 18-102_k141_108649 | Not-determined |
| 18-102_k141_48521 | Not-determined |
| 18-102_k141_108651 | Not-determined |
| 18-102_k141_88759 | Not-determined |
| 18-102_k141_119554 | Not-determined |
| 18-102_k141_88760 | Not-determined |
| 18-102_k141_119555 | Low-quality |
| 18-102_k141_191830 | Low-quality |
| 18-102_k141_140392 | Not-determined |
| 18-102_k141_191833 | Low-quality |
| 18-102_k141_108662 | Not-determined |
| 18-102_k141_78162 | Not-determined |
| 18-102_k141_48532 | Not-determined |
| 18-102_k141_57565 | Not-determined |
| 18-102_k141_140407 | Low-quality |
| 18-102_k141_48539 | Low-quality |
| 18-102_k141_140408 | Not-determined |
| 18-102_k141_191835 | Not-determined |
| 18-102_k141_88766 | Not-determined |
| 18-102_k141_4683 | Not-determined |
| 18-102_k141_140415 | Not-determined |
| 18-102_k141_221913 | Not-determined |
| 18-102_k141_221915 | Not-determined |
| 18-102_k141_201090 | Not-determined |
| 18-102_k141_67385 | Low-quality |
| 18-102_k141_221918 | Not-determined |
| 18-102_k141_88770 | Not-determined |
| 18-102_k141_160320 | Not-determined |
| 18-102_k141_221928 | Not-determined |
| 18-102_k141_171843 | Not-determined |
| 18-102_k141_171849 | Not-determined |
| 18-102_k141_221935 | Low-quality |
| 18-102_k141_128824 | Not-determined |
| 18-102_k141_221937 | Not-determined |
| 18-102_k141_181924 | Not-determined |
| 18-102_k141_48569 | Not-determined |
| 18-102_k141_128825 | Low-quality |
| 18-102_k141_78187 | Not-determined |
| 18-102_k141_233761 | Low-quality |
| 18-102_k141_140426 | Not-determined |
| 18-102_k141_150488 | Not-determined |
| 18-102_k141_160331 | Not-determined |
| 18-102_k141_233770 | Not-determined |
| 18-102_k141_15852 | Not-determined |
| 18-102_k141_160334 | Not-determined |
| 18-102_k141_191843 | Not-determined |
| 18-102_k141_108697 | Not-determined |
| 18-102_k141_160339 | Low-quality |
| 18-102_k141_150497 | Not-determined |
| 18-102_k141_201102 | Low-quality |
| 18-102_k141_99035 | Low-quality |
| 18-102_k141_201106 | Not-determined |
| 18-102_k141_48584 | Not-determined |
| 18-102_k141_150509 | Not-determined |
| 18-102_k141_201112 | Not-determined |
| 18-102_k141_201116 | Not-determined |
| 18-102_k141_201119 | Not-determined |
| 18-102_k141_150513 | Not-determined |
| 18-102_k141_221951 | Not-determined |
| 18-102_k141_181957 | Not-determined |
| 18-102_k141_36984 | Not-determined |
| 18-102_k141_221952 | Not-determined |
| 18-102_k141_57592 | Not-determined |
| 18-102_k141_150517 | Not-determined |
| 18-102_k141_233784 | Not-determined |
| 18-102_k141_181963 | Not-determined |
| 18-102_k141_15860 | Not-determined |
| 18-102_k141_15862 | Not-determined |
| 18-102_k141_15867 | Not-determined |
| 18-102_k141_15870 | Not-determined |
| 18-102_k141_140453 | Not-determined |
| 18-102_k141_99062 | Not-determined |
| 18-102_k141_191885 | Not-determined |
| 18-102_k141_201128 | Not-determined |
| 18-102_k141_191890 | Not-determined |
| 18-102_k141_99073 | Not-determined |
| 18-102_k141_201130 | Not-determined |
| 18-102_k141_108712 | Not-determined |
| 18-102_k141_48597 | Not-determined |
| 18-102_k141_181984 | Not-determined |
| 18-102_k141_140464 | Not-determined |
| 18-102_k141_171900 | Not-determined |
| 18-102_k141_99079 | Low-quality |
| 18-102_k141_150524 | Not-determined |
| 18-102_k141_78215 | Not-determined |
| 18-102_k141_212159 | Not-determined |
| 18-102_k141_99080 | Not-determined |
| 18-102_k141_15894 | Not-determined |
| 18-102_k141_140474 | Not-determined |
| 18-102_k141_15898 | Not-determined |
| 18-102_k141_99083 | Not-determined |
| 18-102_k141_201151 | Not-determined |
| 18-102_k141_233820 | Not-determined |
| 18-102_k141_119625 | Not-determined |
| 18-102_k141_140484 | Not-determined |
| 18-102_k141_48621 | Not-determined |
| 18-102_k141_15910 | Not-determined |
| 18-102_k141_242760 | Not-determined |
| 18-102_k141_15914 | Not-determined |
| 18-102_k141_4687 | Not-determined |
| 18-102_k141_99098 | Not-determined |
| 18-102_k141_15918 | Not-determined |
| 18-102_k141_99100 | Not-determined |
| 18-102_k141_119633 | Not-determined |
| 18-102_k141_171910 | Not-determined |
| 18-102_k141_119637 | Low-quality |
| 18-102_k141_99103 | Low-quality |
| 18-102_k141_119645 | Not-determined |
| 18-102_k141_182003 | Not-determined |
| 18-102_k141_201169 | Not-determined |
| 18-102_k141_191906 | Not-determined |
| 18-102_k141_150532 | Low-quality |
| 18-102_k141_233838 | Not-determined |
| 18-102_k141_119654 | Not-determined |
| 18-102_k141_140501 | Not-determined |
| 18-102_k141_212182 | Not-determined |
| 18-102_k141_140512 | Low-quality |
| 18-102_k141_171946 | Not-determined |
| 18-102_k141_99117 | Not-determined |
| 18-102_k141_140515 | Not-determined |
| 18-102_k141_4703 | Not-determined |
| 18-102_k141_233860 | Not-determined |
| 18-102_k141_4704 | Not-determined |
| 18-102_k141_108774 | Not-determined |
| 18-102_k141_233865 | Not-determined |
| 18-102_k141_108778 | Not-determined |
| 18-102_k141_212200 | Not-determined |
| 18-102_k141_99125 | Not-determined |
| 18-102_k141_221991 | Not-determined |
| 18-110-final-viral-combined_k141_40673\|\|0_partial | Medium-quality |
| 18-102_k141_171959 | Not-determined |
| 18-102_k141_191912 | Not-determined |
| 18-102_k141_150551 | Not-determined |
| 18-102_k141_4711 | Not-determined |
| 18-102_k141_4716 | Not-determined |
| 18-102_k141_150557 | Not-determined |
| 18-102_k141_182021 | Not-determined |
| 18-102_k141_191922 | Not-determined |
| 18-102_k141_182023 | Not-determined |
| 18-102_k141_119685 | Not-determined |
| 18-102_k141_119688 | Not-determined |
| 18-102_k141_36995 | Not-determined |
| 18-102_k141_36997 | Low-quality |
| 18-102_k141_201195 | Not-determined |
| 18-102_k141_48647 | Not-determined |
| 18-102_k141_128863 | Low-quality |
| 18-102_k141_119696 | Low-quality |
| 18-102_k141_191933 | Not-determined |
| 18-102_k141_171973 | Not-determined |
| 18-102_k141_57611 | Not-determined |
| 18-102_k141_212219 | Not-determined |
| 18-102_k141_57613 | Not-determined |
| 18-102_k141_57618 | Not-determined |
| 18-102_k141_88777 | Not-determined |
| 18-102_k141_212228 | Not-determined |
| 18-102_k141_57626 | Not-determined |
| 18-102_k141_48665 | Not-determined |
| 18-102_k141_88778 | Not-determined |
| 18-102_k141_48668 | Not-determined |
| 18-102_k141_171978 | Not-determined |
| 18-102_k141_128885 | Not-determined |
| 18-102_k141_78244 | Not-determined |
| 18-102_k141_128887 | Not-determined |
| 18-102_k141_171984 | Not-determined |
| 18-102_k141_67424 | Not-determined |
| 18-102_k141_37031 | Not-determined |
| 18-102_k141_191950 | Not-determined |
| 18-102_k141_15935 | Not-determined |
| 18-102_k141_57641 | Not-determined |
| 18-102_k141_242814 | Not-determined |
| 18-102_k141_182053 | Not-determined |
| 18-102_k141_48688 | Not-determined |
| 18-102_k141_48691 | Not-determined |
| 18-102_k141_128909 | Low-quality |
| 18-102_k141_191957 | Low-quality |
| 18-102_k141_78271 | Not-determined |
| 18-102_k141_57648 | Not-determined |
| 18-102_k141_242825 | Not-determined |
| 18-102_k141_128918 | Low-quality |
| 18-102_k141_48700 | Not-determined |
| 18-102_k141_119712 | Not-determined |
| 18-102_k141_37059 | Not-determined |
| 18-102_k141_37061 | Not-determined |
| 18-102_k141_48707 | Not-determined |
| 18-102_k141_48708 | Not-determined |
| 18-102_k141_15944 | Not-determined |
| 18-102_k141_78288 | Not-determined |
| 18-102_k141_119720 | Not-determined |
| 18-102_k141_119722 | Not-determined |
| 18-102_k141_128939 | Low-quality |
| 18-102_k141_150570 | Not-determined |
| 18-102_k141_150574 | Not-determined |
| 18-102_k141_150578 | Low-quality |
| 18-102_k141_242851 | Not-determined |
| 18-102_k141_78296 | Not-determined |
| 18-102_k141_48722 | Not-determined |
| 18-102_k141_15959 | Not-determined |
| 18-102_k141_48726 | Not-determined |
| 18-102_k141_15960 | Not-determined |
| 18-102_k141_150598 | Not-determined |
| 18-102_k141_57666 | Not-determined |
| 18-102_k141_140547 | Not-determined |
| 18-102_k141_172063 | Not-determined |
| 18-102_k141_37076 | Not-determined |
| 18-102_k141_88831 | Not-determined |
| 18-102_k141_15971 | Not-determined |
| 18-102_k141_48737 | Not-determined |
| 18-102_k141_172072 | Not-determined |
| 18-102_k141_88841 | Not-determined |
| 18-102_k141_182116 | Not-determined |
| 18-102_k141_15982 | Not-determined |
| 18-102_k141_88843 | Not-determined |
| 18-102_k141_48750 | Not-determined |
| 18-102_k141_119739 | Low-quality |
| 18-102_k141_150614 | Not-determined |
| 18-102_k141_150615 | Not-determined |
| 18-102_k141_37091 | Not-determined |
| 18-102_k141_182126 | Not-determined |
| 18-102_k141_140572 | Not-determined |
| 18-102_k141_140575 | Not-determined |
| 18-102_k141_150620 | Not-determined |
| 18-102_k141_99155 | Low-quality |
| 18-102_k141_67467 | Low-quality |
| 18-102_k141_48772 | Not-determined |
| 18-102_k141_88860 | Not-determined |
| 18-102_k141_48774 | Not-determined |
| 18-102_k141_78322 | Not-determined |
| 18-102_k141_78323 | Not-determined |
| 18-102_k141_140588 | Not-determined |
| 18-102_k141_191980 | Not-determined |
| 18-102_k141_191982 | Not-determined |
| 18-102_k141_48787 | Not-determined |
| 18-102_k141_140594 | Low-quality |
| 18-102_k141_172090 | Not-determined |
| 18-102_k141_67491 | Not-determined |
| 18-102_k141_233885 | Not-determined |
| 18-102_k141_150653 | Not-determined |
| 18-102_k141_78357 | Not-determined |
| 18-102_k141_119759 | Not-determined |
| 18-102_k141_48798 | Not-determined |
| 18-102_k141_48799 | Not-determined |
| 18-102_k141_140604 | Not-determined |
| 18-102_k141_191990 | Low-quality |
| 18-102_k141_48801 | Not-determined |
| 18-102_k141_182158 | Not-determined |
| 18-102_k141_191993 | Not-determined |
| 18-102_k141_119772 | Not-determined |
| 18-102_k141_140613 | Not-determined |
| 18-102_k141_25559 | Not-determined |
| 18-102_k141_182169 | Not-determined |
| 18-102_k141_67498 | Not-determined |
| 18-102_k141_48828 | Low-quality |
| 18-102_k141_16037 | Not-determined |
| 18-102_k141_172107 | Not-determined |
| 18-102_k141_48832 | Not-determined |
| 18-102_k141_78385 | Not-determined |
| 18-102_k141_88903 | Not-determined |
| 18-102_k141_16050 | Not-determined |
| 18-102_k141_242899 | Not-determined |
| 18-102_k141_108851 | Not-determined |
| 18-102_k141_48838 | Not-determined |
| 18-102_k141_160392 | Not-determined |
| 18-102_k141_119797 | Not-determined |
| 18-102_k141_78393 | Not-determined |
| 18-102_k141_88916 | Not-determined |
| 18-102_k141_88921 | Not-determined |
| 18-102_k141_4742 | Not-determined |
| 18-102_k141_160406 | Not-determined |
| 18-102_k141_119801 | Not-determined |
| 18-102_k141_67526 | Not-determined |
| 18-102_k141_192005 | Not-determined |
| 18-102_k141_160411 | Not-determined |
| 18-102_k141_48850 | Not-determined |
| 18-102_k141_48851 | Not-determined |
| 18-102_k141_201247 | Not-determined |
| 18-102_k141_67532 | Not-determined |
| 18-102_k141_201249 | Not-determined |
| 18-102_k141_67540 | Not-determined |
| 18-102_k141_25568 | Not-determined |
| 18-102_k141_99189 | Not-determined |
| 18-102_k141_37118 | Not-determined |
| 18-102_k141_4755 | Not-determined |
| 18-102_k141_108885 | Low-quality |
| 18-102_k141_37127 | Low-quality |
| 18-102_k141_48876 | Not-determined |
| 18-102_k141_150714 | Not-determined |
| 18-102_k141_99194 | Low-quality |
| 18-102_k141_140653 | Low-quality |
| 18-102_k141_99198 | Not-determined |
| 18-102_k141_37142 | Low-quality |
| 18-102_k141_48889 | Not-determined |
| 18-102_k141_160441 | Low-quality |
| 18-102_k141_37149 | Not-determined |
| 18-102_k141_99215 | Not-determined |
| 18-102_k141_37154 | Not-determined |
| 18-102_k141_37160 | Not-determined |
| 18-102_k141_37161 | Not-determined |
| 18-102_k141_78415 | Not-determined |
| 18-102_k141_150741 | Not-determined |
| 18-102_k141_119832 | Not-determined |
| 18-102_k141_119843 | Low-quality |
| 18-102_k141_57713 | Not-determined |
| 18-102_k141_99228 | Not-determined |
| 18-102_k141_140679 | Not-determined |
| 18-106-final-viral-combined_k141_128972\|\|full | Medium-quality |
| 18-102_k141_99233 | Not-determined |
| 18-102_k141_48928 | Not-determined |
| 18-102_k141_99241 | Not-determined |
| 18-102_k141_99243 | Not-determined |
| 18-102_k141_37178 | Not-determined |
| 18-102_k141_140697 | Low-quality |
| 18-102_k141_182238 | Not-determined |
| 18-102_k141_88965 | Not-determined |
| 18-102_k141_182244 | Not-determined |
| 18-102_k141_108911 | Not-determined |
| 18-102_k141_150758 | Not-determined |
| 18-102_k141_48942 | Low-quality |
| 18-102_k141_108915 | Not-determined |
| 18-102_k141_119866 | Low-quality |
| 18-102_k141_37197 | Not-determined |
| 18-102_k141_48946 | Not-determined |
| 18-102_k141_48949 | Not-determined |
| 18-102_k141_88977 | Not-determined |
| 18-102_k141_99250 | Not-determined |
| 18-102_k141_48953 | Not-determined |
| 18-102_k141_140718 | Not-determined |
| 18-102_k141_150765 | Not-determined |
| 18-102_k141_16062 | Not-determined |
| 18-102_k141_233952 | Not-determined |
| 18-102_k141_160486 | Not-determined |
| 18-102_k141_233953 | Low-quality |
| 18-102_k141_160487 | Not-determined |
| 18-102_k141_48961 | Not-determined |
| 18-102_k141_233955 | Not-determined |
| 18-102_k141_160490 | Not-determined |
| 18-102_k141_160491 | Not-determined |
| 18-102_k141_150774 | Not-determined |
| 18-102_k141_160493 | Not-determined |
| 18-102_k141_78443 | Not-determined |
| 18-102_k141_150781 | Not-determined |
| 18-102_k141_99277 | Low-quality |
| 18-102_k141_233963 | Not-determined |
| 18-102_k141_172173 | Not-determined |
| 18-102_k141_172177 | Not-determined |
| 18-102_k141_233971 | Not-determined |
| 18-102_k141_119884 | Not-determined |
| 18-102_k141_48981 | Not-determined |
| 18-102_k141_192027 | Not-determined |
| 18-102_k141_140749 | Not-determined |
| 18-102_k141_48987 | Not-determined |
| 18-102_k141_160517 | Low-quality |
| 18-102_k141_140752 | Not-determined |
| 18-102_k141_160520 | Not-determined |
| 18-102_k141_140757 | Not-determined |
| 18-102_k141_129017 | Not-determined |
| 18-102_k141_140762 | Low-quality |
| 18-102_k141_140764 | Low-quality |
| 18-102_k141_78470 | Not-determined |
| 18-102_k141_108963 | Not-determined |

| | |
|---|---|
| 18-102_k141_140768 | Not-determined |
| 18-102_k141_160528 | Not-determined |
| 18-102_k141_150799 | Low-quality |
| 18-102_k141_129042 | Not-determined |
| 18-102_k141_129043 | Not-determined |
| 18-102_k141_140770 | Low-quality |
| 18-102_k141_49020 | Not-determined |
| 18-102_k141_150820 | Not-determined |
| 18-102_k141_233997 | Not-determined |
| 18-102_k141_192052 | Not-determined |
| 18-102_k141_119898 | Not-determined |
| 18-102_k141_212313 | Not-determined |
| 18-102_k141_57787 | Low-quality |
| 18-102_k141_57789 | Not-determined |
| 18-102_k141_160548 | Not-determined |
| 18-102_k141_57795 | Not-determined |
| 18-102_k141_192066 | Not-determined |
| 18-102_k141_192067 | Not-determined |
| 18-102_k141_160554 | Not-determined |
| 18-102_k141_160555 | Not-determined |
| 18-102_k141_57798 | Low-quality |
| 18-102_k141_182322 | Not-determined |
| 18-102_k141_99323 | Not-determined |
| 18-102_k141_129055 | Not-determined |
| 18-102_k141_160559 | Not-determined |
| 18-102_k141_129058 | Not-determined |
| 18-102_k141_108986 | Not-determined |
| 18-102_k141_57801 | Low-quality |
| 18-102_k141_212334 | Not-determined |
| 18-102_k141_160565 | Not-determined |
| 18-102_k141_99327 | Not-determined |
| 18-102_k141_99330 | Not-determined |
| 18-102_k141_129064 | Not-determined |
| 18-102_k141_89049 | Not-determined |
| 18-102_k141_150845 | Not-determined |
| 18-102_k141_89051 | Not-determined |
| 18-102_k141_108994 | Not-determined |
| 18-102_k141_108996 | Not-determined |
| 18-102_k141_140780 | Not-determined |
| 18-102_k141_140782 | Low-quality |
| 18-102_k141_160570 | Not-determined |
| 18-102_k141_37248 | Not-determined |
| 18-102_k141_140785 | Not-determined |
| 18-102_k141_201306 | Not-determined |
| 18-102_k141_150850 | Not-determined |
| 18-102_k141_172236 | Not-determined |
| 18-102_k141_89064 | Not-determined |
| 18-102_k141_109011 | Low-quality |
| 18-102_k141_89073 | Not-determined |
| 18-102_k141_140798 | Not-determined |
| 18-102_k141_160588 | Not-determined |
| 18-102_k141_150858 | Not-determined |
| 18-102_k141_109015 | Not-determined |
| 18-102_k141_37285 | Not-determined |
| 18-102_k141_172247 | Not-determined |
| 18-102_k141_37287 | Not-determined |
| 18-102_k141_37288 | Not-determined |
| 18-102_k141_201324 | Not-determined |
| 18-102_k141_201325 | Not-determined |
| 18-102_k141_37293 | Not-determined |
| 18-102_k141_201327 | Not-determined |
| 18-102_k141_4793 | Not-determined |
| 18-102_k141_172257 | Not-determined |
| 18-102_k141_37297 | Not-determined |
| 18-102_k141_89081 | Low-quality |
| 18-102_k141_242951 | Low-quality |
| 18-102_k141_201336 | Not-determined |
| 18-102_k141_78499 | Low-quality |
| 18-102_k141_172260 | Not-determined |
| 18-102_k141_78501 | Not-determined |
| 18-102_k141_109021 | Not-determined |
| 18-102_k141_150888 | Not-determined |
| 18-102_k141_150895 | Not-determined |
| 18-102_k141_140842 | Not-determined |
| 18-102_k141_89104 | Not-determined |
| 18-102_k141_150901 | Low-quality |
| 18-102_k141_140845 | Not-determined |
| 18-102_k141_57825 | Low-quality |
| 18-102_k141_57828 | Not-determined |
| 18-102_k141_57829 | Not-determined |
| 18-102_k141_99367 | Not-determined |
| 18-102_k141_172264 | Low-quality |
| 18-102_k141_119950 | Not-determined |
| 18-102_k141_37322 | Not-determined |
| 18-102_k141_172274 | Not-determined |
| 18-102_k141_140865 | Not-determined |
| 18-102_k141_49036 | Not-determined |
| 18-102_k141_119951 | Low-quality |
| 18-102_k141_78523 | Not-determined |
| 18-102_k141_119953 | Not-determined |
| 18-102_k141_119955 | Not-determined |
| 18-102_k141_201361 | Not-determined |
| 18-102_k141_109055 | Not-determined |
| 18-102_k141_119958 | Not-determined |
| 18-102_k141_49046 | Not-determined |
| 18-102_k141_49048 | Not-determined |
| 18-102_k141_242980 | Not-determined |
| 18-102_k141_16125 | Not-determined |
| 18-102_k141_99375 | Not-determined |
| 18-102_k141_89123 | Not-determined |
| 18-102_k141_16133 | Not-determined |
| 18-102_k141_119979 | Not-determined |
| 18-102_k141_201367 | Low-quality |
| 18-102_k141_109071 | Not-determined |
| 18-102_k141_57844 | Not-determined |
| 18-102_k141_57849 | Not-determined |
| 18-102_k141_89134 | Not-determined |
| 18-102_k141_242992 | Not-determined |
| 18-102_k141_89137 | Not-determined |
| 18-102_k141_57855 | Not-determined |
| 18-102_k141_49068 | Not-determined |
| 18-102_k141_49069 | Low-quality |
| 18-102_k141_57860 | Not-determined |
| 18-102_k141_57861 | Not-determined |
| 18-102_k141_201383 | Not-determined |
| 18-102_k141_212369 | Not-determined |
| 18-102_k141_57864 | Not-determined |
| 18-102_k141_89149 | Not-determined |
| 18-102_k141_16156 | Not-determined |
| 18-102_k141_192090 | Not-determined |
| 18-102_k141_109094 | Not-determined |
| 18-102_k141_67614 | Not-determined |
| 18-102_k141_172303 | Not-determined |
| 18-102_k141_192100 | Not-determined |
| 18-102_k141_16166 | Not-determined |
| 18-102_k141_57881 | Not-determined |
| 18-102_k141_49084 | Low-quality |
| 18-102_k141_201389 | Not-determined |
| 18-102_k141_49087 | Not-determined |
| 18-102_k141_172309 | Not-determined |
| 18-102_k141_192113 | Not-determined |
| 18-102_k141_172310 | Not-determined |
| 18-102_k141_192119 | Not-determined |
| 18-102_k141_16178 | Low-quality |
| 18-102_k141_192124 | Not-determined |
| 18-102_k141_89174 | Not-determined |
| 18-102_k141_192129 | Low-quality |
| 18-102_k141_78550 | Not-determined |
| 18-102_k141_89183 | Not-determined |
| 18-102_k141_243012 | Not-determined |
| 18-102_k141_99408 | Not-determined |
| 18-102_k141_109125 | Not-determined |
| 18-102_k141_89189 | Not-determined |
| 18-102_k141_234019 | Not-determined |
| 18-102_k141_109126 | Low-quality |
| 18-102_k141_25605 | Not-determined |
| 18-102_k141_89199 | Not-determined |
| 18-102_k141_172335 | Not-determined |
| 18-102_k141_109133 | Low-quality |
| 18-102_k141_120000 | Not-determined |
| 18-102_k141_109141 | Not-determined |
| 18-102_k141_172344 | Not-determined |
| 18-102_k141_67634 | Not-determined |
| 18-102_k141_78554 | Not-determined |
| 18-102_k141_67639 | Not-determined |
| 18-102_k141_160645 | Not-determined |
| 18-102_k141_234025 | Not-determined |
| 18-102_k141_120008 | Not-determined |
| 18-102_k141_234029 | Not-determined |
| 18-102_k141_99426 | Not-determined |
| 18-102_k141_16212 | Not-determined |
| 18-102_k141_99431 | Not-determined |
| 18-102_k141_49108 | Not-determined |
| 18-102_k141_99434 | Not-determined |
| 18-102_k141_172367 | Not-determined |
| 18-102_k141_172368 | Low-quality |
| 18-102_k141_109152 | Not-determined |
| 18-102_k141_109154 | Not-determined |
| 18-102_k141_234042 | Not-determined |
| 18-102_k141_160682 | Not-determined |
| 18-102_k141_192184 | Not-determined |
| 18-102_k141_120035 | Not-determined |
| 18-102_k141_4817 | Not-determined |
| 18-102_k141_37351 | Not-determined |
| 18-102_k141_25638 | Not-determined |
| 18-102_k141_37372 | Low-quality |
| 18-102_k141_67697 | Low-quality |
| 18-102_k141_4827 | Not-determined |
| 18-102_k141_182375 | Low-quality |
| 18-102_k141_120042 | Not-determined |
| 18-102_k141_172402 | Not-determined |
| 18-102_k141_222001 | Not-determined |
| 18-102_k141_201418 | Low-quality |
| 18-102_k141_37380 | Not-determined |
| 18-102_k141_78579 | Not-determined |
| 18-102_k141_120055 | Not-determined |
| 18-102_k141_120057 | Low-quality |
| 18-102_k141_172413 | Not-determined |
| 18-102_k141_57903 | Not-determined |
| 18-102_k141_160696 | Not-determined |
| 18-102_k141_222008 | Not-determined |
| 18-102_k141_25650 | Low-quality |
| 18-102_k141_49153 | Low-quality |
| 18-102_k141_49156 | Not-determined |
| 18-102_k141_25655 | Not-determined |
| 18-102_k141_192194 | Not-determined |
| 18-102_k141_192196 | Not-determined |
| 18-102_k141_37395 | Not-determined |
| 18-102_k141_37396 | Low-quality |
| 18-102_k141_172434 | Not-determined |
| 18-102_k141_25659 | Not-determined |
| 18-102_k141_37401 | Not-determined |
| 18-102_k141_201433 | Not-determined |
| 18-102_k141_78593 | Not-determined |
| 18-102_k141_57911 | Not-determined |
| 18-102_k141_25662 | Not-determined |
| 18-102_k141_37412 | Not-determined |
| 18-102_k141_234050 | Low-quality |
| 18-102_k141_78608 | Not-determined |
| 18-102_k141_109188 | Not-determined |
| 18-102_k141_109190 | Not-determined |
| 18-102_k141_201447 | Not-determined |
| 18-102_k141_160717 | Not-determined |
| 18-102_k141_25671 | Not-determined |
| 18-102_k141_37418 | Not-determined |
| 18-102_k141_25676 | Not-determined |
| 18-102_k141_234068 | Not-determined |
| 18-102_k141_49180 | Not-determined |
| 18-102_k141_120070 | Not-determined |
| 18-102_k141_25678 | Low-quality |
| 18-102_k141_182428 | Not-determined |
| 18-102_k141_37434 | Not-determined |
| 18-102_k141_57920 | Not-determined |
| 18-102_k141_57921 | Not-determined |
| 18-102_k141_57925 | Not-determined |
| 18-102_k141_172463 | Not-determined |
| 18-102_k141_192201 | Not-determined |
| 18-102_k141_57928 | Low-quality |
| 18-102_k141_182435 | Not-determined |
| 18-102_k141_67751 | Not-determined |
| 18-102_k141_182438 | Low-quality |
| 18-102_k141_67756 | Not-determined |
| 18-102_k141_57931 | Low-quality |
| 18-102_k141_192208 | Not-determined |
| 18-102_k141_182447 | Not-determined |
| 18-102_k141_120079 | Not-determined |
| 18-102_k141_49192 | Not-determined |
| 18-102_k141_182453 | Not-determined |
| 18-102_k141_37454 | Not-determined |
| 18-102_k141_234081 | Not-determined |
| 18-102_k141_172474 | Not-determined |
| 18-102_k141_234083 | Not-determined |
| 18-102_k141_67774 | Not-determined |
| 18-102_k141_49198 | Not-determined |
| 18-102_k141_67777 | Not-determined |
| 18-102_k141_243028 | Not-determined |
| 18-102_k141_67787 | Not-determined |
| 18-102_k141_243049 | Not-determined |
| 18-102_k141_182467 | Not-determined |
| 18-102_k141_201474 | Not-determined |
| 18-102_k141_234109 | Not-determined |
| 18-102_k141_201477 | Not-determined |
| 18-102_k141_25699 | Not-determined |
| 18-102_k141_243057 | Not-determined |
| 18-102_k141_67803 | Not-determined |
| 18-102_k141_67807 | Not-determined |
| 18-102_k141_67808 | Not-determined |
| 18-102_k141_243060 | Not-determined |
| 18-102_k141_120117 | Not-determined |
| 18-102_k141_222055 | Not-determined |
| 18-102_k141_25713 | Not-determined |
| 18-102_k141_67814 | Not-determined |
| 18-102_k141_192225 | Not-determined |
| 18-102_k141_192227 | Not-determined |
| 18-102_k141_89208 | Not-determined |
| 18-102_k141_222062 | Not-determined |
| 18-102_k141_78634 | Low-quality |
| 18-102_k141_234123 | Not-determined |
| 18-102_k141_234124 | Not-determined |
| 18-102_k141_25726 | Not-determined |
| 18-102_k141_160736 | Not-determined |
| 18-102_k141_99450 | Not-determined |
| 18-102_k141_151006 | Low-quality |
| 18-102_k141_151007 | Not-determined |
| 18-102_k141_120132 | Not-determined |
| 18-102_k141_201494 | Not-determined |
| 18-102_k141_151010 | Not-determined |
| 18-102_k141_192237 | Not-determined |
| 18-102_k141_234139 | Not-determined |
| 18-102_k141_234140 | Not-determined |
| 18-102_k141_120139 | Not-determined |
| 18-102_k141_234149 | Not-determined |
| 18-101-final-viral-combined_k141_212892\|\|full | Medium-quality |
| 18-102_k141_120143 | Not-determined |
| 18-102_k141_243078 | Not-determined |
| 18-102_k141_201501 | Low-quality |
| 18-102_k141_212405 | Not-determined |
| 18-102_k141_120150 | Not-determined |
| 18-102_k141_89252 | Not-determined |
| 18-102_k141_99473 | Not-determined |
| 18-102_k141_234163 | Not-determined |
| 18-102_k141_192253 | Not-determined |
| 18-102_k141_182508 | Not-determined |
| 18-102_k141_172556 | Not-determined |
| 18-102_k141_49279 | Not-determined |
| 18-102_k141_172561 | Not-determined |
| 18-102_k141_172563 | Not-determined |
| 18-102_k141_160762 | Low-quality |
| 18-102_k141_49281 | Not-determined |
| 18-102_k141_234184 | Not-determined |
| 18-102_k141_99489 | Not-determined |
| 18-102_k141_49285 | Not-determined |
| 18-102_k141_160766 | Low-quality |
| 18-102_k141_129083 | Not-determined |
| 18-102_k141_160769 | Not-determined |
| 18-102_k141_89273 | Low-quality |
| 18-102_k141_160774 | Not-determined |
| 18-102_k141_16244 | Not-determined |
| 18-102_k141_78643 | Not-determined |
| 18-102_k141_120179 | Not-determined |
| 18-102_k141_212434 | Not-determined |
| 18-102_k141_151028 | Not-determined |
| 18-102_k141_172580 | Low-quality |
| 18-102_k141_140970 | Not-determined |
| 18-102_k141_57950 | Not-determined |
| 18-102_k141_16255 | Not-determined |
| 18-102_k141_140974 | Low-quality |
| 18-102_k141_16257 | Not-determined |
| 18-102_k141_16258 | Not-determined |
| 18-102_k141_99512 | Not-determined |
| 18-102_k141_192275 | Low-quality |
| 18-102_k141_99515 | Low-quality |
| 18-102_k141_151046 | Low-quality |
| 18-102_k141_140982 | Not-determined |
| 18-102_k141_151050 | Not-determined |
| 18-102_k141_16268 | Low-quality |
| 18-102_k141_4833 | Not-determined |
| 18-102_k141_192288 | Not-determined |
| 18-102_k141_99520 | Not-determined |
| 18-102_k141_212457 | Not-determined |
| 18-102_k141_16269 | Not-determined |
| 18-102_k141_212459 | Not-determined |
| 18-102_k141_222098 | Low-quality |
| 18-102_k141_172600 | Not-determined |
| 18-102_k141_49305 | Not-determined |
| 18-102_k141_4843 | Not-determined |
| 18-102_k141_49306 | Not-determined |
| 18-102_k141_222099 | Low-quality |
| 18-102_k141_243131 | Not-determined |
| 18-102_k141_67874 | Low-quality |
| 18-102_k141_129108 | Not-determined |
| 18-102_k141_16283 | Not-determined |
| 18-102_k141_192301 | Not-determined |
| 18-102_k141_49317 | Not-determined |
| 18-102_k141_89309 | Not-determined |
| 18-102_k141_67885 | Not-determined |
| 18-102_k141_57975 | Not-determined |
| 18-102_k141_89318 | Not-determined |
| 18-102_k141_67893 | Not-determined |
| 18-102_k141_67894 | Not-determined |
| 18-102_k141_129120 | Not-determined |
| 18-102_k141_212471 | Not-determined |
| 18-102_k141_89327 | Not-determined |
| 18-102_k141_234213 | Not-determined |
| 18-102_k141_234222 | Not-determined |
| 18-102_k141_109214 | Not-determined |
| 18-102_k141_99545 | Not-determined |
| 18-102_k141_212481 | Not-determined |
| 18-102_k141_49333 | Not-determined |
| 18-102_k141_243155 | Not-determined |
| 18-102_k141_243156 | Not-determined |
| 18-102_k141_129137 | Not-determined |
| 18-102_k141_201545 | Not-determined |
| 18-102_k141_109220 | Not-determined |
| 18-102_k141_234232 | Not-determined |
| 18-102_k141_172672 | Not-determined |
| 18-102_k141_172673 | Not-determined |
| 18-102_k141_49347 | Not-determined |
| 18-102_k141_172674 | Low-quality |
| 18-102_k141_201555 | Not-determined |
| 18-102_k141_243162 | Not-determined |
| 18-102_k141_222112 | Not-determined |
| 18-102_k141_234249 | Not-determined |
| 18-102_k141_243167 | Low-quality |
| 18-102_k141_234252 | Not-determined |
| 18-102_k141_99560 | Low-quality |
| 18-102_k141_234255 | Not-determined |
| 18-102_k141_109232 | Not-determined |
| 18-102_k141_129141 | Not-determined |
| 18-102_k141_129143 | Not-determined |
| 18-102_k141_99573 | Not-determined |
| 18-102_k141_109242 | Not-determined |
| 18-102_k141_109244 | Not-determined |
| 18-102_k141_172687 | Not-determined |
| 18-102_k141_129150 | Not-determined |
| 18-102_k141_67940 | Not-determined |
| 18-102_k141_234267 | Not-determined |
| 18-102_k141_234269 | Not-determined |
| 18-102_k141_99577 | Low-quality |
| 18-102_k141_109246 | Not-determined |
| 18-102_k141_99587 | Not-determined |
| 18-102_k141_243176 | Not-determined |
| 18-102_k141_222137 | Not-determined |
| 18-102_k141_243178 | Not-determined |
| 18-102_k141_172710 | Not-determined |
| 18-102_k141_212491 | Not-determined |
| 18-102_k141_212492 | Not-determined |
| 18-102_k141_212493 | Low-quality |
| 18-102_k141_234289 | Low-quality |
| 18-102_k141_16319 | Not-determined |
| 18-102_k141_234291 | Not-determined |
| 18-102_k141_109254 | Not-determined |
| 18-102_k141_201570 | Not-determined |
| 18-102_k141_16327 | Low-quality |
| 18-102_k141_16329 | Not-determined |
| 18-102_k141_129165 | Not-determined |
| 18-102_k141_4855 | Low-quality |
| 18-102_k141_222153 | Not-determined |
| 18-102_k141_222156 | Not-determined |
| 18-102_k141_4858 | Low-quality |
| 18-102_k141_109257 | Low-quality |
| 18-102_k141_222157 | Not-determined |
| 18-102_k141_120242 | Low-quality |
| 18-102_k141_243204 | Not-determined |
| 18-102_k141_212509 | Not-determined |
| 18-102_k141_243205 | Not-determined |
| 18-102_k141_109263 | Not-determined |
| 18-102_k141_243207 | Not-determined |
| 18-102_k141_243208 | Not-determined |
| 18-102_k141_89381 | Not-determined |
| 18-102_k141_89382 | Not-determined |
| 18-102_k141_222162 | Not-determined |
| 18-102_k141_89383 | Not-determined |
| 18-102_k141_192308 | Not-determined |
| 18-102_k141_222170 | Not-determined |
| 18-102_k141_25759 | Not-determined |
| 18-102_k141_151188 | Not-determined |
| 18-102_k141_16354 | Not-determined |
| 18-102_k141_99609 | Not-determined |
| 18-102_k141_151191 | Not-determined |
| 18-102_k141_201602 | Not-determined |
| 18-102_k141_16360 | Not-determined |
| 18-102_k141_222173 | Not-determined |
| 18-102_k141_212515 | Low-quality |
| 18-102_k141_89391 | Not-determined |
| 18-102_k141_25769 | Not-determined |
| 18-102_k141_99620 | Not-determined |
| 18-102_k141_99625 | Not-determined |
| 18-102_k141_25773 | Not-determined |
| 18-102_k141_16373 | Not-determined |
| 18-102_k141_212531 | Not-determined |
| 18-102_k141_25783 | Not-determined |
| 18-102_k141_25790 | Not-determined |
| 18-102_k141_234319 | Not-determined |
| 18-102_k141_99628 | Not-determined |
| 18-102_k141_99629 | Not-determined |
| 18-102_k141_120248 | Not-determined |
| 18-102_k141_109303 | Not-determined |
| 18-102_k141_99632 | Not-determined |
| 18-102_k141_192315 | Not-determined |
| 18-102_k141_234332 | Not-determined |
| 18-102_k141_67965 | Not-determined |
| 18-102_k141_192318 | Not-determined |
| 18-102_k141_192319 | Low-quality |
| 18-102_k141_212553 | Not-determined |
| 18-102_k141_192324 | Not-determined |
| 18-102_k141_160833 | Not-determined |
| 18-102_k141_160838 | Not-determined |
| 18-102_k141_25811 | Not-determined |
| 18-102_k141_172718 | Not-determined |
| 18-102_k141_58081 | Not-determined |
| 18-102_k141_212568 | Not-determined |
| 18-102_k141_212571 | Not-determined |
| 18-102_k141_67973 | Not-determined |
| 18-102_k141_182604 | Not-determined |
| 18-102_k141_4876 | Not-determined |
| 18-102_k141_212575 | Low-quality |
| 18-102_k141_212579 | Not-determined |
| 18-102_k141_99648 | Not-determined |
| 18-102_k141_58094 | Not-determined |
| 18-102_k141_99649 | Not-determined |
| 18-102_k141_160867 | Not-determined |
| 18-102_k141_160868 | Not-determined |
| 18-102_k141_182618 | Low-quality |
| 18-102_k141_99665 | Not-determined |
| 18-102_k141_89432 | Not-determined |
| 18-102_k141_129198 | Low-quality |
| 18-102_k141_243231 | Low-quality |
| 18-102_k141_182632 | Not-determined |
| 18-102_k141_234364 | Not-determined |
| 18-102_k141_201655 | Not-determined |
| 18-102_k141_4899 | Not-determined |
| 18-102_k141_182645 | Not-determined |
| 18-102_k141_192332 | Low-quality |
| 18-102_k141_234367 | Not-determined |
| 18-102_k141_67987 | Low-quality |
| 18-102_k141_192333 | Not-determined |
| 18-102_k141_172753 | Not-determined |
| 18-102_k141_67996 | Not-determined |
| 18-102_k141_58105 | Low-quality |
| 18-102_k141_109350 | Not-determined |
| 18-102_k141_109353 | Not-determined |
| 18-102_k141_99692 | Not-determined |
| 18-102_k141_160906 | Not-determined |
| 18-102_k141_172769 | Not-determined |
| 18-102_k141_68017 | Not-determined |
| 18-102_k141_109359 | Not-determined |
| 18-102_k141_109365 | Not-determined |
| 18-102_k141_172779 | Low-quality |
| 18-102_k141_37467 | Low-quality |
| 18-102_k141_16392 | Not-determined |
| 18-102_k141_201683 | Not-determined |
| 18-102_k141_182650 | Not-determined |
| 18-102_k141_172784 | Not-determined |
| 18-102_k141_49425 | Not-determined |
| 18-102_k141_201686 | Low-quality |
| 18-102_k141_49430 | Not-determined |
| 18-102_k141_234387 | Not-determined |
| 18-102_k141_109374 | Not-determined |
| 18-102_k141_151235 | Low-quality |
| 18-102_k141_192360 | Not-determined |
| 18-102_k141_182662 | Not-determined |
| 18-102_k141_182664 | Not-determined |
| 18-102_k141_192361 | Low-quality |
| 18-102_k141_120259 | Not-determined |
| 18-102_k141_68036 | Not-determined |
| 18-102_k141_68037 | Not-determined |
| 18-102_k141_49442 | Not-determined |
| 18-102_k141_192369 | Not-determined |
| 18-102_k141_234399 | Not-determined |
| 18-102_k141_192375 | Not-determined |
| 18-102_k141_151249 | Low-quality |
| 18-102_k141_37479 | Low-quality |
| 18-102_k141_120267 | Not-determined |
| 18-102_k141_120268 | Not-determined |
| 18-102_k141_120270 | Not-determined |
| 18-102_k141_172822 | Not-determined |
| 18-102_k141_16426 | Not-determined |
| 18-102_k141_120273 | Not-determined |
| 18-102_k141_68048 | Low-quality |
| 18-102_k141_16429 | Not-determined |
| 18-102_k141_37486 | Not-determined |
| 18-102_k141_222227 | Not-determined |
| 18-102_k141_37491 | Not-determined |
| 18-102_k141_25856 | Not-determined |
| 18-102_k141_151283 | Not-determined |
| 18-102_k141_49478 | Not-determined |
| 18-102_k141_234422 | Low-quality |
| 18-102_k141_37497 | Not-determined |
| 18-102_k141_151288 | Not-determined |
| 18-102_k141_234430 | Not-determined |
| 18-102_k141_109406 | Low-quality |
| 18-102_k141_37506 | Not-determined |
| 18-102_k141_222234 | Low-quality |
| 18-102_k141_89470 | Low-quality |
| 18-102_k141_109412 | Not-determined |
| 18-102_k141_78731 | Not-determined |
| 18-102_k141_222236 | Not-determined |
| 18-102_k141_25872 | Not-determined |
| 18-102_k141_222239 | Not-determined |
| 18-102_k141_172832 | Not-determined |
| 18-102_k141_49494 | Not-determined |
| 18-102_k141_222243 | Low-quality |
| 18-102_k141_151312 | Not-determined |
| 18-102_k141_234450 | Not-determined |
| 18-102_k141_99718 | Not-determined |
| 18-102_k141_172843 | Not-determined |
| 18-102_k141_99722 | Not-determined |
| 18-102_k141_201703 | Not-determined |
| 18-102_k141_49503 | Not-determined |
| 18-102_k141_49504 | Low-quality |
| 18-102_k141_129218 | Low-quality |
| 18-102_k141_49505 | Not-determined |
| 18-102_k141_58123 | Not-determined |
| 18-102_k141_160918 | Low-quality |
| 18-102_k141_25876 | Not-determined |
| 18-102_k141_37517 | Not-determined |
| 18-102_k141_151321 | Not-determined |
| 18-102_k141_37520 | Not-determined |
| 18-102_k141_129221 | Not-determined |
| 18-102_k141_222258 | Not-determined |
| 18-102_k141_172859 | Not-determined |
| 18-102_k141_212584 | Not-determined |
| 18-102_k141_68082 | Not-determined |
| 18-102_k141_68083 | Not-determined |
| 18-102_k141_78763 | Not-determined |
| 18-102_k141_68086 | Not-determined |
| 18-102_k141_68087 | Not-determined |
| 18-102_k141_78766 | Low-quality |
| 18-102_k141_78768 | Not-determined |
| 18-102_k141_129239 | Not-determined |
| 18-102_k141_78771 | Not-determined |
| 18-102_k141_192414 | Not-determined |
| 18-102_k141_25887 | Not-determined |
| 18-102_k141_49534 | Not-determined |
| 18-102_k141_25894 | Not-determined |
| 18-102_k141_25898 | Not-determined |
| 18-102_k141_25900 | Not-determined |
| 18-102_k141_25901 | Not-determined |

| | |
|---|---|
| 18-102_k141_4906 | Not-determined |
| 18-102_k141_151351 | Not-determined |
| 18-102_k141_234479 | Not-determined |
| 18-102_k141_49537 | Not-determined |
| 18-102_k141_68094 | Not-determined |
| 18-102_k141_234485 | Not-determined |
| 18-102_k141_222266 | Low-quality |
| 18-102_k141_222268 | Not-determined |
| 18-102_k141_151352 | Not-determined |
| 18-102_k141_4923 | Not-determined |
| 18-102_k141_212590 | Not-determined |
| 18-102_k141_151361 | Not-determined |
| 18-102_k141_37535 | Not-determined |
| 18-102_k141_172893 | Not-determined |
| 18-102_k141_25918 | Not-determined |
| 18-102_k141_37539 | Low-quality |
| 18-102_k141_172898 | Not-determined |
| 18-102_k141_160948 | Not-determined |
| 18-102_k141_222285 | Not-determined |
| 18-102_k141_222286 | Not-determined |
| 18-102_k141_58166 | Not-determined |
| 18-102_k141_151374 | Not-determined |
| 18-102_k141_151375 | Not-determined |
| 18-102_k141_4931 | Not-determined |
| 18-102_k141_16504 | Low-quality |
| 18-102_k141_192437 | Not-determined |
| 18-102_k141_172925 | Not-determined |
| 18-102_k141_172926 | Low-quality |
| 18-102_k141_172928 | Low-quality |
| 18-102_k141_4945 | Not-determined |
| 18-102_k141_172932 | Not-determined |
| 18-102_k141_49584 | Not-determined |
| 18-102_k141_25955 | Not-determined |
| 18-102_k141_89525 | Low-quality |
| 18-102_k141_234514 | Not-determined |
| 18-102_k141_89531 | Not-determined |
| 18-102_k141_172944 | Not-determined |
| 18-102_k141_222299 | Not-determined |
| 18-102_k141_58196 | Not-determined |
| 18-102_k141_151384 | Not-determined |
| 18-102_k141_58205 | Not-determined |
| 18-102_k141_120322 | Not-determined |
| 18-102_k141_201769 | Not-determined |
| 18-102_k141_4954 | Not-determined |
| 18-102_k141_49608 | Not-determined |
| 18-102_k141_99770 | Not-determined |
| 18-102_k141_49612 | Low-quality |
| 18-102_k141_182739 | Not-determined |
| 18-102_k141_68121 | Not-determined |
| 18-102_k141_141029 | Not-determined |
| 18-102_k141_89546 | Low-quality |
| 18-102_k141_16534 | Not-determined |
| 18-102_k141_49615 | Not-determined |
| 18-102_k141_192442 | Not-determined |
| 18-102_k141_192445 | Low-quality |
| 18-102_k141_222309 | Not-determined |
| 18-102_k141_37573 | Not-determined |
| 18-102_k141_16542 | Not-determined |
| 18-102_k141_212613 | Not-determined |
| 18-102_k141_16550 | Not-determined |
| 18-102_k141_172978 | Low-quality |
| 18-102_k141_16554 | Not-determined |
| 18-102_k141_141044 | Not-determined |
| 18-102_k141_120353 | Not-determined |
| 18-102_k141_89571 | Not-determined |
| 18-102_k141_16560 | Low-quality |
| 18-102_k141_58214 | Not-determined |
| 18-102_k141_161003 | Not-determined |
| 18-102_k141_243296 | Not-determined |
| 18-102_k141_212617 | Low-quality |
| 18-102_k141_89582 | Not-determined |
| 18-102_k141_89584 | Low-quality |
| 18-102_k141_212623 | Not-determined |
| 18-102_k141_68145 | Not-determined |
| 18-102_k141_234546 | Not-determined |
| 18-102_k141_222335 | Not-determined |
| 18-102_k141_68148 | Not-determined |
| 18-102_k141_89591 | Not-determined |
| 18-102_k141_58220 | Not-determined |
| 18-102_k141_109446 | Low-quality |
| 18-102_k141_234550 | Low-quality |
| 18-102_k141_151401 | Not-determined |
| 18-102_k141_141072 | Not-determined |
| 18-102_k141_212638 | Not-determined |
| 18-102_k141_129282 | Not-determined |
| 18-102_k141_243318 | Not-determined |
| 18-102_k141_109455 | Not-determined |
| 18-102_k141_4970 | Not-determined |
| 18-102_k141_161022 | Not-determined |
| 18-102_k141_37622 | Not-determined |
| 18-102_k141_212651 | Not-determined |
| 18-102_k141_151412 | Not-determined |
| 18-102_k141_89615 | Not-determined |
| 18-102_k141_192490 | Low-quality |
| 18-102_k141_4983 | Not-determined |
| 18-102_k141_4984 | Not-determined |
| 18-102_k141_4986 | Not-determined |
| 18-102_k141_201793 | Not-determined |
| 18-102_k141_173053 | Not-determined |
| 18-102_k141_192501 | Not-determined |
| 18-102_k141_173057 | Not-determined |
| 18-102_k141_141098 | Low-quality |
| 18-102_k141_212668 | Not-determined |
| 18-102_k141_141101 | Not-determined |
| 18-102_k141_68170 | Not-determined |
| 18-102_k141_234573 | Low-quality |
| 18-102_k141_173064 | Not-determined |
| 18-102_k141_161035 | Low-quality |
| 18-102_k141_16587 | Not-determined |
| 18-102_k141_37652 | Not-determined |
| 18-102_k141_234579 | Not-determined |
| 18-102_k141_201810 | Not-determined |
| 18-102_k141_16595 | Low-quality |
| 18-102_k141_243335 | Low-quality |
| 18-102_k141_243336 | Not-determined |
| 18-102_k141_37661 | Not-determined |
| 18-102_k141_99820 | Not-determined |
| 18-102_k141_109473 | Low-quality |
| 18-102_k141_173067 | Not-determined |
| 18-102_k141_89661 | Not-determined |
| 18-102_k141_234593 | Not-determined |
| 18-102_k141_173073 | Not-determined |
| 18-102_k141_201818 | Low-quality |
| 18-102_k141_222359 | Not-determined |
| 18-102_k141_173076 | Not-determined |
| 18-102_k141_234601 | Not-determined |
| 18-102_k141_129302 | Not-determined |
| 18-102_k141_151459 | Low-quality |
| 18-102_k141_141130 | Not-determined |
| 18-102_k141_129315 | Not-determined |
| 18-102_k141_89670 | Not-determined |
| 18-102_k141_192532 | Not-determined |
| 18-102_k141_129325 | Not-determined |
| 18-102_k141_222375 | Not-determined |
| 18-102_k141_78799 | Not-determined |
| 18-102_k141_16607 | Not-determined |
| 18-102_k141_16610 | Not-determined |
| 18-102_k141_120413 | Not-determined |
| 18-102_k141_201847 | Not-determined |
| 18-102_k141_212672 | Not-determined |
| 18-102_k141_201851 | Not-determined |
| 18-102_k141_16619 | Not-determined |
| 18-102_k141_129342 | Not-determined |
| 18-102_k141_16625 | Not-determined |
| 18-102_k141_78810 | Not-determined |
| 18-102_k141_89696 | Not-determined |
| 18-102_k141_89697 | Not-determined |
| 18-102_k141_129349 | Not-determined |
| 18-102_k141_78817 | Not-determined |
| 18-102_k141_49617 | Not-determined |
| 18-102_k141_129360 | Not-determined |
| 18-102_k141_49625 | Not-determined |
| 18-102_k141_49626 | Not-determined |
| 18-102_k141_49628 | Not-determined |
| 18-102_k141_49629 | Not-determined |
| 18-102_k141_234654 | Not-determined |
| 18-102_k141_129370 | Low-quality |
| 18-102_k141_161047 | Not-determined |
| 18-102_k141_16640 | Low-quality |
| 18-102_k141_78836 | Not-determined |
| 18-102_k141_68207 | Not-determined |
| 18-102_k141_68208 | Low-quality |
| 18-102_k141_161057 | Not-determined |
| 18-102_k141_25991 | Not-determined |
| 18-102_k141_173110 | Not-determined |
| 18-102_k141_173111 | Not-determined |
| 18-102_k141_161063 | Not-determined |
| 18-102_k141_182754 | Not-determined |
| 18-102_k141_182756 | Not-determined |
| 18-102_k141_173123 | Not-determined |
| 18-102_k141_5030 | Low-quality |
| 18-102_k141_5036 | Not-determined |
| 18-102_k141_201865 | Not-determined |
| 18-102_k141_201867 | Not-determined |
| 18-102_k141_5039 | Low-quality |
| 18-102_k141_161081 | Not-determined |
| 18-102_k141_161082 | Not-determined |
| 18-102_k141_49636 | Not-determined |
| 18-102_k141_26001 | Not-determined |
| 18-102_k141_151481 | Not-determined |
| 18-102_k141_173138 | Low-quality |
| 18-102_k141_201876 | Not-determined |
| 18-102_k141_212724 | Not-determined |
| 18-102_k141_26004 | Low-quality |
| 18-102_k141_182785 | Not-determined |
| 18-102_k141_161089 | Not-determined |
| 18-102_k141_173146 | Not-determined |
| 18-102_k141_129422 | Not-determined |
| 18-102_k141_49653 | Not-determined |
| 18-102_k141_120422 | Low-quality |
| 18-102_k141_120425 | Not-determined |
| 18-102_k141_26021 | Not-determined |
| 18-102_k141_222395 | Not-determined |
| 18-102_k141_49665 | Not-determined |
| 18-102_k141_173156 | Not-determined |
| 18-102_k141_49668 | Not-determined |
| 18-102_k141_182792 | Not-determined |
| 18-102_k141_182795 | Not-determined |
| 18-102_k141_234687 | Not-determined |
| 18-102_k141_99892 | Not-determined |
| 18-102_k141_234691 | Not-determined |
| 18-102_k141_120438 | Not-determined |
| 18-102_k141_161104 | Not-determined |
| 18-102_k141_234693 | Not-determined |
| 18-102_k141_109523 | Low-quality |
| 18-102_k141_109526 | Not-determined |
| 18-102_k141_234701 | Low-quality |
| 18-102_k141_161110 | Not-determined |
| 18-102_k141_173178 | Not-determined |
| 18-102_k141_120444 | Not-determined |
| 18-102_k141_182802 | Not-determined |
| 18-102_k141_120453 | Not-determined |
| 18-102_k141_78851 | Not-determined |
| 18-102_k141_37668 | Not-determined |
| 18-102_k141_37673 | Not-determined |
| 18-102_k141_182812 | Not-determined |
| 18-102_k141_161136 | Not-determined |
| 18-102_k141_201903 | Not-determined |
| 18-102_k141_161139 | Not-determined |
| 18-102_k141_120459 | Not-determined |
| 18-102_k141_120464 | Not-determined |
| 18-102_k141_212766 | Not-determined |
| 18-102_k141_109545 | Not-determined |
| 18-102_k141_182829 | Not-determined |
| 18-102_k141_182830 | Not-determined |
| 18-102_k141_49699 | Not-determined |
| 18-102_k141_78869 | Not-determined |
| 18-102_k141_109546 | Low-quality |
| 18-102_k141_16672 | Not-determined |
| 18-102_k141_109548 | Not-determined |
| 18-102_k141_212775 | Low-quality |
| 18-102_k141_222417 | Not-determined |
| 18-102_k141_234735 | Not-determined |
| 18-102_k141_201915 | Low-quality |
| 18-102_k141_5055 | Not-determined |
| 18-102_k141_161153 | Not-determined |
| 18-102_k141_212780 | Not-determined |
| 18-102_k141_129478 | Not-determined |
| 18-102_k141_161158 | Not-determined |
| 18-102_k141_161159 | Not-determined |
| 18-102_k141_120486 | Not-determined |
| 18-102_k141_222427 | Low-quality |
| 18-102_k141_5070 | Not-determined |
| 18-102_k141_120492 | Not-determined |
| 18-102_k141_182846 | Not-determined |
| 18-102_k141_120499 | Low-quality |
| 18-102_k141_5071 | Not-determined |
| 18-102_k141_120504 | Not-determined |
| 18-102_k141_192553 | Not-determined |
| 18-102_k141_129481 | Not-determined |
| 18-102_k141_16701 | Not-determined |
| 18-102_k141_201925 | Not-determined |
| 18-102_k141_192561 | Not-determined |
| 18-102_k141_120517 | Not-determined |
| 18-102_k141_16709 | Not-determined |
| 18-102_k141_192563 | Not-determined |
| 18-102_k141_201932 | Not-determined |
| 18-102_k141_161185 | Not-determined |
| 18-102_k141_49724 | Not-determined |
| 18-102_k141_49726 | Not-determined |
| 18-102_k141_16716 | Not-determined |
| 18-102_k141_49727 | Not-determined |
| 18-102_k141_151555 | Not-determined |
| 18-102_k141_222462 | Not-determined |
| 18-102_k141_151556 | Not-determined |
| 18-102_k141_120520 | Not-determined |
| 18-102_k141_192579 | Not-determined |
| 18-102_k141_49733 | Not-determined |
| 18-102_k141_129485 | Low-quality |
| 18-102_k141_5098 | Not-determined |
| 18-102_k141_201947 | Not-determined |
| 18-102_k141_161199 | Not-determined |
| 18-102_k141_5103 | Not-determined |
| 18-102_k141_192589 | Not-determined |
| 18-102_k141_99960 | Low-quality |
| 18-102_k141_222477 | Not-determined |
| 18-102_k141_5108 | Not-determined |
| 18-102_k141_49745 | Not-determined |
| 18-102_k141_201959 | Not-determined |
| 18-102_k141_201963 | Not-determined |
| 18-102_k141_173183 | Not-determined |
| 18-102_k141_201964 | Not-determined |
| 18-102_k141_37736 | Not-determined |
| 18-102_k141_243367 | Not-determined |
| 18-102_k141_151561 | Not-determined |
| 18-102_k141_16743 | Not-determined |
| 18-102_k141_151563 | Not-determined |
| 18-102_k141_161215 | Not-determined |
| 18-102_k141_182861 | Not-determined |
| 18-102_k141_58279 | Low-quality |
| 18-102_k141_37751 | Low-quality |
| 18-102_k141_243372 | Not-determined |
| 18-102_k141_16749 | Not-determined |
| 18-102_k141_120548 | Low-quality |
| 18-102_k141_5124 | Low-quality |
| 18-102_k141_173210 | Not-determined |
| 18-102_k141_120552 | Low-quality |
| 18-102_k141_234793 | Not-determined |
| 18-102_k141_151590 | Not-determined |
| 18-102_k141_5137 | Not-determined |
| 18-102_k141_49782 | Not-determined |
| 18-102_k141_68232 | Not-determined |
| 18-102_k141_151603 | Not-determined |
| 18-102_k141_161234 | Not-determined |
| 18-102_k141_182876 | Not-determined |
| 18-102_k141_151606 | Low-quality |
| 18-102_k141_68237 | Not-determined |
| 18-102_k141_68240 | Not-determined |
| 18-102_k141_151608 | Not-determined |
| 18-102_k141_212849 | Not-determined |
| 18-102_k141_212854 | Not-determined |
| 18-102_k141_201975 | Not-determined |
| 18-102_k141_89740 | Not-determined |
| 18-102_k141_161252 | Not-determined |
| 18-102_k141_151621 | Not-determined |
| 18-102_k141_120580 | Not-determined |
| 18-102_k141_49806 | Low-quality |
| 18-102_k141_58320 | Not-determined |
| 18-102_k141_120583 | Not-determined |
| 18-102_k141_151626 | Not-determined |
| 18-102_k141_49809 | Not-determined |
| 18-102_k141_173243 | Not-determined |
| 18-102_k141_243398 | Not-determined |
| 18-102_k141_16772 | Not-determined |
| 18-102_k141_173245 | Not-determined |
| 18-102_k141_120594 | Not-determined |
| 18-102_k141_243400 | Low-quality |
| 18-102_k141_161268 | Low-quality |
| 18-102_k141_182917 | Not-determined |
| 18-102_k141_120601 | Not-determined |
| 18-102_k141_161273 | Not-determined |
| 18-102_k141_5175 | Low-quality |
| 18-102_k141_222489 | Low-quality |
| 18-102_k141_89744 | Not-determined |
| 18-102_k141_109579 | Not-determined |
| 18-102_k141_151669 | Not-determined |
| 18-102_k141_212894 | Not-determined |
| 18-102_k141_109589 | Not-determined |
| 18-102_k141_141152 | Not-determined |
| 18-102_k141_89772 | Low-quality |
| 18-102_k141_5210 | Not-determined |
| 18-102_k141_109603 | Not-determined |
| 18-102_k141_202012 | Low-quality |
| 18-102_k141_16815 | Not-determined |
| 18-102_k141_212912 | Not-determined |
| 18-102_k141_161301 | Not-determined |
| 18-102_k141_26064 | Not-determined |
| 18-102_k141_68277 | Not-determined |
| 18-102_k141_161305 | Not-determined |
| 18-102_k141_182951 | Not-determined |
| 18-102_k141_151694 | Not-determined |
| 18-102_k141_212922 | Not-determined |
| 18-102_k141_243421 | Not-determined |
| 18-102_k141_129565 | Not-determined |
| 18-102_k141_49862 | Low-quality |
| 18-102_k141_129574 | Not-determined |
| 18-102_k141_192630 | Low-quality |
| 18-102_k141_26086 | Not-determined |
| 18-102_k141_78906 | Not-determined |
| 18-102_k141_151707 | Low-quality |
| 18-102_k141_78908 | Not-determined |
| 18-102_k141_151711 | Low-quality |
| 18-102_k141_202014 | Low-quality |
| 18-102_k141_129585 | Not-determined |
| 18-102_k141_78910 | Not-determined |
| 18-102_k141_16844 | Not-determined |
| 18-102_k141_78912 | Not-determined |
| 18-102_k141_182966 | Low-quality |
| 18-102_k141_202021 | Not-determined |
| 18-102_k141_16847 | Not-determined |
| 18-102_k141_26098 | Not-determined |
| 18-102_k141_99992 | Not-determined |
| 18-102_k141_16851 | Not-determined |
| 18-102_k141_68301 | Not-determined |
| 18-102_k141_120634 | Not-determined |
| 18-102_k141_109610 | Not-determined |
| 18-102_k141_109613 | Low-quality |
| 18-102_k141_129595 | Not-determined |
| 18-102_k141_58354 | Not-determined |
| 18-102_k141_49882 | Not-determined |
| 18-102_k141_49885 | Not-determined |
| 18-102_k141_173268 | Not-determined |
| 18-102_k141_49888 | Not-determined |
| 18-102_k141_192643 | Not-determined |
| 18-102_k141_129601 | Low-quality |
| 18-102_k141_192645 | Not-determined |
| 18-102_k141_100004 | Low-quality |
| 18-102_k141_202038 | Not-determined |
| 18-102_k141_202040 | Not-determined |
| 18-102_k141_78922 | Not-determined |
| 18-102_k141_243426 | Not-determined |
| 18-102_k141_192655 | Not-determined |
| 18-102_k141_49896 | Not-determined |
| 18-102_k141_89804 | Not-determined |
| 18-102_k141_49903 | Not-determined |
| 18-102_k141_78927 | Not-determined |
| 18-102_k141_49905 | Not-determined |
| 18-102_k141_243429 | Not-determined |
| 18-102_k141_58375 | Low-quality |
| 18-102_k141_120655 | Low-quality |
| 18-102_k141_49911 | Not-determined |
| 18-102_k141_141177 | Not-determined |
| 18-102_k141_78935 | Not-determined |
| 18-102_k141_100030 | Not-determined |
| 18-102_k141_173282 | Not-determined |
| 18-102_k141_68305 | Not-determined |
| 18-102_k141_49918 | Low-quality |
| 18-102_k141_141183 | Not-determined |
| 18-102_k141_49923 | Not-determined |
| 18-102_k141_68310 | Not-determined |
| 18-102_k141_100041 | Not-determined |
| 18-102_k141_243438 | Not-determined |
| 18-102_k141_120667 | Not-determined |
| 18-102_k141_49927 | Not-determined |
| 18-102_k141_89811 | Not-determined |
| 18-102_k141_100051 | Not-determined |
| 18-102_k141_26104 | Not-determined |
| 18-102_k141_120673 | Not-determined |
| 18-102_k141_161367 | Not-determined |
| 18-102_k141_243439 | Not-determined |
| 18-102_k141_161369 | Not-determined |
| 18-102_k141_243442 | Not-determined |
| 18-102_k141_89819 | Not-determined |
| 18-102_k141_151728 | Low-quality |
| 18-102_k141_212974 | Not-determined |
| 18-102_k141_151733 | Not-determined |
| 18-102_k141_49938 | Not-determined |
| 18-102_k141_49941 | Not-determined |
| 18-102_k141_89826 | Not-determined |
| 18-102_k141_202061 | Low-quality |
| 18-102_k141_49944 | Not-determined |
| 18-102_k141_202063 | Not-determined |
| 17-109-final-viral-combined_k141_383851\|\|full | Medium-quality |
| 18-102_k141_173322 | Not-determined |
| 18-102_k141_173326 | Not-determined |
| 18-102_k141_161402 | Not-determined |
| 18-102_k141_173333 | Not-determined |
| 18-102_k141_141216 | Not-determined |
| 18-102_k141_173341 | Not-determined |
| 18-102_k141_16869 | Not-determined |
| 18-102_k141_68322 | Not-determined |
| 18-102_k141_16873 | Not-determined |
| 18-102_k141_192686 | Not-determined |
| 18-102_k141_16878 | Not-determined |
| 18-102_k141_192687 | Not-determined |
| 18-102_k141_68329 | Not-determined |
| 18-102_k141_192689 | Not-determined |
| 18-102_k141_212981 | Not-determined |
| 18-102_k141_89847 | Not-determined |
| 18-102_k141_16881 | Not-determined |
| 18-102_k141_192704 | Not-determined |
| 18-102_k141_222561 | Not-determined |
| 18-102_k141_161423 | Not-determined |
| 18-102_k141_151764 | Not-determined |
| 18-102_k141_89859 | Not-determined |
| 18-102_k141_100066 | Not-determined |
| 18-102_k141_151767 | Not-determined |
| 18-102_k141_58449 | Not-determined |
| 18-102_k141_161433 | Not-determined |
| 18-102_k141_222562 | Not-determined |
| 18-102_k141_58452 | Not-determined |
| 18-102_k141_192715 | Low-quality |
| 18-102_k141_151772 | Not-determined |
| 18-102_k141_58459 | Not-determined |
| 18-102_k141_173357 | Not-determined |
| 18-102_k141_58462 | Not-determined |
| 18-102_k141_37809 | Not-determined |
| 18-102_k141_222575 | Not-determined |
| 18-102_k141_151783 | Not-determined |
| 18-102_k141_16908 | Not-determined |
| 18-102_k141_212994 | Not-determined |
| 18-102_k141_173362 | Not-determined |
| 18-102_k141_212996 | Not-determined |
| 18-102_k141_192731 | Not-determined |
| 18-102_k141_37816 | Low-quality |
| 18-102_k141_89869 | Not-determined |
| 18-102_k141_222579 | Not-determined |
| 18-102_k141_49973 | Not-determined |
| 18-102_k141_173372 | Not-determined |
| 18-102_k141_109691 | Not-determined |
| 18-102_k141_173373 | Not-determined |
| 18-102_k141_78953 | Not-determined |
| 18-102_k141_213011 | Low-quality |
| 18-102_k141_213013 | Not-determined |
| 18-102_k141_16937 | Not-determined |
| 18-102_k141_222583 | Not-determined |
| 18-102_k141_151801 | Not-determined |
| 18-102_k141_173394 | Not-determined |
| 18-102_k141_234809 | Not-determined |
| 18-102_k141_161447 | Not-determined |
| 18-102_k141_37829 | Not-determined |
| 18-102_k141_37830 | Not-determined |
| 18-102_k141_26106 | Low-quality |
| 18-102_k141_213029 | Not-determined |
| 18-102_k141_89888 | Not-determined |
| 18-102_k141_161452 | Not-determined |
| 18-102_k141_213031 | Low-quality |
| 18-102_k141_26110 | Not-determined |
| 18-102_k141_109706 | Not-determined |
| 18-102_k141_109707 | Not-determined |
| 18-102_k141_161458 | Not-determined |
| 18-102_k141_173408 | Not-determined |
| 18-102_k141_173410 | Not-determined |
| 18-102_k141_213038 | Not-determined |
| 18-102_k141_173412 | Not-determined |
| 18-102_k141_151826 | Low-quality |
| 18-102_k141_213042 | Not-determined |
| 18-102_k141_141275 | Not-determined |
| 18-102_k141_173425 | Not-determined |
| 18-102_k141_16984 | Not-determined |
| 18-102_k141_89912 | Not-determined |
| 18-102_k141_100114 | Not-determined |
| 18-102_k141_5226 | Not-determined |
| 18-102_k141_151836 | Not-determined |
| 18-102_k141_129629 | Not-determined |
| 18-102_k141_141285 | Not-determined |
| 18-102_k141_109717 | Not-determined |
| 18-102_k141_109724 | Low-quality |
| 18-102_k141_213056 | Low-quality |
| 18-102_k141_234834 | Not-determined |
| 18-102_k141_141305 | Not-determined |
| 18-102_k141_141306 | Not-determined |
| 18-102_k141_89929 | Not-determined |
| 18-102_k141_213060 | Not-determined |
| 18-102_k141_213066 | Low-quality |
| 18-102_k141_151850 | Low-quality |
| 18-102_k141_151851 | Not-determined |
| 18-102_k141_192737 | Low-quality |
| 18-102_k141_17000 | Low-quality |
| 18-102_k141_129646 | Not-determined |
| 18-102_k141_213076 | Not-determined |
| 18-102_k141_78993 | Not-determined |
| 18-102_k141_222637 | Not-determined |
| 18-102_k141_222643 | Not-determined |
| 18-102_k141_89945 | Not-determined |
| 18-102_k141_234846 | Not-determined |
| 18-102_k141_141326 | Not-determined |
| 18-102_k141_202079 | Not-determined |
| 18-102_k141_234851 | Not-determined |
| 18-102_k141_50009 | Not-determined |
| 18-102_k141_234853 | Not-determined |
| 18-102_k141_89953 | Not-determined |
| 18-102_k141_37865 | Not-determined |
| 18-102_k141_120724 | Not-determined |
| 18-102_k141_234863 | Not-determined |
| 18-102_k141_234866 | Not-determined |
| 18-102_k141_161472 | Not-determined |
| 18-102_k141_202083 | Low-quality |
| 18-102_k141_37881 | Not-determined |
| 18-102_k141_120729 | Not-determined |
| 18-102_k141_109774 | Not-determined |
| 18-102_k141_173461 | Not-determined |
| 18-102_k141_222650 | Not-determined |
| 18-102_k141_109777 | Not-determined |
| 18-102_k141_120740 | Not-determined |
| 18-102_k141_68367 | Not-determined |
| 18-102_k141_120742 | Not-determined |
| 18-102_k141_120744 | Not-determined |
| 18-102_k141_234886 | Not-determined |
| 18-102_k141_109785 | Not-determined |
| 18-102_k141_222660 | Not-determined |
| 18-102_k141_234892 | Not-determined |
| 18-102_k141_173474 | Not-determined |
| 18-102_k141_222664 | Not-determined |
| 18-102_k141_68385 | Not-determined |
| 18-102_k141_234896 | Not-determined |
| 18-102_k141_68386 | Low-quality |
| 18-102_k141_222672 | Not-determined |
| 18-102_k141_243469 | Low-quality |
| 18-102_k141_100134 | Not-determined |
| 18-102_k141_5268 | Not-determined |
| 18-102_k141_100135 | Not-determined |
| 18-102_k141_50046 | Not-determined |
| 18-102_k141_173491 | Not-determined |
| 18-102_k141_89982 | Not-determined |
| 18-102_k141_120757 | Not-determined |
| 18-102_k141_182997 | Not-determined |
| 18-102_k141_17023 | Not-determined |
| 18-102_k141_161505 | Not-determined |
| 18-102_k141_243478 | Not-determined |
| 18-102_k141_26138 | Not-determined |
| 18-102_k141_50063 | Not-determined |
| 18-102_k141_192761 | Not-determined |
| 18-102_k141_192763 | Not-determined |
| 18-102_k141_89986 | Not-determined |
| 18-102_k141_50072 | Not-determined |
| 18-102_k141_192768 | Not-determined |
| 18-102_k141_243481 | Low-quality |
| 18-102_k141_202118 | Not-determined |

| | |
|---|---|
| 18-102_k141_17027 | Not-determined |
| 18-102_k141_50075 | Not-determined |
| 18-102_k141_202121 | Not-determined |
| 18-102_k141_68411 | Low-quality |
| 18-102_k141_161519 | Not-determined |
| 18-102_k141_89995 | Low-quality |
| 18-102_k141_173503 | Low-quality |
| 18-102_k141_173505 | Not-determined |
| 18-102_k141_37898 | Not-determined |
| 18-102_k141_173509 | Not-determined |
| 18-102_k141_202128 | Not-determined |
| 18-102_k141_79033 | Not-determined |
| 18-102_k141_90003 | Not-determined |
| 18-102_k141_202129 | Not-determined |
| 18-102_k141_173512 | Low-quality |
| 18-102_k141_202131 | Not-determined |
| 18-102_k141_120772 | Not-determined |
| 18-102_k141_50090 | Not-determined |
| 18-102_k141_173517 | Not-determined |
| 18-102_k141_68420 | Not-determined |
| 18-102_k141_68421 | Not-determined |
| 18-102_k141_120779 | Low-quality |
| 18-102_k141_120781 | Not-determined |
| 18-102_k141_37907 | Not-determined |
| 18-102_k141_202137 | Not-determined |
| 18-102_k141_243510 | Not-determined |
| 18-102_k141_68435 | Not-determined |
| 18-102_k141_68439 | Not-determined |
| 18-102_k141_192788 | Not-determined |
| 18-102_k141_68443 | Not-determined |
| 18-102_k141_17056 | Not-determined |
| 18-102_k141_79065 | Not-determined |
| 18-102_k141_120802 | Low-quality |
| 18-102_k141_17057 | Not-determined |
| 18-102_k141_151872 | Not-determined |
| 18-102_k141_222701 | Not-determined |
| 18-102_k141_192798 | Not-determined |
| 18-102_k141_58529 | Not-determined |
| 18-102_k141_222705 | Not-determined |
| 18-102_k141_202173 | Not-determined |
| 18-102_k141_17063 | Not-determined |
| 18-102_k141_58536 | Not-determined |
| 18-102_k141_243521 | Not-determined |
| 18-102_k141_100201 | Not-determined |
| 18-102_k141_202176 | Not-determined |
| 18-102_k141_129670 | Not-determined |
| 18-102_k141_37926 | Not-determined |
| 18-102_k141_68470 | Low-quality |
| 18-102_k141_58540 | Low-quality |
| 18-102_k141_151897 | Not-determined |
| 18-102_k141_129674 | Not-determined |
| 18-102_k141_151898 | Not-determined |
| 18-102_k141_192811 | Not-determined |
| 18-102_k141_202186 | Not-determined |
| 18-102_k141_213095 | Low-quality |
| 18-102_k141_58552 | Not-determined |
| 18-102_k141_161557 | Not-determined |
| 18-102_k141_202190 | Low-quality |
| 18-102_k141_129679 | Not-determined |
| 18-102_k141_37943 | Not-determined |
| 18-102_k141_213103 | Not-determined |
| 18-102_k141_17081 | Low-quality |
| 18-102_k141_141405 | Not-determined |
| 18-102_k141_161566 | Not-determined |
| 18-102_k141_213107 | Not-determined |
| 18-102_k141_173557 | Not-determined |
| 18-102_k141_213110 | Not-determined |
| 18-102_k141_222726 | Not-determined |
| 18-102_k141_37963 | Not-determined |
| 18-102_k141_90030 | Low-quality |
| 18-102_k141_161572 | Not-determined |
| 18-102_k141_192822 | Low-quality |
| 18-102_k141_234936 | Not-determined |
| 18-102_k141_192828 | Not-determined |
| 18-102_k141_173573 | Not-determined |
| 18-102_k141_192829 | Not-determined |
| 18-102_k141_173575 | Not-determined |
| 18-102_k141_37978 | Not-determined |
| 18-102_k141_161576 | Low-quality |
| 18-102_k141_213117 | Not-determined |
| 18-102_k141_202209 | Not-determined |
| 18-102_k141_213122 | Not-determined |
| 18-102_k141_129701 | Not-determined |
| 18-102_k141_37996 | Not-determined |
| 18-102_k141_234960 | Not-determined |
| 18-102_k141_50127 | Not-determined |
| 18-102_k141_141416 | Not-determined |
| 18-102_k141_141422 | Not-determined |
| 18-102_k141_202224 | Not-determined |
| 18-102_k141_243574 | Not-determined |
| 18-102_k141_17132 | Not-determined |
| 18-102_k141_202232 | Not-determined |
| 18-102_k141_50143 | Not-determined |
| 18-102_k141_50147 | Low-quality |
| 18-102_k141_17141 | Not-determined |
| 18-102_k141_50149 | Not-determined |
| 18-102_k141_50150 | Not-determined |
| 18-102_k141_202235 | Not-determined |
| 18-102_k141_151930 | Not-determined |
| 18-102_k141_129723 | Not-determined |
| 18-102_k141_120836 | Not-determined |
| 18-102_k141_129725 | Not-determined |
| 18-102_k141_213143 | Not-determined |
| 18-102_k141_129729 | Not-determined |
| 18-102_k141_173606 | Not-determined |
| 18-102_k141_129732 | Low-quality |
| 18-102_k141_234985 | Not-determined |
| 18-102_k141_151937 | Low-quality |
| 18-102_k141_38034 | Not-determined |
| 18-102_k141_173617 | Low-quality |
| 18-102_k141_120854 | Not-determined |
| 18-102_k141_17150 | Not-determined |
| 18-102_k141_5295 | Not-determined |
| 18-102_k141_120862 | Not-determined |
| 18-102_k141_120863 | Not-determined |
| 18-102_k141_129752 | Not-determined |
| 18-102_k141_151943 | Not-determined |
| 18-102_k141_151944 | Not-determined |
| 18-102_k141_202247 | Not-determined |
| 18-102_k141_90053 | Not-determined |
| 18-102_k141_235004 | Not-determined |
| 18-102_k141_141448 | Not-determined |
| 18-102_k141_192879 | Not-determined |
| 18-102_k141_129766 | Low-quality |
| 18-102_k141_141464 | Low-quality |
| 18-102_k141_192882 | Not-determined |
| 18-102_k141_173629 | Not-determined |
| 18-102_k141_243609 | Not-determined |
| 18-102_k141_173633 | Not-determined |
| 18-102_k141_213172 | Not-determined |
| 18-102_k141_50182 | Not-determined |
| 18-102_k141_120870 | Low-quality |
| 18-102_k141_243613 | Not-determined |
| 18-102_k141_58578 | Not-determined |
| 18-102_k141_129777 | Not-determined |
| 18-102_k141_141476 | Low-quality |
| 18-102_k141_192891 | Not-determined |
| 18-102_k141_243624 | Not-determined |
| 18-102_k141_68521 | Not-determined |
| 18-102_k141_90081 | Not-determined |
| 18-102_k141_243626 | Not-determined |
| 18-102_k141_243628 | Not-determined |
| 18-102_k141_243630 | Not-determined |
| 18-102_k141_50202 | Not-determined |
| 18-102_k141_129793 | Not-determined |
| 18-102_k141_5323 | Low-quality |
| 18-102_k141_90092 | Not-determined |
| 18-102_k141_5324 | Not-determined |
| 18-102_k141_100244 | Not-determined |
| 18-102_k141_235028 | Not-determined |
| 18-102_k141_90095 | Not-determined |
| 18-102_k141_79101 | Not-determined |
| 18-102_k141_79104 | Not-determined |
| 18-102_k141_141489 | Not-determined |
| 18-102_k141_79105 | Not-determined |
| 18-102_k141_79107 | Not-determined |
| 18-102_k141_100248 | Not-determined |
| 18-102_k141_38063 | Not-determined |
| 18-102_k141_100251 | Not-determined |
| 18-102_k141_213202 | Not-determined |
| 18-102_k141_202288 | Not-determined |
| 18-102_k141_202289 | Not-determined |
| 18-102_k141_152002 | Not-determined |
| 18-102_k141_152008 | Low-quality |
| 18-102_k141_202294 | Not-determined |
| 18-102_k141_120884 | Not-determined |
| 18-102_k141_120885 | Not-determined |
| 18-102_k141_202296 | Not-determined |
| 18-102_k141_38087 | Not-determined |
| 18-102_k141_100265 | Not-determined |
| 18-102_k141_68550 | Not-determined |
| 18-102_k141_235055 | Not-determined |
| 18-102_k141_129795 | Not-determined |
| 18-102_k141_90140 | Not-determined |
| 18-102_k141_192922 | Not-determined |
| 18-102_k141_120908 | Not-determined |
| 18-102_k141_235066 | Not-determined |
| 18-102_k141_222784 | Not-determined |
| 18-102_k141_50212 | Not-determined |
| 18-102_k141_79146 | Not-determined |
| 18-102_k141_68571 | Not-determined |
| 18-102_k141_129814 | Low-quality |
| 18-102_k141_222790 | Not-determined |
| 18-102_k141_183045 | Low-quality |
| 18-102_k141_50215 | Not-determined |
| 18-102_k141_235075 | Not-determined |
| 18-102_k141_79157 | Not-determined |
| 18-102_k141_5360 | Not-determined |
| 18-102_k141_100274 | Not-determined |
| 18-102_k141_5363 | Not-determined |
| 18-102_k141_38118 | Low-quality |
| 18-102_k141_222796 | Not-determined |
| 18-102_k141_5365 | Not-determined |
| 18-102_k141_202322 | Not-determined |
| 18-102_k141_120917 | Not-determined |
| 18-102_k141_38123 | Low-quality |
| 18-102_k141_243666 | Not-determined |
| 18-102_k141_26179 | Not-determined |
| 18-102_k141_26181 | Not-determined |
| 18-102_k141_38134 | Not-determined |
| 18-102_k141_26184 | Not-determined |
| 18-102_k141_192950 | Not-determined |
| 18-102_k141_79159 | Low-quality |
| 18-102_k141_222803 | Not-determined |
| 18-102_k141_58643 | Not-determined |
| 18-102_k141_38141 | Not-determined |
| 18-102_k141_79166 | Not-determined |
| 18-102_k141_243670 | Not-determined |
| 18-102_k141_202342 | Low-quality |
| 18-102_k141_38142 | Low-quality |
| 18-102_k141_58647 | Low-quality |
| 18-102_k141_222805 | Not-determined |
| 18-102_k141_100280 | Not-determined |
| 18-102_k141_222807 | Not-determined |
| 18-102_k141_141503 | Low-quality |
| 18-102_k141_183076 | Not-determined |
| 18-102_k141_38150 | Not-determined |
| 18-102_k141_58656 | Not-determined |
| 18-102_k141_50249 | Not-determined |
| 18-102_k141_38157 | Not-determined |
| 18-102_k141_243674 | Not-determined |
| 18-102_k141_183082 | Not-determined |
| 18-102_k141_68585 | Not-determined |
| 18-102_k141_243676 | Not-determined |
| 18-102_k141_183087 | Not-determined |
| 18-102_k141_235090 | Low-quality |
| 18-102_k141_100300 | Not-determined |
| 18-102_k141_5386 | Not-determined |
| 18-102_k141_38167 | Not-determined |
| 18-102_k141_100304 | Not-determined |
| 18-102_k141_235094 | Low-quality |
| 18-102_k141_192980 | Not-determined |
| 18-102_k141_120942 | Not-determined |
| 18-102_k141_120947 | Not-determined |
| 18-102_k141_100313 | Not-determined |
| 18-102_k141_38174 | Low-quality |
| 18-102_k141_100318 | Not-determined |
| 18-102_k141_183103 | Not-determined |
| 18-102_k141_183105 | Not-determined |
| 18-102_k141_183106 | Not-determined |
| 18-102_k141_161589 | Not-determined |
| 18-102_k141_26194 | Not-determined |
| 18-102_k141_141526 | Not-determined |
| 18-102_k141_173645 | Not-determined |
| 18-102_k141_173646 | Not-determined |
| 18-102_k141_202367 | Not-determined |
| 18-102_k141_100324 | Not-determined |
| 18-102_k141_183118 | Low-quality |
| 18-102_k141_90194 | Not-determined |
| 18-102_k141_50289 | Not-determined |
| 18-102_k141_90201 | Not-determined |
| 18-102_k141_202380 | Not-determined |
| 18-102_k141_26216 | Low-quality |
| 18-102_k141_79209 | Not-determined |
| 18-102_k141_90206 | Not-determined |
| 18-102_k141_68621 | Not-determined |
| 18-102_k141_141558 | Not-determined |
| 18-102_k141_26218 | Low-quality |
| 18-102_k141_152043 | Not-determined |
| 18-102_k141_58674 | Not-determined |
| 18-102_k141_58675 | Not-determined |
| 18-102_k141_58676 | Not-determined |
| 18-102_k141_141565 | Not-determined |
| 18-102_k141_193011 | Not-determined |
| 18-102_k141_183132 | Not-determined |
| 18-102_k141_79226 | Not-determined |
| 18-102_k141_90219 | Not-determined |
| 18-102_k141_152050 | Low-quality |
| 18-102_k141_183139 | Not-determined |
| 18-102_k141_90222 | Not-determined |
| 18-102_k141_141572 | Not-determined |
| 18-102_k141_5401 | Low-quality |
| 18-102_k141_129869 | Not-determined |
| 18-102_k141_90230 | Not-determined |
| 18-102_k141_68648 | Not-determined |
| 18-102_k141_26229 | Not-determined |
| 18-102_k141_121000 | Not-determined |
| 18-102_k141_26234 | Not-determined |
| 18-102_k141_193020 | Low-quality |
| 18-102_k141_193021 | Not-determined |
| 18-102_k141_50326 | Not-determined |
| 18-102_k141_79254 | Not-determined |
| 18-102_k141_141584 | Low-quality |
| 18-102_k141_152065 | Not-determined |
| 18-102_k141_58688 | Not-determined |
| 18-102_k141_222861 | Not-determined |
| 18-102_k141_235124 | Not-determined |
| 18-102_k141_79258 | Not-determined |
| 18-102_k141_58692 | Not-determined |
| 18-102_k141_141589 | Low-quality |
| 18-102_k141_235129 | Not-determined |
| 18-102_k141_121007 | Not-determined |
| 18-102_k141_68672 | Not-determined |
| 18-102_k141_213247 | Not-determined |
| 18-102_k141_152081 | Not-determined |
| 18-102_k141_26246 | Low-quality |
| 18-102_k141_213250 | Not-determined |
| 18-102_k141_38210 | Not-determined |
| 18-102_k141_38211 | Not-determined |
| 18-102_k141_121017 | Not-determined |
| 18-102_k141_100392 | Low-quality |
| 18-102_k141_183176 | Not-determined |
| 18-102_k141_152096 | Not-determined |
| 18-102_k141_38217 | Not-determined |
| 18-102_k141_193031 | Not-determined |
| 18-102_k141_141602 | Low-quality |
| 18-102_k141_121035 | Not-determined |
| 18-102_k141_235144 | Not-determined |
| 18-102_k141_141603 | Not-determined |
| 18-102_k141_193033 | Not-determined |
| 18-102_k141_222879 | Not-determined |
| 18-102_k141_38221 | Not-determined |
| 18-102_k141_173691 | Not-determined |
| 18-102_k141_121042 | Not-determined |
| 18-102_k141_222881 | Not-determined |
| 18-102_k141_38224 | Not-determined |
| 18-102_k141_50361 | Not-determined |
| 18-102_k141_68694 | Not-determined |
| 18-102_k141_222891 | Low-quality |
| 18-102_k141_183201 | Not-determined |
| 18-102_k141_213264 | Not-determined |
| 18-102_k141_50370 | Not-determined |
| 18-102_k141_38232 | Not-determined |
| 18-102_k141_121065 | Not-determined |
| 18-102_k141_38238 | Not-determined |
| 18-102_k141_183207 | Not-determined |
| 18-102_k141_213282 | Not-determined |
| 18-102_k141_50375 | Not-determined |
| 18-102_k141_50376 | Not-determined |
| 18-102_k141_235169 | Not-determined |
| 18-102_k141_213287 | Not-determined |
| 18-102_k141_222903 | Not-determined |
| 18-102_k141_129880 | Not-determined |
| 18-102_k141_68716 | Not-determined |
| 18-102_k141_235173 | Not-determined |
| 18-102_k141_17186 | Not-determined |
| 18-102_k141_100428 | Not-determined |
| 18-102_k141_235174 | Not-determined |
| 18-102_k141_58712 | Not-determined |
| 18-102_k141_173705 | Not-determined |
| 18-102_k141_38253 | Not-determined |
| 18-102_k141_213293 | Not-determined |
| 18-102_k141_26294 | Not-determined |
| 18-102_k141_68725 | Not-determined |
| 18-102_k141_38262 | Not-determined |
| 18-102_k141_5444 | Not-determined |
| 18-102_k141_5447 | Not-determined |
| 18-102_k141_68737 | Low-quality |
| 18-102_k141_222927 | Not-determined |
| 18-102_k141_5452 | Not-determined |
| 18-102_k141_213308 | Not-determined |
| 18-102_k141_68744 | Not-determined |
| 18-102_k141_68745 | Not-determined |
| 18-102_k141_213309 | Not-determined |
| 18-109-final-viral-combined_k141_156351\|\|full | Medium-quality |
| 18-102_k141_193045 | Not-determined |
| 18-102_k141_79307 | Not-determined |
| 18-102_k141_68754 | Not-determined |
| 18-102_k141_38273 | Not-determined |
| 18-102_k141_26311 | Not-determined |
| 18-102_k141_5464 | Not-determined |
| 18-102_k141_90322 | Not-determined |
| 18-102_k141_100457 | Not-determined |
| 18-102_k141_68763 | Not-determined |
| 18-102_k141_129918 | Not-determined |
| 18-102_k141_213328 | Not-determined |
| 18-102_k141_68768 | Not-determined |
| 18-102_k141_213330 | Not-determined |
| 18-102_k141_100471 | Not-determined |
| 18-102_k141_50433 | Not-determined |
| 18-102_k141_50437 | Not-determined |
| 18-102_k141_235204 | Not-determined |
| 18-102_k141_213336 | Not-determined |
| 18-102_k141_79336 | Low-quality |
| 18-102_k141_79339 | Not-determined |
| 18-102_k141_152109 | Not-determined |
| 18-102_k141_129937 | Low-quality |
| 18-102_k141_38285 | Not-determined |
| 18-102_k141_68776 | Not-determined |
| 18-102_k141_90338 | Not-determined |
| 18-102_k141_193068 | Not-determined |
| 18-102_k141_152120 | Not-determined |
| 18-102_k141_121081 | Not-determined |
| 18-102_k141_173772 | Not-determined |
| 18-102_k141_173780 | Not-determined |
| 18-102_k141_193076 | Low-quality |
| 18-102_k141_50464 | Not-determined |
| 18-102_k141_38301 | Not-determined |
| 18-102_k141_121092 | Not-determined |
| 18-102_k141_173800 | Not-determined |
| 18-102_k141_90362 | Not-determined |
| 18-102_k141_173808 | Not-determined |
| 18-102_k141_68812 | Not-determined |
| 18-102_k141_38309 | Not-determined |
| 18-102_k141_68814 | Not-determined |
| 18-102_k141_38311 | Not-determined |
| 18-102_k141_193098 | Not-determined |
| 18-102_k141_68820 | Not-determined |
| 18-102_k141_90371 | Not-determined |
| 18-102_k141_50480 | Not-determined |
| 18-102_k141_213365 | Not-determined |
| 18-102_k141_68822 | Not-determined |
| 18-102_k141_50484 | Low-quality |
| 18-102_k141_68826 | Not-determined |
| 18-102_k141_173822 | Not-determined |
| 18-102_k141_121106 | Low-quality |
| 18-102_k141_50489 | Not-determined |
| 18-102_k141_121109 | Not-determined |
| 18-102_k141_79349 | Not-determined |
| 18-102_k141_38326 | Low-quality |
| 18-102_k141_79361 | Not-determined |
| 18-102_k141_17246 | Not-determined |
| 18-102_k141_79368 | Not-determined |
| 18-102_k141_38334 | Not-determined |
| 18-102_k141_152172 | Not-determined |
| 18-102_k141_79376 | Not-determined |
| 18-102_k141_79377 | Not-determined |
| 18-102_k141_17258 | Not-determined |
| 18-102_k141_38340 | Not-determined |
| 18-102_k141_90393 | Not-determined |
| 18-102_k141_141615 | Not-determined |
| 18-102_k141_141616 | Low-quality |
| 18-102_k141_129989 | Not-determined |
| 18-102_k141_109844 | Not-determined |
| 18-102_k141_202474 | Low-quality |
| 18-102_k141_129993 | Not-determined |
| 18-102_k141_90404 | Not-determined |
| 18-102_k141_141626 | Not-determined |
| 18-102_k141_38349 | Not-determined |
| 18-102_k141_243683 | Not-determined |
| 18-102_k141_202481 | Not-determined |
| 18-102_k141_100497 | Not-determined |
| 18-102_k141_129998 | Not-determined |
| 18-102_k141_79406 | Not-determined |
| 18-102_k141_141639 | Not-determined |
| 18-102_k141_38354 | Not-determined |
| 18-102_k141_68863 | Not-determined |
| 18-102_k141_202482 | Low-quality |
| 18-102_k141_79410 | Not-determined |
| 18-102_k141_141644 | Not-determined |
| 18-102_k141_68870 | Low-quality |
| 18-102_k141_79417 | Not-determined |
| 18-102_k141_79418 | Not-determined |
| 18-102_k141_79419 | Not-determined |
| 18-102_k141_121143 | Not-determined |
| 18-102_k141_173860 | Low-quality |
| 18-102_k141_121148 | Low-quality |
| 18-102_k141_38363 | Not-determined |
| 18-102_k141_68876 | Not-determined |
| 18-102_k141_193143 | Not-determined |
| 18-102_k141_202493 | Not-determined |
| 18-102_k141_68878 | Not-determined |
| 18-102_k141_121160 | Not-determined |
| 18-102_k141_193147 | Not-determined |
| 18-102_k141_68881 | Not-determined |
| 18-102_k141_100506 | Not-determined |
| 18-102_k141_235283 | Not-determined |
| 18-102_k141_68899 | Not-determined |
| 18-102_k141_90429 | Not-determined |
| 18-102_k141_79433 | Not-determined |
| 18-102_k141_90430 | Low-quality |
| 18-102_k141_193163 | Not-determined |
| 18-102_k141_109866 | Low-quality |
| 18-102_k141_109870 | Not-determined |
| 18-102_k141_161670 | Not-determined |
| 18-102_k141_235297 | Not-determined |
| 18-102_k141_17344 | Not-determined |
| 18-102_k141_141677 | Not-determined |
| 18-102_k141_17348 | Not-determined |
| 18-102_k141_58740 | Not-determined |
| 18-102_k141_161676 | Not-determined |
| 18-102_k141_68911 | Not-determined |
| 18-102_k141_193170 | Not-determined |
| 18-102_k141_109886 | Low-quality |
| 18-102_k141_50515 | Not-determined |
| 18-102_k141_68914 | Not-determined |
| 18-102_k141_161683 | Not-determined |
| 18-102_k141_68921 | Not-determined |
| 18-102_k141_141682 | Not-determined |
| 18-102_k141_235309 | Not-determined |
| 18-102_k141_50528 | Not-determined |
| 18-102_k141_183247 | Not-determined |
| 18-102_k141_79449 | Not-determined |
| 18-102_k141_58749 | Low-quality |
| 18-102_k141_121177 | Not-determined |
| 18-102_k141_68936 | Not-determined |
| 18-102_k141_152239 | Not-determined |
| 18-102_k141_183252 | Low-quality |
| 18-102_k141_121181 | Not-determined |
| 18-102_k141_109893 | Not-determined |
| 18-102_k141_58754 | Not-determined |
| 18-102_k141_38402 | Not-determined |
| 18-102_k141_38403 | Low-quality |
| 18-102_k141_161695 | Not-determined |
| 18-102_k141_50541 | Not-determined |
| 18-102_k141_38411 | Not-determined |
| 18-102_k141_222931 | Not-determined |
| 18-102_k141_173885 | Not-determined |
| 18-102_k141_222932 | Not-determined |
| 18-102_k141_222934 | Not-determined |
| 18-102_k141_152257 | Not-determined |
| 18-102_k141_222935 | Not-determined |
| 18-102_k141_222941 | Not-determined |
| 18-102_k141_58761 | Not-determined |
| 18-102_k141_222948 | Not-determined |
| 18-102_k141_141693 | Not-determined |
| 18-102_k141_68954 | Not-determined |
| 18-102_k141_152270 | Not-determined |
| 18-102_k141_79482 | Not-determined |
| 18-102_k141_222956 | Not-determined |
| 18-102_k141_79486 | Not-determined |
| 18-102_k141_173903 | Not-determined |
| 18-102_k141_100520 | Not-determined |
| 18-102_k141_222958 | Not-determined |
| 18-102_k141_109916 | Not-determined |
| 18-102_k141_130022 | Low-quality |
| 18-102_k141_130023 | Not-determined |
| 18-102_k141_222960 | Not-determined |
| 18-102_k141_100529 | Not-determined |
| 18-102_k141_79488 | Low-quality |
| 18-102_k141_100531 | Not-determined |
| 18-102_k141_121197 | Low-quality |
| 18-102_k141_68959 | Not-determined |
| 18-102_k141_193196 | Not-determined |
| 18-102_k141_243700 | Not-determined |
| 18-102_k141_202535 | Not-determined |
| 18-102_k141_202538 | Not-determined |
| 18-102_k141_141703 | Not-determined |
| 18-102_k141_100538 | Not-determined |
| 18-102_k141_26369 | Low-quality |
| 18-102_k141_161731 | Low-quality |
| 18-102_k141_141710 | Not-determined |
| 18-102_k141_161733 | Not-determined |
| 18-102_k141_173928 | Not-determined |
| 18-102_k141_213478 | Not-determined |
| 18-102_k141_68973 | Not-determined |
| 18-102_k141_213481 | Not-determined |
| 18-102_k141_173937 | Not-determined |
| 18-102_k141_173938 | Not-determined |
| 18-102_k141_109942 | Low-quality |
| 18-102_k141_109944 | Not-determined |
| 18-102_k141_193217 | Not-determined |
| 18-102_k141_235329 | Not-determined |
| 18-102_k141_130035 | Not-determined |
| 18-102_k141_130038 | Not-determined |
| 18-102_k141_141722 | Not-determined |
| 18-102_k141_222979 | Not-determined |
| 18-102_k141_202541 | Not-determined |
| 18-102_k141_222984 | Not-determined |
| 18-102_k141_235336 | Not-determined |
| 18-102_k141_161744 | Not-determined |
| 18-102_k141_161745 | Low-quality |
| 18-102_k141_109950 | Not-determined |
| 18-102_k141_161749 | Not-determined |
| 18-102_k141_161751 | Not-determined |
| 18-102_k141_222990 | Low-quality |
| 18-102_k141_5481 | Not-determined |
| 18-102_k141_161753 | Not-determined |
| 18-102_k141_130050 | Not-determined |
| 18-102_k141_100570 | Not-determined |
| 18-102_k141_183284 | Not-determined |
| 18-102_k141_69003 | Not-determined |
| 18-102_k141_152311 | Not-determined |
| 18-102_k141_5495 | Not-determined |
| 18-102_k141_202555 | Not-determined |
| 18-102_k141_235346 | Not-determined |
| 18-102_k141_235348 | Not-determined |
| 18-102_k141_100584 | Not-determined |
| 18-102_k141_100588 | Not-determined |
| 18-102_k141_58802 | Not-determined |
| 18-102_k141_58803 | Not-determined |
| 18-102_k141_109973 | Not-determined |
| 18-102_k141_223013 | Not-determined |
| 18-102_k141_38492 | Not-determined |
| 18-102_k141_58809 | Low-quality |
| 18-102_k141_193237 | Not-determined |
| 18-102_k141_109983 | Not-determined |
| 18-102_k141_109984 | Not-determined |
| 18-102_k141_5512 | Not-determined |
| 18-102_k141_26425 | Not-determined |
| 18-102_k141_26426 | Not-determined |
| 18-102_k141_50573 | Not-determined |
| 18-102_k141_50575 | Not-determined |
| 18-102_k141_50576 | Not-determined |
| 18-102_k141_130078 | Not-determined |
| 18-102_k141_26432 | Not-determined |
| 18-102_k141_235371 | Not-determined |
| 18-102_k141_130080 | Not-determined |
| 18-102_k141_121218 | Low-quality |

| | |
|---|---|
| 18-102_k141_58816 | Not-determined |
| 18-102_k141_79512 | Low-quality |
| 18-102_k141_69031 | Not-determined |
| 18-102_k141_121223 | Low-quality |
| 18-102_k141_121228 | Not-determined |
| 18-102_k141_223026 | Not-determined |
| 18-102_k141_235375 | Low-quality |
| 18-102_k141_100600 | Not-determined |
| 18-102_k141_141768 | Low-quality |
| 18-102_k141_100606 | Low-quality |
| 18-102_k141_100608 | Low-quality |
| 18-102_k141_38507 | Not-determined |
| 18-102_k141_26453 | Not-determined |
| 18-102_k141_50603 | Low-quality |
| 18-102_k141_26455 | Not-determined |
| 18-102_k141_121241 | Low-quality |
| 18-102_k141_141781 | Not-determined |
| 18-102_k141_235386 | Not-determined |
| 18-102_k141_141784 | Not-determined |
| 18-102_k141_152353 | Not-determined |
| 18-102_k141_26461 | Not-determined |
| 18-102_k141_17376 | Not-determined |
| 18-102_k141_17379 | Not-determined |
| 18-102_k141_223051 | Not-determined |
| 18-102_k141_17385 | Not-determined |
| 18-102_k141_121254 | Not-determined |
| 18-102_k141_223057 | Not-determined |
| 18-102_k141_130094 | Not-determined |
| 18-102_k141_79526 | Low-quality |
| 18-102_k141_121260 | Not-determined |
| 18-102_k141_213515 | Not-determined |
| 18-102_k141_121262 | Not-determined |
| 18-102_k141_121264 | Not-determined |
| 18-102_k141_5560 | Low-quality |
| 18-102_k141_243736 | Not-determined |
| 18-102_k141_235413 | Not-determined |
| 18-102_k141_58838 | Not-determined |
| 18-102_k141_141801 | Not-determined |
| 18-102_k141_100644 | Not-determined |
| 18-102_k141_5561 | Not-determined |
| 18-102_k141_121280 | Not-determined |
| 18-102_k141_235424 | Not-determined |
| 18-102_k141_223075 | Low-quality |
| 18-102_k141_50652 | Low-quality |
| 18-102_k141_121282 | Not-determined |
| 18-102_k141_50655 | Not-determined |
| 18-102_k141_17410 | Not-determined |
| 18-102_k141_121296 | Not-determined |
| 18-102_k141_121301 | Not-determined |
| 18-102_k141_152370 | Not-determined |
| 18-102_k141_38576 | Not-determined |
| 18-102_k141_121307 | Not-determined |
| 18-102_k141_58841 | Not-determined |
| 18-102_k141_79554 | Not-determined |
| 18-102_k141_243759 | Low-quality |
| 18-102_k141_243760 | Not-determined |
| 18-102_k141_100677 | Not-determined |
| 18-102_k141_173990 | Not-determined |
| 18-102_k141_5581 | Not-determined |
| 18-102_k141_141827 | Not-determined |
| 18-102_k141_152384 | Not-determined |
| 18-102_k141_243772 | Not-determined |
| 18-102_k141_152387 | Not-determined |
| 18-102_k141_235455 | Not-determined |
| 18-102_k141_50687 | Not-determined |
| 18-102_k141_17429 | Not-determined |
| 18-102_k141_223093 | Not-determined |
| 18-102_k141_243781 | Not-determined |
| 18-102_k141_152399 | Not-determined |
| 18-102_k141_17449 | Not-determined |
| 18-102_k141_193255 | Not-determined |
| 18-102_k141_152401 | Not-determined |
| 18-102_k141_38606 | Not-determined |
| 18-102_k141_17451 | Not-determined |
| 18-102_k141_174024 | Not-determined |
| 18-102_k141_193259 | Not-determined |
| 18-102_k141_193260 | Not-determined |
| 18-102_k141_130128 | Not-determined |
| 18-102_k141_193262 | Not-determined |
| 18-102_k141_58860 | Not-determined |
| 18-102_k141_243801 | Not-determined |
| 18-102_k141_38618 | Not-determined |
| 18-102_k141_38621 | Low-quality |
| 18-102_k141_243804 | Not-determined |
| 18-102_k141_5612 | Not-determined |
| 18-102_k141_38624 | Not-determined |
| 18-102_k141_243811 | Not-determined |
| 18-102_k141_58872 | Not-determined |
| 18-102_k141_90472 | Not-determined |
| 18-102_k141_17460 | Not-determined |
| 18-102_k141_183288 | Not-determined |
| 18-102_k141_5615 | Not-determined |
| 18-102_k141_121325 | Not-determined |
| 18-102_k141_193284 | Not-determined |
| 18-102_k141_193286 | Not-determined |
| 18-102_k141_243824 | Not-determined |
| 18-102_k141_90480 | Not-determined |
| 18-102_k141_213551 | Not-determined |
| 18-102_k141_213552 | Not-determined |
| 18-102_k141_141854 | Not-determined |
| 18-102_k141_193296 | Low-quality |
| 18-102_k141_58884 | Not-determined |
| 18-102_k141_17471 | Not-determined |
| 18-102_k141_17472 | Not-determined |
| 18-102_k141_121334 | Not-determined |
| 18-102_k141_213554 | Not-determined |
| 18-102_k141_174062 | Not-determined |
| 18-102_k141_50727 | Not-determined |
| 18-102_k141_174063 | Not-determined |
| 18-102_k141_213560 | Not-determined |
| 18-102_k141_243834 | Not-determined |
| 18-102_k141_121344 | Low-quality |
| 18-102_k141_50732 | Not-determined |
| 18-102_k141_100724 | Not-determined |
| 18-102_k141_79570 | Not-determined |
| 18-102_k141_243838 | Not-determined |
| 18-102_k141_50736 | Not-determined |
| 18-102_k141_243839 | Not-determined |
| 18-102_k141_130152 | Not-determined |
| 18-102_k141_58900 | Not-determined |
| 18-102_k141_213567 | Not-determined |
| 18-102_k141_121362 | Not-determined |
| 18-102_k141_100729 | Not-determined |
| 18-102_k141_235476 | Not-determined |
| 18-102_k141_50748 | Not-determined |
| 18-102_k141_243862 | Not-determined |
| 18-102_k141_223112 | Not-determined |
| 18-102_k141_100747 | Not-determined |
| 18-102_k141_38630 | Low-quality |
| 18-102_k141_130188 | Not-determined |
| 18-102_k141_193326 | Not-determined |
| 18-102_k141_130190 | Not-determined |
| 18-102_k141_174101 | Not-determined |
| 18-102_k141_202589 | Not-determined |
| 18-102_k141_141887 | Not-determined |
| 18-102_k141_50762 | Not-determined |
| 18-102_k141_141889 | Not-determined |
| 18-102_k141_174105 | Not-determined |
| 18-102_k141_152420 | Not-determined |
| 18-102_k141_235490 | Not-determined |
| 18-102_k141_130201 | Low-quality |
| 18-102_k141_130204 | Not-determined |
| 18-102_k141_90505 | Not-determined |
| 18-102_k141_141899 | Not-determined |
| 18-102_k141_90515 | Not-determined |
| 18-102_k141_50780 | Not-determined |
| 18-102_k141_235496 | Not-determined |
| 18-102_k141_152439 | Not-determined |
| 18-102_k141_174114 | Not-determined |
| 18-102_k141_50784 | Low-quality |
| 18-102_k141_50789 | Not-determined |
| 18-102_k141_183327 | Low-quality |
| 18-102_k141_235505 | Not-determined |
| 18-102_k141_17508 | Not-determined |
| 18-102_k141_161799 | Not-determined |
| 18-102_k141_38654 | Not-determined |
| 18-102_k141_69072 | Not-determined |
| 18-102_k141_17511 | Low-quality |
| 18-102_k141_69077 | Not-determined |
| 18-102_k141_100800 | Not-determined |
| 18-102_k141_235521 | Not-determined |
| 18-102_k141_100802 | Not-determined |
| 18-102_k141_121384 | Not-determined |
| 18-102_k141_5636 | Not-determined |
| 18-102_k141_121389 | Not-determined |
| 18-102_k141_58937 | Not-determined |
| 18-102_k141_235530 | Not-determined |
| 18-102_k141_58944 | Not-determined |
| 18-102_k141_100824 | Not-determined |
| 18-102_k141_235536 | Low-quality |
| 18-102_k141_152464 | Low-quality |
| 18-102_k141_243914 | Not-determined |
| 18-102_k141_5647 | Not-determined |
| 18-102_k141_100829 | Not-determined |
| 18-102_k141_100835 | Not-determined |
| 18-102_k141_50804 | Not-determined |
| 18-102_k141_141925 | Not-determined |
| 18-102_k141_183363 | Not-determined |
| 18-102_k141_183364 | Not-determined |
| 18-102_k141_50809 | Not-determined |
| 18-102_k141_183369 | Not-determined |
| 18-102_k141_5672 | Not-determined |
| 18-102_k141_17523 | Not-determined |
| 18-102_k141_38688 | Not-determined |
| 18-102_k141_183378 | Not-determined |
| 18-102_k141_223170 | Not-determined |
| 18-102_k141_50837 | Not-determined |
| 18-102_k141_50838 | Not-determined |
| 18-102_k141_110054 | Not-determined |
| 18-102_k141_202661 | Not-determined |
| 18-102_k141_5681 | Not-determined |
| 18-102_k141_174168 | Not-determined |
| 18-102_k141_183382 | Not-determined |
| 18-102_k141_50858 | Not-determined |
| 18-102_k141_141962 | Not-determined |
| 18-102_k141_193390 | Not-determined |
| 18-102_k141_141965 | Not-determined |
| 18-102_k141_223177 | Not-determined |
| 18-102_k141_130220 | Low-quality |
| 18-102_k141_58962 | Low-quality |
| 18-102_k141_174175 | Low-quality |
| 18-102_k141_202674 | Low-quality |
| 18-102_k141_38704 | Not-determined |
| 18-102_k141_183390 | Low-quality |
| 18-102_k141_38717 | Not-determined |
| 18-102_k141_213613 | Not-determined |
| 18-102_k141_100844 | Not-determined |
| 18-102_k141_193409 | Not-determined |
| 18-102_k141_202695 | Not-determined |
| 18-102_k141_213619 | Not-determined |
| 18-102_k141_202696 | Not-determined |
| 18-102_k141_38737 | Not-determined |
| 18-102_k141_152486 | Low-quality |
| 18-102_k141_90544 | Not-determined |
| 18-102_k141_141987 | Not-determined |
| 18-102_k141_141988 | Not-determined |
| 18-102_k141_223196 | Not-determined |
| 18-102_k141_202712 | Not-determined |
| 18-102_k141_243961 | Low-quality |
| 18-102_k141_152491 | Low-quality |
| 18-102_k141_79597 | Not-determined |
| 18-102_k141_202715 | Low-quality |
| 18-102_k141_90547 | Not-determined |
| 18-102_k141_202718 | Not-determined |
| 18-102_k141_90548 | Not-determined |
| 18-102_k141_152499 | Low-quality |
| 18-102_k141_100872 | Low-quality |
| 18-102_k141_5702 | Low-quality |
| 18-102_k141_17568 | Not-determined |
| 18-102_k141_161808 | Not-determined |
| 18-102_k141_5707 | Not-determined |
| 18-102_k141_50898 | Not-determined |
| 18-102_k141_5708 | Not-determined |
| 18-102_k141_161815 | Not-determined |
| 18-102_k141_213632 | Low-quality |
| 18-102_k141_90561 | Not-determined |
| 18-102_k141_223215 | Not-determined |
| 18-102_k141_174212 | Not-determined |
| 18-102_k141_213638 | Not-determined |
| 18-102_k141_5725 | Not-determined |
| 18-102_k141_142025 | Not-determined |
| 18-102_k141_90570 | Low-quality |
| 18-102_k141_100900 | Not-determined |
| 18-102_k141_202727 | Not-determined |
| 18-102_k141_90573 | Not-determined |
| 18-102_k141_152525 | Not-determined |
| 18-102_k141_90578 | Not-determined |
| 18-102_k141_193431 | Not-determined |
| 18-102_k141_193433 | Not-determined |
| 18-102_k141_121408 | Not-determined |
| 18-102_k141_223229 | Not-determined |
| 18-102_k141_243972 | Not-determined |
| 18-102_k141_243973 | Not-determined |
| 18-102_k141_90589 | Not-determined |
| 18-102_k141_90592 | Not-determined |
| 18-102_k141_223235 | Not-determined |
| 18-102_k141_152546 | Low-quality |
| 18-102_k141_58999 | Not-determined |
| 18-102_k141_90597 | Not-determined |
| 18-102_k141_223238 | Not-determined |
| 18-102_k141_79609 | Not-determined |
| 18-102_k141_90600 | Not-determined |
| 18-102_k141_183400 | Not-determined |
| 18-102_k141_59004 | Not-determined |
| 18-102_k141_59008 | Not-determined |
| 18-102_k141_183403 | Not-determined |
| 18-102_k141_59009 | Not-determined |
| 18-102_k141_79618 | Not-determined |
| 18-102_k141_121420 | Not-determined |
| 18-102_k141_17598 | Not-determined |
| 18-102_k141_193457 | Not-determined |
| 18-102_k141_121424 | Not-determined |
| 18-102_k141_202744 | Not-determined |
| 18-102_k141_121428 | Low-quality |
| 18-102_k141_17599 | Not-determined |
| 18-102_k141_79631 | Not-determined |
| 18-102_k141_17602 | Low-quality |
| 18-102_k141_183426 | Low-quality |
| 18-102_k141_38770 | Not-determined |
| 18-102_k141_38772 | Not-determined |
| 18-102_k141_213670 | Not-determined |
| 18-102_k141_183438 | Not-determined |
| 18-102_k141_223259 | Not-determined |
| 18-102_k141_17630 | Not-determined |
| 18-102_k141_17631 | Not-determined |
| 18-102_k141_142063 | Not-determined |
| 18-102_k141_223262 | Not-determined |
| 18-102_k141_202772 | Not-determined |
| 18-102_k141_79644 | Not-determined |
| 18-102_k141_79645 | Not-determined |
| 18-102_k141_202774 | Not-determined |
| 18-102_k141_59020 | Not-determined |
| 18-102_k141_17640 | Not-determined |
| 18-102_k141_223268 | Not-determined |
| 18-102_k141_50942 | Not-determined |
| 18-102_k141_50943 | Not-determined |
| 18-102_k141_69125 | Low-quality |
| 18-102_k141_235635 | Not-determined |
| 18-102_k141_110116 | Not-determined |
| 18-102_k141_243985 | Low-quality |
| 18-102_k141_213684 | Not-determined |
| 18-102_k141_213685 | Not-determined |
| 18-102_k141_69137 | Not-determined |
| 18-102_k141_223282 | Not-determined |
| 18-102_k141_223283 | Not-determined |
| 18-102_k141_213696 | Not-determined |
| 18-102_k141_183457 | Not-determined |
| 18-102_k141_235642 | Not-determined |
| 18-102_k141_213705 | Not-determined |
| 18-102_k141_17677 | Not-determined |
| 18-102_k141_223294 | Not-determined |
| 18-102_k141_59037 | Not-determined |
| 18-102_k141_130251 | Not-determined |
| 18-102_k141_26494 | Not-determined |
| 18-102_k141_110126 | Not-determined |
| 18-102_k141_130259 | Not-determined |
| 18-102_k141_79664 | Not-determined |
| 18-102_k141_235652 | Not-determined |
| 18-102_k141_183467 | Not-determined |
| 18-102_k141_79670 | Low-quality |
| 18-102_k141_183472 | Not-determined |
| 18-102_k141_79677 | Not-determined |
| 18-102_k141_79678 | Not-determined |
| 18-102_k141_121461 | Low-quality |
| 18-102_k141_213712 | Not-determined |
| 18-102_k141_243991 | Not-determined |
| 18-102_k141_213720 | Not-determined |
| 18-102_k141_213724 | Not-determined |
| 18-102_k141_79681 | Low-quality |
| 18-102_k141_110151 | Not-determined |
| 18-102_k141_5752 | Not-determined |
| 18-102_k141_235675 | Low-quality |
| 18-102_k141_183489 | Not-determined |
| 18-102_k141_174250 | Not-determined |
| 18-102_k141_213738 | Not-determined |
| 18-102_k141_152588 | Not-determined |
| 18-102_k141_79692 | Not-determined |
| 18-102_k141_121480 | Not-determined |
| 18-102_k141_26511 | Not-determined |
| 18-102_k141_152599 | Not-determined |
| 18-102_k141_174273 | Not-determined |
| 18-102_k141_193496 | Not-determined |
| 18-102_k141_38811 | Not-determined |
| 18-102_k141_213749 | Not-determined |
| 18-102_k141_244014 | Not-determined |
| 18-102_k141_152603 | Not-determined |
| 18-102_k141_69183 | Not-determined |
| 18-102_k141_69190 | Not-determined |
| 18-102_k141_202829 | Not-determined |
| 18-102_k141_59058 | Low-quality |
| 18-102_k141_69196 | Not-determined |
| 18-102_k141_110162 | Not-determined |
| 18-102_k141_161847 | Not-determined |
| 18-102_k141_161848 | Low-quality |
| 18-102_k141_202837 | Not-determined |
| 18-102_k141_193524 | Not-determined |
| 18-102_k141_213761 | Not-determined |
| 18-102_k141_244030 | Low-quality |
| 18-102_k141_69226 | Not-determined |
| 18-102_k141_235717 | Low-quality |
| 18-102_k141_69233 | Low-quality |
| 18-102_k141_161862 | Not-determined |
| 18-102_k141_161863 | Not-determined |
| 18-102_k141_244041 | Not-determined |
| 18-102_k141_59094 | Not-determined |
| 18-102_k141_183515 | Not-determined |
| 18-102_k141_223342 | Not-determined |
| 18-102_k141_213781 | Not-determined |
| 18-102_k141_69242 | Low-quality |
| 18-102_k141_130323 | Not-determined |
| 18-102_k141_235745 | Not-determined |
| 18-102_k141_244059 | Not-determined |
| 18-102_k141_79713 | Not-determined |
| 18-102_k141_244064 | Not-determined |
| 18-102_k141_183535 | Not-determined |
| 18-102_k141_202858 | Not-determined |
| 18-102_k141_244065 | Not-determined |
| 18-102_k141_235755 | Not-determined |
| 18-102_k141_161872 | Not-determined |
| 18-102_k141_235759 | Not-determined |
| 18-102_k141_69256 | Not-determined |
| 18-102_k141_50959 | Not-determined |
| 18-102_k141_174302 | Low-quality |
| 18-102_k141_79723 | Low-quality |
| 18-102_k141_79726 | Not-determined |
| 18-102_k141_50964 | Not-determined |
| 18-102_k141_59125 | Not-determined |
| 18-102_k141_110214 | Low-quality |
| 18-102_k141_59130 | Not-determined |
| 18-102_k141_142074 | Not-determined |
| 18-102_k141_142079 | Not-determined |
| 18-102_k141_152652 | Not-determined |
| 18-102_k141_121536 | Not-determined |
| 18-102_k141_59135 | Not-determined |
| 18-102_k141_142084 | Not-determined |
| 18-102_k141_244088 | Not-determined |
| 18-102_k141_79737 | Not-determined |
| 18-102_k141_244090 | Not-determined |
| 18-102_k141_244092 | Not-determined |
| 18-102_k141_223359 | Not-determined |
| 18-102_k141_59156 | Not-determined |
| 18-102_k141_161879 | Not-determined |
| 18-102_k141_193576 | Not-determined |
| 18-102_k141_223364 | Not-determined |
| 18-102_k141_26522 | Not-determined |
| 18-102_k141_183546 | Not-determined |
| 18-102_k141_142111 | Low-quality |
| 18-102_k141_174314 | Low-quality |
| 18-102_k141_50993 | Not-determined |
| 18-102_k141_193593 | Not-determined |
| 18-102_k141_223379 | Not-determined |
| 18-102_k141_193594 | Not-determined |
| 18-102_k141_244100 | Not-determined |
| 18-102_k141_174321 | Not-determined |
| 18-102_k141_26530 | Low-quality |
| 18-102_k141_79762 | Not-determined |
| 18-102_k141_223388 | Not-determined |
| 18-102_k141_244109 | Not-determined |
| 18-102_k141_26535 | Not-determined |
| 18-102_k141_142139 | Not-determined |
| 18-102_k141_79768 | Not-determined |
| 18-102_k141_223394 | Low-quality |
| 18-102_k141_174330 | Not-determined |
| 18-102_k141_223395 | Not-determined |
| 18-102_k141_183560 | Not-determined |
| 18-102_k141_142142 | Not-determined |
| 18-102_k141_183565 | Not-determined |
| 18-102_k141_202879 | Low-quality |
| 18-102_k141_202880 | Not-determined |
| 18-102_k141_69284 | Low-quality |
| 18-102_k141_223406 | Not-determined |
| 18-102_k141_174353 | Not-determined |
| 18-102_k141_223411 | Not-determined |
| 18-102_k141_174354 | Not-determined |
| 18-102_k141_223412 | Low-quality |
| 18-102_k141_202898 | Not-determined |
| 18-102_k141_142158 | Not-determined |
| 18-102_k141_79783 | Not-determined |
| 18-102_k141_202906 | Not-determined |
| 18-102_k141_223414 | Not-determined |
| 18-102_k141_100994 | Not-determined |
| 18-102_k141_223418 | Not-determined |
| 18-102_k141_223423 | Not-determined |
| 18-102_k141_51044 | Not-determined |
| 18-102_k141_121560 | Not-determined |
| 18-102_k141_38836 | Not-determined |
| 18-102_k141_101001 | Not-determined |
| 18-102_k141_17717 | Not-determined |
| 18-102_k141_213799 | Not-determined |
| 18-102_k141_79802 | Not-determined |
| 18-102_k141_244147 | Not-determined |
| 18-102_k141_213807 | Not-determined |
| 18-102_k141_38845 | Not-determined |
| 18-102_k141_223431 | Not-determined |
| 18-102_k141_69318 | Not-determined |
| 18-102_k141_59179 | Not-determined |
| 18-102_k141_193615 | Not-determined |
| 18-102_k141_121564 | Not-determined |
| 18-102_k141_101016 | Not-determined |
| 18-102_k141_223443 | Not-determined |
| 18-102_k141_101017 | Not-determined |
| 18-102_k141_121569 | Low-quality |
| 18-102_k141_142178 | Not-determined |
| 18-102_k141_79816 | Not-determined |
| 18-102_k141_121570 | Low-quality |
| 18-102_k141_17724 | Not-determined |
| 18-102_k141_101019 | Not-determined |
| 18-102_k141_235851 | Not-determined |
| 18-102_k141_193624 | Not-determined |
| 18-102_k141_17730 | Not-determined |
| 18-102_k141_121571 | Not-determined |
| 18-102_k141_142185 | Not-determined |
| 18-102_k141_183587 | Not-determined |
| 18-102_k141_193635 | Not-determined |
| 18-102_k141_183589 | Not-determined |
| 18-102_k141_121577 | Not-determined |
| 18-102_k141_174397 | Not-determined |
| 18-102_k141_17740 | Not-determined |
| 18-102_k141_38865 | Low-quality |
| 18-102_k141_38866 | Not-determined |
| 18-102_k141_38867 | Not-determined |
| 18-102_k141_223449 | Not-determined |
| 18-102_k141_223450 | Not-determined |
| 18-102_k141_69349 | Not-determined |
| 18-102_k141_101045 | Not-determined |
| 18-102_k141_213822 | Not-determined |
| 18-102_k141_38874 | Low-quality |
| 18-102_k141_38876 | Not-determined |
| 18-102_k141_235869 | Low-quality |
| 18-102_k141_51076 | Not-determined |
| 18-102_k141_51077 | Not-determined |
| 18-102_k141_121586 | Not-determined |
| 18-102_k141_69354 | Low-quality |
| 18-102_k141_183594 | Not-determined |
| 18-102_k141_38884 | Not-determined |
| 18-102_k141_174416 | Not-determined |
| 18-102_k141_235876 | Not-determined |
| 18-102_k141_38894 | Not-determined |
| 18-102_k141_101075 | Low-quality |
| 18-102_k141_174422 | Not-determined |
| 18-102_k141_183611 | Not-determined |
| 18-102_k141_235883 | Not-determined |
| 18-102_k141_26572 | Not-determined |
| 18-102_k141_235886 | Not-determined |
| 18-102_k141_193654 | Not-determined |
| 18-102_k141_101090 | Low-quality |
| 18-102_k141_69368 | Not-determined |
| 18-102_k141_183617 | Not-determined |
| 18-102_k141_193656 | Not-determined |
| 18-102_k141_193658 | Low-quality |
| 18-102_k141_174435 | Not-determined |
| 18-102_k141_101091 | Not-determined |
| 18-102_k141_174437 | Not-determined |
| 18-102_k141_174440 | Not-determined |
| 18-102_k141_174441 | Not-determined |
| 18-102_k141_213828 | Not-determined |
| 18-102_k141_174444 | Not-determined |
| 18-102_k141_121599 | Not-determined |
| 18-102_k141_174446 | Not-determined |
| 18-102_k141_101096 | Low-quality |
| 18-102_k141_213832 | Not-determined |
| 18-102_k141_69373 | Not-determined |
| 18-102_k141_59205 | Not-determined |
| 18-102_k141_69375 | Not-determined |
| 18-102_k141_101103 | Not-determined |
| 18-102_k141_69381 | Not-determined |
| 18-102_k141_101105 | Not-determined |
| 18-102_k141_235899 | Not-determined |
| 18-102_k141_51082 | Not-determined |
| 18-102_k141_51084 | Low-quality |
| 18-102_k141_69391 | Not-determined |
| 18-102_k141_202967 | Not-determined |
| 18-102_k141_121613 | Not-determined |
| 18-102_k141_59220 | Not-determined |
| 18-102_k141_213845 | Not-determined |
| 18-102_k141_121615 | Low-quality |
| 18-102_k141_183633 | Not-determined |
| 18-102_k141_59223 | Not-determined |
| 18-102_k141_213849 | Not-determined |
| 18-102_k141_213850 | Low-quality |
| 18-102_k141_193691 | Low-quality |
| 18-102_k141_193695 | Not-determined |
| 18-102_k141_121626 | Not-determined |
| 18-102_k141_51101 | Not-determined |
| 18-102_k141_244160 | Not-determined |
| 18-102_k141_213864 | Not-determined |
| 18-102_k141_110243 | Not-determined |
| 18-102_k141_223485 | Not-determined |
| 18-102_k141_223489 | Not-determined |
| 18-102_k141_202978 | Low-quality |
| 18-102_k141_17763 | Not-determined |
| 18-102_k141_130346 | Not-determined |
| 18-102_k141_213869 | Not-determined |
| 18-102_k141_202982 | Not-determined |
| 18-102_k141_142205 | Low-quality |
| 18-102_k141_69422 | Not-determined |
| 18-102_k141_17772 | Low-quality |
| 18-102_k141_183644 | Not-determined |
| 18-102_k141_69429 | Not-determined |
| 18-102_k141_69434 | Not-determined |
| 18-102_k141_59255 | Not-determined |
| 18-102_k141_26604 | Not-determined |
| 18-102_k141_235931 | Not-determined |
| 18-102_k141_51103 | Not-determined |
| 18-102_k141_202992 | Not-determined |
| 18-102_k141_38936 | Not-determined |
| 18-102_k141_5811 | Not-determined |
| 18-102_k141_142214 | Not-determined |
| 18-102_k141_59270 | Not-determined |
| 18-102_k141_142221 | Not-determined |
| 18-102_k141_130370 | Not-determined |
| 18-102_k141_235933 | Not-determined |
| 18-102_k141_161933 | Not-determined |
| 18-102_k141_235934 | Not-determined |
| 18-102_k141_161935 | Not-determined |
| 18-102_k141_38944 | Not-determined |
| 18-102_k141_38945 | Not-determined |
| 18-102_k141_38949 | Not-determined |
| 18-102_k141_235936 | Not-determined |
| 18-102_k141_59285 | Not-determined |
| 18-102_k141_193733 | Not-determined |
| 18-102_k141_38953 | Not-determined |
| 18-102_k141_59289 | Not-determined |
| 18-102_k141_69451 | Not-determined |
| 18-102_k141_26629 | Low-quality |
| 18-102_k141_161946 | Not-determined |
| 18-102_k141_38968 | Not-determined |
| 18-102_k141_5824 | Not-determined |
| 18-102_k141_59308 | Not-determined |

| | |
|---|---|
| 18-102_k141_38969 | Not-determined |
| 18-102_k141_59311 | Not-determined |
| 18-102_k141_5826 | Low-quality |
| 18-102_k141_244187 | Not-determined |
| 18-102_k141_161956 | Not-determined |
| 18-102_k141_161957 | Not-determined |
| 18-102_k141_235954 | Not-determined |
| 18-102_k141_193737 | Not-determined |
| 18-102_k141_51113 | Not-determined |
| 18-102_k141_69470 | Low-quality |
| 18-102_k141_193739 | Not-determined |
| 18-102_k141_17792 | Not-determined |
| 18-102_k141_193741 | Low-quality |
| 18-102_k141_17795 | Low-quality |
| 18-102_k141_38985 | Not-determined |
| 18-102_k141_183656 | Not-determined |
| 18-102_k141_17798 | Not-determined |
| 18-102_k141_90655 | Not-determined |
| 18-102_k141_174453 | Not-determined |
| 18-102_k141_90660 | Low-quality |
| 18-102_k141_193751 | Not-determined |
| 18-102_k141_235975 | Not-determined |
| 18-102_k141_203019 | Not-determined |
| 18-102_k141_203021 | Not-determined |
| 18-102_k141_51127 | Not-determined |
| 18-102_k141_38995 | Not-determined |
| 18-102_k141_110286 | Not-determined |
| 18-102_k141_174464 | Not-determined |
| 18-102_k141_161977 | Not-determined |
| 18-102_k141_235989 | Not-determined |
| 18-102_k141_193765 | Not-determined |
| 18-102_k141_130393 | Not-determined |
| 18-102_k141_130400 | Not-determined |
| 18-102_k141_17805 | Low-quality |
| 18-102_k141_51143 | Not-determined |
| 18-102_k141_203035 | Low-quality |
| 18-102_k141_152726 | Not-determined |
| 18-102_k141_223549 | Low-quality |
| 18-102_k141_69500 | Not-determined |
| 18-102_k141_59331 | Not-determined |
| 18-102_k141_90679 | Not-determined |
| 18-102_k141_90681 | Not-determined |
| 18-102_k141_130414 | Not-determined |
| 18-102_k141_203038 | Not-determined |
| 18-102_k141_203039 | Not-determined |
| 18-102_k141_183693 | Not-determined |
| 18-102_k141_17832 | Low-quality |
| 18-102_k141_59338 | Not-determined |
| 18-102_k141_17833 | Not-determined |
| 18-102_k141_130420 | Not-determined |
| 18-102_k141_223565 | Not-determined |
| 18-102_k141_183698 | Not-determined |
| 18-102_k141_236000 | Low-quality |
| 18-102_k141_90687 | Low-quality |
| 18-102_k141_236005 | Not-determined |
| 18-102_k141_142237 | Not-determined |
| 18-102_k141_121691 | Not-determined |
| 18-102_k141_90700 | Not-determined |
| 18-102_k141_17863 | Not-determined |
| 18-102_k141_17869 | Not-determined |
| 18-102_k141_183721 | Not-determined |
| 18-102_k141_69538 | Not-determined |
| 18-102_k141_152764 | Not-determined |
| 18-102_k141_223576 | Not-determined |
| 18-102_k141_193811 | Not-determined |
| 18-102_k141_223580 | Not-determined |
| 18-102_k141_69547 | Not-determined |
| 18-102_k141_90727 | Not-determined |
| 18-109_k141_123954 | Medium-quality |
| 18-102_k141_121721 | Low-quality |
| 18-102_k141_26653 | Not-determined |
| 18-102_k141_236015 | Low-quality |
| 18-102_k141_121729 | Low-quality |
| 18-102_k141_17874 | Not-determined |
| 18-102_k141_121732 | Not-determined |
| 18-102_k141_152791 | Not-determined |
| 18-102_k141_236023 | Not-determined |
| 18-102_k141_213923 | Not-determined |
| 18-102_k141_59366 | Not-determined |
| 18-102_k141_59371 | Low-quality |
| 18-102_k141_17880 | Not-determined |
| 18-102_k141_223609 | Not-determined |
| 18-102_k141_51168 | Not-determined |
| 18-102_k141_213937 | Not-determined |
| 18-102_k141_213939 | Not-determined |
| 18-102_k141_236036 | Not-determined |
| 18-102_k141_223616 | Not-determined |
| 18-102_k141_39031 | Not-determined |
| 18-102_k141_223620 | Not-determined |
| 18-102_k141_51180 | Not-determined |
| 18-102_k141_244202 | Not-determined |
| 18-102_k141_213957 | Not-determined |
| 18-102_k141_51186 | Not-determined |
| 18-102_k141_152795 | Not-determined |
| 18-102_k141_244205 | Not-determined |
| 18-102_k141_26683 | Not-determined |
| 18-102_k141_26684 | Low-quality |
| 18-102_k141_152803 | Not-determined |
| 18-102_k141_244208 | Not-determined |
| 18-102_k141_203073 | Not-determined |
| 18-102_k141_51195 | Low-quality |
| 18-102_k141_39046 | Not-determined |
| 18-102_k141_244213 | Not-determined |
| 18-102_k141_39051 | Not-determined |
| 18-102_k141_130437 | Not-determined |
| 18-102_k141_101179 | Not-determined |
| 18-102_k141_203080 | Not-determined |
| 18-102_k141_51201 | Low-quality |
| 18-102_k141_183774 | Low-quality |
| 18-102_k141_213969 | Not-determined |
| 18-102_k141_183775 | Not-determined |
| 18-102_k141_110310 | Low-quality |
| 18-102_k141_69597 | Not-determined |
| 18-102_k141_69599 | Not-determined |
| 18-102_k141_244224 | Low-quality |
| 18-102_k141_69606 | Not-determined |
| 18-102_k141_110315 | Not-determined |
| 18-102_k141_79875 | Not-determined |
| 18-102_k141_152827 | Not-determined |
| 18-102_k141_121793 | Not-determined |
| 18-102_k141_121794 | Not-determined |
| 18-102_k141_142261 | Not-determined |
| 18-102_k141_213981 | Not-determined |
| 18-102_k141_51209 | Not-determined |
| 18-102_k141_142263 | Not-determined |
| 18-102_k141_110320 | Not-determined |
| 18-102_k141_110321 | Not-determined |
| 18-102_k141_213986 | Not-determined |
| 18-102_k141_213991 | Low-quality |
| 18-102_k141_213992 | Not-determined |
| 18-102_k141_203096 | Not-determined |
| 18-102_k141_79887 | Not-determined |
| 18-102_k141_110336 | Not-determined |
| 18-102_k141_110339 | Not-determined |
| 18-102_k141_110340 | Not-determined |
| 18-102_k141_101205 | Not-determined |
| 18-102_k141_214006 | Not-determined |
| 18-102_k141_244241 | Not-determined |
| 18-102_k141_244242 | Not-determined |
| 18-102_k141_193854 | Not-determined |
| 18-102_k141_101210 | Not-determined |
| 18-102_k141_193860 | Not-determined |
| 18-102_k141_39089 | Not-determined |
| 18-102_k141_193862 | Not-determined |
| 18-102_k141_214013 | Not-determined |
| 18-102_k141_51237 | Not-determined |
| 18-102_k141_130472 | Not-determined |
| 18-102_k141_51243 | Not-determined |
| 18-102_k141_193871 | Not-determined |
| 18-102_k141_130478 | Low-quality |
| 18-102_k141_90746 | Low-quality |
| 18-102_k141_90748 | Not-determined |
| 18-102_k141_142294 | Not-determined |
| 18-102_k141_193876 | Not-determined |
| 18-102_k141_214028 | Not-determined |
| 18-102_k141_142295 | Not-determined |
| 18-102_k141_101222 | Low-quality |
| 18-102_k141_152856 | Not-determined |
| 18-102_k141_39108 | Not-determined |
| 18-102_k141_59394 | Not-determined |
| 18-102_k141_130496 | Not-determined |
| 18-102_k141_214034 | Not-determined |
| 18-102_k141_193889 | Low-quality |
| 18-102_k141_214039 | Not-determined |
| 18-102_k141_17903 | Not-determined |
| 18-102_k141_193894 | Not-determined |
| 18-102_k141_214042 | Not-determined |
| 18-102_k141_59402 | Not-determined |
| 18-102_k141_183790 | Not-determined |
| 18-102_k141_152872 | Low-quality |
| 18-102_k141_183796 | Not-determined |
| 18-102_k141_142308 | Not-determined |
| 18-102_k141_193911 | Not-determined |
| 18-102_k141_69659 | Not-determined |
| 18-102_k141_214051 | Not-determined |
| 18-102_k141_183804 | Not-determined |
| 18-102_k141_110387 | Not-determined |
| 18-102_k141_17914 | Not-determined |
| 18-102_k141_69666 | Not-determined |
| 18-102_k141_214058 | Not-determined |
| 18-102_k141_110390 | Not-determined |
| 18-102_k141_214065 | Not-determined |
| 18-102_k141_193921 | Not-determined |
| 18-102_k141_5864 | Not-determined |
| 18-102_k141_214076 | Low-quality |
| 18-102_k141_244257 | Not-determined |
| 18-102_k141_214079 | Not-determined |
| 18-102_k141_17926 | Not-determined |
| 18-102_k141_152920 | Not-determined |
| 18-102_k141_130517 | Not-determined |
| 18-102_k141_142329 | Not-determined |
| 18-102_k141_51283 | Not-determined |
| 18-102_k141_214084 | Low-quality |
| 18-102_k141_223642 | Low-quality |
| 18-102_k141_183818 | Not-determined |
| 18-102_k141_223643 | Not-determined |
| 18-102_k141_90783 | Not-determined |
| 18-102_k141_130532 | Not-determined |
| 18-102_k141_39134 | Not-determined |
| 18-102_k141_130534 | Not-determined |
| 18-102_k141_142330 | Low-quality |
| 18-102_k141_59443 | Not-determined |
| 18-102_k141_142333 | Not-determined |
| 18-102_k141_214086 | Not-determined |
| 18-102_k141_59447 | Not-determined |
| 18-102_k141_39139 | Not-determined |
| 18-102_k141_69699 | Not-determined |
| 18-102_k141_69700 | Not-determined |
| 18-102_k141_203139 | Not-determined |
| 18-102_k141_174510 | Not-determined |
| 18-102_k141_39143 | Not-determined |
| 18-102_k141_59459 | Not-determined |
| 18-102_k141_183853 | Not-determined |
| 18-102_k141_59464 | Not-determined |
| 18-102_k141_59466 | Not-determined |
| 18-102_k141_244268 | Not-determined |
| 18-101-final-viral-combined_k141_308848\|\|full | Medium-quality |
| 18-102_k141_223661 | Not-determined |
| 18-102_k141_152951 | Not-determined |
| 18-102_k141_130553 | Not-determined |
| 18-102_k141_152954 | Low-quality |
| 18-102_k141_152962 | Not-determined |
| 18-102_k141_39172 | Not-determined |
| 18-102_k141_39176 | Not-determined |
| 18-102_k141_152971 | Not-determined |
| 18-102_k141_223675 | Not-determined |
| 18-102_k141_223676 | Not-determined |
| 18-102_k141_59488 | Not-determined |
| 18-102_k141_130581 | Not-determined |
| 18-102_k141_59492 | Not-determined |
| 18-102_k141_130582 | Not-determined |
| 18-102_k141_69711 | Not-determined |
| 18-102_k141_193944 | Not-determined |
| 18-102_k141_183878 | Low-quality |
| 18-102_k141_193948 | Not-determined |
| 18-102_k141_51318 | Not-determined |
| 18-102_k141_51320 | Not-determined |
| 18-102_k141_223693 | Low-quality |
| 18-102_k141_193951 | Not-determined |
| 18-102_k141_174537 | Not-determined |
| 18-102_k141_244285 | Not-determined |
| 18-102_k141_223698 | Not-determined |
| 18-102_k141_152977 | Not-determined |
| 18-102_k141_223709 | Not-determined |
| 18-102_k141_90800 | Not-determined |
| 18-102_k141_223714 | Low-quality |
| 18-102_k141_223719 | Not-determined |
| 18-102_k141_152995 | Not-determined |
| 18-102_k141_51339 | Not-determined |
| 18-102_k141_5896 | Not-determined |
| 18-102_k141_5906 | Not-determined |
| 18-102_k141_101261 | Not-determined |
| 18-102_k141_193982 | Not-determined |
| 18-102_k141_39207 | Not-determined |
| 18-102_k141_130619 | Not-determined |
| 18-102_k141_130621 | Not-determined |
| 18-102_k141_51344 | Not-determined |
| 18-102_k141_26688 | Not-determined |
| 18-102_k141_236113 | Not-determined |
| 18-102_k141_59519 | Not-determined |
| 18-102_k141_51350 | Not-determined |
| 18-102_k141_101265 | Not-determined |
| 18-102_k141_59522 | Not-determined |
| 18-102_k141_59523 | Not-determined |
| 18-102_k141_59524 | Not-determined |
| 18-102_k141_223740 | Not-determined |
| 18-102_k141_110443 | Not-determined |
| 18-102_k141_110447 | Not-determined |
| 18-102_k141_194002 | Not-determined |
| 18-102_k141_203169 | Not-determined |
| 18-102_k141_223745 | Not-determined |
| 18-102_k141_101276 | Not-determined |
| 18-102_k141_174563 | Not-determined |
| 18-102_k141_101278 | Not-determined |
| 18-102_k141_236129 | Not-determined |
| 18-102_k141_236133 | Not-determined |
| 18-102_k141_5930 | Not-determined |
| 18-102_k141_174576 | Not-determined |
| 18-102_k141_51378 | Not-determined |
| 18-102_k141_174577 | Not-determined |
| 18-102_k141_161994 | Not-determined |
| 18-102_k141_244292 | Not-determined |
| 18-102_k141_203181 | Not-determined |
| 18-102_k141_5933 | Not-determined |
| 18-102_k141_203183 | Not-determined |
| 18-102_k141_153046 | Not-determined |
| 18-102_k141_121861 | Not-determined |
| 18-102_k141_5940 | Low-quality |
| 18-102_k141_194016 | Not-determined |
| 18-102_k141_183953 | Not-determined |
| 18-102_k141_194018 | Low-quality |
| 18-102_k141_183955 | Not-determined |
| 18-102_k141_153048 | Low-quality |
| 18-102_k141_5943 | Not-determined |
| 18-102_k141_194021 | Low-quality |
| 18-102_k141_153050 | Not-determined |
| 18-102_k141_5946 | Not-determined |
| 18-102_k141_5948 | Low-quality |
| 18-102_k141_194029 | Not-determined |
| 18-102_k141_214134 | Not-determined |
| 18-102_k141_174589 | Not-determined |
| 18-102_k141_39215 | Not-determined |
| 18-102_k141_236140 | Not-determined |
| 18-102_k141_244304 | Not-determined |
| 18-102_k141_203195 | Not-determined |
| 18-102_k141_59556 | Not-determined |
| 18-102_k141_39222 | Not-determined |
| 18-102_k141_59557 | Not-determined |
| 18-102_k141_130663 | Not-determined |
| 18-102_k141_101311 | Not-determined |
| 18-102_k141_59564 | Not-determined |
| 18-102_k141_236153 | Not-determined |
| 18-102_k141_223783 | Not-determined |
| 18-102_k141_203200 | Not-determined |
| 18-102_k141_130672 | Not-determined |
| 18-102_k141_174602 | Not-determined |
| 18-102_k141_244314 | Not-determined |
| 18-102_k141_244315 | Low-quality |
| 18-102_k141_194052 | Not-determined |
| 18-102_k141_130678 | Not-determined |
| 18-102_k141_69747 | Not-determined |
| 18-102_k141_183993 | Not-determined |
| 18-102_k141_194058 | Not-determined |
| 18-102_k141_110500 | Not-determined |
| 18-102_k141_194059 | Not-determined |
| 18-102_k141_184002 | Not-determined |
| 18-102_k141_184005 | Not-determined |
| 18-102_k141_5975 | Not-determined |
| 18-102_k141_101330 | Not-determined |
| 18-102_k141_80011 | Low-quality |
| 18-102_k141_51398 | Not-determined |
| 18-102_k141_26709 | Not-determined |
| 18-102_k141_69769 | Not-determined |
| 18-102_k141_17986 | Not-determined |
| 18-102_k141_194078 | Not-determined |
| 18-102_k141_174618 | Not-determined |
| 18-102_k141_101339 | Low-quality |
| 18-102_k141_101344 | Not-determined |
| 18-102_k141_194093 | Not-determined |
| 18-102_k141_80028 | Not-determined |
| 18-102_k141_214162 | Not-determined |
| 18-102_k141_90839 | Not-determined |
| 18-102_k141_39249 | Not-determined |
| 18-102_k141_39253 | Not-determined |
| 18-102_k141_194101 | Not-determined |
| 18-102_k141_203242 | Not-determined |
| 18-102_k141_80042 | Not-determined |
| 18-102_k141_162066 | Not-determined |
| 18-102_k141_80051 | Not-determined |
| 18-102_k141_39267 | Not-determined |
| 18-102_k141_174639 | Not-determined |
| 18-102_k141_51408 | Not-determined |
| 18-102_k141_80060 | Not-determined |
| 18-102_k141_69789 | Not-determined |
| 18-102_k141_18012 | Not-determined |
| 18-102_k141_223801 | Not-determined |
| 18-102_k141_80067 | Not-determined |
| 18-102_k141_174647 | Not-determined |
| 18-102_k141_162076 | Low-quality |
| 18-102_k141_90849 | Not-determined |
| 18-102_k141_162077 | Not-determined |
| 18-102_k141_26716 | Not-determined |
| 18-102_k141_26717 | Not-determined |
| 18-102_k141_203249 | Not-determined |
| 18-102_k141_39276 | Low-quality |
| 18-102_k141_101379 | Not-determined |
| 18-102_k141_184028 | Not-determined |
| 18-102_k141_69800 | Low-quality |
| 18-102_k141_214190 | Not-determined |
| 18-102_k141_51429 | Not-determined |
| 18-102_k141_203269 | Not-determined |
| 18-102_k141_174670 | Not-determined |
| 18-102_k141_69804 | Not-determined |
| 18-102_k141_101398 | Low-quality |
| 18-102_k141_174673 | Not-determined |
| 18-102_k141_110547 | Not-determined |
| 18-102_k141_236198 | Not-determined |
| 18-102_k141_69811 | Not-determined |
| 18-102_k141_26730 | Not-determined |
| 18-102_k141_6052 | Not-determined |
| 18-102_k141_184047 | Low-quality |
| 18-102_k141_184049 | Not-determined |
| 18-102_k141_110554 | Low-quality |
| 18-102_k141_223836 | Not-determined |
| 18-102_k141_6057 | Not-determined |
| 18-102_k141_39289 | Not-determined |
| 18-102_k141_223840 | Not-determined |
| 18-102_k141_184059 | Low-quality |
| 18-102_k141_69825 | Not-determined |
| 18-102_k141_69828 | Not-determined |
| 18-102_k141_184065 | Not-determined |
| 18-102_k141_162119 | Not-determined |
| 18-102_k141_203278 | Not-determined |
| 18-102_k141_162121 | Not-determined |
| 18-102_k141_203282 | Not-determined |
| 18-102_k141_6073 | Not-determined |
| 18-102_k141_39301 | Not-determined |
| 18-102_k141_26736 | Low-quality |
| 18-110-final-viral-combined_k141_23371\|\|full | Medium-quality |
| 18-102_k141_69851 | Not-determined |
| 18-102_k141_39312 | Not-determined |
| 18-102_k141_153095 | Not-determined |
| 18-102_k141_39316 | Not-determined |
| 18-102_k141_69856 | Not-determined |
| 18-102_k141_101429 | Not-determined |
| 18-102_k141_90896 | Low-quality |
| 18-102_k141_184084 | Not-determined |
| 18-102_k141_6077 | Not-determined |
| 18-102_k141_236205 | Not-determined |
| 18-102_k141_18073 | Not-determined |
| 18-102_k141_236206 | Not-determined |
| 18-102_k141_110583 | Not-determined |
| 18-102_k141_39325 | Not-determined |
| 18-102_k141_69862 | Low-quality |
| 18-102_k141_236212 | Not-determined |
| 18-102_k141_162146 | Not-determined |
| 18-102_k141_142358 | Not-determined |
| 18-102_k141_90904 | Not-determined |
| 18-102_k141_142359 | Not-determined |
| 18-102_k141_153109 | Not-determined |
| 18-102_k141_90910 | Not-determined |
| 18-102_k141_236227 | Not-determined |
| 18-102_k141_153119 | Not-determined |
| 18-102_k141_174727 | Not-determined |
| 18-102_k141_236231 | Not-determined |
| 18-102_k141_142362 | Not-determined |
| 18-102_k141_236232 | Not-determined |
| 18-102_k141_26753 | Not-determined |
| 18-102_k141_39340 | Low-quality |
| 18-102_k141_69881 | Not-determined |
| 18-102_k141_69885 | Not-determined |
| 18-102_k141_39358 | Not-determined |
| 18-102_k141_236242 | Not-determined |
| 18-102_k141_236244 | Not-determined |
| 18-102_k141_184121 | Not-determined |
| 18-102_k141_90929 | Low-quality |
| 18-102_k141_121891 | Low-quality |
| 18-102_k141_162151 | Not-determined |
| 18-102_k141_236251 | Not-determined |
| 18-102_k141_121894 | Not-determined |
| 18-102_k141_162155 | Not-determined |
| 18-102_k141_39368 | Low-quality |
| 18-102_k141_184136 | Not-determined |
| 18-102_k141_51442 | Not-determined |
| 18-102_k141_142370 | Not-determined |
| 18-102_k141_18117 | Not-determined |
| 18-102_k141_236263 | Not-determined |
| 18-102_k141_90953 | Not-determined |
| 18-102_k141_130702 | Low-quality |
| 18-102_k141_223853 | Not-determined |
| 18-102_k141_184153 | Not-determined |
| 18-102_k141_130705 | Not-determined |
| 18-102_k141_6094 | Not-determined |
| 18-102_k141_121915 | Not-determined |
| 18-102_k141_69912 | Not-determined |
| 18-102_k141_142388 | Not-determined |
| 18-102_k141_69913 | Not-determined |
| 18-102_k141_184171 | Not-determined |
| 18-102_k141_6099 | Not-determined |
| 18-102_k141_69923 | Low-quality |
| 18-102_k141_214234 | Low-quality |
| 18-102_k141_174770 | Not-determined |
| 18-102_k141_69925 | Not-determined |
| 18-102_k141_174773 | Not-determined |
| 18-102_k141_51450 | Not-determined |
| 18-102_k141_101479 | Not-determined |
| 18-102_k141_69929 | Not-determined |
| 18-102_k141_153177 | Not-determined |
| 18-102_k141_39388 | Not-determined |
| 18-102_k141_130730 | Not-determined |
| 18-102_k141_214249 | Not-determined |
| 18-102_k141_39396 | Not-determined |
| 18-102_k141_142407 | Low-quality |
| 18-102_k141_51458 | Not-determined |
| 18-102_k141_59623 | Not-determined |
| 18-102_k141_69936 | Not-determined |
| 18-102_k141_184190 | Not-determined |
| 18-102_k141_59626 | Not-determined |
| 18-102_k141_184202 | Not-determined |
| 18-102_k141_174787 | Not-determined |
| 18-102_k141_214256 | Not-determined |
| 18-102_k141_101491 | Not-determined |
| 18-102_k141_90985 | Low-quality |
| 18-102_k141_6104 | Not-determined |
| 18-102_k141_121937 | Not-determined |
| 18-102_k141_101496 | Not-determined |
| 18-102_k141_110614 | Not-determined |
| 18-102_k141_51474 | Not-determined |
| 18-102_k141_110615 | Not-determined |
| 18-102_k141_51476 | Not-determined |
| 18-102_k141_214269 | Not-determined |
| 18-102_k141_130743 | Not-determined |
| 18-102_k141_162177 | Not-determined |
| 18-102_k141_6107 | Not-determined |
| 18-102_k141_162181 | Not-determined |
| 18-102_k141_130746 | Not-determined |
| 18-102_k141_121950 | Low-quality |
| 18-102_k141_90999 | Not-determined |
| 18-102_k141_214274 | Not-determined |
| 18-102_k141_121953 | Not-determined |
| 18-102_k141_184223 | Not-determined |
| 18-102_k141_121956 | Not-determined |
| 18-102_k141_194122 | Not-determined |
| 18-102_k141_142428 | Not-determined |
| 18-102_k141_101509 | Not-determined |
| 18-102_k141_142436 | Not-determined |
| 18-102_k141_174802 | Low-quality |
| 18-102_k141_236273 | Low-quality |
| 18-102_k141_214285 | Not-determined |
| 18-102_k141_142443 | Not-determined |
| 18-102_k141_110645 | Not-determined |
| 18-102_k141_214288 | Not-determined |
| 18-102_k141_236280 | Not-determined |
| 18-102_k141_142446 | Not-determined |
| 18-102_k141_110650 | Not-determined |
| 18-102_k141_39431 | Not-determined |
| 18-102_k141_101512 | Low-quality |
| 18-102_k141_39433 | Not-determined |
| 18-102_k141_91020 | Not-determined |
| 18-102_k141_214302 | Not-determined |
| 18-102_k141_59634 | Not-determined |
| 18-102_k141_236296 | Not-determined |
| 18-102_k141_59637 | Not-determined |
| 18-102_k141_18167 | Not-determined |
| 18-102_k141_184239 | Low-quality |
| 18-102_k141_110667 | Not-determined |
| 18-102_k141_26839 | Not-determined |
| 18-102_k141_153193 | Not-determined |
| 18-102_k141_174836 | Not-determined |
| 18-102_k141_142469 | Not-determined |
| 18-102_k141_121985 | Not-determined |
| 18-102_k141_174841 | Not-determined |
| 18-102_k141_130777 | Not-determined |
| 18-102_k141_142476 | Not-determined |
| 18-102_k141_121992 | Not-determined |
| 18-102_k141_59647 | Not-determined |
| 18-102_k141_153204 | Not-determined |
| 18-102_k141_162207 | Not-determined |
| 18-102_k141_194145 | Not-determined |
| 18-102_k141_153212 | Not-determined |
| 18-102_k141_101542 | Low-quality |
| 18-102_k141_110684 | Not-determined |
| 18-102_k141_110688 | Not-determined |
| 18-102_k141_184259 | Low-quality |
| 18-102_k141_130786 | Not-determined |
| 18-102_k141_162217 | Not-determined |
| 18-102_k141_142489 | Low-quality |
| 18-102_k141_91048 | Not-determined |
| 18-102_k141_91050 | Not-determined |
| 18-102_k141_174857 | Not-determined |
| 18-102_k141_39466 | Not-determined |
| 18-102_k141_194167 | Not-determined |
| 18-102_k141_101555 | Not-determined |
| 18-102_k141_39468 | Not-determined |
| 18-102_k141_26852 | Not-determined |
| 18-102_k141_39470 | Not-determined |
| 18-102_k141_130789 | Low-quality |
| 18-102_k141_184275 | Not-determined |
| 18-102_k141_6167 | Not-determined |
| 18-102_k141_18218 | Not-determined |
| 18-102_k141_223892 | Not-determined |
| 18-102_k141_18219 | Not-determined |
| 18-102_k141_130797 | Low-quality |
| 18-102_k141_110706 | Not-determined |
| 18-102_k141_51557 | Not-determined |
| 18-102_k141_184283 | Not-determined |
| 18-102_k141_6180 | Not-determined |
| 18-102_k141_80078 | Not-determined |
| 18-102_k141_6185 | Not-determined |
| 18-102_k141_184293 | Not-determined |
| 18-102_k141_6188 | Not-determined |
| 18-102_k141_39492 | Not-determined |
| 18-102_k141_59676 | Not-determined |
| 18-102_k141_39494 | Low-quality |
| 18-102_k141_18242 | Not-determined |
| 18-102_k141_184304 | Not-determined |
| 18-102_k141_142525 | Not-determined |
| 18-102_k141_142527 | Low-quality |
| 18-102_k141_184310 | Not-determined |
| 18-102_k141_142530 | Not-determined |
| 18-102_k141_130824 | Not-determined |
| 18-102_k141_91106 | Not-determined |
| 18-102_k141_101596 | Not-determined |
| 18-102_k141_162263 | Not-determined |
| 18-102_k141_122054 | Not-determined |
| 18-102_k141_18259 | Not-determined |
| 18-102_k141_91117 | Not-determined |
| 18-102_k141_59702 | Not-determined |
| 18-102_k141_184326 | Not-determined |

| | |
|---|---|
| 18-102_k141_101600 | Not-determined |
| 18-102_k141_18265 | Not-determined |
| 18-102_k141_130832 | Not-determined |
| 18-102_k141_184335 | Not-determined |
| 18-102_k141_162275 | Not-determined |
| 18-102_k141_18271 | Not-determined |
| 18-102_k141_153238 | Not-determined |
| 18-102_k141_18277 | Not-determined |
| 18-102_k141_142546 | Not-determined |
| 18-102_k141_80118 | Not-determined |
| 18-102_k141_39526 | Not-determined |
| 18-102_k141_91132 | Not-determined |
| 18-102_k141_6226 | Not-determined |
| 18-102_k141_194182 | Not-determined |
| 18-102_k141_6232 | Not-determined |
| 18-102_k141_194188 | Not-determined |
| 18-102_k141_184360 | Not-determined |
| 18-102_k141_6238 | Not-determined |
| 18-102_k141_142583 | Not-determined |
| 18-102_k141_162289 | Not-determined |
| 18-102_k141_6242 | Not-determined |
| 18-102_k141_101621 | Not-determined |
| 18-102_k141_101623 | Not-determined |
| 18-102_k141_162294 | Not-determined |
| 18-102_k141_39541 | Not-determined |
| 18-102_k141_130858 | Low-quality |
| 18-102_k141_39544 | Not-determined |
| 18-102_k141_6254 | Low-quality |
| 18-102_k141_6257 | Not-determined |
| 18-102_k141_174930 | Not-determined |
| 18-102_k141_6261 | Not-determined |
| 18-102_k141_26907 | Not-determined |
| 18-102_k141_153271 | Not-determined |
| 18-102_k141_162306 | Not-determined |
| 18-102_k141_6263 | Low-quality |
| 18-102_k141_39554 | Not-determined |
| 18-102_k141_153275 | Not-determined |
| 18-102_k141_174942 | Low-quality |
| 18-102_k141_174944 | Not-determined |
| 18-102_k141_236362 | Not-determined |
| 18-102_k141_223912 | Low-quality |
| 18-102_k141_184376 | Not-determined |
| 18-102_k141_101632 | Not-determined |
| 18-102_k141_6275 | Not-determined |
| 18-102_k141_214313 | Not-determined |
| 18-102_k141_223915 | Not-determined |
| 18-102_k141_194226 | Not-determined |
| 18-102_k141_101633 | Low-quality |
| 18-102_k141_91153 | Not-determined |
| 18-102_k141_130890 | Not-determined |
| 18-102_k141_101636 | Low-quality |
| 18-102_k141_26928 | Not-determined |
| 18-102_k141_91157 | Not-determined |
| 18-102_k141_184389 | Not-determined |
| 18-102_k141_39592 | Not-determined |
| 18-102_k141_39593 | Not-determined |
| 18-102_k141_194235 | Not-determined |
| 18-102_k141_80144 | Low-quality |
| 18-102_k141_80147 | Not-determined |
| 18-102_k141_174968 | Not-determined |
| 18-102_k141_39597 | Not-determined |
| 18-102_k141_39598 | Not-determined |
| 18-102_k141_214323 | Not-determined |
| 18-102_k141_184397 | Not-determined |
| 18-102_k141_130914 | Low-quality |
| 18-102_k141_130916 | Low-quality |
| 18-102_k141_184401 | Not-determined |
| 17-109-final-viral-combined_k141_376780\|\|full | Medium-quality |
| 18-102_k141_194250 | Not-determined |
| 18-102_k141_214329 | Not-determined |
| 18-102_k141_59741 | Not-determined |
| 18-102_k141_122120 | Not-determined |
| 18-102_k141_174977 | Not-determined |
| 18-102_k141_214332 | Not-determined |
| 18-102_k141_142599 | Not-determined |
| 18-102_k141_59749 | Not-determined |
| 18-102_k141_214340 | Not-determined |
| 18-102_k141_142603 | Not-determined |
| 18-102_k141_110768 | Not-determined |
| 18-102_k141_184415 | Not-determined |
| 18-102_k141_174991 | Not-determined |
| 18-102_k141_18306 | Not-determined |
| 18-102_k141_153322 | Low-quality |
| 18-102_k141_80164 | Not-determined |
| 18-102_k141_153326 | Not-determined |
| 18-102_k141_51582 | Not-determined |
| 18-102_k141_194269 | Not-determined |
| 18-102_k141_153331 | Not-determined |
| 18-102_k141_26965 | Not-determined |
| 18-102_k141_175005 | Not-determined |
| 18-102_k141_194276 | Not-determined |
| 18-102_k141_26971 | Not-determined |
| 18-102_k141_122135 | Low-quality |
| 18-102_k141_26978 | Not-determined |
| 18-102_k141_69957 | Not-determined |
| 18-102_k141_69958 | Not-determined |
| 18-102_k141_194283 | Not-determined |
| 18-102_k141_130937 | Not-determined |
| 18-102_k141_101656 | Not-determined |
| 18-102_k141_80168 | Low-quality |
| 18-102_k141_91173 | Not-determined |
| 18-102_k141_59786 | Low-quality |
| 18-102_k141_142628 | Not-determined |
| 18-102_k141_142632 | Not-determined |
| 18-102_k141_26992 | Not-determined |
| 18-102_k141_122155 | Not-determined |
| 18-102_k141_153351 | Not-determined |
| 18-102_k141_153353 | Not-determined |
| 18-102_k141_184432 | Not-determined |
| 18-102_k141_91196 | Not-determined |
| 18-102_k141_91197 | Not-determined |
| 18-102_k141_39638 | Not-determined |
| 18-102_k141_184437 | Not-determined |
| 18-102_k141_51591 | Not-determined |
| 18-102_k141_27002 | Not-determined |
| 18-102_k141_80193 | Not-determined |
| 18-102_k141_80194 | Not-determined |
| 18-102_k141_153361 | Not-determined |
| 18-102_k141_153365 | Not-determined |
| 18-102_k141_6290 | Not-determined |
| 18-102_k141_101680 | Not-determined |
| 18-102_k141_184448 | Not-determined |
| 18-102_k141_162336 | Not-determined |
| 18-102_k141_6293 | Not-determined |
| 18-102_k141_91202 | Not-determined |
| 18-102_k141_18332 | Not-determined |
| 18-102_k141_6296 | Not-determined |
| 18-102_k141_153380 | Low-quality |
| 18-102_k141_80209 | Not-determined |
| 18-102_k141_122167 | Not-determined |
| 18-102_k141_236423 | Not-determined |
| 18-102_k141_214350 | Not-determined |
| 18-102_k141_162349 | Not-determined |
| 18-102_k141_91214 | Not-determined |
| 18-102_k141_91216 | Not-determined |
| 18-102_k141_122173 | Not-determined |
| 18-102_k141_6302 | Not-determined |
| 18-102_k141_6305 | Not-determined |
| 18-102_k141_223960 | Not-determined |
| 18-102_k141_27034 | Not-determined |
| 18-102_k141_91229 | Not-determined |
| 18-102_k141_223963 | Not-determined |
| 18-102_k141_194299 | Not-determined |
| 18-102_k141_70017 | Not-determined |
| 18-102_k141_91242 | Not-determined |
| 18-102_k141_91244 | Not-determined |
| 18-102_k141_101701 | Not-determined |
| 18-102_k141_51618 | Not-determined |
| 18-102_k141_244338 | Not-determined |
| 18-102_k141_244340 | Not-determined |
| 18-102_k141_70022 | Low-quality |
| 18-102_k141_244342 | Not-determined |
| 18-102_k141_101705 | Not-determined |
| 18-102_k141_91252 | Not-determined |
| 18-102_k141_101708 | Not-determined |
| 18-102_k141_162371 | Not-determined |
| 18-102_k141_6322 | Not-determined |
| 18-102_k141_244348 | Not-determined |
| 18-102_k141_153425 | Not-determined |
| 18-102_k141_153426 | Not-determined |
| 18-102_k141_142652 | Not-determined |
| 18-102_k141_6326 | Not-determined |
| 18-102_k141_91260 | Not-determined |
| 18-102_k141_39667 | Not-determined |
| 18-102_k141_236452 | Not-determined |
| 18-102_k141_39671 | Not-determined |
| 18-102_k141_142660 | Not-determined |
| 18-102_k141_101717 | Not-determined |
| 18-102_k141_110772 | Not-determined |
| 18-102_k141_70037 | Not-determined |
| 18-102_k141_244360 | Not-determined |
| 18-102_k141_236457 | Not-determined |
| 18-102_k141_70043 | Not-determined |
| 18-102_k141_153446 | Not-determined |
| 18-102_k141_236461 | Not-determined |
| 18-102_k141_110780 | Not-determined |
| 18-102_k141_101726 | Not-determined |
| 18-102_k141_130947 | Not-determined |
| 18-102_k141_130949 | Not-determined |
| 18-102_k141_101729 | Not-determined |
| 18-102_k141_142680 | Not-determined |
| 18-102_k141_162385 | Not-determined |
| 18-102_k141_223995 | Not-determined |
| 18-102_k141_142682 | Not-determined |
| 18-102_k141_153459 | Not-determined |
| 18-102_k141_153460 | Low-quality |
| 18-102_k141_39680 | Not-determined |
| 18-102_k141_153463 | Not-determined |
| 18-102_k141_162390 | Not-determined |
| 18-102_k141_122212 | Not-determined |
| 18-102_k141_153465 | Not-determined |
| 18-102_k141_162394 | Low-quality |
| 18-102_k141_51641 | Not-determined |
| 18-102_k141_184461 | Not-determined |
| 18-102_k141_110793 | Not-determined |
| 18-102_k141_51645 | Low-quality |
| 18-102_k141_142686 | Not-determined |
| 18-102_k141_236480 | Not-determined |
| 18-102_k141_142694 | Not-determined |
| 18-102_k141_70073 | Not-determined |
| 18-102_k141_91279 | Not-determined |
| 18-102_k141_51657 | Not-determined |
| 18-102_k141_130958 | Not-determined |
| 18-102_k141_162401 | Not-determined |
| 18-102_k141_39705 | Not-determined |
| 18-102_k141_101754 | Not-determined |
| 18-102_k141_184471 | Not-determined |
| 18-102_k141_51669 | Not-determined |
| 18-102_k141_130964 | Not-determined |
| 18-102_k141_122224 | Not-determined |
| 18-102_k141_91282 | Not-determined |
| 18-102_k141_142704 | Not-determined |
| 18-102_k141_39718 | Not-determined |
| 18-102_k141_162410 | Not-determined |
| 18-102_k141_142706 | Not-determined |
| 18-102_k141_194333 | Not-determined |
| 18-102_k141_130970 | Low-quality |
| 18-102_k141_91290 | Not-determined |
| 18-102_k141_18376 | Low-quality |
| 18-102_k141_130974 | Not-determined |
| 18-102_k141_175034 | Not-determined |
| 18-102_k141_184492 | Not-determined |
| 18-102_k141_162413 | Not-determined |
| 18-102_k141_130980 | Not-determined |
| 18-102_k141_27076 | Not-determined |
| 18-102_k141_18378 | Not-determined |
| 18-102_k141_142722 | Not-determined |
| 18-102_k141_80272 | Not-determined |
| 18-102_k141_101769 | Not-determined |
| 18-102_k141_27081 | Not-determined |
| 18-102_k141_162420 | Not-determined |
| 18-102_k141_80276 | Low-quality |
| 18-102_k141_122242 | Low-quality |
| 18-102_k141_101772 | Not-determined |
| 18-102_k141_162425 | Not-determined |
| 18-102_k141_6331 | Not-determined |
| 18-102_k141_101778 | Not-determined |
| 18-102_k141_6334 | Not-determined |
| 18-102_k141_142733 | Not-determined |
| 18-102_k141_6336 | Not-determined |
| 18-102_k141_27086 | Not-determined |
| 18-102_k141_18399 | Not-determined |
| 18-102_k141_51692 | Not-determined |
| 18-102_k141_51695 | Not-determined |
| 18-102_k141_130993 | Not-determined |
| 18-102_k141_194362 | Low-quality |
| 18-102_k141_130995 | Not-determined |
| 18-102_k141_110823 | Not-determined |
| 18-102_k141_130998 | Not-determined |
| 18-102_k141_110825 | Not-determined |
| 18-102_k141_131000 | Not-determined |
| 18-102_k141_101792 | Not-determined |
| 18-102_k141_175041 | Not-determined |
| 18-102_k141_224021 | Not-determined |
| 18-102_k141_101796 | Not-determined |
| 18-102_k141_131004 | Not-determined |
| 18-102_k141_70115 | Not-determined |
| 18-102_k141_101801 | Not-determined |
| 18-102_k141_236521 | Not-determined |
| 18-102_k141_27097 | Not-determined |
| 18-102_k141_101806 | Not-determined |
| 18-102_k141_51705 | Not-determined |
| 18-102_k141_224027 | Not-determined |
| 18-102_k141_236528 | Not-determined |
| 18-102_k141_122251 | Not-determined |
| 18-102_k141_59828 | Not-determined |
| 18-102_k141_224031 | Not-determined |
| 18-102_k141_80313 | Not-determined |
| 18-102_k141_224032 | Not-determined |
| 18-102_k141_59835 | Not-determined |
| 18-102_k141_162428 | Not-determined |
| 18-102_k141_70154 | Not-determined |
| 18-102_k141_27110 | Low-quality |
| 18-102_k141_6351 | Not-determined |
| 18-102_k141_27112 | Not-determined |
| 18-102_k141_110840 | Not-determined |
| 18-102_k141_51724 | Not-determined |
| 18-102_k141_6358 | Not-determined |
| 18-102_k141_59854 | Not-determined |
| 18-102_k141_59855 | Not-determined |
| 18-102_k141_162438 | Not-determined |
| 18-102_k141_236548 | Not-determined |
| 18-102_k141_51738 | Not-determined |
| 18-102_k141_80343 | Low-quality |
| 18-102_k141_27126 | Low-quality |
| 18-102_k141_39781 | Not-determined |
| 18-102_k141_18429 | Not-determined |
| 18-102_k141_131024 | Not-determined |
| 18-102_k141_70158 | Low-quality |
| 18-102_k141_51746 | Not-determined |
| 18-102_k141_18435 | Not-determined |
| 18-102_k141_122280 | Low-quality |
| 18-102_k141_110856 | Not-determined |
| 18-102_k141_175099 | Low-quality |
| 18-102_k141_6361 | Not-determined |
| 18-102_k141_184541 | Not-determined |
| 18-102_k141_175101 | Not-determined |
| 18-102_k141_131043 | Not-determined |
| 18-102_k141_142741 | Not-determined |
| 18-102_k141_175109 | Low-quality |
| 18-102_k141_244417 | Not-determined |
| 18-102_k141_27141 | Not-determined |
| 18-102_k141_175114 | Not-determined |
| 18-102_k141_162465 | Low-quality |
| 18-102_k141_110870 | Low-quality |
| 18-102_k141_175117 | Not-determined |
| 18-102_k141_110873 | Low-quality |
| 18-102_k141_70181 | Not-determined |
| 18-102_k141_214411 | Low-quality |
| 18-102_k141_162473 | Not-determined |
| 18-102_k141_194432 | Not-determined |
| 18-102_k141_51764 | Not-determined |
| 18-102_k141_244440 | Low-quality |
| 18-102_k141_142758 | Not-determined |
| 18-102_k141_236553 | Not-determined |
| 18-102_k141_51771 | Not-determined |
| 18-102_k141_244446 | Not-determined |
| 18-102_k141_122319 | Not-determined |
| 18-102_k141_194452 | Not-determined |
| 18-102_k141_162492 | Not-determined |
| 18-102_k141_175141 | Low-quality |
| 18-102_k141_131052 | Not-determined |
| 18-102_k141_224071 | Not-determined |
| 18-102_k141_224074 | Not-determined |
| 18-102_k141_224075 | Not-determined |
| 18-102_k141_224080 | Not-determined |
| 18-102_k141_142764 | Not-determined |
| 18-102_k141_59879 | Not-determined |
| 18-102_k141_59883 | Not-determined |
| 18-102_k141_131057 | Not-determined |
| 18-102_k141_39807 | Not-determined |
| 18-102_k141_59887 | Not-determined |
| 18-102_k141_39809 | Not-determined |
| 18-102_k141_70196 | Not-determined |
| 18-102_k141_175152 | Not-determined |
| 18-102_k141_194462 | Not-determined |
| 18-102_k141_39821 | Not-determined |
| 18-102_k141_70200 | Not-determined |
| 18-102_k141_236570 | Not-determined |
| 18-102_k141_91335 | Not-determined |
| 18-102_k141_91337 | Not-determined |
| 18-102_k141_70209 | Not-determined |
| 18-102_k141_162503 | Not-determined |
| 18-102_k141_91344 | Not-determined |
| 18-102_k141_214449 | Not-determined |
| 18-102_k141_175176 | Not-determined |
| 18-102_k141_214450 | Not-determined |
| 18-102_k141_70227 | Not-determined |
| 18-102_k141_59901 | Not-determined |
| 18-102_k141_194479 | Not-determined |
| 18-102_k141_18460 | Low-quality |
| 18-102_k141_18462 | Not-determined |
| 18-102_k141_91347 | Low-quality |
| 18-102_k141_110906 | Not-determined |
| 18-102_k141_194492 | Not-determined |
| 18-102_k141_162514 | Low-quality |
| 18-102_k141_80360 | Not-determined |
| 18-102_k141_110913 | Not-determined |
| 18-102_k141_142804 | Not-determined |
| 18-102_k141_91357 | Not-determined |
| 18-102_k141_110924 | Not-determined |
| 18-102_k141_224116 | Not-determined |
| 18-102_k141_162533 | Not-determined |
| 18-102_k141_91363 | Not-determined |
| 18-102_k141_27163 | Not-determined |
| 18-102_k141_214473 | Not-determined |
| 18-102_k141_214475 | Not-determined |
| 18-102_k141_91367 | Not-determined |
| 18-102_k141_59926 | Not-determined |
| 18-102_k141_236615 | Not-determined |
| 18-102_k141_6403 | Not-determined |
| 18-102_k141_39854 | Not-determined |
| 18-102_k141_194510 | Not-determined |
| 18-102_k141_236617 | Not-determined |
| 18-102_k141_59928 | Not-determined |
| 18-102_k141_236618 | Not-determined |
| 18-102_k141_59930 | Not-determined |
| 18-102_k141_224123 | Not-determined |
| 18-102_k141_224127 | Not-determined |
| 18-102_k141_175217 | Not-determined |
| 18-102_k141_236627 | Not-determined |
| 18-102_k141_101832 | Not-determined |
| 18-102_k141_59935 | Not-determined |
| 18-102_k141_236629 | Not-determined |
| 18-102_k141_91378 | Not-determined |
| 18-102_k141_214488 | Not-determined |
| 18-102_k141_142829 | Not-determined |
| 18-102_k141_80366 | Not-determined |
| 18-102_k141_39880 | Not-determined |
| 18-102_k141_91388 | Not-determined |
| 18-102_k141_142833 | Not-determined |
| 18-102_k141_110944 | Not-determined |
| 18-102_k141_162539 | Not-determined |
| 18-102_k141_91395 | Not-determined |
| 18-102_k141_142840 | Not-determined |
| 18-102_k141_59947 | Not-determined |
| 18-102_k141_101837 | Low-quality |
| 18-102_k141_80374 | Not-determined |
| 18-102_k141_142845 | Not-determined |
| 18-102_k141_101840 | Not-determined |
| 18-102_k141_59950 | Not-determined |
| 18-102_k141_80379 | Not-determined |
| 18-102_k141_214502 | Not-determined |
| 18-102_k141_59955 | Not-determined |
| 18-102_k141_224156 | Not-determined |
| 18-102_k141_175239 | Not-determined |
| 18-102_k141_101859 | Not-determined |
| 18-102_k141_39905 | Not-determined |
| 18-102_k141_80401 | Low-quality |
| 18-102_k141_122370 | Not-determined |
| 18-102_k141_39919 | Not-determined |
| 18-102_k141_194572 | Not-determined |
| 18-102_k141_122375 | Not-determined |
| 18-102_k141_39931 | Not-determined |
| 18-102_k141_122379 | Not-determined |
| 18-102_k141_194578 | Not-determined |
| 18-102_k141_142872 | Not-determined |
| 18-102_k141_39934 | Not-determined |
| 18-102_k141_18526 | Not-determined |
| 18-102_k141_122383 | Not-determined |
| 18-102_k141_110966 | Not-determined |
| 18-102_k141_214521 | Not-determined |
| 18-102_k141_122386 | Low-quality |
| 18-102_k141_80430 | Not-determined |
| 18-102_k141_39942 | Not-determined |
| 18-102_k141_80433 | Not-determined |
| 18-102_k141_80438 | Not-determined |
| 18-102_k141_6414 | Not-determined |
| 18-102_k141_203333 | Not-determined |
| 18-102_k141_6416 | Not-determined |
| 18-102_k141_70285 | Not-determined |
| 18-102_k141_142882 | Low-quality |
| 18-102_k141_18547 | Low-quality |
| 18-102_k141_18548 | Not-determined |
| 18-102_k141_142886 | Not-determined |
| 18-102_k141_203346 | Not-determined |
| 18-102_k141_110994 | Not-determined |
| 18-102_k141_6421 | Not-determined |
| 18-102_k141_60001 | Not-determined |
| 18-102_k141_60003 | Not-determined |
| 18-102_k141_203353 | Not-determined |
| 18-102_k141_18565 | Low-quality |
| 18-102_k141_194607 | Not-determined |
| 18-102_k141_39957 | Not-determined |
| 18-102_k141_91469 | Not-determined |
| 18-102_k141_91473 | Not-determined |
| 18-102_k141_91479 | Not-determined |
| 18-102_k141_70293 | Not-determined |
| 18-102_k141_101866 | Not-determined |
| 18-102_k141_18580 | Low-quality |
| 18-102_k141_101873 | Not-determined |
| 18-102_k141_60010 | Not-determined |
| 18-102_k141_70302 | Not-determined |
| 18-102_k141_27179 | Not-determined |
| 18-102_k141_194623 | Low-quality |
| 18-102_k141_101879 | Not-determined |
| 18-102_k141_194625 | Not-determined |
| 18-102_k141_39986 | Not-determined |
| 18-102_k141_18594 | Not-determined |
| 18-102_k141_194627 | Not-determined |
| 18-102_k141_70315 | Not-determined |
| 18-102_k141_101881 | Not-determined |
| 18-102_k141_111019 | Not-determined |
| 18-102_k141_111020 | Not-determined |
| 18-102_k141_122423 | Low-quality |
| 18-102_k141_122426 | Not-determined |
| 18-102_k141_214565 | Not-determined |
| 18-102_k141_91497 | Low-quality |
| 18-102_k141_91503 | Low-quality |
| 18-102_k141_111037 | Not-determined |
| 18-102_k141_40014 | Not-determined |
| 18-102_k141_194642 | Not-determined |
| 18-102_k141_60046 | Not-determined |
| 18-102_k141_60047 | Not-determined |
| 18-102_k141_203373 | Not-determined |
| 18-102_k141_60049 | Not-determined |
| 18-102_k141_175258 | Not-determined |
| 18-102_k141_60055 | Not-determined |
| 18-102_k141_80470 | Not-determined |
| 18-102_k141_122444 | Not-determined |
| 18-102_k141_214581 | Not-determined |
| 18-102_k141_175273 | Not-determined |
| 18-102_k141_91520 | Not-determined |
| 18-102_k141_214590 | Not-determined |
| 18-102_k141_175280 | Not-determined |
| 18-102_k141_91529 | Not-determined |
| 18-102_k141_101922 | Not-determined |
| 18-102_k141_70332 | Low-quality |
| 18-102_k141_142924 | Not-determined |
| 18-102_k141_111044 | Not-determined |
| 18-102_k141_80495 | Not-determined |
| 18-102_k141_70349 | Low-quality |
| 18-102_k141_236652 | Not-determined |
| 18-102_k141_101940 | Not-determined |
| 18-102_k141_194672 | Not-determined |
| 18-102_k141_214618 | Not-determined |
| 18-102_k141_91555 | Not-determined |
| 18-102_k141_91557 | Not-determined |
| 18-102_k141_70358 | Not-determined |
| 18-102_k141_91561 | Not-determined |
| 18-102_k141_60097 | Not-determined |
| 18-102_k141_40039 | Low-quality |
| 18-102_k141_194676 | Not-determined |
| 18-102_k141_27254 | Not-determined |
| 18-102_k141_203399 | Not-determined |
| 18-102_k141_203403 | Low-quality |
| 18-102_k141_142962 | Not-determined |
| 18-102_k141_80501 | Not-determined |
| 18-101-final-viral-combined_k141_107073\|\|full | Medium-quality |
| 18-102_k141_175306 | Not-determined |
| 18-102_k141_142971 | Not-determined |
| 18-102_k141_60111 | Not-determined |
| 18-102_k141_27277 | Not-determined |
| 18-102_k141_18633 | Not-determined |
| 18-102_k141_194694 | Not-determined |
| 18-102_k141_80515 | Not-determined |
| 18-102_k141_175318 | Not-determined |
| 18-102_k141_60120 | Not-determined |
| 18-102_k141_40051 | Not-determined |
| 18-102_k141_6433 | Low-quality |
| 18-102_k141_224188 | Not-determined |
| 18-102_k141_91573 | Not-determined |
| 18-102_k141_142979 | Not-determined |
| 18-102_k141_224192 | Not-determined |
| 18-102_k141_18656 | Not-determined |
| 18-102_k141_142983 | Not-determined |
| 18-102_k141_236655 | Low-quality |
| 18-102_k141_175332 | Not-determined |
| 18-102_k141_142997 | Not-determined |
| 18-102_k141_175336 | Not-determined |
| 18-102_k141_194720 | Not-determined |
| 18-102_k141_80542 | Not-determined |
| 18-102_k141_27310 | Low-quality |
| 18-102_k141_40071 | Not-determined |
| 18-102_k141_80545 | Not-determined |
| 18-102_k141_214678 | Not-determined |
| 18-102_k141_60140 | Not-determined |
| 18-102_k141_122486 | Not-determined |
| 18-102_k141_101995 | Not-determined |
| 18-102_k141_203449 | Not-determined |
| 18-102_k141_175340 | Not-determined |
| 18-102_k141_6443 | Not-determined |
| 18-102_k141_40081 | Not-determined |
| 18-102_k141_60150 | Not-determined |
| 18-102_k141_203459 | Not-determined |
| 18-102_k141_40090 | Not-determined |
| 18-102_k141_224218 | Not-determined |
| 18-102_k141_175362 | Not-determined |
| 18-102_k141_60162 | Not-determined |
| 18-102_k141_40095 | Not-determined |
| 18-102_k141_102030 | Not-determined |
| 18-102_k141_18673 | Not-determined |
| 18-102_k141_27329 | Not-determined |
| 18-102_k141_27330 | Not-determined |
| 18-102_k141_143010 | Not-determined |
| 18-102_k141_203467 | Not-determined |
| 18-102_k141_18677 | Not-determined |
| 18-102_k141_214699 | Not-determined |
| 18-102_k141_143012 | Not-determined |
| 18-102_k141_162589 | Not-determined |
| 18-102_k141_91596 | Not-determined |
| 18-102_k141_60173 | Not-determined |
| 18-102_k141_162592 | Low-quality |
| 18-102_k141_236679 | Not-determined |
| 18-102_k141_27333 | Low-quality |
| 18-102_k141_60178 | Not-determined |
| 18-102_k141_18682 | Not-determined |
| 18-102_k141_143016 | Not-determined |
| 18-102_k141_143017 | Not-determined |
| 18-102_k141_194772 | Not-determined |
| 18-102_k141_143024 | Not-determined |
| 18-102_k141_27345 | Not-determined |
| 18-102_k141_214704 | Not-determined |
| 18-102_k141_184586 | Not-determined |
| 18-102_k141_236694 | Not-determined |
| 18-102_k141_184593 | Not-determined |
| 18-102_k141_224231 | Not-determined |
| 18-102_k141_91631 | Not-determined |
| 18-102_k141_6480 | Not-determined |
| 18-102_k141_18713 | Not-determined |
| 18-102_k141_143046 | Not-determined |
| 18-102_k141_224242 | Not-determined |
| 18-102_k141_143050 | Not-determined |
| 18-102_k141_143055 | Not-determined |
| 18-102_k141_175389 | Low-quality |
| 18-102_k141_18723 | Not-determined |
| 18-102_k141_244456 | Not-determined |
| 18-102_k141_6486 | Not-determined |
| 18-102_k141_175390 | Not-determined |

| | |
|---|---|
| 18-102_k141_102054 | Not-determined |
| 18-102_k141_60206 | Not-determined |
| 18-102_k141_131118 | Low-quality |
| 18-102_k141_244458 | Not-determined |
| 18-102_k141_131121 | Not-determined |
| 18-102_k141_91658 | Not-determined |
| 18-102_k141_194793 | Not-determined |
| 18-102_k141_236699 | Not-determined |
| 18-102_k141_236703 | Not-determined |
| 18-102_k141_131126 | Not-determined |
| 18-102_k141_131127 | Not-determined |
| 18-102_k141_184627 | Not-determined |
| 18-102_k141_40152 | Not-determined |
| 18-102_k141_194802 | Not-determined |
| 18-102_k141_162613 | Not-determined |
| 18-102_k141_214713 | Not-determined |
| 18-102_k141_184637 | Not-determined |
| 18-102_k141_6496 | Low-quality |
| 18-102_k141_236718 | Not-determined |
| 18-102_k141_102073 | Not-determined |
| 18-102_k141_27370 | Not-determined |
| 18-102_k141_236724 | Not-determined |
| 18-102_k141_18761 | Not-determined |
| 18-102_k141_70436 | Not-determined |
| 18-102_k141_102086 | Low-quality |
| 18-102_k141_91675 | Not-determined |
| 18-102_k141_6514 | Not-determined |
| 18-102_k141_18766 | Low-quality |
| 18-102_k141_27388 | Not-determined |
| 18-102_k141_236748 | Not-determined |
| 18-102_k141_194826 | Not-determined |
| 18-102_k141_40178 | Not-determined |
| 18-102_k141_131135 | Not-determined |
| 18-102_k141_131137 | Not-determined |
| 18-102_k141_236770 | Not-determined |
| 18-102_k141_236771 | Not-determined |
| 18-102_k141_131140 | Not-determined |
| 18-102_k141_194834 | Not-determined |
| 18-102_k141_214738 | Not-determined |
| 18-102_k141_194841 | Not-determined |
| 18-102_k141_80562 | Not-determined |
| 18-102_k141_80564 | Not-determined |
| 18-102_k141_91695 | Not-determined |
| 18-102_k141_60239 | Not-determined |
| 18-102_k141_80571 | Not-determined |
| 18-102_k141_18782 | Not-determined |
| 18-102_k141_244482 | Not-determined |
| 18-102_k141_70463 | Low-quality |
| 18-102_k141_91704 | Not-determined |
| 18-102_k141_91708 | Not-determined |
| 18-102_k141_70471 | Not-determined |
| 18-102_k141_224254 | Not-determined |
| 18-102_k141_18793 | Low-quality |
| 18-102_k141_102096 | Not-determined |
| 18-102_k141_91718 | Not-determined |
| 18-102_k141_40187 | Not-determined |
| 18-102_k141_91723 | Not-determined |
| 18-102_k141_184678 | Not-determined |
| 18-102_k141_162651 | Not-determined |
| 18-102_k141_184686 | Low-quality |
| 18-102_k141_40195 | Not-determined |
| 18-102_k141_175422 | Not-determined |
| 18-102_k141_184689 | Not-determined |
| 18-102_k141_131153 | Not-determined |
| 18-102_k141_175426 | Not-determined |
| 18-102_k141_224276 | Not-determined |
| 18-102_k141_60268 | Not-determined |
| 18-102_k141_18809 | Not-determined |
| 18-102_k141_184700 | Not-determined |
| 18-102_k141_111073 | Not-determined |
| 18-102_k141_131168 | Not-determined |
| 18-102_k141_224286 | Low-quality |
| 18-102_k141_102125 | Not-determined |
| 18-102_k141_91751 | Not-determined |
| 18-102_k141_224294 | Not-determined |
| 18-102_k141_214773 | Not-determined |
| 18-102_k141_224296 | Not-determined |
| 18-102_k141_102134 | Not-determined |
| 18-102_k141_184716 | Low-quality |
| 18-102_k141_80588 | Not-determined |
| 18-102_k141_131189 | Not-determined |
| 18-102_k141_131193 | Not-determined |
| 18-102_k141_70496 | Not-determined |
| 18-102_k141_175449 | Not-determined |
| 18-102_k141_131198 | Not-determined |
| 18-102_k141_175456 | Not-determined |
| 18-102_k141_91782 | Not-determined |
| 18-102_k141_60292 | Not-determined |
| 18-102_k141_102146 | Not-determined |
| 18-102_k141_175472 | Not-determined |
| 18-102_k141_214798 | Low-quality |
| 18-102_k141_131208 | Not-determined |
| 18-102_k141_143069 | Not-determined |
| 18-102_k141_70501 | Not-determined |
| 18-102_k141_6530 | Low-quality |
| 18-102_k141_91800 | Not-determined |
| 18-102_k141_244548 | Not-determined |
| 18-102_k141_6535 | Not-determined |
| 18-102_k141_236830 | Not-determined |
| 18-102_k141_175495 | Not-determined |
| 18-102_k141_122549 | Low-quality |
| 18-102_k141_175498 | Not-determined |
| 18-102_k141_143075 | Low-quality |
| 18-102_k141_236839 | Low-quality |
| 18-102_k141_40261 | Not-determined |
| 18-102_k141_224322 | Not-determined |
| 18-102_k141_143080 | Not-determined |
| 18-102_k141_143084 | Not-determined |
| 18-102_k141_175505 | Not-determined |
| 18-102_k141_175506 | Low-quality |
| 18-102_k141_70513 | Not-determined |
| 18-102_k141_102174 | Not-determined |
| 18-102_k141_102176 | Not-determined |
| 18-102_k141_214826 | Not-determined |
| 18-102_k141_214835 | Not-determined |
| 18-102_k141_102190 | Not-determined |
| 18-102_k141_80624 | Not-determined |
| 18-102_k141_175509 | Low-quality |
| 18-102_k141_162664 | Not-determined |
| 18-102_k141_143109 | Not-determined |
| 18-102_k141_214839 | Not-determined |
| 18-102_k141_175513 | Not-determined |
| 18-102_k141_40294 | Not-determined |
| 18-102_k141_102194 | Not-determined |
| 18-102_k141_214843 | Not-determined |
| 18-102_k141_111091 | Low-quality |
| 18-102_k141_236872 | Low-quality |
| 18-102_k141_111092 | Not-determined |
| 18-102_k141_143120 | Not-determined |
| 18-102_k141_122559 | Not-determined |
| 18-102_k141_162676 | Not-determined |
| 18-102_k141_40297 | Not-determined |
| 18-102_k141_184767 | Not-determined |
| 18-102_k141_184770 | Not-determined |
| 18-102_k141_70530 | Low-quality |
| 18-102_k141_122567 | Not-determined |
| 18-102_k141_91838 | Not-determined |
| 18-102_k141_70534 | Not-determined |
| 18-102_k141_60348 | Not-determined |
| 18-102_k141_111107 | Not-determined |
| 18-102_k141_6566 | Not-determined |
| 18-102_k141_111118 | Not-determined |
| 18-102_k141_111121 | Not-determined |
| 18-102_k141_6569 | Not-determined |
| 18-102_k141_203536 | Not-determined |
| 18-102_k141_131222 | Low-quality |
| 18-102_k141_70545 | Not-determined |
| 18-102_k141_203538 | Not-determined |
| 18-102_k141_203539 | Not-determined |
| 18-102_k141_70549 | Not-determined |
| 18-102_k141_184789 | Not-determined |
| 18-102_k141_102244 | Not-determined |
| 18-102_k141_236895 | Not-determined |
| 18-102_k141_102251 | Not-determined |
| 18-102_k141_80638 | Not-determined |
| 18-102_k141_60377 | Not-determined |
| 18-102_k141_6591 | Not-determined |
| 18-102_k141_214865 | Low-quality |
| 18-102_k141_102255 | Not-determined |
| 18-102_k141_102256 | Not-determined |
| 18-102_k141_236908 | Not-determined |
| 18-102_k141_214867 | Not-determined |
| 18-102_k141_162692 | Not-determined |
| 18-102_k141_224347 | Not-determined |
| 18-102_k141_91858 | Not-determined |
| 18-102_k141_6598 | Not-determined |
| 18-102_k141_91860 | Not-determined |
| 18-102_k141_162707 | Low-quality |
| 18-102_k141_224350 | Not-determined |
| 18-102_k141_162709 | Not-determined |
| 18-102_k141_162712 | Not-determined |
| 18-102_k141_224360 | Not-determined |
| 18-102_k141_102285 | Not-determined |
| 18-102_k141_131251 | Not-determined |
| 18-102_k141_60417 | Not-determined |
| 18-102_k141_102288 | Not-determined |
| 18-102_k141_60420 | Not-determined |
| 18-102_k141_60422 | Not-determined |
| 18-102_k141_224366 | Not-determined |
| 18-102_k141_175541 | Not-determined |
| 18-102_k141_80669 | Low-quality |
| 18-102_k141_184831 | Not-determined |
| 18-102_k141_60431 | Not-determined |
| 18-102_k141_175549 | Not-determined |
| 18-102_k141_131258 | Not-determined |
| 18-102_k141_194867 | Not-determined |
| 18-102_k141_244570 | Not-determined |
| 18-102_k141_131261 | Not-determined |
| 18-102_k141_244574 | Not-determined |
| 18-102_k141_184841 | Not-determined |
| 18-102_k141_162735 | Not-determined |
| 18-102_k141_131269 | Not-determined |
| 18-102_k141_143172 | Not-determined |
| 18-102_k141_224383 | Not-determined |
| 18-102_k141_224384 | Not-determined |
| 18-102_k141_214876 | Not-determined |
| 18-102_k141_214877 | Not-determined |
| 18-102_k141_175569 | Not-determined |
| 18-102_k141_143178 | Not-determined |
| 18-102_k141_244585 | Low-quality |
| 18-102_k141_184869 | Not-determined |
| 18-102_k141_60463 | Not-determined |
| 18-102_k141_194898 | Not-determined |
| 18-102_k141_244591 | Not-determined |
| 18-102_k141_102303 | Not-determined |
| 18-102_k141_203562 | Not-determined |
| 18-102_k141_111167 | Not-determined |
| 18-102_k141_143189 | Low-quality |
| 18-102_k141_70621 | Not-determined |
| 18-102_k141_122604 | Not-determined |
| 18-102_k141_122605 | Not-determined |
| 18-102_k141_153479 | Low-quality |
| 18-102_k141_194903 | Not-determined |
| 18-102_k141_111176 | Not-determined |
| 18-102_k141_203567 | Not-determined |
| 18-102_k141_70630 | Not-determined |
| 18-102_k141_70633 | Not-determined |
| 18-102_k141_153483 | Low-quality |
| 18-102_k141_153484 | Not-determined |
| 18-102_k141_143200 | Low-quality |
| 18-102_k141_214895 | Not-determined |
| 18-102_k141_27415 | Not-determined |
| 18-102_k141_143204 | Not-determined |
| 18-102_k141_143205 | Not-determined |
| 18-102_k141_236962 | Not-determined |
| 18-102_k141_244598 | Not-determined |
| 18-102_k141_175588 | Not-determined |
| 18-102_k141_91930 | Not-determined |
| 18-102_k141_111183 | Not-determined |
| 18-102_k141_60491 | Not-determined |
| 18-102_k141_70648 | Not-determined |
| 18-102_k141_60498 | Not-determined |
| 18-102_k141_236979 | Not-determined |
| 18-102_k141_194921 | Not-determined |
| 18-102_k141_214914 | Not-determined |
| 18-102_k141_236983 | Not-determined |
| 18-102_k141_91950 | Not-determined |
| 18-102_k141_60508 | Not-determined |
| 18-102_k141_203590 | Not-determined |
| 18-102_k141_236986 | Not-determined |
| 18-102_k141_214920 | Not-determined |
| 18-102_k141_40341 | Not-determined |
| 18-102_k141_111196 | Not-determined |
| 18-102_k141_27429 | Low-quality |
| 18-102_k141_244627 | Not-determined |
| 18-102_k141_203595 | Not-determined |
| 18-102_k141_162780 | Not-determined |
| 18-102_k141_143213 | Not-determined |
| 18-102_k141_122626 | Not-determined |
| 18-102_k141_131293 | Not-determined |
| 18-102_k141_27443 | Not-determined |
| 18-102_k141_224421 | Not-determined |
| 18-102_k141_60538 | Not-determined |
| 18-102_k141_131295 | Not-determined |
| 18-102_k141_102315 | Not-determined |
| 18-102_k141_102316 | Not-determined |
| 18-102_k141_102318 | Not-determined |
| 18-102_k141_214932 | Not-determined |
| 18-102_k141_203607 | Not-determined |
| 18-102_k141_70680 | Not-determined |
| 18-102_k141_80695 | Not-determined |
| 18-102_k141_80702 | Not-determined |
| 18-102_k141_143233 | Not-determined |
| 18-102_k141_214941 | Not-determined |
| 18-102_k141_40352 | Not-determined |
| 18-102_k141_214943 | Not-determined |
| 18-102_k141_244644 | Not-determined |
| 18-102_k141_80719 | Not-determined |
| 18-102_k141_143244 | Not-determined |
| 18-102_k141_27450 | Not-determined |
| 18-102_k141_203625 | Not-determined |
| 18-102_k141_27453 | Not-determined |
| 18-102_k141_143247 | Not-determined |
| 18-102_k141_143249 | Not-determined |
| 18-102_k141_91964 | Not-determined |
| 18-102_k141_143251 | Not-determined |
| 18-102_k141_203635 | Not-determined |
| 18-102_k141_162809 | Not-determined |
| 18-102_k141_6612 | Low-quality |
| 18-102_k141_102341 | Not-determined |
| 18-102_k141_244662 | Not-determined |
| 18-102_k141_203640 | Not-determined |
| 18-102_k141_244663 | Low-quality |
| 18-102_k141_162811 | Not-determined |
| 18-102_k141_214965 | Not-determined |
| 18-102_k141_162812 | Not-determined |
| 18-102_k141_237020 | Not-determined |
| 18-102_k141_244668 | Not-determined |
| 18-102_k141_237021 | Not-determined |
| 18-102_k141_143252 | Not-determined |
| 18-102_k141_203645 | Low-quality |
| 18-102_k141_244672 | Not-determined |
| 18-102_k141_224451 | Not-determined |
| 18-102_k141_91973 | Not-determined |
| 18-102_k141_162822 | Not-determined |
| 18-102_k141_224453 | Not-determined |
| 18-102_k141_143261 | Not-determined |
| 18-102_k141_162824 | Low-quality |
| 18-102_k141_102364 | Not-determined |
| 18-102_k141_237035 | Not-determined |
| 18-102_k141_80749 | Not-determined |
| 18-102_k141_153502 | Low-quality |
| 18-102_k141_237039 | Low-quality |
| 18-102_k141_237055 | Low-quality |
| 18-102_k141_237058 | Not-determined |
| 18-102_k141_237059 | Low-quality |
| 18-102_k141_153519 | Not-determined |
| 18-102_k141_91995 | Not-determined |
| 18-102_k141_70737 | Not-determined |
| 18-102_k141_224461 | Not-determined |
| 18-102_k141_131347 | Not-determined |
| 18-102_k141_91998 | Not-determined |
| 18-102_k141_80766 | Not-determined |
| 18-102_k141_80767 | Not-determined |
| 18-102_k141_60569 | Not-determined |
| 18-102_k141_111202 | Not-determined |
| 18-102_k141_111204 | Not-determined |
| 18-102_k141_203657 | Not-determined |
| 18-102_k141_203659 | Not-determined |
| 18-102_k141_122666 | Not-determined |
| 18-102_k141_224471 | Not-determined |
| 18-102_k141_237081 | Not-determined |
| 18-102_k141_92009 | Not-determined |
| 18-102_k141_111215 | Not-determined |
| 18-102_k141_122670 | Not-determined |
| 18-102_k141_214973 | Not-determined |
| 18-102_k141_175614 | Low-quality |
| 18-102_k141_143280 | Not-determined |
| 18-102_k141_175624 | Not-determined |
| 18-102_k141_162833 | Not-determined |
| 18-102_k141_70760 | Not-determined |
| 18-102_k141_237091 | Not-determined |
| 18-102_k141_102385 | Not-determined |
| 18-102_k141_214987 | Not-determined |
| 18-102_k141_214989 | Not-determined |
| 18-102_k141_143302 | Not-determined |
| 18-102_k141_40386 | Not-determined |
| 18-102_k141_143310 | Not-determined |
| 18-102_k141_131371 | Not-determined |
| 18-102_k141_215003 | Not-determined |
| 18-102_k141_162856 | Not-determined |
| 18-102_k141_92058 | Not-determined |
| 18-102_k141_143322 | Not-determined |
| 18-102_k141_60607 | Not-determined |
| 18-102_k141_40402 | Not-determined |
| 18-102_k141_18900 | Not-determined |
| 18-102_k141_194942 | Not-determined |
| 18-102_k141_143336 | Not-determined |
| 18-102_k141_194944 | Not-determined |
| 18-102_k141_92082 | Low-quality |
| 18-102_k141_224512 | Not-determined |
| 18-102_k141_40416 | Low-quality |
| 18-102_k141_111257 | Not-determined |
| 18-102_k141_194951 | Not-determined |
| 18-102_k141_162867 | Not-determined |
| 18-102_k141_237113 | Not-determined |
| 18-102_k141_237114 | Not-determined |
| 18-102_k141_175652 | Not-determined |
| 18-102_k141_153546 | Not-determined |
| 18-102_k141_92087 | Not-determined |
| 18-102_k141_153548 | Not-determined |
| 18-102_k141_40425 | Low-quality |
| 18-102_k141_215030 | Not-determined |
| 18-102_k141_237126 | Not-determined |
| 18-102_k141_111268 | Low-quality |
| 18-102_k141_6664 | Not-determined |
| 18-102_k141_153563 | Not-determined |
| 18-102_k141_244725 | Not-determined |
| 18-102_k141_175673 | Not-determined |
| 18-102_k141_60619 | Low-quality |
| 18-102_k141_18915 | Not-determined |
| 18-102_k141_175676 | Not-determined |
| 18-102_k141_194961 | Not-determined |
| 18-102_k141_224530 | Not-determined |
| 18-102_k141_92098 | Not-determined |
| 18-102_k141_92099 | Not-determined |
| 18-102_k141_122722 | Low-quality |
| 18-102_k141_6670 | Not-determined |
| 18-102_k141_122727 | Not-determined |
| 18-102_k141_60635 | Not-determined |
| 18-102_k141_70807 | Not-determined |
| 18-102_k141_143377 | Not-determined |
| 18-102_k141_175689 | Not-determined |
| 18-102_k141_143386 | Not-determined |
| 18-102_k141_131388 | Not-determined |
| 18-102_k141_111300 | Not-determined |
| 18-102_k141_60645 | Not-determined |
| 18-102_k141_18949 | Not-determined |
| 18-102_k141_60650 | Not-determined |
| 18-102_k141_70822 | Not-determined |
| 18-102_k141_143396 | Low-quality |
| 18-102_k141_27496 | Not-determined |
| 18-102_k141_111308 | Not-determined |
| 18-102_k141_237167 | Not-determined |
| 18-102_k141_70829 | Not-determined |
| 18-102_k141_131392 | Not-determined |
| 18-102_k141_18958 | Low-quality |
| 18-102_k141_92120 | Low-quality |
| 18-102_k141_92121 | Low-quality |
| 18-102_k141_18960 | Not-determined |
| 18-102_k141_131397 | Not-determined |
| 18-102_k141_6694 | Not-determined |
| 18-102_k141_6695 | Not-determined |
| 18-102_k141_131402 | Not-determined |
| 18-102_k141_153601 | Not-determined |
| 18-102_k141_70845 | Not-determined |
| 18-102_k141_153602 | Not-determined |
| 18-102_k141_203685 | Not-determined |
| 18-102_k141_244747 | Not-determined |
| 18-102_k141_122753 | Not-determined |
| 18-102_k141_244750 | Not-determined |
| 18-102_k141_111311 | Not-determined |
| 18-102_k141_111318 | Not-determined |
| 18-102_k141_215050 | Not-determined |
| 18-102_k141_131410 | Low-quality |
| 18-102_k141_60665 | Not-determined |
| 18-102_k141_153619 | Not-determined |
| 18-102_k141_237220 | Not-determined |
| 18-102_k141_184887 | Not-determined |
| 18-102_k141_60673 | Not-determined |
| 18-102_k141_153620 | Not-determined |
| 18-102_k141_215059 | Not-determined |
| 18-102_k141_244756 | Not-determined |
| 18-102_k141_203702 | Not-determined |
| 18-102_k141_111335 | Not-determined |
| 18-102_k141_92145 | Low-quality |
| 18-102_k141_92146 | Not-determined |
| 18-102_k141_60699 | Not-determined |
| 18-102_k141_143450 | Not-determined |
| 18-102_k141_215076 | Not-determined |
| 18-102_k141_27520 | Not-determined |
| 18-102_k141_131456 | Not-determined |
| 18-102_k141_40480 | Not-determined |
| 18-102_k141_215086 | Low-quality |
| 18-102_k141_60707 | Not-determined |
| 18-102_k141_203713 | Not-determined |
| 18-102_k141_40490 | Not-determined |
| 18-102_k141_153645 | Not-determined |
| 18-102_k141_215096 | Not-determined |
| 18-102_k141_175717 | Not-determined |
| 18-102_k141_175721 | Not-determined |
| 18-102_k141_215109 | Not-determined |
| 18-102_k141_237245 | Not-determined |
| 18-102_k141_215111 | Not-determined |
| 18-102_k141_131473 | Not-determined |
| 18-102_k141_102405 | Not-determined |
| 18-102_k141_102406 | Not-determined |
| 18-102_k141_162889 | Low-quality |
| 18-102_k141_215114 | Not-determined |
| 18-102_k141_92160 | Not-determined |
| 18-102_k141_6726 | Not-determined |
| 18-102_k141_215116 | Not-determined |
| 18-102_k141_175728 | Not-determined |
| 18-102_k141_102416 | Not-determined |
| 18-102_k141_60716 | Not-determined |
| 18-102_k141_122819 | Not-determined |
| 18-102_k141_143472 | Not-determined |
| 18-102_k141_162897 | Not-determined |
| 18-102_k141_18973 | Not-determined |
| 18-102_k141_175735 | Not-determined |
| 18-102_k141_153668 | Not-determined |
| 18-102_k141_215123 | Not-determined |
| 18-102_k141_80829 | Not-determined |
| 18-102_k141_27537 | Not-determined |
| 18-102_k141_203745 | Not-determined |
| 18-102_k141_215127 | Not-determined |
| 18-102_k141_153673 | Not-determined |
| 18-102_k141_153674 | Not-determined |
| 18-102_k141_195006 | Low-quality |
| 18-102_k141_244776 | Not-determined |
| 18-102_k141_175743 | Low-quality |
| 18-102_k141_143482 | Not-determined |
| 18-102_k141_175744 | Not-determined |
| 18-102_k141_143483 | Not-determined |
| 18-102_k141_162911 | Not-determined |
| 18-102_k141_203755 | Not-determined |
| 18-102_k141_92195 | Not-determined |
| 18-102_k141_244780 | Low-quality |
| 18-102_k141_203762 | Not-determined |
| 18-102_k141_195013 | Not-determined |
| 18-102_k141_80859 | Not-determined |
| 18-102_k141_244789 | Not-determined |
| 18-102_k141_111348 | Not-determined |
| 18-102_k141_111349 | Not-determined |
| 18-102_k141_102427 | Not-determined |
| 18-102_k141_60745 | Not-determined |
| 18-102_k141_111352 | Not-determined |
| 18-102_k141_237268 | Not-determined |
| 18-102_k141_162936 | Not-determined |
| 18-102_k141_184922 | Not-determined |
| 18-102_k141_224544 | Not-determined |
| 18-102_k141_70865 | Not-determined |
| 18-102_k141_153729 | Not-determined |
| 18-102_k141_111361 | Not-determined |
| 18-102_k141_80876 | Low-quality |
| 18-102_k141_111367 | Not-determined |
| 18-102_k141_70871 | Not-determined |
| 18-102_k141_60751 | Not-determined |
| 18-102_k141_195030 | Not-determined |
| 18-102_k141_6762 | Not-determined |
| 18-102_k141_70874 | Not-determined |
| 18-102_k141_215182 | Not-determined |
| 18-102_k141_175778 | Not-determined |
| 18-102_k141_92229 | Not-determined |
| 18-102_k141_122845 | Not-determined |
| 18-102_k141_237286 | Not-determined |
| 18-102_k141_175779 | Not-determined |
| 18-102_k141_224549 | Not-determined |
| 18-102_k141_70881 | Not-determined |
| 18-102_k141_19004 | Not-determined |
| 18-102_k141_70887 | Not-determined |
| 18-102_k141_215190 | Not-determined |
| 18-102_k141_215193 | Not-determined |
| 18-102_k141_122855 | Not-determined |
| 18-102_k141_215195 | Not-determined |
| 18-102_k141_203789 | Not-determined |
| 18-102_k141_244827 | Not-determined |
| 18-102_k141_224557 | Not-determined |
| 18-102_k141_215201 | Not-determined |
| 18-102_k141_162960 | Not-determined |
| 18-102_k141_27569 | Not-determined |
| 18-102_k141_60778 | Not-determined |
| 18-102_k141_80903 | Low-quality |
| 18-102_k141_6781 | Not-determined |
| 18-102_k141_80906 | Not-determined |
| 18-102_k141_195043 | Not-determined |
| 18-102_k141_80911 | Not-determined |
| 18-102_k141_102465 | Not-determined |
| 18-102_k141_19024 | Not-determined |
| 18-102_k141_143505 | Not-determined |
| 18-102_k141_19029 | Not-determined |
| 18-102_k141_40531 | Not-determined |
| 18-102_k141_27583 | Not-determined |
| 18-102_k141_19031 | Not-determined |
| 18-102_k141_175817 | Low-quality |
| 18-102_k141_175818 | Low-quality |
| 18-102_k141_195049 | Not-determined |
| 18-102_k141_80922 | Not-determined |
| 18-102_k141_215218 | Not-determined |
| 18-102_k141_175820 | Not-determined |
| 18-102_k141_175821 | Not-determined |
| 18-102_k141_143515 | Not-determined |
| 18-102_k141_215224 | Not-determined |
| 18-102_k141_184960 | Not-determined |
| 18-102_k141_111400 | Low-quality |
| 18-102_k141_237309 | Low-quality |
| 18-102_k141_237311 | Not-determined |
| 18-102_k141_175828 | Not-determined |
| 18-102_k141_203802 | Not-determined |
| 18-102_k141_19037 | Low-quality |
| 18-102_k141_184973 | Not-determined |
| 18-102_k141_60792 | Not-determined |
| 18-102_k141_102489 | Not-determined |
| 18-102_k141_40553 | Not-determined |
| 18-102_k141_237319 | Low-quality |
| 18-102_k141_80953 | Not-determined |
| 18-102_k141_60808 | Not-determined |
| 18-102_k141_102497 | Not-determined |
| 18-102_k141_80955 | Not-determined |
| 18-102_k141_92256 | Not-determined |
| 18-102_k141_80958 | Not-determined |
| 18-102_k141_80962 | Not-determined |
| 18-102_k141_195076 | Not-determined |
| 18-102_k141_92266 | Not-determined |
| 18-102_k141_40558 | Not-determined |
| 18-102_k141_111410 | Not-determined |
| 18-102_k141_102504 | Not-determined |
| 18-102_k141_215252 | Not-determined |
| 18-102_k141_131484 | Low-quality |
| 18-102_k141_111417 | Not-determined |
| 18-102_k141_111418 | Not-determined |
| 18-102_k141_131487 | Not-determined |
| 18-102_k141_237343 | Not-determined |
| 18-102_k141_131488 | Not-determined |
| 18-102_k141_40575 | Not-determined |
| 18-102_k141_60831 | Not-determined |
| 18-102_k141_70927 | Not-determined |
| 18-102_k141_60835 | Not-determined |
| 18-102_k141_70928 | Not-determined |
| 18-102_k141_60836 | Not-determined |
| 18-102_k141_143561 | Not-determined |
| 18-102_k141_80985 | Not-determined |
| 18-102_k141_143564 | Not-determined |
| 18-102_k141_143572 | Not-determined |
| 18-102_k141_60841 | Not-determined |
| 18-102_k141_60845 | Not-determined |
| 18-102_k141_19077 | Low-quality |
| 18-102_k141_184997 | Not-determined |
| 18-102_k141_237364 | Low-quality |

| | |
|---|---|
| 18-102_k141_237365 | Low-quality |
| 18-102_k141_70942 | Not-determined |
| 18-102_k141_215285 | Not-determined |
| 18-102_k141_122885 | Not-determined |
| 18-102_k141_40578 | Not-determined |
| 18-102_k141_81005 | Not-determined |
| 18-102_k141_60850 | Not-determined |
| 18-102_k141_111436 | Not-determined |
| 18-102_k141_102551 | Not-determined |
| 18-102_k141_19088 | Not-determined |
| 18-102_k141_40583 | Not-determined |
| 18-102_k141_111449 | Not-determined |
| 18-102_k141_102566 | Not-determined |
| 18-102_k141_102568 | Not-determined |
| 18-102_k141_111454 | Not-determined |
| 18-102_k141_224570 | Not-determined |
| 18-102_k141_153784 | Not-determined |
| 18-102_k141_237381 | Not-determined |
| 18-102_k141_153786 | Not-determined |
| 18-102_k141_102574 | Not-determined |
| 18-102_k141_153790 | Not-determined |
| 18-102_k141_111461 | Not-determined |
| 18-102_k141_237389 | Not-determined |
| 18-102_k141_122919 | Not-determined |
| 18-102_k141_122920 | Not-determined |
| 18-102_k141_122921 | Low-quality |
| 18-102_k141_19118 | Not-determined |
| 18-102_k141_244849 | Not-determined |
| 18-102_k141_195143 | Not-determined |
| 18-102_k141_215308 | Low-quality |
| 18-102_k141_111473 | Low-quality |
| 18-102_k141_175841 | Not-determined |
| 18-102_k141_40610 | Not-determined |
| 18-102_k141_224582 | Not-determined |
| 18-102_k141_131520 | Not-determined |
| 18-102_k141_131522 | Not-determined |
| 18-102_k141_175851 | Low-quality |
| 18-102_k141_153808 | Low-quality |
| 18-102_k141_195158 | Not-determined |
| 18-102_k141_153810 | Not-determined |
| 18-102_k141_102601 | Not-determined |
| 18-102_k141_102602 | Not-determined |
| 18-102_k141_153813 | Not-determined |
| 18-102_k141_153814 | Not-determined |
| 18-102_k141_131524 | Not-determined |
| 18-102_k141_143593 | Not-determined |
| 18-102_k141_237418 | Not-determined |
| 18-102_k141_237419 | Not-determined |
| 18-102_k141_185030 | Not-determined |
| 18-102_k141_60876 | Not-determined |
| 18-102_k141_175863 | Not-determined |
| 18-102_k141_195170 | Low-quality |
| 18-102_k141_131551 | Not-determined |
| 18-102_k141_131556 | Not-determined |
| 18-102_k141_131562 | Not-determined |
| 18-102_k141_185039 | Not-determined |
| 18-102_k141_102619 | Not-determined |
| 18-102_k141_143597 | Not-determined |
| 18-102_k141_215334 | Not-determined |
| 18-102_k141_203879 | Not-determined |
| 18-102_k141_203888 | Not-determined |
| 18-102_k141_185049 | Not-determined |
| 18-102_k141_203890 | Not-determined |
| 18-102_k141_203891 | Not-determined |
| 18-102_k141_102635 | Not-determined |
| 18-102_k141_175879 | Low-quality |
| 18-102_k141_102642 | Not-determined |
| 18-102_k141_203901 | Not-determined |
| 18-102_k141_60910 | Not-determined |
| 18-102_k141_203907 | Not-determined |
| 18-102_k141_40654 | Not-determined |
| 18-102_k141_40655 | Not-determined |
| 18-102_k141_102647 | Not-determined |
| 18-102_k141_224601 | Not-determined |
| 18-102_k141_92291 | Not-determined |
| 18-102_k141_175901 | Not-determined |
| 18-102_k141_70970 | Not-determined |
| 18-102_k141_203918 | Not-determined |
| 18-102_k141_203919 | Not-determined |
| 18-102_k141_70973 | Not-determined |
| 18-102_k141_70978 | Not-determined |
| 18-102_k141_70979 | Not-determined |
| 18-102_k141_92299 | Not-determined |
| 18-102_k141_70986 | Not-determined |
| 18-102_k141_203931 | Not-determined |
| 18-102_k141_175911 | Not-determined |
| 18-102_k141_237475 | Not-determined |
| 18-102_k141_237477 | Not-determined |
| 18-102_k141_102673 | Not-determined |
| 18-102_k141_215363 | Not-determined |
| 18-102_k141_175917 | Not-determined |
| 18-102_k141_224619 | Low-quality |
| 18-102_k141_19195 | Not-determined |
| 18-102_k141_111481 | Not-determined |
| 18-102_k141_215372 | Not-determined |
| 18-102_k141_203940 | Low-quality |
| 18-102_k141_203943 | Not-determined |
| 18-102_k141_237486 | Not-determined |
| 18-102_k141_224624 | Not-determined |
| 18-102_k141_111490 | Not-determined |
| 18-102_k141_102688 | Not-determined |
| 18-102_k141_175924 | Low-quality |
| 18-102_k141_40693 | Not-determined |
| 18-102_k141_102695 | Not-determined |
| 18-102_k141_60928 | Not-determined |
| 18-102_k141_195179 | Not-determined |
| 18-102_k141_185074 | Not-determined |
| 18-102_k141_153822 | Low-quality |
| 18-102_k141_195189 | Not-determined |
| 18-102_k141_81096 | Not-determined |
| 18-102_k141_60935 | Not-determined |
| 18-102_k141_215389 | Not-determined |
| 18-102_k141_153835 | Not-determined |
| 18-102_k141_185086 | Not-determined |
| 18-102_k141_195199 | Not-determined |
| 18-102_k141_71015 | Not-determined |
| 18-102_k141_40715 | Not-determined |
| 18-102_k141_6814 | Not-determined |
| 18-102_k141_153849 | Not-determined |
| 18-102_k141_195211 | Not-determined |
| 18-102_k141_71024 | Not-determined |
| 18-102_k141_27637 | Not-determined |
| 18-102_k141_40718 | Not-determined |
| 18-102_k141_19219 | Not-determined |
| 18-102_k141_215402 | Not-determined |
| 18-102_k141_195215 | Not-determined |
| 18-102_k141_19220 | Not-determined |
| 18-102_k141_40722 | Not-determined |
| 18-102_k141_19222 | Not-determined |
| 18-102_k141_215405 | Not-determined |
| 18-102_k141_71034 | Not-determined |
| 18-102_k141_19226 | Not-determined |
| 18-102_k141_224660 | Low-quality |
| 18-102_k141_27645 | Not-determined |
| 18-102_k141_81113 | Not-determined |
| 18-102_k141_6833 | Low-quality |
| 18-102_k141_195238 | Not-determined |
| 18-102_k141_81118 | Not-determined |
| 18-102_k141_195239 | Not-determined |
| 18-102_k141_203958 | Not-determined |
| 18-102_k141_40738 | Not-determined |
| 18-102_k141_163014 | Not-determined |
| 18-102_k141_6844 | Not-determined |
| 18-102_k141_19236 | Not-determined |
| 18-102_k141_195247 | Not-determined |
| 18-102_k141_71066 | Not-determined |
| 18-102_k141_153864 | Low-quality |
| 18-102_k141_27658 | Not-determined |
| 18-102_k141_143675 | Not-determined |
| 18-102_k141_185115 | Low-quality |
| 18-102_k141_203975 | Not-determined |
| 18-102_k141_224678 | Not-determined |
| 18-102_k141_237518 | Not-determined |
| 18-102_k141_237522 | Not-determined |
| 18-102_k141_6860 | Not-determined |
| 18-102_k141_244905 | Not-determined |
| 18-102_k141_185119 | Low-quality |
| 18-102_k141_215429 | Not-determined |
| 18-102_k141_215430 | Not-determined |
| 18-102_k141_185121 | Not-determined |
| 18-102_k141_143686 | Not-determined |
| 18-102_k141_185137 | Not-determined |
| 18-102_k141_81139 | Not-determined |
| 18-102_k141_19259 | Not-determined |
| 18-102_k141_163035 | Not-determined |
| 18-102_k141_224686 | Not-determined |
| 18-102_k141_92324 | Not-determined |
| 18-102_k141_71074 | Not-determined |
| 18-102_k141_92327 | Not-determined |
| 18-102_k141_163039 | Not-determined |
| 18-102_k141_203996 | Not-determined |
| 18-102_k141_102731 | Not-determined |
| 18-102_k141_163044 | Not-determined |
| 18-102_k141_143704 | Not-determined |
| 18-102_k141_185164 | Low-quality |
| 18-102_k141_71092 | Not-determined |
| 18-102_k141_224701 | Not-determined |
| 18-102_k141_153916 | Not-determined |
| 18-102_k141_224703 | Not-determined |
| 18-102_k141_6878 | Not-determined |
| 18-102_k141_71099 | Not-determined |
| 18-102_k141_81171 | Not-determined |
| 18-102_k141_195302 | Not-determined |
| 18-102_k141_224711 | Not-determined |
| 18-102_k141_153923 | Not-determined |
| 18-102_k141_163060 | Not-determined |
| 18-102_k141_102751 | Not-determined |
| 18-102_k141_6885 | Not-determined |
| 18-102_k141_6892 | Not-determined |
| 18-102_k141_81189 | Low-quality |
| 18-102_k141_224720 | Not-determined |
| 18-102_k141_224726 | Not-determined |
| 18-102_k141_237563 | Not-determined |
| 18-102_k141_204009 | Not-determined |
| 18-102_k141_81193 | Not-determined |
| 18-102_k141_111511 | Not-determined |
| 18-102_k141_224733 | Not-determined |
| 18-102_k141_81200 | Not-determined |
| 18-102_k141_102766 | Not-determined |
| 18-102_k141_40816 | Not-determined |
| 18-102_k141_102768 | Not-determined |
| 18-102_k141_111515 | Not-determined |
| 18-102_k141_81205 | Low-quality |
| 18-102_k141_81208 | Not-determined |
| 18-102_k141_71137 | Not-determined |
| 18-102_k141_195331 | Not-determined |
| 18-102_k141_153955 | Not-determined |
| 18-102_k141_81213 | Not-determined |
| 18-102_k141_153958 | Not-determined |
| 18-102_k141_237573 | Low-quality |
| 18-102_k141_153959 | Not-determined |
| 18-102_k141_195335 | Not-determined |
| 18-102_k141_81221 | Not-determined |
| 18-102_k141_153960 | Not-determined |
| 18-102_k141_237579 | Not-determined |
| 18-102_k141_131576 | Not-determined |
| 18-102_k141_237583 | Not-determined |
| 18-102_k141_195341 | Not-determined |
| 18-102_k141_163078 | Not-determined |
| 18-102_k141_27698 | Not-determined |
| 18-102_k141_204024 | Not-determined |
| 18-102_k141_6928 | Not-determined |
| 18-102_k141_40824 | Not-determined |
| 18-102_k141_143725 | Not-determined |
| 18-102_k141_143726 | Not-determined |
| 18-102_k141_111534 | Not-determined |
| 18-102_k141_204027 | Not-determined |
| 18-102_k141_204028 | Not-determined |
| 18-102_k141_111536 | Not-determined |
| 18-102_k141_81241 | Not-determined |
| 18-102_k141_111539 | Not-determined |
| 18-102_k141_185220 | Not-determined |
| 18-102_k141_40827 | Not-determined |
| 18-102_k141_40828 | Low-quality |
| 18-102_k141_71144 | Low-quality |
| 18-102_k141_244931 | Low-quality |
| 18-102_k141_153973 | Not-determined |
| 18-102_k141_102794 | Not-determined |
| 18-102_k141_153981 | Not-determined |
| 18-102_k141_131602 | Not-determined |
| 18-102_k141_81259 | Not-determined |
| 18-102_k141_19327 | Not-determined |
| 18-102_k141_215496 | Not-determined |
| 18-102_k141_204050 | Not-determined |
| 18-102_k141_71155 | Not-determined |
| 18-102_k141_163107 | Low-quality |
| 18-102_k141_163111 | Not-determined |
| 18-102_k141_224784 | Not-determined |
| 18-102_k141_163116 | Not-determined |
| 18-102_k141_81276 | Not-determined |
| 18-102_k141_185241 | Not-determined |
| 18-102_k141_185242 | Not-determined |
| 18-102_k141_111545 | Not-determined |
| 18-102_k141_154001 | Not-determined |
| 18-102_k141_185254 | Not-determined |
| 18-102_k141_92356 | Not-determined |
| 18-102_k141_185259 | Not-determined |
| 18-102_k141_163130 | Low-quality |
| 18-102_k141_244947 | Not-determined |
| 18-102_k141_6970 | Not-determined |
| 18-102_k141_224795 | Not-determined |
| 18-102_k141_154009 | Not-determined |
| 18-102_k141_215524 | Not-determined |
| 18-102_k141_224799 | Low-quality |
| 18-102_k141_154017 | Not-determined |
| 18-102_k141_195373 | Not-determined |
| 18-102_k141_154018 | Not-determined |
| 18-102_k141_81286 | Not-determined |
| 18-102_k141_122972 | Not-determined |
| 18-102_k141_224806 | Not-determined |
| 18-102_k141_154021 | Not-determined |
| 18-102_k141_195377 | Not-determined |
| 18-102_k141_19368 | Not-determined |
| 18-102_k141_204067 | Low-quality |
| 18-102_k141_195382 | Not-determined |
| 18-102_k141_102842 | Not-determined |
| 18-102_k141_71197 | Not-determined |
| 18-102_k141_71199 | Not-determined |
| 18-102_k141_19380 | Low-quality |
| 18-102_k141_71204 | Not-determined |
| 18-102_k141_61021 | Low-quality |
| 18-102_k141_71207 | Not-determined |
| 18-102_k141_131647 | Not-determined |
| 18-102_k141_61025 | Not-determined |
| 18-102_k141_61028 | Not-determined |
| 18-102_k141_204076 | Not-determined |
| 18-102_k141_185272 | Not-determined |
| 18-102_k141_102857 | Not-determined |
| 18-102_k141_19394 | Not-determined |
| 18-102_k141_195404 | Not-determined |
| 18-102_k141_195408 | Not-determined |
| 18-102_k141_195410 | Not-determined |
| 18-102_k141_237607 | Not-determined |
| 18-102_k141_71219 | Not-determined |
| 18-102_k141_71221 | Not-determined |
| 18-102_k141_215541 | Low-quality |
| 18-102_k141_163173 | Not-determined |
| 18-102_k141_237611 | Not-determined |
| 18-102_k141_215544 | Not-determined |
| 18-102_k141_163174 | Not-determined |
| 18-102_k141_27728 | Not-determined |
| 18-102_k141_163185 | Not-determined |
| 18-102_k141_92420 | Not-determined |
| 18-102_k141_111614 | Not-determined |
| 18-102_k141_215557 | Not-determined |
| 18-102_k141_92422 | Not-determined |
| 18-102_k141_92424 | Not-determined |
| 18-102_k141_27738 | Not-determined |
| 18-102_k141_215559 | Not-determined |
| 18-102_k141_81312 | Not-determined |
| 18-102_k141_111618 | Not-determined |
| 18-102_k141_163197 | Not-determined |
| 18-102_k141_81315 | Not-determined |
| 18-102_k141_163198 | Not-determined |
| 18-102_k141_204104 | Not-determined |
| 18-102_k141_244981 | Not-determined |
| 18-102_k141_204105 | Not-determined |
| 18-102_k141_27744 | Not-determined |
| 18-102_k141_111627 | Not-determined |
| 18-102_k141_131657 | Not-determined |
| 18-102_k141_195442 | Not-determined |
| 18-102_k141_27752 | Not-determined |
| 18-102_k141_185283 | Not-determined |
| 18-102_k141_27756 | Not-determined |
| 18-102_k141_163204 | Not-determined |
| 18-102_k141_61035 | Not-determined |
| 18-102_k141_27765 | Not-determined |
| 18-102_k141_176012 | Not-determined |
| 18-102_k141_81347 | Not-determined |
| 18-102_k141_61043 | Not-determined |
| 18-102_k141_61044 | Not-determined |
| 18-102_k141_81351 | Not-determined |
| 18-102_k141_176018 | Not-determined |
| 18-102_k141_176022 | Not-determined |
| 18-102_k141_81354 | Not-determined |
| 18-102_k141_81357 | Not-determined |
| 18-102_k141_27773 | Not-determined |
| 18-102_k141_176041 | Not-determined |
| 18-102_k141_111651 | Not-determined |
| 18-102_k141_163225 | Not-determined |
| 18-102_k141_176045 | Not-determined |
| 18-102_k141_27781 | Not-determined |
| 18-102_k141_163231 | Low-quality |
| 18-102_k141_61066 | Not-determined |
| 18-102_k141_27792 | Not-determined |
| 18-102_k141_163241 | Not-determined |
| 18-102_k141_224853 | Not-determined |
| 18-102_k141_176049 | Not-determined |
| 18-102_k141_176053 | Not-determined |
| 18-102_k141_71293 | Not-determined |
| 18-102_k141_27803 | Not-determined |
| 18-102_k141_204136 | Not-determined |
| 18-102_k141_131684 | Not-determined |
| 18-102_k141_27809 | Not-determined |
| 18-102_k141_102878 | Not-determined |
| 18-102_k141_131688 | Not-determined |
| 18-102_k141_224858 | Not-determined |
| 18-102_k141_71304 | Not-determined |
| 18-102_k141_204145 | Not-determined |
| 18-102_k141_163248 | Not-determined |
| 18-102_k141_204146 | Not-determined |
| 18-102_k141_163249 | Not-determined |
| 18-102_k141_27820 | Not-determined |
| 18-102_k141_111660 | Not-determined |
| 18-102_k141_185306 | Not-determined |
| 18-102_k141_81390 | Not-determined |
| 18-102_k141_224877 | Not-determined |
| 18-102_k141_81394 | Not-determined |
| 18-102_k141_123049 | Not-determined |
| 18-102_k141_204154 | Not-determined |
| 18-102_k141_27827 | Not-determined |
| 18-102_k141_204157 | Not-determined |
| 18-102_k141_143748 | Not-determined |
| 18-102_k141_143751 | Not-determined |
| 18-102_k141_71321 | Not-determined |
| 18-102_k141_204159 | Not-determined |
| 18-102_k141_143755 | Not-determined |
| 18-102_k141_27831 | Not-determined |
| 18-102_k141_71326 | Not-determined |
| 18-102_k141_27833 | Not-determined |
| 18-102_k141_131725 | Low-quality |
| 18-102_k141_204168 | Not-determined |
| 18-102_k141_131727 | Low-quality |
| 18-102_k141_71337 | Not-determined |
| 18-102_k141_185332 | Not-determined |
| 18-102_k141_81426 | Not-determined |
| 18-102_k141_204188 | Not-determined |
| 18-102_k141_111676 | Not-determined |
| 18-102_k141_204191 | Not-determined |
| 18-102_k141_40846 | Not-determined |
| 18-102_k141_40848 | Not-determined |
| 18-102_k141_143765 | Not-determined |
| 18-102_k141_40850 | Not-determined |
| 18-102_k141_154044 | Not-determined |
| 18-102_k141_185342 | Not-determined |
| 18-102_k141_111684 | Low-quality |
| 18-102_k141_154048 | Not-determined |
| 18-102_k141_111685 | Not-determined |
| 18-102_k141_143772 | Not-determined |
| 18-102_k141_61135 | Not-determined |
| 18-102_k141_245042 | Not-determined |
| 18-102_k141_185351 | Not-determined |
| 18-102_k141_143775 | Not-determined |
| 18-102_k141_123063 | Not-determined |
| 18-102_k141_224916 | Not-determined |
| 18-102_k141_40860 | Not-determined |
| 18-102_k141_123067 | Not-determined |
| 18-102_k141_111698 | Not-determined |
| 18-102_k141_224921 | Not-determined |
| 18-102_k141_224928 | Not-determined |
| 18-102_k141_154071 | Not-determined |
| 18-102_k141_71366 | Low-quality |
| 18-102_k141_81440 | Not-determined |
| 18-102_k141_195563 | Not-determined |
| 18-102_k141_111703 | Not-determined |
| 18-102_k141_40879 | Not-determined |
| 18-102_k141_204205 | Not-determined |
| 18-102_k141_102916 | Not-determined |
| 18-102_k141_204210 | Not-determined |
| 18-102_k141_102917 | Not-determined |
| 18-102_k141_102921 | Not-determined |
| 18-102_k141_163259 | Not-determined |
| 18-102_k141_61175 | Low-quality |
| 18-102_k141_195586 | Not-determined |
| 18-102_k141_185375 | Not-determined |
| 18-102_k141_195590 | Not-determined |
| 18-102_k141_61178 | Not-determined |
| 18-102_k141_61180 | Low-quality |
| 18-102_k141_61181 | Not-determined |
| 18-102_k141_224945 | Not-determined |
| 18-102_k141_195593 | Not-determined |
| 18-102_k141_102929 | Not-determined |
| 18-102_k141_245088 | Not-determined |
| 18-102_k141_123084 | Low-quality |
| 18-102_k141_111728 | Not-determined |
| 18-102_k141_40903 | Not-determined |
| 18-102_k141_245092 | Not-determined |
| 18-102_k141_111729 | Not-determined |
| 18-102_k141_61199 | Not-determined |
| 18-102_k141_102939 | Not-determined |
| 18-102_k141_40906 | Not-determined |
| 18-102_k141_245102 | Low-quality |
| 18-102_k141_40913 | Not-determined |
| 18-102_k141_111738 | Not-determined |
| 18-102_k141_224970 | Not-determined |
| 18-102_k141_123097 | Not-determined |
| 18-102_k141_154092 | Low-quality |
| 18-102_k141_204236 | Not-determined |
| 18-102_k141_102946 | Not-determined |
| 18-102_k141_245117 | Not-determined |
| 18-102_k141_245118 | Not-determined |
| 18-102_k141_71394 | Not-determined |
| 18-102_k141_131772 | Not-determined |
| 18-102_k141_92426 | Not-determined |
| 18-102_k141_92428 | Not-determined |
| 18-102_k141_204251 | Not-determined |
| 18-102_k141_71411 | Not-determined |
| 18-102_k141_71412 | Not-determined |
| 18-102_k141_195645 | Low-quality |
| 18-102_k141_143821 | Not-determined |
| 18-102_k141_237666 | Not-determined |
| 18-102_k141_61220 | Not-determined |
| 18-102_k141_71414 | Not-determined |
| 18-102_k141_204257 | Not-determined |
| 18-102_k141_185419 | Not-determined |
| 18-102_k141_195650 | Low-quality |
| 18-102_k141_245138 | Not-determined |
| 18-102_k141_185422 | Not-determined |
| 18-102_k141_111765 | Not-determined |
| 18-102_k141_61228 | Not-determined |
| 18-102_k141_163289 | Not-determined |
| 18-102_k141_71422 | Not-determined |
| 18-102_k141_71423 | Low-quality |
| 18-102_k141_7041 | Not-determined |
| 18-102_k141_204260 | Low-quality |
| 18-102_k141_154123 | Not-determined |
| 18-102_k141_185429 | Not-determined |
| 18-102_k141_92453 | Not-determined |
| 18-102_k141_143839 | Not-determined |
| 18-102_k141_154127 | Not-determined |
| 18-102_k141_143844 | Low-quality |
| 18-102_k141_185447 | Not-determined |
| 18-102_k141_215680 | Not-determined |
| 18-102_k141_111777 | Not-determined |
| 18-102_k141_245151 | Low-quality |
| 18-102_k141_102983 | Not-determined |
| 18-102_k141_154141 | Not-determined |
| 18-102_k141_131800 | Not-determined |
| 18-102_k141_237672 | Not-determined |
| 18-102_k141_131801 | Low-quality |
| 18-102_k141_185464 | Not-determined |
| 18-102_k141_215681 | Not-determined |
| 18-102_k141_102998 | Not-determined |
| 18-102_k141_123146 | Not-determined |
| 18-102_k141_123147 | Not-determined |
| 18-102_k141_215689 | Not-determined |
| 18-102_k141_111798 | Not-determined |
| 18-102_k141_103002 | Not-determined |
| 18-102_k141_237684 | Low-quality |
| 18-102_k141_92477 | Not-determined |
| 18-102_k141_195681 | Not-determined |
| 18-102_k141_111809 | Not-determined |
| 18-102_k141_195692 | Not-determined |
| 18-102_k141_237698 | Low-quality |
| 18-102_k141_103020 | Not-determined |
| 18-102_k141_131834 | Not-determined |
| 18-102_k141_92495 | Not-determined |
| 18-102_k141_131837 | Not-determined |
| 18-102_k141_27858 | Not-determined |
| 18-102_k141_195702 | Not-determined |
| 18-102_k141_92498 | Low-quality |
| 18-102_k141_215707 | Not-determined |
| 18-102_k141_195706 | Not-determined |
| 18-102_k141_131844 | Not-determined |
| 18-102_k141_92503 | Not-determined |
| 18-102_k141_215720 | Not-determined |
| 18-102_k141_131852 | Not-determined |
| 18-102_k141_19437 | Not-determined |
| 18-102_k141_131854 | Not-determined |
| 18-102_k141_131855 | Not-determined |
| 18-102_k141_7073 | Low-quality |
| 18-102_k141_185474 | Not-determined |
| 18-102_k141_185477 | Not-determined |
| 18-102_k141_7082 | Not-determined |
| 18-102_k141_185478 | Not-determined |
| 18-102_k141_185480 | Not-determined |
| 18-102_k141_143851 | Not-determined |
| 18-102_k141_123174 | Not-determined |
| 18-102_k141_143852 | Not-determined |
| 18-102_k141_123179 | Not-determined |
| 18-102_k141_27868 | Not-determined |
| 18-102_k141_143862 | Not-determined |
| 18-102_k141_131864 | Not-determined |
| 18-102_k141_27881 | Not-determined |
| 18-102_k141_224997 | Not-determined |
| 18-102_k141_224999 | Not-determined |
| 18-102_k141_225000 | Not-determined |
| 18-102_k141_237737 | Not-determined |
| 18-102_k141_163296 | Low-quality |
| 18-102_k141_19473 | Not-determined |
| 18-102_k141_19474 | Not-determined |
| 18-102_k141_123197 | Not-determined |
| 18-102_k141_19475 | Not-determined |
| 18-102_k141_225011 | Not-determined |
| 18-102_k141_81498 | Not-determined |
| 18-102_k141_225013 | Not-determined |
| 18-102_k141_154199 | Not-determined |
| 18-102_k141_123200 | Not-determined |
| 18-102_k141_225016 | Not-determined |
| 18-102_k141_27896 | Not-determined |
| 18-102_k141_215756 | Not-determined |
| 18-102_k141_19480 | Not-determined |
| 18-102_k141_61313 | Not-determined |
| 18-102_k141_19481 | Not-determined |
| 18-102_k141_154205 | Not-determined |
| 18-102_k141_19482 | Not-determined |
| 18-102_k141_19483 | Not-determined |
| 18-102_k141_154206 | Not-determined |
| 18-102_k141_225021 | Not-determined |
| 18-102_k141_237746 | Not-determined |
| 18-102_k141_61314 | Not-determined |
| 18-102_k141_225023 | Not-determined |
| 18-102_k141_225024 | Not-determined |
| 18-102_k141_92549 | Low-quality |
| 18-102_k141_225025 | Not-determined |
| 18-102_k141_61322 | Not-determined |
| 18-102_k141_143890 | Not-determined |
| 18-102_k141_154219 | Not-determined |
| 18-102_k141_237752 | Not-determined |
| 18-102_k141_195744 | Not-determined |
| 18-102_k141_143893 | Not-determined |
| 18-102_k141_225036 | Not-determined |
| 18-102_k141_81517 | Not-determined |
| 18-102_k141_61332 | Not-determined |
| 18-102_k141_92560 | Not-determined |
| 18-102_k141_81521 | Not-determined |
| 18-102_k141_61336 | Not-determined |
| 18-102_k141_215771 | Not-determined |
| 18-102_k141_40953 | Not-determined |
| 18-102_k141_19495 | Not-determined |
| 18-102_k141_123217 | Not-determined |
| 18-102_k141_215776 | Not-determined |
| 18-102_k141_215778 | Not-determined |
| 18-102_k141_154226 | Not-determined |
| 18-102_k141_215781 | Not-determined |
| 18-102_k141_103046 | Not-determined |
| 18-102_k141_225044 | Not-determined |
| 18-102_k141_237761 | Not-determined |
| 18-102_k141_123222 | Not-determined |

| | |
|---|---|
| 18-102_k141_163335 | Not-determined |
| 18-102_k141_19506 | Not-determined |
| 18-102_k141_19507 | Not-determined |
| 18-102_k141_81533 | Not-determined |
| 18-102_k141_154241 | Not-determined |
| 18-102_k141_19512 | Not-determined |
| 18-102_k141_143916 | Not-determined |
| 18-102_k141_27913 | Not-determined |
| 18-102_k141_27920 | Not-determined |
| 18-102_k141_19531 | Not-determined |
| 18-102_k141_195786 | Not-determined |
| 18-102_k141_103050 | Not-determined |
| 18-102_k141_195788 | Not-determined |
| 18-102_k141_19540 | Not-determined |
| 18-102_k141_27930 | Not-determined |
| 18-102_k141_143934 | Not-determined |
| 18-102_k141_195804 | Not-determined |
| 18-102_k141_143941 | Not-determined |
| 18-102_k141_195808 | Not-determined |
| 18-102_k141_154268 | Not-determined |
| 18-102_k141_103064 | Not-determined |
| 18-102_k141_71445 | Not-determined |
| 18-102_k141_92602 | Not-determined |
| 18-102_k141_92603 | Not-determined |
| 18-102_k141_81568 | Not-determined |
| 18-102_k141_237812 | Not-determined |
| 18-102_k141_143957 | Not-determined |
| 18-102_k141_27954 | Not-determined |
| 18-102_k141_143958 | Not-determined |
| 18-102_k141_71468 | Not-determined |
| 18-102_k141_154279 | Not-determined |
| 18-102_k141_19562 | Not-determined |
| 18-102_k141_154282 | Not-determined |
| 18-102_k141_131895 | Not-determined |
| 18-102_k141_154284 | Not-determined |
| 18-102_k141_27958 | Not-determined |
| 18-102_k141_154287 | Not-determined |
| 18-102_k141_27961 | Not-determined |
| 18-102_k141_163368 | Not-determined |
| 18-102_k141_81572 | Not-determined |
| 18-102_k141_143977 | Low-quality |
| 18-102_k141_7132 | Not-determined |
| 18-102_k141_7133 | Not-determined |
| 18-102_k141_81579 | Not-determined |
| 18-102_k141_7139 | Not-determined |
| 18-102_k141_143988 | Not-determined |
| 18-102_k141_7142 | Not-determined |
| 18-102_k141_7143 | Not-determined |
| 18-102_k141_71490 | Not-determined |
| 18-102_k141_41003 | Not-determined |
| 18-102_k141_71492 | Not-determined |
| 18-102_k141_71493 | Not-determined |
| 18-102_k141_123246 | Not-determined |
| 18-102_k141_143997 | Not-determined |
| 18-102_k141_71495 | Low-quality |
| 18-102_k141_19582 | Not-determined |
| 18-102_k141_103099 | Not-determined |
| 18-102_k141_41008 | Not-determined |
| 18-102_k141_71501 | Not-determined |
| 18-102_k141_163387 | Not-determined |
| 18-102_k141_144005 | Not-determined |
| 18-102_k141_71504 | Not-determined |
| 18-102_k141_237836 | Not-determined |
| 18-102_k141_19592 | Not-determined |
| 18-102_k141_111847 | Not-determined |
| 18-102_k141_27977 | Not-determined |
| 18-102_k141_215826 | Low-quality |
| 18-102_k141_103105 | Not-determined |
| 18-102_k141_71513 | Not-determined |
| 18-102_k141_71514 | Not-determined |
| 18-102_k141_103106 | Not-determined |
| 18-102_k141_27981 | Not-determined |
| 18-102_k141_71516 | Low-quality |
| 18-102_k141_131916 | Not-determined |
| 18-102_k141_195855 | Not-determined |
| 18-102_k141_41013 | Not-determined |
| 18-102_k141_131919 | Not-determined |
| 18-102_k141_41020 | Not-determined |
| 18-102_k141_195859 | Not-determined |
| 18-102_k141_144027 | Low-quality |
| 18-102_k141_144028 | Not-determined |
| 18-102_k141_237851 | Not-determined |
| 18-102_k141_111864 | Not-determined |
| 18-102_k141_61378 | Not-determined |
| 18-102_k141_7163 | Not-determined |
| 18-102_k141_225111 | Not-determined |
| 18-102_k141_154340 | Not-determined |
| 18-102_k141_7165 | Not-determined |
| 18-102_k141_61381 | Not-determined |
| 18-102_k141_103125 | Low-quality |
| 18-102_k141_19601 | Not-determined |
| 18-102_k141_81635 | Not-determined |
| 18-102_k141_81641 | Not-determined |
| 18-102_k141_144035 | Not-determined |
| 18-102_k141_131928 | Not-determined |
| 18-102_k141_61394 | Not-determined |
| 18-102_k141_71537 | Not-determined |
| 18-102_k141_225128 | Not-determined |
| 18-102_k141_144059 | Not-determined |
| 18-102_k141_225129 | Not-determined |
| 18-102_k141_225130 | Not-determined |
| 18-102_k141_81651 | Not-determined |
| 18-102_k141_144066 | Low-quality |
| 18-102_k141_71546 | Not-determined |
| 18-102_k141_71548 | Not-determined |
| 18-102_k141_41046 | Low-quality |
| 18-102_k141_144073 | Not-determined |
| 18-102_k141_163429 | Not-determined |
| 18-102_k141_154357 | Not-determined |
| 18-102_k141_154359 | Not-determined |
| 18-102_k141_131954 | Not-determined |
| 18-102_k141_225147 | Not-determined |
| 18-102_k141_154364 | Not-determined |
| 18-102_k141_163440 | Not-determined |
| 18-102_k141_81671 | Not-determined |
| 18-102_k141_81674 | Not-determined |
| 18-102_k141_81675 | Not-determined |
| 18-102_k141_41058 | Not-determined |
| 18-102_k141_225154 | Low-quality |
| 18-102_k141_154372 | Not-determined |
| 18-102_k141_41072 | Not-determined |
| 18-102_k141_81694 | Not-determined |
| 18-102_k141_225163 | Not-determined |
| 18-102_k141_71570 | Not-determined |
| 18-102_k141_103141 | Low-quality |
| 18-102_k141_103150 | Not-determined |
| 18-102_k141_225172 | Not-determined |
| 18-102_k141_41078 | Not-determined |
| 18-102_k141_41079 | Not-determined |
| 18-102_k141_123254 | Not-determined |
| 18-102_k141_144090 | Not-determined |
| 18-102_k141_71588 | Not-determined |
| 18-102_k141_144093 | Not-determined |
| 18-102_k141_41082 | Not-determined |
| 18-102_k141_225177 | Not-determined |
| 18-102_k141_144094 | Not-determined |
| 18-102_k141_103152 | Not-determined |
| 18-102_k141_225182 | Not-determined |
| 18-102_k141_225183 | Not-determined |
| 18-102_k141_245201 | Not-determined |
| 18-102_k141_123269 | Not-determined |
| 18-102_k141_7221 | Low-quality |
| 18-102_k141_28012 | Not-determined |
| 18-102_k141_7224 | Not-determined |
| 18-102_k141_144110 | Not-determined |
| 18-102_k141_92621 | Not-determined |
| 18-102_k141_7229 | Not-determined |
| 18-102_k141_28018 | Not-determined |
| 18-102_k141_7231 | Not-determined |
| 18-102_k141_19640 | Not-determined |
| 18-102_k141_41106 | Not-determined |
| 18-102_k141_245220 | Not-determined |
| 18-102_k141_41108 | Low-quality |
| 18-102_k141_28021 | Low-quality |
| 18-102_k141_61427 | Not-determined |
| 18-102_k141_81725 | Not-determined |
| 18-102_k141_71616 | Not-determined |
| 18-102_k141_81730 | Not-determined |
| 18-102_k141_195906 | Not-determined |
| 18-102_k141_61430 | Not-determined |
| 18-102_k141_225192 | Not-determined |
| 18-102_k141_28027 | Not-determined |
| 18-102_k141_19658 | Not-determined |
| 18-102_k141_41128 | Not-determined |
| 18-102_k141_7239 | Not-determined |
| 18-102_k141_92641 | Not-determined |
| 18-102_k141_154413 | Not-determined |
| 18-102_k141_195909 | Not-determined |
| 18-102_k141_81754 | Not-determined |
| 18-102_k141_144126 | Not-determined |
| 18-102_k141_195912 | Not-determined |
| 18-102_k141_195913 | Not-determined |
| 18-102_k141_195914 | Not-determined |
| 18-102_k141_195918 | Low-quality |
| 18-102_k141_81761 | Not-determined |
| 18-102_k141_144131 | Not-determined |
| 18-102_k141_28037 | Not-determined |
| 18-102_k141_81762 | Not-determined |
| 18-102_k141_144133 | Not-determined |
| 18-102_k141_144135 | Not-determined |
| 18-102_k141_195930 | Low-quality |
| 18-102_k141_7256 | Not-determined |
| 18-102_k141_28043 | Low-quality |
| 18-102_k141_132012 | Not-determined |
| 18-102_k141_132020 | Not-determined |
| 18-102_k141_154429 | Not-determined |
| 18-102_k141_195935 | Not-determined |
| 18-102_k141_144143 | Low-quality |
| 18-102_k141_204341 | Not-determined |
| 18-102_k141_19701 | Not-determined |
| 18-102_k141_92670 | Not-determined |
| 18-102_k141_144156 | Not-determined |
| 18-102_k141_225237 | Not-determined |
| 18-102_k141_61461 | Not-determined |
| 18-102_k141_163481 | Not-determined |
| 18-102_k141_132036 | Not-determined |
| 18-102_k141_163487 | Not-determined |
| 18-102_k141_195950 | Not-determined |
| 18-102_k141_41163 | Not-determined |
| 18-102_k141_225252 | Not-determined |
| 18-102_k141_225253 | Not-determined |
| 18-102_k141_225257 | Not-determined |
| 18-102_k141_225261 | Not-determined |
| 18-102_k141_19722 | Not-determined |
| 18-110_k141_86758 | Medium-quality |
| 18-102_k141_19723 | Not-determined |
| 18-102_k141_225270 | Not-determined |
| 18-102_k141_225271 | Not-determined |
| 18-102_k141_92705 | Low-quality |
| 18-102_k141_195975 | Not-determined |
| 18-102_k141_195978 | Not-determined |
| 18-102_k141_195979 | Not-determined |
| 18-102_k141_225273 | Not-determined |
| 18-102_k141_92724 | Not-determined |
| 18-102_k141_163508 | Not-determined |
| 18-102_k141_163515 | Not-determined |
| 18-102_k141_185531 | Low-quality |
| 18-102_k141_185532 | Not-determined |
| 18-102_k141_28082 | Not-determined |
| 18-102_k141_185534 | Not-determined |
| 18-102_k141_28083 | Not-determined |
| 18-102_k141_81778 | Not-determined |
| 18-102_k141_144196 | Low-quality |
| 18-102_k141_237892 | Low-quality |
| 18-102_k141_245252 | Not-determined |
| 18-102_k141_71640 | Not-determined |
| 18-102_k141_163534 | Not-determined |
| 18-102_k141_19746 | Not-determined |
| 18-102_k141_71647 | Not-determined |
| 18-102_k141_103169 | Not-determined |
| 18-102_k141_103170 | Not-determined |
| 18-102_k141_103171 | Not-determined |
| 18-102_k141_245258 | Not-determined |
| 18-102_k141_41192 | Not-determined |
| 18-102_k141_163544 | Low-quality |
| 18-102_k141_28092 | Low-quality |
| 18-102_k141_92744 | Low-quality |
| 18-102_k141_28094 | Not-determined |
| 18-102_k141_144210 | Not-determined |
| 18-102_k141_245264 | Not-determined |
| 18-102_k141_111899 | Not-determined |
| 18-102_k141_111905 | Low-quality |
| 18-102_k141_111909 | Not-determined |
| 18-102_k141_92752 | Not-determined |
| 18-102_k141_215856 | Not-determined |
| 18-102_k141_19762 | Not-determined |
| 18-102_k141_215860 | Not-determined |
| 18-102_k141_215864 | Not-determined |
| 18-102_k141_163568 | Low-quality |
| 18-102_k141_19765 | Not-determined |
| 18-102_k141_61514 | Not-determined |
| 18-102_k141_196050 | Not-determined |
| 18-102_k141_19771 | Not-determined |
| 18-102_k141_41212 | Not-determined |
| 18-102_k141_19772 | Not-determined |
| 18-102_k141_41213 | Not-determined |
| 18-102_k141_154494 | Not-determined |
| 18-102_k141_154497 | Not-determined |
| 18-102_k141_163579 | Not-determined |
| 18-102_k141_19781 | Not-determined |
| 18-102_k141_81830 | Not-determined |
| 18-102_k141_144227 | Not-determined |
| 18-102_k141_144231 | Not-determined |
| 18-102_k141_123348 | Not-determined |
| 18-102_k141_196072 | Not-determined |
| 18-102_k141_196073 | Not-determined |
| 18-102_k141_163586 | Not-determined |
| 18-102_k141_196074 | Not-determined |
| 18-102_k141_111917 | Low-quality |
| 18-102_k141_41219 | Not-determined |
| 18-102_k141_163591 | Not-determined |
| 18-102_k141_215882 | Not-determined |
| 18-102_k141_81845 | Not-determined |
| 18-102_k141_111919 | Not-determined |
| 18-102_k141_196082 | Low-quality |
| 18-102_k141_154515 | Low-quality |
| 18-102_k141_111932 | Not-determined |
| 18-102_k141_28152 | Not-determined |
| 18-102_k141_61530 | Not-determined |
| 18-102_k141_19807 | Not-determined |
| 18-102_k141_237927 | Not-determined |
| 18-102_k141_144249 | Not-determined |
| 18-102_k141_215894 | Not-determined |
| 18-102_k141_7303 | Not-determined |
| 18-102_k141_215897 | Not-determined |
| 18-102_k141_144251 | Not-determined |
| 18-102_k141_237929 | Not-determined |
| 18-102_k141_215899 | Not-determined |
| 18-102_k141_111946 | Low-quality |
| 18-102_k141_196100 | Not-determined |
| 18-102_k141_111949 | Not-determined |
| 18-102_k141_215911 | Not-determined |
| 18-102_k141_123367 | Not-determined |
| 18-102_k141_215914 | Not-determined |
| 18-102_k141_215915 | Not-determined |
| 18-102_k141_196108 | Not-determined |
| 18-102_k141_41238 | Not-determined |
| 18-102_k141_154538 | Not-determined |
| 18-102_k141_144268 | Not-determined |
| 18-102_k141_225380 | Not-determined |
| 18-102_k141_111962 | Not-determined |
| 18-102_k141_111963 | Not-determined |
| 18-102_k141_111964 | Not-determined |
| 18-102_k141_196122 | Not-determined |
| 18-102_k141_215928 | Not-determined |
| 18-102_k141_71658 | Not-determined |
| 18-102_k141_196128 | Not-determined |
| 18-102_k141_215930 | Not-determined |
| 18-102_k141_19830 | Low-quality |
| 18-102_k141_144276 | Not-determined |
| 18-102_k141_144279 | Not-determined |
| 18-102_k141_71662 | Not-determined |
| 18-102_k141_28177 | Not-determined |
| 18-102_k141_196138 | Not-determined |
| 18-102_k141_123391 | Not-determined |
| 18-102_k141_154560 | Not-determined |
| 18-102_k141_154563 | Not-determined |
| 18-102_k141_154565 | Not-determined |
| 18-102_k141_196147 | Not-determined |
| 18-102_k141_154567 | Not-determined |
| 18-102_k141_132043 | Not-determined |
| 18-102_k141_111992 | Not-determined |
| 18-102_k141_111996 | Not-determined |
| 18-102_k141_111997 | Not-determined |
| 18-102_k141_28195 | Not-determined |
| 18-102_k141_154575 | Not-determined |
| 18-102_k141_92778 | Not-determined |
| 18-102_k141_71687 | Not-determined |
| 18-102_k141_28200 | Not-determined |
| 18-102_k141_215959 | Not-determined |
| 18-102_k141_154580 | Low-quality |
| 18-102_k141_81877 | Not-determined |
| 18-102_k141_71695 | Low-quality |
| 18-102_k141_81885 | Not-determined |
| 18-102_k141_41265 | Not-determined |
| 18-102_k141_144290 | Not-determined |
| 18-102_k141_71698 | Not-determined |
| 18-102_k141_132053 | Not-determined |
| 18-102_k141_196166 | Not-determined |
| 18-102_k141_81891 | Not-determined |
| 18-102_k141_41272 | Not-determined |
| 18-102_k141_28214 | Not-determined |
| 18-102_k141_163626 | Not-determined |
| 18-102_k141_196170 | Not-determined |
| 18-102_k141_71699 | Not-determined |
| 18-102_k141_81896 | Not-determined |
| 18-102_k141_81900 | Low-quality |
| 18-102_k141_132063 | Not-determined |
| 18-102_k141_196177 | Low-quality |
| 18-102_k141_215983 | Not-determined |
| 18-102_k141_28234 | Not-determined |
| 18-102_k141_154605 | Not-determined |
| 18-102_k141_28240 | Not-determined |
| 18-102_k141_215997 | Not-determined |
| 18-102_k141_28244 | Not-determined |
| 18-102_k141_163644 | Not-determined |
| 18-102_k141_41284 | Not-determined |
| 18-102_k141_132087 | Not-determined |
| 18-102_k141_144306 | Not-determined |
| 18-102_k141_163654 | Not-determined |
| 18-102_k141_196185 | Not-determined |
| 18-102_k141_112021 | Not-determined |
| 18-102_k141_163656 | Not-determined |
| 18-102_k141_132093 | Not-determined |
| 18-102_k141_112028 | Not-determined |
| 18-102_k141_132099 | Not-determined |
| 18-102_k141_196200 | Not-determined |
| 18-102_k141_132101 | Low-quality |
| 18-102_k141_7358 | Not-determined |
| 18-102_k141_132105 | Not-determined |
| 18-102_k141_112034 | Not-determined |
| 18-102_k141_112037 | Not-determined |
| 18-102_k141_216035 | Not-determined |
| 18-102_k141_163678 | Not-determined |
| 18-102_k141_112041 | Not-determined |
| 18-102_k141_71766 | Low-quality |
| 18-102_k141_163684 | Not-determined |
| 18-102_k141_7370 | Not-determined |
| 18-102_k141_81943 | Not-determined |
| 18-102_k141_28275 | Not-determined |
| 18-102_k141_163694 | Not-determined |
| 18-102_k141_7377 | Not-determined |
| 18-102_k141_81946 | Not-determined |
| 18-102_k141_92813 | Not-determined |
| 18-102_k141_144328 | Not-determined |
| 18-102_k141_81949 | Not-determined |
| 18-102_k141_144332 | Not-determined |
| 18-102_k141_204380 | Not-determined |
| 18-102_k141_7387 | Not-determined |
| 18-102_k141_28287 | Not-determined |
| 18-102_k141_28288 | Not-determined |
| 18-102_k141_132128 | Not-determined |
| 18-102_k141_71789 | Not-determined |
| 18-102_k141_71790 | Not-determined |
| 18-102_k141_204390 | Not-determined |
| 18-102_k141_225416 | Not-determined |
| 18-102_k141_28293 | Not-determined |
| 18-102_k141_225418 | Not-determined |
| 18-102_k141_204392 | Not-determined |
| 18-102_k141_41288 | Not-determined |
| 18-102_k141_196221 | Not-determined |
| 18-102_k141_144342 | Not-determined |
| 18-102_k141_144343 | Not-determined |
| 18-102_k141_71798 | Low-quality |
| 18-102_k141_225423 | Low-quality |
| 18-102_k141_92823 | Low-quality |
| 18-102_k141_144348 | Not-determined |
| 18-102_k141_185669 | Not-determined |
| 18-102_k141_41293 | Not-determined |
| 18-102_k141_196234 | Not-determined |
| 18-102_k141_163704 | Not-determined |
| 18-102_k141_196236 | Not-determined |
| 18-102_k141_163707 | Not-determined |
| 18-102_k141_163708 | Not-determined |
| 18-102_k141_92838 | Not-determined |
| 18-102_k141_7417 | Not-determined |
| 18-102_k141_41301 | Not-determined |
| 18-102_k141_132148 | Not-determined |
| 18-102_k141_237978 | Not-determined |
| 18-102_k141_245285 | Not-determined |
| 18-102_k141_132151 | Not-determined |
| 18-102_k141_196243 | Not-determined |
| 18-102_k141_132153 | Not-determined |
| 18-102_k141_163712 | Not-determined |
| 18-102_k141_103216 | Not-determined |
| 18-102_k141_185684 | Not-determined |
| 18-102_k141_245291 | Not-determined |
| 18-102_k141_7429 | Not-determined |
| 18-102_k141_154613 | Not-determined |
| 18-102_k141_103218 | Not-determined |
| 18-102_k141_7432 | Not-determined |
| 18-102_k141_103223 | Not-determined |
| 18-102_k141_123396 | Not-determined |
| 18-102_k141_237991 | Low-quality |
| 18-102_k141_237993 | Not-determined |
| 18-102_k141_28305 | Not-determined |
| 18-102_k141_144365 | Not-determined |
| 18-102_k141_61601 | Not-determined |
| 18-102_k141_132155 | Not-determined |
| 18-102_k141_154625 | Not-determined |
| 18-102_k141_71818 | Not-determined |
| 18-102_k141_71819 | Not-determined |
| 18-102_k141_196259 | Not-determined |
| 18-102_k141_92858 | Not-determined |
| 18-102_k141_144372 | Not-determined |
| 18-102_k141_144377 | Low-quality |
| 18-102_k141_144380 | Not-determined |
| 18-102_k141_245313 | Not-determined |
| 18-102_k141_245316 | Not-determined |
| 18-102_k141_245317 | Not-determined |
| 18-102_k141_61617 | Not-determined |
| 18-102_k141_61619 | Not-determined |
| 18-102_k141_61621 | Not-determined |
| 18-102_k141_103229 | Not-determined |
| 18-102_k141_132170 | Not-determined |
| 18-102_k141_154651 | Not-determined |
| 18-102_k141_154655 | Not-determined |
| 18-102_k141_28323 | Not-determined |
| 18-102_k141_28325 | Low-quality |
| 18-102_k141_132176 | Not-determined |
| 18-102_k141_28329 | Low-quality |
| 18-102_k141_245325 | Not-determined |
| 18-102_k141_245326 | Low-quality |
| 18-102_k141_238007 | Not-determined |
| 18-102_k141_225457 | Not-determined |
| 18-102_k141_123428 | Not-determined |
| 18-102_k141_132187 | Not-determined |
| 18-102_k141_132188 | Not-determined |
| 18-102_k141_154672 | Not-determined |
| 18-102_k141_41335 | Low-quality |
| 18-102_k141_144394 | Not-determined |
| 18-102_k141_132196 | Not-determined |
| 18-102_k141_185723 | Not-determined |
| 18-102_k141_185724 | Not-determined |
| 18-102_k141_92878 | Not-determined |
| 18-102_k141_123449 | Not-determined |
| 18-102_k141_71875 | Not-determined |
| 18-102_k141_185741 | Not-determined |
| 18-102_k141_71879 | Not-determined |
| 18-102_k141_7455 | Not-determined |
| 18-102_k141_216096 | Not-determined |
| 18-102_k141_216099 | Low-quality |
| 18-102_k141_81957 | Not-determined |
| 18-102_k141_123467 | Not-determined |
| 18-102_k141_123468 | Not-determined |
| 18-102_k141_123469 | Low-quality |
| 18-102_k141_28359 | Not-determined |
| 18-102_k141_71910 | Low-quality |
| 18-102_k141_123486 | Not-determined |
| 18-102_k141_123489 | Not-determined |
| 18-102_k141_7468 | Not-determined |
| 18-102_k141_7469 | Not-determined |
| 18-102_k141_154716 | Not-determined |
| 18-102_k141_7474 | Not-determined |
| 18-102_k141_123502 | Not-determined |
| 18-102_k141_71927 | Not-determined |
| 18-102_k141_19935 | Not-determined |
| 18-102_k141_92898 | Not-determined |
| 18-102_k141_28374 | Not-determined |
| 18-102_k141_7485 | Not-determined |
| 18-102_k141_132224 | Not-determined |
| 18-102_k141_204432 | Low-quality |
| 18-102_k141_154726 | Not-determined |
| 18-102_k141_61665 | Not-determined |
| 18-102_k141_61666 | Not-determined |
| 18-102_k141_144452 | Low-quality |
| 18-102_k141_216105 | Not-determined |
| 18-102_k141_216106 | Not-determined |
| 18-102_k141_154732 | Not-determined |
| 18-102_k141_19946 | Not-determined |
| 18-102_k141_92918 | Not-determined |
| 18-102_k141_19953 | Not-determined |
| 18-102_k141_103260 | Low-quality |
| 18-102_k141_92924 | Not-determined |
| 18-102_k141_61685 | Not-determined |
| 18-102_k141_154746 | Not-determined |
| 18-102_k141_41347 | Not-determined |
| 18-102_k141_163739 | Not-determined |
| 18-102_k141_28388 | Not-determined |
| 18-102_k141_28395 | Low-quality |
| 18-102_k141_238038 | Low-quality |
| 18-102_k141_19976 | Not-determined |
| 18-102_k141_132252 | Not-determined |
| 18-102_k141_132258 | Not-determined |
| 18-102_k141_204471 | Not-determined |
| 18-102_k141_61699 | Not-determined |
| 18-102_k141_28411 | Not-determined |
| 18-102_k141_19987 | Not-determined |
| 18-102_k141_163750 | Not-determined |
| 18-102_k141_238055 | Not-determined |
| 18-102_k141_238056 | Low-quality |
| 18-102_k141_144466 | Low-quality |
| 18-102_k141_19991 | Not-determined |
| 18-102_k141_196272 | Not-determined |
| 18-102_k141_61714 | Low-quality |
| 18-102_k141_225501 | Not-determined |
| 18-102_k141_144471 | Not-determined |
| 18-102_k141_61716 | Not-determined |
| 18-102_k141_196277 | Not-determined |
| 18-102_k141_196280 | Not-determined |
| 18-102_k141_123540 | Not-determined |
| 18-102_k141_19994 | Not-determined |
| 18-102_k141_132286 | Not-determined |
| 18-102_k141_61722 | Not-determined |
| 18-102_k141_112104 | Low-quality |
| 18-102_k141_144474 | Low-quality |
| 18-102_k141_123550 | Not-determined |
| 18-102_k141_92951 | Not-determined |
| 18-102_k141_225519 | Not-determined |
| 18-102_k141_185810 | Not-determined |
| 18-102_k141_71988 | Not-determined |
| 18-102_k141_82013 | Not-determined |
| 18-102_k141_61745 | Not-determined |
| 18-102_k141_163781 | Not-determined |
| 18-102_k141_185823 | Not-determined |
| 18-102_k141_7501 | Low-quality |
| 18-102_k141_103311 | Not-determined |
| 18-102_k141_103312 | Low-quality |
| 18-102_k141_103313 | Not-determined |
| 18-102_k141_71997 | Not-determined |
| 18-102_k141_204500 | Not-determined |
| 18-102_k141_185833 | Not-determined |
| 18-102_k141_92969 | Not-determined |
| 18-102_k141_144506 | Not-determined |
| 18-102_k141_72002 | Not-determined |
| 18-102_k141_216162 | Not-determined |
| 18-102_k141_132306 | Not-determined |
| 18-102_k141_185839 | Not-determined |
| 18-102_k141_123584 | Not-determined |
| 18-102_k141_185841 | Not-determined |
| 18-102_k141_123585 | Not-determined |
| 18-102_k141_163803 | Not-determined |
| 18-102_k141_185849 | Not-determined |
| 18-102_k141_154801 | Not-determined |
| 18-102_k141_61772 | Not-determined |
| 18-102_k141_123589 | Not-determined |
| 18-102_k141_61774 | Not-determined |
| 18-102_k141_144521 | Not-determined |
| 18-102_k141_144522 | Not-determined |
| 18-102_k141_61777 | Not-determined |
| 18-102_k141_28433 | Not-determined |
| 18-102_k141_185855 | Not-determined |
| 18-102_k141_123595 | Not-determined |
| 18-102_k141_245365 | Not-determined |
| 18-102_k141_7533 | Not-determined |
| 18-102_k141_92976 | Not-determined |
| 18-102_k141_154817 | Not-determined |
| 18-102_k141_103322 | Not-determined |
| 18-102_k141_7535 | Not-determined |

| | |
|---|---|
| 18-102_k141_28441 | Not-determined |
| 18-102_k141_216168 | Not-determined |
| 18-102_k141_225527 | Not-determined |
| 18-102_k141_163825 | Not-determined |
| 18-102_k141_103330 | Low-quality |
| 18-102_k141_185871 | Not-determined |
| 18-102_k141_28452 | Not-determined |
| 18-102_k141_185877 | Not-determined |
| 18-102_k141_28455 | Not-determined |
| 18-102_k141_185882 | Not-determined |
| 18-102_k141_82068 | Not-determined |
| 18-102_k141_20038 | Not-determined |
| 18-102_k141_204529 | Not-determined |
| 18-102_k141_82079 | Not-determined |
| 18-102_k141_72029 | Not-determined |
| 18-102_k141_82081 | Not-determined |
| 18-102_k141_20041 | Not-determined |
| 18-102_k141_204533 | Not-determined |
| 18-102_k141_72035 | Not-determined |
| 18-102_k141_154857 | Not-determined |
| 18-102_k141_154859 | Not-determined |
| 18-102_k141_204537 | Not-determined |
| 18-102_k141_245374 | Not-determined |
| 18-102_k141_225540 | Not-determined |
| 18-102_k141_112142 | Not-determined |
| 18-102_k141_225545 | Not-determined |
| 18-102_k141_225548 | Not-determined |
| 18-102_k141_82108 | Not-determined |
| 18-102_k141_7567 | Not-determined |
| 18-102_k141_245381 | Not-determined |
| 18-102_k141_225553 | Not-determined |
| 18-102_k141_216204 | Not-determined |
| 18-102_k141_82112 | Not-determined |
| 18-102_k141_238087 | Not-determined |
| 18-102_k141_238089 | Not-determined |
| 18-102_k141_238090 | Not-determined |
| 18-102_k141_216214 | Not-determined |
| 18-102_k141_93005 | Not-determined |
| 18-102_k141_196288 | Not-determined |
| 18-102_k141_245398 | Not-determined |
| 18-102_k141_216223 | Not-determined |
| 18-102_k141_93011 | Not-determined |
| 18-102_k141_216224 | Not-determined |
| 18-102_k141_123618 | Not-determined |
| 18-102_k141_123621 | Not-determined |
| 18-102_k141_238111 | Not-determined |
| 18-102_k141_20054 | Not-determined |
| 18-102_k141_204568 | Not-determined |
| 18-102_k141_154878 | Not-determined |
| 18-102_k141_196311 | Not-determined |
| 18-102_k141_61817 | Not-determined |
| 18-102_k141_196312 | Not-determined |
| 18-102_k141_204570 | Not-determined |
| 18-102_k141_185952 | Not-determined |
| 18-102_k141_61819 | Not-determined |
| 18-102_k141_7595 | Not-determined |
| 18-102_k141_72061 | Not-determined |
| 18-102_k141_112179 | Not-determined |
| 18-102_k141_72068 | Not-determined |
| 18-102_k141_185961 | Not-determined |
| 18-102_k141_238126 | Not-determined |
| 18-102_k141_112184 | Not-determined |
| 18-102_k141_72075 | Not-determined |
| 18-102_k141_112196 | Not-determined |
| 18-102_k141_20072 | Not-determined |
| 18-102_k141_112198 | Not-determined |
| 18-102_k141_216255 | Not-determined |
| 18-102_k141_103407 | Not-determined |
| 18-102_k141_196334 | Not-determined |
| 18-102_k141_61839 | Low-quality |
| 18-102_k141_216264 | Not-determined |
| 18-102_k141_132349 | Not-determined |
| 18-102_k141_225584 | Not-determined |
| 18-102_k141_112215 | Not-determined |
| 18-102_k141_93034 | Not-determined |
| 18-102_k141_185974 | Not-determined |
| 18-102_k141_103411 | Not-determined |
| 18-102_k141_72095 | Not-determined |
| 18-102_k141_61862 | Not-determined |
| 18-102_k141_238161 | Not-determined |
| 18-102_k141_196359 | Not-determined |
| 18-102_k141_112225 | Not-determined |
| 18-102_k141_238164 | Not-determined |
| 18-102_k141_112227 | Not-determined |
| 18-102_k141_72103 | Not-determined |
| 18-102_k141_225606 | Not-determined |
| 18-102_k141_93042 | Not-determined |
| 18-102_k141_123668 | Not-determined |
| 18-102_k141_238175 | Not-determined |
| 18-102_k141_82173 | Not-determined |
| 18-102_k141_186009 | Not-determined |
| 18-102_k141_123684 | Not-determined |
| 18-102_k141_123686 | Not-determined |
| 18-102_k141_7636 | Not-determined |
| 18-102_k141_163916 | Not-determined |
| 18-102_k141_225631 | Not-determined |
| 18-102_k141_225632 | Not-determined |
| 18-102_k141_93053 | Not-determined |
| 18-102_k141_163921 | Not-determined |
| 18-102_k141_144539 | Not-determined |
| 18-102_k141_132407 | Not-determined |
| 18-102_k141_93062 | Not-determined |
| 18-102_k141_123702 | Not-determined |
| 18-102_k141_82198 | Not-determined |
| 18-102_k141_123706 | Not-determined |
| 18-102_k141_144545 | Not-determined |
| 18-102_k141_225644 | Not-determined |
| 18-102_k141_144549 | Not-determined |
| 18-102_k141_144552 | Not-determined |
| 18-102_k141_225649 | Low-quality |
| 18-102_k141_123709 | Not-determined |
| 18-102_k141_7664 | Not-determined |
| 18-102_k141_72129 | Not-determined |
| 18-102_k141_132421 | Not-determined |
| 18-102_k141_72133 | Low-quality |
| 18-102_k141_132423 | Not-determined |
| 18-102_k141_72139 | Low-quality |
| 18-102_k141_82232 | Not-determined |
| 18-102_k141_196418 | Low-quality |
| 18-102_k141_132430 | Low-quality |
| 18-102_k141_163939 | Not-determined |
| 18-102_k141_196423 | Not-determined |
| 18-102_k141_93075 | Not-determined |
| 18-102_k141_72156 | Not-determined |
| 18-102_k141_204592 | Not-determined |
| 18-102_k141_112288 | Low-quality |
| 18-102_k141_123730 | Not-determined |
| 18-102_k141_204594 | Not-determined |
| 18-102_k141_163946 | Low-quality |
| 18-102_k141_196435 | Not-determined |
| 18-102_k141_82250 | Not-determined |
| 18-102_k141_93080 | Not-determined |
| 18-102_k141_144601 | Not-determined |
| 18-102_k141_196442 | Not-determined |
| 18-102_k141_7678 | Not-determined |
| 18-102_k141_132449 | Not-determined |
| 18-102_k141_196456 | Not-determined |
| 18-102_k141_112304 | Not-determined |
| 18-102_k141_112307 | Not-determined |
| 18-102_k141_123745 | Not-determined |
| 18-102_k141_204618 | Not-determined |
| 18-102_k141_225689 | Not-determined |
| 18-102_k141_163974 | Not-determined |
| 18-102_k141_112325 | Not-determined |
| 18-102_k141_103428 | Not-determined |
| 18-102_k141_123756 | Not-determined |
| 18-102_k141_204629 | Not-determined |
| 18-102_k141_132468 | Not-determined |
| 18-102_k141_103436 | Not-determined |
| 18-102_k141_61895 | Not-determined |
| 18-102_k141_204631 | Not-determined |
| 18-102_k141_132475 | Low-quality |
| 18-102_k141_82299 | Not-determined |
| 18-102_k141_225706 | Not-determined |
| 18-102_k141_154938 | Not-determined |
| 18-102_k141_163989 | Not-determined |
| 18-102_k141_204638 | Not-determined |
| 18-102_k141_112329 | Not-determined |
| 18-102_k141_7700 | Not-determined |
| 18-102_k141_82314 | Not-determined |
| 18-102_k141_82318 | Not-determined |
| 18-102_k141_112338 | Not-determined |
| 18-102_k141_225723 | Not-determined |
| 18-102_k141_7712 | Not-determined |
| 18-102_k141_225729 | Low-quality |
| 18-102_k141_144645 | Not-determined |
| 18-102_k141_112348 | Not-determined |
| 18-102_k141_144647 | Not-determined |
| 18-102_k141_225740 | Not-determined |
| 18-102_k141_245434 | Not-determined |
| 18-102_k141_132499 | Not-determined |
| 18-102_k141_112366 | Not-determined |
| 18-102_k141_112369 | Not-determined |
| 18-102_k141_7724 | Not-determined |
| 18-102_k141_132505 | Low-quality |
| 18-102_k141_7727 | Low-quality |
| 18-102_k141_7732 | Not-determined |
| 18-102_k141_132510 | Low-quality |
| 18-102_k141_245447 | Not-determined |
| 18-102_k141_72197 | Not-determined |
| 18-102_k141_72208 | Not-determined |
| 18-102_k141_132519 | Not-determined |
| 18-102_k141_154959 | Not-determined |
| 18-102_k141_132525 | Not-determined |
| 18-102_k141_132526 | Not-determined |
| 18-102_k141_164011 | Not-determined |
| 18-102_k141_144662 | Low-quality |
| 18-102_k141_144663 | Not-determined |
| 18-102_k141_216316 | Low-quality |
| 18-102_k141_103481 | Not-determined |
| 18-102_k141_103482 | Not-determined |
| 18-102_k141_72221 | Not-determined |
| 18-102_k141_144667 | Not-determined |
| 18-102_k141_154980 | Not-determined |
| 18-102_k141_154982 | Not-determined |
| 18-102_k141_186057 | Not-determined |
| 18-102_k141_225762 | Not-determined |
| 18-102_k141_225763 | Not-determined |
| 18-102_k141_225766 | Not-determined |
| 18-102_k141_186066 | Not-determined |
| 18-102_k141_28495 | Not-determined |
| 18-102_k141_112413 | Not-determined |
| 18-102_k141_245472 | Not-determined |
| 18-102_k141_28504 | Not-determined |
| 18-102_k141_28507 | Not-determined |
| 18-102_k141_132562 | Not-determined |
| 18-102_k141_72239 | Low-quality |
| 18-102_k141_204680 | Not-determined |
| 18-102_k141_72243 | Not-determined |
| 18-102_k141_245481 | Not-determined |
| 18-102_k141_132566 | Low-quality |
| 18-102_k141_204688 | Not-determined |
| 18-102_k141_72262 | Not-determined |
| 18-102_k141_196491 | Not-determined |
| 18-102_k141_28526 | Low-quality |
| 18-102_k141_144682 | Low-quality |
| 18-102_k141_204699 | Low-quality |
| 18-102_k141_72269 | Not-determined |
| 18-102_k141_204700 | Not-determined |
| 18-102_k141_155014 | Not-determined |
| 18-102_k141_196501 | Not-determined |
| 18-102_k141_28539 | Not-determined |
| 18-102_k141_196505 | Not-determined |
| 18-102_k141_245489 | Low-quality |
| 18-102_k141_155017 | Not-determined |
| 18-102_k141_144694 | Not-determined |
| 18-102_k141_155024 | Not-determined |
| 18-102_k141_28552 | Low-quality |
| 18-102_k141_123782 | Not-determined |
| 18-102_k141_196516 | Not-determined |
| 18-102_k141_245496 | Not-determined |
| 18-102_k141_216357 | Not-determined |
| 18-102_k141_132588 | Low-quality |
| 18-102_k141_204721 | Not-determined |
| 18-102_k141_123792 | Not-determined |
| 18-102_k141_123794 | Not-determined |
| 18-102_k141_28565 | Not-determined |
| 18-102_k141_93102 | Low-quality |
| 18-102_k141_28570 | Not-determined |
| 18-102_k141_186106 | Not-determined |
| 18-102_k141_204733 | Not-determined |
| 18-102_k141_132603 | Not-determined |
| 18-102_k141_164031 | Not-determined |
| 18-102_k141_20119 | Not-determined |
| 18-102_k141_225799 | Not-determined |
| 18-102_k141_225803 | Not-determined |
| 18-102_k141_61941 | Not-determined |
| 18-102_k141_20125 | Not-determined |
| 18-102_k141_186115 | Not-determined |
| 18-102_k141_132612 | Not-determined |
| 18-102_k141_72284 | Not-determined |
| 18-102_k141_20137 | Not-determined |
| 18-102_k141_164046 | Not-determined |
| 18-102_k141_132616 | Not-determined |
| 18-102_k141_155040 | Not-determined |
| 18-102_k141_196551 | Low-quality |
| 18-102_k141_196552 | Not-determined |
| 18-102_k141_186133 | Not-determined |
| 18-102_k141_28598 | Not-determined |
| 18-102_k141_186135 | Not-determined |
| 18-102_k141_132629 | Low-quality |
| 18-102_k141_123810 | Not-determined |
| 18-102_k141_225817 | Not-determined |
| 18-102_k141_164060 | Not-determined |
| 18-102_k141_225821 | Not-determined |
| 18-102_k141_132635 | Not-determined |
| 18-102_k141_186147 | Low-quality |
| 18-102_k141_123815 | Low-quality |
| 18-102_k141_204764 | Low-quality |
| 18-102_k141_225833 | Not-determined |
| 18-102_k141_61962 | Not-determined |
| 18-102_k141_132645 | Not-determined |
| 18-102_k141_186162 | Not-determined |
| 18-102_k141_196570 | Not-determined |
| 18-102_k141_82392 | Not-determined |
| 18-102_k141_28624 | Not-determined |
| 18-102_k141_132658 | Not-determined |
| 18-102_k141_225850 | Not-determined |
| 18-102_k141_196575 | Not-determined |
| 18-102_k141_61973 | Not-determined |
| 18-102_k141_82407 | Not-determined |
| 18-102_k141_225858 | Not-determined |
| 18-102_k141_72295 | Not-determined |
| 18-102_k141_123828 | Not-determined |
| 18-102_k141_155070 | Not-determined |
| 18-102_k141_196596 | Not-determined |
| 18-102_k141_61987 | Not-determined |
| 18-102_k141_72302 | Not-determined |
| 18-102_k141_225870 | Not-determined |
| 18-102_k141_144725 | Not-determined |
| 18-102_k141_72310 | Not-determined |
| 18-102_k141_82412 | Not-determined |
| 18-102_k141_164076 | Not-determined |
| 18-102_k141_225878 | Not-determined |
| 18-102_k141_82414 | Not-determined |
| 18-102_k141_245533 | Not-determined |
| 18-102_k141_204786 | Not-determined |
| 18-102_k141_112429 | Not-determined |
| 18-102_k141_72325 | Not-determined |
| 18-102_k141_72332 | Low-quality |
| 18-102_k141_196616 | Not-determined |
| 18-102_k141_62009 | Not-determined |
| 18-102_k141_82422 | Not-determined |
| 18-102_k141_132680 | Not-determined |
| 18-102_k141_204794 | Not-determined |
| 18-102_k141_245549 | Not-determined |
| 18-102_k141_204796 | Not-determined |
| 18-102_k141_144732 | Not-determined |
| 18-102_k141_144735 | Not-determined |
| 18-102_k141_62012 | Not-determined |
| 18-102_k141_216379 | Low-quality |
| 18-102_k141_123856 | Not-determined |
| 18-102_k141_20200 | Not-determined |
| 18-102_k141_132694 | Low-quality |
| 18-102_k141_123859 | Not-determined |
| 18-102_k141_204806 | Low-quality |
| 18-102_k141_164094 | Not-determined |
| 18-102_k141_245563 | Not-determined |
| 18-102_k141_20214 | Not-determined |
| 18-102_k141_20216 | Not-determined |
| 18-102_k141_132712 | Not-determined |
| 18-102_k141_132713 | Not-determined |
| 18-102_k141_132716 | Not-determined |
| 18-102_k141_132720 | Not-determined |
| 18-102_k141_204818 | Not-determined |
| 18-102_k141_216388 | Not-determined |
| 18-102_k141_7748 | Not-determined |
| 18-102_k141_196631 | Not-determined |
| 18-102_k141_82444 | Not-determined |
| 18-102_k141_225903 | Not-determined |
| 18-102_k141_196633 | Low-quality |
| 18-102_k141_132733 | Not-determined |
| 18-102_k141_72362 | Not-determined |
| 18-102_k141_155087 | Not-determined |
| 18-102_k141_72370 | Low-quality |
| 18-102_k141_62048 | Low-quality |
| 18-102_k141_132746 | Not-determined |
| 18-102_k141_20237 | Not-determined |
| 18-102_k141_216406 | Not-determined |
| 18-102_k141_72373 | Not-determined |
| 18-102_k141_62057 | Not-determined |
| 18-102_k141_20247 | Not-determined |
| 18-102_k141_62061 | Not-determined |
| 18-102_k141_132766 | Not-determined |
| 18-102_k141_20251 | Low-quality |
| 18-102_k141_62062 | Not-determined |
| 18-102_k141_186192 | Not-determined |
| 18-102_k141_20256 | Not-determined |
| 18-102_k141_204855 | Not-determined |
| 18-102_k141_196664 | Not-determined |
| 18-102_k141_204860 | Not-determined |
| 18-102_k141_164119 | Not-determined |
| 18-102_k141_196666 | Not-determined |
| 18-102_k141_186199 | Not-determined |
| 18-102_k141_204863 | Not-determined |
| 18-102_k141_62067 | Not-determined |
| 18-102_k141_155120 | Not-determined |
| 18-102_k141_62068 | Not-determined |
| 18-102_k141_62070 | Low-quality |
| 18-102_k141_216447 | Not-determined |
| 18-102_k141_155130 | Not-determined |
| 18-102_k141_103551 | Not-determined |
| 18-102_k141_245569 | Low-quality |
| 18-102_k141_93128 | Not-determined |
| 18-102_k141_28703 | Not-determined |
| 18-102_k141_216459 | Not-determined |
| 18-102_k141_123889 | Not-determined |
| 18-102_k141_186210 | Not-determined |
| 18-102_k141_164131 | Not-determined |
| 18-102_k141_72434 | Not-determined |
| 18-102_k141_7754 | Not-determined |
| 18-102_k141_245586 | Low-quality |
| 18-102_k141_28719 | Not-determined |
| 18-102_k141_245588 | Not-determined |
| 18-102_k141_62106 | Not-determined |
| 18-102_k141_62107 | Not-determined |
| 18-102_k141_132836 | Low-quality |
| 18-102_k141_28731 | Not-determined |
| 18-102_k141_28733 | Low-quality |
| 18-102_k141_93147 | Not-determined |
| 18-102_k141_28734 | Low-quality |
| 18-102_k141_132845 | Low-quality |
| 18-102_k141_164139 | Not-determined |
| 18-102_k141_93161 | Not-determined |
| 18-102_k141_62116 | Not-determined |
| 18-102_k141_72456 | Not-determined |
| 18-102_k141_216497 | Not-determined |
| 18-102_k141_112470 | Not-determined |
| 18-102_k141_245604 | Not-determined |
| 18-102_k141_245605 | Low-quality |
| 18-102_k141_132851 | Not-determined |
| 18-102_k141_112481 | Not-determined |
| 18-102_k141_245620 | Not-determined |
| 18-102_k141_72466 | Not-determined |
| 18-102_k141_164162 | Not-determined |
| 18-102_k141_216521 | Low-quality |
| 18-102_k141_216526 | Not-determined |
| 18-102_k141_72476 | Not-determined |
| 18-102_k141_245629 | Not-determined |
| 18-102_k141_93193 | Not-determined |
| 18-102_k141_20332 | Not-determined |
| 18-102_k141_20336 | Not-determined |
| 18-102_k141_28746 | Not-determined |
| 18-102_k141_93199 | Not-determined |
| 18-102_k141_132871 | Not-determined |
| 18-102_k141_20339 | Not-determined |
| 18-102_k141_132876 | Not-determined |
| 18-102_k141_112496 | Not-determined |
| 18-102_k141_216540 | Not-determined |
| 18-102_k141_245636 | Not-determined |
| 18-102_k141_132878 | Not-determined |
| 18-102_k141_155190 | Not-determined |
| 18-102_k141_204884 | Low-quality |
| 18-102_k141_196723 | Not-determined |
| 18-102_k141_123935 | Not-determined |
| 18-102_k141_72490 | Not-determined |
| 18-102_k141_204889 | Not-determined |
| 18-102_k141_155198 | Not-determined |
| 18-102_k141_245652 | Not-determined |
| 18-102_k141_164187 | Low-quality |
| 18-102_k141_216552 | Not-determined |
| 18-102_k141_132896 | Not-determined |
| 18-102_k141_112511 | Low-quality |
| 18-102_k141_245654 | Not-determined |
| 18-102_k141_28776 | Low-quality |
| 18-102_k141_204903 | Not-determined |
| 18-102_k141_132905 | Low-quality |
| 18-102_k141_82469 | Not-determined |
| 18-102_k141_28788 | Not-determined |
| 18-102_k141_20375 | Low-quality |
| 18-102_k141_28795 | Not-determined |
| 18-102_k141_204922 | Not-determined |
| 18-102_k141_20382 | Low-quality |
| 18-102_k141_20383 | Not-determined |
| 18-102_k141_112524 | Not-determined |
| 18-102_k141_155215 | Not-determined |
| 18-102_k141_28802 | Not-determined |
| 18-102_k141_144773 | Not-determined |
| 18-102_k141_186224 | Low-quality |
| 18-102_k141_204944 | Not-determined |
| 18-102_k141_204946 | Not-determined |
| 18-102_k141_186229 | Not-determined |
| 18-102_k141_155241 | Not-determined |
| 18-102_k141_186235 | Not-determined |
| 18-102_k141_82519 | Not-determined |
| 18-102_k141_186237 | Not-determined |
| 18-102_k141_28807 | Not-determined |
| 18-102_k141_216584 | Not-determined |
| 18-102_k141_28817 | Not-determined |
| 18-102_k141_155256 | Not-determined |
| 18-102_k141_186252 | Not-determined |
| 18-102_k141_245689 | Not-determined |
| 18-102_k141_28823 | Not-determined |
| 18-102_k141_216595 | Low-quality |
| 18-102_k141_245694 | Low-quality |
| 18-102_k141_132929 | Not-determined |
| 18-102_k141_204974 | Low-quality |
| 18-102_k141_112536 | Not-determined |
| 18-102_k141_204977 | Not-determined |
| 18-102_k141_112546 | Not-determined |
| 18-102_k141_112550 | Not-determined |
| 18-102_k141_164210 | Not-determined |
| 18-102_k141_132948 | Not-determined |
| 18-102_k141_28837 | Low-quality |
| 18-102_k141_186269 | Low-quality |
| 18-102_k141_164213 | Not-determined |
| 18-102_k141_155289 | Not-determined |
| 18-102_k141_204992 | Not-determined |
| 18-102_k141_20399 | Low-quality |
| 18-102_k141_186283 | Not-determined |
| 18-102_k141_28850 | Not-determined |
| 18-102_k141_186286 | Not-determined |
| 18-102_k141_123962 | Low-quality |
| 18-102_k141_186288 | Not-determined |
| 18-102_k141_20411 | Not-determined |
| 18-102_k141_155303 | Low-quality |
| 18-102_k141_28854 | Not-determined |
| 18-102_k141_20418 | Not-determined |
| 18-102_k141_155308 | Low-quality |
| 18-102_k141_28861 | Not-determined |
| 18-102_k141_28862 | Not-determined |
| 18-102_k141_28865 | Not-determined |
| 18-102_k141_112556 | Low-quality |
| 18-102_k141_112560 | Not-determined |
| 18-102_k141_196747 | Not-determined |
| 18-102_k141_144819 | Not-determined |
| 18-102_k141_155319 | Not-determined |
| 18-102_k141_144826 | Not-determined |
| 18-102_k141_216634 | Not-determined |
| 18-102_k141_186291 | Not-determined |
| 18-102_k141_123973 | Not-determined |
| 18-102_k141_186294 | Not-determined |
| 18-102_k141_196756 | Not-determined |
| 18-102_k141_245712 | Not-determined |
| 18-102_k141_196757 | Not-determined |
| 18-102_k141_123986 | Not-determined |
| 18-102_k141_155325 | Not-determined |
| 18-102_k141_155329 | Not-determined |
| 18-102_k141_28907 | Not-determined |
| 18-102_k141_186306 | Not-determined |
| 18-102_k141_144833 | Not-determined |
| 18-102_k141_123994 | Not-determined |
| 18-102_k141_112594 | Not-determined |
| 18-102_k141_155336 | Not-determined |
| 18-102_k141_28918 | Not-determined |
| 18-102_k141_144853 | Not-determined |
| 18-102_k141_216670 | Not-determined |
| 18-102_k141_216678 | Not-determined |
| 18-102_k141_124014 | Not-determined |
| 18-102_k141_144859 | Not-determined |
| 18-102_k141_186336 | Not-determined |
| 18-102_k141_186337 | Low-quality |
| 18-102_k141_112604 | Not-determined |
| 18-102_k141_112605 | Not-determined |
| 18-102_k141_20440 | Low-quality |
| 18-102_k141_20451 | Not-determined |
| 18-102_k141_205030 | Not-determined |
| 18-102_k141_28931 | Not-determined |
| 18-102_k141_216705 | Not-determined |
| 18-102_k141_186366 | Not-determined |
| 18-102_k141_245753 | Not-determined |
| 18-102_k141_82545 | Low-quality |
| 18-102_k141_205054 | Not-determined |
| 18-102_k141_20465 | Not-determined |
| 18-102_k141_245760 | Not-determined |
| 18-102_k141_112641 | Not-determined |
| 18-102_k141_20467 | Not-determined |
| 18-102_k141_216724 | Not-determined |
| 18-102_k141_186398 | Not-determined |
| 18-102_k141_245763 | Not-determined |
| 18-102_k141_112650 | Not-determined |
| 18-102_k141_245771 | Not-determined |
| 18-102_k141_245773 | Not-determined |
| 18-102_k141_20477 | Not-determined |
| 18-102_k141_20480 | Not-determined |
| 18-102_k141_20484 | Not-determined |
| 18-102_k141_144882 | Not-determined |
| 18-102_k141_144883 | Low-quality |
| 18-102_k141_20489 | Not-determined |
| 18-102_k141_20492 | Not-determined |
| 18-102_k141_216751 | Not-determined |
| 18-102_k141_112681 | Not-determined |
| 18-102_k141_82575 | Not-determined |
| 18-102_k141_124059 | Low-quality |
| 18-102_k141_82591 | Not-determined |
| 18-102_k141_225959 | Not-determined |
| 18-102_k141_225960 | Not-determined |
| 18-102_k141_186410 | Not-determined |
| 18-102_k141_82605 | Not-determined |
| 18-102_k141_20507 | Not-determined |
| 18-102_k141_216813 | Not-determined |
| 18-102_k141_20511 | Not-determined |
| 18-102_k141_216814 | Not-determined |
| 18-102_k141_82606 | Low-quality |
| 18-102_k141_216819 | Not-determined |
| 18-102_k141_225974 | Not-determined |
| 18-102_k141_216827 | Low-quality |
| 18-102_k141_20530 | Low-quality |
| 18-102_k141_225993 | Not-determined |
| 18-102_k141_164240 | Not-determined |
| 18-102_k141_164246 | Not-determined |
| 18-102_k141_225997 | Low-quality |
| 18-102_k141_164258 | Not-determined |
| 18-102_k141_144896 | Not-determined |
| 18-102_k141_82627 | Not-determined |
| 18-102_k141_164278 | Not-determined |
| 18-102_k141_164279 | Not-determined |
| 18-102_k141_144912 | Not-determined |
| 18-102_k141_82635 | Not-determined |
| 18-102_k141_20570 | Not-determined |
| 18-102_k141_164285 | Not-determined |
| 18-102_k141_164288 | Low-quality |
| 18-102_k141_112750 | Not-determined |
| 18-102_k141_144919 | Not-determined |
| 18-102_k141_112767 | Not-determined |
| 18-102_k141_82649 | Not-determined |
| 18-102_k141_216898 | Not-determined |
| 18-102_k141_82654 | Not-determined |
| 18-102_k141_82655 | Not-determined |
| 18-102_k141_20625 | Not-determined |
| 18-102_k141_20626 | Not-determined |
| 18-102_k141_20628 | Not-determined |
| 18-102_k141_28952 | Not-determined |
| 18-102_k141_124080 | Not-determined |
| 18-102_k141_205111 | Not-determined |
| 18-102_k141_216924 | Not-determined |
| 18-102_k141_245795 | Not-determined |
| 18-102_k141_205117 | Not-determined |
| 18-102_k141_82672 | Low-quality |

| | |
|---|---|
| 18-102_k141_205118 | Not-determined |
| 18-102_k141_205125 | Not-determined |
| 18-102_k141_164336 | Not-determined |
| 18-102_k141_205130 | Not-determined |
| 18-102_k141_82685 | Not-determined |
| 18-102_k141_20656 | Not-determined |
| 18-102_k141_226020 | Low-quality |
| 18-102_k141_20666 | Not-determined |
| 18-102_k141_205147 | Not-determined |
| 18-102_k141_205149 | Not-determined |
| 18-102_k141_20670 | Not-determined |
| 18-102_k141_205150 | Not-determined |
| 18-102_k141_164360 | Not-determined |
| 18-102_k141_226030 | Not-determined |
| 18-102_k141_82702 | Not-determined |
| 18-102_k141_144963 | Not-determined |
| 18-102_k141_124123 | Not-determined |
| 18-102_k141_20686 | Not-determined |
| 18-102_k141_82704 | Not-determined |
| 18-102_k141_82709 | Not-determined |
| 18-102_k141_144967 | Not-determined |
| 18-102_k141_28978 | Not-determined |
| 18-102_k141_144969 | Not-determined |
| 18-102_k141_245821 | Not-determined |
| 18-102_k141_124146 | Low-quality |
| 18-102_k141_226069 | Not-determined |
| 18-102_k141_164371 | Not-determined |
| 18-102_k141_28988 | Not-determined |
| 18-102_k141_82717 | Not-determined |
| 18-102_k141_82718 | Not-determined |
| 18-102_k141_82720 | Not-determined |
| 18-102_k141_226077 | Not-determined |
| 18-102_k141_205183 | Not-determined |
| 18-102_k141_29006 | Not-determined |
| 18-102_k141_124169 | Not-determined |
| 18-102_k141_205194 | Not-determined |
| 18-102_k141_20699 | Not-determined |
| 18-102_k141_20705 | Not-determined |
| 18-102_k141_245830 | Not-determined |
| 18-102_k141_205215 | Not-determined |
| 18-102_k141_205217 | Not-determined |
| 18-102_k141_216945 | Not-determined |
| 18-102_k141_226105 | Not-determined |
| 18-102_k141_245835 | Not-determined |
| 18-102_k141_205225 | Not-determined |
| 18-102_k141_205231 | Not-determined |
| 18-102_k141_216967 | Not-determined |
| 18-102_k141_216970 | Not-determined |
| 18-102_k141_245848 | Not-determined |
| 18-102_k141_245851 | Not-determined |
| 18-102_k141_164405 | Low-quality |
| 18-102_k141_164409 | Not-determined |
| 18-102_k141_245857 | Not-determined |
| 18-102_k141_124183 | Not-determined |
| 18-102_k141_226120 | Not-determined |
| 18-102_k141_245859 | Not-determined |
| 18-102_k141_29037 | Not-determined |
| 18-102_k141_29038 | Not-determined |
| 18-102_k141_226125 | Not-determined |
| 18-102_k141_82763 | Not-determined |
| 18-102_k141_82764 | Not-determined |
| 18-102_k141_112869 | Not-determined |
| 18-102_k141_112873 | Low-quality |
| 18-102_k141_124208 | Not-determined |
| 18-102_k141_112877 | Low-quality |
| 18-102_k141_132994 | Not-determined |
| 18-102_k141_132995 | Not-determined |
| 18-102_k141_205275 | Not-determined |
| 18-102_k141_112888 | Not-determined |
| 18-102_k141_226143 | Low-quality |
| 18-102_k141_226148 | Not-determined |
| 18-102_k141_226150 | Not-determined |
| 18-102_k141_245891 | Not-determined |
| 18-102_k141_124235 | Not-determined |
| 18-102_k141_205294 | Not-determined |
| 18-102_k141_112909 | Not-determined |
| 18-102_k141_226161 | Not-determined |
| 18-102_k141_226166 | Not-determined |
| 18-102_k141_133007 | Not-determined |
| 18-102_k141_133011 | Not-determined |
| 18-102_k141_245909 | Not-determined |
| 18-102_k141_226173 | Not-determined |
| 18-102_k141_245913 | Not-determined |
| 18-102_k141_226175 | Low-quality |
| 18-102_k141_245915 | Not-determined |
| 18-102_k141_205307 | Not-determined |
| 18-102_k141_205310 | Not-determined |
| 18-102_k141_226180 | Not-determined |
| 18-102_k141_7783 | Not-determined |
| 18-102_k141_226184 | Not-determined |
| 18-102_k141_82789 | Not-determined |
| 18-102_k141_82791 | Not-determined |
| 18-102_k141_216986 | Not-determined |
| 18-102_k141_82795 | Not-determined |
| 18-102_k141_133034 | Not-determined |
| 18-102_k141_226188 | Not-determined |
| 18-102_k141_226190 | Not-determined |
| 18-102_k141_7787 | Low-quality |
| 18-102_k141_217002 | Low-quality |
| 18-102_k141_7790 | Not-determined |
| 18-102_k141_7798 | Low-quality |
| 18-102_k141_205332 | Not-determined |
| 18-102_k141_226206 | Not-determined |
| 18-102_k141_7802 | Not-determined |
| 18-102_k141_226217 | Not-determined |
| 18-102_k141_226218 | Not-determined |
| 18-102_k141_82804 | Low-quality |
| 18-102_k141_133078 | Not-determined |
| 18-102_k141_82805 | Not-determined |
| 18-102_k141_226224 | Not-determined |
| 18-102_k141_112917 | Not-determined |
| 18-102_k141_112918 | Not-determined |
| 18-102_k141_226231 | Not-determined |
| 18-102_k141_82819 | Not-determined |
| 18-102_k141_29106 | Not-determined |
| 18-102_k141_226236 | Not-determined |
| 18-102_k141_205354 | Not-determined |
| 18-102_k141_82831 | Not-determined |
| 18-102_k141_226247 | Not-determined |
| 18-102_k141_226256 | Not-determined |
| 18-102_k141_133099 | Not-determined |
| 18-102_k141_217025 | Not-determined |
| 18-102_k141_7824 | Not-determined |
| 18-102_k141_82845 | Not-determined |
| 18-102_k141_133118 | Not-determined |
| 18-102_k141_29130 | Not-determined |
| 18-102_k141_29134 | Not-determined |
| 18-102_k141_133127 | Not-determined |
| 18-102_k141_133130 | Low-quality |
| 18-102_k141_29136 | Not-determined |
| 18-102_k141_217048 | Not-determined |
| 18-102_k141_217052 | Low-quality |
| 18-102_k141_217054 | Low-quality |
| 18-102_k141_217064 | Not-determined |
| 18-102_k141_7833 | Not-determined |
| 18-102_k141_133140 | Not-determined |
| 18-102_k141_133141 | Not-determined |
| 18-102_k141_226294 | Low-quality |
| 18-102_k141_217087 | Not-determined |
| 18-102_k141_133156 | Not-determined |
| 18-102_k141_217091 | Not-determined |
| 18-102_k141_7842 | Not-determined |
| 18-102_k141_226322 | Not-determined |
| 18-102_k141_133171 | Not-determined |
| 18-102_k141_226337 | Not-determined |
| 18-102_k141_29168 | Not-determined |
| 18-102_k141_226360 | Low-quality |
| 18-102_k141_245992 | Not-determined |
| 18-102_k141_245993 | Not-determined |
| 18-102_k141_133187 | Low-quality |
| 18-102_k141_217124 | Not-determined |
| 18-102_k141_217127 | Not-determined |
| 18-102_k141_133192 | Low-quality |
| 18-102_k141_164522 | Not-determined |
| 18-102_k141_133211 | Not-determined |
| 18-102_k141_246008 | Not-determined |
| 18-102_k141_133218 | Not-determined |
| 18-102_k141_164550 | Not-determined |
| 18-102_k141_112948 | Not-determined |
| 18-102_k141_217152 | Not-determined |
| 18-102_k141_226393 | Not-determined |
| 18-102_k141_226398 | Not-determined |
| 18-102_k141_217159 | Low-quality |
| 18-102_k141_205386 | Not-determined |
| 18-102_k141_217165 | Not-determined |
| 18-102_k141_7857 | Not-determined |
| 18-102_k141_246038 | Not-determined |
| 18-102_k141_7860 | Not-determined |
| 18-102_k141_205401 | Low-quality |
| 18-102_k141_217176 | Not-determined |
| 18-102_k141_217178 | Not-determined |
| 18-102_k141_246047 | Not-determined |
| 18-102_k141_133265 | Not-determined |
| 18-102_k141_112963 | Not-determined |
| 18-102_k141_133278 | Not-determined |
| 18-102_k141_133284 | Not-determined |
| 18-102_k141_7881 | Not-determined |
| 18-102_k141_112973 | Low-quality |
| 18-102_k141_112978 | Not-determined |
| 18-102_k141_112982 | Not-determined |
| 18-102_k141_133296 | Not-determined |
| 18-102_k141_246052 | Not-determined |
| 18-102_k141_205418 | Not-determined |
| 18-102_k141_133307 | Low-quality |
| 18-102_k141_246059 | Not-determined |
| 18-102_k141_7889 | Not-determined |
| 18-102_k141_205424 | Not-determined |
| 18-102_k141_164605 | Not-determined |
| 18-102_k141_217191 | Not-determined |
| 18-102_k141_205434 | Not-determined |
| 18-102_k141_164618 | Not-determined |
| 18-102_k141_226428 | Not-determined |
| 18-102_k141_7910 | Not-determined |
| 18-102_k141_7912 | Low-quality |
| 18-102_k141_246066 | Not-determined |
| 18-102_k141_246072 | Not-determined |
| 18-102_k141_112997 | Not-determined |
| 18-102_k141_29197 | Not-determined |
| 18-102_k141_205455 | Not-determined |
| 18-102_k141_29202 | Not-determined |
| 18-102_k141_217197 | Not-determined |
| 18-102_k141_113008 | Low-quality |
| 18-102_k141_113011 | Low-quality |
| 18-102_k141_226462 | Not-determined |
| 18-102_k141_113022 | Not-determined |
| 18-102_k141_133335 | Not-determined |
| 18-102_k141_164647 | Not-determined |
| 18-102_k141_164659 | Low-quality |
| 18-102_k141_113038 | Not-determined |
| 18-102_k141_113043 | Not-determined |
| 18-102_k141_29222 | Low-quality |
| 18-102_k141_29223 | Low-quality |
| 18-102_k141_164676 | Not-determined |
| 18-102_k141_133340 | Not-determined |
| 18-102_k141_133344 | Not-determined |
| 18-102_k141_226488 | Not-determined |
| 18-102_k141_217224 | Not-determined |
| 18-102_k141_226501 | Not-determined |
| 18-102_k141_133361 | Not-determined |
| 18-102_k141_113066 | Not-determined |
| 18-102_k141_133363 | Not-determined |
| 18-102_k141_133367 | Not-determined |
| 18-102_k141_164695 | Not-determined |
| 18-102_k141_164696 | Not-determined |
| 18-102_k141_164707 | Not-determined |
| 18-102_k141_29254 | Not-determined |
| 18-102_k141_113100 | Not-determined |
| 18-102_k141_29258 | Low-quality |
| 18-102_k141_29264 | Not-determined |
| 18-102_k141_113112 | Not-determined |
| 18-102_k141_113118 | Not-determined |
| 18-102_k141_113120 | Not-determined |
| 18-102_k141_226507 | Not-determined |
| 18-102_k141_164753 | Not-determined |
| 18-102_k141_246108 | Not-determined |
| 18-102_k141_29282 | Not-determined |
| 18-102_k141_29285 | Not-determined |
| 18-102_k141_246118 | Not-determined |
| 18-102_k141_226531 | Not-determined |
| 18-102_k141_113153 | Not-determined |
| 18-102_k141_164790 | Not-determined |
| 18-102_k141_246132 | Not-determined |
| 18-102_k141_217264 | Not-determined |
| 18-102_k141_246139 | Not-determined |
| 18-102_k141_246141 | Low-quality |
| 18-102_k141_226539 | Low-quality |
| 18-102_k141_164797 | Not-determined |
| 18-102_k141_113175 | Not-determined |
| 18-102_k141_113178 | Not-determined |
| 18-102_k141_113179 | Not-determined |
| 18-102_k141_205531 | Not-determined |
| 18-102_k141_226567 | Not-determined |
| 18-102_k141_205536 | Not-determined |
| 18-102_k141_113187 | Not-determined |
| 18-102_k141_7949 | Not-determined |
| 18-102_k141_217296 | Not-determined |
| 18-102_k141_226575 | Not-determined |
| 18-102_k141_7951 | Not-determined |
| 18-102_k141_226577 | Not-determined |
| 18-102_k141_246162 | Not-determined |
| 18-102_k141_164804 | Not-determined |
| 18-102_k141_217302 | Not-determined |
| 18-102_k141_7954 | Not-determined |
| 18-102_k141_29310 | Not-determined |
| 18-102_k141_164810 | Not-determined |
| 18-102_k141_7957 | Low-quality |
| 18-102_k141_217310 | Not-determined |
| 18-102_k141_7962 | Not-determined |
| 18-102_k141_205553 | Not-determined |
| 18-102_k141_246183 | Not-determined |
| 18-102_k141_246184 | Not-determined |
| 18-102_k141_113213 | Not-determined |
| 18-102_k141_205557 | Not-determined |
| 18-102_k141_226598 | Not-determined |
| 18-102_k141_246200 | Not-determined |
| 18-102_k141_113229 | Not-determined |
| 18-102_k141_217333 | Not-determined |
| 18-102_k141_164858 | Not-determined |
| 18-102_k141_7988 | Not-determined |
| 18-102_k141_217341 | Not-determined |
| 18-102_k141_205580 | Not-determined |
| 18-102_k141_164866 | Not-determined |
| 18-102_k141_7994 | Low-quality |
| 18-102_k141_7995 | Not-determined |
| 18-102_k141_133382 | Low-quality |
| 18-102_k141_7998 | Not-determined |
| 18-102_k141_205591 | Not-determined |
| 18-102_k141_164885 | Not-determined |
| 18-102_k141_164889 | Not-determined |
| 18-102_k141_217356 | Not-determined |
| 18-102_k141_226623 | Not-determined |
| 18-102_k141_133404 | Not-determined |
| 18-102_k141_113267 | Not-determined |
| 18-102_k141_205600 | Not-determined |
| 18-102_k141_205604 | Not-determined |
| 18-102_k141_205606 | Not-determined |
| 18-102_k141_8020 | Not-determined |
| 18-102_k141_113276 | Not-determined |
| 18-102_k141_29343 | Not-determined |
| 18-102_k141_29344 | Not-determined |
| 18-102_k141_8022 | Low-quality |
| 18-102_k141_113284 | Not-determined |
| 18-102_k141_113286 | Not-determined |
| 18-102_k141_226655 | Not-determined |
| 18-102_k141_29358 | Not-determined |
| 18-102_k141_226661 | Not-determined |
| 18-102_k141_226669 | Not-determined |
| 18-102_k141_133437 | Not-determined |
| 18-102_k141_133440 | Low-quality |
| 18-102_k141_113316 | Not-determined |
| 18-102_k141_164908 | Not-determined |
| 18-102_k141_164909 | Not-determined |
| 18-102_k141_205653 | Not-determined |
| 18-102_k141_246227 | Not-determined |
| 18-102_k141_133455 | Not-determined |
| 18-102_k141_164924 | Not-determined |
| 18-102_k141_29380 | Not-determined |
| 18-102_k141_226702 | Not-determined |
| 18-102_k141_133475 | Not-determined |
| 18-102_k141_217391 | Low-quality |
| 18-102_k141_29392 | Not-determined |
| 18-102_k141_164942 | Not-determined |
| 18-102_k141_226727 | Not-determined |
| 18-102_k141_217404 | Not-determined |
| 18-102_k141_164946 | Low-quality |
| 18-102_k141_164948 | Not-determined |
| 18-102_k141_164956 | Not-determined |
| 18-102_k141_8063 | Not-determined |
| 18-102_k141_133497 | Low-quality |
| 18-102_k141_8066 | Not-determined |
| 18-102_k141_164961 | Low-quality |
| 18-102_k141_164967 | Not-determined |
| 18-102_k141_133506 | Not-determined |
| 18-102_k141_246254 | Low-quality |
| 18-102_k141_164988 | Not-determined |
| 18-102_k141_226769 | Low-quality |
| 18-102_k141_246269 | Low-quality |
| 18-102_k141_226773 | Not-determined |
| 18-102_k141_133529 | Not-determined |
| 18-102_k141_165012 | Not-determined |
| 18-102_k141_165016 | Not-determined |
| 18-102_k141_226788 | Not-determined |
| 18-102_k141_165021 | Low-quality |
| 18-102_k141_29444 | Not-determined |
| 18-102_k141_133558 | Not-determined |
| 18-102_k141_205700 | Not-determined |
| 18-102_k141_29450 | Not-determined |
| 18-102_k141_226798 | Not-determined |
| 18-102_k141_205705 | Not-determined |
| 18-102_k141_165030 | Not-determined |
| 18-102_k141_205710 | Not-determined |
| 18-102_k141_133592 | Not-determined |
| 18-102_k141_246293 | Not-determined |
| 18-102_k141_8096 | Not-determined |
| 18-102_k141_29486 | Low-quality |
| 18-102_k141_8106 | Not-determined |
| 18-102_k141_8107 | Not-determined |
| 18-102_k141_205731 | Not-determined |
| 18-102_k141_29503 | Not-determined |
| 18-102_k141_165069 | Not-determined |
| 18-102_k141_8121 | Not-determined |
| 18-102_k141_165079 | Not-determined |
| 18-102_k141_8127 | Not-determined |
| 18-102_k141_165084 | Low-quality |
| 18-102_k141_29514 | Not-determined |
| 18-102_k141_8136 | Low-quality |
| 18-102_k141_113356 | Not-determined |
| 18-102_k141_113357 | Not-determined |
| 18-102_k141_217414 | Not-determined |
| 18-102_k141_165099 | Not-determined |
| 18-102_k141_165102 | Not-determined |
| 18-102_k141_8159 | Low-quality |
| 18-102_k141_226839 | Not-determined |
| 18-102_k141_226840 | Not-determined |
| 18-102_k141_217430 | Not-determined |
| 18-102_k141_113369 | Not-determined |
| 18-102_k141_226847 | Low-quality |
| 18-102_k141_8162 | Not-determined |
| 18-110-final-viral-combined_k141_86758\|\|full | Medium-quality |
| 18-102_k141_8163 | Not-determined |
| 18-102_k141_217451 | Not-determined |
| 18-102_k141_205765 | Not-determined |
| 18-102_k141_29550 | Low-quality |
| 18-102_k141_165130 | Low-quality |
| 18-102_k141_226862 | Low-quality |
| 18-102_k141_226874 | Not-determined |
| 18-102_k141_29560 | Not-determined |
| 18-102_k141_205784 | Not-determined |
| 18-102_k141_246321 | Not-determined |
| 18-102_k141_8201 | Not-determined |
| 18-102_k141_8213 | Not-determined |
| 18-102_k141_8214 | Low-quality |
| 18-102_k141_29578 | Not-determined |
| 18-102_k141_8220 | Not-determined |
| 18-102_k141_113386 | Not-determined |
| 18-102_k141_226903 | Not-determined |
| 18-102_k141_165174 | Low-quality |
| 18-102_k141_113398 | Not-determined |
| 18-102_k141_113411 | Low-quality |
| 18-102_k141_165194 | Not-determined |
| 18-102_k141_226927 | Not-determined |
| 18-102_k141_226936 | Low-quality |
| 18-102_k141_205809 | Not-determined |
| 18-102_k141_165216 | Not-determined |
| 18-102_k141_205817 | Not-determined |
| 18-102_k141_205819 | Not-determined |
| 18-102_k141_226938 | Not-determined |
| 18-102_k141_226943 | Not-determined |
| 18-102_k141_246351 | Not-determined |
| 18-102_k141_217471 | Not-determined |
| 18-102_k141_246354 | Not-determined |
| 18-102_k141_165224 | Not-determined |
| 18-102_k141_217474 | Not-determined |
| 18-102_k141_205830 | Not-determined |
| 18-102_k141_165230 | Not-determined |
| 18-102_k141_113438 | Low-quality |
| 18-102_k141_113446 | Not-determined |
| 18-102_k141_165244 | Not-determined |
| 18-102_k141_205854 | Not-determined |
| 18-102_k141_205857 | Not-determined |
| 18-102_k141_246359 | Not-determined |
| 18-102_k141_165258 | Not-determined |
| 18-102_k141_165264 | Not-determined |
| 18-102_k141_226965 | Not-determined |
| 18-102_k141_226976 | Not-determined |
| 18-102_k141_113460 | Not-determined |
| 18-102_k141_205875 | Not-determined |
| 18-102_k141_113462 | Not-determined |
| 18-102_k141_226985 | Not-determined |
| 18-102_k141_205890 | Not-determined |
| 18-102_k141_165283 | Low-quality |
| 18-102_k141_165288 | Not-determined |
| 18-102_k141_227036 | Not-determined |
| 18-102_k141_165298 | Not-determined |
| 18-102_k141_165301 | Not-determined |
| 18-102_k141_165302 | Not-determined |
| 18-102_k141_246397 | Not-determined |
| 18-102_k141_227050 | Not-determined |
| 18-102_k141_29603 | Not-determined |
| 18-102_k141_165319 | Low-quality |
| 18-102_k141_165332 | Not-determined |
| 18-102_k141_246414 | Not-determined |
| 18-102_k141_133603 | Not-determined |
| 18-102_k141_165353 | Low-quality |
| 18-102_k141_113509 | Not-determined |
| 18-102_k141_8282 | Not-determined |
| 18-102_k141_205922 | Not-determined |
| 18-102_k141_205929 | Not-determined |
| 18-102_k141_8296 | Not-determined |
| 18-102_k141_205949 | Not-determined |
| 18-102_k141_165372 | Not-determined |
| 18-102_k141_205955 | Not-determined |
| 18-102_k141_113525 | Not-determined |
| 18-102_k141_29669 | Low-quality |
| 18-102_k141_227087 | Not-determined |
| 18-102_k141_227095 | Not-determined |
| 18-102_k141_165387 | Not-determined |
| 18-102_k141_165391 | Low-quality |
| 18-102_k141_227103 | Low-quality |
| 18-102_k141_113534 | Not-determined |
| 18-102_k141_8310 | Not-determined |
| 18-102_k141_29703 | Not-determined |
| 18-102_k141_133641 | Not-determined |
| 18-102_k141_133642 | Not-determined |
| 18-102_k141_133645 | Not-determined |
| 18-102_k141_113546 | Not-determined |
| 18-102_k141_206008 | Not-determined |
| 18-102_k141_165416 | Not-determined |
| 18-102_k141_165417 | Not-determined |
| 18-102_k141_246475 | Not-determined |
| 18-102_k141_133659 | Not-determined |
| 18-102_k141_113573 | Not-determined |
| 18-102_k141_133661 | Low-quality |
| 18-102_k141_165443 | Not-determined |
| 18-102_k141_227152 | Not-determined |
| 18-102_k141_246481 | Not-determined |
| 18-102_k141_165450 | Low-quality |
| 18-102_k141_133669 | Not-determined |
| 18-102_k141_246485 | Not-determined |
| 18-102_k141_165458 | Not-determined |
| 18-102_k141_133677 | Not-determined |
| 18-102_k141_165464 | Not-determined |
| 18-102_k141_8337 | Not-determined |
| 18-102_k141_113605 | Not-determined |
| 18-102_k141_165470 | Not-determined |
| 18-102_k141_113609 | Not-determined |
| 18-102_k141_206058 | Not-determined |
| 18-102_k141_29759 | Not-determined |
| 18-102_k141_113611 | Not-determined |
| 18-102_k141_206066 | Not-determined |
| 18-102_k141_29768 | Not-determined |
| 18-102_k141_133704 | Not-determined |
| 18-102_k141_206076 | Not-determined |
| 18-102_k141_206077 | Not-determined |
| 18-102_k141_29777 | Not-determined |
| 18-102_k141_206082 | Not-determined |
| 18-102_k141_206083 | Not-determined |
| 18-102_k141_133710 | Not-determined |
| 18-102_k141_133717 | Not-determined |
| 18-102_k141_133725 | Not-determined |
| 18-102_k141_8377 | Not-determined |
| 18-102_k141_227159 | Not-determined |
| 18-102_k141_227163 | Not-determined |
| 18-102_k141_29805 | Not-determined |
| 18-102_k141_246549 | Not-determined |
| 18-102_k141_227198 | Not-determined |
| 18-102_k141_227203 | Not-determined |
| 18-102_k141_227206 | Not-determined |
| 18-102_k141_246564 | Not-determined |
| 18-102_k141_246571 | Not-determined |
| 18-102_k141_8395 | Not-determined |
| 18-102_k141_246578 | Not-determined |
| 18-102_k141_113653 | Not-determined |
| 18-102_k141_227224 | Not-determined |
| 18-102_k141_165522 | Not-determined |
| 18-102_k141_8399 | Not-determined |
| 18-102_k141_113659 | Not-determined |
| 18-102_k141_113660 | Not-determined |
| 18-102_k141_165532 | Not-determined |
| 18-102_k141_246599 | Not-determined |
| 18-102_k141_246600 | Not-determined |
| 18-102_k141_246603 | Not-determined |
| 18-102_k141_227246 | Not-determined |
| 18-102_k141_227250 | Not-determined |
| 18-102_k141_246610 | Low-quality |
| 18-102_k141_113669 | Not-determined |
| 18-102_k141_246618 | Not-determined |
| 18-102_k141_246622 | Not-determined |
| 18-102_k141_206112 | Not-determined |
| 18-102_k141_227272 | Not-determined |
| 18-102_k141_133750 | Not-determined |
| 18-102_k141_133757 | Not-determined |
| 18-102_k141_206120 | Low-quality |
| 18-102_k141_165558 | Not-determined |
| 18-102_k141_165565 | Low-quality |
| 18-102_k141_227297 | Not-determined |
| 18-102_k141_165570 | Not-determined |
| 18-102_k141_206126 | Not-determined |
| 18-102_k141_206127 | Not-determined |
| 18-102_k141_165596 | Not-determined |
| 18-102_k141_227316 | Not-determined |
| 18-102_k141_206149 | Not-determined |
| 18-102_k141_246675 | Not-determined |
| 18-102_k141_227329 | Not-determined |
| 18-102_k141_227332 | Not-determined |
| 18-102_k141_206175 | Not-determined |
| 18-102_k141_133821 | Low-quality |
| 18-102_k141_113684 | Not-determined |
| 18-102_k141_206177 | Not-determined |
| 18-102_k141_206178 | Not-determined |
| 18-102_k141_206179 | Not-determined |
| 18-102_k141_133824 | Not-determined |
| 18-102_k141_133825 | Low-quality |
| 18-102_k141_133828 | Not-determined |
| 18-102_k141_246698 | Not-determined |
| 18-102_k141_133830 | Not-determined |
| 18-102_k141_246701 | Not-determined |
| 18-102_k141_246703 | Not-determined |
| 18-102_k141_113711 | Not-determined |
| 18-102_k141_113718 | Low-quality |
| 18-102_k141_133858 | Not-determined |
| 18-102_k141_246717 | Not-determined |
| 18-102_k141_29894 | Not-determined |
| 18-102_k141_29904 | Not-determined |
| 18-102_k141_206207 | Low-quality |
| 18-102_k141_29907 | Not-determined |
| 18-102_k141_206213 | Not-determined |
| 18-102_k141_29913 | Not-determined |
| 18-102_k141_8471 | Not-determined |
| 18-102_k141_206219 | Not-determined |
| 18-102_k141_29929 | Not-determined |
| 18-102_k141_246735 | Not-determined |
| 18-102_k141_206242 | Not-determined |
| 18-102_k141_206248 | Not-determined |
| 18-102_k141_133905 | Low-quality |
| 18-102_k141_113761 | Not-determined |
| 18-102_k141_113764 | Not-determined |
| 18-102_k141_227368 | Not-determined |
| 18-102_k141_113781 | Not-determined |
| 18-102_k141_113784 | Not-determined |
| 18-102_k141_8512 | Low-quality |
| 18-102_k141_133938 | Not-determined |
| 18-102_k141_206264 | Not-determined |
| 18-102_k141_246759 | Not-determined |
| 18-102_k141_206267 | Not-determined |
| 18-102_k141_133948 | Not-determined |
| 18-102_k141_113791 | Not-determined |
| 18-102_k141_206269 | Not-determined |
| 18-102_k141_29969 | Not-determined |

18-102_k141_246768 Not-determined
18-102_k141_133984 Not-determined
18-102_k141_30005 Not-determined
18-102_k141_227397 Not-determined
18-102_k141_246785 Not-determined
18-102_k141_206297 Low-quality
18-102_k141_246793 Not-determined
18-102_k141_165611 Low-quality
18-102_k141_246805 Not-determined
18-102_k141_8542 Not-determined
18-102_k141_8548 Not-determined
18-102_k141_30015 Low-quality
18-102_k141_113816 Not-determined
18-102_k141_165625 Not-determined
18-102_k141_30027 Not-determined
18-102_k141_30035 Not-determined
18-102_k141_30038 Not-determined
18-102_k141_30040 Not-determined
18-102_k141_134056 Not-determined
18-102_k141_30059 Not-determined
18-102_k141_30061 Low-quality
18-102_k141_206316 Not-determined
18-102_k141_206317 Not-determined
18-102_k141_246843 Not-determined
18-102_k141_30078 Not-determined
18-102_k141_227418 Not-determined
18-102_k141_30080 Not-determined
18-102_k141_134076 Low-quality
18-102_k141_30099 Not-determined
18-102_k141_246848 Not-determined
18-102_k141_134089 Low-quality
18-102_k141_165648 Low-quality
18-102_k141_246862 Not-determined
18-102_k141_30108 Not-determined
18-102_k141_206337 Not-determined
18-102_k141_165654 Not-determined
18-102_k141_165656 Not-determined
18-102_k141_165657 Not-determined
18-102_k141_246878 Not-determined
18-102_k141_134126 Not-determined
18-102_k141_165662 Not-determined
18-102_k141_165665 Not-determined
18-102_k141_134129 Low-quality
18-102_k141_165668 Not-determined
18-102_k141_165669 Not-determined
18-102_k141_165671 Not-determined
18-102_k141_165672 Not-determined
18-102_k141_246886 Not-determined
18-102_k141_113836 Not-determined
18-102_k141_134143 Not-determined
18-102_k141_30114 Not-determined
18-102_k141_165679 Not-determined
18-102_k141_246898 Not-determined
18-102_k141_206342 Not-determined
18-102_k141_206343 Not-determined
18-102_k141_134159 Not-determined
18-102_k141_165687 Not-determined
18-102_k141_206353 Low-quality
18-102_k141_227428 Low-quality
18-102_k141_134182 Not-determined
18-102_k141_165691 Low-quality
18-102_k141_246910 Low-quality
18-102_k141_206382 Low-quality
18-102_k141_206388 Not-determined
18-102_k141_113849 Not-determined
18-102_k141_227462 Not-determined
18-102_k141_227472 Not-determined
18-102_k141_113865 Not-determined
18-102_k141_113868 Not-determined
18-102_k141_30162 Not-determined
18-102_k141_30166 Not-determined
18-102_k141_30169 Not-determined
18-102_k141_30178 Not-determined
18-102_k141_227490 Low-quality
18-102_k141_227500 Not-determined
18-102_k141_30206 Low-quality
18-102_k141_227506 Not-determined
18-102_k141_246926 Not-determined
18-102_k141_246927 Not-determined
18-102_k141_206421 Not-determined
18-102_k141_246937 Not-determined
18-102_k141_113881 Not-determined
18-102_k141_206434 Not-determined
18-102_k141_227533 Low-quality
18-102_k141_246944 Not-determined
18-102_k141_227541 Not-determined
18-102_k141_227543 Not-determined
18-102_k141_113893 Low-quality
18-102_k141_206457 Low-quality
18-102_k141_246951 Not-determined
18-102_k141_227557 Not-determined
18-102_k141_227565 Not-determined
18-102_k141_246974 Not-determined
18-102_k141_246981 Not-determined
18-102_k141_206484 Not-determined
18-102_k141_134232 Not-determined
18-102_k141_30303 Not-determined
18-102_k141_206494 Not-determined
18-102_k141_246995 Not-determined
18-102_k141_246997 Not-determined
18-102_k141_247004 Not-determined
18-102_k141_30320 Not-determined
18-102_k141_247008 Not-determined
18-102_k141_247009 Low-quality
18-102_k141_206511 Not-determined
18-102_k141_30346 Not-determined
18-102_k141_134279 Not-determined
18-102_k141_134282 Not-determined
18-102_k141_227571 Not-determined
18-102_k141_227580 Not-determined
18-102_k141_134306 Low-quality
18-102_k141_247041 Not-determined
18-102_k141_134314 Not-determined
18-102_k141_227593 Not-determined
18-102_k141_247055 Not-determined
18-102_k141_227611 Low-quality
18-102_k141_227613 Low-quality
18-102_k141_227619 Not-determined
18-102_k141_206518 Not-determined
18-102_k141_227624 Not-determined
18-102_k141_134341 Not-determined
18-102_k141_227627 Low-quality
18-102_k141_206536 Not-determined
18-102_k141_227631 Low-quality
18-102_k141_134357 Low-quality
18-102_k141_134361 Not-determined
18-102_k141_134362 Not-determined
18-102_k141_30377 Not-determined
18-102_k141_30381 Not-determined
18-102_k141_227638 Low-quality
18-102_k141_134367 Not-determined
18-102_k141_227666 Not-determined
18-102_k141_227671 Not-determined
18-102_k141_30401 Low-quality
18-102_k141_30404 Not-determined
18-102_k141_227696 Not-determined
18-102_k141_227697 Not-determined
18-102_k141_206566 Not-determined
18-102_k141_247090 Not-determined
18-102_k141_134382 Not-determined
18-102_k141_206577 Low-quality
18-102_k141_134386 Not-determined
18-102_k141_247104 Not-determined
18-102_k141_206579 Not-determined
18-102_k141_206583 Not-determined
18-102_k141_206585 Not-determined
18-102_k141_134402 Not-determined
18-102_k141_134406 Not-determined
18-102_k141_30457 Low-quality
18-102_k141_227707 Not-determined
18-102_k141_227708 Not-determined
18-102_k141_206594 Not-determined
18-102_k141_206597 Not-determined
18-102_k141_227711 Not-determined
18-102_k141_227716 Not-determined
18-102_k141_134417 Not-determined
18-102_k141_227721 Not-determined
18-102_k141_227722 Not-determined
18-102_k141_227725 Not-determined
18-102_k141_227726 Not-determined
18-102_k141_134438 Not-determined
18-102_k141_227739 Low-quality
18-102_k141_30476 Not-determined
18-102_k141_30477 Not-determined
18-102_k141_227749 Not-determined
18-102_k141_30491 Not-determined
18-102_k141_8615 Not-determined
18-102_k141_30500 Not-determined
18-102_k141_206628 Not-determined
18-102_k141_206643 Not-determined
18-102_k141_8648 Not-determined
18-102_k141_134473 Not-determined
18-102_k141_30528 Not-determined
18-102_k141_30530 Not-determined
18-102_k141_134475 Low-quality
18-102_k141_30557 Not-determined
18-102_k141_30560 Not-determined
18-102_k141_206674 Not-determined
18-102_k141_206680 Not-determined
18-102_k141_134494 Not-determined
18-102_k141_206688 Not-determined
18-102_k141_8672 Not-determined
18-102_k141_8680 Not-determined
18-102_k141_8686 Not-determined
18-102_k141_227789 Not-determined
18-102_k141_134497 Low-quality
18-102_k141_134500 Not-determined
18-102_k141_8713 Not-determined
18-102_k141_30593 Not-determined
18-102_k141_134518 Not-determined
18-102_k141_134524 Low-quality
18-102_k141_8733 Not-determined
18-102_k141_134551 Not-determined
18-102_k141_134554 Not-determined
18-102_k141_134555 Not-determined
18-102_k141_227805 Not-determined
18-102_k141_134576 Low-quality
18-102_k141_30628 Low-quality
18-102_k141_30650 Not-determined
18-102_k141_134593 Not-determined
18-102_k141_30655 Not-determined
18-102_k141_134605 Not-determined
18-102_k141_30666 Not-determined
18-102_k141_134607 Low-quality
18-102_k141_30667 Not-determined
18-102_k141_30700 Low-quality
18-102_k141_134619 Not-determined
18-102_k141_134623 Low-quality
18-102_k141_30740 Not-determined
18-102_k141_247115 Not-determined
18-102_k141_8825 Not-determined
18-102_k141_8830 Low-quality
18-102_k141_247127 Not-determined
18-102_k141_247130 Not-determined
18-102_k141_247131 Not-determined
18-102_k141_247140 Not-determined
18-102_k141_247145 Not-determined
18-102_k141_206721 Low-quality
18-102_k141_247165 Not-determined
18-102_k141_8884 Low-quality
18-102_k141_8888 Not-determined
18-102_k141_206728 Not-determined
18-102_k141_206729 Not-determined
18-102_k141_247201 Not-determined
18-102_k141_8903 Not-determined
18-102_k141_206738 Low-quality
18-102_k141_8920 Low-quality
18-102_k141_247215 Not-determined
18-102_k141_206746 Not-determined
18-102_k141_8926 Not-determined
18-102_k141_247220 Not-determined
18-102_k141_206776 Not-determined
18-102_k141_30764 Not-determined
18-102_k141_8944 Not-determined
18-102_k141_30770 Not-determined
18-102_k141_30771 Low-quality
18-102_k141_30772 Low-quality
18-102_k141_206789 Not-determined
18-102_k141_206791 Not-determined
18-102_k141_206795 Not-determined
18-102_k141_8949 Not-determined
18-102_k141_30792 Low-quality
18-102_k141_30793 Not-determined
18-102_k141_247243 Not-determined
18-102_k141_247251 Not-determined
18-102_k141_8982 Not-determined
18-102_k141_247269 Not-determined
18-102_k141_30797 Not-determined
18-102_k141_247274 Not-determined
18-102_k141_9000 Not-determined
18-102_k141_9002 Not-determined
18-102_k141_30807 Not-determined
18-102_k141_30819 Not-determined
18-102_k141_247288 Not-determined
18-102_k141_247295 Not-determined
18-102_k141_9010 Not-determined
18-102_k141_247298 Low-quality
18-102_k141_206816 Not-determined
18-102_k141_30861 Low-quality
18-102_k141_247322 Not-determined
18-102_k141_247328 Not-determined
18-102_k141_247340 Low-quality
18-102_k141_247350 Not-determined
18-102_k141_30891 Not-determined
18-102_k141_9062 Not-determined
18-102_k141_247387 Not-determined
18-102_k141_247398 Low-quality
18-102_k141_247408 Not-determined
18-102_k141_9090 Not-determined
18-102_k141_9095 Low-quality
18-102_k141_30920 Not-determined
18-102_k141_9099 Not-determined
18-102_k141_247422 Not-determined
18-102_k141_9114 Low-quality
18-102_k141_247424 Not-determined
18-102_k141_9118 Low-quality
18-102_k141_9125 Not-determined
18-102_k141_247429 Not-determined
18-102_k141_247433 Low-quality
18-102_k141_247434 Low-quality
18-102_k141_30962 Low-quality
18-102_k141_247446 Not-determined
18-102_k141_247454 Not-determined
18-102_k141_30975 Not-determined
18-102_k141_30976 Not-determined
18-102_k141_30978 Not-determined
18-102_k141_9138 Not-determined
18-102_k141_9142 Not-determined
18-102_k141_31004 Not-determined
18-102_k141_247478 Not-determined
18-102_k141_247480 Not-determined
18-102_k141_247490 Not-determined
18-102_k141_247491 Low-quality
18-102_k141_247498 Not-determined
18-102_k141_31050 Not-determined
18-102_k141_247505 Not-determined
18-102_k141_31057 Not-determined
18-102_k141_247518 Not-determined
18-102_k141_247536 Not-determined
18-102_k141_9167 Not-determined
18-102_k141_9171 Not-determined
18-102_k141_9179 Not-determined
18-102_k141_9184 Not-determined
18-102_k141_247551 Not-determined
18-102_k141_206837 Not-determined
18-102_k141_247567 Not-determined
18-102_k141_247571 Not-determined
18-102_k141_206843 Not-determined
18-102_k141_9275 Not-determined
18-102_k141_206857 Not-determined
18-102_k141_206860 Low-quality
18-102_k141_9278 Not-determined
18-102_k141_206867 Not-determined
18-102_k141_247581 Not-determined
18-102_k141_206887 Not-determined
18-102_k141_206892 Not-determined
18-102_k141_9291 Not-determined
18-102_k141_247607 Not-determined
18-102_k141_247617 Not-determined
18-102_k141_9315 Not-determined
18-102_k141_9321 Low-quality
18-102_k141_247638 Not-determined
18-102_k141_206937 Not-determined
18-102_k141_247643 Not-determined
18-102_k141_206953 Not-determined
18-102_k141_247669 Low-quality
18-102_k141_247704 Not-determined
18-102_k141_206983 Not-determined
18-102_k141_247739 Low-quality
18-102_k141_247745 Not-determined
18-102_k141_247756 Not-determined
18-102_k141_247763 Not-determined
18-102_k141_9348 Low-quality
18-102_k141_9355 Not-determined
18-102_k141_9359 Not-determined
18-102_k141_247796 Not-determined
18-102_k141_247799 Not-determined
18-102_k141_9367 Not-determined
18-102_k141_247812 Not-determined
18-102_k141_9370 Not-determined
18-102_k141_9375 Not-determined
18-102_k141_247821 Not-determined
18-102_k141_9377 Not-determined
18-102_k141_247839 Not-determined
18-102_k141_9403 Not-determined
18-102_k141_9405 Not-determined
18-102_k141_247850 Not-determined
18-102_k141_247865 Not-determined
18-102_k141_9447 Not-determined
18-102_k141_9455 Not-determined
18-102_k141_9487 Low-quality
18-102_k141_247918 Not-determined
18-102_k141_9508 Low-quality
18-102_k141_9509 Not-determined
18-102_k141_9512 Not-determined
18-102_k141_247930 Not-determined
18-102_k141_207004 Not-determined
18-102_k141_207011 Low-quality
18-102_k141_207021 Low-quality
18-102_k141_207026 Not-determined
18-102_k141_247968 Low-quality
18-102_k141_247972 Not-determined
18-102_k141_247984 Not-determined
18-102_k141_247991 Not-determined
18-102_k141_247992 Not-determined
18-102_k141_247994 Not-determined
18-102_k141_248008 Not-determined
18-102_k141_248012 Not-determined
18-102_k141_207046 Not-determined
18-102_k141_248024 Not-determined
18-102_k141_248031 Not-determined
18-102_k141_248043 Low-quality
18-102_k141_248047 Not-determined
18-102_k141_207062 Low-quality
18-102_k141_207065 Not-determined
18-102_k141_248053 Not-determined
18-102_k141_248055 Not-determined
18-102_k141_9560 Not-determined
18-102_k141_207096 Not-determined
18-102_k141_248084 Not-determined
18-102_k141_248094 Not-determined
18-102_k141_248102 Not-determined
18-102_k141_9574 Not-determined
18-102_k141_248115 Low-quality
18-102_k141_9591 Not-determined
18-102_k141_248125 Not-determined
18-102_k141_248140 Not-determined
18-102_k141_248149 Not-determined
18-102_k141_248152 Not-determined
18-102_k141_248154 Low-quality
18-102_k141_248163 Not-determined
18-102_k141_248173 Not-determined
18-102_k141_248177 Not-determined
18-102_k141_248184 Not-determined
18-102_k141_9603 Not-determined
18-102_k141_248189 Not-determined
18-102_k141_9606 Not-determined
18-102_k141_248195 Not-determined
18-102_k141_248198 Low-quality
18-102_k141_248210 Low-quality
18-102_k141_9634 Not-determined
18-102_k141_248224 Not-determined
18-102_k141_248232 Not-determined
18-102_k141_248236 Not-determined
18-102_k141_248237 Not-determined
18-102_k141_248242 Not-determined
18-102_k141_9652 Not-determined
18-102_k141_248251 Not-determined
18-102_k141_248252 Not-determined
18-102_k141_9655 Not-determined
18-102_k141_9711 Not-determined
17-108-final-viral-combined_k141_326318||full Medium-quality
18-102_k141_9719 Not-determined
18-102_k141_9727 Not-determined
18-102_k141_9745 Not-determined
18-102_k141_248322 Not-determined
18-102_k141_9780 Not-determined
18-102_k141_9790 Not-determined
18-102_k141_9792 Not-determined
18-109-final-viral-combined_k141_123954||full Medium-quality
18-102_k141_9813 Low-quality
18-101-final-viral-combined_k141_159544||full Medium-quality
18-102_k141_248328 Low-quality
18-102_k141_9831 Not-determined
18-102_k141_9838 Not-determined
18-102_k141_248333 Not-determined
18-102_k141_248347 Not-determined
18-102_k141_248351 Low-quality
18-102_k141_248368 Not-determined
18-102_k141_248375 Not-determined
18-102_k141_248381 Not-determined
18-102_k141_248391 Not-determined
18-102_k141_248399 Low-quality
18-102_k141_248402 Not-determined
18-102_k141_248407 Not-determined
18-102_k141_248416 Not-determined
18-102_k141_248423 Not-determined
18-102_k141_9860 Not-determined
18-102_k141_248427 Not-determined
18-102_k141_248432 Not-determined
18-102_k141_9863 Not-determined
18-102_k141_248436 Not-determined
18-102_k141_248439 Not-determined
18-102_k141_9871 Not-determined
18-102_k141_248451 Not-determined
18-102_k141_9880 Not-determined
18-102_k141_9882 Low-quality
18-102_k141_248465 Not-determined
18-102_k141_248466 Not-determined
18-102_k141_248471 Not-determined
18-102_k141_248472 Not-determined
18-102_k141_248476 Not-determined
18-102_k141_248479 Not-determined
18-102_k141_248481 Not-determined
18-102_k141_248486 Low-quality
18-102_k141_248487 Not-determined
18-102_k141_248492 Not-determined
18-102_k141_248494 Not-determined
18-102_k141_248496 Not-determined
18-102_k141_248497 Not-determined
18-102_k141_248501 Not-determined
18-102_k141_248502 Not-determined
18-102_k141_248505 Not-determined
18-102_k141_248508 Not-determined
18-102_k141_248514 Not-determined
18-102_k141_248522 Not-determined
18-102_k141_248523 Not-determined
18-102_k141_248524 Not-determined
18-102_k141_248525 Low-quality
18-102_k141_248533 Not-determined
18-102_k141_248538 Not-determined
18-102_k141_248539 Not-determined
18-102_k141_248540 Not-determined
18-102_k141_248545 Not-determined
18-102_k141_248546 Not-determined
18-102_k141_248547 Not-determined
18-102_k141_9903 Low-quality
18-102_k141_9906 Not-determined
18-102_k141_9934 Not-determined
18-102_k141_9935 Not-determined
18-102_k141_9936 Not-determined
18-102_k141_10006 Not-determined
18-102_k141_10029 Not-determined
18-102_k141_10035 Not-determined
18-102_k141_10038 Not-determined
18-102_k141_10056 Not-determined
18-102_k141_10081 Not-determined
18-102_k141_10083 Not-determined
18-102_k141_10129 Not-determined
18-102_k141_10186 Not-determined
18-102_k141_10202 Not-determined
18-102_k141_10205 Not-determined
18-102_k141_10214 Not-determined
18-102_k141_10226 Not-determined
18-102_k141_10236 Not-determined
18-102_k141_10253 Not-determined
18-102_k141_10254 Not-determined
18-102_k141_10260 Not-determined
18-102_k141_10262 Not-determined
18-102_k141_10278 Not-determined
18-102_k141_10281 Not-determined
18-102_k141_10314 Not-determined
18-102_k141_10316 Not-determined
18-104-final-viral-combined_k141_69815||full Low-quality
18-104-final-viral-combined_k141_49566||full Not-determined
18-104-final-viral-combined_k141_140983||full Low-quality
18-104-final-viral-combined_k141_10417||full Not-determined
18-104-final-viral-combined_k141_143935||full Not-determined
18-104-final-viral-combined_k141_82321||full Not-determined
18-104-final-viral-combined_k141_26994||full Not-determined
18-104-final-viral-combined_k141_167013||full Not-determined
18-104-final-viral-combined_k141_139314||full Not-determined
18-104-final-viral-combined_k141_160214||full Low-quality
18-104-final-viral-combined_k141_48768||full Not-determined
18-104-final-viral-combined_k141_34844||full Low-quality
18-104-final-viral-combined_k141_20921||full Low-quality
18-104-final-viral-combined_k141_146307||full Not-determined
18-104-final-viral-combined_k141_20923||full Low-quality
18-104-final-viral-combined_k141_48774||full Low-quality
18-104-final-viral-combined_k141_48777||full Not-determined
18-104-final-viral-combined_k141_146310||full Low-quality
18-104-final-viral-combined_k141_48787||full Low-quality
18-104-final-viral-combined_k141_90588||full Low-quality
18-104-final-viral-combined_k141_90589||full Not-determined
18-104-final-viral-combined_k141_34881||full Low-quality
18-104-final-viral-combined_k141_104527||full Not-determined
18-104-final-viral-combined_k141_34883||full Not-determined
18-104-final-viral-combined_k141_139357||full Low-quality
18-104-final-viral-combined_k141_34888||full Low-quality
18-104-final-viral-combined_k141_49||full Low-quality
18-104-final-viral-combined_k141_118441||full Not-determined
18-104-final-viral-combined_k141_20951||full Not-determined
18-104-final-viral-combined_k141_125403||full Low-quality
18-104-final-viral-combined_k141_69721||full Not-determined
18-104-final-viral-combined_k141_41824||full Not-determined
18-104-final-viral-combined_k141_132417||full Low-quality
18-104-final-viral-combined_k141_20958||full Not-determined
18-104-final-viral-combined_k141_62769||full Low-quality
18-104-final-viral-combined_k141_118449||full Not-determined
18-104-final-viral-combined_k141_125409||full Low-quality
18-104-final-viral-combined_k141_90621||full Not-determined
18-104-final-viral-combined_k141_41833||full Not-determined
18-104-final-viral-combined_k141_118451||full Not-determined
18-104-final-viral-combined_k141_69730||full Low-quality
18-104-final-viral-combined_k141_153301||full Not-determined
18-104-final-viral-combined_k141_125412||full Low-quality
18-104-final-viral-combined_k141_20970||full Low-quality
18-104-final-viral-combined_k141_48816||full Low-quality
18-104-final-viral-combined_k141_69738||full Low-quality
18-104-final-viral-combined_k141_27927||full Not-determined
18-104-final-viral-combined_k141_153311||full Not-determined
18-104-final-viral-combined_k141_27932||full Low-quality
18-104-final-viral-combined_k141_20982||full Low-quality
18-104-final-viral-combined_k141_125428||full Not-determined
18-104-final-viral-combined_k141_139388||full Not-determined
18-104-final-viral-combined_k141_97566||full Low-quality
18-104-final-viral-combined_k141_153320||full Not-determined
18-104-final-viral-combined_k141_83655||full Not-determined
18-104-final-viral-combined_k141_111522||full Not-determined
18-104-final-viral-combined_k141_132459||full Not-determined
18-104-final-viral-combined_k141_125460||full Low-quality
18-104-final-viral-combined_k141_34944||full Low-quality
18-104-final-viral-combined_k141_41872||full Not-determined
18-104-final-viral-combined_k141_160274||full Low-quality
18-104-final-viral-combined_k141_111541||full Low-quality
18-104-final-viral-combined_k141_139||full Low-quality
18-104-final-viral-combined_k141_62813||full Not-determined
18-104-final-viral-combined_k141_125490||full Not-determined
18-104-final-viral-combined_k141_160297||full Not-determined
18-104-final-viral-combined_k141_104604||full Low-quality
18-104-final-viral-combined_k141_76767||full Low-quality
18-104-final-viral-combined_k141_69810||full Low-quality
18-104-final-viral-combined_k141_152||full Not-determined
18-104-final-viral-combined_k141_90704||full Not-determined
18-104-final-viral-combined_k141_62826||full Low-quality
18-104-final-viral-combined_k141_139458||full Not-determined
18-104-final-viral-combined_k141_111586||full Low-quality
18-104-final-viral-combined_k141_146354||full Not-determined
18-104-final-viral-combined_k141_90720||full Low-quality
18-104-final-viral-combined_k141_69820||full Low-quality
18-104-final-viral-combined_k141_83700||full Low-quality
18-104-final-viral-combined_k141_69822||full Low-quality
18-104-final-viral-combined_k141_69827||full Not-determined
18-104-final-viral-combined_k141_90727||full Low-quality
18-104-final-viral-combined_k141_104646||full Not-determined
18-104-final-viral-combined_k141_7027||full Low-quality
18-104-final-viral-combined_k141_175||full Not-determined
18-104-final-viral-combined_k141_21036||full Not-determined
18-104-final-viral-combined_k141_177||full Low-quality

18-104-final-viral-combined_k141_139493||full Not-determined
18-104-final-viral-combined_k141_27969||full Not-determined
18-104-final-viral-combined_k141_76784||full Low-quality
18-104-final-viral-combined_k141_14051||full Low-quality
18-104-final-viral-combined_k141_14053||full Low-quality
18-104-final-viral-combined_k141_14054||full Not-determined
18-104-final-viral-combined_k141_76787||full Not-determined
18-104-final-viral-combined_k141_62860||full Not-determined
18-104-final-viral-combined_k141_76791||full Not-determined
18-104-final-viral-combined_k141_27992||full Low-quality
18-104-final-viral-combined_k141_7062||full Not-determined
18-104-final-viral-combined_k141_90756||full Low-quality
18-104-final-viral-combined_k141_76797||full Low-quality
18-104-final-viral-combined_k141_90758||full Not-determined
18-104-final-viral-combined_k141_48878||full Not-determined
18-104-final-viral-combined_k141_62885||full Not-determined
18-104-final-viral-combined_k141_55806||full Not-determined
18-104-final-viral-combined_k141_139520||full Not-determined
18-104-final-viral-combined_k141_14074||full Low-quality
18-104-final-viral-combined_k141_90772||full Not-determined
18-104-final-viral-combined_k141_132515||full Low-quality
18-104-final-viral-combined_k141_28003||full Low-quality
18-104-final-viral-combined_k141_55824||full Not-determined
18-104-final-viral-combined_k141_111678||full Not-determined
18-104-final-viral-combined_k141_83770||full Not-determined
18-104-final-viral-combined_k141_125562||full Low-quality
18-104-final-viral-combined_k141_160361||full Not-determined
18-104-final-viral-combined_k141_125563||full Not-determined
18-104-final-viral-combined_k141_28017||full Not-determined
18-104-final-viral-combined_k141_132532||full Not-determined
18-109_k141_80255 Medium-quality
18-104-final-viral-combined_k141_153396||full Low-quality
18-104-final-viral-combined_k141_7135||full Low-quality
18-104-final-viral-combined_k141_14104||full Not-determined
18-104-final-viral-combined_k141_274||full Low-quality
18-104-final-viral-combined_k141_62922||full Not-determined
18-104-final-viral-combined_k141_83789||full Low-quality
18-104-final-viral-combined_k141_160394||full Low-quality
18-104-final-viral-combined_k141_62926||full Not-determined
18-104-final-viral-combined_k141_153419||full Not-determined
18-104-final-viral-combined_k141_7146||full Not-determined
18-104-final-viral-combined_k141_139554||full Low-quality
18-104-final-viral-combined_k141_146388||full Low-quality
18-104-final-viral-combined_k141_69955||full Not-determined
18-104-final-viral-combined_k141_7155||full Low-quality
18-104-final-viral-combined_k141_97733||full Not-determined
18-104-final-viral-combined_k141_125578||full Not-determined
18-104-final-viral-combined_k141_125584||full Not-determined
18-104-final-viral-combined_k141_41997||full Low-quality
18-104-final-viral-combined_k141_111719||full Low-quality
18-104-final-viral-combined_k141_69975||full Low-quality
18-104-final-viral-combined_k141_111723||full Not-determined
18-104-final-viral-combined_k141_321||full Low-quality
18-104-final-viral-combined_k141_28035||full Not-determined
18-104-final-viral-combined_k141_83831||full Not-determined
18-104-final-viral-combined_k141_69991||full Not-determined
18-104-final-viral-combined_k141_62966||full Low-quality
18-104-final-viral-combined_k141_338||full Not-determined
18-104-final-viral-combined_k141_104762||full Low-quality
18-104-final-viral-combined_k141_104763||full Not-determined
18-104-final-viral-combined_k141_48940||full Low-quality
18-104-final-viral-combined_k141_42026||full Low-quality
18-104-final-viral-combined_k141_125636||full Not-determined
18-104-final-viral-combined_k141_146449||full Not-determined
18-104-final-viral-combined_k141_139600||full Low-quality
18-104-final-viral-combined_k141_42029||full Not-determined
18-104-final-viral-combined_k141_365||full Not-determined
18-104-final-viral-combined_k141_146468||full Not-determined
18-104-final-viral-combined_k141_118602||full Not-determined
18-104-final-viral-combined_k141_97785||full Low-quality
18-104-final-viral-combined_k141_21132||full Low-quality
18-104-final-viral-combined_k141_28077||full Not-determined
18-104-final-viral-combined_k141_160450||full Not-determined
18-104-final-viral-combined_k141_28099||full Low-quality
18-104-final-viral-combined_k141_104799||full Low-quality
18-104-final-viral-combined_k141_28101||full Not-determined
18-104-final-viral-combined_k141_14157||full Low-quality
18-104-final-viral-combined_k141_160459||full Low-quality
18-104-final-viral-combined_k141_70030||full Not-determined
18-104-final-viral-combined_k141_90914||full Not-determined
18-104-final-viral-combined_k141_55932||full Low-quality
18-104-final-viral-combined_k141_42052||full Low-quality
18-104-final-viral-combined_k141_48977||full Not-determined
18-104-final-viral-combined_k141_63016||full Low-quality
18-104-final-viral-combined_k141_42055||full Low-quality
18-104-final-viral-combined_k141_14176||full Low-quality
18-104-final-viral-combined_k141_146506||full Low-quality
18-104-final-viral-combined_k141_139654||full Not-determined
18-104-final-viral-combined_k141_118651||full Not-determined
18-104-final-viral-combined_k141_55954||full Not-determined
18-104-final-viral-combined_k141_63034||full Not-determined
18-104-final-viral-combined_k141_139665||full Low-quality
18-104-final-viral-combined_k141_63041||full Not-determined
18-104-final-viral-combined_k141_139669||full Low-quality
18-104-final-viral-combined_k141_83897||full Low-quality
18-104-final-viral-combined_k141_83898||full Not-determined
18-104-final-viral-combined_k141_104841||full Not-determined
18-104-final-viral-combined_k141_76931||full Not-determined
18-104-final-viral-combined_k141_14212||full Low-quality
18-104-final-viral-combined_k141_14223||full Low-quality
18-104-final-viral-combined_k141_104855||full Not-determined
18-104-final-viral-combined_k141_63059||full Low-quality
18-104-final-viral-combined_k141_76970||full Not-determined
18-104-final-viral-combined_k141_139687||full Low-quality
18-104-final-viral-combined_k141_146537||full Low-quality
18-104-final-viral-combined_k141_63065||full Not-determined
18-104-final-viral-combined_k141_28188||full Not-determined
18-104-final-viral-combined_k141_28189||full Not-determined
18-104-final-viral-combined_k141_465||full Not-determined
18-104-final-viral-combined_k141_76979||full Low-quality
18-104-final-viral-combined_k141_111878||full Not-determined
18-104-final-viral-combined_k141_56005||full Not-determined
18-104-final-viral-combined_k141_139695||full Not-determined
18-104-final-viral-combined_k141_104868||full Not-determined
18-104-final-viral-combined_k141_83960||full Low-quality
18-104-final-viral-combined_k141_56019||full Not-determined
18-104-final-viral-combined_k141_7310||full Not-determined
18-104-final-viral-combined_k141_14276||full Not-determined
18-104-final-viral-combined_k141_28204||full Not-determined
18-104-final-viral-combined_k141_63078||full Not-determined
18-104-final-viral-combined_k141_91024||full Not-determined
18-104-final-viral-combined_k141_97909||full Low-quality
18-104-final-viral-combined_k141_42153||full Not-determined
18-104-final-viral-combined_k141_28218||full Not-determined
18-104-final-viral-combined_k141_139730||full Low-quality
18-104-final-viral-combined_k141_42159||full Low-quality
18-104-final-viral-combined_k141_77010||full Low-quality
18-104-final-viral-combined_k141_146589||full Low-quality
18-104-final-viral-combined_k141_56055||full Low-quality
18-104-final-viral-combined_k141_104885||full Not-determined
18-104-final-viral-combined_k141_7317||full Low-quality
18-104-final-viral-combined_k141_500||full Not-determined
18-104-final-viral-combined_k141_104886||full Not-determined
18-104-final-viral-combined_k141_91040||full Not-determined
18-104-final-viral-combined_k141_42181||full Not-determined
18-104-final-viral-combined_k141_42189||full Not-determined
18-104-final-viral-combined_k141_91041||full Low-quality
18-104-final-viral-combined_k141_104892||full Not-determined
18-104-final-viral-combined_k141_501||full Low-quality
18-104-final-viral-combined_k141_104903||full Not-determined
18-104-final-viral-combined_k141_104904||full Low-quality
17-108-final-viral-combined_k141_49632||full Medium-quality
18-104-final-viral-combined_k141_160564||full Not-determined
18-104-final-viral-combined_k141_146615||full Not-determined
18-104-final-viral-combined_k141_49045||full Low-quality
18-104-final-viral-combined_k141_511||full Low-quality
18-104-final-viral-combined_k141_97923||full Low-quality
18-104-final-viral-combined_k141_125813||full Low-quality
18-104-final-viral-combined_k141_160603||full Low-quality
18-104-final-viral-combined_k141_146645||full Not-determined
18-104-final-viral-combined_k141_42225||full Not-determined
18-104-final-viral-combined_k141_42227||full Low-quality
18-104-final-viral-combined_k141_70134||full Not-determined
18-104-final-viral-combined_k141_104933||full Low-quality
18-104-final-viral-combined_k141_104937||full Not-determined
18-104-final-viral-combined_k141_84028||full Not-determined
18-104-final-viral-combined_k141_70148||full Not-determined
18-104-final-viral-combined_k141_536||full Not-determined
18-104-final-viral-combined_k141_132705||full Low-quality
18-104-final-viral-combined_k141_160636||full Low-quality
18-104-final-viral-combined_k141_97939||full Not-determined
18-104-final-viral-combined_k141_146651||full Low-quality
18-104-final-viral-combined_k141_21298||full Not-determined
18-104-final-viral-combined_k141_97943||full Not-determined
18-104-final-viral-combined_k141_118714||full Low-quality
18-104-final-viral-combined_k141_139792||full Not-determined
18-104-final-viral-combined_k141_132708||full Low-quality
18-104-final-viral-combined_k141_139794||full Not-determined
18-104-final-viral-combined_k141_28303||full Low-quality
18-104-final-viral-combined_k141_132714||full Not-determined
18-104-final-viral-combined_k141_111985||full Not-determined
18-104-final-viral-combined_k141_91095||full Not-determined
18-104-final-viral-combined_k141_91096||full Low-quality
18-104-final-viral-combined_k141_97961||full Low-quality
18-104-final-viral-combined_k141_125860||full Low-quality
18-104-final-viral-combined_k141_42236||full Not-determined
18-104-final-viral-combined_k141_125861||full Low-quality
18-104-final-viral-combined_k141_118730||full Low-quality
18-104-final-viral-combined_k141_28315||full Not-determined
18-104-final-viral-combined_k141_104970||full Low-quality
18-104-final-viral-combined_k141_84043||full Low-quality
18-104-final-viral-combined_k141_570||full Low-quality
18-104-final-viral-combined_k141_63174||full Not-determined
18-104-final-viral-combined_k141_572||full Low-quality
18-104-final-viral-combined_k141_146670||full Low-quality
18-104-final-viral-combined_k141_77135||full Low-quality
18-104-final-viral-combined_k141_132725||full Low-quality
18-104-final-viral-combined_k141_125871||full Low-quality
18-104-final-viral-combined_k141_63181||full Not-determined
18-104-final-viral-combined_k141_21317||full Low-quality
18-104-final-viral-combined_k141_56157||full Not-determined
18-104-final-viral-combined_k141_84054||full Not-determined
18-104-final-viral-combined_k141_70200||full Not-determined
18-104-final-viral-combined_k141_160656||full Low-quality
18-104-final-viral-combined_k141_112010||full Not-determined
18-104-final-viral-combined_k141_21329||full Not-determined
18-104-final-viral-combined_k141_112011||full Not-determined
18-104-final-viral-combined_k141_125882||full Not-determined
18-104-final-viral-combined_k141_118753||full Not-determined
18-104-final-viral-combined_k141_132741||full Low-quality
18-104-final-viral-combined_k141_118755||full Low-quality
18-104-final-viral-combined_k141_7350||full Low-quality
18-104-final-viral-combined_k141_125902||full Low-quality
18-104-final-viral-combined_k141_125908||full Not-determined
18-104-final-viral-combined_k141_160678||full Low-quality
18-104-final-viral-combined_k141_118767||full Not-determined
18-104-final-viral-combined_k141_104993||full Low-quality
18-104-final-viral-combined_k141_42289||full Not-determined
18-104-final-viral-combined_k141_139830||full Low-quality
18-104-final-viral-combined_k141_70222||full Not-determined
18-104-final-viral-combined_k141_42300||full Low-quality
18-104-final-viral-combined_k141_98019||full Not-determined
18-104-final-viral-combined_k141_118772||full Not-determined
18-104-final-viral-combined_k141_84127||full Low-quality
18-104-final-viral-combined_k141_125961||full Not-determined
18-104-final-viral-combined_k141_617||full Low-quality
18-104-final-viral-combined_k141_160702||full Low-quality
18-104-final-viral-combined_k141_105011||full Not-determined
18-104-final-viral-combined_k141_21357||full Not-determined
18-104-final-viral-combined_k141_153625||full Not-determined
18-104-final-viral-combined_k141_91182||full Not-determined
18-104-final-viral-combined_k141_125962||full Not-determined
18-104-final-viral-combined_k141_21360||full Not-determined
18-104-final-viral-combined_k141_70253||full Low-quality
18-104-final-viral-combined_k141_105020||full Low-quality
18-104-final-viral-combined_k141_70266||full Low-quality
18-104-final-viral-combined_k141_77220||full Not-determined
18-104-final-viral-combined_k141_70268||full Not-determined
18-104-final-viral-combined_k141_633||full Low-quality
18-104-final-viral-combined_k141_139878||full Not-determined
18-104-final-viral-combined_k141_146711||full Not-determined
18-104-final-viral-combined_k141_643||full Not-determined
18-104-final-viral-combined_k141_105037||full Low-quality
18-104-final-viral-combined_k141_84170||full Not-determined
18-104-final-viral-combined_k141_91198||full Low-quality
18-104-final-viral-combined_k141_125980||full Low-quality
18-104-final-viral-combined_k141_91203||full Not-determined
18-104-final-viral-combined_k141_42353||full Low-quality
18-104-final-viral-combined_k141_14409||full Not-determined
18-104-final-viral-combined_k141_77255||full Low-quality
18-104-final-viral-combined_k141_91219||full Low-quality
18-104-final-viral-combined_k141_91221||full Low-quality
18-104-final-viral-combined_k141_153681||full Not-determined
18-104-final-viral-combined_k141_98092||full Low-quality
18-104-final-viral-combined_k141_687||full Low-quality
18-104-final-viral-combined_k141_21413||full Low-quality
18-104-final-viral-combined_k141_91231||full Not-determined
18-104-final-viral-combined_k141_70316||full Low-quality
18-104-final-viral-combined_k141_160743||full Low-quality
18-104-final-viral-combined_k141_146753||full Low-quality
18-104-final-viral-combined_k141_694||full Not-determined
18-104-final-viral-combined_k141_153699||full Low-quality
18-104-final-viral-combined_k141_160754||full Low-quality
18-104-final-viral-combined_k141_702||full Not-determined
18-104-final-viral-combined_k141_98112||full Not-determined
18-104-final-viral-combined_k141_105080||full Low-quality
18-104-final-viral-combined_k141_112079||full Not-determined
18-104-final-viral-combined_k141_77280||full Not-determined
18-104-final-viral-combined_k141_28444||full Low-quality
18-104-final-viral-combined_k141_118833||full Low-quality
18-110-final-viral-combined_k141_66195||full Medium-quality
18-104-final-viral-combined_k141_70346||full Not-determined
18-104-final-viral-combined_k141_118842||full Low-quality
18-104-final-viral-combined_k141_112091||full Not-determined
18-104-final-viral-combined_k141_132828||full Low-quality
18-104-final-viral-combined_k141_49209||full Not-determined
18-104-final-viral-combined_k141_84214||full Not-determined
18-104-final-viral-combined_k141_139951||full Low-quality
18-104-final-viral-combined_k141_14441||full Low-quality
18-104-final-viral-combined_k141_118848||full Not-determined
18-104-final-viral-combined_k141_139961||full Low-quality
18-104-final-viral-combined_k141_14446||full Not-determined
18-104-final-viral-combined_k141_49221||full Not-determined
18-104-final-viral-combined_k141_77319||full Not-determined
18-109-final-viral-combined_k141_80255||full Medium-quality
18-104-final-viral-combined_k141_118877||full Not-determined
18-104-final-viral-combined_k141_49245||full Not-determined
18-104-final-viral-combined_k141_118879||full Not-determined
18-104-final-viral-combined_k141_70365||full Not-determined
18-104-final-viral-combined_k141_160823||full Low-quality
18-104-final-viral-combined_k141_91333||full Low-quality
18-104-final-viral-combined_k141_160831||full Low-quality
18-104-final-viral-combined_k141_146845||full Low-quality
18-104-final-viral-combined_k141_132852||full Low-quality
18-104-final-viral-combined_k141_42406||full Low-quality
18-104-final-viral-combined_k141_140003||full Low-quality
18-104-final-viral-combined_k141_42418||full Low-quality
18-104-final-viral-combined_k141_21501||full Not-determined
18-104-final-viral-combined_k141_49269||full Not-determined
18-104-final-viral-combined_k141_70410||full Low-quality
18-104-final-viral-combined_k141_56278||full Not-determined
18-104-final-viral-combined_k141_126140||full Low-quality
18-104-final-viral-combined_k141_140014||full Low-quality
18-104-final-viral-combined_k141_140017||full Low-quality
18-104-final-viral-combined_k141_21512||full Not-determined
18-104-final-viral-combined_k141_118922||full Not-determined
18-104-final-viral-combined_k141_91370||full Not-determined
18-104-final-viral-combined_k141_35282||full Not-determined
18-104-final-viral-combined_k141_35284||full Low-quality
18-104-final-viral-combined_k141_35287||full Low-quality
18-104-final-viral-combined_k141_84318||full Low-quality
18-104-final-viral-combined_k141_91374||full Not-determined
18-104-final-viral-combined_k141_118932||full Low-quality
18-104-final-viral-combined_k141_14486||full Not-determined
18-104-final-viral-combined_k141_105180||full Not-determined
18-104-final-viral-combined_k141_7486||full Low-quality
18-104-final-viral-combined_k141_112144||full Low-quality
18-104-final-viral-combined_k141_778||full Not-determined
18-104-final-viral-combined_k141_781||full Not-determined
18-104-final-viral-combined_k141_21539||full Low-quality
18-104-final-viral-combined_k141_49308||full Not-determined
18-104-final-viral-combined_k141_84355||full Not-determined
18-104-final-viral-combined_k141_70459||full Not-determined
18-104-final-viral-combined_k141_14514||full Low-quality
18-104-final-viral-combined_k141_105213||full Not-determined
18-104-final-viral-combined_k141_56340||full Low-quality
18-104-final-viral-combined_k141_787||full Not-determined
18-104-final-viral-combined_k141_35296||full Not-determined
18-104-final-viral-combined_k141_42486||full Low-quality
18-104-final-viral-combined_k141_28555||full Not-determined
18-104-final-viral-combined_k141_56345||full Low-quality
18-104-final-viral-combined_k141_105247||full Not-determined
18-104-final-viral-combined_k141_91416||full Low-quality
18-104-final-viral-combined_k141_132918||full Not-determined
18-104-final-viral-combined_k141_63345||full Not-determined
18-104-final-viral-combined_k141_63347||full Not-determined
18-104-final-viral-combined_k141_815||full Not-determined
18-104-final-viral-combined_k141_21590||full Low-quality
18-104-final-viral-combined_k141_140082||full Not-determined
18-104-final-viral-combined_k141_91432||full Low-quality
18-104-final-viral-combined_k141_153871||full Not-determined
18-104-final-viral-combined_k141_140086||full Not-determined
18-104-final-viral-combined_k141_21592||full Low-quality
18-104-final-viral-combined_k141_98266||full Low-quality
18-104-final-viral-combined_k141_21599||full Low-quality
18-104-final-viral-combined_k141_153881||full Low-quality
18-104-final-viral-combined_k141_105288||full Low-quality
18-104-final-viral-combined_k141_153883||full Not-determined
18-104-final-viral-combined_k141_146944||full Not-determined
18-104-final-viral-combined_k141_105301||full Low-quality
18-104-final-viral-combined_k141_105304||full Low-quality
18-104-final-viral-combined_k141_98272||full Not-determined
18-104-final-viral-combined_k141_42548||full Not-determined
18-104-final-viral-combined_k141_42562||full Not-determined
18-104-final-viral-combined_k141_63394||full Low-quality
18-104-final-viral-combined_k141_140113||full Not-determined
18-104-final-viral-combined_k141_56378||full Low-quality
18-104-final-viral-combined_k141_146970||full Low-quality
18-104-final-viral-combined_k141_105327||full Low-quality
18-104-final-viral-combined_k141_63399||full Low-quality
18-104-final-viral-combined_k141_867||full Not-determined
18-104-final-viral-combined_k141_42587||full Low-quality
18-104-final-viral-combined_k141_49342||full Not-determined
18-104-final-viral-combined_k141_7614||full Not-determined
18-104-final-viral-combined_k141_105356||full Not-determined
18-104-final-viral-combined_k141_70537||full Low-quality
18-104-final-viral-combined_k141_126313||full Low-quality
18-104-final-viral-combined_k141_112248||full Not-determined
18-104-final-viral-combined_k141_56417||full Not-determined
18-104-final-viral-combined_k141_70561||full Low-quality
18-104-final-viral-combined_k141_91472||full Not-determined
18-104-final-viral-combined_k141_70572||full Low-quality
18-104-final-viral-combined_k141_63447||full Low-quality
18-104-final-viral-combined_k141_91481||full Not-determined
18-104-final-viral-combined_k141_28625||full Not-determined
18-104-final-viral-combined_k141_98332||full Low-quality
18-104-final-viral-combined_k141_63455||full Not-determined
18-104-final-viral-combined_k141_14536||full Low-quality
18-104-final-viral-combined_k141_35396||full Not-determined
18-104-final-viral-combined_k141_119041||full Low-quality
18-104-final-viral-combined_k141_28632||full Not-determined
18-104-final-viral-combined_k141_91488||full Not-determined
18-104-final-viral-combined_k141_28635||full Low-quality
18-104-final-viral-combined_k141_119048||full Not-determined
18-104-final-viral-combined_k141_77436||full Low-quality
18-104-final-viral-combined_k141_890||full Low-quality
18-104-final-viral-combined_k141_132997||full Low-quality
18-104-final-viral-combined_k141_147027||full Low-quality
18-104-final-viral-combined_k141_21661||full Low-quality
18-104-final-viral-combined_k141_35418||full Not-determined
18-104-final-viral-combined_k141_153992||full Not-determined
18-104-final-viral-combined_k141_70629||full Low-quality
18-104-final-viral-combined_k141_91546||full Low-quality
18-104-final-viral-combined_k141_56476||full Not-determined
18-104-final-viral-combined_k141_126371||full Not-determined
18-104-final-viral-combined_k141_154002||full Low-quality
18-104-final-viral-combined_k141_14592||full Not-determined
18-104-final-viral-combined_k141_70643||full Low-quality
18-104-final-viral-combined_k141_147053||full Low-quality
18-104-final-viral-combined_k141_140185||full Low-quality
18-104-final-viral-combined_k141_28679||full Low-quality
18-110-final-viral-combined_k141_180957||full Medium-quality
18-104-final-viral-combined_k141_915||full Not-determined
18-104-final-viral-combined_k141_35450||full Not-determined
18-104-final-viral-combined_k141_28694||full Not-determined
18-104-final-viral-combined_k141_77496||full Not-determined
18-104-final-viral-combined_k141_56532||full Low-quality
18-104-final-viral-combined_k141_140199||full Low-quality
18-104-final-viral-combined_k141_7651||full Low-quality
18-104-final-viral-combined_k141_133055||full Not-determined
18-104-final-viral-combined_k141_28720||full Low-quality
18-104-final-viral-combined_k141_935||full Low-quality
18-104-final-viral-combined_k141_119089||full Not-determined
18-104-final-viral-combined_k141_14616||full Low-quality
18-104-final-viral-combined_k141_77530||full Not-determined
18-104-final-viral-combined_k141_35472||full Low-quality
18-104-final-viral-combined_k141_147089||full Not-determined
18-104-final-viral-combined_k141_154051||full Low-quality
18-104-final-viral-combined_k141_105505||full Not-determined
18-104-final-viral-combined_k141_147104||full Not-determined
18-104-final-viral-combined_k141_140224||full Low-quality
18-104-final-viral-combined_k141_42681||full Low-quality
18-104-final-viral-combined_k141_961||full Not-determined
18-104-final-viral-combined_k141_84562||full Low-quality
18-104-final-viral-combined_k141_105507||full Low-quality
18-104-final-viral-combined_k141_77551||full Low-quality
18-104-final-viral-combined_k141_70657||full Not-determined
18-104-final-viral-combined_k141_119125||full Not-determined
18-104-final-viral-combined_k141_140245||full Not-determined
18-104-final-viral-combined_k141_160940||full Not-determined
18-104-final-viral-combined_k141_70673||full Not-determined
18-104-final-viral-combined_k141_154077||full Low-quality
18-104-final-viral-combined_k141_133103||full Not-determined
18-104-final-viral-combined_k141_49445||full Not-determined
18-104-final-viral-combined_k141_105544||full Not-determined
18-104-final-viral-combined_k141_140275||full Not-determined
18-104-final-viral-combined_k141_91658||full Not-determined
18-104-final-viral-combined_k141_140280||full Low-quality
18-104-final-viral-combined_k141_21708||full Not-determined
18-104-final-viral-combined_k141_56574||full Low-quality
18-104-final-viral-combined_k141_84606||full Low-quality
18-104-final-viral-combined_k141_91674||full Low-quality
18-104-final-viral-combined_k141_35552||full Low-quality
18-104-final-viral-combined_k141_133134||full Not-determined
18-104-final-viral-combined_k141_112441||full Not-determined
18-104-final-viral-combined_k141_126572||full Not-determined
18-104-final-viral-combined_k141_154113||full Low-quality
18-104-final-viral-combined_k141_91701||full Not-determined
18-104-final-viral-combined_k141_98421||full Low-quality
18-104-final-viral-combined_k141_112463||full Low-quality
18-104-final-viral-combined_k141_21756||full Low-quality
18-104-final-viral-combined_k141_1019||full Low-quality
18-104-final-viral-combined_k141_1020||full Low-quality
18-104-final-viral-combined_k141_21761||full Low-quality
18-104-final-viral-combined_k141_133165||full Low-quality
18-104-final-viral-combined_k141_160993||full Low-quality
18-104-final-viral-combined_k141_147200||full Low-quality
18-104-final-viral-combined_k141_126606||full Not-determined
18-104-final-viral-combined_k141_112496||full Not-determined
18-104-final-viral-combined_k141_98435||full Low-quality
18-104-final-viral-combined_k141_21779||full Not-determined
18-104-final-viral-combined_k141_7728||full Low-quality
18-104-final-viral-combined_k141_7730||full Low-quality
18-104-final-viral-combined_k141_7738||full Not-determined
18-104-final-viral-combined_k141_77650||full Not-determined
18-104-final-viral-combined_k141_119224||full Low-quality
18-104-final-viral-combined_k141_28891||full Not-determined
18-104-final-viral-combined_k141_35605||full Not-determined
18-104-final-viral-combined_k141_14721||full Low-quality
18-104-final-viral-combined_k141_70715||full Low-quality
18-104-final-viral-combined_k141_77670||full Low-quality
18-104-final-viral-combined_k141_42751||full Not-determined
18-104-final-viral-combined_k141_119236||full Not-determined
18-104-final-viral-combined_k141_154216||full Low-quality
18-104-final-viral-combined_k141_28913||full Not-determined
18-104-final-viral-combined_k141_140375||full Not-determined
18-104-final-viral-combined_k141_98469||full Not-determined
18-104-final-viral-combined_k141_77682||full Not-determined
18-104-final-viral-combined_k141_119246||full Low-quality
18-104-final-viral-combined_k141_84667||full Not-determined
18-104-final-viral-combined_k141_1058||full Low-quality
18-104-final-viral-combined_k141_98475||full Not-determined
18-104-final-viral-combined_k141_161050||full Low-quality
18-104-final-viral-combined_k141_56598||full Low-quality
18-104-final-viral-combined_k141_42772||full Low-quality
18-104-final-viral-combined_k141_147263||full Not-determined
18-104-final-viral-combined_k141_154230||full Not-determined
18-104-final-viral-combined_k141_147266||full Not-determined
18-104-final-viral-combined_k141_14741||full Low-quality
18-104-final-viral-combined_k141_98495||full Not-determined
18-104-final-viral-combined_k141_105648||full Not-determined
18-104-final-viral-combined_k141_147268||full Low-quality
18-104-final-viral-combined_k141_84691||full Not-determined
18-104-final-viral-combined_k141_63720||full Not-determined
18-104-final-viral-combined_k141_98497||full Low-quality
18-104-final-viral-combined_k141_42788||full Low-quality
18-104-final-viral-combined_k141_42789||full Not-determined
18-104-final-viral-combined_k141_140395||full Low-quality
18-104-final-viral-combined_k141_119294||full Not-determined
18-104-final-viral-combined_k141_140403||full Not-determined
18-104-final-viral-combined_k141_1069||full Not-determined
18-104-final-viral-combined_k141_112546||full Not-determined
18-104-final-viral-combined_k141_28977||full Not-determined
18-104-final-viral-combined_k141_140412||full Not-determined
18-104-final-viral-combined_k141_105681||full Not-determined
18-104-final-viral-combined_k141_1074||full Not-determined
18-104-final-viral-combined_k141_140413||full Not-determined
18-104-final-viral-combined_k141_84713||full Not-determined
18-104-final-viral-combined_k141_49474||full Low-quality
18-104-final-viral-combined_k141_112547||full Low-quality
18-104-final-viral-combined_k141_112548||full Not-determined
18-104-final-viral-combined_k141_7817||full Not-determined
18-104-final-viral-combined_k141_35660||full Low-quality
18-104-final-viral-combined_k141_28988||full Not-determined
18-104-final-viral-combined_k141_56661||full Low-quality
18-104-final-viral-combined_k141_56663||full Not-determined
18-104-final-viral-combined_k141_140440||full Low-quality
18-104-final-viral-combined_k141_7823||full Not-determined
18-104-final-viral-combined_k141_49489||full Not-determined
18-104-final-viral-combined_k141_112568||full Low-quality
18-104-final-viral-combined_k141_49493||full Low-quality
18-104-final-viral-combined_k141_7832||full Low-quality
18-104-final-viral-combined_k141_91788||full Not-determined
18-104-final-viral-combined_k141_161129||full Not-determined
18-104-final-viral-combined_k141_49501||full Low-quality
18-104-final-viral-combined_k141_29007||full Low-quality
18-104-final-viral-combined_k141_119354||full Not-determined
18-104-final-viral-combined_k141_35689||full Not-determined
18-104-final-viral-combined_k141_126705||full Low-quality
18-104-final-viral-combined_k141_56690||full Low-quality
18-104-final-viral-combined_k141_140461||full Not-determined
18-104-final-viral-combined_k141_126707||full Low-quality
18-104-final-viral-combined_k141_63772||full Not-determined
18-104-final-viral-combined_k141_140469||full Low-quality
18-104-final-viral-combined_k141_126715||full Not-determined
18-104-final-viral-combined_k141_42871||full Low-quality
18-104-final-viral-combined_k141_98558||full Low-quality
18-104-final-viral-combined_k141_56720||full Not-determined
18-104-final-viral-combined_k141_112627||full Low-quality
18-104-final-viral-combined_k141_91816||full Not-determined
18-104-final-viral-combined_k141_133348||full Not-determined
18-104-final-viral-combined_k141_77810||full Not-determined
18-104-final-viral-combined_k141_112641||full Low-quality
18-104-final-viral-combined_k141_14756||full Low-quality
18-104-final-viral-combined_k141_21837||full Not-determined
18-104-final-viral-combined_k141_42896||full Not-determined
18-104-final-viral-combined_k141_133357||full Low-quality
18-104-final-viral-combined_k141_91844||full Low-quality
18-104-final-viral-combined_k141_98590||full Low-quality
18-104-final-viral-combined_k141_133363||full Not-determined
18-104-final-viral-combined_k141_154321||full Not-determined
18-104-final-viral-combined_k141_1185||full Not-determined
18-104-final-viral-combined_k141_70807||full Low-quality
18-104-final-viral-combined_k141_91866||full Not-determined
18-104-final-viral-combined_k141_91869||full Not-determined
18-104-final-viral-combined_k141_140510||full Low-quality
18-104-final-viral-combined_k141_91886||full Low-quality
18-104-final-viral-combined_k141_140517||full Not-determined
18-104-final-viral-combined_k141_49510||full Not-determined
18-104-final-viral-combined_k141_29071||full Not-determined
18-104-final-viral-combined_k141_63845||full Not-determined
18-104-final-viral-combined_k141_29074||full Low-quality
18-104-final-viral-combined_k141_140527||full Not-determined
18-104-final-viral-combined_k141_91905||full Low-quality
18-104-final-viral-combined_k141_77914||full Low-quality
18-104-final-viral-combined_k141_35785||full Low-quality
18-104-final-viral-combined_k141_112716||full Not-determined
18-104-final-viral-combined_k141_21904||full Low-quality
18-104-final-viral-combined_k141_84810||full Not-determined
18-104-final-viral-combined_k141_133417||full Low-quality
18-104-final-viral-combined_k141_126798||full Not-determined
18-104-final-viral-combined_k141_91922||full Not-determined
18-104-final-viral-combined_k141_35801||full Not-determined
18-104-final-viral-combined_k141_112742||full Not-determined
18-104-final-viral-combined_k141_21925||full Low-quality
18-104-final-viral-combined_k141_147411||full Not-determined
18-104-final-viral-combined_k141_140552||full Not-determined
18-104-final-viral-combined_k141_133437||full Not-determined
18-104-final-viral-combined_k141_7966||full Low-quality
18-104-final-viral-combined_k141_119507||full Low-quality
18-104-final-viral-combined_k141_98639||full Low-quality
18-104-final-viral-combined_k141_49527||full Low-quality
18-104-final-viral-combined_k141_7973||full Low-quality
18-104-final-viral-combined_k141_147422||full Low-quality
18-104-final-viral-combined_k141_29132||full Low-quality
18-104-final-viral-combined_k141_140565||full Not-determined
18-104-final-viral-combined_k141_70836||full Low-quality
18-104-final-viral-combined_k141_98644||full Low-quality
18-104-final-viral-combined_k141_77976||full Not-determined
18-104-final-viral-combined_k141_119517||full Not-determined
18-104-final-viral-combined_k141_98654||full Not-determined
18-104-final-viral-combined_k141_70849||full Low-quality
18-104-final-viral-combined_k141_112761||full Low-quality
18-104-final-viral-combined_k141_14852||full Low-quality
18-104-final-viral-combined_k141_147440||full Low-quality
18-104-final-viral-combined_k141_119529||full Not-determined
18-105-final-viral-combined_k141_73301||full Medium-quality
18-104-final-viral-combined_k141_105849||full Low-quality
18-104-final-viral-combined_k141_70867||full Low-quality

| | |
|---|---|
| 18-104-final-viral-combined_k141_105852\|\|full | Low-quality |
| 18-104-final-viral-combined_k141_112772\|\|full | Low-quality |
| 18-104-final-viral-combined_k141_21981\|\|full | Not-determined |
| 18-104-final-viral-combined_k141_56863\|\|full | Not-determined |
| 18-104-final-viral-combined_k141_119558\|\|full | Low-quality |
| 18-104-final-viral-combined_k141_78022\|\|full | Not-determined |
| 18-104-final-viral-combined_k141_154378\|\|full | Low-quality |
| 18-104-final-viral-combined_k141_35839\|\|full | Not-determined |
| 18-104-final-viral-combined_k141_126854\|\|full | Not-determined |
| 18-104-final-viral-combined_k141_140625\|\|full | Not-determined |
| 18-104-final-viral-combined_k141_8047\|\|full | Low-quality |
| 18-104-final-viral-combined_k141_112780\|\|full | Not-determined |
| 18-104-final-viral-combined_k141_105904\|\|full | Low-quality |
| 18-104-final-viral-combined_k141_1287\|\|full | Low-quality |
| 18-104-final-viral-combined_k141_126868\|\|full | Not-determined |
| 18-104-final-viral-combined_k141_105913\|\|full | Not-determined |
| 18-104-final-viral-combined_k141_14895\|\|full | Not-determined |
| 18-104-final-viral-combined_k141_8086\|\|full | Not-determined |
| 18-104-final-viral-combined_k141_98692\|\|full | Not-determined |
| 18-101-final-viral-combined_k141_21398\|\|full | Medium-quality |
| 18-104-final-viral-combined_k141_63968\|\|full | Low-quality |
| 18-109-final-viral-combined_k141_69024\|\|full | Medium-quality |
| 18-104-final-viral-combined_k141_154452\|\|full | Not-determined |
| 18-104-final-viral-combined_k141_126893\|\|full | Low-quality |
| 18-104-final-viral-combined_k141_154453\|\|full | Low-quality |
| 18-104-final-viral-combined_k141_92014\|\|full | Low-quality |
| 18-104-final-viral-combined_k141_98712\|\|full | Not-determined |
| 18-104-final-viral-combined_k141_154461\|\|full | Not-determined |
| 18-104-final-viral-combined_k141_63995\|\|full | Not-determined |
| 18-104-final-viral-combined_k141_147514\|\|full | Not-determined |
| 18-104-final-viral-combined_k141_154475\|\|full | Not-determined |
| 18-104-final-viral-combined_k141_29223\|\|full | Not-determined |
| 18-104-final-viral-combined_k141_8139\|\|full | Not-determined |
| 18-104-final-viral-combined_k141_78106\|\|full | Low-quality |
| 18-104-final-viral-combined_k141_35902\|\|full | Low-quality |
| 18-104-final-viral-combined_k141_14964\|\|full | Not-determined |
| 18-104-final-viral-combined_k141_147536\|\|full | Low-quality |
| 18-104-final-viral-combined_k141_161277\|\|full | Low-quality |
| 18-104-final-viral-combined_k141_56922\|\|full | Not-determined |
| 18-104-final-viral-combined_k141_49656\|\|full | Low-quality |
| 18-104-final-viral-combined_k141_22099\|\|full | Low-quality |
| 18-104-final-viral-combined_k141_70984\|\|full | Low-quality |
| 18-104-final-viral-combined_k141_78140\|\|full | Low-quality |
| 18-104-final-viral-combined_k141_92076\|\|full | Not-determined |
| 18-104-final-viral-combined_k141_29280\|\|full | Low-quality |
| 18-104-final-viral-combined_k141_29283\|\|full | Not-determined |
| 18-104-final-viral-combined_k141_84935\|\|full | Not-determined |
| 18-104-final-viral-combined_k141_84940\|\|full | Not-determined |
| 18-104-final-viral-combined_k141_92098\|\|full | Low-quality |
| 18-104-final-viral-combined_k141_15020\|\|full | Low-quality |
| 18-104-final-viral-combined_k141_1378\|\|full | Not-determined |
| 18-104-final-viral-combined_k141_64035\|\|full | Low-quality |
| 18-104-final-viral-combined_k141_147573\|\|full | Low-quality |
| 18-104-final-viral-combined_k141_119632\|\|full | Not-determined |
| 18-104-final-viral-combined_k141_15052\|\|full | Not-determined |
| 18-104-final-viral-combined_k141_1397\|\|full | Not-determined |
| 18-104-final-viral-combined_k141_43028\|\|full | Low-quality |
| 18-104-final-viral-combined_k141_98741\|\|full | Not-determined |
| 18-104-final-viral-combined_k141_147584\|\|full | Not-determined |
| 18-104-final-viral-combined_k141_154535\|\|full | Not-determined |
| 18-104-final-viral-combined_k141_105935\|\|full | Low-quality |
| 18-104-final-viral-combined_k141_147586\|\|full | Not-determined |
| 18-104-final-viral-combined_k141_15055\|\|full | Low-quality |
| 18-104-final-viral-combined_k141_35966\|\|full | Not-determined |
| 18-104-final-viral-combined_k141_29297\|\|full | Not-determined |
| 18-104-final-viral-combined_k141_147591\|\|full | Low-quality |
| 18-104-final-viral-combined_k141_119646\|\|full | Low-quality |
| 18-104-final-viral-combined_k141_147592\|\|full | Not-determined |
| 18-104-final-viral-combined_k141_140809\|\|full | Low-quality |
| 18-104-final-viral-combined_k141_8187\|\|full | Not-determined |
| 18-104-final-viral-combined_k141_29306\|\|full | Low-quality |
| 18-104-final-viral-combined_k141_84989\|\|full | Not-determined |
| 18-104-final-viral-combined_k141_49709\|\|full | Not-determined |
| 18-104-final-viral-combined_k141_98759\|\|full | Not-determined |
| 18-104-final-viral-combined_k141_78182\|\|full | Low-quality |
| 18-104-final-viral-combined_k141_92145\|\|full | Not-determined |
| 18-104-final-viral-combined_k141_29315\|\|full | Not-determined |
| 18-104-final-viral-combined_k141_85002\|\|full | Not-determined |
| 18-104-final-viral-combined_k141_49713\|\|full | Low-quality |
| 18-104-final-viral-combined_k141_29328\|\|full | Low-quality |
| 18-104-final-viral-combined_k141_57003\|\|full | Not-determined |
| 18-104-final-viral-combined_k141_57004\|\|full | Not-determined |
| 18-104-final-viral-combined_k141_98777\|\|full | Not-determined |
| 18-104-final-viral-combined_k141_85008\|\|full | Low-quality |
| 18-104-final-viral-combined_k141_92156\|\|full | Not-determined |
| 18-104-final-viral-combined_k141_85018\|\|full | Low-quality |
| 18-104-final-viral-combined_k141_105979\|\|full | Not-determined |
| 18-104-final-viral-combined_k141_1430\|\|full | Not-determined |
| 18-104-final-viral-combined_k141_92157\|\|full | Not-determined |
| 18-104-final-viral-combined_k141_154599\|\|full | Not-determined |
| 18-104-final-viral-combined_k141_105987\|\|full | Low-quality |
| 18-104-final-viral-combined_k141_8219\|\|full | Not-determined |
| 18-104-final-viral-combined_k141_49733\|\|full | Not-determined |
| 18-104-final-viral-combined_k141_57025\|\|full | Not-determined |
| 18-104-final-viral-combined_k141_64074\|\|full | Low-quality |
| 18-104-final-viral-combined_k141_112823\|\|full | Low-quality |
| 18-104-final-viral-combined_k141_8224\|\|full | Low-quality |
| 18-104-final-viral-combined_k141_98801\|\|full | Low-quality |
| 18-104-final-viral-combined_k141_36023\|\|full | Low-quality |
| 18-104-final-viral-combined_k141_119712\|\|full | Low-quality |
| 18-104-final-viral-combined_k141_57028\|\|full | Low-quality |
| 18-104-final-viral-combined_k141_8227\|\|full | Not-determined |
| 18-104-final-viral-combined_k141_105996\|\|full | Not-determined |
| 18-104-final-viral-combined_k141_161371\|\|full | Not-determined |
| 18-104-final-viral-combined_k141_57044\|\|full | Low-quality |
| 18-104-final-viral-combined_k141_161379\|\|full | Not-determined |
| 18-104-final-viral-combined_k141_78236\|\|full | Low-quality |
| 18-104-final-viral-combined_k141_147666\|\|full | Not-determined |
| 18-104-final-viral-combined_k141_85054\|\|full | Not-determined |
| 18-104-final-viral-combined_k141_106017\|\|full | Low-quality |
| 18-104-final-viral-combined_k141_43122\|\|full | Low-quality |
| 18-104-final-viral-combined_k141_43123\|\|full | Low-quality |
| 18-104-final-viral-combined_k141_119739\|\|full | Not-determined |
| 18-104-final-viral-combined_k141_154622\|\|full | Not-determined |
| 18-104-final-viral-combined_k141_8272\|\|full | Not-determined |
| 18-104-final-viral-combined_k141_98827\|\|full | Not-determined |
| 18-104-final-viral-combined_k141_36069\|\|full | Low-quality |
| 18-104-final-viral-combined_k141_112866\|\|full | Low-quality |
| 18-104-final-viral-combined_k141_1456\|\|full | Low-quality |
| 18-104-final-viral-combined_k141_15124\|\|full | Not-determined |
| 18-104-final-viral-combined_k141_127047\|\|full | Not-determined |
| 18-104-final-viral-combined_k141_15128\|\|full | Low-quality |
| 18-104-final-viral-combined_k141_29385\|\|full | Not-determined |
| 18-104-final-viral-combined_k141_43128\|\|full | Low-quality |
| 18-104-final-viral-combined_k141_49780\|\|full | Not-determined |
| 18-104-final-viral-combined_k141_49781\|\|full | Low-quality |
| 18-104-final-viral-combined_k141_8280\|\|full | Low-quality |
| 18-104-final-viral-combined_k141_154643\|\|full | Low-quality |
| 18-104-final-viral-combined_k141_106061\|\|full | Not-determined |
| 18-104-final-viral-combined_k141_36085\|\|full | Not-determined |
| 18-104-final-viral-combined_k141_43147\|\|full | Not-determined |
| 18-104-final-viral-combined_k141_78284\|\|full | Not-determined |
| 18-104-final-viral-combined_k141_29409\|\|full | Low-quality |
| 18-104-final-viral-combined_k141_15149\|\|full | Not-determined |
| 18-104-final-viral-combined_k141_43151\|\|full | Not-determined |
| 18-104-final-viral-combined_k141_29411\|\|full | Low-quality |
| 18-103_k141_94494 | Medium-quality |
| 18-104-final-viral-combined_k141_43157\|\|full | Not-determined |
| 18-104-final-viral-combined_k141_154655\|\|full | Low-quality |
| 18-104-final-viral-combined_k141_98877\|\|full | Not-determined |
| 18-104-final-viral-combined_k141_64133\|\|full | Low-quality |
| 18-104-final-viral-combined_k141_127103\|\|full | Not-determined |
| 18-104-final-viral-combined_k141_106080\|\|full | Not-determined |
| 18-104-final-viral-combined_k141_29431\|\|full | Low-quality |
| 18-104-final-viral-combined_k141_57112\|\|full | Low-quality |
| 18-104-final-viral-combined_k141_43172\|\|full | Not-determined |
| 18-104-final-viral-combined_k141_49822\|\|full | Not-determined |
| 18-104-final-viral-combined_k141_127107\|\|full | Not-determined |
| 18-104-final-viral-combined_k141_15176\|\|full | Not-determined |
| 18-104-final-viral-combined_k141_8326\|\|full | Low-quality |
| 18-104-final-viral-combined_k141_43173\|\|full | Not-determined |
| 18-104-final-viral-combined_k141_106090\|\|full | Not-determined |
| 18-104-final-viral-combined_k141_22189\|\|full | Not-determined |
| 18-104-final-viral-combined_k141_43177\|\|full | Low-quality |
| 18-104-final-viral-combined_k141_140923\|\|full | Not-determined |
| 18-104-final-viral-combined_k141_133764\|\|full | Low-quality |
| 18-104-final-viral-combined_k141_43190\|\|full | Low-quality |
| 18-104-final-viral-combined_k141_8346\|\|full | Not-determined |
| 18-104-final-viral-combined_k141_161488\|\|full | Not-determined |
| 18-104-final-viral-combined_k141_106116\|\|full | Low-quality |
| 18-104-final-viral-combined_k141_71182\|\|full | Not-determined |
| 18-104-final-viral-combined_k141_147722\|\|full | Low-quality |
| 18-104-final-viral-combined_k141_98920\|\|full | Not-determined |
| 18-104-final-viral-combined_k141_71184\|\|full | Low-quality |
| 18-104-final-viral-combined_k141_98925\|\|full | Not-determined |
| 18-104-final-viral-combined_k141_140946\|\|full | Not-determined |
| 18-104-final-viral-combined_k141_49856\|\|full | Low-quality |
| 18-104-final-viral-combined_k141_49866\|\|full | Not-determined |
| 18-104-final-viral-combined_k141_98935\|\|full | Low-quality |
| 18-104-final-viral-combined_k141_1553\|\|full | Not-determined |
| 17-109-final-viral-combined_k141_223043\|\|full | Medium-quality |
| 18-104-final-viral-combined_k141_98947\|\|full | Not-determined |
| 18-104-final-viral-combined_k141_64211\|\|full | Not-determined |
| 18-104-final-viral-combined_k141_29503\|\|full | Not-determined |
| 18-104-final-viral-combined_k141_1562\|\|full | Low-quality |
| 18-104-final-viral-combined_k141_1565\|\|full | Not-determined |
| 18-104-final-viral-combined_k141_71204\|\|full | Not-determined |
| 18-104-final-viral-combined_k141_85163\|\|full | Not-determined |
| 18-104-final-viral-combined_k141_127132\|\|full | Not-determined |
| 18-104-final-viral-combined_k141_141015\|\|full | Low-quality |
| 18-104-final-viral-combined_k141_57163\|\|full | Low-quality |
| 18-104-final-viral-combined_k141_8373\|\|full | Not-determined |
| 18-104-final-viral-combined_k141_98976\|\|full | Not-determined |
| 18-104-final-viral-combined_k141_98977\|\|full | Not-determined |
| 18-104-final-viral-combined_k141_57173\|\|full | Not-determined |
| 18-104-final-viral-combined_k141_147780\|\|full | Low-quality |
| 18-104-final-viral-combined_k141_64246\|\|full | Low-quality |
| 18-104-final-viral-combined_k141_1589\|\|full | Not-determined |
| 18-104-final-viral-combined_k141_8382\|\|full | Not-determined |
| 18-104-final-viral-combined_k141_147790\|\|full | Low-quality |
| 18-104-final-viral-combined_k141_29535\|\|full | Low-quality |
| 18-104-final-viral-combined_k141_15260\|\|full | Not-determined |
| 18-104-final-viral-combined_k141_154774\|\|full | Not-determined |
| 18-104-final-viral-combined_k141_71221\|\|full | Not-determined |
| 18-104-final-viral-combined_k141_29538\|\|full | Low-quality |
| 18-104-final-viral-combined_k141_98994\|\|full | Low-quality |
| 18-104-final-viral-combined_k141_29546\|\|full | Low-quality |
| 18-104-final-viral-combined_k141_154784\|\|full | Not-determined |
| 18-104-final-viral-combined_k141_161566\|\|full | Low-quality |
| 18-104-final-viral-combined_k141_106188\|\|full | Not-determined |
| 18-104-final-viral-combined_k141_133853\|\|full | Not-determined |
| 18-104-final-viral-combined_k141_29549\|\|full | Low-quality |
| 18-104-final-viral-combined_k141_127161\|\|full | Not-determined |
| 18-104-final-viral-combined_k141_92278\|\|full | Not-determined |
| 18-104-final-viral-combined_k141_154793\|\|full | Low-quality |
| 18-104-final-viral-combined_k141_36123\|\|full | Low-quality |
| 18-104-final-viral-combined_k141_8387\|\|full | Not-determined |
| 18-104-final-viral-combined_k141_127167\|\|full | Not-determined |
| 18-104-final-viral-combined_k141_22286\|\|full | Low-quality |
| 18-104-final-viral-combined_k141_78417\|\|full | Not-determined |
| 18-104-final-viral-combined_k141_57208\|\|full | Not-determined |
| 18-104-final-viral-combined_k141_106199\|\|full | Low-quality |
| 18-104-final-viral-combined_k141_78418\|\|full | Low-quality |
| 18-104-final-viral-combined_k141_127177\|\|full | Not-determined |
| 18-104-final-viral-combined_k141_1622\|\|full | Not-determined |
| 18-104-final-viral-combined_k141_57210\|\|full | Low-quality |
| 18-104-final-viral-combined_k141_8407\|\|full | Low-quality |
| 18-104-final-viral-combined_k141_71258\|\|full | Low-quality |
| 18-104-final-viral-combined_k141_127185\|\|full | Low-quality |
| 18-104-final-viral-combined_k141_85224\|\|full | Not-determined |
| 18-104-final-viral-combined_k141_133881\|\|full | Low-quality |
| 18-104-final-viral-combined_k141_57222\|\|full | Not-determined |
| 18-104-final-viral-combined_k141_133893\|\|full | Not-determined |
| 18-104-final-viral-combined_k141_78438\|\|full | Not-determined |
| 18-104-final-viral-combined_k141_133897\|\|full | Not-determined |
| 18-104-final-viral-combined_k141_29609\|\|full | Low-quality |
| 18-104-final-viral-combined_k141_64284\|\|full | Not-determined |
| 18-104-final-viral-combined_k141_106236\|\|full | Not-determined |
| 18-104-final-viral-combined_k141_133904\|\|full | Low-quality |
| 18-104-final-viral-combined_k141_64286\|\|full | Not-determined |
| 18-104-final-viral-combined_k141_141060\|\|full | Not-determined |
| 18-104-final-viral-combined_k141_119857\|\|full | Not-determined |
| 18-104-final-viral-combined_k141_78451\|\|full | Low-quality |
| 18-104-final-viral-combined_k141_119859\|\|full | Low-quality |
| 18-104-final-viral-combined_k141_119867\|\|full | Not-determined |
| 18-104-final-viral-combined_k141_154830\|\|full | Not-determined |
| 18-104-final-viral-combined_k141_99068\|\|full | Not-determined |
| 18-104-final-viral-combined_k141_36170\|\|full | Low-quality |
| 18-104-final-viral-combined_k141_15316\|\|full | Not-determined |
| 18-104-final-viral-combined_k141_29627\|\|full | Low-quality |
| 18-104-final-viral-combined_k141_99070\|\|full | Not-determined |
| 18-104-final-viral-combined_k141_1653\|\|full | Not-determined |
| 18-104-final-viral-combined_k141_43329\|\|full | Low-quality |
| 18-104-final-viral-combined_k141_99074\|\|full | Low-quality |
| 18-104-final-viral-combined_k141_85244\|\|full | Not-determined |
| 18-104-final-viral-combined_k141_127225\|\|full | Low-quality |
| 18-104-final-viral-combined_k141_15325\|\|full | Not-determined |
| 18-104-final-viral-combined_k141_1665\|\|full | Low-quality |
| 18-104-final-viral-combined_k141_29636\|\|full | Low-quality |
| 18-104-final-viral-combined_k141_92326\|\|full | Low-quality |
| 18-104-final-viral-combined_k141_147851\|\|full | Low-quality |
| 18-104-final-viral-combined_k141_113085\|\|full | Not-determined |
| 18-104-final-viral-combined_k141_154845\|\|full | Not-determined |
| 18-104-final-viral-combined_k141_127259\|\|full | Low-quality |
| 18-104-final-viral-combined_k141_8479\|\|full | Low-quality |
| 18-104-final-viral-combined_k141_127268\|\|full | Not-determined |
| 18-104-final-viral-combined_k141_8493\|\|full | Not-determined |
| 18-104-final-viral-combined_k141_119890\|\|full | Not-determined |
| 18-104-final-viral-combined_k141_78494\|\|full | Not-determined |
| 18-104-final-viral-combined_k141_154863\|\|full | Not-determined |
| 18-104-final-viral-combined_k141_78497\|\|full | Not-determined |
| 18-104-final-viral-combined_k141_64320\|\|full | Not-determined |
| 18-104-final-viral-combined_k141_36218\|\|full | Low-quality |
| 18-104-final-viral-combined_k141_43382\|\|full | Low-quality |
| 18-104-final-viral-combined_k141_161648\|\|full | Low-quality |
| 18-104-final-viral-combined_k141_119911\|\|full | Not-determined |
| 18-104-final-viral-combined_k141_113161\|\|full | Not-determined |
| 18-104-final-viral-combined_k141_92371\|\|full | Low-quality |
| 18-104-final-viral-combined_k141_154886\|\|full | Low-quality |
| 18-104-final-viral-combined_k141_8521\|\|full | Low-quality |
| 18-104-final-viral-combined_k141_92384\|\|full | Low-quality |
| 18-104-final-viral-combined_k141_36248\|\|full | Low-quality |
| 18-104-final-viral-combined_k141_36250\|\|full | Low-quality |
| 18-104-final-viral-combined_k141_71356\|\|full | Not-determined |
| 18-104-final-viral-combined_k141_106333\|\|full | Low-quality |
| 18-104-final-viral-combined_k141_71359\|\|full | Not-determined |
| 18-104-final-viral-combined_k141_85356\|\|full | Not-determined |
| 18-104-final-viral-combined_k141_15403\|\|full | Not-determined |
| 18-104-final-viral-combined_k141_133941\|\|full | Low-quality |
| 18-104-final-viral-combined_k141_43403\|\|full | Low-quality |
| 18-104-final-viral-combined_k141_85359\|\|full | Low-quality |
| 18-104-final-viral-combined_k141_36258\|\|full | Not-determined |
| 18-104-final-viral-combined_k141_8536\|\|full | Not-determined |
| 18-104-final-viral-combined_k141_106350\|\|full | Not-determined |
| 18-104-final-viral-combined_k141_29693\|\|full | Low-quality |
| 18-104-final-viral-combined_k141_50001\|\|full | Low-quality |
| 18-104-final-viral-combined_k141_106357\|\|full | Low-quality |
| 18-104-final-viral-combined_k141_43419\|\|full | Low-quality |
| 18-104-final-viral-combined_k141_64343\|\|full | Low-quality |
| 18-104-final-viral-combined_k141_161683\|\|full | Not-determined |
| 18-104-final-viral-combined_k141_29720\|\|full | Not-determined |
| 18-104-final-viral-combined_k141_85378\|\|full | Not-determined |
| 18-104-final-viral-combined_k141_71395\|\|full | Not-determined |
| 18-104-final-viral-combined_k141_133956\|\|full | Low-quality |
| 18-104-final-viral-combined_k141_113216\|\|full | Not-determined |
| 18-104-final-viral-combined_k141_15433\|\|full | Low-quality |
| 18-104-final-viral-combined_k141_106363\|\|full | Not-determined |
| 18-104-final-viral-combined_k141_15440\|\|full | Not-determined |
| 18-104-final-viral-combined_k141_29728\|\|full | Not-determined |
| 18-104-final-viral-combined_k141_85407\|\|full | Low-quality |
| 18-104-final-viral-combined_k141_127291\|\|full | Not-determined |
| 18-104-final-viral-combined_k141_29740\|\|full | Not-determined |
| 18-104-final-viral-combined_k141_1774\|\|full | Not-determined |
| 18-104-final-viral-combined_k141_29744\|\|full | Not-determined |
| 18-104-final-viral-combined_k141_36291\|\|full | Not-determined |
| 18-104-final-viral-combined_k141_36301\|\|full | Low-quality |
| 18-104-final-viral-combined_k141_113234\|\|full | Not-determined |
| 18-104-final-viral-combined_k141_147927\|\|full | Not-determined |
| 18-104-final-viral-combined_k141_71418\|\|full | Not-determined |
| 18-104-final-viral-combined_k141_106387\|\|full | Not-determined |
| 18-104-final-viral-combined_k141_113238\|\|full | Not-determined |
| 18-104-final-viral-combined_k141_113250\|\|full | Low-quality |
| 18-104-final-viral-combined_k141_8591\|\|full | Low-quality |
| 18-104-final-viral-combined_k141_147942\|\|full | Not-determined |
| 18-104-final-viral-combined_k141_64450\|\|full | Not-determined |
| 18-104-final-viral-combined_k141_92455\|\|full | Not-determined |
| 18-104-final-viral-combined_k141_113259\|\|full | Not-determined |
| 18-104-final-viral-combined_k141_29801\|\|full | Not-determined |
| 18-104-final-viral-combined_k141_154972\|\|full | Low-quality |
| 18-104-final-viral-combined_k141_141227\|\|full | Low-quality |
| 18-104-final-viral-combined_k141_36347\|\|full | Low-quality |
| 18-104-final-viral-combined_k141_15461\|\|full | Low-quality |
| 18-104-final-viral-combined_k141_161792\|\|full | Not-determined |
| 18-104-final-viral-combined_k141_127363\|\|full | Low-quality |
| 18-104-final-viral-combined_k141_43499\|\|full | Not-determined |
| 18-104-final-viral-combined_k141_127371\|\|full | Not-determined |
| 18-104-final-viral-combined_k141_36365\|\|full | Not-determined |
| 18-104-final-viral-combined_k141_155005\|\|full | Low-quality |
| 18-104-final-viral-combined_k141_113293\|\|full | Not-determined |
| 18-104-final-viral-combined_k141_127385\|\|full | Not-determined |
| 18-104-final-viral-combined_k141_22390\|\|full | Low-quality |
| 18-104-final-viral-combined_k141_36376\|\|full | Low-quality |
| 18-104-final-viral-combined_k141_161822\|\|full | Low-quality |
| 18-104-final-viral-combined_k141_22396\|\|full | Low-quality |
| 18-104-final-viral-combined_k141_113296\|\|full | Low-quality |
| 18-104-final-viral-combined_k141_155028\|\|full | Not-determined |
| 18-104-final-viral-combined_k141_161832\|\|full | Not-determined |
| 18-104-final-viral-combined_k141_29833\|\|full | Low-quality |
| 18-104-final-viral-combined_k141_1864\|\|full | Low-quality |
| 18-104-final-viral-combined_k141_8656\|\|full | Not-determined |
| 18-104-final-viral-combined_k141_71496\|\|full | Not-determined |
| 18-104-final-viral-combined_k141_134114\|\|full | Low-quality |
| 18-104-final-viral-combined_k141_113307\|\|full | Low-quality |
| 18-104-final-viral-combined_k141_155051\|\|full | Not-determined |
| 18-104-final-viral-combined_k141_57451\|\|full | Low-quality |
| 18-104-final-viral-combined_k141_22434\|\|full | Not-determined |
| 18-104-final-viral-combined_k141_64527\|\|full | Low-quality |
| 18-104-final-viral-combined_k141_50123\|\|full | Low-quality |
| 18-104-final-viral-combined_k141_57467\|\|full | Low-quality |
| 18-104-final-viral-combined_k141_22444\|\|full | Not-determined |
| 18-104-final-viral-combined_k141_36396\|\|full | Low-quality |
| 18-104-final-viral-combined_k141_50136\|\|full | Low-quality |
| 18-104-final-viral-combined_k141_50139\|\|full | Not-determined |
| 18-104-final-viral-combined_k141_22454\|\|full | Not-determined |
| 18-104-final-viral-combined_k141_29879\|\|full | Not-determined |
| 18-104-final-viral-combined_k141_15551\|\|full | Not-determined |
| 18-104-final-viral-combined_k141_78672\|\|full | Not-determined |
| 18-104-final-viral-combined_k141_22465\|\|full | Not-determined |
| 18-104-final-viral-combined_k141_36422\|\|full | Low-quality |
| 18-104-final-viral-combined_k141_85521\|\|full | Not-determined |
| 18-104-final-viral-combined_k141_22468\|\|full | Low-quality |
| 18-104-final-viral-combined_k141_57503\|\|full | Not-determined |
| 18-104-final-viral-combined_k141_113338\|\|full | Not-determined |
| 18-104-final-viral-combined_k141_161914\|\|full | Not-determined |
| 18-104-final-viral-combined_k141_71522\|\|full | Not-determined |
| 18-104-final-viral-combined_k141_8713\|\|full | Not-determined |
| 18-104-final-viral-combined_k141_43602\|\|full | Low-quality |
| 18-104-final-viral-combined_k141_57521\|\|full | Not-determined |
| 18-104-final-viral-combined_k141_141267\|\|full | Not-determined |
| 18-104-final-viral-combined_k141_64557\|\|full | Low-quality |
| 18-104-final-viral-combined_k141_71531\|\|full | Low-quality |
| 18-104-final-viral-combined_k141_106505\|\|full | Not-determined |
| 18-104-final-viral-combined_k141_161936\|\|full | Not-determined |
| 18-104-final-viral-combined_k141_85541\|\|full | Not-determined |
| 18-104-final-viral-combined_k141_78721\|\|full | Not-determined |
| 18-104-final-viral-combined_k141_71537\|\|full | Not-determined |
| 18-104-final-viral-combined_k141_92582\|\|full | Low-quality |
| 18-104-final-viral-combined_k141_134187\|\|full | Not-determined |
| 18-104-final-viral-combined_k141_92588\|\|full | Not-determined |
| 18-104-final-viral-combined_k141_57542\|\|full | Not-determined |
| 18-104-final-viral-combined_k141_113346\|\|full | Not-determined |
| 18-104-final-viral-combined_k141_127479\|\|full | Low-quality |
| 18-104-final-viral-combined_k141_78733\|\|full | Not-determined |
| 18-104-final-viral-combined_k141_8739\|\|full | Low-quality |
| 18-104-final-viral-combined_k141_106545\|\|full | Low-quality |
| 18-104-final-viral-combined_k141_161963\|\|full | Low-quality |
| 18-104-final-viral-combined_k141_22531\|\|full | Low-quality |
| 18-104-final-viral-combined_k141_99390\|\|full | Low-quality |
| 18-104-final-viral-combined_k141_43660\|\|full | Low-quality |
| 18-104-final-viral-combined_k141_85548\|\|full | Low-quality |
| 18-104-final-viral-combined_k141_22543\|\|full | Not-determined |
| 18-104-final-viral-combined_k141_134204\|\|full | Low-quality |
| 18-104-final-viral-combined_k141_106565\|\|full | Low-quality |
| 18-104-final-viral-combined_k141_113376\|\|full | Low-quality |
| 18-104-final-viral-combined_k141_92619\|\|full | Low-quality |
| 18-104-final-viral-combined_k141_92621\|\|full | Low-quality |
| 18-104-final-viral-combined_k141_1954\|\|full | Low-quality |
| 18-104-final-viral-combined_k141_99406\|\|full | Not-determined |
| 18-104-final-viral-combined_k141_134211\|\|full | Not-determined |
| 18-104-final-viral-combined_k141_92624\|\|full | Not-determined |
| 18-104-final-viral-combined_k141_106568\|\|full | Not-determined |
| 18-104-final-viral-combined_k141_134224\|\|full | Not-determined |
| 18-104-final-viral-combined_k141_106571\|\|full | Low-quality |
| 18-104-final-viral-combined_k141_99415\|\|full | Not-determined |
| 18-104-final-viral-combined_k141_1967\|\|full | Not-determined |
| 18-104-final-viral-combined_k141_141296\|\|full | Not-determined |
| 18-104-final-viral-combined_k141_22581\|\|full | Not-determined |
| 18-104-final-viral-combined_k141_127515\|\|full | Low-quality |
| 18-104-final-viral-combined_k141_50248\|\|full | Not-determined |
| 18-104-final-viral-combined_k141_36489\|\|full | Not-determined |
| 18-104-final-viral-combined_k141_85573\|\|full | Not-determined |
| 18-104-final-viral-combined_k141_29981\|\|full | Not-determined |
| 18-104-final-viral-combined_k141_50252\|\|full | Not-determined |
| 18-104-final-viral-combined_k141_43715\|\|full | Low-quality |
| 18-104-final-viral-combined_k141_36502\|\|full | Not-determined |
| 18-104-final-viral-combined_k141_134260\|\|full | Not-determined |
| 18-104-final-viral-combined_k141_134262\|\|full | Not-determined |
| 18-104-final-viral-combined_k141_50258\|\|full | Not-determined |
| 18-104-final-viral-combined_k141_99446\|\|full | Not-determined |
| 18-104-final-viral-combined_k141_50259\|\|full | Not-determined |
| 18-104-final-viral-combined_k141_43721\|\|full | Low-quality |
| 18-104-final-viral-combined_k141_127538\|\|full | Not-determined |
| 18-104-final-viral-combined_k141_78806\|\|full | Low-quality |
| 18-104-final-viral-combined_k141_71614\|\|full | Low-quality |
| 18-104-final-viral-combined_k141_155184\|\|full | Low-quality |
| 18-104-final-viral-combined_k141_43729\|\|full | Low-quality |
| 18-104-final-viral-combined_k141_162013\|\|full | Not-determined |
| 18-104-final-viral-combined_k141_15691\|\|full | Not-determined |
| 18-104-final-viral-combined_k141_148152\|\|full | Low-quality |
| 18-104-final-viral-combined_k141_50277\|\|full | Not-determined |
| 18-104-final-viral-combined_k141_57616\|\|full | Low-quality |
| 18-104-final-viral-combined_k141_162029\|\|full | Not-determined |
| 18-104-final-viral-combined_k141_148163\|\|full | Not-determined |
| 18-104-final-viral-combined_k141_71631\|\|full | Not-determined |
| 18-104-final-viral-combined_k141_22621\|\|full | Low-quality |
| 18-104-final-viral-combined_k141_141336\|\|full | Low-quality |
| 18-104-final-viral-combined_k141_141337\|\|full | Low-quality |
| 18-104-final-viral-combined_k141_71645\|\|full | Low-quality |
| 18-104-final-viral-combined_k141_134298\|\|full | Not-determined |
| 18-104-final-viral-combined_k141_71651\|\|full | Low-quality |
| 18-104-final-viral-combined_k141_162053\|\|full | Not-determined |
| 18-104-final-viral-combined_k141_15722\|\|full | Not-determined |
| KM-final-viral-combined_k141_16309\|\|full | Medium-quality |
| 18-104-final-viral-combined_k141_155195\|\|full | Low-quality |
| 18-104-final-viral-combined_k141_2019\|\|full | Low-quality |
| 18-104-final-viral-combined_k141_36543\|\|full | Low-quality |
| 18-104-final-viral-combined_k141_2024\|\|full | Low-quality |
| 18-104-final-viral-combined_k141_43777\|\|full | Not-determined |
| 18-104-final-viral-combined_k141_15734\|\|full | Low-quality |
| 18-104-final-viral-combined_k141_2042\|\|full | Not-determined |
| 18-104-final-viral-combined_k141_85644\|\|full | Not-determined |
| 18-104-final-viral-combined_k141_106614\|\|full | Not-determined |
| 18-104-final-viral-combined_k141_15742\|\|full | Not-determined |
| 18-104-final-viral-combined_k141_85649\|\|full | Not-determined |
| 18-104-final-viral-combined_k141_127595\|\|full | Low-quality |
| 18-104-final-viral-combined_k141_162101\|\|full | Low-quality |
| 18-104-final-viral-combined_k141_15755\|\|full | Low-quality |
| 18-104-final-viral-combined_k141_99499\|\|full | Not-determined |
| 18-104-final-viral-combined_k141_36561\|\|full | Not-determined |
| 18-104-final-viral-combined_k141_43796\|\|full | Not-determined |
| 18-104-final-viral-combined_k141_8843\|\|full | Not-determined |
| 18-104-final-viral-combined_k141_127604\|\|full | Not-determined |
| 18-104-final-viral-combined_k141_155203\|\|full | Low-quality |
| 18-104-final-viral-combined_k141_8847\|\|full | Not-determined |
| 18-104-final-viral-combined_k141_57633\|\|full | Not-determined |
| 18-104-final-viral-combined_k141_106632\|\|full | Low-quality |
| 18-104-final-viral-combined_k141_50369\|\|full | Not-determined |
| 18-104-final-viral-combined_k141_127624\|\|full | Low-quality |
| 18-104-final-viral-combined_k141_162136\|\|full | Low-quality |
| 18-104-final-viral-combined_k141_50377\|\|full | Low-quality |
| 18-104-final-viral-combined_k141_8874\|\|full | Not-determined |
| 18-104-final-viral-combined_k141_8877\|\|full | Low-quality |
| 18-104-final-viral-combined_k141_50380\|\|full | Low-quality |
| 18-104-final-viral-combined_k141_64683\|\|full | Not-determined |
| 18-104-final-viral-combined_k141_50393\|\|full | Low-quality |
| 18-104-final-viral-combined_k141_22707\|\|full | Low-quality |
| 18-104-final-viral-combined_k141_162147\|\|full | Not-determined |
| 18-104-final-viral-combined_k141_134382\|\|full | Not-determined |
| 18-104-final-viral-combined_k141_71721\|\|full | Low-quality |
| 18-104-final-viral-combined_k141_85696\|\|full | Not-determined |
| 18-104-final-viral-combined_k141_30088\|\|full | Low-quality |
| 18-104-final-viral-combined_k141_148264\|\|full | Low-quality |
| 18-104-final-viral-combined_k141_22726\|\|full | Not-determined |
| 18-104-final-viral-combined_k141_71734\|\|full | Low-quality |
| 18-104-final-viral-combined_k141_162162\|\|full | Not-determined |
| 18-104-final-viral-combined_k141_106677\|\|full | Low-quality |
| 18-104-final-viral-combined_k141_85715\|\|full | Not-determined |
| 18-104-final-viral-combined_k141_148282\|\|full | Not-determined |
| 18-104-final-viral-combined_k141_8911\|\|full | Low-quality |
| 18-104-final-viral-combined_k141_22737\|\|full | Low-quality |
| 18-104-final-viral-combined_k141_162184\|\|full | Low-quality |
| 18-104-final-viral-combined_k141_43829\|\|full | Low-quality |
| 18-104-final-viral-combined_k141_141415\|\|full | Not-determined |
| 18-104-final-viral-combined_k141_162190\|\|full | Low-quality |
| 18-104-final-viral-combined_k141_141423\|\|full | Low-quality |
| 18-104-final-viral-combined_k141_134429\|\|full | Low-quality |
| 18-104-final-viral-combined_k141_141426\|\|full | Low-quality |
| 18-104-final-viral-combined_k141_57680\|\|full | Low-quality |
| 18-104-final-viral-combined_k141_85757\|\|full | Not-determined |
| 18-104-final-viral-combined_k141_162198\|\|full | Not-determined |
| 18-104-final-viral-combined_k141_78976\|\|full | Low-quality |
| 18-104-final-viral-combined_k141_85766\|\|full | Low-quality |
| 18-104-final-viral-combined_k141_162201\|\|full | Not-determined |
| 18-104-final-viral-combined_k141_155307\|\|full | Not-determined |
| 18-104-final-viral-combined_k141_2135\|\|full | Low-quality |
| 18-104-final-viral-combined_k141_85774\|\|full | Low-quality |
| 18-104-final-viral-combined_k141_113465\|\|full | Not-determined |
| 18-104-final-viral-combined_k141_43845\|\|full | Low-quality |
| 18-104-final-viral-combined_k141_85784\|\|full | Not-determined |
| 18-104-final-viral-combined_k141_22764\|\|full | Not-determined |
| 18-104-final-viral-combined_k141_134450\|\|full | Not-determined |
| 18-104-final-viral-combined_k141_78999\|\|full | Not-determined |
| 18-104-final-viral-combined_k141_119987\|\|full | Low-quality |
| 18-104-final-viral-combined_k141_79008\|\|full | Not-determined |
| 18-104-final-viral-combined_k141_57711\|\|full | Not-determined |
| 18-104-final-viral-combined_k141_106746\|\|full | Low-quality |
| 18-104-final-viral-combined_k141_43868\|\|full | Not-determined |
| 18-104-final-viral-combined_k141_2156\|\|full | Not-determined |
| 18-104-final-viral-combined_k141_155347\|\|full | Not-determined |
| 18-104-final-viral-combined_k141_162239\|\|full | Not-determined |
| 18-104-final-viral-combined_k141_106760\|\|full | Low-quality |
| 18-104-final-viral-combined_k141_148313\|\|full | Not-determined |
| 18-104-final-viral-combined_k141_162245\|\|full | Low-quality |
| 18-104-final-viral-combined_k141_92906\|\|full | Low-quality |
| 18-104-final-viral-combined_k141_113497\|\|full | Low-quality |
| 18-104-final-viral-combined_k141_43893\|\|full | Not-determined |
| 18-104-final-viral-combined_k141_162258\|\|full | Not-determined |
| 18-104-final-viral-combined_k141_22818\|\|full | Low-quality |
| 18-104-final-viral-combined_k141_36669\|\|full | Not-determined |
| 18-105-final-viral-combined_k141_2195\|\|full | Not-determined |
| 18-104-final-viral-combined_k141_36674\|\|full | Not-determined |
| 18-104-final-viral-combined_k141_36688\|\|full | Low-quality |
| 18-104-final-viral-combined_k141_22838\|\|full | Not-determined |
| 18-104-final-viral-combined_k141_127725\|\|full | Low-quality |
| 18-104-final-viral-combined_k141_22847\|\|full | Not-determined |
| 18-104-final-viral-combined_k141_79080\|\|full | Not-determined |
| 18-104-final-viral-combined_k141_9017\|\|full | Not-determined |
| 18-104-final-viral-combined_k141_36701\|\|full | Not-determined |
| 18-104-final-viral-combined_k141_120062\|\|full | Not-determined |
| 18-104-final-viral-combined_k141_57793\|\|full | Low-quality |
| 18-104-final-viral-combined_k141_43916\|\|full | Low-quality |
| 18-104-final-viral-combined_k141_9027\|\|full | Not-determined |
| 18-104-final-viral-combined_k141_92965\|\|full | Low-quality |
| 18-104-final-viral-combined_k141_71792\|\|full | Low-quality |
| 18-104-final-viral-combined_k141_71801\|\|full | Not-determined |
| 18-104-final-viral-combined_k141_57816\|\|full | Not-determined |
| 18-104-final-viral-combined_k141_113518\|\|full | Low-quality |
| 18-104-final-viral-combined_k141_2218\|\|full | Not-determined |
| 18-104-final-viral-combined_k141_57823\|\|full | Low-quality |
| 18-104-final-viral-combined_k141_106783\|\|full | Not-determined |
| 18-104-final-viral-combined_k141_9044\|\|full | Low-quality |
| 18-104-final-viral-combined_k141_22903\|\|full | Not-determined |
| 18-104-final-viral-combined_k141_16004\|\|full | Not-determined |
| 18-104-final-viral-combined_k141_43955\|\|full | Low-quality |
| 18-104-final-viral-combined_k141_64848\|\|full | Not-determined |
| 18-104-final-viral-combined_k141_162311\|\|full | Low-quality |
| 18-104-final-viral-combined_k141_162313\|\|full | Low-quality |
| 18-104-final-viral-combined_k141_127765\|\|full | Low-quality |
| 18-104-final-viral-combined_k141_141541\|\|full | Not-determined |
| 18-104-final-viral-combined_k141_113554\|\|full | Low-quality |
| 18-104-final-viral-combined_k141_57861\|\|full | Not-determined |
| 18-104-final-viral-combined_k141_64857\|\|full | Not-determined |
| 18-104-final-viral-combined_k141_36754\|\|full | Not-determined |
| 18-105_k141_210460 | Medium-quality |
| 18-104-final-viral-combined_k141_148437\|\|full | Not-determined |
| 18-104-final-viral-combined_k141_162327\|\|full | Not-determined |
| 18-104-final-viral-combined_k141_64863\|\|full | Not-determined |
| 18-104-final-viral-combined_k141_113567\|\|full | Not-determined |
| 18-104-final-viral-combined_k141_9087\|\|full | Not-determined |
| 18-104-final-viral-combined_k141_43979\|\|full | Not-determined |
| 18-104-final-viral-combined_k141_64864\|\|full | Low-quality |
| 18-104-final-viral-combined_k141_30157\|\|full | Low-quality |
| 18-104-final-viral-combined_k141_22957\|\|full | Not-determined |
| 18-104-final-viral-combined_k141_43994\|\|full | Low-quality |
| 18-104-final-viral-combined_k141_134603\|\|full | Low-quality |
| 18-104-final-viral-combined_k141_141571\|\|full | Not-determined |
| 18-104-final-viral-combined_k141_36792\|\|full | Low-quality |
| 18-104-final-viral-combined_k141_127794\|\|full | Low-quality |
| 18-104-final-viral-combined_k141_113591\|\|full | Low-quality |
| 18-104-final-viral-combined_k141_141582\|\|full | Low-quality |
| 18-104-final-viral-combined_k141_16050\|\|full | Not-determined |
| 18-104-final-viral-combined_k141_106839\|\|full | Not-determined |
| 18-104-final-viral-combined_k141_50501\|\|full | Not-determined |
| 18-104-final-viral-combined_k141_127807\|\|full | Low-quality |
| 18-104-final-viral-combined_k141_113593\|\|full | Low-quality |
| 18-104-final-viral-combined_k141_79169\|\|full | Low-quality |
| 18-104-final-viral-combined_k141_85896\|\|full | Not-determined |
| 18-104-final-viral-combined_k141_79171\|\|full | Low-quality |
| 18-104-final-viral-combined_k141_120130\|\|full | Low-quality |

| | |
|---|---|
| 18-104-final-viral-combined_k141_79175\|\|full | Not-determined |
| 18-104-final-viral-combined_k141_9124\|\|full | Not-determined |
| 18-104-final-viral-combined_k141_134635\|\|full | Low-quality |
| 18-104-final-viral-combined_k141_85906\|\|full | Not-determined |
| 18-104-final-viral-combined_k141_106855\|\|full | Low-quality |
| 18-104-final-viral-combined_k141_134640\|\|full | Low-quality |
| 18-104-final-viral-combined_k141_50505\|\|full | Not-determined |
| 18-104-final-viral-combined_k141_79194\|\|full | Not-determined |
| 18-104-final-viral-combined_k141_79197\|\|full | Low-quality |
| 18-104-final-viral-combined_k141_30202\|\|full | Low-quality |
| 18-104-final-viral-combined_k141_113621\|\|full | Low-quality |
| 18-104-final-viral-combined_k141_148498\|\|full | Low-quality |
| 18-104-final-viral-combined_k141_155487\|\|full | Not-determined |
| 18-104-final-viral-combined_k141_120168\|\|full | Low-quality |
| 18-104-final-viral-combined_k141_9174\|\|full | Not-determined |
| 18-104-final-viral-combined_k141_113649\|\|full | Low-quality |
| 18-104-final-viral-combined_k141_120179\|\|full | Low-quality |
| 18-105-final-viral-combined_k141_210460\|\|full | Medium-quality |
| 18-104-final-viral-combined_k141_44054\|\|full | Low-quality |
| 18-104-final-viral-combined_k141_93092\|\|full | Not-determined |
| 18-104-final-viral-combined_k141_57954\|\|full | Low-quality |
| 18-104-final-viral-combined_k141_2243\|\|full | Low-quality |
| 18-104-final-viral-combined_k141_71916\|\|full | Low-quality |
| 18-104-final-viral-combined_k141_71917\|\|full | Not-determined |
| 18-104-final-viral-combined_k141_9211\|\|full | Not-determined |
| 18-104-final-viral-combined_k141_155514\|\|full | Low-quality |
| 18-104-final-viral-combined_k141_44077\|\|full | Not-determined |
| 18-104-final-viral-combined_k141_2247\|\|full | Low-quality |
| 18-104-final-viral-combined_k141_162394\|\|full | Low-quality |
| 18-104-final-viral-combined_k141_162395\|\|full | Low-quality |
| 18-104-final-viral-combined_k141_162399\|\|full | Not-determined |
| 18-104-final-viral-combined_k141_134679\|\|full | Low-quality |
| 18-104-final-viral-combined_k141_148536\|\|full | Low-quality |
| 18-104-final-viral-combined_k141_79259\|\|full | Not-determined |
| 18-104-final-viral-combined_k141_2273\|\|full | Low-quality |
| 18-104-final-viral-combined_k141_50577\|\|full | Low-quality |
| 18-104-final-viral-combined_k141_79279\|\|full | Not-determined |
| 18-104-final-viral-combined_k141_23088\|\|full | Low-quality |
| 18-104-final-viral-combined_k141_120200\|\|full | Low-quality |
| 18-104-final-viral-combined_k141_106914\|\|full | Low-quality |
| 18-104-final-viral-combined_k141_155544\|\|full | Low-quality |
| 18-104-final-viral-combined_k141_148548\|\|full | Not-determined |
| 18-104-final-viral-combined_k141_2300\|\|full | Not-determined |
| 18-104-final-viral-combined_k141_44123\|\|full | Not-determined |
| 18-104-final-viral-combined_k141_120218\|\|full | Not-determined |
| 18-104-final-viral-combined_k141_85997\|\|full | Not-determined |
| 18-104-final-viral-combined_k141_9273\|\|full | Not-determined |
| 18-104-final-viral-combined_k141_30262\|\|full | Not-determined |
| 18-104-final-viral-combined_k141_16124\|\|full | Not-determined |
| 18-104-final-viral-combined_k141_86013\|\|full | Low-quality |
| 18-104-final-viral-combined_k141_30277\|\|full | Not-determined |
| 18-104-final-viral-combined_k141_23100\|\|full | Not-determined |
| 18-104-final-viral-combined_k141_36952\|\|full | Not-determined |
| 18-104-final-viral-combined_k141_93146\|\|full | Low-quality |
| 18-101-final-viral-combined_k141_136097\|\|0_partial | Medium-quality |
| 18-104-final-viral-combined_k141_23107\|\|full | Not-determined |
| 18-104-final-viral-combined_k141_93155\|\|full | Low-quality |
| 18-104-final-viral-combined_k141_72011\|\|full | Low-quality |
| 18-104-final-viral-combined_k141_30296\|\|full | Low-quality |
| 18-104-final-viral-combined_k141_86034\|\|full | Low-quality |
| 18-104-final-viral-combined_k141_16164\|\|full | Low-quality |
| 18-104-final-viral-combined_k141_134748\|\|full | Low-quality |
| 18-104-final-viral-combined_k141_106961\|\|full | Low-quality |
| 18-104-final-viral-combined_k141_127868\|\|full | Not-determined |
| 18-104-final-viral-combined_k141_36983\|\|full | Low-quality |
| 18-104-final-viral-combined_k141_134762\|\|full | Low-quality |
| 18-104-final-viral-combined_k141_113738\|\|full | Not-determined |
| 18-104-final-viral-combined_k141_134772\|\|full | Not-determined |
| 18-104-final-viral-combined_k141_106993\|\|full | Low-quality |
| 18-104-final-viral-combined_k141_141725\|\|full | Not-determined |
| 18-104-final-viral-combined_k141_72058\|\|full | Low-quality |
| 18-104-final-viral-combined_k141_141731\|\|full | Not-determined |
| 18-104-final-viral-combined_k141_113762\|\|full | Low-quality |
| 18-104-final-viral-combined_k141_86101\|\|full | Low-quality |
| 18-104-final-viral-combined_k141_9358\|\|full | Not-determined |
| 18-104-final-viral-combined_k141_9369\|\|full | Not-determined |
| 18-104-final-viral-combined_k141_113778\|\|full | Low-quality |
| 18-104-final-viral-combined_k141_93177\|\|full | Not-determined |
| 18-104-final-viral-combined_k141_23135\|\|full | Low-quality |
| 18-104-final-viral-combined_k141_155596\|\|full | Not-determined |
| 18-104-final-viral-combined_k141_23138\|\|full | Low-quality |
| 18-104-final-viral-combined_k141_16221\|\|full | Low-quality |
| 18-104-final-viral-combined_k141_16225\|\|full | Low-quality |
| 18-104-final-viral-combined_k141_2384\|\|full | Not-determined |
| 18-104-final-viral-combined_k141_127925\|\|full | Not-determined |
| 18-104-final-viral-combined_k141_79404\|\|full | Low-quality |
| 18-104-final-viral-combined_k141_148607\|\|full | Low-quality |
| 18-104-final-viral-combined_k141_44212\|\|full | Low-quality |
| 18-104-final-viral-combined_k141_99572\|\|full | Not-determined |
| 18-104-final-viral-combined_k141_127930\|\|full | Not-determined |
| 18-104-final-viral-combined_k141_30377\|\|full | Not-determined |
| 18-104-final-viral-combined_k141_65119\|\|full | Low-quality |
| 18-104-final-viral-combined_k141_50633\|\|full | Low-quality |
| 18-104-final-viral-combined_k141_44221\|\|full | Not-determined |
| 18-104-final-viral-combined_k141_86166\|\|full | Not-determined |
| 18-104-final-viral-combined_k141_86167\|\|full | Low-quality |
| 18-104-final-viral-combined_k141_30395\|\|full | Not-determined |
| 18-104-final-viral-combined_k141_50644\|\|full | Not-determined |
| 18-104-final-viral-combined_k141_107057\|\|full | Not-determined |
| 18-104-final-viral-combined_k141_79416\|\|full | Low-quality |
| 18-104-final-viral-combined_k141_50645\|\|full | Not-determined |
| 18-104-final-viral-combined_k141_2439\|\|full | Not-determined |
| 18-104-final-viral-combined_k141_37057\|\|full | Low-quality |
| 18-104-final-viral-combined_k141_72178\|\|full | Not-determined |
| 18-104-final-viral-combined_k141_120392\|\|full | Low-quality |
| 18-104-final-viral-combined_k141_107073\|\|full | Not-determined |
| 18-104-final-viral-combined_k141_155633\|\|full | Not-determined |
| 18-104-final-viral-combined_k141_93204\|\|full | Low-quality |
| 18-104-final-viral-combined_k141_148666\|\|full | Not-determined |
| 18-104-final-viral-combined_k141_155640\|\|full | Not-determined |
| 18-104-final-viral-combined_k141_50682\|\|full | Low-quality |
| 18-104-final-viral-combined_k141_155652\|\|full | Low-quality |
| 18-104-final-viral-combined_k141_72196\|\|full | Not-determined |
| 18-104-final-viral-combined_k141_30432\|\|full | Not-determined |
| 17-109-final-viral-combined_k141_264845\|\|full | High-quality |
| 18-104-final-viral-combined_k141_2496\|\|full | Low-quality |
| 18-104-final-viral-combined_k141_148685\|\|full | Not-determined |
| 18-104-final-viral-combined_k141_134860\|\|full | Not-determined |
| 18-104-final-viral-combined_k141_50697\|\|full | Low-quality |
| 18-104-final-viral-combined_k141_148688\|\|full | Low-quality |
| 18-104-final-viral-combined_k141_2510\|\|full | Not-determined |
| 18-104-final-viral-combined_k141_86193\|\|full | Not-determined |
| 18-104-final-viral-combined_k141_44293\|\|full | Not-determined |
| 18-104-final-viral-combined_k141_148690\|\|full | Low-quality |
| 18-104-final-viral-combined_k141_127992\|\|full | Not-determined |
| 18-104-final-viral-combined_k141_65202\|\|full | Low-quality |
| 18-104-final-viral-combined_k141_107107\|\|full | Not-determined |
| 18-104-final-viral-combined_k141_86203\|\|full | Not-determined |
| 18-104-final-viral-combined_k141_9470\|\|full | Low-quality |
| 18-104-final-viral-combined_k141_23217\|\|full | Not-determined |
| 18-104-final-viral-combined_k141_148701\|\|full | Low-quality |
| 18-104-final-viral-combined_k141_65211\|\|full | Low-quality |
| 18-104-final-viral-combined_k141_37098\|\|full | Not-determined |
| 18-104-final-viral-combined_k141_113872\|\|full | Low-quality |
| 18-104-final-viral-combined_k141_79529\|\|full | Not-determined |
| 18-104-final-viral-combined_k141_72234\|\|full | Low-quality |
| 18-104-final-viral-combined_k141_65217\|\|full | Low-quality |
| 18-104-final-viral-combined_k141_107120\|\|full | Low-quality |
| 18-104-final-viral-combined_k141_65222\|\|full | Low-quality |
| 18-104-final-viral-combined_k141_65225\|\|full | Not-determined |
| 18-104-final-viral-combined_k141_57994\|\|full | Low-quality |
| 18-104-final-viral-combined_k141_99665\|\|full | Not-determined |
| 18-104-final-viral-combined_k141_148711\|\|full | Not-determined |
| 18-104-final-viral-combined_k141_141902\|\|full | Not-determined |
| 18-104-final-viral-combined_k141_37121\|\|full | Not-determined |
| 18-104-final-viral-combined_k141_113901\|\|full | Low-quality |
| 18-104-final-viral-combined_k141_30487\|\|full | Low-quality |
| 18-104-final-viral-combined_k141_148727\|\|full | Low-quality |
| 18-104-final-viral-combined_k141_148728\|\|full | Not-determined |
| 18-104-final-viral-combined_k141_23260\|\|full | Not-determined |
| 18-104-final-viral-combined_k141_134911\|\|full | Low-quality |
| 18-104-final-viral-combined_k141_44332\|\|full | Low-quality |
| 18-104-final-viral-combined_k141_16337\|\|full | Low-quality |
| 18-104-final-viral-combined_k141_120456\|\|full | Low-quality |
| 18-104-final-viral-combined_k141_155724\|\|full | Not-determined |
| 18-104-final-viral-combined_k141_37132\|\|full | Low-quality |
| 18-104-final-viral-combined_k141_30499\|\|full | Not-determined |
| 18-104-final-viral-combined_k141_134926\|\|full | Low-quality |
| 18-104-final-viral-combined_k141_93251\|\|full | Low-quality |
| 18-104-final-viral-combined_k141_37136\|\|full | Not-determined |
| 18-104-final-viral-combined_k141_162596\|\|full | Low-quality |
| 18-104-final-viral-combined_k141_30509\|\|full | Low-quality |
| 18-104-final-viral-combined_k141_155739\|\|full | Low-quality |
| 18-104-final-viral-combined_k141_72288\|\|full | Not-determined |
| 18-104-final-viral-combined_k141_44348\|\|full | Not-determined |
| 18-104-final-viral-combined_k141_162604\|\|full | Not-determined |
| 18-104-final-viral-combined_k141_44350\|\|full | Low-quality |
| 18-104-final-viral-combined_k141_120484\|\|full | Not-determined |
| 18-104-final-viral-combined_k141_120485\|\|full | Low-quality |
| 18-104-final-viral-combined_k141_141952\|\|full | Not-determined |
| 18-104-final-viral-combined_k141_58066\|\|full | Not-determined |
| 18-104-final-viral-combined_k141_155766\|\|full | Not-determined |
| 18-104-final-viral-combined_k141_86285\|\|full | Low-quality |
| 18-104-final-viral-combined_k141_155770\|\|full | Low-quality |
| 18-104-final-viral-combined_k141_79583\|\|full | Low-quality |
| 18-104-final-viral-combined_k141_86293\|\|full | Low-quality |
| 18-104-final-viral-combined_k141_148760\|\|full | Low-quality |
| 18-104-final-viral-combined_k141_93282\|\|full | Not-determined |
| 18-104-final-viral-combined_k141_2632\|\|full | Low-quality |
| 18-104-final-viral-combined_k141_30561\|\|full | Not-determined |
| 18-104-final-viral-combined_k141_72335\|\|full | Not-determined |
| 18-104-final-viral-combined_k141_155808\|\|full | Low-quality |
| 18-104-final-viral-combined_k141_44378\|\|full | Not-determined |
| 18-104-final-viral-combined_k141_9549\|\|full | Not-determined |
| 18-104-final-viral-combined_k141_37182\|\|full | Low-quality |
| 18-104-final-viral-combined_k141_79615\|\|full | Not-determined |
| 18-104-final-viral-combined_k141_155817\|\|full | Not-determined |
| 18-104-final-viral-combined_k141_30585\|\|full | Not-determined |
| 18-104-final-viral-combined_k141_120537\|\|full | Not-determined |
| 18-104-final-viral-combined_k141_37190\|\|full | Not-determined |
| 18-104-final-viral-combined_k141_79625\|\|full | Not-determined |
| 18-104-final-viral-combined_k141_99748\|\|full | Not-determined |
| 17-108-final-viral-combined_k141_112147\|\|full | Medium-quality |
| 18-104-final-viral-combined_k141_93335\|\|full | Not-determined |
| 18-104-final-viral-combined_k141_16397\|\|full | Not-determined |
| 18-104-final-viral-combined_k141_99761\|\|full | Not-determined |
| 18-104-final-viral-combined_k141_148799\|\|full | Not-determined |
| 18-104-final-viral-combined_k141_58157\|\|full | Low-quality |
| 18-104-final-viral-combined_k141_162637\|\|full | Not-determined |
| 18-104-final-viral-combined_k141_58159\|\|full | Low-quality |
| 18-104-final-viral-combined_k141_155859\|\|full | Not-determined |
| 18-104-final-viral-combined_k141_44419\|\|full | Not-determined |
| 18-104-final-viral-combined_k141_23405\|\|full | Low-quality |
| 18-104-final-viral-combined_k141_107227\|\|full | Not-determined |
| 18-104-final-viral-combined_k141_44423\|\|full | Low-quality |
| 18-104-final-viral-combined_k141_44437\|\|full | Low-quality |
| 18-104-final-viral-combined_k141_93375\|\|full | Low-quality |
| 18-104-final-viral-combined_k141_148847\|\|full | Not-determined |
| 18-104-final-viral-combined_k141_113969\|\|full | Not-determined |
| 18-104-final-viral-combined_k141_50860\|\|full | Low-quality |
| 18-104-final-viral-combined_k141_58191\|\|full | Low-quality |
| 18-104-final-viral-combined_k141_155873\|\|full | Low-quality |
| 18-104-final-viral-combined_k141_2729\|\|full | Low-quality |
| 18-104-final-viral-combined_k141_72415\|\|full | Low-quality |
| 18-104-final-viral-combined_k141_99791\|\|full | Low-quality |
| 18-104-final-viral-combined_k141_86408\|\|full | Not-determined |
| 18-104-final-viral-combined_k141_148860\|\|full | Not-determined |
| 18-104-final-viral-combined_k141_30678\|\|full | Low-quality |
| 18-104-final-viral-combined_k141_50873\|\|full | Low-quality |
| 18-104-final-viral-combined_k141_113983\|\|full | Low-quality |
| 18-104-final-viral-combined_k141_113984\|\|full | Not-determined |
| 18-104-final-viral-combined_k141_99803\|\|full | Low-quality |
| 18-104-final-viral-combined_k141_2748\|\|full | Not-determined |
| 18-104-final-viral-combined_k141_23462\|\|full | Low-quality |
| 18-104-final-viral-combined_k141_30690\|\|full | Low-quality |
| 18-104-final-viral-combined_k141_2750\|\|full | Low-quality |
| 18-104-final-viral-combined_k141_162711\|\|full | Low-quality |
| 18-104-final-viral-combined_k141_148887\|\|full | Low-quality |
| 18-104-final-viral-combined_k141_148888\|\|full | Not-determined |
| 18-104-final-viral-combined_k141_93396\|\|full | Low-quality |
| 18-104-final-viral-combined_k141_162724\|\|full | Low-quality |
| 18-104-final-viral-combined_k141_50903\|\|full | Low-quality |
| 18-104-final-viral-combined_k141_30707\|\|full | Not-determined |
| 18-104-final-viral-combined_k141_120642\|\|full | Low-quality |
| 18-104-final-viral-combined_k141_142112\|\|full | Not-determined |
| 18-104-final-viral-combined_k141_30719\|\|full | Not-determined |
| 18-104-final-viral-combined_k141_65413\|\|full | Low-quality |
| 18-104-final-viral-combined_k141_72466\|\|full | Not-determined |
| 18-104-final-viral-combined_k141_30732\|\|full | Low-quality |
| 18-104-final-viral-combined_k141_37268\|\|full | Not-determined |
| 18-104-final-viral-combined_k141_120657\|\|full | Not-determined |
| 18-104-final-viral-combined_k141_107258\|\|full | Not-determined |
| 18-104-final-viral-combined_k141_37278\|\|full | Not-determined |
| 18-104-final-viral-combined_k141_120668\|\|full | Low-quality |
| 18-104-final-viral-combined_k141_23531\|\|full | Low-quality |
| 18-104-final-viral-combined_k141_120669\|\|full | Not-determined |
| 18-104-final-viral-combined_k141_162794\|\|full | Low-quality |
| 18-104-final-viral-combined_k141_65441\|\|full | Low-quality |
| 18-104-final-viral-combined_k141_148952\|\|full | Not-determined |
| 18-104-final-viral-combined_k141_58237\|\|full | Low-quality |
| 18-104-final-viral-combined_k141_155987\|\|full | Not-determined |
| 18-104-final-viral-combined_k141_142156\|\|full | Low-quality |
| 18-104-final-viral-combined_k141_135099\|\|full | Not-determined |
| 18-104-final-viral-combined_k141_120692\|\|full | Not-determined |
| 18-104-final-viral-combined_k141_93450\|\|full | Not-determined |
| 18-104-final-viral-combined_k141_156017\|\|full | Low-quality |
| 18-104-final-viral-combined_k141_162829\|\|full | Low-quality |
| 18-104-final-viral-combined_k141_114046\|\|full | Not-determined |
| 18-104-final-viral-combined_k141_156029\|\|full | Low-quality |
| 18-104-final-viral-combined_k141_9637\|\|full | Low-quality |
| 18-104-final-viral-combined_k141_162843\|\|full | Not-determined |
| 18-104-final-viral-combined_k141_2868\|\|full | Not-determined |
| 18-104-final-viral-combined_k141_156049\|\|full | Not-determined |
| 18-104-final-viral-combined_k141_65482\|\|full | Not-determined |
| 18-104-final-viral-combined_k141_51009\|\|full | Not-determined |
| 18-104-final-viral-combined_k141_99943\|\|full | Not-determined |
| 18-104-final-viral-combined_k141_30840\|\|full | Low-quality |
| 18-104-final-viral-combined_k141_37331\|\|full | Low-quality |
| 18-104-final-viral-combined_k141_58287\|\|full | Low-quality |
| 18-104-final-viral-combined_k141_99949\|\|full | Low-quality |
| 18-104-final-viral-combined_k141_99950\|\|full | Not-determined |
| 18-104-final-viral-combined_k141_93492\|\|full | Low-quality |
| 18-104-final-viral-combined_k141_142220\|\|full | Not-determined |
| 18-104-final-viral-combined_k141_72546\|\|full | Not-determined |
| 18-104-final-viral-combined_k141_2882\|\|full | Low-quality |
| 18-104-final-viral-combined_k141_72548\|\|full | Low-quality |
| 18-104-final-viral-combined_k141_58293\|\|full | Low-quality |
| 18-104-final-viral-combined_k141_120754\|\|full | Low-quality |
| 18-104-final-viral-combined_k141_2897\|\|full | Low-quality |
| 18-104-final-viral-combined_k141_142235\|\|full | Not-determined |
| 18-104-final-viral-combined_k141_162879\|\|full | Low-quality |
| 18-104-final-viral-combined_k141_86501\|\|full | Low-quality |
| 18-104-final-viral-combined_k141_156091\|\|full | Low-quality |
| 18-104-final-viral-combined_k141_156098\|\|full | Low-quality |
| 18-104-final-viral-combined_k141_16582\|\|full | Low-quality |
| 18-104-final-viral-combined_k141_58328\|\|full | Not-determined |
| 18-104-final-viral-combined_k141_2922\|\|full | Low-quality |
| 18-104-final-viral-combined_k141_107367\|\|full | Not-determined |
| 18-104-final-viral-combined_k141_107369\|\|full | Low-quality |
| 18-104-final-viral-combined_k141_79777\|\|full | Low-quality |
| 18-104-final-viral-combined_k141_72591\|\|full | Not-determined |
| 18-104-final-viral-combined_k141_23610\|\|full | Low-quality |
| 18-104-final-viral-combined_k141_2931\|\|full | Not-determined |
| 18-104-final-viral-combined_k141_23614\|\|full | Not-determined |
| 18-104-final-viral-combined_k141_142278\|\|full | Not-determined |
| 18-104-final-viral-combined_k141_162908\|\|full | Not-determined |
| 18-104-final-viral-combined_k141_65527\|\|full | Low-quality |
| 18-104-final-viral-combined_k141_100013\|\|full | Low-quality |
| 18-104-final-viral-combined_k141_142293\|\|full | Not-determined |
| 18-104-final-viral-combined_k141_79805\|\|full | Not-determined |
| 18-104-final-viral-combined_k141_30912\|\|full | Not-determined |
| 18-104-final-viral-combined_k141_9728\|\|full | Low-quality |
| 18-104-final-viral-combined_k141_135207\|\|full | Low-quality |
| 18-104-final-viral-combined_k141_100023\|\|full | Not-determined |
| 18-104-final-viral-combined_k141_128049\|\|full | Low-quality |
| 18-104-final-viral-combined_k141_156136\|\|full | Not-determined |
| 18-104-final-viral-combined_k141_86578\|\|full | Not-determined |
| 18-104-final-viral-combined_k141_86583\|\|full | Not-determined |
| 18-104-final-viral-combined_k141_23644\|\|full | Not-determined |
| 18-104-final-viral-combined_k141_30944\|\|full | Low-quality |
| 18-104-final-viral-combined_k141_107416\|\|full | Not-determined |
| 18-104-final-viral-combined_k141_79827\|\|full | Not-determined |
| 18-104-final-viral-combined_k141_149072\|\|full | Not-determined |
| 18-104-final-viral-combined_k141_156157\|\|full | Low-quality |
| 18-104-final-viral-combined_k141_58387\|\|full | Not-determined |
| 18-104-final-viral-combined_k141_107422\|\|full | Not-determined |
| 18-104-final-viral-combined_k141_149080\|\|full | Not-determined |
| 18-104-final-viral-combined_k141_37417\|\|full | Not-determined |
| 18-104-final-viral-combined_k141_9748\|\|full | Low-quality |
| 18-109-final-viral-combined_k141_90393\|\|full | Medium-quality |
| 18-104-final-viral-combined_k141_114127\|\|full | Not-determined |
| 18-104-final-viral-combined_k141_156163\|\|full | Not-determined |
| 18-104-final-viral-combined_k141_162918\|\|full | Low-quality |
| 18-104-final-viral-combined_k141_72639\|\|full | Not-determined |
| 18-104-final-viral-combined_k141_135233\|\|full | Not-determined |
| 18-104-final-viral-combined_k141_107445\|\|full | Not-determined |
| 18-104-final-viral-combined_k141_58408\|\|full | Not-determined |
| 18-104-final-viral-combined_k141_128108\|\|full | Not-determined |
| 18-104-final-viral-combined_k141_58414\|\|full | Low-quality |
| 18-104-final-viral-combined_k141_51096\|\|full | Not-determined |
| 18-104-final-viral-combined_k141_114151\|\|full | Low-quality |
| 18-104-final-viral-combined_k141_128131\|\|full | Not-determined |
| 18-104-final-viral-combined_k141_51105\|\|full | Not-determined |
| 18-104-final-viral-combined_k141_149125\|\|full | Not-determined |
| 18-104-final-viral-combined_k141_107476\|\|full | Low-quality |
| 18-104-final-viral-combined_k141_149137\|\|full | Not-determined |
| 18-104-final-viral-combined_k141_149138\|\|full | Not-determined |
| 18-104-final-viral-combined_k141_156204\|\|full | Low-quality |
| 18-104-final-viral-combined_k141_162948\|\|full | Low-quality |
| 18-104-final-viral-combined_k141_142386\|\|full | Low-quality |
| 18-104-final-viral-combined_k141_162950\|\|full | Not-determined |
| 18-104-final-viral-combined_k141_100107\|\|full | Low-quality |
| 18-104-final-viral-combined_k141_135256\|\|full | Not-determined |
| 18-104-final-viral-combined_k141_9842\|\|full | Not-determined |
| 18-104-final-viral-combined_k141_51119\|\|full | Low-quality |
| 18-104-final-viral-combined_k141_107506\|\|full | Not-determined |
| 18-104-final-viral-combined_k141_142401\|\|full | Low-quality |
| 18-104-final-viral-combined_k141_100117\|\|full | Low-quality |
| 18-104-final-viral-combined_k141_86639\|\|full | Low-quality |
| 18-104-final-viral-combined_k141_65602\|\|full | Not-determined |
| 18-104-final-viral-combined_k141_86644\|\|full | Not-determined |
| 18-104-final-viral-combined_k141_3031\|\|full | Not-determined |
| 18-104-final-viral-combined_k141_114177\|\|full | Low-quality |
| 18-104-final-viral-combined_k141_93550\|\|full | Low-quality |
| 18-104-final-viral-combined_k141_23744\|\|full | Low-quality |
| 18-104-final-viral-combined_k141_31015\|\|full | Not-determined |
| 18-104-final-viral-combined_k141_156252\|\|full | Not-determined |
| 18-104-final-viral-combined_k141_65629\|\|full | Not-determined |
| 18-104-final-viral-combined_k141_86666\|\|full | Low-quality |
| 18-104-final-viral-combined_k141_72656\|\|full | Low-quality |
| 18-104-final-viral-combined_k141_31037\|\|full | Not-determined |
| 18-104-final-viral-combined_k141_156279\|\|full | Low-quality |
| 18-104-final-viral-combined_k141_149208\|\|full | Low-quality |
| 18-104-final-viral-combined_k141_156300\|\|full | Not-determined |
| 18-104-final-viral-combined_k141_120921\|\|full | Low-quality |
| 18-104-final-viral-combined_k141_107547\|\|full | Not-determined |
| 18-104-final-viral-combined_k141_163002\|\|full | Not-determined |
| 18-104-final-viral-combined_k141_93576\|\|full | Low-quality |
| 18-104-final-viral-combined_k141_142456\|\|full | Not-determined |
| 18-104-final-viral-combined_k141_142462\|\|full | Not-determined |
| 18-104-final-viral-combined_k141_16708\|\|full | Not-determined |
| 18-104-final-viral-combined_k141_65694\|\|full | Low-quality |
| 18-104-final-viral-combined_k141_51188\|\|full | Low-quality |
| 18-104-final-viral-combined_k141_114194\|\|full | Not-determined |
| 18-104-final-viral-combined_k141_128264\|\|full | Low-quality |
| 18-104-final-viral-combined_k141_65706\|\|full | Not-determined |
| 18-104-final-viral-combined_k141_163010\|\|full | Not-determined |
| 18-104-final-viral-combined_k141_93600\|\|full | Not-determined |
| 18-104-final-viral-combined_k141_156322\|\|full | Not-determined |
| 18-104-final-viral-combined_k141_44768\|\|full | Not-determined |
| 18-104-final-viral-combined_k141_65732\|\|full | Low-quality |
| 18-104-final-viral-combined_k141_16731\|\|full | Low-quality |
| 18-104-final-viral-combined_k141_135290\|\|full | Low-quality |
| 18-104-final-viral-combined_k141_107573\|\|full | Not-determined |
| 18-104-final-viral-combined_k141_163035\|\|full | Not-determined |
| 18-104-final-viral-combined_k141_163038\|\|full | Not-determined |
| 18-104-final-viral-combined_k141_16737\|\|full | Low-quality |
| 18-104-final-viral-combined_k141_16739\|\|full | Low-quality |
| 18-104-final-viral-combined_k141_3074\|\|full | Low-quality |
| 18-104-final-viral-combined_k141_79979\|\|full | Not-determined |
| 18-104-final-viral-combined_k141_58524\|\|full | Low-quality |
| 18-104-final-viral-combined_k141_163042\|\|full | Not-determined |
| 18-104-final-viral-combined_k141_163043\|\|full | Not-determined |
| 18-104-final-viral-combined_k141_3092\|\|full | Not-determined |
| 18-104-final-viral-combined_k141_120958\|\|full | Not-determined |
| 18-104-final-viral-combined_k141_156371\|\|full | Not-determined |
| 18-104-final-viral-combined_k141_9949\|\|full | Low-quality |
| 18-104-final-viral-combined_k141_44816\|\|full | Not-determined |
| 18-104-final-viral-combined_k141_156381\|\|full | Not-determined |
| 18-104-final-viral-combined_k141_79997\|\|full | Low-quality |
| 18-104-final-viral-combined_k141_86808\|\|full | Not-determined |
| 18-104-final-viral-combined_k141_114301\|\|full | Low-quality |
| 18-104-final-viral-combined_k141_107616\|\|full | Not-determined |
| 18-104-final-viral-combined_k141_163089\|\|full | Not-determined |
| 18-104-final-viral-combined_k141_23794\|\|full | Low-quality |
| 18-104-final-viral-combined_k141_44821\|\|full | Low-quality |
| 18-104-final-viral-combined_k141_80019\|\|full | Not-determined |
| 18-104-final-viral-combined_k141_114312\|\|full | Low-quality |
| 18-104-final-viral-combined_k141_9997\|\|full | Low-quality |
| 18-104-final-viral-combined_k141_128331\|\|full | Low-quality |
| 18-104-final-viral-combined_k141_37571\|\|full | Low-quality |
| 18-104-final-viral-combined_k141_107661\|\|full | Low-quality |
| 18-104-final-viral-combined_k141_31156\|\|full | Not-determined |
| 18-104-final-viral-combined_k141_107662\|\|full | Low-quality |
| 18-104-final-viral-combined_k141_80025\|\|full | Low-quality |
| 18-104-final-viral-combined_k141_37586\|\|full | Not-determined |
| 18-104-final-viral-combined_k141_128353\|\|full | Not-determined |
| 18-104-final-viral-combined_k141_142594\|\|full | Low-quality |
| 18-104-final-viral-combined_k141_128360\|\|full | Low-quality |
| 18-104-final-viral-combined_k141_31175\|\|full | Not-determined |
| 18-104-final-viral-combined_k141_23845\|\|full | Not-determined |
| 18-104-final-viral-combined_k141_149285\|\|full | Low-quality |
| 18-104-final-viral-combined_k141_142612\|\|full | Not-determined |
| 18-104-final-viral-combined_k141_10046\|\|full | Not-determined |
| 18-104-final-viral-combined_k141_72761\|\|full | Not-determined |
| 18-104-final-viral-combined_k141_37645\|\|full | Low-quality |
| 18-104-final-viral-combined_k141_93764\|\|full | Low-quality |
| 18-104-final-viral-combined_k141_86841\|\|full | Low-quality |
| 18-104-final-viral-combined_k141_149308\|\|full | Not-determined |
| 18-104-final-viral-combined_k141_156443\|\|full | Not-determined |
| 18-104-final-viral-combined_k141_142639\|\|full | Low-quality |
| 18-104-final-viral-combined_k141_163189\|\|full | Low-quality |
| 18-104-final-viral-combined_k141_149317\|\|full | Low-quality |
| 18-104-final-viral-combined_k141_37661\|\|full | Low-quality |
| 18-104-final-viral-combined_k141_114360\|\|full | Not-determined |
| 18-104-final-viral-combined_k141_37693\|\|full | Not-determined |
| 18-104-final-viral-combined_k141_107770\|\|full | Not-determined |
| 18-104-final-viral-combined_k141_51277\|\|full | Low-quality |
| 18-104-final-viral-combined_k141_72798\|\|full | Not-determined |
| 18-104-final-viral-combined_k141_120999\|\|full | Not-determined |
| 18-104-final-viral-combined_k141_72799\|\|full | Low-quality |
| 18-104-final-viral-combined_k141_121007\|\|full | Low-quality |
| 18-104-final-viral-combined_k141_37713\|\|full | Not-determined |
| 18-104-final-viral-combined_k141_156476\|\|full | Low-quality |
| 18-104-final-viral-combined_k141_107810\|\|full | Low-quality |
| 18-104-final-viral-combined_k141_44983\|\|full | Low-quality |
| 18-104-final-viral-combined_k141_156488\|\|full | Not-determined |
| 18-104-final-viral-combined_k141_149379\|\|full | Not-determined |
| 18-104-final-viral-combined_k141_156491\|\|full | Not-determined |
| 18-104-final-viral-combined_k141_86889\|\|full | Not-determined |
| 18-104-final-viral-combined_k141_51313\|\|full | Not-determined |
| 18-104-final-viral-combined_k141_37737\|\|full | Low-quality |
| 18-104-final-viral-combined_k141_72861\|\|full | Not-determined |
| 18-104-final-viral-combined_k141_23940\|\|full | Low-quality |
| 18-104-final-viral-combined_k141_37740\|\|full | Not-determined |
| 18-104-final-viral-combined_k141_86900\|\|full | Not-determined |
| 18-104-final-viral-combined_k141_58563\|\|full | Low-quality |
| 18-104-final-viral-combined_k141_45025\|\|full | Low-quality |
| 18-104-final-viral-combined_k141_58565\|\|full | Not-determined |
| 18-104-final-viral-combined_k141_51328\|\|full | Not-determined |
| 18-104-final-viral-combined_k141_142705\|\|full | Low-quality |
| 18-104-final-viral-combined_k141_100394\|\|full | Not-determined |
| 18-104-final-viral-combined_k141_142711\|\|full | Not-determined |
| 18-104-final-viral-combined_k141_114409\|\|full | Not-determined |
| 18-104-final-viral-combined_k141_156523\|\|full | Not-determined |
| 18-104-final-viral-combined_k141_128475\|\|full | Not-determined |
| 18-104-final-viral-combined_k141_100403\|\|full | Not-determined |
| 18-104-final-viral-combined_k141_10152\|\|full | Not-determined |
| 18-104-final-viral-combined_k141_37764\|\|full | Low-quality |
| 18-104-final-viral-combined_k141_10165\|\|full | Not-determined |
| 18-104-final-viral-combined_k141_135508\|\|full | Not-determined |
| 18-104-final-viral-combined_k141_45073\|\|full | Not-determined |
| 18-104-final-viral-combined_k141_93820\|\|full | Not-determined |
| 18-104-final-viral-combined_k141_10175\|\|full | Not-determined |
| 18-104-final-viral-combined_k141_149420\|\|full | Not-determined |
| 18-104-final-viral-combined_k141_156536\|\|full | Low-quality |
| 18-104-final-viral-combined_k141_37778\|\|full | Low-quality |
| 18-104-final-viral-combined_k141_51357\|\|full | Low-quality |
| 18-104-final-viral-combined_k141_114431\|\|full | Low-quality |
| 18-104-final-viral-combined_k141_121106\|\|full | Not-determined |
| 18-104-final-viral-combined_k141_58639\|\|full | Not-determined |
| 18-104-final-viral-combined_k141_121108\|\|full | Not-determined |
| 18-104-final-viral-combined_k141_10199\|\|full | Not-determined |
| 18-104-final-viral-combined_k141_23997\|\|full | Not-determined |
| 18-104-final-viral-combined_k141_107856\|\|full | Low-quality |
| 18-104-final-viral-combined_k141_51381\|\|full | Low-quality |
| 18-104-final-viral-combined_k141_156566\|\|full | Not-determined |
| 18-104-final-viral-combined_k141_65953\|\|full | Low-quality |
| 18-104-final-viral-combined_k141_24018\|\|full | Not-determined |
| 18-104-final-viral-combined_k141_107877\|\|full | Low-quality |
| 18-104-final-viral-combined_k141_142769\|\|full | Low-quality |
| 18-104-final-viral-combined_k141_149453\|\|full | Low-quality |
| 18-104-final-viral-combined_k141_45126\|\|full | Not-determined |
| 18-104-final-viral-combined_k141_149457\|\|full | Low-quality |
| 18-104-final-viral-combined_k141_72963\|\|full | Low-quality |
| 18-104-final-viral-combined_k141_86948\|\|full | Not-determined |
| 18-104-final-viral-combined_k141_65961\|\|full | Not-determined |
| 18-104-final-viral-combined_k141_58682\|\|full | Not-determined |
| 18-104-final-viral-combined_k141_80254\|\|full | Not-determined |
| 18-104-final-viral-combined_k141_86961\|\|full | Not-determined |
| 18-104-final-viral-combined_k141_24034\|\|full | Low-quality |
| 18-104-final-viral-combined_k141_51418\|\|full | Not-determined |
| 18-104-final-viral-combined_k141_24050\|\|full | Not-determined |
| 18-104-final-viral-combined_k141_128571\|\|full | Not-determined |
| 18-104-final-viral-combined_k141_114521\|\|full | Not-determined |
| 18-104-final-viral-combined_k141_135585\|\|full | Low-quality |
| 18-104-final-viral-combined_k141_10239\|\|full | Low-quality |
| 18-104-final-viral-combined_k141_10245\|\|full | Low-quality |
| 18-104-final-viral-combined_k141_31382\|\|full | Low-quality |
| 18-110_k141_113693 | Medium-quality |
| 18-104-final-viral-combined_k141_93854\|\|full | Low-quality |
| 18-104-final-viral-combined_k141_121132\|\|full | Low-quality |
| 18-104-final-viral-combined_k141_58722\|\|full | Low-quality |
| 18-104-final-viral-combined_k141_121149\|\|full | Not-determined |
| 18-104-final-viral-combined_k141_10264\|\|full | Not-determined |
| 18-104-final-viral-combined_k141_24088\|\|full | Low-quality |
| 18-104-final-viral-combined_k141_156635\|\|full | Low-quality |
| 18-104-final-viral-combined_k141_163352\|\|full | Not-determined |
| 18-104-final-viral-combined_k141_149512\|\|full | Not-determined |
| 18-104-final-viral-combined_k141_45136\|\|full | Not-determined |
| 18-104-final-viral-combined_k141_73034\|\|full | Not-determined |
| 18-104-final-viral-combined_k141_114534\|\|full | Not-determined |
| 18-104-final-viral-combined_k141_31438\|\|full | Low-quality |
| 18-104-final-viral-combined_k141_86982\|\|full | Not-determined |
| 18-104-final-viral-combined_k141_121166\|\|full | Low-quality |
| 18-104-final-viral-combined_k141_31446\|\|full | Low-quality |
| 18-104-final-viral-combined_k141_73062\|\|full | Not-determined |
| 18-104-final-viral-combined_k141_31447\|\|full | Not-determined |
| 18-104-final-viral-combined_k141_149556\|\|full | Low-quality |
| 18-104-final-viral-combined_k141_121178\|\|full | Low-quality |
| 18-104-final-viral-combined_k141_58771\|\|full | Not-determined |
| 18-104-final-viral-combined_k141_10304\|\|full | Low-quality |
| 18-104-final-viral-combined_k141_3343\|\|full | Not-determined |
| 18-104-final-viral-combined_k141_10307\|\|full | Low-quality |
| 18-104-final-viral-combined_k141_149565\|\|full | Not-determined |
| 18-104-final-viral-combined_k141_100566\|\|full | Not-determined |
| 18-104-final-viral-combined_k141_65992\|\|full | Low-quality |
| 18-104-final-viral-combined_k141_163381\|\|full | Low-quality |
| 18-104-final-viral-combined_k141_58795\|\|full | Not-determined |
| 18-104-final-viral-combined_k141_93941\|\|full | Not-determined |
| 18-104-final-viral-combined_k141_107996\|\|full | Low-quality |
| 18-104-final-viral-combined_k141_37874\|\|full | Not-determined |
| 18-104-final-viral-combined_k141_45195\|\|full | Not-determined |
| 18-104-final-viral-combined_k141_3366\|\|full | Low-quality |
| 18-104-final-viral-combined_k141_149590\|\|full | Not-determined |
| 18-104-final-viral-combined_k141_58798\|\|full | Not-determined |
| 18-104-final-viral-combined_k141_121187\|\|full | Not-determined |
| 18-104-final-viral-combined_k141_163409\|\|full | Low-quality |
| 18-104-final-viral-combined_k141_80275\|\|full | Low-quality |
| 18-104-final-viral-combined_k141_93961\|\|full | Not-determined |
| 18-104-final-viral-combined_k141_10366\|\|full | Low-quality |
| 18-104-final-viral-combined_k141_65998\|\|full | Not-determined |
| 18-104-final-viral-combined_k141_135673\|\|full | Not-determined |
| 18-104-final-viral-combined_k141_66000\|\|full | Not-determined |
| 18-104-final-viral-combined_k141_45222\|\|full | Not-determined |
| 18-104-final-viral-combined_k141_45223\|\|full | Low-quality |
| 18-104-final-viral-combined_k141_66004\|\|full | Low-quality |
| 18-104-final-viral-combined_k141_73110\|\|full | Not-determined |
| 18-104-final-viral-combined_k141_163416\|\|full | Not-determined |
| 18-104-final-viral-combined_k141_3381\|\|full | Not-determined |
| 18-104-final-viral-combined_k141_128646\|\|full | Not-determined |
| 18-104-final-viral-combined_k141_10394\|\|full | Low-quality |
| 18-104-final-viral-combined_k141_31514\|\|full | Low-quality |
| 18-104-final-viral-combined_k141_73128\|\|full | Low-quality |
| 18-104-final-viral-combined_k141_45250\|\|full | Low-quality |
| 18-104-final-viral-combined_k141_135686\|\|full | Not-determined |
| 18-104-final-viral-combined_k141_163434\|\|full | Not-determined |
| 18-104-final-viral-combined_k141_10406\|\|full | Low-quality |
| 18-104-final-viral-combined_k141_66022\|\|full | Low-quality |
| 18-104-final-viral-combined_k141_87080\|\|full | Not-determined |
| 18-104-final-viral-combined_k141_10414\|\|full | Not-determined |
| 18-104-final-viral-combined_k141_108063\|\|full | Not-determined |
| 18-104-final-viral-combined_k141_156689\|\|full | Low-quality |
| 18-104-final-viral-combined_k141_156690\|\|full | Low-quality |
| 18-104-final-viral-combined_k141_45269\|\|full | Low-quality |
| 18-104-final-viral-combined_k141_45270\|\|full | Low-quality |
| 18-104-final-viral-combined_k141_87092\|\|full | Low-quality |
| 18-104-final-viral-combined_k141_37948\|\|full | Low-quality |
| 18-104-final-viral-combined_k141_87124\|\|full | Low-quality |
| 18-104-final-viral-combined_k141_66051\|\|full | Not-determined |
| 18-104-final-viral-combined_k141_149653\|\|full | Low-quality |
| 18-104-final-viral-combined_k141_142891\|\|full | Low-quality |
| 18-104-final-viral-combined_k141_108092\|\|full | Not-determined |
| 18-104-final-viral-combined_k141_135741\|\|full | Not-determined |
| 18-104-final-viral-combined_k141_80368\|\|full | Not-determined |
| 18-104-final-viral-combined_k141_121278\|\|full | Low-quality |
| 18-104-final-viral-combined_k141_163450\|\|full | Not-determined |
| 18-104-final-viral-combined_k141_45316\|\|full | Low-quality |
| 18-104-final-viral-combined_k141_149674\|\|full | Not-determined |

| | |
|---|---|
| 18-104-final-viral-combined_k141_163458\|\|full | Low-quality |
| 18-104-final-viral-combined_k141_16953\|\|full | Not-determined |
| 18-104-final-viral-combined_k141_73226\|\|full | Not-determined |
| 18-104-final-viral-combined_k141_114651\|\|full | Low-quality |
| 18-104-final-viral-combined_k141_142924\|\|full | Not-determined |
| 18-104-final-viral-combined_k141_121300\|\|full | Low-quality |
| 18-104-final-viral-combined_k141_128668\|\|full | Not-determined |
| 18-104-final-viral-combined_k141_114656\|\|full | Low-quality |
| 18-104-final-viral-combined_k141_24218\|\|full | Low-quality |
| 18-104-final-viral-combined_k141_45340\|\|full | Low-quality |
| 18-104-final-viral-combined_k141_163478\|\|full | Not-determined |
| 18-104-final-viral-combined_k141_16991\|\|full | Low-quality |
| 18-104-final-viral-combined_k141_156784\|\|full | Low-quality |
| 18-104-final-viral-combined_k141_121304\|\|full | Low-quality |
| 18-104-final-viral-combined_k141_66110\|\|full | Low-quality |
| 18-104-final-viral-combined_k141_66114\|\|full | Not-determined |
| 18-104-final-viral-combined_k141_128690\|\|full | Low-quality |
| 18-104-final-viral-combined_k141_94046\|\|full | Low-quality |
| 18-104-final-viral-combined_k141_100742\|\|full | Low-quality |
| 18-104-final-viral-combined_k141_10461\|\|full | Not-determined |
| 18-104-final-viral-combined_k141_45347\|\|full | Low-quality |
| 18-104-final-viral-combined_k141_149706\|\|full | Not-determined |
| 18-104-final-viral-combined_k141_51569\|\|full | Not-determined |
| 18-104-final-viral-combined_k141_10469\|\|full | Not-determined |
| 18-104-final-viral-combined_k141_73257\|\|full | Low-quality |
| 18-104-final-viral-combined_k141_58949\|\|full | Not-determined |
| 18-104-final-viral-combined_k141_121346\|\|full | Low-quality |
| 18-104-final-viral-combined_k141_114724\|\|full | Low-quality |
| 18-104-final-viral-combined_k141_114731\|\|full | Not-determined |
| 18-104-final-viral-combined_k141_17033\|\|full | Low-quality |
| 18-104-final-viral-combined_k141_58973\|\|full | Not-determined |
| 18-104-final-viral-combined_k141_156839\|\|full | Low-quality |
| 18-104-final-viral-combined_k141_45389\|\|full | Low-quality |
| 18-104-final-viral-combined_k141_73298\|\|full | Not-determined |
| 18-104-final-viral-combined_k141_163530\|\|full | Low-quality |
| 18-104-final-viral-combined_k141_58981\|\|full | Not-determined |
| 18-104-final-viral-combined_k141_100768\|\|full | Low-quality |
| 18-104-final-viral-combined_k141_31553\|\|full | Low-quality |
| 18-104-final-viral-combined_k141_17046\|\|full | Low-quality |
| 18-104-final-viral-combined_k141_121387\|\|full | Low-quality |
| 18-104-final-viral-combined_k141_94077\|\|full | Low-quality |
| 18-104-final-viral-combined_k141_80415\|\|full | Low-quality |
| 18-104-final-viral-combined_k141_3548\|\|full | Not-determined |
| 18-104-final-viral-combined_k141_73312\|\|full | Not-determined |
| 18-104-final-viral-combined_k141_31559\|\|full | Not-determined |
| 18-104-final-viral-combined_k141_94097\|\|full | Low-quality |
| 18-104-final-viral-combined_k141_3551\|\|full | Low-quality |
| 18-104-final-viral-combined_k141_94099\|\|full | Low-quality |
| 18-104-final-viral-combined_k141_142965\|\|full | Not-determined |
| 18-104-final-viral-combined_k141_108238\|\|full | Not-determined |
| 18-104-final-viral-combined_k141_10555\|\|full | Low-quality |
| 18-104-final-viral-combined_k141_94101\|\|full | Not-determined |
| 18-104-final-viral-combined_k141_51578\|\|full | Not-determined |
| 18-104-final-viral-combined_k141_142967\|\|full | Low-quality |
| 18-104-final-viral-combined_k141_100777\|\|full | Low-quality |
| 18-104-final-viral-combined_k141_94104\|\|full | Not-determined |
| 18-104-final-viral-combined_k141_108251\|\|full | Not-determined |
| 18-104-final-viral-combined_k141_38034\|\|full | Not-determined |
| 18-104-final-viral-combined_k141_24315\|\|full | Not-determined |
| 18-104-final-viral-combined_k141_17073\|\|full | Low-quality |
| 18-104-final-viral-combined_k141_51591\|\|full | Not-determined |
| 18-104-final-viral-combined_k141_114774\|\|full | Low-quality |
| 18-104-final-viral-combined_k141_24321\|\|full | Low-quality |
| 18-104-final-viral-combined_k141_51595\|\|full | Low-quality |
| 18-104-final-viral-combined_k141_80454\|\|full | Not-determined |
| 18-104-final-viral-combined_k141_149782\|\|full | Not-determined |
| 18-104-final-viral-combined_k141_108271\|\|full | Low-quality |
| 18-104-final-viral-combined_k141_128748\|\|full | Not-determined |
| 18-104-final-viral-combined_k141_80462\|\|full | Not-determined |
| 18-104-final-viral-combined_k141_51607\|\|full | Low-quality |
| 18-104-final-viral-combined_k141_73346\|\|full | Not-determined |
| 18-104-final-viral-combined_k141_143017\|\|full | Low-quality |
| 18-104-final-viral-combined_k141_51609\|\|full | Low-quality |
| 18-104-final-viral-combined_k141_135883\|\|full | Low-quality |
| 18-104-final-viral-combined_k141_59036\|\|full | Low-quality |
| 18-104-final-viral-combined_k141_94128\|\|full | Low-quality |
| 18-104-final-viral-combined_k141_31600\|\|full | Low-quality |
| 18-104-final-viral-combined_k141_149808\|\|full | Low-quality |
| 18-104-final-viral-combined_k141_121434\|\|full | Not-determined |
| 18-104-final-viral-combined_k141_80480\|\|full | Not-determined |
| 18-104-final-viral-combined_k141_149816\|\|full | Not-determined |
| 18-104-final-viral-combined_k141_135908\|\|full | Low-quality |
| 18-104-final-viral-combined_k141_38073\|\|full | Low-quality |
| 18-104-final-viral-combined_k141_87206\|\|full | Not-determined |
| 18-104-final-viral-combined_k141_108311\|\|full | Not-determined |
| 18-104-final-viral-combined_k141_24368\|\|full | Low-quality |
| 18-104-final-viral-combined_k141_38078\|\|full | Low-quality |
| 18-104-final-viral-combined_k141_45487\|\|full | Not-determined |
| 18-104-final-viral-combined_k141_10619\|\|full | Not-determined |
| 18-104-final-viral-combined_k141_59083\|\|full | Not-determined |
| 18-104-final-viral-combined_k141_66285\|\|full | Low-quality |
| 18-104-final-viral-combined_k141_80525\|\|full | Not-determined |
| 18-104-final-viral-combined_k141_17143\|\|full | Not-determined |
| 18-104-final-viral-combined_k141_157003\|\|full | Low-quality |
| 18-104-final-viral-combined_k141_51653\|\|full | Low-quality |
| 18-104-final-viral-combined_k141_10630\|\|full | Not-determined |
| 18-104-final-viral-combined_k141_66301\|\|full | Low-quality |
| 18-104-final-viral-combined_k141_149870\|\|full | Low-quality |
| 18-104-final-viral-combined_k141_100887\|\|full | Low-quality |
| 18-104-final-viral-combined_k141_80541\|\|full | Not-determined |
| 18-104-final-viral-combined_k141_10645\|\|full | Not-determined |
| 18-104-final-viral-combined_k141_121491\|\|full | Low-quality |
| 18-104-final-viral-combined_k141_73457\|\|full | Low-quality |
| 18-104-final-viral-combined_k141_108322\|\|full | Not-determined |
| 18-104-final-viral-combined_k141_163664\|\|full | Not-determined |
| 18-104-final-viral-combined_k141_143116\|\|full | Not-determined |
| 18-104-final-viral-combined_k141_51670\|\|full | Not-determined |
| 18-104-final-viral-combined_k141_135983\|\|full | Low-quality |
| 18-104-final-viral-combined_k141_66324\|\|full | Not-determined |
| 18-104-final-viral-combined_k141_114850\|\|full | Low-quality |
| 18-104-final-viral-combined_k141_143127\|\|full | Not-determined |
| 18-104-final-viral-combined_k141_100904\|\|full | Not-determined |
| 18-104-final-viral-combined_k141_51685\|\|full | Low-quality |
| 18-104-final-viral-combined_k141_149900\|\|full | Not-determined |
| 18-104-final-viral-combined_k141_59135\|\|full | Low-quality |
| 18-104-final-viral-combined_k141_66328\|\|full | Low-quality |
| 18-104-final-viral-combined_k141_87262\|\|full | Not-determined |
| 18-104-final-viral-combined_k141_100926\|\|full | Not-determined |
| 18-104-final-viral-combined_k141_128856\|\|full | Not-determined |
| 18-104-final-viral-combined_k141_10701\|\|full | Low-quality |
| 18-104-final-viral-combined_k141_59152\|\|full | Not-determined |
| 18-104-final-viral-combined_k141_80593\|\|full | Low-quality |
| 18-104-final-viral-combined_k141_38187\|\|full | Low-quality |
| 18-104-final-viral-combined_k141_66344\|\|full | Not-determined |
| 18-104-final-viral-combined_k141_114887\|\|full | Not-determined |
| 18-104-final-viral-combined_k141_3657\|\|full | Low-quality |
| 18-104-final-viral-combined_k141_31681\|\|full | Not-determined |
| 18-104-final-viral-combined_k141_108391\|\|full | Not-determined |
| 18-104-final-viral-combined_k141_31688\|\|full | Low-quality |
| 18-104-final-viral-combined_k141_31694\|\|full | Low-quality |
| 18-104-final-viral-combined_k141_108399\|\|full | Not-determined |
| 18-104-final-viral-combined_k141_38218\|\|full | Low-quality |
| 18-104-final-viral-combined_k141_3674\|\|full | Not-determined |
| 18-104-final-viral-combined_k141_31695\|\|full | Not-determined |
| 18-104-final-viral-combined_k141_114901\|\|full | Low-quality |
| 18-104-final-viral-combined_k141_24500\|\|full | Low-quality |
| 18-104-final-viral-combined_k141_157108\|\|full | Low-quality |
| 18-104-final-viral-combined_k141_163689\|\|full | Low-quality |
| 18-104-final-viral-combined_k141_121544\|\|full | Low-quality |
| 18-104-final-viral-combined_k141_149966\|\|full | Not-determined |
| 18-104-final-viral-combined_k141_10744\|\|full | Low-quality |
| 18-104-final-viral-combined_k141_121549\|\|full | Not-determined |
| 18-104-final-viral-combined_k141_143225\|\|full | Not-determined |
| 18-104-final-viral-combined_k141_73521\|\|full | Not-determined |
| 18-104-final-viral-combined_k141_128877\|\|full | Low-quality |
| 18-104-final-viral-combined_k141_143232\|\|full | Low-quality |
| 18-104-final-viral-combined_k141_108423\|\|full | Low-quality |
| 18-104-final-viral-combined_k141_59177\|\|full | Low-quality |
| 18-104-final-viral-combined_k141_45560\|\|full | Low-quality |
| 18-104-final-viral-combined_k141_143247\|\|full | Not-determined |
| 18-104-final-viral-combined_k141_45573\|\|full | Not-determined |
| 18-104-final-viral-combined_k141_87315\|\|full | Low-quality |
| 18-104-final-viral-combined_k141_121580\|\|full | Not-determined |
| 18-104-final-viral-combined_k141_114930\|\|full | Not-determined |
| 18-104-final-viral-combined_k141_136094\|\|full | Low-quality |
| 18-104-final-viral-combined_k141_66433\|\|full | Not-determined |
| 18-104-final-viral-combined_k141_94319\|\|full | Not-determined |
| 18-104-final-viral-combined_k141_73558\|\|full | Low-quality |
| 18-104-final-viral-combined_k141_114937\|\|full | Not-determined |
| 18-104-final-viral-combined_k141_17223\|\|full | Not-determined |
| 18-104-final-viral-combined_k141_80617\|\|full | Not-determined |
| 18-104-final-viral-combined_k141_51751\|\|full | Low-quality |
| 18-104-final-viral-combined_k141_157148\|\|full | Not-determined |
| 18-104-final-viral-combined_k141_59218\|\|full | Not-determined |
| 18-104-final-viral-combined_k141_31782\|\|full | Not-determined |
| 18-104-final-viral-combined_k141_94351\|\|full | Low-quality |
| 18-104-final-viral-combined_k141_87349\|\|full | Not-determined |
| 18-104-final-viral-combined_k141_143298\|\|full | Low-quality |
| 18-104-final-viral-combined_k141_143302\|\|full | Low-quality |
| 18-104-final-viral-combined_k141_150017\|\|full | Low-quality |
| 18-104-final-viral-combined_k141_17244\|\|full | Not-determined |
| 18-104-final-viral-combined_k141_3729\|\|full | Low-quality |
| 18-104-final-viral-combined_k141_114974\|\|full | Low-quality |
| 18-104-final-viral-combined_k141_3733\|\|full | Low-quality |
| 18-104-final-viral-combined_k141_150034\|\|full | Not-determined |
| 18-104-final-viral-combined_k141_87371\|\|full | Not-determined |
| 18-104-final-viral-combined_k141_136134\|\|full | Low-quality |
| 18-104-final-viral-combined_k141_128952\|\|full | Not-determined |
| 18-104-final-viral-combined_k141_108481\|\|full | Not-determined |
| 18-104-final-viral-combined_k141_114998\|\|full | Low-quality |
| 18-104-final-viral-combined_k141_80651\|\|full | Low-quality |
| 18-104-final-viral-combined_k141_136140\|\|full | Not-determined |
| 18-104-final-viral-combined_k141_94416\|\|full | Low-quality |
| 18-101-final-viral-combined_k141_374219\|\|full | Medium-quality |
| 18-104-final-viral-combined_k141_45686\|\|full | Low-quality |
| 18-104-final-viral-combined_k141_73638\|\|full | Low-quality |
| 18-104-final-viral-combined_k141_143334\|\|full | Low-quality |
| 18-104-final-viral-combined_k141_3783\|\|full | Low-quality |
| 18-104-final-viral-combined_k141_45697\|\|full | Not-determined |
| 18-104-final-viral-combined_k141_73654\|\|full | Low-quality |
| 18-104-final-viral-combined_k141_51805\|\|full | Not-determined |
| 18-104-final-viral-combined_k141_94428\|\|full | Not-determined |
| 18-104-final-viral-combined_k141_129003\|\|full | Not-determined |
| 18-104-final-viral-combined_k141_129016\|\|full | Not-determined |
| 18-104-final-viral-combined_k141_121681\|\|full | Low-quality |
| 18-104-final-viral-combined_k141_51832\|\|full | Not-determined |
| 18-104-final-viral-combined_k141_80699\|\|full | Not-determined |
| 18-104-final-viral-combined_k141_129031\|\|full | Low-quality |
| 18-104-final-viral-combined_k141_59304\|\|full | Low-quality |
| 18-104-final-viral-combined_k141_24586\|\|full | Low-quality |
| 18-104-final-viral-combined_k141_51843\|\|full | Not-determined |
| 18-104-final-viral-combined_k141_38394\|\|full | Not-determined |
| 18-104-final-viral-combined_k141_66569\|\|full | Not-determined |
| 18-104-final-viral-combined_k141_101079\|\|full | Low-quality |
| 18-104-final-viral-combined_k141_73684\|\|full | Low-quality |
| 18-104-final-viral-combined_k141_45757\|\|full | Not-determined |
| 18-104-final-viral-combined_k141_38403\|\|full | Not-determined |
| 18-104-final-viral-combined_k141_94468\|\|full | Low-quality |
| 18-104-final-viral-combined_k141_115086\|\|full | Not-determined |
| 18-104-final-viral-combined_k141_10817\|\|full | Not-determined |
| 18-104-final-viral-combined_k141_59314\|\|full | Not-determined |
| 18-104-final-viral-combined_k141_101094\|\|full | Low-quality |
| 18-104-final-viral-combined_k141_80720\|\|full | Low-quality |
| 18-104-final-viral-combined_k141_129071\|\|full | Not-determined |
| 18-104-final-viral-combined_k141_101101\|\|full | Low-quality |
| 18-104-final-viral-combined_k141_163835\|\|full | Low-quality |
| 18-104-final-viral-combined_k141_143403\|\|full | Low-quality |
| 18-104-final-viral-combined_k141_51888\|\|full | Not-determined |
| 18-104-final-viral-combined_k141_136195\|\|full | Not-determined |
| 18-102-final-viral-combined_k141_126134\|\|full | Medium-quality |
| 18-104-final-viral-combined_k141_51892\|\|full | Not-determined |
| 18-104-final-viral-combined_k141_150097\|\|full | Not-determined |
| 18-104-final-viral-combined_k141_66600\|\|full | Low-quality |
| 18-104-final-viral-combined_k141_143421\|\|full | Low-quality |
| 18-104-final-viral-combined_k141_143423\|\|full | Not-determined |
| 18-104-final-viral-combined_k141_163860\|\|full | Not-determined |
| 18-104-final-viral-combined_k141_163862\|\|full | Low-quality |
| 18-104-final-viral-combined_k141_51904\|\|full | Not-determined |
| 18-104-final-viral-combined_k141_115111\|\|full | Low-quality |
| 18-104-final-viral-combined_k141_129103\|\|full | Low-quality |
| 18-104-final-viral-combined_k141_38442\|\|full | Low-quality |
| 18-104-final-viral-combined_k141_121686\|\|full | Not-determined |
| 18-104-final-viral-combined_k141_136223\|\|full | Low-quality |
| 18-104-final-viral-combined_k141_73708\|\|full | Not-determined |
| 18-104-final-viral-combined_k141_108577\|\|full | Not-determined |
| 18-104-final-viral-combined_k141_80780\|\|full | Not-determined |
| KM-final-viral-combined_k141_79709\|\|full | Medium-quality |
| 18-104-final-viral-combined_k141_51939\|\|full | Not-determined |
| 18-104-final-viral-combined_k141_31921\|\|full | Not-determined |
| 18-104-final-viral-combined_k141_31922\|\|full | Not-determined |
| 18-104-final-viral-combined_k141_38466\|\|full | Low-quality |
| 18-104-final-viral-combined_k141_10876\|\|full | Low-quality |
| 18-104-final-viral-combined_k141_66663\|\|full | Low-quality |
| 18-104-final-viral-combined_k141_163906\|\|full | Low-quality |
| 18-104-final-viral-combined_k141_87470\|\|full | Not-determined |
| 18-104-final-viral-combined_k141_38479\|\|full | Low-quality |
| 18-104-final-viral-combined_k141_101181\|\|full | Not-determined |
| 18-104-final-viral-combined_k141_150151\|\|full | Not-determined |
| 18-104-final-viral-combined_k141_121732\|\|full | Low-quality |
| 18-104-final-viral-combined_k141_31946\|\|full | Not-determined |
| 18-104-final-viral-combined_k141_136262\|\|full | Low-quality |
| 18-104-final-viral-combined_k141_136265\|\|full | Low-quality |
| 18-104-final-viral-combined_k141_150160\|\|full | Not-determined |
| 18-104-final-viral-combined_k141_143468\|\|full | Low-quality |
| 18-104-final-viral-combined_k141_59405\|\|full | Not-determined |
| 18-104-final-viral-combined_k141_143474\|\|full | Not-determined |
| 18-104-final-viral-combined_k141_80850\|\|full | Low-quality |
| 18-104-final-viral-combined_k141_66699\|\|full | Low-quality |
| 18-104-final-viral-combined_k141_38502\|\|full | Low-quality |
| 18-104-final-viral-combined_k141_115171\|\|full | Low-quality |
| 18-104-final-viral-combined_k141_3980\|\|full | Not-determined |
| 18-104-final-viral-combined_k141_59429\|\|full | Not-determined |
| 18-104-final-viral-combined_k141_59435\|\|full | Low-quality |
| 18-104-final-viral-combined_k141_108662\|\|full | Low-quality |
| 18-104-final-viral-combined_k141_59447\|\|full | Low-quality |
| 18-104-final-viral-combined_k141_66718\|\|full | Not-determined |
| 18-104-final-viral-combined_k141_73789\|\|full | Not-determined |
| 18-104-final-viral-combined_k141_108675\|\|full | Not-determined |
| 18-104-final-viral-combined_k141_17321\|\|full | Low-quality |
| 18-104-final-viral-combined_k141_31975\|\|full | Low-quality |
| 18-104-final-viral-combined_k141_164003\|\|full | Low-quality |
| 18-104-final-viral-combined_k141_24783\|\|full | Low-quality |
| 18-104-final-viral-combined_k141_129210\|\|full | Low-quality |
| 18-104-final-viral-combined_k141_51995\|\|full | Not-determined |
| 18-104-final-viral-combined_k141_66733\|\|full | Low-quality |
| 18-104-final-viral-combined_k141_31986\|\|full | Low-quality |
| 18-104-final-viral-combined_k141_129219\|\|full | Low-quality |
| 18-104-final-viral-combined_k141_66743\|\|full | Not-determined |
| 18-104-final-viral-combined_k141_31990\|\|full | Low-quality |
| 18-104-final-viral-combined_k141_38581\|\|full | Not-determined |
| 18-104-final-viral-combined_k141_73831\|\|full | Not-determined |
| 18-104-final-viral-combined_k141_108726\|\|full | Low-quality |
| 18-104-final-viral-combined_k141_143554\|\|full | Not-determined |
| 18-104-final-viral-combined_k141_66750\|\|full | Low-quality |
| 18-104-final-viral-combined_k141_66752\|\|full | Not-determined |
| 18-104-final-viral-combined_k141_143555\|\|full | Not-determined |
| 18-104-final-viral-combined_k141_129236\|\|full | Not-determined |
| 18-104-final-viral-combined_k141_80974\|\|full | Low-quality |
| 18-104-final-viral-combined_k141_52002\|\|full | Not-determined |
| 18-104-final-viral-combined_k141_10967\|\|full | Not-determined |
| 18-104-final-viral-combined_k141_129251\|\|full | Not-determined |
| 18-104-final-viral-combined_k141_10986\|\|full | Low-quality |
| 18-104-final-viral-combined_k141_52032\|\|full | Not-determined |
| 18-104-final-viral-combined_k141_108761\|\|full | Low-quality |
| 18-104-final-viral-combined_k141_4097\|\|full | Low-quality |
| 18-104-final-viral-combined_k141_121821\|\|full | Not-determined |
| 18-104-final-viral-combined_k141_94668\|\|full | Not-determined |
| 18-104-final-viral-combined_k141_73867\|\|full | Low-quality |
| 18-104-final-viral-combined_k141_157333\|\|full | Not-determined |
| 18-104-final-viral-combined_k141_94671\|\|full | Low-quality |
| 18-104-final-viral-combined_k141_81006\|\|full | Not-determined |
| 18-104-final-viral-combined_k141_24854\|\|full | Not-determined |
| 18-104-final-viral-combined_k141_143595\|\|full | Low-quality |
| 18-104-final-viral-combined_k141_115277\|\|full | Not-determined |
| 18-104-final-viral-combined_k141_24858\|\|full | Not-determined |
| 18-104-final-viral-combined_k141_143599\|\|full | Low-quality |
| 18-104-final-viral-combined_k141_81011\|\|full | Low-quality |
| 18-104-final-viral-combined_k141_52055\|\|full | Not-determined |
| 18-104-final-viral-combined_k141_143608\|\|full | Not-determined |
| 18-104-final-viral-combined_k141_157340\|\|full | Low-quality |
| 18-104-final-viral-combined_k141_24872\|\|full | Not-determined |
| 18-104-final-viral-combined_k141_164096\|\|full | Not-determined |
| 18-104-final-viral-combined_k141_94704\|\|full | Not-determined |
| 18-104-final-viral-combined_k141_121837\|\|full | Low-quality |
| 18-104-final-viral-combined_k141_101352\|\|full | Not-determined |
| 18-104-final-viral-combined_k141_46006\|\|full | Not-determined |
| 18-104-final-viral-combined_k141_32053\|\|full | Low-quality |
| 18-104-final-viral-combined_k141_101360\|\|full | Low-quality |
| 18-104-final-viral-combined_k141_46013\|\|full | Low-quality |
| 18-104-final-viral-combined_k141_73904\|\|full | Not-determined |
| 18-104-final-viral-combined_k141_52094\|\|full | Low-quality |
| 18-104-final-viral-combined_k141_164127\|\|full | Not-determined |
| 18-104-final-viral-combined_k141_38640\|\|full | Low-quality |
| 18-104-final-viral-combined_k141_81029\|\|full | Not-determined |
| 18-104-final-viral-combined_k141_94739\|\|full | Low-quality |
| 18-104-final-viral-combined_k141_11082\|\|full | Low-quality |
| 18-104-final-viral-combined_k141_73918\|\|full | Low-quality |
| 18-104-final-viral-combined_k141_164140\|\|full | Not-determined |
| 18-104-final-viral-combined_k141_17373\|\|full | Low-quality |
| 18-104-final-viral-combined_k141_101397\|\|full | Not-determined |
| 18-104-final-viral-combined_k141_94768\|\|full | Not-determined |
| 18-104-final-viral-combined_k141_11116\|\|full | Not-determined |
| 18-104-final-viral-combined_k141_38658\|\|full | Not-determined |
| 18-104-final-viral-combined_k141_46062\|\|full | Low-quality |
| 18-104-final-viral-combined_k141_129361\|\|full | Not-determined |
| 18-104-final-viral-combined_k141_121919\|\|full | Not-determined |
| 18-104-final-viral-combined_k141_73971\|\|full | Low-quality |
| 18-104-final-viral-combined_k141_24925\|\|full | Low-quality |
| 18-104-final-viral-combined_k141_81106\|\|full | Not-determined |
| 18-104-final-viral-combined_k141_164207\|\|full | Low-quality |
| 18-104-final-viral-combined_k141_59553\|\|full | Low-quality |
| 18-104-final-viral-combined_k141_143647\|\|full | Low-quality |
| 18-104-final-viral-combined_k141_150331\|\|full | Low-quality |
| 18-104-final-viral-combined_k141_59567\|\|full | Not-determined |
| 18-104-final-viral-combined_k141_38695\|\|full | Not-determined |
| 18-104-final-viral-combined_k141_87631\|\|full | Not-determined |
| 18-104-final-viral-combined_k141_136411\|\|full | Low-quality |
| 18-104-final-viral-combined_k141_66929\|\|full | Not-determined |
| 18-104-final-viral-combined_k141_24970\|\|full | Not-determined |
| 18-104-final-viral-combined_k141_17490\|\|full | Low-quality |
| 18-104-final-viral-combined_k141_11206\|\|full | Low-quality |
| 18-104-final-viral-combined_k141_59601\|\|full | Low-quality |
| 18-104-final-viral-combined_k141_101477\|\|full | Not-determined |
| 18-104-final-viral-combined_k141_11211\|\|full | Low-quality |
| 18-104-final-viral-combined_k141_87641\|\|full | Not-determined |
| 18-104-final-viral-combined_k141_143683\|\|full | Not-determined |
| 18-104-final-viral-combined_k141_4171\|\|full | Not-determined |
| 18-104-final-viral-combined_k141_32115\|\|full | Low-quality |
| 18-104-final-viral-combined_k141_101485\|\|full | Not-determined |
| 18-104-final-viral-combined_k141_66954\|\|full | Not-determined |
| 18-104-final-viral-combined_k141_157426\|\|full | Not-determined |
| 18-104-final-viral-combined_k141_66956\|\|full | Low-quality |
| 18-104-final-viral-combined_k141_150396\|\|full | Not-determined |
| 18-104-final-viral-combined_k141_136464\|\|full | Low-quality |
| 18-104-final-viral-combined_k141_129461\|\|full | Low-quality |
| 18-104-final-viral-combined_k141_129464\|\|full | Not-determined |
| 18-104-final-viral-combined_k141_59628\|\|full | Not-determined |
| 18-104-final-viral-combined_k141_66976\|\|full | Low-quality |
| 18-104-final-viral-combined_k141_66977\|\|full | Low-quality |
| 18-104-final-viral-combined_k141_52258\|\|full | Not-determined |
| 18-102-final-viral-combined_k141_156305\|\|full | Medium-quality |
| 18-104-final-viral-combined_k141_164283\|\|full | Not-determined |
| 18-104-final-viral-combined_k141_11241\|\|full | Not-determined |
| 18-104-final-viral-combined_k141_136485\|\|full | Not-determined |
| 18-104-final-viral-combined_k141_11242\|\|full | Not-determined |
| 18-104-final-viral-combined_k141_32175\|\|full | Not-determined |
| 18-104-final-viral-combined_k141_150425\|\|full | Not-determined |
| 18-104-final-viral-combined_k141_150430\|\|full | Low-quality |
| 18-104-final-viral-combined_k141_157451\|\|full | Not-determined |
| 18-104-final-viral-combined_k141_164301\|\|full | Low-quality |
| 18-104-final-viral-combined_k141_67003\|\|full | Not-determined |
| 18-104-final-viral-combined_k141_11243\|\|full | Low-quality |
| 18-104-final-viral-combined_k141_81160\|\|full | Low-quality |
| 18-104-final-viral-combined_k141_101534\|\|full | Low-quality |
| 18-104-final-viral-combined_k141_143748\|\|full | Low-quality |
| 18-104-final-viral-combined_k141_38786\|\|full | Not-determined |
| 18-104-final-viral-combined_k141_108998\|\|full | Low-quality |
| 18-104-final-viral-combined_k141_94812\|\|full | Not-determined |
| 18-104-final-viral-combined_k141_87714\|\|full | Low-quality |
| 18-104-final-viral-combined_k141_109016\|\|full | Low-quality |
| 18-104-final-viral-combined_k141_101540\|\|full | Low-quality |
| 18-104-final-viral-combined_k141_4222\|\|full | Low-quality |
| 18-104-final-viral-combined_k141_25049\|\|full | Not-determined |
| 18-104-final-viral-combined_k141_81187\|\|full | Not-determined |
| 18-104-final-viral-combined_k141_157484\|\|full | Low-quality |
| 18-104-final-viral-combined_k141_46192\|\|full | Not-determined |
| 18-104-final-viral-combined_k141_150442\|\|full | Low-quality |
| 18-104-final-viral-combined_k141_38825\|\|full | Not-determined |
| 18-104-final-viral-combined_k141_136536\|\|full | Low-quality |
| 18-104-final-viral-combined_k141_59693\|\|full | Not-determined |
| 18-104-final-viral-combined_k141_109045\|\|full | Low-quality |
| 18-104-final-viral-combined_k141_164359\|\|full | Low-quality |
| 18-104-final-viral-combined_k141_109055\|\|full | Low-quality |
| 18-104-final-viral-combined_k141_81231\|\|full | Low-quality |
| 18-104-final-viral-combined_k141_94878\|\|full | Not-determined |
| 18-104-final-viral-combined_k141_94882\|\|full | Low-quality |
| 18-104-final-viral-combined_k141_129553\|\|full | Low-quality |
| 18-104-final-viral-combined_k141_67066\|\|full | Low-quality |
| 18-104-final-viral-combined_k141_101570\|\|full | Not-determined |
| 18-104-final-viral-combined_k141_150488\|\|full | Not-determined |
| 18-104-final-viral-combined_k141_164380\|\|full | Not-determined |
| 18-104-final-viral-combined_k141_87790\|\|full | Not-determined |
| 18-104-final-viral-combined_k141_136592\|\|full | Not-determined |
| 18-104-final-viral-combined_k141_59731\|\|full | Not-determined |
| 18-104-final-viral-combined_k141_129563\|\|full | Low-quality |
| 18-110-final-viral-combined_k141_113693\|\|full | Medium-quality |
| 18-104-final-viral-combined_k141_164391\|\|full | Low-quality |
| 18-104-final-viral-combined_k141_4291\|\|full | Not-determined |
| 18-104-final-viral-combined_k141_32288\|\|full | Not-determined |
| 18-104-final-viral-combined_k141_4295\|\|full | Low-quality |
| 18-104-final-viral-combined_k141_4296\|\|full | Low-quality |
| 18-104-final-viral-combined_k141_101599\|\|full | Not-determined |
| 18-104-final-viral-combined_k141_164401\|\|full | Low-quality |
| 18-104-final-viral-combined_k141_101603\|\|full | Low-quality |
| 18-104-final-viral-combined_k141_136614\|\|full | Low-quality |
| 18-104-final-viral-combined_k141_164405\|\|full | Not-determined |
| 18-104-final-viral-combined_k141_74123\|\|full | Low-quality |
| 18-104-final-viral-combined_k141_143826\|\|full | Not-determined |
| 18-104-final-viral-combined_k141_32302\|\|full | Not-determined |
| 18-104-final-viral-combined_k141_121996\|\|full | Not-determined |
| 18-104-final-viral-combined_k141_32306\|\|full | Not-determined |
| 18-104-final-viral-combined_k141_52370\|\|full | Not-determined |
| 18-104-final-viral-combined_k141_150514\|\|full | Low-quality |
| 18-104-final-viral-combined_k141_25139\|\|full | Not-determined |
| 18-104-final-viral-combined_k141_4306\|\|full | Low-quality |
| 18-104-final-viral-combined_k141_46299\|\|full | Not-determined |
| 18-104-final-viral-combined_k141_101622\|\|full | Not-determined |
| 18-104-final-viral-combined_k141_46301\|\|full | Not-determined |
| 18-104-final-viral-combined_k141_157589\|\|full | Low-quality |
| 18-104-final-viral-combined_k141_59766\|\|full | Low-quality |
| 18-104-final-viral-combined_k141_38902\|\|full | Low-quality |
| 18-104-final-viral-combined_k141_87831\|\|full | Not-determined |
| 18-104-final-viral-combined_k141_4318\|\|full | Low-quality |
| 18-104-final-viral-combined_k141_109126\|\|full | Low-quality |
| 18-104-final-viral-combined_k141_136635\|\|full | Low-quality |
| 18-104-final-viral-combined_k141_157597\|\|full | Low-quality |
| 18-104-final-viral-combined_k141_52383\|\|full | Not-determined |
| 18-104-final-viral-combined_k141_115463\|\|full | Not-determined |
| 18-104-final-viral-combined_k141_164421\|\|full | Low-quality |
| 18-104-final-viral-combined_k141_46327\|\|full | Low-quality |
| 18-104-final-viral-combined_k141_11334\|\|full | Low-quality |
| 18-104-final-viral-combined_k141_81329\|\|full | Low-quality |
| 18-104-final-viral-combined_k141_150543\|\|full | Not-determined |
| 18-104-final-viral-combined_k141_11347\|\|full | Not-determined |
| 18-104-final-viral-combined_k141_59787\|\|full | Not-determined |
| 18-104-final-viral-combined_k141_38945\|\|full | Not-determined |
| 18-104-final-viral-combined_k141_4320\|\|full | Low-quality |
| 18-104-final-viral-combined_k141_38953\|\|full | Low-quality |
| 18-104-final-viral-combined_k141_25182\|\|full | Low-quality |
| 18-104-final-viral-combined_k141_157634\|\|full | Low-quality |
| 18-104-final-viral-combined_k141_59802\|\|full | Low-quality |
| 18-104-final-viral-combined_k141_122067\|\|full | Low-quality |
| 18-104-final-viral-combined_k141_59811\|\|full | Low-quality |
| 18-104-final-viral-combined_k141_115498\|\|full | Low-quality |
| 18-104-final-viral-combined_k141_59819\|\|full | Low-quality |
| 18-104-final-viral-combined_k141_157658\|\|full | Low-quality |
| 18-104-final-viral-combined_k141_46371\|\|full | Low-quality |
| 18-104-final-viral-combined_k141_109173\|\|full | Not-determined |
| 18-104-final-viral-combined_k141_122099\|\|full | Not-determined |
| 18-104-final-viral-combined_k141_4344\|\|full | Not-determined |
| 18-104-final-viral-combined_k141_52423\|\|full | Not-determined |
| 18-104-final-viral-combined_k141_109177\|\|full | Low-quality |
| 18-104-final-viral-combined_k141_67185\|\|full | Not-determined |
| 18-104-final-viral-combined_k141_136700\|\|full | Not-determined |
| 18-104-final-viral-combined_k141_38995\|\|full | Not-determined |
| 18-104-final-viral-combined_k141_87860\|\|full | Low-quality |
| 18-104-final-viral-combined_k141_115516\|\|full | Not-determined |
| 18-104-final-viral-combined_k141_109188\|\|full | Not-determined |
| 18-104-final-viral-combined_k141_67197\|\|full | Not-determined |
| 18-104-final-viral-combined_k141_136711\|\|full | Low-quality |
| 18-104-final-viral-combined_k141_17597\|\|full | Not-determined |
| 18-104-final-viral-combined_k141_74254\|\|full | Not-determined |
| 18-104-final-viral-combined_k141_17602\|\|full | Not-determined |
| 18-104-final-viral-combined_k141_136734\|\|full | Not-determined |
| 18-104-final-viral-combined_k141_122112\|\|full | Not-determined |
| 18-104-final-viral-combined_k141_4396\|\|full | Low-quality |
| 18-101-final-viral-combined_k141_166392\|\|full | Medium-quality |
| 18-104-final-viral-combined_k141_32396\|\|full | Low-quality |
| 18-104-final-viral-combined_k141_17605\|\|full | Low-quality |
| 18-104-final-viral-combined_k141_17608\|\|full | Not-determined |
| 18-104-final-viral-combined_k141_157726\|\|full | Low-quality |
| 18-104-final-viral-combined_k141_32415\|\|full | Low-quality |
| 18-104-final-viral-combined_k141_25210\|\|full | Low-quality |
| 18-104-final-viral-combined_k141_81430\|\|full | Not-determined |
| 18-104-final-viral-combined_k141_122136\|\|full | Not-determined |
| 18-104-final-viral-combined_k141_25217\|\|full | Not-determined |
| 18-104-final-viral-combined_k141_52508\|\|full | Not-determined |
| 18-104-final-viral-combined_k141_150632\|\|full | Not-determined |
| 18-104-final-viral-combined_k141_129664\|\|full | Not-determined |
| 18-104-final-viral-combined_k141_164551\|\|full | Not-determined |
| 18-104-final-viral-combined_k141_11456\|\|full | Not-determined |
| 18-104-final-viral-combined_k141_122166\|\|full | Low-quality |
| 18-104-final-viral-combined_k141_59900\|\|full | Not-determined |
| 18-104-final-viral-combined_k141_109260\|\|full | Low-quality |
| 18-104-final-viral-combined_k141_11479\|\|full | Not-determined |
| 18-104-final-viral-combined_k141_25242\|\|full | Not-determined |
| 18-104-final-viral-combined_k141_67277\|\|full | Low-quality |
| 18-104-final-viral-combined_k141_32458\|\|full | Not-determined |
| 18-104-final-viral-combined_k141_101770\|\|full | Low-quality |
| 18-104-final-viral-combined_k141_25246\|\|full | Not-determined |
| 18-104-final-viral-combined_k141_95060\|\|full | Not-determined |
| 18-104-final-viral-combined_k141_32461\|\|full | Low-quality |
| 18-104-final-viral-combined_k141_59912\|\|full | Low-quality |
| 18-104-final-viral-combined_k141_39066\|\|full | Low-quality |
| 18-104-final-viral-combined_k141_59914\|\|full | Not-determined |
| 18-104-final-viral-combined_k141_81470\|\|full | Low-quality |
| 18-104-final-viral-combined_k141_164575\|\|full | Not-determined |
| 18-104-final-viral-combined_k141_95067\|\|full | Not-determined |
| 18-104-final-viral-combined_k141_115556\|\|full | Low-quality |
| 18-104-final-viral-combined_k141_25262\|\|full | Low-quality |
| 18-104-final-viral-combined_k141_122191\|\|full | Low-quality |
| 18-104-final-viral-combined_k141_115560\|\|full | Low-quality |
| 18-104-final-viral-combined_k141_95072\|\|full | Not-determined |
| 18-104-final-viral-combined_k141_95074\|\|full | Low-quality |
| 18-104-final-viral-combined_k141_81480\|\|full | Not-determined |
| 18-104-final-viral-combined_k141_52551\|\|full | Not-determined |
| 18-104-final-viral-combined_k141_101808\|\|full | Low-quality |
| 18-104-final-viral-combined_k141_87960\|\|full | Low-quality |
| 18-104-final-viral-combined_k141_87965\|\|full | Not-determined |
| 18-104-final-viral-combined_k141_25282\|\|full | Not-determined |
| 18-104-final-viral-combined_k141_46513\|\|full | Low-quality |
| 18-104-final-viral-combined_k141_17708\|\|full | Low-quality |
| 18-104-final-viral-combined_k141_150663\|\|full | Low-quality |
| 18-104-final-viral-combined_k141_46518\|\|full | Low-quality |
| 18-104-final-viral-combined_k141_157811\|\|full | Not-determined |
| 18-104-final-viral-combined_k141_122206\|\|full | Not-determined |
| 18-104-final-viral-combined_k141_109314\|\|full | Not-determined |
| 18-104-final-viral-combined_k141_11548\|\|full | Low-quality |
| 18-104-final-viral-combined_k141_136807\|\|full | Not-determined |
| 18-104-final-viral-combined_k141_81516\|\|full | Not-determined |
| 18-104-final-viral-combined_k141_17727\|\|full | Low-quality |
| 18-104-final-viral-combined_k141_74383\|\|full | Not-determined |
| 18-104-final-viral-combined_k141_32497\|\|full | Not-determined |
| 18-104-final-viral-combined_k141_52587\|\|full | Not-determined |
| 18-104-final-viral-combined_k141_74385\|\|full | Low-quality |
| 18-104-final-viral-combined_k141_150686\|\|full | Low-quality |
| 18-104-final-viral-combined_k141_157843\|\|full | Not-determined |
| 18-104-final-viral-combined_k141_74390\|\|full | Not-determined |
| 18-104-final-viral-combined_k141_122233\|\|full | Not-determined |
| 18-104-final-viral-combined_k141_87997\|\|full | Not-determined |
| 18-104-final-viral-combined_k141_46552\|\|full | Not-determined |
| 18-104-final-viral-combined_k141_46556\|\|full | Not-determined |
| 18-104-final-viral-combined_k141_81533\|\|full | Not-determined |
| 18-104-final-viral-combined_k141_74403\|\|full | Not-determined |
| 18-104-final-viral-combined_k141_115646\|\|full | Not-determined |
| 18-104-final-viral-combined_k141_25324\|\|full | Low-quality |
| 18-104-final-viral-combined_k141_136824\|\|full | Low-quality |
| 18-104-final-viral-combined_k141_32520\|\|full | Low-quality |
| 18-104-final-viral-combined_k141_59948\|\|full | Not-determined |
| 18-104-final-viral-combined_k141_136830\|\|full | Not-determined |
| 18-104-final-viral-combined_k141_122263\|\|full | Low-quality |
| 18-104-final-viral-combined_k141_25335\|\|full | Not-determined |
| 18-104-final-viral-combined_k141_157866\|\|full | Not-determined |
| 18-104-final-viral-combined_k141_39139\|\|full | Not-determined |
| 18-104-final-viral-combined_k141_164636\|\|full | Not-determined |
| 18-104-final-viral-combined_k141_67341\|\|full | Not-determined |
| 18-104-final-viral-combined_k141_101860\|\|full | Low-quality |
| 18-104-final-viral-combined_k141_164639\|\|full | Low-quality |
| 18-104-final-viral-combined_k141_95121\|\|full | Not-determined |
| 18-104-final-viral-combined_k141_115676\|\|full | Not-determined |
| 18-104-final-viral-combined_k141_101866\|\|full | Not-determined |
| 18-104-final-viral-combined_k141_32549\|\|full | Not-determined |
| 18-104-final-viral-combined_k141_52635\|\|full | Not-determined |
| 18-104-final-viral-combined_k141_164644\|\|full | Low-quality |
| 18-104-final-viral-combined_k141_32550\|\|full | Low-quality |
| 18-104-final-viral-combined_k141_136853\|\|full | Low-quality |
| 18-104-final-viral-combined_k141_59972\|\|full | Not-determined |
| 18-104-final-viral-combined_k141_25356\|\|full | Not-determined |
| 18-104-final-viral-combined_k141_81591\|\|full | Low-quality |
| 18-104-final-viral-combined_k141_129710\|\|full | Not-determined |
| 18-104-final-viral-combined_k141_136867\|\|full | Low-quality |
| 18-104-final-viral-combined_k141_74425\|\|full | Low-quality |
| 18-104-final-viral-combined_k141_17780\|\|full | Low-quality |
| 18-104-final-viral-combined_k141_129730\|\|full | Not-determined |
| 18-104-final-viral-combined_k141_52671\|\|full | Low-quality |
| 18-104-final-viral-combined_k141_39175\|\|full | Not-determined |
| 18-104-final-viral-combined_k141_59997\|\|full | Low-quality |
| 18-104-final-viral-combined_k141_136882\|\|full | Not-determined |
| 18-104-final-viral-combined_k141_46614\|\|full | Low-quality |

| | |
|---|---|
| 18-104-final-viral-combined_k141_115717\|\|full | Not-determined |
| 18-104-final-viral-combined_k141_32576\|\|full | Not-determined |
| 18-104-final-viral-combined_k141_74442\|\|full | Low-quality |
| 18-104-final-viral-combined_k141_129750\|\|full | Not-determined |
| 18-104-final-viral-combined_k141_81628\|\|full | Low-quality |
| 18-104-final-viral-combined_k141_164686\|\|full | Low-quality |
| 18-104-final-viral-combined_k141_81630\|\|full | Low-quality |
| 18-104-final-viral-combined_k141_17790\|\|full | Not-determined |
| 18-104-final-viral-combined_k141_67386\|\|full | Not-determined |
| 18-104-final-viral-combined_k141_144112\|\|full | Not-determined |
| 18-104-final-viral-combined_k141_109403\|\|full | Not-determined |
| 18-104-final-viral-combined_k141_4491\|\|full | Not-determined |
| 18-104-final-viral-combined_k141_144115\|\|full | Not-determined |
| 18-104-final-viral-combined_k141_25431\|\|full | Not-determined |
| 18-104-final-viral-combined_k141_39193\|\|full | Low-quality |
| 18-104-final-viral-combined_k141_101945\|\|full | Not-determined |
| 18-104-final-viral-combined_k141_46634\|\|full | Not-determined |
| 18-104-final-viral-combined_k141_46635\|\|full | Low-quality |
| 18-104-final-viral-combined_k141_67396\|\|full | Low-quality |
| 18-104-final-viral-combined_k141_129778\|\|full | Not-determined |
| 18-104-final-viral-combined_k141_25436\|\|full | Low-quality |
| 18-104-final-viral-combined_k141_17812\|\|full | Low-quality |
| 18-104-final-viral-combined_k141_32604\|\|full | Low-quality |
| 18-104-final-viral-combined_k141_46642\|\|full | Low-quality |
| 18-104-final-viral-combined_k141_157937\|\|full | Low-quality |
| 18-104-final-viral-combined_k141_52731\|\|full | Not-determined |
| 18-104-final-viral-combined_k141_150758\|\|full | Low-quality |
| 18-104-final-viral-combined_k141_25453\|\|full | Low-quality |
| 18-104-final-viral-combined_k141_4514\|\|full | Not-determined |
| 18-104-final-viral-combined_k141_52734\|\|full | Not-determined |
| 18-104-final-viral-combined_k141_88095\|\|full | Low-quality |
| 18-104-final-viral-combined_k141_115765\|\|full | Not-determined |
| 18-104-final-viral-combined_k141_115766\|\|full | Not-determined |
| 18-104-final-viral-combined_k141_39236\|\|full | Not-determined |
| 18-104-final-viral-combined_k141_95173\|\|full | Not-determined |
| 18-104-final-viral-combined_k141_67442\|\|full | Not-determined |
| 18-104-final-viral-combined_k141_136943\|\|full | Not-determined |
| 18-104-final-viral-combined_k141_115787\|\|full | Not-determined |
| 18-104-final-viral-combined_k141_150777\|\|full | Not-determined |
| 18-104-final-viral-combined_k141_60071\|\|full | Low-quality |
| 18-104-final-viral-combined_k141_32628\|\|full | Not-determined |
| 18-104-final-viral-combined_k141_150781\|\|full | Not-determined |
| 18-104-final-viral-combined_k141_157979\|\|full | Not-determined |
| 18-104-final-viral-combined_k141_150784\|\|full | Not-determined |
| 18-104-final-viral-combined_k141_46666\|\|full | Low-quality |
| 18-104-final-viral-combined_k141_88132\|\|full | Not-determined |
| 18-104-final-viral-combined_k141_11710\|\|full | Low-quality |
| 18-104-final-viral-combined_k141_88135\|\|full | Not-determined |
| 18-104-final-viral-combined_k141_88138\|\|full | Low-quality |
| 18-104-final-viral-combined_k141_4559\|\|full | Low-quality |
| 18-104-final-viral-combined_k141_32648\|\|full | Not-determined |
| 18-104-final-viral-combined_k141_11726\|\|full | Low-quality |
| 18-104-final-viral-combined_k141_115823\|\|full | Low-quality |
| 18-104-final-viral-combined_k141_158011\|\|full | Low-quality |
| 18-104-final-viral-combined_k141_102007\|\|full | Not-determined |
| 18-104-final-viral-combined_k141_115830\|\|full | Not-determined |
| 18-104-final-viral-combined_k141_122412\|\|full | Low-quality |
| 18-104-final-viral-combined_k141_150820\|\|full | Low-quality |
| 18-104-final-viral-combined_k141_109474\|\|full | Not-determined |
| 18-104-final-viral-combined_k141_137001\|\|full | Not-determined |
| 18-104-final-viral-combined_k141_122418\|\|full | Low-quality |
| 18-104-final-viral-combined_k141_115839\|\|full | Not-determined |
| 18-104-final-viral-combined_k141_17865\|\|full | Low-quality |
| 18-104-final-viral-combined_k141_4595\|\|full | Low-quality |
| 18-104-final-viral-combined_k141_88180\|\|full | Not-determined |
| 18-104-final-viral-combined_k141_67513\|\|full | Not-determined |
| 18-104-final-viral-combined_k141_144225\|\|full | Low-quality |
| 18-104-final-viral-combined_k141_60148\|\|full | Not-determined |
| 18-104-final-viral-combined_k141_67523\|\|full | Low-quality |
| 18-104-final-viral-combined_k141_150867\|\|full | Low-quality |
| 18-104-final-viral-combined_k141_88197\|\|full | Low-quality |
| 18-104-final-viral-combined_k141_25525\|\|full | Low-quality |
| 18-104-final-viral-combined_k141_158084\|\|full | Not-determined |
| 18-104-final-viral-combined_k141_144267\|\|full | Not-determined |
| 18-104-final-viral-combined_k141_81774\|\|full | Not-determined |
| 18-104-final-viral-combined_k141_17893\|\|full | Low-quality |
| 18-104-final-viral-combined_k141_32688\|\|full | Low-quality |
| 18-104-final-viral-combined_k141_60160\|\|full | Not-determined |
| 18-104-final-viral-combined_k141_122456\|\|full | Low-quality |
| 18-104-final-viral-combined_k141_60163\|\|full | Low-quality |
| 18-104-final-viral-combined_k141_52861\|\|full | Low-quality |
| 18-104-final-viral-combined_k141_150907\|\|full | Not-determined |
| 18-104-final-viral-combined_k141_32702\|\|full | Not-determined |
| 18-104-final-viral-combined_k141_74630\|\|full | Not-determined |
| 18-104-final-viral-combined_k141_115929\|\|full | Not-determined |
| 18-104-final-viral-combined_k141_88251\|\|full | Low-quality |
| 18-104-final-viral-combined_k141_102034\|\|full | Low-quality |
| 18-104-final-viral-combined_k141_158122\|\|full | Low-quality |
| 18-104-final-viral-combined_k141_81820\|\|full | Not-determined |
| 18-104-final-viral-combined_k141_25585\|\|full | Low-quality |
| 18-104-final-viral-combined_k141_4697\|\|full | Low-quality |
| 18-104-final-viral-combined_k141_95322\|\|full | Not-determined |
| 18-104-final-viral-combined_k141_88262\|\|full | Not-determined |
| 18-104-final-viral-combined_k141_32729\|\|full | Not-determined |
| 18-104-final-viral-combined_k141_144299\|\|full | Low-quality |
| 18-104-final-viral-combined_k141_81824\|\|full | Not-determined |
| 18-104-final-viral-combined_k141_129923\|\|full | Low-quality |
| 18-104-final-viral-combined_k141_115953\|\|full | Low-quality |
| 18-104-final-viral-combined_k141_52887\|\|full | Low-quality |
| 18-104-final-viral-combined_k141_11854\|\|full | Not-determined |
| 18-104-final-viral-combined_k141_164880\|\|full | Not-determined |
| 18-104-final-viral-combined_k141_144304\|\|full | Not-determined |
| 18-104-final-viral-combined_k141_102069\|\|full | Not-determined |
| 18-104-final-viral-combined_k141_11864\|\|full | Not-determined |
| 18-104-final-viral-combined_k141_95345\|\|full | Low-quality |
| 18-104-final-viral-combined_k141_11866\|\|full | Low-quality |
| 18-104-final-viral-combined_k141_109638\|\|full | Not-determined |
| 18-104-final-viral-combined_k141_74662\|\|full | Not-determined |
| 18-104-final-viral-combined_k141_115975\|\|full | Low-quality |
| 18-104-final-viral-combined_k141_137106\|\|full | Low-quality |
| 18-104-final-viral-combined_k141_164898\|\|full | Not-determined |
| 18-104-final-viral-combined_k141_25620\|\|full | Low-quality |
| 18-104-final-viral-combined_k141_81863\|\|full | Not-determined |
| 18-104-final-viral-combined_k141_32761\|\|full | Low-quality |
| 18-104-final-viral-combined_k141_122504\|\|full | Low-quality |
| 18-104-final-viral-combined_k141_102075\|\|full | Not-determined |
| 18-104-final-viral-combined_k141_137113\|\|full | Low-quality |
| 18-104-final-viral-combined_k141_25627\|\|full | Not-determined |
| 18-104-final-viral-combined_k141_164909\|\|full | Not-determined |
| 18-104-final-viral-combined_k141_122507\|\|full | Low-quality |
| 18-104-final-viral-combined_k141_74688\|\|full | Low-quality |
| 18-104-final-viral-combined_k141_25637\|\|full | Low-quality |
| 18-104-final-viral-combined_k141_60240\|\|full | Low-quality |
| 18-104-final-viral-combined_k141_116015\|\|full | Not-determined |
| 18-104-final-viral-combined_k141_46778\|\|full | Not-determined |
| 18-104-final-viral-combined_k141_122525\|\|full | Not-determined |
| 18-104-final-viral-combined_k141_151007\|\|full | Not-determined |
| 18-104-final-viral-combined_k141_144342\|\|full | Not-determined |
| 18-104-final-viral-combined_k141_109677\|\|full | Not-determined |
| 18-104-final-viral-combined_k141_11895\|\|full | Not-determined |
| 18-104-final-viral-combined_k141_46783\|\|full | Not-determined |
| 18-104-final-viral-combined_k141_46786\|\|full | Low-quality |
| 18-104-final-viral-combined_k141_81898\|\|full | Low-quality |
| 18-104-final-viral-combined_k141_158205\|\|full | Not-determined |
| 18-104-final-viral-combined_k141_18022\|\|full | Low-quality |
| 18-104-final-viral-combined_k141_81907\|\|full | Not-determined |
| 18-104-final-viral-combined_k141_122555\|\|full | Not-determined |
| 18-104-final-viral-combined_k141_109689\|\|full | Not-determined |
| 18-104-final-viral-combined_k141_88335\|\|full | Low-quality |
| 18-104-final-viral-combined_k141_46794\|\|full | Low-quality |
| 18-104-final-viral-combined_k141_46803\|\|full | Not-determined |
| 18-104-final-viral-combined_k141_25684\|\|full | Low-quality |
| 18-104-final-viral-combined_k141_46807\|\|full | Low-quality |
| 18-104-final-viral-combined_k141_116046\|\|full | Low-quality |
| 18-104-final-viral-combined_k141_144385\|\|full | Low-quality |
| 18-104-final-viral-combined_k141_60284\|\|full | Not-determined |
| 18-104-final-viral-combined_k141_116057\|\|full | Not-determined |
| 18-104-final-viral-combined_k141_74758\|\|full | Low-quality |
| 18-104-final-viral-combined_k141_144395\|\|full | Not-determined |
| 18-104-final-viral-combined_k141_164972\|\|full | Low-quality |
| 18-104-final-viral-combined_k141_74761\|\|full | Low-quality |
| 18-104-final-viral-combined_k141_25695\|\|full | Low-quality |
| 18-104-final-viral-combined_k141_95450\|\|full | Not-determined |
| 18-104-final-viral-combined_k141_25699\|\|full | Low-quality |
| 18-104-final-viral-combined_k141_130066\|\|full | Not-determined |
| 18-104-final-viral-combined_k141_46817\|\|full | Not-determined |
| 18-104-final-viral-combined_k141_144408\|\|full | Low-quality |
| 18-104-final-viral-combined_k141_130072\|\|full | Low-quality |
| 18-104-final-viral-combined_k141_137191\|\|full | Low-quality |
| 18-104-final-viral-combined_k141_52994\|\|full | Low-quality |
| 18-104-final-viral-combined_k141_60314\|\|full | Low-quality |
| 18-104-final-viral-combined_k141_39491\|\|full | Low-quality |
| 18-104-final-viral-combined_k141_144428\|\|full | Low-quality |
| 18-104-final-viral-combined_k141_109734\|\|full | Low-quality |
| 18-104-final-viral-combined_k141_46849\|\|full | Not-determined |
| 18-104-final-viral-combined_k141_144431\|\|full | Low-quality |
| 18-104-final-viral-combined_k141_158296\|\|full | Not-determined |
| 18-104-final-viral-combined_k141_102196\|\|full | Low-quality |
| 18-104-final-viral-combined_k141_18089\|\|full | Not-determined |
| 18-104-final-viral-combined_k141_130109\|\|full | Not-determined |
| 18-104-final-viral-combined_k141_25751\|\|full | Not-determined |
| 18-104-final-viral-combined_k141_53008\|\|full | Not-determined |
| 18-104-final-viral-combined_k141_18100\|\|full | Not-determined |
| 18-104-final-viral-combined_k141_39521\|\|full | Not-determined |
| 18-104-final-viral-combined_k141_67725\|\|full | Low-quality |
| 18-104-final-viral-combined_k141_39525\|\|full | Not-determined |
| 18-104-final-viral-combined_k141_116115\|\|full | Low-quality |
| 18-104-final-viral-combined_k141_95493\|\|full | Not-determined |
| 18-104-final-viral-combined_k141_67726\|\|full | Low-quality |
| 18-104-final-viral-combined_k141_81970\|\|full | Low-quality |
| 18-104-final-viral-combined_k141_151054\|\|full | Low-quality |
| 18-104-final-viral-combined_k141_67740\|\|full | Low-quality |
| 18-104-final-viral-combined_k141_151061\|\|full | Low-quality |
| 18-104-final-viral-combined_k141_53033\|\|full | Not-determined |
| 18-104-final-viral-combined_k141_122663\|\|full | Not-determined |
| 18-104-final-viral-combined_k141_109786\|\|full | Low-quality |
| 18-104-final-viral-combined_k141_95505\|\|full | Not-determined |
| 18-104-final-viral-combined_k141_81978\|\|full | Not-determined |
| 18-104-final-viral-combined_k141_95512\|\|full | Not-determined |
| 18-104-final-viral-combined_k141_144488\|\|full | Not-determined |
| 18-104-final-viral-combined_k141_109797\|\|full | Not-determined |
| 18-104-final-viral-combined_k141_60361\|\|full | Not-determined |
| 18-104-final-viral-combined_k141_102271\|\|full | Not-determined |
| 18-104-final-viral-combined_k141_122679\|\|full | Low-quality |
| 18-104-final-viral-combined_k141_4875\|\|full | Not-determined |
| 18-104-final-viral-combined_k141_165078\|\|full | Low-quality |
| 18-104-final-viral-combined_k141_67770\|\|full | Not-determined |
| 18-104-final-viral-combined_k141_60373\|\|full | Low-quality |
| 18-104-final-viral-combined_k141_116159\|\|full | Not-determined |
| 18-104-final-viral-combined_k141_116160\|\|full | Low-quality |
| 18-104-final-viral-combined_k141_165096\|\|full | Not-determined |
| 18-104-final-viral-combined_k141_116173\|\|full | Not-determined |
| 18-104-final-viral-combined_k141_46916\|\|full | Not-determined |
| 18-104-final-viral-combined_k141_60384\|\|full | Not-determined |
| 18-104-final-viral-combined_k141_130158\|\|full | Low-quality |
| 18-104-final-viral-combined_k141_102287\|\|full | Low-quality |
| 18-104-final-viral-combined_k141_74858\|\|full | Low-quality |
| 18-104-final-viral-combined_k141_116196\|\|full | Low-quality |
| 18-104-final-viral-combined_k141_25808\|\|full | Low-quality |
| 18-104-final-viral-combined_k141_95534\|\|full | Not-determined |
| 18-104-final-viral-combined_k141_122714\|\|full | Low-quality |
| 18-104-final-viral-combined_k141_53068\|\|full | Not-determined |
| 18-104-final-viral-combined_k141_60399\|\|full | Not-determined |
| 18-104-final-viral-combined_k141_122718\|\|full | Not-determined |
| 18-104-final-viral-combined_k141_25826\|\|full | Low-quality |
| 18-104-final-viral-combined_k141_4918\|\|full | Not-determined |
| 18-104-final-viral-combined_k141_116212\|\|full | Not-determined |
| 18-104-final-viral-combined_k141_137300\|\|full | Not-determined |
| 18-104-final-viral-combined_k141_116219\|\|full | Low-quality |
| 18-104-final-viral-combined_k141_109841\|\|full | Not-determined |
| 18-104-final-viral-combined_k141_116223\|\|full | Low-quality |
| 18-104-final-viral-combined_k141_82060\|\|full | Low-quality |
| 18-104-final-viral-combined_k141_144579\|\|full | Not-determined |
| 18-104-final-viral-combined_k141_18213\|\|full | Not-determined |
| 18-104-final-viral-combined_k141_67828\|\|full | Not-determined |
| 18-104-final-viral-combined_k141_137318\|\|full | Low-quality |
| 18-104-final-viral-combined_k141_32959\|\|full | Low-quality |
| 18-104-final-viral-combined_k141_46974\|\|full | Not-determined |
| 18-104-final-viral-combined_k141_39591\|\|full | Not-determined |
| 18-104-final-viral-combined_k141_53101\|\|full | Low-quality |
| 18-104-final-viral-combined_k141_67837\|\|full | Low-quality |
| 18-104-final-viral-combined_k141_4959\|\|full | Not-determined |
| 18-104-final-viral-combined_k141_39594\|\|full | Not-determined |
| 18-104-final-viral-combined_k141_53112\|\|full | Not-determined |
| 18-104-final-viral-combined_k141_151176\|\|full | Not-determined |
| 18-104-final-viral-combined_k141_82074\|\|full | Low-quality |
| 18-104-final-viral-combined_k141_130215\|\|full | Low-quality |
| 18-104-final-viral-combined_k141_158480\|\|full | Not-determined |
| 18-104-final-viral-combined_k141_137341\|\|full | Not-determined |
| 18-104-final-viral-combined_k141_158482\|\|full | Low-quality |
| 18-104-final-viral-combined_k141_158486\|\|full | Not-determined |
| 18-104-final-viral-combined_k141_4978\|\|full | Low-quality |
| 18-104-final-viral-combined_k141_53134\|\|full | Low-quality |
| 18-104-final-viral-combined_k141_74958\|\|full | Not-determined |
| 18-104-final-viral-combined_k141_102367\|\|full | Low-quality |
| 18-104-final-viral-combined_k141_46987\|\|full | Low-quality |
| 18-104-final-viral-combined_k141_144619\|\|full | Not-determined |
| 18-104-final-viral-combined_k141_109889\|\|full | Not-determined |
| 18-104-final-viral-combined_k141_109891\|\|full | Low-quality |
| 18-104-final-viral-combined_k141_144628\|\|full | Not-determined |
| 18-104-final-viral-combined_k141_33001\|\|full | Not-determined |
| 18-104-final-viral-combined_k141_4988\|\|full | Low-quality |
| 18-104-final-viral-combined_k141_18266\|\|full | Not-determined |
| 18-104-final-viral-combined_k141_95634\|\|full | Low-quality |
| 18-104-final-viral-combined_k141_144631\|\|full | Not-determined |
| 18-104-final-viral-combined_k141_67853\|\|full | Not-determined |
| 18-104-final-viral-combined_k141_158524\|\|full | Low-quality |
| 18-104-final-viral-combined_k141_53172\|\|full | Low-quality |
| 18-104-final-viral-combined_k141_116281\|\|full | Not-determined |
| 18-104-final-viral-combined_k141_25906\|\|full | Not-determined |
| 18-104-final-viral-combined_k141_12085\|\|full | Not-determined |
| 18-104-final-viral-combined_k141_102402\|\|full | Not-determined |
| 18-104-final-viral-combined_k141_18294\|\|full | Low-quality |
| 18-104-final-viral-combined_k141_39629\|\|full | Not-determined |
| 18-104-final-viral-combined_k141_116290\|\|full | Not-determined |
| 18-104-final-viral-combined_k141_12095\|\|full | Not-determined |
| 18-104-final-viral-combined_k141_82096\|\|full | Not-determined |
| 18-104-final-viral-combined_k141_12096\|\|full | Low-quality |
| 18-104-final-viral-combined_k141_67889\|\|full | Low-quality |
| 18-104-final-viral-combined_k141_102428\|\|full | Low-quality |
| 18-104-final-viral-combined_k141_165258\|\|full | Not-determined |
| 18-104-final-viral-combined_k141_165264\|\|full | Not-determined |
| 18-104-final-viral-combined_k141_18340\|\|full | Low-quality |
| 18-104-final-viral-combined_k141_95659\|\|full | Not-determined |
| 18-104-final-viral-combined_k141_95660\|\|full | Not-determined |
| 18-104-final-viral-combined_k141_67915\|\|full | Low-quality |
| 18-104-final-viral-combined_k141_18353\|\|full | Low-quality |
| 18-104-final-viral-combined_k141_151264\|\|full | Not-determined |
| 18-104-final-viral-combined_k141_151269\|\|full | Low-quality |
| 18-104-final-viral-combined_k141_102460\|\|full | Low-quality |
| 18-104-final-viral-combined_k141_122815\|\|full | Low-quality |
| 18-104-final-viral-combined_k141_67937\|\|full | Not-determined |
| 18-104-final-viral-combined_k141_18363\|\|full | Low-quality |
| 18-104-final-viral-combined_k141_82144\|\|full | Not-determined |
| 18-104-final-viral-combined_k141_137375\|\|full | Low-quality |
| 18-104-final-viral-combined_k141_5001\|\|full | Not-determined |
| 18-104-final-viral-combined_k141_39675\|\|full | Low-quality |
| 18-104-final-viral-combined_k141_144693\|\|full | Low-quality |
| 18-104-final-viral-combined_k141_102486\|\|full | Low-quality |
| 18-104-final-viral-combined_k141_109975\|\|full | Low-quality |
| 18-104-final-viral-combined_k141_144697\|\|full | Low-quality |
| 18-104-final-viral-combined_k141_53240\|\|full | Not-determined |
| 18-104-final-viral-combined_k141_130277\|\|full | Low-quality |
| 18-104-final-viral-combined_k141_39697\|\|full | Not-determined |
| 18-104-final-viral-combined_k141_39699\|\|full | Not-determined |
| 18-104-final-viral-combined_k141_151281\|\|full | Low-quality |
| 18-104-final-viral-combined_k141_33065\|\|full | Low-quality |
| 18-104-final-viral-combined_k141_53252\|\|full | Not-determined |
| 18-104-final-viral-combined_k141_25982\|\|full | Low-quality |
| 18-104-final-viral-combined_k141_109994\|\|full | Low-quality |
| 18-104-final-viral-combined_k141_122866\|\|full | Not-determined |
| 18-104-final-viral-combined_k141_165320\|\|full | Not-determined |
| 18-104-final-viral-combined_k141_33073\|\|full | Not-determined |
| 18-104-final-viral-combined_k141_158663\|\|full | Not-determined |
| 18-104-final-viral-combined_k141_158664\|\|full | Low-quality |
| 18-104-final-viral-combined_k141_88641\|\|full | Not-determined |
| 18-104-final-viral-combined_k141_39721\|\|full | Not-determined |
| 18-104-final-viral-combined_k141_122879\|\|full | Low-quality |
| 18-104-final-viral-combined_k141_158682\|\|full | Low-quality |
| 18-104-final-viral-combined_k141_165341\|\|full | Not-determined |
| 18-104-final-viral-combined_k141_102546\|\|full | Low-quality |
| 18-104-final-viral-combined_k141_33086\|\|full | Low-quality |
| 18-104-final-viral-combined_k141_88659\|\|full | Low-quality |
| 18-104-final-viral-combined_k141_39737\|\|full | Low-quality |
| 18-104-final-viral-combined_k141_158687\|\|full | Low-quality |
| 18-104-final-viral-combined_k141_130344\|\|full | Not-determined |
| 18-104-final-viral-combined_k141_33102\|\|full | Low-quality |
| 18-104-final-viral-combined_k141_18439\|\|full | Not-determined |
| 18-104-final-viral-combined_k141_12202\|\|full | Not-determined |
| 18-104-final-viral-combined_k141_18445\|\|full | Low-quality |
| 18-104-final-viral-combined_k141_67986\|\|full | Not-determined |
| 18-104-final-viral-combined_k141_18448\|\|full | Low-quality |
| 18-104-final-viral-combined_k141_53281\|\|full | Low-quality |
| 18-104-final-viral-combined_k141_67991\|\|full | Low-quality |
| 18-104-final-viral-combined_k141_110061\|\|full | Low-quality |
| 18-104-final-viral-combined_k141_60551\|\|full | Low-quality |
| 18-104-final-viral-combined_k141_53297\|\|full | Low-quality |
| 18-104-final-viral-combined_k141_144783\|\|full | Low-quality |
| 18-104-final-viral-combined_k141_110083\|\|full | Not-determined |
| 18-104-final-viral-combined_k141_144787\|\|full | Low-quality |
| 18-104-final-viral-combined_k141_88695\|\|full | Low-quality |
| 18-104-final-viral-combined_k141_26048\|\|full | Not-determined |
| 18-104-final-viral-combined_k141_110092\|\|full | Low-quality |
| 18-104-final-viral-combined_k141_110095\|\|full | Low-quality |
| 18-104-final-viral-combined_k141_53313\|\|full | Not-determined |
| 18-104-final-viral-combined_k141_144801\|\|full | Not-determined |
| 18-104-final-viral-combined_k141_110097\|\|full | Not-determined |
| 18-104-final-viral-combined_k141_53315\|\|full | Low-quality |
| 18-104-final-viral-combined_k141_130407\|\|full | Low-quality |
| 18-104-final-viral-combined_k141_53319\|\|full | Not-determined |
| 18-104-final-viral-combined_k141_151378\|\|full | Low-quality |
| 18-104-final-viral-combined_k141_33150\|\|full | Not-determined |
| 18-104-final-viral-combined_k141_75094\|\|full | Low-quality |
| 18-104-final-viral-combined_k141_88718\|\|full | Low-quality |
| 18-104-final-viral-combined_k141_151381\|\|full | Not-determined |
| 18-104-final-viral-combined_k141_75096\|\|full | Not-determined |
| 18-104-final-viral-combined_k141_18480\|\|full | Not-determined |
| 18-104-final-viral-combined_k141_82218\|\|full | Low-quality |
| 18-104-final-viral-combined_k141_144819\|\|full | Low-quality |
| 18-104-final-viral-combined_k141_151398\|\|full | Not-determined |
| 18-104-final-viral-combined_k141_82224\|\|full | Not-determined |
| 18-104-final-viral-combined_k141_95825\|\|full | Low-quality |
| 18-104-final-viral-combined_k141_110136\|\|full | Low-quality |
| 18-104-final-viral-combined_k141_151412\|\|full | Low-quality |
| 18-104-final-viral-combined_k141_5074\|\|full | Low-quality |
| 18-104-final-viral-combined_k141_130443\|\|full | Low-quality |
| 18-104-final-viral-combined_k141_110143\|\|full | Not-determined |
| 18-104-final-viral-combined_k141_130448\|\|full | Low-quality |
| 18-104-final-viral-combined_k141_39856\|\|full | Not-determined |
| 18-104-final-viral-combined_k141_18506\|\|full | Not-determined |
| 18-104-final-viral-combined_k141_151424\|\|full | Low-quality |
| 18-104-final-viral-combined_k141_88763\|\|full | Low-quality |
| 18-104-final-viral-combined_k141_158797\|\|full | Low-quality |
| 18-104-final-viral-combined_k141_144830\|\|full | Low-quality |
| 18-104-final-viral-combined_k141_82241\|\|full | Not-determined |
| 18-104-final-viral-combined_k141_151432\|\|full | Not-determined |
| 18-104-final-viral-combined_k141_47159\|\|full | Low-quality |
| 18-104-final-viral-combined_k141_144841\|\|full | Low-quality |
| 18-104-final-viral-combined_k141_158818\|\|full | Low-quality |
| 18-104-final-viral-combined_k141_39875\|\|full | Not-determined |
| 18-104-final-viral-combined_k141_75133\|\|full | Not-determined |
| 18-104-final-viral-combined_k141_123014\|\|full | Low-quality |
| 18-104-final-viral-combined_k141_82253\|\|full | Not-determined |
| 18-104-final-viral-combined_k141_33225\|\|full | Low-quality |
| 18-104-final-viral-combined_k141_110204\|\|full | Not-determined |
| 18-104-final-viral-combined_k141_144863\|\|full | Not-determined |
| 18-104-final-viral-combined_k141_151446\|\|full | Not-determined |
| 18-104-final-viral-combined_k141_88801\|\|full | Low-quality |
| 18-104-final-viral-combined_k141_137471\|\|full | Not-determined |
| 18-104-final-viral-combined_k141_33237\|\|full | Low-quality |
| 18-104-final-viral-combined_k141_26140\|\|full | Not-determined |
| 18-104-final-viral-combined_k141_5096\|\|full | Not-determined |
| 18-104-final-viral-combined_k141_75154\|\|full | Not-determined |
| 18-104-final-viral-combined_k141_82279\|\|full | Not-determined |
| 18-104-final-viral-combined_k141_47213\|\|full | Not-determined |
| 18-104-final-viral-combined_k141_33246\|\|full | Not-determined |
| 18-104-final-viral-combined_k141_151465\|\|full | Not-determined |
| 18-104-final-viral-combined_k141_5102\|\|full | Not-determined |
| 18-104-final-viral-combined_k141_116569\|\|full | Not-determined |
| 18-104-final-viral-combined_k141_18559\|\|full | Not-determined |
| 18-104-final-viral-combined_k141_110227\|\|full | Not-determined |
| 18-104-final-viral-combined_k141_123043\|\|full | Low-quality |
| 18-104-final-viral-combined_k141_82284\|\|full | Not-determined |
| 18-104-final-viral-combined_k141_12364\|\|full | Low-quality |
| 18-104-final-viral-combined_k141_110239\|\|full | Low-quality |
| 18-104-final-viral-combined_k141_151477\|\|full | Low-quality |
| 18-104-final-viral-combined_k141_75191\|\|full | Low-quality |
| 18-104-final-viral-combined_k141_88822\|\|full | Not-determined |
| 18-104-final-viral-combined_k141_82299\|\|full | Not-determined |
| 18-104-final-viral-combined_k141_53437\|\|full | Low-quality |
| 18-104-final-viral-combined_k141_82303\|\|full | Not-determined |
| 18-104-final-viral-combined_k141_33293\|\|full | Not-determined |
| 18-104-final-viral-combined_k141_53447\|\|full | Not-determined |
| 18-104-final-viral-combined_k141_95964\|\|full | Not-determined |
| 18-104-final-viral-combined_k141_123075\|\|full | Low-quality |
| 18-104-final-viral-combined_k141_5146\|\|full | Low-quality |
| 18-104-final-viral-combined_k141_137534\|\|full | Not-determined |
| 18-104-final-viral-combined_k141_68138\|\|full | Not-determined |
| 18-104-final-viral-combined_k141_110268\|\|full | Not-determined |
| 18-104-final-viral-combined_k141_47260\|\|full | Not-determined |
| 18-104-final-viral-combined_k141_151509\|\|full | Low-quality |
| 18-104-final-viral-combined_k141_39970\|\|full | Low-quality |
| 18-104-final-viral-combined_k141_68142\|\|full | Not-determined |
| 18-104-final-viral-combined_k141_60683\|\|full | Low-quality |
| 18-104-final-viral-combined_k141_75237\|\|full | Not-determined |
| 18-104-final-viral-combined_k141_123084\|\|full | Low-quality |
| 18-104-final-viral-combined_k141_47273\|\|full | Low-quality |
| 18-104-final-viral-combined_k141_144958\|\|full | Low-quality |
| 18-104-final-viral-combined_k141_12414\|\|full | Low-quality |
| 18-104-final-viral-combined_k141_123086\|\|full | Not-determined |
| 18-104-final-viral-combined_k141_110295\|\|full | Not-determined |
| 18-104-final-viral-combined_k141_88855\|\|full | Not-determined |
| 18-104-final-viral-combined_k141_18649\|\|full | Low-quality |
| 18-104-final-viral-combined_k141_158890\|\|full | Low-quality |
| 18-104-final-viral-combined_k141_158891\|\|full | Low-quality |
| 18-104-final-viral-combined_k141_96024\|\|full | Low-quality |
| 18-104-final-viral-combined_k141_130575\|\|full | Low-quality |
| 18-104-final-viral-combined_k141_137570\|\|full | Not-determined |
| 18-104-final-viral-combined_k141_151552\|\|full | Low-quality |
| 18-104-final-viral-combined_k141_88864\|\|full | Low-quality |
| 18-104-final-viral-combined_k141_151554\|\|full | Low-quality |
| 18-104-final-viral-combined_k141_144978\|\|full | Low-quality |
| 18-104-final-viral-combined_k141_33350\|\|full | Not-determined |
| 18-104-final-viral-combined_k141_47298\|\|full | Low-quality |
| 18-104-final-viral-combined_k141_144981\|\|full | Not-determined |
| 18-104-final-viral-combined_k141_39997\|\|full | Not-determined |
| 18-103-final-viral-combined_k141_45305\|\|full | Medium-quality |
| 18-104-final-viral-combined_k141_18683\|\|full | Not-determined |
| 18-104-final-viral-combined_k141_123135\|\|full | Low-quality |
| 18-104-final-viral-combined_k141_12426\|\|full | Not-determined |
| 18-104-final-viral-combined_k141_123138\|\|full | Low-quality |
| 18-104-final-viral-combined_k141_116636\|\|full | Low-quality |
| 18-104-final-viral-combined_k141_18696\|\|full | Not-determined |
| 18-104-final-viral-combined_k141_40014\|\|full | Low-quality |
| 18-104-final-viral-combined_k141_110329\|\|full | Not-determined |
| 18-104-final-viral-combined_k141_151586\|\|full | Low-quality |
| 18-104-final-viral-combined_k141_145005\|\|full | Low-quality |
| 18-104-final-viral-combined_k141_130626\|\|full | Low-quality |
| 18-104-final-viral-combined_k141_88906\|\|full | Low-quality |
| 18-104-final-viral-combined_k141_88914\|\|full | Not-determined |
| 18-104-final-viral-combined_k141_130632\|\|full | Not-determined |
| 18-104-final-viral-combined_k141_96088\|\|full | Low-quality |
| 18-104-final-viral-combined_k141_110344\|\|full | Low-quality |
| 18-104-final-viral-combined_k141_82360\|\|full | Low-quality |
| 18-104-final-viral-combined_k141_158992\|\|full | Low-quality |
| 18-104-final-viral-combined_k141_40052\|\|full | Low-quality |
| 18-104-final-viral-combined_k141_130646\|\|full | Low-quality |
| 18-104-final-viral-combined_k141_82367\|\|full | Not-determined |
| 18-104-final-viral-combined_k141_137650\|\|full | Not-determined |
| 18-104-final-viral-combined_k141_75306\|\|full | Low-quality |
| 18-104-final-viral-combined_k141_110381\|\|full | Low-quality |
| 18-104-final-viral-combined_k141_130661\|\|full | Not-determined |
| 18-104-final-viral-combined_k141_47373\|\|full | Low-quality |
| 18-104-final-viral-combined_k141_5309\|\|full | Low-quality |
| 18-104-final-viral-combined_k141_68277\|\|full | Not-determined |
| 18-104-final-viral-combined_k141_40084\|\|full | Low-quality |
| 18-104-final-viral-combined_k141_130670\|\|full | Low-quality |
| 18-104-final-viral-combined_k141_145061\|\|full | Low-quality |
| 18-104-final-viral-combined_k141_53582\|\|full | Not-determined |
| 18-104-final-viral-combined_k141_47388\|\|full | Low-quality |
| 18-104-final-viral-combined_k141_40094\|\|full | Low-quality |
| 18-104-final-viral-combined_k141_82411\|\|full | Low-quality |
| 18-104-final-viral-combined_k141_26308\|\|full | Not-determined |
| 18-104-final-viral-combined_k141_116721\|\|full | Not-determined |
| 18-104-final-viral-combined_k141_26312\|\|full | Not-determined |
| 18-104-final-viral-combined_k141_18774\|\|full | Not-determined |
| 18-104-final-viral-combined_k141_47407\|\|full | Low-quality |
| 18-104-final-viral-combined_k141_26325\|\|full | Low-quality |
| 18-104-final-viral-combined_k141_145077\|\|full | Low-quality |
| 18-104-final-viral-combined_k141_33424\|\|full | Not-determined |
| 18-104-final-viral-combined_k141_18784\|\|full | Not-determined |
| 18-104-final-viral-combined_k141_110429\|\|full | Not-determined |
| 18-104-final-viral-combined_k141_123238\|\|full | Low-quality |
| 18-104-final-viral-combined_k141_102708\|\|full | Low-quality |
| 18-104-final-viral-combined_k141_60856\|\|full | Not-determined |
| 18-104-final-viral-combined_k141_12514\|\|full | Low-quality |
| 18-104-final-viral-combined_k141_40140\|\|full | Low-quality |
| 18-104-final-viral-combined_k141_130685\|\|full | Low-quality |
| 18-104-final-viral-combined_k141_40149\|\|full | Low-quality |
| 18-104-final-viral-combined_k141_151707\|\|full | Not-determined |
| 18-104-final-viral-combined_k141_40153\|\|full | Low-quality |
| 18-104-final-viral-combined_k141_102732\|\|full | Low-quality |
| 18-104-final-viral-combined_k141_130693\|\|full | Not-determined |
| 18-104-final-viral-combined_k141_96192\|\|full | Not-determined |
| 18-104-final-viral-combined_k141_123259\|\|full | Low-quality |
| 18-104-final-viral-combined_k141_130698\|\|full | Not-determined |
| 18-104-final-viral-combined_k141_96200\|\|full | Low-quality |
| 18-104-final-viral-combined_k141_68324\|\|full | Not-determined |
| 18-104-final-viral-combined_k141_116745\|\|full | Not-determined |
| 18-104-final-viral-combined_k141_137733\|\|full | Not-determined |
| 18-104-final-viral-combined_k141_53616\|\|full | Not-determined |
| 18-104-final-viral-combined_k141_123264\|\|full | Not-determined |
| 18-104-final-viral-combined_k141_82479\|\|full | Low-quality |
| 18-104-final-viral-combined_k141_102752\|\|full | Low-quality |
| 18-104-final-viral-combined_k141_40183\|\|full | Low-quality |
| 18-104-final-viral-combined_k141_137737\|\|full | Not-determined |
| 18-104-final-viral-combined_k141_40184\|\|full | Not-determined |
| 18-104-final-viral-combined_k141_123285\|\|full | Not-determined |
| 18-104-final-viral-combined_k141_5424\|\|full | Low-quality |
| 18-104-final-viral-combined_k141_53630\|\|full | Not-determined |
| 18-104-final-viral-combined_k141_33501\|\|full | Low-quality |
| 18-104-final-viral-combined_k141_116765\|\|full | Low-quality |
| 18-104-final-viral-combined_k141_33512\|\|full | Not-determined |
| 18-104-final-viral-combined_k141_96247\|\|full | Low-quality |
| 18-104-final-viral-combined_k141_130742\|\|full | Not-determined |
| 18-104-final-viral-combined_k141_151738\|\|full | Not-determined |
| 18-104-final-viral-combined_k141_75388\|\|full | Low-quality |
| 18-104-final-viral-combined_k141_26399\|\|full | Low-quality |
| 18-104-final-viral-combined_k141_75392\|\|full | Not-determined |
| 18-104-final-viral-combined_k141_12595\|\|full | Low-quality |
| 18-104-final-viral-combined_k141_89083\|\|full | Low-quality |
| 18-104-final-viral-combined_k141_151766\|\|full | Low-quality |
| 18-104-final-viral-combined_k141_159098\|\|full | Low-quality |
| 18-104-final-viral-combined_k141_116800\|\|full | Not-determined |
| 18-104-final-viral-combined_k141_18867\|\|full | Not-determined |
| 18-104-final-viral-combined_k141_18868\|\|full | Low-quality |
| 18-104-final-viral-combined_k141_145180\|\|full | Low-quality |
| 18-104-final-viral-combined_k141_89113\|\|full | Not-determined |
| 18-104-final-viral-combined_k141_75418\|\|full | Not-determined |
| 18-104-final-viral-combined_k141_26442\|\|full | Low-quality |
| 18-104-final-viral-combined_k141_5473\|\|full | Low-quality |
| 18-104-final-viral-combined_k141_116818\|\|full | Low-quality |
| 18-104-final-viral-combined_k141_5479\|\|full | Low-quality |
| 18-104-final-viral-combined_k141_145190\|\|full | Low-quality |
| 18-104-final-viral-combined_k141_47479\|\|full | Not-determined |
| 18-104-final-viral-combined_k141_96296\|\|full | Low-quality |
| 18-104-final-viral-combined_k141_102863\|\|full | Low-quality |
| 18-104-final-viral-combined_k141_159128\|\|full | Low-quality |
| 18-104-final-viral-combined_k141_116824\|\|full | Low-quality |
| 18-104-final-viral-combined_k141_26450\|\|full | Not-determined |
| 18-104-final-viral-combined_k141_145207\|\|full | Low-quality |
| 18-104-final-viral-combined_k141_5505\|\|full | Not-determined |
| 18-104-final-viral-combined_k141_60984\|\|full | Not-determined |
| 18-104-final-viral-combined_k141_18920\|\|full | Not-determined |
| 18-104-final-viral-combined_k141_116838\|\|full | Not-determined |
| 18-104-final-viral-combined_k141_26463\|\|full | Not-determined |
| 18-104-final-viral-combined_k141_89146\|\|full | Low-quality |
| 18-104-final-viral-combined_k141_89148\|\|full | Not-determined |
| 18-104-final-viral-combined_k141_5522\|\|full | Not-determined |
| 18-104-final-viral-combined_k141_60997\|\|full | Low-quality |
| 18-104-final-viral-combined_k141_110565\|\|full | Low-quality |
| 18-104-final-viral-combined_k141_5523\|\|full | Low-quality |
| 18-104-final-viral-combined_k141_33601\|\|full | Not-determined |
| 18-104-final-viral-combined_k141_47494\|\|full | Not-determined |
| 18-104-final-viral-combined_k141_18938\|\|full | Low-quality |
| 18-104-final-viral-combined_k141_33604\|\|full | Low-quality |
| 18-104-final-viral-combined_k141_130767\|\|full | Low-quality |
| 18-104-final-viral-combined_k141_75455\|\|full | Low-quality |
| 18-104-final-viral-combined_k141_145219\|\|full | Low-quality |
| 18-104-final-viral-combined_k141_40313\|\|full | Not-determined |
| 18-104-final-viral-combined_k141_159157\|\|full | Low-quality |
| 18-104-final-viral-combined_k141_33611\|\|full | Not-determined |
| 18-104-final-viral-combined_k141_53727\|\|full | Low-quality |
| 18-104-final-viral-combined_k141_12684\|\|full | Not-determined |
| 18-104-final-viral-combined_k141_116863\|\|full | Low-quality |
| 18-104-final-viral-combined_k141_18951\|\|full | Low-quality |
| 18-104-final-viral-combined_k141_159162\|\|full | Not-determined |
| 18-104-final-viral-combined_k141_61011\|\|full | Not-determined |
| 18-104-final-viral-combined_k141_18958\|\|full | Not-determined |
| 18-104-final-viral-combined_k141_110592\|\|full | Low-quality |
| 18-104-final-viral-combined_k141_82676\|\|full | Not-determined |
| 18-104-final-viral-combined_k141_145244\|\|full | Not-determined |
| 18-104-final-viral-combined_k141_75487\|\|full | Low-quality |
| 18-104-final-viral-combined_k141_26475\|\|full | Not-determined |
| 18-104-final-viral-combined_k141_40340\|\|full | Low-quality |
| 18-104-final-viral-combined_k141_61032\|\|full | Low-quality |
| 18-104-final-viral-combined_k141_89191\|\|full | Not-determined |
| 18-104-final-viral-combined_k141_145249\|\|full | Not-determined |
| 18-104-final-viral-combined_k141_159187\|\|full | Not-determined |
| 18-104-final-viral-combined_k141_47548\|\|full | Not-determined |
| 18-104-final-viral-combined_k141_26484\|\|full | Not-determined |
| 18-104-final-viral-combined_k141_26491\|\|full | Low-quality |
| 18-104-final-viral-combined_k141_110617\|\|full | Low-quality |
| 18-104-final-viral-combined_k141_123459\|\|full | Low-quality |
| 18-104-final-viral-combined_k141_5543\|\|full | Low-quality |
| 18-104-final-viral-combined_k141_145265\|\|full | Not-determined |
| 18-104-final-viral-combined_k141_40353\|\|full | Low-quality |
| 18-104-final-viral-combined_k141_145266\|\|full | Low-quality |
| 18-104-final-viral-combined_k141_33651\|\|full | Low-quality |

| | |
|---|---|
| 18-104-final-viral-combined_k141_96371\|\|full | Low-quality |
| 18-104-final-viral-combined_k141_165568\|\|full | Low-quality |
| 18-104-final-viral-combined_k141_130821\|\|full | Not-determined |
| 18-104-final-viral-combined_k141_130823\|\|full | Not-determined |
| 18-104-final-viral-combined_k141_89215\|\|full | Low-quality |
| 18-104-final-viral-combined_k141_89221\|\|full | Low-quality |
| 18-104-final-viral-combined_k141_165580\|\|full | Not-determined |
| 18-104-final-viral-combined_k141_75526\|\|full | Low-quality |
| 18-104-final-viral-combined_k141_116918\|\|full | Low-quality |
| 18-104-final-viral-combined_k141_89228\|\|full | Low-quality |
| 18-104-final-viral-combined_k141_68466\|\|full | Not-determined |
| 18-104-final-viral-combined_k141_137887\|\|full | Low-quality |
| 18-104-final-viral-combined_k141_151826\|\|full | Not-determined |
| 18-104-final-viral-combined_k141_68479\|\|full | Low-quality |
| 18-105_k141_49310 | Medium-quality |
| 18-104-final-viral-combined_k141_123490\|\|full | Not-determined |
| 18-104-final-viral-combined_k141_68496\|\|full | Not-determined |
| 18-104-final-viral-combined_k141_116952\|\|full | Low-quality |
| 18-104-final-viral-combined_k141_47574\|\|full | Low-quality |
| 18-104-final-viral-combined_k141_33686\|\|full | Low-quality |
| 18-104-final-viral-combined_k141_47575\|\|full | Not-determined |
| 18-104-final-viral-combined_k141_110667\|\|full | Not-determined |
| 18-104-final-viral-combined_k141_145321\|\|full | Low-quality |
| 18-104-final-viral-combined_k141_159222\|\|full | Not-determined |
| 18-104-final-viral-combined_k141_137914\|\|full | Low-quality |
| 18-104-final-viral-combined_k141_26564\|\|full | Low-quality |
| 18-104-final-viral-combined_k141_159233\|\|full | Low-quality |
| 18-104-final-viral-combined_k141_159234\|\|full | Low-quality |
| 18-104-final-viral-combined_k141_47594\|\|full | Not-determined |
| 18-104-final-viral-combined_k141_116984\|\|full | Not-determined |
| 18-104-final-viral-combined_k141_19054\|\|full | Low-quality |
| 18-104-final-viral-combined_k141_96406\|\|full | Not-determined |
| 18-104-final-viral-combined_k141_110691\|\|full | Not-determined |
| 18-104-final-viral-combined_k141_33711\|\|full | Not-determined |
| 18-104-final-viral-combined_k141_89288\|\|full | Low-quality |
| 18-104-final-viral-combined_k141_75600\|\|full | Low-quality |
| 18-104-final-viral-combined_k141_82784\|\|full | Not-determined |
| 18-104-final-viral-combined_k141_89292\|\|full | Not-determined |
| 18-104-final-viral-combined_k141_110695\|\|full | Low-quality |
| 18-104-final-viral-combined_k141_130909\|\|full | Low-quality |
| 18-104-final-viral-combined_k141_151856\|\|full | Low-quality |
| 18-104-final-viral-combined_k141_26574\|\|full | Not-determined |
| 18-104-final-viral-combined_k141_116996\|\|full | Not-determined |
| 18-104-final-viral-combined_k141_82796\|\|full | Not-determined |
| 18-104-final-viral-combined_k141_40441\|\|full | Low-quality |
| 18-104-final-viral-combined_k141_40443\|\|full | Not-determined |
| 18-104-final-viral-combined_k141_159263\|\|full | Not-determined |
| 18-104-final-viral-combined_k141_103028\|\|full | Not-determined |
| 18-104-final-viral-combined_k141_53823\|\|full | Low-quality |
| 18-104-final-viral-combined_k141_5646\|\|full | Not-determined |
| 18-104-final-viral-combined_k141_40458\|\|full | Not-determined |
| 18-104-final-viral-combined_k141_165657\|\|full | Not-determined |
| 18-104-final-viral-combined_k141_40460\|\|full | Low-quality |
| 18-104-final-viral-combined_k141_33743\|\|full | Low-quality |
| 18-104-final-viral-combined_k141_47662\|\|full | Not-determined |
| 18-104-final-viral-combined_k141_110715\|\|full | Low-quality |
| 18-104-final-viral-combined_k141_75661\|\|full | Low-quality |
| 18-104-final-viral-combined_k141_82837\|\|full | Low-quality |
| 18-104-final-viral-combined_k141_40476\|\|full | Not-determined |
| 18-104-final-viral-combined_k141_110725\|\|full | Not-determined |
| 18-104-final-viral-combined_k141_165672\|\|full | Low-quality |
| 18-104-final-viral-combined_k141_145411\|\|full | Low-quality |
| 18-104-final-viral-combined_k141_61176\|\|full | Not-determined |
| 18-104-final-viral-combined_k141_12747\|\|full | Not-determined |
| 18-104-final-viral-combined_k141_82843\|\|full | Not-determined |
| 18-104-final-viral-combined_k141_19125\|\|full | Low-quality |
| 18-104-final-viral-combined_k141_123563\|\|full | Not-determined |
| 18-104-final-viral-combined_k141_151902\|\|full | Not-determined |
| 18-104-final-viral-combined_k141_159288\|\|full | Low-quality |
| 18-104-final-viral-combined_k141_96445\|\|full | Low-quality |
| 18-104-final-viral-combined_k141_19148\|\|full | Low-quality |
| 18-104-final-viral-combined_k141_12772\|\|full | Low-quality |
| 18-104-final-viral-combined_k141_68618\|\|full | Low-quality |
| 18-104-final-viral-combined_k141_33764\|\|full | Not-determined |
| 18-104-final-viral-combined_k141_33769\|\|full | Not-determined |
| 18-104-final-viral-combined_k141_130964\|\|full | Not-determined |
| 18-104-final-viral-combined_k141_12805\|\|full | Not-determined |
| 18-104-final-viral-combined_k141_130968\|\|full | Low-quality |
| 18-104-final-viral-combined_k141_145453\|\|full | Not-determined |
| 18-104-final-viral-combined_k141_159325\|\|full | Not-determined |
| 18-104-final-viral-combined_k141_53879\|\|full | Not-determined |
| 18-104-final-viral-combined_k141_47732\|\|full | Not-determined |
| 18-104-final-viral-combined_k141_165717\|\|full | Low-quality |
| 18-104-final-viral-combined_k141_137991\|\|full | Not-determined |
| 18-104-final-viral-combined_k141_75723\|\|full | Low-quality |
| 18-104-final-viral-combined_k141_103131\|\|full | Low-quality |
| 18-104-final-viral-combined_k141_19200\|\|full | Not-determined |
| 18-104-final-viral-combined_k141_103138\|\|full | Not-determined |
| 18-104-final-viral-combined_k141_145500\|\|full | Not-determined |
| 18-104-final-viral-combined_k141_103143\|\|full | Not-determined |
| 18-104-final-viral-combined_k141_33810\|\|full | Not-determined |
| 18-104-final-viral-combined_k141_131008\|\|full | Low-quality |
| 18-104-final-viral-combined_k141_82902\|\|full | Low-quality |
| 18-104-final-viral-combined_k141_165742\|\|full | Not-determined |
| 18-104-final-viral-combined_k141_151985\|\|full | Not-determined |
| 18-104-final-viral-combined_k141_96518\|\|full | Low-quality |
| 18-104-final-viral-combined_k141_131026\|\|full | Low-quality |
| 18-104-final-viral-combined_k141_12879\|\|full | Not-determined |
| 18-104-final-viral-combined_k141_152002\|\|full | Not-determined |
| 18-104-final-viral-combined_k141_89457\|\|full | Not-determined |
| 18-104-final-viral-combined_k141_12881\|\|full | Not-determined |
| 18-104-final-viral-combined_k141_53923\|\|full | Low-quality |
| 18-104-final-viral-combined_k141_53927\|\|full | Low-quality |
| 18-104-final-viral-combined_k141_19239\|\|full | Not-determined |
| 18-104-final-viral-combined_k141_145545\|\|full | Not-determined |
| 18-104-final-viral-combined_k141_165768\|\|full | Not-determined |
| 18-104-final-viral-combined_k141_89473\|\|full | Not-determined |
| 18-104-final-viral-combined_k141_53938\|\|full | Not-determined |
| 18-104-final-viral-combined_k141_82960\|\|full | Low-quality |
| 18-104-final-viral-combined_k141_33847\|\|full | Low-quality |
| 18-104-final-viral-combined_k141_5747\|\|full | Low-quality |
| 18-104-final-viral-combined_k141_103198\|\|full | Low-quality |
| 18-104-final-viral-combined_k141_82971\|\|full | Low-quality |
| 18-104-final-viral-combined_k141_68742\|\|full | Low-quality |
| 18-104-final-viral-combined_k141_47790\|\|full | Low-quality |
| 18-104-final-viral-combined_k141_5764\|\|full | Low-quality |
| 18-104-final-viral-combined_k141_47792\|\|full | Not-determined |
| 18-104-final-viral-combined_k141_82991\|\|full | Not-determined |
| 18-104-final-viral-combined_k141_152020\|\|full | Low-quality |
| 18-104-final-viral-combined_k141_138056\|\|full | Not-determined |
| 18-104-final-viral-combined_k141_131090\|\|full | Not-determined |
| 18-104-final-viral-combined_k141_26632\|\|full | Low-quality |
| 18-104-final-viral-combined_k141_165795\|\|full | Low-quality |
| 18-104-final-viral-combined_k141_110887\|\|full | Not-determined |
| 18-104-final-viral-combined_k141_138078\|\|full | Low-quality |
| 18-104-final-viral-combined_k141_103219\|\|full | Not-determined |
| 18-104-final-viral-combined_k141_145609\|\|full | Not-determined |
| 18-104-final-viral-combined_k141_33890\|\|full | Low-quality |
| 18-104-final-viral-combined_k141_89541\|\|full | Low-quality |
| 18-104-final-viral-combined_k141_75783\|\|full | Not-determined |
| 18-104-final-viral-combined_k141_103232\|\|full | Not-determined |
| 18-104-final-viral-combined_k141_138100\|\|full | Low-quality |
| 18-104-final-viral-combined_k141_68792\|\|full | Not-determined |
| 18-104-final-viral-combined_k141_83037\|\|full | Not-determined |
| 18-104-final-viral-combined_k141_165814\|\|full | Not-determined |
| 18-104-final-viral-combined_k141_138105\|\|full | Low-quality |
| 18-104-final-viral-combined_k141_40606\|\|full | Not-determined |
| 18-104-final-viral-combined_k141_103244\|\|full | Not-determined |
| 18-104-final-viral-combined_k141_5797\|\|full | Not-determined |
| 18-104-final-viral-combined_k141_54007\|\|full | Not-determined |
| 18-104-final-viral-combined_k141_159484\|\|full | Not-determined |
| 18-104-final-viral-combined_k141_138114\|\|full | Not-determined |
| 18-104-final-viral-combined_k141_33934\|\|full | Low-quality |
| 18-104-final-viral-combined_k141_19262\|\|full | Not-determined |
| 18-104-final-viral-combined_k141_165832\|\|full | Low-quality |
| 18-104-final-viral-combined_k141_159494\|\|full | Not-determined |
| 18-104-final-viral-combined_k141_165838\|\|full | Not-determined |
| 18-104-final-viral-combined_k141_117215\|\|full | Low-quality |
| 18-104-final-viral-combined_k141_5820\|\|full | Not-determined |
| 18-104-final-viral-combined_k141_83079\|\|full | Not-determined |
| 18-104-final-viral-combined_k141_75838\|\|full | Low-quality |
| 18-104-final-viral-combined_k141_138146\|\|full | Not-determined |
| 18-104-final-viral-combined_k141_5826\|\|full | Not-determined |
| 18-104-final-viral-combined_k141_138153\|\|full | Not-determined |
| 18-104-final-viral-combined_k141_103258\|\|full | Low-quality |
| 18-104-final-viral-combined_k141_138160\|\|full | Low-quality |
| 18-104-final-viral-combined_k141_40668\|\|full | Not-determined |
| 18-104-final-viral-combined_k141_145687\|\|full | Low-quality |
| 18-104-final-viral-combined_k141_103275\|\|full | Low-quality |
| 18-104-final-viral-combined_k141_19287\|\|full | Not-determined |
| 18-104-final-viral-combined_k141_5855\|\|full | Low-quality |
| 18-104-final-viral-combined_k141_13000\|\|full | Low-quality |
| 18-104-final-viral-combined_k141_96656\|\|full | Not-determined |
| 18-104-final-viral-combined_k141_54047\|\|full | Low-quality |
| 18-104-final-viral-combined_k141_145697\|\|full | Low-quality |
| 18-104-final-viral-combined_k141_131146\|\|full | Not-determined |
| 18-104-final-viral-combined_k141_103288\|\|full | Low-quality |
| 18-104-final-viral-combined_k141_5867\|\|full | Low-quality |
| 18-104-final-viral-combined_k141_61383\|\|full | Low-quality |
| 18-104-final-viral-combined_k141_103291\|\|full | Low-quality |
| 18-104-final-viral-combined_k141_159559\|\|full | Not-determined |
| 18-104-final-viral-combined_k141_138201\|\|full | Low-quality |
| 18-104-final-viral-combined_k141_5881\|\|full | Not-determined |
| 18-104-final-viral-combined_k141_123792\|\|full | Low-quality |
| 18-104-final-viral-combined_k141_54079\|\|full | Low-quality |
| 18-104-final-viral-combined_k141_103308\|\|full | Not-determined |
| 18-104-final-viral-combined_k141_110997\|\|full | Low-quality |
| 18-104-final-viral-combined_k141_83139\|\|full | Low-quality |
| 18-104-final-viral-combined_k141_75904\|\|full | Not-determined |
| 18-104-final-viral-combined_k141_83150\|\|full | Not-determined |
| 18-104-final-viral-combined_k141_117289\|\|full | Not-determined |
| 18-104-final-viral-combined_k141_111003\|\|full | Low-quality |
| 18-104-final-viral-combined_k141_54104\|\|full | Low-quality |
| 18-104-final-viral-combined_k141_117294\|\|full | Not-determined |
| 18-104-final-viral-combined_k141_138226\|\|full | Low-quality |
| 18-104-final-viral-combined_k141_5893\|\|full | Not-determined |
| 18-104-final-viral-combined_k141_152156\|\|full | Low-quality |
| 18-104-final-viral-combined_k141_19331\|\|full | Low-quality |
| 18-104-final-viral-combined_k141_26725\|\|full | Low-quality |
| 18-104-final-viral-combined_k141_83166\|\|full | Not-determined |
| 18-104-final-viral-combined_k141_68905\|\|full | Not-determined |
| 18-104-final-viral-combined_k141_26736\|\|full | Low-quality |
| 18-104-final-viral-combined_k141_47987\|\|full | Low-quality |
| 18-104-final-viral-combined_k141_83180\|\|full | Not-determined |
| 18-104-final-viral-combined_k141_68919\|\|full | Low-quality |
| 18-104-final-viral-combined_k141_159586\|\|full | Low-quality |
| 18-104-final-viral-combined_k141_61427\|\|full | Not-determined |
| 18-104-final-viral-combined_k141_13050\|\|full | Not-determined |
| 18-104-final-viral-combined_k141_117329\|\|full | Low-quality |
| 18-104-final-viral-combined_k141_75956\|\|full | Low-quality |
| 18-104-final-viral-combined_k141_117331\|\|full | Not-determined |
| 18-104-final-viral-combined_k141_75963\|\|full | Not-determined |
| 18-104-final-viral-combined_k141_117336\|\|full | Not-determined |
| 18-104-final-viral-combined_k141_111061\|\|full | Not-determined |
| 18-104-final-viral-combined_k141_34046\|\|full | Not-determined |
| 18-104-final-viral-combined_k141_89583\|\|full | Not-determined |
| 18-104-final-viral-combined_k141_75982\|\|full | Low-quality |
| 18-104-final-viral-combined_k141_13081\|\|full | Not-determined |
| 18-104-final-viral-combined_k141_48002\|\|full | Not-determined |
| 18-104-final-viral-combined_k141_54158\|\|full | Not-determined |
| 18-104-final-viral-combined_k141_26792\|\|full | Not-determined |
| 18-104-final-viral-combined_k141_152218\|\|full | Not-determined |
| 18-104-final-viral-combined_k141_138294\|\|full | Low-quality |
| 18-104-final-viral-combined_k141_145834\|\|full | Not-determined |
| 18-104-final-viral-combined_k141_61454\|\|full | Not-determined |
| 18-104-final-viral-combined_k141_165974\|\|full | Not-determined |
| 18-104-final-viral-combined_k141_40824\|\|full | Not-determined |
| 18-104-final-viral-combined_k141_34068\|\|full | Low-quality |
| 18-104-final-viral-combined_k141_165978\|\|full | Low-quality |
| 18-104-final-viral-combined_k141_111106\|\|full | Low-quality |
| 18-104-final-viral-combined_k141_40840\|\|full | Low-quality |
| 18-104-final-viral-combined_k141_123886\|\|full | Low-quality |
| 18-104-final-viral-combined_k141_131179\|\|full | Low-quality |
| 18-104-final-viral-combined_k141_61466\|\|full | Not-determined |
| 18-104-final-viral-combined_k141_5977\|\|full | Not-determined |
| 18-104-final-viral-combined_k141_61472\|\|full | Not-determined |
| 18-104-final-viral-combined_k141_54202\|\|full | Not-determined |
| 18-104-final-viral-combined_k141_40865\|\|full | Not-determined |
| 18-104-final-viral-combined_k141_117392\|\|full | Low-quality |
| 18-104-final-viral-combined_k141_111133\|\|full | Low-quality |
| 18-104-final-viral-combined_k141_5992\|\|full | Not-determined |
| 18-104-final-viral-combined_k141_138309\|\|full | Not-determined |
| 18-104-final-viral-combined_k141_34100\|\|full | Low-quality |
| 18-104-final-viral-combined_k141_83307\|\|full | Not-determined |
| 18-104-final-viral-combined_k141_89626\|\|full | Not-determined |
| 18-104-final-viral-combined_k141_111149\|\|full | Not-determined |
| 18-104-final-viral-combined_k141_61506\|\|full | Not-determined |
| 18-104-final-viral-combined_k141_76034\|\|full | Not-determined |
| 18-104-final-viral-combined_k141_117407\|\|full | Not-determined |
| 18-104-final-viral-combined_k141_138318\|\|full | Low-quality |
| 18-104-final-viral-combined_k141_61507\|\|full | Low-quality |
| 18-104-final-viral-combined_k141_145844\|\|full | Not-determined |
| 18-104-final-viral-combined_k141_26854\|\|full | Low-quality |
| 18-104-final-viral-combined_k141_145846\|\|full | Not-determined |
| 18-104-final-viral-combined_k141_40903\|\|full | Not-determined |
| 18-104-final-viral-combined_k141_76036\|\|full | Low-quality |
| 18-104-final-viral-combined_k141_40907\|\|full | Low-quality |
| 18-104-final-viral-combined_k141_48096\|\|full | Low-quality |
| 18-104-final-viral-combined_k141_68963\|\|full | Low-quality |
| 18-104-final-viral-combined_k141_131248\|\|full | Not-determined |
| 18-104-final-viral-combined_k141_13177\|\|full | Not-determined |
| 18-104-final-viral-combined_k141_138339\|\|full | Not-determined |
| 18-104-final-viral-combined_k141_117416\|\|full | Low-quality |
| 18-104-final-viral-combined_k141_48109\|\|full | Low-quality |
| 18-104-final-viral-combined_k141_34137\|\|full | Low-quality |
| 18-104-final-viral-combined_k141_76061\|\|full | Not-determined |
| 18-104-final-viral-combined_k141_76065\|\|full | Low-quality |
| 18-104-final-viral-combined_k141_145859\|\|full | Not-determined |
| 18-104-final-viral-combined_k141_13186\|\|full | Low-quality |
| 18-104-final-viral-combined_k141_54264\|\|full | Low-quality |
| 18-104-final-viral-combined_k141_96774\|\|full | Low-quality |
| 18-104-final-viral-combined_k141_76073\|\|full | Low-quality |
| 18-104-final-viral-combined_k141_138352\|\|full | Low-quality |
| 18-104-final-viral-combined_k141_48124\|\|full | Low-quality |
| 18-104-final-viral-combined_k141_103353\|\|full | Not-determined |
| 18-104-final-viral-combined_k141_123953\|\|full | Not-determined |
| 18-104-final-viral-combined_k141_111180\|\|full | Not-determined |
| 18-104-final-viral-combined_k141_145880\|\|full | Low-quality |
| 18-104-final-viral-combined_k141_6074\|\|full | Low-quality |
| 18-104-final-viral-combined_k141_131273\|\|full | Not-determined |
| 18-104-final-viral-combined_k141_117460\|\|full | Low-quality |
| 18-104-final-viral-combined_k141_152244\|\|full | Not-determined |
| 18-104-final-viral-combined_k141_48146\|\|full | Not-determined |
| 18-104-final-viral-combined_k141_123982\|\|full | Not-determined |
| 18-104-final-viral-combined_k141_152247\|\|full | Not-determined |
| 18-104-final-viral-combined_k141_34168\|\|full | Low-quality |
| 18-104-final-viral-combined_k141_76079\|\|full | Not-determined |
| 18-104-final-viral-combined_k141_34170\|\|full | Low-quality |
| 18-104-final-viral-combined_k141_26901\|\|full | Not-determined |
| 18-104-final-viral-combined_k141_48165\|\|full | Low-quality |
| 18-104-final-viral-combined_k141_34188\|\|full | Not-determined |
| 18-104-final-viral-combined_k141_96815\|\|full | Not-determined |
| 18-104-final-viral-combined_k141_103413\|\|full | Not-determined |
| 18-104-final-viral-combined_k141_152259\|\|full | Low-quality |
| 18-104-final-viral-combined_k141_54309\|\|full | Not-determined |
| 18-104-final-viral-combined_k141_124007\|\|full | Not-determined |
| 18-104-final-viral-combined_k141_111204\|\|full | Not-determined |
| 18-104-final-viral-combined_k141_69045\|\|full | Not-determined |
| 18-104-final-viral-combined_k141_69046\|\|full | Not-determined |
| 18-104-final-viral-combined_k141_138407\|\|full | Not-determined |
| 18-104-final-viral-combined_k141_19514\|\|full | Not-determined |
| 18-104-final-viral-combined_k141_152272\|\|full | Low-quality |
| 18-104-final-viral-combined_k141_117505\|\|full | Not-determined |
| 18-104-final-viral-combined_k141_19530\|\|full | Low-quality |
| 18-104-final-viral-combined_k141_145917\|\|full | Low-quality |
| 18-104-final-viral-combined_k141_34209\|\|full | Low-quality |
| 18-104-final-viral-combined_k141_48186\|\|full | Not-determined |
| 18-104-final-viral-combined_k141_26946\|\|full | Low-quality |
| 18-104-final-viral-combined_k141_19533\|\|full | Not-determined |
| 18-104-final-viral-combined_k141_19534\|\|full | Not-determined |
| 18-104-final-viral-combined_k141_54341\|\|full | Low-quality |
| 18-104-final-viral-combined_k141_26951\|\|full | Low-quality |
| 18-104-final-viral-combined_k141_69081\|\|full | Low-quality |
| 18-104-final-viral-combined_k141_124053\|\|full | Not-determined |
| 18-104-final-viral-combined_k141_54354\|\|full | Low-quality |
| 18-104-final-viral-combined_k141_103471\|\|full | Not-determined |
| 18-104-final-viral-combined_k141_117539\|\|full | Low-quality |
| 18-104-final-viral-combined_k141_89763\|\|full | Not-determined |
| 18-104-final-viral-combined_k141_6144\|\|full | Low-quality |
| 18-104-final-viral-combined_k141_89765\|\|full | Not-determined |
| 18-104-final-viral-combined_k141_103485\|\|full | Low-quality |
| 18-104-final-viral-combined_k141_61645\|\|full | Not-determined |
| 18-104-final-viral-combined_k141_152335\|\|full | Not-determined |
| 18-104-final-viral-combined_k141_48223\|\|full | Not-determined |
| 18-104-final-viral-combined_k141_6156\|\|full | Not-determined |
| 18-104-final-viral-combined_k141_117557\|\|full | Not-determined |
| 18-104-final-viral-combined_k141_19558\|\|full | Not-determined |
| 18-104-final-viral-combined_k141_166146\|\|full | Not-determined |
| 18-104-final-viral-combined_k141_124091\|\|full | Low-quality |
| 18-104-final-viral-combined_k141_76185\|\|full | Low-quality |
| 18-104-final-viral-combined_k141_152349\|\|full | Not-determined |
| 18-104-final-viral-combined_k141_124102\|\|full | Low-quality |
| 18-104-final-viral-combined_k141_159776\|\|full | Low-quality |
| 18-104-final-viral-combined_k141_76200\|\|full | Low-quality |
| 18-104-final-viral-combined_k141_83400\|\|full | Low-quality |
| 18-104-final-viral-combined_k141_41039\|\|full | Low-quality |
| 18-104-final-viral-combined_k141_41048\|\|full | Not-determined |
| 18-104-final-viral-combined_k141_96883\|\|full | Not-determined |
| 18-104-final-viral-combined_k141_89820\|\|full | Not-determined |
| 18-104-final-viral-combined_k141_159804\|\|full | Not-determined |
| 18-104-final-viral-combined_k141_166184\|\|full | Low-quality |
| 18-104-final-viral-combined_k141_166199\|\|full | Not-determined |
| 18-104-final-viral-combined_k141_138521\|\|full | Low-quality |
| 18-104-final-viral-combined_k141_145984\|\|full | Not-determined |
| 18-104-final-viral-combined_k141_48282\|\|full | Low-quality |
| 18-104-final-viral-combined_k141_54466\|\|full | Low-quality |
| 18-104-final-viral-combined_k141_34272\|\|full | Low-quality |
| 18-104-final-viral-combined_k141_166208\|\|full | Not-determined |
| 18-104-final-viral-combined_k141_131383\|\|full | Low-quality |
| 18-104-final-viral-combined_k141_48288\|\|full | Not-determined |
| 18-104-final-viral-combined_k141_41073\|\|full | Not-determined |
| 18-104-final-viral-combined_k141_41074\|\|full | Not-determined |
| 18-104-final-viral-combined_k141_61698\|\|full | Low-quality |
| 18-104-final-viral-combined_k141_138535\|\|full | Low-quality |
| 18-104-final-viral-combined_k141_34284\|\|full | Low-quality |
| 18-104-final-viral-combined_k141_41078\|\|full | Not-determined |
| 18-104-final-viral-combined_k141_41079\|\|full | Low-quality |
| 18-104-final-viral-combined_k141_111278\|\|full | Low-quality |
| 18-104-final-viral-combined_k141_27042\|\|full | Low-quality |
| 18-104-final-viral-combined_k141_61715\|\|full | Not-determined |
| 18-104-final-viral-combined_k141_41086\|\|full | Not-determined |
| 18-104-final-viral-combined_k141_103540\|\|full | Not-determined |
| 18-104-final-viral-combined_k141_27047\|\|full | Not-determined |
| 18-104-final-viral-combined_k141_146031\|\|full | Low-quality |
| 18-104-final-viral-combined_k141_41092\|\|full | Not-determined |
| 18-104-final-viral-combined_k141_34294\|\|full | Not-determined |
| 18-104-final-viral-combined_k141_34298\|\|full | Low-quality |
| 18-104-final-viral-combined_k141_146037\|\|full | Low-quality |
| 18-104-final-viral-combined_k141_48319\|\|full | Low-quality |
| 18-104-final-viral-combined_k141_124204\|\|full | Not-determined |
| 18-104-final-viral-combined_k141_6242\|\|full | Low-quality |
| 18-104-final-viral-combined_k141_69195\|\|full | Low-quality |
| 18-104-final-viral-combined_k141_117648\|\|full | Not-determined |
| 18-104-final-viral-combined_k141_96955\|\|full | Low-quality |
| 18-104-final-viral-combined_k141_166272\|\|full | Low-quality |
| 18-104-final-viral-combined_k141_117655\|\|full | Low-quality |
| 18-104-final-viral-combined_k141_6262\|\|full | Not-determined |
| 18-104-final-viral-combined_k141_166281\|\|full | Not-determined |
| 18-104-final-viral-combined_k141_27080\|\|full | Low-quality |
| 18-104-final-viral-combined_k141_19652\|\|full | Not-determined |
| 18-104-final-viral-combined_k141_138585\|\|full | Low-quality |
| 18-104-final-viral-combined_k141_83476\|\|full | Not-determined |
| 18-104-final-viral-combined_k141_41135\|\|full | Low-quality |
| 18-104-final-viral-combined_k141_111316\|\|full | Low-quality |
| 18-104-final-viral-combined_k141_83485\|\|full | Not-determined |
| 18-104-final-viral-combined_k141_6281\|\|full | Low-quality |
| 18-104-final-viral-combined_k141_27095\|\|full | Low-quality |
| 18-104-final-viral-combined_k141_61805\|\|full | Low-quality |
| 18-104-final-viral-combined_k141_117697\|\|full | Low-quality |
| 18-104-final-viral-combined_k141_117700\|\|full | Not-determined |
| 18-104-final-viral-combined_k141_103586\|\|full | Low-quality |
| 18-104-final-viral-combined_k141_124257\|\|full | Not-determined |
| 18-104-final-viral-combined_k141_124258\|\|full | Low-quality |
| 18-104-final-viral-combined_k141_159905\|\|full | Not-determined |
| 18-104-final-viral-combined_k141_61823\|\|full | Low-quality |
| 18-104-final-viral-combined_k141_124275\|\|full | Low-quality |
| 18-104-final-viral-combined_k141_159913\|\|full | Low-quality |
| 18-104-final-viral-combined_k141_159914\|\|full | Low-quality |
| 18-104-final-viral-combined_k141_97029\|\|full | Low-quality |
| 18-104-final-viral-combined_k141_48383\|\|full | Not-determined |
| 18-104-final-viral-combined_k141_27116\|\|full | Not-determined |
| 18-109-final-viral-combined_k141_224523\|\|full | Medium-quality |
| 18-104-final-viral-combined_k141_27129\|\|full | Not-determined |
| 18-104-final-viral-combined_k141_19716\|\|full | Low-quality |
| 18-104-final-viral-combined_k141_61860\|\|full | Not-determined |
| 18-104-final-viral-combined_k141_69250\|\|full | Low-quality |
| 18-104-final-viral-combined_k141_117754\|\|full | Not-determined |
| 18-104-final-viral-combined_k141_27134\|\|full | Low-quality |
| 18-104-final-viral-combined_k141_152452\|\|full | Not-determined |
| 18-104-final-viral-combined_k141_138662\|\|full | Low-quality |
| 18-104-final-viral-combined_k141_117761\|\|full | Low-quality |
| 18-104-final-viral-combined_k141_111378\|\|full | Low-quality |
| 18-104-final-viral-combined_k141_27148\|\|full | Low-quality |
| 18-104-final-viral-combined_k141_166310\|\|full | Low-quality |
| 18-104-final-viral-combined_k141_6344\|\|full | Low-quality |
| 18-104-final-viral-combined_k141_61897\|\|full | Low-quality |
| 18-104-final-viral-combined_k141_19743\|\|full | Not-determined |
| 18-104-final-viral-combined_k141_117781\|\|full | Low-quality |
| 18-104-final-viral-combined_k141_27156\|\|full | Low-quality |
| 18-104-final-viral-combined_k141_89950\|\|full | Not-determined |
| 18-104-final-viral-combined_k141_19754\|\|full | Not-determined |
| 18-104-final-viral-combined_k141_117799\|\|full | Low-quality |
| 18-104-final-viral-combined_k141_13535\|\|full | Not-determined |
| 18-104-final-viral-combined_k141_41214\|\|full | Not-determined |
| 18-104-final-viral-combined_k141_152488\|\|full | Not-determined |
| 18-104-final-viral-combined_k141_103678\|\|full | Low-quality |
| 18-104-final-viral-combined_k141_166360\|\|full | Low-quality |
| 18-104-final-viral-combined_k141_41225\|\|full | Low-quality |
| 18-104-final-viral-combined_k141_61946\|\|full | Low-quality |
| 18-104-final-viral-combined_k141_13559\|\|full | Low-quality |
| 18-104-final-viral-combined_k141_111418\|\|full | Not-determined |
| 18-104-final-viral-combined_k141_166369\|\|full | Low-quality |
| 18-104-final-viral-combined_k141_146222\|\|full | Low-quality |
| 18-104-final-viral-combined_k141_160034\|\|full | Not-determined |
| 18-104-final-viral-combined_k141_146228\|\|full | Not-determined |
| 18-104-final-viral-combined_k141_131533\|\|full | Not-determined |
| 18-104-final-viral-combined_k141_76405\|\|full | Low-quality |
| 18-104-final-viral-combined_k141_160043\|\|full | Not-determined |
| 18-104-final-viral-combined_k141_160046\|\|full | Low-quality |
| 18-104-final-viral-combined_k141_41243\|\|full | Not-determined |
| 18-104-final-viral-combined_k141_27227\|\|full | Low-quality |
| 18-104-final-viral-combined_k141_54707\|\|full | Low-quality |
| 18-104-final-viral-combined_k141_103718\|\|full | Low-quality |
| 18-104-final-viral-combined_k141_160047\|\|full | Low-quality |
| 18-104-final-viral-combined_k141_69355\|\|full | Low-quality |
| 18-104-final-viral-combined_k141_166402\|\|full | Not-determined |
| 18-104-final-viral-combined_k141_97070\|\|full | Not-determined |
| 18-104-final-viral-combined_k141_117827\|\|full | Low-quality |
| 18-104-final-viral-combined_k141_124371\|\|full | Low-quality |
| 18-104-final-viral-combined_k141_166412\|\|full | Low-quality |
| 18-104-final-viral-combined_k141_138751\|\|full | Not-determined |
| 18-104-final-viral-combined_k141_124376\|\|full | Not-determined |
| 18-104-final-viral-combined_k141_27264\|\|full | Not-determined |
| 18-104-final-viral-combined_k141_41250\|\|full | Low-quality |
| 18-104-final-viral-combined_k141_166434\|\|full | Low-quality |
| 18-104-final-viral-combined_k141_124392\|\|full | Not-determined |
| 18-104-final-viral-combined_k141_166436\|\|full | Not-determined |
| 18-104-final-viral-combined_k141_61999\|\|full | Low-quality |
| 18-104-final-viral-combined_k141_6469\|\|full | Not-determined |
| 18-104-final-viral-combined_k141_19837\|\|full | Not-determined |
| 18-104-final-viral-combined_k141_97111\|\|full | Not-determined |
| 18-104-final-viral-combined_k141_160086\|\|full | Not-determined |
| 18-104-final-viral-combined_k141_166447\|\|full | Not-determined |
| 18-104-final-viral-combined_k141_69435\|\|full | Not-determined |
| 18-104-final-viral-combined_k141_62016\|\|full | Low-quality |
| 18-104-final-viral-combined_k141_160099\|\|full | Low-quality |
| 18-104-final-viral-combined_k141_97117\|\|full | Not-determined |
| 18-104-final-viral-combined_k141_62020\|\|full | Not-determined |
| 18-104-final-viral-combined_k141_27307\|\|full | Low-quality |
| 18-104-final-viral-combined_k141_69450\|\|full | Not-determined |
| 18-104-final-viral-combined_k141_124412\|\|full | Low-quality |
| 18-104-final-viral-combined_k141_90029\|\|full | Low-quality |
| 18-104-final-viral-combined_k141_166457\|\|full | Low-quality |
| 18-104-final-viral-combined_k141_90036\|\|full | Not-determined |
| 18-104-final-viral-combined_k141_124426\|\|full | Not-determined |
| 18-104-final-viral-combined_k141_90042\|\|full | Not-determined |
| 18-104-final-viral-combined_k141_138837\|\|full | Low-quality |
| 18-104-final-viral-combined_k141_97169\|\|full | Not-determined |
| 18-104-final-viral-combined_k141_97171\|\|full | Not-determined |
| 18-104-final-viral-combined_k141_54808\|\|full | Not-determined |
| 18-104-final-viral-combined_k141_62071\|\|full | Not-determined |
| 18-104-final-viral-combined_k141_166488\|\|full | Not-determined |
| 18-104-final-viral-combined_k141_69502\|\|full | Not-determined |
| 18-104-final-viral-combined_k141_97198\|\|full | Not-determined |
| 18-104-final-viral-combined_k141_6543\|\|full | Low-quality |
| 18-104-final-viral-combined_k141_13647\|\|full | Not-determined |
| 18-104-final-viral-combined_k141_160175\|\|full | Low-quality |
| 18-104-final-viral-combined_k141_131636\|\|full | Low-quality |
| 18-104-final-viral-combined_k141_76500\|\|full | Not-determined |
| 18-104-final-viral-combined_k141_13658\|\|full | Low-quality |
| 18-104-final-viral-combined_k141_69529\|\|full | Low-quality |
| 18-104-final-viral-combined_k141_62121\|\|full | Low-quality |
| 18-104-final-viral-combined_k141_34429\|\|full | Low-quality |
| 18-104-final-viral-combined_k141_19920\|\|full | Not-determined |
| 18-104-final-viral-combined_k141_131655\|\|full | Not-determined |
| 18-104-final-viral-combined_k141_124503\|\|full | Low-quality |
| 18-104-final-viral-combined_k141_117869\|\|full | Not-determined |
| 18-104-final-viral-combined_k141_69546\|\|full | Low-quality |
| 18-104-final-viral-combined_k141_90122\|\|full | Low-quality |
| 18-104-final-viral-combined_k141_166529\|\|full | Low-quality |
| 18-104-final-viral-combined_k141_54883\|\|full | Not-determined |
| 18-104-final-viral-combined_k141_166539\|\|full | Not-determined |
| 18-104-final-viral-combined_k141_19929\|\|full | Not-determined |
| 18-104-final-viral-combined_k141_54893\|\|full | Not-determined |
| 18-104-final-viral-combined_k141_138929\|\|full | Not-determined |
| 18-104-final-viral-combined_k141_19937\|\|full | Not-determined |
| 18-104-final-viral-combined_k141_41362\|\|full | Not-determined |
| 18-104-final-viral-combined_k141_6619\|\|full | Low-quality |
| 18-104-final-viral-combined_k141_19940\|\|full | Low-quality |
| 18-104-final-viral-combined_k141_62152\|\|full | Not-determined |
| 18-104-final-viral-combined_k141_6622\|\|full | Low-quality |
| 18-104-final-viral-combined_k141_6627\|\|full | Not-determined |
| 18-104-final-viral-combined_k141_166564\|\|full | Low-quality |
| 18-104-final-viral-combined_k141_34481\|\|full | Not-determined |
| 18-104-final-viral-combined_k141_41389\|\|full | Not-determined |
| 18-104-final-viral-combined_k141_166576\|\|full | Not-determined |
| 18-104-final-viral-combined_k141_13691\|\|full | Low-quality |
| 18-104-final-viral-combined_k141_19955\|\|full | Not-determined |
| 18-104-final-viral-combined_k141_62182\|\|full | Not-determined |
| 18-104-final-viral-combined_k141_69575\|\|full | Not-determined |
| 18-104-final-viral-combined_k141_6646\|\|full | Low-quality |
| 18-104-final-viral-combined_k141_138984\|\|full | Low-quality |
| 18-104-final-viral-combined_k141_97248\|\|full | Low-quality |
| 18-104-final-viral-combined_k141_48625\|\|full | Not-determined |
| 18-104-final-viral-combined_k141_97251\|\|full | Low-quality |
| 18-104-final-viral-combined_k141_54941\|\|full | Not-determined |
| 18-104-final-viral-combined_k141_27422\|\|full | Low-quality |
| 18-104-final-viral-combined_k141_131729\|\|full | Not-determined |
| 18-104-final-viral-combined_k141_13723\|\|full | Not-determined |
| 18-104-final-viral-combined_k141_166615\|\|full | Low-quality |
| 18-104-final-viral-combined_k141_131741\|\|full | Low-quality |
| 18-104-final-viral-combined_k141_90201\|\|full | Not-determined |
| 18-104-final-viral-combined_k141_166630\|\|full | Not-determined |
| 18-104-final-viral-combined_k141_139026\|\|full | Not-determined |
| 18-104-final-viral-combined_k141_152675\|\|full | Low-quality |
| 18-104-final-viral-combined_k141_27452\|\|full | Low-quality |
| 18-104-final-viral-combined_k141_117952\|\|full | Low-quality |
| 18-104-final-viral-combined_k141_90218\|\|full | Not-determined |
| 18-104-final-viral-combined_k141_117956\|\|full | Low-quality |
| 18-104-final-viral-combined_k141_6681\|\|full | Low-quality |
| 18-104-final-viral-combined_k141_62240\|\|full | Low-quality |
| 18-104-final-viral-combined_k141_20006\|\|full | Low-quality |
| 18-104-final-viral-combined_k141_48654\|\|full | Not-determined |
| 18-104-final-viral-combined_k141_166660\|\|full | Not-determined |
| 18-104-final-viral-combined_k141_34582\|\|full | Not-determined |
| 18-104-final-viral-combined_k141_139068\|\|full | Not-determined |
| 18-104-final-viral-combined_k141_117977\|\|full | Not-determined |
| 18-104-final-viral-combined_k141_152701\|\|full | Low-quality |
| 18-104-final-viral-combined_k141_34599\|\|full | Not-determined |
| 18-104-final-viral-combined_k141_124600\|\|full | Low-quality |
| 18-104-final-viral-combined_k141_48678\|\|full | Low-quality |
| 18-104-final-viral-combined_k141_20025\|\|full | Low-quality |
| 18-104-final-viral-combined_k141_69601\|\|full | Low-quality |
| 18-104-final-viral-combined_k141_13770\|\|full | Not-determined |
| 18-104-final-viral-combined_k141_118000\|\|full | Not-determined |
| 18-104-final-viral-combined_k141_41468\|\|full | Not-determined |
| 18-104-final-viral-combined_k141_166683\|\|full | Low-quality |
| 18-104-final-viral-combined_k141_124614\|\|full | Low-quality |
| 18-104-final-viral-combined_k141_152734\|\|full | Low-quality |
| 18-104-final-viral-combined_k141_103831\|\|full | Low-quality |
| 18-104-final-viral-combined_k141_13785\|\|full | Low-quality |
| 18-104-final-viral-combined_k141_34614\|\|full | Not-determined |
| 18-104-final-viral-combined_k141_139092\|\|full | Not-determined |
| 18-104-final-viral-combined_k141_69618\|\|full | Low-quality |
| 18-104-final-viral-combined_k141_103842\|\|full | Not-determined |
| 18-104-final-viral-combined_k141_103847\|\|full | Not-determined |
| 18-104-final-viral-combined_k141_62283\|\|full | Not-determined |
| 18-104-final-viral-combined_k141_118013\|\|full | Not-determined |
| 18-104-final-viral-combined_k141_166701\|\|full | Not-determined |
| 18-104-final-viral-combined_k141_34632\|\|full | Low-quality |
| 18-104-final-viral-combined_k141_90316\|\|full | Not-determined |
| 18-104-final-viral-combined_k141_48709\|\|full | Not-determined |
| 18-104-final-viral-combined_k141_48718\|\|full | Not-determined |
| 18-104-final-viral-combined_k141_103877\|\|full | Low-quality |
| 18-104-final-viral-combined_k141_34662\|\|full | Low-quality |
| 18-104-final-viral-combined_k141_34663\|\|full | Low-quality |
| 18-104-final-viral-combined_k141_103879\|\|full | Not-determined |
| 18-104-final-viral-combined_k141_103881\|\|full | Not-determined |
| 18-104-final-viral-combined_k141_124693\|\|full | Low-quality |
| 18-104-final-viral-combined_k141_97368\|\|full | Low-quality |
| 18-104-final-viral-combined_k141_69655\|\|full | Low-quality |
| 18-104-final-viral-combined_k141_34676\|\|full | Low-quality |
| 18-104-final-viral-combined_k141_62335\|\|full | Low-quality |
| 18-104-final-viral-combined_k141_103896\|\|full | Not-determined |
| 18-104-final-viral-combined_k141_90346\|\|full | Low-quality |
| 18-104-final-viral-combined_k141_97391\|\|full | Low-quality |
| 18-104-final-viral-combined_k141_55020\|\|full | Not-determined |
| 18-104-final-viral-combined_k141_131844\|\|full | Not-determined |
| 18-104-final-viral-combined_k141_90355\|\|full | Low-quality |
| 18-104-final-viral-combined_k141_6764\|\|full | Not-determined |
| 18-104-final-viral-combined_k141_34715\|\|full | Not-determined |
| 18-104-final-viral-combined_k141_139192\|\|full | Not-determined |
| 18-104-final-viral-combined_k141_13842\|\|full | Not-determined |
| 18-104-final-viral-combined_k141_90385\|\|full | Low-quality |
| 18-105-final-viral-combined_k141_49310\|\|full | Medium-quality |
| 18-104-final-viral-combined_k141_62378\|\|full | Not-determined |
| 18-104-final-viral-combined_k141_34740\|\|full | Not-determined |
| 18-104-final-viral-combined_k141_6803\|\|full | Not-determined |
| 18-104-final-viral-combined_k141_103968\|\|full | Not-determined |

| | |
|---|---|
| 18-104-final-viral-combined_k141_6806\|\|full | Low-quality |
| 18-104-final-viral-combined_k141_166794\|\|full | Not-determined |
| 18-104-final-viral-combined_k141_124761\|\|full | Low-quality |
| 18-104-final-viral-combined_k141_27626\|\|full | Not-determined |
| 18-104-final-viral-combined_k141_152897\|\|full | Low-quality |
| 18-104-final-viral-combined_k141_118144\|\|full | Not-determined |
| 18-104-final-viral-combined_k141_90412\|\|full | Low-quality |
| 18-104-final-viral-combined_k141_27636\|\|full | Low-quality |
| 18-104-final-viral-combined_k141_62410\|\|full | Not-determined |
| 18-104-final-viral-combined_k141_27644\|\|full | Low-quality |
| 18-104-final-viral-combined_k141_34787\|\|full | Not-determined |
| 18-104-final-viral-combined_k141_55031\|\|full | Low-quality |
| 18-104-final-viral-combined_k141_139264\|\|full | Not-determined |
| 18-104-final-viral-combined_k141_55034\|\|full | Not-determined |
| 18-104-final-viral-combined_k141_41561\|\|full | Not-determined |
| 18-104-final-viral-combined_k141_6850\|\|full | Low-quality |
| 18-104-final-viral-combined_k141_27668\|\|full | Low-quality |
| 18-104-final-viral-combined_k141_124841\|\|full | Low-quality |
| 18-104-final-viral-combined_k141_34815\|\|full | Not-determined |
| 18-104-final-viral-combined_k141_62445\|\|full | Low-quality |
| 18-104-final-viral-combined_k141_34820\|\|full | Low-quality |
| 18-104-final-viral-combined_k141_152974\|\|full | Low-quality |
| 18-104-final-viral-combined_k141_55048\|\|full | Not-determined |
| 18-104-final-viral-combined_k141_90475\|\|full | Low-quality |
| 18-104-final-viral-combined_k141_6854\|\|full | Not-determined |
| 18-104-final-viral-combined_k141_27686\|\|full | Not-determined |
| 18-104-final-viral-combined_k141_97453\|\|full | Not-determined |
| 18-104-final-viral-combined_k141_166858\|\|full | Not-determined |
| 18-104-final-viral-combined_k141_27687\|\|full | Low-quality |
| 18-104-final-viral-combined_k141_97457\|\|full | Low-quality |
| 18-104-final-viral-combined_k141_41584\|\|full | Low-quality |
| 18-104-final-viral-combined_k141_152999\|\|full | Low-quality |
| 18-104-final-viral-combined_k141_20220\|\|full | Not-determined |
| 18-104-final-viral-combined_k141_153011\|\|full | Not-determined |
| 18-104-final-viral-combined_k141_97471\|\|full | Low-quality |
| 18-104-final-viral-combined_k141_6870\|\|full | Not-determined |
| 18-104-final-viral-combined_k141_166884\|\|full | Not-determined |
| 18-104-final-viral-combined_k141_55099\|\|full | Not-determined |
| 18-104-final-viral-combined_k141_20278\|\|full | Low-quality |
| 18-104-final-viral-combined_k141_118272\|\|full | Low-quality |
| 18-104-final-viral-combined_k141_124930\|\|full | Low-quality |
| 18-104-final-viral-combined_k141_6902\|\|full | Not-determined |
| 18-104-final-viral-combined_k141_124936\|\|full | Low-quality |
| 18-104-final-viral-combined_k141_6906\|\|full | Not-determined |
| 18-104-final-viral-combined_k141_41632\|\|full | Not-determined |
| 18-104-final-viral-combined_k141_41634\|\|full | Low-quality |
| 18-104-final-viral-combined_k141_124939\|\|full | Not-determined |
| 18-104-final-viral-combined_k141_132027\|\|full | Low-quality |
| 18-104-final-viral-combined_k141_104120\|\|full | Low-quality |
| 18-104-final-viral-combined_k141_20307\|\|full | Low-quality |
| 18-104-final-viral-combined_k141_153050\|\|full | Not-determined |
| 18-104-final-viral-combined_k141_55102\|\|full | Low-quality |
| 18-104-final-viral-combined_k141_55108\|\|full | Not-determined |
| 18-104-final-viral-combined_k141_27792\|\|full | Not-determined |
| 18-104-final-viral-combined_k141_62522\|\|full | Low-quality |
| 18-104-final-viral-combined_k141_124979\|\|full | Low-quality |
| 18-104-final-viral-combined_k141_20335\|\|full | Not-determined |
| 18-104-final-viral-combined_k141_20339\|\|full | Not-determined |
| 18-104-final-viral-combined_k141_104180\|\|full | Low-quality |
| 18-104-final-viral-combined_k141_132100\|\|full | Low-quality |
| 18-104-final-viral-combined_k141_153077\|\|full | Not-determined |
| 18-104-final-viral-combined_k141_55137\|\|full | Low-quality |
| 18-104-final-viral-combined_k141_55140\|\|full | Not-determined |
| 18-104-final-viral-combined_k141_62557\|\|full | Low-quality |
| 18-104-final-viral-combined_k141_132131\|\|full | Low-quality |
| 18-104-final-viral-combined_k141_55171\|\|full | Low-quality |
| 18-104-final-viral-combined_k141_153097\|\|full | Low-quality |
| 18-104-final-viral-combined_k141_132169\|\|full | Not-determined |
| 18-104-final-viral-combined_k141_153121\|\|full | Low-quality |
| 18-104-final-viral-combined_k141_27848\|\|full | Low-quality |
| 18-104-final-viral-combined_k141_118397\|\|full | Low-quality |
| 18-104-final-viral-combined_k141_62625\|\|full | Low-quality |
| 18-104-final-viral-combined_k141_118408\|\|full | Low-quality |
| 18-104-final-viral-combined_k141_104233\|\|full | Low-quality |
| 18-104-final-viral-combined_k141_55197\|\|full | Not-determined |
| 18-104-final-viral-combined_k141_125081\|\|full | Not-determined |
| 18-104-final-viral-combined_k141_125086\|\|full | Low-quality |
| 18-104-final-viral-combined_k141_20404\|\|full | Not-determined |
| 18-104-final-viral-combined_k141_125094\|\|full | Low-quality |
| 18-104-final-viral-combined_k141_132224\|\|full | Not-determined |
| 18-104-final-viral-combined_k141_125106\|\|full | Not-determined |
| 18-105_k141_10783 | High-quality |
| 18-104-final-viral-combined_k141_132257\|\|full | Not-determined |
| 18-104-final-viral-combined_k141_132259\|\|full | Low-quality |
| 18-104-final-viral-combined_k141_41788\|\|full | Low-quality |
| 18-104-final-viral-combined_k141_104298\|\|full | Low-quality |
| 18-104-final-viral-combined_k141_132281\|\|full | Not-determined |
| 18-104-final-viral-combined_k141_153178\|\|full | Low-quality |
| 18-104-final-viral-combined_k141_62652\|\|full | Low-quality |
| 18-104-final-viral-combined_k141_125109\|\|full | Low-quality |
| 18-104-final-viral-combined_k141_62656\|\|full | Low-quality |
| 18-104-final-viral-combined_k141_125116\|\|full | Not-determined |
| 18-104-final-viral-combined_k141_55296\|\|full | Not-determined |
| 18-104-final-viral-combined_k141_132283\|\|full | Low-quality |
| 18-104-final-viral-combined_k141_153213\|\|full | Not-determined |
| 18-104-final-viral-combined_k141_62673\|\|full | Not-determined |
| 18-104-final-viral-combined_k141_132304\|\|full | Not-determined |
| 18-104-final-viral-combined_k141_104351\|\|full | Not-determined |
| 18-104-final-viral-combined_k141_104357\|\|full | Not-determined |
| 18-104-final-viral-combined_k141_55317\|\|full | Not-determined |
| 18-104-final-viral-combined_k141_125180\|\|full | Not-determined |
| 18-104-final-viral-combined_k141_20504\|\|full | Low-quality |
| 18-104-final-viral-combined_k141_55369\|\|full | Low-quality |
| 18-104-final-viral-combined_k141_20513\|\|full | Not-determined |
| 18-104-final-viral-combined_k141_125193\|\|full | Not-determined |
| 18-104-final-viral-combined_k141_125205\|\|full | Low-quality |
| 18-104-final-viral-combined_k141_20535\|\|full | Not-determined |
| 18-104-final-viral-combined_k141_125221\|\|full | Not-determined |
| 18-104-final-viral-combined_k141_125223\|\|full | Not-determined |
| 18-104-final-viral-combined_k141_55404\|\|full | Low-quality |
| 18-104-final-viral-combined_k141_132345\|\|full | Low-quality |
| 18-104-final-viral-combined_k141_55448\|\|full | Not-determined |
| 18-104-final-viral-combined_k141_20631\|\|full | Not-determined |
| 18-104-final-viral-combined_k141_125295\|\|full | Not-determined |
| 18-104-final-viral-combined_k141_55512\|\|full | Not-determined |
| 18-104-final-viral-combined_k141_125313\|\|full | Not-determined |
| 18-104-final-viral-combined_k141_20726\|\|full | Not-determined |
| 18-104-final-viral-combined_k141_55697\|\|full | Low-quality |
| 18-104-final-viral-combined_k141_55706\|\|full | Low-quality |
| 18-104-final-viral-combined_k141_20807\|\|full | Low-quality |
| 18-104-final-viral-combined_k141_55720\|\|full | Low-quality |
| 18-104-final-viral-combined_k141_20823\|\|full | Not-determined |
| 18-104-final-viral-combined_k141_20824\|\|full | Low-quality |
| 18-104-final-viral-combined_k141_20833\|\|full | Not-determined |
| 18-104-final-viral-combined_k141_20852\|\|full | Not-determined |
| 18-104-final-viral-combined_k141_20858\|\|full | Not-determined |
| 18-104-final-viral-combined_k141_132506\|\|0_partial | Not-determined |
| 18-104-final-viral-combined_k141_36148\|\|0_partial | Low-quality |
| 18-104-final-viral-combined_k141_57313\|\|0_partial | Not-determined |
| 18-104-final-viral-combined_k141_85444\|\|0_partial | Low-quality |
| 18-104-final-viral-combined_k141_30070\|\|0_partial | Low-quality |
| 18-101_k141_208875 | Medium-quality |
| 18-104-final-viral-combined_k141_23755\|\|0_partial | Not-determined |
| 18-104-final-viral-combined_k141_93625\|\|0_partial | Low-quality |
| 18-104-final-viral-combined_k141_65995\|\|0_partial | Low-quality |
| 18-104-final-viral-combined_k141_87187\|\|0_partial | Not-determined |
| 18-104-final-viral-combined_k141_150126\|\|0_partial | Not-determined |
| 18-101-final-viral-combined_k141_208875\|\|full | Medium-quality |
| 18-104-final-viral-combined_k141_19257\|\|0_partial | Not-determined |
| 18-104-final-viral-combined_k141_131178\|\|0_partial | Low-quality |
| 18-104-final-viral-combined_k141_103336\|\|0_partial | Not-determined |
| 18-104-final-viral-combined_k141_152228\|\|0_partial | Low-quality |
| 18-104-final-viral-combined_k141_19767\|\|0_partial | Low-quality |
| 18-104-final-viral-combined_k141_6948\|\|0_partial | Low-quality |
| 18-104-final-viral-combined_k141_104496\|\|lt2gene | Not-determined |
| 18-104-final-viral-combined_k141_76635\|\|lt2gene | Low-quality |
| 18-104-final-viral-combined_k141_125397\|\|lt2gene | Low-quality |
| 18-104-final-viral-combined_k141_153257\|\|lt2gene | Low-quality |
| 18-104-final-viral-combined_k141_62740\|\|lt2gene | Low-quality |
| 18-104-final-viral-combined_k141_132402\|\|lt2gene | Low-quality |
| 18-104-final-viral-combined_k141_13988\|\|lt2gene | Low-quality |
| 18-104-final-viral-combined_k141_111504\|\|lt2gene | Low-quality |
| 18-104-final-viral-combined_k141_83685\|\|lt2gene | Low-quality |
| 18-104-final-viral-combined_k141_83708\|\|lt2gene | Not-determined |
| 18-104-final-viral-combined_k141_41905\|\|lt2gene | Not-determined |
| 18-104-final-viral-combined_k141_139503\|\|lt2gene | Low-quality |
| 18-104-final-viral-combined_k141_14056\|\|lt2gene | Not-determined |
| 18-104-final-viral-combined_k141_48863\|\|lt2gene | Low-quality |
| 18-104-final-viral-combined_k141_62865\|\|lt2gene | Low-quality |
| 18-104-final-viral-combined_k141_21072\|\|lt2gene | Low-quality |
| 18-104-final-viral-combined_k141_28004\|\|lt2gene | Not-determined |
| 18-104-final-viral-combined_k141_132523\|\|lt2gene | Low-quality |
| 18-104-final-viral-combined_k141_14082\|\|lt2gene | Not-determined |
| 18-104-final-viral-combined_k141_55845\|\|lt2gene | Low-quality |
| 18-104-final-viral-combined_k141_160390\|\|lt2gene | Low-quality |
| 18-104-final-viral-combined_k141_153421\|\|lt2gene | Low-quality |
| 18-104-final-viral-combined_k141_55895\|\|lt2gene | Not-determined |
| 18-104-final-viral-combined_k141_83804\|\|lt2gene | Low-quality |
| 18-104-final-viral-combined_k141_146398\|\|lt2gene | Low-quality |
| 18-104-final-viral-combined_k141_118552\|\|lt2gene | Low-quality |
| 18-104-final-viral-combined_k141_111755\|\|lt2gene | Low-quality |
| 18-104-final-viral-combined_k141_28061\|\|lt2gene | Low-quality |
| 18-104-final-viral-combined_k141_118577\|\|lt2gene | Low-quality |
| 18-104-final-viral-combined_k141_375\|\|lt2gene | Low-quality |
| 18-104-final-viral-combined_k141_28085\|\|lt2gene | Not-determined |
| 18-104-final-viral-combined_k141_21136\|\|lt2gene | Not-determined |
| 18-104-final-viral-combined_k141_62998\|\|lt2gene | Not-determined |
| 18-104-final-viral-combined_k141_55929\|\|lt2gene | Low-quality |
| 18-104-final-viral-combined_k141_139637\|\|lt2gene | Low-quality |
| 18-104-final-viral-combined_k141_48978\|\|lt2gene | Low-quality |
| 18-104-final-viral-combined_k141_76900\|\|lt2gene | Low-quality |
| 18-104-final-viral-combined_k141_125714\|\|lt2gene | Low-quality |
| 18-104-final-viral-combined_k141_97842\|\|lt2gene | Not-determined |
| 18-104-final-viral-combined_k141_153516\|\|lt2gene | Low-quality |
| 18-104-final-viral-combined_k141_76941\|\|lt2gene | Low-quality |
| 18-104-final-viral-combined_k141_153537\|\|lt2gene | Low-quality |
| 18-104-final-viral-combined_k141_42117\|\|lt2gene | Not-determined |
| 18-104-final-viral-combined_k141_42125\|\|lt2gene | Low-quality |
| 18-104-final-viral-combined_k141_125759\|\|lt2gene | Low-quality |
| 18-104-final-viral-combined_k141_77001\|\|lt2gene | Not-determined |
| 18-104-final-viral-combined_k141_125798\|\|lt2gene | Low-quality |
| 18-104-final-viral-combined_k141_49047\|\|lt2gene | Low-quality |
| 18-104-final-viral-combined_k141_63109\|\|lt2gene | Not-determined |
| 18-104-final-viral-combined_k141_111977\|\|lt2gene | Low-quality |
| 18-104-final-viral-combined_k141_118743\|\|lt2gene | Low-quality |
| 18-104-final-viral-combined_k141_91112\|\|lt2gene | Low-quality |
| 18-104-final-viral-combined_k141_84067\|\|lt2gene | Low-quality |
| 18-104-final-viral-combined_k141_97987\|\|lt2gene | Low-quality |
| 18-104-final-viral-combined_k141_49098\|\|lt2gene | Not-determined |
| 18-104-final-viral-combined_k141_28351\|\|lt2gene | Low-quality |
| 18-104-final-viral-combined_k141_28355\|\|lt2gene | Not-determined |
| 18-104-final-viral-combined_k141_42288\|\|lt2gene | Low-quality |
| 18-104-final-viral-combined_k141_84115\|\|lt2gene | Not-determined |
| 18-104-final-viral-combined_k141_146695\|\|lt2gene | Low-quality |
| 18-104-final-viral-combined_k141_21350\|\|lt2gene | Not-determined |
| 18-104-final-viral-combined_k141_21354\|\|lt2gene | Low-quality |
| 18-104-final-viral-combined_k141_153622\|\|lt2gene | Low-quality |
| 18-104-final-viral-combined_k141_105009\|\|lt2gene | Low-quality |
| 18-104-final-viral-combined_k141_641\|\|lt2gene | Low-quality |
| 18-104-final-viral-combined_k141_21388\|\|lt2gene | Low-quality |
| 18-104-final-viral-combined_k141_63222\|\|lt2gene | Low-quality |
| 18-104-final-viral-combined_k141_153669\|\|lt2gene | Low-quality |
| 18-104-final-viral-combined_k141_63238\|\|lt2gene | Low-quality |
| 18-104-final-viral-combined_k141_160753\|\|lt2gene | Not-determined |
| 18-104-final-viral-combined_k141_126025\|\|lt2gene | Not-determined |
| 18-104-final-viral-combined_k141_84207\|\|lt2gene | Low-quality |
| 18-104-final-viral-combined_k141_84218\|\|lt2gene | Low-quality |
| 18-104-final-viral-combined_k141_35236\|\|lt2gene | Not-determined |
| 18-104-final-viral-combined_k141_21448\|\|lt2gene | Low-quality |
| 18-104-final-viral-combined_k141_91313\|\|lt2gene | Low-quality |
| 18-104-final-viral-combined_k141_14455\|\|lt2gene | Not-determined |
| 18-104-final-viral-combined_k141_105111\|\|lt2gene | Low-quality |
| 18-104-final-viral-combined_k141_112096\|\|lt2gene | Low-quality |
| 18-104-final-viral-combined_k141_21479\|\|lt2gene | Not-determined |
| 18-104-final-viral-combined_k141_28486\|\|lt2gene | Low-quality |
| 18-104-final-viral-combined_k141_63250\|\|lt2gene | Low-quality |
| 18-104-final-viral-combined_k141_105135\|\|lt2gene | Low-quality |
| 18-104-final-viral-combined_k141_118909\|\|lt2gene | Low-quality |
| 18-104-final-viral-combined_k141_28522\|\|lt2gene | Low-quality |
| 18-104-final-viral-combined_k141_146882\|\|lt2gene | Low-quality |
| 18-104-final-viral-combined_k141_105174\|\|lt2gene | Not-determined |
| 18-104-final-viral-combined_k141_112135\|\|lt2gene | Low-quality |
| 18-104-final-viral-combined_k141_84223\|\|lt2gene | Low-quality |
| 18-104-final-viral-combined_k141_105188\|\|lt2gene | Not-determined |
| 18-104-final-viral-combined_k141_146898\|\|lt2gene | Not-determined |
| 18-104-final-viral-combined_k141_112152\|\|lt2gene | Low-quality |
| 18-104-final-viral-combined_k141_126197\|\|lt2gene | Low-quality |
| 18-104-final-viral-combined_k141_42472\|\|lt2gene | Not-determined |
| 18-104-final-viral-combined_k141_42490\|\|lt2gene | Low-quality |
| 18-104-final-viral-combined_k141_63335\|\|lt2gene | Not-determined |
| 18-104-final-viral-combined_k141_140070\|\|lt2gene | Low-quality |
| 18-104-final-viral-combined_k141_84392\|\|lt2gene | Low-quality |
| 18-104-final-viral-combined_k141_105267\|\|lt2gene | Low-quality |
| 18-104-final-viral-combined_k141_98259\|\|lt2gene | Low-quality |
| 18-104-final-viral-combined_k141_91436\|\|lt2gene | Low-quality |
| 18-104-final-viral-combined_k141_42550\|\|lt2gene | Low-quality |
| 18-104-final-viral-combined_k141_140107\|\|lt2gene | Low-quality |
| 18-104-final-viral-combined_k141_35349\|\|lt2gene | Low-quality |
| 18-104-final-viral-combined_k141_70579\|\|lt2gene | Low-quality |
| 18-104-final-viral-combined_k141_91492\|\|lt2gene | Low-quality |
| 18-104-final-viral-combined_k141_7624\|\|lt2gene | Low-quality |
| 18-104-final-viral-combined_k141_56454\|\|lt2gene | Not-determined |
| 18-104-final-viral-combined_k141_28688\|\|lt2gene | Low-quality |
| 18-104-final-viral-combined_k141_154025\|\|lt2gene | Not-determined |
| 18-104-final-viral-combined_k141_126400\|\|lt2gene | Low-quality |
| 18-104-final-viral-combined_k141_91595\|\|lt2gene | Low-quality |
| 18-104-final-viral-combined_k141_84540\|\|lt2gene | Low-quality |
| 18-104-final-viral-combined_k141_28776\|\|lt2gene | Low-quality |
| 18-104-final-viral-combined_k141_42693\|\|lt2gene | Low-quality |
| 18-104-final-viral-combined_k141_112391\|\|lt2gene | Low-quality |
| 18-104-final-viral-combined_k141_154066\|\|lt2gene | Low-quality |
| 18-104-final-viral-combined_k141_49436\|\|lt2gene | Not-determined |
| 18-104-final-viral-combined_k141_84589\|\|lt2gene | Low-quality |
| 18-104-final-viral-combined_k141_56563\|\|lt2gene | Low-quality |
| 18-104-final-viral-combined_k141_147152\|\|lt2gene | Not-determined |
| 18-104-final-viral-combined_k141_133117\|\|lt2gene | Low-quality |
| 18-104-final-viral-combined_k141_77601\|\|lt2gene | Low-quality |
| 18-104-final-viral-combined_k141_35560\|\|lt2gene | Low-quality |
| 18-104-final-viral-combined_k141_35562\|\|lt2gene | Low-quality |
| 18-104-final-viral-combined_k141_105597\|\|lt2gene | Not-determined |
| 18-104-final-viral-combined_k141_140308\|\|lt2gene | Low-quality |
| 18-104-final-viral-combined_k141_140332\|\|lt2gene | Low-quality |
| 18-104-final-viral-combined_k141_126618\|\|lt2gene | Low-quality |
| 18-104-final-viral-combined_k141_7753\|\|lt2gene | Low-quality |
| 18-104-final-viral-combined_k141_154210\|\|lt2gene | Low-quality |
| 18-104-final-viral-combined_k141_28920\|\|lt2gene | Low-quality |
| 18-104-final-viral-combined_k141_14739\|\|lt2gene | Low-quality |
| 18-104-final-viral-combined_k141_133233\|\|lt2gene | Low-quality |
| 18-104-final-viral-combined_k141_133240\|\|lt2gene | Not-determined |
| 18-104-final-viral-combined_k141_140387\|\|lt2gene | Low-quality |
| 18-104-final-viral-combined_k141_133251\|\|lt2gene | Not-determined |
| 18-104-final-viral-combined_k141_119304\|\|lt2gene | Low-quality |
| 18-104-final-viral-combined_k141_42816\|\|lt2gene | Low-quality |
| 18-104-final-viral-combined_k141_77739\|\|lt2gene | Not-determined |
| 18-104-final-viral-combined_k141_112560\|\|lt2gene | Not-determined |
| 18-104-final-viral-combined_k141_1097\|\|lt2gene | Not-determined |
| 18-104-final-viral-combined_k141_42830\|\|lt2gene | Not-determined |
| 18-104-final-viral-combined_k141_1134\|\|lt2gene | Low-quality |
| 18-104-final-viral-combined_k141_42867\|\|lt2gene | Low-quality |
| 18-104-final-viral-combined_k141_154305\|\|lt2gene | Low-quality |
| 18-104-final-viral-combined_k141_98584\|\|lt2gene | Not-determined |
| 18-104-final-viral-combined_k141_70805\|\|lt2gene | Low-quality |
| 18-104-final-viral-combined_k141_63817\|\|lt2gene | Low-quality |
| 18-104-final-viral-combined_k141_112694\|\|lt2gene | Not-determined |
| 18-104-final-viral-combined_k141_126761\|\|lt2gene | Low-quality |
| 18-104-final-viral-combined_k141_42956\|\|lt2gene | Not-determined |
| 18-104-final-viral-combined_k141_42960\|\|lt2gene | Not-determined |
| 18-104-final-viral-combined_k141_7913\|\|lt2gene | Not-determined |
| 18-104-final-viral-combined_k141_42988\|\|lt2gene | Low-quality |
| 18-104-final-viral-combined_k141_112728\|\|lt2gene | Not-determined |
| 18-104-final-viral-combined_k141_42999\|\|lt2gene | Not-determined |
| 18-104-final-viral-combined_k141_161229\|\|lt2gene | Low-quality |
| 18-104-final-viral-combined_k141_140584\|\|lt2gene | Not-determined |
| 18-104-final-viral-combined_k141_70848\|\|lt2gene | Not-determined |
| 18-104-final-viral-combined_k141_147434\|\|lt2gene | Low-quality |
| 18-104-final-viral-combined_k141_8023\|\|lt2gene | Not-determined |
| 18-104-final-viral-combined_k141_105877\|\|lt2gene | Low-quality |
| 18-104-final-viral-combined_k141_78029\|\|lt2gene | Low-quality |
| 18-104-final-viral-combined_k141_154434\|\|lt2gene | Low-quality |
| 18-104-final-viral-combined_k141_22017\|\|lt2gene | Low-quality |
| 18-104-final-viral-combined_k141_8101\|\|lt2gene | Low-quality |
| 18-104-final-viral-combined_k141_29243\|\|lt2gene | Not-determined |
| 18-104-final-viral-combined_k141_35915\|\|lt2gene | Low-quality |
| 18-104-final-viral-combined_k141_49645\|\|lt2gene | Low-quality |
| 18-104-final-viral-combined_k141_84912\|\|lt2gene | Low-quality |
| 18-104-final-viral-combined_k141_22100\|\|lt2gene | Low-quality |
| 18-104-final-viral-combined_k141_49695\|\|lt2gene | Not-determined |
| 18-104-final-viral-combined_k141_57013\|\|lt2gene | Low-quality |
| 18-104-final-viral-combined_k141_36038\|\|lt2gene | Not-determined |
| 18-104-final-viral-combined_k141_112840\|\|lt2gene | Low-quality |
| 18-104-final-viral-combined_k141_29359\|\|lt2gene | Low-quality |
| 18-104-final-viral-combined_k141_161382\|\|lt2gene | Not-determined |
| 18-104-final-viral-combined_k141_85056\|\|lt2gene | Low-quality |
| 18-104-final-viral-combined_k141_133668\|\|lt2gene | Low-quality |
| 18-104-final-viral-combined_k141_8265\|\|lt2gene | Low-quality |
| 18-104-final-viral-combined_k141_127026\|\|lt2gene | Low-quality |
| 18-104-final-viral-combined_k141_127030\|\|lt2gene | Low-quality |
| 18-104-final-viral-combined_k141_15130\|\|lt2gene | Not-determined |
| 18-104-final-viral-combined_k141_36097\|\|lt2gene | Low-quality |
| 18-104-final-viral-combined_k141_49799\|\|lt2gene | Low-quality |
| 18-104-final-viral-combined_k141_133732\|\|lt2gene | Low-quality |
| 18-104-final-viral-combined_k141_64142\|\|lt2gene | Low-quality |
| 18-104-final-viral-combined_k141_154712\|\|lt2gene | Low-quality |
| 18-104-final-viral-combined_k141_22194\|\|lt2gene | Not-determined |
| 18-104-final-viral-combined_k141_154716\|\|lt2gene | Low-quality |
| 18-104-final-viral-combined_k141_29464\|\|lt2gene | Low-quality |
| 18-104-final-viral-combined_k141_154732\|\|lt2gene | Not-determined |
| 18-104-final-viral-combined_k141_140937\|\|lt2gene | Low-quality |
| 18-104-final-viral-combined_k141_147729\|\|lt2gene | Low-quality |
| 18-104-final-viral-combined_k141_140965\|\|lt2gene | Low-quality |
| 18-104-final-viral-combined_k141_71212\|\|lt2gene | Low-quality |
| 18-104-final-viral-combined_k141_106157\|\|lt2gene | Low-quality |
| 18-104-final-viral-combined_k141_141024\|\|lt2gene | Not-determined |
| 18-104-final-viral-combined_k141_127154\|\|lt2gene | Not-determined |
| 18-104-final-viral-combined_k141_49929\|\|lt2gene | Low-quality |
| 18-104-final-viral-combined_k141_57189\|\|lt2gene | Not-determined |
| 18-104-final-viral-combined_k141_8388\|\|lt2gene | Low-quality |
| 18-104-final-viral-combined_k141_99029\|\|lt2gene | Low-quality |
| 18-104-final-viral-combined_k141_57217\|\|lt2gene | Not-determined |
| 18-104-final-viral-combined_k141_1633\|\|lt2gene | Low-quality |
| 18-104-final-viral-combined_k141_8420\|\|lt2gene | Low-quality |
| 18-104-final-viral-combined_k141_71266\|\|lt2gene | Low-quality |
| 18-104-final-viral-combined_k141_113047\|\|lt2gene | Low-quality |
| 18-104-final-viral-combined_k141_106271\|\|lt2gene | Low-quality |
| 18-104-final-viral-combined_k141_43340\|\|lt2gene | Low-quality |
| 18-104-final-viral-combined_k141_113073\|\|lt2gene | Low-quality |
| 18-104-final-viral-combined_k141_92323\|\|lt2gene | Low-quality |
| 18-104-final-viral-combined_k141_92344\|\|lt2gene | Low-quality |
| 18-104-final-viral-combined_k141_92358\|\|lt2gene | Low-quality |
| 18-104-final-viral-combined_k141_78489\|\|lt2gene | Low-quality |
| 18-104-final-viral-combined_k141_85323\|\|lt2gene | Not-determined |
| 18-104-final-viral-combined_k141_113141\|\|lt2gene | Low-quality |
| 18-104-final-viral-combined_k141_161639\|\|lt2gene | Low-quality |
| 18-104-final-viral-combined_k141_36236\|\|lt2gene | Low-quality |
| 18-104-final-viral-combined_k141_36238\|\|lt2gene | Low-quality |
| 18-104-final-viral-combined_k141_29672\|\|lt2gene | Low-quality |
| 18-104-final-viral-combined_k141_29683\|\|lt2gene | Low-quality |
| 18-104-final-viral-combined_k141_113191\|\|lt2gene | Low-quality |
| 18-104-final-viral-combined_k141_147895\|\|lt2gene | Not-determined |
| 18-104-final-viral-combined_k141_71399\|\|lt2gene | Not-determined |
| 18-104-final-viral-combined_k141_99178\|\|lt2gene | Low-quality |
| 18-104-final-viral-combined_k141_64387\|\|lt2gene | Not-determined |
| 18-104-final-viral-combined_k141_154935\|\|lt2gene | Low-quality |
| 18-104-final-viral-combined_k141_154957\|\|lt2gene | Low-quality |
| 18-104-final-viral-combined_k141_99242\|\|lt2gene | Not-determined |
| 18-104-final-viral-combined_k141_141215\|\|lt2gene | Not-determined |
| 18-104-final-viral-combined_k141_106409\|\|lt2gene | Low-quality |
| 18-104-final-viral-combined_k141_134025\|\|lt2gene | Not-determined |
| 18-104-final-viral-combined_k141_147954\|\|lt2gene | Low-quality |
| 18-104-final-viral-combined_k141_36331\|\|lt2gene | Low-quality |
| 18-104-final-viral-combined_k141_29811\|\|lt2gene | Low-quality |
| 18-104-final-viral-combined_k141_36353\|\|lt2gene | Low-quality |
| 18-104-final-viral-combined_k141_113285\|\|lt2gene | Low-quality |
| 18-104-final-viral-combined_k141_71491\|\|lt2gene | Not-determined |
| 18-104-final-viral-combined_k141_155033\|\|lt2gene | Low-quality |
| 18-104-final-viral-combined_k141_57446\|\|lt2gene | Low-quality |
| 18-104-final-viral-combined_k141_127417\|\|lt2gene | Low-quality |
| 18-104-final-viral-combined_k141_99316\|\|lt2gene | Low-quality |
| 18-104-final-viral-combined_k141_8673\|\|lt2gene | Not-determined |
| 18-104-final-viral-combined_k141_22456\|\|lt2gene | Low-quality |
| 18-104-final-viral-combined_k141_22457\|\|lt2gene | Low-quality |
| 18-104-final-viral-combined_k141_85513\|\|lt2gene | Not-determined |
| 18-104-final-viral-combined_k141_161890\|\|lt2gene | Low-quality |
| 18-104-final-viral-combined_k141_43584\|\|lt2gene | Low-quality |
| 18-104-final-viral-combined_k141_78680\|\|lt2gene | Low-quality |
| 18-104-final-viral-combined_k141_106490\|\|lt2gene | Low-quality |
| 18-104-final-viral-combined_k141_43598\|\|lt2gene | Low-quality |
| 18-104-final-viral-combined_k141_155103\|\|lt2gene | Low-quality |
| 18-104-final-viral-combined_k141_43609\|\|lt2gene | Low-quality |
| 18-104-final-viral-combined_k141_1919\|\|lt2gene | Low-quality |
| 18-104-final-viral-combined_k141_99380\|\|lt2gene | Not-determined |
| 18-104-final-viral-combined_k141_29950\|\|lt2gene | Low-quality |
| 18-104-final-viral-combined_k141_22530\|\|lt2gene | Low-quality |
| 18-104-final-viral-combined_k141_22535\|\|lt2gene | Not-determined |
| 18-104-final-viral-combined_k141_78750\|\|lt2gene | Not-determined |
| 18-104-final-viral-combined_k141_1949\|\|lt2gene | Not-determined |
| 18-104-final-viral-combined_k141_64611\|\|lt2gene | Not-determined |
| 18-104-final-viral-combined_k141_29962\|\|lt2gene | Low-quality |
| 18-104-final-viral-combined_k141_43674\|\|lt2gene | Low-quality |
| 18-104-final-viral-combined_k141_50255\|\|lt2gene | Low-quality |
| 18-104-final-viral-combined_k141_1988\|\|lt2gene | Low-quality |
| 18-104-final-viral-combined_k141_92655\|\|lt2gene | Low-quality |
| 18-104-final-viral-combined_k141_85607\|\|lt2gene | Low-quality |
| 18-104-final-viral-combined_k141_8832\|\|lt2gene | Low-quality |
| 18-104-final-viral-combined_k141_106607\|\|lt2gene | Not-determined |
| 18-104-final-viral-combined_k141_36552\|\|lt2gene | Low-quality |
| 18-104-final-viral-combined_k141_22642\|\|lt2gene | Low-quality |
| 18-104-final-viral-combined_k141_22673\|\|lt2gene | Low-quality |
| 18-104-final-viral-combined_k141_134338\|\|lt2gene | Low-quality |
| 18-104-final-viral-combined_k141_99519\|\|lt2gene | Not-determined |
| 18-104-final-viral-combined_k141_50374\|\|lt2gene | Low-quality |
| 18-104-final-viral-combined_k141_50396\|\|lt2gene | Low-quality |
| 18-104-final-viral-combined_k141_85702\|\|lt2gene | Not-determined |
| 18-104-final-viral-combined_k141_22739\|\|lt2gene | Low-quality |
| 18-104-final-viral-combined_k141_64732\|\|lt2gene | Low-quality |
| 18-104-final-viral-combined_k141_127692\|\|lt2gene | Low-quality |
| 18-104-final-viral-combined_k141_162197\|\|lt2gene | Not-determined |
| 18-104-final-viral-combined_k141_113443\|\|lt2gene | Low-quality |
| 18-104-final-viral-combined_k141_57690\|\|lt2gene | Not-determined |
| 18-104-final-viral-combined_k141_85790\|\|lt2gene | Low-quality |
| 18-104-final-viral-combined_k141_15896\|\|lt2gene | Low-quality |
| 18-104-final-viral-combined_k141_162247\|\|lt2gene | Low-quality |
| 18-104-final-viral-combined_k141_43904\|\|lt2gene | Not-determined |
| 18-104-final-viral-combined_k141_36691\|\|lt2gene | Not-determined |
| 18-104-final-viral-combined_k141_148369\|\|lt2gene | Low-quality |
| 18-104-final-viral-combined_k141_71788\|\|lt2gene | Low-quality |
| 18-104-final-viral-combined_k141_36732\|\|lt2gene | Low-quality |
| 18-104-final-viral-combined_k141_155395\|\|lt2gene | Not-determined |
| 18-104-final-viral-combined_k141_43949\|\|lt2gene | Low-quality |
| 18-104-final-viral-combined_k141_85876\|\|lt2gene | Not-determined |
| 18-104-final-viral-combined_k141_106803\|\|lt2gene | Low-quality |
| 18-104-final-viral-combined_k141_113543\|\|lt2gene | Not-determined |
| 18-104-final-viral-combined_k141_148438\|\|lt2gene | Not-determined |
| 18-104-final-viral-combined_k141_36773\|\|lt2gene | Low-quality |
| 18-104-final-viral-combined_k141_106826\|\|lt2gene | Not-determined |
| 18-104-final-viral-combined_k141_22959\|\|lt2gene | Low-quality |
| 18-104-final-viral-combined_k141_148458\|\|lt2gene | Low-quality |
| 18-104-final-viral-combined_k141_30161\|\|lt2gene | Not-determined |
| 18-104-final-viral-combined_k141_120115\|\|lt2gene | Not-determined |
| 18-104-final-viral-combined_k141_44008\|\|lt2gene | Low-quality |
| 18-104-final-viral-combined_k141_155464\|\|lt2gene | Low-quality |
| 18-104-final-viral-combined_k141_155468\|\|lt2gene | Not-determined |
| 18-104-final-viral-combined_k141_16059\|\|lt2gene | Not-determined |
| 18-104-final-viral-combined_k141_44040\|\|lt2gene | Not-determined |
| 18-104-final-viral-combined_k141_64940\|\|lt2gene | Not-determined |
| 18-104-final-viral-combined_k141_148510\|\|lt2gene | Low-quality |
| 18-104-final-viral-combined_k141_113661\|\|lt2gene | Not-determined |
| 18-104-final-viral-combined_k141_9196\|\|lt2gene | Not-determined |
| 18-104-final-viral-combined_k141_50562\|\|lt2gene | Low-quality |
| 18-104-final-viral-combined_k141_9226\|\|lt2gene | Low-quality |
| 18-104-final-viral-combined_k141_79269\|\|lt2gene | Low-quality |
| 18-104-final-viral-combined_k141_79277\|\|lt2gene | Low-quality |
| 18-104-final-viral-combined_k141_113709\|\|lt2gene | Not-determined |
| 18-104-final-viral-combined_k141_9254\|\|lt2gene | Low-quality |
| 18-104-final-viral-combined_k141_23092\|\|lt2gene | Low-quality |
| 18-104-final-viral-combined_k141_23093\|\|lt2gene | Not-determined |
| 18-104-final-viral-combined_k141_36911\|\|lt2gene | Low-quality |
| 18-104-final-viral-combined_k141_16117\|\|lt2gene | Not-determined |
| 18-104-final-viral-combined_k141_106929\|\|lt2gene | Low-quality |
| 18-104-final-viral-combined_k141_36928\|\|lt2gene | Low-quality |
| 18-104-final-viral-combined_k141_2311\|\|lt2gene | Not-determined |
| 18-104-final-viral-combined_k141_36956\|\|lt2gene | Low-quality |
| 18-104-final-viral-combined_k141_65042\|\|lt2gene | Not-determined |
| 18-104-final-viral-combined_k141_72035\|\|lt2gene | Low-quality |
| 18-104-final-viral-combined_k141_134773\|\|lt2gene | Not-determined |
| 18-104-final-viral-combined_k141_86067\|\|lt2gene | Low-quality |
| 18-104-final-viral-combined_k141_141747\|\|lt2gene | Low-quality |
| 18-104-final-viral-combined_k141_86107\|\|lt2gene | Low-quality |
| 18-104-final-viral-combined_k141_155606\|\|lt2gene | Low-quality |
| 18-104-final-viral-combined_k141_44202\|\|lt2gene | Low-quality |
| 18-104-final-viral-combined_k141_79401\|\|lt2gene | Low-quality |
| 18-104-final-viral-combined_k141_72129\|\|lt2gene | Not-determined |
| 18-104-final-viral-combined_k141_50640\|\|lt2gene | Low-quality |
| 18-104-final-viral-combined_k141_79417\|\|lt2gene | Not-determined |
| 18-104-final-viral-combined_k141_37062\|\|lt2gene | Not-determined |
| 18-104-final-viral-combined_k141_50659\|\|lt2gene | Low-quality |
| 18-104-final-viral-combined_k141_2456\|\|lt2gene | Low-quality |
| 18-104-final-viral-combined_k141_2460\|\|lt2gene | Low-quality |
| 18-104-final-viral-combined_k141_155649\|\|lt2gene | Low-quality |
| 18-104-final-viral-combined_k141_141860\|\|lt2gene | Low-quality |
| 18-104-final-viral-combined_k141_107094\|\|lt2gene | Low-quality |
| 18-104-final-viral-combined_k141_86192\|\|lt2gene | Not-determined |
| 18-104-final-viral-combined_k141_30438\|\|lt2gene | Low-quality |
| 18-104-final-viral-combined_k141_93232\|\|lt2gene | Not-determined |
| 18-104-final-viral-combined_k141_148696\|\|lt2gene | Not-determined |
| 18-104-final-viral-combined_k141_148708\|\|lt2gene | Low-quality |
| 18-104-final-viral-combined_k141_57995\|\|lt2gene | Low-quality |
| 18-104-final-viral-combined_k141_86232\|\|lt2gene | Low-quality |
| 18-104-final-viral-combined_k141_58024\|\|lt2gene | Low-quality |
| 18-104-final-viral-combined_k141_120465\|\|lt2gene | Low-quality |
| 18-104-final-viral-combined_k141_134959\|\|lt2gene | Low-quality |
| 18-104-final-viral-combined_k141_2597\|\|lt2gene | Low-quality |
| 18-104-final-viral-combined_k141_113924\|\|lt2gene | Low-quality |
| 18-104-final-viral-combined_k141_58072\|\|lt2gene | Low-quality |
| 18-104-final-viral-combined_k141_107160\|\|lt2gene | Low-quality |
| 18-104-final-viral-combined_k141_141983\|\|lt2gene | Not-determined |
| 18-104-final-viral-combined_k141_9559\|\|lt2gene | Low-quality |
| 18-104-final-viral-combined_k141_37183\|\|lt2gene | Not-determined |
| 18-104-final-viral-combined_k141_23385\|\|lt2gene | Low-quality |
| 18-104-final-viral-combined_k141_155855\|\|lt2gene | Not-determined |
| 18-104-final-viral-combined_k141_99776\|\|lt2gene | Not-determined |
| 18-104-final-viral-combined_k141_107243\|\|lt2gene | Low-quality |
| 18-104-final-viral-combined_k141_44455\|\|lt2gene | Low-quality |
| 18-104-final-viral-combined_k141_2730\|\|lt2gene | Low-quality |
| 18-104-final-viral-combined_k141_58198\|\|lt2gene | Low-quality |
| 18-104-final-viral-combined_k141_162698\|\|lt2gene | Not-determined |
| 18-104-final-viral-combined_k141_120620\|\|lt2gene | Not-determined |
| 18-104-final-viral-combined_k141_37244\|\|lt2gene | Not-determined |
| 18-104-final-viral-combined_k141_2772\|\|lt2gene | Low-quality |
| 18-104-final-viral-combined_k141_37253\|\|lt2gene | Not-determined |
| 18-104-final-viral-combined_k141_72490\|\|lt2gene | Low-quality |
| 18-104-final-viral-combined_k141_2804\|\|lt2gene | Not-determined |
| 18-104-final-viral-combined_k141_142127\|\|lt2gene | Not-determined |
| 18-104-final-viral-combined_k141_148939\|\|lt2gene | Low-quality |
| 18-104-final-viral-combined_k141_107272\|\|lt2gene | Low-quality |
| 18-104-final-viral-combined_k141_16481\|\|lt2gene | Not-determined |
| 18-104-final-viral-combined_k141_162798\|\|lt2gene | Low-quality |
| 18-104-final-viral-combined_k141_107286\|\|lt2gene | Not-determined |
| 18-104-final-viral-combined_k141_2832\|\|lt2gene | Not-determined |
| 18-104-final-viral-combined_k141_44539\|\|lt2gene | Low-quality |
| 18-104-final-viral-combined_k141_30823\|\|lt2gene | Low-quality |
| 18-104-final-viral-combined_k141_9646\|\|lt2gene | Not-determined |
| 18-104-final-viral-combined_k141_99940\|\|lt2gene | Not-determined |
| 18-104-final-viral-combined_k141_135126\|\|lt2gene | Not-determined |
| 18-104-final-viral-combined_k141_72564\|\|lt2gene | Low-quality |
| 18-104-final-viral-combined_k141_86504\|\|lt2gene | Low-quality |
| 18-104-final-viral-combined_k141_79757\|\|lt2gene | Not-determined |
| 18-104-final-viral-combined_k141_65507\|\|lt2gene | Low-quality |
| 18-104-final-viral-combined_k141_86513\|\|lt2gene | Low-quality |
| 18-104-final-viral-combined_k141_142256\|\|lt2gene | Low-quality |
| 18-104-final-viral-combined_k141_107360\|\|lt2gene | Not-determined |
| 18-104-final-viral-combined_k141_58330\|\|lt2gene | Low-quality |
| 18-104-final-viral-combined_k141_120797\|\|lt2gene | Not-determined |
| 18-104-final-viral-combined_k141_16607\|\|lt2gene | Not-determined |
| 18-104-final-viral-combined_k141_9701\|\|lt2gene | Low-quality |
| 18-104-final-viral-combined_k141_9711\|\|lt2gene | Not-determined |
| 18-104-final-viral-combined_k141_23621\|\|lt2gene | Low-quality |
| 18-104-final-viral-combined_k141_149035\|\|lt2gene | Low-quality |
| 18-104-final-viral-combined_k141_51068\|\|lt2gene | Not-determined |
| 18-104-final-viral-combined_k141_2950\|\|lt2gene | Low-quality |
| 18-104-final-viral-combined_k141_16634\|\|lt2gene | Low-quality |
| 18-104-final-viral-combined_k141_107405\|\|lt2gene | Low-quality |
| 18-104-final-viral-combined_k141_120832\|\|lt2gene | Low-quality |
| 18-104-final-viral-combined_k141_100055\|\|lt2gene | Not-determined |
| 18-104-final-viral-combined_k141_16639\|\|lt2gene | Low-quality |
| 18-104-final-viral-combined_k141_58399\|\|lt2gene | Low-quality |
| 18-104-final-viral-combined_k141_128113\|\|lt2gene | Low-quality |
| 18-104-final-viral-combined_k141_142349\|\|lt2gene | Low-quality |
| 18-104-final-viral-combined_k141_128116\|\|lt2gene | Low-quality |
| 18-104-final-viral-combined_k141_156193\|\|lt2gene | Low-quality |
| 18-104-final-viral-combined_k141_16649\|\|lt2gene | Not-determined |
| 18-104-final-viral-combined_k141_162935\|\|lt2gene | Low-quality |
| 18-104-final-viral-combined_k141_23704\|\|lt2gene | Low-quality |
| 18-104-final-viral-combined_k141_114164\|\|lt2gene | Low-quality |
| 18-104-final-viral-combined_k141_142385\|\|lt2gene | Low-quality |
| 18-104-final-viral-combined_k141_107509\|\|lt2gene | Not-determined |
| 18-104-final-viral-combined_k141_3042\|\|lt2gene | Low-quality |
| 18-104-final-viral-combined_k141_93544\|\|lt2gene | Not-determined |
| 18-104-final-viral-combined_k141_93553\|\|lt2gene | Low-quality |
| 18-104-final-viral-combined_k141_51168\|\|lt2gene | Low-quality |
| 18-104-final-viral-combined_k141_9881\|\|lt2gene | Low-quality |
| 18-104-final-viral-combined_k141_58482\|\|lt2gene | Low-quality |
| 18-104-final-viral-combined_k141_86723\|\|lt2gene | Low-quality |
| 18-104-final-viral-combined_k141_86725\|\|lt2gene | Low-quality |
| 18-104-final-viral-combined_k141_51193\|\|lt2gene | Not-determined |
| 18-104-final-viral-combined_k141_86762\|\|lt2gene | Low-quality |
| 18-104-final-viral-combined_k141_86764\|\|lt2gene | Low-quality |
| 18-104-final-viral-combined_k141_65750\|\|lt2gene | Low-quality |
| 18-104-final-viral-combined_k141_23756\|\|lt2gene | Not-determined |
| 18-104-final-viral-combined_k141_9947\|\|lt2gene | Low-quality |
| 18-104-final-viral-combined_k141_156382\|\|lt2gene | Not-determined |
| 18-104-final-viral-combined_k141_23795\|\|lt2gene | Low-quality |
| 18-104-final-viral-combined_k141_107642\|\|lt2gene | Low-quality |
| 18-104-final-viral-combined_k141_37590\|\|lt2gene | Low-quality |
| 18-104-final-viral-combined_k141_37591\|\|lt2gene | Low-quality |
| 18-104-final-viral-combined_k141_31172\|\|lt2gene | Low-quality |
| 18-104-final-viral-combined_k141_10036\|\|lt2gene | Low-quality |
| 18-104-final-viral-combined_k141_10044\|\|lt2gene | Not-determined |
| 18-104-final-viral-combined_k141_156441\|\|lt2gene | Not-determined |

18-104-final-viral-combined_k141_149326|||lt2gene Low-quality
18-104-final-viral-combined_k141_37702|||lt2gene Low-quality
18-104-final-viral-combined_k141_37708|||lt2gene Low-quality
18-104-final-viral-combined_k141_142693|||lt2gene Low-quality
18-104-final-viral-combined_k141_100354|||lt2gene Low-quality
18-104-final-viral-combined_k141_135451|||lt2gene Low-quality
18-104-final-viral-combined_k141_107820|||lt2gene Low-quality
18-104-final-viral-combined_k141_3246|||lt2gene Low-quality
18-104-final-viral-combined_k141_51326|||lt2gene Low-quality
18-104-final-viral-combined_k141_23973|||lt2gene Not-determined
18-104-final-viral-combined_k141_31299|||lt2gene Not-determined
18-104-final-viral-combined_k141_51364|||lt2gene Not-determined
18-104-final-viral-combined_k141_58636|||lt2gene Low-quality
18-104-final-viral-combined_k141_156553|||lt2gene Low-quality
18-104-final-viral-combined_k141_24009|||lt2gene Low-quality
18-104-final-viral-combined_k141_135538|||lt2gene Low-quality
18-104-final-viral-combined_k141_128553|||lt2gene Low-quality
18-104-final-viral-combined_k141_86950|||lt2gene Low-quality
18-104-final-viral-combined_k141_142792|||lt2gene Low-quality
18-104-final-viral-combined_k141_72989|||lt2gene Not-determined
18-104-final-viral-combined_k141_37816|||lt2gene Low-quality
18-104-final-viral-combined_k141_149476|||lt2gene Low-quality
18-104-final-viral-combined_k141_100509|||lt2gene Low-quality
18-104-final-viral-combined_k141_149485|||lt2gene Low-quality
18-104-final-viral-combined_k141_121137|||lt2gene Low-quality
18-104-final-viral-combined_k141_135617|||lt2gene Low-quality
18-104-final-viral-combined_k141_31413|||lt2gene Low-quality
18-104-final-viral-combined_k141_93870|||lt2gene Not-determined
18-104-final-viral-combined_k141_93879|||lt2gene Low-quality
18-104-final-viral-combined_k141_16828|||lt2gene Low-quality
18-104-final-viral-combined_k141_128633|||lt2gene Not-determined
18-104-final-viral-combined_k141_73056|||lt2gene Low-quality
18-104-final-viral-combined_k141_100561|||lt2gene Not-determined
18-104-final-viral-combined_k141_58783|||lt2gene Low-quality
18-104-final-viral-combined_k141_16862|||lt2gene Not-determined
18-104-final-viral-combined_k141_100591|||lt2gene Low-quality
18-104-final-viral-combined_k141_163399|||lt2gene Low-quality
18-104-final-viral-combined_k141_142890|||lt2gene Low-quality
18-104-final-viral-combined_k141_73179|||lt2gene Low-quality
18-104-final-viral-combined_k141_66041|||lt2gene Low-quality
18-104-final-viral-combined_k141_87110|||lt2gene Low-quality
18-104-final-viral-combined_k141_3444|||lt2gene Low-quality
18-104-final-viral-combined_k141_45293|||lt2gene Low-quality
18-104-final-viral-combined_k141_87137|||lt2gene Not-determined
18-104-final-viral-combined_k141_100736|||lt2gene Low-quality
18-104-final-viral-combined_k141_128683|||lt2gene Low-quality
18-104-final-viral-combined_k141_108163|||lt2gene Not-determined
18-104-final-viral-combined_k141_156790|||lt2gene Low-quality
18-104-final-viral-combined_k141_58926|||lt2gene Low-quality
18-104-final-viral-combined_k141_114680|||lt2gene Low-quality
18-104-final-viral-combined_k141_66128|||lt2gene Low-quality
18-104-final-viral-combined_k141_37997|||lt2gene Low-quality
18-104-final-viral-combined_k141_58959|||lt2gene Low-quality
18-104-final-viral-combined_k141_10510|||lt2gene Not-determined
18-104-final-viral-combined_k141_121369|||lt2gene Not-determined
18-104-final-viral-combined_k141_10515|||lt2gene Low-quality
18-104-final-viral-combined_k141_121379|||lt2gene Low-quality
18-104-final-viral-combined_k141_58989|||lt2gene Low-quality
18-104-final-viral-combined_k141_142955|||lt2gene Low-quality
18-104-final-viral-combined_k141_10580|||lt2gene Not-determined
18-104-final-viral-combined_k141_51615|||lt2gene Low-quality
18-104-final-viral-combined_k141_135911|||lt2gene Low-quality
18-104-final-viral-combined_k141_128781|||lt2gene Low-quality
18-104-final-viral-combined_k141_156973|||lt2gene Low-quality
18-104-final-viral-combined_k141_135940|||lt2gene Low-quality
18-104-final-viral-combined_k141_24398|||lt2gene Low-quality
18-104-final-viral-combined_k141_80536|||lt2gene Not-determined
18-104-final-viral-combined_k141_128798|||lt2gene Low-quality
18-104-final-viral-combined_k141_157019|||lt2gene Low-quality
18-104-final-viral-combined_k141_59105|||lt2gene Low-quality
18-104-final-viral-combined_k141_66322|||lt2gene Low-quality
18-104-final-viral-combined_k141_135996|||lt2gene Low-quality
18-104-final-viral-combined_k141_80585|||lt2gene Low-quality
18-104-final-viral-combined_k141_87248|||lt2gene Not-determined
18-104-final-viral-combined_k141_100922|||lt2gene Low-quality
18-104-final-viral-combined_k141_108376|||lt2gene Low-quality
18-104-final-viral-combined_k141_59143|||lt2gene Low-quality
18-104-final-viral-combined_k141_100935|||lt2gene Low-quality
18-104-final-viral-combined_k141_114900|||lt2gene Low-quality
18-104-final-viral-combined_k141_108422|||lt2gene Low-quality
18-104-final-viral-combined_k141_51719|||lt2gene Low-quality
18-104-final-viral-combined_k141_108438|||lt2gene Low-quality
18-104-final-viral-combined_k141_73544|||lt2gene Not-determined
18-104-final-viral-combined_k141_136087|||lt2gene Low-quality
18-104-final-viral-combined_k141_128919|||lt2gene Not-determined
18-104-final-viral-combined_k141_121588|||lt2gene Low-quality
18-104-final-viral-combined_k141_136097|||lt2gene Not-determined
18-104-final-viral-combined_k141_114945|||lt2gene Low-quality
18-104-final-viral-combined_k141_108452|||lt2gene Not-determined
18-104-final-viral-combined_k141_45624|||lt2gene Low-quality
18-104-final-viral-combined_k141_143304|||lt2gene Low-quality
18-104-final-viral-combined_k141_94368|||lt2gene Low-quality
18-104-final-viral-combined_k141_66487|||lt2gene Low-quality
18-104-final-viral-combined_k141_114996|||lt2gene Not-determined
18-104-final-viral-combined_k141_157184|||lt2gene Low-quality
18-104-final-viral-combined_k141_59272|||lt2gene Low-quality
18-104-final-viral-combined_k141_108532|||lt2gene Low-quality
18-104-final-viral-combined_k141_73683|||lt2gene Low-quality
18-104-final-viral-combined_k141_59354|||lt2gene Low-quality
18-104-final-viral-combined_k141_150114|||lt2gene Not-determined
18-104-final-viral-combined_k141_80777|||lt2gene Low-quality
18-104-final-viral-combined_k141_31916|||lt2gene Not-determined
18-104-final-viral-combined_k141_45800|||lt2gene Not-determined
18-104-final-viral-combined_k141_108592|||lt2gene Low-quality
18-104-final-viral-combined_k141_73752|||lt2gene Not-determined
18-104-final-viral-combined_k141_31960|||lt2gene Low-quality
18-104-final-viral-combined_k141_129148|||lt2gene Not-determined
18-104-final-viral-combined_k141_10885|||lt2gene Not-determined
18-104-final-viral-combined_k141_115207|||lt2gene Not-determined
18-104-final-viral-combined_k141_80926|||lt2gene Low-quality
18-104-final-viral-combined_k141_73815|||lt2gene Low-quality
18-104-final-viral-combined_k141_10937|||lt2gene Not-determined
18-104-final-viral-combined_k141_94625|||lt2gene Low-quality
18-104-final-viral-combined_k141_45919|||lt2gene Not-determined
18-104-final-viral-combined_k141_108709|||lt2gene Not-determined
18-104-final-viral-combined_k141_108710|||lt2gene Not-determined
18-104-final-viral-combined_k141_73827|||lt2gene Low-quality
18-104-final-viral-combined_k141_10979|||lt2gene Low-quality
18-104-final-viral-combined_k141_101292|||lt2gene Not-determined
18-104-final-viral-combined_k141_24839|||lt2gene Not-determined
18-104-final-viral-combined_k141_52033|||lt2gene Not-determined
18-104-final-viral-combined_k141_101338|||lt2gene Not-determined
18-104-final-viral-combined_k141_32039|||lt2gene Low-quality
18-104-final-viral-combined_k141_66802|||lt2gene Low-quality
18-104-final-viral-combined_k141_101353|||lt2gene Low-quality
18-104-final-viral-combined_k141_46015|||lt2gene Low-quality
18-104-final-viral-combined_k141_94742|||lt2gene Low-quality
18-104-final-viral-combined_k141_108842|||lt2gene Not-determined
18-104-final-viral-combined_k141_108847|||lt2gene Not-determined
18-104-final-viral-combined_k141_121890|||lt2gene Low-quality
18-104-final-viral-combined_k141_101420|||lt2gene Low-quality
18-104-final-viral-combined_k141_52147|||lt2gene Low-quality
18-104-final-viral-combined_k141_129351|||lt2gene Low-quality
18-104-final-viral-combined_k141_81063|||lt2gene Not-determined
18-104-final-viral-combined_k141_17425|||lt2gene Low-quality
18-104-final-viral-combined_k141_108881|||lt2gene Not-determined
18-104-final-viral-combined_k141_11144|||lt2gene Low-quality
18-104-final-viral-combined_k141_143627|||lt2gene Low-quality
18-104-final-viral-combined_k141_164219|||lt2gene Low-quality
18-104-final-viral-combined_k141_108915|||lt2gene Low-quality
18-104-final-viral-combined_k141_59561|||lt2gene Low-quality
18-104-final-viral-combined_k141_46074|||lt2gene Not-determined
18-104-final-viral-combined_k141_17467|||lt2gene Not-determined
18-104-final-viral-combined_k141_17476|||lt2gene Low-quality
18-104-final-viral-combined_k141_66944|||lt2gene Low-quality
18-104-final-viral-combined_k141_66946|||lt2gene Low-quality
18-104-final-viral-combined_k141_129440|||lt2gene Low-quality
18-104-final-viral-combined_k141_129456|||lt2gene Low-quality
18-104-final-viral-combined_k141_101499|||lt2gene Low-quality
18-104-final-viral-combined_k141_38734|||lt2gene Low-quality
18-104-final-viral-combined_k141_32145|||lt2gene Low-quality
18-104-final-viral-combined_k141_101525|||lt2gene Low-quality
18-104-final-viral-combined_k141_38817|||lt2gene Low-quality
18-104-final-viral-combined_k141_74040|||lt2gene Low-quality
18-104-final-viral-combined_k141_143776|||lt2gene Low-quality
18-104-final-viral-combined_k141_87748|||lt2gene Not-determined
18-104-final-viral-combined_k141_59704|||lt2gene Low-quality
18-104-final-viral-combined_k141_74069|||lt2gene Low-quality
18-104-final-viral-combined_k141_143824|||lt2gene Not-determined
18-104-final-viral-combined_k141_136634|||lt2gene Low-quality
18-104-final-viral-combined_k141_11329|||lt2gene Not-determined
18-104-final-viral-combined_k141_25157|||lt2gene Not-determined
18-104-final-viral-combined_k141_46330|||lt2gene Low-quality
18-104-final-viral-combined_k141_11360|||lt2gene Low-quality
18-104-final-viral-combined_k141_74175|||lt2gene Low-quality
18-104-final-viral-combined_k141_150565|||lt2gene Low-quality
18-104-final-viral-combined_k141_81342|||lt2gene Not-determined
18-104-final-viral-combined_k141_67160|||lt2gene Low-quality
18-104-final-viral-combined_k141_67166|||lt2gene Low-quality
18-104-final-viral-combined_k141_101633|||lt2gene Not-determined
18-104-final-viral-combined_k141_4351|||lt2gene Low-quality
18-104-final-viral-combined_k141_95019|||lt2gene Low-quality
18-104-final-viral-combined_k141_150601|||lt2gene Not-determined
18-104-final-viral-combined_k141_74253|||lt2gene Low-quality
18-104-final-viral-combined_k141_101686|||lt2gene Low-quality
18-104-final-viral-combined_k141_59855|||lt2gene Low-quality
18-104-final-viral-combined_k141_87873|||lt2gene Not-determined
18-104-final-viral-combined_k141_59861|||lt2gene Not-determined
18-104-final-viral-combined_k141_52481|||lt2gene Low-quality
18-104-final-viral-combined_k141_67246|||lt2gene Low-quality
18-104-final-viral-combined_k141_164531|||lt2gene Not-determined
18-104-final-viral-combined_k141_122144|||lt2gene Not-determined
18-104-final-viral-combined_k141_87892|||lt2gene Low-quality
18-104-final-viral-combined_k141_52517|||lt2gene Low-quality
18-104-final-viral-combined_k141_17629|||lt2gene Low-quality
18-104-final-viral-combined_k141_157783|||lt2gene Low-quality
18-104-final-viral-combined_k141_87937|||lt2gene Low-quality
18-104-final-viral-combined_k141_144001|||lt2gene Not-determined
18-104-final-viral-combined_k141_115592|||lt2gene Not-determined
18-104-final-viral-combined_k141_164599|||lt2gene Low-quality
18-104-final-viral-combined_k141_115616|||lt2gene Not-determined
18-104-final-viral-combined_k141_52601|||lt2gene Not-determined
18-104-final-viral-combined_k141_157854|||lt2gene Low-quality
18-104-final-viral-combined_k141_52637|||lt2gene Low-quality
18-104-final-viral-combined_k141_101875|||lt2gene Low-quality
18-104-final-viral-combined_k141_144079|||lt2gene Not-determined
18-104-final-viral-combined_k141_122289|||lt2gene Low-quality
18-104-final-viral-combined_k141_88055|||lt2gene Low-quality
18-104-final-viral-combined_k141_32575|||lt2gene Low-quality
18-104-final-viral-combined_k141_122309|||lt2gene Low-quality
18-104-final-viral-combined_k141_39192|||lt2gene Low-quality
18-104-final-viral-combined_k141_109395|||lt2gene Not-determined
18-104-final-viral-combined_k141_25430|||lt2gene Low-quality
18-104-final-viral-combined_k141_17805|||lt2gene Not-determined
18-104-final-viral-combined_k141_157932|||lt2gene Low-quality
18-104-final-viral-combined_k141_115745|||lt2gene Low-quality
18-104-final-viral-combined_k141_4527|||lt2gene Low-quality
18-104-final-viral-combined_k141_109431|||lt2gene Low-quality
18-104-final-viral-combined_k141_150802|||lt2gene Low-quality
18-104-final-viral-combined_k141_109467|||lt2gene Not-determined
18-104-final-viral-combined_k141_164801|||lt2gene Low-quality
18-104-final-viral-combined_k141_74548|||lt2gene Low-quality
18-104-final-viral-combined_k141_74556|||lt2gene Low-quality
18-104-final-viral-combined_k141_52794|||lt2gene Low-quality
18-104-final-viral-combined_k141_122425|||lt2gene Not-determined
18-104-final-viral-combined_k141_17882|||lt2gene Low-quality
18-104-final-viral-combined_k141_11790|||lt2gene Not-determined
18-104-final-viral-combined_k141_46683|||lt2gene Low-quality
18-104-final-viral-combined_k141_39334|||lt2gene Not-determined
18-104-final-viral-combined_k141_11825|||lt2gene Low-quality
18-104-final-viral-combined_k141_150928|||lt2gene Not-determined
18-104-final-viral-combined_k141_122488|||lt2gene Not-determined
18-104-final-viral-combined_k141_115958|||lt2gene Not-determined
18-104-final-viral-combined_k141_88283|||lt2gene Not-determined
18-104-final-viral-combined_k141_74663|||lt2gene Low-quality
18-104-final-viral-combined_k141_39418|||lt2gene Low-quality
18-104-final-viral-combined_k141_109664|||lt2gene Low-quality
18-104-final-viral-combined_k141_115988|||lt2gene Low-quality
18-104-final-viral-combined_k141_74697|||lt2gene Low-quality
18-104-final-viral-combined_k141_67641|||lt2gene Low-quality
18-104-final-viral-combined_k141_52939|||lt2gene Low-quality
18-104-final-viral-combined_k141_130046|||lt2gene Low-quality
18-104-final-viral-combined_k141_88360|||lt2gene Low-quality
18-104-final-viral-combined_k141_52969|||lt2gene Not-determined
18-104-final-viral-combined_k141_116065|||lt2gene Low-quality
18-104-final-viral-combined_k141_122590|||lt2gene Not-determined
18-104-final-viral-combined_k141_18075|||lt2gene Low-quality
18-104-final-viral-combined_k141_60313|||lt2gene Not-determined
18-104-final-viral-combined_k141_137201|||lt2gene Low-quality
18-104-final-viral-combined_k141_88416|||lt2gene Low-quality
18-104-final-viral-combined_k141_60323|||lt2gene Not-determined
18-104-final-viral-combined_k141_88421|||lt2gene Low-quality
18-104-final-viral-combined_k141_122660|||lt2gene Low-quality
18-104-final-viral-combined_k141_144476|||lt2gene Low-quality
18-104-final-viral-combined_k141_165110|||lt2gene Low-quality
18-104-final-viral-combined_k141_74897|||lt2gene Low-quality
18-104-final-viral-combined_k141_109833|||lt2gene Not-determined
18-104-final-viral-combined_k141_137310|||lt2gene Not-determined
18-104-final-viral-combined_k141_116224|||lt2gene Not-determined
18-104-final-viral-combined_k141_88511|||lt2gene Low-quality
18-104-final-viral-combined_k141_165170|||lt2gene Not-determined
18-104-final-viral-combined_k141_151162|||lt2gene Low-quality
18-104-final-viral-combined_k141_109865|||lt2gene Low-quality
18-104-final-viral-combined_k141_158481|||lt2gene Low-quality
18-104-final-viral-combined_k141_102368|||lt2gene Not-determined
18-104-final-viral-combined_k141_53150|||lt2gene Low-quality
18-104-final-viral-combined_k141_39611|||lt2gene Low-quality
18-104-final-viral-combined_k141_102421|||lt2gene Low-quality
18-104-final-viral-combined_k141_151226|||lt2gene Not-determined
18-104-final-viral-combined_k141_151245|||lt2gene Not-determined
18-104-final-viral-combined_k141_102440|||lt2gene Low-quality
18-104-final-viral-combined_k141_67912|||lt2gene Not-determined
18-104-final-viral-combined_k141_116350|||lt2gene Not-determined
18-104-final-viral-combined_k141_144663|||lt2gene Not-determined
18-104-final-viral-combined_k141_116354|||lt2gene Not-determined
18-104-final-viral-combined_k141_60406|||lt2gene Low-quality
18-104-final-viral-combined_k141_158606|||lt2gene Not-determined
18-104-final-viral-combined_k141_122811|||lt2gene Not-determined
18-104-final-viral-combined_k141_95684|||lt2gene Low-quality
18-104-final-viral-combined_k141_122818|||lt2gene Low-quality
18-104-final-viral-combined_k141_39684|||lt2gene Low-quality
18-104-final-viral-combined_k141_158634|||lt2gene Low-quality
18-104-final-viral-combined_k141_12184|||lt2gene Not-determined
18-104-final-viral-combined_k141_60481|||lt2gene Not-determined
18-104-final-viral-combined_k141_95728|||lt2gene Low-quality
18-104-final-viral-combined_k141_95731|||lt2gene Low-quality
18-104-final-viral-combined_k141_144741|||lt2gene Not-determined
18-104-final-viral-combined_k141_116441|||lt2gene Not-determined
18-104-final-viral-combined_k141_39749|||lt2gene Low-quality
18-104-final-viral-combined_k141_144767|||lt2gene Low-quality
18-104-final-viral-combined_k141_68037|||lt2gene Low-quality
18-104-final-viral-combined_k141_102607|||lt2gene Low-quality
18-104-final-viral-combined_k141_122922|||lt2gene Low-quality
18-104-final-viral-combined_k141_110100|||lt2gene Low-quality
18-104-final-viral-combined_k141_75098|||lt2gene Low-quality
18-104-final-viral-combined_k141_110128|||lt2gene Low-quality
18-104-final-viral-combined_k141_158762|||lt2gene Low-quality
18-104-final-viral-combined_k141_12297|||lt2gene Low-quality
18-104-final-viral-combined_k141_122998|||lt2gene Low-quality
18-104-final-viral-combined_k141_110186|||lt2gene Low-quality
18-104-final-viral-combined_k141_18534|||lt2gene Not-determined
18-104-final-viral-combined_k141_130476|||lt2gene Not-determined
18-104-final-viral-combined_k141_53411|||lt2gene Low-quality
18-104-final-viral-combined_k141_144891|||lt2gene Not-determined
18-104-final-viral-combined_k141_60633|||lt2gene Low-quality
18-104-final-viral-combined_k141_33254|||lt2gene Low-quality
18-104-final-viral-combined_k141_12365|||lt2gene Not-determined
18-104-final-viral-combined_k141_33284|||lt2gene Not-determined
18-104-final-viral-combined_k141_144926|||lt2gene Low-quality
18-104-final-viral-combined_k141_60674|||lt2gene Low-quality
18-104-final-viral-combined_k141_18655|||lt2gene Low-quality
18-104-final-viral-combined_k141_5195|||lt2gene Low-quality
18-104-final-viral-combined_k141_96038|||lt2gene Low-quality
18-104-final-viral-combined_k141_60715|||lt2gene Low-quality
18-104-final-viral-combined_k141_123125|||lt2gene Low-quality
18-104-final-viral-combined_k141_102651|||lt2gene Not-determined
18-104-final-viral-combined_k141_158929|||lt2gene Low-quality
18-104-final-viral-combined_k141_96062|||lt2gene Low-quality
18-104-final-viral-combined_k141_123141|||lt2gene Low-quality
18-104-final-viral-combined_k141_60769|||lt2gene Low-quality
18-104-final-viral-combined_k141_18742|||lt2gene Low-quality
18-104-final-viral-combined_k141_165407|||lt2gene Not-determined
18-104-final-viral-combined_k141_5312|||lt2gene Low-quality
18-104-final-viral-combined_k141_47410|||lt2gene Low-quality
18-104-final-viral-combined_k141_5377|||lt2gene Low-quality
18-104-final-viral-combined_k141_53610|||lt2gene Not-determined
18-104-final-viral-combined_k141_137716|||lt2gene Not-determined
18-104-final-viral-combined_k141_116735|||lt2gene Low-quality
18-104-final-viral-combined_k141_130700|||lt2gene Low-quality
18-104-final-viral-combined_k141_159065|||lt2gene Low-quality
18-104-final-viral-combined_k141_18838|||lt2gene Low-quality
18-104-final-viral-combined_k141_82533|||lt2gene Low-quality
18-104-final-viral-combined_k141_116793|||lt2gene Low-quality
18-104-final-viral-combined_k141_102810|||lt2gene Low-quality
18-104-final-viral-combined_k141_40246|||lt2gene Low-quality
18-104-final-viral-combined_k141_40264|||lt2gene Low-quality
18-104-final-viral-combined_k141_5477|||lt2gene Low-quality
18-104-final-viral-combined_k141_26451|||lt2gene Low-quality
18-104-final-viral-combined_k141_82650|||lt2gene Low-quality
18-104-final-viral-combined_k141_47507|||lt2gene Not-determined
18-104-final-viral-combined_k141_53754|||lt2gene Low-quality
18-104-final-viral-combined_k141_18972|||lt2gene Not-determined
18-104-final-viral-combined_k141_26483|||lt2gene Not-determined
18-104-final-viral-combined_k141_96347|||lt2gene Not-determined
18-104-final-viral-combined_k141_151819|||lt2gene Low-quality
18-104-final-viral-combined_k141_75530|||lt2gene Low-quality
18-104-final-viral-combined_k141_61086|||lt2gene Low-quality
18-104-final-viral-combined_k141_33687|||lt2gene Low-quality
18-104-final-viral-combined_k141_130887|||lt2gene Low-quality
18-104-final-viral-combined_k141_151852|||lt2gene Low-quality
18-104-final-viral-combined_k141_33730|||lt2gene Low-quality
18-104-final-viral-combined_k141_68555|||lt2gene Not-determined
18-104-final-viral-combined_k141_123536|||lt2gene Low-quality
18-104-final-viral-combined_k141_103071|||lt2gene Low-quality
18-104-final-viral-combined_k141_82863|||lt2gene Not-determined
18-104-final-viral-combined_k141_75685|||lt2gene Not-determined
18-104-final-viral-combined_k141_96455|||lt2gene Low-quality
18-104-final-viral-combined_k141_19151|||lt2gene Low-quality
18-104-final-viral-combined_k141_53852|||lt2gene Low-quality
18-104-final-viral-combined_k141_53871|||lt2gene Low-quality
18-104-final-viral-combined_k141_117074|||lt2gene Low-quality
18-104-final-viral-combined_k141_68667|||lt2gene Not-determined
18-104-final-viral-combined_k141_47740|||lt2gene Low-quality
18-104-final-viral-combined_k141_151998|||lt2gene Low-quality
18-104-final-viral-combined_k141_33841|||lt2gene Not-determined
18-104-final-viral-combined_k141_53952|||lt2gene Low-quality
18-104-final-viral-combined_k141_61249|||lt2gene Low-quality
18-104-final-viral-combined_k141_110870|||lt2gene Low-quality
18-104-final-viral-combined_k141_96559|||lt2gene Low-quality
18-104-final-viral-combined_k141_68775|||lt2gene Low-quality
18-104-final-viral-combined_k141_61322|||lt2gene Low-quality
18-104-final-viral-combined_k141_145667|||lt2gene Low-quality
18-104-final-viral-combined_k141_47920|||lt2gene Not-determined
18-104-final-viral-combined_k141_61372|||lt2gene Low-quality
18-104-final-viral-combined_k141_61374|||lt2gene Low-quality
18-104-final-viral-combined_k141_75852|||lt2gene Low-quality
18-104-final-viral-combined_k141_54045|||lt2gene Low-quality
18-104-final-viral-combined_k141_131153|||lt2gene Low-quality
18-104-final-viral-combined_k141_96684|||lt2gene Low-quality
18-104-final-viral-combined_k141_83142|||lt2gene Low-quality
18-104-final-viral-combined_k141_123827|||lt2gene Low-quality
18-104-final-viral-combined_k141_40748|||lt2gene Low-quality
18-104-final-viral-combined_k141_61422|||lt2gene Low-quality
18-104-final-viral-combined_k141_152196|||lt2gene Low-quality
18-104-final-viral-combined_k141_96736|||lt2gene Low-quality
18-104-final-viral-combined_k141_111063|||lt2gene Not-determined
18-104-final-viral-combined_k141_138276|||lt2gene Low-quality
18-104-final-viral-combined_k141_54173|||lt2gene Not-determined
18-104-final-viral-combined_k141_165990|||lt2gene Low-quality
18-104-final-viral-combined_k141_123893|||lt2gene Not-determined
18-104-final-viral-combined_k141_131184|||lt2gene Low-quality
18-104-final-viral-combined_k141_111129|||lt2gene Low-quality
18-104-final-viral-combined_k141_19397|||lt2gene Not-determined
18-104-final-viral-combined_k141_19430|||lt2gene Low-quality
18-104-final-viral-combined_k141_19446|||lt2gene Low-quality
18-104-final-viral-combined_k141_89693|||lt2gene Low-quality
18-104-final-viral-combined_k141_19465|||lt2gene Low-quality
18-104-final-viral-combined_k141_6073|||lt2gene Low-quality
18-104-final-viral-combined_k141_117455|||lt2gene Low-quality
18-104-final-viral-combined_k141_26886|||lt2gene Not-determined
18-104-final-viral-combined_k141_76116|||lt2gene Not-determined
18-104-final-viral-combined_k141_111214|||lt2gene Not-determined
18-104-final-viral-combined_k141_34204|||lt2gene Not-determined
18-104-final-viral-combined_k141_96834|||lt2gene Low-quality
18-104-final-viral-combined_k141_152282|||lt2gene Low-quality
18-104-final-viral-combined_k141_145938|||lt2gene Not-determined
18-104-final-viral-combined_k141_69091|||lt2gene Low-quality
18-104-final-viral-combined_k141_19543|||lt2gene Low-quality
18-104-final-viral-combined_k141_89766|||lt2gene Low-quality
18-104-final-viral-combined_k141_103495|||lt2gene Low-quality
18-104-final-viral-combined_k141_159783|||lt2gene Low-quality
18-104-final-viral-combined_k141_138513|||lt2gene Low-quality
18-104-final-viral-combined_k141_124140|||lt2gene Low-quality
18-104-final-viral-combined_k141_76239|||lt2gene Not-determined
18-104-final-viral-combined_k141_19614|||lt2gene Not-determined
18-104-final-viral-combined_k141_145998|||lt2gene Low-quality
18-104-final-viral-combined_k141_27030|||lt2gene Low-quality
18-104-final-viral-combined_k141_111271|||lt2gene Not-determined
18-104-final-viral-combined_k141_124174|||lt2gene Low-quality
18-104-final-viral-combined_k141_146036|||lt2gene Low-quality
18-104-final-viral-combined_k141_61742|||lt2gene Low-quality
18-104-final-viral-combined_k141_124212|||lt2gene Not-determined
18-104-final-viral-combined_k141_159881|||lt2gene Low-quality
18-104-final-viral-combined_k141_152413|||lt2gene Low-quality
18-104-final-viral-combined_k141_96994|||lt2gene Not-determined
18-104-final-viral-combined_k141_83488|||lt2gene Low-quality
18-104-final-viral-combined_k141_152423|||lt2gene Not-determined
18-104-final-viral-combined_k141_97020|||lt2gene Low-quality
18-104-final-viral-combined_k141_103610|||lt2gene Low-quality
18-104-final-viral-combined_k141_146128|||lt2gene Low-quality
18-104-final-viral-combined_k141_19706|||lt2gene Low-quality
18-104-final-viral-combined_k141_13487|||lt2gene Not-determined
18-104-final-viral-combined_k141_159937|||lt2gene Low-quality
18-104-final-viral-combined_k141_6343|||lt2gene Low-quality
18-104-final-viral-combined_k141_83518|||lt2gene Low-quality
18-104-final-viral-combined_k141_138678|||lt2gene Not-determined
18-104-final-viral-combined_k141_83558|||lt2gene Not-determined
18-104-final-viral-combined_k141_41211|||lt2gene Not-determined
18-104-final-viral-combined_k141_61938|||lt2gene Not-determined
18-104-final-viral-combined_k141_48429|||lt2gene Low-quality
18-104-final-viral-combined_k141_152507|||lt2gene Not-determined
18-104-final-viral-combined_k141_41231|||lt2gene Low-quality
18-104-final-viral-combined_k141_166406|||lt2gene Not-determined
18-104-final-viral-combined_k141_54729|||lt2gene Not-determined
18-104-final-viral-combined_k141_62003|||lt2gene Low-quality
18-104-final-viral-combined_k141_97137|||lt2gene Low-quality
18-104-final-viral-combined_k141_97141|||lt2gene Low-quality
18-104-final-viral-combined_k141_41277|||lt2gene Low-quality
18-104-final-viral-combined_k141_54793|||lt2gene Low-quality
18-104-final-viral-combined_k141_90076|||lt2gene Not-determined
18-104-final-viral-combined_k141_62079|||lt2gene Low-quality
18-104-final-viral-combined_k141_62100|||lt2gene Not-determined
18-104-final-viral-combined_k141_69519|||lt2gene Low-quality
18-104-final-viral-combined_k141_76512|||lt2gene Low-quality
18-104-final-viral-combined_k141_97225|||lt2gene Low-quality
18-104-final-viral-combined_k141_69552|||lt2gene Not-determined
18-104-final-viral-combined_k141_41346|||lt2gene Low-quality
18-104-final-viral-combined_k141_138954|||lt2gene Low-quality
18-104-final-viral-combined_k141_131703|||lt2gene Low-quality
18-104-final-viral-combined_k141_124556|||lt2gene Low-quality
18-104-final-viral-combined_k141_124572|||lt2gene Low-quality
18-104-final-viral-combined_k141_62186|||lt2gene Low-quality
18-104-final-viral-combined_k141_41402|||lt2gene Low-quality
18-104-final-viral-combined_k141_138985|||lt2gene Low-quality
18-104-final-viral-combined_k141_139002|||lt2gene Low-quality
18-104-final-viral-combined_k141_152649|||lt2gene Low-quality
18-104-final-viral-combined_k141_139013|||lt2gene Low-quality
18-104-final-viral-combined_k141_27465|||lt2gene Low-quality
18-104-final-viral-combined_k141_34579|||lt2gene Low-quality
18-104-final-viral-combined_k141_20014|||lt2gene Not-determined
18-104-final-viral-combined_k141_27490|||lt2gene Low-quality
18-104-final-viral-combined_k141_90288|||lt2gene Low-quality
18-104-final-viral-combined_k141_139134|||lt2gene Low-quality
18-104-final-viral-combined_k141_118066|||lt2gene Low-quality
18-104-final-viral-combined_k141_131860|||lt2gene Low-quality
18-104-final-viral-combined_k141_166782|||lt2gene Not-determined
18-104-final-viral-combined_k141_103976|||lt2gene Low-quality
18-104-final-viral-combined_k141_41524|||lt2gene Low-quality
18-104-final-viral-combined_k141_124798|||lt2gene Low-quality
18-104-final-viral-combined_k141_13906|||lt2gene Not-determined
18-104-final-viral-combined_k141_6835|||lt2gene Not-determined
18-104-final-viral-combined_k141_6837|||lt2gene Low-quality
18-104-final-viral-combined_k141_139284|||lt2gene Low-quality
18-104-final-viral-combined_k141_55045|||lt2gene Not-determined
18-104-final-viral-combined_k141_104068|||lt2gene Low-quality
18-104-final-viral-combined_k141_62496|||lt2gene Low-quality
18-104-final-viral-combined_k141_166915|||lt2gene Low-quality
18-104-final-viral-combined_k141_166917|||lt2gene Low-quality
18-104-final-viral-combined_k141_153037|||lt2gene Not-determined
18-104-final-viral-combined_k141_55109|||lt2gene Low-quality
18-104-final-viral-combined_k141_90538|||lt2gene Low-quality
18-104-final-viral-combined_k141_90546|||lt2gene Not-determined
18-104-final-viral-combined_k141_124980|||lt2gene Low-quality
18-104-final-viral-combined_k141_153066|||lt2gene Low-quality
18-104-final-viral-combined_k141_118368|||lt2gene Low-quality
18-104-final-viral-combined_k141_132144|||lt2gene Low-quality
18-104-final-viral-combined_k141_132148|||lt2gene Low-quality
18-104-final-viral-combined_k141_125023|||lt2gene Low-quality
18-104-final-viral-combined_k141_125057|||lt2gene Not-determined
18-104-final-viral-combined_k141_104254|||lt2gene Low-quality
18-104-final-viral-combined_k141_55242|||lt2gene Low-quality
18-104-final-viral-combined_k141_62637|||lt2gene Not-determined
18-104-final-viral-combined_k141_41782|||lt2gene Not-determined
18-104-final-viral-combined_k141_41790|||lt2gene Not-determined
18-104-final-viral-combined_k141_62687|||lt2gene Low-quality
18-104-final-viral-combined_k141_104370|||lt2gene Low-quality
18-104-final-viral-combined_k141_20477|||lt2gene Low-quality
18-104-final-viral-combined_k141_20515|||lt2gene Not-determined
18-104-final-viral-combined_k141_55430|||lt2gene Not-determined
18-104-final-viral-combined_k141_125358|||lt2gene Low-quality
18-104_k141_27865 Not-determined
18-104_k141_104489 Not-determined
18-104_k141_6968 Not-determined
18-104_k141_146282 Not-determined
18-104_k141_160212 Not-determined
18-104_k141_160214 Low-quality
18-104_k141_41805 Not-determined
18-104_k141_76630 Not-determined
18-104_k141_160218 Not-determined
18-104_k141_48765 Not-determined
18-104_k141_55741 Not-determined
18-104_k141_6977 Not-determined
18-104_k141_125390 Not-determined
18-104_k141_83605 Not-determined
18-104_k141_20911 Low-quality
18-104_k141_62707 Not-determined
18-104_k141_55744 Not-determined
18-104_k141_55745 Low-quality
18-104_k141_160220 Not-determined
18-104_k141_34849 Not-determined
18-104_k141_69670 Not-determined
18-104_k141_104502 Not-determined
18-104_k141_27874 Not-determined
18-104_k141_69674 Not-determined
18-104_k141_132356 Not-determined
18-104_k141_34853 Not-determined
18-104_k141_6983 Not-determined
18-104_k141_97540 Not-determined
18-104_k141_55756 Not-determined
18-104_k141_118423 Not-determined
18-104_k141_97543 Not-determined
18-104_k141_139330 Not-determined
18-104_k141_139331 Not-determined
18-104_k141_132365 Not-determined
18-104_k141_160231 Not-determined
18-104_k141_132367 Not-determined
18-104_k141_62717 Not-determined
18-104_k141_34861 Not-determined
18-104_k141_69681 Not-determined
18-104_k141_27889 Not-determined
18-104_k141_48784 Not-determined
18-104_k141_132372 Not-determined
18-104_k141_90581 Not-determined
18-104_k141_160233 Not-determined
18-104_k141_48787 Low-quality
18-104_k141_160234 Not-determined
18-104_k141_69691 Not-determined
18-104_k141_23 Not-determined
18-104_k141_20936 Not-determined
18-104_k141_153266 Not-determined
18-104_k141_69696 Not-determined
18-104_k141_20937 Not-determined
18-104_k141_20939 Not-determined
18-104_k141_76663 Not-determined
18-104_k141_76664 Not-determined
18-104_k141_139352 Not-determined
18-104_k141_69702 Not-determined
18-104_k141_76665 Not-determined
18-104_k141_160242 Not-determined
18-104_k141_35 Not-determined
18-104_k141_69703 Not-determined
18-104_k141_62736 Not-determined
18-104_k141_90591 Not-determined
18-104_k141_34881 Low-quality
18-104_k141_62739 Not-determined
18-104_k141_76674 Not-determined
18-104_k141_48797 Not-determined
18-104_k141_146315 Not-determined
18-104_k141_69708 Not-determined
18-104_k141_43 Low-quality
18-104_k141_76678 Not-determined
18-104_k141_132390 Low-quality
18-104_k141_104531 Not-determined
18-104_k141_83638 Not-determined
18-104_k141_132395 Not-determined
18-104_k141_139357 Low-quality
18-104_k141_62753 Not-determined
18-104_k141_160255 Not-determined
18-104_k141_90604 Not-determined
18-104_k141_125403 Low-quality
18-104_k141_118442 Not-determined
18-104_k141_20956 Not-determined
18-104_k141_34900 Not-determined
18-104_k141_90611 Not-determined
18-104_k141_153291 Not-determined
18-104_k141_83652 Not-determined
18-104_k141_153293 Not-determined
18-104_k141_41824 Not-determined
18-104_k141_125407 Not-determined

| | |
|---|---|
| 18-104_k141_13979 | Not-determined |
| 18-104_k141_48810 | Not-determined |
| 18-104_k141_62772 | Not-determined |
| 18-104_k141_34905 | Not-determined |
| 18-104_k141_13981 | Not-determined |
| 18-104_k141_90626 | Not-determined |
| 18-104_k141_13988 | Low-quality |
| 18-104_k141_125413 | Not-determined |
| 18-104_k141_41842 | Not-determined |
| 18-104_k141_62775 | Not-determined |
| 18-104_k141_20972 | Low-quality |
| 18-104_k141_76704 | Not-determined |
| 18-104_k141_97553 | Not-determined |
| 18-104_k141_27927 | Not-determined |
| 18-104_k141_139378 | Not-determined |
| 18-104_k141_79 | Not-determined |
| 18-104_k141_139380 | Not-determined |
| 18-104_k141_27932 | Low-quality |
| 18-104_k141_146327 | Not-determined |
| 18-104_k141_76712 | Not-determined |
| 18-104_k141_85 | Not-determined |
| 18-104_k141_132428 | Not-determined |
| 18-104_k141_14003 | Not-determined |
| 18-104_k141_20978 | Not-determined |
| 18-104_k141_90640 | Not-determined |
| 18-104_k141_90641 | Not-determined |
| 18-104_k141_90642 | Not-determined |
| 18-104_k141_139383 | Not-determined |
| 18-104_k141_125427 | Low-quality |
| 18-104_k141_104544 | Not-determined |
| 18-104_k141_104545 | Not-determined |
| 18-104_k141_139387 | Low-quality |
| 18-104_k141_111506 | Not-determined |
| 18-104_k141_118457 | Not-determined |
| 18-104_k141_139392 | Not-determined |
| 18-104_k141_153317 | Not-determined |
| 18-104_k141_69754 | Not-determined |
| 18-104_k141_132435 | Not-determined |
| 18-104_k141_41848 | Not-determined |
| 18-104_k141_118464 | Not-determined |
| 18-104_k141_97571 | Not-determined |
| 18-104_k141_69761 | Not-determined |
| 18-104_k141_69764 | Not-determined |
| 18-104_k141_104559 | Not-determined |
| 18-104_k141_139405 | Not-determined |
| 18-104_k141_20997 | Not-determined |
| 18-104_k141_69769 | Not-determined |
| 18-104_k141_132447 | Not-determined |
| 18-104_k141_90670 | Not-determined |
| 18-104_k141_69772 | Not-determined |
| 18-104_k141_125443 | Not-determined |
| 18-104_k141_132453 | Not-determined |
| 18-104_k141_34941 | Not-determined |
| 18-104_k141_132457 | Not-determined |
| 18-104_k141_34943 | Not-determined |
| 18-104_k141_69784 | Low-quality |
| 18-104_k141_41862 | Not-determined |
| 18-104_k141_41863 | Not-determined |
| 18-104_k141_83665 | Not-determined |
| 18-104_k141_69790 | Low-quality |
| 18-104_k141_120 | Not-determined |
| 18-104_k141_62785 | Not-determined |
| 18-104_k141_76727 | Not-determined |
| 18-104_k141_122 | Not-determined |
| 18-104_k141_128 | Not-determined |
| 18-104_k141_111531 | Not-determined |
| 18-104_k141_62793 | Not-determined |
| 18-104_k141_129 | Not-determined |
| 18-104_k141_153340 | Not-determined |
| 18-104_k141_130 | Not-determined |
| 18-104_k141_132473 | Not-determined |
| 18-104_k141_131 | Not-determined |
| 18-104_k141_153343 | Not-determined |
| 18-104_k141_160276 | Not-determined |
| 18-104_k141_90682 | Not-determined |
| 18-104_k141_76742 | Not-determined |
| 18-104_k141_138 | Not-determined |
| 18-104_k141_111547 | Not-determined |
| 18-104_k141_160280 | Not-determined |
| 18-104_k141_160282 | Not-determined |
| 18-104_k141_55764 | Not-determined |
| 18-104_k141_34954 | Not-determined |
| 18-104_k141_55766 | Not-determined |
| 18-104_k141_55768 | Not-determined |
| 18-104_k141_55772 | Not-determined |
| 18-104_k141_160290 | Not-determined |
| 18-104_k141_55776 | Not-determined |
| 18-104_k141_139433 | Not-determined |
| 18-104_k141_97599 | Not-determined |
| 18-104_k141_97600 | Not-determined |
| 18-104_k141_125492 | Not-determined |
| 18-104_k141_111555 | Not-determined |
| 18-104_k141_111558 | Not-determined |
| 18-104_k141_14034 | Not-determined |
| 18-104_k141_76766 | Not-determined |
| 18-104_k141_125499 | Not-determined |
| 18-104_k141_139439 | Not-determined |
| 18-104_k141_76767 | Low-quality |
| 18-104_k141_90701 | Not-determined |
| 18-104_k141_48834 | Not-determined |
| 18-104_k141_83688 | Not-determined |
| 18-104_k141_83689 | Not-determined |
| 18-104_k141_111565 | Not-determined |
| 18-104_k141_97608 | Not-determined |
| 18-104_k141_111571 | Low-quality |
| 18-104_k141_160307 | Low-quality |
| 18-104_k141_90708 | Not-determined |
| 18-104_k141_153367 | Not-determined |
| 18-104_k141_159 | Not-determined |
| 18-104_k141_62826 | Low-quality |
| 18-104_k141_160314 | Not-determined |
| 18-104_k141_69815 | Low-quality |
| 18-104_k141_97617 | Not-determined |
| 18-104_k141_62830 | Not-determined |
| 18-104_k141_69819 | Not-determined |
| 18-104_k141_41888 | Not-determined |
| 18-104_k141_83697 | Not-determined |
| 18-104_k141_118518 | Not-determined |
| 18-104_k141_139468 | Not-determined |
| 18-104_k141_62837 | Not-determined |
| 18-104_k141_62838 | Low-quality |
| 18-104_k141_118522 | Not-determined |
| 18-104_k141_139471 | Not-determined |
| 18-104_k141_118524 | Not-determined |
| 18-104_k141_118525 | Not-determined |
| 18-104_k141_97626 | Not-determined |
| 18-104_k141_111593 | Not-determined |
| 18-104_k141_153383 | Not-determined |
| 18-104_k141_153386 | Not-determined |
| 18-104_k141_146365 | Not-determined |
| 18-104_k141_160328 | Not-determined |
| 18-104_k141_160329 | Not-determined |
| 18-104_k141_83708 | Not-determined |
| 18-104_k141_139478 | Not-determined |
| 18-104_k141_69830 | Not-determined |
| 18-104_k141_62850 | Not-determined |
| 18-104_k141_41905 | Not-determined |
| 18-104_k141_41906 | Not-determined |
| 18-104_k141_90727 | Low-quality |
| 18-104_k141_41907 | Not-determined |
| 18-104_k141_111605 | Not-determined |
| 18-104_k141_174 | Low-quality |
| 18-104_k141_48848 | Not-determined |
| 18-104_k141_69839 | Not-determined |
| 18-104_k141_139491 | Not-determined |
| 18-104_k141_186 | Not-determined |
| 18-104_k141_48854 | Not-determined |
| 18-104_k141_21042 | Not-determined |
| 18-104_k141_69843 | Not-determined |
| 18-104_k141_104655 | Not-determined |
| 18-104_k141_97632 | Not-determined |
| 18-104_k141_188 | Low-quality |
| 18-104_k141_41919 | Not-determined |
| 18-104_k141_139493 | Not-determined |
| 18-104_k141_27968 | Not-determined |
| 18-104_k141_7039 | Not-determined |
| 18-104_k141_104662 | Not-determined |
| 18-104_k141_111620 | Not-determined |
| 18-104_k141_14045 | Not-determined |
| 18-104_k141_97643 | Not-determined |
| 18-104_k141_76784 | Low-quality |
| 18-104_k141_104668 | Low-quality |
| 18-104_k141_69856 | Not-determined |
| 18-104_k141_90737 | Not-determined |
| 18-104_k141_69861 | Not-determined |
| 18-104_k141_41930 | Not-determined |
| 18-104_k141_69863 | Not-determined |
| 18-104_k141_41931 | Not-determined |
| 18-104_k141_21055 | Not-determined |
| 18-104_k141_111633 | Not-determined |
| 18-104_k141_7049 | Not-determined |
| 18-104_k141_90741 | Not-determined |
| 18-104_k141_97653 | Not-determined |
| 18-104_k141_55785 | Not-determined |
| 18-104_k141_7051 | Not-determined |
| 18-104_k141_111640 | Not-determined |
| 18-104_k141_21061 | Not-determined |
| 18-104_k141_41936 | Not-determined |
| 18-104_k141_41937 | Not-determined |
| 18-104_k141_160348 | Not-determined |
| 18-104_k141_7060 | Not-determined |
| 18-104_k141_62871 | Low-quality |
| 18-104_k141_90754 | Not-determined |
| 18-104_k141_206 | Not-determined |
| 18-104_k141_7061 | Not-determined |
| 18-104_k141_62874 | Not-determined |
| 18-104_k141_62875 | Not-determined |
| 18-104_k141_125511 | Not-determined |
| 18-104_k141_21065 | Not-determined |
| 18-104_k141_62876 | Low-quality |
| 18-104_k141_76798 | Not-determined |
| 18-104_k141_62882 | Not-determined |
| 18-104_k141_111648 | Not-determined |
| 18-104_k141_139514 | Not-determined |
| 18-104_k141_69886 | Not-determined |
| 18-104_k141_69887 | Not-determined |
| 18-104_k141_97676 | Not-determined |
| 18-104_k141_41950 | Not-determined |
| 18-104_k141_132511 | Not-determined |
| 18-104_k141_48883 | Not-determined |
| 18-104_k141_48884 | Not-determined |
| 18-104_k141_111656 | Not-determined |
| 18-104_k141_111657 | Not-determined |
| 18-104_k141_69893 | Not-determined |
| 18-104_k141_146368 | Low-quality |
| 18-104_k141_90766 | Low-quality |
| 18-104_k141_90767 | Not-determined |
| 18-104_k141_111661 | Low-quality |
| 18-104_k141_223 | Low-quality |
| 18-104_k141_14073 | Not-determined |
| 18-104_k141_41951 | Not-determined |
| 18-104_k141_41953 | Not-determined |
| 18-104_k141_83746 | Not-determined |
| 18-104_k141_97683 | Not-determined |
| 18-104_k141_90771 | Not-determined |
| 18-104_k141_55817 | Not-determined |
| 18-104_k141_14074 | Low-quality |
| 18-104_k141_21072 | Low-quality |
| 18-104_k141_111667 | Not-determined |
| 18-104_k141_7074 | Not-determined |
| 18-104_k141_146378 | Not-determined |
| 18-104_k141_21077 | Not-determined |
| 18-104_k141_83747 | Not-determined |
| 18-104_k141_55824 | Not-determined |
| 18-104_k141_28004 | Not-determined |
| 18-104_k141_104698 | Not-determined |
| 18-104_k141_104700 | Not-determined |
| 18-104_k141_125546 | Not-determined |
| 18-104_k141_132522 | Not-determined |
| 18-104_k141_90778 | Not-determined |
| 18-104_k141_125549 | Not-determined |
| 18-104_k141_104712 | Not-determined |
| 18-104_k141_55835 | Low-quality |
| 18-104_k141_7098 | Not-determined |
| 18-104_k141_248 | Not-determined |
| 18-104_k141_55840 | Not-determined |
| 18-104_k141_90783 | Not-determined |
| 18-104_k141_62893 | Not-determined |
| 18-104_k141_254 | Not-determined |
| 18-104_k141_7115 | Not-determined |
| 18-104_k141_55848 | Low-quality |
| 18-104_k141_14087 | Not-determined |
| 18-104_k141_69923 | Not-determined |
| 18-104_k141_48888 | Not-determined |
| 18-104_k141_111685 | Not-determined |
| 18-104_k141_14089 | Not-determined |
| 18-104_k141_160366 | Not-determined |
| 18-104_k141_104726 | Not-determined |
| 18-104_k141_264 | Not-determined |
| 18-104_k141_265 | Not-determined |
| 18-104_k141_111689 | Not-determined |
| 18-104_k141_69929 | Not-determined |
| 18-104_k141_21086 | Not-determined |
| 18-104_k141_268 | Not-determined |
| 18-104_k141_269 | Not-determined |
| 18-104_k141_160376 | Not-determined |
| 18-104_k141_153403 | Not-determined |
| 18-104_k141_69936 | Not-determined |
| 18-104_k141_14103 | Not-determined |
| 18-104_k141_97715 | Not-determined |
| 18-104_k141_97716 | Not-determined |
| 18-104_k141_273 | Not-determined |
| 18-104_k141_55878 | Not-determined |
| 18-104_k141_14106 | Not-determined |
| 18-104_k141_97721 | Not-determined |
| 18-104_k141_41972 | Not-determined |
| 18-104_k141_125567 | Not-determined |
| 18-104_k141_69943 | Not-determined |
| 18-104_k141_160394 | Low-quality |
| 18-104_k141_125570 | Not-determined |
| 18-104_k141_83793 | Not-determined |
| 18-104_k141_104745 | Not-determined |
| 18-104_k141_7143 | Not-determined |
| 18-104_k141_69948 | Not-determined |
| 18-104_k141_35046 | Not-determined |
| 18-104_k141_283 | Not-determined |
| 18-104_k141_132559 | Not-determined |
| 18-104_k141_21106 | Not-determined |
| 18-104_k141_35052 | Not-determined |
| 18-104_k141_62931 | Not-determined |
| 18-104_k141_153423 | Not-determined |
| 18-104_k141_41979 | Not-determined |
| 18-104_k141_76831 | Not-determined |
| 18-104_k141_139556 | Not-determined |
| 18-104_k141_118545 | Not-determined |
| 18-104_k141_160398 | Not-determined |
| 18-104_k141_14122 | Not-determined |
| 18-104_k141_160401 | Not-determined |
| 18-104_k141_76837 | Not-determined |
| 18-104_k141_132568 | Not-determined |
| 18-104_k141_21112 | Low-quality |
| 18-104_k141_62940 | Not-determined |
| 18-104_k141_294 | Not-determined |
| 18-104_k141_139563 | Not-determined |
| 18-104_k141_14132 | Not-determined |
| 18-104_k141_146394 | Not-determined |
| 18-104_k141_21118 | Not-determined |
| 18-104_k141_83804 | Low-quality |
| 18-104_k141_69955 | Not-determined |
| 18-104_k141_83805 | Not-determined |
| 18-104_k141_90805 | Not-determined |
| 18-104_k141_35063 | Not-determined |
| 18-104_k141_69956 | Low-quality |
| 18-104_k141_298 | Not-determined |
| 18-104_k141_90808 | Low-quality |
| 18-104_k141_7161 | Not-determined |
| 18-104_k141_132577 | Not-determined |
| 18-104_k141_125581 | Not-determined |
| 18-104_k141_125583 | Not-determined |
| 18-104_k141_14145 | Not-determined |
| 18-104_k141_14146 | Low-quality |
| 18-104_k141_69963 | Not-determined |
| 18-104_k141_69966 | Not-determined |
| 18-104_k141_146405 | Low-quality |
| 18-104_k141_21130 | Not-determined |
| 18-104_k141_304 | Not-determined |
| 18-104_k141_153442 | Not-determined |
| 18-104_k141_305 | Not-determined |
| 18-104_k141_21131 | Not-determined |
| 18-104_k141_7171 | Not-determined |
| 18-104_k141_139584 | Not-determined |
| 18-104_k141_97743 | Not-determined |
| 18-104_k141_35074 | Low-quality |
| 18-104_k141_111723 | Not-determined |
| 18-104_k141_111724 | Not-determined |
| 18-104_k141_111726 | Not-determined |
| 18-104_k141_111731 | Not-determined |
| 18-104_k141_118552 | Low-quality |
| 18-104_k141_62953 | Not-determined |
| 18-104_k141_321 | Low-quality |
| 18-104_k141_97749 | Not-determined |
| 18-104_k141_97750 | Not-determined |
| 18-104_k141_48922 | Not-determined |
| 18-104_k141_153452 | Not-determined |
| 18-104_k141_326 | Not-determined |
| 18-104_k141_97754 | Not-determined |
| 18-104_k141_160412 | Not-determined |
| 18-104_k141_28036 | Not-determined |
| 18-104_k141_83831 | Not-determined |
| 18-104_k141_160415 | Not-determined |
| 18-104_k141_62960 | Not-determined |
| 18-104_k141_104750 | Not-determined |
| 18-104_k141_146421 | Not-determined |
| 18-104_k141_28041 | Not-determined |
| 18-104_k141_111749 | Low-quality |
| 18-104_k141_111752 | Not-determined |
| 18-104_k141_335 | Low-quality |
| 18-104_k141_90835 | Not-determined |
| 18-104_k141_146430 | Not-determined |
| 18-104_k141_90840 | Not-determined |
| 18-104_k141_62966 | Low-quality |
| 18-104_k141_7188 | Not-determined |
| 18-104_k141_76852 | Not-determined |
| 18-104_k141_97760 | Not-determined |
| 18-104_k141_7191 | Not-determined |
| 18-104_k141_339 | Not-determined |
| 18-104_k141_28055 | Not-determined |
| 18-104_k141_125618 | Low-quality |
| 18-104_k141_7196 | Not-determined |
| 18-104_k141_146436 | Not-determined |
| 18-104_k141_42020 | Not-determined |
| 18-104_k141_35094 | Not-determined |
| 18-104_k141_35095 | Not-determined |
| 18-104_k141_7203 | Not-determined |
| 18-104_k141_90856 | Not-determined |
| 18-104_k141_125627 | Low-quality |
| 18-104_k141_48940 | Low-quality |
| 18-104_k141_355 | Not-determined |
| 18-104_k141_28063 | Not-determined |
| 18-104_k141_7209 | Not-determined |
| 18-104_k141_125636 | Not-determined |
| 18-104_k141_104765 | Low-quality |
| 18-104_k141_48942 | Low-quality |
| 18-104_k141_146453 | Not-determined |
| 18-104_k141_28072 | Not-determined |
| 18-104_k141_42029 | Not-determined |
| 18-104_k141_104772 | Not-determined |
| 18-104_k141_118586 | Not-determined |
| 18-104_k141_48949 | Not-determined |
| 18-104_k141_76867 | Not-determined |
| 18-104_k141_62988 | Not-determined |
| 18-104_k141_62989 | Not-determined |
| 18-104_k141_118591 | Not-determined |
| 18-104_k141_35108 | Not-determined |
| 18-104_k141_146464 | Low-quality |
| 18-104_k141_42036 | Not-determined |
| 18-104_k141_42037 | Not-determined |
| 18-104_k141_373 | Low-quality |
| 18-104_k141_374 | Not-determined |
| 18-104_k141_125661 | Not-determined |
| 18-104_k141_55906 | Not-determined |
| 18-104_k141_104781 | Not-determined |
| 18-104_k141_146470 | Not-determined |
| 18-104_k141_125663 | Not-determined |
| 18-104_k141_146472 | Not-determined |
| 18-104_k141_48955 | Not-determined |
| 18-104_k141_139617 | Low-quality |
| 18-104_k141_384 | Not-determined |
| 18-104_k141_35113 | Not-determined |
| 18-104_k141_14152 | Low-quality |
| 18-104_k141_55916 | Not-determined |
| 18-104_k141_139621 | Not-determined |
| 18-104_k141_48959 | Not-determined |
| 18-104_k141_139623 | Not-determined |
| 18-104_k141_48961 | Not-determined |
| 18-104_k141_97791 | Not-determined |
| 18-104_k141_391 | Not-determined |
| 18-104_k141_83852 | Not-determined |
| 18-104_k141_28087 | Not-determined |
| 18-104_k141_35117 | Low-quality |
| 18-104_k141_83856 | Low-quality |
| 18-104_k141_160450 | Not-determined |
| 18-104_k141_42046 | Not-determined |
| 18-104_k141_28100 | Not-determined |
| 18-104_k141_70021 | Low-quality |
| 18-104_k141_146484 | Not-determined |
| 18-104_k141_146485 | Not-determined |
| 18-104_k141_111795 | Low-quality |
| 18-104_k141_104803 | Not-determined |
| 18-104_k141_125676 | Not-determined |
| 18-104_k141_125677 | Not-determined |
| 18-104_k141_70027 | Not-determined |
| 18-104_k141_111798 | Not-determined |
| 18-104_k141_125681 | Not-determined |
| 18-104_k141_63008 | Not-determined |
| 18-104_k141_42049 | Low-quality |
| 18-104_k141_139635 | Not-determined |
| 18-104_k141_90914 | Not-determined |
| 18-104_k141_55932 | Low-quality |
| 18-104_k141_118629 | Not-determined |
| 18-104_k141_125685 | Not-determined |
| 18-104_k141_97800 | Not-determined |
| 18-104_k141_21143 | Not-determined |
| 18-104_k141_48978 | Low-quality |
| 18-104_k141_97804 | Not-determined |
| 18-104_k141_42054 | Not-determined |
| 18-104_k141_63017 | Not-determined |
| 18-104_k141_70038 | Not-determined |
| 18-104_k141_90925 | Not-determined |
| 18-104_k141_70041 | Not-determined |
| 18-104_k141_76887 | Not-determined |
| 18-104_k141_70043 | Not-determined |
| 18-104_k141_63021 | Not-determined |
| 18-104_k141_146504 | Not-determined |
| 18-104_k141_118644 | Low-quality |
| 18-104_k141_83873 | Not-determined |
| 18-104_k141_63022 | Not-determined |
| 18-104_k141_139652 | Not-determined |
| 18-104_k141_153499 | Not-determined |
| 18-104_k141_97812 | Not-determined |
| 18-104_k141_97813 | Not-determined |
| 18-104_k141_63026 | Not-determined |
| 18-104_k141_90934 | Not-determined |
| 18-104_k141_63029 | Not-determined |
| 18-104_k141_7226 | Not-determined |
| 18-104_k141_97817 | Not-determined |
| 18-104_k141_21149 | Not-determined |
| 18-104_k141_83877 | Not-determined |
| 18-104_k141_14176 | Low-quality |
| 18-104_k141_48987 | Low-quality |
| 18-104_k141_132609 | Not-determined |
| 18-104_k141_146506 | Low-quality |
| 18-104_k141_21150 | Not-determined |
| 18-104_k141_21151 | Not-determined |
| 18-104_k141_104820 | Not-determined |
| 18-104_k141_55952 | Low-quality |
| 18-104_k141_97820 | Not-determined |
| 18-104_k141_76899 | Not-determined |
| 18-104_k141_55955 | Not-determined |
| 18-104_k141_104830 | Not-determined |
| 18-104_k141_146512 | Low-quality |
| 18-104_k141_63039 | Not-determined |
| 18-104_k141_118653 | Not-determined |
| 18-104_k141_70046 | Not-determined |
| 18-104_k141_42068 | Not-determined |
| 18-104_k141_42070 | Not-determined |
| 18-104_k141_421 | Not-determined |
| 18-104_k141_83890 | Not-determined |
| 18-104_k141_28139 | Not-determined |
| 18-104_k141_97828 | Not-determined |
| 18-104_k141_21164 | Not-determined |
| 18-104_k141_76913 | Not-determined |
| 18-104_k141_139670 | Not-determined |
| 18-104_k141_76917 | Not-determined |
| 18-104_k141_132615 | Not-determined |
| 18-104_k141_76919 | Not-determined |
| 18-104_k141_146515 | Low-quality |
| 18-104_k141_97836 | Not-determined |
| 18-104_k141_433 | Not-determined |
| 18-104_k141_14202 | Not-determined |
| 18-104_k141_146517 | Not-determined |
| 18-104_k141_125725 | Low-quality |
| 18-104_k141_146521 | Not-determined |
| 18-104_k141_111838 | Not-determined |
| 18-104_k141_21165 | Low-quality |
| 18-104_k141_35131 | Not-determined |
| 18-104_k141_14210 | Not-determined |
| 18-104_k141_90958 | Not-determined |
| 18-104_k141_28152 | Not-determined |
| 18-104_k141_104841 | Not-determined |
| 18-104_k141_111846 | Not-determined |
| 18-104_k141_104844 | Not-determined |
| 18-104_k141_125727 | Not-determined |
| 18-104_k141_125729 | Not-determined |
| 18-104_k141_139679 | Not-determined |
| 18-104_k141_14217 | Not-determined |
| 18-104_k141_83912 | Not-determined |
| 18-104_k141_139681 | Not-determined |
| 18-104_k141_83913 | Low-quality |
| 18-104_k141_21183 | Low-quality |
| 18-104_k141_28162 | Not-determined |
| 18-104_k141_125735 | Not-determined |
| 18-104_k141_55974 | Not-determined |
| 18-104_k141_118673 | Not-determined |
| 18-104_k141_153521 | Not-determined |
| 18-104_k141_76939 | Not-determined |
| 18-104_k141_146528 | Not-determined |
| 18-104_k141_76942 | Not-determined |
| 18-104_k141_76948 | Low-quality |
| 18-104_k141_90975 | Not-determined |
| 18-104_k141_436 | Not-determined |
| 18-104_k141_14231 | Not-determined |
| 18-104_k141_146531 | Not-determined |
| 18-104_k141_132630 | Low-quality |
| 18-104_k141_28174 | Not-determined |
| 18-104_k141_438 | Not-determined |
| 18-104_k141_7282 | Not-determined |
| 18-104_k141_90984 | Not-determined |
| 18-104_k141_97876 | Not-determined |
| 18-104_k141_160509 | Not-determined |
| 18-104_k141_21190 | Not-determined |
| 18-104_k141_153541 | Not-determined |
| 18-104_k141_63054 | Not-determined |
| 18-104_k141_125748 | Not-determined |
| 18-104_k141_153543 | Low-quality |
| 18-104_k141_55993 | Low-quality |
| 18-104_k141_42117 | Not-determined |
| 18-104_k141_21193 | Not-determined |
| 18-104_k141_76967 | Not-determined |
| 18-104_k141_76974 | Not-determined |
| 18-104_k141_132656 | Not-determined |
| 18-104_k141_111867 | Not-determined |
| 18-104_k141_139688 | Not-determined |
| 18-104_k141_146537 | Low-quality |
| 18-104_k141_42124 | Not-determined |
| 18-104_k141_146539 | Not-determined |
| 18-104_k141_14258 | Not-determined |
| 18-104_k141_42126 | Not-determined |
| 18-104_k141_111875 | Not-determined |
| 18-104_k141_76979 | Low-quality |
| 18-104_k141_132664 | Not-determined |
| 18-104_k141_42132 | Not-determined |
| 18-104_k141_76980 | Not-determined |
| 18-104_k141_125759 | Low-quality |
| 18-104_k141_111880 | Not-determined |
| 18-104_k141_48997 | Not-determined |
| 18-104_k141_91009 | Not-determined |
| 18-104_k141_56010 | Not-determined |
| 18-104_k141_14266 | Not-determined |
| 18-104_k141_56014 | Not-determined |
| 18-104_k141_139702 | Not-determined |
| 18-104_k141_125764 | Low-quality |
| 18-104_k141_146556 | Low-quality |
| 18-104_k141_7309 | Not-determined |
| 18-104_k141_104878 | Not-determined |
| 18-104_k141_49007 | Not-determined |
| 18-104_k141_14274 | Not-determined |
| 18-104_k141_91015 | Not-determined |
| 18-104_k141_83969 | Not-determined |
| 18-104_k141_56022 | Not-determined |
| 18-104_k141_139710 | Not-determined |
| 18-104_k141_28204 | Not-determined |
| 18-104_k141_118687 | Not-determined |
| 18-104_k141_160517 | Low-quality |
| 18-104_k141_91019 | Not-determined |
| 18-104_k141_28207 | Not-determined |
| 18-104_k141_111884 | Not-determined |
| 18-104_k141_160521 | Not-determined |
| 18-104_k141_160522 | Not-determined |
| 18-104_k141_132687 | Not-determined |
| 18-104_k141_42148 | Not-determined |
| 18-104_k141_139719 | Not-determined |
| 18-104_k141_146568 | Not-determined |
| 18-104_k141_76996 | Not-determined |
| 18-104_k141_160525 | Not-determined |
| 18-104_k141_160526 | Not-determined |
| 18-104_k141_160528 | Not-determined |
| 18-104_k141_97909 | Low-quality |
| 18-104_k141_28212 | Not-determined |

| | |
|---|---|
| 18-104_k141_104884 | Not-determined |
| 18-104_k141_56035 | Not-determined |
| 18-104_k141_63089 | Not-determined |
| 18-104_k141_14296 | Not-determined |
| 18-104_k141_483 | Not-determined |
| 18-104_k141_28219 | Not-determined |
| 18-104_k141_132696 | Not-determined |
| 18-104_k141_70074 | Low-quality |
| 18-104_k141_83987 | Low-quality |
| 18-104_k141_56044 | Not-determined |
| 18-104_k141_28224 | Not-determined |
| 18-104_k141_70078 | Not-determined |
| 18-104_k141_77008 | Not-determined |
| 18-104_k141_63092 | Not-determined |
| 18-104_k141_63095 | Not-determined |
| 18-104_k141_153571 | Low-quality |
| 18-104_k141_91026 | Not-determined |
| 18-104_k141_91029 | Not-determined |
| 18-104_k141_111905 | Low-quality |
| 18-104_k141_125783 | Not-determined |
| 18-104_k141_42173 | Not-determined |
| 18-104_k141_146590 | Not-determined |
| 18-104_k141_146591 | Not-determined |
| 18-104_k141_91033 | Not-determined |
| 18-104_k141_91034 | Not-determined |
| 18-104_k141_56058 | Not-determined |
| 18-104_k141_21220 | Not-determined |
| 18-104_k141_28227 | Not-determined |
| 18-104_k141_7317 | Low-quality |
| 18-104_k141_139744 | Not-determined |
| 18-104_k141_160538 | Not-determined |
| 18-104_k141_111915 | Not-determined |
| 18-104_k141_153583 | Not-determined |
| 18-104_k141_56065 | Not-determined |
| 18-104_k141_56067 | Not-determined |
| 18-104_k141_160543 | Low-quality |
| 18-104_k141_28229 | Not-determined |
| 18-104_k141_153585 | Not-determined |
| 18-104_k141_56069 | Not-determined |
| 18-104_k141_139756 | Not-determined |
| 18-104_k141_111925 | Not-determined |
| 18-104_k141_21235 | Not-determined |
| 18-104_k141_56070 | Not-determined |
| 18-104_k141_14311 | Not-determined |
| 18-104_k141_153592 | Not-determined |
| 18-104_k141_83991 | Not-determined |
| 18-104_k141_70094 | Low-quality |
| 18-104_k141_21241 | Not-determined |
| 18-104_k141_77034 | Not-determined |
| 18-104_k141_501 | Low-quality |
| 18-104_k141_160554 | Not-determined |
| 18-104_k141_56080 | Not-determined |
| 18-104_k141_83996 | Not-determined |
| 18-104_k141_160556 | Not-determined |
| 18-104_k141_56086 | Not-determined |
| [illegible] | [illegible] |

| | |
|---|---|
| 18-104_k141_14496 | Not-determined |
| 18-104_k141_105197 | Low-quality |
| 18-104_k141_126191 | Not-determined |
| 18-104_k141_7499 | Not-determined |
| 18-104_k141_14500 | Not-determined |
| 18-104_k141_49306 | Not-determined |
| 18-104_k141_112152 | Low-quality |
| 18-104_k141_140043 | Not-determined |
| 18-104_k141_105201 | Not-determined |
| 18-104_k141_153796 | Not-determined |
| 18-104_k141_77414 | Not-determined |
| 18-104_k141_126199 | Not-determined |
| 18-104_k141_126200 | Not-determined |
| 18-104_k141_105203 | Not-determined |
| 18-104_k141_132874 | Not-determined |
| 18-104_k141_98213 | Not-determined |
| 18-104_k141_126202 | Not-determined |
| 18-104_k141_132876 | Not-determined |
| 18-104_k141_49308 | Not-determined |
| 18-104_k141_63293 | Not-determined |
| 18-104_k141_132878 | Not-determined |
| 18-104_k141_153800 | Not-determined |
| 18-104_k141_21543 | Not-determined |
| 18-104_k141_63296 | Low-quality |
| 18-104_k141_14512 | Not-determined |
| 18-104_k141_112161 | Not-determined |
| 18-104_k141_91396 | Not-determined |
| 18-104_k141_35295 | Not-determined |
| 18-104_k141_118955 | Not-determined |
| 18-104_k141_91400 | Not-determined |
| 18-104_k141_126205 | Not-determined |
| 18-104_k141_84358 | Not-determined |
| 18-104_k141_63305 | Not-determined |
| 18-104_k141_112167 | Not-determined |
| 18-104_k141_56334 | Not-determined |
| 18-104_k141_14514 | Low-quality |
| 18-104_k141_91401 | Not-determined |
| 18-104_k141_7520 | Not-determined |
| 18-104_k141_105215 | Not-determined |
| 18-104_k141_28540 | Not-determined |
| 18-104_k141_63311 | Not-determined |
| 18-104_k141_28541 | Not-determined |
| 18-104_k141_21545 | Not-determined |
| 18-104_k141_132896 | Not-determined |
| 18-104_k141_28546 | Low-quality |
| 18-104_k141_56341 | Not-determined |
| 18-104_k141_28547 | Not-determined |
| 18-104_k141_28548 | Not-determined |
| 18-104_k141_132899 | Not-determined |
| 18-104_k141_105224 | Not-determined |
| 18-104_k141_28550 | Not-determined |
| 18-104_k141_14521 | Not-determined |
| 18-104_k141_35302 | Not-determined |
| 18-104_k141_14522 | Not-determined |
| 18-104_k141_35306 | Not-determined |
| 18-104_k141_28561 | Not-determined |
| 18-104_k141_146911 | Not-determined |
| 18-104_k141_28567 | Not-determined |
| 18-104_k141_118962 | Not-determined |
| 18-104_k141_126227 | Not-determined |
| 18-104_k141_7530 | Not-determined |
| 18-104_k141_84376 | Not-determined |
| 18-104_k141_146916 | Not-determined |
| 18-104_k141_105234 | Not-determined |
| 18-104_k141_42497 | Not-determined |
| 18-104_k141_84378 | Not-determined |
| 18-104_k141_153833 | Not-determined |
| 18-104_k141_105236 | Not-determined |
| 18-104_k141_21568 | Not-determined |
| 18-104_k141_153838 | Not-determined |
| 18-104_k141_112174 | Not-determined |
| 18-104_k141_793 | Not-determined |
| 18-104_k141_63340 | Not-determined |
| 18-104_k141_112177 | Not-determined |
| 18-104_k141_795 | Not-determined |
| 18-104_k141_91415 | Not-determined |
| 18-104_k141_112181 | Not-determined |
| 18-104_k141_140071 | Not-determined |
| 18-104_k141_21575 | Not-determined |
| 18-104_k141_70487 | Not-determined |
| 18-104_k141_153853 | Not-determined |
| 18-104_k141_112188 | Not-determined |
| 18-104_k141_42509 | Not-determined |
| 18-104_k141_84385 | Not-determined |
| 18-104_k141_126244 | Not-determined |
| 18-104_k141_21583 | Not-determined |
| 18-104_k141_84390 | Not-determined |
| 18-104_k141_98249 | Not-determined |
| 18-104_k141_84391 | Not-determined |
| 18-104_k141_84392 | Low-quality |
| 18-104_k141_70502 | Low-quality |
| 18-104_k141_132928 | Not-determined |
| 18-104_k141_7539 | Low-quality |
| 18-104_k141_70505 | Not-determined |
| 18-104_k141_118977 | Not-determined |
| 18-104_k141_140077 | Not-determined |
| 18-104_k141_42519 | Not-determined |
| 18-104_k141_118980 | Not-determined |
| 18-104_k141_153866 | Not-determined |
| 18-104_k141_105272 | Not-determined |
| 18-104_k141_98259 | Low-quality |
| 18-104_k141_153868 | Not-determined |
| 18-104_k141_126254 | Not-determined |
| 18-104_k141_84412 | Not-determined |
| 18-104_k141_91436 | Low-quality |
| 18-104_k141_126257 | Not-determined |
| 18-104_k141_49314 | Not-determined |
| 18-104_k141_821 | Not-determined |
| 18-104_k141_98263 | Not-determined |
| 18-104_k141_84419 | Low-quality |
| 18-104_k141_140087 | Low-quality |
| 18-104_k141_7556 | Low-quality |
| 18-104_k141_98264 | Not-determined |
| 18-104_k141_153876 | Not-determined |
| 18-104_k141_84422 | Not-determined |
| 18-104_k141_105281 | Not-determined |
| 18-104_k141_126263 | Low-quality |
| 18-104_k141_42531 | Not-determined |
| 18-104_k141_126265 | Not-determined |
| 18-104_k141_84424 | Low-quality |
| 18-104_k141_828 | Not-determined |
| 18-104_k141_84428 | Low-quality |
| 18-104_k141_831 | Not-determined |
| 18-104_k141_28573 | Not-determined |
| 18-104_k141_28575 | Low-quality |
| 18-104_k141_91444 | Not-determined |
| 18-104_k141_28576 | Not-determined |
| 18-104_k141_70518 | Not-determined |
| 18-104_k141_84435 | Not-determined |
| 18-104_k141_146941 | Low-quality |
| 18-104_k141_21601 | Not-determined |
| 18-104_k141_118996 | Low-quality |
| 18-104_k141_140094 | Not-determined |
| 18-104_k141_126277 | Not-determined |
| 18-104_k141_105295 | Not-determined |
| 18-104_k141_21607 | Not-determined |
| 18-104_k141_140099 | Not-determined |
| 18-104_k141_35339 | Not-determined |
| 18-104_k141_146944 | Not-determined |
| 18-104_k141_105299 | Not-determined |
| 18-104_k141_35340 | Not-determined |
| 18-104_k141_63380 | Not-determined |
| 18-104_k141_91451 | Not-determined |
| 18-104_k141_35345 | Not-determined |
| 18-104_k141_105307 | Not-determined |
| 18-104_k141_42550 | Low-quality |
| 18-104_k141_42551 | Not-determined |
| 18-104_k141_42553 | Not-determined |
| 18-104_k141_140107 | Low-quality |
| 18-104_k141_119012 | Not-determined |
| 18-104_k141_858 | Not-determined |
| 18-104_k141_7597 | Not-determined |
| 18-104_k141_146961 | Not-determined |
| 18-104_k141_140111 | Not-determined |
| 18-104_k141_63394 | Low-quality |
| 18-104_k141_35358 | Not-determined |
| 18-104_k141_56379 | Not-determined |
| 18-104_k141_84465 | Not-determined |
| 18-104_k141_146969 | Not-determined |
| 18-104_k141_42576 | Not-determined |
| 18-104_k141_42578 | Not-determined |
| 18-104_k141_63398 | Not-determined |
| 18-104_k141_28600 | Not-determined |
| 18-104_k141_126296 | Not-determined |
| 18-104_k141_35361 | Not-determined |
| 18-104_k141_84474 | Not-determined |
| 18-104_k141_153909 | Not-determined |
| 18-104_k141_874 | Not-determined |
| 18-104_k141_84482 | Not-determined |
| 18-104_k141_876 | Not-determined |
| 18-104_k141_132949 | Not-determined |
| 18-104_k141_132950 | Not-determined |
| 18-104_k141_84486 | Not-determined |
| 18-104_k141_140131 | Low-quality |
| 18-104_k141_140132 | Low-quality |
| 18-104_k141_49353 | Low-quality |
| 18-104_k141_42597 | Not-determined |
| 18-104_k141_7612 | Not-determined |
| 18-104_k141_42600 | Low-quality |
| 18-104_k141_105346 | Not-determined |
| 18-104_k141_153915 | Not-determined |
| 18-104_k141_42602 | Not-determined |
| 18-104_k141_91461 | Not-determined |
| 18-104_k141_140135 | Not-determined |
| 18-104_k141_70530 | Low-quality |
| 18-104_k141_105352 | Not-determined |
| 18-104_k141_21615 | Not-determined |
| 18-104_k141_56403 | Not-determined |
| 18-104_k141_84506 | Not-determined |
| 18-104_k141_49365 | Low-quality |
| 18-104_k141_153926 | Not-determined |
| 18-104_k141_56405 | Low-quality |
| 18-104_k141_21620 | Not-determined |
| 18-104_k141_28612 | Not-determined |
| 18-104_k141_63412 | Not-determined |
| 18-104_k141_105361 | Not-determined |
| 18-104_k141_132976 | Not-determined |
| 18-104_k141_153934 | Not-determined |
| 18-104_k141_140144 | Not-determined |
| 18-104_k141_140145 | Not-determined |
| 18-104_k141_140148 | Not-determined |
| 18-104_k141_49373 | Not-determined |
| 18-104_k141_28623 | Low-quality |
| 18-104_k141_132983 | Low-quality |
| 18-104_k141_49377 | Not-determined |
| 18-104_k141_153944 | Low-quality |
| 18-104_k141_153945 | Low-quality |
| 18-104_k141_112258 | Not-determined |
| 18-104_k141_42606 | Not-determined |
| 18-104_k141_105387 | Not-determined |
| 18-104_k141_70555 | Not-determined |
| 18-104_k141_49380 | Not-determined |
| 18-104_k141_56415 | Not-determined |
| 18-104_k141_126330 | Not-determined |
| 18-104_k141_126332 | Not-determined |
| 18-104_k141_153952 | Not-determined |
| 18-104_k141_35379 | Not-determined |
| 18-104_k141_146994 | Not-determined |
| 18-104_k141_49385 | Low-quality |
| 18-104_k141_112261 | Low-quality |
| 18-104_k141_70563 | Not-determined |
| 18-104_k141_21633 | Not-determined |
| 18-104_k141_98325 | Not-determined |
| 18-104_k141_77420 | Not-determined |
| 18-104_k141_105407 | Not-determined |
| 18-104_k141_42614 | Low-quality |
| 18-104_k141_42617 | Low-quality |
| 18-104_k141_105410 | Not-determined |
| 18-104_k141_63449 | Not-determined |
| 18-104_k141_91478 | Not-determined |
| 18-104_k141_49389 | Not-determined |
| 18-104_k141_14536 | Low-quality |
| 18-104_k141_112271 | Not-determined |
| 18-104_k141_98336 | Not-determined |
| 18-104_k141_63458 | Not-determined |
| 18-104_k141_28630 | Not-determined |
| 18-104_k141_63459 | Not-determined |
| 18-104_k141_153965 | Not-determined |
| 18-104_k141_84523 | Not-determined |
| 18-104_k141_42635 | Not-determined |
| 18-104_k141_91492 | Low-quality |
| 18-104_k141_119047 | Not-determined |
| 18-104_k141_77432 | Not-determined |
| 18-104_k141_63468 | Not-determined |
| 18-104_k141_14551 | Not-determined |
| 18-104_k141_14552 | Low-quality |
| 18-104_k141_14554 | Not-determined |
| 18-104_k141_28636 | Not-determined |
| 18-104_k141_56441 | Not-determined |
| 18-104_k141_42643 | Not-determined |
| 18-104_k141_77440 | Not-determined |
| 18-104_k141_14558 | Not-determined |
| 18-104_k141_119051 | Not-determined |
| 18-104_k141_91506 | Not-determined |
| 18-104_k141_7625 | Not-determined |
| 18-104_k141_112293 | Not-determined |
| 18-104_k141_14561 | Not-determined |
| 18-104_k141_77444 | Not-determined |
| 18-104_k141_21646 | Not-determined |
| 18-104_k141_112298 | Not-determined |
| 18-104_k141_112299 | Not-determined |
| 18-104_k141_56443 | Not-determined |
| 18-104_k141_91513 | Not-determined |
| 18-104_k141_21650 | Not-determined |
| 18-104_k141_56444 | Not-determined |
| 18-104_k141_91515 | Not-determined |
| 18-104_k141_56447 | Not-determined |
| 18-104_k141_891 | Not-determined |
| 18-104_k141_91519 | Not-determined |
| 18-104_k141_119061 | Not-determined |
| 18-104_k141_140169 | Not-determined |
| 18-104_k141_147027 | Low-quality |
| 18-104_k141_91525 | Not-determined |
| 18-104_k141_112308 | Not-determined |
| 18-104_k141_91528 | Not-determined |
| 18-104_k141_140173 | Not-determined |
| 18-104_k141_56463 | Not-determined |
| 18-104_k141_70616 | Not-determined |
| 18-104_k141_133003 | Not-determined |
| 18-104_k141_147033 | Not-determined |
| 18-104_k141_56465 | Not-determined |
| 18-104_k141_70621 | Not-determined |
| 18-104_k141_147037 | Not-determined |
| 18-104_k141_153988 | Not-determined |
| 18-104_k141_28659 | Low-quality |
| 18-104_k141_56469 | Not-determined |
| 18-104_k141_21671 | Not-determined |
| 18-104_k141_133015 | Not-determined |
| 18-104_k141_153994 | Not-determined |
| 18-104_k141_70627 | Not-determined |
| 18-104_k141_126369 | Not-determined |
| 18-104_k141_133019 | Not-determined |
| 18-104_k141_56471 | Not-determined |
| 18-104_k141_147045 | Not-determined |
| 18-104_k141_112318 | Not-determined |
| 18-104_k141_14587 | Not-determined |
| 18-104_k141_112319 | Not-determined |
| 18-104_k141_153996 | Not-determined |
| 18-104_k141_70635 | Not-determined |
| 18-104_k141_153997 | Not-determined |
| 18-104_k141_56478 | Low-quality |
| 18-104_k141_70639 | Not-determined |
| 18-104_k141_126372 | Not-determined |
| 18-104_k141_28677 | Not-determined |
| 18-104_k141_105443 | Not-determined |
| 18-104_k141_126375 | Not-determined |
| 18-104_k141_898 | Not-determined |
| 18-104_k141_70643 | Low-quality |
| 18-104_k141_105447 | Not-determined |
| 18-104_k141_56489 | Not-determined |
| 18-104_k141_140185 | Low-quality |
| 18-104_k141_105449 | Not-determined |
| 18-104_k141_906 | Not-determined |
| 18-104_k141_28679 | Low-quality |
| 18-104_k141_154013 | Not-determined |
| 18-104_k141_140191 | Not-determined |
| 18-104_k141_910 | Not-determined |
| 18-104_k141_63485 | Not-determined |
| 18-104_k141_140192 | Not-determined |
| 18-104_k141_56502 | Not-determined |
| 18-104_k141_56503 | Not-determined |
| 18-104_k141_14601 | Not-determined |
| 18-104_k141_91561 | Not-determined |
| 18-104_k141_133026 | Not-determined |
| 18-104_k141_77485 | Not-determined |
| 18-104_k141_133035 | Not-determined |
| 18-104_k141_7629 | Low-quality |
| 18-104_k141_154024 | Not-determined |
| 18-104_k141_14607 | Low-quality |
| 18-104_k141_119071 | Not-determined |
| 18-104_k141_126389 | Not-determined |
| 18-104_k141_147060 | Not-determined |
| 18-104_k141_147061 | Not-determined |
| 18-104_k141_35451 | Not-determined |
| 18-104_k141_119077 | Not-determined |
| 18-104_k141_119079 | Not-determined |
| 18-104_k141_21684 | Not-determined |
| 18-104_k141_21687 | Low-quality |
| 18-104_k141_77495 | Not-determined |
| 18-104_k141_63501 | Not-determined |
| 18-104_k141_147066 | Not-determined |
| 18-104_k141_56531 | Not-determined |
| 18-104_k141_112347 | Not-determined |
| 18-104_k141_147068 | Not-determined |
| 18-104_k141_56532 | Low-quality |
| 18-104_k141_77501 | Not-determined |
| 18-104_k141_28705 | Not-determined |
| 18-104_k141_63513 | Not-determined |
| 18-104_k141_126414 | Not-determined |
| 18-104_k141_91596 | Not-determined |
| 18-104_k141_126418 | Not-determined |
| 18-104_k141_14615 | Not-determined |
| 18-104_k141_63518 | Low-quality |
| 18-104_k141_28716 | Not-determined |
| 18-104_k141_112351 | Low-quality |
| 18-104_k141_140202 | Not-determined |
| 18-104_k141_133055 | Not-determined |
| 18-104_k141_154047 | Not-determined |
| 18-104_k141_105478 | Not-determined |
| 18-104_k141_147076 | Not-determined |
| 18-104_k141_154049 | Not-determined |
| 18-104_k141_112358 | Not-determined |
| 18-104_k141_63526 | Not-determined |
| 18-104_k141_91602 | Not-determined |
| 18-104_k141_112361 | Not-determined |
| 18-104_k141_28730 | Not-determined |
| 18-104_k141_77529 | Not-determined |
| 18-104_k141_147083 | Not-determined |
| 18-104_k141_160909 | Low-quality |
| 18-104_k141_91606 | Not-determined |
| 18-104_k141_28734 | Low-quality |
| 18-104_k141_56539 | Not-determined |
| 18-104_k141_91612 | Low-quality |
| 18-104_k141_28741 | Not-determined |
| 18-104_k141_42666 | Not-determined |
| 18-104_k141_133068 | Not-determined |
| 18-104_k141_119104 | Not-determined |
| 18-104_k141_35472 | Low-quality |
| 18-104_k141_56544 | Low-quality |
| 18-104_k141_133070 | Not-determined |
| 18-104_k141_942 | Low-quality |
| 18-104_k141_42673 | Not-determined |
| 18-104_k141_140215 | Not-determined |
| 18-104_k141_56547 | Not-determined |
| 18-104_k141_945 | Not-determined |
| 18-104_k141_133075 | Not-determined |
| 18-104_k141_112374 | Not-determined |
| 18-104_k141_147097 | Not-determined |
| 18-104_k141_119111 | Not-determined |
| 18-104_k141_105497 | Not-determined |
| 18-104_k141_953 | Not-determined |
| 18-104_k141_954 | Not-determined |
| 18-104_k141_133078 | Not-determined |
| 18-104_k141_77540 | Not-determined |
| 18-104_k141_63534 | Not-determined |
| 18-104_k141_140218 | Not-determined |
| 18-104_k141_958 | Not-determined |
| 18-104_k141_105500 | Not-determined |
| 18-104_k141_63536 | Not-determined |
| 18-104_k141_42676 | Not-determined |
| 18-104_k141_35484 | Not-determined |
| 18-104_k141_77542 | Not-determined |
| 18-104_k141_126461 | Low-quality |
| 18-104_k141_140223 | Not-determined |
| 18-104_k141_119120 | Not-determined |
| 18-104_k141_84549 | Not-determined |
| 18-104_k141_126465 | Not-determined |
| 18-104_k141_112378 | Not-determined |
| 18-104_k141_28762 | Not-determined |
| 18-104_k141_35493 | Not-determined |
| 18-104_k141_42681 | Low-quality |
| 18-104_k141_63542 | Low-quality |
| 18-104_k141_49416 | Not-determined |
| 18-104_k141_28768 | Not-determined |
| 18-104_k141_49417 | Not-determined |
| 18-104_k141_49419 | Not-determined |
| 18-104_k141_63546 | Not-determined |
| 18-104_k141_7682 | Not-determined |
| 18-104_k141_70655 | Not-determined |
| 18-104_k141_63549 | Not-determined |
| 18-104_k141_147113 | Not-determined |
| 18-104_k141_126477 | Not-determined |
| 18-104_k141_49425 | Not-determined |
| 18-104_k141_84562 | Low-quality |
| 18-104_k141_112386 | Not-determined |
| 18-104_k141_140234 | Not-determined |
| 18-104_k141_126480 | Not-determined |
| 18-104_k141_77553 | Not-determined |
| 18-104_k141_160936 | Not-determined |
| 18-104_k141_105509 | Not-determined |
| 18-104_k141_35513 | Low-quality |
| 18-104_k141_98352 | Not-determined |
| 18-104_k141_112391 | Low-quality |
| 18-104_k141_126483 | Not-determined |
| 18-104_k141_84570 | Not-determined |
| 18-104_k141_140240 | Not-determined |
| 18-104_k141_35518 | Not-determined |
| 18-104_k141_84573 | Not-determined |
| 18-104_k141_35519 | Not-determined |
| 18-104_k141_98358 | Not-determined |
| 18-104_k141_140245 | Not-determined |
| 18-104_k141_126493 | Not-determined |
| 18-104_k141_126494 | Not-determined |
| 18-104_k141_126495 | Not-determined |
| 18-104_k141_14641 | Not-determined |
| 18-104_k141_133095 | Not-determined |
| 18-104_k141_133097 | Not-determined |
| 18-104_k141_154069 | Not-determined |
| 18-104_k141_77561 | Not-determined |
| 18-104_k141_49436 | Not-determined |
| 18-104_k141_63561 | Low-quality |
| 18-104_k141_49439 | Not-determined |
| 18-104_k141_140256 | Not-determined |
| 18-104_k141_154071 | Not-determined |
| 18-104_k141_91645 | Not-determined |
| 18-104_k141_14651 | Not-determined |
| 18-104_k141_84583 | Not-determined |
| 18-104_k141_154074 | Not-determined |
| 18-104_k141_147133 | Not-determined |
| 18-104_k141_147136 | Low-quality |
| 18-104_k141_147139 | Not-determined |
| 18-104_k141_112413 | Not-determined |
| 18-104_k141_126501 | Not-determined |
| 18-104_k141_160949 | Not-determined |
| 18-104_k141_98382 | Not-determined |
| 18-104_k141_160950 | Low-quality |
| 18-104_k141_21694 | Not-determined |
| 18-104_k141_35538 | Not-determined |
| 18-104_k141_14664 | Not-determined |
| 18-104_k141_63576 | Not-determined |
| 18-104_k141_70687 | Not-determined |
| 18-104_k141_119150 | Low-quality |
| 18-104_k141_105541 | Not-determined |
| 18-104_k141_77573 | Not-determined |
| 18-104_k141_91652 | Not-determined |
| 18-104_k141_28802 | Low-quality |
| 18-104_k141_21696 | Not-determined |
| 18-104_k141_91654 | Not-determined |
| 18-104_k141_63581 | Not-determined |
| 18-104_k141_84597 | Not-determined |
| 18-104_k141_77578 | Not-determined |
| 18-104_k141_140275 | Not-determined |
| 18-104_k141_56568 | Not-determined |
| 18-104_k141_112423 | Not-determined |
| 18-104_k141_119162 | Low-quality |
| 18-104_k141_140277 | Not-determined |
| 18-104_k141_21701 | Not-determined |
| 18-104_k141_49448 | Not-determined |
| 18-104_k141_35547 | Not-determined |
| 18-104_k141_126528 | Not-determined |
| 18-104_k141_28807 | Low-quality |
| 18-104_k141_63596 | Not-determined |
| 18-104_k141_91670 | Not-determined |
| 18-104_k141_77592 | Not-determined |
| 18-104_k141_84603 | Not-determined |
| 18-104_k141_77595 | Low-quality |
| 18-104_k141_147162 | Not-determined |
| 18-104_k141_28814 | Low-quality |
| 18-104_k141_77603 | Not-determined |
| 18-104_k141_21717 | Not-determined |
| 18-104_k141_14684 | Not-determined |
| 18-104_k141_63616 | Not-determined |
| 18-104_k141_91677 | Not-determined |
| 18-104_k141_133125 | Not-determined |
| 18-104_k141_119184 | Not-determined |
| 18-104_k141_126554 | Not-determined |
| 18-104_k141_28819 | Not-determined |
| 18-104_k141_98404 | Not-determined |
| 18-104_k141_126561 | Not-determined |
| 18-104_k141_133133 | Low-quality |
| 18-104_k141_21723 | Not-determined |
| 18-104_k141_7703 | Not-determined |
| 18-104_k141_21726 | Not-determined |
| 18-104_k141_35552 | Low-quality |
| 18-104_k141_91690 | Not-determined |
| 18-104_k141_119194 | Not-determined |
| 18-104_k141_28830 | Not-determined |
| 18-104_k141_154099 | Not-determined |
| 18-104_k141_105568 | Not-determined |
| 18-104_k141_84624 | Not-determined |
| 18-104_k141_154103 | Not-determined |
| 18-104_k141_28833 | Not-determined |
| 18-104_k141_21734 | Not-determined |
| 18-104_k141_126568 | Not-determined |
| 18-104_k141_77614 | Not-determined |
| 18-104_k141_35560 | Low-quality |
| 18-104_k141_119207 | Not-determined |
| 18-104_k141_84631 | Not-determined |
| 18-104_k141_154110 | Not-determined |
| 18-104_k141_35562 | Low-quality |
| 18-104_k141_14702 | Not-determined |
| 18-104_k141_77615 | Not-determined |
| 18-104_k141_154112 | Not-determined |
| 18-104_k141_28834 | Not-determined |
| 18-104_k141_21741 | Not-determined |
| 18-104_k141_140292 | Not-determined |
| 18-104_k141_112451 | Not-determined |
| 18-104_k141_126576 | Not-determined |
| 18-104_k141_14710 | Not-determined |
| 18-104_k141_21753 | Not-determined |
| 18-104_k141_98431 | Not-determined |
| 18-104_k141_154135 | Not-determined |
| 18-104_k141_7708 | Not-determined |
| 18-104_k141_21757 | Low-quality |
| 18-104_k141_160979 | Not-determined |
| 18-104_k141_133157 | Not-determined |
| 18-104_k141_21760 | Low-quality |
| 18-104_k141_91710 | Not-determined |
| 18-104_k141_119221 | Not-determined |
| 18-104_k141_7710 | Not-determined |
| 18-104_k141_105596 | Not-determined |
| 18-104_k141_28853 | Not-determined |
| 18-104_k141_126589 | Not-determined |
| 18-104_k141_1020 | Low-quality |
| 18-104_k141_63638 | Not-determined |
| 18-104_k141_28858 | Not-determined |
| 18-104_k141_105602 | Not-determined |
| 18-104_k141_140303 | Not-determined |
| 18-104_k141_112474 | Not-determined |
| 18-104_k141_28866 | Not-determined |
| 18-104_k141_133165 | Low-quality |
| 18-104_k141_147195 | Not-determined |
| 18-104_k141_140312 | Not-determined |
| 18-104_k141_105612 | Low-quality |
| 18-104_k141_28876 | Not-determined |
| 18-104_k141_112485 | Not-determined |
| 18-104_k141_28878 | Not-determined |
| 18-104_k141_28879 | Not-determined |
| 18-104_k141_140319 | Not-determined |
| 18-104_k141_28882 | Not-determined |
| 18-104_k141_154161 | Not-determined |
| 18-104_k141_63643 | Not-determined |
| 18-104_k141_147206 | Low-quality |
| 18-104_k141_1039 | Not-determined |
| 18-104_k141_147209 | Not-determined |
| 18-104_k141_154171 | Not-determined |
| 18-104_k141_21779 | Not-determined |
| 18-104_k141_140334 | Not-determined |
| 18-104_k141_7728 | Low-quality |
| 18-104_k141_7729 | Not-determined |
| 18-104_k141_7730 | Low-quality |
| 18-104_k141_21782 | Not-determined |
| 18-104_k141_77647 | Low-quality |
| 18-104_k141_21785 | Not-determined |
| 18-104_k141_21786 | Not-determined |
| 18-104_k141_21787 | Not-determined |
| 18-104_k141_7737 | Not-determined |
| 18-104_k141_147220 | Not-determined |
| 18-104_k141_147224 | Not-determined |

| | |
|---|---|
| 18-104_k141_7740 | Not-determined |
| 18-104_k141_147226 | Not-determined |
| 18-104_k141_161012 | Not-determined |
| 18-104_k141_77651 | Not-determined |
| 18-104_k141_7742 | Not-determined |
| 18-104_k141_161013 | Not-determined |
| 18-104_k141_63654 | Not-determined |
| 18-104_k141_21797 | Not-determined |
| 18-104_k141_154182 | Low-quality |
| 18-104_k141_126609 | Not-determined |
| 18-104_k141_63664 | Not-determined |
| 18-104_k141_140351 | Not-determined |
| 18-104_k141_70708 | Not-determined |
| 18-104_k141_154192 | Not-determined |
| 18-104_k141_126615 | Not-determined |
| 18-104_k141_147228 | Not-determined |
| 18-104_k141_35613 | Not-determined |
| 18-104_k141_28899 | Low-quality |
| 18-104_k141_140360 | Not-determined |
| 18-104_k141_42745 | Not-determined |
| 18-104_k141_63681 | Not-determined |
| 18-104_k141_77670 | Low-quality |
| 18-104_k141_161020 | Not-determined |
| 18-104_k141_133203 | Not-determined |
| 18-104_k141_35617 | Not-determined |
| 18-104_k141_98460 | Not-determined |
| 18-104_k141_112509 | Not-determined |
| 18-104_k141_112511 | Not-determined |
| 18-104_k141_126627 | Not-determined |
| 18-104_k141_63688 | Not-determined |
| 18-104_k141_56589 | Low-quality |
| 18-104_k141_56590 | Low-quality |
| 18-104_k141_14725 | Not-determined |
| 18-104_k141_119237 | Not-determined |
| 18-104_k141_119238 | Not-determined |
| 18-104_k141_56593 | Low-quality |
| 18-104_k141_7763 | Not-determined |
| 18-104_k141_98469 | Not-determined |
| 18-104_k141_119242 | Low-quality |
| 18-104_k141_63696 | Not-determined |
| 18-104_k141_1057 | Not-determined |
| 18-104_k141_161040 | Not-determined |
| 18-104_k141_126637 | Not-determined |
| 18-104_k141_84669 | Not-determined |
| 18-104_k141_7769 | Not-determined |
| 18-104_k141_126638 | Not-determined |
| 18-104_k141_14730 | Not-determined |
| 18-104_k141_147252 | Not-determined |
| 18-104_k141_63704 | Not-determined |
| 18-104_k141_28921 | Not-determined |
| 18-104_k141_112518 | Not-determined |
| 18-104_k141_63706 | Not-determined |
| 18-104_k141_77691 | Not-determined |
| 18-104_k141_112520 | Not-determined |
| 18-104_k141_56597 | Not-determined |
| 18-104_k141_161054 | Not-determined |
| 18-104_k141_119258 | Not-determined |
| 18-104_k141_84674 | Not-determined |
| 18-104_k141_63712 | Not-determined |
| 18-104_k141_77700 | Not-determined |
| 18-104_k141_28930 | Not-determined |
| 18-104_k141_77701 | Not-determined |
| 18-104_k141_56602 | Not-determined |
| 18-104_k141_63717 | Not-determined |
| 18-104_k141_77702 | Not-determined |
| 18-104_k141_84677 | Not-determined |
| 18-104_k141_77704 | Not-determined |
| 18-104_k141_119265 | Not-determined |
| 18-104_k141_77705 | Not-determined |
| 18-104_k141_56605 | Not-determined |
| 18-104_k141_112527 | Not-determined |
| 18-104_k141_84682 | Not-determined |
| 18-104_k141_77710 | Not-determined |
| 18-104_k141_56614 | Not-determined |
| 18-104_k141_42781 | Not-determined |
| 18-104_k141_84687 | Not-determined |
| 18-104_k141_28939 | Not-determined |
| 18-104_k141_56616 | Not-determined |
| 18-104_k141_147268 | Low-quality |
| 18-104_k141_161068 | Not-determined |
| 18-104_k141_119278 | Not-determined |
| 18-104_k141_112538 | Not-determined |
| 18-104_k141_133241 | Not-determined |
| 18-104_k141_161071 | Not-determined |
| 18-104_k141_154240 | Not-determined |
| 18-104_k141_77717 | Not-determined |
| 18-104_k141_119288 | Not-determined |
| 18-104_k141_56627 | Not-determined |
| 18-104_k141_98503 | Not-determined |
| 18-104_k141_140390 | Low-quality |
| 18-104_k141_63728 | Not-determined |
| 18-104_k141_147275 | Low-quality |
| 18-104_k141_161083 | Not-determined |
| 18-104_k141_133249 | Not-determined |
| 18-104_k141_161084 | Not-determined |
| 18-104_k141_91751 | Not-determined |
| 18-104_k141_161085 | Not-determined |
| 18-104_k141_133252 | Not-determined |
| 18-104_k141_161086 | Not-determined |
| 18-104_k141_133253 | Not-determined |
| 18-104_k141_28963 | Not-determined |
| 18-104_k141_140402 | Not-determined |
| 18-104_k141_98509 | Not-determined |
| 18-104_k141_42797 | Not-determined |
| 18-104_k141_14752 | Not-determined |
| 18-104_k141_7794 | Low-quality |
| 18-104_k141_147283 | Not-determined |
| 18-104_k141_84701 | Not-determined |
| 18-104_k141_119300 | Not-determined |
| 18-104_k141_91763 | Not-determined |
| 18-104_k141_1072 | Not-determined |
| 18-104_k141_119303 | Not-determined |
| 18-104_k141_98513 | Not-determined |
| 18-104_k141_119304 | Low-quality |
| 18-104_k141_7800 | Not-determined |
| 18-104_k141_140409 | Not-determined |
| 18-104_k141_56646 | Not-determined |
| 18-104_k141_7801 | Not-determined |
| 18-104_k141_105676 | Not-determined |
| 18-104_k141_126655 | Not-determined |
| 18-104_k141_105677 | Not-determined |
| 18-104_k141_49466 | Not-determined |
| 18-104_k141_126659 | Not-determined |
| 18-104_k141_77723 | Not-determined |
| 18-104_k141_147290 | Not-determined |
| 18-104_k141_126661 | Not-determined |
| 18-104_k141_133266 | Not-determined |
| 18-104_k141_147292 | Not-determined |
| 18-104_k141_70743 | Not-determined |
| 18-104_k141_77729 | Not-determined |
| 18-104_k141_63744 | Not-determined |
| 18-104_k141_7815 | Not-determined |
| 18-104_k141_42816 | Low-quality |
| 18-104_k141_42818 | Not-determined |
| 18-104_k141_112547 | Low-quality |
| 18-104_k141_35656 | Not-determined |
| 18-104_k141_28984 | Low-quality |
| 18-104_k141_77733 | Not-determined |
| 18-104_k141_119328 | Not-determined |
| 18-104_k141_147299 | Not-determined |
| 18-104_k141_1083 | Not-determined |
| 18-104_k141_154264 | Not-determined |
| 18-104_k141_112549 | Not-determined |
| 18-104_k141_140428 | Not-determined |
| 18-104_k141_77735 | Not-determined |
| 18-104_k141_7818 | Not-determined |
| 18-104_k141_147303 | Not-determined |
| 18-104_k141_35660 | Low-quality |
| 18-104_k141_77741 | Not-determined |
| 18-104_k141_63749 | Not-determined |
| 18-104_k141_28990 | Not-determined |
| 18-104_k141_1090 | Not-determined |
| 18-104_k141_1092 | Not-determined |
| 18-104_k141_140442 | Not-determined |
| 18-104_k141_133294 | Not-determined |
| 18-104_k141_112563 | Not-determined |
| 18-104_k141_49489 | Not-determined |
| 18-104_k141_147312 | Not-determined |
| 18-104_k141_70756 | Not-determined |
| 18-104_k141_161108 | Low-quality |
| 18-104_k141_140446 | Not-determined |
| 18-104_k141_91775 | Not-determined |
| 18-104_k141_133301 | Low-quality |
| 18-104_k141_119346 | Not-determined |
| 18-104_k141_91779 | Not-determined |
| 18-104_k141_70759 | Not-determined |
| 18-104_k141_147316 | Not-determined |
| 18-104_k141_91780 | Low-quality |
| 18-104_k141_77760 | Not-determined |
| 18-104_k141_161116 | Not-determined |
| 18-104_k141_105713 | Not-determined |
| 18-104_k141_98528 | Not-determined |
| 18-104_k141_105715 | Not-determined |
| 18-104_k141_42840 | Not-determined |
| 18-104_k141_49497 | Not-determined |
| 18-104_k141_91787 | Not-determined |
| 18-104_k141_126697 | Not-determined |
| 18-104_k141_98534 | Not-determined |
| 18-104_k141_98536 | Not-determined |
| 18-104_k141_112581 | Not-determined |
| 18-104_k141_77769 | Not-determined |
| 18-104_k141_77771 | Not-determined |
| 18-104_k141_112583 | Not-determined |
| 18-104_k141_98539 | Not-determined |
| 18-104_k141_42847 | Not-determined |
| 18-104_k141_1118 | Not-determined |
| 18-104_k141_112590 | Not-determined |
| 18-104_k141_126704 | Not-determined |
| 18-104_k141_105724 | Not-determined |
| 18-104_k141_63760 | Not-determined |
| 18-104_k141_63761 | Not-determined |
| 18-104_k141_35684 | Not-determined |
| 18-104_k141_77781 | Not-determined |
| 18-104_k141_63764 | Not-determined |
| 18-104_k141_1122 | Not-determined |
| 18-104_k141_42857 | Not-determined |
| 18-104_k141_1125 | Not-determined |
| 18-104_k141_77784 | Low-quality |
| 18-104_k141_119356 | Not-determined |
| 18-104_k141_56690 | Low-quality |
| 18-104_k141_29010 | Not-determined |
| 18-104_k141_126706 | Not-determined |
| 18-104_k141_119358 | Not-determined |
| 18-104_k141_1127 | Not-determined |
| 18-104_k141_70766 | Not-determined |
| 18-104_k141_1129 | Not-determined |
| 18-104_k141_84751 | Not-determined |
| 18-104_k141_105738 | Low-quality |
| 18-104_k141_126707 | Low-quality |
| 18-104_k141_91800 | Low-quality |
| 18-104_k141_56696 | Not-determined |
| 18-104_k141_84755 | Not-determined |
| 18-104_k141_1131 | Not-determined |
| 18-104_k141_1134 | Low-quality |
| 18-104_k141_56698 | Not-determined |
| 18-104_k141_133316 | Not-determined |
| 18-104_k141_154293 | Not-determined |
| 18-104_k141_63777 | Not-determined |
| 18-104_k141_119378 | Not-determined |
| 18-104_k141_70771 | Not-determined |
| 18-104_k141_42867 | Low-quality |
| 18-104_k141_105756 | Not-determined |
| 18-104_k141_133325 | Not-determined |
| 18-104_k141_105757 | Not-determined |
| 18-104_k141_126718 | Not-determined |
| 18-104_k141_35701 | Not-determined |
| 18-104_k141_133328 | Not-determined |
| 18-104_k141_35706 | Not-determined |
| 18-104_k141_133334 | Not-determined |
| 18-104_k141_77790 | Low-quality |
| 18-104_k141_98556 | Not-determined |
| 18-104_k141_133341 | Not-determined |
| 18-104_k141_77795 | Not-determined |
| 18-104_k141_70780 | Not-determined |
| 18-104_k141_35714 | Not-determined |
| 18-104_k141_77798 | Not-determined |
| 18-104_k141_105764 | Not-determined |
| 18-104_k141_112622 | Not-determined |
| 18-104_k141_112626 | Not-determined |
| 18-104_k141_105768 | Low-quality |
| 18-104_k141_119416 | Not-determined |
| 18-104_k141_112636 | Not-determined |
| 18-104_k141_56736 | Not-determined |
| 18-104_k141_56737 | Not-determined |
| 18-104_k141_112641 | Low-quality |
| 18-104_k141_91820 | Not-determined |
| 18-104_k141_35728 | Not-determined |
| 18-104_k141_35729 | Not-determined |
| 18-104_k141_105782 | Low-quality |
| 18-104_k141_14754 | Not-determined |
| 18-104_k141_77819 | Not-determined |
| 18-104_k141_42890 | Low-quality |
| 18-104_k141_42891 | Not-determined |
| 18-104_k141_98576 | Low-quality |
| 18-104_k141_98578 | Not-determined |
| 18-104_k141_84773 | Not-determined |
| 18-104_k141_112651 | Not-determined |
| 18-104_k141_161145 | Not-determined |
| 18-104_k141_119421 | Not-determined |
| 18-104_k141_21838 | Low-quality |
| 18-104_k141_84781 | Not-determined |
| 18-104_k141_161148 | Not-determined |
| 18-104_k141_21841 | Low-quality |
| 18-104_k141_105795 | Not-determined |
| 18-104_k141_1174 | Not-determined |
| 18-104_k141_91842 | Not-determined |
| 18-104_k141_126732 | Not-determined |
| 18-104_k141_21844 | Not-determined |
| 18-104_k141_154312 | Not-determined |
| 18-104_k141_14763 | Not-determined |
| 18-104_k141_77827 | Not-determined |
| 18-104_k141_56753 | Low-quality |
| 18-104_k141_7849 | Low-quality |
| 18-104_k141_161161 | Not-determined |
| 18-104_k141_1177 | Not-determined |
| 18-104_k141_112671 | Not-determined |
| 18-104_k141_77834 | Not-determined |
| 18-104_k141_112672 | Not-determined |
| 18-104_k141_77838 | Not-determined |
| 18-104_k141_147341 | Low-quality |
| 18-104_k141_161171 | Not-determined |
| 18-104_k141_29043 | Not-determined |
| 18-104_k141_21857 | Not-determined |
| 18-104_k141_14780 | Low-quality |
| 18-104_k141_161173 | Not-determined |
| 18-104_k141_14782 | Not-determined |
| 18-104_k141_14784 | Not-determined |
| 18-104_k141_91856 | Not-determined |
| 18-104_k141_29048 | Not-determined |
| 18-104_k141_77850 | Not-determined |
| 18-104_k141_98598 | Not-determined |
| 18-104_k141_7862 | Not-determined |
| 18-104_k141_21866 | Not-determined |
| 18-104_k141_77852 | Not-determined |
| 18-104_k141_112688 | Low-quality |
| 18-104_k141_77853 | Not-determined |
| 18-104_k141_63811 | Not-determined |
| 18-104_k141_77854 | Not-determined |
| 18-104_k141_1191 | Low-quality |
| 18-104_k141_14797 | Not-determined |
| 18-104_k141_154329 | Not-determined |
| 18-104_k141_70811 | Not-determined |
| 18-104_k141_140498 | Not-determined |
| 18-104_k141_126753 | Not-determined |
| 18-104_k141_63818 | Not-determined |
| 18-104_k141_1195 | Not-determined |
| 18-104_k141_126756 | Not-determined |
| 18-104_k141_161189 | Not-determined |
| 18-104_k141_154337 | Not-determined |
| 18-104_k141_126764 | Not-determined |
| 18-104_k141_63825 | Not-determined |
| 18-104_k141_29054 | Not-determined |
| 18-104_k141_1206 | Not-determined |
| 18-104_k141_147364 | Not-determined |
| 18-104_k141_63827 | Not-determined |
| 18-104_k141_140513 | Not-determined |
| 18-104_k141_42933 | Not-determined |
| 18-104_k141_35748 | Low-quality |
| 18-104_k141_91881 | Not-determined |
| 18-104_k141_91882 | Not-determined |
| 18-104_k141_84794 | Not-determined |
| 18-104_k141_91887 | Not-determined |
| 18-104_k141_140518 | Not-determined |
| 18-104_k141_7872 | Not-determined |
| 18-104_k141_126768 | Not-determined |
| 18-104_k141_77890 | Not-determined |
| 18-104_k141_35761 | Not-determined |
| 18-104_k141_147375 | Not-determined |
| 18-104_k141_98610 | Not-determined |
| 18-104_k141_77894 | Not-determined |
| 18-104_k141_42946 | Not-determined |
| 18-104_k141_29071 | Not-determined |
| 18-104_k141_133387 | Not-determined |
| 18-104_k141_133390 | Not-determined |
| 18-104_k141_126776 | Not-determined |
| 18-104_k141_7883 | Not-determined |
| 18-104_k141_126777 | Not-determined |
| 18-104_k141_35767 | Not-determined |
| 18-104_k141_35768 | Not-determined |
| 18-104_k141_35769 | Not-determined |
| 18-104_k141_7888 | Not-determined |
| 18-104_k141_126782 | Not-determined |
| 18-104_k141_140528 | Not-determined |
| 18-104_k141_70829 | Not-determined |
| 18-104_k141_133399 | Low-quality |
| 18-104_k141_42966 | Not-determined |
| 18-104_k141_49516 | Not-determined |
| 18-104_k141_29078 | Not-determined |
| 18-104_k141_56776 | Not-determined |
| 18-104_k141_140532 | Not-determined |
| 18-104_k141_119476 | Not-determined |
| 18-104_k141_133410 | Not-determined |
| 18-104_k141_14816 | Not-determined |
| 18-104_k141_140541 | Not-determined |
| 18-104_k141_133415 | Not-determined |
| 18-104_k141_29091 | Not-determined |
| 18-104_k141_161222 | Not-determined |
| 18-104_k141_42988 | Low-quality |
| 18-104_k141_84814 | Not-determined |
| 18-104_k141_112726 | Low-quality |
| 18-104_k141_42992 | Not-determined |
| 18-104_k141_29098 | Not-determined |
| 18-104_k141_126798 | Not-determined |
| 18-104_k141_21911 | Not-determined |
| 18-104_k141_119492 | Not-determined |
| 18-104_k141_42999 | Not-determined |
| 18-104_k141_91930 | Not-determined |
| 18-104_k141_21913 | Not-determined |
| 18-104_k141_161225 | Not-determined |
| 18-104_k141_161226 | Not-determined |
| 18-104_k141_84826 | Not-determined |
| 18-104_k141_91934 | Not-determined |
| 18-104_k141_43003 | Not-determined |
| 18-104_k141_21918 | Not-determined |
| 18-104_k141_161229 | Low-quality |
| 18-104_k141_161230 | Not-determined |
| 18-104_k141_147398 | Not-determined |
| 18-104_k141_7943 | Not-determined |
| 18-104_k141_56795 | Not-determined |
| 18-104_k141_7944 | Not-determined |
| 18-104_k141_112732 | Not-determined |
| 18-104_k141_112735 | Not-determined |
| 18-104_k141_147403 | Not-determined |
| 18-104_k141_161234 | Not-determined |
| 18-104_k141_147405 | Not-determined |
| 18-104_k141_133431 | Not-determined |
| 18-104_k141_91942 | Not-determined |
| 18-104_k141_63875 | Not-determined |
| 18-104_k141_91946 | Not-determined |
| 18-104_k141_21925 | Low-quality |
| 18-104_k141_77936 | Not-determined |
| 18-104_k141_98638 | Not-determined |
| 18-104_k141_29113 | Not-determined |
| 18-104_k141_147411 | Not-determined |
| 18-104_k141_147415 | Not-determined |
| 18-104_k141_161249 | Low-quality |
| 18-104_k141_43017 | Not-determined |
| 18-104_k141_140554 | Not-determined |
| 18-104_k141_140555 | Not-determined |
| 18-104_k141_7966 | Low-quality |
| 18-104_k141_133441 | Not-determined |
| 18-104_k141_35805 | Not-determined |
| 18-104_k141_43022 | Not-determined |
| 18-104_k141_56816 | Not-determined |
| 18-104_k141_35807 | Not-determined |
| 18-104_k141_98639 | Low-quality |
| 18-104_k141_140561 | Low-quality |
| 18-104_k141_119508 | Not-determined |
| 18-104_k141_56819 | Not-determined |
| 18-104_k141_147420 | Not-determined |
| 18-104_k141_77957 | Not-determined |
| 18-104_k141_56820 | Not-determined |
| 18-104_k141_147421 | Low-quality |
| 18-104_k141_1225 | Not-determined |
| 18-104_k141_49528 | Not-determined |
| 18-104_k141_29132 | Low-quality |
| 18-104_k141_147424 | Not-determined |
| 18-104_k141_133443 | Low-quality |
| 18-104_k141_63887 | Not-determined |
| 18-104_k141_35821 | Not-determined |
| 18-104_k141_140570 | Not-determined |
| 18-104_k141_133447 | Not-determined |
| 18-104_k141_7979 | Not-determined |
| 18-104_k141_133450 | Not-determined |
| 18-104_k141_7981 | Not-determined |
| 18-104_k141_77969 | Not-determined |
| 18-104_k141_77975 | Low-quality |
| 18-104_k141_112753 | Low-quality |
| 18-104_k141_56831 | Not-determined |
| 18-104_k141_98647 | Not-determined |
| 18-104_k141_77979 | Not-determined |
| 18-104_k141_77982 | Not-determined |
| 18-104_k141_7992 | Not-determined |
| 18-104_k141_14842 | Not-determined |
| 18-104_k141_63897 | Not-determined |
| 18-104_k141_63899 | Not-determined |
| 18-104_k141_29142 | Not-determined |
| 18-104_k141_63901 | Not-determined |
| 18-104_k141_21959 | Not-determined |
| 18-104_k141_29144 | Not-determined |
| 18-104_k141_14845 | Not-determined |
| 18-104_k141_91960 | Not-determined |
| 18-104_k141_133458 | Not-determined |
| 18-104_k141_154361 | Not-determined |
| 18-104_k141_119521 | Not-determined |
| 18-104_k141_154363 | Not-determined |
| 18-104_k141_140595 | Not-determined |
| 18-104_k141_91965 | Not-determined |
| 18-104_k141_56848 | Not-determined |
| 18-104_k141_63903 | Low-quality |
| 18-104_k141_21962 | Not-determined |
| 18-104_k141_119527 | Not-determined |
| 18-104_k141_126835 | Not-determined |
| 18-104_k141_77998 | Not-determined |
| 18-104_k141_112762 | Not-determined |
| 18-104_k141_63906 | Not-determined |
| 18-104_k141_140603 | Not-determined |
| 18-104_k141_154371 | Not-determined |
| 18-104_k141_56853 | Not-determined |
| 18-104_k141_147442 | Not-determined |
| 18-104_k141_70862 | Not-determined |
| 18-104_k141_1247 | Not-determined |
| 18-104_k141_70864 | Not-determined |
| 18-104_k141_147446 | Not-determined |
| 18-104_k141_126846 | Not-determined |
| 18-104_k141_119536 | Not-determined |
| 18-104_k141_147453 | Not-determined |
| 18-104_k141_29165 | Not-determined |
| 18-104_k141_112768 | Low-quality |
| 18-104_k141_126849 | Not-determined |
| 18-104_k141_70870 | Not-determined |
| 18-104_k141_56857 | Not-determined |
| 18-104_k141_119545 | Not-determined |
| 18-104_k141_119546 | Low-quality |
| 18-104_k141_98658 | Not-determined |
| 18-104_k141_84866 | Not-determined |
| 18-104_k141_78011 | Not-determined |
| 18-104_k141_8026 | Not-determined |
| 18-104_k141_91977 | Not-determined |
| 18-104_k141_56866 | Not-determined |
| 18-104_k141_105868 | Not-determined |
| 18-104_k141_133468 | Not-determined |
| 18-104_k141_119559 | Not-determined |
| 18-104_k141_105871 | Low-quality |
| 18-104_k141_154383 | Low-quality |
| 18-104_k141_14862 | Not-determined |
| 18-104_k141_35839 | Not-determined |
| 18-104_k141_49557 | Not-determined |
| 18-104_k141_1267 | Not-determined |
| 18-104_k141_35842 | Not-determined |
| 18-104_k141_147465 | Not-determined |
| 18-104_k141_21989 | Not-determined |
| 18-104_k141_78037 | Not-determined |
| 18-104_k141_56888 | Not-determined |
| 18-104_k141_8045 | Not-determined |
| 18-104_k141_84886 | Not-determined |
| 18-104_k141_91986 | Low-quality |
| 18-104_k141_98676 | Not-determined |
| 18-104_k141_8049 | Not-determined |
| 18-104_k141_22003 | Not-determined |
| 18-104_k141_105892 | Not-determined |
| 18-104_k141_140641 | Not-determined |
| 18-104_k141_78051 | Not-determined |
| 18-104_k141_35851 | Not-determined |
| 18-104_k141_1276 | Not-determined |
| 18-104_k141_154406 | Low-quality |
| 18-104_k141_35852 | Not-determined |
| 18-104_k141_140647 | Not-determined |
| 18-104_k141_133490 | Not-determined |
| 18-104_k141_84899 | Not-determined |
| 18-104_k141_14871 | Not-determined |
| 18-104_k141_49566 | Not-determined |
| 18-104_k141_126866 | Not-determined |
| 18-104_k141_147472 | Not-determined |
| 18-104_k141_154415 | Low-quality |
| 18-104_k141_63946 | Not-determined |
| 18-104_k141_154416 | Not-determined |
| 18-104_k141_49569 | Not-determined |
| 18-104_k141_133502 | Not-determined |
| 18-104_k141_70887 | Low-quality |
| 18-104_k141_49577 | Not-determined |
| 18-104_k141_133507 | Not-determined |
| 18-104_k141_63949 | Not-determined |
| 18-104_k141_154428 | Not-determined |
| 18-104_k141_154431 | Not-determined |
| 18-104_k141_154432 | Not-determined |
| 18-104_k141_112800 | Not-determined |
| 18-104_k141_56910 | Low-quality |
| 18-104_k141_112805 | Not-determined |
| 18-104_k141_8083 | Not-determined |
| 18-104_k141_14891 | Not-determined |
| 18-104_k141_154440 | Not-determined |
| 18-104_k141_98687 | Low-quality |
| 18-104_k141_63962 | Not-determined |
| 18-104_k141_126883 | Not-determined |
| 18-104_k141_147481 | Not-determined |
| 18-104_k141_14901 | Not-determined |
| 18-104_k141_78075 | Not-determined |
| 18-104_k141_1299 | Not-determined |
| 18-104_k141_112816 | Not-determined |
| 18-104_k141_1300 | Not-determined |
| 18-104_k141_1301 | Not-determined |
| 18-104_k141_154446 | Not-determined |
| 18-104_k141_8093 | Not-determined |
| 18-104_k141_14908 | Not-determined |
| 18-104_k141_126891 | Not-determined |
| 18-104_k141_133511 | Not-determined |
| 18-106-final-viral-combined_k141_118484\|\|full | Medium-quality |
| 18-104_k141_126892 | Not-determined |
| 18-104_k141_154448 | Low-quality |
| 18-104_k141_14913 | Not-determined |
| 18-104_k141_98696 | Not-determined |
| 18-104_k141_133514 | Not-determined |
| 18-104_k141_22017 | Low-quality |
| 18-104_k141_1307 | Not-determined |
| 18-104_k141_63977 | Not-determined |
| 18-104_k141_154453 | Low-quality |
| 18-104_k141_63979 | Not-determined |
| 18-104_k141_22024 | Not-determined |
| 18-104_k141_22025 | Not-determined |
| 18-104_k141_154455 | Not-determined |
| 18-104_k141_154457 | Not-determined |
| 18-104_k141_56920 | Low-quality |
| 18-104_k141_29193 | Not-determined |
| 18-104_k141_8106 | Not-determined |
| 18-104_k141_1315 | Not-determined |
| 18-104_k141_140700 | Low-quality |
| 18-104_k141_14923 | Not-determined |
| 18-104_k141_1319 | Not-determined |
| 18-104_k141_70923 | Not-determined |
| 18-104_k141_140707 | Not-determined |
| 18-104_k141_140712 | Not-determined |
| 18-104_k141_8121 | Not-determined |
| 18-104_k141_8124 | Not-determined |
| 18-104_k141_29208 | Not-determined |
| 18-104_k141_1332 | Not-determined |
| 18-104_k141_63994 | Not-determined |
| 18-104_k141_92024 | Not-determined |
| 18-104_k141_22050 | Not-determined |
| 18-104_k141_92025 | Low-quality |
| 18-104_k141_140718 | Not-determined |
| 18-104_k141_78085 | Not-determined |
| 18-104_k141_35870 | Not-determined |
| 18-104_k141_35872 | Not-determined |
| 18-104_k141_140725 | Not-determined |
| 18-104_k141_22053 | Not-determined |
| 18-104_k141_92033 | Not-determined |
| 18-104_k141_154470 | Low-quality |
| 18-104_k141_35877 | Not-determined |

| | |
|---|---|
| 18-104_k141_14949 | Not-determined |
| 18-104_k141_133534 | Not-determined |
| 18-104_k141_49628 | Not-determined |
| 18-104_k141_78094 | Low-quality |
| 18-104_k141_29224 | Not-determined |
| 18-104_k141_49634 | Not-determined |
| 18-104_k141_49635 | Not-determined |
| 18-104_k141_133544 | Not-determined |
| 18-104_k141_29229 | Not-determined |
| 18-104_k141_161260 | Low-quality |
| 18-104_k141_154485 | Not-determined |
| 18-104_k141_35893 | Not-determined |
| 18-104_k141_78104 | Not-determined |
| 18-104_k141_1358 | Low-quality |
| 18-104_k141_161264 | Not-determined |
| 18-104_k141_35897 | Low-quality |
| 18-104_k141_35898 | Not-determined |
| 18-104_k141_8141 | Low-quality |
| 18-104_k141_8144 | Not-determined |
| 18-104_k141_8146 | Not-determined |
| 18-104_k141_8151 | Not-determined |
| 18-104_k141_147537 | Not-determined |
| 18-104_k141_49641 | Not-determined |
| 18-104_k141_14968 | Low-quality |
| 18-104_k141_35914 | Not-determined |
| 18-104_k141_126897 | Not-determined |
| 18-104_k141_22085 | Not-determined |
| 18-104_k141_14974 | Not-determined |
| 18-104_k141_161279 | Not-determined |
| 18-104_k141_119590 | Not-determined |
| 18-104_k141_133560 | Not-determined |
| 18-104_k141_126900 | Not-determined |
| 18-104_k141_161283 | Not-determined |
| 18-104_k141_70958 | Not-determined |
| 18-104_k141_84908 | Not-determined |
| 18-104_k141_84910 | Not-determined |
| 18-104_k141_35920 | Not-determined |
| 18-104_k141_70961 | Not-determined |
| 18-104_k141_35923 | Not-determined |
| 18-104_k141_84916 | Not-determined |
| 18-104_k141_84917 | Not-determined |
| 18-104_k141_49651 | Not-determined |
| 18-104_k141_161286 | Not-determined |
| 18-104_k141_49654 | Not-determined |
| 18-104_k141_29253 | Not-determined |
| 18-104_k141_14990 | Not-determined |
| 18-104_k141_84921 | Not-determined |
| 18-104_k141_14995 | Not-determined |
| 18-104_k141_161294 | Low-quality |
| 18-104_k141_140759 | Low-quality |
| 18-104_k141_147553 | Not-determined |
| 18-104_k141_22101 | Not-determined |
| 18-104_k141_70971 | Not-determined |
| 18-104_k141_92067 | Not-determined |
| 18-104_k141_92069 | Not-determined |
| 18-104_k141_56933 | Not-determined |
| 18-104_k141_70984 | Low-quality |
| 18-104_k141_56935 | Not-determined |
| 18-104_k141_64012 | Not-determined |
| 18-104_k141_133570 | Not-determined |
| 18-104_k141_84933 | Not-determined |
| 18-104_k141_22107 | Not-determined |
| 18-104_k141_126908 | Low-quality |
| 18-104_k141_92078 | Not-determined |
| 18-104_k141_78142 | Not-determined |
| 18-104_k141_92083 | Not-determined |
| 18-104_k141_70997 | Not-determined |
| 18-104_k141_147559 | Not-determined |
| 18-104_k141_15012 | Low-quality |
| 18-104_k141_126918 | Not-determined |
| 18-104_k141_147565 | Not-determined |
| 18-104_k141_84939 | Low-quality |
| 18-104_k141_126921 | Not-determined |
| 18-104_k141_126922 | Low-quality |
| 18-104_k141_1376 | Not-determined |
| 18-104_k141_15016 | Not-determined |
| 18-104_k141_92096 | Not-determined |
| 18-104_k141_71005 | Not-determined |
| 18-104_k141_49669 | Not-determined |
| 18-104_k141_49670 | Not-determined |
| 18-104_k141_84946 | Not-determined |
| 18-104_k141_49677 | Not-determined |
| 18-104_k141_84950 | Not-determined |
| 18-104_k141_71008 | Low-quality |
| 18-104_k141_119610 | Not-determined |
| 18-104_k141_49678 | Not-determined |
| 18-104_k141_71009 | Not-determined |
| 18-104_k141_64029 | Low-quality |
| 18-104_k141_56950 | Low-quality |
| 18-104_k141_84953 | Not-determined |
| 18-104_k141_84954 | Not-determined |
| 18-104_k141_15031 | Not-determined |
| 18-104_k141_119614 | Not-determined |
| 18-104_k141_161308 | Not-determined |
| 18-104_k141_56955 | Low-quality |
| 18-104_k141_161310 | Not-determined |
| 18-104_k141_147570 | Not-determined |
| 18-104_k141_161314 | Not-determined |
| 18-104_k141_119626 | Not-determined |
| 18-104_k141_126940 | Not-determined |
| 18-104_k141_161322 | Not-determined |
| 18-104_k141_56960 | Not-determined |
| 18-104_k141_49692 | Not-determined |
| 18-104_k141_84961 | Not-determined |
| 18-104_k141_154525 | Not-determined |
| 18-104_k141_71021 | Not-determined |
| 18-104_k141_43025 | Not-determined |
| 18-104_k141_154527 | Not-determined |
| 18-104_k141_154528 | Not-determined |
| 18-104_k141_15046 | Not-determined |
| 18-104_k141_105925 | Not-determined |
| 18-104_k141_154530 | Not-determined |
| 18-104_k141_98738 | Not-determined |
| 18-104_k141_92114 | Not-determined |
| 18-104_k141_140791 | Not-determined |
| 18-104_k141_140792 | Not-determined |
| 18-104_k141_92116 | Not-determined |
| 18-104_k141_1396 | Not-determined |
| 18-104_k141_29291 | Low-quality |
| 18-104_k141_56965 | Not-determined |
| 18-104_k141_56966 | Low-quality |
| 18-104_k141_105935 | Low-quality |
| 18-104_k141_43035 | Not-determined |
| 18-104_k141_84966 | Not-determined |
| 18-104_k141_92126 | Not-determined |
| 18-104_k141_43036 | Not-determined |
| 18-104_k141_43038 | Not-determined |
| 18-104_k141_64055 | Not-determined |
| 18-104_k141_15057 | Not-determined |
| 18-104_k141_154541 | Not-determined |
| 18-104_k141_29295 | Not-determined |
| 18-104_k141_64059 | Not-determined |
| 18-104_k141_126953 | Not-determined |
| 18-104_k141_147591 | Low-quality |
| 18-104_k141_84976 | Not-determined |
| 18-104_k141_8181 | Not-determined |
| 18-104_k141_147599 | Not-determined |
| 18-104_k141_43041 | Not-determined |
| 18-104_k141_147601 | Not-determined |
| 18-104_k141_140809 | Low-quality |
| 18-104_k141_92132 | Not-determined |
| 18-104_k141_140813 | Not-determined |
| 18-104_k141_35975 | Not-determined |
| 18-104_k141_154558 | Not-determined |
| 18-104_k141_154559 | Not-determined |
| 18-104_k141_140815 | Not-determined |
| 18-104_k141_35978 | Not-determined |
| 18-104_k141_56987 | Not-determined |
| 18-104_k141_49708 | Not-determined |
| 18-104_k141_119655 | Not-determined |
| 18-104_k141_71042 | Not-determined |
| 18-104_k141_29309 | Low-quality |
| 18-104_k141_49711 | Not-determined |
| 18-104_k141_84994 | Not-determined |
| 18-104_k141_105952 | Not-determined |
| 18-104_k141_84995 | Not-determined |
| 18-104_k141_1404 | Not-determined |
| 18-104_k141_147604 | Low-quality |
| 18-104_k141_119669 | Not-determined |
| 18-104_k141_85001 | Low-quality |
| 18-104_k141_43063 | Not-determined |
| 18-104_k141_154565 | Not-determined |
| 18-104_k141_98767 | Low-quality |
| 18-104_k141_126968 | Low-quality |
| 18-104_k141_126972 | Not-determined |
| 18-104_k141_126973 | Not-determined |
| 18-104_k141_140839 | Not-determined |
| 18-104_k141_126978 | Not-determined |
| 18-104_k141_35998 | Not-determined |
| 18-104_k141_154581 | Not-determined |
| 18-104_k141_140844 | Low-quality |
| 18-104_k141_8197 | Not-determined |
| 18-104_k141_43072 | Not-determined |
| 18-104_k141_119690 | Not-determined |
| 18-104_k141_36000 | Not-determined |
| 18-104_k141_85009 | Not-determined |
| 18-104_k141_1420 | Not-determined |
| 18-104_k141_85010 | Not-determined |
| 18-104_k141_105976 | Not-determined |
| 18-104_k141_85015 | Not-determined |
| 18-104_k141_154585 | Not-determined |
| 18-104_k141_154586 | Not-determined |
| 18-104_k141_154589 | Not-determined |
| 18-104_k141_43087 | Not-determined |
| 18-104_k141_22137 | Not-determined |
| 18-104_k141_1426 | Not-determined |
| 18-104_k141_71057 | Not-determined |
| 18-104_k141_43090 | Not-determined |
| 18-104_k141_154597 | Not-determined |
| 18-104_k141_29336 | Not-determined |
| 18-104_k141_22143 | Not-determined |
| 18-104_k141_78198 | Not-determined |
| 18-104_k141_85022 | Not-determined |
| 18-104_k141_8210 | Not-determined |
| 18-104_k141_78203 | Not-determined |
| 18-104_k141_22148 | Not-determined |
| 18-104_k141_85026 | Not-determined |
| 18-104_k141_29342 | Not-determined |
| 18-104_k141_78206 | Low-quality |
| 18-104_k141_1432 | Not-determined |
| 18-104_k141_36012 | Not-determined |
| 18-104_k141_105983 | Not-determined |
| 18-104_k141_8215 | Not-determined |
| 18-104_k141_92162 | Not-determined |
| 18-104_k141_78214 | Not-determined |
| 18-104_k141_147636 | Not-determined |
| 18-104_k141_78215 | Not-determined |
| 18-104_k141_8218 | Not-determined |
| 18-104_k141_147637 | Not-determined |
| 18-104_k141_98793 | Not-determined |
| 18-104_k141_154604 | Not-determined |
| 18-104_k141_92166 | Not-determined |
| 18-104_k141_36021 | Not-determined |
| 18-104_k141_154606 | Not-determined |
| 18-104_k141_105993 | Not-determined |
| 18-104_k141_119709 | Not-determined |
| 18-104_k141_71068 | Not-determined |
| 18-104_k141_57027 | Not-determined |
| 18-104_k141_78222 | Not-determined |
| 18-104_k141_43103 | Not-determined |
| 18-104_k141_15074 | Not-determined |
| 18-104_k141_57028 | Low-quality |
| 18-104_k141_85038 | Not-determined |
| 18-104_k141_119715 | Not-determined |
| 18-104_k141_140864 | Not-determined |
| 18-104_k141_126999 | Not-determined |
| 18-104_k141_119717 | Not-determined |
| 18-104_k141_119720 | Low-quality |
| 18-104_k141_49742 | Not-determined |
| 18-104_k141_78226 | Not-determined |
| 18-104_k141_22158 | Not-determined |
| 18-104_k141_106000 | Not-determined |
| 18-104_k141_36032 | Not-determined |
| 18-104_k141_1448 | Low-quality |
| 18-104_k141_71080 | Not-determined |
| 18-104_k141_57043 | Not-determined |
| 18-104_k141_36033 | Not-determined |
| 18-104_k141_161374 | Not-determined |
| 18-104_k141_36036 | Not-determined |
| 18-104_k141_29357 | Not-determined |
| 18-104_k141_161378 | Not-determined |
| 18-104_k141_29360 | Low-quality |
| 18-104_k141_119724 | Low-quality |
| 18-104_k141_36047 | Not-determined |
| 18-104_k141_71096 | Not-determined |
| 18-104_k141_78236 | Low-quality |
| 18-104_k141_15095 | Not-determined |
| 18-104_k141_8250 | Not-determined |
| 18-104_k141_127015 | Not-determined |
| 18-104_k141_15097 | Not-determined |
| 18-104_k141_112843 | Not-determined |
| 18-104_k141_154610 | Low-quality |
| 18-104_k141_57060 | Not-determined |
| 18-104_k141_154612 | Not-determined |
| 18-104_k141_64084 | Not-determined |
| 18-104_k141_64086 | Not-determined |
| 18-104_k141_133670 | Not-determined |
| 18-104_k141_112847 | Not-determined |
| 18-104_k141_57067 | Not-determined |
| 18-104_k141_57069 | Not-determined |
| 18-104_k141_57070 | Not-determined |
| 18-104_k141_36060 | Not-determined |
| 18-104_k141_43123 | Low-quality |
| 18-104_k141_49754 | Not-determined |
| 18-104_k141_49756 | Low-quality |
| 18-104_k141_29371 | Not-determined |
| 18-104_k141_78248 | Not-determined |
| 18-104_k141_15108 | Not-determined |
| 18-104_k141_154627 | Not-determined |
| 18-104_k141_49757 | Not-determined |
| 18-104_k141_127028 | Not-determined |
| 18-104_k141_71118 | Not-determined |
| 18-104_k141_85064 | Not-determined |
| 18-104_k141_147673 | Not-determined |
| 18-104_k141_8275 | Not-determined |
| 18-104_k141_36069 | Low-quality |
| 18-104_k141_1457 | Not-determined |
| 18-104_k141_133688 | Not-determined |
| 18-104_k141_127038 | Not-determined |
| 18-104_k141_1459 | Not-determined |
| 18-104_k141_78259 | Not-determined |
| 18-104_k141_98834 | Not-determined |
| 18-104_k141_57077 | Not-determined |
| 18-104_k141_127041 | Not-determined |
| 18-104_k141_161422 | Low-quality |
| 18-104_k141_106033 | Not-determined |
| 18-104_k141_161423 | Not-determined |
| 18-104_k141_15122 | Not-determined |
| 18-104_k141_15125 | Not-determined |
| 18-104_k141_127045 | Not-determined |
| 18-104_k141_106037 | Not-determined |
| 18-104_k141_57080 | Not-determined |
| 18-104_k141_49774 | Not-determined |
| 18-104_k141_49777 | Not-determined |
| 18-104_k141_1462 | Not-determined |
| 18-104_k141_29390 | Not-determined |
| 18-104_k141_1465 | Low-quality |
| 18-104_k141_78268 | Not-determined |
| 18-104_k141_15130 | Not-determined |
| 18-104_k141_71131 | Not-determined |
| 18-104_k141_161435 | Not-determined |
| 18-104_k141_154638 | Not-determined |
| 18-104_k141_127057 | Not-determined |
| 18-104_k141_78272 | Not-determined |
| 18-104_k141_22182 | Low-quality |
| 18-104_k141_85071 | Not-determined |
| 18-104_k141_140877 | Low-quality |
| 18-104_k141_127070 | Not-determined |
| 18-104_k141_71137 | Not-determined |
| 18-104_k141_29407 | Not-determined |
| 18-104_k141_127076 | Not-determined |
| 18-104_k141_140887 | Not-determined |
| 18-104_k141_140889 | Not-determined |
| 18-104_k141_64106 | Not-determined |
| 18-104_k141_15146 | Not-determined |
| 18-104_k141_133714 | Not-determined |
| 18-104_k141_36085 | Not-determined |
| 18-104_k141_78283 | Not-determined |
| 18-104_k141_36088 | Low-quality |
| 18-104_k141_36089 | Not-determined |
| 18-104_k141_71146 | Not-determined |
| 18-104_k141_78287 | Not-determined |
| 18-104_k141_98857 | Not-determined |
| 18-104_k141_119767 | Not-determined |
| 18-104_k141_98859 | Not-determined |
| 18-104_k141_57095 | Not-determined |
| 18-104_k141_43152 | Not-determined |
| 18-104_k141_64118 | Not-determined |
| 18-101-final-viral-combined_k141_32878\|\|full | Medium-quality |
| 18-104_k141_36097 | Low-quality |
| 18-104_k141_98865 | Not-determined |
| 18-104_k141_8286 | Not-determined |
| 18-104_k141_57099 | Not-determined |
| 18-104_k141_1471 | Not-determined |
| 18-104_k141_1473 | Not-determined |
| 18-104_k141_133721 | Not-determined |
| 18-104_k141_8290 | Not-determined |
| 18-104_k141_8291 | Not-determined |
| 18-104_k141_29419 | Not-determined |
| 18-104_k141_15155 | Not-determined |
| 18-104_k141_1481 | Not-determined |
| 18-104_k141_140895 | Not-determined |
| 18-104_k141_8300 | Not-determined |
| 18-104_k141_127095 | Not-determined |
| 18-104_k141_78303 | Not-determined |
| 18-104_k141_133735 | Not-determined |
| 18-104_k141_78307 | Not-determined |
| 18-104_k141_147692 | Not-determined |
| 18-104_k141_133737 | Not-determined |
| 18-104_k141_161471 | Not-determined |
| 18-104_k141_1493 | Not-determined |
| 18-104_k141_8313 | Not-determined |
| 18-104_k141_29431 | Low-quality |
| 18-104_k141_154674 | Not-determined |
| 18-104_k141_112903 | Not-determined |
| 18-104_k141_78316 | Not-determined |
| 18-104_k141_29433 | Not-determined |
| 18-104_k141_29435 | Not-determined |
| 18-104_k141_140907 | Not-determined |
| 18-104_k141_154680 | Not-determined |
| 18-104_k141_154682 | Not-determined |
| 18-104_k141_71163 | Not-determined |
| 18-104_k141_78325 | Not-determined |
| 18-104_k141_85116 | Not-determined |
| 18-104_k141_57118 | Not-determined |
| 18-104_k141_1506 | Not-determined |
| 18-104_k141_78335 | Not-determined |
| 18-104_k141_57119 | Not-determined |
| 18-104_k141_64142 | Low-quality |
| 18-104_k141_85121 | Not-determined |
| 18-104_k141_106088 | Not-determined |
| 18-104_k141_64148 | Not-determined |
| 18-104_k141_154696 | Low-quality |
| 18-104_k141_43176 | Not-determined |
| 18-104_k141_85122 | Not-determined |
| 18-104_k141_43177 | Low-quality |
| 18-104_k141_78339 | Not-determined |
| 18-104_k141_29443 | Not-determined |
| 18-104_k141_64156 | Not-determined |
| 18-104_k141_43185 | Not-determined |
| 18-104_k141_1512 | Not-determined |
| 18-104_k141_140921 | Not-determined |
| 18-104_k141_64165 | Not-determined |
| 18-104_k141_49833 | Not-determined |
| 18-104_k141_133761 | Not-determined |
| 18-104_k141_64167 | Not-determined |
| 18-104_k141_154710 | Not-determined |
| 18-104_k141_140924 | Not-determined |
| 18-104_k141_22193 | Not-determined |
| 18-104_k141_29454 | Low-quality |
| 18-104_k141_22194 | Not-determined |
| 18-104_k141_112912 | Not-determined |
| 18-104_k141_98898 | Not-determined |
| 18-104_k141_127127 | Not-determined |
| 18-104_k141_154718 | Not-determined |
| 18-104_k141_98900 | Not-determined |
| 18-104_k141_147708 | Not-determined |
| 18-104_k141_140928 | Not-determined |
| 18-104_k141_22198 | Not-determined |
| 18-104_k141_49839 | Not-determined |
| 18-104_k141_112920 | Not-determined |
| 18-104_k141_8342 | Not-determined |
| 18-104_k141_147709 | Not-determined |
| 18-104_k141_78352 | Not-determined |
| 18-104_k141_98906 | Low-quality |
| 18-104_k141_106106 | Not-determined |
| 18-104_k141_78355 | Not-determined |
| 18-104_k141_64179 | Low-quality |
| 18-104_k141_98910 | Not-determined |
| 18-104_k141_22203 | Not-determined |
| 18-104_k141_133775 | Not-determined |
| 18-104_k141_133777 | Not-determined |
| 18-104_k141_133782 | Not-determined |
| 18-104_k141_112925 | Not-determined |
| 18-104_k141_154735 | Low-quality |
| 18-104_k141_49846 | Not-determined |
| 18-104_k141_161489 | Not-determined |
| 18-104_k141_49849 | Not-determined |
| 18-104_k141_1529 | Not-determined |
| 18-104_k141_43205 | Not-determined |
| 18-104_k141_154737 | Not-determined |
| 18-104_k141_154738 | Not-determined |
| 18-104_k141_64192 | Not-determined |
| 18-104_k141_161497 | Not-determined |
| 18-104_k141_15211 | Not-determined |
| 18-104_k141_133788 | Not-determined |
| 18-104_k141_43212 | Not-determined |
| 18-104_k141_43214 | Not-determined |
| 18-104_k141_29486 | Not-determined |
| 18-104_k141_133793 | Not-determined |
| 18-104_k141_161504 | Not-determined |
| 18-104_k141_49856 | Low-quality |
| 18-104_k141_98934 | Not-determined |
| 18-104_k141_78368 | Not-determined |
| 18-104_k141_133799 | Not-determined |
| 18-104_k141_78369 | Not-determined |
| 18-104_k141_112951 | Not-determined |
| 18-104_k141_112952 | Not-determined |
| 18-104_k141_161506 | Not-determined |
| 18-104_k141_71195 | Not-determined |
| 18-104_k141_133804 | Not-determined |
| 18-104_k141_161512 | Low-quality |
| 18-104_k141_43220 | Not-determined |
| 18-104_k141_112957 | Not-determined |
| 18-104_k141_133809 | Not-determined |
| 18-104_k141_15226 | Not-determined |
| 18-104_k141_64206 | Not-determined |
| 18-104_k141_49874 | Not-determined |
| 18-104_k141_98946 | Not-determined |
| 18-104_k141_78386 | Not-determined |
| 18-104_k141_57124 | Not-determined |
| 18-104_k141_106140 | Not-determined |
| 18-104_k141_98959 | Not-determined |
| 18-104_k141_78401 | Not-determined |
| 18-104_k141_140988 | Not-determined |
| 18-104_k141_98963 | Not-determined |
| 18-104_k141_85147 | Not-determined |
| 18-104_k141_1562 | Low-quality |
| 18-104_k141_147765 | Not-determined |
| 18-104_k141_92207 | Not-determined |
| 18-104_k141_15256 | Not-determined |
| 18-104_k141_92210 | Not-determined |
| 18-104_k141_49897 | Not-determined |
| 18-104_k141_29515 | Not-determined |
| 18-104_k141_154759 | Not-determined |
| 18-104_k141_43230 | Not-determined |
| 18-104_k141_64213 | Not-determined |
| 18-104_k141_112984 | Not-determined |
| 18-104_k141_43232 | Not-determined |
| 18-104_k141_49901 | Not-determined |
| 18-104_k141_161540 | Not-determined |
| 18-104_k141_29521 | Not-determined |
| 18-104_k141_140999 | Not-determined |
| 18-104_k141_64218 | Not-determined |
| 18-104_k141_147766 | Low-quality |
| 18-104_k141_92217 | Not-determined |
| 18-104_k141_22222 | Not-determined |
| 18-104_k141_147768 | Not-determined |
| 18-104_k141_119801 | Not-determined |
| 18-104_k141_78405 | Not-determined |
| 18-104_k141_1572 | Not-determined |
| 18-104_k141_85155 | Not-determined |
| 18-104_k141_141009 | Not-determined |
| 18-104_k141_64228 | Not-determined |
| 18-104_k141_141010 | Not-determined |
| 18-104_k141_22229 | Not-determined |
| 18-104_k141_92222 | Not-determined |
| 18-104_k141_22232 | Not-determined |
| 18-104_k141_57162 | Not-determined |
| 18-104_k141_43247 | Not-determined |
| 18-104_k141_92232 | Not-determined |
| 18-104_k141_57164 | Not-determined |
| 18-104_k141_85169 | Not-determined |
| 18-104_k141_106157 | Low-quality |
| 18-104_k141_1585 | Not-determined |
| 18-104_k141_92246 | Low-quality |
| 18-104_k141_161550 | Not-determined |
| 18-104_k141_64250 | Low-quality |
| 18-104_k141_8376 | Not-determined |
| 18-104_k141_8377 | Not-determined |
| 18-104_k141_133830 | Not-determined |
| 18-104_k141_64253 | Not-determined |
| 18-104_k141_22254 | Not-determined |
| 18-104_k141_1590 | Not-determined |
| 18-104_k141_1591 | Low-quality |
| 18-104_k141_92255 | Not-determined |
| 18-104_k141_154769 | Not-determined |
| 18-104_k141_29536 | Not-determined |
| 18-104_k141_154770 | Not-determined |
| 18-104_k141_64257 | Not-determined |
| 18-104_k141_92259 | Low-quality |
| 18-104_k141_15261 | Not-determined |
| 18-104_k141_49925 | Not-determined |
| 18-104_k141_154776 | Not-determined |
| 18-104_k141_8385 | Not-determined |
| 18-104_k141_112995 | Low-quality |
| 18-104_k141_29539 | Not-determined |
| 18-104_k141_141028 | Not-determined |
| 18-104_k141_106182 | Not-determined |
| 18-104_k141_49928 | Not-determined |
| 18-104_k141_49929 | Low-quality |
| 18-104_k141_154779 | Not-determined |
| 18-104_k141_22266 | Not-determined |
| 18-104_k141_154782 | Not-determined |
| 18-104_k141_106184 | Low-quality |
| 18-104_k141_57193 | Not-determined |
| 18-104_k141_71223 | Low-quality |
| 18-104_k141_133848 | Not-determined |
| 18-104_k141_133849 | Not-determined |
| 18-104_k141_57195 | Not-determined |
| 18-104_k141_147795 | Not-determined |
| 18-104_k141_29547 | Not-determined |
| 18-104_k141_71227 | Not-determined |
| 18-104_k141_154786 | Not-determined |
| 18-104_k141_127159 | Not-determined |
| 18-104_k141_29548 | Not-determined |
| 18-104_k141_161569 | Not-determined |
| 18-104_k141_92275 | Not-determined |
| 18-104_k141_106192 | Not-determined |
| 18-104_k141_36119 | Not-determined |
| 18-104_k141_85204 | Not-determined |
| 18-104_k141_49940 | Not-determined |
| 18-104_k141_43258 | Not-determined |
| 18-104_k141_29551 | Not-determined |
| 18-104_k141_22271 | Not-determined |
| 18-104_k141_127164 | Not-determined |
| 18-104_k141_71238 | Not-determined |
| 18-104_k141_36124 | Not-determined |
| 18-104_k141_8388 | Low-quality |
| 18-104_k141_85208 | Not-determined |
| 18-104_k141_1616 | Not-determined |
| 18-104_k141_99008 | Not-determined |
| 18-104_k141_161581 | Low-quality |
| 18-104_k141_113017 | Not-determined |
| 18-104_k141_119838 | Not-determined |
| 18-104_k141_57204 | Not-determined |
| 18-104_k141_147804 | Not-determined |
| 18-104_k141_161584 | Not-determined |
| 18-104_k141_8396 | Not-determined |
| 18-104_k141_29564 | Not-determined |
| 18-104_k141_147807 | Not-determined |
| 18-104_k141_99012 | Not-determined |
| 18-104_k141_161586 | Not-determined |
| 18-104_k141_99015 | Low-quality |
| 18-104_k141_36136 | Not-determined |
| 18-104_k141_154803 | Not-determined |
| 18-104_k141_29570 | Not-determined |
| 18-104_k141_113027 | Not-determined |
| 18-104_k141_92288 | Not-determined |
| 18-104_k141_119848 | Not-determined |
| 18-104_k141_1622 | Not-determined |
| 18-104_k141_8407 | Low-quality |
| 18-104_k141_1625 | Not-determined |
| 18-104_k141_133877 | Not-determined |
| 18-104_k141_15277 | Low-quality |
| 18-104_k141_99029 | Low-quality |
| 18-104_k141_15279 | Not-determined |
| 18-104_k141_99032 | Not-determined |
| 18-104_k141_8412 | Not-determined |
| 18-104_k141_85227 | Not-determined |
| 18-104_k141_49959 | Low-quality |
| 18-104_k141_133883 | Low-quality |
| 18-104_k141_57218 | Not-determined |
| 18-104_k141_113038 | Low-quality |
| 18-104_k141_1633 | Low-quality |
| 18-104_k141_147823 | Not-determined |
| 18-104_k141_133889 | Not-determined |
| 18-104_k141_133890 | Not-determined |
| 18-104_k141_57223 | Not-determined |

| | |
|---|---|
| 18-104_k141_43285 | Not-determined |
| 18-104_k141_161617 | Low-quality |
| 18-104_k141_147826 | Not-determined |
| 18-104_k141_43289 | Not-determined |
| 18-104_k141_43290 | Not-determined |
| 18-104_k141_57230 | Not-determined |
| 18-104_k141_57231 | Not-determined |
| 18-104_k141_147829 | Not-determined |
| 18-104_k141_15294 | Not-determined |
| 18-104_k141_141057 | Not-determined |
| 18-104_k141_29605 | Not-determined |
| 18-104_k141_133900 | Not-determined |
| 18-104_k141_113055 | Not-determined |
| 18-104_k141_141063 | Not-determined |
| 18-104_k141_29616 | Not-determined |
| 18-104_k141_1641 | Not-determined |
| 18-104_k141_78451 | Low-quality |
| 18-104_k141_29618 | Not-determined |
| 18-104_k141_119861 | Not-determined |
| 18-104_k141_141069 | Not-determined |
| 18-104_k141_141070 | Not-determined |
| 18-104_k141_36162 | Not-determined |
| 18-104_k141_49987 | Low-quality |
| 18-104_k141_113061 | Not-determined |
| 18-104_k141_119863 | Not-determined |
| 18-104_k141_43308 | Not-determined |
| 18-104_k141_49989 | Low-quality |
| 18-104_k141_8444 | Not-determined |
| 18-104_k141_36167 | Not-determined |
| 18-104_k141_127212 | Not-determined |
| 18-104_k141_43311 | Not-determined |
| 18-104_k141_29620 | Not-determined |
| 18-104_k141_57244 | Not-determined |
| 18-104_k141_8449 | Not-determined |
| 18-104_k141_8451 | Not-determined |
| 18-104_k141_22318 | Not-determined |
| 18-104_k141_15312 | Not-determined |
| 18-104_k141_15315 | Not-determined |
| 18-104_k141_127217 | Not-determined |
| 18-104_k141_99068 | Not-determined |
| 18-104_k141_57247 | Low-quality |
| 18-104_k141_1649 | Not-determined |
| 18-104_k141_57249 | Not-determined |
| 18-104_k141_1652 | Low-quality |
| 18-104_k141_43327 | Not-determined |
| 18-104_k141_147835 | Not-determined |
| 18-104_k141_99072 | Not-determined |
| 18-104_k141_29632 | Low-quality |
| 18-104_k141_43330 | Not-determined |
| 18-104_k141_99075 | Not-determined |
| 18-104_k141_22332 | Not-determined |
| 18-104_k141_78472 | Not-determined |
| 18-104_k141_141090 | Not-determined |
| 18-104_k141_92310 | Not-determined |
| 18-104_k141_99078 | Not-determined |
| 18-104_k141_49992 | Not-determined |
| 18-104_k141_36176 | Not-determined |
| 18-104_k141_71275 | Not-determined |
| 18-104_k141_57264 | Not-determined |
| 18-104_k141_43343 | Not-determined |
| 18-104_k141_127240 | Not-determined |
| 18-104_k141_119873 | Not-determined |
| 18-104_k141_99100 | Not-determined |
| 18-104_k141_99101 | Not-determined |
| 18-104_k141_29643 | Not-determined |
| 18-104_k141_99102 | Not-determined |
| 18-104_k141_127244 | Not-determined |
| 18-104_k141_141107 | Not-determined |
| 18-104_k141_85270 | Not-determined |
| 18-104_k141_147860 | Not-determined |
| 18-104_k141_1670 | Not-determined |
| 18-104_k141_43351 | Not-determined |
| 18-104_k141_92332 | Not-determined |
| 18-104_k141_85272 | Not-determined |
| 18-104_k141_8461 | Not-determined |
| 18-104_k141_154838 | Not-determined |
| 18-104_k141_127252 | Not-determined |
| 18-104_k141_127254 | Low-quality |
| 18-104_k141_106287 | Not-determined |
| 18-104_k141_127256 | Not-determined |
| 18-104_k141_106291 | Not-determined |
| 18-104_k141_92338 | Not-determined |
| 18-104_k141_15342 | Not-determined |
| 18-104_k141_85283 | Not-determined |
| 18-104_k141_43360 | Not-determined |
| 18-104_k141_92344 | Low-quality |
| 18-104_k141_147869 | Low-quality |
| 18-104_k141_106292 | Not-determined |
| 18-104_k141_15344 | Not-determined |
| 18-104_k141_127259 | Not-determined |
| 18-104_k141_141110 | Not-determined |
| 18-104_k141_57272 | Not-determined |
| 18-104_k141_1672 | Not-determined |
| 18-104_k141_8478 | Not-determined |
| 18-104_k141_113109 | Not-determined |
| 18-104_k141_36186 | Low-quality |
| 18-104_k141_57278 | Not-determined |
| 18-104_k141_36188 | Low-quality |
| 18-104_k141_1682 | Not-determined |
| 18-104_k141_127264 | Not-determined |
| 18-104_k141_71313 | Not-determined |
| 18-104_k141_92351 | Not-determined |
| 18-104_k141_15360 | Not-determined |
| 18-104_k141_147877 | Low-quality |
| 18-104_k141_64311 | Not-determined |
| 18-104_k141_64312 | Not-determined |
| 18-104_k141_1685 | Not-determined |
| 18-104_k141_85306 | Not-determined |
| 18-104_k141_1686 | Not-determined |
| 18-104_k141_71318 | Not-determined |
| 18-104_k141_119882 | Not-determined |
| 18-104_k141_57291 | Not-determined |
| 18-104_k141_141127 | Low-quality |
| 18-104_k141_71324 | Not-determined |
| 18-104_k141_57293 | Not-determined |
| 18-104_k141_15367 | Not-determined |
| 18-104_k141_141129 | Low-quality |
| 18-104_k141_154855 | Not-determined |
| 18-104_k141_85317 | Not-determined |
| 18-104_k141_113130 | Not-determined |
| 18-104_k141_78489 | Low-quality |
| 18-104_k141_57295 | Not-determined |
| 18-104_k141_113133 | Not-determined |
| 18-104_k141_71333 | Not-determined |
| 18-104_k141_71334 | Not-determined |
| 18-104_k141_36212 | Not-determined |
| 18-104_k141_154862 | Low-quality |
| 18-104_k141_113138 | Not-determined |
| 18-104_k141_64313 | Not-determined |
| 18-104_k141_119895 | Not-determined |
| 18-104_k141_154866 | Not-determined |
| 18-104_k141_154867 | Not-determined |
| 18-104_k141_64317 | Low-quality |
| 18-104_k141_64323 | Not-determined |
| 18-104_k141_161635 | Not-determined |
| 18-104_k141_119897 | Not-determined |
| 18-104_k141_57307 | Not-determined |
| 18-104_k141_15375 | Not-determined |
| 18-104_k141_1701 | Not-determined |
| 18-104_k141_154880 | Not-determined |
| 18-104_k141_85331 | Not-determined |
| 18-104_k141_85333 | Not-determined |
| 18-104_k141_36227 | Not-determined |
| 18-104_k141_43382 | Low-quality |
| 18-104_k141_43383 | Low-quality |
| 18-104_k141_64326 | Not-determined |
| 18-104_k141_113146 | Not-determined |
| 18-104_k141_36230 | Not-determined |
| 18-104_k141_106325 | Not-determined |
| 18-104_k141_15379 | Not-determined |
| 18-104_k141_78513 | Not-determined |
| 18-104_k141_36235 | Not-determined |
| 18-104_k141_36236 | Low-quality |
| 18-104_k141_71345 | Not-determined |
| 18-104_k141_99153 | Not-determined |
| 18-104_k141_78520 | Not-determined |
| 18-104_k141_161647 | Not-determined |
| 18-104_k141_113159 | Not-determined |
| 18-104_k141_8518 | Not-determined |
| 18-104_k141_29647 | Not-determined |
| 18-104_k141_78524 | Not-determined |
| 18-104_k141_78525 | Not-determined |
| 18-104_k141_85344 | Not-determined |
| 18-104_k141_85346 | Not-determined |
| 18-104_k141_15390 | Not-determined |
| 18-104_k141_113167 | Not-determined |
| 18-104_k141_1717 | Not-determined |
| 18-104_k141_154886 | Low-quality |
| 18-104_k141_36245 | Not-determined |
| 18-104_k141_92384 | Low-quality |
| 18-104_k141_36250 | Low-quality |
| 18-104_k141_119928 | Not-determined |
| 18-104_k141_113178 | Not-determined |
| 18-104_k141_154889 | Not-determined |
| 18-104_k141_106333 | Low-quality |
| 18-104_k141_29670 | Not-determined |
| 18-104_k141_29672 | Low-quality |
| 18-104_k141_1722 | Low-quality |
| 18-104_k141_1724 | Not-determined |
| 18-104_k141_133937 | Low-quality |
| 18-104_k141_8531 | Not-determined |
| 18-104_k141_8533 | Not-determined |
| 18-104_k141_99158 | Not-determined |
| 18-104_k141_29676 | Not-determined |
| 18-104_k141_106340 | Not-determined |
| 18-104_k141_71365 | Not-determined |
| 18-104_k141_106347 | Not-determined |
| 18-104_k141_106348 | Not-determined |
| 18-104_k141_36257 | Low-quality |
| 18-104_k141_57315 | Not-determined |
| 18-104_k141_113189 | Not-determined |
| 18-104_k141_57319 | Not-determined |
| 18-104_k141_57325 | Not-determined |
| 18-104_k141_133954 | Not-determined |
| 18-104_k141_57327 | Not-determined |
| 18-104_k141_154891 | Not-determined |
| 18-104_k141_43415 | Low-quality |
| 18-104_k141_57329 | Low-quality |
| 18-104_k141_64340 | Not-determined |
| 18-104_k141_141132 | Not-determined |
| 18-104_k141_43420 | Not-determined |
| 18-104_k141_85370 | Not-determined |
| 18-104_k141_29707 | Not-determined |
| 18-104_k141_29708 | Not-determined |
| 18-104_k141_64346 | Not-determined |
| 18-104_k141_15419 | Not-determined |
| 18-104_k141_29710 | Not-determined |
| 18-104_k141_161678 | Not-determined |
| 18-104_k141_29711 | Not-determined |
| 18-104_k141_85375 | Not-determined |
| 18-104_k141_147894 | Not-determined |
| 18-104_k141_8547 | Not-determined |
| 18-104_k141_85377 | Not-determined |
| 18-104_k141_43431 | Not-determined |
| 18-104_k141_141149 | Not-determined |
| 18-104_k141_147902 | Not-determined |
| 18-104_k141_36279 | Not-determined |
| 18-104_k141_71398 | Not-determined |
| 18-104_k141_141155 | Not-determined |
| 18-104_k141_161689 | Not-determined |
| 18-104_k141_50016 | Not-determined |
| 18-104_k141_8565 | Not-determined |
| 18-104_k141_71410 | Not-determined |
| 18-104_k141_64380 | Not-determined |
| 18-104_k141_57347 | Not-determined |
| 18-104_k141_161698 | Not-determined |
| 18-104_k141_57350 | Low-quality |
| 18-104_k141_106366 | Not-determined |
| 18-104_k141_64387 | Not-determined |
| 18-104_k141_15443 | Not-determined |
| 18-104_k141_133972 | Not-determined |
| 18-104_k141_161709 | Not-determined |
| 18-104_k141_64402 | Not-determined |
| 18-104_k141_141158 | Not-determined |
| 18-104_k141_57361 | Not-determined |
| 18-104_k141_133978 | Not-determined |
| 18-104_k141_154912 | Not-determined |
| 18-104_k141_57366 | Not-determined |
| 18-104_k141_106369 | Not-determined |
| 18-104_k141_15455 | Not-determined |
| 18-104_k141_133984 | Not-determined |
| 18-104_k141_154919 | Not-determined |
| 18-104_k141_29748 | Not-determined |
| 18-104_k141_133990 | Not-determined |
| 18-104_k141_154921 | Not-determined |
| 18-104_k141_147914 | Not-determined |
| 18-104_k141_99208 | Low-quality |
| 18-104_k141_154924 | Not-determined |
| 18-104_k141_147917 | Not-determined |
| 18-104_k141_154928 | Low-quality |
| 18-104_k141_57371 | Not-determined |
| 18-104_k141_99212 | Not-determined |
| 18-104_k141_36293 | Low-quality |
| 18-104_k141_57373 | Not-determined |
| 18-104_k141_64419 | Low-quality |
| 18-104_k141_161727 | Not-determined |
| 18-104_k141_154935 | Low-quality |
| 18-104_k141_127307 | Not-determined |
| 18-104_k141_57378 | Not-determined |
| 18-104_k141_29754 | Not-determined |
| 18-104_k141_154941 | Not-determined |
| 18-104_k141_29761 | Not-determined |
| 18-104_k141_43451 | Not-determined |
| 18-104_k141_64431 | Not-determined |
| 18-104_k141_99221 | Not-determined |
| 18-104_k141_92438 | Not-determined |
| 18-104_k141_85429 | Not-determined |
| 18-104_k141_106385 | Not-determined |
| 18-104_k141_1791 | Low-quality |
| 18-104_k141_92443 | Not-determined |
| 18-104_k141_29768 | Not-determined |
| 18-104_k141_57396 | Not-determined |
| 18-104_k141_85430 | Not-determined |
| 18-104_k141_36309 | Not-determined |
| 18-104_k141_106387 | Not-determined |
| 18-104_k141_1797 | Not-determined |
| 18-104_k141_127326 | Low-quality |
| 18-104_k141_99229 | Low-quality |
| 18-104_k141_141193 | Not-determined |
| 18-104_k141_57398 | Not-determined |
| 18-104_k141_64439 | Not-determined |
| 18-104_k141_85436 | Not-determined |
| 18-104_k141_85438 | Not-determined |
| 18-104_k141_161749 | Not-determined |
| 18-104_k141_154953 | Not-determined |
| 18-104_k141_113250 | Low-quality |
| 18-104_k141_85442 | Not-determined |
| 18-104_k141_50036 | Not-determined |
| 18-104_k141_50037 | Not-determined |
| 18-104_k141_36323 | Not-determined |
| 18-104_k141_127342 | Not-determined |
| 18-104_k141_127344 | Not-determined |
| 18-104_k141_8591 | Low-quality |
| 18-104_k141_127346 | Not-determined |
| 18-104_k141_154956 | Not-determined |
| 18-104_k141_127348 | Not-determined |
| 18-104_k141_8594 | Not-determined |
| 18-104_k141_8595 | Not-determined |
| 18-104_k141_22354 | Low-quality |
| 18-104_k141_8597 | Not-determined |
| 18-104_k141_29784 | Not-determined |
| 18-104_k141_78554 | Not-determined |
| 18-104_k141_147945 | Not-determined |
| 18-104_k141_78562 | Not-determined |
| 18-104_k141_141215 | Not-determined |
| 18-104_k141_71440 | Not-determined |
| 18-104_k141_43460 | Not-determined |
| 18-104_k141_99248 | Not-determined |
| 18-104_k141_141219 | Not-determined |
| 18-104_k141_106412 | Not-determined |
| 18-104_k141_22359 | Not-determined |
| 18-104_k141_29798 | Not-determined |
| 18-104_k141_147954 | Low-quality |
| 18-104_k141_154964 | Not-determined |
| 18-104_k141_8611 | Not-determined |
| 18-104_k141_64461 | Not-determined |
| 18-104_k141_154968 | Not-determined |
| 18-104_k141_134035 | Not-determined |
| 18-104_k141_154970 | Not-determined |
| 18-104_k141_71455 | Not-determined |
| 18-104_k141_29802 | Not-determined |
| 18-104_k141_141223 | Low-quality |
| 18-104_k141_134036 | Not-determined |
| 18-104_k141_29803 | Not-determined |
| 18-104_k141_43470 | Not-determined |
| 18-104_k141_78571 | Not-determined |
| 18-104_k141_8613 | Low-quality |
| 18-104_k141_8614 | Not-determined |
| 18-104_k141_134040 | Not-determined |
| 18-104_k141_141228 | Not-determined |
| 18-104_k141_134044 | Not-determined |
| 18-104_k141_161777 | Low-quality |
| 18-104_k141_71461 | Not-determined |
| 18-104_k141_43478 | Not-determined |
| 18-104_k141_106419 | Not-determined |
| 18-104_k141_134046 | Not-determined |
| 18-104_k141_36347 | Low-quality |
| 18-104_k141_92467 | Not-determined |
| 18-104_k141_29814 | Not-determined |
| 18-104_k141_147962 | Not-determined |
| 18-104_k141_154984 | Not-determined |
| 18-104_k141_50054 | Not-determined |
| 18-104_k141_1834 | Not-determined |
| 18-104_k141_99256 | Not-determined |
| 18-104_k141_127361 | Not-determined |
| 18-104_k141_64470 | Not-determined |
| 18-104_k141_8622 | Low-quality |
| 18-104_k141_15468 | Not-determined |
| 18-104_k141_36353 | Low-quality |
| 18-104_k141_113276 | Not-determined |
| 18-104_k141_64475 | Not-determined |
| 18-104_k141_36354 | Not-determined |
| 18-104_k141_113278 | Not-determined |
| 18-104_k141_154990 | Not-determined |
| 18-104_k141_99262 | Not-determined |
| 18-104_k141_1840 | Not-determined |
| 18-104_k141_71482 | Low-quality |
| 18-104_k141_78590 | Not-determined |
| 18-104_k141_71483 | Not-determined |
| 18-104_k141_8625 | Not-determined |
| 18-104_k141_127364 | Not-determined |
| 18-104_k141_154994 | Low-quality |
| 18-104_k141_92486 | Not-determined |
| 18-104_k141_161800 | Not-determined |
| 18-104_k141_127366 | Not-determined |
| 18-104_k141_1847 | Not-determined |
| 18-104_k141_64485 | Not-determined |
| 18-104_k141_113285 | Low-quality |
| 18-104_k141_1849 | Not-determined |
| 18-104_k141_8628 | Not-determined |
| 18-104_k141_71491 | Not-determined |
| 18-104_k141_113288 | Not-determined |
| 18-104_k141_43502 | Not-determined |
| 18-104_k141_78595 | Not-determined |
| 18-104_k141_155003 | Not-determined |
| 18-104_k141_92497 | Not-determined |
| 18-104_k141_57432 | Not-determined |
| 18-104_k141_85453 | Not-determined |
| 18-104_k141_127381 | Not-determined |
| 18-104_k141_78598 | Not-determined |
| 18-104_k141_85454 | Not-determined |
| 18-104_k141_43511 | Not-determined |
| 18-104_k141_155005 | Low-quality |
| 18-104_k141_155008 | Not-determined |
| 18-104_k141_106447 | Not-determined |
| 18-104_k141_155009 | Not-determined |
| 18-104_k141_99281 | Not-determined |
| 18-104_k141_127386 | Low-quality |
| 18-104_k141_85460 | Not-determined |
| 18-104_k141_85461 | Not-determined |
| 18-104_k141_64493 | Not-determined |
| 18-104_k141_15492 | Not-determined |
| 18-104_k141_106452 | Not-determined |
| 18-104_k141_99286 | Not-determined |
| 18-104_k141_22396 | Low-quality |
| 18-104_k141_15495 | Not-determined |
| 18-104_k141_113298 | Not-determined |
| 18-104_k141_113300 | Not-determined |
| 18-104_k141_15498 | Not-determined |
| 18-104_k141_15499 | Not-determined |
| 18-104_k141_50102 | Not-determined |
| 18-104_k141_15502 | Not-determined |
| 18-104_k141_85472 | Not-determined |
| 18-104_k141_43532 | Not-determined |
| 18-104_k141_22401 | Not-determined |
| 18-104_k141_1859 | Not-determined |
| 18-104_k141_155029 | Not-determined |
| 18-104_k141_155030 | Not-determined |
| 18-104_k141_147979 | Not-determined |
| 18-104_k141_43534 | Not-determined |
| 18-104_k141_1861 | Not-determined |
| 18-104_k141_99301 | Not-determined |
| 18-104_k141_43542 | Not-determined |
| 18-104_k141_147981 | Not-determined |
| 18-104_k141_155038 | Not-determined |
| 18-104_k141_155039 | Not-determined |
| 18-104_k141_50117 | Low-quality |
| 18-104_k141_29842 | Not-determined |
| 18-104_k141_15516 | Not-determined |
| 18-104_k141_106472 | Not-determined |
| 18-104_k141_64508 | Not-determined |
| 18-104_k141_71498 | Not-determined |
| 18-104_k141_155047 | Not-determined |
| 18-104_k141_155048 | Not-determined |
| 18-104_k141_92516 | Not-determined |
| 18-104_k141_106474 | Not-determined |
| 18-104_k141_22424 | Not-determined |
| 18-104_k141_134120 | Not-determined |
| 18-104_k141_92524 | Not-determined |
| 18-104_k141_64518 | Not-determined |
| 18-104_k141_64519 | Not-determined |
| 18-104_k141_147989 | Not-determined |
| 18-104_k141_1867 | Not-determined |
| 18-104_k141_147993 | Not-determined |
| 18-104_k141_147994 | Not-determined |
| 18-104_k141_161860 | Low-quality |
| 18-104_k141_134127 | Not-determined |
| 18-104_k141_57459 | Not-determined |
| 18-104_k141_161862 | Not-determined |
| 18-104_k141_85497 | Not-determined |
| 18-104_k141_85498 | Not-determined |
| 18-104_k141_50125 | Not-determined |
| 18-104_k141_148003 | Not-determined |
| 18-104_k141_57464 | Not-determined |
| 18-104_k141_1878 | Not-determined |
| 18-104_k141_50130 | Not-determined |
| 18-104_k141_64531 | Not-determined |
| 18-104_k141_161872 | Not-determined |
| 18-104_k141_1883 | Not-determined |
| 18-104_k141_57474 | Not-determined |
| 18-104_k141_64533 | Not-determined |
| 18-104_k141_22445 | Not-determined |
| 18-104_k141_50136 | Low-quality |
| 18-104_k141_64534 | Not-determined |
| 18-104_k141_36398 | Not-determined |
| 18-104_k141_155075 | Not-determined |
| 18-104_k141_36399 | Low-quality |
| 18-104_k141_155078 | Not-determined |
| 18-104_k141_1892 | Not-determined |
| 18-104_k141_161877 | Low-quality |
| 18-104_k141_155080 | Not-determined |
| 18-104_k141_148010 | Not-determined |
| 18-104_k141_155081 | Not-determined |
| 18-104_k141_99321 | Not-determined |
| 18-104_k141_161882 | Not-determined |
| 18-104_k141_64541 | Not-determined |
| 18-104_k141_78640 | Not-determined |
| 18-104_k141_78642 | Not-determined |
| 18-104_k141_127427 | Not-determined |
| 18-104_k141_99325 | Not-determined |
| 18-104_k141_161885 | Not-determined |
| 18-104_k141_99329 | Not-determined |
| 18-104_k141_134141 | Not-determined |
| 18-104_k141_8678 | Not-determined |
| 18-104_k141_127433 | Not-determined |
| 18-104_k141_8681 | Not-determined |
| 18-104_k141_57493 | Not-determined |
| 18-104_k141_50161 | Not-determined |
| 18-104_k141_141234 | Not-determined |
| 18-104_k141_85520 | Not-determined |
| 18-104_k141_99339 | Not-determined |
| 18-104_k141_148023 | Not-determined |
| 18-104_k141_134148 | Not-determined |
| 18-104_k141_134149 | Not-determined |
| 18-104_k141_29880 | Not-determined |
| 18-104_k141_29881 | Not-determined |
| 18-104_k141_155085 | Not-determined |
| 18-104_k141_36412 | Not-determined |
| 18-104_k141_57496 | Not-determined |
| 18-104_k141_50167 | Not-determined |
| 18-104_k141_1901 | Not-determined |
| 18-104_k141_113317 | Not-determined |
| 18-104_k141_1904 | Not-determined |
| 18-104_k141_50174 | Not-determined |
| 18-104_k141_29893 | Not-determined |
| 18-104_k141_78669 | Not-determined |
| 18-104_k141_15558 | Not-determined |
| 18-104_k141_8696 | Not-determined |
| 18-104_k141_29900 | Not-determined |
| 18-104_k141_71505 | Not-determined |
| 18-104_k141_92533 | Not-determined |
| 18-104_k141_141244 | Not-determined |
| 18-104_k141_22468 | Low-quality |
| 18-104_k141_29905 | Not-determined |
| 18-104_k141_141246 | Not-determined |
| 18-104_k141_22471 | Not-determined |
| 18-104_k141_113326 | Not-determined |
| 18-104_k141_29909 | Not-determined |
| 18-104_k141_43587 | Not-determined |
| 18-104_k141_78680 | Low-quality |
| 18-104_k141_71513 | Not-determined |
| 18-104_k141_113333 | Not-determined |
| 18-104_k141_71514 | Not-determined |
| 18-104_k141_127457 | Not-determined |
| 18-104_k141_127458 | Not-determined |
| 18-104_k141_106487 | Not-determined |
| 18-104_k141_29918 | Not-determined |
| 18-104_k141_71519 | Not-determined |
| 18-104_k141_29919 | Not-determined |
| 18-104_k141_92553 | Not-determined |
| 18-104_k141_127462 | Not-determined |
| 18-104_k141_36432 | Not-determined |
| 18-104_k141_15579 | Not-determined |
| 18-104_k141_141260 | Not-determined |
| 18-104_k141_29922 | Not-determined |
| 18-104_k141_29926 | Not-determined |
| 18-104_k141_64550 | Not-determined |
| 18-104_k141_113340 | Not-determined |
| 18-104_k141_106500 | Not-determined |
| 18-104_k141_155103 | Low-quality |
| 18-104_k141_71529 | Not-determined |
| 18-104_k141_106501 | Not-determined |
| 18-104_k141_161921 | Not-determined |
| 18-104_k141_22485 | Not-determined |
| 18-104_k141_155107 | Not-determined |
| 18-104_k141_148051 | Not-determined |
| 18-104_k141_85527 | Not-determined |
| 18-104_k141_43606 | Not-determined |
| 18-104_k141_141268 | Not-determined |
| 18-104_k141_64558 | Not-determined |
| 18-104_k141_99357 | Not-determined |
| 18-104_k141_71531 | Low-quality |
| 18-104_k141_141271 | Not-determined |
| 18-104_k141_106505 | Not-determined |
| 18-104_k141_92571 | Not-determined |
| 18-104_k141_78719 | Not-determined |
| 18-104_k141_43619 | Not-determined |
| 18-104_k141_92576 | Not-determined |
| 18-104_k141_127467 | Not-determined |
| 18-104_k141_22500 | Not-determined |
| 18-104_k141_161946 | Not-determined |
| 18-104_k141_155108 | Not-determined |
| 18-104_k141_155109 | Low-quality |
| 18-104_k141_106515 | Not-determined |
| 18-104_k141_141284 | Not-determined |
| 18-104_k141_92580 | Not-determined |
| 18-104_k141_148063 | Not-determined |
| 18-104_k141_50187 | Not-determined |
| 18-104_k141_78722 | Not-determined |
| 18-104_k141_8729 | Not-determined |
| 18-104_k141_50191 | Not-determined |
| 18-104_k141_148065 | Not-determined |
| 18-104_k141_106522 | Not-determined |
| 18-104_k141_43635 | Not-determined |
| 18-104_k141_127473 | Not-determined |
| 18-104_k141_43636 | Not-determined |
| 18-104_k141_43637 | Not-determined |
| 18-104_k141_71540 | Not-determined |
| 18-104_k141_15602 | Not-determined |
| 18-104_k141_106529 | Not-determined |
| 18-104_k141_113346 | Not-determined |
| 18-104_k141_92589 | Low-quality |
| 18-104_k141_36449 | Not-determined |
| 18-104_k141_78729 | Not-determined |
| 18-104_k141_22507 | Not-determined |
| 18-104_k141_29932 | Not-determined |
| 18-104_k141_29933 | Not-determined |
| 18-104_k141_106531 | Not-determined |
| 18-104_k141_43646 | Not-determined |
| 18-104_k141_36453 | Not-determined |
| 18-104_k141_99379 | Not-determined |
| 18-104_k141_57553 | Not-determined |
| 18-104_k141_148077 | Not-determined |
| 18-104_k141_36454 | Not-determined |
| 18-104_k141_64584 | Not-determined |
| 18-104_k141_50207 | Low-quality |
| 18-104_k141_29940 | Not-determined |
| 18-104_k141_1928 | Not-determined |
| 18-104_k141_36458 | Not-determined |

| | |
|---|---|
| 18-104_k141_1930 | Not-determined |
| 18-104_k141_113353 | Not-determined |
| 18-104_k141_78741 | Not-determined |
| 18-104_k141_22521 | Not-determined |
| 18-104_k141_99387 | Not-determined |
| 18-104_k141_92602 | Not-determined |
| 18-104_k141_43657 | Not-determined |
| 18-104_k141_161956 | Not-determined |
| 18-104_k141_36461 | Not-determined |
| 18-104_k141_78744 | Not-determined |
| 18-104_k141_15617 | Not-determined |
| 18-104_k141_8748 | Low-quality |
| 18-104_k141_8750 | Not-determined |
| 18-104_k141_161963 | Low-quality |
| 18-104_k141_85547 | Not-determined |
| 18-104_k141_106557 | Not-determined |
| 18-104_k141_15629 | Not-determined |
| 18-104_k141_78751 | Not-determined |
| 18-104_k141_106561 | Not-determined |
| 18-104_k141_85551 | Not-determined |
| 18-104_k141_99394 | Not-determined |
| 18-104_k141_15632 | Not-determined |
| 18-104_k141_106565 | Low-quality |
| 18-104_k141_78754 | Low-quality |
| 18-104_k141_78755 | Not-determined |
| 18-104_k141_148089 | Not-determined |
| 18-104_k141_113377 | Not-determined |
| 18-104_k141_15636 | Not-determined |
| 18-104_k141_1951 | Not-determined |
| 18-104_k141_155136 | Not-determined |
| 18-104_k141_92621 | Low-quality |
| 18-104_k141_15640 | Not-determined |
| 18-104_k141_15641 | Not-determined |
| 18-104_k141_78760 | Not-determined |
| 18-104_k141_113382 | Not-determined |
| 18-104_k141_1954 | Low-quality |
| 18-104_k141_1955 | Not-determined |
| 18-104_k141_134211 | Not-determined |
| 18-104_k141_71557 | Not-determined |
| 18-104_k141_127497 | Not-determined |
| 18-104_k141_1957 | Low-quality |
| 18-104_k141_64620 | Not-determined |
| 18-104_k141_78767 | Not-determined |
| 18-104_k141_36468 | Not-determined |
| 18-104_k141_92626 | Low-quality |
| 18-104_k141_99413 | Not-determined |
| 18-104_k141_134228 | Not-determined |
| 18-104_k141_57576 | Not-determined |
| 18-104_k141_127504 | Not-determined |
| 18-104_k141_22573 | Not-determined |
| 18-104_k141_50243 | Not-determined |
| 18-104_k141_71575 | Not-determined |
| 18-104_k141_78775 | Not-determined |
| 18-104_k141_148118 | Not-determined |
| 18-104_k141_8766 | Not-determined |
| 18-104_k141_43687 | Not-determined |
| 18-104_k141_92636 | Not-determined |
| 18-104_k141_43691 | Not-determined |
| 18-104_k141_155152 | Not-determined |
| 18-104_k141_78785 | Not-determined |
| 18-104_k141_141296 | Not-determined |
| 18-104_k141_22580 | Not-determined |
| 18-104_k141_57589 | Not-determined |
| 18-104_k141_113400 | Low-quality |
| 18-104_k141_148127 | Not-determined |
| 18-104_k141_134236 | Not-determined |
| 18-104_k141_141300 | Not-determined |
| 18-104_k141_148130 | Not-determined |
| 18-104_k141_43701 | Not-determined |
| 18-104_k141_43702 | Not-determined |
| 18-104_k141_92642 | Not-determined |
| 18-104_k141_8776 | Not-determined |
| 18-104_k141_141308 | Not-determined |
| 18-104_k141_134242 | Not-determined |
| 18-104_k141_8779 | Not-determined |
| 18-104_k141_36486 | Not-determined |
| 18-104_k141_92650 | Low-quality |
| 18-104_k141_50250 | Not-determined |
| 18-104_k141_134247 | Not-determined |
| 18-104_k141_8784 | Not-determined |
| 18-104_k141_155164 | Not-determined |
| 18-104_k141_155165 | Not-determined |
| 18-104_k141_161990 | Not-determined |
| 18-104_k141_127525 | Not-determined |
| 18-104_k141_36492 | Not-determined |
| 18-104_k141_8788 | Low-quality |
| 18-104_k141_71596 | Not-determined |
| 18-104_k141_155168 | Not-determined |
| 18-104_k141_71597 | Not-determined |
| 18-104_k141_161993 | Not-determined |
| 18-104_k141_141314 | Not-determined |
| 18-104_k141_134254 | Not-determined |
| 18-104_k141_8792 | Low-quality |
| 18-104_k141_127531 | Not-determined |
| 18-104_k141_36501 | Not-determined |
| 18-104_k141_50255 | Low-quality |
| 18-104_k141_36502 | Not-determined |
| 18-104_k141_36503 | Not-determined |
| 18-104_k141_99439 | Not-determined |
| 18-104_k141_85578 | Not-determined |
| 18-104_k141_15670 | Not-determined |
| 18-104_k141_134259 | Not-determined |
| 18-104_k141_85580 | Not-determined |
| 18-104_k141_162003 | Not-determined |
| 18-104_k141_43717 | Not-determined |
| 18-104_k141_8794 | Not-determined |
| 18-104_k141_148142 | Not-determined |
| 18-104_k141_36507 | Not-determined |
| 18-104_k141_106593 | Not-determined |
| 18-104_k141_29997 | Low-quality |
| 18-104_k141_15678 | Not-determined |
| 18-104_k141_15679 | Not-determined |
| 18-104_k141_78799 | Not-determined |
| 18-104_k141_78801 | Not-determined |
| 18-104_k141_141322 | Not-determined |
| 18-104_k141_99446 | Low-quality |
| 18-104_k141_141324 | Not-determined |
| 18-104_k141_141326 | Not-determined |
| 18-104_k141_43721 | Low-quality |
| 18-104_k141_30006 | Not-determined |
| 18-104_k141_30007 | Not-determined |
| 18-104_k141_141331 | Not-determined |
| 18-104_k141_43725 | Not-determined |
| 18-104_k141_43728 | Not-determined |
| 18-104_k141_127544 | Not-determined |
| 18-104_k141_99458 | Low-quality |
| 18-104_k141_43729 | Low-quality |
| 18-104_k141_148147 | Not-determined |
| 18-104_k141_50267 | Low-quality |
| 18-104_k141_2008 | Not-determined |
| 18-104_k141_43733 | Not-determined |
| 18-104_k141_78814 | Not-determined |
| 18-104_k141_162017 | Not-determined |
| 18-104_k141_78819 | Not-determined |
| 18-104_k141_71619 | Not-determined |
| 18-104_k141_15692 | Not-determined |
| 18-104_k141_148152 | Low-quality |
| 18-104_k141_57614 | Not-determined |
| 18-104_k141_43747 | Not-determined |
| 18-104_k141_50278 | Not-determined |
| 18-104_k141_148155 | Not-determined |
| 18-104_k141_134281 | Not-determined |
| 18-104_k141_50279 | Not-determined |
| 18-104_k141_50280 | Not-determined |
| 18-104_k141_15697 | Not-determined |
| 18-104_k141_15700 | Not-determined |
| 18-104_k141_148160 | Not-determined |
| 18-104_k141_148161 | Not-determined |
| 18-104_k141_134289 | Not-determined |
| 18-104_k141_134290 | Not-determined |
| 18-104_k141_127550 | Not-determined |
| 18-104_k141_22614 | Not-determined |
| 18-104_k141_127555 | Low-quality |
| 18-104_k141_162034 | Not-determined |
| 18-104_k141_162035 | Not-determined |
| 18-104_k141_30030 | Not-determined |
| 18-104_k141_71637 | Not-determined |
| 18-104_k141_148171 | Not-determined |
| 18-104_k141_30032 | Not-determined |
| 18-104_k141_148173 | Not-determined |
| 18-104_k141_22621 | Low-quality |
| 18-104_k141_30038 | Not-determined |
| 18-104_k141_141336 | Low-quality |
| 18-104_k141_50295 | Not-determined |
| 18-104_k141_78842 | Not-determined |
| 18-104_k141_141340 | Not-determined |
| 18-104_k141_30048 | Not-determined |
| 18-104_k141_15716 | Not-determined |
| 18-104_k141_141348 | Not-determined |
| 18-104_k141_155187 | Not-determined |
| 18-104_k141_141354 | Not-determined |
| 18-104_k141_15723 | Not-determined |
| 18-104_k141_64633 | Not-determined |
| 18-104_k141_127568 | Not-determined |
| 18-104_k141_127571 | Not-determined |
| 18-104_k141_78855 | Not-determined |
| 18-104_k141_36537 | Not-determined |
| 18-104_k141_127575 | Not-determined |
| 18-104_k141_30053 | Not-determined |
| 18-104_k141_64642 | Low-quality |
| 18-104_k141_155197 | Not-determined |
| 18-104_k141_15730 | Not-determined |
| 18-104_k141_155199 | Not-determined |
| 18-104_k141_8833 | Not-determined |
| 18-104_k141_15731 | Low-quality |
| 18-104_k141_71654 | Not-determined |
| 18-104_k141_127578 | Not-determined |
| 18-104_k141_127579 | Not-determined |
| 18-104_k141_134318 | Not-determined |
| 18-104_k141_71660 | Not-determined |
| 18-104_k141_8838 | Low-quality |
| 18-104_k141_148182 | Not-determined |
| 18-104_k141_71662 | Not-determined |
| 18-104_k141_2038 | Not-determined |
| 18-104_k141_127587 | Not-determined |
| 18-104_k141_148188 | Not-determined |
| 18-104_k141_148189 | Not-determined |
| 18-104_k141_106604 | Low-quality |
| 18-104_k141_78869 | Not-determined |
| 18-104_k141_71671 | Not-determined |
| 18-104_k141_106610 | Not-determined |
| 18-104_k141_106613 | Not-determined |
| 18-104_k141_15740 | Not-determined |
| 18-104_k141_43785 | Not-determined |
| 18-104_k141_43786 | Not-determined |
| 18-104_k141_50335 | Not-determined |
| 18-104_k141_148209 | Not-determined |
| 18-104_k141_127594 | Not-determined |
| 18-104_k141_36554 | Not-determined |
| 18-104_k141_15746 | Not-determined |
| 18-104_k141_36557 | Not-determined |
| 18-104_k141_92735 | Not-determined |
| 18-104_k141_43789 | Not-determined |
| 18-104_k141_127596 | Low-quality |
| 18-104_k141_22640 | Not-determined |
| 18-104_k141_162105 | Not-determined |
| 18-104_k141_85658 | Not-determined |
| 18-104_k141_57629 | Not-determined |
| 18-104_k141_22642 | Low-quality |
| 18-104_k141_148222 | Not-determined |
| 18-104_k141_8844 | Not-determined |
| 18-104_k141_134326 | Not-determined |
| 18-104_k141_141363 | Not-determined |
| 18-104_k141_36565 | Not-determined |
| 18-104_k141_22650 | Not-determined |
| 18-104_k141_155204 | Not-determined |
| 18-104_k141_22652 | Not-determined |
| 18-104_k141_141369 | Not-determined |
| 18-104_k141_15763 | Not-determined |
| 18-104_k141_36573 | Not-determined |
| 18-104_k141_148234 | Not-determined |
| 18-104_k141_155209 | Not-determined |
| 18-104_k141_22666 | Not-determined |
| 18-104_k141_85664 | Not-determined |
| 18-104_k141_127612 | Not-determined |
| 18-104_k141_92764 | Low-quality |
| 18-104_k141_127613 | Not-determined |
| 18-104_k141_57635 | Not-determined |
| 18-104_k141_78893 | Not-determined |
| 18-104_k141_85667 | Not-determined |
| 18-104_k141_155213 | Not-determined |
| 18-104_k141_92770 | Not-determined |
| 18-104_k141_15778 | Not-determined |
| 18-104_k141_50362 | Not-determined |
| 18-104_k141_99514 | Low-quality |
| 18-104_k141_148245 | Not-determined |
| 18-104_k141_99516 | Low-quality |
| 18-104_k141_155217 | Not-determined |
| 18-104_k141_8865 | Not-determined |
| 18-104_k141_99518 | Not-determined |
| 18-104_k141_99520 | Not-determined |
| 18-104_k141_2084 | Not-determined |
| 18-104_k141_92787 | Not-determined |
| 18-104_k141_50372 | Not-determined |
| 18-104_k141_127623 | Low-quality |
| 18-104_k141_78907 | Not-determined |
| 18-104_k141_127624 | Low-quality |
| 18-104_k141_127625 | Not-determined |
| 18-104_k141_127629 | Not-determined |
| 18-104_k141_106649 | Not-determined |
| 18-104_k141_134354 | Low-quality |
| 18-104_k141_71693 | Not-determined |
| 18-104_k141_134355 | Not-determined |
| 18-104_k141_22693 | Not-determined |
| 18-104_k141_134361 | Not-determined |
| 18-104_k141_2086 | Not-determined |
| 18-104_k141_2089 | Not-determined |
| 18-104_k141_71703 | Not-determined |
| 18-104_k141_78929 | Not-determined |
| 18-104_k141_141378 | Not-determined |
| 18-104_k141_78933 | Not-determined |
| 18-104_k141_155240 | Not-determined |
| 18-104_k141_64689 | Not-determined |
| 18-104_k141_155245 | Low-quality |
| 18-104_k141_71713 | Low-quality |
| 18-104_k141_162148 | Not-determined |
| 18-104_k141_30073 | Not-determined |
| 18-104_k141_134381 | Not-determined |
| 18-104_k141_141389 | Not-determined |
| 18-104_k141_162150 | Low-quality |
| 18-104_k141_155248 | Low-quality |
| 18-104_k141_64694 | Not-determined |
| 18-104_k141_127656 | Not-determined |
| 18-104_k141_2103 | Not-determined |
| 18-104_k141_134384 | Not-determined |
| 18-104_k141_30078 | Not-determined |
| 18-104_k141_50405 | Not-determined |
| 18-104_k141_71719 | Not-determined |
| 18-104_k141_50406 | Not-determined |
| 18-104_k141_50407 | Not-determined |
| 18-104_k141_15809 | Not-determined |
| 18-104_k141_162157 | Not-determined |
| 18-104_k141_127659 | Not-determined |
| 18-104_k141_155254 | Not-determined |
| 18-104_k141_162158 | Not-determined |
| 18-104_k141_127662 | Not-determined |
| 18-104_k141_162159 | Low-quality |
| 18-104_k141_22725 | Not-determined |
| 18-104_k141_85706 | Not-determined |
| 18-104_k141_148264 | Low-quality |
| 18-104_k141_155264 | Not-determined |
| 18-104_k141_127669 | Not-determined |
| 18-104_k141_15817 | Not-determined |
| 18-104_k141_162162 | Not-determined |
| 18-104_k141_64710 | Not-determined |
| 18-104_k141_155272 | Not-determined |
| 18-104_k141_71737 | Not-determined |
| 18-104_k141_119952 | Low-quality |
| 18-104_k141_57659 | Not-determined |
| 18-104_k141_78953 | Not-determined |
| 18-104_k141_15829 | Low-quality |
| 18-104_k141_78956 | Not-determined |
| 18-104_k141_155275 | Not-determined |
| 18-104_k141_43810 | Not-determined |
| 18-104_k141_134406 | Not-determined |
| 18-104_k141_71746 | Not-determined |
| 18-104_k141_134410 | Not-determined |
| 18-104_k141_155277 | Not-determined |
| 18-104_k141_36600 | Not-determined |
| 18-104_k141_22735 | Not-determined |
| 18-104_k141_43814 | Not-determined |
| 18-104_k141_92813 | Not-determined |
| 18-104_k141_113425 | Not-determined |
| 18-104_k141_8911 | Low-quality |
| 18-104_k141_8912 | Not-determined |
| 18-104_k141_148284 | Not-determined |
| 18-104_k141_22737 | Low-quality |
| 18-104_k141_43822 | Not-determined |
| 18-104_k141_36610 | Not-determined |
| 18-104_k141_36612 | Not-determined |
| 18-104_k141_85731 | Not-determined |
| 18-104_k141_78962 | Not-determined |
| 18-104_k141_43829 | Low-quality |
| 18-104_k141_92824 | Not-determined |
| 18-104_k141_119961 | Not-determined |
| 18-104_k141_141419 | Not-determined |
| 18-104_k141_22745 | Not-determined |
| 18-104_k141_127693 | Not-determined |
| 18-104_k141_36628 | Not-determined |
| 18-104_k141_50428 | Low-quality |
| 18-104_k141_141424 | Not-determined |
| 18-104_k141_50431 | Not-determined |
| 18-104_k141_134429 | Low-quality |
| 18-104_k141_119969 | Not-determined |
| 18-104_k141_50437 | Not-determined |
| 18-104_k141_50438 | Not-determined |
| 18-104_k141_85752 | Not-determined |
| 18-104_k141_15857 | Not-determined |
| 18-104_k141_119970 | Not-determined |
| 18-104_k141_141432 | Not-determined |
| 18-104_k141_57680 | Low-quality |
| 18-104_k141_64747 | Not-determined |
| 18-104_k141_92841 | Not-determined |
| 18-104_k141_113443 | Low-quality |
| 18-104_k141_15863 | Not-determined |
| 18-104_k141_113447 | Low-quality |
| 18-104_k141_141440 | Not-determined |
| 18-104_k141_57689 | Not-determined |
| 18-104_k141_22752 | Not-determined |
| 18-104_k141_113452 | Low-quality |
| 18-104_k141_92847 | Not-determined |
| 18-104_k141_106724 | Not-determined |
| 18-104_k141_162203 | Not-determined |
| 18-104_k141_113459 | Not-determined |
| 18-104_k141_106730 | Not-determined |
| 18-104_k141_162211 | Not-determined |
| 18-104_k141_22763 | Not-determined |
| 18-104_k141_8959 | Not-determined |
| 18-104_k141_162214 | Not-determined |
| 18-104_k141_155323 | Not-determined |
| 18-104_k141_141454 | Not-determined |
| 18-104_k141_57707 | Not-determined |
| 18-104_k141_43852 | Not-determined |
| 18-104_k141_43857 | Not-determined |
| 18-104_k141_148305 | Not-determined |
| 18-104_k141_30108 | Low-quality |
| 18-104_k141_162221 | Not-determined |
| 18-104_k141_141465 | Not-determined |
| 18-104_k141_85791 | Not-determined |
| 18-104_k141_134455 | Not-determined |
| 18-104_k141_22775 | Not-determined |
| 18-104_k141_22778 | Not-determined |
| 18-104_k141_22781 | Not-determined |
| 18-104_k141_36649 | Not-determined |
| 18-104_k141_43873 | Not-determined |
| 18-104_k141_92877 | Not-determined |
| 18-104_k141_43875 | Not-determined |
| 18-104_k141_92878 | Not-determined |
| 18-104_k141_155337 | Not-determined |
| 18-104_k141_141474 | Low-quality |
| 18-104_k141_119990 | Not-determined |
| 18-104_k141_57730 | Not-determined |
| 18-104_k141_113488 | Not-determined |
| 18-104_k141_2157 | Not-determined |
| 18-104_k141_50460 | Not-determined |
| 18-104_k141_43879 | Low-quality |
| 18-104_k141_22792 | Not-determined |
| 18-104_k141_15904 | Not-determined |
| 18-104_k141_92893 | Not-determined |
| 18-104_k141_50467 | Not-determined |
| 18-104_k141_134480 | Not-determined |
| 18-104_k141_43884 | Not-determined |
| 18-104_k141_162239 | Not-determined |
| 18-104_k141_148312 | Low-quality |
| 18-104_k141_106760 | Low-quality |
| 18-104_k141_141481 | Low-quality |
| 18-104_k141_79024 | Not-determined |
| 18-104_k141_50471 | Not-determined |
| 18-104_k141_50472 | Not-determined |
| 18-104_k141_92900 | Low-quality |
| 18-104_k141_2169 | Not-determined |
| 18-104_k141_22801 | Not-determined |
| 18-104_k141_162254 | Not-determined |
| 18-104_k141_155355 | Not-determined |
| 18-104_k141_15919 | Not-determined |
| 18-104_k141_148330 | Not-determined |
| 18-104_k141_134487 | Not-determined |
| 18-104_k141_57743 | Not-determined |
| 18-104_k141_134491 | Not-determined |
| 18-104_k141_64773 | Not-determined |
| 18-104_k141_148335 | Not-determined |
| 18-104_k141_22813 | Not-determined |
| 18-104_k141_113506 | Not-determined |
| 18-104_k141_79042 | Not-determined |
| 18-104_k141_120024 | Not-determined |
| 18-104_k141_8984 | Not-determined |
| 18-104_k141_36661 | Not-determined |
| 18-104_k141_8988 | Not-determined |
| 18-104_k141_162262 | Not-determined |
| 18-104_k141_120030 | Low-quality |
| 18-104_k141_36666 | Not-determined |
| 18-104_k141_92908 | Not-determined |
| 18-104_k141_64785 | Not-determined |
| 18-104_k141_57759 | Not-determined |
| 18-104_k141_8996 | Not-determined |
| 18-104_k141_22824 | Not-determined |
| 18-104_k141_92913 | Not-determined |
| 18-104_k141_71761 | Not-determined |
| 18-104_k141_134505 | Low-quality |
| 18-104_k141_92915 | Not-determined |
| 18-104_k141_85830 | Not-determined |
| 18-104_k141_22829 | Not-determined |
| 18-104_k141_134510 | Not-determined |
| 18-104_k141_64794 | Not-determined |
| 18-104_k141_134514 | Not-determined |
| 18-104_k141_79066 | Not-determined |
| 18-104_k141_36687 | Not-determined |
| 18-104_k141_92923 | Not-determined |
| 18-104_k141_162278 | Not-determined |
| 18-104_k141_134516 | Not-determined |
| 18-104_k141_9001 | Not-determined |
| 18-104_k141_64802 | Not-determined |
| 18-104_k141_134522 | Not-determined |
| 18-104_k141_134527 | Not-determined |
| 18-104_k141_9012 | Not-determined |
| 18-104_k141_79082 | Not-determined |
| 18-104_k141_71781 | Not-determined |
| 18-104_k141_9016 | Not-determined |
| 18-104_k141_71782 | Not-determined |
| 18-104_k141_64810 | Not-determined |
| 18-104_k141_9025 | Not-determined |
| 18-104_k141_79091 | Not-determined |
| 18-104_k141_43916 | Low-quality |
| 18-104_k141_148364 | Not-determined |
| 18-104_k141_36708 | Not-determined |
| 18-104_k141_92953 | Not-determined |
| 18-104_k141_57795 | Not-determined |
| 18-104_k141_71784 | Not-determined |
| 18-104_k141_22861 | Not-determined |
| 18-104_k141_22864 | Not-determined |
| 18-104_k141_141503 | Not-determined |
| 18-104_k141_148369 | Low-quality |
| 18-104_k141_57806 | Not-determined |
| 18-104_k141_22874 | Not-determined |
| 18-104_k141_148379 | Not-determined |
| 18-104_k141_15961 | Not-determined |
| 18-104_k141_22877 | Not-determined |
| 18-104_k141_36726 | Not-determined |
| 18-104_k141_71792 | Low-quality |
| 18-104_k141_106769 | Not-determined |
| 18-104_k141_64825 | Not-determined |
| 18-104_k141_71798 | Not-determined |
| 18-104_k141_106773 | Not-determined |
| 18-104_k141_64828 | Not-determined |
| 18-104_k141_162307 | Not-determined |
| 18-104_k141_141518 | Not-determined |
| 18-104_k141_43927 | Not-determined |
| 18-104_k141_36727 | Not-determined |
| 18-104_k141_127745 | Not-determined |
| 18-104_k141_113516 | Not-determined |
| 18-104_k141_79109 | Not-determined |
| 18-104_k141_15976 | Not-determined |
| 18-104_k141_2212 | Not-determined |
| 18-104_k141_148393 | Not-determined |
| 18-104_k141_2215 | Not-determined |
| 18-104_k141_113523 | Low-quality |
| 18-104_k141_30119 | Not-determined |
| 18-104_k141_127752 | Not-determined |
| 18-104_k141_2231 | Not-determined |
| 18-104_k141_57836 | Not-determined |
| 18-104_k141_43944 | Not-determined |
| 18-104_k141_15999 | Not-determined |
| 18-104_k141_79116 | Not-determined |
| 18-104_k141_16002 | Not-determined |
| 18-104_k141_141532 | Not-determined |
| 18-104_k141_79118 | Low-quality |
| 18-104_k141_22907 | Not-determined |
| 18-104_k141_148412 | Not-determined |
| 18-104_k141_79120 | Not-determined |
| 18-104_k141_22908 | Not-determined |
| 18-104_k141_127761 | Not-determined |
| 18-104_k141_141536 | Not-determined |
| 18-104_k141_30122 | Not-determined |
| 18-104_k141_64844 | Not-determined |
| 18-104_k141_106798 | Not-determined |
| 18-104_k141_106799 | Not-determined |
| 18-104_k141_79129 | Not-determined |
| 18-104_k141_79131 | Not-determined |
| 18-104_k141_79132 | Not-determined |
| 18-104_k141_64850 | Not-determined |
| 18-104_k141_30127 | Not-determined |
| 18-104_k141_134559 | Not-determined |
| 18-104_k141_9054 | Low-quality |
| 18-104_k141_79141 | Not-determined |
| 18-104_k141_22918 | Low-quality |
| 18-104_k141_22919 | Not-determined |
| 18-104_k141_71810 | Low-quality |
| 18-104_k141_155411 | Not-determined |
| 18-104_k141_127765 | Low-quality |
| 18-104_k141_79146 | Not-determined |
| 18-104_k141_36745 | Not-determined |
| 18-104_k141_134573 | Not-determined |
| 18-104_k141_85885 | Not-determined |
| 18-104_k141_30137 | Not-determined |
| 18-104_k141_113560 | Not-determined |
| 18-104_k141_64859 | Not-determined |
| 18-104_k141_134583 | Not-determined |
| 17-109-final-viral-combined_k141_156667\|\|full | Medium-quality |
| 18-104_k141_134588 | Not-determined |
| 18-104_k141_50476 | Not-determined |
| 18-104_k141_148439 | Not-determined |
| 18-104_k141_50482 | Not-determined |
| 18-104_k141_134594 | Not-determined |
| 18-104_k141_71829 | Low-quality |
| 18-104_k141_9087 | Not-determined |
| 18-104_k141_36769 | Not-determined |
| 18-104_k141_120099 | Not-determined |
| 18-104_k141_43981 | Not-determined |
| 18-104_k141_36776 | Not-determined |
| 18-104_k141_134602 | Not-determined |
| 18-104_k141_93007 | Not-determined |
| 18-104_k141_162341 | Not-determined |
| 18-104_k141_93009 | Not-determined |
| 18-104_k141_30159 | Not-determined |
| 18-104_k141_71844 | Not-determined |
| 18-104_k141_43994 | Low-quality |
| 18-104_k141_93010 | Not-determined |
| 18-104_k141_22959 | Low-quality |
| 18-104_k141_43997 | Not-determined |
| 18-104_k141_134604 | Low-quality |
| 18-104_k141_43998 | Not-determined |
| 18-104_k141_162342 | Not-determined |
| 18-104_k141_162343 | Not-determined |
| 18-104_k141_93015 | Not-determined |
| 18-104_k141_106833 | Not-determined |
| 18-104_k141_120112 | Not-determined |
| 18-104_k141_148465 | Not-determined |
| 18-104_k141_141575 | Not-determined |
| 18-104_k141_120115 | Not-determined |
| 18-104_k141_71854 | Not-determined |
| 18-104_k141_71856 | Not-determined |
| 18-104_k141_36792 | Low-quality |
| 18-104_k141_9106 | Not-determined |
| 18-104_k141_148470 | Not-determined |
| 18-104_k141_9107 | Not-determined |
| 18-104_k141_44006 | Low-quality |
| 18-104_k141_127796 | Not-determined |
| 18-104_k141_71858 | Not-determined |
| 18-104_k141_113591 | Low-quality |
| 18-104_k141_127799 | Not-determined |
| 18-104_k141_127802 | Not-determined |
| 18-104_k141_64878 | Not-determined |
| 18-104_k141_155444 | Not-determined |
| 18-104_k141_36805 | Not-determined |
| 18-104_k141_155446 | Not-determined |
| 18-104_k141_30168 | Not-determined |
| 18-104_k141_106838 | Not-determined |
| 18-104_k141_36806 | Not-determined |
| 18-104_k141_30170 | Not-determined |

| | |
|---|---|
| 18-104_k141_155447 | Not-determined |
| 18-104_k141_16050 | Not-determined |
| 18-104_k141_44013 | Not-determined |
| 18-104_k141_71862 | Not-determined |
| 18-104_k141_113593 | Low-quality |
| 18-104_k141_79169 | Low-quality |
| 18-104_k141_16053 | Not-determined |
| 18-104_k141_127808 | Not-determined |
| 18-104_k141_120129 | Not-determined |
| 18-104_k141_57896 | Not-determined |
| 18-104_k141_64888 | Not-determined |
| 18-104_k141_9120 | Not-determined |
| 18-104_k141_30180 | Not-determined |
| 18-104_k141_93027 | Not-determined |
| 18-104_k141_106845 | Not-determined |
| 18-104_k141_155454 | Not-determined |
| 18-104_k141_9123 | Not-determined |
| 18-104_k141_127810 | Not-determined |
| 18-104_k141_30189 | Not-determined |
| 18-104_k141_85903 | Not-determined |
| 18-104_k141_134633 | Not-determined |
| 18-104_k141_85904 | Not-determined |
| 18-104_k141_134634 | Not-determined |
| 18-104_k141_64896 | Low-quality |
| 18-104_k141_57898 | Not-determined |
| 18-104_k141_64897 | Not-determined |
| 18-104_k141_93035 | Not-determined |
| 18-104_k141_30192 | Not-determined |
| 18-104_k141_106851 | Not-determined |
| 18-104_k141_93036 | Not-determined |
| 18-104_k141_30194 | Not-determined |
| 18-104_k141_141606 | Not-determined |
| 18-104_k141_113608 | Not-determined |
| 18-104_k141_127816 | Not-determined |
| 18-104_k141_9130 | Not-determined |
| 18-104_k141_155468 | Not-determined |
| 18-104_k141_64902 | Not-determined |
| 18-104_k141_64903 | Not-determined |
| 18-104_k141_57903 | Low-quality |
| 18-104_k141_148483 | Not-determined |
| 18-104_k141_79193 | Not-determined |
| 18-104_k141_9141 | Not-determined |
| 18-104_k141_127822 | Not-determined |
| 18-104_k141_127823 | Low-quality |
| 18-104_k141_79195 | Not-determined |
| 18-104_k141_113619 | Not-determined |
| 18-104_k141_22992 | Not-determined |
| 18-104_k141_30202 | Low-quality |
| 18-104_k141_155476 | Not-determined |
| 18-104_k141_148493 | Not-determined |
| 18-104_k141_127835 | Not-determined |
| 18-104_k141_9150 | Not-determined |
| 18-104_k141_93058 | Not-determined |
| 18-104_k141_44033 | Not-determined |
| 18-104_k141_93061 | Not-determined |
| 18-104_k141_134646 | Not-determined |
| 18-104_k141_9155 | Not-determined |
| 18-104_k141_50518 | Not-determined |
| 18-104_k141_50521 | Not-determined |
| 18-104_k141_23003 | Not-determined |
| 18-104_k141_30213 | Not-determined |
| 18-104_k141_23004 | Not-determined |
| 18-104_k141_50523 | Not-determined |
| 18-104_k141_93071 | Not-determined |
| 18-104_k141_120163 | Not-determined |
| 18-104_k141_93075 | Not-determined |
| 18-104_k141_93077 | Not-determined |
| 18-104_k141_79215 | Not-determined |
| 18-104_k141_155488 | Not-determined |
| 18-104_k141_9172 | Not-determined |
| 18-104_k141_113633 | Low-quality |
| 18-104_k141_50531 | Not-determined |
| 18-104_k141_120167 | Not-determined |
| 18-104_k141_120168 | Low-quality |
| 18-104_k141_79222 | Not-determined |
| 18-104_k141_64926 | Low-quality |
| 18-104_k141_120170 | Not-determined |
| 18-104_k141_155492 | Not-determined |
| 18-104_k141_57933 | Not-determined |
| 18-104_k141_44047 | Not-determined |
| 18-104_k141_44049 | Not-determined |
| 18-104_k141_50537 | Not-determined |
| 18-104_k141_106871 | Low-quality |
| 18-104_k141_134661 | Not-determined |
| 18-104_k141_106872 | Not-determined |
| 18-104_k141_120176 | Not-determined |
| 18-104_k141_36848 | Not-determined |
| 18-104_k141_85930 | Not-determined |
| 18-104_k141_64935 | Not-determined |
| 18-104_k141_85931 | Not-determined |
| 18-104_k141_50539 | Not-determined |
| 18-104_k141_36850 | Not-determined |
| 18-104_k141_120180 | Not-determined |
| 18-104_k141_93091 | Not-determined |
| 17-109-final-viral-combined_k141_210942\|\|full | High-quality |
| 18-104_k141_148503 | Not-determined |
| 18-104_k141_9181 | Not-determined |
| 18-104_k141_120184 | Not-determined |
| 18-104_k141_9185 | Not-determined |
| 18-104_k141_9186 | Not-determined |
| 18-104_k141_93094 | Not-determined |
| 18-104_k141_113660 | Not-determined |
| 18-104_k141_93095 | Not-determined |
| 18-104_k141_44058 | Not-determined |
| 18-104_k141_71910 | Not-determined |
| 18-104_k141_9190 | Not-determined |
| 18-104_k141_106880 | Low-quality |
| 18-104_k141_71913 | Not-determined |
| 18-104_k141_57954 | Low-quality |
| 18-104_k141_162377 | Not-determined |
| 18-104_k141_106881 | Not-determined |
| 18-104_k141_79239 | Not-determined |
| 18-104_k141_79240 | Not-determined |
| 18-104_k141_93102 | Not-determined |
| 18-104_k141_85938 | Not-determined |
| 18-104_k141_23025 | Not-determined |
| 18-104_k141_2243 | Low-quality |
| 18-104_k141_113666 | Not-determined |
| 18-104_k141_120188 | Not-determined |
| 18-104_k141_93103 | Low-quality |
| 18-104_k141_71916 | Low-quality |
| 18-104_k141_9203 | Not-determined |
| 18-104_k141_106894 | Not-determined |
| 18-104_k141_85945 | Not-determined |
| 18-104_k141_2247 | Low-quality |
| 18-104_k141_113678 | Not-determined |
| 18-104_k141_64957 | Not-determined |
| 18-104_k141_134671 | Not-determined |
| 18-104_k141_36880 | Not-determined |
| 18-104_k141_44083 | Not-determined |
| 18-104_k141_64963 | Not-determined |
| 18-104_k141_120192 | Not-determined |
| 18-104_k141_64966 | Not-determined |
| 18-104_k141_79246 | Not-determined |
| 18-104_k141_134679 | Low-quality |
| 18-104_k141_50557 | Not-determined |
| 18-104_k141_85961 | Not-determined |
| 18-104_k141_148538 | Not-determined |
| 18-104_k141_93106 | Low-quality |
| 18-104_k141_71939 | Not-determined |
| 18-104_k141_113692 | Not-determined |
| 18-104_k141_71942 | Not-determined |
| 18-104_k141_2269 | Not-determined |
| 18-104_k141_93112 | Not-determined |
| 18-104_k141_2270 | Not-determined |
| 18-104_k141_2272 | Not-determined |
| 18-104_k141_79263 | Not-determined |
| 18-104_k141_36898 | Not-determined |
| 18-104_k141_93117 | Not-determined |
| 18-104_k141_23068 | Not-determined |
| 18-104_k141_93120 | Not-determined |
| 18-104_k141_36901 | Not-determined |
| 18-104_k141_36902 | Not-determined |
| 18-104_k141_85976 | Not-determined |
| 18-104_k141_2273 | Low-quality |
| 18-104_k141_79272 | Not-determined |
| 18-104_k141_9236 | Not-determined |
| 18-104_k141_162416 | Not-determined |
| 18-104_k141_134700 | Low-quality |
| 18-104_k141_50583 | Not-determined |
| 18-104_k141_134701 | Not-determined |
| 18-104_k141_79278 | Not-determined |
| 18-104_k141_155526 | Not-determined |
| 18-104_k141_71951 | Not-determined |
| 18-104_k141_134703 | Not-determined |
| 18-104_k141_23083 | Not-determined |
| 18-104_k141_85987 | Low-quality |
| 18-104_k141_71956 | Not-determined |
| 18-104_k141_36905 | Not-determined |
| 18-104_k141_9252 | Not-determined |
| 18-104_k141_23088 | Low-quality |
| 18-104_k141_16109 | Not-determined |
| 18-104_k141_30245 | Not-determined |
| 18-104_k141_113713 | Not-determined |
| 18-104_k141_30246 | Not-determined |
| 18-104_k141_113716 | Not-determined |
| 18-104_k141_127852 | Not-determined |
| 18-104_k141_79293 | Not-determined |
| 18-104_k141_162430 | Not-determined |
| 18-104_k141_106917 | Not-determined |
| 18-104_k141_79300 | Not-determined |
| 18-104_k141_85992 | Not-determined |
| 18-104_k141_2296 | Not-determined |
| 18-104_k141_44116 | Not-determined |
| 18-104_k141_120212 | Not-determined |
| 18-104_k141_2299 | Not-determined |
| 18-104_k141_120214 | Not-determined |
| 18-104_k141_79306 | Not-determined |
| 18-104_k141_155548 | Not-determined |
| 18-104_k141_16119 | Not-determined |
| 18-104_k141_155550 | Not-determined |
| 18-104_k141_9269 | Not-determined |
| 18-104_k141_148554 | Not-determined |
| 18-104_k141_30259 | Not-determined |
| 18-104_k141_36929 | Not-determined |
| 18-104_k141_120225 | Not-determined |
| 18-104_k141_71979 | Not-determined |
| 18-104_k141_155557 | Not-determined |
| 18-104_k141_36932 | Not-determined |
| 18-104_k141_79311 | Not-determined |
| 18-104_k141_134718 | Not-determined |
| 18-104_k141_71981 | Not-determined |
| 18-104_k141_93141 | Not-determined |
| 18-104_k141_120228 | Low-quality |
| 18-104_k141_93143 | Not-determined |
| 18-104_k141_2310 | Not-determined |
| 18-104_k141_148558 | Not-determined |
| 18-104_k141_36936 | Not-determined |
| 18-104_k141_30268 | Not-determined |
| 18-104_k141_64999 | Not-determined |
| 18-104_k141_141665 | Not-determined |
| 18-104_k141_86005 | Not-determined |
| 18-104_k141_106937 | Not-determined |
| 18-104_k141_65001 | Not-determined |
| 18-104_k141_106939 | Not-determined |
| 18-104_k141_36941 | Not-determined |
| 18-104_k141_106942 | Low-quality |
| 18-104_k141_148567 | Not-determined |
| 18-104_k141_36947 | Low-quality |
| 18-104_k141_16140 | Not-determined |
| 18-104_k141_2329 | Not-determined |
| 18-104_k141_23103 | Not-determined |
| 18-104_k141_30285 | Not-determined |
| 18-104_k141_71998 | Not-determined |
| 18-104_k141_141682 | Not-determined |
| 18-104_k141_86032 | Not-determined |
| 18-104_k141_16147 | Not-determined |
| 18-104_k141_16149 | Not-determined |
| 18-104_k141_93147 | Not-determined |
| 18-104_k141_30292 | Not-determined |
| 18-104_k141_127857 | Not-determined |
| 18-104_k141_141683 | Not-determined |
| 18-104_k141_9290 | Not-determined |
| 18-104_k141_2343 | Not-determined |
| 18-104_k141_16162 | Not-determined |
| 18-104_k141_16163 | Not-determined |
| 18-104_k141_134740 | Not-determined |
| 18-104_k141_30296 | Low-quality |
| 18-104_k141_2349 | Not-determined |
| 18-104_k141_127862 | Not-determined |
| 18-104_k141_155567 | Not-determined |
| 18-104_k141_141697 | Not-determined |
| 18-104_k141_36976 | Not-determined |
| 18-104_k141_106961 | Low-quality |
| 18-104_k141_72020 | Not-determined |
| 18-104_k141_36980 | Not-determined |
| 18-104_k141_36983 | Low-quality |
| 18-104_k141_50588 | Not-determined |
| 18-104_k141_106972 | Not-determined |
| 18-104_k141_65029 | Not-determined |
| 18-104_k141_23112 | Low-quality |
| 18-104_k141_106976 | Not-determined |
| 18-104_k141_113737 | Not-determined |
| 18-104_k141_106979 | Not-determined |
| 18-104_k141_127872 | Not-determined |
| 18-104_k141_155576 | Low-quality |
| 18-104_k141_50599 | Not-determined |
| 18-104_k141_79346 | Not-determined |
| 18-104_k141_72034 | Not-determined |
| 18-104_k141_23126 | Low-quality |
| 18-104_k141_113738 | Not-determined |
| 18-104_k141_79348 | Not-determined |
| 18-104_k141_16178 | Not-determined |
| 18-104_k141_30313 | Not-determined |
| 18-104_k141_86059 | Not-determined |
| 18-104_k141_113743 | Not-determined |
| 18-104_k141_141716 | Not-determined |
| 18-104_k141_134766 | Not-determined |
| 18-104_k141_16185 | Not-determined |
| 18-104_k141_141719 | Not-determined |
| 18-104_k141_134768 | Not-determined |
| 18-104_k141_16188 | Not-determined |
| 18-104_k141_72047 | Not-determined |
| 18-104_k141_44148 | Not-determined |
| 18-104_k141_65051 | Not-determined |
| 18-104_k141_134770 | Not-determined |
| 18-104_k141_72049 | Not-determined |
| 18-104_k141_44152 | Not-determined |
| 18-104_k141_99539 | Not-determined |
| 18-104_k141_16189 | Not-determined |
| 18-104_k141_106994 | Not-determined |
| 18-104_k141_106997 | Not-determined |
| 18-104_k141_65059 | Not-determined |
| 18-104_k141_106998 | Not-determined |
| 18-104_k141_113755 | Not-determined |
| 18-104_k141_86075 | Not-determined |
| 18-104_k141_79366 | Not-determined |
| 18-104_k141_50613 | Not-determined |
| 18-104_k141_72058 | Low-quality |
| 18-104_k141_44163 | Not-determined |
| 18-104_k141_44167 | Not-determined |
| 18-104_k141_134775 | Low-quality |
| 18-104_k141_86086 | Not-determined |
| 18-104_k141_107006 | Not-determined |
| 18-104_k141_79376 | Not-determined |
| 18-104_k141_72068 | Low-quality |
| 18-104_k141_141733 | Not-determined |
| 18-104_k141_72069 | Not-determined |
| 18-104_k141_141734 | Not-determined |
| 18-104_k141_86095 | Not-determined |
| 18-104_k141_2352 | Not-determined |
| 18-104_k141_72075 | Not-determined |
| 18-104_k141_72076 | Not-determined |
| 18-104_k141_79380 | Not-determined |
| 18-104_k141_86097 | Not-determined |
| 18-104_k141_9347 | Not-determined |
| 18-104_k141_30336 | Not-determined |
| 18-104_k141_36985 | Not-determined |
| 18-104_k141_148596 | Low-quality |
| 18-104_k141_79385 | Not-determined |
| 18-104_k141_9351 | Not-determined |
| 18-104_k141_9354 | Not-determined |
| 18-104_k141_44179 | Not-determined |
| 18-104_k141_16197 | Not-determined |
| 18-104_k141_72086 | Not-determined |
| 18-104_k141_44181 | Not-determined |
| 18-104_k141_141749 | Not-determined |
| 18-104_k141_120313 | Not-determined |
| 18-104_k141_72093 | Not-determined |
| 18-104_k141_86108 | Not-determined |
| 18-104_k141_141759 | Not-determined |
| 18-104_k141_107021 | Low-quality |
| 18-104_k141_23131 | Not-determined |
| 18-104_k141_162464 | Not-determined |
| 18-104_k141_72099 | Not-determined |
| 18-104_k141_107026 | Not-determined |
| 18-104_k141_134784 | Not-determined |
| 18-104_k141_113780 | Not-determined |
| 18-104_k141_72103 | Not-determined |
| 18-104_k141_16207 | Not-determined |
| 18-104_k141_44191 | Not-determined |
| 18-104_k141_16208 | Not-determined |
| 18-104_k141_162467 | Low-quality |
| 18-104_k141_93178 | Not-determined |
| 18-104_k141_36994 | Low-quality |
| 18-104_k141_134792 | Low-quality |
| 18-104_k141_162471 | Low-quality |
| 18-104_k141_113785 | Not-determined |
| 18-104_k141_162475 | Not-determined |
| 18-104_k141_113787 | Not-determined |
| 18-104_k141_23136 | Not-determined |
| 18-104_k141_155599 | Not-determined |
| 18-104_k141_162477 | Not-determined |
| 18-104_k141_134795 | Low-quality |
| 18-104_k141_72116 | Not-determined |
| 18-104_k141_44199 | Not-determined |
| 18-104_k141_2378 | Not-determined |
| 18-104_k141_72118 | Not-determined |
| 18-104_k141_44200 | Not-determined |
| 18-104_k141_141769 | Not-determined |
| 18-104_k141_162480 | Not-determined |
| 18-104_k141_44201 | Not-determined |
| 18-104_k141_72122 | Not-determined |
| 18-104_k141_79397 | Not-determined |
| 18-104_k141_23138 | Low-quality |
| 18-104_k141_16221 | Low-quality |
| 18-104_k141_44202 | Low-quality |
| 18-104_k141_16222 | Not-determined |
| 18-104_k141_44203 | Not-determined |
| 18-104_k141_44205 | Not-determined |
| 18-104_k141_9400 | Not-determined |
| 18-104_k141_65106 | Not-determined |
| 18-104_k141_72125 | Low-quality |
| 18-104_k141_79405 | Not-determined |
| 18-104_k141_30366 | Not-determined |
| 18-104_k141_127926 | Not-determined |
| 18-104_k141_127927 | Not-determined |
| 18-104_k141_155612 | Not-determined |
| 18-104_k141_37010 | Not-determined |
| 18-104_k141_99571 | Not-determined |
| 18-104_k141_37011 | Not-determined |
| 18-104_k141_16237 | Not-determined |
| 18-104_k141_86140 | Not-determined |
| 18-104_k141_120330 | Not-determined |
| 18-104_k141_155615 | Not-determined |
| 18-104_k141_30371 | Not-determined |
| 18-104_k141_23149 | Not-determined |
| 18-104_k141_134809 | Low-quality |
| 18-104_k141_9408 | Low-quality |
| 18-104_k141_16239 | Not-determined |
| 18-104_k141_23152 | Not-determined |
| 18-104_k141_16240 | Not-determined |
| 18-104_k141_37017 | Not-determined |
| 18-104_k141_127931 | Not-determined |
| 18-104_k141_86146 | Not-determined |
| 18-104_k141_148615 | Not-determined |
| 18-104_k141_127935 | Not-determined |
| 18-104_k141_9414 | Not-determined |
| 18-104_k141_148617 | Not-determined |
| 18-104_k141_155622 | Not-determined |
| 18-104_k141_99579 | Not-determined |
| 18-104_k141_65119 | Low-quality |
| 18-104_k141_37033 | Not-determined |
| 18-104_k141_16254 | Low-quality |
| 18-104_k141_44217 | Not-determined |
| 18-104_k141_113809 | Not-determined |
| 18-104_k141_30389 | Not-determined |
| 18-104_k141_9421 | Not-determined |
| 18-104_k141_50639 | Not-determined |
| 18-104_k141_30393 | Low-quality |
| 18-104_k141_44229 | Not-determined |
| 18-104_k141_134822 | Not-determined |
| 18-104_k141_44231 | Not-determined |
| 18-104_k141_16269 | Not-determined |
| 18-104_k141_134826 | Not-determined |
| 18-104_k141_148627 | Low-quality |
| 18-104_k141_99598 | Not-determined |
| 18-104_k141_2425 | Not-determined |
| 18-104_k141_134831 | Not-determined |
| 18-104_k141_2428 | Not-determined |
| 18-104_k141_44237 | Not-determined |
| 18-104_k141_65145 | Not-determined |
| 18-104_k141_148640 | Not-determined |
| 18-104_k141_2433 | Not-determined |
| 18-104_k141_141806 | Not-determined |
| 18-104_k141_134837 | Not-determined |
| 18-104_k141_162508 | Not-determined |
| 18-104_k141_2436 | Not-determined |
| 18-104_k141_134838 | Not-determined |
| 18-104_k141_127941 | Not-determined |
| 18-104_k141_120376 | Not-determined |
| 18-104_k141_134841 | Not-determined |
| 18-104_k141_37055 | Not-determined |
| 18-104_k141_127944 | Low-quality |
| 18-104_k141_134845 | Not-determined |
| 18-104_k141_127946 | Not-determined |
| 18-104_k141_120385 | Not-determined |
| 18-104_k141_50657 | Not-determined |
| 18-104_k141_37061 | Not-determined |
| 18-104_k141_37063 | Not-determined |
| 18-104_k141_50659 | Low-quality |
| 18-104_k141_141822 | Not-determined |
| 18-104_k141_120389 | Not-determined |
| 18-104_k141_93190 | Not-determined |
| 18-104_k141_120391 | Low-quality |
| 18-104_k141_155627 | Not-determined |
| 18-104_k141_93193 | Not-determined |
| 18-104_k141_113840 | Not-determined |
| 18-104_k141_93194 | Not-determined |
| 18-104_k141_127958 | Not-determined |
| 18-104_k141_148659 | Not-determined |
| 18-104_k141_50663 | Low-quality |
| 18-104_k141_120397 | Not-determined |
| 18-104_k141_141827 | Not-determined |
| 18-104_k141_65154 | Not-determined |
| 18-104_k141_141828 | Not-determined |
| 18-104_k141_141831 | Not-determined |
| 18-104_k141_2459 | Not-determined |
| 18-104_k141_23173 | Not-determined |
| 18-104_k141_113845 | Not-determined |
| 18-104_k141_23175 | Not-determined |
| 18-104_k141_113846 | Not-determined |
| 18-104_k141_127971 | Low-quality |
| 18-104_k141_65161 | Not-determined |
| 18-104_k141_2464 | Not-determined |
| 18-104_k141_50670 | Not-determined |
| 18-104_k141_141840 | Not-determined |
| 18-104_k141_72183 | Not-determined |
| 18-104_k141_50672 | Not-determined |
| 18-104_k141_37079 | Not-determined |
| 18-104_k141_65168 | Not-determined |
| 18-104_k141_9435 | Not-determined |
| 18-104_k141_65170 | Not-determined |
| 18-104_k141_44264 | Not-determined |
| 18-104_k141_9440 | Not-determined |
| 18-104_k141_148670 | Not-determined |
| 18-104_k141_155644 | Not-determined |
| 18-104_k141_155645 | Not-determined |
| 18-104_k141_113855 | Not-determined |
| 18-104_k141_65172 | Not-determined |
| 18-104_k141_162513 | Not-determined |
| 18-104_k141_93215 | Not-determined |
| 18-104_k141_30427 | Not-determined |
| 18-104_k141_79469 | Not-determined |
| 18-104_k141_113861 | Not-determined |
| 18-104_k141_72198 | Low-quality |
| 18-104_k141_65180 | Low-quality |
| 18-104_k141_134856 | Not-determined |
| 18-104_k141_2486 | Not-determined |
| 18-104_k141_30429 | Not-determined |
| 18-104_k141_37086 | Not-determined |
| 18-104_k141_30431 | Not-determined |
| 18-104_k141_107088 | Not-determined |
| 18-104_k141_148677 | Not-determined |
| 18-104_k141_127983 | Not-determined |
| 18-104_k141_65188 | Not-determined |
| 18-104_k141_107094 | Low-quality |
| 18-104_k141_50689 | Not-determined |
| 18-104_k141_141864 | Not-determined |
| 18-104_k141_107097 | Not-determined |
| 18-104_k141_79481 | Not-determined |
| 18-104_k141_107099 | Not-determined |
| 18-104_k141_65195 | Not-determined |
| 18-104_k141_2496 | Low-quality |
| 18-104_k141_86178 | Not-determined |
| 18-104_k141_44286 | Not-determined |
| 18-104_k141_79487 | Not-determined |
| 18-104_k141_44288 | Not-determined |
| 18-104_k141_162529 | Not-determined |
| 18-104_k141_9456 | Not-determined |
| 18-104_k141_148688 | Low-quality |
| 18-104_k141_127987 | Not-determined |
| 18-104_k141_134869 | Not-determined |
| 18-104_k141_127988 | Not-determined |
| 18-104_k141_9460 | Not-determined |
| 18-104_k141_155669 | Not-determined |
| 18-104_k141_99640 | Not-determined |
| 18-104_k141_9462 | Not-determined |
| 18-104_k141_50701 | Not-determined |
| 18-104_k141_57966 | Not-determined |
| 18-104_k141_79501 | Not-determined |
| 18-104_k141_86193 | Not-determined |
| 18-104_k141_134872 | Not-determined |
| 18-104_k141_57968 | Not-determined |
| 18-104_k141_99646 | Not-determined |
| 18-104_k141_23204 | Not-determined |
| 18-104_k141_57970 | Not-determined |
| 18-104_k141_93229 | Not-determined |
| 18-104_k141_107107 | Not-determined |
| 18-104_k141_9469 | Not-determined |
| 18-104_k141_57977 | Not-determined |
| 18-104_k141_127999 | Low-quality |
| 18-104_k141_86204 | Not-determined |
| 18-104_k141_93234 | Not-determined |
| 18-104_k141_113863 | Not-determined |
| 18-104_k141_79510 | Not-determined |
| 18-104_k141_99656 | Not-determined |
| 18-104_k141_128003 | Not-determined |
| 18-104_k141_86206 | Not-determined |
| 18-104_k141_44296 | Not-determined |
| 18-104_k141_50705 | Not-determined |
| 18-104_k141_79515 | Not-determined |
| 18-104_k141_99660 | Not-determined |
| 18-104_k141_57979 | Not-determined |
| 18-104_k141_57980 | Not-determined |
| 18-104_k141_113868 | Not-determined |
| 18-104_k141_148699 | Not-determined |
| 18-104_k141_79519 | Not-determined |
| 18-104_k141_30456 | Not-determined |
| 18-104_k141_113871 | Not-determined |
| 18-104_k141_148701 | Not-determined |
| 18-104_k141_148704 | Not-determined |
| 18-104_k141_65213 | Not-determined |
| 18-104_k141_113873 | Not-determined |
| 18-104_k141_37100 | Not-determined |
| 18-104_k141_128015 | Not-determined |
| 18-104_k141_72235 | Not-determined |
| 18-104_k141_44310 | Not-determined |
| 18-104_k141_128019 | Not-determined |
| 18-104_k141_57989 | Not-determined |
| 18-104_k141_79532 | Not-determined |
| 18-104_k141_86212 | Not-determined |
| 18-104_k141_44314 | Not-determined |
| 18-104_k141_44316 | Not-determined |
| 18-104_k141_107125 | Not-determined |
| 18-104_k141_57994 | Low-quality |
| 18-104_k141_50719 | Low-quality |
| 18-104_k141_72243 | Not-determined |
| 18-104_k141_2538 | Low-quality |
| 18-104_k141_50720 | Not-determined |
| 18-104_k141_57998 | Not-determined |
| 18-104_k141_107133 | Low-quality |
| 18-104_k141_141903 | Not-determined |
| 18-104_k141_86225 | Not-determined |
| 18-104_k141_128027 | Not-determined |
| 18-104_k141_23239 | Low-quality |
| 18-104_k141_155699 | Not-determined |
| 18-104_k141_23243 | Not-determined |
| 18-104_k141_141904 | Not-determined |
| 18-104_k141_155703 | Not-determined |
| 18-104_k141_65241 | Not-determined |
| 18-104_k141_30483 | Not-determined |
| 18-104_k141_50732 | Low-quality |
| 18-104_k141_16322 | Not-determined |
| 18-104_k141_128041 | Not-determined |
| 18-104_k141_58008 | Not-determined |
| 18-104_k141_162571 | Low-quality |
| 18-104_k141_107136 | Not-determined |
| 18-104_k141_23254 | Low-quality |
| 18-104_k141_113903 | Not-determined |
| 18-104_k141_65248 | Not-determined |
| 18-104_k141_37125 | Not-determined |
| 18-104_k141_162576 | Not-determined |
| 18-104_k141_141920 | Not-determined |
| 18-104_k141_141923 | Not-determined |
| 18-104_k141_16327 | Not-determined |
| 18-104_k141_37129 | Not-determined |
| 18-104_k141_72259 | Not-determined |
| 18-104_k141_16330 | Not-determined |

| | |
|---|---|
| 18-104_k141_65253 | Not-determined |
| 18-104_k141_134911 | Low-quality |
| 18-104_k141_58024 | Low-quality |
| 18-104_k141_2553 | Not-determined |
| 18-104_k141_37131 | Not-determined |
| 18-104_k141_72266 | Not-determined |
| 18-104_k141_99678 | Not-determined |
| 18-104_k141_107143 | Not-determined |
| 18-104_k141_134917 | Not-determined |
| 18-104_k141_120455 | Not-determined |
| 18-104_k141_134918 | Not-determined |
| 18-104_k141_99683 | Not-determined |
| 18-104_k141_107148 | Not-determined |
| 18-104_k141_44333 | Low-quality |
| 18-104_k141_37132 | Low-quality |
| 18-104_k141_72274 | Not-determined |
| 18-104_k141_50743 | Low-quality |
| 18-104_k141_58035 | Not-determined |
| 18-104_k141_93249 | Not-determined |
| 18-104_k141_9486 | Not-determined |
| 18-104_k141_134927 | Not-determined |
| 18-104_k141_50746 | Not-determined |
| 18-104_k141_50747 | Not-determined |
| 18-104_k141_162587 | Not-determined |
| 18-104_k141_155731 | Low-quality |
| 18-104_k141_16340 | Not-determined |
| 18-104_k141_30502 | Not-determined |
| 18-104_k141_86251 | Not-determined |
| 18-104_k141_58038 | Not-determined |
| 18-104_k141_134933 | Not-determined |
| 18-104_k141_72283 | Not-determined |
| 18-104_k141_50752 | Not-determined |
| 18-104_k141_30508 | Not-determined |
| 18-104_k141_30510 | Not-determined |
| 18-104_k141_162600 | Not-determined |
| 18-104_k141_120459 | Not-determined |
| 18-104_k141_44344 | Not-determined |
| 18-104_k141_79549 | Not-determined |
| 18-104_k141_120465 | Low-quality |
| 18-104_k141_44348 | Not-determined |
| 18-104_k141_9502 | Not-determined |
| 18-104_k141_2587 | Not-determined |
| 18-104_k141_120474 | Not-determined |
| 18-104_k141_9508 | Not-determined |
| 18-104_k141_155746 | Not-determined |
| 18-104_k141_9509 | Not-determined |
| 18-104_k141_2589 | Not-determined |
| 18-104_k141_134955 | Not-determined |
| 18-104_k141_58054 | Not-determined |
| 18-104_k141_65269 | Not-determined |
| 18-104_k141_23271 | Not-determined |
| 18-104_k141_93264 | Not-determined |
| 18-104_k141_9515 | Not-determined |
| 18-104_k141_93265 | Not-determined |
| 18-104_k141_162609 | Not-determined |
| 18-104_k141_162610 | Not-determined |
| 18-104_k141_141944 | Not-determined |
| 18-104_k141_30516 | Not-determined |
| 18-104_k141_79560 | Not-determined |
| 18-104_k141_9520 | Not-determined |
| 18-104_k141_65276 | Not-determined |
| 18-104_k141_120478 | Not-determined |
| 18-104_k141_65278 | Not-determined |
| 18-104_k141_162615 | Not-determined |
| 18-104_k141_120480 | Not-determined |
| 18-104_k141_65281 | Not-determined |
| 18-104_k141_72293 | Low-quality |
| 18-104_k141_155752 | Not-determined |
| 18-104_k141_65283 | Not-determined |
| 18-104_k141_44362 | Not-determined |
| 18-104_k141_99692 | Not-determined |
| 18-104_k141_23289 | Not-determined |
| 18-104_k141_155755 | Not-determined |
| 18-104_k141_155757 | Not-determined |
| 18-104_k141_2599 | Low-quality |
| 18-104_k141_86277 | Not-determined |
| 18-104_k141_86280 | Not-determined |
| 18-104_k141_155760 | Not-determined |
| 18-104_k141_79569 | Not-determined |
| 18-104_k141_2603 | Not-determined |
| 18-104_k141_72296 | Not-determined |
| 18-104_k141_113918 | Not-determined |
| 18-104_k141_30531 | Not-determined |
| 18-104_k141_141956 | Not-determined |
| 18-104_k141_44367 | Not-determined |
| 18-104_k141_99706 | Not-determined |
| 18-104_k141_99708 | Not-determined |
| 18-104_k141_50774 | Not-determined |
| 18-104_k141_16380 | Low-quality |
| 18-104_k141_79582 | Not-determined |
| 18-104_k141_155771 | Not-determined |
| 18-104_k141_65291 | Low-quality |
| 18-104_k141_162627 | Not-determined |
| 18-104_k141_65296 | Not-determined |
| 18-104_k141_58072 | Low-quality |
| 18-104_k141_30547 | Not-determined |
| 18-104_k141_30548 | Not-determined |
| 18-104_k141_30549 | Low-quality |
| 18-104_k141_155781 | Not-determined |
| 18-104_k141_99712 | Not-determined |
| 18-104_k141_23307 | Not-determined |
| 18-104_k141_58079 | Not-determined |
| 18-104_k141_2614 | Not-determined |
| 18-104_k141_23308 | Not-determined |
| 18-104_k141_86297 | Not-determined |
| 18-104_k141_58080 | Not-determined |
| 18-104_k141_58083 | Not-determined |
| 18-104_k141_23312 | Not-determined |
| 18-104_k141_79588 | Not-determined |
| 18-104_k141_2617 | Not-determined |
| 18-104_k141_148766 | Not-determined |
| 18-104_k141_58090 | Not-determined |
| 18-104_k141_72321 | Not-determined |
| 18-104_k141_162632 | Not-determined |
| 18-104_k141_148775 | Not-determined |
| 18-104_k141_99721 | Low-quality |
| 18-104_k141_120513 | Not-determined |
| 18-104_k141_134998 | Not-determined |
| 18-104_k141_141971 | Low-quality |
| 18-104_k141_58092 | Not-determined |
| 18-104_k141_9537 | Not-determined |
| 18-104_k141_23324 | Not-determined |
| 18-104_k141_2629 | Not-determined |
| 18-104_k141_148785 | Not-determined |
| 18-104_k141_79600 | Not-determined |
| 18-104_k141_72328 | Not-determined |
| 18-104_k141_99731 | Not-determined |
| 18-104_k141_72334 | Not-determined |
| 18-104_k141_135005 | Low-quality |
| 18-104_k141_113929 | Not-determined |
| 18-104_k141_2637 | Not-determined |
| 18-104_k141_30561 | Not-determined |
| 18-104_k141_30562 | Not-determined |
| 18-104_k141_155804 | Not-determined |
| 18-104_k141_141983 | Not-determined |
| 18-104_k141_50796 | Not-determined |
| 18-104_k141_86325 | Not-determined |
| 18-104_k141_141984 | Not-determined |
| 18-104_k141_44374 | Not-determined |
| 18-104_k141_155806 | Not-determined |
| 18-104_k141_93303 | Low-quality |
| 18-104_k141_141987 | Not-determined |
| 18-104_k141_58120 | Not-determined |
| 18-104_k141_65314 | Not-determined |
| 18-104_k141_65315 | Not-determined |
| 18-104_k141_9549 | Not-determined |
| 18-104_k141_135013 | Not-determined |
| 18-104_k141_135014 | Not-determined |
| 18-104_k141_23338 | Not-determined |
| 18-104_k141_86332 | Not-determined |
| 18-104_k141_72346 | Not-determined |
| 18-104_k141_107180 | Not-determined |
| 18-104_k141_86334 | Not-determined |
| 18-104_k141_86336 | Not-determined |
| 18-104_k141_65324 | Not-determined |
| 18-104_k141_30574 | Not-determined |
| 18-104_k141_23348 | Not-determined |
| 18-104_k141_65326 | Not-determined |
| 18-104_k141_120534 | Not-determined |
| 18-104_k141_23351 | Not-determined |
| 18-104_k141_44388 | Low-quality |
| 18-104_k141_135018 | Not-determined |
| 18-104_k141_9560 | Not-determined |
| 18-104_k141_44389 | Not-determined |
| 18-104_k141_86345 | Not-determined |
| 18-104_k141_37182 | Low-quality |
| 18-104_k141_50803 | Not-determined |
| 18-104_k141_23357 | Not-determined |
| 18-104_k141_58130 | Not-determined |
| 18-104_k141_79617 | Not-determined |
| 18-104_k141_30580 | Not-determined |
| 18-104_k141_30584 | Not-determined |
| 18-104_k141_72358 | Not-determined |
| 18-104_k141_155822 | Not-determined |
| 18-104_k141_44400 | Low-quality |
| 18-104_k141_120539 | Not-determined |
| 18-104_k141_16384 | Not-determined |
| 18-104_k141_155825 | Low-quality |
| 18-104_k141_2662 | Not-determined |
| 18-104_k141_155828 | Not-determined |
| 18-104_k141_79627 | Not-determined |
| 18-104_k141_72367 | Not-determined |
| 18-104_k141_2669 | Not-determined |
| 18-104_k141_9579 | Not-determined |
| 18-104_k141_79628 | Not-determined |
| 18-104_k141_93331 | Not-determined |
| 18-101-final-viral-combined_k141_271164\|\|full | Medium-quality |
| 18-104_k141_30596 | Not-determined |
| 18-104_k141_9589 | Not-determined |
| 18-104_k141_30598 | Not-determined |
| 18-104_k141_23385 | Low-quality |
| 18-104_k141_107188 | Not-determined |
| 18-104_k141_93337 | Not-determined |
| 18-104_k141_58151 | Not-determined |
| 18-104_k141_30601 | Not-determined |
| 18-104_k141_99763 | Not-determined |
| 18-104_k141_86348 | Not-determined |
| 18-104_k141_107190 | Not-determined |
| 18-104_k141_162636 | Low-quality |
| 18-104_k141_99768 | Not-determined |
| 18-104_k141_142016 | Not-determined |
| 18-104_k141_107195 | Not-determined |
| 18-104_k141_148812 | Not-determined |
| 18-104_k141_148814 | Not-determined |
| 18-104_k141_79644 | Low-quality |
| 18-104_k141_50828 | Not-determined |
| 18-104_k141_72388 | Not-determined |
| 18-104_k141_2686 | Not-determined |
| 18-104_k141_148815 | Not-determined |
| 18-104_k141_120554 | Not-determined |
| 18-104_k141_30618 | Not-determined |
| 18-104_k141_50833 | Not-determined |
| 18-104_k141_120562 | Not-determined |
| 18-104_k141_135026 | Not-determined |
| 18-104_k141_120567 | Not-determined |
| 18-104_k141_93346 | Not-determined |
| 18-104_k141_148822 | Not-determined |
| 18-104_k141_142042 | Not-determined |
| 18-104_k141_30627 | Not-determined |
| 18-104_k141_135031 | Not-determined |
| 18-104_k141_148824 | Not-determined |
| 18-104_k141_2700 | Not-determined |
| 18-104_k141_148827 | Not-determined |
| 18-104_k141_2702 | Low-quality |
| 18-104_k141_72401 | Not-determined |
| 18-104_k141_107223 | Not-determined |
| 18-104_k141_86379 | Not-determined |
| 18-104_k141_30637 | Not-determined |
| 18-104_k141_135042 | Not-determined |
| 18-104_k141_120570 | Not-determined |
| 18-104_k141_93362 | Not-determined |
| 18-104_k141_23410 | Not-determined |
| 18-104_k141_2717 | Not-determined |
| 18-104_k141_44426 | Not-determined |
| 18-104_k141_142056 | Not-determined |
| 18-104_k141_23412 | Not-determined |
| 18-104_k141_142057 | Not-determined |
| 18-104_k141_148833 | Not-determined |
| 18-104_k141_148834 | Not-determined |
| 18-104_k141_155865 | Low-quality |
| 18-104_k141_93370 | Low-quality |
| 18-104_k141_155866 | Not-determined |
| 18-104_k141_142060 | Not-determined |
| 18-104_k141_50846 | Not-determined |
| 18-104_k141_155870 | Low-quality |
| 18-104_k141_120576 | Not-determined |
| 18-104_k141_162655 | Not-determined |
| 18-104_k141_30647 | Not-determined |
| 18-104_k141_30648 | Low-quality |
| 18-104_k141_44440 | Not-determined |
| 18-104_k141_148843 | Not-determined |
| 18-104_k141_162661 | Not-determined |
| 18-104_k141_50851 | Not-determined |
| 18-104_k141_50853 | Not-determined |
| 18-104_k141_30652 | Not-determined |
| 18-104_k141_44449 | Not-determined |
| 18-104_k141_9625 | Not-determined |
| 18-104_k141_30656 | Not-determined |
| 18-104_k141_79669 | Not-determined |
| 18-104_k141_93382 | Not-determined |
| 18-104_k141_23431 | Not-determined |
| 18-104_k141_50860 | Low-quality |
| 18-104_k141_72413 | Not-determined |
| 18-104_k141_155873 | Low-quality |
| 18-104_k141_44456 | Not-determined |
| 18-104_k141_44457 | Not-determined |
| 18-104_k141_93387 | Not-determined |
| 18-104_k141_2729 | Low-quality |
| 18-104_k141_148854 | Not-determined |
| 18-104_k141_2730 | Low-quality |
| 18-104_k141_148856 | Not-determined |
| 18-104_k141_155877 | Not-determined |
| 18-104_k141_58198 | Low-quality |
| 18-104_k141_155880 | Low-quality |
| 18-104_k141_50866 | Not-determined |
| 18-104_k141_162671 | Not-determined |
| 18-104_k141_23437 | Not-determined |
| 18-104_k141_65373 | Not-determined |
| 18-104_k141_113979 | Not-determined |
| 18-104_k141_99794 | Not-determined |
| 18-104_k141_135074 | Low-quality |
| 18-104_k141_148861 | Not-determined |
| 18-104_k141_99795 | Not-determined |
| 18-104_k141_142083 | Not-determined |
| 18-104_k141_30677 | Not-determined |
| 18-104_k141_135076 | Not-determined |
| 18-104_k141_2734 | Not-determined |
| 18-104_k141_135078 | Not-determined |
| 18-104_k141_37195 | Not-determined |
| 18-104_k141_37199 | Not-determined |
| 18-104_k141_30688 | Not-determined |
| 18-104_k141_99803 | Low-quality |
| 18-104_k141_142094 | Not-determined |
| 18-104_k141_162690 | Not-determined |
| 18-104_k141_120604 | Not-determined |
| 18-104_k141_44481 | Not-determined |
| 18-104_k141_2747 | Not-determined |
| 18-104_k141_50877 | Not-determined |
| 18-104_k141_135082 | Not-determined |
| 18-104_k141_162696 | Not-determined |
| 18-104_k141_30691 | Not-determined |
| 18-104_k141_162704 | Not-determined |
| 18-104_k141_86412 | Not-determined |
| 18-104_k141_162707 | Not-determined |
| 18-104_k141_23472 | Not-determined |
| 18-104_k141_155896 | Not-determined |
| 18-104_k141_86418 | Low-quality |
| 18-104_k141_72438 | Low-quality |
| 18-104_k141_162711 | Low-quality |
| 18-104_k141_162712 | Not-determined |
| 18-104_k141_155902 | Not-determined |
| 18-104_k141_155909 | Not-determined |
| 18-104_k141_2760 | Not-determined |
| 18-104_k141_37230 | Not-determined |
| 18-104_k141_37231 | Not-determined |
| 18-104_k141_30700 | Not-determined |
| 18-104_k141_37233 | Not-determined |
| 18-104_k141_2763 | Not-determined |
| 18-104_k141_93396 | Low-quality |
| 18-104_k141_86421 | Not-determined |
| 18-104_k141_93399 | Not-determined |
| 18-104_k141_93405 | Not-determined |
| 18-104_k141_23481 | Not-determined |
| 18-104_k141_65395 | Not-determined |
| 18-104_k141_86427 | Not-determined |
| 18-104_k141_65396 | Not-determined |
| 18-104_k141_50903 | Low-quality |
| 18-104_k141_120642 | Low-quality |
| 18-104_k141_30709 | Not-determined |
| 18-104_k141_142109 | Not-determined |
| 18-104_k141_2771 | Not-determined |
| 18-104_k141_30717 | Not-determined |
| 18-104_k141_50906 | Not-determined |
| 18-104_k141_16437 | Not-determined |
| 18-104_k141_162738 | Not-determined |
| 18-104_k141_50908 | Not-determined |
| 18-104_k141_86435 | Not-determined |
| 18-104_k141_50912 | Not-determined |
| 18-104_k141_2778 | Not-determined |
| 18-104_k141_114001 | Not-determined |
| 18-104_k141_37258 | Not-determined |
| 18-104_k141_50913 | Not-determined |
| 18-104_k141_155938 | Low-quality |
| 18-104_k141_120650 | Not-determined |
| 18-104_k141_162745 | Not-determined |
| 18-104_k141_2780 | Not-determined |
| 18-104_k141_148908 | Low-quality |
| 18-104_k141_86439 | Not-determined |
| 18-104_k141_142115 | Not-determined |
| 18-104_k141_93419 | Not-determined |
| 18-104_k141_30724 | Not-determined |
| 18-104_k141_16450 | Not-determined |
| 18-104_k141_16454 | Not-determined |
| 18-104_k141_2793 | Not-determined |
| 18-104_k141_99852 | Not-determined |
| 18-104_k141_162761 | Not-determined |
| 18-104_k141_107249 | Not-determined |
| 18-104_k141_107250 | Not-determined |
| 18-104_k141_93431 | Not-determined |
| 18-104_k141_23497 | Not-determined |
| 18-104_k141_93434 | Not-determined |
| 18-104_k141_93435 | Not-determined |
| 18-104_k141_30732 | Low-quality |
| 18-104_k141_93438 | Not-determined |
| 18-104_k141_86450 | Not-determined |
| 18-104_k141_93439 | Not-determined |
| 18-104_k141_72488 | Not-determined |
| 18-104_k141_72489 | Not-determined |
| 18-104_k141_58221 | Not-determined |
| 18-104_k141_162769 | Not-determined |
| 18-104_k141_23508 | Not-determined |
| 18-104_k141_114007 | Not-determined |
| 18-104_k141_58224 | Not-determined |
| 18-104_k141_50947 | Not-determined |
| 18-104_k141_30747 | Not-determined |
| 18-104_k141_72495 | Not-determined |
| 18-104_k141_30751 | Not-determined |
| 18-104_k141_99872 | Not-determined |
| 18-104_k141_107263 | Not-determined |
| 18-104_k141_162774 | Not-determined |
| 18-104_k141_30752 | Not-determined |
| 18-104_k141_148931 | Not-determined |
| 18-104_k141_114018 | Not-determined |
| 18-104_k141_30754 | Not-determined |
| 18-104_k141_142132 | Not-determined |
| 18-104_k141_30756 | Not-determined |
| 18-104_k141_23519 | Not-determined |
| 18-104_k141_99876 | Not-determined |
| 18-104_k141_107268 | Not-determined |
| 18-104_k141_162776 | Not-determined |
| 18-104_k141_162778 | Not-determined |
| 18-104_k141_148939 | Low-quality |
| 18-104_k141_72505 | Not-determined |
| 18-104_k141_107272 | Low-quality |
| 18-104_k141_30763 | Not-determined |
| 18-104_k141_65437 | Not-determined |
| 18-104_k141_148943 | Not-determined |
| 18-104_k141_99886 | Not-determined |
| 18-104_k141_50955 | Not-determined |
| 18-104_k141_23528 | Low-quality |
| 18-104_k141_72513 | Not-determined |
| 18-104_k141_142142 | Not-determined |
| 18-104_k141_162787 | Low-quality |
| 18-104_k141_30770 | Low-quality |
| 18-104_k141_72517 | Low-quality |
| 18-104_k141_99896 | Not-determined |
| 18-104_k141_23534 | Not-determined |
| 18-104_k141_16484 | Not-determined |
| 18-104_k141_16485 | Not-determined |
| 18-104_k141_162794 | Low-quality |
| 18-104_k141_155976 | Not-determined |
| 18-104_k141_120676 | Not-determined |
| 18-104_k141_72523 | Not-determined |
| 18-104_k141_16488 | Not-determined |
| 18-104_k141_162798 | Low-quality |
| 18-104_k141_148952 | Not-determined |
| 18-104_k141_162799 | Not-determined |
| 18-104_k141_30781 | Not-determined |
| 18-104_k141_72528 | Not-determined |
| 18-104_k141_120681 | Not-determined |
| 18-104_k141_99906 | Not-determined |
| 18-104_k141_155991 | Not-determined |
| 18-104_k141_99907 | Not-determined |
| 18-104_k141_50956 | Low-quality |
| 18-104_k141_148962 | Not-determined |
| 18-104_k141_72532 | Not-determined |
| 18-104_k141_2823 | Not-determined |
| 18-104_k141_148964 | Not-determined |
| 18-104_k141_30795 | Not-determined |
| 18-104_k141_16500 | Not-determined |
| 18-104_k141_16501 | Not-determined |
| 18-104_k141_58246 | Not-determined |
| 18-104_k141_155995 | Not-determined |
| 18-104_k141_72537 | Not-determined |
| 18-104_k141_120684 | Low-quality |
| 18-104_k141_16504 | Not-determined |
| 18-104_k141_148966 | Not-determined |
| 18-104_k141_155997 | Low-quality |
| 18-104_k141_44502 | Not-determined |
| 18-104_k141_16510 | Not-determined |
| 18-104_k141_44504 | Not-determined |
| 18-104_k141_155999 | Not-determined |
| 18-104_k141_135108 | Low-quality |
| 18-104_k141_44508 | Not-determined |
| 18-104_k141_114026 | Not-determined |
| 18-104_k141_120690 | Not-determined |
| 18-104_k141_114027 | Not-determined |
| 18-104_k141_162807 | Not-determined |
| 18-104_k141_50968 | Not-determined |
| 18-104_k141_148971 | Not-determined |
| 18-104_k141_120693 | Not-determined |
| 18-104_k141_2834 | Not-determined |
| 18-104_k141_156007 | Not-determined |
| 18-104_k141_23542 | Not-determined |
| 18-104_k141_142168 | Not-determined |
| 18-104_k141_44513 | Not-determined |
| 18-104_k141_156012 | Not-determined |
| 18-104_k141_16516 | Not-determined |
| 18-104_k141_99917 | Not-determined |
| 18-104_k141_142173 | Not-determined |
| 18-104_k141_142174 | Not-determined |
| 18-104_k141_107295 | Low-quality |
| 18-104_k141_2836 | Not-determined |
| 18-104_k141_156013 | Low-quality |
| 18-104_k141_2839 | Not-determined |
| 18-104_k141_107302 | Not-determined |
| 18-104_k141_79679 | Not-determined |
| 18-104_k141_148988 | Not-determined |
| 18-104_k141_148989 | Not-determined |
| 18-104_k141_30802 | Not-determined |
| 18-104_k141_30803 | Not-determined |
| 18-104_k141_2845 | Low-quality |
| 18-104_k141_93457 | Not-determined |
| 18-104_k141_58253 | Not-determined |
| 18-104_k141_2847 | Not-determined |
| 18-104_k141_44520 | Not-determined |
| 18-104_k141_79688 | Not-determined |
| 18-104_k141_93460 | Not-determined |
| 18-104_k141_135114 | Not-determined |
| 18-104_k141_99924 | Not-determined |
| 18-104_k141_162831 | Not-determined |
| 18-104_k141_37305 | Not-determined |
| 18-104_k141_9629 | Not-determined |
| 18-104_k141_93462 | Not-determined |
| 18-104_k141_9632 | Not-determined |
| 18-104_k141_79696 | Not-determined |
| 18-104_k141_16527 | Not-determined |
| 18-104_k141_86492 | Not-determined |
| 18-104_k141_44533 | Not-determined |
| 18-104_k141_50998 | Low-quality |
| 18-104_k141_16533 | Not-determined |
| 18-104_k141_37312 | Not-determined |
| 18-104_k141_114054 | Not-determined |
| 18-104_k141_99933 | Not-determined |
| 18-104_k141_120725 | Not-determined |
| 18-104_k141_162845 | Not-determined |
| 18-104_k141_156038 | Not-determined |
| 18-104_k141_142191 | Not-determined |
| 18-104_k141_30818 | Not-determined |
| 18-104_k141_23564 | Not-determined |
| 18-104_k141_30822 | Not-determined |
| 18-104_k141_58263 | Not-determined |
| 18-104_k141_142193 | Not-determined |
| 18-104_k141_44550 | Not-determined |
| 18-104_k141_162855 | Low-quality |
| 18-104_k141_9646 | Not-determined |
| 18-104_k141_142195 | Not-determined |
| 18-104_k141_120730 | Not-determined |
| 18-104_k141_44555 | Not-determined |
| 18-104_k141_135125 | Not-determined |
| 18-104_k141_9652 | Not-determined |
| 18-104_k141_162865 | Not-determined |
| 18-104_k141_114067 | Not-determined |
| 18-104_k141_114069 | Not-determined |
| 18-104_k141_79704 | Not-determined |
| 18-104_k141_30838 | Not-determined |
| 18-104_k141_9658 | Not-determined |
| 18-104_k141_16543 | Not-determined |
| 18-104_k141_162871 | Not-determined |
| 18-104_k141_79707 | Not-determined |
| 18-104_k141_30840 | Low-quality |
| 18-104_k141_93474 | Not-determined |
| 18-104_k141_79709 | Not-determined |
| 18-104_k141_156062 | Not-determined |
| 18-104_k141_142206 | Not-determined |
| 18-104_k141_16546 | Low-quality |
| 18-104_k141_156066 | Not-determined |
| 18-104_k141_30846 | Not-determined |
| 18-104_k141_107333 | Not-determined |
| 18-104_k141_37338 | Not-determined |
| 18-104_k141_107337 | Not-determined |
| 18-104_k141_58289 | Not-determined |
| 18-104_k141_135128 | Not-determined |
| 18-104_k141_58290 | Not-determined |
| 18-104_k141_135130 | Not-determined |
| 18-104_k141_107341 | Not-determined |
| 18-104_k141_37345 | Low-quality |
| 18-104_k141_120744 | Low-quality |
| 18-104_k141_142214 | Not-determined |
| 18-104_k141_93483 | Not-determined |
| 18-104_k141_30850 | Low-quality |
| 18-104_k141_37349 | Not-determined |
| 18-104_k141_114075 | Not-determined |
| 18-104_k141_30851 | Low-quality |
| 18-104_k141_9669 | Not-determined |
| 18-104_k141_9671 | Low-quality |
| 18-104_k141_107346 | Low-quality |
| 18-104_k141_65492 | Not-determined |
| 18-104_k141_114080 | Not-determined |
| 18-104_k141_114081 | Not-determined |
| 18-104_k141_51024 | Not-determined |
| 18-104_k141_114082 | Not-determined |
| 18-104_k141_2885 | Not-determined |
| 18-104_k141_142227 | Not-determined |
| 18-104_k141_2888 | Not-determined |
| 18-104_k141_30854 | Not-determined |
| 18-104_k141_30856 | Not-determined |
| 18-104_k141_2894 | Not-determined |
| 18-104_k141_135153 | Low-quality |
| 18-104_k141_99963 | Not-determined |
| 18-104_k141_162875 | Not-determined |
| 18-104_k141_65498 | Not-determined |
| 18-104_k141_37356 | Not-determined |
| 18-104_k141_44565 | Not-determined |
| 18-104_k141_120758 | Not-determined |
| 18-104_k141_156082 | Not-determined |
| 18-104_k141_142237 | Not-determined |
| 18-104_k141_72560 | Not-determined |
| 18-104_k141_58307 | Not-determined |
| 18-104_k141_72567 | Not-determined |
| 18-104_k141_79754 | Not-determined |
| 18-104_k141_120772 | Not-determined |
| 18-104_k141_99973 | Not-determined |
| 18-104_k141_72569 | Not-determined |
| 18-104_k141_142251 | Not-determined |
| 18-104_k141_72571 | Not-determined |
| 18-104_k141_93511 | Not-determined |
| 18-104_k141_135166 | Not-determined |
| 18-104_k141_30864 | Not-determined |
| 18-104_k141_23591 | Not-determined |
| 18-104_k141_16582 | Low-quality |
| 18-104_k141_44588 | Not-determined |
| 18-104_k141_135172 | Not-determined |
| 18-104_k141_37372 | Low-quality |
| 18-104_k141_135176 | Not-determined |
| 18-104_k141_37373 | Not-determined |
| 18-104_k141_86527 | Not-determined |
| 18-104_k141_162882 | Not-determined |
| 18-104_k141_9697 | Not-determined |

| | |
|---|---|
| 18-104_k141_86531 | Not-determined |
| 18-104_k141_30876 | Not-determined |
| 18-104_k141_16591 | Not-determined |
| 18-104_k141_99992 | Not-determined |
| 18-104_k141_79768 | Not-determined |
| 18-104_k141_30878 | Not-determined |
| 18-104_k141_120800 | Not-determined |
| 18-104_k141_16596 | Not-determined |
| 18-104_k141_120802 | Not-determined |
| 18-104_k141_79778 | Not-determined |
| 18-104_k141_23610 | Low-quality |
| 18-104_k141_2937 | Not-determined |
| 18-104_k141_2942 | Not-determined |
| 18-104_k141_135190 | Not-determined |
| 18-104_k141_156120 | Not-determined |
| 18-104_k141_51052 | Not-determined |
| 18-104_k141_135191 | Not-determined |
| 18-104_k141_120818 | Not-determined |
| 18-104_k141_16615 | Not-determined |
| 18-104_k141_9712 | Not-determined |
| 18-104_k141_79791 | Not-determined |
| 18-104_k141_58339 | Not-determined |
| 18-104_k141_44614 | Not-determined |
| 18-104_k141_107383 | Not-determined |
| 18-104_k141_9714 | Not-determined |
| 18-104_k141_16618 | Not-determined |
| 18-104_k141_79796 | Not-determined |
| 18-104_k141_23620 | Not-determined |
| 18-104_k141_114101 | Not-determined |
| 18-104_k141_9718 | Not-determined |
| 18-104_k141_51065 | Not-determined |
| 18-104_k141_65527 | Low-quality |
| 18-104_k141_23624 | Not-determined |
| 18-104_k141_142289 | Not-determined |
| 18-104_k141_2952 | Not-determined |
| 18-104_k141_44623 | Not-determined |
| 18-104_k141_72607 | Not-determined |
| 18-104_k141_100013 | Low-quality |
| 18-104_k141_100014 | Low-quality |
| 18-104_k141_86558 | Not-determined |
| 18-104_k141_44628 | Not-determined |
| 18-104_k141_93532 | Low-quality |
| 18-104_k141_128044 | Low-quality |
| 18-104_k141_142298 | Not-determined |
| 18-104_k141_93536 | Not-determined |
| 18-104_k141_128045 | Not-determined |
| 18-104_k141_37392 | Not-determined |
| 18-104_k141_9734 | Not-determined |
| 18-104_k141_37394 | Low-quality |
| 18-104_k141_72618 | Not-determined |
| 18-104_k141_86569 | Not-determined |
| 18-104_k141_23631 | Not-determined |
| 18-104_k141_156136 | Not-determined |
| 18-104_k141_156139 | Not-determined |
| 18-104_k141_30931 | Not-determined |
| 18-104_k141_16634 | Low-quality |
| 18-104_k141_23636 | Not-determined |
| 18-104_k141_23637 | Not-determined |
| 18-104_k141_128056 | Not-determined |
| 18-104_k141_86580 | Not-determined |
| 18-104_k141_44645 | Not-determined |
| 18-104_k141_149056 | Not-determined |
| 18-104_k141_107408 | Not-determined |
| 18-104_k141_142308 | Not-determined |
| 18-104_k141_128062 | Not-determined |
| 18-104_k141_65552 | Not-determined |
| 18-104_k141_30940 | Not-determined |
| 18-104_k141_128066 | Not-determined |
| 18-104_k141_37405 | Low-quality |
| 18-104_k141_30944 | Low-quality |
| 18-104_k141_128072 | Not-determined |
| 18-104_k141_58380 | Not-determined |
| 18-104_k141_2967 | Not-determined |
| 18-104_k141_128076 | Not-determined |
| 18-104_k141_37414 | Not-determined |
| 18-104_k141_156157 | Low-quality |
| 18-104_k141_2971 | Not-determined |
| 18-104_k141_72633 | Not-determined |
| 18-104_k141_23656 | Not-determined |
| 18-104_k141_2974 | Not-determined |
| 18-104_k141_44661 | Not-determined |
| 18-104_k141_86590 | Not-determined |
| 18-104_k141_9748 | Low-quality |
| 18-101-final-viral-combined_k141_227917\|\|full | Medium-quality |
| 18-104_k141_2979 | Not-determined |
| 18-104_k141_9750 | Not-determined |
| 18-104_k141_120841 | Not-determined |
| 18-104_k141_100063 | Not-determined |
| 18-104_k141_58391 | Not-determined |
| 18-104_k141_58392 | Not-determined |
| 18-104_k141_23666 | Not-determined |
| 18-104_k141_100065 | Not-determined |
| 18-104_k141_58395 | Not-determined |
| 18-104_k141_156163 | Not-determined |
| 18-104_k141_156164 | Not-determined |
| 18-104_k141_107433 | Not-determined |
| 18-104_k141_2985 | Not-determined |
| 18-104_k141_156167 | Not-determined |
| 18-104_k141_2986 | Not-determined |
| 18-104_k141_44665 | Not-determined |
| 18-104_k141_135224 | Not-determined |
| 18-104_k141_156173 | Not-determined |
| 18-104_k141_107437 | Not-determined |
| 18-104_k141_142329 | Not-determined |
| 18-104_k141_58399 | Low-quality |
| 18-104_k141_23674 | Not-determined |
| 18-104_k141_156177 | Not-determined |
| 18-104_k141_72639 | Not-determined |
| 18-104_k141_156182 | Not-determined |
| 18-104_k141_23681 | Not-determined |
| 18-104_k141_107444 | Not-determined |
| 18-104_k141_30966 | Not-determined |
| 18-104_k141_107445 | Not-determined |
| 18-104_k141_107446 | Not-determined |
| 18-104_k141_100080 | Not-determined |
| 18-104_k141_30969 | Low-quality |
| 18-104_k141_2999 | Not-determined |
| 18-104_k141_51081 | Not-determined |
| 18-104_k141_100083 | Not-determined |
| 18-104_k141_79849 | Low-quality |
| 18-104_k141_9768 | Not-determined |
| 18-104_k141_58409 | Not-determined |
| 18-104_k141_30974 | Not-determined |
| 18-104_k141_30976 | Not-determined |
| 18-104_k141_114136 | Not-determined |
| 18-104_k141_44669 | Not-determined |
| 18-104_k141_100087 | Not-determined |
| 18-104_k141_107452 | Not-determined |
| 18-104_k141_79856 | Not-determined |
| 18-104_k141_72649 | Not-determined |
| 18-104_k141_135241 | Not-determined |
| 18-104_k141_114142 | Not-determined |
| 18-104_k141_107454 | Not-determined |
| 18-104_k141_162923 | Not-determined |
| 18-104_k141_58416 | Not-determined |
| 18-104_k141_107457 | Not-determined |
| 18-104_k141_51094 | Not-determined |
| 18-104_k141_9782 | Not-determined |
| 18-104_k141_30986 | Not-determined |
| 18-104_k141_9783 | Not-determined |
| 18-104_k141_9784 | Not-determined |
| 18-104_k141_142357 | Not-determined |
| 18-104_k141_114146 | Not-determined |
| 18-104_k141_51097 | Not-determined |
| 18-104_k141_23693 | Not-determined |
| 18-104_k141_86605 | Not-determined |
| 18-104_k141_128128 | Not-determined |
| 18-104_k141_44677 | Not-determined |
| 18-104_k141_107464 | Not-determined |
| 18-104_k141_16654 | Not-determined |
| 18-104_k141_107465 | Not-determined |
| 18-104_k141_44679 | Not-determined |
| 18-104_k141_44682 | Not-determined |
| 18-104_k141_9796 | Not-determined |
| 18-104_k141_44683 | Not-determined |
| 18-104_k141_128133 | Not-determined |
| 18-104_k141_51102 | Not-determined |
| 18-104_k141_114156 | Not-determined |
| 18-104_k141_79869 | Low-quality |
| 18-104_k141_114160 | Not-determined |
| 18-104_k141_100097 | Not-determined |
| 18-104_k141_142370 | Not-determined |
| 18-104_k141_65556 | Not-determined |
| 18-104_k141_44693 | Not-determined |
| 18-104_k141_86617 | Not-determined |
| 18-104_k141_107476 | Low-quality |
| 18-104_k141_128142 | Low-quality |
| 18-104_k141_37425 | Not-determined |
| 18-104_k141_128147 | Not-determined |
| 18-104_k141_37429 | Not-determined |
| 18-104_k141_107481 | Not-determined |
| 18-104_k141_128152 | Low-quality |
| 18-104_k141_31001 | Not-determined |
| 18-104_k141_16656 | Not-determined |
| 18-104_k141_37435 | Low-quality |
| 18-104_k141_107489 | Not-determined |
| 18-104_k141_128155 | Not-determined |
| 18-104_k141_31008 | Not-determined |
| 18-104_k141_162946 | Not-determined |
| 18-104_k141_156206 | Not-determined |
| 18-104_k141_9818 | Not-determined |
| 18-104_k141_149141 | Not-determined |
| 18-104_k141_128160 | Not-determined |
| 18-104_k141_9821 | Not-determined |
| 18-104_k141_107497 | Not-determined |
| 18-104_k141_58448 | Not-determined |
| 18-104_k141_65577 | Not-determined |
| 18-104_k141_149150 | Not-determined |
| 18-104_k141_128169 | Not-determined |
| 18-104_k141_162950 | Not-determined |
| 18-104_k141_128170 | Not-determined |
| 18-104_k141_9836 | Not-determined |
| 18-104_k141_9838 | Not-determined |
| 18-104_k141_120881 | Not-determined |
| 18-104_k141_162954 | Not-determined |
| 18-104_k141_128176 | Not-determined |
| 18-104_k141_142398 | Not-determined |
| 18-104_k141_142399 | Not-determined |
| 18-104_k141_100111 | Not-determined |
| 18-104_k141_128181 | Not-determined |
| 18-104_k141_135262 | Not-determined |
| 18-104_k141_65591 | Not-determined |
| 18-104_k141_3017 | Not-determined |
| 18-104_k141_9850 | Not-determined |
| 18-104_k141_156225 | Not-determined |
| 18-104_k141_100115 | Not-determined |
| 18-104_k141_142401 | Low-quality |
| 18-104_k141_100117 | Low-quality |
| 18-104_k141_86639 | Low-quality |
| 18-104_k141_149170 | Low-quality |
| 18-104_k141_162960 | Not-determined |
| 18-104_k141_9854 | Low-quality |
| 18-104_k141_3030 | Not-determined |
| 18-104_k141_107522 | Not-determined |
| 18-104_k141_156239 | Not-determined |
| 18-104_k141_149180 | Low-quality |
| 18-104_k141_142416 | Low-quality |
| 18-104_k141_3042 | Low-quality |
| 18-104_k141_128185 | Low-quality |
| 18-104_k141_149187 | Not-determined |
| 18-104_k141_128186 | Not-determined |
| 18-104_k141_100130 | Not-determined |
| 18-104_k141_65620 | Low-quality |
| 18-104_k141_114177 | Low-quality |
| 18-104_k141_23741 | Not-determined |
| 18-104_k141_100135 | Not-determined |
| 18-104_k141_93553 | Low-quality |
| 18-104_k141_128195 | Not-determined |
| 18-104_k141_23747 | Not-determined |
| 18-104_k141_128199 | Not-determined |
| 18-104_k141_128202 | Not-determined |
| 18-104_k141_44706 | Not-determined |
| 18-104_k141_44708 | Not-determined |
| 18-104_k141_135275 | Not-determined |
| 18-104_k141_120893 | Not-determined |
| 18-104_k141_44710 | Not-determined |
| 18-104_k141_23751 | Not-determined |
| 18-104_k141_149199 | Not-determined |
| 18-104_k141_114192 | Not-determined |
| 18-104_k141_114193 | Not-determined |
| 18-104_k141_86673 | Not-determined |
| 18-104_k141_86674 | Not-determined |
| 18-104_k141_79889 | Not-determined |
| 18-104_k141_65638 | Not-determined |
| 18-104_k141_51142 | Not-determined |
| 18-104_k141_128212 | Low-quality |
| 18-104_k141_79894 | Not-determined |
| 18-104_k141_79895 | Not-determined |
| 18-104_k141_162983 | Not-determined |
| 18-104_k141_86692 | Not-determined |
| 18-104_k141_128221 | Not-determined |
| 18-104_k141_86696 | Not-determined |
| 18-104_k141_162993 | Not-determined |
| 18-104_k141_120907 | Not-determined |
| 18-104_k141_156279 | Low-quality |
| 18-104_k141_156281 | Not-determined |
| 18-104_k141_156283 | Not-determined |
| 18-104_k141_31042 | Not-determined |
| 18-104_k141_65668 | Not-determined |
| 18-104_k141_86706 | Not-determined |
| 18-104_k141_16672 | Not-determined |
| 18-104_k141_128235 | Not-determined |
| 18-104_k141_142431 | Not-determined |
| 18-104_k141_93564 | Not-determined |
| 18-104_k141_142432 | Not-determined |
| 18-104_k141_156297 | Not-determined |
| 18-104_k141_156299 | Not-determined |
| 18-104_k141_93568 | Not-determined |
| 18-104_k141_128247 | Low-quality |
| 18-104_k141_51172 | Not-determined |
| 18-104_k141_142446 | Not-determined |
| 18-104_k141_31057 | Not-determined |
| 18-104_k141_120925 | Not-determined |
| 18-104_k141_51180 | Not-determined |
| 18-104_k141_51181 | Not-determined |
| 18-104_k141_163005 | Low-quality |
| 18-104_k141_65683 | Not-determined |
| 18-104_k141_142457 | Not-determined |
| 18-104_k141_156309 | Not-determined |
| 18-104_k141_9886 | Not-determined |
| 18-104_k141_44747 | Not-determined |
| 18-104_k141_142467 | Not-determined |
| 18-104_k141_79925 | Not-determined |
| 18-104_k141_58482 | Low-quality |
| 18-104_k141_65697 | Not-determined |
| 18-104_k141_58484 | Not-determined |
| 18-104_k141_142469 | Not-determined |
| 18-104_k141_142471 | Low-quality |
| 18-104_k141_163007 | Not-determined |
| 18-104_k141_51191 | Not-determined |
| 18-104_k141_37469 | Not-determined |
| 18-104_k141_16717 | Not-determined |
| 18-104_k141_31063 | Low-quality |
| 18-104_k141_93589 | Not-determined |
| 18-104_k141_100148 | Not-determined |
| 18-104_k141_93592 | Not-determined |
| 18-104_k141_128264 | Low-quality |
| 18-104_k141_51193 | Not-determined |
| 18-104_k141_163011 | Not-determined |
| 18-104_k141_79939 | Not-determined |
| 18-104_k141_93597 | Low-quality |
| 18-104_k141_58502 | Not-determined |
| 18-104_k141_31071 | Not-determined |
| 18-104_k141_86737 | Not-determined |
| 18-104_k141_79944 | Not-determined |
| 18-104_k141_51201 | Not-determined |
| 18-104_k141_156324 | Low-quality |
| 18-104_k141_107556 | Not-determined |
| 18-104_k141_51205 | Low-quality |
| 18-104_k141_58508 | Low-quality |
| 18-104_k141_128270 | Low-quality |
| 18-104_k141_114214 | Not-determined |
| 18-104_k141_156334 | Not-determined |
| 18-104_k141_9898 | Not-determined |
| 18-104_k141_31085 | Not-determined |
| 18-104_k141_93616 | Not-determined |
| 18-104_k141_44775 | Not-determined |
| 18-104_k141_9903 | Not-determined |
| 18-104_k141_31088 | Not-determined |
| 18-104_k141_100164 | Not-determined |
| 18-104_k141_9906 | Not-determined |
| 18-104_k141_37491 | Not-determined |
| 18-104_k141_128284 | Not-determined |
| 18-104_k141_65733 | Not-determined |
| 18-104_k141_44784 | Not-determined |
| 18-104_k141_72679 | Not-determined |
| 18-104_k141_86752 | Low-quality |
| 18-104_k141_65738 | Not-determined |
| 18-104_k141_86753 | Not-determined |
| 18-104_k141_65739 | Not-determined |
| 18-104_k141_79960 | Not-determined |
| 18-104_k141_16729 | Low-quality |
| 18-104_k141_51216 | Not-determined |
| 18-104_k141_65742 | Not-determined |
| 18-104_k141_114241 | Not-determined |
| 18-104_k141_86761 | Not-determined |
| 18-104_k141_16732 | Not-determined |
| 18-104_k141_156344 | Not-determined |
| 18-104_k141_72686 | Not-determined |
| 18-104_k141_100167 | Not-determined |
| 18-104_k141_107575 | Not-determined |
| 18-104_k141_9920 | Not-determined |
| 18-104_k141_37503 | Not-determined |
| 18-104_k141_65747 | Not-determined |
| 18-104_k141_135293 | Not-determined |
| 18-104_k141_65750 | Low-quality |
| 18-104_k141_72689 | Not-determined |
| 18-104_k141_3063 | Not-determined |
| 18-104_k141_16738 | Not-determined |
| 18-104_k141_79967 | Not-determined |
| 18-104_k141_3066 | Not-determined |
| 18-104_k141_86773 | Not-determined |
| 18-104_k141_44795 | Not-determined |
| 18-104_k141_128298 | Not-determined |
| 18-104_k141_114251 | Not-determined |
| 18-104_k141_9927 | Not-determined |
| 18-104_k141_16747 | Not-determined |
| 18-104_k141_163040 | Not-determined |
| 18-104_k141_9932 | Not-determined |
| 18-104_k141_3078 | Not-determined |
| 18-104_k141_16749 | Not-determined |
| 18-104_k141_3081 | Not-determined |
| 18-104_k141_79981 | Not-determined |
| 18-104_k141_58524 | Low-quality |
| 18-104_k141_16752 | Not-determined |
| 18-104_k141_44801 | Not-determined |
| 18-104_k141_16755 | Not-determined |
| 18-104_k141_16756 | Not-determined |
| 18-104_k141_114266 | Not-determined |
| 18-104_k141_72703 | Not-determined |
| 18-104_k141_163042 | Not-determined |
| 18-104_k141_120953 | Not-determined |
| 18-104_k141_9942 | Not-determined |
| 18-104_k141_58531 | Not-determined |
| 18-104_k141_100177 | Not-determined |
| 18-104_k141_114275 | Not-determined |
| 18-104_k141_58533 | Not-determined |
| 18-104_k141_37523 | Not-determined |
| 18-104_k141_100179 | Not-determined |
| 18-104_k141_100183 | Not-determined |
| 18-104_k141_44812 | Not-determined |
| 18-104_k141_142541 | Not-determined |
| 18-104_k141_156370 | Not-determined |
| 18-104_k141_44815 | Not-determined |
| 18-104_k141_163056 | Not-determined |
| 18-104_k141_142544 | Not-determined |
| 18-104_k141_58535 | Not-determined |
| 18-104_k141_72714 | Not-determined |
| 18-104_k141_9949 | Low-quality |
| 18-104_k141_23762 | Not-determined |
| 18-104_k141_135325 | Not-determined |
| 18-104_k141_72719 | Not-determined |
| 18-104_k141_156373 | Not-determined |
| 18-104_k141_9956 | Not-determined |
| 18-104_k141_72720 | Not-determined |
| 18-104_k141_107587 | Not-determined |
| 18-104_k141_3100 | Not-determined |
| 18-104_k141_163064 | Not-determined |
| 18-104_k141_3101 | Not-determined |
| 18-104_k141_3102 | Not-determined |
| 18-104_k141_3103 | Not-determined |
| 18-104_k141_163066 | Not-determined |
| 18-104_k141_156378 | Not-determined |
| 18-104_k141_58539 | Not-determined |
| 18-104_k141_86796 | Not-determined |
| 18-104_k141_149224 | Low-quality |
| 18-104_k141_107599 | Not-determined |
| 18-104_k141_9967 | Not-determined |
| 18-104_k141_120963 | Low-quality |
| 18-104_k141_163071 | Not-determined |
| 18-104_k141_58546 | Not-determined |
| 18-104_k141_156382 | Not-determined |
| 18-104_k141_100199 | Not-determined |
| 18-104_k141_107606 | Not-determined |
| 18-104_k141_120969 | Not-determined |
| 18-104_k141_114295 | Not-determined |
| 18-104_k141_23782 | Not-determined |
| 18-104_k141_156390 | Not-determined |
| 18-104_k141_100205 | Low-quality |
| 18-104_k141_149230 | Not-determined |
| 18-104_k141_65757 | Not-determined |
| 18-104_k141_51240 | Not-determined |
| 18-104_k141_142555 | Not-determined |
| 18-104_k141_65759 | Not-determined |
| 18-104_k141_51242 | Not-determined |
| 18-104_k141_37546 | Not-determined |
| 18-104_k141_9984 | Not-determined |
| 18-104_k141_156392 | Low-quality |
| 18-104_k141_80003 | Not-determined |
| 18-104_k141_86815 | Not-determined |
| 18-104_k141_65765 | Not-determined |
| 18-104_k141_72727 | Not-determined |
| 18-104_k141_80005 | Not-determined |
| 18-104_k141_80006 | Not-determined |
| 18-104_k141_114310 | Not-determined |
| 18-104_k141_16768 | Not-determined |
| 18-104_k141_120981 | Not-determined |
| 18-104_k141_72731 | Not-determined |
| 18-104_k141_149242 | Not-determined |
| 18-104_k141_149243 | Not-determined |
| 18-104_k141_135344 | Not-determined |
| 18-104_k141_3132 | Not-determined |
| 18-104_k141_135346 | Not-determined |
| 18-104_k141_100221 | Not-determined |
| 18-104_k141_37553 | Not-determined |
| 18-104_k141_100222 | Not-determined |
| 18-104_k141_31144 | Not-determined |
| 18-104_k141_107630 | Not-determined |
| 18-104_k141_23795 | Low-quality |
| 18-104_k141_9992 | Not-determined |
| 18-104_k141_44821 | Low-quality |
| 18-104_k141_163100 | Not-determined |
| 18-104_k141_93660 | Not-determined |
| 18-104_k141_23804 | Not-determined |
| 18-104_k141_135355 | Not-determined |
| 18-104_k141_23805 | Not-determined |
| 18-104_k141_93661 | Not-determined |
| 18-104_k141_107642 | Low-quality |
| 18-104_k141_156402 | Not-determined |
| 18-104_k141_93664 | Not-determined |
| 18-104_k141_3139 | Not-determined |
| 18-104_k141_114317 | Not-determined |
| 18-104_k141_65785 | Not-determined |
| 18-104_k141_128329 | Not-determined |
| 18-104_k141_37564 | Not-determined |
| 18-104_k141_93673 | Not-determined |
| 18-104_k141_128331 | Low-quality |
| 18-104_k141_107660 | Not-determined |
| 18-104_k141_120990 | Not-determined |
| 18-104_k141_37572 | Not-determined |
| 18-104_k141_135369 | Not-determined |
| 18-104_k141_37573 | Not-determined |
| 18-104_k141_44833 | Not-determined |
| 18-104_k141_23822 | Not-determined |
| 18-104_k141_51268 | Not-determined |
| 18-104_k141_10001 | Not-determined |
| 18-104_k141_51273 | Not-determined |
| 18-104_k141_80029 | Not-determined |
| 18-104_k141_16781 | Not-determined |
| 18-104_k141_80030 | Not-determined |
| 18-104_k141_128347 | Not-determined |
| 18-104_k141_80033 | Not-determined |
| 18-104_k141_65794 | Not-determined |
| 18-104_k141_142582 | Not-determined |
| 18-104_k141_16788 | Not-determined |
| 18-104_k141_142583 | Not-determined |
| 18-104_k141_142584 | Not-determined |
| 18-104_k141_93687 | Low-quality |
| 18-104_k141_44841 | Not-determined |
| 18-104_k141_65796 | Low-quality |
| 18-104_k141_142588 | Not-determined |
| 18-104_k141_107682 | Not-determined |
| 18-104_k141_31169 | Not-determined |
| 18-104_k141_107686 | Not-determined |
| 18-104_k141_100251 | Not-determined |
| 18-104_k141_128356 | Not-determined |
| 18-104_k141_107688 | Not-determined |
| 18-104_k141_10011 | Low-quality |
| 18-104_k141_37593 | Not-determined |
| 18-104_k141_10014 | Not-determined |
| 18-104_k141_163124 | Not-determined |
| 18-104_k141_65809 | Not-determined |
| 18-104_k141_100257 | Not-determined |
| 18-104_k141_37596 | Not-determined |
| 18-104_k141_37600 | Not-determined |
| 18-104_k141_44855 | Not-determined |
| 18-104_k141_10021 | Not-determined |
| 18-104_k141_163133 | Not-determined |
| 18-104_k141_93717 | Low-quality |
| 18-104_k141_86823 | Not-determined |
| 18-104_k141_31175 | Not-determined |
| 18-104_k141_93718 | Not-determined |
| 18-104_k141_86826 | Low-quality |
| 18-104_k141_3152 | Not-determined |
| 18-104_k141_156420 | Not-determined |
| 18-104_k141_44861 | Not-determined |
| 18-104_k141_142604 | Not-determined |
| 18-104_k141_135390 | Not-determined |
| 18-104_k141_80068 | Not-determined |
| 18-104_k141_37606 | Not-determined |
| 18-104_k141_107699 | Low-quality |
| 18-104_k141_44869 | Not-determined |
| 18-104_k141_23839 | Not-determined |
| 18-104_k141_135392 | Not-determined |
| 18-104_k141_135394 | Not-determined |
| 18-104_k141_31181 | Not-determined |
| 18-104_k141_44871 | Not-determined |
| 18-104_k141_93733 | Not-determined |
| 18-104_k141_23842 | Not-determined |
| 18-104_k141_114335 | Not-determined |
| 18-104_k141_135397 | Not-determined |
| 18-104_k141_163149 | Low-quality |
| 18-104_k141_3158 | Not-determined |
| 18-104_k141_114336 | Not-determined |
| 18-104_k141_156428 | Not-determined |
| 18-104_k141_65842 | Not-determined |
| 18-104_k141_44883 | Not-determined |
| 18-104_k141_44884 | Not-determined |
| 18-104_k141_31193 | Low-quality |
| 18-104_k141_135409 | Not-determined |
| 18-104_k141_93748 | Not-determined |
| 18-104_k141_3167 | Not-determined |
| 18-104_k141_163160 | Not-determined |
| 18-104_k141_10040 | Not-determined |
| 18-104_k141_163163 | Not-determined |
| 18-104_k141_135415 | Not-determined |
| 18-104_k141_107715 | Not-determined |
| 18-104_k141_10043 | Not-determined |
| 18-104_k141_44894 | Not-determined |
| 18-104_k141_135421 | Not-determined |
| 18-104_k141_149300 | Not-determined |
| 18-104_k141_135422 | Not-determined |
| 18-104_k141_135423 | Not-determined |
| 18-104_k141_3179 | Not-determined |
| 18-104_k141_135424 | Not-determined |
| 18-104_k141_100294 | Not-determined |
| 18-104_k141_163176 | Not-determined |
| 18-104_k141_3186 | Not-determined |
| 18-104_k141_3187 | Not-determined |
| 18-104_k141_100300 | Not-determined |
| 18-104_k141_3189 | Not-determined |
| 18-104_k141_44904 | Not-determined |
| 18-104_k141_142630 | Not-determined |
| 18-104_k141_100304 | Not-determined |
| 18-104_k141_93764 | Low-quality |
| 18-104_k141_72769 | Not-determined |
| 18-104_k141_86844 | Not-determined |
| 18-104_k141_44909 | Not-determined |
| 18-104_k141_3202 | Not-determined |
| 18-104_k141_10062 | Not-determined |
| 18-104_k141_128379 | Not-determined |
| 18-104_k141_128382 | Low-quality |
| 18-104_k141_107746 | Not-determined |
| 18-104_k141_37664 | Not-determined |
| 18-104_k141_23859 | Not-determined |
| 18-104_k141_23861 | Not-determined |
| 18-104_k141_149323 | Not-determined |
| 18-104_k141_149324 | Not-determined |
| 18-104_k141_163196 | Not-determined |
| 18-104_k141_80096 | Not-determined |
| 18-104_k141_10077 | Low-quality |
| 18-104_k141_93772 | Not-determined |
| 18-104_k141_128395 | Not-determined |
| 18-104_k141_3214 | Not-determined |
| 18-104_k141_37678 | Not-determined |
| 18-104_k141_107754 | Not-determined |
| 18-104_k141_100310 | Not-determined |
| 18-104_k141_93783 | Not-determined |
| 18-104_k141_86856 | Not-determined |

| | |
|---|---|
| 18-104_k141_93784 | Not-determined |
| 18-104_k141_149338 | Not-determined |
| 18-104_k141_100312 | Not-determined |
| 18-104_k141_156459 | Not-determined |
| 18-104_k141_93785 | Not-determined |
| 18-104_k141_86860 | Not-determined |
| 18-104_k141_3229 | Not-determined |
| 18-104_k141_93787 | Not-determined |
| 18-104_k141_93788 | Not-determined |
| 18-104_k141_37687 | Not-determined |
| 18-104_k141_128403 | Not-determined |
| 18-104_k141_23880 | Not-determined |
| 18-104_k141_10081 | Low-quality |
| 18-104_k141_10082 | Not-determined |
| 18-104_k141_23886 | Not-determined |
| 18-104_k141_65876 | Not-determined |
| 18-104_k141_128407 | Not-determined |
| 18-104_k141_10087 | Not-determined |
| 18-104_k141_23891 | Not-determined |
| 18-104_k141_107771 | Not-determined |
| 18-104_k141_107775 | Not-determined |
| 18-104_k141_86879 | Not-determined |
| 18-104_k141_121001 | Not-determined |
| 18-104_k141_107779 | Not-determined |
| 18-104_k141_149349 | Not-determined |
| 18-104_k141_107782 | Not-determined |
| 18-104_k141_51282 | Not-determined |
| 18-104_k141_72802 | Not-determined |
| 18-104_k141_93809 | Not-determined |
| 18-104_k141_93811 | Not-determined |
| 18-104_k141_149362 | Not-determined |
| 18-104_k141_128416 | Not-determined |
| 18-104_k141_107795 | Low-quality |
| 18-104_k141_163209 | Not-determined |
| 18-104_k141_107797 | Not-determined |
| 18-104_k141_142679 | Not-determined |
| 18-104_k141_114363 | Not-determined |
| 18-104_k141_163213 | Not-determined |
| 18-104_k141_121014 | Not-determined |
| 18-104_k141_163216 | Not-determined |
| 18-104_k141_114366 | Not-determined |
| 18-104_k141_114368 | Not-determined |
| 18-104_k141_51287 | Not-determined |
| 18-104_k141_51288 | Not-determined |
| 18-104_k141_128432 | Not-determined |
| 18-104_k141_142686 | Not-determined |
| 18-104_k141_10097 | Not-determined |
| 18-104_k141_72815 | Not-determined |
| 18-104_k141_163223 | Not-determined |
| 18-104_k141_80135 | Not-determined |
| 18-104_k141_37708 | Low-quality |
| 18-104_k141_156472 | Not-determined |
| 18-104_k141_72819 | Not-determined |
| 18-104_k141_128438 | Not-determined |
| 18-104_k141_44978 | Not-determined |
| 18-104_k141_23914 | Not-determined |
| 18-104_k141_72823 | Not-determined |
| 18-104_k141_72824 | Not-determined |
| 18-104_k141_44980 | Low-quality |
| 18-104_k141_23916 | Not-determined |
| 18-104_k141_142698 | Not-determined |
| 18-104_k141_163231 | Not-determined |
| 18-104_k141_23919 | Not-determined |
| 18-104_k141_37716 | Not-determined |
| 18-104_k141_37717 | Not-determined |
| 18-104_k141_3232 | Not-determined |
| 18-104_k141_10110 | Low-quality |
| 18-104_k141_3235 | Not-determined |
| 18-104_k141_149366 | Not-determined |
| 18-104_k141_114384 | Low-quality |
| 18-104_k141_107812 | Not-determined |
| 18-104_k141_149368 | Not-determined |
| 18-104_k141_72839 | Not-determined |
| 18-104_k141_163236 | Low-quality |
| 18-104_k141_128451 | Not-determined |
| 18-104_k141_149369 | Not-determined |
| 18-104_k141_107818 | Not-determined |
| 18-104_k141_149373 | Not-determined |
| 18-104_k141_10125 | Not-determined |
| 18-104_k141_3246 | Low-quality |
| 18-104_k141_72847 | Not-determined |
| 18-104_k141_65919 | Not-determined |
| 18-104_k141_23933 | Not-determined |
| 18-104_k141_128457 | Not-determined |
| 18-104_k141_10128 | Low-quality |
| 18-104_k141_45007 | Not-determined |
| 18-104_k141_45008 | Not-determined |
| 18-104_k141_3251 | Not-determined |
| 18-104_k141_135462 | Not-determined |
| 18-104_k141_51317 | Not-determined |
| 18-104_k141_72862 | Not-determined |
| 18-104_k141_128464 | Not-determined |
| 18-104_k141_107829 | Not-determined |
| 18-104_k141_86899 | Low-quality |
| 18-104_k141_114400 | Not-determined |
| 18-104_k141_45023 | Not-determined |
| 18-104_k141_114402 | Not-determined |
| 18-104_k141_10143 | Not-determined |
| 18-104_k141_121060 | Not-determined |
| 18-104_k141_121061 | Not-determined |
| 18-104_k141_3264 | Not-determined |
| 18-104_k141_3267 | Not-determined |
| 18-104_k141_142703 | Not-determined |
| 18-104_k141_45032 | Not-determined |
| 18-104_k141_45036 | Not-determined |
| 18-104_k141_58575 | Not-determined |
| 18-104_k141_58577 | Not-determined |
| 18-104_k141_156513 | Low-quality |
| 18-104_k141_135467 | Not-determined |
| 18-104_k141_156517 | Not-determined |
| 18-104_k141_23950 | Not-determined |
| 18-104_k141_58582 | Not-determined |
| 18-104_k141_156520 | Not-determined |
| 18-104_k141_23954 | Not-determined |
| 18-104_k141_3280 | Low-quality |
| 18-104_k141_58589 | Not-determined |
| 18-104_k141_121072 | Not-determined |
| 18-104_k141_80165 | Not-determined |
| 18-104_k141_58591 | Not-determined |
| 18-104_k141_37757 | Not-determined |
| 18-104_k141_23959 | Not-determined |
| 18-104_k141_23963 | Not-determined |
| 18-104_k141_114413 | Low-quality |
| 18-104_k141_80174 | Not-determined |
| 18-104_k141_114414 | Not-determined |
| 18-104_k141_72872 | Low-quality |
| 18-104_k141_37761 | Not-determined |
| 18-104_k141_23971 | Not-determined |
| 18-104_k141_31277 | Not-determined |
| 18-104_k141_163281 | Not-determined |
| 18-104_k141_163282 | Not-determined |
| 18-104_k141_31280 | Not-determined |
| 18-104_k141_142722 | Not-determined |
| 18-104_k141_114422 | Not-determined |
| 18-104_k141_80182 | Not-determined |
| 18-104_k141_86913 | Not-determined |
| 18-104_k141_23980 | Not-determined |
| 18-104_k141_80187 | Not-determined |
| 18-104_k141_72888 | Not-determined |
| 18-104_k141_142726 | Not-determined |
| 18-104_k141_72889 | Not-determined |
| 18-104_k141_86915 | Not-determined |
| 18-104_k141_100421 | Not-determined |
| 18-104_k141_100425 | Not-determined |
| 18-104_k141_58615 | Not-determined |
| 18-104_k141_31294 | Low-quality |
| 18-104_k141_86917 | Not-determined |
| 18-104_k141_45062 | Not-determined |
| 18-104_k141_72890 | Low-quality |
| 18-104_k141_45066 | Not-determined |
| 18-104_k141_58621 | Not-determined |
| 18-104_k141_45069 | Not-determined |
| 18-104_k141_135507 | Low-quality |
| 18-104_k141_31302 | Not-determined |
| 18-104_k141_51342 | Not-determined |
| 18-104_k141_100434 | Not-determined |
| 18-104_k141_58624 | Not-determined |
| 18-104_k141_72898 | Not-determined |
| 18-104_k141_72900 | Not-determined |
| 18-104_k141_142738 | Not-determined |
| 18-104_k141_51347 | Not-determined |
| 18-104_k141_72903 | Low-quality |
| 18-104_k141_80205 | Low-quality |
| 18-104_k141_128494 | Not-determined |
| 18-104_k141_135515 | Not-determined |
| 18-104_k141_10177 | Low-quality |
| 18-104_k141_128496 | Not-determined |
| 18-104_k141_142750 | Not-determined |
| 18-104_k141_149423 | Not-determined |
| 18-104_k141_86921 | Not-determined |
| 18-104_k141_121102 | Not-determined |
| 18-104_k141_80209 | Not-determined |
| 18-104_k141_128499 | Not-determined |
| 18-104_k141_10185 | Not-determined |
| 18-104_k141_51358 | Not-determined |
| 18-104_k141_86924 | Not-determined |
| 18-104_k141_114432 | Not-determined |
| 18-104_k141_10194 | Low-quality |
| 18-104_k141_156549 | Low-quality |
| 18-104_k141_45086 | Not-determined |
| 18-104_k141_10198 | Not-determined |
| 18-104_k141_156557 | Not-determined |
| 18-104_k141_114439 | Not-determined |
| 18-104_k141_100444 | Not-determined |
| 18-104_k141_72927 | Not-determined |
| 18-104_k141_86932 | Not-determined |
| 18-104_k141_24002 | Not-determined |
| 18-104_k141_163294 | Not-determined |
| 18-104_k141_86934 | Not-determined |
| 18-104_k141_24005 | Not-determined |
| 18-104_k141_24006 | Low-quality |
| 18-104_k141_128514 | Not-determined |
| 18-104_k141_163298 | Not-determined |
| 18-104_k141_65940 | Not-determined |
| 18-104_k141_65942 | Not-determined |
| 18-104_k141_128519 | Not-determined |
| 18-104_k141_163305 | Not-determined |
| 18-104_k141_156571 | Not-determined |
| 18-104_k141_114449 | Not-determined |
| 18-104_k141_24015 | Not-determined |
| 18-104_k141_114451 | Not-determined |
| 18-104_k141_142755 | Not-determined |
| 18-104_k141_45109 | Not-determined |
| 18-104_k141_135533 | Low-quality |
| 18-104_k141_114460 | Not-determined |
| 18-104_k141_128526 | Not-determined |
| 18-104_k141_114466 | Not-determined |
| 18-104_k141_142763 | Not-determined |
| 18-104_k141_51390 | Low-quality |
| 18-104_k141_51394 | Not-determined |
| 18-104_k141_135538 | Low-quality |
| 18-104_k141_58660 | Not-determined |
| 18-104_k141_45123 | Not-determined |
| 18-104_k141_142769 | Low-quality |
| 18-104_k141_156588 | Not-determined |
| 18-104_k141_58662 | Not-determined |
| 18-104_k141_80237 | Not-determined |
| 18-104_k141_72958 | Not-determined |
| 18-104_k141_114479 | Not-determined |
| 18-104_k141_58667 | Not-determined |
| 18-104_k141_149453 | Low-quality |
| 18-104_k141_114483 | Not-determined |
| 18-104_k141_100455 | Not-determined |
| 18-104_k141_142783 | Not-determined |
| 18-104_k141_58672 | Not-determined |
| 18-104_k141_58673 | Low-quality |
| 18-104_k141_114489 | Not-determined |
| 18-104_k141_58675 | Not-determined |
| 18-104_k141_135551 | Not-determined |
| 18-104_k141_107899 | Low-quality |
| 18-104_k141_163322 | Not-determined |
| 18-104_k141_128556 | Not-determined |
| 18-104_k141_72967 | Not-determined |
| 18-104_k141_128559 | Not-determined |
| 18-104_k141_114500 | Not-determined |
| 18-104_k141_72972 | Not-determined |
| 18-104_k141_80252 | Low-quality |
| 18-104_k141_114504 | Not-determined |
| 18-104_k141_65965 | Not-determined |
| 18-104_k141_51413 | Low-quality |
| 18-104_k141_100464 | Low-quality |
| 18-104_k141_65966 | Not-determined |
| 18-104_k141_100465 | Not-determined |
| 18-104_k141_10221 | Not-determined |
| 18-104_k141_51416 | Not-determined |
| 18-104_k141_135561 | Not-determined |
| 18-104_k141_142803 | Not-determined |
| 18-104_k141_51418 | Not-determined |
| 18-104_k141_128565 | Not-determined |
| 18-104_k141_114510 | Not-determined |
| 18-104_k141_107911 | Not-determined |
| 18-104_k141_24038 | Not-determined |
| 18-104_k141_107914 | Not-determined |
| 18-104_k141_31367 | Not-determined |
| 18-104_k141_31369 | Not-determined |
| 18-104_k141_114515 | Not-determined |
| 18-104_k141_72987 | Not-determined |
| 18-104_k141_107919 | Not-determined |
| 18-104_k141_135570 | Not-determined |
| 18-104_k141_72989 | Not-determined |
| 18-104_k141_135571 | Not-determined |
| 18-104_k141_135572 | Low-quality |
| 18-104_k141_58695 | Not-determined |
| 18-104_k141_135573 | Not-determined |
| 18-104_k141_37806 | Not-determined |
| 18-104_k141_24050 | Not-determined |
| 18-104_k141_37808 | Not-determined |
| 18-104_k141_51427 | Not-determined |
| 18-104_k141_107922 | Low-quality |
| 18-104_k141_24054 | Not-determined |
| 18-104_k141_114518 | Not-determined |
| 18-104_k141_72999 | Not-determined |
| 18-104_k141_142813 | Not-determined |
| 18-104_k141_37816 | Low-quality |
| 18-104_k141_149476 | Low-quality |
| 18-104_k141_114520 | Not-determined |
| 18-104_k141_37817 | Not-determined |
| 18-104_k141_149481 | Not-determined |
| 18-104_k141_142819 | Not-determined |
| 18-104_k141_135587 | Low-quality |
| 18-104_k141_142820 | Not-determined |
| 18-104_k141_10244 | Not-determined |
| 18-104_k141_100499 | Not-determined |
| 18-104_k141_142824 | Not-determined |
| 18-104_k141_31382 | Low-quality |
| 18-104_k141_135592 | Not-determined |
| 18-104_k141_142827 | Not-determined |
| 18-104_k141_142828 | Not-determined |
| 18-104_k141_24070 | Not-determined |
| 18-104_k141_128580 | Not-determined |
| 18-104_k141_100509 | Low-quality |
| 18-104_k141_128581 | Low-quality |
| 18-104_k141_135602 | Low-quality |
| 18-104_k141_58713 | Not-determined |
| 18-104_k141_37829 | Not-determined |
| 18-110_k141_126814 | Medium-quality |
| 18-104_k141_45129 | Not-determined |
| 18-104_k141_10251 | Not-determined |
| 18-104_k141_135608 | Not-determined |
| 18-104_k141_93853 | Not-determined |
| 18-104_k141_10253 | Not-determined |
| 18-104_k141_163331 | Low-quality |
| 18-104_k141_45132 | Not-determined |
| 18-104_k141_163332 | Not-determined |
| 18-104_k141_142846 | Not-determined |
| 18-104_k141_100519 | Not-determined |
| 18-104_k141_31404 | Not-determined |
| 18-104_k141_135610 | Not-determined |
| 18-104_k141_93854 | Low-quality |
| 18-104_k141_107934 | Not-determined |
| 18-104_k141_121134 | Not-determined |
| 18-104_k141_107937 | Not-determined |
| 18-104_k141_121139 | Low-quality |
| 18-104_k141_107942 | Not-determined |
| 18-104_k141_37839 | Not-determined |
| 18-104_k141_163341 | Not-determined |
| 18-104_k141_93860 | Not-determined |
| 18-104_k141_121141 | Not-determined |
| 18-104_k141_73026 | Not-determined |
| 18-104_k141_128596 | Not-determined |
| 18-104_k141_163344 | Not-determined |
| 18-104_k141_135618 | Not-determined |
| 18-104_k141_24087 | Not-determined |
| 18-104_k141_100528 | Not-determined |
| 18-104_k141_24091 | Not-determined |
| 18-104_k141_73032 | Not-determined |
| 18-104_k141_149509 | Low-quality |
| 18-104_k141_58734 | Not-determined |
| 18-104_k141_128609 | Not-determined |
| 18-104_k141_135625 | Not-determined |
| 18-104_k141_10270 | Not-determined |
| 18-104_k141_163358 | Not-determined |
| 18-104_k141_58740 | Not-determined |
| 18-104_k141_31428 | Low-quality |
| 18-104_k141_114526 | Not-determined |
| 18-104_k141_114530 | Not-determined |
| 18-104_k141_93874 | Not-determined |
| 18-104_k141_135630 | Not-determined |
| 18-104_k141_24096 | Low-quality |
| 18-104_k141_16821 | Low-quality |
| 18-104_k141_107957 | Not-determined |
| 18-104_k141_100544 | Not-determined |
| 18-104_k141_163366 | Not-determined |
| 18-104_k141_86983 | Not-determined |
| 18-104_k141_73039 | Not-determined |
| 18-104_k141_128625 | Not-determined |
| 18-104_k141_93885 | Not-determined |
| 18-104_k141_121157 | Not-determined |
| 18-104_k141_16828 | Low-quality |
| 18-104_k141_16830 | Not-determined |
| 18-104_k141_45149 | Not-determined |
| 18-104_k141_73046 | Not-determined |
| 18-104_k141_16832 | Not-determined |
| 18-104_k141_16833 | Not-determined |
| 18-104_k141_31444 | Not-determined |
| 18-104_k141_73049 | Not-determined |
| 18-104_k141_121166 | Low-quality |
| 18-104_k141_107967 | Not-determined |
| 18-104_k141_73052 | Not-determined |
| 18-104_k141_73053 | Not-determined |
| 18-104_k141_10300 | Low-quality |
| 18-104_k141_149546 | Not-determined |
| 18-104_k141_128637 | Not-determined |
| 18-104_k141_100555 | Not-determined |
| 18-104_k141_100556 | Not-determined |
| 18-104_k141_149554 | Not-determined |
| 18-104_k141_31447 | Not-determined |
| 18-104_k141_149556 | Low-quality |
| 18-104_k141_121178 | Low-quality |
| 18-104_k141_121179 | Not-determined |
| 18-104_k141_100565 | Not-determined |
| 18-104_k141_114566 | Not-determined |
| 18-104_k141_45174 | Low-quality |
| 18-104_k141_114570 | Not-determined |
| 18-104_k141_163375 | Not-determined |
| 18-104_k141_58778 | Not-determined |
| 18-104_k141_10313 | Not-determined |
| 18-104_k141_93929 | Not-determined |
| 18-104_k141_100572 | Not-determined |
| 18-104_k141_107990 | Not-determined |
| 18-104_k141_31466 | Not-determined |
| 18-104_k141_107991 | Not-determined |
| 18-104_k141_10320 | Not-determined |
| 18-104_k141_65994 | Not-determined |
| 18-104_k141_3349 | Not-determined |
| 18-104_k141_163382 | Not-determined |
| 18-104_k141_107994 | Not-determined |
| 18-104_k141_3351 | Not-determined |
| 18-104_k141_100586 | Not-determined |
| 18-104_k141_100588 | Not-determined |
| 18-104_k141_10328 | Not-determined |
| 18-104_k141_114579 | Not-determined |
| 18-104_k141_37869 | Not-determined |
| 18-104_k141_114581 | Not-determined |
| 18-104_k141_16868 | Low-quality |
| 18-104_k141_149584 | Not-determined |
| 18-104_k141_37875 | Not-determined |
| 18-104_k141_142876 | Not-determined |
| 18-104_k141_37876 | Not-determined |
| 18-104_k141_45196 | Not-determined |
| 18-104_k141_93944 | Not-determined |
| 18-104_k141_10341 | Not-determined |
| 18-104_k141_114586 | Low-quality |
| 18-104_k141_73081 | Not-determined |
| 18-104_k141_163400 | Low-quality |
| 18-104_k141_149590 | Not-determined |
| 18-104_k141_100610 | Not-determined |
| 18-104_k141_73087 | Not-determined |
| 18-104_k141_114589 | Not-determined |
| 18-104_k141_100614 | Not-determined |
| 18-104_k141_3370 | Not-determined |
| 18-104_k141_51450 | Not-determined |
| 18-104_k141_24126 | Not-determined |
| 18-104_k141_87024 | Low-quality |
| 18-104_k141_16892 | Not-determined |
| 18-104_k141_108004 | Not-determined |
| 18-104_k141_58801 | Not-determined |
| 18-104_k141_100618 | Low-quality |
| 18-104_k141_10358 | Not-determined |
| 18-104_k141_16897 | Not-determined |
| 18-104_k141_73093 | Low-quality |
| 18-104_k141_108008 | Not-determined |
| 18-104_k141_93958 | Not-determined |
| 18-104_k141_163404 | Not-determined |
| 18-104_k141_58809 | Not-determined |
| 18-104_k141_135665 | Not-determined |
| 18-104_k141_87034 | Not-determined |
| 18-104_k141_51457 | Not-determined |
| 18-104_k141_10365 | Not-determined |
| 18-104_k141_45213 | Not-determined |
| 18-104_k141_51461 | Not-determined |
| 18-104_k141_58816 | Not-determined |
| 18-104_k141_58818 | Not-determined |
| 18-104_k141_37883 | Not-determined |
| 18-104_k141_135670 | Not-determined |
| 18-104_k141_58822 | Not-determined |
| 18-104_k141_37888 | Not-determined |
| 18-104_k141_93961 | Not-determined |
| 18-104_k141_24136 | Not-determined |
| 18-104_k141_51469 | Not-determined |
| 18-104_k141_10371 | Not-determined |
| 18-104_k141_51470 | Not-determined |
| 18-104_k141_87044 | Not-determined |
| 18-104_k141_149595 | Not-determined |
| 18-104_k141_10376 | Not-determined |
| 18-104_k141_31496 | Not-determined |
| 18-104_k141_37897 | Not-determined |
| 18-104_k141_37898 | Not-determined |
| 18-104_k141_10379 | Low-quality |
| 18-104_k141_66004 | Low-quality |
| 18-104_k141_149600 | Not-determined |
| 18-104_k141_163416 | Not-determined |
| 18-104_k141_87052 | Not-determined |
| 18-104_k141_149604 | Not-determined |
| 18-104_k141_93976 | Not-determined |
| 18-104_k141_87053 | Not-determined |
| 18-104_k141_114595 | Not-determined |
| 18-104_k141_37907 | Not-determined |
| 18-104_k141_45232 | Not-determined |
| 18-104_k141_66014 | Not-determined |
| 18-104_k141_58829 | Low-quality |
| 18-104_k141_163421 | Not-determined |
| 18-104_k141_163424 | Not-determined |
| 18-104_k141_87060 | Not-determined |
| 18-104_k141_73119 | Not-determined |
| 18-104_k141_24152 | Not-determined |
| 18-104_k141_128644 | Not-determined |
| 18-104_k141_58832 | Not-determined |
| 18-104_k141_24153 | Not-determined |
| 18-104_k141_45244 | Not-determined |
| 18-104_k141_10394 | Low-quality |
| 18-104_k141_128648 | Not-determined |
| 18-104_k141_51482 | Not-determined |
| 18-104_k141_3386 | Not-determined |
| 18-104_k141_100642 | Not-determined |
| 18-104_k141_114613 | Not-determined |
| 18-104_k141_114614 | Not-determined |
| 18-104_k141_114615 | Not-determined |
| 18-104_k141_121204 | Not-determined |
| 18-104_k141_45255 | Not-determined |
| 18-104_k141_3395 | Not-determined |
| 18-104_k141_114619 | Not-determined |
| 18-104_k141_108055 | Not-determined |
| 18-104_k141_114621 | Not-determined |
| 18-104_k141_149614 | Not-determined |
| 18-104_k141_73140 | Low-quality |
| 18-104_k141_45262 | Not-determined |
| 18-104_k141_121215 | Not-determined |
| 18-104_k141_142883 | Not-determined |
| 18-104_k141_87078 | Not-determined |
| 18-104_k141_73143 | Not-determined |
| 18-104_k141_94009 | Not-determined |
| 18-104_k141_80314 | Not-determined |
| 18-104_k141_94013 | Not-determined |
| 18-104_k141_73152 | Not-determined |
| 18-104_k141_80319 | Not-determined |
| 18-104_k141_94018 | Not-determined |
| 18-104_k141_73154 | Not-determined |
| 18-104_k141_31518 | Not-determined |
| 18-104_k141_94020 | Not-determined |
| 18-104_k141_3416 | Not-determined |
| 18-104_k141_73160 | Not-determined |
| 18-104_k141_16913 | Not-determined |
| 18-104_k141_121226 | Not-determined |
| 18-104_k141_73169 | Not-determined |
| 18-104_k141_149633 | Not-determined |
| 18-104_k141_10430 | Not-determined |
| 18-104_k141_73173 | Not-determined |
| 18-104_k141_51489 | Not-determined |
| 18-104_k141_80338 | Not-determined |
| 18-104_k141_87098 | Not-determined |
| 18-104_k141_3433 | Not-determined |
| 18-104_k141_66032 | Not-determined |
| 18-104_k141_73175 | Not-determined |
| 18-104_k141_121243 | Not-determined |
| 18-104_k141_80340 | Not-determined |
| 18-104_k141_135716 | Not-determined |
| 18-104_k141_87102 | Not-determined |
| 18-104_k141_149639 | Not-determined |
| 18-104_k141_121247 | Not-determined |
| 18-104_k141_156701 | Not-determined |
| 18-104_k141_121248 | Not-determined |
| 18-104_k141_3438 | Not-determined |
| 18-104_k141_135718 | Not-determined |
| 18-104_k141_73181 | Not-determined |
| 18-104_k141_94033 | Low-quality |
| 18-104_k141_58864 | Not-determined |
| 18-104_k141_3444 | Low-quality |
| 18-104_k141_156704 | Not-determined |
| 18-104_k141_66044 | Not-determined |
| 18-104_k141_37943 | Low-quality |
| 18-104_k141_3445 | Not-determined |
| 18-104_k141_66047 | Not-determined |
| 18-104_k141_108085 | Not-determined |
| 18-104_k141_37948 | Low-quality |
| 18-104_k141_45282 | Not-determined |
| 18-104_k141_16918 | Not-determined |
| 18-104_k141_37950 | Not-determined |
| 18-104_k141_87123 | Not-determined |
| 18-104_k141_156708 | Not-determined |
| 18-104_k141_45284 | Not-determined |
| 18-104_k141_135733 | Not-determined |
| 18-104_k141_156713 | Not-determined |
| 18-104_k141_121263 | Not-determined |
| 18-104_k141_156716 | Not-determined |
| 18-104_k141_80356 | Not-determined |
| 18-104_k141_45293 | Low-quality |
| 18-104_k141_73190 | Not-determined |
| 18-104_k141_37959 | Not-determined |
| 18-104_k141_87130 | Not-determined |
| 18-104_k141_16928 | Not-determined |
| 18-104_k141_51508 | Not-determined |
| 18-104_k141_156722 | Not-determined |
| 18-104_k141_73194 | Not-determined |
| 18-104_k141_87132 | Not-determined |
| 18-104_k141_66055 | Not-determined |
| 18-104_k141_66056 | Not-determined |
| 18-104_k141_149658 | Not-determined |
| 18-104_k141_121273 | Not-determined |
| 18-104_k141_156727 | Not-determined |
| 18-104_k141_87138 | Not-determined |
| 18-104_k141_114641 | Low-quality |
| 18-104_k141_87141 | Not-determined |
| 18-104_k141_142899 | Not-determined |
| 18-104_k141_135746 | Not-determined |
| 18-104_k141_87143 | Not-determined |
| 18-104_k141_24166 | Not-determined |
| 18-104_k141_121278 | Low-quality |
| 18-104_k141_121282 | Low-quality |
| 18-104_k141_58872 | Not-determined |
| 18-104_k141_87157 | Not-determined |
| 18-104_k141_66063 | Not-determined |
| 18-104_k141_45308 | Not-determined |
| 18-104_k141_121290 | Low-quality |
| 18-104_k141_51517 | Not-determined |
| 18-104_k141_73213 | Not-determined |
| 18-104_k141_51519 | Not-determined |
| 18-104_k141_51521 | Not-determined |
| 18-104_k141_149670 | Not-determined |
| 18-104_k141_51523 | Not-determined |
| 18-104_k141_73216 | Low-quality |
| 18-104_k141_45311 | Not-determined |
| 18-104_k141_135748 | Low-quality |
| 18-104_k141_58879 | Not-determined |
| 18-104_k141_51527 | Not-determined |
| 18-104_k141_3465 | Not-determined |
| 18-104_k141_51529 | Not-determined |
| 18-104_k141_45314 | Not-determined |
| 18-104_k141_45316 | Low-quality |

| | |
|---|---|
| 18-104_k141_58882 | Not-determined |
| 18-104_k141_73224 | Not-determined |
| 18-104_k141_24194 | Not-determined |
| 18-104_k141_142915 | Not-determined |
| 18-104_k141_45320 | Low-quality |
| 18-104_k141_156747 | Not-determined |
| 18-104_k141_45321 | Not-determined |
| 18-104_k141_163458 | Low-quality |
| 18-104_k141_149679 | Not-determined |
| 18-104_k141_73228 | Not-determined |
| 18-104_k141_51533 | Not-determined |
| 18-104_k141_100704 | Not-determined |
| 18-104_k141_163465 | Not-determined |
| 18-104_k141_3475 | Not-determined |
| 18-104_k141_114651 | Low-quality |
| 18-104_k141_58895 | Low-quality |
| 18-104_k141_100710 | Not-determined |
| 18-104_k141_128663 | Not-determined |
| 18-104_k141_58896 | Not-determined |
| 18-104_k141_58897 | Not-determined |
| 18-104_k141_3480 | Low-quality |
| 18-104_k141_58901 | Not-determined |
| 18-104_k141_135769 | Not-determined |
| 18-104_k141_142928 | Not-determined |
| 18-104_k141_80381 | Not-determined |
| 18-104_k141_24210 | Not-determined |
| 18-104_k141_114656 | Low-quality |
| 18-104_k141_24215 | Not-determined |
| 18-104_k141_163475 | Not-determined |
| 18-104_k141_58905 | Not-determined |
| 18-104_k141_114661 | Not-determined |
| 18-104_k141_66092 | Not-determined |
| 18-104_k141_100733 | Not-determined |
| 18-104_k141_100734 | Not-determined |
| 18-104_k141_135780 | Not-determined |
| 18-104_k141_114664 | Low-quality |
| 18-104_k141_16989 | Not-determined |
| 18-104_k141_66098 | Not-determined |
| 18-104_k141_45344 | Not-determined |
| 18-104_k141_66101 | Not-determined |
| 18-104_k141_10448 | Not-determined |
| 18-104_k141_24223 | Not-determined |
| 18-104_k141_58912 | Not-determined |
| 18-104_k141_149688 | Not-determined |
| 18-104_k141_51544 | Not-determined |
| 18-104_k141_100736 | Low-quality |
| 18-104_k141_3489 | Not-determined |
| 18-104_k141_10451 | Not-determined |
| 18-104_k141_24226 | Not-determined |
| 18-104_k141_128681 | Low-quality |
| 18-104_k141_100741 | Not-determined |
| 18-104_k141_149694 | Not-determined |
| 18-104_k141_121304 | Low-quality |
| 18-104_k141_128685 | Not-determined |
| 18-104_k141_108160 | Low-quality |
| 18-104_k141_108164 | Not-determined |
| 18-104_k141_17001 | Not-determined |
| 18-104_k141_149701 | Not-determined |
| 18-104_k141_3503 | Not-determined |
| 18-104_k141_58924 | Not-determined |
| 18-104_k141_51554 | Not-determined |
| 18-104_k141_163494 | Not-determined |
| 18-104_k141_10459 | Not-determined |
| 18-104_k141_149703 | Not-determined |
| 18-104_k141_156793 | Not-determined |
| 18-104_k141_135786 | Not-determined |
| 18-104_k141_156794 | Not-determined |
| 18-104_k141_37978 | Not-determined |
| 18-104_k141_94048 | Not-determined |
| 18-104_k141_58930 | Not-determined |
| 18-104_k141_66121 | Not-determined |
| 18-104_k141_37981 | Not-determined |
| 18-104_k141_114684 | Not-determined |
| 18-104_k141_45347 | Low-quality |
| 18-104_k141_128698 | Not-determined |
| 18-104_k141_24243 | Not-determined |
| 18-104_k141_114688 | Not-determined |
| 18-104_k141_108170 | Not-determined |
| 18-104_k141_114689 | Not-determined |
| 18-104_k141_73250 | Not-determined |
| 18-104_k141_135791 | Not-determined |
| 18-104_k141_37990 | Not-determined |
| 18-104_k141_45352 | Not-determined |
| 18-104_k141_114697 | Not-determined |
| 18-104_k141_114698 | Not-determined |
| 18-104_k141_108176 | Not-determined |
| 18-104_k141_121328 | Not-determined |
| 18-104_k141_87179 | Not-determined |
| 18-104_k141_45354 | Not-determined |
| 18-104_k141_149716 | Not-determined |
| 18-104_k141_45355 | Not-determined |
| 18-104_k141_114701 | Not-determined |
| 18-104_k141_128706 | Not-determined |
| 18-104_k141_37997 | Low-quality |
| 18-104_k141_37998 | Not-determined |
| 18-104_k141_37999 | Not-determined |
| 18-104_k141_108179 | Not-determined |
| 18-104_k141_149724 | Not-determined |
| 18-104_k141_80394 | Not-determined |
| 18-104_k141_135802 | Not-determined |
| 18-104_k141_121335 | Not-determined |
| 18-104_k141_135805 | Not-determined |
| 18-104_k141_45362 | Not-determined |
| 18-104_k141_3519 | Not-determined |
| 18-104_k141_135806 | Not-determined |
| 18-104_k141_135807 | Not-determined |
| 18-104_k141_108183 | Not-determined |
| 18-104_k141_149728 | Not-determined |
| 18-104_k141_135809 | Not-determined |
| 18-104_k141_73263 | Not-determined |
| 18-104_k141_17022 | Not-determined |
| 18-104_k141_10482 | Not-determined |
| 18-104_k141_73271 | Not-determined |
| 18-104_k141_10484 | Not-determined |
| 18-104_k141_142948 | Not-determined |
| 18-104_k141_10491 | Not-determined |
| 18-104_k141_114723 | Not-determined |
| 18-104_k141_66153 | Not-determined |
| 18-104_k141_94065 | Not-determined |
| 18-104_k141_114724 | Low-quality |
| 18-104_k141_135825 | Low-quality |
| 18-104_k141_24263 | Low-quality |
| 18-104_k141_94068 | Not-determined |
| 18-104_k141_80413 | Not-determined |
| 18-104_k141_114728 | Not-determined |
| 18-104_k141_66161 | Not-determined |
| 18-104_k141_58965 | Not-determined |
| 18-104_k141_121356 | Not-determined |
| 18-104_k141_100754 | Not-determined |
| 18-104_k141_100755 | Not-determined |
| 18-104_k141_10503 | Not-determined |
| 18-104_k141_128724 | Not-determined |
| 18-104_k141_108207 | Not-determined |
| 18-104_k141_114737 | Low-quality |
| 18-104_k141_10510 | Not-determined |
| 18-104_k141_58979 | Not-determined |
| 18-104_k141_121376 | Low-quality |
| 18-104_k141_10520 | Not-determined |
| 18-104_k141_73300 | Not-determined |
| 18-104_k141_24279 | Not-determined |
| 18-104_k141_10521 | Not-determined |
| 18-104_k141_121381 | Not-determined |
| 18-104_k141_17040 | Not-determined |
| 18-104_k141_73303 | Not-determined |
| 18-104_k141_156842 | Low-quality |
| 18-104_k141_31552 | Not-determined |
| 18-104_k141_31553 | Low-quality |
| 18-104_k141_10527 | Not-determined |
| 18-104_k141_114756 | Not-determined |
| 18-104_k141_45398 | Not-determined |
| 18-104_k141_58991 | Not-determined |
| 18-104_k141_114760 | Not-determined |
| 18-104_k141_66192 | Not-determined |
| 18-104_k141_80418 | Not-determined |
| 18-104_k141_17055 | Not-determined |
| 18-104_k141_38025 | Not-determined |
| 18-104_k141_66195 | Not-determined |
| 18-104_k141_108232 | Not-determined |
| 18-104_k141_3549 | Not-determined |
| 18-104_k141_73313 | Not-determined |
| 18-104_k141_163544 | Not-determined |
| 18-104_k141_10546 | Not-determined |
| 18-104_k141_73316 | Not-determined |
| 18-104_k141_142962 | Not-determined |
| 18-104_k141_163547 | Not-determined |
| 18-104_k141_163548 | Not-determined |
| 18-104_k141_24290 | Not-determined |
| 18-104_k141_3551 | Low-quality |
| 18-104_k141_45407 | Not-determined |
| 18-104_k141_10550 | Not-determined |
| 18-104_k141_163554 | Not-determined |
| 18-104_k141_10552 | Not-determined |
| 18-104_k141_45410 | Not-determined |
| 18-104_k141_149757 | Low-quality |
| 18-104_k141_156858 | Not-determined |
| 18-104_k141_31566 | Not-determined |
| 18-104_k141_108245 | Not-determined |
| 18-104_k141_38031 | Not-determined |
| 18-104_k141_135850 | Not-determined |
| 18-104_k141_45418 | Not-determined |
| 18-104_k141_17063 | Not-determined |
| 18-104_k141_156865 | Not-determined |
| 18-104_k141_17064 | Not-determined |
| 18-104_k141_31570 | Not-determined |
| 18-104_k141_17068 | Not-determined |
| 18-104_k141_59007 | Low-quality |
| 18-104_k141_149762 | Not-determined |
| 18-104_k141_24312 | Not-determined |
| 18-104_k141_149765 | Not-determined |
| 18-104_k141_156872 | Not-determined |
| 18-104_k141_163561 | Not-determined |
| 18-104_k141_121401 | Not-determined |
| 18-104_k141_135863 | Not-determined |
| 18-104_k141_142976 | Not-determined |
| 18-104_k141_121405 | Low-quality |
| 18-104_k141_156880 | Not-determined |
| 18-104_k141_121407 | Not-determined |
| 18-104_k141_114772 | Not-determined |
| 18-104_k141_156882 | Not-determined |
| 18-104_k141_135868 | Not-determined |
| 18-104_k141_3573 | Low-quality |
| 18-104_k141_24317 | Not-determined |
| 18-104_k141_38040 | Not-determined |
| 18-104_k141_142989 | Not-determined |
| 18-104_k141_142991 | Not-determined |
| 18-104_k141_100790 | Not-determined |
| 18-104_k141_163570 | Not-determined |
| 18-104_k141_24321 | Low-quality |
| 18-104_k141_156887 | Not-determined |
| 18-104_k141_100792 | Not-determined |
| 18-104_k141_80452 | Not-determined |
| 18-104_k141_80453 | Not-determined |
| 18-104_k141_142995 | Not-determined |
| 18-104_k141_121419 | Not-determined |
| 18-104_k141_149780 | Not-determined |
| 18-104_k141_38052 | Not-determined |
| 18-104_k141_59025 | Not-determined |
| 18-104_k141_163586 | Not-determined |
| 18-104_k141_66216 | Low-quality |
| 18-104_k141_10590 | Not-determined |
| 18-104_k141_114783 | Not-determined |
| 18-104_k141_128743 | Not-determined |
| 18-104_k141_143007 | Not-determined |
| 18-104_k141_114784 | Not-determined |
| 18-104_k141_73340 | Not-determined |
| 18-104_k141_143012 | Not-determined |
| 18-104_k141_149788 | Not-determined |
| 18-104_k141_108270 | Not-determined |
| 18-104_k141_94118 | Not-determined |
| 18-104_k141_24336 | Not-determined |
| 18-104_k141_108274 | Not-determined |
| 18-104_k141_94121 | Not-determined |
| 18-104_k141_51607 | Low-quality |
| 18-104_k141_128752 | Low-quality |
| 18-104_k141_31590 | Not-determined |
| 18-104_k141_135882 | Low-quality |
| 18-104_k141_66224 | Not-determined |
| 18-104_k141_66226 | Not-determined |
| 18-104_k141_17087 | Not-determined |
| 18-104_k141_128755 | Not-determined |
| 18-104_k141_31596 | Not-determined |
| 18-104_k141_51610 | Not-determined |
| 18-104_k141_66231 | Not-determined |
| 18-104_k141_100811 | Not-determined |
| 18-104_k141_24352 | Not-determined |
| 18-104_k141_51612 | Not-determined |
| 18-104_k141_94129 | Not-determined |
| 18-104_k141_108284 | Not-determined |
| 18-104_k141_31603 | Not-determined |
| 18-104_k141_31604 | Not-determined |
| 18-104_k141_80476 | Not-determined |
| 18-104_k141_108287 | Low-quality |
| 18-104_k141_51616 | Not-determined |
| 18-104_k141_73360 | Not-determined |
| 18-104_k141_135889 | Low-quality |
| 18-104_k141_59043 | Not-determined |
| 18-104_k141_80479 | Not-determined |
| 18-104_k141_149808 | Low-quality |
| 18-104_k141_73364 | Not-determined |
| 18-104_k141_114799 | Not-determined |
| 18-104_k141_73366 | Not-determined |
| 18-104_k141_114800 | Not-determined |
| 18-104_k141_59048 | Not-determined |
| 18-104_k141_66247 | Not-determined |
| 18-104_k141_114802 | Not-determined |
| 18-104_k141_80482 | Not-determined |
| 18-104_k141_73372 | Not-determined |
| 18-104_k141_66251 | Not-determined |
| 18-104_k141_45451 | Not-determined |
| 18-104_k141_156926 | Not-determined |
| 18-104_k141_156927 | Not-determined |
| 18-104_k141_156929 | Low-quality |
| 18-104_k141_156936 | Not-determined |
| 18-104_k141_80487 | Not-determined |
| 18-104_k141_3600 | Not-determined |
| 18-104_k141_94137 | Low-quality |
| 18-104_k141_3602 | Not-determined |
| 18-104_k141_94140 | Low-quality |
| 18-104_k141_128774 | Not-determined |
| 18-104_k141_87199 | Not-determined |
| 18-104_k141_149818 | Not-determined |
| 18-104_k141_128776 | Not-determined |
| 18-104_k141_17100 | Not-determined |
| 18-104_k141_108309 | Not-determined |
| 18-104_k141_149820 | Low-quality |
| 18-104_k141_100835 | Not-determined |
| 18-104_k141_143050 | Not-determined |
| 18-104_k141_143052 | Not-determined |
| 18-104_k141_143054 | Not-determined |
| 18-104_k141_10612 | Not-determined |
| 18-104_k141_3612 | Not-determined |
| 18-104_k141_149827 | Not-determined |
| 18-104_k141_143056 | Not-determined |
| 18-104_k141_24365 | Not-determined |
| 18-104_k141_156954 | Not-determined |
| 18-104_k141_73385 | Not-determined |
| 18-104_k141_156956 | Not-determined |
| 18-104_k141_10615 | Not-determined |
| 18-104_k141_10616 | Not-determined |
| 18-104_k141_73389 | Not-determined |
| 18-104_k141_100840 | Not-determined |
| 18-104_k141_87211 | Low-quality |
| 18-104_k141_17116 | Not-determined |
| 18-104_k141_45486 | Not-determined |
| 18-104_k141_143064 | Not-determined |
| 18-104_k141_38081 | Not-determined |
| 18-104_k141_87218 | Not-determined |
| 18-104_k141_143067 | Not-determined |
| 18-104_k141_17119 | Not-determined |
| 18-104_k141_94167 | Low-quality |
| 18-104_k141_80501 | Not-determined |
| 18-104_k141_31619 | Not-determined |
| 18-104_k141_45491 | Not-determined |
| 18-104_k141_80502 | Not-determined |
| 18-104_k141_31621 | Not-determined |
| 18-104_k141_59073 | Not-determined |
| 18-104_k141_100853 | Not-determined |
| 18-104_k141_156966 | Not-determined |
| 18-104_k141_17125 | Not-determined |
| 18-104_k141_51629 | Not-determined |
| 18-104_k141_17126 | Not-determined |
| 18-104_k141_100862 | Not-determined |
| 18-104_k141_17130 | Low-quality |
| 18-104_k141_143085 | Not-determined |
| 18-104_k141_121456 | Not-determined |
| 18-104_k141_17133 | Not-determined |
| 18-104_k141_163609 | Not-determined |
| 18-104_k141_163612 | Not-determined |
| 18-104_k141_156983 | Not-determined |
| 18-104_k141_31632 | Not-determined |
| 18-104_k141_24386 | Low-quality |
| 18-104_k141_135942 | Not-determined |
| 18-104_k141_59077 | Not-determined |
| 18-104_k141_24392 | Not-determined |
| 18-104_k141_128786 | Not-determined |
| 18-104_k141_163624 | Not-determined |
| 18-104_k141_24396 | Not-determined |
| 18-104_k141_24397 | Not-determined |
| 18-104_k141_94189 | Not-determined |
| 18-104_k141_100880 | Not-determined |
| 18-104_k141_80527 | Low-quality |
| 18-104_k141_38106 | Not-determined |
| 18-104_k141_73427 | Not-determined |
| 18-104_k141_121468 | Not-determined |
| 18-104_k141_114813 | Not-determined |
| 18-104_k141_31638 | Not-determined |
| 18-104_k141_108317 | Not-determined |
| 18-104_k141_31640 | Not-determined |
| 18-104_k141_114816 | Not-determined |
| 18-104_k141_108318 | Not-determined |
| 18-104_k141_73434 | Not-determined |
| 18-104_k141_51652 | Not-determined |
| 18-104_k141_38111 | Not-determined |
| 18-104_k141_121471 | Low-quality |
| 18-104_k141_114818 | Not-determined |
| 18-104_k141_17144 | Not-determined |
| 18-104_k141_135959 | Low-quality |
| 18-104_k141_17149 | Not-determined |
| 18-104_k141_80536 | Not-determined |
| 18-104_k141_38115 | Not-determined |
| 18-104_k141_157003 | Low-quality |
| 18-104_k141_24400 | Not-determined |
| 18-104_k141_80539 | Not-determined |
| 18-104_k141_149869 | Not-determined |
| 18-104_k141_38124 | Not-determined |
| 18-104_k141_128807 | Low-quality |
| 18-104_k141_94197 | Not-determined |
| 18-104_k141_10639 | Not-determined |
| 18-104_k141_143103 | Not-determined |
| 18-104_k141_73445 | Not-determined |
| 18-104_k141_51658 | Not-determined |
| 18-104_k141_128814 | Not-determined |
| 18-104_k141_45513 | Not-determined |
| 18-104_k141_73447 | Low-quality |
| 18-104_k141_59100 | Not-determined |
| 18-104_k141_135971 | Not-determined |
| 18-104_k141_10643 | Not-determined |
| 18-104_k141_45516 | Not-determined |
| 18-104_k141_163654 | Not-determined |
| 18-104_k141_143109 | Not-determined |
| 18-104_k141_128818 | Not-determined |
| 18-104_k141_114833 | Not-determined |
| 18-104_k141_80544 | Not-determined |
| 18-104_k141_59104 | Not-determined |
| 18-104_k141_128820 | Not-determined |
| 18-104_k141_114834 | Not-determined |
| 18-104_k141_17155 | Not-determined |
| 18-104_k141_94204 | Not-determined |
| 18-104_k141_51662 | Not-determined |
| 18-104_k141_73457 | Low-quality |
| 18-104_k141_163657 | Not-determined |
| 18-104_k141_128828 | Not-determined |
| 18-104_k141_31648 | Not-determined |
| 18-104_k141_114838 | Not-determined |
| 18-104_k141_135978 | Not-determined |
| 18-104_k141_114840 | Not-determined |
| 18-104_k141_108323 | Not-determined |
| 18-104_k141_128833 | Not-determined |
| 18-104_k141_163665 | Not-determined |
| 18-104_k141_66323 | Not-determined |
| 18-104_k141_45531 | Not-determined |
| 18-104_k141_80564 | Not-determined |
| 18-104_k141_51669 | Not-determined |
| 18-104_k141_51670 | Not-determined |
| 18-104_k141_17169 | Not-determined |
| 18-104_k141_45533 | Not-determined |
| 18-104_k141_135983 | Low-quality |
| 18-104_k141_163672 | Not-determined |
| 18-104_k141_135985 | Not-determined |
| 18-104_k141_114847 | Low-quality |
| 18-104_k141_108329 | Not-determined |
| 18-104_k141_135986 | Not-determined |
| 18-104_k141_80568 | Not-determined |
| 18-104_k141_143119 | Not-determined |
| 18-104_k141_114849 | Not-determined |
| 18-104_k141_80569 | Not-determined |
| 18-104_k141_108331 | Not-determined |
| 18-104_k141_149877 | Not-determined |
| 18-104_k141_135990 | Not-determined |
| 18-104_k141_114851 | Not-determined |
| 18-104_k141_59114 | Not-determined |
| 18-104_k141_10661 | Not-determined |
| 18-104_k141_108335 | Not-determined |
| 18-104_k141_24429 | Not-determined |
| 18-104_k141_59115 | Not-determined |
| 18-104_k141_114854 | Low-quality |
| 18-104_k141_51676 | Not-determined |
| 18-104_k141_94223 | Not-determined |
| 18-104_k141_94225 | Not-determined |
| 18-104_k141_135995 | Not-determined |
| 18-104_k141_135996 | Low-quality |
| 18-104_k141_59119 | Not-determined |
| 18-104_k141_100900 | Not-determined |
| 18-104_k141_100901 | Not-determined |
| 18-104_k141_143130 | Not-determined |
| 18-104_k141_108342 | Not-determined |
| 18-104_k141_80578 | Not-determined |
| 18-104_k141_136000 | Not-determined |
| 18-104_k141_157044 | Low-quality |
| 18-104_k141_45540 | Not-determined |
| 18-104_k141_157045 | Not-determined |
| 18-104_k141_31667 | Not-determined |
| 18-104_k141_136001 | Not-determined |
| 18-104_k141_24442 | Low-quality |
| 18-104_k141_10670 | Not-determined |
| 18-104_k141_136004 | Not-determined |
| 18-104_k141_114863 | Not-determined |
| 18-104_k141_121502 | Not-determined |
| 18-104_k141_114866 | Not-determined |
| 18-104_k141_51683 | Not-determined |
| 18-104_k141_59134 | Not-determined |
| 18-104_k141_108364 | Not-determined |
| 18-104_k141_157056 | Not-determined |
| 18-104_k141_143141 | Not-determined |
| 18-104_k141_3628 | Not-determined |
| 18-104_k141_24458 | Not-determined |
| 18-104_k141_100919 | Low-quality |
| 18-104_k141_136008 | Not-determined |
| 18-104_k141_24460 | Not-determined |
| 18-104_k141_100923 | Not-determined |
| 18-104_k141_143149 | Not-determined |
| 18-104_k141_108377 | Not-determined |
| 18-104_k141_66329 | Not-determined |
| 18-104_k141_66331 | Not-determined |
| 18-104_k141_59143 | Low-quality |
| 18-104_k141_143152 | Not-determined |
| 18-104_k141_149915 | Not-determined |
| 18-104_k141_157070 | Not-determined |
| 18-104_k141_10694 | Not-determined |
| 18-104_k141_136015 | Not-determined |
| 18-104_k141_3636 | Not-determined |
| 18-104_k141_24465 | Not-determined |
| 18-104_k141_136018 | Not-determined |
| 18-104_k141_24469 | Not-determined |
| 18-104_k141_38173 | Not-determined |
| 18-104_k141_24472 | Not-determined |
| 18-104_k141_108384 | Low-quality |
| 18-104_k141_114885 | Not-determined |
| 18-104_k141_38182 | Not-determined |
| 18-104_k141_17187 | Not-determined |
| 18-104_k141_87266 | Not-determined |
| 18-104_k141_38188 | Not-determined |
| 18-104_k141_100935 | Low-quality |
| 18-104_k141_17190 | Not-determined |
| 18-104_k141_38189 | Not-determined |
| 18-104_k141_3654 | Not-determined |
| 18-104_k141_38191 | Not-determined |
| 18-104_k141_66347 | Not-determined |
| 18-104_k141_143178 | Not-determined |
| 18-104_k141_87275 | Not-determined |
| 18-104_k141_108391 | Not-determined |
| 18-104_k141_143185 | Not-determined |
| 18-104_k141_136042 | Not-determined |
| 18-104_k141_143191 | Not-determined |
| 18-104_k141_66358 | Not-determined |
| 18-104_k141_143192 | Not-determined |
| 18-104_k141_100941 | Not-determined |
| 18-104_k141_136046 | Not-determined |
| 18-104_k141_157092 | Not-determined |
| 18-104_k141_100944 | Not-determined |
| 18-104_k141_3665 | Not-determined |
| 18-104_k141_100945 | Not-determined |
| 18-104_k141_87279 | Low-quality |
| 18-104_k141_136049 | Not-determined |
| 18-104_k141_121528 | Not-determined |
| 18-104_k141_66369 | Not-determined |
| 18-104_k141_31694 | Low-quality |
| 18-104_k141_163678 | Not-determined |
| 18-104_k141_121533 | Not-determined |
| 18-104_k141_157098 | Not-determined |
| 18-104_k141_157100 | Low-quality |
| 18-104_k141_143200 | Not-determined |
| 18-104_k141_136054 | Not-determined |
| 18-104_k141_149956 | Not-determined |
| 18-104_k141_38222 | Not-determined |
| 18-104_k141_143206 | Not-determined |
| 18-104_k141_128863 | Not-determined |
| 18-104_k141_31700 | Not-determined |
| 18-104_k141_38226 | Not-determined |
| 18-104_k141_108404 | Low-quality |
| 18-104_k141_143210 | Not-determined |
| 18-104_k141_73505 | Not-determined |
| 18-104_k141_94264 | Not-determined |
| 18-104_k141_121542 | Not-determined |
| 18-104_k141_73509 | Not-determined |
| 18-104_k141_100954 | Not-determined |
| 18-104_k141_66385 | Not-determined |
| 18-104_k141_157112 | Low-quality |
| 18-104_k141_149966 | Not-determined |
| 18-104_k141_87291 | Not-determined |
| 18-104_k141_51709 | Not-determined |
| 18-104_k141_114906 | Not-determined |
| 18-104_k141_38234 | Not-determined |
| 18-104_k141_45554 | Not-determined |
| 18-104_k141_87298 | Not-determined |
| 18-104_k141_149978 | Not-determined |
| 18-104_k141_10753 | Not-determined |
| 18-104_k141_114912 | Not-determined |
| 18-104_k141_108418 | Not-determined |
| 18-104_k141_10754 | Not-determined |
| 18-104_k141_143228 | Not-determined |
| 18-104_k141_128877 | Low-quality |
| 18-104_k141_114913 | Not-determined |
| 18-104_k141_108420 | Not-determined |
| 18-104_k141_94279 | Not-determined |
| 18-104_k141_128880 | Not-determined |
| 18-104_k141_114917 | Not-determined |
| 18-104_k141_38244 | Not-determined |
| 18-104_k141_143235 | Not-determined |
| 18-104_k141_121557 | Not-determined |
| 18-104_k141_114922 | Not-determined |
| 18-104_k141_59176 | Not-determined |
| 18-104_k141_108424 | Not-determined |
| 18-104_k141_51719 | Low-quality |
| 18-104_k141_94283 | Not-determined |
| 18-104_k141_94284 | Not-determined |
| 18-104_k141_3686 | Not-determined |
| 18-104_k141_45568 | Not-determined |
| 18-104_k141_45570 | Not-determined |
| 18-104_k141_66412 | Not-determined |
| 18-104_k141_100967 | Not-determined |
| 18-104_k141_73530 | Not-determined |
| 18-104_k141_66414 | Not-determined |
| 18-104_k141_3688 | Not-determined |
| 18-104_k141_73533 | Not-determined |
| 18-104_k141_136075 | Not-determined |
| 18-104_k141_3690 | Not-determined |
| 18-104_k141_24521 | Not-determined |
| 18-104_k141_108438 | Low-quality |
| 18-104_k141_100974 | Not-determined |
| 18-104_k141_128902 | Not-determined |
| 18-104_k141_87315 | Low-quality |
| 18-104_k141_143253 | Not-determined |
| 18-104_k141_94299 | Not-determined |
| 18-104_k141_38256 | Not-determined |
| 18-104_k141_143257 | Not-determined |
| 18-104_k141_128908 | Not-determined |
| 18-104_k141_73546 | Not-determined |
| 18-104_k141_59193 | Not-determined |
| 18-104_k141_45576 | Not-determined |
| 18-104_k141_157132 | Not-determined |
| 18-104_k141_121582 | Not-determined |
| 18-104_k141_87325 | Not-determined |
| 18-104_k141_51731 | Not-determined |
| 18-104_k141_157136 | Not-determined |
| 18-104_k141_24529 | Not-determined |
| 18-104_k141_143268 | Low-quality |
| 18-104_k141_100986 | Low-quality |
| 18-104_k141_136092 | Not-determined |

| | |
|---|---|
| 18-104_k141_121588 | Low-quality |
| 18-104_k141_94311 | Not-determined |
| 18-104_k141_136094 | Low-quality |
| 18-104_k141_31752 | Not-determined |
| 18-104_k141_51737 | Not-determined |
| 18-104_k141_94318 | Not-determined |
| 18-104_k141_157145 | Not-determined |
| 18-104_k141_31757 | Not-determined |
| 18-104_k141_66433 | Not-determined |
| 18-104_k141_73557 | Not-determined |
| 18-104_k141_59202 | Low-quality |
| 18-104_k141_45586 | Not-determined |
| 18-104_k141_17211 | Not-determined |
| 18-104_k141_45588 | Not-determined |
| 18-104_k141_38284 | Low-quality |
| 18-104_k141_17214 | Not-determined |
| 18-104_k141_94323 | Not-determined |
| 18-104_k141_17217 | Not-determined |
| 18-104_k141_17219 | Not-determined |
| 18-104_k141_94325 | Not-determined |
| 18-104_k141_149997 | Not-determined |
| 18-104_k141_143283 | Not-determined |
| 18-104_k141_31775 | Not-determined |
| 18-104_k141_128940 | Not-determined |
| 18-104_k141_31776 | Not-determined |
| 18-104_k141_121597 | Not-determined |
| 18-104_k141_51756 | Not-determined |
| 18-104_k141_121598 | Not-determined |
| 18-104_k141_163702 | Not-determined |
| 18-104_k141_73565 | Low-quality |
| 18-104_k141_38291 | Not-determined |
| 18-104_k141_163705 | Not-determined |
| 18-104_k141_121602 | Not-determined |
| 18-104_k141_163708 | Not-determined |
| 18-104_k141_114952 | Not-determined |
| 18-104_k141_94347 | Not-determined |
| 18-104_k141_114956 | Not-determined |
| 18-104_k141_45607 | Not-determined |
| 18-104_k141_17239 | Not-determined |
| 18-104_k141_45608 | Low-quality |
| 18-104_k141_45609 | Not-determined |
| 18-104_k141_121611 | Not-determined |
| 18-104_k141_121612 | Low-quality |
| 18-104_k141_3718 | Low-quality |
| 18-104_k141_66463 | Not-determined |
| 18-104_k141_136119 | Not-determined |
| 18-104_k141_87350 | Not-determined |
| 18-104_k141_66467 | Not-determined |
| 18-104_k141_45616 | Not-determined |
| 18-104_k141_143298 | Low-quality |
| 18-104_k141_163722 | Not-determined |
| 18-104_k141_24552 | Not-determined |
| 18-104_k141_24553 | Not-determined |
| 18-104_k141_150016 | Not-determined |
| 18-104_k141_3726 | Not-determined |
| 18-104_k141_136124 | Not-determined |
| 18-104_k141_150017 | Low-quality |
| 18-104_k141_87357 | Not-determined |
| 18-104_k141_17244 | Not-determined |
| 18-104_k141_31795 | Low-quality |
| 18-104_k141_45624 | Low-quality |
| 18-104_k141_45625 | Not-determined |
| 18-104_k141_150020 | Not-determined |
| 18-104_k141_3731 | Not-determined |
| 18-104_k141_157155 | Not-determined |
| 18-104_k141_45634 | Not-determined |
| 18-104_k141_80624 | Not-determined |
| 18-104_k141_121631 | Not-determined |
| 18-104_k141_143312 | Not-determined |
| 18-104_k141_45639 | Not-determined |
| 18-104_k141_143313 | Not-determined |
| 18-104_k141_163737 | Not-determined |
| 18-104_k141_87368 | Not-determined |
| 18-104_k141_143316 | Not-determined |
| 18-104_k141_3733 | Low-quality |
| 18-104_k141_45644 | Not-determined |
| 18-104_k141_94382 | Not-determined |
| 18-104_k141_108479 | Not-determined |
| 18-104_k141_45651 | Low-quality |
| 18-104_k141_163738 | Not-determined |
| 18-104_k141_94390 | Low-quality |
| 18-104_k141_128954 | Not-determined |
| 18-104_k141_66482 | Low-quality |
| 18-104_k141_51774 | Not-determined |
| 18-104_k141_45655 | Not-determined |
| 18-104_k141_38308 | Not-determined |
| 18-104_k141_80641 | Not-determined |
| 18-104_k141_66489 | Not-determined |
| 18-104_k141_163748 | Not-determined |
| 18-104_k141_108485 | Not-determined |
| 18-104_k141_38314 | Not-determined |
| 18-104_k141_66492 | Not-determined |
| 18-104_k141_128961 | Low-quality |
| 18-104_k141_163754 | Not-determined |
| 18-104_k141_38319 | Not-determined |
| 18-104_k141_73602 | Not-determined |
| 18-104_k141_45663 | Not-determined |
| 18-104_k141_115003 | Not-determined |
| 18-104_k141_45668 | Not-determined |
| 18-104_k141_115007 | Not-determined |
| 18-104_k141_66500 | Not-determined |
| 18-104_k141_38327 | Not-determined |
| 18-104_k141_157189 | Not-determined |
| 18-104_k141_73617 | Not-determined |
| 18-104_k141_73618 | Not-determined |
| 18-104_k141_73619 | Not-determined |
| 18-104_k141_51776 | Not-determined |
| 18-104_k141_59266 | Low-quality |
| 18-104_k141_87387 | Low-quality |
| 18-104_k141_115021 | Not-determined |
| 18-104_k141_128970 | Not-determined |
| 18-104_k141_38340 | Not-determined |
| 18-104_k141_108503 | Not-determined |
| 18-104_k141_66513 | Not-determined |
| 18-104_k141_73634 | Not-determined |
| 18-104_k141_38343 | Not-determined |
| 18-104_k141_94416 | Low-quality |
| 18-104_k141_121653 | Not-determined |
| 18-104_k141_121655 | Not-determined |
| 18-104_k141_3764 | Not-determined |
| 18-104_k141_51785 | Not-determined |
| 18-104_k141_10764 | Not-determined |
| 18-104_k141_45691 | Not-determined |
| 18-104_k141_45692 | Not-determined |
| 18-104_k141_163766 | Not-determined |
| 18-104_k141_3771 | Low-quality |
| 18-104_k141_10768 | Not-determined |
| 18-104_k141_115037 | Not-determined |
| 18-104_k141_73644 | Not-determined |
| 18-104_k141_128982 | Not-determined |
| 18-104_k141_143331 | Not-determined |
| 18-104_k141_136151 | Not-determined |
| 18-104_k141_128985 | Not-determined |
| 18-104_k141_66523 | Not-determined |
| 18-104_k141_128986 | Not-determined |
| 18-104_k141_38356 | Not-determined |
| 18-104_k141_38357 | Not-determined |
| 18-104_k141_128990 | Not-determined |
| 18-104_k141_80662 | Not-determined |
| 18-104_k141_94421 | Not-determined |
| 18-104_k141_45698 | Not-determined |
| 18-104_k141_45699 | Low-quality |
| 18-104_k141_163778 | Not-determined |
| 18-104_k141_31849 | Not-determined |
| 18-104_k141_51801 | Not-determined |
| 18-104_k141_128997 | Low-quality |
| 18-104_k141_51804 | Not-determined |
| 18-104_k141_121668 | Not-determined |
| 18-104_k141_10781 | Not-determined |
| 18-104_k141_73656 | Not-determined |
| 18-104_k141_129001 | Not-determined |
| 18-104_k141_80667 | Not-determined |
| 18-104_k141_31855 | Not-determined |
| 18-104_k141_24567 | Not-determined |
| 18-104_k141_143342 | Not-determined |
| 18-104_k141_108523 | Not-determined |
| 18-104_k141_73660 | Not-determined |
| 18-104_k141_143343 | Not-determined |
| 18-104_k141_87414 | Not-determined |
| 18-104_k141_94430 | Not-determined |
| 18-104_k141_38367 | Not-determined |
| 18-104_k141_121676 | Not-determined |
| 18-104_k141_45713 | Not-determined |
| 18-104_k141_38369 | Not-determined |
| 18-104_k141_10787 | Not-determined |
| 18-104_k141_10789 | Not-determined |
| 18-104_k141_10791 | Not-determined |
| 18-104_k141_38377 | Low-quality |
| 18-104_k141_45721 | Low-quality |
| 18-104_k141_10793 | Not-determined |
| 18-104_k141_51811 | Not-determined |
| 18-104_k141_150048 | Not-determined |
| 18-104_k141_10795 | Not-determined |
| 18-104_k141_80680 | Not-determined |
| 18-104_k141_73676 | Not-determined |
| 18-104_k141_129014 | Not-determined |
| 18-104_k141_80681 | Not-determined |
| 18-104_k141_129015 | Not-determined |
| 18-104_k141_87429 | Not-determined |
| 18-104_k141_51820 | Not-determined |
| 18-104_k141_45726 | Low-quality |
| 18-104_k141_157232 | Not-determined |
| 18-104_k141_115056 | Not-determined |
| 18-104_k141_51823 | Not-determined |
| 18-104_k141_143366 | Not-determined |
| 18-104_k141_51825 | Not-determined |
| 18-104_k141_129018 | Not-determined |
| 18-104_k141_150055 | Not-determined |
| 18-104_k141_66551 | Not-determined |
| 18-104_k141_45730 | Not-determined |
| 18-104_k141_80693 | Not-determined |
| 18-104_k141_115062 | Low-quality |
| 18-104_k141_150058 | Not-determined |
| 18-104_k141_80694 | Not-determined |
| 18-104_k141_115063 | Not-determined |
| 18-104_k141_150059 | Not-determined |
| 18-104_k141_87441 | Not-determined |
| 18-104_k141_3813 | Not-determined |
| 18-104_k141_80698 | Low-quality |
| 18-104_k141_115066 | Not-determined |
| 18-104_k141_115068 | Not-determined |
| 18-104_k141_24581 | Not-determined |
| 18-104_k141_163801 | Not-determined |
| 18-104_k141_129032 | Not-determined |
| 18-104_k141_59304 | Low-quality |
| 18-104_k141_129034 | Not-determined |
| 18-104_k141_150061 | Not-determined |
| 18-104_k141_150063 | Not-determined |
| 18-104_k141_101069 | Not-determined |
| 18-104_k141_108547 | Not-determined |
| 18-104_k141_150064 | Not-determined |
| 18-104_k141_129041 | Not-determined |
| 18-104_k141_3831 | Not-determined |
| 18-104_k141_163810 | Not-determined |
| 18-104_k141_129049 | Not-determined |
| 18-104_k141_163813 | Not-determined |
| 18-104_k141_136166 | Not-determined |
| 18-104_k141_80704 | Not-determined |
| 18-104_k141_45755 | Not-determined |
| 18-104_k141_94453 | Not-determined |
| 18-104_k141_59310 | Not-determined |
| 18-104_k141_101077 | Not-determined |
| 18-104_k141_51851 | Not-determined |
| 18-104_k141_73683 | Low-quality |
| 18-104_k141_129053 | Not-determined |
| 18-104_k141_157241 | Low-quality |
| 18-104_k141_80712 | Not-determined |
| 18-104_k141_101079 | Low-quality |
| 18-104_k141_94459 | Not-determined |
| 18-104_k141_163816 | Not-determined |
| 18-104_k141_45758 | Not-determined |
| 18-104_k141_101085 | Not-determined |
| 18-104_k141_94463 | Not-determined |
| 18-104_k141_38401 | Not-determined |
| 18-104_k141_101087 | Not-determined |
| 18-104_k141_94464 | Not-determined |
| 18-104_k141_24601 | Not-determined |
| 18-104_k141_24602 | Not-determined |
| 18-104_k141_115085 | Not-determined |
| 18-104_k141_115088 | Not-determined |
| 18-104_k141_115091 | Not-determined |
| 18-104_k141_115092 | Not-determined |
| 18-104_k141_150080 | Not-determined |
| 18-104_k141_10829 | Not-determined |
| 18-104_k141_163824 | Not-determined |
| 18-104_k141_115094 | Not-determined |
| 18-104_k141_150084 | Low-quality |
| 18-104_k141_45767 | Not-determined |
| 18-104_k141_163827 | Not-determined |
| 18-104_k141_163828 | Low-quality |
| 18-104_k141_24621 | Not-determined |
| 18-104_k141_115095 | Not-determined |
| 18-104_k141_143400 | Not-determined |
| 18-104_k141_38424 | Not-determined |
| 18-104_k141_51880 | Not-determined |
| 18-104_k141_51883 | Not-determined |
| 18-104_k141_66588 | Not-determined |
| 18-104_k141_3870 | Not-determined |
| 18-104_k141_163839 | Not-determined |
| 18-104_k141_59323 | Not-determined |
| 18-104_k141_129082 | Not-determined |
| 18-104_k141_59324 | Not-determined |
| 18-104_k141_59325 | Low-quality |
| 18-104_k141_163846 | Not-determined |
| 18-104_k141_51892 | Not-determined |
| 18-104_k141_136197 | Not-determined |
| 18-104_k141_163854 | Not-determined |
| 18-104_k141_87458 | Not-determined |
| 18-104_k141_87459 | Not-determined |
| 18-104_k141_59338 | Not-determined |
| 18-104_k141_59346 | Not-determined |
| 18-104_k141_150102 | Not-determined |
| 18-104_k141_143421 | Low-quality |
| 18-104_k141_24637 | Not-determined |
| 18-104_k141_51898 | Not-determined |
| 18-104_k141_150107 | Not-determined |
| 18-104_k141_66606 | Not-determined |
| 18-104_k141_136207 | Not-determined |
| 18-104_k141_51900 | Low-quality |
| 18-104_k141_59354 | Low-quality |
| 18-104_k141_80746 | Not-determined |
| 18-104_k141_150112 | Not-determined |
| 18-104_k141_101119 | Not-determined |
| 18-104_k141_24642 | Not-determined |
| 18-104_k141_24643 | Not-determined |
| 18-104_k141_59355 | Not-determined |
| 18-104_k141_24646 | Not-determined |
| 18-104_k141_129098 | Low-quality |
| 18-104_k141_129100 | Not-determined |
| 18-104_k141_115107 | Not-determined |
| 18-104_k141_31878 | Not-determined |
| 18-104_k141_80752 | Not-determined |
| 18-104_k141_59360 | Not-determined |
| 18-104_k141_66614 | Not-determined |
| 18-104_k141_136213 | Not-determined |
| 18-104_k141_87468 | Not-determined |
| 18-104_k141_51905 | Not-determined |
| 18-104_k141_31888 | Not-determined |
| 18-104_k141_66619 | Not-determined |
| 18-104_k141_108556 | Not-determined |
| 18-104_k141_163868 | Not-determined |
| 18-104_k141_115111 | Low-quality |
| 18-104_k141_3896 | Not-determined |
| 18-104_k141_38441 | Not-determined |
| 18-104_k141_129108 | Low-quality |
| 18-104_k141_66625 | Not-determined |
| 18-104_k141_38442 | Low-quality |
| 18-104_k141_101128 | Not-determined |
| 18-104_k141_51919 | Not-determined |
| 18-104_k141_73706 | Not-determined |
| 18-104_k141_163880 | Not-determined |
| 18-104_k141_115124 | Not-determined |
| 18-104_k141_66626 | Not-determined |
| 18-104_k141_3905 | Not-determined |
| 18-104_k141_163883 | Not-determined |
| 18-104_k141_45783 | Not-determined |
| 18-104_k141_143431 | Not-determined |
| 18-104_k141_143434 | Not-determined |
| 18-104_k141_38446 | Not-determined |
| 18-104_k141_51931 | Not-determined |
| 18-104_k141_51933 | Not-determined |
| 18-104_k141_121698 | Low-quality |
| 18-104_k141_51934 | Not-determined |
| 18-104_k141_3912 | Not-determined |
| 18-104_k141_59374 | Not-determined |
| 18-104_k141_51935 | Low-quality |
| 18-104_k141_66635 | Not-determined |
| 18-104_k141_101135 | Not-determined |
| 18-104_k141_80786 | Not-determined |
| 18-104_k141_31911 | Not-determined |
| 18-104_k141_101137 | Not-determined |
| 18-104_k141_3915 | Not-determined |
| 18-104_k141_51940 | Not-determined |
| 18-104_k141_80787 | Not-determined |
| 18-104_k141_31916 | Not-determined |
| 18-104_k141_59378 | Not-determined |
| 18-104_k141_80792 | Not-determined |
| 18-104_k141_115139 | Not-determined |
| 18-104_k141_121705 | Not-determined |
| 18-104_k141_115143 | Not-determined |
| 18-104_k141_3923 | Not-determined |
| 18-104_k141_101145 | Not-determined |
| 18-104_k141_101146 | Not-determined |
| 18-104_k141_66647 | Not-determined |
| 18-104_k141_38465 | Low-quality |
| 18-104_k141_10868 | Not-determined |
| 18-104_k141_115148 | Not-determined |
| 18-104_k141_121708 | Not-determined |
| 18-104_k141_163894 | Not-determined |
| 18-104_k141_157268 | Not-determined |
| 18-104_k141_163895 | Not-determined |
| 18-104_k141_108583 | Not-determined |
| 18-104_k141_136235 | Not-determined |
| 18-104_k141_129121 | Not-determined |
| 18-104_k141_121712 | Not-determined |
| 18-104_k141_66657 | Not-determined |
| 18-104_k141_108587 | Not-determined |
| 18-104_k141_66662 | Not-determined |
| 18-104_k141_3935 | Not-determined |
| 18-104_k141_38476 | Not-determined |
| 18-104_k141_3937 | Not-determined |
| 18-104_k141_94510 | Not-determined |
| 18-104_k141_150131 | Not-determined |
| 18-104_k141_121718 | Not-determined |
| 18-104_k141_24678 | Not-determined |
| 18-104_k141_101173 | Not-determined |
| 18-104_k141_51954 | Not-determined |
| 18-104_k141_136243 | Not-determined |
| 18-104_k141_163906 | Low-quality |
| 18-104_k141_101178 | Not-determined |
| 18-104_k141_163908 | Not-determined |
| 18-104_k141_163909 | Not-determined |
| 18-104_k141_121724 | Not-determined |
| 18-104_k141_101180 | Not-determined |
| 18-104_k141_51959 | Not-determined |
| 18-104_k141_129135 | Low-quality |
| 18-104_k141_136251 | Low-quality |
| 18-104_k141_38480 | Not-determined |
| 18-104_k141_121728 | Not-determined |
| 18-104_k141_24686 | Not-determined |
| 18-104_k141_136256 | Not-determined |
| 18-104_k141_87477 | Not-determined |
| 18-104_k141_24692 | Not-determined |
| 18-104_k141_150149 | Not-determined |
| 18-104_k141_73739 | Not-determined |
| 18-104_k141_108597 | Low-quality |
| 18-104_k141_3957 | Not-determined |
| 18-104_k141_24695 | Not-determined |
| 18-104_k141_66675 | Not-determined |
| 18-104_k141_80827 | Not-determined |
| 18-104_k141_66677 | Not-determined |
| 18-104_k141_150156 | Not-determined |
| 18-104_k141_150159 | Not-determined |
| 18-104_k141_136265 | Low-quality |
| 18-104_k141_143467 | Not-determined |
| 18-104_k141_94527 | Not-determined |
| 18-104_k141_108609 | Not-determined |
| 18-104_k141_73746 | Not-determined |
| 18-104_k141_66687 | Not-determined |
| 18-104_k141_3968 | Not-determined |
| 18-104_k141_24706 | Low-quality |
| 18-104_k141_73751 | Not-determined |
| 18-104_k141_45827 | Not-determined |
| 18-104_k141_66689 | Not-determined |
| 18-104_k141_163918 | Not-determined |
| 18-104_k141_73757 | Not-determined |
| 18-104_k141_94537 | Low-quality |
| 18-104_k141_136281 | Not-determined |
| 18-104_k141_94539 | Low-quality |
| 18-104_k141_163928 | Not-determined |
| 18-104_k141_73763 | Not-determined |
| 18-104_k141_136289 | Not-determined |
| 18-104_k141_150177 | Not-determined |
| 18-104_k141_163937 | Not-determined |
| 18-104_k141_163938 | Not-determined |
| 18-104_k141_66697 | Not-determined |
| 18-104_k141_157295 | Not-determined |
| 18-104_k141_45839 | Not-determined |
| 18-104_k141_80856 | Not-determined |
| 18-104_k141_38487 | Not-determined |
| 18-104_k141_150185 | Not-determined |
| 18-104_k141_24727 | Not-determined |
| 18-104_k141_129147 | Not-determined |
| 18-104_k141_59409 | Not-determined |
| 18-104_k141_150189 | Not-determined |
| 18-104_k141_108636 | Not-determined |
| 18-104_k141_80866 | Not-determined |
| 18-104_k141_45845 | Not-determined |
| 18-104_k141_94560 | Not-determined |
| 18-104_k141_150191 | Not-determined |
| 18-104_k141_94561 | Not-determined |
| 18-104_k141_94562 | Not-determined |
| 18-104_k141_108641 | Low-quality |
| 18-104_k141_94563 | Not-determined |
| 18-104_k141_80872 | Not-determined |
| 18-104_k141_24738 | Not-determined |
| 18-104_k141_66706 | Not-determined |
| 18-104_k141_73768 | Not-determined |
| 18-104_k141_31964 | Not-determined |
| 18-104_k141_129157 | Not-determined |
| 18-104_k141_31966 | Not-determined |
| 18-104_k141_10893 | Not-determined |
| 18-104_k141_10894 | Not-determined |
| 18-104_k141_31969 | Not-determined |
| 18-104_k141_45852 | Not-determined |
| 18-104_k141_129160 | Not-determined |
| 18-104_k141_150199 | Not-determined |
| 18-104_k141_3979 | Low-quality |
| 18-104_k141_108648 | Not-determined |
| 18-104_k141_143490 | Not-determined |
| 18-104_k141_101207 | Not-determined |
| 18-104_k141_163965 | Not-determined |
| 18-104_k141_10901 | Not-determined |
| 18-104_k141_80885 | Not-determined |
| 18-104_k141_101210 | Not-determined |
| 18-104_k141_101213 | Not-determined |
| 18-104_k141_163972 | Not-determined |
| 18-104_k141_163974 | Not-determined |
| 18-104_k141_80892 | Not-determined |
| 18-104_k141_59435 | Low-quality |
| 18-104_k141_66712 | Not-determined |
| 18-104_k141_10908 | Not-determined |
| 18-104_k141_94572 | Not-determined |
| 18-104_k141_150215 | Not-determined |
| 18-104_k141_163983 | Not-determined |
| 18-104_k141_143503 | Not-determined |
| 18-104_k141_129173 | Not-determined |
| 18-104_k141_94575 | Not-determined |
| 18-104_k141_163986 | Low-quality |
| 18-104_k141_115184 | Not-determined |
| 18-104_k141_80899 | Not-determined |
| 18-104_k141_38516 | Not-determined |
| 18-104_k141_66720 | Not-determined |
| 18-104_k141_38518 | Not-determined |
| 18-104_k141_45879 | Low-quality |
| 18-104_k141_24758 | Not-determined |
| 18-104_k141_143514 | Not-determined |
| 18-104_k141_115188 | Not-determined |
| 18-104_k141_24759 | Not-determined |
| 18-104_k141_24760 | Not-determined |
| 18-104_k141_115192 | Not-determined |
| 18-104_k141_163993 | Low-quality |
| 18-104_k141_143515 | Not-determined |
| 18-104_k141_45886 | Not-determined |
| 18-104_k141_24766 | Not-determined |
| 18-104_k141_80903 | Not-determined |
| 18-104_k141_4002 | Not-determined |
| 18-104_k141_101238 | Not-determined |
| 18-104_k141_121771 | Not-determined |
| 18-104_k141_24776 | Not-determined |
| 18-104_k141_121774 | Not-determined |
| 18-104_k141_136338 | Not-determined |
| 18-104_k141_136340 | Not-determined |
| 18-104_k141_94599 | Not-determined |
| 18-104_k141_164003 | Low-quality |
| 18-104_k141_164009 | Not-determined |
| 18-104_k141_164010 | Not-determined |
| 18-104_k141_164012 | Not-determined |
| 18-104_k141_73807 | Not-determined |
| 18-104_k141_136345 | Not-determined |
| 18-104_k141_17331 | Not-determined |
| 18-104_k141_45905 | Not-determined |
| 18-104_k141_94607 | Not-determined |
| 18-104_k141_38559 | Not-determined |
| 18-104_k141_31979 | Not-determined |
| 18-104_k141_24783 | Low-quality |
| 18-104_k141_51988 | Not-determined |
| 18-104_k141_66723 | Not-determined |
| 18-104_k141_31982 | Not-determined |
| 18-104_k141_94609 | Not-determined |
| 18-104_k141_129198 | Not-determined |
| 18-104_k141_73813 | Not-determined |
| 18-104_k141_164019 | Low-quality |
| 18-104_k141_17338 | Not-determined |
| 18-104_k141_38563 | Not-determined |
| 18-104_k141_38564 | Not-determined |
| 18-104_k141_129207 | Not-determined |
| 18-104_k141_94618 | Not-determined |
| 18-104_k141_129209 | Low-quality |
| 18-104_k141_121784 | Not-determined |
| 18-104_k141_38568 | Not-determined |
| 18-104_k141_10935 | Not-determined |
| 18-104_k141_129210 | Low-quality |
| 18-104_k141_73819 | Low-quality |
| 18-104_k141_115214 | Not-determined |
| 18-104_k141_73820 | Not-determined |
| 18-104_k141_121786 | Low-quality |
| 18-104_k141_10937 | Not-determined |
| 18-104_k141_10938 | Not-determined |
| 18-104_k141_80939 | Not-determined |
| 18-104_k141_164027 | Not-determined |
| 18-104_k141_4032 | Not-determined |
| 18-104_k141_94629 | Not-determined |
| 18-104_k141_164029 | Not-determined |
| 18-104_k141_94630 | Not-determined |
| 18-104_k141_108709 | Not-determined |
| 18-104_k141_38576 | Not-determined |
| 18-104_k141_108710 | Not-determined |
| 18-104_k141_108715 | Low-quality |
| 18-104_k141_164034 | Not-determined |
| 18-104_k141_66739 | Not-determined |
| 18-104_k141_164039 | Not-determined |
| 18-104_k141_24795 | Not-determined |
| 18-104_k141_87481 | Not-determined |
| 18-104_k141_66742 | Not-determined |
| 18-104_k141_59455 | Not-determined |
| 18-104_k141_115222 | Not-determined |
| 18-104_k141_66743 | Not-determined |
| 18-104_k141_80954 | Not-determined |
| 18-104_k141_164042 | Low-quality |
| 18-104_k141_87482 | Not-determined |
| 18-104_k141_10948 | Not-determined |
| 18-104_k141_80956 | Not-determined |
| 18-104_k141_94633 | Not-determined |
| 18-104_k141_10950 | Not-determined |
| 18-104_k141_10952 | Not-determined |
| 18-104_k141_87485 | Not-determined |
| 18-104_k141_108727 | Not-determined |
| 18-104_k141_38585 | Not-determined |
| 18-104_k141_115231 | Not-determined |
| 18-104_k141_45939 | Low-quality |
| 18-104_k141_94643 | Not-determined |
| 18-104_k141_31996 | Not-determined |
| 18-104_k141_4051 | Not-determined |
| 18-104_k141_10957 | Not-determined |
| 18-104_k141_10958 | Low-quality |
| 18-104_k141_4056 | Not-determined |
| 18-104_k141_101278 | Not-determined |
| 18-104_k141_32004 | Not-determined |
| 18-104_k141_121800 | Not-determined |
| 18-104_k141_66754 | Not-determined |
| 18-104_k141_143559 | Not-determined |
| 18-104_k141_108736 | Not-determined |
| 18-104_k141_80977 | Not-determined |
| 18-104_k141_121805 | Not-determined |
| 18-104_k141_94654 | Not-determined |
| 18-104_k141_94655 | Not-determined |
| 18-104_k141_10969 | Not-determined |
| 18-104_k141_115253 | Not-determined |
| 18-104_k141_24814 | Low-quality |
| 18-104_k141_4073 | Not-determined |
| 18-104_k141_52018 | Not-determined |
| 18-104_k141_164064 | Not-determined |
| 18-104_k141_101290 | Not-determined |
| 18-104_k141_73851 | Low-quality |
| 18-104_k141_94667 | Not-determined |
| 18-104_k141_108753 | Not-determined |
| 18-104_k141_101293 | Not-determined |
| 18-104_k141_10987 | Not-determined |

| | |
|---|---|
| 18-104_k141_4089 | Not-determined |
| 18-104_k141_32019 | Not-determined |
| 18-104_k141_45949 | Not-determined |
| 18-104_k141_24837 | Not-determined |
| 18-104_k141_101305 | Low-quality |
| 18-104_k141_157322 | Not-determined |
| 18-104_k141_143573 | Not-determined |
| 18-104_k141_45960 | Not-determined |
| 18-104_k141_32026 | Not-determined |
| 18-104_k141_66763 | Not-determined |
| 18-104_k141_80993 | Not-determined |
| 18-104_k141_80994 | Low-quality |
| 18-104_k141_87513 | Not-determined |
| 18-104_k141_66767 | Not-determined |
| 18-104_k141_80996 | Not-determined |
| 18-104_k141_164070 | Not-determined |
| 18-104_k141_73858 | Not-determined |
| 18-104_k141_73859 | Not-determined |
| 18-104_k141_45967 | Not-determined |
| 18-104_k141_94670 | Low-quality |
| 18-104_k141_164078 | Not-determined |
| 18-104_k141_157332 | Not-determined |
| 18-104_k141_4102 | Not-determined |
| 18-104_k141_4104 | Not-determined |
| 18-104_k141_4105 | Not-determined |
| 18-104_k141_101325 | Not-determined |
| 18-104_k141_38595 | Not-determined |
| 18-104_k141_81002 | Not-determined |
| 18-104_k141_52043 | Low-quality |
| 18-104_k141_32027 | Not-determined |
| 18-104_k141_150218 | Not-determined |
| 18-104_k141_32028 | Not-determined |
| 18-104_k141_73877 | Not-determined |
| 18-104_k141_164087 | Not-determined |
| 18-104_k141_129289 | Not-determined |
| 18-104_k141_66782 | Not-determined |
| 18-104_k141_164089 | Low-quality |
| 18-104_k141_4113 | Not-determined |
| 18-104_k141_52054 | Not-determined |
| 18-104_k141_164091 | Not-determined |
| 18-104_k141_115281 | Not-determined |
| 18-104_k141_17363 | Not-determined |
| 18-104_k141_38605 | Not-determined |
| 18-104_k141_32035 | Not-determined |
| 18-104_k141_66792 | Not-determined |
| 18-104_k141_38610 | Not-determined |
| 18-104_k141_17367 | Not-determined |
| 18-104_k141_32040 | Not-determined |
| 18-104_k141_11026 | Not-determined |
| 18-104_k141_73889 | Low-quality |
| 18-104_k141_66800 | Not-determined |
| 18-104_k141_164097 | Not-determined |
| 18-104_k141_66802 | Low-quality |
| 18-104_k141_4129 | Not-determined |
| 18-104_k141_59484 | Not-determined |
| 18-104_k141_45998 | Not-determined |
| 18-104_k141_59485 | Not-determined |
| 18-104_k141_4136 | Not-determined |
| 18-104_k141_66811 | Not-determined |
| 18-104_k141_101350 | Not-determined |
| 18-104_k141_32049 | Not-determined |
| 18-104_k141_101351 | Not-determined |
| 18-104_k141_11036 | Not-determined |
| 18-104_k141_46000 | Not-determined |
| 18-104_k141_81017 | Not-determined |
| 18-104_k141_121837 | Low-quality |
| 18-104_k141_24887 | Not-determined |
| 18-104_k141_108811 | Not-determined |
| 18-104_k141_87568 | Not-determined |
| 18-104_k141_157345 | Not-determined |
| 18-104_k141_46006 | Not-determined |
| 18-104_k141_59487 | Not-determined |
| 18-104_k141_52083 | Not-determined |
| 18-104_k141_121844 | Not-determined |
| 18-104_k141_121846 | Low-quality |
| 18-104_k141_121852 | Not-determined |
| 18-104_k141_59497 | Not-determined |
| 18-104_k141_150250 | Not-determined |
| 18-104_k141_46013 | Low-quality |
| 18-104_k141_87578 | Not-determined |
| 18-104_k141_94721 | Not-determined |
| 18-104_k141_101374 | Not-determined |
| 18-104_k141_66821 | Low-quality |
| 18-104_k141_52099 | Not-determined |
| 18-104_k141_66822 | Not-determined |
| 18-104_k141_46015 | Low-quality |
| 18-104_k141_101379 | Not-determined |
| 18-104_k141_164128 | Not-determined |
| 18-104_k141_150259 | Not-determined |
| 18-104_k141_52103 | Not-determined |
| 18-104_k141_46019 | Not-determined |
| 18-104_k141_94732 | Not-determined |
| 18-104_k141_11067 | Not-determined |
| 18-104_k141_59506 | Not-determined |
| 18-104_k141_59508 | Not-determined |
| 18-104_k141_87588 | Not-determined |
| 18-104_k141_150267 | Not-determined |
| 18-104_k141_164135 | Not-determined |
| 18-104_k141_52109 | Not-determined |
| 18-104_k141_52110 | Not-determined |
| 18-104_k141_129301 | Low-quality |
| 18-104_k141_73913 | Not-determined |
| 18-104_k141_66839 | Not-determined |
| 18-104_k141_150271 | Not-determined |
| 18-104_k141_121873 | Not-determined |
| 18-104_k141_101387 | Not-determined |
| 18-104_k141_121875 | Not-determined |
| 18-104_k141_81033 | Not-determined |
| 18-104_k141_108841 | Not-determined |
| 18-104_k141_101393 | Not-determined |
| 18-104_k141_52118 | Not-determined |
| 18-104_k141_17373 | Low-quality |
| 18-104_k141_108847 | Not-determined |
| 18-104_k141_121883 | Not-determined |
| 18-104_k141_73925 | Not-determined |
| 18-104_k141_87598 | Not-determined |
| 18-104_k141_101394 | Not-determined |
| 18-104_k141_73926 | Not-determined |
| 18-104_k141_129310 | Not-determined |
| 18-104_k141_108852 | Low-quality |
| 18-104_k141_32064 | Not-determined |
| 18-104_k141_32065 | Not-determined |
| 18-104_k141_157375 | Not-determined |
| 18-104_k141_108855 | Not-determined |
| 18-104_k141_46037 | Not-determined |
| 18-104_k141_121889 | Low-quality |
| 18-104_k141_17379 | Not-determined |
| 18-104_k141_87606 | Not-determined |
| 18-104_k141_121890 | Low-quality |
| 18-104_k141_121891 | Not-determined |
| 18-104_k141_17386 | Not-determined |
| 18-104_k141_157380 | Not-determined |
| 18-104_k141_73937 | Not-determined |
| 18-104_k141_59516 | Not-determined |
| 18-104_k141_157383 | Not-determined |
| 18-104_k141_32074 | Not-determined |
| 18-104_k141_17397 | Not-determined |
| 18-104_k141_81051 | Not-determined |
| 18-104_k141_81054 | Not-determined |
| 18-104_k141_101421 | Not-determined |
| 18-104_k141_73950 | Not-determined |
| 18-104_k141_32081 | Not-determined |
| 18-104_k141_164153 | Not-determined |
| 18-104_k141_11117 | Not-determined |
| 18-104_k141_38657 | Not-determined |
| 18-104_k141_129341 | Not-determined |
| 18-104_k141_150285 | Not-determined |
| 18-104_k141_11120 | Not-determined |
| 18-104_k141_52141 | Not-determined |
| 18-104_k141_24913 | Not-determined |
| 18-104_k141_46059 | Not-determined |
| 18-104_k141_129352 | Not-determined |
| 18-104_k141_121905 | Not-determined |
| 18-104_k141_66890 | Not-determined |
| 18-104_k141_17420 | Not-determined |
| 18-104_k141_46062 | Low-quality |
| 18-104_k141_164166 | Not-determined |
| 18-104_k141_66894 | Not-determined |
| 18-104_k141_136360 | Not-determined |
| 18-104_k141_121910 | Not-determined |
| 18-104_k141_73960 | Not-determined |
| 18-104_k141_164173 | Not-determined |
| 18-104_k141_46067 | Not-determined |
| 18-104_k141_150305 | Not-determined |
| 18-104_k141_121915 | Not-determined |
| 18-104_k141_121916 | Not-determined |
| 18-104_k141_121917 | Not-determined |
| 18-104_k141_108883 | Not-determined |
| 18-104_k141_11147 | Not-determined |
| 18-104_k141_59527 | Not-determined |
| 18-104_k141_81084 | Low-quality |
| 18-104_k141_143627 | Low-quality |
| 18-104_k141_94783 | Not-determined |
| 18-104_k141_11149 | Not-determined |
| 18-104_k141_38664 | Not-determined |
| 18-104_k141_143632 | Not-determined |
| 18-104_k141_59530 | Not-determined |
| 18-104_k141_32096 | Not-determined |
| 18-104_k141_11157 | Not-determined |
| 18-104_k141_73971 | Low-quality |
| 18-104_k141_121923 | Not-determined |
| 18-104_k141_24921 | Not-determined |
| 18-104_k141_136374 | Not-determined |
| 18-104_k141_32103 | Not-determined |
| 18-104_k141_150311 | Not-determined |
| 18-104_k141_52166 | Not-determined |
| 18-104_k141_121932 | Not-determined |
| 18-104_k141_38674 | Not-determined |
| 18-104_k141_24928 | Not-determined |
| 18-104_k141_11169 | Not-determined |
| 18-104_k141_87619 | Not-determined |
| 18-104_k141_59545 | Not-determined |
| 18-104_k141_87621 | Not-determined |
| 18-104_k141_59546 | Not-determined |
| 18-104_k141_59547 | Low-quality |
| 18-104_k141_164203 | Low-quality |
| 18-104_k141_59551 | Not-determined |
| 18-104_k141_143646 | Not-determined |
| 18-104_k141_11170 | Not-determined |
| 18-104_k141_11171 | Not-determined |
| 18-104_k141_115323 | Low-quality |
| 18-104_k141_11174 | Not-determined |
| 18-104_k141_73982 | Not-determined |
| 18-104_k141_164212 | Not-determined |
| 18-104_k141_73984 | Not-determined |
| 18-104_k141_94801 | Not-determined |
| 18-104_k141_136402 | Not-determined |
| 18-104_k141_73986 | Not-determined |
| 18-104_k141_136404 | Not-determined |
| 18-104_k141_150331 | Low-quality |
| 18-104_k141_52176 | Not-determined |
| 18-104_k141_87625 | Not-determined |
| 18-104_k141_11181 | Not-determined |
| 18-104_k141_101438 | Not-determined |
| 18-104_k141_115325 | Not-determined |
| 18-104_k141_164222 | Not-determined |
| 18-104_k141_38688 | Not-determined |
| 18-104_k141_157400 | Not-determined |
| 18-104_k141_59561 | Low-quality |
| 18-104_k141_129411 | Not-determined |
| 18-104_k141_115337 | Low-quality |
| 18-104_k141_115338 | Not-determined |
| 18-104_k141_38693 | Not-determined |
| 18-104_k141_143665 | Not-determined |
| 18-104_k141_108922 | Not-determined |
| 18-104_k141_24943 | Not-determined |
| 18-104_k141_164225 | Not-determined |
| 18-104_k141_17467 | Not-determined |
| 18-104_k141_150342 | Not-determined |
| 18-104_k141_164228 | Not-determined |
| 18-104_k141_121942 | Not-determined |
| 18-104_k141_59579 | Not-determined |
| 18-104_k141_52201 | Not-determined |
| 18-104_k141_38695 | Not-determined |
| 18-104_k141_108934 | Not-determined |
| 18-104_k141_121945 | Not-determined |
| 18-104_k141_164230 | Not-determined |
| 18-104_k141_115352 | Not-determined |
| 18-104_k141_46090 | Not-determined |
| 18-104_k141_115353 | Not-determined |
| 18-104_k141_38700 | Not-determined |
| 18-104_k141_108939 | Not-determined |
| 18-104_k141_59586 | Low-quality |
| 18-104_k141_66926 | Not-determined |
| 18-104_k141_52210 | Not-determined |
| 18-104_k141_17481 | Not-determined |
| 18-104_k141_11192 | Not-determined |
| 18-104_k141_150351 | Not-determined |
| 18-104_k141_136411 | Low-quality |
| 18-104_k141_52213 | Not-determined |
| 18-104_k141_52215 | Low-quality |
| 18-104_k141_59591 | Not-determined |
| 18-104_k141_24971 | Not-determined |
| 18-104_k141_11195 | Not-determined |
| 18-104_k141_101463 | Not-determined |
| 18-104_k141_11197 | Not-determined |
| 18-104_k141_4159 | Not-determined |
| 18-104_k141_17487 | Low-quality |
| 18-104_k141_11199 | Not-determined |
| 18-104_k141_121959 | Not-determined |
| 18-104_k141_115365 | Low-quality |
| 18-104_k141_66935 | Not-determined |
| 18-104_k141_101467 | Not-determined |
| 18-104_k141_24974 | Not-determined |
| 18-104_k141_59594 | Not-determined |
| 18-104_k141_11204 | Not-determined |
| 18-104_k141_150362 | Not-determined |
| 18-104_k141_108944 | Not-determined |
| 18-104_k141_121963 | Not-determined |
| 18-104_k141_164249 | Low-quality |
| 18-104_k141_136416 | Not-determined |
| 18-104_k141_52230 | Not-determined |
| 18-104_k141_121969 | Not-determined |
| 18-104_k141_164254 | Not-determined |
| 18-104_k141_66946 | Low-quality |
| 18-104_k141_121971 | Not-determined |
| 18-104_k141_136421 | Not-determined |
| 18-104_k141_143681 | Not-determined |
| 18-104_k141_46110 | Not-determined |
| 18-104_k141_38713 | Not-determined |
| 18-104_k141_11211 | Low-quality |
| 18-104_k141_59607 | Not-determined |
| 18-104_k141_52238 | Not-determined |
| 18-104_k141_108955 | Not-determined |
| 18-104_k141_17503 | Not-determined |
| 18-104_k141_101479 | Low-quality |
| 18-104_k141_121975 | Not-determined |
| 18-104_k141_136432 | Not-determined |
| 18-104_k141_129442 | Low-quality |
| 18-104_k141_52246 | Not-determined |
| 18-104_k141_108969 | Not-determined |
| 18-104_k141_52248 | Not-determined |
| 18-104_k141_32115 | Low-quality |
| 18-104_k141_143685 | Low-quality |
| 18-104_k141_101484 | Not-determined |
| 18-104_k141_136438 | Not-determined |
| 18-104_k141_87645 | Not-determined |
| 18-104_k141_136443 | Not-determined |
| 18-104_k141_38721 | Low-quality |
| 18-104_k141_150391 | Not-determined |
| 18-104_k141_157426 | Not-determined |
| 18-104_k141_32126 | Not-determined |
| 18-104_k141_66956 | Low-quality |
| 18-104_k141_46124 | Not-determined |
| 18-104_k141_108985 | Not-determined |
| 18-104_k141_46128 | Low-quality |
| 18-104_k141_24996 | Not-determined |
| 18-104_k141_150398 | Not-determined |
| 18-104_k141_101496 | Not-determined |
| 18-104_k141_38729 | Not-determined |
| 18-104_k141_108992 | Not-determined |
| 18-104_k141_157432 | Low-quality |
| 18-104_k141_150403 | Not-determined |
| 18-104_k141_32138 | Not-determined |
| 18-104_k141_101502 | Low-quality |
| 18-104_k141_143702 | Not-determined |
| 18-104_k141_66969 | Not-determined |
| 18-104_k141_101506 | Not-determined |
| 18-104_k141_129457 | Not-determined |
| 18-104_k141_101508 | Low-quality |
| 18-104_k141_66975 | Not-determined |
| 18-104_k141_32149 | Not-determined |
| 18-104_k141_157439 | Not-determined |
| 18-104_k141_32155 | Not-determined |
| 18-104_k141_164270 | Not-determined |
| 18-104_k141_32159 | Not-determined |
| 18-104_k141_32160 | Not-determined |
| 18-104_k141_52258 | Not-determined |
| 18-104_k141_66979 | Not-determined |
| 18-104_k141_164272 | Not-determined |
| 18-104_k141_87670 | Low-quality |
| 18-104_k141_150411 | Not-determined |
| 18-104_k141_129470 | Not-determined |
| 18-104_k141_81135 | Not-determined |
| 18-104_k141_11230 | Not-determined |
| 18-104_k141_101522 | Not-determined |
| 18-104_k141_101525 | Low-quality |
| 18-110-final-viral-combined_k141_126814\|\|full | Medium-quality |
| 18-104_k141_11235 | Not-determined |
| 18-104_k141_25023 | Not-determined |
| 18-104_k141_38747 | Not-determined |
| 18-104_k141_52272 | Not-determined |
| 18-104_k141_74003 | Low-quality |
| 18-104_k141_129483 | Not-determined |
| 18-104_k141_81144 | Not-determined |
| 18-104_k141_25026 | Not-determined |
| 18-104_k141_129486 | Not-determined |
| 18-104_k141_66992 | Not-determined |
| 18-104_k141_164287 | Not-determined |
| 18-104_k141_38755 | Not-determined |
| 18-104_k141_143719 | Not-determined |
| 18-104_k141_32178 | Not-determined |
| 18-104_k141_74019 | Not-determined |
| 18-104_k141_46168 | Not-determined |
| 18-104_k141_74020 | Not-determined |
| 18-104_k141_32180 | Not-determined |
| 18-104_k141_81152 | Not-determined |
| 18-104_k141_143728 | Not-determined |
| 18-104_k141_157452 | Not-determined |
| 18-104_k141_67002 | Not-determined |
| 18-104_k141_87694 | Not-determined |
| 18-104_k141_164299 | Not-determined |
| 18-104_k141_164301 | Low-quality |
| 18-104_k141_32188 | Not-determined |
| 18-104_k141_115400 | Not-determined |
| 18-104_k141_32192 | Not-determined |
| 18-104_k141_38778 | Not-determined |
| 18-104_k141_32195 | Not-determined |
| 18-104_k141_81160 | Low-quality |
| 18-104_k141_81162 | Not-determined |
| 18-104_k141_108994 | Not-determined |
| 18-104_k141_115405 | Not-determined |
| 18-104_k141_115406 | Not-determined |
| 18-104_k141_115407 | Not-determined |
| 18-104_k141_164306 | Not-determined |
| 18-104_k141_32201 | Not-determined |
| 18-104_k141_32202 | Not-determined |
| 18-104_k141_115411 | Not-determined |
| 18-104_k141_11249 | Not-determined |
| 18-104_k141_115412 | Not-determined |
| 18-104_k141_136501 | Not-determined |
| 18-104_k141_164310 | Low-quality |
| 18-104_k141_143755 | Not-determined |
| 18-104_k141_94805 | Not-determined |
| 18-104_k141_59647 | Not-determined |
| 18-104_k141_143756 | Not-determined |
| 18-104_k141_11257 | Not-determined |
| 18-104_k141_136503 | Not-determined |
| 18-104_k141_52293 | Not-determined |
| 18-104_k141_136508 | Not-determined |
| 18-104_k141_59651 | Not-determined |
| 18-104_k141_81173 | Not-determined |
| 18-104_k141_136510 | Not-determined |
| 18-104_k141_52297 | Not-determined |
| 18-104_k141_157467 | Not-determined |
| 18-104_k141_164312 | Not-determined |
| 18-104_k141_109013 | Not-determined |
| 18-104_k141_157474 | Not-determined |
| 18-104_k141_59657 | Not-determined |
| 18-104_k141_109016 | Low-quality |
| 18-104_k141_32218 | Not-determined |
| 18-104_k141_136517 | Not-determined |
| 18-104_k141_94827 | Low-quality |
| 18-104_k141_109018 | Not-determined |
| 18-104_k141_94830 | Not-determined |
| 18-104_k141_109022 | Not-determined |
| 18-104_k141_38812 | Not-determined |
| 18-104_k141_81186 | Not-determined |
| 18-104_k141_94832 | Not-determined |
| 18-104_k141_46185 | Not-determined |
| 18-104_k141_74025 | Not-determined |
| 18-104_k141_81188 | Low-quality |
| 18-104_k141_38820 | Not-determined |
| 18-104_k141_74027 | Not-determined |
| 18-104_k141_94836 | Not-determined |
| 18-104_k141_164323 | Not-determined |
| 18-104_k141_25055 | Not-determined |
| 18-104_k141_150435 | Not-determined |
| 18-104_k141_4234 | Not-determined |
| 18-104_k141_74031 | Not-determined |
| 18-104_k141_4235 | Not-determined |
| 18-104_k141_74035 | Not-determined |
| 18-104_k141_157486 | Not-determined |
| 18-104_k141_94837 | Not-determined |
| 18-104_k141_59677 | Low-quality |
| 18-104_k141_150440 | Not-determined |
| 18-104_k141_136527 | Not-determined |
| 18-104_k141_25063 | Not-determined |
| 18-104_k141_157490 | Not-determined |
| 18-104_k141_136533 | Not-determined |
| 18-104_k141_25068 | Low-quality |
| 18-104_k141_129512 | Not-determined |
| 18-104_k141_32229 | Not-determined |
| 18-104_k141_94844 | Not-determined |
| 18-104_k141_136534 | Not-determined |
| 18-104_k141_109030 | Not-determined |
| 18-104_k141_25070 | Not-determined |
| 18-104_k141_150443 | Low-quality |
| 18-104_k141_150447 | Not-determined |
| 18-104_k141_109033 | Not-determined |
| 18-104_k141_150449 | Not-determined |
| 18-104_k141_143776 | Low-quality |
| 18-104_k141_150450 | Not-determined |
| 18-104_k141_32230 | Not-determined |
| 18-104_k141_67043 | Not-determined |
| 18-104_k141_46203 | Not-determined |
| 18-104_k141_143779 | Not-determined |
| 18-104_k141_143780 | Not-determined |
| 18-104_k141_87746 | Not-determined |
| 18-104_k141_59694 | Low-quality |
| 18-104_k141_11281 | Not-determined |
| 18-104_k141_59696 | Not-determined |
| 18-104_k141_32236 | Not-determined |
| 18-104_k141_32240 | Not-determined |
| 18-104_k141_109039 | Not-determined |
| 18-104_k141_38833 | Low-quality |
| 18-104_k141_157506 | Not-determined |
| 18-104_k141_157507 | Not-determined |
| 18-104_k141_94857 | Not-determined |
| 18-104_k141_129519 | Not-determined |
| 18-104_k141_164352 | Not-determined |
| 18-104_k141_25081 | Not-determined |
| 18-104_k141_150459 | Not-determined |
| 18-104_k141_94859 | Not-determined |
| 18-104_k141_150462 | Not-determined |
| 18-104_k141_32254 | Low-quality |
| 18-104_k141_115433 | Not-determined |
| 18-104_k141_25088 | Not-determined |
| 18-104_k141_32258 | Not-determined |
| 18-104_k141_32261 | Not-determined |
| 18-104_k141_74047 | Not-determined |
| 18-104_k141_109051 | Not-determined |
| 18-104_k141_38847 | Not-determined |
| 18-104_k141_94868 | Not-determined |
| 18-104_k141_164357 | Not-determined |
| 18-104_k141_94871 | Not-determined |
| 18-104_k141_94872 | Not-determined |
| 18-104_k141_150466 | Not-determined |
| 18-104_k141_46218 | Not-determined |
| 18-104_k141_4258 | Not-determined |
| 18-104_k141_74050 | Not-determined |
| 18-104_k141_81230 | Not-determined |
| 18-104_k141_32262 | Not-determined |
| 18-104_k141_136558 | Low-quality |
| 18-104_k141_74054 | Not-determined |
| 18-104_k141_150470 | Not-determined |
| 18-104_k141_109055 | Low-quality |
| 18-104_k141_143803 | Not-determined |
| 18-104_k141_25096 | Low-quality |
| 18-104_k141_109057 | Not-determined |
| 18-104_k141_129534 | Not-determined |
| 18-104_k141_109059 | Not-determined |
| 18-104_k141_81231 | Low-quality |
| 18-104_k141_129536 | Not-determined |
| 18-104_k141_59704 | Low-quality |
| 18-104_k141_74062 | Not-determined |
| 18-104_k141_4269 | Not-determined |
| 18-104_k141_164368 | Not-determined |
| 18-104_k141_94880 | Not-determined |
| 18-104_k141_67063 | Not-determined |
| 18-104_k141_94882 | Low-quality |
| 18-104_k141_109063 | Not-determined |
| 18-104_k141_129545 | Not-determined |
| 18-104_k141_74069 | Low-quality |
| 18-104_k141_157535 | Not-determined |
| 18-104_k141_101557 | Not-determined |
| 18-104_k141_32269 | Not-determined |
| 18-104_k141_101560 | Not-determined |
| 18-104_k141_136570 | Not-determined |
| 18-104_k141_46234 | Not-determined |
| 18-104_k141_38856 | Not-determined |
| 18-104_k141_81240 | Not-determined |
| 18-104_k141_52337 | Not-determined |
| 18-104_k141_129551 | Not-determined |
| 18-104_k141_81241 | Not-determined |
| 18-104_k141_74073 | Not-determined |
| 18-104_k141_136574 | Not-determined |
| 18-104_k141_150482 | Not-determined |
| 18-104_k141_115441 | Low-quality |
| 18-104_k141_129555 | Not-determined |
| 18-104_k141_59716 | Not-determined |
| 18-104_k141_101566 | Not-determined |
| 18-104_k141_87780 | Not-determined |
| 18-104_k141_38861 | Not-determined |
| 18-104_k141_164376 | Not-determined |
| 18-104_k141_25114 | Not-determined |
| 18-104_k141_115446 | Not-determined |
| 18-104_k141_143815 | Not-determined |
| 18-104_k141_25122 | Low-quality |
| 18-104_k141_164380 | Not-determined |
| 18-104_k141_87785 | Low-quality |
| 18-104_k141_101578 | Not-determined |
| 18-104_k141_11319 | Not-determined |
| 18-104_k141_143819 | Not-determined |
| 18-104_k141_87790 | Not-determined |
| 18-104_k141_59729 | Not-determined |
| 18-104_k141_81264 | Not-determined |
| 18-104_k141_74094 | Low-quality |
| 18-104_k141_59734 | Not-determined |
| 18-104_k141_109072 | Not-determined |
| 18-104_k141_129568 | Not-determined |
| 18-104_k141_164392 | Not-determined |
| 18-104_k141_74098 | Low-quality |
| 18-104_k141_46257 | Not-determined |
| 18-104_k141_74099 | Not-determined |
| 18-104_k141_74104 | Not-determined |
| 18-104_k141_74105 | Not-determined |
| 18-104_k141_46266 | Not-determined |
| 18-104_k141_17519 | Not-determined |
| 18-104_k141_164397 | Not-determined |
| 18-104_k141_136605 | Not-determined |
| 18-104_k141_157557 | Not-determined |
| 18-104_k141_17521 | Not-determined |
| 18-104_k141_101596 | Not-determined |
| 18-104_k141_101597 | Not-determined |
| 18-104_k141_74111 | Low-quality |
| 18-104_k141_150492 | Not-determined |
| 18-104_k141_87801 | Not-determined |
| 18-104_k141_121991 | Not-determined |
| 18-104_k141_129582 | Not-determined |
| 18-104_k141_129583 | Not-determined |
| 18-104_k141_59742 | Not-determined |
| 18-104_k141_46272 | Not-determined |
| 18-104_k141_52361 | Not-determined |
| 18-104_k141_32295 | Not-determined |
| 18-104_k141_74118 | Not-determined |
| 18-104_k141_94914 | Not-determined |
| 18-104_k141_94915 | Not-determined |
| 18-104_k141_32300 | Not-determined |
| 18-104_k141_101606 | Not-determined |
| 18-104_k141_46277 | Not-determined |
| 18-104_k141_74121 | Not-determined |
| 18-104_k141_59747 | Not-determined |
| 18-104_k141_17524 | Not-determined |
| 18-104_k141_136618 | Not-determined |
| 18-104_k141_52364 | Not-determined |
| 18-104_k141_143824 | Not-determined |
| 18-104_k141_115460 | Not-determined |
| 18-104_k141_115461 | Not-determined |
| 18-104_k141_136619 | Not-determined |
| 18-104_k141_109090 | Not-determined |
| 18-104_k141_81281 | Low-quality |
| 18-104_k141_109099 | Low-quality |
| 18-104_k141_4305 | Not-determined |
| 18-104_k141_94925 | Not-determined |
| 18-104_k141_122001 | Low-quality |
| 18-104_k141_109105 | Not-determined |
| 18-104_k141_81293 | Not-determined |
| 18-104_k141_81294 | Not-determined |

| | |
|---|---|
| 18-104_k141_157579 | Not-determined |
| 18-104_k141_46290 | Not-determined |
| 18-104_k141_129604 | Not-determined |
| 18-104_k141_157580 | Not-determined |
| 18-104_k141_67093 | Not-determined |
| 18-104_k141_109111 | Not-determined |
| 18-104_k141_74137 | Not-determined |
| 18-104_k141_52371 | Not-determined |
| 18-104_k141_32310 | Low-quality |
| 18-104_k141_52374 | Not-determined |
| 18-104_k141_17534 | Not-determined |
| 18-104_k141_4309 | Not-determined |
| 18-104_k141_46298 | Not-determined |
| 18-104_k141_101621 | Not-determined |
| 18-104_k141_157588 | Not-determined |
| 18-104_k141_38899 | Not-determined |
| 18-104_k141_25144 | Not-determined |
| 18-104_k141_122018 | Not-determined |
| 18-104_k141_67104 | Not-determined |
| 18-104_k141_81303 | Not-determined |
| 18-104_k141_109121 | Not-determined |
| 18-104_k141_17538 | Not-determined |
| 18-104_k141_164419 | Not-determined |
| 18-104_k141_59770 | Not-determined |
| 18-104_k141_67109 | Not-determined |
| 18-104_k141_81309 | Not-determined |
| 18-104_k141_17540 | Not-determined |
| 18-104_k141_17542 | Low-quality |
| 18-104_k141_59772 | Not-determined |
| 18-104_k141_122021 | Not-determined |
| 18-104_k141_17544 | Not-determined |
| 18-104_k141_25148 | Not-determined |
| 18-104_k141_67111 | Not-determined |
| 18-104_k141_143841 | Low-quality |
| 18-104_k141_46315 | Not-determined |
| 18-104_k141_109126 | Low-quality |
| 18-104_k141_38911 | Not-determined |
| 18-104_k141_136635 | Low-quality |
| 18-104_k141_74146 | Not-determined |
| 18-104_k141_143845 | Not-determined |
| 18-104_k141_17551 | Not-determined |
| 18-104_k141_67118 | Not-determined |
| 18-104_k141_46321 | Not-determined |
| 18-104_k141_46323 | Not-determined |
| 18-104_k141_46324 | Not-determined |
| 18-104_k141_129617 | Not-determined |
| 18-104_k141_52382 | Not-determined |
| 18-104_k141_94950 | Low-quality |
| 18-104_k141_81324 | Not-determined |
| 18-104_k141_157603 | Not-determined |
| 18-104_k141_52385 | Not-determined |
| 18-104_k141_74154 | Not-determined |
| 18-104_k141_25157 | Not-determined |
| 18-104_k141_46328 | Not-determined |
| 18-104_k141_46329 | Not-determined |
| 18-104_k141_38924 | Not-determined |
| 18-104_k141_74156 | Not-determined |
| 18-104_k141_150536 | Low-quality |
| 18-104_k141_74158 | Not-determined |
| 18-104_k141_67121 | Not-determined |
| 18-104_k141_11338 | Not-determined |
| 18-104_k141_122028 | Not-determined |
| 18-104_k141_150537 | Not-determined |
| 18-104_k141_94960 | Not-determined |
| 18-104_k141_122030 | Low-quality |
| 18-104_k141_122031 | Not-determined |
| 18-104_k141_32328 | Not-determined |
| 18-104_k141_74160 | Low-quality |
| 18-104_k141_46337 | Not-determined |
| 18-104_k141_94970 | Not-determined |
| 18-104_k141_38934 | Not-determined |
| 18-104_k141_150548 | Not-determined |
| 18-104_k141_109131 | Not-determined |
| 18-104_k141_25170 | Not-determined |
| 18-104_k141_11357 | Not-determined |
| 18-104_k141_164424 | Not-determined |
| 18-104_k141_164426 | Not-determined |
| 18-104_k141_109135 | Not-determined |
| 18-104_k141_109137 | Not-determined |
| 18-104_k141_74169 | Not-determined |
| 18-104_k141_157622 | Low-quality |
| 18-104_k141_109142 | Not-determined |
| 18-104_k141_129629 | Not-determined |
| 18-104_k141_94980 | Not-determined |
| 18-104_k141_157626 | Not-determined |
| 18-104_k141_129632 | Not-determined |
| 18-104_k141_164433 | Not-determined |
| 18-104_k141_94983 | Not-determined |
| 18-104_k141_150559 | Not-determined |
| 18-104_k141_59796 | Not-determined |
| 18-104_k141_122055 | Not-determined |
| 18-104_k141_11368 | Not-determined |
| 18-104_k141_74179 | Not-determined |
| 18-104_k141_157634 | Low-quality |
| 18-104_k141_157635 | Not-determined |
| 18-104_k141_74181 | Not-determined |
| 18-104_k141_94987 | Low-quality |
| 18-104_k141_143873 | Not-determined |
| 18-104_k141_129637 | Not-determined |
| 18-104_k141_164440 | Not-determined |
| 18-104_k141_157640 | Not-determined |
| 18-104_k141_150570 | Not-determined |
| 18-104_k141_136668 | Not-determined |
| 18-104_k141_17576 | Not-determined |
| 18-104_k141_59811 | Low-quality |
| 18-104_k141_150575 | Not-determined |
| 18-104_k141_109161 | Not-determined |
| 18-104_k141_25192 | Not-determined |
| 18-104_k141_81336 | Not-determined |
| 18-104_k141_52403 | Not-determined |
| 18-104_k141_115498 | Low-quality |
| 18-104_k141_59819 | Low-quality |
| 18-104_k141_52406 | Not-determined |
| 18-104_k141_136675 | Not-determined |
| 18-104_k141_157656 | Not-determined |
| 18-104_k141_101632 | Low-quality |
| 18-104_k141_52408 | Not-determined |
| 18-104_k141_101633 | Not-determined |
| 18-104_k141_4325 | Not-determined |
| 18-104_k141_122093 | Not-determined |
| 18-104_k141_136683 | Not-determined |
| 18-104_k141_4333 | Not-determined |
| 18-104_k141_4335 | Not-determined |
| 18-104_k141_157666 | Low-quality |
| 18-104_k141_157667 | Not-determined |
| 18-104_k141_157669 | Low-quality |
| 18-104_k141_4340 | Not-determined |
| 18-104_k141_4342 | Not-determined |
| 18-104_k141_32345 | Not-determined |
| 18-104_k141_115509 | Not-determined |
| 18-104_k141_164467 | Not-determined |
| 18-104_k141_81362 | Not-determined |
| 18-104_k141_87849 | Low-quality |
| 18-104_k141_150590 | Not-determined |
| 18-104_k141_4349 | Not-determined |
| 18-104_k141_109179 | Not-determined |
| 18-104_k141_164472 | Not-determined |
| 18-104_k141_157677 | Not-determined |
| 18-104_k141_143880 | Not-determined |
| 18-104_k141_11404 | Not-determined |
| 18-104_k141_87859 | Not-determined |
| 18-104_k141_11405 | Not-determined |
| 18-104_k141_38998 | Not-determined |
| 18-104_k141_95014 | Not-determined |
| 18-104_k141_81374 | Not-determined |
| 18-104_k141_95015 | Not-determined |
| 18-104_k141_109184 | Not-determined |
| 18-104_k141_32361 | Not-determined |
| 18-104_k141_115516 | Not-determined |
| 18-104_k141_46383 | Not-determined |
| 18-104_k141_101661 | Not-determined |
| 18-104_k141_67194 | Not-determined |
| 18-104_k141_46384 | Not-determined |
| 18-104_k141_74226 | Not-determined |
| 18-104_k141_101663 | Not-determined |
| 18-104_k141_52427 | Not-determined |
| 18-104_k141_101667 | Not-determined |
| 18-104_k141_46386 | Not-determined |
| 18-104_k141_122104 | Not-determined |
| 18-104_k141_11421 | Not-determined |
| 18-104_k141_11422 | Not-determined |
| 18-104_k141_74235 | Not-determined |
| 18-104_k141_95025 | Low-quality |
| 18-104_k141_52429 | Not-determined |
| 18-104_k141_164486 | Not-determined |
| 18-104_k141_95028 | Not-determined |
| 18-104_k141_109199 | Not-determined |
| 18-104_k141_101669 | Not-determined |
| 18-104_k141_46389 | Not-determined |
| 18-104_k141_74240 | Not-determined |
| 18-104_k141_115534 | Not-determined |
| 18-104_k141_136715 | Not-determined |
| 18-104_k141_136717 | Not-determined |
| 18-104_k141_143900 | Not-determined |
| 18-104_k141_52440 | Low-quality |
| 18-104_k141_4380 | Not-determined |
| 18-104_k141_136722 | Not-determined |
| 18-104_k141_157707 | Low-quality |
| 18-104_k141_74248 | Not-determined |
| 18-104_k141_32388 | Not-determined |
| 18-104_k141_115536 | Not-determined |
| 18-104_k141_74252 | Not-determined |
| 18-104_k141_74253 | Low-quality |
| 18-104_k141_67219 | Not-determined |
| 18-104_k141_101688 | Not-determined |
| 18-104_k141_17600 | Not-determined |
| 18-104_k141_74254 | Not-determined |
| 18-104_k141_101691 | Not-determined |
| 18-104_k141_11438 | Not-determined |
| 18-104_k141_164504 | Not-determined |
| 18-104_k141_101694 | Not-determined |
| 18-104_k141_52462 | Not-determined |
| 18-104_k141_59853 | Not-determined |
| 18-104_k141_136734 | Not-determined |
| 18-104_k141_52463 | Not-determined |
| 18-104_k141_39008 | Low-quality |
| 18-104_k141_143917 | Not-determined |
| 18-104_k141_143918 | Not-determined |
| 18-104_k141_101698 | Not-determined |
| 18-104_k141_81407 | Not-determined |
| 18-104_k141_74259 | Not-determined |
| 18-104_k141_32393 | Not-determined |
| 18-104_k141_67227 | Not-determined |
| 18-104_k141_109216 | Not-determined |
| 18-104_k141_101702 | Not-determined |
| 18-104_k141_122117 | Not-determined |
| 18-104_k141_164516 | Not-determined |
| 18-104_k141_109218 | Not-determined |
| 18-104_k141_74263 | Not-determined |
| 18-104_k141_109220 | Not-determined |
| 18-104_k141_74267 | Not-determined |
| 18-104_k141_109221 | Not-determined |
| 18-104_k141_122122 | Not-determined |
| 18-104_k141_59866 | Not-determined |
| 18-104_k141_52481 | Low-quality |
| 18-104_k141_81419 | Not-determined |
| 18-104_k141_25204 | Not-determined |
| 18-104_k141_25205 | Not-determined |
| 18-104_k141_101708 | Low-quality |
| 18-104_k141_101711 | Not-determined |
| 18-104_k141_143932 | Not-determined |
| 18-104_k141_157726 | Low-quality |
| 18-104_k141_52486 | Not-determined |
| 18-104_k141_74275 | Not-determined |
| 18-104_k141_101718 | Not-determined |
| 18-104_k141_122129 | Not-determined |
| 18-104_k141_122130 | Not-determined |
| 18-104_k141_46418 | Not-determined |
| 18-104_k141_95044 | Not-determined |
| 18-104_k141_74279 | Not-determined |
| 18-104_k141_129654 | Not-determined |
| 18-104_k141_164530 | Not-determined |
| 18-104_k141_32415 | Low-quality |
| 18-104_k141_32416 | Not-determined |
| 18-104_k141_46427 | Not-determined |
| 18-104_k141_52499 | Not-determined |
| 18-104_k141_25212 | Not-determined |
| 18-104_k141_52501 | Not-determined |
| 18-104_k141_17613 | Not-determined |
| 18-104_k141_52506 | Not-determined |
| 18-104_k141_32420 | Not-determined |
| 18-104_k141_17614 | Not-determined |
| 18-104_k141_39033 | Not-determined |
| 18-104_k141_67252 | Not-determined |
| 18-104_k141_67254 | Not-determined |
| 18-104_k141_17616 | Not-determined |
| 18-104_k141_74289 | Not-determined |
| 18-104_k141_81435 | Not-determined |
| 18-104_k141_136747 | Not-determined |
| 18-104_k141_4417 | Not-determined |
| 18-104_k141_129659 | Not-determined |
| 18-104_k141_164539 | Not-determined |
| 18-104_k141_109254 | Not-determined |
| 18-104_k141_129661 | Not-determined |
| 18-104_k141_136749 | Not-determined |
| 18-104_k141_164543 | Not-determined |
| 18-104_k141_150627 | Not-determined |
| 18-104_k141_81443 | Not-determined |
| 18-104_k141_11448 | Not-determined |
| 18-104_k141_25227 | Not-determined |
| 18-104_k141_150630 | Not-determined |
| 18-104_k141_11451 | Not-determined |
| 18-104_k141_122157 | Not-determined |
| 18-104_k141_122159 | Not-determined |
| 18-104_k141_74301 | Not-determined |
| 18-104_k141_11453 | Not-determined |
| 18-104_k141_122161 | Low-quality |
| 18-104_k141_59891 | Not-determined |
| 18-104_k141_39041 | Not-determined |
| 18-104_k141_136755 | Not-determined |
| 18-104_k141_59893 | Not-determined |
| 18-104_k141_11455 | Not-determined |
| 18-104_k141_52518 | Not-determined |
| 18-104_k141_17632 | Not-determined |
| 18-104_k141_59895 | Not-determined |
| 18-104_k141_46450 | Not-determined |
| 18-104_k141_129670 | Not-determined |
| 18-104_k141_87900 | Not-determined |
| 18-104_k141_32437 | Not-determined |
| 18-104_k141_129672 | Not-determined |
| 18-104_k141_164555 | Not-determined |
| 18-104_k141_143943 | Low-quality |
| 18-104_k141_25239 | Not-determined |
| 18-104_k141_59898 | Not-determined |
| 18-104_k141_157747 | Not-determined |
| 18-104_k141_122166 | Low-quality |
| 18-104_k141_143947 | Not-determined |
| 18-104_k141_11461 | Not-determined |
| 18-104_k141_109260 | Low-quality |
| 18-104_k141_32443 | Not-determined |
| 18-104_k141_109262 | Not-determined |
| 18-104_k141_109263 | Not-determined |
| 18-104_k141_129678 | Not-determined |
| 18-104_k141_129679 | Not-determined |
| 18-104_k141_157753 | Not-determined |
| 18-104_k141_32451 | Low-quality |
| 18-104_k141_11470 | Not-determined |
| 18-104_k141_157760 | Not-determined |
| 18-104_k141_11472 | Not-determined |
| 18-104_k141_143961 | Not-determined |
| 18-104_k141_46464 | Not-determined |
| 18-104_k141_157768 | Not-determined |
| 18-104_k141_136768 | Not-determined |
| 18-104_k141_11475 | Not-determined |
| 18-104_k141_11476 | Not-determined |
| 18-104_k141_101765 | Not-determined |
| 18-104_k141_95054 | Low-quality |
| 18-104_k141_95056 | Not-determined |
| 18-104_k141_101769 | Not-determined |
| 18-104_k141_122178 | Not-determined |
| 18-104_k141_81455 | Low-quality |
| 18-104_k141_101770 | Low-quality |
| 18-104_k141_25246 | Not-determined |
| 18-104_k141_143967 | Not-determined |
| 18-104_k141_101773 | Not-determined |
| 18-104_k141_87927 | Not-determined |
| 18-104_k141_17660 | Not-determined |
| 18-104_k141_101779 | Not-determined |
| 18-104_k141_122182 | Not-determined |
| 18-104_k141_39062 | Not-determined |
| 18-104_k141_87929 | Not-determined |
| 18-104_k141_67281 | Not-determined |
| 18-104_k141_67282 | Not-determined |
| 18-104_k141_25250 | Not-determined |
| 18-104_k141_17662 | Not-determined |
| 18-104_k141_164568 | Not-determined |
| 18-104_k141_95063 | Not-determined |
| 18-104_k141_59913 | Not-determined |
| 18-104_k141_87933 | Not-determined |
| 18-104_k141_87934 | Not-determined |
| 18-104_k141_157789 | Not-determined |
| 18-104_k141_59914 | Not-determined |
| 18-104_k141_59916 | Not-determined |
| 18-104_k141_143988 | Not-determined |
| 18-104_k141_157791 | Not-determined |
| 18-104_k141_32473 | Not-determined |
| 18-104_k141_25255 | Not-determined |
| 18-104_k141_101789 | Not-determined |
| 18-104_k141_87938 | Not-determined |
| 18-104_k141_164575 | Not-determined |
| 18-104_k141_87941 | Not-determined |
| 18-104_k141_136773 | Not-determined |
| 18-104_k141_164577 | Not-determined |
| 18-104_k141_143996 | Not-determined |
| 18-104_k141_115556 | Low-quality |
| 18-104_k141_59922 | Not-determined |
| 18-104_k141_67295 | Not-determined |
| 18-104_k141_87946 | Not-determined |
| 18-104_k141_67297 | Not-determined |
| 18-104_k141_67298 | Not-determined |
| 18-104_k141_122192 | Not-determined |
| 18-104_k141_74343 | Not-determined |
| 18-104_k141_67300 | Not-determined |
| 18-104_k141_157792 | Low-quality |
| 18-104_k141_52539 | Not-determined |
| 18-104_k141_109278 | Not-determined |
| 18-104_k141_39079 | Low-quality |
| 18-104_k141_39080 | Not-determined |
| 18-104_k141_32484 | Not-determined |
| 18-104_k141_25264 | Not-determined |
| 18-104_k141_74351 | Not-determined |
| 18-104_k141_46497 | Low-quality |
| 18-104_k141_52545 | Low-quality |
| 18-104_k141_150635 | Low-quality |
| 18-104_k141_136782 | Not-determined |
| 18-104_k141_17690 | Not-determined |
| 18-104_k141_39084 | Not-determined |
| 18-104_k141_150637 | Not-determined |
| 18-104_k141_32489 | Low-quality |
| 18-104_k141_122201 | Not-determined |
| 18-104_k141_17692 | Not-determined |
| 18-104_k141_17693 | Not-determined |
| 18-104_k141_109287 | Low-quality |
| 18-104_k141_52549 | Not-determined |
| 18-104_k141_59932 | Not-determined |
| 18-104_k141_52551 | Not-determined |
| 18-104_k141_52552 | Not-determined |
| 18-104_k141_67314 | Not-determined |
| 18-104_k141_87960 | Low-quality |
| 18-104_k141_101811 | Not-determined |
| 18-104_k141_17700 | Not-determined |
| 18-104_k141_81486 | Not-determined |
| 18-104_k141_115580 | Not-determined |
| 18-104_k141_150651 | Not-determined |
| 18-104_k141_164591 | Not-determined |
| 18-104_k141_25286 | Not-determined |
| 18-104_k141_101816 | Not-determined |
| 18-104_k141_17707 | Not-determined |
| 18-104_k141_109305 | Not-determined |
| 18-104_k141_157804 | Not-determined |
| 18-104_k141_150664 | Not-determined |
| 18-104_k141_157810 | Not-determined |
| 18-104_k141_115592 | Not-determined |
| 18-104_k141_87972 | Low-quality |
| 18-104_k141_81500 | Not-determined |
| 18-104_k141_87976 | Not-determined |
| 18-104_k141_39102 | Low-quality |
| 18-104_k141_11548 | Low-quality |
| 18-104_k141_144018 | Low-quality |
| 18-104_k141_144022 | Not-determined |
| 18-104_k141_74373 | Not-determined |
| 18-104_k141_39107 | Not-determined |
| 18-104_k141_122212 | Not-determined |
| 18-104_k141_157821 | Not-determined |
| 18-104_k141_115606 | Not-determined |
| 18-104_k141_115609 | Not-determined |
| 18-104_k141_144029 | Not-determined |
| 18-104_k141_17725 | Not-determined |
| 18-104_k141_17727 | Low-quality |
| 18-104_k141_11558 | Not-determined |
| 18-104_k141_122221 | Not-determined |
| 18-104_k141_157830 | Not-determined |
| 18-104_k141_46540 | Low-quality |
| 18-104_k141_46541 | Not-determined |
| 18-104_k141_74385 | Low-quality |
| 18-104_k141_46544 | Not-determined |
| 18-104_k141_101825 | Not-determined |
| 18-104_k141_157836 | Not-determined |
| 18-104_k141_157838 | Not-determined |
| 18-104_k141_115629 | Not-determined |
| 18-104_k141_136820 | Not-determined |
| 18-104_k141_101830 | Not-determined |
| 18-104_k141_32503 | Not-determined |
| 18-104_k141_157843 | Not-determined |
| 18-104_k141_150694 | Not-determined |
| 18-104_k141_17743 | Low-quality |
| 18-104_k141_157846 | Not-determined |
| 18-104_k141_122236 | Low-quality |
| 18-104_k141_32507 | Not-determined |
| 18-104_k141_122238 | Not-determined |
| 18-104_k141_109328 | Not-determined |
| 18-104_k141_32510 | Not-determined |
| 18-104_k141_74399 | Not-determined |
| 18-104_k141_4444 | Not-determined |
| 18-104_k141_4445 | Not-determined |
| 18-104_k141_4448 | Not-determined |
| 18-104_k141_157854 | Low-quality |
| 18-104_k141_101841 | Not-determined |
| 18-104_k141_144039 | Not-determined |
| 18-104_k141_109337 | Not-determined |
| 18-104_k141_52604 | Not-determined |
| 18-104_k141_4456 | Not-determined |
| 18-104_k141_157863 | Not-determined |
| 18-104_k141_25323 | Not-determined |
| 18-104_k141_157865 | Not-determined |
| 18-104_k141_115648 | Not-determined |
| 18-104_k141_4463 | Not-determined |
| 18-104_k141_109349 | Not-determined |
| 18-104_k141_81540 | Not-determined |
| 18-104_k141_144044 | Not-determined |
| 18-104_k141_11573 | Not-determined |
| 18-104_k141_52610 | Not-determined |
| 18-104_k141_136824 | Low-quality |
| 18-104_k141_52611 | Not-determined |
| 18-104_k141_136825 | Not-determined |
| 18-104_k141_4466 | Not-determined |
| 18-104_k141_32517 | Not-determined |
| 18-104_k141_52613 | Not-determined |
| 18-104_k141_46567 | Not-determined |
| 18-104_k141_115656 | Not-determined |
| 18-104_k141_46569 | Not-determined |
| 18-104_k141_101850 | Not-determined |
| 18-104_k141_11582 | Not-determined |
| 18-104_k141_109362 | Not-determined |
| 18-104_k141_115659 | Not-determined |
| 18-104_k141_88019 | Not-determined |
| 18-104_k141_32526 | Not-determined |
| 18-104_k141_88020 | Not-determined |
| 18-104_k141_32527 | Not-determined |
| 18-104_k141_25334 | Not-determined |
| 18-104_k141_164631 | Not-determined |
| 18-104_k141_39137 | Not-determined |
| 18-104_k141_150701 | Not-determined |
| 18-104_k141_59952 | Not-determined |
| 18-104_k141_136838 | Not-determined |
| 18-104_k141_144052 | Not-determined |
| 18-104_k141_32534 | Not-determined |
| 18-104_k141_59955 | Not-determined |
| 18-104_k141_46577 | Not-determined |
| 18-104_k141_144055 | Not-determined |
| 18-104_k141_109372 | Not-determined |
| 18-104_k141_144057 | Not-determined |
| 18-104_k141_136843 | Not-determined |
| 18-104_k141_144060 | Not-determined |
| 18-104_k141_25347 | Not-determined |
| 18-104_k141_17756 | Not-determined |
| 18-104_k141_59960 | Not-determined |
| 18-104_k141_11598 | Not-determined |
| 18-104_k141_115674 | Not-determined |
| 18-104_k141_101863 | Not-determined |
| 18-104_k141_122270 | Not-determined |
| 18-104_k141_129696 | Not-determined |
| 18-104_k141_101866 | Not-determined |
| 18-104_k141_59968 | Not-determined |
| 18-104_k141_101871 | Not-determined |
| 18-104_k141_74422 | Not-determined |
| 18-104_k141_67351 | Not-determined |
| 18-104_k141_164644 | Low-quality |
| 18-104_k141_32550 | Low-quality |
| 18-104_k141_81574 | Not-determined |
| 18-104_k141_101874 | Low-quality |
| 18-104_k141_157885 | Low-quality |
| 18-104_k141_101875 | Low-quality |
| 18-104_k141_67361 | Not-determined |
| 18-104_k141_164646 | Not-determined |
| 18-104_k141_136860 | Low-quality |
| 18-104_k141_164650 | Not-determined |
| 18-104_k141_52647 | Not-determined |
| 18-104_k141_81587 | Not-determined |
| 18-104_k141_129704 | Not-determined |
| 18-104_k141_52650 | Not-determined |
| 18-104_k141_17766 | Low-quality |
| 18-104_k141_59979 | Not-determined |
| 18-104_k141_39150 | Low-quality |
| 18-104_k141_88041 | Not-determined |
| 18-104_k141_88042 | Not-determined |
| 18-104_k141_122288 | Not-determined |
| 18-104_k141_32563 | Not-determined |
| 18-104_k141_39157 | Not-determined |
| 18-104_k141_144080 | Low-quality |
| 18-104_k141_32564 | Not-determined |
| 18-104_k141_136870 | Not-determined |
| 18-104_k141_81599 | Not-determined |
| 18-104_k141_150716 | Not-determined |
| 18-104_k141_46597 | Not-determined |
| 18-104_k141_39161 | Low-quality |
| 18-104_k141_74427 | Low-quality |
| 18-104_k141_129721 | Not-determined |
| 18-104_k141_25371 | Not-determined |
| 18-104_k141_81604 | Not-determined |
| 18-104_k141_39163 | Not-determined |
| 18-104_k141_129723 | Not-determined |
| 18-104_k141_122297 | Not-determined |
| 18-104_k141_39167 | Not-determined |
| 18-104_k141_129727 | Not-determined |
| 18-104_k141_115692 | Not-determined |
| 18-104_k141_67375 | Low-quality |
| 18-104_k141_115697 | Not-determined |
| 18-104_k141_150726 | Low-quality |
| 18-104_k141_164662 | Not-determined |
| 18-104_k141_101890 | Not-determined |
| 18-104_k141_25376 | Not-determined |
| 18-104_k141_164663 | Not-determined |
| 18-104_k141_157893 | Not-determined |
| 18-104_k141_115701 | Not-determined |
| 18-104_k141_52665 | Not-determined |
| 18-104_k141_59995 | Not-determined |
| 18-104_k141_74440 | Not-determined |
| 18-104_k141_25382 | Low-quality |
| 18-104_k141_52673 | Not-determined |
| 18-104_k141_136881 | Not-determined |
| 18-104_k141_81617 | Not-determined |
| 18-104_k141_25383 | Not-determined |
| 18-104_k141_136883 | Not-determined |
| 18-104_k141_39176 | Low-quality |
| 18-104_k141_52679 | Not-determined |
| 18-104_k141_136889 | Not-determined |
| 18-104_k141_46609 | Not-determined |
| 18-104_k141_144099 | Not-determined |
| 18-104_k141_52682 | Not-determined |
| 18-104_k141_39187 | Not-determined |
| 18-104_k141_32571 | Not-determined |
| 18-104_k141_25395 | Not-determined |
| 18-104_k141_144104 | Not-determined |
| 18-104_k141_122309 | Low-quality |
| 18-104_k141_129747 | Not-determined |
| 18-104_k141_60013 | Not-determined |
| 18-104_k141_115727 | Not-determined |
| 18-104_k141_164684 | Not-determined |
| 18-104_k141_32585 | Not-determined |
| 18-104_k141_157911 | Not-determined |
| 18-104_k141_60025 | Not-determined |
| 18-104_k141_122316 | Not-determined |
| 18-104_k141_52701 | Not-determined |
| 18-104_k141_101916 | Not-determined |
| 18-104_k141_25408 | Not-determined |
| 18-104_k141_67386 | Not-determined |
| 18-104_k141_136906 | Not-determined |
| 18-104_k141_11638 | Not-determined |
| 18-104_k141_74451 | Not-determined |
| 18-104_k141_101922 | Low-quality |
| 18-104_k141_129766 | Not-determined |
| 18-104_k141_122322 | Not-determined |
| 18-104_k141_52711 | Not-determined |
| 18-104_k141_136909 | Not-determined |
| 18-104_k141_88076 | Not-determined |
| 18-104_k141_25414 | Not-determined |
| 18-104_k141_46623 | Not-determined |
| 18-104_k141_74455 | Not-determined |

| | |
|---|---|
| 18-104_k141_74461 | Not-determined |
| 18-104_k141_11647 | Not-determined |
| 18-104_k141_74462 | Not-determined |
| 18-104_k141_101936 | Not-determined |
| 18-104_k141_101938 | Not-determined |
| 18-104_k141_157923 | Not-determined |
| 18-104_k141_74467 | Not-determined |
| 18-104_k141_17799 | Not-determined |
| 18-104_k141_11654 | Not-determined |
| 18-104_k141_122339 | Not-determined |
| 18-104_k141_74472 | Not-determined |
| 18-104_k141_11658 | Not-determined |
| 18-104_k141_67396 | Low-quality |
| 18-104_k141_157929 | Not-determined |
| 18-104_k141_150747 | Not-determined |
| 18-104_k141_11661 | Not-determined |
| 18-104_k141_115741 | Not-determined |
| 18-104_k141_115744 | Not-determined |
| 18-104_k141_32606 | Not-determined |
| 18-104_k141_60038 | Not-determined |
| 18-104_k141_115752 | Not-determined |
| 18-104_k141_95147 | Low-quality |
| 18-104_k141_46646 | Not-determined |
| 18-104_k141_95149 | Not-determined |
| 18-104_k141_95150 | Not-determined |
| 18-104_k141_95151 | Not-determined |
| 18-104_k141_46648 | Not-determined |
| 18-104_k141_46650 | Not-determined |
| 18-104_k141_46652 | Not-determined |
| 18-104_k141_150759 | Not-determined |
| 18-104_k141_115756 | Not-determined |
| 18-104_k141_109411 | Low-quality |
| 18-104_k141_74478 | Not-determined |
| 18-104_k141_115757 | Not-determined |
| 18-104_k141_95156 | Not-determined |
| 18-104_k141_150761 | Not-determined |
| 18-104_k141_4509 | Not-determined |
| 18-104_k141_157947 | Not-determined |
| 18-104_k141_4510 | Not-determined |
| 18-104_k141_101963 | Not-determined |
| 18-104_k141_17819 | Not-determined |
| 18-104_k141_109418 | Not-determined |
| 18-104_k141_81668 | Not-determined |
| 18-104_k141_81669 | Not-determined |
| 18-104_k141_95164 | Not-determined |
| 18-104_k141_81671 | Not-determined |
| 18-104_k141_95167 | Not-determined |
| 18-104_k141_11679 | Not-determined |
| 18-104_k141_52736 | Not-determined |
| 18-104_k141_157953 | Not-determined |
| 18-104_k141_157955 | Not-determined |
| 18-104_k141_17828 | Not-determined |
| 18-104_k141_88099 | Not-determined |
| 18-104_k141_88106 | Not-determined |
| 18-104_k141_164719 | Not-determined |
| 18-104_k141_39228 | Not-determined |
| 18-104_k141_101972 | Not-determined |
| 18-104_k141_144142 | Not-determined |
| 18-104_k141_25466 | Not-determined |
| 18-104_k141_74481 | Not-determined |
| 18-104_k141_4531 | Not-determined |
| 18-104_k141_136930 | Not-determined |
| 18-104_k141_52748 | Not-determined |
| 18-104_k141_39233 | Not-determined |
| 18-104_k141_81685 | Not-determined |
| 18-104_k141_74485 | Not-determined |
| 18-104_k141_144146 | Not-determined |
| 18-104_k141_67426 | Not-determined |
| 18-104_k141_52752 | Not-determined |
| 18-104_k141_109431 | Low-quality |
| 18-104_k141_164729 | Not-determined |
| 18-104_k141_60058 | Not-determined |
| 18-104_k141_81687 | Not-determined |
| 18-104_k141_32623 | Not-determined |
| 18-104_k141_150773 | Not-determined |
| 18-104_k141_39240 | Not-determined |
| 18-104_k141_25472 | Not-determined |
| 18-104_k141_109433 | Not-determined |
| 18-104_k141_67437 | Not-determined |
| 18-104_k141_67439 | Not-determined |
| 18-104_k141_46658 | Not-determined |
| 18-104_k141_109438 | Not-determined |
| 18-104_k141_11693 | Not-determined |
| 18-104_k141_4540 | Low-quality |
| 18-104_k141_88116 | Not-determined |
| 18-104_k141_122368 | Not-determined |
| 18-104_k141_74502 | Not-determined |
| 18-104_k141_52762 | Not-determined |
| 18-104_k141_11694 | Not-determined |
| 18-104_k141_25481 | Not-determined |
| 18-104_k141_52765 | Not-determined |
| 18-104_k141_150779 | Not-determined |
| 18-104_k141_52768 | Not-determined |
| 18-104_k141_11700 | Not-determined |
| 18-104_k141_115797 | Not-determined |
| 18-104_k141_115800 | Not-determined |
| 18-104_k141_157979 | Not-determined |
| 18-104_k141_67448 | Not-determined |
| 18-104_k141_67449 | Not-determined |
| 18-104_k141_4549 | Not-determined |
| 18-104_k141_164748 | Not-determined |
| 18-104_k141_88131 | Not-determined |
| 18-104_k141_164750 | Not-determined |
| 18-104_k141_67454 | Not-determined |
| 18-104_k141_46668 | Not-determined |
| 18-104_k141_95183 | Low-quality |
| 18-104_k141_164754 | Not-determined |
| 18-104_k141_95185 | Not-determined |
| 18-104_k141_101992 | Low-quality |
| 18-104_k141_46671 | Not-determined |
| 18-104_k141_88133 | Not-determined |
| 18-104_k141_67460 | Not-determined |
| 18-104_k141_109454 | Not-determined |
| 18-104_k141_95190 | Not-determined |
| 18-104_k141_46674 | Not-determined |
| 18-104_k141_67463 | Not-determined |
| 18-104_k141_46677 | Not-determined |
| 18-104_k141_25497 | Not-determined |
| 18-104_k141_157987 | Not-determined |
| 18-104_k141_109461 | Not-determined |
| 18-104_k141_25500 | Not-determined |
| 18-104_k141_39265 | Not-determined |
| 18-104_k141_144165 | Not-determined |
| 18-104_k141_60082 | Not-determined |
| 18-104_k141_32645 | Low-quality |
| 18-104_k141_136973 | Not-determined |
| 18-104_k141_60085 | Not-determined |
| 18-104_k141_81717 | Not-determined |
| 18-104_k141_122389 | Not-determined |
| 18-104_k141_150799 | Not-determined |
| 18-104_k141_60088 | Low-quality |
| 18-104_k141_122391 | Not-determined |
| 18-104_k141_136978 | Not-determined |
| 18-104_k141_81722 | Not-determined |
| 18-104_k141_39278 | Not-determined |
| 18-104_k141_150803 | Not-determined |
| 18-104_k141_52777 | Low-quality |
| 18-104_k141_158006 | Not-determined |
| 18-104_k141_95198 | Not-determined |
| 18-104_k141_11730 | Not-determined |
| 18-104_k141_136984 | Not-determined |
| 18-104_k141_136985 | Not-determined |
| 18-104_k141_11735 | Not-determined |
| 18-104_k141_136987 | Not-determined |
| 18-104_k141_164779 | Not-determined |
| 18-104_k141_4574 | Not-determined |
| 18-104_k141_122402 | Not-determined |
| 18-104_k141_129800 | Not-determined |
| 18-104_k141_164784 | Not-determined |
| 18-104_k141_164785 | Not-determined |
| 18-104_k141_88161 | Not-determined |
| 18-104_k141_144183 | Not-determined |
| 18-104_k141_150816 | Not-determined |
| 18-104_k141_4578 | Not-determined |
| 18-104_k141_74538 | Not-determined |
| 18-104_k141_164791 | Not-determined |
| 18-104_k141_129811 | Not-determined |
| 18-104_k141_129812 | Not-determined |
| 18-104_k141_17854 | Low-quality |
| 18-104_k141_158017 | Not-determined |
| 18-104_k141_67489 | Low-quality |
| 18-104_k141_74545 | Not-determined |
| 18-104_k141_11745 | Not-determined |
| 18-104_k141_81736 | Low-quality |
| 18-104_k141_74548 | Low-quality |
| 18-104_k141_17861 | Not-determined |
| 18-104_k141_11750 | Not-determined |
| 18-104_k141_158033 | Not-determined |
| 18-104_k141_60115 | Not-determined |
| 18-104_k141_122422 | Low-quality |
| 18-104_k141_150828 | Not-determined |
| 18-104_k141_60119 | Not-determined |
| 18-104_k141_137008 | Not-determined |
| 18-104_k141_74556 | Low-quality |
| 18-104_k141_60122 | Not-determined |
| 18-104_k141_129826 | Low-quality |
| 18-104_k141_144210 | Not-determined |
| 18-104_k141_158040 | Not-determined |
| 18-104_k141_115842 | Not-determined |
| 18-104_k141_67503 | Not-determined |
| 18-104_k141_102024 | Not-determined |
| 18-104_k141_67505 | Not-determined |
| 18-104_k141_158045 | Not-determined |
| 18-104_k141_81756 | Not-determined |
| 18-104_k141_88182 | Not-determined |
| 18-104_k141_81757 | Not-determined |
| 18-104_k141_52804 | Not-determined |
| 18-104_k141_60135 | Not-determined |
| 18-104_k141_88184 | Not-determined |
| 18-104_k141_150844 | Not-determined |
| 18-104_k141_95214 | Not-determined |
| 18-104_k141_115854 | Not-determined |
| 18-104_k141_74563 | Not-determined |
| 18-104_k141_39308 | Not-determined |
| 18-104_k141_95218 | Not-determined |
| 18-104_k141_144225 | Low-quality |
| 18-104_k141_122432 | Not-determined |
| 18-104_k141_11776 | Not-determined |
| 18-104_k141_88190 | Not-determined |
| 18-104_k141_52820 | Not-determined |
| 18-104_k141_88192 | Not-determined |
| 18-104_k141_122435 | Not-determined |
| 18-104_k141_129845 | Not-determined |
| 18-104_k141_17878 | Not-determined |
| 18-104_k141_32652 | Not-determined |
| 18-104_k141_137026 | Not-determined |
| 18-104_k141_144234 | Not-determined |
| 18-104_k141_95232 | Not-determined |
| 18-104_k141_129849 | Not-determined |
| 18-104_k141_95236 | Not-determined |
| 18-104_k141_95239 | Not-determined |
| 18-104_k141_122447 | Not-determined |
| 18-104_k141_95241 | Not-determined |
| 18-104_k141_158067 | Not-determined |
| 18-104_k141_164826 | Not-determined |
| 18-104_k141_67528 | Not-determined |
| 18-104_k141_95242 | Not-determined |
| 18-104_k141_115860 | Not-determined |
| 18-104_k141_25523 | Not-determined |
| 18-104_k141_122450 | Not-determined |
| 18-104_k141_60154 | Not-determined |
| 18-104_k141_11795 | Not-determined |
| 18-104_k141_17890 | Not-determined |
| 18-104_k141_25530 | Not-determined |
| 18-104_k141_46680 | Not-determined |
| 18-104_k141_129862 | Not-determined |
| 18-104_k141_11803 | Not-determined |
| 18-104_k141_32672 | Not-determined |
| 18-104_k141_25538 | Not-determined |
| 18-104_k141_88207 | Not-determined |
| 18-104_k141_74599 | Not-determined |
| 18-104_k141_67549 | Not-determined |
| 18-104_k141_11812 | Not-determined |
| 18-104_k141_88209 | Not-determined |
| 18-104_k141_74600 | Not-determined |
| 18-104_k141_129868 | Not-determined |
| 18-104_k141_46692 | Not-determined |
| 18-104_k141_52845 | Not-determined |
| 18-104_k141_25541 | Not-determined |
| 18-104_k141_67553 | Not-determined |
| 18-104_k141_67555 | Not-determined |
| 18-104_k141_11821 | Not-determined |
| 18-104_k141_32684 | Low-quality |
| 18-104_k141_88216 | Not-determined |
| 18-104_k141_39334 | Not-determined |
| 18-104_k141_95264 | Not-determined |
| 18-104_k141_81781 | Low-quality |
| 18-104_k141_81782 | Not-determined |
| 18-104_k141_81784 | Not-determined |
| 18-104_k141_46704 | Not-determined |
| 18-104_k141_88222 | Not-determined |
| 18-104_k141_67569 | Not-determined |
| 18-104_k141_150898 | Not-determined |
| 18-104_k141_4650 | Low-quality |
| 18-104_k141_81791 | Low-quality |
| 18-104_k141_150902 | Not-determined |
| 18-104_k141_60163 | Low-quality |
| 18-104_k141_52861 | Low-quality |
| 18-104_k141_4654 | Not-determined |
| 18-104_k141_4655 | Not-determined |
| 18-104_k141_4656 | Not-determined |
| 18-104_k141_32696 | Not-determined |
| 18-104_k141_81799 | Not-determined |
| 18-104_k141_122464 | Not-determined |
| 18-104_k141_60164 | Not-determined |
| 18-104_k141_39356 | Not-determined |
| 18-104_k141_67575 | Not-determined |
| 18-104_k141_4659 | Not-determined |
| 18-104_k141_4660 | Not-determined |
| 18-104_k141_4661 | Not-determined |
| 18-104_k141_129889 | Not-determined |
| 18-104_k141_150909 | Not-determined |
| 18-104_k141_74614 | Not-determined |
| 18-104_k141_4665 | Not-determined |
| 18-104_k141_109556 | Not-determined |
| 18-104_k141_25573 | Not-determined |
| 18-104_k141_17916 | Not-determined |
| 18-104_k141_109560 | Not-determined |
| 18-104_k141_115909 | Not-determined |
| 18-104_k141_164845 | Not-determined |
| 18-104_k141_4674 | Not-determined |
| 18-104_k141_46719 | Not-determined |
| 18-104_k141_88226 | Not-determined |
| 18-104_k141_4676 | Not-determined |
| 18-104_k141_46723 | Not-determined |
| 18-104_k141_150918 | Not-determined |
| 18-104_k141_88229 | Not-determined |
| 18-104_k141_158106 | Not-determined |
| 18-104_k141_74621 | Not-determined |
| 18-104_k141_95291 | Not-determined |
| 18-104_k141_17928 | Not-determined |
| 18-104_k141_109571 | Not-determined |
| 18-104_k141_32706 | Not-determined |
| 18-104_k141_25580 | Not-determined |
| 18-104_k141_32707 | Not-determined |
| 18-104_k141_17934 | Not-determined |
| 18-104_k141_109576 | Not-determined |
| 18-104_k141_32711 | Low-quality |
| 18-104_k141_109578 | Not-determined |
| 18-104_k141_164854 | Not-determined |
| 18-104_k141_115923 | Low-quality |
| 18-104_k141_32714 | Not-determined |
| 18-104_k141_39373 | Not-determined |
| 18-104_k141_32718 | Not-determined |
| 18-104_k141_4689 | Not-determined |
| 18-104_k141_32721 | Not-determined |
| 18-104_k141_39378 | Not-determined |
| 18-104_k141_150938 | Not-determined |
| 18-104_k141_115933 | Not-determined |
| 18-104_k141_88251 | Low-quality |
| 18-104_k141_129907 | Not-determined |
| 18-104_k141_109589 | Not-determined |
| 18-104_k141_102034 | Low-quality |
| 18-104_k141_88258 | Not-determined |
| 18-104_k141_115938 | Not-determined |
| 18-104_k141_122483 | Not-determined |
| 18-104_k141_102040 | Not-determined |
| 18-104_k141_102042 | Not-determined |
| 18-104_k141_115944 | Not-determined |
| 18-104_k141_95323 | Not-determined |
| 18-104_k141_25590 | Not-determined |
| 18-104_k141_67594 | Not-determined |
| 18-104_k141_129917 | Not-determined |
| 18-104_k141_144299 | Low-quality |
| 18-104_k141_137070 | Not-determined |
| 18-104_k141_11845 | Not-determined |
| 18-104_k141_74645 | Not-determined |
| 18-104_k141_137073 | Not-determined |
| 18-104_k141_102052 | Not-determined |
| 18-104_k141_25596 | Not-determined |
| 18-104_k141_137075 | Low-quality |
| 18-104_k141_137077 | Not-determined |
| 18-104_k141_102056 | Not-determined |
| 18-104_k141_32736 | Not-determined |
| 18-104_k141_129923 | Low-quality |
| 18-104_k141_17954 | Not-determined |
| 18-104_k141_129924 | Not-determined |
| 18-104_k141_74656 | Not-determined |
| 18-104_k141_129925 | Not-determined |
| 18-104_k141_95341 | Not-determined |
| 18-104_k141_39399 | Not-determined |
| 18-104_k141_4718 | Not-determined |
| 18-104_k141_52887 | Low-quality |
| 18-104_k141_164881 | Not-determined |
| 18-104_k141_95343 | Not-determined |
| 18-104_k141_17966 | Not-determined |
| 18-104_k141_158140 | Not-determined |
| 18-104_k141_129942 | Not-determined |
| 18-104_k141_46727 | Not-determined |
| 18-104_k141_137092 | Not-determined |
| 18-104_k141_60208 | Not-determined |
| 18-104_k141_4731 | Not-determined |
| 18-104_k141_17974 | Not-determined |
| 18-104_k141_32749 | Not-determined |
| 18-104_k141_95347 | Not-determined |
| 18-104_k141_115974 | Not-determined |
| 18-104_k141_95350 | Not-determined |
| 18-104_k141_88280 | Not-determined |
| 18-104_k141_39412 | Not-determined |
| 18-104_k141_46736 | Not-determined |
| 18-104_k141_74662 | Not-determined |
| 18-104_k141_137096 | Not-determined |
| 18-104_k141_137098 | Low-quality |
| 18-104_k141_137099 | Not-determined |
| 18-104_k141_129959 | Not-determined |
| 18-104_k141_122495 | Low-quality |
| 18-104_k141_122496 | Not-determined |
| 18-104_k141_88290 | Not-determined |
| 18-104_k141_17983 | Not-determined |
| 18-104_k141_74669 | Not-determined |
| 18-104_k141_137105 | Not-determined |
| 18-104_k141_4745 | Not-determined |
| 18-104_k141_95359 | Not-determined |
| 18-104_k141_39418 | Low-quality |
| 18-104_k141_4746 | Not-determined |
| 18-104_k141_164893 | Not-determined |
| 18-104_k141_109645 | Not-determined |
| 18-104_k141_17985 | Not-determined |
| 18-104_k141_67614 | Not-determined |
| 18-104_k141_129965 | Not-determined |
| 18-104_k141_60224 | Not-determined |
| 18-104_k141_95368 | Not-determined |
| 18-104_k141_46748 | Not-determined |
| 18-104_k141_109650 | Not-determined |
| 18-104_k141_74681 | Not-determined |
| 18-104_k141_67622 | Not-determined |
| 18-104_k141_102075 | Not-determined |
| 18-104_k141_46754 | Low-quality |
| 18-104_k141_109655 | Not-determined |
| 18-104_k141_88302 | Not-determined |
| 18-104_k141_95377 | Not-determined |
| 18-104_k141_102079 | Not-determined |
| 18-104_k141_67629 | Not-determined |
| 18-104_k141_52897 | Not-determined |
| 18-104_k141_102082 | Not-determined |
| 18-104_k141_46758 | Not-determined |
| 18-104_k141_88306 | Not-determined |
| 18-104_k141_52900 | Not-determined |
| 18-104_k141_109664 | Low-quality |
| 18-104_k141_115988 | Low-quality |
| 18-104_k141_4766 | Not-determined |
| 18-104_k141_4767 | Not-determined |
| 18-104_k141_150981 | Not-determined |
| 18-104_k141_81875 | Not-determined |
| 18-104_k141_144325 | Not-determined |
| 18-104_k141_150982 | Not-determined |
| 18-104_k141_4770 | Not-determined |
| 18-104_k141_74688 | Low-quality |
| 18-104_k141_4772 | Not-determined |
| 18-104_k141_144328 | Not-determined |
| 18-104_k141_95390 | Not-determined |
| 18-104_k141_122512 | Not-determined |
| 18-104_k141_39422 | Not-determined |
| 18-104_k141_25642 | Not-determined |
| 18-104_k141_60243 | Not-determined |
| 18-104_k141_95394 | Not-determined |
| 18-104_k141_102093 | Not-determined |
| 18-104_k141_74694 | Not-determined |
| 18-104_k141_158187 | Not-determined |
| 18-104_k141_81890 | Not-determined |
| 18-104_k141_74696 | Not-determined |
| 18-104_k141_74697 | Low-quality |
| 18-104_k141_52908 | Low-quality |
| 18-104_k141_129984 | Not-determined |
| 18-104_k141_129985 | Not-determined |
| 18-104_k141_144341 | Not-determined |
| 18-104_k141_25657 | Not-determined |
| 18-104_k141_102106 | Not-determined |
| 18-104_k141_52912 | Not-determined |
| 18-104_k141_150999 | Not-determined |
| 18-104_k141_158197 | Not-determined |
| 18-104_k141_122519 | Not-determined |
| 18-104_k141_88315 | Not-determined |
| 18-104_k141_158203 | Not-determined |
| 18-104_k141_25661 | Low-quality |
| 18-104_k141_4776 | Not-determined |
| 18-104_k141_102113 | Not-determined |
| 18-104_k141_32802 | Not-determined |
| 18-104_k141_52916 | Not-determined |
| 18-104_k141_39445 | Not-determined |
| 18-104_k141_102118 | Not-determined |
| 18-104_k141_137129 | Not-determined |
| 18-104_k141_137130 | Not-determined |
| 18-104_k141_39449 | Not-determined |
| 18-104_k141_32805 | Not-determined |
| 18-104_k141_137133 | Not-determined |
| 18-104_k141_164934 | Low-quality |
| 18-104_k141_116027 | Not-determined |
| 18-104_k141_11902 | Not-determined |
| 18-104_k141_144352 | Not-determined |
| 18-104_k141_144353 | Not-determined |
| 18-104_k141_88325 | Low-quality |
| 18-104_k141_46785 | Not-determined |
| 18-104_k141_116031 | Not-determined |
| 18-104_k141_67641 | Low-quality |
| 18-104_k141_52926 | Not-determined |
| 18-104_k141_137140 | Not-determined |
| 18-104_k141_116036 | Not-determined |
| 18-104_k141_130014 | Not-determined |
| 18-104_k141_116038 | Not-determined |
| 18-104_k141_130015 | Not-determined |
| 18-104_k141_88331 | Not-determined |
| 18-104_k141_18017 | Not-determined |
| 18-104_k141_102132 | Not-determined |
| 18-104_k141_60258 | Not-determined |
| 18-104_k141_102134 | Low-quality |
| 18-104_k141_81903 | Not-determined |
| 18-104_k141_109688 | Not-determined |
| 18-104_k141_95420 | Not-determined |
| 18-104_k141_130019 | Not-determined |
| 18-104_k141_4786 | Not-determined |
| 18-104_k141_60261 | Not-determined |
| 18-104_k141_46792 | Not-determined |
| 18-104_k141_164944 | Not-determined |
| 18-104_k141_60264 | Not-determined |
| 18-104_k141_4790 | Not-determined |
| 18-104_k141_122552 | Not-determined |
| 18-104_k141_18027 | Not-determined |
| 18-104_k141_46793 | Not-determined |
| 18-104_k141_137146 | Not-determined |
| 18-104_k141_52934 | Not-determined |
| 18-104_k141_137147 | Not-determined |
| 18-104_k141_52935 | Not-determined |
| 18-104_k141_122556 | Not-determined |
| 18-104_k141_52937 | Not-determined |
| 18-104_k141_109689 | Not-determined |
| 18-104_k141_18038 | Not-determined |
| 18-104_k141_81919 | Not-determined |
| 18-104_k141_158223 | Not-determined |
| 18-104_k141_116045 | Low-quality |
| 18-104_k141_137151 | Not-determined |
| 18-104_k141_18040 | Not-determined |
| 18-104_k141_18041 | Not-determined |
| 18-104_k141_18043 | Not-determined |
| 18-104_k141_164958 | Not-determined |
| 18-104_k141_81925 | Not-determined |
| 18-104_k141_46798 | Not-determined |
| 18-104_k141_46799 | Not-determined |
| 18-104_k141_130038 | Not-determined |
| 18-104_k141_88341 | Not-determined |
| 18-104_k141_144374 | Not-determined |
| 18-104_k141_144375 | Not-determined |
| 18-104_k141_102167 | Not-determined |
| 18-104_k141_88346 | Not-determined |
| 18-104_k141_4805 | Not-determined |
| 18-104_k141_81938 | Not-determined |
| 18-104_k141_74739 | Not-determined |
| 18-104_k141_60274 | Not-determined |
| 18-104_k141_46808 | Not-determined |
| 18-104_k141_81940 | Not-determined |
| 18-104_k141_11930 | Not-determined |
| 18-104_k141_158238 | Not-determined |
| 18-104_k141_158242 | Not-determined |
| 18-104_k141_102175 | Not-determined |
| 18-104_k141_67656 | Not-determined |
| 18-104_k141_116048 | Not-determined |
| 18-104_k141_158247 | Not-determined |
| 18-104_k141_130053 | Not-determined |
| 18-104_k141_18056 | Not-determined |
| 18-104_k141_109702 | Not-determined |
| 18-104_k141_11936 | Not-determined |
| 18-104_k141_144391 | Not-determined |
| 18-104_k141_88362 | Not-determined |
| 18-104_k141_102182 | Low-quality |
| 18-104_k141_102184 | Not-determined |
| 18-104_k141_102185 | Not-determined |
| 18-104_k141_109706 | Not-determined |
| 18-104_k141_52969 | Not-determined |
| 18-104_k141_109708 | Not-determined |
| 18-104_k141_95442 | Not-determined |
| 18-104_k141_116060 | Not-determined |
| 18-104_k141_52970 | Not-determined |
| 18-104_k141_144399 | Not-determined |
| 18-104_k141_39466 | Low-quality |
| 18-104_k141_18064 | Not-determined |
| 18-104_k141_109714 | Not-determined |
| 18-104_k141_25694 | Low-quality |
| 18-104_k141_164975 | Not-determined |
| 18-104_k141_164978 | Not-determined |
| 18-104_k141_67672 | Low-quality |
| 18-104_k141_116065 | Low-quality |
| 18-104_k141_164979 | Not-determined |
| 18-104_k141_137175 | Not-determined |
| 18-104_k141_102193 | Not-determined |
| 18-104_k141_122590 | Not-determined |
| 18-104_k141_32809 | Not-determined |
| 18-104_k141_144407 | Not-determined |
| 18-104_k141_122593 | Not-determined |
| 18-104_k141_67680 | Not-determined |
| 18-104_k141_158267 | Not-determined |
| 18-104_k141_137178 | Not-determined |
| 18-104_k141_130072 | Low-quality |
| 18-104_k141_60299 | Not-determined |
| 18-104_k141_46823 | Not-determined |
| 18-104_k141_122597 | Not-determined |
| 18-104_k141_116070 | Not-determined |
| 18-104_k141_158281 | Not-determined |
| 18-104_k141_81945 | Low-quality |
| 18-104_k141_25715 | Not-determined |
| 18-104_k141_52992 | Not-determined |
| 18-104_k141_74767 | Not-determined |
| 18-104_k141_46830 | Not-determined |
| 18-104_k141_39480 | Not-determined |
| 18-104_k141_25723 | Not-determined |
| 18-104_k141_60305 | Not-determined |
| 18-104_k141_158290 | Not-determined |
| 18-104_k141_151011 | Not-determined |
| 18-104_k141_25731 | Not-determined |
| 18-104_k141_60312 | Not-determined |
| 18-104_k141_18079 | Not-determined |
| 18-104_k141_164993 | Not-determined |
| 18-104_k141_122610 | Not-determined |
| 18-104_k141_88402 | Not-determined |
| 18-104_k141_95477 | Not-determined |
| 18-104_k141_4832 | Not-determined |
| 18-104_k141_88405 | Not-determined |
| 18-104_k141_137202 | Not-determined |
| 18-104_k141_130101 | Not-determined |
| 18-104_k141_88408 | Not-determined |
| 18-104_k141_130102 | Not-determined |
| 18-104_k141_116086 | Not-determined |
| 18-104_k141_60316 | Not-determined |
| 18-104_k141_39492 | Not-determined |
| 18-104_k141_39494 | Not-determined |
| 18-104_k141_165002 | Not-determined |
| 18-104_k141_39495 | Not-determined |
| 18-104_k141_116091 | Not-determined |
| 18-104_k141_144431 | Low-quality |
| 18-104_k141_165005 | Not-determined |

| | |
|---|---|
| 18-104_k141_4840 | Not-determined |
| 18-104_k141_4841 | Not-determined |
| 18-104_k141_18089 | Not-determined |
| 18-104_k141_144433 | Not-determined |
| 18-104_k141_165011 | Not-determined |
| 18-104_k141_32818 | Not-determined |
| 18-104_k141_39503 | Not-determined |
| 18-104_k141_39505 | Not-determined |
| 18-104_k141_60323 | Not-determined |
| 18-104_k141_46856 | Not-determined |
| 18-104_k141_39510 | Not-determined |
| 18-104_k141_46861 | Not-determined |
| 18-104_k141_144439 | Not-determined |
| 18-104_k141_88424 | Low-quality |
| 18-104_k141_11946 | Not-determined |
| 18-104_k141_151035 | Not-determined |
| 18-104_k141_165026 | Not-determined |
| 18-104_k141_95491 | Not-determined |
| 18-104_k141_74784 | Not-determined |
| 18-104_k141_39523 | Not-determined |
| 18-104_k141_39524 | Not-determined |
| 18-104_k141_4851 | Not-determined |
| 18-104_k141_74791 | Not-determined |
| 18-104_k141_102217 | Not-determined |
| 18-104_k141_130119 | Not-determined |
| 18-104_k141_60340 | Not-determined |
| 18-104_k141_60341 | Not-determined |
| 18-104_k141_67724 | Not-determined |
| 18-104_k141_18108 | Not-determined |
| 18-104_k141_67725 | Low-quality |
| 18-104_k141_122645 | Not-determined |
| 18-104_k141_122646 | Not-determined |
| 18-104_k141_116117 | Not-determined |
| 18-104_k141_109761 | Not-determined |
| 18-104_k141_32841 | Not-determined |
| 18-104_k141_67728 | Not-determined |
| 18-104_k141_95497 | Not-determined |
| 18-104_k141_158313 | Not-determined |
| 18-104_k141_116120 | Not-determined |
| 18-104_k141_158315 | Not-determined |
| 18-104_k141_11954 | Not-determined |
| 18-104_k141_67736 | Not-determined |
| 18-104_k141_151057 | Not-determined |
| 18-104_k141_165042 | Not-determined |
| 18-104_k141_53028 | Not-determined |
| 18-104_k141_67742 | Not-determined |
| 18-104_k141_11966 | Not-determined |
| 18-104_k141_88445 | Low-quality |
| 18-104_k141_165052 | Not-determined |
| 18-104_k141_116129 | Not-determined |
| 18-104_k141_130138 | Not-determined |
| 18-104_k141_11967 | Not-determined |
| 18-104_k141_144476 | Low-quality |
| 18-104_k141_116134 | Not-determined |
| 18-104_k141_18125 | Not-determined |
| 18-104_k141_11968 | Not-determined |
| 18-104_k141_60348 | Not-determined |
| 18-104_k141_109782 | Not-determined |
| 18-104_k141_67752 | Not-determined |
| 18-104_k141_46879 | Not-determined |
| 18-104_k141_109786 | Low-quality |
| 18-104_k141_81975 | Not-determined |
| 18-104_k141_18131 | Low-quality |
| 18-104_k141_46881 | Not-determined |
| 18-104_k141_25769 | Not-determined |
| 18-104_k141_165060 | Not-determined |
| 18-104_k141_95506 | Not-determined |
| 18-104_k141_151080 | Not-determined |
| 18-104_k141_144485 | Not-determined |
| 18-104_k141_165062 | Not-determined |
| 18-104_k141_158333 | Low-quality |
| 18-104_k141_60351 | Not-determined |
| 18-104_k141_25773 | Not-determined |
| 18-104_k141_95510 | Not-determined |
| 18-104_k141_60357 | Low-quality |
| 18-104_k141_11979 | Not-determined |
| 18-104_k141_67767 | Not-determined |
| 18-104_k141_144488 | Not-determined |
| 18-104_k141_102268 | Not-determined |
| 18-104_k141_46890 | Not-determined |
| 18-104_k141_116150 | Not-determined |
| 18-104_k141_60363 | Not-determined |
| 18-104_k141_122673 | Not-determined |
| 18-104_k141_116153 | Not-determined |
| 18-104_k141_165072 | Low-quality |
| 18-104_k141_60366 | Not-determined |
| 18-104_k141_11989 | Not-determined |
| 18-104_k141_25788 | Low-quality |
| 18-104_k141_158358 | Not-determined |
| 18-104_k141_53054 | Not-determined |
| 18-104_k141_60369 | Low-quality |
| 18-104_k141_60371 | Not-determined |
| 18-104_k141_88465 | Not-determined |
| 18-104_k141_81995 | Not-determined |
| 18-104_k141_158362 | Not-determined |
| 18-104_k141_81997 | Not-determined |
| 18-104_k141_122682 | Not-determined |
| 18-104_k141_102275 | Not-determined |
| 18-104_k141_4878 | Not-determined |
| 18-104_k141_151099 | Not-determined |
| 18-104_k141_116160 | Low-quality |
| 18-104_k141_137252 | Not-determined |
| 18-104_k141_116161 | Not-determined |
| 18-104_k141_137253 | Not-determined |
| 18-104_k141_32854 | Not-determined |
| 18-104_k141_18156 | Not-determined |
| 18-104_k141_32856 | Not-determined |
| 18-104_k141_144510 | Low-quality |
| 18-104_k141_88477 | Not-determined |
| 18-104_k141_46906 | Low-quality |
| 18-104_k141_116169 | Not-determined |
| 18-104_k141_82010 | Not-determined |
| 18-104_k141_82012 | Not-determined |
| 18-104_k141_137266 | Not-determined |
| 18-104_k141_60380 | Not-determined |
| 18-104_k141_137270 | Not-determined |
| 18-104_k141_32873 | Not-determined |
| 18-104_k141_4888 | Not-determined |
| 18-104_k141_122692 | Not-determined |
| 18-104_k141_102281 | Not-determined |
| 18-104_k141_32884 | Not-determined |
| 18-104_k141_102287 | Low-quality |
| 18-104_k141_151124 | Not-determined |
| 18-104_k141_95522 | Not-determined |
| 18-104_k141_95527 | Not-determined |
| 18-104_k141_109810 | Not-determined |
| 18-104_k141_32895 | Not-determined |
| 18-104_k141_102294 | Low-quality |
| 18-104_k141_74868 | Not-determined |
| 18-104_k141_116194 | Not-determined |
| 18-104_k141_95531 | Not-determined |
| 18-104_k141_130182 | Low-quality |
| 18-104_k141_102295 | Not-determined |
| 18-104_k141_18176 | Not-determined |
| 18-104_k141_74870 | Not-determined |
| 18-104_k141_116197 | Not-determined |
| 18-104_k141_32902 | Not-determined |
| 18-104_k141_158394 | Not-determined |
| 18-104_k141_82047 | Not-determined |
| 18-104_k141_158395 | Not-determined |
| 18-104_k141_130190 | Not-determined |
| 18-104_k141_95534 | Not-determined |
| 18-104_k141_95535 | Not-determined |
| 18-104_k141_95538 | Not-determined |
| 18-104_k141_109821 | Not-determined |
| 18-104_k141_116205 | Not-determined |
| 18-104_k141_74881 | Not-determined |
| 18-104_k141_60392 | Not-determined |
| 18-104_k141_95540 | Not-determined |
| 18-104_k141_137278 | Not-determined |
| 18-104_k141_32914 | Not-determined |
| 18-104_k141_95544 | Not-determined |
| 18-104_k141_88503 | Not-determined |
| 18-104_k141_60394 | Not-determined |
| 18-104_k141_137283 | Not-determined |
| 18-104_k141_67800 | Not-determined |
| 18-104_k141_60395 | Low-quality |
| 18-104_k141_158410 | Not-determined |
| 18-104_k141_122714 | Low-quality |
| 18-104_k141_144545 | Not-determined |
| 18-104_k141_60398 | Not-determined |
| 18-104_k141_95548 | Not-determined |
| 18-104_k141_95550 | Not-determined |
| 18-104_k141_165115 | Not-determined |
| 18-104_k141_18197 | Not-determined |
| 18-104_k141_109828 | Not-determined |
| 18-104_k141_39546 | Not-determined |
| 18-104_k141_18199 | Not-determined |
| 18-104_k141_25821 | Not-determined |
| 18-104_k141_158419 | Not-determined |
| 18-104_k141_137290 | Low-quality |
| 18-104_k141_32925 | Not-determined |
| 18-104_k141_137294 | Not-determined |
| 18-104_k141_67813 | Not-determined |
| 18-104_k141_130207 | Not-determined |
| 18-104_k141_116215 | Not-determined |
| 18-104_k141_25829 | Not-determined |
| 18-104_k141_165125 | Not-determined |
| 18-104_k141_12012 | Not-determined |
| 18-104_k141_109840 | Not-determined |
| 18-104_k141_95574 | Not-determined |
| 18-104_k141_144565 | Low-quality |
| 18-104_k141_95578 | Not-determined |
| 18-104_k141_82052 | Not-determined |
| 18-104_k141_102315 | Not-determined |
| 18-104_k141_144569 | Low-quality |
| 18-104_k141_165146 | Not-determined |
| 18-104_k141_158458 | Not-determined |
| 18-104_k141_158459 | Not-determined |
| 18-104_k141_144576 | Low-quality |
| 18-104_k141_151149 | Not-determined |
| 18-104_k141_4941 | Not-determined |
| 18-104_k141_144578 | Not-determined |
| 18-104_k141_151152 | Not-determined |
| 18-104_k141_18212 | Not-determined |
| 18-104_k141_67827 | Not-determined |
| 18-104_k141_12027 | Not-determined |
| 18-104_k141_46957 | Not-determined |
| 18-104_k141_12028 | Not-determined |
| 18-104_k141_122742 | Not-determined |
| 18-104_k141_32950 | Not-determined |
| 18-104_k141_25852 | Not-determined |
| 18-104_k141_122744 | Not-determined |
| 18-104_k141_151153 | Not-determined |
| 18-104_k141_12030 | Not-determined |
| 18-104_k141_122745 | Not-determined |
| 18-104_k141_137314 | Not-determined |
| 18-104_k141_116231 | Not-determined |
| 18-104_k141_95596 | Not-determined |
| 18-104_k141_25854 | Low-quality |
| 18-104_k141_18220 | Not-determined |
| 18-104_k141_137318 | Low-quality |
| 18-104_k141_95600 | Not-determined |
| 18-104_k141_109849 | Not-determined |
| 18-104_k141_74922 | Not-determined |
| 18-104_k141_137324 | Not-determined |
| 18-104_k141_74926 | Not-determined |
| 18-104_k141_18224 | Not-determined |
| 18-104_k141_165165 | Not-determined |
| 18-104_k141_25863 | Not-determined |
| 18-104_k141_102336 | Low-quality |
| 18-104_k141_144595 | Not-determined |
| 18-104_k141_116240 | Not-determined |
| 18-104_k141_102338 | Not-determined |
| 18-104_k141_165171 | Not-determined |
| 18-104_k141_53101 | Low-quality |
| 18-104_k141_151167 | Not-determined |
| 18-104_k141_116249 | Not-determined |
| 18-104_k141_102345 | Not-determined |
| 18-104_k141_32974 | Not-determined |
| 18-104_k141_165181 | Not-determined |
| 18-104_k141_53109 | Not-determined |
| 18-104_k141_102347 | Not-determined |
| 18-104_k141_95611 | Not-determined |
| 18-104_k141_144599 | Not-determined |
| 18-104_k141_74943 | Not-determined |
| 18-104_k141_74945 | Not-determined |
| 18-104_k141_18234 | Not-determined |
| 18-104_k141_4967 | Not-determined |
| 18-104_k141_165193 | Not-determined |
| 18-104_k141_158481 | Low-quality |
| 18-104_k141_67843 | Not-determined |
| 18-104_k141_151182 | Not-determined |
| 18-104_k141_39598 | Not-determined |
| 18-104_k141_137344 | Not-determined |
| 18-104_k141_116256 | Not-determined |
| 18-104_k141_158487 | Not-determined |
| 18-104_k141_25873 | Not-determined |
| 18-104_k141_158489 | Not-determined |
| 18-104_k141_95629 | Not-determined |
| 18-104_k141_137349 | Not-determined |
| 18-104_k141_116263 | Not-determined |
| 18-104_k141_4978 | Low-quality |
| 18-104_k141_53134 | Low-quality |
| 18-104_k141_18248 | Not-determined |
| 18-104_k141_165218 | Not-determined |
| 18-104_k141_102360 | Not-determined |
| 18-104_k141_32996 | Not-determined |
| 18-104_k141_116271 | Not-determined |
| 18-104_k141_151203 | Not-determined |
| 18-104_k141_144622 | Not-determined |
| 18-104_k141_53141 | Not-determined |
| 18-104_k141_67846 | Not-determined |
| 18-104_k141_151207 | Not-determined |
| 18-104_k141_53145 | Low-quality |
| 18-104_k141_18261 | Low-quality |
| 18-104_k141_158514 | Not-determined |
| 18-104_k141_46994 | Not-determined |
| 18-104_k141_4988 | Low-quality |
| 18-104_k141_67850 | Not-determined |
| 18-104_k141_67851 | Not-determined |
| 18-104_k141_33004 | Not-determined |
| 18-104_k141_122761 | Not-determined |
| 18-104_k141_144631 | Not-determined |
| 18-104_k141_33006 | Not-determined |
| 18-104_k141_18273 | Not-determined |
| 18-104_k141_122763 | Not-determined |
| 18-104_k141_144632 | Not-determined |
| 18-104_k141_122764 | Not-determined |
| 18-104_k141_122767 | Not-determined |
| 18-104_k141_33013 | Not-determined |
| 18-104_k141_102386 | Not-determined |
| 18-104_k141_18275 | Not-determined |
| 18-104_k141_158520 | Low-quality |
| 18-104_k141_33017 | Not-determined |
| 18-104_k141_39616 | Not-determined |
| 18-104_k141_158522 | Not-determined |
| 18-104_k141_158523 | Not-determined |
| 18-104_k141_102392 | Not-determined |
| 18-104_k141_67865 | Not-determined |
| 18-104_k141_88538 | Not-determined |
| 18-104_k141_33026 | Not-determined |
| 18-104_k141_67871 | Not-determined |
| 18-104_k141_33030 | Not-determined |
| 18-104_k141_158528 | Not-determined |
| 18-104_k141_39628 | Low-quality |
| 18-104_k141_102400 | Not-determined |
| 18-104_k141_116285 | Not-determined |
| 18-104_k141_53174 | Low-quality |
| 18-104_k141_116286 | Low-quality |
| 18-104_k141_95639 | Not-determined |
| 18-104_k141_116293 | Not-determined |
| 18-104_k141_39631 | Not-determined |
| 18-104_k141_130257 | Not-determined |
| 18-104_k141_102410 | Not-determined |
| 18-104_k141_39632 | Not-determined |
| 18-104_k141_82093 | Not-determined |
| 18-104_k141_102421 | Low-quality |
| 18-104_k141_47011 | Not-determined |
| 18-104_k141_67883 | Not-determined |
| 18-104_k141_151233 | Not-determined |
| 18-104_k141_122773 | Low-quality |
| 18-104_k141_158560 | Not-determined |
| 18-104_k141_67893 | Low-quality |
| 18-104_k141_82107 | Not-determined |
| 18-104_k141_158566 | Not-determined |
| 18-104_k141_102431 | Not-determined |
| 18-104_k141_151244 | Not-determined |
| 18-104_k141_53187 | Not-determined |
| 18-104_k141_102433 | Not-determined |
| 18-104_k141_122779 | Not-determined |
| 18-104_k141_158575 | Not-determined |
| 18-104_k141_47036 | Not-determined |
| 18-104_k141_88577 | Not-determined |
| 18-104_k141_18331 | Not-determined |
| 18-104_k141_95644 | Not-determined |
| 18-104_k141_18332 | Not-determined |
| 18-104_k141_67901 | Not-determined |
| 18-104_k141_144657 | Not-determined |
| 18-104_k141_102440 | Low-quality |
| 18-104_k141_88581 | Not-determined |
| 18-104_k141_18334 | Not-determined |
| 18-104_k141_95652 | Not-determined |
| 18-104_k141_18336 | Not-determined |
| 18-104_k141_158580 | Not-determined |
| 18-104_k141_47045 | Not-determined |
| 18-104_k141_102448 | Not-determined |
| 18-104_k141_122788 | Not-determined |
| 18-104_k141_18340 | Low-quality |
| 18-104_k141_82125 | Not-determined |
| 18-104_k141_53198 | Not-determined |
| 18-104_k141_109946 | Not-determined |
| 18-104_k141_67912 | Not-determined |
| 18-104_k141_67913 | Not-determined |
| 18-104_k141_102457 | Not-determined |
| 18-104_k141_88590 | Not-determined |
| 18-104_k141_109949 | Not-determined |
| 18-104_k141_25923 | Low-quality |
| 18-104_k141_18347 | Not-determined |
| 18-104_k141_102459 | Not-determined |
| 18-104_k141_95662 | Not-determined |
| 18-104_k141_165271 | Not-determined |
| 18-104_k141_25927 | Not-determined |
| 18-104_k141_116352 | Not-determined |
| 18-104_k141_60403 | Not-determined |
| 18-104_k141_53209 | Not-determined |
| 18-104_k141_122802 | Not-determined |
| 18-104_k141_158600 | Not-determined |
| 18-104_k141_53211 | Low-quality |
| 18-104_k141_47053 | Not-determined |
| 18-104_k141_67928 | Not-determined |
| 18-104_k141_39653 | Not-determined |
| 18-104_k141_95676 | Not-determined |
| 18-104_k141_88606 | Not-determined |
| 18-104_k141_109961 | Not-determined |
| 18-104_k141_144677 | Not-determined |
| 18-104_k141_165276 | Not-determined |
| 18-104_k141_122812 | Not-determined |
| 18-104_k141_39659 | Not-determined |
| 18-104_k141_67935 | Not-determined |
| 18-104_k141_39661 | Not-determined |
| 18-104_k141_25941 | Not-determined |
| 18-104_k141_67939 | Not-determined |
| 18-104_k141_39665 | Not-determined |
| 18-104_k141_12156 | Not-determined |
| 18-104_k141_12158 | Not-determined |
| 18-104_k141_67941 | Not-determined |
| 18-104_k141_95689 | Not-determined |
| 18-104_k141_122822 | Not-determined |
| 18-104_k141_137372 | Not-determined |
| 18-104_k141_88618 | Not-determined |
| 18-104_k141_82146 | Not-determined |
| 18-104_k141_88621 | Not-determined |
| 18-104_k141_165293 | Not-determined |
| 18-104_k141_109972 | Not-determined |
| 18-104_k141_144691 | Not-determined |
| 18-104_k141_25955 | Not-determined |
| 18-104_k141_95696 | Not-determined |
| 18-104_k141_82148 | Not-determined |
| 18-104_k141_95697 | Not-determined |
| 18-104_k141_102481 | Not-determined |
| 18-104_k141_39675 | Low-quality |
| 18-104_k141_158626 | Not-determined |
| 18-104_k141_158627 | Not-determined |
| 18-104_k141_102484 | Not-determined |
| 18-104_k141_18370 | Not-determined |
| 18-104_k141_165297 | Not-determined |
| 18-104_k141_88628 | Not-determined |
| 18-104_k141_25960 | Not-determined |
| 18-104_k141_82153 | Not-determined |
| 18-104_k141_33037 | Not-determined |
| 18-104_k141_74979 | Not-determined |
| 18-104_k141_102486 | Low-quality |
| 18-104_k141_25965 | Not-determined |
| 18-104_k141_39684 | Low-quality |
| 18-104_k141_25966 | Not-determined |
| 18-104_k141_109976 | Low-quality |
| 18-104_k141_82157 | Not-determined |
| 18-104_k141_25968 | Not-determined |
| 18-104_k141_95702 | Not-determined |
| 18-104_k141_5004 | Not-determined |
| 18-104_k141_60455 | Not-determined |
| 18-104_k141_137387 | Not-determined |
| 18-104_k141_60456 | Low-quality |
| 18-104_k141_137388 | Not-determined |
| 18-104_k141_74983 | Not-determined |
| 18-104_k141_5006 | Not-determined |
| 18-104_k141_25976 | Not-determined |
| 18-104_k141_53239 | Not-determined |
| 18-104_k141_82168 | Not-determined |
| 18-104_k141_53240 | Not-determined |
| 18-104_k141_12170 | Not-determined |
| 18-104_k141_130277 | Low-quality |
| 18-104_k141_18375 | Not-determined |
| 18-104_k141_158633 | Not-determined |
| 18-104_k141_53245 | Not-determined |
| 18-104_k141_60460 | Not-determined |
| 18-104_k141_60461 | Not-determined |
| 18-104_k141_130286 | Not-determined |
| 18-104_k141_18384 | Not-determined |
| 18-104_k141_5018 | Not-determined |
| 18-104_k141_102511 | Not-determined |
| 18-104_k141_165315 | Not-determined |
| 18-104_k141_102513 | Not-determined |
| 18-104_k141_122859 | Not-determined |
| 18-104_k141_18387 | Not-determined |
| 18-104_k141_33061 | Not-determined |
| 18-104_k141_12186 | Low-quality |
| 18-104_k141_102521 | Not-determined |
| 18-104_k141_39707 | Not-determined |
| 18-104_k141_158659 | Not-determined |
| 18-104_k141_74986 | Low-quality |
| 18-104_k141_33067 | Not-determined |
| 18-104_k141_18394 | Not-determined |
| 18-104_k141_67950 | Not-determined |
| 18-104_k141_116392 | Not-determined |
| 18-104_k141_116394 | Not-determined |
| 18-104_k141_25984 | Not-determined |
| 18-104_k141_130296 | Not-determined |
| 18-104_k141_109996 | Not-determined |
| 18-104_k141_18402 | Not-determined |
| 18-104_k141_95721 | Not-determined |
| 18-104_k141_74997 | Not-determined |
| 18-104_k141_82179 | Low-quality |
| 18-104_k141_122873 | Not-determined |
| 18-104_k141_110001 | Not-determined |
| 18-104_k141_102541 | Not-determined |
| 18-104_k141_95728 | Low-quality |
| 18-104_k141_158678 | Not-determined |
| 18-104_k141_33084 | Not-determined |
| 18-104_k141_60487 | Not-determined |
| 18-104_k141_144732 | Not-determined |
| 18-104_k141_110006 | Low-quality |
| 18-104_k141_25998 | Not-determined |
| 18-104_k141_95731 | Low-quality |
| 18-104_k141_88649 | Not-determined |
| 18-104_k141_158682 | Low-quality |
| 18-104_k141_18421 | Not-determined |
| 18-104_k141_130321 | Not-determined |
| 18-104_k141_75002 | Low-quality |
| 18-104_k141_75003 | Not-determined |
| 18-104_k141_102546 | Low-quality |
| 18-104_k141_88654 | Not-determined |
| 18-104_k141_39728 | Not-determined |
| 18-104_k141_110023 | Not-determined |
| 18-104_k141_165349 | Not-determined |
| 18-104_k141_53265 | Not-determined |
| 18-104_k141_60493 | Low-quality |
| 18-104_k141_165352 | Not-determined |
| 18-104_k141_67968 | Not-determined |
| 18-104_k141_60500 | Not-determined |
| 18-104_k141_130333 | Not-determined |
| 18-104_k141_60503 | Not-determined |
| 18-104_k141_47074 | Not-determined |
| 18-104_k141_47075 | Not-determined |
| 18-104_k141_165357 | Not-determined |
| 18-104_k141_95748 | Not-determined |
| 18-104_k141_18431 | Low-quality |
| 18-104_k141_18432 | Not-determined |
| 18-104_k141_88665 | Not-determined |
| 18-104_k141_67972 | Not-determined |
| 18-104_k141_47080 | Not-determined |
| 18-104_k141_144746 | Not-determined |
| 18-104_k141_47083 | Not-determined |
| 18-104_k141_158687 | Low-quality |
| 18-104_k141_95756 | Not-determined |
| 18-104_k141_47086 | Not-determined |
| 18-104_k141_75021 | Not-determined |
| 18-104_k141_88674 | Not-determined |
| 18-104_k141_122895 | Not-determined |
| 18-104_k141_67977 | Not-determined |
| 18-104_k141_47089 | Not-determined |
| 18-104_k141_39741 | Not-determined |
| 18-104_k141_158691 | Not-determined |
| 18-104_k141_67980 | Not-determined |
| 18-104_k141_102564 | Not-determined |
| 18-104_k141_12204 | Not-determined |
| 18-104_k141_158699 | Not-determined |
| 18-104_k141_110051 | Not-determined |
| 18-104_k141_130357 | Not-determined |
| 18-104_k141_110054 | Not-determined |
| 18-104_k141_151336 | Not-determined |
| 18-104_k141_33116 | Not-determined |
| 18-104_k141_102577 | Not-determined |
| 18-104_k141_110060 | Not-determined |
| 18-104_k141_67991 | Low-quality |
| 18-104_k141_110061 | Low-quality |
| 18-104_k141_60535 | Not-determined |
| 18-104_k141_110062 | Not-determined |
| 18-104_k141_26027 | Not-determined |
| 18-104_k141_144765 | Not-determined |
| 18-104_k141_151342 | Not-determined |
| 18-104_k141_33124 | Not-determined |
| 18-104_k141_110069 | Not-determined |
| 18-104_k141_151346 | Not-determined |
| 18-104_k141_110072 | Not-determined |
| 18-104_k141_82185 | Not-determined |
| 18-104_k141_110075 | Not-determined |
| 18-104_k141_102584 | Not-determined |
| 18-104_k141_144771 | Not-determined |
| 18-104_k141_130376 | Not-determined |
| 18-104_k141_39770 | Not-determined |
| 18-104_k141_116462 | Not-determined |
| 18-104_k141_39771 | Not-determined |
| 18-104_k141_60551 | Low-quality |
| 18-104_k141_88685 | Not-determined |
| 18-104_k141_39773 | Not-determined |
| 18-104_k141_130379 | Not-determined |
| 18-104_k141_39774 | Not-determined |
| 18-104_k141_12220 | Not-determined |
| 18-104_k141_95785 | Not-determined |
| 18-104_k141_75051 | Not-determined |
| 18-104_k141_26042 | Not-determined |
| 18-104_k141_26043 | Not-determined |
| 18-104_k141_39778 | Not-determined |
| 18-104_k141_68012 | Not-determined |
| 18-104_k141_95787 | Not-determined |
| 18-104_k141_102591 | Not-determined |
| 18-104_k141_144783 | Low-quality |
| 18-104_k141_144784 | Low-quality |
| 18-104_k141_53301 | Not-determined |
| 18-104_k141_12224 | Not-determined |
| 18-104_k141_68018 | Not-determined |
| 18-104_k141_12227 | Not-determined |
| 18-104_k141_47107 | Low-quality |
| 18-104_k141_53306 | Not-determined |
| 18-104_k141_144787 | Low-quality |
| 18-104_k141_12228 | Not-determined |
| 18-104_k141_68024 | Low-quality |
| 18-104_k141_130388 | Not-determined |
| 18-104_k141_88695 | Low-quality |
| 18-104_k141_60572 | Not-determined |
| 18-104_k141_68028 | Not-determined |
| 18-104_k141_12234 | Not-determined |
| 18-104_k141_82209 | Not-determined |
| 18-104_k141_39788 | Not-determined |
| 18-104_k141_122912 | Not-determined |
| 18-104_k141_151366 | Low-quality |
| 18-104_k141_102600 | Not-determined |
| 18-104_k141_88703 | Not-determined |
| 18-104_k141_60581 | Not-determined |
| 18-104_k141_144799 | Not-determined |
| 18-104_k141_122917 | Not-determined |
| 18-104_k141_110092 | Low-quality |
| 18-104_k141_116477 | Not-determined |
| 18-104_k141_12241 | Not-determined |
| 18-104_k141_39794 | Not-determined |
| 18-104_k141_33134 | Not-determined |
| 18-104_k141_26058 | Not-determined |
| 18-104_k141_144803 | Low-quality |
| 18-104_k141_102609 | Low-quality |
| 18-104_k141_39799 | Not-determined |
| 18-104_k141_60589 | Not-determined |
| 18-104_k141_75080 | Not-determined |
| 18-104_k141_151372 | Not-determined |
| 18-104_k141_130411 | Not-determined |
| 18-104_k141_75082 | Not-determined |
| 18-104_k141_116490 | Not-determined |
| 18-104_k141_130413 | Not-determined |

| | |
|---|---|
| 18-104_k141_39803 | Not-determined |
| 18-104_k141_18461 | Not-determined |
| 18-104_k141_122929 | Not-determined |
| 18-104_k141_68047 | Not-determined |
| 18-104_k141_18466 | Not-determined |
| 18-104_k141_39807 | Not-determined |
| 18-104_k141_12260 | Not-determined |
| 18-104_k141_68051 | Not-determined |
| 18-104_k141_151380 | Not-determined |
| 18-104_k141_33151 | Not-determined |
| 18-104_k141_53328 | Not-determined |
| 18-104_k141_33152 | Not-determined |
| 18-104_k141_33153 | Not-determined |
| 18-104_k141_88718 | Low-quality |
| 18-104_k141_144813 | Not-determined |
| 18-104_k141_33156 | Not-determined |
| 18-104_k141_95812 | Not-determined |
| 18-104_k141_95813 | Not-determined |
| 18-104_k141_33157 | Not-determined |
| 18-104_k141_110121 | Not-determined |
| 18-104_k141_122933 | Not-determined |
| 18-104_k141_116503 | Not-determined |
| 18-104_k141_95816 | Not-determined |
| 18-104_k141_53336 | Not-determined |
| 18-104_k141_95817 | Not-determined |
| 18-104_k141_33164 | Not-determined |
| 18-104_k141_116506 | Not-determined |
| 18-104_k141_60610 | Not-determined |
| 18-104_k141_26068 | Not-determined |
| 18-104_k141_151389 | Not-determined |
| 18-104_k141_47118 | Not-determined |
| 18-104_k141_82217 | Low-quality |
| 18-104_k141_151391 | Not-determined |
| 18-104_k141_39819 | Not-determined |
| 18-104_k141_116508 | Not-determined |
| 18-104_k141_151394 | Not-determined |
| 18-104_k141_12262 | Not-determined |
| 18-104_k141_39824 | Not-determined |
| 18-104_k141_33177 | Not-determined |
| 18-104_k141_116510 | Low-quality |
| 18-104_k141_88736 | Not-determined |
| 18-104_k141_33179 | Not-determined |
| 18-104_k141_110128 | Low-quality |
| 18-104_k141_68072 | Low-quality |
| 18-104_k141_137412 | Not-determined |
| 18-104_k141_88742 | Not-determined |
| 18-104_k141_158761 | Not-determined |
| 18-104_k141_122944 | Low-quality |
| 18-104_k141_53349 | Not-determined |
| 18-104_k141_158763 | Not-determined |
| 18-104_k141_82226 | Not-determined |
| 18-104_k141_39838 | Not-determined |
| 18-104_k141_5062 | Not-determined |
| 18-104_k141_110134 | Not-determined |
| 18-104_k141_116518 | Low-quality |
| 18-104_k141_26084 | Not-determined |
| 18-104_k141_158766 | Low-quality |
| 18-104_k141_88755 | Not-determined |
| 18-104_k141_53355 | Not-determined |
| 18-104_k141_68087 | Not-determined |
| 18-104_k141_122958 | Not-determined |
| 18-104_k141_158770 | Not-determined |
| 18-104_k141_53361 | Not-determined |
| 18-104_k141_47130 | Not-determined |
| 18-104_k141_60619 | Not-determined |
| 18-104_k141_53365 | Not-determined |
| 18-104_k141_53366 | Not-determined |
| 18-104_k141_95846 | Not-determined |
| 18-104_k141_122976 | Not-determined |
| 18-104_k141_130448 | Low-quality |
| 18-104_k141_130449 | Not-determined |
| 18-104_k141_110148 | Low-quality |
| 18-104_k141_122980 | Not-determined |
| 18-104_k141_158789 | Not-determined |
| 18-104_k141_110154 | Not-determined |
| 18-104_k141_110155 | Not-determined |
| 18-104_k141_110157 | Not-determined |
| 18-104_k141_137424 | Not-determined |
| 18-104_k141_158797 | Low-quality |
| 18-104_k141_95863 | Not-determined |
| 18-104_k141_12295 | Not-determined |
| 18-104_k141_47142 | Not-determined |
| 18-104_k141_18514 | Not-determined |
| 18-104_k141_47144 | Not-determined |
| 18-104_k141_144831 | Not-determined |
| 18-104_k141_12297 | Low-quality |
| 18-104_k141_137429 | Not-determined |
| 18-104_k141_75122 | Not-determined |
| 18-104_k141_158799 | Not-determined |
| 18-104_k141_158802 | Not-determined |
| 18-104_k141_116523 | Not-determined |
| 18-104_k141_82242 | Not-determined |
| 18-104_k141_158806 | Not-determined |
| 18-104_k141_47149 | Not-determined |
| 18-104_k141_75125 | Not-determined |
| 18-104_k141_95870 | Not-determined |
| 18-104_k141_95871 | Not-determined |
| 18-104_k141_144838 | Not-determined |
| 18-104_k141_47159 | Low-quality |
| 18-104_k141_123003 | Not-determined |
| 18-104_k141_39873 | Not-determined |
| 18-104_k141_158818 | Low-quality |
| 18-104_k141_12309 | Not-determined |
| 18-104_k141_144845 | Low-quality |
| 18-104_k141_47170 | Not-determined |
| 18-104_k141_47171 | Not-determined |
| 18-104_k141_144848 | Not-determined |
| 18-104_k141_88784 | Not-determined |
| 18-104_k141_123013 | Not-determined |
| 18-104_k141_18522 | Not-determined |
| 18-104_k141_88789 | Not-determined |
| 18-104_k141_123014 | Low-quality |
| 18-104_k141_123015 | Not-determined |
| 18-104_k141_47179 | Not-determined |
| 18-104_k141_39882 | Not-determined |
| 18-104_k141_33223 | Not-determined |
| 18-104_k141_82252 | Not-determined |
| 18-104_k141_26121 | Not-determined |
| 18-104_k141_158829 | Not-determined |
| 18-104_k141_123017 | Low-quality |
| 18-104_k141_12330 | Not-determined |
| 18-104_k141_137455 | Low-quality |
| 18-104_k141_75138 | Low-quality |
| 18-104_k141_18536 | Not-determined |
| 18-104_k141_130480 | Not-determined |
| 18-104_k141_116542 | Not-determined |
| 18-104_k141_158832 | Not-determined |
| 18-104_k141_116544 | Not-determined |
| 18-104_k141_137465 | Low-quality |
| 18-104_k141_158835 | Not-determined |
| 18-104_k141_82266 | Not-determined |
| 18-104_k141_5089 | Not-determined |
| 18-104_k141_47198 | Not-determined |
| 18-104_k141_39905 | Not-determined |
| 18-104_k141_47201 | Low-quality |
| 18-104_k141_53413 | Not-determined |
| 18-104_k141_39909 | Not-determined |
| 18-104_k141_12344 | Not-determined |
| 18-104_k141_47204 | Not-determined |
| 18-104_k141_47206 | Not-determined |
| 18-104_k141_53418 | Not-determined |
| 18-104_k141_110212 | Not-determined |
| 18-104_k141_33238 | Not-determined |
| 18-104_k141_39916 | Not-determined |
| 18-104_k141_47211 | Not-determined |
| 18-104_k141_39918 | Not-determined |
| 18-104_k141_144881 | Not-determined |
| 18-104_k141_82278 | Not-determined |
| 18-104_k141_137481 | Not-determined |
| 18-104_k141_47214 | Not-determined |
| 18-104_k141_144884 | Not-determined |
| 18-104_k141_60622 | Not-determined |
| 18-104_k141_130495 | Low-quality |
| 18-104_k141_151464 | Not-determined |
| 18-104_k141_95912 | Not-determined |
| 18-104_k141_123035 | Not-determined |
| 18-104_k141_144887 | Not-determined |
| 18-104_k141_5101 | Not-determined |
| 18-104_k141_144890 | Not-determined |
| 18-104_k141_130499 | Not-determined |
| 18-104_k141_5104 | Not-determined |
| 18-104_k141_158840 | Not-determined |
| 18-104_k141_53420 | Not-determined |
| 18-104_k141_33251 | Not-determined |
| 18-104_k141_110226 | Not-determined |
| 18-104_k141_60636 | Not-determined |
| 18-104_k141_33258 | Not-determined |
| 18-104_k141_95924 | Not-determined |
| 18-104_k141_18561 | Not-determined |
| 18-104_k141_5112 | Low-quality |
| 18-104_k141_33259 | Not-determined |
| 18-104_k141_95925 | Not-determined |
| 18-104_k141_12360 | Not-determined |
| 18-104_k141_12362 | Not-determined |
| 18-104_k141_144903 | Not-determined |
| 18-104_k141_88817 | Not-determined |
| 18-104_k141_82285 | Not-determined |
| 18-104_k141_60645 | Not-determined |
| 18-104_k141_110241 | Not-determined |
| 18-104_k141_82286 | Low-quality |
| 18-104_k141_33267 | Low-quality |
| 18-104_k141_144910 | Not-determined |
| 18-104_k141_123052 | Not-determined |
| 18-104_k141_88819 | Low-quality |
| 18-104_k141_12373 | Not-determined |
| 18-104_k141_75191 | Low-quality |
| 18-104_k141_53432 | Not-determined |
| 18-104_k141_68106 | Not-determined |
| 18-104_k141_95949 | Not-determined |
| 18-104_k141_75196 | Not-determined |
| 18-104_k141_33281 | Not-determined |
| 18-104_k141_144921 | Not-determined |
| 18-104_k141_88826 | Not-determined |
| 18-104_k141_33283 | Not-determined |
| 18-104_k141_95956 | Not-determined |
| 18-104_k141_137514 | Not-determined |
| 18-104_k141_5131 | Low-quality |
| 18-104_k141_5135 | Not-determined |
| 18-104_k141_18590 | Not-determined |
| 18-104_k141_75207 | Not-determined |
| 18-104_k141_137518 | Not-determined |
| 18-104_k141_33294 | Low-quality |
| 18-104_k141_88839 | Not-determined |
| 18-104_k141_39947 | Not-determined |
| 18-104_k141_18596 | Not-determined |
| 18-104_k141_88840 | Not-determined |
| 18-104_k141_26152 | Not-determined |
| 18-104_k141_33305 | Not-determined |
| 18-104_k141_75222 | Not-determined |
| 18-104_k141_123068 | Not-determined |
| 18-104_k141_33309 | Not-determined |
| 18-104_k141_47252 | Not-determined |
| 18-104_k141_110263 | Not-determined |
| 18-104_k141_151508 | Not-determined |
| 18-104_k141_39961 | Not-determined |
| 18-104_k141_68137 | Not-determined |
| 18-104_k141_123079 | Not-determined |
| 18-104_k141_137537 | Not-determined |
| 18-104_k141_5151 | Not-determined |
| 18-104_k141_39965 | Not-determined |
| 18-104_k141_110269 | Not-determined |
| 18-104_k141_53460 | Not-determined |
| 18-104_k141_68140 | Not-determined |
| 18-104_k141_5155 | Not-determined |
| 18-104_k141_95992 | Not-determined |
| 18-104_k141_137542 | Not-determined |
| 18-104_k141_110271 | Not-determined |
| 18-104_k141_18619 | Not-determined |
| 18-104_k141_110274 | Not-determined |
| 18-104_k141_95997 | Not-determined |
| 18-104_k141_60683 | Low-quality |
| 18-104_k141_18623 | Not-determined |
| 18-104_k141_47270 | Not-determined |
| 18-104_k141_158864 | Low-quality |
| 18-104_k141_18626 | Not-determined |
| 18-104_k141_130555 | Not-determined |
| 18-104_k141_88842 | Not-determined |
| 18-104_k141_39977 | Not-determined |
| 18-104_k141_123084 | Low-quality |
| 18-104_k141_75242 | Not-determined |
| 18-104_k141_75243 | Not-determined |
| 18-104_k141_5171 | Not-determined |
| 18-104_k141_60692 | Not-determined |
| 18-104_k141_88847 | Low-quality |
| 18-104_k141_53481 | Not-determined |
| 18-104_k141_165393 | Low-quality |
| 18-104_k141_53483 | Not-determined |
| 18-104_k141_18638 | Not-determined |
| 18-104_k141_158878 | Not-determined |
| 18-104_k141_75250 | Not-determined |
| 18-104_k141_130570 | Not-determined |
| 18-104_k141_151524 | Not-determined |
| 18-104_k141_5181 | Not-determined |
| 18-104_k141_5183 | Not-determined |
| 18-104_k141_137561 | Not-determined |
| 18-104_k141_123092 | Not-determined |
| 18-104_k141_123094 | Not-determined |
| 18-104_k141_88854 | Not-determined |
| 18-104_k141_102626 | Not-determined |
| 18-104_k141_96018 | Not-determined |
| 18-104_k141_26194 | Not-determined |
| 18-104_k141_123100 | Not-determined |
| 18-104_k141_47289 | Not-determined |
| 18-104_k141_26196 | Not-determined |
| 18-104_k141_68165 | Not-determined |
| 18-104_k141_110300 | Not-determined |
| 18-104_k141_18650 | Not-determined |
| 18-104_k141_158890 | Low-quality |
| 18-104_k141_123105 | Not-determined |
| 18-104_k141_137568 | Not-determined |
| 18-104_k141_26199 | Not-determined |
| 18-104_k141_110302 | Not-determined |
| 18-104_k141_39982 | Not-determined |
| 18-104_k141_144974 | Not-determined |
| 18-104_k141_151543 | Not-determined |
| 18-104_k141_158893 | Not-determined |
| 18-104_k141_144977 | Not-determined |
| 18-104_k141_60707 | Not-determined |
| 18-104_k141_39984 | Not-determined |
| 18-104_k141_39985 | Low-quality |
| 18-104_k141_88861 | Not-determined |
| 18-104_k141_53503 | Not-determined |
| 18-104_k141_151550 | Not-determined |
| 18-104_k141_110310 | Not-determined |
| 18-104_k141_116599 | Not-determined |
| 18-104_k141_18665 | Not-determined |
| 18-104_k141_88865 | Not-determined |
| 18-104_k141_151554 | Low-quality |
| 18-104_k141_68184 | Not-determined |
| 18-104_k141_53510 | Not-determined |
| 18-104_k141_158906 | Not-determined |
| 18-104_k141_68185 | Not-determined |
| 18-104_k141_68186 | Not-determined |
| 18-104_k141_110317 | Low-quality |
| 18-104_k141_144978 | Low-quality |
| 18-104_k141_75278 | Not-determined |
| 18-104_k141_96038 | Low-quality |
| 18-104_k141_130583 | Not-determined |
| 18-104_k141_123118 | Not-determined |
| 18-104_k141_39988 | Low-quality |
| 18-104_k141_151566 | Not-determined |
| 18-104_k141_33352 | Not-determined |
| 18-104_k141_47298 | Low-quality |
| 18-104_k141_18676 | Low-quality |
| 18-104_k141_47299 | Not-determined |
| 18-104_k141_102649 | Not-determined |
| 18-104_k141_123125 | Low-quality |
| 18-104_k141_60719 | Not-determined |
| 18-104_k141_26218 | Not-determined |
| 18-104_k141_68196 | Not-determined |
| 18-104_k141_158914 | Not-determined |
| 18-104_k141_123128 | Not-determined |
| 18-104_k141_158916 | Not-determined |
| 18-104_k141_123129 | Not-determined |
| 18-104_k141_53526 | Not-determined |
| 18-104_k141_5205 | Not-determined |
| 18-104_k141_39996 | Not-determined |
| 18-104_k141_26225 | Not-determined |
| 18-104_k141_47300 | Not-determined |
| 18-104_k141_5208 | Not-determined |
| 18-104_k141_68202 | Not-determined |
| 18-104_k141_158922 | Not-determined |
| 18-104_k141_68203 | Not-determined |
| 18-104_k141_12425 | Not-determined |
| 18-104_k141_75284 | Not-determined |
| 18-104_k141_137583 | Not-determined |
| 18-104_k141_26230 | Not-determined |
| 18-104_k141_75288 | Low-quality |
| 18-104_k141_68209 | Not-determined |
| 18-104_k141_60722 | Not-determined |
| 18-104_k141_116625 | Not-determined |
| 18-104_k141_116627 | Not-determined |
| 18-104_k141_82321 | Not-determined |
| 18-104_k141_12427 | Not-determined |
| 18-104_k141_53540 | Not-determined |
| 18-104_k141_40005 | Not-determined |
| 18-104_k141_116632 | Not-determined |
| 18-104_k141_102660 | Not-determined |
| 18-104_k141_158935 | Not-determined |
| 18-104_k141_96059 | Not-determined |
| 18-104_k141_68213 | Not-determined |
| 18-104_k141_102662 | Not-determined |
| 18-104_k141_88881 | Not-determined |
| 18-104_k141_5217 | Not-determined |
| 18-104_k141_110324 | Not-determined |
| 18-104_k141_130603 | Not-determined |
| 18-104_k141_68216 | Not-determined |
| 18-104_k141_26242 | Not-determined |
| 18-104_k141_60731 | Not-determined |
| 18-104_k141_47314 | Not-determined |
| 18-104_k141_82332 | Not-determined |
| 18-104_k141_123149 | Not-determined |
| 18-104_k141_116641 | Not-determined |
| 18-104_k141_137592 | Not-determined |
| 18-104_k141_137593 | Not-determined |
| 18-104_k141_68223 | Low-quality |
| 18-104_k141_123154 | Not-determined |
| 18-104_k141_5234 | Not-determined |
| 18-104_k141_26248 | Not-determined |
| 18-104_k141_88893 | Not-determined |
| 18-104_k141_40011 | Not-determined |
| 18-104_k141_82338 | Not-determined |
| 18-104_k141_137603 | Not-determined |
| 18-104_k141_40014 | Low-quality |
| 18-104_k141_12451 | Not-determined |
| 18-104_k141_96065 | Not-determined |
| 18-104_k141_88897 | Not-determined |
| 18-104_k141_82344 | Not-determined |
| 18-104_k141_12457 | Not-determined |
| 18-104_k141_151591 | Not-determined |
| 18-104_k141_96074 | Low-quality |
| 18-104_k141_82347 | Not-determined |
| 18-104_k141_158964 | Low-quality |
| 18-104_k141_151594 | Not-determined |
| 18-104_k141_5247 | Not-determined |
| 18-104_k141_18703 | Not-determined |
| 18-104_k141_88909 | Not-determined |
| 18-104_k141_60742 | Not-determined |
| 18-104_k141_123168 | Not-determined |
| 18-104_k141_110337 | Not-determined |
| 18-104_k141_47336 | Low-quality |
| 18-104_k141_12462 | Not-determined |
| 18-104_k141_26267 | Not-determined |
| 18-104_k141_60746 | Not-determined |
| 18-104_k141_116658 | Not-determined |
| 18-104_k141_123174 | Not-determined |
| 18-104_k141_82352 | Not-determined |
| 18-104_k141_116660 | Not-determined |
| 18-104_k141_116663 | Not-determined |
| 18-104_k141_137620 | Not-determined |
| 18-104_k141_40029 | Not-determined |
| 18-104_k141_18712 | Not-determined |
| 18-104_k141_53557 | Not-determined |
| 18-104_k141_40035 | Not-determined |
| 18-104_k141_137624 | Not-determined |
| 18-104_k141_137626 | Not-determined |
| 18-104_k141_82355 | Not-determined |
| 18-104_k141_145020 | Not-determined |
| 18-104_k141_110347 | Not-determined |
| 18-104_k141_158988 | Not-determined |
| 18-104_k141_40042 | Not-determined |
| 18-104_k141_116672 | Not-determined |
| 18-104_k141_123180 | Not-determined |
| 18-104_k141_60757 | Not-determined |
| 18-104_k141_110348 | Not-determined |
| 18-104_k141_123182 | Not-determined |
| 18-104_k141_40046 | Not-determined |
| 18-104_k141_145029 | Low-quality |
| 18-104_k141_145031 | Not-determined |
| 18-104_k141_82360 | Low-quality |
| 18-104_k141_158992 | Low-quality |
| 18-104_k141_40049 | Not-determined |
| 18-104_k141_5273 | Not-determined |
| 18-104_k141_40052 | Low-quality |
| 18-104_k141_137635 | Not-determined |
| 18-104_k141_151610 | Not-determined |
| 18-104_k141_110356 | Not-determined |
| 18-104_k141_82366 | Not-determined |
| 18-104_k141_151615 | Not-determined |
| 18-104_k141_88942 | Not-determined |
| 18-104_k141_110360 | Not-determined |
| 18-104_k141_47358 | Not-determined |
| 18-104_k141_75297 | Not-determined |
| 18-104_k141_137641 | Not-determined |
| 18-104_k141_18730 | Low-quality |
| 18-104_k141_145037 | Low-quality |
| 18-104_k141_145038 | Not-determined |
| 18-104_k141_40070 | Not-determined |
| 18-104_k141_145039 | Not-determined |
| 18-104_k141_145040 | Not-determined |
| 18-104_k141_18733 | Low-quality |
| 18-104_k141_60767 | Not-determined |
| 18-104_k141_88950 | Not-determined |
| 18-104_k141_137648 | Not-determined |
| 18-104_k141_68254 | Not-determined |
| 18-104_k141_40073 | Not-determined |
| 18-104_k141_96132 | Not-determined |
| 18-104_k141_145044 | Not-determined |
| 18-104_k141_82372 | Not-determined |
| 18-104_k141_5288 | Not-determined |
| 18-104_k141_145046 | Not-determined |
| 18-104_k141_68263 | Not-determined |
| 18-104_k141_75306 | Low-quality |
| 18-104_k141_130658 | Not-determined |
| 18-104_k141_75307 | Not-determined |
| 18-104_k141_102677 | Not-determined |
| 18-104_k141_137660 | Not-determined |
| 18-104_k141_151624 | Not-determined |
| 18-104_k141_123224 | Not-determined |
| 18-104_k141_5296 | Not-determined |
| 18-104_k141_96142 | Not-determined |
| 18-104_k141_137662 | Not-determined |
| 18-104_k141_116683 | Not-determined |
| 18-104_k141_82380 | Not-determined |
| 18-104_k141_123228 | Not-determined |
| 18-104_k141_165404 | Not-determined |
| 18-104_k141_137663 | Not-determined |
| 18-104_k141_60796 | Not-determined |
| 18-104_k141_165408 | Not-determined |
| 18-104_k141_82386 | Not-determined |
| 18-104_k141_60800 | Not-determined |
| 18-104_k141_18749 | Not-determined |
| 18-104_k141_145059 | Not-determined |
| 18-104_k141_102690 | Not-determined |
| 18-104_k141_130670 | Low-quality |
| 18-104_k141_47385 | Not-determined |
| 18-104_k141_82399 | Not-determined |
| 18-104_k141_53584 | Not-determined |
| 18-104_k141_53585 | Not-determined |
| 18-104_k141_5326 | Not-determined |
| 18-104_k141_53586 | Not-determined |
| 18-104_k141_116707 | Not-determined |
| 18-104_k141_145062 | Not-determined |
| 18-104_k141_53587 | Not-determined |
| 18-104_k141_116710 | Not-determined |
| 18-104_k141_47390 | Not-determined |
| 18-104_k141_116712 | Not-determined |
| 18-104_k141_82407 | Not-determined |
| 18-104_k141_40092 | Not-determined |
| 18-104_k141_165422 | Not-determined |
| 18-104_k141_110403 | Not-determined |
| 18-104_k141_60811 | Not-determined |
| 18-104_k141_151651 | Not-determined |
| 18-104_k141_165423 | Not-determined |
| 18-104_k141_60817 | Not-determined |
| 18-104_k141_60818 | Not-determined |
| 18-104_k141_137677 | Not-determined |
| 18-104_k141_159013 | Not-determined |
| 18-104_k141_82411 | Low-quality |
| 18-104_k141_40099 | Not-determined |
| 18-104_k141_68299 | Low-quality |
| 18-104_k141_96171 | Not-determined |
| 18-104_k141_12493 | Not-determined |
| 18-104_k141_82414 | Not-determined |
| 18-104_k141_60831 | Not-determined |
| 18-104_k141_40104 | Not-determined |
| 18-104_k141_137683 | Low-quality |
| 18-104_k141_110417 | Not-determined |
| 18-104_k141_5349 | Not-determined |
| 18-104_k141_5351 | Not-determined |
| 18-104_k141_12496 | Not-determined |
| 18-104_k141_159027 | Not-determined |
| 18-104_k141_82421 | Not-determined |
| 18-104_k141_159029 | Not-determined |
| 18-104_k141_159032 | Not-determined |
| 18-104_k141_165443 | Not-determined |
| 18-104_k141_33423 | Not-determined |
| 18-104_k141_40116 | Low-quality |
| 18-104_k141_116725 | Not-determined |
| 18-104_k141_96180 | Not-determined |
| 18-104_k141_18776 | Not-determined |
| 18-104_k141_110422 | Not-determined |
| 18-104_k141_145082 | Low-quality |
| 18-104_k141_40134 | Not-determined |
| 18-104_k141_82435 | Not-determined |
| 18-104_k141_88986 | Not-determined |
| 18-104_k141_60850 | Low-quality |
| 18-104_k141_40138 | Not-determined |
| 18-104_k141_47415 | Not-determined |
| 18-104_k141_33437 | Not-determined |
| 18-104_k141_53603 | Not-determined |
| 18-104_k141_102708 | Low-quality |
| 18-104_k141_82442 | Not-determined |
| 18-104_k141_151691 | Not-determined |
| 18-104_k141_110432 | Not-determined |
| 18-104_k141_53606 | Not-determined |
| 18-104_k141_75329 | Low-quality |
| 18-104_k141_60861 | Not-determined |
| 18-104_k141_26346 | Not-determined |
| 18-104_k141_165449 | Not-determined |
| 18-104_k141_75331 | Low-quality |
| 18-104_k141_40143 | Not-determined |
| 18-104_k141_60863 | Not-determined |
| 18-104_k141_40144 | Not-determined |
| 18-104_k141_151700 | Not-determined |
| 18-104_k141_151702 | Not-determined |
| 18-104_k141_88994 | Not-determined |
| 18-104_k141_40148 | Low-quality |
| 18-104_k141_151706 | Not-determined |
| 18-104_k141_47428 | Not-determined |
| 18-104_k141_47431 | Not-determined |
| 18-104_k141_18792 | Not-determined |
| 18-104_k141_102722 | Low-quality |
| 18-104_k141_151709 | Not-determined |
| 18-104_k141_12527 | Not-determined |
| 18-104_k141_18793 | Not-determined |
| 18-104_k141_75337 | Not-determined |
| 18-104_k141_151711 | Not-determined |
| 18-104_k141_26356 | Not-determined |
| 18-104_k141_33442 | Not-determined |
| 18-104_k141_60875 | Not-determined |
| 18-104_k141_26358 | Not-determined |
| 18-104_k141_33443 | Not-determined |
| 18-104_k141_151714 | Low-quality |
| 18-104_k141_47437 | Not-determined |
| 18-104_k141_89007 | Not-determined |
| 18-104_k141_75341 | Not-determined |
| 18-104_k141_116727 | Not-determined |
| 18-104_k141_5392 | Not-determined |
| 18-104_k141_102728 | Not-determined |
| 18-104_k141_5393 | Not-determined |
| 18-104_k141_40157 | Not-determined |
| 18-104_k141_33448 | Not-determined |
| 18-104_k141_82461 | Not-determined |
| 18-104_k141_12532 | Not-determined |
| 18-104_k141_82462 | Not-determined |
| 18-104_k141_82465 | Not-determined |
| 18-104_k141_96193 | Not-determined |
| 18-104_k141_5399 | Not-determined |
| 18-104_k141_75346 | Not-determined |
| 18-104_k141_5400 | Not-determined |
| 18-104_k141_123257 | Not-determined |
| 18-104_k141_89010 | Not-determined |
| 18-104_k141_40166 | Not-determined |
| 18-104_k141_12537 | Not-determined |
| 18-104_k141_60884 | Not-determined |
| 18-104_k141_82470 | Not-determined |
| 18-104_k141_130697 | Not-determined |
| 18-104_k141_60886 | Not-determined |
| 18-104_k141_33456 | Not-determined |
| 18-104_k141_47448 | Not-determined |
| 18-104_k141_116744 | Not-determined |
| 18-104_k141_123259 | Low-quality |
| 18-104_k141_40175 | Not-determined |
| 18-104_k141_12540 | Not-determined |
| 18-104_k141_102747 | Not-determined |
| 18-104_k141_102751 | Not-determined |
| 18-104_k141_123268 | Not-determined |
| 18-104_k141_47452 | Not-determined |

| | |
|---|---|
| 18-104_k141_33464 | Not-determined |
| 18-104_k141_68334 | Not-determined |
| 18-104_k141_130704 | Not-determined |
| 18-104_k141_123270 | Not-determined |
| 18-104_k141_96206 | Not-determined |
| 18-104_k141_40183 | Low-quality |
| 18-104_k141_130708 | Not-determined |
| 18-104_k141_68343 | Not-determined |
| 18-104_k141_75361 | Not-determined |
| 18-104_k141_75363 | Not-determined |
| 18-104_k141_165465 | Not-determined |
| 18-104_k141_33480 | Not-determined |
| 18-104_k141_5414 | Not-determined |
| 18-104_k141_18810 | Not-determined |
| 18-104_k141_60912 | Not-determined |
| 18-104_k141_96222 | Low-quality |
| 18-104_k141_96223 | Low-quality |
| 18-104_k141_60913 | Low-quality |
| 18-104_k141_26375 | Not-determined |
| 18-104_k141_116755 | Not-determined |
| 18-104_k141_145119 | Not-determined |
| 18-104_k141_68351 | Not-determined |
| 18-104_k141_26380 | Not-determined |
| 18-104_k141_68352 | Not-determined |
| 18-104_k141_75373 | Not-determined |
| 18-104_k141_145124 | Not-determined |
| 18-104_k141_145125 | Not-determined |
| 18-104_k141_18819 | Not-determined |
| 18-104_k141_26387 | Not-determined |
| 18-104_k141_18822 | Not-determined |
| 18-104_k141_68360 | Not-determined |
| 18-104_k141_123296 | Not-determined |
| 18-104_k141_96236 | Not-determined |
| 18-104_k141_96238 | Not-determined |
| 18-104_k141_102772 | Not-determined |
| 18-104_k141_130737 | Not-determined |
| 18-104_k141_151722 | Not-determined |
| 18-104_k141_40206 | Not-determined |
| 18-104_k141_102774 | Not-determined |
| 18-104_k141_116765 | Low-quality |
| 18-104_k141_116766 | Low-quality |
| 18-104_k141_151724 | Not-determined |
| 18-104_k141_82511 | Not-determined |
| 18-104_k141_116767 | Not-determined |
| 18-104_k141_96245 | Not-determined |
| 18-104_k141_53640 | Not-determined |
| 18-104_k141_102782 | Not-determined |
| 18-104_k141_110461 | Not-determined |
| 18-104_k141_123300 | Low-quality |
| 18-104_k141_123301 | Not-determined |
| 18-104_k141_123302 | Not-determined |
| 18-104_k141_137748 | Not-determined |
| 18-104_k141_130741 | Not-determined |
| 18-104_k141_110465 | Not-determined |
| 18-104_k141_12586 | Low-quality |
| 18-104_k141_96249 | Not-determined |
| 18-104_k141_116776 | Not-determined |
| 18-104_k141_82522 | Not-determined |
| 18-104_k141_82523 | Not-determined |
| 18-104_k141_12589 | Not-determined |
| 18-104_k141_89064 | Not-determined |
| 18-104_k141_159080 | Not-determined |
| 18-104_k141_53647 | Not-determined |
| 18-104_k141_159083 | Not-determined |
| 18-104_k141_123309 | Not-determined |
| 18-104_k141_40220 | Not-determined |
| 18-104_k141_82531 | Low-quality |
| 18-104_k141_130747 | Not-determined |
| 18-104_k141_82533 | Low-quality |
| 18-104_k141_130748 | Low-quality |
| 18-104_k141_123314 | Not-determined |
| 18-104_k141_116789 | Not-determined |
| 18-104_k141_116791 | Not-determined |
| 18-104_k141_33525 | Not-determined |
| 18-104_k141_110477 | Not-determined |
| 18-104_k141_18855 | Not-determined |
| 18-104_k141_40232 | Not-determined |
| 18-104_k141_60921 | Not-determined |
| 18-104_k141_110482 | Not-determined |
| 18-104_k141_33532 | Not-determined |
| 18-104_k141_123317 | Not-determined |
| 18-104_k141_75399 | Not-determined |
| 18-104_k141_33537 | Not-determined |
| 18-104_k141_26403 | Not-determined |
| 18-104_k141_33539 | Not-determined |
| 18-104_k141_137764 | Low-quality |
| 18-104_k141_5446 | Not-determined |
| 18-104_k141_89083 | Low-quality |
| 18-104_k141_12598 | Not-determined |
| 18-104_k141_33547 | Not-determined |
| 18-104_k141_89087 | Not-determined |
| 18-104_k141_102816 | Not-determined |
| 18-104_k141_82560 | Not-determined |
| 18-104_k141_102818 | Not-determined |
| 18-104_k141_82561 | Not-determined |
| 18-104_k141_89097 | Not-determined |
| 18-104_k141_151772 | Not-determined |
| 18-104_k141_145161 | Not-determined |
| 18-104_k141_53681 | Not-determined |
| 18-104_k141_40253 | Not-determined |
| 18-104_k141_12605 | Not-determined |
| 18-104_k141_137776 | Not-determined |
| 18-104_k141_33559 | Not-determined |
| 18-104_k141_145165 | Not-determined |
| 18-104_k141_123331 | Not-determined |
| 18-104_k141_40264 | Low-quality |
| 18-104_k141_159108 | Not-determined |
| 18-104_k141_110507 | Not-determined |
| 18-104_k141_102839 | Not-determined |
| 18-104_k141_137783 | Not-determined |
| 18-104_k141_12611 | Not-determined |
| 18-104_k141_137784 | Not-determined |
| 18-104_k141_40268 | Not-determined |
| 18-104_k141_47468 | Not-determined |
| 18-104_k141_96275 | Not-determined |
| 18-104_k141_145174 | Not-determined |
| 18-104_k141_40274 | Not-determined |
| 18-104_k141_110512 | Not-determined |
| 18-104_k141_96276 | Not-determined |
| 18-104_k141_110514 | Not-determined |
| 18-104_k141_110516 | Not-determined |
| 18-104_k141_5461 | Not-determined |
| 18-104_k141_68374 | Not-determined |
| 18-104_k141_18880 | Low-quality |
| 18-104_k141_82588 | Not-determined |
| 18-104_k141_5465 | Not-determined |
| 18-104_k141_89112 | Not-determined |
| 18-104_k141_96284 | Not-determined |
| 18-104_k141_53694 | Not-determined |
| 18-104_k141_123348 | Not-determined |
| 18-104_k141_145180 | Low-quality |
| 18-104_k141_82597 | Not-determined |
| 18-104_k141_75417 | Not-determined |
| 18-104_k141_82599 | Not-determined |
| 18-104_k141_26442 | Low-quality |
| 18-104_k141_82603 | Not-determined |
| 18-104_k141_116811 | Not-determined |
| 18-104_k141_5473 | Low-quality |
| 18-104_k141_116813 | Not-determined |
| 18-104_k141_18889 | Not-determined |
| 18-104_k141_96290 | Not-determined |
| 18-104_k141_12627 | Low-quality |
| 18-104_k141_116817 | Not-determined |
| 18-104_k141_89121 | Not-determined |
| 18-104_k141_40291 | Not-determined |
| 18-104_k141_53702 | Not-determined |
| 18-104_k141_116818 | Low-quality |
| 18-104_k141_40295 | Not-determined |
| 18-104_k141_47480 | Not-determined |
| 18-104_k141_53705 | Not-determined |
| 18-104_k141_130757 | Not-determined |
| 18-104_k141_33582 | Not-determined |
| 18-104_k141_75425 | Not-determined |
| 18-104_k141_5487 | Not-determined |
| 18-104_k141_96298 | Not-determined |
| 18-104_k141_12642 | Not-determined |
| 18-104_k141_159127 | Not-determined |
| 18-104_k141_5490 | Not-determined |
| 18-104_k141_53711 | Not-determined |
| 18-104_k141_75427 | Not-determined |
| 18-104_k141_145199 | Not-determined |
| 18-104_k141_159128 | Low-quality |
| 18-104_k141_89131 | Not-determined |
| 18-104_k141_60970 | Not-determined |
| 18-104_k141_159131 | Not-determined |
| 18-104_k141_116832 | Not-determined |
| 18-104_k141_145207 | Low-quality |
| 18-104_k141_96310 | Not-determined |
| 18-104_k141_89138 | Not-determined |
| 18-104_k141_123382 | Not-determined |
| 18-104_k141_12660 | Not-determined |
| 18-104_k141_102870 | Not-determined |
| 18-104_k141_75439 | Not-determined |
| 18-104_k141_75443 | Not-determined |
| 18-104_k141_145211 | Not-determined |
| 18-104_k141_96319 | Not-determined |
| 18-104_k141_18920 | Not-determined |
| 18-104_k141_5516 | Not-determined |
| 18-104_k141_60990 | Not-determined |
| 18-104_k141_5518 | Not-determined |
| 18-104_k141_47487 | Not-determined |
| 18-104_k141_60995 | Not-determined |
| 18-104_k141_53722 | Not-determined |
| 18-104_k141_110561 | Not-determined |
| 18-104_k141_33600 | Not-determined |
| 18-104_k141_18930 | Not-determined |
| 18-104_k141_89149 | Not-determined |
| 18-104_k141_47491 | Not-determined |
| 18-104_k141_18932 | Not-determined |
| 18-104_k141_102887 | Not-determined |
| 18-104_k141_159142 | Not-determined |
| 18-104_k141_159143 | Not-determined |
| 18-104_k141_116846 | Not-determined |
| 18-104_k141_102889 | Not-determined |
| 18-104_k141_96332 | Not-determined |
| 18-104_k141_96333 | Not-determined |
| 18-104_k141_5523 | Low-quality |
| 18-104_k141_165534 | Not-determined |
| 18-104_k141_151794 | Not-determined |
| 18-104_k141_165535 | Not-determined |
| 18-104_k141_116850 | Not-determined |
| 18-104_k141_18937 | Not-determined |
| 18-104_k141_123403 | Not-determined |
| 18-104_k141_165539 | Low-quality |
| 18-104_k141_116853 | Not-determined |
| 18-104_k141_165542 | Low-quality |
| 18-104_k141_116857 | Not-determined |
| 18-104_k141_61006 | Not-determined |
| 18-104_k141_102894 | Not-determined |
| 18-104_k141_102895 | Not-determined |
| 18-104_k141_33607 | Not-determined |
| 18-104_k141_116859 | Not-determined |
| 18-104_k141_68421 | Not-determined |
| 18-104_k141_82660 | Not-determined |
| 18-104_k141_40313 | Not-determined |
| 18-104_k141_47512 | Not-determined |
| 18-104_k141_40315 | Not-determined |
| 18-104_k141_159157 | Low-quality |
| 18-104_k141_123417 | Not-determined |
| 18-104_k141_102904 | Not-determined |
| 18-104_k141_102905 | Not-determined |
| 18-104_k141_18950 | Not-determined |
| 18-104_k141_130781 | Not-determined |
| 18-104_k141_12685 | Not-determined |
| 18-104_k141_137818 | Not-determined |
| 18-104_k141_68427 | Not-determined |
| 18-104_k141_89171 | Not-determined |
| 18-104_k141_145227 | Not-determined |
| 18-104_k141_89173 | Not-determined |
| 18-104_k141_47525 | Not-determined |
| 18-104_k141_53745 | Low-quality |
| 18-104_k141_40326 | Not-determined |
| 18-104_k141_82674 | Not-determined |
| 18-104_k141_47528 | Not-determined |
| 18-104_k141_68435 | Not-determined |
| 18-104_k141_159170 | Not-determined |
| 18-104_k141_145241 | Not-determined |
| 18-104_k141_102914 | Not-determined |
| 18-104_k141_61018 | Not-determined |
| 18-104_k141_61022 | Low-quality |
| 18-104_k141_159174 | Not-determined |
| 18-104_k141_53754 | Low-quality |
| 18-104_k141_145246 | Not-determined |
| 18-104_k141_89187 | Not-determined |
| 18-104_k141_102923 | Not-determined |
| 18-104_k141_47538 | Not-determined |
| 18-104_k141_18968 | Not-determined |
| 18-104_k141_68446 | Not-determined |
| 18-104_k141_33621 | Not-determined |
| 18-104_k141_40335 | Not-determined |
| 18-104_k141_110605 | Not-determined |
| 18-104_k141_33623 | Not-determined |
| 18-104_k141_159183 | Not-determined |
| 18-104_k141_26480 | Not-determined |
| 18-104_k141_96344 | Not-determined |
| 18-104_k141_40342 | Not-determined |
| 18-104_k141_145249 | Not-determined |
| 18-104_k141_82689 | Not-determined |
| 18-104_k141_33626 | Not-determined |
| 18-104_k141_137838 | Not-determined |
| 18-104_k141_75496 | Not-determined |
| 18-104_k141_75497 | Not-determined |
| 18-104_k141_151814 | Not-determined |
| 18-104_k141_116874 | Not-determined |
| 18-104_k141_137841 | Not-determined |
| 18-104_k141_26484 | Not-determined |
| 18-104_k141_96352 | Not-determined |
| 18-104_k141_110610 | Not-determined |
| 18-104_k141_82695 | Not-determined |
| 18-104_k141_137843 | Not-determined |
| 18-104_k141_82696 | Not-determined |
| 18-104_k141_47552 | Not-determined |
| 18-104_k141_53768 | Not-determined |
| 18-104_k141_47554 | Not-determined |
| 18-104_k141_151822 | Not-determined |
| 18-104_k141_40345 | Not-determined |
| 18-104_k141_116886 | Not-determined |
| 18-104_k141_33643 | Not-determined |
| 18-104_k141_75502 | Not-determined |
| 18-104_k141_96365 | Not-determined |
| 18-104_k141_26495 | Not-determined |
| 18-104_k141_130815 | Low-quality |
| 18-104_k141_145262 | Not-determined |
| 18-104_k141_116896 | Not-determined |
| 18-104_k141_137854 | Not-determined |
| 18-104_k141_130817 | Not-determined |
| 18-104_k141_145264 | Not-determined |
| 18-104_k141_102930 | Not-determined |
| 18-104_k141_75515 | Not-determined |
| 18-104_k141_26498 | Not-determined |
| 18-104_k141_165563 | Low-quality |
| 18-104_k141_137859 | Not-determined |
| 18-104_k141_116904 | Not-determined |
| 18-104_k141_26501 | Not-determined |
| 18-104_k141_137862 | Not-determined |
| 18-104_k141_26502 | Not-determined |
| 18-104_k141_96371 | Low-quality |
| 18-104_k141_102935 | Not-determined |
| 18-104_k141_18988 | Not-determined |
| 18-104_k141_102936 | Not-determined |
| 18-104_k141_82716 | Not-determined |
| 18-104_k141_82717 | Not-determined |
| 18-104_k141_53788 | Not-determined |
| 18-104_k141_61055 | Not-determined |
| 18-104_k141_130823 | Not-determined |
| 18-104_k141_116910 | Not-determined |
| 18-104_k141_26507 | Not-determined |
| 18-104_k141_123471 | Not-determined |
| 18-104_k141_26509 | Not-determined |
| 18-104_k141_116912 | Not-determined |
| 18-104_k141_116913 | Not-determined |
| 18-104_k141_53792 | Not-determined |
| 18-104_k141_123475 | Not-determined |
| 18-104_k141_165577 | Not-determined |
| 18-104_k141_12700 | Not-determined |
| 18-104_k141_12701 | Not-determined |
| 18-104_k141_145276 | Not-determined |
| 18-104_k141_159204 | Not-determined |
| 18-104_k141_145283 | Not-determined |
| 18-104_k141_40369 | Not-determined |
| 18-104_k141_123479 | Not-determined |
| 18-104_k141_137882 | Not-determined |
| 18-104_k141_137883 | Not-determined |
| 18-104_k141_40371 | Not-determined |
| 18-104_k141_5559 | Not-determined |
| 18-104_k141_40373 | Low-quality |
| 18-104_k141_40374 | Not-determined |
| 18-104_k141_130835 | Low-quality |
| 18-104_k141_151824 | Not-determined |
| 18-104_k141_82731 | Not-determined |
| 18-104_k141_82734 | Not-determined |
| 18-104_k141_89239 | Low-quality |
| 18-104_k141_26520 | Not-determined |
| 18-104_k141_82735 | Not-determined |
| 18-104_k141_5568 | Not-determined |
| 18-104_k141_33661 | Not-determined |
| 18-104_k141_33662 | Not-determined |
| 18-104_k141_26528 | Not-determined |
| 18-104_k141_68484 | Not-determined |
| 18-104_k141_33664 | Low-quality |
| 18-104_k141_26530 | Not-determined |
| 18-104_k141_5575 | Low-quality |
| 18-104_k141_75544 | Not-determined |
| 18-104_k141_61074 | Not-determined |
| 18-104_k141_5579 | Not-determined |
| 18-104_k141_40385 | Not-determined |
| 18-104_k141_116939 | Not-determined |
| 18-104_k141_151832 | Not-determined |
| 18-104_k141_68491 | Not-determined |
| 18-104_k141_26535 | Not-determined |
| 18-104_k141_40388 | Not-determined |
| 18-104_k141_102966 | Not-determined |
| 18-104_k141_137904 | Low-quality |
| 18-104_k141_61081 | Low-quality |
| 18-104_k141_102968 | Low-quality |
| 18-104_k141_123495 | Not-determined |
| 18-104_k141_110650 | Not-determined |
| 18-104_k141_26539 | Not-determined |
| 18-104_k141_110651 | Not-determined |
| 18-104_k141_110654 | Not-determined |
| 18-104_k141_5590 | Not-determined |
| 18-104_k141_110655 | Not-determined |
| 18-104_k141_102974 | Not-determined |
| 18-104_k141_110657 | Not-determined |
| 18-104_k141_26545 | Not-determined |
| 18-104_k141_130854 | Not-determined |
| 18-104_k141_130855 | Not-determined |
| 18-104_k141_33676 | Not-determined |
| 18-104_k141_82749 | Low-quality |
| 18-104_k141_151835 | Not-determined |
| 18-104_k141_145310 | Not-determined |
| 18-104_k141_75553 | Not-determined |
| 18-104_k141_47574 | Low-quality |
| 18-104_k141_61090 | Low-quality |
| 18-104_k141_61091 | Not-determined |
| 18-104_k141_26549 | Not-determined |
| 18-104_k141_145314 | Not-determined |
| 18-104_k141_102981 | Low-quality |
| 18-104_k141_75559 | Not-determined |
| 18-104_k141_33686 | Low-quality |
| 18-104_k141_33687 | Low-quality |
| 18-104_k141_61095 | Not-determined |
| 18-104_k141_165588 | Not-determined |
| 18-104_k141_47576 | Not-determined |
| 18-104_k141_110668 | Not-determined |
| 18-104_k141_137910 | Not-determined |
| 18-104_k141_130868 | Not-determined |
| 18-104_k141_12723 | Not-determined |
| 18-104_k141_40400 | Not-determined |
| 18-104_k141_61104 | Not-determined |
| 18-104_k141_40405 | Not-determined |
| 18-104_k141_19036 | Not-determined |
| 18-104_k141_159229 | Not-determined |
| 18-104_k141_137915 | Not-determined |
| 18-104_k141_82769 | Not-determined |
| 18-104_k141_47584 | Not-determined |
| 18-104_k141_130881 | Not-determined |
| 18-104_k141_145326 | Not-determined |
| 18-104_k141_130883 | Not-determined |
| 18-104_k141_89280 | Low-quality |
| 18-104_k141_110678 | Not-determined |
| 18-104_k141_103005 | Not-determined |
| 18-104_k141_110679 | Not-determined |
| 18-104_k141_110682 | Not-determined |
| 18-104_k141_82775 | Not-determined |
| 18-104_k141_130887 | Low-quality |
| 18-104_k141_89283 | Not-determined |
| 18-104_k141_103007 | Not-determined |
| 18-104_k141_89285 | Not-determined |
| 18-104_k141_19049 | Not-determined |
| 18-104_k141_110689 | Not-determined |
| 18-104_k141_75580 | Not-determined |
| 18-104_k141_33703 | Not-determined |
| 18-104_k141_159233 | Low-quality |
| 18-104_k141_165602 | Low-quality |
| 18-104_k141_159236 | Not-determined |
| 18-104_k141_96408 | Not-determined |
| 18-104_k141_130904 | Not-determined |
| 18-104_k141_110692 | Not-determined |
| 18-104_k141_145348 | Not-determined |
| 18-104_k141_123527 | Not-determined |
| 18-104_k141_159248 | Not-determined |
| 18-104_k141_61125 | Not-determined |
| 18-104_k141_68520 | Not-determined |
| 18-104_k141_75600 | Low-quality |
| 18-104_k141_47604 | Not-determined |
| 18-104_k141_53796 | Not-determined |
| 18-104_k141_151850 | Not-determined |
| 18-104_k141_33721 | Not-determined |
| 18-104_k141_68521 | Not-determined |
| 18-104_k141_75604 | Not-determined |
| 18-104_k141_130909 | Low-quality |
| 18-104_k141_151852 | Low-quality |
| 18-104_k141_75605 | Not-determined |
| 18-104_k141_130919 | Not-determined |
| 18-104_k141_5617 | Not-determined |
| 18-104_k141_165627 | Not-determined |
| 18-104_k141_68531 | Not-determined |
| 18-104_k141_33730 | Low-quality |
| 18-104_k141_5624 | Not-determined |
| 18-104_k141_47620 | Not-determined |
| 18-104_k141_19070 | Not-determined |
| 18-104_k141_68540 | Not-determined |
| 18-104_k141_116997 | Not-determined |
| 18-104_k141_61138 | Not-determined |
| 18-104_k141_40441 | Low-quality |
| 18-104_k141_165635 | Not-determined |
| 18-104_k141_47622 | Not-determined |
| 18-104_k141_89310 | Not-determined |
| 18-104_k141_151872 | Not-determined |
| 18-104_k141_5630 | Low-quality |
| 18-104_k141_151874 | Not-determined |
| 18-104_k141_159259 | Not-determined |
| 18-104_k141_159262 | Not-determined |
| 18-104_k141_19078 | Not-determined |
| 18-104_k141_53816 | Not-determined |
| 18-104_k141_151877 | Not-determined |
| 18-104_k141_82806 | Not-determined |
| 18-104_k141_53817 | Not-determined |
| 18-104_k141_5638 | Not-determined |
| 18-104_k141_82813 | Not-determined |
| 18-104_k141_145383 | Low-quality |
| 18-104_k141_130930 | Low-quality |
| 18-104_k141_47635 | Not-determined |
| 18-104_k141_5642 | Not-determined |
| 18-104_k141_19092 | Not-determined |
| 18-104_k141_61142 | Not-determined |
| 18-104_k141_53822 | Not-determined |
| 18-104_k141_145391 | Low-quality |
| 18-104_k141_130934 | Not-determined |
| 18-104_k141_145392 | Low-quality |
| 18-104_k141_68555 | Not-determined |
| 18-104_k141_159270 | Not-determined |
| 18-104_k141_47647 | Low-quality |
| 18-104_k141_117016 | Not-determined |
| 18-104_k141_130939 | Not-determined |
| 18-104_k141_165654 | Not-determined |
| 18-104_k141_61151 | Not-determined |
| 18-104_k141_68568 | Not-determined |
| 18-104_k141_61155 | Not-determined |
| 18-104_k141_61156 | Not-determined |
| 18-104_k141_145400 | Not-determined |
| 18-104_k141_75647 | Not-determined |
| 18-104_k141_103048 | Not-determined |
| 18-104_k141_40464 | Not-determined |
| 18-104_k141_89326 | Not-determined |
| 18-104_k141_19107 | Not-determined |
| 18-104_k141_89327 | Low-quality |
| 18-104_k141_47661 | Not-determined |
| 18-104_k141_89332 | Not-determined |
| 18-104_k141_145405 | Not-determined |
| 18-104_k141_75658 | Not-determined |
| 18-104_k141_47664 | Not-determined |
| 18-104_k141_40472 | Low-quality |
| 18-104_k141_68577 | Not-determined |
| 18-104_k141_68580 | Not-determined |
| 18-104_k141_40473 | Not-determined |
| 18-104_k141_68586 | Not-determined |
| 18-104_k141_103054 | Not-determined |
| 18-104_k141_68588 | Not-determined |
| 18-104_k141_12741 | Not-determined |
| 18-104_k141_68589 | Not-determined |
| 18-104_k141_103057 | Not-determined |
| 18-104_k141_68598 | Not-determined |
| 18-104_k141_110729 | Not-determined |
| 18-104_k141_110731 | Not-determined |
| 18-104_k141_61182 | Not-determined |
| 18-104_k141_19126 | Not-determined |
| 18-104_k141_82846 | Not-determined |
| 18-104_k141_103068 | Not-determined |
| 18-104_k141_12758 | Not-determined |
| 18-104_k141_68603 | Low-quality |
| 18-104_k141_82851 | Not-determined |
| 18-104_k141_103071 | Low-quality |
| 18-104_k141_96427 | Not-determined |
| 18-104_k141_123570 | Not-determined |
| 18-104_k141_47683 | Not-determined |
| 18-104_k141_151902 | Not-determined |
| 18-104_k141_40485 | Not-determined |
| 18-104_k141_53831 | Not-determined |
| 18-104_k141_165689 | Not-determined |
| 18-104_k141_19135 | Not-determined |
| 18-104_k141_96434 | Not-determined |
| 18-104_k141_68607 | Not-determined |
| 18-104_k141_19138 | Not-determined |
| 18-104_k141_82854 | Not-determined |
| 18-104_k141_96437 | Not-determined |
| 18-104_k141_68609 | Not-determined |
| 18-104_k141_75680 | Not-determined |
| 18-104_k141_103083 | Not-determined |
| 18-104_k141_151907 | Not-determined |
| 18-104_k141_33763 | Not-determined |
| 18-104_k141_103085 | Not-determined |
| 18-104_k141_165699 | Not-determined |
| 18-104_k141_96445 | Low-quality |
| 18-104_k141_82859 | Not-determined |
| 18-104_k141_165702 | Not-determined |
| 18-104_k141_5690 | Not-determined |
| 18-104_k141_82863 | Not-determined |
| 18-104_k141_12772 | Low-quality |
| 18-104_k141_61199 | Low-quality |
| 18-104_k141_47694 | Low-quality |
| 18-104_k141_89368 | Not-determined |
| 18-104_k141_61201 | Not-determined |
| 18-104_k141_137949 | Not-determined |
| 18-104_k141_75685 | Not-determined |
| 18-104_k141_145439 | Not-determined |
| 18-104_k141_137950 | Not-determined |
| 18-104_k141_89371 | Not-determined |
| 18-104_k141_12775 | Not-determined |
| 18-104_k141_137951 | Not-determined |
| 18-104_k141_89372 | Not-determined |
| 18-104_k141_19150 | Not-determined |
| 18-104_k141_96455 | Low-quality |
| 18-104_k141_165705 | Not-determined |
| 18-104_k141_12787 | Not-determined |
| 18-104_k141_151918 | Not-determined |
| 18-104_k141_47700 | Not-determined |
| 18-104_k141_40502 | Not-determined |
| 18-104_k141_123598 | Not-determined |
| 18-104_k141_130958 | Not-determined |
| 18-104_k141_75695 | Not-determined |
| 18-104_k141_68630 | Not-determined |
| 18-104_k141_75696 | Not-determined |
| 18-104_k141_47714 | Not-determined |
| 18-104_k141_151933 | Not-determined |
| 18-104_k141_33774 | Not-determined |
| 18-104_k141_68640 | Not-determined |
| 18-104_k141_110751 | Not-determined |
| 18-104_k141_53862 | Low-quality |
| 18-104_k141_117049 | Not-determined |
| 18-104_k141_68642 | Low-quality |
| 18-104_k141_75709 | Not-determined |
| 18-104_k141_137960 | Not-determined |
| 18-104_k141_165714 | Not-determined |
| 18-104_k141_103106 | Not-determined |
| 18-104_k141_117058 | Not-determined |
| 18-104_k141_123602 | Low-quality |
| 18-104_k141_159320 | Not-determined |
| 18-104_k141_123604 | Not-determined |
| 18-104_k141_82869 | Low-quality |
| 18-104_k141_61218 | Not-determined |
| 18-104_k141_33789 | Not-determined |
| 18-104_k141_103113 | Not-determined |
| 18-104_k141_117065 | Not-determined |
| 18-104_k141_151945 | Low-quality |
| 18-104_k141_47725 | Not-determined |
| 18-104_k141_53873 | Low-quality |
| 18-104_k141_89414 | Not-determined |

| | |
|---|---|
| 18-104_k141_33797 | Low-quality |
| 18-104_k141_110763 | Not-determined |
| 18-104_k141_12830 | Not-determined |
| 18-104_k141_68653 | Not-determined |
| 18-104_k141_103122 | Not-determined |
| 18-104_k141_159332 | Not-determined |
| 18-104_k141_19184 | Not-determined |
| 18-104_k141_103124 | Not-determined |
| 18-104_k141_117082 | Not-determined |
| 18-104_k141_137988 | Not-determined |
| 18-104_k141_96492 | Not-determined |
| 18-104_k141_145469 | Low-quality |
| 18-104_k141_159344 | Not-determined |
| 18-104_k141_19192 | Not-determined |
| 18-104_k141_159345 | Not-determined |
| 18-104_k141_117086 | Not-determined |
| 18-104_k141_123620 | Low-quality |
| 18-104_k141_12845 | Not-determined |
| 18-104_k141_117087 | Not-determined |
| 18-104_k141_75724 | Not-determined |
| 18-104_k141_137995 | Not-determined |
| 18-104_k141_117093 | Not-determined |
| 18-104_k141_75726 | Low-quality |
| 18-104_k141_130984 | Not-determined |
| 18-104_k141_89433 | Not-determined |
| 18-104_k141_137998 | Not-determined |
| 18-104_k141_75728 | Not-determined |
| 18-104_k141_117104 | Not-determined |
| 18-104_k141_47743 | Not-determined |
| 18-104_k141_19200 | Not-determined |
| 18-104_k141_61229 | Not-determined |
| 18-104_k141_138003 | Not-determined |
| 18-104_k141_75735 | Not-determined |
| 18-104_k141_82889 | Not-determined |
| 18-104_k141_145499 | Not-determined |
| 18-104_k141_68669 | Not-determined |
| 18-104_k141_33804 | Not-determined |
| 18-104_k141_123646 | Not-determined |
| 18-104_k141_159358 | Not-determined |
| 18-104_k141_82897 | Not-determined |
| 18-104_k141_117108 | Not-determined |
| 18-104_k141_165737 | Not-determined |
| 18-104_k141_159364 | Not-determined |
| 18-104_k141_5725 | Not-determined |
| 18-104_k141_33811 | Not-determined |
| 18-104_k141_19211 | Not-determined |
| 18-104_k141_5730 | Not-determined |
| 18-104_k141_117116 | Not-determined |
| 18-104_k141_131006 | Low-quality |
| 18-104_k141_165741 | Not-determined |
| 18-104_k141_103151 | Not-determined |
| 18-104_k141_159371 | Not-determined |
| 18-104_k141_145508 | Not-determined |
| 18-104_k141_82902 | Low-quality |
| 18-104_k141_165743 | Not-determined |
| 18-104_k141_12862 | Not-determined |
| 18-104_k141_165745 | Not-determined |
| 18-104_k141_68686 | Low-quality |
| 18-104_k141_75753 | Not-determined |
| 18-104_k141_103157 | Not-determined |
| 18-104_k141_159376 | Not-determined |
| 18-104_k141_131014 | Not-determined |
| 18-104_k141_117125 | Not-determined |
| 18-104_k141_151988 | Not-determined |
| 18-104_k141_103163 | Not-determined |
| 18-104_k141_103167 | Not-determined |
| 18-104_k141_68694 | Not-determined |
| 18-104_k141_131023 | Not-determined |
| 18-104_k141_53914 | Not-determined |
| 18-104_k141_82915 | Not-determined |
| 18-104_k141_33817 | Not-determined |
| 18-104_k141_151999 | Not-determined |
| 18-104_k141_152000 | Not-determined |
| 18-104_k141_89453 | Not-determined |
| 18-104_k141_145528 | Not-determined |
| 18-104_k141_165753 | Not-determined |
| 18-104_k141_165754 | Not-determined |
| 18-104_k141_68698 | Not-determined |
| 18-104_k141_19225 | Not-determined |
| 18-104_k141_165755 | Not-determined |
| 18-104_k141_131031 | Not-determined |
| 18-104_k141_19226 | Not-determined |
| 18-104_k141_159387 | Not-determined |
| 18-104_k141_47753 | Not-determined |
| 18-104_k141_26591 | Not-determined |
| 18-104_k141_145535 | Low-quality |
| 18-104_k141_53921 | Not-determined |
| 18-104_k141_12882 | Not-determined |
| 18-104_k141_103179 | Not-determined |
| 18-104_k141_89460 | Low-quality |
| 18-104_k141_26595 | Not-determined |
| 18-104_k141_53923 | Low-quality |
| 18-104_k141_159396 | Not-determined |
| 18-104_k141_152008 | Not-determined |
| 18-104_k141_19234 | Not-determined |
| 18-104_k141_131041 | Not-determined |
| 18-104_k141_131042 | Not-determined |
| 18-104_k141_19235 | Not-determined |
| 18-104_k141_33834 | Not-determined |
| 18-104_k141_12887 | Not-determined |
| 18-104_k141_159399 | Not-determined |
| 18-104_k141_165762 | Not-determined |
| 18-104_k141_33838 | Not-determined |
| 18-104_k141_152016 | Not-determined |
| 18-104_k141_123672 | Not-determined |
| 18-104_k141_123673 | Not-determined |
| 18-104_k141_123675 | Not-determined |
| 18-104_k141_19247 | Not-determined |
| 18-104_k141_19248 | Not-determined |
| 18-104_k141_53931 | Not-determined |
| 18-104_k141_159412 | Not-determined |
| 18-104_k141_40520 | Not-determined |
| 18-104_k141_47766 | Low-quality |
| 18-104_k141_165770 | Not-determined |
| 18-104_k141_53936 | Not-determined |
| 18-104_k141_159417 | Not-determined |
| 18-104_k141_110847 | Not-determined |
| 18-104_k141_145555 | Low-quality |
| 18-104_k141_82960 | Low-quality |
| 18-104_k141_117144 | Not-determined |
| 18-104_k141_40533 | Not-determined |
| 18-104_k141_68727 | Not-determined |
| 18-104_k141_117146 | Not-determined |
| 18-104_k141_40535 | Not-determined |
| 18-104_k141_33851 | Not-determined |
| 18-104_k141_82969 | Not-determined |
| 18-104_k141_40539 | Not-determined |
| 18-104_k141_12905 | Not-determined |
| 18-104_k141_145566 | Not-determined |
| 18-104_k141_159430 | Not-determined |
| 18-104_k141_165781 | Not-determined |
| 18-104_k141_138041 | Not-determined |
| 18-104_k141_40542 | Not-determined |
| 18-104_k141_47786 | Not-determined |
| 18-104_k141_5749 | Not-determined |
| 18-104_k141_131068 | Not-determined |
| 18-104_k141_165782 | Not-determined |
| 18-104_k141_12909 | Not-determined |
| 18-104_k141_82977 | Not-determined |
| 18-104_k141_159436 | Not-determined |
| 18-104_k141_131074 | Not-determined |
| 18-104_k141_103205 | Not-determined |
| 18-104_k141_145571 | Not-determined |
| 18-104_k141_138048 | Not-determined |
| 18-104_k141_61249 | Low-quality |
| 18-104_k141_40549 | Not-determined |
| 18-104_k141_61253 | Not-determined |
| 18-104_k141_145585 | Not-determined |
| 18-104_k141_96548 | Not-determined |
| 18-104_k141_89511 | Not-determined |
| 18-104_k141_131083 | Not-determined |
| 18-104_k141_103212 | Not-determined |
| 18-104_k141_145589 | Not-determined |
| 18-104_k141_110878 | Not-determined |
| 18-104_k141_159445 | Not-determined |
| 18-104_k141_47798 | Not-determined |
| 18-104_k141_33876 | Not-determined |
| 18-104_k141_33878 | Not-determined |
| 18-104_k141_165790 | Not-determined |
| 18-104_k141_138067 | Not-determined |
| 18-104_k141_138070 | Not-determined |
| 18-104_k141_68762 | Not-determined |
| 18-104_k141_131092 | Not-determined |
| 18-104_k141_138072 | Not-determined |
| 18-104_k141_103215 | Not-determined |
| 18-104_k141_165794 | Low-quality |
| 18-104_k141_53972 | Not-determined |
| 18-104_k141_131095 | Not-determined |
| 18-104_k141_117165 | Not-determined |
| 18-104_k141_117169 | Not-determined |
| 18-104_k141_33886 | Not-determined |
| 18-104_k141_68769 | Not-determined |
| 18-104_k141_145606 | Not-determined |
| 18-104_k141_145608 | Not-determined |
| 18-104_k141_47818 | Not-determined |
| 18-104_k141_61275 | Not-determined |
| 18-104_k141_47820 | Not-determined |
| 18-104_k141_89530 | Not-determined |
| 18-104_k141_89532 | Not-determined |
| 18-104_k141_159461 | Not-determined |
| 18-104_k141_165802 | Not-determined |
| 18-104_k141_103223 | Not-determined |
| 18-104_k141_117182 | Not-determined |
| 18-104_k141_61280 | Low-quality |
| 18-104_k141_152049 | Not-determined |
| 18-104_k141_103227 | Not-determined |
| 18-104_k141_131117 | Not-determined |
| 18-104_k141_138086 | Not-determined |
| 18-104_k141_53985 | Not-determined |
| 18-104_k141_89541 | Low-quality |
| 18-104_k141_53987 | Not-determined |
| 18-104_k141_159471 | Not-determined |
| 18-104_k141_47830 | Low-quality |
| 18-104_k141_89542 | Not-determined |
| 18-104_k141_68784 | Not-determined |
| 18-104_k141_47835 | Not-determined |
| 18-104_k141_12930 | Not-determined |
| 18-104_k141_96568 | Not-determined |
| 18-104_k141_75791 | Not-determined |
| 18-104_k141_165809 | Not-determined |
| 18-104_k141_68789 | Low-quality |
| 18-104_k141_40595 | Not-determined |
| 18-104_k141_138099 | Not-determined |
| 18-104_k141_47840 | Not-determined |
| 18-104_k141_47843 | Not-determined |
| 18-104_k141_68794 | Not-determined |
| 18-104_k141_40602 | Not-determined |
| 18-104_k141_47845 | Low-quality |
| 18-104_k141_75798 | Not-determined |
| 18-104_k141_159474 | Not-determined |
| 18-104_k141_47852 | Not-determined |
| 18-104_k141_165817 | Not-determined |
| 18-104_k141_165819 | Not-determined |
| 18-104_k141_47856 | Not-determined |
| 18-104_k141_33912 | Low-quality |
| 18-104_k141_138108 | Low-quality |
| 18-104_k141_145631 | Not-determined |
| 18-104_k141_138111 | Not-determined |
| 18-104_k141_152078 | Not-determined |
| 18-104_k141_61303 | Not-determined |
| 18-104_k141_96595 | Not-determined |
| 18-104_k141_75811 | Not-determined |
| 18-104_k141_165824 | Not-determined |
| 18-104_k141_5801 | Not-determined |
| 18-104_k141_96598 | Not-determined |
| 18-104_k141_26640 | Not-determined |
| 18-104_k141_75813 | Not-determined |
| 18-104_k141_40611 | Not-determined |
| 18-104_k141_26642 | Not-determined |
| 18-104_k141_26643 | Not-determined |
| 18-104_k141_68811 | Not-determined |
| 18-104_k141_152085 | Not-determined |
| 18-104_k141_152086 | Not-determined |
| 18-104_k141_61311 | Not-determined |
| 18-104_k141_61313 | Low-quality |
| 18-104_k141_165831 | Not-determined |
| 18-104_k141_40619 | Not-determined |
| 18-104_k141_54012 | Not-determined |
| 18-104_k141_5809 | Not-determined |
| 18-104_k141_40621 | Not-determined |
| 18-104_k141_40624 | Not-determined |
| 18-104_k141_117200 | Not-determined |
| 18-104_k141_123702 | Not-determined |
| 18-104_k141_123704 | Not-determined |
| 18-104_k141_145639 | Not-determined |
| 18-104_k141_138120 | Not-determined |
| 18-104_k141_138121 | Not-determined |
| 18-104_k141_75828 | Low-quality |
| 18-104_k141_123706 | Not-determined |
| 18-104_k141_54023 | Not-determined |
| 18-104_k141_83064 | Not-determined |
| 18-104_k141_54026 | Not-determined |
| 18-104_k141_61323 | Not-determined |
| 18-104_k141_5819 | Not-determined |
| 18-104_k141_83070 | Not-determined |
| 18-104_k141_110914 | Not-determined |
| 18-104_k141_165837 | Not-determined |
| 18-104_k141_68824 | Not-determined |
| 18-104_k141_12956 | Not-determined |
| 18-104_k141_152104 | Not-determined |
| 18-104_k141_152105 | Not-determined |
| 18-104_k141_83071 | Not-determined |
| 18-104_k141_68827 | Low-quality |
| 18-104_k141_123724 | Not-determined |
| 18-104_k141_33945 | Not-determined |
| 18-104_k141_12959 | Not-determined |
| 18-104_k141_83076 | Not-determined |
| 18-104_k141_159505 | Not-determined |
| 18-104_k141_159507 | Not-determined |
| 18-104_k141_12964 | Not-determined |
| 18-104_k141_26651 | Not-determined |
| 18-104_k141_110919 | Not-determined |
| 18-104_k141_159510 | Not-determined |
| 18-104_k141_12970 | Not-determined |
| 18-104_k141_75834 | Not-determined |
| 18-104_k141_159514 | Not-determined |
| 18-104_k141_165848 | Not-determined |
| 18-104_k141_12974 | Not-determined |
| 18-104_k141_83081 | Not-determined |
| 18-104_k141_145667 | Low-quality |
| 18-104_k141_47903 | Not-determined |
| 18-104_k141_83085 | Not-determined |
| 18-104_k141_75841 | Not-determined |
| 18-104_k141_138151 | Not-determined |
| 18-104_k141_19277 | Not-determined |
| 18-104_k141_96629 | Not-determined |
| 18-104_k141_40657 | Not-determined |
| 18-104_k141_117226 | Not-determined |
| 18-104_k141_61350 | Not-determined |
| 18-104_k141_47911 | Not-determined |
| 18-104_k141_103259 | Not-determined |
| 18-104_k141_19282 | Not-determined |
| 18-104_k141_61357 | Low-quality |
| 18-104_k141_5841 | Not-determined |
| 18-104_k141_68857 | Not-determined |
| 18-104_k141_110944 | Not-determined |
| 18-104_k141_165861 | Not-determined |
| 18-104_k141_138166 | Not-determined |
| 18-104_k141_96645 | Not-determined |
| 18-104_k141_103265 | Not-determined |
| 18-104_k141_40670 | Not-determined |
| 18-104_k141_61374 | Low-quality |
| 18-104_k141_138175 | Not-determined |
| 18-104_k141_26682 | Not-determined |
| 18-104_k141_96651 | Not-determined |
| 18-104_k141_12986 | Not-determined |
| 18-104_k141_110947 | Low-quality |
| 18-104_k141_159537 | Not-determined |
| 18-104_k141_12988 | Not-determined |
| 18-104_k141_159539 | Not-determined |
| 18-104_k141_26685 | Not-determined |
| 18-104_k141_5850 | Not-determined |
| 18-104_k141_26688 | Not-determined |
| 18-104_k141_5854 | Not-determined |
| 18-104_k141_165877 | Not-determined |
| 18-104_k141_123766 | Not-determined |
| 18-104_k141_40685 | Not-determined |
| 18-104_k141_5855 | Low-quality |
| 18-104_k141_12999 | Not-determined |
| 18-104_k141_159543 | Low-quality |
| 18-104_k141_75862 | Low-quality |
| 18-104_k141_40689 | Not-determined |
| 18-104_k141_13000 | Low-quality |
| 18-104_k141_5858 | Not-determined |
| 18-104_k141_110955 | Not-determined |
| 18-104_k141_138179 | Not-determined |
| 18-104_k141_117235 | Not-determined |
| 18-104_k141_110956 | Not-determined |
| 18-104_k141_75865 | Not-determined |
| 18-104_k141_54047 | Low-quality |
| 18-104_k141_145695 | Not-determined |
| 18-104_k141_47938 | Not-determined |
| 18-104_k141_5864 | Not-determined |
| 18-104_k141_61381 | Not-determined |
| 18-104_k141_54050 | Not-determined |
| 18-104_k141_47942 | Not-determined |
| 18-104_k141_131144 | Not-determined |
| 18-104_k141_131146 | Not-determined |
| 18-104_k141_54052 | Not-determined |
| 18-104_k141_75870 | Not-determined |
| 18-104_k141_110969 | Not-determined |
| 18-104_k141_26694 | Not-determined |
| 18-104_k141_131148 | Not-determined |
| 18-104_k141_61383 | Low-quality |
| 18-104_k141_26695 | Not-determined |
| 18-104_k141_110972 | Not-determined |
| 18-104_k141_131150 | Not-determined |
| 18-104_k141_159553 | Not-determined |
| 18-104_k141_96664 | Not-determined |
| 18-104_k141_145704 | Not-determined |
| 18-104_k141_54058 | Not-determined |
| 18-104_k141_75878 | Not-determined |
| 18-104_k141_40702 | Not-determined |
| 18-104_k141_5874 | Not-determined |
| 18-104_k141_152117 | Not-determined |
| 18-104_k141_33963 | Not-determined |
| 18-104_k141_61389 | Low-quality |
| 18-104_k141_13013 | Not-determined |
| 18-104_k141_117258 | Not-determined |
| 18-104_k141_13019 | Not-determined |
| 18-104_k141_75880 | Not-determined |
| 18-104_k141_68879 | Not-determined |
| 18-104_k141_165893 | Not-determined |
| 18-104_k141_123790 | Not-determined |
| 18-104_k141_145716 | Not-determined |
| 18-104_k141_117264 | Not-determined |
| 18-104_k141_123792 | Low-quality |
| 18-104_k141_131156 | Not-determined |
| 18-104_k141_54077 | Not-determined |
| 18-104_k141_138208 | Low-quality |
| 18-104_k141_83124 | Low-quality |
| 18-104_k141_117266 | Not-determined |
| 18-104_k141_152133 | Not-determined |
| 18-104_k141_123802 | Not-determined |
| 18-104_k141_68882 | Not-determined |
| 18-104_k141_123805 | Not-determined |
| 18-104_k141_68885 | Not-determined |
| 18-104_k141_145724 | Not-determined |
| 18-104_k141_96680 | Not-determined |
| 18-104_k141_68888 | Not-determined |
| 18-104_k141_103309 | Not-determined |
| 18-104_k141_110996 | Not-determined |
| 18-104_k141_33975 | Not-determined |
| 18-104_k141_131161 | Not-determined |
| 18-104_k141_96681 | Not-determined |
| 18-104_k141_131163 | Not-determined |
| 18-104_k141_83132 | Not-determined |
| 18-104_k141_117282 | Not-determined |
| 18-104_k141_152143 | Not-determined |
| 18-104_k141_165901 | Not-determined |
| 18-104_k141_54095 | Not-determined |
| 18-104_k141_68892 | Not-determined |
| 18-104_k141_83142 | Low-quality |
| 18-104_k141_111000 | Not-determined |
| 18-104_k141_83144 | Low-quality |
| 18-104_k141_83146 | Not-determined |
| 18-104_k141_145740 | Not-determined |
| 18-104_k141_145741 | Not-determined |
| 18-104_k141_40744 | Not-determined |
| 18-104_k141_123827 | Low-quality |
| 18-104_k141_83153 | Not-determined |
| 18-104_k141_96698 | Not-determined |
| 18-104_k141_5887 | Not-determined |
| 18-104_k141_111004 | Not-determined |
| 18-104_k141_5889 | Low-quality |
| 18-104_k141_96700 | Not-determined |
| 18-104_k141_111006 | Not-determined |
| 18-104_k141_111007 | Low-quality |
| 18-104_k141_19329 | Not-determined |
| 18-104_k141_131177 | Not-determined |
| 18-104_k141_54107 | Not-determined |
| 18-104_k141_117295 | Not-determined |
| 18-104_k141_26723 | Not-determined |
| 18-104_k141_138226 | Low-quality |
| 18-104_k141_138228 | Not-determined |
| 18-104_k141_33993 | Not-determined |
| 18-104_k141_165905 | Not-determined |
| 18-104_k141_19334 | Not-determined |
| 18-104_k141_138231 | Not-determined |
| 18-104_k141_165907 | Not-determined |
| 18-104_k141_138236 | Not-determined |
| 18-104_k141_159567 | Not-determined |
| 18-104_k141_152165 | Low-quality |
| 18-104_k141_40757 | Not-determined |
| 18-104_k141_145760 | Not-determined |
| 18-104_k141_138239 | Not-determined |
| 18-104_k141_165910 | Not-determined |
| 18-104_k141_159570 | Not-determined |
| 18-104_k141_33999 | Not-determined |
| 18-104_k141_159573 | Not-determined |
| 18-104_k141_165914 | Not-determined |
| 18-104_k141_68907 | Not-determined |
| 18-104_k141_26737 | Not-determined |
| 18-104_k141_123855 | Not-determined |
| 18-104_k141_152172 | Not-determined |
| 18-104_k141_123857 | Not-determined |
| 18-104_k141_123860 | Not-determined |
| 18-104_k141_111033 | Not-determined |
| 18-104_k141_75930 | Not-determined |
| 18-104_k141_165920 | Not-determined |
| 18-104_k141_61419 | Not-determined |
| 18-104_k141_111034 | Not-determined |
| 18-104_k141_40766 | Not-determined |
| 18-104_k141_26747 | Not-determined |
| 18-104_k141_68919 | Low-quality |
| 18-104_k141_34010 | Not-determined |
| 18-104_k141_34011 | Not-determined |
| 18-104_k141_152183 | Not-determined |
| 18-104_k141_61422 | Low-quality |
| 18-104_k141_61425 | Not-determined |
| 18-104_k141_34020 | Not-determined |
| 18-104_k141_145782 | Not-determined |
| 18-104_k141_152191 | Not-determined |
| 18-104_k141_117322 | Not-determined |
| 18-104_k141_152193 | Not-determined |
| 18-104_k141_13050 | Not-determined |
| 18-104_k141_83197 | Not-determined |
| 18-104_k141_61433 | Not-determined |
| 18-104_k141_75948 | Low-quality |
| 18-104_k141_40781 | Not-determined |
| 18-104_k141_34029 | Not-determined |
| 18-104_k141_26763 | Not-determined |
| 18-104_k141_145790 | Not-determined |
| 18-104_k141_145791 | Not-determined |
| 18-104_k141_152198 | Not-determined |
| 18-104_k141_159594 | Low-quality |
| 18-104_k141_13053 | Not-determined |
| 18-104_k141_75956 | Low-quality |
| 18-104_k141_145797 | Not-determined |
| 18-104_k141_165949 | Not-determined |
| 18-104_k141_5929 | Not-determined |
| 18-104_k141_96736 | Low-quality |
| 18-104_k141_145803 | Not-determined |
| 18-104_k141_54141 | Not-determined |
| 18-104_k141_54143 | Not-determined |
| 18-104_k141_117334 | Not-determined |
| 18-104_k141_26773 | Low-quality |
| 18-104_k141_40794 | Not-determined |
| 18-104_k141_145810 | Not-determined |
| 18-104_k141_68931 | Low-quality |
| 18-104_k141_117338 | Not-determined |
| 18-104_k141_159613 | Low-quality |
| 18-104_k141_75968 | Not-determined |
| 18-104_k141_138274 | Not-determined |
| 18-104_k141_5934 | Not-determined |
| 18-104_k141_83218 | Not-determined |
| 18-104_k141_13070 | Not-determined |
| 18-104_k141_75978 | Not-determined |
| 18-104_k141_68937 | Not-determined |
| 18-104_k141_159620 | Not-determined |
| 18-104_k141_145819 | Not-determined |
| 18-104_k141_5940 | Not-determined |
| 18-104_k141_111068 | Not-determined |
| 18-104_k141_117344 | Not-determined |
| 18-104_k141_13073 | Not-determined |
| 18-104_k141_75982 | Low-quality |
| 18-104_k141_138279 | Not-determined |
| 18-104_k141_111071 | Not-determined |
| 18-104_k141_13077 | Not-determined |
| 18-104_k141_61440 | Not-determined |
| 18-104_k141_165962 | Not-determined |
| 18-104_k141_145824 | Not-determined |
| 18-104_k141_61445 | Not-determined |
| 18-104_k141_47999 | Not-determined |
| 18-104_k141_145830 | Not-determined |
| 18-104_k141_68951 | Not-determined |
| 18-104_k141_75985 | Not-determined |
| 18-104_k141_75990 | Not-determined |
| 18-104_k141_75991 | Not-determined |
| 18-104_k141_48003 | Not-determined |
| 18-104_k141_117354 | Not-determined |
| 18-104_k141_68955 | Not-determined |
| 18-104_k141_117358 | Not-determined |
| 18-104_k141_83237 | Not-determined |
| 18-104_k141_61451 | Not-determined |
| 18-104_k141_123869 | Not-determined |
| 18-104_k141_89601 | Low-quality |
| 18-104_k141_40822 | Not-determined |
| 18-104_k141_138294 | Low-quality |
| 18-104_k141_48006 | Not-determined |
| 18-104_k141_117365 | Not-determined |
| 18-104_k141_111093 | Not-determined |
| 18-104_k141_48008 | Not-determined |
| 18-104_k141_152223 | Not-determined |
| 18-104_k141_54174 | Not-determined |
| 18-104_k141_111096 | Not-determined |
| 18-104_k141_117370 | Not-determined |
| 18-104_k141_13111 | Not-determined |
| 18-104_k141_34075 | Low-quality |
| 18-104_k141_19346 | Low-quality |
| 18-104_k141_13116 | Not-determined |
| 18-104_k141_54183 | Not-determined |
| 18-104_k141_117372 | Not-determined |
| 18-104_k141_40839 | Not-determined |
| 18-104_k141_34077 | Not-determined |
| 18-104_k141_34078 | Not-determined |
| 18-104_k141_83248 | Not-determined |
| 18-104_k141_5971 | Not-determined |
| 18-104_k141_83259 | Not-determined |
| 18-104_k141_83261 | Not-determined |
| 18-104_k141_40840 | Low-quality |
| 18-104_k141_40842 | Not-determined |
| 18-104_k141_117383 | Not-determined |
| 18-104_k141_13130 | Not-determined |
| 18-104_k141_48031 | Not-determined |
| 18-104_k141_13132 | Not-determined |
| 18-104_k141_48034 | Not-determined |
| 18-104_k141_13134 | Not-determined |
| 18-104_k141_131179 | Low-quality |
| 18-104_k141_89611 | Not-determined |
| 18-104_k141_19368 | Not-determined |
| 18-104_k141_5979 | Not-determined |
| 18-104_k141_5980 | Not-determined |
| 18-104_k141_48045 | Not-determined |
| 18-104_k141_131194 | Not-determined |
| 18-104_k141_111123 | Not-determined |
| 18-104_k141_40864 | Not-determined |
| 18-104_k141_117387 | Not-determined |
| 18-104_k141_26834 | Not-determined |
| 18-104_k141_40866 | Not-determined |
| 18-104_k141_166015 | Not-determined |
| 18-104_k141_26838 | Low-quality |
| 18-104_k141_111131 | Not-determined |
| 18-104_k141_54210 | Not-determined |
| 18-104_k141_48058 | Not-determined |
| 18-104_k141_54213 | Not-determined |
| 18-104_k141_26840 | Not-determined |
| 18-104_k141_83298 | Not-determined |
| 18-104_k141_19386 | Not-determined |
| 18-104_k141_54221 | Not-determined |
| 18-104_k141_54222 | Not-determined |
| 18-104_k141_138308 | Not-determined |
| 18-104_k141_40882 | Not-determined |
| 18-104_k141_117404 | Not-determined |
| 18-104_k141_61498 | Low-quality |
| 18-104_k141_61501 | Not-determined |
| 18-104_k141_166029 | Not-determined |
| 18-104_k141_54229 | Not-determined |
| 18-104_k141_61502 | Low-quality |
| 18-104_k141_48077 | Not-determined |
| 18-104_k141_19392 | Not-determined |
| 18-104_k141_89624 | Not-determined |
| 18-104_k141_13162 | Not-determined |
| 18-104_k141_138317 | Not-determined |
| 18-104_k141_76028 | Not-determined |
| 18-104_k141_19398 | Not-determined |
| 18-104_k141_54234 | Not-determined |
| 18-104_k141_40892 | Not-determined |
| 18-104_k141_123905 | Not-determined |

| | |
|---|---|
| 18-104_k141_123908 | Not-determined |
| 18-104_k141_34112 | Low-quality |
| 18-104_k141_6009 | Not-determined |
| 18-104_k141_123914 | Not-determined |
| 18-104_k141_89636 | Not-determined |
| 18-104_k141_138321 | Low-quality |
| 18-104_k141_117411 | Not-determined |
| 18-104_k141_123917 | Not-determined |
| 18-104_k141_123920 | Not-determined |
| 18-104_k141_89639 | Not-determined |
| 18-104_k141_138326 | Not-determined |
| 18-104_k141_131242 | Not-determined |
| 18-104_k141_40905 | Not-determined |
| 18-104_k141_96751 | Not-determined |
| 18-104_k141_6019 | Not-determined |
| 18-104_k141_138332 | Not-determined |
| 18-104_k141_83320 | Low-quality |
| 18-104_k141_76045 | Not-determined |
| 18-104_k141_54246 | Not-determined |
| 18-104_k141_131247 | Not-determined |
| 18-104_k141_68963 | Low-quality |
| 18-104_k141_34119 | Not-determined |
| 18-104_k141_48098 | Not-determined |
| 18-104_k141_123931 | Not-determined |
| 18-104_k141_89655 | Not-determined |
| 18-104_k141_34123 | Not-determined |
| 18-104_k141_123934 | Low-quality |
| 18-104_k141_34129 | Not-determined |
| 18-104_k141_48105 | Not-determined |
| 18-104_k141_111154 | Low-quality |
| 18-104_k141_34132 | Not-determined |
| 18-104_k141_13180 | Not-determined |
| 18-104_k141_145851 | Not-determined |
| 18-104_k141_166038 | Not-determined |
| 18-104_k141_54258 | Not-determined |
| 18-104_k141_34136 | Not-determined |
| 18-104_k141_96761 | Not-determined |
| 18-104_k141_111160 | Not-determined |
| 18-104_k141_111162 | Not-determined |
| 18-104_k141_145857 | Not-determined |
| 18-104_k141_40932 | Not-determined |
| 18-104_k141_19439 | Not-determined |
| 18-104_k141_13186 | Low-quality |
| 18-104_k141_145860 | Not-determined |
| 18-104_k141_19442 | Not-determined |
| 18-104_k141_34147 | Not-determined |
| 18-104_k141_96766 | Not-determined |
| 18-104_k141_96767 | Not-determined |
| 18-104_k141_68980 | Not-determined |
| 18-104_k141_103351 | Not-determined |
| 18-104_k141_54264 | Low-quality |
| 18-104_k141_61524 | Not-determined |
| 18-104_k141_6044 | Not-determined |
| 18-104_k141_40940 | Not-determined |
| 18-104_k141_68987 | Not-determined |
| 18-104_k141_76073 | Low-quality |
| 18-104_k141_48124 | Low-quality |
| 18-104_k141_89681 | Not-determined |
| 18-104_k141_19451 | Not-determined |
| 18-104_k141_89687 | Low-quality |
| 18-104_k141_61538 | Not-determined |
| 18-104_k141_89688 | Not-determined |
| 18-104_k141_13196 | Not-determined |
| 18-104_k141_19455 | Not-determined |
| 18-104_k141_103362 | Not-determined |
| 18-104_k141_19457 | Not-determined |
| 18-104_k141_123959 | Not-determined |
| 18-104_k141_13199 | Not-determined |
| 18-104_k141_13200 | Not-determined |
| 18-104_k141_19461 | Not-determined |
| 18-104_k141_103364 | Not-determined |
| 18-104_k141_69003 | Not-determined |
| 18-104_k141_19464 | Not-determined |
| 18-104_k141_61550 | Low-quality |
| 18-104_k141_131263 | Not-determined |
| 18-104_k141_103368 | Not-determined |
| 18-104_k141_145877 | Not-determined |
| 18-104_k141_19467 | Not-determined |
| 18-104_k141_48135 | Low-quality |
| 18-104_k141_48136 | Not-determined |
| 18-104_k141_6062 | Low-quality |
| 18-104_k141_103371 | Not-determined |
| 18-104_k141_152239 | Not-determined |
| 18-104_k141_131268 | Not-determined |
| 18-104_k141_138369 | Low-quality |
| 18-104_k141_61556 | Not-determined |
| 18-104_k141_103378 | Not-determined |
| 18-104_k141_69009 | Not-determined |
| 18-104_k141_6072 | Not-determined |
| 18-104_k141_69013 | Not-determined |
| 18-104_k141_131274 | Not-determined |
| 18-104_k141_54294 | Low-quality |
| 18-104_k141_19473 | Not-determined |
| 18-104_k141_138377 | Not-determined |
| 18-104_k141_54298 | Not-determined |
| 18-104_k141_96800 | Not-determined |
| 18-104_k141_26883 | Not-determined |
| 18-104_k141_145892 | Not-determined |
| 18-104_k141_138382 | Not-determined |
| 18-104_k141_13230 | Not-determined |
| 18-104_k141_54301 | Not-determined |
| 18-104_k141_19476 | Not-determined |
| 18-104_k141_19482 | Not-determined |
| 18-104_k141_123994 | Not-determined |
| 18-104_k141_96808 | Not-determined |
| 18-104_k141_96809 | Not-determined |
| 18-104_k141_131293 | Not-determined |
| 18-104_k141_138386 | Not-determined |
| 18-104_k141_26894 | Not-determined |
| 18-104_k141_159681 | Not-determined |
| 18-104_k141_6078 | Low-quality |
| 18-104_k141_138390 | Not-determined |
| 18-104_k141_159682 | Not-determined |
| 18-104_k141_159683 | Not-determined |
| 18-104_k141_6081 | Not-determined |
| 18-104_k141_159685 | Not-determined |
| 18-104_k141_166061 | Not-determined |
| 18-104_k141_117471 | Not-determined |
| 18-104_k141_96814 | Not-determined |
| 18-104_k141_117474 | Not-determined |
| 18-104_k141_34187 | Not-determined |
| 18-104_k141_61576 | Low-quality |
| 18-104_k141_69032 | Not-determined |
| 18-104_k141_48165 | Low-quality |
| 18-104_k141_48166 | Low-quality |
| 18-104_k141_83342 | Not-determined |
| 18-104_k141_124001 | Not-determined |
| 18-104_k141_48169 | Not-determined |
| 18-104_k141_145895 | Not-determined |
| 18-104_k141_83346 | Not-determined |
| 18-104_k141_48170 | Low-quality |
| 18-104_k141_124002 | Not-determined |
| 18-104_k141_166071 | Not-determined |
| 18-104_k141_13252 | Not-determined |
| 18-104_k141_48172 | Not-determined |
| 18-104_k141_19501 | Not-determined |
| 18-104_k141_76095 | Not-determined |
| 18-104_k141_111202 | Not-determined |
| 18-104_k141_40956 | Not-determined |
| 18-104_k141_69042 | Not-determined |
| 18-104_k141_152259 | Low-quality |
| 18-104_k141_159690 | Not-determined |
| 18-104_k141_13257 | Not-determined |
| 18-104_k141_40957 | Not-determined |
| 18-104_k141_48176 | Not-determined |
| 18-104_k141_117490 | Not-determined |
| 18-104_k141_96824 | Not-determined |
| 18-104_k141_166079 | Low-quality |
| 18-104_k141_76104 | Not-determined |
| 18-104_k141_48182 | Not-determined |
| 18-104_k141_159696 | Low-quality |
| 18-104_k141_6099 | Not-determined |
| 18-104_k141_111207 | Not-determined |
| 18-104_k141_83362 | Not-determined |
| 18-104_k141_26924 | Not-determined |
| 18-104_k141_26925 | Not-determined |
| 18-104_k141_13263 | Low-quality |
| 18-104_k141_19508 | Not-determined |
| 18-104_k141_166084 | Not-determined |
| 18-104_k141_13266 | Not-determined |
| 18-104_k141_13267 | Not-determined |
| 18-104_k141_83372 | Not-determined |
| 18-104_k141_13269 | Not-determined |
| 18-104_k141_76112 | Low-quality |
| 18-104_k141_76116 | Not-determined |
| 18-104_k141_89727 | Not-determined |
| 18-104_k141_6107 | Not-determined |
| 18-104_k141_40970 | Low-quality |
| 18-104_k141_111213 | Not-determined |
| 18-104_k141_54331 | Not-determined |
| 18-104_k141_19519 | Not-determined |
| 18-104_k141_152274 | Not-determined |
| 18-104_k141_34203 | Not-determined |
| 18-104_k141_54334 | Not-determined |
| 18-104_k141_69063 | Not-determined |
| 18-104_k141_19526 | Not-determined |
| 18-104_k141_166092 | Not-determined |
| 18-104_k141_69069 | Not-determined |
| 18-104_k141_6115 | Not-determined |
| 18-104_k141_6117 | Not-determined |
| 18-104_k141_69072 | Not-determined |
| 18-104_k141_40986 | Not-determined |
| 18-104_k141_96834 | Low-quality |
| 18-104_k141_48184 | Low-quality |
| 18-104_k141_13287 | Not-determined |
| 18-104_k141_89744 | Not-determined |
| 18-104_k141_145917 | Low-quality |
| 18-104_k141_34209 | Low-quality |
| 18-104_k141_145921 | Not-determined |
| 18-104_k141_26950 | Not-determined |
| 18-104_k141_152293 | Not-determined |
| 18-104_k141_159720 | Not-determined |
| 18-104_k141_89760 | Not-determined |
| 18-104_k141_69082 | Not-determined |
| 18-104_k141_76131 | Not-determined |
| 18-104_k141_152300 | Not-determined |
| 18-104_k141_166105 | Not-determined |
| 18-104_k141_34214 | Not-determined |
| 18-104_k141_103462 | Not-determined |
| 18-104_k141_96849 | Not-determined |
| 18-104_k141_6131 | Not-determined |
| 18-104_k141_103467 | Not-determined |
| 18-104_k141_131337 | Not-determined |
| 18-104_k141_103468 | Not-determined |
| 18-104_k141_131339 | Not-determined |
| 18-104_k141_48197 | Low-quality |
| 18-104_k141_34218 | Not-determined |
| 18-104_k141_61629 | Not-determined |
| 18-104_k141_152316 | Not-determined |
| 18-104_k141_61632 | Low-quality |
| 18-104_k141_13302 | Not-determined |
| 18-104_k141_6141 | Not-determined |
| 18-104_k141_76150 | Not-determined |
| 18-104_k141_117539 | Low-quality |
| 18-104_k141_13304 | Not-determined |
| 18-104_k141_152319 | Not-determined |
| 18-104_k141_19543 | Low-quality |
| 18-104_k141_48210 | Not-determined |
| 18-104_k141_13309 | Not-determined |
| 18-104_k141_131347 | Not-determined |
| 18-104_k141_124062 | Not-determined |
| 18-104_k141_124065 | Not-determined |
| 18-104_k141_145949 | Not-determined |
| 18-104_k141_124066 | Not-determined |
| 18-104_k141_76160 | Not-determined |
| 18-104_k141_131352 | Not-determined |
| 18-104_k141_6144 | Low-quality |
| 18-104_k141_13314 | Not-determined |
| 18-104_k141_159752 | Low-quality |
| 18-104_k141_34231 | Not-determined |
| 18-104_k141_13315 | Not-determined |
| 18-104_k141_166127 | Not-determined |
| 18-104_k141_41013 | Not-determined |
| 18-104_k141_166131 | Not-determined |
| 18-104_k141_145954 | Not-determined |
| 18-104_k141_111223 | Not-determined |
| 18-104_k141_124073 | Not-determined |
| 18-104_k141_117552 | Low-quality |
| 18-104_k141_54383 | Low-quality |
| 18-104_k141_69105 | Not-determined |
| 18-104_k141_145958 | Not-determined |
| 18-104_k141_152331 | Not-determined |
| 18-104_k141_34241 | Not-determined |
| 18-104_k141_69111 | Not-determined |
| 18-104_k141_83389 | Not-determined |
| 18-104_k141_83391 | Not-determined |
| 18-104_k141_6161 | Not-determined |
| 18-104_k141_54390 | Not-determined |
| 18-104_k141_166141 | Not-determined |
| 18-104_k141_124077 | Not-determined |
| 18-104_k141_103492 | Not-determined |
| 18-104_k141_124079 | Not-determined |
| 18-104_k141_13318 | Not-determined |
| 18-104_k141_103494 | Low-quality |
| 18-104_k141_76178 | Not-determined |
| 18-104_k141_13322 | Not-determined |
| 18-104_k141_54400 | Low-quality |
| 18-104_k141_26998 | Not-determined |
| 18-104_k141_124091 | Low-quality |
| 18-104_k141_19565 | Not-determined |
| 18-104_k141_96858 | Not-determined |
| 18-104_k141_111253 | Not-determined |
| 18-104_k141_138497 | Not-determined |
| 18-104_k141_124098 | Not-determined |
| 18-104_k141_48238 | Not-determined |
| 18-104_k141_159771 | Not-determined |
| 18-104_k141_76193 | Not-determined |
| 18-104_k141_96869 | Not-determined |
| 18-104_k141_166162 | Not-determined |
| 18-104_k141_48244 | Not-determined |
| 18-104_k141_54410 | Not-determined |
| 18-104_k141_124104 | Not-determined |
| 18-104_k141_89800 | Not-determined |
| 18-104_k141_159779 | Not-determined |
| 18-104_k141_13336 | Not-determined |
| 18-104_k141_83398 | Not-determined |
| 18-104_k141_54415 | Not-determined |
| 18-104_k141_159783 | Low-quality |
| 18-104_k141_41038 | Not-determined |
| 18-104_k141_19585 | Not-determined |
| 18-104_k141_89807 | Not-determined |
| 18-104_k141_13340 | Not-determined |
| 18-104_k141_54418 | Not-determined |
| 18-104_k141_159786 | Not-determined |
| 18-104_k141_103513 | Not-determined |
| 18-104_k141_83405 | Not-determined |
| 18-104_k141_131369 | Not-determined |
| 18-104_k141_117559 | Not-determined |
| 18-104_k141_13346 | Not-determined |
| 18-104_k141_34251 | Not-determined |
| 18-104_k141_61679 | Not-determined |
| 18-104_k141_103518 | Not-determined |
| 18-104_k141_6183 | Low-quality |
| 18-104_k141_34253 | Not-determined |
| 18-104_k141_124125 | Not-determined |
| 18-104_k141_54428 | Not-determined |
| 18-104_k141_19591 | Not-determined |
| 18-104_k141_166180 | Not-determined |
| 18-104_k141_54435 | Not-determined |
| 18-104_k141_124130 | Low-quality |
| 18-104_k141_6185 | Not-determined |
| 18-104_k141_138513 | Low-quality |
| 18-104_k141_6188 | Low-quality |
| 18-104_k141_166184 | Low-quality |
| 18-104_k141_19605 | Not-determined |
| 18-104_k141_6193 | Low-quality |
| 18-104_k141_138519 | Not-determined |
| 18-104_k141_166190 | Not-determined |
| 18-104_k141_152371 | Not-determined |
| 18-104_k141_76217 | Not-determined |
| 18-104_k141_76218 | Not-determined |
| 18-104_k141_76219 | Low-quality |
| 18-104_k141_13355 | Low-quality |
| 18-104_k141_103535 | Not-determined |
| 18-104_k141_6198 | Low-quality |
| 18-104_k141_34268 | Not-determined |
| 18-104_k141_166194 | Not-determined |
| 18-104_k141_124143 | Not-determined |
| 18-104_k141_131374 | Not-determined |
| 18-104_k141_48273 | Not-determined |
| 18-104_k141_131376 | Not-determined |
| 18-104_k141_41057 | Not-determined |
| 18-104_k141_76226 | Not-determined |
| 18-104_k141_48278 | Not-determined |
| 18-104_k141_76227 | Not-determined |
| 18-104_k141_6214 | Not-determined |
| 18-104_k141_96896 | Not-determined |
| 18-104_k141_159815 | Not-determined |
| 18-104_k141_138528 | Not-determined |
| 18-104_k141_6220 | Not-determined |
| 18-104_k141_124157 | Not-determined |
| 18-104_k141_19610 | Not-determined |
| 18-104_k141_131383 | Low-quality |
| 18-104_k141_124160 | Not-determined |
| 18-104_k141_34277 | Not-determined |
| 18-104_k141_13372 | Not-determined |
| 18-104_k141_117597 | Not-determined |
| 18-104_k141_145999 | Not-determined |
| 18-104_k141_152383 | Not-determined |
| 18-104_k141_54477 | Not-determined |
| 18-104_k141_48294 | Low-quality |
| 18-104_k141_48298 | Not-determined |
| 18-104_k141_111265 | Not-determined |
| 18-104_k141_146003 | Not-determined |
| 18-104_k141_61703 | Not-determined |
| 18-104_k141_76251 | Not-determined |
| 18-104_k141_166214 | Low-quality |
| 18-104_k141_27030 | Low-quality |
| 18-104_k141_166216 | Not-determined |
| 18-104_k141_166218 | Not-determined |
| 18-104_k141_48302 | Not-determined |
| 18-104_k141_146005 | Not-determined |
| 18-104_k141_138538 | Not-determined |
| 18-104_k141_138539 | Not-determined |
| 18-104_k141_89827 | Not-determined |
| 18-104_k141_111274 | Not-determined |
| 18-104_k141_146010 | Not-determined |
| 18-104_k141_166229 | Not-determined |
| 18-104_k141_69158 | Not-determined |
| 18-104_k141_111278 | Low-quality |
| 18-104_k141_13390 | Not-determined |
| 18-104_k141_124176 | Not-determined |
| 18-104_k141_13391 | Not-determined |
| 18-104_k141_89836 | Not-determined |
| 18-104_k141_13394 | Not-determined |
| 18-104_k141_124179 | Not-determined |
| 18-104_k141_146020 | Not-determined |
| 18-104_k141_159833 | Not-determined |
| 18-104_k141_96916 | Not-determined |
| 18-104_k141_159834 | Not-determined |
| 18-104_k141_69162 | Not-determined |
| 18-104_k141_13399 | Not-determined |
| 18-104_k141_96919 | Not-determined |
| 18-104_k141_117610 | Not-determined |
| 18-104_k141_117612 | Not-determined |
| 18-104_k141_61720 | Not-determined |
| 18-104_k141_69167 | Not-determined |
| 18-104_k141_61722 | Not-determined |
| 18-104_k141_124185 | Not-determined |
| 18-104_k141_111283 | Not-determined |
| 18-104_k141_146031 | Low-quality |
| 18-104_k141_61727 | Not-determined |
| 18-104_k141_111286 | Not-determined |
| 18-104_k141_103543 | Not-determined |
| 18-104_k141_111288 | Not-determined |
| 18-104_k141_152403 | Not-determined |
| 18-104_k141_96926 | Not-determined |
| 18-104_k141_69175 | Not-determined |
| 18-104_k141_96927 | Not-determined |
| 18-104_k141_54511 | Not-determined |
| 18-104_k141_41092 | Not-determined |
| 18-104_k141_138552 | Not-determined |
| 18-104_k141_124196 | Not-determined |
| 18-104_k141_54516 | Not-determined |
| 18-104_k141_6235 | Not-determined |
| 18-104_k141_111297 | Not-determined |
| 18-104_k141_111298 | Not-determined |
| 18-104_k141_166243 | Not-determined |
| 18-104_k141_89855 | Not-determined |
| 18-104_k141_83446 | Not-determined |
| 18-104_k141_96932 | Not-determined |
| 18-104_k141_34303 | Not-determined |
| 18-104_k141_117627 | Not-determined |
| 18-104_k141_41099 | Not-determined |
| 18-104_k141_48320 | Not-determined |
| 18-104_k141_69183 | Not-determined |
| 18-104_k141_13419 | Not-determined |
| 18-104_k141_34309 | Not-determined |
| 18-104_k141_76284 | Not-determined |
| 18-104_k141_159860 | Not-determined |
| 18-104_k141_96935 | Not-determined |
| 18-104_k141_69186 | Not-determined |
| 18-104_k141_138569 | Not-determined |
| 18-104_k141_89861 | Not-determined |
| 18-104_k141_61749 | Not-determined |
| 18-104_k141_13425 | Not-determined |
| 18-104_k141_76287 | Low-quality |
| 18-104_k141_69192 | Not-determined |
| 18-104_k141_89865 | Not-determined |
| 18-104_k141_83454 | Not-determined |
| 18-104_k141_96943 | Low-quality |
| 18-104_k141_166255 | Not-determined |
| 18-104_k141_166256 | Not-determined |
| 18-104_k141_89870 | Not-determined |
| 18-104_k141_138575 | Not-determined |
| 18-104_k141_13431 | Not-determined |
| 18-104_k141_159869 | Not-determined |
| 18-104_k141_54539 | Not-determined |
| 18-104_k141_138578 | Not-determined |
| 18-104_k141_6252 | Not-determined |
| 18-104_k141_166260 | Not-determined |
| 18-104_k141_76294 | Not-determined |
| 18-104_k141_146054 | Not-determined |
| 18-104_k141_61769 | Not-determined |
| 18-104_k141_96960 | Not-determined |
| 18-104_k141_96962 | Not-determined |
| 18-104_k141_159874 | Not-determined |
| 18-104_k141_146059 | Not-determined |
| 18-104_k141_117652 | Not-determined |
| 18-104_k141_41113 | Not-determined |
| 18-104_k141_111310 | Not-determined |
| 18-104_k141_152407 | Not-determined |
| 18-104_k141_131399 | Not-determined |
| 18-104_k141_89880 | Not-determined |
| 18-104_k141_166282 | Not-determined |
| 18-104_k141_27080 | Low-quality |
| 18-104_k141_61783 | Not-determined |
| 18-104_k141_117667 | Not-determined |
| 18-104_k141_61785 | Not-determined |
| 18-104_k141_117672 | Not-determined |
| 18-104_k141_159889 | Not-determined |
| 18-104_k141_146074 | Not-determined |
| 18-104_k141_146075 | Not-determined |
| 18-104_k141_61788 | Not-determined |
| 18-104_k141_89886 | Not-determined |
| 18-104_k141_27082 | Not-determined |
| 18-104_k141_27083 | Not-determined |
| 18-104_k141_89888 | Not-determined |
| 18-104_k141_69218 | Not-determined |
| 18-104_k141_27085 | Low-quality |
| 18-104_k141_76304 | Not-determined |
| 18-104_k141_54562 | Not-determined |
| 18-104_k141_54565 | Not-determined |
| 18-104_k141_83482 | Not-determined |
| 18-104_k141_61796 | Low-quality |
| 18-104_k141_54569 | Not-determined |
| 18-104_k141_96990 | Not-determined |
| 18-104_k141_117684 | Not-determined |
| 18-104_k141_103564 | Not-determined |
| 18-104_k141_96992 | Low-quality |
| 18-104_k141_54574 | Low-quality |
| 18-104_k141_13451 | Not-determined |
| 18-104_k141_27089 | Not-determined |
| 18-104_k141_138594 | Not-determined |
| 18-104_k141_54577 | Not-determined |
| 18-104_k141_27092 | Not-determined |
| 18-104_k141_27093 | Not-determined |
| 18-104_k141_96998 | Not-determined |
| 18-104_k141_83488 | Low-quality |
| 18-104_k141_111326 | Not-determined |
| 18-104_k141_117692 | Not-determined |
| 18-104_k141_103577 | Not-determined |
| 18-104_k141_117696 | Not-determined |
| 18-104_k141_146093 | Not-determined |
| 18-104_k141_76314 | Not-determined |
| 18-104_k141_138599 | Not-determined |
| 18-104_k141_76316 | Low-quality |
| 18-104_k141_97012 | Not-determined |
| 18-104_k141_41150 | Not-determined |
| 18-104_k141_41151 | Not-determined |
| 18-104_k141_13461 | Not-determined |
| 18-104_k141_131415 | Not-determined |
| 18-104_k141_111338 | Not-determined |
| 18-104_k141_19667 | Not-determined |
| 18-104_k141_138601 | Not-determined |
| 18-104_k141_76323 | Not-determined |
| 18-104_k141_146097 | Not-determined |
| 18-104_k141_111341 | Not-determined |
| 18-104_k141_146098 | Not-determined |
| 18-104_k141_146100 | Not-determined |
| 18-104_k141_124254 | Not-determined |
| 18-104_k141_54590 | Low-quality |
| 18-104_k141_117707 | Not-determined |
| 18-104_k141_146106 | Not-determined |
| 18-104_k141_159902 | Low-quality |
| 18-104_k141_48377 | Not-determined |
| 18-104_k141_48378 | Not-determined |
| 18-104_k141_89909 | Not-determined |
| 18-104_k141_117715 | Not-determined |
| 18-104_k141_124261 | Not-determined |
| 18-104_k141_13467 | Not-determined |
| 18-104_k141_89915 | Not-determined |
| 18-104_k141_124267 | Not-determined |
| 18-104_k141_54602 | Not-determined |
| 18-104_k141_54605 | Not-determined |
| 18-104_k141_103592 | Not-determined |
| 18-104_k141_138612 | Not-determined |
| 18-104_k141_54608 | Not-determined |
| 18-104_k141_19685 | Not-determined |
| 18-104_k141_117723 | Not-determined |
| 18-104_k141_146123 | Not-determined |
| 18-104_k141_69240 | Not-determined |
| 18-104_k141_97028 | Not-determined |
| 18-104_k141_61830 | Not-determined |
| 18-104_k141_13477 | Not-determined |
| 18-104_k141_131434 | Not-determined |
| 18-104_k141_138624 | Not-determined |
| 18-104_k141_61838 | Not-determined |
| 18-104_k141_27116 | Not-determined |
| 18-104_k141_61841 | Not-determined |
| 18-104_k141_131437 | Not-determined |
| 18-104_k141_146124 | Not-determined |
| 18-104_k141_13482 | Low-quality |
| 18-104_k141_61843 | Not-determined |
| 18-104_k141_146129 | Not-determined |
| 18-104_k141_111360 | Not-determined |
| 18-104_k141_6315 | Not-determined |
| 18-104_k141_27121 | Not-determined |
| 18-104_k141_27122 | Not-determined |
| 18-104_k141_6320 | Not-determined |
| 18-104_k141_124281 | Not-determined |
| 18-104_k141_61852 | Not-determined |
| 18-104_k141_166301 | Not-determined |
| 18-104_k141_117743 | Not-determined |
| 18-104_k141_13486 | Not-determined |
| 18-104_k141_117747 | Not-determined |
| 18-104_k141_131453 | Low-quality |
| 18-104_k141_117748 | Not-determined |
| 18-104_k141_103626 | Not-determined |
| 18-104_k141_76347 | Not-determined |
| 18-104_k141_103631 | Not-determined |
| 18-104_k141_61863 | Not-determined |
| 18-104_k141_69250 | Low-quality |
| 18-104_k141_61866 | Not-determined |
| 18-104_k141_124289 | Low-quality |
| 18-104_k141_6327 | Not-determined |
| 18-104_k141_6328 | Not-determined |
| 18-104_k141_83498 | Not-determined |
| 18-104_k141_138654 | Not-determined |
| 18-104_k141_41158 | Not-determined |
| 18-104_k141_138660 | Not-determined |
| 18-104_k141_69259 | Not-determined |
| 18-104_k141_146152 | Not-determined |
| 18-104_k141_69261 | Not-determined |
| 18-104_k141_117760 | Low-quality |
| 18-104_k141_41165 | Not-determined |
| 18-104_k141_159945 | Not-determined |
| 18-104_k141_27140 | Not-determined |
| 18-104_k141_159946 | Not-determined |
| 18-104_k141_89930 | Not-determined |
| 18-104_k141_146156 | Not-determined |
| 18-104_k141_76364 | Not-determined |
| 18-104_k141_76366 | Not-determined |
| 18-104_k141_83512 | Low-quality |
| 18-104_k141_89936 | Not-determined |
| 18-104_k141_41175 | Not-determined |
| 18-104_k141_6343 | Low-quality |
| 18-104_k141_54630 | Not-determined |
| 18-104_k141_19735 | Not-determined |
| 18-104_k141_166313 | Not-determined |
| 18-104_k141_117773 | Not-determined |
| 18-104_k141_138672 | Not-determined |
| 18-104_k141_41179 | Not-determined |
| 18-104_k141_6349 | Not-determined |
| 18-104_k141_124309 | Not-determined |
| 18-104_k141_54635 | Not-determined |
| 18-104_k141_19743 | Not-determined |
| 18-104_k141_124316 | Not-determined |
| 18-104_k141_6359 | Not-determined |

| | |
|---|---|
| 18-104_k141_166320 | Not-determined |
| 18-104_k141_69276 | Not-determined |
| 18-104_k141_159977 | Not-determined |
| 18-104_k141_13514 | Not-determined |
| 18-104_k141_131483 | Low-quality |
| 18-104_k141_61905 | Not-determined |
| 18-104_k141_124322 | Not-determined |
| 18-104_k141_61906 | Not-determined |
| 18-104_k141_61907 | Not-determined |
| 18-104_k141_54649 | Not-determined |
| 18-104_k141_83537 | Not-determined |
| 18-104_k141_83539 | Not-determined |
| 18-104_k141_159986 | Not-determined |
| 18-104_k141_13527 | Not-determined |
| 18-104_k141_69286 | Not-determined |
| 18-104_k141_166330 | Not-determined |
| 18-104_k141_6374 | Not-determined |
| 18-104_k141_54659 | Not-determined |
| 18-104_k141_124330 | Not-determined |
| 18-104_k141_111387 | Not-determined |
| 18-104_k141_69289 | Not-determined |
| 18-104_k141_13531 | Not-determined |
| 18-104_k141_13532 | Not-determined |
| 18-104_k141_146176 | Not-determined |
| 18-104_k141_6377 | Low-quality |
| 18-104_k141_159993 | Not-determined |
| 18-104_k141_6378 | Not-determined |
| 18-104_k141_19759 | Not-determined |
| 18-104_k141_6379 | Not-determined |
| 18-104_k141_83551 | Not-determined |
| 18-104_k141_76380 | Not-determined |
| 18-104_k141_103659 | Not-determined |
| 18-104_k141_146180 | Not-determined |
| 18-104_k141_103660 | Not-determined |
| 18-104_k141_152471 | Not-determined |
| 18-104_k141_146184 | Not-determined |
| 18-104_k141_166340 | Not-determined |
| 18-104_k141_13534 | Not-determined |
| 18-104_k141_61913 | Not-determined |
| 18-104_k141_48403 | Not-determined |
| 18-104_k141_166345 | Not-determined |
| 18-104_k141_152477 | Not-determined |
| 18-104_k141_124341 | Not-determined |
| 18-104_k141_48409 | Not-determined |
| 18-104_k141_152481 | Not-determined |
| 18-104_k141_160004 | Not-determined |
| 18-104_k141_48413 | Not-determined |
| 18-104_k141_13540 | Not-determined |
| 18-104_k141_124346 | Low-quality |
| 18-104_k141_48417 | Not-determined |
| 18-104_k141_152486 | Not-determined |
| 18-104_k141_124349 | Not-determined |
| 18-104_k141_103674 | Not-determined |
| 18-104_k141_48420 | Not-determined |
| 18-104_k141_131509 | Not-determined |
| 18-104_k141_117809 | Not-determined |
| 18-104_k141_41220 | Not-determined |
| 18-104_k141_138714 | Not-determined |
| 18-104_k141_41224 | Not-determined |
| 18-104_k141_83575 | Not-determined |
| 18-104_k141_131513 | Not-determined |
| 18-104_k141_124353 | Not-determined |
| 18-104_k141_6399 | Low-quality |
| 18-104_k141_166363 | Not-determined |
| 18-104_k141_166364 | Not-determined |
| 18-104_k141_146207 | Not-determined |
| 18-104_k141_152499 | Not-determined |
| 18-104_k141_124360 | Not-determined |
| 18-104_k141_48434 | Not-determined |
| 18-104_k141_13557 | Not-determined |
| 18-104_k141_111416 | Not-determined |
| 18-104_k141_152502 | Not-determined |
| 18-104_k141_146213 | Not-determined |
| 18-104_k141_117817 | Not-determined |
| 18-104_k141_76391 | Not-determined |
| 18-104_k141_146216 | Low-quality |
| 18-104_k141_76393 | Not-determined |
| 18-104_k141_146218 | Low-quality |
| 18-104_k141_166369 | Low-quality |
| 18-104_k141_76399 | Not-determined |
| 18-104_k141_146222 | Low-quality |
| 18-104_k141_54685 | Not-determined |
| 18-104_k141_131527 | Not-determined |
| 18-104_k141_41231 | Low-quality |
| 18-104_k141_27217 | Not-determined |
| 18-104_k141_97046 | Not-determined |
| 18-104_k141_89966 | Not-determined |
| 18-104_k141_152518 | Not-determined |
| 18-104_k141_48451 | Not-determined |
| 18-104_k141_27225 | Not-determined |
| 18-104_k141_48452 | Not-determined |
| 18-104_k141_48454 | Not-determined |
| 18-104_k141_41245 | Not-determined |
| 18-104_k141_89972 | Not-determined |
| 18-104_k141_152524 | Not-determined |
| 18-104_k141_69336 | Low-quality |
| 18-104_k141_19770 | Not-determined |
| 18-104_k141_48461 | Not-determined |
| 18-104_k141_54708 | Low-quality |
| 18-104_k141_131546 | Not-determined |
| 18-104_k141_131549 | Not-determined |
| 18-104_k141_54711 | Not-determined |
| 18-104_k141_19779 | Not-determined |
| 18-104_k141_103720 | Not-determined |
| 18-104_k141_160050 | Not-determined |
| 18-104_k141_103721 | Not-determined |
| 18-104_k141_166391 | Not-determined |
| 18-104_k141_19784 | Not-determined |
| 18-104_k141_111443 | Not-determined |
| 18-104_k141_166396 | Low-quality |
| 18-104_k141_54715 | Not-determined |
| 18-104_k141_97063 | Not-determined |
| 18-104_k141_166400 | Not-determined |
| 18-104_k141_69355 | Low-quality |
| 18-104_k141_160057 | Not-determined |
| 18-104_k141_34311 | Not-determined |
| 18-104_k141_160060 | Not-determined |
| 18-104_k141_34313 | Not-determined |
| 18-104_k141_48466 | Not-determined |
| 18-104_k141_111451 | Not-determined |
| 18-104_k141_69365 | Not-determined |
| 18-104_k141_27261 | Not-determined |
| 18-104_k141_97078 | Not-determined |
| 18-104_k141_48471 | Not-determined |
| 18-104_k141_97079 | Low-quality |
| 18-104_k141_97085 | Not-determined |
| 18-104_k141_19795 | Not-determined |
| 18-104_k141_146254 | Not-determined |
| 18-104_k141_146255 | Not-determined |
| 18-104_k141_13595 | Not-determined |
| 18-104_k141_117836 | Low-quality |
| 18-104_k141_69379 | Low-quality |
| 18-104_k141_166415 | Not-determined |
| 18-104_k141_117838 | Not-determined |
| 18-104_k141_19802 | Low-quality |
| 18-104_k141_97089 | Not-determined |
| 18-104_k141_131564 | Not-determined |
| 18-104_k141_138753 | Not-determined |
| 18-104_k141_6439 | Low-quality |
| 18-104_k141_69388 | Not-determined |
| 18-104_k141_69389 | Not-determined |
| 18-104_k141_61991 | Not-determined |
| 18-104_k141_138759 | Not-determined |
| 18-104_k141_6448 | Not-determined |
| 18-104_k141_6450 | Not-determined |
| 18-104_k141_41253 | Not-determined |
| 18-104_k141_76424 | Not-determined |
| 18-104_k141_166431 | Not-determined |
| 18-104_k141_166432 | Not-determined |
| 18-104_k141_124387 | Not-determined |
| 18-104_k141_69401 | Not-determined |
| 18-104_k141_6461 | Not-determined |
| 18-104_k141_146267 | Not-determined |
| 18-104_k141_54756 | Not-determined |
| 18-104_k141_90004 | Not-determined |
| 18-104_k141_138773 | Not-determined |
| 18-104_k141_69421 | Not-determined |
| 18-104_k141_62000 | Not-determined |
| 18-104_k141_54763 | Not-determined |
| 18-104_k141_27295 | Not-determined |
| 18-104_k141_6471 | Low-quality |
| 18-104_k141_48497 | Not-determined |
| 18-104_k141_27296 | Not-determined |
| 18-104_k141_48498 | Not-determined |
| 18-104_k141_97112 | Not-determined |
| 18-104_k141_138786 | Not-determined |
| 18-104_k141_76442 | Not-determined |
| 18-104_k141_19848 | Not-determined |
| 18-104_k141_27301 | Not-determined |
| 18-104_k141_138794 | Not-determined |
| 18-104_k141_6479 | Not-determined |
| 18-104_k141_138796 | Not-determined |
| 18-104_k141_62020 | Not-determined |
| 18-104_k141_62021 | Not-determined |
| 18-104_k141_6485 | Not-determined |
| 18-104_k141_19853 | Not-determined |
| 18-104_k141_97122 | Not-determined |
| 18-104_k141_19855 | Not-determined |
| 18-104_k141_48512 | Not-determined |
| 18-104_k141_62025 | Not-determined |
| 18-104_k141_138802 | Low-quality |
| 18-104_k141_146275 | Not-determined |
| 18-104_k141_19858 | Not-determined |
| 18-104_k141_90015 | Not-determined |
| 18-104_k141_160117 | Not-determined |
| 18-104_k141_54767 | Not-determined |
| 18-104_k141_138810 | Not-determined |
| 18-104_k141_41267 | Not-determined |
| 18-104_k141_62034 | Not-determined |
| 18-104_k141_76464 | Not-determined |
| 18-104_k141_97134 | Not-determined |
| 18-104_k141_124412 | Low-quality |
| 18-104_k141_90028 | Not-determined |
| 18-104_k141_160124 | Not-determined |
| 18-104_k141_138816 | Not-determined |
| 18-104_k141_90032 | Not-determined |
| 18-104_k141_54775 | Not-determined |
| 18-104_k141_19867 | Not-determined |
| 18-104_k141_54777 | Low-quality |
| 18-104_k141_34348 | Not-determined |
| 18-104_k141_34356 | Not-determined |
| 18-104_k141_54787 | Not-determined |
| 18-104_k141_6500 | Not-determined |
| 18-104_k141_90039 | Not-determined |
| 18-104_k141_90040 | Low-quality |
| 18-104_k141_48536 | Not-determined |
| 18-104_k141_54791 | Not-determined |
| 18-104_k141_27317 | Not-determined |
| 18-104_k141_138825 | Not-determined |
| 18-104_k141_34362 | Not-determined |
| 18-104_k141_76475 | Low-quality |
| 18-104_k141_97155 | Not-determined |
| 18-104_k141_76477 | Not-determined |
| 18-104_k141_138836 | Low-quality |
| 18-104_k141_6513 | Not-determined |
| 18-104_k141_69474 | Not-determined |
| 18-104_k141_34373 | Low-quality |
| 18-104_k141_97161 | Not-determined |
| 18-104_k141_62050 | Not-determined |
| 18-104_k141_19877 | Low-quality |
| 18-104_k141_138843 | Not-determined |
| 18-104_k141_124444 | Not-determined |
| 18-104_k141_97170 | Not-determined |
| 18-104_k141_62065 | Not-determined |
| 18-104_k141_62067 | Not-determined |
| 18-104_k141_62070 | Not-determined |
| 18-104_k141_160142 | Low-quality |
| 18-104_k141_13631 | Low-quality |
| 18-104_k141_160145 | Not-determined |
| 18-104_k141_54815 | Not-determined |
| 18-104_k141_34379 | Not-determined |
| 18-104_k141_27326 | Not-determined |
| 18-104_k141_69496 | Not-determined |
| 18-104_k141_160155 | Not-determined |
| 18-104_k141_41299 | Not-determined |
| 18-104_k141_138856 | Not-determined |
| 18-104_k141_124469 | Not-determined |
| 18-104_k141_27329 | Not-determined |
| 18-104_k141_41305 | Not-determined |
| 18-104_k141_6535 | Not-determined |
| 18-104_k141_62087 | Not-determined |
| 18-104_k141_6538 | Not-determined |
| 18-104_k141_160160 | Not-determined |
| 18-104_k141_160167 | Not-determined |
| 18-104_k141_62101 | Not-determined |
| 18-104_k141_34404 | Not-determined |
| 18-104_k141_13643 | Low-quality |
| 18-104_k141_97202 | Not-determined |
| 18-104_k141_41320 | Not-determined |
| 18-104_k141_34407 | Not-determined |
| 18-104_k141_62104 | Not-determined |
| 18-104_k141_34410 | Low-quality |
| 18-104_k141_62107 | Not-determined |
| 18-104_k141_54841 | Not-determined |
| 18-104_k141_6553 | Not-determined |
| 18-104_k141_138869 | Low-quality |
| 18-104_k141_160172 | Not-determined |
| 18-104_k141_166500 | Not-determined |
| 18-104_k141_166501 | Not-determined |
| 18-104_k141_166502 | Not-determined |
| 18-104_k141_76501 | Not-determined |
| 18-104_k141_6557 | Low-quality |
| 18-104_k141_6558 | Not-determined |
| 18-104_k141_13651 | Not-determined |
| 18-104_k141_6560 | Not-determined |
| 18-104_k141_76508 | Not-determined |
| 18-104_k141_138880 | Not-determined |
| 18-104_k141_41323 | Not-determined |
| 18-104_k141_160182 | Not-determined |
| 18-104_k141_152545 | Not-determined |
| 18-104_k141_19916 | Not-determined |
| 18-104_k141_166513 | Not-determined |
| 18-104_k141_48581 | Low-quality |
| 18-104_k141_97225 | Low-quality |
| 18-104_k141_124492 | Not-determined |
| 18-104_k141_69533 | Not-determined |
| 18-104_k141_34430 | Not-determined |
| 18-104_k141_54866 | Not-determined |
| 18-104_k141_97229 | Not-determined |
| 18-104_k141_160193 | Not-determined |
| 18-104_k141_160195 | Not-determined |
| 18-104_k141_124497 | Not-determined |
| 18-104_k141_6580 | Not-determined |
| 18-104_k141_160203 | Not-determined |
| 18-104_k141_160204 | Not-determined |
| 18-104_k141_131656 | Not-determined |
| 18-104_k141_131660 | Not-determined |
| 18-104_k141_34446 | Low-quality |
| 18-104_k141_69545 | Low-quality |
| 18-104_k141_48585 | Not-determined |
| 18-104_k141_41341 | Not-determined |
| 18-104_k141_76525 | Not-determined |
| 18-104_k141_54869 | Not-determined |
| 18-104_k141_166530 | Not-determined |
| 18-104_k141_54874 | Not-determined |
| 18-104_k141_138916 | Not-determined |
| 18-104_k141_124518 | Not-determined |
| 18-104_k141_138917 | Not-determined |
| 18-104_k141_54876 | Low-quality |
| 18-104_k141_6587 | Not-determined |
| 18-104_k141_124520 | Not-determined |
| 18-104_k141_54879 | Not-determined |
| 18-104_k141_54880 | Not-determined |
| 18-104_k141_54881 | Not-determined |
| 18-104_k141_62137 | Not-determined |
| 18-104_k141_69564 | Not-determined |
| 18-104_k141_34463 | Not-determined |
| 18-104_k141_34464 | Not-determined |
| 18-104_k141_48606 | Not-determined |
| 18-104_k141_34465 | Not-determined |
| 18-104_k141_138922 | Not-determined |
| 18-104_k141_90141 | Not-determined |
| 18-104_k141_124525 | Not-determined |
| 18-104_k141_131680 | Not-determined |
| 18-104_k141_34468 | Not-determined |
| 18-104_k141_54887 | Not-determined |
| 18-104_k141_41348 | Not-determined |
| 18-104_k141_76538 | Not-determined |
| 18-104_k141_76539 | Low-quality |
| 18-104_k141_117891 | Not-determined |
| 18-104_k141_6607 | Not-determined |
| 18-104_k141_76543 | Not-determined |
| 18-104_k141_117894 | Not-determined |
| 18-104_k141_152586 | Not-determined |
| 18-104_k141_41353 | Not-determined |
| 18-104_k141_117897 | Not-determined |
| 18-104_k141_6615 | Not-determined |
| 18-104_k141_27392 | Not-determined |
| 18-104_k141_76551 | Not-determined |
| 18-104_k141_62147 | Not-determined |
| 18-104_k141_6620 | Not-determined |
| 18-104_k141_62148 | Not-determined |
| 18-104_k141_76560 | Not-determined |
| 18-104_k141_166555 | Not-determined |
| 18-104_k141_103724 | Not-determined |
| 18-104_k141_34476 | Not-determined |
| 18-104_k141_62152 | Not-determined |
| 18-104_k141_124541 | Not-determined |
| 18-104_k141_152602 | Not-determined |
| 18-104_k141_41372 | Not-determined |
| 18-104_k141_13679 | Not-determined |
| 18-104_k141_138945 | Not-determined |
| 18-104_k141_124548 | Not-determined |
| 18-104_k141_41386 | Not-determined |
| 18-104_k141_131697 | Not-determined |
| 18-104_k141_34482 | Not-determined |
| 18-104_k141_6633 | Not-determined |
| 18-104_k141_124552 | Not-determined |
| 18-104_k141_166569 | Not-determined |
| 18-104_k141_13687 | Not-determined |
| 18-104_k141_62165 | Not-determined |
| 18-104_k141_34487 | Not-determined |
| 18-104_k141_13689 | Not-determined |
| 18-104_k141_62171 | Not-determined |
| 18-104_k141_138960 | Not-determined |
| 18-104_k141_76580 | Not-determined |
| 18-104_k141_103739 | Not-determined |
| 18-104_k141_124562 | Not-determined |
| 18-104_k141_166578 | Not-determined |
| 18-104_k141_54920 | Low-quality |
| 18-104_k141_13691 | Low-quality |
| 18-104_k141_166579 | Not-determined |
| 18-104_k141_13693 | Not-determined |
| 18-104_k141_13694 | Not-determined |
| 18-104_k141_103743 | Not-determined |
| 18-104_k141_76586 | Low-quality |
| 18-104_k141_19957 | Not-determined |
| 18-104_k141_138971 | Not-determined |
| 18-104_k141_124573 | Not-determined |
| 18-104_k141_19961 | Not-determined |
| 18-104_k141_166587 | Not-determined |
| 18-104_k141_13701 | Not-determined |
| 18-104_k141_6639 | Low-quality |
| 18-104_k141_54931 | Not-determined |
| 18-104_k141_54934 | Not-determined |
| 18-104_k141_90175 | Not-determined |
| 18-104_k141_13703 | Not-determined |
| 18-104_k141_166592 | Not-determined |
| 18-104_k141_41401 | Not-determined |
| 18-104_k141_152612 | Not-determined |
| 18-104_k141_62194 | Low-quality |
| 18-104_k141_166595 | Not-determined |
| 18-104_k141_152614 | Not-determined |
| 18-104_k141_19967 | Not-determined |
| 18-104_k141_152618 | Not-determined |
| 18-104_k141_166599 | Not-determined |
| 18-104_k141_34518 | Not-determined |
| 18-104_k141_97247 | Not-determined |
| 18-104_k141_54938 | Not-determined |
| 18-104_k141_138993 | Not-determined |
| 18-104_k141_166604 | Not-determined |
| 18-104_k141_54940 | Not-determined |
| 18-104_k141_97249 | Not-determined |
| 18-104_k141_152632 | Not-determined |
| 18-104_k141_62197 | Not-determined |
| 18-104_k141_166607 | Not-determined |
| 18-104_k141_6660 | Not-determined |
| 18-104_k141_76614 | Not-determined |
| 18-104_k141_97254 | Not-determined |
| 18-104_k141_62199 | Not-determined |
| 18-104_k141_27422 | Low-quality |
| 18-104_k141_139003 | Not-determined |
| 18-104_k141_97260 | Not-determined |
| 18-104_k141_27426 | Not-determined |
| 18-104_k141_62202 | Not-determined |
| 18-104_k141_34531 | Not-determined |
| 18-104_k141_27429 | Not-determined |
| 18-104_k141_34534 | Not-determined |
| 18-104_k141_62204 | Not-determined |
| 18-104_k141_166618 | Not-determined |
| 18-104_k141_103764 | Not-determined |
| 18-104_k141_152652 | Not-determined |
| 18-104_k141_27439 | Not-determined |
| 18-104_k141_139014 | Not-determined |
| 18-104_k141_97274 | Not-determined |
| 18-104_k141_139020 | Not-determined |
| 18-104_k141_62212 | Low-quality |
| 18-104_k141_6669 | Low-quality |
| 18-104_k141_62214 | Not-determined |
| 18-104_k141_13739 | Not-determined |
| 18-104_k141_62217 | Not-determined |
| 18-104_k141_19981 | Not-determined |
| 18-104_k141_90201 | Not-determined |
| 18-104_k141_97278 | Not-determined |
| 18-104_k141_90202 | Not-determined |
| 18-104_k141_41406 | Not-determined |
| 18-104_k141_166632 | Not-determined |
| 18-104_k141_6676 | Not-determined |
| 18-104_k141_103785 | Not-determined |
| 18-104_k141_19991 | Not-determined |
| 18-104_k141_34549 | Not-determined |
| 18-104_k141_54946 | Not-determined |
| 18-104_k141_90211 | Not-determined |
| 18-104_k141_6678 | Not-determined |
| 18-104_k141_117950 | Not-determined |
| 18-104_k141_34554 | Not-determined |
| 18-104_k141_27452 | Low-quality |
| 18-104_k141_34555 | Not-determined |
| 18-104_k141_48637 | Not-determined |
| 18-104_k141_34557 | Not-determined |
| 18-104_k141_34558 | Not-determined |
| 18-104_k141_117954 | Not-determined |
| 18-104_k141_48642 | Not-determined |
| 18-104_k141_97282 | Not-determined |
| 18-104_k141_54948 | Not-determined |
| 18-104_k141_54949 | Low-quality |
| 18-104_k141_166646 | Not-determined |
| 18-104_k141_90219 | Not-determined |
| 18-104_k141_13743 | Not-determined |
| 18-104_k141_41418 | Not-determined |
| 18-104_k141_90221 | Not-determined |
| 18-104_k141_90223 | Not-determined |
| 18-104_k141_6681 | Low-quality |
| 18-104_k141_13748 | Not-determined |
| 18-104_k141_6685 | Not-determined |
| 18-104_k141_48652 | Not-determined |
| 18-104_k141_48653 | Not-determined |
| 18-104_k141_27465 | Low-quality |
| 18-104_k141_41429 | Not-determined |
| 18-104_k141_41430 | Not-determined |
| 18-104_k141_6690 | Not-determined |
| 18-104_k141_41432 | Not-determined |
| 18-104_k141_34576 | Not-determined |
| 18-104_k141_97293 | Not-determined |
| 18-104_k141_20006 | Low-quality |
| 18-104_k141_90233 | Not-determined |
| 18-104_k141_13752 | Not-determined |
| 18-104_k141_13754 | Not-determined |
| 18-104_k141_34581 | Not-determined |
| 18-104_k141_41441 | Not-determined |
| 18-104_k141_103814 | Not-determined |
| 18-104_k141_90240 | Not-determined |
| 18-104_k141_62247 | Not-determined |
| 18-104_k141_117972 | Not-determined |
| 18-104_k141_13761 | Not-determined |
| 18-104_k141_20016 | Not-determined |
| 18-104_k141_62255 | Not-determined |
| 18-104_k141_13764 | Not-determined |
| 18-104_k141_62256 | Not-determined |
| 18-104_k141_34591 | Low-quality |
| 18-104_k141_69593 | Not-determined |
| 18-104_k141_54956 | Not-determined |
| 18-104_k141_34598 | Not-determined |
| 18-104_k141_117987 | Not-determined |
| 18-104_k141_69597 | Not-determined |
| 18-104_k141_41453 | Not-determined |
| 18-104_k141_124600 | Low-quality |
| 18-104_k141_131760 | Not-determined |
| 18-104_k141_41459 | Not-determined |
| 18-104_k141_69599 | Not-determined |
| 18-104_k141_152714 | Not-determined |
| 18-104_k141_152716 | Not-determined |
| 18-104_k141_20026 | Not-determined |
| 18-104_k141_152717 | Not-determined |
| 18-104_k141_117994 | Not-determined |
| 18-104_k141_41461 | Not-determined |
| 18-104_k141_27483 | Not-determined |
| 18-104_k141_41464 | Not-determined |
| 18-104_k141_69601 | Low-quality |
| 18-104_k141_166677 | Not-determined |
| 18-104_k141_139084 | Low-quality |
| 18-104_k141_54974 | Low-quality |
| 18-104_k141_124611 | Not-determined |
| 18-104_k141_166682 | Low-quality |
| 18-104_k141_13776 | Not-determined |
| 18-104_k141_152730 | Not-determined |
| 18-104_k141_69610 | Not-determined |
| 18-104_k141_48689 | Not-determined |
| 18-104_k141_34608 | Low-quality |
| 18-104_k141_103831 | Low-quality |
| 18-104_k141_27490 | Low-quality |
| 18-104_k141_90258 | Not-determined |
| 18-104_k141_34615 | Not-determined |
| 18-104_k141_131788 | Low-quality |
| 18-104_k141_131789 | Not-determined |
| 18-104_k141_62281 | Not-determined |
| 18-104_k141_13789 | Not-determined |
| 18-104_k141_34618 | Not-determined |
| 18-104_k141_41482 | Not-determined |
| 18-104_k141_20048 | Not-determined |
| 18-104_k141_13794 | Not-determined |
| 18-104_k141_27497 | Not-determined |
| 18-104_k141_139090 | Not-determined |
| 18-104_k141_166695 | Not-determined |
| 18-104_k141_131796 | Not-determined |
| 18-104_k141_124636 | Not-determined |
| 18-104_k141_69621 | Not-determined |
| 18-104_k141_103844 | Not-determined |
| 18-104_k141_124641 | Not-determined |
| 18-104_k141_27507 | Not-determined |
| 18-104_k141_139105 | Not-determined |
| 18-104_k141_34631 | Not-determined |
| 18-104_k141_90279 | Not-determined |
| 18-104_k141_131807 | Not-determined |
| 18-104_k141_13804 | Not-determined |
| 18-104_k141_152756 | Not-determined |
| 18-104_k141_90283 | Not-determined |
| 18-104_k141_20064 | Not-determined |
| 18-104_k141_6712 | Not-determined |
| 18-104_k141_90284 | Not-determined |
| 18-104_k141_103855 | Not-determined |
| 18-104_k141_103856 | Not-determined |
| 18-104_k141_34632 | Low-quality |
| 18-104_k141_118026 | Low-quality |
| 18-104_k141_152763 | Not-determined |
| 18-104_k141_131821 | Not-determined |
| 18-104_k141_139116 | Not-determined |
| 18-104_k141_20072 | Not-determined |
| 18-104_k141_90303 | Not-determined |
| 18-104_k141_20077 | Not-determined |
| 18-104_k141_90308 | Not-determined |
| 18-104_k141_90310 | Not-determined |
| 18-104_k141_90311 | Not-determined |
| 18-104_k141_152769 | Not-determined |
| 18-104_k141_152770 | Not-determined |
| 18-104_k141_118042 | Not-determined |
| 18-104_k141_48700 | Not-determined |
| 18-104_k141_48701 | Not-determined |
| 18-104_k141_152775 | Not-determined |
| 18-104_k141_48702 | Not-determined |
| 18-104_k141_118045 | Not-determined |
| 18-104_k141_20090 | Not-determined |
| 18-104_k141_48710 | Not-determined |
| 18-104_k141_166721 | Not-determined |
| 18-104_k141_48712 | Not-determined |
| 18-104_k141_20096 | Low-quality |
| 18-104_k141_118050 | Not-determined |
| 18-104_k141_118051 | Not-determined |
| 18-104_k141_166724 | Low-quality |
| 18-104_k141_90326 | Not-determined |
| 18-104_k141_34660 | Not-determined |
| 18-104_k141_118060 | Not-determined |
| 18-104_k141_55015 | Not-determined |
| 18-104_k141_139129 | Not-determined |
| 18-104_k141_13822 | Not-determined |
| 18-104_k141_13823 | Low-quality |
| 18-104_k141_90334 | Not-determined |
| 18-104_k141_103880 | Not-determined |
| 18-104_k141_124687 | Not-determined |
| 18-104_k141_69649 | Not-determined |
| 18-104_k141_152791 | Not-determined |
| 18-104_k141_124690 | Not-determined |
| 18-104_k141_62307 | Not-determined |
| 18-104_k141_62309 | Not-determined |
| 18-104_k141_90341 | Not-determined |
| 18-104_k141_20101 | Not-determined |
| 18-104_k141_34675 | Not-determined |
| 18-104_k141_69656 | Not-determined |
| 18-104_k141_152799 | Not-determined |

| | |
|---|---|
| 18-104_k141_69657 | Not-determined |
| 18-104_k141_152801 | Not-determined |
| 18-104_k141_166750 | Low-quality |
| 18-104_k141_97381 | Not-determined |
| 18-104_k141_27559 | Not-determined |
| 18-104_k141_166755 | Not-determined |
| 18-104_k141_97387 | Not-determined |
| 18-104_k141_131838 | Not-determined |
| 18-104_k141_97389 | Not-determined |
| 18-104_k141_62332 | Not-determined |
| 18-104_k141_131840 | Not-determined |
| 18-104_k141_13835 | Low-quality |
| 18-104_k141_6754 | Low-quality |
| 18-104_k141_20113 | Not-determined |
| 18-104_k141_103894 | Not-determined |
| 18-104_k141_166763 | Not-determined |
| 18-104_k141_6756 | Not-determined |
| 18-104_k141_139163 | Not-determined |
| 18-104_k141_34688 | Not-determined |
| 18-104_k141_48739 | Not-determined |
| 18-104_k141_103898 | Not-determined |
| 18-104_k141_27569 | Not-determined |
| 18-104_k141_34691 | Not-determined |
| 18-104_k141_152829 | Not-determined |
| 18-104_k141_20124 | Not-determined |
| 18-104_k141_103913 | Not-determined |
| 18-104_k141_90360 | Low-quality |
| 18-104_k141_6765 | Not-determined |
| 18-104_k141_103915 | Not-determined |
| 18-104_k141_118085 | Not-determined |
| 18-104_k141_62345 | Not-determined |
| 18-104_k141_62346 | Not-determined |
| 18-104_k141_62347 | Not-determined |
| 18-104_k141_103920 | Not-determined |
| 18-104_k141_166776 | Not-determined |
| 18-104_k141_103924 | Not-determined |
| 18-104_k141_55028 | Not-determined |
| 18-104_k141_103925 | Not-determined |
| 18-104_k141_118094 | Not-determined |
| 18-104_k141_34710 | Not-determined |
| 18-104_k141_152833 | Not-determined |
| 18-104_k141_118098 | Not-determined |
| 18-104_k141_103927 | Not-determined |
| 18-104_k141_152839 | Low-quality |
| 18-104_k141_131860 | Low-quality |
| 18-104_k141_131861 | Not-determined |
| 18-104_k141_90363 | Not-determined |
| 18-104_k141_20138 | Not-determined |
| 18-104_k141_90369 | Not-determined |
| 18-104_k141_62360 | Not-determined |
| 18-104_k141_20148 | Not-determined |
| 18-104_k141_6782 | Not-determined |
| 18-104_k141_6783 | Not-determined |
| 18-104_k141_48759 | Not-determined |
| 18-104_k141_34725 | Not-determined |
| 18-104_k141_103944 | Not-determined |
| 18-104_k141_103946 | Not-determined |
| 18-104_k141_62364 | Not-determined |
| 18-104_k141_103948 | Not-determined |
| 18-104_k141_124724 | Not-determined |
| 18-104_k141_139196 | Not-determined |
| 18-104_k141_118111 | Not-determined |
| 18-104_k141_41491 | Not-determined |
| 18-104_k141_34732 | Low-quality |
| 18-104_k141_34733 | Not-determined |
| 18-104_k141_152869 | Not-determined |
| 18-104_k141_62381 | Not-determined |
| 18-104_k141_62382 | Not-determined |
| 18-104_k141_62383 | Not-determined |
| 18-104_k141_34742 | Not-determined |
| 18-104_k141_27622 | Not-determined |
| 18-104_k141_124747 | Not-determined |
| 18-104_k141_124749 | Not-determined |
| 18-104_k141_124750 | Not-determined |
| 18-104_k141_124758 | Not-determined |
| 18-104_k141_13865 | Not-determined |
| 18-104_k141_152883 | Not-determined |
| 18-104_k141_62389 | Not-determined |
| 18-104_k141_13867 | Not-determined |
| 18-104_k141_62390 | Not-determined |
| 18-104_k141_62392 | Not-determined |
| 18-104_k141_124761 | Low-quality |
| 18-104_k141_13870 | Not-determined |
| 18-104_k141_27626 | Not-determined |
| 18-104_k141_152887 | Not-determined |
| 18-104_k141_103975 | Not-determined |
| 18-104_k141_6820 | Not-determined |
| 18-104_k141_103976 | Low-quality |
| 18-104_k141_90404 | Not-determined |
| 18-104_k141_152893 | Not-determined |
| 18-104_k141_152895 | Not-determined |
| 18-104_k141_131883 | Not-determined |
| 18-104_k141_131884 | Not-determined |
| 18-104_k141_166797 | Not-determined |
| 18-104_k141_41515 | Not-determined |
| 18-104_k141_41516 | Not-determined |
| 18-104_k141_152902 | Not-determined |
| 18-104_k141_124778 | Not-determined |
| 18-104_k141_41524 | Low-quality |
| 18-104_k141_118147 | Not-determined |
| 18-104_k141_124780 | Not-determined |
| 18-104_k141_13888 | Not-determined |
| 18-104_k141_139242 | Not-determined |
| 18-104_k141_166807 | Not-determined |
| 18-104_k141_118154 | Not-determined |
| 18-104_k141_131892 | Not-determined |
| 18-104_k141_118155 | Not-determined |
| 18-104_k141_131894 | Not-determined |
| 18-104_k141_62404 | Not-determined |
| 18-104_k141_90424 | Not-determined |
| 18-104_k141_152906 | Not-determined |
| 18-104_k141_27636 | Low-quality |
| 18-104_k141_118163 | Not-determined |
| 18-104_k141_62412 | Not-determined |
| 18-104_k141_62413 | Not-determined |
| 18-104_k141_118170 | Not-determined |
| 18-104_k141_20165 | Not-determined |
| 18-104_k141_90432 | Not-determined |
| 18-104_k141_41529 | Not-determined |
| 18-104_k141_34772 | Not-determined |
| 18-104_k141_152924 | Not-determined |
| 18-104_k141_41531 | Not-determined |
| 18-104_k141_20171 | Not-determined |
| 18-104_k141_13911 | Not-determined |
| 18-104_k141_62421 | Not-determined |
| 18-104_k141_13912 | Not-determined |
| 18-104_k141_139249 | Not-determined |
| 18-104_k141_41542 | Not-determined |
| 18-104_k141_41544 | Not-determined |
| 18-104_k141_6829 | Not-determined |
| 18-104_k141_131908 | Not-determined |
| 18-104_k141_124815 | Not-determined |
| 18-104_k141_131912 | Not-determined |
| 18-104_k141_139256 | Not-determined |
| 18-104_k141_131914 | Low-quality |
| 18-104_k141_27653 | Not-determined |
| 18-104_k141_139259 | Not-determined |
| 18-104_k141_62426 | Not-determined |
| 18-104_k141_55031 | Low-quality |
| 18-104_k141_97443 | Not-determined |
| 18-104_k141_20182 | Not-determined |
| 18-104_k141_55041 | Not-determined |
| 18-104_k141_34807 | Not-determined |
| 18-104_k141_27666 | Not-determined |
| 18-104_k141_6850 | Low-quality |
| 18-104_k141_27668 | Low-quality |
| 18-104_k141_6851 | Not-determined |
| 18-104_k141_124841 | Low-quality |
| 18-104_k141_41572 | Not-determined |
| 18-104_k141_90462 | Not-determined |
| 18-104_k141_152969 | Not-determined |
| 18-104_k141_139282 | Not-determined |
| 18-104_k141_34819 | Not-determined |
| 18-104_k141_131933 | Not-determined |
| 18-104_k141_166837 | Not-determined |
| 18-104_k141_104004 | Not-determined |
| 18-104_k141_139290 | Not-determined |
| 18-104_k141_27681 | Not-determined |
| 18-104_k141_20201 | Not-determined |
| 18-104_k141_20203 | Not-determined |
| 18-104_k141_104011 | Not-determined |
| 18-104_k141_104012 | Not-determined |
| 18-104_k141_27683 | Not-determined |
| 18-104_k141_55049 | Not-determined |
| 18-104_k141_131941 | Not-determined |
| 18-104_k141_166845 | Not-determined |
| 18-104_k141_152982 | Not-determined |
| 18-104_k141_152983 | Low-quality |
| 18-104_k141_139293 | Not-determined |
| 18-104_k141_124865 | Not-determined |
| 18-104_k141_166852 | Not-determined |
| 18-104_k141_62461 | Not-determined |
| 18-104_k141_104019 | Not-determined |
| 18-104_k141_55057 | Not-determined |
| 18-104_k141_124880 | Not-determined |
| 18-104_k141_131967 | Not-determined |
| 18-104_k141_104028 | Not-determined |
| 18-104_k141_166859 | Not-determined |
| 18-104_k141_152996 | Low-quality |
| 18-104_k141_90490 | Not-determined |
| 18-104_k141_104032 | Not-determined |
| 18-104_k141_131969 | Not-determined |
| 18-104_k141_118218 | Not-determined |
| 18-104_k141_97457 | Low-quality |
| 18-104_k141_90494 | Not-determined |
| 18-104_k141_55063 | Not-determined |
| 18-104_k141_62473 | Not-determined |
| 18-104_k141_152999 | Low-quality |
| 18-104_k141_62476 | Not-determined |
| 18-104_k141_55068 | Not-determined |
| 18-104_k141_153004 | Not-determined |
| 18-104_k141_62479 | Not-determined |
| 18-104_k141_20221 | Not-determined |
| 18-104_k141_41586 | Not-determined |
| 18-104_k141_131988 | Not-determined |
| 18-104_k141_27703 | Not-determined |
| 18-104_k141_104050 | Not-determined |
| 18-104_k141_55076 | Not-determined |
| 18-104_k141_41596 | Low-quality |
| 18-104_k141_62488 | Low-quality |
| 18-104_k141_118228 | Not-determined |
| 18-104_k141_6867 | Not-determined |
| 18-104_k141_20234 | Not-determined |
| 18-104_k141_104058 | Not-determined |
| 18-104_k141_6869 | Not-determined |
| 18-104_k141_20235 | Not-determined |
| 18-104_k141_90515 | Not-determined |
| 18-104_k141_124886 | Not-determined |
| 18-104_k141_41606 | Not-determined |
| 18-104_k141_124887 | Not-determined |
| 18-104_k141_104064 | Not-determined |
| 18-104_k141_27712 | Not-determined |
| 18-104_k141_20242 | Not-determined |
| 18-104_k141_27714 | Low-quality |
| 18-104_k141_41610 | Not-determined |
| 18-104_k141_6879 | Not-determined |
| 18-104_k141_55086 | Not-determined |
| 18-104_k141_27717 | Low-quality |
| 18-104_k141_118242 | Not-determined |
| 18-104_k141_124896 | Not-determined |
| 18-104_k141_97474 | Not-determined |
| 18-104_k141_118245 | Not-determined |
| 18-104_k141_27720 | Low-quality |
| 18-104_k141_97476 | Not-determined |
| 18-104_k141_118250 | Not-determined |
| 18-104_k141_166874 | Not-determined |
| 18-104_k141_124907 | Not-determined |
| 18-104_k141_104078 | Not-determined |
| 18-104_k141_124908 | Not-determined |
| 18-104_k141_90530 | Not-determined |
| 18-104_k141_97481 | Not-determined |
| 18-104_k141_97482 | Not-determined |
| 18-104_k141_27727 | Not-determined |
| 18-104_k141_20270 | Not-determined |
| 18-104_k141_118258 | Not-determined |
| 18-104_k141_118260 | Not-determined |
| 18-104_k141_118261 | Not-determined |
| 18-104_k141_132005 | Not-determined |
| 18-104_k141_27741 | Not-determined |
| 18-104_k141_166891 | Not-determined |
| 18-104_k141_20283 | Not-determined |
| 18-104_k141_97497 | Not-determined |
| 18-104_k141_41626 | Not-determined |
| 18-104_k141_104100 | Not-determined |
| 18-104_k141_118283 | Low-quality |
| 18-104_k141_132023 | Low-quality |
| 18-104_k141_6909 | Not-determined |
| 18-104_k141_124942 | Not-determined |
| 18-104_k141_132028 | Not-determined |
| 18-104_k141_132031 | Not-determined |
| 18-104_k141_104113 | Not-determined |
| 18-104_k141_20304 | Not-determined |
| 18-104_k141_6917 | Not-determined |
| 18-104_k141_166915 | Low-quality |
| 18-104_k141_41640 | Not-determined |
| 18-104_k141_41643 | Not-determined |
| 18-104_k141_118298 | Not-determined |
| 18-104_k141_104122 | Low-quality |
| 18-104_k141_118300 | Low-quality |
| 18-104_k141_62508 | Not-determined |
| 18-104_k141_20314 | Not-determined |
| 18-104_k141_153033 | Not-determined |
| 18-104_k141_62512 | Not-determined |
| 18-104_k141_27781 | Not-determined |
| 18-104_k141_104144 | Not-determined |
| 18-104_k141_27784 | Not-determined |
| 18-104_k141_20322 | Not-determined |
| 18-104_k141_55103 | Low-quality |
| 18-104_k141_6923 | Low-quality |
| 18-104_k141_41659 | Not-determined |
| 18-104_k141_55114 | Not-determined |
| 18-104_k141_118303 | Not-determined |
| 18-104_k141_41660 | Not-determined |
| 18-104_k141_118304 | Not-determined |
| 18-104_k141_41663 | Not-determined |
| 18-104_k141_118309 | Not-determined |
| 18-104_k141_41665 | Not-determined |
| 18-104_k141_90539 | Not-determined |
| 18-104_k141_104169 | Not-determined |
| 18-104_k141_132090 | Not-determined |
| 18-104_k141_124980 | Low-quality |
| 18-104_k141_153065 | Not-determined |
| 18-104_k141_27805 | Not-determined |
| 18-104_k141_27806 | Not-determined |
| 18-104_k141_90549 | Not-determined |
| 18-104_k141_153070 | Not-determined |
| 18-104_k141_166945 | Not-determined |
| 18-104_k141_90552 | Not-determined |
| 18-104_k141_27810 | Low-quality |
| 18-104_k141_132096 | Not-determined |
| 18-104_k141_132098 | Not-determined |
| 18-104_k141_104181 | Not-determined |
| 18-104_k141_20344 | Not-determined |
| 18-104_k141_55128 | Not-determined |
| 18-104_k141_132102 | Not-determined |
| 18-104_k141_153080 | Not-determined |
| 18-104_k141_166960 | Not-determined |
| 18-104_k141_62548 | Not-determined |
| 18-104_k141_166964 | Not-determined |
| 18-104_k141_62550 | Not-determined |
| 18-104_k141_62551 | Not-determined |
| 18-104_k141_62552 | Not-determined |
| 18-104_k141_118313 | Not-determined |
| 18-104_k141_97511 | Low-quality |
| 18-104_k141_132117 | Not-determined |
| 18-104_k141_62558 | Not-determined |
| 18-104_k141_55142 | Not-determined |
| 18-104_k141_118320 | Not-determined |
| 18-104_k141_97516 | Not-determined |
| 18-104_k141_153089 | Low-quality |
| 18-104_k141_97519 | Not-determined |
| 18-104_k141_97521 | Not-determined |
| 18-104_k141_132124 | Not-determined |
| 18-104_k141_41695 | Low-quality |
| 18-104_k141_62570 | Not-determined |
| 18-104_k141_41702 | Not-determined |
| 18-104_k141_41703 | Not-determined |
| 18-104_k141_27840 | Not-determined |
| 18-104_k141_132137 | Not-determined |
| 18-104_k141_132140 | Not-determined |
| 18-104_k141_132141 | Not-determined |
| 18-104_k141_132142 | Low-quality |
| 18-104_k141_153096 | Not-determined |
| 18-104_k141_132144 | Low-quality |
| 18-104_k141_125002 | Not-determined |
| 18-104_k141_153098 | Not-determined |
| 18-104_k141_62592 | Not-determined |
| 18-104_k141_125007 | Not-determined |
| 18-104_k141_118375 | Not-determined |
| 18-104_k141_125009 | Not-determined |
| 18-104_k141_125010 | Not-determined |
| 18-104_k141_132166 | Not-determined |
| 18-104_k141_153112 | Not-determined |
| 18-104_k141_118391 | Not-determined |
| 18-104_k141_104214 | Not-determined |
| 18-104_k141_166970 | Not-determined |
| 18-104_k141_118394 | Not-determined |
| 18-104_k141_132175 | Not-determined |
| 18-104_k141_153124 | Not-determined |
| 18-104_k141_132182 | Not-determined |
| 18-104_k141_104224 | Not-determined |
| 18-104_k141_104225 | Not-determined |
| 18-104_k141_41732 | Not-determined |
| 18-104_k141_104232 | Not-determined |
| 18-104_k141_20377 | Not-determined |
| 18-104_k141_125042 | Not-determined |
| 18-104_k141_153137 | Low-quality |
| 18-104_k141_153138 | Not-determined |
| 18-104_k141_166981 | Not-determined |
| 18-104_k141_125051 | Not-determined |
| 18-104_k141_125057 | Not-determined |
| 18-104_k141_62633 | Not-determined |
| 18-104_k141_125064 | Not-determined |
| 18-104_k141_55186 | Not-determined |
| 18-104_k141_125073 | Not-determined |
| 18-104_k141_41749 | Not-determined |
| 18-104_k141_153154 | Not-determined |
| 18-104_k141_132185 | Not-determined |
| 18-104_k141_41754 | Not-determined |
| 18-104_k141_132191 | Not-determined |
| 18-104_k141_55197 | Not-determined |
| 18-104_k141_55198 | Low-quality |
| 18-104_k141_118416 | Not-determined |
| 18-104_k141_55203 | Not-determined |
| 18-104_k141_153160 | Not-determined |
| 18-104_k141_104244 | Not-determined |
| 18-104_k141_125082 | Not-determined |
| 18-104_k141_20400 | Not-determined |
| 18-104_k141_20403 | Not-determined |
| 18-104_k141_20404 | Not-determined |
| 18-104_k141_153165 | Not-determined |
| 18-104_k141_55210 | Not-determined |
| 18-104_k141_125094 | Low-quality |
| 18-104_k141_55218 | Not-determined |
| 18-104_k141_55220 | Not-determined |
| 18-104_k141_153173 | Not-determined |
| 18-104_k141_104261 | Not-determined |
| 18-104_k141_20425 | Not-determined |
| 18-104_k141_132237 | Not-determined |
| 18-104_k141_104278 | Not-determined |
| 18-104_k141_166990 | Not-determined |
| 18-101-final-viral-combined_k141_354899\|\|full | Medium-quality |
| 18-104_k141_62637 | Not-determined |
| 18-104_k141_41788 | Low-quality |
| 18-104_k141_41794 | Not-determined |
| 18-104_k141_55273 | Not-determined |
| 18-104_k141_104296 | Not-determined |
| 18-104_k141_55280 | Not-determined |
| 18-104_k141_55281 | Not-determined |
| 18-104_k141_132280 | Not-determined |
| 18-104_k141_153178 | Low-quality |
| 18-104_k141_104303 | Not-determined |
| 18-104_k141_104304 | Not-determined |
| 18-104_k141_167002 | Not-determined |
| 18-104_k141_104313 | Not-determined |
| 18-104_k141_104318 | Not-determined |
| 18-104_k141_62656 | Low-quality |
| 18-104_k141_20430 | Not-determined |
| 18-104_k141_62660 | Not-determined |
| 18-104_k141_153196 | Not-determined |
| 18-104_k141_153197 | Not-determined |
| 18-104_k141_104329 | Not-determined |
| 18-104_k141_6963 | Not-determined |
| 18-104_k141_167018 | Not-determined |
| 18-104_k141_153206 | Not-determined |
| 18-104_k141_167020 | Not-determined |
| 18-104_k141_167026 | Not-determined |
| 18-104_k141_167029 | Not-determined |
| 18-104_k141_167031 | Not-determined |
| 18-104_k141_132285 | Not-determined |
| 18-104_k141_153215 | Not-determined |
| 18-104_k141_125121 | Not-determined |
| 18-104_k141_153219 | Not-determined |
| 18-104_k141_167039 | Not-determined |
| 18-104_k141_62675 | Not-determined |
| 18-104_k141_167044 | Not-determined |
| 18-104_k141_62677 | Low-quality |
| 18-104_k141_20441 | Not-determined |
| 18-104_k141_104345 | Not-determined |
| 18-104_k141_125134 | Not-determined |
| 18-104_k141_167051 | Not-determined |
| 18-104_k141_167056 | Not-determined |
| 18-104_k141_104350 | Not-determined |
| 18-104_k141_167063 | Not-determined |
| 18-104_k141_153241 | Not-determined |
| 18-104_k141_125142 | Not-determined |
| 18-104_k141_55306 | Low-quality |
| 18-104_k141_62686 | Not-determined |
| 18-104_k141_55314 | Not-determined |
| 18-104_k141_55316 | Not-determined |
| 18-104_k141_167075 | Not-determined |
| 18-104_k141_167077 | Not-determined |
| 18-104_k141_167087 | Not-determined |
| 18-104_k141_125154 | Not-determined |
| 18-104_k141_104370 | Low-quality |
| 18-104_k141_167091 | Not-determined |
| 18-104_k141_104373 | Not-determined |
| 18-104_k141_167100 | Not-determined |
| 18-104_k141_167104 | Not-determined |
| 18-104_k141_167106 | Not-determined |
| 18-104_k141_167107 | Not-determined |
| 18-104_k141_20472 | Not-determined |
| 18-104_k141_55320 | Not-determined |
| 18-104_k141_167114 | Not-determined |
| 18-104_k141_125165 | Not-determined |
| 18-104_k141_167116 | Not-determined |
| 18-104_k141_125166 | Low-quality |
| 18-104_k141_167124 | Not-determined |
| 18-104_k141_55333 | Low-quality |
| 18-104_k141_167127 | Not-determined |
| 18-104_k141_125170 | Not-determined |
| 18-104_k141_167130 | Not-determined |
| 18-104_k141_125172 | Not-determined |
| 18-104_k141_167133 | Not-determined |
| 18-104_k141_167136 | Not-determined |
| 18-104_k141_167140 | Not-determined |
| 18-104_k141_167143 | Not-determined |
| 18-104_k141_167146 | Not-determined |
| 18-104_k141_167150 | Not-determined |
| 18-104_k141_167154 | Not-determined |
| 18-104_k141_125180 | Not-determined |
| 18-104_k141_55345 | Not-determined |
| 18-104_k141_167164 | Not-determined |
| 18-104_k141_167167 | Not-determined |
| 18-104_k141_104401 | Not-determined |
| 18-104_k141_55365 | Not-determined |
| 18-104_k141_20506 | Not-determined |
| 18-104_k141_20509 | Not-determined |
| 18-104_k141_132330 | Not-determined |
| 18-104_k141_104419 | Not-determined |
| 18-104_k141_20519 | Not-determined |
| 18-104_k141_20523 | Not-determined |
| 18-104_k141_20529 | Not-determined |
| 18-104_k141_125205 | Low-quality |
| 18-104_k141_132336 | Not-determined |
| 18-104_k141_125216 | Not-determined |
| 18-104_k141_104441 | Low-quality |
| 18-104_k141_20542 | Low-quality |
| 18-104_k141_104447 | Low-quality |
| 18-104_k141_125227 | Not-determined |
| 18-104_k141_125232 | Not-determined |
| 18-104_k141_104460 | Low-quality |
| 18-104_k141_20569 | Not-determined |
| 18-104_k141_55402 | Not-determined |
| 18-104_k141_20571 | Not-determined |
| 18-104_k141_20573 | Not-determined |
| 18-104_k141_125241 | Not-determined |
| 18-104_k141_132342 | Not-determined |
| 18-104_k141_20588 | Not-determined |
| 18-104_k141_55415 | Not-determined |
| 18-104_k141_20599 | Not-determined |
| 18-104_k141_104468 | Low-quality |
| 18-104_k141_104470 | Not-determined |
| 18-104_k141_104471 | Not-determined |
| 18-104_k141_132345 | Low-quality |
| 18-104_k141_20616 | Not-determined |
| 18-104_k141_104482 | Not-determined |
| 18-104_k141_104484 | Not-determined |
| 18-104_k141_55448 | Not-determined |
| 18-104_k141_104487 | Not-determined |
| 18-104_k141_125258 | Not-determined |
| 18-104_k141_125260 | Not-determined |
| 18-104_k141_125261 | Low-quality |
| 18-104_k141_125266 | Not-determined |
| 18-104_k141_20629 | Not-determined |
| 18-104_k141_55476 | Not-determined |
| 18-104_k141_125277 | Not-determined |
| 18-104_k141_55487 | Not-determined |
| 18-104_k141_55489 | Not-determined |
| 18-104_k141_55490 | Not-determined |
| 18-104_k141_55492 | Not-determined |
| 18-104_k141_125288 | Low-quality |
| 18-104_k141_55504 | Low-quality |
| 18-104_k141_20667 | Not-determined |
| 18-104_k141_20671 | Not-determined |
| 18-104_k141_55507 | Not-determined |
| 18-104_k141_20673 | Not-determined |
| 18-104_k141_125302 | Not-determined |
| 18-104_k141_20681 | Not-determined |
| 18-104_k141_125310 | Not-determined |
| 18-104_k141_55511 | Not-determined |
| 18-104_k141_55516 | Not-determined |
| 18-104_k141_55529 | Not-determined |
| 18-104_k141_20705 | Not-determined |
| 18-104_k141_20707 | Not-determined |
| 18-104_k141_20708 | Not-determined |
| 18-104_k141_55552 | Not-determined |
| 18-104_k141_55556 | Not-determined |
| 18-104_k141_20716 | Not-determined |
| 18-104_k141_55563 | Not-determined |
| 18-104_k141_55585 | Not-determined |
| 18-104_k141_55591 | Not-determined |
| 18-104_k141_55604 | Not-determined |
| 18-104_k141_55610 | Not-determined |
| 18-104_k141_20750 | Not-determined |
| 18-104_k141_55611 | Not-determined |
| 18-104_k141_20755 | Not-determined |
| 18-104_k141_55625 | Not-determined |
| 18-104_k141_55634 | Not-determined |
| 18-104_k141_55640 | Not-determined |
| 18-104_k141_55662 | Not-determined |
| 18-104_k141_55664 | Not-determined |
| 18-104_k141_55665 | Not-determined |
| 18-104_k141_20772 | Not-determined |
| 18-104_k141_20775 | Not-determined |
| 18-104_k141_55677 | Not-determined |
| 18-104_k141_125348 | Not-determined |
| 18-104_k141_125352 | Not-determined |
| 18-104_k141_20789 | Not-determined |
| 18-104_k141_55685 | Not-determined |
| 18-104_k141_125358 | Low-quality |
| 18-104_k141_55702 | Not-determined |
| 18-104_k141_55715 | Low-quality |
| 18-104_k141_55716 | Not-determined |
| 18-104_k141_55722 | Not-determined |
| 18-104_k141_20830 | Not-determined |
| 18-104_k141_20841 | Not-determined |
| 18-104_k141_20848 | Not-determined |
| 18-104_k141_20863 | Not-determined |
| 18-104_k141_20865 | Not-determined |
| 18-104_k141_20873 | Not-determined |
| 18-106-final-viral-combined_k141_58886\|\|full | Not-determined |
| KM-final-viral-combined_k141_179363\|\|full | Medium-quality |
| 18-106-final-viral-combined_k141_6450\|\|full | Not-determined |
| 18-106-final-viral-combined_k141_136044\|\|full | Not-determined |
| 18-106-final-viral-combined_k141_14387\|\|full | Not-determined |
| 18-106-final-viral-combined_k141_14416\|\|full | Not-determined |
| 18-106-final-viral-combined_k141_55841\|\|full | Not-determined |
| 18-105-final-viral-combined_k141_147993\|\|full | Medium-quality |
| 18-106-final-viral-combined_k141_139327\|\|full | Not-determined |
| 18-104-final-viral-combined_k141_92005\|\|full | Medium-quality |
| 18-106-final-viral-combined_k141_140770\|\|full | Low-quality |
| 18-106-final-viral-combined_k141_140808\|\|full | Low-quality |
| 18-106-final-viral-combined_k141_129167\|\|full | Not-determined |
| 18-106-final-viral-combined_k141_123301\|\|full | Not-determined |
| 18-106-final-viral-combined_k141_64598\|\|full | Not-determined |
| 18-106-final-viral-combined_k141_11753\|\|full | Not-determined |
| 18-106-final-viral-combined_k141_35241\|\|full | Not-determined |
| 18-106-final-viral-combined_k141_123304\|\|full | Not-determined |
| 18-106-final-viral-combined_k141_64605\|\|full | Low-quality |
| 18-106-final-viral-combined_k141_29377\|\|full | Not-determined |
| 18-106-final-viral-combined_k141_129179\|\|full | Low-quality |
| 18-106-final-viral-combined_k141_58737\|\|full | Low-quality |
| 18-106-final-viral-combined_k141_5888\|\|full | Low-quality |
| 18-106-final-viral-combined_k141_23502\|\|full | Not-determined |
| 18-106-final-viral-combined_k141_23503\|\|full | Not-determined |
| 18-106-final-viral-combined_k141_93968\|\|full | Not-determined |
| 18-106-final-viral-combined_k141_64607\|\|full | Low-quality |
| 18-106-final-viral-combined_k141_82240\|\|full | Not-determined |

| | |
|---|---|
| 18-106-final-viral-combined_k141_11771\|\|full | Low-quality |
| 18-106-final-viral-combined_k141_105721\|\|full | Low-quality |
| 18-106-final-viral-combined_k141_135061\|\|full | Not-determined |
| 18-106-final-viral-combined_k141_99848\|\|full | Low-quality |
| 18-106-final-viral-combined_k141_82256\|\|full | Not-determined |
| 18-106-final-viral-combined_k141_60\|\|full | Not-determined |
| 18-106-final-viral-combined_k141_23525\|\|full | Not-determined |
| 18-106-final-viral-combined_k141_11789\|\|full | Not-determined |
| 18-106-final-viral-combined_k141_11791\|\|full | Low-quality |
| 18-106-final-viral-combined_k141_52862\|\|full | Not-determined |
| 18-106-final-viral-combined_k141_105738\|\|full | Not-determined |
| 18-106-final-viral-combined_k141_88114\|\|full | Not-determined |
| 18-106-final-viral-combined_k141_123339\|\|full | Low-quality |
| 18-106-final-viral-combined_k141_52870\|\|full | Low-quality |
| 18-106-final-viral-combined_k141_76381\|\|full | Not-determined |
| 18-106-final-viral-combined_k141_17667\|\|full | Low-quality |
| 18-106-final-viral-combined_k141_88\|\|full | Not-determined |
| 18-106-final-viral-combined_k141_17670\|\|full | Not-determined |
| 18-106-final-viral-combined_k141_41169\|\|full | Low-quality |
| 18-106-final-viral-combined_k141_123345\|\|full | Low-quality |
| 18-106-final-viral-combined_k141_58768\|\|full | Low-quality |
| 18-106-final-viral-combined_k141_117447\|\|full | Not-determined |
| 18-106-final-viral-combined_k141_117448\|\|full | Not-determined |
| 18-106-final-viral-combined_k141_76398\|\|full | Low-quality |
| 18-106-final-viral-combined_k141_58774\|\|full | Low-quality |
| 18-106-final-viral-combined_k141_46890\|\|full | Not-determined |
| 18-106-final-viral-combined_k141_17690\|\|full | Not-determined |
| 18-106-final-viral-combined_k141_117461\|\|full | Not-determined |
| 18-106-final-viral-combined_k141_41192\|\|full | Low-quality |
| 18-106-final-viral-combined_k141_76411\|\|full | Low-quality |
| 18-106-final-viral-combined_k141_94018\|\|full | Low-quality |
| 18-106-final-viral-combined_k141_105760\|\|full | Not-determined |
| 18-106-final-viral-combined_k141_88170\|\|full | Not-determined |
| 18-106-final-viral-combined_k141_29463\|\|full | Not-determined |
| 18-106-final-viral-combined_k141_129255\|\|full | Low-quality |
| 18-106-final-viral-combined_k141_35319\|\|full | Not-determined |
| 18-106-final-viral-combined_k141_111649\|\|full | Low-quality |
| 18-106-final-viral-combined_k141_129259\|\|full | Low-quality |
| 18-106-final-viral-combined_k141_123370\|\|full | Low-quality |
| 18-106-final-viral-combined_k141_111656\|\|full | Not-determined |
| 18-106-final-viral-combined_k141_17730\|\|full | Not-determined |
| 18-106-final-viral-combined_k141_130\|\|full | Low-quality |
| 18-106-final-viral-combined_k141_41208\|\|full | Low-quality |
| 18-106-final-viral-combined_k141_129263\|\|full | Not-determined |
| 18-106-final-viral-combined_k141_111657\|\|full | Not-determined |
| 18-106-final-viral-combined_k141_111658\|\|full | Low-quality |
| 18-106-final-viral-combined_k141_94058\|\|full | Not-determined |
| 18-106-final-viral-combined_k141_123381\|\|full | Low-quality |
| 18-106-final-viral-combined_k141_111661\|\|full | Not-determined |
| 18-106-final-viral-combined_k141_135156\|\|full | Not-determined |
| 18-106-final-viral-combined_k141_11853\|\|full | Low-quality |
| 18-106-final-viral-combined_k141_123395\|\|full | Low-quality |
| 18-106-final-viral-combined_k141_64661\|\|full | Low-quality |
| 18-106-final-viral-combined_k141_165\|\|full | Not-determined |
| 18-106-final-viral-combined_k141_88220\|\|full | Low-quality |
| 18-106-final-viral-combined_k141_64669\|\|full | Not-determined |
| 18-106-final-viral-combined_k141_117500\|\|full | Not-determined |
| 18-106-final-viral-combined_k141_111708\|\|full | Low-quality |
| 18-106-final-viral-combined_k141_123414\|\|full | Not-determined |
| 18-106-final-viral-combined_k141_88242\|\|full | Not-determined |
| 18-106-final-viral-combined_k141_111714\|\|full | Not-determined |
| 18-106-final-viral-combined_k141_129328\|\|full | Not-determined |
| 18-106-final-viral-combined_k141_41236\|\|full | Low-quality |
| 18-106-final-viral-combined_k141_190\|\|full | Low-quality |
| 18-106-final-viral-combined_k141_41237\|\|full | Not-determined |
| 18-106-final-viral-combined_k141_23568\|\|full | Not-determined |
| 18-106-final-viral-combined_k141_201\|\|full | Low-quality |
| 18-106-final-viral-combined_k141_123420\|\|full | Not-determined |
| 18-106-final-viral-combined_k141_99923\|\|full | Low-quality |
| 18-106-final-viral-combined_k141_76431\|\|full | Low-quality |
| 18-106-final-viral-combined_k141_5934\|\|full | Low-quality |
| 18-106-final-viral-combined_k141_64713\|\|full | Not-determined |
| 18-106-final-viral-combined_k141_88267\|\|full | Not-determined |
| 18-106-final-viral-combined_k141_58884\|\|full | Low-quality |
| 18-106-final-viral-combined_k141_11900\|\|full | Not-determined |
| 18-106-final-viral-combined_k141_111742\|\|full | Low-quality |
| 18-106-final-viral-combined_k141_64715\|\|full | Not-determined |
| 18-106-final-viral-combined_k141_17795\|\|full | Low-quality |
| 18-106-final-viral-combined_k141_105845\|\|full | Not-determined |
| 18-106-final-viral-combined_k141_23580\|\|full | Not-determined |
| 18-106-final-viral-combined_k141_58906\|\|full | Not-determined |
| 18-106-final-viral-combined_k141_5962\|\|full | Low-quality |
| 18-106-final-viral-combined_k141_5963\|\|full | Low-quality |
| 18-106-final-viral-combined_k141_105868\|\|full | Not-determined |
| 18-106-final-viral-combined_k141_255\|\|full | Not-determined |
| 18-106-final-viral-combined_k141_129383\|\|full | Not-determined |
| 18-104_k141_92005 | Medium-quality |
| 18-106-final-viral-combined_k141_94135\|\|full | Low-quality |
| 18-106-final-viral-combined_k141_76479\|\|full | Low-quality |
| 18-106-final-viral-combined_k141_76480\|\|full | Not-determined |
| 18-106-final-viral-combined_k141_88297\|\|full | Not-determined |
| 18-106-final-viral-combined_k141_129386\|\|full | Low-quality |
| 18-106-final-viral-combined_k141_111796\|\|full | Not-determined |
| 18-106-final-viral-combined_k141_76496\|\|full | Low-quality |
| 18-106-final-viral-combined_k141_123461\|\|full | Not-determined |
| 18-106-final-viral-combined_k141_64769\|\|full | Not-determined |
| 18-106-final-viral-combined_k141_105914\|\|full | Low-quality |
| 18-106-final-viral-combined_k141_117574\|\|full | Low-quality |
| 18-106-final-viral-combined_k141_35394\|\|full | Not-determined |
| 18-106-final-viral-combined_k141_41263\|\|full | Not-determined |
| 18-106-final-viral-combined_k141_129400\|\|full | Low-quality |
| 18-106-final-viral-combined_k141_88339\|\|full | Not-determined |
| 18-106-final-viral-combined_k141_94186\|\|full | Low-quality |
| 18-106-final-viral-combined_k141_123476\|\|full | Low-quality |
| 18-106-final-viral-combined_k141_11963\|\|full | Not-determined |
| 18-106-final-viral-combined_k141_111823\|\|full | Not-determined |
| 18-106-final-viral-combined_k141_129415\|\|full | Not-determined |
| 18-106-final-viral-combined_k141_99955\|\|full | Low-quality |
| 18-106-final-viral-combined_k141_123490\|\|full | Not-determined |
| 18-106-final-viral-combined_k141_70637\|\|full | Not-determined |
| 18-106-final-viral-combined_k141_11983\|\|full | Low-quality |
| 18-106-final-viral-combined_k141_94215\|\|full | Not-determined |
| 18-106-final-viral-combined_k141_58969\|\|full | Low-quality |
| 18-106-final-viral-combined_k141_64810\|\|full | Not-determined |
| 18-106-final-viral-combined_k141_70651\|\|full | Low-quality |
| 18-106-final-viral-combined_k141_64824\|\|full | Not-determined |
| 18-106-final-viral-combined_k141_35442\|\|full | Not-determined |
| 18-106-final-viral-combined_k141_325\|\|full | Low-quality |
| 18-106-final-viral-combined_k141_123521\|\|full | Not-determined |
| 18-106-final-viral-combined_k141_41320\|\|full | Low-quality |
| 18-106-final-viral-combined_k141_58998\|\|full | Not-determined |
| 18-106-final-viral-combined_k141_76592\|\|full | Not-determined |
| 18-106-final-viral-combined_k141_12028\|\|full | Not-determined |
| 18-106-final-viral-combined_k141_64861\|\|full | Low-quality |
| 18-106-final-viral-combined_k141_106003\|\|full | Not-determined |
| 18-106-final-viral-combined_k141_94249\|\|full | Low-quality |
| 18-106-final-viral-combined_k141_12059\|\|full | Not-determined |
| 18-106-final-viral-combined_k141_23646\|\|full | Not-determined |
| 18-106-final-viral-combined_k141_371\|\|full | Not-determined |
| 18-106-final-viral-combined_k141_106030\|\|full | Not-determined |
| 18-106-final-viral-combined_k141_6044\|\|full | Not-determined |
| 18-106-final-viral-combined_k141_64885\|\|full | Not-determined |
| 18-106-final-viral-combined_k141_12080\|\|full | Low-quality |
| 18-106-final-viral-combined_k141_111905\|\|full | Low-quality |
| 18-106-final-viral-combined_k141_59044\|\|full | Not-determined |
| 18-106-final-viral-combined_k141_76640\|\|full | Low-quality |
| 18-106-final-viral-combined_k141_35522\|\|full | Not-determined |
| 18-106-final-viral-combined_k141_117727\|\|full | Low-quality |
| 18-106-final-viral-combined_k141_59075\|\|full | Low-quality |
| 18-106-final-viral-combined_k141_41365\|\|full | Not-determined |
| 18-106-final-viral-combined_k141_123573\|\|full | Low-quality |
| 18-106-final-viral-combined_k141_12127\|\|full | Not-determined |
| 17-108-final-viral-combined_k141_268967\|\|full | Medium-quality |
| 18-106-final-viral-combined_k141_59082\|\|full | Not-determined |
| 18-106-final-viral-combined_k141_41390\|\|full | Not-determined |
| 18-106-final-viral-combined_k141_29549\|\|full | Not-determined |
| 18-106-final-viral-combined_k141_29550\|\|full | Not-determined |
| 18-106-final-viral-combined_k141_135237\|\|full | Low-quality |
| 18-106-final-viral-combined_k141_82370\|\|full | Low-quality |
| 18-106-final-viral-combined_k141_70799\|\|full | Low-quality |
| 18-106-final-viral-combined_k141_76684\|\|full | Low-quality |
| 18-102-final-viral-combined_k141_55495\|\|full | Medium-quality |
| 18-106-final-viral-combined_k141_70812\|\|full | Not-determined |
| 18-106-final-viral-combined_k141_6065\|\|full | Low-quality |
| 18-106-final-viral-combined_k141_29590\|\|full | Low-quality |
| 18-106-final-viral-combined_k141_454\|\|full | Low-quality |
| 18-106-final-viral-combined_k141_59093\|\|full | Not-determined |
| 18-106-final-viral-combined_k141_94344\|\|full | Not-determined |
| 18-106-final-viral-combined_k141_12162\|\|full | Not-determined |
| 18-106-final-viral-combined_k141_123640\|\|full | Not-determined |
| 18-106-final-viral-combined_k141_100034\|\|full | Not-determined |
| 18-106-final-viral-combined_k141_70848\|\|full | Low-quality |
| 18-105-final-viral-combined_k141_10783\|\|full | High-quality |
| 18-106-final-viral-combined_k141_88422\|\|full | Not-determined |
| 18-106-final-viral-combined_k141_111980\|\|full | Low-quality |
| 18-106-final-viral-combined_k141_94367\|\|full | Not-determined |
| 18-106-final-viral-combined_k141_6096\|\|full | Not-determined |
| 18-106-final-viral-combined_k141_82444\|\|full | Not-determined |
| 18-106-final-viral-combined_k141_59157\|\|full | Not-determined |
| 18-106-final-viral-combined_k141_23773\|\|full | Not-determined |
| 18-106-final-viral-combined_k141_82446\|\|full | Low-quality |
| 18-106-final-viral-combined_k141_82447\|\|full | Low-quality |
| 18-106-final-viral-combined_k141_100066\|\|full | Low-quality |
| 18-106-final-viral-combined_k141_17839\|\|full | Low-quality |
| 18-106-final-viral-combined_k141_41465\|\|full | Not-determined |
| 18-106-final-viral-combined_k141_59177\|\|full | Not-determined |
| 18-106-final-viral-combined_k141_76701\|\|full | Not-determined |
| 18-106-final-viral-combined_k141_17841\|\|full | Low-quality |
| 18-106-final-viral-combined_k141_6122\|\|full | Not-determined |
| 18-106-final-viral-combined_k141_522\|\|full | Not-determined |
| 18-106-final-viral-combined_k141_88474\|\|full | Low-quality |
| 18-106-final-viral-combined_k141_23802\|\|full | Low-quality |
| 18-106-final-viral-combined_k141_94427\|\|full | Not-determined |
| 18-106-final-viral-combined_k141_29714\|\|full | Not-determined |
| 18-106-final-viral-combined_k141_6156\|\|full | Low-quality |
| 18-106-final-viral-combined_k141_70921\|\|full | Low-quality |
| 18-106-final-viral-combined_k141_29722\|\|full | Not-determined |
| 18-106-final-viral-combined_k141_29733\|\|full | Low-quality |
| 18-106-final-viral-combined_k141_41527\|\|full | Low-quality |
| 18-106-final-viral-combined_k141_106241\|\|full | Not-determined |
| 18-106-final-viral-combined_k141_41535\|\|full | Low-quality |
| 18-106-final-viral-combined_k141_41549\|\|full | Not-determined |
| 18-106-final-viral-combined_k141_82494\|\|full | Low-quality |
| 18-106-final-viral-combined_k141_88548\|\|full | Not-determined |
| 18-106-final-viral-combined_k141_76715\|\|full | Not-determined |
| 18-106-final-viral-combined_k141_106244\|\|full | Low-quality |
| 18-106-final-viral-combined_k141_12272\|\|full | Not-determined |
| 18-106-final-viral-combined_k141_82513\|\|full | Not-determined |
| 18-106-final-viral-combined_k141_123743\|\|full | Not-determined |
| 18-106-final-viral-combined_k141_88570\|\|full | Low-quality |
| 18-106-final-viral-combined_k141_47267\|\|full | Low-quality |
| 18-106-final-viral-combined_k141_52926\|\|full | Low-quality |
| 18-106-final-viral-combined_k141_47273\|\|full | Low-quality |
| 18-106-final-viral-combined_k141_100106\|\|full | Low-quality |
| 18-106-final-viral-combined_k141_6219\|\|full | Not-determined |
| 18-106-final-viral-combined_k141_59262\|\|full | Low-quality |
| 18-106-final-viral-combined_k141_29791\|\|full | Low-quality |
| 18-106-final-viral-combined_k141_41577\|\|full | Low-quality |
| 18-106-final-viral-combined_k141_52935\|\|full | Not-determined |
| 18-106-final-viral-combined_k141_100116\|\|full | Not-determined |
| 18-106-final-viral-combined_k141_41601\|\|full | Not-determined |
| 18-106-final-viral-combined_k141_52967\|\|full | Low-quality |
| 18-106-final-viral-combined_k141_76763\|\|full | Low-quality |
| 18-106-final-viral-combined_k141_88619\|\|full | Low-quality |
| 18-106-final-viral-combined_k141_70983\|\|full | Low-quality |
| 18-106-final-viral-combined_k141_590\|\|full | Not-determined |
| 18-106-final-viral-combined_k141_88628\|\|full | Not-determined |
| 18-106-final-viral-combined_k141_601\|\|full | Not-determined |
| 18-106-final-viral-combined_k141_135250\|\|full | Low-quality |
| 18-106-final-viral-combined_k141_123834\|\|full | Not-determined |
| 18-106-final-viral-combined_k141_94551\|\|full | Low-quality |
| 18-106-final-viral-combined_k141_52995\|\|full | Not-determined |
| 18-106-final-viral-combined_k141_602\|\|full | Low-quality |
| 18-106-final-viral-combined_k141_123859\|\|full | Not-determined |
| 18-106-final-viral-combined_k141_94567\|\|full | Not-determined |
| 18-106-final-viral-combined_k141_100160\|\|full | Low-quality |
| 18-106-final-viral-combined_k141_88664\|\|full | Low-quality |
| 18-106-final-viral-combined_k141_624\|\|full | Low-quality |
| 18-106-final-viral-combined_k141_6274\|\|full | Not-determined |
| 18-106-final-viral-combined_k141_29894\|\|full | Not-determined |
| 18-106-final-viral-combined_k141_629\|\|full | Low-quality |
| 18-106-final-viral-combined_k141_12337\|\|full | Low-quality |
| 18-106-final-viral-combined_k141_117852\|\|full | Not-determined |
| 18-106-final-viral-combined_k141_117873\|\|full | Not-determined |
| 18-106-final-viral-combined_k141_47386\|\|full | Low-quality |
| 18-106-final-viral-combined_k141_117875\|\|full | Not-determined |
| 18-106-final-viral-combined_k141_117881\|\|full | Not-determined |
| 18-106-final-viral-combined_k141_112081\|\|full | Not-determined |
| 18-106-final-viral-combined_k141_676\|\|full | Not-determined |
| 18-106-final-viral-combined_k141_100235\|\|full | Low-quality |
| 18-106-final-viral-combined_k141_88696\|\|full | Not-determined |
| 18-106-final-viral-combined_k141_106381\|\|full | Low-quality |
| 18-106-final-viral-combined_k141_135357\|\|full | Low-quality |
| 18-106-final-viral-combined_k141_123931\|\|full | Not-determined |
| 18-106-final-viral-combined_k141_53111\|\|full | Not-determined |
| 18-106-final-viral-combined_k141_71023\|\|full | Not-determined |
| 18-106-final-viral-combined_k141_88710\|\|full | Low-quality |
| 18-106-final-viral-combined_k141_59333\|\|full | Not-determined |
| 18-106-final-viral-combined_k141_117915\|\|full | Low-quality |
| 18-106-final-viral-combined_k141_704\|\|full | Low-quality |
| 18-106-final-viral-combined_k141_65092\|\|full | Low-quality |
| 18-106-final-viral-combined_k141_47467\|\|full | Not-determined |
| 18-106-final-viral-combined_k141_53141\|\|full | Low-quality |
| 18-106-final-viral-combined_k141_112133\|\|full | Low-quality |
| 18-106-final-viral-combined_k141_718\|\|full | Low-quality |
| 18-106-final-viral-combined_k141_135394\|\|full | Low-quality |
| 18-106-final-viral-combined_k141_106410\|\|full | Low-quality |
| 18-106-final-viral-combined_k141_47485\|\|full | Low-quality |
| 18-106-final-viral-combined_k141_106419\|\|full | Low-quality |
| 18-106-final-viral-combined_k141_47487\|\|full | Not-determined |
| 18-106-final-viral-combined_k141_47490\|\|full | Low-quality |
| 18-106-final-viral-combined_k141_123985\|\|full | Not-determined |
| 18-106-final-viral-combined_k141_47495\|\|full | Not-determined |
| 18-106-final-viral-combined_k141_88773\|\|full | Low-quality |
| 18-106-final-viral-combined_k141_94664\|\|full | Not-determined |
| 18-106-final-viral-combined_k141_124003\|\|full | Not-determined |
| 18-106-final-viral-combined_k141_117967\|\|full | Not-determined |
| 18-106-final-viral-combined_k141_41774\|\|full | Low-quality |
| 18-106-final-viral-combined_k141_124004\|\|full | Low-quality |
| 18-106-final-viral-combined_k141_6399\|\|full | Low-quality |
| 18-106-final-viral-combined_k141_135431\|\|full | Low-quality |
| 18-106-final-viral-combined_k141_135434\|\|full | Low-quality |
| 18-106-final-viral-combined_k141_106449\|\|full | Low-quality |
| 18-106-final-viral-combined_k141_47525\|\|full | Not-determined |
| 18-106-final-viral-combined_k141_71067\|\|full | Not-determined |
| 18-106-final-viral-combined_k141_135452\|\|full | Not-determined |
| 18-106-final-viral-combined_k141_94701\|\|full | Not-determined |
| 18-106-final-viral-combined_k141_129788\|\|full | Not-determined |
| 18-101-final-viral-combined_k141_252439\|\|full | Medium-quality |
| 18-106-final-viral-combined_k141_135470\|\|full | Low-quality |
| 18-106-final-viral-combined_k141_6446\|\|full | Low-quality |
| 18-106-final-viral-combined_k141_6449\|\|full | Not-determined |
| 18-106-final-viral-combined_k141_12391\|\|full | Not-determined |
| 18-106-final-viral-combined_k141_53168\|\|full | Not-determined |
| 18-106-final-viral-combined_k141_88800\|\|full | Not-determined |
| 18-106-final-viral-combined_k141_6467\|\|full | Low-quality |
| 18-106-final-viral-combined_k141_135502\|\|full | Low-quality |
| 18-106-final-viral-combined_k141_6479\|\|full | Not-determined |
| 18-106-final-viral-combined_k141_59372\|\|full | Not-determined |
| 18-106-final-viral-combined_k141_65184\|\|full | Low-quality |
| 18-106-final-viral-combined_k141_88823\|\|full | Not-determined |
| 18-106-final-viral-combined_k141_94758\|\|full | Low-quality |
| 18-106-final-viral-combined_k141_88836\|\|full | Not-determined |
| 18-106-final-viral-combined_k141_23970\|\|full | Not-determined |
| 18-106-final-viral-combined_k141_124080\|\|full | Not-determined |
| 18-106-final-viral-combined_k141_94772\|\|full | Not-determined |
| 18-106-final-viral-combined_k141_65208\|\|full | Not-determined |
| 18-106-final-viral-combined_k141_100353\|\|full | Not-determined |
| 18-106-final-viral-combined_k141_41787\|\|full | Low-quality |
| 18-106-final-viral-combined_k141_88854\|\|full | Not-determined |
| 18-106-final-viral-combined_k141_124105\|\|full | Low-quality |
| 18-106-final-viral-combined_k141_803\|\|full | Low-quality |
| 18-106-final-viral-combined_k141_59403\|\|full | Not-determined |
| 18-106-final-viral-combined_k141_805\|\|full | Low-quality |
| 18-106-final-viral-combined_k141_808\|\|full | Not-determined |
| 18-110_k141_244525 | Medium-quality |
| 18-102-final-viral-combined_k141_199384\|\|full | Medium-quality |
| 18-106-final-viral-combined_k141_106495\|\|full | Low-quality |
| 18-106-final-viral-combined_k141_47611\|\|full | Not-determined |
| 18-106-final-viral-combined_k141_100378\|\|full | Not-determined |
| 18-106-final-viral-combined_k141_841\|\|full | Low-quality |
| 18-106-final-viral-combined_k141_24006\|\|full | Not-determined |
| 18-106-final-viral-combined_k141_100388\|\|full | Not-determined |
| 18-106-final-viral-combined_k141_41848\|\|full | Not-determined |
| 18-106-final-viral-combined_k141_12485\|\|full | Low-quality |
| 18-106-final-viral-combined_k141_41863\|\|full | Not-determined |
| 18-106-final-viral-combined_k141_24032\|\|full | Not-determined |
| 18-106-final-viral-combined_k141_53229\|\|full | Not-determined |
| 18-106-final-viral-combined_k141_12493\|\|full | Low-quality |
| 18-106-final-viral-combined_k141_24033\|\|full | Not-determined |
| 18-106-final-viral-combined_k141_12515\|\|full | Not-determined |
| 18-106-final-viral-combined_k141_129848\|\|full | Not-determined |
| 18-106-final-viral-combined_k141_24056\|\|full | Low-quality |
| 18-106-final-viral-combined_k141_82661\|\|full | Not-determined |
| 18-106-final-viral-combined_k141_53259\|\|full | Not-determined |
| 18-106-final-viral-combined_k141_82692\|\|full | Not-determined |
| 18-106-final-viral-combined_k141_65322\|\|full | Not-determined |
| 18-106-final-viral-combined_k141_6606\|\|full | Not-determined |
| 18-106-final-viral-combined_k141_82717\|\|full | Low-quality |
| 18-106-final-viral-combined_k141_94874\|\|full | Not-determined |
| 18-106-final-viral-combined_k141_47683\|\|full | Low-quality |
| 18-106-final-viral-combined_k141_956\|\|full | Not-determined |
| 18-106-final-viral-combined_k141_24094\|\|full | Low-quality |
| 18-106-final-viral-combined_k141_82734\|\|full | Not-determined |
| 18-106-final-viral-combined_k141_30081\|\|full | Low-quality |
| 18-106-final-viral-combined_k141_41946\|\|full | Low-quality |
| 18-106-final-viral-combined_k141_12575\|\|full | Low-quality |
| 18-106-final-viral-combined_k141_6625\|\|full | Low-quality |
| 18-106-final-viral-combined_k141_129901\|\|full | Low-quality |
| 18-106-final-viral-combined_k141_76955\|\|full | Not-determined |
| 18-106-final-viral-combined_k141_129903\|\|full | Low-quality |
| 18-106-final-viral-combined_k141_17983\|\|full | Not-determined |
| 18-106-final-viral-combined_k141_94898\|\|full | Low-quality |
| 18-106-final-viral-combined_k141_35635\|\|full | Not-determined |
| 18-106-final-viral-combined_k141_53309\|\|full | Not-determined |
| 18-106-final-viral-combined_k141_89021\|\|full | Low-quality |
| 18-106-final-viral-combined_k141_65385\|\|full | Not-determined |
| 18-106-final-viral-combined_k141_30098\|\|full | Low-quality |
| 18-106-final-viral-combined_k141_65398\|\|full | Not-determined |
| 18-106-final-viral-combined_k141_6666\|\|full | Low-quality |
| 18-106-final-viral-combined_k141_6670\|\|full | Not-determined |
| 18-106-final-viral-combined_k141_71264\|\|full | Low-quality |
| 18-106-final-viral-combined_k141_89045\|\|full | Low-quality |
| 18-106-final-viral-combined_k141_1040\|\|full | Low-quality |
| 18-106-final-viral-combined_k141_94931\|\|full | Low-quality |
| 18-106-final-viral-combined_k141_59501\|\|full | Low-quality |
| 18-106-final-viral-combined_k141_94939\|\|full | Not-determined |
| 18-106-final-viral-combined_k141_59505\|\|full | Low-quality |
| 18-106-final-viral-combined_k141_6705\|\|full | Not-determined |
| 18-106-final-viral-combined_k141_76992\|\|full | Low-quality |
| 18-106-final-viral-combined_k141_30162\|\|full | Not-determined |
| 18-106-final-viral-combined_k141_59531\|\|full | Low-quality |
| 18-106-final-viral-combined_k141_77001\|\|full | Not-determined |
| 18-106-final-viral-combined_k141_94975\|\|full | Low-quality |
| 18-106-final-viral-combined_k141_82844\|\|full | Low-quality |
| 18-106-final-viral-combined_k141_47715\|\|full | Not-determined |
| 18-106-final-viral-combined_k141_135593\|\|full | Not-determined |
| 18-106-final-viral-combined_k141_65485\|\|full | Low-quality |
| 18-106-final-viral-combined_k141_89180\|\|full | Not-determined |
| 18-106-final-viral-combined_k141_89181\|\|full | Not-determined |
| 18-106-final-viral-combined_k141_1151\|\|full | Not-determined |
| 18-106-final-viral-combined_k141_24196\|\|full | Not-determined |
| 18-106-final-viral-combined_k141_42017\|\|full | Low-quality |
| 18-106-final-viral-combined_k141_89185\|\|full | Low-quality |
| 18-106-final-viral-combined_k141_42022\|\|full | Not-determined |
| 18-106-final-viral-combined_k141_65516\|\|full | Not-determined |
| 18-106-final-viral-combined_k141_71292\|\|full | Not-determined |
| 18-106-final-viral-combined_k141_35780\|\|full | Low-quality |
| 18-106-final-viral-combined_k141_18072\|\|full | Not-determined |
| 18-106-final-viral-combined_k141_30238\|\|full | Low-quality |
| 18-106-final-viral-combined_k141_89197\|\|full | Not-determined |
| 18-106-final-viral-combined_k141_129972\|\|full | Not-determined |
| 18-106-final-viral-combined_k141_95071\|\|full | Low-quality |
| 18-106-final-viral-combined_k141_65535\|\|full | Low-quality |
| 18-106-final-viral-combined_k141_18094\|\|full | Low-quality |
| 18-106-final-viral-combined_k141_89206\|\|full | Low-quality |
| 18-106-final-viral-combined_k141_18101\|\|full | Low-quality |
| 18-106-final-viral-combined_k141_89208\|\|full | Not-determined |
| 18-104_k141_107182 | Medium-quality |
| 18-106-final-viral-combined_k141_95083\|\|full | Not-determined |
| 18-106-final-viral-combined_k141_65554\|\|full | Not-determined |
| 18-106-final-viral-combined_k141_18102\|\|full | Low-quality |
| 18-106-final-viral-combined_k141_129984\|\|full | Low-quality |
| 18-106-final-viral-combined_k141_65555\|\|full | Not-determined |
| 18-106-final-viral-combined_k141_12613\|\|full | Not-determined |
| 18-106-final-viral-combined_k141_24230\|\|full | Not-determined |
| 18-106-final-viral-combined_k141_89233\|\|full | Not-determined |
| 18-106-final-viral-combined_k141_42088\|\|full | Low-quality |
| 18-106-final-viral-combined_k141_18118\|\|full | Not-determined |
| 18-106-final-viral-combined_k141_89236\|\|full | Not-determined |
| 18-106-final-viral-combined_k141_89241\|\|full | Not-determined |
| 18-106-final-viral-combined_k141_18127\|\|full | Low-quality |
| 18-106-final-viral-combined_k141_30297\|\|full | Not-determined |
| 18-106-final-viral-combined_k141_124188\|\|full | Not-determined |
| 18-106-final-viral-combined_k141_1245\|\|full | Low-quality |
| 18-106-final-viral-combined_k141_12629\|\|full | Not-determined |
| 18-106-final-viral-combined_k141_35840\|\|full | Not-determined |
| 18-106-final-viral-combined_k141_89261\|\|full | Not-determined |
| 18-106-final-viral-combined_k141_6857\|\|full | Not-determined |
| 18-106-final-viral-combined_k141_82926\|\|full | Not-determined |
| 18-106-final-viral-combined_k141_82931\|\|full | Low-quality |
| 18-106-final-viral-combined_k141_53380\|\|full | Low-quality |
| 18-106-final-viral-combined_k141_106585\|\|full | Low-quality |
| 18-106-final-viral-combined_k141_59688\|\|full | Low-quality |
| 18-106-final-viral-combined_k141_124203\|\|full | Not-determined |
| 18-106-final-viral-combined_k141_35870\|\|full | Low-quality |
| 18-106-final-viral-combined_k141_35874\|\|full | Low-quality |
| 18-106-final-viral-combined_k141_130033\|\|full | Low-quality |
| 18-106-final-viral-combined_k141_65621\|\|full | Not-determined |
| 18-106-final-viral-combined_k141_130037\|\|full | Not-determined |
| 18-106-final-viral-combined_k141_89314\|\|full | Low-quality |
| 18-106-final-viral-combined_k141_77130\|\|full | Low-quality |
| 18-106-final-viral-combined_k141_135649\|\|full | Not-determined |
| 18-106-final-viral-combined_k141_124232\|\|full | Not-determined |
| 18-106-final-viral-combined_k141_18168\|\|full | Low-quality |
| 18-106-final-viral-combined_k141_47857\|\|full | Low-quality |
| 18-106-final-viral-combined_k141_42141\|\|full | Not-determined |
| 18-106-final-viral-combined_k141_18172\|\|full | Not-determined |
| 18-106-final-viral-combined_k141_100515\|\|full | Low-quality |
| 18-106-final-viral-combined_k141_106648\|\|full | Not-determined |
| 18-106-final-viral-combined_k141_100516\|\|full | Not-determined |
| 18-106-final-viral-combined_k141_82977\|\|full | Not-determined |
| 18-106-final-viral-combined_k141_18178\|\|full | Not-determined |
| 18-106-final-viral-combined_k141_59743\|\|full | Not-determined |
| 18-106-final-viral-combined_k141_59745\|\|full | Not-determined |
| 18-106-final-viral-combined_k141_112260\|\|full | Low-quality |
| 18-106-final-viral-combined_k141_130059\|\|full | Low-quality |
| 18-106-final-viral-combined_k141_1321\|\|full | Low-quality |
| 18-106-final-viral-combined_k141_71444\|\|full | Not-determined |
| 18-106-final-viral-combined_k141_47901\|\|full | Not-determined |
| 18-106-final-viral-combined_k141_1325\|\|full | Not-determined |
| 18-106-final-viral-combined_k141_130079\|\|full | Not-determined |
| 18-106-final-viral-combined_k141_112290\|\|full | Low-quality |
| 18-106-final-viral-combined_k141_47913\|\|full | Not-determined |
| 18-106-final-viral-combined_k141_24254\|\|full | Low-quality |
| 18-106-final-viral-combined_k141_82987\|\|full | Not-determined |
| 18-106-final-viral-combined_k141_12662\|\|full | Low-quality |
| 18-106-final-viral-combined_k141_1339\|\|full | Low-quality |
| 18-106-final-viral-combined_k141_30386\|\|full | Low-quality |
| 18-106-final-viral-combined_k141_6944\|\|full | Not-determined |
| 18-106-final-viral-combined_k141_35961\|\|full | Low-quality |
| 18-106-final-viral-combined_k141_6951\|\|full | Low-quality |
| 18-106-final-viral-combined_k141_106685\|\|full | Low-quality |
| 18-106-final-viral-combined_k141_59800\|\|full | Not-determined |
| 18-106-final-viral-combined_k141_130095\|\|full | Low-quality |
| 18-106-final-viral-combined_k141_112335\|\|full | Not-determined |
| 18-106-final-viral-combined_k141_12678\|\|full | Not-determined |
| 18-106-final-viral-combined_k141_24269\|\|full | Low-quality |
| 18-106-final-viral-combined_k141_112337\|\|full | Not-determined |
| 18-106-final-viral-combined_k141_89390\|\|full | Low-quality |
| 18-106-final-viral-combined_k141_83007\|\|full | Not-determined |
| 18-106-final-viral-combined_k141_30410\|\|full | Low-quality |
| 18-106-final-viral-combined_k141_112339\|\|full | Not-determined |
| 18-106-final-viral-combined_k141_106691\|\|full | Not-determined |
| 18-106-final-viral-combined_k141_6972\|\|full | Low-quality |
| 18-106-final-viral-combined_k141_6978\|\|full | Low-quality |
| 18-106-final-viral-combined_k141_83030\|\|full | Not-determined |
| 18-106-final-viral-combined_k141_6985\|\|full | Not-determined |
| 18-106-final-viral-combined_k141_89406\|\|full | Low-quality |
| 18-106-final-viral-combined_k141_1370\|\|full | Low-quality |
| 18-106-final-viral-combined_k141_6990\|\|full | Low-quality |
| 18-106-final-viral-combined_k141_100626\|\|full | Low-quality |
| 18-106-final-viral-combined_k141_18272\|\|full | Not-determined |
| 18-106-final-viral-combined_k141_47990\|\|full | Not-determined |
| 18-106-final-viral-combined_k141_53430\|\|full | Not-determined |
| 18-106-final-viral-combined_k141_100634\|\|full | Not-determined |
| 18-106-final-viral-combined_k141_59833\|\|full | Low-quality |
| 18-106-final-viral-combined_k141_36000\|\|full | Not-determined |
| 18-106-final-viral-combined_k141_106727\|\|full | Not-determined |
| 18-106-final-viral-combined_k141_71492\|\|full | Low-quality |
| 18-106-final-viral-combined_k141_36021\|\|full | Not-determined |
| 18-106-final-viral-combined_k141_1423\|\|full | Low-quality |
| 18-106-final-viral-combined_k141_135684\|\|full | Low-quality |
| 18-106-final-viral-combined_k141_106754\|\|full | Not-determined |
| 18-106-final-viral-combined_k141_18293\|\|full | Not-determined |
| 18-106-final-viral-combined_k141_106758\|\|full | Low-quality |
| 18-106-final-viral-combined_k141_30485\|\|full | Not-determined |
| 18-106-final-viral-combined_k141_124326\|\|full | Not-determined |
| 18-106-final-viral-combined_k141_1445\|\|full | Not-determined |
| 18-106-final-viral-combined_k141_24374\|\|full | Low-quality |
| 18-106-final-viral-combined_k141_48055\|\|full | Low-quality |
| 18-106-final-viral-combined_k141_36056\|\|full | Low-quality |
| 18-106-final-viral-combined_k141_71541\|\|full | Not-determined |
| 18-106-final-viral-combined_k141_77220\|\|full | Not-determined |
| 18-106-final-viral-combined_k141_36070\|\|full | Not-determined |
| 18-106-final-viral-combined_k141_71546\|\|full | Not-determined |
| 18-106-final-viral-combined_k141_77222\|\|full | Not-determined |
| 18-106-final-viral-combined_k141_106788\|\|full | Low-quality |
| 18-106-final-viral-combined_k141_118159\|\|full | Not-determined |
| 18-106-final-viral-combined_k141_118163\|\|full | Not-determined |
| 18-106-final-viral-combined_k141_106795\|\|full | Low-quality |
| 18-106-final-viral-combined_k141_89441\|\|full | Low-quality |
| 18-106-final-viral-combined_k141_18333\|\|full | Not-determined |
| 18-106-final-viral-combined_k141_77244\|\|full | Not-determined |
| 18-106-final-viral-combined_k141_18340\|\|full | Low-quality |
| 18-106-final-viral-combined_k141_7023\|\|full | Not-determined |
| 18-106-final-viral-combined_k141_112448\|\|full | Not-determined |
| 18-106-final-viral-combined_k141_48106\|\|full | Low-quality |
| 18-106-final-viral-combined_k141_124365\|\|full | Not-determined |
| 18-106-final-viral-combined_k141_18360\|\|full | Low-quality |
| 18-106-final-viral-combined_k141_36114\|\|full | Low-quality |
| 18-106-final-viral-combined_k141_124395\|\|full | Low-quality |
| 18-106-final-viral-combined_k141_12734\|\|full | Not-determined |
| 18-106-final-viral-combined_k141_42290\|\|full | Low-quality |
| 18-106-final-viral-combined_k141_12738\|\|full | Not-determined |
| 18-106-final-viral-combined_k141_112475\|\|full | Not-determined |
| 18-106-final-viral-combined_k141_1516\|\|full | Not-determined |
| 18-106-final-viral-combined_k141_12742\|\|full | Not-determined |
| 18-106-final-viral-combined_k141_18372\|\|full | Low-quality |
| 18-106-final-viral-combined_k141_30510\|\|full | Not-determined |
| 18-106-final-viral-combined_k141_83161\|\|full | Low-quality |
| 18-106-final-viral-combined_k141_124410\|\|full | Not-determined |
| 18-106-final-viral-combined_k141_112480\|\|full | Not-determined |
| 18-106-final-viral-combined_k141_112484\|\|full | Not-determined |
| 18-106-final-viral-combined_k141_59888\|\|full | Not-determined |
| 18-106-final-viral-combined_k141_18384\|\|full | Not-determined |
| 18-106-final-viral-combined_k141_95237\|\|full | Not-determined |
| 18-104-final-viral-combined_k141_107182\|\|full | Medium-quality |
| 18-106-final-viral-combined_k141_95243\|\|full | Low-quality |
| 18-106-final-viral-combined_k141_30548\|\|full | Low-quality |
| 18-106-final-viral-combined_k141_95251\|\|full | Low-quality |
| 18-106-final-viral-combined_k141_30553\|\|full | Not-determined |
| 18-106-final-viral-combined_k141_95259\|\|full | Not-determined |
| 18-106-final-viral-combined_k141_48173\|\|full | Low-quality |
| 18-106-final-viral-combined_k141_77296\|\|full | Not-determined |
| 18-106-final-viral-combined_k141_135772\|\|full | Low-quality |
| 18-106-final-viral-combined_k141_83183\|\|full | Not-determined |
| 18-106-final-viral-combined_k141_112521\|\|full | Not-determined |
| 18-106-final-viral-combined_k141_12786\|\|full | Low-quality |
| 18-106-final-viral-combined_k141_48187\|\|full | Low-quality |
| 18-106-final-viral-combined_k141_30577\|\|full | Not-determined |
| 18-106-final-viral-combined_k141_48188\|\|full | Not-determined |
| 18-106-final-viral-combined_k141_53504\|\|full | Not-determined |
| 18-106-final-viral-combined_k141_83196\|\|full | Not-determined |
| 18-106-final-viral-combined_k141_124476\|\|full | Not-determined |
| 18-106-final-viral-combined_k141_124477\|\|full | Not-determined |
| 18-106-final-viral-combined_k141_135796\|\|full | Low-quality |
| 18-106-final-viral-combined_k141_77325\|\|full | Not-determined |
| 18-106-final-viral-combined_k141_124491\|\|full | Not-determined |
| 18-106-final-viral-combined_k141_48207\|\|full | Not-determined |
| 18-106-final-viral-combined_k141_36207\|\|full | Low-quality |
| 18-106-final-viral-combined_k141_112536\|\|full | Not-determined |
| 18-106-final-viral-combined_k141_1599\|\|full | Not-determined |
| 18-106-final-viral-combined_k141_24426\|\|full | Not-determined |
| 18-106-final-viral-combined_k141_118282\|\|full | Low-quality |
| 18-106-final-viral-combined_k141_24427\|\|full | Low-quality |
| 18-106-final-viral-combined_k141_59955\|\|full | Not-determined |
| 18-106-final-viral-combined_k141_77355\|\|full | Not-determined |
| 18-106-final-viral-combined_k141_36230\|\|full | Not-determined |
| 18-106-final-viral-combined_k141_100770\|\|full | Not-determined |
| 18-106-final-viral-combined_k141_42401\|\|full | Not-determined |
| 18-106-final-viral-combined_k141_59967\|\|full | Not-determined |
| 18-106-final-viral-combined_k141_1613\|\|full | Low-quality |
| 18-106-final-viral-combined_k141_89503\|\|full | Low-quality |
| 18-106-final-viral-combined_k141_18448\|\|full | Low-quality |
| 18-106-final-viral-combined_k141_36238\|\|full | Not-determined |
| 18-106-final-viral-combined_k141_18449\|\|full | Not-determined |
| 18-106-final-viral-combined_k141_48215\|\|full | Not-determined |
| 18-106-final-viral-combined_k141_42413\|\|full | Not-determined |
| 18-106-final-viral-combined_k141_100776\|\|full | Low-quality |
| 18-106-final-viral-combined_k141_36247\|\|full | Low-quality |
| 18-106-final-viral-combined_k141_59984\|\|full | Not-determined |
| 18-106-final-viral-combined_k141_1652\|\|full | Not-determined |
| 18-106-final-viral-combined_k141_95299\|\|full | Low-quality |
| 18-106-final-viral-combined_k141_59995\|\|full | Low-quality |
| 18-106-final-viral-combined_k141_42454\|\|full | Low-quality |
| 18-106-final-viral-combined_k141_59999\|\|full | Low-quality |
| 18-106-final-viral-combined_k141_12824\|\|full | Not-determined |
| 18-106-final-viral-combined_k141_89520\|\|full | Not-determined |
| 18-106-final-viral-combined_k141_30622\|\|full | Not-determined |
| 18-106-final-viral-combined_k141_135837\|\|full | Not-determined |
| 18-106-final-viral-combined_k141_12825\|\|full | Low-quality |
| 18-106-final-viral-combined_k141_48253\|\|full | Not-determined |
| 18-106-final-viral-combined_k141_60003\|\|full | Low-quality |
| 18-106-final-viral-combined_k141_95325\|\|full | Not-determined |
| 18-106-final-viral-combined_k141_42480\|\|full | Not-determined |
| 18-106-final-viral-combined_k141_77409\|\|full | Low-quality |
| 18-106-final-viral-combined_k141_83250\|\|full | Not-determined |
| 18-106-final-viral-combined_k141_7128\|\|full | Low-quality |

| | |
|---|---|
| 18-106-final-viral-combined_k141_95340\|\|full | Not-determined |
| 18-106-final-viral-combined_k141_65897\|\|full | Low-quality |
| 18-106-final-viral-combined_k141_12842\|\|full | Low-quality |
| 18-106-final-viral-combined_k141_100814\|\|full | Not-determined |
| 18-106-final-viral-combined_k141_53517\|\|full | Not-determined |
| 18-106-final-viral-combined_k141_42489\|\|full | Low-quality |
| 18-106-final-viral-combined_k141_130271\|\|full | Low-quality |
| 18-106-final-viral-combined_k141_24453\|\|full | Low-quality |
| 18-106-final-viral-combined_k141_53526\|\|full | Low-quality |
| 18-106-final-viral-combined_k141_83283\|\|full | Low-quality |
| 18-106-final-viral-combined_k141_106955\|\|full | Not-determined |
| 18-106-final-viral-combined_k141_12863\|\|full | Not-determined |
| 18-106-final-viral-combined_k141_118357\|\|full | Low-quality |
| 18-106-final-viral-combined_k141_89559\|\|full | Low-quality |
| 18-106-final-viral-combined_k141_83297\|\|full | Not-determined |
| 18-106-final-viral-combined_k141_77448\|\|full | Low-quality |
| 18-106-final-viral-combined_k141_36338\|\|full | Low-quality |
| 18-106-final-viral-combined_k141_12877\|\|full | Not-determined |
| 18-106-final-viral-combined_k141_18507\|\|full | Not-determined |
| 18-106-final-viral-combined_k141_1682\|\|full | Not-determined |
| 18-106-final-viral-combined_k141_60077\|\|full | Not-determined |
| 18-106-final-viral-combined_k141_60082\|\|full | Not-determined |
| 18-106-final-viral-combined_k141_18534\|\|full | Low-quality |
| 18-106-final-viral-combined_k141_42519\|\|full | Low-quality |
| 18-106-final-viral-combined_k141_48323\|\|full | Not-determined |
| 18-106-final-viral-combined_k141_36349\|\|full | Not-determined |
| 18-106-final-viral-combined_k141_130310\|\|full | Not-determined |
| 18-106-final-viral-combined_k141_65938\|\|full | Not-determined |
| 18-106-final-viral-combined_k141_7195\|\|full | Low-quality |
| 18-106-final-viral-combined_k141_112635\|\|full | Low-quality |
| 18-106-final-viral-combined_k141_135866\|\|full | Not-determined |
| 18-106-final-viral-combined_k141_1700\|\|full | Low-quality |
| 18-106-final-viral-combined_k141_65946\|\|full | Low-quality |
| 18-106-final-viral-combined_k141_60099\|\|full | Not-determined |
| 18-106-final-viral-combined_k141_48340\|\|full | Low-quality |
| 18-106-final-viral-combined_k141_24496\|\|full | Not-determined |
| 18-106-final-viral-combined_k141_1706\|\|full | Low-quality |
| 18-106-final-viral-combined_k141_83344\|\|full | Not-determined |
| 18-106-final-viral-combined_k141_30707\|\|full | Low-quality |
| 18-106-final-viral-combined_k141_60112\|\|full | Not-determined |
| 18-106-final-viral-combined_k141_42536\|\|full | Not-determined |
| 18-106-final-viral-combined_k141_53584\|\|full | Not-determined |
| 18-106-final-viral-combined_k141_18593\|\|full | Low-quality |
| 18-106-final-viral-combined_k141_95422\|\|full | Not-determined |
| 18-106-final-viral-combined_k141_83395\|\|full | Not-determined |
| 18-106-final-viral-combined_k141_24533\|\|full | Not-determined |
| 18-106-final-viral-combined_k141_89679\|\|full | Not-determined |
| 18-106-final-viral-combined_k141_77483\|\|full | Not-determined |
| 18-106-final-viral-combined_k141_77485\|\|full | Not-determined |
| 18-106-final-viral-combined_k141_18620\|\|full | Not-determined |
| 18-106-final-viral-combined_k141_12953\|\|full | Low-quality |
| 18-106-final-viral-combined_k141_7261\|\|full | Not-determined |
| 18-106-final-viral-combined_k141_77486\|\|full | Low-quality |
| 18-106-final-viral-combined_k141_18628\|\|full | Low-quality |
| 18-106-final-viral-combined_k141_77492\|\|full | Not-determined |
| 18-106-final-viral-combined_k141_18645\|\|full | Not-determined |
| 18-106-final-viral-combined_k141_65998\|\|full | Low-quality |
| 18-106-final-viral-combined_k141_7280\|\|full | Not-determined |
| 18-106-final-viral-combined_k141_7283\|\|full | Low-quality |
| 18-106-final-viral-combined_k141_100905\|\|full | Not-determined |
| 18-106-final-viral-combined_k141_1762\|\|full | Not-determined |
| 18-106-final-viral-combined_k141_89727\|\|full | Not-determined |
| 18-106-final-viral-combined_k141_30750\|\|full | Low-quality |
| 18-106-final-viral-combined_k141_24592\|\|full | Not-determined |
| 18-106-final-viral-combined_k141_66026\|\|full | Not-determined |
| 18-106-final-viral-combined_k141_42641\|\|full | Low-quality |
| 18-106-final-viral-combined_k141_77524\|\|full | Low-quality |
| 18-106-final-viral-combined_k141_42644\|\|full | Not-determined |
| 18-106-final-viral-combined_k141_100921\|\|full | Low-quality |
| 18-106-final-viral-combined_k141_107135\|\|full | Not-determined |
| 18-106-final-viral-combined_k141_42645\|\|full | Low-quality |
| 18-106-final-viral-combined_k141_83454\|\|full | Not-determined |
| 18-106-final-viral-combined_k141_18691\|\|full | Low-quality |
| 18-106-final-viral-combined_k141_77548\|\|full | Not-determined |
| 18-106-final-viral-combined_k141_42659\|\|full | Low-quality |
| 18-106-final-viral-combined_k141_66055\|\|full | Low-quality |
| 18-106-final-viral-combined_k141_95471\|\|full | Low-quality |
| 18-106-final-viral-combined_k141_42663\|\|full | Low-quality |
| 18-106-final-viral-combined_k141_107152\|\|full | Low-quality |
| 18-106-final-viral-combined_k141_53673\|\|full | Not-determined |
| 18-106-final-viral-combined_k141_30797\|\|full | Not-determined |
| 18-106-final-viral-combined_k141_30800\|\|full | Not-determined |
| 18-106-final-viral-combined_k141_95491\|\|full | Not-determined |
| 18-106-final-viral-combined_k141_1786\|\|full | Low-quality |
| 18-106-final-viral-combined_k141_135952\|\|full | Not-determined |
| 18-106-final-viral-combined_k141_7312\|\|full | Low-quality |
| 18-106-final-viral-combined_k141_130412\|\|full | Not-determined |
| 18-106-final-viral-combined_k141_53694\|\|full | Not-determined |
| 18-106-final-viral-combined_k141_42688\|\|full | Low-quality |
| 18-106-final-viral-combined_k141_53698\|\|full | Low-quality |
| 18-106-final-viral-combined_k141_53712\|\|full | Low-quality |
| 18-106-final-viral-combined_k141_83515\|\|full | Low-quality |
| 18-106-final-viral-combined_k141_100943\|\|full | Not-determined |
| 18-106-final-viral-combined_k141_71717\|\|full | Not-determined |
| 18-106-final-viral-combined_k141_60284\|\|full | Low-quality |
| 18-106-final-viral-combined_k141_107203\|\|full | Low-quality |
| 18-106-final-viral-combined_k141_71719\|\|full | Low-quality |
| 18-106-final-viral-combined_k141_1810\|\|full | Not-determined |
| 18-106-final-viral-combined_k141_30821\|\|full | Not-determined |
| 18-106-final-viral-combined_k141_60287\|\|full | Low-quality |
| 18-106-final-viral-combined_k141_1825\|\|full | Not-determined |
| 18-106-final-viral-combined_k141_83526\|\|full | Not-determined |
| 18-106-final-viral-combined_k141_18771\|\|full | Not-determined |
| 18-106-final-viral-combined_k141_18775\|\|full | Not-determined |
| 18-106-final-viral-combined_k141_89744\|\|full | Low-quality |
| 18-106-final-viral-combined_k141_71736\|\|full | Low-quality |
| 18-106-final-viral-combined_k141_130452\|\|full | Not-determined |
| 18-106-final-viral-combined_k141_95535\|\|full | Not-determined |
| 18-106-final-viral-combined_k141_107217\|\|full | Not-determined |
| 18-106-final-viral-combined_k141_36376\|\|full | Not-determined |
| 18-106-final-viral-combined_k141_83556\|\|full | Low-quality |
| 18-106-final-viral-combined_k141_89761\|\|full | Low-quality |
| 18-106-final-viral-combined_k141_71757\|\|full | Low-quality |
| 18-106-final-viral-combined_k141_53747\|\|full | Not-determined |
| 18-106-final-viral-combined_k141_12993\|\|full | Low-quality |
| 18-102_k141_79304 | Medium-quality |
| 18-106-final-viral-combined_k141_136017\|\|full | Low-quality |
| 18-106-final-viral-combined_k141_1898\|\|full | Not-determined |
| 18-106-final-viral-combined_k141_112725\|\|full | Not-determined |
| 18-106-final-viral-combined_k141_136029\|\|full | Not-determined |
| 18-106-final-viral-combined_k141_48562\|\|full | Low-quality |
| 18-106-final-viral-combined_k141_107260\|\|full | Low-quality |
| 18-106-final-viral-combined_k141_66123\|\|full | Low-quality |
| 18-106-final-viral-combined_k141_112731\|\|full | Low-quality |
| 18-106-final-viral-combined_k141_1923\|\|full | Not-determined |
| 18-106-final-viral-combined_k141_118466\|\|full | Low-quality |
| 18-106-final-viral-combined_k141_89825\|\|full | Not-determined |
| 18-106-final-viral-combined_k141_130510\|\|full | Not-determined |
| 18-106-final-viral-combined_k141_66138\|\|full | Not-determined |
| 18-106-final-viral-combined_k141_136070\|\|full | Low-quality |
| 18-106-final-viral-combined_k141_53771\|\|full | Not-determined |
| 18-106-final-viral-combined_k141_36441\|\|full | Low-quality |
| 18-106-final-viral-combined_k141_18796\|\|full | Not-determined |
| 18-106-final-viral-combined_k141_89835\|\|full | Low-quality |
| 18-106-final-viral-combined_k141_66146\|\|full | Not-determined |
| 18-106-final-viral-combined_k141_30931\|\|full | Low-quality |
| 18-106-final-viral-combined_k141_53787\|\|full | Not-determined |
| 18-106-final-viral-combined_k141_30935\|\|full | Not-determined |
| 18-106-final-viral-combined_k141_66158\|\|full | Not-determined |
| 18-106-final-viral-combined_k141_42763\|\|full | Low-quality |
| 18-106-final-viral-combined_k141_83614\|\|full | Low-quality |
| 18-106-final-viral-combined_k141_53805\|\|full | Low-quality |
| 18-106-final-viral-combined_k141_13071\|\|full | Not-determined |
| 18-102-final-viral-combined_k141_79304\|\|full | Medium-quality |
| 18-106-final-viral-combined_k141_95641\|\|full | Low-quality |
| 18-106-final-viral-combined_k141_66172\|\|full | Low-quality |
| 18-106-final-viral-combined_k141_36472\|\|full | Low-quality |
| 18-106-final-viral-combined_k141_77591\|\|full | Low-quality |
| 18-106-final-viral-combined_k141_13091\|\|full | Not-determined |
| 18-106-final-viral-combined_k141_42788\|\|full | Low-quality |
| 18-106-final-viral-combined_k141_30988\|\|full | Not-determined |
| 18-106-final-viral-combined_k141_42809\|\|full | Not-determined |
| 18-106-final-viral-combined_k141_130587\|\|full | Not-determined |
| 18-106-final-viral-combined_k141_53834\|\|full | Low-quality |
| 18-106-final-viral-combined_k141_66184\|\|full | Low-quality |
| 18-106-final-viral-combined_k141_42814\|\|full | Low-quality |
| 18-106-final-viral-combined_k141_60457\|\|full | Low-quality |
| 18-106-final-viral-combined_k141_124662\|\|full | Not-determined |
| 18-106-final-viral-combined_k141_118557\|\|full | Low-quality |
| 18-106-final-viral-combined_k141_95692\|\|full | Low-quality |
| 18-106-final-viral-combined_k141_24682\|\|full | Low-quality |
| 18-106-final-viral-combined_k141_130612\|\|full | Not-determined |
| 18-106-final-viral-combined_k141_124685\|\|full | Not-determined |
| 18-106-final-viral-combined_k141_130615\|\|full | Not-determined |
| 18-106-final-viral-combined_k141_130616\|\|full | Low-quality |
| 18-106-final-viral-combined_k141_18812\|\|full | Low-quality |
| 18-106-final-viral-combined_k141_112764\|\|full | Low-quality |
| 18-106-final-viral-combined_k141_2061\|\|full | Not-determined |
| 18-106-final-viral-combined_k141_83659\|\|full | Low-quality |
| 18-106-final-viral-combined_k141_112781\|\|full | Not-determined |
| 18-106-final-viral-combined_k141_36539\|\|full | Not-determined |
| 18-106-final-viral-combined_k141_24723\|\|full | Not-determined |
| 18-106-final-viral-combined_k141_83673\|\|full | Not-determined |
| 18-106-final-viral-combined_k141_112786\|\|full | Low-quality |
| 18-106-final-viral-combined_k141_2064\|\|full | Not-determined |
| 18-106-final-viral-combined_k141_130653\|\|full | Not-determined |
| 18-106-final-viral-combined_k141_2076\|\|full | Not-determined |
| 18-106-final-viral-combined_k141_130658\|\|full | Not-determined |
| 18-106-final-viral-combined_k141_42867\|\|full | Not-determined |
| 18-106-final-viral-combined_k141_36557\|\|full | Low-quality |
| 18-106-final-viral-combined_k141_71850\|\|full | Not-determined |
| 18-106-final-viral-combined_k141_31051\|\|full | Not-determined |
| 18-106-final-viral-combined_k141_95739\|\|full | Not-determined |
| 18-106-final-viral-combined_k141_24739\|\|full | Not-determined |
| 18-106-final-viral-combined_k141_31067\|\|full | Not-determined |
| 18-106-final-viral-combined_k141_36578\|\|full | Low-quality |
| 18-106-final-viral-combined_k141_130668\|\|full | Low-quality |
| 18-106-final-viral-combined_k141_60522\|\|full | Low-quality |
| 18-106-final-viral-combined_k141_89939\|\|full | Not-determined |
| 18-106-final-viral-combined_k141_53903\|\|full | Low-quality |
| 18-106-final-viral-combined_k141_36596\|\|full | Low-quality |
| 18-106-final-viral-combined_k141_36600\|\|full | Not-determined |
| 18-106-final-viral-combined_k141_77728\|\|full | Not-determined |
| 18-106-final-viral-combined_k141_24748\|\|full | Low-quality |
| 18-106-final-viral-combined_k141_107422\|\|full | Not-determined |
| 18-106-final-viral-combined_k141_13275\|\|full | Not-determined |
| 18-106-final-viral-combined_k141_71875\|\|full | Not-determined |
| 18-106-final-viral-combined_k141_31093\|\|full | Not-determined |
| 18-106-final-viral-combined_k141_77729\|\|full | Not-determined |
| 18-106-final-viral-combined_k141_42922\|\|full | Low-quality |
| 18-106-final-viral-combined_k141_71880\|\|full | Not-determined |
| 18-106-final-viral-combined_k141_24766\|\|full | Low-quality |
| 18-106-final-viral-combined_k141_60561\|\|full | Not-determined |
| 18-106-final-viral-combined_k141_118600\|\|full | Not-determined |
| 18-106-final-viral-combined_k141_60564\|\|full | Not-determined |
| 18-106-final-viral-combined_k141_13286\|\|full | Not-determined |
| 18-106-final-viral-combined_k141_136228\|\|full | Low-quality |
| 18-106-final-viral-combined_k141_31096\|\|full | Low-quality |
| 18-106-final-viral-combined_k141_60587\|\|full | Not-determined |
| 18-106-final-viral-combined_k141_77772\|\|full | Not-determined |
| 18-106-final-viral-combined_k141_53986\|\|full | Not-determined |
| 18-106-final-viral-combined_k141_2095\|\|full | Low-quality |
| 18-106-final-viral-combined_k141_60590\|\|full | Low-quality |
| 18-106-final-viral-combined_k141_48804\|\|full | Not-determined |
| 18-106-final-viral-combined_k141_130736\|\|full | Not-determined |
| 18-106-final-viral-combined_k141_107437\|\|full | Not-determined |
| 18-106-final-viral-combined_k141_112879\|\|full | Low-quality |
| 18-106-final-viral-combined_k141_130749\|\|full | Low-quality |
| 18-106-final-viral-combined_k141_107440\|\|full | Not-determined |
| 18-106-final-viral-combined_k141_90009\|\|full | Not-determined |
| 18-106-final-viral-combined_k141_136237\|\|full | Not-determined |
| 18-106-final-viral-combined_k141_36699\|\|full | Not-determined |
| 18-106-final-viral-combined_k141_118611\|\|full | Not-determined |
| 18-106-final-viral-combined_k141_36703\|\|full | Low-quality |
| 18-106-final-viral-combined_k141_136245\|\|full | Not-determined |
| 18-106-final-viral-combined_k141_13297\|\|full | Low-quality |
| 18-106-final-viral-combined_k141_112897\|\|full | Low-quality |
| 18-106-final-viral-combined_k141_107454\|\|full | Not-determined |
| 18-106-final-viral-combined_k141_107461\|\|full | Not-determined |
| 18-106-final-viral-combined_k141_13308\|\|full | Not-determined |
| 18-106-final-viral-combined_k141_136267\|\|full | Low-quality |
| 18-106-final-viral-combined_k141_24834\|\|full | Low-quality |
| 18-106-final-viral-combined_k141_2153\|\|full | Low-quality |
| 18-106-final-viral-combined_k141_112931\|\|full | Low-quality |
| 18-106-final-viral-combined_k141_124804\|\|full | Low-quality |
| 18-106-final-viral-combined_k141_2158\|\|full | Low-quality |
| 18-106-final-viral-combined_k141_13344\|\|full | Low-quality |
| 18-106-final-viral-combined_k141_13347\|\|full | Not-determined |
| 18-106-final-viral-combined_k141_71940\|\|full | Not-determined |
| 18-106-final-viral-combined_k141_130792\|\|full | Not-determined |
| 18-106-final-viral-combined_k141_31193\|\|full | Not-determined |
| 18-106-final-viral-combined_k141_13366\|\|full | Low-quality |
| 18-106-final-viral-combined_k141_118684\|\|full | Not-determined |
| 18-106-final-viral-combined_k141_24874\|\|full | Not-determined |
| 18-106-final-viral-combined_k141_77779\|\|full | Not-determined |
| 18-106-final-viral-combined_k141_95829\|\|full | Low-quality |
| 18-106-final-viral-combined_k141_18832\|\|full | Low-quality |
| 18-106-final-viral-combined_k141_130814\|\|full | Not-determined |
| 18-106-final-viral-combined_k141_2232\|\|full | Not-determined |
| 18-106-final-viral-combined_k141_2238\|\|full | Low-quality |
| 18-106-final-viral-combined_k141_66383\|\|full | Not-determined |
| 18-106-final-viral-combined_k141_95846\|\|full | Not-determined |
| 18-106-final-viral-combined_k141_71987\|\|full | Not-determined |
| 18-106-final-viral-combined_k141_130826\|\|full | Low-quality |
| 18-106-final-viral-combined_k141_112998\|\|full | Low-quality |
| 18-106-final-viral-combined_k141_90125\|\|full | Low-quality |
| 18-106-final-viral-combined_k141_48910\|\|full | Low-quality |
| 18-106-final-viral-combined_k141_43001\|\|full | Low-quality |
| 18-106-final-viral-combined_k141_31242\|\|full | Low-quality |
| 18-106-final-viral-combined_k141_36870\|\|full | Low-quality |
| 18-106-final-viral-combined_k141_136373\|\|full | Not-determined |
| 18-106-final-viral-combined_k141_18868\|\|full | Low-quality |
| 18-106-final-viral-combined_k141_13408\|\|full | Low-quality |
| 18-106-final-viral-combined_k141_130851\|\|full | Low-quality |
| 18-106-final-viral-combined_k141_43031\|\|full | Not-determined |
| 18-106-final-viral-combined_k141_130853\|\|full | Low-quality |
| 18-106-final-viral-combined_k141_124845\|\|full | Low-quality |
| 18-106-final-viral-combined_k141_83850\|\|full | Not-determined |
| 18-106-final-viral-combined_k141_113027\|\|full | Low-quality |
| 18-106-final-viral-combined_k141_60712\|\|full | Low-quality |
| 18-106-final-viral-combined_k141_13423\|\|full | Not-determined |
| 18-106-final-viral-combined_k141_43060\|\|full | Not-determined |
| 18-106-final-viral-combined_k141_77813\|\|full | Not-determined |
| 18-106-final-viral-combined_k141_13438\|\|full | Low-quality |
| 18-106-final-viral-combined_k141_43078\|\|full | Low-quality |
| 18-106-final-viral-combined_k141_77826\|\|full | Low-quality |
| 18-106-final-viral-combined_k141_36943\|\|full | Not-determined |
| 18-106-final-viral-combined_k141_18909\|\|full | Not-determined |
| 18-106-final-viral-combined_k141_124894\|\|full | Low-quality |
| 18-106-final-viral-combined_k141_136388\|\|full | Low-quality |
| 18-106-final-viral-combined_k141_113057\|\|full | Not-determined |
| 18-106-final-viral-combined_k141_18911\|\|full | Not-determined |
| 18-106-final-viral-combined_k141_13487\|\|full | Low-quality |
| 18-106-final-viral-combined_k141_77843\|\|full | Low-quality |
| 18-106-final-viral-combined_k141_136409\|\|full | Low-quality |
| 18-106-final-viral-combined_k141_136416\|\|full | Low-quality |
| 18-106-final-viral-combined_k141_13494\|\|full | Not-determined |
| 18-106-final-viral-combined_k141_24927\|\|full | Not-determined |
| 18-106-final-viral-combined_k141_72082\|\|full | Not-determined |
| 18-106-final-viral-combined_k141_60749\|\|full | Not-determined |
| 18-106-final-viral-combined_k141_43139\|\|full | Not-determined |
| 18-106-final-viral-combined_k141_136430\|\|full | Not-determined |
| 18-106-final-viral-combined_k141_2327\|\|full | Not-determined |
| 18-106-final-viral-combined_k141_136434\|\|full | Not-determined |
| 18-106-final-viral-combined_k141_24945\|\|full | Not-determined |
| 18-106-final-viral-combined_k141_7554\|\|full | Low-quality |
| 18-106-final-viral-combined_k141_95884\|\|full | Not-determined |
| 18-106-final-viral-combined_k141_95885\|\|full | Low-quality |
| 18-106-final-viral-combined_k141_2357\|\|full | Not-determined |
| 18-106-final-viral-combined_k141_72130\|\|full | Low-quality |
| 18-106-final-viral-combined_k141_113105\|\|full | Low-quality |
| 18-106-final-viral-combined_k141_136465\|\|full | Not-determined |
| 18-106-final-viral-combined_k141_66503\|\|full | Low-quality |
| 18-106-final-viral-combined_k141_49003\|\|full | Not-determined |
| 18-106-final-viral-combined_k141_95908\|\|full | Not-determined |
| 18-106-final-viral-combined_k141_113118\|\|full | Low-quality |
| 18-106-final-viral-combined_k141_66514\|\|full | Low-quality |
| 18-106-final-viral-combined_k141_136490\|\|full | Not-determined |
| 18-106-final-viral-combined_k141_113131\|\|full | Not-determined |
| 18-106-final-viral-combined_k141_113138\|\|full | Not-determined |
| 18-106-final-viral-combined_k141_95919\|\|full | Not-determined |
| 18-106-final-viral-combined_k141_31357\|\|full | Low-quality |
| 18-106-final-viral-combined_k141_124973\|\|full | Not-determined |
| 18-106-final-viral-combined_k141_7596\|\|full | Not-determined |
| 18-106-final-viral-combined_k141_113147\|\|full | Not-determined |
| 18-106-final-viral-combined_k141_124989\|\|full | Not-determined |
| 18-106-final-viral-combined_k141_124994\|\|full | Low-quality |
| 18-106-final-viral-combined_k141_31371\|\|full | Not-determined |
| 18-106-final-viral-combined_k141_31378\|\|full | Not-determined |
| 18-106-final-viral-combined_k141_25004\|\|full | Low-quality |
| 18-106-final-viral-combined_k141_25009\|\|full | Not-determined |
| 18-106-final-viral-combined_k141_136551\|\|full | Not-determined |
| 18-106-final-viral-combined_k141_49080\|\|full | Low-quality |
| 18-106-final-viral-combined_k141_36964\|\|full | Not-determined |
| 18-106-final-viral-combined_k141_113167\|\|full | Not-determined |
| 18-106-final-viral-combined_k141_31383\|\|full | Low-quality |
| 18-106-final-viral-combined_k141_7631\|\|full | Not-determined |
| 18-106-final-viral-combined_k141_2477\|\|full | Not-determined |
| 18-106-final-viral-combined_k141_77888\|\|full | Not-determined |
| 18-106-final-viral-combined_k141_113175\|\|full | Low-quality |
| 18-106-final-viral-combined_k141_66565\|\|full | Not-determined |
| 18-106-final-viral-combined_k141_72190\|\|full | Not-determined |
| 18-106-final-viral-combined_k141_43303\|\|full | Not-determined |
| 18-106-final-viral-combined_k141_125086\|\|full | Low-quality |
| 18-106-final-viral-combined_k141_31418\|\|full | Not-determined |
| 18-106-final-viral-combined_k141_95942\|\|full | Not-determined |
| 18-106-final-viral-combined_k141_113203\|\|full | Not-determined |
| 18-106-final-viral-combined_k141_2525\|\|full | Not-determined |
| 18-106-final-viral-combined_k141_77932\|\|full | Not-determined |
| 18-106-final-viral-combined_k141_136621\|\|full | Not-determined |
| 18-106-final-viral-combined_k141_113231\|\|full | Not-determined |
| 18-106-final-viral-combined_k141_2537\|\|full | Not-determined |
| 18-106-final-viral-combined_k141_101045\|\|full | Not-determined |
| 18-106-final-viral-combined_k141_66639\|\|full | Not-determined |
| 18-106-final-viral-combined_k141_130980\|\|full | Not-determined |
| 18-106-final-viral-combined_k141_136635\|\|full | Not-determined |
| 18-106-final-viral-combined_k141_2552\|\|full | Low-quality |
| 18-106-final-viral-combined_k141_136636\|\|full | Not-determined |
| 18-106-final-viral-combined_k141_31459\|\|full | Low-quality |
| 18-106-final-viral-combined_k141_113241\|\|full | Low-quality |
| 18-106-final-viral-combined_k141_101058\|\|full | Not-determined |
| 18-106-final-viral-combined_k141_77954\|\|full | Not-determined |
| 18-106-final-viral-combined_k141_95972\|\|full | Not-determined |
| 18-106-final-viral-combined_k141_37025\|\|full | Not-determined |
| 18-106-final-viral-combined_k141_37030\|\|full | Low-quality |
| 18-106-final-viral-combined_k141_43381\|\|full | Low-quality |
| 18-106-final-viral-combined_k141_118858\|\|full | Not-determined |
| 18-106-final-viral-combined_k141_95994\|\|full | Low-quality |
| 18-106-final-viral-combined_k141_66692\|\|full | Low-quality |
| 18-106-final-viral-combined_k141_43386\|\|full | Low-quality |
| 18-106-final-viral-combined_k141_54138\|\|full | Not-determined |
| 18-106-final-viral-combined_k141_13680\|\|full | Low-quality |
| 18-106-final-viral-combined_k141_83909\|\|full | Not-determined |
| 18-106-final-viral-combined_k141_136692\|\|full | Low-quality |
| 18-106-final-viral-combined_k141_113301\|\|full | Not-determined |
| 18-106-final-viral-combined_k141_31509\|\|full | Not-determined |
| 18-106-final-viral-combined_k141_66711\|\|full | Low-quality |
| 18-106-final-viral-combined_k141_77992\|\|full | Not-determined |
| 18-106-final-viral-combined_k141_7713\|\|full | Not-determined |
| 18-106-final-viral-combined_k141_136722\|\|full | Not-determined |
| 18-106-final-viral-combined_k141_31536\|\|full | Low-quality |
| 18-106-final-viral-combined_k141_54178\|\|full | Not-determined |
| 18-106-final-viral-combined_k141_37124\|\|full | Low-quality |
| 18-106-final-viral-combined_k141_96052\|\|full | Not-determined |
| 18-106-final-viral-combined_k141_107657\|\|full | Low-quality |
| 18-109_k141_53338 | Medium-quality |
| 18-106-final-viral-combined_k141_83923\|\|full | Not-determined |
| 18-110-final-viral-combined_k141_244525\|\|full | Medium-quality |
| 18-106-final-viral-combined_k141_96063\|\|full | Not-determined |
| 18-106-final-viral-combined_k141_101102\|\|full | Not-determined |
| 18-106-final-viral-combined_k141_131003\|\|full | Low-quality |
| 18-106-final-viral-combined_k141_72231\|\|full | Not-determined |
| 18-106-final-viral-combined_k141_2636\|\|full | Not-determined |
| 18-106-final-viral-combined_k141_107671\|\|full | Not-determined |
| 18-106-final-viral-combined_k141_96075\|\|full | Not-determined |
| 18-106-final-viral-combined_k141_37159\|\|full | Low-quality |
| 18-106-final-viral-combined_k141_125246\|\|full | Low-quality |
| 18-106-final-viral-combined_k141_78058\|\|full | Low-quality |
| 18-106-final-viral-combined_k141_96082\|\|full | Low-quality |
| 18-106-final-viral-combined_k141_43423\|\|full | Low-quality |
| 18-106-final-viral-combined_k141_54208\|\|full | Not-determined |
| 18-106-final-viral-combined_k141_7753\|\|full | Not-determined |
| 18-106-final-viral-combined_k141_136772\|\|full | Low-quality |
| 18-106-final-viral-combined_k141_72284\|\|full | Low-quality |
| 18-106-final-viral-combined_k141_60952\|\|full | Not-determined |
| 18-106-final-viral-combined_k141_43443\|\|full | Not-determined |
| 18-106-final-viral-combined_k141_136794\|\|full | Not-determined |
| 18-106-final-viral-combined_k141_13725\|\|full | Low-quality |
| 18-106-final-viral-combined_k141_13726\|\|full | Low-quality |
| 18-106-final-viral-combined_k141_54248\|\|full | Not-determined |
| 18-106-final-viral-combined_k141_113354\|\|full | Low-quality |
| 18-106-final-viral-combined_k141_37217\|\|full | Low-quality |
| 18-106-final-viral-combined_k141_101171\|\|full | Not-determined |
| 18-106-final-viral-combined_k141_125299\|\|full | Not-determined |
| 18-106-final-viral-combined_k141_96129\|\|full | Low-quality |
| 18-106-final-viral-combined_k141_54264\|\|full | Low-quality |
| 18-106-final-viral-combined_k141_101175\|\|full | Not-determined |
| 18-106-final-viral-combined_k141_66833\|\|full | Not-determined |
| 18-106-final-viral-combined_k141_131064\|\|full | Not-determined |
| 18-106-final-viral-combined_k141_60987\|\|full | Not-determined |
| 18-106-final-viral-combined_k141_107761\|\|full | Not-determined |
| 18-106-final-viral-combined_k141_43494\|\|full | Low-quality |
| 18-106-final-viral-combined_k141_96157\|\|full | Low-quality |
| 18-106-final-viral-combined_k141_118996\|\|full | Not-determined |
| 18-106-final-viral-combined_k141_2725\|\|full | Not-determined |
| 18-106-final-viral-combined_k141_66866\|\|full | Not-determined |
| 18-106-final-viral-combined_k141_136848\|\|full | Low-quality |
| 18-106-final-viral-combined_k141_136849\|\|full | Low-quality |
| 18-106-final-viral-combined_k141_43498\|\|full | Not-determined |
| 18-106-final-viral-combined_k141_2735\|\|full | Not-determined |
| 18-106-final-viral-combined_k141_19053\|\|full | Not-determined |
| 18-106-final-viral-combined_k141_96165\|\|full | Low-quality |
| 18-106-final-viral-combined_k141_78103\|\|full | Low-quality |
| 18-106-final-viral-combined_k141_84033\|\|full | Not-determined |
| 18-106-final-viral-combined_k141_66880\|\|full | Low-quality |
| 18-106-final-viral-combined_k141_107776\|\|full | Low-quality |
| 18-106-final-viral-combined_k141_13752\|\|full | Low-quality |
| 18-106-final-viral-combined_k141_31609\|\|full | Not-determined |
| 18-106-final-viral-combined_k141_54294\|\|full | Not-determined |
| 18-106-final-viral-combined_k141_37283\|\|full | Not-determined |
| 18-106-final-viral-combined_k141_31618\|\|full | Low-quality |
| 18-106-final-viral-combined_k141_84052\|\|full | Not-determined |
| 18-106-final-viral-combined_k141_31625\|\|full | Low-quality |
| 18-106-final-viral-combined_k141_25177\|\|full | Low-quality |
| 18-106-final-viral-combined_k141_61027\|\|full | Low-quality |
| 18-106-final-viral-combined_k141_31636\|\|full | Not-determined |
| 18-106-final-viral-combined_k141_31639\|\|full | Low-quality |
| 18-106-final-viral-combined_k141_2766\|\|full | Not-determined |
| 18-109_k141_239833 | Medium-quality |
| 18-106-final-viral-combined_k141_37330\|\|full | Not-determined |
| 18-106-final-viral-combined_k141_61041\|\|full | Not-determined |
| 18-106-final-viral-combined_k141_31647\|\|full | Low-quality |
| 18-106-final-viral-combined_k141_54323\|\|full | Not-determined |
| 18-106-final-viral-combined_k141_113449\|\|full | Not-determined |
| 18-106-final-viral-combined_k141_78112\|\|full | Low-quality |
| 18-106-final-viral-combined_k141_119027\|\|full | Low-quality |
| 18-106-final-viral-combined_k141_125389\|\|full | Low-quality |
| 18-106-final-viral-combined_k141_31686\|\|full | Low-quality |
| 18-106-final-viral-combined_k141_2822\|\|full | Low-quality |
| 18-106-final-viral-combined_k141_119045\|\|full | Low-quality |
| 18-106-final-viral-combined_k141_2829\|\|full | Low-quality |
| 18-106-final-viral-combined_k141_66929\|\|full | Low-quality |
| 18-106-final-viral-combined_k141_31700\|\|full | Not-determined |
| 18-106-final-viral-combined_k141_13779\|\|full | Not-determined |
| 18-106-final-viral-combined_k141_78114\|\|full | Not-determined |
| 18-106-final-viral-combined_k141_78116\|\|full | Low-quality |
| 18-106-final-viral-combined_k141_125401\|\|full | Low-quality |
| 18-106-final-viral-combined_k141_31708\|\|full | Low-quality |
| 18-106-final-viral-combined_k141_66939\|\|full | Not-determined |
| 18-106-final-viral-combined_k141_49254\|\|full | Low-quality |
| 18-106-final-viral-combined_k141_72399\|\|full | Low-quality |
| 18-106-final-viral-combined_k141_66941\|\|full | Low-quality |
| 18-106-final-viral-combined_k141_25253\|\|full | Not-determined |
| 18-106-final-viral-combined_k141_66944\|\|full | Low-quality |
| 18-106-final-viral-combined_k141_49264\|\|full | Not-determined |
| 18-106-final-viral-combined_k141_25258\|\|full | Low-quality |
| 18-106-final-viral-combined_k141_43571\|\|full | Not-determined |
| 18-106-final-viral-combined_k141_19105\|\|full | Not-determined |
| 18-106-final-viral-combined_k141_125450\|\|full | Low-quality |
| 18-106-final-viral-combined_k141_101258\|\|full | Low-quality |
| 18-106-final-viral-combined_k141_78135\|\|full | Not-determined |
| 18-106-final-viral-combined_k141_78144\|\|full | Not-determined |
| 18-106-final-viral-combined_k141_131181\|\|full | Low-quality |
| 18-106-final-viral-combined_k141_7958\|\|full | Not-determined |
| 18-106-final-viral-combined_k141_37414\|\|full | Low-quality |
| 18-106-final-viral-combined_k141_49304\|\|full | Low-quality |
| 18-106-final-viral-combined_k141_13865\|\|full | Low-quality |
| 18-106-final-viral-combined_k141_31752\|\|full | Not-determined |
| 18-106-final-viral-combined_k141_125496\|\|full | Low-quality |
| 18-106-final-viral-combined_k141_78159\|\|full | Not-determined |
| 18-106-final-viral-combined_k141_19143\|\|full | Low-quality |
| 18-106-final-viral-combined_k141_25336\|\|full | Low-quality |
| 18-106-final-viral-combined_k141_19149\|\|full | Not-determined |
| 18-106-final-viral-combined_k141_101333\|\|full | Low-quality |
| 18-106-final-viral-combined_k141_43612\|\|full | Low-quality |
| 18-106-final-viral-combined_k141_8002\|\|full | Not-determined |
| 18-106-final-viral-combined_k141_25350\|\|full | Low-quality |
| 18-106-final-viral-combined_k141_101340\|\|full | Not-determined |
| 18-106-final-viral-combined_k141_84085\|\|full | Low-quality |
| 18-106-final-viral-combined_k141_2914\|\|full | Not-determined |
| 18-106-final-viral-combined_k141_25361\|\|full | Low-quality |
| 18-106-final-viral-combined_k141_72460\|\|full | Not-determined |
| 18-106-final-viral-combined_k141_66964\|\|full | Not-determined |
| 18-106-final-viral-combined_k141_90340\|\|full | Not-determined |
| 18-106-final-viral-combined_k141_101364\|\|full | Low-quality |
| 18-106-final-viral-combined_k141_8027\|\|full | Not-determined |
| 18-106-final-viral-combined_k141_90346\|\|full | Low-quality |
| 18-106-final-viral-combined_k141_72462\|\|full | Not-determined |
| 18-106-final-viral-combined_k141_90348\|\|full | Low-quality |
| 18-106-final-viral-combined_k141_96307\|\|full | Not-determined |
| 18-106-final-viral-combined_k141_25388\|\|full | Low-quality |
| 18-106-final-viral-combined_k141_25389\|\|full | Not-determined |
| 18-106-final-viral-combined_k141_90358\|\|full | Low-quality |
| 18-106-final-viral-combined_k141_96317\|\|full | Low-quality |
| 18-106-final-viral-combined_k141_119116\|\|full | Low-quality |
| 18-106-final-viral-combined_k141_2968\|\|full | Not-determined |
| 18-106-final-viral-combined_k141_72468\|\|full | Not-determined |
| 18-106-final-viral-combined_k141_61132\|\|full | Low-quality |
| 18-106-final-viral-combined_k141_96320\|\|full | Not-determined |
| 18-106-final-viral-combined_k141_107947\|\|full | Not-determined |
| 18-106-final-viral-combined_k141_90377\|\|full | Not-determined |
| 18-106-final-viral-combined_k141_61139\|\|full | Low-quality |
| 18-106-final-viral-combined_k141_78232\|\|full | Not-determined |
| 18-106-final-viral-combined_k141_2999\|\|full | Not-determined |
| 18-106-final-viral-combined_k141_125541\|\|full | Low-quality |
| 18-106-final-viral-combined_k141_90388\|\|full | Not-determined |
| 18-106-final-viral-combined_k141_72511\|\|full | Low-quality |
| 18-106-final-viral-combined_k141_107981\|\|full | Low-quality |
| 18-106-final-viral-combined_k141_107985\|\|full | Low-quality |
| 18-106-final-viral-combined_k141_90405\|\|full | Low-quality |
| 18-106-final-viral-combined_k141_113502\|\|full | Not-determined |
| 18-106-final-viral-combined_k141_19272\|\|full | Low-quality |
| 18-106-final-viral-combined_k141_8089\|\|full | Low-quality |
| 18-106-final-viral-combined_k141_84172\|\|full | Low-quality |
| 18-106-final-viral-combined_k141_37573\|\|full | Low-quality |
| 18-106-final-viral-combined_k141_37577\|\|full | Low-quality |
| 18-106-final-viral-combined_k141_101429\|\|full | Low-quality |
| 18-106-final-viral-combined_k141_25454\|\|full | Not-determined |
| 18-106-final-viral-combined_k141_3027\|\|full | Low-quality |
| 18-106-final-viral-combined_k141_19288\|\|full | Low-quality |
| 18-106-final-viral-combined_k141_72558\|\|full | Not-determined |
| 18-106-final-viral-combined_k141_131243\|\|full | Low-quality |
| 18-106-final-viral-combined_k141_37583\|\|full | Not-determined |
| 18-106-final-viral-combined_k141_25465\|\|full | Low-quality |
| 18-106-final-viral-combined_k141_84194\|\|full | Not-determined |
| 18-106-final-viral-combined_k141_113528\|\|full | Low-quality |
| 18-106-final-viral-combined_k141_90408\|\|full | Not-determined |
| 18-106-final-viral-combined_k141_84203\|\|full | Low-quality |
| 18-102_k141_53881 | Medium-quality |
| 18-106-final-viral-combined_k141_125565\|\|full | Not-determined |
| 18-106-final-viral-combined_k141_37604\|\|full | Not-determined |
| 18-106-final-viral-combined_k141_19308\|\|full | Low-quality |
| 18-106-final-viral-combined_k141_72577\|\|full | Not-determined |
| 18-106-final-viral-combined_k141_14016\|\|full | Low-quality |
| 18-106-final-viral-combined_k141_72578\|\|full | Low-quality |
| 18-106-final-viral-combined_k141_3071\|\|full | Not-determined |
| 18-106-final-viral-combined_k141_37625\|\|full | Low-quality |
| 18-106-final-viral-combined_k141_72582\|\|full | Low-quality |
| 18-106-final-viral-combined_k141_96396\|\|full | Not-determined |
| 18-106-final-viral-combined_k141_119146\|\|full | Not-determined |
| 18-106-final-viral-combined_k141_67072\|\|full | Low-quality |
| 18-106-final-viral-combined_k141_54459\|\|full | Not-determined |
| 18-106-final-viral-combined_k141_49480\|\|full | Not-determined |
| 18-106-final-viral-combined_k141_113571\|\|full | Not-determined |
| 18-106-final-viral-combined_k141_108059\|\|full | Not-determined |
| 18-106-final-viral-combined_k141_113573\|\|full | Low-quality |
| 18-106-final-viral-combined_k141_67093\|\|full | Low-quality |
| 18-106-final-viral-combined_k141_90492\|\|full | Low-quality |
| 18-106-final-viral-combined_k141_78340\|\|full | Not-determined |
| 18-106-final-viral-combined_k141_108064\|\|full | Not-determined |
| 18-106-final-viral-combined_k141_19348\|\|full | Low-quality |
| 18-106-final-viral-combined_k141_84259\|\|full | Low-quality |
| 18-106-final-viral-combined_k141_84278\|\|full | Not-determined |
| 18-106-final-viral-combined_k141_67138\|\|full | Low-quality |
| 18-106-final-viral-combined_k141_14065\|\|full | Not-determined |
| 18-106-final-viral-combined_k141_125617\|\|full | Low-quality |
| 18-106-final-viral-combined_k141_19364\|\|full | Not-determined |
| 18-106-final-viral-combined_k141_125620\|\|full | Low-quality |
| 18-106-final-viral-combined_k141_14070\|\|full | Low-quality |
| 18-106-final-viral-combined_k141_96437\|\|full | Not-determined |
| 18-106-final-viral-combined_k141_3121\|\|full | Low-quality |
| 18-106-final-viral-combined_k141_90524\|\|full | Not-determined |
| 18-106-final-viral-combined_k141_37690\|\|full | Not-determined |
| 18-106-final-viral-combined_k141_78377\|\|full | Low-quality |
| 18-106-final-viral-combined_k141_108096\|\|full | Not-determined |
| 18-106-final-viral-combined_k141_61196\|\|full | Not-determined |
| 18-106-final-viral-combined_k141_31854\|\|full | Not-determined |
| 18-106-final-viral-combined_k141_3182\|\|full | Low-quality |
| 18-106-final-viral-combined_k141_25642\|\|full | Low-quality |
| 18-106-final-viral-combined_k141_84322\|\|full | Low-quality |
| 18-106-final-viral-combined_k141_37754\|\|full | Not-determined |
| 18-106-final-viral-combined_k141_84324\|\|full | Low-quality |
| 18-106-final-viral-combined_k141_108143\|\|full | Not-determined |
| 18-106-final-viral-combined_k141_72664\|\|full | Not-determined |
| 18-106-final-viral-combined_k141_125705\|\|full | Not-determined |
| 18-106-final-viral-combined_k141_96502\|\|full | Low-quality |
| 18-106-final-viral-combined_k141_125711\|\|full | Not-determined |
| 18-106-final-viral-combined_k141_90564\|\|full | Not-determined |
| 18-106-final-viral-combined_k141_72681\|\|full | Low-quality |

| | |
|---|---|
| 18-106-final-viral-combined_k141_49582\|\|full | Not-determined |
| 18-106-final-viral-combined_k141_108151\|\|full | Low-quality |
| 18-106-final-viral-combined_k141_72697\|\|full | Not-determined |
| 18-106-final-viral-combined_k141_101557\|\|full | Not-determined |
| 18-106-final-viral-combined_k141_72704\|\|full | Low-quality |
| 18-106-final-viral-combined_k141_72706\|\|full | Low-quality |
| 18-106-final-viral-combined_k141_84361\|\|full | Not-determined |
| 18-106-final-viral-combined_k141_61269\|\|full | Low-quality |
| 18-106-final-viral-combined_k141_113680\|\|full | Low-quality |
| 18-106-final-viral-combined_k141_49624\|\|full | Not-determined |
| 18-106-final-viral-combined_k141_108175\|\|full | Low-quality |
| 18-106-final-viral-combined_k141_72718\|\|full | Not-determined |
| 18-106-final-viral-combined_k141_19466\|\|full | Not-determined |
| 18-106-final-viral-combined_k141_119330\|\|full | Not-determined |
| 18-109-final-viral-combined_k141_239833\|\|full | Medium-quality |
| 18-106-final-viral-combined_k141_131302\|\|full | Low-quality |
| 18-106-final-viral-combined_k141_96542\|\|full | Low-quality |
| 18-106-final-viral-combined_k141_119352\|\|full | Low-quality |
| 18-106-final-viral-combined_k141_90657\|\|full | Not-determined |
| 18-106-final-viral-combined_k141_3298\|\|full | Not-determined |
| 18-106-final-viral-combined_k141_43894\|\|full | Low-quality |
| 18-106-final-viral-combined_k141_108189\|\|full | Low-quality |
| 18-106-final-viral-combined_k141_119373\|\|full | Low-quality |
| 18-106-final-viral-combined_k141_25708\|\|full | Not-determined |
| 18-106-final-viral-combined_k141_108198\|\|full | Not-determined |
| 18-106-final-viral-combined_k141_119382\|\|full | Not-determined |
| 18-106-final-viral-combined_k141_90686\|\|full | Not-determined |
| 18-106-final-viral-combined_k141_25729\|\|full | Low-quality |
| 18-106-final-viral-combined_k141_131364\|\|full | Low-quality |
| 18-106-final-viral-combined_k141_37874\|\|full | Low-quality |
| 18-106-final-viral-combined_k141_19532\|\|full | Low-quality |
| 18-106-final-viral-combined_k141_19534\|\|full | Not-determined |
| 18-106-final-viral-combined_k141_49692\|\|full | Not-determined |
| 18-106-final-viral-combined_k141_101641\|\|full | Not-determined |
| 18-106-final-viral-combined_k141_19541\|\|full | Low-quality |
| 18-106-final-viral-combined_k141_43949\|\|full | Not-determined |
| 18-106-final-viral-combined_k141_78542\|\|full | Not-determined |
| 18-106-final-viral-combined_k141_131393\|\|full | Not-determined |
| 18-106-final-viral-combined_k141_8217\|\|full | Not-determined |
| 18-106-final-viral-combined_k141_31966\|\|full | Low-quality |
| 18-106-final-viral-combined_k141_78548\|\|full | Low-quality |
| 18-106-final-viral-combined_k141_137031\|\|full | Not-determined |
| 18-106-final-viral-combined_k141_49740\|\|full | Low-quality |
| 18-106-final-viral-combined_k141_125878\|\|full | Low-quality |
| 18-106-final-viral-combined_k141_67371\|\|full | Not-determined |
| 18-106-final-viral-combined_k141_131432\|\|full | Low-quality |
| 18-106-final-viral-combined_k141_101691\|\|full | Not-determined |
| 18-106-final-viral-combined_k141_125884\|\|full | Not-determined |
| 18-106-final-viral-combined_k141_78591\|\|full | Not-determined |
| 18-106-final-viral-combined_k141_108259\|\|full | Low-quality |
| 18-106-final-viral-combined_k141_72796\|\|full | Low-quality |
| 18-106-final-viral-combined_k141_54693\|\|full | Not-determined |
| 18-106-final-viral-combined_k141_25833\|\|full | Not-determined |
| 18-106-final-viral-combined_k141_90754\|\|full | Not-determined |
| 18-106-final-viral-combined_k141_96691\|\|full | Low-quality |
| 18-106-final-viral-combined_k141_54706\|\|full | Not-determined |
| 18-106-final-viral-combined_k141_14166\|\|full | Not-determined |
| 18-106-final-viral-combined_k141_90759\|\|full | Low-quality |
| 18-106-final-viral-combined_k141_108285\|\|full | Low-quality |
| 18-106-final-viral-combined_k141_37960\|\|full | Not-determined |
| 18-106-final-viral-combined_k141_54708\|\|full | Not-determined |
| 18-106-final-viral-combined_k141_49778\|\|full | Low-quality |
| 18-106-final-viral-combined_k141_84496\|\|full | Low-quality |
| 18-106-final-viral-combined_k141_72833\|\|full | Low-quality |
| 18-106-final-viral-combined_k141_72839\|\|full | Not-determined |
| 18-106-final-viral-combined_k141_49802\|\|full | Low-quality |
| 18-106-final-viral-combined_k141_37966\|\|full | Low-quality |
| 18-106-final-viral-combined_k141_14174\|\|full | Not-determined |
| 18-106-final-viral-combined_k141_131507\|\|full | Not-determined |
| 18-106-final-viral-combined_k141_84528\|\|full | Low-quality |
| 18-106-final-viral-combined_k141_67437\|\|full | Low-quality |
| 18-106-final-viral-combined_k141_119495\|\|full | Low-quality |
| 18-106-final-viral-combined_k141_90791\|\|full | Not-determined |
| 18-106-final-viral-combined_k141_38009\|\|full | Low-quality |
| 18-106-final-viral-combined_k141_137152\|\|full | Low-quality |
| 18-106-final-viral-combined_k141_14217\|\|full | Low-quality |
| 18-106-final-viral-combined_k141_72898\|\|full | Low-quality |
| 18-106-final-viral-combined_k141_101739\|\|full | Not-determined |
| 18-106-final-viral-combined_k141_78645\|\|full | Not-determined |
| 18-106-final-viral-combined_k141_32058\|\|full | Low-quality |
| 18-106-final-viral-combined_k141_19629\|\|full | Not-determined |
| 18-106-final-viral-combined_k141_32065\|\|full | Low-quality |
| 18-106-final-viral-combined_k141_90819\|\|full | Not-determined |
| 18-106-final-viral-combined_k141_137173\|\|full | Not-determined |
| 18-106-final-viral-combined_k141_32069\|\|full | Low-quality |
| 18-106-final-viral-combined_k141_32071\|\|full | Not-determined |
| 18-106-final-viral-combined_k141_3454\|\|full | Low-quality |
| 18-106-final-viral-combined_k141_61496\|\|full | Not-determined |
| 18-106-final-viral-combined_k141_101758\|\|full | Low-quality |
| 18-106-final-viral-combined_k141_38046\|\|full | Not-determined |
| 18-106-final-viral-combined_k141_14261\|\|full | Low-quality |
| 18-106-final-viral-combined_k141_32094\|\|full | Not-determined |
| 18-106-final-viral-combined_k141_72969\|\|full | Not-determined |
| 18-106-final-viral-combined_k141_137203\|\|full | Not-determined |
| 18-106-final-viral-combined_k141_101781\|\|full | Low-quality |
| 18-106-final-viral-combined_k141_61508\|\|full | Low-quality |
| 18-106-final-viral-combined_k141_90852\|\|full | Not-determined |
| 18-106-final-viral-combined_k141_119537\|\|full | Low-quality |
| 18-106-final-viral-combined_k141_84575\|\|full | Low-quality |
| 18-106-final-viral-combined_k141_113903\|\|full | Low-quality |
| 18-109-final-viral-combined_k141_53338\|\|full | Medium-quality |
| 18-106-final-viral-combined_k141_73019\|\|full | Low-quality |
| 18-105_k141_33855 | Medium-quality |
| 18-106-final-viral-combined_k141_25846\|\|full | Not-determined |
| 18-106-final-viral-combined_k141_137244\|\|full | Not-determined |
| 18-106-final-viral-combined_k141_84584\|\|full | Not-determined |
| 18-106-final-viral-combined_k141_78658\|\|full | Not-determined |
| 18-106-final-viral-combined_k141_90874\|\|full | Low-quality |
| 18-106-final-viral-combined_k141_113912\|\|full | Low-quality |
| 18-106-final-viral-combined_k141_78660\|\|full | Low-quality |
| 18-106-final-viral-combined_k141_38090\|\|full | Low-quality |
| 18-106-final-viral-combined_k141_8320\|\|full | Low-quality |
| 18-106-final-viral-combined_k141_119572\|\|full | Not-determined |
| 18-106-final-viral-combined_k141_101819\|\|full | Low-quality |
| 18-106-final-viral-combined_k141_67545\|\|full | Not-determined |
| 18-106-final-viral-combined_k141_96859\|\|full | Low-quality |
| 18-106-final-viral-combined_k141_137271\|\|full | Not-determined |
| 18-106-final-viral-combined_k141_54858\|\|full | Low-quality |
| 18-106-final-viral-combined_k141_101827\|\|full | Not-determined |
| 18-106-final-viral-combined_k141_113942\|\|full | Low-quality |
| 18-106-final-viral-combined_k141_84623\|\|full | Low-quality |
| 18-106-final-viral-combined_k141_19719\|\|full | Not-determined |
| 18-106-final-viral-combined_k141_19721\|\|full | Not-determined |
| 18-106-final-viral-combined_k141_38123\|\|full | Not-determined |
| 18-106-final-viral-combined_k141_90936\|\|full | Not-determined |
| 18-106-final-viral-combined_k141_61556\|\|full | Not-determined |
| 18-106-final-viral-combined_k141_96895\|\|full | Not-determined |
| 18-106-final-viral-combined_k141_67580\|\|full | Low-quality |
| 18-106-final-viral-combined_k141_108321\|\|full | Low-quality |
| 18-106-final-viral-combined_k141_19737\|\|full | Low-quality |
| 18-106-final-viral-combined_k141_54916\|\|full | Low-quality |
| 18-106-final-viral-combined_k141_3558\|\|full | Low-quality |
| 18-106-final-viral-combined_k141_108325\|\|full | Low-quality |
| 18-106-final-viral-combined_k141_44205\|\|full | Not-determined |
| 18-106-final-viral-combined_k141_108336\|\|full | Not-determined |
| 18-106-final-viral-combined_k141_54930\|\|full | Low-quality |
| 18-106-final-viral-combined_k141_14359\|\|full | Not-determined |
| 18-106-final-viral-combined_k141_14367\|\|full | Not-determined |
| 18-106-final-viral-combined_k141_108363\|\|full | Not-determined |
| 18-106-final-viral-combined_k141_61608\|\|full | Low-quality |
| 18-106-final-viral-combined_k141_131652\|\|full | Not-determined |
| 18-106-final-viral-combined_k141_84654\|\|full | Low-quality |
| 18-102-final-viral-combined_k141_233107\|\|full | Medium-quality |
| 18-106-final-viral-combined_k141_78739\|\|full | Low-quality |
| 18-106-final-viral-combined_k141_3578\|\|full | Not-determined |
| 18-106-final-viral-combined_k141_96933\|\|full | Low-quality |
| 18-106-final-viral-combined_k141_32183\|\|full | Not-determined |
| 18-106-final-viral-combined_k141_44235\|\|full | Not-determined |
| 18-106-final-viral-combined_k141_67639\|\|full | Low-quality |
| 18-106-final-viral-combined_k141_108388\|\|full | Low-quality |
| 18-106-final-viral-combined_k141_3584\|\|full | Not-determined |
| 18-106-final-viral-combined_k141_44253\|\|full | Not-determined |
| 18-106-final-viral-combined_k141_73150\|\|full | Not-determined |
| 18-106-final-viral-combined_k141_96941\|\|full | Not-determined |
| 18-106-final-viral-combined_k141_131685\|\|full | Low-quality |
| 18-106-final-viral-combined_k141_67663\|\|full | Not-determined |
| 18-106-final-viral-combined_k141_38218\|\|full | Not-determined |
| 18-106-final-viral-combined_k141_137318\|\|full | Low-quality |
| 18-106-final-viral-combined_k141_73167\|\|full | Low-quality |
| 18-106-final-viral-combined_k141_44276\|\|full | Not-determined |
| 18-106-final-viral-combined_k141_131696\|\|full | Low-quality |
| 18-106-final-viral-combined_k141_38224\|\|full | Not-determined |
| 18-106-final-viral-combined_k141_125916\|\|full | Not-determined |
| 18-106-final-viral-combined_k141_32221\|\|full | Not-determined |
| 18-106-final-viral-combined_k141_8378\|\|full | Low-quality |
| 18-106-final-viral-combined_k141_32231\|\|full | Low-quality |
| 18-106-final-viral-combined_k141_44287\|\|full | Not-determined |
| 18-106-final-viral-combined_k141_96982\|\|full | Not-determined |
| 18-106-final-viral-combined_k141_32236\|\|full | Not-determined |
| 18-106-final-viral-combined_k141_131726\|\|full | Low-quality |
| 18-106-final-viral-combined_k141_19855\|\|full | Low-quality |
| 18-106-final-viral-combined_k141_50048\|\|full | Not-determined |
| 18-106-final-viral-combined_k141_14428\|\|full | Low-quality |
| 18-106-final-viral-combined_k141_44297\|\|full | Low-quality |
| 18-106-final-viral-combined_k141_3634\|\|full | Not-determined |
| 18-106-final-viral-combined_k141_125932\|\|full | Low-quality |
| 18-106-final-viral-combined_k141_19883\|\|full | Not-determined |
| 18-106-final-viral-combined_k141_32261\|\|full | Not-determined |
| 18-106-final-viral-combined_k141_102018\|\|full | Not-determined |
| 18-106-final-viral-combined_k141_14441\|\|full | Not-determined |
| 18-106-final-viral-combined_k141_84739\|\|full | Not-determined |
| 18-106-final-viral-combined_k141_125940\|\|full | Low-quality |
| 18-106-final-viral-combined_k141_78806\|\|full | Low-quality |
| 18-106-final-viral-combined_k141_119653\|\|full | Low-quality |
| 18-106-final-viral-combined_k141_67737\|\|full | Low-quality |
| 18-106-final-viral-combined_k141_137407\|\|full | Low-quality |
| 18-106-final-viral-combined_k141_78823\|\|full | Low-quality |
| 18-106-final-viral-combined_k141_67746\|\|full | Not-determined |
| 18-106-final-viral-combined_k141_84753\|\|full | Not-determined |
| 18-106-final-viral-combined_k141_14470\|\|full | Low-quality |
| 18-106-final-viral-combined_k141_3674\|\|full | Low-quality |
| 18-106-final-viral-combined_k141_84775\|\|full | Not-determined |
| 18-106-final-viral-combined_k141_32284\|\|full | Not-determined |
| 18-106-final-viral-combined_k141_67761\|\|full | Low-quality |
| 18-106-final-viral-combined_k141_97073\|\|full | Low-quality |
| 18-106-final-viral-combined_k141_137438\|\|full | Not-determined |
| 18-106-final-viral-combined_k141_61699\|\|full | Not-determined |
| 18-106-final-viral-combined_k141_131828\|\|full | Low-quality |
| 18-106-final-viral-combined_k141_114069\|\|full | Not-determined |
| 18-106-final-viral-combined_k141_19933\|\|full | Low-quality |
| 18-106-final-viral-combined_k141_73252\|\|full | Not-determined |
| 18-106-final-viral-combined_k141_38337\|\|full | Low-quality |
| 18-106-final-viral-combined_k141_3728\|\|full | Low-quality |
| 18-106-final-viral-combined_k141_108515\|\|full | Low-quality |
| 18-106-final-viral-combined_k141_131860\|\|full | Not-determined |
| 18-106-final-viral-combined_k141_44340\|\|full | Low-quality |
| 18-106-final-viral-combined_k141_119695\|\|full | Low-quality |
| 18-106-final-viral-combined_k141_55071\|\|full | Low-quality |
| 18-106-final-viral-combined_k141_114107\|\|full | Not-determined |
| 18-106-final-viral-combined_k141_131879\|\|full | Low-quality |
| 18-106-final-viral-combined_k141_102116\|\|full | Not-determined |
| 18-106-final-viral-combined_k141_131883\|\|full | Low-quality |
| 18-106-final-viral-combined_k141_44352\|\|full | Not-determined |
| 18-106-final-viral-combined_k141_73259\|\|full | Not-determined |
| 18-106-final-viral-combined_k141_73260\|\|full | Not-determined |
| 18-106-final-viral-combined_k141_32338\|\|full | Low-quality |
| 18-106-final-viral-combined_k141_119713\|\|full | Not-determined |
| 18-106-final-viral-combined_k141_84820\|\|full | Low-quality |
| 18-106-final-viral-combined_k141_114133\|\|full | Low-quality |
| 18-106-final-viral-combined_k141_55090\|\|full | Not-determined |
| 18-106-final-viral-combined_k141_73290\|\|full | Low-quality |
| 18-106-final-viral-combined_k141_14512\|\|full | Low-quality |
| 18-106-final-viral-combined_k141_14513\|\|full | Not-determined |
| 18-106-final-viral-combined_k141_114146\|\|full | Low-quality |
| 18-106-final-viral-combined_k141_8452\|\|full | Not-determined |
| 18-106-final-viral-combined_k141_126064\|\|full | Low-quality |
| 18-106-final-viral-combined_k141_131931\|\|full | Low-quality |
| 18-106-final-viral-combined_k141_38400\|\|full | Not-determined |
| 18-106-final-viral-combined_k141_102124\|\|full | Low-quality |
| 18-106-final-viral-combined_k141_73324\|\|full | Not-determined |
| 18-106-final-viral-combined_k141_97147\|\|full | Not-determined |
| 18-106-final-viral-combined_k141_3811\|\|full | Low-quality |
| 18-106-final-viral-combined_k141_102138\|\|full | Not-determined |
| 18-106-final-viral-combined_k141_108580\|\|full | Low-quality |
| 18-106-final-viral-combined_k141_73339\|\|full | Not-determined |
| 18-106-final-viral-combined_k141_119768\|\|full | Not-determined |
| 18-106-final-viral-combined_k141_67898\|\|full | Not-determined |
| 18-106-final-viral-combined_k141_61796\|\|full | Low-quality |
| 18-106-final-viral-combined_k141_3851\|\|full | Low-quality |
| 18-106-final-viral-combined_k141_32435\|\|full | Low-quality |
| 18-106-final-viral-combined_k141_14564\|\|full | Not-determined |
| 18-106-final-viral-combined_k141_114244\|\|full | Not-determined |
| 18-106-final-viral-combined_k141_44423\|\|full | Low-quality |
| 18-106-final-viral-combined_k141_32441\|\|full | Not-determined |
| 18-106-final-viral-combined_k141_131949\|\|full | Not-determined |
| 18-106-final-viral-combined_k141_19978\|\|full | Low-quality |
| 18-106-final-viral-combined_k141_84887\|\|full | Not-determined |
| 18-106-final-viral-combined_k141_126174\|\|full | Not-determined |
| 18-106-final-viral-combined_k141_97227\|\|full | Not-determined |
| 18-106-final-viral-combined_k141_19993\|\|full | Not-determined |
| 18-106-final-viral-combined_k141_108679\|\|full | Low-quality |
| 18-106-final-viral-combined_k141_119820\|\|full | Low-quality |
| 18-106-final-viral-combined_k141_97256\|\|full | Not-determined |
| 18-106-final-viral-combined_k141_26037\|\|full | Not-determined |
| 18-106-final-viral-combined_k141_20013\|\|full | Low-quality |
| 18-106-final-viral-combined_k141_55210\|\|full | Not-determined |
| 18-106-final-viral-combined_k141_119835\|\|full | Not-determined |
| 18-106-final-viral-combined_k141_55216\|\|full | Not-determined |
| 18-106-final-viral-combined_k141_137542\|\|full | Low-quality |
| 18-106-final-viral-combined_k141_119848\|\|full | Not-determined |
| 18-109-final-viral-combined_k141_225260\|\|full | High-quality |
| 18-106-final-viral-combined_k141_32485\|\|full | Low-quality |
| 18-106-final-viral-combined_k141_97286\|\|full | Low-quality |
| 18-106-final-viral-combined_k141_126206\|\|full | Low-quality |
| 18-106-final-viral-combined_k141_84926\|\|full | Not-determined |
| 18-106-final-viral-combined_k141_20044\|\|full | Not-determined |
| 18-106-final-viral-combined_k141_78828\|\|full | Not-determined |
| 18-106-final-viral-combined_k141_68002\|\|full | Not-determined |
| 18-106-final-viral-combined_k141_97296\|\|full | Not-determined |
| 18-106-final-viral-combined_k141_119859\|\|full | Low-quality |
| 18-106-final-viral-combined_k141_132009\|\|full | Low-quality |
| 18-106-final-viral-combined_k141_32506\|\|full | Not-determined |
| 18-106-final-viral-combined_k141_3948\|\|full | Low-quality |
| 18-106-final-viral-combined_k141_137577\|\|full | Not-determined |
| 18-106-final-viral-combined_k141_102250\|\|full | Low-quality |
| 18-106-final-viral-combined_k141_84966\|\|full | Not-determined |
| 18-106-final-viral-combined_k141_84969\|\|full | Low-quality |
| 18-106-final-viral-combined_k141_97310\|\|full | Low-quality |
| 18-106-final-viral-combined_k141_3949\|\|full | Not-determined |
| 18-106-final-viral-combined_k141_102269\|\|full | Not-determined |
| 18-106-final-viral-combined_k141_55258\|\|full | Not-determined |
| 18-106-final-viral-combined_k141_3967\|\|full | Not-determined |
| 18-106-final-viral-combined_k141_8561\|\|full | Not-determined |
| 18-106-final-viral-combined_k141_26119\|\|full | Low-quality |
| 18-109_k141_157264 | Medium-quality |
| 18-106-final-viral-combined_k141_44531\|\|full | Not-determined |
| 18-106-final-viral-combined_k141_26124\|\|full | Low-quality |
| 18-106-final-viral-combined_k141_132082\|\|full | Low-quality |
| 18-106-final-viral-combined_k141_108787\|\|full | Not-determined |
| 18-106-final-viral-combined_k141_119925\|\|full | Not-determined |
| 18-106-final-viral-combined_k141_84993\|\|full | Not-determined |
| 18-106-final-viral-combined_k141_38499\|\|full | Not-determined |
| 18-106-final-viral-combined_k141_137610\|\|full | Low-quality |
| 18-106-final-viral-combined_k141_55298\|\|full | Not-determined |
| 18-106-final-viral-combined_k141_14630\|\|full | Low-quality |
| 18-106-final-viral-combined_k141_20065\|\|full | Low-quality |
| 18-106-final-viral-combined_k141_137615\|\|full | Not-determined |
| 18-101-final-viral-combined_k141_32863\|\|full | Medium-quality |
| 18-106-final-viral-combined_k141_26151\|\|full | Not-determined |
| 18-102-final-viral-combined_k141_53881\|\|full | Medium-quality |
| 18-106-final-viral-combined_k141_73455\|\|full | Not-determined |
| 18-106-final-viral-combined_k141_137626\|\|full | Low-quality |
| 18-106-final-viral-combined_k141_73461\|\|full | Low-quality |
| 18-106-final-viral-combined_k141_102339\|\|full | Not-determined |
| 18-106-final-viral-combined_k141_20089\|\|full | Not-determined |
| 18-106-final-viral-combined_k141_97353\|\|full | Not-determined |
| 18-106-final-viral-combined_k141_126266\|\|full | Not-determined |
| 18-106-final-viral-combined_k141_61976\|\|full | Low-quality |
| 18-106-final-viral-combined_k141_50262\|\|full | Low-quality |
| 18-106-final-viral-combined_k141_32553\|\|full | Low-quality |
| 18-106-final-viral-combined_k141_4038\|\|full | Low-quality |
| 18-106-final-viral-combined_k141_38555\|\|full | Low-quality |
| 18-106-final-viral-combined_k141_38557\|\|full | Low-quality |
| 18-106-final-viral-combined_k141_132139\|\|full | Not-determined |
| 18-106-final-viral-combined_k141_102367\|\|full | Low-quality |
| 18-106-final-viral-combined_k141_108833\|\|full | Low-quality |
| 18-106-final-viral-combined_k141_44568\|\|full | Low-quality |
| 18-106-final-viral-combined_k141_85070\|\|full | Not-determined |
| 18-106-final-viral-combined_k141_126319\|\|full | Not-determined |
| 18-106-final-viral-combined_k141_91044\|\|full | Low-quality |
| 18-106-final-viral-combined_k141_20143\|\|full | Not-determined |
| 18-106-final-viral-combined_k141_55342\|\|full | Not-determined |
| 18-106-final-viral-combined_k141_14703\|\|full | Low-quality |
| 18-106-final-viral-combined_k141_108878\|\|full | Not-determined |
| 18-106-final-viral-combined_k141_126336\|\|full | Low-quality |
| 18-106-final-viral-combined_k141_78995\|\|full | Not-determined |
| 18-106-final-viral-combined_k141_38637\|\|full | Not-determined |
| 18-106-final-viral-combined_k141_108911\|\|full | Low-quality |
| 18-106-final-viral-combined_k141_126359\|\|full | Not-determined |
| 18-106-final-viral-combined_k141_4097\|\|full | Not-determined |
| 18-106-final-viral-combined_k141_79034\|\|full | Low-quality |
| 18-106-final-viral-combined_k141_102424\|\|full | Not-determined |
| 18-106-final-viral-combined_k141_62057\|\|full | Low-quality |
| 18-106-final-viral-combined_k141_126383\|\|full | Low-quality |
| 18-106-final-viral-combined_k141_132215\|\|full | Not-determined |
| 18-106-final-viral-combined_k141_20158\|\|full | Low-quality |
| 18-106-final-viral-combined_k141_20164\|\|full | Not-determined |
| 18-106-final-viral-combined_k141_108928\|\|full | Not-determined |
| 18-106-final-viral-combined_k141_132224\|\|full | Not-determined |
| 18-106-final-viral-combined_k141_55357\|\|full | Low-quality |
| 18-106-final-viral-combined_k141_97502\|\|full | Not-determined |
| 18-106-final-viral-combined_k141_26204\|\|full | Low-quality |
| 18-106-final-viral-combined_k141_68148\|\|full | Low-quality |
| 18-106-final-viral-combined_k141_120043\|\|full | Not-determined |
| 18-106-final-viral-combined_k141_91059\|\|full | Not-determined |
| 18-106-final-viral-combined_k141_32608\|\|full | Low-quality |
| 18-106-final-viral-combined_k141_97515\|\|full | Not-determined |
| 18-106-final-viral-combined_k141_50310\|\|full | Low-quality |
| 18-106-final-viral-combined_k141_62073\|\|full | Low-quality |
| 18-106-final-viral-combined_k141_137734\|\|full | Low-quality |
| 18-106-final-viral-combined_k141_38689\|\|full | Low-quality |
| 18-106-final-viral-combined_k141_68167\|\|full | Not-determined |
| 18-106-final-viral-combined_k141_102462\|\|full | Not-determined |
| 18-106-final-viral-combined_k141_68175\|\|full | Low-quality |
| 18-106-final-viral-combined_k141_68176\|\|full | Low-quality |
| 18-106-final-viral-combined_k141_26237\|\|full | Not-determined |
| 18-106-final-viral-combined_k141_32632\|\|full | Not-determined |
| 18-106-final-viral-combined_k141_20199\|\|full | Not-determined |
| 18-106-final-viral-combined_k141_97544\|\|full | Not-determined |
| 18-106-final-viral-combined_k141_132260\|\|full | Not-determined |
| 18-106-final-viral-combined_k141_120066\|\|full | Not-determined |
| 18-106-final-viral-combined_k141_26255\|\|full | Low-quality |
| 18-106-final-viral-combined_k141_73621\|\|full | Not-determined |
| 18-106-final-viral-combined_k141_38737\|\|full | Low-quality |
| 18-106-final-viral-combined_k141_68229\|\|full | Not-determined |
| 18-106-final-viral-combined_k141_62139\|\|full | Not-determined |
| 18-106-final-viral-combined_k141_120110\|\|full | Not-determined |
| 18-106-final-viral-combined_k141_79106\|\|full | Not-determined |
| 18-102-final-viral-combined_k141_221944\|\|full | High-quality |
| 18-106-final-viral-combined_k141_4205\|\|full | Low-quality |
| 18-106-final-viral-combined_k141_55402\|\|full | Low-quality |
| 18-106-final-viral-combined_k141_120124\|\|full | Low-quality |
| 18-106-final-viral-combined_k141_120126\|\|full | Low-quality |
| 18-106-final-viral-combined_k141_137785\|\|full | Low-quality |
| 18-106-final-viral-combined_k141_44630\|\|full | Low-quality |
| 18-106-final-viral-combined_k141_126482\|\|full | Not-determined |
| 18-106-final-viral-combined_k141_20271\|\|full | Low-quality |
| 18-106-final-viral-combined_k141_44659\|\|full | Low-quality |
| 18-106-final-viral-combined_k141_62217\|\|full | Not-determined |
| 18-104_k141_161443 | Medium-quality |
| 18-106-final-viral-combined_k141_102549\|\|full | Low-quality |
| 18-106-final-viral-combined_k141_32657\|\|full | Not-determined |
| 18-106-final-viral-combined_k141_102553\|\|full | Low-quality |
| 18-106-final-viral-combined_k141_44679\|\|full | Not-determined |
| 18-106-final-viral-combined_k141_137806\|\|full | Not-determined |
| 18-106-final-viral-combined_k141_79170\|\|full | Low-quality |
| 18-106-final-viral-combined_k141_8694\|\|full | Low-quality |
| 18-106-final-viral-combined_k141_102573\|\|full | Low-quality |
| 18-106-final-viral-combined_k141_126522\|\|full | Not-determined |
| 18-106-final-viral-combined_k141_38770\|\|full | Low-quality |
| 18-106-final-viral-combined_k141_4275\|\|full | Not-determined |
| 18-106-final-viral-combined_k141_38777\|\|full | Low-quality |
| 18-106-final-viral-combined_k141_8718\|\|full | Not-determined |
| 18-106-final-viral-combined_k141_38778\|\|full | Not-determined |
| 18-106-final-viral-combined_k141_8727\|\|full | Not-determined |
| 18-106-final-viral-combined_k141_32705\|\|full | Low-quality |
| 18-106-final-viral-combined_k141_20299\|\|full | Low-quality |
| 18-106-final-viral-combined_k141_137846\|\|full | Not-determined |
| 18-106-final-viral-combined_k141_68339\|\|full | Low-quality |
| 18-106-final-viral-combined_k141_62300\|\|full | Low-quality |
| 18-106-final-viral-combined_k141_50321\|\|full | Low-quality |
| 18-106-final-viral-combined_k141_73644\|\|full | Low-quality |
| 18-106-final-viral-combined_k141_102643\|\|full | Low-quality |
| 18-106-final-viral-combined_k141_102644\|\|full | Not-determined |
| 18-106-final-viral-combined_k141_50341\|\|full | Low-quality |
| 18-106-final-viral-combined_k141_55470\|\|full | Low-quality |
| 18-106-final-viral-combined_k141_85275\|\|full | Not-determined |
| 18-106-final-viral-combined_k141_26406\|\|full | Not-determined |
| 18-106-final-viral-combined_k141_137863\|\|full | Low-quality |
| 18-106-final-viral-combined_k141_73674\|\|full | Not-determined |
| 18-106-final-viral-combined_k141_91243\|\|full | Low-quality |
| 18-106-final-viral-combined_k141_91249\|\|full | Low-quality |
| 18-106-final-viral-combined_k141_44715\|\|full | Low-quality |
| 18-106-final-viral-combined_k141_97616\|\|full | Not-determined |
| 18-106-final-viral-combined_k141_20369\|\|full | Not-determined |
| 18-106-final-viral-combined_k141_137884\|\|full | Low-quality |
| 18-106-final-viral-combined_k141_126584\|\|full | Not-determined |
| 18-106-final-viral-combined_k141_79244\|\|full | Low-quality |
| 18-106-final-viral-combined_k141_32743\|\|full | Not-determined |
| 18-106-final-viral-combined_k141_91261\|\|full | Not-determined |
| 18-106-final-viral-combined_k141_132342\|\|full | Not-determined |
| 18-106-final-viral-combined_k141_8845\|\|full | Not-determined |
| 18-106-final-viral-combined_k141_4382\|\|full | Low-quality |
| 18-106-final-viral-combined_k141_44767\|\|full | Low-quality |
| 18-106-final-viral-combined_k141_132360\|\|full | Not-determined |
| 18-106-final-viral-combined_k141_4387\|\|full | Low-quality |
| 18-106-final-viral-combined_k141_79276\|\|full | Not-determined |
| 18-106-final-viral-combined_k141_91275\|\|full | Low-quality |
| 18-106-final-viral-combined_k141_32751\|\|full | Low-quality |
| 18-106-final-viral-combined_k141_132374\|\|full | Low-quality |
| 18-106-final-viral-combined_k141_126591\|\|full | Low-quality |
| 18-106-final-viral-combined_k141_62351\|\|full | Low-quality |
| 18-106-final-viral-combined_k141_4408\|\|full | Not-determined |
| 18-106-final-viral-combined_k141_44770\|\|full | Not-determined |
| 18-106-final-viral-combined_k141_132393\|\|full | Low-quality |
| 18-106-final-viral-combined_k141_44774\|\|full | Not-determined |
| 18-106-final-viral-combined_k141_8887\|\|full | Low-quality |
| 18-106-final-viral-combined_k141_62383\|\|full | Low-quality |
| 18-106-final-viral-combined_k141_55534\|\|full | Low-quality |
| 18-106-final-viral-combined_k141_26481\|\|full | Not-determined |
| 18-106-final-viral-combined_k141_73732\|\|full | Not-determined |
| 18-106-final-viral-combined_k141_62392\|\|full | Low-quality |
| 18-106-final-viral-combined_k141_91318\|\|full | Low-quality |
| 18-106-final-viral-combined_k141_132416\|\|full | Low-quality |
| 18-106-final-viral-combined_k141_44792\|\|full | Low-quality |
| 18-106-final-viral-combined_k141_50471\|\|full | Low-quality |
| 18-106-final-viral-combined_k141_137951\|\|full | Not-determined |
| 18-106-final-viral-combined_k141_62410\|\|full | Not-determined |
| 18-106-final-viral-combined_k141_132432\|\|full | Not-determined |
| 18-106-final-viral-combined_k141_91348\|\|full | Not-determined |
| 18-106-final-viral-combined_k141_132436\|\|full | Low-quality |
| 18-106-final-viral-combined_k141_50479\|\|full | Not-determined |
| 18-106-final-viral-combined_k141_79331\|\|full | Low-quality |
| 18-106-final-viral-combined_k141_8931\|\|full | Not-determined |
| 18-106-final-viral-combined_k141_97686\|\|full | Not-determined |
| 18-106-final-viral-combined_k141_55585\|\|full | Not-determined |
| 18-106-final-viral-combined_k141_62428\|\|full | Low-quality |
| 18-106-final-viral-combined_k141_132470\|\|full | Low-quality |
| 18-106-final-viral-combined_k141_26523\|\|full | Not-determined |
| 18-106-final-viral-combined_k141_73764\|\|full | Not-determined |
| 18-106-final-viral-combined_k141_38969\|\|full | Low-quality |
| 18-106-final-viral-combined_k141_38971\|\|full | Low-quality |
| 18-106-final-viral-combined_k141_79354\|\|full | Not-determined |
| 18-106-final-viral-combined_k141_8936\|\|full | Low-quality |
| 18-106-final-viral-combined_k141_114753\|\|full | Low-quality |
| 18-106-final-viral-combined_k141_114754\|\|full | Not-determined |
| 18-106-final-viral-combined_k141_55615\|\|full | Not-determined |
| 18-106-final-viral-combined_k141_50513\|\|full | Not-determined |
| 18-106-final-viral-combined_k141_73791\|\|full | Not-determined |
| 18-106-final-viral-combined_k141_102734\|\|full | Low-quality |
| 18-106-final-viral-combined_k141_38998\|\|full | Low-quality |
| 18-106-final-viral-combined_k141_44845\|\|full | Low-quality |
| 18-106-final-viral-combined_k141_79367\|\|full | Low-quality |
| 18-106-final-viral-combined_k141_102749\|\|full | Low-quality |
| 18-106-final-viral-combined_k141_39003\|\|full | Not-determined |
| 18-106-final-viral-combined_k141_126668\|\|full | Not-determined |
| 18-106-final-viral-combined_k141_91394\|\|full | Low-quality |
| 18-106-final-viral-combined_k141_55639\|\|full | Not-determined |
| 18-106-final-viral-combined_k141_73802\|\|full | Not-determined |
| 18-106-final-viral-combined_k141_114792\|\|full | Not-determined |
| 18-106-final-viral-combined_k141_73805\|\|full | Not-determined |
| 18-106-final-viral-combined_k141_85453\|\|full | Low-quality |
| 18-106-final-viral-combined_k141_73808\|\|full | Not-determined |
| 18-106-final-viral-combined_k141_73817\|\|full | Not-determined |
| 18-106-final-viral-combined_k141_91425\|\|full | Not-determined |
| 18-106-final-viral-combined_k141_132547\|\|full | Not-determined |
| 18-106-final-viral-combined_k141_68562\|\|full | Low-quality |
| 18-106-final-viral-combined_k141_138004\|\|full | Low-quality |
| 18-106-final-viral-combined_k141_97788\|\|full | Not-determined |
| 18-106-final-viral-combined_k141_132551\|\|full | Low-quality |
| 18-106-final-viral-combined_k141_102804\|\|full | Low-quality |
| 18-106-final-viral-combined_k141_55675\|\|full | Not-determined |
| 18-106-final-viral-combined_k141_9002\|\|full | Not-determined |
| 18-106-final-viral-combined_k141_9005\|\|full | Low-quality |
| 18-106-final-viral-combined_k141_20462\|\|full | Low-quality |
| 18-106-final-viral-combined_k141_120387\|\|full | Low-quality |
| 18-106-final-viral-combined_k141_79421\|\|full | Not-determined |
| 18-106-final-viral-combined_k141_97800\|\|full | Not-determined |
| 18-106-final-viral-combined_k141_9015\|\|full | Low-quality |
| 18-106-final-viral-combined_k141_120402\|\|full | Not-determined |
| 18-106-final-viral-combined_k141_73845\|\|full | Not-determined |
| 18-106-final-viral-combined_k141_102837\|\|full | Not-determined |
| 18-106-final-viral-combined_k141_15051\|\|full | Low-quality |
| 18-106-final-viral-combined_k141_97810\|\|full | Not-determined |
| 18-106-final-viral-combined_k141_132581\|\|full | Low-quality |
| 18-106-final-viral-combined_k141_102854\|\|full | Not-determined |
| 18-106-final-viral-combined_k141_126715\|\|full | Low-quality |
| 18-106-final-viral-combined_k141_85521\|\|full | Low-quality |
| 18-106-final-viral-combined_k141_85523\|\|full | Not-determined |
| 18-106-final-viral-combined_k141_62462\|\|full | Not-determined |
| 18-106-final-viral-combined_k141_44970\|\|full | Low-quality |
| 18-106-final-viral-combined_k141_109109\|\|full | Low-quality |
| 18-106-final-viral-combined_k141_102872\|\|full | Not-determined |
| 18-104-final-viral-combined_k141_161443\|\|full | Medium-quality |
| 18-106-final-viral-combined_k141_79429\|\|full | Low-quality |
| 18-106-final-viral-combined_k141_85550\|\|full | Not-determined |
| 18-106-final-viral-combined_k141_79432\|\|full | Not-determined |
| 18-106-final-viral-combined_k141_62504\|\|full | Not-determined |
| 18-106-final-viral-combined_k141_39029\|\|full | Low-quality |
| 18-106-final-viral-combined_k141_126735\|\|full | Not-determined |
| 18-106-final-viral-combined_k141_50646\|\|full | Not-determined |
| 18-106-final-viral-combined_k141_62526\|\|full | Low-quality |
| 18-106-final-viral-combined_k141_85567\|\|full | Low-quality |
| 18-106-final-viral-combined_k141_45014\|\|full | Low-quality |
| 18-106-final-viral-combined_k141_138089\|\|full | Low-quality |
| 18-106-final-viral-combined_k141_85570\|\|full | Not-determined |
| 18-106-final-viral-combined_k141_79466\|\|full | Low-quality |
| 18-109-final-viral-combined_k141_157264\|\|full | Medium-quality |
| 18-106-final-viral-combined_k141_32934\|\|full | Not-determined |
| 18-106-final-viral-combined_k141_132593\|\|full | Low-quality |
| 18-106-final-viral-combined_k141_4567\|\|full | Not-determined |
| 18-106-final-viral-combined_k141_132608\|\|full | Not-determined |
| 18-106-final-viral-combined_k141_102876\|\|full | Low-quality |
| 18-106-final-viral-combined_k141_26645\|\|full | Low-quality |
| 18-106-final-viral-combined_k141_120411\|\|full | Not-determined |
| 18-106-final-viral-combined_k141_114850\|\|full | Not-determined |
| 18-106-final-viral-combined_k141_50662\|\|full | Not-determined |
| 18-106-final-viral-combined_k141_68611\|\|full | Not-determined |
| 18-106-final-viral-combined_k141_50671\|\|full | Low-quality |
| 18-106-final-viral-combined_k141_15069\|\|full | Low-quality |
| 18-106-final-viral-combined_k141_62576\|\|full | Not-determined |
| 18-106-final-viral-combined_k141_114859\|\|full | Not-determined |
| 18-106-final-viral-combined_k141_50688\|\|full | Low-quality |
| 18-106-final-viral-combined_k141_85634\|\|full | Low-quality |
| 18-106-final-viral-combined_k141_50707\|\|full | Low-quality |
| 18-106-final-viral-combined_k141_79497\|\|full | Not-determined |
| 18-106-final-viral-combined_k141_120461\|\|full | Not-determined |
| 18-106-final-viral-combined_k141_20576\|\|full | Low-quality |
| 18-106-final-viral-combined_k141_120469\|\|full | Not-determined |
| 18-101-final-viral-combined_k141_373308\|\|full | High-quality |
| 18-106-final-viral-combined_k141_114894\|\|full | Low-quality |
| 18-106-final-viral-combined_k141_126817\|\|full | Low-quality |
| 18-106-final-viral-combined_k141_120472\|\|full | Not-determined |
| 18-106-final-viral-combined_k141_97900\|\|full | Not-determined |
| 18-106-final-viral-combined_k141_45088\|\|full | Not-determined |
| 18-106-final-viral-combined_k141_102908\|\|full | Low-quality |
| 18-106-final-viral-combined_k141_126826\|\|full | Low-quality |
| 18-106-final-viral-combined_k141_79544\|\|full | Not-determined |
| 18-106-final-viral-combined_k141_26715\|\|full | Low-quality |
| 18-106-final-viral-combined_k141_39108\|\|full | Low-quality |
| 18-106-final-viral-combined_k141_102917\|\|full | Low-quality |
| 18-106-final-viral-combined_k141_132658\|\|full | Not-determined |
| 18-106-final-viral-combined_k141_26725\|\|full | Not-determined |
| 18-106-final-viral-combined_k141_45102\|\|full | Low-quality |
| 18-106-final-viral-combined_k141_45111\|\|full | Not-determined |
| 18-106-final-viral-combined_k141_20586\|\|full | Low-quality |
| 18-106-final-viral-combined_k141_32969\|\|full | Not-determined |
| 18-106-final-viral-combined_k141_62660\|\|full | Not-determined |
| 18-106-final-viral-combined_k141_68687\|\|full | Low-quality |
| 18-106-final-viral-combined_k141_20612\|\|full | Not-determined |
| 18-106-final-viral-combined_k141_91631\|\|full | Not-determined |
| 18-106-final-viral-combined_k141_91632\|\|full | Not-determined |
| 18-106-final-viral-combined_k141_68693\|\|full | Not-determined |
| 18-106-final-viral-combined_k141_20616\|\|full | Low-quality |
| 18-106-final-viral-combined_k141_26746\|\|full | Low-quality |
| 18-106-final-viral-combined_k141_20618\|\|full | Not-determined |
| 18-106-final-viral-combined_k141_85682\|\|full | Low-quality |
| 18-106-final-viral-combined_k141_39148\|\|full | Not-determined |
| 18-106-final-viral-combined_k141_4709\|\|full | Not-determined |
| 18-106-final-viral-combined_k141_26770\|\|full | Not-determined |
| 18-106-final-viral-combined_k141_97978\|\|full | Low-quality |
| 18-106-final-viral-combined_k141_85702\|\|full | Not-determined |
| 18-106-final-viral-combined_k141_55890\|\|full | Not-determined |
| 18-106-final-viral-combined_k141_114971\|\|full | Not-determined |
| 18-106-final-viral-combined_k141_97993\|\|full | Low-quality |
| 18-106-final-viral-combined_k141_26802\|\|full | Low-quality |
| 18-106-final-viral-combined_k141_98028\|\|full | Low-quality |
| 18-106-final-viral-combined_k141_20665\|\|full | Low-quality |
| 18-106-final-viral-combined_k141_26825\|\|full | Low-quality |

| | |
|---|---|
| 18-106-final-viral-combined_k141_26829||full | Not-determined |
| 18-106-final-viral-combined_k141_15247||full | Low-quality |
| 18-106-final-viral-combined_k141_91708||full | Not-determined |
| 18-106-final-viral-combined_k141_20685||full | Low-quality |
| 18-105-final-viral-combined_k141_7292||full | Medium-quality |
| 18-106-final-viral-combined_k141_15258||full | Low-quality |
| 18-106-final-viral-combined_k141_91711||full | Not-determined |
| 18-106-final-viral-combined_k141_91725||full | Not-determined |
| 18-106-final-viral-combined_k141_20689||full | Low-quality |
| 18-106-final-viral-combined_k141_91733||full | Not-determined |
| 18-106-final-viral-combined_k141_115004||full | Low-quality |
| 18-106-final-viral-combined_k141_91735||full | Low-quality |
| 18-106-final-viral-combined_k141_62794||full | Low-quality |
| 18-106-final-viral-combined_k141_55952||full | Low-quality |
| 18-106-final-viral-combined_k141_62798||full | Low-quality |
| 18-106-final-viral-combined_k141_9147||full | Low-quality |
| 18-106-final-viral-combined_k141_109378||full | Low-quality |
| 18-106-final-viral-combined_k141_73953||full | Low-quality |
| 18-106-final-viral-combined_k141_91761||full | Low-quality |
| 18-106-final-viral-combined_k141_39195||full | Not-determined |
| 18-106-final-viral-combined_k141_68782||full | Low-quality |
| 18-106-final-viral-combined_k141_91768||full | Not-determined |
| 18-106-final-viral-combined_k141_98092||full | Low-quality |
| 18-106-final-viral-combined_k141_4881||full | Low-quality |
| 18-106-final-viral-combined_k141_4884||full | Not-determined |
| 18-106-final-viral-combined_k141_115058||full | Not-determined |
| 18-106-final-viral-combined_k141_120567||full | Low-quality |
| 18-106-final-viral-combined_k141_79660||full | Low-quality |
| 18-106-final-viral-combined_k141_126917||full | Low-quality |
| 18-106-final-viral-combined_k141_79664||full | Not-determined |
| 18-106-final-viral-combined_k141_9175||full | Low-quality |
| 18-106-final-viral-combined_k141_68810||full | Low-quality |
| 18-106-final-viral-combined_k141_103086||full | Not-determined |
| 18-106-final-viral-combined_k141_98112||full | Low-quality |
| 18-106-final-viral-combined_k141_50782||full | Not-determined |
| 18-106-final-viral-combined_k141_62862||full | Low-quality |
| 18-106-final-viral-combined_k141_85853||full | Low-quality |
| 18-106-final-viral-combined_k141_109432||full | Low-quality |
| 18-106-final-viral-combined_k141_85854||full | Not-determined |
| 18-106-final-viral-combined_k141_74023||full | Not-determined |
| 18-106-final-viral-combined_k141_26952||full | Not-determined |
| 18-106-final-viral-combined_k141_74027||full | Not-determined |
| 18-106-final-viral-combined_k141_103108||full | Not-determined |
| 18-106-final-viral-combined_k141_115112||full | Low-quality |
| 18-106-final-viral-combined_k141_50784||full | Low-quality |
| 18-106-final-viral-combined_k141_85874||full | Not-determined |
| 18-106-final-viral-combined_k141_20704||full | Low-quality |
| 18-106-final-viral-combined_k141_126936||full | Not-determined |
| 18-106-final-viral-combined_k141_20713||full | Low-quality |
| 18-106-final-viral-combined_k141_85883||full | Not-determined |
| 18-106-final-viral-combined_k141_15389||full | Low-quality |
| 18-106-final-viral-combined_k141_62908||full | Low-quality |
| 18-106-final-viral-combined_k141_91861||full | Low-quality |
| 18-106-final-viral-combined_k141_120656||full | Low-quality |
| 18-106-final-viral-combined_k141_4953||full | Low-quality |
| 18-106-final-viral-combined_k141_115131||full | Not-determined |
| 18-106-final-viral-combined_k141_85897||full | Low-quality |
| 18-106-final-viral-combined_k141_55974||full | Low-quality |
| 18-106-final-viral-combined_k141_26973||full | Not-determined |
| 18-106-final-viral-combined_k141_20737||full | Low-quality |
| 18-106-final-viral-combined_k141_85901||full | Not-determined |
| 18-106-final-viral-combined_k141_115138||full | Not-determined |
| 18-106-final-viral-combined_k141_33025||full | Low-quality |
| 18-106-final-viral-combined_k141_62924||full | Not-determined |
| 18-106-final-viral-combined_k141_115149||full | Low-quality |
| 18-106-final-viral-combined_k141_109473||full | Low-quality |
| 18-106-final-viral-combined_k141_45267||full | Low-quality |
| 18-106-final-viral-combined_k141_26999||full | Low-quality |
| 18-106-final-viral-combined_k141_4963||full | Not-determined |
| 18-106-final-viral-combined_k141_45284||full | Not-determined |
| 18-106-final-viral-combined_k141_27001||full | Not-determined |
| 18-106-final-viral-combined_k141_120696||full | Low-quality |
| 18-106-final-viral-combined_k141_109485||full | Low-quality |
| 18-106-final-viral-combined_k141_74076||full | Low-quality |
| 18-106-final-viral-combined_k141_56012||full | Not-determined |
| 18-106-final-viral-combined_k141_103188||full | Not-determined |
| 18-106-final-viral-combined_k141_103191||full | Low-quality |
| 18-106-final-viral-combined_k141_45287||full | Not-determined |
| 18-106-final-viral-combined_k141_109496||full | Low-quality |
| 18-106-final-viral-combined_k141_74096||full | Not-determined |
| 18-106-final-viral-combined_k141_45289||full | Not-determined |
| 18-106-final-viral-combined_k141_126986||full | Low-quality |
| 18-106-final-viral-combined_k141_33068||full | Not-determined |
| 18-106-final-viral-combined_k141_109504||full | Low-quality |
| 18-106-final-viral-combined_k141_45307||full | Low-quality |
| 18-106-final-viral-combined_k141_74117||full | Not-determined |
| 18-106-final-viral-combined_k141_79776||full | Not-determined |
| 18-106-final-viral-combined_k141_20786||full | Low-quality |
| 18-106-final-viral-combined_k141_39320||full | Not-determined |
| 18-106-final-viral-combined_k141_33080||full | Not-determined |
| 18-106-final-viral-combined_k141_98201||full | Not-determined |
| 18-106-final-viral-combined_k141_85936||full | Low-quality |
| 18-106-final-viral-combined_k141_85943||full | Not-determined |
| 18-106-final-viral-combined_k141_39341||full | Not-determined |
| 18-106-final-viral-combined_k141_56034||full | Not-determined |
| 18-106-final-viral-combined_k141_5029||full | Low-quality |
| 18-106-final-viral-combined_k141_91924||full | Not-determined |
| 18-106-final-viral-combined_k141_127000||full | Low-quality |
| 18-106-final-viral-combined_k141_120754||full | Not-determined |
| 18-106-final-viral-combined_k141_109578||full | Not-determined |
| 18-106-final-viral-combined_k141_79812||full | Not-determined |
| 18-106-final-viral-combined_k141_56054||full | Not-determined |
| 18-106-final-viral-combined_k141_56066||full | Low-quality |
| 18-106-final-viral-combined_k141_39375||full | Low-quality |
| 18-106-final-viral-combined_k141_103232||full | Not-determined |
| 18-106-final-viral-combined_k141_127027||full | Low-quality |
| 18-106-final-viral-combined_k141_33132||full | Not-determined |
| 18-106-final-viral-combined_k141_138381||full | Low-quality |
| 18-106-final-viral-combined_k141_15520||full | Not-determined |
| 18-106-final-viral-combined_k141_103238||full | Low-quality |
| 18-106-final-viral-combined_k141_103249||full | Low-quality |
| 18-106-final-viral-combined_k141_68988||full | Low-quality |
| 18-106-final-viral-combined_k141_15543||full | Not-determined |
| 18-106-final-viral-combined_k141_91949||full | Low-quality |
| 18-106-final-viral-combined_k141_56097||full | Low-quality |
| 18-106-final-viral-combined_k141_103277||full | Not-determined |
| 18-106-final-viral-combined_k141_39441||full | Low-quality |
| 18-106-final-viral-combined_k141_109617||full | Low-quality |
| 18-106-final-viral-combined_k141_109618||full | Low-quality |
| 18-106-final-viral-combined_k141_27095||full | Low-quality |
| 18-106-final-viral-combined_k141_127058||full | Not-determined |
| 18-106-final-viral-combined_k141_27114||full | Not-determined |
| 18-106-final-viral-combined_k141_127060||full | Not-determined |
| 18-106-final-viral-combined_k141_74204||full | Not-determined |
| 18-106-final-viral-combined_k141_39459||full | Not-determined |
| 18-106-final-viral-combined_k141_138435||full | Low-quality |
| 18-106-final-viral-combined_k141_138437||full | Not-determined |
| 18-106-final-viral-combined_k141_127065||full | Low-quality |
| 18-106-final-viral-combined_k141_20900||full | Low-quality |
| 18-106-final-viral-combined_k141_103298||full | Low-quality |
| 18-106-final-viral-combined_k141_86063||full | Not-determined |
| 18-106-final-viral-combined_k141_115325||full | Not-determined |
| 18-106-final-viral-combined_k141_69024||full | Not-determined |
| 18-106-final-viral-combined_k141_51009||full | Not-determined |
| 18-106-final-viral-combined_k141_27146||full | Low-quality |
| 18-106-final-viral-combined_k141_138467||full | Low-quality |
| 18-106-final-viral-combined_k141_39484||full | Not-determined |
| 18-106-final-viral-combined_k141_115350||full | Not-determined |
| 18-106-final-viral-combined_k141_74236||full | Low-quality |
| 18-106-final-viral-combined_k141_39494||full | Low-quality |
| 18-106-final-viral-combined_k141_103335||full | Not-determined |
| 18-106-final-viral-combined_k141_33277||full | Low-quality |
| 18-106-final-viral-combined_k141_74245||full | Not-determined |
| 18-106-final-viral-combined_k141_127118||full | Not-determined |
| 18-106-final-viral-combined_k141_86121||full | Low-quality |
| 18-106-final-viral-combined_k141_138482||full | Low-quality |
| 18-106-final-viral-combined_k141_63180||full | Not-determined |
| 18-106-final-viral-combined_k141_115374||full | Low-quality |
| 18-106-final-viral-combined_k141_63188||full | Low-quality |
| 18-106-final-viral-combined_k141_27200||full | Low-quality |
| 18-106-final-viral-combined_k141_9462||full | Not-determined |
| 18-106-final-viral-combined_k141_20943||full | Not-determined |
| 18-106-final-viral-combined_k141_115387||full | Low-quality |
| 18-106-final-viral-combined_k141_63201||full | Not-determined |
| 18-106-final-viral-combined_k141_138523||full | Not-determined |
| 18-106-final-viral-combined_k141_98354||full | Low-quality |
| 18-106-final-viral-combined_k141_9482||full | Low-quality |
| 18-106-final-viral-combined_k141_103390||full | Not-determined |
| 18-106-final-viral-combined_k141_51052||full | Low-quality |
| 18-106-final-viral-combined_k141_45485||full | Low-quality |
| 18-106-final-viral-combined_k141_103411||full | Not-determined |
| 18-106-final-viral-combined_k141_9507||full | Not-determined |
| 18-106-final-viral-combined_k141_33359||full | Not-determined |
| 18-106-final-viral-combined_k141_63235||full | Not-determined |
| 18-106-final-viral-combined_k141_51081||full | Low-quality |
| 18-106-final-viral-combined_k141_86179||full | Low-quality |
| 18-106-final-viral-combined_k141_27244||full | Low-quality |
| 18-106-final-viral-combined_k141_33378||full | Low-quality |
| 18-106-final-viral-combined_k141_138566||full | Low-quality |
| 18-106-final-viral-combined_k141_33383||full | Low-quality |
| 18-106-final-viral-combined_k141_86188||full | Low-quality |
| 18-106-final-viral-combined_k141_5274||full | Low-quality |
| 18-106-final-viral-combined_k141_74318||full | Low-quality |
| 18-106-final-viral-combined_k141_33406||full | Not-determined |
| 18-106-final-viral-combined_k141_63259||full | Not-determined |
| 18-106-final-viral-combined_k141_92028||full | Low-quality |
| 18-106-final-viral-combined_k141_138583||full | Low-quality |
| 18-106-final-viral-combined_k141_127219||full | Low-quality |
| 18-106-final-viral-combined_k141_33415||full | Not-determined |
| 18-106-final-viral-combined_k141_98410||full | Low-quality |
| 18-106-final-viral-combined_k141_120901||full | Not-determined |
| 18-106-final-viral-combined_k141_92039||full | Low-quality |
| 18-106-final-viral-combined_k141_120921||full | Not-determined |
| 18-106-final-viral-combined_k141_33427||full | Not-determined |
| 18-106-final-viral-combined_k141_27282||full | Low-quality |
| 18-106-final-viral-combined_k141_27283||full | Low-quality |
| 18-106-final-viral-combined_k141_9587||full | Low-quality |
| 18-106-final-viral-combined_k141_15741||full | Not-determined |
| 18-106-final-viral-combined_k141_120948||full | Low-quality |
| 18-106-final-viral-combined_k141_120952||full | Low-quality |
| 18-106-final-viral-combined_k141_109701||full | Not-determined |
| 18-106-final-viral-combined_k141_86244||full | Not-determined |
| 18-106-final-viral-combined_k141_9595||full | Not-determined |
| 18-106-final-viral-combined_k141_69074||full | Low-quality |
| 18-106-final-viral-combined_k141_86248||full | Low-quality |
| 18-106-final-viral-combined_k141_9600||full | Low-quality |
| 18-106-final-viral-combined_k141_109709||full | Not-determined |
| 18-106-final-viral-combined_k141_79976||full | Not-determined |
| 18-106-final-viral-combined_k141_74375||full | Not-determined |
| 18-106-final-viral-combined_k141_127250||full | Low-quality |
| 18-106-final-viral-combined_k141_109713||full | Low-quality |
| 18-106-final-viral-combined_k141_69101||full | Low-quality |
| 18-106-final-viral-combined_k141_33479||full | Not-determined |
| 18-106-final-viral-combined_k141_79983||full | Low-quality |
| 18-106-final-viral-combined_k141_51172||full | Low-quality |
| 18-106-final-viral-combined_k141_120988||full | Low-quality |
| 18-106-final-viral-combined_k141_9638||full | Not-determined |
| 18-106-final-viral-combined_k141_132846||full | Not-determined |
| 18-106-final-viral-combined_k141_45557||full | Low-quality |
| 18-106-final-viral-combined_k141_120999||full | Low-quality |
| 18-106-final-viral-combined_k141_69103||full | Not-determined |
| 18-106-final-viral-combined_k141_121003||full | Low-quality |
| 18-106-final-viral-combined_k141_45569||full | Low-quality |
| 18-106-final-viral-combined_k141_86281||full | Not-determined |
| 18-106-final-viral-combined_k141_109738||full | Low-quality |
| 18-106-final-viral-combined_k141_51203||full | Low-quality |
| 18-106-final-viral-combined_k141_45586||full | Not-determined |
| 18-106-final-viral-combined_k141_74416||full | Not-determined |
| 18-106-final-viral-combined_k141_39668||full | Not-determined |
| 18-106-final-viral-combined_k141_98488||full | Not-determined |
| 18-106-final-viral-combined_k141_27339||full | Low-quality |
| 18-106-final-viral-combined_k141_45601||full | Low-quality |
| 18-106-final-viral-combined_k141_33513||full | Low-quality |
| 18-106-final-viral-combined_k141_27347||full | Low-quality |
| 18-106-final-viral-combined_k141_109778||full | Not-determined |
| 18-106-final-viral-combined_k141_98512||full | Low-quality |
| 18-106-final-viral-combined_k141_27355||full | Not-determined |
| 18-106-final-viral-combined_k141_5412||full | Low-quality |
| 18-106-final-viral-combined_k141_21011||full | Not-determined |
| 18-106-final-viral-combined_k141_74427||full | Low-quality |
| 18-106-final-viral-combined_k141_92124||full | Low-quality |
| 18-106-final-viral-combined_k141_5419||full | Not-determined |
| 18-106-final-viral-combined_k141_98539||full | Low-quality |
| 18-106-final-viral-combined_k141_15869||full | Not-determined |
| 18-106-final-viral-combined_k141_80069||full | Low-quality |
| 18-106-final-viral-combined_k141_15880||full | Low-quality |
| 18-106-final-viral-combined_k141_121049||full | Not-determined |
| 18-106-final-viral-combined_k141_98544||full | Low-quality |
| 18-106-final-viral-combined_k141_45666||full | Not-determined |
| 18-106-final-viral-combined_k141_9682||full | Low-quality |
| 18-106-final-viral-combined_k141_27402||full | Low-quality |
| 18-106-final-viral-combined_k141_5465||full | Not-determined |
| 18-106-final-viral-combined_k141_121057||full | Not-determined |
| 18-106-final-viral-combined_k141_86352||full | Low-quality |
| 18-106-final-viral-combined_k141_80100||full | Not-determined |
| 18-106-final-viral-combined_k141_15908||full | Low-quality |
| 18-106-final-viral-combined_k141_15920||full | Low-quality |
| 18-106-final-viral-combined_k141_27429||full | Not-determined |
| 18-106-final-viral-combined_k141_92145||full | Not-determined |
| 18-106-final-viral-combined_k141_80124||full | Not-determined |
| 18-106-final-viral-combined_k141_51356||full | Not-determined |
| 18-106-final-viral-combined_k141_109862||full | Not-determined |
| 18-106-final-viral-combined_k141_15922||full | Not-determined |
| 18-106-final-viral-combined_k141_80126||full | Low-quality |
| 18-106-final-viral-combined_k141_109873||full | Low-quality |
| 18-106-final-viral-combined_k141_69164||full | Low-quality |
| 18-106-final-viral-combined_k141_27438||full | Not-determined |
| 18-106-final-viral-combined_k141_9718||full | Not-determined |
| 18-106-final-viral-combined_k141_56206||full | Not-determined |
| 18-106-final-viral-combined_k141_15941||full | Not-determined |
| 18-106-final-viral-combined_k141_45707||full | Not-determined |
| 18-106-final-viral-combined_k141_56214||full | Low-quality |
| 18-106-final-viral-combined_k141_121086||full | Low-quality |
| 18-106-final-viral-combined_k141_63420||full | Not-determined |
| 18-106-final-viral-combined_k141_127335||full | Low-quality |
| 18-106-final-viral-combined_k141_69201||full | Not-determined |
| 18-106-final-viral-combined_k141_80146||full | Low-quality |
| 18-106-final-viral-combined_k141_15969||full | Not-determined |
| 18-106-final-viral-combined_k141_115656||full | Not-determined |
| 18-106-final-viral-combined_k141_109910||full | Low-quality |
| 18-106-final-viral-combined_k141_132973||full | Low-quality |
| 18-106-final-viral-combined_k141_138695||full | Low-quality |
| 18-106-final-viral-combined_k141_5473||full | Not-determined |
| 18-106-final-viral-combined_k141_51386||full | Low-quality |
| 18-106-final-viral-combined_k141_63442||full | Low-quality |
| 18-106-final-viral-combined_k141_115664||full | Low-quality |
| 18-106-final-viral-combined_k141_80166||full | Not-determined |
| 18-106-final-viral-combined_k141_74514||full | Not-determined |
| 18-106-final-viral-combined_k141_15970||full | Low-quality |
| 18-106-final-viral-combined_k141_127364||full | Low-quality |
| 18-106-final-viral-combined_k141_80177||full | Not-determined |
| 18-106-final-viral-combined_k141_45763||full | Low-quality |
| 18-106-final-viral-combined_k141_138698||full | Low-quality |
| 18-106-final-viral-combined_k141_80182||full | Low-quality |
| 18-106-final-viral-combined_k141_39815||full | Not-determined |
| 18-106-final-viral-combined_k141_103577||full | Not-determined |
| 18-106-final-viral-combined_k141_103579||full | Low-quality |
| 18-106-final-viral-combined_k141_51407||full | Low-quality |
| 18-106-final-viral-combined_k141_56285||full | Not-determined |
| 18-106-final-viral-combined_k141_9746||full | Low-quality |
| 18-106-final-viral-combined_k141_21147||full | Not-determined |
| 18-106-final-viral-combined_k141_16010||full | Not-determined |
| 18-106-final-viral-combined_k141_27534||full | Low-quality |
| 18-106-final-viral-combined_k141_39834||full | Low-quality |
| 18-106-final-viral-combined_k141_98643||full | Low-quality |
| 18-106-final-viral-combined_k141_21155||full | Low-quality |
| 18-106-final-viral-combined_k141_109939||full | Not-determined |
| 18-106-final-viral-combined_k141_9771||full | Low-quality |
| 18-106-final-viral-combined_k141_115701||full | Low-quality |
| 18-106-final-viral-combined_k141_56322||full | Not-determined |
| 18-106-final-viral-combined_k141_92239||full | Low-quality |
| 18-106-final-viral-combined_k141_51431||full | Not-determined |
| 18-106-final-viral-combined_k141_74554||full | Low-quality |
| 18-106-final-viral-combined_k141_16028||full | Not-determined |
| 18-106-final-viral-combined_k141_51439||full | Not-determined |
| 18-106-final-viral-combined_k141_45801||full | Low-quality |
| 18-106-final-viral-combined_k141_5510||full | Low-quality |
| 18-106-final-viral-combined_k141_56336||full | Not-determined |
| 18-106-final-viral-combined_k141_133048||full | Not-determined |
| 18-106-final-viral-combined_k141_9790||full | Not-determined |
| 18-102-final-viral-combined_k141_4685||0_partial | High-quality |
| 18-106-final-viral-combined_k141_27537||full | Not-determined |
| 18-106-final-viral-combined_k141_27539||full | Low-quality |
| 18-106-final-viral-combined_k141_92264||full | Not-determined |
| 18-106-final-viral-combined_k141_115722||full | Not-determined |
| 18-106-final-viral-combined_k141_109983||full | Not-determined |
| 18-106-final-viral-combined_k141_69294||full | Not-determined |
| 18-106-final-viral-combined_k141_39886||full | Low-quality |
| 18-106-final-viral-combined_k141_110001||full | Not-determined |
| 18-106-final-viral-combined_k141_138759||full | Not-determined |
| 18-106-final-viral-combined_k141_21192||full | Low-quality |
| 18-106-final-viral-combined_k141_69307||full | Low-quality |
| 18-106-final-viral-combined_k141_98705||full | Low-quality |
| 18-106-final-viral-combined_k141_103601||full | Not-determined |
| 18-106-final-viral-combined_k141_74577||full | Low-quality |
| 18-106-final-viral-combined_k141_27575||full | Low-quality |
| 18-106-final-viral-combined_k141_45893||full | Not-determined |
| 18-106-final-viral-combined_k141_16087||full | Not-determined |
| 18-106-final-viral-combined_k141_39916||full | Low-quality |
| 18-106-final-viral-combined_k141_21228||full | Not-determined |
| 18-106-final-viral-combined_k141_115776||full | Not-determined |
| 18-106-final-viral-combined_k141_92307||full | Low-quality |
| 18-106-final-viral-combined_k141_51490||full | Not-determined |
| 18-106-final-viral-combined_k141_138795||full | Low-quality |
| 18-106-final-viral-combined_k141_74601||full | Not-determined |
| 18-106-final-viral-combined_k141_56382||full | Low-quality |
| 18-106-final-viral-combined_k141_133126||full | Not-determined |
| 18-106-final-viral-combined_k141_74607||full | Not-determined |
| 18-106-final-viral-combined_k141_69337||full | Not-determined |
| 18-106-final-viral-combined_k141_133133||full | Not-determined |
| 18-106-final-viral-combined_k141_138815||full | Not-determined |
| 18-106-final-viral-combined_k141_133134||full | Low-quality |
| 18-106-final-viral-combined_k141_45908||full | Low-quality |
| 18-106-final-viral-combined_k141_80288||full | Not-determined |
| 18-106-final-viral-combined_k141_98757||full | Low-quality |
| 18-106-final-viral-combined_k141_115805||full | Not-determined |
| 18-106-final-viral-combined_k141_27601||full | Low-quality |
| 18-106-final-viral-combined_k141_121166||full | Low-quality |
| 18-106-final-viral-combined_k141_80295||full | Low-quality |
| 18-106-final-viral-combined_k141_21255||full | Low-quality |
| 18-106-final-viral-combined_k141_121178||full | Low-quality |
| 18-106-final-viral-combined_k141_27606||full | Low-quality |
| 18-106-final-viral-combined_k141_51529||full | Not-determined |
| 18-106-final-viral-combined_k141_138842||full | Low-quality |
| 18-106-final-viral-combined_k141_74650||full | Low-quality |
| 18-106-final-viral-combined_k141_92365||full | Low-quality |
| 18-106-final-viral-combined_k141_51542||full | Not-determined |
| 18-106-final-viral-combined_k141_115837||full | Low-quality |
| 18-106-final-viral-combined_k141_63644||full | Not-determined |
| 18-106-final-viral-combined_k141_51548||full | Not-determined |
| 18-106-final-viral-combined_k141_92377||full | Not-determined |
| 18-106-final-viral-combined_k141_138880||full | Low-quality |
| 18-106-final-viral-combined_k141_45959||full | Low-quality |
| 18-106-final-viral-combined_k141_98802||full | Not-determined |
| 18-106-final-viral-combined_k141_98805||full | Not-determined |
| 18-106-final-viral-combined_k141_45970||full | Not-determined |
| 18-106-final-viral-combined_k141_115867||full | Low-quality |
| 18-106-final-viral-combined_k141_138907||full | Low-quality |
| 18-106-final-viral-combined_k141_45974||full | Not-determined |
| 18-106-final-viral-combined_k141_27675||full | Low-quality |
| 18-106-final-viral-combined_k141_121217||full | Low-quality |
| 18-106-final-viral-combined_k141_98832||full | Not-determined |
| 18-106-final-viral-combined_k141_63675||full | Not-determined |
| 18-106-final-viral-combined_k141_16195||full | Low-quality |
| 18-106-final-viral-combined_k141_33515||full | Not-determined |
| 18-106-final-viral-combined_k141_33528||full | Low-quality |
| 18-106-final-viral-combined_k141_127460||full | Not-determined |
| 18-106-final-viral-combined_k141_133189||full | Not-determined |
| 18-106-final-viral-combined_k141_74730||full | Low-quality |
| 18-106-final-viral-combined_k141_86472||full | Not-determined |
| 18-106-final-viral-combined_k141_133191||full | Not-determined |
| 18-106-final-viral-combined_k141_21305||full | Not-determined |
| 17-109_k141_373775 | High-quality |
| 18-106-final-viral-combined_k141_92486||full | Low-quality |
| 18-106-final-viral-combined_k141_110152||full | Low-quality |
| 18-106-final-viral-combined_k141_21323||full | Not-determined |
| 18-106-final-viral-combined_k141_110162||full | Low-quality |
| 18-106-final-viral-combined_k141_27750||full | Low-quality |
| 18-106-final-viral-combined_k141_9960||full | Low-quality |
| 18-106-final-viral-combined_k141_74780||full | Not-determined |
| 18-106-final-viral-combined_k141_16246||full | Low-quality |
| 18-106-final-viral-combined_k141_5739||full | Not-determined |
| 18-106-final-viral-combined_k141_56419||full | Low-quality |
| 18-106-final-viral-combined_k141_98888||full | Not-determined |
| 18-106-final-viral-combined_k141_98890||full | Low-quality |
| 18-106-final-viral-combined_k141_133226||full | Low-quality |
| 18-106-final-viral-combined_k141_138986||full | Low-quality |
| 18-106-final-viral-combined_k141_46001||full | Low-quality |
| 18-106-final-viral-combined_k141_46006||full | Low-quality |
| 18-106-final-viral-combined_k141_127498||full | Low-quality |
| 18-106-final-viral-combined_k141_121313||full | Not-determined |
| 18-106-final-viral-combined_k141_92525||full | Low-quality |
| 18-106-final-viral-combined_k141_27785||full | Not-determined |
| 18-106-final-viral-combined_k141_63726||full | Not-determined |
| 18-106-final-viral-combined_k141_121325||full | Low-quality |
| 18-106-final-viral-combined_k141_110207||full | Not-determined |
| 18-106-final-viral-combined_k141_63732||full | Not-determined |
| 18-106-final-viral-combined_k141_63742||full | Not-determined |
| 18-106-final-viral-combined_k141_74847||full | Not-determined |
| 18-106-final-viral-combined_k141_127513||full | Not-determined |
| 18-106-final-viral-combined_k141_33563||full | Low-quality |
| 18-106-final-viral-combined_k141_40095||full | Low-quality |
| 18-106-final-viral-combined_k141_127517||full | Low-quality |
| 18-106-final-viral-combined_k141_40097||full | Low-quality |
| 18-106-final-viral-combined_k141_86564||full | Not-determined |
| 18-106-final-viral-combined_k141_110243||full | Not-determined |
| 18-106-final-viral-combined_k141_74882||full | Low-quality |
| 17-109-final-viral-combined_k141_373775||full | High-quality |
| 18-106-final-viral-combined_k141_33619||full | Not-determined |
| 18-106-final-viral-combined_k141_92577||full | Not-determined |
| 18-106-final-viral-combined_k141_98966||full | Not-determined |
| 18-106-final-viral-combined_k141_92584||full | Low-quality |
| 18-106-final-viral-combined_k141_16284||full | Low-quality |
| 18-106-final-viral-combined_k141_98972||full | Low-quality |
| 18-106-final-viral-combined_k141_21356||full | Low-quality |
| 18-106-final-viral-combined_k141_92592||full | Low-quality |
| 18-106-final-viral-combined_k141_92595||full | Not-determined |
| 18-106-final-viral-combined_k141_92601||full | Not-determined |
| 18-106-final-viral-combined_k141_40149||full | Low-quality |
| 18-106-final-viral-combined_k141_92622||full | Low-quality |
| 18-106-final-viral-combined_k141_63824||full | Not-determined |
| 18-106-final-viral-combined_k141_115961||full | Not-determined |
| 18-106-final-viral-combined_k141_121398||full | Low-quality |
| 18-106-final-viral-combined_k141_92637||full | Not-determined |
| 18-106-final-viral-combined_k141_99005||full | Low-quality |
| 18-106-final-viral-combined_k141_110309||full | Low-quality |
| 18-106-final-viral-combined_k141_133382||full | Not-determined |
| 18-106-final-viral-combined_k141_133385||full | Not-determined |
| 18-106-final-viral-combined_k141_133390||full | Low-quality |
| 18-106-final-viral-combined_k141_51695||full | Not-determined |
| 18-106-final-viral-combined_k141_127600||full | Low-quality |
| 18-106-final-viral-combined_k141_133394||full | Not-determined |
| 18-106-final-viral-combined_k141_133397||full | Low-quality |
| 18-106-final-viral-combined_k141_51721||full | Low-quality |
| 18-106-final-viral-combined_k141_103750||full | Low-quality |
| 18-106-final-viral-combined_k141_9984||full | Not-determined |
| 18-106-final-viral-combined_k141_16372||full | Not-determined |
| 18-106-final-viral-combined_k141_92679||full | Low-quality |
| 18-106-final-viral-combined_k141_16378||full | Low-quality |
| 18-106-final-viral-combined_k141_9987||full | Low-quality |
| 18-106-final-viral-combined_k141_21410||full | Low-quality |
| 18-106-final-viral-combined_k141_40182||full | Low-quality |
| 18-106-final-viral-combined_k141_33755||full | Not-determined |
| 18-106-final-viral-combined_k141_33766||full | Not-determined |
| 18-106-final-viral-combined_k141_99091||full | Not-determined |
| 18-106-final-viral-combined_k141_27926||full | Low-quality |
| 18-106-final-viral-combined_k141_27931||full | Low-quality |
| 18-106-final-viral-combined_k141_46228||full | Not-determined |
| 18-106-final-viral-combined_k141_63914||full | Not-determined |
| 18-106-final-viral-combined_k141_116041||full | Not-determined |
| 18-106-final-viral-combined_k141_10008||full | Low-quality |
| 18-106-final-viral-combined_k141_16436||full | Low-quality |
| 18-106-final-viral-combined_k141_127700||full | Not-determined |
| 18-106-final-viral-combined_k141_139084||full | Not-determined |
| 18-106-final-viral-combined_k141_46245||full | Low-quality |
| 18-106-final-viral-combined_k141_51782||full | Low-quality |
| 18-106-final-viral-combined_k141_10027||full | Not-determined |
| 18-106-final-viral-combined_k141_116070||full | Low-quality |
| 18-106-final-viral-combined_k141_127712||full | Low-quality |
| 18-106-final-viral-combined_k141_74934||full | Not-determined |
| 18-106-final-viral-combined_k141_33797||full | Low-quality |
| 18-106-final-viral-combined_k141_21478||full | Not-determined |
| 18-106-final-viral-combined_k141_51798||full | Not-determined |
| 18-106-final-viral-combined_k141_86740||full | Not-determined |
| 18-106-final-viral-combined_k141_116092||full | Low-quality |
| 18-106-final-viral-combined_k141_86744||full | Not-determined |
| 18-106-final-viral-combined_k141_69573||full | Low-quality |
| 18-106-final-viral-combined_k141_69576||full | Not-determined |
| 18-106-final-viral-combined_k141_127719||full | Low-quality |
| 18-106-final-viral-combined_k141_51819||full | Low-quality |
| 18-106-final-viral-combined_k141_86752||full | Not-determined |
| 18-106-final-viral-combined_k141_99161||full | Low-quality |
| 18-106-final-viral-combined_k141_116108||full | Not-determined |
| 18-106-final-viral-combined_k141_63946||full | Not-determined |
| 18-106-final-viral-combined_k141_56566||full | Low-quality |
| 18-106-final-viral-combined_k141_127741||full | Not-determined |
| 18-106-final-viral-combined_k141_27996||full | Not-determined |
| 18-106-final-viral-combined_k141_99177||full | Not-determined |
| 18-106-final-viral-combined_k141_16484||full | Not-determined |
| 18-106-final-viral-combined_k141_10094||full | Not-determined |
| 18-106-final-viral-combined_k141_121419||full | Low-quality |
| 18-106-final-viral-combined_k141_28014||full | Low-quality |
| 18-106-final-viral-combined_k141_139132||full | Not-determined |
| 18-106-final-viral-combined_k141_74991||full | Not-determined |
| 18-106-final-viral-combined_k141_28020||full | Low-quality |
| 18-106-final-viral-combined_k141_10100||full | Low-quality |
| 18-106-final-viral-combined_k141_139142||full | Low-quality |
| 18-106-final-viral-combined_k141_40325||full | Low-quality |
| 18-106-final-viral-combined_k141_46325||full | Not-determined |
| 18-106-final-viral-combined_k141_16512||full | Low-quality |
| 18-106-final-viral-combined_k141_74999||full | Low-quality |
| 18-106-final-viral-combined_k141_16514||full | Low-quality |
| 18-106-final-viral-combined_k141_139168||full | Not-determined |
| 18-106-final-viral-combined_k141_16516||full | Not-determined |
| 18-106-final-viral-combined_k141_16518||full | Not-determined |
| 18-106-final-viral-combined_k141_21578||full | Not-determined |
| 18-106-final-viral-combined_k141_69672||full | Low-quality |
| 18-106-final-viral-combined_k141_21583||full | Not-determined |
| 18-106-final-viral-combined_k141_127778||full | Low-quality |
| 18-106-final-viral-combined_k141_16537||full | Low-quality |
| 18-106-final-viral-combined_k141_16541||full | Low-quality |
| 18-106-final-viral-combined_k141_133522||full | Low-quality |
| 18-106-final-viral-combined_k141_99232||full | Low-quality |
| 18-106-final-viral-combined_k141_133530||full | Low-quality |
| 18-106-final-viral-combined_k141_64047||full | Not-determined |
| 18-106-final-viral-combined_k141_116214||full | Not-determined |
| 18-106-final-viral-combined_k141_127811||full | Low-quality |
| 18-106-final-viral-combined_k141_40376||full | Not-determined |
| 18-106-final-viral-combined_k141_51871||full | Not-determined |
| 18-106-final-viral-combined_k141_139215||full | Not-determined |
| 18-106-final-viral-combined_k141_133531||full | Not-determined |
| 18-106-final-viral-combined_k141_133540||full | Not-determined |
| 18-106-final-viral-combined_k141_16570||full | Low-quality |
| 18-106-final-viral-combined_k141_40395||full | Not-determined |
| 18-106-final-viral-combined_k141_75063||full | Not-determined |
| 18-106-final-viral-combined_k141_75064||full | Low-quality |
| 18-106-final-viral-combined_k141_80629||full | Low-quality |
| 18-106-final-viral-combined_k141_121463||full | Low-quality |
| 18-106-final-viral-combined_k141_64081||full | Not-determined |
| 18-106-final-viral-combined_k141_51903||full | Low-quality |
| 18-106-final-viral-combined_k141_51920||full | Low-quality |
| 18-106-final-viral-combined_k141_121484||full | Not-determined |
| 18-106-final-viral-combined_k141_56685||full | Low-quality |
| 18-106-final-viral-combined_k141_80653||full | Low-quality |
| 18-106-final-viral-combined_k141_127850||full | Low-quality |
| 18-106-final-viral-combined_k141_103804||full | Low-quality |
| 18-106-final-viral-combined_k141_99266||full | Not-determined |
| 18-106-final-viral-combined_k141_133575||full | Low-quality |
| 18-106-final-viral-combined_k141_56689||full | Not-determined |
| 18-106-final-viral-combined_k141_51936||full | Low-quality |
| 18-106-final-viral-combined_k141_99273||full | Not-determined |
| 18-106-final-viral-combined_k141_33894||full | Low-quality |
| 18-106-final-viral-combined_k141_92841||full | Not-determined |
| 18-106-final-viral-combined_k141_121493||full | Not-determined |
| 18-106-final-viral-combined_k141_99281||full | Not-determined |
| 18-106-final-viral-combined_k141_86874||full | Low-quality |
| 18-106-final-viral-combined_k141_103839||full | Low-quality |
| 18-106-final-viral-combined_k141_92852||full | Low-quality |
| 18-106-final-viral-combined_k141_99284||full | Not-determined |
| 18-106-final-viral-combined_k141_51965||full | Low-quality |
| 18-106-final-viral-combined_k141_139254||full | Not-determined |
| 18-106-final-viral-combined_k141_16643||full | Low-quality |
| 18-106-final-viral-combined_k141_56714||full | Not-determined |
| 18-106-final-viral-combined_k141_121539||full | Low-quality |
| 18-106-final-viral-combined_k141_16644||full | Not-determined |
| 18-106-final-viral-combined_k141_46387||full | Low-quality |
| 18-106-final-viral-combined_k141_139263||full | Low-quality |
| 18-106-final-viral-combined_k141_133626||full | Not-determined |
| 18-106-final-viral-combined_k141_33968||full | Low-quality |
| 18-106-final-viral-combined_k141_64144||full | Low-quality |
| 18-106-final-viral-combined_k141_121571||full | Not-determined |
| 18-106-final-viral-combined_k141_139280||full | Low-quality |
| 18-106-final-viral-combined_k141_28193||full | Low-quality |
| 18-106-final-viral-combined_k141_80713||full | Not-determined |
| 18-106-final-viral-combined_k141_10230||full | Not-determined |
| 18-106-final-viral-combined_k141_121578||full | Not-determined |
| 18-106-final-viral-combined_k141_116305||full | Low-quality |
| 18-106-final-viral-combined_k141_64165||full | Not-determined |
| 18-106-final-viral-combined_k141_46417||full | Not-determined |
| 18-106-final-viral-combined_k141_64168||full | Not-determined |
| 18-106-final-viral-combined_k141_110543||full | Low-quality |
| 18-106-final-viral-combined_k141_40478||full | Not-determined |
| 18-106-final-viral-combined_k141_64173||full | Not-determined |
| 18-106-final-viral-combined_k141_133667||full | Low-quality |
| 18-106-final-viral-combined_k141_21719||full | Not-determined |
| 18-106-final-viral-combined_k141_64181||full | Not-determined |
| 18-106-final-viral-combined_k141_69875||full | Not-determined |
| 18-106-final-viral-combined_k141_64198||full | Low-quality |
| 18-106-final-viral-combined_k141_99327||full | Not-determined |
| 18-106-final-viral-combined_k141_86986||full | Low-quality |
| 18-106-final-viral-combined_k141_86993||full | Low-quality |
| 18-106-final-viral-combined_k141_10291||full | Low-quality |
| 18-106-final-viral-combined_k141_133715||full | Not-determined |
| 18-106-final-viral-combined_k141_139360||full | Low-quality |
| 18-106-final-viral-combined_k141_52102||full | Low-quality |
| 18-106-final-viral-combined_k141_69919||full | Not-determined |
| 18-106-final-viral-combined_k141_121619||full | Not-determined |
| 18-106-final-viral-combined_k141_139364||full | Not-determined |
| 18-106-final-viral-combined_k141_21781||full | Low-quality |

| | |
|---|---|
| 18-106-final-viral-combined_k141_99376\|\|full | Low-quality |
| 18-106-final-viral-combined_k141_103961\|\|full | Not-determined |
| 18-106-final-viral-combined_k141_128000\|\|full | Not-determined |
| 18-106-final-viral-combined_k141_99380\|\|full | Low-quality |
| 18-106-final-viral-combined_k141_69940\|\|full | Low-quality |
| 18-106-final-viral-combined_k141_34011\|\|full | Not-determined |
| 18-106-final-viral-combined_k141_75167\|\|full | Low-quality |
| 18-106-final-viral-combined_k141_116402\|\|full | Not-determined |
| 18-106-final-viral-combined_k141_69972\|\|full | Not-determined |
| 18-106-final-viral-combined_k141_110648\|\|full | Not-determined |
| 18-106-final-viral-combined_k141_139419\|\|full | Not-determined |
| 18-106-final-viral-combined_k141_10343\|\|full | Not-determined |
| 18-106-final-viral-combined_k141_139422\|\|full | Not-determined |
| 18-106-final-viral-combined_k141_34068\|\|full | Low-quality |
| 18-106-final-viral-combined_k141_87038\|\|full | Not-determined |
| 18-106-final-viral-combined_k141_99443\|\|full | Low-quality |
| 18-106-final-viral-combined_k141_52129\|\|full | Not-determined |
| 18-106-final-viral-combined_k141_92867\|\|full | Low-quality |
| 18-106-final-viral-combined_k141_80841\|\|full | Low-quality |
| 18-106-final-viral-combined_k141_133800\|\|full | Not-determined |
| 18-106-final-viral-combined_k141_70036\|\|full | Low-quality |
| 18-106-final-viral-combined_k141_21832\|\|full | Low-quality |
| 18-106-final-viral-combined_k141_133804\|\|full | Not-determined |
| 18-106-final-viral-combined_k141_104030\|\|full | Not-determined |
| 18-106-final-viral-combined_k141_133813\|\|full | Not-determined |
| 18-106-final-viral-combined_k141_10371\|\|full | Not-determined |
| 18-106-final-viral-combined_k141_99471\|\|full | Not-determined |
| 18-106-final-viral-combined_k141_34122\|\|full | Low-quality |
| 18-106-final-viral-combined_k141_116450\|\|full | Not-determined |
| 18-106-final-viral-combined_k141_21855\|\|full | Low-quality |
| 18-106-final-viral-combined_k141_16847\|\|full | Low-quality |
| 18-106-final-viral-combined_k141_128091\|\|full | Low-quality |
| 18-106-final-viral-combined_k141_46451\|\|full | Not-determined |
| 18-106-final-viral-combined_k141_70073\|\|full | Not-determined |
| 18-106-final-viral-combined_k141_52149\|\|full | Low-quality |
| 18-106-final-viral-combined_k141_139467\|\|full | Low-quality |
| 18-106-final-viral-combined_k141_99514\|\|full | Low-quality |
| 18-106-final-viral-combined_k141_110710\|\|full | Not-determined |
| 18-106-final-viral-combined_k141_34166\|\|full | Low-quality |
| 18-106-final-viral-combined_k141_133869\|\|full | Not-determined |
| 18-106-final-viral-combined_k141_64398\|\|full | Low-quality |
| 18-106-final-viral-combined_k141_40668\|\|full | Low-quality |
| 18-106-final-viral-combined_k141_133870\|\|full | Not-determined |
| 18-106-final-viral-combined_k141_64404\|\|full | Not-determined |
| 18-106-final-viral-combined_k141_87141\|\|full | Low-quality |
| 18-106-final-viral-combined_k141_87146\|\|full | Low-quality |
| 18-106-final-viral-combined_k141_128136\|\|full | Low-quality |
| 18-106-final-viral-combined_k141_40688\|\|full | Not-determined |
| 18-106-final-viral-combined_k141_28366\|\|full | Not-determined |
| 18-106-final-viral-combined_k141_10442\|\|full | Not-determined |
| 18-106-final-viral-combined_k141_21890\|\|full | Low-quality |
| 18-106-final-viral-combined_k141_110757\|\|full | Not-determined |
| 18-106-final-viral-combined_k141_87179\|\|full | Not-determined |
| 18-106-final-viral-combined_k141_40722\|\|full | Not-determined |
| 18-106-final-viral-combined_k141_92980\|\|full | Not-determined |
| 18-106-final-viral-combined_k141_128166\|\|full | Low-quality |
| 18-106-final-viral-combined_k141_40727\|\|full | Low-quality |
| 18-106-final-viral-combined_k141_10463\|\|full | Not-determined |
| 18-106-final-viral-combined_k141_121776\|\|full | Not-determined |
| 18-106-final-viral-combined_k141_10466\|\|full | Not-determined |
| 18-106-final-viral-combined_k141_133918\|\|full | Not-determined |
| 18-106-final-viral-combined_k141_75261\|\|full | Not-determined |
| 18-106-final-viral-combined_k141_56893\|\|full | Not-determined |
| 18-106-final-viral-combined_k141_10472\|\|full | Low-quality |
| 18-106-final-viral-combined_k141_93000\|\|full | Not-determined |
| 18-106-final-viral-combined_k141_10474\|\|full | Low-quality |
| 18-106-final-viral-combined_k141_80886\|\|full | Not-determined |
| 18-106-final-viral-combined_k141_139518\|\|full | Not-determined |
| 18-106-final-viral-combined_k141_93006\|\|full | Low-quality |
| 18-106-final-viral-combined_k141_110817\|\|full | Not-determined |
| 18-106-final-viral-combined_k141_93014\|\|full | Not-determined |
| 18-106-final-viral-combined_k141_139533\|\|full | Not-determined |
| 18-106-final-viral-combined_k141_56938\|\|full | Not-determined |
| 18-106-final-viral-combined_k141_70158\|\|full | Low-quality |
| 18-106-final-viral-combined_k141_128200\|\|full | Low-quality |
| 18-106-final-viral-combined_k141_93024\|\|full | Low-quality |
| 18-106-final-viral-combined_k141_46563\|\|full | Low-quality |
| 18-106-final-viral-combined_k141_116539\|\|full | Low-quality |
| 18-106-final-viral-combined_k141_10489\|\|full | Low-quality |
| 18-106-final-viral-combined_k141_40774\|\|full | Low-quality |
| 18-106-final-viral-combined_k141_128213\|\|full | Low-quality |
| 18-106-final-viral-combined_k141_16968\|\|full | Low-quality |
| 18-106-final-viral-combined_k141_34232\|\|full | Low-quality |
| 18-106-final-viral-combined_k141_93046\|\|full | Not-determined |
| 18-106-final-viral-combined_k141_10511\|\|full | Not-determined |
| 18-106-final-viral-combined_k141_110836\|\|full | Low-quality |
| 18-106-final-viral-combined_k141_139569\|\|full | Low-quality |
| 18-106-final-viral-combined_k141_40792\|\|full | Not-determined |
| 18-106-final-viral-combined_k141_110854\|\|full | Low-quality |
| 18-106-final-viral-combined_k141_116578\|\|full | Low-quality |
| 18-106-final-viral-combined_k141_104225\|\|full | Low-quality |
| 18-106-final-viral-combined_k141_128260\|\|full | Low-quality |
| 18-106-final-viral-combined_k141_110875\|\|full | Low-quality |
| 18-106-final-viral-combined_k141_34268\|\|full | Not-determined |
| 18-106-final-viral-combined_k141_110879\|\|full | Low-quality |
| 18-106-final-viral-combined_k141_46628\|\|full | Not-determined |
| 18-106-final-viral-combined_k141_52269\|\|full | Low-quality |
| 18-106-final-viral-combined_k141_128282\|\|full | Low-quality |
| 18-106-final-viral-combined_k141_99600\|\|full | Not-determined |
| 18-106-final-viral-combined_k141_121863\|\|full | Not-determined |
| 18-106-final-viral-combined_k141_17004\|\|full | Not-determined |
| 18-106-final-viral-combined_k141_99604\|\|full | Not-determined |
| 18-106-final-viral-combined_k141_87305\|\|full | Low-quality |
| 18-106-final-viral-combined_k141_110894\|\|full | Low-quality |
| 18-106-final-viral-combined_k141_104252\|\|full | Not-determined |
| 18-106-final-viral-combined_k141_17019\|\|full | Low-quality |
| 18-106-final-viral-combined_k141_57036\|\|full | Low-quality |
| 18-106-final-viral-combined_k141_116630\|\|full | Not-determined |
| 18-106-final-viral-combined_k141_139652\|\|full | Not-determined |
| 18-106-final-viral-combined_k141_57048\|\|full | Low-quality |
| 18-106-final-viral-combined_k141_46672\|\|full | Not-determined |
| 18-106-final-viral-combined_k141_57050\|\|full | Low-quality |
| 18-106-final-viral-combined_k141_116651\|\|full | Low-quality |
| 18-106-final-viral-combined_k141_116652\|\|full | Low-quality |
| 18-106-final-viral-combined_k141_52305\|\|full | Low-quality |
| 18-106-final-viral-combined_k141_80991\|\|full | Not-determined |
| 18-106-final-viral-combined_k141_57055\|\|full | Low-quality |
| 18-106-final-viral-combined_k141_116655\|\|full | Not-determined |
| 18-106-final-viral-combined_k141_99657\|\|full | Not-determined |
| 18-106-final-viral-combined_k141_133965\|\|full | Low-quality |
| 18-106-final-viral-combined_k141_52327\|\|full | Not-determined |
| 18-106-final-viral-combined_k141_139679\|\|full | Low-quality |
| 18-106-final-viral-combined_k141_28535\|\|full | Low-quality |
| 18-106-final-viral-combined_k141_57071\|\|full | Low-quality |
| 18-106-final-viral-combined_k141_52334\|\|full | Low-quality |
| 18-106-final-viral-combined_k141_116687\|\|full | Not-determined |
| 18-106-final-viral-combined_k141_57089\|\|full | Not-determined |
| 18-106-final-viral-combined_k141_22140\|\|full | Low-quality |
| 18-106-final-viral-combined_k141_52355\|\|full | Not-determined |
| 18-106-final-viral-combined_k141_104329\|\|full | Low-quality |
| 18-106-final-viral-combined_k141_128356\|\|full | Low-quality |
| 18-106-final-viral-combined_k141_121898\|\|full | Low-quality |
| 18-106-final-viral-combined_k141_99683\|\|full | Low-quality |
| 18-106-final-viral-combined_k141_75414\|\|full | Low-quality |
| 18-106-final-viral-combined_k141_139738\|\|full | Not-determined |
| 18-106-final-viral-combined_k141_128371\|\|full | Low-quality |
| 18-106-final-viral-combined_k141_57138\|\|full | Not-determined |
| 18-106-final-viral-combined_k141_57140\|\|full | Not-determined |
| 18-106-final-viral-combined_k141_10638\|\|full | Not-determined |
| 18-106-final-viral-combined_k141_139759\|\|full | Not-determined |
| 18-106-final-viral-combined_k141_57142\|\|full | Low-quality |
| 18-106-final-viral-combined_k141_87423\|\|full | Not-determined |
| 18-106-final-viral-combined_k141_99729\|\|full | Low-quality |
| 18-106-final-viral-combined_k141_104381\|\|full | Low-quality |
| 18-106-final-viral-combined_k141_22208\|\|full | Low-quality |
| 18-106-final-viral-combined_k141_121910\|\|full | Low-quality |
| 18-106-final-viral-combined_k141_22212\|\|full | Not-determined |
| 18-106-final-viral-combined_k141_134033\|\|full | Not-determined |
| 18-106-final-viral-combined_k141_64554\|\|full | Not-determined |
| 18-106-final-viral-combined_k141_139780\|\|full | Not-determined |
| 18-106-final-viral-combined_k141_93203\|\|full | Low-quality |
| 18-106-final-viral-combined_k141_121919\|\|full | Not-determined |
| 18-106-final-viral-combined_k141_46770\|\|full | Low-quality |
| 18-106-final-viral-combined_k141_22223\|\|full | Not-determined |
| 18-106-final-viral-combined_k141_128383\|\|full | Not-determined |
| 18-106-final-viral-combined_k141_134039\|\|full | Low-quality |
| 18-106-final-viral-combined_k141_87449\|\|full | Not-determined |
| 18-106-final-viral-combined_k141_17119\|\|full | Low-quality |
| 18-106_k141_137591 | Complete |
| 18-106-final-viral-combined_k141_34327\|\|full | Not-determined |
| 18-106-final-viral-combined_k141_22231\|\|full | Low-quality |
| 18-106-final-viral-combined_k141_10678\|\|full | Not-determined |
| 18-106-final-viral-combined_k141_139823\|\|full | Not-determined |
| 18-106-final-viral-combined_k141_93258\|\|full | Not-determined |
| 18-106-final-viral-combined_k141_99789\|\|full | Low-quality |
| 18-106-final-viral-combined_k141_75520\|\|full | Low-quality |
| 18-106-final-viral-combined_k141_40934\|\|full | Low-quality |
| 18-106-final-viral-combined_k141_22260\|\|full | Not-determined |
| 18-106-final-viral-combined_k141_104441\|\|full | Not-determined |
| 18-106-final-viral-combined_k141_121985\|\|full | Not-determined |
| 18-106-final-viral-combined_k141_93280\|\|full | Not-determined |
| 18-106-final-viral-combined_k141_40957\|\|full | Not-determined |
| 18-106-final-viral-combined_k141_57252\|\|full | Low-quality |
| 18-106-final-viral-combined_k141_116799\|\|full | Not-determined |
| 18-106-final-viral-combined_k141_139875\|\|full | Low-quality |
| 18-106-final-viral-combined_k141_17153\|\|full | Low-quality |
| 18-106-final-viral-combined_k141_87494\|\|full | Low-quality |
| 18-106-final-viral-combined_k141_139881\|\|full | Not-determined |
| 18-106-final-viral-combined_k141_104460\|\|full | Not-determined |
| 18-106-final-viral-combined_k141_87499\|\|full | Low-quality |
| 18-106-final-viral-combined_k141_110981\|\|full | Low-quality |
| 18-106-final-viral-combined_k141_128463\|\|full | Low-quality |
| 18-106-final-viral-combined_k141_22306\|\|full | Not-determined |
| 18-106-final-viral-combined_k141_34400\|\|full | Low-quality |
| 18-106-final-viral-combined_k141_81122\|\|full | Not-determined |
| 18-106-final-viral-combined_k141_22316\|\|full | Not-determined |
| 18-106-final-viral-combined_k141_57279\|\|full | Not-determined |
| 18-106-final-viral-combined_k141_52545\|\|full | Not-determined |
| 18-106-final-viral-combined_k141_134134\|\|full | Not-determined |
| 18-106-final-viral-combined_k141_93340\|\|full | Not-determined |
| 18-106-final-viral-combined_k141_34431\|\|full | Low-quality |
| 18-106-final-viral-combined_k141_52550\|\|full | Not-determined |
| 18-106-final-viral-combined_k141_75586\|\|full | Not-determined |
| 18-106-final-viral-combined_k141_81163\|\|full | Not-determined |
| 18-106-final-viral-combined_k141_34446\|\|full | Not-determined |
| 18-106-final-viral-combined_k141_52555\|\|full | Not-determined |
| 18-106-final-viral-combined_k141_116851\|\|full | Low-quality |
| 18-106-final-viral-combined_k141_57301\|\|full | Low-quality |
| 18-106-final-viral-combined_k141_134173\|\|full | Low-quality |
| 18-106-final-viral-combined_k141_111009\|\|full | Low-quality |
| 18-106-final-viral-combined_k141_70451\|\|full | Not-determined |
| 18-106-final-viral-combined_k141_116867\|\|full | Low-quality |
| 18-106-final-viral-combined_k141_93374\|\|full | Low-quality |
| 18-106-final-viral-combined_k141_122058\|\|full | Not-determined |
| 18-106-final-viral-combined_k141_116874\|\|full | Not-determined |
| 18-106-final-viral-combined_k141_134190\|\|full | Not-determined |
| 18-106-final-viral-combined_k141_93380\|\|full | Not-determined |
| 18-106-final-viral-combined_k141_52602\|\|full | Not-determined |
| 18-106-final-viral-combined_k141_87597\|\|full | Not-determined |
| 18-106-final-viral-combined_k141_10694\|\|full | Low-quality |
| 18-106-final-viral-combined_k141_93390\|\|full | Low-quality |
| 18-106-final-viral-combined_k141_34480\|\|full | Low-quality |
| 18-106-final-viral-combined_k141_128555\|\|full | Low-quality |
| 18-106-final-viral-combined_k141_17219\|\|full | Low-quality |
| 18-106-final-viral-combined_k141_34493\|\|full | Low-quality |
| 18-106-final-viral-combined_k141_10722\|\|full | Not-determined |
| 18-106-final-viral-combined_k141_93399\|\|full | Low-quality |
| 18-106-final-viral-combined_k141_10733\|\|full | Low-quality |
| 18-106-final-viral-combined_k141_57347\|\|full | Low-quality |
| 18-106-final-viral-combined_k141_104554\|\|full | Low-quality |
| 18-106-final-viral-combined_k141_17227\|\|full | Not-determined |
| 18-106-final-viral-combined_k141_104558\|\|full | Not-determined |
| 18-106-final-viral-combined_k141_10749\|\|full | Low-quality |
| 18-106-final-viral-combined_k141_28644\|\|full | Not-determined |
| 18-106-final-viral-combined_k141_134242\|\|full | Low-quality |
| 18-106-final-viral-combined_k141_104565\|\|full | Not-determined |
| 18-106-final-viral-combined_k141_10753\|\|full | Not-determined |
| 18-106-final-viral-combined_k141_34515\|\|full | Not-determined |
| 18-106-final-viral-combined_k141_46841\|\|full | Not-determined |
| 18-106-final-viral-combined_k141_111112\|\|full | Not-determined |
| 18-106-final-viral-combined_k141_10783\|\|full | Low-quality |
| 18-106-final-viral-combined_k141_41052\|\|full | Not-determined |
| 18-106-final-viral-combined_k141_52711\|\|full | Not-determined |
| 18-106-final-viral-combined_k141_34542\|\|full | Low-quality |
| 18-106-final-viral-combined_k141_22347\|\|full | Not-determined |
| 18-106-final-viral-combined_k141_41071\|\|full | Low-quality |
| 18-106-final-viral-combined_k141_52718\|\|full | Low-quality |
| 18-106-final-viral-combined_k141_122140\|\|full | Not-determined |
| 18-106-final-viral-combined_k141_17281\|\|full | Not-determined |
| 18-106-final-viral-combined_k141_46900\|\|full | Low-quality |
| 18-106-final-viral-combined_k141_122146\|\|full | Not-determined |
| 18-106-final-viral-combined_k141_87628\|\|full | Not-determined |
| 18-106-final-viral-combined_k141_81283\|\|full | Low-quality |
| 18-106-final-viral-combined_k141_111148\|\|full | Low-quality |
| 18-106-final-viral-combined_k141_128604\|\|full | Not-determined |
| 18-106-final-viral-combined_k141_104616\|\|full | Not-determined |
| 18-106-final-viral-combined_k141_10817\|\|full | Low-quality |
| 18-106-final-viral-combined_k141_117017\|\|full | Not-determined |
| 18-106-final-viral-combined_k141_34577\|\|full | Not-determined |
| 18-106-final-viral-combined_k141_22388\|\|full | Not-determined |
| 18-106-final-viral-combined_k141_10821\|\|full | Low-quality |
| 18-106-final-viral-combined_k141_139992\|\|full | Low-quality |
| 18-106-final-viral-combined_k141_139996\|\|full | Low-quality |
| 18-106-final-viral-combined_k141_87657\|\|full | Not-determined |
| 18-106-final-viral-combined_k141_134228\|\|full | Low-quality |
| 18-106-final-viral-combined_k141_81312\|\|full | Low-quality |
| 18-106-final-viral-combined_k141_122216\|\|full | Not-determined |
| 18-106-final-viral-combined_k141_128656\|\|full | Low-quality |
| 18-106-final-viral-combined_k141_52736\|\|full | Not-determined |
| 18-106-final-viral-combined_k141_134340\|\|full | Low-quality |
| 18-106-final-viral-combined_k141_93468\|\|full | Not-determined |
| 18-106-final-viral-combined_k141_128669\|\|full | Low-quality |
| 18-106-final-viral-combined_k141_10843\|\|full | Low-quality |
| 18-106-final-viral-combined_k141_75740\|\|full | Low-quality |
| 18-106-final-viral-combined_k141_117069\|\|full | Low-quality |
| 18-106-final-viral-combined_k141_57416\|\|full | Not-determined |
| 18-106-final-viral-combined_k141_134369\|\|full | Low-quality |
| 18-106-final-viral-combined_k141_134371\|\|full | Low-quality |
| 18-106-final-viral-combined_k141_10860\|\|full | Low-quality |
| 18-106-final-viral-combined_k141_10864\|\|full | Low-quality |
| 18-106-final-viral-combined_k141_134380\|\|full | Not-determined |
| 18-106-final-viral-combined_k141_104691\|\|full | Not-determined |
| 18-106-final-viral-combined_k141_81357\|\|full | Low-quality |
| 18-106-final-viral-combined_k141_93525\|\|full | Not-determined |
| 18-106-final-viral-combined_k141_10867\|\|full | Not-determined |
| 18-106-final-viral-combined_k141_128716\|\|full | Not-determined |
| 18-106-final-viral-combined_k141_81365\|\|full | Low-quality |
| 18-106-final-viral-combined_k141_134397\|\|full | Low-quality |
| 18-106-final-viral-combined_k141_57436\|\|full | Low-quality |
| 18-106-final-viral-combined_k141_10891\|\|full | Low-quality |
| 18-106-final-viral-combined_k141_57456\|\|full | Low-quality |
| 18-106-final-viral-combined_k141_140076\|\|full | Not-determined |
| 18-106-final-viral-combined_k141_93550\|\|full | Low-quality |
| 18-106-final-viral-combined_k141_17375\|\|full | Low-quality |
| 18-106-final-viral-combined_k141_22486\|\|full | Not-determined |
| 18-106-final-viral-combined_k141_140087\|\|full | Low-quality |
| 18-106-final-viral-combined_k141_22506\|\|full | Not-determined |
| 18-106-final-viral-combined_k141_17399\|\|full | Not-determined |
| 18-106-final-viral-combined_k141_93580\|\|full | Low-quality |
| 18-106-final-viral-combined_k141_28819\|\|full | Not-determined |
| 18-106-final-viral-combined_k141_17418\|\|full | Low-quality |
| 18-106-final-viral-combined_k141_17428\|\|full | Low-quality |
| 18-106-final-viral-combined_k141_57505\|\|full | Low-quality |
| 18-106-final-viral-combined_k141_104813\|\|full | Not-determined |
| 18-106-final-viral-combined_k141_104815\|\|full | Low-quality |
| 18-106-final-viral-combined_k141_28835\|\|full | Not-determined |
| 18-106-final-viral-combined_k141_111303\|\|full | Low-quality |
| 18-106-final-viral-combined_k141_52812\|\|full | Low-quality |
| 18-106-final-viral-combined_k141_17452\|\|full | Not-determined |
| 18-106-final-viral-combined_k141_52816\|\|full | Low-quality |
| 18-106-final-viral-combined_k141_28846\|\|full | Not-determined |
| 18-106-final-viral-combined_k141_134508\|\|full | Not-determined |
| 18-106-final-viral-combined_k141_128836\|\|full | Low-quality |
| 18-106-final-viral-combined_k141_111313\|\|full | Not-determined |
| 18-106-final-viral-combined_k141_87811\|\|full | Not-determined |
| 18-106-final-viral-combined_k141_111318\|\|full | Low-quality |
| 18-106-final-viral-combined_k141_34629\|\|full | Not-determined |
| 18-106-final-viral-combined_k141_10981\|\|full | Not-determined |
| 18-106-final-viral-combined_k141_75828\|\|full | Low-quality |
| 18-106-final-viral-combined_k141_87822\|\|full | Not-determined |
| 18-106-final-viral-combined_k141_22573\|\|full | Not-determined |
| 18-106-final-viral-combined_k141_17484\|\|full | Low-quality |
| 18-106-final-viral-combined_k141_75834\|\|full | Not-determined |
| 18-106-final-viral-combined_k141_81527\|\|full | Not-determined |
| 18-106-final-viral-combined_k141_10988\|\|full | Not-determined |
| 18-106-final-viral-combined_k141_111331\|\|full | Not-determined |
| 18-106-final-viral-combined_k141_104867\|\|full | Not-determined |
| 18-106-final-viral-combined_k141_128858\|\|full | Not-determined |
| 18-106-final-viral-combined_k141_104871\|\|full | Low-quality |
| 18-106-final-viral-combined_k141_134549\|\|full | Not-determined |
| 18-106-final-viral-combined_k141_57559\|\|full | Low-quality |
| 18-106-final-viral-combined_k141_22600\|\|full | Low-quality |
| 18-106-final-viral-combined_k141_34649\|\|full | Not-determined |
| 18-106-final-viral-combined_k141_11013\|\|full | Low-quality |
| 18-106-final-viral-combined_k141_140150\|\|full | Low-quality |
| 18-106-final-viral-combined_k141_104907\|\|full | Low-quality |
| 18-106-final-viral-combined_k141_140152\|\|full | Low-quality |
| 18-106-final-viral-combined_k141_93681\|\|full | Low-quality |
| 18-106-final-viral-combined_k141_87862\|\|full | Not-determined |
| 18-106-final-viral-combined_k141_140162\|\|full | Low-quality |
| 18-106-final-viral-combined_k141_81573\|\|full | Low-quality |
| 18-106-final-viral-combined_k141_87867\|\|full | Low-quality |
| 18-106-final-viral-combined_k141_87870\|\|full | Not-determined |
| 18-106-final-viral-combined_k141_93692\|\|full | Low-quality |
| 18-106-final-viral-combined_k141_104928\|\|full | Not-determined |
| 18-106-final-viral-combined_k141_57587\|\|full | Not-determined |
| 18-106-final-viral-combined_k141_111358\|\|full | Not-determined |
| 18-106-final-viral-combined_k141_122429\|\|full | Low-quality |
| 18-106-final-viral-combined_k141_128893\|\|full | Low-quality |
| 18-106-final-viral-combined_k141_34688\|\|full | Low-quality |
| 18-106-final-viral-combined_k141_93720\|\|full | Low-quality |
| 18-106-final-viral-combined_k141_140197\|\|full | Low-quality |
| 18-106-final-viral-combined_k141_22627\|\|full | Low-quality |
| 18-106-final-viral-combined_k141_34698\|\|full | Low-quality |
| 18-106-final-viral-combined_k141_134624\|\|full | Low-quality |
| 18-106-final-viral-combined_k141_128922\|\|full | Not-determined |
| 18-106-final-viral-combined_k141_34715\|\|full | Not-determined |
| 18-106-final-viral-combined_k141_140203\|\|full | Not-determined |
| 18-106-final-viral-combined_k141_87896\|\|full | Not-determined |
| 18-106-final-viral-combined_k141_81655\|\|full | Not-determined |
| 18-106-final-viral-combined_k141_122480\|\|full | Low-quality |
| 18-106-final-viral-combined_k141_140230\|\|full | Not-determined |
| 18-106-final-viral-combined_k141_134694\|\|full | Not-determined |
| 18-106-final-viral-combined_k141_57653\|\|full | Low-quality |
| 18-106-final-viral-combined_k141_81672\|\|full | Not-determined |
| 18-106-final-viral-combined_k141_93777\|\|full | Not-determined |
| 18-106-final-viral-combined_k141_22704\|\|full | Not-determined |
| 18-106-final-viral-combined_k141_134718\|\|full | Low-quality |
| 18-106-final-viral-combined_k141_22708\|\|full | Not-determined |
| 18-106-final-viral-combined_k141_117275\|\|full | Low-quality |
| 18-106-final-viral-combined_k141_22723\|\|full | Low-quality |
| 18-106-final-viral-combined_k141_28937\|\|full | Low-quality |
| 18-106-final-viral-combined_k141_140272\|\|full | Low-quality |
| 18-106-final-viral-combined_k141_164485\|\|full | Medium-quality |
| 18-106-final-viral-combined_k141_34780\|\|full | Not-determined |
| 18-106-final-viral-combined_k141_140296\|\|full | Low-quality |
| 18-106-final-viral-combined_k141_28974\|\|full | Not-determined |
| 18-106-final-viral-combined_k141_28980\|\|full | Not-determined |
| 18-106-final-viral-combined_k141_81723\|\|full | Low-quality |
| 18-106-final-viral-combined_k141_140335\|\|full | Not-determined |
| 18-106-final-viral-combined_k141_11083\|\|full | Low-quality |
| 18-106-final-viral-combined_k141_140344\|\|full | Low-quality |
| 18-106-final-viral-combined_k141_122573\|\|full | Low-quality |
| 18-106-final-viral-combined_k141_29027\|\|full | Low-quality |
| 18-106-final-viral-combined_k141_88001\|\|full | Low-quality |
| 18-106-final-viral-combined_k141_117347\|\|full | Not-determined |
| 18-106-final-viral-combined_k141_75923\|\|full | Not-determined |
| 18-106-final-viral-combined_k141_93876\|\|full | Low-quality |
| 18-106-final-viral-combined_k141_29031\|\|full | Low-quality |
| 18-106-final-viral-combined_k141_129039\|\|full | Not-determined |
| 18-106-final-viral-combined_k141_140364\|\|full | Low-quality |
| 18-106-final-viral-combined_k141_75943\|\|full | Low-quality |
| 18-106-final-viral-combined_k141_140369\|\|full | Low-quality |
| 18-106-final-viral-combined_k141_93898\|\|full | Not-determined |
| 18-106-final-viral-combined_k141_57772\|\|full | Not-determined |
| 18-106-final-viral-combined_k141_34821\|\|full | Low-quality |
| 18-106-final-viral-combined_k141_22869\|\|full | Not-determined |
| 18-106-final-viral-combined_k141_57797\|\|full | Not-determined |
| 18-106-final-viral-combined_k141_34854\|\|full | Not-determined |
| 18-106-final-viral-combined_k141_122647\|\|full | Not-determined |
| 18-106-final-viral-combined_k141_34858\|\|full | Not-determined |
| 18-106-final-viral-combined_k141_129103\|\|full | Low-quality |
| 18-106-final-viral-combined_k141_76011\|\|full | Low-quality |
| 18-106-final-viral-combined_k141_34865\|\|full | Not-determined |
| 18-106-final-viral-combined_k141_11151\|\|full | Low-quality |
| 18-106-final-viral-combined_k141_11154\|\|full | Not-determined |
| 18-106-final-viral-combined_k141_57833\|\|full | Low-quality |
| 18-106-final-viral-combined_k141_140460\|\|full | Not-determined |
| 18-106-final-viral-combined_k141_76033\|\|full | Not-determined |
| 18-106-final-viral-combined_k141_105086\|\|full | Not-determined |
| 18-106-final-viral-combined_k141_122776\|\|full | Not-determined |
| 18-106-final-viral-combined_k141_76046\|\|full | Low-quality |
| 18-106-final-viral-combined_k141_88057\|\|full | Low-quality |
| 18-106-final-viral-combined_k141_11212\|\|full | Not-determined |
| 18-106-final-viral-combined_k141_134838\|\|full | Low-quality |
| 18-106-final-viral-combined_k141_11223\|\|full | Not-determined |
| 18-106-final-viral-combined_k141_122831\|\|full | Low-quality |
| 18-106-final-viral-combined_k141_134863\|\|full | Low-quality |
| 18-106-final-viral-combined_k141_111470\|\|full | Not-determined |
| 18-106-final-viral-combined_k141_134869\|\|full | Not-determined |
| 18-106-final-viral-combined_k141_11280\|\|full | Not-determined |
| 18-106-final-viral-combined_k141_34987\|\|full | Not-determined |
| 18-106-final-viral-combined_k141_57906\|\|full | Low-quality |
| 18-106-final-viral-combined_k141_81841\|\|full | Low-quality |
| 18-106-final-viral-combined_k141_105124\|\|full | Low-quality |
| 18-106-final-viral-combined_k141_122869\|\|full | Low-quality |
| 18-106-final-viral-combined_k141_35019\|\|full | Low-quality |
| 18-106-final-viral-combined_k141_81858\|\|full | Not-determined |
| 18-106-final-viral-combined_k141_140565\|\|full | Low-quality |
| 18-106-final-viral-combined_k141_35035\|\|full | Low-quality |
| 18-106-final-viral-combined_k141_81864\|\|full | Not-determined |
| 18-106-final-viral-combined_k141_29267\|\|full | Not-determined |
| 18-106-final-viral-combined_k141_57955\|\|full | Low-quality |
| 18-106-final-viral-combined_k141_81872\|\|full | Not-determined |
| 18-106-final-viral-combined_k141_29278\|\|full | Not-determined |
| 18-106-final-viral-combined_k141_122920\|\|full | Not-determined |
| 18-106-final-viral-combined_k141_105188\|\|full | Not-determined |
| 18-106-final-viral-combined_k141_29291\|\|full | Not-determined |
| 18-106-final-viral-combined_k141_81879\|\|full | Not-determined |
| 18-106-final-viral-combined_k141_122927\|\|full | Low-quality |
| 18-106-final-viral-combined_k141_81899\|\|full | Not-determined |
| 18-106-final-viral-combined_k141_140622\|\|full | Low-quality |
| 18-106-final-viral-combined_k141_105200\|\|full | Low-quality |
| 18-106-final-viral-combined_k141_134905\|\|full | Low-quality |
| 18-106-final-viral-combined_k141_35086\|\|full | Low-quality |
| 18-106-final-viral-combined_k141_76148\|\|full | Not-determined |
| 18-106-final-viral-combined_k141_35107\|\|full | Not-determined |
| 18-106-final-viral-combined_k141_105241\|\|full | Not-determined |
| 18-106-final-viral-combined_k141_35124\|\|full | Not-determined |
| 18-106-final-viral-combined_k141_105252\|\|full | Not-determined |
| 18-106-final-viral-combined_k141_81971\|\|full | Not-determined |
| 18-106-final-viral-combined_k141_122976\|\|full | Not-determined |
| 18-106-final-viral-combined_k141_81986\|\|full | Not-determined |
| 18-106-final-viral-combined_k141_140702\|\|full | Not-determined |
| 18-106-final-viral-combined_k141_76211\|\|full | Low-quality |
| 18-106-final-viral-combined_k141_35161\|\|full | Low-quality |
| 18-106-final-viral-combined_k141_123017\|\|full | Low-quality |
| 18-106-final-viral-combined_k141_105350\|\|full | Not-determined |
| 18-106-final-viral-combined_k141_76238\|\|full | Not-determined |
| 18-106-final-viral-combined_k141_82057\|\|full | Not-determined |
| 18-106-final-viral-combined_k141_135021\|\|full | Not-determined |
| 18-102_k141_23562 | High-quality |
| 18-106-final-viral-combined_k141_35200\|\|full | Low-quality |
| 18-106-final-viral-combined_k141_82075\|\|full | Low-quality |
| 18-106-final-viral-combined_k141_76252\|\|full | Low-quality |
| 18-106-final-viral-combined_k141_123089\|\|full | Not-determined |
| 18-106-final-viral-combined_k141_123098\|\|full | Low-quality |
| 18-106-final-viral-combined_k141_123102\|\|full | Low-quality |
| 18-106-final-viral-combined_k141_82089\|\|full | Not-determined |
| 18-106-final-viral-combined_k141_123116\|\|full | Not-determined |
| 18-106-final-viral-combined_k141_23313\|\|full | Not-determined |
| 18-106-final-viral-combined_k141_123121\|\|full | Not-determined |
| 18-106-final-viral-combined_k141_123140\|\|full | Not-determined |
| 18-106-final-viral-combined_k141_23352\|\|full | Not-determined |
| 18-106-final-viral-combined_k141_58215\|\|full | Not-determined |
| 18-106-final-viral-combined_k141_105483\|\|full | Not-determined |
| 18-106-final-viral-combined_k141_58243\|\|full | Not-determined |
| 18-106-final-viral-combined_k141_105509\|\|full | Low-quality |
| 18-106-final-viral-combined_k141_58255\|\|full | Low-quality |
| 18-106-final-viral-combined_k141_105533\|\|full | Low-quality |
| 18-106-final-viral-combined_k141_23402\|\|full | Low-quality |
| 18-106-final-viral-combined_k141_123233\|\|full | Low-quality |
| 18-106-final-viral-combined_k141_23461\|\|full | Low-quality |
| 18-106-final-viral-combined_k141_23465\|\|full | Not-determined |
| 18-106-final-viral-combined_k141_105611\|\|full | Low-quality |
| 18-106-final-viral-combined_k141_105630\|\|full | Low-quality |
| 18-106-final-viral-combined_k141_105649\|\|full | Not-determined |
| 18-106-final-viral-combined_k141_58410\|\|full | Not-determined |
| 18-106-final-viral-combined_k141_58426\|\|full | Not-determined |
| 18-106-final-viral-combined_k141_23486\|\|full | Not-determined |
| 18-106-final-viral-combined_k141_11469\|\|full | Not-determined |
| 18-106-final-viral-combined_k141_11480\|\|full | Not-determined |
| 18-106-final-viral-combined_k141_58560\|\|full | Low-quality |
| 18-106-final-viral-combined_k141_58662\|\|full | Low-quality |
| 18-106-final-viral-combined_k141_58691\|\|full | Low-quality |
| 18-106-final-viral-combined_k141_11645\|\|full | Not-determined |
| 18-106-final-viral-combined_k141_11662\|\|full | Low-quality |
| 18-106-final-viral-combined_k141_11671\|\|full | Not-determined |
| 18-106-final-viral-combined_k141_11675\|\|full | Low-quality |
| 18-106-final-viral-combined_k141_11714\|\|full | Not-determined |
| 18-106-final-viral-combined_k141_11723\|\|full | Low-quality |
| 18-106-final-viral-combined_k141_23614\|\|0_partial | Not-determined |
| 18-106-final-viral-combined_k141_29467\|\|0_partial | Low-quality |
| 18-106-final-viral-combined_k141_76704\|\|0_partial | Low-quality |
| 18-106-final-viral-combined_k141_117996\|\|0_partial | Low-quality |
| 18-106-final-viral-combined_k141_35594\|\|0_partial | Low-quality |
| 18-106-final-viral-combined_k141_118037\|\|0_partial | Not-determined |
| 18-106-final-viral-combined_k141_12636\|\|0_partial | Not-determined |
| 18-106-final-viral-combined_k141_95143\|\|0_partial | Low-quality |
| 18-106-final-viral-combined_k141_12728\|\|0_partial | Not-determined |
| 18-106-final-viral-combined_k141_107641\|\|0_partial | Not-determined |
| 18-106-final-viral-combined_k141_107641\|\|1_partial | Low-quality |
| 18-106-final-viral-combined_k141_49236\|\|0_partial | Low-quality |
| 18-106-final-viral-combined_k141_49241\|\|0_partial | Not-determined |
| 18-106-final-viral-combined_k141_61052\|\|0_partial | Low-quality |
| 18-106-final-viral-combined_k141_151708\|\|full | Medium-quality |
| 18-106-final-viral-combined_k141_31764\|\|0_partial | Not-determined |
| 18-106-final-viral-combined_k141_78615\|\|0_partial | Not-determined |
| 18-106-final-viral-combined_k141_125904\|\|0_partial | Not-determined |
| 18-106-final-viral-combined_k141_3824\|\|0_partial | Not-determined |
| KM-final-viral-combined_k141_146850\|\|full | Medium-quality |
| 18-106-final-viral-combined_k141_8614\|\|0_partial | Low-quality |
| 18-106-final-viral-combined_k141_109628\|\|0_partial | Not-determined |
| 18-106-final-viral-combined_k141_103573\|\|0_partial | Low-quality |
| 18-106-final-viral-combined_k141_69378\|\|0_partial | Not-determined |
| 18-106-final-viral-combined_k141_34000\|\|0_partial | Not-determined |
| 18-106-final-viral-combined_k141_46418\|\|0_partial | Not-determined |
| 18-106-final-viral-combined_k141_34613\|\|0_partial | Low-quality |
| 18-106-final-viral-combined_k141_17626\|\|lt2gene | Not-determined |
| 18-106-final-viral-combined_k141_93952\|\|lt2gene | Not-determined |
| 18-106-final-viral-combined_k141_88096\|\|lt2gene | Low-quality |
| 18-106-final-viral-combined_k141_23510\|\|lt2gene | Low-quality |
| 18-106-final-viral-combined_k141_29405\|\|lt2gene | Low-quality |
| 18-106-final-viral-combined_k141_123325\|\|lt2gene | Low-quality |
| 18-106-final-viral-combined_k141_35266\|\|lt2gene | Low-quality |
| 18-106-final-viral-combined_k141_99860\|\|lt2gene | Low-quality |
| 18-106-final-viral-combined_k141_35273\|\|lt2gene | Low-quality |
| 18-106-final-viral-combined_k141_92\|\|lt2gene | Low-quality |
| 18-106-final-viral-combined_k141_76401\|\|lt2gene | Low-quality |
| 18-106-final-viral-combined_k141_117462\|\|lt2gene | Low-quality |
| 18-106-final-viral-combined_k141_123365\|\|lt2gene | Low-quality |
| 18-106-final-viral-combined_k141_35336\|\|lt2gene | Low-quality |
| 18-106-final-viral-combined_k141_135161\|\|lt2gene | Low-quality |
| 18-106-final-viral-combined_k141_11868\|\|lt2gene | Low-quality |
| 18-106-final-viral-combined_k141_129302\|\|lt2gene | Not-determined |
| 18-106-final-viral-combined_k141_117502\|\|lt2gene | Not-determined |
| 18-106-final-viral-combined_k141_99915\|\|lt2gene | Low-quality |
| 18-106-final-viral-combined_k141_41243\|\|lt2gene | Low-quality |
| 18-106-final-viral-combined_k141_47107\|\|lt2gene | Not-determined |
| 18-106-final-viral-combined_k141_88262\|\|lt2gene | Low-quality |
| 18-106-final-viral-combined_k141_111727\|\|lt2gene | Low-quality |
| 18-106-final-viral-combined_k141_123437\|\|lt2gene | Low-quality |
| 18-106-final-viral-combined_k141_58900\|\|lt2gene | Low-quality |
| 18-106-final-viral-combined_k141_64738\|\|lt2gene | Not-determined |
| 18-106-final-viral-combined_k141_105853\|\|lt2gene | Low-quality |
| 18-106-final-viral-combined_k141_70621\|\|lt2gene | Low-quality |
| 18-106-final-viral-combined_k141_99932\|\|lt2gene | Low-quality |
| 18-106-final-viral-combined_k141_64789\|\|lt2gene | Low-quality |
| 18-106-final-viral-combined_k141_117601\|\|lt2gene | Low-quality |
| 18-106-final-viral-combined_k141_35414\|\|lt2gene | Low-quality |
| 18-106-final-viral-combined_k141_105940\|\|lt2gene | Low-quality |
| 18-106-final-viral-combined_k141_117629\|\|lt2gene | Low-quality |
| 18-106-final-viral-combined_k141_111842\|\|lt2gene | Not-determined |
| 18-106-final-viral-combined_k141_70665\|\|lt2gene | Low-quality |
| 18-106-final-viral-combined_k141_339\|\|lt2gene | Low-quality |
| 18-106-final-viral-combined_k141_41325\|\|lt2gene | Not-determined |
| 18-106-final-viral-combined_k141_105985\|\|lt2gene | Low-quality |
| 18-106-final-viral-combined_k141_6017\|\|lt2gene | Not-determined |
| 18-106-final-viral-combined_k141_105993\|\|lt2gene | Low-quality |
| 18-106-final-viral-combined_k141_117684\|\|lt2gene | Not-determined |
| 18-106-final-viral-combined_k141_47242\|\|lt2gene | Low-quality |
| 18-106-final-viral-combined_k141_94272\|\|lt2gene | Not-determined |
| 18-106-final-viral-combined_k141_94288\|\|lt2gene | Not-determined |
| 18-106-final-viral-combined_k141_111913\|\|lt2gene | Low-quality |
| 18-106-final-viral-combined_k141_59071\|\|lt2gene | Low-quality |
| 18-106-final-viral-combined_k141_426\|\|lt2gene | Not-determined |
| 18-106-final-viral-combined_k141_35548\|\|lt2gene | Low-quality |
| 18-106-final-viral-combined_k141_123603\|\|lt2gene | Low-quality |
| 18-106-final-viral-combined_k141_100004\|\|lt2gene | Not-determined |
| 18-106-final-viral-combined_k141_12175\|\|lt2gene | Low-quality |
| 18-106-final-viral-combined_k141_23734\|\|lt2gene | Low-quality |
| 18-106-final-viral-combined_k141_70844\|\|lt2gene | Low-quality |
| 18-106-final-viral-combined_k141_106172\|\|lt2gene | Low-quality |
| 18-106-final-viral-combined_k141_70875\|\|lt2gene | Low-quality |
| 18-106-final-viral-combined_k141_17831\|\|lt2gene | Not-determined |
| 18-106-final-viral-combined_k141_129522\|\|lt2gene | Low-quality |
| 18-106-final-viral-combined_k141_514\|\|lt2gene | Low-quality |
| 18-106-final-viral-combined_k141_94403\|\|lt2gene | Low-quality |
| 18-106-final-viral-combined_k141_70897\|\|lt2gene | Not-determined |
| 18-106-final-viral-combined_k141_88557\|\|lt2gene | Not-determined |
| 18-106-final-viral-combined_k141_59254\|\|lt2gene | Not-determined |
| 18-106-final-viral-combined_k141_129601\|\|lt2gene | Not-determined |
| 18-106-final-viral-combined_k141_47285\|\|lt2gene | Low-quality |
| 18-106-final-viral-combined_k141_41625\|\|lt2gene | Low-quality |
| 18-106-final-viral-combined_k141_53005\|\|lt2gene | Low-quality |
| 18-106-final-viral-combined_k141_610\|\|lt2gene | Low-quality |
| 18-106-final-viral-combined_k141_100173\|\|lt2gene | Not-determined |
| 18-106-final-viral-combined_k141_135305\|\|lt2gene | Low-quality |
| 18-106-final-viral-combined_k141_47380\|\|lt2gene | Low-quality |

18-106-final-viral-combined_k141_100185||lt2gene Not-determined
18-106-final-viral-combined_k141_94588||lt2gene Low-quality
18-106-final-viral-combined_k141_65049||lt2gene Low-quality
18-106-final-viral-combined_k141_129661||lt2gene Low-quality
18-106-final-viral-combined_k141_23869||lt2gene Low-quality
18-106-final-viral-combined_k141_122930||lt2gene Low-quality
18-106-final-viral-combined_k141_100249||lt2gene Low-quality
18-106-final-viral-combined_k141_117910||lt2gene Low-quality
18-106-final-viral-combined_k141_129710||lt2gene Low-quality
18-106-final-viral-combined_k141_122965||lt2gene Not-determined
18-106-final-viral-combined_k141_135389||lt2gene Low-quality
18-106-final-viral-combined_k141_6355||lt2gene Low-quality
18-106-final-viral-combined_k141_41753||lt2gene Low-quality
18-106-final-viral-combined_k141_135451||lt2gene Not-determined
18-106-final-viral-combined_k141_106465||lt2gene Low-quality
18-106-final-viral-combined_k141_88795||lt2gene Low-quality
18-106-final-viral-combined_k141_112198||lt2gene Not-determined
18-106-final-viral-combined_k141_53164||lt2gene Not-determined
18-106-final-viral-combined_k141_94740||lt2gene Low-quality
18-106-final-viral-combined_k141_6496||lt2gene Low-quality
18-106-final-viral-combined_k141_59390||lt2gene Not-determined
18-106-final-viral-combined_k141_94774||lt2gene Low-quality
18-106-final-viral-combined_k141_47561||lt2gene Low-quality
18-106-final-viral-combined_k141_135523||lt2gene Low-quality
18-106-final-viral-combined_k141_12435||lt2gene Not-determined
18-106-final-viral-combined_k141_59409||lt2gene Not-determined
18-106-final-viral-combined_k141_71190||lt2gene Not-determined
18-106-final-viral-combined_k141_100376||lt2gene Not-determined
18-106-final-viral-combined_k141_82622||lt2gene Low-quality
18-106-final-viral-combined_k141_65238||lt2gene Not-determined
18-106-final-viral-combined_k141_76872||lt2gene Low-quality
18-106-final-viral-combined_k141_88898||lt2gene Low-quality
18-106-final-viral-combined_k141_100395||lt2gene Low-quality
18-106-final-viral-combined_k141_6504||lt2gene Low-quality
18-106-final-viral-combined_k141_94792||lt2gene Low-quality
18-106-final-viral-combined_k141_76933||lt2gene Not-determined
18-106-final-viral-combined_k141_41870||lt2gene Not-determined
18-106-final-viral-combined_k141_6573||lt2gene Not-determined
18-106-final-viral-combined_k141_129864||lt2gene Low-quality
18-106-final-viral-combined_k141_47662||lt2gene Low-quality
18-106-final-viral-combined_k141_41933||lt2gene Low-quality
18-106-final-viral-combined_k141_35627||lt2gene Not-determined
18-106-final-viral-combined_k141_76961||lt2gene Low-quality
18-106-final-viral-combined_k141_993||lt2gene Low-quality
18-106-final-viral-combined_k141_35644||lt2gene Low-quality
18-106-final-viral-combined_k141_6667||lt2gene Low-quality
18-106-final-viral-combined_k141_89041||lt2gene Low-quality
18-106-final-viral-combined_k141_53377||lt2gene Not-determined
18-106-final-viral-combined_k141_35701||lt2gene Low-quality
18-106-final-viral-combined_k141_82804||lt2gene Low-quality
18-106-final-viral-combined_k141_1096||lt2gene Low-quality
18-106-final-viral-combined_k141_89090||lt2gene Low-quality
18-106-final-viral-combined_k141_6719||lt2gene Not-determined
18-106-final-viral-combined_k141_82824||lt2gene Low-quality
18-106-final-viral-combined_k141_30183||lt2gene Low-quality
18-106-final-viral-combined_k141_18039||lt2gene Low-quality
18-106-final-viral-combined_k141_95008||lt2gene Low-quality
18-106-final-viral-combined_k141_95009||lt2gene Not-determined
18-106-final-viral-combined_k141_18055||lt2gene Low-quality
18-106-final-viral-combined_k141_95038||lt2gene Not-determined
18-106-final-viral-combined_k141_6791||lt2gene Low-quality
18-106-final-viral-combined_k141_71302||lt2gene Low-quality
18-106-final-viral-combined_k141_95052||lt2gene Not-determined
18-106-final-viral-combined_k141_35791||lt2gene Low-quality
18-106-final-viral-combined_k141_77081||lt2gene Not-determined
18-106-final-viral-combined_k141_24231||lt2gene Not-determined
18-106-final-viral-combined_k141_1223||lt2gene Low-quality
18-106-final-viral-combined_k141_18116||lt2gene Low-quality
18-106-final-viral-combined_k141_1235||lt2gene Not-determined
18-106-final-viral-combined_k141_47822||lt2gene Not-determined
18-106-final-viral-combined_k141_6885||lt2gene Not-determined
18-106-final-viral-combined_k141_106645||lt2gene Low-quality
18-106-final-viral-combined_k141_77144||lt2gene Low-quality
18-106-final-viral-combined_k141_71433||lt2gene Not-determined
18-106-final-viral-combined_k141_100526||lt2gene Low-quality
18-106-final-viral-combined_k141_6930||lt2gene Low-quality
18-106-final-viral-combined_k141_6945||lt2gene Low-quality
18-106-final-viral-combined_k141_59790||lt2gene Not-determined
18-106-final-viral-combined_k141_30397||lt2gene Low-quality
18-106-final-viral-combined_k141_42183||lt2gene Low-quality
18-106-final-viral-combined_k141_112340||lt2gene Low-quality
18-106-final-viral-combined_k141_24304||lt2gene Low-quality
18-106-final-viral-combined_k141_1389||lt2gene Low-quality
18-106-final-viral-combined_k141_1415||lt2gene Not-determined
18-106-final-viral-combined_k141_53459||lt2gene Low-quality
18-106-final-viral-combined_k141_30469||lt2gene Low-quality
18-106-final-viral-combined_k141_42226||lt2gene Low-quality
18-106-final-viral-combined_k141_106767||lt2gene Not-determined
18-106-final-viral-combined_k141_112430||lt2gene Low-quality
18-106-final-viral-combined_k141_1457||lt2gene Not-determined
18-106-final-viral-combined_k141_100654||lt2gene Low-quality
18-106-final-viral-combined_k141_1469||lt2gene Low-quality
18-106-final-viral-combined_k141_83115||lt2gene Low-quality
18-106-final-viral-combined_k141_106815||lt2gene Not-determined
18-106-final-viral-combined_k141_36108||lt2gene Not-determined
18-106-final-viral-combined_k141_42274||lt2gene Not-determined
18-106-final-viral-combined_k141_118238||lt2gene Low-quality
18-106-final-viral-combined_k141_106856||lt2gene Low-quality
18-106-final-viral-combined_k141_30528||lt2gene Low-quality
18-106-final-viral-combined_k141_59896||lt2gene Low-quality
18-106-final-viral-combined_k141_53494||lt2gene Low-quality
18-106-final-viral-combined_k141_24421||lt2gene Not-determined
18-106-final-viral-combined_k141_59952||lt2gene Not-determined
18-106-final-viral-combined_k141_1605||lt2gene Low-quality
18-106-final-viral-combined_k141_118293||lt2gene Not-determined
18-106-final-viral-combined_k141_1631||lt2gene Not-determined
18-106-final-viral-combined_k141_124562||lt2gene Low-quality
18-106-final-viral-combined_k141_83242||lt2gene Low-quality
18-106-final-viral-combined_k141_77422||lt2gene Not-determined
18-106-final-viral-combined_k141_36303||lt2gene Low-quality
18-106-final-viral-combined_k141_89553||lt2gene Low-quality
18-106-final-viral-combined_k141_95363||lt2gene Low-quality
18-106-final-viral-combined_k141_30671||lt2gene Not-determined
18-106-final-viral-combined_k141_30678||lt2gene Not-determined
18-106-final-viral-combined_k141_42525||lt2gene Low-quality
18-106-final-viral-combined_k141_135865||lt2gene Low-quality
18-106-final-viral-combined_k141_77466||lt2gene Low-quality
18-106-final-viral-combined_k141_60118||lt2gene Not-determined
18-106-final-viral-combined_k141_30717||lt2gene Low-quality
18-106-final-viral-combined_k141_107056||lt2gene Low-quality
18-106-final-viral-combined_k141_60196||lt2gene Low-quality
18-106-final-viral-combined_k141_83442||lt2gene Low-quality
18-106-final-viral-combined_k141_89737||lt2gene Low-quality
18-106-final-viral-combined_k141_95477||lt2gene Not-determined
18-106-final-viral-combined_k141_95490||lt2gene Low-quality
18-106-final-viral-combined_k141_1776||lt2gene Low-quality
18-106-final-viral-combined_k141_48445||lt2gene Not-determined
18-106-final-viral-combined_k141_1848||lt2gene Low-quality
18-106-final-viral-combined_k141_30846||lt2gene Not-determined
18-106-final-viral-combined_k141_42724||lt2gene Low-quality
18-106-final-viral-combined_k141_30862||lt2gene Low-quality
18-106-final-viral-combined_k141_83583||lt2gene Low-quality
18-106-final-viral-combined_k141_60393||lt2gene Not-determined
18-106-final-viral-combined_k141_60408||lt2gene Low-quality
18-106-final-viral-combined_k141_30926||lt2gene Low-quality
18-106-final-viral-combined_k141_77568||lt2gene Low-quality
18-106-final-viral-combined_k141_124621||lt2gene Low-quality
18-106-final-viral-combined_k141_77589||lt2gene Low-quality
18-106-final-viral-combined_k141_13095||lt2gene Low-quality
18-106-final-viral-combined_k141_89894||lt2gene Low-quality
18-106-final-viral-combined_k141_95661||lt2gene Low-quality
18-106-final-viral-combined_k141_130583||lt2gene Low-quality
18-106-final-viral-combined_k141_107355||lt2gene Low-quality
18-106-final-viral-combined_k141_66187||lt2gene Not-determined
18-106-final-viral-combined_k141_60453||lt2gene Not-determined
18-106-final-viral-combined_k141_124678||lt2gene Low-quality
18-106-final-viral-combined_k141_13145||lt2gene Not-determined
18-106-final-viral-combined_k141_2052||lt2gene Low-quality
18-106-final-viral-combined_k141_7473||lt2gene Low-quality
18-106-final-viral-combined_k141_36555||lt2gene Not-determined
18-106-final-viral-combined_k141_112806||lt2gene Not-determined
18-106-final-viral-combined_k141_136165||lt2gene Low-quality
18-106-final-viral-combined_k141_48749||lt2gene Low-quality
18-106-final-viral-combined_k141_36611||lt2gene Low-quality
18-106-final-viral-combined_k141_42930||lt2gene Low-quality
18-106-final-viral-combined_k141_60576||lt2gene Low-quality
18-106-final-viral-combined_k141_36659||lt2gene Low-quality
18-106-final-viral-combined_k141_124777||lt2gene Low-quality
18-106-final-viral-combined_k141_42981||lt2gene Low-quality
18-106-final-viral-combined_k141_112888||lt2gene Low-quality
18-106-final-viral-combined_k141_13299||lt2gene Low-quality
18-106-final-viral-combined_k141_83747||lt2gene Low-quality
18-106-final-viral-combined_k141_36739||lt2gene Not-determined
18-106-final-viral-combined_k141_95784||lt2gene Not-determined
18-106-final-viral-combined_k141_136290||lt2gene Not-determined
18-106-final-viral-combined_k141_60660||lt2gene Low-quality
18-106-final-viral-combined_k141_112941||lt2gene Low-quality
18-106-final-viral-combined_k141_2166||lt2gene Low-quality
18-106-final-viral-combined_k141_31203||lt2gene Low-quality
18-106-final-viral-combined_k141_130834||lt2gene Low-quality
18-106-final-viral-combined_k141_36893||lt2gene Low-quality
18-106-final-viral-combined_k141_130905||lt2gene Not-determined
18-106-final-viral-combined_k141_130910||lt2gene Not-determined
18-106-final-viral-combined_k141_66440||lt2gene Low-quality
18-106-final-viral-combined_k141_136433||lt2gene Low-quality
18-106-final-viral-combined_k141_2344||lt2gene Not-determined
18-106-final-viral-combined_k141_43170||lt2gene Low-quality
18-106-final-viral-combined_k141_113109||lt2gene Low-quality
18-106-final-viral-combined_k141_13545||lt2gene Low-quality
18-106-final-viral-combined_k141_60807||lt2gene Not-determined
18-106-final-viral-combined_k141_136557||lt2gene Low-quality
18-106-final-viral-combined_k141_36967||lt2gene Low-quality
18-106-final-viral-combined_k141_7628||lt2gene Low-quality
18-106-final-viral-combined_k141_118779||lt2gene Low-quality
18-106-final-viral-combined_k141_2473||lt2gene Not-determined
18-106-final-viral-combined_k141_7636||lt2gene Not-determined
18-106-final-viral-combined_k141_66586||lt2gene Not-determined
18-106-final-viral-combined_k141_7643||lt2gene Low-quality
18-106-final-viral-combined_k141_13607||lt2gene Low-quality
18-106-final-viral-combined_k141_72208||lt2gene Low-quality
18-106-final-viral-combined_k141_13612||lt2gene Low-quality
18-106-final-viral-combined_k141_37004||lt2gene Low-quality
18-106-final-viral-combined_k141_113197||lt2gene Low-quality
18-106-final-viral-combined_k141_43340||lt2gene Low-quality
18-106-final-viral-combined_k141_136616||lt2gene Low-quality
18-106-final-viral-combined_k141_130983||lt2gene Low-quality
18-106-final-viral-combined_k141_125124||lt2gene Low-quality
18-106-final-viral-combined_k141_43362||lt2gene Low-quality
18-106-final-viral-combined_k141_125149||lt2gene Low-quality
18-106-final-viral-combined_k141_25048||lt2gene Low-quality
18-106-final-viral-combined_k141_7682||lt2gene Not-determined
18-106-final-viral-combined_k141_66689||lt2gene Low-quality
18-106-final-viral-combined_k141_13667||lt2gene Low-quality
18-106-final-viral-combined_k141_136681||lt2gene Low-quality
18-106-final-viral-combined_k141_66698||lt2gene Low-quality
18-106-final-viral-combined_k141_125172||lt2gene Low-quality
18-106-final-viral-combined_k141_125182||lt2gene Not-determined
18-106-final-viral-combined_k141_113300||lt2gene Low-quality
18-106-final-viral-combined_k141_66715||lt2gene Low-quality
18-106-final-viral-combined_k141_31527||lt2gene Low-quality
18-106-final-viral-combined_k141_101095||lt2gene Not-determined
18-106-final-viral-combined_k141_60881||lt2gene Low-quality
18-106-final-viral-combined_k141_72234||lt2gene Low-quality
18-106-final-viral-combined_k141_13695||lt2gene Low-quality
18-106-final-viral-combined_k141_107708||lt2gene Not-determined
18-106-final-viral-combined_k141_72278||lt2gene Not-determined
18-106-final-viral-combined_k141_83946||lt2gene Not-determined
18-106-final-viral-combined_k141_113335||lt2gene Low-quality
18-106-final-viral-combined_k141_43442||lt2gene Low-quality
18-106-final-viral-combined_k141_66803||lt2gene Low-quality
18-106-final-viral-combined_k141_60977||lt2gene Low-quality
18-106-final-viral-combined_k141_101170||lt2gene Low-quality
18-106-final-viral-combined_k141_84005||lt2gene Not-determined
18-106-final-viral-combined_k141_43492||lt2gene Low-quality
18-106-final-viral-combined_k141_118993||lt2gene Not-determined
18-106-final-viral-combined_k141_54276||lt2gene Low-quality
18-106-final-viral-combined_k141_72348||lt2gene Low-quality
18-106-final-viral-combined_k141_113411||lt2gene Low-quality
18-106-final-viral-combined_k141_113424||lt2gene Low-quality
18-106-final-viral-combined_k141_131103||lt2gene Low-quality
18-106-final-viral-combined_k141_136906||lt2gene Not-determined
18-106-final-viral-combined_k141_101257||lt2gene Low-quality
18-106-final-viral-combined_k141_107838||lt2gene Not-determined
18-106-final-viral-combined_k141_7898||lt2gene Not-determined
18-106-final-viral-combined_k141_54347||lt2gene Not-determined
18-106-final-viral-combined_k141_78119||lt2gene Not-determined
18-106-final-viral-combined_k141_54366||lt2gene Low-quality
18-106-final-viral-combined_k141_72400||lt2gene Low-quality
18-106-final-viral-combined_k141_125435||lt2gene Low-quality
18-106-final-viral-combined_k141_25273||lt2gene Low-quality
18-106-final-viral-combined_k141_101268||lt2gene Low-quality
18-106-final-viral-combined_k141_61062||lt2gene Low-quality
18-106-final-viral-combined_k141_125490||lt2gene Low-quality
18-106-final-viral-combined_k141_49359||lt2gene Not-determined
18-106-final-viral-combined_k141_25355||lt2gene Low-quality
18-106-final-viral-combined_k141_25366||lt2gene Low-quality
18-106-final-viral-combined_k141_101366||lt2gene Not-determined
18-106-final-viral-combined_k141_107922||lt2gene Low-quality
18-106-final-viral-combined_k141_37501||lt2gene Low-quality
18-106-final-viral-combined_k141_84120||lt2gene Low-quality
18-106-final-viral-combined_k141_107940||lt2gene Low-quality
18-106-final-viral-combined_k141_8044||lt2gene Not-determined
18-106-final-viral-combined_k141_78212||lt2gene Not-determined
18-106-final-viral-combined_k141_107956||lt2gene Not-determined
18-106-final-viral-combined_k141_96347||lt2gene Low-quality
18-106-final-viral-combined_k141_37531||lt2gene Not-determined
18-106-final-viral-combined_k141_25426||lt2gene Low-quality
18-106-final-viral-combined_k141_72501||lt2gene Low-quality
18-106-final-viral-combined_k141_61154||lt2gene Not-determined
18-106-final-viral-combined_k141_101415||lt2gene Not-determined
18-106-final-viral-combined_k141_113492||lt2gene Low-quality
18-106-final-viral-combined_k141_101423||lt2gene Low-quality
18-106-final-viral-combined_k141_101427||lt2gene Low-quality
18-106-final-viral-combined_k141_84198||lt2gene Low-quality
18-106-final-viral-combined_k141_14003||lt2gene Low-quality
18-106-final-viral-combined_k141_125559||lt2gene Low-quality
18-106-final-viral-combined_k141_90442||lt2gene Low-quality
18-106-final-viral-combined_k141_113557||lt2gene Low-quality
18-106-final-viral-combined_k141_90467||lt2gene Not-determined
18-106-final-viral-combined_k141_54458||lt2gene Low-quality
18-106-final-viral-combined_k141_8163||lt2gene Not-determined
18-106-final-viral-combined_k141_96415||lt2gene Low-quality
18-106-final-viral-combined_k141_25521||lt2gene Low-quality
18-106-final-viral-combined_k141_119175||lt2gene Low-quality
18-106-final-viral-combined_k141_67136||lt2gene Low-quality
18-106-final-viral-combined_k141_37678||lt2gene Not-determined
18-106-final-viral-combined_k141_54505||lt2gene Low-quality
18-106-final-viral-combined_k141_61195||lt2gene Low-quality
18-106-final-viral-combined_k141_119234||lt2gene Not-determined
18-106-final-viral-combined_k141_25625||lt2gene Not-determined
18-106-final-viral-combined_k141_25627||lt2gene Low-quality
18-106-final-viral-combined_k141_43876||lt2gene Not-determined
18-106-final-viral-combined_k141_3222||lt2gene Low-quality
18-106-final-viral-combined_k141_119275||lt2gene Low-quality
18-106-final-viral-combined_k141_96511||lt2gene Low-quality
18-106-final-viral-combined_k141_84350||lt2gene Low-quality
18-106-final-viral-combined_k141_37775||lt2gene Low-quality
18-106-final-viral-combined_k141_37800||lt2gene Not-determined
18-106-final-viral-combined_k141_131306||lt2gene Low-quality
18-106-final-viral-combined_k141_54593||lt2gene Low-quality
18-106-final-viral-combined_k141_113716||lt2gene Low-quality
18-106-final-viral-combined_k141_61298||lt2gene Low-quality
18-106-final-viral-combined_k141_78479||lt2gene Low-quality
18-106-final-viral-combined_k141_136979||lt2gene Low-quality
18-106-final-viral-combined_k141_125839||lt2gene Low-quality
18-106-final-viral-combined_k141_108203||lt2gene Not-determined
18-106-final-viral-combined_k141_25720||lt2gene Low-quality
18-106-final-viral-combined_k141_84478||lt2gene Not-determined
18-106-final-viral-combined_k141_19567||lt2gene Low-quality
18-106-final-viral-combined_k141_37909||lt2gene Low-quality
18-106-final-viral-combined_k141_31972||lt2gene Low-quality
18-106-final-viral-combined_k141_108236||lt2gene Low-quality
18-106-final-viral-combined_k141_78561||lt2gene Low-quality
18-106-final-viral-combined_k141_19601||lt2gene Low-quality
18-106-final-viral-combined_k141_43989||lt2gene Not-determined
18-106-final-viral-combined_k141_90732||lt2gene Low-quality
18-106-final-viral-combined_k141_78614||lt2gene Not-determined
18-106-final-viral-combined_k141_90756||lt2gene Low-quality
18-106-final-viral-combined_k141_44026||lt2gene Not-determined
18-106-final-viral-combined_k141_44037||lt2gene Low-quality
18-106-final-viral-combined_k141_44045||lt2gene Not-determined
18-106-final-viral-combined_k141_49799||lt2gene Low-quality
18-106-final-viral-combined_k141_44051||lt2gene Low-quality
18-106-final-viral-combined_k141_37975||lt2gene Low-quality
18-106-final-viral-combined_k141_49833||lt2gene Low-quality
18-106-final-viral-combined_k141_113825||lt2gene Low-quality
18-106-final-viral-combined_k141_137155||lt2gene Low-quality
18-106-final-viral-combined_k141_96752||lt2gene Not-determined
18-106-final-viral-combined_k141_84550||lt2gene Low-quality
18-106-final-viral-combined_k141_8284||lt2gene Low-quality
18-106-final-viral-combined_k141_14269||lt2gene Not-determined
18-106-final-viral-combined_k141_73011||lt2gene Not-determined
18-106-final-viral-combined_k141_3493||lt2gene Low-quality
18-106-final-viral-combined_k141_119546||lt2gene Not-determined
18-106-final-viral-combined_k141_49911||lt2gene Not-determined
18-106-final-viral-combined_k141_90876||lt2gene Low-quality
18-106-final-viral-combined_k141_96871||lt2gene Not-determined
18-106-final-viral-combined_k141_44156||lt2gene Low-quality
18-106-final-viral-combined_k141_49933||lt2gene Low-quality
18-106-final-viral-combined_k141_67559||lt2gene Low-quality
18-106-final-viral-combined_k141_73070||lt2gene Low-quality
18-106-final-viral-combined_k141_14345||lt2gene Low-quality
18-106-final-viral-combined_k141_73075||lt2gene Low-quality
18-106-final-viral-combined_k141_54892||lt2gene Low-quality
18-106-final-viral-combined_k141_25861||lt2gene Low-quality
18-106-final-viral-combined_k141_101896||lt2gene Low-quality
18-106-final-viral-combined_k141_137290||lt2gene Low-quality
18-106-final-viral-combined_k141_101905||lt2gene Not-determined
18-106-final-viral-combined_k141_67631||lt2gene Low-quality
18-106-final-viral-combined_k141_44242||lt2gene Low-quality
18-106-final-viral-combined_k141_38198||lt2gene Not-determined
18-106-final-viral-combined_k141_19820||lt2gene Not-determined
18-106-final-viral-combined_k141_119588||lt2gene Low-quality
18-106-final-viral-combined_k141_101970||lt2gene Low-quality
18-106-final-viral-combined_k141_101981||lt2gene Low-quality
18-106-final-viral-combined_k141_125922||lt2gene Not-determined
18-106-final-viral-combined_k141_55010||lt2gene Not-determined
18-106-final-viral-combined_k141_97012||lt2gene Low-quality
18-106-final-viral-combined_k141_114018||lt2gene Low-quality
18-106-final-viral-combined_k141_137416||lt2gene Not-determined
18-106-final-viral-combined_k141_44313||lt2gene Low-quality
18-106-final-viral-combined_k141_84790||lt2gene Not-determined
18-106-final-viral-combined_k141_67790||lt2gene Not-determined
18-106-final-viral-combined_k141_8396||lt2gene Low-quality
18-106-final-viral-combined_k141_3708||lt2gene Low-quality
18-106-final-viral-combined_k141_32313||lt2gene Low-quality
18-106-final-viral-combined_k141_19947||lt2gene Low-quality
18-106-final-viral-combined_k141_102091||lt2gene Low-quality
18-106-final-viral-combined_k141_97131||lt2gene Low-quality
18-106-final-viral-combined_k141_137513||lt2gene Low-quality
18-106-final-viral-combined_k141_25922||lt2gene Low-quality
18-106-final-viral-combined_k141_97155||lt2gene Not-determined
18-106-final-viral-combined_k141_73331||lt2gene Not-determined
18-106-final-viral-combined_k141_25954||lt2gene Low-quality
18-106-final-viral-combined_k141_84833||lt2gene Low-quality
18-106-final-viral-combined_k141_32404||lt2gene Not-determined
18-106-final-viral-combined_k141_84838||lt2gene Low-quality
18-106-final-viral-combined_k141_108624||lt2gene Not-determined
18-106-final-viral-combined_k141_67902||lt2gene Low-quality
18-106-final-viral-combined_k141_84881||lt2gene Low-quality
18-106-final-viral-combined_k141_50175||lt2gene Low-quality
18-106-final-viral-combined_k141_14571||lt2gene Not-determined
18-106-final-viral-combined_k141_73370||lt2gene Not-determined
18-106-final-viral-combined_k141_73389||lt2gene Low-quality
18-106-final-viral-combined_k141_26030||lt2gene Low-quality
18-106-final-viral-combined_k141_84917||lt2gene Low-quality
18-106-final-viral-combined_k141_26041||lt2gene Low-quality
18-106-final-viral-combined_k141_55215||lt2gene Low-quality
18-106-final-viral-combined_k141_102237||lt2gene Not-determined
18-106-final-viral-combined_k141_8510||lt2gene Not-determined
18-106-final-viral-combined_k141_126217||lt2gene Not-determined
18-106-final-viral-combined_k141_114343||lt2gene Low-quality
18-106-final-viral-combined_k141_102260||lt2gene Low-quality
18-106-final-viral-combined_k141_8548||lt2gene Low-quality
18-106-final-viral-combined_k141_50214||lt2gene Low-quality
18-106-final-viral-combined_k141_119923||lt2gene Low-quality
18-106-final-viral-combined_k141_26129||lt2gene Low-quality
18-106-final-viral-combined_k141_26136||lt2gene Low-quality
18-106-final-viral-combined_k141_38483||lt2gene Not-determined
18-106-final-viral-combined_k141_38507||lt2gene Low-quality
18-106-final-viral-combined_k141_55304||lt2gene Low-quality
18-106-final-viral-combined_k141_38512||lt2gene Not-determined
18-106-final-viral-combined_k141_97341||lt2gene Low-quality
18-106-final-viral-combined_k141_102343||lt2gene Low-quality
18-106-final-viral-combined_k141_20091||lt2gene Not-determined
18-106-final-viral-combined_k141_14656||lt2gene Low-quality
18-106-final-viral-combined_k141_20101||lt2gene Low-quality
18-106-final-viral-combined_k141_114409||lt2gene Low-quality
18-106-final-viral-combined_k141_20157||lt2gene Low-quality
18-106-final-viral-combined_k141_97430||lt2gene Not-determined
18-106-final-viral-combined_k141_132216||lt2gene Not-determined
18-106-final-viral-combined_k141_32600||lt2gene Low-quality
18-106-final-viral-combined_k141_97524||lt2gene Not-determined
18-106-final-viral-combined_k141_26225||lt2gene Low-quality
18-106-final-viral-combined_k141_68169||lt2gene Low-quality
18-106-final-viral-combined_k141_26230||lt2gene Not-determined
18-106-final-viral-combined_k141_85119||lt2gene Low-quality
18-106-final-viral-combined_k141_73617||lt2gene Low-quality
18-106-final-viral-combined_k141_32645||lt2gene Not-determined
18-106-final-viral-combined_k141_126445||lt2gene Low-quality
18-106-final-viral-combined_k141_44601||lt2gene Low-quality
18-106-final-viral-combined_k141_137761||lt2gene Not-determined
18-106-final-viral-combined_k141_20255||lt2gene Low-quality
18-106-final-viral-combined_k141_44648||lt2gene Not-determined
18-106-final-viral-combined_k141_120152||lt2gene Low-quality
18-106-final-viral-combined_k141_62244||lt2gene Not-determined
18-106-final-viral-combined_k141_120161||lt2gene Not-determined
18-106-final-viral-combined_k141_32680||lt2gene Low-quality
18-106-final-viral-combined_k141_14868||lt2gene Low-quality
18-106-final-viral-combined_k141_137837||lt2gene Low-quality
18-106-final-viral-combined_k141_137839||lt2gene Low-quality
18-106-final-viral-combined_k141_102627||lt2gene Not-determined
18-106-final-viral-combined_k141_102639||lt2gene Low-quality
18-106-final-viral-combined_k141_120181||lt2gene Low-quality
18-106-final-viral-combined_k141_50365||lt2gene Low-quality
18-106-final-viral-combined_k141_114635||lt2gene Low-quality
18-106-final-viral-combined_k141_50389||lt2gene Low-quality
18-106-final-viral-combined_k141_26423||lt2gene Not-determined
18-106-final-viral-combined_k141_79271||lt2gene Low-quality
18-106-final-viral-combined_k141_26441||lt2gene Low-quality
18-106-final-viral-combined_k141_4390||lt2gene Low-quality
18-106-final-viral-combined_k141_38911||lt2gene Not-determined
18-106-final-viral-combined_k141_120236||lt2gene Low-quality
18-106-final-viral-combined_k141_85326||lt2gene Not-determined
18-106-final-viral-combined_k141_85339||lt2gene Low-quality
18-106-final-viral-combined_k141_32764||lt2gene Not-determined
18-106-final-viral-combined_k141_126628||lt2gene Low-quality
18-106-final-viral-combined_k141_4445||lt2gene Low-quality
18-106-final-viral-combined_k141_32777||lt2gene Low-quality
18-106-final-viral-combined_k141_32787||lt2gene Low-quality
18-106-final-viral-combined_k141_102674||lt2gene Not-determined
18-106-final-viral-combined_k141_114730||lt2gene Low-quality
18-106-final-viral-combined_k141_137957||lt2gene Low-quality
18-106-final-viral-combined_k141_68499||lt2gene Not-determined
18-106-final-viral-combined_k141_120319||lt2gene Not-determined
18-106-final-viral-combined_k141_91361||lt2gene Not-determined
18-106-final-viral-combined_k141_109023||lt2gene Not-determined
18-106-final-viral-combined_k141_109027||lt2gene Not-determined
18-106-final-viral-combined_k141_8961||lt2gene Not-determined
18-106-final-viral-combined_k141_91399||lt2gene Low-quality
18-106-final-viral-combined_k141_44854||lt2gene Low-quality
18-106-final-viral-combined_k141_15033||lt2gene Low-quality
18-106-final-viral-combined_k141_50563||lt2gene Low-quality
18-106-final-viral-combined_k141_102823||lt2gene Low-quality
18-106-final-viral-combined_k141_102831||lt2gene Low-quality
18-106-final-viral-combined_k141_44954||lt2gene Not-determined
18-106-final-viral-combined_k141_20491||lt2gene Not-determined
18-106-final-viral-combined_k141_138064||lt2gene Low-quality
18-106-final-viral-combined_k141_97847||lt2gene Low-quality
18-106-final-viral-combined_k141_109118||lt2gene Not-determined
18-106-final-viral-combined_k141_109126||lt2gene Low-quality
18-106-final-viral-combined_k141_132617||lt2gene Low-quality
18-106-final-viral-combined_k141_45070||lt2gene Not-determined
18-106-final-viral-combined_k141_91551||lt2gene Low-quality
18-106-final-viral-combined_k141_50709||lt2gene Not-determined
18-106-final-viral-combined_k141_109241||lt2gene Low-quality
18-106-final-viral-combined_k141_4641||lt2gene Low-quality
18-106-final-viral-combined_k141_55816||lt2gene Not-determined
18-106-final-viral-combined_k141_97936||lt2gene Low-quality
18-106-final-viral-combined_k141_20595||lt2gene Low-quality
18-106-final-viral-combined_k141_114936||lt2gene Not-determined
18-106-final-viral-combined_k141_138223||lt2gene Low-quality
18-106-final-viral-combined_k141_20621||lt2gene Low-quality
18-106-final-viral-combined_k141_45158||lt2gene Low-quality
18-106-final-viral-combined_k141_62686||lt2gene Low-quality
18-106-final-viral-combined_k141_132702||lt2gene Low-quality
18-106-final-viral-combined_k141_98017||lt2gene Low-quality
18-106-final-viral-combined_k141_91702||lt2gene Not-determined
18-106-final-viral-combined_k141_98061||lt2gene Not-determined
18-106-final-viral-combined_k141_50723||lt2gene Low-quality
18-106-final-viral-combined_k141_109382||lt2gene Low-quality
18-106-final-viral-combined_k141_73960||lt2gene Not-determined
18-106-final-viral-combined_k141_126921||lt2gene Low-quality
18-106-final-viral-combined_k141_68816||lt2gene Low-quality
18-106-final-viral-combined_k141_79686||lt2gene Not-determined
18-106-final-viral-combined_k141_91822||lt2gene Not-determined
18-106-final-viral-combined_k141_120627||lt2gene Low-quality
18-106-final-viral-combined_k141_9209||lt2gene Low-quality
18-106-final-viral-combined_k141_120644||lt2gene Not-determined
18-106-final-viral-combined_k141_50816||lt2gene Not-determined
18-106-final-viral-combined_k141_39252||lt2gene Low-quality
18-106-final-viral-combined_k141_103151||lt2gene Not-determined
18-106-final-viral-combined_k141_20745||lt2gene Low-quality
18-106-final-viral-combined_k141_4979||lt2gene Low-quality
18-106-final-viral-combined_k141_50866||lt2gene Not-determined
18-106-final-viral-combined_k141_62974||lt2gene Not-determined
18-106-final-viral-combined_k141_15459||lt2gene Low-quality
18-106-final-viral-combined_k141_5010||lt2gene Low-quality
18-106-final-viral-combined_k141_85954||lt2gene Not-determined
18-106-final-viral-combined_k141_27052||lt2gene Low-quality
18-106-final-viral-combined_k141_9334||lt2gene Low-quality
18-106-final-viral-combined_k141_138395||lt2gene Low-quality
18-106-final-viral-combined_k141_115253||lt2gene Low-quality
18-106-final-viral-combined_k141_63065||lt2gene Low-quality
18-106-final-viral-combined_k141_79865||lt2gene Low-quality
18-106-final-viral-combined_k141_50962||lt2gene Low-quality
18-106-final-viral-combined_k141_56106||lt2gene Not-determined
18-106-final-viral-combined_k141_5123||lt2gene Low-quality
18-106-final-viral-combined_k141_120820||lt2gene Low-quality
18-106-final-viral-combined_k141_50993||lt2gene Low-quality
18-106-final-viral-combined_k141_5169||lt2gene Low-quality
18-106-final-viral-combined_k141_86095||lt2gene Low-quality
18-106-final-viral-combined_k141_127091||lt2gene Not-determined
18-106-final-viral-combined_k141_15610||lt2gene Not-determined
18-106-final-viral-combined_k141_33275||lt2gene Low-quality
18-106-final-viral-combined_k141_15641||lt2gene Low-quality
18-106-final-viral-combined_k141_63185||lt2gene Not-determined
18-106-final-viral-combined_k141_63191||lt2gene Not-determined
18-106-final-viral-combined_k141_45456||lt2gene Low-quality
18-106-final-viral-combined_k141_63208||lt2gene Low-quality
18-106-final-viral-combined_k141_109665||lt2gene Not-determined
18-106-final-viral-combined_k141_109666||lt2gene Low-quality
18-106-final-viral-combined_k141_15667||lt2gene Not-determined
18-106-final-viral-combined_k141_33337||lt2gene Low-quality
18-106-final-viral-combined_k141_5265||lt2gene Low-quality
18-106-final-viral-combined_k141_138559||lt2gene Low-quality
18-106-final-viral-combined_k141_33377||lt2gene Low-quality
18-106-final-viral-combined_k141_45496||lt2gene Not-determined
18-106-final-viral-combined_k141_9523||lt2gene Low-quality
18-106-final-viral-combined_k141_138568||lt2gene Low-quality
18-106-final-viral-combined_k141_9529||lt2gene Not-determined
18-106-final-viral-combined_k141_103430||lt2gene Low-quality
18-106-final-viral-combined_k141_103437||lt2gene Low-quality
18-106-final-viral-combined_k141_120898||lt2gene Low-quality
18-106-final-viral-combined_k141_27266||lt2gene Low-quality
18-106-final-viral-combined_k141_103447||lt2gene Not-determined
18-106-final-viral-combined_k141_51126||lt2gene Not-determined
18-106-final-viral-combined_k141_115456||lt2gene Not-determined
18-106-final-viral-combined_k141_86260||lt2gene Not-determined
18-106-final-viral-combined_k141_5350||lt2gene Low-quality
18-106-final-viral-combined_k141_63337||lt2gene Low-quality
18-106-final-viral-combined_k141_103513||lt2gene Low-quality
18-106-final-viral-combined_k141_15839||lt2gene Low-quality
18-106-final-viral-combined_k141_45619||lt2gene Low-quality
18-106-final-viral-combined_k141_121047||lt2gene Low-quality
18-106-final-viral-combined_k141_98536||lt2gene Low-quality
18-106-final-viral-combined_k141_63354||lt2gene Not-determined
18-106-final-viral-combined_k141_80068||lt2gene Low-quality
18-106-final-viral-combined_k141_27387||lt2gene Low-quality
18-106-final-viral-combined_k141_86345||lt2gene Not-determined
18-106-final-viral-combined_k141_109838||lt2gene Low-quality
18-106-final-viral-combined_k141_115619||lt2gene Low-quality
18-106-final-viral-combined_k141_15929||lt2gene Low-quality
18-106-final-viral-combined_k141_92152||lt2gene Low-quality
18-106-final-viral-combined_k141_74481||lt2gene Low-quality
18-106-final-viral-combined_k141_9720||lt2gene Low-quality
18-106-final-viral-combined_k141_121082||lt2gene Not-determined
18-106-final-viral-combined_k141_9736||lt2gene Not-determined
18-106-final-viral-combined_k141_98586||lt2gene Not-determined
18-106-final-viral-combined_k141_80170||lt2gene Not-determined
18-106-final-viral-combined_k141_80189||lt2gene Low-quality
18-106-final-viral-combined_k141_133005||lt2gene Low-quality
18-106-final-viral-combined_k141_63469||lt2gene Low-quality
18-106-final-viral-combined_k141_56296||lt2gene Not-determined
18-106-final-viral-combined_k141_21158||lt2gene Low-quality
18-106-final-viral-combined_k141_69262||lt2gene Low-quality
18-106-final-viral-combined_k141_56324||lt2gene Low-quality
18-106-final-viral-combined_k141_45799||lt2gene Not-determined
18-106-final-viral-combined_k141_16041||lt2gene Low-quality
18-106-final-viral-combined_k141_92261||lt2gene Low-quality
18-106-final-viral-combined_k141_98691||lt2gene Not-determined
18-106-final-viral-combined_k141_45846||lt2gene Low-quality
18-106-final-viral-combined_k141_103592||lt2gene Low-quality
18-106-final-viral-combined_k141_110016||lt2gene Low-quality
18-106-final-viral-combined_k141_51511||lt2gene Low-quality
18-106-final-viral-combined_k141_9841||lt2gene Low-quality
18-106-final-viral-combined_k141_138821||lt2gene Low-quality
18-106-final-viral-combined_k141_69363||lt2gene Not-determined
18-106-final-viral-combined_k141_80308||lt2gene Not-determined
18-106-final-viral-combined_k141_45929||lt2gene Low-quality
18-106-final-viral-combined_k141_127396||lt2gene Not-determined
18-106-final-viral-combined_k141_103668||lt2gene Not-determined
18-106-final-viral-combined_k141_92393||lt2gene Not-determined
18-106-final-viral-combined_k141_21258||lt2gene Not-determined
18-106-final-viral-combined_k141_86439||lt2gene Low-quality
18-106-final-viral-combined_k141_86440||lt2gene Not-determined
18-106-final-viral-combined_k141_121232||lt2gene Not-determined
18-106-final-viral-combined_k141_27707||lt2gene Not-determined
18-106-final-viral-combined_k141_5704||lt2gene Not-determined

| | |
|---|---|
| 18-106-final-viral-combined_k141_33542||lt2gene | Not-determined |
| 18-106-final-viral-combined_k141_127474||lt2gene | Not-determined |
| 18-106-final-viral-combined_k141_133201||lt2gene | Not-determined |
| 18-106-final-viral-combined_k141_39985||lt2gene | Low-quality |
| 18-106-final-viral-combined_k141_103726||lt2gene | Low-quality |
| 18-106-final-viral-combined_k141_74788||lt2gene | Not-determined |
| 18-106-final-viral-combined_k141_80423||lt2gene | Low-quality |
| 18-106-final-viral-combined_k141_74807||lt2gene | Low-quality |
| 18-106-final-viral-combined_k141_40049||lt2gene | Low-quality |
| 18-106-final-viral-combined_k141_139037||lt2gene | Low-quality |
| 18-106-final-viral-combined_k141_80471||lt2gene | Low-quality |
| 18-106-final-viral-combined_k141_127529||lt2gene | Not-determined |
| 18-106-final-viral-combined_k141_46106||lt2gene | Not-determined |
| 18-106-final-viral-combined_k141_51655||lt2gene | Low-quality |
| 18-106-final-viral-combined_k141_86593||lt2gene | Not-determined |
| 18-106-final-viral-combined_k141_69427||lt2gene | Low-quality |
| 18-106-final-viral-combined_k141_46133||lt2gene | Low-quality |
| 18-106-final-viral-combined_k141_40138||lt2gene | Low-quality |
| 18-106-final-viral-combined_k141_121394||lt2gene | Low-quality |
| 18-106-final-viral-combined_k141_56491||lt2gene | Low-quality |
| 18-106-final-viral-combined_k141_46182||lt2gene | Low-quality |
| 18-106-final-viral-combined_k141_27874||lt2gene | Low-quality |
| 18-106-final-viral-combined_k141_27884||lt2gene | Low-quality |
| 18-106-final-viral-combined_k141_40167||lt2gene | Low-quality |
| 18-106-final-viral-combined_k141_63880||lt2gene | Not-determined |
| 18-106-final-viral-combined_k141_33748||lt2gene | Low-quality |
| 18-106-final-viral-combined_k141_33758||lt2gene | Low-quality |
| 18-106-final-viral-combined_k141_27930||lt2gene | Not-determined |
| 18-106-final-viral-combined_k141_139071||lt2gene | Low-quality |
| 18-106-final-viral-combined_k141_51776||lt2gene | Not-determined |
| 18-106-final-viral-combined_k141_110373||lt2gene | Low-quality |
| 18-106-final-viral-combined_k141_16467||lt2gene | Not-determined |
| 18-106-final-viral-combined_k141_63959||lt2gene | Low-quality |
| 18-106-final-viral-combined_k141_116140||lt2gene | Low-quality |
| 18-106-final-viral-combined_k141_10090||lt2gene | Not-determined |
| 18-106-final-viral-combined_k141_92697||lt2gene | Low-quality |
| 18-106-final-viral-combined_k141_40332||lt2gene | Low-quality |
| 18-106-final-viral-combined_k141_10127||lt2gene | Not-determined |
| 18-106-final-viral-combined_k141_116177||lt2gene | Low-quality |
| 18-106-final-viral-combined_k141_56630||lt2gene | Low-quality |
| 18-106-final-viral-combined_k141_133523||lt2gene | Not-determined |
| 18-106-final-viral-combined_k141_86800||lt2gene | Not-determined |
| 18-106-final-viral-combined_k141_139220||lt2gene | Low-quality |
| 18-106-final-viral-combined_k141_16573||lt2gene | Low-quality |
| 18-106-final-viral-combined_k141_133553||lt2gene | Low-quality |
| 18-106-final-viral-combined_k141_56670||lt2gene | Not-determined |
| 18-106-final-viral-combined_k141_64084||lt2gene | Not-determined |
| 18-106-final-viral-combined_k141_51931||lt2gene | Low-quality |
| 18-106-final-viral-combined_k141_64107||lt2gene | Not-determined |
| 18-106-final-viral-combined_k141_51953||lt2gene | Low-quality |
| 18-106-final-viral-combined_k141_51985||lt2gene | Low-quality |
| 18-106-final-viral-combined_k141_121552||lt2gene | Not-determined |
| 18-106-final-viral-combined_k141_21681||lt2gene | Low-quality |
| 18-106-final-viral-combined_k141_21695||lt2gene | Low-quality |
| 18-106-final-viral-combined_k141_133644||lt2gene | Low-quality |
| 18-106-final-viral-combined_k141_69854||lt2gene | Not-determined |
| 18-106-final-viral-combined_k141_64187||lt2gene | Low-quality |
| 18-106-final-viral-combined_k141_139316||lt2gene | Low-quality |
| 18-106-final-viral-combined_k141_103936||lt2gene | Not-determined |
| 18-106-final-viral-combined_k141_121616||lt2gene | Low-quality |
| 18-106-final-viral-combined_k141_40550||lt2gene | Low-quality |
| 18-106-final-viral-combined_k141_110624||lt2gene | Low-quality |
| 18-106-final-viral-combined_k141_116420||lt2gene | Low-quality |
| 18-106-final-viral-combined_k141_121691||lt2gene | Not-determined |
| 18-106-final-viral-combined_k141_121699||lt2gene | Not-determined |
| 18-106-final-viral-combined_k141_40607||lt2gene | Low-quality |
| 18-106-final-viral-combined_k141_128051||lt2gene | Not-determined |
| 18-106-final-viral-combined_k141_40637||lt2gene | Not-determined |
| 18-106-final-viral-combined_k141_34120||lt2gene | Low-quality |
| 18-106-final-viral-combined_k141_10380||lt2gene | Not-determined |
| 18-106-final-viral-combined_k141_133828||lt2gene | Low-quality |
| 18-106-final-viral-combined_k141_46459||lt2gene | Not-determined |
| 18-106-final-viral-combined_k141_87107||lt2gene | Not-determined |
| 18-106-final-viral-combined_k141_133844||lt2gene | Low-quality |
| 18-106-final-viral-combined_k141_10387||lt2gene | Not-determined |
| 18-106-final-viral-combined_k141_34153||lt2gene | Low-quality |
| 18-106-final-viral-combined_k141_110717||lt2gene | Low-quality |
| 18-106-final-viral-combined_k141_46493||lt2gene | Low-quality |
| 18-106-final-viral-combined_k141_10428||lt2gene | Not-determined |
| 18-106-final-viral-combined_k141_104109||lt2gene | Low-quality |
| 18-106-final-viral-combined_k141_56872||lt2gene | Low-quality |
| 18-106-final-viral-combined_k141_87181||lt2gene | Not-determined |
| 18-106-final-viral-combined_k141_64422||lt2gene | Low-quality |
| 18-106-final-viral-combined_k141_128162||lt2gene | Low-quality |
| 18-106-final-viral-combined_k141_52227||lt2gene | Low-quality |
| 18-106-final-viral-combined_k141_110775||lt2gene | Not-determined |
| 18-106-final-viral-combined_k141_56886||lt2gene | Not-determined |
| 18-106-final-viral-combined_k141_87220||lt2gene | Low-quality |
| 18-106-final-viral-combined_k141_28422||lt2gene | Low-quality |
| 18-106-final-viral-combined_k141_128184||lt2gene | Low-quality |
| 18-106-final-viral-combined_k141_75292||lt2gene | Not-determined |
| 18-106-final-viral-combined_k141_116544||lt2gene | Low-quality |
| 18-106-final-viral-combined_k141_116550||lt2gene | Low-quality |
| 18-106-final-viral-combined_k141_128252||lt2gene | Not-determined |
| 18-106-final-viral-combined_k141_56995||lt2gene | Not-determined |
| 18-106-final-viral-combined_k141_70208||lt2gene | Low-quality |
| 18-106-final-viral-combined_k141_99584||lt2gene | Not-determined |
| 18-106-final-viral-combined_k141_57017||lt2gene | Not-determined |
| 18-106-final-viral-combined_k141_22042||lt2gene | Low-quality |
| 18-106-final-viral-combined_k141_99608||lt2gene | Not-determined |
| 18-106-final-viral-combined_k141_57034||lt2gene | Low-quality |
| 18-106-final-viral-combined_k141_22068||lt2gene | Low-quality |
| 18-106-final-viral-combined_k141_10521||lt2gene | Not-determined |
| 18-106-final-viral-combined_k141_22077||lt2gene | Low-quality |
| 18-106-final-viral-combined_k141_10555||lt2gene | Low-quality |
| 18-106-final-viral-combined_k141_87346||lt2gene | Not-determined |
| 18-106-final-viral-combined_k141_128378||lt2gene | Low-quality |
| 18-106-final-viral-combined_k141_57181||lt2gene | Low-quality |
| 18-106-final-viral-combined_k141_104408||lt2gene | Not-determined |
| 18-106-final-viral-combined_k141_139803||lt2gene | Not-determined |
| 18-106-final-viral-combined_k141_81072||lt2gene | Low-quality |
| 18-106-final-viral-combined_k141_139811||lt2gene | Not-determined |
| 18-106-final-viral-combined_k141_70387||lt2gene | Not-determined |
| 18-106-final-viral-combined_k141_116807||lt2gene | Low-quality |
| 18-106-final-viral-combined_k141_87514||lt2gene | Low-quality |
| 18-106-final-viral-combined_k141_93319||lt2gene | Not-determined |
| 18-106-final-viral-combined_k141_122045||lt2gene | Low-quality |
| 18-106-final-viral-combined_k141_134141||lt2gene | Not-determined |
| 18-106-final-viral-combined_k141_34441||lt2gene | Not-determined |
| 18-106-final-viral-combined_k141_116842||lt2gene | Not-determined |
| 18-106-final-viral-combined_k141_134172||lt2gene | Low-quality |
| 18-106-final-viral-combined_k141_93372||lt2gene | Not-determined |
| 18-106-final-viral-combined_k141_122062||lt2gene | Low-quality |
| 18-106-final-viral-combined_k141_81180||lt2gene | Low-quality |
| 18-106-final-viral-combined_k141_34467||lt2gene | Low-quality |
| 18-106-final-viral-combined_k141_104508||lt2gene | Low-quality |
| 18-106-final-viral-combined_k141_57322||lt2gene | Low-quality |
| 18-106-final-viral-combined_k141_52623||lt2gene | Not-determined |
| 18-106-final-viral-combined_k141_10727||lt2gene | Low-quality |
| 18-106-final-viral-combined_k141_34514||lt2gene | Low-quality |
| 18-106-final-viral-combined_k141_116932||lt2gene | Not-determined |
| 18-106-final-viral-combined_k141_46855||lt2gene | Low-quality |
| 18-106-final-viral-combined_k141_122136||lt2gene | Not-determined |
| 18-106-final-viral-combined_k141_128607||lt2gene | Not-determined |
| 18-106-final-viral-combined_k141_41080||lt2gene | Low-quality |
| 18-106-final-viral-combined_k141_34592||lt2gene | Not-determined |
| 18-106-final-viral-combined_k141_134339||lt2gene | Low-quality |
| 18-106-final-viral-combined_k141_134345||lt2gene | Low-quality |
| 18-106-final-viral-combined_k141_134366||lt2gene | Not-determined |
| 18-106-final-viral-combined_k141_104696||lt2gene | Not-determined |
| 18-106-final-viral-combined_k141_22440||lt2gene | Low-quality |
| 18-106-final-viral-combined_k141_81398||lt2gene | Not-determined |
| 18-106-final-viral-combined_k141_140062||lt2gene | Not-determined |
| 18-106-final-viral-combined_k141_10918||lt2gene | Low-quality |
| 18-106-final-viral-combined_k141_134451||lt2gene | Not-determined |
| 18-106-final-viral-combined_k141_128773||lt2gene | Low-quality |
| 18-106-final-viral-combined_k141_104778||lt2gene | Low-quality |
| 18-106-final-viral-combined_k141_17404||lt2gene | Low-quality |
| 18-106-final-viral-combined_k141_128819||lt2gene | Low-quality |
| 18-106-final-viral-combined_k141_111309||lt2gene | Low-quality |
| 18-106-final-viral-combined_k141_122338||lt2gene | Not-determined |
| 18-106-final-viral-combined_k141_28880||lt2gene | Low-quality |
| 18-106-final-viral-combined_k141_81556||lt2gene | Low-quality |
| 18-106-final-viral-combined_k141_140148||lt2gene | Not-determined |
| 18-106-final-viral-combined_k141_122396||lt2gene | Low-quality |
| 18-106-final-viral-combined_k141_11034||lt2gene | Low-quality |
| 18-106-final-viral-combined_k141_93698||lt2gene | Low-quality |
| 18-106-final-viral-combined_k141_134597||lt2gene | Low-quality |
| 18-106-final-viral-combined_k141_134607||lt2gene | Not-determined |
| 18-106-final-viral-combined_k141_117211||lt2gene | Not-determined |
| 18-106-final-viral-combined_k141_117227||lt2gene | Low-quality |
| 18-106-final-viral-combined_k141_134658||lt2gene | Low-quality |
| 18-106-final-viral-combined_k141_22692||lt2gene | Low-quality |
| 18-106-final-viral-combined_k141_22699||lt2gene | Not-determined |
| 18-106-final-viral-combined_k141_22706||lt2gene | Low-quality |
| 18-106-final-viral-combined_k141_140311||lt2gene | Not-determined |
| 18-106-final-viral-combined_k141_140349||lt2gene | Low-quality |
| 18-106-final-viral-combined_k141_11099||lt2gene | Low-quality |
| 18-106-final-viral-combined_k141_22820||lt2gene | Low-quality |
| 18-106-final-viral-combined_k141_29041||lt2gene | Not-determined |
| 18-106-final-viral-combined_k141_134792||lt2gene | Not-determined |
| 18-106-final-viral-combined_k141_34813||lt2gene | Low-quality |
| 18-106-final-viral-combined_k141_93929||lt2gene | Low-quality |
| 18-106-final-viral-combined_k141_29087||lt2gene | Not-determined |
| 18-106-final-viral-combined_k141_29091||lt2gene | Low-quality |
| 18-106-final-viral-combined_k141_29093||lt2gene | Low-quality |
| 18-106-final-viral-combined_k141_122695||lt2gene | Not-determined |
| 18-106-final-viral-combined_k141_11163||lt2gene | Low-quality |
| 18-106-final-viral-combined_k141_76021||lt2gene | Not-determined |
| 18-106-final-viral-combined_k141_76043||lt2gene | Not-determined |
| 18-106-final-viral-combined_k141_140481||lt2gene | Not-determined |
| 18-106-final-viral-combined_k141_122771||lt2gene | Low-quality |
| 18-106-final-viral-combined_k141_76071||lt2gene | Low-quality |
| 18-106-final-viral-combined_k141_122808||lt2gene | Low-quality |
| 18-106-final-viral-combined_k141_11258||lt2gene | Low-quality |
| 18-106-final-viral-combined_k141_111493||lt2gene | Low-quality |
| 18-106-final-viral-combined_k141_122940||lt2gene | Low-quality |
| 18-106-final-viral-combined_k141_81898||lt2gene | Low-quality |
| 18-106-final-viral-combined_k141_105221||lt2gene | Low-quality |
| 18-106-final-viral-combined_k141_134930||lt2gene | Low-quality |
| 18-106-final-viral-combined_k141_105255||lt2gene | Low-quality |
| 18-106-final-viral-combined_k141_81995||lt2gene | Not-determined |
| 18-106-final-viral-combined_k141_81996||lt2gene | Low-quality |
| 18-106-final-viral-combined_k141_140718||lt2gene | Low-quality |
| 18-106-final-viral-combined_k141_105306||lt2gene | Low-quality |
| 18-106-final-viral-combined_k141_35169||lt2gene | Not-determined |
| 18-106-final-viral-combined_k141_23319||lt2gene | Low-quality |
| 18-106-final-viral-combined_k141_105447||lt2gene | Low-quality |
| 18-106-final-viral-combined_k141_58272||lt2gene | Low-quality |
| 18-106-final-viral-combined_k141_58318||lt2gene | Low-quality |
| 18-106-final-viral-combined_k141_23444||lt2gene | Low-quality |
| 18-106-final-viral-combined_k141_58417||lt2gene | Not-determined |
| 18-106-final-viral-combined_k141_105673||lt2gene | Not-determined |
| 18-106-final-viral-combined_k141_11468||lt2gene | Low-quality |
| 18-106-final-viral-combined_k141_58486||lt2gene | Low-quality |
| 18-106-final-viral-combined_k141_58509||lt2gene | Low-quality |
| 18-106-final-viral-combined_k141_11579||lt2gene | Low-quality |
| 18-106-final-viral-combined_k141_11707||lt2gene | Low-quality |
| 18-106_k141_0 | Not-determined |
| 18-106_k141_2 | Not-determined |
| 18-106_k141_46973 | Not-determined |
| 18-106_k141_11748 | Not-determined |
| 18-106_k141_11749 | Not-determined |
| 18-106_k141_35237 | Not-determined |
| 18-106_k141_82212 | Not-determined |
| 18-106_k141_29365 | Not-determined |
| 18-106_k141_41111 | Not-determined |
| 18-106_k141_41112 | Not-determined |
| 18-106_k141_82215 | Not-determined |
| 18-106_k141_105689 | Not-determined |
| 18-106_k141_129172 | Not-determined |
| 18-106_k141_117439 | Not-determined |
| 18-106_k141_88079 | Not-determined |
| 18-106_k141_11762 | Not-determined |
| 18-106_k141_76334 | Not-determined |
| 18-106_k141_35247 | Not-determined |
| 18-106_k141_70474 | Not-determined |
| 18-106_k141_35249 | Not-determined |
| 18-106_k141_111563 | Not-determined |
| 18-106_k141_40 | Not-determined |
| 18-106_k141_111569 | Not-determined |
| 18-106_k141_5889 | Not-determined |
| 18-106_k141_99839 | Not-determined |
| 18-106_k141_70492 | Not-determined |
| 18-106_k141_105717 | Not-determined |
| 18-106_k141_5892 | Not-determined |
| 18-106_k141_129185 | Not-determined |
| 18-106_k141_58742 | Not-determined |
| 18-106_k141_23506 | Not-determined |
| 18-106_k141_29391 | Not-determined |
| 18-106_k141_23510 | Low-quality |
| 18-106_k141_129189 | Not-determined |
| 18-106_k141_105721 | Low-quality |
| 18-106_k141_129191 | Not-determined |
| 18-106_k141_105723 | Not-determined |
| 18-106_k141_129193 | Not-determined |
| 18-106_k141_64619 | Not-determined |
| 18-106_k141_29405 | Low-quality |
| 18-106_k141_58746 | Low-quality |
| 18-106_k141_135065 | Not-determined |
| 18-106_k141_123324 | Not-determined |
| 18-106_k141_11784 | Not-determined |
| 18-106_k141_52848 | Not-determined |
| 18-106_k141_76351 | Not-determined |
| 18-106_k141_5912 | Low-quality |
| 18-106_k141_41161 | Not-determined |
| 18-106_k141_135077 | Not-determined |
| 18-106_k141_129206 | Not-determined |
| 18-106_k141_35275 | Not-determined |
| 18-106_k141_135084 | Low-quality |
| 18-106_k141_99869 | Not-determined |
| 18-106_k141_78 | Not-determined |
| 18-106_k141_135097 | Not-determined |
| 18-106_k141_76378 | Not-determined |
| 18-106_k141_111612 | Not-determined |
| 18-106_k141_35292 | Not-determined |
| 18-106_k141_35295 | Not-determined |
| 18-106_k141_64635 | Not-determined |
| 18-106_k141_76387 | Not-determined |
| 18-106_k141_64637 | Not-determined |
| 18-106_k141_64639 | Not-determined |
| 18-106_k141_35300 | Not-determined |
| 18-106_k141_17675 | Low-quality |
| 18-106_k141_105751 | Not-determined |
| 18-106_k141_46986 | Not-determined |
| 18-106_k141_129221 | Not-determined |
| 18-106_k141_135112 | Not-determined |
| 18-106_k141_41182 | Not-determined |
| 18-106_k141_76398 | Low-quality |
| 18-106_k141_41184 | Not-determined |
| 18-106_k141_17687 | Not-determined |
| 18-106_k141_129223 | Low-quality |
| 18-106_k141_11812 | Not-determined |
| 18-106_k141_58780 | Not-determined |
| 18-106_k141_29441 | Not-determined |
| 18-106_k141_17693 | Not-determined |
| 18-106_k141_41191 | Low-quality |
| 18-106_k141_94014 | Not-determined |
| 18-106_k141_47005 | Not-determined |
| 18-106_k141_112 | Not-determined |
| 18-106_k141_135115 | Not-determined |
| 18-106_k141_88157 | Not-determined |
| 18-106_k141_114 | Not-determined |
| 18-106_k141_64645 | Not-determined |
| 18-106_k141_58799 | Not-determined |
| 18-106_k141_135123 | Not-determined |
| 18-106_k141_129242 | Not-determined |
| 18-106_k141_58804 | Not-determined |
| 18-106_k141_94027 | Not-determined |
| 18-106_k141_88168 | Not-determined |
| 18-106_k141_88169 | Not-determined |
| 18-106_k141_122 | Not-determined |
| 18-106_k141_123356 | Not-determined |
| 18-106_k141_41203 | Not-determined |
| 18-106_k141_82313 | Not-determined |
| 18-106_k141_58812 | Not-determined |
| 18-106_k141_117468 | Not-determined |
| 18-106_k141_58815 | Not-determined |
| 18-106_k141_135136 | Not-determined |
| 18-106_k141_94036 | Not-determined |
| 18-106_k141_111635 | Not-determined |
| 18-106_k141_82317 | Low-quality |
| 18-106_k141_29462 | Not-determined |
| 18-106_k141_111644 | Not-determined |
| 18-106_k141_11840 | Not-determined |
| 18-106_k141_29463 | Not-determined |
| 18-106_k141_94047 | Not-determined |
| 18-106_k141_17725 | Not-determined |
| 18-106_k141_17732 | Not-determined |
| 18-106_k141_135146 | Not-determined |
| 18-106_k141_88185 | Not-determined |
| 18-106_k141_58840 | Not-determined |
| 18-106_k141_47048 | Not-determined |
| 18-106_k141_94059 | Low-quality |
| 18-106_k141_11847 | Not-determined |
| 18-106_k141_58849 | Not-determined |
| 18-106_k141_88204 | Not-determined |
| 18-106_k141_23542 | Not-determined |
| 18-106_k141_135162 | Not-determined |
| 18-106_k141_11858 | Not-determined |
| 18-106_k141_105785 | Not-determined |
| 18-106_k141_11862 | Not-determined |
| 18-106_k141_88208 | Not-determined |
| 18-106_k141_117475 | Not-determined |
| 18-106_k141_11867 | Not-determined |
| 18-106_k141_17743 | Not-determined |
| 18-106_k141_47063 | Not-determined |
| 18-106_k141_135179 | Low-quality |
| 18-106_k141_88220 | Low-quality |
| 18-106_k141_123402 | Not-determined |
| 18-106_k141_123403 | Not-determined |
| 18-106_k141_129306 | Not-determined |
| 18-106_k141_99892 | Not-determined |
| 18-106_k141_70528 | Not-determined |
| 18-106_k141_129311 | Not-determined |
| 18-106_k141_129312 | Not-determined |
| 18-106_k141_41220 | Not-determined |
| 18-106_k141_70533 | Not-determined |
| 18-106_k141_64683 | Not-determined |
| 18-106_k141_41224 | Not-determined |
| 18-106_k141_41232 | Not-determined |
| 18-106_k141_94090 | Not-determined |
| 18-106_k141_135188 | Not-determined |
| 18-106_k141_188 | Low-quality |
| 18-106_k141_5926 | Not-determined |
| 18-106_k141_190 | Low-quality |
| 18-106_k141_47098 | Not-determined |
| 18-106_k141_5929 | Not-determined |
| 18-106_k141_99916 | Not-determined |
| 18-106_k141_76425 | Not-determined |
| 18-106_k141_41244 | Not-determined |
| 18-106_k141_41245 | Not-determined |
| 18-106_k141_117515 | Not-determined |
| 18-106_k141_88251 | Not-determined |
| 18-106_k141_117517 | Not-determined |
| 18-106_k141_117519 | Not-determined |
| 18-106_k141_105830 | Low-quality |
| 18-106_k141_88253 | Not-determined |
| 18-106_k141_123425 | Not-determined |
| 18-106_k141_117523 | Not-determined |
| 18-106_k141_11890 | Not-determined |
| 18-106_k141_209 | Not-determined |
| 18-106_k141_76431 | Low-quality |
| 18-106_k141_123430 | Not-determined |
| 18-106_k141_99926 | Low-quality |
| 18-106_k141_123431 | Not-determined |
| 18-106_k141_123432 | Not-determined |
| 18-106_k141_64708 | Not-determined |
| 18-106_k141_47112 | Not-determined |
| 18-106_k141_11898 | Not-determined |
| 18-106_k141_111728 | Not-determined |
| 18-106_k141_111730 | Not-determined |
| 18-106_k141_129355 | Not-determined |
| 18-106_k141_88268 | Not-determined |
| 18-106_k141_70558 | Not-determined |
| 18-106_k141_58884 | Low-quality |
| 18-106_k141_58885 | Not-determined |
| 18-106_k141_135204 | Not-determined |
| 18-106_k141_129361 | Not-determined |
| 18-106_k141_123444 | Low-quality |
| 18-106_k141_111742 | Low-quality |
| 18-106_k141_135206 | Not-determined |
| 18-106_k141_233 | Not-determined |
| 18-106_k141_111746 | Not-determined |
| 18-106_k141_236 | Not-determined |
| 18-106_k141_47120 | Not-determined |
| 18-106_k141_47121 | Not-determined |
| 18-106_k141_117550 | Not-determined |
| 18-106_k141_64719 | Not-determined |
| 18-106_k141_239 | Not-determined |
| 18-106_k141_76447 | Not-determined |
| 18-106_k141_11921 | Low-quality |
| 18-106_k141_5955 | Not-determined |
| 18-106_k141_17795 | Low-quality |
| 18-106_k141_94113 | Not-determined |
| 18-106_k141_105847 | Not-determined |
| 18-106_k141_70574 | Not-determined |
| 18-106_k141_94118 | Not-determined |
| 18-106_k141_70578 | Not-determined |
| 18-106_k141_94121 | Not-determined |
| 18-106_k141_70579 | Not-determined |
| 18-106_k141_76461 | Not-determined |
| 18-106_k141_23588 | Not-determined |
| 18-106_k141_70585 | Not-determined |
| 18-106_k141_5968 | Not-determined |
| 18-106_k141_94126 | Low-quality |
| 18-106_k141_94134 | Not-determined |
| 18-106_k141_64750 | Not-determined |
| 18-106_k141_76478 | Not-determined |
| 18-106_k141_58920 | Not-determined |
| 18-106_k141_105875 | Not-determined |
| 18-106_k141_94139 | Not-determined |
| 18-106_k141_64753 | Not-determined |
| 18-106_k141_64755 | Not-determined |
| 18-106_k141_111778 | Not-determined |
| 18-106_k141_76487 | Not-determined |
| 18-106_k141_105884 | Not-determined |
| 18-106_k141_58927 | Not-determined |
| 18-106_k141_94143 | Not-determined |
| 18-106_k141_105887 | Not-determined |
| 18-106_k141_88300 | Not-determined |
| 18-106_k141_76489 | Not-determined |
| 18-106_k141_76493 | Not-determined |
| 18-106_k141_88305 | Not-determined |
| 18-106_k141_35385 | Not-determined |
| 18-106_k141_76503 | Low-quality |
| 18-106_k141_58938 | Not-determined |
| 18-106_k141_105903 | Not-determined |
| 18-106_k141_11928 | Not-determined |
| 18-106_k141_278 | Not-determined |
| 18-106_k141_47169 | Not-determined |
| 18-106_k141_88311 | Not-determined |
| 18-106_k141_47171 | Not-determined |
| 18-106_k141_88312 | Not-determined |
| 18-106_k141_76511 | Not-determined |
| 18-106_k141_76512 | Not-determined |
| 18-106_k141_123463 | Not-determined |
| 18-106_k141_11942 | Not-determined |
| 18-106_k141_64769 | Not-determined |
| 18-106_k141_123466 | Not-determined |
| 18-106_k141_41262 | Not-determined |
| 18-106_k141_35394 | Not-determined |
| 18-106_k141_35397 | Not-determined |
| 18-106_k141_117583 | Low-quality |
| 18-106_k141_64778 | Not-determined |
| 18-106_k141_88330 | Not-determined |
| 18-106_k141_41263 | Not-determined |
| 18-106_k141_64780 | Low-quality |
| 18-106_k141_117588 | Not-determined |
| 18-106_k141_70623 | Not-determined |
| 18-106_k141_41269 | Low-quality |
| 18-106_k141_94181 | Not-determined |
| 18-106_k141_70626 | Not-determined |
| 18-106_k141_123470 | Not-determined |
| 18-106_k141_94185 | Low-quality |
| 18-106_k141_294 | Not-determined |
| 18-106_k141_123473 | Not-determined |
| 18-106_k141_117596 | Not-determined |
| 18-106_k141_88342 | Not-determined |
| 18-106_k141_94188 | Not-determined |
| 18-106_k141_11963 | Not-determined |
| 18-106_k141_123479 | Low-quality |
| 18-106_k141_11964 | Not-determined |
| 18-106_k141_35409 | Not-determined |
| 18-106_k141_64788 | Not-determined |
| 18-106_k141_64790 | Not-determined |
| 18-106_k141_58945 | Not-determined |
| 18-106_k141_76538 | Not-determined |
| 18-106_k141_76539 | Not-determined |
| 18-106_k141_41280 | Not-determined |
| 18-106_k141_94207 | Not-determined |
| 18-106_k141_70637 | Not-determined |
| 18-106_k141_117615 | Not-determined |
| 18-106_k141_64799 | Not-determined |
| 18-106_k141_76551 | Not-determined |
| 18-106_k141_117623 | Not-determined |
| 18-106_k141_76555 | Not-determined |
| 18-106_k141_123493 | Not-determined |
| 18-106_k141_117629 | Low-quality |
| 18-106_k141_111842 | Not-determined |
| 18-106_k141_117630 | Not-determined |
| 18-106_k141_76557 | Not-determined |
| 18-106_k141_76562 | Not-determined |
| 18-106_k141_105955 | Not-determined |
| 18-106_k141_41296 | Not-determined |
| 18-106_k141_64811 | Not-determined |
| 18-106_k141_47204 | Not-determined |
| 18-106_k141_64812 | Not-determined |
| 18-106_k141_12004 | Not-determined |
| 18-106_k141_117644 | Not-determined |
| 18-106_k141_76571 | Not-determined |
| 18-106_k141_111852 | Not-determined |
| 18-106_k141_117651 | Not-determined |
| 18-106_k141_317 | Not-determined |
| 18-106_k141_64826 | Not-determined |
| 18-106_k141_12013 | Not-determined |
| 18-106_k141_322 | Not-determined |
| 18-106_k141_324 | Not-determined |
| 18-106_k141_325 | Low-quality |
| 18-106_k141_35447 | Not-determined |
| 18-106_k141_123520 | Not-determined |
| 18-106_k141_64838 | Not-determined |
| 18-106_k141_58996 | Not-determined |
| 18-106_k141_35450 | Low-quality |
| 18-106_k141_70665 | Low-quality |
| 18-106_k141_99978 | Not-determined |
| 18-106_k141_41320 | Low-quality |
| 18-106_k141_23618 | Not-determined |
| 18-106_k141_23619 | Not-determined |
| 18-106_k141_23623 | Low-quality |
| 18-106_k141_343 | Not-determined |
| 18-106_k141_23626 | Not-determined |
| 18-106_k141_105985 | Low-quality |
| 18-106_k141_94236 | Not-determined |
| 18-106_k141_347 | Not-determined |
| 18-106_k141_94240 | Not-determined |
| 18-106_k141_64859 | Not-determined |
| 18-106_k141_12029 | Not-determined |
| 18-106_k141_12033 | Not-determined |
| 18-106_k141_105995 | Not-determined |
| 18-106_k141_35467 | Not-determined |
| 18-106_k141_117672 | Not-determined |
| 18-106_k141_105996 | Not-determined |
| 18-106_k141_99995 | Not-determined |
| 18-106_k141_59005 | Not-determined |
| 18-106_k141_59006 | Not-determined |
| 18-106_k141_117675 | Not-determined |
| 18-106_k141_47214 | Not-determined |
| 18-106_k141_70688 | Not-determined |
| 18-106_k141_35477 | Not-determined |
| 18-106_k141_76605 | Not-determined |
| 18-106_k141_106008 | Not-determined |
| 18-106_k141_59015 | Low-quality |
| 18-106_k141_117685 | Not-determined |
| 18-106_k141_35488 | Not-determined |
| 18-106_k141_12053 | Not-determined |
| 18-106_k141_106018 | Low-quality |
| 18-106_k141_117691 | Not-determined |
| 18-106_k141_94258 | Not-determined |
| 18-106_k141_59028 | Not-determined |
| 18-106_k141_23648 | Not-determined |
| 18-106_k141_106027 | Not-determined |
| 18-106_k141_6033 | Not-determined |
| 18-106_k141_117697 | Not-determined |
| 18-106_k141_76629 | Low-quality |
| 18-106_k141_70699 | Not-determined |
| 18-106_k141_41343 | Not-determined |
| 18-106_k141_6038 | Not-determined |
| 18-106_k141_70703 | Not-determined |
| 18-106_k141_12068 | Not-determined |
| 18-106_k141_35504 | Not-determined |
| 18-106_k141_41351 | Not-determined |
| 18-106_k141_88385 | Not-determined |
| 18-106_k141_70706 | Not-determined |
| 18-106_k141_47244 | Not-determined |
| 18-106_k141_6052 | Not-determined |
| 18-106_k141_94278 | Not-determined |
| 18-106_k141_106046 | Not-determined |
| 18-106_k141_111903 | Not-determined |
| 18-106_k141_117717 | Low-quality |
| 18-106_k141_70734 | Not-determined |
| 18-106_k141_23664 | Not-determined |
| 18-106_k141_123547 | Low-quality |
| 18-106_k141_111919 | Not-determined |
| 18-106_k141_29512 | Not-determined |
| 18-106_k141_123554 | Not-determined |
| 18-106_k141_59062 | Not-determined |
| 18-106_k141_117725 | Not-determined |
| 18-106_k141_59063 | Not-determined |
| 18-106_k141_23673 | Not-determined |
| 18-106_k141_35533 | Not-determined |
| 18-106_k141_111943 | Not-determined |
| 18-106_k141_70745 | Not-determined |
| 18-106_k141_23686 | Not-determined |
| 18-106_k141_111947 | Not-determined |
| 18-106_k141_70748 | Not-determined |
| 18-106_k141_123573 | Low-quality |
| 18-106_k141_76665 | Not-determined |
| 18-106_k141_35541 | Not-determined |

| | |
|---|---|
| 18-106_k141_35542 | Low-quality |
| 18-106_k141_41375 | Not-determined |
| 18-106_k141_412 | Not-determined |
| 18-106_k141_23690 | Not-determined |
| 18-106_k141_135225 | Not-determined |
| 18-106_k141_12137 | Not-determined |
| 18-106_k141_70770 | Low-quality |
| 18-106_k141_70774 | Not-determined |
| 18-106_k141_70775 | Not-determined |
| 18-106_k141_12142 | Not-determined |
| 18-106_k141_41393 | Low-quality |
| 18-106_k141_41395 | Not-determined |
| 18-106_k141_70780 | Low-quality |
| 18-106_k141_123589 | Not-determined |
| 18-106_k141_123593 | Low-quality |
| 18-106_k141_29549 | Not-determined |
| 18-106_k141_123608 | Not-determined |
| 18-106_k141_444 | Not-determined |
| 18-106_k141_70799 | Low-quality |
| 18-106_k141_123619 | Not-determined |
| 18-106_k141_94317 | Not-determined |
| 18-106_k141_82384 | Not-determined |
| 18-106_k141_450 | Not-determined |
| 18-106_k141_6063 | Not-determined |
| 18-106_k141_23701 | Low-quality |
| 18-101-final-viral-combined_k141_313851\|\|full | High-quality |
| 18-106_k141_70812 | Not-determined |
| 18-106_k141_6065 | Low-quality |
| 18-106_k141_94335 | Not-determined |
| 18-106_k141_29598 | Not-determined |
| 18-106_k141_129441 | Not-determined |
| 18-106_k141_106124 | Low-quality |
| 18-106_k141_59098 | Not-determined |
| 18-106_k141_106128 | Not-determined |
| 18-106_k141_29619 | Not-determined |
| 18-106_k141_12165 | Not-determined |
| 18-106_k141_12166 | Not-determined |
| 18-106_k141_129456 | Not-determined |
| 18-106_k141_129458 | Low-quality |
| 18-106_k141_35563 | Not-determined |
| 18-106_k141_29624 | Not-determined |
| 18-106_k141_129459 | Not-determined |
| 18-106_k141_17800 | Not-determined |
| 18-106_k141_100019 | Not-determined |
| 18-106_k141_23727 | Not-determined |
| 18-106_k141_59105 | Not-determined |
| 18-106_k141_59106 | Not-determined |
| 18-106_k141_129470 | Not-determined |
| 18-106_k141_23733 | Not-determined |
| 18-106_k141_6087 | Not-determined |
| 18-106_k141_23737 | Not-determined |
| 18-106_k141_106139 | Not-determined |
| 18-106_k141_106141 | Not-determined |
| 18-106_k141_129483 | Not-determined |
| 18-106_k141_35583 | Not-determined |
| 18-106_k141_12184 | Not-determined |
| 18-106_k141_35588 | Not-determined |
| 18-106_k141_35590 | Not-determined |
| 18-106_k141_29643 | Not-determined |
| 18-106_k141_106153 | Not-determined |
| 18-106_k141_17812 | Not-determined |
| 18-106_k141_476 | Not-determined |
| 18-106_k141_29651 | Not-determined |
| 18-106_k141_59144 | Not-determined |
| 18-106_k141_23763 | Not-determined |
| 18-106_k141_100047 | Not-determined |
| 18-106_k141_12200 | Low-quality |
| 18-106_k141_88423 | Not-determined |
| 18-106_k141_100052 | Not-determined |
| 18-106_k141_17824 | Not-determined |
| 18-106_k141_100055 | Not-determined |
| 18-106_k141_100056 | Not-determined |
| 18-106_k141_29672 | Not-determined |
| 18-106_k141_17830 | Not-determined |
| 18-106_k141_100066 | Low-quality |
| 18-106_k141_64906 | Not-determined |
| 18-106_k141_64907 | Not-determined |
| 18-106_k141_59172 | Not-determined |
| 18-106_k141_70885 | Not-determined |
| 18-106_k141_100076 | Not-determined |
| 18-106_k141_17841 | Low-quality |
| 18-106_k141_59180 | Not-determined |
| 18-106_k141_29680 | Not-determined |
| 18-106_k141_41475 | Not-determined |
| 18-106_k141_112013 | Not-determined |
| 18-106_k141_17847 | Not-determined |
| 18-106_k141_100090 | Not-determined |
| 18-106_k141_106186 | Not-determined |
| 18-106_k141_64932 | Not-determined |
| 18-106_k141_70897 | Not-determined |
| 18-106_k141_41490 | Not-determined |
| 18-106_k141_100097 | Not-determined |
| 18-106_k141_17856 | Not-determined |
| 18-106_k141_12219 | Not-determined |
| 18-106_k141_6130 | Not-determined |
| 18-106_k141_17858 | Not-determined |
| 18-106_k141_88465 | Not-determined |
| 18-106_k141_64945 | Not-determined |
| 18-106_k141_17860 | Not-determined |
| 18-106_k141_88473 | Not-determined |
| 18-106_k141_533 | Low-quality |
| 18-106_k141_64957 | Low-quality |
| 18-106_k141_123669 | Not-determined |
| 18-106_k141_41511 | Not-determined |
| 18-106_k141_41515 | Not-determined |
| 18-106_k141_17877 | Not-determined |
| 18-106_k141_29712 | Not-determined |
| 18-106_k141_129540 | Not-determined |
| 18-106_k141_12234 | Not-determined |
| 18-106_k141_70915 | Low-quality |
| 18-106_k141_41523 | Low-quality |
| 18-106_k141_129544 | Low-quality |
| 18-106_k141_123691 | Not-determined |
| 18-106_k141_117763 | Not-determined |
| 18-106_k141_542 | Not-determined |
| 18-106_k141_29722 | Not-determined |
| 18-106_k141_6161 | Not-determined |
| 18-106_k141_6166 | Not-determined |
| 18-106_k141_12253 | Not-determined |
| 18-106_k141_88508 | Not-determined |
| 18-106_k141_123701 | Not-determined |
| 18-106_k141_6172 | Not-determined |
| 18-106_k141_106225 | Not-determined |
| 18-106_k141_123710 | Not-determined |
| 18-106_k141_6180 | Not-determined |
| 18-106_k141_106238 | Not-determined |
| 18-106_k141_88529 | Not-determined |
| 18-106_k141_94446 | Not-determined |
| 18-106_k141_41533 | Not-determined |
| 18-106_k141_29743 | Not-determined |
| 18-106_k141_41534 | Not-determined |
| 18-106_k141_117795 | Not-determined |
| 18-106_k141_41535 | Low-quality |
| 18-106_k141_29748 | Not-determined |
| 18-106_k141_29750 | Not-determined |
| 18-106_k141_41539 | Low-quality |
| 18-106_k141_41540 | Not-determined |
| 18-106_k141_70932 | Not-determined |
| 18-106_k141_76711 | Not-determined |
| 18-106_k141_59225 | Not-determined |
| 18-106_k141_76712 | Not-determined |
| 18-106_k141_88538 | Not-determined |
| 18-106_k141_82498 | Not-determined |
| 18-106_k141_94460 | Not-determined |
| 18-106_k141_94461 | Not-determined |
| 18-106_k141_52881 | Not-determined |
| 18-106_k141_106244 | Low-quality |
| 18-106_k141_106246 | Not-determined |
| 18-106_k141_94465 | Not-determined |
| 18-106_k141_59238 | Not-determined |
| 18-106_k141_94473 | Not-determined |
| 18-106_k141_52890 | Not-determined |
| 18-106_k141_52891 | Not-determined |
| 18-106_k141_29774 | Not-determined |
| 18-106_k141_59250 | Not-determined |
| 18-106_k141_123741 | Not-determined |
| 18-106_k141_82512 | Not-determined |
| 18-106_k141_12278 | Not-determined |
| 18-106_k141_6205 | Not-determined |
| 18-106_k141_52920 | Not-determined |
| 18-106_k141_6209 | Not-determined |
| 18-106_k141_129588 | Not-determined |
| 18-106_k141_52926 | Low-quality |
| 18-106_k141_129591 | Not-determined |
| 18-106_k141_94482 | Not-determined |
| 18-106_k141_129593 | Not-determined |
| 18-106_k141_129596 | Not-determined |
| 18-106_k141_123758 | Not-determined |
| 18-106_k141_129599 | Not-determined |
| 18-106_k141_41570 | Low-quality |
| 18-106_k141_94490 | Low-quality |
| 18-106_k141_123760 | Not-determined |
| 18-106_k141_6224 | Not-determined |
| 18-106_k141_59262 | Low-quality |
| 18-106_k141_59266 | Not-determined |
| 18-106_k141_123770 | Not-determined |
| 18-106_k141_41581 | Not-determined |
| 18-106_k141_123774 | Not-determined |
| 18-106_k141_82535 | Not-determined |
| 18-106_k141_41591 | Low-quality |
| 18-106_k141_76747 | Not-determined |
| 18-106_k141_555 | Not-determined |
| 18-106_k141_12310 | Not-determined |
| 18-106_k141_559 | Not-determined |
| 18-106_k141_94516 | Not-determined |
| 18-106_k141_12315 | Not-determined |
| 18-106_k141_41605 | Not-determined |
| 18-106_k141_94531 | Not-determined |
| 18-106_k141_41607 | Not-determined |
| 18-106_k141_70972 | Not-determined |
| 18-106_k141_6234 | Not-determined |
| 18-106_k141_76758 | Not-determined |
| 18-106_k141_47311 | Not-determined |
| 18-106_k141_571 | Low-quality |
| 18-106_k141_88611 | Not-determined |
| 18-106_k141_70980 | Not-determined |
| 18-106_k141_94543 | Not-determined |
| 18-106_k141_29813 | Not-determined |
| 18-106_k141_29815 | Not-determined |
| 18-106_k141_106303 | Not-determined |
| 18-106_k141_41625 | Low-quality |
| 18-106_k141_64964 | Not-determined |
| 18-106_k141_585 | Not-determined |
| 18-106_k141_29825 | Not-determined |
| 18-106_k141_41638 | Not-determined |
| 18-106_k141_123833 | Not-determined |
| 18-106_k141_47327 | Not-determined |
| 18-106_k141_135249 | Not-determined |
| 18-106_k141_135256 | Not-determined |
| 18-106_k141_123837 | Not-determined |
| 18-106_k141_135258 | Not-determined |
| 18-106_k141_64977 | Not-determined |
| 18-106_k141_41649 | Not-determined |
| 18-106_k141_29839 | Not-determined |
| 18-106_k141_29843 | Not-determined |
| 18-106_k141_94551 | Low-quality |
| 18-106_k141_41655 | Not-determined |
| 18-106_k141_135268 | Not-determined |
| 18-106_k141_41657 | Low-quality |
| 18-106_k141_41663 | Not-determined |
| 18-106_k141_106330 | Not-determined |
| 18-106_k141_112039 | Not-determined |
| 18-106_k141_17909 | Not-determined |
| 18-106_k141_100148 | Not-determined |
| 18-106_k141_17913 | Not-determined |
| 18-106_k141_71001 | Not-determined |
| 18-106_k141_100149 | Not-determined |
| 18-106_k141_94556 | Not-determined |
| 18-106_k141_64983 | Not-determined |
| 18-106_k141_94558 | Not-determined |
| 18-106_k141_135279 | Not-determined |
| 18-106_k141_135280 | Not-determined |
| 18-106_k141_29863 | Not-determined |
| 18-106_k141_12328 | Not-determined |
| 18-106_k141_100152 | Not-determined |
| 18-106_k141_88649 | Not-determined |
| 18-106_k141_82573 | Not-determined |
| 18-106_k141_41677 | Not-determined |
| 18-106_k141_53012 | Not-determined |
| 18-106_k141_6270 | Not-determined |
| 18-106_k141_29880 | Not-determined |
| 18-106_k141_41689 | Not-determined |
| 18-106_k141_88664 | Low-quality |
| 18-106_k141_88667 | Not-determined |
| 18-106_k141_76784 | Not-determined |
| 18-106_k141_12334 | Not-determined |
| 18-106_k141_53013 | Not-determined |
| 18-106_k141_53015 | Not-determined |
| 18-106_k141_625 | Low-quality |
| 18-106_k141_82588 | Not-determined |
| 18-106_k141_29893 | Not-determined |
| 18-106_k141_88674 | Not-determined |
| 18-106_k141_117831 | Not-determined |
| 18-106_k141_88675 | Low-quality |
| 18-106_k141_65007 | Not-determined |
| 18-106_k141_29898 | Not-determined |
| 18-106_k141_6277 | Not-determined |
| 18-106_k141_6278 | Not-determined |
| 18-106_k141_117835 | Not-determined |
| 18-106_k141_82590 | Not-determined |
| 18-106_k141_135293 | Not-determined |
| 18-106_k141_117841 | Not-determined |
| 18-106_k141_117846 | Not-determined |
| 18-106_k141_135298 | Not-determined |
| 18-106_k141_112072 | Not-determined |
| 18-106_k141_29909 | Not-determined |
| 18-106_k141_123871 | Not-determined |
| 18-106_k141_123872 | Not-determined |
| 18-106_k141_637 | Low-quality |
| 18-106_k141_135301 | Not-determined |
| 18-106_k141_82592 | Not-determined |
| 18-106_k141_640 | Not-determined |
| 18-106_k141_65019 | Not-determined |
| 18-106_k141_53035 | Not-determined |
| 18-106_k141_135308 | Low-quality |
| 18-106_k141_129618 | Not-determined |
| 18-106_k141_47380 | Low-quality |
| 18-106_k141_123883 | Not-determined |
| 18-106_k141_643 | Not-determined |
| 18-106_k141_29925 | Not-determined |
| 18-106_k141_76814 | Not-determined |
| 18-106_k141_53043 | Not-determined |
| 18-106_k141_76817 | Not-determined |
| 18-106_k141_29927 | Low-quality |
| 18-106_k141_123890 | Not-determined |
| 18-106_k141_29929 | Not-determined |
| 18-106_k141_100192 | Not-determined |
| 18-106_k141_129628 | Not-determined |
| 18-106_k141_88685 | Not-determined |
| 18-106_k141_29938 | Low-quality |
| 18-106_k141_88686 | Not-determined |
| 18-106_k141_76826 | Not-determined |
| 18-106_k141_41702 | Not-determined |
| 18-106_k141_129639 | Low-quality |
| 18-106_k141_41703 | Not-determined |
| 18-106_k141_76835 | Not-determined |
| 18-106_k141_53075 | Not-determined |
| 18-106_k141_76836 | Low-quality |
| 18-106_k141_100218 | Not-determined |
| 18-106_k141_123910 | Not-determined |
| 18-106_k141_94600 | Low-quality |
| 18-106_k141_94602 | Not-determined |
| 18-106_k141_94604 | Not-determined |
| 18-106_k141_65048 | Not-determined |
| 18-106_k141_23852 | Low-quality |
| 18-106_k141_65049 | Low-quality |
| 18-106_k141_123915 | Not-determined |
| 18-106_k141_65051 | Not-determined |
| 18-106_k141_23854 | Not-determined |
| 18-106_k141_41732 | Not-determined |
| 18-106_k141_53090 | Not-determined |
| 18-106_k141_41734 | Low-quality |
| 18-106_k141_59293 | Not-determined |
| 18-106_k141_135340 | Not-determined |
| 18-106_k141_29958 | Not-determined |
| 18-106_k141_65061 | Not-determined |
| 18-106_k141_47411 | Low-quality |
| 18-106_k141_47415 | Not-determined |
| 18-106_k141_129659 | Not-determined |
| 18-106_k141_47420 | Not-determined |
| 18-106_k141_17954 | Not-determined |
| 18-106_k141_106378 | Not-determined |
| 18-106_k141_112093 | Not-determined |
| 18-106_k141_23875 | Not-determined |
| 18-106_k141_123930 | Low-quality |
| 18-106_k141_12355 | Not-determined |
| 18-106_k141_59308 | Not-determined |
| 18-106_k141_100247 | Not-determined |
| 18-106_k141_106394 | Not-determined |
| 18-106_k141_59313 | Not-determined |
| 18-106_k141_117900 | Not-determined |
| 18-106_k141_59316 | Not-determined |
| 18-106_k141_53116 | Not-determined |
| 18-106_k141_100254 | Not-determined |
| 18-106_k141_117906 | Not-determined |
| 18-106_k141_112111 | Not-determined |
| 18-106_k141_29965 | Not-determined |
| 18-106_k141_100262 | Low-quality |
| 18-106_k141_23884 | Not-determined |
| 18-106_k141_135364 | Not-determined |
| 18-106_k141_704 | Low-quality |
| 18-106_k141_117919 | Not-determined |
| 18-106_k141_135368 | Not-determined |
| 18-106_k141_117920 | Not-determined |
| 18-106_k141_135369 | Not-determined |
| 18-106_k141_53137 | Not-determined |
| 18-106_k141_88726 | Not-determined |
| 18-106_k141_117928 | Not-determined |
| 18-106_k141_6344 | Not-determined |
| 18-106_k141_117932 | Not-determined |
| 18-106_k141_59344 | Not-determined |
| 18-106_k141_123958 | Not-determined |
| 18-106_k141_88737 | Not-determined |
| 18-106_k141_47473 | Low-quality |
| 18-106_k141_117938 | Not-determined |
| 18-106_k141_94632 | Not-determined |
| 18-106_k141_100282 | Not-determined |
| 18-106_k141_129720 | Not-determined |
| 18-106_k141_88746 | Not-determined |
| 18-106_k141_94635 | Not-determined |
| 18-106_k141_88750 | Not-determined |
| 18-106_k141_88753 | Not-determined |
| 18-106_k141_23910 | Not-determined |
| 18-106_k141_100295 | Not-determined |
| 18-106_k141_106405 | Not-determined |
| 18-106_k141_6358 | Not-determined |
| 18-106_k141_123979 | Low-quality |
| 18-106_k141_733 | Not-determined |
| 18-106_k141_735 | Not-determined |
| 18-106_k141_737 | Not-determined |
| 18-106_k141_6363 | Not-determined |
| 18-106_k141_738 | Not-determined |
| 18-106_k141_23916 | Not-determined |
| 18-106_k141_23920 | Not-determined |
| 18-106_k141_100305 | Not-determined |
| 18-106_k141_100306 | Not-determined |
| 18-106_k141_94656 | Not-determined |
| 18-106_k141_23927 | Low-quality |
| 18-106_k141_112153 | Not-determined |
| 18-106_k141_41764 | Not-determined |
| 18-106_k141_129738 | Not-determined |
| 18-106_k141_47495 | Not-determined |
| 18-106_k141_88773 | Low-quality |
| 18-106_k141_41771 | Not-determined |
| 18-106_k141_88775 | Not-determined |
| 18-106_k141_41773 | Not-determined |
| 18-106_k141_94666 | Not-determined |
| 18-106_k141_6389 | Not-determined |
| 18-106_k141_100317 | Low-quality |
| 18-106_k141_47504 | Not-determined |
| 18-106_k141_94675 | Not-determined |
| 18-106_k141_94676 | Not-determined |
| 18-106_k141_94678 | Not-determined |
| 18-106_k141_124004 | Low-quality |
| 18-106_k141_129745 | Low-quality |
| 18-106_k141_129746 | Low-quality |
| 18-106_k141_47512 | Not-determined |
| 18-106_k141_6399 | Low-quality |
| 18-106_k141_135431 | Low-quality |
| 18-106_k141_29996 | Not-determined |
| 18-106_k141_47518 | Not-determined |
| 18-106_k141_94690 | Not-determined |
| 18-106_k141_47522 | Not-determined |
| 18-106_k141_129769 | Not-determined |
| 18-106_k141_47524 | Not-determined |
| 18-106_k141_71065 | Not-determined |
| 18-106_k141_30012 | Not-determined |
| 18-106_k141_30013 | Not-determined |
| 18-106_k141_23954 | Not-determined |
| 18-106_k141_124025 | Not-determined |
| 18-106_k141_30017 | Not-determined |
| 18-106_k141_88788 | Low-quality |
| 18-106_k141_112166 | Not-determined |
| 18-106_k141_88789 | Not-determined |
| 18-106_k141_6416 | Not-determined |
| 18-106_k141_12384 | Not-determined |
| 18-106_k141_112179 | Not-determined |
| 18-106_k141_106460 | Not-determined |
| 18-106_k141_746 | Low-quality |
| 18-106_k141_135466 | Not-determined |
| 18-106_k141_106467 | Not-determined |
| 18-106_k141_754 | Not-determined |
| 18-106_k141_112201 | Not-determined |
| 18-106_k141_94723 | Low-quality |
| 18-106_k141_124048 | Not-determined |
| 18-106_k141_65152 | Not-determined |
| 18-106_k141_65159 | Not-determined |
| 18-106_k141_12393 | Not-determined |
| 18-106_k141_12397 | Not-determined |
| 18-106_k141_65170 | Not-determined |
| 18-106_k141_65174 | Not-determined |
| 18-106_k141_88820 | Not-determined |
| 18-106_k141_100334 | Not-determined |
| 18-106_k141_88821 | Not-determined |
| 18-106_k141_6490 | Not-determined |
| 18-106_k141_100337 | Not-determined |
| 18-106_k141_94745 | Not-determined |
| 18-106_k141_12412 | Not-determined |
| 18-106_k141_112217 | Not-determined |
| 18-106_k141_23964 | Not-determined |
| 18-106_k141_23968 | Not-determined |
| 18-106_k141_94758 | Low-quality |
| 18-106_k141_71115 | Not-determined |
| 18-106_k141_65195 | Not-determined |
| 18-106_k141_12422 | Low-quality |
| 18-106_k141_23978 | Not-determined |
| 18-106_k141_94769 | Not-determined |
| 18-106_k141_775 | Not-determined |
| 18-106_k141_71131 | Not-determined |
| 18-106_k141_88844 | Not-determined |
| 18-106_k141_65211 | Low-quality |
| 18-106_k141_71133 | Low-quality |
| 18-106_k141_124089 | Not-determined |
| 18-106_k141_71136 | Not-determined |
| 18-106_k141_786 | Not-determined |
| 18-106_k141_65216 | Not-determined |
| 18-106_k141_135509 | Not-determined |
| 18-106_k141_124094 | Not-determined |
| 18-106_k141_65218 | Not-determined |
| 18-106_k141_41787 | Low-quality |
| 18-106_k141_41790 | Not-determined |
| 18-106_k141_41791 | Not-determined |
| 18-106_k141_798 | Low-quality |
| 18-106_k141_135512 | Not-determined |
| 18-106_k141_799 | Not-determined |
| 18-106_k141_71159 | Not-determined |
| 18-106_k141_124105 | Low-quality |
| 18-106_k141_802 | Not-determined |
| 18-106_k141_135519 | Not-determined |
| 18-106_k141_71169 | Not-determined |
| 18-106_k141_41803 | Not-determined |
| 18-106_k141_47581 | Not-determined |
| 18-106_k141_41806 | Not-determined |
| 18-106_k141_59409 | Not-determined |
| 18-106_k141_71181 | Not-determined |
| 18-106_k141_41812 | Not-determined |
| 18-106_k141_41820 | Not-determined |
| 18-106_k141_41821 | Not-determined |
| 18-106_k141_827 | Not-determined |
| 18-106_k141_88880 | Low-quality |
| 18-106_k141_88881 | Not-determined |
| 18-106_k141_82612 | Not-determined |
| 18-106_k141_106498 | Not-determined |
| 18-106_k141_65238 | Not-determined |
| 18-106_k141_23999 | Not-determined |
| 18-106_k141_839 | Low-quality |
| 18-106_k141_65241 | Not-determined |
| 18-106_k141_76872 | Low-quality |
| 18-106_k141_12458 | Not-determined |
| 18-106_k141_106508 | Not-determined |
| 18-106_k141_24005 | Not-determined |
| 18-106_k141_106515 | Not-determined |
| 18-106_k141_76882 | Not-determined |
| 18-106_k141_100387 | Not-determined |
| 18-106_k141_129834 | Not-determined |
| 18-106_k141_47616 | Low-quality |
| 18-106_k141_59412 | Not-determined |
| 18-106_k141_12469 | Not-determined |
| 18-106_k141_106534 | Not-determined |
| 18-106_k141_88912 | Not-determined |
| 18-106_k141_76892 | Not-determined |
| 18-106_k141_859 | Not-determined |
| 18-106_k141_59418 | Not-determined |
| 18-106_k141_41838 | Not-determined |
| 18-106_k141_76896 | Not-determined |
| 18-106_k141_117990 | Not-determined |
| 18-106_k141_65250 | Not-determined |
| 18-106_k141_41851 | Not-determined |
| 18-106_k141_24022 | Not-determined |
| 18-106_k141_865 | Not-determined |
| 18-106_k141_76910 | Not-determined |
| 18-106_k141_59432 | Not-determined |
| 18-106_k141_41856 | Not-determined |
| 18-106_k141_878 | Not-determined |
| 18-106_k141_12485 | Low-quality |
| 18-106_k141_6522 | Not-determined |
| 18-106_k141_53226 | Not-determined |
| 18-106_k141_24032 | Not-determined |
| 18-106_k141_59443 | Not-determined |
| 18-106_k141_12496 | Not-determined |
| 18-106_k141_12499 | Not-determined |
| 18-106_k141_65278 | Low-quality |
| 18-106_k141_12502 | Not-determined |
| 18-106_k141_88953 | Not-determined |
| 18-106_k141_24038 | Not-determined |
| 18-106_k141_6528 | Not-determined |
| 18-106_k141_88956 | Not-determined |
| 18-106_k141_53236 | Not-determined |
| 18-106_k141_12511 | Not-determined |
| 18-106_k141_88964 | Not-determined |
| 18-106_k141_65281 | Not-determined |
| 18-106_k141_82653 | Not-determined |
| 18-106_k141_100423 | Not-determined |
| 18-106_k141_82658 | Not-determined |
| 18-106_k141_65294 | Low-quality |
| 18-106_k141_24066 | Not-determined |
| 18-106_k141_88973 | Not-determined |
| 18-106_k141_24071 | Not-determined |
| 18-106_k141_6565 | Not-determined |
| 18-106_k141_41897 | Not-determined |
| 18-106_k141_94820 | Low-quality |
| 18-106_k141_898 | Not-determined |
| 18-106_k141_88983 | Not-determined |
| 18-106_k141_30029 | Not-determined |
| 18-106_k141_65306 | Not-determined |
| 18-106_k141_82674 | Not-determined |
| 18-106_k141_76948 | Not-determined |
| 18-106_k141_59452 | Not-determined |
| 18-106_k141_59454 | Not-determined |
| 18-106_k141_30045 | Not-determined |
| 18-106_k141_30046 | Not-determined |
| 18-106_k141_12550 | Not-determined |
| 18-106_k141_30050 | Low-quality |
| 18-106_k141_12552 | Low-quality |
| 18-106_k141_94850 | Not-determined |
| 18-106_k141_65318 | Low-quality |
| 18-106_k141_82701 | Not-determined |
| 18-106_k141_129871 | Not-determined |
| 18-106_k141_24079 | Not-determined |
| 18-106_k141_59472 | Not-determined |
| 18-106_k141_47655 | Not-determined |
| 18-106_k141_47658 | Not-determined |
| 18-106_k141_100463 | Not-determined |
| 18-106_k141_129875 | Not-determined |
| 18-106_k141_65326 | Not-determined |
| 18-106_k141_41919 | Not-determined |
| 18-106_k141_94874 | Not-determined |
| 18-106_k141_41923 | Not-determined |
| 18-106_k141_47684 | Not-determined |
| 18-106_k141_30063 | Not-determined |
| 18-106_k141_41927 | Not-determined |
| 18-106_k141_41929 | Not-determined |
| 18-106_k141_953 | Not-determined |
| 18-106_k141_118025 | Low-quality |
| 18-106_k141_30069 | Not-determined |
| 18-106_k141_12571 | Not-determined |
| 18-106_k141_30077 | Not-determined |
| 18-106_k141_65341 | Not-determined |
| 18-106_k141_41944 | Not-determined |
| 18-106_k141_24102 | Not-determined |
| 18-106_k141_65347 | Not-determined |
| 18-106_k141_100485 | Not-determined |
| 18-106_k141_35615 | Not-determined |
| 18-106_k141_41948 | Low-quality |
| 18-106_k141_974 | Not-determined |
| 18-106_k141_975 | Not-determined |
| 18-106_k141_47697 | Low-quality |
| 18-106_k141_35618 | Not-determined |
| 18-106_k141_94885 | Not-determined |

| | |
|---|---|
| 18-106_k141_100488 | Not-determined |
| 18-106_k141_53290 | Not-determined |
| 18-106_k141_129900 | Not-determined |
| 18-106_k141_6626 | Not-determined |
| 18-106_k141_35627 | Not-determined |
| 18-106_k141_89008 | Low-quality |
| 18-106_k141_6630 | Low-quality |
| 18-106_k141_129908 | Not-determined |
| 18-106_k141_12585 | Not-determined |
| 18-106_k141_982 | Not-determined |
| 18-106_k141_129911 | Not-determined |
| 18-106_k141_94898 | Low-quality |
| 18-106_k141_17988 | Not-determined |
| 18-106_k141_53306 | Not-determined |
| 18-106_k141_65363 | Not-determined |
| 18-106_k141_94903 | Not-determined |
| 18-106_k141_71226 | Not-determined |
| 18-106_k141_129923 | Not-determined |
| 18-106_k141_76969 | Not-determined |
| 18-106_k141_89026 | Not-determined |
| 18-106_k141_6649 | Not-determined |
| 18-106_k141_53320 | Not-determined |
| 18-106_k141_999 | Low-quality |
| 18-106_k141_53326 | Not-determined |
| 18-106_k141_53332 | Not-determined |
| 18-106_k141_35646 | Not-determined |
| 18-106_k141_35649 | Not-determined |
| 18-106_k141_35651 | Not-determined |
| 18-106_k141_129928 | Not-determined |
| 18-106_k141_71243 | Not-determined |
| 18-106_k141_30092 | Not-determined |
| 18-106_k141_53338 | Not-determined |
| 18-106_k141_35663 | Not-determined |
| 18-106_k141_24117 | Low-quality |
| 18-106_k141_71251 | Not-determined |
| 18-106_k141_24118 | Not-determined |
| 18-106_k141_12591 | Not-determined |
| 18-106_k141_71253 | Not-determined |
| 18-106_k141_30097 | Not-determined |
| 18-106_k141_53342 | Not-determined |
| 18-106_k141_6661 | Not-determined |
| 18-106_k141_24126 | Not-determined |
| 18-106_k141_65402 | Not-determined |
| 18-106_k141_6669 | Not-determined |
| 18-106_k141_65407 | Not-determined |
| 18-106_k141_53356 | Not-determined |
| 18-106_k141_89042 | Low-quality |
| 18-106_k141_35679 | Not-determined |
| 18-106_k141_41967 | Not-determined |
| 18-106_k141_1039 | Not-determined |
| 18-106_k141_41970 | Not-determined |
| 18-106_k141_53367 | Low-quality |
| 18-106_k141_1040 | Low-quality |
| 18-106_k141_59497 | Not-determined |
| 18-106_k141_6684 | Not-determined |
| 18-106_k141_1048 | Not-determined |
| 18-106_k141_65425 | Not-determined |
| 18-106_k141_59501 | Low-quality |
| 18-106_k141_41987 | Not-determined |
| 18-106_k141_71283 | Not-determined |
| 18-106_k141_41995 | Not-determined |
| 18-106_k141_30133 | Not-determined |
| 18-106_k141_65431 | Not-determined |
| 18-106_k141_135534 | Not-determined |
| 18-106_k141_30146 | Not-determined |
| 18-106_k141_30148 | Not-determined |
| 18-106_k141_18015 | Not-determined |
| 18-106_k141_135552 | Not-determined |
| 18-106_k141_59518 | Not-determined |
| 18-106_k141_35701 | Low-quality |
| 18-106_k141_30161 | Not-determined |
| 18-106_k141_18018 | Not-determined |
| 18-106_k141_1085 | Not-determined |
| 18-106_k141_89083 | Not-determined |
| 18-106_k141_135558 | Not-determined |
| 18-106_k141_135562 | Not-determined |
| 18-106_k141_89087 | Not-determined |
| 18-106_k141_18025 | Not-determined |
| 18-106_k141_89093 | Not-determined |
| 18-106_k141_77011 | Not-determined |
| 18-106_k141_24140 | Not-determined |
| 18-106_k141_89097 | Not-determined |
| 18-106_k141_82822 | Low-quality |
| 18-106_k141_59544 | Not-determined |
| 18-106_k141_89109 | Not-determined |
| 18-106_k141_59558 | Not-determined |
| 18-106_k141_89114 | Not-determined |
| 18-106_k141_82841 | Not-determined |
| 18-106_k141_65472 | Not-determined |
| 18-106_k141_35730 | Not-determined |
| 18-106_k141_135591 | Not-determined |
| 18-106_k141_59569 | Not-determined |
| 18-106_k141_24164 | Low-quality |
| 18-106_k141_65479 | Not-determined |
| 18-106_k141_89135 | Not-determined |
| 18-106_k141_59583 | Not-determined |
| 18-106_k141_89152 | Not-determined |
| 18-106_k141_24175 | Not-determined |
| 18-106_k141_1129 | Not-determined |
| 18-106_k141_18045 | Low-quality |
| 18-106_k141_18048 | Not-determined |
| 18-106_k141_89171 | Not-determined |
| 18-106_k141_89172 | Not-determined |
| 18-106_k141_24180 | Not-determined |
| 18-106_k141_95018 | Not-determined |
| 18-106_k141_30207 | Not-determined |
| 18-106_k141_59600 | Not-determined |
| 18-106_k141_59602 | Not-determined |
| 18-106_k141_95023 | Not-determined |
| 18-106_k141_65494 | Not-determined |
| 18-106_k141_65495 | Not-determined |
| 18-106_k141_18059 | Not-determined |
| 18-106_k141_24191 | Not-determined |
| 18-106_k141_30214 | Not-determined |
| 18-106_k141_89182 | Not-determined |
| 18-106_k141_77043 | Not-determined |
| 18-106_k141_6769 | Not-determined |
| 18-106_k141_6778 | Not-determined |
| 18-106_k141_77057 | Not-determined |
| 18-106_k141_59624 | Not-determined |
| 18-106_k141_35766 | Not-determined |
| 18-106_k141_124129 | Not-determined |
| 18-106_k141_6786 | Not-determined |
| 18-106_k141_35773 | Not-determined |
| 18-106_k141_1173 | Not-determined |
| 18-106_k141_124133 | Not-determined |
| 18-106_k141_6792 | Not-determined |
| 18-106_k141_47750 | Not-determined |
| 18-106_k141_47756 | Not-determined |
| 18-106_k141_82863 | Not-determined |
| 18-106_k141_24207 | Not-determined |
| 18-106_k141_82868 | Not-determined |
| 18-106_k141_124143 | Not-determined |
| 18-106_k141_30238 | Low-quality |
| 18-106_k141_124144 | Not-determined |
| 18-106_k141_95046 | Not-determined |
| 18-106_k141_6805 | Not-determined |
| 18-106_k141_59640 | Not-determined |
| 18-106_k141_47763 | Not-determined |
| 18-106_k141_35791 | Low-quality |
| 18-106_k141_1190 | Not-determined |
| 18-106_k141_82883 | Not-determined |
| 18-106_k141_124150 | Not-determined |
| 18-106_k141_42049 | Not-determined |
| 18-106_k141_71318 | Not-determined |
| 18-106_k141_124156 | Not-determined |
| 18-106_k141_124157 | Not-determined |
| 18-106_k141_71320 | Not-determined |
| 18-106_k141_6816 | Not-determined |
| 18-106_k141_6819 | Not-determined |
| 18-106_k141_24221 | Not-determined |
| 18-106_k141_124161 | Not-determined |
| 18-106_k141_24225 | Not-determined |
| 18-106_k141_59661 | Not-determined |
| 18-106_k141_95072 | Not-determined |
| 18-106_k141_124168 | Not-determined |
| 18-106_k141_35808 | Not-determined |
| 18-106_k141_65545 | Not-determined |
| 18-106_k141_124175 | Not-determined |
| 18-106_k141_30268 | Not-determined |
| 18-106_k141_82894 | Not-determined |
| 18-106_k141_18102 | Low-quality |
| 18-106_k141_1208 | Not-determined |
| 18-106_k141_30284 | Not-determined |
| 18-106_k141_18106 | Not-determined |
| 18-106_k141_35823 | Not-determined |
| 18-106_k141_71344 | Not-determined |
| 18-106_k141_89228 | Not-determined |
| 18-106_k141_89229 | Not-determined |
| 18-106_k141_6832 | Not-determined |
| 18-106_k141_82907 | Not-determined |
| 18-106_k141_42092 | Not-determined |
| 18-106_k141_89237 | Not-determined |
| 18-106_k141_89239 | Not-determined |
| 18-106_k141_12616 | Not-determined |
| 18-106_k141_129992 | Not-determined |
| 18-106_k141_65574 | Not-determined |
| 18-106_k141_12618 | Not-determined |
| 18-106_k141_1242 | Not-determined |
| 18-106_k141_12623 | Not-determined |
| 18-106_k141_47802 | Not-determined |
| 18-106_k141_12628 | Not-determined |
| 18-106_k141_42098 | Not-determined |
| 18-106_k141_124191 | Not-determined |
| 18-106_k141_42108 | Not-determined |
| 18-106_k141_82919 | Not-determined |
| 18-106_k141_47811 | Low-quality |
| 18-106_k141_65589 | Not-determined |
| 18-106_k141_82925 | Not-determined |
| 18-106_k141_47815 | Not-determined |
| 18-106_k141_71368 | Not-determined |
| 18-106_k141_65600 | Not-determined |
| 18-106_k141_89278 | Not-determined |
| 18-106_k141_89283 | Not-determined |
| 18-106_k141_82933 | Not-determined |
| 18-106_k141_82935 | Low-quality |
| 18-106_k141_77104 | Low-quality |
| 18-106_k141_82940 | Not-determined |
| 18-106_k141_124199 | Low-quality |
| 18-106_k141_130016 | Not-determined |
| 18-106_k141_6862 | Low-quality |
| 18-106_k141_106590 | Low-quality |
| 18-106_k141_106595 | Not-determined |
| 18-106_k141_18133 | Not-determined |
| 18-106_k141_6866 | Not-determined |
| 18-106_k141_35861 | Low-quality |
| 18-106_k141_106597 | Not-determined |
| 18-106_k141_130026 | Not-determined |
| 18-106_k141_130029 | Not-determined |
| 18-106_k141_106608 | Not-determined |
| 18-106_k141_42120 | Not-determined |
| 18-106_k141_71392 | Not-determined |
| 18-106_k141_82966 | Not-determined |
| 18-106_k141_65620 | Not-determined |
| 18-106_k141_1271 | Not-determined |
| 18-106_k141_77114 | Not-determined |
| 18-106_k141_42127 | Not-determined |
| 18-106_k141_89308 | Low-quality |
| 18-106_k141_89309 | Not-determined |
| 18-106_k141_59718 | Not-determined |
| 18-106_k141_89312 | Not-determined |
| 18-106_k141_65636 | Not-determined |
| 18-106_k141_135637 | Not-determined |
| 18-106_k141_106631 | Low-quality |
| 18-106_k141_30330 | Not-determined |
| 18-106_k141_89317 | Not-determined |
| 18-106_k141_65648 | Not-determined |
| 18-106_k141_135648 | Not-determined |
| 18-106_k141_42140 | Not-determined |
| 18-106_k141_1295 | Low-quality |
| 18-106_k141_53404 | Not-determined |
| 18-106_k141_18168 | Low-quality |
| 18-106_k141_124239 | Not-determined |
| 18-106_k141_47857 | Low-quality |
| 18-106_k141_6903 | Not-determined |
| 18-106_k141_95097 | Not-determined |
| 18-106_k141_35907 | Not-determined |
| 18-106_k141_89324 | Low-quality |
| 18-106_k141_124241 | Not-determined |
| 18-106_k141_47862 | Not-determined |
| 18-106_k141_65657 | Not-determined |
| 18-106_k141_71429 | Not-determined |
| 18-106_k141_82971 | Not-determined |
| 18-106_k141_47867 | Not-determined |
| 18-106_k141_30338 | Not-determined |
| 18-106_k141_42148 | Not-determined |
| 18-106_k141_135660 | Not-determined |
| 18-106_k141_100515 | Low-quality |
| 18-106_k141_47872 | Low-quality |
| 18-106_k141_130057 | Not-determined |
| 18-106_k141_95109 | Not-determined |
| 18-106_k141_65663 | Not-determined |
| 18-106_k141_30349 | Not-determined |
| 18-106_k141_112257 | Not-determined |
| 18-106_k141_30353 | Not-determined |
| 18-106_k141_82983 | Low-quality |
| 18-106_k141_47884 | Not-determined |
| 18-106_k141_124254 | Not-determined |
| 18-106_k141_95112 | Not-determined |
| 18-106_k141_47885 | Not-determined |
| 18-106_k141_89332 | Not-determined |
| 18-106_k141_118051 | Low-quality |
| 18-106_k141_135674 | Not-determined |
| 18-106_k141_100526 | Low-quality |
| 18-106_k141_82986 | Not-determined |
| 18-106_k141_130066 | Not-determined |
| 18-106_k141_71437 | Not-determined |
| 18-106_k141_12637 | Not-determined |
| 18-106_k141_118055 | Not-determined |
| 18-106_k141_18194 | Not-determined |
| 18-106_k141_112274 | Not-determined |
| 18-106_k141_1321 | Low-quality |
| 18-106_k141_100533 | Not-determined |
| 18-106_k141_89343 | Not-determined |
| 18-106_k141_35937 | Low-quality |
| 18-106_k141_106668 | Not-determined |
| 18-106_k141_35940 | Not-determined |
| 18-106_k141_6914 | Not-determined |
| 18-106_k141_59761 | Not-determined |
| 18-106_k141_59764 | Not-determined |
| 18-106_k141_24246 | Not-determined |
| 18-106_k141_71445 | Not-determined |
| 18-106_k141_24247 | Not-determined |
| 18-106_k141_42165 | Not-determined |
| 18-106_k141_65671 | Not-determined |
| 18-106_k141_124271 | Not-determined |
| 18-106_k141_42168 | Not-determined |
| 18-106_k141_12652 | Not-determined |
| 18-106_k141_6926 | Low-quality |
| 18-106_k141_65673 | Not-determined |
| 18-106_k141_12655 | Not-determined |
| 18-106_k141_12658 | Low-quality |
| 18-106_k141_47914 | Not-determined |
| 18-106_k141_47916 | Not-determined |
| 18-106_k141_35945 | Low-quality |
| 18-106_k141_1336 | Not-determined |
| 18-106_k141_112294 | Not-determined |
| 18-106_k141_65677 | Not-determined |
| 18-106_k141_12663 | Not-determined |
| 18-106_k141_59782 | Not-determined |
| 18-106_k141_65680 | Not-determined |
| 18-106_k141_12665 | Not-determined |
| 18-106_k141_71460 | Not-determined |
| 18-106_k141_124282 | Not-determined |
| 18-106_k141_118080 | Not-determined |
| 18-106_k141_100553 | Not-determined |
| 18-106_k141_124286 | Not-determined |
| 18-106_k141_124288 | Not-determined |
| 18-106_k141_35962 | Not-determined |
| 18-106_k141_118084 | Not-determined |
| 18-106_k141_59789 | Not-determined |
| 18-106_k141_118086 | Not-determined |
| 18-106_k141_18214 | Not-determined |
| 18-106_k141_30395 | Not-determined |
| 18-106_k141_18216 | Not-determined |
| 18-106_k141_100563 | Not-determined |
| 18-106_k141_89381 | Not-determined |
| 18-106_k141_112320 | Not-determined |
| 18-106_k141_65689 | Not-determined |
| 18-106_k141_1350 | Not-determined |
| 18-106_k141_106684 | Not-determined |
| 18-106_k141_65692 | Not-determined |
| 18-106_k141_24262 | Not-determined |
| 18-106_k141_89386 | Low-quality |
| 18-106_k141_130096 | Not-determined |
| 18-106_k141_130097 | Not-determined |
| 18-106_k141_89389 | Not-determined |
| 18-106_k141_47947 | Not-determined |
| 18-106_k141_30410 | Low-quality |
| 18-106_k141_42183 | Low-quality |
| 18-106_k141_83020 | Not-determined |
| 18-106_k141_47960 | Not-determined |
| 18-106_k141_47962 | Not-determined |
| 18-106_k141_59820 | Low-quality |
| 18-106_k141_112353 | Not-determined |
| 18-106_k141_130100 | Not-determined |
| 18-106_k141_89402 | Not-determined |
| 18-106_k141_65729 | Not-determined |
| 18-106_k141_89405 | Not-determined |
| 18-106_k141_6984 | Not-determined |
| 18-106_k141_100606 | Not-determined |
| 18-106_k141_100608 | Low-quality |
| 18-106_k141_106702 | Not-determined |
| 18-106_k141_83035 | Not-determined |
| 18-106_k141_112363 | Not-determined |
| 18-106_k141_130115 | Not-determined |
| 18-106_k141_53416 | Not-determined |
| 18-106_k141_118107 | Not-determined |
| 18-106_k141_47972 | Not-determined |
| 18-106_k141_18254 | Not-determined |
| 18-106_k141_118112 | Not-determined |
| 18-106_k141_30443 | Not-determined |
| 18-106_k141_71485 | Not-determined |
| 18-106_k141_71487 | Low-quality |
| 18-106_k141_118115 | Not-determined |
| 18-106_k141_77178 | Not-determined |
| 18-106_k141_118117 | Not-determined |
| 18-106_k141_83039 | Low-quality |
| 18-106_k141_83042 | Not-determined |
| 18-106_k141_118124 | Not-determined |
| 18-106_k141_130135 | Not-determined |
| 18-106_k141_6996 | Not-determined |
| 18-106_k141_18268 | Not-determined |
| 18-106_k141_1381 | Not-determined |
| 18-106_k141_112380 | Low-quality |
| 18-106_k141_83046 | Not-determined |
| 18-106_k141_77191 | Not-determined |
| 18-106_k141_100630 | Not-determined |
| 18-106_k141_35998 | Not-determined |
| 18-106_k141_106717 | Not-determined |
| 18-106_k141_18273 | Not-determined |
| 18-106_k141_7005 | Not-determined |
| 18-106_k141_59829 | Low-quality |
| 18-106_k141_1390 | Not-determined |
| 18-106_k141_47989 | Not-determined |
| 18-106_k141_112388 | Not-determined |
| 18-106_k141_106726 | Not-determined |
| 18-106_k141_47992 | Not-determined |
| 18-106_k141_59834 | Low-quality |
| 18-106_k141_36005 | Not-determined |
| 18-106_k141_106730 | Not-determined |
| 18-106_k141_112397 | Not-determined |
| 18-106_k141_24338 | Not-determined |
| 18-106_k141_71493 | Not-determined |
| 18-106_k141_106733 | Not-determined |
| 18-106_k141_106735 | Not-determined |
| 18-106_k141_112402 | Not-determined |
| 18-106_k141_30452 | Not-determined |
| 18-106_k141_1412 | Not-determined |
| 18-106_k141_48012 | Not-determined |
| 18-106_k141_77199 | Not-determined |
| 18-106_k141_65755 | Low-quality |
| 18-106_k141_59837 | Not-determined |
| 18-106_k141_30455 | Not-determined |
| 18-106_k141_59838 | Not-determined |
| 18-106_k141_59839 | Not-determined |
| 18-106_k141_130155 | Not-determined |
| 18-106_k141_130156 | Not-determined |
| 18-106_k141_1420 | Not-determined |
| 18-106_k141_124322 | Low-quality |
| 18-106_k141_106746 | Not-determined |
| 18-106_k141_48033 | Not-determined |
| 18-106_k141_106753 | Not-determined |
| 18-106_k141_12705 | Not-determined |
| 18-106_k141_106754 | Not-determined |
| 18-106_k141_135686 | Not-determined |
| 18-106_k141_59845 | Not-determined |
| 18-106_k141_1431 | Not-determined |
| 18-106_k141_95156 | Not-determined |
| 18-106_k141_30478 | Not-determined |
| 18-106_k141_36044 | Not-determined |
| 18-106_k141_36046 | Not-determined |
| 18-106_k141_18304 | Not-determined |
| 18-106_k141_36049 | Not-determined |
| 18-106_k141_42222 | Not-determined |
| 18-106_k141_112428 | Not-determined |
| 18-106_k141_65780 | Not-determined |
| 18-106_k141_83090 | Low-quality |
| 18-106_k141_36056 | Low-quality |
| 18-106_k141_118150 | Not-determined |
| 18-106_k141_1457 | Not-determined |
| 18-106_k141_124337 | Low-quality |
| 18-106_k141_36061 | Not-determined |
| 18-106_k141_36063 | Not-determined |
| 18-106_k141_100644 | Not-determined |
| 18-106_k141_36066 | Low-quality |
| 18-106_k141_42241 | Low-quality |
| 18-106_k141_18320 | Not-determined |
| 18-106_k141_83102 | Not-determined |
| 18-106_k141_106785 | Not-determined |
| 18-106_k141_65791 | Not-determined |
| 18-106_k141_135704 | Not-determined |
| 18-106_k141_18328 | Not-determined |
| 18-106_k141_59871 | Not-determined |
| 18-106_k141_100658 | Not-determined |
| 18-106_k141_36079 | Not-determined |
| 18-106_k141_100665 | Low-quality |
| 18-106_k141_100668 | Low-quality |
| 18-106_k141_100669 | Not-determined |
| 18-106_k141_83108 | Not-determined |
| 18-106_k141_118164 | Not-determined |
| 18-106_k141_89441 | Low-quality |
| 18-106_k141_118176 | Not-determined |
| 18-106_k141_36083 | Not-determined |
| 18-106_k141_124363 | Low-quality |
| 18-106_k141_36085 | Not-determined |
| 18-106_k141_65802 | Not-determined |
| 18-106_k141_106815 | Not-determined |
| 18-106_k141_118184 | Not-determined |
| 18-106_k141_124364 | Not-determined |
| 18-106_k141_24401 | Not-determined |
| 18-106_k141_48095 | Not-determined |
| 18-106_k141_48096 | Not-determined |
| 18-106_k141_48097 | Not-determined |
| 18-106_k141_7027 | Not-determined |
| 18-106_k141_83124 | Not-determined |
| 18-106_k141_89453 | Not-determined |
| 18-106_k141_7032 | Not-determined |
| 18-106_k141_77253 | Not-determined |
| 18-106_k141_106835 | Not-determined |
| 18-106_k141_135726 | Not-determined |
| 18-106_k141_48111 | Not-determined |
| 18-106_k141_36110 | Not-determined |
| 18-106_k141_65816 | Not-determined |
| 18-106_k141_124368 | Low-quality |
| 18-106_k141_18351 | Not-determined |
| 18-106_k141_42268 | Not-determined |
| 18-106_k141_124374 | Not-determined |
| 18-106_k141_36113 | Not-determined |
| 18-106_k141_124375 | Not-determined |
| 18-106_k141_42275 | Not-determined |
| 18-106_k141_77270 | Not-determined |
| 18-106_k141_18359 | Not-determined |
| 18-106_k141_124384 | Not-determined |
| 18-106_k141_95198 | Not-determined |
| 18-106_k141_83147 | Not-determined |
| 18-106_k141_65834 | Not-determined |
| 18-106_k141_1507 | Not-determined |
| 18-106_k141_24406 | Not-determined |
| 18-106_k141_36116 | Not-determined |
| 18-106_k141_118231 | Not-determined |
| 18-106_k141_36119 | Not-determined |
| 18-106_k141_12733 | Not-determined |
| 18-106_k141_36124 | Not-determined |
| 18-106_k141_30508 | Not-determined |
| 18-106_k141_48135 | Not-determined |
| 18-106_k141_42294 | Not-determined |
| 18-106_k141_71580 | Not-determined |
| 18-106_k141_18369 | Not-determined |
| 18-106_k141_106856 | Low-quality |
| 18-106_k141_18372 | Low-quality |
| 18-106_k141_118244 | Not-determined |
| 18-106_k141_48138 | Not-determined |
| 18-106_k141_106860 | Low-quality |
| 18-106_k141_30511 | Low-quality |
| 18-106_k141_118249 | Not-determined |
| 18-106_k141_53470 | Not-determined |
| 18-106_k141_95218 | Not-determined |
| 18-106_k141_36135 | Not-determined |
| 18-106_k141_71585 | Low-quality |
| 18-106_k141_118252 | Not-determined |
| 18-106_k141_53477 | Not-determined |
| 18-106_k141_135755 | Not-determined |
| 18-106_k141_124422 | Not-determined |
| 18-106_k141_124427 | Not-determined |
| 18-106_k141_30529 | Not-determined |
| 18-106_k141_30531 | Not-determined |
| 18-106_k141_12764 | Not-determined |
| 18-106_k141_95239 | Not-determined |
| 18-106_k141_65844 | Not-determined |
| 18-106_k141_48152 | Not-determined |
| 18-106_k141_48154 | Not-determined |
| 18-106_k141_1542 | Not-determined |
| 18-106_k141_48157 | Not-determined |
| 18-106_k141_124443 | Not-determined |
| 18-106_k141_12770 | Not-determined |
| 18-106_k141_30547 | Not-determined |
| 18-106_k141_1549 | Not-determined |
| 18-106_k141_124448 | Low-quality |
| 18-106_k141_65848 | Not-determined |
| 18-106_k141_1551 | Not-determined |
| 18-106_k141_7066 | Not-determined |
| 18-106_k141_77282 | Not-determined |
| 18-106_k141_59905 | Not-determined |
| 18-106_k141_118274 | Not-determined |
| 18-106_k141_12776 | Not-determined |
| 18-106_k141_18409 | Not-determined |
| 18-106_k141_48172 | Not-determined |
| 18-106_k141_77286 | Not-determined |
| 18-106_k141_59909 | Not-determined |
| 18-106_k141_18415 | Not-determined |
| 18-106_k141_77292 | Not-determined |
| 18-106_k141_1558 | Not-determined |
| 18-106_k141_1559 | Not-determined |
| 18-106_k141_48173 | Low-quality |
| 18-106_k141_1560 | Not-determined |
| 18-106_k141_7085 | Not-determined |
| 18-106_k141_7086 | Not-determined |
| 18-106_k141_30566 | Not-determined |
| 18-106_k141_77300 | Not-determined |
| 18-106_k141_12781 | Not-determined |
| 18-106_k141_12784 | Not-determined |
| 18-106_k141_112519 | Low-quality |
| 18-106_k141_112520 | Not-determined |
| 18-106_k141_124469 | Not-determined |
| 18-106_k141_30575 | Not-determined |
| 18-106_k141_59926 | Not-determined |
| 18-106_k141_1571 | Not-determined |
| 18-106_k141_48186 | Not-determined |
| 18-106_k141_18428 | Not-determined |
| 18-106_k141_36193 | Not-determined |
| 18-106_k141_30579 | Not-determined |
| 18-106_k141_36195 | Not-determined |
| 18-106_k141_42356 | Not-determined |
| 18-106_k141_53506 | Not-determined |
| 18-106_k141_100743 | Not-determined |
| 18-106_k141_112529 | Not-determined |
| 18-106_k141_100745 | Not-determined |
| 18-106_k141_59936 | Not-determined |
| 18-106_k141_12789 | Not-determined |
| 18-106_k141_1580 | Not-determined |
| 18-106_k141_83208 | Not-determined |
| 18-106_k141_24413 | Not-determined |
| 18-106_k141_18434 | Low-quality |
| 18-106_k141_124486 | Low-quality |
| 18-106_k141_1593 | Not-determined |
| 18-106_k141_36201 | Not-determined |
| 18-106_k141_100757 | Not-determined |
| 18-106_k141_59947 | Not-determined |
| 18-106_k141_124499 | Not-determined |
| 18-106_k141_12809 | Not-determined |
| 18-106_k141_71594 | Not-determined |
| 18-106_k141_124502 | Not-determined |
| 18-106_k141_59951 | Not-determined |
| 18-106_k141_77344 | Not-determined |
| 18-106_k141_112546 | Not-determined |
| 18-106_k141_89494 | Not-determined |
| 18-106_k141_36225 | Not-determined |
| 18-106_k141_24433 | Not-determined |
| 18-106_k141_42390 | Not-determined |
| 18-106_k141_77356 | Not-determined |
| 18-106_k141_77358 | Not-determined |
| 18-106_k141_42393 | Not-determined |
| 18-106_k141_130172 | Not-determined |
| 18-106_k141_124527 | Not-determined |
| 18-106_k141_118290 | Low-quality |
| 18-106_k141_42402 | Not-determined |
| 18-106_k141_89503 | Low-quality |
| 18-106_k141_18447 | Not-determined |

| | |
|---|---|
| 18-106_k141_100775 | Not-determined |
| 18-106_k141_1624 | Not-determined |
| 18-106_k141_42412 | Low-quality |
| 18-106_k141_18452 | Not-determined |
| 18-106_k141_1631 | Not-determined |
| 18-106_k141_1634 | Not-determined |
| 18-106_k141_59975 | Not-determined |
| 18-106_k141_135809 | Not-determined |
| 18-106_k141_124545 | Not-determined |
| 18-106_k141_77373 | Not-determined |
| 18-106_k141_95284 | Not-determined |
| 18-106_k141_95285 | Not-determined |
| 18-106_k141_42430 | Not-determined |
| 18-106_k141_36247 | Low-quality |
| 18-106_k141_135812 | Not-determined |
| 18-106_k141_118323 | Not-determined |
| 18-106_k141_77375 | Not-determined |
| 18-106_k141_59987 | Not-determined |
| 18-106_k141_130198 | Not-determined |
| 18-106_k141_77377 | Not-determined |
| 18-106_k141_7117 | Not-determined |
| 18-106_k141_42443 | Not-determined |
| 18-106_k141_1655 | Not-determined |
| 18-106_k141_130205 | Not-determined |
| 18-106_k141_59992 | Not-determined |
| 18-106_k141_48231 | Not-determined |
| 18-106_k141_48235 | Not-determined |
| 18-106_k141_48236 | Not-determined |
| 18-106_k141_95307 | Not-determined |
| 18-106_k141_30614 | Not-determined |
| 18-106_k141_59999 | Low-quality |
| 18-106_k141_48240 | Not-determined |
| 18-106_k141_48241 | Not-determined |
| 18-106_k141_48247 | Not-determined |
| 18-106_k141_42463 | Not-determined |
| 18-106_k141_48248 | Not-determined |
| 18-106_k141_124582 | Not-determined |
| 18-106_k141_124583 | Not-determined |
| 18-106_k141_130228 | Not-determined |
| 18-106_k141_89530 | Not-determined |
| 18-106_k141_42470 | Not-determined |
| 18-106_k141_130231 | Not-determined |
| 18-106_k141_24441 | Not-determined |
| 18-106_k141_48258 | Not-determined |
| 18-106_k141_106946 | Not-determined |
| 18-106_k141_42480 | Not-determined |
| 18-106_k141_95327 | Not-determined |
| 18-106_k141_36285 | Not-determined |
| 18-106_k141_65888 | Low-quality |
| 18-106_k141_42483 | Not-determined |
| 18-106_k141_95336 | Not-determined |
| 18-106_k141_135849 | Not-determined |
| 18-106_k141_77420 | Not-determined |
| 18-106_k141_18488 | Not-determined |
| 18-106_k141_48279 | Not-determined |
| 18-106_k141_95342 | Not-determined |
| 18-106_k141_36301 | Not-determined |
| 18-106_k141_130261 | Not-determined |
| 18-106_k141_36303 | Low-quality |
| 18-106_k141_77427 | Not-determined |
| 18-106_k141_95344 | Not-determined |
| 18-106_k141_77429 | Not-determined |
| 18-106_k141_30644 | Not-determined |
| 18-106_k141_100825 | Not-determined |
| 18-106_k141_77430 | Not-determined |
| 18-106_k141_77431 | Not-determined |
| 18-106_k141_7143 | Not-determined |
| 18-106_k141_24456 | Not-determined |
| 18-106_k141_100841 | Not-determined |
| 18-106_k141_48297 | Not-determined |
| 18-106_k141_106960 | Not-determined |
| 18-106_k141_42502 | Not-determined |
| 18-106_k141_89558 | Not-determined |
| 18-106_k141_118357 | Low-quality |
| 18-106_k141_118359 | Not-determined |
| 18-106_k141_130282 | Not-determined |
| 18-106_k141_60046 | Not-determined |
| 18-106_k141_53539 | Not-determined |
| 18-106_k141_65903 | Not-determined |
| 18-106_k141_83294 | Not-determined |
| 18-106_k141_65907 | Not-determined |
| 18-106_k141_36329 | Not-determined |
| 18-106_k141_60055 | Low-quality |
| 18-106_k141_95366 | Low-quality |
| 18-106_k141_12874 | Not-determined |
| 18-106_k141_118361 | Low-quality |
| 18-106_k141_18499 | Low-quality |
| 18-106_k141_112627 | Not-determined |
| 18-106_k141_60062 | Not-determined |
| 18-106_k141_118366 | Not-determined |
| 18-106_k141_7163 | Not-determined |
| 18-106_k141_89573 | Not-determined |
| 18-106_k141_30673 | Not-determined |
| 18-106_k141_1676 | Not-determined |
| 18-106_k141_83299 | Not-determined |
| 18-106_k141_24464 | Not-determined |
| 18-106_k141_83302 | Not-determined |
| 18-106_k141_89576 | Not-determined |
| 18-106_k141_53553 | Not-determined |
| 18-106_k141_7172 | Not-determined |
| 18-106_k141_95367 | Not-determined |
| 18-106_k141_60069 | Not-determined |
| 18-106_k141_106994 | Not-determined |
| 18-106_k141_83311 | Not-determined |
| 18-106_k141_65917 | Not-determined |
| 18-106_k141_89587 | Not-determined |
| 18-106_k141_89588 | Not-determined |
| 18-106_k141_7184 | Not-determined |
| 18-106_k141_18533 | Not-determined |
| 18-106_k141_77461 | Not-determined |
| 18-106_k141_65924 | Not-determined |
| 18-106_k141_107007 | Not-determined |
| 18-106_k141_18534 | Low-quality |
| 18-106_k141_60089 | Not-determined |
| 18-106_k141_12893 | Low-quality |
| 18-106_k141_95384 | Not-determined |
| 18-106_k141_89609 | Not-determined |
| 18-106_k141_1689 | Not-determined |
| 18-106_k141_95387 | Not-determined |
| 18-106_k141_36350 | Low-quality |
| 18-106_k141_42525 | Low-quality |
| 18-106_k141_7195 | Low-quality |
| 18-106_k141_135863 | Not-determined |
| 18-106_k141_48327 | Not-determined |
| 18-106_k141_118415 | Not-determined |
| 18-106_k141_12895 | Not-determined |
| 18-106_k141_135867 | Low-quality |
| 18-106_k141_53561 | Not-determined |
| 18-106_k141_18557 | Not-determined |
| 18-106_k141_130314 | Not-determined |
| 18-106_k141_24494 | Not-determined |
| 18-106_k141_135872 | Not-determined |
| 18-106_k141_65952 | Not-determined |
| 18-106_k141_112637 | Not-determined |
| 18-106_k141_77466 | Low-quality |
| 18-106_k141_18564 | Not-determined |
| 18-106_k141_60109 | Low-quality |
| 18-106_k141_60110 | Not-determined |
| 18-106_k141_18570 | Not-determined |
| 18-106_k141_83349 | Low-quality |
| 18-106_k141_65956 | Not-determined |
| 18-106_k141_30709 | Not-determined |
| 18-106_k141_24505 | Not-determined |
| 18-106_k141_1716 | Not-determined |
| 18-106_k141_7196 | Not-determined |
| 18-106_k141_83355 | Not-determined |
| 18-106_k141_89643 | Low-quality |
| 18-106_k141_1722 | Not-determined |
| 18-106_k141_77472 | Not-determined |
| 18-106_k141_135877 | Not-determined |
| 18-106_k141_77473 | Not-determined |
| 18-106_k141_42545 | Not-determined |
| 18-106_k141_107043 | Not-determined |
| 18-106_k141_7204 | Not-determined |
| 18-106_k141_30717 | Low-quality |
| 18-106_k141_30719 | Not-determined |
| 18-106_k141_12918 | Not-determined |
| 18-106_k141_95410 | Not-determined |
| 18-106_k141_89646 | Not-determined |
| 18-106_k141_53584 | Not-determined |
| 18-106_k141_53585 | Not-determined |
| 18-106_k141_30726 | Not-determined |
| 18-106_k141_65968 | Not-determined |
| 18-106_k141_89648 | Not-determined |
| 18-106_k141_7215 | Not-determined |
| 18-106_k141_7216 | Not-determined |
| 18-106_k141_42557 | Not-determined |
| 18-106_k141_18589 | Not-determined |
| 18-106_k141_83377 | Not-determined |
| 18-106_k141_7222 | Not-determined |
| 18-106_k141_30732 | Not-determined |
| 18-106_k141_30733 | Not-determined |
| 18-106_k141_135882 | Not-determined |
| 18-106_k141_60137 | Not-determined |
| 18-106_k141_12925 | Not-determined |
| 18-106_k141_18598 | Not-determined |
| 18-106_k141_53595 | Not-determined |
| 18-106_k141_89662 | Not-determined |
| 18-106_k141_83391 | Low-quality |
| 18-106_k141_24522 | Not-determined |
| 18-106_k141_71623 | Not-determined |
| 18-106_k141_42571 | Not-determined |
| 18-106_k141_24533 | Not-determined |
| 18-106_k141_83404 | Not-determined |
| 18-106_k141_135896 | Not-determined |
| 18-106_k141_83409 | Low-quality |
| 18-106_k141_24540 | Not-determined |
| 18-106_k141_12953 | Low-quality |
| 18-106_k141_107095 | Not-determined |
| 18-106_k141_135906 | Not-determined |
| 18-106_k141_18627 | Not-determined |
| 18-106_k141_77486 | Low-quality |
| 18-106_k141_18628 | Low-quality |
| 18-106_k141_89691 | Not-determined |
| 18-106_k141_112687 | Not-determined |
| 18-106_k141_60172 | Low-quality |
| 18-106_k141_12957 | Not-determined |
| 18-106_k141_95430 | Not-determined |
| 18-106_k141_83433 | Not-determined |
| 18-106_k141_42600 | Not-determined |
| 18-106_k141_71636 | Not-determined |
| 18-106_k141_12967 | Not-determined |
| 18-106_k141_135919 | Not-determined |
| 18-106_k141_7270 | Not-determined |
| 18-106_k141_7271 | Not-determined |
| 18-106_k141_24570 | Not-determined |
| 18-106_k141_77503 | Not-determined |
| 18-106_k141_130357 | Not-determined |
| 18-106_k141_24578 | Not-determined |
| 18-106_k141_18654 | Not-determined |
| 18-106_k141_18656 | Not-determined |
| 18-106_k141_53620 | Not-determined |
| 18-106_k141_130361 | Not-determined |
| 18-106_k141_66014 | Not-determined |
| 18-106_k141_60199 | Not-determined |
| 18-106_k141_7284 | Not-determined |
| 18-106_k141_42626 | Not-determined |
| 18-106_k141_77513 | Not-determined |
| 18-106_k141_30741 | Not-determined |
| 18-106_k141_83437 | Not-determined |
| 18-106_k141_1762 | Not-determined |
| 18-106_k141_60207 | Not-determined |
| 18-106_k141_30746 | Not-determined |
| 18-106_k141_53631 | Not-determined |
| 18-106_k141_48393 | Low-quality |
| 18-106_k141_18672 | Not-determined |
| 18-106_k141_83447 | Low-quality |
| 18-106_k141_71655 | Not-determined |
| 18-106_k141_24593 | Not-determined |
| 18-106_k141_107115 | Not-determined |
| 18-106_k141_60213 | Not-determined |
| 18-106_k141_42635 | Not-determined |
| 18-106_k141_95435 | Not-determined |
| 18-106_k141_77517 | Not-determined |
| 18-106_k141_66032 | Not-determined |
| 18-106_k141_18687 | Not-determined |
| 18-106_k141_66036 | Low-quality |
| 18-106_k141_7290 | Not-determined |
| 18-106_k141_7291 | Not-determined |
| 18-106_k141_95453 | Not-determined |
| 18-106_k141_135931 | Not-determined |
| 18-106_k141_100921 | Low-quality |
| 18-106_k141_130378 | Not-determined |
| 18-106_k141_66041 | Low-quality |
| 18-106_k141_107138 | Not-determined |
| 18-106_k141_66044 | Not-determined |
| 18-106_k141_112700 | Not-determined |
| 18-106_k141_18691 | Low-quality |
| 18-106_k141_130385 | Low-quality |
| 18-106_k141_135943 | Not-determined |
| 18-106_k141_83461 | Not-determined |
| 18-106_k141_60223 | Not-determined |
| 18-106_k141_18702 | Not-determined |
| 18-106_k141_18704 | Not-determined |
| 18-106_k141_100931 | Not-determined |
| 18-106_k141_95472 | Not-determined |
| 18-106_k141_107140 | Not-determined |
| 18-106_k141_71679 | Not-determined |
| 18-106_k141_95477 | Not-determined |
| 18-106_k141_53667 | Not-determined |
| 18-106_k141_95480 | Not-determined |
| 18-106_k141_30795 | Not-determined |
| 18-106_k141_71689 | Not-determined |
| 18-106_k141_1775 | Not-determined |
| 18-106_k141_71692 | Not-determined |
| 18-106_k141_71694 | Not-determined |
| 18-106_k141_95491 | Not-determined |
| 18-106_k141_53686 | Not-determined |
| 18-106_k141_130397 | Not-determined |
| 18-106_k141_42674 | Not-determined |
| 18-106_k141_107164 | Low-quality |
| 18-106_k141_53689 | Not-determined |
| 18-106_k141_66074 | Low-quality |
| 18-106_k141_66077 | Not-determined |
| 18-106_k141_42678 | Not-determined |
| 18-106_k141_66078 | Not-determined |
| 18-106_k141_66080 | Not-determined |
| 18-106_k141_66082 | Not-determined |
| 18-106_k141_95499 | Not-determined |
| 18-106_k141_107173 | Not-determined |
| 18-106_k141_107175 | Not-determined |
| 18-106_k141_7316 | Not-determined |
| 18-106_k141_7317 | Not-determined |
| 18-106_k141_1795 | Low-quality |
| 18-106_k141_95512 | Not-determined |
| 18-106_k141_95513 | Not-determined |
| 18-106_k141_60261 | Low-quality |
| 18-106_k141_95514 | Not-determined |
| 18-106_k141_12988 | Not-determined |
| 18-106_k141_60265 | Not-determined |
| 18-106_k141_53698 | Low-quality |
| 18-106_k141_18754 | Not-determined |
| 18-106_k141_71704 | Not-determined |
| 18-106_k141_42695 | Not-determined |
| 18-106_k141_71708 | Not-determined |
| 18-106_k141_135969 | Not-determined |
| 18-106_k141_18761 | Not-determined |
| 18-106_k141_100942 | Not-determined |
| 18-106_k141_107203 | Low-quality |
| 18-106_k141_71719 | Low-quality |
| 18-106_k141_53716 | Not-determined |
| 18-106_k141_7324 | Not-determined |
| 18-106_k141_1816 | Not-determined |
| 18-106_k141_100951 | Not-determined |
| 18-106_k141_71732 | Low-quality |
| 18-106_k141_130435 | Not-determined |
| 18-106_k141_60302 | Not-determined |
| 18-106_k141_83530 | Low-quality |
| 18-106_k141_83531 | Not-determined |
| 18-106_k141_100965 | Not-determined |
| 18-106_k141_42699 | Not-determined |
| 18-106_k141_48483 | Not-determined |
| 18-106_k141_130441 | Not-determined |
| 18-106_k141_36352 | Not-determined |
| 18-106_k141_30837 | Not-determined |
| 18-106_k141_1857 | Not-determined |
| 18-106_k141_30839 | Low-quality |
| 18-106_k141_83552 | Not-determined |
| 18-106_k141_36358 | Not-determined |
| 18-106_k141_95538 | Not-determined |
| 18-106_k141_1866 | Not-determined |
| 18-106_k141_107206 | Not-determined |
| 18-106_k141_42717 | Low-quality |
| 18-106_k141_71741 | Not-determined |
| 18-106_k141_89755 | Not-determined |
| 18-106_k141_53724 | Not-determined |
| 18-106_k141_30850 | Not-determined |
| 18-106_k141_42723 | Not-determined |
| 18-106_k141_42726 | Not-determined |
| 18-106_k141_71744 | Not-determined |
| 18-106_k141_71746 | Not-determined |
| 18-106_k141_107218 | Not-determined |
| 18-106_k141_83556 | Low-quality |
| 18-106_k141_71750 | Not-determined |
| 18-106_k141_95560 | Not-determined |
| 18-106_k141_42738 | Not-determined |
| 18-106_k141_83565 | Not-determined |
| 18-106_k141_30865 | Not-determined |
| 18-106_k141_7358 | Not-determined |
| 18-106_k141_136008 | Not-determined |
| 18-106_k141_7365 | Not-determined |
| 18-106_k141_12993 | Low-quality |
| 18-106_k141_36388 | Not-determined |
| 18-106_k141_95563 | Not-determined |
| 18-106_k141_60335 | Not-determined |
| 18-106_k141_1895 | Low-quality |
| 18-106_k141_89777 | Low-quality |
| 18-106_k141_83578 | Not-determined |
| 18-106_k141_89779 | Low-quality |
| 18-106_k141_36398 | Not-determined |
| 18-106_k141_7374 | Not-determined |
| 18-106_k141_13004 | Not-determined |
| 18-106_k141_7377 | Not-determined |
| 18-106_k141_30883 | Not-determined |
| 18-106_k141_71781 | Not-determined |
| 18-106_k141_13010 | Not-determined |
| 18-106_k141_7381 | Not-determined |
| 18-106_k141_136033 | Not-determined |
| 18-106_k141_107255 | Not-determined |
| 18-106_k141_36412 | Not-determined |
| 18-106_k141_112729 | Not-determined |
| 18-106_k141_48562 | Low-quality |
| 18-106_k141_71786 | Not-determined |
| 18-106_k141_60356 | Not-determined |
| 18-106_k141_118449 | Not-determined |
| 18-106_k141_36421 | Not-determined |
| 18-106_k141_13016 | Not-determined |
| 18-106_k141_66120 | Not-determined |
| 18-106_k141_130501 | Low-quality |
| 18-106_k141_60375 | Not-determined |
| 18-106_k141_13026 | Not-determined |
| 18-106_k141_66123 | Low-quality |
| 18-106_k141_130506 | Not-determined |
| 18-106_k141_112731 | Low-quality |
| 18-106_k141_95596 | Not-determined |
| 18-106_k141_1925 | Not-determined |
| 18-106_k141_60398 | Not-determined |
| 18-106_k141_18790 | Low-quality |
| 18-106_k141_42762 | Not-determined |
| 18-106_k141_60400 | Not-determined |
| 18-106_k141_48597 | Not-determined |
| 18-106_k141_1935 | Not-determined |
| 18-106_k141_36442 | Not-determined |
| 18-106_k141_60407 | Not-determined |
| 18-106_k141_18796 | Low-quality |
| 18-106_k141_7397 | Not-determined |
| 18-106_k141_1942 | Not-determined |
| 18-106_k141_89835 | Low-quality |
| 18-106_k141_13051 | Not-determined |
| 18-106_k141_30930 | Not-determined |
| 18-106_k141_7411 | Not-determined |
| 18-106_k141_95619 | Low-quality |
| 18-106_k141_95620 | Not-determined |
| 18-106_k141_13061 | Not-determined |
| 18-106_k141_66152 | Not-determined |
| 18-106_k141_71805 | Not-determined |
| 18-106_k141_83610 | Not-determined |
| 18-106_k141_30935 | Not-determined |
| 18-106_k141_53797 | Not-determined |
| 18-106_k141_77568 | Low-quality |
| 18-106_k141_136093 | Not-determined |
| 18-106_k141_42770 | Not-determined |
| 18-106_k141_13072 | Not-determined |
| 18-106_k141_107283 | Not-determined |
| 18-106_k141_60425 | Not-determined |
| 18-106_k141_1958 | Not-determined |
| 18-106_k141_60426 | Low-quality |
| 18-106_k141_77581 | Not-determined |
| 18-106_k141_95639 | Not-determined |
| 18-106_k141_48633 | Not-determined |
| 18-106_k141_89866 | Not-determined |
| 18-106_k141_83621 | Not-determined |
| 18-106_k141_136106 | Not-determined |
| 18-106_k141_66172 | Low-quality |
| 18-106_k141_83623 | Not-determined |
| 18-106_k141_24616 | Not-determined |
| 18-106_k141_66175 | Not-determined |
| 18-106_k141_30960 | Not-determined |
| 18-106_k141_89872 | Not-determined |
| 18-106_k141_30961 | Not-determined |
| 18-106_k141_42778 | Not-determined |
| 18-106_k141_83628 | Not-determined |
| 18-106_k141_89877 | Not-determined |
| 18-106_k141_53825 | Not-determined |
| 18-106_k141_24625 | Not-determined |
| 18-106_k141_95645 | Not-determined |
| 18-106_k141_7420 | Not-determined |
| 18-106_k141_107309 | Not-determined |
| 18-106_k141_36476 | Not-determined |
| 18-106_k141_77591 | Low-quality |
| 18-106_k141_107321 | Not-determined |
| 18-106_k141_107324 | Not-determined |
| 18-106_k141_77595 | Not-determined |
| 18-106_k141_136116 | Not-determined |
| 18-106_k141_77608 | Not-determined |
| 18-106_k141_130573 | Not-determined |
| 18-106_k141_118518 | Not-determined |
| 18-106_k141_24633 | Not-determined |
| 18-106_k141_60448 | Not-determined |
| 18-106_k141_30981 | Low-quality |
| 18-106_k141_107339 | Not-determined |
| 18-106_k141_30982 | Not-determined |
| 18-106_k141_95659 | Not-determined |
| 18-106_k141_83650 | Not-determined |
| 18-106_k141_118527 | Not-determined |
| 18-106_k141_136122 | Not-determined |
| 18-106_k141_30986 | Not-determined |
| 18-106_k141_36499 | Not-determined |
| 18-106_k141_107351 | Not-determined |
| 18-106_k141_136127 | Not-determined |
| 18-106_k141_136128 | Not-determined |
| 18-106_k141_124650 | Not-determined |
| 18-106_k141_107355 | Low-quality |
| 18-106_k141_42809 | Not-determined |
| 18-106_k141_2011 | Not-determined |
| 18-106_k141_95677 | Not-determined |
| 18-106_k141_13110 | Not-determined |
| 18-106_k141_118540 | Not-determined |
| 18-106_k141_53838 | Not-determined |
| 18-106_k141_13120 | Not-determined |
| 18-106_k141_77628 | Not-determined |
| 18-106_k141_42825 | Not-determined |
| 18-106_k141_136141 | Not-determined |
| 18-106_k141_124668 | Not-determined |
| 18-106_k141_118557 | Low-quality |
| 18-106_k141_130597 | Not-determined |
| 18-106_k141_2030 | Not-determined |
| 18-106_k141_95699 | Not-determined |
| 18-106_k141_77639 | Not-determined |
| 18-106_k141_124672 | Not-determined |
| 18-106_k141_130602 | Not-determined |
| 18-106_k141_42833 | Not-determined |
| 18-106_k141_77646 | Not-determined |
| 18-106_k141_130607 | Not-determined |
| 18-106_k141_30997 | Not-determined |
| 18-106_k141_24685 | Not-determined |
| 18-106_k141_42839 | Not-determined |
| 18-106_k141_24687 | Not-determined |
| 18-106_k141_95701 | Not-determined |
| 18-106_k141_66200 | Not-determined |
| 18-106_k141_77655 | Not-determined |
| 18-106_k141_2044 | Not-determined |
| 18-106_k141_77657 | Not-determined |
| 18-106_k141_2045 | Not-determined |
| 18-106_k141_95704 | Low-quality |
| 18-106_k141_13152 | Not-determined |
| 18-106_k141_107380 | Not-determined |
| 18-106_k141_18803 | Not-determined |
| 18-106_k141_107382 | Not-determined |
| 18-106_k141_112759 | Not-determined |
| 18-106_k141_18808 | Not-determined |
| 18-106_k141_36522 | Not-determined |
| 18-106_k141_24707 | Not-determined |
| 18-106_k141_18813 | Not-determined |
| 18-106_k141_24708 | Not-determined |
| 18-106_k141_2054 | Not-determined |
| 18-106_k141_18814 | Not-determined |
| 18-106_k141_2057 | Not-determined |
| 18-106_k141_112777 | Not-determined |
| 18-106_k141_112779 | Not-determined |
| 18-106_k141_31009 | Not-determined |
| 18-106_k141_118590 | Low-quality |
| 18-106_k141_83659 | Low-quality |
| 18-106_k141_83660 | Not-determined |
| 18-106_k141_107400 | Not-determined |
| 18-106_k141_130642 | Not-determined |
| 18-106_k141_24725 | Not-determined |
| 18-106_k141_112786 | Low-quality |
| 18-106_k141_31018 | Not-determined |
| 18-106_k141_130648 | Not-determined |
| 18-106_k141_95726 | Not-determined |
| 18-106_k141_60503 | Not-determined |
| 18-106_k141_13205 | Not-determined |
| 18-106_k141_48711 | Not-determined |
| 18-106_k141_7464 | Not-determined |
| 18-106_k141_7466 | Not-determined |
| 18-106_k141_42851 | Not-determined |
| 18-106_k141_7467 | Not-determined |
| 18-106_k141_71832 | Not-determined |
| 18-106_k141_36554 | Not-determined |
| 18-106_k141_112813 | Not-determined |
| 18-106_k141_136171 | Not-determined |
| 18-106_k141_77688 | Not-determined |
| 18-106_k141_42871 | Not-determined |
| 18-106_k141_95736 | Not-determined |
| 18-106_k141_77694 | Not-determined |
| 18-106_k141_83693 | Not-determined |
| 18-106_k141_48737 | Not-determined |
| 18-106_k141_13248 | Not-determined |
| 18-106_k141_77706 | Not-determined |
| 18-106_k141_77709 | Not-determined |
| 18-106_k141_89926 | Not-determined |
| 18-106_k141_36575 | Not-determined |
| 18-106_k141_112826 | Not-determined |
| 18-106_k141_77712 | Low-quality |
| 18-106_k141_112830 | Not-determined |
| 18-106_k141_60518 | Not-determined |
| 18-106_k141_31082 | Not-determined |
| 18-106_k141_95745 | Not-determined |
| 18-106_k141_36596 | Low-quality |
| 18-106_k141_31092 | Not-determined |
| 18-106_k141_42900 | Not-determined |
| 18-106_k141_60527 | Not-determined |
| 18-106_k141_2080 | Not-determined |
| 18-106_k141_24742 | Low-quality |
| 18-106_k141_36603 | Not-determined |
| 18-106_k141_107416 | Not-determined |
| 18-106_k141_42910 | Not-determined |
| 18-106_k141_130688 | Not-determined |
| 18-106_k141_77728 | Not-determined |
| 18-106_k141_112844 | Not-determined |
| 18-106_k141_36614 | Not-determined |
| 18-106_k141_130700 | Not-determined |
| 18-106_k141_36619 | Not-determined |
| 18-106_k141_112850 | Not-determined |
| 18-106_k141_71883 | Not-determined |
| 18-106_k141_36626 | Not-determined |
| 18-106_k141_24760 | Not-determined |
| 18-106_k141_53950 | Not-determined |
| 18-106_k141_89963 | Not-determined |
| 18-106_k141_77745 | Not-determined |
| 18-106_k141_136214 | Not-determined |
| 18-106_k141_36638 | Not-determined |
| 18-106_k141_66224 | Not-determined |
| 18-106_k141_36641 | Not-determined |
| 18-106_k141_48768 | Not-determined |
| 18-106_k141_118594 | Not-determined |
| 18-106_k141_130711 | Not-determined |
| 18-106_k141_36646 | Not-determined |
| 18-106_k141_124759 | Not-determined |
| 18-106_k141_124761 | Not-determined |
| 18-106_k141_48774 | Low-quality |
| 18-106_k141_118597 | Low-quality |
| 18-106_k141_66236 | Low-quality |
| 18-106_k141_118599 | Not-determined |
| 18-106_k141_124770 | Not-determined |
| 18-106_k141_13285 | Not-determined |
| 18-106_k141_60566 | Not-determined |
| 18-106_k141_130721 | Not-determined |
| 18-106_k141_13288 | Not-determined |
| 18-106_k141_48792 | Not-determined |
| 18-106_k141_36652 | Not-determined |
| 18-106_k141_36653 | Low-quality |
| 18-106_k141_48797 | Not-determined |
| 18-106_k141_42942 | Not-determined |
| 18-106_k141_118610 | Low-quality |
| 18-106_k141_66268 | Not-determined |
| 18-106_k141_112870 | Not-determined |
| 18-106_k141_36672 | Not-determined |

| | |
|---|---|
| 18-106_k141_48799 | Low-quality |
| 18-106_k141_60590 | Low-quality |
| 18-106_k141_112874 | Not-determined |
| 18-106_k141_42955 | Not-determined |
| 18-106_k141_48806 | Not-determined |
| 18-106_k141_48807 | Not-determined |
| 18-106_k141_24799 | Not-determined |
| 18-106_k141_89986 | Low-quality |
| 18-106_k141_124785 | Not-determined |
| 18-106_k141_2109 | Not-determined |
| 18-106_k141_124791 | Not-determined |
| 18-106_k141_48814 | Not-determined |
| 18-106_k141_130742 | Not-determined |
| 18-106_k141_31112 | Low-quality |
| 18-106_k141_60609 | Not-determined |
| 18-106_k141_107437 | Not-determined |
| 18-106_k141_36697 | Not-determined |
| 18-106_k141_130749 | Low-quality |
| 18-106_k141_107441 | Not-determined |
| 18-106_k141_90010 | Not-determined |
| 18-106_k141_112888 | Low-quality |
| 18-106_k141_118617 | Not-determined |
| 18-106_k141_90015 | Not-determined |
| 18-106_k141_118620 | Not-determined |
| 18-106_k141_83741 | Not-determined |
| 18-106_k141_13299 | Low-quality |
| 18-106_k141_130762 | Not-determined |
| 18-106_k141_83747 | Low-quality |
| 18-106_k141_90024 | Not-determined |
| 18-106_k141_48838 | Not-determined |
| 18-106_k141_66287 | Not-determined |
| 18-106_k141_112908 | Not-determined |
| 18-106_k141_60639 | Not-determined |
| 18-106_k141_112914 | Not-determined |
| 18-106_k141_2133 | Not-determined |
| 18-106_k141_2134 | Not-determined |
| 18-106_k141_66295 | Not-determined |
| 18-106_k141_66298 | Not-determined |
| 18-106_k141_66300 | Not-determined |
| 18-106_k141_2140 | Not-determined |
| 18-106_k141_136271 | Not-determined |
| 18-106_k141_36732 | Not-determined |
| 18-106_k141_107472 | Not-determined |
| 18-106_k141_36736 | Not-determined |
| 18-106_k141_31129 | Not-determined |
| 18-106_k141_107476 | Not-determined |
| 18-106_k141_31130 | Not-determined |
| 18-106_k141_66301 | Low-quality |
| 18-106_k141_31136 | Not-determined |
| 18-106_k141_24842 | Not-determined |
| 18-106_k141_36753 | Not-determined |
| 18-106_k141_112931 | Low-quality |
| 18-106_k141_31143 | Not-determined |
| 18-106_k141_31146 | Not-determined |
| 18-106_k141_13327 | Not-determined |
| 18-106_k141_31156 | Not-determined |
| 18-106_k141_124806 | Not-determined |
| 18-106_k141_124810 | Not-determined |
| 18-106_k141_60658 | Not-determined |
| 18-106_k141_66330 | Not-determined |
| 18-106_k141_66332 | Not-determined |
| 18-106_k141_112941 | Low-quality |
| 18-106_k141_36783 | Not-determined |
| 18-106_k141_31161 | Not-determined |
| 18-106_k141_95798 | Not-determined |
| 18-106_k141_90048 | Not-determined |
| 18-106_k141_136301 | Not-determined |
| 18-106_k141_90051 | Not-determined |
| 18-106_k141_13347 | Not-determined |
| 18-106_k141_130785 | Low-quality |
| 18-106_k141_112944 | Not-determined |
| 18-106_k141_60669 | Not-determined |
| 18-106_k141_107480 | Not-determined |
| 18-106_k141_136308 | Not-determined |
| 18-106_k141_13352 | Not-determined |
| 18-106_k141_95811 | Low-quality |
| 18-106_k141_130790 | Not-determined |
| 18-106_k141_31173 | Not-determined |
| 18-106_k141_136312 | Not-determined |
| 18-106_k141_48875 | Not-determined |
| 18-106_k141_90063 | Not-determined |
| 18-106_k141_136319 | Not-determined |
| 18-106_k141_71952 | Low-quality |
| 18-106_k141_101016 | Not-determined |
| 18-106_k141_71954 | Not-determined |
| 18-106_k141_95819 | Low-quality |
| 18-106_k141_95821 | Not-determined |
| 18-106_k141_90072 | Not-determined |
| 18-106_k141_24867 | Not-determined |
| 18-106_k141_124835 | Not-determined |
| 18-106_k141_90079 | Not-determined |
| 18-106_k141_18827 | Not-determined |
| 18-106_k141_90081 | Not-determined |
| [illegible] | [illegible] |
| 18-106_k141_7547 | Low-quality |
| 18-106_k141_95872 | Not-determined |
| 18-106_k141_60752 | Not-determined |
| 18-106_k141_18958 | Not-determined |
| 18-106_k141_18960 | Not-determined |
| 18-106_k141_95878 | Not-determined |
| 18-106_k141_18963 | Not-determined |
| 18-106_k141_31331 | Not-determined |
| 18-106_k141_66468 | Not-determined |
| 18-106_k141_130940 | Not-determined |
| 18-106_k141_24944 | Not-determined |
| 18-106_k141_60765 | Not-determined |
| 18-106_k141_43161 | Not-determined |
| 18-106_k141_95883 | Not-determined |
| 18-106_k141_48969 | Not-determined |
| 18-106_k141_66476 | Low-quality |
| 18-106_k141_130950 | Not-determined |
| 18-106_k141_48979 | Low-quality |
| 18-106_k141_2350 | Not-determined |
| 18-106_k141_130952 | Not-determined |
| 18-106_k141_24957 | Not-determined |
| 18-106_k141_2353 | Not-determined |
| 18-106_k141_60780 | Not-determined |
| 18-106_k141_7562 | Not-determined |
| 18-106_k141_18971 | Not-determined |
| 18-106_k141_60783 | Not-determined |
| 18-106_k141_113105 | Low-quality |
| 18-106_k141_18977 | Not-determined |
| 18-106_k141_13518 | Not-determined |
| 18-106_k141_24966 | Low-quality |
| 18-106_k141_124943 | Not-determined |
| 18-106_k141_43187 | Not-determined |
| 18-106_k141_2369 | Not-determined |
| 18-106_k141_95894 | Not-determined |
| 18-106_k141_13527 | Not-determined |
| 18-106_k141_13530 | Not-determined |
| 18-106_k141_95902 | Not-determined |
| 18-106_k141_136478 | Not-determined |
| 18-106_k141_2376 | Not-determined |
| 18-106_k141_24970 | Not-determined |
| 18-106_k141_95909 | Not-determined |
| 18-106_k141_43209 | Not-determined |
| 18-106_k141_2387 | Not-determined |
| 18-106_k141_130970 | Not-determined |
| 18-106_k141_136491 | Not-determined |
| 18-106_k141_113131 | Not-determined |
| 18-106_k141_7579 | Not-determined |
| 18-106_k141_66521 | Low-quality |
| 18-106_k141_113143 | Not-determined |
| 18-106_k141_124972 | Not-determined |
| 18-106_k141_2392 | Low-quality |
| 18-106_k141_66533 | Not-determined |
| 18-106_k141_124977 | Not-determined |
| 18-106_k141_124979 | Not-determined |
| 18-106_k141_136501 | Low-quality |
| 18-106_k141_13550 | Not-determined |
| 18-106_k141_31370 | Not-determined |
| 18-106_k141_136504 | Not-determined |
| 18-106_k141_49061 | Not-determined |
| 18-106_k141_13569 | Not-determined |
| 18-106_k141_72141 | Not-determined |
| 18-106_k141_13576 | Not-determined |
| 18-106_k141_2408 | Not-determined |
| 18-106_k141_2409 | Not-determined |
| 18-106_k141_136532 | Not-determined |
| 18-106_k141_136535 | Not-determined |
| 18-106_k141_13583 | Not-determined |
| 18-106_k141_136543 | Low-quality |
| 18-106_k141_113154 | Not-determined |
| 18-106_k141_113156 | Not-determined |
| 18-106_k141_136547 | Not-determined |
| 18-106_k141_13590 | Not-determined |
| 18-106_k141_13595 | Not-determined |
| 18-106_k141_2441 | Not-determined |
| 18-106_k141_13598 | Not-determined |
| 18-106_k141_43247 | Not-determined |
| 18-106_k141_136563 | Not-determined |
| 18-106_k141_118772 | Not-determined |
| 18-106_k141_77869 | Not-determined |
| 18-106_k141_43257 | Not-determined |
| 18-106_k141_49089 | Not-determined |
| 18-106_k141_36967 | Low-quality |
| 18-106_k141_7624 | Not-determined |
| 18-106_k141_113171 | Not-determined |
| 18-106_k141_7628 | Low-quality |
| 18-106_k141_43269 | Not-determined |
| 18-106_k141_125036 | Low-quality |
| 18-106_k141_36977 | Low-quality |
| 18-106_k141_60834 | Not-determined |
| 18-106_k141_7631 | Not-determined |
| 18-106_k141_43273 | Not-determined |
| 18-106_k141_7636 | Not-determined |
| 18-106_k141_43276 | Not-determined |
| 18-106_k141_31389 | Not-determined |
| 18-106_k141_72158 | Not-determined |
| 18-106_k141_43280 | Not-determined |
| 18-106_k141_43282 | Not-determined |
| 18-106_k141_118786 | Not-determined |
| 18-106_k141_77892 | Not-determined |
| 18-106_k141_118789 | Not-determined |
| 18-106_k141_125054 | Not-determined |
| 18-106_k141_49106 | Not-determined |
| 18-106_k141_66554 | Not-determined |
| 18-106_k141_43292 | Not-determined |
| 18-106_k141_66560 | Not-determined |
| 18-106_k141_2490 | Not-determined |
| 18-106_k141_49113 | Not-determined |
| 18-106_k141_136588 | Low-quality |
| 18-106_k141_43300 | Not-determined |
| 18-106_k141_54003 | Not-determined |
| 18-106_k141_49122 | Not-determined |
| [illegible] | [illegible] |

| | |
|---|---|
| 18-106_k141_107758 | Not-determined |
| 18-106_k141_136845 | Not-determined |
| 18-106_k141_125308 | Not-determined |
| 18-106_k141_131080 | Not-determined |
| 18-106_k141_66862 | Not-determined |
| 18-106_k141_96157 | Low-quality |
| 18-106_k141_118996 | Not-determined |
| 18-106_k141_2727 | Not-determined |
| 18-106_k141_101199 | Not-determined |
| 18-106_k141_113385 | Low-quality |
| 18-106_k141_19051 | Not-determined |
| 18-106_k141_113387 | Not-determined |
| 18-106_k141_37243 | Not-determined |
| 18-106_k141_2741 | Not-determined |
| 18-106_k141_72330 | Not-determined |
| 18-106_k141_96167 | Not-determined |
| 18-106_k141_96170 | Not-determined |
| 18-106_k141_136852 | Not-determined |
| 18-106_k141_131093 | Not-determined |
| 18-106_k141_72335 | Not-determined |
| 18-106_k141_78106 | Not-determined |
| 18-106_k141_7829 | Not-determined |
| 18-106_k141_125315 | Not-determined |
| 18-106_k141_25163 | Not-determined |
| 18-106_k141_31592 | Not-determined |
| 18-106_k141_31593 | Not-determined |
| 18-106_k141_37272 | Low-quality |
| 18-106_k141_107776 | Low-quality |
| 18-106_k141_31606 | Not-determined |
| 18-106_k141_125328 | Not-determined |
| 18-106_k141_43517 | Not-determined |
| 18-106_k141_72348 | Low-quality |
| 18-106_k141_31611 | Not-determined |
| 18-106_k141_7842 | Low-quality |
| 18-106_k141_136877 | Not-determined |
| 18-106_k141_107788 | Not-determined |
| 18-106_k141_72357 | Not-determined |
| 18-106_k141_61023 | Not-determined |
| 18-106_k141_96206 | Not-determined |
| 18-106_k141_72362 | Not-determined |
| 18-106_k141_72363 | Not-determined |
| 18-106_k141_37304 | Not-determined |
| 18-106_k141_2763 | Low-quality |
| 18-106_k141_72366 | Not-determined |
| 18-106_k141_72370 | Not-determined |
| 18-106_k141_113409 | Not-determined |
| 18-106_k141_31638 | Not-determined |
| 18-106_k141_84063 | Not-determined |
| 18-106_k141_37312 | Not-determined |
| 18-106_k141_136882 | Not-determined |
| 18-106_k141_31645 | Not-determined |
| 18-106_k141_7859 | Not-determined |
| 18-106_k141_25197 | Not-determined |
| 18-106_k141_61042 | Not-determined |
| 18-106_k141_101241 | Not-determined |
| 18-106_k141_66910 | Not-determined |
| 18-106_k141_96213 | Not-determined |
| 18-106_k141_61051 | Not-determined |
| 18-106_k141_2785 | Not-determined |
| 18-106_k141_7876 | Not-determined |
| 18-106_k141_37341 | Not-determined |
| 18-106_k141_107811 | Not-determined |
| 18-106_k141_125368 | Not-determined |
| 18-106_k141_7884 | Not-determined |
| 18-106_k141_107812 | Not-determined |
| 18-106_k141_113438 | Not-determined |
| 18-106_k141_113439 | Not-determined |
| 18-106_k141_125374 | Not-determined |
| 18-106_k141_136922 | Low-quality |
| 18-106_k141_2795 | Not-determined |
| 18-106_k141_96221 | Not-determined |
| 18-106_k141_131110 | Not-determined |
| 18-106_k141_125377 | Not-determined |
| 18-106_k141_119023 | Not-determined |
| 18-106_k141_119024 | Not-determined |
| 18-106_k141_101254 | Not-determined |
| 18-106_k141_13766 | Not-determined |
| 18-106_k141_119027 | Low-quality |
| 18-106_k141_54333 | Not-determined |
| 18-106_k141_119029 | Not-determined |
| 18-106_k141_54337 | Not-determined |
| 18-106_k141_37347 | Not-determined |
| 18-106_k141_125387 | Not-determined |
| 18-106_k141_131111 | Not-determined |
| 18-106_k141_19060 | Not-determined |
| 18-106_k141_107838 | Not-determined |
| 18-106_k141_125390 | Not-determined |
| 18-106_k141_37349 | Low-quality |
| 18-106_k141_7896 | Not-determined |
| 18-106_k141_2826 | Not-determined |
| 18-106_k141_25231 | Not-determined |
| 18-106_k141_19064 | Low-quality |
| 18-106_k141_113453 | Not-determined |
| 18-106_k141_2837 | Not-determined |
| 18-106_k141_113457 | Not-determined |
| 18-106_k141_13784 | Not-determined |
| 18-106_k141_43531 | Not-determined |
| 18-106_k141_2845 | Low-quality |
| 18-106_k141_90199 | Not-determined |
| 18-106_k141_54359 | Not-determined |
| 18-106_k141_78117 | Low-quality |
| 18-106_k141_78119 | Not-determined |
| 18-106_k141_125400 | Not-determined |
| 18-106_k141_96237 | Not-determined |
| 18-106_k141_7911 | Not-determined |
| 18-106_k141_131129 | Not-determined |
| 18-106_k141_7913 | Not-determined |
| 18-106_k141_31711 | Not-determined |
| 18-106_k141_25246 | Not-determined |
| 18-106_k141_19090 | Low-quality |
| 18-106_k141_31714 | Not-determined |
| 18-106_k141_90211 | Not-determined |
| 18-106_k141_90214 | Not-determined |
| 18-106_k141_54373 | Not-determined |
| 18-106_k141_49254 | Low-quality |
| 18-106_k141_72399 | Low-quality |
| 18-106_k141_107867 | Not-determined |
| 18-106_k141_43552 | Not-determined |
| 18-106_k141_43554 | Not-determined |
| 18-106_k141_25249 | Not-determined |
| 18-106_k141_31716 | Not-determined |
| 18-106_k141_31717 | Not-determined |
| 18-106_k141_72411 | Not-determined |
| 18-106_k141_125422 | Not-determined |
| 18-106_k141_107881 | Not-determined |
| 18-106_k141_131147 | Not-determined |
| 18-106_k141_131150 | Not-determined |
| 18-106_k141_66944 | Low-quality |
| 18-106_k141_66946 | Not-determined |
| 18-106_k141_13812 | Low-quality |
| 18-106_k141_49264 | Not-determined |
| 18-106_k141_25258 | Low-quality |
| 18-106_k141_49269 | Not-determined |
| 18-106_k141_90242 | Low-quality |
| 18-106_k141_119051 | Not-determined |
| 18-106_k141_31727 | Not-determined |
| 18-106_k141_131159 | Not-determined |
| 18-106_k141_125448 | Not-determined |
| 18-106_k141_131160 | Not-determined |
| 18-106_k141_25268 | Not-determined |
| 18-106_k141_49277 | Not-determined |
| 18-106_k141_101258 | Low-quality |
| 18-106_k141_78136 | Not-determined |
| 18-106_k141_78139 | Not-determined |
| 18-106_k141_25283 | Not-determined |
| 18-106_k141_7948 | Not-determined |
| 18-106_k141_72446 | Not-determined |
| 18-106_k141_25286 | Low-quality |
| 18-106_k141_125461 | Not-determined |
| 18-106_k141_54387 | Not-determined |
| 18-106_k141_37404 | Not-determined |
| 18-106_k141_37406 | Not-determined |
| 18-106_k141_2874 | Not-determined |
| 18-106_k141_25293 | Not-determined |
| 18-106_k141_72459 | Not-determined |
| 18-106_k141_49292 | Not-determined |
| 18-106_k141_54403 | Not-determined |
| 18-106_k141_49298 | Not-determined |
| 18-106_k141_61062 | Low-quality |
| 18-106_k141_78152 | Not-determined |
| 18-106_k141_31742 | Low-quality |
| 18-106_k141_37414 | Low-quality |
| 18-106_k141_37415 | Low-quality |
| 18-106_k141_119071 | Not-determined |
| 18-106_k141_31749 | Not-determined |
| 18-106_k141_90265 | Not-determined |
| 18-106_k141_37420 | Not-determined |
| 18-106_k141_49307 | Not-determined |
| 18-106_k141_125478 | Not-determined |
| 18-106_k141_31752 | Not-determined |
| 18-106_k141_78158 | Not-determined |
| 18-106_k141_90274 | Not-determined |
| 18-106_k141_31757 | Not-determined |
| 18-106_k141_37430 | Not-determined |
| 18-106_k141_125488 | Low-quality |
| 18-106_k141_125495 | Not-determined |
| 18-106_k141_78164 | Not-determined |
| 18-106_k141_37439 | Not-determined |
| 18-106_k141_43581 | Low-quality |
| 18-106_k141_13885 | Not-determined |
| 18-106_k141_19148 | Not-determined |
| 18-106_k141_25334 | Not-determined |
| 18-106_k141_90287 | Not-determined |
| 18-106_k141_78172 | Low-quality |
| 18-106_k141_49330 | Not-determined |
| 18-106_k141_78178 | Not-determined |
| 18-106_k141_131196 | Not-determined |
| 18-106_k141_49340 | Not-determined |
| 18-106_k141_78186 | Not-determined |
| 18-106_k141_96274 | Not-determined |
| 18-106_k141_49347 | Not-determined |
| 18-106_k141_13899 | Not-determined |
| 18-106_k141_61078 | Not-determined |
| 18-106_k141_101339 | Not-determined |
| 18-106_k141_90310 | Low-quality |
| 18-106_k141_61080 | Not-determined |
| 18-106_k141_8008 | Low-quality |
| 18-106_k141_61084 | Not-determined |
| 18-106_k141_90322 | Not-determined |
| 18-106_k141_84087 | Not-determined |
| 18-106_k141_8020 | Not-determined |
| 18-106_k141_25354 | Not-determined |
| 18-106_k141_90327 | Not-determined |
| 18-106_k141_84092 | Not-determined |
| 18-106_k141_107908 | Not-determined |
| 18-106_k141_19180 | Not-determined |
| 18-106_k141_84094 | Not-determined |
| 18-106_k141_90332 | Not-determined |
| 18-106_k141_84097 | Not-determined |
| 18-106_k141_125499 | Not-determined |
| 18-106_k141_125503 | Not-determined |
| 18-106_k141_66963 | Not-determined |
| 18-106_k141_125507 | Not-determined |
| 18-106_k141_84099 | Not-determined |
| 18-106_k141_90340 | Not-determined |
| 18-106_k141_49375 | Not-determined |
| 18-106_k141_13922 | Not-determined |
| 18-106_k141_84103 | Not-determined |
| 18-106_k141_49378 | Not-determined |
| 18-106_k141_90345 | Not-determined |
| 18-106_k141_61110 | Not-determined |
| 18-106_k141_8027 | Not-determined |
| 18-106_k141_25385 | Not-determined |
| 18-106_k141_8029 | Not-determined |
| 18-106_k141_49389 | Not-determined |
| 18-106_k141_61118 | Not-determined |
| 18-106_k141_37497 | Not-determined |
| 18-106_k141_54410 | Not-determined |
| 18-106_k141_84113 | Not-determined |
| 18-106_k141_25388 | Low-quality |
| 18-106_k141_96313 | Not-determined |
| 18-106_k141_37510 | Not-determined |
| 18-106_k141_37512 | Not-determined |
| 18-106_k141_119107 | Not-determined |
| 18-106_k141_107934 | Not-determined |
| 18-106_k141_90358 | Low-quality |
| 18-106_k141_78201 | Not-determined |
| 18-106_k141_8041 | Not-determined |
| 18-106_k141_54419 | Not-determined |
| 18-106_k141_25396 | Not-determined |
| 18-106_k141_25401 | Not-determined |
| 18-106_k141_101382 | Not-determined |
| 18-106_k141_19224 | Not-determined |
| 18-106_k141_19227 | Not-determined |
| 18-106_k141_107946 | Not-determined |
| 18-106_k141_107948 | Not-determined |
| 18-106_k141_37522 | Not-determined |
| 18-106_k141_96327 | Not-determined |
| 18-106_k141_96328 | Not-determined |
| 18-106_k141_78220 | Not-determined |
| 18-106_k141_2984 | Not-determined |
| 18-106_k141_125531 | Not-determined |
| 18-106_k141_90379 | Not-determined |
| 18-106_k141_8059 | Not-determined |
| 18-106_k141_54431 | Not-determined |
| 18-106_k141_8060 | Not-determined |
| 18-106_k141_96342 | Not-determined |
| 18-106_k141_25420 | Not-determined |
| 18-106_k141_54445 | Not-determined |
| 18-106_k141_61139 | Low-quality |
| 18-106_k141_96354 | Not-determined |
| 18-106_k141_125537 | Not-determined |
| 18-106_k141_61146 | Not-determined |
| 18-106_k141_72500 | Not-determined |
| 18-106_k141_61154 | Not-determined |
| 18-106_k141_90394 | Not-determined |
| 18-106_k141_107977 | Not-determined |
| 18-106_k141_72512 | Not-determined |
| 18-106_k141_19253 | Low-quality |
| 18-106_k141_72519 | Not-determined |
| 18-106_k141_131214 | Not-determined |
| 18-106_k141_8079 | Not-determined |
| 18-106_k141_131217 | Not-determined |
| 18-106_k141_19262 | Low-quality |
| 18-106_k141_25441 | Not-determined |
| 18-106_k141_90407 | Not-determined |
| 18-106_k141_125554 | Low-quality |
| 18-106_k141_131223 | Not-determined |
| 18-106_k141_25443 | Not-determined |
| 18-106_k141_113492 | Low-quality |
| 18-106_k141_84162 | Not-determined |
| 18-106_k141_84163 | Not-determined |
| 18-106_k141_19264 | Not-determined |
| 18-106_k141_84165 | Not-determined |
| 18-106_k141_84167 | Not-determined |
| 18-106_k141_72539 | Not-determined |
| 18-106_k141_72542 | Not-determined |
| 18-106_k141_67028 | Low-quality |
| 18-106_k141_67029 | Not-determined |
| 18-106_k141_72544 | Not-determined |
| 18-106_k141_13968 | Not-determined |
| 18-106_k141_13969 | Not-determined |
| 18-106_k141_84172 | Low-quality |
| 18-106_k141_101421 | Not-determined |
| 18-106_k141_131234 | Not-determined |
| 18-106_k141_13975 | Not-determined |
| 18-106_k141_67044 | Not-determined |
| 18-106_k141_37578 | Not-determined |
| 18-106_k141_84186 | Not-determined |
| 18-106_k141_19285 | Not-determined |
| 18-106_k141_113519 | Not-determined |
| 18-106_k141_3027 | Low-quality |
| 18-106_k141_13979 | Not-determined |
| 18-106_k141_43714 | Not-determined |
| 18-106_k141_19289 | Not-determined |
| 18-106_k141_25462 | Not-determined |
| 18-106_k141_19291 | Not-determined |
| 18-106_k141_84190 | Not-determined |
| 18-106_k141_13994 | Not-determined |
| 18-106_k141_101437 | Not-determined |
| 18-106_k141_113528 | Low-quality |
| 18-106_k141_49452 | Not-determined |
| 18-106_k141_131250 | Not-determined |
| 18-106_k141_19302 | Not-determined |
| 18-106_k141_14002 | Not-determined |
| 18-106_k141_101449 | Not-determined |
| 18-106_k141_8112 | Not-determined |
| 18-106_k141_67046 | Not-determined |
| 18-106_k141_8115 | Not-determined |
| 18-106_k141_37599 | Not-determined |
| 18-106_k141_25482 | Not-determined |
| 18-106_k141_131257 | Not-determined |
| 18-106_k141_43730 | Not-determined |
| 18-106_k141_84211 | Not-determined |
| 18-106_k141_3059 | Not-determined |
| 18-106_k141_78291 | Not-determined |
| 18-106_k141_3060 | Not-determined |
| 18-106_k141_8122 | Not-determined |
| 18-106_k141_113545 | Not-determined |
| 18-106_k141_8123 | Not-determined |
| 18-106_k141_43734 | Not-determined |
| 18-106_k141_78294 | Low-quality |
| 18-106_k141_19307 | Not-determined |
| 18-106_k141_25486 | Not-determined |
| 18-106_k141_8130 | Not-determined |
| 18-106_k141_67062 | Not-determined |
| 18-106_k141_101464 | Not-determined |
| 18-106_k141_108023 | Not-determined |
| 18-106_k141_72575 | Not-determined |
| 18-106_k141_14018 | Not-determined |
| 18-106_k141_113559 | Not-determined |
| 18-106_k141_90448 | Not-determined |
| 18-106_k141_19318 | Not-determined |
| 18-106_k141_8144 | Not-determined |
| 18-106_k141_8150 | Not-determined |
| 18-106_k141_8152 | Not-determined |
| 18-106_k141_67069 | Not-determined |
| 18-106_k141_78314 | Not-determined |
| 18-106_k141_14032 | Not-determined |
| 18-106_k141_19326 | Low-quality |
| 18-106_k141_19327 | Low-quality |
| 18-106_k141_54453 | Not-determined |
| 18-106_k141_96397 | Not-determined |
| 18-106_k141_14035 | Not-determined |
| 18-106_k141_101491 | Not-determined |
| 18-106_k141_14037 | Not-determined |
| 18-106_k141_119147 | Not-determined |
| 18-106_k141_72584 | Not-determined |
| 18-106_k141_19333 | Not-determined |
| 18-106_k141_67073 | Not-determined |
| 18-106_k141_108049 | Not-determined |
| 18-106_k141_19334 | Not-determined |
| 18-106_k141_19335 | Not-determined |
| 18-106_k141_78327 | Not-determined |
| 18-106_k141_19336 | Not-determined |
| 18-106_k141_72588 | Not-determined |
| 18-106_k141_49478 | Not-determined |
| 18-106_k141_90476 | Not-determined |
| 18-106_k141_19338 | Not-determined |
| 18-106_k141_119156 | Not-determined |
| 18-106_k141_37657 | Not-determined |
| 18-106_k141_113573 | Low-quality |
| 18-106_k141_125577 | Not-determined |
| 18-106_k141_101507 | Not-determined |
| 18-106_k141_67091 | Not-determined |
| 18-106_k141_96410 | Low-quality |
| 18-106_k141_67093 | Low-quality |
| 18-106_k141_119163 | Not-determined |
| 18-106_k141_67102 | Not-determined |
| 18-106_k141_119170 | Not-determined |
| 18-106_k141_96419 | Not-determined |
| 18-106_k141_67106 | Not-determined |
| 18-106_k141_96421 | Not-determined |
| 18-106_k141_31773 | Not-determined |
| 18-106_k141_25521 | Low-quality |
| 18-106_k141_84258 | Not-determined |
| 18-106_k141_67113 | Not-determined |
| 18-106_k141_3084 | Not-determined |
| 18-106_k141_72606 | Not-determined |
| 18-106_k141_49507 | Not-determined |
| 18-106_k141_78354 | Not-determined |
| 18-106_k141_43790 | Not-determined |
| 18-106_k141_19363 | Not-determined |
| 18-106_k141_49515 | Not-determined |
| 18-106_k141_90519 | Not-determined |
| 18-106_k141_67122 | Not-determined |
| 18-106_k141_125600 | Not-determined |
| 18-106_k141_67130 | Low-quality |
| 18-106_k141_125607 | Not-determined |
| 18-106_k141_119185 | Not-determined |
| 18-106_k141_43795 | Not-determined |
| 18-106_k141_84283 | Not-determined |
| 18-106_k141_3103 | Not-determined |
| 18-106_k141_67138 | Low-quality |
| 18-106_k141_31795 | Not-determined |
| 18-106_k141_54493 | Low-quality |
| 18-106_k141_125617 | Low-quality |
| 18-106_k141_19364 | Not-determined |
| 18-106_k141_113601 | Not-determined |
| 18-106_k141_43811 | Not-determined |
| 18-106_k141_3112 | Not-determined |
| 18-106_k141_67145 | Not-determined |
| 18-106_k141_108081 | Not-determined |
| 18-106_k141_108082 | Not-determined |
| 18-106_k141_101547 | Not-determined |
| 18-106_k141_96440 | Not-determined |
| 18-106_k141_43825 | Not-determined |
| 18-106_k141_14074 | Not-determined |
| 18-106_k141_3124 | Not-determined |
| 18-106_k141_90528 | Not-determined |
| 18-106_k141_90529 | Not-determined |
| 18-106_k141_19375 | Not-determined |
| 18-106_k141_72622 | Not-determined |
| 18-106_k141_119194 | Not-determined |
| 18-106_k141_25573 | Not-determined |
| 18-106_k141_14079 | Not-determined |
| 18-106_k141_90533 | Low-quality |
| 18-106_k141_113617 | Not-determined |
| 18-106_k141_43839 | Not-determined |
| 18-106_k141_125648 | Not-determined |
| 18-106_k141_125651 | Not-determined |
| 18-106_k141_96443 | Not-determined |
| 18-106_k141_113624 | Not-determined |
| 18-106_k141_119208 | Not-determined |
| 18-106_k141_14088 | Not-determined |
| 18-106_k141_19394 | Not-determined |
| 18-106_k141_3136 | Not-determined |
| 18-106_k141_49524 | Not-determined |
| 18-106_k141_108093 | Not-determined |
| 18-106_k141_96452 | Not-determined |
| 18-106_k141_54517 | Not-determined |
| 18-106_k141_108099 | Not-determined |
| 18-106_k141_43851 | Not-determined |
| 18-106_k141_31849 | Not-determined |
| 18-106_k141_14093 | Low-quality |
| 18-106_k141_3151 | Not-determined |
| 18-106_k141_67165 | Not-determined |
| 18-106_k141_61202 | Not-determined |
| 18-106_k141_125674 | Not-determined |
| 18-106_k141_78390 | Not-determined |
| 18-106_k141_25613 | Not-determined |
| 18-106_k141_54527 | Not-determined |
| 18-106_k141_49545 | Not-determined |
| 18-106_k141_37729 | Not-determined |
| 18-106_k141_113638 | Not-determined |
| 18-106_k141_43854 | Not-determined |
| 18-106_k141_78413 | Not-determined |
| 18-106_k141_96474 | Not-determined |
| 18-106_k141_108125 | Not-determined |
| 18-106_k141_78414 | Not-determined |
| 18-106_k141_49564 | Low-quality |
| 18-106_k141_25636 | Not-determined |
| 18-106_k141_3192 | Not-determined |
| 18-106_k141_125691 | Not-determined |
| 18-106_k141_49567 | Low-quality |
| 18-106_k141_54534 | Not-determined |
| 18-106_k141_54541 | Not-determined |
| 18-106_k141_119251 | Not-determined |
| 18-106_k141_54545 | Not-determined |
| 18-106_k141_119254 | Not-determined |
| 18-106_k141_119258 | Not-determined |
| 18-106_k141_96492 | Not-determined |
| 18-106_k141_84323 | Not-determined |
| 18-106_k141_108142 | Not-determined |
| 18-106_k141_67200 | Not-determined |
| 18-106_k141_19407 | Low-quality |
| 18-106_k141_119262 | Not-determined |
| 18-106_k141_37759 | Not-determined |
| 18-106_k141_72667 | Low-quality |
| 18-106_k141_3222 | Low-quality |
| 18-106_k141_61240 | Not-determined |
| 18-106_k141_78426 | Not-determined |
| 18-106_k141_19426 | Not-determined |
| 18-106_k141_108151 | Low-quality |
| 18-106_k141_67214 | Not-determined |
| 18-106_k141_31901 | Not-determined |
| 18-106_k141_78431 | Not-determined |
| 18-106_k141_54574 | Not-determined |
| 18-106_k141_25673 | Not-determined |
| 18-106_k141_125726 | Not-determined |
| 18-106_k141_19431 | Not-determined |
| 18-106_k141_90577 | Not-determined |
| 18-106_k141_37775 | Low-quality |
| 18-106_k141_37776 | Not-determined |
| 18-106_k141_3237 | Low-quality |
| 18-106_k141_113667 | Not-determined |
| 18-106_k141_119291 | Not-determined |
| 18-106_k141_96517 | Not-determined |
| 18-106_k141_96518 | Not-determined |
| 18-106_k141_101565 | Not-determined |
| 18-106_k141_61259 | Not-determined |
| 18-106_k141_125744 | Not-determined |
| 18-106_k141_37790 | Not-determined |
| 18-106_k141_19452 | Not-determined |
| 18-106_k141_49601 | Not-determined |
| 18-106_k141_37795 | Not-determined |
| 18-106_k141_84368 | Not-determined |
| 18-106_k141_90589 | Not-determined |
| 18-106_k141_90590 | Not-determined |
| 18-106_k141_37801 | Not-determined |
| 18-106_k141_119307 | Not-determined |
| 18-106_k141_125757 | Not-determined |
| 18-106_k141_61273 | Not-determined |
| 18-106_k141_125760 | Not-determined |
| 18-106_k141_37806 | Not-determined |
| 18-106_k141_119316 | Not-determined |
| 18-106_k141_125769 | Not-determined |
| 18-106_k141_119319 | Not-determined |
| 18-106_k141_119324 | Low-quality |
| 18-106_k141_96534 | Not-determined |
| 18-106_k141_25690 | Not-determined |
| 18-106_k141_37815 | Not-determined |
| 18-106_k141_131288 | Not-determined |
| 18-106_k141_113695 | Not-determined |
| 18-106_k141_49635 | Not-determined |
| 18-102-final-viral-combined_k141_23562\|\|full | High-quality |
| 18-106_k141_119337 | Not-determined |
| 18-106_k141_54584 | Not-determined |
| 18-106_k141_131297 | Not-determined |
| 18-106_k141_78463 | Not-determined |
| 18-106_k141_131302 | Low-quality |
| 18-106_k141_84405 | Not-determined |
| 18-106_k141_131317 | Not-determined |
| 18-106_k141_67262 | Not-determined |
| 18-106_k141_25695 | Low-quality |
| 18-106_k141_125814 | Low-quality |
| 18-106_k141_67277 | Not-determined |
| 18-106_k141_3287 | Not-determined |
| 18-106_k141_125820 | Not-determined |
| 18-106_k141_3289 | Not-determined |
| 18-106_k141_19492 | Not-determined |
| 18-106_k141_113720 | Not-determined |
| 18-106_k141_67283 | Not-determined |
| 18-106_k141_3291 | Not-determined |
| 18-106_k141_54606 | Not-determined |
| 18-106_k141_119361 | Not-determined |
| 18-106_k141_67285 | Not-determined |
| 18-106_k141_96563 | Not-determined |
| 18-106_k141_3301 | Not-determined |
| 18-106_k141_78470 | Not-determined |
| 18-106_k141_131329 | Not-determined |
| 18-106_k141_54616 | Not-determined |
| 18-106_k141_43894 | Low-quality |
| 18-106_k141_78471 | Not-determined |
| 18-106_k141_108183 | Not-determined |
| 18-106_k141_101603 | Not-determined |
| 18-106_k141_84434 | Not-determined |
| 18-106_k141_90668 | Low-quality |
| 18-106_k141_131337 | Not-determined |
| 18-106_k141_90669 | Not-determined |
| 18-106_k141_19511 | Not-determined |
| 18-106_k141_37839 | Low-quality |
| 18-106_k141_67304 | Not-determined |
| 18-106_k141_67305 | Not-determined |
| 18-106_k141_78480 | Not-determined |
| 18-106_k141_3323 | Not-determined |
| 18-106_k141_119373 | Low-quality |
| 18-106_k141_43908 | Not-determined |
| 18-106_k141_108191 | Not-determined |
| 18-106_k141_43914 | Not-determined |
| 18-106_k141_43915 | Not-determined |
| 18-106_k141_49672 | Not-determined |
| 18-106_k141_101620 | Not-determined |
| 18-106_k141_67320 | Not-determined |
| 18-106_k141_8198 | Not-determined |
| 18-106_k141_54639 | Not-determined |
| 18-106_k141_37851 | Not-determined |
| 18-106_k141_8202 | Not-determined |
| 18-106_k141_90683 | Not-determined |
| 18-106_k141_90685 | Not-determined |
| 18-106_k141_125848 | Not-determined |
| 18-106_k141_49683 | Not-determined |
| 18-106_k141_67329 | Not-determined |
| 18-106_k141_131354 | Not-determined |
| 18-106_k141_3334 | Not-determined |
| 18-106_k141_125849 | Not-determined |
| 18-106_k141_90688 | Not-determined |
| 18-106_k141_101634 | Low-quality |
| 18-106_k141_125857 | Not-determined |

| | |
|---|---|
| 18-106_k141_101635 | Not-determined |
| 18-106_k141_96615 | Not-determined |
| 18-106_k141_137009 | Not-determined |
| 18-106_k141_131368 | Not-determined |
| 18-106_k141_90698 | Not-determined |
| 18-106_k141_131375 | Low-quality |
| 18-106_k141_37876 | Not-determined |
| 18-106_k141_119399 | Not-determined |
| 18-106_k141_49696 | Not-determined |
| 18-106_k141_37878 | Not-determined |
| 18-106_k141_78522 | Not-determined |
| 18-106_k141_67355 | Not-determined |
| 18-106_k141_3356 | Not-determined |
| 18-106_k141_78531 | Not-determined |
| 18-106_k141_49713 | Not-determined |
| 18-106_k141_61348 | Not-determined |
| 18-106_k141_19555 | Not-determined |
| 18-106_k141_78540 | Not-determined |
| 18-106_k141_78542 | Not-determined |
| 18-106_k141_19561 | Not-determined |
| 18-106_k141_31957 | Not-determined |
| 18-106_k141_37898 | Not-determined |
| 18-106_k141_125863 | Not-determined |
| 18-106_k141_49720 | Not-determined |
| 18-106_k141_125868 | Not-determined |
| 18-106_k141_37901 | Not-determined |
| 18-106_k141_37902 | Not-determined |
| 18-106_k141_49726 | Not-determined |
| 18-106_k141_72756 | Not-determined |
| 18-106_k141_43961 | Not-determined |
| 18-106_k141_43963 | Low-quality |
| 18-106_k141_78546 | Not-determined |
| 18-106_k141_72757 | Not-determined |
| 18-106_k141_131400 | Not-determined |
| 18-106_k141_72761 | Not-determined |
| 18-106_k141_37911 | Not-determined |
| 18-106_k141_131403 | Not-determined |
| 18-106_k141_78550 | Not-determined |
| 18-106_k141_72770 | Not-determined |
| 18-106_k141_108231 | Not-determined |
| 18-106_k141_61355 | Not-determined |
| 18-106_k141_61357 | Not-determined |
| 18-106_k141_61359 | Not-determined |
| 18-106_k141_61361 | Not-determined |
| 18-106_k141_19589 | Not-determined |
| 18-106_k141_78562 | Not-determined |
| 18-106_k141_72778 | Not-determined |
| 18-106_k141_108239 | Not-determined |
| 18-106_k141_14144 | Not-determined |
| 18-106_k141_108242 | Not-determined |
| 18-106_k141_119444 | Not-determined |
| 18-106_k141_49750 | Not-determined |
| 18-106_k141_67373 | Not-determined |
| 18-106_k141_101684 | Low-quality |
| 18-106_k141_19601 | Low-quality |
| 18-106_k141_90710 | Not-determined |
| 18-106_k141_108251 | Not-determined |
| 18-106_k141_78584 | Not-determined |
| 18-106_k141_101688 | Not-determined |
| 18-106_k141_43986 | Not-determined |
| 18-106_k141_108256 | Not-determined |
| 18-106_k141_125888 | Not-determined |
| 18-106_k141_25808 | Not-determined |
| 18-106_k141_14153 | Not-determined |
| 18-106_k141_14156 | Not-determined |
| 18-106_k141_113772 | Not-determined |
| 18-106_k141_101706 | Not-determined |
| 18-106_k141_61376 | Not-determined |
| 18-106_k141_25809 | Low-quality |
| 18-106_k141_61386 | Not-determined |
| 18-106_k141_96663 | Not-determined |
| 18-106_k141_96665 | Not-determined |
| 18-106_k141_25811 | Not-determined |
| 18-106_k141_119461 | Not-determined |
| 18-106_k141_72798 | Not-determined |
| 18-106_k141_14164 | Not-determined |
| 18-106_k141_37959 | Not-determined |
| 18-106_k141_72807 | Not-determined |
| 18-106_k141_25824 | Not-determined |
| 18-106_k141_44016 | Not-determined |
| 18-106_k141_25830 | Not-determined |
| 18-106_k141_49763 | Low-quality |
| 18-106_k141_108275 | Not-determined |
| 18-106_k141_84490 | Not-determined |
| 18-106_k141_90756 | Low-quality |
| 18-106_k141_96696 | Not-determined |
| 18-106_k141_131484 | Not-determined |
| 18-106_k141_137087 | Not-determined |
| 18-106_k141_137088 | Not-determined |
| 18-106_k141_113789 | Not-determined |
| 18-106_k141_96704 | Low-quality |
| 18-106_k141_14169 | Not-determined |
| 18-106_k141_113793 | Not-determined |
| 18-106_k141_72830 | Not-determined |
| 18-106_k141_54709 | Not-determined |
| 18-106_k141_49778 | Low-quality |
| 18-106_k141_54713 | Not-determined |
| 18-106_k141_96713 | Not-determined |
| 18-106_k141_101716 | Not-determined |
| 18-106_k141_49789 | Not-determined |
| 18-106_k141_137102 | Not-determined |
| 18-106_k141_101722 | Not-determined |
| 18-106_k141_67403 | Not-determined |
| 18-106_k141_84515 | Not-determined |
| 18-106_k141_67404 | Not-determined |
| 18-106_k141_72845 | Not-determined |
| 18-106_k141_119473 | Not-determined |
| 18-106_k141_49805 | Not-determined |
| 18-106_k141_49808 | Not-determined |
| 18-106_k141_19612 | Not-determined |
| 18-106_k141_90776 | Not-determined |
| 18-106_k141_54733 | Not-determined |
| 18-106_k141_119479 | Not-determined |
| 18-106_k141_3418 | Not-determined |
| 18-106_k141_44053 | Not-determined |
| 18-106_k141_72853 | Not-determined |
| 18-106_k141_119486 | Not-determined |
| 18-106_k141_137123 | Not-determined |
| 18-106_k141_54745 | Not-determined |
| 18-106_k141_49820 | Not-determined |
| 18-106_k141_37984 | Not-determined |
| 18-106_k141_49821 | Not-determined |
| 18-106_k141_137128 | Not-determined |
| 18-106_k141_137129 | Not-determined |
| 18-106_k141_67424 | Not-determined |
| 18-106_k141_72864 | Not-determined |
| 18-106_k141_3431 | Not-determined |
| 18-106_k141_14187 | Not-determined |
| 18-106_k141_3432 | Not-determined |
| 18-106_k141_84522 | Not-determined |
| 18-106_k141_67430 | Not-determined |
| 18-106_k141_96738 | Not-determined |
| 18-106_k141_90790 | Not-determined |
| 18-106_k141_119493 | Not-determined |
| 18-106_k141_3446 | Not-determined |
| 18-106_k141_137147 | Not-determined |
| 18-106_k141_72883 | Not-determined |
| 18-106_k141_137152 | Low-quality |
| 18-106_k141_14213 | Not-determined |
| 18-106_k141_84537 | Not-determined |
| 18-106_k141_14216 | Not-determined |
| 18-106_k141_3449 | Not-determined |
| 18-106_k141_137156 | Not-determined |
| 18-106_k141_137157 | Not-determined |
| 18-106_k141_84539 | Not-determined |
| 18-106_k141_78624 | Not-determined |
| 18-106_k141_96757 | Not-determined |
| 18-106_k141_119505 | Not-determined |
| 18-106_k141_84546 | Not-determined |
| 18-106_k141_49850 | Not-determined |
| 18-106_k141_84549 | Not-determined |
| 18-106_k141_3452 | Not-determined |
| 18-106_k141_78634 | Not-determined |
| 18-106_k141_8234 | Not-determined |
| 18-106_k141_72918 | Not-determined |
| 18-106_k141_38020 | Not-determined |
| 18-106_k141_14235 | Not-determined |
| 18-106_k141_54779 | Low-quality |
| 18-106_k141_113850 | Not-determined |
| 18-106_k141_101748 | Not-determined |
| 18-106_k141_61486 | Not-determined |
| 18-106_k141_101754 | Not-determined |
| 18-106_k141_32069 | Low-quality |
| 18-106_k141_90826 | Not-determined |
| 18-106_k141_54789 | Not-determined |
| 18-106_k141_90827 | Not-determined |
| 18-106_k141_137179 | Not-determined |
| 18-106_k141_49880 | Not-determined |
| 18-106_k141_96770 | Not-determined |
| 18-106_k141_54795 | Low-quality |
| 18-106_k141_131550 | Not-determined |
| 18-106_k141_54796 | Not-determined |
| 18-106_k141_113859 | Not-determined |
| 18-106_k141_8263 | Not-determined |
| 18-106_k141_78651 | Not-determined |
| 18-106_k141_137190 | Not-determined |
| 18-106_k141_78654 | Not-determined |
| 18-106_k141_96780 | Not-determined |
| 18-106_k141_54808 | Not-determined |
| 18-106_k141_137192 | Not-determined |
| 18-106_k141_119518 | Not-determined |
| 18-106_k141_119519 | Not-determined |
| 18-106_k141_8273 | Not-determined |
| 18-106_k141_54814 | Not-determined |
| 18-106_k141_96788 | Not-determined |
| 18-106_k141_38042 | Not-determined |
| 18-106_k141_96790 | Not-determined |
| 18-106_k141_101758 | Low-quality |
| 18-106_k141_32084 | Not-determined |
| 18-106_k141_96797 | Not-determined |
| 18-106_k141_67487 | Not-determined |
| 18-106_k141_54820 | Not-determined |
| 18-106_k141_32097 | Not-determined |
| 18-106_k141_101767 | Not-determined |
| 18-106_k141_38052 | Not-determined |
| 18-106_k141_90833 | Not-determined |
| 18-106_k141_113872 | Not-determined |
| 18-106_k141_96803 | Not-determined |
| 18-106_k141_54831 | Not-determined |
| 18-106_k141_96808 | Low-quality |
| 18-106_k141_101777 | Not-determined |
| 18-106_k141_101779 | Not-determined |
| 18-106_k141_72982 | Not-determined |
| 18-106_k141_19664 | Not-determined |
| 18-106_k141_19666 | Not-determined |
| 18-106_k141_72985 | Not-determined |
| 18-106_k141_67513 | Not-determined |
| 18-106_k141_38058 | Not-determined |
| 18-106_k141_8287 | Not-determined |
| 18-106_k141_61511 | Not-determined |
| 18-106_k141_61513 | Not-determined |
| 18-106_k141_38064 | Not-determined |
| 18-106_k141_101794 | Not-determined |
| 18-106_k141_84563 | Not-determined |
| 18-106_k141_3479 | Not-determined |
| 18-106_k141_14286 | Low-quality |
| 18-106_k141_119537 | Low-quality |
| 18-106_k141_113899 | Not-determined |
| 18-106_k141_137237 | Not-determined |
| 18-106_k141_137238 | Not-determined |
| 18-106_k141_32107 | Not-determined |
| 18-106_k141_25842 | Not-determined |
| 18-106_k141_113905 | Not-determined |
| 18-106_k141_113909 | Not-determined |
| 18-106_k141_78656 | Not-determined |
| 18-106_k141_8308 | Not-determined |
| 18-106_k141_3512 | Not-determined |
| 18-106_k141_90874 | Low-quality |
| 18-106_k141_8309 | Not-determined |
| 18-106_k141_137251 | Not-determined |
| 18-106_k141_119569 | Not-determined |
| 18-106_k141_38090 | Low-quality |
| 18-106_k141_14304 | Low-quality |
| 18-106_k141_96852 | Not-determined |
| 18-106_k141_84600 | Not-determined |
| 18-106_k141_78663 | Not-determined |
| 18-106_k141_73032 | Not-determined |
| 18-106_k141_73033 | Not-determined |
| 18-106_k141_84602 | Not-determined |
| 18-106_k141_38097 | Not-determined |
| 18-106_k141_19694 | Not-determined |
| 18-106_k141_73035 | Not-determined |
| 18-106_k141_96858 | Not-determined |
| 18-106_k141_137264 | Not-determined |
| 18-106_k141_49919 | Low-quality |
| 18-106_k141_137267 | Not-determined |
| 18-106_k141_113920 | Not-determined |
| 18-106_k141_19703 | Not-determined |
| 18-106_k141_19708 | Not-determined |
| 18-106_k141_54856 | Not-determined |
| 18-106_k141_96866 | Not-determined |
| 18-106_k141_90896 | Not-determined |
| 18-106_k141_54858 | Low-quality |
| 18-106_k141_90901 | Not-determined |
| 18-106_k141_8347 | Not-determined |
| 18-106_k141_113939 | Low-quality |
| 18-106_k141_90908 | Low-quality |
| 18-106_k141_90913 | Not-determined |
| 18-106_k141_73058 | Not-determined |
| 18-106_k141_113945 | Not-determined |
| 18-106_k141_19718 | Not-determined |
| 18-106_k141_78684 | Not-determined |
| 18-106_k141_84613 | Not-determined |
| 18-106_k141_90918 | Not-determined |
| 18-106_k141_73059 | Not-determined |
| 18-106_k141_113948 | Not-determined |
| 18-106_k141_90923 | Not-determined |
| 18-106_k141_131561 | Not-determined |
| 18-106_k141_54870 | Not-determined |
| 18-106_k141_101839 | Not-determined |
| 18-106_k141_101840 | Not-determined |
| 18-106_k141_90927 | Not-determined |
| 18-106_k141_3532 | Not-determined |
| 18-106_k141_67573 | Not-determined |
| 18-106_k141_19721 | Not-determined |
| 18-106_k141_3534 | Not-determined |
| 18-106_k141_90934 | Not-determined |
| 18-106_k141_3537 | Not-determined |
| 18-106_k141_78686 | Not-determined |
| 18-106_k141_54887 | Low-quality |
| 18-106_k141_84631 | Not-determined |
| 18-106_k141_101850 | Not-determined |
| 18-106_k141_131577 | Not-determined |
| 18-106_k141_108310 | Low-quality |
| 18-106_k141_73077 | Not-determined |
| 18-106_k141_101853 | Not-determined |
| 18-106_k141_61557 | Not-determined |
| 18-106_k141_44181 | Not-determined |
| 18-106_k141_44182 | Not-determined |
| 18-106_k141_54906 | Not-determined |
| 18-106_k141_54911 | Not-determined |
| 18-106_k141_113972 | Not-determined |
| 18-106_k141_90944 | Not-determined |
| 18-106_k141_44193 | Not-determined |
| 18-106_k141_131601 | Not-determined |
| 18-106_k141_14353 | Not-determined |
| 18-106_k141_67584 | Not-determined |
| 18-106_k141_73093 | Not-determined |
| 18-106_k141_84639 | Not-determined |
| 18-106_k141_3556 | Not-determined |
| 18-106_k141_90950 | Not-determined |
| 18-106_k141_73096 | Not-determined |
| 18-106_k141_84643 | Low-quality |
| 18-106_k141_49965 | Not-determined |
| 18-106_k141_38144 | Not-determined |
| 18-106_k141_44203 | Not-determined |
| 18-106_k141_67595 | Not-determined |
| 18-106_k141_14354 | Not-determined |
| 18-106_k141_32138 | Not-determined |
| 18-106_k141_19753 | Not-determined |
| 18-106_k141_131619 | Not-determined |
| 18-106_k141_108336 | Not-determined |
| 18-106_k141_44212 | Not-determined |
| 18-106_k141_96920 | Not-determined |
| 18-106_k141_19762 | Low-quality |
| 18-106_k141_101887 | Not-determined |
| 18-106_k141_96926 | Not-determined |
| 18-106_k141_73107 | Not-determined |
| 18-106_k141_44222 | Not-determined |
| 18-106_k141_67614 | Not-determined |
| 18-106_k141_101889 | Not-determined |
| 18-106_k141_25866 | Not-determined |
| 18-106_k141_101898 | Not-determined |
| 18-106_k141_14372 | Low-quality |
| 18-106_k141_19785 | Not-determined |
| 18-106_k141_14378 | Not-determined |
| 18-106_k141_101906 | Not-determined |
| 18-106_k141_19793 | Not-determined |
| 18-106_k141_137297 | Not-determined |
| 18-106_k141_54937 | Not-determined |
| 18-106_k141_19803 | Not-determined |
| 18-106_k141_67631 | Low-quality |
| 18-106_k141_113992 | Not-determined |
| 18-106_k141_38177 | Not-determined |
| 18-106_k141_84668 | Not-determined |
| 18-106_k141_84669 | Not-determined |
| 18-106_k141_61629 | Not-determined |
| 18-106_k141_78746 | Not-determined |
| 18-106_k141_25879 | Not-determined |
| 18-106_k141_84672 | Not-determined |
| 18-106_k141_108386 | Not-determined |
| 18-106_k141_131668 | Not-determined |
| 18-106_k141_78754 | Not-determined |
| 18-106_k141_119577 | Not-determined |
| 18-106_k141_101929 | Not-determined |
| 18-106_k141_73147 | Not-determined |
| 18-106_k141_3591 | Not-determined |
| 18-106_k141_108394 | Not-determined |
| 18-106_k141_108396 | Not-determined |
| 18-106_k141_67651 | Low-quality |
| 18-106_k141_101935 | Not-determined |
| 18-106_k141_38198 | Not-determined |
| 18-106_k141_3600 | Not-determined |
| 18-106_k141_44254 | Not-determined |
| 18-106_k141_101942 | Not-determined |
| 18-106_k141_38204 | Not-determined |
| 18-106_k141_44257 | Not-determined |
| 18-106_k141_67657 | Not-determined |
| 18-106_k141_38206 | Low-quality |
| 18-106_k141_73152 | Not-determined |
| 18-106_k141_131685 | Low-quality |
| 18-106_k141_50005 | Not-determined |
| 18-106_k141_96949 | Not-determined |
| 18-106_k141_108412 | Not-determined |
| 18-106_k141_119589 | Not-determined |
| 18-106_k141_73158 | Not-determined |
| 18-106_k141_32209 | Not-determined |
| 18-106_k141_50012 | Not-determined |
| 18-106_k141_54967 | Low-quality |
| 18-106_k141_50013 | Not-determined |
| 18-106_k141_19823 | Not-determined |
| 18-106_k141_73161 | Not-determined |
| 18-106_k141_84703 | Not-determined |
| 18-106_k141_90991 | Not-determined |
| 18-106_k141_3615 | Not-determined |
| 18-106_k141_38223 | Not-determined |
| 18-106_k141_96967 | Not-determined |
| 18-106_k141_32218 | Not-determined |
| 18-106_k141_91007 | Not-determined |
| 18-106_k141_91009 | Not-determined |
| 18-106_k141_101966 | Not-determined |
| 18-106_k141_73174 | Not-determined |
| 18-106_k141_67681 | Not-determined |
| 18-106_k141_14392 | Not-determined |
| 18-106_k141_19832 | Not-determined |
| 18-106_k141_44285 | Not-determined |
| 18-106_k141_54982 | Not-determined |
| 18-106_k141_19837 | Not-determined |
| 18-106_k141_84720 | Not-determined |
| 18-106_k141_84722 | Not-determined |
| 18-106_k141_137345 | Not-determined |
| 18-106_k141_38229 | Not-determined |
| 18-106_k141_91031 | Not-determined |
| 18-106_k141_78778 | Not-determined |
| 18-106_k141_131715 | Not-determined |
| 18-106_k141_91034 | Not-determined |
| 18-106_k141_96984 | Not-determined |
| 18-106_k141_101978 | Not-determined |
| 18-106_k141_61635 | Not-determined |
| 18-106_k141_44294 | Not-determined |
| 18-106_k141_38236 | Not-determined |
| 18-106_k141_137354 | Not-determined |
| 18-106_k141_50033 | Not-determined |
| 18-106_k141_78781 | Not-determined |
| 18-106_k141_84733 | Low-quality |
| 18-106_k141_67692 | Not-determined |
| 18-106_k141_14416 | Not-determined |
| 18-106_k141_14417 | Not-determined |
| 18-106_k141_131739 | Not-determined |
| 18-106_k141_137369 | Not-determined |
| 18-106_k141_32245 | Not-determined |
| 18-106_k141_101998 | Not-determined |
| 18-106_k141_131743 | Low-quality |
| 18-106_k141_91038 | Not-determined |
| 18-106_k141_137373 | Not-determined |
| 18-106_k141_19869 | Not-determined |
| 18-106_k141_50053 | Not-determined |
| 18-106_k141_137375 | Not-determined |
| 18-106_k141_108449 | Not-determined |
| 18-106_k141_19873 | Not-determined |
| 18-106_k141_44298 | Not-determined |
| 18-106_k141_61648 | Not-determined |
| 18-106_k141_3633 | Not-determined |
| 18-106_k141_137381 | Low-quality |
| 18-106_k141_3636 | Not-determined |
| 18-106_k141_32259 | Not-determined |
| 18-106_k141_67704 | Not-determined |
| 18-106_k141_108455 | Low-quality |
| 18-106_k141_19884 | Not-determined |
| 18-106_k141_32261 | Not-determined |
| 18-106_k141_67711 | Not-determined |
| 18-106_k141_119642 | Not-determined |
| 18-106_k141_97007 | Not-determined |
| 18-106_k141_119643 | Not-determined |
| 18-106_k141_125936 | Not-determined |
| 18-106_k141_3655 | Not-determined |
| 18-106_k141_38266 | Not-determined |
| 18-106_k141_131769 | Not-determined |
| 18-106_k141_119646 | Not-determined |
| 18-106_k141_108472 | Not-determined |
| 18-106_k141_78810 | Not-determined |
| 18-106_k141_114018 | Low-quality |
| 18-106_k141_61672 | Not-determined |
| 18-106_k141_50088 | Not-determined |
| 18-106_k141_61677 | Low-quality |
| 18-106_k141_102035 | Not-determined |
| 18-106_k141_84753 | Not-determined |
| 18-106_k141_125954 | Not-determined |
| 18-106_k141_61683 | Not-determined |
| 18-106_k141_97047 | Not-determined |
| 18-106_k141_97048 | Not-determined |
| 18-106_k141_137416 | Not-determined |
| 18-106_k141_131797 | Not-determined |
| 18-106_k141_114036 | Not-determined |
| 18-106_k141_114037 | Not-determined |
| 18-106_k141_84770 | Not-determined |
| 18-106_k141_131799 | Not-determined |
| 18-106_k141_102046 | Not-determined |
| 18-106_k141_125970 | Not-determined |
| 18-106_k141_114045 | Not-determined |
| 18-106_k141_44305 | Not-determined |
| 18-106_k141_32275 | Not-determined |
| 18-106_k141_38300 | Not-determined |
| 18-106_k141_19896 | Not-determined |
| 18-106_k141_61697 | Not-determined |
| 18-106_k141_114052 | Not-determined |
| 18-106_k141_114053 | Not-determined |
| 18-106_k141_131812 | Not-determined |
| 18-106_k141_137430 | Low-quality |
| 18-106_k141_114055 | Not-determined |
| 18-106_k141_131817 | Not-determined |
| 18-106_k141_73241 | Not-determined |
| 18-106_k141_97072 | Not-determined |
| 18-106_k141_84782 | Not-determined |
| 18-106_k141_38305 | Low-quality |
| 18-106_k141_38306 | Low-quality |
| 18-106_k141_97074 | Not-determined |
| 18-106_k141_44314 | Not-determined |
| 18-106_k141_67763 | Not-determined |
| 18-106_k141_131828 | Low-quality |
| 18-106_k141_19917 | Not-determined |
| 18-106_k141_38321 | Not-determined |
| 18-106_k141_19922 | Not-determined |
| 18-106_k141_61724 | Not-determined |
| 18-106_k141_102074 | Not-determined |
| 18-106_k141_3704 | Not-determined |
| 18-106_k141_44326 | Not-determined |
| 18-106_k141_3708 | Low-quality |
| 18-106_k141_44330 | Not-determined |
| 18-106_k141_97104 | Not-determined |
| 18-106_k141_125988 | Not-determined |
| 18-106_k141_131850 | Not-determined |
| 18-106_k141_32313 | Low-quality |
| 18-106_k141_131853 | Not-determined |
| 18-106_k141_32315 | Not-determined |
| 18-106_k141_32319 | Not-determined |
| 18-106_k141_61733 | Not-determined |
| 18-106_k141_55044 | Not-determined |
| 18-106_k141_102083 | Not-determined |
| 18-106_k141_3718 | Not-determined |
| 18-106_k141_19955 | Low-quality |
| 18-106_k141_3722 | Not-determined |
| 18-106_k141_119694 | Not-determined |
| 18-106_k141_126008 | Not-determined |
| 18-106_k141_108518 | Not-determined |
| 18-106_k141_126013 | Not-determined |
| 18-106_k141_44338 | Not-determined |
| 18-106_k141_3732 | Not-determined |
| 18-106_k141_102108 | Not-determined |
| 18-106_k141_119695 | Low-quality |
| 18-106_k141_102113 | Low-quality |
| 18-106_k141_119697 | Low-quality |
| 18-106_k141_108528 | Not-determined |
| 18-106_k141_126018 | Not-determined |
| 18-106_k141_114108 | Not-determined |
| 18-106_k141_131878 | Not-determined |
| 18-106_k141_3746 | Not-determined |
| 18-106_k141_126026 | Not-determined |
| 18-106_k141_119703 | Not-determined |
| 18-106_k141_50108 | Not-determined |
| 18-106_k141_73261 | Not-determined |
| 18-106_k141_32338 | Low-quality |
| 18-106_k141_67824 | Not-determined |
| 18-106_k141_32342 | Not-determined |
| 18-106_k141_114119 | Not-determined |
| 18-106_k141_67827 | Not-determined |
| 18-106_k141_131910 | Not-determined |
| 18-106_k141_67831 | Not-determined |
| 18-106_k141_73282 | Not-determined |
| 18-106_k141_97127 | Not-determined |
| 18-106_k141_67837 | Not-determined |
| 18-106_k141_8434 | Not-determined |
| 18-106_k141_3775 | Not-determined |
| 18-106_k141_8436 | Not-determined |
| 18-106_k141_114138 | Not-determined |
| 18-106_k141_3779 | Low-quality |
| 18-106_k141_14499 | Not-determined |
| 18-106_k141_73293 | Not-determined |
| 18-106_k141_14503 | Not-determined |
| 18-106_k141_14504 | Low-quality |
| 18-106_k141_14508 | Not-determined |
| 18-106_k141_14509 | Not-determined |
| 18-106_k141_32364 | Not-determined |
| 18-106_k141_131927 | Not-determined |
| 18-106_k141_55109 | Not-determined |
| 18-106_k141_3787 | Not-determined |
| 18-106_k141_8458 | Not-determined |
| 18-106_k141_73313 | Not-determined |
| 18-106_k141_126066 | Not-determined |
| 18-106_k141_44365 | Not-determined |
| 18-106_k141_38402 | Not-determined |
| 18-106_k141_38403 | Not-determined |
| 18-106_k141_55116 | Not-determined |
| 18-106_k141_55117 | Not-determined |
| 18-106_k141_3797 | Not-determined |
| 18-106_k141_32384 | Not-determined |
| 18-106_k141_14528 | Not-determined |
| 18-106_k141_73318 | Not-determined |
| 18-106_k141_8473 | Not-determined |
| 18-106_k141_102122 | Not-determined |
| 18-106_k141_119726 | Not-determined |
| 18-106_k141_102129 | Not-determined |
| 18-106_k141_61751 | Not-determined |
| 18-106_k141_114177 | Not-determined |
| 18-106_k141_114178 | Not-determined |
| 18-106_k141_14533 | Not-determined |
| 18-106_k141_50127 | Not-determined |
| 18-106_k141_14536 | Not-determined |
| 18-106_k141_8487 | Not-determined |
| 18-106_k141_8488 | Not-determined |
| 18-106_k141_131939 | Not-determined |
| 18-106_k141_119738 | Low-quality |
| 18-106_k141_126078 | Not-determined |
| 18-106_k141_131944 | Not-determined |
| 18-106_k141_131945 | Not-determined |
| 18-106_k141_8494 | Not-determined |
| 18-106_k141_61763 | Not-determined |
| 18-106_k141_25952 | Not-determined |
| 18-106_k141_8499 | Not-determined |
| 18-106_k141_108576 | Not-determined |
| 18-106_k141_126092 | Not-determined |
| 18-106_k141_108577 | Not-determined |
| 18-106_k141_55142 | Not-determined |
| 18-106_k141_67862 | Not-determined |
| 18-106_k141_73334 | Not-determined |
| 18-106_k141_44373 | Not-determined |
| 18-106_k141_32393 | Not-determined |
| 18-106_k141_8500 | Not-determined |
| 18-106_k141_126104 | Not-determined |

| | |
|---|---|
| 18-106_k141_73343 | Not-determined |
| 18-106_k141_97167 | Not-determined |
| 18-106_k141_119755 | Not-determined |
| 18-106_k141_119757 | Not-determined |
| 18-106_k141_126118 | Not-determined |
| 18-106_k141_119759 | Not-determined |
| 18-106_k141_25970 | Not-determined |
| 18-106_k141_126122 | Not-determined |
| 18-106_k141_102146 | Not-determined |
| 18-106_k141_114207 | Not-determined |
| 18-106_k141_32419 | Not-determined |
| 18-106_k141_3824 | Not-determined |
| 18-106_k141_119766 | Not-determined |
| 18-106_k141_61790 | Not-determined |
| 18-106_k141_73354 | Not-determined |
| 18-106_k141_102156 | Not-determined |
| 18-106_k141_44387 | Not-determined |
| 18-106_k141_97173 | Not-determined |
| 18-106_k141_108629 | Not-determined |
| 18-106_k141_32427 | Not-determined |
| 18-106_k141_114227 | Not-determined |
| 18-106_k141_55182 | Not-determined |
| 18-106_k141_3839 | Not-determined |
| 18-106_k141_44395 | Not-determined |
| 18-106_k141_25983 | Not-determined |
| 18-106_k141_108636 | Low-quality |
| 18-106_k141_25989 | Low-quality |
| 18-106_k141_44406 | Not-determined |
| 18-106_k141_114238 | Not-determined |
| 18-106_k141_32435 | Low-quality |
| 18-106_k141_114243 | Not-determined |
| 18-106_k141_102173 | Not-determined |
| 18-106_k141_3856 | Not-determined |
| 18-106_k141_67932 | Not-determined |
| 18-106_k141_97194 | Not-determined |
| 18-106_k141_32443 | Not-determined |
| 18-106_k141_84871 | Not-determined |
| 18-106_k141_3861 | Not-determined |
| 18-106_k141_44427 | Not-determined |
| 18-106_k141_26001 | Not-determined |
| 18-106_k141_97201 | Not-determined |
| 18-106_k141_61817 | Not-determined |
| 18-106_k141_67941 | Not-determined |
| 18-106_k141_61819 | Not-determined |
| 18-106_k141_32450 | Not-determined |
| 18-106_k141_108658 | Not-determined |
| 18-106_k141_67948 | Not-determined |
| 18-106_k141_3871 | Not-determined |
| 18-106_k141_3874 | Not-determined |
| 18-106_k141_119792 | Not-determined |
| 18-106_k141_102188 | Not-determined |
| 18-106_k141_97210 | Not-determined |
| 18-106_k141_97211 | Not-determined |
| 18-106_k141_84890 | Not-determined |
| 18-106_k141_84891 | Not-determined |
| 18-106_k141_14573 | Not-determined |
| 18-106_k141_97215 | Not-determined |
| 18-106_k141_3886 | Not-determined |
| 18-106_k141_102198 | Not-determined |
| 18-106_k141_131960 | Not-determined |
| 18-106_k141_102199 | Not-determined |
| 18-106_k141_3889 | Not-determined |
| 18-106_k141_55197 | Not-determined |
| 18-106_k141_114277 | Not-determined |
| 18-106_k141_67968 | Low-quality |
| 18-106_k141_108668 | Not-determined |
| 18-106_k141_97227 | Not-determined |
| 18-106_k141_14577 | Not-determined |
| 18-106_k141_84905 | Not-determined |
| 18-106_k141_19995 | Not-determined |
| 18-106_k141_67973 | Not-determined |
| 18-106_k141_114280 | Not-determined |
| 18-106_k141_67977 | Not-determined |
| 18-106_k141_32456 | Not-determined |
| 18-106_k141_26024 | Not-determined |
| 18-106_k141_119813 | Not-determined |
| 18-106_k141_108679 | Low-quality |
| 18-106_k141_20009 | Not-determined |
| 18-106_k141_26029 | Low-quality |
| 18-106_k141_119820 | Low-quality |
| 18-106_k141_131980 | Not-determined |
| 18-106_k141_131985 | Low-quality |
| 18-106_k141_61849 | Not-determined |
| 18-106_k141_131988 | Low-quality |
| 18-106_k141_97259 | Not-determined |
| 18-106_k141_97260 | Low-quality |
| 18-106_k141_108688 | Not-determined |
| 18-106_k141_3913 | Not-determined |
| 18-106_k141_97263 | Not-determined |
| 18-106_k141_61855 | Not-determined |
| 18-106_k141_20013 | Low-quality |
| 18-106_k141_55200 | Not-determined |
| 18-106_k141_32474 | Not-determined |
| 18-106_k141_97270 | Not-determined |
| 18-106_k141_55207 | Not-determined |
| 18-106_k141_55209 | Not-determined |
| 18-106_k141_67981 | Not-determined |
| 18-106_k141_131998 | Not-determined |
| 18-106_k141_108704 | Not-determined |
| 18-106_k141_132003 | Not-determined |
| 18-106_k141_26062 | Not-determined |
| 18-106_k141_126190 | Low-quality |
| 18-106_k141_102233 | Not-determined |
| 18-106_k141_44461 | Not-determined |
| 18-106_k141_126194 | Not-determined |
| 18-106_k141_20034 | Not-determined |
| 18-106_k141_126203 | Not-determined |
| 18-106_k141_3935 | Not-determined |
| 18-106_k141_137542 | Low-quality |
| 18-106_k141_20040 | Not-determined |
| 18-106_k141_3938 | Not-determined |
| 18-106_k141_8509 | Low-quality |
| 18-106_k141_108723 | Not-determined |
| 18-106_k141_3943 | Not-determined |
| 18-106_k141_20049 | Not-determined |
| 18-106_k141_78829 | Not-determined |
| 18-106_k141_119854 | Not-determined |
| 18-106_k141_8518 | Not-determined |
| 18-106_k141_119856 | Not-determined |
| 18-106_k141_68010 | Low-quality |
| 18-106_k141_44483 | Not-determined |
| 18-106_k141_84940 | Not-determined |
| 18-106_k141_84941 | Not-determined |
| 18-106_k141_32496 | Not-determined |
| 18-106_k141_114310 | Not-determined |
| 18-106_k141_32505 | Not-determined |
| 18-106_k141_114316 | Not-determined |
| 18-106_k141_137561 | Not-determined |
| 18-106_k141_114317 | Not-determined |
| 18-106_k141_114321 | Not-determined |
| 18-106_k141_38427 | Not-determined |
| 18-106_k141_61902 | Not-determined |
| 18-106_k141_44501 | Not-determined |
| 18-106_k141_84958 | Not-determined |
| 18-106_k141_38431 | Not-determined |
| 18-106_k141_38436 | Not-determined |
| 18-106_k141_38439 | Not-determined |
| 18-106_k141_137575 | Not-determined |
| 18-106_k141_44515 | Not-determined |
| 18-106_k141_44516 | Not-determined |
| 18-106_k141_38455 | Not-determined |
| 18-106_k141_84966 | Not-determined |
| 18-106_k141_55234 | Not-determined |
| 18-106_k141_78876 | Not-determined |
| 18-106_k141_26100 | Not-determined |
| 18-106_k141_84969 | Low-quality |
| 18-106_k141_97309 | Not-determined |
| 18-106_k141_8537 | Not-determined |
| 18-106_k141_61921 | Not-determined |
| 18-106_k141_102260 | Low-quality |
| 18-106_k141_8540 | Not-determined |
| 18-106_k141_137587 | Not-determined |
| 18-106_k141_78890 | Not-determined |
| 18-106_k141_132045 | Not-determined |
| 18-106_k141_119894 | Not-determined |
| 18-106_k141_8550 | Not-determined |
| 18-106_k141_61933 | Not-determined |
| 18-106_k141_108761 | Not-determined |
| 18-106_k141_26113 | Not-determined |
| 18-106_k141_108762 | Not-determined |
| 18-106_k141_55261 | Not-determined |
| 18-106_k141_97317 | Not-determined |
| 18-106_k141_97318 | Not-determined |
| 18-106_k141_55268 | Not-determined |
| 18-106_k141_119901 | Not-determined |
| 18-106_k141_132059 | Not-determined |
| 18-106_k141_102278 | Not-determined |
| 18-106_k141_84979 | Not-determined |
| 18-106_k141_26118 | Not-determined |
| 18-106_k141_8561 | Not-determined |
| 18-106_k141_108785 | Not-determined |
| 17-109-final-viral-combined_k141_63649\|\|full | Medium-quality |
| 18-106_k141_50213 | Not-determined |
| 18-106_k141_26126 | Not-determined |
| 18-106_k141_26129 | Low-quality |
| 18-106_k141_26130 | Not-determined |
| 18-106_k141_55291 | Not-determined |
| 18-106_k141_73419 | Not-determined |
| 18-106_k141_8572 | Not-determined |
| 18-106_k141_8574 | Not-determined |
| 18-106_k141_38486 | Not-determined |
| 18-106_k141_3981 | Not-determined |
| 18-106_k141_3986 | Not-determined |
| 18-106_k141_3987 | Not-determined |
| 18-106_k141_38495 | Not-determined |
| 18-106_k141_20061 | Not-determined |
| 18-106_k141_3990 | Low-quality |
| 18-106_k141_26145 | Low-quality |
| 18-106_k141_14631 | Low-quality |
| 18-106_k141_102316 | Not-determined |
| 18-106_k141_38503 | Not-determined |
| 18-106_k141_55302 | Not-determined |
| 18-106_k141_55308 | Not-determined |
| 18-106_k141_38512 | Not-determined |
| 18-106_k141_132093 | Not-determined |
| 18-106_k141_137619 | Not-determined |
| 18-106_k141_102331 | Not-determined |
| 18-106_k141_132096 | Not-determined |
| 18-106_k141_20071 | Not-determined |
| 18-106_k141_108810 | Not-determined |
| 18-106_k141_137625 | Not-determined |
| 18-106_k141_78912 | Not-determined |
| 18-106_k141_38520 | Not-determined |
| 18-106_k141_132104 | Not-determined |
| 18-106_k141_20082 | Not-determined |
| 18-106_k141_50249 | Not-determined |
| 18-106_k141_78918 | Not-determined |
| 18-106_k141_97347 | Not-determined |
| 18-106_k141_20087 | Not-determined |
| 18-106_k141_137635 | Not-determined |
| 18-106_k141_119935 | Not-determined |
| 18-106_k141_14653 | Not-determined |
| 18-106_k141_4030 | Low-quality |
| 18-106_k141_126256 | Not-determined |
| 18-106_k141_97349 | Not-determined |
| 18-106_k141_126257 | Not-determined |
| 18-106_k141_137639 | Not-determined |
| 18-106_k141_102348 | Not-determined |
| 18-106_k141_50259 | Not-determined |
| 18-106_k141_108828 | Not-determined |
| 18-106_k141_44541 | Not-determined |
| 18-106_k141_97360 | Low-quality |
| 18-106_k141_8595 | Not-determined |
| 18-106_k141_114391 | Not-determined |
| 18-106_k141_132126 | Not-determined |
| 18-106_k141_32555 | Not-determined |
| 18-106_k141_114393 | Not-determined |
| 18-106_k141_102352 | Not-determined |
| 18-106_k141_20094 | Not-determined |
| 18-106_k141_44545 | Not-determined |
| 18-106_k141_85038 | Not-determined |
| 18-106_k141_14658 | Not-determined |
| 18-106_k141_8603 | Not-determined |
| 18-106_k141_20099 | Not-determined |
| 18-106_k141_102362 | Not-determined |
| 18-106_k141_137659 | Not-determined |
| 18-106_k141_126286 | Not-determined |
| 18-106_k141_137661 | Not-determined |
| 18-106_k141_126287 | Not-determined |
| 18-106_k141_68036 | Not-determined |
| 18-106_k141_8612 | Low-quality |
| 18-106_k141_132142 | Not-determined |
| 18-106_k141_61993 | Not-determined |
| 18-106_k141_44560 | Not-determined |
| 18-106_k141_126291 | Not-determined |
| 18-106_k141_97370 | Not-determined |
| 18-106_k141_44565 | Low-quality |
| 18-106_k141_78947 | Not-determined |
| 18-106_k141_55320 | Not-determined |
| 18-106_k141_137671 | Not-determined |
| 18-106_k141_61999 | Not-determined |
| 18-106_k141_4064 | Not-determined |
| 18-106_k141_85061 | Not-determined |
| 18-106_k141_108849 | Not-determined |
| 18-106_k141_62006 | Not-determined |
| 18-106_k141_14678 | Not-determined |
| 18-106_k141_78961 | Low-quality |
| 18-106_k141_68042 | Not-determined |
| 18-106_k141_97386 | Not-determined |
| 18-106_k141_137688 | Not-determined |
| 18-106_k141_55328 | Not-determined |
| 18-106_k141_68049 | Not-determined |
| 18-106_k141_126315 | Not-determined |
| 18-106_k141_126316 | Not-determined |
| 18-106_k141_4067 | Not-determined |
| 18-106_k141_14683 | Not-determined |
| 18-106_k141_68051 | Not-determined |
| 18-106_k141_126319 | Not-determined |
| 18-106_k141_132149 | Not-determined |
| 18-106_k141_137697 | Not-determined |
| 18-106_k141_91043 | Not-determined |
| 18-106_k141_20138 | Not-determined |
| 18-106_k141_91048 | Not-determined |
| 18-106_k141_62017 | Not-determined |
| 18-106_k141_20143 | Not-determined |
| 18-106_k141_55343 | Not-determined |
| 18-106_k141_20144 | Not-determined |
| 18-106_k141_68069 | Not-determined |
| 18-106_k141_38600 | Not-determined |
| 18-106_k141_102400 | Not-determined |
| 18-106_k141_68077 | Not-determined |
| 18-106_k141_78986 | Not-determined |
| 18-106_k141_108892 | Low-quality |
| 18-106_k141_20150 | Not-determined |
| 18-106_k141_78995 | Not-determined |
| 18-106_k141_14713 | Not-determined |
| 18-106_k141_79003 | Not-determined |
| 18-106_k141_38619 | Not-determined |
| 18-106_k141_14721 | Not-determined |
| 18-106_k141_14725 | Not-determined |
| 18-106_k141_4090 | Not-determined |
| 18-106_k141_62047 | Not-determined |
| 18-106_k141_4094 | Not-determined |
| 18-106_k141_120013 | Not-determined |
| 18-106_k141_73508 | Not-determined |
| 18-106_k141_14728 | Low-quality |
| 18-106_k141_79013 | Not-determined |
| 18-106_k141_73511 | Not-determined |
| 18-106_k141_108911 | Low-quality |
| 18-106_k141_97433 | Not-determined |
| 18-106_k141_73515 | Not-determined |
| 18-106_k141_114455 | Not-determined |
| 18-106_k141_50284 | Not-determined |
| 18-106_k141_114457 | Not-determined |
| 18-106_k141_79028 | Not-determined |
| 18-106_k141_79030 | Not-determined |
| 18-106_k141_4101 | Not-determined |
| 18-106_k141_132197 | Not-determined |
| 18-106_k141_97438 | Not-determined |
| 18-106_k141_79034 | Low-quality |
| 18-106_k141_14739 | Not-determined |
| 18-106_k141_126374 | Not-determined |
| 18-106_k141_14740 | Not-determined |
| 18-106_k141_97445 | Not-determined |
| 18-106_k141_14744 | Not-determined |
| 18-106_k141_14747 | Not-determined |
| 18-106_k141_4110 | Not-determined |
| 18-106_k141_97454 | Not-determined |
| 18-106_k141_73534 | Not-determined |
| 18-106_k141_4115 | Not-determined |
| 18-106_k141_73537 | Not-determined |
| 18-106_k141_137700 | Not-determined |
| 18-106_k141_4116 | Not-determined |
| 18-106_k141_120024 | Not-determined |
| 18-106_k141_120025 | Not-determined |
| 18-106_k141_132215 | Not-determined |
| 18-106_k141_85080 | Not-determined |
| 18-106_k141_32583 | Not-determined |
| 18-106_k141_73560 | Not-determined |
| 18-106_k141_132221 | Not-determined |
| 18-106_k141_26184 | Not-determined |
| 18-106_k141_79041 | Not-determined |
| 18-106_k141_79042 | Not-determined |
| 18-106_k141_26186 | Not-determined |
| 18-106_k141_50306 | Not-determined |
| 18-106_k141_85087 | Not-determined |
| 18-106_k141_68126 | Low-quality |
| 18-106_k141_108931 | Not-determined |
| 18-106_k141_137712 | Not-determined |
| 18-106_k141_97489 | Not-determined |
| 18-106_k141_38664 | Not-determined |
| 18-106_k141_108940 | Not-determined |
| 18-106_k141_114490 | Not-determined |
| 18-106_k141_38669 | Not-determined |
| 18-106_k141_126416 | Not-determined |
| 18-106_k141_126420 | Not-determined |
| 18-106_k141_38675 | Not-determined |
| 18-106_k141_38677 | Not-determined |
| 18-106_k141_20180 | Not-determined |
| 18-106_k141_102449 | Not-determined |
| 18-106_k141_68153 | Low-quality |
| 18-106_k141_32600 | Low-quality |
| 18-106_k141_126429 | Not-determined |
| 18-106_k141_20186 | Not-determined |
| 18-106_k141_32605 | Not-determined |
| 18-106_k141_102456 | Not-determined |
| 18-106_k141_32615 | Not-determined |
| 18-106_k141_137734 | Low-quality |
| 18-106_k141_97521 | Not-determined |
| 18-106_k141_26222 | Not-determined |
| 18-106_k141_97524 | Not-determined |
| 18-106_k141_68169 | Low-quality |
| 18-106_k141_68170 | Not-determined |
| 18-106_k141_62088 | Not-determined |
| 18-106_k141_132246 | Not-determined |
| 18-106_k141_26236 | Not-determined |
| 18-106_k141_38707 | Not-determined |
| 18-106_k141_68180 | Not-determined |
| 18-106_k141_120051 | Not-determined |
| 18-106_k141_120053 | Not-determined |
| 18-106_k141_32631 | Not-determined |
| 18-106_k141_132252 | Not-determined |
| 18-106_k141_26238 | Not-determined |
| 18-106_k141_38710 | Not-determined |
| 18-106_k141_120060 | Not-determined |
| 18-106_k141_68192 | Not-determined |
| 18-106_k141_55379 | Not-determined |
| 18-106_k141_32638 | Not-determined |
| 18-106_k141_26245 | Not-determined |
| 18-106_k141_120065 | Not-determined |
| 18-106_k141_68200 | Not-determined |
| 18-106_k141_108974 | Not-determined |
| 18-106_k141_38719 | Not-determined |
| 18-106_k141_120071 | Not-determined |
| 18-106_k141_91079 | Not-determined |
| 18-106_k141_120076 | Not-determined |
| 18-106_k141_73624 | Not-determined |
| 18-106_k141_73625 | Not-determined |
| 18-106_k141_132275 | Not-determined |
| 18-106_k141_114502 | Low-quality |
| 18-106_k141_85146 | Not-determined |
| 18-106_k141_114505 | Not-determined |
| 18-106_k141_20218 | Not-determined |
| 18-106_k141_73630 | Not-determined |
| 18-106_k141_44590 | Not-determined |
| 18-106_k141_62122 | Not-determined |
| 18-106_k141_132289 | Low-quality |
| 18-106_k141_62128 | Not-determined |
| 18-106_k141_126445 | Low-quality |
| 18-106_k141_126453 | Not-determined |
| 18-106_k141_132308 | Low-quality |
| 18-106_k141_20239 | Not-determined |
| 18-106_k141_120107 | Not-determined |
| 18-106_k141_85161 | Not-determined |
| 18-106_k141_132316 | Not-determined |
| 18-106_k141_102495 | Not-determined |
| 18-106_k141_132321 | Not-determined |
| 18-106_k141_137771 | Not-determined |
| 18-106_k141_137773 | Not-determined |
| 18-106_k141_126471 | Not-determined |
| 18-106_k141_44614 | Low-quality |
| 18-106_k141_20251 | Not-determined |
| 18-106_k141_132329 | Not-determined |
| 18-106_k141_14789 | Not-determined |
| 18-106_k141_62179 | Not-determined |
| 18-106_k141_68260 | Not-determined |
| 18-106_k141_38753 | Not-determined |
| 18-106_k141_126480 | Not-determined |
| 18-106_k141_20268 | Not-determined |
| 18-106_k141_62192 | Not-determined |
| 18-106_k141_114531 | Not-determined |
| 18-106_k141_8630 | Not-determined |
| 18-106_k141_44631 | Not-determined |
| 18-106_k141_14795 | Not-determined |
| 18-106_k141_4232 | Not-determined |
| 18-106_k141_68268 | Not-determined |
| 18-106_k141_8633 | Not-determined |
| 18-106_k141_4236 | Not-determined |
| 18-106_k141_79129 | Not-determined |
| 18-106_k141_14800 | Not-determined |
| 18-106_k141_8641 | Not-determined |
| 18-106_k141_62199 | Low-quality |
| 18-106_k141_85182 | Not-determined |
| 18-106_k141_20275 | Not-determined |
| 18-106_k141_20276 | Not-determined |
| 18-106_k141_79147 | Not-determined |
| 18-106_k141_132336 | Not-determined |
| 18-106_k141_14811 | Low-quality |
| 18-106_k141_79149 | Not-determined |
| 18-106_k141_91133 | Not-determined |
| 18-106_k141_14818 | Not-determined |
| 18-106_k141_68293 | Not-determined |
| 18-106_k141_126496 | Not-determined |
| 18-106_k141_91138 | Not-determined |
| 18-106_k141_14825 | Not-determined |
| 18-106_k141_97575 | Not-determined |
| 18-106_k141_97576 | Not-determined |
| 18-106_k141_97578 | Not-determined |
| 18-106_k141_114550 | Not-determined |
| 18-106_k141_4245 | Not-determined |
| 18-106_k141_4246 | Not-determined |
| 18-106_k141_4247 | Not-determined |
| 18-106_k141_102548 | Not-determined |
| 18-106_k141_14827 | Low-quality |
| 18-106_k141_62240 | Not-determined |
| 18-106_k141_8677 | Not-determined |
| 18-106_k141_14836 | Not-determined |
| 18-106_k141_32662 | Not-determined |
| 18-106_k141_68305 | Not-determined |
| 18-106_k141_79168 | Not-determined |
| 18-106_k141_137809 | Not-determined |
| 18-106_k141_137811 | Not-determined |
| 18-106_k141_137819 | Not-determined |
| 18-106_k141_68316 | Not-determined |
| 18-106_k141_102569 | Not-determined |
| 18-106_k141_62252 | Not-determined |
| 18-106_k141_85211 | Not-determined |
| 18-106_k141_62257 | Not-determined |
| 18-106_k141_8700 | Not-determined |
| 18-106_k141_91168 | Not-determined |
| 18-106_k141_102574 | Not-determined |
| 18-106_k141_32680 | Low-quality |
| 18-106_k141_8703 | Low-quality |
| 18-106_k141_32683 | Not-determined |
| 18-106_k141_8715 | Not-determined |
| 18-106_k141_38777 | Low-quality |
| 18-106_k141_8718 | Not-determined |
| 18-106_k141_14866 | Not-determined |
| 18-106_k141_14868 | Low-quality |
| 18-106_k141_38782 | Not-determined |
| 18-106_k141_38783 | Not-determined |
| 18-106_k141_8727 | Not-determined |
| 18-106_k141_62271 | Not-determined |
| 18-106_k141_32708 | Not-determined |
| 18-106_k141_137830 | Not-determined |
| 18-106_k141_91187 | Not-determined |
| 18-106_k141_126538 | Not-determined |
| 18-106_k141_14874 | Not-determined |
| 18-106_k141_102601 | Not-determined |
| 18-106_k141_14875 | Not-determined |
| 18-106_k141_8735 | Not-determined |
| 18-106_k141_102603 | Low-quality |
| 18-106_k141_126542 | Not-determined |
| 18-106_k141_91191 | Not-determined |
| 18-106_k141_20299 | Low-quality |
| 18-106_k141_137844 | Not-determined |
| 18-106_k141_85255 | Not-determined |
| 18-106_k141_55438 | Not-determined |
| 18-106_k141_62285 | Low-quality |
| 18-106_k141_126558 | Not-determined |
| 18-106_k141_85259 | Not-determined |
| 18-106_k141_85261 | Not-determined |
| 18-106_k141_85263 | Not-determined |
| 18-106_k141_79218 | Not-determined |
| 18-106_k141_68343 | Not-determined |
| 18-106_k141_26390 | Not-determined |
| 18-106_k141_91212 | Not-determined |
| 18-106_k141_8776 | Not-determined |
| 18-106_k141_20321 | Not-determined |
| 18-106_k141_50325 | Not-determined |
| 18-106_k141_85272 | Not-determined |
| 18-106_k141_38820 | Low-quality |
| 18-106_k141_4294 | Not-determined |
| 18-106_k141_120175 | Low-quality |
| 18-106_k141_55470 | Low-quality |
| 18-106_k141_97595 | Not-determined |
| 18-106_k141_85277 | Not-determined |
| 18-106_k141_4308 | Not-determined |
| 18-106_k141_8785 | Not-determined |
| 18-106_k141_137855 | Not-determined |
| 18-106_k141_50349 | Not-determined |
| 18-106_k141_8786 | Not-determined |
| 18-106_k141_137857 | Not-determined |
| 18-106_k141_26407 | Not-determined |
| 18-106_k141_137862 | Not-determined |
| 18-106_k141_73672 | Not-determined |
| 18-106_k141_8792 | Not-determined |
| 18-106_k141_26412 | Not-determined |
| 18-106_k141_4315 | Not-determined |
| 18-106_k141_26413 | Not-determined |
| 18-106_k141_44695 | Not-determined |
| 18-106_k141_114617 | Not-determined |
| 18-106_k141_8794 | Not-determined |
| 18-106_k141_114618 | Not-determined |
| 18-106_k141_120191 | Not-determined |
| 18-106_k141_50362 | Low-quality |
| 18-106_k141_44698 | Not-determined |
| 18-106_k141_50365 | Low-quality |
| 18-106_k141_114619 | Not-determined |
| 18-106_k141_4319 | Not-determined |
| 18-106_k141_32719 | Not-determined |
| 18-106_k141_114626 | Not-determined |
| 18-106_k141_50376 | Not-determined |
| 18-106_k141_44700 | Not-determined |
| 18-106_k141_114631 | Not-determined |
| 18-106_k141_114637 | Not-determined |
| 18-106_k141_20362 | Not-determined |
| 18-106_k141_44711 | Not-determined |
| 18-106_k141_8796 | Not-determined |
| 18-106_k141_126579 | Not-determined |
| 18-106_k141_73687 | Not-determined |
| 18-106_k141_68392 | Not-determined |
| 18-106_k141_8801 | Not-determined |
| 18-106_k141_79239 | Not-determined |
| 18-106_k141_55503 | Not-determined |
| 18-106_k141_126582 | Not-determined |
| 18-106_k141_68400 | Not-determined |
| 18-106_k141_38872 | Not-determined |
| 18-106_k141_73693 | Not-determined |
| 18-106_k141_68407 | Not-determined |
| 18-106_k141_26421 | Not-determined |
| 18-106_k141_68412 | Not-determined |
| 18-106_k141_38880 | Not-determined |
| 18-106_k141_91257 | Not-determined |
| 18-106_k141_8826 | Not-determined |
| 18-106_k141_62316 | Not-determined |
| 18-106_k141_20380 | Not-determined |
| 18-106_k141_44738 | Not-determined |
| 18-106_k141_137897 | Not-determined |
| 18-106_k141_38889 | Not-determined |
| 18-106_k141_97634 | Not-determined |
| 18-106_k141_44744 | Not-determined |
| 18-106_k141_44746 | Not-determined |
| 18-106_k141_4363 | Not-determined |
| 18-106_k141_4371 | Not-determined |
| 18-106_k141_8843 | Not-determined |
| 18-106_k141_132344 | Not-determined |
| 18-106_k141_132350 | Not-determined |
| 18-106_k141_114659 | Not-determined |
| 18-106_k141_62324 | Not-determined |
| 18-106_k141_132355 | Not-determined |
| 18-106_k141_132356 | Not-determined |
| 18-106_k141_73703 | Low-quality |
| 18-106_k141_50420 | Not-determined |
| 18-106_k141_91270 | Not-determined |
| 18-106_k141_50422 | Not-determined |
| 18-106_k141_4382 | Low-quality |
| 18-106_k141_20398 | Not-determined |
| 18-106_k141_85315 | Not-determined |

| | |
|---|---|
| 18-106_k141_91273 | Not-determined |
| 18-106_k141_73708 | Not-determined |
| 18-106_k141_120220 | Not-determined |
| 18-106_k141_79273 | Not-determined |
| 18-106_k141_132362 | Not-determined |
| 18-106_k141_132363 | Not-determined |
| 18-106_k141_73714 | Not-determined |
| 18-106_k141_62338 | Not-determined |
| 18-106_k141_38911 | Not-determined |
| 18-106_k141_68444 | Not-determined |
| 18-106_k141_50438 | Not-determined |
| 18-106_k141_120238 | Low-quality |
| 18-106_k141_55509 | Not-determined |
| 18-106_k141_126588 | Not-determined |
| 18-106_k141_62346 | Not-determined |
| 18-106_k141_126598 | Low-quality |
| 18-106_k141_137917 | Not-determined |
| 18-106_k141_4414 | Not-determined |
| 18-106_k141_91295 | Not-determined |
| 18-106_k141_120256 | Not-determined |
| 18-106_k141_120257 | Not-determined |
| 18-106_k141_91300 | Not-determined |
| 18-106_k141_132394 | Not-determined |
| 18-106_k141_91305 | Not-determined |
| 18-106_k141_8874 | Not-determined |
| 18-106_k141_137925 | Not-determined |
| 18-106_k141_114709 | Not-determined |
| 18-106_k141_26466 | Not-determined |
| 18-106_k141_44776 | Not-determined |
| 18-106_k141_120266 | Not-determined |
| 18-106_k141_79292 | Not-determined |
| 18-106_k141_114712 | Not-determined |
| 18-106_k141_102650 | Not-determined |
| 18-106_k141_79299 | Not-determined |
| 18-106_k141_68460 | Not-determined |
| 18-106_k141_14939 | Not-determined |
| 18-106_k141_73730 | Not-determined |
| 18-106_k141_126620 | Not-determined |
| 18-106_k141_137945 | Not-determined |
| 18-106_k141_68465 | Not-determined |
| 18-106_k141_32770 | Not-determined |
| 18-106_k141_102666 | Not-determined |
| 18-106_k141_85344 | Not-determined |
| 18-106_k141_102667 | Not-determined |
| 18-106_k141_97639 | Not-determined |
| 18-106_k141_50468 | Not-determined |
| 18-106_k141_120284 | Low-quality |
| 18-106_k141_85348 | Low-quality |
| 18-106_k141_79317 | Not-determined |
| 18-106_k141_73734 | Not-determined |
| 18-106_k141_91330 | Not-determined |
| 18-106_k141_62402 | Not-determined |
| 18-106_k141_91332 | Not-determined |
| 18-106_k141_8908 | Not-determined |
| 18-106_k141_32792 | Not-determined |
| 18-106_k141_132428 | Not-determined |
| 18-106_k141_120301 | Not-determined |
| 18-106_k141_126638 | Not-determined |
| 18-106_k141_8923 | Not-determined |
| 18-106_k141_114737 | Not-determined |
| 18-106_k141_38948 | Not-determined |
| 18-106_k141_97668 | Not-determined |
| 18-106_k141_55564 | Not-determined |
| 18-106_k141_132443 | Not-determined |
| 18-106_k141_79334 | Low-quality |
| 18-106_k141_91355 | Not-determined |
| 18-106_k141_102704 | Not-determined |
| 18-106_k141_79336 | Not-determined |
| 18-106_k141_137971 | Not-determined |
| 18-106_k141_55573 | Not-determined |
| 18-106_k141_97678 | Low-quality |
| 18-106_k141_102707 | Not-determined |
| 18-106_k141_132452 | Not-determined |
| 18-106_k141_79340 | Not-determined |
| 18-106_k141_38957 | Not-determined |
| 18-106_k141_14989 | Not-determined |
| 18-106_k141_4462 | Not-determined |
| 18-106_k141_4463 | Not-determined |
| 18-106_k141_62428 | Low-quality |
| 18-106_k141_68506 | Low-quality |
| 18-106_k141_79353 | Not-determined |
| 18-106_k141_97704 | Not-determined |
| 18-106_k141_85396 | Low-quality |
| 18-106_k141_55603 | Not-determined |
| 18-106_k141_62450 | Not-determined |
| 18-106_k141_108997 | Not-determined |
| 18-106_k141_97715 | Not-determined |
| 18-106_k141_73772 | Not-determined |
| 18-106_k141_8940 | Not-determined |
| 18-106_k141_97721 | Not-determined |
| 18-106_k141_14993 | Low-quality |
| 18-106_k141_114753 | Low-quality |
| 18-106_k141_132485 | Not-determined |
| 18-106_k141_126657 | Not-determined |
| 18-106_k141_85402 | Not-determined |
| 18-106_k141_14998 | Not-determined |
| 18-106_k141_15000 | Not-determined |
| 18-106_k141_102730 | Not-determined |
| 18-106_k141_26536 | Not-determined |
| 18-106_k141_20401 | Not-determined |
| 18-106_k141_38986 | Not-determined |
| 18-106_k141_132491 | Low-quality |
| 18-106_k141_20408 | Not-determined |
| 18-106_k141_73787 | Not-determined |
| 18-106_k141_85411 | Not-determined |
| 18-106_k141_73790 | Not-determined |
| 18-106_k141_73792 | Not-determined |
| 18-106_k141_44844 | Not-determined |
| 18-106_k141_20412 | Not-determined |
| 18-106_k141_38999 | Not-determined |
| 18-106_k141_4468 | Not-determined |
| 18-106_k141_102741 | Not-determined |
| 18-106_k141_39001 | Not-determined |
| 18-106_k141_97742 | Not-determined |
| 18-106_k141_50524 | Not-determined |
| 18-106_k141_85422 | Low-quality |
| 18-106_k141_15019 | Not-determined |
| 18-106_k141_26543 | Not-determined |
| 18-106_k141_15022 | Not-determined |
| 18-106_k141_20423 | Not-determined |
| 18-106_k141_15024 | Not-determined |
| 18-106_k141_114773 | Not-determined |
| 18-106_k141_8967 | Not-determined |
| 18-106_k141_15029 | Not-determined |
| 18-106_k141_79375 | Not-determined |
| 18-106_k141_55644 | Not-determined |
| 18-106_k141_39016 | Not-determined |
| 18-106_k141_39017 | Not-determined |
| 18-106_k141_26545 | Low-quality |
| 18-106_k141_15033 | Low-quality |
| 18-106_k141_85444 | Not-determined |
| 18-106_k141_15034 | Not-determined |
| 18-106_k141_85445 | Not-determined |
| 18-106_k141_114784 | Not-determined |
| 18-106_k141_26554 | Not-determined |
| 18-106_k141_8978 | Low-quality |
| 18-106_k141_102770 | Not-determined |
| 18-106_k141_50538 | Not-determined |
| 18-106_k141_120357 | Not-determined |
| 18-106_k141_97766 | Not-determined |
| 18-106_k141_32824 | Not-determined |
| 18-106_k141_32825 | Not-determined |
| 18-106_k141_102775 | Not-determined |
| 18-106_k141_4478 | Not-determined |
| 18-106_k141_132527 | Not-determined |
| 18-106_k141_120364 | Not-determined |
| 18-106_k141_20444 | Not-determined |
| 18-106_k141_85459 | Not-determined |
| 18-106_k141_85462 | Not-determined |
| 18-106_k141_91425 | Not-determined |
| 18-106_k141_8988 | Not-determined |
| 18-106_k141_44901 | Not-determined |
| 18-106_k141_32850 | Not-determined |
| 18-106_k141_85470 | Not-determined |
| 18-106_k141_126708 | Low-quality |
| 18-106_k141_73821 | Not-determined |
| 18-106_k141_68565 | Not-determined |
| 18-106_k141_79399 | Not-determined |
| 18-106_k141_55672 | Not-determined |
| 18-106_k141_79401 | Not-determined |
| 18-106_k141_85473 | Not-determined |
| 18-106_k141_68571 | Not-determined |
| 18-106_k141_55678 | Not-determined |
| 18-106_k141_102808 | Not-determined |
| 18-106_k141_9004 | Not-determined |
| 18-106_k141_85481 | Not-determined |
| 18-106_k141_79419 | Not-determined |
| 18-106_k141_132564 | Not-determined |
| 18-106_k141_32865 | Not-determined |
| 18-106_k141_120396 | Not-determined |
| 18-106_k141_55686 | Not-determined |
| 18-106_k141_138029 | Not-determined |
| 18-106_k141_4491 | Not-determined |
| 18-106_k141_73846 | Not-determined |
| 18-106_k141_4495 | Not-determined |
| 18-106_k141_73852 | Not-determined |
| 18-106_k141_9023 | Low-quality |
| 18-106_k141_44931 | Not-determined |
| 18-106_k141_85504 | Not-determined |
| 18-106_k141_44938 | Not-determined |
| 18-106_k141_32879 | Not-determined |
| 18-106_k141_85515 | Low-quality |
| 18-106_k141_85517 | Not-determined |
| 18-106_k141_9037 | Not-determined |
| 18-106_k141_102861 | Not-determined |
| 18-106_k141_126718 | Not-determined |
| 18-106_k141_26581 | Not-determined |
| 18-106_k141_109089 | Not-determined |
| 18-106_k141_138059 | Low-quality |
| 18-106_k141_114803 | Not-determined |
| 18-106_k141_20491 | Not-determined |
| 18-106_k141_114812 | Not-determined |
| 18-106_k141_138066 | Not-determined |
| 18-106_k141_26598 | Not-determined |
| 18-106_k141_97848 | Not-determined |
| 18-106_k141_32901 | Not-determined |
| 18-106_k141_26608 | Low-quality |
| 18-106_k141_39023 | Not-determined |
| 18-106_k141_138072 | Not-determined |
| 18-106_k141_114818 | Not-determined |
| 18-106_k141_32902 | Not-determined |
| 18-106_k141_97854 | Not-determined |
| 18-106_k141_4532 | Not-determined |
| 18-106_k141_138082 | Not-determined |
| 18-106_k141_79435 | Not-determined |
| 18-106_k141_4538 | Not-determined |
| 18-106_k141_45003 | Not-determined |
| 18-106_k141_50641 | Not-determined |
| 18-106_k141_45005 | Not-determined |
| 18-106_k141_45007 | Not-determined |
| 18-106_k141_55729 | Not-determined |
| 18-106_k141_109141 | Not-determined |
| 18-106_k141_32910 | Not-determined |
| 18-106_k141_109148 | Not-determined |
| 18-106_k141_50646 | Not-determined |
| 18-106_k141_79449 | Not-determined |
| 18-106_k141_79450 | Not-determined |
| 18-106_k141_32920 | Not-determined |
| 18-106_k141_91480 | Not-determined |
| 18-106_k141_45014 | Low-quality |
| 18-106_k141_109152 | Not-determined |
| 18-106_k141_91488 | Low-quality |
| 18-106_k141_79456 | Not-determined |
| 18-106_k141_79458 | Not-determined |
| 18-106_k141_45016 | Not-determined |
| 18-106_k141_109155 | Not-determined |
| 18-106_k141_109156 | Not-determined |
| 18-106_k141_62535 | Not-determined |
| 18-106_k141_4551 | Not-determined |
| 18-106_k141_79465 | Not-determined |
| 18-106_k141_32933 | Not-determined |
| 18-106_k141_109166 | Not-determined |
| 18-106-final-viral-combined_k141_137591\|\|full | High-quality |
| 18-106_k141_32934 | Not-determined |
| 18-106_k141_20546 | Not-determined |
| 18-106_k141_138098 | Not-determined |
| 18-106_k141_32935 | Not-determined |
| 18-106_k141_85590 | Not-determined |
| 18-106_k141_50651 | Not-determined |
| 18-106_k141_4557 | Not-determined |
| 18-106_k141_50653 | Not-determined |
| 18-106_k141_109174 | Not-determined |
| 18-106_k141_50657 | Not-determined |
| 18-106_k141_97881 | Not-determined |
| 18-106_k141_97884 | Not-determined |
| 18-106_k141_132604 | Not-determined |
| 18-106_k141_15056 | Not-determined |
| 18-106_k141_79475 | Not-determined |
| 18-106_k141_4570 | Not-determined |
| 18-106_k141_32954 | Not-determined |
| 18-106_k141_85608 | Not-determined |
| 18-106_k141_55753 | Not-determined |
| 18-106_k141_62563 | Not-determined |
| 18-106_k141_26645 | Low-quality |
| 18-106_k141_45047 | Not-determined |
| 18-106_k141_126791 | Not-determined |
| 18-106_k141_109201 | Not-determined |
| 18-106_k141_68603 | Not-determined |
| 18-106_k141_138109 | Not-determined |
| 18-106_k141_26648 | Not-determined |
| 18-106_k141_120416 | Not-determined |
| 18-106_k141_50662 | Not-determined |
| 18-106_k141_55763 | Not-determined |
| 18-106_k141_50666 | Not-determined |
| 18-106_k141_85617 | Not-determined |
| 18-106_k141_85619 | Not-determined |
| 18-106_k141_4595 | Not-determined |
| 18-106_k141_39047 | Not-determined |
| 18-106_k141_15067 | Not-determined |
| 18-106_k141_132623 | Not-determined |
| 18-106_k141_15070 | Not-determined |
| 18-106_k141_50674 | Not-determined |
| 18-106_k141_55770 | Not-determined |
| 18-106_k141_55771 | Not-determined |
| 18-106_k141_4602 | Not-determined |
| 18-106_k141_50676 | Not-determined |
| 18-106_k141_102880 | Not-determined |
| 18-106_k141_91539 | Not-determined |
| 18-106_k141_26669 | Not-determined |
| 18-106_k141_50689 | Not-determined |
| 18-106_k141_138116 | Not-determined |
| 18-106_k141_91543 | Not-determined |
| 18-106_k141_138120 | Not-determined |
| 18-106_k141_102888 | Not-determined |
| 18-106_k141_26676 | Not-determined |
| 18-106_k141_4618 | Not-determined |
| 18-106_k141_15097 | Not-determined |
| 18-106_k141_26687 | Not-determined |
| 18-106_k141_85643 | Not-determined |
| 18-106_k141_4624 | Not-determined |
| 18-106_k141_50712 | Not-determined |
| 18-106_k141_4625 | Not-determined |
| 18-106_k141_114873 | Not-determined |
| 18-106_k141_55803 | Low-quality |
| 18-106_k141_26699 | Not-determined |
| 18-106_k141_114878 | Not-determined |
| 18-106_k141_91565 | Not-determined |
| 18-106_k141_26702 | Not-determined |
| 18-106_k141_68641 | Not-determined |
| 18-106_k141_55807 | Not-determined |
| 18-106_k141_62591 | Not-determined |
| 18-106_k141_109252 | Not-determined |
| 18-106_k141_4645 | Not-determined |
| 18-106_k141_39088 | Not-determined |
| 18-106_k141_109256 | Not-determined |
| 18-106_k141_114894 | Low-quality |
| 18-106_k141_138166 | Not-determined |
| 18-106_k141_55822 | Low-quality |
| 18-106_k141_114895 | Not-determined |
| 18-106_k141_138167 | Not-determined |
| 18-106_k141_9059 | Not-determined |
| 18-106_k141_79528 | Low-quality |
| 18-106_k141_9061 | Not-determined |
| 18-106_k141_138171 | Not-determined |
| 18-106_k141_62604 | Not-determined |
| 18-106_k141_4660 | Not-determined |
| 18-106_k141_91579 | Low-quality |
| 18-106_k141_62610 | Not-determined |
| 18-106_k141_9065 | Not-determined |
| 18-106_k141_97902 | Low-quality |
| 18-106_k141_85657 | Not-determined |
| 18-106_k141_114906 | Not-determined |
| 18-106_k141_85664 | Not-determined |
| 18-106_k141_126828 | Not-determined |
| 18-106_k141_4672 | Not-determined |
| 18-106_k141_79545 | Not-determined |
| 18-106_k141_120476 | Not-determined |
| 18-106_k141_120477 | Not-determined |
| 18-106_k141_79547 | Not-determined |
| 18-106_k141_85671 | Not-determined |
| 18-106_k141_138190 | Not-determined |
| 18-106_k141_73859 | Not-determined |
| 18-106_k141_132657 | Not-determined |
| 18-106_k141_109274 | Not-determined |
| 18-106_k141_68661 | Not-determined |
| 18-106_k141_9088 | Not-determined |
| 18-106_k141_9089 | Not-determined |
| 18-106_k141_9090 | Low-quality |
| 18-106_k141_68663 | Not-determined |
| 18-106_k141_62635 | Not-determined |
| 18-106_k141_138194 | Not-determined |
| 18-106_k141_26733 | Not-determined |
| 18-106_k141_138196 | Not-determined |
| 18-106_k141_26735 | Not-determined |
| 18-106_k141_62642 | Not-determined |
| 18-106_k141_97934 | Not-determined |
| 18-106_k141_39123 | Not-determined |
| 18-106_k141_32965 | Not-determined |
| 18-106_k141_9100 | Not-determined |
| 18-106_k141_32970 | Not-determined |
| 18-106_k141_9104 | Not-determined |
| 18-106_k141_126864 | Not-determined |
| 18-106_k141_91619 | Not-determined |
| 18-106_k141_91622 | Low-quality |
| 18-106_k141_68687 | Low-quality |
| 18-106_k141_62664 | Not-determined |
| 18-106_k141_55835 | Not-determined |
| 18-106_k141_45133 | Not-determined |
| 18-106_k141_45134 | Not-determined |
| 18-106_k141_97949 | Not-determined |
| 18-106_k141_138214 | Not-determined |
| 18-106_k141_26742 | Not-determined |
| 18-106_k141_45136 | Not-determined |
| 18-106_k141_114942 | Not-determined |
| 18-106_k141_45142 | Not-determined |
| 18-106_k141_4698 | Not-determined |
| 18-106_k141_20616 | Low-quality |
| 18-106_k141_79553 | Not-determined |
| 18-106_k141_109313 | Not-determined |
| 18-106_k141_132682 | Not-determined |
| 18-106_k141_68706 | Not-determined |
| 18-106_k141_20618 | Not-determined |
| 18-106_k141_32992 | Not-determined |
| 18-106_k141_85686 | Not-determined |
| 18-106_k141_97954 | Not-determined |
| 18-106_k141_39147 | Not-determined |
| 18-106_k141_132692 | Not-determined |
| 18-106_k141_26759 | Not-determined |
| 18-106_k141_73878 | Not-determined |
| 18-106_k141_55849 | Not-determined |
| 18-106_k141_91646 | Not-determined |
| 18-106_k141_20627 | Low-quality |
| 18-106_k141_55851 | Not-determined |
| 18-106_k141_4716 | Not-determined |
| 18-106_k141_26769 | Not-determined |
| 18-106_k141_55857 | Not-determined |
| 18-106_k141_102965 | Not-determined |
| 18-106_k141_4720 | Not-determined |
| 18-106_k141_26775 | Low-quality |
| 18-106_k141_79567 | Not-determined |
| 18-106_k141_85697 | Not-determined |
| 18-106_k141_79571 | Not-determined |
| 18-106_k141_55880 | Not-determined |
| 18-106_k141_132712 | Not-determined |
| 18-106_k141_26790 | Not-determined |
| 18-106_k141_132715 | Not-determined |
| 18-106_k141_68727 | Not-determined |
| 18-106_k141_114967 | Not-determined |
| 18-106_k141_102968 | Low-quality |
| 18-106_k141_39173 | Not-determined |
| 18-106_k141_4730 | Not-determined |
| 18-106_k141_85704 | Not-determined |
| 18-106_k141_79584 | Not-determined |
| 18-106_k141_9121 | Not-determined |
| 18-106_k141_9122 | Not-determined |
| 18-106_k141_4735 | Not-determined |
| 18-106_k141_85708 | Not-determined |
| 18-106_k141_132724 | Not-determined |
| 18-106_k141_132728 | Not-determined |
| 18-106_k141_132730 | Not-determined |
| 18-106_k141_73901 | Not-determined |
| 18-106_k141_97987 | Not-determined |
| 18-106_k141_4739 | Not-determined |
| 18-106_k141_73906 | Not-determined |
| 18-106_k141_15185 | Not-determined |
| 18-106_k141_97998 | Not-determined |
| 18-106_k141_85722 | Not-determined |
| 18-106_k141_79592 | Not-determined |
| 18-106_k141_98006 | Not-determined |
| 18-106_k141_26796 | Not-determined |
| 18-106_k141_98009 | Not-determined |
| 18-106_k141_26805 | Not-determined |
| 18-106_k141_102975 | Not-determined |
| 18-106_k141_98017 | Low-quality |
| 18-106_k141_15203 | Not-determined |
| 18-106_k141_68738 | Not-determined |
| 18-106_k141_79606 | Not-determined |
| 18-106_k141_55908 | Not-determined |
| 18-106_k141_73921 | Not-determined |
| 18-106_k141_55915 | Not-determined |
| 18-106_k141_26815 | Not-determined |
| 18-106_k141_85741 | Not-determined |
| 18-106_k141_73927 | Low-quality |
| 18-106_k141_4778 | Not-determined |
| 18-106_k141_102996 | Not-determined |
| 18-106_k141_102998 | Not-determined |
| 18-106_k141_4780 | Not-determined |
| 18-106_k141_26822 | Not-determined |
| 18-106_k141_62746 | Not-determined |
| 18-106_k141_132762 | Not-determined |
| 18-106_k141_4788 | Not-determined |
| 18-106_k141_109320 | Not-determined |
| 18-106_k141_138241 | Not-determined |
| 18-106_k141_20671 | Not-determined |
| 18-106_k141_68762 | Not-determined |
| 18-106_k141_26831 | Low-quality |
| 18-106_k141_26836 | Not-determined |
| 18-106_k141_91698 | Not-determined |
| 18-106_k141_109336 | Not-determined |
| 18-106_k141_45197 | Not-determined |
| 18-106_k141_114982 | Not-determined |
| 18-106_k141_138254 | Not-determined |
| 18-106_k141_120518 | Not-determined |
| 18-106_k141_26845 | Not-determined |
| 18-106_k141_9135 | Not-determined |
| 18-106_k141_45200 | Not-determined |
| 18-106_k141_91713 | Not-determined |
| 18-106_k141_85759 | Not-determined |
| 18-106_k141_55932 | Not-determined |
| 18-106_k141_4808 | Not-determined |
| 18-106_k141_62768 | Not-determined |
| 18-106_k141_62773 | Not-determined |
| 18-106_k141_20688 | Not-determined |
| 18-106_k141_50728 | Not-determined |
| 18-106_k141_4820 | Not-determined |
| 18-106_k141_15274 | Not-determined |
| 18-106_k141_91733 | Not-determined |
| 18-106_k141_9142 | Not-determined |
| 18-106_k141_4831 | Not-determined |
| 18-106_k141_55945 | Not-determined |
| 18-106_k141_120543 | Low-quality |
| 18-106_k141_91738 | Low-quality |
| 18-106_k141_55951 | Not-determined |
| 18-106_k141_85773 | Not-determined |
| 18-106_k141_91741 | Not-determined |
| 18-106_k141_55954 | Not-determined |
| 18-106_k141_91744 | Not-determined |
| 18-106_k141_109374 | Not-determined |
| 18-106_k141_138272 | Low-quality |
| 18-106_k141_50736 | Not-determined |
| 18-106_k141_85786 | Not-determined |
| 18-106_k141_62801 | Not-determined |
| 18-106_k141_50739 | Not-determined |
| 18-106_k141_103045 | Not-determined |
| 18-106_k141_115022 | Not-determined |
| 18-106_k141_91751 | Not-determined |
| 18-106_k141_138283 | Not-determined |
| 18-106_k141_109378 | Low-quality |
| 18-106_k141_62811 | Not-determined |
| 18-106_k141_39189 | Not-determined |
| 18-106_k141_39191 | Not-determined |
| 18-106_k141_109390 | Not-determined |
| 18-106_k141_39195 | Not-determined |
| 18-106_k141_26881 | Not-determined |
| 18-106_k141_115040 | Not-determined |
| 18-106_k141_138301 | Not-determined |
| 18-106_k141_138302 | Not-determined |
| 18-106_k141_85799 | Not-determined |
| 18-106_k141_4877 | Not-determined |
| 18-106_k141_126894 | Not-determined |
| 18-106_k141_91770 | Not-determined |
| 18-106_k141_91772 | Not-determined |
| 18-106_k141_120559 | Not-determined |
| 18-106_k141_68791 | Not-determined |
| 18-106_k141_50765 | Not-determined |
| 18-106_k141_26887 | Not-determined |
| 18-106_k141_50766 | Not-determined |
| 18-106_k141_62828 | Not-determined |
| 18-106_k141_45231 | Not-determined |
| 18-106_k141_91777 | Not-determined |
| 18-106_k141_45233 | Not-determined |
| 18-106_k141_39212 | Not-determined |
| 18-106_k141_4885 | Not-determined |
| 18-106_k141_39215 | Not-determined |
| 18-106_k141_39218 | Low-quality |
| 18-106_k141_120567 | Low-quality |
| 18-106_k141_91782 | Not-determined |
| 18-106_k141_9169 | Not-determined |
| 18-106_k141_9170 | Not-determined |
| 18-106_k141_62835 | Not-determined |
| 18-106_k141_62838 | Not-determined |
| 18-106_k141_73982 | Not-determined |
| 18-106_k141_73983 | Not-determined |
| 18-106_k141_26907 | Not-determined |
| 18-106_k141_91789 | Not-determined |
| 18-106_k141_79665 | Not-determined |
| 18-106_k141_26909 | Not-determined |
| 18-106_k141_91791 | Not-determined |
| 18-106_k141_103078 | Not-determined |
| 18-106_k141_26910 | Not-determined |
| 18-106_k141_26913 | Not-determined |
| 18-106_k141_109411 | Not-determined |
| 18-106_k141_62848 | Not-determined |
| 18-106_k141_91799 | Not-determined |
| 18-106_k141_109416 | Not-determined |
| 18-106_k141_85837 | Not-determined |
| 18-106_k141_98111 | Not-determined |
| 18-106_k141_85840 | Not-determined |
| 18-106_k141_74003 | Not-determined |
| 18-106_k141_138321 | Not-determined |
| 18-106_k141_26923 | Not-determined |
| 18-106_k141_138325 | Not-determined |
| 18-106_k141_68819 | Not-determined |
| 18-106_k141_138326 | Not-determined |
| 18-106_k141_79686 | Not-determined |
| 18-106_k141_50780 | Not-determined |
| 18-106_k141_103101 | Not-determined |
| 18-106_k141_62859 | Not-determined |
| 18-106_k141_115083 | Not-determined |
| 18-106_k141_26929 | Not-determined |
| 18-106_k141_109427 | Not-determined |
| 18-106_k141_62862 | Low-quality |
| 18-106_k141_62863 | Not-determined |
| 18-106_k141_138331 | Not-determined |
| 18-106_k141_26936 | Not-determined |
| 18-106_k141_138332 | Not-determined |
| 18-106_k141_115090 | Not-determined |
| 18-106_k141_115091 | Not-determined |
| 18-106_k141_138333 | Not-determined |
| 18-106_k141_91815 | Low-quality |
| 18-106_k141_68837 | Not-determined |
| 18-106_k141_62870 | Low-quality |
| 18-106_k141_15329 | Not-determined |
| 18-106_k141_115105 | Not-determined |
| 18-106_k141_4911 | Not-determined |
| 18-106_k141_4913 | Not-determined |
| 18-106_k141_120621 | Not-determined |
| 18-106_k141_15341 | Not-determined |
| 18-106_k141_62880 | Not-determined |
| 18-106_k141_74036 | Not-determined |
| 18-106_k141_115112 | Low-quality |
| 18-106_k141_62884 | Not-determined |
| 18-106_k141_9201 | Not-determined |
| 18-106_k141_109446 | Not-determined |
| 18-106_k141_20696 | Not-determined |
| 18-106_k141_91831 | Not-determined |
| 18-106_k141_85864 | Not-determined |
| 18-106_k141_15354 | Not-determined |
| 18-106_k141_120632 | Not-determined |
| 18-106_k141_103115 | Not-determined |
| 18-106_k141_15356 | Not-determined |
| 18-106_k141_15358 | Not-determined |
| 18-106_k141_50801 | Not-determined |
| 18-106_k141_50803 | Not-determined |
| 18-106_k141_74048 | Not-determined |
| 18-106_k141_120636 | Not-determined |
| 18-106_k141_74052 | Not-determined |
| 18-106_k141_120637 | Not-determined |
| 18-106_k141_103127 | Not-determined |

| | |
|---|---|
| 18-106_k141_15371 | Not-determined |
| 18-106_k141_120642 | Not-determined |
| 18-106_k141_9210 | Not-determined |
| 18-106_k141_20707 | Not-determined |
| 18-106_k141_62903 | Not-determined |
| 18-106_k141_15377 | Not-determined |
| 18-106_k141_98127 | Not-determined |
| 18-106_k141_50816 | Not-determined |
| 18-106_k141_15380 | Not-determined |
| 18-106_k141_91856 | Not-determined |
| 18-106_k141_126942 | Not-determined |
| 18-106_k141_126944 | Not-determined |
| 18-106_k141_50819 | Not-determined |
| 18-106_k141_120654 | Not-determined |
| 18-106_k141_15387 | Not-determined |
| 18-106_k141_50822 | Not-determined |
| 18-106_k141_15394 | Not-determined |
| 18-106_k141_15396 | Not-determined |
| 18-106_k141_39251 | Not-determined |
| 18-106_k141_62908 | Low-quality |
| 18-106_k141_85889 | Not-determined |
| 18-106_k141_62910 | Not-determined |
| 18-106_k141_85894 | Not-determined |
| 18-106_k141_33008 | Not-determined |
| 18-106_k141_39259 | Not-determined |
| 18-106_k141_39262 | Not-determined |
| 18-106_k141_91869 | Not-determined |
| 18-106_k141_126955 | Not-determined |
| 18-106_k141_98146 | Not-determined |
| 18-106_k141_26980 | Not-determined |
| 18-106_k141_26982 | Not-determined |
| 18-106_k141_33023 | Low-quality |
| 18-106_k141_120680 | Not-determined |
| 18-106_k141_115144 | Not-determined |
| 18-106_k141_39275 | Not-determined |
| 18-106_k141_115147 | Not-determined |
| 18-106_k141_39277 | Not-determined |
| 18-106_k141_9234 | Not-determined |
| 18-106_k141_9236 | Not-determined |
| 18-106_k141_109475 | Not-determined |
| 18-106_k141_45267 | Low-quality |
| 18-106_k141_85911 | Not-determined |
| 18-106_k141_33035 | Not-determined |
| 18-106_k141_98165 | Low-quality |
| 18-106_k141_15426 | Not-determined |
| 18-106_k141_120684 | Not-determined |
| 18-106_k141_9246 | Not-determined |
| 18-106_k141_62944 | Not-determined |
| 18-106_k141_62951 | Not-determined |
| 18-106_k141_98174 | Not-determined |
| 18-106_k141_103181 | Not-determined |
| 18-106_k141_98177 | Not-determined |
| 18-106_k141_4985 | Not-determined |
| 18-106_k141_98181 | Not-determined |
| 18-106_k141_98184 | Not-determined |
| 18-106_k141_50867 | Not-determined |
| 18-106_k141_109495 | Not-determined |
| 18-106_k141_39292 | Not-determined |
| 18-106_k141_103192 | Not-determined |
| 18-106_k141_68890 | Not-determined |
| 18-106_k141_120704 | Not-determined |
| 18-106_k141_4993 | Not-determined |
| 18-106_k141_74095 | Not-determined |
| 18-106_k141_33061 | Not-determined |
| 18-106_k141_109497 | Not-determined |
| 18-106_k141_62978 | Not-determined |
| 18-106_k141_45290 | Not-determined |
| 18-106_k141_20779 | Not-determined |
| 18-106_k141_109502 | Not-determined |
| 18-106_k141_45303 | Not-determined |
| 18-106_k141_33073 | Low-quality |
| 18-106_k141_120720 | Not-determined |
| 18-106_k141_120722 | Not-determined |
| 18-106_k141_115201 | Not-determined |
| 18-106_k141_68904 | Not-determined |
| 18-106_k141_85934 | Not-determined |
| 18-106_k141_85935 | Not-determined |
| 18-106_k141_79773 | Not-determined |
| 18-106_k141_15456 | Not-determined |
| 18-106_k141_15459 | Low-quality |
| 18-106_k141_91905 | Not-determined |
| 18-106_k141_50878 | Not-determined |
| 18-106_k141_20789 | Not-determined |
| 18-106_k141_15466 | Not-determined |
| 18-106_k141_68911 | Not-determined |
| 18-106_k141_103214 | Not-determined |
| 18-106_k141_15470 | Not-determined |
| 18-106_k141_20792 | Not-determined |
| 18-106_k141_33088 | Not-determined |
| 18-106_k141_9261 | Not-determined |
| 18-106_k141_62995 | Low-quality |
| 18-106_k141_9266 | Not-determined |
| 18-106_k141_9267 | Not-determined |
| 18-106_k141_33095 | Not-determined |
| 18-106_k141_62998 | Not-determined |
| 18-106_k141_109536 | Not-determined |
| 18-106_k141_56017 | Not-determined |
| 18-106_k141_85943 | Not-determined |
| 18-106_k141_85944 | Not-determined |
| 18-106_k141_109542 | Not-determined |
| 18-106_k141_74139 | Not-determined |
| 18-106_k141_85951 | Not-determined |
| 18-106_k141_109545 | Not-determined |
| 18-106_k141_56022 | Not-determined |
| 18-106_k141_109547 | Not-determined |
| 17-109-final-viral-combined_k141_351847\|\|full | Medium-quality |
| 18-106_k141_63010 | Not-determined |
| 18-106_k141_68941 | Not-determined |
| 18-106_k141_63013 | Not-determined |
| 18-106_k141_39347 | Not-determined |
| 18-106_k141_56033 | Not-determined |
| 18-106_k141_126996 | Not-determined |
| 18-106_k141_63019 | Not-determined |
| 18-106_k141_56034 | Not-determined |
| 18-106_k141_68946 | Low-quality |
| 18-106_k141_127000 | Low-quality |
| 18-106_k141_115225 | Not-determined |
| 18-106_k141_15491 | Low-quality |
| 18-106_k141_50902 | Not-determined |
| 18-106_k141_120754 | Not-determined |
| 18-106_k141_74148 | Not-determined |
| 18-106_k141_85963 | Not-determined |
| 18-106_k141_91927 | Not-determined |
| 18-106_k141_79808 | Not-determined |
| 18-106_k141_15501 | Not-determined |
| 18-106_k141_98240 | Low-quality |
| 18-106_k141_33111 | Not-determined |
| 18-106_k141_79809 | Not-determined |
| 18-106_k141_9298 | Not-determined |
| 18-106_k141_33114 | Not-determined |
| 18-106_k141_56046 | Not-determined |
| 18-106_k141_33115 | Not-determined |
| 18-106_k141_39351 | Not-determined |
| 18-106_k141_120763 | Not-determined |
| 18-106_k141_56054 | Not-determined |
| 18-106_k141_85986 | Not-determined |
| 18-106_k141_9310 | Not-determined |
| 18-106_k141_5049 | Not-determined |
| 18-106_k141_9311 | Not-determined |
| 18-106_k141_33130 | Not-determined |
| 18-106_k141_9315 | Not-determined |
| 18-106_k141_138373 | Not-determined |
| 18-106_k141_50910 | Low-quality |
| 18-106_k141_50911 | Not-determined |
| 18-106_k141_68974 | Not-determined |
| 18-106_k141_27052 | Low-quality |
| 18-106_k141_115241 | Not-determined |
| 18-106_k141_68978 | Not-determined |
| 18-106_k141_27056 | Not-determined |
| 18-106_k141_63042 | Not-determined |
| 18-106_k141_50917 | Not-determined |
| 18-106_k141_79827 | Low-quality |
| 18-106_k141_109594 | Not-determined |
| 18-106_k141_91938 | Not-determined |
| 18-106_k141_27059 | Not-determined |
| 18-106_k141_74168 | Not-determined |
| 18-106_k141_39388 | Not-determined |
| 18-106_k141_109601 | Not-determined |
| 18-106_k141_9340 | Not-determined |
| 18-106_k141_39395 | Not-determined |
| 18-106_k141_15534 | Not-determined |
| 18-106_k141_138405 | Not-determined |
| 18-106_k141_45329 | Not-determined |
| 18-106_k141_109608 | Not-determined |
| 18-106_k141_20833 | Not-determined |
| 18-106_k141_79844 | Low-quality |
| 18-106_k141_15544 | Not-determined |
| 18-106_k141_5074 | Not-determined |
| 18-106_k141_27071 | Not-determined |
| 18-106_k141_33175 | Not-determined |
| 18-106_k141_39403 | Not-determined |
| 18-106_k141_33178 | Not-determined |
| 18-106_k141_79854 | Not-determined |
| 18-106_k141_33179 | Low-quality |
| 18-106_k141_138421 | Not-determined |
| 18-106_k141_91955 | Not-determined |
| 18-106_k141_39412 | Not-determined |
| 18-106_k141_91958 | Not-determined |
| 18-106_k141_5091 | Not-determined |
| 18-106_k141_115282 | Not-determined |
| 18-106_k141_98306 | Not-determined |
| 18-106_k141_20850 | Not-determined |
| 18-106_k141_127037 | Not-determined |
| 18-106_k141_50962 | Low-quality |
| 18-106_k141_120779 | Not-determined |
| 18-106_k141_120780 | Not-determined |
| 18-106_k141_45354 | Not-determined |
| 18-106_k141_56097 | Low-quality |
| 18-106_k141_50963 | Not-determined |
| 18-106_k141_68995 | Not-determined |
| 18-106_k141_56107 | Not-determined |
| 18-106_k141_5109 | Not-determined |
| 18-106_k141_56108 | Not-determined |
| 18-106_k141_33204 | Not-determined |
| 18-106_k141_5110 | Not-determined |
| 18-106_k141_120787 | Not-determined |
| 18-106_k141_120795 | Not-determined |
| 18-106_k141_9379 | Not-determined |
| 18-106_k141_39445 | Not-determined |
| 18-106_k141_20859 | Not-determined |
| 18-106_k141_115294 | Not-determined |
| 18-106_k141_69005 | Not-determined |
| 18-106_k141_27101 | Not-determined |
| 18-106_k141_120798 | Not-determined |
| 18-106_k141_20870 | Not-determined |
| 18-106_k141_86025 | Not-determined |
| 18-106_k141_69010 | Not-determined |
| 18-106_k141_109627 | Low-quality |
| 18-106_k141_115301 | Not-determined |
| 18-106_k141_20880 | Not-determined |
| 18-106_k141_33214 | Low-quality |
| 18-106_k141_27115 | Not-determined |
| 18-106_k141_27118 | Not-determined |
| 18-106_k141_103285 | Not-determined |
| 18-106_k141_86038 | Not-determined |
| 18-106_k141_50988 | Not-determined |
| 18-106_k141_138440 | Not-determined |
| 18-106_k141_138441 | Not-determined |
| 18-106_k141_103295 | Not-determined |
| 18-106_k141_20898 | Not-determined |
| 18-106_k141_120813 | Not-determined |
| 18-106_k141_27133 | Not-determined |
| 18-106_k141_103302 | Not-determined |
| 18-106_k141_15579 | Not-determined |
| 18-106_k141_63105 | Not-determined |
| 18-106_k141_9423 | Not-determined |
| 18-106_k141_63112 | Not-determined |
| 18-106_k141_5156 | Not-determined |
| 18-106_k141_5157 | Not-determined |
| 18-106_k141_45387 | Not-determined |
| 18-106_k141_9428 | Not-determined |
| 18-106_k141_127079 | Not-determined |
| 18-106_k141_127080 | Not-determined |
| 18-106_k141_63119 | Not-determined |
| 18-106_k141_86070 | Low-quality |
| 18-106_k141_86071 | Not-determined |
| 18-106_k141_127081 | Not-determined |
| 18-106_k141_63125 | Not-determined |
| 18-106_k141_127084 | Not-determined |
| 18-106_k141_63127 | Not-determined |
| 18-106_k141_39476 | Not-determined |
| 18-106_k141_127086 | Not-determined |
| 18-106_k141_74217 | Not-determined |
| 18-106_k141_79910 | Low-quality |
| 18-106_k141_15599 | Not-determined |
| 18-106_k141_120854 | Not-determined |
| 18-106_k141_39481 | Not-determined |
| 18-106_k141_115343 | Not-determined |
| 18-106_k141_103310 | Not-determined |
| 18-106_k141_86094 | Not-determined |
| 18-106_k141_103312 | Not-determined |
| 18-106_k141_45415 | Not-determined |
| 18-106_k141_98314 | Not-determined |
| 18-106_k141_69041 | Not-determined |
| 18-106_k141_127099 | Not-determined |
| 18-106_k141_69046 | Not-determined |
| 18-106_k141_138474 | Not-determined |
| 18-106_k141_103316 | Not-determined |
| 18-106_k141_20915 | Low-quality |
| 18-106_k141_86105 | Not-determined |
| 18-106_k141_120869 | Not-determined |
| 18-106_k141_15618 | Not-determined |
| 18-106_k141_15619 | Not-determined |
| 18-106_k141_86115 | Not-determined |
| 18-106_k141_63161 | Not-determined |
| 18-106_k141_115357 | Not-determined |
| 18-106_k141_9434 | Not-determined |
| 18-106_k141_9435 | Low-quality |
| 18-106_k141_15632 | Not-determined |
| 18-106_k141_63166 | Not-determined |
| 18-106_k141_27174 | Not-determined |
| 18-106_k141_33275 | Low-quality |
| 18-106_k141_103340 | Not-determined |
| 18-106_k141_15639 | Not-determined |
| 18-106_k141_15640 | Not-determined |
| 18-106_k141_86121 | Low-quality |
| 18-106_k141_15641 | Low-quality |
| 18-106_k141_74247 | Not-determined |
| 18-106_k141_33285 | Not-determined |
| 18-106_k141_79934 | Not-determined |
| 18-106_k141_103352 | Not-determined |
| 18-106_k141_86138 | Not-determined |
| 18-106_k141_74256 | Not-determined |
| 18-106_k141_127135 | Not-determined |
| 18-106_k141_115368 | Not-determined |
| 18-106_k141_15658 | Not-determined |
| 18-106_k141_103363 | Not-determined |
| 18-106_k141_86151 | Not-determined |
| 18-106_k141_39534 | Not-determined |
| 18-106_k141_138512 | Not-determined |
| 18-106_k141_39536 | Not-determined |
| 18-106_k141_27205 | Not-determined |
| 18-106_k141_103373 | Not-determined |
| 18-106_k141_63199 | Not-determined |
| 18-106_k141_109647 | Not-determined |
| 18-106_k141_51051 | Not-determined |
| 18-106_k141_9467 | Not-determined |
| 18-106_k141_109654 | Not-determined |
| 18-106_k141_103377 | Not-determined |
| 18-106_k141_115389 | Not-determined |
| 18-106_k141_103378 | Not-determined |
| 18-106_k141_74269 | Not-determined |
| 18-106_k141_39539 | Not-determined |
| 18-106_k141_127159 | Not-determined |
| 18-106_k141_127160 | Not-determined |
| 18-106_k141_115399 | Not-determined |
| 18-106_k141_9482 | Low-quality |
| 18-106_k141_33322 | Not-determined |
| 18-106_k141_20949 | Not-determined |
| 18-106_k141_33325 | Not-determined |
| 18-106_k141_98361 | Not-determined |
| 18-106_k141_63219 | Not-determined |
| 18-106_k141_63222 | Not-determined |
| 18-106_k141_98365 | Not-determined |
| 18-106_k141_138525 | Not-determined |
| 18-106_k141_5235 | Not-determined |
| 18-106_k141_9497 | Not-determined |
| 18-106_k141_115407 | Not-determined |
| 18-106_k141_115408 | Not-determined |
| 18-106_k141_5244 | Not-determined |
| 18-106_k141_9500 | Not-determined |
| 18-106_k141_138535 | Not-determined |
| 18-106_k141_27219 | Not-determined |
| 18-106_k141_27221 | Not-determined |
| 18-106_k141_9503 | Not-determined |
| 18-106_k141_5255 | Not-determined |
| 18-106_k141_91983 | Not-determined |
| 18-106_k141_5259 | Not-determined |
| 18-106_k141_33353 | Not-determined |
| 18-106_k141_39564 | Not-determined |
| 18-106_k141_63232 | Not-determined |
| 18-106_k141_51079 | Not-determined |
| 18-106_k141_33362 | Not-determined |
| 18-106_k141_15674 | Not-determined |
| 18-106_k141_109685 | Not-determined |
| 18-106_k141_15678 | Not-determined |
| 18-106_k141_9522 | Not-determined |
| 18-106_k141_33378 | Low-quality |
| 18-106_k141_15684 | Not-determined |
| 18-106_k141_15687 | Not-determined |
| 18-106_k141_127199 | Not-determined |
| 18-106_k141_127204 | Not-determined |
| 18-106_k141_15689 | Not-determined |
| 18-106_k141_103433 | Not-determined |
| 18-106_k141_27253 | Not-determined |
| 18-106_k141_127208 | Not-determined |
| 18-106_k141_127210 | Not-determined |
| 18-106_k141_98387 | Not-determined |
| 18-106_k141_74313 | Not-determined |
| 18-106_k141_5280 | Not-determined |
| 18-106_k141_86196 | Low-quality |
| 18-106_k141_86198 | Not-determined |
| 18-106_k141_86199 | Not-determined |
| 18-106_k141_98395 | Low-quality |
| 18-106_k141_98398 | Not-determined |
| 18-106_k141_98400 | Not-determined |
| 18-106_k141_45515 | Not-determined |
| 18-106_k141_120889 | Not-determined |
| 18-106_k141_9553 | Not-determined |
| 18-106_k141_86206 | Not-determined |
| 18-106_k141_15714 | Not-determined |
| 18-106_k141_9557 | Not-determined |
| 18-106_k141_138585 | Not-determined |
| 18-106_k141_5301 | Not-determined |
| 18-106_k141_86214 | Not-determined |
| 18-106_k141_9562 | Low-quality |
| 18-106_k141_33417 | Not-determined |
| 18-106_k141_98410 | Low-quality |
| 18-106_k141_92035 | Not-determined |
| 18-106_k141_120903 | Not-determined |
| 18-106_k141_120905 | Not-determined |
| 18-106_k141_138593 | Low-quality |
| 18-106_k141_63265 | Not-determined |
| 18-106_k141_120915 | Not-determined |
| 18-106_k141_74333 | Not-determined |
| 18-106_k141_115431 | Not-determined |
| 18-106_k141_115435 | Not-determined |
| 18-106_k141_120925 | Low-quality |
| 18-106_k141_63280 | Not-determined |
| 18-106_k141_138614 | Not-determined |
| 18-106_k141_63284 | Not-determined |
| 18-106_k141_138618 | Not-determined |
| 18-106_k141_15737 | Not-determined |
| 18-106_k141_63285 | Not-determined |
| 18-106_k141_138624 | Not-determined |
| 18-106_k141_9587 | Low-quality |
| 18-106_k141_132783 | Low-quality |
| 18-106_k141_15742 | Not-determined |
| 18-106_k141_109699 | Not-determined |
| 18-106_k141_27291 | Not-determined |
| 18-106_k141_33440 | Not-determined |
| 18-106_k141_74357 | Not-determined |
| 18-106_k141_98432 | Not-determined |
| 18-106_k141_103459 | Not-determined |
| 18-106_k141_63298 | Not-determined |
| 18-106_k141_39621 | Not-determined |
| 18-106_k141_79958 | Not-determined |
| 18-106_k141_33447 | Not-determined |
| 18-106_k141_132805 | Low-quality |
| 18-106_k141_86249 | Not-determined |
| 18-106_k141_74360 | Low-quality |
| 18-106_k141_115453 | Not-determined |
| 18-106_k141_98439 | Not-determined |
| 18-106_k141_63313 | Not-determined |
| 18-106_k141_51154 | Not-determined |
| 18-106_k141_9604 | Low-quality |
| 18-106_k141_33456 | Not-determined |
| 18-106_k141_79971 | Not-determined |
| 18-106_k141_63317 | Not-determined |
| 18-106_k141_109708 | Not-determined |
| 18-106_k141_9612 | Not-determined |
| 18-106_k141_74375 | Not-determined |
| 18-106_k141_120986 | Not-determined |
| 18-106_k141_15779 | Not-determined |
| 18-106_k141_63321 | Not-determined |
| 18-106_k141_39632 | Not-determined |
| 18-106_k141_45543 | Not-determined |
| 18-106_k141_5349 | Not-determined |
| 18-106_k141_15783 | Not-determined |
| 18-106_k141_69095 | Not-determined |
| 18-106_k141_5353 | Not-determined |
| 18-106_k141_109722 | Not-determined |
| 18-106_k141_98449 | Not-determined |
| 18-106_k141_33477 | Not-determined |
| 18-106_k141_51168 | Not-determined |
| 18-106_k141_74389 | Not-determined |
| 18-106_k141_15792 | Low-quality |
| 18-106_k141_103483 | Not-determined |
| 18-106_k141_20966 | Not-determined |
| 18-106_k141_20967 | Not-determined |
| 18-106_k141_138652 | Not-determined |
| 18-106_k141_5357 | Not-determined |
| 18-106_k141_9639 | Not-determined |
| 18-106_k141_79989 | Low-quality |
| 18-106_k141_79990 | Not-determined |
| 18-106_k141_127269 | Not-determined |
| 18-106_k141_5361 | Not-determined |
| 18-106_k141_92089 | Not-determined |
| 18-106_k141_27328 | Low-quality |
| 18-106_k141_92095 | Not-determined |
| 18-106_k141_45569 | Low-quality |
| 18-106_k141_9652 | Not-determined |
| 18-106_k141_92097 | Not-determined |
| 18-106_k141_27333 | Not-determined |
| 18-106_k141_39658 | Not-determined |
| 18-106_k141_39659 | Not-determined |
| 18-106_k141_80002 | Not-determined |
| 18-106_k141_86291 | Not-determined |
| 18-106_k141_15800 | Low-quality |
| 18-106_k141_98476 | Not-determined |
| 18-106_k141_109741 | Not-determined |
| 18-106_k141_20989 | Not-determined |
| 18-106_k141_51199 | Low-quality |
| 18-106_k141_15803 | Not-determined |
| 18-106_k141_109744 | Not-determined |
| 18-106_k141_63339 | Not-determined |
| 18-106_k141_109747 | Not-determined |
| 18-106_k141_115503 | Not-determined |
| 18-106_k141_56151 | Not-determined |
| 18-106_k141_27334 | Not-determined |
| 18-106_k141_115505 | Not-determined |
| 18-106_k141_33499 | Not-determined |
| 18-106_k141_121017 | Not-determined |
| 18-106_k141_15814 | Not-determined |
| 18-106_k141_39672 | Low-quality |
| 18-106_k141_33504 | Not-determined |
| 18-106_k141_127300 | Not-determined |
| 18-106_k141_5386 | Not-determined |
| 18-106_k141_115512 | Not-determined |
| 18-106_k141_127307 | Not-determined |
| 18-106_k141_56164 | Not-determined |
| 18-106_k141_127309 | Not-determined |
| 18-106_k141_27347 | Low-quality |
| 18-106_k141_98503 | Not-determined |
| 18-106_k141_109774 | Not-determined |
| 18-106_k141_103532 | Not-determined |
| 18-106_k141_121030 | Not-determined |
| 18-106_k141_69115 | Not-determined |
| 18-106_k141_21002 | Not-determined |
| 18-106_k141_98509 | Not-determined |
| 18-106_k141_5402 | Not-determined |
| 18-106_k141_109779 | Not-determined |
| 18-106_k141_121035 | Not-determined |
| 18-106_k141_45614 | Not-determined |
| 18-106_k141_80023 | Not-determined |
| 18-106_k141_132885 | Not-determined |
| 18-106_k141_132887 | Not-determined |
| 18-106_k141_45618 | Not-determined |
| 18-106_k141_103534 | Not-determined |
| 18-106_k141_127315 | Not-determined |
| 18-106_k141_92101 | Not-determined |
| 18-106_k141_45619 | Low-quality |
| 18-106_k141_51238 | Low-quality |
| 18-106_k141_27358 | Low-quality |
| 18-106_k141_103543 | Not-determined |
| 18-106_k141_5412 | Low-quality |
| 18-106_k141_103552 | Not-determined |
| 18-106_k141_45640 | Not-determined |
| 18-106_k141_74427 | Low-quality |
| 18-106_k141_92120 | Low-quality |
| 18-106_k141_27372 | Not-determined |
| 18-106_k141_92127 | Not-determined |
| 18-106_k141_115564 | Not-determined |
| 18-106_k141_51263 | Not-determined |
| 18-106_k141_109804 | Not-determined |
| 18-106_k141_80054 | Not-determined |
| 18-106_k141_51275 | Not-determined |
| 18-106_k141_5431 | Not-determined |
| 18-106_k141_21029 | Not-determined |
| 18-106_k141_5435 | Not-determined |
| 18-106_k141_86311 | Not-determined |
| 18-106_k141_5436 | Not-determined |
| 18-106_k141_27381 | Not-determined |
| 18-106_k141_51281 | Not-determined |
| 18-106_k141_51287 | Low-quality |
| 18-106_k141_39710 | Not-determined |
| 18-106_k141_86319 | Not-determined |
| 18-106_k141_63351 | Not-determined |
| 18-106_k141_132912 | Not-determined |
| 18-106_k141_80066 | Low-quality |
| 18-106_k141_86321 | Not-determined |
| 18-106_k141_15875 | Not-determined |
| 18-106_k141_15879 | Not-determined |
| 18-106_k141_39720 | Not-determined |
| 18-106_k141_39725 | Not-determined |
| 18-106_k141_132920 | Not-determined |
| 18-106_k141_63361 | Not-determined |
| 18-106_k141_86336 | Not-determined |
| 18-106_k141_80081 | Not-determined |
| 18-106_k141_45668 | Not-determined |
| 18-106_k141_86338 | Not-determined |
| 18-106_k141_63367 | Not-determined |
| 18-106_k141_115596 | Not-determined |
| 18-106_k141_9690 | Not-determined |
| 18-106_k141_51308 | Not-determined |
| 18-106_k141_63370 | Not-determined |
| 18-106_k141_9693 | Not-determined |
| 18-106_k141_63373 | Not-determined |
| 18-106_k141_27402 | Low-quality |
| 18-106_k141_74433 | Not-determined |
| 18-106_k141_74434 | Low-quality |
| 18-106_k141_21053 | Not-determined |
| 18-106_k141_115609 | Not-determined |
| 18-106_k141_121057 | Not-determined |
| 18-106_k141_74440 | Not-determined |
| 18-106_k141_98568 | Not-determined |
| 18-106_k141_86352 | Low-quality |
| 18-106_k141_86353 | Not-determined |
| 18-106_k141_121060 | Not-determined |
| 18-106_k141_115617 | Not-determined |
| 18-106_k141_9702 | Low-quality |
| 18-106_k141_115619 | Low-quality |
| 18-106_k141_51328 | Low-quality |
| 18-106_k141_27414 | Not-determined |
| 18-106_k141_80107 | Not-determined |
| 18-106_k141_27424 | Not-determined |
| 18-106_k141_45682 | Not-determined |
| 18-106_k141_45685 | Not-determined |
| 18-106_k141_92141 | Not-determined |
| 18-106_k141_21073 | Not-determined |
| 18-106_k141_69148 | Not-determined |
| 18-106_k141_80125 | Not-determined |
| 18-106_k141_21077 | Not-determined |
| 18-106_k141_21080 | Not-determined |
| 18-106_k141_21081 | Not-determined |
| 18-106_k141_15923 | Not-determined |
| 18-106_k141_45688 | Low-quality |
| 18-106_k141_39761 | Not-determined |
| 18-106_k141_21084 | Not-determined |
| 18-106_k141_63401 | Not-determined |
| 18-106_k141_63402 | Not-determined |
| 18-106_k141_15932 | Not-determined |
| 18-106_k141_69165 | Not-determined |
| 18-106_k141_138675 | Not-determined |
| 18-106_k141_92156 | Not-determined |
| 18-106_k141_69175 | Not-determined |
| 18-106_k141_74481 | Low-quality |
| 18-106_k141_56208 | Not-determined |
| 18-106_k141_115633 | Low-quality |
| 18-106_k141_92161 | Not-determined |
| 18-106_k141_45714 | Not-determined |
| 18-106_k141_109892 | Not-determined |
| 18-106_k141_51365 | Not-determined |
| 18-106_k141_45716 | Not-determined |
| 18-106_k141_15951 | Not-determined |
| 18-106_k141_15952 | Not-determined |
| 18-106_k141_86396 | Not-determined |
| 18-106_k141_109901 | Not-determined |
| 18-106_k141_9737 | Not-determined |

| | |
|---|---|
| 18-106_k141_45725 | Not-determined |
| 18-106_k141_92177 | Not-determined |
| 18-106_k141_121091 | Not-determined |
| 18-106_k141_15962 | Not-determined |
| 18-106_k141_132957 | Not-determined |
| 18-106_k141_127335 | Low-quality |
| 18-106_k141_51379 | Not-determined |
| 18-106_k141_109907 | Not-determined |
| 18-106_k141_80145 | Not-determined |
| 18-106_k141_92192 | Not-determined |
| 18-106_k141_109909 | Not-determined |
| 18-106_k141_132964 | Not-determined |
| 18-106_k141_45735 | Not-determined |
| 18-106_k141_51384 | Not-determined |
| 18-106_k141_86409 | Not-determined |
| 18-106_k141_86410 | Low-quality |
| 18-106_k141_132970 | Not-determined |
| 18-106_k141_98590 | Not-determined |
| 18-106_k141_63441 | Not-determined |
| 18-106_k141_98597 | Not-determined |
| 18-106_k141_109918 | Not-determined |
| 18-106_k141_27479 | Not-determined |
| 18-106_k141_92202 | Not-determined |
| 18-106_k141_109921 | Not-determined |
| 18-106_k141_51388 | Not-determined |
| 18-106_k141_127364 | Low-quality |
| 18-106_k141_27489 | Not-determined |
| 18-106_k141_15975 | Not-determined |
| 18-106_k141_15979 | Not-determined |
| 18-106_k141_132992 | Not-determined |
| 18-106_k141_92213 | Not-determined |
| 18-106_k141_74525 | Not-determined |
| 18-106_k141_74530 | Not-determined |
| 18-106_k141_92223 | Not-determined |
| 18-106_k141_74532 | Not-determined |
| 18-106_k141_115671 | Low-quality |
| 18-106_k141_92226 | Not-determined |
| 18-106_k141_27508 | Not-determined |
| 18-106_k141_69219 | Not-determined |
| 18-106_k141_56274 | Not-determined |
| 18-106_k141_98638 | Not-determined |
| 18-106_k141_74541 | Not-determined |
| 18-106_k141_63463 | Not-determined |
| 18-106_k141_56285 | Not-determined |
| 18-106_k141_21134 | Not-determined |
| 18-106_k141_9748 | Not-determined |
| 18-106_k141_21136 | Not-determined |
| 18-106_k141_63477 | Not-determined |
| 18-106_k141_21138 | Not-determined |
| 18-106_k141_86426 | Not-determined |
| 18-106_k141_63482 | Not-determined |
| 18-106_k141_9758 | Not-determined |
| 18-106_k141_21147 | Not-determined |
| 18-106_k141_9761 | Not-determined |
| 18-106_k141_56301 | Not-determined |
| 18-106_k141_115697 | Not-determined |
| 18-106_k141_21150 | Not-determined |
| 18-106_k141_9764 | Not-determined |
| 18-106_k141_39834 | Low-quality |
| 18-106_k141_98643 | Low-quality |
| 18-106_k141_92234 | Not-determined |
| 18-106_k141_16013 | Not-determined |
| 18-106_k141_92236 | Not-determined |
| 18-106_k141_21156 | Not-determined |
| 18-106_k141_98649 | Not-determined |
| 18-106_k141_16016 | Not-determined |
| 18-106_k141_69262 | Low-quality |
| 18-106_k141_133032 | Not-determined |
| 18-106_k141_115701 | Low-quality |
| 18-106_k141_9781 | Not-determined |
| 18-106_k141_69263 | Not-determined |
| 18-106_k141_115705 | Not-determined |
| 18-106_k141_92239 | Low-quality |
| 18-106_k141_63498 | Not-determined |
| 18-106_k141_45799 | Not-determined |
| 18-106_k141_133045 | Not-determined |
| 18-106_k141_39854 | Not-determined |
| 18-106_k141_56329 | Not-determined |
| 18-106_k141_63504 | Not-determined |
| 18-106_k141_80240 | Not-determined |
| 18-106_k141_80242 | Low-quality |
| 18-106_k141_98664 | Not-determined |
| 18-106_k141_92254 | Not-determined |
| 18-106_k141_80246 | Not-determined |
| 18-106_k141_63514 | Not-determined |
| 18-106_k141_51448 | Not-determined |
| 18-106_k141_63516 | Not-determined |
| 18-106_k141_16035 | Not-determined |
| 18-106_k141_9792 | Not-determined |
| 18-106_k141_121117 | Not-determined |
| 18-106_k141_9794 | Not-determined |
| 18-106_k141_69280 | Not-determined |
| 18-106_k141_92259 | Not-determined |
| 18-106_k141_121122 | Not-determined |
| 18-106_k141_109971 | Not-determined |
| 18-106_k141_121124 | Not-determined |
| 18-106_k141_109975 | Not-determined |
| 18-106_k141_16049 | Not-determined |
| 18-106_k141_16051 | Not-determined |
| 18-106_k141_45821 | Not-determined |
| 18-106_k141_92266 | Not-determined |
| 18-106_k141_27545 | Not-determined |
| 18-106_k141_98681 | Not-determined |
| 18-106_k141_98684 | Not-determined |
| 18-106_k141_39878 | Not-determined |
| 18-106_k141_9806 | Not-determined |
| 18-106_k141_9807 | Not-determined |
| 18-106_k141_133061 | Not-determined |
| 18-106_k141_80262 | Not-determined |
| 18-106_k141_39884 | Not-determined |
| 18-106_k141_39885 | Not-determined |
| 18-106_k141_133063 | Not-determined |
| 18-106_k141_133065 | Not-determined |
| 18-106_k141_109985 | Not-determined |
| 18-106_k141_138733 | Not-determined |
| 18-106_k141_21174 | Not-determined |
| 18-106_k141_9815 | Not-determined |
| 18-106_k141_45842 | Not-determined |
| 18-106_k141_63542 | Not-determined |
| 18-106_k141_103583 | Not-determined |
| 18-106_k141_39886 | Low-quality |
| 18-106_k141_69297 | Not-determined |
| 18-106_k141_63558 | Low-quality |
| 18-106_k141_92276 | Not-determined |
| 18-106_k141_80280 | Not-determined |
| 18-106_k141_121131 | Not-determined |
| 18-106_k141_63563 | Not-determined |
| 18-106_k141_21192 | Low-quality |
| 18-106_k141_110006 | Not-determined |
| 18-106_k141_69303 | Not-determined |
| 18-106_k141_138766 | Not-determined |
| 18-106_k141_45864 | Not-determined |
| 18-106_k141_110011 | Not-determined |
| 18-106_k141_98705 | Low-quality |
| 18-106_k141_103595 | Not-determined |
| 18-106_k141_74560 | Not-determined |
| 18-106_k141_138774 | Low-quality |
| 18-106_k141_56349 | Not-determined |
| 18-106_k141_74563 | Not-determined |
| 18-106_k141_9826 | Not-determined |
| 18-106_k141_56355 | Not-determined |
| 18-106_k141_16063 | Not-determined |
| 18-106_k141_74570 | Not-determined |
| 18-106_k141_5576 | Not-determined |
| 18-106_k141_98717 | Not-determined |
| 18-106_k141_92285 | Not-determined |
| 18-106_k141_45877 | Not-determined |
| 18-106_k141_110020 | Not-determined |
| 18-106_k141_63588 | Not-determined |
| 18-106_k141_51477 | Not-determined |
| 18-106_k141_92300 | Not-determined |
| 18-106_k141_92302 | Not-determined |
| 18-106_k141_16076 | Not-determined |
| 18-106_k141_133114 | Not-determined |
| 18-106_k141_16078 | Not-determined |
| 18-106_k141_5594 | Not-determined |
| 18-106_k141_21224 | Not-determined |
| 18-106_k141_39916 | Low-quality |
| 18-106_k141_39917 | Not-determined |
| 18-106_k141_92307 | Low-quality |
| 18-106_k141_5600 | Not-determined |
| 18-106_k141_63595 | Not-determined |
| 18-106_k141_5607 | Not-determined |
| 18-106_k141_27580 | Not-determined |
| 18-106_k141_98738 | Not-determined |
| 18-106_k141_74598 | Not-determined |
| 18-106_k141_63602 | Not-determined |
| 18-106_k141_16096 | Not-determined |
| 18-106_k141_115779 | Not-determined |
| 18-106_k141_56377 | Not-determined |
| 18-106_k141_56378 | Not-determined |
| 18-106_k141_16106 | Not-determined |
| 18-106_k141_16107 | Not-determined |
| 18-106_k141_45907 | Not-determined |
| 18-106_k141_110063 | Not-determined |
| 18-106_k141_51501 | Not-determined |
| 18-106_k141_133124 | Not-determined |
| 18-106_k141_9834 | Not-determined |
| 18-106_k141_56386 | Not-determined |
| 18-106_k141_21243 | Not-determined |
| 18-106_k141_121159 | Not-determined |
| 18-106_k141_121160 | Not-determined |
| 18-106_k141_110069 | Not-determined |
| 18-106_k141_51513 | Not-determined |
| 18-106_k141_9841 | Low-quality |
| 18-106_k141_110079 | Not-determined |
| 18-106_k141_92331 | Not-determined |
| 18-106_k141_45908 | Low-quality |
| 18-106_k141_74622 | Not-determined |
| 18-106_k141_110085 | Not-determined |
| 18-106_k141_69353 | Not-determined |
| 18-106_k141_92336 | Not-determined |
| 18-106_k141_92340 | Not-determined |
| 18-106_k141_121171 | Not-determined |
| 18-106_k141_115809 | Not-determined |
| 18-106_k141_121176 | Not-determined |
| 18-106_k141_69363 | Not-determined |
| 18-106_k141_45921 | Low-quality |
| 18-106_k141_74639 | Low-quality |
| 18-106_k141_80305 | Not-determined |
| 18-106_k141_80307 | Low-quality |
| 18-106_k141_80308 | Not-determined |
| 18-106_k141_80309 | Low-quality |
| 18-106_k141_51539 | Not-determined |
| 18-106_k141_69373 | Not-determined |
| 18-106_k141_63632 | Not-determined |
| 18-106_k141_69375 | Not-determined |
| [illegible] | [illegible] |
| 18-106_k141_27625 | Not-determined |
| 18-106_k141_98780 | Low-quality |
| 18-106_k141_138854 | Not-determined |
| 18-106_k141_51542 | Not-determined |
| 18-106_k141_74654 | Not-determined |
| 18-106_k141_115837 | Low-quality |
| 18-106_k141_74656 | Not-determined |
| 18-106_k141_45948 | Not-determined |
| 18-106_k141_74667 | Not-determined |
| 18-106_k141_121208 | Not-determined |
| 18-106_k141_74669 | Not-determined |
| 18-106_k141_138870 | Low-quality |
| 18-106_k141_74673 | Not-determined |
| 18-106_k141_92376 | Not-determined |
| 18-106_k141_45957 | Not-determined |
| 18-106_k141_5655 | Not-determined |
| 18-106_k141_80330 | Not-determined |
| 18-106_k141_27647 | Not-determined |
| 18-106_k141_103670 | Not-determined |
| 18-106_k141_16155 | Not-determined |
| 18-106_k141_5660 | Not-determined |
| 18-106_k141_5661 | Not-determined |
| 18-106_k141_138883 | Not-determined |
| 18-106_k141_121215 | Not-determined |
| 18-106_k141_51563 | Not-determined |
| 18-106_k141_74681 | Not-determined |
| 18-106_k141_92389 | Not-determined |
| 18-106_k141_5670 | Not-determined |
| 18-106_k141_127409 | Not-determined |
| 18-106_k141_92400 | Not-determined |
| 18-106_k141_92405 | Not-determined |
| 18-106_k141_80337 | Not-determined |
| 18-106_k141_103685 | Not-determined |
| 18-106_k141_98809 | Not-determined |
| 18-106_k141_16163 | Not-determined |
| 18-106_k141_115864 | Not-determined |
| 18-106_k141_138894 | Not-determined |
| 18-106_k141_27670 | Not-determined |
| 18-106_k141_45970 | Not-determined |
| 18-106_k141_103697 | Not-determined |
| 18-106_k141_80347 | Not-determined |
| 18-106_k141_16167 | Not-determined |
| 18-106_k141_98818 | Not-determined |
| 18-106_k141_121217 | Low-quality |
| 18-106_k141_127434 | Not-determined |
| 18-106_k141_115874 | Not-determined |
| 18-106_k141_16176 | Not-determined |
| 18-106_k141_16180 | Not-determined |
| 18-106_k141_9893 | Not-determined |
| 18-106_k141_9894 | Not-determined |
| 18-106_k141_39938 | Not-determined |
| 18-106_k141_138920 | Not-determined |
| 18-106_k141_121235 | Not-determined |
| 18-106_k141_21278 | Not-determined |
| 18-106_k141_110130 | Not-determined |
| 18-106_k141_9904 | Not-determined |
| 18-106_k141_5677 | Not-determined |
| 18-106_k141_63678 | Not-determined |
| 18-106_k141_86447 | Not-determined |
| 18-106_k141_9906 | Not-determined |
| 18-106_k141_121240 | Not-determined |
| 18-106_k141_16200 | Not-determined |
| 18-106_k141_127450 | Low-quality |
| 18-106_k141_27695 | Low-quality |
| 18-106_k141_86452 | Not-determined |
| 18-106_k141_74716 | Not-determined |
| 18-106_k141_121253 | Not-determined |
| 18-106_k141_74719 | Not-determined |
| 18-106_k141_74720 | Not-determined |
| 18-106_k141_33531 | Not-determined |
| 18-106_k141_21284 | Not-determined |
| 18-106_k141_92448 | Not-determined |
| 18-106_k141_92453 | Not-determined |
| 18-106_k141_115909 | Not-determined |
| 18-106_k141_80357 | Not-determined |
| 18-106_k141_74733 | Not-determined |
| 18-106_k141_5711 | Not-determined |
| 18-106_k141_39977 | Not-determined |
| 18-106_k141_138950 | Not-determined |
| 18-106_k141_39979 | Low-quality |
| 18-106_k141_127469 | Not-determined |
| 18-106_k141_33534 | Not-determined |
| 18-106_k141_33535 | Not-determined |
| 18-106_k141_9926 | Not-determined |
| 18-106_k141_27718 | Not-determined |
| 18-106_k141_115926 | Not-determined |
| 18-106_k141_133203 | Not-determined |
| 18-106_k141_110141 | Not-determined |
| 18-106_k141_27727 | Not-determined |
| 18-106_k141_103710 | Not-determined |
| 18-106_k141_110145 | Not-determined |
| 18-106_k141_80382 | Not-determined |
| 18-106_k141_27730 | Not-determined |
| 18-106_k141_9940 | Not-determined |
| 18-106_k141_92477 | Not-determined |
| 18-106_k141_5724 | Not-determined |
| 18-106_k141_27736 | Not-determined |
| 18-106_k141_98870 | Not-determined |
| 18-106_k141_80393 | Not-determined |
| 18-106_k141_98872 | Not-determined |
| 18-106_k141_56394 | Not-determined |
| [illegible] | [illegible] |
| 18-106_k141_103726 | Low-quality |
| 18-106_k141_21323 | Not-determined |
| 18-106_k141_9953 | Not-determined |
| 18-106_k141_138962 | Not-determined |
| 18-106_k141_5738 | Not-determined |
| 18-106_k141_56411 | Low-quality |
| 18-106_k141_40017 | Not-determined |
| 18-106_k141_74777 | Not-determined |
| 18-106_k141_110164 | Not-determined |
| 18-106_k141_74779 | Not-determined |
| 18-106_k141_121293 | Not-determined |
| 18-106_k141_110173 | Not-determined |
| 18-106_k141_74785 | Not-determined |
| 18-106_k141_16249 | Not-determined |
| 18-106_k141_92496 | Not-determined |
| 18-106_k141_80412 | Not-determined |
| 18-106_k141_127485 | Not-determined |
| 18-106_k141_80415 | Not-determined |
| 18-106_k141_56419 | Low-quality |
| 18-106_k141_45998 | Not-determined |
| 18-106_k141_110180 | Not-determined |
| 18-106_k141_92510 | Low-quality |
| 18-106_k141_74800 | Not-determined |
| 18-106_k141_40038 | Not-determined |
| 18-106_k141_56426 | Not-determined |
| 18-106_k141_138979 | Not-determined |
| 18-106_k141_133219 | Not-determined |
| 18-106_k141_74809 | Not-determined |
| 18-106_k141_46001 | Low-quality |
| 18-106_k141_9967 | Not-determined |
| 18-106_k141_40055 | Low-quality |
| 18-106_k141_56450 | Not-determined |
| 18-106_k141_121312 | Not-determined |
| 18-106_k141_74820 | Not-determined |
| 18-106_k141_80434 | Not-determined |
| 18-106_k141_86522 | Not-determined |
| 18-106_k141_63718 | Not-determined |
| 18-106_k141_121315 | Not-determined |
| 18-106_k141_80435 | Not-determined |
| 18-106_k141_63720 | Not-determined |
| 18-106_k141_133245 | Not-determined |
| 18-106_k141_121318 | Low-quality |
| 18-106_k141_133246 | Not-determined |
| 18-106_k141_139005 | Not-determined |
| 18-106_k141_121323 | Not-determined |
| 18-106_k141_139010 | Not-determined |
| 18-106_k141_63728 | Low-quality |
| 18-106_k141_46024 | Not-determined |
| 18-106_k141_92537 | Not-determined |
| 18-106_k141_40078 | Not-determined |
| 18-106_k141_5774 | Not-determined |
| 18-106_k141_46025 | Not-determined |
| 18-106_k141_40080 | Not-determined |
| 18-106_k141_98916 | Not-determined |
| 18-106_k141_139015 | Not-determined |
| 18-106_k141_127501 | Not-determined |
| 18-106_k141_46028 | Not-determined |
| 18-106_k141_51608 | Not-determined |
| 18-106_k141_27791 | Not-determined |
| 18-106_k141_5782 | Not-determined |
| 18-106_k141_56458 | Not-determined |
| 18-106_k141_98925 | Not-determined |
| 18-106_k141_51615 | Not-determined |
| 18-106_k141_74846 | Not-determined |
| 18-106_k141_40085 | Not-determined |
| 18-106_k141_127512 | Not-determined |
| 18-106_k141_74853 | Not-determined |
| 18-106_k141_40086 | Not-determined |
| 18-106_k141_46048 | Not-determined |
| 18-106_k141_139035 | Not-determined |
| 18-106_k141_33568 | Not-determined |
| 18-106_k141_74863 | Not-determined |
| 18-106_k141_5816 | Not-determined |
| 18-106_k141_139048 | Not-determined |
| 18-106_k141_139049 | Not-determined |
| 18-106_k141_5822 | Not-determined |
| 18-106_k141_92547 | Not-determined |
| 18-106_k141_69381 | Not-determined |
| 18-106_k141_74874 | Not-determined |
| 18-106_k141_127527 | Not-determined |
| 18-106_k141_16263 | Not-determined |
| 18-106_k141_127532 | Not-determined |
| 18-106_k141_33597 | Not-determined |
| 18-106_k141_86559 | Not-determined |
| 18-106_k141_33599 | Not-determined |
| 18-106_k141_92557 | Not-determined |
| 18-106_k141_92559 | Low-quality |
| 18-106_k141_74878 | Not-determined |
| 18-106_k141_33604 | Not-determined |
| 18-106_k141_5837 | Not-determined |
| 18-106_k141_127540 | Low-quality |
| 18-106_k141_69392 | Not-determined |
| 18-106_k141_69393 | Low-quality |
| 18-106_k141_86576 | Not-determined |
| 18-106_k141_69400 | Not-determined |
| 18-106_k141_40126 | Not-determined |
| 18-106_k141_86586 | Not-determined |
| 18-106_k141_46107 | Not-determined |
| 18-106_k141_86587 | Not-determined |
| 18-106_k141_16271 | Not-determined |
| 18-106_k141_21343 | Not-determined |
| 18-106_k141_121334 | Low-quality |
| 18-106_k141_27803 | Not-determined |
| 18-106_k141_46115 | Not-determined |
| 18-106_k141_121338 | Not-determined |
| 18-106_k141_16280 | Not-determined |
| 18-106_k141_21345 | Not-determined |
| 18-106_k141_121340 | Not-determined |
| 18-106_k141_121341 | Not-determined |
| 18-106_k141_133325 | Not-determined |
| 18-106_k141_5861 | Not-determined |
| 18-106_k141_5863 | Not-determined |
| 18-106_k141_51661 | Low-quality |
| 18-106_k141_16287 | Not-determined |
| 18-106_k141_98972 | Low-quality |
| 18-106_k141_69422 | Not-determined |
| 18-106_k141_21354 | Not-determined |
| 18-106_k141_27812 | Not-determined |
| 18-106_k141_121360 | Not-determined |
| 18-106_k141_27816 | Not-determined |
| 18-106_k141_86605 | Not-determined |
| 18-106_k141_98980 | Not-determined |
| 18-106_k141_121371 | Not-determined |
| 18-106_k141_121373 | Not-determined |
| 18-106_k141_63792 | Not-determined |
| 18-106_k141_27827 | Not-determined |
| 18-106_k141_92596 | Not-determined |
| 18-106_k141_63794 | Not-determined |
| 18-106_k141_56470 | Not-determined |
| 18-106_k141_92597 | Not-determined |
| 18-106_k141_121375 | Not-determined |
| 18-106_k141_33646 | Low-quality |
| 18-106_k141_69439 | Not-determined |
| 18-106_k141_46140 | Not-determined |
| 18-106_k141_46142 | Not-determined |
| 18-106_k141_33654 | Not-determined |
| 18-106_k141_46146 | Not-determined |
| 18-106_k141_63802 | Not-determined |
| 18-106_k141_110288 | Not-determined |
| 18-106_k141_110289 | Not-determined |
| 18-106_k141_69448 | Not-determined |
| 18-106_k141_121383 | Not-determined |
| 18-106_k141_46155 | Not-determined |
| 18-106_k141_133366 | Low-quality |
| 18-106_k141_33673 | Low-quality |
| 18-106_k141_121387 | Not-determined |
| 18-106_k141_63814 | Not-determined |
| 18-106_k141_46159 | Not-determined |
| 18-106_k141_121389 | Not-determined |
| 18-106_k141_33683 | Low-quality |
| 18-106_k141_133371 | Not-determined |
| 18-106_k141_115967 | Not-determined |
| 18-106_k141_80481 | Not-determined |
| 18-106_k141_56491 | Low-quality |
| 18-106_k141_63834 | Not-determined |
| 18-106_k141_127581 | Not-determined |
| 18-106_k141_56494 | Not-determined |
| 18-106_k141_99007 | Not-determined |
| 18-106_k141_115975 | Not-determined |
| 18-106_k141_110309 | Low-quality |
| 18-106_k141_110312 | Not-determined |
| 18-106_k141_133386 | Low-quality |
| 18-106_k141_115984 | Not-determined |
| 18-106_k141_27863 | Not-determined |
| 18-106_k141_33699 | Not-determined |
| 18-106_k141_127589 | Not-determined |
| 18-106_k141_86651 | Not-determined |
| 18-106_k141_92654 | Not-determined |
| 18-106_k141_46171 | Not-determined |
| 18-106_k141_21368 | Not-determined |
| 18-106_k141_80494 | Not-determined |
| 18-106_k141_21374 | Not-determined |
| 18-106_k141_51699 | Low-quality |
| 18-106_k141_51700 | Not-determined |
| 18-106_k141_115994 | Not-determined |
| 18-106_k141_127605 | Not-determined |
| 18-106_k141_33702 | Not-determined |
| 18-106_k141_116003 | Not-determined |
| 18-106_k141_51709 | Not-determined |
| 18-106_k141_99037 | Not-determined |
| 18-106_k141_69487 | Not-determined |
| 18-106_k141_69489 | Not-determined |
| [illegible] | [illegible] |
| 18-106_k141_16390 | Not-determined |
| 18-106_k141_63882 | Not-determined |
| 18-106_k141_99074 | Not-determined |
| 18-106_k141_86680 | Not-determined |
| 18-106_k141_9994 | Not-determined |
| 18-106_k141_92690 | Not-determined |
| 18-106_k141_9998 | Not-determined |
| 18-106_k141_63886 | Not-determined |
| 18-106_k141_63889 | Not-determined |
| 18-106_k141_40182 | Low-quality |
| 18-106_k141_40187 | Not-determined |
| 18-106_k141_33765 | Not-determined |
| 18-106_k141_86691 | Not-determined |
| 18-106_k141_86692 | Not-determined |
| 18-106_k141_116028 | Not-determined |
| 18-106_k141_21428 | Not-determined |
| 18-106_k141_74899 | Low-quality |
| 18-106_k141_46221 | Not-determined |
| 18-106_k141_21436 | Not-determined |
| 18-106_k141_127679 | Not-determined |
| 18-106_k141_86701 | Not-determined |
| 18-106_k141_116037 | Not-determined |
| 18-106_k141_86703 | Not-determined |
| 18-106_k141_16430 | Not-determined |
| 18-106_k141_127681 | Not-determined |
| 18-106_k141_116040 | Not-determined |
| 18-106_k141_86709 | Not-determined |
| 18-106_k141_74914 | Not-determined |
| 18-106_k141_99107 | Low-quality |
| 18-106_k141_127685 | Not-determined |
| 18-106_k141_127687 | Not-determined |
| 18-106_k141_139078 | Not-determined |
| 18-106_k141_46235 | Not-determined |
| 18-106_k141_139083 | Not-determined |
| 18-106_k141_40215 | Not-determined |
| 18-106_k141_10013 | Not-determined |
| 18-106_k141_10016 | Not-determined |
| 18-106_k141_69532 | Not-determined |
| 18-106_k141_110373 | Low-quality |
| 18-106_k141_69535 | Not-determined |
| 18-106_k141_16444 | Low-quality |
| 18-106_k141_21461 | Low-quality |
| 18-106_k141_127704 | Low-quality |
| 18-106_k141_40228 | Not-determined |
| 18-106_k141_110378 | Not-determined |
| 18-106_k141_74926 | Not-determined |
| 18-106_k141_16450 | Not-determined |
| 18-106_k141_33794 | Not-determined |
| 18-106_k141_139090 | Not-determined |
| 18-106_k141_74928 | Not-determined |
| 18-106_k141_116068 | Not-determined |
| 18-106_k141_40234 | Not-determined |
| 18-106_k141_99127 | Not-determined |
| 18-106_k141_116069 | Not-determined |
| 18-106_k141_139094 | Not-determined |
| 18-106_k141_10029 | Not-determined |
| 18-106_k141_127711 | Not-determined |
| 18-106_k141_21473 | Not-determined |
| 18-106_k141_51790 | Not-determined |
| 18-106_k141_40247 | Not-determined |
| 18-106_k141_40249 | Not-determined |
| 18-106_k141_40251 | Not-determined |
| 18-106_k141_110384 | Not-determined |
| 18-106_k141_116082 | Not-determined |
| 18-106_k141_16458 | Not-determined |

| | |
|---|---|
| 18-106_k141_51796 | Not-determined |
| 18-106_k141_56529 | Not-determined |
| 18-106_k141_99138 | Not-determined |
| 18-106_k141_99140 | Not-determined |
| 18-106_k141_56538 | Not-determined |
| 18-106_k141_110387 | Low-quality |
| 18-106_k141_21489 | Not-determined |
| 18-106_k141_21490 | Not-determined |
| 18-106_k141_99148 | Not-determined |
| 18-106_k141_86742 | Not-determined |
| 18-106_k141_110392 | Low-quality |
| 18-106_k141_133410 | Not-determined |
| 18-106_k141_110394 | Not-determined |
| 18-106_k141_51812 | Not-determined |
| 18-106_k141_116102 | Not-determined |
| 18-106_k141_110402 | Not-determined |
| 18-106_k141_51818 | Not-determined |
| 18-106_k141_40272 | Not-determined |
| 18-106_k141_99158 | Not-determined |
| 18-106_k141_139111 | Not-determined |
| 18-106_k141_27986 | Not-determined |
| 18-106_k141_46268 | Low-quality |
| 18-106_k141_139113 | Not-determined |
| 18-106_k141_51828 | Not-determined |
| 18-106_k141_86758 | Not-determined |
| 18-106_k141_86762 | Not-determined |
| 18-106_k141_40283 | Not-determined |
| 18-106_k141_110420 | Not-determined |
| 18-106_k141_56560 | Not-determined |
| 18-106_k141_63947 | Not-determined |
| 18-106_k141_80563 | Not-determined |
| 18-106_k141_110434 | Not-determined |
| 18-106_k141_21497 | Not-determined |
| 18-106_k141_80568 | Not-determined |
| 18-106_k141_21500 | Not-determined |
| 18-106_k141_69618 | Not-determined |
| 18-106_k141_46298 | Low-quality |
| 18-106_k141_116131 | Not-determined |
| 18-106_k141_133457 | Not-determined |
| 18-106_k141_27992 | Not-determined |
| 18-106_k141_133459 | Not-determined |
| 18-106_k141_116135 | Not-determined |
| 18-106_k141_116137 | Not-determined |
| 18-106_k141_10085 | Not-determined |
| 18-106_k141_116142 | Not-determined |
| 18-106_k141_28003 | Not-determined |
| 18-106_k141_51842 | Not-determined |
| 18-106_k141_92698 | Not-determined |
| 18-106_k141_21519 | Not-determined |
| 18-106_k141_110451 | Not-determined |
| 18-106_k141_127763 | Not-determined |
| 18-106_k141_21526 | Not-determined |
| 18-106_k141_40323 | Not-determined |
| 18-106_k141_16487 | Low-quality |
| 18-106_k141_21541 | Not-determined |
| 18-106_k141_69651 | Not-determined |
| 18-106_k141_56593 | Not-determined |
| 18-106_k141_139142 | Low-quality |
| 18-106_k141_46312 | Not-determined |
| 18-106_k141_40325 | Low-quality |
| 18-106_k141_92711 | Not-determined |
| 18-106_k141_10106 | Not-determined |
| 18-106_k141_69660 | Not-determined |
| 18-106_k141_63993 | Not-determined |
| 18-106_k141_46316 | Not-determined |
| 18-106_k141_69664 | Not-determined |
| 18-106_k141_133486 | Not-determined |
| 18-106_k141_92724 | Not-determined |
| 18-106_k141_92725 | Not-determined |
| 18-106_k141_99202 | Not-determined |
| 18-106_k141_51868 | Not-determined |
| 18-106_k141_10116 | Not-determined |
| 18-106_k141_121446 | Not-determined |
| 18-106_k141_133490 | Not-determined |
| 18-106_k141_139156 | Not-determined |
| 18-106_k141_16508 | Not-determined |
| 18-106_k141_28038 | Not-determined |
| 18-106_k141_139164 | Not-determined |
| 18-106_k141_133505 | Not-determined |
| 18-106_k141_99207 | Not-determined |
| 18-106_k141_21571 | Not-determined |
| 18-106_k141_110490 | Not-determined |
| 18-106_k141_21574 | Not-determined |
| 18-106_k141_139178 | Not-determined |
| 18-106_k141_116178 | Not-determined |
| 18-106_k141_116181 | Not-determined |
| 18-106_k141_64019 | Not-determined |
| 18-106_k141_40344 | Not-determined |
| 18-106_k141_64022 | Not-determined |
| 18-106_k141_133515 | Not-determined |
| 18-106_k141_28058 | Not-determined |
| 18-106_k141_116182 | Not-determined |
| 18-106_k141_99220 | Not-determined |
| 18-106_k141_28066 | Not-determined |
| 18-106_k141_92760 | Not-determined |
| 18-106_k141_69675 | Not-determined |
| 18-106_k141_116194 | Not-determined |
| 18-106_k141_139198 | Not-determined |
| 18-106_k141_127778 | Low-quality |
| 18-106_k141_69688 | Not-determined |
| 18-106_k141_127788 | Not-determined |
| 18-106_k141_21601 | Not-determined |
| 18-106_k141_127790 | Not-determined |
| 18-106_k141_21605 | Not-determined |
| 18-106_k141_86773 | Not-determined |
| 18-106_k141_86774 | Not-determined |
| 18-106_k141_75046 | Not-determined |
| 18-106_k141_16550 | Low-quality |
| 18-106_k141_80618 | Not-determined |
| 18-106_k141_40366 | Not-determined |
| 18-106_k141_92790 | Not-determined |
| 18-106_k141_40370 | Not-determined |
| 18-106_k141_33827 | Not-determined |
| 18-106_k141_127810 | Not-determined |
| 18-106_k141_133530 | Low-quality |
| 18-106_k141_69706 | Not-determined |
| 18-106_k141_16562 | Not-determined |
| 18-106_k141_116214 | Not-determined |
| 18-106_k141_33831 | Not-determined |
| 18-106_k141_92803 | Not-determined |
| 18-106_k141_64051 | Low-quality |
| 18-106_k141_69720 | Not-determined |
| 18-106_k141_69722 | Not-determined |
| 18-106_k141_40390 | Not-determined |
| 18-106_k141_86810 | Not-determined |
| 18-106_k141_139220 | Low-quality |
| 18-106_k141_86812 | Not-determined |
| 18-106_k141_64066 | Not-determined |
| 18-106_k141_133537 | Not-determined |
| 18-106_k141_69728 | Not-determined |
| 18-106_k141_51885 | Not-determined |
| 18-106_k141_133539 | Not-determined |
| 18-106_k141_16569 | Not-determined |
| 18-106_k141_69731 | Not-determined |
| 18-106_k141_56660 | Low-quality |
| 18-106_k141_33856 | Not-determined |
| 18-106_k141_80627 | Not-determined |
| 18-106_k141_40399 | Not-determined |
| 18-106_k141_75066 | Not-determined |
| 18-106_k141_21634 | Not-determined |
| 18-106_k141_121460 | Not-determined |
| 18-106_k141_127814 | Not-determined |
| 18-106_k141_80631 | Not-determined |
| 18-106_k141_80632 | Not-determined |
| 18-106_k141_127818 | Low-quality |
| 18-106_k141_103764 | Not-determined |
| 18-106_k141_127819 | Not-determined |
| 18-106_k141_69747 | Not-determined |
| 18-106_k141_127836 | Low-quality |
| 18-106_k141_121474 | Not-determined |
| 18-106_k141_64086 | Not-determined |
| 18-106_k141_86848 | Not-determined |
| 18-106_k141_80649 | Not-determined |
| 18-106_k141_80651 | Not-determined |
| 18-106_k141_103788 | Not-determined |
| 18-106_k141_33876 | Not-determined |
| 18-106_k141_103793 | Not-determined |
| 18-106_k141_64096 | Not-determined |
| 18-106_k141_69764 | Not-determined |
| 18-106_k141_121491 | Not-determined |
| 18-106_k141_28115 | Not-determined |
| 18-106_k141_69771 | Not-determined |
| 18-106_k141_80665 | Not-determined |
| 18-106_k141_92829 | Not-determined |
| 18-106_k141_127853 | Not-determined |
| 18-106_k141_10179 | Not-determined |
| 18-106_k141_127856 | Not-determined |
| 18-106_k141_69774 | Not-determined |
| 18-106_k141_51936 | Low-quality |
| 18-106_k141_69777 | Not-determined |
| 18-106_k141_116244 | Not-determined |
| 18-106_k141_99269 | Not-determined |
| 18-106_k141_103817 | Not-determined |
| 18-106_k141_28128 | Not-determined |
| 18-106_k141_103819 | Not-determined |
| 18-106_k141_51945 | Not-determined |
| 18-106_k141_10184 | Not-determined |
| 18-106_k141_103822 | Not-determined |
| 18-106_k141_99274 | Not-determined |
| 18-106_k141_33898 | Not-determined |
| 18-106_k141_51950 | Not-determined |
| 18-106_k141_33901 | Low-quality |
| 18-106_k141_21655 | Not-determined |
| 18-106_k141_103836 | Not-determined |
| 18-106_k141_21659 | Not-determined |
| 18-106_k141_64114 | Not-determined |
| 18-106_k141_21661 | Low-quality |
| 18-106_k141_80677 | Not-determined |
| 18-106_k141_116268 | Not-determined |
| 18-106_k141_33912 | Not-determined |
| 18-106_k141_99282 | Not-determined |
| 18-106_k141_86876 | Not-determined |
| 18-106_k141_103841 | Low-quality |
| 18-106_k141_116272 | Not-determined |
| 18-106_k141_103844 | Not-determined |
| 18-106_k141_56694 | Not-determined |
| 18-106_k141_64123 | Not-determined |
| 18-106_k141_51963 | Not-determined |
| 18-106_k141_133612 | Not-determined |
| 18-106_k141_56698 | Not-determined |
| 18-106_k141_86895 | Low-quality |
| 18-106_k141_33927 | Low-quality |
| 18-106_k141_121530 | Not-determined |
| 18-106_k141_127865 | Not-determined |
| 18-106_k141_28166 | Not-determined |
| 18-106_k141_121540 | Not-determined |
| 18-106_k141_86911 | Not-determined |
| 18-106_k141_86912 | Not-determined |
| 18-106_k141_40422 | Not-determined |
| 18-106_k141_10226 | Not-determined |
| 18-106_k141_121553 | Not-determined |
| 18-106_k141_121555 | Not-determined |
| 18-106_k141_86921 | Not-determined |
| 18-106_k141_86923 | Not-determined |
| 18-106_k141_33965 | Not-determined |
| 18-106_k141_21681 | Low-quality |
| 18-106_k141_28189 | Low-quality |
| 18-106_k141_16676 | Not-determined |
| 18-106_k141_33970 | Not-determined |
| 18-106_k141_21689 | Not-determined |
| 18-106_k141_69820 | Not-determined |
| 18-106_k141_21691 | Not-determined |
| 18-106_k141_69821 | Not-determined |
| 18-106_k141_51996 | Not-determined |
| 18-106_k141_80708 | Not-determined |
| 18-106_k141_110508 | Not-determined |
| 18-106_k141_139279 | Not-determined |
| 18-106_k141_80712 | Not-determined |
| 18-106_k141_28193 | Low-quality |
| 18-106_k141_75072 | Low-quality |
| 18-106_k141_121583 | Not-determined |
| 18-106_k141_33982 | Not-determined |
| 18-106_k141_110520 | Not-determined |
| 18-106_k141_133644 | Low-quality |
| 18-106_k141_139293 | Low-quality |
| 18-106_k141_133645 | Not-determined |
| 18-106_k141_40452 | Not-determined |
| 18-106_k141_46407 | Not-determined |
| 18-106_k141_16700 | Not-determined |
| 18-106_k141_133652 | Low-quality |
| 18-106_k141_10242 | Not-determined |
| 18-106_k141_80729 | Not-determined |
| 18-106_k141_16703 | Not-determined |
| 18-106_k141_64162 | Not-determined |
| 18-106_k141_16708 | Not-determined |
| 18-106_k141_40465 | Not-determined |
| 18-106_k141_64166 | Not-determined |
| 18-106_k141_133653 | Not-determined |
| 18-106_k141_21712 | Not-determined |
| 18-106_k141_86956 | Low-quality |
| 18-106_k141_110544 | Not-determined |
| 18-106_k141_64170 | Not-determined |
| 18-106_k141_64171 | Low-quality |
| 18-106_k141_16720 | Not-determined |
| 18-106_k141_127908 | Not-determined |
| 18-106_k141_52032 | Not-determined |
| 18-106_k141_133666 | Not-determined |
| 18-106_k141_64175 | Not-determined |
| 18-106_k141_28218 | Not-determined |
| 18-106_k141_80746 | Not-determined |
| 18-106_k141_21719 | Not-determined |
| 18-106_k141_80747 | Low-quality |
| 18-106_k141_86969 | Not-determined |
| 18-106_k141_16729 | Not-determined |
| 18-106_k141_21724 | Not-determined |
| 18-106_k141_21728 | Not-determined |
| 18-106_k141_64183 | Not-determined |
| 18-106_k141_69872 | Not-determined |
| 18-106_k141_64186 | Not-determined |
| 18-106_k141_16738 | Not-determined |
| 18-106_k141_10264 | Not-determined |
| 18-106_k141_75092 | Not-determined |
| 18-106_k141_64191 | Not-determined |
| 18-106_k141_139316 | Low-quality |
| 18-106_k141_116322 | Not-determined |
| 18-106_k141_28230 | Not-determined |
| 18-106_k141_21742 | Not-determined |
| 18-106_k141_52062 | Not-determined |
| 18-106_k141_133689 | Not-determined |
| 18-106_k141_116326 | Not-determined |
| 18-106_k141_139325 | Not-determined |
| 18-106_k141_40496 | Not-determined |
| 18-106_k141_52072 | Low-quality |
| 18-106_k141_64202 | Not-determined |
| 18-106_k141_10277 | Not-determined |
| 18-106_k141_75107 | Low-quality |
| 18-106_k141_139332 | Not-determined |
| 18-106_k141_52084 | Not-determined |
| 18-106_k141_52086 | Not-determined |
| 18-106_k141_75117 | Not-determined |
| 18-106_k141_56730 | Low-quality |
| 18-106_k141_10287 | Not-determined |
| 18-106_k141_56731 | Not-determined |
| 18-106_k141_80770 | Not-determined |
| 18-106_k141_64227 | Not-determined |
| 18-106_k141_56734 | Not-determined |
| 18-106_k141_69904 | Not-determined |
| 18-106_k141_139348 | Not-determined |
| 18-106_k141_21763 | Low-quality |
| 18-106_k141_64233 | Not-determined |
| 18-106_k141_121606 | Not-determined |
| 18-106_k141_116354 | Not-determined |
| 18-106_k141_133715 | Not-determined |
| 18-106_k141_121612 | Not-determined |
| 18-106_k141_10295 | Not-determined |
| 18-106_k141_52102 | Low-quality |
| 18-106_k141_64240 | Low-quality |
| 18-106_k141_110590 | Not-determined |
| 18-106_k141_116363 | Not-determined |
| 18-106_k141_10298 | Not-determined |
| 18-106_k141_99374 | Not-determined |
| 18-106_k141_64244 | Not-determined |
| 18-106_k141_103950 | Not-determined |
| 18-106_k141_16779 | Low-quality |
| 18-106_k141_127992 | Not-determined |
| 18-106_k141_21780 | Low-quality |
| 18-106_k141_133739 | Not-determined |
| 18-106_k141_133740 | Not-determined |
| 18-106_k141_69928 | Low-quality |
| 18-106_k141_40537 | Not-determined |
| 18-106_k141_133744 | Not-determined |
| 18-106_k141_121627 | Not-determined |
| 18-106_k141_16784 | Not-determined |
| 18-106_k141_16786 | Not-determined |
| 18-106_k141_133754 | Not-determined |
| 18-106_k141_75152 | Not-determined |
| 18-106_k141_121638 | Not-determined |
| 18-106_k141_40547 | Not-determined |
| 18-106_k141_128002 | Not-determined |
| 18-106_k141_139375 | Not-determined |
| 18-106_k141_121645 | Not-determined |
| 18-106_k141_10323 | Not-determined |
| 18-106_k141_64279 | Not-determined |
| 18-106_k141_128007 | Not-determined |
| 18-106_k141_116390 | Not-determined |
| 18-106_k141_110610 | Not-determined |
| 18-106_k141_75167 | Low-quality |
| 18-106_k141_34015 | Not-determined |
| 18-106_k141_133766 | Not-determined |
| 18-106_k141_110612 | Not-determined |
| 18-106_k141_40571 | Not-determined |
| 18-106_k141_34021 | Not-determined |
| 18-106_k141_103986 | Not-determined |
| 18-106_k141_34025 | Not-determined |
| 18-106_k141_40572 | Not-determined |
| 18-106_k141_110617 | Not-determined |
| 18-106_k141_69953 | Not-determined |
| 18-106_k141_128025 | Not-determined |
| 18-106_k141_99402 | Not-determined |
| 18-106_k141_121662 | Not-determined |
| 18-106_k141_116401 | Not-determined |
| 18-106_k141_110623 | Not-determined |
| 18-106_k141_116403 | Not-determined |
| 18-106_k141_56764 | Not-determined |
| 18-106_k141_75186 | Not-determined |
| 18-106_k141_116408 | Not-determined |
| 18-106_k141_139404 | Not-determined |
| 18-106_k141_16798 | Not-determined |
| 18-106_k141_34042 | Not-determined |
| 18-106_k141_116414 | Not-determined |
| 18-106_k141_34043 | Not-determined |
| 18-106_k141_75188 | Not-determined |
| 18-106_k141_139405 | Low-quality |
| 18-106_k141_116419 | Not-determined |
| 18-106_k141_75192 | Not-determined |
| 18-106_k141_10327 | Not-determined |
| 18-106_k141_116422 | Not-determined |
| 18-106_k141_99424 | Not-determined |
| 18-106_k141_139414 | Not-determined |
| 18-106_k141_99426 | Not-determined |
| 18-106_k141_99430 | Low-quality |
| 18-106_k141_10342 | Not-determined |
| 18-106_k141_56774 | Not-determined |
| 18-106_k141_56775 | Not-determined |
| 18-106_k141_64331 | Low-quality |
| 18-106_k141_121697 | Not-determined |
| 18-106_k141_121699 | Not-determined |
| 18-106_k141_70011 | Not-determined |
| 18-106_k141_110664 | Not-determined |
| 18-106_k141_56788 | Not-determined |
| 18-106_k141_87046 | Not-determined |
| 18-106_k141_70014 | Not-determined |
| 18-106_k141_70016 | Not-determined |
| 18-106_k141_139433 | Not-determined |
| 18-106_k141_70019 | Not-determined |
| 18-106_k141_70020 | Not-determined |
| 18-106_k141_128040 | Not-determined |
| 18-106_k141_87057 | Not-determined |
| 18-106_k141_28270 | Not-determined |
| 18-106_k141_92856 | Not-determined |
| 18-106_k141_139446 | Not-determined |
| 18-106_k141_121717 | Not-determined |
| 18-106_k141_99451 | Not-determined |
| 18-106_k141_56809 | Not-determined |
| 18-106_k141_80826 | Low-quality |
| 18-106_k141_92863 | Not-determined |
| 18-106_k141_34083 | Not-determined |
| 18-106_k141_80829 | Not-determined |
| 18-106_k141_128052 | Not-determined |
| 18-106_k141_110671 | Not-determined |
| 18-106_k141_99455 | Not-determined |
| 18-106_k141_56812 | Not-determined |
| 18-106_k141_28280 | Not-determined |
| 18-106_k141_92867 | Low-quality |
| 18-106_k141_87066 | Not-determined |
| 18-106_k141_34090 | Not-determined |
| 18-106_k141_34091 | Not-determined |
| 18-106_k141_87069 | Not-determined |
| 18-106_k141_34094 | Not-determined |
| 18-106_k141_40626 | Not-determined |
| 18-106_k141_133801 | Not-determined |
| 18-106_k141_34098 | Not-determined |
| 18-106_k141_70036 | Low-quality |
| 18-106_k141_64362 | Not-determined |
| 18-106_k141_64363 | Not-determined |
| 18-106_k141_10365 | Not-determined |
| 18-106_k141_92885 | Not-determined |
| 18-106_k141_128059 | Not-determined |
| 18-106_k141_28294 | Not-determined |
| 18-106_k141_121728 | Not-determined |
| 18-106_k141_28296 | Not-determined |
| 18-106_k141_70044 | Not-determined |
| 18-106_k141_64369 | Not-determined |
| 18-106_k141_104033 | Not-determined |
| 18-106_k141_34114 | Not-determined |
| 18-106_k141_116447 | Not-determined |
| 18-106_k141_70054 | Not-determined |
| 18-106_k141_40637 | Not-determined |
| 18-106_k141_21842 | Not-determined |
| 18-106_k141_34120 | Low-quality |
| 18-106_k141_99476 | Not-determined |
| 18-106_k141_34122 | Low-quality |
| 18-106_k141_133824 | Not-determined |
| 18-106_k141_16840 | Not-determined |
| 18-106_k141_128079 | Not-determined |
| 18-106_k141_87094 | Not-determined |
| 18-106_k141_21851 | Not-determined |
| 18-106_k141_128083 | Not-determined |
| 18-106_k141_99485 | Not-determined |
| 18-106_k141_56847 | Not-determined |
| 18-106_k141_104050 | Not-determined |
| 18-106_k141_34131 | Not-determined |
| 18-106_k141_46447 | Not-determined |
| 18-106_k141_128090 | Not-determined |
| 18-106_k141_34134 | Not-determined |
| 18-106_k141_16851 | Not-determined |
| 18-106_k141_28319 | Not-determined |
| 18-106_k141_46453 | Not-determined |
| 18-106_k141_70075 | Not-determined |
| 18-106_k141_75215 | Not-determined |
| 18-106_k141_10388 | Not-determined |
| 18-106_k141_75220 | Not-determined |
| 18-106_k141_99510 | Not-determined |
| 18-106_k141_133853 | Not-determined |
| 18-106_k141_10397 | Low-quality |
| 18-106_k141_28330 | Not-determined |
| 18-106_k141_104077 | Not-determined |
| 18-106_k141_10400 | Low-quality |
| 18-106_k141_128100 | Low-quality |
| 18-106_k141_52158 | Not-determined |
| 18-106_k141_116464 | Not-determined |
| 18-106_k141_34162 | Not-determined |
| 18-106_k141_75232 | Not-determined |
| 18-106_k141_87123 | Not-determined |
| 18-106_k141_34166 | Low-quality |
| 18-106_k141_133867 | Not-determined |
| 18-106_k141_121732 | Not-determined |
| 18-106_k141_104087 | Not-determined |
| 18-106_k141_28335 | Not-determined |
| 18-106_k141_87129 | Not-determined |
| 18-106_k141_110720 | Not-determined |
| 18-106_k141_40668 | Low-quality |
| 18-106_k141_128117 | Not-determined |
| 18-106_k141_64400 | Not-determined |
| 18-106_k141_46486 | Not-determined |
| 18-106_k141_10421 | Not-determined |
| 18-106_k141_128123 | Not-determined |
| 18-106_k141_28350 | Not-determined |
| 18-106_k141_21871 | Not-determined |
| 18-106_k141_10424 | Not-determined |
| 18-106_k141_121743 | Not-determined |
| 18-106_k141_46492 | Not-determined |
| 18-106_k141_28354 | Not-determined |
| 18-106_k141_21873 | Not-determined |
| 18-106_k141_40675 | Not-determined |
| 18-106_k141_87141 | Low-quality |
| 18-106_k141_128132 | Not-determined |
| 18-106_k141_52188 | Not-determined |
| 18-106_k141_40685 | Low-quality |
| 18-106_k141_104101 | Not-determined |
| 18-106_k141_87151 | Not-determined |
| 18-106_k141_133881 | Not-determined |
| 18-106_k141_16891 | Not-determined |
| 18-106_k141_104109 | Low-quality |
| 18-106_k141_52195 | Not-determined |
| 18-106_k141_110746 | Not-determined |
| 18-106_k141_52196 | Not-determined |
| 18-106_k141_87158 | Not-determined |
| 18-106_k141_16898 | Not-determined |
| 18-106_k141_16899 | Not-determined |
| 18-106_k141_139495 | Not-determined |
| 18-106_k141_40695 | Not-determined |
| 18-106_k141_46501 | Not-determined |
| 18-106_k141_139500 | Not-determined |
| 18-106_k141_52202 | Not-determined |
| 18-106_k141_116483 | Not-determined |
| 18-106_k141_104110 | Not-determined |
| 18-106_k141_92960 | Not-determined |
| 18-106_k141_34197 | Not-determined |
| 18-106_k141_92963 | Not-determined |
| 18-106_k141_28376 | Not-determined |
| 18-106_k141_21883 | Low-quality |
| 18-106_k141_40707 | Not-determined |
| 18-106_k141_121747 | Not-determined |
| 18-106_k141_56874 | Not-determined |
| 18-106_k141_10446 | Not-determined |
| 18-106_k141_121751 | Not-determined |
| 18-106_k141_110756 | Not-determined |
| 18-106_k141_121755 | Not-determined |
| 18-106_k141_121756 | Low-quality |
| 18-106_k141_34207 | Not-determined |
| 18-106_k141_110758 | Not-determined |
| 18-106_k141_56877 | Not-determined |
| 18-106_k141_10452 | Not-determined |
| 18-106_k141_110761 | Not-determined |
| 18-106_k141_52219 | Not-determined |
| 18-106_k141_87186 | Not-determined |
| 18-106_k141_64423 | Not-determined |
| 18-106_k141_121766 | Not-determined |
| 18-106_k141_92978 | Not-determined |
| 18-106_k141_10458 | Not-determined |
| 18-106_k141_80874 | Not-determined |
| 18-106_k141_40722 | Not-determined |
| 18-106_k141_10461 | Not-determined |
| 18-106_k141_128160 | Not-determined |
| 18-106_k141_87193 | Not-determined |
| 18-106_k141_128162 | Low-quality |
| 18-106_k141_34220 | Not-determined |
| 18-106_k141_75246 | Not-determined |
| 18-106_k141_21909 | Not-determined |
| 18-106_k141_121773 | Not-determined |
| 18-106_k141_52228 | Not-determined |
| 18-106_k141_128169 | Not-determined |
| 18-106_k141_92982 | Not-determined |
| 18-106_k141_46516 | Not-determined |
| 18-106_k141_87205 | Not-determined |
| 18-106_k141_56890 | Not-determined |
| 18-106_k141_64437 | Not-determined |
| 18-106_k141_104127 | Not-determined |
| 18-106_k141_87212 | Not-determined |
| 18-106_k141_28410 | Not-determined |
| 18-106_k141_46519 | Not-determined |
| 18-106_k141_116496 | Not-determined |
| 18-106_k141_104140 | Not-determined |
| 18-106_k141_128174 | Not-determined |
| 18-106_k141_46522 | Not-determined |
| 18-106_k141_87223 | Not-determined |
| 18-106_k141_40740 | Not-determined |
| 18-106_k141_28417 | Not-determined |
| 18-106_k141_128177 | Not-determined |
| 18-106_k141_28418 | Low-quality |
| 18-106_k141_56899 | Not-determined |
| 18-106_k141_46528 | Not-determined |
| 18-106_k141_139516 | Not-determined |
| 18-106_k141_75276 | Not-determined |
| 18-106_k141_116516 | Not-determined |
| 18-106_k141_70134 | Not-determined |
| 18-106_k141_139517 | Not-determined |
| 18-106_k141_75283 | Not-determined |
| 18-106_k141_80886 | Not-determined |
| 18-106_k141_16941 | Not-determined |
| 18-106_k141_80888 | Not-determined |
| 18-106_k141_21936 | Not-determined |
| 18-106_k141_139518 | Not-determined |
| 18-106_k141_104159 | Not-determined |
| 18-106_k141_139520 | Not-determined |
| 18-106_k141_46550 | Not-determined |
| 18-106_k141_121786 | Not-determined |
| 18-106_k141_46553 | Not-determined |
| 18-106_k141_56929 | Not-determined |
| 18-106_k141_46555 | Not-determined |
| 18-106_k141_121789 | Not-determined |
| 18-106_k141_70152 | Not-determined |
| 18-106_k141_116520 | Not-determined |
| 18-106_k141_110820 | Not-determined |
| 18-106_k141_139528 | Not-determined |
| 18-106_k141_139530 | Not-determined |
| 18-106_k141_28453 | Low-quality |
| 18-106_k141_139532 | Not-determined |

| | |
|---|---|
| 18-106_k141_16948 | Not-determined |
| 18-106_k141_64463 | Not-determined |
| 18-106_k141_16950 | Not-determined |
| 18-106_k141_75299 | Not-determined |
| 18-106_k141_139536 | Not-determined |
| 18-106_k141_21965 | Not-determined |
| 18-106_k141_64471 | Not-determined |
| 18-106_k141_70161 | Not-determined |
| 18-106_k141_139540 | Not-determined |
| 18-106_k141_21969 | Not-determined |
| 18-106_k141_80915 | Not-determined |
| 18-106_k141_128207 | Not-determined |
| 18-106_k141_104185 | Not-determined |
| 18-106_k141_10489 | Low-quality |
| 18-106_k141_75304 | Not-determined |
| 18-106_k141_46571 | Not-determined |
| 18-106_k141_16960 | Not-determined |
| 18-106_k141_116550 | Low-quality |
| 18-106_k141_40780 | Not-determined |
| 18-106_k141_21980 | Not-determined |
| 18-106_k141_87248 | Not-determined |
| 18-106_k141_21986 | Not-determined |
| 18-106_k141_139552 | Not-determined |
| 18-106_k141_56960 | Not-determined |
| 18-106_k141_10505 | Not-determined |
| 18-106_k141_46590 | Not-determined |
| 18-106_k141_52245 | Not-determined |
| 18-106_k141_139564 | Not-determined |
| 18-106_k141_10510 | Not-determined |
| 18-106_k141_56970 | Not-determined |
| 18-106_k141_104213 | Not-determined |
| 18-106_k141_10514 | Not-determined |
| 18-106_k141_99560 | Not-determined |
| 18-106_k141_21999 | Not-determined |
| 18-106_k141_128233 | Not-determined |
| 18-106_k141_87256 | Not-determined |
| 18-106_k141_121797 | Not-determined |
| 18-106_k141_139568 | Not-determined |
| 18-106_k141_87262 | Not-determined |
| 18-106_k141_46601 | Not-determined |
| 18-106_k141_110854 | Low-quality |
| 18-106_k141_99572 | Not-determined |
| 18-106_k141_139578 | Not-determined |
| 18-106_k141_87269 | Not-determined |
| 18-106_k141_139582 | Not-determined |
| 18-106_k141_128252 | Not-determined |
| 18-106_k141_40801 | Not-determined |
| 18-106_k141_128254 | Not-determined |
| 18-106_k141_56990 | Not-determined |
| 18-106_k141_80931 | Not-determined |
| 18-106_k141_80933 | Not-determined |
| 18-106_k141_28488 | Not-determined |
| 18-106_k141_139594 | Not-determined |
| 18-106_k141_56991 | Not-determined |
| 18-106_k141_110871 | Not-determined |
| 18-106_k141_28495 | Not-determined |
| 18-106_k141_56998 | Not-determined |
| 18-106_k141_46618 | Not-determined |
| 18-106_k141_121828 | Not-determined |
| 18-106_k141_70201 | Not-determined |
| 18-106_k141_116590 | Not-determined |
| 18-106_k141_104237 | Not-determined |
| 18-106_k141_139604 | Not-determined |
| 18-106_k141_121833 | Not-determined |
| 18-106_k141_93075 | Not-determined |
| 18-106_k141_121843 | Not-determined |
| 18-106_k141_121844 | Not-determined |
| 18-106_k141_22033 | Not-determined |
| 18-106_k141_139608 | Not-determined |
| 18-106_k141_87287 | Not-determined |
| 18-106_k141_87289 | Low-quality |
| 18-106_k141_64494 | Not-determined |
| 18-106_k141_110882 | Not-determined |
| 18-106_k141_99591 | Not-determined |
| 18-106_k141_121853 | Not-determined |
| 18-106_k141_99592 | Not-determined |
| 18-106_k141_139610 | Not-determined |
| 18-106_k141_139611 | Not-determined |
| 18-106_k141_116606 | Not-determined |
| 18-106_k141_64499 | Low-quality |
| 18-106_k141_22040 | Not-determined |
| 18-106_k141_40821 | Not-determined |
| 18-106_k141_87298 | Not-determined |
| 18-106_k141_99600 | Not-determined |
| 18-106_k141_87301 | Not-determined |
| 18-106_k141_110889 | Not-determined |
| 18-106_k141_22043 | Not-determined |
| 18-106_k141_22045 | Not-determined |
| 18-106_k141_116621 | Not-determined |
| 18-106_k141_139624 | Not-determined |
| 18-106_k141_87304 | Not-determined |
| 18-106_k141_22048 | Not-determined |
| 18-106_k141_99608 | Not-determined |
| 18-106_k141_87305 | Low-quality |
| 18-106_k141_128284 | Not-determined |
| 18-106_k141_22050 | Not-determined |
| 18-106_k141_80966 | Not-determined |
| 18-106_k141_110894 | Low-quality |
| 18-106_k141_46652 | Not-determined |
| 18-106_k141_22053 | Not-determined |
| 18-106_k141_34304 | Not-determined |
| 18-106_k141_34305 | Not-determined |
| 18-106_k141_110899 | Not-determined |
| 18-106_k141_139632 | Not-determined |
| 18-106_k141_64524 | Not-determined |
| 18-106_k141_57038 | Not-determined |
| 18-106_k141_139636 | Not-determined |
| 18-106_k141_17024 | Not-determined |
| 18-106_k141_28503 | Not-determined |
| 18-106_k141_28506 | Not-determined |
| 18-106_k141_116630 | Not-determined |
| 18-106_k141_22069 | Not-determined |
| 18-106_k141_34313 | Not-determined |
| 18-106_k141_116635 | Not-determined |
| 18-106_k141_57047 | Not-determined |
| 18-106_k141_40851 | Not-determined |
| 18-106_k141_87334 | Not-determined |
| 18-106_k141_10524 | Not-determined |
| 18-106_k141_99633 | Not-determined |
| 18-106_k141_99634 | Not-determined |
| 18-106_k141_75354 | Low-quality |
| 18-106_k141_87339 | Not-determined |
| 18-106_k141_99638 | Not-determined |
| 18-106_k141_70254 | Not-determined |
| 18-106_k141_40864 | Not-determined |
| 18-106_k141_52297 | Not-determined |
| 18-106_k141_70256 | Not-determined |
| 18-106_k141_80984 | Not-determined |
| 18-106_k141_80985 | Not-determined |
| 18-106_k141_70263 | Not-determined |
| 18-106_k141_80990 | Not-determined |
| 18-106_k141_10533 | Not-determined |
| 18-106_k141_10534 | Low-quality |
| 18-106_k141_133951 | Not-determined |
| 18-106_k141_80991 | Not-determined |
| 18-106_k141_46686 | Not-determined |
| 18-106_k141_75367 | Low-quality |
| 18-106_k141_70271 | Not-determined |
| 18-106_k141_139663 | Not-determined |
| 18-106_k141_70274 | Not-determined |
| 18-106_k141_28522 | Not-determined |
| 18-106_k141_116655 | Not-determined |
| 18-106_k141_116658 | Not-determined |
| 18-106_k141_116662 | Not-determined |
| 18-106_k141_22103 | Not-determined |
| 18-106_k141_52325 | Not-determined |
| 18-106_k141_22110 | Not-determined |
| 18-106_k141_133965 | Low-quality |
| 18-106_k141_139676 | Not-determined |
| 18-106_k141_81001 | Not-determined |
| 18-106_k141_22112 | Not-determined |
| 18-106_k141_139678 | Not-determined |
| 18-106_k141_75374 | Not-determined |
| 18-106_k141_139679 | Low-quality |
| 18-106_k141_128308 | Not-determined |
| 18-106_k141_22116 | Not-determined |
| 18-106_k141_46695 | Not-determined |
| 18-106_k141_128316 | Not-determined |
| 18-106_k141_52334 | Low-quality |
| 18-106_k141_93104 | Not-determined |
| 18-106_k141_40884 | Not-determined |
| 18-106_k141_87357 | Not-determined |
| 18-106_k141_40887 | Not-determined |
| 18-106_k141_10574 | Not-determined |
| 18-106_k141_75391 | Not-determined |
| 18-106_k141_52343 | Not-determined |
| 18-106_k141_139693 | Low-quality |
| 18-106_k141_81029 | Not-determined |
| 18-106_k141_22125 | Low-quality |
| 18-106_k141_104313 | Not-determined |
| 18-106_k141_133975 | Not-determined |
| 18-106_k141_40899 | Not-determined |
| 18-106_k141_139701 | Not-determined |
| 18-106_k141_121883 | Not-determined |
| 18-106_k141_128332 | Not-determined |
| 18-106_k141_139705 | Not-determined |
| 18-106_k141_104319 | Not-determined |
| 18-106_k141_93114 | Low-quality |
| 18-106_k141_133978 | Low-quality |
| 18-106_k141_10590 | Not-determined |
| 18-106_k141_46725 | Not-determined |
| 18-106_k141_128339 | Not-determined |
| 18-106_k141_133981 | Not-determined |
| 18-106_k141_128341 | Not-determined |
| 18-106_k141_46728 | Not-determined |
| 18-106_k141_70318 | Not-determined |
| 18-106_k141_93126 | Not-determined |
| 18-106_k141_70321 | Not-determined |
| 18-106_k141_116697 | Not-determined |
| 18-106_k141_128346 | Not-determined |
| 18-106_k141_17063 | Not-determined |
| 18-106_k141_104329 | Low-quality |
| 18-106_k141_81048 | Not-determined |
| 18-106_k141_99674 | Not-determined |
| 18-106_k141_10608 | Not-determined |
| 18-106_k141_17069 | Not-determined |
| 18-106_k141_110907 | Not-determined |
| 18-106_k141_46738 | Not-determined |
| 18-106_k141_99683 | Low-quality |
| 18-106_k141_57114 | Not-determined |
| 18-106_k141_110911 | Not-determined |
| 18-106_k141_75420 | Not-determined |
| 18-106_k141_87393 | Not-determined |
| 18-106_k141_116724 | Low-quality |
| 18-106_k141_22166 | Not-determined |
| 18-106_k141_139734 | Not-determined |
| 18-106_k141_104342 | Not-determined |
| 18-106_k141_139735 | Low-quality |
| 18-106_k141_116725 | Low-quality |
| 18-106_k141_139748 | Low-quality |
| 18-106_k141_10631 | Not-determined |
| 18-106_k141_128365 | Not-determined |
| 18-106_k141_116732 | Not-determined |
| 18-106_k141_10633 | Not-determined |
| 18-106_k141_99713 | Not-determined |
| 18-106_k141_75443 | Not-determined |
| 18-106_k141_75447 | Not-determined |
| 18-106_k141_116737 | Not-determined |
| 18-106_k141_87415 | Not-determined |
| 18-106_k141_17104 | Not-determined |
| 18-106_k141_110929 | Not-determined |
| 18-106_k141_87421 | Not-determined |
| 18-106_k141_52383 | Not-determined |
| 18-106_k141_10640 | Not-determined |
| 18-106_k141_17105 | Not-determined |
| 18-106_k141_116746 | Not-determined |
| 18-106_k141_46759 | Not-determined |
| 18-106_k141_110941 | Not-determined |
| 18-106_k141_93172 | Not-determined |
| 18-106_k141_116750 | Not-determined |
| 18-106_k141_57149 | Not-determined |
| 18-106_k141_99729 | Low-quality |
| 18-106_k141_87434 | Not-determined |
| 18-106_k141_10644 | Not-determined |
| 18-106_k141_10645 | Not-determined |
| 18-106_k141_121909 | Not-determined |
| 18-106_k141_134022 | Not-determined |
| 18-106_k141_10646 | Low-quality |
| 18-106_k141_10648 | Not-determined |
| 18-106_k141_104389 | Not-determined |
| 18-106_k141_22210 | Not-determined |
| 18-106_k141_121911 | Not-determined |
| 18-106_k141_22212 | Not-determined |
| 18-106_k141_121914 | Not-determined |
| 18-106_k141_116762 | Low-quality |
| 18-106_k141_22216 | Not-determined |
| 18-106_k141_93201 | Not-determined |
| 18-106_k141_57166 | Not-determined |
| 18-106_k141_52401 | Not-determined |
| 18-106_k141_93208 | Not-determined |
| 18-106_k141_116773 | Not-determined |
| 18-106_k141_64561 | Not-determined |
| 18-106_k141_121926 | Not-determined |
| 18-106_k141_46770 | Low-quality |
| 18-106_k141_121927 | Not-determined |
| 18-106_k141_75487 | Not-determined |
| 18-106_k141_93215 | Not-determined |
| 18-106_k141_121929 | Not-determined |
| 18-106_k141_104402 | Not-determined |
| 18-106_k141_104403 | Not-determined |
| 18-106_k141_10661 | Not-determined |
| 18-106_k141_93220 | Not-determined |
| 18-106_k141_10664 | Not-determined |
| 18-106_k141_99756 | Not-determined |
| 18-106_k141_22225 | Not-determined |
| 18-106_k141_139795 | Not-determined |
| 18-106_k141_64569 | Not-determined |
| 18-106_k141_64570 | Not-determined |
| 18-106_k141_57190 | Not-determined |
| 18-106_k141_10666 | Not-determined |
| 18-106_k141_104416 | Not-determined |
| 18-106_k141_57195 | Not-determined |
| 18-106_k141_70346 | Not-determined |
| 18-106_k141_104423 | Not-determined |
| 18-106_k141_139804 | Not-determined |
| 18-106_k141_70351 | Not-determined |
| 18-106_k141_52429 | Not-determined |
| 18-106_k141_64572 | Low-quality |
| 18-106_k141_121954 | Not-determined |
| 18-106_k141_87453 | Not-determined |
| 18-106_k141_93245 | Low-quality |
| 18-106_k141_81074 | Not-determined |
| 18-106_k141_121956 | Not-determined |
| 18-106_k141_128408 | Not-determined |
| 18-106_k141_22237 | Not-determined |
| 18-106_k141_10678 | Not-determined |
| 18-106_k141_139822 | Not-determined |
| 18-106_k141_40927 | Not-determined |
| 18-106_k141_134050 | Low-quality |
| 18-106_k141_134051 | Not-determined |
| 18-106_k141_128412 | Low-quality |
| 18-106_k141_87460 | Not-determined |
| 18-106_k141_17123 | Not-determined |
| 18-106_k141_87464 | Not-determined |
| 18-106_k141_87465 | Not-determined |
| 18-106_k141_99787 | Not-determined |
| 18-106_k141_57218 | Not-determined |
| 18-106_k141_134060 | Not-determined |
| 18-106_k141_46782 | Not-determined |
| 18-106_k141_52463 | Low-quality |
| 18-106_k141_70362 | Not-determined |
| 18-106_k141_139838 | Not-determined |
| 18-106_k141_116780 | Not-determined |
| 18-106_k141_28607 | Not-determined |
| 18-106_k141_75520 | Low-quality |
| 18-106_k141_134067 | Not-determined |
| 18-106_k141_134068 | Not-determined |
| 18-106_k141_28611 | Not-determined |
| 18-106_k141_22253 | Not-determined |
| 18-106_k141_121977 | Not-determined |
| 18-106_k141_52475 | Not-determined |
| 18-106_k141_70373 | Not-determined |
| 18-106_k141_128431 | Not-determined |
| 18-106_k141_139847 | Not-determined |
| 18-106_k141_34344 | Not-determined |
| 18-106_k141_139848 | Not-determined |
| 18-106_k141_134075 | Not-determined |
| 18-106_k141_46788 | Low-quality |
| 18-106_k141_134079 | Not-determined |
| 18-106_k141_28625 | Not-determined |
| 18-106_k141_93276 | Not-determined |
| 18-106_k141_116783 | Not-determined |
| 18-106_k141_93278 | Not-determined |
| 18-106_k141_81095 | Not-determined |
| 18-106_k141_40949 | Not-determined |
| 18-106_k141_22276 | Not-determined |
| 18-106_k141_81100 | Not-determined |
| 18-106_k141_116796 | Not-determined |
| 18-106_k141_139864 | Not-determined |
| 18-106_k141_139869 | Low-quality |
| 18-106_k141_57252 | Low-quality |
| 18-106_k141_57255 | Not-determined |
| 18-106_k141_116800 | Not-determined |
| 18-106_k141_122007 | Not-determined |
| 18-106_k141_104459 | Not-determined |
| 18-106_k141_34384 | Not-determined |
| 18-106_k141_57269 | Not-determined |
| 18-106_k141_52522 | Not-determined |
| 18-106_k141_70402 | Not-determined |
| 18-106_k141_52523 | Not-determined |
| 18-106_k141_122020 | Not-determined |
| 18-106_k141_57276 | Not-determined |
| 18-106_k141_34402 | Not-determined |
| 18-106_k141_75563 | Not-determined |
| 18-106_k141_134113 | Not-determined |
| 18-106_k141_87508 | Not-determined |
| 18-106_k141_110994 | Low-quality |
| 18-106_k141_52529 | Not-determined |
| 18-106_k141_17169 | Not-determined |
| 18-106_k141_34410 | Not-determined |
| 18-106_k141_22324 | Not-determined |
| 18-106_k141_110996 | Not-determined |
| 18-106_k141_70427 | Not-determined |
| 18-106_k141_110998 | Low-quality |
| 18-106_k141_81126 | Not-determined |
| 18-106_k141_122031 | Not-determined |
| 18-106_k141_52535 | Not-determined |
| 18-106_k141_134123 | Low-quality |
| 18-106_k141_87520 | Not-determined |
| 18-106_k141_87522 | Low-quality |
| 18-106_k141_122036 | Low-quality |
| 18-106_k141_40988 | Not-determined |
| 18-106_k141_75573 | Not-determined |
| 18-106_k141_34417 | Not-determined |
| 18-106_k141_57290 | Not-determined |
| 18-106_k141_81141 | Not-determined |
| 18-106_k141_122047 | Not-determined |
| 18-106_k141_116832 | Not-determined |
| 18-106_k141_116834 | Not-determined |
| 18-106_k141_139906 | Not-determined |
| 18-106_k141_104475 | Not-determined |
| 18-106_k141_139910 | Not-determined |
| 18-106_k141_93342 | Not-determined |
| 18-106_k141_17182 | Not-determined |
| 18-106_k141_87546 | Not-determined |
| 18-106_k141_93352 | Not-determined |
| 18-106_k141_134144 | Not-determined |
| 18-106_k141_17184 | Low-quality |
| 18-106_k141_116841 | Not-determined |
| 18-106_k141_134146 | Not-determined |
| 18-106_k141_81163 | Not-determined |
| 18-106_k141_93362 | Not-determined |
| 18-106_k141_52561 | Not-determined |
| 18-106_k141_41023 | Not-determined |
| 18-106_k141_139939 | Not-determined |
| 18-106_k141_93366 | Not-determined |
| 18-106_k141_75609 | Not-determined |
| 18-106_k141_111007 | Not-determined |
| 18-106_k141_128511 | Not-determined |
| 18-106_k141_134174 | Not-determined |
| 18-106_k141_57305 | Not-determined |
| 18-106_k141_57307 | Not-determined |
| 18-106_k141_111015 | Low-quality |
| 18-106_k141_111017 | Not-determined |
| 18-106_k141_87580 | Not-determined |
| 18-106_k141_75626 | Not-determined |
| 18-106_k141_52589 | Not-determined |
| 18-106_k141_87585 | Not-determined |
| 18-106_k141_122067 | Not-determined |
| 18-106_k141_104502 | Not-determined |
| 18-106_k141_134196 | Not-determined |
| 18-106_k141_111026 | Not-determined |
| 18-106_k141_81184 | Not-determined |
| 18-106_k141_34465 | Not-determined |
| 18-106_k141_111031 | Not-determined |
| 18-106_k141_111032 | Not-determined |
| 18-106_k141_87594 | Not-determined |
| 18-106_k141_87596 | Not-determined |
| 18-106_k141_111035 | Not-determined |
| 18-106_k141_128526 | Low-quality |
| 18-106_k141_17202 | Not-determined |
| 18-106_k141_122074 | Not-determined |
| 18-106_k141_104522 | Not-determined |
| 18-106_k141_57317 | Not-determined |
| 18-106_k141_134207 | Low-quality |
| 18-106_k141_122079 | Not-determined |
| 18-106_k141_134211 | Low-quality |
| 18-106_k141_57322 | Low-quality |
| 18-106_k141_52628 | Not-determined |
| 18-106_k141_81205 | Not-determined |
| 18-106_k141_111052 | Not-determined |
| 18-106_k141_17219 | Low-quality |
| 18-106_k141_81214 | Not-determined |
| 18-106_k141_111059 | Not-determined |
| 18-106_k141_116899 | Not-determined |
| 18-106_k141_52647 | Not-determined |
| 18-106_k141_81222 | Not-determined |
| 18-106_k141_57337 | Not-determined |
| 18-106_k141_116901 | Not-determined |
| 18-106_k141_93406 | Not-determined |
| 18-106_k141_57345 | Not-determined |
| 18-106_k141_111076 | Not-determined |
| 18-106_k141_116913 | Not-determined |
| 18-106_k141_57353 | Not-determined |
| 18-106_k141_116915 | Not-determined |
| 18-106_k141_111080 | Not-determined |
| 18-106_k141_34506 | Not-determined |
| 18-106_k141_93420 | Not-determined |
| 18-106_k141_57361 | Not-determined |
| 18-106_k141_134232 | Not-determined |
| 18-106_k141_52657 | Low-quality |
| 18-106_k141_111088 | Low-quality |
| 18-106_k141_10747 | Not-determined |
| 18-106_k141_116925 | Not-determined |
| 18-106_k141_10753 | Not-determined |
| 18-106_k141_139956 | Not-determined |
| 18-106_k141_111092 | Not-determined |
| 18-106_k141_17240 | Not-determined |
| 18-106_k141_75644 | Not-determined |
| 18-106_k141_104579 | Not-determined |
| 18-106_k141_52682 | Not-determined |
| 18-106_k141_17257 | Not-determined |
| 18-106_k141_52688 | Low-quality |
| 18-106_k141_10772 | Not-determined |
| 18-106_k141_139974 | Not-determined |
| 18-106_k141_75657 | Not-determined |
| 18-106_k141_17262 | Not-determined |
| 18-106_k141_134251 | Not-determined |
| 18-106_k141_46846 | Not-determined |
| 18-106_k141_41039 | Not-determined |
| 18-106_k141_116938 | Not-determined |
| 18-106_k141_93447 | Not-determined |
| 18-106_k141_81259 | Not-determined |
| 18-106_k141_116944 | Not-determined |
| 18-106_k141_46857 | Not-determined |
| 18-106_k141_46863 | Not-determined |
| 18-106_k141_46868 | Not-determined |
| 18-106_k141_116953 | Not-determined |
| 18-106_k141_41053 | Not-determined |
| 18-106_k141_46875 | Not-determined |
| 18-106_k141_28682 | Not-determined |
| 18-106_k141_41059 | Low-quality |
| 18-106_k141_116970 | Not-determined |
| 18-106_k141_28686 | Not-determined |
| 18-106_k141_57392 | Not-determined |
| 18-106_k141_87614 | Not-determined |
| 18-106_k141_41073 | Not-determined |
| 18-106_k141_34551 | Not-determined |
| 18-106_k141_22349 | Not-determined |
| 18-106_k141_46894 | Not-determined |
| 18-106_k141_116982 | Not-determined |
| 18-106_k141_22354 | Not-determined |
| 18-106_k141_75684 | Not-determined |
| 18-106_k141_93457 | Not-determined |
| 18-106_k141_111141 | Not-determined |
| 18-106_k141_116988 | Not-determined |
| 18-106_k141_93458 | Not-determined |
| 18-106_k141_22359 | Not-determined |
| 18-106_k141_116993 | Not-determined |
| 18-106_k141_17295 | Not-determined |
| 18-106_k141_81271 | Not-determined |
| 18-106_k141_81275 | Not-determined |
| 18-106_k141_104603 | Not-determined |
| 18-106_k141_104606 | Not-determined |
| 18-106_k141_134303 | Not-determined |
| 18-106_k141_75700 | Low-quality |
| 18-106_k141_104613 | Not-determined |
| 18-106_k141_128606 | Not-determined |
| 18-106_k141_104617 | Not-determined |
| 18-106_k141_17305 | Not-determined |
| 18-106_k141_28723 | Not-determined |
| 18-106_k141_22375 | Not-determined |
| 18-106_k141_134320 | Not-determined |
| 18-106_k141_87643 | Not-determined |
| 18-106_k141_46920 | Not-determined |
| 18-106_k141_111162 | Not-determined |
| 18-106_k141_10818 | Not-determined |
| 18-106_k141_122185 | Not-determined |
| 18-106_k141_104645 | Not-determined |
| 18-106_k141_81300 | Not-determined |
| 18-106_k141_111176 | Not-determined |
| 18-106_k141_46934 | Not-determined |
| 18-106_k141_28733 | Not-determined |
| 18-106_k141_117024 | Not-determined |
| 18-106_k141_117026 | Not-determined |
| 18-106_k141_134328 | Low-quality |
| 18-106_k141_128645 | Not-determined |
| 18-106_k141_117028 | Low-quality |
| 18-106_k141_128649 | Not-determined |
| 18-106_k141_134336 | Not-determined |
| 18-106_k141_22402 | Not-determined |
| 18-106_k141_140004 | Not-determined |
| 18-106_k141_140005 | Not-determined |
| 18-106_k141_81320 | Not-determined |
| 18-106_k141_140008 | Not-determined |
| 18-106_k141_128656 | Low-quality |
| 18-106_k141_87666 | Low-quality |
| 18-106_k141_117037 | Not-determined |
| 18-106_k141_104678 | Not-determined |
| 18-106_k141_10830 | Not-determined |
| 18-106_k141_117038 | Not-determined |
| 18-106_k141_128662 | Low-quality |
| 18-106_k141_140015 | Not-determined |
| 18-106_k141_134340 | Low-quality |
| 18-106_k141_93468 | Not-determined |
| 18-106_k141_134344 | Not-determined |
| 18-106_k141_52743 | Not-determined |
| 18-106_k141_52744 | Not-determined |
| 18-106_k141_87677 | Low-quality |
| 18-106_k141_75723 | Not-determined |
| 18-106_k141_140023 | Low-quality |
| 18-106_k141_111206 | Not-determined |
| 18-106_k141_122234 | Not-determined |
| 18-106_k141_117055 | Not-determined |
| 18-106_k141_134351 | Not-determined |
| 18-106_k141_93478 | Not-determined |
| 18-106_k141_128676 | Not-determined |
| 18-106_k141_52757 | Not-determined |
| 18-106_k141_111213 | Low-quality |
| 18-106_k141_93480 | Not-determined |
| 18-106_k141_93486 | Not-determined |
| 18-106_k141_93487 | Not-determined |
| 18-106_k141_52769 | Low-quality |
| 18-106_k141_93504 | Not-determined |
| 18-106_k141_140031 | Not-determined |
| 18-106_k141_81343 | Not-determined |
| 18-106_k141_128692 | Not-determined |
| 18-106_k141_128695 | Not-determined |
| 18-106_k141_10861 | Not-determined |
| 18-106_k141_122261 | Not-determined |
| 18-106_k141_81352 | Not-determined |
| 18-106_k141_75754 | Not-determined |
| 18-106_k141_134378 | Not-determined |
| 18-106_k141_128702 | Not-determined |
| 18-106_k141_117080 | Low-quality |
| 18-106_k141_134382 | Not-determined |
| 18-106_k141_128708 | Not-determined |
| 18-106_k141_140047 | Not-determined |
| 18-106_k141_117086 | Not-determined |
| 18-106_k141_104700 | Not-determined |
| 18-106_k141_122266 | Not-determined |
| 18-106_k141_128716 | Not-determined |
| 18-106_k141_52775 | Not-determined |
| 18-106_k141_81365 | Low-quality |
| 18-106_k141_81366 | Not-determined |
| 18-106_k141_57436 | Low-quality |
| 18-106_k141_10883 | Not-determined |
| 18-106_k141_104713 | Not-determined |
| 18-106_k141_22457 | Not-determined |
| 18-106_k141_28762 | Not-determined |
| 18-106_k141_52786 | Not-determined |
| 18-106_k141_28764 | Not-determined |
| 18-106_k141_134405 | Not-determined |
| 18-106_k141_128729 | Not-determined |
| 18-106_k141_52791 | Not-determined |
| 18-106_k141_134413 | Not-determined |
| 18-106_k141_93535 | Not-determined |
| 18-106_k141_17344 | Not-determined |
| 18-106_k141_128731 | Not-determined |

| | |
|---|---|
| 18-106_k141_81380 | Not-determined |
| 18-106_k141_104728 | Not-determined |
| 18-106_k141_10893 | Not-determined |
| 18-106_k141_87724 | Not-determined |
| 18-106_k141_57443 | Not-determined |
| 18-106_k141_57449 | Not-determined |
| 18-106_k141_104740 | Not-determined |
| 18-106_k141_87729 | Not-determined |
| 18-106_k141_57451 | Not-determined |
| 18-106_k141_81395 | Low-quality |
| 18-106_k141_57455 | Not-determined |
| 18-106_k141_104742 | Not-determined |
| 18-106_k141_122297 | Not-determined |
| 18-106_k141_93545 | Low-quality |
| 18-106_k141_28783 | Not-determined |
| 18-106_k141_75783 | Not-determined |
| 18-106_k141_140074 | Not-determined |
| 18-106_k141_22475 | Not-determined |
| 18-106_k141_28788 | Not-determined |
| 18-106_k141_140076 | Not-determined |
| 18-106_k141_140078 | Not-determined |
| 18-106_k141_28794 | Low-quality |
| 18-106_k141_134449 | Not-determined |
| 18-106_k141_122306 | Not-determined |
| 18-106_k141_111256 | Not-determined |
| 18-106_k141_140081 | Not-determined |
| 18-106_k141_104761 | Not-determined |
| 18-106_k141_93550 | Low-quality |
| 18-106_k141_93551 | Not-determined |
| 18-106_k141_128762 | Not-determined |
| 18-106_k141_111261 | Not-determined |
| 18-106_k141_134454 | Low-quality |
| 18-106_k141_10936 | Not-determined |
| 18-106_k141_17379 | Not-determined |
| 18-106_k141_17382 | Not-determined |
| 18-106_k141_81428 | Not-determined |
| 18-106_k141_128773 | Low-quality |
| 18-106_k141_10945 | Not-determined |
| 18-106_k141_117126 | Not-determined |
| 18-106_k141_81436 | Not-determined |
| 18-106_k141_28803 | Not-determined |
| 18-106_k141_128790 | Not-determined |
| 18-106_k141_17394 | Not-determined |
| 18-106_k141_10954 | Not-determined |
| 18-106_k141_17398 | Not-determined |
| 18-106_k141_57479 | Not-determined |
| 18-106_k141_28814 | Low-quality |
| 18-106_k141_22507 | Not-determined |
| 18-106_k141_17402 | Low-quality |
| 18-106_k141_22513 | Not-determined |
| 18-106_k141_134470 | Not-determined |
| 18-106_k141_17418 | Low-quality |
| 18-106_k141_57494 | Not-determined |
| 18-106_k141_81443 | Not-determined |
| 18-106_k141_128815 | Not-determined |
| 18-106_k141_75797 | Not-determined |
| 18-106_k141_104804 | Not-determined |
| 18-106_k141_87781 | Not-determined |
| 18-106_k141_104810 | Low-quality |
| 18-106_k141_75806 | Not-determined |
| 18-106_k141_17433 | Low-quality |
| 18-106_k141_128827 | Not-determined |
| 18-106_k141_87790 | Not-determined |
| 18-106_k141_140105 | Not-determined |
| 18-106_k141_134496 | Not-determined |
| 18-106_k141_22537 | Not-determined |
| 18-106_k141_117155 | Not-determined |
| 18-106_k141_57513 | Not-determined |
| 18-106_k141_104818 | Low-quality |
| 18-106_k141_93596 | Not-determined |
| 18-106_k141_17448 | Not-determined |
| 18-106_k141_52812 | Low-quality |
| 18-106_k141_22549 | Not-determined |
| 18-106_k141_81490 | Not-determined |
| 18-106_k141_93605 | Not-determined |
| 18-106_k141_122323 | Low-quality |
| 18-106_k141_81495 | Not-determined |
| 18-106_k141_122327 | Not-determined |
| 18-106_k141_34619 | Not-determined |
| 18-106_k141_81500 | Not-determined |
| 18-106_k141_17460 | Not-determined |
| 18-106_k141_87806 | Not-determined |
| 18-106_k141_81506 | Not-determined |
| 18-106_k141_134510 | Not-determined |
| 18-106_k141_17463 | Not-determined |
| 18-106_k141_134518 | Not-determined |
| 18-106_k141_117177 | Not-determined |
| 18-106_k141_104846 | Not-determined |
| 18-106_k141_81515 | Not-determined |
| 18-106_k141_17472 | Not-determined |
| 18-106_k141_122337 | Not-determined |
| 18-106_k141_34628 | Not-determined |
| 18-106_k141_75828 | Low-quality |
| 18-106_k141_81523 | Not-determined |
| 18-106_k141_10986 | Low-quality |
| 18-106_k141_93631 | Not-determined |
| 18-106_k141_134533 | Not-determined |
| 18-106_k141_122349 | Not-determined |
| 18-106_k141_128849 | Not-determined |
| 18-106_k141_122350 | Not-determined |
| 18-106_k141_17484 | Low-quality |
| 18-106_k141_117183 | Not-determined |
| 18-106_k141_87835 | Not-determined |
| 18-106_k141_28868 | Not-determined |
| 18-106_k141_104865 | Not-determined |
| 18-106_k141_140139 | Low-quality |
| 18-106_k141_75845 | Not-determined |
| 18-106_k141_75847 | Not-determined |
| 18-106_k141_81534 | Not-determined |
| 18-106_k141_57548 | Not-determined |
| 18-106_k141_134550 | Not-determined |
| 18-106_k141_28884 | Low-quality |
| 18-106_k141_134561 | Not-determined |
| 18-106_k141_11009 | Not-determined |
| 18-106_k141_87847 | Not-determined |
| 18-106_k141_128871 | Low-quality |
| 18-106_k141_122383 | Not-determined |
| 18-106_k141_75863 | Not-determined |
| 18-106_k141_140148 | Not-determined |
| 18-106_k141_17526 | Not-determined |
| 18-106_k141_104910 | Not-determined |
| 18-106_k141_17531 | Not-determined |
| 18-106_k141_140162 | Low-quality |
| 18-106_k141_122403 | Not-determined |
| 18-106_k141_81574 | Not-determined |
| 18-106_k141_28901 | Not-determined |
| 18-106_k141_57574 | Low-quality |
| 18-106_k141_134584 | Not-determined |
| 18-106_k141_104921 | Not-determined |
| 18-106_k141_57575 | Not-determined |
| 18-106_k141_81580 | Not-determined |
| 18-106_k141_104924 | Not-determined |
| 18-106_k141_11031 | Not-determined |
| 18-106_k141_34670 | Not-determined |
| 18-106_k141_81586 | Not-determined |
| 18-106_k141_93693 | Not-determined |
| 18-106_k141_22616 | Not-determined |
| 18-106_k141_11040 | Not-determined |
| 18-106_k141_11041 | Not-determined |
| 18-106_k141_134594 | Not-determined |
| 18-106_k141_81597 | Not-determined |
| 18-106_k141_134595 | Not-determined |
| 18-106_k141_11047 | Not-determined |
| 18-106_k141_81602 | Low-quality |
| 18-106_k141_117197 | Not-determined |
| 18-106_k141_140184 | Not-determined |
| 18-106_k141_104945 | Not-determined |
| 18-106_k141_17552 | Not-determined |
| 18-106_k141_93710 | Not-determined |
| 18-106_k141_93712 | Not-determined |
| 18-106_k141_134606 | Not-determined |
| 18-106_k141_128894 | Not-determined |
| 18-106_k141_17554 | Not-determined |
| 18-106_k141_17555 | Not-determined |
| 18-106_k141_122436 | Not-determined |
| 18-106_k141_34688 | Low-quality |
| 18-106_k141_128897 | Not-determined |
| 18-106_k141_134613 | Not-determined |
| 18-106_k141_128905 | Not-determined |
| 18-106_k141_22627 | Low-quality |
| 18-106_k141_57605 | Low-quality |
| 18-106_k141_134633 | Not-determined |
| 18-106_k141_81636 | Not-determined |
| 18-106_k141_134640 | Not-determined |
| 18-106_k141_111384 | Not-determined |
| 18-106_k141_111386 | Not-determined |
| 18-106_k141_34714 | Not-determined |
| 18-106_k141_17591 | Not-determined |
| 18-106_k141_128925 | Not-determined |
| 18-106_k141_111389 | Not-determined |
| 18-106_k141_128929 | Not-determined |
| 18-106_k141_128930 | Not-determined |
| 18-106_k141_134645 | Not-determined |
| 18-106_k141_134647 | Not-determined |
| 18-106_k141_117220 | Not-determined |
| 18-106_k141_140208 | Not-determined |
| 18-106_k141_93737 | Low-quality |
| 18-106_k141_128933 | Not-determined |
| 18-106_k141_57617 | Not-determined |
| 18-106_k141_17604 | Not-determined |
| 18-106_k141_134664 | Not-determined |
| 18-106_k141_17615 | Not-determined |
| 18-106_k141_87890 | Not-determined |
| 18-106_k141_122474 | Not-determined |
| 18-106_k141_87893 | Not-determined |
| 18-106_k141_57619 | Not-determined |
| 18-106_k141_134674 | Low-quality |
| 18-106_k141_134680 | Low-quality |
| 18-106_k141_57630 | Not-determined |
| 18-106_k141_22668 | Not-determined |
| 18-106_k141_134682 | Not-determined |
| 18-106_k141_57636 | Not-determined |
| 18-106_k141_87900 | Not-determined |
| 18-106_k141_122483 | Not-determined |
| 18-106_k141_93761 | Not-determined |
| 18-106_k141_134687 | Not-determined |
| 18-106_k141_117254 | Not-determined |
| 18-106_k141_134697 | Not-determined |
| 18-106_k141_22692 | Low-quality |
| 18-106_k141_128963 | Not-determined |
| 18-106_k141_134701 | Not-determined |
| 18-106_k141_87914 | Not-determined |
| 18-106_k141_22700 | Not-determined |
| 18-106_k141_93780 | Not-determined |
| 18-106_k141_22710 | Not-determined |
| 18-106_k141_34751 | Not-determined |
| 18-106_k141_122518 | Not-determined |
| 18-106_k141_57681 | Not-determined |
| 18-106_k141_87938 | Not-determined |
| 18-106_k141_28934 | Not-determined |
| 18-106_k141_140272 | Low-quality |
| 18-106_k141_140275 | Not-determined |
| 18-106_k141_34772 | Not-determined |
| 18-106_k141_87947 | Not-determined |
| 18-106_k141_117293 | Not-determined |
| 18-106_k141_87949 | Not-determined |
| 18-106_k141_128974 | Not-determined |
| 18-106_k141_34777 | Low-quality |
| 18-106_k141_28951 | Not-determined |
| 18-106_k141_140286 | Not-determined |
| 18-106_k141_22739 | Not-determined |
| 18-106_k141_140289 | Not-determined |
| 18-106_k141_34781 | Not-determined |
| 18-106_k141_81705 | Not-determined |
| 18-106_k141_117296 | Not-determined |
| 18-106_k141_140303 | Not-determined |
| 18-106_k141_122539 | Not-determined |
| 18-106_k141_140308 | Low-quality |
| 18-106_k141_28975 | Not-determined |
| 18-106_k141_140310 | Not-determined |
| 18-106_k141_57720 | Not-determined |
| 18-106_k141_122544 | Not-determined |
| 18-106_k141_57729 | Not-determined |
| 18-106_k141_117324 | Not-determined |
| 18-106_k141_28992 | Low-quality |
| 18-106_k141_28999 | Not-determined |
| 18-106_k141_122555 | Not-determined |
| 18-106_k141_122560 | Not-determined |
| 18-106_k141_140338 | Not-determined |
| 18-106_k141_129012 | Not-determined |
| 18-106_k141_93848 | Low-quality |
| 18-106_k141_117338 | Not-determined |
| 18-106_k141_29019 | Not-determined |
| 18-106_k141_22781 | Not-determined |
| 18-106_k141_29020 | Not-determined |
| 18-106_k141_29023 | Not-determined |
| 18-106_k141_22790 | Not-determined |
| 18-106_k141_29025 | Not-determined |
| 18-106_k141_87993 | Not-determined |
| 18-106_k141_11093 | Not-determined |
| 18-106_k141_88000 | Not-determined |
| 18-106_k141_129032 | Not-determined |
| 18-106_k141_93875 | Not-determined |
| 18-106_k141_122587 | Not-determined |
| 18-106_k141_81758 | Not-determined |
| 18-106_k141_11096 | Not-determined |
| 18-106_k141_11099 | Low-quality |
| 18-106_k141_129043 | Not-determined |
| 18-106_k141_104978 | Not-determined |
| 18-106_k141_104979 | Not-determined |
| 18-106_k141_117357 | Not-determined |
| 18-106_k141_81774 | Not-determined |
| 18-106_k141_93896 | Not-determined |
| 18-106_k141_104996 | Not-determined |
| 18-106_k141_88005 | Not-determined |
| 18-106_k141_11107 | Not-determined |
| 18-106_k141_93900 | Not-determined |
| 18-106_k141_22838 | Not-determined |
| 18-106_k141_122609 | Not-determined |
| 18-106_k141_129066 | Not-determined |
| 18-106_k141_22846 | Not-determined |
| 18-106_k141_11118 | Not-determined |
| 18-106_k141_129072 | Not-determined |
| 18-106_k141_11119 | Not-determined |
| 18-106_k141_22863 | Not-determined |
| 18-106_k141_140389 | Not-determined |
| 18-106_k141_122628 | Not-determined |
| 18-106_k141_75966 | Not-determined |
| 18-106_k141_22869 | Not-determined |
| 18-106_k141_93918 | Not-determined |
| 18-106_k141_34830 | Not-determined |
| 18-106_k141_22871 | Not-determined |
| 18-106_k141_29061 | Not-determined |
| 18-106_k141_93925 | Not-determined |
| 18-106_k141_122639 | Not-determined |
| 18-106_k141_122640 | Not-determined |
| 18-106_k141_122643 | Not-determined |
| 18-106_k141_117396 | Not-determined |
| 18-106_k141_57808 | Not-determined |
| 18-106_k141_88043 | Not-determined |
| 18-106_k141_34852 | Not-determined |
| 18-106_k141_29075 | Not-determined |
| 18-106_k141_57816 | Not-determined |
| 18-106_k141_29083 | Not-determined |
| 18-106_k141_29084 | Not-determined |
| 18-106_k141_129096 | Not-determined |
| 18-106_k141_140417 | Not-determined |
| 18-106_k141_105039 | Low-quality |
| 18-106_k141_122669 | Not-determined |
| 18-106_k141_129101 | Not-determined |
| 18-106_k141_22885 | Not-determined |
| 18-106_k141_76011 | Low-quality |
| 18-106_k141_76015 | Not-determined |
| 18-106_k141_129117 | Not-determined |
| 18-106_k141_29119 | Not-determined |
| 18-106_k141_29120 | Not-determined |
| 18-106_k141_129120 | Not-determined |
| 18-106_k141_11148 | Not-determined |
| 18-106_k141_129123 | Not-determined |
| 18-106_k141_140436 | Not-determined |
| 18-106_k141_129125 | Not-determined |
| 18-106_k141_140438 | Not-determined |
| 18-106_k141_34884 | Not-determined |
| 18-106_k141_122707 | Not-determined |
| 18-106_k141_122709 | Not-determined |
| 18-106_k141_122710 | Not-determined |
| 18-106_k141_140445 | Not-determined |
| 18-106_k141_140450 | Not-determined |
| 18-106_k141_122743 | Not-determined |
| 18-106_k141_122747 | Not-determined |
| 18-106_k141_117415 | Not-determined |
| 18-106_k141_76037 | Not-determined |
| 18-106_k141_140469 | Not-determined |
| 18-106_k141_122753 | Not-determined |
| 18-106_k141_134815 | Not-determined |
| 18-106_k141_34911 | Not-determined |
| 18-106_k141_122773 | Not-determined |
| 18-106_k141_11188 | Not-determined |
| 18-106_k141_134824 | Not-determined |
| 18-106_k141_29156 | Not-determined |
| 18-106_k141_34923 | Not-determined |
| 18-106_k141_134827 | Not-determined |
| 18-106_k141_57846 | Not-determined |
| 18-106_k141_122778 | Not-determined |
| 18-106_k141_22961 | Not-determined |
| 18-106_k141_105093 | Not-determined |
| 18-106_k141_111396 | Low-quality |
| 18-106_k141_29164 | Not-determined |
| 18-106_k141_140496 | Not-determined |
| 18-106_k141_88055 | Not-determined |
| 18-106_k141_140502 | Not-determined |
| 18-106_k141_76064 | Not-determined |
| 18-106_k141_57861 | Not-determined |
| 18-106_k141_140505 | Not-determined |
| 18-106_k141_29177 | Not-determined |
| 18-106_k141_76076 | Not-determined |
| 18-106_k141_11219 | Not-determined |
| 18-106_k141_11220 | Not-determined |
| 18-106_k141_134840 | Not-determined |
| 18-106_k141_22994 | Not-determined |
| 18-106_k141_111420 | Not-determined |
| 18-106_k141_122812 | Not-determined |
| 18-106_k141_88067 | Not-determined |
| 18-106_k141_11229 | Not-determined |
| 18-106_k141_34949 | Not-determined |
| 18-106_k141_29185 | Not-determined |
| 18-106_k141_81800 | Low-quality |
| 18-106_k141_140514 | Not-determined |
| 18-106_k141_134844 | Not-determined |
| 18-106_k141_76083 | Not-determined |
| 18-106_k141_11247 | Not-determined |
| 18-106_k141_34968 | Not-determined |
| 18-106_k141_111443 | Not-determined |
| 18-106_k141_111445 | Low-quality |
| 18-106_k141_111448 | Not-determined |
| 18-106_k141_81808 | Not-determined |
| 18-106_k141_122835 | Low-quality |
| 18-106_k141_81814 | Not-determined |
| 18-106_k141_81821 | Low-quality |
| 18-106_k141_81822 | Not-determined |
| 18-106_k141_105110 | Not-determined |
| 18-106_k141_29211 | Not-determined |
| 18-106_k141_76091 | Not-determined |
| 18-106_k141_23017 | Not-determined |
| 18-106_k141_23018 | Not-determined |
| 18-106_k141_134866 | Not-determined |
| 18-106_k141_111468 | Not-determined |
| 18-106_k141_23024 | Not-determined |
| 18-106_k141_23027 | Not-determined |
| 18-106_k141_34977 | Not-determined |
| 18-106_k141_76110 | Not-determined |
| 18-106_k141_11286 | Not-determined |
| 18-106_k141_29221 | Not-determined |
| 18-106_k141_34992 | Not-determined |
| 18-106_k141_81839 | Low-quality |
| 18-106_k141_81841 | Low-quality |
| 18-106_k141_122865 | Not-determined |
| 18-106_k141_57918 | Not-determined |
| 18-106_k141_140558 | Not-determined |
| 18-106_k141_140561 | Not-determined |
| 18-106_k141_140563 | Not-determined |
| 18-106_k141_29241 | Not-determined |
| 18-106_k141_57926 | Not-determined |
| 18-106_k141_111492 | Not-determined |
| 18-106_k141_57928 | Not-determined |
| 18-106_k141_11309 | Not-determined |
| 18-106_k141_105138 | Not-determined |
| 18-106_k141_105139 | Not-determined |
| 18-106_k141_122885 | Not-determined |
| 18-106_k141_134881 | Not-determined |
| 18-106_k141_35023 | Not-determined |
| 18-106_k141_134884 | Not-determined |
| 18-106_k141_23073 | Not-determined |
| 18-106_k141_29257 | Low-quality |
| 18-106_k141_57946 | Not-determined |
| 18-106_k141_35034 | Not-determined |
| 18-106_k141_11332 | Not-determined |
| 18-106_k141_11333 | Not-determined |
| 18-106_k141_11336 | Not-determined |
| 18-106_k141_81867 | Not-determined |
| 18-106_k141_134898 | Not-determined |
| 18-106_k141_140577 | Low-quality |
| 18-106_k141_134902 | Not-determined |
| 18-106_k141_57954 | Not-determined |
| 18-106_k141_29273 | Not-determined |
| 18-106_k141_105164 | Not-determined |
| 18-106_k141_122906 | Not-determined |
| 18-106_k141_23080 | Not-determined |
| 18-106_k141_105166 | Not-determined |
| 18-106_k141_35054 | Not-determined |
| 18-106_k141_11345 | Low-quality |
| 18-106_k141_23083 | Not-determined |
| 18-106_k141_122909 | Not-determined |
| 18-106_k141_23085 | Not-determined |
| 18-106_k141_140598 | Not-determined |
| 18-106_k141_57955 | Low-quality |
| 18-106_k141_122911 | Not-determined |
| 18-106_k141_105173 | Not-determined |
| 18-106_k141_57962 | Not-determined |
| 18-106_k141_105177 | Not-determined |
| 18-106_k141_57964 | Low-quality |
| 18-106_k141_29287 | Not-determined |
| 18-106_k141_105182 | Not-determined |
| 18-106_k141_105194 | Low-quality |
| 18-106_k141_57978 | Not-determined |
| 18-106_k141_140613 | Not-determined |
| 18-106_k141_122936 | Not-determined |
| 18-106_k141_11362 | Not-determined |
| 18-106_k141_122944 | Not-determined |
| 18-106_k141_11366 | Not-determined |
| 18-106_k141_29312 | Not-determined |
| 18-106_k141_29316 | Not-determined |
| 18-106_k141_29319 | Not-determined |
| 18-106_k141_23109 | Not-determined |
| 18-106_k141_111551 | Not-determined |
| 18-106_k141_23115 | Not-determined |
| 18-106_k141_29325 | Not-determined |
| 18-106_k141_23121 | Not-determined |
| 18-106_k141_11382 | Not-determined |
| 18-106_k141_140630 | Not-determined |
| 18-106_k141_140637 | Not-determined |
| 18-106_k141_11397 | Not-determined |
| 18-106_k141_35099 | Not-determined |
| 18-106_k141_105235 | Not-determined |
| 18-106_k141_134925 | Not-determined |
| 18-106_k141_29340 | Not-determined |
| 18-106_k141_35109 | Not-determined |
| 18-106_k141_35112 | Not-determined |
| 18-106_k141_57997 | Not-determined |
| 18-106_k141_29353 | Not-determined |
| 18-106_k141_134943 | Low-quality |
| 18-106_k141_81942 | Not-determined |
| 18-106_k141_81954 | Not-determined |
| 18-106_k141_140689 | Not-determined |
| 18-106_k141_81960 | Not-determined |
| 18-106_k141_81961 | Low-quality |
| 18-106_k141_76178 | Not-determined |
| 18-106_k141_23183 | Not-determined |
| 18-106_k141_81974 | Not-determined |
| 18-106_k141_35127 | Not-determined |
| 18-106_k141_35130 | Not-determined |
| 18-106_k141_134966 | Not-determined |
| 18-106_k141_81985 | Not-determined |
| 18-106_k141_140695 | Not-determined |
| 18-106_k141_122981 | Not-determined |
| 18-106_k141_81987 | Not-determined |
| 18-106_k141_76195 | Not-determined |
| 18-106_k141_81997 | Not-determined |
| 18-106_k141_35148 | Not-determined |
| 18-106_k141_105289 | Not-determined |
| 18-106_k141_35154 | Not-determined |
| 18-106_k141_35155 | Not-determined |
| 18-106_k141_76220 | Not-determined |
| 18-106_k141_35158 | Not-determined |
| 18-106_k141_35159 | Not-determined |
| 18-106_k141_105301 | Not-determined |
| 18-106_k141_35167 | Not-determined |
| 18-106_k141_82014 | Not-determined |
| 18-106_k141_134991 | Not-determined |
| 18-106_k141_140728 | Not-determined |
| 18-106_k141_123016 | Not-determined |
| 18-106_k141_23228 | Not-determined |
| 18-106_k141_23233 | Not-determined |
| 18-106_k141_123025 | Not-determined |
| 18-106_k141_134996 | Low-quality |
| 18-106_k141_105330 | Not-determined |
| 18-106_k141_105335 | Not-determined |
| 18-106_k141_23240 | Not-determined |
| 18-106_k141_105342 | Not-determined |
| 18-106_k141_23244 | Not-determined |
| 18-106_k141_123032 | Not-determined |
| 18-106_k141_23251 | Low-quality |
| 18-106_k141_23253 | Not-determined |
| 18-106_k141_82047 | Not-determined |
| 18-106_k141_135012 | Not-determined |
| 18-106_k141_11421 | Not-determined |
| 18-106_k141_135015 | Not-determined |
| 18-106_k141_123051 | Not-determined |
| 18-106_k141_58067 | Not-determined |
| 18-106_k141_23271 | Not-determined |
| 18-106_k141_76249 | Not-determined |
| 18-106_k141_135024 | Not-determined |
| 18-106_k141_58068 | Not-determined |
| 18-106_k141_123067 | Not-determined |
| 18-106_k141_123071 | Not-determined |
| 18-106_k141_123074 | Not-determined |
| 18-106_k141_123077 | Not-determined |
| 18-106_k141_23282 | Not-determined |
| 18-106_k141_35203 | Not-determined |
| 18-106_k141_23285 | Not-determined |
| 18-106_k141_23293 | Not-determined |
| 18-106_k141_23296 | Not-determined |
| 18-106_k141_123087 | Not-determined |
| 18-106_k141_76257 | Not-determined |
| 18-106_k141_105381 | Not-determined |
| 18-106_k141_123100 | Not-determined |
| 18-106_k141_58106 | Not-determined |
| 18-106_k141_58109 | Not-determined |
| 18-106_k141_58111 | Not-determined |
| 18-106_k141_58130 | Not-determined |
| 18-106_k141_82096 | Not-determined |
| 18-106_k141_82099 | Not-determined |
| 18-106_k141_105396 | Low-quality |
| 18-106_k141_105425 | Not-determined |
| 18-106_k141_82135 | Not-determined |
| 18-106_k141_58170 | Not-determined |
| 18-106_k141_105433 | Not-determined |
| 18-106_k141_82138 | Not-determined |
| 18-106_k141_105449 | Not-determined |
| 18-106_k141_105450 | Not-determined |
| 18-106_k141_58194 | Not-determined |
| 18-106_k141_123151 | Not-determined |
| 18-106_k141_123155 | Low-quality |
| 18-106_k141_58204 | Not-determined |
| 18-106_k141_76278 | Not-determined |
| 18-106_k141_23360 | Not-determined |
| 18-106_k141_76280 | Not-determined |
| 18-106_k141_123165 | Not-determined |
| 18-106_k141_76282 | Not-determined |
| 18-106_k141_82164 | Not-determined |
| 18-106_k141_23369 | Not-determined |
| 18-106_k141_76289 | Not-determined |
| 18-106_k141_105473 | Not-determined |
| 18-106_k141_82170 | Not-determined |
| 18-106_k141_58227 | Not-determined |
| 18-106_k141_76295 | Not-determined |
| 18-106_k141_76306 | Not-determined |
| 18-106_k141_123179 | Not-determined |
| 18-106_k141_23372 | Not-determined |
| 18-106_k141_58241 | Low-quality |
| 18-106_k141_105498 | Not-determined |
| 18-106_k141_82178 | Not-determined |
| 18-106_k141_82181 | Not-determined |
| 18-106_k141_105509 | Low-quality |
| 18-106_k141_58252 | Not-determined |
| 18-106_k141_58254 | Low-quality |
| 18-106_k141_58257 | Not-determined |
| 18-106_k141_123203 | Not-determined |
| 18-106_k141_58258 | Not-determined |
| 18-106_k141_82195 | Low-quality |
| 18-106_k141_123206 | Not-determined |
| 18-106_k141_58266 | Low-quality |
| 18-106_k141_105526 | Not-determined |
| 18-106_k141_123210 | Not-determined |
| 18-106_k141_58269 | Not-determined |
| 18-106_k141_58272 | Low-quality |
| 18-106_k141_123217 | Not-determined |
| 18-106_k141_58291 | Not-determined |
| 18-106_k141_23407 | Not-determined |
| 18-106_k141_105580 | Not-determined |
| 18-106_k141_105592 | Not-determined |
| 18-106_k141_123242 | Not-determined |
| 18-106_k141_105598 | Not-determined |
| 18-106_k141_58327 | Not-determined |
| 18-106_k141_58329 | Not-determined |
| 18-106_k141_58337 | Not-determined |
| 18-106_k141_123262 | Not-determined |
| 18-106_k141_23433 | Not-determined |
| 18-106_k141_23461 | Low-quality |

| | |
|---|---|
| 18-106_k141_58379 | Not-determined |
| 18-106_k141_58384 | Not-determined |
| 18-106_k141_58387 | Not-determined |
| 18-106_k141_105616 | Not-determined |
| 18-106_k141_123284 | Not-determined |
| 18-106_k141_105618 | Not-determined |
| 18-106_k141_58394 | Not-determined |
| 18-106_k141_58401 | Not-determined |
| 18-106_k141_58408 | Not-determined |
| 18-106_k141_105651 | Not-determined |
| 18-106_k141_23487 | Not-determined |
| 18-106_k141_58476 | Not-determined |
| 18-106_k141_58498 | Not-determined |
| 18-106_k141_58508 | Not-determined |
| 18-106_k141_58546 | Not-determined |
| 18-106_k141_58566 | Not-determined |
| 18-106_k141_58571 | Not-determined |
| 18-106_k141_58578 | Not-determined |
| 18-106_k141_11500 | Low-quality |
| 18-106_k141_11516 | Low-quality |
| 18-106_k141_11526 | Not-determined |
| 18-101-final-viral-combined_k141_231411\|\|full | Medium-quality |
| 18-106_k141_11543 | Not-determined |
| 18-106_k141_140744 | Not-determined |
| 18-106_k141_11557 | Not-determined |
| 18-106_k141_11563 | Not-determined |
| 18-106_k141_11564 | Low-quality |
| 18-106_k141_140752 | Not-determined |
| 18-106_k141_140757 | Not-determined |
| 18-106_k141_140759 | Not-determined |
| 18-106_k141_11569 | Not-determined |
| 18-106_k141_11586 | Not-determined |
| 18-106_k141_140782 | Not-determined |
| 18-106_k141_140801 | Not-determined |
| 18-106_k141_140817 | Not-determined |
| 18-106_k141_140824 | Not-determined |
| 18-106_k141_140826 | Not-determined |
| 18-106_k141_140834 | Not-determined |
| 18-106_k141_140837 | Not-determined |
| 18-106_k141_140847 | Not-determined |
| 18-106_k141_140850 | Not-determined |
| 18-106_k141_140883 | Not-determined |
| 18-106_k141_140894 | Not-determined |
| 18-106_k141_11602 | Low-quality |
| 18-106_k141_11648 | Not-determined |
| 18-106_k141_11654 | Low-quality |
| 18-106_k141_11676 | Not-determined |
| 18-106_k141_11677 | Not-determined |
| 18-106_k141_11711 | Not-determined |
| 18-106_k141_11717 | Not-determined |
| 18-106_k141_11724 | Not-determined |
| 18-106_k141_11735 | Not-determined |
| 18-106_k141_11738 | Not-determined |
| 18-110-final-viral-combined_k141_103929\|\|full | Not-determined |
| 18-110-final-viral-combined_k141_233147\|\|full | Low-quality |
| 18-110-final-viral-combined_k141_141557\|\|full | Low-quality |
| 18-110-final-viral-combined_k141_268426\|\|full | Not-determined |
| 17-109_k141_17950 | Medium-quality |
| 18-110-final-viral-combined_k141_86965\|\|full | Not-determined |
| 18-110-final-viral-combined_k141_168300\|\|full | Low-quality |
| 17-109-final-viral-combined_k141_17950\|\|full | Medium-quality |
| 18-110-final-viral-combined_k141_192515\|\|full | Not-determined |
| 18-110-final-viral-combined_k141_124471\|\|full | Low-quality |
| 17-109-final-viral-combined_k141_393703\|\|full | High-quality |
| 18-110-final-viral-combined_k141_276202\|\|full | Not-determined |
| 17-109-final-viral-combined_k141_47214\|\|full | Medium-quality |
| KM-final-viral-combined_k141_103269\|\|full | Medium-quality |
| 18-110-final-viral-combined_k141_276212\|\|full | Not-determined |
| 18-110-final-viral-combined_k141_276246\|\|full | Not-determined |
| 18-110-final-viral-combined_k141_276313\|\|full | Low-quality |
| 18-110-final-viral-combined_k141_172791\|\|full | Not-determined |
| 18-110-final-viral-combined_k141_115210\|\|full | Low-quality |
| 18-110-final-viral-combined_k141_138251\|\|full | Not-determined |
| 18-110-final-viral-combined_k141_149766\|\|full | Low-quality |
| 18-110-final-viral-combined_k141_218886\|\|full | Not-determined |
| 18-110-final-viral-combined_k141_230425\|\|full | Low-quality |
| 18-110-final-viral-combined_k141_126740\|\|full | Low-quality |
| 18-110-final-viral-combined_k141_253427\|\|full | Not-determined |
| 18-110-final-viral-combined_k141_126744\|\|full | Not-determined |
| 18-110-final-viral-combined_k141_241916\|\|full | Low-quality |
| 18-110-final-viral-combined_k141_11540\|\|full | Not-determined |
| 18-110-final-viral-combined_k141_149780\|\|full | Not-determined |
| 18-110-final-viral-combined_k141_218896\|\|full | Low-quality |
| 18-110-final-viral-combined_k141_230439\|\|full | Not-determined |
| 18-110-final-viral-combined_k141_46104\|\|full | Low-quality |
| 18-110-final-viral-combined_k141_34589\|\|full | Not-determined |
| 18-110-final-viral-combined_k141_241935\|\|full | Low-quality |
| 18-110-final-viral-combined_k141_80639\|\|full | Low-quality |
| 18-110-final-viral-combined_k141_253456\|\|full | Not-determined |
| 18-110-final-viral-combined_k141_207372\|\|full | Not-determined |
| 18-110-final-viral-combined_k141_253459\|\|full | Low-quality |
| 18-110-final-viral-combined_k141_138323\|\|full | Not-determined |
| 18-110-final-viral-combined_k141_80651\|\|full | Low-quality |
| 18-110-final-viral-combined_k141_265009\|\|full | Low-quality |
| 18-110-final-viral-combined_k141_241952\|\|full | Low-quality |
| 18-110-final-viral-combined_k141_253463\|\|full | Low-quality |
| 18-110-final-viral-combined_k141_184372\|\|full | Low-quality |
| 18-110-final-viral-combined_k141_253464\|\|full | Not-determined |
| 18-110-final-viral-combined_k141_23053\|\|full | Not-determined |
| 18-110-final-viral-combined_k141_57621\|\|full | Not-determined |
| 18-110-final-viral-combined_k141_149812\|\|full | Low-quality |
| 18-110-final-viral-combined_k141_80657\|\|full | Low-quality |
| 18-110-final-viral-combined_k141_161289\|\|full | Low-quality |
| 18-110-final-viral-combined_k141_265058\|\|full | Not-determined |
| 18-110-final-viral-combined_k141_34653\|\|full | Not-determined |
| 18-110-final-viral-combined_k141_230510\|\|full | Not-determined |
| 18-110-final-viral-combined_k141_115288\|\|full | Not-determined |
| 18-110-final-viral-combined_k141_92229\|\|full | Not-determined |
| 18-110-final-viral-combined_k141_103711\|\|full | Not-determined |
| 18-110-final-viral-combined_k141_138381\|\|full | Not-determined |
| 18-110-final-viral-combined_k141_149833\|\|full | Low-quality |
| 18-110-final-viral-combined_k141_207406\|\|full | Not-determined |
| 18-110-final-viral-combined_k141_149834\|\|full | Low-quality |
| 18-110-final-viral-combined_k141_103718\|\|full | Not-determined |
| 18-110-final-viral-combined_k141_265064\|\|full | Not-determined |
| 18-110-final-viral-combined_k141_218941\|\|full | Not-determined |
| 18-110-final-viral-combined_k141_207414\|\|full | Not-determined |
| 18-110-final-viral-combined_k141_230540\|\|full | Low-quality |
| 18-110-final-viral-combined_k141_34683\|\|full | Not-determined |
| 18-110-final-viral-combined_k141_115289\|\|full | Low-quality |
| 18-110-final-viral-combined_k141_34684\|\|full | Low-quality |
| 18-110-final-viral-combined_k141_113\|\|full | Not-determined |
| 18-110-final-viral-combined_k141_149844\|\|full | Not-determined |
| 18-110-final-viral-combined_k141_195921\|\|full | Low-quality |
| 18-110-final-viral-combined_k141_126782\|\|full | Not-determined |
| 18-110-final-viral-combined_k141_218953\|\|full | Not-determined |
| 18-110-final-viral-combined_k141_207428\|\|full | Low-quality |
| 18-110-final-viral-combined_k141_242003\|\|full | Low-quality |
| 18-110-final-viral-combined_k141_23119\|\|full | Not-determined |
| 18-110-final-viral-combined_k141_184391\|\|full | Low-quality |
| 18-110-final-viral-combined_k141_135\|\|full | Not-determined |
| 18-110-final-viral-combined_k141_92296\|\|full | Not-determined |
| 18-110-final-viral-combined_k141_184397\|\|full | Low-quality |
| 18-110-final-viral-combined_k141_242016\|\|full | Low-quality |
| 18-110-final-viral-combined_k141_242017\|\|full | Low-quality |
| 18-110-final-viral-combined_k141_46215\|\|full | Low-quality |
| 18-110-final-viral-combined_k141_242024\|\|full | Not-determined |
| 18-110-final-viral-combined_k141_184410\|\|full | Low-quality |
| 18-110-final-viral-combined_k141_149891\|\|full | Low-quality |
| 18-110-final-viral-combined_k141_23149\|\|full | Low-quality |
| 18-110-final-viral-combined_k141_184415\|\|full | Not-determined |
| 18-110-final-viral-combined_k141_172899\|\|full | Not-determined |
| 18-110-final-viral-combined_k141_138413\|\|full | Not-determined |
| 18-104_k141_114120 | High-quality |
| 18-110-final-viral-combined_k141_138422\|\|full | Not-determined |
| 18-110-final-viral-combined_k141_242079\|\|full | Not-determined |
| 18-110-final-viral-combined_k141_115299\|\|full | Low-quality |
| 18-110-final-viral-combined_k141_138445\|\|full | Low-quality |
| 18-110-final-viral-combined_k141_242096\|\|full | Not-determined |
| 18-110-final-viral-combined_k141_57747\|\|full | Low-quality |
| 18-110-final-viral-combined_k141_11568\|\|full | Low-quality |
| 18-110-final-viral-combined_k141_265140\|\|full | Low-quality |
| 18-110-final-viral-combined_k141_46267\|\|full | Not-determined |
| 18-110-final-viral-combined_k141_242103\|\|full | Low-quality |
| 18-110-final-viral-combined_k141_69340\|\|full | Low-quality |
| 18-110-final-viral-combined_k141_172941\|\|full | Not-determined |
| 18-110-final-viral-combined_k141_183\|\|full | Not-determined |
| 18-110-final-viral-combined_k141_23224\|\|full | Not-determined |
| 18-110-final-viral-combined_k141_230656\|\|full | Not-determined |
| 18-110-final-viral-combined_k141_207503\|\|full | Low-quality |
| 18-110-final-viral-combined_k141_138485\|\|full | Not-determined |
| 18-110-final-viral-combined_k141_11578\|\|full | Not-determined |
| 18-110-final-viral-combined_k141_184476\|\|full | Low-quality |
| 18-110-final-viral-combined_k141_80885\|\|full | Low-quality |
| 18-110-final-viral-combined_k141_57801\|\|full | Low-quality |
| 18-110-final-viral-combined_k141_11590\|\|full | Not-determined |
| 18-110-final-viral-combined_k141_138502\|\|full | Low-quality |
| 18-110-final-viral-combined_k141_219020\|\|full | Low-quality |
| 18-110-final-viral-combined_k141_23252\|\|full | Low-quality |
| 18-110-final-viral-combined_k141_115360\|\|full | Not-determined |
| 18-110-final-viral-combined_k141_69361\|\|full | Low-quality |
| 18-110-final-viral-combined_k141_138516\|\|full | Low-quality |
| 18-110-final-viral-combined_k141_230695\|\|full | Not-determined |
| 18-110-final-viral-combined_k141_69362\|\|full | Low-quality |
| 18-110-final-viral-combined_k141_195994\|\|full | Not-determined |
| 18-110-final-viral-combined_k141_138520\|\|full | Not-determined |
| 18-110-final-viral-combined_k141_69365\|\|full | Low-quality |
| 18-110-final-viral-combined_k141_80913\|\|full | Not-determined |
| 18-110-final-viral-combined_k141_253554\|\|full | Not-determined |
| 18-110-final-viral-combined_k141_210\|\|full | Low-quality |
| 18-110-final-viral-combined_k141_23271\|\|full | Not-determined |
| 18-110-final-viral-combined_k141_219035\|\|full | Low-quality |
| 18-110-final-viral-combined_k141_213\|\|full | Not-determined |
| 18-110-final-viral-combined_k141_215\|\|full | Low-quality |
| 18-110-final-viral-combined_k141_92396\|\|full | Not-determined |
| 18-110-final-viral-combined_k141_126914\|\|full | Not-determined |
| 18-110-final-viral-combined_k141_196025\|\|full | Not-determined |
| 18-110-final-viral-combined_k141_103744\|\|full | Low-quality |
| 18-110-final-viral-combined_k141_184511\|\|full | Low-quality |
| 18-110-final-viral-combined_k141_69377\|\|full | Low-quality |
| 18-110-final-viral-combined_k141_80954\|\|full | Not-determined |
| 18-110-final-viral-combined_k141_103749\|\|full | Low-quality |
| 18-110-final-viral-combined_k141_80959\|\|full | Not-determined |
| 18-110-final-viral-combined_k141_103754\|\|full | Not-determined |
| 18-110-final-viral-combined_k141_23302\|\|full | Not-determined |
| 18-110-final-viral-combined_k141_57841\|\|full | Low-quality |
| 18-110-final-viral-combined_k141_230731\|\|full | Not-determined |
| 18-110-final-viral-combined_k141_257\|\|full | Not-determined |
| 18-110-final-viral-combined_k141_242171\|\|full | Not-determined |
| 18-110-final-viral-combined_k141_196057\|\|full | Low-quality |
| 18-110-final-viral-combined_k141_242177\|\|full | Low-quality |
| 18-110-final-viral-combined_k141_103785\|\|full | Not-determined |
| 18-110-final-viral-combined_k141_57858\|\|full | Not-determined |
| 18-110-final-viral-combined_k141_242180\|\|full | Low-quality |
| 18-110-final-viral-combined_k141_196060\|\|full | Low-quality |
| 18-110-final-viral-combined_k141_207589\|\|full | Not-determined |
| 18-110-final-viral-combined_k141_103788\|\|full | Not-determined |
| 18-110-final-viral-combined_k141_173053\|\|full | Low-quality |
| 18-110-final-viral-combined_k141_115427\|\|full | Not-determined |
| 18-110-final-viral-combined_k141_161357\|\|full | Not-determined |
| 18-110-final-viral-combined_k141_138596\|\|full | Low-quality |
| 18-110-final-viral-combined_k141_242190\|\|full | Low-quality |
| 18-110-final-viral-combined_k141_46317\|\|full | Not-determined |
| 18-110-final-viral-combined_k141_196078\|\|full | Not-determined |
| 18-110-final-viral-combined_k141_11648\|\|full | Not-determined |
| 18-110-final-viral-combined_k141_80993\|\|full | Not-determined |
| 18-110-final-viral-combined_k141_46323\|\|full | Not-determined |
| 18-110-final-viral-combined_k141_149952\|\|full | Low-quality |
| 18-105_k141_164032 | Medium-quality |
| 18-110-final-viral-combined_k141_11660\|\|full | Not-determined |
| 18-110-final-viral-combined_k141_184559\|\|full | Not-determined |
| 18-110-final-viral-combined_k141_230809\|\|full | Not-determined |
| 18-110-final-viral-combined_k141_46373\|\|full | Low-quality |
| 18-110-final-viral-combined_k141_196128\|\|full | Not-determined |
| 18-110-final-viral-combined_k141_11667\|\|full | Not-determined |
| 18-110-final-viral-combined_k141_23348\|\|full | Low-quality |
| 18-110-final-viral-combined_k141_115447\|\|full | Low-quality |
| 18-110-final-viral-combined_k141_57910\|\|full | Not-determined |
| 18-110-final-viral-combined_k141_230823\|\|full | Not-determined |
| 18-110-final-viral-combined_k141_219084\|\|full | Low-quality |
| 18-110-final-viral-combined_k141_46382\|\|full | Not-determined |
| 18-110-final-viral-combined_k141_23358\|\|full | Low-quality |
| 18-110-final-viral-combined_k141_230837\|\|full | Not-determined |
| 18-110-final-viral-combined_k141_297\|\|full | Low-quality |
| 18-110-final-viral-combined_k141_138659\|\|full | Not-determined |
| 18-110-final-viral-combined_k141_23370\|\|full | Not-determined |
| 18-110-final-viral-combined_k141_207642\|\|full | Low-quality |
| 18-110-final-viral-combined_k141_11700\|\|full | Not-determined |
| 18-110-final-viral-combined_k141_242270\|\|full | Low-quality |
| 18-110-final-viral-combined_k141_138666\|\|full | Not-determined |
| 18-110-final-viral-combined_k141_57952\|\|full | Not-determined |
| 18-110-final-viral-combined_k141_184613\|\|full | Not-determined |
| 18-110-final-viral-combined_k141_69471\|\|full | Low-quality |
| 18-110-final-viral-combined_k141_207644\|\|full | Not-determined |
| 18-110-final-viral-combined_k141_207645\|\|full | Low-quality |
| 18-110-final-viral-combined_k141_69477\|\|full | Not-determined |
| 18-110-final-viral-combined_k141_173152\|\|full | Not-determined |
| 18-110-final-viral-combined_k141_57966\|\|full | Low-quality |
| 18-110-final-viral-combined_k141_115474\|\|full | Low-quality |
| 18-110-final-viral-combined_k141_57967\|\|full | Low-quality |
| 18-110-final-viral-combined_k141_219128\|\|full | Low-quality |
| 18-110-final-viral-combined_k141_103855\|\|full | Not-determined |
| 18-110-final-viral-combined_k141_242285\|\|full | Not-determined |
| 18-110-final-viral-combined_k141_219132\|\|full | Not-determined |
| 18-110-final-viral-combined_k141_161405\|\|full | Not-determined |
| 18-110-final-viral-combined_k141_103873\|\|full | Low-quality |
| 18-110-final-viral-combined_k141_230892\|\|full | Low-quality |
| 18-110-final-viral-combined_k141_242302\|\|full | Low-quality |
| 18-110-final-viral-combined_k141_150037\|\|full | Not-determined |
| 18-110-final-viral-combined_k141_138711\|\|full | Not-determined |
| 18-110-final-viral-combined_k141_196155\|\|full | Not-determined |
| 18-110-final-viral-combined_k141_127042\|\|full | Not-determined |
| 18-110-final-viral-combined_k141_196162\|\|full | Low-quality |
| 18-110-final-viral-combined_k141_196165\|\|full | Not-determined |
| 18-110-final-viral-combined_k141_23405\|\|full | Not-determined |
| 18-110-final-viral-combined_k141_253667\|\|full | Not-determined |
| 18-110-final-viral-combined_k141_265263\|\|full | Not-determined |
| 18-110-final-viral-combined_k141_230909\|\|full | Low-quality |
| 18-110-final-viral-combined_k141_103893\|\|full | Not-determined |
| 18-110-final-viral-combined_k141_46477\|\|full | Low-quality |
| 18-110-final-viral-combined_k141_161443\|\|full | Not-determined |
| 18-110-final-viral-combined_k141_150076\|\|full | Not-determined |
| 18-110-final-viral-combined_k141_230915\|\|full | Low-quality |
| 18-110-final-viral-combined_k141_58020\|\|full | Low-quality |
| 18-110-final-viral-combined_k141_69538\|\|full | Low-quality |
| 18-110-final-viral-combined_k141_196187\|\|full | Not-determined |
| 18-110-final-viral-combined_k141_207684\|\|full | Low-quality |
| 18-110-final-viral-combined_k141_69542\|\|full | Not-determined |
| 18-110-final-viral-combined_k141_11757\|\|full | Low-quality |
| 18-110-final-viral-combined_k141_92542\|\|full | Low-quality |
| 18-110-final-viral-combined_k141_265298\|\|full | Low-quality |
| 18-110-final-viral-combined_k141_81092\|\|full | Not-determined |
| 18-110-final-viral-combined_k141_92546\|\|full | Not-determined |
| 18-110-final-viral-combined_k141_81102\|\|full | Not-determined |
| 18-110-final-viral-combined_k141_115546\|\|full | Low-quality |
| 18-110-final-viral-combined_k141_368\|\|full | Not-determined |
| 18-110-final-viral-combined_k141_23426\|\|full | Not-determined |
| 18-110-final-viral-combined_k141_81125\|\|full | Low-quality |
| 18-110-final-viral-combined_k141_372\|\|full | Low-quality |
| 18-110-final-viral-combined_k141_242362\|\|full | Not-determined |
| 18-110-final-viral-combined_k141_173183\|\|full | Not-determined |
| 18-110-final-viral-combined_k141_127103\|\|full | Not-determined |
| 18-110-final-viral-combined_k141_242373\|\|full | Low-quality |
| 18-110-final-viral-combined_k141_196198\|\|full | Not-determined |
| 18-110-final-viral-combined_k141_242379\|\|full | Low-quality |
| 18-110-final-viral-combined_k141_265301\|\|full | Not-determined |
| 18-110-final-viral-combined_k141_23463\|\|full | Low-quality |
| 18-110-final-viral-combined_k141_253720\|\|full | Not-determined |
| 18-110-final-viral-combined_k141_150102\|\|full | Not-determined |
| 18-110-final-viral-combined_k141_219149\|\|full | Not-determined |
| 18-110-final-viral-combined_k141_92602\|\|full | Not-determined |
| 18-110-final-viral-combined_k141_115565\|\|full | Not-determined |
| 18-110-final-viral-combined_k141_138807\|\|full | Not-determined |
| 18-110-final-viral-combined_k141_196241\|\|full | Low-quality |
| 18-110-final-viral-combined_k141_219190\|\|full | Not-determined |
| 18-110-final-viral-combined_k141_173250\|\|full | Not-determined |
| 18-110-final-viral-combined_k141_46531\|\|full | Low-quality |
| 18-110-final-viral-combined_k141_242447\|\|full | Not-determined |
| 18-110-final-viral-combined_k141_92620\|\|full | Low-quality |
| 18-110-final-viral-combined_k141_253752\|\|full | Low-quality |
| 18-110-final-viral-combined_k141_81153\|\|full | Low-quality |
| 18-110-final-viral-combined_k141_103941\|\|full | Not-determined |
| 18-110-final-viral-combined_k141_11861\|\|full | Not-determined |
| 18-110-final-viral-combined_k141_115605\|\|full | Low-quality |
| 18-110-final-viral-combined_k141_219217\|\|full | Low-quality |
| 18-110-final-viral-combined_k141_115607\|\|full | Not-determined |
| 18-110-final-viral-combined_k141_219218\|\|full | Low-quality |
| 18-110-final-viral-combined_k141_58166\|\|full | Low-quality |
| 18-110-final-viral-combined_k141_265370\|\|full | Low-quality |
| 18-110-final-viral-combined_k141_196277\|\|full | Low-quality |
| 18-110-final-viral-combined_k141_150110\|\|full | Low-quality |
| 18-110-final-viral-combined_k141_207765\|\|full | Not-determined |
| 18-110-final-viral-combined_k141_439\|\|full | Not-determined |
| 18-110-final-viral-combined_k141_115631\|\|full | Not-determined |
| 18-110-final-viral-combined_k141_81194\|\|full | Not-determined |
| 18-110-final-viral-combined_k141_81205\|\|full | Not-determined |
| 18-110-final-viral-combined_k141_447\|\|full | Not-determined |
| 18-110-final-viral-combined_k141_69667\|\|full | Not-determined |
| 18-110-final-viral-combined_k141_115659\|\|full | Low-quality |
| 18-110-final-viral-combined_k141_81209\|\|full | Not-determined |
| 18-110-final-viral-combined_k141_92659\|\|full | Not-determined |
| 18-110-final-viral-combined_k141_161613\|\|full | Not-determined |
| 18-110-final-viral-combined_k141_219255\|\|full | Not-determined |
| 18-110-final-viral-combined_k141_11922\|\|full | Low-quality |
| 18-110-final-viral-combined_k141_81224\|\|full | Not-determined |
| 18-110-final-viral-combined_k141_92678\|\|full | Not-determined |
| 18-110-final-viral-combined_k141_81227\|\|full | Low-quality |
| 18-110-final-viral-combined_k141_23543\|\|full | Low-quality |
| 18-110-final-viral-combined_k141_219279\|\|full | Not-determined |
| 18-110-final-viral-combined_k141_485\|\|full | Low-quality |
| 18-110-final-viral-combined_k141_138918\|\|full | Low-quality |
| 18-110-final-viral-combined_k141_92683\|\|full | Not-determined |
| 18-110-final-viral-combined_k141_150166\|\|full | Low-quality |
| 18-110-final-viral-combined_k141_81241\|\|full | Not-determined |
| 18-110-final-viral-combined_k141_253766\|\|full | Low-quality |
| 18-110-final-viral-combined_k141_69678\|\|full | Low-quality |
| 18-110-final-viral-combined_k141_253770\|\|full | Low-quality |
| 18-110-final-viral-combined_k141_138943\|\|full | Not-determined |
| 18-110-final-viral-combined_k141_253771\|\|full | Not-determined |
| 18-110-final-viral-combined_k141_242563\|\|full | Not-determined |
| 18-110-final-viral-combined_k141_23561\|\|full | Low-quality |
| 18-110-final-viral-combined_k141_265455\|\|full | Not-determined |
| 18-110-final-viral-combined_k141_508\|\|full | Not-determined |
| 18-110-final-viral-combined_k141_196329\|\|full | Not-determined |
| 18-110-final-viral-combined_k141_127137\|\|full | Low-quality |
| 18-110-final-viral-combined_k141_253777\|\|full | Low-quality |
| 18-110-final-viral-combined_k141_253780\|\|full | Not-determined |
| 18-110-final-viral-combined_k141_115710\|\|full | Low-quality |
| 18-110-final-viral-combined_k141_219336\|\|full | Low-quality |
| 18-110-final-viral-combined_k141_196362\|\|full | Not-determined |
| 18-110-final-viral-combined_k141_92747\|\|full | Not-determined |
| 18-110-final-viral-combined_k141_69706\|\|full | Not-determined |
| 18-110-final-viral-combined_k141_196366\|\|full | Not-determined |
| 18-110-final-viral-combined_k141_58200\|\|full | Low-quality |
| 18-110-final-viral-combined_k141_127176\|\|full | Not-determined |
| 18-110-final-viral-combined_k141_242591\|\|full | Not-determined |
| 18-110-final-viral-combined_k141_46601\|\|full | Low-quality |
| 18-110-final-viral-combined_k141_46604\|\|full | Low-quality |
| 18-110-final-viral-combined_k141_230942\|\|full | Not-determined |
| 18-110-final-viral-combined_k141_46621\|\|full | Not-determined |
| 18-110-final-viral-combined_k141_92780\|\|full | Not-determined |
| 18-110-final-viral-combined_k141_219376\|\|full | Not-determined |
| 18-110-final-viral-combined_k141_230948\|\|full | Low-quality |
| 18-110-final-viral-combined_k141_92803\|\|full | Low-quality |
| 18-110-final-viral-combined_k141_58232\|\|full | Low-quality |
| 18-110-final-viral-combined_k141_230983\|\|full | Not-determined |
| 18-110-final-viral-combined_k141_265523\|\|full | Not-determined |
| 18-110-final-viral-combined_k141_242624\|\|full | Not-determined |
| 18-110-final-viral-combined_k141_173408\|\|full | Not-determined |
| 18-110-final-viral-combined_k141_184808\|\|full | Not-determined |
| 18-110-final-viral-combined_k141_34807\|\|full | Not-determined |
| 18-110-final-viral-combined_k141_173430\|\|full | Not-determined |
| 18-110-final-viral-combined_k141_92834\|\|full | Not-determined |
| 18-110-final-viral-combined_k141_34817\|\|full | Low-quality |
| 18-110-final-viral-combined_k141_196429\|\|full | Low-quality |
| 18-110-final-viral-combined_k141_58281\|\|full | Low-quality |
| 18-110-final-viral-combined_k141_115750\|\|full | Not-determined |
| 17-109-final-viral-combined_k141_48648\|\|full | Medium-quality |
| 18-110-final-viral-combined_k141_69768\|\|full | Low-quality |
| 18-105-final-viral-combined_k141_164032\|\|full | Medium-quality |
| 18-110-final-viral-combined_k141_219431\|\|full | Low-quality |
| 18-110-final-viral-combined_k141_196455\|\|full | Not-determined |
| 18-110-final-viral-combined_k141_161703\|\|full | Low-quality |
| 18-110-final-viral-combined_k141_173456\|\|full | Not-determined |
| 18-110-final-viral-combined_k141_161708\|\|full | Not-determined |
| 18-110-final-viral-combined_k141_58308\|\|full | Low-quality |
| 18-110-final-viral-combined_k141_12027\|\|full | Not-determined |
| 18-110-final-viral-combined_k141_196468\|\|full | Not-determined |
| 18-110-final-viral-combined_k141_242661\|\|full | Not-determined |
| 18-110-final-viral-combined_k141_184839\|\|full | Low-quality |
| 18-110-final-viral-combined_k141_81399\|\|full | Low-quality |
| 18-110-final-viral-combined_k141_674\|\|full | Not-determined |
| 18-110-final-viral-combined_k141_173485\|\|full | Low-quality |
| 18-110-final-viral-combined_k141_150247\|\|full | Low-quality |
| 18-110-final-viral-combined_k141_231055\|\|full | Not-determined |
| 18-110-final-viral-combined_k141_678\|\|full | Not-determined |
| 18-110-final-viral-combined_k141_253850\|\|full | Low-quality |
| 18-110-final-viral-combined_k141_150257\|\|full | Not-determined |
| 18-110-final-viral-combined_k141_92892\|\|full | Not-determined |
| 18-110-final-viral-combined_k141_34871\|\|full | Low-quality |
| 18-110-final-viral-combined_k141_231058\|\|full | Low-quality |
| 18-110-final-viral-combined_k141_265574\|\|full | Not-determined |
| 18-110-final-viral-combined_k141_34877\|\|full | Not-determined |
| 18-110-final-viral-combined_k141_58352\|\|full | Not-determined |
| 18-110-final-viral-combined_k141_150283\|\|full | Not-determined |
| 18-110-final-viral-combined_k141_58355\|\|full | Low-quality |
| 18-110-final-viral-combined_k141_685\|\|full | Low-quality |
| 18-110-final-viral-combined_k141_23653\|\|full | Not-determined |
| 18-110-final-viral-combined_k141_688\|\|full | Not-determined |
| 18-110-final-viral-combined_k141_161766\|\|full | Low-quality |
| 18-110-final-viral-combined_k141_242716\|\|full | Low-quality |
| 18-110-final-viral-combined_k141_161775\|\|full | Low-quality |
| 18-110-final-viral-combined_k141_173530\|\|full | Low-quality |
| 18-110-final-viral-combined_k141_12063\|\|full | Not-determined |
| 18-110-final-viral-combined_k141_127279\|\|full | Low-quality |
| 18-110-final-viral-combined_k141_265621\|\|full | Low-quality |
| 18-110-final-viral-combined_k141_34920\|\|full | Low-quality |
| 18-110-final-viral-combined_k141_23687\|\|full | Low-quality |
| 18-110-final-viral-combined_k141_115768\|\|full | Low-quality |
| 18-110-final-viral-combined_k141_34925\|\|full | Low-quality |
| 18-110-final-viral-combined_k141_81409\|\|full | Low-quality |
| 18-110-final-viral-combined_k141_69808\|\|full | Not-determined |
| 18-110-final-viral-combined_k141_161796\|\|full | Not-determined |
| 18-110-final-viral-combined_k141_104223\|\|full | Not-determined |
| 18-110-final-viral-combined_k141_23718\|\|full | Not-determined |
| 18-110-final-viral-combined_k141_92971\|\|full | Not-determined |
| 18-110-final-viral-combined_k141_115790\|\|full | Low-quality |
| 18-110-final-viral-combined_k141_104242\|\|full | Not-determined |
| 18-110-final-viral-combined_k141_115793\|\|full | Low-quality |
| 18-110-final-viral-combined_k141_23724\|\|full | Low-quality |
| 18-110-final-viral-combined_k141_139032\|\|full | Low-quality |
| 18-110-final-viral-combined_k141_208037\|\|full | Low-quality |
| 18-110-final-viral-combined_k141_92983\|\|full | Low-quality |
| 18-110-final-viral-combined_k141_115801\|\|full | Low-quality |
| 18-110-final-viral-combined_k141_69815\|\|full | Low-quality |
| 18-110-final-viral-combined_k141_92997\|\|full | Not-determined |
| 18-110-final-viral-combined_k141_92999\|\|full | Low-quality |
| 18-110-final-viral-combined_k141_115815\|\|full | Not-determined |
| 18-110-final-viral-combined_k141_69829\|\|full | Not-determined |
| 18-110-final-viral-combined_k141_127314\|\|full | Low-quality |
| 18-110-final-viral-combined_k141_46677\|\|full | Not-determined |
| 18-110-final-viral-combined_k141_115821\|\|full | Low-quality |
| 18-110-final-viral-combined_k141_208055\|\|full | Low-quality |
| 18-110-final-viral-combined_k141_46679\|\|full | Low-quality |
| 18-110-final-viral-combined_k141_127331\|\|full | Not-determined |
| 18-110-final-viral-combined_k141_69861\|\|full | Not-determined |
| 18-110-final-viral-combined_k141_752\|\|full | Not-determined |
| 18-110-final-viral-combined_k141_93032\|\|full | Not-determined |
| 18-110-final-viral-combined_k141_184976\|\|full | Not-determined |
| 18-110-final-viral-combined_k141_208094\|\|full | Not-determined |
| 18-110-final-viral-combined_k141_69890\|\|full | Not-determined |
| 18-110-final-viral-combined_k141_69892\|\|full | Low-quality |
| 18-110-final-viral-combined_k141_219544\|\|full | Low-quality |
| 18-110-final-viral-combined_k141_93043\|\|full | Low-quality |
| 18-110-final-viral-combined_k141_69899\|\|full | Not-determined |
| 18-110-final-viral-combined_k141_69903\|\|full | Low-quality |
| 18-110-final-viral-combined_k141_242753\|\|full | Low-quality |
| 18-110-final-viral-combined_k141_253985\|\|full | Not-determined |
| 18-110-final-viral-combined_k141_161889\|\|full | Low-quality |
| 18-110-final-viral-combined_k141_115862\|\|full | Low-quality |
| 18-110-final-viral-combined_k141_161899\|\|full | Not-determined |
| 18-110-final-viral-combined_k141_265748\|\|full | Low-quality |
| 18-110-final-viral-combined_k141_46747\|\|full | Not-determined |
| 18-110-final-viral-combined_k141_196612\|\|full | Low-quality |
| 18-110-final-viral-combined_k141_242759\|\|full | Low-quality |
| 18-110-final-viral-combined_k141_58404\|\|full | Not-determined |
| 18-110-final-viral-combined_k141_46768\|\|full | Low-quality |
| 18-110-final-viral-combined_k141_127423\|\|full | Low-quality |
| 18-110-final-viral-combined_k141_770\|\|full | Low-quality |
| 18-110-final-viral-combined_k141_115875\|\|full | Low-quality |
| 18-110-final-viral-combined_k141_161901\|\|full | Low-quality |
| 18-110-final-viral-combined_k141_81509\|\|full | Low-quality |
| 18-110-final-viral-combined_k141_127426\|\|full | Not-determined |
| 18-110-final-viral-combined_k141_115884\|\|full | Not-determined |
| 18-110-final-viral-combined_k141_12093\|\|full | Low-quality |
| 18-110-final-viral-combined_k141_139116\|\|full | Not-determined |
| 18-110-final-viral-combined_k141_46795\|\|full | Low-quality |
| 18-110-final-viral-combined_k141_93127\|\|full | Not-determined |
| 18-110-final-viral-combined_k141_69971\|\|full | Low-quality |
| 18-110-final-viral-combined_k141_161928\|\|full | Low-quality |
| 18-110-final-viral-combined_k141_127464\|\|full | Not-determined |
| 18-110-final-viral-combined_k141_46812\|\|full | Not-determined |
| 18-110-final-viral-combined_k141_185066\|\|full | Not-determined |
| 18-110-final-viral-combined_k141_173674\|\|full | Low-quality |
| 18-110-final-viral-combined_k141_185072\|\|full | Not-determined |
| 18-110-final-viral-combined_k141_219596\|\|full | Low-quality |
| 18-110-final-viral-combined_k141_185085\|\|full | Not-determined |
| 18-110-final-viral-combined_k141_231213\|\|full | Low-quality |
| 18-110-final-viral-combined_k141_104365\|\|full | Not-determined |
| 18-110-final-viral-combined_k141_127501\|\|full | Low-quality |
| 18-110-final-viral-combined_k141_23869\|\|full | Low-quality |
| 18-110-final-viral-combined_k141_12120\|\|full | Not-determined |
| 18-110-final-viral-combined_k141_70024\|\|full | Low-quality |
| 18-110-final-viral-combined_k141_139152\|\|full | Low-quality |
| 18-110-final-viral-combined_k141_161967\|\|full | Low-quality |
| 18-110-final-viral-combined_k141_231227\|\|full | Not-determined |
| 18-110-final-viral-combined_k141_150522\|\|full | Low-quality |
| 18-110-final-viral-combined_k141_826\|\|full | Not-determined |
| 18-110-final-viral-combined_k141_161971\|\|full | Not-determined |
| 18-110-final-viral-combined_k141_81567\|\|full | Not-determined |
| 18-110-final-viral-combined_k141_35150\|\|full | Low-quality |
| 18-110-final-viral-combined_k141_219628\|\|full | Not-determined |
| 18-110-final-viral-combined_k141_833\|\|full | Not-determined |
| 18-110-final-viral-combined_k141_81580\|\|full | Low-quality |
| 18-110-final-viral-combined_k141_139170\|\|full | Not-determined |
| 18-110-final-viral-combined_k141_254047\|\|full | Not-determined |
| 18-110-final-viral-combined_k141_219637\|\|full | Low-quality |
| 18-110-final-viral-combined_k141_173732\|\|full | Not-determined |
| 18-110-final-viral-combined_k141_115995\|\|full | Not-determined |
| 18-110-final-viral-combined_k141_127521\|\|full | Not-determined |
| 18-110-final-viral-combined_k141_219661\|\|full | Low-quality |
| 18-110-final-viral-combined_k141_161990\|\|full | Low-quality |
| 18-110-final-viral-combined_k141_58490\|\|full | Not-determined |
| 18-110-final-viral-combined_k141_139205\|\|full | Low-quality |
| 18-110-final-viral-combined_k141_23909\|\|full | Low-quality |
| 18-110-final-viral-combined_k141_857\|\|full | Not-determined |
| 18-110-final-viral-combined_k141_162003\|\|full | Not-determined |
| 18-110-final-viral-combined_k141_35168\|\|full | Low-quality |
| 18-110-final-viral-combined_k141_162005\|\|full | Not-determined |
| 18-110-final-viral-combined_k141_46878\|\|full | Not-determined |
| 18-110-final-viral-combined_k141_23928\|\|full | Low-quality |
| 18-110-final-viral-combined_k141_868\|\|full | Not-determined |
| 18-101-final-viral-combined_k141_156637\|\|full | Medium-quality |
| 18-110-final-viral-combined_k141_70067\|\|full | Not-determined |
| 18-110-final-viral-combined_k141_208252\|\|full | Low-quality |
| 18-110-final-viral-combined_k141_70070\|\|full | Not-determined |
| 18-110-final-viral-combined_k141_35208\|\|full | Not-determined |
| 18-101_k141_313991 | Medium-quality |
| 18-110-final-viral-combined_k141_150596\|\|full | Low-quality |
| 18-110-final-viral-combined_k141_885\|\|full | Low-quality |
| 18-110-final-viral-combined_k141_23945\|\|full | Low-quality |
| 18-110-final-viral-combined_k141_254110\|\|full | Not-determined |
| 18-110-final-viral-combined_k141_116015\|\|full | Not-determined |
| 18-110-final-viral-combined_k141_104445\|\|full | Not-determined |
| 18-110-final-viral-combined_k141_185138\|\|full | Low-quality |
| 18-110-final-viral-combined_k141_70101\|\|full | Low-quality |
| 18-110-final-viral-combined_k141_35239\|\|full | Not-determined |
| 18-110-final-viral-combined_k141_162036\|\|full | Not-determined |
| 18-110-final-viral-combined_k141_919\|\|full | Low-quality |
| 18-110-final-viral-combined_k141_208289\|\|full | Low-quality |
| 18-110-final-viral-combined_k141_150622\|\|full | Low-quality |
| 18-110-final-viral-combined_k141_116025\|\|full | Not-determined |
| 18-110-final-viral-combined_k141_196739\|\|full | Not-determined |
| 18-110-final-viral-combined_k141_150630\|\|full | Not-determined |
| 18-110-final-viral-combined_k141_35250\|\|full | Not-determined |
| 18-110-final-viral-combined_k141_70116\|\|full | Low-quality |
| 18-110-final-viral-combined_k141_70125\|\|full | Low-quality |
| 18-110-final-viral-combined_k141_254135\|\|full | Not-determined |
| 18-110-final-viral-combined_k141_196755\|\|full | Not-determined |
| 18-110-final-viral-combined_k141_58542\|\|full | Low-quality |
| 18-110-final-viral-combined_k141_941\|\|full | Low-quality |
| 18-110-final-viral-combined_k141_58561\|\|full | Not-determined |
| 18-110-final-viral-combined_k141_945\|\|full | Not-determined |
| 18-110-final-viral-combined_k141_231410\|\|full | Low-quality |
| 18-110-final-viral-combined_k141_949\|\|full | Not-determined |
| 18-110-final-viral-combined_k141_58563\|\|full | Low-quality |
| 18-110-final-viral-combined_k141_58565\|\|full | Not-determined |
| 18-110-final-viral-combined_k141_254162\|\|full | Low-quality |
| 18-110-final-viral-combined_k141_58568\|\|full | Not-determined |
| 18-110-final-viral-combined_k141_35288\|\|full | Low-quality |
| 18-110-final-viral-combined_k141_127559\|\|full | Not-determined |
| 18-110-final-viral-combined_k141_58577\|\|full | Not-determined |
| 18-110-final-viral-combined_k141_70160\|\|full | Not-determined |
| 18-110-final-viral-combined_k141_231426\|\|full | Not-determined |
| 18-110-final-viral-combined_k141_208348\|\|full | Low-quality |
| 18-110-final-viral-combined_k141_150706\|\|full | Not-determined |
| 18-110-final-viral-combined_k141_150707\|\|full | Low-quality |
| 18-110-final-viral-combined_k141_162098\|\|full | Low-quality |
| 18-110-final-viral-combined_k141_173926\|\|full | Not-determined |
| 18-110-final-viral-combined_k141_12305\|\|full | Low-quality |
| 18-110-final-viral-combined_k141_173930\|\|full | Low-quality |
| 18-110-final-viral-combined_k141_185218\|\|full | Low-quality |
| 18-110-final-viral-combined_k141_162115\|\|full | Low-quality |
| 18-110-final-viral-combined_k141_70161\|\|full | Low-quality |
| 18-110-final-viral-combined_k141_12312\|\|full | Not-determined |
| 18-110-final-viral-combined_k141_46933\|\|full | Low-quality |
| 18-110-final-viral-combined_k141_93191\|\|full | Not-determined |
| 18-110-final-viral-combined_k141_185228\|\|full | Low-quality |
| 18-110-final-viral-combined_k141_162126\|\|full | Low-quality |

18-110-final-viral-combined_k141_70175||full Low-quality
18-110-final-viral-combined_k141_196823||full Not-determined
18-110-final-viral-combined_k141_162141||full Not-determined
18-110-final-viral-combined_k141_231471||full Not-determined
18-110-final-viral-combined_k141_173973||full Low-quality
18-110-final-viral-combined_k141_58647||full Low-quality
18-110-final-viral-combined_k141_81723||full Low-quality
18-110-final-viral-combined_k141_116077||full Not-determined
18-110-final-viral-combined_k141_46948||full Not-determined
18-110-final-viral-combined_k141_231491||full Not-determined
18-110-final-viral-combined_k141_104532||full Low-quality
18-110-final-viral-combined_k141_162152||full Not-determined
18-110-final-viral-combined_k141_70200||full Low-quality
18-110-final-viral-combined_k141_139378||full Not-determined
18-110-final-viral-combined_k141_70205||full Not-determined
18-110-final-viral-combined_k141_231507||full Low-quality
18-110-final-viral-combined_k141_70213||full Low-quality
18-110-final-viral-combined_k141_231512||full Not-determined
18-110-final-viral-combined_k141_93218||full Low-quality
18-110-final-viral-combined_k141_70218||full Low-quality
18-110-final-viral-combined_k141_981||full Not-determined
18-110-final-viral-combined_k141_150781||full Low-quality
18-110-final-viral-combined_k141_208414||full Not-determined
18-110-final-viral-combined_k141_58685||full Not-determined
18-110-final-viral-combined_k141_81758||full Low-quality
18-110-final-viral-combined_k141_196854||full Low-quality
18-110-final-viral-combined_k141_208421||full Not-determined
18-110-final-viral-combined_k141_93252||full Low-quality
18-110-final-viral-combined_k141_70248||full Not-determined
18-110-final-viral-combined_k141_196865||full Not-determined
18-110-final-viral-combined_k141_265880||full Low-quality
18-110-final-viral-combined_k141_116131||full Not-determined
18-110-final-viral-combined_k141_116135||full Low-quality
18-110-final-viral-combined_k141_81782||full Low-quality
18-110-final-viral-combined_k141_243122||full Low-quality
18-110-final-viral-combined_k141_231568||full Low-quality
18-110-final-viral-combined_k141_93293||full Low-quality
18-110-final-viral-combined_k141_185355||full Low-quality
18-110-final-viral-combined_k141_116170||full Low-quality
18-110-final-viral-combined_k141_81804||full Low-quality
18-110-final-viral-combined_k141_243147||full Low-quality
18-110-final-viral-combined_k141_139476||full Not-determined
18-110-final-viral-combined_k141_70295||full Not-determined
18-110-final-viral-combined_k141_185379||full Low-quality
18-110-final-viral-combined_k141_70304||full Low-quality
18-110-final-viral-combined_k141_93325||full Low-quality
18-110-final-viral-combined_k141_127725||full Not-determined
18-110-final-viral-combined_k141_35416||full Low-quality
18-110-final-viral-combined_k141_93337||full Low-quality
18-110-final-viral-combined_k141_81836||full Low-quality
18-110-final-viral-combined_k141_196892||full Low-quality
18-110-final-viral-combined_k141_81840||full Low-quality
18-110-final-viral-combined_k141_231619||full Not-determined
18-110-final-viral-combined_k141_35438||full Low-quality
18-110-final-viral-combined_k141_104645||full Not-determined
18-110-final-viral-combined_k141_127729||full Not-determined
18-110-final-viral-combined_k141_116210||full Low-quality
18-110-final-viral-combined_k141_35476||full Low-quality
18-110-final-viral-combined_k141_231642||full Not-determined
18-110-final-viral-combined_k141_208581||full Not-determined
18-110-final-viral-combined_k141_185466||full Not-determined
18-110-final-viral-combined_k141_12361||full Low-quality
18-110-final-viral-combined_k141_116224||full Low-quality
18-110-final-viral-combined_k141_12369||full Low-quality
18-110-final-viral-combined_k141_24140||full Not-determined
18-110-final-viral-combined_k141_104703||full Not-determined
18-110-final-viral-combined_k141_70385||full Low-quality
18-110-final-viral-combined_k141_35484||full Low-quality
18-110-final-viral-combined_k141_70389||full Low-quality
18-110-final-viral-combined_k141_162260||full Not-determined
18-110-final-viral-combined_k141_162261||full Not-determined
18-110-final-viral-combined_k141_162263||full Not-determined
18-110-final-viral-combined_k141_185495||full Not-determined
18-110-final-viral-combined_k141_93421||full Low-quality
18-110-final-viral-combined_k141_58808||full Not-determined
18-110-final-viral-combined_k141_265932||full Not-determined
18-110-final-viral-combined_k141_104732||full Low-quality
18-110-final-viral-combined_k141_70390||full Low-quality
18-110-final-viral-combined_k141_104734||full Not-determined
18-110-final-viral-combined_k141_47125||full Not-determined
18-110-final-viral-combined_k141_265933||full Not-determined
18-110-final-viral-combined_k141_150949||full Low-quality
18-110-final-viral-combined_k141_219751||full Not-determined
18-110-final-viral-combined_k141_219752||full Low-quality
18-110-final-viral-combined_k141_58819||full Not-determined
18-110-final-viral-combined_k141_47134||full Not-determined
18-110-final-viral-combined_k141_58822||full Low-quality
18-110-final-viral-combined_k141_185526||full Low-quality
18-110-final-viral-combined_k141_127766||full Low-quality
18-110-final-viral-combined_k141_47141||full Low-quality
18-110-final-viral-combined_k141_162276||full Not-determined
18-110-final-viral-combined_k141_104783||full Not-determined
18-110-final-viral-combined_k141_58855||full Not-determined
18-110-final-viral-combined_k141_58860||full Not-determined
18-110-final-viral-combined_k141_24217||full Not-determined
18-110-final-viral-combined_k141_197001||full Not-determined
18-110-final-viral-combined_k141_93486||full Low-quality
18-110-final-viral-combined_k141_104804||full Not-determined
18-110-final-viral-combined_k141_58871||full Not-determined
18-110-final-viral-combined_k141_1090||full Not-determined
18-110-final-viral-combined_k141_24234||full Not-determined
18-110-final-viral-combined_k141_243265||full Low-quality
18-110-final-viral-combined_k141_70435||full Not-determined
18-110-final-viral-combined_k141_197007||full Low-quality
18-110-final-viral-combined_k141_162320||full Low-quality
18-110-final-viral-combined_k141_139581||full Not-determined
18-110-final-viral-combined_k141_243269||full Low-quality
18-110-final-viral-combined_k141_58885||full Not-determined
18-110-final-viral-combined_k141_81946||full Low-quality
18-110-final-viral-combined_k141_185595||full Not-determined
18-110-final-viral-combined_k141_219792||full Low-quality
18-110-final-viral-combined_k141_197019||full Not-determined
18-110-final-viral-combined_k141_139595||full Low-quality
18-110-final-viral-combined_k141_24268||full Low-quality
18-110-final-viral-combined_k141_243281||full Low-quality
18-110-final-viral-combined_k141_47179||full Not-determined
18-110-final-viral-combined_k141_185608||full Low-quality
18-110-final-viral-combined_k141_231689||full Not-determined
18-110-final-viral-combined_k141_81964||full Low-quality
18-110-final-viral-combined_k141_35554||full Low-quality
18-110-final-viral-combined_k141_81981||full Not-determined
18-101-final-viral-combined_k141_313991||full Medium-quality
18-110-final-viral-combined_k141_174057||full Low-quality
18-110-final-viral-combined_k141_243307||full Low-quality
18-110-final-viral-combined_k141_254378||full Not-determined
18-110-final-viral-combined_k141_58939||full Not-determined
18-110-final-viral-combined_k141_1122||full Not-determined
18-110-final-viral-combined_k141_127858||full Not-determined
18-110-final-viral-combined_k141_47196||full Low-quality
18-110-final-viral-combined_k141_82005||full Low-quality
18-110-final-viral-combined_k141_82006||full Low-quality
18-101-final-viral-combined_k141_238898||full Medium-quality
18-110-final-viral-combined_k141_58953||full Low-quality
18-110-final-viral-combined_k141_24299||full Not-determined
18-110-final-viral-combined_k141_127867||full Low-quality
18-110-final-viral-combined_k141_47207||full Not-determined
18-110-final-viral-combined_k141_58962||full Low-quality
18-110-final-viral-combined_k141_35572||full Not-determined
18-110-final-viral-combined_k141_231764||full Low-quality
18-110-final-viral-combined_k141_70527||full Not-determined
18-110-final-viral-combined_k141_58967||full Not-determined
18-110-final-viral-combined_k141_139677||full Low-quality
18-110-final-viral-combined_k141_231786||full Not-determined
18-110-final-viral-combined_k141_70542||full Low-quality
18-110-final-viral-combined_k141_47229||full Low-quality
18-110-final-viral-combined_k141_219813||full Low-quality
18-110-final-viral-combined_k141_82017||full Low-quality
18-110-final-viral-combined_k141_127877||full Not-determined
18-110-final-viral-combined_k141_12400||full Not-determined
18-110-final-viral-combined_k141_219820||full Low-quality
18-110-final-viral-combined_k141_116416||full Low-quality
18-110-final-viral-combined_k141_185652||full Not-determined
18-110-final-viral-combined_k141_82043||full Not-determined
18-110-final-viral-combined_k141_12422||full Low-quality
18-110-final-viral-combined_k141_93624||full Not-determined
18-110-final-viral-combined_k141_162416||full Low-quality
18-110-final-viral-combined_k141_254451||full Low-quality
18-110-final-viral-combined_k141_127915||full Low-quality
18-110-final-viral-combined_k141_47265||full Low-quality
18-110-final-viral-combined_k141_219846||full Not-determined
18-110-final-viral-combined_k141_243360||full Not-determined
18-110-final-viral-combined_k141_70572||full Not-determined
18-110-final-viral-combined_k141_93641||full Low-quality
18-110-final-viral-combined_k141_174146||full Low-quality
18-110-final-viral-combined_k141_162436||full Low-quality
18-110-final-viral-combined_k141_254463||full Not-determined
18-110-final-viral-combined_k141_219854||full Low-quality
18-110-final-viral-combined_k141_185683||full Not-determined
18-110-final-viral-combined_k141_162446||full Not-determined
18-110-final-viral-combined_k141_47286||full Not-determined
18-110-final-viral-combined_k141_174160||full Not-determined
18-110-final-viral-combined_k141_47289||full Low-quality
18-110-final-viral-combined_k141_82084||full Low-quality
18-110-final-viral-combined_k141_139734||full Not-determined
18-110-final-viral-combined_k141_139737||full Not-determined
18-110-final-viral-combined_k141_162463||full Not-determined
18-110-final-viral-combined_k141_12455||full Not-determined
18-110-final-viral-combined_k141_208821||full Low-quality
18-110-final-viral-combined_k141_35692||full Low-quality
18-110-final-viral-combined_k141_208830||full Low-quality
18-110-final-viral-combined_k141_104911||full Not-determined
18-110-final-viral-combined_k141_35695||full Not-determined
18-110-final-viral-combined_k141_35701||full Not-determined
18-110-final-viral-combined_k141_12466||full Low-quality
18-110-final-viral-combined_k141_151071||full Low-quality
18-110-final-viral-combined_k141_254486||full Low-quality
18-110-final-viral-combined_k141_139777||full Not-determined
18-110-final-viral-combined_k141_104938||full Not-determined
18-110-final-viral-combined_k141_151096||full Low-quality
18-110-final-viral-combined_k141_162528||full Low-quality
18-110-final-viral-combined_k141_231895||full Not-determined
18-110-final-viral-combined_k141_139810||full Low-quality
18-110-final-viral-combined_k141_104959||full Low-quality
18-110-final-viral-combined_k141_12497||full Not-determined
18-110-final-viral-combined_k141_266025||full Low-quality
18-110-final-viral-combined_k141_24438||full Not-determined
18-110-final-viral-combined_k141_151117||full Not-determined
18-110-final-viral-combined_k141_35746||full Low-quality
18-110-final-viral-combined_k141_243433||full Not-determined
18-110-final-viral-combined_k141_24466||full Not-determined
18-110-final-viral-combined_k141_35758||full Not-determined
18-110-final-viral-combined_k141_151130||full Low-quality
18-110-final-viral-combined_k141_185768||full Low-quality
18-110-final-viral-combined_k141_162581||full Not-determined
18-110-final-viral-combined_k141_254495||full Not-determined
18-110-final-viral-combined_k141_82150||full Low-quality
18-110-final-viral-combined_k141_231937||full Not-determined
18-110-final-viral-combined_k141_162590||full Low-quality
18-110-final-viral-combined_k141_266073||full Not-determined
18-110-final-viral-combined_k141_24477||full Not-determined
18-110-final-viral-combined_k141_82161||full Not-determined
18-110-final-viral-combined_k141_59194||full Low-quality
18-110-final-viral-combined_k141_151138||full Not-determined
18-110-final-viral-combined_k141_127985||full Low-quality
18-110-final-viral-combined_k141_70717||full Not-determined
18-110-final-viral-combined_k141_266077||full Low-quality
18-110-final-viral-combined_k141_219945||full Not-determined
18-110-final-viral-combined_k141_59222||full Low-quality
18-110-final-viral-combined_k141_12507||full Not-determined
18-110-final-viral-combined_k141_1264||full Not-determined
18-110-final-viral-combined_k141_116481||full Not-determined
18-110-final-viral-combined_k141_185812||full Low-quality
18-110-final-viral-combined_k141_139866||full Low-quality
18-110-final-viral-combined_k141_59247||full Not-determined
18-110-final-viral-combined_k141_82178||full Not-determined
18-110-final-viral-combined_k141_254525||full Not-determined
18-110-final-viral-combined_k141_24503||full Low-quality
18-110-final-viral-combined_k141_105025||full Not-determined
18-110-final-viral-combined_k141_59270||full Low-quality
18-110-final-viral-combined_k141_24512||full Not-determined
18-110-final-viral-combined_k141_208837||full Not-determined
18-110-final-viral-combined_k141_35813||full Not-determined
18-110-final-viral-combined_k141_185837||full Low-quality
18-110-final-viral-combined_k141_185842||full Not-determined
18-110-final-viral-combined_k141_59274||full Not-determined
18-110-final-viral-combined_k141_266120||full Not-determined
18-110-final-viral-combined_k141_174270||full Not-determined
18-110-final-viral-combined_k141_162624||full Not-determined
18-110-final-viral-combined_k141_197047||full Not-determined
18-110-final-viral-combined_k141_254566||full Low-quality
18-110-final-viral-combined_k141_162632||full Not-determined
18-110-final-viral-combined_k141_70801||full Low-quality
18-110-final-viral-combined_k141_139937||full Not-determined
18-110-final-viral-combined_k141_197059||full Not-determined
18-110-final-viral-combined_k141_24556||full Low-quality
18-110-final-viral-combined_k141_266156||full Not-determined
18-110-final-viral-combined_k141_197065||full Low-quality
18-110-final-viral-combined_k141_59304||full Low-quality
18-110-final-viral-combined_k141_24564||full Not-determined
18-110-final-viral-combined_k141_47374||full Low-quality
18-110-final-viral-combined_k141_185890||full Not-determined
18-110-final-viral-combined_k141_174315||full Low-quality
18-110-final-viral-combined_k141_174323||full Not-determined
18-110-final-viral-combined_k141_24596||full Not-determined
18-110-final-viral-combined_k141_93730||full Low-quality
18-110-final-viral-combined_k141_24604||full Low-quality
18-110-final-viral-combined_k141_128083||full Low-quality
18-110-final-viral-combined_k141_128089||full Low-quality
18-110-final-viral-combined_k141_116547||full Low-quality
18-110-final-viral-combined_k141_220095||full Not-determined
18-110-final-viral-combined_k141_59346||full Not-determined
18-110-final-viral-combined_k141_151284||full Not-determined
18-110-final-viral-combined_k141_243579||full Not-determined
18-110-final-viral-combined_k141_93757||full Not-determined
18-110-final-viral-combined_k141_35908||full Not-determined
18-110-final-viral-combined_k141_266214||full Not-determined
18-110-final-viral-combined_k141_139965||full Not-determined
18-110-final-viral-combined_k141_220108||full Low-quality
18-110-final-viral-combined_k141_35910||full Low-quality
18-110-final-viral-combined_k141_139968||full Not-determined
18-110-final-viral-combined_k141_128103||full Low-quality
18-110-final-viral-combined_k141_174368||full Not-determined
18-110-final-viral-combined_k141_93796||full Not-determined
18-110-final-viral-combined_k141_12551||full Low-quality
18-110-final-viral-combined_k141_151303||full Low-quality
18-110-final-viral-combined_k141_185920||full Low-quality
18-110-final-viral-combined_k141_266250||full Not-determined
18-110-final-viral-combined_k141_232066||full Low-quality
18-110-final-viral-combined_k141_59395||full Not-determined
18-110-final-viral-combined_k141_35922||full Not-determined
18-110-final-viral-combined_k141_12563||full Not-determined
18-110-final-viral-combined_k141_35926||full Not-determined
18-110-final-viral-combined_k141_105141||full Not-determined
18-110-final-viral-combined_k141_243625||full Low-quality
18-110-final-viral-combined_k141_35928||full Not-determined
18-110-final-viral-combined_k141_128138||full Low-quality
18-110-final-viral-combined_k141_116569||full Not-determined
18-110-final-viral-combined_k141_93827||full Low-quality
18-110-final-viral-combined_k141_24694||full Low-quality
18-110-final-viral-combined_k141_93840||full Low-quality
18-110-final-viral-combined_k141_162714||full Low-quality
18-110-final-viral-combined_k141_59436||full Not-determined
18-110-final-viral-combined_k141_1473||full Low-quality
18-110-final-viral-combined_k141_70878||full Not-determined
18-110-final-viral-combined_k141_116615||full Not-determined
18-110-final-viral-combined_k141_128178||full Low-quality
18-110-final-viral-combined_k141_140049||full Low-quality
18-110-final-viral-combined_k141_70890||full Low-quality
18-110-final-viral-combined_k141_266285||full Not-determined
18-110-final-viral-combined_k141_197226||full Not-determined
18-110-final-viral-combined_k141_266286||full Low-quality
18-110-final-viral-combined_k141_59469||full Not-determined
18-110-final-viral-combined_k141_82266||full Not-determined
18-110-final-viral-combined_k141_232121||full Low-quality
18-110-final-viral-combined_k141_1485||full Not-determined
18-110-final-viral-combined_k141_174430||full Not-determined
18-110-final-viral-combined_k141_1490||full Not-determined
18-110-final-viral-combined_k141_116647||full Not-determined
18-110-final-viral-combined_k141_105173||full Not-determined
18-110-final-viral-combined_k141_12603||full Not-determined
18-110-final-viral-combined_k141_128208||full Low-quality
18-110-final-viral-combined_k141_232141||full Low-quality
18-110-final-viral-combined_k141_59479||full Low-quality
18-110-final-viral-combined_k141_186014||full Not-determined
17-109-final-viral-combined_k141_401379||full Medium-quality
18-110-final-viral-combined_k141_174450||full Not-determined
18-110-final-viral-combined_k141_128213||full Not-determined
18-110-final-viral-combined_k141_140062||full Low-quality
18-110-final-viral-combined_k141_197278||full Low-quality
18-110-final-viral-combined_k141_243662||full Low-quality
18-110-final-viral-combined_k141_105184||full Not-determined
18-110-final-viral-combined_k141_209051||full Not-determined
18-110-final-viral-combined_k141_140088||full Not-determined
18-101-final-viral-combined_k141_394628||full Medium-quality
18-110-final-viral-combined_k141_140097||full Not-determined
18-110-final-viral-combined_k141_24774||full Not-determined
18-110-final-viral-combined_k141_105200||full Not-determined
18-110-final-viral-combined_k141_128240||full Not-determined
18-110-final-viral-combined_k141_220189||full Not-determined
18-110-final-viral-combined_k141_24795||full Low-quality
18-110-final-viral-combined_k141_59499||full Not-determined
18-110-final-viral-combined_k141_59501||full Not-determined
18-110-final-viral-combined_k141_47418||full Low-quality
18-110-final-viral-combined_k141_266302||full Low-quality
18-110-final-viral-combined_k141_105232||full Not-determined
18-110-final-viral-combined_k141_128286||full Low-quality
18-110-final-viral-combined_k141_116701||full Low-quality
18-110-final-viral-combined_k141_59518||full Low-quality
18-110-final-viral-combined_k141_174518||full Not-determined
18-110-final-viral-combined_k141_151502||full Low-quality
18-110-final-viral-combined_k141_93920||full Not-determined
18-110-final-viral-combined_k141_105244||full Low-quality
18-110-final-viral-combined_k141_174532||full Low-quality
18-110-final-viral-combined_k141_209135||full Not-determined
18-110-final-viral-combined_k141_197324||full Low-quality
18-110-final-viral-combined_k141_93937||full Low-quality
18-102-final-viral-combined_k141_95987||full High-quality
18-110-final-viral-combined_k141_116737||full Not-determined
18-110-final-viral-combined_k141_93941||full Not-determined
18-110-final-viral-combined_k141_197339||full Not-determined
18-110-final-viral-combined_k141_254757||full Not-determined
18-110-final-viral-combined_k141_186067||full Low-quality
18-110-final-viral-combined_k141_232220||full Low-quality
18-110-final-viral-combined_k141_174574||full Not-determined
18-110-final-viral-combined_k141_266343||full Low-quality
18-110-final-viral-combined_k141_140201||full Low-quality
18-110-final-viral-combined_k141_174579||full Low-quality
18-110-final-viral-combined_k141_82288||full Not-determined
18-110-final-viral-combined_k141_162823||full Low-quality
18-110-final-viral-combined_k141_151547||full Not-determined
18-110-final-viral-combined_k141_71014||full Low-quality
18-110-final-viral-combined_k141_1590||full Low-quality
18-110-final-viral-combined_k141_266352||full Low-quality
18-110-final-viral-combined_k141_1594||full Not-determined
18-110-final-viral-combined_k141_243758||full Low-quality
18-110-final-viral-combined_k141_1596||full Not-determined
18-110-final-viral-combined_k141_140224||full Not-determined
18-110-final-viral-combined_k141_243767||full Not-determined
18-110-final-viral-combined_k141_151577||full Not-determined
18-110-final-viral-combined_k141_59569||full Not-determined
18-110-final-viral-combined_k141_116795||full Low-quality
18-110-final-viral-combined_k141_243785||full Low-quality
18-110-final-viral-combined_k141_82320||full Low-quality
18-110-final-viral-combined_k141_12691||full Low-quality
18-110-final-viral-combined_k141_116796||full Low-quality
18-110-final-viral-combined_k141_12696||full Low-quality
18-110-final-viral-combined_k141_186135||full Not-determined
18-110-final-viral-combined_k141_71053||full Not-determined
18-110-final-viral-combined_k141_116802||full Low-quality
18-110-final-viral-combined_k141_82336||full Not-determined
18-110-final-viral-combined_k141_93973||full Not-determined
18-110-final-viral-combined_k141_243791||full Low-quality
18-110-final-viral-combined_k141_93976||full Low-quality
18-110-final-viral-combined_k141_243793||full Not-determined
18-110-final-viral-combined_k141_151633||full Not-determined
18-110-final-viral-combined_k141_232282||full Low-quality
18-110-final-viral-combined_k141_186169||full Low-quality
18-110-final-viral-combined_k141_82355||full Low-quality
18-110-final-viral-combined_k141_36090||full Low-quality
18-110-final-viral-combined_k141_151653||full Low-quality
18-110-final-viral-combined_k141_82373||full Low-quality
18-110-final-viral-combined_k141_140306||full Not-determined
18-110-final-viral-combined_k141_232297||full Not-determined
18-110-final-viral-combined_k141_186183||full Low-quality
18-110-final-viral-combined_k141_36101||full Low-quality
18-110-final-viral-combined_k141_140320||full Low-quality
18-110-final-viral-combined_k141_36104||full Not-determined
18-110-final-viral-combined_k141_12751||full Not-determined
18-110-final-viral-combined_k141_151674||full Not-determined
18-110-final-viral-combined_k141_24950||full Low-quality
18-110-final-viral-combined_k141_105351||full Not-determined
18-110-final-viral-combined_k141_232309||full Low-quality
18-110-final-viral-combined_k141_47539||full Not-determined
18-110-final-viral-combined_k141_162898||full Low-quality
18-110-final-viral-combined_k141_209250||full Not-determined
18-110-final-viral-combined_k141_243838||full Low-quality
18-110-final-viral-combined_k141_71090||full Low-quality
18-110-final-viral-combined_k141_254832||full Low-quality
18-110-final-viral-combined_k141_151700||full Low-quality
18-110-final-viral-combined_k141_266418||full Not-determined
18-110-final-viral-combined_k141_162899||full Low-quality
18-110-final-viral-combined_k141_186211||full Not-determined
18-110-final-viral-combined_k141_266419||full Low-quality
18-110-final-viral-combined_k141_12787||full Low-quality
18-110-final-viral-combined_k141_82406||full Low-quality
18-110-final-viral-combined_k141_105374||full Low-quality
18-110-final-viral-combined_k141_186236||full Not-determined
18-110-final-viral-combined_k141_36142||full Low-quality
18-110-final-viral-combined_k141_116882||full Low-quality
18-110-final-viral-combined_k141_140361||full Not-determined
18-110-final-viral-combined_k141_266458||full Low-quality
18-110-final-viral-combined_k141_197432||full Not-determined
18-110-final-viral-combined_k141_243883||full Low-quality
18-110-final-viral-combined_k141_266463||full Low-quality
18-110-final-viral-combined_k141_220358||full Low-quality
18-110-final-viral-combined_k141_243893||full Not-determined
18-110-final-viral-combined_k141_151735||full Low-quality
18-110-final-viral-combined_k141_12802||full Low-quality
18-110-final-viral-combined_k141_94027||full Not-determined
18-110-final-viral-combined_k141_1671||full Low-quality
18-110-final-viral-combined_k141_209278||full Not-determined
18-110-final-viral-combined_k141_128471||full Not-determined
18-110-final-viral-combined_k141_82461||full Low-quality
18-110-final-viral-combined_k141_151749||full Not-determined
18-110-final-viral-combined_k141_243907||full Low-quality
18-110-final-viral-combined_k141_243909||full Not-determined
18-110-final-viral-combined_k141_186269||full Not-determined
18-110-final-viral-combined_k141_140393||full Low-quality
18-110-final-viral-combined_k141_82485||full Low-quality
18-110-final-viral-combined_k141_162989||full Not-determined
18-110-final-viral-combined_k141_243927||full Low-quality
18-110-final-viral-combined_k141_232357||full Not-determined
18-110-final-viral-combined_k141_163000||full Not-determined
18-110-final-viral-combined_k141_151787||full Not-determined
18-110-final-viral-combined_k141_94092||full Low-quality
18-110-final-viral-combined_k141_266525||full Not-determined
18-110-final-viral-combined_k141_151790||full Not-determined
18-110-final-viral-combined_k141_140423||full Not-determined
18-110-final-viral-combined_k141_232362||full Low-quality
18-110-final-viral-combined_k141_94099||full Low-quality
18-110-final-viral-combined_k141_25055||full Low-quality
18-110-final-viral-combined_k141_232367||full Low-quality
18-110-final-viral-combined_k141_220402||full Low-quality
18-110-final-viral-combined_k141_94116||full Low-quality
18-110-final-viral-combined_k141_266543||full Not-determined
18-110-final-viral-combined_k141_25071||full Low-quality
18-110-final-viral-combined_k141_105451||full Not-determined
18-110-final-viral-combined_k141_94121||full Low-quality
18-110-final-viral-combined_k141_254916||full Low-quality
18-110-final-viral-combined_k141_116991||full Low-quality
18-110-final-viral-combined_k141_220408||full Not-determined
18-110-final-viral-combined_k141_140447||full Low-quality
18-110-final-viral-combined_k141_151812||full Not-determined
18-110-final-viral-combined_k141_117002||full Not-determined
18-110-final-viral-combined_k141_25089||full Low-quality
18-110-final-viral-combined_k141_117021||full Low-quality
18-110-final-viral-combined_k141_254918||full Not-determined
18-110-final-viral-combined_k141_243965||full Not-determined
18-110-final-viral-combined_k141_105475||full Not-determined
18-110-final-viral-combined_k141_254925||full Low-quality
18-110-final-viral-combined_k141_105476||full Low-quality
18-110-final-viral-combined_k141_94130||full Not-determined
18-110-final-viral-combined_k141_254929||full Low-quality
18-110-final-viral-combined_k141_117031||full Not-determined
18-110-final-viral-combined_k141_232430||full Not-determined
18-110-final-viral-combined_k141_71212||full Low-quality
18-110-final-viral-combined_k141_220423||full Not-determined
18-110-final-viral-combined_k141_163066||full Not-determined
18-110-final-viral-combined_k141_209343||full Not-determined
18-110-final-viral-combined_k141_82543||full Not-determined
18-110-final-viral-combined_k141_36235||full Low-quality
18-110-final-viral-combined_k141_186354||full Low-quality
18-110-final-viral-combined_k141_266591||full Not-determined
18-110-final-viral-combined_k141_94162||full Low-quality
18-110-final-viral-combined_k141_163104||full Not-determined
18-110-final-viral-combined_k141_220452||full Not-determined
18-110-final-viral-combined_k141_244012||full Not-determined
18-110-final-viral-combined_k141_1749||full Low-quality
18-110-final-viral-combined_k141_94178||full Low-quality
18-110-final-viral-combined_k141_82574||full Not-determined
18-110-final-viral-combined_k141_163119||full Not-determined
18-110-final-viral-combined_k141_25126||full Not-determined
18-110-final-viral-combined_k141_25129||full Low-quality
18-110-final-viral-combined_k141_25130||full Low-quality
18-110-final-viral-combined_k141_36287||full Not-determined
18-110-final-viral-combined_k141_197583||full Not-determined
18-110-final-viral-combined_k141_71291||full Low-quality
18-110-final-viral-combined_k141_82612||full Not-determined
18-110-final-viral-combined_k141_105501||full Not-determined
18-110-final-viral-combined_k141_244034||full Low-quality
18-110-final-viral-combined_k141_117049||full Not-determined
18-110-final-viral-combined_k141_254940||full Not-determined
18-110-final-viral-combined_k141_47697||full Low-quality
18-110-final-viral-combined_k141_244043||full Not-determined
18-110-final-viral-combined_k141_254950||full Not-determined
18-110-final-viral-combined_k141_140576||full Low-quality
18-110-final-viral-combined_k141_128565||full Low-quality
18-110-final-viral-combined_k141_117065||full Low-quality
18-110-final-viral-combined_k141_174862||full Low-quality
18-110-final-viral-combined_k141_36335||full Not-determined
18-110-final-viral-combined_k141_220497||full Low-quality
18-110-final-viral-combined_k141_232498||full Low-quality
18-110-final-viral-combined_k141_128584||full Not-determined
18-110-final-viral-combined_k141_266646||full Low-quality
18-110-final-viral-combined_k141_1785||full Low-quality
18-110-final-viral-combined_k141_220505||full Low-quality
18-110-final-viral-combined_k141_209441||full Not-determined
18-110-final-viral-combined_k141_25192||full Low-quality
18-110-final-viral-combined_k141_266658||full Low-quality
18-110-final-viral-combined_k141_244096||full Low-quality
18-110-final-viral-combined_k141_151863||full Not-determined
18-104-final-viral-combined_k141_114120||full High-quality
18-110-final-viral-combined_k141_244106||full Low-quality
18-110-final-viral-combined_k141_163175||full Not-determined
18-110-final-viral-combined_k141_1796||full Low-quality
18-110-final-viral-combined_k141_209453||full Low-quality
18-110-final-viral-combined_k141_254969||full Low-quality
18-110-final-viral-combined_k141_244122||full Not-determined
18-110-final-viral-combined_k141_71344||full Low-quality
18-110-final-viral-combined_k141_209458||full Not-determined
18-110-final-viral-combined_k141_71346||full Low-quality
18-110-final-viral-combined_k141_105589||full Low-quality
18-110-final-viral-combined_k141_266693||full Low-quality
18-110-final-viral-combined_k141_105590||full Low-quality
18-110-final-viral-combined_k141_59803||full Not-determined
18-110-final-viral-combined_k141_174928||full Not-determined
18-110-final-viral-combined_k141_105614||full Not-determined
18-110-final-viral-combined_k141_266704||full Not-determined
18-110-final-viral-combined_k141_71356||full Not-determined
18-110-final-viral-combined_k141_174941||full Not-determined
18-110-final-viral-combined_k141_128616||full Low-quality
18-110-final-viral-combined_k141_140657||full Low-quality
18-110-final-viral-combined_k141_163231||full Not-determined
18-110-final-viral-combined_k141_186408||full Low-quality
18-110-final-viral-combined_k141_266710||full Low-quality
18-110-final-viral-combined_k141_174948||full Not-determined
18-110-final-viral-combined_k141_255001||full Not-determined
18-110-final-viral-combined_k141_244157||full Low-quality
18-110-final-viral-combined_k141_59817||full Not-determined
18-110-final-viral-combined_k141_71392||full Not-determined
18-110-final-viral-combined_k141_1859||full Low-quality
18-110-final-viral-combined_k141_25303||full Not-determined
18-110-final-viral-combined_k141_59835||full Not-determined
18-110-final-viral-combined_k141_1863||full Not-determined
18-110-final-viral-combined_k141_117092||full Not-determined
18-110-final-viral-combined_k141_174990||full Low-quality
18-110-final-viral-combined_k141_59849||full Low-quality
18-110-final-viral-combined_k141_186479||full Not-determined
18-110-final-viral-combined_k141_151920||full Not-determined
18-110-final-viral-combined_k141_232562||full Not-determined
18-110-final-viral-combined_k141_47807||full Not-determined
18-110-final-viral-combined_k141_151927||full Not-determined
18-110-final-viral-combined_k141_117112||full Low-quality
18-110-final-viral-combined_k141_59861||full Low-quality
18-110-final-viral-combined_k141_175016||full Not-determined
18-110-final-viral-combined_k141_59862||full Not-determined
18-110-final-viral-combined_k141_197697||full Not-determined
18-110-final-viral-combined_k141_117128||full Low-quality
18-110-final-viral-combined_k141_232589||full Not-determined
18-110-final-viral-combined_k141_47826||full Low-quality
18-110-final-viral-combined_k141_175023||full Not-determined
18-110-final-viral-combined_k141_117132||full Low-quality
18-110-final-viral-combined_k141_163249||full Not-determined
18-110-final-viral-combined_k141_71446||full Low-quality
18-110-final-viral-combined_k141_244196||full Low-quality
18-110-final-viral-combined_k141_197703||full Low-quality
18-110-final-viral-combined_k141_94305||full Not-determined
18-110-final-viral-combined_k141_47847||full Not-determined
18-110-final-viral-combined_k141_232595||full Low-quality
18-110-final-viral-combined_k141_266776||full Low-quality
18-110-final-viral-combined_k141_186488||full Low-quality
18-110-final-viral-combined_k141_128703||full Not-determined
18-110-final-viral-combined_k141_186489||full Not-determined
18-110-final-viral-combined_k141_244219||full Low-quality
18-110-final-viral-combined_k141_71481||full Low-quality
18-110-final-viral-combined_k141_175055||full Low-quality
18-110-final-viral-combined_k141_94320||full Low-quality
18-110-final-viral-combined_k141_266813||full Low-quality
18-110-final-viral-combined_k141_25414||full Not-determined

| | |
|---|---|
| 18-110-final-viral-combined_k141_197749||full | Low-quality |
| 18-110-final-viral-combined_k141_105687||full | Low-quality |
| 18-110-final-viral-combined_k141_209496||full | Low-quality |
| 18-110-final-viral-combined_k141_59889||full | Low-quality |
| 18-110-final-viral-combined_k141_197763||full | Not-determined |
| 18-110-final-viral-combined_k141_128755||full | Not-determined |
| 18-110-final-viral-combined_k141_220595||full | Not-determined |
| 18-110-final-viral-combined_k141_244273||full | Low-quality |
| 18-110-final-viral-combined_k141_232638||full | Low-quality |
| 18-110-final-viral-combined_k141_36444||full | Low-quality |
| 18-110-final-viral-combined_k141_105693||full | Low-quality |
| 18-110-final-viral-combined_k141_105698||full | Low-quality |
| 18-110-final-viral-combined_k141_59906||full | Low-quality |
| 18-110-final-viral-combined_k141_128776||full | Not-determined |
| 18-110-final-viral-combined_k141_220612||full | Low-quality |
| 18-110-final-viral-combined_k141_209529||full | Low-quality |
| 18-110-final-viral-combined_k141_128782||full | Low-quality |
| 18-110-final-viral-combined_k141_175100||full | Not-determined |
| 18-110-final-viral-combined_k141_266855||full | Low-quality |
| 18-110-final-viral-combined_k141_94367||full | Not-determined |
| 18-110-final-viral-combined_k141_163349||full | Not-determined |
| 18-110-final-viral-combined_k141_175110||full | Low-quality |
| 18-110-final-viral-combined_k141_186540||full | Not-determined |
| 18-110-final-viral-combined_k141_266856||full | Not-determined |
| 18-110-final-viral-combined_k141_152035||full | Not-determined |
| 18-110-final-viral-combined_k141_220630||full | Low-quality |
| 18-110-final-viral-combined_k141_244332||full | Low-quality |
| 18-110-final-viral-combined_k141_220634||full | Low-quality |
| 18-110-final-viral-combined_k141_152043||full | Not-determined |
| 18-110-final-viral-combined_k141_117153||full | Low-quality |
| 18-110-final-viral-combined_k141_105731||full | Not-determined |
| 18-110-final-viral-combined_k141_197780||full | Low-quality |
| 18-110-final-viral-combined_k141_117165||full | Not-determined |
| 18-110-final-viral-combined_k141_232691||full | Low-quality |
| 18-110-final-viral-combined_k141_255164||full | Not-determined |
| 18-110-final-viral-combined_k141_175161||full | Not-determined |
| 18-110-final-viral-combined_k141_197804||full | Not-determined |
| 18-110-final-viral-combined_k141_244370||full | Not-determined |
| 18-110-final-viral-combined_k141_209555||full | Not-determined |
| 18-110-final-viral-combined_k141_186574||full | Low-quality |
| 18-110-final-viral-combined_k141_186577||full | Low-quality |
| 18-110-final-viral-combined_k141_197826||full | Low-quality |
| 18-110-final-viral-combined_k141_244375||full | Not-determined |
| 18-110-final-viral-combined_k141_59946||full | Not-determined |
| 18-110-final-viral-combined_k141_152064||full | Not-determined |
| 18-110-final-viral-combined_k141_220705||full | Not-determined |
| 18-110-final-viral-combined_k141_117192||full | Low-quality |
| 18-110-final-viral-combined_k141_59954||full | Low-quality |
| 18-110-final-viral-combined_k141_128847||full | Low-quality |
| 18-110-final-viral-combined_k141_255182||full | Low-quality |
| 18-110-final-viral-combined_k141_71650||full | Low-quality |
| 18-110-final-viral-combined_k141_255186||full | Not-determined |
| 18-110-final-viral-combined_k141_163383||full | Not-determined |
| 18-110-final-viral-combined_k141_71654||full | Not-determined |
| 18-110-final-viral-combined_k141_59969||full | Low-quality |
| 18-110-final-viral-combined_k141_71656||full | Low-quality |
| 18-110-final-viral-combined_k141_117202||full | Low-quality |
| 18-110-final-viral-combined_k141_163399||full | Low-quality |
| 18-110-final-viral-combined_k141_175198||full | Low-quality |
| 18-110-final-viral-combined_k141_220732||full | Low-quality |
| 18-110-final-viral-combined_k141_71674||full | Not-determined |
| 18-110-final-viral-combined_k141_59977||full | Low-quality |
| 18-110-final-viral-combined_k141_117215||full | Low-quality |
| 18-110-final-viral-combined_k141_25514||full | Low-quality |
| 18-110-final-viral-combined_k141_186614||full | Not-determined |
| 18-110-final-viral-combined_k141_94469||full | Low-quality |
| 18-110-final-viral-combined_k141_186630||full | Low-quality |
| 18-110-final-viral-combined_k141_36540||full | Low-quality |
| 18-110-final-viral-combined_k141_25531||full | Low-quality |
| 18-110-final-viral-combined_k141_209603||full | Low-quality |
| 18-110-final-viral-combined_k141_36549||full | Not-determined |
| 18-110-final-viral-combined_k141_82752||full | Not-determined |
| 18-110-final-viral-combined_k141_209612||full | Not-determined |
| 18-110-final-viral-combined_k141_220744||full | Low-quality |
| 18-110-final-viral-combined_k141_163426||full | Low-quality |
| 18-110-final-viral-combined_k141_71686||full | Not-determined |
| 18-110-final-viral-combined_k141_25542||full | Low-quality |
| 18-110-final-viral-combined_k141_36556||full | Low-quality |
| 18-110-final-viral-combined_k141_82781||full | Not-determined |
| 18-110-final-viral-combined_k141_152116||full | Low-quality |
| 18-110-final-viral-combined_k141_209630||full | Not-determined |
| 18-110-final-viral-combined_k141_232796||full | Not-determined |
| 18-110-final-viral-combined_k141_209631||full | Not-determined |
| 18-110-final-viral-combined_k141_220747||full | Low-quality |
| 18-110-final-viral-combined_k141_232809||full | Not-determined |
| 18-110-final-viral-combined_k141_94508||full | Low-quality |
| 18-110-final-viral-combined_k141_244423||full | Low-quality |
| 18-110-final-viral-combined_k141_255214||full | Low-quality |
| 18-110-final-viral-combined_k141_255215||full | Not-determined |
| 18-110-final-viral-combined_k141_94524||full | Low-quality |
| 18-110-final-viral-combined_k141_25585||full | Low-quality |
| 18-110-final-viral-combined_k141_152132||full | Low-quality |
| 18-110-final-viral-combined_k141_1980||full | Not-determined |
| 18-110-final-viral-combined_k141_36583||full | Not-determined |
| 18-110-final-viral-combined_k141_71736||full | Not-determined |
| 18-110-final-viral-combined_k141_197865||full | Low-quality |
| 18-110-final-viral-combined_k141_163460||full | Low-quality |
| 18-110-final-viral-combined_k141_232843||full | Not-determined |
| 18-110-final-viral-combined_k141_244456||full | Low-quality |
| 18-110-final-viral-combined_k141_1996||full | Low-quality |
| 18-110-final-viral-combined_k141_186668||full | Low-quality |
| 18-110-final-viral-combined_k141_197875||full | Low-quality |
| 18-110-final-viral-combined_k141_60069||full | Low-quality |
| 18-110-final-viral-combined_k141_94534||full | Not-determined |
| 18-110-final-viral-combined_k141_60071||full | Not-determined |
| 18-110-final-viral-combined_k141_13061||full | Low-quality |
| 17-106-final-viral-combined_k141_27311||full | Medium-quality |
| 18-110-final-viral-combined_k141_244465||full | Not-determined |
| 18-110-final-viral-combined_k141_232863||full | Low-quality |
| 18-110-final-viral-combined_k141_220802||full | Not-determined |
| 18-110-final-viral-combined_k141_175286||full | Low-quality |
| 18-110-final-viral-combined_k141_152142||full | Not-determined |
| 18-110-final-viral-combined_k141_140792||full | Low-quality |
| 18-110-final-viral-combined_k141_244473||full | Low-quality |
| 18-110-final-viral-combined_k141_128913||full | Low-quality |
| 18-110-final-viral-combined_k141_175296||full | Not-determined |
| 18-110-final-viral-combined_k141_105807||full | Low-quality |
| 18-110-final-viral-combined_k141_140802||full | Not-determined |
| 18-110-final-viral-combined_k141_82852||full | Not-determined |
| 18-110-final-viral-combined_k141_163508||full | Low-quality |
| 18-110-final-viral-combined_k141_186689||full | Low-quality |
| 18-110-final-viral-combined_k141_232885||full | Not-determined |
| 18-110-final-viral-combined_k141_94566||full | Not-determined |
| 18-110-final-viral-combined_k141_197909||full | Low-quality |
| 18-110-final-viral-combined_k141_255279||full | Not-determined |
| 18-110-final-viral-combined_k141_209692||full | Low-quality |
| 18-110-final-viral-combined_k141_163521||full | Low-quality |
| 18-110-final-viral-combined_k141_117307||full | Not-determined |
| 18-110-final-viral-combined_k141_105838||full | Not-determined |
| 18-109-final-viral-combined_k141_123085||full | Medium-quality |
| 18-110-final-viral-combined_k141_60117||full | Low-quality |
| 18-110-final-viral-combined_k141_140822||full | Low-quality |
| 18-110-final-viral-combined_k141_255297||full | Low-quality |
| 18-110-final-viral-combined_k141_140825||full | Not-determined |
| 18-110-final-viral-combined_k141_209715||full | Not-determined |
| 18-110-final-viral-combined_k141_60129||full | Not-determined |
| 18-110-final-viral-combined_k141_94595||full | Low-quality |
| 18-110-final-viral-combined_k141_267086||full | Low-quality |
| 18-102_k141_248327 | Medium-quality |
| 18-110-final-viral-combined_k141_267090||full | Not-determined |
| 18-110-final-viral-combined_k141_117348||full | Not-determined |
| 18-110-final-viral-combined_k141_2053||full | Low-quality |
| 18-110-final-viral-combined_k141_220840||full | Not-determined |
| 18-110-final-viral-combined_k141_117365||full | Not-determined |
| 18-110-final-viral-combined_k141_36667||full | Not-determined |
| 18-110-final-viral-combined_k141_2076||full | Not-determined |
| 18-110-final-viral-combined_k141_220853||full | Not-determined |
| 18-102-final-viral-combined_k141_73327||full | High-quality |
| 18-110-final-viral-combined_k141_2098||full | Not-determined |
| 18-110-final-viral-combined_k141_140891||full | Low-quality |
| 18-110-final-viral-combined_k141_209744||full | Not-determined |
| 18-110-final-viral-combined_k141_71786||full | Low-quality |
| 18-110-final-viral-combined_k141_36715||full | Low-quality |
| 18-110-final-viral-combined_k141_71798||full | Low-quality |
| 18-110-final-viral-combined_k141_197985||full | Not-determined |
| 18-110-final-viral-combined_k141_209772||full | Low-quality |
| 18-110-final-viral-combined_k141_82897||full | Low-quality |
| 18-110-final-viral-combined_k141_186703||full | Not-determined |
| 18-110-final-viral-combined_k141_220890||full | Low-quality |
| 18-110-final-viral-combined_k141_94646||full | Low-quality |
| 18-110-final-viral-combined_k141_209789||full | Low-quality |
| 18-102-final-viral-combined_k141_248327||full | Medium-quality |
| 18-110-final-viral-combined_k141_209790||full | Low-quality |
| 18-110-final-viral-combined_k141_198030||full | Low-quality |
| 18-110-final-viral-combined_k141_244530||full | Not-determined |
| 18-110-final-viral-combined_k141_71845||full | Not-determined |
| 18-110-final-viral-combined_k141_82941||full | Low-quality |
| 18-110-final-viral-combined_k141_13154||full | Low-quality |
| 18-110-final-viral-combined_k141_175414||full | Not-determined |
| 18-110-final-viral-combined_k141_82959||full | Low-quality |
| 18-110-final-viral-combined_k141_13155||full | Not-determined |
| 18-110-final-viral-combined_k141_140929||full | Low-quality |
| 18-110-final-viral-combined_k141_140932||full | Not-determined |
| 18-110-final-viral-combined_k141_2166||full | Not-determined |
| 18-110-final-viral-combined_k141_267152||full | Low-quality |
| 18-110-final-viral-combined_k141_232953||full | Low-quality |
| 18-110-final-viral-combined_k141_220932||full | Low-quality |
| 18-110-final-viral-combined_k141_175441||full | Low-quality |
| 18-110-final-viral-combined_k141_128942||full | Low-quality |
| 18-110-final-viral-combined_k141_244536||full | Not-determined |
| 18-110-final-viral-combined_k141_128945||full | Low-quality |
| 18-110-final-viral-combined_k141_94669||full | Not-determined |
| 18-110-final-viral-combined_k141_140974||full | Low-quality |
| 18-110-final-viral-combined_k141_94670||full | Not-determined |
| 18-110-final-viral-combined_k141_2182||full | Low-quality |
| 18-110-final-viral-combined_k141_36792||full | Low-quality |
| 18-110-final-viral-combined_k141_2189||full | Low-quality |
| 18-110-final-viral-combined_k141_71889||full | Low-quality |
| 18-110-final-viral-combined_k141_244551||full | Low-quality |
| 18-110-final-viral-combined_k141_209841||full | Low-quality |
| 18-110-final-viral-combined_k141_48219||full | Not-determined |
| 18-110-final-viral-combined_k141_186797||full | Not-determined |
| 18-110-final-viral-combined_k141_175448||full | Low-quality |
| 18-110-final-viral-combined_k141_220971||full | Not-determined |
| 18-110-final-viral-combined_k141_128954||full | Low-quality |
| 18-110-final-viral-combined_k141_2214||full | Low-quality |
| 18-110-final-viral-combined_k141_244572||full | Not-determined |
| 18-110-final-viral-combined_k141_220991||full | Low-quality |
| 18-110-final-viral-combined_k141_94719||full | Not-determined |
| 18-110-final-viral-combined_k141_198138||full | Not-determined |
| 18-110-final-viral-combined_k141_25706||full | Not-determined |
| 18-110-final-viral-combined_k141_186819||full | Low-quality |
| 18-110-final-viral-combined_k141_152341||full | Low-quality |
| 18-110-final-viral-combined_k141_105856||full | Low-quality |
| 18-110-final-viral-combined_k141_255370||full | Low-quality |
| 18-110-final-viral-combined_k141_221006||full | Not-determined |
| 18-110-final-viral-combined_k141_105867||full | Not-determined |
| 18-110-final-viral-combined_k141_244610||full | Low-quality |
| 18-110-final-viral-combined_k141_244612||full | Not-determined |
| 18-110-final-viral-combined_k141_94749||full | Low-quality |
| 18-110-final-viral-combined_k141_267198||full | Low-quality |
| 18-110-final-viral-combined_k141_36877||full | Not-determined |
| 18-110-final-viral-combined_k141_105876||full | Not-determined |
| 18-110-final-viral-combined_k141_267208||full | Low-quality |
| 18-110-final-viral-combined_k141_209914||full | Low-quality |
| 18-110-final-viral-combined_k141_221016||full | Not-determined |
| 18-110-final-viral-combined_k141_244643||full | Low-quality |
| 18-110-final-viral-combined_k141_175516||full | Low-quality |
| 18-110-final-viral-combined_k141_152396||full | Low-quality |
| 18-110-final-viral-combined_k141_209925||full | Not-determined |
| 18-110-final-viral-combined_k141_198167||full | Low-quality |
| 18-110-final-viral-combined_k141_198171||full | Not-determined |
| 18-110-final-viral-combined_k141_255409||full | Low-quality |
| 18-110-final-viral-combined_k141_255410||full | Low-quality |
| 18-110-final-viral-combined_k141_186851||full | Not-determined |
| 18-110-final-viral-combined_k141_36912||full | Not-determined |
| 18-110-final-viral-combined_k141_141107||full | Low-quality |
| 18-110-final-viral-combined_k141_233020||full | Low-quality |
| 18-110-final-viral-combined_k141_71938||full | Not-determined |
| 18-110-final-viral-combined_k141_2267||full | Low-quality |
| 18-110-final-viral-combined_k141_48314||full | Not-determined |
| 18-110-final-viral-combined_k141_152425||full | Not-determined |
| 18-110-final-viral-combined_k141_233042||full | Low-quality |
| 18-110-final-viral-combined_k141_175565||full | Not-determined |
| 18-110-final-viral-combined_k141_105882||full | Low-quality |
| 18-110-final-viral-combined_k141_233058||full | Low-quality |
| 18-110-final-viral-combined_k141_163717||full | Not-determined |
| 18-110-final-viral-combined_k141_233064||full | Low-quality |
| 18-110-final-viral-combined_k141_163721||full | Low-quality |
| 18-110-final-viral-combined_k141_2290||full | Not-determined |
| 18-110-final-viral-combined_k141_94823||full | Not-determined |
| 18-110-final-viral-combined_k141_152452||full | Low-quality |
| 18-110-final-viral-combined_k141_175575||full | Low-quality |
| 18-110-final-viral-combined_k141_25814||full | Low-quality |
| 18-110-final-viral-combined_k141_244768||full | Not-determined |
| 18-110-final-viral-combined_k141_94849||full | Low-quality |
| 18-110-final-viral-combined_k141_36975||full | Low-quality |
| 18-110-final-viral-combined_k141_186939||full | Low-quality |
| 18-110-final-viral-combined_k141_129131||full | Not-determined |
| 18-110-final-viral-combined_k141_221097||full | Not-determined |
| 18-110-final-viral-combined_k141_175582||full | Low-quality |
| 18-110-final-viral-combined_k141_163766||full | Low-quality |
| 18-110-final-viral-combined_k141_210025||full | Low-quality |
| 18-110-final-viral-combined_k141_94868||full | Not-determined |
| 18-110-final-viral-combined_k141_233113||full | Not-determined |
| 18-110-final-viral-combined_k141_210029||full | Not-determined |
| 18-110-final-viral-combined_k141_48385||full | Not-determined |
| 18-110-final-viral-combined_k141_255495||full | Not-determined |
| 18-110-final-viral-combined_k141_129143||full | Not-determined |
| 18-110-final-viral-combined_k141_163769||full | Not-determined |
| 18-110-final-viral-combined_k141_186952||full | Not-determined |
| 18-110-final-viral-combined_k141_25843||full | Low-quality |
| 18-110-final-viral-combined_k141_163783||full | Not-determined |
| 18-110-final-viral-combined_k141_233126||full | Low-quality |
| 18-110-final-viral-combined_k141_13267||full | Low-quality |
| 18-110-final-viral-combined_k141_198255||full | Low-quality |
| 18-110-final-viral-combined_k141_94896||full | Not-determined |
| 18-110-final-viral-combined_k141_255510||full | Low-quality |
| 18-110-final-viral-combined_k141_210045||full | Low-quality |
| 18-110-final-viral-combined_k141_255518||full | Low-quality |
| 18-110-final-viral-combined_k141_233135||full | Not-determined |
| 18-110-final-viral-combined_k141_152507||full | Low-quality |
| 18-110-final-viral-combined_k141_175617||full | Not-determined |
| 18-110-final-viral-combined_k141_210048||full | Low-quality |
| 18-110-final-viral-combined_k141_163813||full | Not-determined |
| 18-110-final-viral-combined_k141_13282||full | Not-determined |
| 18-110-final-viral-combined_k141_175622||full | Low-quality |
| 18-110-final-viral-combined_k141_94913||full | Low-quality |
| 18-110-final-viral-combined_k141_25881||full | Low-quality |
| 18-110-final-viral-combined_k141_2366||full | Low-quality |
| 18-110-final-viral-combined_k141_244832||full | Not-determined |
| 18-110-final-viral-combined_k141_129183||full | Not-determined |
| 18-110-final-viral-combined_k141_244838||full | Not-determined |
| 18-110-final-viral-combined_k141_233151||full | Not-determined |
| 18-110-final-viral-combined_k141_255561||full | Low-quality |
| 18-110-final-viral-combined_k141_152549||full | Not-determined |
| 18-110-final-viral-combined_k141_105953||full | Not-determined |
| 18-110-final-viral-combined_k141_105954||full | Low-quality |
| 18-110-final-viral-combined_k141_94950||full | Low-quality |
| 18-110-final-viral-combined_k141_83022||full | Low-quality |
| 18-110-final-viral-combined_k141_141218||full | Not-determined |
| 18-110-final-viral-combined_k141_255582||full | Low-quality |
| 18-110-final-viral-combined_k141_141224||full | Not-determined |
| 18-110-final-viral-combined_k141_37056||full | Low-quality |
| 18-110-final-viral-combined_k141_94965||full | Low-quality |
| 18-110-final-viral-combined_k141_198327||full | Not-determined |
| 18-110-final-viral-combined_k141_152574||full | Low-quality |
| 18-110-final-viral-combined_k141_267314||full | Not-determined |
| 18-110-final-viral-combined_k141_83045||full | Low-quality |
| 18-110-final-viral-combined_k141_37068||full | Low-quality |
| 18-110-final-viral-combined_k141_267322||full | Not-determined |
| 18-110-final-viral-combined_k141_83053||full | Low-quality |
| 18-110-final-viral-combined_k141_267324||full | Low-quality |
| 18-110-final-viral-combined_k141_141241||full | Not-determined |
| 18-110-final-viral-combined_k141_187000||full | Low-quality |
| 18-110-final-viral-combined_k141_267339||full | Not-determined |
| 18-110-final-viral-combined_k141_255595||full | Low-quality |
| 18-110-final-viral-combined_k141_117646||full | Low-quality |
| 18-110-final-viral-combined_k141_48403||full | Not-determined |
| 18-110-final-viral-combined_k141_25954||full | Low-quality |
| 18-110-final-viral-combined_k141_83070||full | Low-quality |
| 18-110-final-viral-combined_k141_244898||full | Low-quality |
| 18-110-final-viral-combined_k141_267360||full | Not-determined |
| 18-110-final-viral-combined_k141_152619||full | Not-determined |
| 18-110-final-viral-combined_k141_187020||full | Not-determined |
| 18-110-final-viral-combined_k141_129264||full | Not-determined |
| 18-110-final-viral-combined_k141_72197||full | Not-determined |
| 18-110-final-viral-combined_k141_13381||full | Not-determined |
| 18-110-final-viral-combined_k141_37087||full | Not-determined |
| 18-110-final-viral-combined_k141_198372||full | Not-determined |
| 18-110-final-viral-combined_k141_129280||full | Low-quality |
| 18-110-final-viral-combined_k141_60286||full | Not-determined |
| 18-110-final-viral-combined_k141_175743||full | Low-quality |
| 18-110-final-viral-combined_k141_2443||full | Low-quality |
| 18-110-final-viral-combined_k141_244932||full | Not-determined |
| 18-110-final-viral-combined_k141_198381||full | Not-determined |
| 18-110-final-viral-combined_k141_37108||full | Low-quality |
| 18-110-final-viral-combined_k141_255638||full | Low-quality |
| 18-110-final-viral-combined_k141_72219||full | Not-determined |
| 18-110-final-viral-combined_k141_244948||full | Not-determined |
| 18-110-final-viral-combined_k141_129285||full | Not-determined |
| 18-110-final-viral-combined_k141_83115||full | Low-quality |
| 18-110-final-viral-combined_k141_117686||full | Low-quality |
| 18-110-final-viral-combined_k141_175764||full | Not-determined |
| 18-110-final-viral-combined_k141_129298||full | Not-determined |
| 18-110-final-viral-combined_k141_198397||full | Low-quality |
| 18-110-final-viral-combined_k141_233257||full | Not-determined |
| 18-110-final-viral-combined_k141_163885||full | Low-quality |
| 18-110-final-viral-combined_k141_175786||full | Not-determined |
| 18-110-final-viral-combined_k141_141331||full | Not-determined |
| 18-110-final-viral-combined_k141_13440||full | Low-quality |
| 18-110-final-viral-combined_k141_198412||full | Not-determined |
| 18-110-final-viral-combined_k141_175794||full | Low-quality |
| 18-110-final-viral-combined_k141_83129||full | Low-quality |
| 18-110-final-viral-combined_k141_60335||full | Low-quality |
| 18-110-final-viral-combined_k141_26019||full | Not-determined |
| 18-110-final-viral-combined_k141_13444||full | Low-quality |
| 18-110-final-viral-combined_k141_26024||full | Not-determined |
| 18-110-final-viral-combined_k141_244965||full | Low-quality |
| 18-110-final-viral-combined_k141_60347||full | Low-quality |
| 18-110-final-viral-combined_k141_72285||full | Low-quality |
| 18-110-final-viral-combined_k141_60351||full | Low-quality |
| 18-110-final-viral-combined_k141_60354||full | Not-determined |
| 18-110-final-viral-combined_k141_37164||full | Low-quality |
| 18-110-final-viral-combined_k141_48449||full | Not-determined |
| 18-110-final-viral-combined_k141_233315||full | Not-determined |
| 18-110-final-viral-combined_k141_244992||full | Not-determined |
| 18-110-final-viral-combined_k141_106052||full | Low-quality |
| 18-110-final-viral-combined_k141_152693||full | Not-determined |
| 18-110-final-viral-combined_k141_245000||full | Not-determined |
| 18-110-final-viral-combined_k141_2482||full | Low-quality |
| 18-110-final-viral-combined_k141_175865||full | Low-quality |
| 18-110-final-viral-combined_k141_141357||full | Not-determined |
| 18-110-final-viral-combined_k141_175867||full | Not-determined |
| 18-110-final-viral-combined_k141_175868||full | Not-determined |
| 18-110-final-viral-combined_k141_267476||full | Low-quality |
| 18-110-final-viral-combined_k141_152707||full | Low-quality |
| 18-110-final-viral-combined_k141_141364||full | Not-determined |
| 18-110-final-viral-combined_k141_37208||full | Not-determined |
| 18-110-final-viral-combined_k141_129368||full | Low-quality |
| 18-110-final-viral-combined_k141_37209||full | Low-quality |
| 18-110-final-viral-combined_k141_175877||full | Not-determined |
| 18-110-final-viral-combined_k141_221201||full | Not-determined |
| 18-110-final-viral-combined_k141_221204||full | Not-determined |
| 18-110-final-viral-combined_k141_129388||full | Not-determined |
| 18-110-final-viral-combined_k141_117697||full | Low-quality |
| 18-110-final-viral-combined_k141_255744||full | Not-determined |
| 18-110-final-viral-combined_k141_37221||full | Not-determined |
| 18-110-final-viral-combined_k141_210160||full | Not-determined |
| 18-110-final-viral-combined_k141_267504||full | Not-determined |
| 18-110-final-viral-combined_k141_106104||full | Not-determined |
| 18-110-final-viral-combined_k141_129400||full | Low-quality |
| 18-110-final-viral-combined_k141_233394||full | Not-determined |
| 18-110-final-viral-combined_k141_13520||full | Low-quality |
| 18-110-final-viral-combined_k141_13522||full | Not-determined |
| 18-110-final-viral-combined_k141_267515||full | Low-quality |
| 18-110-final-viral-combined_k141_13523||full | Low-quality |
| 18-110-final-viral-combined_k141_187123||full | Not-determined |
| 18-110-final-viral-combined_k141_106119||full | Not-determined |
| 18-110-final-viral-combined_k141_2537||full | Low-quality |
| 18-110-final-viral-combined_k141_2539||full | Low-quality |
| 18-110-final-viral-combined_k141_141434||full | Low-quality |
| 18-110-final-viral-combined_k141_233410||full | Low-quality |
| 18-110-final-viral-combined_k141_198423||full | Low-quality |
| 18-110-final-viral-combined_k141_117707||full | Not-determined |
| 18-110-final-viral-combined_k141_72364||full | Low-quality |
| 18-110-final-viral-combined_k141_117709||full | Not-determined |
| 18-110-final-viral-combined_k141_233416||full | Low-quality |
| 18-110-final-viral-combined_k141_152775||full | Not-determined |
| 18-110-final-viral-combined_k141_221225||full | Low-quality |
| 18-110-final-viral-combined_k141_13568||full | Not-determined |
| 18-110-final-viral-combined_k141_175974||full | Not-determined |
| 18-110-final-viral-combined_k141_72385||full | Low-quality |
| 18-110-final-viral-combined_k141_233432||full | Not-determined |
| 18-110-final-viral-combined_k141_210185||full | Low-quality |
| 18-110-final-viral-combined_k141_255774||full | Not-determined |
| 18-110-final-viral-combined_k141_267582||full | Not-determined |
| 18-110-final-viral-combined_k141_95200||full | Low-quality |
| 18-110-final-viral-combined_k141_175984||full | Not-determined |
| 18-110-final-viral-combined_k141_221263||full | Low-quality |
| 18-110-final-viral-combined_k141_106161||full | Not-determined |
| 18-110-final-viral-combined_k141_176008||full | Low-quality |
| 18-110-final-viral-combined_k141_267602||full | Not-determined |
| 18-110-final-viral-combined_k141_221271||full | Not-determined |
| 18-110-final-viral-combined_k141_2595||full | Low-quality |
| 18-110-final-viral-combined_k141_210200||full | Not-determined |
| 18-110-final-viral-combined_k141_176019||full | Not-determined |
| 18-110-final-viral-combined_k141_267605||full | Low-quality |
| 18-110-final-viral-combined_k141_245074||full | Low-quality |
| 18-110-final-viral-combined_k141_176026||full | Not-determined |
| 18-110-final-viral-combined_k141_95224||full | Not-determined |
| 18-110-final-viral-combined_k141_129488||full | Low-quality |
| 18-110-final-viral-combined_k141_129489||full | Low-quality |
| 18-110-final-viral-combined_k141_2610||full | Low-quality |
| 18-110-final-viral-combined_k141_152823||full | Low-quality |
| 18-110-final-viral-combined_k141_129494||full | Low-quality |
| 18-110-final-viral-combined_k141_13623||full | Low-quality |
| 18-110-final-viral-combined_k141_60514||full | Not-determined |
| 18-110-final-viral-combined_k141_72428||full | Not-determined |
| 18-110-final-viral-combined_k141_2632||full | Low-quality |
| 17-109_k141_336780 | Medium-quality |
| 18-110-final-viral-combined_k141_2633||full | Low-quality |
| 18-110-final-viral-combined_k141_245079||full | Low-quality |
| 18-110-final-viral-combined_k141_198490||full | Low-quality |
| 18-110-final-viral-combined_k141_255826||full | Low-quality |
| 18-110-final-viral-combined_k141_255827||full | Low-quality |
| 18-110-final-viral-combined_k141_129539||full | Low-quality |
| 18-110-final-viral-combined_k141_95267||full | Low-quality |
| 18-104-final-viral-combined_k141_24659||full | Medium-quality |
| 18-110-final-viral-combined_k141_2657||full | Low-quality |
| 18-110-final-viral-combined_k141_26123||full | Not-determined |
| 18-110-final-viral-combined_k141_233529||full | Not-determined |
| 18-110-final-viral-combined_k141_141535||full | Not-determined |
| 18-110-final-viral-combined_k141_152836||full | Low-quality |
| 18-110-final-viral-combined_k141_221310||full | Not-determined |
| 18-110-final-viral-combined_k141_152841||full | Low-quality |
| 18-110-final-viral-combined_k141_233549||full | Not-determined |
| 18-110-final-viral-combined_k141_152848||full | Not-determined |
| 18-110-final-viral-combined_k141_106202||full | Low-quality |
| 18-110-final-viral-combined_k141_95325||full | Low-quality |
| 18-110-final-viral-combined_k141_129565||full | Not-determined |
| 18-110-final-viral-combined_k141_267673||full | Not-determined |
| 18-110-final-viral-combined_k141_95334||full | Not-determined |
| 18-110-final-viral-combined_k141_129569||full | Low-quality |
| 18-110-final-viral-combined_k141_163927||full | Low-quality |
| 18-110-final-viral-combined_k141_233567||full | Low-quality |
| 18-110-final-viral-combined_k141_26172||full | Low-quality |
| 18-110-final-viral-combined_k141_163928||full | Not-determined |
| 18-110-final-viral-combined_k141_60591||full | Low-quality |
| 18-110-final-viral-combined_k141_95349||full | Not-determined |
| 18-110-final-viral-combined_k141_129571||full | Low-quality |
| 18-110-final-viral-combined_k141_141590||full | Not-determined |
| 18-110-final-viral-combined_k141_255909||full | Not-determined |
| 18-110-final-viral-combined_k141_26189||full | Not-determined |
| 18-110-final-viral-combined_k141_198557||full | Low-quality |
| 18-110-final-viral-combined_k141_255921||full | Low-quality |
| 18-110-final-viral-combined_k141_163973||full | Not-determined |
| 18-110-final-viral-combined_k141_117828||full | Low-quality |
| 18-110-final-viral-combined_k141_26204||full | Not-determined |
| 18-110-final-viral-combined_k141_267700||full | Low-quality |
| 18-110-final-viral-combined_k141_2702||full | Not-determined |
| 18-110-final-viral-combined_k141_129608||full | Low-quality |
| 18-110-final-viral-combined_k141_221376||full | Not-determined |
| 18-110-final-viral-combined_k141_37330||full | Not-determined |
| 18-110-final-viral-combined_k141_37334||full | Low-quality |
| 18-110-final-viral-combined_k141_255956||full | Not-determined |
| 18-110-final-viral-combined_k141_267727||full | Low-quality |
| 18-110-final-viral-combined_k141_37344||full | Low-quality |
| 18-110-final-viral-combined_k141_176166||full | Low-quality |
| 18-110-final-viral-combined_k141_129632||full | Not-determined |
| 18-110-final-viral-combined_k141_26234||full | Not-determined |
| 18-110-final-viral-combined_k141_267733||full | Not-determined |
| 18-110-final-viral-combined_k141_164020||full | Low-quality |
| 18-110-final-viral-combined_k141_37364||full | Low-quality |
| 18-110-final-viral-combined_k141_152908||full | Low-quality |
| 18-110-final-viral-combined_k141_95435||full | Not-determined |
| 18-110-final-viral-combined_k141_221420||full | Not-determined |
| 18-110-final-viral-combined_k141_95441||full | Low-quality |
| 18-110-final-viral-combined_k141_26268||full | Low-quality |
| 18-110-final-viral-combined_k141_95454||full | Not-determined |
| 18-110-final-viral-combined_k141_72527||full | Low-quality |
| 18-110-final-viral-combined_k141_233666||full | Low-quality |
| 18-110-final-viral-combined_k141_164065||full | Low-quality |
| 18-110-final-viral-combined_k141_48676||full | Low-quality |
| 18-110-final-viral-combined_k141_141687||full | Low-quality |
| 18-110-final-viral-combined_k141_210350||full | Low-quality |
| 18-110-final-viral-combined_k141_233681||full | Not-determined |
| 18-110-final-viral-combined_k141_48682||full | Not-determined |
| 18-110-final-viral-combined_k141_245145||full | Low-quality |
| 18-110-final-viral-combined_k141_233682||full | Low-quality |
| 18-110-final-viral-combined_k141_164078||full | Not-determined |
| 18-110-final-viral-combined_k141_129643||full | Not-determined |
| 18-110-final-viral-combined_k141_245153||full | Low-quality |
| 18-110-final-viral-combined_k141_48704||full | Low-quality |
| 18-110-final-viral-combined_k141_26307||full | Low-quality |
| 18-110-final-viral-combined_k141_37401||full | Low-quality |
| 18-110-final-viral-combined_k141_267803||full | Not-determined |
| 18-110-final-viral-combined_k141_245162||full | Not-determined |
| 18-110-final-viral-combined_k141_255987||full | Not-determined |
| 18-110-final-viral-combined_k141_210374||full | Not-determined |
| 18-110-final-viral-combined_k141_245180||full | Not-determined |
| 18-110-final-viral-combined_k141_245183||full | Low-quality |
| 18-110-final-viral-combined_k141_198641||full | Not-determined |
| 18-110-final-viral-combined_k141_164116||full | Not-determined |
| 18-110-final-viral-combined_k141_2771||full | Not-determined |
| 18-110-final-viral-combined_k141_129693||full | Low-quality |
| 18-110-final-viral-combined_k141_37431||full | Not-determined |
| 18-110-final-viral-combined_k141_245191||full | Not-determined |
| 18-110-final-viral-combined_k141_245197||full | Low-quality |
| 18-110-final-viral-combined_k141_48739||full | Not-determined |
| 18-110-final-viral-combined_k141_164125||full | Low-quality |
| 18-110-final-viral-combined_k141_256013||full | Not-determined |
| 18-110-final-viral-combined_k141_48747||full | Not-determined |
| 18-110-final-viral-combined_k141_198670||full | Low-quality |
| 18-110-final-viral-combined_k141_187326||full | Low-quality |
| 18-110-final-viral-combined_k141_117932||full | Not-determined |
| 18-110-final-viral-combined_k141_129714||full | Not-determined |
| 18-110-final-viral-combined_k141_233743||full | Low-quality |
| 18-110-final-viral-combined_k141_129719||full | Low-quality |
| 18-110-final-viral-combined_k141_256045||full | Low-quality |
| 18-110-final-viral-combined_k141_48764||full | Low-quality |
| 18-110-final-viral-combined_k141_187349||full | Not-determined |
| 18-110-final-viral-combined_k141_48770||full | Not-determined |
| 18-110-final-viral-combined_k141_221515||full | Low-quality |
| 18-110-final-viral-combined_k141_48771||full | Not-determined |
| 18-110-final-viral-combined_k141_267887||full | Low-quality |
| 18-110-final-viral-combined_k141_176357||full | Not-determined |
| 18-110-final-viral-combined_k141_187369||full | Low-quality |
| 18-110-final-viral-combined_k141_72676||full | Not-determined |
| 18-110-final-viral-combined_k141_95486||full | Not-determined |
| 18-110-final-viral-combined_k141_187402||full | Low-quality |
| 18-110-final-viral-combined_k141_72690||full | Low-quality |
| 18-110-final-viral-combined_k141_176371||full | Not-determined |
| 18-110-final-viral-combined_k141_95509||full | Not-determined |
| 18-110-final-viral-combined_k141_245278||full | Not-determined |
| 18-110-final-viral-combined_k141_141852||full | Low-quality |
| 18-110-final-viral-combined_k141_106250||full | Not-determined |
| 18-110-final-viral-combined_k141_37499||full | Not-determined |
| 18-110-final-viral-combined_k141_106254||full | Low-quality |
| 18-110-final-viral-combined_k141_13729||full | Low-quality |
| 18-110-final-viral-combined_k141_141859||full | Low-quality |
| 18-110-final-viral-combined_k141_118035||full | Low-quality |
| 18-110-final-viral-combined_k141_141866||full | Not-determined |
| 18-110-final-viral-combined_k141_245292||full | Low-quality |
| 18-110-final-viral-combined_k141_83167||full | Low-quality |
| 18-110-final-viral-combined_k141_60716||full | Low-quality |
| 18-110-final-viral-combined_k141_245293||full | Not-determined |
| 18-110-final-viral-combined_k141_187442||full | Not-determined |
| 18-110-final-viral-combined_k141_83169||full | Low-quality |
| 18-110-final-viral-combined_k141_129794||full | Not-determined |
| 18-110-final-viral-combined_k141_72722||full | Not-determined |
| 18-110-final-viral-combined_k141_2821||full | Low-quality |
| 18-110-final-viral-combined_k141_37524||full | Not-determined |
| 18-110-final-viral-combined_k141_256083||full | Low-quality |
| 18-110-final-viral-combined_k141_267947||full | Not-determined |
| 18-110-final-viral-combined_k141_141890||full | Low-quality |
| 18-110-final-viral-combined_k141_221596||full | Not-determined |
| 18-110-final-viral-combined_k141_2840||full | Low-quality |
| 18-110-final-viral-combined_k141_233810||full | Not-determined |
| 18-110-final-viral-combined_k141_83207||full | Low-quality |
| 18-110-final-viral-combined_k141_13752||full | Not-determined |
| 18-110-final-viral-combined_k141_221601||full | Not-determined |
| 18-110-final-viral-combined_k141_153016||full | Low-quality |
| 18-110-final-viral-combined_k141_95571||full | Low-quality |
| 18-110-final-viral-combined_k141_118063||full | Not-determined |
| 18-110-final-viral-combined_k141_141901||full | Not-determined |
| 18-110-final-viral-combined_k141_106280||full | Low-quality |
| 18-110-final-viral-combined_k141_129845||full | Not-determined |
| 18-110-final-viral-combined_k141_256103||full | Not-determined |
| 18-110-final-viral-combined_k141_233844||full | Not-determined |
| 18-110-final-viral-combined_k141_95595||full | Not-determined |
| 18-110-final-viral-combined_k141_106296||full | Low-quality |
| 18-110-final-viral-combined_k141_2873||full | Low-quality |
| 18-110-final-viral-combined_k141_83235||full | Not-determined |
| 18-110-final-viral-combined_k141_129857||full | Low-quality |
| 18-110-final-viral-combined_k141_256120||full | Not-determined |
| 18-110-final-viral-combined_k141_37603||full | Not-determined |
| 18-110-final-viral-combined_k141_48868||full | Not-determined |
| 18-110-final-viral-combined_k141_141905||full | Not-determined |

| | |
|---|---|
| 18-110-final-viral-combined_k141_233850\|\|full | Low-quality |
| 18-110-final-viral-combined_k141_129880\|\|full | Low-quality |
| 18-110-final-viral-combined_k141_141912\|\|full | Low-quality |
| 18-110-final-viral-combined_k141_72791\|\|full | Low-quality |
| 18-110-final-viral-combined_k141_267979\|\|full | Low-quality |
| 18-110-final-viral-combined_k141_48886\|\|full | Not-determined |
| 18-110-final-viral-combined_k141_256135\|\|full | Low-quality |
| 18-110-final-viral-combined_k141_37636\|\|full | Low-quality |
| 18-110-final-viral-combined_k141_26391\|\|full | Low-quality |
| 18-110-final-viral-combined_k141_245392\|\|full | Not-determined |
| 18-110-final-viral-combined_k141_129904\|\|full | Not-determined |
| 18-110-final-viral-combined_k141_245400\|\|full | Not-determined |
| 18-110-final-viral-combined_k141_37638\|\|full | Not-determined |
| 18-110-final-viral-combined_k141_153078\|\|full | Not-determined |
| 18-110-final-viral-combined_k141_268022\|\|full | Low-quality |
| 18-110-final-viral-combined_k141_129908\|\|full | Not-determined |
| 18-105_k141_212596 | Medium-quality |
| 18-110-final-viral-combined_k141_106361\|\|full | Not-determined |
| 18-110-final-viral-combined_k141_198797\|\|full | Low-quality |
| 18-110-final-viral-combined_k141_187539\|\|full | Low-quality |
| 18-110-final-viral-combined_k141_245402\|\|full | Low-quality |
| 18-110-final-viral-combined_k141_37654\|\|full | Not-determined |
| 18-110-final-viral-combined_k141_256156\|\|full | Low-quality |
| 18-110-final-viral-combined_k141_141978\|\|full | Not-determined |
| 18-110-final-viral-combined_k141_153089\|\|full | Not-determined |
| 18-110-final-viral-combined_k141_72840\|\|full | Not-determined |
| 18-110-final-viral-combined_k141_118118\|\|full | Not-determined |
| 18-110-final-viral-combined_k141_83316\|\|full | Not-determined |
| 18-110-final-viral-combined_k141_187560\|\|full | Low-quality |
| 18-110-final-viral-combined_k141_106380\|\|full | Low-quality |
| 18-110-final-viral-combined_k141_198803\|\|full | Not-determined |
| 18-110-final-viral-combined_k141_187565\|\|full | Not-determined |
| 18-110-final-viral-combined_k141_256176\|\|full | Low-quality |
| 18-110-final-viral-combined_k141_164316\|\|full | Low-quality |
| 18-110-final-viral-combined_k141_268054\|\|full | Not-determined |
| 18-110-final-viral-combined_k141_129957\|\|full | Low-quality |
| 18-110-final-viral-combined_k141_187569\|\|full | Low-quality |
| 18-110-final-viral-combined_k141_26436\|\|full | Not-determined |
| 18-110-final-viral-combined_k141_118136\|\|full | Not-determined |
| 18-110-final-viral-combined_k141_221690\|\|full | Not-determined |
| 18-110-final-viral-combined_k141_198820\|\|full | Low-quality |
| 18-110-final-viral-combined_k141_95615\|\|full | Not-determined |
| 18-110-final-viral-combined_k141_2975\|\|full | Not-determined |
| 18-110-final-viral-combined_k141_176504\|\|full | Not-determined |
| 18-110-final-viral-combined_k141_60888\|\|full | Low-quality |
| 18-110-final-viral-combined_k141_268090\|\|full | Not-determined |
| 18-110-final-viral-combined_k141_164347\|\|full | Not-determined |
| 18-110-final-viral-combined_k141_2996\|\|full | Not-determined |
| 18-110-final-viral-combined_k141_210542\|\|full | Not-determined |
| 18-110-final-viral-combined_k141_95628\|\|full | Not-determined |
| 18-110-final-viral-combined_k141_72882\|\|full | Low-quality |
| 18-110-final-viral-combined_k141_3002\|\|full | Low-quality |
| 18-105-final-viral-combined_k141_212596\|\|full | Medium-quality |
| 18-110-final-viral-combined_k141_268107\|\|full | Not-determined |
| 18-110-final-viral-combined_k141_106422\|\|full | Low-quality |
| 18-110-final-viral-combined_k141_95637\|\|full | Not-determined |
| 18-110-final-viral-combined_k141_72891\|\|full | Not-determined |
| 18-110-final-viral-combined_k141_187622\|\|full | Not-determined |
| 18-110-final-viral-combined_k141_95641\|\|full | Not-determined |
| 18-110-final-viral-combined_k141_72896\|\|full | Not-determined |
| 18-110-final-viral-combined_k141_245434\|\|full | Low-quality |
| 18-110-final-viral-combined_k141_118177\|\|full | Low-quality |
| 18-110-final-viral-combined_k141_83362\|\|full | Not-determined |
| 18-110-final-viral-combined_k141_3022\|\|full | Low-quality |
| 18-110-final-viral-combined_k141_37709\|\|full | Low-quality |
| 18-110-final-viral-combined_k141_256265\|\|full | Not-determined |
| 18-110-final-viral-combined_k141_118190\|\|full | Low-quality |
| 18-110-final-viral-combined_k141_48939\|\|full | Not-determined |
| 18-110-final-viral-combined_k141_268118\|\|full | Not-determined |
| 18-110-final-viral-combined_k141_268119\|\|full | Not-determined |
| 18-110-final-viral-combined_k141_13803\|\|full | Low-quality |
| 18-110-final-viral-combined_k141_3045\|\|full | Low-quality |
| 18-110-final-viral-combined_k141_13804\|\|full | Not-determined |
| 18-110-final-viral-combined_k141_95675\|\|full | Not-determined |
| 18-110-final-viral-combined_k141_210611\|\|full | Low-quality |
| 18-110-final-viral-combined_k141_210614\|\|full | Not-determined |
| 18-110-final-viral-combined_k141_48968\|\|full | Not-determined |
| 18-110-final-viral-combined_k141_13814\|\|full | Not-determined |
| 18-110-final-viral-combined_k141_118231\|\|full | Not-determined |
| 18-110-final-viral-combined_k141_106496\|\|full | Not-determined |
| 18-110-final-viral-combined_k141_26546\|\|full | Low-quality |
| 18-110-final-viral-combined_k141_153167\|\|full | Not-determined |
| 18-110-final-viral-combined_k141_187671\|\|full | Low-quality |
| 18-110-final-viral-combined_k141_3071\|\|full | Not-determined |
| 18-110-final-viral-combined_k141_245438\|\|full | Low-quality |
| 18-110-final-viral-combined_k141_106502\|\|full | Low-quality |
| 18-110-final-viral-combined_k141_26595\|\|full | Not-determined |
| 18-110-final-viral-combined_k141_153191\|\|full | Low-quality |
| 18-110-final-viral-combined_k141_245448\|\|full | Low-quality |
| 18-110-final-viral-combined_k141_118265\|\|full | Low-quality |
| 18-110-final-viral-combined_k141_176625\|\|full | Low-quality |
| 18-110-final-viral-combined_k141_153198\|\|full | Not-determined |
| 18-110-final-viral-combined_k141_48995\|\|full | Not-determined |
| 18-110-final-viral-combined_k141_106522\|\|full | Low-quality |
| 18-110-final-viral-combined_k141_153209\|\|full | Low-quality |
| 18-110-final-viral-combined_k141_3085\|\|full | Not-determined |
| 18-110-final-viral-combined_k141_106528\|\|full | Low-quality |
| 18-110-final-viral-combined_k141_49009\|\|full | Not-determined |
| 18-110-final-viral-combined_k141_233963\|\|full | Low-quality |
| 18-101-final-viral-combined_k141_120533\|\|full | High-quality |
| 18-110-final-viral-combined_k141_49016\|\|full | Not-determined |
| 18-110-final-viral-combined_k141_106540\|\|full | Low-quality |
| 18-110-final-viral-combined_k141_26650\|\|full | Low-quality |
| 18-110-final-viral-combined_k141_72927\|\|full | Not-determined |
| 18-110-final-viral-combined_k141_26653\|\|full | Low-quality |
| 18-110-final-viral-combined_k141_118325\|\|full | Low-quality |
| 18-110-final-viral-combined_k141_245472\|\|full | Low-quality |
| 18-110-final-viral-combined_k141_49027\|\|full | Not-determined |
| 18-110-final-viral-combined_k141_245480\|\|full | Low-quality |
| 18-110-final-viral-combined_k141_49032\|\|full | Low-quality |
| 18-110-final-viral-combined_k141_26665\|\|full | Low-quality |
| 18-110-final-viral-combined_k141_72942\|\|full | Not-determined |
| 18-110-final-viral-combined_k141_130098\|\|full | Low-quality |
| 18-110-final-viral-combined_k141_37817\|\|full | Not-determined |
| 18-110-final-viral-combined_k141_118353\|\|full | Low-quality |
| 18-110-final-viral-combined_k141_106563\|\|full | Not-determined |
| 18-110-final-viral-combined_k141_3113\|\|full | Low-quality |
| 18-110-final-viral-combined_k141_130107\|\|full | Low-quality |
| 18-110-final-viral-combined_k141_245504\|\|full | Not-determined |
| 18-110-final-viral-combined_k141_130116\|\|full | Not-determined |
| 18-110-final-viral-combined_k141_234012\|\|full | Low-quality |
| 18-110-final-viral-combined_k141_221840\|\|full | Not-determined |
| 18-110-final-viral-combined_k141_72977\|\|full | Not-determined |
| 18-110-final-viral-combined_k141_245524\|\|full | Not-determined |
| 18-110-final-viral-combined_k141_198976\|\|full | Low-quality |
| 18-110-final-viral-combined_k141_187774\|\|full | Low-quality |
| 18-110-final-viral-combined_k141_13906\|\|full | Not-determined |
| 18-110-final-viral-combined_k141_221848\|\|full | Not-determined |
| 18-110-final-viral-combined_k141_187775\|\|full | Low-quality |
| 18-110-final-viral-combined_k141_210674\|\|full | Not-determined |
| 18-110-final-viral-combined_k141_73026\|\|full | Low-quality |
| 18-110-final-viral-combined_k141_49107\|\|full | Not-determined |
| 18-110-final-viral-combined_k141_106634\|\|full | Not-determined |
| 18-110-final-viral-combined_k141_26720\|\|full | Not-determined |
| 18-110-final-viral-combined_k141_49115\|\|full | Low-quality |
| 18-110-final-viral-combined_k141_164489\|\|full | Low-quality |
| 18-110-final-viral-combined_k141_210709\|\|full | Low-quality |
| 18-110-final-viral-combined_k141_130206\|\|full | Not-determined |
| 18-110-final-viral-combined_k141_130212\|\|full | Not-determined |
| 18-110-final-viral-combined_k141_73058\|\|full | Low-quality |
| 18-110-final-viral-combined_k141_164515\|\|full | Not-determined |
| 18-110-final-viral-combined_k141_187829\|\|full | Low-quality |
| 18-110-final-viral-combined_k141_268268\|\|full | Low-quality |
| 18-110-final-viral-combined_k141_210720\|\|full | Not-determined |
| 18-110-final-viral-combined_k141_199013\|\|full | Not-determined |
| 18-110-final-viral-combined_k141_130259\|\|full | Not-determined |
| 18-110-final-viral-combined_k141_268325\|\|full | Not-determined |
| 18-110-final-viral-combined_k141_210736\|\|full | Low-quality |
| 18-110-final-viral-combined_k141_199028\|\|full | Low-quality |
| 18-110-final-viral-combined_k141_187861\|\|full | Not-determined |
| 18-110-final-viral-combined_k141_49187\|\|full | Low-quality |
| 18-110-final-viral-combined_k141_176817\|\|full | Not-determined |
| 18-110-final-viral-combined_k141_187902\|\|full | Low-quality |
| 18-110-final-viral-combined_k141_187903\|\|full | Not-determined |
| 18-110-final-viral-combined_k141_268368\|\|full | Low-quality |
| 18-110-final-viral-combined_k141_210796\|\|full | Not-determined |
| 18-110-final-viral-combined_k141_245647\|\|full | Not-determined |
| 18-110-final-viral-combined_k141_118437\|\|full | Not-determined |
| 18-110-final-viral-combined_k141_37970\|\|full | Not-determined |
| 18-110-final-viral-combined_k141_268379\|\|full | Low-quality |
| 18-110-final-viral-combined_k141_176854\|\|full | Low-quality |
| 18-110-final-viral-combined_k141_95805\|\|full | Not-determined |
| 18-110-final-viral-combined_k141_176857\|\|full | Low-quality |
| 18-110-final-viral-combined_k141_49211\|\|full | Not-determined |
| 18-110-final-viral-combined_k141_142201\|\|full | Low-quality |
| 18-110-final-viral-combined_k141_268400\|\|full | Not-determined |
| 18-110-final-viral-combined_k141_234181\|\|full | Not-determined |
| 18-110-final-viral-combined_k141_176868\|\|full | Not-determined |
| 18-110-final-viral-combined_k141_210830\|\|full | Not-determined |
| 18-110-final-viral-combined_k141_118462\|\|full | Not-determined |
| 18-110-final-viral-combined_k141_210832\|\|full | Not-determined |
| 18-110-final-viral-combined_k141_83546\|\|full | Not-determined |
| 18-110-final-viral-combined_k141_221999\|\|full | Low-quality |
| 18-110-final-viral-combined_k141_130357\|\|full | Not-determined |
| 18-110-final-viral-combined_k141_153405\|\|full | Low-quality |
| 18-110-final-viral-combined_k141_73160\|\|full | Not-determined |
| 18-110-final-viral-combined_k141_14035\|\|full | Low-quality |
| 18-110-final-viral-combined_k141_176896\|\|full | Not-determined |
| 18-110-final-viral-combined_k141_49248\|\|full | Not-determined |
| 18-110-final-viral-combined_k141_26793\|\|full | Low-quality |
| 18-110-final-viral-combined_k141_199113\|\|full | Low-quality |
| 18-110-final-viral-combined_k141_14053\|\|full | Not-determined |
| 18-110-final-viral-combined_k141_3252\|\|full | Low-quality |
| 18-110-final-viral-combined_k141_153417\|\|full | Not-determined |
| 18-110-final-viral-combined_k141_106774\|\|full | Not-determined |
| 18-110-final-viral-combined_k141_38007\|\|full | Low-quality |
| 18-110-final-viral-combined_k141_245718\|\|full | Low-quality |
| 18-110-final-viral-combined_k141_268446\|\|full | Low-quality |
| 18-110-final-viral-combined_k141_210858\|\|full | Not-determined |
| 18-110-final-viral-combined_k141_38011\|\|full | Not-determined |
| 18-110-final-viral-combined_k141_164626\|\|full | Low-quality |
| 18-110-final-viral-combined_k141_14065\|\|full | Not-determined |
| 18-110-final-viral-combined_k141_153431\|\|full | Low-quality |
| 18-110-final-viral-combined_k141_118528\|\|full | Not-determined |
| 18-110-final-viral-combined_k141_164630\|\|full | Low-quality |
| 18-110-final-viral-combined_k141_164633\|\|full | Low-quality |
| 18-110-final-viral-combined_k141_153446\|\|full | Not-determined |
| 18-110-final-viral-combined_k141_199135\|\|full | Low-quality |
| 18-110-final-viral-combined_k141_118538\|\|full | Not-determined |
| 18-110-final-viral-combined_k141_106790\|\|full | Not-determined |
| 18-110-final-viral-combined_k141_130411\|\|full | Low-quality |
| 18-110-final-viral-combined_k141_199148\|\|full | Low-quality |
| 18-110-final-viral-combined_k141_26879\|\|full | Not-determined |
| 18-110-final-viral-combined_k141_187987\|\|full | Low-quality |
| 18-110-final-viral-combined_k141_38067\|\|full | Low-quality |
| 18-110-final-viral-combined_k141_187998\|\|full | Not-determined |
| 18-110-final-viral-combined_k141_61112\|\|full | Not-determined |
| 18-110-final-viral-combined_k141_245753\|\|full | Low-quality |
| 18-110-final-viral-combined_k141_118581\|\|full | Not-determined |
| 18-110-final-viral-combined_k141_95935\|\|full | Low-quality |
| 18-110-final-viral-combined_k141_118586\|\|full | Not-determined |
| 18-110-final-viral-combined_k141_164670\|\|full | Low-quality |
| 18-110-final-viral-combined_k141_142328\|\|full | Low-quality |
| 18-110-final-viral-combined_k141_106856\|\|full | Low-quality |
| 18-110-final-viral-combined_k141_14080\|\|full | Low-quality |
| 18-110-final-viral-combined_k141_142329\|\|full | Low-quality |
| 18-110-final-viral-combined_k141_199165\|\|full | Low-quality |
| 18-110-final-viral-combined_k141_245777\|\|full | Not-determined |
| 18-110-final-viral-combined_k141_83631\|\|full | Low-quality |
| 18-110-final-viral-combined_k141_199183\|\|full | Low-quality |
| 18-110-final-viral-combined_k141_61163\|\|full | Low-quality |
| 18-110-final-viral-combined_k141_106898\|\|full | Low-quality |
| 18-110-final-viral-combined_k141_49349\|\|full | Low-quality |
| 18-110-final-viral-combined_k141_245792\|\|full | Low-quality |
| 18-110-final-viral-combined_k141_268501\|\|full | Not-determined |
| 18-110-final-viral-combined_k141_210884\|\|full | Not-determined |
| 18-110-final-viral-combined_k141_14145\|\|full | Low-quality |
| 18-110-final-viral-combined_k141_153564\|\|full | Not-determined |
| 18-110-final-viral-combined_k141_234307\|\|full | Not-determined |
| 18-110-final-viral-combined_k141_3370\|\|full | Low-quality |
| 18-110-final-viral-combined_k141_245820\|\|full | Not-determined |
| 18-110-final-viral-combined_k141_222152\|\|full | Low-quality |
| 18-110-final-viral-combined_k141_177061\|\|full | Low-quality |
| 18-110-final-viral-combined_k141_245828\|\|full | Low-quality |
| 18-110-final-viral-combined_k141_268542\|\|full | Low-quality |
| 18-110-final-viral-combined_k141_95988\|\|full | Not-determined |
| 18-110-final-viral-combined_k141_26998\|\|full | Not-determined |
| 18-110-final-viral-combined_k141_142404\|\|full | Low-quality |
| 18-110-final-viral-combined_k141_118672\|\|full | Low-quality |
| 18-110-final-viral-combined_k141_83694\|\|full | Not-determined |
| 18-110-final-viral-combined_k141_118688\|\|full | Not-determined |
| 18-110-final-viral-combined_k141_234370\|\|full | Low-quality |
| 18-110-final-viral-combined_k141_234371\|\|full | Low-quality |
| 18-110-final-viral-combined_k141_130462\|\|full | Low-quality |
| 18-110-final-viral-combined_k141_73280\|\|full | Low-quality |
| 18-110-final-viral-combined_k141_38151\|\|full | Low-quality |
| 18-110-final-viral-combined_k141_61239\|\|full | Low-quality |
| 18-110-final-viral-combined_k141_268594\|\|full | Low-quality |
| 18-110-final-viral-combined_k141_164765\|\|full | Not-determined |
| 18-110-final-viral-combined_k141_268601\|\|full | Not-determined |
| 18-110-final-viral-combined_k141_210978\|\|full | Low-quality |
| 18-110-final-viral-combined_k141_106998\|\|full | Low-quality |
| 18-110-final-viral-combined_k141_164769\|\|full | Not-determined |
| 18-110-final-viral-combined_k141_118713\|\|full | Low-quality |
| 18-110-final-viral-combined_k141_245918\|\|full | Not-determined |
| 18-102-final-viral-combined_k141_51446\|\|full | Medium-quality |
| 18-110-final-viral-combined_k141_61281\|\|full | Not-determined |
| 18-110-final-viral-combined_k141_234391\|\|full | Not-determined |
| 18-110-final-viral-combined_k141_210990\|\|full | Low-quality |
| 18-110-final-viral-combined_k141_96055\|\|full | Not-determined |
| 18-110-final-viral-combined_k141_210993\|\|full | Not-determined |
| 18-110-final-viral-combined_k141_164821\|\|full | Not-determined |
| 18-110-final-viral-combined_k141_83756\|\|full | Not-determined |
| 18-110-final-viral-combined_k141_130546\|\|full | Low-quality |
| 18-110-final-viral-combined_k141_177132\|\|full | Not-determined |
| 18-110-final-viral-combined_k141_83765\|\|full | Low-quality |
| 18-110-final-viral-combined_k141_245972\|\|full | Low-quality |
| 18-110-final-viral-combined_k141_164834\|\|full | Low-quality |
| 18-110-final-viral-combined_k141_61302\|\|full | Not-determined |
| 18-110-final-viral-combined_k141_164837\|\|full | Low-quality |
| 18-110-final-viral-combined_k141_164839\|\|full | Not-determined |
| 18-110-final-viral-combined_k141_27115\|\|full | Not-determined |
| 18-110-final-viral-combined_k141_142497\|\|full | Not-determined |
| 18-110-final-viral-combined_k141_14163\|\|full | Low-quality |
| 18-110-final-viral-combined_k141_142513\|\|full | Low-quality |
| 18-104-final-viral-combined_k141_1246\|\|full | Medium-quality |
| 18-110-final-viral-combined_k141_27131\|\|full | Not-determined |
| 18-110-final-viral-combined_k141_245992\|\|full | Low-quality |
| 18-110-final-viral-combined_k141_38250\|\|full | Low-quality |
| 18-110-final-viral-combined_k141_118790\|\|full | Not-determined |
| 18-110-final-viral-combined_k141_245999\|\|full | Low-quality |
| 18-110-final-viral-combined_k141_177173\|\|full | Low-quality |
| 18-110-final-viral-combined_k141_234448\|\|full | Not-determined |
| 18-110-final-viral-combined_k141_234451\|\|full | Not-determined |
| 18-110-final-viral-combined_k141_153683\|\|full | Low-quality |
| 18-110-final-viral-combined_k141_268681\|\|full | Not-determined |
| 18-110-final-viral-combined_k141_246007\|\|full | Not-determined |
| 18-110-final-viral-combined_k141_61328\|\|full | Not-determined |
| 18-110-final-viral-combined_k141_107078\|\|full | Low-quality |
| 18-110-final-viral-combined_k141_222264\|\|full | Not-determined |
| 18-110-final-viral-combined_k141_164893\|\|full | Not-determined |
| 18-110-final-viral-combined_k141_256401\|\|full | Not-determined |
| 18-110-final-viral-combined_k141_246017\|\|full | Low-quality |
| 18-110-final-viral-combined_k141_61341\|\|full | Not-determined |
| 18-110-final-viral-combined_k141_211075\|\|full | Low-quality |
| 18-110-final-viral-combined_k141_61342\|\|full | Not-determined |
| 18-110-final-viral-combined_k141_118818\|\|full | Low-quality |
| 18-110-final-viral-combined_k141_107094\|\|full | Not-determined |
| KM-final-viral-combined_k141_126600\|\|full | High-quality |
| 18-110-final-viral-combined_k141_256407\|\|full | Not-determined |
| 18-110-final-viral-combined_k141_61351\|\|full | Not-determined |
| 18-110-final-viral-combined_k141_3480\|\|full | Low-quality |
| 18-110-final-viral-combined_k141_3482\|\|full | Low-quality |
| 18-110-final-viral-combined_k141_14211\|\|full | Low-quality |
| 18-110-final-viral-combined_k141_27189\|\|full | Not-determined |
| 18-110-final-viral-combined_k141_38289\|\|full | Low-quality |
| 18-110-final-viral-combined_k141_3483\|\|full | Not-determined |
| 18-110-final-viral-combined_k141_234507\|\|full | Not-determined |
| 18-110-final-viral-combined_k141_130623\|\|full | Not-determined |
| 18-110-final-viral-combined_k141_153730\|\|full | Not-determined |
| 18-110-final-viral-combined_k141_199267\|\|full | Not-determined |
| 18-110-final-viral-combined_k141_153736\|\|full | Low-quality |
| 18-110-final-viral-combined_k141_27195\|\|full | Low-quality |
| 18-110-final-viral-combined_k141_188165\|\|full | Low-quality |
| 18-110-final-viral-combined_k141_3511\|\|full | Not-determined |
| 18-110-final-viral-combined_k141_130631\|\|full | Low-quality |
| 18-110-final-viral-combined_k141_234548\|\|full | Not-determined |
| 18-110-final-viral-combined_k141_199279\|\|full | Low-quality |
| 18-110-final-viral-combined_k141_177250\|\|full | Low-quality |
| 18-110-final-viral-combined_k141_83825\|\|full | Low-quality |
| 18-110-final-viral-combined_k141_234549\|\|full | Low-quality |
| 18-110-final-viral-combined_k141_177251\|\|full | Not-determined |
| 18-102_k141_110761 | Medium-quality |
| 18-110-final-viral-combined_k141_107147\|\|full | Low-quality |
| 18-110-final-viral-combined_k141_3527\|\|full | Low-quality |
| 18-110-final-viral-combined_k141_142597\|\|full | Not-determined |
| 18-110-final-viral-combined_k141_96146\|\|full | Low-quality |
| 18-110-final-viral-combined_k141_130665\|\|full | Not-determined |
| 18-110-final-viral-combined_k141_177285\|\|full | Not-determined |
| 18-110-final-viral-combined_k141_222362\|\|full | Not-determined |
| 18-110-final-viral-combined_k141_14279\|\|full | Low-quality |
| 18-110-final-viral-combined_k141_234560\|\|full | Low-quality |
| 18-110-final-viral-combined_k141_73378\|\|full | Not-determined |
| 18-110-final-viral-combined_k141_211140\|\|full | Low-quality |
| 18-110-final-viral-combined_k141_177289\|\|full | Low-quality |
| 18-110-final-viral-combined_k141_234581\|\|full | Not-determined |
| 18-110-final-viral-combined_k141_177294\|\|full | Not-determined |
| 18-110-final-viral-combined_k141_38328\|\|full | Low-quality |
| 18-110-final-viral-combined_k141_199332\|\|full | Not-determined |
| 18-110-final-viral-combined_k141_83868\|\|full | Low-quality |
| 18-110-final-viral-combined_k141_177309\|\|full | Not-determined |
| 18-110-final-viral-combined_k141_234588\|\|full | Low-quality |
| 18-110-final-viral-combined_k141_107216\|\|full | Not-determined |
| 18-110-final-viral-combined_k141_188266\|\|full | Low-quality |
| 18-110-final-viral-combined_k141_3562\|\|full | Low-quality |
| 18-110-final-viral-combined_k141_234606\|\|full | Low-quality |
| 18-110-final-viral-combined_k141_61418\|\|full | Low-quality |
| 18-110-final-viral-combined_k141_177347\|\|full | Not-determined |
| 18-110-final-viral-combined_k141_107235\|\|full | Not-determined |
| 18-110-final-viral-combined_k141_96197\|\|full | Not-determined |
| 18-110-final-viral-combined_k141_153767\|\|full | Not-determined |
| 18-110-final-viral-combined_k141_177367\|\|full | Not-determined |
| 18-110-final-viral-combined_k141_234624\|\|full | Low-quality |
| 18-110-final-viral-combined_k141_83929\|\|full | Not-determined |
| 18-110-final-viral-combined_k141_38359\|\|full | Not-determined |
| 18-110-final-viral-combined_k141_268766\|\|full | Not-determined |
| 18-110-final-viral-combined_k141_61445\|\|full | Not-determined |
| 18-110-final-viral-combined_k141_96216\|\|full | Low-quality |
| 18-110-final-viral-combined_k141_165078\|\|full | Low-quality |
| 18-110-final-viral-combined_k141_199396\|\|full | Not-determined |
| 18-110-final-viral-combined_k141_83957\|\|full | Low-quality |
| 18-110-final-viral-combined_k141_38370\|\|full | Not-determined |
| 18-110-final-viral-combined_k141_256488\|\|full | Low-quality |
| 18-110-final-viral-combined_k141_211255\|\|full | Not-determined |
| 18-110-final-viral-combined_k141_96267\|\|full | Low-quality |
| 18-110-final-viral-combined_k141_268795\|\|full | Low-quality |
| 18-110-final-viral-combined_k141_119005\|\|full | Low-quality |
| 18-110-final-viral-combined_k141_61493\|\|full | Not-determined |
| 18-110-final-viral-combined_k141_49659\|\|full | Not-determined |
| 18-110-final-viral-combined_k141_177418\|\|full | Low-quality |
| 18-110-final-viral-combined_k141_61496\|\|full | Not-determined |
| 18-110-final-viral-combined_k141_83998\|\|full | Not-determined |
| 18-110-final-viral-combined_k141_49663\|\|full | Low-quality |
| 18-110-final-viral-combined_k141_256495\|\|full | Not-determined |
| 18-110-final-viral-combined_k141_222526\|\|full | Low-quality |
| 18-110-final-viral-combined_k141_107278\|\|full | Not-determined |
| 18-110-final-viral-combined_k141_234686\|\|full | Low-quality |
| 18-110-final-viral-combined_k141_61519\|\|full | Low-quality |
| 18-110-final-viral-combined_k141_165130\|\|full | Not-determined |
| 18-110-final-viral-combined_k141_165138\|\|full | Not-determined |
| 18-110-final-viral-combined_k141_107304\|\|full | Not-determined |
| 18-110-final-viral-combined_k141_211287\|\|full | Low-quality |
| 18-110-final-viral-combined_k141_84009\|\|full | Not-determined |
| 18-110-final-viral-combined_k141_165148\|\|full | Low-quality |
| 18-110-final-viral-combined_k141_211297\|\|full | Low-quality |
| 18-110-final-viral-combined_k141_199477\|\|full | Low-quality |
| 18-110-final-viral-combined_k141_3645\|\|full | Not-determined |
| 18-110-final-viral-combined_k141_96360\|\|full | Low-quality |
| 18-110-final-viral-combined_k141_234754\|\|full | Not-determined |
| 18-110-final-viral-combined_k141_222602\|\|full | Low-quality |
| 18-110-final-viral-combined_k141_246152\|\|full | Not-determined |
| 18-110-final-viral-combined_k141_38501\|\|full | Not-determined |
| 18-110-final-viral-combined_k141_61546\|\|full | Low-quality |
| 18-110-final-viral-combined_k141_61547\|\|full | Low-quality |
| 18-110-final-viral-combined_k141_3653\|\|full | Not-determined |
| 18-110-final-viral-combined_k141_61555\|\|full | Low-quality |
| 18-110-final-viral-combined_k141_107326\|\|full | Low-quality |
| 18-110-final-viral-combined_k141_3659\|\|full | Not-determined |
| 18-110-final-viral-combined_k141_153864\|\|full | Not-determined |
| 18-110-final-viral-combined_k141_84046\|\|full | Not-determined |
| 18-110-final-viral-combined_k141_61561\|\|full | Low-quality |
| 18-110-final-viral-combined_k141_107332\|\|full | Low-quality |
| 18-110-final-viral-combined_k141_256576\|\|full | Not-determined |
| 18-110-final-viral-combined_k141_268887\|\|full | Not-determined |
| 18-110-final-viral-combined_k141_142696\|\|full | Low-quality |
| 18-110-final-viral-combined_k141_142697\|\|full | Low-quality |
| 18-110-final-viral-combined_k141_61567\|\|full | Not-determined |
| 18-110-final-viral-combined_k141_268902\|\|full | Not-determined |
| 18-110-final-viral-combined_k141_49724\|\|full | Low-quality |
| 18-110-final-viral-combined_k141_3670\|\|full | Low-quality |
| 18-110-final-viral-combined_k141_73539\|\|full | Not-determined |
| 18-110-final-viral-combined_k141_49726\|\|full | Not-determined |
| 18-110-final-viral-combined_k141_38539\|\|full | Low-quality |
| 18-110-final-viral-combined_k141_73549\|\|full | Not-determined |
| 18-110-final-viral-combined_k141_130819\|\|full | Not-determined |
| 18-110-final-viral-combined_k141_84081\|\|full | Not-determined |
| 18-110-final-viral-combined_k141_49731\|\|full | Low-quality |
| 18-110-final-viral-combined_k141_153892\|\|full | Not-determined |
| 18-110-final-viral-combined_k141_256596\|\|full | Not-determined |
| 18-110-final-viral-combined_k141_211342\|\|full | Low-quality |
| 18-110-final-viral-combined_k141_107363\|\|full | Low-quality |
| 18-110-final-viral-combined_k141_177505\|\|full | Not-determined |
| 18-110-final-viral-combined_k141_177507\|\|full | Not-determined |
| 18-110-final-viral-combined_k141_268949\|\|full | Not-determined |
| 18-110-final-viral-combined_k141_177518\|\|full | Low-quality |
| 18-110-final-viral-combined_k141_130847\|\|full | Not-determined |
| 18-110-final-viral-combined_k141_107374\|\|full | Low-quality |
| 18-110-final-viral-combined_k141_49782\|\|full | Low-quality |
| 18-110-final-viral-combined_k141_107375\|\|full | Low-quality |
| 18-110-final-viral-combined_k141_153912\|\|full | Not-determined |
| 18-110-final-viral-combined_k141_14364\|\|full | Low-quality |
| 18-110-final-viral-combined_k141_165265\|\|full | Low-quality |
| 18-110-final-viral-combined_k141_38572\|\|full | Not-determined |
| 18-110-final-viral-combined_k141_153922\|\|full | Low-quality |
| 18-110-final-viral-combined_k141_119094\|\|full | Not-determined |
| 18-110-final-viral-combined_k141_177526\|\|full | Not-determined |
| 18-110-final-viral-combined_k141_268958\|\|full | Not-determined |
| 18-110-final-viral-combined_k141_130865\|\|full | Low-quality |
| 18-110-final-viral-combined_k141_27461\|\|full | Low-quality |
| 18-110-final-viral-combined_k141_153930\|\|full | Not-determined |
| 18-110-final-viral-combined_k141_130872\|\|full | Not-determined |
| 18-110-final-viral-combined_k141_38592\|\|full | Low-quality |
| 18-110-final-viral-combined_k141_73579\|\|full | Not-determined |
| 18-110-final-viral-combined_k141_268980\|\|full | Not-determined |
| 18-110-final-viral-combined_k141_142745\|\|full | Not-determined |
| 18-110-final-viral-combined_k141_49824\|\|full | Not-determined |
| 18-110-final-viral-combined_k141_73584\|\|full | Low-quality |
| 18-110-final-viral-combined_k141_107424\|\|full | Not-determined |
| 18-110-final-viral-combined_k141_38602\|\|full | Not-determined |
| 18-110-final-viral-combined_k141_188518\|\|full | Low-quality |
| 18-110-final-viral-combined_k141_222710\|\|full | Not-determined |
| 18-110-final-viral-combined_k141_153962\|\|full | Low-quality |
| 18-110-final-viral-combined_k141_61694\|\|full | Not-determined |
| 18-110-final-viral-combined_k141_14433\|\|full | Not-determined |
| 18-110-final-viral-combined_k141_38626\|\|full | Low-quality |
| 18-110-final-viral-combined_k141_107451\|\|full | Not-determined |
| 18-110-final-viral-combined_k141_177548\|\|full | Low-quality |
| 18-110-final-viral-combined_k141_3746\|\|full | Low-quality |
| 18-110-final-viral-combined_k141_130915\|\|full | Low-quality |
| 18-110-final-viral-combined_k141_256653\|\|full | Not-determined |
| 18-110-final-viral-combined_k141_107453\|\|full | Not-determined |
| 18-110-final-viral-combined_k141_61699\|\|full | Not-determined |
| 18-110-final-viral-combined_k141_246208\|\|full | Low-quality |
| 18-110-final-viral-combined_k141_211407\|\|full | Low-quality |
| 18-110-final-viral-combined_k141_234836\|\|full | Low-quality |
| 18-110-final-viral-combined_k141_130932\|\|full | Low-quality |
| 18-110-final-viral-combined_k141_269059\|\|full | Low-quality |
| 18-110-final-viral-combined_k141_84207\|\|full | Not-determined |
| 18-110-final-viral-combined_k141_38688\|\|full | Not-determined |
| 18-110-final-viral-combined_k141_61735\|\|full | Not-determined |
| 18-110-final-viral-combined_k141_107489\|\|full | Not-determined |
| 18-110-final-viral-combined_k141_188566\|\|full | Not-determined |
| 18-110-final-viral-combined_k141_96480\|\|full | Not-determined |
| 18-110-final-viral-combined_k141_107498\|\|full | Not-determined |
| 18-110-final-viral-combined_k141_84222\|\|full | Not-determined |
| 18-110-final-viral-combined_k141_107501\|\|full | Low-quality |
| 18-110-final-viral-combined_k141_269082\|\|full | Not-determined |
| 18-110-final-viral-combined_k141_188585\|\|full | Low-quality |
| 18-110-final-viral-combined_k141_165391\|\|full | Low-quality |
| 18-110-final-viral-combined_k141_211451\|\|full | Not-determined |
| 18-110-final-viral-combined_k141_96488\|\|full | Low-quality |
| 18-110-final-viral-combined_k141_96491\|\|full | Low-quality |
| 18-110-final-viral-combined_k141_234873\|\|full | Not-determined |
| 18-110-final-viral-combined_k141_96493\|\|full | Not-determined |
| 18-110-final-viral-combined_k141_246288\|\|full | Not-determined |
| 18-110-final-viral-combined_k141_49910\|\|full | Low-quality |
| 18-110-final-viral-combined_k141_234885\|\|full | Not-determined |
| 18-110-final-viral-combined_k141_199584\|\|full | Not-determined |
| 18-110-final-viral-combined_k141_130977\|\|full | Low-quality |
| 18-110-final-viral-combined_k141_269106\|\|full | Low-quality |
| 18-110-final-viral-combined_k141_38720\|\|full | Low-quality |
| 18-110-final-viral-combined_k141_154065\|\|full | Not-determined |
| 18-110-final-viral-combined_k141_222829\|\|full | Low-quality |
| 18-110-final-viral-combined_k141_234904\|\|full | Low-quality |
| 18-110-final-viral-combined_k141_154087\|\|full | Not-determined |
| 18-110-final-viral-combined_k141_84247\|\|full | Not-determined |
| 18-110-final-viral-combined_k141_49948\|\|full | Low-quality |
| 18-110-final-viral-combined_k141_154097\|\|full | Not-determined |
| 18-110-final-viral-combined_k141_222840\|\|full | Not-determined |
| 18-110-final-viral-combined_k141_84260\|\|full | Not-determined |
| 18-110-final-viral-combined_k141_177641\|\|full | Not-determined |
| 18-110-final-viral-combined_k141_222848\|\|full | Not-determined |
| 18-110-final-viral-combined_k141_269147\|\|full | Low-quality |
| 18-110-final-viral-combined_k141_38742\|\|full | Not-determined |
| 18-110-final-viral-combined_k141_199623\|\|full | Not-determined |
| 18-110-final-viral-combined_k141_3850\|\|full | Low-quality |
| 18-110-final-viral-combined_k141_49979\|\|full | Low-quality |
| 18-110-final-viral-combined_k141_177668\|\|full | Not-determined |
| 18-110-final-viral-combined_k141_119217\|\|full | Not-determined |
| 18-110-final-viral-combined_k141_177673\|\|full | Not-determined |
| 18-110-final-viral-combined_k141_3871\|\|full | Low-quality |
| 18-110-final-viral-combined_k141_154172\|\|full | Not-determined |
| 18-110-final-viral-combined_k141_96590\|\|full | Not-determined |
| 18-110-final-viral-combined_k141_61822\|\|full | Low-quality |
| 18-110-final-viral-combined_k141_222927\|\|full | Low-quality |
| 18-110-final-viral-combined_k141_27614\|\|full | Not-determined |
| 18-110-final-viral-combined_k141_14592\|\|full | Not-determined |
| 18-110-final-viral-combined_k141_177710\|\|full | Low-quality |
| 18-110-final-viral-combined_k141_27627\|\|full | Not-determined |
| 18-110-final-viral-combined_k141_246316\|\|full | Low-quality |
| 18-110-final-viral-combined_k141_96614\|\|full | Low-quality |
| 18-110-final-viral-combined_k141_256735\|\|full | Low-quality |
| 18-110-final-viral-combined_k141_107537\|\|full | Not-determined |
| 18-110-final-viral-combined_k141_84345\|\|full | Low-quality |
| 18-110-final-viral-combined_k141_84353\|\|full | Low-quality |
| 18-110-final-viral-combined_k141_142916\|\|full | Not-determined |
| 18-110-final-viral-combined_k141_131104\|\|full | Not-determined |
| 18-110-final-viral-combined_k141_14642\|\|full | Low-quality |
| 18-110-final-viral-combined_k141_177766\|\|full | Not-determined |
| 18-110-final-viral-combined_k141_73652\|\|full | Low-quality |
| 18-110-final-viral-combined_k141_119304\|\|full | Not-determined |
| 18-110-final-viral-combined_k141_107581\|\|full | Not-determined |
| 18-110-final-viral-combined_k141_165482\|\|full | Low-quality |
| 18-110-final-viral-combined_k141_177778\|\|full | Low-quality |
| 18-110-final-viral-combined_k141_61916\|\|full | Low-quality |
| 18-110-final-viral-combined_k141_107590\|\|full | Not-determined |
| 18-110-final-viral-combined_k141_256786\|\|full | Not-determined |
| 18-110-final-viral-combined_k141_235003\|\|full | Low-quality |
| 18-110-final-viral-combined_k141_188760\|\|full | Not-determined |
| 18-110-final-viral-combined_k141_3942\|\|full | Low-quality |
| 18-110-final-viral-combined_k141_107601\|\|full | Not-determined |
| 18-110-final-viral-combined_k141_61922\|\|full | Not-determined |
| 18-110-final-viral-combined_k141_211647\|\|full | Low-quality |
| 18-110-final-viral-combined_k141_3950\|\|full | Low-quality |
| 18-110-final-viral-combined_k141_38886\|\|full | Not-determined |
| 18-110-final-viral-combined_k141_73672\|\|full | Low-quality |
| 18-110-final-viral-combined_k141_188790\|\|full | Not-determined |
| 18-110-final-viral-combined_k141_235045\|\|full | Not-determined |
| 18-110-final-viral-combined_k141_235053\|\|full | Low-quality |
| 18-110-final-viral-combined_k141_73700\|\|full | Not-determined |
| 18-110-final-viral-combined_k141_61982\|\|full | Not-determined |
| 18-110-final-viral-combined_k141_256829\|\|full | Low-quality |
| 18-110-final-viral-combined_k141_256831\|\|full | Low-quality |
| 18-110-final-viral-combined_k141_38918\|\|full | Low-quality |
| 18-110-final-viral-combined_k141_119317\|\|full | Not-determined |
| 18-110-final-viral-combined_k141_3974\|\|full | Low-quality |
| 18-110-final-viral-combined_k141_96652\|\|full | Not-determined |
| 18-110-final-viral-combined_k141_256860\|\|full | Not-determined |
| 18-110-final-viral-combined_k141_246353\|\|full | Not-determined |
| 18-110-final-viral-combined_k141_50020\|\|full | Not-determined |
| 18-110-final-viral-combined_k141_165580\|\|full | Low-quality |
| 18-110-final-viral-combined_k141_246374\|\|full | Low-quality |
| 18-110-final-viral-combined_k141_256873\|\|full | Low-quality |
| 18-110-final-viral-combined_k141_73712\|\|full | Low-quality |
| 18-110-final-viral-combined_k141_3986\|\|full | Low-quality |
| 18-110-final-viral-combined_k141_235069\|\|full | Low-quality |
| 18-110-final-viral-combined_k141_256878\|\|full | Low-quality |
| 18-110-final-viral-combined_k141_3995\|\|full | Low-quality |
| 18-110-final-viral-combined_k141_269283\|\|full | Low-quality |
| 18-110-final-viral-combined_k141_107662\|\|full | Not-determined |
| 18-110-final-viral-combined_k141_165596\|\|full | Low-quality |
| 18-110-final-viral-combined_k141_235087\|\|full | Low-quality |
| 18-110-final-viral-combined_k141_165601\|\|full | Low-quality |
| 18-110-final-viral-combined_k141_188883\|\|full | Not-determined |
| 18-110-final-viral-combined_k141_154284\|\|full | Low-quality |
| 18-110-final-viral-combined_k141_188886\|\|full | Low-quality |
| 18-110-final-viral-combined_k141_131219\|\|full | Low-quality |
| 18-110-final-viral-combined_k141_4027\|\|full | Not-determined |
| 18-110-final-viral-combined_k141_131224\|\|full | Not-determined |
| 18-110-final-viral-combined_k141_73767\|\|full | Not-determined |
| 18-110-final-viral-combined_k141_62124\|\|full | Not-determined |
| 18-110-final-viral-combined_k141_107694\|\|full | Low-quality |
| 18-110-final-viral-combined_k141_269324\|\|full | Low-quality |
| 18-110-final-viral-combined_k141_269329\|\|full | Not-determined |
| 18-110-final-viral-combined_k141_223006\|\|full | Low-quality |
| 18-110-final-viral-combined_k141_96731\|\|full | Low-quality |
| 18-110-final-viral-combined_k141_165662\|\|full | Not-determined |
| 18-110-final-viral-combined_k141_14794\|\|full | Not-determined |
| 18-110-final-viral-combined_k141_4046\|\|full | Low-quality |

| | |
|---|---|
| 18-110-final-viral-combined_k141_223018\|\|full | Low-quality |
| 18-110-final-viral-combined_k141_177858\|\|full | Low-quality |
| 18-110-final-viral-combined_k141_107729\|\|full | Not-determined |
| 18-110-final-viral-combined_k141_131283\|\|full | Not-determined |
| 18-110-final-viral-combined_k141_165673\|\|full | Not-determined |
| 18-110-final-viral-combined_k141_62162\|\|full | Low-quality |
| 18-110-final-viral-combined_k141_177882\|\|full | Not-determined |
| 18-110-final-viral-combined_k141_154342\|\|full | Not-determined |
| 18-110-final-viral-combined_k141_96766\|\|full | Low-quality |
| 18-110-final-viral-combined_k141_4060\|\|full | Not-determined |
| 18-110-final-viral-combined_k141_177892\|\|full | Low-quality |
| 18-110-final-viral-combined_k141_131304\|\|full | Low-quality |
| 18-110-final-viral-combined_k141_223047\|\|full | Not-determined |
| 18-110-final-viral-combined_k141_177897\|\|full | Not-determined |
| 18-110-final-viral-combined_k141_269399\|\|full | Low-quality |
| 18-110-final-viral-combined_k141_246425\|\|full | Not-determined |
| 18-110-final-viral-combined_k141_119403\|\|full | Low-quality |
| 18-110-final-viral-combined_k141_14823\|\|full | Low-quality |
| 18-110-final-viral-combined_k141_143135\|\|full | Not-determined |
| 18-110-final-viral-combined_k141_269407\|\|full | Low-quality |
| 18-110-final-viral-combined_k141_96791\|\|full | Low-quality |
| 18-110-final-viral-combined_k141_73826\|\|full | Not-determined |
| 18-110-final-viral-combined_k141_14840\|\|full | Not-determined |
| 18-110-final-viral-combined_k141_246447\|\|full | Not-determined |
| 18-110-final-viral-combined_k141_50172\|\|full | Low-quality |
| 18-110-final-viral-combined_k141_211848\|\|full | Low-quality |
| 18-110-final-viral-combined_k141_14872\|\|full | Not-determined |
| 18-110-final-viral-combined_k141_62226\|\|full | Not-determined |
| 18-110-final-viral-combined_k141_188950\|\|full | Not-determined |
| 18-110-final-viral-combined_k141_235198\|\|full | Not-determined |
| 18-110-final-viral-combined_k141_50192\|\|full | Low-quality |
| 18-110-final-viral-combined_k141_165736\|\|full | Not-determined |
| 18-110-final-viral-combined_k141_235203\|\|full | Low-quality |
| 18-110-final-viral-combined_k141_96821\|\|full | Low-quality |
| 18-110-final-viral-combined_k141_131361\|\|full | Low-quality |
| 18-110-final-viral-combined_k141_235206\|\|full | Low-quality |
| 18-110-final-viral-combined_k141_4111\|\|full | Low-quality |
| 18-110-final-viral-combined_k141_223133\|\|full | Not-determined |
| 18-110-final-viral-combined_k141_131375\|\|full | Low-quality |
| 18-110-final-viral-combined_k141_39136\|\|full | Not-determined |
| 18-110-final-viral-combined_k141_165748\|\|full | Not-determined |
| 18-110-final-viral-combined_k141_107824\|\|full | Low-quality |
| 18-110-final-viral-combined_k141_96838\|\|full | Not-determined |
| 18-110-final-viral-combined_k141_223141\|\|full | Low-quality |
| 18-110-final-viral-combined_k141_96847\|\|full | Not-determined |
| 18-110-final-viral-combined_k141_269476\|\|full | Low-quality |
| 18-110-final-viral-combined_k141_223144\|\|full | Low-quality |
| 18-110-final-viral-combined_k141_39145\|\|full | Not-determined |
| 18-110-final-viral-combined_k141_178001\|\|full | Low-quality |
| 18-110-final-viral-combined_k141_223158\|\|full | Not-determined |
| 18-110-final-viral-combined_k141_39158\|\|full | Not-determined |
| 18-110-final-viral-combined_k141_235236\|\|full | Not-determined |
| 18-110-final-viral-combined_k141_178017\|\|full | Low-quality |
| 18-110-final-viral-combined_k141_165765\|\|full | Low-quality |
| 18-110-final-viral-combined_k141_73965\|\|full | Not-determined |
| 18-110-final-viral-combined_k141_73966\|\|full | Low-quality |
| 18-110-final-viral-combined_k141_84575\|\|full | Low-quality |
| 18-110-final-viral-combined_k141_27750\|\|full | Low-quality |
| 18-110-final-viral-combined_k141_119501\|\|full | Low-quality |
| 18-110-final-viral-combined_k141_119502\|\|full | Low-quality |
| 18-110-final-viral-combined_k141_269525\|\|full | Not-determined |
| 18-110-final-viral-combined_k141_14975\|\|full | Not-determined |
| 18-110-final-viral-combined_k141_269537\|\|full | Not-determined |
| 18-110-final-viral-combined_k141_96917\|\|full | Low-quality |
| 18-110-final-viral-combined_k141_62339\|\|full | Not-determined |
| 18-110-final-viral-combined_k141_84620\|\|full | Low-quality |
| 18-110-final-viral-combined_k141_143206\|\|full | Low-quality |
| 18-110-final-viral-combined_k141_84642\|\|full | Low-quality |
| 18-110-final-viral-combined_k141_74051\|\|full | Not-determined |
| 18-110-final-viral-combined_k141_107953\|\|full | Not-determined |
| 18-110-final-viral-combined_k141_15009\|\|full | Not-determined |
| 18-110-final-viral-combined_k141_143239\|\|full | Not-determined |
| 18-110-final-viral-combined_k141_15011\|\|full | Not-determined |
| 18-110-final-viral-combined_k141_131471\|\|full | Not-determined |
| 18-110-final-viral-combined_k141_119578\|\|full | Low-quality |
| 18-110-final-viral-combined_k141_96982\|\|full | Not-determined |
| 18-110-final-viral-combined_k141_39228\|\|full | Not-determined |
| 18-110-final-viral-combined_k141_223240\|\|full | Low-quality |
| 18-110-final-viral-combined_k141_235372\|\|full | Low-quality |
| 18-110-final-viral-combined_k141_62412\|\|full | Low-quality |
| 18-110-final-viral-combined_k141_178108\|\|full | Not-determined |
| 18-110-final-viral-combined_k141_74085\|\|full | Not-determined |
| 18-110-final-viral-combined_k141_235381\|\|full | Not-determined |
| 18-110-final-viral-combined_k141_62417\|\|full | Not-determined |
| 18-110-final-viral-combined_k141_131502\|\|full | Low-quality |
| 18-110-final-viral-combined_k141_27829\|\|full | Not-determined |
| 18-110-final-viral-combined_k141_223263\|\|full | Not-determined |
| 18-110-final-viral-combined_k141_246593\|\|full | Not-determined |
| 18-110-final-viral-combined_k141_15044\|\|full | Not-determined |
| 18-110-final-viral-combined_k141_235394\|\|full | Not-determined |
| 18-110-final-viral-combined_k141_165807\|\|full | Not-determined |
| 18-110-final-viral-combined_k141_39275\|\|full | Low-quality |
| 18-110-final-viral-combined_k141_27852\|\|full | Low-quality |
| 18-110-final-viral-combined_k141_27854\|\|full | Not-determined |
| 18-110-final-viral-combined_k141_154550\|\|full | Not-determined |
| 18-110-final-viral-combined_k141_108014\|\|full | Not-determined |
| 18-110-final-viral-combined_k141_50396\|\|full | Not-determined |
| 18-110-final-viral-combined_k141_84748\|\|full | Not-determined |
| 18-110-final-viral-combined_k141_119637\|\|full | Low-quality |
| 18-110-final-viral-combined_k141_212056\|\|full | Not-determined |
| 18-110-final-viral-combined_k141_108030\|\|full | Low-quality |
| 18-110-final-viral-combined_k141_212067\|\|full | Low-quality |
| 18-110-final-viral-combined_k141_246658\|\|full | Low-quality |
| 18-110-final-viral-combined_k141_131581\|\|full | Low-quality |
| 18-110-final-viral-combined_k141_235439\|\|full | Not-determined |
| 18-110-final-viral-combined_k141_178162\|\|full | Not-determined |
| 18-110-final-viral-combined_k141_50437\|\|full | Low-quality |
| 18-110-final-viral-combined_k141_246664\|\|full | Not-determined |
| 18-110-final-viral-combined_k141_74205\|\|full | Not-determined |
| 18-110-final-viral-combined_k141_178188\|\|full | Low-quality |
| 18-110-final-viral-combined_k141_189169\|\|full | Low-quality |
| 18-110-final-viral-combined_k141_74207\|\|full | Not-determined |
| 18-110-final-viral-combined_k141_4245\|\|full | Not-determined |
| 18-110-final-viral-combined_k141_74209\|\|full | Not-determined |
| 18-110-final-viral-combined_k141_235459\|\|full | Not-determined |
| 18-110-final-viral-combined_k141_27890\|\|full | Low-quality |
| 18-110-final-viral-combined_k141_4260\|\|full | Not-determined |
| 18-110-final-viral-combined_k141_189197\|\|full | Not-determined |
| 18-110-final-viral-combined_k141_143378\|\|full | Not-determined |
| 18-110-final-viral-combined_k141_15124\|\|full | Not-determined |
| 18-110-final-viral-combined_k141_50488\|\|full | Not-determined |
| 18-110-final-viral-combined_k141_257027\|\|full | Low-quality |
| 18-110-final-viral-combined_k141_131656\|\|full | Low-quality |
| 18-110-final-viral-combined_k141_4281\|\|full | Not-determined |
| 18-110-final-viral-combined_k141_4283\|\|full | Not-determined |
| 18-110-final-viral-combined_k141_257029\|\|full | Low-quality |
| 18-110-final-viral-combined_k141_27919\|\|full | Not-determined |
| 18-110-final-viral-combined_k141_84792\|\|full | Low-quality |
| 18-110-final-viral-combined_k141_143408\|\|full | Low-quality |
| 18-110-final-viral-combined_k141_4296\|\|full | Not-determined |
| 18-110-final-viral-combined_k141_199815\|\|full | Not-determined |
| 18-110-final-viral-combined_k141_108082\|\|full | Low-quality |
| 18-110-final-viral-combined_k141_235514\|\|full | Not-determined |
| 18-110-final-viral-combined_k141_39422\|\|full | Not-determined |
| 18-110-final-viral-combined_k141_189230\|\|full | Low-quality |
| 18-110-final-viral-combined_k141_257045\|\|full | Low-quality |
| 18-110-final-viral-combined_k141_212170\|\|full | Not-determined |
| 18-110-final-viral-combined_k141_212171\|\|full | Not-determined |
| 18-110-final-viral-combined_k141_199834\|\|full | Low-quality |
| 18-110-final-viral-combined_k141_199835\|\|full | Not-determined |
| 18-110-final-viral-combined_k141_178292\|\|full | Low-quality |
| 18-110-final-viral-combined_k141_178299\|\|full | Low-quality |
| 18-110-final-viral-combined_k141_154680\|\|full | Low-quality |
| 18-110-final-viral-combined_k141_257058\|\|full | Not-determined |
| 18-110-final-viral-combined_k141_154684\|\|full | Low-quality |
| 18-110-final-viral-combined_k141_50540\|\|full | Low-quality |
| 18-110-final-viral-combined_k141_257061\|\|full | Not-determined |
| 18-110-final-viral-combined_k141_108147\|\|full | Low-quality |
| 18-110-final-viral-combined_k141_84841\|\|full | Low-quality |
| 18-110-final-viral-combined_k141_97112\|\|full | Not-determined |
| 18-110-final-viral-combined_k141_212193\|\|full | Not-determined |
| 18-110-final-viral-combined_k141_154697\|\|full | Not-determined |
| 18-110-final-viral-combined_k141_165914\|\|full | Not-determined |
| 18-110-final-viral-combined_k141_178349\|\|full | Not-determined |
| 18-110-final-viral-combined_k141_119746\|\|full | Not-determined |
| 18-110-final-viral-combined_k141_143445\|\|full | Not-determined |
| 18-110-final-viral-combined_k141_4374\|\|full | Not-determined |
| 18-110-final-viral-combined_k141_74267\|\|full | Low-quality |
| 18-110-final-viral-combined_k141_62606\|\|full | Not-determined |
| 18-110-final-viral-combined_k141_257081\|\|full | Not-determined |
| 18-110-final-viral-combined_k141_119752\|\|full | Not-determined |
| 18-110-final-viral-combined_k141_235587\|\|full | Not-determined |
| 18-110-final-viral-combined_k141_154721\|\|full | Not-determined |
| 18-110-final-viral-combined_k141_269790\|\|full | Not-determined |
| 18-110-final-viral-combined_k141_246790\|\|full | Low-quality |
| 18-110-final-viral-combined_k141_223383\|\|full | Low-quality |
| 18-110-final-viral-combined_k141_4389\|\|full | Low-quality |
| 18-110-final-viral-combined_k141_27999\|\|full | Low-quality |
| 18-110-final-viral-combined_k141_50598\|\|full | Low-quality |
| 18-110-final-viral-combined_k141_269806\|\|full | Not-determined |
| 18-110-final-viral-combined_k141_97165\|\|full | Low-quality |
| 18-110-final-viral-combined_k141_97167\|\|full | Not-determined |
| 18-110-final-viral-combined_k141_223417\|\|full | Not-determined |
| 18-110-final-viral-combined_k141_189312\|\|full | Low-quality |
| 18-110-final-viral-combined_k141_97179\|\|full | Not-determined |
| 18-110-final-viral-combined_k141_235635\|\|full | Low-quality |
| 18-110-final-viral-combined_k141_15235\|\|full | Not-determined |
| 18-102-final-viral-combined_k141_110761\|\|full | Medium-quality |
| 18-110-final-viral-combined_k141_269829\|\|full | Not-determined |
| 18-110-final-viral-combined_k141_74281\|\|full | Low-quality |
| 18-110-final-viral-combined_k141_131709\|\|full | Low-quality |
| 18-110-final-viral-combined_k141_108190\|\|full | Not-determined |
| 18-110-final-viral-combined_k141_223438\|\|full | Low-quality |
| 18-110-final-viral-combined_k141_62705\|\|full | Not-determined |
| 18-110-final-viral-combined_k141_97205\|\|full | Not-determined |
| 18-110-final-viral-combined_k141_223445\|\|full | Not-determined |
| 18-110-final-viral-combined_k141_212255\|\|full | Low-quality |
| 18-110-final-viral-combined_k141_62724\|\|full | Low-quality |
| 18-110-final-viral-combined_k141_189358\|\|full | Not-determined |
| 18-110-final-viral-combined_k141_212267\|\|full | Not-determined |
| 18-110-final-viral-combined_k141_74309\|\|full | Not-determined |
| 18-110-final-viral-combined_k141_84940\|\|full | Low-quality |
| 18-110-final-viral-combined_k141_119793\|\|full | Low-quality |
| 18-110-final-viral-combined_k141_15286\|\|full | Not-determined |
| 18-110-final-viral-combined_k141_84941\|\|full | Low-quality |
| 18-110-final-viral-combined_k141_212274\|\|full | Low-quality |
| 18-110-final-viral-combined_k141_166057\|\|full | Low-quality |
| 18-110-final-viral-combined_k141_74322\|\|full | Low-quality |
| 18-110-final-viral-combined_k141_246861\|\|full | Not-determined |
| 18-110-final-viral-combined_k141_269876\|\|full | Not-determined |
| 18-110-final-viral-combined_k141_119812\|\|full | Low-quality |
| 18-110-final-viral-combined_k141_235680\|\|full | Not-determined |
| 18-110-final-viral-combined_k141_269885\|\|full | Not-determined |
| 18-110-final-viral-combined_k141_189383\|\|full | Not-determined |
| 18-110-final-viral-combined_k141_50674\|\|full | Low-quality |
| 18-110-final-viral-combined_k141_39572\|\|full | Low-quality |
| 18-110-final-viral-combined_k141_119825\|\|full | Low-quality |
| 18-110-final-viral-combined_k141_15316\|\|full | Not-determined |
| 18-110-final-viral-combined_k141_119826\|\|full | Low-quality |
| 18-110-final-viral-combined_k141_166079\|\|full | Low-quality |
| 18-110-final-viral-combined_k141_50685\|\|full | Not-determined |
| 18-110-final-viral-combined_k141_119838\|\|full | Low-quality |
| 18-110-final-viral-combined_k141_269906\|\|full | Not-determined |
| 18-110-final-viral-combined_k141_131735\|\|full | Not-determined |
| 18-110-final-viral-combined_k141_50700\|\|full | Not-determined |
| 18-110-final-viral-combined_k141_108233\|\|full | Low-quality |
| 18-110-final-viral-combined_k141_223495\|\|full | Not-determined |
| 18-110-final-viral-combined_k141_50713\|\|full | Low-quality |
| 18-110-final-viral-combined_k141_223503\|\|full | Not-determined |
| 18-110-final-viral-combined_k141_189401\|\|full | Not-determined |
| 18-110-final-viral-combined_k141_246908\|\|full | Low-quality |
| 18-110-final-viral-combined_k141_246916\|\|full | Not-determined |
| 18-110-final-viral-combined_k141_189423\|\|full | Not-determined |
| 18-110-final-viral-combined_k141_178503\|\|full | Not-determined |
| 18-110-final-viral-combined_k141_131782\|\|full | Low-quality |
| 18-110-final-viral-combined_k141_189427\|\|full | Not-determined |
| 18-110-final-viral-combined_k141_269955\|\|full | Not-determined |
| 18-110-final-viral-combined_k141_28147\|\|full | Not-determined |
| 18-110-final-viral-combined_k141_62819\|\|full | Low-quality |
| 18-110-final-viral-combined_k141_212303\|\|full | Low-quality |
| 18-110-final-viral-combined_k141_28155\|\|full | Not-determined |
| 18-110-final-viral-combined_k141_108248\|\|full | Not-determined |
| 18-110-final-viral-combined_k141_15368\|\|full | Low-quality |
| 18-110-final-viral-combined_k141_154739\|\|full | Low-quality |
| 18-110-final-viral-combined_k141_50732\|\|full | Low-quality |
| 18-110-final-viral-combined_k141_178515\|\|full | Not-determined |
| 18-110-final-viral-combined_k141_62841\|\|full | Low-quality |
| 18-110-final-viral-combined_k141_178516\|\|full | Low-quality |
| 18-110-final-viral-combined_k141_74369\|\|full | Not-determined |
| 18-110-final-viral-combined_k141_50739\|\|full | Not-determined |
| 18-110-final-viral-combined_k141_257197\|\|full | Low-quality |
| 18-110-final-viral-combined_k141_143567\|\|full | Low-quality |
| 18-110-final-viral-combined_k141_257204\|\|full | Low-quality |
| 18-110-final-viral-combined_k141_108289\|\|full | Not-determined |
| 18-110-final-viral-combined_k141_50744\|\|full | Low-quality |
| 18-110-final-viral-combined_k141_119897\|\|full | Not-determined |
| 18-110-final-viral-combined_k141_199878\|\|full | Low-quality |
| 18-110-final-viral-combined_k141_223549\|\|full | Low-quality |
| 18-110-final-viral-combined_k141_62907\|\|full | Not-determined |
| 18-110-final-viral-combined_k141_247027\|\|full | Not-determined |
| 18-110-final-viral-combined_k141_108344\|\|full | Not-determined |
| 18-110-final-viral-combined_k141_235844\|\|full | Not-determined |
| 18-110-final-viral-combined_k141_28240\|\|full | Not-determined |
| 18-110-final-viral-combined_k141_108354\|\|full | Low-quality |
| 18-110-final-viral-combined_k141_62935\|\|full | Not-determined |
| 18-110-final-viral-combined_k141_247043\|\|full | Low-quality |
| 18-110-final-viral-combined_k141_270025\|\|full | Not-determined |
| 18-110-final-viral-combined_k141_199888\|\|full | Not-determined |
| 18-110-final-viral-combined_k141_15444\|\|full | Not-determined |
| 18-110-final-viral-combined_k141_257256\|\|full | Low-quality |
| 18-110-final-viral-combined_k141_189504\|\|full | Not-determined |
| 18-110-final-viral-combined_k141_108382\|\|full | Not-determined |
| 18-110-final-viral-combined_k141_257262\|\|full | Low-quality |
| 18-110-final-viral-combined_k141_62960\|\|full | Low-quality |
| 18-110-final-viral-combined_k141_199900\|\|full | Not-determined |
| 18-110-final-viral-combined_k141_108391\|\|full | Not-determined |
| 18-110-final-viral-combined_k141_108393\|\|full | Not-determined |
| 18-110-final-viral-combined_k141_178632\|\|full | Not-determined |
| 18-110-final-viral-combined_k141_97267\|\|full | Not-determined |
| 18-110-final-viral-combined_k141_97281\|\|full | Low-quality |
| 18-110-final-viral-combined_k141_189547\|\|full | Not-determined |
| 18-110-final-viral-combined_k141_63002\|\|full | Low-quality |
| 18-110-final-viral-combined_k141_247115\|\|full | Not-determined |
| 18-110-final-viral-combined_k141_97302\|\|full | Not-determined |
| 18-110-final-viral-combined_k141_143633\|\|full | Low-quality |
| 18-110-final-viral-combined_k141_270080\|\|full | Not-determined |
| 18-110-final-viral-combined_k141_199955\|\|full | Not-determined |
| 18-110-final-viral-combined_k141_212425\|\|full | Low-quality |
| 18-110-final-viral-combined_k141_108413\|\|full | Low-quality |
| 18-110-final-viral-combined_k141_270090\|\|full | Not-determined |
| 18-110-final-viral-combined_k141_154873\|\|full | Low-quality |
| 18-110-final-viral-combined_k141_178688\|\|full | Not-determined |
| 18-110-final-viral-combined_k141_212452\|\|full | Not-determined |
| 18-110-final-viral-combined_k141_97336\|\|full | Not-determined |
| 18-110-final-viral-combined_k141_270098\|\|full | Low-quality |
| 18-110-final-viral-combined_k141_50856\|\|full | Not-determined |
| 18-110-final-viral-combined_k141_85211\|\|full | Low-quality |
| 18-110-final-viral-combined_k141_50870\|\|full | Not-determined |
| 18-110-final-viral-combined_k141_166286\|\|full | Low-quality |
| 18-110-final-viral-combined_k141_154907\|\|full | Not-determined |
| 18-110-final-viral-combined_k141_247181\|\|full | Not-determined |
| 18-110-final-viral-combined_k141_97367\|\|full | Not-determined |
| 18-110-final-viral-combined_k141_143694\|\|full | Low-quality |
| 18-110-final-viral-combined_k141_235979\|\|full | Not-determined |
| 18-110-final-viral-combined_k141_85224\|\|full | Low-quality |
| 18-110-final-viral-combined_k141_108464\|\|full | Not-determined |
| 18-110-final-viral-combined_k141_223617\|\|full | Not-determined |
| 18-110-final-viral-combined_k141_212490\|\|full | Not-determined |
| 18-110-final-viral-combined_k141_108470\|\|full | Low-quality |
| 18-110-final-viral-combined_k141_212494\|\|full | Low-quality |
| 18-110-final-viral-combined_k141_257335\|\|full | Not-determined |
| 18-110-final-viral-combined_k141_97388\|\|full | Not-determined |
| 18-110-final-viral-combined_k141_189595\|\|full | Not-determined |
| 18-110-final-viral-combined_k141_257345\|\|full | Not-determined |
| 18-110-final-viral-combined_k141_39788\|\|full | Low-quality |
| 18-110-final-viral-combined_k141_154952\|\|full | Low-quality |
| 18-110-final-viral-combined_k141_28389\|\|full | Not-determined |
| 18-110-final-viral-combined_k141_28390\|\|full | Not-determined |
| 18-110-final-viral-combined_k141_143739\|\|full | Low-quality |
| 18-110-final-viral-combined_k141_4555\|\|full | Not-determined |
| 18-110-final-viral-combined_k141_74508\|\|full | Not-determined |
| 18-110-final-viral-combined_k141_131922\|\|full | Low-quality |
| 18-110-final-viral-combined_k141_97407\|\|full | Low-quality |
| 18-110-final-viral-combined_k141_85259\|\|full | Low-quality |
| 18-110-final-viral-combined_k141_143747\|\|full | Not-determined |
| 18-110-final-viral-combined_k141_200019\|\|full | Not-determined |
| 18-110-final-viral-combined_k141_97416\|\|full | Not-determined |
| 18-110-final-viral-combined_k141_257361\|\|full | Not-determined |
| 18-110-final-viral-combined_k141_120094\|\|full | Not-determined |
| 18-110-final-viral-combined_k141_257362\|\|full | Not-determined |
| 18-110-final-viral-combined_k141_236032\|\|full | Low-quality |
| 18-110-final-viral-combined_k141_257366\|\|full | Low-quality |
| 18-110-final-viral-combined_k141_223673\|\|full | Low-quality |
| 18-110-final-viral-combined_k141_178816\|\|full | Not-determined |
| 18-110-final-viral-combined_k141_223674\|\|full | Not-determined |
| 18-110-final-viral-combined_k141_74529\|\|full | Low-quality |
| 18-110-final-viral-combined_k141_178821\|\|full | Low-quality |
| 18-110-final-viral-combined_k141_131945\|\|full | Not-determined |
| 18-110-final-viral-combined_k141_189651\|\|full | Not-determined |
| 18-110-final-viral-combined_k141_178829\|\|full | Low-quality |
| 18-110-final-viral-combined_k141_131953\|\|full | Not-determined |
| 18-110-final-viral-combined_k141_97437\|\|full | Not-determined |
| 18-110-final-viral-combined_k141_270192\|\|full | Not-determined |
| 18-110-final-viral-combined_k141_85286\|\|full | Low-quality |
| 18-110-final-viral-combined_k141_50919\|\|full | Low-quality |
| 18-110-final-viral-combined_k141_97440\|\|full | Not-determined |
| 18-110-final-viral-combined_k141_28450\|\|full | Low-quality |
| 18-110-final-viral-combined_k141_63118\|\|full | Low-quality |
| 18-110-final-viral-combined_k141_63122\|\|full | Not-determined |
| 18-110-final-viral-combined_k141_236065\|\|full | Not-determined |
| 18-110-final-viral-combined_k141_39868\|\|full | Not-determined |
| 18-110-final-viral-combined_k141_15633\|\|full | Not-determined |
| 18-110-final-viral-combined_k141_131986\|\|full | Low-quality |
| 18-110-final-viral-combined_k141_155026\|\|full | Low-quality |
| 18-110-final-viral-combined_k141_212537\|\|full | Not-determined |
| 18-110-final-viral-combined_k141_155031\|\|full | Low-quality |
| 18-110-final-viral-combined_k141_143807\|\|full | Not-determined |
| 18-110-final-viral-combined_k141_15655\|\|full | Low-quality |
| 18-110-final-viral-combined_k141_189681\|\|full | Low-quality |
| 18-110-final-viral-combined_k141_143836\|\|full | Not-determined |
| 18-110-final-viral-combined_k141_50984\|\|full | Low-quality |
| 18-110-final-viral-combined_k141_132026\|\|full | Low-quality |
| 18-110-final-viral-combined_k141_257421\|\|full | Not-determined |
| 18-110-final-viral-combined_k141_15694\|\|full | Low-quality |
| 18-110-final-viral-combined_k141_39911\|\|full | Not-determined |
| 18-110-final-viral-combined_k141_155052\|\|full | Low-quality |
| 18-110-final-viral-combined_k141_120156\|\|full | Low-quality |
| 18-110-final-viral-combined_k141_39917\|\|full | Not-determined |
| 18-110-final-viral-combined_k141_39922\|\|full | Low-quality |
| 18-110-final-viral-combined_k141_28564\|\|full | Low-quality |
| 18-110-final-viral-combined_k141_74661\|\|full | Low-quality |
| 18-110-final-viral-combined_k141_132046\|\|full | Low-quality |
| 18-110-final-viral-combined_k141_189701\|\|full | Not-determined |
| 18-110-final-viral-combined_k141_4615\|\|full | Low-quality |
| 18-110-final-viral-combined_k141_63182\|\|full | Low-quality |
| 18-110-final-viral-combined_k141_155075\|\|full | Not-determined |
| 18-110-final-viral-combined_k141_143894\|\|full | Low-quality |
| 18-110-final-viral-combined_k141_189706\|\|full | Low-quality |
| 18-110-final-viral-combined_k141_39942\|\|full | Not-determined |
| 18-110-final-viral-combined_k141_236164\|\|full | Not-determined |
| 18-110-final-viral-combined_k141_4633\|\|full | Low-quality |
| 18-110-final-viral-combined_k141_74683\|\|full | Low-quality |
| 18-110-final-viral-combined_k141_15710\|\|full | Low-quality |
| 18-110-final-viral-combined_k141_247407\|\|full | Not-determined |
| 18-110-final-viral-combined_k141_247410\|\|full | Not-determined |
| 18-110-final-viral-combined_k141_132075\|\|full | Low-quality |
| 18-110-final-viral-combined_k141_143923\|\|full | Not-determined |
| 18-110-final-viral-combined_k141_74692\|\|full | Low-quality |
| 18-110-final-viral-combined_k141_28613\|\|full | Not-determined |
| 18-110-final-viral-combined_k141_39979\|\|full | Not-determined |
| 18-110-final-viral-combined_k141_178912\|\|full | Low-quality |
| 18-110-final-viral-combined_k141_270332\|\|full | Not-determined |
| 18-110-final-viral-combined_k141_143942\|\|full | Not-determined |
| 18-110-final-viral-combined_k141_247428\|\|full | Not-determined |
| 18-110-final-viral-combined_k141_166355\|\|full | Not-determined |
| 18-110-final-viral-combined_k141_28620\|\|full | Not-determined |
| 18-110-final-viral-combined_k141_51097\|\|full | Low-quality |
| 18-110-final-viral-combined_k141_143958\|\|full | Not-determined |
| 18-110-final-viral-combined_k141_178934\|\|full | Not-determined |
| 18-110-final-viral-combined_k141_4659\|\|full | Low-quality |
| 18-110-final-viral-combined_k141_247447\|\|full | Not-determined |
| 18-110-final-viral-combined_k141_63247\|\|full | Not-determined |
| 18-110-final-viral-combined_k141_51135\|\|full | Not-determined |
| 18-110-final-viral-combined_k141_4679\|\|full | Low-quality |
| 18-110-final-viral-combined_k141_257494\|\|full | Not-determined |
| 18-110-final-viral-combined_k141_97623\|\|full | Low-quality |
| 18-110-final-viral-combined_k141_74710\|\|full | Low-quality |
| 18-110-final-viral-combined_k141_247463\|\|full | Not-determined |
| 18-110-final-viral-combined_k141_28640\|\|full | Not-determined |
| 18-110-final-viral-combined_k141_85409\|\|full | Not-determined |
| 18-110-final-viral-combined_k141_166375\|\|full | Not-determined |
| 18-110-final-viral-combined_k141_155092\|\|full | Low-quality |
| 18-110-final-viral-combined_k141_155099\|\|full | Low-quality |
| 18-110-final-viral-combined_k141_40045\|\|full | Low-quality |
| 18-110-final-viral-combined_k141_144002\|\|full | Not-determined |
| 18-110-final-viral-combined_k141_247477\|\|full | Not-determined |
| 18-110-final-viral-combined_k141_144004\|\|full | Low-quality |
| 18-101-final-viral-combined_k141_268922\|\|full | Medium-quality |
| 18-110-final-viral-combined_k141_28654\|\|full | Low-quality |
| 18-110-final-viral-combined_k141_189775\|\|full | Not-determined |
| 18-110-final-viral-combined_k141_108581\|\|full | Low-quality |
| 18-110-final-viral-combined_k141_166415\|\|full | Not-determined |
| 18-110-final-viral-combined_k141_108584\|\|full | Low-quality |
| 18-110-final-viral-combined_k141_155135\|\|full | Low-quality |
| 18-110-final-viral-combined_k141_257513\|\|full | Low-quality |
| 18-110-final-viral-combined_k141_97652\|\|full | Low-quality |
| 18-110-final-viral-combined_k141_97656\|\|full | Not-determined |
| 18-110-final-viral-combined_k141_200152\|\|full | Low-quality |
| 18-110-final-viral-combined_k141_74790\|\|full | Low-quality |
| 18-110-final-viral-combined_k141_212617\|\|full | Low-quality |
| 18-110-final-viral-combined_k141_4710\|\|full | Not-determined |
| 18-110-final-viral-combined_k141_85485\|\|full | Not-determined |
| 18-110-final-viral-combined_k141_144052\|\|full | Not-determined |
| 18-110-final-viral-combined_k141_257532\|\|full | Not-determined |
| 18-110-final-viral-combined_k141_28694\|\|full | Low-quality |
| 18-110-final-viral-combined_k141_236227\|\|full | Low-quality |
| 18-110-final-viral-combined_k141_4720\|\|full | Not-determined |
| 18-110-final-viral-combined_k141_212640\|\|full | Not-determined |
| 18-110-final-viral-combined_k141_63343\|\|full | Low-quality |
| 18-110-final-viral-combined_k141_144080\|\|full | Not-determined |
| 18-110-final-viral-combined_k141_189837\|\|full | Low-quality |
| 18-110-final-viral-combined_k141_257546\|\|full | Not-determined |
| 18-110-final-viral-combined_k141_74809\|\|full | Not-determined |
| 18-110-final-viral-combined_k141_108645\|\|full | Low-quality |
| 18-110-final-viral-combined_k141_212657\|\|full | Low-quality |
| 18-110-final-viral-combined_k141_132212\|\|full | Not-determined |
| 18-110-final-viral-combined_k141_85501\|\|full | Not-determined |
| 18-110-final-viral-combined_k141_155200\|\|full | Low-quality |
| 18-110-final-viral-combined_k141_120232\|\|full | Not-determined |
| 18-110-final-viral-combined_k141_28720\|\|full | Low-quality |
| 18-110-final-viral-combined_k141_257551\|\|full | Not-determined |
| 18-110-final-viral-combined_k141_85515\|\|full | Not-determined |
| 18-110-final-viral-combined_k141_189850\|\|full | Not-determined |
| 18-110-final-viral-combined_k141_97668\|\|full | Not-determined |
| 18-110-final-viral-combined_k141_28751\|\|full | Not-determined |
| 18-110-final-viral-combined_k141_179048\|\|full | Low-quality |
| 18-110-final-viral-combined_k141_74832\|\|full | Low-quality |
| 18-110-final-viral-combined_k141_155239\|\|full | Not-determined |
| 18-110-final-viral-combined_k141_179053\|\|full | Low-quality |
| 18-110-final-viral-combined_k141_120244\|\|full | Not-determined |
| 18-110-final-viral-combined_k141_236275\|\|full | Not-determined |
| 18-110-final-viral-combined_k141_257581\|\|full | Not-determined |
| 18-110-final-viral-combined_k141_166485\|\|full | Not-determined |
| 18-110-final-viral-combined_k141_28758\|\|full | Not-determined |
| 18-110-final-viral-combined_k141_247523\|\|full | Low-quality |
| 18-110-final-viral-combined_k141_144141\|\|full | Low-quality |
| 18-110-final-viral-combined_k141_166488\|\|full | Not-determined |
| 18-101-final-viral-combined_k141_17651\|\|full | High-quality |
| 18-110-final-viral-combined_k141_97714\|\|full | Not-determined |
| 18-110-final-viral-combined_k141_4771\|\|full | Low-quality |
| 18-110-final-viral-combined_k141_236296\|\|full | Low-quality |
| 18-110-final-viral-combined_k141_223975\|\|full | Low-quality |
| 18-110-final-viral-combined_k141_4777\|\|full | Not-determined |
| 18-110-final-viral-combined_k141_144162\|\|full | Low-quality |
| 18-110-final-viral-combined_k141_108746\|\|full | Not-determined |
| 18-110-final-viral-combined_k141_51225\|\|full | Low-quality |
| 18-110-final-viral-combined_k141_144193\|\|full | Low-quality |
| 18-110-final-viral-combined_k141_132277\|\|full | Low-quality |
| 18-110-final-viral-combined_k141_85603\|\|full | Not-determined |
| 18-110-final-viral-combined_k141_155288\|\|full | Not-determined |
| 18-110-final-viral-combined_k141_166515\|\|full | Not-determined |
| 18-110-final-viral-combined_k141_85606\|\|full | Not-determined |
| 18-110-final-viral-combined_k141_15816\|\|full | Not-determined |
| 18-110-final-viral-combined_k141_4790\|\|full | Low-quality |
| 17-108-final-viral-combined_k141_200633\|\|full | Medium-quality |
| 18-110-final-viral-combined_k141_247573\|\|full | Low-quality |
| 18-110-final-viral-combined_k141_85627\|\|full | Low-quality |
| 18-110-final-viral-combined_k141_236370\|\|full | Not-determined |
| 18-110-final-viral-combined_k141_28820\|\|full | Low-quality |
| 18-110-final-viral-combined_k141_200178\|\|full | Low-quality |
| 18-110-final-viral-combined_k141_247594\|\|full | Low-quality |
| 18-110-final-viral-combined_k141_97743\|\|full | Not-determined |
| 18-110-final-viral-combined_k141_74869\|\|full | Low-quality |
| 18-110-final-viral-combined_k141_97745\|\|full | Low-quality |
| 18-110-final-viral-combined_k141_40149\|\|full | Low-quality |
| 18-110-final-viral-combined_k141_51264\|\|full | Not-determined |
| 18-110-final-viral-combined_k141_236400\|\|full | Not-determined |
| 18-110-final-viral-combined_k141_97766\|\|full | Low-quality |
| 18-110-final-viral-combined_k141_200194\|\|full | Low-quality |
| 18-110-final-viral-combined_k141_108792\|\|full | Low-quality |
| 18-110-final-viral-combined_k141_270435\|\|full | Not-determined |
| 18-110-final-viral-combined_k141_40166\|\|full | Not-determined |
| 18-110-final-viral-combined_k141_28866\|\|full | Low-quality |
| 18-110-final-viral-combined_k141_40172\|\|full | Low-quality |
| 18-110-final-viral-combined_k141_189982\|\|full | Not-determined |
| 18-110-final-viral-combined_k141_63462\|\|full | Not-determined |
| 18-110-final-viral-combined_k141_200224\|\|full | Not-determined |
| 18-110-final-viral-combined_k141_51302\|\|full | Not-determined |
| 18-110-final-viral-combined_k141_247623\|\|full | Low-quality |
| 18-110-final-viral-combined_k141_270448\|\|full | Not-determined |
| 18-110-final-viral-combined_k141_15871\|\|full | Low-quality |
| 18-110-final-viral-combined_k141_257678\|\|full | Not-determined |
| 18-110-final-viral-combined_k141_40203\|\|full | Low-quality |
| 18-110-final-viral-combined_k141_236430\|\|full | Not-determined |
| 18-110-final-viral-combined_k141_270449\|\|full | Low-quality |
| 18-110-final-viral-combined_k141_179179\|\|full | Not-determined |
| 18-110-final-viral-combined_k141_236433\|\|full | Not-determined |
| 18-110-final-viral-combined_k141_74929\|\|full | Not-determined |
| 18-110-final-viral-combined_k141_270455\|\|full | Not-determined |
| 18-110-final-viral-combined_k141_74933\|\|full | Low-quality |
| 18-110-final-viral-combined_k141_28924\|\|full | Not-determined |
| 18-110-final-viral-combined_k141_166603\|\|full | Not-determined |
| 18-110-final-viral-combined_k141_40226\|\|full | Not-determined |
| 18-110-final-viral-combined_k141_120402\|\|full | Not-determined |
| 18-110-final-viral-combined_k141_85739\|\|full | Low-quality |
| 18-110-final-viral-combined_k141_51325\|\|full | Low-quality |
| 18-110-final-viral-combined_k141_51334\|\|full | Low-quality |
| 18-110-final-viral-combined_k141_4887\|\|full | Not-determined |
| 18-110-final-viral-combined_k141_190049\|\|full | Not-determined |
| 18-110-final-viral-combined_k141_212835\|\|full | Low-quality |
| 18-110-final-viral-combined_k141_51351\|\|full | Low-quality |
| 18-110-final-viral-combined_k141_257723\|\|full | Low-quality |
| 18-110-final-viral-combined_k141_85754\|\|full | Low-quality |
| 18-110-final-viral-combined_k141_74985\|\|full | Not-determined |
| 18-110-final-viral-combined_k141_4896\|\|full | Low-quality |
| 18-110-final-viral-combined_k141_85761\|\|full | Low-quality |
| 18-110-final-viral-combined_k141_63523\|\|full | Low-quality |
| 18-110-final-viral-combined_k141_270470\|\|full | Not-determined |
| 18-110-final-viral-combined_k141_257726\|\|full | Not-determined |
| 18-110-final-viral-combined_k141_224085\|\|full | Low-quality |
| 18-110-final-viral-combined_k141_166651\|\|full | Not-determined |
| 18-110-final-viral-combined_k141_270482\|\|full | Not-determined |
| 18-110-final-viral-combined_k141_74997\|\|full | Low-quality |
| 18-110-final-viral-combined_k141_120468\|\|full | Low-quality |
| 18-110-final-viral-combined_k141_257731\|\|full | Not-determined |
| 18-110-final-viral-combined_k141_4917\|\|full | Low-quality |
| 18-110-final-viral-combined_k141_108824\|\|full | Not-determined |
| 18-110-final-viral-combined_k141_270498\|\|full | Low-quality |
| 18-110-final-viral-combined_k141_224109\|\|full | Low-quality |
| 18-110-final-viral-combined_k141_257741\|\|full | Not-determined |
| 18-110-final-viral-combined_k141_270503\|\|full | Low-quality |
| 18-110-final-viral-combined_k141_51393\|\|full | Low-quality |
| 18-110-final-viral-combined_k141_236488\|\|full | Low-quality |
| 18-110-final-viral-combined_k141_166683\|\|full | Not-determined |
| 18-110-final-viral-combined_k141_224114\|\|full | Not-determined |
| 18-110-final-viral-combined_k141_75022\|\|full | Low-quality |
| 18-110-final-viral-combined_k141_270506\|\|full | Not-determined |
| 18-110-final-viral-combined_k141_224122\|\|full | Low-quality |
| 18-110-final-viral-combined_k141_63568\|\|full | Not-determined |
| 18-110-final-viral-combined_k141_224129\|\|full | Low-quality |
| 18-110-final-viral-combined_k141_120484\|\|full | Not-determined |
| 18-110-final-viral-combined_k141_200357\|\|full | Low-quality |
| 18-110-final-viral-combined_k141_270528\|\|full | Low-quality |
| 18-110-final-viral-combined_k141_63584\|\|full | Not-determined |
| 18-110-final-viral-combined_k141_257762\|\|full | Low-quality |
| 18-110-final-viral-combined_k141_51431\|\|full | Low-quality |
| 18-110-final-viral-combined_k141_51432\|\|full | Not-determined |
| 18-110-final-viral-combined_k141_179266\|\|full | Not-determined |
| 18-110-final-viral-combined_k141_132363\|\|full | Low-quality |
| 18-110-final-viral-combined_k141_270550\|\|full | Low-quality |
| 18-110-final-viral-combined_k141_40312\|\|full | Not-determined |
| 18-110-final-viral-combined_k141_29080\|\|full | Not-determined |
| 18-110-final-viral-combined_k141_108848\|\|full | Not-determined |
| 18-110-final-viral-combined_k141_132391\|\|full | Low-quality |
| 18-110-final-viral-combined_k141_247732\|\|full | Low-quality |
| 18-110-final-viral-combined_k141_224155\|\|full | Low-quality |
| 18-110-final-viral-combined_k141_29095\|\|full | Not-determined |
| 18-110-final-viral-combined_k141_179289\|\|full | Not-determined |
| 18-110-final-viral-combined_k141_166764\|\|full | Not-determined |
| 18-110-final-viral-combined_k141_120497\|\|full | Low-quality |
| 18-105_k141_52566 | High-quality |
| 18-110-final-viral-combined_k141_97849\|\|full | Low-quality |
| 18-110-final-viral-combined_k141_29118\|\|full | Not-determined |
| 18-110-final-viral-combined_k141_200399\|\|full | Low-quality |
| 18-110-final-viral-combined_k141_120509\|\|full | Not-determined |
| 18-110-final-viral-combined_k141_63621\|\|full | Low-quality |
| 18-110-final-viral-combined_k141_236576\|\|full | Not-determined |
| 18-110-final-viral-combined_k141_63626\|\|full | Not-determined |
| 18-110-final-viral-combined_k141_212931\|\|full | Not-determined |
| 18-110-final-viral-combined_k141_97869\|\|full | Low-quality |
| 18-110-final-viral-combined_k141_40364\|\|full | Low-quality |
| 18-110-final-viral-combined_k141_155473\|\|full | Low-quality |
| 18-110-final-viral-combined_k141_5050\|\|full | Low-quality |
| 18-110-final-viral-combined_k141_247778\|\|full | Not-determined |
| 18-110-final-viral-combined_k141_179335\|\|full | Low-quality |
| 18-110-final-viral-combined_k141_108880\|\|full | Low-quality |
| 18-110-final-viral-combined_k141_15983\|\|full | Not-determined |
| 18-110-final-viral-combined_k141_212949\|\|full | Not-determined |
| 18-110-final-viral-combined_k141_155484\|\|full | Low-quality |

18-110-final-viral-combined_k141_120543||full Low-quality
18-110-final-viral-combined_k141_190172||full Not-determined
18-110-final-viral-combined_k141_29165||full Low-quality
18-110-final-viral-combined_k141_120545||full Low-quality
18-110-final-viral-combined_k141_40382||full Not-determined
18-110-final-viral-combined_k141_108888||full Low-quality
18-110-final-viral-combined_k141_224178||full Low-quality
18-110-final-viral-combined_k141_190182||full Not-determined
18-110-final-viral-combined_k141_212951||full Low-quality
18-110-final-viral-combined_k141_29183||full Low-quality
18-110-final-viral-combined_k141_108908||full Low-quality
18-110-final-viral-combined_k141_51530||full Not-determined
18-110-final-viral-combined_k141_236609||full Low-quality
18-110-final-viral-combined_k141_63641||full Low-quality
18-110-final-viral-combined_k141_236616||full Not-determined
18-110-final-viral-combined_k141_155513||full Not-determined
18-110-final-viral-combined_k141_212969||full Not-determined
18-110-final-viral-combined_k141_40398||full Low-quality
18-110-final-viral-combined_k141_270659||full Not-determined
18-110-final-viral-combined_k141_5070||full Low-quality
18-110-final-viral-combined_k141_270665||full Not-determined
18-110-final-viral-combined_k141_155536||full Low-quality
18-110-final-viral-combined_k141_120565||full Not-determined
18-110-final-viral-combined_k141_236635||full Not-determined
18-110-final-viral-combined_k141_120574||full Not-determined
18-110-final-viral-combined_k141_236638||full Low-quality
18-110-final-viral-combined_k141_155563||full Low-quality
18-110-final-viral-combined_k141_190208||full Not-determined
18-110-final-viral-combined_k141_257847||full Not-determined
18-110-final-viral-combined_k141_155572||full Low-quality
18-110-final-viral-combined_k141_16045||full Not-determined
18-110-final-viral-combined_k141_63697||full Low-quality
18-110-final-viral-combined_k141_247827||full Low-quality
18-110-final-viral-combined_k141_16056||full Low-quality
18-110-final-viral-combined_k141_51582||full Not-determined
18-110-final-viral-combined_k141_270728||full Not-determined
18-110-final-viral-combined_k141_155622||full Low-quality
18-110-final-viral-combined_k141_29262||full Not-determined
18-110-final-viral-combined_k141_51590||full Low-quality
18-110-final-viral-combined_k141_120611||full Not-determined
KM_k141_254872 High-quality
18-110-final-viral-combined_k141_270743||full Low-quality
18-110-final-viral-combined_k141_247840||full Not-determined
18-110-final-viral-combined_k141_155637||full Not-determined
18-110-final-viral-combined_k141_270750||full Not-determined
18-110-final-viral-combined_k141_120627||full Not-determined
18-110-final-viral-combined_k141_5151||full Low-quality
18-110-final-viral-combined_k141_85809||full Low-quality
18-110-final-viral-combined_k141_98018||full Not-determined
18-110-final-viral-combined_k141_29282||full Not-determined
18-110-final-viral-combined_k141_120636||full Not-determined
18-110-final-viral-combined_k141_247870||full Low-quality
18-110-final-viral-combined_k141_40508||full Not-determined
18-110-final-viral-combined_k141_51616||full Low-quality
18-110-final-viral-combined_k141_200478||full Not-determined
18-110-final-viral-combined_k141_270783||full Low-quality
18-110-final-viral-combined_k141_51641||full Not-determined
18-110-final-viral-combined_k141_85842||full Low-quality
18-110-final-viral-combined_k141_257915||full Not-determined
18-110-final-viral-combined_k141_40538||full Not-determined
18-110-final-viral-combined_k141_120656||full Low-quality
18-110-final-viral-combined_k141_98085||full Low-quality
18-110-final-viral-combined_k141_190284||full Not-determined
18-110-final-viral-combined_k141_29343||full Low-quality
18-110-final-viral-combined_k141_98100||full Low-quality
18-110-final-viral-combined_k141_155656||full Not-determined
18-110-final-viral-combined_k141_5214||full Low-quality
18-110-final-viral-combined_k141_132540||full Not-determined
18-110-final-viral-combined_k141_75098||full Not-determined
18-110-final-viral-combined_k141_179484||full Not-determined
18-110-final-viral-combined_k141_85918||full Not-determined
18-110-final-viral-combined_k141_155691||full Not-determined
18-110-final-viral-combined_k141_247928||full Low-quality
18-110-final-viral-combined_k141_200556||full Low-quality
18-110-final-viral-combined_k141_270857||full Not-determined
18-110-final-viral-combined_k141_236712||full Not-determined
18-110-final-viral-combined_k141_236715||full Not-determined
18-110-final-viral-combined_k141_75139||full Not-determined
18-110-final-viral-combined_k141_224390||full Not-determined
18-110-final-viral-combined_k141_120720||full Low-quality
18-110-final-viral-combined_k141_190357||full Not-determined
18-110-final-viral-combined_k141_247958||full Low-quality
18-110-final-viral-combined_k141_51753||full Not-determined
18-110-final-viral-combined_k141_270881||full Not-determined
18-110-final-viral-combined_k141_109011||full Low-quality
18-110-final-viral-combined_k141_179503||full Not-determined
18-110-final-viral-combined_k141_155722||full Not-determined
18-110-final-viral-combined_k141_247976||full Not-determined
18-110-final-viral-combined_k141_224413||full Low-quality
18-110-final-viral-combined_k141_109016||full Not-determined
18-110-final-viral-combined_k141_5261||full Not-determined
18-110-final-viral-combined_k141_75183||full Not-determined
18-110-final-viral-combined_k141_224419||full Not-determined
18-110-final-viral-combined_k141_98215||full Low-quality
18-110-final-viral-combined_k141_98217||full Not-determined
18-110-final-viral-combined_k141_247991||full Low-quality
18-110-final-viral-combined_k141_236738||full Not-determined
18-110-final-viral-combined_k141_155748||full Not-determined
18-110-final-viral-combined_k141_63752||full Low-quality
18-110-final-viral-combined_k141_248004||full Low-quality
18-110-final-viral-combined_k141_155762||full Low-quality
18-110-final-viral-combined_k141_132680||full Low-quality
18-110-final-viral-combined_k141_29456||full Low-quality
18-110-final-viral-combined_k141_236755||full Not-determined
18-110-final-viral-combined_k141_75205||full Not-determined
18-110-final-viral-combined_k141_5298||full Low-quality
18-110-final-viral-combined_k141_5303||full Low-quality
18-110-final-viral-combined_k141_224453||full Not-determined
18-110-final-viral-combined_k141_63761||full Low-quality
18-110-final-viral-combined_k141_270936||full Low-quality
18-110-final-viral-combined_k141_155781||full Not-determined
18-110-final-viral-combined_k141_200646||full Low-quality
18-110-final-viral-combined_k141_40640||full Low-quality
18-110-final-viral-combined_k141_29477||full Low-quality
18-110-final-viral-combined_k141_40641||full Low-quality
18-110-final-viral-combined_k141_144236||full Not-determined
18-110-final-viral-combined_k141_63775||full Not-determined
18-110-final-viral-combined_k141_109072||full Low-quality
18-110-final-viral-combined_k141_270944||full Not-determined
18-110-final-viral-combined_k141_270947||full Not-determined
18-110-final-viral-combined_k141_109078||full Not-determined
18-110-final-viral-combined_k141_236782||full Not-determined
18-110-final-viral-combined_k141_40659||full Not-determined
18-110-final-viral-combined_k141_270949||full Not-determined
18-110-final-viral-combined_k141_63789||full Not-determined
18-110-final-viral-combined_k141_29509||full Not-determined
18-110-final-viral-combined_k141_144245||full Low-quality
18-110-final-viral-combined_k141_155802||full Not-determined
18-110-final-viral-combined_k141_16230||full Not-determined
18-110-final-viral-combined_k141_200676||full Low-quality
18-110-final-viral-combined_k141_179586||full Low-quality
18-110-final-viral-combined_k141_63813||full Not-determined
18-110-final-viral-combined_k141_29533||full Not-determined
17-106_k141_128232 Complete
18-110-final-viral-combined_k141_248077||full Low-quality
18-110-final-viral-combined_k141_155816||full Low-quality
18-110-final-viral-combined_k141_86061||full Low-quality
18-110-final-viral-combined_k141_190401||full Low-quality
18-110-final-viral-combined_k141_270960||full Low-quality
18-110-final-viral-combined_k141_144267||full Low-quality
18-110-final-viral-combined_k141_120802||full Low-quality
18-110-final-viral-combined_k141_179612||full Not-determined
18-110-final-viral-combined_k141_155827||full Not-determined
18-110-final-viral-combined_k141_224539||full Not-determined
18-110-final-viral-combined_k141_190417||full Low-quality
18-110-final-viral-combined_k141_179622||full Not-determined
18-110-final-viral-combined_k141_258057||full Low-quality
18-110-final-viral-combined_k141_144269||full Low-quality
18-110-final-viral-combined_k141_29571||full Low-quality
18-110-final-viral-combined_k141_5375||full Low-quality
18-110-final-viral-combined_k141_190439||full Low-quality
18-110-final-viral-combined_k141_248107||full Not-determined
18-110-final-viral-combined_k141_190445||full Low-quality
18-110-final-viral-combined_k141_51869||full Low-quality
18-110-final-viral-combined_k141_236823||full Not-determined
18-110-final-viral-combined_k141_5389||full Low-quality
18-110-final-viral-combined_k141_224572||full Low-quality
18-110-final-viral-combined_k141_5396||full Not-determined
18-110-final-viral-combined_k141_51886||full Not-determined
18-110-final-viral-combined_k141_179655||full Not-determined
18-110-final-viral-combined_k141_167117||full Low-quality
18-110-final-viral-combined_k141_155871||full Not-determined
18-110-final-viral-combined_k141_109159||full Low-quality
18-110-final-viral-combined_k141_120869||full Low-quality
18-110-final-viral-combined_k141_51895||full Not-determined
KM-final-viral-combined_k141_254872||full High-quality
18-110-final-viral-combined_k141_132807||full Low-quality
18-110-final-viral-combined_k141_98288||full Low-quality
18-110-final-viral-combined_k141_109169||full Not-determined
18-110-final-viral-combined_k141_248152||full Low-quality
18-110-final-viral-combined_k141_224605||full Not-determined
18-110-final-viral-combined_k141_179662||full Low-quality
18-110-final-viral-combined_k141_190461||full Low-quality
18-110-final-viral-combined_k141_120889||full Low-quality
18-110-final-viral-combined_k141_75325||full Not-determined
18-110-final-viral-combined_k141_29618||full Low-quality
18-110-final-viral-combined_k141_190470||full Low-quality
18-110-final-viral-combined_k141_167153||full Not-determined
18-110-final-viral-combined_k141_213115||full Not-determined
18-110-final-viral-combined_k141_98336||full Low-quality
18-110-final-viral-combined_k141_29636||full Low-quality
18-110-final-viral-combined_k141_109206||full Low-quality
18-110-final-viral-combined_k141_224634||full Low-quality
18-110-final-viral-combined_k141_144352||full Not-determined
18-110-final-viral-combined_k141_144354||full Low-quality
18-110-final-viral-combined_k141_120920||full Not-determined
18-110-final-viral-combined_k141_144366||full Low-quality
18-110-final-viral-combined_k141_63876||full Not-determined
18-110-final-viral-combined_k141_155955||full Not-determined
18-110-final-viral-combined_k141_86109||full Not-determined
18-110-final-viral-combined_k141_236915||full Not-determined
18-110-final-viral-combined_k141_16385||full Not-determined
18-110-final-viral-combined_k141_271016||full Not-determined
18-110-final-viral-combined_k141_29670||full Low-quality
18-110-final-viral-combined_k141_63894||full Not-determined
18-110-final-viral-combined_k141_224663||full Low-quality
18-110-final-viral-combined_k141_248226||full Not-determined
18-110-final-viral-combined_k141_271025||full Low-quality
18-110-final-viral-combined_k141_29687||full Low-quality
18-110-final-viral-combined_k141_258164||full Low-quality
18-110-final-viral-combined_k141_248242||full Not-determined
18-110-final-viral-combined_k141_271034||full Not-determined
18-110-final-viral-combined_k141_40746||full Low-quality
18-110-final-viral-combined_k141_29692||full Not-determined
18-110-final-viral-combined_k141_63937||full Low-quality
18-110-final-viral-combined_k141_109245||full Low-quality
18-110-final-viral-combined_k141_132855||full Not-determined
18-110-final-viral-combined_k141_51977||full Low-quality
18-110-final-viral-combined_k141_51978||full Not-determined
18-110-final-viral-combined_k141_144418||full Not-determined
18-110-final-viral-combined_k141_155984||full Low-quality
18-110-final-viral-combined_k141_40783||full Low-quality
18-110-final-viral-combined_k141_120990||full Low-quality
18-110-final-viral-combined_k141_16440||full Not-determined
18-110-final-viral-combined_k141_132904||full Not-determined
18-110-final-viral-combined_k141_237026||full Not-determined
18-110-final-viral-combined_k141_190547||full Low-quality
18-110-final-viral-combined_k141_29729||full Not-determined
18-110-final-viral-combined_k141_144457||full Not-determined
18-110-final-viral-combined_k141_86222||full Not-determined
18-110-final-viral-combined_k141_237035||full Low-quality
18-110-final-viral-combined_k141_155998||full Not-determined
18-110-final-viral-combined_k141_179815||full Not-determined
18-110-final-viral-combined_k141_213236||full Not-determined
18-110-final-viral-combined_k141_16455||full Not-determined
18-110-final-viral-combined_k141_29760||full Low-quality
18-110-final-viral-combined_k141_224776||full Not-determined
18-110-final-viral-combined_k141_132950||full Low-quality
18-110-final-viral-combined_k141_213246||full Not-determined
18-110-final-viral-combined_k141_248303||full Not-determined
18-110-final-viral-combined_k141_86237||full Not-determined
18-110-final-viral-combined_k141_190587||full Not-determined
18-110-final-viral-combined_k141_5560||full Low-quality
18-110-final-viral-combined_k141_144509||full Not-determined
18-110-final-viral-combined_k141_271091||full Low-quality
18-110-final-viral-combined_k141_40858||full Not-determined
18-110-final-viral-combined_k141_271099||full Not-determined
18-110-final-viral-combined_k141_5569||full Low-quality
18-110-final-viral-combined_k141_213255||full Low-quality
18-110-final-viral-combined_k141_109360||full Low-quality
18-110-final-viral-combined_k141_121041||full Not-determined
18-110-final-viral-combined_k141_190590||full Low-quality
18-110-final-viral-combined_k141_64043||full Not-determined
18-110-final-viral-combined_k141_224798||full Not-determined
18-110-final-viral-combined_k141_75383||full Not-determined
18-110-final-viral-combined_k141_86261||full Low-quality
18-110-final-viral-combined_k141_190591||full Low-quality
18-110-final-viral-combined_k141_224814||full Low-quality
18-110-final-viral-combined_k141_132977||full Low-quality
18-110-final-viral-combined_k141_5580||full Not-determined
18-110-final-viral-combined_k141_237109||full Low-quality
18-110-final-viral-combined_k141_109385||full Not-determined
18-110-final-viral-combined_k141_190619||full Not-determined
18-110-final-viral-combined_k141_52093||full Low-quality
18-110-final-viral-combined_k141_75433||full Not-determined
18-110-final-viral-combined_k141_237129||full Not-determined
18-110-final-viral-combined_k141_29807||full Low-quality
18-110-final-viral-combined_k141_121096||full Low-quality
18-110-final-viral-combined_k141_64094||full Not-determined
18-110-final-viral-combined_k141_156015||full Low-quality
18-110-final-viral-combined_k141_144561||full Low-quality
18-110-final-viral-combined_k141_52116||full Not-determined
18-110-final-viral-combined_k141_190637||full Low-quality
18-110-final-viral-combined_k141_190638||full Not-determined
18-110-final-viral-combined_k141_52117||full Not-determined
18-110-final-viral-combined_k141_40904||full Not-determined
18-110-final-viral-combined_k141_156022||full Not-determined
18-110-final-viral-combined_k141_156027||full Not-determined
18-110-final-viral-combined_k141_109423||full Not-determined
18-110-final-viral-combined_k141_213332||full Not-determined
18-110-final-viral-combined_k141_167439||full Not-determined
18-110-final-viral-combined_k141_109437||full Low-quality
18-110-final-viral-combined_k141_75485||full Low-quality
18-110-final-viral-combined_k141_190664||full Low-quality
18-110-final-viral-combined_k141_167449||full Not-determined
18-110-final-viral-combined_k141_224898||full Not-determined
18-110-final-viral-combined_k141_133021||full Low-quality
18-110-final-viral-combined_k141_156050||full Not-determined
18-110-final-viral-combined_k141_98450||full Low-quality
18-110-final-viral-combined_k141_75523||full Not-determined
18-110-final-viral-combined_k141_98459||full Not-determined
18-110-final-viral-combined_k141_156068||full Not-determined
18-110-final-viral-combined_k141_248379||full Low-quality
18-110-final-viral-combined_k141_248385||full Low-quality
18-110-final-viral-combined_k141_156096||full Not-determined
18-110-final-viral-combined_k141_167487||full Not-determined
18-110-final-viral-combined_k141_271227||full Low-quality
18-110-final-viral-combined_k141_213405||full Not-determined
18-110-final-viral-combined_k141_200920||full Not-determined
18-110-final-viral-combined_k141_52198||full Low-quality
18-110-final-viral-combined_k141_16485||full Low-quality
18-110-final-viral-combined_k141_41016||full Low-quality
18-110-final-viral-combined_k141_224929||full Not-determined
18-110-final-viral-combined_k141_200928||full Low-quality
18-110-final-viral-combined_k141_167500||full Low-quality
18-110-final-viral-combined_k141_258257||full Not-determined
18-110-final-viral-combined_k141_64209||full Low-quality
18-110-final-viral-combined_k141_52202||full Low-quality
18-110-final-viral-combined_k141_41025||full Not-determined
18-110-final-viral-combined_k141_98500||full Low-quality
18-110-final-viral-combined_k141_98507||full Low-quality
18-110-final-viral-combined_k141_258267||full Low-quality
18-110-final-viral-combined_k141_41027||full Not-determined
18-110-final-viral-combined_k141_213430||full Not-determined
18-110-final-viral-combined_k141_190742||full Low-quality
18-110-final-viral-combined_k141_213437||full Low-quality
18-110-final-viral-combined_k141_52213||full Not-determined
18-110-final-viral-combined_k141_271249||full Not-determined
18-110-final-viral-combined_k141_237195||full Low-quality
18-110-final-viral-combined_k141_200966||full Low-quality
18-110-final-viral-combined_k141_29833||full Not-determined
18-110-final-viral-combined_k141_224968||full Low-quality
18-110-final-viral-combined_k141_121188||full Not-determined
18-110-final-viral-combined_k141_5646||full Not-determined
18-110-final-viral-combined_k141_167525||full Low-quality
18-110-final-viral-combined_k141_41054||full Low-quality
18-110-final-viral-combined_k141_258279||full Low-quality
18-110-final-viral-combined_k141_144656||full Low-quality
18-110-final-viral-combined_k141_200982||full Low-quality
18-110-final-viral-combined_k141_86414||full Not-determined
18-110-final-viral-combined_k141_41071||full Not-determined
18-110-final-viral-combined_k141_64307||full Low-quality
18-110-final-viral-combined_k141_41088||full Low-quality
18-110-final-viral-combined_k141_64310||full Low-quality
17-106-final-viral-combined_k141_128232||full High-quality
18-110-final-viral-combined_k141_190785||full Not-determined
18-110-final-viral-combined_k141_258326||full Not-determined
18-110-final-viral-combined_k141_201046||full Low-quality
18-110-final-viral-combined_k141_52291||full Not-determined
18-110-final-viral-combined_k141_109599||full Low-quality
18-110-final-viral-combined_k141_167570||full Low-quality
18-110-final-viral-combined_k141_258330||full Not-determined
18-110-final-viral-combined_k141_190805||full Not-determined
18-110-final-viral-combined_k141_41121||full Not-determined
18-110-final-viral-combined_k141_190810||full Not-determined
18-110-final-viral-combined_k141_213539||full Low-quality
18-110-final-viral-combined_k141_41157||full Low-quality
18-110-final-viral-combined_k141_201060||full Not-determined
18-110-final-viral-combined_k141_52337||full Low-quality
18-110-final-viral-combined_k141_156238||full Low-quality
18-110-final-viral-combined_k141_190843||full Not-determined
18-110-final-viral-combined_k141_201063||full Not-determined
18-110-final-viral-combined_k141_213580||full Not-determined
18-110-final-viral-combined_k141_180065||full Not-determined
18-110-final-viral-combined_k141_29961||full Low-quality
18-110-final-viral-combined_k141_29974||full Not-determined
18-110-final-viral-combined_k141_64387||full Not-determined
18-110-final-viral-combined_k141_201083||full Not-determined
18-110-final-viral-combined_k141_52389||full Not-determined
18-110-final-viral-combined_k141_248532||full Not-determined
18-110-final-viral-combined_k141_109660||full Not-determined
18-110-final-viral-combined_k141_30003||full Not-determined
18-110-final-viral-combined_k141_213612||full Not-determined
18-110-final-viral-combined_k141_5726||full Not-determined
18-110-final-viral-combined_k141_213617||full Not-determined
18-110-final-viral-combined_k141_237249||full Not-determined
18-110-final-viral-combined_k141_75729||full Low-quality
18-110-final-viral-combined_k141_213621||full Low-quality
18-110-final-viral-combined_k141_201104||full Not-determined
18-110-final-viral-combined_k141_225109||full Not-determined
18-110-final-viral-combined_k141_237255||full Low-quality
18-110-final-viral-combined_k141_271343||full Low-quality
18-110-final-viral-combined_k141_98639||full Not-determined
18-110-final-viral-combined_k141_225113||full Low-quality
18-110-final-viral-combined_k141_64443||full Not-determined
18-110-final-viral-combined_k141_98648||full Not-determined
18-110-final-viral-combined_k141_41246||full Not-determined
18-110-final-viral-combined_k141_5763||full Not-determined
18-110-final-viral-combined_k141_180152||full Not-determined
18-110-final-viral-combined_k141_109685||full Low-quality
18-110-final-viral-combined_k141_144857||full Not-determined
18-110-final-viral-combined_k141_248566||full Not-determined
18-110-final-viral-combined_k141_248567||full Low-quality
18-110-final-viral-combined_k141_248568||full Not-determined
18-110-final-viral-combined_k141_86532||full Low-quality
18-110-final-viral-combined_k141_201130||full Low-quality
18-110-final-viral-combined_k141_109698||full Low-quality
18-110-final-viral-combined_k141_271371||full Not-determined
18-110-final-viral-combined_k141_258423||full Not-determined
18-110-final-viral-combined_k141_75795||full Low-quality
18-110-final-viral-combined_k141_121377||full Not-determined
18-110-final-viral-combined_k141_30067||full Low-quality
18-110-final-viral-combined_k141_180221||full Low-quality
18-109-final-viral-combined_k141_143218||0_partial High-quality
17-106-final-viral-combined_k141_56773||0_partial Medium-quality
18-110-final-viral-combined_k141_271396||full Low-quality
18-110-final-viral-combined_k141_75807||full Low-quality
18-110-final-viral-combined_k141_16712||full Low-quality
18-110-final-viral-combined_k141_16714||full Low-quality
18-110-final-viral-combined_k141_121407||full Low-quality
18-110-final-viral-combined_k141_180235||full Not-determined
18-110-final-viral-combined_k141_86576||full Low-quality
18-110-final-viral-combined_k141_271422||full Not-determined
18-110-final-viral-combined_k141_64539||full Not-determined
18-110-final-viral-combined_k141_5829||full Not-determined
18-110-final-viral-combined_k141_121421||full Low-quality
18-110-final-viral-combined_k141_133333||full Low-quality
18-110-final-viral-combined_k141_237380||full Low-quality
18-110-final-viral-combined_k141_52432||full Low-quality
18-110-final-viral-combined_k141_248616||full Low-quality
18-110-final-viral-combined_k141_64575||full Not-determined
18-110-final-viral-combined_k141_258477||full Not-determined
18-110-final-viral-combined_k141_167716||full Not-determined
18-110-final-viral-combined_k141_121444||full Not-determined
18-110-final-viral-combined_k141_201204||full Not-determined
18-110-final-viral-combined_k141_248637||full Not-determined
18-110-final-viral-combined_k141_201211||full Not-determined
18-110-final-viral-combined_k141_75850||full Not-determined
18-110-final-viral-combined_k141_86603||full Low-quality
18-110-final-viral-combined_k141_121453||full Low-quality
18-110-final-viral-combined_k141_180279||full Not-determined
18-110-final-viral-combined_k141_156301||full Not-determined
18-110-final-viral-combined_k141_75862||full Low-quality
18-110-final-viral-combined_k141_237415||full Not-determined
18-110-final-viral-combined_k141_109759||full Low-quality
18-110-final-viral-combined_k141_86622||full Not-determined
18-110-final-viral-combined_k141_109765||full Not-determined
18-110-final-viral-combined_k141_52477||full Not-determined
18-110-final-viral-combined_k141_16756||full Low-quality
18-110-final-viral-combined_k141_167766||full Not-determined
18-110-final-viral-combined_k141_121477||full Low-quality
18-110-final-viral-combined_k141_144975||full Not-determined
18-110-final-viral-combined_k141_144976||full Not-determined
18-110-final-viral-combined_k141_167767||full Low-quality
18-110-final-viral-combined_k141_5883||full Low-quality
18-110-final-viral-combined_k141_109797||full Low-quality
18-110-final-viral-combined_k141_86661||full Not-determined
18-110-final-viral-combined_k141_156356||full Not-determined
18-110-final-viral-combined_k141_225189||full Low-quality
18-110-final-viral-combined_k141_156358||full Low-quality
18-110-final-viral-combined_k141_52502||full Low-quality
18-110-final-viral-combined_k141_52505||full Not-determined
18-110-final-viral-combined_k141_156366||full Low-quality
18-110-final-viral-combined_k141_167785||full Low-quality
18-110-final-viral-combined_k141_225197||full Low-quality
18-110-final-viral-combined_k141_271521||full Low-quality
18-110-final-viral-combined_k141_271527||full Not-determined
18-110-final-viral-combined_k141_201279||full Low-quality
18-110-final-viral-combined_k141_109826||full Low-quality
18-110-final-viral-combined_k141_271528||full Not-determined
18-110-final-viral-combined_k141_213694||full Not-determined
18-110-final-viral-combined_k141_237494||full Not-determined
18-110-final-viral-combined_k141_41388||full Not-determined
18-110-final-viral-combined_k141_201327||full Not-determined
18-110-final-viral-combined_k141_180382||full Not-determined
18-110-final-viral-combined_k141_30131||full Not-determined
18-110-final-viral-combined_k141_145039||full Low-quality
18-110-final-viral-combined_k141_248719||full Not-determined
18-110-final-viral-combined_k141_41415||full Not-determined
18-110-final-viral-combined_k141_133459||full Not-determined
18-110-final-viral-combined_k141_52559||full Not-determined
18-110-final-viral-combined_k141_258564||full Not-determined
18-110-final-viral-combined_k141_225268||full Not-determined
18-110-final-viral-combined_k141_156453||full Low-quality
18-110-final-viral-combined_k141_201366||full Not-determined
18-110-final-viral-combined_k141_167869||full Not-determined
18-110-final-viral-combined_k141_201367||full Not-determined
18-110-final-viral-combined_k141_258578||full Not-determined
18-110-final-viral-combined_k141_258580||full Low-quality
18-110-final-viral-combined_k141_98826||full Not-determined
18-110-final-viral-combined_k141_133466||full Not-determined
18-110-final-viral-combined_k141_75963||full Not-determined
18-110-final-viral-combined_k141_225295||full Low-quality
18-110-final-viral-combined_k141_225299||full Not-determined
18-110-final-viral-combined_k141_86711||full Not-determined
18-110-final-viral-combined_k141_145084||full Not-determined
18-110-final-viral-combined_k141_201385||full Low-quality
18-110-final-viral-combined_k141_121582||full Low-quality
18-110-final-viral-combined_k141_201390||full Low-quality
18-110-final-viral-combined_k141_109918||full Not-determined
18-110-final-viral-combined_k141_16877||full Low-quality
18-110-final-viral-combined_k141_86727||full Low-quality
18-110-final-viral-combined_k141_6000||full Not-determined
18-110-final-viral-combined_k141_30224||full Not-determined
18-110-final-viral-combined_k141_201400||full Low-quality
18-110-final-viral-combined_k141_109932||full Low-quality
18-110-final-viral-combined_k141_145134||full Low-quality
18-110-final-viral-combined_k141_271615||full Low-quality
18-110-final-viral-combined_k141_145138||full Not-determined
18-110-final-viral-combined_k141_213802||full Not-determined
18-110-final-viral-combined_k141_109945||full Low-quality
18-110-final-viral-combined_k141_271626||full Not-determined
18-110-final-viral-combined_k141_52653||full Low-quality
18-110-final-viral-combined_k141_6037||full Low-quality
18-110-final-viral-combined_k141_248820||full Not-determined
18-110-final-viral-combined_k141_201427||full Low-quality
18-110-final-viral-combined_k141_191048||full Not-determined
17-106-final-viral-combined_k141_3878||full High-quality
18-110-final-viral-combined_k141_41497||full Low-quality
18-110-final-viral-combined_k141_41498||full Low-quality
18-110-final-viral-combined_k141_30234||full Not-determined
18-110-final-viral-combined_k141_133536||full Low-quality
18-110-final-viral-combined_k141_191056||full Low-quality
18-110-final-viral-combined_k141_258661||full Not-determined
18-110-final-viral-combined_k141_237584||full Not-determined
18-110-final-viral-combined_k141_213858||full Not-determined
18-110-final-viral-combined_k141_225399||full Not-determined
18-110-final-viral-combined_k141_201456||full Low-quality
18-110-final-viral-combined_k141_237585||full Not-determined
18-110-final-viral-combined_k141_41528||full Low-quality
18-110-final-viral-combined_k141_109996||full Low-quality
18-110-final-viral-combined_k141_225410||full Not-determined
18-110-final-viral-combined_k141_145180||full Low-quality
18-110-final-viral-combined_k141_121654||full Low-quality
18-110-final-viral-combined_k141_213876||full Not-determined
18-110-final-viral-combined_k141_271684||full Not-determined
18-110-final-viral-combined_k141_41535||full Not-determined
18-110-final-viral-combined_k141_86813||full Not-determined
18-110-final-viral-combined_k141_248890||full Low-quality
18-110-final-viral-combined_k141_64681||full Low-quality
18-110-final-viral-combined_k141_191110||full Not-determined
18-110-final-viral-combined_k141_237607||full Low-quality
18-110-final-viral-combined_k141_98929||full Not-determined
18-110-final-viral-combined_k141_110007||full Not-determined
18-110-final-viral-combined_k141_133581||full Low-quality
18-110-final-viral-combined_k141_110009||full Not-determined
18-110-final-viral-combined_k141_98942||full Low-quality
18-110-final-viral-combined_k141_213910||full Low-quality
18-110-final-viral-combined_k141_133599||full Not-determined
18-110-final-viral-combined_k141_237616||full Not-determined
18-110-final-viral-combined_k141_258722||full Low-quality
18-110-final-viral-combined_k141_168029||full Low-quality
18-110-final-viral-combined_k141_156563||full Not-determined
18-110-final-viral-combined_k141_52757||full Low-quality
18-110-final-viral-combined_k141_201484||full Low-quality
18-110-final-viral-combined_k141_168032||full Not-determined
18-110-final-viral-combined_k141_110037||full Low-quality
18-110-final-viral-combined_k141_191159||full Low-quality
18-110-final-viral-combined_k141_271733||full Not-determined
18-110-final-viral-combined_k141_86842||full Low-quality
18-110-final-viral-combined_k141_237632||full Low-quality
18-110-final-viral-combined_k141_248932||full Low-quality
18-110-final-viral-combined_k141_110046||full Not-determined
18-110-final-viral-combined_k141_110052||full Low-quality
18-110-final-viral-combined_k141_98983||full Low-quality
18-110-final-viral-combined_k141_201513||full Not-determined
18-110-final-viral-combined_k141_86869||full Not-determined
18-110-final-viral-combined_k141_41578||full Low-quality
18-110-final-viral-combined_k141_76073||full Not-determined
18-110-final-viral-combined_k141_6145||full Not-determined
18-110-final-viral-combined_k141_213967||full Not-determined
18-110-final-viral-combined_k141_168056||full Low-quality
18-110-final-viral-combined_k141_6152||full Low-quality
18-110-final-viral-combined_k141_86886||full Not-determined
18-110-final-viral-combined_k141_41580||full Not-determined
18-110-final-viral-combined_k141_30367||full Low-quality
18-110-final-viral-combined_k141_145214||full Low-quality
18-110-final-viral-combined_k141_86891||full Low-quality
18-110-final-viral-combined_k141_76104||full Not-determined
18-110-final-viral-combined_k141_258737||full Low-quality
18-110-final-viral-combined_k141_41597||full Low-quality
18-110-final-viral-combined_k141_258738||full Not-determined
18-110-final-viral-combined_k141_76107||full Not-determined
18-110-final-viral-combined_k141_121710||full Low-quality
18-110-final-viral-combined_k141_191219||full Not-determined
18-110-final-viral-combined_k141_30395||full Not-determined
18-110-final-viral-combined_k141_237676||full Low-quality
18-110-final-viral-combined_k141_258759||full Low-quality
18-110-final-viral-combined_k141_133682||full Low-quality
18-110-final-viral-combined_k141_110078||full Not-determined
18-110-final-viral-combined_k141_168106||full Low-quality
18-110-final-viral-combined_k141_6186||full Not-determined
18-110-final-viral-combined_k141_271786||full Low-quality
18-110-final-viral-combined_k141_133694||full Low-quality
18-110-final-viral-combined_k141_110084||full Not-determined
18-110-final-viral-combined_k141_156700||full Low-quality
18-110-final-viral-combined_k141_121734||full Not-determined
18-110-final-viral-combined_k141_237708||full Low-quality
18-110-final-viral-combined_k141_121737||full Low-quality
18-110-final-viral-combined_k141_156727||full Low-quality
18-110-final-viral-combined_k141_110102||full Not-determined
18-110-final-viral-combined_k141_180607||full Not-determined
18-110-final-viral-combined_k141_225577||full Low-quality
18-110-final-viral-combined_k141_64777||full Low-quality
18-110-final-viral-combined_k141_225586||full Not-determined
18-110-final-viral-combined_k141_64781||full Not-determined
18-110-final-viral-combined_k141_237720||full Low-quality
18-110-final-viral-combined_k141_225590||full Not-determined
18-110-final-viral-combined_k141_41661||full Not-determined
18-110-final-viral-combined_k141_156736||full Not-determined
18-110-final-viral-combined_k141_214041||full Low-quality
18-110-final-viral-combined_k141_110123||full Not-determined
18-110-final-viral-combined_k141_237730||full Not-determined
18-110-final-viral-combined_k141_271824||full Low-quality
18-110-final-viral-combined_k141_271826||full Low-quality
18-110-final-viral-combined_k141_133746||full Low-quality
18-110-final-viral-combined_k141_41683||full Low-quality
18-110-final-viral-combined_k141_121758||full Low-quality
18-110-final-viral-combined_k141_168194||full Not-determined

| | |
|---|---|
| 18-110-final-viral-combined_k141_180632\|\|full | Low-quality |
| 18-110-final-viral-combined_k141_6242\|\|full | Not-determined |
| 18-110-final-viral-combined_k141_99151\|\|full | Not-determined |
| 18-110-final-viral-combined_k141_52779\|\|full | Not-determined |
| 18-110-final-viral-combined_k141_99155\|\|full | Low-quality |
| 18-110-final-viral-combined_k141_180645\|\|full | Low-quality |
| 18-110-final-viral-combined_k141_64824\|\|full | Low-quality |
| 17-109-final-viral-combined_k141_401383\|\|full | High-quality |
| 18-110-final-viral-combined_k141_145326\|\|full | Low-quality |
| 18-110-final-viral-combined_k141_99156\|\|full | Not-determined |
| 18-110-final-viral-combined_k141_271840\|\|full | Low-quality |
| 18-110-final-viral-combined_k141_225621\|\|full | Low-quality |
| 18-110-final-viral-combined_k141_133779\|\|full | Not-determined |
| 18-110-final-viral-combined_k141_145341\|\|full | Low-quality |
| 18-110-final-viral-combined_k141_201589\|\|full | Not-determined |
| 18-110-final-viral-combined_k141_52806\|\|full | Not-determined |
| 18-110-final-viral-combined_k141_41724\|\|full | Not-determined |
| 18-110-final-viral-combined_k141_145352\|\|full | Not-determined |
| 18-110-final-viral-combined_k141_258826\|\|full | Not-determined |
| 18-110-final-viral-combined_k141_52815\|\|full | Not-determined |
| 18-110-final-viral-combined_k141_271876\|\|full | Not-determined |
| 18-110-final-viral-combined_k141_110137\|\|full | Not-determined |
| 18-110-final-viral-combined_k141_99192\|\|full | Not-determined |
| 18-110-final-viral-combined_k141_86981\|\|full | Not-determined |
| 18-110-final-viral-combined_k141_64853\|\|full | Low-quality |
| 18-110-final-viral-combined_k141_64855\|\|full | Low-quality |
| 18-110-final-viral-combined_k141_214092\|\|full | Not-determined |
| 18-110-final-viral-combined_k141_99198\|\|full | Low-quality |
| 18-110-final-viral-combined_k141_145369\|\|full | Not-determined |
| 18-110-final-viral-combined_k141_145375\|\|full | Not-determined |
| 18-110-final-viral-combined_k141_258849\|\|full | Not-determined |
| 18-110-final-viral-combined_k141_76254\|\|full | Low-quality |
| 18-110-final-viral-combined_k141_121839\|\|full | Not-determined |
| 18-110-final-viral-combined_k141_191341\|\|full | Not-determined |
| 18-110-final-viral-combined_k141_145381\|\|full | Not-determined |
| 18-110-final-viral-combined_k141_30540\|\|full | Not-determined |
| 18-110-final-viral-combined_k141_133832\|\|full | Not-determined |
| 18-110-final-viral-combined_k141_17089\|\|full | Low-quality |
| 18-110-final-viral-combined_k141_156843\|\|full | Low-quality |
| 18-110-final-viral-combined_k141_133844\|\|full | Not-determined |
| 18-110-final-viral-combined_k141_64867\|\|full | Low-quality |
| 18-110-final-viral-combined_k141_121856\|\|full | Low-quality |
| 18-110-final-viral-combined_k141_76285\|\|full | Low-quality |
| 18-110-final-viral-combined_k141_225671\|\|full | Not-determined |
| 18-110-final-viral-combined_k141_258886\|\|full | Not-determined |
| 18-110-final-viral-combined_k141_30585\|\|full | Not-determined |
| 18-110-final-viral-combined_k141_121874\|\|full | Low-quality |
| 18-110-final-viral-combined_k141_168306\|\|full | Low-quality |
| 18-110-final-viral-combined_k141_145424\|\|full | Not-determined |
| 18-110-final-viral-combined_k141_237757\|\|full | Not-determined |
| 18-110-final-viral-combined_k141_248993\|\|full | Not-determined |
| 18-110-final-viral-combined_k141_30609\|\|full | Not-determined |
| 18-110-final-viral-combined_k141_248996\|\|full | Not-determined |
| 18-110-final-viral-combined_k141_99216\|\|full | Not-determined |
| 18-110-final-viral-combined_k141_271929\|\|full | Not-determined |
| 18-110-final-viral-combined_k141_99221\|\|full | Low-quality |
| 18-110-final-viral-combined_k141_249008\|\|full | Not-determined |
| 18-110-final-viral-combined_k141_258904\|\|full | Low-quality |
| 18-110-final-viral-combined_k141_110179\|\|full | Low-quality |
| 18-110-final-viral-combined_k141_168325\|\|full | Not-determined |
| 18-110-final-viral-combined_k141_145442\|\|full | Low-quality |
| 18-110-final-viral-combined_k141_214158\|\|full | Not-determined |
| 18-110-final-viral-combined_k141_180757\|\|full | Not-determined |
| 18-110-final-viral-combined_k141_145445\|\|full | Not-determined |
| 18-110-final-viral-combined_k141_168336\|\|full | Not-determined |
| 18-110-final-viral-combined_k141_191417\|\|full | Not-determined |
| 18-110-final-viral-combined_k141_6351\|\|full | Low-quality |
| 18-110-final-viral-combined_k141_30656\|\|full | Not-determined |
| 18-110-final-viral-combined_k141_145455\|\|full | Not-determined |
| 18-110-final-viral-combined_k141_30669\|\|full | Not-determined |
| 18-110-final-viral-combined_k141_249054\|\|full | Low-quality |
| 18-110-final-viral-combined_k141_214183\|\|full | Not-determined |
| 18-110-final-viral-combined_k141_133892\|\|full | Not-determined |
| 18-110-final-viral-combined_k141_214187\|\|full | Low-quality |
| 18-110-final-viral-combined_k141_30683\|\|full | Low-quality |
| 18-110-final-viral-combined_k141_249078\|\|full | Low-quality |
| 18-110-final-viral-combined_k141_237823\|\|full | Not-determined |
| 18-110-final-viral-combined_k141_99265\|\|full | Not-determined |
| 18-110-final-viral-combined_k141_30706\|\|full | Not-determined |
| 18-110-final-viral-combined_k141_237831\|\|full | Not-determined |
| 18-110-final-viral-combined_k141_121932\|\|full | Low-quality |
| 18-110-final-viral-combined_k141_156936\|\|full | Not-determined |
| 18-110-final-viral-combined_k141_110230\|\|full | Not-determined |
| 18-110-final-viral-combined_k141_87156\|\|full | Not-determined |
| 18-110-final-viral-combined_k141_168396\|\|full | Low-quality |
| 18-110-final-viral-combined_k141_214200\|\|full | Not-determined |
| 18-110-final-viral-combined_k141_191505\|\|full | Low-quality |
| 18-110-final-viral-combined_k141_156983\|\|full | Not-determined |
| 18-110-final-viral-combined_k141_191512\|\|full | Not-determined |
| 18-110-final-viral-combined_k141_6389\|\|full | Low-quality |
| 18-110-final-viral-combined_k141_87229\|\|full | Low-quality |
| 18-110-final-viral-combined_k141_110285\|\|full | Not-determined |
| 18-110-final-viral-combined_k141_168430\|\|full | Low-quality |
| 18-110-final-viral-combined_k141_157004\|\|full | Low-quality |
| 18-110-final-viral-combined_k141_53019\|\|full | Low-quality |
| 18-110-final-viral-combined_k141_76422\|\|full | Low-quality |
| 18-110-final-viral-combined_k141_225811\|\|full | Low-quality |
| 18-110-final-viral-combined_k141_237917\|\|full | Not-determined |
| 18-110-final-viral-combined_k141_168450\|\|full | Low-quality |
| 18-110-final-viral-combined_k141_65021\|\|full | Not-determined |
| 18-110-final-viral-combined_k141_87256\|\|full | Not-determined |
| 18-110-final-viral-combined_k141_6434\|\|full | Low-quality |
| 18-110-final-viral-combined_k141_157043\|\|full | Low-quality |
| 18-110-final-viral-combined_k141_157049\|\|full | Low-quality |
| 18-110-final-viral-combined_k141_237943\|\|full | Low-quality |
| 18-110-final-viral-combined_k141_249145\|\|full | Low-quality |
| 18-110-final-viral-combined_k141_180883\|\|full | Low-quality |
| 18-110-final-viral-combined_k141_237950\|\|full | Low-quality |
| 18-110-final-viral-combined_k141_6451\|\|full | Low-quality |
| 18-110-final-viral-combined_k141_30785\|\|full | Not-determined |
| 18-110-final-viral-combined_k141_110338\|\|full | Not-determined |
| 18-110-final-viral-combined_k141_87296\|\|full | Not-determined |
| 18-110-final-viral-combined_k141_134073\|\|full | Low-quality |
| 18-110-final-viral-combined_k141_53056\|\|full | Low-quality |
| 18-110-final-viral-combined_k141_157105\|\|full | Not-determined |
| 18-110-final-viral-combined_k141_76482\|\|full | Low-quality |
| 18-110-final-viral-combined_k141_180913\|\|full | Low-quality |
| 18-110-final-viral-combined_k141_30804\|\|full | Not-determined |
| 18-110-final-viral-combined_k141_191582\|\|full | Low-quality |
| 18-110-final-viral-combined_k141_17247\|\|full | Not-determined |
| 18-110-final-viral-combined_k141_272120\|\|full | Not-determined |
| 18-110-final-viral-combined_k141_99396\|\|full | Not-determined |
| 18-110-final-viral-combined_k141_65100\|\|full | Not-determined |
| 18-110-final-viral-combined_k141_237982\|\|full | Not-determined |
| 18-110-final-viral-combined_k141_99404\|\|full | Not-determined |
| 18-110-final-viral-combined_k141_76526\|\|full | Low-quality |
| 18-110-final-viral-combined_k141_225836\|\|full | Not-determined |
| 18-110-final-viral-combined_k141_191604\|\|full | Not-determined |
| 18-110-final-viral-combined_k141_110362\|\|full | Low-quality |
| 18-110-final-viral-combined_k141_65114\|\|full | Not-determined |
| 18-110-final-viral-combined_k141_17259\|\|full | Not-determined |
| 18-110-final-viral-combined_k141_272127\|\|full | Not-determined |
| 18-110-final-viral-combined_k141_87389\|\|full | Low-quality |
| 18-110-final-viral-combined_k141_180942\|\|full | Not-determined |
| 17-106_k141_64472 | High-quality |
| 18-110-final-viral-combined_k141_122060\|\|full | Low-quality |
| 18-110-final-viral-combined_k141_168476\|\|full | Low-quality |
| 18-110-final-viral-combined_k141_110394\|\|full | Not-determined |
| 18-110-final-viral-combined_k141_65127\|\|full | Not-determined |
| 18-110-final-viral-combined_k141_30868\|\|full | Not-determined |
| 18-110-final-viral-combined_k141_272147\|\|full | Low-quality |
| 18-110-final-viral-combined_k141_87427\|\|full | Low-quality |
| 18-110-final-viral-combined_k141_238031\|\|full | Not-determined |
| 17-109-final-viral-combined_k141_75733\|\|full | High-quality |
| 18-110-final-viral-combined_k141_272158\|\|full | Low-quality |
| 18-110-final-viral-combined_k141_157157\|\|full | Not-determined |
| 18-110-final-viral-combined_k141_41803\|\|full | Not-determined |
| 18-110-final-viral-combined_k141_272160\|\|full | Low-quality |
| 18-110-final-viral-combined_k141_272162\|\|full | Not-determined |
| 18-110-final-viral-combined_k141_134139\|\|full | Not-determined |
| 18-110-final-viral-combined_k141_225890\|\|full | Not-determined |
| 18-110-final-viral-combined_k141_201844\|\|full | Not-determined |
| 18-110-final-viral-combined_k141_110416\|\|full | Low-quality |
| 18-110-final-viral-combined_k141_87458\|\|full | Not-determined |
| 18-110-final-viral-combined_k141_17333\|\|full | Low-quality |
| 18-110-final-viral-combined_k141_110417\|\|full | Low-quality |
| 18-110-final-viral-combined_k141_238061\|\|full | Low-quality |
| 18-110-final-viral-combined_k141_201849\|\|full | Not-determined |
| 18-110-final-viral-combined_k141_30918\|\|full | Not-determined |
| 18-110-final-viral-combined_k141_87465\|\|full | Not-determined |
| 18-110-final-viral-combined_k141_238067\|\|full | Not-determined |
| 18-110-final-viral-combined_k141_272181\|\|full | Low-quality |
| 18-110-final-viral-combined_k141_30926\|\|full | Low-quality |
| 18-110-final-viral-combined_k141_201866\|\|full | Low-quality |
| 18-110-final-viral-combined_k141_157201\|\|full | Low-quality |
| 18-110-final-viral-combined_k141_249245\|\|full | Low-quality |
| 18-110-final-viral-combined_k141_214365\|\|full | Low-quality |
| 18-110-final-viral-combined_k141_157208\|\|full | Low-quality |
| 18-110-final-viral-combined_k141_168547\|\|full | Low-quality |
| 18-110-final-viral-combined_k141_6522\|\|full | Not-determined |
| 18-110-final-viral-combined_k141_53162\|\|full | Low-quality |
| 18-110-final-viral-combined_k141_76613\|\|full | Not-determined |
| 18-110-final-viral-combined_k141_110487\|\|full | Low-quality |
| 18-110-final-viral-combined_k141_110493\|\|full | Low-quality |
| 18-110-final-viral-combined_k141_30994\|\|full | Low-quality |
| 18-110-final-viral-combined_k141_201884\|\|full | Low-quality |
| 18-110-final-viral-combined_k141_122171\|\|full | Low-quality |
| 18-110-final-viral-combined_k141_65204\|\|full | Low-quality |
| 18-110-final-viral-combined_k141_134225\|\|full | Low-quality |
| 18-110-final-viral-combined_k141_134227\|\|full | Low-quality |
| 18-110-final-viral-combined_k141_214398\|\|full | Not-determined |
| 18-110-final-viral-combined_k141_41843\|\|full | Not-determined |
| 18-110-final-viral-combined_k141_65222\|\|full | Not-determined |
| 18-110-final-viral-combined_k141_122179\|\|full | Low-quality |
| 18-110-final-viral-combined_k141_87570\|\|full | Not-determined |
| 18-110-final-viral-combined_k141_87578\|\|full | Low-quality |
| 18-110-final-viral-combined_k141_87581\|\|full | Low-quality |
| 18-110-final-viral-combined_k141_238154\|\|full | Not-determined |
| 18-110-final-viral-combined_k141_122200\|\|full | Not-determined |
| 18-110-final-viral-combined_k141_181120\|\|full | Not-determined |
| 18-110-final-viral-combined_k141_31023\|\|full | Low-quality |
| 18-110-final-viral-combined_k141_168637\|\|full | Not-determined |
| 18-110-final-viral-combined_k141_225941\|\|full | Not-determined |
| 18-110-final-viral-combined_k141_225942\|\|full | Not-determined |
| 18-110-final-viral-combined_k141_238184\|\|full | Not-determined |
| 18-110-final-viral-combined_k141_65269\|\|full | Not-determined |
| 18-110_k141_270 | High-quality |
| 18-110-final-viral-combined_k141_145826\|\|full | Not-determined |
| 18-110-final-viral-combined_k141_214453\|\|full | Not-determined |
| 18-110-final-viral-combined_k141_157283\|\|full | Low-quality |
| 18-110-final-viral-combined_k141_157285\|\|full | Not-determined |
| 18-110-final-viral-combined_k141_41875\|\|full | Low-quality |
| 18-110-final-viral-combined_k141_76670\|\|full | Not-determined |
| 18-110-final-viral-combined_k141_272268\|\|full | Not-determined |
| 18-110-final-viral-combined_k141_214463\|\|full | Low-quality |
| 18-110-final-viral-combined_k141_65280\|\|full | Low-quality |
| 18-110-final-viral-combined_k141_259209\|\|full | Low-quality |
| 18-110-final-viral-combined_k141_65295\|\|full | Not-determined |
| 18-110-final-viral-combined_k141_122252\|\|full | Not-determined |
| 18-110-final-viral-combined_k141_238233\|\|full | Not-determined |
| 18-110-final-viral-combined_k141_145864\|\|full | Not-determined |
| 18-110-final-viral-combined_k141_145872\|\|full | Low-quality |
| 18-110-final-viral-combined_k141_168694\|\|full | Not-determined |
| KM-final-viral-combined_k141_30308\|\|full | Medium-quality |
| 18-110-final-viral-combined_k141_226013\|\|full | Low-quality |
| 18-110-final-viral-combined_k141_134290\|\|full | Low-quality |
| 18-110-final-viral-combined_k141_41922\|\|full | Low-quality |
| 18-110-final-viral-combined_k141_249367\|\|full | Not-determined |
| 18-110-final-viral-combined_k141_76704\|\|full | Not-determined |
| 18-110-final-viral-combined_k141_201961\|\|full | Not-determined |
| 18-110-final-viral-combined_k141_201964\|\|full | Low-quality |
| 18-110-final-viral-combined_k141_17369\|\|full | Not-determined |
| 18-110-final-viral-combined_k141_214510\|\|full | Not-determined |
| 18-110-final-viral-combined_k141_31107\|\|full | Low-quality |
| 18-110-final-viral-combined_k141_201972\|\|full | Low-quality |
| 18-110-final-viral-combined_k141_168738\|\|full | Low-quality |
| 18-110-final-viral-combined_k141_214541\|\|full | Not-determined |
| 18-110-final-viral-combined_k141_157353\|\|full | Low-quality |
| 18-110-final-viral-combined_k141_157358\|\|full | Low-quality |
| 18-110-final-viral-combined_k141_17401\|\|full | Not-determined |
| 18-110-final-viral-combined_k141_157361\|\|full | Low-quality |
| 18-110-final-viral-combined_k141_145924\|\|full | Not-determined |
| 18-110-final-viral-combined_k141_122349\|\|full | Low-quality |
| 18-110-final-viral-combined_k141_168757\|\|full | Not-determined |
| 18-110-final-viral-combined_k141_99637\|\|full | Not-determined |
| 18-110-final-viral-combined_k141_53307\|\|full | Not-determined |
| 18-110-final-viral-combined_k141_168768\|\|full | Not-determined |
| 18-110-final-viral-combined_k141_31179\|\|full | Low-quality |
| 18-110-final-viral-combined_k141_65422\|\|full | Low-quality |
| 18-110-final-viral-combined_k141_168789\|\|full | Not-determined |
| 18-110-final-viral-combined_k141_6630\|\|full | Not-determined |
| 18-110-final-viral-combined_k141_122373\|\|full | Not-determined |
| 18-110-final-viral-combined_k141_202032\|\|full | Not-determined |
| 18-110-final-viral-combined_k141_42026\|\|full | Not-determined |
| 18-110-final-viral-combined_k141_145963\|\|full | Low-quality |
| 18-110-final-viral-combined_k141_226073\|\|full | Not-determined |
| 18-110-final-viral-combined_k141_134413\|\|full | Not-determined |
| 18-110-final-viral-combined_k141_31217\|\|full | Not-determined |
| 18-110-final-viral-combined_k141_168822\|\|full | Low-quality |
| 18-110-final-viral-combined_k141_6645\|\|full | Not-determined |
| 18-110-final-viral-combined_k141_99685\|\|full | Low-quality |
| 18-110-final-viral-combined_k141_238391\|\|full | Not-determined |
| 18-110-final-viral-combined_k141_31228\|\|full | Not-determined |
| 18-110-final-viral-combined_k141_202073\|\|full | Low-quality |
| 18-110-final-viral-combined_k141_53373\|\|full | Low-quality |
| 18-110-final-viral-combined_k141_42075\|\|full | Not-determined |
| 18-110-final-viral-combined_k141_272359\|\|full | Low-quality |
| 18-110-final-viral-combined_k141_6669\|\|full | Low-quality |
| 18-110-final-viral-combined_k141_168831\|\|full | Low-quality |
| 18-110-final-viral-combined_k141_76790\|\|full | Low-quality |
| 18-110-final-viral-combined_k141_146005\|\|full | Not-determined |
| 18-110-final-viral-combined_k141_249448\|\|full | Low-quality |
| 18-110-final-viral-combined_k141_99718\|\|full | Not-determined |
| 18-110-final-viral-combined_k141_134459\|\|full | Low-quality |
| 18-110-final-viral-combined_k141_87689\|\|full | Not-determined |
| 18-110-final-viral-combined_k141_65484\|\|full | Low-quality |
| 18-110-final-viral-combined_k141_270\|\|full | High-quality |
| 18-110-final-viral-combined_k141_87697\|\|full | Not-determined |
| 18-110-final-viral-combined_k141_214626\|\|full | Not-determined |
| 18-110-final-viral-combined_k141_249477\|\|full | Low-quality |
| 18-110-final-viral-combined_k141_202121\|\|full | Low-quality |
| 18-110-final-viral-combined_k141_17468\|\|full | Not-determined |
| 18-110-final-viral-combined_k141_202124\|\|full | Low-quality |
| 18-110-final-viral-combined_k141_259324\|\|full | Not-determined |
| 18-110-final-viral-combined_k141_87701\|\|full | Not-determined |
| 18-110-final-viral-combined_k141_226091\|\|full | Low-quality |
| 18-110-final-viral-combined_k141_134493\|\|full | Not-determined |
| 18-110-final-viral-combined_k141_122449\|\|full | Not-determined |
| 18-110-final-viral-combined_k141_272378\|\|full | Not-determined |
| 18-110-final-viral-combined_k141_31283\|\|full | Not-determined |
| 18-110-final-viral-combined_k141_168887\|\|full | Low-quality |
| 18-110-final-viral-combined_k141_238444\|\|full | Low-quality |
| 18-110-final-viral-combined_k141_214632\|\|full | Low-quality |
| 18-110-final-viral-combined_k141_157464\|\|full | Low-quality |
| 18-110-final-viral-combined_k141_31286\|\|full | Not-determined |
| 18-110-final-viral-combined_k141_259326\|\|full | Low-quality |
| 18-110-final-viral-combined_k141_157467\|\|full | Not-determined |
| 18-110-final-viral-combined_k141_238450\|\|full | Not-determined |
| 18-110-final-viral-combined_k141_31289\|\|full | Low-quality |
| 18-110-final-viral-combined_k141_65487\|\|full | Low-quality |
| 18-110-final-viral-combined_k141_53447\|\|full | Low-quality |
| 18-110-final-viral-combined_k141_272403\|\|full | Low-quality |
| 18-110-final-viral-combined_k141_53450\|\|full | Not-determined |
| 18-110-final-viral-combined_k141_76852\|\|full | Low-quality |
| 18-110-final-viral-combined_k141_110608\|\|full | Not-determined |
| 18-110-final-viral-combined_k141_259341\|\|full | Not-determined |
| 18-110-final-viral-combined_k141_76858\|\|full | Not-determined |
| 18-110-final-viral-combined_k141_259345\|\|full | Low-quality |
| 18-110-final-viral-combined_k141_17509\|\|full | Not-determined |
| 18-110-final-viral-combined_k141_226127\|\|full | Not-determined |
| 18-110-final-viral-combined_k141_168930\|\|full | Not-determined |
| 18-110-final-viral-combined_k141_6751\|\|full | Not-determined |
| 18-110-final-viral-combined_k141_214653\|\|full | Low-quality |
| 18-110-final-viral-combined_k141_53481\|\|full | Low-quality |
| 18-110-final-viral-combined_k141_249529\|\|full | Low-quality |
| 18-110-final-viral-combined_k141_99755\|\|full | Not-determined |
| 18-110-final-viral-combined_k141_259363\|\|full | Not-determined |
| 18-110-final-viral-combined_k141_110641\|\|full | Not-determined |
| 17-106-final-viral-combined_k141_64472\|\|full | Medium-quality |
| 18-110-final-viral-combined_k141_53492\|\|full | Low-quality |
| 18-110-final-viral-combined_k141_31323\|\|full | Not-determined |
| 18-110-final-viral-combined_k141_214665\|\|full | Low-quality |
| 18-110-final-viral-combined_k141_134521\|\|full | Low-quality |
| 18-110-final-viral-combined_k141_249546\|\|full | Low-quality |
| 18-110-final-viral-combined_k141_168952\|\|full | Low-quality |
| 18-110-final-viral-combined_k141_17527\|\|full | Not-determined |
| 18-110-final-viral-combined_k141_214667\|\|full | Not-determined |
| 18-110-final-viral-combined_k141_226147\|\|full | Not-determined |
| 18-110-final-viral-combined_k141_157544\|\|full | Not-determined |
| 18-110-final-viral-combined_k141_157545\|\|full | Low-quality |
| 18-110-final-viral-combined_k141_249548\|\|full | Low-quality |
| 18-110-final-viral-combined_k141_238545\|\|full | Low-quality |
| 18-110-final-viral-combined_k141_226162\|\|full | Not-determined |
| 18-110-final-viral-combined_k141_6781\|\|full | Low-quality |
| 18-110-final-viral-combined_k141_42221\|\|full | Low-quality |
| 18-110-final-viral-combined_k141_6783\|\|full | Not-determined |
| 18-110-final-viral-combined_k141_53529\|\|full | Low-quality |
| 18-110-final-viral-combined_k141_249573\|\|full | Not-determined |
| 18-110-final-viral-combined_k141_76945\|\|full | Not-determined |
| 18-110-final-viral-combined_k141_99792\|\|full | Not-determined |
| 18-110-final-viral-combined_k141_110729\|\|full | Not-determined |
| 18-110-final-viral-combined_k141_259413\|\|full | Not-determined |
| 18-110-final-viral-combined_k141_134547\|\|full | Low-quality |
| 18-110-final-viral-combined_k141_53555\|\|full | Not-determined |
| 18-110-final-viral-combined_k141_99810\|\|full | Not-determined |
| 18-110-final-viral-combined_k141_181378\|\|full | Low-quality |
| 18-110-final-viral-combined_k141_146184\|\|full | Low-quality |
| 18-110-final-viral-combined_k141_17588\|\|full | Low-quality |
| 18-110-final-viral-combined_k141_134565\|\|full | Not-determined |
| 18-110-final-viral-combined_k141_110763\|\|full | Low-quality |
| 18-110-final-viral-combined_k141_249608\|\|full | Not-determined |
| 18-110-final-viral-combined_k141_87800\|\|full | Not-determined |
| 18-110-final-viral-combined_k141_6828\|\|full | Low-quality |
| 18-110-final-viral-combined_k141_65607\|\|full | Low-quality |
| 18-110-final-viral-combined_k141_76996\|\|full | Low-quality |
| 18-110-final-viral-combined_k141_259437\|\|full | Not-determined |
| 18-110-final-viral-combined_k141_31415\|\|full | Low-quality |
| 18-110-final-viral-combined_k141_202246\|\|full | Not-determined |
| 18-110-final-viral-combined_k141_157592\|\|full | Low-quality |
| 18-110-final-viral-combined_k141_134587\|\|full | Not-determined |
| 18-110-final-viral-combined_k141_259438\|\|full | Not-determined |
| 18-110-final-viral-combined_k141_191768\|\|full | Low-quality |
| 18-110-final-viral-combined_k141_110801\|\|full | Not-determined |
| 18-110-final-viral-combined_k141_272493\|\|full | Low-quality |
| 18-110-final-viral-combined_k141_169026\|\|full | Not-determined |
| 18-110-final-viral-combined_k141_157607\|\|full | Not-determined |
| 18-110-final-viral-combined_k141_249627\|\|full | Low-quality |
| 18-110-final-viral-combined_k141_146233\|\|full | Not-determined |
| 18-110-final-viral-combined_k141_110813\|\|full | Low-quality |
| 18-110-final-viral-combined_k141_110814\|\|full | Not-determined |
| 18-110-final-viral-combined_k141_202272\|\|full | Low-quality |
| 18-110-final-viral-combined_k141_65662\|\|full | Not-determined |
| 18-110-final-viral-combined_k141_214780\|\|full | Low-quality |
| 18-110-final-viral-combined_k141_134605\|\|full | Low-quality |
| 18-110-final-viral-combined_k141_169053\|\|full | Not-determined |
| 18-110-final-viral-combined_k141_87847\|\|full | Low-quality |
| 18-110-final-viral-combined_k141_17648\|\|full | Low-quality |
| 18-110-final-viral-combined_k141_87853\|\|full | Low-quality |
| 18-110-final-viral-combined_k141_65683\|\|full | Not-determined |
| 18-110-final-viral-combined_k141_17650\|\|full | Low-quality |
| 18-110-final-viral-combined_k141_65689\|\|full | Not-determined |
| 18-110-final-viral-combined_k141_249665\|\|full | Low-quality |
| 18-110-final-viral-combined_k141_77045\|\|full | Low-quality |
| 18-110-final-viral-combined_k141_146274\|\|full | Not-determined |
| 18-110-final-viral-combined_k141_42281\|\|full | Not-determined |
| 18-110-final-viral-combined_k141_249675\|\|full | Low-quality |
| 18-110-final-viral-combined_k141_77065\|\|full | Not-determined |
| 18-110-final-viral-combined_k141_157672\|\|full | Low-quality |
| 18-110-final-viral-combined_k141_99938\|\|full | Not-determined |
| 18-110-final-viral-combined_k141_157681\|\|full | Not-determined |
| 18-110-final-viral-combined_k141_6955\|\|full | Not-determined |
| 18-110-final-viral-combined_k141_272544\|\|full | Not-determined |
| 18-110-final-viral-combined_k141_214849\|\|full | Not-determined |
| 18-110-final-viral-combined_k141_31516\|\|full | Low-quality |
| 18-110-final-viral-combined_k141_157703\|\|full | Not-determined |
| 18-110-final-viral-combined_k141_226307\|\|full | Low-quality |
| 18-110-final-viral-combined_k141_272563\|\|full | Not-determined |
| 18-110-final-viral-combined_k141_214864\|\|full | Not-determined |
| 18-110-final-viral-combined_k141_31529\|\|full | Low-quality |
| 18-110-final-viral-combined_k141_214866\|\|full | Not-determined |
| 18-110-final-viral-combined_k141_17713\|\|full | Not-determined |
| 18-110-final-viral-combined_k141_6979\|\|full | Low-quality |
| 18-110-final-viral-combined_k141_202354\|\|full | Low-quality |
| 18-110-final-viral-combined_k141_226313\|\|full | Not-determined |
| 18-110-final-viral-combined_k141_202359\|\|full | Not-determined |
| 18-110-final-viral-combined_k141_110899\|\|full | Low-quality |
| 18-110-final-viral-combined_k141_214872\|\|full | Not-determined |
| 18-110-final-viral-combined_k141_99988\|\|full | Not-determined |
| 18-110-final-viral-combined_k141_191921\|\|full | Not-determined |
| 18-110-final-viral-combined_k141_87954\|\|full | Low-quality |
| 18-110-final-viral-combined_k141_259507\|\|full | Not-determined |
| 18-110-final-viral-combined_k141_100009\|\|full | Low-quality |
| 18-110-final-viral-combined_k141_7023\|\|full | Low-quality |
| 18-110-final-viral-combined_k141_53718\|\|full | Low-quality |
| 18-110-final-viral-combined_k141_7030\|\|full | Low-quality |
| 18-110-final-viral-combined_k141_226362\|\|full | Low-quality |
| 18-110-final-viral-combined_k141_249735\|\|full | Low-quality |
| 18-110-final-viral-combined_k141_214910\|\|full | Low-quality |
| 18-110-final-viral-combined_k141_134668\|\|full | Low-quality |
| 18-110-final-viral-combined_k141_31599\|\|full | Not-determined |
| 18-110-final-viral-combined_k141_191981\|\|full | Not-determined |
| 18-110-final-viral-combined_k141_214917\|\|full | Low-quality |
| 18-110-final-viral-combined_k141_7063\|\|full | Not-determined |
| 18-110-final-viral-combined_k141_214925\|\|full | Not-determined |
| 18-110-final-viral-combined_k141_146423\|\|full | Low-quality |
| 18-110-final-viral-combined_k141_146424\|\|full | Low-quality |
| 18-110-final-viral-combined_k141_259568\|\|full | Low-quality |
| 18-110-final-viral-combined_k141_214933\|\|full | Low-quality |
| 18-110-final-viral-combined_k141_87995\|\|full | Low-quality |
| 18-110-final-viral-combined_k141_202428\|\|full | Not-determined |
| 18-110-final-viral-combined_k141_53761\|\|full | Not-determined |
| 18-110-final-viral-combined_k141_157794\|\|full | Not-determined |
| 18-110-final-viral-combined_k141_249769\|\|full | Low-quality |
| 18-110-final-viral-combined_k141_31631\|\|full | Low-quality |
| 18-110-final-viral-combined_k141_169108\|\|full | Not-determined |
| 18-110-final-viral-combined_k141_77098\|\|full | Low-quality |
| 18-110-final-viral-combined_k141_7099\|\|full | Not-determined |
| 18-110-final-viral-combined_k141_238810\|\|full | Not-determined |
| 18-110-final-viral-combined_k141_7101\|\|full | Low-quality |
| 18-110-final-viral-combined_k141_110984\|\|full | Not-determined |
| 18-110-final-viral-combined_k141_272663\|\|full | Not-determined |
| 18-110-final-viral-combined_k141_157826\|\|full | Not-determined |
| 18-110-final-viral-combined_k141_214967\|\|full | Low-quality |
| 18-110-final-viral-combined_k141_226428\|\|full | Low-quality |
| 18-110-final-viral-combined_k141_7111\|\|full | Not-determined |
| 18-110-final-viral-combined_k141_17848\|\|full | Low-quality |
| 18-110-final-viral-combined_k141_157843\|\|full | Low-quality |
| 18-110-final-viral-combined_k141_134754\|\|full | Not-determined |
| 18-110-final-viral-combined_k141_226435\|\|full | Not-determined |
| 18-110-final-viral-combined_k141_169128\|\|full | Not-determined |
| 18-110-final-viral-combined_k141_238854\|\|full | Low-quality |
| 18-110-final-viral-combined_k141_31689\|\|full | Not-determined |
| 18-110-final-viral-combined_k141_146490\|\|full | Not-determined |
| 18-110-final-viral-combined_k141_122795\|\|full | Not-determined |
| 18-110-final-viral-combined_k141_146491\|\|full | Not-determined |
| 18-110-final-viral-combined_k141_157861\|\|full | Low-quality |
| 18-110-final-viral-combined_k141_7133\|\|full | Not-determined |
| 18-110-final-viral-combined_k141_88073\|\|full | Not-determined |
| 18-110-final-viral-combined_k141_111027\|\|full | Not-determined |
| 18-110-final-viral-combined_k141_77169\|\|full | Low-quality |
| 18-110-final-viral-combined_k141_53823\|\|full | Low-quality |
| 18-110-final-viral-combined_k141_238884\|\|full | Not-determined |
| 18-110-final-viral-combined_k141_169159\|\|full | Low-quality |
| 18-110-final-viral-combined_k141_122826\|\|full | Not-determined |
| 18-110-final-viral-combined_k141_77194\|\|full | Low-quality |
| 18-110-final-viral-combined_k141_259653\|\|full | Not-determined |
| 18-110-final-viral-combined_k141_122828\|\|full | Not-determined |
| 18-110-final-viral-combined_k141_42430\|\|full | Not-determined |
| 18-110-final-viral-combined_k141_134781\|\|full | Not-determined |
| 18-110-final-viral-combined_k141_215025\|\|full | Not-determined |
| 18-110-final-viral-combined_k141_226476\|\|full | Not-determined |
| 18-110-final-viral-combined_k141_169175\|\|full | Low-quality |
| 18-110-final-viral-combined_k141_53845\|\|full | Not-determined |
| 18-110-final-viral-combined_k141_42442\|\|full | Not-determined |
| 18-110-final-viral-combined_k141_7145\|\|full | Not-determined |
| 18-110-final-viral-combined_k141_122847\|\|full | Not-determined |
| 18-110-final-viral-combined_k141_259663\|\|full | Not-determined |
| 18-110-final-viral-combined_k141_42451\|\|full | Not-determined |
| 18-110-final-viral-combined_k141_88113\|\|full | Not-determined |
| 18-110-final-viral-combined_k141_122851\|\|full | Low-quality |
| 18-110-final-viral-combined_k141_192109\|\|full | Not-determined |
| 18-110-final-viral-combined_k141_88115\|\|full | Low-quality |
| 18-110-final-viral-combined_k141_31728\|\|full | Not-determined |
| 18-110-final-viral-combined_k141_88119\|\|full | Not-determined |
| 18-110-final-viral-combined_k141_122859\|\|full | Low-quality |
| 18-110-final-viral-combined_k141_134813\|\|full | Low-quality |
| 18-110-final-viral-combined_k141_272718\|\|full | Low-quality |
| 18-110-final-viral-combined_k141_215056\|\|full | Not-determined |
| 18-110-final-viral-combined_k141_100090\|\|full | Not-determined |
| 18-110-final-viral-combined_k141_192125\|\|full | Low-quality |
| 18-110-final-viral-combined_k141_65861\|\|full | Low-quality |
| 18-110-final-viral-combined_k141_181681\|\|full | Low-quality |
| 18-110-final-viral-combined_k141_226502\|\|full | Not-determined |
| 18-110-final-viral-combined_k141_157917\|\|full | Low-quality |
| 18-110-final-viral-combined_k141_249891\|\|full | Low-quality |
| 18-110-final-viral-combined_k141_77249\|\|full | Low-quality |
| 18-110-final-viral-combined_k141_31732\|\|full | Low-quality |
| 18-110-final-viral-combined_k141_65877\|\|full | Not-determined |
| 18-110-final-viral-combined_k141_238934\|\|full | Not-determined |
| 18-110-final-viral-combined_k141_157933\|\|full | Low-quality |
| 18-110-final-viral-combined_k141_100117\|\|full | Not-determined |
| 18-110-final-viral-combined_k141_7195\|\|full | Not-determined |
| 17-109-final-viral-combined_k141_390985\|\|full | High-quality |
| 18-110-final-viral-combined_k141_215088\|\|full | Low-quality |
| 18-110-final-viral-combined_k141_65903\|\|full | Low-quality |
| KM-final-viral-combined_k141_223748\|\|full | Medium-quality |
| 18-110-final-viral-combined_k141_272792\|\|full | Low-quality |
| 18-110-final-viral-combined_k141_122918\|\|full | Not-determined |
| 18-110-final-viral-combined_k141_238977\|\|full | Not-determined |
| 18-110-final-viral-combined_k141_88145\|\|full | Low-quality |
| 18-110-final-viral-combined_k141_88153\|\|full | Low-quality |
| 18-110-final-viral-combined_k141_157977\|\|full | Low-quality |
| 18-110-final-viral-combined_k141_192145\|\|full | Not-determined |
| 18-110-final-viral-combined_k141_31797\|\|full | Low-quality |
| 18-110-final-viral-combined_k141_202482\|\|full | Not-determined |
| 18-110-final-viral-combined_k141_157984\|\|full | Low-quality |
| 18-110-final-viral-combined_k141_202488\|\|full | Low-quality |
| 18-110-final-viral-combined_k141_157988\|\|full | Not-determined |
| 18-110-final-viral-combined_k141_259713\|\|full | Low-quality |
| 18-110-final-viral-combined_k141_157991\|\|full | Low-quality |
| 18-110-final-viral-combined_k141_239027\|\|full | Low-quality |
| 18-110-final-viral-combined_k141_202500\|\|full | Low-quality |
| 18-110-final-viral-combined_k141_17958\|\|full | Low-quality |
| 18-110-final-viral-combined_k141_169312\|\|full | Low-quality |
| 18-110-final-viral-combined_k141_53973\|\|full | Not-determined |
| 18-110-final-viral-combined_k141_192183\|\|full | Not-determined |
| 18-110-final-viral-combined_k141_111114\|\|full | Low-quality |
| 18-110-final-viral-combined_k141_17971\|\|full | Not-determined |
| 18-110-final-viral-combined_k141_65936\|\|full | Low-quality |
| 18-110-final-viral-combined_k141_146599\|\|full | Low-quality |
| 18-110-final-viral-combined_k141_77313\|\|full | Low-quality |
| 18-110-final-viral-combined_k141_31813\|\|full | Not-determined |
| 18-110-final-viral-combined_k141_100173\|\|full | Low-quality |
| 18-110-final-viral-combined_k141_202521\|\|full | Not-determined |
| 18-110-final-viral-combined_k141_54003\|\|full | Not-determined |
| 18-110-final-viral-combined_k141_158014\|\|full | Low-quality |
| 18-110-final-viral-combined_k141_192193\|\|full | Not-determined |
| 18-110-final-viral-combined_k141_111124\|\|full | Low-quality |
| 18-110-final-viral-combined_k141_111127\|\|full | Low-quality |
| 18-110-final-viral-combined_k141_202529\|\|full | Not-determined |
| 18-110-final-viral-combined_k141_272859\|\|full | Not-determined |
| 18-110-final-viral-combined_k141_31823\|\|full | Low-quality |
| 18-110-final-viral-combined_k141_54011\|\|full | Not-determined |
| 18-110-final-viral-combined_k141_239065\|\|full | Not-determined |
| 18-110-final-viral-combined_k141_122979\|\|full | Not-determined |
| 18-110-final-viral-combined_k141_249907\|\|full | Low-quality |
| 18-110-final-viral-combined_k141_202542\|\|full | Low-quality |
| 18-110-final-viral-combined_k141_239070\|\|full | Not-determined |
| 18-110-final-viral-combined_k141_31829\|\|full | Not-determined |
| 18-110-final-viral-combined_k141_18013\|\|full | Low-quality |
| 18-110-final-viral-combined_k141_181844\|\|full | Not-determined |
| 18-110-final-viral-combined_k141_239082\|\|full | Low-quality |
| 18-110-final-viral-combined_k141_202560\|\|full | Not-determined |
| 18-110-final-viral-combined_k141_7245\|\|full | Low-quality |
| 18-110-final-viral-combined_k141_31856\|\|full | Low-quality |
| 18-110-final-viral-combined_k141_239087\|\|full | Not-determined |
| 18-110-final-viral-combined_k141_31859\|\|full | Low-quality |
| 18-110-final-viral-combined_k141_42579\|\|full | Not-determined |
| 18-110-final-viral-combined_k141_192227\|\|full | Not-determined |
| 18-110-final-viral-combined_k141_31864\|\|full | Not-determined |
| 18-110-final-viral-combined_k141_54053\|\|full | Not-determined |
| 18-110-final-viral-combined_k141_192242\|\|full | Low-quality |
| 18-110-final-viral-combined_k141_272928\|\|full | Not-determined |
| 18-110-final-viral-combined_k141_239118\|\|full | Not-determined |
| 18-110-final-viral-combined_k141_7271\|\|full | Low-quality |
| 18-110-final-viral-combined_k141_181882\|\|full | Not-determined |
| 18-110-final-viral-combined_k141_7278\|\|full | Not-determined |
| 18-110-final-viral-combined_k141_239129\|\|full | Not-determined |
| 18-110-final-viral-combined_k141_100214\|\|full | Not-determined |
| 18-110-final-viral-combined_k141_215148\|\|full | Low-quality |
| 18-110-final-viral-combined_k141_272964\|\|full | Not-determined |
| 18-110-final-viral-combined_k141_77446\|\|full | Not-determined |
| 18-110-final-viral-combined_k141_31907\|\|full | Low-quality |
| 18-110-final-viral-combined_k141_7303\|\|full | Not-determined |
| 18-110-final-viral-combined_k141_100221\|\|full | Not-determined |
| 18-110-final-viral-combined_k141_272969\|\|full | Not-determined |
| 18-110-final-viral-combined_k141_272972\|\|full | Low-quality |
| 18-110-final-viral-combined_k141_31913\|\|full | Low-quality |
| 18-110-final-viral-combined_k141_215167\|\|full | Not-determined |
| 18-110-final-viral-combined_k141_54105\|\|full | Not-determined |
| 18-110-final-viral-combined_k141_100233\|\|full | Low-quality |
| 18-110-final-viral-combined_k141_272994\|\|full | Not-determined |
| 18-110-final-viral-combined_k141_123093\|\|full | Low-quality |
| 18-110-final-viral-combined_k141_215190\|\|full | Not-determined |
| 18-110-final-viral-combined_k141_77506\|\|full | Low-quality |
| 18-110-final-viral-combined_k141_123098\|\|full | Low-quality |
| 18-110-final-viral-combined_k141_226645\|\|full | Not-determined |
| 18-110-final-viral-combined_k141_100278\|\|full | Low-quality |
| 18-110-final-viral-combined_k141_42714\|\|full | Not-determined |
| 18-110-final-viral-combined_k141_146759\|\|full | Not-determined |
| 18-110-final-viral-combined_k141_249966\|\|full | Low-quality |
| 18-110-final-viral-combined_k141_135088\|\|full | Not-determined |
| 18-110-final-viral-combined_k141_31954\|\|full | Not-determined |
| 18-110-final-viral-combined_k141_54178\|\|full | Low-quality |
| 18-110-final-viral-combined_k141_273063\|\|full | Not-determined |
| 18-110-final-viral-combined_k141_123165\|\|full | Low-quality |
| 18-110-final-viral-combined_k141_42760\|\|full | Not-determined |
| 18-110-final-viral-combined_k141_77581\|\|full | Low-quality |
| 18-110-final-viral-combined_k141_182015\|\|full | Low-quality |

| | |
|---|---|
| 18-110-final-viral-combined_k141_202649\|\|full | Low-quality |
| 18-110-final-viral-combined_k141_158152\|\|full | Low-quality |
| 18-110-final-viral-combined_k141_215251\|\|full | Low-quality |
| 18-110-final-viral-combined_k141_239313\|\|full | Not-determined |
| 18-110-final-viral-combined_k141_215258\|\|full | Low-quality |
| 18-110-final-viral-combined_k141_42782\|\|full | Low-quality |
| 18-110-final-viral-combined_k141_66042\|\|full | Low-quality |
| 18-110-final-viral-combined_k141_54183\|\|full | Not-determined |
| 18-110-final-viral-combined_k141_158176\|\|full | Low-quality |
| 18-110-final-viral-combined_k141_215263\|\|full | Not-determined |
| 18-110-final-viral-combined_k141_54188\|\|full | Not-determined |
| 18-110-final-viral-combined_k141_239337\|\|full | Not-determined |
| 18-110-final-viral-combined_k141_226695\|\|full | Low-quality |
| 18-110-final-viral-combined_k141_77640\|\|full | Low-quality |
| 18-110-final-viral-combined_k141_239359\|\|full | Not-determined |
| 18-110-final-viral-combined_k141_111298\|\|full | Not-determined |
| 18-110-final-viral-combined_k141_250057\|\|full | Not-determined |
| 18-110-final-viral-combined_k141_182098\|\|full | Low-quality |
| 18-110-final-viral-combined_k141_250071\|\|full | Low-quality |
| 18-110-final-viral-combined_k141_215288\|\|full | Not-determined |
| 18-110-final-viral-combined_k141_77668\|\|full | Not-determined |
| 18-110-final-viral-combined_k141_32038\|\|full | Low-quality |
| 18-110-final-viral-combined_k141_135188\|\|full | Not-determined |
| 18-110-final-viral-combined_k141_250086\|\|full | Not-determined |
| 18-110-final-viral-combined_k141_192339\|\|full | Not-determined |
| 18-110-final-viral-combined_k141_146875\|\|full | Not-determined |
| 18-110-final-viral-combined_k141_66112\|\|full | Not-determined |
| 18-110-final-viral-combined_k141_54260\|\|full | Low-quality |
| 18-110-final-viral-combined_k141_158208\|\|full | Low-quality |
| 18-110-final-viral-combined_k141_250112\|\|full | Low-quality |
| 18-110-final-viral-combined_k141_182150\|\|full | Not-determined |
| 18-110-final-viral-combined_k141_215315\|\|full | Not-determined |
| 18-110-final-viral-combined_k141_111335\|\|full | Not-determined |
| 18-110-final-viral-combined_k141_123296\|\|full | Low-quality |
| 18-110-final-viral-combined_k141_146892\|\|full | Low-quality |
| 18-110-final-viral-combined_k141_123297\|\|full | Not-determined |
| 18-110-final-viral-combined_k141_100404\|\|full | Low-quality |
| 18-110-final-viral-combined_k141_77712\|\|full | Not-determined |
| 18-110-final-viral-combined_k141_215324\|\|full | Low-quality |
| 18-110-final-viral-combined_k141_32090\|\|full | Not-determined |
| 18-110-final-viral-combined_k141_42869\|\|full | Not-determined |
| 18-110-final-viral-combined_k141_182179\|\|full | Low-quality |
| 18-110-final-viral-combined_k141_202688\|\|full | Not-determined |
| 18-110-final-viral-combined_k141_100431\|\|full | Not-determined |
| 18-110-final-viral-combined_k141_202693\|\|full | Not-determined |
| 18-110-final-viral-combined_k141_259859\|\|full | Low-quality |
| 18-110-final-viral-combined_k141_146960\|\|full | Low-quality |
| 18-110-final-viral-combined_k141_111386\|\|full | Not-determined |
| 18-110-final-viral-combined_k141_192410\|\|full | Not-determined |
| 18-110-final-viral-combined_k141_202732\|\|full | Low-quality |
| 18-110-final-viral-combined_k141_18183\|\|full | Low-quality |
| 18-110-final-viral-combined_k141_54304\|\|full | Low-quality |
| 18-110-final-viral-combined_k141_18185\|\|full | Not-determined |
| 18-110-final-viral-combined_k141_54310\|\|full | Low-quality |
| 18-110-final-viral-combined_k141_202757\|\|full | Not-determined |
| 18-110-final-viral-combined_k141_158281\|\|full | Not-determined |
| 18-110-final-viral-combined_k141_111410\|\|full | Not-determined |
| 18-110-final-viral-combined_k141_158286\|\|full | Low-quality |
| 18-110-final-viral-combined_k141_158287\|\|full | Low-quality |
| 18-110-final-viral-combined_k141_18209\|\|full | Low-quality |
| 18-110-final-viral-combined_k141_169688\|\|full | Not-determined |
| 18-110-final-viral-combined_k141_88273\|\|full | Low-quality |
| 18-110-final-viral-combined_k141_32178\|\|full | Low-quality |
| 18-110-final-viral-combined_k141_135268\|\|full | Low-quality |
| 18-110-final-viral-combined_k141_18217\|\|full | Not-determined |
| 18-110-final-viral-combined_k141_239436\|\|full | Not-determined |
| 18-110-final-viral-combined_k141_135273\|\|full | Low-quality |
| 18-110-final-viral-combined_k141_192468\|\|full | Not-determined |
| 18-110-final-viral-combined_k141_215429\|\|full | Low-quality |
| 18-110-final-viral-combined_k141_111438\|\|full | Low-quality |
| 18-110-final-viral-combined_k141_32196\|\|full | Not-determined |
| 18-110-final-viral-combined_k141_42984\|\|full | Not-determined |
| 18-110-final-viral-combined_k141_135314\|\|full | Not-determined |
| 18-110-final-viral-combined_k141_226847\|\|full | Not-determined |
| 18-110-final-viral-combined_k141_215473\|\|full | Low-quality |
| 18-110-final-viral-combined_k141_7342\|\|full | Low-quality |
| 18-110-final-viral-combined_k141_147074\|\|full | Not-determined |
| 18-110-final-viral-combined_k141_169741\|\|full | Not-determined |
| 18-110-final-viral-combined_k141_158340\|\|full | Not-determined |
| 18-110-final-viral-combined_k141_147097\|\|full | Low-quality |
| 18-110-final-viral-combined_k141_169745\|\|full | Not-determined |
| 18-110-final-viral-combined_k141_43022\|\|full | Not-determined |
| 18-110-final-viral-combined_k141_158349\|\|full | Not-determined |
| 18-110-final-viral-combined_k141_43025\|\|full | Low-quality |
| 18-110-final-viral-combined_k141_32235\|\|full | Not-determined |
| 18-110-final-viral-combined_k141_100518\|\|full | Low-quality |
| 18-110-final-viral-combined_k141_215500\|\|full | Low-quality |
| 18-110-final-viral-combined_k141_77866\|\|full | Not-determined |
| 18-110-final-viral-combined_k141_192538\|\|full | Not-determined |
| 18-110-final-viral-combined_k141_273107\|\|full | Not-determined |
| 18-110-final-viral-combined_k141_169768\|\|full | Not-determined |
| 18-110-final-viral-combined_k141_43039\|\|full | Low-quality |
| 18-110-final-viral-combined_k141_273124\|\|full | Low-quality |
| 18-110-final-viral-combined_k141_215506\|\|full | Not-determined |
| 18-110-final-viral-combined_k141_100540\|\|full | Low-quality |
| 18-110-final-viral-combined_k141_111512\|\|full | Not-determined |
| 18-110-final-viral-combined_k141_32278\|\|full | Not-determined |
| 18-110-final-viral-combined_k141_147162\|\|full | Low-quality |
| 18-110-final-viral-combined_k141_182224\|\|full | Low-quality |
| 18-110-final-viral-combined_k141_250239\|\|full | Low-quality |
| 18-110-final-viral-combined_k141_43065\|\|full | Low-quality |
| 18-110-final-viral-combined_k141_158401\|\|full | Low-quality |
| 18-110-final-viral-combined_k141_54404\|\|full | Low-quality |
| 18-110-final-viral-combined_k141_100573\|\|full | Not-determined |
| 18-101-final-viral-combined_k141_169788\|\|full | Medium-quality |
| 18-110-final-viral-combined_k141_18319\|\|full | Low-quality |
| 18-110-final-viral-combined_k141_215534\|\|full | Low-quality |
| 18-110-final-viral-combined_k141_123439\|\|full | Not-determined |
| 18-110-final-viral-combined_k141_226938\|\|full | Not-determined |
| 18-110-final-viral-combined_k141_202874\|\|full | Not-determined |
| 18-110-final-viral-combined_k141_158414\|\|full | Not-determined |
| 18-110-final-viral-combined_k141_239532\|\|full | Not-determined |
| 18-110-final-viral-combined_k141_226939\|\|full | Not-determined |
| 18-110-final-viral-combined_k141_273163\|\|full | Not-determined |
| 18-110-final-viral-combined_k141_259900\|\|full | Low-quality |
| 18-110-final-viral-combined_k141_192626\|\|full | Not-determined |
| 18-110-final-viral-combined_k141_88408\|\|full | Low-quality |
| 18-110-final-viral-combined_k141_32295\|\|full | Not-determined |
| 18-110-final-viral-combined_k141_111566\|\|full | Not-determined |
| 18-110-final-viral-combined_k141_111570\|\|full | Not-determined |
| 18-110-final-viral-combined_k141_239569\|\|full | Low-quality |
| 18-110-final-viral-combined_k141_147204\|\|full | Not-determined |
| 18-110-final-viral-combined_k141_43118\|\|full | Not-determined |
| 18-110-final-viral-combined_k141_7418\|\|full | Not-determined |
| 18-110-final-viral-combined_k141_123493\|\|full | Not-determined |
| 18-110-final-viral-combined_k141_147213\|\|full | Not-determined |
| 18-110-final-viral-combined_k141_239595\|\|full | Low-quality |
| 18-110-final-viral-combined_k141_32326\|\|full | Low-quality |
| 18-110-final-viral-combined_k141_227007\|\|full | Low-quality |
| 18-110-final-viral-combined_k141_7420\|\|full | Low-quality |
| 18-110-final-viral-combined_k141_43149\|\|full | Low-quality |
| 18-110-final-viral-combined_k141_202935\|\|full | Low-quality |
| 18-110-final-viral-combined_k141_250331\|\|full | Not-determined |
| 18-110-final-viral-combined_k141_77966\|\|full | Low-quality |
| 17-106-final-viral-combined_k141_23408\|\|0_partial | High-quality |
| 18-110-final-viral-combined_k141_239614\|\|full | Low-quality |
| 18-109-final-viral-combined_k141_3669\|\|full | Medium-quality |
| 17-108-final-viral-combined_k141_265453\|\|full | High-quality |
| 18-110-final-viral-combined_k141_88440\|\|full | Low-quality |
| 18-110-final-viral-combined_k141_215615\|\|full | Low-quality |
| 18-110-final-viral-combined_k141_43171\|\|full | Low-quality |
| 18-110-final-viral-combined_k141_88442\|\|full | Low-quality |
| 18-110-final-viral-combined_k141_100674\|\|full | Not-determined |
| 18-110-final-viral-combined_k141_111631\|\|full | Not-determined |
| 18-110-final-viral-combined_k141_135482\|\|full | Low-quality |
| 18-110-final-viral-combined_k141_259930\|\|full | Not-determined |
| 18-110-final-viral-combined_k141_227034\|\|full | Low-quality |
| 18-110-final-viral-combined_k141_147277\|\|full | Low-quality |
| 18-110-final-viral-combined_k141_66211\|\|full | Not-determined |
| 18-110-final-viral-combined_k141_273217\|\|full | Low-quality |
| 18-110-final-viral-combined_k141_239658\|\|full | Low-quality |
| 18-110-final-viral-combined_k141_273220\|\|full | Low-quality |
| 18-110-final-viral-combined_k141_239661\|\|full | Low-quality |
| 18-110-final-viral-combined_k141_135505\|\|full | Not-determined |
| 18-110-final-viral-combined_k141_227041\|\|full | Low-quality |
| 18-110-final-viral-combined_k141_123556\|\|full | Low-quality |
| 18-110-final-viral-combined_k141_135523\|\|full | Low-quality |
| 18-110-final-viral-combined_k141_18462\|\|full | Not-determined |
| 18-110-final-viral-combined_k141_192710\|\|full | Low-quality |
| 18-110-final-viral-combined_k141_259954\|\|full | Low-quality |
| 18-110-final-viral-combined_k141_135528\|\|full | Not-determined |
| 18-110-final-viral-combined_k141_111676\|\|full | Not-determined |
| 18-110-final-viral-combined_k141_32372\|\|full | Not-determined |
| 18-110-final-viral-combined_k141_273240\|\|full | Low-quality |
| 18-110-final-viral-combined_k141_111681\|\|full | Low-quality |
| 18-110-final-viral-combined_k141_239707\|\|full | Low-quality |
| 18-110-final-viral-combined_k141_227060\|\|full | Low-quality |
| 18-110-final-viral-combined_k141_100722\|\|full | Low-quality |
| 18-110-final-viral-combined_k141_7500\|\|full | Not-determined |
| 18-110-final-viral-combined_k141_259986\|\|full | Not-determined |
| 18-110-final-viral-combined_k141_100754\|\|full | Low-quality |
| 18-110-final-viral-combined_k141_32403\|\|full | Not-determined |
| 18-110-final-viral-combined_k141_54559\|\|full | Not-determined |
| 18-110-final-viral-combined_k141_192761\|\|full | Low-quality |
| 18-110-final-viral-combined_k141_32411\|\|full | Not-determined |
| 18-110-final-viral-combined_k141_78011\|\|full | Not-determined |
| 18-110-final-viral-combined_k141_192767\|\|full | Low-quality |
| 18-102-final-viral-combined_k141_132966\|\|full | Medium-quality |
| 18-110-final-viral-combined_k141_78030\|\|full | Low-quality |
| 18-110-final-viral-combined_k141_54596\|\|full | Not-determined |
| 18-110-final-viral-combined_k141_18490\|\|full | Not-determined |
| 18-110-final-viral-combined_k141_100775\|\|full | Low-quality |
| 18-110-final-viral-combined_k141_147366\|\|full | Not-determined |
| 18-110-final-viral-combined_k141_192790\|\|full | Low-quality |
| 18-110-final-viral-combined_k141_123652\|\|full | Not-determined |
| 18-110-final-viral-combined_k141_100787\|\|full | Not-determined |
| 18-110-final-viral-combined_k141_54615\|\|full | Low-quality |
| 18-110-final-viral-combined_k141_250345\|\|full | Low-quality |
| 18-110-final-viral-combined_k141_182290\|\|full | Not-determined |
| 18-110-final-viral-combined_k141_43298\|\|full | Not-determined |
| 18-110-final-viral-combined_k141_54625\|\|full | Not-determined |
| 18-110-final-viral-combined_k141_66278\|\|full | Not-determined |
| 18-110-final-viral-combined_k141_7541\|\|full | Not-determined |
| 18-110-final-viral-combined_k141_123687\|\|full | Low-quality |
| 18-110-final-viral-combined_k141_66290\|\|full | Low-quality |
| 18-110-final-viral-combined_k141_273311\|\|full | Low-quality |
| 18-110-final-viral-combined_k141_158482\|\|full | Low-quality |
| 18-110-final-viral-combined_k141_123702\|\|full | Low-quality |
| 18-110-final-viral-combined_k141_260029\|\|full | Not-determined |
| 18-110-final-viral-combined_k141_227149\|\|full | Not-determined |
| 18-110-final-viral-combined_k141_203063\|\|full | Not-determined |
| 18-110-final-viral-combined_k141_250362\|\|full | Low-quality |
| 18-110-final-viral-combined_k141_227151\|\|full | Not-determined |
| 18-110-final-viral-combined_k141_203070\|\|full | Not-determined |
| 18-110-final-viral-combined_k141_88543\|\|full | Low-quality |
| 18-110-final-viral-combined_k141_135665\|\|full | Not-determined |
| 18-110-final-viral-combined_k141_147458\|\|full | Not-determined |
| 18-110-final-viral-combined_k141_158523\|\|full | Not-determined |
| 18-110-final-viral-combined_k141_147460\|\|full | Low-quality |
| 18-110-final-viral-combined_k141_169950\|\|full | Low-quality |
| 18-110-final-viral-combined_k141_100849\|\|full | Low-quality |
| 18-110-final-viral-combined_k141_260052\|\|full | Not-determined |
| 18-110-final-viral-combined_k141_135678\|\|full | Low-quality |
| 18-110-final-viral-combined_k141_7589\|\|full | Low-quality |
| 18-110-final-viral-combined_k141_273353\|\|full | Not-determined |
| 18-110-final-viral-combined_k141_215767\|\|full | Low-quality |
| 18-110-final-viral-combined_k141_192856\|\|full | Low-quality |
| 18-110-final-viral-combined_k141_147482\|\|full | Low-quality |
| 18-110-final-viral-combined_k141_169963\|\|full | Not-determined |
| 18-110-final-viral-combined_k141_43333\|\|full | Not-determined |
| 18-110-final-viral-combined_k141_182332\|\|full | Not-determined |
| 18-110-final-viral-combined_k141_18567\|\|full | Low-quality |
| 18-110-final-viral-combined_k141_100882\|\|full | Low-quality |
| 18-110-final-viral-combined_k141_43349\|\|full | Not-determined |
| 18-110-final-viral-combined_k141_66369\|\|full | Low-quality |
| 18-110-final-viral-combined_k141_135728\|\|full | Low-quality |
| 18-110-final-viral-combined_k141_203134\|\|full | Not-determined |
| 18-110-final-viral-combined_k141_123769\|\|full | Low-quality |
| 18-110-final-viral-combined_k141_18577\|\|full | Not-determined |
| 18-110-final-viral-combined_k141_203141\|\|full | Low-quality |
| 18-110-final-viral-combined_k141_215802\|\|full | Low-quality |
| 18-110-final-viral-combined_k141_169981\|\|full | Low-quality |
| 17-109-final-viral-combined_k141_117447\|\|0_partial | Medium-quality |
| 18-110-final-viral-combined_k141_215803\|\|full | Low-quality |
| 18-110-final-viral-combined_k141_215804\|\|full | Not-determined |
| 18-110-final-viral-combined_k141_18590\|\|full | Low-quality |
| 18-110-final-viral-combined_k141_192886\|\|full | Low-quality |
| 18-110-final-viral-combined_k141_182355\|\|full | Not-determined |
| 18-110-final-viral-combined_k141_123782\|\|full | Not-determined |
| 18-110-final-viral-combined_k141_111834\|\|full | Low-quality |
| 18-110-final-viral-combined_k141_169991\|\|full | Not-determined |
| 18-110-final-viral-combined_k141_158562\|\|full | Low-quality |
| 18-110-final-viral-combined_k141_7635\|\|full | Low-quality |
| 18-110-final-viral-combined_k141_169996\|\|full | Not-determined |
| 18-110-final-viral-combined_k141_18598\|\|full | Low-quality |
| [illegible] | [illegible] |

18-110-final-viral-combined_k141_55332||full Low-quality
18-110-final-viral-combined_k141_148386||full Low-quality
18-110-final-viral-combined_k141_8462||full Not-determined
18-110-final-viral-combined_k141_274183||full Not-determined
18-110-final-viral-combined_k141_33391||full Not-determined
18-110-final-viral-combined_k141_216696||full Low-quality
18-110-final-viral-combined_k141_260643||full Not-determined
18-110-final-viral-combined_k141_251207||full Not-determined
18-110-final-viral-combined_k141_55338||full Not-determined
18-110-final-viral-combined_k141_124584||full Low-quality
18-110-final-viral-combined_k141_251211||full Low-quality
18-110-final-viral-combined_k141_148408||full Not-determined
18-110-final-viral-combined_k141_8490||full Low-quality
18-110-final-viral-combined_k141_203921||full Not-determined
18-110-final-viral-combined_k141_170777||full Low-quality
18-110-final-viral-combined_k141_112653||full Low-quality
18-110-final-viral-combined_k141_228080||full Not-determined
18-110-final-viral-combined_k141_228090||full Low-quality
18-110-final-viral-combined_k141_228091||full Not-determined
18-110-final-viral-combined_k141_240682||full Low-quality
18-110-final-viral-combined_k141_79099||full Low-quality
18-110-final-viral-combined_k141_148461||full Low-quality
18-110-final-viral-combined_k141_33450||full Low-quality
18-110-final-viral-combined_k141_67166||full Not-determined
18-110-final-viral-combined_k141_8516||full Not-determined
18-110-final-viral-combined_k141_124626||full Low-quality
18-110-final-viral-combined_k141_251248||full Low-quality
18-110-final-viral-combined_k141_79120||full Low-quality
18-110-final-viral-combined_k141_228107||full Low-quality
18-110-final-viral-combined_k141_67175||full Low-quality
18-110-final-viral-combined_k141_183467||full Low-quality
18-110-final-viral-combined_k141_193709||full Not-determined
18-110-final-viral-combined_k141_228111||full Low-quality
18-110-final-viral-combined_k141_112705||full Not-determined
18-110-final-viral-combined_k141_183471||full Low-quality
18-110-final-viral-combined_k141_228119||full Low-quality
18-110-final-viral-combined_k141_228120||full Low-quality
18-110-final-viral-combined_k141_136791||full Not-determined
18-110-final-viral-combined_k141_251274||full Not-determined
18-110-final-viral-combined_k141_260701||full Not-determined
18-110-final-viral-combined_k141_148518||full Low-quality
18-110-final-viral-combined_k141_216811||full Not-determined
18-110-final-viral-combined_k141_159466||full Not-determined
18-110-final-viral-combined_k141_148528||full Not-determined
18-110-final-viral-combined_k141_89634||full Low-quality
18-110-final-viral-combined_k141_159498||full Low-quality
18-110-final-viral-combined_k141_79172||full Not-determined
18-110-final-viral-combined_k141_19531||full Low-quality
18-110-final-viral-combined_k141_148556||full Not-determined
18-110-final-viral-combined_k141_79181||full Low-quality
18-110-final-viral-combined_k141_67224||full Not-determined
18-110-final-viral-combined_k141_216867||full Not-determined
18-110-final-viral-combined_k141_170843||full Not-determined
18-110-final-viral-combined_k141_101743||full Low-quality
18-110-final-viral-combined_k141_67259||full Low-quality
18-110-final-viral-combined_k141_55457||full Not-determined
18-110-final-viral-combined_k141_148581||full Not-determined
18-110-final-viral-combined_k141_55462||full Not-determined
18-110-final-viral-combined_k141_183530||full Low-quality
18-110-final-viral-combined_k141_204063||full Not-determined
18-110-final-viral-combined_k141_260770||full Low-quality
18-110-final-viral-combined_k141_240796||full Not-determined
18-110-final-viral-combined_k141_170864||full Not-determined
18-110-final-viral-combined_k141_124718||full Low-quality
18-110-final-viral-combined_k141_67292||full Not-determined
18-110-final-viral-combined_k141_101791||full Low-quality
18-110-final-viral-combined_k141_159576||full Not-determined
18-110-final-viral-combined_k141_148599||full Not-determined
18-110-final-viral-combined_k141_101795||full Low-quality
18-110-final-viral-combined_k141_19632||full Not-determined
18-110-final-viral-combined_k141_260799||full Not-determined
18-110-final-viral-combined_k141_240848||full Not-determined
18-110-final-viral-combined_k141_251332||full Not-determined
18-110-final-viral-combined_k141_101824||full Not-determined
18-110-final-viral-combined_k141_240850||full Not-determined
18-110-final-viral-combined_k141_33584||full Low-quality
18-110-final-viral-combined_k141_19657||full Not-determined
18-110-final-viral-combined_k141_33594||full Low-quality
18-110-final-viral-combined_k141_193866||full Not-determined
18-110-final-viral-combined_k141_55559||full Not-determined
18-110-final-viral-combined_k141_67363||full Not-determined
18-110-final-viral-combined_k141_251346||full Low-quality
18-110-final-viral-combined_k141_33601||full Not-determined
18-110-final-viral-combined_k141_228256||full Not-determined
18-110-final-viral-combined_k141_44569||full Low-quality
18-110-final-viral-combined_k141_44576||full Not-determined
18-110-final-viral-combined_k141_79302||full Not-determined
18-110-final-viral-combined_k141_204130||full Not-determined
18-110-final-viral-combined_k141_44587||full Low-quality
18-110-final-viral-combined_k141_136957||full Not-determined
18-110-final-viral-combined_k141_33645||full Low-quality
18-110-final-viral-combined_k141_101869||full Not-determined
18-110-final-viral-combined_k141_89808||full Low-quality
18-110-final-viral-combined_k141_79337||full Not-determined
18-110-final-viral-combined_k141_79339||full Low-quality
18-110-final-viral-combined_k141_112912||full Low-quality
18-110-final-viral-combined_k141_216926||full Low-quality
18-110-final-viral-combined_k141_19731||full Low-quality
18-110-final-viral-combined_k141_67445||full Low-quality
18-110-final-viral-combined_k141_193918||full Low-quality
18-110-final-viral-combined_k141_148736||full Not-determined
18-110-final-viral-combined_k141_67447||full Not-determined
18-110-final-viral-combined_k141_112930||full Not-determined
18-110-final-viral-combined_k141_193922||full Not-determined
18-110-final-viral-combined_k141_159705||full Not-determined
18-110-final-viral-combined_k141_148749||full Low-quality
18-110-final-viral-combined_k141_251394||full Not-determined
18-110-final-viral-combined_k141_55612||full Not-determined
18-110-final-viral-combined_k141_183673||full Low-quality
18-110-final-viral-combined_k141_79363||full Low-quality
18-110-final-viral-combined_k141_148762||full Low-quality
18-110-final-viral-combined_k141_137010||full Not-determined
18-110-final-viral-combined_k141_124828||full Low-quality
18-110-final-viral-combined_k141_19757||full Low-quality
18-110-final-viral-combined_k141_216963||full Low-quality
18-110-final-viral-combined_k141_251430||full Low-quality
18-110-final-viral-combined_k141_112977||full Low-quality
18-110-final-viral-combined_k141_124844||full Low-quality
18-110-final-viral-combined_k141_193956||full Low-quality
18-110-final-viral-combined_k141_183713||full Not-determined
18-110-final-viral-combined_k141_260898||full Low-quality
18-110-final-viral-combined_k141_67491||full Not-determined
18-110-final-viral-combined_k141_183717||full Low-quality
18-110-final-viral-combined_k141_101972||full Not-determined
18-110-final-viral-combined_k141_159754||full Low-quality
18-110-final-viral-combined_k141_159759||full Low-quality
18-110-final-viral-combined_k141_55654||full Low-quality
18-110-final-viral-combined_k141_44663||full Not-determined
18-110-final-viral-combined_k141_8592||full Low-quality
18-110-final-viral-combined_k141_148799||full Low-quality
18-110-final-viral-combined_k141_228279||full Low-quality
18-110-final-viral-combined_k141_228284||full Low-quality
18-110-final-viral-combined_k141_8595||full Not-determined
18-110-final-viral-combined_k141_124873||full Low-quality
18-110-final-viral-combined_k141_171050||full Low-quality
18-110-final-viral-combined_k141_8608||full Low-quality
18-110-final-viral-combined_k141_251499||full Low-quality
18-110-final-viral-combined_k141_44683||full Low-quality
18-110-final-viral-combined_k141_79453||full Not-determined
18-110-final-viral-combined_k141_89874||full Low-quality
18-110-final-viral-combined_k141_8615||full Low-quality
18-110-final-viral-combined_k141_124919||full Not-determined
18-110-final-viral-combined_k141_33756||full Low-quality
18-110-final-viral-combined_k141_137115||full Not-determined
18-110-final-viral-combined_k141_44698||full Not-determined
18-110-final-viral-combined_k141_194031||full Not-determined
18-110-final-viral-combined_k141_148851||full Not-determined
18-110-final-viral-combined_k141_113053||full Not-determined
18-110-final-viral-combined_k141_171081||full Low-quality
18-110-final-viral-combined_k141_44706||full Not-determined
18-110-final-viral-combined_k141_148854||full Not-determined
18-110-final-viral-combined_k141_124939||full Low-quality
18-110-final-viral-combined_k141_171084||full Low-quality
18-110-final-viral-combined_k141_204194||full Low-quality
18-110-final-viral-combined_k141_183742||full Low-quality
18-110-final-viral-combined_k141_159819||full Low-quality
18-110-final-viral-combined_k141_159824||full Not-determined
18-110-final-viral-combined_k141_55707||full Low-quality
18-110-final-viral-combined_k141_183758||full Low-quality
18-110-final-viral-combined_k141_124965||full Low-quality
18-110-final-viral-combined_k141_183765||full Low-quality
18-110-final-viral-combined_k141_228383||full Low-quality
18-110-final-viral-combined_k141_8668||full Not-determined
18-110-final-viral-combined_k141_44746||full Not-determined
18-110-final-viral-combined_k141_183779||full Not-determined
18-110-final-viral-combined_k141_260998||full Low-quality
18-110-final-viral-combined_k141_19856||full Low-quality
18-110-final-viral-combined_k141_183791||full Not-determined
18-110-final-viral-combined_k141_228389||full Low-quality
18-110-final-viral-combined_k141_102059||full Low-quality
18-110-final-viral-combined_k141_44758||full Not-determined
18-110-final-viral-combined_k141_33824||full Not-determined
18-110-final-viral-combined_k141_124995||full Not-determined
18-110-final-viral-combined_k141_67593||full Not-determined
18-110-final-viral-combined_k141_137167||full Low-quality
18-110-final-viral-combined_k141_67595||full Low-quality
18-110-final-viral-combined_k141_194073||full Low-quality
18-110-final-viral-combined_k141_89952||full Not-determined
18-110-final-viral-combined_k141_8728||full Low-quality
18-110-final-viral-combined_k141_67622||full Low-quality
18-110-final-viral-combined_k141_19916||full Not-determined
18-110-final-viral-combined_k141_8740||full Low-quality
18-110-final-viral-combined_k141_19924||full Not-determined
18-110-final-viral-combined_k141_274284||full Not-determined
18-110-final-viral-combined_k141_241168||full Not-determined
18-110-final-viral-combined_k141_261063||full Low-quality
18-110-final-viral-combined_k141_274295||full Low-quality
18-110-final-viral-combined_k141_89981||full Low-quality
18-110-final-viral-combined_k141_19941||full Low-quality
18-110-final-viral-combined_k141_44812||full Low-quality
18-110-final-viral-combined_k141_274314||full Not-determined
18-110-final-viral-combined_k141_8768||full Not-determined
18-110-final-viral-combined_k141_228464||full Low-quality
18-110-final-viral-combined_k141_216978||full Not-determined
18-110-final-viral-combined_k141_171169||full Low-quality
18-110-final-viral-combined_k141_137259||full Low-quality
18-110-final-viral-combined_k141_90033||full Not-determined
18-110-final-viral-combined_k141_171186||full Low-quality
18-110-final-viral-combined_k141_55789||full Not-determined
18-110-final-viral-combined_k141_79618||full Low-quality
18-110-final-viral-combined_k141_261092||full Low-quality
18-110-final-viral-combined_k141_90048||full Low-quality
18-110-final-viral-combined_k141_228512||full Not-determined
18-110-final-viral-combined_k141_217019||full Not-determined
18-110-final-viral-combined_k141_241198||full Low-quality
18-110-final-viral-combined_k141_274348||full Low-quality
18-110-final-viral-combined_k141_194170||full Low-quality
18-110-final-viral-combined_k141_20010||full Not-determined
18-110-final-viral-combined_k141_228519||full Low-quality
18-110-final-viral-combined_k141_137303||full Not-determined
18-110-final-viral-combined_k141_228522||full Low-quality
18-110-final-viral-combined_k141_194176||full Low-quality
18-110-final-viral-combined_k141_194186||full Not-determined
18-110-final-viral-combined_k141_113216||full Not-determined
18-110-final-viral-combined_k141_149008||full Low-quality
18-110-final-viral-combined_k141_79666||full Not-determined
18-110-final-viral-combined_k141_241224||full Not-determined
18-110-final-viral-combined_k141_274374||full Not-determined
18-110-final-viral-combined_k141_79675||full Low-quality
18-110-final-viral-combined_k141_67729||full Low-quality
18-110-final-viral-combined_k141_183906||full Not-determined
18-110-final-viral-combined_k141_79682||full Low-quality
18-110-final-viral-combined_k141_228550||full Low-quality
18-110-final-viral-combined_k141_20038||full Low-quality
18-110-final-viral-combined_k141_204367||full Not-determined
18-110-final-viral-combined_k141_149016||full Low-quality
18-110-final-viral-combined_k141_33984||full Low-quality
18-110-final-viral-combined_k141_171252||full Not-determined
18-110-final-viral-combined_k141_204371||full Low-quality
18-110-final-viral-combined_k141_228568||full Low-quality
18-110-final-viral-combined_k141_251789||full Low-quality
18-110-final-viral-combined_k141_194199||full Not-determined
18-110-final-viral-combined_k141_113282||full Low-quality
18-110-final-viral-combined_k141_217031||full Low-quality
18-110-final-viral-combined_k141_20056||full Not-determined
18-110-final-viral-combined_k141_149048||full Not-determined
18-110-final-viral-combined_k141_44955||full Not-determined
18-110-final-viral-combined_k141_79718||full Not-determined
18-110-final-viral-combined_k141_102202||full Not-determined
18-110-final-viral-combined_k141_20072||full Not-determined
18-110-final-viral-combined_k141_44988||full Low-quality
18-110-final-viral-combined_k141_159934||full Not-determined
18-110-final-viral-combined_k141_8867||full Not-determined
18-110-final-viral-combined_k141_44991||full Not-determined
18-110-final-viral-combined_k141_204411||full Low-quality
18-110-final-viral-combined_k141_241294||full Low-quality
18-110-final-viral-combined_k141_67818||full Low-quality
18-110-final-viral-combined_k141_274472||full Low-quality
18-110-final-viral-combined_k141_159955||full Not-determined
18-110-final-viral-combined_k141_55935||full Low-quality
18-110-final-viral-combined_k141_241320||full Not-determined
18-110-final-viral-combined_k141_34054||full Not-determined
18-110-final-viral-combined_k141_45035||full Low-quality
18-110-final-viral-combined_k141_79768||full Not-determined
18-110-final-viral-combined_k141_137406||full Not-determined
18-110-final-viral-combined_k141_217079||full Low-quality
18-110-final-viral-combined_k141_241329||full Not-determined
18-110-final-viral-combined_k141_184006||full Not-determined
18-110-final-viral-combined_k141_34068||full Low-quality
18-110-final-viral-combined_k141_149133||full Low-quality
18-110-final-viral-combined_k141_137423||full Not-determined
18-110-final-viral-combined_k141_160009||full Low-quality
18-110-final-viral-combined_k141_113346||full Low-quality
18-110-final-viral-combined_k141_113352||full Low-quality
18-110-final-viral-combined_k141_251886||full Not-determined
18-110-final-viral-combined_k141_274543||full Not-determined
18-110-final-viral-combined_k141_90250||full Low-quality
18-110-final-viral-combined_k141_160024||full Not-determined
18-110-final-viral-combined_k141_102323||full Not-determined
18-110-final-viral-combined_k141_261200||full Not-determined
18-110-final-viral-combined_k141_204473||full Not-determined
18-110-final-viral-combined_k141_125132||full Not-determined
18-110-final-viral-combined_k141_171351||full Low-quality
18-110-final-viral-combined_k141_160036||full Not-determined
18-110-final-viral-combined_k141_113371||full Low-quality
18-110-final-viral-combined_k141_90272||full Not-determined
18-110-final-viral-combined_k141_204486||full Not-determined
18-110-final-viral-combined_k141_113379||full Low-quality
18-110-final-viral-combined_k141_67925||full Low-quality
18-110-final-viral-combined_k141_261213||full Low-quality
18-110-final-viral-combined_k141_79838||full Not-determined
18-110-final-viral-combined_k141_90291||full Not-determined
18-110-final-viral-combined_k141_184015||full Not-determined
18-110-final-viral-combined_k141_204497||full Low-quality
18-110-final-viral-combined_k141_274588||full Low-quality
18-110-final-viral-combined_k141_34114||full Low-quality
18-110-final-viral-combined_k141_171381||full Low-quality
18-110-final-viral-combined_k141_67949||full Low-quality
18-110-final-viral-combined_k141_261239||full Low-quality
18-110-final-viral-combined_k141_137490||full Not-determined
18-110-final-viral-combined_k141_274599||full Low-quality
18-110-final-viral-combined_k141_113414||full Low-quality
18-110-final-viral-combined_k141_184032||full Low-quality
18-110-final-viral-combined_k141_56032||full Not-determined
18-110-final-viral-combined_k141_125176||full Not-determined
18-110-final-viral-combined_k141_90330||full Low-quality
18-110-final-viral-combined_k141_194365||full Not-determined
18-110-final-viral-combined_k141_56044||full Not-determined
18-110-final-viral-combined_k141_204506||full Low-quality
18-110-final-viral-combined_k141_149245||full Low-quality
18-110-final-viral-combined_k141_184052||full Not-determined
18-110-final-viral-combined_k141_194379||full Not-determined
18-110-final-viral-combined_k141_67990||full Not-determined
18-110-final-viral-combined_k141_79910||full Low-quality
18-110-final-viral-combined_k141_217127||full Low-quality
18-110-final-viral-combined_k141_274642||full Not-determined
18-110-final-viral-combined_k141_149277||full Low-quality
18-110-final-viral-combined_k141_113451||full Not-determined
18-110-final-viral-combined_k141_184082||full Low-quality
18-110-final-viral-combined_k141_56067||full Low-quality
18-110-final-viral-combined_k141_68020||full Low-quality
18-110-final-viral-combined_k141_102371||full Low-quality
18-110-final-viral-combined_k141_194397||full Not-determined
18-110-final-viral-combined_k141_45254||full Not-determined
18-110-final-viral-combined_k141_34165||full Not-determined
18-110-final-viral-combined_k141_274657||full Low-quality
18-110-final-viral-combined_k141_194408||full Not-determined
18-110-final-viral-combined_k141_113483||full Low-quality
18-110-final-viral-combined_k141_45281||full Not-determined
18-110-final-viral-combined_k141_56094||full Not-determined
18-110-final-viral-combined_k141_9015||full Low-quality
18-110-final-viral-combined_k141_137584||full Low-quality
18-110-final-viral-combined_k141_113501||full Low-quality
18-110-final-viral-combined_k141_204603||full Low-quality
18-110-final-viral-combined_k141_125256||full Low-quality
18-110-final-viral-combined_k141_137592||full Low-quality
18-110-final-viral-combined_k141_45310||full Low-quality
18-110-final-viral-combined_k141_184166||full Low-quality
18-110-final-viral-combined_k141_160105||full Not-determined
18-110-final-viral-combined_k141_9036||full Not-determined
18-110-final-viral-combined_k141_102406||full Low-quality
18-110-final-viral-combined_k141_56117||full Not-determined
18-110-final-viral-combined_k141_102407||full Not-determined
18-110-final-viral-combined_k141_68039||full Not-determined
18-110-final-viral-combined_k141_217205||full Low-quality
18-110-final-viral-combined_k141_217207||full Low-quality
18-110-final-viral-combined_k141_274665||full Low-quality
18-110-final-viral-combined_k141_68046||full Low-quality
18-110-final-viral-combined_k141_137618||full Not-determined
18-110-final-viral-combined_k141_45319||full Low-quality
18-110-final-viral-combined_k141_274675||full Not-determined
18-110-final-viral-combined_k141_68074||full Low-quality
18-110-final-viral-combined_k141_45336||full Not-determined
18-110-final-viral-combined_k141_125286||full Low-quality
18-110-final-viral-combined_k141_194478||full Low-quality
18-110-final-viral-combined_k141_261330||full Low-quality
18-110-final-viral-combined_k141_160136||full Not-determined
18-110-final-viral-combined_k141_68078||full Not-determined
18-110-final-viral-combined_k141_137659||full Not-determined
18-110-final-viral-combined_k141_90466||full Not-determined
18-110-final-viral-combined_k141_184194||full Not-determined
18-110-final-viral-combined_k141_160163||full Not-determined
18-110-final-viral-combined_k141_171482||full Low-quality
18-110-final-viral-combined_k141_194515||full Low-quality
18-110-final-viral-combined_k141_45371||full Low-quality
18-110-final-viral-combined_k141_9111||full Low-quality
18-110-final-viral-combined_k141_184204||full Low-quality
18-110-final-viral-combined_k141_68105||full Low-quality
18-110-final-viral-combined_k141_194523||full Not-determined
18-110-final-viral-combined_k141_274705||full Low-quality
18-110-final-viral-combined_k141_45387||full Not-determined
18-110-final-viral-combined_k141_56203||full Low-quality
18-110-final-viral-combined_k141_184221||full Low-quality
18-110-final-viral-combined_k141_149454||full Not-determined
18-110-final-viral-combined_k141_80041||full Low-quality
18-110-final-viral-combined_k141_125335||full Low-quality
18-110-final-viral-combined_k141_149467||full Low-quality
18-110-final-viral-combined_k141_204683||full Low-quality
18-110-final-viral-combined_k141_274740||full Low-quality
18-110-final-viral-combined_k141_68155||full Not-determined
18-110-final-viral-combined_k141_194553||full Not-determined
18-110-final-viral-combined_k141_274747||full Not-determined
18-110-final-viral-combined_k141_171540||full Not-determined
18-110-final-viral-combined_k141_90502||full Not-determined
18-110-final-viral-combined_k141_102474||full Low-quality
18-110-final-viral-combined_k141_184270||full Not-determined
18-110-final-viral-combined_k141_9157||full Not-determined
18-110-final-viral-combined_k141_20238||full Low-quality
18-110-final-viral-combined_k141_228890||full Low-quality
18-110-final-viral-combined_k141_204705||full Not-determined
18-110-final-viral-combined_k141_68191||full Not-determined
18-110-final-viral-combined_k141_137749||full Low-quality
18-110-final-viral-combined_k141_204711||full Not-determined
18-110-final-viral-combined_k141_45467||full Low-quality
18-110-final-viral-combined_k141_252017||full Not-determined
18-110-final-viral-combined_k141_90521||full Not-determined
18-110-final-viral-combined_k141_252021||full Not-determined
18-110-final-viral-combined_k141_125369||full Not-determined
18-110-final-viral-combined_k141_204720||full Low-quality
18-110-final-viral-combined_k141_171585||full Not-determined
18-110-final-viral-combined_k141_137774||full Low-quality
18-110-final-viral-combined_k141_9173||full Not-determined
18-110-final-viral-combined_k141_149506||full Not-determined
18-110-final-viral-combined_k141_149513||full Not-determined
18-110-final-viral-combined_k141_204738||full Low-quality
18-110-final-viral-combined_k141_160288||full Not-determined
18-110-final-viral-combined_k141_252040||full Low-quality
18-110-final-viral-combined_k141_204742||full Not-determined
18-110-final-viral-combined_k141_137787||full Low-quality
18-110-final-viral-combined_k141_217350||full Low-quality
18-110-final-viral-combined_k141_45502||full Low-quality
18-110-final-viral-combined_k141_217369||full Not-determined
18-110-final-viral-combined_k141_274815||full Not-determined
18-110-final-viral-combined_k141_9193||full Not-determined
18-110-final-viral-combined_k141_228948||full Not-determined
18-110-final-viral-combined_k141_217377||full Low-quality
18-110-final-viral-combined_k141_34370||full Not-determined
18-110-final-viral-combined_k141_102545||full Low-quality
18-110-final-viral-combined_k141_34380||full Low-quality
18-110-final-viral-combined_k141_261434||full Low-quality
18-110-final-viral-combined_k141_228966||full Low-quality
18-110-final-viral-combined_k141_217390||full Not-determined
18-110-final-viral-combined_k141_20305||full Not-determined
18-110-final-viral-combined_k141_34398||full Low-quality
18-110-final-viral-combined_k141_125445||full Low-quality
18-110-final-viral-combined_k141_261462||full Not-determined
18-110-final-viral-combined_k141_204807||full Not-determined
17-106_k141_104169 Medium-quality
18-110-final-viral-combined_k141_160357||full Low-quality
18-110-final-viral-combined_k141_113701||full Low-quality
18-110-final-viral-combined_k141_125462||full Not-determined
18-110-final-viral-combined_k141_68330||full Not-determined
18-110-final-viral-combined_k141_204839||full Not-determined
18-110-final-viral-combined_k141_274912||full Not-determined
18-110-final-viral-combined_k141_137854||full Not-determined
18-110-final-viral-combined_k141_274917||full Not-determined
18-110-final-viral-combined_k141_20348||full Low-quality
18-110-final-viral-combined_k141_194696||full Not-determined
18-110-final-viral-combined_k141_102600||full Low-quality
18-110-final-viral-combined_k141_229031||full Not-determined
18-110-final-viral-combined_k141_137865||full Low-quality
18-110-final-viral-combined_k141_241539||full Low-quality
18-110-final-viral-combined_k141_68354||full Not-determined
18-110-final-viral-combined_k141_204886||full Low-quality
18-110-final-viral-combined_k141_90674||full Not-determined
18-110-final-viral-combined_k141_204889||full Low-quality
18-110-final-viral-combined_k141_194702||full Low-quality
18-110-final-viral-combined_k141_204892||full Not-determined
18-110-final-viral-combined_k141_217468||full Not-determined
18-110-final-viral-combined_k141_90677||full Not-determined
18-110-final-viral-combined_k141_20362||full Low-quality
18-110-final-viral-combined_k141_102627||full Low-quality
18-110-final-viral-combined_k141_194726||full Not-determined
18-110-final-viral-combined_k141_204902||full Not-determined
18-110-final-viral-combined_k141_45615||full Low-quality
18-110-final-viral-combined_k141_252184||full Not-determined
18-110-final-viral-combined_k141_45620||full Low-quality
18-110-final-viral-combined_k141_171708||full Not-determined
18-110-final-viral-combined_k141_102649||full Low-quality
18-110-final-viral-combined_k141_217509||full Low-quality
18-110-final-viral-combined_k141_20386||full Low-quality
18-110-final-viral-combined_k141_160440||full Low-quality
18-110-final-viral-combined_k141_137929||full Low-quality
18-110-final-viral-combined_k141_241595||full Not-determined
18-110-final-viral-combined_k141_252199||full Low-quality
18-110-final-viral-combined_k141_9324||full Not-determined
18-110-final-viral-combined_k141_171727||full Not-determined
18-110-final-viral-combined_k141_102655||full Low-quality
18-110-final-viral-combined_k141_34475||full Low-quality
18-110-final-viral-combined_k141_274974||full Not-determined
18-110-final-viral-combined_k141_20404||full Not-determined
18-110-final-viral-combined_k141_102656||full Not-determined
18-110-final-viral-combined_k141_160466||full Not-determined
18-110-final-viral-combined_k141_252220||full Low-quality
18-110-final-viral-combined_k141_125547||full Low-quality
18-110-final-viral-combined_k141_90744||full Low-quality
18-110-final-viral-combined_k141_241632||full Not-determined
18-110-final-viral-combined_k141_241635||full Low-quality
18-110-final-viral-combined_k141_149573||full Low-quality
18-110-final-viral-combined_k141_80218||full Low-quality
18-110-final-viral-combined_k141_194757||full Low-quality
18-110-final-viral-combined_k141_194759||full Low-quality
18-110-final-viral-combined_k141_252230||full Not-determined
18-110-final-viral-combined_k141_194761||full Not-determined
18-110-final-viral-combined_k141_194763||full Low-quality
18-110-final-viral-combined_k141_102708||full Low-quality
18-110-final-viral-combined_k141_9346||full Not-determined
18-110-final-viral-combined_k141_20441||full Not-determined
18-110-final-viral-combined_k141_275001||full Not-determined
18-110-final-viral-combined_k141_80235||full Not-determined
18-110-final-viral-combined_k141_102714||full Not-determined
18-110-final-viral-combined_k141_68465||full Low-quality
18-110-final-viral-combined_k141_90781||full Not-determined
18-110-final-viral-combined_k141_45688||full Not-determined
18-110-final-viral-combined_k141_275009||full Low-quality
18-110-final-viral-combined_k141_149604||full Low-quality
18-110-final-viral-combined_k141_241678||full Low-quality
18-110-final-viral-combined_k141_252243||full Low-quality
18-110-final-viral-combined_k141_275013||full Not-determined
18-110-final-viral-combined_k141_68474||full Not-determined
18-110-final-viral-combined_k141_125604||full Low-quality
18-110-final-viral-combined_k141_113787||full Not-determined
18-110-final-viral-combined_k141_252252||full Low-quality
18-110-final-viral-combined_k141_229048||full Not-determined
18-110-final-viral-combined_k141_217599||full Low-quality
18-110-final-viral-combined_k141_34545||full Low-quality
18-110-final-viral-combined_k141_113798||full Low-quality
18-110-final-viral-combined_k141_102766||full Low-quality
18-110-final-viral-combined_k141_138029||full Not-determined
18-110-final-viral-combined_k141_241728||full Low-quality
18-110-final-viral-combined_k141_9410||full Low-quality
18-110-final-viral-combined_k141_102768||full Low-quality
18-110-final-viral-combined_k141_275029||full Not-determined
18-110-final-viral-combined_k141_194864||full Low-quality
18-110-final-viral-combined_k141_252272||full Not-determined
18-110-final-viral-combined_k141_90881||full Not-determined
18-110-final-viral-combined_k141_125646||full Low-quality
18-110-final-viral-combined_k141_261626||full Not-determined
18-110-final-viral-combined_k141_90886||full Not-determined
18-110-final-viral-combined_k141_90894||full Low-quality
18-110-final-viral-combined_k141_125660||full Low-quality
18-110-final-viral-combined_k141_217633||full Not-determined
18-110-final-viral-combined_k141_160530||full Not-determined
18-110-final-viral-combined_k141_194902||full Low-quality
18-110-final-viral-combined_k141_229099||full Not-determined
18-110-final-viral-combined_k141_275061||full Low-quality
18-110-final-viral-combined_k141_229104||full Not-determined
18-110-final-viral-combined_k141_102829||full Not-determined
18-110-final-viral-combined_k141_275068||full Not-determined
18-110-final-viral-combined_k141_241778||full Not-determined
18-110-final-viral-combined_k141_102847||full Low-quality
18-110-final-viral-combined_k141_80303||full Low-quality
18-110-final-viral-combined_k141_125689||full Low-quality
18-110-final-viral-combined_k141_125691||full Not-determined
18-110-final-viral-combined_k141_160554||full Low-quality
18-110-final-viral-combined_k141_252331||full Low-quality
18-110-final-viral-combined_k141_217666||full Not-determined
18-110-final-viral-combined_k141_252334||full Not-determined
18-110-final-viral-combined_k141_56441||full Low-quality
18-110-final-viral-combined_k141_160557||full Low-quality
18-110-final-viral-combined_k141_90951||full Not-determined
18-110-final-viral-combined_k141_9469||full Low-quality
18-110-final-viral-combined_k141_56445||full Low-quality
18-110-final-viral-combined_k141_45772||full Low-quality
18-110-final-viral-combined_k141_102871||full Not-determined
18-110-final-viral-combined_k141_205057||full Not-determined
18-110-final-viral-combined_k141_125715||full Low-quality
18-110-final-viral-combined_k141_125721||full Not-determined
18-110-final-viral-combined_k141_229142||full Low-quality
18-110-final-viral-combined_k141_80327||full Low-quality
18-110-final-viral-combined_k141_160574||full Low-quality
18-110-final-viral-combined_k141_68516||full Low-quality
18-110-final-viral-combined_k141_194964||full Low-quality
18-110-final-viral-combined_k141_149670||full Not-determined
18-110-final-viral-combined_k141_261686||full Low-quality
18-110-final-viral-combined_k141_20559||full Low-quality
18-110-final-viral-combined_k141_229184||full Low-quality
18-110-final-viral-combined_k141_9507||full Not-determined
18-110-final-viral-combined_k141_80364||full Not-determined
18-110-final-viral-combined_k141_45812||full Low-quality
18-110-final-viral-combined_k141_275163||full Not-determined
18-110-final-viral-combined_k141_91029||full Not-determined
18-110-final-viral-combined_k141_229197||full Not-determined
18-110-final-viral-combined_k141_241823||full Not-determined
18-110-final-viral-combined_k141_149715||full Low-quality
18-110-final-viral-combined_k141_138118||full Not-determined
18-110-final-viral-combined_k141_125828||full Low-quality
18-110-final-viral-combined_k141_217780||full Not-determined
18-110-final-viral-combined_k141_261722||full Not-determined
18-110-final-viral-combined_k141_171922||full Low-quality
18-110-final-viral-combined_k141_217791||full Low-quality
18-110-final-viral-combined_k141_138138||full Not-determined
18-110-final-viral-combined_k141_91083||full Not-determined
18-110-final-viral-combined_k141_113913||full Not-determined
18-110-final-viral-combined_k141_56546||full Not-determined
18-110-final-viral-combined_k141_9570||full Low-quality
18-110-final-viral-combined_k141_241858||full Low-quality
18-110-final-viral-combined_k141_91097||full Not-determined
18-110-final-viral-combined_k141_45848||full Not-determined
18-110-final-viral-combined_k141_149733||full Low-quality
18-110-final-viral-combined_k141_195067||full Low-quality
18-110-final-viral-combined_k141_275212||full Low-quality
18-110-final-viral-combined_k141_229273||full Low-quality
18-110-final-viral-combined_k141_113955||full Low-quality
18-110-final-viral-combined_k141_68533||full Low-quality
18-110-final-viral-combined_k141_91116||full Low-quality
18-110-final-viral-combined_k141_217841||full Not-determined
18-110-final-viral-combined_k141_80412||full Low-quality
18-110-final-viral-combined_k141_160646||full Not-determined
18-110-final-viral-combined_k141_149738||full Low-quality
18-110-final-viral-combined_k141_80414||full Low-quality
18-110-final-viral-combined_k141_91132||full Low-quality
18-110-final-viral-combined_k141_56565||full Low-quality
18-110-final-viral-combined_k141_20634||full Not-determined
18-110-final-viral-combined_k141_113966||full Low-quality
18-110-final-viral-combined_k141_229294||full Low-quality
18-110-final-viral-combined_k141_20641||full Low-quality
18-110-final-viral-combined_k141_261791||full Low-quality
18-110-final-viral-combined_k141_68552||full Low-quality

18-110-final-viral-combined_k141_252446||full Not-determined
18-110-final-viral-combined_k141_229308||full Not-determined
18-110-final-viral-combined_k141_160686||full Not-determined
18-110-final-viral-combined_k141_275246||full Low-quality
18-110-final-viral-combined_k141_172064||full Low-quality
18-110-final-viral-combined_k141_20669||full Not-determined
18-110-final-viral-combined_k141_217861||full Low-quality
18-110-final-viral-combined_k141_68570||full Low-quality
18-110-final-viral-combined_k141_20674||full Low-quality
18-110-final-viral-combined_k141_125937||full Low-quality
18-110-final-viral-combined_k141_275263||full Low-quality
18-110-final-viral-combined_k141_113993||full Low-quality
18-110-final-viral-combined_k141_138221||full Low-quality
18-110-final-viral-combined_k141_125948||full Not-determined
18-110-final-viral-combined_k141_160712||full Not-determined
18-110-final-viral-combined_k141_138223||full Not-determined
18-110-final-viral-combined_k141_261809||full Not-determined
18-110-final-viral-combined_k141_114003||full Low-quality
18-110-final-viral-combined_k141_252529||full Low-quality
18-110-final-viral-combined_k141_252531||full Low-quality
18-110-final-viral-combined_k141_229346||full Low-quality
18-110-final-viral-combined_k141_9613||full Low-quality
18-110-final-viral-combined_k141_125971||full Not-determined
18-110-final-viral-combined_k141_125972||full Not-determined
18-110-final-viral-combined_k141_252553||full Not-determined
18-110-final-viral-combined_k141_114046||full Low-quality
18-110-final-viral-combined_k141_114049||full Low-quality
18-110-final-viral-combined_k141_125991||full Low-quality
18-110-final-viral-combined_k141_20734||full Not-determined
17-106-final-viral-combined_k141_104169||full Medium-quality
18-110-final-viral-combined_k141_275323||full Low-quality
18-110-final-viral-combined_k141_126004||full Not-determined
18-110-final-viral-combined_k141_45942||full Low-quality
18-110-final-viral-combined_k141_20748||full Low-quality
18-110-final-viral-combined_k141_205250||full Low-quality
18-110-final-viral-combined_k141_56676||full Low-quality
18-110-final-viral-combined_k141_91209||full Low-quality
18-110-final-viral-combined_k141_91214||full Low-quality
18-110-final-viral-combined_k141_68634||full Not-determined
18-110-final-viral-combined_k141_91221||full Not-determined
18-110-final-viral-combined_k141_275386||full Not-determined
18-110-final-viral-combined_k141_9658||full Not-determined
18-110-final-viral-combined_k141_126033||full Not-determined
18-110-final-viral-combined_k141_275391||full Not-determined
18-110-final-viral-combined_k141_229406||full Not-determined
18-110-final-viral-combined_k141_114092||full Not-determined
18-110-final-viral-combined_k141_252633||full Low-quality
18-110-final-viral-combined_k141_20800||full Not-determined
18-110-final-viral-combined_k141_252638||full Not-determined
18-110-final-viral-combined_k141_205290||full Low-quality
18-110-final-viral-combined_k141_45985||full Not-determined
18-110-final-viral-combined_k141_217911||full Not-determined
18-110-final-viral-combined_k141_160772||full Not-determined
18-110-final-viral-combined_k141_172184||full Low-quality
18-110-final-viral-combined_k141_20816||full Not-determined
18-110-final-viral-combined_k141_9687||full Low-quality
18-110-final-viral-combined_k141_275418||full Not-determined
18-110-final-viral-combined_k141_172201||full Low-quality
18-110-final-viral-combined_k141_205317||full Low-quality
18-110-final-viral-combined_k141_91279||full Low-quality
18-110-final-viral-combined_k141_275448||full Not-determined
18-110-final-viral-combined_k141_195120||full Low-quality
18-110-final-viral-combined_k141_229468||full Not-determined
18-110-final-viral-combined_k141_91296||full Not-determined
18-110-final-viral-combined_k141_252674||full Low-quality
18-110-final-viral-combined_k141_46023||full Low-quality
18-110-final-viral-combined_k141_229477||full Not-determined
18-110-final-viral-combined_k141_252688||full Not-determined
18-110-final-viral-combined_k141_68664||full Low-quality
18-110-final-viral-combined_k141_56783||full Low-quality
18-110-final-viral-combined_k141_205342||full Low-quality
18-110-final-viral-combined_k141_91303||full Not-determined
18-110-final-viral-combined_k141_20869||full Not-determined
18-110-final-viral-combined_k141_160838||full Low-quality
18-110-final-viral-combined_k141_114135||full Not-determined
18-110-final-viral-combined_k141_20882||full Low-quality
18-110-final-viral-combined_k141_46050||full Not-determined
18-110-final-viral-combined_k141_217979||full Low-quality
18-110-final-viral-combined_k141_46058||full Not-determined
18-110-final-viral-combined_k141_80580||full Not-determined
18-110-final-viral-combined_k141_68714||full Not-determined
18-110-final-viral-combined_k141_229529||full Not-determined
18-110-final-viral-combined_k141_218006||full Low-quality
18-110-final-viral-combined_k141_56848||full Not-determined
18-110-final-viral-combined_k141_114150||full Not-determined
18-110-final-viral-combined_k141_218029||full Low-quality
18-110-final-viral-combined_k141_114151||full Low-quality
18-110-final-viral-combined_k141_80617||full Not-determined
18-110-final-viral-combined_k141_126161||full Low-quality
18-110-final-viral-combined_k141_56874||full Not-determined
18-110-final-viral-combined_k141_195215||full Low-quality
18-110-final-viral-combined_k141_56882||full Not-determined
18-110-final-viral-combined_k141_91399||full Not-determined
18-110-final-viral-combined_k141_9762||full Low-quality
18-110-final-viral-combined_k141_80636||full Low-quality
18-110-final-viral-combined_k141_275523||full Low-quality
18-110-final-viral-combined_k141_275535||full Not-determined
18-110-final-viral-combined_k141_103090||full Low-quality
18-110-final-viral-combined_k141_160946||full Not-determined
18-110-final-viral-combined_k141_160950||full Not-determined
18-110-final-viral-combined_k141_9804||full Low-quality
18-110-final-viral-combined_k141_9814||full Low-quality
18-110-final-viral-combined_k141_68750||full Low-quality
18-110-final-viral-combined_k141_9815||full Not-determined
18-110-final-viral-combined_k141_275540||full Low-quality
18-110-final-viral-combined_k141_218130||full Not-determined
18-110-final-viral-combined_k141_103107||full Not-determined
18-110-final-viral-combined_k141_261910||full Low-quality
18-110-final-viral-combined_k141_68778||full Low-quality
18-110-final-viral-combined_k141_160972||full Not-determined
18-110-final-viral-combined_k141_275574||full Low-quality
18-110-final-viral-combined_k141_275577||full Not-determined
18-110-final-viral-combined_k141_218153||full Not-determined
18-110-final-viral-combined_k141_103130||full Not-determined
18-110-final-viral-combined_k141_103134||full Not-determined
18-110-final-viral-combined_k141_91514||full Low-quality
18-110-final-viral-combined_k141_20976||full Low-quality
18-110-final-viral-combined_k141_91518||full Not-determined
18-110-final-viral-combined_k141_126236||full Not-determined
18-110-final-viral-combined_k141_172450||full Low-quality
18-110-final-viral-combined_k141_275609||full Low-quality
18-110-final-viral-combined_k141_161004||full Not-determined
18-110-final-viral-combined_k141_218183||full Not-determined
18-110-final-viral-combined_k141_275615||full Low-quality
18-110-final-viral-combined_k141_126269||full Low-quality
18-110-final-viral-combined_k141_126270||full Low-quality
18-110-final-viral-combined_k141_103160||full Low-quality
18-110-final-viral-combined_k141_20999||full Not-determined
18-110-final-viral-combined_k141_275616||full Low-quality
18-110-final-viral-combined_k141_126278||full Not-determined
18-110-final-viral-combined_k141_261973||full Low-quality
18-110-final-viral-combined_k141_261975||full Not-determined
18-110-final-viral-combined_k141_9897||full Not-determined
18-110-final-viral-combined_k141_103177||full Not-determined
18-110-final-viral-combined_k141_161032||full Low-quality
18-110-final-viral-combined_k141_114293||full Low-quality
18-110-final-viral-combined_k141_218224||full Not-determined
17-109_k141_223235 High-quality
18-110-final-viral-combined_k141_218237||full Not-determined
18-110-final-viral-combined_k141_205633||full Low-quality
18-110-final-viral-combined_k141_161057||full Low-quality
18-110-final-viral-combined_k141_9954||full Not-determined
18-110-final-viral-combined_k141_114361||full Not-determined
18-110-final-viral-combined_k141_229748||full Low-quality
18-110-final-viral-combined_k141_275654||full Low-quality
18-110-final-viral-combined_k141_68811||full Not-determined
18-110-final-viral-combined_k141_9969||full Low-quality
18-110-final-viral-combined_k141_68815||full Low-quality
18-110-final-viral-combined_k141_103247||full Low-quality
18-110-final-viral-combined_k141_195288||full Not-determined
18-110-final-viral-combined_k141_103272||full Not-determined
18-110-final-viral-combined_k141_126412||full Low-quality
18-110-final-viral-combined_k141_57029||full Low-quality
18-110-final-viral-combined_k141_10019||full Low-quality
18-110-final-viral-combined_k141_126429||full Not-determined
18-110-final-viral-combined_k141_91658||full Not-determined
18-110-final-viral-combined_k141_218309||full Not-determined
18-110-final-viral-combined_k141_57049||full Not-determined
18-110-final-viral-combined_k141_68852||full Low-quality
18-110-final-viral-combined_k141_68853||full Low-quality
18-110-final-viral-combined_k141_91689||full Not-determined
18-110-final-viral-combined_k141_91700||full Not-determined
18-110-final-viral-combined_k141_229856||full Low-quality
18-110-final-viral-combined_k141_229858||full Low-quality
18-110-final-viral-combined_k141_252857||full Low-quality
18-110-final-viral-combined_k141_21153||full Not-determined
18-110-final-viral-combined_k141_91759||full Not-determined
18-110-final-viral-combined_k141_252880||full Not-determined
18-110-final-viral-combined_k141_10065||full Not-determined
18-110-final-viral-combined_k141_195391||full Not-determined
18-110-final-viral-combined_k141_10070||full Not-determined
18-110-final-viral-combined_k141_195393||full Not-determined
18-110-final-viral-combined_k141_91775||full Low-quality
18-110-final-viral-combined_k141_103321||full Not-determined
18-110-final-viral-combined_k141_161155||full Low-quality
18-110-final-viral-combined_k141_195400||full Low-quality
18-110-final-viral-combined_k141_161167||full Not-determined
18-110-final-viral-combined_k141_275819||full Low-quality
18-110-final-viral-combined_k141_252927||full Not-determined
18-110-final-viral-combined_k141_114468||full Not-determined
18-110-final-viral-combined_k141_218401||full Not-determined
17-106_k141_10480 High-quality
18-110-final-viral-combined_k141_91828||full Low-quality
18-110-final-viral-combined_k141_218404||full Not-determined
17-106-final-viral-combined_k141_10480||full High-quality
18-110-final-viral-combined_k141_218406||full Not-determined
18-110-final-viral-combined_k141_91846||full Low-quality
18-110-final-viral-combined_k141_229942||full Low-quality
18-110-final-viral-combined_k141_10130||full Low-quality
18-110-final-viral-combined_k141_252977||full Not-determined
18-110-final-viral-combined_k141_103380||full Not-determined
18-110-final-viral-combined_k141_10137||full Not-determined
18-110-final-viral-combined_k141_91870||full Low-quality
18-110-final-viral-combined_k141_21252||full Not-determined
18-110-final-viral-combined_k141_69007||full Low-quality
18-110-final-viral-combined_k141_57173||full Not-determined
18-110-final-viral-combined_k141_21256||full Not-determined
18-110-final-viral-combined_k141_21257||full Low-quality
18-110-final-viral-combined_k141_57192||full Low-quality
18-110-final-viral-combined_k141_91888||full Low-quality
18-110-final-viral-combined_k141_21271||full Low-quality
18-110-final-viral-combined_k141_2174||0_partial High-quality
18-110-final-viral-combined_k141_126550||full Low-quality
18-110-final-viral-combined_k141_218454||full Low-quality
18-110-final-viral-combined_k141_103440||full Not-determined
18-110-final-viral-combined_k141_275884||full Not-determined
18-110-final-viral-combined_k141_21280||full Not-determined
18-110-final-viral-combined_k141_103448||full Low-quality
18-110-final-viral-combined_k141_91917||full Not-determined
18-110-final-viral-combined_k141_262225||full Low-quality
18-110-final-viral-combined_k141_126565||full Not-determined
18-110-final-viral-combined_k141_275908||full Low-quality
18-110-final-viral-combined_k141_57207||full Not-determined
18-110-final-viral-combined_k141_172683||full Not-determined
18-110-final-viral-combined_k141_253009||full Low-quality
18-110-final-viral-combined_k141_275936||full Low-quality
18-110-final-viral-combined_k141_114524||full Low-quality
18-110-final-viral-combined_k141_230057||full Low-quality
18-110-final-viral-combined_k141_172723||full Low-quality
18-110-final-viral-combined_k141_57259||full Low-quality
18-110-final-viral-combined_k141_253039||full Low-quality
18-110-final-viral-combined_k141_218548||full Not-determined
18-110-final-viral-combined_k141_253051||full Not-determined
18-110-final-viral-combined_k141_218558||full Not-determined
18-110-final-viral-combined_k141_262298||full Low-quality
18-110-final-viral-combined_k141_126604||full Low-quality
18-110-final-viral-combined_k141_230095||full Low-quality
18-110-final-viral-combined_k141_21376||full Not-determined
18-110-final-viral-combined_k141_218582||full Low-quality
18-110-final-viral-combined_k141_57316||full Not-determined
18-110-final-viral-combined_k141_10211||full Not-determined
18-110-final-viral-combined_k141_195537||full Low-quality
18-110-final-viral-combined_k141_218611||full Low-quality
18-110-final-viral-combined_k141_10245||full Low-quality
18-110-final-viral-combined_k141_57342||full Not-determined
18-110-final-viral-combined_k141_103563||full Not-determined
18-110-final-viral-combined_k141_57346||full Not-determined
18-110-final-viral-combined_k141_230165||full Low-quality
18-110-final-viral-combined_k141_21434||full Not-determined
18-110-final-viral-combined_k141_10271||full Low-quality
18-110-final-viral-combined_k141_126707||full Not-determined
18-110-final-viral-combined_k141_57362||full Low-quality
18-110-final-viral-combined_k141_275979||full Low-quality
18-110-final-viral-combined_k141_10276||full Low-quality
18-110-final-viral-combined_k141_92007||full Not-determined
18-110-final-viral-combined_k141_275988||full Not-determined
18-110-final-viral-combined_k141_218666||full Low-quality
18-110-final-viral-combined_k141_218676||full Low-quality
18-110-final-viral-combined_k141_262405||full Not-determined
18-110-final-viral-combined_k141_275992||full Not-determined
18-110-final-viral-combined_k141_253211||full Not-determined
18-110-final-viral-combined_k141_195681||full Low-quality
18-110-final-viral-combined_k141_57428||full Low-quality
18-110-final-viral-combined_k141_195693||full Not-determined
18-110-final-viral-combined_k141_206059||full Low-quality
18-110-final-viral-combined_k141_21493||full Not-determined
18-110-final-viral-combined_k141_195702||full Not-determined
18-110-final-viral-combined_k141_253262||full Low-quality
18-110-final-viral-combined_k141_218713||full Not-determined
18-110-final-viral-combined_k141_10320||full Not-determined
18-110-final-viral-combined_k141_276049||full Low-quality
18-110-final-viral-combined_k141_10336||full Low-quality
18-110-final-viral-combined_k141_253293||full Low-quality
18-110-final-viral-combined_k141_206093||full Low-quality
18-110-final-viral-combined_k141_262467||full Not-determined
18-110-final-viral-combined_k141_57494||full Low-quality
18-110-final-viral-combined_k141_21563||full Not-determined
18-110-final-viral-combined_k141_218764||full Low-quality
18-110-final-viral-combined_k141_21574||full Low-quality
18-110-final-viral-combined_k141_218769||full Not-determined
18-110-final-viral-combined_k141_21582||full Not-determined
18-110-final-viral-combined_k141_206195||full Not-determined
18-110-final-viral-combined_k141_230324||full Low-quality
18-110-final-viral-combined_k141_92110||full Low-quality
18-110-final-viral-combined_k141_230336||full Low-quality
18-110-final-viral-combined_k141_114714||full Not-determined
18-110-final-viral-combined_k141_21620||full Not-determined
18-110-final-viral-combined_k141_114716||full Not-determined
18-110-final-viral-combined_k141_21621||full Not-determined
18-110-final-viral-combined_k141_21624||full Not-determined
18-110-final-viral-combined_k141_218823||full Low-quality
18-110-final-viral-combined_k141_92134||full Low-quality
18-110-final-viral-combined_k141_230359||full Not-determined
18-110-final-viral-combined_k141_57558||full Not-determined
18-110-final-viral-combined_k141_10450||full Low-quality
18-110-final-viral-combined_k141_114736||full Low-quality
18-110-final-viral-combined_k141_195726||full Low-quality
18-110-final-viral-combined_k141_10479||full Low-quality
18-110-final-viral-combined_k141_262565||full Low-quality
18-110-final-viral-combined_k141_10480||full Low-quality
18-110-final-viral-combined_k141_21669||full Low-quality
18-110-final-viral-combined_k141_114780||full Not-determined
18-110-final-viral-combined_k141_92148||full Not-determined
18-110-final-viral-combined_k141_10507||full Low-quality
18-110-final-viral-combined_k141_21699||full Low-quality
18-110-final-viral-combined_k141_21708||full Not-determined
18-110-final-viral-combined_k141_195778||full Low-quality
18-110-final-viral-combined_k141_114833||full Low-quality
18-110-final-viral-combined_k141_10553||full Low-quality
18-110-final-viral-combined_k141_21746||full Low-quality
18-110-final-viral-combined_k141_114901||full Low-quality
18-110-final-viral-combined_k141_206338||full Not-determined
18-110-final-viral-combined_k141_10621||full Low-quality
18-110-final-viral-combined_k141_195815||full Low-quality
18-110-final-viral-combined_k141_206372||full Not-determined
18-110-final-viral-combined_k141_206402||full Not-determined
18-110-final-viral-combined_k141_206425||full Low-quality
18-110-final-viral-combined_k141_10691||full Low-quality
18-110-final-viral-combined_k141_206449||full Low-quality
18-110-final-viral-combined_k141_21797||full Not-determined
18-110-final-viral-combined_k141_206474||full Low-quality
18-110-final-viral-combined_k141_262750||full Not-determined
18-110-final-viral-combined_k141_114997||full Not-determined
18-110-final-viral-combined_k141_262763||full Low-quality
18-110-final-viral-combined_k141_10754||full Not-determined
18-110-final-viral-combined_k141_115055||full Not-determined
18-110-final-viral-combined_k141_21844||full Low-quality
18-110-final-viral-combined_k141_262828||full Not-determined
18-110-final-viral-combined_k141_21875||full Not-determined
18-110-final-viral-combined_k141_115101||full Not-determined
18-110-final-viral-combined_k141_206568||full Not-determined
18-110-final-viral-combined_k141_206574||full Not-determined
18-110-final-viral-combined_k141_206578||full Not-determined
18-110-final-viral-combined_k141_262872||full Low-quality
18-110-final-viral-combined_k141_206585||full Low-quality
18-110-final-viral-combined_k141_262896||full Not-determined
18-110-final-viral-combined_k141_115190||full Low-quality
18-110-final-viral-combined_k141_206667||full Not-determined
18-110-final-viral-combined_k141_22007||full Low-quality
18-110-final-viral-combined_k141_206685||full Low-quality
18-110-final-viral-combined_k141_263002||full Not-determined
18-110-final-viral-combined_k141_206698||full Low-quality
18-110-final-viral-combined_k141_263005||full Not-determined
18-110-final-viral-combined_k141_22057||full Low-quality
18-110-final-viral-combined_k141_263026||full Low-quality
18-110-final-viral-combined_k141_22061||full Low-quality
18-110-final-viral-combined_k141_263034||full Low-quality
18-110-final-viral-combined_k141_10960||full Not-determined
18-110-final-viral-combined_k141_10961||full Not-determined
18-110-final-viral-combined_k141_22089||full Not-determined
18-110-final-viral-combined_k141_22107||full Not-determined
18-110-final-viral-combined_k141_10975||full Not-determined
18-110-final-viral-combined_k141_22140||full Low-quality
18-110-final-viral-combined_k141_263126||full Not-determined
18-110-final-viral-combined_k141_263130||full Low-quality
18-110-final-viral-combined_k141_206785||full Low-quality
18-110-final-viral-combined_k141_263142||full Not-determined
18-110-final-viral-combined_k141_22198||full Low-quality
18-110-final-viral-combined_k141_263172||full Low-quality
18-110-final-viral-combined_k141_10986||full Low-quality
18-110-final-viral-combined_k141_206813||full Low-quality
18-110-final-viral-combined_k141_206819||full Not-determined
18-110-final-viral-combined_k141_11018||full Low-quality
18-110-final-viral-combined_k141_11041||full Low-quality
18-110-final-viral-combined_k141_22236||full Low-quality
18-110-final-viral-combined_k141_11070||full Low-quality
18-110-final-viral-combined_k141_22272||full Low-quality
18-110-final-viral-combined_k141_22299||full Low-quality
18-110-final-viral-combined_k141_206861||full Not-determined
18-110-final-viral-combined_k141_11114||full Not-determined
18-110-final-viral-combined_k141_22358||full Low-quality
18-110-final-viral-combined_k141_263340||full Not-determined
18-110-final-viral-combined_k141_22398||full Not-determined
18-110-final-viral-combined_k141_263368||full Low-quality
18-110-final-viral-combined_k141_263419||full Not-determined
18-110-final-viral-combined_k141_22476||full Not-determined
18-110-final-viral-combined_k141_206987||full Not-determined
18-110-final-viral-combined_k141_263459||full Not-determined
18-110-final-viral-combined_k141_263507||full Not-determined
18-110-final-viral-combined_k141_22495||full Low-quality
18-110-final-viral-combined_k141_263568||full Low-quality
18-110-final-viral-combined_k141_263569||full Low-quality
18-110-final-viral-combined_k141_263572||full Not-determined
18-110-final-viral-combined_k141_11130||full Not-determined
18-110-final-viral-combined_k141_207093||full Not-determined
18-110-final-viral-combined_k141_22554||full Low-quality
18-110-final-viral-combined_k141_11165||full Not-determined
18-110-final-viral-combined_k141_11173||full Not-determined
18-110-final-viral-combined_k141_11209||full Low-quality
18-110-final-viral-combined_k141_263684||full Low-quality
18-110-final-viral-combined_k141_22615||full Low-quality
18-110-final-viral-combined_k141_263752||full Not-determined
18-110-final-viral-combined_k141_22654||full Low-quality
18-110-final-viral-combined_k141_22659||full Not-determined
18-110-final-viral-combined_k141_263758||full Not-determined
18-110-final-viral-combined_k141_207258||full Low-quality
18-110-final-viral-combined_k141_207261||full Low-quality
18-110-final-viral-combined_k141_207262||full Low-quality
18-110-final-viral-combined_k141_22706||full Not-determined
18-110-final-viral-combined_k141_22728||full Not-determined
18-110-final-viral-combined_k141_11334||full Not-determined
18-110-final-viral-combined_k141_22739||full Low-quality
18-110-final-viral-combined_k141_207302||full Not-determined
18-110-final-viral-combined_k141_207304||full Low-quality
18-110-final-viral-combined_k141_207308||full Not-determined
18-110-final-viral-combined_k141_11380||full Not-determined
18-110-final-viral-combined_k141_207339||full Not-determined
18-110-final-viral-combined_k141_11420||full Not-determined
18-110-final-viral-combined_k141_22809||full Not-determined
18-110-final-viral-combined_k141_11433||full Low-quality
18-110-final-viral-combined_k141_22883||full Not-determined
18-110-final-viral-combined_k141_264048||full Low-quality
18-110-final-viral-combined_k141_22948||full Not-determined
18-110-final-viral-combined_k141_264111||full Low-quality
18-110-final-viral-combined_k141_264158||full Not-determined
18-110-final-viral-combined_k141_264244||full Low-quality
18-110-final-viral-combined_k141_264319||full Not-determined
18-110-final-viral-combined_k141_264432||full Low-quality
18-110-final-viral-combined_k141_264487||full Low-quality
18-110-final-viral-combined_k141_264558||full Not-determined
18-110-final-viral-combined_k141_264592||full Low-quality
18-110-final-viral-combined_k141_264685||full Not-determined
18-110-final-viral-combined_k141_264687||full Not-determined
18-110-final-viral-combined_k141_264692||full Low-quality
17-109-final-viral-combined_k141_157167||0_partial High-quality
18-110-final-viral-combined_k141_264778||full Low-quality
18-110-final-viral-combined_k141_264865||full Not-determined
18-110-final-viral-combined_k141_212554||0_partial Low-quality
18-110-final-viral-combined_k141_92329||0_partial Low-quality
18-110-final-viral-combined_k141_207717||0_partial Low-quality
18-110-final-viral-combined_k141_46557||0_partial Not-determined
18-110-final-viral-combined_k141_58168||0_partial Low-quality
18-110-final-viral-combined_k141_34759||0_partial Low-quality
18-110-final-viral-combined_k141_34759||1_partial Low-quality
18-110-final-viral-combined_k141_139022||0_partial Low-quality
18-110-final-viral-combined_k141_219482||0_partial Not-determined
18-110-final-viral-combined_k141_185132||0_partial Low-quality
18-110-final-viral-combined_k141_219682||0_partial Low-quality
18-110-final-viral-combined_k141_162592||0_partial Low-quality
18-110-final-viral-combined_k141_12543||0_partial Not-determined
18-110-final-viral-combined_k141_220187||0_partial Low-quality
18-104-final-viral-combined_k141_143879||0_partial High-quality
18-110-final-viral-combined_k141_2174||1_partial Low-quality
18-110-final-viral-combined_k141_2174||2_partial Low-quality
18-110-final-viral-combined_k141_105853||0_partial Not-determined
18-110-final-viral-combined_k141_163586||0_partial Low-quality
18-110-final-viral-combined_k141_60264||0_partial Low-quality
18-110-final-viral-combined_k141_48399||0_partial Low-quality
18-110-final-viral-combined_k141_37125||0_partial Low-quality
18-110-final-viral-combined_k141_106223||0_partial Low-quality
18-110-final-viral-combined_k141_83144||0_partial Low-quality
18-110-final-viral-combined_k141_164484||0_partial Low-quality
18-110-final-viral-combined_k141_268464||0_partial Not-determined
18-110-final-viral-combined_k141_188103||0_partial Low-quality
18-110-final-viral-combined_k141_256295||0_partial Low-quality
18-102-final-viral-combined_k141_48661||full Medium-quality
18-110-final-viral-combined_k141_257014||0_partial Low-quality
18-110-final-viral-combined_k141_223339||0_partial Low-quality
18-110-final-viral-combined_k141_178559||0_partial Low-quality
18-110-final-viral-combined_k141_4428||0_partial Low-quality
18-110-final-viral-combined_k141_212916||0_partial Low-quality
18-110-final-viral-combined_k141_190185||0_partial Not-determined
18-110-final-viral-combined_k141_155650||0_partial Low-quality
18-110-final-viral-combined_k141_63699||0_partial Low-quality
18-102-final-viral-combined_k141_230282||full Medium-quality
18-110-final-viral-combined_k141_259203||0_partial Low-quality
18-110-final-viral-combined_k141_226044||0_partial Low-quality
18-110-final-viral-combined_k141_259282||0_partial Low-quality
18-110-final-viral-combined_k141_169056||0_partial Low-quality
18-110-final-viral-combined_k141_88253||0_partial Low-quality
18-110-final-viral-combined_k141_88389||0_partial Low-quality
18-110-final-viral-combined_k141_273181||0_partial Low-quality
18-110-final-viral-combined_k141_78097||0_partial Not-determined
18-110-final-viral-combined_k141_19380||0_partial Low-quality
18-110-final-viral-combined_k141_216894||0_partial Low-quality
18-110-final-viral-combined_k141_8521||0_partial Low-quality
18-110-final-viral-combined_k141_194197||0_partial Low-quality
18-110-final-viral-combined_k141_228797||0_partial Not-determined
18-110-final-viral-combined_k141_20235||0_partial Not-determined
18-110-final-viral-combined_k141_229038||0_partial Not-determined
18-110-final-viral-combined_k141_160505||0_partial Not-determined
18-110-final-viral-combined_k141_205724||0_partial Low-quality
18-110-final-viral-combined_k141_114651||0_partial Low-quality
18-110-final-viral-combined_k141_92109||0_partial Not-determined
18-110-final-viral-combined_k141_241905||lt2gene Low-quality
18-110-final-viral-combined_k141_230450||lt2gene Low-quality
18-110-final-viral-combined_k141_46111||lt2gene Low-quality
18-110-final-viral-combined_k141_34600||lt2gene Not-determined
18-110-final-viral-combined_k141_207391||lt2gene Low-quality
18-110-final-viral-combined_k141_172833||lt2gene Low-quality
18-110-final-viral-combined_k141_253467||lt2gene Not-determined
18-110-final-viral-combined_k141_69228||lt2gene Low-quality
18-110-final-viral-combined_k141_70||lt2gene Low-quality
18-110-final-viral-combined_k141_115262||lt2gene Low-quality
18-110-final-viral-combined_k141_69241||lt2gene Low-quality
18-110-final-viral-combined_k141_69249||lt2gene Low-quality
18-110-final-viral-combined_k141_149879||lt2gene Not-determined
18-110-final-viral-combined_k141_207437||lt2gene Low-quality
18-110-final-viral-combined_k141_218972||lt2gene Low-quality
18-110-final-viral-combined_k141_92304||lt2gene Low-quality
18-110-final-viral-combined_k141_265093||lt2gene Low-quality
18-110-final-viral-combined_k141_138412||lt2gene Low-quality
18-110-final-viral-combined_k141_23178||lt2gene Not-determined
18-110-final-viral-combined_k141_172911||lt2gene Low-quality
18-110-final-viral-combined_k141_80824||lt2gene Not-determined
18-110-final-viral-combined_k141_115311||lt2gene Low-quality
18-110-final-viral-combined_k141_92351||lt2gene Low-quality
18-110-final-viral-combined_k141_92354||lt2gene Not-determined
18-110-final-viral-combined_k141_138484||lt2gene Low-quality
18-110-final-viral-combined_k141_138497||lt2gene Low-quality
18-110-final-viral-combined_k141_219022||lt2gene Low-quality
18-110-final-viral-combined_k141_11600||lt2gene Low-quality
18-110-final-viral-combined_k141_242119||lt2gene Low-quality
18-110-final-viral-combined_k141_196012||lt2gene Not-determined
18-110-final-viral-combined_k141_196017||lt2gene Low-quality
18-110-final-viral-combined_k141_196018||lt2gene Low-quality
18-110-final-viral-combined_k141_224||lt2gene Low-quality
18-110-final-viral-combined_k141_196029||lt2gene Low-quality
18-110-final-viral-combined_k141_161334||lt2gene Not-determined
18-110-final-viral-combined_k141_161347||lt2gene Low-quality
18-110-final-viral-combined_k141_161348||lt2gene Not-determined
18-110-final-viral-combined_k141_57851||lt2gene Low-quality
18-110-final-viral-combined_k141_69393||lt2gene Low-quality
18-110-final-viral-combined_k141_57863||lt2gene Low-quality
18-110-final-viral-combined_k141_126965||lt2gene Not-determined
18-110-final-viral-combined_k141_149953||lt2gene Low-quality
18-110-final-viral-combined_k141_196081||lt2gene Low-quality
18-110-final-viral-combined_k141_46337||lt2gene Not-determined
18-110-final-viral-combined_k141_81010||lt2gene Low-quality
18-110-final-viral-combined_k141_57914||lt2gene Low-quality
18-110-final-viral-combined_k141_230831||lt2gene Low-quality
18-110-final-viral-combined_k141_230840||lt2gene Low-quality
18-110-final-viral-combined_k141_115468||lt2gene Low-quality
18-110-final-viral-combined_k141_11701||lt2gene Low-quality
18-110-final-viral-combined_k141_303||lt2gene Low-quality
18-110-final-viral-combined_k141_219112||lt2gene Low-quality
18-110-final-viral-combined_k141_314||lt2gene Low-quality
18-110-final-viral-combined_k141_69481||lt2gene Low-quality
18-110-final-viral-combined_k141_150020||lt2gene Low-quality
18-110-final-viral-combined_k141_219138||lt2gene Low-quality
18-110-final-viral-combined_k141_265227||lt2gene Low-quality
18-110-final-viral-combined_k141_184641||lt2gene Not-determined
18-110-final-viral-combined_k141_103890||lt2gene Low-quality
18-110-final-viral-combined_k141_230902||lt2gene Low-quality
18-110-final-viral-combined_k141_92528||lt2gene Not-determined
18-110-final-viral-combined_k141_265286||lt2gene Low-quality
18-110-final-viral-combined_k141_161454||lt2gene Low-quality
18-110-final-viral-combined_k141_23422||lt2gene Low-quality
18-110-final-viral-combined_k141_11789||lt2gene Low-quality
18-110-final-viral-combined_k141_138770||lt2gene Low-quality
18-110-final-viral-combined_k141_92583||lt2gene Low-quality
18-110-final-viral-combined_k141_253728||lt2gene Low-quality
18-110-final-viral-combined_k141_58121||lt2gene Not-determined
18-110-final-viral-combined_k141_173235||lt2gene Low-quality
18-110-final-viral-combined_k141_242424||lt2gene Low-quality
18-110-final-viral-combined_k141_173251||lt2gene Not-determined
18-110-final-viral-combined_k141_92627||lt2gene Not-determined
18-110-final-viral-combined_k141_253756||lt2gene Low-quality
18-110-final-viral-combined_k141_161545||lt2gene Low-quality
18-110-final-viral-combined_k141_138865||lt2gene Not-determined
18-110-final-viral-combined_k141_69646||lt2gene Not-determined
18-110-final-viral-combined_k141_219224||lt2gene Low-quality
18-110-final-viral-combined_k141_92655||lt2gene Low-quality
18-110-final-viral-combined_k141_150142||lt2gene Not-determined
18-110-final-viral-combined_k141_161622||lt2gene Low-quality
18-110-final-viral-combined_k141_23546||lt2gene Low-quality
18-110-final-viral-combined_k141_265424||lt2gene Low-quality
18-110-final-viral-combined_k141_11935||lt2gene Not-determined
18-110-final-viral-combined_k141_489||lt2gene Low-quality
18-110-final-viral-combined_k141_207839||lt2gene Low-quality
18-110-final-viral-combined_k141_81290||lt2gene Not-determined
18-110-final-viral-combined_k141_184719||lt2gene Low-quality
18-110-final-viral-combined_k141_127162||lt2gene Low-quality
18-110-final-viral-combined_k141_138969||lt2gene Not-determined
18-110-final-viral-combined_k141_58199||lt2gene Low-quality
18-110-final-viral-combined_k141_196367||lt2gene Low-quality
18-110-final-viral-combined_k141_242603||lt2gene Not-determined
18-110-final-viral-combined_k141_92776||lt2gene Low-quality
18-110-final-viral-combined_k141_34765||lt2gene Not-determined
18-110-final-viral-combined_k141_242613||lt2gene Low-quality
18-110-final-viral-combined_k141_58233||lt2gene Not-determined
18-110-final-viral-combined_k141_104122||lt2gene Low-quality
18-110-final-viral-combined_k141_34813||lt2gene Not-determined
18-110-final-viral-combined_k141_69747||lt2gene Low-quality
18-110-final-viral-combined_k141_173519||lt2gene Not-determined
18-110-final-viral-combined_k141_23639||lt2gene Low-quality
18-110-final-viral-combined_k141_69780||lt2gene Low-quality
18-110-final-viral-combined_k141_58266||lt2gene Low-quality
18-110-final-viral-combined_k141_231082||lt2gene Low-quality
18-110-final-viral-combined_k141_231123||lt2gene Not-determined
18-110-final-viral-combined_k141_92965||lt2gene Low-quality
18-110-final-viral-combined_k141_196541||lt2gene Low-quality
18-110-final-viral-combined_k141_139023||lt2gene Low-quality
18-110-final-viral-combined_k141_184907||lt2gene Not-determined
18-110-final-viral-combined_k141_208045||lt2gene Low-quality
18-110-final-viral-combined_k141_184943||lt2gene Low-quality
18-110-final-viral-combined_k141_58375||lt2gene Low-quality
18-110-final-viral-combined_k141_184955||lt2gene Low-quality
18-110-final-viral-combined_k141_219495||lt2gene Low-quality

| | |
|---|---|
| 18-110-final-viral-combined_k141_69879\|\|lt2gene | Low-quality |
| 18-110-final-viral-combined_k141_23797\|\|lt2gene | Low-quality |
| 18-110-final-viral-combined_k141_23805\|\|lt2gene | Low-quality |
| 18-110-final-viral-combined_k141_115852\|\|lt2gene | Low-quality |
| 18-110-final-viral-combined_k141_35023\|\|lt2gene | Low-quality |
| 18-110-final-viral-combined_k141_35026\|\|lt2gene | Low-quality |
| 18-110-final-viral-combined_k141_46792\|\|lt2gene | Not-determined |
| 18-110-final-viral-combined_k141_69967\|\|lt2gene | Not-determined |
| 18-110-final-viral-combined_k141_12110\|\|lt2gene | Low-quality |
| 18-110-final-viral-combined_k141_139137\|\|lt2gene | Low-quality |
| 18-110-final-viral-combined_k141_231186\|\|lt2gene | Not-determined |
| 18-110-final-viral-combined_k141_208186\|\|lt2gene | Low-quality |
| 18-110-final-viral-combined_k141_93149\|\|lt2gene | Not-determined |
| 18-110-final-viral-combined_k141_35159\|\|lt2gene | Low-quality |
| 18-110-final-viral-combined_k141_231259\|\|lt2gene | Low-quality |
| 18-110-final-viral-combined_k141_46866\|\|lt2gene | Not-determined |
| 18-110-final-viral-combined_k141_242853\|\|lt2gene | Not-determined |
| 18-110-final-viral-combined_k141_139234\|\|lt2gene | Not-determined |
| 18-110-final-viral-combined_k141_208256\|\|lt2gene | Low-quality |
| 18-110-final-viral-combined_k141_231342\|\|lt2gene | Low-quality |
| 18-110-final-viral-combined_k141_242922\|\|lt2gene | Low-quality |
| 18-110-final-viral-combined_k141_150603\|\|lt2gene | Not-determined |
| 18-110-final-viral-combined_k141_196758\|\|lt2gene | Not-determined |
| 18-110-final-viral-combined_k141_35286\|\|lt2gene | Low-quality |
| 18-110-final-viral-combined_k141_208335\|\|lt2gene | Low-quality |
| 18-110-final-viral-combined_k141_35308\|\|lt2gene | Low-quality |
| 18-110-final-viral-combined_k141_254205\|\|lt2gene | Low-quality |
| 18-110-final-viral-combined_k141_173935\|\|lt2gene | Not-determined |
| 18-110-final-viral-combined_k141_162155\|\|lt2gene | Low-quality |
| 18-110-final-viral-combined_k141_150770\|\|lt2gene | Low-quality |
| 18-110-final-viral-combined_k141_127628\|\|lt2gene | Low-quality |
| 18-110-final-viral-combined_k141_139398\|\|lt2gene | Not-determined |
| 18-110-final-viral-combined_k141_196845\|\|lt2gene | Low-quality |
| 18-110-final-viral-combined_k141_265857\|\|lt2gene | Low-quality |
| 18-110-final-viral-combined_k141_127658\|\|lt2gene | Low-quality |
| 18-110-final-viral-combined_k141_231538\|\|lt2gene | Low-quality |
| 18-110-final-viral-combined_k141_24056\|\|lt2gene | Not-determined |
| 18-110-final-viral-combined_k141_47004\|\|lt2gene | Low-quality |
| 18-110-final-viral-combined_k141_47016\|\|lt2gene | Low-quality |
| 18-110-final-viral-combined_k141_47017\|\|lt2gene | Not-determined |
| 18-110-final-viral-combined_k141_208456\|\|lt2gene | Not-determined |
| 18-110-final-viral-combined_k141_231558\|\|lt2gene | Low-quality |
| 18-110-final-viral-combined_k141_127673\|\|lt2gene | Low-quality |
| 18-110-final-viral-combined_k141_70284\|\|lt2gene | Low-quality |
| 18-110-final-viral-combined_k141_185348\|\|lt2gene | Low-quality |
| 18-110-final-viral-combined_k141_58722\|\|lt2gene | Not-determined |
| 18-110-final-viral-combined_k141_231573\|\|lt2gene | Low-quality |
| 18-110-final-viral-combined_k141_35399\|\|lt2gene | Low-quality |
| 18-110-final-viral-combined_k141_150863\|\|lt2gene | Low-quality |
| 18-110-final-viral-combined_k141_116218\|\|lt2gene | Low-quality |
| 18-110-final-viral-combined_k141_104663\|\|lt2gene | Low-quality |
| 18-110-final-viral-combined_k141_24128\|\|lt2gene | Low-quality |
| 18-110-final-viral-combined_k141_35458\|\|lt2gene | Not-determined |
| 18-110-final-viral-combined_k141_162211\|\|lt2gene | Not-determined |
| 18-110-final-viral-combined_k141_219696\|\|lt2gene | Low-quality |
| 18-110-final-viral-combined_k141_24161\|\|lt2gene | Low-quality |
| 18-110-final-viral-combined_k141_185498\|\|lt2gene | Low-quality |
| 18-110-final-viral-combined_k141_93436\|\|lt2gene | Low-quality |
| 18-110-final-viral-combined_k141_219756\|\|lt2gene | Low-quality |
| 18-110-final-viral-combined_k141_162270\|\|lt2gene | Low-quality |
| 18-110-final-viral-combined_k141_58867\|\|lt2gene | Low-quality |
| 18-110-final-viral-combined_k141_208693\|\|lt2gene | Low-quality |
| 18-110-final-viral-combined_k141_81980\|\|lt2gene | Low-quality |
| 18-110-final-viral-combined_k141_174053\|\|lt2gene | Low-quality |
| 18-110-final-viral-combined_k141_231717\|\|lt2gene | Not-determined |
| 18-110-final-viral-combined_k141_231719\|\|lt2gene | Not-determined |
| 18-110-final-viral-combined_k141_231734\|\|lt2gene | Low-quality |
| 18-110-final-viral-combined_k141_254380\|\|lt2gene | Low-quality |
| 18-110-final-viral-combined_k141_243319\|\|lt2gene | Low-quality |
| 18-110-final-viral-combined_k141_162401\|\|lt2gene | Low-quality |
| 18-110-final-viral-combined_k141_59002\|\|lt2gene | Low-quality |
| 18-110-final-viral-combined_k141_93625\|\|lt2gene | Low-quality |
| 18-110-final-viral-combined_k141_59044\|\|lt2gene | Low-quality |
| 18-110-final-viral-combined_k141_208791\|\|lt2gene | Not-determined |
| 18-110-final-viral-combined_k141_104886\|\|lt2gene | Low-quality |
| 18-110-final-viral-combined_k141_208814\|\|lt2gene | Not-determined |
| 18-110-final-viral-combined_k141_208816\|\|lt2gene | Low-quality |
| 18-110-final-viral-combined_k141_24403\|\|lt2gene | Not-determined |
| 18-110-final-viral-combined_k141_162504\|\|lt2gene | Low-quality |
| 18-110-final-viral-combined_k141_116442\|\|lt2gene | Low-quality |
| 18-110-final-viral-combined_k141_104950\|\|lt2gene | Low-quality |
| 18-110-final-viral-combined_k141_162532\|\|lt2gene | Not-determined |
| 18-110-final-viral-combined_k141_185754\|\|lt2gene | Low-quality |
| 18-110-final-viral-combined_k141_162560\|\|lt2gene | Low-quality |
| 18-110-final-viral-combined_k141_266041\|\|lt2gene | Low-quality |
| 18-110-final-viral-combined_k141_24470\|\|lt2gene | Low-quality |
| 18-110-final-viral-combined_k141_127964\|\|lt2gene | Not-determined |
| 18-110-final-viral-combined_k141_1240\|\|lt2gene | Not-determined |
| 18-110-final-viral-combined_k141_1246\|\|lt2gene | Not-determined |
| 18-110-final-viral-combined_k141_151146\|\|lt2gene | Not-determined |
| 18-110-final-viral-combined_k141_128013\|\|lt2gene | Low-quality |
| 18-110-final-viral-combined_k141_70749\|\|lt2gene | Not-determined |
| 18-110-final-viral-combined_k141_231978\|\|lt2gene | Low-quality |
| 18-110-final-viral-combined_k141_24528\|\|lt2gene | Low-quality |
| 18-110-final-viral-combined_k141_231993\|\|lt2gene | Not-determined |
| 18-110-final-viral-combined_k141_197042\|\|lt2gene | Low-quality |
| 18-110-final-viral-combined_k141_151222\|\|lt2gene | Low-quality |
| 18-110-final-viral-combined_k141_174285\|\|lt2gene | Low-quality |
| 18-110-final-viral-combined_k141_197082\|\|lt2gene | Low-quality |
| 18-110-final-viral-combined_k141_197085\|\|lt2gene | Not-determined |
| 18-110-final-viral-combined_k141_1381\|\|lt2gene | Not-determined |
| 18-110-final-viral-combined_k141_47376\|\|lt2gene | Not-determined |
| 18-110-final-viral-combined_k141_174341\|\|lt2gene | Not-determined |
| 18-110-final-viral-combined_k141_1399\|\|lt2gene | Low-quality |
| 18-110-final-viral-combined_k141_1405\|\|lt2gene | Low-quality |
| 18-110-final-viral-combined_k141_1406\|\|lt2gene | Low-quality |
| 18-110-final-viral-combined_k141_254622\|\|lt2gene | Low-quality |
| 18-110-final-viral-combined_k141_220117\|\|lt2gene | Not-determined |
| 18-110-final-viral-combined_k141_139975\|\|lt2gene | Low-quality |
| 18-110-final-viral-combined_k141_128114\|\|lt2gene | Low-quality |
| 18-110-final-viral-combined_k141_70832\|\|lt2gene | Low-quality |
| 18-110-final-viral-combined_k141_59400\|\|lt2gene | Not-determined |
| 18-110-final-viral-combined_k141_220162\|\|lt2gene | Low-quality |
| 18-110-final-viral-combined_k141_128162\|\|lt2gene | Not-determined |
| 18-110-final-viral-combined_k141_116608\|\|lt2gene | Low-quality |
| 18-110-final-viral-combined_k141_162761\|\|lt2gene | Low-quality |
| 18-110-final-viral-combined_k141_93888\|\|lt2gene | Not-determined |
| 18-110-final-viral-combined_k141_105160\|\|lt2gene | Low-quality |
| 18-110-final-viral-combined_k141_254672\|\|lt2gene | Low-quality |
| 18-110-final-viral-combined_k141_1504\|\|lt2gene | Low-quality |
| 18-110-final-viral-combined_k141_151405\|\|lt2gene | Low-quality |
| 18-110-final-viral-combined_k141_254681\|\|lt2gene | Low-quality |
| 18-110-final-viral-combined_k141_116658\|\|lt2gene | Low-quality |
| 18-110-final-viral-combined_k141_151420\|\|lt2gene | Not-determined |
| 18-110-final-viral-combined_k141_174466\|\|lt2gene | Low-quality |
| 18-110-final-viral-combined_k141_197283\|\|lt2gene | Low-quality |
| 18-110-final-viral-combined_k141_12655\|\|lt2gene | Low-quality |
| 18-110-final-viral-combined_k141_47399\|\|lt2gene | Low-quality |
| 18-110-final-viral-combined_k141_174500\|\|lt2gene | Not-determined |
| 18-110-final-viral-combined_k141_59509\|\|lt2gene | Low-quality |
| 18-110-final-viral-combined_k141_116699\|\|lt2gene | Low-quality |
| 18-110-final-viral-combined_k141_197309\|\|lt2gene | Low-quality |
| 18-110-final-viral-combined_k141_151504\|\|lt2gene | Low-quality |
| 18-110-final-viral-combined_k141_128299\|\|lt2gene | Low-quality |
| 18-110-final-viral-combined_k141_254753\|\|lt2gene | Low-quality |
| 18-110-final-viral-combined_k141_35992\|\|lt2gene | Low-quality |
| 18-110-final-viral-combined_k141_209165\|\|lt2gene | Low-quality |
| 18-110-final-viral-combined_k141_93961\|\|lt2gene | Low-quality |
| 18-110-final-viral-combined_k141_174594\|\|lt2gene | Not-determined |
| 18-110-final-viral-combined_k141_197365\|\|lt2gene | Not-determined |
| 18-110-final-viral-combined_k141_128363\|\|lt2gene | Low-quality |
| 18-110-final-viral-combined_k141_151567\|\|lt2gene | Low-quality |
| 18-110-final-viral-combined_k141_1605\|\|lt2gene | Low-quality |
| 18-110-final-viral-combined_k141_151574\|\|lt2gene | Not-determined |
| 18-110-final-viral-combined_k141_59574\|\|lt2gene | Not-determined |
| 18-110-final-viral-combined_k141_209203\|\|lt2gene | Low-quality |
| 18-110-final-viral-combined_k141_151608\|\|lt2gene | Low-quality |
| 18-110-final-viral-combined_k141_82339\|\|lt2gene | Not-determined |
| 18-110-final-viral-combined_k141_36084\|\|lt2gene | Not-determined |
| 18-110-final-viral-combined_k141_82360\|\|lt2gene | Low-quality |
| 18-110-final-viral-combined_k141_232291\|\|lt2gene | Low-quality |
| 18-110-final-viral-combined_k141_82367\|\|lt2gene | Low-quality |
| 18-110-final-viral-combined_k141_151654\|\|lt2gene | Not-determined |
| 18-110-final-viral-combined_k141_232298\|\|lt2gene | Low-quality |
| 18-110-final-viral-combined_k141_12743\|\|lt2gene | Not-determined |
| 18-110-final-viral-combined_k141_12744\|\|lt2gene | Low-quality |
| 18-110-final-viral-combined_k141_12750\|\|lt2gene | Not-determined |
| 18-110-final-viral-combined_k141_162894\|\|lt2gene | Not-determined |
| 18-110-final-viral-combined_k141_36125\|\|lt2gene | Low-quality |
| 18-110-final-viral-combined_k141_116867\|\|lt2gene | Low-quality |
| 18-110-final-viral-combined_k141_243842\|\|lt2gene | Low-quality |
| 18-110-final-viral-combined_k141_266428\|\|lt2gene | Low-quality |
| 18-110-final-viral-combined_k141_232314\|\|lt2gene | Low-quality |
| 18-110-final-viral-combined_k141_82407\|\|lt2gene | Low-quality |
| 18-110-final-viral-combined_k141_266437\|\|lt2gene | Low-quality |
| 18-110-final-viral-combined_k141_82418\|\|lt2gene | Not-determined |
| 18-110-final-viral-combined_k141_266445\|\|lt2gene | Low-quality |
| 18-110-final-viral-combined_k141_162963\|\|lt2gene | Not-determined |
| 18-110-final-viral-combined_k141_209285\|\|lt2gene | Low-quality |
| 18-110-final-viral-combined_k141_25043\|\|lt2gene | Low-quality |
| 18-110-final-viral-combined_k141_163008\|\|lt2gene | Low-quality |
| 18-110-final-viral-combined_k141_94127\|\|lt2gene | Low-quality |
| 18-110-final-viral-combined_k141_59679\|\|lt2gene | Low-quality |
| 18-110-final-viral-combined_k141_232425\|\|lt2gene | Not-determined |
| 18-110-final-viral-combined_k141_94157\|\|lt2gene | Low-quality |
| 18-110-final-viral-combined_k141_59708\|\|lt2gene | Low-quality |
| 18-110-final-viral-combined_k141_209362\|\|lt2gene | Low-quality |
| 18-110-final-viral-combined_k141_209383\|\|lt2gene | Low-quality |
| 18-110-final-viral-combined_k141_36294\|\|lt2gene | Not-determined |
| 18-110-final-viral-combined_k141_59750\|\|lt2gene | Low-quality |
| 18-110-final-viral-combined_k141_82646\|\|lt2gene | Low-quality |
| 18-110-final-viral-combined_k141_266651\|\|lt2gene | Low-quality |
| 18-110-final-viral-combined_k141_82669\|\|lt2gene | Not-determined |
| 18-110-final-viral-combined_k141_59756\|\|lt2gene | Not-determined |
| 18-110-final-viral-combined_k141_47732\|\|lt2gene | Low-quality |
| 18-110-final-viral-combined_k141_59787\|\|lt2gene | Not-determined |
| 18-110-final-viral-combined_k141_105601\|\|lt2gene | Not-determined |
| 18-110-final-viral-combined_k141_47751\|\|lt2gene | Low-quality |
| 18-110-final-viral-combined_k141_186398\|\|lt2gene | Low-quality |
| 18-110-final-viral-combined_k141_36367\|\|lt2gene | Low-quality |
| 18-110-final-viral-combined_k141_128620\|\|lt2gene | Low-quality |
| 18-110-final-viral-combined_k141_36373\|\|lt2gene | Not-determined |
| 18-110-final-viral-combined_k141_255003\|\|lt2gene | Low-quality |
| 18-110-final-viral-combined_k141_140668\|\|lt2gene | Not-determined |
| 18-110-final-viral-combined_k141_36385\|\|lt2gene | Not-determined |
| 18-110-final-viral-combined_k141_174959\|\|lt2gene | Low-quality |
| 18-110-final-viral-combined_k141_128637\|\|lt2gene | Low-quality |
| 18-110-final-viral-combined_k141_94270\|\|lt2gene | Low-quality |
| 18-110-final-viral-combined_k141_232552\|\|lt2gene | Low-quality |
| 18-110-final-viral-combined_k141_59851\|\|lt2gene | Low-quality |
| 18-110-final-viral-combined_k141_174998\|\|lt2gene | Low-quality |
| 18-110-final-viral-combined_k141_232563\|\|lt2gene | Low-quality |
| 18-110-final-viral-combined_k141_71444\|\|lt2gene | Not-determined |
| 18-110-final-viral-combined_k141_220564\|\|lt2gene | Low-quality |
| 18-110-final-viral-combined_k141_117130\|\|lt2gene | Low-quality |
| 18-110-final-viral-combined_k141_25354\|\|lt2gene | Low-quality |
| 18-110-final-viral-combined_k141_128674\|\|lt2gene | Low-quality |
| 18-110-final-viral-combined_k141_255088\|\|lt2gene | Low-quality |
| 18-110-final-viral-combined_k141_94336\|\|lt2gene | Low-quality |
| 18-110-final-viral-combined_k141_244254\|\|lt2gene | Low-quality |
| 18-110-final-viral-combined_k141_197754\|\|lt2gene | Low-quality |
| 18-110-final-viral-combined_k141_59903\|\|lt2gene | Low-quality |
| 18-110-final-viral-combined_k141_152003\|\|lt2gene | Low-quality |
| 18-110-final-viral-combined_k141_220605\|\|lt2gene | Not-determined |
| 18-110-final-viral-combined_k141_105706\|\|lt2gene | Not-determined |
| 18-110-final-viral-combined_k141_175099\|\|lt2gene | Low-quality |
| 18-110-final-viral-combined_k141_128792\|\|lt2gene | Low-quality |
| 18-110-final-viral-combined_k141_255126\|\|lt2gene | Not-determined |
| 18-110-final-viral-combined_k141_232695\|\|lt2gene | Low-quality |
| 18-110-final-viral-combined_k141_266907\|\|lt2gene | Not-determined |
| 18-110-final-viral-combined_k141_232706\|\|lt2gene | Low-quality |
| 18-110-final-viral-combined_k141_244369\|\|lt2gene | Low-quality |
| 18-110-final-viral-combined_k141_140740\|\|lt2gene | Low-quality |
| 18-110-final-viral-combined_k141_266935\|\|lt2gene | Not-determined |
| 18-110-final-viral-combined_k141_94440\|\|lt2gene | Low-quality |
| 18-110-final-viral-combined_k141_175201\|\|lt2gene | Low-quality |
| 18-110-final-viral-combined_k141_163413\|\|lt2gene | Not-determined |
| 18-110-final-viral-combined_k141_47999\|\|lt2gene | Low-quality |
| 18-110-final-viral-combined_k141_25523\|\|lt2gene | Low-quality |
| 18-110-final-viral-combined_k141_60007\|\|lt2gene | Low-quality |
| 18-110-final-viral-combined_k141_117258\|\|lt2gene | Not-determined |
| 18-110-final-viral-combined_k141_232790\|\|lt2gene | Low-quality |
| 18-110-final-viral-combined_k141_175274\|\|lt2gene | Low-quality |
| 18-110-final-viral-combined_k141_82846\|\|lt2gene | Low-quality |
| 18-110-final-viral-combined_k141_220812\|\|lt2gene | Low-quality |
| 18-110-final-viral-combined_k141_60139\|\|lt2gene | Low-quality |
| 18-110-final-viral-combined_k141_197951\|\|lt2gene | Low-quality |
| 18-110-final-viral-combined_k141_2086\|\|lt2gene | Not-determined |
| 18-110-final-viral-combined_k141_197971\|\|lt2gene | Low-quality |
| 18-110-final-viral-combined_k141_220875\|\|lt2gene | Not-determined |
| 18-110-final-viral-combined_k141_48169\|\|lt2gene | Low-quality |
| 18-110-final-viral-combined_k141_186737\|\|lt2gene | Low-quality |
| 18-110-final-viral-combined_k141_71875\|\|lt2gene | Low-quality |
| 18-110-final-viral-combined_k141_140948\|\|lt2gene | Low-quality |
| 18-110-final-viral-combined_k141_117429\|\|lt2gene | Low-quality |
| 18-110-final-viral-combined_k141_25682\|\|lt2gene | Low-quality |
| 18-110-final-viral-combined_k141_36788\|\|lt2gene | Low-quality |
| 18-110-final-viral-combined_k141_13219\|\|lt2gene | Low-quality |
| 18-110-final-viral-combined_k141_117454\|\|lt2gene | Not-determined |
| 18-110-final-viral-combined_k141_105854\|\|lt2gene | Low-quality |
| 18-110-final-viral-combined_k141_267187\|\|lt2gene | Not-determined |
| 18-110-final-viral-combined_k141_163594\|\|lt2gene | Low-quality |
| 18-110-final-viral-combined_k141_220990\|\|lt2gene | Low-quality |
| 18-110-final-viral-combined_k141_105863\|\|lt2gene | Low-quality |
| 18-110-final-viral-combined_k141_128988\|\|lt2gene | Low-quality |
| 18-110-final-viral-combined_k141_209904\|\|lt2gene | Low-quality |
| 18-110-final-viral-combined_k141_244625\|\|lt2gene | Low-quality |
| 18-110-final-viral-combined_k141_186827\|\|lt2gene | Low-quality |
| 18-110-final-viral-combined_k141_186828\|\|lt2gene | Low-quality |
| 18-110-final-viral-combined_k141_71917\|\|lt2gene | Low-quality |
| 18-110-final-viral-combined_k141_94779\|\|lt2gene | Low-quality |
| 18-110-final-viral-combined_k141_244666\|\|lt2gene | Not-determined |
| 18-110-final-viral-combined_k141_175550\|\|lt2gene | Not-determined |
| 18-110-final-viral-combined_k141_94806\|\|lt2gene | Low-quality |
| 18-110-final-viral-combined_k141_129069\|\|lt2gene | Low-quality |
| 18-110-final-viral-combined_k141_209957\|\|lt2gene | Not-determined |
| 18-110-final-viral-combined_k141_94818\|\|lt2gene | Low-quality |
| 18-110-final-viral-combined_k141_186911\|\|lt2gene | Not-determined |
| 18-110-final-viral-combined_k141_94848\|\|lt2gene | Not-determined |
| 18-110-final-viral-combined_k141_221077\|\|lt2gene | Low-quality |
| 18-110-final-viral-combined_k141_36985\|\|lt2gene | Low-quality |
| 18-110-final-viral-combined_k141_255501\|\|lt2gene | Not-determined |
| 18-110-final-viral-combined_k141_198240\|\|lt2gene | Not-determined |
| 18-110-final-viral-combined_k141_25849\|\|lt2gene | Low-quality |
| 18-110-final-viral-combined_k141_152503\|\|lt2gene | Low-quality |
| 18-110-final-viral-combined_k141_163800\|\|lt2gene | Low-quality |
| 18-110-final-viral-combined_k141_198260\|\|lt2gene | Not-determined |
| 18-110-final-viral-combined_k141_210053\|\|lt2gene | Low-quality |
| 18-110-final-viral-combined_k141_129175\|\|lt2gene | Not-determined |
| 18-110-final-viral-combined_k141_198278\|\|lt2gene | Not-determined |
| 18-110-final-viral-combined_k141_186988\|\|lt2gene | Low-quality |
| 18-110-final-viral-combined_k141_152570\|\|lt2gene | Low-quality |
| 18-110-final-viral-combined_k141_94977\|\|lt2gene | Low-quality |
| 18-110-final-viral-combined_k141_233205\|\|lt2gene | Low-quality |
| 18-110-final-viral-combined_k141_2397\|\|lt2gene | Not-determined |
| 18-110-final-viral-combined_k141_255588\|\|lt2gene | Low-quality |
| 18-110-final-viral-combined_k141_255604\|\|lt2gene | Low-quality |
| 18-110-final-viral-combined_k141_83104\|\|lt2gene | Low-quality |
| 18-110-final-viral-combined_k141_233263\|\|lt2gene | Low-quality |
| 18-110-final-viral-combined_k141_2454\|\|lt2gene | Not-determined |
| 18-110-final-viral-combined_k141_13434\|\|lt2gene | Low-quality |
| 18-110-final-viral-combined_k141_106025\|\|lt2gene | Low-quality |
| 18-110-final-viral-combined_k141_129345\|\|lt2gene | Low-quality |
| 18-110-final-viral-combined_k141_141362\|\|lt2gene | Low-quality |
| 18-110-final-viral-combined_k141_152708\|\|lt2gene | Not-determined |
| 18-110-final-viral-combined_k141_13497\|\|lt2gene | Low-quality |
| 18-110-final-viral-combined_k141_106091\|\|lt2gene | Low-quality |
| 18-110-final-viral-combined_k141_37232\|\|lt2gene | Low-quality |
| 18-110-final-viral-combined_k141_60425\|\|lt2gene | Not-determined |
| 18-110-final-viral-combined_k141_2558\|\|lt2gene | Low-quality |
| 18-110-final-viral-combined_k141_198450\|\|lt2gene | Not-determined |
| 18-110-final-viral-combined_k141_13626\|\|lt2gene | Low-quality |
| 18-110-final-viral-combined_k141_26088\|\|lt2gene | Not-determined |
| 18-110-final-viral-combined_k141_255824\|\|lt2gene | Low-quality |
| 18-110-final-viral-combined_k141_26106\|\|lt2gene | Low-quality |
| 18-110-final-viral-combined_k141_2653\|\|lt2gene | Not-determined |
| 18-110-final-viral-combined_k141_255853\|\|lt2gene | Low-quality |
| 18-110-final-viral-combined_k141_152846\|\|lt2gene | Low-quality |
| 18-110-final-viral-combined_k141_255858\|\|lt2gene | Low-quality |
| 18-110-final-viral-combined_k141_60582\|\|lt2gene | Low-quality |
| 18-110-final-viral-combined_k141_95351\|\|lt2gene | Low-quality |
| 18-110-final-viral-combined_k141_233584\|\|lt2gene | Low-quality |
| 18-110-final-viral-combined_k141_163942\|\|lt2gene | Low-quality |
| 18-110-final-viral-combined_k141_117815\|\|lt2gene | Not-determined |
| 18-110-final-viral-combined_k141_255933\|\|lt2gene | Not-determined |
| 18-110-final-viral-combined_k141_48656\|\|lt2gene | Low-quality |
| 18-110-final-viral-combined_k141_152924\|\|lt2gene | Low-quality |
| 18-110-final-viral-combined_k141_26300\|\|lt2gene | Low-quality |
| 18-110-final-viral-combined_k141_129659\|\|lt2gene | Not-determined |
| 18-110-final-viral-combined_k141_48710\|\|lt2gene | Not-determined |
| 18-110-final-viral-combined_k141_245188\|\|lt2gene | Low-quality |
| 18-110-final-viral-combined_k141_129694\|\|lt2gene | Low-quality |
| 18-110-final-viral-combined_k141_2775\|\|lt2gene | Low-quality |
| 18-110-final-viral-combined_k141_37437\|\|lt2gene | Low-quality |
| 18-110-final-viral-combined_k141_2789\|\|lt2gene | Low-quality |
| 18-110-final-viral-combined_k141_187331\|\|lt2gene | Not-determined |
| 18-110-final-viral-combined_k141_233741\|\|lt2gene | Not-determined |
| 18-110-final-viral-combined_k141_141792\|\|lt2gene | Not-determined |
| 18-110-final-viral-combined_k141_26323\|\|lt2gene | Not-determined |
| 18-110-final-viral-combined_k141_245233\|\|lt2gene | Low-quality |
| 18-110-final-viral-combined_k141_37496\|\|lt2gene | Low-quality |
| 18-110-final-viral-combined_k141_118022\|\|lt2gene | Not-determined |
| 18-110-final-viral-combined_k141_129782\|\|lt2gene | Not-determined |
| 18-110-final-viral-combined_k141_267938\|\|lt2gene | Not-determined |
| 18-110-final-viral-combined_k141_83164\|\|lt2gene | Not-determined |
| 18-110-final-viral-combined_k141_13749\|\|lt2gene | Low-quality |
| 18-110-final-viral-combined_k141_221580\|\|lt2gene | Low-quality |
| 18-110-final-viral-combined_k141_256084\|\|lt2gene | Low-quality |
| 18-110-final-viral-combined_k141_153000\|\|lt2gene | Low-quality |
| 18-110-final-viral-combined_k141_129818\|\|lt2gene | Not-determined |
| 18-110-final-viral-combined_k141_256093\|\|lt2gene | Low-quality |
| 18-110-final-viral-combined_k141_118073\|\|lt2gene | Low-quality |
| 18-110-final-viral-combined_k141_268001\|\|lt2gene | Low-quality |
| 18-110-final-viral-combined_k141_48912\|\|lt2gene | Low-quality |
| 18-110-final-viral-combined_k141_187553\|\|lt2gene | Not-determined |
| 18-110-final-viral-combined_k141_72844\|\|lt2gene | Not-determined |
| 18-110-final-viral-combined_k141_26431\|\|lt2gene | Low-quality |
| 18-110-final-viral-combined_k141_129962\|\|lt2gene | Not-determined |
| 18-110-final-viral-combined_k141_176507\|\|lt2gene | Low-quality |
| 18-110-final-viral-combined_k141_268108\|\|lt2gene | Low-quality |
| 18-110-final-viral-combined_k141_164368\|\|lt2gene | Low-quality |
| 18-110-final-viral-combined_k141_198867\|\|lt2gene | Low-quality |
| 18-110-final-viral-combined_k141_187630\|\|lt2gene | Low-quality |
| 18-110-final-viral-combined_k141_60946\|\|lt2gene | Not-determined |
| 18-110-final-viral-combined_k141_118202\|\|lt2gene | Low-quality |
| 18-110-final-viral-combined_k141_3050\|\|lt2gene | Low-quality |
| 18-110-final-viral-combined_k141_37737\|\|lt2gene | Low-quality |
| 18-110-final-viral-combined_k141_37739\|\|lt2gene | Not-determined |
| 18-110-final-viral-combined_k141_26562\|\|lt2gene | Low-quality |
| 18-110-final-viral-combined_k141_164406\|\|lt2gene | Low-quality |
| 18-110-final-viral-combined_k141_130050\|\|lt2gene | Low-quality |
| 18-110-final-viral-combined_k141_198920\|\|lt2gene | Low-quality |
| 18-110-final-viral-combined_k141_37767\|\|lt2gene | Low-quality |
| 18-110-final-viral-combined_k141_198924\|\|lt2gene | Low-quality |
| 18-110-final-viral-combined_k141_187690\|\|lt2gene | Low-quality |
| 18-110-final-viral-combined_k141_245455\|\|lt2gene | Not-determined |
| 18-110-final-viral-combined_k141_233960\|\|lt2gene | Not-determined |
| 18-110-final-viral-combined_k141_3102\|\|lt2gene | Low-quality |
| 18-110-final-viral-combined_k141_245502\|\|lt2gene | Low-quality |
| 18-110-final-viral-combined_k141_130113\|\|lt2gene | Low-quality |
| 18-110-final-viral-combined_k141_72959\|\|lt2gene | Low-quality |
| 18-110-final-viral-combined_k141_72997\|\|lt2gene | Not-determined |
| 18-110-final-viral-combined_k141_210657\|\|lt2gene | Low-quality |
| 18-110-final-viral-combined_k141_176712\|\|lt2gene | Low-quality |
| 18-110-final-viral-combined_k141_176714\|\|lt2gene | Not-determined |
| 18-110-final-viral-combined_k141_13926\|\|lt2gene | Low-quality |
| 18-110-final-viral-combined_k141_176751\|\|lt2gene | Low-quality |
| 18-110-final-viral-combined_k141_130201\|\|lt2gene | Low-quality |
| 18-110-final-viral-combined_k141_187810\|\|lt2gene | Low-quality |
| 18-110-final-viral-combined_k141_83451\|\|lt2gene | Low-quality |
| 18-110-final-viral-combined_k141_73078\|\|lt2gene | Low-quality |
| 18-110-final-viral-combined_k141_118420\|\|lt2gene | Low-quality |
| 18-110-final-viral-combined_k141_130324\|\|lt2gene | Not-determined |
| 18-110-final-viral-combined_k141_106714\|\|lt2gene | Not-determined |
| 18-110-final-viral-combined_k141_210806\|\|lt2gene | Low-quality |
| 18-110-final-viral-combined_k141_268396\|\|lt2gene | Low-quality |
| 18-110-final-viral-combined_k141_95827\|\|lt2gene | Not-determined |
| 18-110-final-viral-combined_k141_245689\|\|lt2gene | Not-determined |
| 18-110-final-viral-combined_k141_106749\|\|lt2gene | Low-quality |
| 18-110-final-viral-combined_k141_37984\|\|lt2gene | Low-quality |
| 18-110-final-viral-combined_k141_153408\|\|lt2gene | Low-quality |
| 18-110-final-viral-combined_k141_26810\|\|lt2gene | Low-quality |
| 18-110-final-viral-combined_k141_26822\|\|lt2gene | Not-determined |
| 18-110-final-viral-combined_k141_187999\|\|lt2gene | Not-determined |
| 18-110-final-viral-combined_k141_153514\|\|lt2gene | Low-quality |
| 18-110-final-viral-combined_k141_245772\|\|lt2gene | Low-quality |
| 18-110-final-viral-combined_k141_153535\|\|lt2gene | Low-quality |
| 18-110-final-viral-combined_k141_49339\|\|lt2gene | Low-quality |
| 18-110-final-viral-combined_k141_268468\|\|lt2gene | Low-quality |
| 18-110-final-viral-combined_k141_14107\|\|lt2gene | Not-determined |
| 18-110-final-viral-combined_k141_61145\|\|lt2gene | Not-determined |
| 18-110-final-viral-combined_k141_49344\|\|lt2gene | Low-quality |
| 18-110-final-viral-combined_k141_188041\|\|lt2gene | Low-quality |
| 18-110-final-viral-combined_k141_153561\|\|lt2gene | Not-determined |
| 18-110-final-viral-combined_k141_106904\|\|lt2gene | Low-quality |
| 18-110-final-viral-combined_k141_164698\|\|lt2gene | Not-determined |
| 18-110-final-viral-combined_k141_188067\|\|lt2gene | Low-quality |
| 18-110-final-viral-combined_k141_153598\|\|lt2gene | Low-quality |
| 18-110-final-viral-combined_k141_234369\|\|lt2gene | Low-quality |
| 18-110-final-viral-combined_k141_245932\|\|lt2gene | Not-determined |
| 18-110-final-viral-combined_k141_130527\|\|lt2gene | Low-quality |
| 18-110-final-viral-combined_k141_268623\|\|lt2gene | Low-quality |
| 18-110-final-viral-combined_k141_130531\|\|lt2gene | Not-determined |
| 18-110-final-viral-combined_k141_256330\|\|lt2gene | Low-quality |
| 18-110-final-viral-combined_k141_27091\|\|lt2gene | Not-determined |
| 18-110-final-viral-combined_k141_164835\|\|lt2gene | Low-quality |
| 18-110-final-viral-combined_k141_49421\|\|lt2gene | Not-determined |
| 18-110-final-viral-combined_k141_96064\|\|lt2gene | Low-quality |
| 18-110-final-viral-combined_k141_256373\|\|lt2gene | Low-quality |
| 18-110-final-viral-combined_k141_118795\|\|lt2gene | Low-quality |
| 18-110-final-viral-combined_k141_3463\|\|lt2gene | Not-determined |
| 18-110-final-viral-combined_k141_256396\|\|lt2gene | Not-determined |
| 18-110-final-viral-combined_k141_118801\|\|lt2gene | Not-determined |
| 18-110-final-viral-combined_k141_14198\|\|lt2gene | Not-determined |
| 18-110-final-viral-combined_k141_14200\|\|lt2gene | Not-determined |
| 18-110-final-viral-combined_k141_234489\|\|lt2gene | Low-quality |
| 18-110-final-viral-combined_k141_164922\|\|lt2gene | Low-quality |
| 18-110-final-viral-combined_k141_153726\|\|lt2gene | Not-determined |
| 18-110-final-viral-combined_k141_199270\|\|lt2gene | Low-quality |
| 18-110-final-viral-combined_k141_3514\|\|lt2gene | Low-quality |
| 18-110-final-viral-combined_k141_222336\|\|lt2gene | Not-determined |
| 18-110-final-viral-combined_k141_130653\|\|lt2gene | Low-quality |
| 18-110-final-viral-combined_k141_130669\|\|lt2gene | Low-quality |
| 18-110-final-viral-combined_k141_73363\|\|lt2gene | Low-quality |
| 18-110-final-viral-combined_k141_83841\|\|lt2gene | Not-determined |
| 18-110-final-viral-combined_k141_142624\|\|lt2gene | Not-determined |
| 18-110-final-viral-combined_k141_130696\|\|lt2gene | Not-determined |
| 18-110-final-viral-combined_k141_211149\|\|lt2gene | Not-determined |
| 18-110-final-viral-combined_k141_3571\|\|lt2gene | Low-quality |
| 18-110-final-viral-combined_k141_142643\|\|lt2gene | Low-quality |
| 18-110-final-viral-combined_k141_268760\|\|lt2gene | Low-quality |
| 18-110-final-viral-combined_k141_246100\|\|lt2gene | Low-quality |
| 18-110-final-viral-combined_k141_118986\|\|lt2gene | Low-quality |
| 18-110-final-viral-combined_k141_83970\|\|lt2gene | Low-quality |
| 18-110-final-viral-combined_k141_83999\|\|lt2gene | Low-quality |
| 18-110-final-viral-combined_k141_107289\|\|lt2gene | Low-quality |
| 18-110-final-viral-combined_k141_61531\|\|lt2gene | Low-quality |
| 18-110-final-viral-combined_k141_222567\|\|lt2gene | Low-quality |
| 18-110-final-viral-combined_k141_38467\|\|lt2gene | Not-determined |
| 18-110-final-viral-combined_k141_199481\|\|lt2gene | Low-quality |
| 18-110-final-viral-combined_k141_96381\|\|lt2gene | Low-quality |
| 18-110-final-viral-combined_k141_3666\|\|lt2gene | Low-quality |
| 18-110-final-viral-combined_k141_142698\|\|lt2gene | Low-quality |
| 18-110-final-viral-combined_k141_177488\|\|lt2gene | Low-quality |
| 18-110-final-viral-combined_k141_153896\|\|lt2gene | Not-determined |
| 18-110-final-viral-combined_k141_84100\|\|lt2gene | Low-quality |
| 18-110-final-viral-combined_k141_27459\|\|lt2gene | Not-determined |
| 18-110-final-viral-combined_k141_49807\|\|lt2gene | Low-quality |
| 18-110-final-viral-combined_k141_38611\|\|lt2gene | Low-quality |
| 18-110-final-viral-combined_k141_165336\|\|lt2gene | Low-quality |
| 18-110-final-viral-combined_k141_142783\|\|lt2gene | Low-quality |
| 18-110-final-viral-combined_k141_38633\|\|lt2gene | Low-quality |
| 18-110-final-viral-combined_k141_14448\|\|lt2gene | Low-quality |
| 18-110-final-viral-combined_k141_165365\|\|lt2gene | Not-determined |
| 18-110-final-viral-combined_k141_107490\|\|lt2gene | Not-determined |
| 18-110-final-viral-combined_k141_14485\|\|lt2gene | Not-determined |
| 18-110-final-viral-combined_k141_130955\|\|lt2gene | Not-determined |
| 18-110-final-viral-combined_k141_49888\|\|lt2gene | Not-determined |
| 18-110-final-viral-combined_k141_49891\|\|lt2gene | Low-quality |
| 18-110-final-viral-combined_k141_211462\|\|lt2gene | Low-quality |
| 18-110-final-viral-combined_k141_199583\|\|lt2gene | Low-quality |
| 18-110-final-viral-combined_k141_130982\|\|lt2gene | Low-quality |
| 18-110-final-viral-combined_k141_177626\|\|lt2gene | Low-quality |
| 18-110-final-viral-combined_k141_49930\|\|lt2gene | Low-quality |
| 18-110-final-viral-combined_k141_84261\|\|lt2gene | Low-quality |
| 18-110-final-viral-combined_k141_119186\|\|lt2gene | Low-quality |
| 18-110-final-viral-combined_k141_119208\|\|lt2gene | Low-quality |
| 18-110-final-viral-combined_k141_165453\|\|lt2gene | Low-quality |
| 18-110-final-viral-combined_k141_119232\|\|lt2gene | Low-quality |
| 18-110-final-viral-combined_k141_96598\|\|lt2gene | Not-determined |
| 18-110-final-viral-combined_k141_84333\|\|lt2gene | Not-determined |
| 18-110-final-viral-combined_k141_96611\|\|lt2gene | Low-quality |
| 18-110-final-viral-combined_k141_256734\|\|lt2gene | Not-determined |
| 18-110-final-viral-combined_k141_3908\|\|lt2gene | Not-determined |
| 18-110-final-viral-combined_k141_27644\|\|lt2gene | Not-determined |
| 18-110-final-viral-combined_k141_246326\|\|lt2gene | Low-quality |
| 18-110-final-viral-combined_k141_211618\|\|lt2gene | Low-quality |
| 18-110-final-viral-combined_k141_131146\|\|lt2gene | Low-quality |
| 18-110-final-viral-combined_k141_38878\|\|lt2gene | Low-quality |
| 18-110-final-viral-combined_k141_188803\|\|lt2gene | Low-quality |
| 18-110-final-viral-combined_k141_119318\|\|lt2gene | Low-quality |
| 18-110-final-viral-combined_k141_119335\|\|lt2gene | Low-quality |
| 18-110-final-viral-combined_k141_131164\|\|lt2gene | Not-determined |
| 18-110-final-viral-combined_k141_165592\|\|lt2gene | Low-quality |
| 18-110-final-viral-combined_k141_62079\|\|lt2gene | Not-determined |
| 18-110-final-viral-combined_k141_62122\|\|lt2gene | Low-quality |
| 18-110-final-viral-combined_k141_165626\|\|lt2gene | Low-quality |
| 18-110-final-viral-combined_k141_39055\|\|lt2gene | Low-quality |
| 18-110-final-viral-combined_k141_119351\|\|lt2gene | Not-determined |
| 18-110-final-viral-combined_k141_50114\|\|lt2gene | Low-quality |
| 18-110-final-viral-combined_k141_84508\|\|lt2gene | Not-determined |
| 18-110-final-viral-combined_k141_14803\|\|lt2gene | Not-determined |
| 18-110-final-viral-combined_k141_165688\|\|lt2gene | Low-quality |
| 18-110-final-viral-combined_k141_211797\|\|lt2gene | Low-quality |
| 18-110-final-viral-combined_k141_119409\|\|lt2gene | Low-quality |
| 18-110-final-viral-combined_k141_50175\|\|lt2gene | Low-quality |
| 18-110-final-viral-combined_k141_50186\|\|lt2gene | Low-quality |
| 18-110-final-viral-combined_k141_154383\|\|lt2gene | Low-quality |
| 18-110-final-viral-combined_k141_131366\|\|lt2gene | Low-quality |
| 18-110-final-viral-combined_k141_4112\|\|lt2gene | Not-determined |
| 18-110-final-viral-combined_k141_269459\|\|lt2gene | Low-quality |
| 18-110-final-viral-combined_k141_14904\|\|lt2gene | Low-quality |
| 18-110-final-viral-combined_k141_50208\|\|lt2gene | Not-determined |
| 18-110-final-viral-combined_k141_223139\|\|lt2gene | Low-quality |
| 18-110-final-viral-combined_k141_188989\|\|lt2gene | Low-quality |
| 18-110-final-viral-combined_k141_62253\|\|lt2gene | Not-determined |
| 18-110-final-viral-combined_k141_269488\|\|lt2gene | Low-quality |
| 18-110-final-viral-combined_k141_189016\|\|lt2gene | Low-quality |
| 18-110-final-viral-combined_k141_165771\|\|lt2gene | Not-determined |
| 18-110-final-viral-combined_k141_189040\|\|lt2gene | Not-determined |
| 18-110-final-viral-combined_k141_119505\|\|lt2gene | Low-quality |
| 18-110-final-viral-combined_k141_235278\|\|lt2gene | Low-quality |
| 18-110-final-viral-combined_k141_50275\|\|lt2gene | Low-quality |
| 18-110-final-viral-combined_k141_62320\|\|lt2gene | Not-determined |
| 18-110-final-viral-combined_k141_119521\|\|lt2gene | Low-quality |
| 18-110-final-viral-combined_k141_119526\|\|lt2gene | Low-quality |
| 18-110-final-viral-combined_k141_107943\|\|lt2gene | Low-quality |
| 18-110-final-viral-combined_k141_178060\|\|lt2gene | Low-quality |
| 18-110-final-viral-combined_k141_211937\|\|lt2gene | Not-determined |
| 18-110-final-viral-combined_k141_27797\|\|lt2gene | Not-determined |
| 18-110-final-viral-combined_k141_178106\|\|lt2gene | Low-quality |
| 18-110-final-viral-combined_k141_27809\|\|lt2gene | Low-quality |
| 18-110-final-viral-combined_k141_154500\|\|lt2gene | Not-determined |
| 18-110-final-viral-combined_k141_269602\|\|lt2gene | Low-quality |
| 18-110-final-viral-combined_k141_84733\|\|lt2gene | Not-determined |
| 18-110-final-viral-combined_k141_74133\|\|lt2gene | Low-quality |
| 18-110-final-viral-combined_k141_246633\|\|lt2gene | Low-quality |
| 18-110-final-viral-combined_k141_143320\|\|lt2gene | Not-determined |
| 18-110-final-viral-combined_k141_62498\|\|lt2gene | Low-quality |
| 18-110-final-viral-combined_k141_235453\|\|lt2gene | Low-quality |
| 18-110-final-viral-combined_k141_246670\|\|lt2gene | Not-determined |
| 18-110-final-viral-combined_k141_178186\|\|lt2gene | Low-quality |
| 18-110-final-viral-combined_k141_62520\|\|lt2gene | Low-quality |
| 18-110-final-viral-combined_k141_235472\|\|lt2gene | Low-quality |
| 18-110-final-viral-combined_k141_131636\|\|lt2gene | Low-quality |
| 18-110-final-viral-combined_k141_178254\|\|lt2gene | Low-quality |
| 18-110-final-viral-combined_k141_108105\|\|lt2gene | Not-determined |
| 18-110-final-viral-combined_k141_108107\|\|lt2gene | Low-quality |
| 18-110-final-viral-combined_k141_27950\|\|lt2gene | Not-determined |
| 18-110-final-viral-combined_k141_4338\|\|lt2gene | Not-determined |
| 18-110-final-viral-combined_k141_189259\|\|lt2gene | Low-quality |
| 18-110-final-viral-combined_k141_97129\|\|lt2gene | Low-quality |
| 18-110-final-viral-combined_k141_74274\|\|lt2gene | Not-determined |
| 18-110-final-viral-combined_k141_178362\|\|lt2gene | Low-quality |
| 18-110-final-viral-combined_k141_165934\|\|lt2gene | Low-quality |
| 18-110-final-viral-combined_k141_235592\|\|lt2gene | Low-quality |
| 18-110-final-viral-combined_k141_108157\|\|lt2gene | Low-quality |
| 18-110-final-viral-combined_k141_97173\|\|lt2gene | Low-quality |
| 18-110-final-viral-combined_k141_15224\|\|lt2gene | Low-quality |
| 18-110-final-viral-combined_k141_15236\|\|lt2gene | Not-determined |
| 18-110-final-viral-combined_k141_4410\|\|lt2gene | Low-quality |
| 18-110-final-viral-combined_k141_166030\|\|lt2gene | Low-quality |
| 18-110-final-viral-combined_k141_212262\|\|lt2gene | Low-quality |
| 18-110-final-viral-combined_k141_62744\|\|lt2gene | Low-quality |
| 18-110-final-viral-combined_k141_39567\|\|lt2gene | Not-determined |
| 18-110-final-viral-combined_k141_74346\|\|lt2gene | Not-determined |
| 18-110-final-viral-combined_k141_246927\|\|lt2gene | Low-quality |
| 18-110-final-viral-combined_k141_62836\|\|lt2gene | Not-determined |
| 18-110-final-viral-combined_k141_235760\|\|lt2gene | Low-quality |
| 18-110-final-viral-combined_k141_178523\|\|lt2gene | Low-quality |
| 18-110-final-viral-combined_k141_39689\|\|lt2gene | Low-quality |
| 18-110-final-viral-combined_k141_257212\|\|lt2gene | Low-quality |
| 18-110-final-viral-combined_k141_143587\|\|lt2gene | Low-quality |
| 18-110-final-viral-combined_k141_28198\|\|lt2gene | Low-quality |
| 18-110-final-viral-combined_k141_74413\|\|lt2gene | Low-quality |
| 18-110-final-viral-combined_k141_257220\|\|lt2gene | Not-determined |
| 18-110-final-viral-combined_k141_166189\|\|lt2gene | Low-quality |
| 18-110-final-viral-combined_k141_85080\|\|lt2gene | Low-quality |
| 18-110-final-viral-combined_k141_166203\|\|lt2gene | Not-determined |
| 18-110-final-viral-combined_k141_62914\|\|lt2gene | Not-determined |
| 18-110-final-viral-combined_k141_28245\|\|lt2gene | Low-quality |
| 18-110-final-viral-combined_k141_4432\|\|lt2gene | Low-quality |
| 18-110-final-viral-combined_k141_189515\|\|lt2gene | Not-determined |
| 18-110-final-viral-combined_k141_62982\|\|lt2gene | Low-quality |
| 18-110-final-viral-combined_k141_97279\|\|lt2gene | Low-quality |
| 18-110-final-viral-combined_k141_199932\|\|lt2gene | Low-quality |
| 18-110-final-viral-combined_k141_257310\|\|lt2gene | Low-quality |
| 18-110-final-viral-combined_k141_63006\|\|lt2gene | Not-determined |
| 18-110-final-viral-combined_k141_120004\|\|lt2gene | Not-determined |

| | |
|---|---|
| 18-110-final-viral-combined_k141_131900\|\|lt2gene | Low-quality |
| 18-110-final-viral-combined_k141_235945\|\|lt2gene | Low-quality |
| 18-110-final-viral-combined_k141_50868\|\|lt2gene | Low-quality |
| 18-110-final-viral-combined_k141_39740\|\|lt2gene | Low-quality |
| 18-110-final-viral-combined_k141_257330\|\|lt2gene | Low-quality |
| 18-110-final-viral-combined_k141_143726\|\|lt2gene | Not-determined |
| 18-110-final-viral-combined_k141_178765\|\|lt2gene | Not-determined |
| 18-110-final-viral-combined_k141_200010\|\|lt2gene | Low-quality |
| 18-110-final-viral-combined_k141_4556\|\|lt2gene | Low-quality |
| 18-110-final-viral-combined_k141_120090\|\|lt2gene | Low-quality |
| 18-110-final-viral-combined_k141_131940\|\|lt2gene | Not-determined |
| 18-110-final-viral-combined_k141_200028\|\|lt2gene | Not-determined |
| 18-110-final-viral-combined_k141_39831\|\|lt2gene | Low-quality |
| 18-110-final-viral-combined_k141_257374\|\|lt2gene | Not-determined |
| 18-110-final-viral-combined_k141_143770\|\|lt2gene | Low-quality |
| 18-110-final-viral-combined_k141_236070\|\|lt2gene | Not-determined |
| 18-110-final-viral-combined_k141_236072\|\|lt2gene | Not-determined |
| 18-110-final-viral-combined_k141_257384\|\|lt2gene | Low-quality |
| 18-110-final-viral-combined_k141_108522\|\|lt2gene | Not-determined |
| 18-110-final-viral-combined_k141_15635\|\|lt2gene | Not-determined |
| 18-110-final-viral-combined_k141_212528\|\|lt2gene | Low-quality |
| 18-110-final-viral-combined_k141_257394\|\|lt2gene | Low-quality |
| 18-110-final-viral-combined_k141_39880\|\|lt2gene | Low-quality |
| 18-110-final-viral-combined_k141_28503\|\|lt2gene | Low-quality |
| 18-110-final-viral-combined_k141_15657\|\|lt2gene | Low-quality |
| 18-110-final-viral-combined_k141_236107\|\|lt2gene | Low-quality |
| 18-110-final-viral-combined_k141_257416\|\|lt2gene | Not-determined |
| 18-110-final-viral-combined_k141_155059\|\|lt2gene | Low-quality |
| 18-110-final-viral-combined_k141_200067\|\|lt2gene | Not-determined |
| 18-110-final-viral-combined_k141_74695\|\|lt2gene | Not-determined |
| 18-110-final-viral-combined_k141_200072\|\|lt2gene | Low-quality |
| 18-110-final-viral-combined_k141_28614\|\|lt2gene | Low-quality |
| 18-110-final-viral-combined_k141_178954\|\|lt2gene | Low-quality |
| 18-110-final-viral-combined_k141_97632\|\|lt2gene | Low-quality |
| 18-110-final-viral-combined_k141_40058\|\|lt2gene | Low-quality |
| 18-110-final-viral-combined_k141_132136\|\|lt2gene | Not-determined |
| 18-110-final-viral-combined_k141_144046\|\|lt2gene | Not-determined |
| 18-110-final-viral-combined_k141_155154\|\|lt2gene | Low-quality |
| 18-110-final-viral-combined_k141_85471\|\|lt2gene | Low-quality |
| 18-110-final-viral-combined_k141_257533\|\|lt2gene | Low-quality |
| 18-110-final-viral-combined_k141_144072\|\|lt2gene | Low-quality |
| 18-110-final-viral-combined_k141_236246\|\|lt2gene | Low-quality |
| 18-110-final-viral-combined_k141_144120\|\|lt2gene | Not-determined |
| 18-110-final-viral-combined_k141_108716\|\|lt2gene | Not-determined |
| 18-110-final-viral-combined_k141_120254\|\|lt2gene | Not-determined |
| 18-110-final-viral-combined_k141_155277\|\|lt2gene | Low-quality |
| 18-110-final-viral-combined_k141_28793\|\|lt2gene | Not-determined |
| 18-110-final-viral-combined_k141_179112\|\|lt2gene | Low-quality |
| 18-110-final-viral-combined_k141_120292\|\|lt2gene | Low-quality |
| 18-110-final-viral-combined_k141_270395\|\|lt2gene | Low-quality |
| 18-110-final-viral-combined_k141_270424\|\|lt2gene | Low-quality |
| 18-110-final-viral-combined_k141_247587\|\|lt2gene | Low-quality |
| 18-110-final-viral-combined_k141_51259\|\|lt2gene | Low-quality |
| 18-110-final-viral-combined_k141_28842\|\|lt2gene | Low-quality |
| 18-110-final-viral-combined_k141_236397\|\|lt2gene | Low-quality |
| 18-110-final-viral-combined_k141_85666\|\|lt2gene | Not-determined |
| 18-110-final-viral-combined_k141_51279\|\|lt2gene | Low-quality |
| 18-110-final-viral-combined_k141_63466\|\|lt2gene | Low-quality |
| 18-110-final-viral-combined_k141_247630\|\|lt2gene | Low-quality |
| 18-110-final-viral-combined_k141_4852\|\|lt2gene | Not-determined |
| 18-110-final-viral-combined_k141_85702\|\|lt2gene | Low-quality |
| 18-110-final-viral-combined_k141_97827\|\|lt2gene | Low-quality |
| 18-110-final-viral-combined_k141_224038\|\|lt2gene | Not-determined |
| 18-110-final-viral-combined_k141_74931\|\|lt2gene | Low-quality |
| 18-110-final-viral-combined_k141_15879\|\|lt2gene | Low-quality |
| 18-110-final-viral-combined_k141_51370\|\|lt2gene | Low-quality |
| 18-110-final-viral-combined_k141_212883\|\|lt2gene | Not-determined |
| 18-110-final-viral-combined_k141_212884\|\|lt2gene | Not-determined |
| 18-110-final-viral-combined_k141_15901\|\|lt2gene | Low-quality |
| 18-110-final-viral-combined_k141_4941\|\|lt2gene | Not-determined |
| 18-110-final-viral-combined_k141_179253\|\|lt2gene | Low-quality |
| 18-110-final-viral-combined_k141_132380\|\|lt2gene | Low-quality |
| 18-110-final-viral-combined_k141_132388\|\|lt2gene | Low-quality |
| 18-110-final-viral-combined_k141_51473\|\|lt2gene | Not-determined |
| 18-110-final-viral-combined_k141_270610\|\|lt2gene | Low-quality |
| 18-110-final-viral-combined_k141_29127\|\|lt2gene | Not-determined |
| 18-110-final-viral-combined_k141_190181\|\|lt2gene | Not-determined |
| 18-110-final-viral-combined_k141_270646\|\|lt2gene | Low-quality |
| 18-110-final-viral-combined_k141_212979\|\|lt2gene | Not-determined |
| 18-110-final-viral-combined_k141_29214\|\|lt2gene | Not-determined |
| 18-110-final-viral-combined_k141_51548\|\|lt2gene | Low-quality |
| 18-110-final-viral-combined_k141_166858\|\|lt2gene | Low-quality |
| 18-110-final-viral-combined_k141_224221\|\|lt2gene | Low-quality |
| [illegible] | [illegible] |
| 18-110-final-viral-combined_k141_54875\|\|lt2gene | Low-quality |
| 18-110-final-viral-combined_k141_54882\|\|lt2gene | Not-determined |
| 18-110-final-viral-combined_k141_88712\|\|lt2gene | Low-quality |
| 18-110-final-viral-combined_k141_260252\|\|lt2gene | Low-quality |
| 18-110-final-viral-combined_k141_88721\|\|lt2gene | Low-quality |
| 18-110-final-viral-combined_k141_170081\|\|lt2gene | Low-quality |
| 18-110-final-viral-combined_k141_88737\|\|lt2gene | Low-quality |
| 18-110-final-viral-combined_k141_203295\|\|lt2gene | Low-quality |
| 18-110-final-viral-combined_k141_135965\|\|lt2gene | Low-quality |
| 18-110-final-viral-combined_k141_182596\|\|lt2gene | Low-quality |
| 18-110-final-viral-combined_k141_260275\|\|lt2gene | Low-quality |
| 18-110-final-viral-combined_k141_135995\|\|lt2gene | Low-quality |
| 18-110-final-viral-combined_k141_112017\|\|lt2gene | Not-determined |
| 18-110-final-viral-combined_k141_273615\|\|lt2gene | Low-quality |
| 18-110-final-viral-combined_k141_193023\|\|lt2gene | Not-determined |
| 18-110-final-viral-combined_k141_88798\|\|lt2gene | Low-quality |
| 18-110-final-viral-combined_k141_193025\|\|lt2gene | Not-determined |
| 18-110-final-viral-combined_k141_239967\|\|lt2gene | Low-quality |
| 18-110-final-viral-combined_k141_216081\|\|lt2gene | Not-determined |
| 18-110-final-viral-combined_k141_18827\|\|lt2gene | Not-determined |
| 18-110-final-viral-combined_k141_250707\|\|lt2gene | Low-quality |
| 18-110-final-viral-combined_k141_43699\|\|lt2gene | Low-quality |
| 18-110-final-viral-combined_k141_136087\|\|lt2gene | Low-quality |
| 18-110-final-viral-combined_k141_250727\|\|lt2gene | Low-quality |
| 18-110-final-viral-combined_k141_182752\|\|lt2gene | Not-determined |
| 18-110-final-viral-combined_k141_182756\|\|lt2gene | Low-quality |
| 18-110-final-viral-combined_k141_158876\|\|lt2gene | Low-quality |
| 18-110-final-viral-combined_k141_158877\|\|lt2gene | Low-quality |
| 18-110-final-viral-combined_k141_250765\|\|lt2gene | Not-determined |
| 18-110-final-viral-combined_k141_32854\|\|lt2gene | Not-determined |
| 18-110-final-viral-combined_k141_112134\|\|lt2gene | Low-quality |
| 18-110-final-viral-combined_k141_112147\|\|lt2gene | Not-determined |
| 18-110-final-viral-combined_k141_193177\|\|lt2gene | Low-quality |
| 18-110-final-viral-combined_k141_240058\|\|lt2gene | Low-quality |
| 18-110-final-viral-combined_k141_203456\|\|lt2gene | Low-quality |
| 18-110-final-viral-combined_k141_18934\|\|lt2gene | Low-quality |
| 18-110-final-viral-combined_k141_216227\|\|lt2gene | Low-quality |
| 18-110-final-viral-combined_k141_89009\|\|lt2gene | Low-quality |
| 18-110-final-viral-combined_k141_136226\|\|lt2gene | Low-quality |
| 18-110-final-viral-combined_k141_18967\|\|lt2gene | Not-determined |
| 18-110-final-viral-combined_k141_273835\|\|lt2gene | Low-quality |
| 18-110-final-viral-combined_k141_78532\|\|lt2gene | Not-determined |
| 18-110-final-viral-combined_k141_89061\|\|lt2gene | Low-quality |
| 18-110-final-viral-combined_k141_124107\|\|lt2gene | Low-quality |
| 18-110-final-viral-combined_k141_19020\|\|lt2gene | Not-determined |
| 18-110-final-viral-combined_k141_260442\|\|lt2gene | Not-determined |
| 18-110-final-viral-combined_k141_8065\|\|lt2gene | Low-quality |
| 18-110-final-viral-combined_k141_260450\|\|lt2gene | Not-determined |
| 18-110-final-viral-combined_k141_227703\|\|lt2gene | Not-determined |
| 18-110-final-viral-combined_k141_227712\|\|lt2gene | Not-determined |
| 18-110-final-viral-combined_k141_43958\|\|lt2gene | Low-quality |
| 18-110-final-viral-combined_k141_136358\|\|lt2gene | Low-quality |
| 18-110-final-viral-combined_k141_203579\|\|lt2gene | Not-determined |
| 18-110-final-viral-combined_k141_136370\|\|lt2gene | Low-quality |

| | |
|---|---|
| 18-110-final-viral-combined_k141_112316\|\|lt2gene | Low-quality |
| 18-110-final-viral-combined_k141_148023\|\|lt2gene | Not-determined |
| 18-110-final-viral-combined_k141_193367\|\|lt2gene | Not-determined |
| 18-110-final-viral-combined_k141_193373\|\|lt2gene | Not-determined |
| 18-110-final-viral-combined_k141_250937\|\|lt2gene | Low-quality |
| 18-110-final-viral-combined_k141_44016\|\|lt2gene | Low-quality |
| 18-110-final-viral-combined_k141_193391\|\|lt2gene | Low-quality |
| 18-110-final-viral-combined_k141_78683\|\|lt2gene | Low-quality |
| 18-110-final-viral-combined_k141_8154\|\|lt2gene | Low-quality |
| 18-110-final-viral-combined_k141_112398\|\|lt2gene | Low-quality |
| 18-110-final-viral-combined_k141_203657\|\|lt2gene | Low-quality |
| 18-110-final-viral-combined_k141_44048\|\|lt2gene | Not-determined |
| 18-110-final-viral-combined_k141_136460\|\|lt2gene | Low-quality |
| 18-110-final-viral-combined_k141_136461\|\|lt2gene | Low-quality |
| 18-110-final-viral-combined_k141_170538\|\|lt2gene | Low-quality |
| 18-110-final-viral-combined_k141_227802\|\|lt2gene | Low-quality |
| 18-110-final-viral-combined_k141_159084\|\|lt2gene | Not-determined |
| 18-110-final-viral-combined_k141_78759\|\|lt2gene | Low-quality |
| 18-110-final-viral-combined_k141_44098\|\|lt2gene | Low-quality |
| 18-110-final-viral-combined_k141_136499\|\|lt2gene | Not-determined |
| 18-110-final-viral-combined_k141_193464\|\|lt2gene | Not-determined |
| 18-110-final-viral-combined_k141_124314\|\|lt2gene | Low-quality |
| 18-110-final-viral-combined_k141_203728\|\|lt2gene | Low-quality |
| 18-110-final-viral-combined_k141_170554\|\|lt2gene | Low-quality |
| 18-110-final-viral-combined_k141_216484\|\|lt2gene | Not-determined |
| 18-110-final-viral-combined_k141_159158\|\|lt2gene | Not-determined |
| 18-110-final-viral-combined_k141_78868\|\|lt2gene | Not-determined |
| 18-110-final-viral-combined_k141_227884\|\|lt2gene | Low-quality |
| 18-110-final-viral-combined_k141_136590\|\|lt2gene | Not-determined |
| 18-110-final-viral-combined_k141_19296\|\|lt2gene | Low-quality |
| 18-110-final-viral-combined_k141_216563\|\|lt2gene | Low-quality |
| 18-110-final-viral-combined_k141_78906\|\|lt2gene | Not-determined |
| 18-110-final-viral-combined_k141_19341\|\|lt2gene | Low-quality |
| 18-110-final-viral-combined_k141_148257\|\|lt2gene | Not-determined |
| 18-110-final-viral-combined_k141_55258\|\|lt2gene | Low-quality |
| 18-110-final-viral-combined_k141_89420\|\|lt2gene | Low-quality |
| 18-110-final-viral-combined_k141_203872\|\|lt2gene | Low-quality |
| 18-110-final-viral-combined_k141_8351\|\|lt2gene | Low-quality |
| 18-110-final-viral-combined_k141_183275\|\|lt2gene | Low-quality |
| 18-110-final-viral-combined_k141_227994\|\|lt2gene | Not-determined |
| 18-110-final-viral-combined_k141_8380\|\|lt2gene | Not-determined |
| 18-110-final-viral-combined_k141_8450\|\|lt2gene | Not-determined |
| 18-110-final-viral-combined_k141_19408\|\|lt2gene | Low-quality |
| 18-110-final-viral-combined_k141_33382\|\|lt2gene | Low-quality |
| 18-110-final-viral-combined_k141_240660\|\|lt2gene | Low-quality |
| 18-110-final-viral-combined_k141_136744\|\|lt2gene | Low-quality |
| 18-110-final-viral-combined_k141_183418\|\|lt2gene | Low-quality |
| 18-110-final-viral-combined_k141_203929\|\|lt2gene | Not-determined |
| 18-110-final-viral-combined_k141_228079\|\|lt2gene | Low-quality |
| 18-110-final-viral-combined_k141_216775\|\|lt2gene | Low-quality |
| 18-110-final-viral-combined_k141_251241\|\|lt2gene | Low-quality |
| 18-110-final-viral-combined_k141_240702\|\|lt2gene | Low-quality |
| 18-110-final-viral-combined_k141_55375\|\|lt2gene | Low-quality |
| 18-110-final-viral-combined_k141_67180\|\|lt2gene | Low-quality |
| 18-110-final-viral-combined_k141_251275\|\|lt2gene | Low-quality |
| 18-110-final-viral-combined_k141_89642\|\|lt2gene | Low-quality |
| 18-110-final-viral-combined_k141_193762\|\|lt2gene | Low-quality |
| 18-110-final-viral-combined_k141_19551\|\|lt2gene | Low-quality |
| 18-110-final-viral-combined_k141_19604\|\|lt2gene | Low-quality |
| 18-110-final-viral-combined_k141_228226\|\|lt2gene | Low-quality |
| 18-110-final-viral-combined_k141_204085\|\|lt2gene | Low-quality |
| 18-110-final-viral-combined_k141_55497\|\|lt2gene | Low-quality |
| 18-110-final-viral-combined_k141_55501\|\|lt2gene | Not-determined |
| 18-110-final-viral-combined_k141_148609\|\|lt2gene | Low-quality |
| 18-110-final-viral-combined_k141_79267\|\|lt2gene | Low-quality |
| 18-110-final-viral-combined_k141_55538\|\|lt2gene | Not-determined |
| 18-110-final-viral-combined_k141_44524\|\|lt2gene | Low-quality |
| 18-110-final-viral-combined_k141_44527\|\|lt2gene | Low-quality |
| 18-110-final-viral-combined_k141_55552\|\|lt2gene | Low-quality |
| 18-110-final-viral-combined_k141_148659\|\|lt2gene | Low-quality |
| 18-110-final-viral-combined_k141_67395\|\|lt2gene | Low-quality |
| 18-110-final-viral-combined_k141_148688\|\|lt2gene | Not-determined |
| 18-110-final-viral-combined_k141_101883\|\|lt2gene | Not-determined |
| 18-110-final-viral-combined_k141_89843\|\|lt2gene | Low-quality |
| 18-110-final-viral-combined_k141_251403\|\|lt2gene | Not-determined |
| 18-110-final-viral-combined_k141_101942\|\|lt2gene | Low-quality |
| [illegible] | [illegible] |
| 18-110-final-viral-combined_k141_103662\|\|lt2gene | Low-quality |
| 18-110-final-viral-combined_k141_230328\|\|lt2gene | Not-determined |
| 18-110-final-viral-combined_k141_230329\|\|lt2gene | Not-determined |
| 18-110-final-viral-combined_k141_57560\|\|lt2gene | Low-quality |
| 18-110-final-viral-combined_k141_195745\|\|lt2gene | Low-quality |
| 18-110-final-viral-combined_k141_114873\|\|lt2gene | Low-quality |
| 18-110-final-viral-combined_k141_262650\|\|lt2gene | Low-quality |
| 18-110-final-viral-combined_k141_206393\|\|lt2gene | Not-determined |
| 18-110-final-viral-combined_k141_206452\|\|lt2gene | Not-determined |
| 18-110-final-viral-combined_k141_262813\|\|lt2gene | Low-quality |
| 18-110-final-viral-combined_k141_262908\|\|lt2gene | Low-quality |
| 18-110-final-viral-combined_k141_10929\|\|lt2gene | Low-quality |
| 18-110-final-viral-combined_k141_22051\|\|lt2gene | Low-quality |
| 18-110-final-viral-combined_k141_263075\|\|lt2gene | Low-quality |
| 18-110-final-viral-combined_k141_263114\|\|lt2gene | Low-quality |
| 18-110-final-viral-combined_k141_22211\|\|lt2gene | Low-quality |
| 18-110-final-viral-combined_k141_22282\|\|lt2gene | Low-quality |
| 18-110-final-viral-combined_k141_263239\|\|lt2gene | Not-determined |
| 18-110-final-viral-combined_k141_263253\|\|lt2gene | Low-quality |
| 18-110-final-viral-combined_k141_22340\|\|lt2gene | Low-quality |
| 18-110-final-viral-combined_k141_263272\|\|lt2gene | Not-determined |
| 18-110-final-viral-combined_k141_263354\|\|lt2gene | Low-quality |
| 18-110-final-viral-combined_k141_207018\|\|lt2gene | Low-quality |
| 18-110-final-viral-combined_k141_207041\|\|lt2gene | Not-determined |
| 18-110-final-viral-combined_k141_11332\|\|lt2gene | Low-quality |
| 18-110-final-viral-combined_k141_11443\|\|lt2gene | Low-quality |
| 18-110-final-viral-combined_k141_11503\|\|lt2gene | Low-quality |
| 18-110-final-viral-combined_k141_11512\|\|lt2gene | Not-determined |
| 18-110-final-viral-combined_k141_11514\|\|lt2gene | Not-determined |
| 18-110-final-viral-combined_k141_22912\|\|lt2gene | Low-quality |
| 18-110-final-viral-combined_k141_264113\|\|lt2gene | Low-quality |
| 18-110-final-viral-combined_k141_264162\|\|lt2gene | Not-determined |
| 18-110-final-viral-combined_k141_264188\|\|lt2gene | Low-quality |
| 18-110_k141_69118 | Not-determined |
| 18-110_k141_126715 | Not-determined |
| 18-110_k141_184314 | Not-determined |
| 18-110_k141_241905 | Low-quality |
| 18-110_k141_103685 | Not-determined |
| 18-110_k141_34565 | Low-quality |
| 18-110_k141_195833 | Not-determined |
| 18-110_k141_149757 | Not-determined |
| 18-110_k141_115207 | Not-determined |
| 18-110_k141_230399 | Not-determined |
| 18-110_k141_172794 | Not-determined |
| 18-110_k141_6 | Not-determined |
| 18-110_k141_92163 | Not-determined |
| 18-110_k141_230410 | Not-determined |
| 18-110_k141_46088 | Not-determined |
| 18-110_k141_92172 | Not-determined |
| 18-110_k141_69137 | Not-determined |
| 18-110_k141_92185 | Not-determined |
| 18-110_k141_138257 | Not-determined |
| 18-110_k141_138258 | Not-determined |
| 18-110_k141_92195 | Not-determined |
| 18-110_k141_92196 | Not-determined |
| 18-110_k141_195851 | Not-determined |
| 18-110_k141_230430 | Not-determined |
| 18-110_k141_264968 | Not-determined |
| 18-110_k141_126740 | Low-quality |
| 18-110_k141_253427 | Not-determined |
| 18-110_k141_34573 | Not-determined |
| 18-110_k141_172812 | Not-determined |
| 18-110_k141_11542 | Not-determined |
| 18-110_k141_138273 | Not-determined |
| 18-110_k141_34585 | Not-determined |
| 18-110_k141_253436 | Not-determined |
| 18-110_k141_207357 | Not-determined |
| 18-110_k141_92203 | Not-determined |
| 18-110_k141_92207 | Not-determined |
| 18-110_k141_241930 | Not-determined |
| 18-110_k141_46109 | Not-determined |
| 18-110_k141_138290 | Not-determined |
| 18-110_k141_34592 | Not-determined |
| 18-110_k141_46111 | Low-quality |
| 18-110_k141_195857 | Not-determined |
| 18-110_k141_195859 | Not-determined |
| 18-110_k141_34598 | Not-determined |
| 18-110_k141_138303 | Not-determined |
| 18-110_k141_218920 | Not-determined |
| 18-110_k141_230457 | Not-determined |
| 18-110_k141_57614 | Not-determined |
| 18-110_k141_218923 | Not-determined |
| 18-110_k141_172819 | Not-determined |
| 18-110_k141_34607 | Not-determined |
| 18-110_k141_34609 | Not-determined |
| 18-110_k141_149793 | Not-determined |
| 18-110_k141_195862 | Not-determined |
| 18-110_k141_264990 | Not-determined |
| 18-110_k141_195867 | Not-determined |
| 18-110_k141_172822 | Not-determined |
| 18-110_k141_184359 | Not-determined |
| 18-110_k141_264997 | Not-determined |
| 18-110_k141_149800 | Not-determined |
| 18-110_k141_46130 | Not-determined |
| 18-110_k141_230460 | Not-determined |
| 18-110_k141_172824 | Low-quality |
| 18-110_k141_195871 | Not-determined |
| 18-110_k141_138320 | Not-determined |
| 18-110_k141_92218 | Low-quality |
| 18-110_k141_69187 | Not-determined |
| 18-110_k141_80650 | Not-determined |
| 18-110_k141_138324 | Not-determined |
| 18-110_k141_265005 | Not-determined |
| 18-110_k141_230470 | Not-determined |
| 18-110_k141_69188 | Not-determined |
| 18-110_k141_184368 | Not-determined |
| 18-110_k141_46138 | Not-determined |
| 18-110_k141_184373 | Not-determined |
| 18-110_k141_149808 | Not-determined |
| 18-110_k141_34628 | Not-determined |
| 18-110_k141_161281 | Not-determined |
| 18-110_k141_69217 | Not-determined |
| 18-110_k141_195888 | Low-quality |
| 18-110_k141_265025 | Not-determined |
| 18-110_k141_161283 | Not-determined |
| 18-110_k141_115253 | Not-determined |
| 18-110_k141_46155 | Not-determined |
| 18-110_k141_265034 | Not-determined |
| 18-110_k141_57632 | Not-determined |
| 18-110_k141_218936 | Not-determined |
| 18-110_k141_230494 | Not-determined |
| 18-110_k141_138358 | Not-determined |
| 18-110_k141_149824 | Not-determined |
| 18-110_k141_138362 | Not-determined |
| 18-110_k141_80665 | Not-determined |
| 18-110_k141_34639 | Not-determined |
| 18-110_k141_80666 | Not-determined |
| 18-110_k141_115265 | Not-determined |
| 18-110_k141_230505 | Not-determined |
| 18-110_k141_138369 | Not-determined |
| 18-110_k141_253482 | Not-determined |
| 18-110_k141_80680 | Not-determined |
| 18-110_k141_69235 | Not-determined |
| 18-110_k141_115279 | Not-determined |
| 18-110_k141_86 | Not-determined |
| 18-110_k141_11557 | Not-determined |
| 18-110_k141_46165 | Not-determined |
| 18-110_k141_241986 | Not-determined |
| 18-110_k141_103705 | Low-quality |
| 18-110_k141_265062 | Not-determined |
| 18-110_k141_138377 | Not-determined |
| 18-110_k141_69244 | Low-quality |
| 18-110_k141_57648 | Not-determined |
| 18-110_k141_103707 | Not-determined |
| 18-110_k141_57652 | Not-determined |
| 18-110_k141_230523 | Not-determined |
| 18-110_k141_138384 | Not-determined |
| 18-110_k141_149836 | Low-quality |
| 18-110_k141_149837 | Low-quality |
| 18-110_k141_265065 | Not-determined |
| 18-110_k141_80695 | Not-determined |
| 18-110_k141_172877 | Not-determined |
| 18-110_k141_195915 | Not-determined |
| 18-110_k141_34683 | Not-determined |
| [illegible] | [illegible] |

| | |
|---|---|
| 18-110_k141_115354 | Not-determined |
| 18-110_k141_92377 | Not-determined |
| 18-110_k141_138515 | Not-determined |
| 18-110_k141_149909 | Not-determined |
| 18-110_k141_11600 | Low-quality |
| 18-110_k141_80904 | Not-determined |
| 18-110_k141_92382 | Not-determined |
| 18-110_k141_149913 | Not-determined |
| 18-110_k141_138516 | Low-quality |
| 18-110_k141_69365 | Low-quality |
| 18-110_k141_138522 | Not-determined |
| 18-110_k141_23268 | Not-determined |
| 18-110_k141_80916 | Not-determined |
| 18-110_k141_219029 | Not-determined |
| 18-110_k141_196004 | Not-determined |
| 18-110_k141_230703 | Not-determined |
| 18-110_k141_138525 | Not-determined |
| 18-110_k141_219032 | Not-determined |
| 18-110_k141_172995 | Not-determined |
| 18-110_k141_92393 | Not-determined |
| 18-110_k141_242119 | Low-quality |
| 18-110_k141_23277 | Not-determined |
| 18-110_k141_173003 | Not-determined |
| 18-110_k141_92395 | Low-quality |
| 18-110_k141_196014 | Not-determined |
| 18-110_k141_173007 | Not-determined |
| 18-110_k141_23280 | Not-determined |
| 18-110_k141_138536 | Not-determined |
| 18-110_k141_92400 | Not-determined |
| 18-110_k141_253564 | Low-quality |
| 18-110_k141_196021 | Not-determined |
| 18-110_k141_222 | Not-determined |
| 18-110_k141_46290 | Not-determined |
| 18-110_k141_173010 | Not-determined |
| 18-110_k141_242131 | Not-determined |
| 18-110_k141_253567 | Not-determined |
| 18-110_k141_242133 | Not-determined |
| 18-110_k141_92403 | Not-determined |
| 18-110_k141_242134 | Not-determined |
| 18-110_k141_242142 | Not-determined |
| 18-110_k141_80942 | Not-determined |
| 18-110_k141_196026 | Not-determined |
| 18-110_k141_242144 | Not-determined |
| 18-110_k141_229 | Not-determined |
| 18-110_k141_115388 | Not-determined |
| 18-110_k141_115390 | Not-determined |
| 18-110_k141_23294 | Not-determined |
| 18-110_k141_46301 | Not-determined |
| 18-110_k141_196034 | Not-determined |
| 18-110_k141_236 | Not-determined |
| 18-110_k141_196036 | Not-determined |
| 18-110_k141_240 | Not-determined |
| 18-110_k141_184518 | Not-determined |
| 18-110_k141_184519 | Not-determined |
| 18-110_k141_184520 | Not-determined |
| 18-110_k141_230721 | Not-determined |
| 18-110_k141_242 | Not-determined |
| 18-110_k141_184522 | Not-determined |
| 18-110_k141_138552 | Not-determined |
| 18-110_k141_57838 | Not-determined |
| 18-110_k141_138553 | Not-determined |
| 18-110_k141_242156 | Not-determined |
| 18-110_k141_207566 | Not-determined |
| 18-110_k141_173027 | Not-determined |
| 18-110_k141_161339 | Not-determined |
| 18-110_k141_173030 | Not-determined |
| 18-110_k141_69381 | Not-determined |
| 18-110_k141_173031 | Not-determined |
| 18-110_k141_184533 | Not-determined |
| 18-110_k141_23313 | Not-determined |
| 18-110_k141_69385 | Not-determined |
| 18-110_k141_230739 | Not-determined |
| 18-110_k141_260 | Not-determined |
| 18-110_k141_230741 | Not-determined |
| 18-110_k141_69394 | Not-determined |
| 18-110_k141_69396 | Not-determined |
| 18-110_k141_242169 | Not-determined |
| 18-110_k141_207580 | Not-determined |
| 18-110_k141_138581 | Not-determined |
| 18-110_k141_265 | Low-quality |
| 18-110_k141_242175 | Not-determined |
| 18-110_k141_242176 | Not-determined |
| 18-110_k141_138589 | Not-determined |
| 18-110_k141_126945 | Not-determined |
| 18-110_k141_138594 | Not-determined |
| 18-110_k141_230753 | Not-determined |
| 18-110_k141_230754 | Not-determined |
| 18-110_k141_46312 | Not-determined |
| 18-110_k141_207596 | Not-determined |
| 18-110_k141_46316 | Not-determined |
| 18-110_k141_196069 | Not-determined |
| 18-110_k141_149929 | Not-determined |
| 18-110_k141_242189 | Not-determined |
| 18-110_k141_92410 | Not-determined |
| 18-110_k141_46317 | Not-determined |
| 18-110_k141_242199 | Not-determined |
| 18-110_k141_80992 | Not-determined |
| 18-110_k141_138605 | Not-determined |
| 18-110_k141_103801 | Not-determined |
| 18-110_k141_69423 | Not-determined |
| 18-110_k141_126962 | Not-determined |
| 18-110_k141_242205 | Not-determined |
| 18-110_k141_115438 | Not-determined |
| 18-110_k141_196081 | Low-quality |
| 18-110_k141_46333 | Not-determined |
| 18-110_k141_242215 | Not-determined |
| 18-110_k141_46337 | Not-determined |
| 18-110_k141_149962 | Not-determined |
| 18-110_k141_230781 | Not-determined |
| 18-110_k141_57883 | Not-determined |
| 18-110_k141_138615 | Not-determined |
| 17-109-final-viral-combined_k141_223235\|\|full | High-quality |
| 18-110_k141_219067 | Not-determined |
| 18-110_k141_271 | Low-quality |
| 18-110_k141_92426 | Not-determined |
| 18-110_k141_138618 | Not-determined |
| 18-110_k141_57889 | Not-determined |
| 18-110_k141_81005 | Not-determined |
| 18-110_k141_138620 | Not-determined |
| 18-110_k141_57892 | Not-determined |
| 18-110_k141_126981 | Not-determined |
| 18-110_k141_265179 | Not-determined |
| 18-110_k141_46353 | Not-determined |
| 18-110_k141_265181 | Not-determined |
| 18-110_k141_253589 | Not-determined |
| 18-110_k141_46361 | Not-determined |
| 18-110_k141_103826 | Not-determined |
| 18-110_k141_184560 | Not-determined |
| 18-110_k141_103828 | Low-quality |
| 18-110_k141_127006 | Not-determined |
| 18-110_k141_173100 | Not-determined |
| 18-110_k141_253591 | Not-determined |
| 18-110_k141_23350 | Not-determined |
| 18-110_k141_115448 | Not-determined |
| 18-110_k141_173103 | Not-determined |
| 18-110_k141_69451 | Not-determined |
| 18-110_k141_219082 | Not-determined |
| 18-110_k141_23354 | Not-determined |
| 18-110_k141_149987 | Not-determined |
| 18-110_k141_253600 | Not-determined |
| 18-110_k141_92449 | Not-determined |
| 18-110_k141_219084 | Low-quality |
| 18-110_k141_242246 | Not-determined |
| 18-110_k141_219085 | Not-determined |
| 18-110_k141_92456 | Not-determined |
| 18-110_k141_219089 | Not-determined |
| 18-110_k141_297 | Low-quality |
| 18-110_k141_149992 | Not-determined |
| 18-110_k141_253608 | Not-determined |
| 18-110_k141_127022 | Not-determined |
| 18-110_k141_173134 | Low-quality |
| 18-110_k141_253611 | Low-quality |
| 18-110_k141_149998 | Not-determined |
| 18-110_k141_138660 | Not-determined |
| 18-110_k141_57935 | Not-determined |
| 18-110_k141_57938 | Not-determined |
| 18-110_k141_57940 | Not-determined |
| 18-110_k141_184603 | Not-determined |
| 18-110_k141_115471 | Not-determined |
| 18-110_k141_230842 | Not-determined |
| 18-110_k141_57948 | Not-determined |
| 18-110_k141_69471 | Low-quality |
| 18-110_k141_23384 | Low-quality |
| 18-110_k141_253623 | Not-determined |
| 18-110_k141_196140 | Not-determined |
| 18-110_k141_184623 | Not-determined |
| 18-110_k141_207660 | Not-determined |
| 18-110_k141_184627 | Low-quality |
| 18-110_k141_207661 | Not-determined |
| 18-110_k141_103853 | Not-determined |
| 18-110_k141_115474 | Low-quality |
| 18-110_k141_230868 | Not-determined |
| 18-110_k141_69492 | Not-determined |
| 18-110_k141_184634 | Not-determined |
| 18-110_k141_115486 | Not-determined |
| 18-110_k141_173160 | Not-determined |
| 18-110_k141_69498 | Not-determined |
| 18-110_k141_196146 | Low-quality |
| 18-110_k141_265213 | Not-determined |
| 18-110_k141_150014 | Not-determined |
| 18-110_k141_161397 | Not-determined |
| 18-110_k141_207670 | Not-determined |
| 18-110_k141_230876 | Not-determined |
| 18-110_k141_57982 | Not-determined |
| 18-110_k141_173163 | Not-determined |
| 18-110_k141_138692 | Not-determined |
| 18-110_k141_161402 | Not-determined |
| 18-110_k141_115500 | Not-determined |
| 18-110_k141_161404 | Not-determined |
| 18-110_k141_219136 | Not-determined |
| 18-110_k141_69507 | Not-determined |
| 18-110_k141_103870 | Not-determined |
| 18-110_k141_242300 | Not-determined |
| 18-110_k141_46444 | Not-determined |
| 18-110_k141_219142 | Not-determined |
| 18-110_k141_138697 | Not-determined |
| 18-110_k141_242302 | Low-quality |
| 18-110_k141_184644 | Not-determined |
| 18-110_k141_184646 | Not-determined |
| 18-110_k141_23392 | Not-determined |
| 18-110_k141_138702 | Not-determined |
| 18-110_k141_242305 | Not-determined |
| 18-110_k141_150036 | Low-quality |
| 18-110_k141_150038 | Low-quality |
| 18-110_k141_81021 | Not-determined |
| 18-110_k141_81027 | Not-determined |
| 18-110_k141_92504 | Not-determined |
| 18-110_k141_150051 | Not-determined |
| 18-110_k141_161424 | Not-determined |
| 18-110_k141_265254 | Low-quality |
| 18-110_k141_161427 | Not-determined |
| 18-110_k141_103889 | Not-determined |
| 18-110_k141_81045 | Not-determined |
| 18-110_k141_230905 | Not-determined |
| 18-110_k141_81049 | Not-determined |
| 18-110_k141_253667 | Not-determined |
| 18-110_k141_265264 | Not-determined |
| 18-110_k141_11756 | Not-determined |
| 18-110_k141_81052 | Not-determined |
| 18-110_k141_150072 | Not-determined |
| 18-110_k141_115516 | Not-determined |
| 18-110_k141_161441 | Not-determined |
| 18-110_k141_346 | Not-determined |
| 18-110_k141_127048 | Not-determined |
| 18-110_k141_230914 | Not-determined |
| 18-110_k141_161445 | Not-determined |
| 18-110_k141_46479 | Not-determined |
| 18-110_k141_207678 | Not-determined |
| 18-110_k141_253681 | Not-determined |
| 18-110_k141_115523 | Not-determined |
| 18-110_k141_138720 | Not-determined |
| 18-110_k141_58020 | Low-quality |
| 18-110_k141_351 | Not-determined |
| 18-110_k141_265282 | Low-quality |
| 18-110_k141_58021 | Not-determined |
| 18-110_k141_265284 | Not-determined |
| 18-110_k141_207684 | Low-quality |
| 18-110_k141_265290 | Not-determined |
| 18-110_k141_207689 | Not-determined |
| 18-110_k141_11757 | Low-quality |
| 18-110_k141_103909 | Not-determined |
| 18-110_k141_11759 | Not-determined |
| 18-110_k141_242348 | Not-determined |
| 18-110_k141_69551 | Not-determined |
| 18-110_k141_11768 | Not-determined |
| 18-110_k141_115538 | Not-determined |
| 18-110_k141_127075 | Not-determined |
| 18-110_k141_161449 | Not-determined |
| 18-110_k141_127078 | Not-determined |
| 18-110_k141_11772 | Not-determined |
| 18-110_k141_161455 | Not-determined |
| 18-110_k141_184667 | Not-determined |
| 18-110_k141_127087 | Not-determined |
| 18-110_k141_127089 | Not-determined |
| 18-110_k141_92560 | Not-determined |
| 18-110_k141_207711 | Not-determined |
| 18-110_k141_369 | Not-determined |
| 18-110_k141_161458 | Low-quality |
| 18-110_k141_161459 | Not-determined |
| 18-110_k141_81125 | Low-quality |
| 18-110_k141_46516 | Not-determined |
| 18-110_k141_11782 | Not-determined |
| 18-110_k141_127092 | Not-determined |
| 18-110_k141_242358 | Not-determined |
| 18-110_k141_173180 | Not-determined |
| 18-110_k141_58053 | Not-determined |
| 18-110_k141_103929 | Not-determined |
| 18-110_k141_23441 | Not-determined |
| 18-110_k141_69567 | Not-determined |
| 18-110_k141_242371 | Not-determined |
| 18-110_k141_11801 | Not-determined |
| 18-110_k141_127105 | Low-quality |
| 18-110_k141_127106 | Low-quality |
| 18-110_k141_196198 | Not-determined |
| 18-110_k141_58090 | Not-determined |
| 18-110_k141_184688 | Not-determined |
| 18-110_k141_173207 | Not-determined |
| 18-110_k141_11823 | Not-determined |
| 18-110_k141_11826 | Not-determined |
| 18-110_k141_242395 | Not-determined |
| 18-110_k141_389 | Not-determined |
| 18-110_k141_253717 | Not-determined |
| 18-110_k141_58105 | Not-determined |
| 18-110_k141_161463 | Not-determined |
| 18-110_k141_253721 | Not-determined |
| 18-110_k141_207734 | Not-determined |
| 18-110_k141_150104 | Not-determined |
| 18-110_k141_161467 | Not-determined |
| 18-110_k141_397 | Not-determined |
| 18-110_k141_196224 | Low-quality |
| 18-110_k141_69589 | Low-quality |
| 18-110_k141_196228 | Not-determined |
| 18-110_k141_161476 | Not-determined |
| 18-110_k141_219155 | Not-determined |
| 18-110_k141_253729 | Not-determined |
| 18-110_k141_161479 | Not-determined |
| 18-110_k141_58113 | Low-quality |
| 18-110_k141_58116 | Not-determined |
| 18-110_k141_161485 | Not-determined |
| 18-110_k141_138797 | Not-determined |
| 18-110_k141_265323 | Not-determined |
| 18-110_k141_219163 | Not-determined |
| 18-110_k141_138800 | Not-determined |
| 18-110_k141_115556 | Not-determined |
| 18-110_k141_196236 | Not-determined |
| 18-110_k141_58123 | Not-determined |
| 18-110_k141_242412 | Not-determined |
| 18-110_k141_173228 | Not-determined |
| 18-110_k141_58125 | Not-determined |
| 18-110_k141_265332 | Not-determined |
| 18-110_k141_58128 | Not-determined |
| 18-110_k141_23475 | Not-determined |
| 18-110_k141_173235 | Low-quality |
| 18-110_k141_161504 | Not-determined |
| 18-110_k141_219177 | Not-determined |
| 18-110_k141_161510 | Not-determined |
| 18-110_k141_161511 | Not-determined |
| 18-110_k141_253737 | Not-determined |
| 18-110_k141_92615 | Not-determined |
| 18-110_k141_23483 | Not-determined |
| 18-110_k141_92617 | Not-determined |
| 18-110_k141_138813 | Not-determined |
| 18-110_k141_196243 | Not-determined |
| 18-110_k141_207744 | Not-determined |
| 18-110_k141_58141 | Not-determined |
| 18-110_k141_207747 | Not-determined |
| 18-110_k141_196246 | Not-determined |
| 18-110_k141_219194 | Not-determined |
| 18-110_k141_196247 | Not-determined |
| 18-110_k141_253745 | Not-determined |
| 18-110_k141_242448 | Not-determined |
| 18-110_k141_115584 | Not-determined |
| 18-110_k141_58154 | Not-determined |
| 18-110_k141_265347 | Low-quality |
| 18-110_k141_92628 | Not-determined |
| 18-110_k141_161533 | Not-determined |
| 18-110_k141_196262 | Not-determined |
| 18-110_k141_103945 | Low-quality |
| 18-110_k141_103946 | Not-determined |
| 18-110_k141_430 | Not-determined |
| 18-110_k141_138835 | Not-determined |
| 18-110_k141_81162 | Not-determined |
| 18-110_k141_138836 | Not-determined |
| 18-110_k141_103948 | Not-determined |
| 18-110_k141_58157 | Not-determined |
| 18-110_k141_219214 | Not-determined |
| 18-110_k141_23519 | Not-determined |
| 18-110_k141_23521 | Not-determined |
| 18-110_k141_138847 | Not-determined |
| 18-110_k141_138848 | Not-determined |
| 18-110_k141_11862 | Not-determined |
| 18-110_k141_161553 | Not-determined |
| 18-110_k141_196272 | Not-determined |
| 18-110_k141_196273 | Not-determined |
| 18-110_k141_138852 | Not-determined |
| 18-110_k141_103953 | Not-determined |
| 18-110_k141_265363 | Not-determined |
| 18-110_k141_11870 | Not-determined |
| 18-110_k141_265364 | Not-determined |
| 18-110_k141_138855 | Not-determined |
| 18-110_k141_58166 | Low-quality |
| 18-110_k141_150110 | Low-quality |
| 18-110_k141_196282 | Not-determined |
| 18-110_k141_138867 | Not-determined |
| 18-110_k141_81186 | Not-determined |
| 18-110_k141_207769 | Low-quality |
| 18-110_k141_103972 | Not-determined |
| 18-110_k141_161568 | Not-determined |
| 18-110_k141_173291 | Not-determined |
| 18-110_k141_81191 | Not-determined |
| 18-110_k141_443 | Not-determined |
| 18-110_k141_207780 | Not-determined |
| 18-110_k141_69642 | Not-determined |
| 18-110_k141_69643 | Not-determined |
| 18-110_k141_11885 | Not-determined |
| 18-110_k141_161582 | Not-determined |
| 18-110_k141_103986 | Not-determined |
| 18-110_k141_69651 | Not-determined |
| 18-110_k141_103989 | Not-determined |
| 18-110_k141_161590 | Not-determined |
| 18-110_k141_265379 | Not-determined |
| 18-110_k141_81203 | Not-determined |
| 18-110_k141_115639 | Not-determined |
| 18-110_k141_115644 | Not-determined |
| 18-110_k141_115645 | Not-determined |
| 18-110_k141_161592 | Not-determined |
| 18-110_k141_81205 | Not-determined |
| 18-110_k141_196303 | Not-determined |
| 18-110_k141_92646 | Not-determined |
| 18-110_k141_161596 | Not-determined |
| 18-110_k141_11902 | Not-determined |
| 18-110_k141_11903 | Not-determined |
| 18-110_k141_454 | Not-determined |
| 18-110_k141_207800 | Not-determined |
| 18-110_k141_11913 | Not-determined |
| 18-110_k141_92660 | Not-determined |
| 18-110_k141_11915 | Low-quality |
| 18-110_k141_104000 | Not-determined |
| 18-110_k141_115664 | Not-determined |
| 18-110_k141_173323 | Not-determined |
| 18-110_k141_242521 | Not-determined |
| 18-110_k141_242522 | Not-determined |
| 18-110_k141_104001 | Not-determined |
| 18-110_k141_115667 | Not-determined |
| 18-110_k141_219251 | Not-determined |
| 18-110_k141_104005 | Not-determined |
| 18-110_k141_173329 | Not-determined |
| 18-110_k141_138896 | Not-determined |
| 18-110_k141_161618 | Not-determined |
| 18-110_k141_23537 | Not-determined |
| 18-110_k141_207813 | Not-determined |
| 18-110_k141_150145 | Low-quality |
| 18-110_k141_460 | Not-determined |
| 18-110_k141_242531 | Not-determined |
| 18-110_k141_207818 | Not-determined |
| 18-110_k141_219258 | Not-determined |
| 18-110_k141_469 | Not-determined |
| 18-110_k141_471 | Low-quality |
| 18-110_k141_265422 | Not-determined |
| 18-110_k141_150157 | Not-determined |
| 18-110_k141_242543 | Not-determined |
| 18-110_k141_219281 | Low-quality |
| 18-110_k141_219282 | Not-determined |
| 18-110_k141_150166 | Low-quality |
| 18-110_k141_207824 | Not-determined |
| 18-110_k141_92693 | Not-determined |
| 18-110_k141_500 | Not-determined |
| 18-110_k141_69677 | Not-determined |
| 18-110_k141_265445 | Low-quality |
| 18-110_k141_138928 | Not-determined |
| 18-110_k141_69678 | Low-quality |
| 18-110_k141_138934 | Not-determined |
| 18-110_k141_138936 | Not-determined |
| 18-110_k141_138943 | Not-determined |
| 18-110_k141_115695 | Not-determined |
| 18-110_k141_219299 | Not-determined |
| 18-110_k141_115700 | Low-quality |
| 18-110_k141_242570 | Not-determined |
| 18-110_k141_127139 | Not-determined |
| 18-110_k141_23569 | Not-determined |
| 18-110_k141_242575 | Not-determined |
| 18-110_k141_58179 | Not-determined |
| 18-110_k141_127143 | Low-quality |
| 18-110_k141_219318 | Not-determined |
| 18-110_k141_219319 | Not-determined |
| 18-110_k141_161637 | Not-determined |
| 18-110_k141_92731 | Not-determined |
| 18-110_k141_92732 | Not-determined |
| 18-110_k141_184715 | Not-determined |
| 18-110_k141_11954 | Not-determined |
| 18-110_k141_242582 | Not-determined |
| 18-110_k141_127149 | Not-determined |
| 18-110_k141_253777 | Low-quality |
| 18-110_k141_161647 | Not-determined |
| 18-110_k141_127155 | Not-determined |
| 18-110_k141_92737 | Low-quality |
| 18-110_k141_161654 | Not-determined |
| 18-110_k141_161656 | Not-determined |
| 18-110_k141_196350 | Not-determined |
| 18-110_k141_150173 | Not-determined |
| 18-110_k141_253780 | Not-determined |
| 18-110_k141_184727 | Not-determined |
| 18-110_k141_69693 | Not-determined |
| 18-110_k141_115712 | Not-determined |
| 18-110_k141_11962 | Not-determined |
| 18-110_k141_138972 | Not-determined |
| 18-110_k141_69697 | Not-determined |
| 18-110_k141_69700 | Not-determined |
| 18-110_k141_207867 | Not-determined |
| 18-110_k141_253785 | Not-determined |
| 18-110_k141_184742 | Not-determined |
| 18-110_k141_58199 | Low-quality |
| 18-110_k141_92757 | Low-quality |
| 18-110_k141_207878 | Not-determined |
| 18-110_k141_138993 | Not-determined |
| 18-110_k141_104037 | Not-determined |
| 18-110_k141_58200 | Low-quality |
| 18-110_k141_536 | Not-determined |
| 18-110_k141_139000 | Not-determined |
| 18-110_k141_230934 | Not-determined |
| 18-110_k141_219359 | Not-determined |
| 18-110_k141_546 | Not-determined |
| 18-110_k141_548 | Not-determined |
| 18-110_k141_207885 | Low-quality |
| 18-110_k141_230943 | Not-determined |
| 18-110_k141_253799 | Not-determined |
| 18-110_k141_219373 | Not-determined |
| 18-110_k141_150187 | Not-determined |
| 18-110_k141_127185 | Not-determined |
| 18-110_k141_265474 | Not-determined |
| 18-110_k141_219376 | Not-determined |
| 18-110_k141_104062 | Not-determined |
| 18-110_k141_46629 | Low-quality |
| 18-110_k141_104065 | Not-determined |
| 18-110_k141_219379 | Not-determined |
| 18-110_k141_219384 | Low-quality |
| 18-110_k141_573 | Not-determined |
| 18-110_k141_219387 | Not-determined |
| 18-110_k141_150197 | Not-determined |
| 18-110_k141_58212 | Not-determined |
| 18-110_k141_575 | Not-determined |
| 18-110_k141_196382 | Not-determined |
| 18-110_k141_577 | Not-determined |
| 18-110_k141_58214 | Not-determined |
| 18-110_k141_578 | Not-determined |
| 18-110_k141_150199 | Not-determined |
| 18-110_k141_580 | Low-quality |
| 18-110_k141_150203 | Not-determined |
| 18-110_k141_196385 | Not-determined |
| 18-110_k141_265501 | Not-determined |
| 18-110_k141_58219 | Not-determined |
| 18-110_k141_196389 | Not-determined |
| 18-110_k141_81305 | Not-determined |
| 18-110_k141_207897 | Not-determined |
| 18-110_k141_104090 | Not-determined |
| 18-110_k141_196394 | Not-determined |
| 18-110_k141_81311 | Not-determined |
| 18-110_k141_58232 | Low-quality |
| 18-110_k141_265518 | Not-determined |
| 18-110_k141_173401 | Not-determined |
| 18-110_k141_196398 | Not-determined |
| 18-110_k141_81318 | Not-determined |
| 18-110_k141_173406 | Not-determined |
| 18-110_k141_161672 | Not-determined |
| 18-110_k141_34787 | Not-determined |
| 18-110_k141_34788 | Not-determined |
| 18-110_k141_242626 | Not-determined |
| 18-110_k141_265528 | Not-determined |
| 18-110_k141_58246 | Not-determined |
| 18-110_k141_265530 | Not-determined |
| 18-110_k141_184790 | Not-determined |
| 18-110_k141_196406 | Not-determined |
| 18-110_k141_265533 | Not-determined |
| 18-110_k141_614 | Not-determined |
| 18-110_k141_219404 | Not-determined |
| 18-110_k141_46636 | Not-determined |
| 18-110_k141_173421 | Not-determined |
| 18-110_k141_12006 | Not-determined |
| 18-110_k141_69734 | Not-determined |
| 18-110_k141_161685 | Not-determined |
| 18-110_k141_92827 | Not-determined |
| 18-110_k141_265541 | Low-quality |
| 18-110_k141_58260 | Not-determined |
| 18-110_k141_115744 | Not-determined |
| 18-110_k141_265543 | Not-determined |
| 18-110_k141_34809 | Not-determined |
| 18-110_k141_173430 | Not-determined |
| 18-110_k141_242633 | Low-quality |
| 18-110_k141_81347 | Not-determined |
| 18-110_k141_219410 | Not-determined |
| 18-110_k141_69753 | Not-determined |
| 18-110_k141_34822 | Not-determined |
| 18-110_k141_92842 | Not-determined |
| 18-110_k141_34824 | Not-determined |
| 18-110_k141_69761 | Not-determined |
| 18-110_k141_81354 | Low-quality |
| 18-110_k141_92847 | Not-determined |
| 18-110-final-viral-combined_k141_264711\|\|full | Medium-quality |
| 18-110_k141_196434 | Not-determined |
| 18-110_k141_69768 | Low-quality |
| 18-110_k141_196440 | Not-determined |
| 18-110_k141_23587 | Not-determined |
| 18-110_k141_631 | Not-determined |
| 18-110_k141_173453 | Not-determined |
| 18-110_k141_150236 | Not-determined |
| 18-110_k141_115755 | Not-determined |
| 18-110_k141_161702 | Low-quality |
| 18-110_k141_81371 | Not-determined |
| 18-110_k141_173457 | Not-determined |
| 18-110_k141_173459 | Not-determined |
| 18-110_k141_231032 | Not-determined |
| 18-110_k141_652 | Not-determined |
| 18-110_k141_253845 | Not-determined |
| 18-110_k141_253846 | Not-determined |
| 18-110_k141_265553 | Low-quality |
| 18-110_k141_231043 | Not-determined |
| 18-110_k141_34850 | Not-determined |
| 18-110_k141_184825 | Not-determined |
| 18-110_k141_161716 | Not-determined |
| 18-110_k141_242650 | Not-determined |
| 18-110_k141_58307 | Not-determined |
| 18-110_k141_656 | Not-determined |
| 18-110_k141_231044 | Not-determined |
| 18-110_k141_242656 | Not-determined |
| 18-110_k141_207975 | Not-determined |
| 18-110_k141_161723 | Not-determined |
| 18-110_k141_34867 | Not-determined |
| 18-110_k141_58310 | Not-determined |
| 18-110_k141_81395 | Not-determined |

| | |
|---|---|
| 18-110_k141_219445 | Not-determined |
| 18-110_k141_242660 | Not-determined |
| 18-110_k141_58324 | Not-determined |
| 18-110_k141_92869 | Low-quality |
| 18-110_k141_58327 | Not-determined |
| 18-110_k141_92871 | Not-determined |
| 18-110_k141_242672 | Not-determined |
| 18-110_k141_265566 | Not-determined |
| 18-110_k141_81399 | Low-quality |
| 18-110_k141_150248 | Not-determined |
| 18-110_k141_161741 | Not-determined |
| 18-110_k141_253853 | Not-determined |
| 18-110_k141_104145 | Not-determined |
| 18-110_k141_104149 | Not-determined |
| 18-110_k141_104150 | Not-determined |
| 18-110_k141_196486 | Not-determined |
| 18-110_k141_23623 | Not-determined |
| 18-110_k141_173502 | Not-determined |
| 18-110_k141_23625 | Not-determined |
| 18-110_k141_161747 | Not-determined |
| 18-110_k141_34872 | Not-determined |
| 18-110_k141_150266 | Not-determined |
| 18-110_k141_265574 | Not-determined |
| 18-110_k141_184858 | Not-determined |
| 18-110_k141_150271 | Not-determined |
| 18-110_k141_150272 | Not-determined |
| 18-110_k141_196496 | Not-determined |
| 18-110_k141_242683 | Not-determined |
| 18-110_k141_242685 | Not-determined |
| 18-110_k141_184862 | Not-determined |
| 18-110_k141_92916 | Not-determined |
| 18-110_k141_12053 | Not-determined |
| 18-110_k141_253857 | Not-determined |
| 18-110_k141_219470 | Not-determined |
| 18-110_k141_34884 | Not-determined |
| 18-110_k141_184871 | Not-determined |
| 18-110_k141_23645 | Not-determined |
| 18-110_k141_242697 | Not-determined |
| 18-110_k141_92927 | Not-determined |
| 18-110_k141_161763 | Not-determined |
| 18-110_k141_265587 | Not-determined |
| 18-110_k141_92931 | Not-determined |
| 18-110_k141_58362 | Not-determined |
| 18-110_k141_104162 | Not-determined |
| 18-110_k141_104165 | Not-determined |
| 18-110_k141_34896 | Not-determined |
| 18-110_k141_253868 | Not-determined |
| 18-110_k141_58366 | Low-quality |
| 18-110_k141_127254 | Not-determined |
| 18-110_k141_207991 | Not-determined |
| 18-110_k141_242711 | Low-quality |
| 18-110_k141_694 | Not-determined |
| 18-110_k141_161770 | Not-determined |
| 18-110_k141_104173 | Not-determined |
| 18-110_k141_104178 | Not-determined |
| 18-110_k141_34898 | Not-determined |
| 18-110_k141_242719 | Not-determined |
| 18-110_k141_242721 | Not-determined |
| 18-110_k141_104188 | Not-determined |
| 18-110_k141_127274 | Low-quality |
| 18-110_k141_58371 | Not-determined |
| 18-110_k141_58372 | Not-determined |
| 18-110_k141_23668 | Not-determined |
| 18-110_k141_265614 | Not-determined |
| 18-110_k141_34916 | Not-determined |
| 18-110_k141_242734 | Not-determined |
| 18-110_k141_265619 | Not-determined |
| 18-110_k141_265620 | Not-determined |
| 18-110_k141_92944 | Not-determined |
| 18-110_k141_231110 | Not-determined |
| 18-110_k141_173550 | Not-determined |
| 18-110_k141_173554 | Low-quality |
| 18-110_k141_265627 | Not-determined |
| 18-110_k141_115768 | Low-quality |
| 18-110_k141_265632 | Not-determined |
| 18-110_k141_265633 | Not-determined |
| 18-110_k141_69794 | Not-determined |
| 18-110_k141_104211 | Not-determined |
| 18-110_k141_196514 | Not-determined |
| 18-110_k141_265639 | Not-determined |
| 18-110_k141_150291 | Not-determined |
| 18-110_k141_34925 | Low-quality |
| 18-110_k141_150292 | Not-determined |
| 18-110_k141_92961 | Not-determined |
| 18-110_k141_253905 | Not-determined |
| 18-110_k141_196517 | Not-determined |
| 18-110_k141_81409 | Low-quality |
| 18-110_k141_127290 | Not-determined |
| 18-110_k141_23709 | Not-determined |
| 18-110_k141_92965 | Low-quality |
| 18-110_k141_92966 | Not-determined |
| 18-110_k141_161797 | Not-determined |
| 18-110_k141_196530 | Not-determined |
| 18-110_k141_161803 | Not-determined |
| 18-110_k141_253918 | Not-determined |
| 18-110_k141_81423 | Not-determined |
| 18-110_k141_196536 | Not-determined |
| 18-110_k141_208016 | Not-determined |
| 18-110_k141_253927 | Not-determined |
| 18-110_k141_150316 | Not-determined |
| 18-110_k141_104236 | Not-determined |
| 18-110_k141_184903 | Low-quality |
| 18-110_k141_265670 | Not-determined |
| 18-110_k141_104240 | Low-quality |
| 18-110_k141_161812 | Not-determined |
| 18-110_k141_115790 | Low-quality |
| 18-110_k141_196542 | Low-quality |
| 18-110_k141_139028 | Not-determined |
| 18-110_k141_265682 | Not-determined |
| 18-110_k141_115795 | Not-determined |
| 18-110_k141_23724 | Low-quality |
| 18-110_k141_12075 | Not-determined |
| 18-110_k141_92979 | Not-determined |
| 18-110_k141_196549 | Not-determined |
| 18-110_k141_127309 | Not-determined |
| 18-110_k141_23731 | Not-determined |
| 18-110_k141_92985 | Not-determined |
| 18-110_k141_115801 | Low-quality |
| 18-110_k141_115802 | Not-determined |
| 18-110_k141_92989 | Not-determined |
| 18-110_k141_69815 | Low-quality |
| 18-110_k141_725 | Not-determined |
| 18-110_k141_161821 | Not-determined |
| 18-110_k141_196555 | Not-determined |
| 18-110_k141_69821 | Low-quality |
| 18-110_k141_115812 | Not-determined |
| 18-110_k141_208045 | Low-quality |
| 18-110_k141_208046 | Not-determined |
| 18-110_k141_139044 | Not-determined |
| 18-110_k141_150338 | Not-determined |
| 18-110_k141_196564 | Not-determined |
| 18-110_k141_81456 | Not-determined |
| 18-110_k141_92999 | Low-quality |
| 18-110_k141_139055 | Not-determined |
| 18-110_k141_93007 | Not-determined |
| 18-110_k141_150355 | Not-determined |
| 18-110_k141_81462 | Not-determined |
| 18-110_k141_93010 | Not-determined |
| 18-110_k141_104253 | Not-determined |
| 18-110_k141_208053 | Not-determined |
| 18-110_k141_69846 | Not-determined |
| 18-110_k141_23748 | Not-determined |
| 18-110_k141_265699 | Not-determined |
| 18-110_k141_139067 | Not-determined |
| 18-110_k141_127336 | Not-determined |
| 18-110_k141_231156 | Not-determined |
| 18-110_k141_231157 | Not-determined |
| 18-110_k141_265705 | Low-quality |
| 18-110_k141_219484 | Not-determined |
| 18-110_k141_69858 | Low-quality |
| 18-110_k141_58375 | Low-quality |
| 18-110_k141_23756 | Not-determined |
| 18-110_k141_139075 | Not-determined |
| 18-110_k141_208068 | Not-determined |
| 18-110_k141_23758 | Not-determined |
| 18-110_k141_219496 | Not-determined |
| 18-110_k141_184958 | Not-determined |
| 18-110_k141_69863 | Not-determined |
| 18-110_k141_58385 | Low-quality |
| 18-110_k141_751 | Not-determined |
| 18-110_k141_184967 | Not-determined |
| 18-110_k141_161865 | Not-determined |
| 18-110_k141_161868 | Not-determined |
| 18-110_k141_184973 | Not-determined |
| 18-110_k141_219512 | Not-determined |
| 18-110_k141_219514 | Not-determined |
| 18-110_k141_173605 | Low-quality |
| 18-110_k141_127359 | Not-determined |
| 18-110_k141_93032 | Not-determined |
| 18-110_k141_208084 | Not-determined |
| 18-110_k141_93038 | Not-determined |
| 18-110_k141_173609 | Not-determined |
| 18-110_k141_208086 | Not-determined |
| 18-110_k141_23795 | Not-determined |
| 18-110_k141_23796 | Not-determined |
| 18-110_k141_219523 | Not-determined |
| 18-110_k141_184980 | Not-determined |
| 18-110_k141_115846 | Low-quality |
| 18-110_k141_69888 | Not-determined |
| 18-110_k141_173621 | Not-determined |
| 18-110_k141_115850 | Not-determined |
| 18-110_k141_115854 | Not-determined |
| 18-110_k141_69893 | Not-determined |
| 18-110_k141_104271 | Not-determined |
| 18-110_k141_150395 | Not-determined |
| 18-110_k141_150396 | Not-determined |
| 18-110_k141_46723 | Not-determined |
| 18-110_k141_104276 | Not-determined |
| 18-110_k141_173645 | Not-determined |
| 18-110_k141_35006 | Not-determined |
| 18-110_k141_253973 | Not-determined |
| 18-110_k141_242745 | Not-determined |
| 18-110_k141_69906 | Not-determined |
| 18-110_k141_35013 | Not-determined |
| 18-110_k141_161887 | Not-determined |
| 18-110_k141_196591 | Not-determined |
| 18-110_k141_196592 | Not-determined |
| 18-110_k141_208126 | Not-determined |
| 18-110_k141_208127 | Not-determined |
| 18-110_k141_208131 | Not-determined |
| 18-110_k141_104292 | Not-determined |
| 18-110_k141_35023 | Low-quality |
| 18-110_k141_196601 | Not-determined |
| 18-110_k141_196602 | Not-determined |
| 18-110_k141_69917 | Not-determined |
| 18-110_k141_150421 | Not-determined |
| 18-110_k141_93072 | Not-determined |
| 18-110_k141_127388 | Not-determined |
| 18-110_k141_104301 | Not-determined |
| 18-110_k141_127396 | Not-determined |
| 18-110_k141_93087 | Not-determined |
| 18-110_k141_161895 | Not-determined |
| 18-110_k141_196606 | Not-determined |
| 18-110_k141_161896 | Not-determined |
| 18-110_k141_69929 | Not-determined |
| 18-110_k141_253991 | Not-determined |
| 18-110_k141_208135 | Not-determined |
| 18-110_k141_35042 | Not-determined |
| 18-110_k141_81489 | Not-determined |
| 18-110_k141_253995 | Not-determined |
| 18-110_k141_115867 | Not-determined |
| 18-110_k141_253997 | Not-determined |
| 18-110_k141_46762 | Not-determined |
| 18-110_k141_242761 | Not-determined |
| 18-110_k141_127415 | Low-quality |
| 18-110_k141_208154 | Not-determined |
| 18-110_k141_196621 | Not-determined |
| 18-110_k141_196625 | Not-determined |
| 18-110_k141_150465 | Not-determined |
| 18-110_k141_127423 | Low-quality |
| 18-110_k141_161901 | Low-quality |
| 18-110_k141_35076 | Not-determined |
| 18-110_k141_139093 | Not-determined |
| 18-110_k141_35080 | Not-determined |
| 18-110_k141_12081 | Not-determined |
| 18-110_k141_58419 | Low-quality |
| 18-110_k141_93103 | Not-determined |
| 18-110_k141_231161 | Not-determined |
| 18-110_k141_93105 | Not-determined |
| 18-110_k141_231164 | Not-determined |
| 18-110_k141_150478 | Not-determined |
| 18-110_k141_139105 | Not-determined |
| 18-110_k141_139106 | Not-determined |
| 18-110_k141_219567 | Not-determined |
| 18-110_k141_93110 | Low-quality |
| 18-110_k141_219570 | Not-determined |
| 18-110_k141_779 | Low-quality |
| 18-110_k141_69951 | Not-determined |
| 18-110_k141_93112 | Not-determined |
| 18-110_k141_115897 | Not-determined |
| 18-110_k141_139112 | Not-determined |
| 18-110_k141_115898 | Not-determined |
| 18-110_k141_185031 | Not-determined |
| 18-110_k141_185032 | Not-determined |
| 18-110_k141_219572 | Not-determined |
| 18-110_k141_69956 | Not-determined |
| 18-110_k141_127434 | Not-determined |
| 18-110_k141_127444 | Not-determined |
| 18-110_k141_127446 | Not-determined |
| 18-110_k141_93134 | Not-determined |
| 18-110_k141_242783 | Not-determined |
| 18-110_k141_254010 | Low-quality |
| 18-110_k141_12107 | Not-determined |
| 18-110_k141_173662 | Not-determined |
| 18-110_k141_104331 | Not-determined |
| 18-110_k141_12110 | Low-quality |
| 18-110_k141_150487 | Not-determined |
| 18-110_k141_185046 | Not-determined |
| 18-110_k141_185047 | Not-determined |
| 18-110_k141_139137 | Low-quality |
| 18-110_k141_150491 | Not-determined |
| 18-110_k141_150493 | Not-determined |
| 18-110_k141_150495 | Not-determined |
| 18-110_k141_127465 | Low-quality |
| 18-110_k141_242796 | Not-determined |
| 18-110_k141_127476 | Not-determined |
| 18-110_k141_242799 | Not-determined |
| 18-110_k141_115920 | Not-determined |
| 18-110_k141_35099 | Not-determined |
| 18-110_k141_93149 | Not-determined |
| 18-110_k141_69991 | Not-determined |
| 18-110_k141_115921 | Not-determined |
| 18-110_k141_127482 | Not-determined |
| 18-110_k141_81528 | Not-determined |
| 18-110_k141_173675 | Not-determined |
| 18-110_k141_23842 | Not-determined |
| 18-110_k141_208200 | Not-determined |
| 18-110_k141_173683 | Not-determined |
| 18-110_k141_23850 | Not-determined |
| 18-110_k141_35116 | Not-determined |
| 18-110_k141_104346 | Not-determined |
| 18-110_k141_23855 | Not-determined |
| 18-110_k141_69998 | Not-determined |
| 18-110_k141_69999 | Not-determined |
| 18-110_k141_115940 | Not-determined |
| 18-110_k141_104352 | Not-determined |
| 18-110_k141_70003 | Not-determined |
| 18-110_k141_46835 | Not-determined |
| 18-110_k141_46838 | Not-determined |
| 18-110_k141_127497 | Not-determined |
| 18-110_k141_196658 | Low-quality |
| 18-110_k141_127500 | Not-determined |
| 18-110_k141_35127 | Not-determined |
| 18-110_k141_196662 | Not-determined |
| 18-110_k141_185088 | Not-determined |
| 18-110_k141_115952 | Not-determined |
| 18-110_k141_185091 | Low-quality |
| 18-110_k141_35133 | Not-determined |
| 18-110_k141_70009 | Not-determined |
| 18-110_k141_23867 | Low-quality |
| 18-110_k141_58438 | Not-determined |
| 18-110_k141_127501 | Low-quality |
| 18-110_k141_231222 | Not-determined |
| 18-110_k141_173711 | Not-determined |
| 18-110_k141_161967 | Low-quality |
| 18-110_k141_58448 | Not-determined |
| 18-110_k141_173713 | Low-quality |
| 18-110_k141_150529 | Not-determined |
| 18-110_k141_161973 | Not-determined |
| 18-110_k141_231238 | Not-determined |
| 18-110_k141_35151 | Not-determined |
| 18-110_k141_104375 | Not-determined |
| 18-110_k141_231242 | Not-determined |
| 18-110_k141_173723 | Not-determined |
| 18-110_k141_231247 | Not-determined |
| 18-110_k141_254043 | Not-determined |
| 18-110_k141_104382 | Not-determined |
| 18-110_k141_12144 | Not-determined |
| 18-110_k141_139167 | Not-determined |
| 18-110_k141_254045 | Not-determined |
| 18-110_k141_23891 | Not-determined |
| 18-110_k141_173730 | Not-determined |
| 18-110_k141_81584 | Not-determined |
| 18-110_k141_139175 | Not-determined |
| 18-110_k141_242813 | Not-determined |
| 18-110_k141_81589 | Not-determined |
| 18-110_k141_219643 | Not-determined |
| 18-110_k141_242816 | Not-determined |
| 18-110_k141_161982 | Low-quality |
| 18-110_k141_150552 | Not-determined |
| 18-110_k141_219648 | Not-determined |
| 18-110_k141_219650 | Not-determined |
| 18-110_k141_219652 | Not-determined |
| 18-110_k141_231264 | Not-determined |
| 18-110_k141_231267 | Not-determined |
| 18-110_k141_70043 | Not-determined |
| 18-110_k141_231271 | Not-determined |
| 18-110_k141_70048 | Not-determined |
| 18-110_k141_150560 | Not-determined |
| 18-110_k141_161991 | Not-determined |
| 18-110_k141_231285 | Not-determined |
| 18-110_k141_139204 | Not-determined |
| 18-110_k141_219672 | Not-determined |
| 18-110_k141_231287 | Not-determined |
| 18-110_k141_231292 | Not-determined |
| 18-110_k141_196669 | Not-determined |
| 18-110_k141_150568 | Not-determined |
| 18-110_k141_162003 | Not-determined |
| 18-110_k141_219675 | Not-determined |
| 18-110_k141_208240 | Not-determined |
| 18-110_k141_219681 | Low-quality |
| 18-110_k141_23920 | Not-determined |
| 18-110_k141_23927 | Not-determined |
| 18-110_k141_173746 | Not-determined |
| 18-110_k141_70057 | Not-determined |
| 18-110_k141_242849 | Not-determined |
| 18-110_k141_81600 | Not-determined |
| 18-110_k141_162016 | Low-quality |
| 18-110_k141_242853 | Not-determined |
| 18-110_k141_104396 | Not-determined |
| 18-110_k141_150573 | Not-determined |
| 18-110_k141_104400 | Not-determined |
| 18-110_k141_254091 | Not-determined |
| 18-110_k141_242864 | Not-determined |
| 18-110_k141_139232 | Not-determined |
| 18-110_k141_231322 | Not-determined |
| 18-110_k141_196691 | Not-determined |
| 18-110_k141_242873 | Not-determined |
| 18-110_k141_208257 | Not-determined |
| 18-110_k141_23938 | Not-determined |
| 18-110_k141_242884 | Not-determined |
| 18-110_k141_254095 | Not-determined |
| 18-110_k141_265770 | Not-determined |
| 18-110_k141_139240 | Not-determined |
| 18-110_k141_254100 | Not-determined |
| 18-110_k141_46897 | Low-quality |
| 18-110_k141_231333 | Not-determined |
| 18-110_k141_265774 | Not-determined |
| 18-110_k141_162026 | Not-determined |
| 18-110_k141_242901 | Not-determined |
| 18-110_k141_196707 | Not-determined |
| 18-110_k141_150592 | Not-determined |
| 18-110_k141_265777 | Not-determined |
| 18-110_k141_104431 | Low-quality |
| 18-110_k141_242911 | Not-determined |
| 18-110_k141_173791 | Not-determined |
| 18-102-final-viral-combined_k141_181095\|\|0_partial | Medium-quality |
| 18-110_k141_231342 | Low-quality |
| 18-110_k141_231343 | Not-determined |
| 18-110_k141_265787 | Not-determined |
| 18-110_k141_242917 | Not-determined |
| 18-110_k141_265789 | Not-determined |
| 18-110_k141_139260 | Not-determined |
| 18-110_k141_58520 | Not-determined |
| 18-110_k141_231345 | Low-quality |
| 18-110_k141_139261 | Not-determined |
| 18-110_k141_104435 | Not-determined |
| 18-110_k141_231347 | Not-determined |
| 18-110_k141_35216 | Not-determined |
| 18-110_k141_231351 | Not-determined |
| 18-110_k141_35218 | Not-determined |
| 18-110_k141_139266 | Low-quality |
| 18-110_k141_104443 | Not-determined |
| 18-110_k141_12227 | Not-determined |
| 18-110_k141_70096 | Not-determined |
| 18-110_k141_58527 | Not-determined |
| 18-110_k141_173820 | Low-quality |
| 18-110_k141_173821 | Not-determined |
| 18-110_k141_35236 | Not-determined |
| 18-110_k141_231357 | Not-determined |
| 18-110_k141_254124 | Not-determined |
| 18-110_k141_150616 | Not-determined |
| 18-110_k141_116020 | Not-determined |
| 18-110_k141_150619 | Not-determined |
| 18-110_k141_208287 | Not-determined |
| 18-110_k141_139286 | Not-determined |
| 18-110_k141_116023 | Not-determined |
| 18-110_k141_139289 | Not-determined |
| 18-110_k141_46902 | Not-determined |
| 18-110_k141_231373 | Not-determined |
| 18-110_k141_924 | Not-determined |
| 18-110_k141_929 | Low-quality |
| 18-110_k141_208298 | Not-determined |
| 18-110_k141_231385 | Not-determined |
| 18-110_k141_185159 | Not-determined |
| 18-110_k141_242952 | Not-determined |
| 18-110_k141_127532 | Low-quality |
| 18-110_k141_185162 | Not-determined |
| 18-110_k141_116034 | Not-determined |
| 18-110_k141_937 | Not-determined |
| 18-110_k141_58543 | Not-determined |
| 18-110_k141_35274 | Not-determined |
| 18-110_k141_208311 | Not-determined |
| 18-110_k141_150663 | Not-determined |
| 18-110_k141_231398 | Not-determined |
| 18-110_k141_35276 | Not-determined |
| 18-110_k141_173847 | Not-determined |
| 18-110_k141_231401 | Not-determined |
| 18-110_k141_104459 | Not-determined |
| 18-110_k141_254142 | Not-determined |
| 18-110_k141_58556 | Not-determined |
| 18-110_k141_173849 | Not-determined |
| 18-110_k141_104477 | Not-determined |
| 18-110_k141_196774 | Not-determined |
| 18-110_k141_231410 | Low-quality |
| 18-110_k141_70144 | Not-determined |
| 18-110_k141_70145 | Not-determined |
| 18-110_k141_173862 | Not-determined |
| 18-110_k141_254160 | Not-determined |
| 18-110_k141_104483 | Not-determined |
| 18-110_k141_35286 | Low-quality |
| 18-110_k141_104485 | Not-determined |
| 18-110_k141_173869 | Low-quality |
| 18-110_k141_81657 | Not-determined |
| 18-110_k141_185183 | Not-determined |
| 18-110_k141_139310 | Not-determined |
| 18-110_k141_185184 | Not-determined |
| 18-110_k141_81660 | Not-determined |
| 18-110_k141_242968 | Not-determined |
| 18-110_k141_173887 | Not-determined |
| 18-110_k141_70156 | Not-determined |
| 18-110_k141_208330 | Not-determined |
| 18-110_k141_173892 | Not-determined |
| 18-110_k141_173894 | Not-determined |
| 18-110_k141_104505 | Not-determined |
| 18-110_k141_58587 | Not-determined |
| 18-110_k141_208335 | Low-quality |
| 18-110_k141_242989 | Not-determined |
| 18-110_k141_196788 | Not-determined |
| 18-110_k141_12287 | Not-determined |
| 18-110_k141_12289 | Not-determined |
| 18-110_k141_162080 | Not-determined |
| 18-110_k141_35304 | Low-quality |
| 18-110_k141_173906 | Not-determined |
| 18-110_k141_150705 | Not-determined |
| 18-110_k141_81691 | Not-determined |
| 18-110_k141_231432 | Not-determined |
| 18-110_k141_208350 | Not-determined |
| 18-110_k141_23966 | Not-determined |
| 18-110_k141_12302 | Not-determined |
| 18-110_k141_185215 | Not-determined |
| 18-110_k141_173925 | Not-determined |
| 18-110_k141_150712 | Not-determined |
| 18-110_k141_58600 | Not-determined |
| 18-110_k141_231439 | Not-determined |
| 18-110_k141_58602 | Not-determined |
| 18-110_k141_196801 | Not-determined |
| 18-110_k141_150718 | Not-determined |
| 18-110_k141_58609 | Not-determined |
| 18-110_k141_231447 | Not-determined |
| 18-110_k141_231448 | Not-determined |
| 18-110_k141_58611 | Not-determined |
| 18-110_k141_243007 | Not-determined |
| 18-110_k141_93158 | Not-determined |
| 18-110_k141_81703 | Not-determined |
| 18-110_k141_243012 | Not-determined |
| 18-110_k141_254201 | Not-determined |
| 18-110_k141_81708 | Not-determined |
| 18-110_k141_46921 | Not-determined |
| 18-110_k141_46923 | Low-quality |
| 18-110_k141_12313 | Not-determined |
| 18-110_k141_116050 | Not-determined |
| 18-110_k141_231462 | Not-determined |
| 18-110_k141_265816 | Low-quality |
| 18-110_k141_70167 | Not-determined |
| 18-110_k141_58633 | Not-determined |
| 18-110_k141_46930 | Low-quality |
| 18-110_k141_58636 | Not-determined |
| 18-110_k141_139331 | Not-determined |
| 18-110_k141_127587 | Not-determined |
| 18-110_k141_139333 | Not-determined |
| 18-110_k141_265825 | Not-determined |
| 18-110_k141_93199 | Not-determined |
| 18-110_k141_173957 | Not-determined |
| 18-110_k141_23999 | Not-determined |
| 18-110_k141_243037 | Not-determined |
| 18-110_k141_46943 | Not-determined |
| 18-110_k141_231473 | Not-determined |
| 18-110_k141_265850 | Not-determined |
| 18-110_k141_127595 | Not-determined |
| 18-110_k141_139355 | Not-determined |
| 18-110_k141_243040 | Not-determined |
| 18-110_k141_12324 | Not-determined |
| 18-110_k141_127601 | Not-determined |
| 18-110_k141_243046 | Not-determined |
| 18-110_k141_127602 | Not-determined |
| 18-110_k141_81723 | Low-quality |
| 18-110_k141_231483 | Not-determined |
| 18-110_k141_104526 | Not-determined |
| 18-110_k141_173983 | Low-quality |
| 18-110_k141_70196 | Not-determined |
| 18-110_k141_81733 | Not-determined |
| 18-110_k141_70198 | Not-determined |
| 18-110_k141_116080 | Not-determined |
| 18-110_k141_81735 | Not-determined |
| 18-110_k141_104534 | Not-determined |
| 18-110_k141_162155 | Low-quality |
| 18-110_k141_116086 | Not-determined |
| 18-110_k141_173991 | Not-determined |
| 18-110_k141_139373 | Not-determined |
| 18-110_k141_24016 | Not-determined |
| 18-110_k141_35313 | Not-determined |
| 18-110_k141_173993 | Not-determined |
| 18-110_k141_58660 | Not-determined |
| 18-110_k141_116090 | Not-determined |
| 18-110_k141_104545 | Not-determined |
| 18-110_k141_127626 | Not-determined |
| 18-110_k141_24031 | Low-quality |
| 18-110_k141_70212 | Not-determined |
| 18-110_k141_127631 | Not-determined |
| 18-110_k141_243063 | Not-determined |
| 18-110_k141_70217 | Not-determined |
| 18-110_k141_254255 | Low-quality |
| 18-110_k141_35327 | Not-determined |
| 18-110_k141_139391 | Not-determined |
| 18-110_k141_46969 | Not-determined |
| 18-110_k141_254257 | Not-determined |
| 18-110_k141_162176 | Not-determined |
| 18-110_k141_139393 | Not-determined |
| 18-110_k141_70220 | Not-determined |
| 18-110_k141_185282 | Not-determined |
| 18-110_k141_254262 | Not-determined |
| 18-110_k141_185285 | Not-determined |
| 18-110_k141_254264 | Not-determined |
| 18-110_k141_127641 | Low-quality |
| 18-110_k141_208394 | Not-determined |
| 18-110_k141_104563 | Not-determined |
| 18-110_k141_243079 | Not-determined |
| 18-110_k141_127648 | Not-determined |
| 18-110_k141_208401 | Not-determined |
| 18-110_k141_162187 | Not-determined |
| 18-110_k141_989 | Not-determined |
| 18-110_k141_196846 | Not-determined |
| 18-110_k141_58675 | Not-determined |
| 18-110_k141_208410 | Low-quality |
| 18-110_k141_231530 | Not-determined |
| 18-110_k141_150794 | Not-determined |
| 18-110_k141_46979 | Not-determined |
| 18-110_k141_243089 | Not-determined |

| | |
|---|---|
| 18-110_k141_231533 | Low-quality |
| 18-110_k141_243092 | Not-determined |
| 18-110_k141_139420 | Not-determined |
| 18-110_k141_139421 | Not-determined |
| 18-110_k141_35349 | Not-determined |
| 18-110_k141_46992 | Not-determined |
| 18-110_k141_104575 | Not-determined |
| 18-110_k141_70240 | Not-determined |
| 18-110_k141_70242 | Not-determined |
| 18-110_k141_58695 | Not-determined |
| 18-110_k141_116118 | Low-quality |
| 18-110_k141_208430 | Not-determined |
| 18-110_k141_185316 | Not-determined |
| 18-110_k141_81779 | Not-determined |
| 18-110_k141_1002 | Not-determined |
| 18-110_k141_254290 | Not-determined |
| 18-110_k141_116126 | Not-determined |
| 18-110_k141_150834 | Low-quality |
| 18-110_k141_150835 | Not-determined |
| 18-110_k141_208447 | Not-determined |
| 18-110_k141_24078 | Not-determined |
| 18-110_k141_208448 | Not-determined |
| 18-110_k141_1008 | Not-determined |
| 18-110_k141_116133 | Not-determined |
| 18-110_k141_243106 | Low-quality |
| 18-110_k141_47022 | Not-determined |
| 18-110_k141_1012 | Not-determined |
| 18-110_k141_35380 | Not-determined |
| 18-110_k141_265882 | Not-determined |
| 18-110_k141_185329 | Not-determined |
| 18-110_k141_35383 | Not-determined |
| 18-110_k141_47026 | Not-determined |
| 18-110_k141_104586 | Not-determined |
| 18-110_k141_81785 | Not-determined |
| 18-110_k141_47034 | Low-quality |
| 18-110_k141_243118 | Not-determined |
| 18-110_k141_265886 | Not-determined |
| 18-110_k141_139464 | Not-determined |
| 18-110_k141_243120 | Not-determined |
| 18-110_k141_265889 | Not-determined |
| 18-110_k141_231557 | Not-determined |
| 18-110_k141_139466 | Low-quality |
| 18-110_k141_243122 | Low-quality |
| 18-110_k141_196883 | Low-quality |
| 18-110_k141_127678 | Not-determined |
| 18-110_k141_243126 | Not-determined |
| 18-110_k141_231565 | Not-determined |
| 18-110_k141_265904 | Not-determined |
| 18-110_k141_127682 | Not-determined |
| 18-110_k141_58716 | Not-determined |
| 18-110_k141_81799 | Not-determined |
| 18-110_k141_93292 | Not-determined |
| 18-110_k141_116168 | Not-determined |
| 18-110_k141_93295 | Not-determined |
| 18-110_k141_139475 | Not-determined |
| 18-110_k141_208494 | Not-determined |
| 18-110_k141_243144 | Not-determined |
| 18-110_k141_231580 | Not-determined |
| 18-110_k141_116176 | Not-determined |
| 18-110_k141_104596 | Not-determined |
| 18-110_k141_231583 | Low-quality |
| 18-110_k141_185365 | Not-determined |
| 18-110_k141_58732 | Not-determined |
| 18-110_k141_58733 | Not-determined |
| 18-110_k141_81807 | Not-determined |
| 18-110_k141_116179 | Not-determined |
| 18-110_k141_254310 | Not-determined |
| 18-110_k141_116180 | Not-determined |
| 18-110_k141_104600 | Not-determined |
| 18-110_k141_116184 | Low-quality |
| 18-110_k141_231601 | Not-determined |
| 18-110_k141_185383 | Not-determined |
| 18-110_k141_127711 | Not-determined |
| 18-110_k141_127713 | Not-determined |
| 18-110_k141_58755 | Not-determined |
| 18-110_k141_208507 | Low-quality |
| 18-110_k141_265910 | Low-quality |
| 18-110_k141_127718 | Not-determined |
| 18-110_k141_127719 | Not-determined |
| 18-110_k141_265918 | Not-determined |
| 18-110_k141_265919 | Not-determined |
| 18-110_k141_1021 | Not-determined |
| 18-110_k141_58762 | Not-determined |
| 18-110_k141_1022 | Not-determined |
| 18-110_k141_265923 | Not-determined |
| 18-110_k141_127726 | Not-determined |
| 18-110_k141_93329 | Not-determined |
| 18-110_k141_35419 | Low-quality |
| 18-110_k141_231615 | Low-quality |
| 18-110_k141_58770 | Not-determined |
| 18-110_k141_208534 | Not-determined |
| 18-110_k141_81841 | Not-determined |
| 18-110_k141_70326 | Not-determined |
| 18-110_k141_231621 | Not-determined |
| 18-110_k141_231623 | Not-determined |
| 18-110_k141_231624 | Not-determined |
| 18-110_k141_196896 | Not-determined |
| 18-110_k141_208539 | Not-determined |
| 18-110_k141_35435 | Not-determined |
| 18-110_k141_104642 | Not-determined |
| 18-110_k141_58779 | Not-determined |
| 18-110_k141_116210 | Low-quality |
| 18-110_k141_127732 | Not-determined |
| 18-110_k141_116212 | Not-determined |
| 18-110_k141_150883 | Not-determined |
| 18-110_k141_243191 | Not-determined |
| 18-110_k141_231632 | Low-quality |
| 18-110_k141_81858 | Low-quality |
| 18-110_k141_47058 | Not-determined |
| 18-110_k141_219690 | Not-determined |
| 18-110_k141_116220 | Not-determined |
| 18-110_k141_70339 | Not-determined |
| 18-110_k141_104659 | Not-determined |
| 18-110_k141_58792 | Not-determined |
| 18-110_k141_104660 | Low-quality |
| 18-110_k141_35454 | Not-determined |
| 18-110_k141_1032 | Low-quality |
| 18-110_k141_185442 | Not-determined |
| 18-110_k141_150887 | Low-quality |
| 18-110_k141_35456 | Not-determined |
| 18-110_k141_24131 | Not-determined |
| 18-110_k141_12347 | Not-determined |
| 18-110_k141_93357 | Low-quality |
| 18-110_k141_81872 | Not-determined |
| 18-110_k141_243204 | Not-determined |
| 18-110_k141_35475 | Low-quality |
| 18-110_k141_162211 | Not-determined |
| 18-110_k141_104667 | Low-quality |
| 18-110_k141_185462 | Not-determined |
| 18-110_k141_93371 | Not-determined |
| 18-110_k141_104673 | Not-determined |
| 18-110_k141_1059 | Not-determined |
| 18-110_k141_243213 | Not-determined |
| 18-110_k141_243216 | Low-quality |
| 18-110_k141_150905 | Not-determined |
| 18-110_k141_243224 | Not-determined |
| 18-110_k141_70365 | Not-determined |
| 18-110_k141_70366 | Not-determined |
| 18-110_k141_12363 | Not-determined |
| 18-110_k141_93380 | Not-determined |
| 18-110_k141_93381 | Not-determined |
| 18-110_k141_196910 | Not-determined |
| 18-110_k141_70371 | Not-determined |
| 18-110_k141_104689 | Not-determined |
| 18-110_k141_116225 | Not-determined |
| 18-110_k141_162234 | Not-determined |
| 18-110_k141_196915 | Low-quality |
| 18-110_k141_219701 | Not-determined |
| 18-110_k141_47088 | Not-determined |
| 18-110_k141_162235 | Not-determined |
| 18-110_k141_47089 | Not-determined |
| 18-110_k141_162239 | Not-determined |
| 18-110_k141_219705 | Not-determined |
| 18-110_k141_12371 | Not-determined |
| 18-110_k141_196922 | Low-quality |
| 18-110_k141_150920 | Not-determined |
| 18-110_k141_104698 | Low-quality |
| 18-110_k141_185474 | Not-determined |
| 18-110_k141_196926 | Low-quality |
| 18-110_k141_185476 | Not-determined |
| 18-110_k141_208614 | Not-determined |
| 18-110_k141_70377 | Low-quality |
| 18-110_k141_139510 | Not-determined |
| 18-110_k141_1073 | Not-determined |
| 18-110_k141_116229 | Not-determined |
| 18-110_k141_1077 | Not-determined |
| 18-110_k141_150924 | Not-determined |
| 18-110_k141_219724 | Not-determined |
| 18-110_k141_196931 | Not-determined |
| 18-110_k141_104714 | Not-determined |
| 18-110_k141_24160 | Not-determined |
| 18-110_k141_35490 | Not-determined |
| 18-110_k141_58795 | Not-determined |
| 18-110_k141_243227 | Not-determined |
| 18-110_k141_12387 | Not-determined |
| 18-110_k141_104719 | Not-determined |
| 18-110_k141_196944 | Not-determined |
| 18-110_k141_104722 | Low-quality |
| 18-110_k141_139536 | Not-determined |
| 18-110_k141_93425 | Not-determined |
| 18-110_k141_104727 | Not-determined |
| 18-110_k141_150940 | Not-determined |
| 18-110_k141_35500 | Not-determined |
| 18-110_k141_243234 | Not-determined |
| 18-110_k141_93432 | Not-determined |
| 18-110_k141_150942 | Not-determined |
| 18-110_k141_196959 | Not-determined |
| 18-110_k141_219745 | Not-determined |
| 18-110_k141_47126 | Not-determined |
| 18-110_k141_185508 | Not-determined |
| 18-110_k141_93441 | Low-quality |
| 18-110_k141_208639 | Not-determined |
| 18-110_k141_243240 | Not-determined |
| 18-110_k141_24179 | Not-determined |
| 18-110_k141_196964 | Low-quality |
| 18-110_k141_243246 | Not-determined |
| 18-110_k141_116247 | Low-quality |
| 18-110_k141_58818 | Not-determined |
| 18-110_k141_35513 | Not-determined |
| 18-110_k141_35519 | Not-determined |
| 18-110_k141_70398 | Not-determined |
| 18-110_k141_70401 | Not-determined |
| 18-110_k141_104766 | Not-determined |
| 18-110_k141_35527 | Not-determined |
| 18-110_k141_104767 | Not-determined |
| 18-110_k141_208647 | Not-determined |
| 18-110_k141_219763 | Not-determined |
| 18-110_k141_104774 | Not-determined |
| 18-110_k141_185532 | Low-quality |
| 18-110_k141_265958 | Not-determined |
| 18-110_k141_254317 | Not-determined |
| 18-110_k141_127774 | Not-determined |
| 18-110_k141_70416 | Not-determined |
| 18-110_k141_93470 | Not-determined |
| 18-110_k141_93471 | Not-determined |
| 18-110_k141_254326 | Not-determined |
| 18-110_k141_162284 | Not-determined |
| 18-110_k141_196989 | Not-determined |
| 18-110_k141_127784 | Not-determined |
| 18-110_k141_162289 | Not-determined |
| 18-110_k141_185552 | Not-determined |
| 18-110_k141_58859 | Not-determined |
| 18-110_k141_93477 | Not-determined |
| 18-110_k141_243262 | Not-determined |
| 18-110_k141_93479 | Not-determined |
| 18-110_k141_196999 | Not-determined |
| 18-110_k141_162298 | Not-determined |
| 18-110_k141_254330 | Not-determined |
| 18-110_k141_208682 | Not-determined |
| 18-110_k141_116278 | Low-quality |
| 18-110_k141_127803 | Not-determined |
| 18-110_k141_139567 | Not-determined |
| 18-110_k141_104801 | Not-determined |
| 18-110_k141_185568 | Not-determined |
| 18-110_k141_197003 | Not-determined |
| 18-110_k141_162307 | Low-quality |
| 18-110_k141_24232 | Not-determined |
| 18-110_k141_93486 | Low-quality |
| 18-110_k141_127805 | Not-determined |
| 18-110_k141_162312 | Not-determined |
| 18-110_k141_162316 | Not-determined |
| 18-110_k141_24234 | Not-determined |
| 18-110_k141_197004 | Low-quality |
| 18-110_k141_197007 | Low-quality |
| 18-110_k141_127810 | Not-determined |
| 18-110_k141_254339 | Not-determined |
| 18-110_k141_24241 | Not-determined |
| 18-110_k141_208700 | Not-determined |
| 18-110_k141_104824 | Not-determined |
| 18-110_k141_243270 | Not-determined |
| 18-110_k141_139584 | Low-quality |
| 18-110_k141_243272 | Not-determined |
| 18-110_k141_219785 | Not-determined |
| 18-110_k141_127826 | Not-determined |
| 18-110_k141_127828 | Not-determined |
| 18-110_k141_219787 | Not-determined |
| 18-110_k141_139590 | Not-determined |
| 18-110_k141_35551 | Not-determined |
| 18-110_k141_243278 | Not-determined |
| 18-110_k141_243279 | Not-determined |
| 18-110_k141_208713 | Not-determined |
| 18-110_k141_208716 | Not-determined |
| 18-110_k141_24262 | Not-determined |
| 18-110_k141_139595 | Low-quality |
| 18-110_k141_24268 | Low-quality |
| 18-110_k141_174021 | Not-determined |
| 18-110_k141_254341 | Not-determined |
| 18-110_k141_243282 | Not-determined |
| 18-110_k141_93527 | Not-determined |
| 18-110_k141_139603 | Not-determined |
| 18-110_k141_70456 | Not-determined |
| 18-110_k141_243287 | Not-determined |
| 18-110_k141_208722 | Not-determined |
| 18-110_k141_116318 | Not-determined |
| 18-110_k141_254349 | Not-determined |
| 18-110_k141_139610 | Not-determined |
| 18-110_k141_139612 | Not-determined |
| 18-110_k141_70462 | Not-determined |
| 18-110_k141_254355 | Not-determined |
| 18-110_k141_35554 | Low-quality |
| 18-110_k141_81967 | Not-determined |
| 18-110_k141_104843 | Not-determined |
| 18-110_k141_70470 | Not-determined |
| 18-110_k141_70471 | Not-determined |
| 18-110_k141_81973 | Not-determined |
| 18-110_k141_231702 | Not-determined |
| 18-110_k141_58921 | Not-determined |
| 18-110_k141_58923 | Not-determined |
| 18-110_k141_139617 | Not-determined |
| 18-110_k141_231708 | Not-determined |
| 18-110_k141_231709 | Not-determined |
| 18-110_k141_139624 | Not-determined |
| 18-110_k141_116337 | Not-determined |
| 18-110_k141_254366 | Not-determined |
| 18-110_k141_116341 | Not-determined |
| 18-110_k141_231717 | Not-determined |
| 18-110_k141_70479 | Not-determined |
| 18-110_k141_174056 | Not-determined |
| 18-110_k141_208746 | Not-determined |
| 18-110_k141_208747 | Not-determined |
| 18-110_k141_70481 | Not-determined |
| 18-110_k141_116347 | Not-determined |
| 18-110_k141_231729 | Not-determined |
| 18-110_k141_231730 | Not-determined |
| 18-110_k141_70486 | Not-determined |
| 18-110_k141_208750 | Not-determined |
| 18-110_k141_139640 | Low-quality |
| 18-110_k141_93552 | Not-determined |
| 18-110_k141_139641 | Not-determined |
| 18-110_k141_243309 | Not-determined |
| 18-110_k141_70491 | Not-determined |
| 18-110_k141_58938 | Not-determined |
| 18-110_k141_243310 | Not-determined |
| 18-110_k141_254381 | Not-determined |
| 18-110_k141_116359 | Not-determined |
| 18-110_k141_254383 | Not-determined |
| 18-110_k141_70495 | Not-determined |
| 18-110_k141_70496 | Low-quality |
| 18-110_k141_93558 | Not-determined |
| 18-110_k141_81997 | Not-determined |
| 18-110_k141_58946 | Not-determined |
| 18-110_k141_93561 | Not-determined |
| 18-110_k141_58948 | Not-determined |
| 18-110_k141_162344 | Not-determined |
| 18-110_k141_70503 | Not-determined |
| 18-110_k141_162345 | Not-determined |
| 18-110_k141_70504 | Not-determined |
| 18-110_k141_231750 | Not-determined |
| 18-110_k141_47202 | Not-determined |
| 18-110_k141_162360 | Not-determined |
| 18-110_k141_116370 | Not-determined |
| 18-110_k141_70523 | Not-determined |
| 18-110_k141_174092 | Not-determined |
| 18-110_k141_174093 | Not-determined |
| 18-110_k141_139674 | Not-determined |
| 18-110_k141_35581 | Not-determined |
| 18-110_k141_150959 | Not-determined |
| 18-110_k141_185617 | Not-determined |
| 18-110_k141_254397 | Not-determined |
| 18-110_k141_35586 | Not-determined |
| 18-110_k141_93582 | Not-determined |
| 18-110_k141_70538 | Not-determined |
| 18-110_k141_47212 | Not-determined |
| 18-110_k141_231773 | Not-determined |
| 18-110_k141_231774 | Not-determined |
| 18-110_k141_116383 | Not-determined |
| 18-110_k141_116386 | Low-quality |
| 18-110_k141_24316 | Not-determined |
| 18-110_k141_93595 | Not-determined |
| 18-110_k141_116393 | Not-determined |
| 18-110_k141_35595 | Not-determined |
| 18-110_k141_150966 | Not-determined |
| 18-110_k141_174110 | Not-determined |
| 18-110_k141_70544 | Not-determined |
| 18-110_k141_243322 | Low-quality |
| 18-110_k141_116398 | Not-determined |
| 18-110_k141_208765 | Not-determined |
| 18-110_k141_47229 | Low-quality |
| 18-110_k141_162392 | Not-determined |
| 18-110_k141_162393 | Not-determined |
| 18-110_k141_185644 | Not-determined |
| 18-110_k141_254428 | Not-determined |
| 18-110_k141_185647 | Not-determined |
| 18-110_k141_47244 | Not-determined |
| 18-110_k141_12403 | Not-determined |
| 18-110_k141_231806 | Not-determined |
| 18-110_k141_219825 | Not-determined |
| 18-110_k141_35625 | Not-determined |
| 18-110_k141_162397 | Not-determined |
| 18-110_k141_12417 | Not-determined |
| 18-110_k141_59003 | Not-determined |
| 18-110_k141_150998 | Not-determined |
| 18-110_k141_127895 | Not-determined |
| 18-110_k141_254445 | Not-determined |
| 18-110_k141_162410 | Not-determined |
| 18-110_k141_59008 | Not-determined |
| 18-110_k141_12422 | Low-quality |
| 18-110_k141_70565 | Not-determined |
| 18-110_k141_265986 | Not-determined |
| 18-110_k141_70567 | Not-determined |
| 18-110_k141_162416 | Low-quality |
| 18-110_k141_93629 | Not-determined |
| 18-110_k141_35645 | Low-quality |
| 18-110_k141_127912 | Not-determined |
| 18-110_k141_59017 | Low-quality |
| 18-110_k141_151010 | Not-determined |
| 18-110_k141_47268 | Not-determined |
| 18-110_k141_219847 | Not-determined |
| 18-110_k141_12434 | Not-determined |
| 18-110_k141_35655 | Not-determined |
| 18-110_k141_59020 | Not-determined |
| 18-110_k141_93641 | Low-quality |
| 18-110_k141_219850 | Not-determined |
| 18-110_k141_70574 | Not-determined |
| 18-110_k141_70578 | Not-determined |
| 18-110_k141_162433 | Not-determined |
| 18-110_k141_254458 | Not-determined |
| 18-110_k141_243369 | Not-determined |
| 18-110_k141_59027 | Not-determined |
| 18-110_k141_243380 | Not-determined |
| 18-110_k141_59032 | Not-determined |
| 18-110_k141_59034 | Not-determined |
| 18-110_k141_12443 | Not-determined |
| 18-110_k141_82070 | Not-determined |
| 18-110_k141_47281 | Low-quality |
| 18-110_k141_139730 | Not-determined |
| 18-110_k141_243382 | Not-determined |
| 18-110_k141_231821 | Not-determined |
| 18-110_k141_70593 | Low-quality |
| 18-110_k141_265996 | Not-determined |
| 18-110_k141_243395 | Not-determined |
| 18-110_k141_185693 | Not-determined |
| 18-110_k141_243396 | Not-determined |
| 18-110_k141_208791 | Not-determined |
| 18-110_k141_59047 | Not-determined |
| 18-110_k141_139732 | Not-determined |
| 18-110_k141_35662 | Not-determined |
| 18-110_k141_185695 | Not-determined |
| 18-110_k141_174166 | Not-determined |
| 18-110_k141_35666 | Not-determined |
| 18-110_k141_243402 | Not-determined |
| 18-110_k141_59053 | Not-determined |
| 18-110_k141_59056 | Not-determined |
| 18-110_k141_151048 | Low-quality |
| 18-110_k141_162460 | Not-determined |
| 18-110_k141_35676 | Not-determined |
| 18-110_k141_1194 | Not-determined |
| 18-110_k141_59062 | Not-determined |
| 18-110_k141_231838 | Not-determined |
| 18-110_k141_174173 | Low-quality |
| 18-110_k141_24367 | Not-determined |
| 18-110_k141_116425 | Not-determined |
| 18-110_k141_70612 | Not-determined |
| 18-110_k141_70613 | Low-quality |
| 18-110_k141_24372 | Not-determined |
| 18-110_k141_243420 | Not-determined |
| 18-110_k141_231842 | Low-quality |
| 18-110_k141_208817 | Not-determined |
| 18-110_k141_59070 | Not-determined |
| 18-110_k141_24379 | Not-determined |
| 18-110_k141_59073 | Not-determined |
| 18-110_k141_70625 | Not-determined |
| 18-110_k141_47298 | Not-determined |
| 18-110_k141_24384 | Not-determined |
| 18-110_k141_59074 | Low-quality |
| 18-110_k141_59075 | Not-determined |
| 18-110_k141_151058 | Not-determined |
| 18-110_k141_35690 | Not-determined |
| 18-110_k141_35692 | Low-quality |
| 18-110_k141_219881 | Not-determined |
| 18-110_k141_219882 | Not-determined |
| 18-110_k141_219883 | Not-determined |
| 18-110_k141_231853 | Not-determined |
| 18-110_k141_1211 | Low-quality |
| 18-110_k141_47316 | Not-determined |
| 18-110_k141_47319 | Low-quality |
| 18-110_k141_185716 | Not-determined |
| 18-110_k141_82098 | Not-determined |
| 18-110_k141_1217 | Not-determined |
| 18-110_k141_174200 | Not-determined |
| 18-110_k141_35703 | Not-determined |
| 18-110_k141_162492 | Not-determined |
| 18-110_k141_174203 | Not-determined |
| 18-110_k141_139769 | Not-determined |
| 18-110_k141_174204 | Not-determined |
| 18-110_k141_162494 | Not-determined |
| 18-110_k141_82101 | Not-determined |
| 18-110_k141_70628 | Low-quality |
| 18-110_k141_162495 | Not-determined |
| 18-110_k141_174207 | Not-determined |
| 18-110_k141_59103 | Low-quality |
| 18-110_k141_47322 | Not-determined |
| 18-110_k141_231860 | Not-determined |
| 18-110_k141_151076 | Not-determined |
| 18-110_k141_104937 | Not-determined |
| 18-110_k141_70640 | Not-determined |
| 18-110_k141_12475 | Not-determined |
| 18-110_k141_139778 | Not-determined |
| 18-110_k141_24423 | Not-determined |
| 18-110_k141_266008 | Not-determined |
| 18-110_k141_219911 | Not-determined |
| 18-110_k141_266010 | Not-determined |
| 18-110_k141_59117 | Not-determined |
| 18-110_k141_70646 | Low-quality |
| 18-110_k141_151092 | Not-determined |
| 18-110_k141_70648 | Not-determined |
| 18-110_k141_162522 | Not-determined |
| 18-110_k141_231873 | Not-determined |
| 18-110_k141_162524 | Not-determined |
| 18-110_k141_104941 | Not-determined |
| 18-110_k141_35721 | Not-determined |
| 18-110_k141_266014 | Not-determined |
| 18-110_k141_70657 | Low-quality |
| 18-110_k141_139800 | Not-determined |
| 18-110_k141_139801 | Not-determined |
| 18-110_k141_12493 | Not-determined |
| 18-110_k141_12495 | Not-determined |
| 18-110_k141_1230 | Not-determined |
| 18-110_k141_162540 | Not-determined |
| 18-110_k141_162541 | Not-determined |
| 18-110_k141_82127 | Not-determined |
| 18-110_k141_82128 | Not-determined |
| 18-110_k141_59149 | Not-determined |
| 18-110_k141_104964 | Not-determined |
| 18-110_k141_104966 | Not-determined |
| 18-110_k141_139826 | Not-determined |
| 18-110_k141_231908 | Not-determined |
| 18-110_k141_59168 | Not-determined |
| 18-110_k141_24445 | Not-determined |
| 18-110_k141_266041 | Low-quality |
| 18-110_k141_162564 | Not-determined |
| 18-110_k141_162567 | Not-determined |
| 18-110_k141_82145 | Not-determined |
| 18-110_k141_24463 | Not-determined |
| 18-110_k141_243434 | Not-determined |
| 18-110_k141_162573 | Not-determined |
| 18-110_k141_231918 | Not-determined |
| 18-110_k141_104974 | Not-determined |
| 18-110_k141_266056 | Not-determined |
| 18-110_k141_162576 | Not-determined |
| 18-110_k141_231922 | Not-determined |
| 18-110_k141_162580 | Not-determined |
| 18-110_k141_70707 | Not-determined |
| 18-110_k141_70708 | Not-determined |
| 18-110_k141_24471 | Not-determined |
| 18-110_k141_162583 | Not-determined |
| 18-110_k141_162585 | Not-determined |
| 18-110_k141_82154 | Not-determined |
| 18-110_k141_266070 | Not-determined |
| 18-110_k141_185773 | Not-determined |
| 18-110_k141_243446 | Not-determined |
| 18-110_k141_104982 | Not-determined |
| 18-110_k141_139841 | Not-determined |
| 18-110_k141_139842 | Not-determined |
| 18-110_k141_254499 | Not-determined |
| 18-110_k141_127987 | Not-determined |
| 18-110_k141_139845 | Not-determined |
| 18-110_k141_82165 | Not-determined |
| 18-110_k141_219941 | Not-determined |
| 18-110_k141_185789 | Not-determined |
| 18-110_k141_266078 | Not-determined |
| 18-110_k141_116464 | Not-determined |
| 18-110_k141_35787 | Not-determined |
| 18-110_k141_59220 | Low-quality |
| 18-110_k141_219947 | Not-determined |
| 18-110_k141_1258 | Not-determined |
| 18-110_k141_116473 | Not-determined |
| 18-110_k141_266085 | Not-determined |
| 18-110_k141_151152 | Not-determined |
| 18-110_k141_128002 | Low-quality |
| 18-110_k141_104989 | Not-determined |
| 18-110_k141_104993 | Not-determined |
| 18-110_k141_151156 | Not-determined |
| 18-110_k141_185806 | Not-determined |
| 18-110_k141_219963 | Not-determined |
| 18-110_k141_1271 | Not-determined |
| 18-110_k141_1272 | Not-determined |
| 18-110_k141_219968 | Not-determined |
| 18-110_k141_24491 | Not-determined |
| 18-110_k141_116485 | Not-determined |
| 18-110_k141_219974 | Not-determined |
| 18-110_k141_35809 | Not-determined |
| 18-110_k141_151176 | Not-determined |
| 18-110_k141_82175 | Not-determined |
| 18-110_k141_1278 | Not-determined |
| 18-110_k141_105009 | Not-determined |
| 18-110_k141_174264 | Not-determined |
| 18-110_k141_266095 | Low-quality |
| 18-110_k141_151185 | Not-determined |
| 18-110_k141_105011 | Not-determined |
| 18-110_k141_105017 | Not-determined |
| 18-110_k141_59260 | Not-determined |
| 18-110_k141_1292 | Not-determined |
| 18-110_k141_70759 | Not-determined |
| 18-110_k141_231965 | Low-quality |
| 18-110_k141_59270 | Low-quality |
| 18-110_k141_139891 | Low-quality |
| 18-110_k141_24513 | Not-determined |
| 18-110_k141_231974 | Not-determined |
| 18-110_k141_243482 | Not-determined |
| 18-110_k141_185839 | Low-quality |
| 18-110_k141_231977 | Not-determined |
| 18-110_k141_254535 | Not-determined |
| 18-110_k141_70769 | Not-determined |
| 18-110_k141_70773 | Not-determined |
| 18-110_k141_162594 | Not-determined |
| 18-110_k141_185848 | Not-determined |
| 18-110_k141_243503 | Not-determined |
| 18-110_k141_70779 | Not-determined |

| | |
|---|---|
| 18-110_k141_1309 | Not-determined |
| 18-110_k141_220011 | Not-determined |
| 18-110_k141_162604 | Not-determined |
| 18-110_k141_35829 | Not-determined |
| 18-110_k141_70784 | Low-quality |
| 18-110_k141_35832 | Not-determined |
| 18-110_k141_243512 | Not-determined |
| 18-110_k141_220020 | Not-determined |
| 18-110_k141_162614 | Not-determined |
| 18-110_k141_116512 | Not-determined |
| 18-110_k141_70787 | Low-quality |
| 18-110_k141_266126 | Not-determined |
| 18-110_k141_151219 | Not-determined |
| 18-110_k141_162616 | Not-determined |
| 18-110_k141_105060 | Not-determined |
| 18-110_k141_105061 | Not-determined |
| 18-110_k141_139923 | Not-determined |
| 18-110_k141_105062 | Not-determined |
| 18-110_k141_208865 | Not-determined |
| 18-110_k141_35842 | Not-determined |
| 18-110_k141_162629 | Not-determined |
| 18-110_k141_151234 | Not-determined |
| 18-110_k141_162631 | Not-determined |
| 18-110_k141_47350 | Not-determined |
| 18-110_k141_185862 | Not-determined |
| 18-110_k141_128063 | Not-determined |
| 18-110_k141_243540 | Not-determined |
| 18-110_k141_70798 | Not-determined |
| 18-110_k141_197053 | Not-determined |
| 18-110_k141_185867 | Not-determined |
| 18-110_k141_35852 | Not-determined |
| 18-110_k141_185868 | Not-determined |
| 18-110_k141_254570 | Not-determined |
| 18-110_k141_197055 | Not-determined |
| 18-110_k141_139933 | Not-determined |
| 18-110_k141_174285 | Low-quality |
| 18-110_k141_1342 | Not-determined |
| 18-110_k141_162644 | Not-determined |
| 18-110_k141_1352 | Not-determined |
| 18-110_k141_1354 | Not-determined |
| 18-110_k141_35867 | Low-quality |
| 18-110_k141_35869 | Not-determined |
| 18-110_k141_59304 | Low-quality |
| 18-110_k141_266160 | Not-determined |
| 18-110_k141_254581 | Not-determined |
| 18-110_k141_24559 | Not-determined |
| 18-110_k141_208886 | Low-quality |
| 18-110_k141_162649 | Not-determined |
| 18-110_k141_266163 | Not-determined |
| 18-110_k141_266164 | Not-determined |
| 18-110_k141_93701 | Not-determined |
| 18-110_k141_162654 | Not-determined |
| 18-110_k141_162656 | Not-determined |
| 18-110_k141_59308 | Low-quality |
| 18-110_k141_93706 | Not-determined |
| 18-110_k141_254583 | Not-determined |
| 18-110_k141_35879 | Not-determined |
| 18-110_k141_93708 | Not-determined |
| 18-110_k141_35881 | Not-determined |
| 18-110_k141_12520 | Not-determined |
| 18-110_k141_24572 | Not-determined |
| 18-110_k141_208904 | Not-determined |
| 18-110_k141_12524 | Not-determined |
| 18-110_k141_174314 | Not-determined |
| 18-110_k141_208908 | Not-determined |
| 18-110_k141_197088 | Not-determined |
| 18-110_k141_197089 | Not-determined |
| 18-110_k141_12528 | Low-quality |
| 18-110_k141_197092 | Not-determined |
| 18-110_k141_174316 | Not-determined |
| 18-110_k141_266169 | Not-determined |
| 18-110_k141_24581 | Not-determined |
| 18-110_k141_35890 | Not-determined |
| 18-110_k141_254591 | Not-determined |
| 18-110_k141_24588 | Not-determined |
| 18-110_k141_254592 | Low-quality |
| 18-110_k141_220084 | Not-determined |
| 18-110_k141_220085 | Not-determined |
| 18-110_k141_59329 | Not-determined |
| 18-110_k141_151277 | Not-determined |
| 18-110_k141_243564 | Not-determined |
| 18-110_k141_232026 | Not-determined |
| 18-110_k141_243565 | Not-determined |
| 18-110_k141_266188 | Not-determined |
| 18-110_k141_116539 | Not-determined |
| 18-110_k141_35899 | Not-determined |
| 18-110_k141_35900 | Not-determined |
| 18-110_k141_254600 | Not-determined |
| 18-110_k141_243569 | Not-determined |
| 18-110_k141_128087 | Not-determined |
| 18-110_k141_266192 | Not-determined |
| 18-110_k141_35903 | Not-determined |
| 18-110_k141_254603 | Not-determined |
| 18-110_k141_232029 | Not-determined |
| 18-110_k141_266200 | Not-determined |
| 18-110_k141_105122 | Low-quality |
| 18-110_k141_208923 | Low-quality |
| 18-110_k141_1390 | Not-determined |
| 18-110_k141_59354 | Not-determined |
| 18-110_k141_24624 | Not-determined |
| 18-110_k141_208926 | Not-determined |
| 18-110_k141_59357 | Not-determined |
| 18-110_k141_116556 | Not-determined |
| 18-110_k141_24629 | Not-determined |
| 18-110_k141_1398 | Not-determined |
| 18-110_k141_220099 | Not-determined |
| 18-110_k141_1402 | Not-determined |
| 18-110_k141_162669 | Not-determined |
| 18-110_k141_139954 | Not-determined |
| 18-110_k141_162675 | Not-determined |
| 18-110_k141_232046 | Not-determined |
| 18-110_k141_174356 | Not-determined |
| 18-110_k141_254620 | Not-determined |
| 18-110_k141_151302 | Not-determined |
| 18-110_k141_254621 | Not-determined |
| 18-110_k141_162679 | Not-determined |
| 18-110_k141_243592 | Not-determined |
| 18-110_k141_59367 | Not-determined |
| 18-110_k141_220109 | Not-determined |
| 18-110_k141_243596 | Not-determined |
| 18-110_k141_220111 | Not-determined |
| 18-110_k141_35910 | Low-quality |
| 18-110_k141_254630 | Not-determined |
| 18-110_k141_24655 | Not-determined |
| 18-110_k141_139969 | Not-determined |
| 18-110_k141_162690 | Not-determined |
| 18-110_k141_162691 | Not-determined |
| 18-110_k141_208937 | Not-determined |
| 18-110_k141_197129 | Not-determined |
| 18-110_k141_197130 | Not-determined |
| 18-110_k141_266230 | Not-determined |
| 18-110_k141_70813 | Not-determined |
| 18-110_k141_139975 | Low-quality |
| 18-110_k141_139977 | Not-determined |
| 18-110_k141_139978 | Not-determined |
| 18-110_k141_220132 | Not-determined |
| 18-110_k141_243617 | Not-determined |
| 18-110_k141_12553 | Not-determined |
| 18-110_k141_59387 | Not-determined |
| 18-110_k141_139981 | Not-determined |
| 18-110_k141_128126 | Low-quality |
| 18-110_k141_151305 | Low-quality |
| 18-110_k141_59388 | Not-determined |
| 18-110_k141_35917 | Not-determined |
| 18-110_k141_220148 | Not-determined |
| 18-110_k141_128132 | Low-quality |
| 18-110_k141_70830 | Not-determined |
| 18-110_k141_151317 | Low-quality |
| 18-110_k141_59399 | Not-determined |
| 18-110_k141_220158 | Not-determined |
| 18-110_k141_59401 | Low-quality |
| 18-110_k141_139996 | Not-determined |
| 18-110_k141_162708 | Not-determined |
| 18-110_k141_59405 | Not-determined |
| 18-110_k141_105141 | Not-determined |
| 18-110_k141_116567 | Not-determined |
| 18-110_k141_151323 | Not-determined |
| 18-110_k141_151325 | Not-determined |
| 18-110_k141_128137 | Not-determined |
| 18-110_k141_128138 | Low-quality |
| 18-110_k141_93830 | Not-determined |
| 18-110_k141_128144 | Not-determined |
| 18-110_k141_93831 | Not-determined |
| 18-110_k141_93834 | Not-determined |
| 18-110_k141_185934 | Not-determined |
| 18-110_k141_1456 | Not-determined |
| 18-110_k141_208949 | Not-determined |
| 18-110_k141_266274 | Not-determined |
| 18-110_k141_220180 | Low-quality |
| 18-110_k141_266278 | Not-determined |
| 18-110_k141_59429 | Not-determined |
| 18-110_k141_243648 | Not-determined |
| 18-110_k141_232090 | Not-determined |
| 18-110_k141_208958 | Not-determined |
| 18-110_k141_208960 | Not-determined |
| 18-110_k141_140027 | Not-determined |
| 18-110_k141_232096 | Not-determined |
| 18-110_k141_140028 | Low-quality |
| 18-110_k141_162744 | Low-quality |
| 18-110_k141_70868 | Not-determined |
| 18-110_k141_243651 | Not-determined |
| 18-110_k141_140043 | Not-determined |
| 18-110_k141_116600 | Not-determined |
| 18-110_k141_140044 | Not-determined |
| 18-110_k141_116604 | Not-determined |
| 18-110_k141_174387 | Not-determined |
| 18-110_k141_151338 | Not-determined |
| 18-110_k141_243657 | Not-determined |
| 18-110_k141_197221 | Not-determined |
| 18-110_k141_151342 | Not-determined |
| 18-110_k141_116617 | Not-determined |
| 18-110_k141_243659 | Not-determined |
| 18-110_k141_59455 | Not-determined |
| 18-110_k141_266282 | Not-determined |
| 18-110_k141_116621 | Low-quality |
| 18-110_k141_140050 | Low-quality |
| 18-110_k141_174394 | Not-determined |
| 18-110_k141_70891 | Not-determined |
| 18-110_k141_174401 | Not-determined |
| 18-110_k141_266286 | Low-quality |
| 18-110_k141_208985 | Not-determined |
| 18-110_k141_174404 | Not-determined |
| 18-110_k141_59471 | Not-determined |
| 18-110_k141_185971 | Not-determined |
| 18-110_k141_254647 | Not-determined |
| 18-110_k141_151365 | Not-determined |
| 18-110_k141_174409 | Not-determined |
| 18-110_k141_174419 | Low-quality |
| 18-110_k141_35934 | Not-determined |
| 18-110_k141_93887 | Not-determined |
| 18-110_k141_12585 | Low-quality |
| 18-110_k141_116634 | Not-determined |
| 18-110_k141_70911 | Not-determined |
| 18-110_k141_232128 | Not-determined |
| 18-110_k141_232129 | Not-determined |
| 18-110_k141_162773 | Not-determined |
| 18-110_k141_185991 | Not-determined |
| 18-110_k141_105166 | Not-determined |
| 18-110_k141_209005 | Not-determined |
| 18-110_k141_105169 | Not-determined |
| 18-110_k141_1497 | Low-quality |
| 18-110_k141_254663 | Not-determined |
| 18-110_k141_162777 | Not-determined |
| 18-110_k141_35953 | Not-determined |
| 18-110_k141_254672 | Low-quality |
| 18-110_k141_254674 | Not-determined |
| 18-110_k141_59478 | Not-determined |
| 18-110_k141_70926 | Not-determined |
| 18-110_k141_151405 | Low-quality |
| 18-110_k141_151407 | Not-determined |
| 18-110_k141_12616 | Not-determined |
| 18-110_k141_254678 | Low-quality |
| 18-110_k141_232153 | Not-determined |
| 18-110_k141_254680 | Not-determined |
| 18-110_k141_254681 | Low-quality |
| 18-110_k141_151418 | Not-determined |
| 18-110_k141_116656 | Not-determined |
| 18-110_k141_197272 | Not-determined |
| 18-110_k141_12631 | Not-determined |
| 18-110_k141_140067 | Not-determined |
| 18-110_k141_140068 | Not-determined |
| 18-110_k141_116670 | Not-determined |
| 18-110_k141_47380 | Not-determined |
| 18-110_k141_116674 | Not-determined |
| 18-110_k141_128221 | Low-quality |
| 18-110_k141_47381 | Not-determined |
| 18-110_k141_140081 | Not-determined |
| 18-110_k141_151434 | Not-determined |
| 18-110_k141_186040 | Not-determined |
| 18-110_k141_243672 | Not-determined |
| 18-110_k141_162792 | Not-determined |
| 18-110_k141_12645 | Not-determined |
| 18-110_k141_186043 | Not-determined |
| 18-110_k141_243675 | Not-determined |
| 18-110_k141_47391 | Not-determined |
| 18-110_k141_162797 | Not-determined |
| 18-110_k141_151438 | Not-determined |
| 18-110_k141_254695 | Not-determined |
| 18-110_k141_186049 | Not-determined |
| 18-110_k141_12655 | Low-quality |
| 18-110_k141_116680 | Not-determined |
| 18-110_k141_47395 | Not-determined |
| 18-110_k141_174481 | Not-determined |
| 18-110_k141_140106 | Not-determined |
| 18-110_k141_93904 | Not-determined |
| 18-110_k141_105200 | Not-determined |
| 18-110_k141_140109 | Not-determined |
| 18-110_k141_47399 | Low-quality |
| 18-110_k141_174484 | Not-determined |
| 18-110_k141_140113 | Not-determined |
| 18-110_k141_116684 | Low-quality |
| 18-110_k141_116690 | Not-determined |
| 18-110_k141_1513 | Not-determined |
| 18-110_k141_116692 | Not-determined |
| 18-110_k141_105214 | Not-determined |
| 18-110_k141_128262 | Not-determined |
| 18-110_k141_151468 | Not-determined |
| 18-110_k141_151469 | Not-determined |
| 18-110_k141_105223 | Not-determined |
| 18-110_k141_220193 | Low-quality |
| 18-110_k141_24808 | Not-determined |
| 18-110_k141_59506 | Low-quality |
| 18-110_k141_220196 | Not-determined |
| 18-110_k141_254721 | Not-determined |
| 18-110_k141_24813 | Not-determined |
| 18-110_k141_105227 | Not-determined |
| 18-110_k141_140138 | Not-determined |
| 18-110_k141_220203 | Not-determined |
| 18-110_k141_24822 | Not-determined |
| 18-110_k141_266314 | Not-determined |
| 18-110_k141_209098 | Not-determined |
| 18-110_k141_128286 | Low-quality |
| 18-110_k141_243707 | Not-determined |
| 18-110_k141_59515 | Not-determined |
| 18-110_k141_151495 | Not-determined |
| 18-110_k141_59520 | Not-determined |
| 18-110_k141_254733 | Low-quality |
| 18-110_k141_151501 | Not-determined |
| 18-110_k141_24842 | Not-determined |
| 18-110_k141_93919 | Not-determined |
| 18-110_k141_47430 | Not-determined |
| 18-110_k141_47431 | Not-determined |
| 18-110_k141_59522 | Not-determined |
| 18-110_k141_209123 | Not-determined |
| 18-110_k141_197320 | Not-determined |
| 18-110_k141_116727 | Not-determined |
| 18-110_k141_209126 | Not-determined |
| 18-110_k141_220212 | Not-determined |
| 18-110_k141_105252 | Not-determined |
| 18-110_k141_151518 | Not-determined |
| 18-110_k141_24854 | Low-quality |
| 18-110_k141_82280 | Not-determined |
| 18-110_k141_24859 | Not-determined |
| 18-110_k141_162808 | Not-determined |
| 18-110_k141_162809 | Not-determined |
| 18-110_k141_24861 | Not-determined |
| 18-110_k141_174543 | Not-determined |
| 18-110_k141_128312 | Not-determined |
| 18-110_k141_128314 | Not-determined |
| 18-110_k141_1546 | Not-determined |
| 18-110_k141_151523 | Not-determined |
| 18-110_k141_116739 | Low-quality |
| 18-110_k141_105262 | Not-determined |
| 18-110_k141_151528 | Not-determined |
| 18-110_k141_1552 | Not-determined |
| 18-110_k141_174557 | Not-determined |
| 18-110_k141_128318 | Not-determined |
| 18-110_k141_1557 | Not-determined |
| 18-110_k141_151531 | Not-determined |
| 18-110_k141_197339 | Not-determined |
| 18-110_k141_36014 | Not-determined |
| 18-110_k141_174566 | Not-determined |
| 18-110_k141_243740 | Not-determined |
| 18-110_k141_128331 | Not-determined |
| 18-110_k141_140190 | Not-determined |
| 18-110_k141_128332 | Not-determined |
| 18-110_k141_232221 | Not-determined |
| 18-110_k141_116759 | Not-determined |
| 18-110_k141_93957 | Not-determined |
| 18-110_k141_93959 | Low-quality |
| 18-110_k141_174577 | Not-determined |
| 18-110_k141_24893 | Not-determined |
| 18-110_k141_197353 | Low-quality |
| 18-110_k141_209170 | Not-determined |
| 18-110_k141_174581 | Not-determined |
| 18-110_k141_174582 | Not-determined |
| 18-110_k141_162823 | Low-quality |
| 18-110_k141_105281 | Not-determined |
| 18-110_k141_174589 | Not-determined |
| 18-110_k141_174590 | Not-determined |
| 18-110_k141_105283 | Not-determined |
| 18-110_k141_105284 | Not-determined |
| 18-110_k141_174594 | Not-determined |
| 18-110_k141_59559 | Not-determined |
| 18-110_k141_186090 | Not-determined |
| 18-110_k141_12667 | Not-determined |
| 18-110_k141_266353 | Not-determined |
| 18-110_k141_266358 | Not-determined |
| 18-110_k141_209175 | Not-determined |
| 18-110_k141_186103 | Not-determined |
| 18-110_k141_151561 | Not-determined |
| 18-110_k141_243757 | Not-determined |
| 18-110_k141_140216 | Not-determined |
| 18-110_k141_24897 | Not-determined |
| 18-110_k141_266362 | Not-determined |
| 18-110_k141_174597 | Not-determined |
| 18-110_k141_128369 | Not-determined |
| 18-110_k141_82295 | Not-determined |
| 18-110_k141_24905 | Not-determined |
| 18-110_k141_128372 | Not-determined |
| 18-110_k141_140223 | Not-determined |
| 18-110_k141_243758 | Low-quality |
| 18-110_k141_12677 | Not-determined |
| 18-110_k141_266372 | Not-determined |
| 18-110_k141_12679 | Not-determined |
| 18-110_k141_24911 | Not-determined |
| 18-110_k141_24913 | Not-determined |
| 18-110_k141_197381 | Not-determined |
| 18-110_k141_1608 | Not-determined |
| 18-110_k141_59568 | Not-determined |
| 18-110_k141_36046 | Not-determined |
| 18-110_k141_174622 | Not-determined |
| 18-110_k141_243770 | Not-determined |
| 18-110_k141_162854 | Not-determined |
| 18-110_k141_243772 | Not-determined |
| 18-110_k141_116777 | Not-determined |
| 18-110_k141_140237 | Not-determined |
| 18-110_k141_1620 | Not-determined |
| 18-110_k141_174629 | Not-determined |
| 18-110_k141_71041 | Not-determined |
| 18-110_k141_243782 | Not-determined |
| 18-110_k141_162866 | Not-determined |
| 18-110_k141_59572 | Not-determined |
| 18-110_k141_140250 | Not-determined |
| 18-110_k141_82320 | Low-quality |
| 18-110_k141_140252 | Not-determined |
| 18-110_k141_12691 | Low-quality |
| 18-110_k141_186127 | Not-determined |
| 18-110_k141_220228 | Not-determined |
| 18-110_k141_186128 | Not-determined |
| 18-110_k141_116797 | Not-determined |
| 18-110_k141_266382 | Not-determined |
| 18-110_k141_232253 | Not-determined |
| 18-110_k141_243787 | Not-determined |
| 18-110_k141_220236 | Not-determined |
| 18-110_k141_186139 | Not-determined |
| 18-110_k141_209196 | Not-determined |
| 18-110_k141_232264 | Not-determined |
| 18-110_k141_254797 | Not-determined |
| 18-110_k141_59582 | Not-determined |
| 18-110_k141_71056 | Not-determined |
| 18-110_k141_232265 | Not-determined |
| 18-110_k141_59588 | Not-determined |
| 18-110_k141_82331 | Not-determined |
| 18-110_k141_128386 | Not-determined |
| 18-110_k141_232266 | Not-determined |
| 18-110_k141_151614 | Not-determined |
| 18-110_k141_254813 | Not-determined |
| 18-110_k141_140264 | Not-determined |
| 18-110_k141_220254 | Not-determined |
| 18-110_k141_24932 | Not-determined |
| 18-110_k141_59599 | Not-determined |
| 18-110_k141_243791 | Low-quality |
| 18-110_k141_24938 | Low-quality |
| 18-110_k141_105324 | Not-determined |
| 18-110_k141_12722 | Not-determined |
| 18-110_k141_105329 | Not-determined |
| 18-110_k141_128404 | Not-determined |
| 18-110_k141_116822 | Low-quality |
| 18-110_k141_105333 | Not-determined |
| 18-110_k141_82356 | Not-determined |
| 18-110_k141_116828 | Not-determined |
| 18-110_k141_59610 | Not-determined |
| 18-110_k141_47495 | Not-determined |
| 18-110_k141_36084 | Not-determined |
| 18-110_k141_151647 | Not-determined |
| 18-110_k141_47499 | Not-determined |
| 18-110_k141_232287 | Low-quality |
| 18-110_k141_36088 | Low-quality |
| 18-110_k141_59617 | Not-determined |
| 18-110_k141_47501 | Not-determined |
| 18-110_k141_243808 | Not-determined |
| 18-110_k141_82362 | Not-determined |
| 18-110_k141_82366 | Not-determined |
| 18-110_k141_59629 | Not-determined |
| 18-110_k141_151655 | Not-determined |
| 18-110_k141_82374 | Not-determined |
| 18-110_k141_59630 | Not-determined |
| 18-110_k141_82375 | Not-determined |
| 18-110_k141_82381 | Not-determined |
| 18-110_k141_220287 | Not-determined |
| 18-110_k141_140312 | Not-determined |
| 18-110_k141_186183 | Low-quality |
| 18-110_k141_59640 | Not-determined |
| 18-110_k141_174640 | Not-determined |
| 18-110_k141_82387 | Low-quality |
| 18-110_k141_174645 | Not-determined |
| 18-110_k141_140317 | Not-determined |
| 18-110_k141_220296 | Not-determined |
| 18-110_k141_82392 | Not-determined |
| 18-110_k141_116840 | Not-determined |
| 18-110_k141_162876 | Not-determined |
| 18-110_k141_174656 | Not-determined |
| 18-110_k141_12748 | Not-determined |
| 18-110_k141_116841 | Not-determined |
| 18-110_k141_94008 | Not-determined |
| 18-110_k141_151684 | Not-determined |
| 18-110_k141_128431 | Not-determined |
| 18-110_k141_36111 | Not-determined |
| 18-110_k141_232309 | Low-quality |
| 18-110_k141_116847 | Not-determined |
| 18-110_k141_186197 | Not-determined |
| 18-110_k141_24956 | Not-determined |
| 18-110_k141_162898 | Low-quality |
| 18-110_k141_36114 | Not-determined |
| 18-110_k141_254828 | Not-determined |
| 18-110_k141_140329 | Not-determined |
| 18-110_k141_24961 | Not-determined |
| 18-110_k141_151690 | Low-quality |
| 18-110_k141_151691 | Not-determined |
| 18-110_k141_140336 | Not-determined |
| 18-110_k141_12773 | Not-determined |
| 18-110_k141_209255 | Not-determined |
| 18-110_k141_186210 | Not-determined |
| 18-110_k141_24971 | Not-determined |
| 18-110_k141_254834 | Not-determined |
| 18-110_k141_151705 | Not-determined |
| 18-110_k141_36133 | Not-determined |
| 18-110_k141_36136 | Not-determined |
| 18-110_k141_186215 | Not-determined |
| 18-110_k141_36138 | Not-determined |
| 18-110_k141_266424 | Not-determined |
| 18-110_k141_36139 | Low-quality |
| 18-110_k141_243846 | Not-determined |
| 18-110_k141_243847 | Not-determined |
| 18-110_k141_71115 | Not-determined |
| 18-110_k141_186229 | Not-determined |
| 18-110_k141_266428 | Low-quality |
| 18-110_k141_243856 | Low-quality |
| 18-110_k141_232313 | Not-determined |
| 18-110_k141_82407 | Low-quality |
| 18-110_k141_266437 | Low-quality |
| 18-110_k141_266440 | Not-determined |
| 18-110_k141_266441 | Not-determined |
| 18-110_k141_82412 | Low-quality |
| 18-110_k141_36142 | Low-quality |
| 18-110_k141_186238 | Not-determined |
| 18-110_k141_140354 | Not-determined |
| 18-110_k141_82418 | Not-determined |
| 18-110_k141_82419 | Not-determined |
| 18-110_k141_36147 | Not-determined |
| 18-110_k141_1643 | Not-determined |
| 18-110_k141_162937 | Not-determined |
| 18-110_k141_36153 | Not-determined |
| 18-110_k141_1645 | Not-determined |
| 18-110_k141_209263 | Not-determined |
| 18-110_k141_116891 | Not-determined |
| 18-110_k141_197430 | Not-determined |
| 18-110_k141_82430 | Not-determined |
| 18-110_k141_243878 | Not-determined |
| 18-110_k141_128440 | Not-determined |
| 18-110_k141_82434 | Not-determined |
| 18-110_k141_24986 | Not-determined |
| 18-110_k141_1659 | Low-quality |
| 18-110_k141_243888 | Low-quality |
| 18-110_k141_116898 | Not-determined |
| 18-110_k141_220360 | Not-determined |
| 18-110_k141_128450 | Not-determined |
| 18-110_k141_266468 | Low-quality |
| 18-110_k141_128453 | Not-determined |
| 18-110_k141_82447 | Not-determined |
| 18-110_k141_128454 | Not-determined |
| 18-110_k141_162961 | Not-determined |
| 18-110_k141_116906 | Not-determined |
| 18-110_k141_243896 | Not-determined |
| 18-110_k141_128463 | Not-determined |
| 18-110_k141_25001 | Not-determined |
| 18-110_k141_197441 | Not-determined |
| 18-110_k141_162970 | Not-determined |
| 18-110_k141_162971 | Not-determined |
| 18-110_k141_105405 | Not-determined |
| 18-110_k141_266486 | Not-determined |
| 18-110_k141_36179 | Not-determined |
| 18-110_k141_94036 | Not-determined |
| 18-110_k141_140376 | Low-quality |
| 18-110_k141_197445 | Not-determined |
| 18-110_k141_94045 | Not-determined |
| 18-110_k141_82461 | Low-quality |
| 18-110_k141_94048 | Not-determined |
| 18-110_k141_12818 | Not-determined |
| 18-110_k141_94052 | Not-determined |
| 18-110_k141_151748 | Not-determined |
| 18-110_k141_197450 | Not-determined |
| 18-110_k141_151749 | Not-determined |
| 18-110_k141_105417 | Low-quality |
| 18-110_k141_197451 | Not-determined |
| 18-110_k141_94055 | Not-determined |
| 18-110_k141_140384 | Not-determined |
| 18-110_k141_82472 | Not-determined |
| 18-110_k141_243909 | Low-quality |
| 18-110_k141_82474 | Not-determined |
| 18-110_k141_116922 | Not-determined |
| 18-110_k141_140388 | Not-determined |
| 18-110_k141_71126 | Not-determined |
| 18-110_k141_105426 | Not-determined |
| 18-110_k141_116928 | Not-determined |
| 18-110_k141_186274 | Not-determined |
| 18-110_k141_25031 | Not-determined |
| 18-110_k141_197462 | Not-determined |
| 18-110_k141_197463 | Not-determined |
| 18-110_k141_209290 | Not-determined |
| 18-110_k141_266494 | Not-determined |
| 18-110_k141_47589 | Not-determined |
| 18-110_k141_25039 | Not-determined |
| 18-110_k141_47594 | Low-quality |
| 18-110_k141_266499 | Not-determined |
| 18-110_k141_186285 | Not-determined |
| 18-110_k141_162987 | Not-determined |
| 18-110_k141_116946 | Not-determined |
| 18-110_k141_186289 | Not-determined |
| 18-110_k141_82485 | Low-quality |
| 18-110_k141_151773 | Not-determined |
| 18-110_k141_94086 | Not-determined |
| 18-110_k141_47605 | Not-determined |
| 18-110_k141_174702 | Not-determined |
| 18-110_k141_82490 | Not-determined |
| 18-110_k141_116954 | Not-determined |
| 18-110_k141_162993 | Low-quality |
| 18-110_k141_243930 | Not-determined |

| | |
|---|---|
| 18-110_k141_266518 | Not-determined |
| 18-110_k141_94091 | Not-determined |
| 18-110_k141_140415 | Not-determined |
| 18-110_k141_116962 | Not-determined |
| 18-110_k141_186307 | Low-quality |
| 18-110_k141_197475 | Not-determined |
| 18-110_k141_105432 | Not-determined |
| 18-110_k141_220396 | Not-determined |
| 18-110_k141_82507 | Not-determined |
| 18-110_k141_243935 | Not-determined |
| 18-110_k141_243936 | Not-determined |
| 18-110_k141_243937 | Not-determined |
| 18-110_k141_116967 | Not-determined |
| 18-110_k141_25055 | Low-quality |
| 18-110_k141_220400 | Not-determined |
| 18-110_k141_220405 | Not-determined |
| 18-110_k141_232372 | Not-determined |
| 18-110_k141_266538 | Not-determined |
| 18-110_k141_186321 | Not-determined |
| 18-110_k141_232375 | Not-determined |
| 18-110_k141_186324 | Not-determined |
| 18-110_k141_140440 | Not-determined |
| 18-110_k141_232377 | Not-determined |
| 18-110_k141_243945 | Not-determined |
| 18-110_k141_197498 | Not-determined |
| 18-110_k141_174719 | Not-determined |
| 18-110_k141_209296 | Not-determined |
| 18-110_k141_209297 | Not-determined |
| 18-110_k141_105454 | Not-determined |
| 18-110_k141_197503 | Not-determined |
| 18-110_k141_128493 | Not-determined |
| 18-110_k141_105457 | Not-determined |
| 18-110_k141_105459 | Not-determined |
| 18-110_k141_116996 | Not-determined |
| 18-110_k141_140445 | Not-determined |
| 18-110_k141_1732 | Low-quality |
| 18-110_k141_243950 | Not-determined |
| 18-110_k141_266555 | Not-determined |
| 18-110_k141_71164 | Low-quality |
| 18-110_k141_71166 | Not-determined |
| 18-110_k141_232405 | Not-determined |
| 18-110_k141_71171 | Not-determined |
| 18-110_k141_151827 | Not-determined |
| 18-110_k141_71172 | Not-determined |
| 18-110_k141_197513 | Low-quality |
| 18-110_k141_151828 | Not-determined |
| 18-110_k141_163042 | Low-quality |
| 18-110_k141_71174 | Not-determined |
| 18-110_k141_105470 | Not-determined |
| 18-110_k141_163047 | Not-determined |
| 18-110_k141_209321 | Not-determined |
| 18-110_k141_197523 | Not-determined |
| 18-110_k141_140462 | Not-determined |
| 18-110_k141_105474 | Not-determined |
| 18-110_k141_266569 | Not-determined |
| 18-110_k141_266573 | Not-determined |
| 18-110_k141_266574 | Not-determined |
| 18-110_k141_266577 | Not-determined |
| 18-110_k141_105476 | Low-quality |
| 18-110_k141_71191 | Low-quality |
| 18-110_k141_128519 | Not-determined |
| 18-110_k141_140475 | Not-determined |
| 18-110_k141_105482 | Not-determined |
| 18-110_k141_163060 | Not-determined |
| 18-110_k141_140480 | Not-determined |
| 18-110_k141_163063 | Not-determined |
| 18-110_k141_243981 | Not-determined |
| 18-110_k141_232425 | Not-determined |
| 18-110_k141_36214 | Not-determined |
| 18-110_k141_220419 | Low-quality |
| 18-110_k141_151854 | Not-determined |
| 18-110_k141_197542 | Not-determined |
| 18-110_k141_59686 | Low-quality |
| 18-110_k141_94151 | Not-determined |
| 18-110_k141_82524 | Not-determined |
| 18-110_k141_140485 | Not-determined |
| 18-110_k141_232435 | Not-determined |
| 18-110_k141_94157 | Low-quality |
| 18-110_k141_82532 | Not-determined |
| 18-110_k141_174736 | Not-determined |
| 18-110_k141_59700 | Not-determined |
| 18-110_k141_243997 | Not-determined |
| 18-110_k141_186349 | Not-determined |
| 18-110_k141_71232 | Not-determined |
| 18-110_k141_36239 | Not-determined |
| 18-110_k141_232449 | Not-determined |
| 18-110_k141_232454 | Not-determined |
| 18-110_k141_140508 | Not-determined |
| 18-110_k141_36248 | Not-determined |
| 18-110_k141_71248 | Not-determined |
| 18-110_k141_197553 | Not-determined |
| 18-110_k141_220450 | Not-determined |
| 18-110_k141_174759 | Not-determined |
| 18-110_k141_140514 | Not-determined |
| 18-110_k141_244010 | Not-determined |
| 18-110_k141_94168 | Not-determined |
| 18-110_k141_47671 | Not-determined |
| 18-110_k141_140519 | Not-determined |
| 18-110_k141_47677 | Not-determined |
| 18-110_k141_47678 | Low-quality |
| 18-110_k141_36263 | Not-determined |
| 18-110_k141_47679 | Not-determined |
| 18-110_k141_220459 | Not-determined |
| 18-110_k141_220461 | Not-determined |
| 18-110_k141_71260 | Not-determined |
| 18-110_k141_59730 | Not-determined |
| 18-110_k141_71267 | Not-determined |
| 18-110_k141_244014 | Not-determined |
| 18-110_k141_36276 | Not-determined |
| 18-110_k141_174784 | Not-determined |
| 18-110_k141_94181 | Not-determined |
| 18-110_k141_209369 | Not-determined |
| 18-110_k141_36284 | Not-determined |
| 18-110_k141_244023 | Not-determined |
| 18-110_k141_163135 | Not-determined |
| 18-110_k141_266602 | Low-quality |
| 18-110_k141_140539 | Not-determined |
| 18-110_k141_82594 | Not-determined |
| 18-110_k141_82596 | Not-determined |
| 18-110_k141_82599 | Not-determined |
| 18-110_k141_266613 | Not-determined |
| 18-110_k141_117045 | Not-determined |
| 18-110_k141_82607 | Not-determined |
| 18-110_k141_25146 | Not-determined |
| 18-110_k141_174824 | Not-determined |
| 18-110_k141_82609 | Not-determined |
| 18-110_k141_105499 | Not-determined |
| 18-110_k141_105502 | Not-determined |
| 18-110_k141_266626 | Not-determined |
| 18-110_k141_59748 | Not-determined |
| 18-110_k141_59751 | Not-determined |
| 18-110_k141_128562 | Not-determined |
| 18-110_k141_140563 | Not-determined |
| 18-110_k141_36305 | Not-determined |
| 18-110_k141_71307 | Not-determined |
| 18-110_k141_25163 | Not-determined |
| 18-110_k141_232469 | Not-determined |
| 18-110_k141_36308 | Not-determined |
| 18-110_k141_232473 | Not-determined |
| 18-110_k141_254941 | Not-determined |
| 18-110_k141_254943 | Not-determined |
| 18-110_k141_209400 | Not-determined |
| 18-110_k141_82631 | Not-determined |
| 18-110_k141_82635 | Low-quality |
| 18-110_k141_140574 | Not-determined |
| 18-110_k141_244050 | Not-determined |
| 18-110_k141_82642 | Not-determined |
| 18-110_k141_209416 | Not-determined |
| 18-110_k141_36326 | Not-determined |
| 18-110_k141_117062 | Not-determined |
| 18-110_k141_174856 | Not-determined |
| 18-110_k141_163167 | Low-quality |
| 18-110_k141_244061 | Not-determined |
| 18-110_k141_209425 | Not-determined |
| 18-110_k141_244064 | Not-determined |
| 18-110_k141_220493 | Low-quality |
| 18-110_k141_209427 | Not-determined |
| 18-110_k141_36333 | Not-determined |
| 18-110_k141_36334 | Not-determined |
| 18-110_k141_244075 | Not-determined |
| 18-110_k141_117075 | Not-determined |
| 18-110_k141_244076 | Not-determined |
| 18-110_k141_117076 | Not-determined |
| 18-110_k141_266646 | Low-quality |
| 18-110_k141_82664 | Not-determined |
| 18-110_k141_266651 | Low-quality |
| 18-110_k141_220505 | Low-quality |
| 18-110_k141_82669 | Not-determined |
| 18-110_k141_209449 | Not-determined |
| 18-110_k141_266659 | Not-determined |
| 18-110_k141_82674 | Not-determined |
| 18-110_k141_174877 | Not-determined |
| 18-110_k141_174878 | Not-determined |
| 18-110_k141_232529 | Low-quality |
| 18-110_k141_266667 | Not-determined |
| 18-110_k141_105552 | Not-determined |
| 18-110_k141_82682 | Not-determined |
| 18-110_k141_82684 | Not-determined |
| 18-110_k141_82685 | Not-determined |
| 18-105_k141_123499 | Medium-quality |
| 18-110_k141_94207 | Not-determined |
| 18-110_k141_140614 | Low-quality |
| 18-110_k141_105559 | Not-determined |
| 18-110_k141_232546 | Not-determined |
| 18-110_k141_254967 | Not-determined |
| 18-110_k141_71341 | Not-determined |
| 18-110_k141_244114 | Not-determined |
| 18-110_k141_25216 | Not-determined |
| 18-110_k141_140623 | Not-determined |
| 18-110_k141_140624 | Not-determined |
| 18-110_k141_174895 | Not-determined |
| 18-110_k141_47717 | Not-determined |
| 18-110_k141_266678 | Not-determined |
| 18-110_k141_82692 | Not-determined |
| 18-110_k141_244131 | Not-determined |
| 18-110_k141_71346 | Low-quality |
| 18-110_k141_25227 | Not-determined |
| 18-110_k141_25228 | Not-determined |
| 18-110_k141_254978 | Not-determined |
| 18-110_k141_105585 | Not-determined |
| 18-110_k141_82702 | Not-determined |
| 18-110_k141_71353 | Not-determined |
| 18-110_k141_25231 | Not-determined |
| 18-110_k141_82704 | Not-determined |
| 18-110_k141_59787 | Not-determined |
| 18-110_k141_128591 | Not-determined |
| 18-110_k141_140646 | Not-determined |
| 18-110_k141_140647 | Not-determined |
| 18-110_k141_244143 | Not-determined |
| 18-110_k141_59793 | Not-determined |
| 18-110_k141_266696 | Not-determined |
| 18-110_k141_128597 | Not-determined |
| 18-110_k141_47744 | Not-determined |
| 18-110_k141_128599 | Not-determined |
| 18-110_k141_105598 | Not-determined |
| 18-110_k141_197599 | Not-determined |
| 18-110_k141_25254 | Not-determined |
| 18-110_k141_163228 | Low-quality |
| 18-110_k141_47755 | Low-quality |
| 18-110_k141_47756 | Not-determined |
| 18-110_k141_186401 | Not-determined |
| 18-110_k141_244152 | Not-determined |
| 18-110_k141_1826 | Not-determined |
| 18-110_k141_36357 | Not-determined |
| 18-110_k141_47759 | Not-determined |
| 18-110_k141_36361 | Not-determined |
| 18-110_k141_36362 | Not-determined |
| 18-110_k141_36364 | Not-determined |
| 18-110_k141_186408 | Low-quality |
| 18-110_k141_197621 | Not-determined |
| 18-110_k141_36367 | Low-quality |
| 18-110_k141_186409 | Not-determined |
| 18-110_k141_128620 | Low-quality |
| 18-110_k141_1837 | Not-determined |
| 18-110_k141_105620 | Not-determined |
| 18-110_k141_117080 | Not-determined |
| 18-110_k141_71370 | Not-determined |
| 18-110_k141_244155 | Not-determined |
| 18-110_k141_174955 | Not-determined |
| 18-110_k141_36378 | Not-determined |
| 18-110_k141_105631 | Not-determined |
| 18-110_k141_255003 | Low-quality |
| 18-110_k141_244160 | Not-determined |
| 18-110_k141_1850 | Not-determined |
| 18-110_k141_128626 | Not-determined |
| 18-110_k141_117090 | Low-quality |
| 18-110_k141_105635 | Low-quality |
| 18-110_k141_105637 | Not-determined |
| 18-110_k141_186433 | Not-determined |
| 18-110_k141_25287 | Not-determined |
| 18-110_k141_94240 | Not-determined |
| 18-110_k141_25289 | Not-determined |
| 18-110_k141_47788 | Not-determined |
| 18-110_k141_244177 | Not-determined |
| 18-110_k141_197635 | Not-determined |
| 18-110_k141_186438 | Not-determined |
| 18-110_k141_174964 | Not-determined |
| 18-110_k141_244181 | Low-quality |
| 18-110_k141_266719 | Not-determined |
| 18-110_k141_1859 | Low-quality |
| 18-110_k141_266722 | Not-determined |
| 18-110_k141_151884 | Not-determined |
| 18-110_k141_266727 | Not-determined |
| 18-110_k141_94266 | Not-determined |
| 18-110_k141_266735 | Not-determined |
| 18-110_k141_94272 | Not-determined |
| 18-110_k141_47800 | Low-quality |
| 18-110_k141_117096 | Not-determined |
| 18-110_k141_266739 | Not-determined |
| 18-110_k141_94275 | Not-determined |
| 18-110_k141_266742 | Not-determined |
| 18-110_k141_25316 | Not-determined |
| 18-110_k141_186462 | Not-determined |
| 18-110_k141_25319 | Not-determined |
| 18-110_k141_174988 | Not-determined |
| 18-110_k141_197663 | Not-determined |
| 18-110_k141_151914 | Not-determined |
| 18-110_k141_105649 | Not-determined |
| 18-110_k141_266751 | Not-determined |
| 18-110_k141_220551 | Not-determined |
| 18-110_k141_174998 | Low-quality |
| 18-110_k141_163234 | Not-determined |
| 18-110_k141_220555 | Not-determined |
| 18-110_k141_59855 | Not-determined |
| 18-110_k141_220558 | Low-quality |
| 18-110_k141_47813 | Not-determined |
| 18-110_k141_175010 | Not-determined |
| 18-110_k141_232576 | Not-determined |
| 18-110_k141_232578 | Not-determined |
| 18-110_k141_25345 | Not-determined |
| 18-110_k141_1882 | Not-determined |
| 18-110_k141_25347 | Not-determined |
| 18-110_k141_151938 | Not-determined |
| 18-110_k141_128664 | Not-determined |
| 18-110_k141_232585 | Low-quality |
| 18-110_k141_128667 | Not-determined |
| 18-110_k141_94292 | Not-determined |
| 18-110_k141_117128 | Low-quality |
| 18-110_k141_47826 | Low-quality |
| 18-110_k141_117130 | Low-quality |
| 18-110_k141_175023 | Not-determined |
| 18-110_k141_232591 | Not-determined |
| 18-110_k141_25354 | Low-quality |
| 18-110_k141_117132 | Low-quality |
| 18-110_k141_25359 | Not-determined |
| 18-110_k141_94302 | Not-determined |
| 18-110_k141_163257 | Not-determined |
| 18-110_k141_244206 | Not-determined |
| 18-110_k141_47839 | Not-determined |
| 18-110_k141_220579 | Not-determined |
| 18-110_k141_59866 | Not-determined |
| 18-110_k141_59867 | Not-determined |
| 18-110_k141_140676 | Low-quality |
| 18-110_k141_59871 | Not-determined |
| 18-110_k141_128690 | Not-determined |
| 18-110_k141_244214 | Not-determined |
| 18-110_k141_128693 | Not-determined |
| 18-110_k141_209463 | Not-determined |
| 18-110_k141_209467 | Not-determined |
| 18-110_k141_175043 | Not-determined |
| 18-110_k141_232597 | Not-determined |
| 18-110_k141_71472 | Not-determined |
| 18-110_k141_105660 | Not-determined |
| 18-110_k141_232600 | Low-quality |
| 18-110_k141_140689 | Not-determined |
| 18-110_k141_140690 | Low-quality |
| 18-110_k141_266783 | Not-determined |
| 18-110_k141_47862 | Not-determined |
| 18-110_k141_197733 | Not-determined |
| 18-110_k141_232608 | Not-determined |
| 18-110_k141_94320 | Low-quality |
| 18-110_k141_151971 | Not-determined |
| 18-110_k141_94325 | Not-determined |
| 18-110_k141_255072 | Low-quality |
| 18-110_k141_105674 | Not-determined |
| 18-110_k141_71499 | Low-quality |
| 18-110_k141_232610 | Not-determined |
| 18-110_k141_163290 | Not-determined |
| 18-110_k141_47877 | Low-quality |
| 18-110_k141_128723 | Not-determined |
| 18-110_k141_232620 | Not-determined |
| 18-110_k141_266808 | Not-determined |
| 18-110_k141_232623 | Low-quality |
| 18-110_k141_94339 | Not-determined |
| 18-110_k141_140708 | Not-determined |
| 18-110_k141_197747 | Not-determined |
| 18-110_k141_140710 | Not-determined |
| 18-110_k141_128737 | Not-determined |
| 18-110_k141_47888 | Not-determined |
| 18-110_k141_197752 | Not-determined |
| 18-110_k141_244259 | Not-determined |
| 18-110_k141_209500 | Not-determined |
| 18-110_k141_47904 | Low-quality |
| 18-110_k141_220592 | Not-determined |
| 18-110_k141_197760 | Not-determined |
| 18-110_k141_94351 | Not-determined |
| 18-110_k141_128759 | Not-determined |
| 18-110_k141_244273 | Low-quality |
| 18-110_k141_47909 | Not-determined |
| 18-110_k141_232642 | Not-determined |
| 18-110_k141_105695 | Not-determined |
| 18-110_k141_152006 | Not-determined |
| 18-110_k141_25442 | Not-determined |
| 18-110_k141_266829 | Not-determined |
| 18-110_k141_266830 | Not-determined |
| 18-110_k141_47918 | Not-determined |
| 18-110_k141_152013 | Not-determined |
| 18-110_k141_163318 | Not-determined |
| 18-110_k141_244281 | Not-determined |
| 18-110_k141_163322 | Not-determined |
| 18-110_k141_128775 | Not-determined |
| 18-110_k141_175093 | Not-determined |
| 18-110_k141_128777 | Not-determined |
| 18-110_k141_71550 | Not-determined |
| 18-110_k141_266844 | Low-quality |
| 18-110_k141_59915 | Not-determined |
| 18-110_k141_220619 | Not-determined |
| 18-110_k141_128784 | Not-determined |
| 18-110_k141_71560 | Not-determined |
| 18-110_k141_36455 | Not-determined |
| 18-110_k141_59918 | Not-determined |
| 18-110_k141_163341 | Not-determined |
| 18-110_k141_163345 | Not-determined |
| 18-110_k141_94368 | Not-determined |
| 18-110_k141_12908 | Not-determined |
| 18-110_k141_36460 | Not-determined |
| 18-110_k141_128797 | Not-determined |
| 18-110_k141_244316 | Not-determined |
| 18-110_k141_232652 | Low-quality |
| 18-110_k141_71570 | Not-determined |
| 18-110_k141_128801 | Not-determined |
| 18-110_k141_12916 | Not-determined |
| 18-110_k141_232657 | Not-determined |
| 18-110_k141_128804 | Low-quality |
| 18-110_k141_244325 | Low-quality |
| 18-110_k141_94378 | Not-determined |
| 18-110_k141_128807 | Not-determined |
| 18-110_k141_232664 | Not-determined |
| 18-110_k141_71581 | Not-determined |
| 18-110_k141_152041 | Not-determined |
| 18-110_k141_12920 | Not-determined |
| 18-110_k141_232666 | Not-determined |
| 18-110_k141_266863 | Not-determined |
| 18-110_k141_220634 | Low-quality |
| 18-110_k141_163366 | Not-determined |
| 18-110_k141_220639 | Not-determined |
| 18-110_k141_117153 | Low-quality |
| 18-110_k141_117154 | Not-determined |
| 18-110_k141_36467 | Not-determined |
| 18-110_k141_266875 | Low-quality |
| 18-110_k141_47929 | Not-determined |
| 18-110_k141_197779 | Not-determined |
| 18-110_k141_152050 | Not-determined |
| 18-110_k141_255135 | Not-determined |
| 18-110_k141_105746 | Not-determined |
| 18-110_k141_117160 | Not-determined |
| 18-110_k141_244351 | Not-determined |
| 18-110_k141_47936 | Not-determined |
| 18-110_k141_128822 | Not-determined |
| 18-110_k141_232683 | Not-determined |
| 18-110_k141_220657 | Not-determined |
| 18-110_k141_266891 | Not-determined |
| 18-110_k141_255148 | Low-quality |
| 18-110_k141_128828 | Not-determined |
| 18-110_k141_175146 | Not-determined |
| 18-110_k141_197792 | Not-determined |
| 18-110_k141_36485 | Not-determined |
| 18-110_k141_266901 | Not-determined |
| 18-110_k141_209547 | Not-determined |
| 18-110_k141_71609 | Not-determined |
| 18-110_k141_232695 | Low-quality |
| 18-110_k141_117177 | Not-determined |
| 18-110_k141_266914 | Not-determined |
| 18-110_k141_232704 | Low-quality |
| 18-110_k141_244364 | Not-determined |
| 18-110_k141_25460 | Not-determined |
| 18-110_k141_209554 | Not-determined |
| 18-110_k141_117187 | Not-determined |
| 18-110_k141_220677 | Not-determined |
| 18-110_k141_71619 | Not-determined |
| 18-110_k141_186566 | Low-quality |
| 18-110_k141_220681 | Not-determined |
| 18-110_k141_140735 | Low-quality |
| 18-110_k141_59935 | Not-determined |
| 18-110_k141_197807 | Not-determined |
| 18-110_k141_232716 | Not-determined |
| 18-110_k141_220686 | Not-determined |
| 18-110_k141_59938 | Not-determined |
| 18-110_k141_266930 | Not-determined |
| 18-110_k141_244370 | Not-determined |
| 18-110_k141_266932 | Not-determined |
| 18-110_k141_186573 | Low-quality |
| 18-110_k141_71629 | Not-determined |
| 18-110_k141_232720 | Low-quality |
| 18-110_k141_71631 | Not-determined |
| 18-110_k141_94421 | Not-determined |
| 18-110_k141_197820 | Not-determined |
| 18-110_k141_244372 | Not-determined |
| 18-110_k141_186574 | Low-quality |
| 18-110_k141_220698 | Not-determined |
| 18-110_k141_140750 | Not-determined |
| 18-110_k141_36506 | Not-determined |
| 18-110_k141_25473 | Not-determined |
| 18-110_k141_25476 | Not-determined |
| 18-110_k141_36509 | Not-determined |
| 18-110_k141_197829 | Not-determined |
| 18-110_k141_140756 | Not-determined |
| 18-110_k141_36512 | Low-quality |
| 18-110_k141_140758 | Not-determined |
| 18-110_k141_94436 | Not-determined |
| 18-110_k141_266950 | Not-determined |
| 18-110_k141_25484 | Not-determined |
| 18-110_k141_140759 | Not-determined |
| 18-110_k141_255179 | Not-determined |
| 18-110_k141_266952 | Not-determined |
| 18-110_k141_209570 | Not-determined |
| 18-110_k141_128847 | Low-quality |
| 18-110_k141_59958 | Not-determined |
| 18-110_k141_232740 | Low-quality |
| 18-110_k141_128851 | Not-determined |
| 18-110_k141_59963 | Not-determined |
| 18-110_k141_232744 | Not-determined |
| 18-110_k141_232745 | Not-determined |
| 18-110_k141_128855 | Not-determined |
| 18-110_k141_255186 | Not-determined |
| 18-110_k141_232750 | Not-determined |
| 18-110_k141_186589 | Not-determined |
| 18-110_k141_25500 | Not-determined |
| 18-110_k141_36530 | Not-determined |
| 18-110_k141_94442 | Not-determined |
| 18-110_k141_220715 | Not-determined |
| 18-110_k141_152073 | Not-determined |
| 18-110_k141_255191 | Not-determined |
| 18-110_k141_59969 | Low-quality |
| 18-110_k141_255192 | Not-determined |
| 18-110_k141_59970 | Not-determined |
| 18-110_k141_220718 | Not-determined |
| 18-110_k141_220719 | Not-determined |
| 18-110_k141_12973 | Not-determined |
| 18-110_k141_47978 | Not-determined |
| 18-110_k141_232758 | Not-determined |
| 18-110_k141_163394 | Not-determined |
| 18-110_k141_71659 | Not-determined |
| 18-110_k141_163395 | Not-determined |
| 18-110_k141_255195 | Not-determined |
| 18-110_k141_232762 | Not-determined |
| 18-110_k141_152079 | Not-determined |
| 18-110_k141_94454 | Not-determined |
| 18-110_k141_117209 | Not-determined |
| 18-110_k141_266964 | Not-determined |
| 18-110_k141_47994 | Not-determined |
| 18-110_k141_117214 | Not-determined |
| 18-110_k141_152085 | Not-determined |
| 18-110_k141_175200 | Not-determined |
| 18-110_k141_266972 | Not-determined |
| 18-110_k141_186613 | Not-determined |
| 18-110_k141_71675 | Not-determined |
| 18-110_k141_94467 | Not-determined |
| 18-110_k141_94468 | Not-determined |
| 18-110_k141_25514 | Low-quality |
| 18-110_k141_82729 | Low-quality |
| 18-110_k141_94469 | Low-quality |
| 18-110_k141_48009 | Not-determined |
| 18-110_k141_266986 | Not-determined |
| 18-110_k141_105784 | Not-determined |
| 18-110_k141_1917 | Not-determined |
| 18-110_k141_71682 | Not-determined |
| 18-110_k141_186620 | Not-determined |
| 18-110_k141_209598 | Not-determined |
| 18-110_k141_186628 | Not-determined |
| 18-110_k141_186630 | Low-quality |
| 18-110_k141_1930 | Not-determined |
| 18-110_k141_1931 | Not-determined |
| 18-110_k141_59990 | Not-determined |
| 18-110_k141_59995 | Not-determined |
| 18-110_k141_105792 | Not-determined |
| 18-110_k141_36551 | Not-determined |
| 18-110_k141_60002 | Not-determined |
| 18-110_k141_244386 | Not-determined |
| 18-110_k141_1950 | Not-determined |
| 18-110_k141_94484 | Not-determined |
| 18-110_k141_244388 | Not-determined |
| 18-110_k141_163426 | Low-quality |
| 18-110_k141_267002 | Low-quality |
| 18-110_k141_12995 | Not-determined |
| 18-110_k141_152107 | Not-determined |
| 18-110_k141_12998 | Not-determined |
| 18-110_k141_117234 | Not-determined |
| 18-110_k141_152113 | Not-determined |
| 18-110_k141_94488 | Not-determined |
| 18-110_k141_71690 | Not-determined |
| 18-110_k141_117242 | Not-determined |
| 18-110_k141_232774 | Not-determined |
| 18-110_k141_71695 | Not-determined |
| 18-110_k141_94497 | Not-determined |
| 18-110_k141_175250 | Low-quality |
| 18-110_k141_94498 | Not-determined |
| 18-110_k141_25548 | Not-determined |
| 18-110_k141_175251 | Not-determined |
| 18-110_k141_244399 | Not-determined |
| 18-110_k141_25550 | Not-determined |
| 18-110_k141_60023 | Not-determined |
| 18-110_k141_25552 | Not-determined |
| 18-110_k141_186645 | Low-quality |
| 18-110_k141_117256 | Not-determined |
| 18-110_k141_13015 | Low-quality |
| 18-110_k141_186652 | Not-determined |
| 18-110_k141_82783 | Not-determined |
| 18-110_k141_128890 | Low-quality |
| 18-110_k141_105802 | Not-determined |
| 18-110_k141_71707 | Not-determined |
| 18-110_k141_232799 | Not-determined |
| 18-110_k141_232801 | Not-determined |
| 18-110_k141_117266 | Not-determined |
| 18-110_k141_71716 | Low-quality |
| 18-110_k141_220758 | Not-determined |
| 18-110_k141_209641 | Not-determined |
| 18-110_k141_197844 | Not-determined |
| 18-110_k141_163455 | Not-determined |
| 18-110_k141_71721 | Not-determined |
| 18-110_k141_244422 | Not-determined |
| 18-110_k141_197849 | Low-quality |
| 18-110_k141_244423 | Low-quality |
| 18-110_k141_244427 | Not-determined |
| 18-110_k141_94520 | Low-quality |
| 18-110_k141_220769 | Not-determined |
| 18-110_k141_94525 | Not-determined |
| 18-110_k141_25578 | Not-determined |
| 18-110_k141_175274 | Low-quality |

| | |
|---|---|
| 18-110_k141_255223 | Not-determined |
| 18-110_k141_25582 | Not-determined |
| 18-110_k141_175277 | Not-determined |
| 18-110_k141_232827 | Low-quality |
| 18-110_k141_232828 | Not-determined |
| 18-110_k141_220774 | Low-quality |
| 18-110_k141_232830 | Not-determined |
| 18-110_k141_197857 | Low-quality |
| 18-110_k141_13046 | Not-determined |
| 18-110_k141_152129 | Not-determined |
| 18-110_k141_197859 | Not-determined |
| 18-110_k141_25585 | Low-quality |
| 18-110_k141_152131 | Not-determined |
| 18-110_k141_244434 | Not-determined |
| 18-110_k141_36577 | Not-determined |
| 18-110_k141_36578 | Not-determined |
| 18-110_k141_1978 | Not-determined |
| 18-110_k141_36580 | Not-determined |
| 18-110_k141_244442 | Not-determined |
| 18-110_k141_267034 | Not-determined |
| 18-110_k141_13051 | Not-determined |
| 18-110_k141_60058 | Not-determined |
| 18-110_k141_82812 | Low-quality |
| 18-110_k141_244450 | Not-determined |
| 18-110_k141_48043 | Not-determined |
| 18-110_k141_60065 | Not-determined |
| 18-110_k141_209670 | Not-determined |
| 18-110_k141_48045 | Not-determined |
| 18-110_k141_209673 | Not-determined |
| 18-110_k141_232847 | Not-determined |
| 18-110_k141_197873 | Not-determined |
| 18-110_k141_163464 | Not-determined |
| 18-110_k141_255245 | Not-determined |
| 18-110_k141_186668 | Low-quality |
| 18-110_k141_209680 | Not-determined |
| 18-110_k141_36600 | Not-determined |
| 18-110_k141_244460 | Not-determined |
| 18-110_k141_186669 | Not-determined |
| 18-110_k141_163468 | Not-determined |
| 18-110_k141_82831 | Not-determined |
| 18-110_k141_152135 | Not-determined |
| 18-110_k141_163471 | Not-determined |
| 18-110_k141_232854 | Not-determined |
| 18-110_k141_163472 | Low-quality |
| 18-110_k141_163473 | Not-determined |
| 18-110_k141_232857 | Not-determined |
| 18-110_k141_186673 | Not-determined |
| 18-110_k141_2000 | Not-determined |
| 18-110_k141_2001 | Not-determined |
| 18-110_k141_13061 | Low-quality |
| 18-110_k141_60073 | Not-determined |
| 18-110_k141_94536 | Not-determined |
| 18-110_k141_140783 | Not-determined |
| 18-110_k141_82837 | Not-determined |
| 18-110_k141_232866 | Not-determined |
| 18-110_k141_232868 | Not-determined |
| 18-110_k141_152140 | Not-determined |
| 18-110_k141_163477 | Not-determined |
| 18-110_k141_94544 | Low-quality |
| 18-110_k141_140790 | Not-determined |
| 18-110_k141_13076 | Low-quality |
| 18-110_k141_13077 | Not-determined |
| 18-110_k141_2010 | Not-determined |
| 18-110_k141_152152 | Not-determined |
| 18-110_k141_82841 | Low-quality |
| 18-110_k141_244474 | Not-determined |
| 18-110_k141_255263 | Not-determined |
| 18-110_k141_244476 | Not-determined |
| 18-110_k141_152154 | Not-determined |
| 18-110_k141_128910 | Not-determined |
| 18-110_k141_244477 | Not-determined |
| 18-110_k141_163493 | Not-determined |
| 18-110_k141_128915 | Not-determined |
| 18-110_k141_232872 | Not-determined |
| 18-110_k141_232873 | Not-determined |
| 18-110_k141_244483 | Not-determined |
| 18-110_k141_163501 | Not-determined |
| 18-110_k141_186686 | Not-determined |
| 18-110_k141_186687 | Not-determined |
| 18-110_k141_94562 | Not-determined |
| 18-110_k141_175300 | Not-determined |
| 18-110_k141_267036 | Not-determined |
| 18-110_k141_244486 | Not-determined |
| 18-110_k141_232884 | Not-determined |
| 18-110_k141_197900 | Not-determined |
| 18-110_k141_13091 | Not-determined |
| 18-110_k141_267039 | Not-determined |
| 18-110_k141_152167 | Not-determined |
| 18-110_k141_82855 | Not-determined |
| 18-110_k141_152169 | Not-determined |
| 18-110_k141_152171 | Not-determined |
| 18-110_k141_232885 | Not-determined |
| 18-110_k141_232886 | Not-determined |
| 18-110_k141_255270 | Low-quality |
| 18-110_k141_117293 | Not-determined |
| 18-110_k141_117294 | Not-determined |
| 18-110_k141_36627 | Not-determined |
| 18-110_k141_197909 | Low-quality |
| 18-110_k141_255279 | Not-determined |
| 18-110_k141_255281 | Not-determined |
| 18-110_k141_60105 | Not-determined |
| 18-110_k141_255282 | Not-determined |
| 18-110_k141_197914 | Not-determined |
| 18-110_k141_94573 | Not-determined |
| 18-110_k141_267053 | Not-determined |
| 18-110_k141_117308 | Not-determined |
| 18-110_k141_2030 | Not-determined |
| 18-110_k141_117313 | Not-determined |
| 18-110_k141_117314 | Not-determined |
| 18-110_k141_267060 | Not-determined |
| 18-110_k141_209703 | Not-determined |
| 18-110_k141_25614 | Not-determined |
| 18-110_k141_105847 | Not-determined |
| 18-110_k141_197929 | Not-determined |
| 18-110_k141_197931 | Not-determined |
| 18-110_k141_267069 | Not-determined |
| 18-110_k141_175335 | Not-determined |
| 18-110_k141_60126 | Not-determined |
| 18-110_k141_94585 | Not-determined |
| 18-110_k141_152200 | Not-determined |
| 18-110_k141_94590 | Not-determined |
| 18-110_k141_267073 | Not-determined |
| 18-110_k141_25624 | Not-determined |
| 18-110_k141_163552 | Not-determined |
| 18-110_k141_25629 | Not-determined |
| 18-110_k141_267083 | Not-determined |
| 18-110_k141_267086 | Low-quality |
| 18-110_k141_232898 | Not-determined |
| 18-110_k141_25630 | Not-determined |
| 18-110_k141_232900 | Not-determined |
| 18-110_k141_60139 | Low-quality |
| 18-110_k141_2049 | Not-determined |
| 18-110_k141_267090 | Not-determined |
| 18-110_k141_13106 | Not-determined |
| 18-110_k141_220830 | Not-determined |
| 18-110_k141_36654 | Not-determined |
| 18-110_k141_267093 | Not-determined |
| 18-110_k141_163562 | Not-determined |
| 18-110_k141_152206 | Not-determined |
| 18-110_k141_13117 | Not-determined |
| 18-110_k141_71771 | Not-determined |
| 18-110_k141_267098 | Not-determined |
| 18-110_k141_163568 | Not-determined |
| 18-110_k141_267103 | Not-determined |
| 18-110_k141_2056 | Not-determined |
| 18-110_k141_117359 | Not-determined |
| 18-110_k141_220839 | Not-determined |
| 18-110_k141_140855 | Not-determined |
| 18-110_k141_209726 | Not-determined |
| 18-110_k141_152223 | Not-determined |
| 18-110_k141_140857 | Low-quality |
| 18-110_k141_267109 | Not-determined |
| 18-110_k141_140862 | Not-determined |
| 18-110_k141_2073 | Not-determined |
| 18-110_k141_2075 | Not-determined |
| 18-110_k141_197949 | Low-quality |
| 18-110_k141_209732 | Not-determined |
| 18-110_k141_140874 | Not-determined |
| 18-110_k141_94607 | Not-determined |
| 18-110_k141_117373 | Not-determined |
| 18-110_k141_2078 | Not-determined |
| 18-110_k141_48066 | Not-determined |
| 18-110_k141_2082 | Not-determined |
| 18-110_k141_140879 | Not-determined |
| 18-110_k141_36679 | Not-determined |
| 18-110_k141_48072 | Not-determined |
| 18-110_k141_2086 | Not-determined |
| 18-110_k141_2095 | Not-determined |
| 18-110_k141_36688 | Not-determined |
| 18-105-final-viral-combined_k141_123499\|\|full | Medium-quality |
| 18-110_k141_60169 | Not-determined |
| 18-110_k141_82886 | Not-determined |
| 18-110_k141_36697 | Not-determined |
| 18-110_k141_60174 | Not-determined |
| 18-110_k141_36699 | Not-determined |
| 18-110_k141_48090 | Not-determined |
| 18-110_k141_197967 | Not-determined |
| 18-110_k141_197971 | Low-quality |
| 18-110_k141_152262 | Not-determined |
| 18-110_k141_48104 | Not-determined |
| 18-110_k141_48109 | Not-determined |
| 18-110_k141_197994 | Not-determined |
| 18-110_k141_209773 | Not-determined |
| 18-110_k141_244523 | Not-determined |
| 18-110_k141_2101 | Not-determined |
| 18-110_k141_197999 | Low-quality |
| 18-110_k141_71808 | Not-determined |
| 18-110_k141_71815 | Low-quality |
| 18-110_k141_82901 | Not-determined |
| 18-110_k141_175362 | Not-determined |
| 18-110_k141_140918 | Not-determined |
| 18-110_k141_140920 | Not-determined |
| 18-110_k141_82906 | Not-determined |
| 18-110_k141_82907 | Not-determined |
| 18-110_k141_94642 | Not-determined |
| 18-110_k141_13141 | Not-determined |
| 18-110_k141_36736 | Not-determined |
| 18-110_k141_94644 | Not-determined |
| 18-110_k141_175370 | Not-determined |
| 18-110_k141_232907 | Not-determined |
| 18-110_k141_94646 | Low-quality |
| 18-110_k141_232912 | Not-determined |
| 18-110_k141_82917 | Not-determined |
| 18-110_k141_48144 | Not-determined |
| 17-109-final-viral-combined_k141_133608\|\|0_partial | Medium-quality |
| 18-110_k141_36745 | Not-determined |
| 18-110_k141_82919 | Not-determined |
| 18-110_k141_232917 | Not-determined |
| 18-110_k141_48150 | Not-determined |
| 18-110_k141_36753 | Not-determined |
| 18-110_k141_186716 | Not-determined |
| 18-110_k141_2132 | Not-determined |
| 18-110_k141_198042 | Low-quality |
| 18-110_k141_198044 | Not-determined |
| 18-110_k141_60185 | Low-quality |
| 18-110_k141_198046 | Not-determined |
| 18-110_k141_36758 | Not-determined |
| 18-110_k141_48160 | Not-determined |
| 18-110_k141_82939 | Not-determined |
| 18-110_k141_48164 | Not-determined |
| 18-110_k141_198048 | Not-determined |
| 18-110_k141_209806 | Not-determined |
| 18-110_k141_13150 | Not-determined |
| 18-110_k141_71863 | Not-determined |
| 18-110_k141_71865 | Not-determined |
| 18-110_k141_48174 | Not-determined |
| 18-110_k141_82954 | Not-determined |
| 18-110_k141_232929 | Not-determined |
| 18-110_k141_128929 | Not-determined |
| 18-110_k141_94658 | Not-determined |
| 18-110_k141_186740 | Not-determined |
| 18-110_k141_255307 | Not-determined |
| 18-110_k141_60198 | Not-determined |
| 18-110_k141_82962 | Not-determined |
| 18-110_k141_175419 | Not-determined |
| 18-110_k141_82964 | Not-determined |
| 18-110_k141_13165 | Not-determined |
| 18-110_k141_117412 | Not-determined |
| 18-110_k141_186748 | Not-determined |
| 18-110_k141_13168 | Not-determined |
| 18-110_k141_48192 | Not-determined |
| 18-110_k141_2162 | Not-determined |
| 18-110_k141_36773 | Not-determined |
| 18-110_k141_2166 | Not-determined |
| 18-110_k141_13179 | Low-quality |
| 18-110_k141_60222 | Low-quality |
| 18-110_k141_71873 | Not-determined |
| 18-110_k141_232945 | Not-determined |
| 18-110_k141_267151 | Not-determined |
| 18-110_k141_232947 | Not-determined |
| 18-110_k141_117419 | Not-determined |
| 18-110_k141_36776 | Not-determined |
| 18-110_k141_255329 | Not-determined |
| 18-110_k141_13189 | Not-determined |
| 18-110_k141_232952 | Not-determined |
| 18-110_k141_267156 | Not-determined |
| 18-110_k141_140950 | Low-quality |
| 18-110_k141_2171 | Not-determined |
| 18-110_k141_186767 | Not-determined |
| 18-110_k141_117429 | Low-quality |
| 18-110_k141_186770 | Not-determined |
| 18-110_k141_152288 | Not-determined |
| 18-110_k141_117433 | Not-determined |
| 18-110_k141_232961 | Not-determined |
| 18-110_k141_152294 | Low-quality |
| 18-110_k141_232966 | Not-determined |
| 18-110_k141_13206 | Not-determined |
| 18-110_k141_198076 | Not-determined |
| 18-110_k141_2175 | Not-determined |
| 18-110_k141_198079 | Not-determined |
| 18-110_k141_175446 | Not-determined |
| 18-110_k141_36794 | Not-determined |
| 18-110_k141_36795 | Not-determined |
| 18-110_k141_2187 | Not-determined |
| 18-110_k141_140986 | Not-determined |
| 18-110_k141_198093 | Not-determined |
| 18-110_k141_220964 | Not-determined |
| 18-110_k141_152308 | Not-determined |
| 18-110_k141_140998 | Not-determined |
| 18-110_k141_220970 | Not-determined |
| 18-110_k141_2191 | Low-quality |
| 18-110_k141_255357 | Low-quality |
| 18-110_k141_48218 | Not-determined |
| 18-110_k141_198103 | Not-determined |
| 18-110_k141_209841 | Low-quality |
| 18-110_k141_48220 | Not-determined |
| 18-110_k141_2198 | Not-determined |
| 18-110_k141_152312 | Not-determined |
| 18-110_k141_244562 | Not-determined |
| 18-110_k141_233000 | Not-determined |
| 18-110_k141_152315 | Not-determined |
| 18-110_k141_209852 | Not-determined |
| 18-110_k141_220979 | Not-determined |
| 18-110_k141_2211 | Not-determined |
| 18-110_k141_244571 | Not-determined |
| 18-110_k141_2214 | Low-quality |
| 18-110_k141_141020 | Low-quality |
| 18-110_k141_141027 | Not-determined |
| 18-110_k141_175451 | Not-determined |
| 18-110_k141_163600 | Not-determined |
| 18-110_k141_48234 | Not-determined |
| 18-110_k141_244581 | Not-determined |
| 18-110_k141_36838 | Not-determined |
| 18-110_k141_36843 | Not-determined |
| 18-110_k141_198141 | Not-determined |
| 18-110_k141_163620 | Not-determined |
| 18-110_k141_128970 | Not-determined |
| 18-110_k141_255363 | Not-determined |
| 18-110_k141_117474 | Not-determined |
| 18-110_k141_220999 | Low-quality |
| 18-110_k141_141048 | Not-determined |
| 18-110_k141_117476 | Not-determined |
| 18-110_k141_82999 | Not-determined |
| 18-110_k141_105856 | Low-quality |
| 18-110_k141_152346 | Not-determined |
| 18-110_k141_94731 | Not-determined |
| 18-110_k141_163637 | Low-quality |
| 18-110_k141_2219 | Not-determined |
| 18-110_k141_128983 | Not-determined |
| 18-110_k141_209887 | Not-determined |
| 18-110_k141_163646 | Not-determined |
| 18-110_k141_105864 | Not-determined |
| 18-110_k141_152360 | Not-determined |
| 18-110_k141_175475 | Not-determined |
| 18-110_k141_255375 | Not-determined |
| 18-110_k141_152363 | Not-determined |
| 18-110_k141_175477 | Low-quality |
| 18-110_k141_163653 | Not-determined |
| 18-110_k141_152364 | Not-determined |
| 18-110_k141_2230 | Not-determined |
| 18-110_k141_128994 | Not-determined |
| 18-110_k141_128995 | Not-determined |
| 18-110_k141_209899 | Not-determined |
| 18-110_k141_128996 | Not-determined |
| 18-110_k141_152370 | Not-determined |
| 18-110_k141_163655 | Low-quality |
| 18-110_k141_244615 | Not-determined |
| 18-110_k141_198157 | Not-determined |
| 18-110_k141_94749 | Low-quality |
| 18-110_k141_129002 | Not-determined |
| 18-110_k141_25729 | Not-determined |
| 18-110_k141_117479 | Not-determined |
| 18-110_k141_129008 | Low-quality |
| 18-110_k141_2238 | Not-determined |
| 18-110_k141_117484 | Not-determined |
| 18-110_k141_255384 | Not-determined |
| 18-110_k141_244625 | Low-quality |
| 18-110_k141_48257 | Not-determined |
| 18-110_k141_48259 | Not-determined |
| 18-110_k141_141071 | Not-determined |
| 18-110_k141_175501 | Not-determined |
| 18-110_k141_36886 | Not-determined |
| 18-110_k141_255393 | Not-determined |
| 18-110_k141_209919 | Not-determined |
| 18-110_k141_25745 | Not-determined |
| 18-110_k141_141083 | Not-determined |
| 18-110_k141_152384 | Not-determined |
| 18-110_k141_175515 | Not-determined |
| 18-110_k141_255400 | Not-determined |
| 18-110_k141_175521 | Not-determined |
| 18-110_k141_255404 | Not-determined |
| 18-110_k141_267227 | Not-determined |
| 18-110_k141_2253 | Not-determined |
| 18-110_k141_48288 | Not-determined |
| 18-110_k141_36900 | Not-determined |
| 18-110_k141_48290 | Not-determined |
| 18-110_k141_186841 | Not-determined |
| 18-110_k141_48294 | Not-determined |
| 18-110_k141_2259 | Not-determined |
| 18-110_k141_48296 | Not-determined |
| 18-110_k141_36907 | Not-determined |
| 18-110_k141_267234 | Not-determined |
| 18-110_k141_198174 | Not-determined |
| 18-110_k141_267238 | Not-determined |
| 18-110_k141_152411 | Not-determined |
| 18-110_k141_152412 | Not-determined |
| 18-110_k141_129045 | Not-determined |
| 18-110_k141_198178 | Not-determined |
| 18-110_k141_255410 | Low-quality |
| 18-110_k141_25757 | Not-determined |
| 18-110_k141_186852 | Not-determined |
| 18-110_k141_244666 | Not-determined |
| 18-110_k141_186856 | Not-determined |
| 18-110_k141_25760 | Not-determined |
| 18-110_k141_48308 | Not-determined |
| 18-110_k141_152419 | Low-quality |
| 18-110_k141_152421 | Not-determined |
| 18-110_k141_25762 | Not-determined |
| 18-110_k141_233015 | Not-determined |
| 18-110_k141_94789 | Not-determined |
| 18-110_k141_244682 | Not-determined |
| 18-110_k141_94796 | Not-determined |
| 18-110_k141_255420 | Not-determined |
| 18-110_k141_209937 | Not-determined |
| 18-110_k141_186864 | Not-determined |
| 18-110_k141_244692 | Not-determined |
| 18-110_k141_209944 | Not-determined |
| 18-110_k141_129062 | Not-determined |
| 18-110_k141_186874 | Low-quality |
| 18-110_k141_141116 | Not-determined |
| 18-110_k141_71944 | Not-determined |
| 18-110_k141_117493 | Not-determined |
| 18-110_k141_244704 | Not-determined |
| 18-110_k141_71947 | Low-quality |
| 18-110_k141_152426 | Not-determined |
| 18-110_k141_163688 | Not-determined |
| 18-110_k141_71949 | Not-determined |
| 18-110_k141_221043 | Not-determined |
| 18-110_k141_186887 | Not-determined |
| 18-110_k141_186896 | Not-determined |
| 18-110_k141_255437 | Not-determined |
| 18-110_k141_186899 | Low-quality |
| 18-110_k141_163704 | Not-determined |
| 18-110_k141_152442 | Not-determined |
| 18-110_k141_175559 | Not-determined |
| 18-110_k141_255439 | Low-quality |
| 18-110_k141_175562 | Not-determined |
| 18-110_k141_152444 | Not-determined |
| 18-110_k141_13228 | Not-determined |
| 18-110_k141_152446 | Not-determined |
| 18-110_k141_13232 | Not-determined |
| 18-110_k141_13233 | Not-determined |
| 18-110_k141_244718 | Not-determined |
| 18-110_k141_186911 | Not-determined |
| 18-110_k141_152450 | Not-determined |
| 18-110_k141_2281 | Not-determined |
| 18-110_k141_186918 | Not-determined |
| 18-110_k141_198200 | Not-determined |
| 18-110_k141_36943 | Not-determined |
| 18-110_k141_117504 | Not-determined |
| 18-110_k141_255450 | Not-determined |
| 18-110_k141_117506 | Not-determined |
| 18-110_k141_175567 | Not-determined |
| 18-110_k141_175569 | Not-determined |
| 18-110_k141_71977 | Not-determined |
| 18-110_k141_71978 | Not-determined |
| 18-110_k141_221055 | Not-determined |
| 18-110_k141_233072 | Not-determined |
| 18-110_k141_244751 | Not-determined |
| 18-110_k141_71989 | Not-determined |
| 18-110_k141_94828 | Not-determined |
| 18-110_k141_163732 | Not-determined |
| 18-110_k141_198221 | Not-determined |
| 18-110_k141_244755 | Not-determined |
| 18-110_k141_36956 | Not-determined |
| 18-110_k141_209981 | Not-determined |
| 18-110_k141_117520 | Not-determined |
| 18-110_k141_244758 | Not-determined |
| 18-110_k141_209989 | Not-determined |
| 18-110_k141_175579 | Not-determined |
| 18-110_k141_255469 | Not-determined |
| 18-110_k141_244764 | Not-determined |
| 18-110_k141_244765 | Not-determined |
| 18-110_k141_244766 | Not-determined |
| 18-110_k141_13246 | Not-determined |
| 18-110_k141_210000 | Not-determined |
| 18-110_k141_48365 | Not-determined |
| 18-110_k141_36963 | Not-determined |
| 18-110_k141_233081 | Low-quality |
| 18-110_k141_210002 | Not-determined |
| 18-110_k141_105915 | Not-determined |
| 18-110_k141_48370 | Not-determined |
| 18-110_k141_255471 | Not-determined |
| 18-110_k141_221077 | Low-quality |
| 18-110_k141_129115 | Low-quality |
| 18-110_k141_221079 | Not-determined |
| 18-110_k141_2299 | Not-determined |
| 18-110_k141_163750 | Not-determined |
| 18-110_k141_71995 | Not-determined |
| 18-110_k141_186938 | Not-determined |
| 18-110_k141_105920 | Not-determined |
| 18-110_k141_221081 | Not-determined |
| 18-110_k141_117543 | Not-determined |
| 18-110_k141_72001 | Not-determined |
| 18-110_k141_221087 | Not-determined |
| 18-110_k141_36980 | Not-determined |
| 18-110_k141_233100 | Not-determined |
| 18-110_k141_221094 | Not-determined |
| 18-110_k141_163760 | Low-quality |
| 18-110_k141_186941 | Not-determined |
| 18-110_k141_36987 | Not-determined |
| 18-110_k141_36994 | Not-determined |
| 18-110_k141_210026 | Not-determined |
| 18-110_k141_152463 | Not-determined |
| 18-110_k141_36999 | Not-determined |
| 18-110_k141_233113 | Not-determined |
| 18-110_k141_2321 | Not-determined |
| 18-110_k141_233114 | Not-determined |
| 18-110_k141_244794 | Not-determined |
| 18-110_k141_48386 | Not-determined |
| 18-110_k141_2324 | Not-determined |
| 18-110_k141_198236 | Not-determined |
| 18-110_k141_141165 | Not-determined |
| 18-110_k141_152474 | Not-determined |
| 18-110_k141_94877 | Low-quality |
| 18-110_k141_2331 | Not-determined |
| 18-110_k141_233125 | Not-determined |
| 18-110_k141_152483 | Not-determined |
| 18-110_k141_72026 | Low-quality |
| 18-110_k141_198244 | Not-determined |
| 18-110_k141_48395 | Not-determined |
| 18-110_k141_117555 | Not-determined |
| 18-110_k141_141175 | Not-determined |
| 18-110_k141_94884 | Not-determined |
| 18-110_k141_175600 | Not-determined |
| 18-110_k141_2342 | Not-determined |
| 18-110_k141_152491 | Not-determined |
| 18-110_k141_163778 | Not-determined |
| 18-110_k141_94887 | Not-determined |
| 18-110_k141_198253 | Not-determined |
| 18-110_k141_233126 | Low-quality |
| 18-110_k141_72039 | Not-determined |
| 18-110_k141_163788 | Not-determined |
| 18-110_k141_129160 | Not-determined |
| 18-110_k141_72041 | Not-determined |
| 18-110_k141_129164 | Low-quality |
| 18-110_k141_94898 | Not-determined |
| 18-110_k141_2355 | Not-determined |
| 18-110_k141_141186 | Not-determined |
| 18-110_k141_186971 | Not-determined |
| 18-110_k141_255517 | Not-determined |
| 18-110_k141_141188 | Not-determined |
| 18-110_k141_13275 | Not-determined |
| 18-110_k141_255524 | Not-determined |
| 18-110_k141_152512 | Not-determined |
| 18-110_k141_233140 | Not-determined |
| 18-110_k141_25879 | Not-determined |
| 18-110_k141_94911 | Not-determined |
| 18-110_k141_105940 | Not-determined |
| 18-110_k141_210057 | Not-determined |
| 18-110_k141_175624 | Not-determined |
| 18-110_k141_152519 | Not-determined |
| 18-110_k141_255533 | Not-determined |
| 18-110_k141_72062 | Not-determined |
| 18-110_k141_141201 | Low-quality |
| 18-110_k141_72064 | Not-determined |
| 18-110_k141_25881 | Low-quality |
| 18-110_k141_163821 | Not-determined |
| 18-110_k141_141204 | Not-determined |
| 18-110_k141_255539 | Not-determined |
| 18-110_k141_94921 | Not-determined |
| 18-110_k141_2368 | Not-determined |
| 18-110_k141_198269 | Not-determined |
| 18-110_k141_244834 | Low-quality |
| 18-110_k141_2374 | Not-determined |
| 18-110_k141_255546 | Not-determined |
| 18-110_k141_129181 | Not-determined |
| 18-110_k141_255547 | Not-determined |
| 18-110_k141_163823 | Not-determined |
| 18-110_k141_175633 | Not-determined |
| 18-110_k141_117573 | Not-determined |
| 18-110_k141_2384 | Not-determined |
| 18-110_k141_117576 | Not-determined |
| 18-110_k141_255552 | Low-quality |
| 18-110_k141_198292 | Not-determined |
| 18-110_k141_152545 | Not-determined |
| 18-110_k141_105948 | Not-determined |
| 18-110_k141_25888 | Not-determined |
| 18-110_k141_117588 | Not-determined |
| 18-110_k141_152547 | Not-determined |
| 18-110_k141_210082 | Not-determined |
| 18-110_k141_163842 | Not-determined |
| 18-110_k141_267258 | Not-determined |
| 18-110_k141_244852 | Not-determined |
| 18-110_k141_255566 | Low-quality |
| 18-110_k141_72094 | Not-determined |
| 18-110_k141_37042 | Not-determined |
| 18-110_k141_267263 | Not-determined |
| 18-110_k141_255570 | Not-determined |
| 18-110_k141_13310 | Not-determined |
| 18-110_k141_255574 | Not-determined |
| 18-110_k141_152555 | Not-determined |
| 18-110_k141_152556 | Not-determined |
| 18-110_k141_83017 | Low-quality |
| 18-110_k141_198314 | Not-determined |
| 18-110_k141_186984 | Not-determined |
| 18-110_k141_152563 | Not-determined |
| 18-110_k141_210097 | Not-determined |
| 18-110_k141_117600 | Not-determined |
| 18-110_k141_175657 | Not-determined |
| 18-110_k141_267276 | Not-determined |
| 18-110_k141_94953 | Not-determined |
| 18-110_k141_210100 | Not-determined |
| 18-110_k141_198318 | Not-determined |
| 18-110_k141_152570 | Low-quality |
| 18-110_k141_210104 | Low-quality |
| 18-110_k141_267288 | Not-determined |
| 18-110_k141_198325 | Not-determined |
| 18-110_k141_72117 | Not-determined |

| | |
|---|---|
| 18-110_k141_267291 | Not-determined |
| 18-110_k141_94963 | Not-determined |
| 18-110_k141_267300 | Not-determined |
| 18-110_k141_94967 | Not-determined |
| 18-110_k141_83037 | Not-determined |
| 18-110_k141_233192 | Not-determined |
| 18-110_k141_152574 | Low-quality |
| 18-110_k141_94973 | Low-quality |
| 18-110_k141_83041 | Not-determined |
| 18-110_k141_117617 | Not-determined |
| 18-110_k141_13332 | Not-determined |
| 18-110_k141_13334 | Not-determined |
| 18-110_k141_117619 | Not-determined |
| 18-110_k141_25925 | Not-determined |
| 18-110_k141_37066 | Not-determined |
| 18-110_k141_72131 | Not-determined |
| 18-110_k141_233200 | Not-determined |
| 18-110_k141_83048 | Not-determined |
| 18-110_k141_186995 | Not-determined |
| 18-110_k141_83049 | Not-determined |
| 18-110_k141_163854 | Not-determined |
| 18-110_k141_198336 | Not-determined |
| 18-110_k141_141236 | Not-determined |
| 18-110_k141_25935 | Not-determined |
| 18-110_k141_175689 | Not-determined |
| 18-110_k141_37068 | Low-quality |
| 18-110_k141_37069 | Not-determined |
| 18-110_k141_255585 | Low-quality |
| 18-110_k141_267327 | Not-determined |
| 18-110_k141_175705 | Not-determined |
| 18-110_k141_267335 | Not-determined |
| 18-110_k141_117628 | Low-quality |
| 18-110_k141_152592 | Not-determined |
| 18-110_k141_72156 | Not-determined |
| 18-110_k141_244888 | Not-determined |
| 18-110_k141_117634 | Not-determined |
| 18-110_k141_141251 | Low-quality |
| 18-110_k141_13366 | Not-determined |
| 18-110_k141_267345 | Not-determined |
| 18-110_k141_117637 | Not-determined |
| 18-110_k141_255599 | Not-determined |
| 18-110_k141_37078 | Not-determined |
| 18-110_k141_25951 | Not-determined |
| 18-110_k141_267356 | Not-determined |
| 18-110_k141_37080 | Not-determined |
| 18-110_k141_2414 | Not-determined |
| 18-110_k141_129244 | Not-determined |
| 18-110_k141_255606 | Not-determined |
| 18-110_k141_117643 | Not-determined |
| 18-110_k141_152611 | Not-determined |
| 18-110_k141_83070 | Low-quality |
| 18-110_k141_141267 | Not-determined |
| 18-110_k141_105977 | Not-determined |
| 18-110_k141_105978 | Not-determined |
| 18-110_k141_141276 | Not-determined |
| 18-110_k141_187022 | Not-determined |
| 18-110_k141_25967 | Not-determined |
| 18-110_k141_198364 | Not-determined |
| 18-110_k141_117656 | Not-determined |
| 18-110_k141_141284 | Not-determined |
| 18-110_k141_117658 | Not-determined |
| 18-110_k141_60280 | Not-determined |
| 18-110_k141_187029 | Not-determined |
| 18-110_k141_152627 | Not-determined |
| 18-110_k141_2437 | Not-determined |
| 18-110_k141_244916 | Not-determined |
| 18-110_k141_152629 | Not-determined |
| 18-110_k141_244918 | Not-determined |
| 18-110_k141_60282 | Low-quality |
| 18-110_k141_244921 | Not-determined |
| 18-110_k141_25973 | Not-determined |
| 18-110_k141_117663 | Not-determined |
| 18-110_k141_152633 | Not-determined |
| 18-110_k141_72206 | Not-determined |
| 18-110_k141_175741 | Not-determined |
| 18-110_k141_117664 | Not-determined |
| 18-110_k141_13395 | Not-determined |
| 18-110_k141_255636 | Not-determined |
| 18-110_k141_37099 | Not-determined |
| 18-110_k141_48427 | Not-determined |
| 18-110_k141_37102 | Not-determined |
| 18-110_k141_48441 | Low-quality |
| 18-110_k141_175756 | Not-determined |
| 18-110_k141_233251 | Not-determined |
| 18-110_k141_37108 | Low-quality |
| 18-110_k141_105995 | Not-determined |
| 18-110_k141_210133 | Low-quality |
| 18-110_k141_233254 | Not-determined |
| 18-110_k141_198391 | Not-determined |
| 18-110_k141_163869 | Not-determined |
| 18-110_k141_163870 | Not-determined |
| 18-110_k141_25996 | Not-determined |
| 18-110_k141_117686 | Low-quality |
| 18-110_k141_267401 | Not-determined |
| 18-110_k141_255645 | Low-quality |
| 18-110_k141_267405 | Not-determined |
| 18-110_k141_37122 | Not-determined |
| 18-110_k141_210136 | Not-determined |
| 18-110_k141_72231 | Not-determined |
| 18-110_k141_255649 | Not-determined |
| 18-110_k141_198399 | Not-determined |
| 18-110_k141_198400 | Not-determined |
| 18-110_k141_72236 | Not-determined |
| 18-110_k141_267414 | Not-determined |
| 18-110_k141_37126 | Low-quality |
| 18-110_k141_72239 | Not-determined |
| 18-110_k141_95012 | Not-determined |
| 18-110_k141_72248 | Not-determined |
| 18-110_k141_106013 | Not-determined |
| 18-110_k141_163887 | Not-determined |
| 18-110_k141_2453 | Not-determined |
| 18-110_k141_141321 | Not-determined |
| 18-110_k141_2455 | Not-determined |
| 18-110_k141_83121 | Not-determined |
| 18-110_k141_141330 | Not-determined |
| 18-110_k141_267426 | Not-determined |
| 18-110_k141_106017 | Low-quality |
| 18-110_k141_244952 | Not-determined |
| 18-110_k141_187038 | Not-determined |
| 18-110_k141_2464 | Not-determined |
| 18-110_k141_2466 | Not-determined |
| 18-110_k141_60330 | Not-determined |
| 18-110_k141_233274 | Not-determined |
| 18-110_k141_60337 | Not-determined |
| 18-110_k141_129320 | Not-determined |
| 18-110_k141_244962 | Not-determined |
| 18-110_k141_13449 | Not-determined |
| 18-110_k141_267442 | Not-determined |
| 18-110_k141_26028 | Not-determined |
| 18-110_k141_175806 | Not-determined |
| 18-110_k141_187053 | Not-determined |
| 18-110_k141_13456 | Not-determined |
| 18-110_k141_37156 | Not-determined |
| 18-110_k141_233295 | Not-determined |
| 18-110_k141_244979 | Not-determined |
| 18-110_k141_95043 | Not-determined |
| 18-110_k141_244986 | Not-determined |
| 18-110_k141_60352 | Not-determined |
| 18-110_k141_129340 | Not-determined |
| 18-110_k141_244987 | Not-determined |
| 18-110_k141_175825 | Not-determined |
| 18-110_k141_129345 | Low-quality |
| 18-110_k141_106042 | Low-quality |
| 18-110_k141_95057 | Low-quality |
| 18-110_k141_60354 | Not-determined |
| 18-110_k141_221171 | Not-determined |
| 18-110_k141_233313 | Not-determined |
| 18-110_k141_221174 | Not-determined |
| 18-110_k141_72295 | Not-determined |
| 18-110_k141_37171 | Not-determined |
| 18-110_k141_13476 | Not-determined |
| 18-110_k141_95068 | Not-determined |
| 18-110_k141_255696 | Not-determined |
| 18-110_k141_72306 | Not-determined |
| 18-110_k141_267463 | Not-determined |
| 18-110_k141_175845 | Not-determined |
| 18-110_k141_106056 | Not-determined |
| 18-110_k141_106057 | Low-quality |
| 18-110_k141_245005 | Not-determined |
| 18-110_k141_187064 | Not-determined |
| 18-110_k141_106063 | Not-determined |
| 18-110_k141_141352 | Not-determined |
| 18-110_k141_221178 | Not-determined |
| 18-110_k141_175861 | Not-determined |
| 18-110_k141_37196 | Not-determined |
| 18-110_k141_37198 | Not-determined |
| 18-110_k141_13483 | Not-determined |
| 18-110_k141_129363 | Not-determined |
| 18-110_k141_152706 | Low-quality |
| 18-110_k141_267477 | Not-determined |
| 18-110_k141_233362 | Not-determined |
| 18-110_k141_37206 | Low-quality |
| 18-110_k141_267480 | Not-determined |
| 18-110_k141_233366 | Not-determined |
| 18-110_k141_83142 | Not-determined |
| 18-110_k141_255718 | Not-determined |
| 18-110_k141_255721 | Not-determined |
| 18-110_k141_221198 | Not-determined |
| 18-110_k141_255725 | Not-determined |
| 18-110_k141_60378 | Not-determined |
| 18-110_k141_141376 | Not-determined |
| 18-110_k141_13504 | Not-determined |
| 18-110_k141_13506 | Not-determined |
| 18-110_k141_13507 | Not-determined |
| 18-110_k141_267489 | Not-determined |
| 18-110_k141_141385 | Not-determined |
| 18-110_k141_152730 | Not-determined |
| 18-110_k141_152731 | Not-determined |
| 18-110_k141_255734 | Not-determined |
| 18-110_k141_48492 | Not-determined |
| 18-110_k141_255736 | Low-quality |
| 18-110_k141_129390 | Not-determined |
| 18-110_k141_95118 | Not-determined |
| 18-110_k141_141401 | Not-determined |
| 18-110_k141_141405 | Not-determined |
| 18-110_k141_13516 | Not-determined |
| 18-110_k141_267499 | Not-determined |
| 18-110_k141_2521 | Not-determined |
| 18-110_k141_141408 | Not-determined |
| 18-110_k141_267500 | Not-determined |
| 18-110_k141_267502 | Not-determined |
| 18-110_k141_233393 | Not-determined |
| 18-110_k141_129400 | Low-quality |
| 18-110_k141_26044 | Not-determined |
| 18-110_k141_26050 | Not-determined |
| 18-110_k141_26051 | Not-determined |
| 18-110_k141_13521 | Not-determined |
| 18-110_k141_26053 | Not-determined |
| 18-110_k141_2524 | Not-determined |
| 18-110_k141_2527 | Not-determined |
| 18-110_k141_221215 | Not-determined |
| 18-110_k141_221216 | Not-determined |
| 18-110_k141_95135 | Not-determined |
| 18-110_k141_95139 | Not-determined |
| 18-110_k141_175914 | Not-determined |
| 18-110_k141_26066 | Not-determined |
| 18-110_k141_60417 | Not-determined |
| 18-110_k141_221218 | Not-determined |
| 18-110_k141_2534 | Not-determined |
| 18-110_k141_95145 | Not-determined |
| 18-110_k141_26074 | Not-determined |
| 18-110_k141_2537 | Low-quality |
| 18-110_k141_187133 | Not-determined |
| 18-110_k141_187134 | Not-determined |
| 18-110_k141_37233 | Not-determined |
| 18-110_k141_37235 | Not-determined |
| 18-110_k141_60427 | Not-determined |
| 18-110_k141_198414 | Not-determined |
| 18-110_k141_198418 | Not-determined |
| 18-110_k141_187142 | Not-determined |
| 18-110_k141_221221 | Not-determined |
| 18-110_k141_233412 | Not-determined |
| 18-110_k141_187146 | Not-determined |
| 18-110_k141_175937 | Low-quality |
| 18-110_k141_175938 | Not-determined |
| 18-110_k141_2549 | Not-determined |
| 18-110_k141_198430 | Low-quality |
| 18-110_k141_95172 | Not-determined |
| 18-110_k141_233419 | Not-determined |
| 18-110_k141_255759 | Not-determined |
| 18-110_k141_13558 | Not-determined |
| 18-110_k141_245030 | Not-determined |
| 18-110_k141_210181 | Not-determined |
| 18-110_k141_48513 | Not-determined |
| 18-110_k141_48514 | Not-determined |
| 18-110_k141_2570 | Not-determined |
| 18-110_k141_60450 | Not-determined |
| 18-110_k141_129452 | Not-determined |
| 18-110_k141_198447 | Not-determined |
| 18-110_k141_48517 | Not-determined |
| 18-110_k141_95192 | Not-determined |
| 18-110_k141_198450 | Not-determined |
| 18-110_k141_60455 | Low-quality |
| 18-110_k141_48525 | Not-determined |
| 18-110_k141_48529 | Not-determined |
| 18-110_k141_245046 | Not-determined |
| 18-110_k141_72391 | Not-determined |
| 18-110_k141_106145 | Not-determined |
| 18-110_k141_72392 | Not-determined |
| 18-110_k141_187188 | Not-determined |
| 18-110_k141_187189 | Not-determined |
| 18-110_k141_175985 | Not-determined |
| 18-110_k141_60469 | Not-determined |
| 18-110_k141_245050 | Not-determined |
| 18-110_k141_175989 | Not-determined |
| 18-110_k141_141467 | Not-determined |
| 18-110_k141_210194 | Not-determined |
| 18-110_k141_48542 | Not-determined |
| 18-110_k141_267586 | Not-determined |
| 18-110_k141_106158 | Not-determined |
| 18-110_k141_233449 | Not-determined |
| 18-110_k141_175997 | Not-determined |
| 18-110_k141_48549 | Not-determined |
| 18-110_k141_13591 | Not-determined |
| 18-110_k141_255786 | Not-determined |
| 18-110_k141_245063 | Not-determined |
| 18-110_k141_152796 | Not-determined |
| 18-110_k141_37260 | Low-quality |
| 18-110_k141_95205 | Not-determined |
| 18-110_k141_2595 | Low-quality |
| 18-110_k141_221274 | Not-determined |
| 18-110_k141_60491 | Not-determined |
| 18-110_k141_13600 | Not-determined |
| 18-110_k141_187205 | Not-determined |
| 18-110_k141_187206 | Not-determined |
| 18-110_k141_37264 | Not-determined |
| 18-110_k141_233463 | Not-determined |
| 18-110_k141_245071 | Not-determined |
| 18-110_k141_245072 | Not-determined |
| 18-110_k141_152808 | Low-quality |
| 18-110_k141_255794 | Not-determined |
| 18-110_k141_2603 | Not-determined |
| 18-110_k141_13608 | Not-determined |
| 18-110_k141_13609 | Not-determined |
| 18-110_k141_152813 | Not-determined |
| 18-110_k141_106177 | Not-determined |
| 18-110_k141_60504 | Not-determined |
| 18-110_k141_245075 | Low-quality |
| 18-110_k141_210209 | Not-determined |
| 18-110_k141_233482 | Not-determined |
| 18-110_k141_48576 | Not-determined |
| 18-110_k141_117731 | Not-determined |
| 18-110_k141_48578 | Not-determined |
| 18-110_k141_2610 | Low-quality |
| 18-110_k141_141474 | Not-determined |
| 18-110_k141_72412 | Not-determined |
| 18-110_k141_152823 | Low-quality |
| 18-110_k141_129494 | Low-quality |
| 18-110_k141_95228 | Not-determined |
| 18-110_k141_187214 | Not-determined |
| 18-110_k141_187215 | Not-determined |
| 18-110_k141_152825 | Not-determined |
| 18-110_k141_129500 | Not-determined |
| 18-110_k141_152827 | Not-determined |
| 18-110_k141_198484 | Not-determined |
| 18-110_k141_129507 | Not-determined |
| 18-110_k141_129510 | Not-determined |
| 18-110_k141_60517 | Not-determined |
| 18-110_k141_267622 | Not-determined |
| 18-110_k141_95240 | Not-determined |
| 18-110_k141_129513 | Not-determined |
| 18-110_k141_95243 | Not-determined |
| 18-110_k141_233503 | Not-determined |
| 18-110_k141_267626 | Not-determined |
| 18-110_k141_2633 | Low-quality |
| 18-110_k141_267634 | Not-determined |
| 18-110_k141_117750 | Not-determined |
| 18-110_k141_245079 | Low-quality |
| 18-110_k141_210228 | Not-determined |
| 18-110_k141_48592 | Not-determined |
| 18-110_k141_95250 | Not-determined |
| 18-110_k141_245082 | Not-determined |
| 18-110_k141_95254 | Not-determined |
| 18-110_k141_255818 | Low-quality |
| 18-110_k141_129529 | Not-determined |
| 18-110_k141_26095 | Not-determined |
| 18-110_k141_198492 | Not-determined |
| 18-110_k141_13648 | Not-determined |
| 18-110_k141_117762 | Not-determined |
| 18-110_k141_255824 | Low-quality |
| 18-110_k141_117767 | Not-determined |
| 18-110_k141_141517 | Not-determined |
| 18-110_k141_141518 | Not-determined |
| 18-110_k141_198502 | Not-determined |
| 18-110_k141_210243 | Not-determined |
| 18-110_k141_198503 | Not-determined |
| 18-110_k141_163892 | Not-determined |
| 18-110_k141_233525 | Not-determined |
| 18-110_k141_267644 | Low-quality |
| 18-110_k141_198514 | Not-determined |
| 18-101_k141_388292 | Medium-quality |
| 18-110_k141_187246 | Not-determined |
| 18-110_k141_26117 | Not-determined |
| 18-110_k141_13654 | Not-determined |
| 18-110_k141_187252 | Not-determined |
| 18-110_k141_221301 | Not-determined |
| 18-110_k141_2657 | Low-quality |
| 18-110_k141_221302 | Not-determined |
| 18-110_k141_95277 | Low-quality |
| 18-110_k141_26123 | Not-determined |
| 18-110_k141_233530 | Not-determined |
| 18-110_k141_245107 | Low-quality |
| 18-110_k141_60547 | Not-determined |
| 18-110_k141_233533 | Not-determined |
| 18-110_k141_245109 | Not-determined |
| 18-110_k141_117770 | Not-determined |
| 18-110_k141_245110 | Low-quality |
| 18-110_k141_60552 | Not-determined |
| 18-110_k141_48628 | Not-determined |
| 18-110_k141_141538 | Not-determined |
| 18-110_k141_60553 | Not-determined |
| 18-110_k141_48629 | Not-determined |
| 18-110_k141_152836 | Low-quality |
| 18-110_k141_117777 | Not-determined |
| 18-110_k141_95287 | Not-determined |
| 18-110_k141_60557 | Not-determined |
| 18-110_k141_221311 | Not-determined |
| 18-110_k141_221312 | Not-determined |
| 18-110_k141_60559 | Low-quality |
| 18-110_k141_245115 | Not-determined |
| 18-110_k141_255845 | Not-determined |
| 18-110_k141_163912 | Not-determined |
| 18-110_k141_117789 | Not-determined |
| 18-110_k141_152842 | Not-determined |
| 18-110_k141_48642 | Not-determined |
| 18-110_k141_267656 | Not-determined |
| 18-110_k141_117790 | Not-determined |
| 18-110_k141_26141 | Low-quality |
| 18-110_k141_117791 | Not-determined |
| 18-110_k141_117793 | Low-quality |
| 18-110_k141_117796 | Not-determined |
| 18-110_k141_267658 | Not-determined |
| 18-110_k141_48648 | Not-determined |
| 18-110_k141_267665 | Not-determined |
| 18-110_k141_60573 | Not-determined |
| 18-110_k141_60576 | Not-determined |
| 18-110_k141_95317 | Not-determined |
| 18-110_k141_60579 | Not-determined |
| 18-110_k141_198530 | Not-determined |
| 18-110_k141_187273 | Not-determined |
| 18-110_k141_95320 | Not-determined |
| 18-110_k141_187276 | Not-determined |
| 18-110_k141_129561 | Not-determined |
| 18-110_k141_187277 | Not-determined |
| 18-110_k141_95325 | Low-quality |
| 18-110_k141_95328 | Not-determined |
| 18-110_k141_187281 | Low-quality |
| 18-110_k141_95332 | Not-determined |
| 18-110_k141_26162 | Low-quality |
| 18-110_k141_255874 | Not-determined |
| 18-110_k141_95336 | Not-determined |
| 18-110_k141_26164 | Not-determined |
| 18-110_k141_255879 | Not-determined |
| 18-110_k141_210266 | Not-determined |
| 18-110_k141_176093 | Low-quality |
| 18-110_k141_60593 | Low-quality |
| 18-110_k141_95340 | Low-quality |
| 18-110_k141_233573 | Not-determined |
| 18-110_k141_198535 | Not-determined |
| 18-110_k141_13686 | Low-quality |
| 18-110_k141_95351 | Low-quality |
| 18-110_k141_72433 | Not-determined |
| 18-110_k141_221326 | Not-determined |
| 18-110_k141_129573 | Not-determined |
| 18-110_k141_255899 | Not-determined |
| 18-110_k141_37267 | Not-determined |
| 18-110_k141_141588 | Not-determined |
| 18-110_k141_129577 | Not-determined |
| 18-110_k141_72444 | Not-determined |
| 18-110_k141_255904 | Not-determined |
| 18-110_k141_37274 | Low-quality |
| 18-110_k141_233597 | Not-determined |
| 18-110_k141_72445 | Not-determined |
| 18-110_k141_37278 | Not-determined |
| 18-110_k141_163947 | Not-determined |
| 18-110_k141_95361 | Not-determined |
| 18-110_k141_72449 | Not-determined |
| 18-110_k141_255909 | Not-determined |
| 18-110_k141_163953 | Not-determined |
| 18-110_k141_152876 | Not-determined |
| 18-110_k141_255914 | Not-determined |
| 18-110_k141_95368 | Not-determined |
| 18-110_k141_95370 | Not-determined |
| 18-110_k141_163961 | Not-determined |
| 18-110_k141_37288 | Not-determined |
| 18-110_k141_245123 | Not-determined |
| 18-110_k141_72465 | Not-determined |
| 18-110_k141_72466 | Not-determined |
| 18-110_k141_198557 | Low-quality |
| 18-110_k141_221337 | Not-determined |
| 18-110_k141_163970 | Not-determined |
| 18-110_k141_117821 | Not-determined |
| 18-110_k141_233610 | Not-determined |
| 18-110_k141_95379 | Not-determined |
| 18-110_k141_233612 | Not-determined |
| 18-110_k141_163972 | Not-determined |
| 18-110_k141_141623 | Not-determined |
| 18-110_k141_141627 | Not-determined |
| 18-110_k141_2691 | Not-determined |
| 18-110_k141_129594 | Not-determined |
| 18-110_k141_255936 | Not-determined |
| 18-110_k141_95389 | Not-determined |
| 18-110_k141_176134 | Not-determined |
| 18-110_k141_233621 | Not-determined |
| 18-110_k141_2700 | Not-determined |
| 18-110_k141_176145 | Not-determined |
| 18-110_k141_2704 | Not-determined |
| 18-110_k141_267706 | Not-determined |
| 18-110_k141_37324 | Low-quality |
| 18-110_k141_37325 | Not-determined |
| 18-110_k141_129616 | Not-determined |
| 18-110_k141_129619 | Not-determined |
| 18-110_k141_221377 | Not-determined |
| 18-110_k141_221379 | Not-determined |
| 18-110_k141_37330 | Low-quality |
| 18-110_k141_26224 | Not-determined |
| 18-110_k141_117854 | Not-determined |
| 18-110_k141_267718 | Not-determined |
| 18-110_k141_26230 | Not-determined |
| 18-110_k141_198583 | Not-determined |
| 18-110_k141_2720 | Not-determined |
| 18-110_k141_267725 | Not-determined |
| 18-110_k141_129626 | Not-determined |
| 18-110_k141_210286 | Not-determined |
| 18-110_k141_37340 | Not-determined |
| 18-110_k141_210294 | Not-determined |
| 18-110_k141_141647 | Not-determined |
| 18-110_k141_267732 | Not-determined |
| 18-110_k141_117868 | Low-quality |
| 18-110_k141_164018 | Not-determined |
| 18-110_k141_210301 | Not-determined |
| 18-110_k141_164020 | Low-quality |
| 18-110_k141_26240 | Not-determined |
| 18-110_k141_37358 | Not-determined |
| 18-110_k141_37362 | Not-determined |
| 18-110_k141_72505 | Not-determined |
| 18-110_k141_37369 | Not-determined |
| 18-110_k141_26247 | Not-determined |
| 18-110_k141_176179 | Not-determined |
| 18-110_k141_176180 | Not-determined |
| 18-110_k141_152908 | Low-quality |
| 18-110_k141_187317 | Not-determined |
| 18-110_k141_48656 | Low-quality |
| 18-110_k141_141657 | Not-determined |
| 18-110_k141_198609 | Not-determined |
| 18-110_k141_152917 | Not-determined |
| 18-110_k141_255979 | Not-determined |
| 18-110_k141_141662 | Not-determined |
| 18-110_k141_198621 | Not-determined |
| 18-110_k141_141666 | Not-determined |
| 18-110_k141_176202 | Not-determined |
| 18-110_k141_26257 | Not-determined |
| 18-110_k141_176205 | Not-determined |
| 18-110_k141_152930 | Not-determined |
| 18-110_k141_141670 | Not-determined |
| 18-110_k141_164050 | Not-determined |
| 18-110_k141_152932 | Not-determined |
| 18-110_k141_255983 | Not-determined |
| 18-110_k141_233658 | Not-determined |
| 18-110_k141_72527 | Low-quality |
| 18-110_k141_233663 | Not-determined |
| 18-110_k141_141682 | Not-determined |
| 18-110_k141_267760 | Not-determined |
| 18-110_k141_233666 | Low-quality |
| 18-110_k141_152942 | Not-determined |
| 18-110_k141_141692 | Not-determined |
| 18-110_k141_267774 | Not-determined |
| 18-110_k141_233677 | Not-determined |
| 18-110_k141_176223 | Not-determined |
| 18-110_k141_26291 | Not-determined |
| 18-110_k141_152958 | Low-quality |
| 18-110_k141_152959 | Not-determined |
| 18-110_k141_129638 | Not-determined |
| 18-110_k141_72551 | Not-determined |
| 18-110_k141_152965 | Not-determined |
| 18-110_k141_245145 | Low-quality |
| 18-110_k141_176226 | Not-determined |
| 18-110_k141_48690 | Not-determined |
| 18-110_k141_176228 | Low-quality |
| 18-110_k141_129645 | Not-determined |
| 18-110_k141_60595 | Not-determined |
| 18-110_k141_60596 | Not-determined |
| 18-110_k141_267790 | Not-determined |
| 18-110_k141_176251 | Not-determined |
| 18-110_k141_141729 | Not-determined |
| 18-110_k141_141732 | Not-determined |
| 18-110_k141_221451 | Not-determined |
| 18-110_k141_2737 | Not-determined |
| 18-110_k141_267805 | Not-determined |
| 18-110_k141_164084 | Not-determined |
| 18-110_k141_164085 | Not-determined |
| 18-110_k141_221455 | Low-quality |
| 18-110_k141_233704 | Not-determined |
| 18-110_k141_176267 | Low-quality |
| 18-110_k141_2743 | Not-determined |
| 18-110_k141_233707 | Not-determined |
| 18-110_k141_255988 | Not-determined |
| 18-110_k141_255991 | Not-determined |
| 18-110_k141_72574 | Low-quality |
| 18-110_k141_72575 | Not-determined |
| 18-110_k141_72579 | Not-determined |
| 18-110_k141_48712 | Not-determined |
| 18-110_k141_245177 | Not-determined |
| 18-110_k141_2761 | Not-determined |
| 18-110_k141_129667 | Not-determined |
| 18-110_k141_117889 | Low-quality |
| 18-110_k141_72585 | Not-determined |
| 18-110_k141_198633 | Not-determined |
| 18-110_k141_221468 | Not-determined |
| 18-110_k141_164112 | Not-determined |
| 18-110_k141_72588 | Not-determined |
| 18-110_k141_267814 | Low-quality |
| 18-110_k141_129675 | Not-determined |
| 18-110_k141_245183 | Low-quality |
| 18-110_k141_245185 | Not-determined |
| 18-110_k141_117899 | Not-determined |
| 18-110_k141_72593 | Not-determined |
| 18-110_k141_245186 | Not-determined |
| 18-110_k141_141746 | Not-determined |
| 18-110_k141_210382 | Not-determined |
| 18-110_k141_141753 | Not-determined |
| 18-110_k141_72599 | Not-determined |
| 18-110_k141_72600 | Not-determined |
| 18-110_k141_198646 | Not-determined |
| 18-110_k141_129689 | Not-determined |
| 18-110_k141_117909 | Not-determined |
| 18-110_k141_198649 | Not-determined |
| 18-110_k141_141757 | Low-quality |
| 18-110_k141_117913 | Not-determined |

| | |
|---|---|
| 18-110_k141_176293 | Not-determined |
| 18-110_k141_72609 | Not-determined |
| 18-110_k141_176294 | Not-determined |
| 18-110_k141_72610 | Not-determined |
| 18-110_k141_129692 | Not-determined |
| 18-110_k141_256005 | Not-determined |
| 18-110_k141_129693 | Low-quality |
| 18-110_k141_37432 | Not-determined |
| 18-110_k141_48734 | Not-determined |
| 18-110_k141_48735 | Not-determined |
| 18-110_k141_48738 | Not-determined |
| 18-110_k141_37439 | Low-quality |
| 18-110_k141_2784 | Not-determined |
| 18-110_k141_141769 | Not-determined |
| 18-110_k141_48747 | Not-determined |
| 18-110_k141_256014 | Not-determined |
| 18-110_k141_60618 | Not-determined |
| 18-110_k141_37452 | Not-determined |
| 18-110_k141_72636 | Not-determined |
| 18-110_k141_72638 | Not-determined |
| 18-110_k141_187326 | Low-quality |
| 18-110_k141_37461 | Not-determined |
| 18-110_k141_106205 | Not-determined |
| 18-110_k141_233741 | Not-determined |
| 18-110_k141_256031 | Not-determined |
| 18-110_k141_106210 | Low-quality |
| 18-110_k141_256033 | Not-determined |
| 18-110_k141_60641 | Not-determined |
| 18-110_k141_106212 | Not-determined |
| 18-110_k141_198688 | Not-determined |
| 18-110_k141_256037 | Not-determined |
| 18-110_k141_176332 | Not-determined |
| 18-110_k141_256039 | Not-determined |
| 18-110_k141_60653 | Not-determined |
| 18-110_k141_164138 | Not-determined |
| 18-110_k141_245221 | Not-determined |
| 18-110_k141_60660 | Not-determined |
| 18-110_k141_141792 | Not-determined |
| 18-110_k141_26322 | Not-determined |
| 18-110_k141_48761 | Not-determined |
| 18-110_k141_26324 | Not-determined |
| 18-110_k141_256048 | Low-quality |
| 18-110_k141_48767 | Not-determined |
| 18-110_k141_198696 | Not-determined |
| 18-110_k141_256054 | Not-determined |
| 18-110_k141_221515 | Low-quality |
| 18-110_k141_95470 | Not-determined |
| 18-110_k141_198704 | Not-determined |
| 18-110_k141_233758 | Not-determined |
| 18-110_k141_26340 | Not-determined |
| 18-110_k141_176352 | Not-determined |
| 18-110_k141_176353 | Not-determined |
| 18-110_k141_117960 | Not-determined |
| 18-110_k141_256065 | Not-determined |
| 18-110_k141_256066 | Not-determined |
| 18-110_k141_210441 | Not-determined |
| 18-110_k141_176359 | Not-determined |
| 18-110_k141_26355 | Not-determined |
| 18-110_k141_95486 | Not-determined |
| 18-110_k141_26361 | Not-determined |
| 18-110_k141_37477 | Not-determined |
| 18-110_k141_210448 | Not-determined |
| 18-110_k141_37481 | Low-quality |
| 18-110_k141_187386 | Not-determined |
| 18-110_k141_117982 | Not-determined |
| 18-110_k141_198735 | Not-determined |
| 18-110_k141_233785 | Not-determined |
| 18-110_k141_37490 | Not-determined |
| 18-110_k141_210455 | Low-quality |
| 18-110_k141_176363 | Low-quality |
| 18-110_k141_72685 | Not-determined |
| 18-110_k141_233787 | Not-determined |
| 18-110_k141_176365 | Not-determined |
| 18-110_k141_117995 | Not-determined |
| 18-110_k141_141843 | Not-determined |
| 18-110_k141_72690 | Low-quality |
| 18-110_k141_118005 | Not-determined |
| 18-110_k141_245276 | Not-determined |
| 18-110_k141_164155 | Low-quality |
| 18-110_k141_164156 | Low-quality |
| 18-110_k141_106249 | Not-determined |
| 18-110_k141_245281 | Not-determined |
| 18-110_k141_176377 | Not-determined |
| 18-110_k141_48798 | Not-determined |
| 18-110_k141_267929 | Not-determined |
| 18-110_k141_141855 | Not-determined |
| 18-110_k141_106253 | Not-determined |
| 18-110_k141_2802 | Not-determined |
| 18-110_k141_129777 | Not-determined |
| 18-110_k141_221563 | Not-determined |
| 18-110_k141_118027 | Not-determined |
| 18-110_k141_37501 | Not-determined |
| 18-110_k141_95529 | Low-quality |
| 18-110_k141_164179 | Not-determined |
| 18-110_k141_83160 | Low-quality |
| 18-110_k141_2809 | Not-determined |
| 18-110_k141_83164 | Not-determined |
| 18-110_k141_2811 | Not-determined |
| 18-110_k141_106266 | Not-determined |
| 18-110_k141_152969 | Not-determined |
| 18-110_k141_245292 | Low-quality |
| 18-110_k141_106268 | Low-quality |
| 18-110_k141_118040 | Not-determined |
| 18-110_k141_106271 | Not-determined |
| 18-110_k141_60716 | Low-quality |
| 18-110_k141_245294 | Not-determined |
| 18-110_k141_141877 | Not-determined |
| 18-110_k141_48810 | Not-determined |
| 18-110_k141_48811 | Not-determined |
| 18-110_k141_72721 | Not-determined |
| 18-110_k141_60724 | Not-determined |
| 18-110_k141_83174 | Not-determined |
| 18-110_k141_164201 | Not-determined |
| 18-110_k141_48818 | Not-determined |
| 18-110_k141_187453 | Not-determined |
| 18-110_k141_221576 | Not-determined |
| 18-110_k141_129794 | Not-determined |
| 18-110_k141_221580 | Low-quality |
| 18-110_k141_221581 | Not-determined |
| 18-110_k141_221583 | Not-determined |
| 18-110_k141_2823 | Not-determined |
| 18-110_k141_141885 | Not-determined |
| 18-110_k141_48824 | Not-determined |
| 18-110_k141_141887 | Low-quality |
| 18-110_k141_141890 | Low-quality |
| 18-110_k141_129810 | Not-determined |
| 18-110_k141_164211 | Not-determined |
| 18-110_k141_2834 | Not-determined |
| 18-110_k141_83194 | Not-determined |
| 18-110_k141_95557 | Not-determined |
| 18-110_k141_152999 | Not-determined |
| 18-110_k141_210502 | Not-determined |
| 18-110_k141_210504 | Not-determined |
| 18-110_k141_233809 | Not-determined |
| 18-110_k141_221595 | Not-determined |
| 18-110_k141_267959 | Not-determined |
| 18-110_k141_83201 | Not-determined |
| 18-110_k141_95561 | Not-determined |
| 18-110_k141_233810 | Not-determined |
| 18-110_k141_118052 | Not-determined |
| 18-110_k141_164220 | Not-determined |
| 18-110_k141_233812 | Not-determined |
| 18-110_k141_176421 | Not-determined |
| 18-110_k141_164223 | Not-determined |
| 18-110_k141_267963 | Low-quality |
| 18-110_k141_37552 | Not-determined |
| 18-110_k141_267964 | Not-determined |
| 18-110_k141_129830 | Not-determined |
| 18-110_k141_164227 | Not-determined |
| 18-110_k141_153016 | Low-quality |
| 18-110_k141_210524 | Not-determined |
| 18-110_k141_2858 | Not-determined |
| 18-110_k141_37562 | Not-determined |
| 18-110_k141_2862 | Not-determined |
| 18-110_k141_83214 | Not-determined |
| 18-110_k141_129840 | Not-determined |
| 18-110_k141_60767 | Not-determined |
| 18-110_k141_83221 | Not-determined |
| 18-110_k141_2867 | Not-determined |
| 18-110_k141_233832 | Not-determined |
| 18-110_k141_245320 | Not-determined |
| 18-110_k141_176435 | Not-determined |
| 18-110_k141_256103 | Not-determined |
| 18-110_k141_245325 | Not-determined |
| 18-110_k141_256107 | Not-determined |
| 18-110_k141_256110 | Not-determined |
| 18-110_k141_210535 | Not-determined |
| 18-110_k141_106292 | Not-determined |
| 18-110_k141_83232 | Not-determined |
| 18-110_k141_153044 | Not-determined |
| 18-110_k141_2873 | Low-quality |
| 18-110_k141_60784 | Low-quality |
| 18-110_k141_129857 | Low-quality |
| 18-110_k141_2878 | Not-determined |
| 18-110_k141_256125 | Not-determined |
| 18-110_k141_37608 | Not-determined |
| 18-110_k141_48868 | Not-determined |
| 18-110_k141_72789 | Not-determined |
| 18-110_k141_129872 | Not-determined |
| 18-110_k141_141906 | Not-determined |
| 18-110_k141_83249 | Not-determined |
| 18-110_k141_13791 | Not-determined |
| 18-110_k141_233850 | Low-quality |
| 18-110_k141_118083 | Not-determined |
| 18-110_k141_164278 | Not-determined |
| 18-110_k141_141913 | Not-determined |
| 18-110_k141_72791 | Low-quality |
| 18-110_k141_37612 | Not-determined |
| 18-110_k141_37613 | Not-determined |
| 18-110_k141_164281 | Not-determined |
| 18-110_k141_141915 | Not-determined |
| 18-110_k141_187513 | Not-determined |
| 18-110_k141_118094 | Not-determined |
| 18-110_k141_198761 | Not-determined |
| 18-110_k141_2886 | Not-determined |
| 18-110_k141_164288 | Not-determined |
| 18-110_k141_245360 | Not-determined |
| 18-110_k141_83270 | Not-determined |
| 18-110_k141_245364 | Not-determined |
| 18-110_k141_72802 | Not-determined |
| 18-110_k141_267997 | Not-determined |
| 18-110_k141_60818 | Not-determined |
| 18-110_k141_106325 | Not-determined |
| 18-110_k141_221654 | Not-determined |
| 18-110_k141_176480 | Not-determined |
| 18-110_k141_106327 | Not-determined |
| 18-110_k141_176481 | Not-determined |
| 18-110_k141_129902 | Not-determined |
| 18-110_k141_83275 | Not-determined |
| 18-110_k141_245374 | Not-determined |
| 18-110_k141_2908 | Not-determined |
| 18-110_k141_221660 | Not-determined |
| 18-110_k141_141941 | Not-determined |
| 18-110_k141_48899 | Not-determined |
| 18-110_k141_106335 | Not-determined |
| 18-110_k141_106337 | Not-determined |
| 18-110_k141_60832 | Not-determined |
| 18-110_k141_72817 | Not-determined |
| 18-110_k141_106339 | Not-determined |
| 18-110_k141_153072 | Not-determined |
| 18-110_k141_60840 | Not-determined |
| 18-110_k141_245395 | Not-determined |
| 18-110_k141_141950 | Not-determined |
| 18-110_k141_106345 | Not-determined |
| 18-110_k141_48925 | Not-determined |
| 18-110_k141_26403 | Not-determined |
| 18-110_k141_37640 | Not-determined |
| 18-110_k141_106354 | Not-determined |
| 18-110_k141_37642 | Not-determined |
| 18-110_k141_2935 | Low-quality |
| 18-110_k141_2937 | Not-determined |
| 18-110_k141_2938 | Not-determined |
| 18-110_k141_141970 | Not-determined |
| 18-110_k141_26410 | Not-determined |
| 18-110_k141_164292 | Not-determined |
| 18-110_k141_268033 | Not-determined |
| 18-110_k141_37648 | Not-determined |
| 18-110_k141_187539 | Low-quality |
| 18-110_k141_198801 | Not-determined |
| 18-110_k141_256155 | Not-determined |
| 18-110_k141_141982 | Not-determined |
| 18-110_k141_26418 | Not-determined |
| 18-110_k141_2948 | Not-determined |
| 18-110_k141_26420 | Not-determined |
| 18-110_k141_129931 | Not-determined |
| 18-110_k141_245408 | Low-quality |
| 18-110_k141_60844 | Not-determined |
| 18-110_k141_256164 | Not-determined |
| 18-110_k141_268048 | Not-determined |
| 18-110_k141_256167 | Low-quality |
| 18-110_k141_37666 | Not-determined |
| 18-110_k141_72850 | Not-determined |
| 18-110_k141_106377 | Not-determined |
| 18-110_k141_245409 | Not-determined |
| 18-110_k141_106380 | Low-quality |
| 18-110_k141_129952 | Not-determined |
| 18-110_k141_187564 | Not-determined |
| 18-110_k141_118135 | Not-determined |
| 18-110_k141_72861 | Not-determined |
| 18-110_k141_256176 | Low-quality |
| 18-110_k141_60864 | Not-determined |
| 18-110_k141_164324 | Not-determined |
| 18-110_k141_187569 | Low-quality |
| 18-110_k141_72869 | Not-determined |
| 18-110_k141_72870 | Not-determined |
| 18-110_k141_129964 | Not-determined |
| 18-110_k141_268065 | Not-determined |
| 18-110_k141_164329 | Not-determined |
| 18-110_k141_153120 | Not-determined |
| 18-110_k141_26446 | Not-determined |
| 18-110_k141_2970 | Low-quality |
| 18-110_k141_83346 | Not-determined |
| 18-110_k141_164330 | Low-quality |
| 18-110_k141_60884 | Not-determined |
| 18-110_k141_198831 | Not-determined |
| 18-110_k141_176505 | Not-determined |
| 18-110_k141_164332 | Not-determined |
| 18-110_k141_268082 | Not-determined |
| 18-110_k141_153126 | Not-determined |
| 18-110_k141_187582 | Not-determined |
| 18-110_k141_153129 | Not-determined |
| 18-110_k141_198841 | Not-determined |
| 18-110_k141_198842 | Not-determined |
| 18-110_k141_37672 | Not-determined |
| 18-110_k141_141999 | Not-determined |
| 18-110_k141_187586 | Not-determined |
| 18-110_k141_268091 | Not-determined |
| 18-110_k141_106405 | Not-determined |
| 18-110_k141_106407 | Not-determined |
| 18-110_k141_187596 | Not-determined |
| 18-110_k141_72879 | Not-determined |
| 18-110_k141_268105 | Not-determined |
| 18-110-final-viral-combined_k141_234473\|\|full | Medium-quality |
| 18-110_k141_221715 | Not-determined |
| 18-110_k141_106421 | Low-quality |
| 18-110_k141_210552 | Not-determined |
| 18-110_k141_72885 | Not-determined |
| 18-110_k141_198856 | Not-determined |
| 18-110_k141_164372 | Not-determined |
| 18-110_k141_37687 | Not-determined |
| 18-110_k141_256229 | Not-determined |
| 18-110_k141_164376 | Not-determined |
| 18-110_k141_210563 | Not-determined |
| 18-110_k141_130012 | Not-determined |
| 18-110_k141_256235 | Not-determined |
| 18-110_k141_176537 | Not-determined |
| 18-110_k141_245432 | Not-determined |
| 18-110_k141_37699 | Not-determined |
| 18-110_k141_164381 | Not-determined |
| 18-110_k141_83358 | Not-determined |
| 18-110_k141_60922 | Not-determined |
| 18-110_k141_95649 | Not-determined |
| 18-110_k141_187630 | Low-quality |
| 18-110_k141_3017 | Not-determined |
| 18-110_k141_118177 | Low-quality |
| 18-110_k141_106443 | Not-determined |
| 18-110_k141_3023 | Not-determined |
| 18-110_k141_256257 | Not-determined |
| 18-110_k141_118185 | Not-determined |
| 18-110_k141_106450 | Not-determined |
| 18-110_k141_268114 | Not-determined |
| 18-110_k141_3032 | Not-determined |
| 18-110_k141_37712 | Not-determined |
| 18-110_k141_118190 | Low-quality |
| 18-110_k141_106460 | Not-determined |
| 18-110_k141_95661 | Not-determined |
| 18-110_k141_118194 | Not-determined |
| 18-110_k141_13796 | Not-determined |
| 18-110_k141_210591 | Not-determined |
| 18-110_k141_118198 | Not-determined |
| 18-110_k141_3039 | Low-quality |
| 18-110_k141_233886 | Not-determined |
| 18-110_k141_176561 | Not-determined |
| 18-110_k141_176562 | Not-determined |
| 18-110_k141_176563 | Not-determined |
| 18-110_k141_26520 | Not-determined |
| 18-110_k141_48950 | Not-determined |
| 18-110_k141_83383 | Not-determined |
| 18-110_k141_60957 | Not-determined |
| 18-110_k141_233898 | Not-determined |
| 18-110_k141_233900 | Not-determined |
| 18-110_k141_48957 | Low-quality |
| 18-110_k141_83391 | Not-determined |
| 18-110_k141_221753 | Not-determined |
| 18-110_k141_198878 | Not-determined |
| 18-110_k141_256285 | Not-determined |
| 18-110_k141_118220 | Not-determined |
| 18-110_k141_106486 | Not-determined |
| 18-110_k141_268129 | Not-determined |
| 18-110_k141_268130 | Not-determined |
| 18-110_k141_106489 | Not-determined |
| 18-110_k141_26535 | Not-determined |
| 18-110_k141_48966 | Not-determined |
| 18-110_k141_60976 | Not-determined |
| 18-110_k141_233913 | Not-determined |
| 18-110_k141_153150 | Not-determined |
| 18-110_k141_48972 | Not-determined |
| 18-110_k141_210623 | Not-determined |
| 18-110_k141_221763 | Not-determined |
| 18-110_k141_26547 | Not-determined |
| 18-110_k141_210629 | Not-determined |
| 18-110_k141_153158 | Not-determined |
| 18-110_k141_210636 | Not-determined |
| 18-110_k141_268137 | Not-determined |
| 18-110_k141_210637 | Not-determined |
| 18-110_k141_187658 | Not-determined |
| 18-110_k141_153166 | Not-determined |
| 18-110_k141_83406 | Not-determined |
| 18-110_k141_176596 | Not-determined |
| 18-110_k141_118242 | Not-determined |
| 18-110_k141_268139 | Not-determined |
| 18-110_k141_37745 | Low-quality |
| 18-110_k141_268146 | Not-determined |
| 18-110_k141_37748 | Not-determined |
| 18-110_k141_60983 | Not-determined |
| 18-110_k141_142019 | Low-quality |
| 18-110_k141_60986 | Not-determined |
| 18-110_k141_153181 | Not-determined |
| 18-110_k141_13849 | Not-determined |
| 18-110_k141_118247 | Not-determined |
| 18-110_k141_198908 | Not-determined |
| 18-110_k141_142026 | Not-determined |
| 18-110_k141_130042 | Not-determined |
| 18-110_k141_26584 | Not-determined |
| 18-110_k141_118255 | Not-determined |
| 18-110_k141_106499 | Not-determined |
| 18-110_k141_37760 | Not-determined |
| 18-110_k141_245436 | Not-determined |
| 18-110_k141_26591 | Not-determined |
| 18-110_k141_130051 | Not-determined |
| 18-110_k141_221782 | Not-determined |
| 18-110_k141_164414 | Not-determined |
| 18-110_k141_37765 | Not-determined |
| 18-110_k141_37767 | Low-quality |
| 18-110_k141_187676 | Not-determined |
| 18-110_k141_37771 | Not-determined |
| 18-110_k141_48987 | Not-determined |
| 18-110_k141_176627 | Not-determined |
| 18-110_k141_187683 | Not-determined |
| 18-110_k141_153197 | Not-determined |
| 18-110_k141_106519 | Not-determined |
| 18-110_k141_142060 | Not-determined |
| 18-110_k141_37778 | Not-determined |
| 18-110_k141_187688 | Not-determined |
| 18-110_k141_153203 | Not-determined |
| 18-110_k141_176642 | Low-quality |
| 18-110_k141_142064 | Not-determined |
| 18-110_k141_106522 | Low-quality |
| 18-110_k141_245455 | Not-determined |
| 18-110_k141_13877 | Not-determined |
| 18-110_k141_118288 | Not-determined |
| 18-110_k141_106526 | Not-determined |
| 18-110_k141_233954 | Not-determined |
| 18-110_k141_106528 | Low-quality |
| 18-110_k141_118297 | Not-determined |
| 18-110_k141_233958 | Not-determined |
| 18-110_k141_245461 | Not-determined |
| 18-110_k141_118300 | Not-determined |
| 18-110_k141_153214 | Not-determined |
| 18-110_k141_118303 | Not-determined |
| 18-110_k141_49006 | Not-determined |
| 18-110_k141_164452 | Not-determined |
| 18-110_k141_61035 | Not-determined |
| 18-110_k141_61038 | Not-determined |
| 18-110_k141_37786 | Not-determined |
| 18-110_k141_233964 | Not-determined |
| 18-110_k141_118314 | Not-determined |
| 18-110_k141_118319 | Not-determined |
| 18-110_k141_130073 | Not-determined |
| 18-110_k141_49018 | Not-determined |
| 18-110_k141_164465 | Not-determined |
| 18-110_k141_61047 | Not-determined |
| 18-110_k141_26643 | Not-determined |
| 18-110_k141_72918 | Not-determined |
| 18-110_k141_142084 | Not-determined |
| 18-110_k141_37795 | Not-determined |
| 18-110_k141_118324 | Low-quality |
| 18-110_k141_164471 | Not-determined |
| 18-110_k141_268170 | Not-determined |
| 18-110_k141_268171 | Not-determined |
| 18-110_k141_26653 | Low-quality |
| 18-110_k141_245472 | Low-quality |
| 18-110_k141_118327 | Not-determined |
| 18-110_k141_176659 | Not-determined |
| 18-110_k141_26657 | Not-determined |
| 18-110_k141_130081 | Not-determined |
| 18-110_k141_268182 | Not-determined |
| 18-110_k141_187708 | Not-determined |
| 18-110_k141_26665 | Low-quality |
| 18-110_k141_106557 | Not-determined |
| 18-110_k141_49036 | Not-determined |
| 18-110_k141_268199 | Not-determined |
| 18-110_k141_233990 | Low-quality |
| 18-110_k141_130100 | Not-determined |
| 18-110_k141_221822 | Low-quality |
| 18-110_k141_198957 | Not-determined |
| 18-110_k141_245495 | Not-determined |
| 18-110_k141_3113 | Low-quality |
| 18-110_k141_106569 | Low-quality |
| 18-110_k141_187721 | Not-determined |
| 18-110_k141_72953 | Not-determined |
| 18-110_k141_37824 | Not-determined |
| 18-110_k141_49048 | Not-determined |
| 18-110_k141_187725 | Not-determined |
| 18-110_k141_187726 | Not-determined |
| 18-110_k141_72959 | Low-quality |
| 18-110_k141_49056 | Not-determined |
| 18-110_k141_49057 | Not-determined |
| 18-110_k141_49058 | Not-determined |
| 18-110_k141_49060 | Not-determined |
| 18-110_k141_106581 | Not-determined |
| 18-110_k141_26683 | Not-determined |
| 18-110_k141_176687 | Not-determined |
| 18-110_k141_130131 | Not-determined |
| 18-110_k141_245514 | Not-determined |
| 18-110_k141_245516 | Not-determined |
| 18-110_k141_3134 | Not-determined |
| 18-110_k141_187750 | Not-determined |
| 18-110_k141_37856 | Not-determined |
| 18-110_k141_37857 | Not-determined |
| 18-110_k141_245524 | Not-determined |
| 18-110_k141_37858 | Low-quality |
| 18-110_k141_245528 | Not-determined |
| 18-110_k141_198979 | Not-determined |
| 18-110_k141_210650 | Not-determined |
| 18-110_k141_234035 | Not-determined |
| 18-110_k141_187769 | Not-determined |
| 18-110_k141_221844 | Not-determined |
| 18-110_k141_176697 | Not-determined |
| 18-110_k141_72997 | Not-determined |
| 18-110_k141_234047 | Not-determined |
| 18-110_k141_210660 | Not-determined |
| 18-110_k141_73004 | Low-quality |
| 18-110_k141_13916 | Not-determined |
| 18-110_k141_13917 | Not-determined |
| 18-110_k141_73008 | Not-determined |
| 18-110_k141_49092 | Not-determined |
| 18-110_k141_245553 | Low-quality |
| 18-110_k141_268204 | Not-determined |
| 18-110_k141_268205 | Not-determined |
| 18-110_k141_130150 | Not-determined |
| 18-110_k141_153227 | Not-determined |
| 18-110_k141_73018 | Not-determined |
| 18-110_k141_130155 | Not-determined |
| 18-110_k141_142124 | Not-determined |
| 18-110_k141_49104 | Not-determined |
| 18-110_k141_210677 | Not-determined |
| 18-110_k141_73026 | Low-quality |
| 18-110_k141_73027 | Not-determined |
| 18-110_k141_245569 | Not-determined |
| 18-110_k141_130169 | Not-determined |
| 18-110_k141_26700 | Not-determined |
| 18-110_k141_106615 | Not-determined |
| 18-110_k141_245570 | Not-determined |
| 18-110_k141_130171 | Not-determined |
| 18-110_k141_245573 | Not-determined |
| 18-110_k141_106624 | Not-determined |
| 18-110_k141_268213 | Not-determined |
| 18-110_k141_268215 | Not-determined |
| 18-110_k141_210693 | Not-determined |
| 18-110_k141_26713 | Not-determined |
| 18-110_k141_221878 | Not-determined |
| 18-110_k141_118374 | Not-determined |
| 18-110_k141_37892 | Not-determined |
| 18-110_k141_49111 | Not-determined |
| 18-110_k141_268225 | Not-determined |
| 18-110_k141_153260 | Not-determined |
| 18-110_k141_26722 | Not-determined |
| 18-110_k141_234083 | Not-determined |
| 18-110_k141_130182 | Not-determined |
| 18-110_k141_26727 | Not-determined |
| 18-110_k141_268231 | Not-determined |
| 18-110_k141_187799 | Not-determined |
| 18-110_k141_176742 | Not-determined |
| 18-110_k141_142142 | Not-determined |
| 18-110_k141_164503 | Not-determined |
| 18-110_k141_164507 | Not-determined |
| 18-110_k141_187813 | Not-determined |
| 18-110_k141_268240 | Not-determined |
| 18-110_k141_187814 | Low-quality |
| 18-110_k141_221907 | Low-quality |
| 18-110_k141_49128 | Not-determined |
| 18-110_k141_210714 | Not-determined |
| 18-110_k141_130216 | Not-determined |
| 18-110_k141_130219 | Low-quality |
| 18-110_k141_3179 | Not-determined |
| 18-110_k141_130222 | Not-determined |
| 18-110_k141_3183 | Not-determined |
| 18-110_k141_234115 | Not-determined |
| 18-110_k141_37908 | Not-determined |
| 18-110_k141_37909 | Not-determined |
| 18-110_k141_176772 | Not-determined |
| 18-110_k141_13968 | Not-determined |
| 18-110_k141_187827 | Not-determined |
| 18-110_k141_73071 | Not-determined |
| 18-110_k141_106655 | Not-determined |
| 18-110_k141_83472 | Not-determined |
| 18-110_k141_130231 | Not-determined |
| 18-110_k141_234125 | Not-determined |
| 18-110_k141_106657 | Not-determined |
| 18-110_k141_73074 | Not-determined |
| 18-110_k141_95733 | Not-determined |
| 18-110_k141_49148 | Not-determined |
| 18-110_k141_73081 | Not-determined |
| 18-110_k141_73082 | Not-determined |
| 18-110_k141_164536 | Not-determined |
| 18-110_k141_73084 | Not-determined |
| 18-110_k141_95739 | Not-determined |
| 18-110_k141_268277 | Not-determined |
| 18-110_k141_187837 | Not-determined |
| 18-110_k141_95743 | Not-determined |
| 18-110_k141_37911 | Not-determined |
| 18-110_k141_153293 | Low-quality |
| 18-110_k141_268287 | Not-determined |
| 18-110_k141_3205 | Not-determined |
| 18-110_k141_49168 | Not-determined |
| 18-110_k141_49169 | Not-determined |
| 18-110_k141_164543 | Not-determined |
| 18-110_k141_268297 | Low-quality |
| 18-110_k141_234133 | Not-determined |
| 18-110_k141_164545 | Not-determined |
| 18-110_k141_130243 | Not-determined |
| 18-110_k141_164549 | Not-determined |
| 18-110_k141_106677 | Not-determined |
| 18-110_k141_268305 | Not-determined |
| 18-110_k141_106680 | Not-determined |
| 18-110_k141_106681 | Not-determined |
| 18-110_k141_176789 | Not-determined |
| 18-110_k141_37927 | Not-determined |

| | |
|---|---|
| 18-110_k141_37929 | Not-determined |
| 18-110_k141_83490 | Not-determined |
| 18-110_k141_130254 | Not-determined |
| 18-110_k141_234141 | Not-determined |
| 18-110_k141_210729 | Not-determined |
| 18-110_k141_106691 | Not-determined |
| 18-110_k141_130257 | Not-determined |
| 18-110_k141_37932 | Not-determined |
| 18-110_k141_130258 | Not-determined |
| 18-110_k141_37935 | Not-determined |
| 18-110_k141_83495 | Not-determined |
| 18-110_k141_187848 | Not-determined |
| 18-110_k141_106695 | Not-determined |
| 18-110_k141_83502 | Not-determined |
| 18-110_k141_37939 | Not-determined |
| 18-110_k141_187859 | Not-determined |
| 18-110_k141_164566 | Not-determined |
| 18-110_k141_210739 | Not-determined |
| 18-110_k141_164568 | Not-determined |
| 18-110_k141_49179 | Low-quality |
| 18-110_k141_49181 | Not-determined |
| 18-110_k141_245599 | Not-determined |
| 18-110_k141_118417 | Not-determined |
| 18-110_k141_210744 | Not-determined |
| 18-110_k141_245607 | Not-determined |
| 18-110_k141_130276 | Not-determined |
| 18-110_k141_37947 | Not-determined |
| 18-110_k141_49186 | Not-determined |
| 18-110_k141_130282 | Not-determined |
| 18-110_k141_199042 | Not-determined |
| 18-110_k141_130288 | Not-determined |
| 18-110_k141_37956 | Not-determined |
| 18-110_k141_199044 | Not-determined |
| 18-110_k141_245614 | Not-determined |
| 18-110_k141_153327 | Not-determined |
| 18-110_k141_73100 | Not-determined |
| 18-110_k141_164575 | Not-determined |
| 18-110_k141_176818 | Not-determined |
| 18-110_k141_245617 | Not-determined |
| 18-110_k141_187879 | Not-determined |
| 18-110_k141_187886 | Not-determined |
| 18-110_k141_268342 | Not-determined |
| 18-110_k141_37963 | Not-determined |
| 18-110_k141_176824 | Not-determined |
| 18-110_k141_187889 | Low-quality |
| 18-110_k141_106704 | Not-determined |
| 18-110_k141_268352 | Not-determined |
| 18-110_k141_26738 | Not-determined |
| 18-110_k141_199068 | Not-determined |
| 18-110_k141_187896 | Not-determined |
| 18-110_k141_210781 | Not-determined |
| 18-110_k141_187902 | Low-quality |
| 18-110_k141_234174 | Not-determined |
| 18-110_k141_118434 | Not-determined |
| 18-110_k141_187911 | Not-determined |
| 18-110_k141_199078 | Not-determined |
| 18-110_k141_268374 | Not-determined |
| 18-110_k141_176845 | Not-determined |
| 18-110_k141_164583 | Low-quality |
| 18-110_k141_245650 | Not-determined |
| 18-110_k141_199083 | Not-determined |
| 18-110_k141_83525 | Not-determined |
| 18-110_k141_187920 | Not-determined |
| 18-110_k141_37973 | Not-determined |
| 18-110_k141_268379 | Low-quality |
| 18-110_k141_83532 | Not-determined |
| 18-110_k141_210819 | Not-determined |
| 18-110_k141_187935 | Not-determined |
| 18-110_k141_153367 | Not-determined |
| 18-110_k141_49214 | Low-quality |
| 18-110_k141_187937 | Not-determined |
| 18-110_k141_176861 | Not-determined |
| 18-110_k141_210825 | Not-determined |
| 18-110_k141_106720 | Not-determined |
| 18-110_k141_95809 | Not-determined |
| 18-110_k141_142199 | Not-determined |
| 18-110_k141_118454 | Not-determined |
| 18-110_k141_118458 | Not-determined |
| 18-110_k141_142207 | Not-determined |
| 18-110_k141_95818 | Not-determined |
| 18-110_k141_106733 | Not-determined |
| 18-110_k141_83543 | Not-determined |
| 18-110_k141_153381 | Not-determined |
| 18-110_k141_153383 | Low-quality |
| 18-110_k141_176877 | Not-determined |
| 18-110_k141_26766 | Low-quality |
| 18-110_k141_221998 | Not-determined |
| 18-110_k141_234187 | Low-quality |
| 18-110_k141_3237 | Not-determined |
| 18-110_k141_268412 | Not-determined |
| 18-110_k141_153396 | Not-determined |
| 18-110_k141_210833 | Not-determined |
| 18-110_k141_245690 | Not-determined |
| 18-110_k141_210835 | Not-determined |
| 18-110_k141_245691 | Not-determined |
| 18-110_k141_118474 | Not-determined |
| 18-110_k141_234191 | Not-determined |
| 18-110_k141_153401 | Not-determined |
| 18-110_k141_118475 | Not-determined |
| 18-110_k141_245694 | Not-determined |
| 18-110_k141_164594 | Not-determined |
| 18-110_k141_268422 | Not-determined |
| 18-110_k141_106752 | Not-determined |
| 18-110_k141_234196 | Not-determined |
| 18-110_k141_199102 | Not-determined |
| 18-110_k141_234199 | Not-determined |
| 18-110_k141_142217 | Not-determined |
| 18-110_k141_26780 | Not-determined |
| 18-110_k141_199110 | Not-determined |
| 18-110_k141_118498 | Not-determined |
| 18-110_k141_3250 | Not-determined |
| 18-110_k141_37988 | Not-determined |
| 18-110_k141_142221 | Not-determined |
| 18-110_k141_95852 | Not-determined |
| 18-110_k141_222019 | Not-determined |
| 18-110_k141_142226 | Not-determined |
| 18-110_k141_222020 | Not-determined |
| 18-110_k141_37994 | Not-determined |
| 18-110_k141_106768 | Low-quality |
| 18-110_k141_37995 | Not-determined |
| 18-110_k141_268432 | Not-determined |
| 18-110_k141_118510 | Low-quality |
| 18-110_k141_118511 | Not-determined |
| 18-110_k141_26787 | Not-determined |
| 18-110_k141_26789 | Not-determined |
| 18-110_k141_118515 | Not-determined |
| 18-110_k141_49246 | Not-determined |
| 18-110_k141_176899 | Not-determined |
| 18-110_k141_95860 | Not-determined |
| 18-110_k141_142238 | Not-determined |
| 18-110_k141_176901 | Not-determined |
| 18-110_k141_38002 | Not-determined |
| 18-110_k141_153408 | Low-quality |
| 18-110_k141_153410 | Not-determined |
| 18-110_k141_49259 | Not-determined |
| 18-110_k141_142242 | Not-determined |
| 18-110_k141_153412 | Not-determined |
| 18-110_k141_26796 | Low-quality |
| 18-110_k141_245712 | Not-determined |
| 18-110_k141_38006 | Not-determined |
| 18-110_k141_95868 | Not-determined |
| 18-110_k141_164613 | Not-determined |
| 18-110_k141_222043 | Not-determined |
| 18-110_k141_49267 | Not-determined |
| 18-110_k141_164615 | Not-determined |
| 18-110_k141_210853 | Not-determined |
| 18-110_k141_3254 | Not-determined |
| 18-110_k141_164620 | Not-determined |
| 18-110_k141_164627 | Not-determined |
| 18-110_k141_130384 | Low-quality |
| 18-110_k141_153431 | Low-quality |
| 18-110_k141_38014 | Not-determined |
| 18-110_k141_153434 | Not-determined |
| 18-110_k141_118528 | Not-determined |
| 18-110_k141_153435 | Not-determined |
| 18-110_k141_199126 | Not-determined |
| 18-110_k141_222056 | Not-determined |
| 18-110_k141_106778 | Not-determined |
| 18-110_k141_130391 | Not-determined |
| 18-110_k141_38023 | Not-determined |
| 18-110_k141_3274 | Not-determined |
| 18-110_k141_38024 | Not-determined |
| 18-110_k141_106786 | Not-determined |
| 18-110_k141_130401 | Not-determined |
| 18-110_k141_153448 | Not-determined |
| 18-110_k141_95904 | Low-quality |
| 18-110_k141_95906 | Low-quality |
| 18-110_k141_142279 | Not-determined |
| 18-110_k141_153456 | Not-determined |
| 18-110_k141_106792 | Not-determined |
| 18-110_k141_187978 | Not-determined |
| 18-110_k141_164645 | Not-determined |
| 18-110_k141_73169 | Not-determined |
| 18-110_k141_176934 | Not-determined |
| 18-110_k141_153460 | Not-determined |
| 18-110_k141_26852 | Not-determined |
| 18-110_k141_130412 | Not-determined |
| 18-110_k141_26863 | Not-determined |
| 18-110_k141_199147 | Not-determined |
| 18-110_k141_118551 | Not-determined |
| 18-110_k141_106805 | Low-quality |
| 18-110_k141_118554 | Not-determined |
| 18-110_k141_222076 | Not-determined |
| 18-110_k141_106810 | Not-determined |
| 18-110_k141_38047 | Not-determined |
| 18-110_k141_142296 | Not-determined |
| 18-110_k141_199148 | Low-quality |
| 18-110_k141_106816 | Not-determined |
| 18-110_k141_26878 | Not-determined |
| 18-110_k141_83606 | Not-determined |
| 18-110_k141_83607 | Not-determined |
| 18-110_k141_187987 | Low-quality |
| 18-110_k141_187988 | Not-determined |
| 18-110_k141_176964 | Not-determined |
| 18-110_k141_73188 | Not-determined |
| 18-110_k141_49291 | Not-determined |
| 18-110_k141_118566 | Not-determined |
| 18-110_k141_234243 | Not-determined |
| 18-110_k141_61099 | Not-determined |
| 18-110_k141_142305 | Not-determined |
| 18-110_k141_14076 | Not-determined |
| 18-110_k141_234246 | Not-determined |
| 18-110_k141_187994 | Not-determined |
| 18-110_k141_153489 | Not-determined |
| 18-110_k141_3302 | Not-determined |
| 18-110_k141_3304 | Not-determined |
| 18-110_k141_153496 | Not-determined |
| 18-110_k141_26893 | Not-determined |
| 18-110_k141_106836 | Not-determined |
| 18-110_k141_176972 | Not-determined |
| 18-110_k141_26897 | Not-determined |
| 18-110_k141_199155 | Not-determined |
| 18-110_k141_245750 | Not-determined |
| 18-110_k141_234259 | Not-determined |
| 18-110_k141_176979 | Not-determined |
| 18-110_k141_118576 | Not-determined |
| 18-110_k141_83618 | Not-determined |
| 18-110_k141_95935 | Low-quality |
| 18-110_k141_49309 | Not-determined |
| 18-110_k141_245759 | Not-determined |
| 18-110_k141_153518 | Not-determined |
| 18-110_k141_245763 | Not-determined |
| 18-110_k141_153520 | Not-determined |
| 18-110_k141_234276 | Not-determined |
| 18-110_k141_245765 | Not-determined |
| 18-110_k141_245766 | Not-determined |
| 18-110_k141_95939 | Not-determined |
| 18-110_k141_61127 | Not-determined |
| 18-110_k141_95941 | Not-determined |
| 18-110_k141_38080 | Not-determined |
| 18-110_k141_199162 | Low-quality |
| 18-110_k141_199163 | Low-quality |
| 18-110_k141_106866 | Not-determined |
| 18-110_k141_49319 | Not-determined |
| 18-110_k141_49320 | Not-determined |
| 18-110_k141_234284 | Not-determined |
| 18-110_k141_3324 | Not-determined |
| 18-110_k141_118601 | Not-determined |
| 18-110_k141_14087 | Not-determined |
| 18-110_k141_3330 | Not-determined |
| 18-110_k141_188022 | Not-determined |
| 18-110_k141_26923 | Not-determined |
| 18-110_k141_95962 | Not-determined |
| 18-110_k141_38089 | Not-determined |
| 18-110_k141_26934 | Not-determined |
| 18-110_k141_222088 | Not-determined |
| 18-110_k141_61137 | Not-determined |
| 18-110_k141_61138 | Not-determined |
| 18-110_k141_188037 | Not-determined |
| 18-110_k141_38097 | Not-determined |
| 18-110_k141_142365 | Not-determined |
| 18-110_k141_26942 | Low-quality |
| 18-110_k141_14107 | Not-determined |
| 18-110_k141_61146 | Not-determined |
| 18-110_k141_188041 | Low-quality |
| 18-110_k141_177019 | Not-determined |
| 18-110_k141_188044 | Not-determined |
| 18-110_k141_153542 | Not-determined |
| 18-110_k141_14115 | Not-determined |
| 18-110_k141_26949 | Not-determined |
| 18-110_k141_268480 | Not-determined |
| 18-110_k141_14121 | Not-determined |
| 18-110_k141_14124 | Not-determined |
| 18-110_k141_268482 | Not-determined |
| 18-110_k141_61162 | Not-determined |
| 18-110_k141_177032 | Not-determined |
| 18-110_k141_26962 | Not-determined |
| 18-110_k141_268488 | Not-determined |
| 18-110_k141_199190 | Not-determined |
| 18-110_k141_177037 | Not-determined |
| 18-110_k141_234294 | Not-determined |
| 18-110_k141_199193 | Not-determined |
| 18-110_k141_245781 | Not-determined |
| 18-110_k141_164688 | Not-determined |
| 18-110_k141_245782 | Not-determined |
| 18-110_k141_210874 | Not-determined |
| 18-110_k141_210878 | Not-determined |
| 18-110_k141_73227 | Low-quality |
| 18-110_k141_26964 | Not-determined |
| 18-110_k141_245792 | Low-quality |
| 18-110_k141_268500 | Not-determined |
| 18-110_k141_106907 | Not-determined |
| 18-110_k141_61179 | Not-determined |
| 18-110_k141_245796 | Not-determined |
| 18-110_k141_142376 | Not-determined |
| 18-110_k141_245799 | Not-determined |
| 18-110_k141_245800 | Not-determined |
| 18-110_k141_142377 | Not-determined |
| 18-110_k141_73236 | Not-determined |
| 18-110_k141_61183 | Not-determined |
| 18-110_k141_245804 | Not-determined |
| 18-110_k141_177039 | Low-quality |
| 18-110_k141_14145 | Low-quality |
| 18-110_k141_61191 | Not-determined |
| 18-110_k141_142385 | Not-determined |
| 18-110_k141_38136 | Not-determined |
| 18-110_k141_142391 | Not-determined |
| 18-110_k141_234309 | Not-determined |
| 18-110_k141_142394 | Not-determined |
| 18-110_k141_188072 | Not-determined |
| 18-110_k141_188075 | Not-determined |
| 18-110_k141_118646 | Not-determined |
| 18-110_k141_164722 | Not-determined |
| 18-110_k141_26985 | Not-determined |
| 18-110_k141_73248 | Not-determined |
| 18-110_k141_26988 | Not-determined |
| 18-110_k141_177059 | Not-determined |
| 18-110_k141_268530 | Not-determined |
| 18-110_k141_268531 | Low-quality |
| 18-110_k141_188085 | Not-determined |
| 18-110_k141_222152 | Low-quality |
| 18-110_k141_73257 | Not-determined |
| 18-110_k141_177062 | Low-quality |
| 18-110_k141_210907 | Not-determined |
| 18-110_k141_73260 | Not-determined |
| 18-110_k141_210908 | Not-determined |
| 18-110_k141_106934 | Not-determined |
| 18-110_k141_95987 | Not-determined |
| 18-110_k141_118655 | Not-determined |
| 18-110_k141_188095 | Not-determined |
| 18-110_k141_61211 | Not-determined |
| 18-110_k141_164736 | Not-determined |
| 18-110_k141_130441 | Not-determined |
| 18-110_k141_95991 | Not-determined |
| 18-110_k141_49362 | Not-determined |
| 18-110_k141_130447 | Low-quality |
| 18-110_k141_73270 | Not-determined |
| 18-110_k141_153578 | Not-determined |
| 18-110_k141_234333 | Not-determined |
| 18-110_k141_142401 | Low-quality |
| 18-110_k141_268557 | Not-determined |
| 18-110_k141_222157 | Not-determined |
| 18-110_k141_49365 | Not-determined |
| 18-110_k141_268562 | Not-determined |
| 18-110_k141_118664 | Not-determined |
| 18-110_k141_96001 | Not-determined |
| 18-110_k141_118665 | Not-determined |
| 18-110_k141_222169 | Not-determined |
| 18-110_k141_27009 | Not-determined |
| 18-110_k141_153589 | Not-determined |
| 18-110_k141_118672 | Low-quality |
| 18-110_k141_210939 | Low-quality |
| 18-110_k141_118674 | Not-determined |
| 18-110_k141_14153 | Not-determined |
| 18-110_k141_27022 | Not-determined |
| 18-110_k141_142409 | Not-determined |
| 18-110_k141_118680 | Not-determined |
| 18-110_k141_118681 | Not-determined |
| 18-110_k141_245861 | Low-quality |
| 18-110_k141_222183 | Not-determined |
| 18-110_k141_61218 | Not-determined |
| 18-110_k141_96028 | Low-quality |
| 18-110_k141_142415 | Not-determined |
| 18-110_k141_268585 | Not-determined |
| 18-110_k141_210953 | Not-determined |
| 18-110_k141_38143 | Not-determined |
| 18-110_k141_222194 | Not-determined |
| 18-110_k141_27032 | Low-quality |
| 18-110_k141_73280 | Low-quality |
| 18-110_k141_27034 | Not-determined |
| 18-110_k141_234378 | Low-quality |
| 18-110_k141_130467 | Not-determined |
| 18-110_k141_130469 | Not-determined |
| 18-110_k141_73285 | Not-determined |
| 18-110_k141_130477 | Not-determined |
| 18-110_k141_106978 | Not-determined |
| 18-110_k141_210968 | Not-determined |
| 18-110_k141_38157 | Not-determined |
| 18-110_k141_268599 | Not-determined |
| 18-110_k141_38159 | Not-determined |
| 18-110_k141_83708 | Not-determined |
| 18-110_k141_83709 | Low-quality |
| 18-110_k141_83712 | Not-determined |
| 18-110_k141_245906 | Low-quality |
| 18-110_k141_38170 | Not-determined |
| 18-110_k141_96044 | Not-determined |
| 18-110_k141_222202 | Not-determined |
| 18-110_k141_3383 | Not-determined |
| 18-110_k141_27041 | Not-determined |
| 18-110_k141_61260 | Not-determined |
| 18-110_k141_118714 | Not-determined |
| 18-110_k141_130498 | Not-determined |
| 18-110_k141_118715 | Not-determined |
| 18-110_k141_222211 | Not-determined |
| 18-110_k141_164771 | Low-quality |
| 18-110_k141_27048 | Not-determined |
| 18-110_k141_164774 | Not-determined |
| 18-110_k141_142454 | Not-determined |
| 18-110_k141_142455 | Not-determined |
| 18-110_k141_83719 | Not-determined |
| 18-110_k141_142464 | Not-determined |
| 18-110_k141_256307 | Not-determined |
| 18-110_k141_130510 | Not-determined |
| 18-110_k141_130511 | Not-determined |
| 18-110_k141_142469 | Not-determined |
| 18-110_k141_107017 | Low-quality |
| 18-110_k141_118733 | Not-determined |
| 18-110_k141_38184 | Not-determined |
| 18-110_k141_49384 | Not-determined |
| 18-110_k141_61288 | Not-determined |
| 18-110_k141_83735 | Not-determined |
| 18-110_k141_234394 | Not-determined |
| 18-110_k141_210992 | Not-determined |
| 18-110_k141_61292 | Not-determined |
| 18-110_k141_61293 | Not-determined |
| 18-110_k141_27073 | Not-determined |
| 18-110_k141_177110 | Low-quality |
| 18-110_k141_245948 | Not-determined |
| 18-110_k141_38196 | Not-determined |
| 18-110_k141_3397 | Not-determined |
| 18-110_k141_83755 | Not-determined |
| 18-110_k141_268633 | Not-determined |
| 18-110_k141_3403 | Not-determined |
| 18-110_k141_245954 | Not-determined |
| 18-110_k141_177126 | Not-determined |
| 18-110_k141_245961 | Not-determined |
| 18-110_k141_107020 | Not-determined |
| 18-110_k141_83765 | Low-quality |
| 18-110_k141_118778 | Not-determined |
| 18-110_k141_83772 | Not-determined |
| 18-110_k141_38207 | Not-determined |
| 18-110_k141_211018 | Not-determined |
| 18-110_k141_211020 | Not-determined |
| 18-110_k141_245980 | Low-quality |
| 18-110_k141_142490 | Not-determined |
| 18-110_k141_142495 | Not-determined |
| 18-110_k141_130565 | Not-determined |
| 18-110_k141_130567 | Not-determined |
| 18-110_k141_211035 | Not-determined |
| 18-110_k141_164843 | Not-determined |
| 18-110_k141_130576 | Not-determined |
| 18-110_k141_96067 | Not-determined |
| 18-110_k141_38233 | Not-determined |
| 18-110_k141_96071 | Not-determined |
| 18-110_k141_268646 | Not-determined |
| 18-110_k141_49440 | Not-determined |
| 18-110_k141_268649 | Not-determined |
| 18-101-final-viral-combined_k141_357373\|\|full | Medium-quality |
| 18-110_k141_49446 | Not-determined |
| 18-110_k141_118780 | Not-determined |
| 18-110_k141_49447 | Not-determined |
| 18-110_k141_96079 | Not-determined |
| 18-110_k141_38244 | Not-determined |
| 18-110_k141_73306 | Not-determined |
| 18-110_k141_245989 | Not-determined |
| 18-110_k141_73307 | Low-quality |
| 18-110_k141_234436 | Not-determined |
| 18-110_k141_73310 | Not-determined |
| 18-110_k141_73316 | Low-quality |
| 18-110_k141_107060 | Not-determined |
| 18-110_k141_130592 | Not-determined |
| 18-110_k141_142530 | Not-determined |
| 18-110_k141_130595 | Not-determined |
| 18-110_k141_177165 | Not-determined |
| 18-110_k141_118792 | Low-quality |
| 18-110_k141_61317 | Not-determined |
| 18-110_k141_177170 | Not-determined |
| 18-110_k141_153679 | Not-determined |
| 18-110_k141_177173 | Low-quality |
| 18-110_k141_153681 | Not-determined |
| 18-110_k141_73324 | Not-determined |
| 18-110_k141_211053 | Not-determined |
| 18-110_k141_83792 | Not-determined |
| 18-110_k141_96099 | Not-determined |
| 18-110_k141_246004 | Not-determined |
| 18-110_k141_3455 | Not-determined |
| 18-110_k141_83797 | Not-determined |
| 18-110_k141_38258 | Not-determined |
| 18-110_k141_188120 | Not-determined |
| 18-110_k141_153686 | Not-determined |
| 18-110_k141_49474 | Not-determined |
| 18-110_k141_83801 | Not-determined |
| 18-110_k141_222263 | Low-quality |
| 18-110_k141_14185 | Not-determined |
| 18-110_k141_164885 | Not-determined |
| 18-110_k141_256396 | Not-determined |
| 18-110_k141_188131 | Not-determined |
| 18-110_k141_246011 | Not-determined |
| 18-110_k141_164891 | Not-determined |
| 18-110_k141_164894 | Not-determined |
| 18-110_k141_27172 | Low-quality |
| 18-110_k141_27173 | Not-determined |
| 18-110_k141_3467 | Not-determined |
| 18-110_k141_118813 | Not-determined |
| 18-110_k141_153705 | Not-determined |
| 18-110_k141_246022 | Not-determined |
| 18-110_k141_234469 | Not-determined |
| 18-110_k141_256404 | Not-determined |
| 18-110_k141_188146 | Not-determined |
| 18-110_k141_14203 | Not-determined |
| 18-110_k141_256405 | Not-determined |
| 18-110_k141_118823 | Not-determined |
| 18-110_k141_38272 | Not-determined |
| 18-110_k141_61350 | Not-determined |
| 18-110_k141_177206 | Not-determined |
| 18-110_k141_107098 | Not-determined |
| 18-110_k141_27180 | Not-determined |
| 18-110_k141_3480 | Low-quality |
| 18-110_k141_27184 | Not-determined |
| 18-110_k141_107102 | Not-determined |
| 18-110_k141_49486 | Low-quality |
| 18-110_k141_107105 | Not-determined |
| 18-110_k141_234485 | Not-determined |
| 18-110_k141_107108 | Not-determined |
| 18-110_k141_188159 | Low-quality |
| 18-110_k141_211098 | Low-quality |
| 18-110_k141_14215 | Not-determined |
| 18-110_k141_256416 | Not-determined |
| 18-110_k141_256417 | Not-determined |
| 18-110_k141_211105 | Not-determined |
| 18-110_k141_234492 | Not-determined |
| 18-110_k141_234494 | Not-determined |
| 18-110_k141_268722 | Not-determined |
| 18-110_k141_3484 | Not-determined |
| 18-110_k141_268731 | Low-quality |
| 18-110_k141_3486 | Not-determined |
| 18-110_k141_256429 | Not-determined |
| 18-110_k141_130618 | Not-determined |
| 18-110_k141_3490 | Not-determined |
| 18-110_k141_142570 | Not-determined |
| 18-110_k141_153719 | Not-determined |
| 18-110_k141_234506 | Not-determined |
| 18-110_k141_222294 | Not-determined |
| 18-110_k141_14228 | Not-determined |
| 18-110_k141_14229 | Not-determined |
| 18-110_k141_164925 | Not-determined |
| 18-110_k141_177217 | Not-determined |
| 18-110_k141_177219 | Not-determined |
| 18-110_k141_3500 | Not-determined |
| 18-110_k141_234509 | Not-determined |
| 18-110_k141_222305 | Not-determined |
| 18-110_k141_234511 | Low-quality |
| 18-110_k141_14235 | Not-determined |
| 18-110_k141_142572 | Not-determined |
| 18-110_k141_3503 | Not-determined |
| 18-110_k141_142575 | Not-determined |
| 18-110_k141_3504 | Not-determined |
| 18-110_k141_246059 | Not-determined |
| 18-110_k141_177225 | Not-determined |
| 18-110_k141_246060 | Not-determined |
| 18-110_k141_14240 | Low-quality |
| 18-110_k141_177227 | Not-determined |
| 18-110_k141_199272 | Not-determined |
| 18-110_k141_222313 | Not-determined |
| 18-110_k141_83815 | Not-determined |
| 18-110_k141_96130 | Not-determined |
| 18-110_k141_222314 | Not-determined |
| 18-110_k141_222319 | Not-determined |
| 18-110_k141_164942 | Low-quality |
| 18-110_k141_164943 | Not-determined |
| 18-110_k141_83816 | Not-determined |
| 18-110_k141_27198 | Not-determined |
| 18-110_k141_142586 | Not-determined |
| 18-110_k141_49514 | Not-determined |
| 18-110_k141_188168 | Not-determined |
| 18-110_k141_199278 | Not-determined |
| 18-110_k141_222334 | Not-determined |
| 18-110_k141_49519 | Not-determined |
| 18-110_k141_130636 | Not-determined |
| 18-110_k141_199286 | Not-determined |
| 18-110_k141_188184 | Not-determined |
| 18-110_k141_49529 | Not-determined |
| 18-110_k141_199294 | Not-determined |
| 18-110_k141_177255 | Low-quality |
| 18-110_k141_142593 | Not-determined |
| 18-110_k141_38302 | Not-determined |
| 18-110_k141_130652 | Low-quality |
| 18-110_k141_177265 | Not-determined |
| 18-110_k141_14274 | Not-determined |
| 18-110_k141_222350 | Not-determined |
| 18-110_k141_164975 | Not-determined |
| 18-110_k141_3528 | Not-determined |
| 18-110_k141_3530 | Not-determined |
| 18-110_k141_107160 | Not-determined |
| 18-110_k141_211129 | Not-determined |
| 18-110_k141_73359 | Not-determined |
| 18-110_k141_38318 | Not-determined |
| 18-110_k141_96152 | Low-quality |
| 18-110_k141_222360 | Low-quality |
| 18-110_k141_14279 | Low-quality |
| 18-110_k141_222365 | Not-determined |
| 18-110_k141_199312 | Not-determined |
| 18-110_k141_211137 | Not-determined |
| 18-110_k141_96164 | Not-determined |
| 18-110_k141_14282 | Not-determined |
| 18-110_k141_49549 | Not-determined |
| 18-110_k141_107175 | Not-determined |
| 18-110_k141_234558 | Not-determined |
| 18-110_k141_118873 | Not-determined |

| | |
|---|---|
| 18-110_k141_118874 | Not-determined |
| 18-110_k141_118875 | Not-determined |
| 18-110_k141_142618 | Not-determined |
| 18-110_k141_188229 | Not-determined |
| 18-110_k141_61366 | Low-quality |
| 18-110_k141_61367 | Not-determined |
| 18-110_k141_142623 | Not-determined |
| 18-110_k141_73380 | Not-determined |
| 18-110_k141_3553 | Not-determined |
| 18-110_k141_211143 | Not-determined |
| 18-110_k141_83845 | Not-determined |
| 18-110_k141_164984 | Not-determined |
| 18-110_k141_199318 | Not-determined |
| 18-110_k141_164992 | Low-quality |
| 18-110_k141_118895 | Not-determined |
| 18-110_k141_27261 | Not-determined |
| 18-110_k141_118897 | Not-determined |
| 18-110_k141_83853 | Not-determined |
| 18-110_k141_27263 | Not-determined |
| 18-110_k141_107205 | Not-determined |
| 18-110_k141_188250 | Not-determined |
| 18-110_k141_27275 | Not-determined |
| 18-110_k141_27276 | Not-determined |
| 18-110_k141_188252 | Not-determined |
| 18-110_k141_107212 | Low-quality |
| 18-110_k141_188253 | Not-determined |
| 18-110_k141_177305 | Not-determined |
| 18-110_k141_199335 | Not-determined |
| 18-110_k141_199338 | Not-determined |
| 18-110_k141_165000 | Not-determined |
| 18-110_k141_118920 | Not-determined |
| 18-110_k141_234588 | Low-quality |
| 18-110_k141_199342 | Not-determined |
| 18-110_k141_199344 | Not-determined |
| 18-110_k141_61403 | Not-determined |
| 18-110_k141_256455 | Not-determined |
| 18-110_k141_165005 | Not-determined |
| 18-110_k141_61409 | Low-quality |
| 18-110_k141_61411 | Not-determined |
| 18-110_k141_211179 | Not-determined |
| 18-110_k141_118932 | Not-determined |
| 18-110_k141_118934 | Not-determined |
| 18-110_k141_118935 | Not-determined |
| 18-110_k141_165023 | Not-determined |
| 18-110_k141_3561 | Not-determined |
| 18-110_k141_199357 | Not-determined |
| 18-110_k141_222422 | Not-determined |
| 18-110_k141_107229 | Not-determined |
| 18-110_k141_107233 | Not-determined |
| 18-110_k141_165036 | Not-determined |
| 18-110_k141_118951 | Not-determined |
| 18-110_k141_165044 | Not-determined |
| 18-110_k141_256483 | Not-determined |
| 18-110_k141_165046 | Not-determined |
| 18-110_k141_49559 | Not-determined |
| 18-110_k141_49560 | Not-determined |
| 18-110_k141_96195 | Not-determined |
| 18-110_k141_107242 | Not-determined |
| 18-110_k141_177355 | Not-determined |
| 18-110_k141_107244 | Not-determined |
| 18-110_k141_153758 | Not-determined |
| 18-110_k141_153759 | Not-determined |
| 18-110_k141_153760 | Not-determined |
| 18-110_k141_83914 | Not-determined |
| 18-110_k141_211195 | Not-determined |
| 18-110_k141_118960 | Not-determined |
| 18-110_k141_199372 | Not-determined |
| 18-110_k141_153764 | Not-determined |
| 18-110_k141_177359 | Not-determined |
| 18-110_k141_153766 | Not-determined |
| 18-110_k141_199375 | Not-determined |
| 18-110_k141_49573 | Not-determined |
| 18-110_k141_222458 | Not-determined |
| 18-110_k141_118963 | Not-determined |
| 18-110_k141_222460 | Not-determined |
| 18-110_k141_222462 | Not-determined |
| 18-110_k141_188278 | Not-determined |
| 18-110_k141_177367 | Not-determined |
| 18-110_k141_188280 | Not-determined |
| 18-110_k141_49584 | Not-determined |
| 18-110_k141_234625 | Not-determined |
| 18-110_k141_153776 | Not-determined |
| 18-110_k141_153779 | Not-determined |
| 18-110_k141_83947 | Not-determined |
| 18-110_k141_38359 | Not-determined |
| 18-110_k141_83949 | Not-determined |
| 18-110_k141_211220 | Not-determined |
| 18-110_k141_211221 | Not-determined |
| 18-110_k141_268764 | Not-determined |
| 18-110_k141_27338 | Not-determined |
| 18-110_k141_222474 | Not-determined |
| 18-110_k141_27339 | Not-determined |
| 18-110_k141_188298 | Not-determined |
| 18-110_k141_188301 | Not-determined |
| 18-110_k141_27343 | Not-determined |
| 18-110_k141_96218 | Not-determined |
| 18-110_k141_96221 | Not-determined |
| 18-110_k141_211234 | Not-determined |
| 18-110_k141_199396 | Not-determined |
| 18-110_k141_199397 | Low-quality |
| 18-110_k141_234651 | Low-quality |
| 18-110_k141_199404 | Not-determined |
| 18-110_k141_49612 | Not-determined |
| 18-110_k141_61458 | Not-determined |
| 18-110_k141_246113 | Not-determined |
| 18-110_k141_199409 | Not-determined |
| 18-110_k141_246116 | Not-determined |
| 18-110_k141_83960 | Not-determined |
| 18-110_k141_199414 | Not-determined |
| 18-110_k141_211243 | Not-determined |
| 18-110_k141_73456 | Not-determined |
| 18-110_k141_96248 | Low-quality |
| 18-110_k141_61466 | Low-quality |
| 18-110_k141_27353 | Not-determined |
| 18-110_k141_96250 | Not-determined |
| 18-110_k141_211248 | Not-determined |
| 18-110_k141_96251 | Not-determined |
| 18-110_k141_107273 | Not-determined |
| 18-110_k141_83963 | Not-determined |
| 18-110_k141_38380 | Not-determined |
| 18-110_k141_83966 | Not-determined |
| 18-110_k141_188327 | Not-determined |
| 18-110_k141_96255 | Not-determined |
| 18-110_k141_49621 | Not-determined |
| 18-110_k141_96263 | Not-determined |
| 18-110_k141_188337 | Not-determined |
| 18-110_k141_246122 | Not-determined |
| 18-110_k141_268790 | Not-determined |
| 18-110_k141_3629 | Not-determined |
| 18-110_k141_246130 | Not-determined |
| 18-110_k141_38394 | Low-quality |
| 18-110_k141_246134 | Not-determined |
| 18-110_k141_49636 | Not-determined |
| 18-110_k141_268804 | Not-determined |
| 18-110_k141_49639 | Not-determined |
| 18-110_k141_49645 | Not-determined |
| 18-110_k141_96278 | Not-determined |
| 18-110_k141_96279 | Not-determined |
| 18-110_k141_83993 | Not-determined |
| 18-110_k141_27388 | Not-determined |
| 18-110_k141_119001 | Not-determined |
| 18-110_k141_61491 | Not-determined |
| 18-110_k141_119008 | Not-determined |
| 18-110_k141_165103 | Not-determined |
| 18-110_k141_119011 | Not-determined |
| 18-110_k141_222512 | Not-determined |
| 18-110_k141_234671 | Not-determined |
| 18-110_k141_130751 | Not-determined |
| 18-110_k141_27397 | Not-determined |
| 18-110_k141_268821 | Not-determined |
| 18-110_k141_96282 | Not-determined |
| 18-110_k141_38429 | Not-determined |
| 18-110_k141_222519 | Not-determined |
| 18-110_k141_234675 | Not-determined |
| 18-110_k141_61504 | Not-determined |
| 18-110_k141_130755 | Not-determined |
| 18-110_k141_96290 | Not-determined |
| 18-110_k141_119023 | Low-quality |
| 18-110_k141_256506 | Not-determined |
| 18-110_k141_61510 | Not-determined |
| 18-110_k141_119030 | Low-quality |
| 18-110_k141_222531 | Not-determined |
| 18-110_k141_96300 | Not-determined |
| 18-110_k141_119037 | Not-determined |
| 18-110_k141_177433 | Not-determined |
| 18-110_k141_61519 | Low-quality |
| 18-110_k141_199449 | Not-determined |
| 18-110_k141_107289 | Low-quality |
| 18-110_k141_222543 | Not-determined |
| 18-110_k141_222544 | Not-determined |
| 18-110_k141_96310 | Not-determined |
| 18-110_k141_188382 | Not-determined |
| 18-110_k141_165136 | Low-quality |
| 18-110_k141_234702 | Not-determined |
| 18-110_k141_234703 | Not-determined |
| 18-110_k141_107298 | Not-determined |
| 18-110_k141_96320 | Not-determined |
| 18-110_k141_73487 | Not-determined |
| 18-110_k141_107301 | Not-determined |
| 18-110_k141_73488 | Not-determined |
| 18-110_k141_234711 | Not-determined |
| 18-110_k141_188392 | Not-determined |
| 18-110_k141_130783 | Not-determined |
| 18-110_k141_107304 | Not-determined |
| 18-110_k141_222557 | Not-determined |
| 18-110_k141_38447 | Not-determined |
| 18-110_k141_256527 | Low-quality |
| 18-110_k141_199467 | Not-determined |
| 18-110_k141_165148 | Low-quality |
| 18-110_k141_268845 | Not-determined |
| 18-110_k141_14295 | Not-determined |
| 18-110_k141_177456 | Not-determined |
| 18-110_k141_153809 | Not-determined |
| 18-110_k141_211298 | Not-determined |
| 18-110_k141_234735 | Not-determined |
| 18-110_k141_256540 | Not-determined |
| 18-110_k141_222569 | Not-determined |
| 18-110_k141_234738 | Not-determined |
| 18-110_k141_222572 | Not-determined |
| 18-110_k141_96353 | Not-determined |
| 18-110_k141_3646 | Not-determined |
| 18-110_k141_211310 | Not-determined |
| 18-110_k141_256544 | Low-quality |
| 18-110_k141_222581 | Not-determined |
| 18-110_k141_142685 | Not-determined |
| 18-110_k141_153827 | Not-determined |
| 18-110_k141_38486 | Not-determined |
| 18-110_k141_38487 | Not-determined |
| 18-110_k141_188424 | Not-determined |
| 18-110_k141_222590 | Not-determined |
| 18-110_k141_153834 | Not-determined |
| 18-110_k141_49690 | Not-determined |
| 18-110_k141_268866 | Not-determined |
| 18-110_k141_234749 | Not-determined |
| 18-110_k141_38490 | Not-determined |
| 18-110_k141_49694 | Not-determined |
| 18-110_k141_222598 | Low-quality |
| 18-110_k141_268873 | Not-determined |
| 18-110_k141_199488 | Not-determined |
| 18-110_k141_38493 | Not-determined |
| 18-110_k141_256557 | Not-determined |
| 18-110_k141_165185 | Not-determined |
| 18-110_k141_38499 | Not-determined |
| 18-110_k141_38500 | Not-determined |
| 18-110_k141_73507 | Low-quality |
| 18-110_k141_73509 | Not-determined |
| 18-110_k141_61543 | Not-determined |
| 18-110_k141_14300 | Not-determined |
| 18-110_k141_3650 | Not-determined |
| 18-110_k141_107319 | Not-determined |
| 18-110_k141_73516 | Not-determined |
| 18-110_k141_73518 | Not-determined |
| 18-110_k141_256565 | Not-determined |
| 18-110_k141_153856 | Not-determined |
| 18-110_k141_153858 | Not-determined |
| 18-110_k141_222616 | Not-determined |
| 18-110_k141_199503 | Not-determined |
| 18-110_k141_107327 | Not-determined |
| 18-110_k141_107329 | Not-determined |
| 18-110_k141_222622 | Not-determined |
| 18-110_k141_211325 | Not-determined |
| 18-110_k141_246179 | Low-quality |
| 18-110_k141_268888 | Not-determined |
| 18-110_k141_38522 | Not-determined |
| 18-110_k141_268891 | Not-determined |
| 18-110_k141_142690 | Not-determined |
| 18-110_k141_142691 | Not-determined |
| 18-110_k141_38525 | Not-determined |
| 18-110_k141_268892 | Not-determined |
| 18-110_k141_142692 | Not-determined |
| 18-110_k141_165198 | Not-determined |
| 18-110_k141_222637 | Not-determined |
| 18-110_k141_49715 | Not-determined |
| 18-110_k141_27415 | Not-determined |
| 18-110_k141_188443 | Not-determined |
| 18-110_k141_199516 | Not-determined |
| 18-110_k141_38534 | Not-determined |
| 18-110_k141_222642 | Not-determined |
| 18-110_k141_49723 | Not-determined |
| 18-110_k141_14317 | Not-determined |
| 18-110_k141_38538 | Not-determined |
| 18-110_k141_199523 | Not-determined |
| 18-110_k141_130817 | Not-determined |
| 18-110_k141_73547 | Not-determined |
| 18-110_k141_199528 | Not-determined |
| 18-110_k141_130819 | Not-determined |
| 18-110_k141_14325 | Not-determined |
| 18-110_k141_153887 | Not-determined |
| 18-110_k141_3676 | Not-determined |
| 18-110_k141_84082 | Not-determined |
| 18-110_k141_153892 | Not-determined |
| 18-110_k141_177484 | Low-quality |
| 18-110_k141_130821 | Not-determined |
| 18-110_k141_49738 | Not-determined |
| 18-110_k141_119052 | Not-determined |
| 18-110_k141_199539 | Not-determined |
| 18-110_k141_84095 | Not-determined |
| 18-110_k141_27433 | Not-determined |
| 18-110_k141_73556 | Not-determined |
| 18-110_k141_3691 | Not-determined |
| 18-110_k141_268915 | Not-determined |
| 18-110_k141_268918 | Not-determined |
| 18-110_k141_211345 | Not-determined |
| 18-110_k141_96407 | Not-determined |
| 18-110_k141_73559 | Not-determined |
| 18-110_k141_268921 | Not-determined |
| 18-110_k141_165216 | Not-determined |
| 18-110_k141_211348 | Not-determined |
| 18-110_k141_119058 | Not-determined |
| 18-110_k141_256606 | Not-determined |
| 18-110_k141_211352 | Not-determined |
| 18-110_k141_130827 | Low-quality |
| 18-110_k141_107365 | Not-determined |
| 18-110_k141_268927 | Low-quality |
| 18-110_k141_3713 | Low-quality |
| 18-110_k141_130832 | Not-determined |
| 18-110_k141_49753 | Not-determined |
| 18-110_k141_61608 | Not-determined |
| 18-110_k141_211362 | Not-determined |
| 18-110_k141_268934 | Not-determined |
| 18-110_k141_211370 | Not-determined |
| 18-110_k141_61619 | Not-determined |
| 18-110_k141_222676 | Not-determined |
| 18-110_k141_268940 | Not-determined |
| 18-110_k141_61625 | Not-determined |
| 18-110_k141_119063 | Not-determined |
| 18-110_k141_268945 | Not-determined |
| 18-110_k141_49777 | Low-quality |
| 18-110_k141_177518 | Low-quality |
| 18-110_k141_27438 | Not-determined |
| 18-110_k141_188468 | Not-determined |
| 18-110_k141_268951 | Not-determined |
| 18-110_k141_268953 | Not-determined |
| 18-110_k141_165262 | Not-determined |
| 18-110_k141_27443 | Not-determined |
| 18-110_k141_165264 | Not-determined |
| 18-110_k141_107379 | Not-determined |
| 18-110_k141_268956 | Not-determined |
| 18-110_k141_153920 | Not-determined |
| 18-110_k141_84136 | Low-quality |
| 18-110_k141_14377 | Low-quality |
| 18-110_k141_14379 | Not-determined |
| 18-110_k141_188484 | Not-determined |
| 18-110_k141_96434 | Not-determined |
| 18-110_k141_119102 | Low-quality |
| 18-110_k141_142726 | Not-determined |
| 18-110_k141_165281 | Not-determined |
| 18-110_k141_84139 | Low-quality |
| 18-110_k141_177533 | Not-determined |
| 18-110_k141_14386 | Not-determined |
| 18-110_k141_188491 | Not-determined |
| 18-110_k141_14389 | Not-determined |
| 18-110_k141_119107 | Not-determined |
| 18-110_k141_14391 | Not-determined |
| 18-110_k141_165288 | Low-quality |
| 18-110_k141_268973 | Not-determined |
| 18-110_k141_27479 | Not-determined |
| 18-110_k141_27486 | Not-determined |
| 18-110_k141_27488 | Not-determined |
| 18-110_k141_38597 | Not-determined |
| 18-110_k141_27491 | Not-determined |
| 18-110_k141_61665 | Not-determined |
| 18-110_k141_49813 | Not-determined |
| 18-110_k141_27495 | Not-determined |
| 18-110_k141_256642 | Not-determined |
| 18-110_k141_49818 | Not-determined |
| 18-110_k141_49822 | Not-determined |
| 18-110_k141_61674 | Low-quality |
| 18-110_k141_222699 | Not-determined |
| 18-110_k141_73583 | Not-determined |
| 18-110_k141_61678 | Not-determined |
| 18-110_k141_61679 | Not-determined |
| 18-110_k141_165303 | Not-determined |
| 18-110_k141_222704 | Not-determined |
| 18-110_k141_268985 | Not-determined |
| 18-110_k141_234800 | Not-determined |
| 18-110_k141_73586 | Not-determined |
| 18-110_k141_130885 | Not-determined |
| 18-110_k141_222709 | Not-determined |
| 18-110_k141_38605 | Not-determined |
| 18-110_k141_188521 | Not-determined |
| 18-110_k141_142758 | Not-determined |
| 18-110_k141_268994 | Not-determined |
| 18-110_k141_142761 | Not-determined |
| 18-110_k141_269005 | Not-determined |
| 18-110_k141_234815 | Not-determined |
| 18-110_k141_222720 | Not-determined |
| 18-110_k141_234817 | Not-determined |
| 18-110_k141_84181 | Low-quality |
| 18-110_k141_234820 | Not-determined |
| 18-110_k141_165330 | Not-determined |
| 18-110_k141_142773 | Not-determined |
| 18-110_k141_269019 | Not-determined |
| 18-110_k141_269022 | Not-determined |
| 18-110_k141_222730 | Not-determined |
| 18-110_k141_130912 | Not-determined |
| 18-110_k141_84190 | Not-determined |
| 18-110_k141_211385 | Low-quality |
| 18-110_k141_256648 | Low-quality |
| 18-110_k141_14434 | Not-determined |
| 18-110_k141_234835 | Not-determined |
| 18-110_k141_107449 | Not-determined |
| 18-110_k141_246189 | Not-determined |
| 18-110_k141_3741 | Not-determined |
| 18-110_k141_84192 | Not-determined |
| 18-110_k141_177542 | Low-quality |
| 18-110_k141_84195 | Not-determined |
| 18-110_k141_27503 | Not-determined |
| 18-110_k141_38631 | Not-determined |
| 18-110_k141_84200 | Not-determined |
| 18-110_k141_3746 | Low-quality |
| 18-110_k141_14443 | Low-quality |
| 18-110_k141_142795 | Not-determined |
| 18-110_k141_165358 | Not-determined |
| 18-110_k141_130919 | Not-determined |
| 18-110_k141_96441 | Not-determined |
| 18-110_k141_256656 | Not-determined |
| 18-110_k141_246202 | Not-determined |
| 18-110_k141_130921 | Not-determined |
| 18-110_k141_38648 | Not-determined |
| 18-110_k141_165363 | Not-determined |
| 18-110_k141_14450 | Not-determined |
| 18-110_k141_130925 | Not-determined |
| 18-110_k141_27519 | Low-quality |
| 18-110_k141_246213 | Not-determined |
| 18-110_k141_107467 | Not-determined |
| 18-110_k141_269046 | Not-determined |
| 18-110_k141_3763 | Not-determined |
| 18-110_k141_234838 | Not-determined |
| 18-110_k141_269050 | Not-determined |
| 18-110_k141_153998 | Not-determined |
| 18-110_k141_107473 | Not-determined |
| 18-110_k141_14459 | Low-quality |
| 18-110_k141_61712 | Not-determined |
| 18-110_k141_96460 | Not-determined |
| 18-110_k141_256669 | Not-determined |
| 18-110_k141_130932 | Low-quality |
| 18-110_k141_211422 | Low-quality |
| 18-110_k141_177585 | Not-determined |
| 18-110_k141_96461 | Not-determined |
| 18-110_k141_142816 | Not-determined |
| 18-110_k141_154007 | Not-determined |
| 18-110_k141_188563 | Not-determined |
| 18-110_k141_154008 | Not-determined |
| 18-110_k141_130936 | Not-determined |
| 18-110_k141_61725 | Not-determined |
| 18-110_k141_38682 | Not-determined |
| 18-110_k141_130941 | Not-determined |
| 18-110_k141_38684 | Not-determined |
| 18-110_k141_234854 | Not-determined |
| 18-110_k141_96471 | Low-quality |
| 18-110_k141_154018 | Not-determined |
| 18-110_k141_234857 | Not-determined |
| 18-110_k141_130944 | Not-determined |
| 18-110_k141_142830 | Low-quality |
| 18-110_k141_142831 | Not-determined |
| 18-110_k141_154026 | Not-determined |
| 18-110_k141_27555 | Not-determined |
| 18-110_k141_27556 | Not-determined |
| 18-110_k141_27557 | Not-determined |
| 18-110_k141_27558 | Low-quality |
| 18-110_k141_234864 | Not-determined |
| 18-110_k141_27560 | Not-determined |
| 18-110_k141_49876 | Not-determined |
| 18-110_k141_14490 | Not-determined |
| 18-110_k141_84221 | Not-determined |
| 18-110_k141_211441 | Not-determined |
| 18-110_k141_199552 | Not-determined |
| 18-110_k141_234868 | Not-determined |
| 18-110_k141_165383 | Not-determined |
| 18-110_k141_211444 | Not-determined |
| 18-110_k141_269079 | Not-determined |
| 18-110_k141_211445 | Not-determined |
| 18-110_k141_38691 | Not-determined |
| 18-110_k141_269080 | Not-determined |
| 18-110_k141_222794 | Not-determined |
| 18-110_k141_199557 | Not-determined |
| 18-110_k141_177604 | Not-determined |
| 18-110_k141_188586 | Not-determined |
| 18-110_k141_269084 | Low-quality |
| 18-110_k141_49887 | Not-determined |
| 18-110_k141_49888 | Not-determined |
| 18-110_k141_130963 | Not-determined |
| 18-110_k141_3787 | Not-determined |
| 18-110_k141_177611 | Not-determined |
| 18-110_k141_107508 | Not-determined |
| 18-110_k141_177612 | Not-determined |
| 18-110_k141_38697 | Low-quality |
| 18-110_k141_154046 | Not-determined |
| 18-110_k141_188600 | Not-determined |
| 18-110_k141_199568 | Not-determined |
| 18-110_k141_246278 | Not-determined |
| 18-110_k141_211463 | Not-determined |
| 18-110_k141_246281 | Not-determined |
| 18-110_k141_49900 | Not-determined |
| 18-110_k141_246283 | Not-determined |
| 18-110_k141_211469 | Not-determined |
| 18-110_k141_211471 | Not-determined |
| 18-110_k141_38709 | Not-determined |
| 18-110_k141_96502 | Low-quality |
| 18-110_k141_269098 | Not-determined |
| 18-110_k141_165408 | Not-determined |
| 18-110_k141_154053 | Not-determined |
| 18-110_k141_269104 | Not-determined |
| 18-110_k141_49917 | Not-determined |
| 18-110_k141_256703 | Not-determined |
| 18-110_k141_154062 | Not-determined |
| 18-110_k141_14519 | Not-determined |
| 18-110_k141_3813 | Not-determined |
| 18-110_k141_188603 | Not-determined |
| 18-110_k141_256707 | Not-determined |
| 18-110_k141_119158 | Not-determined |
| 18-110_k141_246298 | Not-determined |
| 18-110_k141_49924 | Not-determined |
| 18-110_k141_49928 | Not-determined |
| 18-110_k141_49930 | Low-quality |
| 18-110_k141_130990 | Not-determined |
| 18-110_k141_84238 | Not-determined |
| 18-110_k141_130992 | Not-determined |
| 18-110_k141_199595 | Not-determined |
| 18-110_k141_154074 | Not-determined |
| 18-110_k141_269120 | Not-determined |
| 18-110_k141_61757 | Not-determined |
| 18-110_k141_130998 | Not-determined |
| 18-110_k141_96516 | Not-determined |
| 18-110_k141_96517 | Not-determined |
| 18-110_k141_61762 | Not-determined |
| 18-110_k141_234908 | Not-determined |
| 18-110_k141_269132 | Not-determined |
| 18-110_k141_188625 | Not-determined |
| 18-110_k141_49942 | Low-quality |
| 18-110_k141_188629 | Not-determined |
| 18-110_k141_177635 | Not-determined |
| 18-110_k141_188634 | Not-determined |
| 18-110_k141_269141 | Low-quality |
| 18-110_k141_84257 | Not-determined |
| 18-110_k141_199612 | Not-determined |
| 18-110_k141_49950 | Not-determined |
| 18-110_k141_49951 | Not-determined |
| 18-110_k141_177638 | Not-determined |
| 18-110_k141_61768 | Not-determined |
| 18-110_k141_234923 | Not-determined |
| 18-110_k141_154105 | Not-determined |
| 18-110_k141_61771 | Not-determined |
| 18-110_k141_211535 | Not-determined |
| 18-110_k141_142840 | Low-quality |
| 18-110_k141_61774 | Not-determined |
| 18-110_k141_234930 | Not-determined |
| 18-110_k141_269148 | Not-determined |
| 18-110_k141_199620 | Low-quality |
| 18-110_k141_211540 | Not-determined |
| 18-110_k141_199621 | Low-quality |
| 18-110_k141_222861 | Not-determined |
| 18-110_k141_131015 | Not-determined |
| 18-110_k141_38742 | Not-determined |
| 18-110_k141_3839 | Not-determined |
| 18-110_k141_84273 | Not-determined |
| 18-110_k141_96548 | Not-determined |
| 18-110_k141_154119 | Not-determined |
| 18-110_k141_142855 | Not-determined |
| 18-110_k141_154122 | Not-determined |
| 18-110_k141_188641 | Not-determined |
| 18-110_k141_119187 | Not-determined |
| 18-110_k141_84279 | Not-determined |
| 18-110_k141_154125 | Not-determined |
| 18-110_k141_96554 | Not-determined |
| 18-110_k141_154129 | Not-determined |
| 18-110_k141_269166 | Not-determined |
| 18-110_k141_14556 | Low-quality |
| 18-110_k141_177660 | Not-determined |
| 18-110_k141_269168 | Low-quality |
| 18-110_k141_154135 | Not-determined |
| 18-110_k141_119196 | Not-determined |
| 18-110_k141_49970 | Not-determined |
| 18-110_k141_38755 | Not-determined |
| 18-110_k141_222880 | Not-determined |
| 18-110_k141_96561 | Not-determined |
| 18-110_k141_49972 | Not-determined |
| 18-110_k141_61800 | Not-determined |
| 18-110_k141_222888 | Not-determined |
| 18-110_k141_188664 | Not-determined |
| 18-110_k141_119208 | Low-quality |
| 18-110_k141_222896 | Not-determined |
| 18-110_k141_119215 | Not-determined |
| 18-110_k141_177669 | Not-determined |
| 18-110_k141_38780 | Not-determined |
| 18-110_k141_269191 | Not-determined |
| 18-110_k141_119224 | Not-determined |
| 18-110_k141_96573 | Low-quality |
| 18-110_k141_177674 | Not-determined |
| 18-110_k141_131050 | Not-determined |
| 18-110_k141_177681 | Not-determined |
| 18-110_k141_188686 | Not-determined |
| 18-110_k141_222919 | Not-determined |
| 18-110_k141_38799 | Not-determined |
| 18-110_k141_154166 | Not-determined |
| 18-110_k141_3871 | Low-quality |
| 18-110_k141_188690 | Not-determined |
| 18-110_k141_119228 | Not-determined |
| 18-110_k141_165449 | Not-determined |
| 18-110_k141_3884 | Not-determined |
| 18-110_k141_27613 | Not-determined |
| 18-110_k141_96591 | Not-determined |
| 18-110_k141_73613 | Not-determined |
| 18-110_k141_84314 | Low-quality |
| 18-110_k141_84315 | Not-determined |
| 18-110_k141_165457 | Not-determined |
| 18-110_k141_222931 | Not-determined |
| 18-110_k141_38806 | Not-determined |

| | |
|---|---|
| 18-110_k141_73622 | Low-quality |
| 18-110_k141_14600 | Not-determined |
| 18-110_k141_3889 | Not-determined |
| 18-110_k141_119244 | Not-determined |
| 18-110_k141_84317 | Not-determined |
| 18-110_k141_38814 | Not-determined |
| 18-110_k141_27626 | Not-determined |
| 18-110_k141_84324 | Low-quality |
| 18-110_k141_119250 | Not-determined |
| 18-110_k141_84332 | Not-determined |
| 18-110_k141_246316 | Low-quality |
| 18-110_k141_14617 | Low-quality |
| 18-110_k141_96610 | Not-determined |
| 18-110_k141_96612 | Not-determined |
| 18-110_k141_27635 | Not-determined |
| 18-110_k141_142895 | Not-determined |
| 18-110_k141_177720 | Low-quality |
| 18-110_k141_256736 | Not-determined |
| 18-110_k141_256738 | Not-determined |
| 18-110_k141_256740 | Not-determined |
| 18-110_k141_107538 | Not-determined |
| 18-110_k141_38830 | Not-determined |
| 18-110_k141_84343 | Not-determined |
| 18-110_k141_119266 | Not-determined |
| 18-110_k141_269233 | Low-quality |
| 18-110_k141_211569 | Low-quality |
| 18-110_k141_256749 | Not-determined |
| 18-110_k141_211581 | Not-determined |
| 18-110_k141_14629 | Not-determined |
| 18-110_k141_142908 | Not-determined |
| 18-110_k141_38847 | Not-determined |
| 18-110_k141_14636 | Not-determined |
| 18-110_k141_119273 | Not-determined |
| 18-110_k141_73636 | Not-determined |
| 18-110_k141_119278 | Not-determined |
| 18-110_k141_84363 | Not-determined |
| 18-110_k141_177750 | Low-quality |
| 18-110_k141_119280 | Not-determined |
| 18-110_k141_211598 | Not-determined |
| 18-110_k141_199650 | Not-determined |
| 18-110_k141_234961 | Not-determined |
| 18-110_k141_142928 | Low-quality |
| 18-110_k141_107557 | Not-determined |
| 18-110_k141_14645 | Not-determined |
| 18-110_k141_61880 | Low-quality |
| 18-110_k141_73644 | Not-determined |
| 18-110_k141_73645 | Not-determined |
| 18-110_k141_73646 | Not-determined |
| 18-110_k141_142935 | Not-determined |
| 18-110_k141_73651 | Not-determined |
| 18-110_k141_119297 | Not-determined |
| 18-110_k141_165465 | Not-determined |
| 18-110_k141_142949 | Not-determined |
| 18-110_k141_142950 | Not-determined |
| 18-110_k141_142951 | Not-determined |
| 18-110_k141_14658 | Not-determined |
| 18-110_k141_73652 | Low-quality |
| 18-110_k141_211624 | Not-determined |
| 18-110_k141_107580 | Not-determined |
| 18-110_k141_119304 | Not-determined |
| 18-110_k141_234978 | Not-determined |
| 18-110_k141_165485 | Not-determined |
| 18-110_k141_142958 | Not-determined |
| 18-110_k141_38860 | Not-determined |
| 18-110_k141_84400 | Not-determined |
| 18-110_k141_199673 | Not-determined |
| 18-110_k141_177781 | Not-determined |
| 18-110_k141_14665 | Not-determined |
| 18-110_k141_154199 | Not-determined |
| 18-110_k141_84414 | Not-determined |
| 18-110_k141_188764 | Not-determined |
| 18-110_k141_235006 | Not-determined |
| 18-110_k141_177805 | Not-determined |
| 18-110_k141_14680 | Not-determined |
| 18-110_k141_73665 | Not-determined |
| 18-110_k141_14682 | Not-determined |
| 18-110_k141_188775 | Not-determined |
| 18-110_k141_165510 | Not-determined |
| 18-110_k141_14686 | Not-determined |
| 18-110_k141_235018 | Not-determined |
| 18-110_k141_61933 | Low-quality |
| 18-110_k141_84432 | Not-determined |
| 18-110_k141_211645 | Not-determined |
| 18-110_k141_235024 | Not-determined |
| 18-110_k141_38874 | Not-determined |
| 18-110_k141_165516 | Not-determined |
| 18-110_k141_222946 | Not-determined |
| 18-110_k141_38876 | Not-determined |
| 18-110_k141_131148 | Not-determined |
| 18-110_k141_61950 | Not-determined |
| 18-110_k141_84444 | Not-determined |
| 18-110_k141_222955 | Not-determined |
| 18-110_k141_222957 | Not-determined |
| 18-110_k141_222958 | Not-determined |
| 18-110_k141_38895 | Low-quality |
| 18-110_k141_211663 | Not-determined |
| 18-110_k141_211665 | Not-determined |
| 18-110_k141_73674 | Not-determined |
| 18-110_k141_84449 | Not-determined |
| 18-110_k141_61964 | Not-determined |
| 18-110_k141_38900 | Not-determined |
| 18-110_k141_84453 | Not-determined |
| 18-110_k141_50007 | Not-determined |
| 18-110_k141_165532 | Not-determined |
| 18-110_k141_73685 | Not-determined |
| 18-110_k141_84470 | Not-determined |
| 18-110_k141_73697 | Not-determined |
| 18-110_k141_188803 | Low-quality |
| 18-110_k141_73700 | Not-determined |
| 18-110_k141_235055 | Not-determined |
| 18-110_k141_199681 | Not-determined |
| 18-110_k141_73706 | Not-determined |
| 18-110_k141_61991 | Not-determined |
| 18-110_k141_222975 | Not-determined |
| 18-110_k141_188818 | Not-determined |
| 18-110_k141_96624 | Not-determined |
| 18-110_k141_188820 | Not-determined |
| 18-110_k141_211680 | Low-quality |
| 18-110_k141_256832 | Not-determined |
| 18-110_k141_61996 | Not-determined |
| 18-110_k141_84490 | Not-determined |
| 18-110_k141_84491 | Not-determined |
| 18-110_k141_96639 | Not-determined |
| 18-110_k141_84493 | Not-determined |
| 18-110_k141_211696 | Not-determined |
| 18-110_k141_119318 | Low-quality |
| 18-110_k141_222989 | Not-determined |
| 18-110_k141_96647 | Not-determined |
| 18-110_k141_211708 | Not-determined |
| 18-110_k141_62010 | Not-determined |
| 18-110_k141_165565 | Not-determined |
| 18-110_k141_246348 | Not-determined |
| 18-110_k141_188833 | Low-quality |
| 18-110_k141_62026 | Not-determined |
| 18-110_k141_62034 | Not-determined |
| 18-110_k141_96663 | Not-determined |
| 18-110_k141_165574 | Not-determined |
| 18-110_k141_211724 | Not-determined |
| 18-110_k141_269243 | Not-determined |
| 18-110_k141_131164 | Not-determined |
| 18-110_k141_62038 | Low-quality |
| 18-110_k141_246363 | Not-determined |
| 18-110_k141_3983 | Not-determined |
| 18-110_k141_211730 | Not-determined |
| 18-110_k141_62043 | Not-determined |
| 18-110_k141_165580 | Low-quality |
| 18-110_k141_235064 | Not-determined |
| 18-110_k141_188846 | Not-determined |
| 18-110_k141_165584 | Not-determined |
| 18-110_k141_188848 | Not-determined |
| 18-110_k141_188850 | Not-determined |
| 18-110_k141_62049 | Not-determined |
| 18-110_k141_154236 | Not-determined |
| 18-110_k141_119345 | Not-determined |
| 18-110_k141_154241 | Not-determined |
| 18-110_k141_269256 | Not-determined |
| 18-110_k141_246380 | Low-quality |
| 18-110_k141_165587 | Not-determined |
| 18-110_k141_3986 | Low-quality |
| 18-110_k141_211752 | Not-determined |
| 18-110_k141_3987 | Not-determined |
| 18-110_k141_235069 | Low-quality |
| 18-110_k141_73720 | Not-determined |
| 18-110_k141_3991 | Not-determined |
| 18-110_k141_246387 | Not-determined |
| 18-110_k141_256880 | Not-determined |
| 18-110_k141_131192 | Not-determined |
| 18-110_k141_235076 | Not-determined |
| 18-110_k141_107650 | Not-determined |
| 18-110_k141_50031 | Not-determined |
| 18-110_k141_269279 | Not-determined |
| 18-110_k141_256888 | Not-determined |
| 18-110_k141_73745 | Not-determined |
| 18-110_k141_131204 | Not-determined |
| 18-110_k141_73746 | Not-determined |
| 18-110_k141_269294 | Not-determined |
| 18-110_k141_107667 | Low-quality |
| 18-110_k141_143012 | Low-quality |
| 18-110_k141_107672 | Not-determined |
| 18-110_k141_131215 | Not-determined |
| 18-110_k141_107676 | Not-determined |
| 18-110_k141_14769 | Not-determined |
| 18-110_k141_269303 | Not-determined |
| 18-110_k141_256896 | Not-determined |
| 18-110_k141_14773 | Not-determined |
| 18-110_k141_188879 | Low-quality |
| 18-110_k141_96719 | Not-determined |
| 18-110_k141_4018 | Not-determined |
| 18-110_k141_143042 | Not-determined |
| 18-110_k141_62106 | Low-quality |
| 18-110_k141_177827 | Not-determined |
| 18-110_k141_256903 | Not-determined |
| 18-110_k141_269310 | Low-quality |
| 18-110_k141_14776 | Low-quality |
| 18-110_k141_143049 | Not-determined |
| 18-110_k141_96726 | Not-determined |
| 18-110_k141_188890 | Not-determined |
| 18-110_k141_50073 | Not-determined |
| 18-110_k141_39013 | Not-determined |
| 18-110_k141_131219 | Low-quality |
| 18-110_k141_50078 | Not-determined |
| 18-110_k141_50083 | Not-determined |
| 18-110_k141_256914 | Low-quality |
| 18-110_k141_143062 | Not-determined |
| 18-110_k141_107686 | Low-quality |
| 18-110_k141_246392 | Not-determined |
| 18-110_k141_73770 | Not-determined |
| 18-110_k141_269320 | Not-determined |
| 18-110_k141_143069 | Not-determined |
| 18-110_k141_39035 | Not-determined |
| 18-110_k141_154304 | Not-determined |
| 18-110_k141_143073 | Not-determined |
| 18-110_k141_39039 | Not-determined |
| 18-110_k141_143075 | Not-determined |
| 18-110_k141_73777 | Not-determined |
| 18-110_k141_222998 | Not-determined |
| 18-110_k141_165641 | Not-determined |
| 18-110_k141_131240 | Not-determined |
| 18-110_k141_107698 | Not-determined |
| 18-110_k141_165645 | Not-determined |
| 18-110_k141_199699 | Not-determined |
| 18-110_k141_131242 | Not-determined |
| 18-110_k141_165648 | Not-determined |
| 18-110_k141_4038 | Not-determined |
| 18-110_k141_269332 | Not-determined |
| 18-110_k141_165655 | Not-determined |
| 18-110_k141_96733 | Not-determined |
| 18-110_k141_62147 | Not-determined |
| 18-110_k141_96740 | Not-determined |
| 18-110_k141_96741 | Not-determined |
| 18-110_k141_131255 | Not-determined |
| 18-110_k141_4046 | Low-quality |
| 18-110_k141_154320 | Not-determined |
| 18-110_k141_154322 | Not-determined |
| 18-110_k141_50115 | Not-determined |
| 18-110_k141_269347 | Not-determined |
| 18-110_k141_154324 | Not-determined |
| 18-110_k141_73811 | Not-determined |
| 18-110_k141_50119 | Not-determined |
| 18-110_k141_154329 | Low-quality |
| 18-110_k141_246403 | Not-determined |
| 18-110_k141_39075 | Not-determined |
| 18-110_k141_269358 | Not-determined |
| 18-110_k141_246405 | Not-determined |
| 18-110_k141_119364 | Not-determined |
| 18-110_k141_39081 | Not-determined |
| 18-110_k141_119365 | Low-quality |
| 18-110_k141_235160 | Not-determined |
| 18-110_k141_4051 | Not-determined |
| 18-110_k141_211773 | Not-determined |
| 18-110_k141_4052 | Not-determined |
| 18-110_k141_143107 | Not-determined |
| 18-110_k141_131284 | Not-determined |
| 18-110_k141_235169 | Not-determined |
| 18-110_k141_143109 | Not-determined |
| 18-110_k141_211782 | Not-determined |
| 18-110_k141_131290 | Not-determined |
| 18-110_k141_131291 | Not-determined |
| 18-110_k141_119371 | Not-determined |
| 18-110_k141_165677 | Low-quality |
| 18-110_k141_269372 | Not-determined |
| 18-110_k141_84511 | Low-quality |
| 18-110_k141_131295 | Not-determined |
| 18-110_k141_119374 | Not-determined |
| 18-110_k141_39099 | Not-determined |
| 18-110_k141_269377 | Not-determined |
| 18-110_k141_107736 | Not-determined |
| 18-110_k141_4062 | Not-determined |
| 18-110_k141_107739 | Not-determined |
| 18-110_k141_177892 | Low-quality |
| 18-110_k141_119388 | Not-determined |
| 18-110_k141_154351 | Not-determined |
| 18-110_k141_84533 | Not-determined |
| 18-110_k141_269389 | Not-determined |
| 18-110_k141_256952 | Not-determined |
| 18-110_k141_119395 | Not-determined |
| 18-110_k141_131312 | Not-determined |
| 18-110_k141_223044 | Not-determined |
| 18-110_k141_256953 | Not-determined |
| 18-110_k141_154358 | Not-determined |
| 18-110_k141_131317 | Not-determined |
| 18-110_k141_107745 | Not-determined |
| 18-110_k141_269395 | Not-determined |
| 18-110_k141_165692 | Not-determined |
| 18-110_k141_256957 | Not-determined |
| 18-110_k141_73820 | Low-quality |
| 18-110_k141_269400 | Not-determined |
| 18-110_k141_84543 | Not-determined |
| 18-110_k141_62190 | Not-determined |
| 18-110_k141_107747 | Not-determined |
| 18-110_k141_269404 | Not-determined |
| 18-110_k141_211796 | Not-determined |
| 18-110_k141_211797 | Low-quality |
| 18-110_k141_107752 | Not-determined |
| 18-110_k141_177906 | Not-determined |
| 18-110_k141_165702 | Low-quality |
| 18-110_k141_223060 | Low-quality |
| 18-110_k141_223062 | Not-determined |
| 18-110_k141_269412 | Not-determined |
| 18-110_k141_131332 | Not-determined |
| 18-110_k141_177911 | Not-determined |
| 18-110_k141_131333 | Not-determined |
| 18-110_k141_119412 | Not-determined |
| 18-110_k141_131336 | Not-determined |
| 18-110_k141_154373 | Not-determined |
| 18-110_k141_62209 | Not-determined |
| 18-110_k141_131342 | Not-determined |
| 18-110_k141_119424 | Not-determined |
| 18-110_k141_177919 | Not-determined |
| 18-110_k141_73845 | Not-determined |
| 18-110_k141_211819 | Not-determined |
| 18-110_k141_50159 | Not-determined |
| 18-110_k141_107775 | Not-determined |
| 18-110_k141_211824 | Not-determined |
| 18-110_k141_235180 | Not-determined |
| 18-110_k141_188941 | Not-determined |
| 18-110_k141_50166 | Low-quality |
| 18-110_k141_223083 | Not-determined |
| 18-110_k141_14862 | Not-determined |
| 18-110_k141_14866 | Not-determined |
| 18-110_k141_131358 | Not-determined |
| 18-110_k141_143149 | Not-determined |
| 18-110_k141_143150 | Not-determined |
| 18-110_k141_50175 | Low-quality |
| 18-110_k141_4085 | Not-determined |
| 18-110_k141_177943 | Not-determined |
| 18-110_k141_96799 | Not-determined |
| 18-110_k141_256969 | Not-determined |
| 18-110_k141_188946 | Not-determined |
| 18-110_k141_177947 | Low-quality |
| 18-110_k141_177949 | Not-determined |
| 18-110_k141_119456 | Low-quality |
| 18-110_k141_165728 | Not-determined |
| 18-110_k141_96811 | Not-determined |
| 18-110_k141_14875 | Not-determined |
| 18-110_k141_165733 | Not-determined |
| 18-110_k141_188956 | Not-determined |
| 18-110_k141_50188 | Not-determined |
| 18-110_k141_96817 | Not-determined |
| 18-110_k141_223099 | Not-determined |
| 18-110_k141_4101 | Not-determined |
| 18-110_k141_188964 | Not-determined |
| 18-110_k141_131361 | Low-quality |
| 18-110_k141_211867 | Not-determined |
| 18-110_k141_235210 | Not-determined |
| 18-110_k141_14889 | Not-determined |
| 18-110_k141_154385 | Not-determined |
| 18-110_k141_188971 | Not-determined |
| 18-110_k141_235217 | Not-determined |
| 18-110_k141_39121 | Not-determined |
| 18-110_k141_4114 | Not-determined |
| 18-110_k141_73895 | Not-determined |
| 18-110_k141_256995 | Not-determined |
| 18-110_k141_269450 | Not-determined |
| 18-110_k141_223131 | Not-determined |
| 18-110_k141_50197 | Not-determined |
| 18-110_k141_269456 | Low-quality |
| 18-110_k141_269457 | Not-determined |
| 18-110_k141_131376 | Not-determined |
| 18-110_k141_269460 | Not-determined |
| 18-110_k141_107823 | Not-determined |
| 18-110_k141_27682 | Not-determined |
| 18-110_k141_188989 | Low-quality |
| 18-110_k141_107830 | Not-determined |
| 18-110_k141_73917 | Not-determined |
| 18-110_k141_73918 | Not-determined |
| 18-110_k141_62243 | Not-determined |
| 18-110_k141_62244 | Not-determined |
| 18-110_k141_223144 | Low-quality |
| 18-110_k141_165758 | Low-quality |
| 18-110_k141_107844 | Not-determined |
| 18-110_k141_178002 | Not-determined |
| 18-110_k141_246480 | Not-determined |
| 18-110_k141_27715 | Low-quality |
| 18-110_k141_62255 | Not-determined |
| 18-110_k141_27722 | Not-determined |
| 18-110_k141_189009 | Not-determined |
| 18-110_k141_178006 | Low-quality |
| 18-110_k141_73939 | Not-determined |
| 18-110_k141_246485 | Not-determined |
| 18-110_k141_119480 | Not-determined |
| 18-110_k141_4154 | Not-determined |
| 18-110_k141_154399 | Not-determined |
| 18-110_k141_84556 | Not-determined |
| 18-110_k141_189019 | Not-determined |
| 18-110_k141_269491 | Not-determined |
| 18-110_k141_96870 | Not-determined |
| 18-110_k141_154401 | Not-determined |
| 18-110_k141_189026 | Not-determined |
| 18-110_k141_269495 | Not-determined |
| 18-110_k141_27736 | Not-determined |
| 18-110_k141_189030 | Not-determined |
| 18-110_k141_189031 | Not-determined |
| 18-110_k141_235243 | Not-determined |
| 18-110_k141_4166 | Not-determined |
| 18-110_k141_223164 | Not-determined |
| 18-110_k141_4168 | Not-determined |
| 18-110_k141_4169 | Not-determined |
| 18-110_k141_178025 | Low-quality |
| 18-110_k141_96878 | Not-determined |
| 18-110_k141_4171 | Not-determined |
| 18-110_k141_50247 | Not-determined |
| 18-110_k141_4175 | Low-quality |
| 18-110_k141_73964 | Low-quality |
| 18-110_k141_96886 | Not-determined |
| 18-110_k141_14937 | Not-determined |
| 18-110_k141_50254 | Not-determined |
| 18-110_k141_96887 | Not-determined |
| 18-110_k141_84568 | Not-determined |
| 18-110_k141_73971 | Not-determined |
| 18-110_k141_84572 | Not-determined |
| 18-110_k141_143177 | Not-determined |
| 18-110_k141_246526 | Not-determined |
| 18-110_k141_14955 | Not-determined |
| 18-110_k141_84575 | Low-quality |
| 18-110_k141_14957 | Not-determined |
| 18-110_k141_143185 | Not-determined |
| 18-110_k141_119496 | Not-determined |
| 18-110_k141_131401 | Not-determined |
| 18-110_k141_84585 | Not-determined |
| 18-110_k141_246533 | Not-determined |
| 18-110_k141_269517 | Not-determined |
| 18-110_k141_107882 | Low-quality |
| 18-110_k141_96914 | Not-determined |
| 18-110_k141_235264 | Not-determined |
| 18-110_k141_119502 | Low-quality |
| 18-110_k141_73997 | Not-determined |
| 18-110_k141_39184 | Not-determined |
| 18-110_k141_269526 | Not-determined |
| 18-110_k141_154436 | Not-determined |
| 18-110_k141_74003 | Not-determined |
| 18-110_k141_211893 | Not-determined |
| 18-110_k141_269534 | Not-determined |
| 18-110_k141_235280 | Not-determined |
| 18-110_k141_223191 | Not-determined |
| 18-110_k141_154444 | Not-determined |
| 18-110_k141_131426 | Not-determined |
| 18-110_k141_269539 | Not-determined |
| 18-110_k141_131428 | Not-determined |
| 18-110_k141_39199 | Not-determined |
| 18-110_k141_84597 | Not-determined |
| 18-110_k141_269543 | Not-determined |
| 18-110_k141_189059 | Not-determined |
| 18-110_k141_14989 | Not-determined |
| 18-110_k141_84605 | Not-determined |
| 18-110_k141_84613 | Not-determined |
| 18-110_k141_223196 | Not-determined |
| 18-110_k141_131435 | Not-determined |
| 18-110_k141_223200 | Not-determined |
| 18-110_k141_131438 | Not-determined |
| 18-110_k141_131440 | Low-quality |
| 18-110_k141_131443 | Low-quality |
| 18-110_k141_119528 | Not-determined |
| 18-110_k141_119530 | Not-determined |
| 18-110_k141_131445 | Not-determined |
| 18-110_k141_235298 | Not-determined |
| 18-110_k141_235299 | Not-determined |
| 18-110_k141_246570 | Not-determined |
| 18-110_k141_84618 | Not-determined |
| 18-110_k141_211909 | Not-determined |
| 18-110_k141_96930 | Not-determined |
| 18-110_k141_178053 | Low-quality |
| 18-110_k141_62342 | Not-determined |
| 18-110_k141_189074 | Not-determined |
| 18-110_k141_211912 | Not-determined |
| 18-110_k141_131461 | Not-determined |
| 18-110_k141_96941 | Not-determined |
| 18-110_k141_131462 | Not-determined |
| 18-110_k141_74034 | Not-determined |
| 18-110_k141_235318 | Not-determined |
| 18-110_k141_143216 | Not-determined |
| 18-110_k141_84633 | Low-quality |
| 18-110_k141_96948 | Not-determined |
| 18-110_k141_62352 | Not-determined |
| 18-110_k141_62354 | Not-determined |
| 18-110_k141_74037 | Not-determined |
| 18-110_k141_199745 | Not-determined |
| 18-110_k141_62356 | Not-determined |
| 18-110_k141_143222 | Not-determined |
| 18-110_k141_199748 | Not-determined |
| 18-110_k141_211924 | Not-determined |
| 18-110_k141_211925 | Not-determined |
| 18-110_k141_74044 | Not-determined |
| 18-110_k141_107943 | Low-quality |
| 18-110_k141_74049 | Not-determined |
| 18-110_k141_154452 | Not-determined |
| 18-110_k141_154454 | Not-determined |
| 18-110_k141_154455 | Not-determined |
| 18-110_k141_189099 | Not-determined |
| 18-110_k141_143234 | Not-determined |
| 18-110_k141_211939 | Not-determined |
| 18-110_k141_211940 | Not-determined |
| 18-110_k141_84647 | Not-determined |
| 18-110_k141_96968 | Not-determined |
| 18-110_k141_15002 | Not-determined |
| 18-110_k141_84650 | Not-determined |
| 18-110_k141_223221 | Not-determined |
| 18-110_k141_15010 | Not-determined |
| 18-110_k141_107969 | Low-quality |
| 18-110_k141_27775 | Not-determined |
| 18-110_k141_107971 | Not-determined |
| 18-110_k141_235341 | Not-determined |
| 18-110_k141_189118 | Not-determined |
| 18-110_k141_119567 | Not-determined |
| 18-110_k141_223228 | Low-quality |
| 18-110_k141_84664 | Not-determined |
| 18-110_k141_223229 | Low-quality |
| 18-110_k141_269564 | Not-determined |
| 18-110_k141_84674 | Not-determined |
| 18-110_k141_143251 | Not-determined |
| 18-110_k141_74065 | Not-determined |
| 18-110_k141_211951 | Not-determined |
| 18-110_k141_223233 | Not-determined |
| 18-110_k141_223235 | Not-determined |
| 18-110_k141_62393 | Not-determined |
| 18-110_k141_131484 | Not-determined |
| 18-110_k141_235355 | Not-determined |
| 18-110_k141_84683 | Not-determined |
| 18-110_k141_178094 | Not-determined |
| 18-110_k141_96992 | Not-determined |
| 18-110_k141_235360 | Not-determined |
| 18-110_k141_235362 | Not-determined |
| 18-110_k141_269572 | Not-determined |
| 18-110_k141_235364 | Not-determined |
| 18-110_k141_50342 | Not-determined |
| 18-110_k141_74066 | Low-quality |
| 18-110_k141_211962 | Not-determined |
| 18-110_k141_62402 | Not-determined |
| 18-110_k141_211967 | Not-determined |
| 18-110_k141_62408 | Not-determined |
| 18-110_k141_15033 | Low-quality |
| 18-110_k141_15034 | Not-determined |
| 18-110_k141_189144 | Not-determined |
| 18-110_k141_235375 | Not-determined |
| 18-110_k141_211973 | Low-quality |
| 18-110_k141_62411 | Not-determined |
| 18-110_k141_97012 | Low-quality |
| 18-110_k141_235380 | Not-determined |
| 18-110_k141_211976 | Not-determined |
| 18-110_k141_189153 | Not-determined |
| 18-110_k141_131501 | Not-determined |
| 18-110_k141_97016 | Not-determined |
| 18-110_k141_97021 | Not-determined |
| 18-110_k141_39238 | Not-determined |
| 18-110_k141_131502 | Low-quality |
| 18-110_k141_269581 | Not-determined |
| 18-110_k141_154502 | Not-determined |
| 18-110_k141_50362 | Not-determined |
| 18-110_k141_211985 | Not-determined |
| 18-110_k141_97025 | Not-determined |
| 18-110_k141_97027 | Not-determined |
| 18-110_k141_235392 | Not-determined |
| 18-110_k141_211992 | Not-determined |
| 18-110_k141_211994 | Not-determined |
| 18-110_k141_62422 | Not-determined |
| 18-110_k141_189166 | Not-determined |
| 18-110_k141_27836 | Not-determined |
| 18-110_k141_84722 | Not-determined |
| 18-110_k141_154519 | Not-determined |
| 18-110_k141_39250 | Low-quality |
| 18-110_k141_74100 | Not-determined |
| 18-110_k141_223272 | Low-quality |
| 18-110_k141_143280 | Not-determined |
| 18-110_k141_165790 | Not-determined |
| 18-110_k141_119607 | Not-determined |
| 18-110_k141_119608 | Not-determined |
| 18-110_k141_143282 | Not-determined |
| 18-110_k141_74109 | Not-determined |
| 18-110_k141_165806 | Not-determined |
| 18-110_k141_74115 | Not-determined |
| 18-110_k141_269615 | Not-determined |
| 18-110_k141_62442 | Not-determined |
| 18-110_k141_212018 | Not-determined |
| 18-110_k141_131544 | Not-determined |
| 18-110_k141_212021 | Not-determined |
| 18-110_k141_39273 | Not-determined |
| 18-110_k141_131547 | Not-determined |
| 18-110_k141_62448 | Not-determined |
| 18-110_k141_165817 | Not-determined |
| 18-110_k141_39275 | Low-quality |
| 18-110_k141_74129 | Not-determined |
| 18-110_k141_212031 | Not-determined |
| 18-110_k141_131553 | Not-determined |
| 18-110_k141_62457 | Not-determined |
| 18-110_k141_143290 | Not-determined |
| 18-110_k141_119620 | Not-determined |
| 18-110_k141_212037 | Low-quality |
| 18-110_k141_143294 | Not-determined |
| 18-110_k141_62462 | Not-determined |

| | |
|---|---|
| 18-110_k141_108012 | Not-determined |
| 18-110_k141_143297 | Not-determined |
| 18-110_k141_74145 | Not-determined |
| 18-110_k141_39287 | Not-determined |
| 18-110_k141_178156 | Not-determined |
| 18-110_k141_246634 | Not-determined |
| 18-110_k141_246636 | Not-determined |
| 18-110_k141_84744 | Not-determined |
| 18-110_k141_50398 | Not-determined |
| 18-110_k141_269635 | Not-determined |
| 18-110_k141_39298 | Not-determined |
| 18-110_k141_84754 | Not-determined |
| 18-110_k141_108029 | Low-quality |
| 18-110_k141_131575 | Not-determined |
| 18-110_k141_269645 | Not-determined |
| 18-110_k141_108030 | Low-quality |
| 18-110_k141_4206 | Low-quality |
| 18-110_k141_4207 | Not-determined |
| 18-110_k141_143322 | Not-determined |
| 18-110_k141_84757 | Not-determined |
| 18-110_k141_246654 | Not-determined |
| 18-110_k141_223327 | Not-determined |
| 18-110_k141_84759 | Not-determined |
| 18-110_k141_74173 | Not-determined |
| 18-110_k141_223329 | Not-determined |
| 18-110_k141_4212 | Not-determined |
| 18-110_k141_235430 | Low-quality |
| 18-110_k141_212067 | Low-quality |
| 18-110_k141_131581 | Low-quality |
| 18-110_k141_246659 | Not-determined |
| 18-110_k141_74179 | Not-determined |
| 18-110_k141_269661 | Not-determined |
| 18-110_k141_223336 | Not-determined |
| 18-110_k141_223337 | Not-determined |
| 18-110_k141_108053 | Not-determined |
| 18-110_k141_97040 | Not-determined |
| 18-110_k141_154578 | Not-determined |
| 18-110_k141_50429 | Not-determined |
| 18-110_k141_212081 | Not-determined |
| 18-110_k141_4230 | Not-determined |
| 18-110_k141_50437 | Low-quality |
| 18-110_k141_39348 | Not-determined |
| 18-110_k141_143341 | Not-determined |
| 18-110_k141_235453 | Low-quality |
| 18-110_k141_50442 | Not-determined |
| 18-110_k141_15090 | Low-quality |
| 18-110_k141_50449 | Not-determined |
| 18-110_k141_62507 | Not-determined |
| 18-110_k141_154595 | Not-determined |
| 18-110_k141_62510 | Low-quality |
| 18-110_k141_131593 | Not-determined |
| 18-110_k141_15099 | Not-determined |
| 18-110_k141_131599 | Not-determined |
| 18-110_k141_131600 | Not-determined |
| 18-110_k141_97065 | Not-determined |
| 18-110_k141_189180 | Not-determined |
| 18-110_k141_39376 | Not-determined |
| 18-110_k141_235456 | Not-determined |
| 18-110_k141_4253 | Not-determined |
| 18-110_k141_131614 | Not-determined |
| 18-110_k141_39380 | Not-determined |
| 18-110_k141_15111 | Not-determined |
| 18-110_k141_246682 | Not-determined |
| 18-110_k141_15113 | Not-determined |
| 18-110_k141_50467 | Not-determined |
| 18-110_k141_4263 | Not-determined |
| 18-110_k141_39393 | Not-determined |
| 18-110_k141_143377 | Not-determined |
| 18-110_k141_131638 | Not-determined |
| 18-110_k141_15121 | Not-determined |
| 18-110_k141_269704 | Low-quality |
| 18-110_k141_119687 | Not-determined |
| 18-110_k141_27911 | Not-determined |
| 18-110_k141_199805 | Not-determined |
| 18-110_k141_15123 | Not-determined |
| 18-110_k141_143388 | Not-determined |
| 18-110_k141_4269 | Not-determined |
| 18-110_k141_257015 | Not-determined |
| 18-110_k141_131643 | Not-determined |
| 18-110_k141_246700 | Not-determined |
| 18-110_k141_15128 | Not-determined |
| 18-110_k141_131646 | Not-determined |
| 18-110_k141_246703 | Not-determined |
| 18-110_k141_178229 | Not-determined |
| 18-110_k141_212134 | Low-quality |
| 18-110_k141_131650 | Not-determined |
| 18-110_k141_4274 | Low-quality |
| 18-110_k141_119699 | Not-determined |
| 18-110_k141_178230 | Not-determined |
| 18-110_k141_119701 | Not-determined |
| 18-110_k141_246710 | Not-determined |
| 18-110_k141_119703 | Not-determined |
| 18-110_k141_15140 | Not-determined |
| 18-110_k141_246720 | Not-determined |
| 18-110_k141_15141 | Not-determined |
| 18-110_k141_15144 | Not-determined |
| 18-110_k141_199813 | Not-determined |
| 18-110_k141_84784 | Not-determined |
| 18-110_k141_235488 | Not-determined |
| 18-110_k141_269728 | Not-determined |
| 18-110_k141_50493 | Not-determined |
| 18-110_k141_269731 | Not-determined |
| 18-110_k141_15151 | Not-determined |
| 18-110_k141_27918 | Not-determined |
| 18-110_k141_143405 | Not-determined |
| 18-110_k141_212138 | Not-determined |
| 18-110_k141_189219 | Not-determined |
| 18-110_k141_27919 | Not-determined |
| 18-110_k141_84793 | Not-determined |
| 18-110_k141_84794 | Not-determined |
| 18-110_k141_84795 | Not-determined |
| 18-110_k141_131680 | Not-determined |
| 18-110_k141_108082 | Low-quality |
| 18-110_k141_108086 | Not-determined |
| 18-110_k141_62575 | Not-determined |
| 18-110_k141_84796 | Not-determined |
| 18-110_k141_27929 | Low-quality |
| 18-110_k141_235515 | Low-quality |
| 18-110_k141_27933 | Not-determined |
| 18-110_k141_50502 | Not-determined |
| 18-110_k141_178267 | Not-determined |
| 18-110_k141_27939 | Not-determined |
| 18-110_k141_178273 | Not-determined |
| 18-110_k141_246757 | Not-determined |
| 18-110_k141_212167 | Not-determined |
| 18-110_k141_269756 | Not-determined |
| 18-110_k141_235526 | Not-determined |
| 18-110_k141_246762 | Not-determined |
| 18-110_k141_257049 | Low-quality |
| 18-110_k141_189235 | Not-determined |
| 18-110_k141_189236 | Not-determined |
| 18-110_k141_143432 | Not-determined |
| 18-110_k141_84811 | Not-determined |
| 18-110_k141_212174 | Not-determined |
| 18-110_k141_154658 | Not-determined |
| 18-110_k141_178281 | Not-determined |
| 18-110_k141_178283 | Not-determined |
| 18-110_k141_62580 | Not-determined |
| 18-110_k141_108113 | Not-determined |
| 18-110_k141_39444 | Not-determined |
| 18-110_k141_4323 | Not-determined |
| 18-110_k141_4327 | Not-determined |
| 18-110_k141_223350 | Not-determined |
| 18-110_k141_212177 | Not-determined |
| 18-110_k141_39449 | Not-determined |
| 18-110_k141_154671 | Not-determined |
| 18-110_k141_235540 | Low-quality |
| 18-110_k141_178299 | Low-quality |
| 18-110_k141_84830 | Not-determined |
| 18-110_k141_97100 | Not-determined |
| 18-110_k141_257059 | Not-determined |
| 18-110_k141_165908 | Low-quality |
| 18-110_k141_4345 | Not-determined |
| 18-110_k141_74248 | Not-determined |
| 18-110_k141_39454 | Not-determined |
| 18-110_k141_119738 | Low-quality |
| 18-110_k141_178317 | Not-determined |
| 18-110_k141_4350 | Not-determined |
| 18-110_k141_27975 | Not-determined |
| 18-110_k141_97108 | Not-determined |
| 18-110_k141_84837 | Not-determined |
| 18-110_k141_235561 | Not-determined |
| 18-110_k141_84839 | Not-determined |
| 18-110_k141_235563 | Not-determined |
| 18-110_k141_235564 | Not-determined |
| 18-110_k141_212188 | Not-determined |
| 18-110_k141_39459 | Not-determined |
| 18-110_k141_235567 | Not-determined |
| 18-110_k141_189244 | Not-determined |
| 18-110_k141_257072 | Not-determined |
| 18-110_k141_212194 | Not-determined |
| 18-110_k141_97118 | Not-determined |
| 18-110_k141_4363 | Not-determined |
| 18-110_k141_50567 | Not-determined |
| 18-110_k141_27981 | Not-determined |
| 18-110_k141_154704 | Not-determined |
| 18-110_k141_97125 | Not-determined |
| 18-110_k141_39465 | Not-determined |
| 18-110_k141_4370 | Not-determined |
| 18-110_k141_15177 | Not-determined |
| 18-110_k141_178353 | Not-determined |
| 18-110_k141_74271 | Not-determined |
| 18-110_k141_257084 | Not-determined |
| 18-110_k141_257087 | Not-determined |
| 18-110_k141_189275 | Not-determined |
| 18-110_k141_165930 | Not-determined |
| 18-110_k141_15191 | Not-determined |
| 18-110_k141_154718 | Not-determined |
| 18-110_k141_119762 | Low-quality |
| 18-110_k141_165935 | Not-determined |
| 18-110_k141_178371 | Not-determined |
| 18-110_k141_269788 | Not-determined |
| 18-110_k141_246782 | Not-determined |
| 18-110_k141_143460 | Not-determined |
| 18-110_k141_189283 | Not-determined |
| 18-110_k141_257094 | Not-determined |
| 18-110_k141_39491 | Not-determined |
| 18-110_k141_39492 | Not-determined |
| 18-110_k141_189287 | Not-determined |
| 18-110_k141_27997 | Not-determined |
| 18-110_k141_62625 | Not-determined |
| 18-110_k141_223383 | Low-quality |
| 18-110_k141_223384 | Not-determined |
| 18-110_k141_39502 | Not-determined |
| 18-110_k141_235595 | Low-quality |
| 18-110_k141_4391 | Not-determined |
| 18-110_k141_235602 | Not-determined |
| 18-110_k141_269799 | Not-determined |
| 18-110_k141_246799 | Not-determined |
| 18-110_k141_131697 | Not-determined |
| 18-110_k141_50601 | Not-determined |
| 18-110_k141_97163 | Not-determined |
| 18-110_k141_62643 | Not-determined |
| 18-110_k141_269804 | Not-determined |
| 18-110_k141_246806 | Not-determined |
| 18-110_k141_62645 | Not-determined |
| 18-110_k141_143487 | Not-determined |
| 18-110_k141_143488 | Not-determined |
| 18-110_k141_143492 | Not-determined |
| 18-110_k141_223405 | Not-determined |
| 18-110_k141_62647 | Not-determined |
| 18-110_k141_97166 | Not-determined |
| 18-110_k141_15215 | Not-determined |
| 18-110_k141_223412 | Not-determined |
| 18-110_k141_269813 | Not-determined |
| 18-110_k141_108156 | Not-determined |
| 18-110_k141_28010 | Not-determined |
| 18-110_k141_178403 | Low-quality |
| 18-110_k141_108159 | Not-determined |
| 18-110_k141_108160 | Not-determined |
| 18-110_k141_212216 | Not-determined |
| 18-110_k141_165981 | Not-determined |
| 18-110_k141_108165 | Not-determined |
| 18-110_k141_178406 | Not-determined |
| 18-110_k141_246819 | Not-determined |
| 18-110_k141_235627 | Not-determined |
| 18-110_k141_246821 | Not-determined |
| 18-110_k141_257133 | Not-determined |
| 18-110_k141_223422 | Not-determined |
| 18-110_k141_62663 | Not-determined |
| 18-110_k141_178412 | Not-determined |
| 18-110_k141_62669 | Not-determined |
| 18-110_k141_269824 | Not-determined |
| 18-110_k141_50607 | Not-determined |
| 18-110_k141_257148 | Not-determined |
| 18-110_k141_62674 | Not-determined |
| 18-110_k141_165993 | Not-determined |
| 18-110_k141_84909 | Not-determined |
| 18-110_k141_15242 | Not-determined |
| 18-110_k141_15243 | Low-quality |
| 18-110_k141_15246 | Not-determined |
| 18-110_k141_212228 | Not-determined |
| 18-110_k141_212239 | Not-determined |
| 18-110_k141_4402 | Not-determined |
| 18-110_k141_74289 | Not-determined |
| 18-110_k141_97201 | Not-determined |
| 18-110_k141_50627 | Not-determined |
| 18-110_k141_235651 | Not-determined |
| 18-110_k141_62704 | Not-determined |
| 18-110_k141_235652 | Not-determined |
| 18-110_k141_50628 | Not-determined |
| 18-110_k141_28046 | Low-quality |
| 18-110_k141_131719 | Not-determined |
| 18-110_k141_131721 | Not-determined |
| 18-110_k141_39527 | Not-determined |
| 18-110_k141_15266 | Not-determined |
| 18-110_k141_178428 | Not-determined |
| 18-110_k141_212245 | Not-determined |
| 18-110_k141_74294 | Not-determined |
| 18-110_k141_246844 | Not-determined |
| 18-110_k141_246846 | Not-determined |
| 18-110_k141_50643 | Not-determined |
| 18-110_k141_235662 | Low-quality |
| 18-110_k141_15270 | Not-determined |
| 18-110_k141_212251 | Not-determined |
| 18-110_k141_119777 | Not-determined |
| 18-110_k141_246853 | Not-determined |
| 18-110_k141_246857 | Low-quality |
| 18-110_k141_119779 | Not-determined |
| 18-110_k141_212258 | Not-determined |
| 18-110_k141_246859 | Not-determined |
| 18-110_k141_166043 | Not-determined |
| 18-110_k141_189356 | Not-determined |
| 18-110_k141_269854 | Not-determined |
| 18-110_k141_50657 | Not-determined |
| 18-110_k141_212263 | Not-determined |
| 18-110_k141_119786 | Not-determined |
| 18-110_k141_50662 | Not-determined |
| 18-110_k141_189358 | Not-determined |
| 18-110_k141_223456 | Not-determined |
| 18-110_k141_97229 | Low-quality |
| 18-110_k141_15288 | Not-determined |
| 18-110_k141_189363 | Not-determined |
| 18-110_k141_269868 | Not-determined |
| 18-110_k141_74316 | Not-determined |
| 18-110_k141_84944 | Not-determined |
| 18-110_k141_166057 | Low-quality |
| 18-110_k141_269873 | Not-determined |
| 18-110_k141_84948 | Not-determined |
| 18-110_k141_223463 | Not-determined |
| 18-110_k141_189372 | Not-determined |
| 18-110_k141_62753 | Not-determined |
| 18-110_k141_15297 | Not-determined |
| 18-110_k141_108215 | Not-determined |
| 18-110_k141_15304 | Not-determined |
| 18-110_k141_235680 | Not-determined |
| 18-110_k141_235681 | Not-determined |
| 18-110_k141_15306 | Not-determined |
| 18-110_k141_15307 | Not-determined |
| 18-110_k141_50671 | Not-determined |
| 18-110_k141_223475 | Not-determined |
| 18-110_k141_166072 | Not-determined |
| 18-110_k141_74334 | Not-determined |
| 18-110_k141_119824 | Not-determined |
| 18-110_k141_246875 | Low-quality |
| 18-110_k141_39576 | Not-determined |
| 18-110_k141_119825 | Low-quality |
| 18-110_k141_246879 | Not-determined |
| 18-110_k141_50684 | Not-determined |
| 18-110_k141_246881 | Not-determined |
| 18-110_k141_28078 | Not-determined |
| 18-110_k141_223478 | Not-determined |
| 18-110_k141_269904 | Not-determined |
| 18-110_k141_269905 | Not-determined |
| 18-110_k141_235693 | Not-determined |
| 18-110_k141_131729 | Not-determined |
| 18-110_k141_166082 | Not-determined |
| 18-110_k141_50695 | Not-determined |
| 18-110_k141_28096 | Not-determined |
| 18-110_k141_269914 | Not-determined |
| 18-110_k141_223486 | Not-determined |
| 18-110_k141_131739 | Not-determined |
| 18-110_k141_257177 | Not-determined |
| 18-110_k141_269917 | Not-determined |
| 18-110_k141_235707 | Not-determined |
| 18-110_k141_257179 | Not-determined |
| 18-110_k141_235712 | Low-quality |
| 18-110_k141_50709 | Not-determined |
| 18-110_k141_108238 | Not-determined |
| 18-110_k141_84995 | Not-determined |
| 18-110_k141_62782 | Not-determined |
| 18-110_k141_166095 | Not-determined |
| 18-110_k141_84997 | Not-determined |
| 18-110_k141_235726 | Not-determined |
| 18-110_k141_74345 | Low-quality |
| 18-110_k141_246891 | Not-determined |
| 18-110_k141_246896 | Not-determined |
| 18-110_k141_178494 | Not-determined |
| 18-110_k141_85006 | Not-determined |
| 18-110_k141_85010 | Not-determined |
| 18-110_k141_189397 | Not-determined |
| 18-110_k141_28131 | Not-determined |
| 18-110_k141_189403 | Not-determined |
| 18-110_k141_85015 | Not-determined |
| 18-110_k141_131771 | Not-determined |
| 18-110_k141_85017 | Not-determined |
| 18-110_k141_62795 | Not-determined |
| 18-110_k141_74357 | Low-quality |
| 18-110_k141_269945 | Not-determined |
| 18-110_k141_119862 | Not-determined |
| 18-110_k141_62801 | Not-determined |
| 18-110_k141_246918 | Not-determined |
| 18-110_k141_39617 | Not-determined |
| 18-110_k141_223524 | Not-determined |
| 18-110_k141_166128 | Not-determined |
| 18-110_k141_178513 | Not-determined |
| 18-110_k141_28149 | Not-determined |
| 18-110_k141_62819 | Not-determined |
| 18-110_k141_15358 | Not-determined |
| 18-110_k141_85028 | Not-determined |
| 18-110_k141_50719 | Not-determined |
| 18-110_k141_28155 | Not-determined |
| 18-110_k141_50721 | Not-determined |
| 18-110_k141_143531 | Not-determined |
| 18-110_k141_39629 | Not-determined |
| 18-110_k141_166139 | Not-determined |
| 18-110_k141_257186 | Not-determined |
| 18-110_k141_257188 | Not-determined |
| 18-110_k141_15365 | Not-determined |
| 18-110_k141_143539 | Not-determined |
| 18-110_k141_235760 | Low-quality |
| 18-110_k141_15371 | Not-determined |
| 18-110_k141_85039 | Not-determined |
| 18-110_k141_269980 | Not-determined |
| 18-110_k141_235762 | Not-determined |
| 18-110_k141_154742 | Not-determined |
| 18-110_k141_235763 | Not-determined |
| 18-110_k141_74361 | Not-determined |
| 18-110_k141_15379 | Not-determined |
| 18-110_k141_143555 | Not-determined |
| 18-110_k141_15382 | Not-determined |
| 18-110_k141_39665 | Not-determined |
| 18-110_k141_143561 | Low-quality |
| 18-110_k141_257194 | Not-determined |
| 18-110_k141_189436 | Low-quality |
| 18-110_k141_39668 | Not-determined |
| 18-110_k141_119868 | Not-determined |
| 18-110_k141_39671 | Not-determined |
| 18-110_k141_62847 | Not-determined |
| 18-110_k141_178526 | Not-determined |
| 18-110_k141_154756 | Not-determined |
| 18-110_k141_178527 | Not-determined |
| 18-110_k141_269992 | Not-determined |
| 18-110_k141_131799 | Not-determined |
| 18-110_k141_235777 | Not-determined |
| 18-110_k141_74379 | Not-determined |
| 18-110_k141_189455 | Not-determined |
| 18-110_k141_189457 | Not-determined |
| 18-110_k141_119878 | Not-determined |
| 18-110_k141_189463 | Not-determined |
| 18-110_k141_212334 | Not-determined |
| 18-110_k141_39688 | Not-determined |
| 18-110_k141_154778 | Not-determined |
| 18-110_k141_212343 | Not-determined |
| 18-110_k141_257212 | Low-quality |
| 18-110_k141_178552 | Not-determined |
| 18-110_k141_154783 | Not-determined |
| 18-110_k141_39697 | Not-determined |
| 18-110_k141_178555 | Not-determined |
| 18-110_k141_212352 | Not-determined |
| 18-110_k141_62870 | Not-determined |
| 18-110_k141_74395 | Not-determined |
| 18-110_k141_143595 | Not-determined |
| 18-110_k141_143596 | Not-determined |
| 18-110_k141_62872 | Not-determined |
| 18-110_k141_74400 | Not-determined |
| 18-110_k141_166157 | Not-determined |
| 18-110_k141_62875 | Not-determined |
| 18-110_k141_246985 | Not-determined |
| 18-110_k141_74413 | Low-quality |
| 18-110_k141_166167 | Not-determined |
| 18-110_k141_62882 | Not-determined |
| 18-110_k141_108302 | Low-quality |
| 18-110_k141_166173 | Not-determined |
| 18-110_k141_108303 | Not-determined |
| 18-110_k141_108307 | Not-determined |
| 18-110_k141_235817 | Not-determined |
| 18-110_k141_85065 | Not-determined |
| 18-110_k141_50746 | Not-determined |
| 18-110_k141_119896 | Not-determined |
| 18-110_k141_166182 | Not-determined |
| 18-110_k141_199869 | Not-determined |
| 18-110_k141_50751 | Not-determined |
| 18-110_k141_166186 | Not-determined |
| 18-110_k141_199872 | Low-quality |
| 18-110_k141_235828 | Not-determined |
| 18-110_k141_223539 | Not-determined |
| 18-110_k141_189495 | Not-determined |
| 18-110_k141_178562 | Not-determined |
| 18-110_k141_62892 | Not-determined |
| 18-110_k141_223544 | Not-determined |
| 18-110_k141_247009 | Not-determined |
| 18-110_k141_189500 | Not-determined |
| 18-110_k141_223547 | Not-determined |
| 18-110_k141_166198 | Not-determined |
| 18-110_k141_257235 | Not-determined |
| 18-110_k141_223548 | Low-quality |
| 18-110_k141_85080 | Low-quality |
| 18-110_k141_235840 | Not-determined |
| 18-110_k141_257239 | Not-determined |
| 18-110_k141_223549 | Low-quality |
| 18-110_k141_257243 | Low-quality |
| 18-110_k141_257247 | Not-determined |
| 18-110_k141_247016 | Not-determined |
| 18-110_k141_143611 | Low-quality |
| 18-110_k141_62911 | Not-determined |
| 18-110_k141_143613 | Not-determined |
| 18-110_k141_108338 | Not-determined |
| 18-110_k141_270019 | Not-determined |
| 18-110_k141_247026 | Not-determined |
| 18-110_k141_223562 | Not-determined |
| 18-110_k141_50778 | Not-determined |
| 18-110_k141_62921 | Not-determined |
| 18-110_k141_15429 | Not-determined |
| 18-110_k141_247031 | Not-determined |
| 18-110_k141_247037 | Not-determined |
| 18-110_k141_15434 | Not-determined |
| 18-110_k141_85094 | Not-determined |
| 18-110_k141_50788 | Low-quality |
| 18-110_k141_15439 | Not-determined |
| 18-110_k141_85100 | Not-determined |
| 18-110_k141_235847 | Low-quality |
| 18-110_k141_62935 | Not-determined |
| 18-110_k141_154794 | Not-determined |
| 18-110_k141_257256 | Low-quality |
| 18-110_k141_119928 | Not-determined |
| 18-110_k141_178603 | Low-quality |
| 18-110_k141_154800 | Not-determined |
| 18-110_k141_199893 | Not-determined |
| 18-110_k141_199895 | Not-determined |
| 18-110_k141_4432 | Low-quality |
| 18-110_k141_178610 | Not-determined |
| 18-110_k141_166237 | Not-determined |
| 18-110_k141_85113 | Not-determined |
| 18-110_k141_235854 | Not-determined |
| 18-110_k141_166240 | Not-determined |
| 18-110_k141_4436 | Not-determined |
| 18-110_k141_62954 | Not-determined |
| 18-110_k141_108384 | Not-determined |
| 18-110_k141_247059 | Not-determined |
| 18-110_k141_166246 | Not-determined |
| 18-110_k141_212386 | Not-determined |
| 18-110_k141_131857 | Not-determined |
| 18-110_k141_154811 | Not-determined |
| 18-110_k141_131859 | Not-determined |
| 18-110_k141_189519 | Low-quality |
| 18-110_k141_178619 | Not-determined |
| 18-110_k141_178621 | Not-determined |
| 18-110_k141_15466 | Not-determined |
| 18-110_k141_212391 | Not-determined |
| 18-110_k141_199908 | Not-determined |
| 18-110_k141_178626 | Not-determined |
| 18-110_k141_257272 | Not-determined |
| 18-110_k141_119950 | Not-determined |
| 18-110_k141_178629 | Not-determined |
| 18-110_k141_4452 | Not-determined |
| 18-110_k141_28270 | Not-determined |
| 18-110_k141_97264 | Not-determined |
| 18-110_k141_257279 | Low-quality |
| 18-110_k141_97265 | Not-determined |
| 18-110_k141_235872 | Not-determined |
| 18-110_k141_257282 | Not-determined |
| 18-110_k141_62979 | Not-determined |
| 18-110_k141_62981 | Not-determined |
| 18-110_k141_235880 | Low-quality |
| 18-110_k141_15470 | Not-determined |
| 18-110_k141_119960 | Not-determined |
| 18-110_k141_4465 | Not-determined |
| 18-110_k141_131881 | Not-determined |
| 18-110_k141_247088 | Not-determined |
| 18-110_k141_270046 | Not-determined |
| 18-110_k141_108395 | Low-quality |
| 18-110_k141_270052 | Not-determined |
| 18-110_k141_247099 | Not-determined |
| 18-110_k141_85149 | Not-determined |
| 18-110_k141_85150 | Not-determined |
| 18-110_k141_235900 | Low-quality |
| 18-110_k141_85157 | Not-determined |
| 18-110_k141_189555 | Not-determined |
| 18-110_k141_62987 | Not-determined |
| 18-110_k141_247109 | Not-determined |
| 18-110_k141_270062 | Not-determined |
| 18-110_k141_154843 | Not-determined |
| 18-110_k141_50815 | Not-determined |
| 18-110_k141_50816 | Not-determined |
| 18-110_k141_97296 | Low-quality |
| 18-110_k141_247114 | Not-determined |
| 18-110_k141_143621 | Not-determined |
| 18-110_k141_85162 | Not-determined |
| 18-110_k141_63002 | Low-quality |
| 18-110_k141_270069 | Not-determined |
| 18-110_k141_63004 | Not-determined |
| 18-110_k141_143627 | Not-determined |
| 18-110_k141_63008 | Not-determined |
| 18-110_k141_28314 | Not-determined |
| 18-110_k141_74429 | Not-determined |
| 18-110_k141_63009 | Low-quality |
| 18-110_k141_143632 | Not-determined |
| 18-110_k141_63012 | Low-quality |
| 18-110_k141_270081 | Not-determined |
| 18-110_k141_15499 | Not-determined |
| 18-110_k141_85172 | Not-determined |
| 18-110_k141_143637 | Not-determined |
| 18-110_k141_74438 | Not-determined |
| 18-110_k141_199963 | Not-determined |
| 18-110_k141_143644 | Not-determined |
| 18-110_k141_154856 | Not-determined |
| 18-110_k141_270089 | Not-determined |
| 18-110_k141_223589 | Not-determined |
| 18-110_k141_63025 | Not-determined |
| 18-110_k141_154862 | Not-determined |
| 18-110_k141_50826 | Not-determined |
| 18-110_k141_223592 | Not-determined |
| 18-110_k141_50830 | Not-determined |
| 18-110_k141_178684 | Not-determined |
| 18-110_k141_28319 | Not-determined |
| 18-110_k141_50834 | Not-determined |
| 18-110_k141_247140 | Not-determined |
| 18-110_k141_131901 | Not-determined |
| 18-110_k141_15514 | Not-determined |
| 18-110_k141_247144 | Not-determined |
| 18-110_k141_50842 | Not-determined |
| 18-110_k141_85190 | Not-determined |
| 18-110_k141_50844 | Not-determined |
| 18-110_k141_50847 | Not-determined |
| 18-110_k141_154873 | Low-quality |
| 18-110_k141_212445 | Not-determined |
| 18-110_k141_223602 | Not-determined |
| 18-110_k141_143661 | Low-quality |

| | |
|---|---|
| 18-110_k141_154875 | Not-determined |
| 18-110_k141_223610 | Not-determined |
| 18-110_k141_235936 | Not-determined |
| 18-110_k141_131911 | Not-determined |
| 18-110_k141_85198 | Not-determined |
| 18-110_k141_39704 | Not-determined |
| 18-110_k141_223613 | Not-determined |
| 18-110_k141_28335 | Not-determined |
| 18-110_k141_178696 | Not-determined |
| 18-110_k141_212455 | Not-determined |
| 18-110_k141_199971 | Not-determined |
| 18-110_k141_120016 | Not-determined |
| 18-110_k141_143676 | Not-determined |
| 18-110_k141_270098 | Low-quality |
| 18-110_k141_74492 | Not-determined |
| 18-110_k141_154898 | Not-determined |
| 18-110_k141_39733 | Not-determined |
| 18-110_k141_120035 | Not-determined |
| 18-110_k141_4513 | Not-determined |
| 18-110_k141_50872 | Not-determined |
| 18-110_k141_235972 | Not-determined |
| 18-110_k141_28367 | Not-determined |
| 18-110_k141_154910 | Not-determined |
| 18-110_k141_257321 | Not-determined |
| 18-110_k141_108456 | Not-determined |
| 18-110_k141_257322 | Low-quality |
| 18-110_k141_15539 | Low-quality |
| 18-110_k141_39749 | Not-determined |
| 18-110_k141_257325 | Not-determined |
| 18-110_k141_235984 | Not-determined |
| 18-110_k141_223617 | Not-determined |
| 18-110_k141_63033 | Not-determined |
| 18-110_k141_97377 | Low-quality |
| 18-110_k141_154923 | Not-determined |
| 18-110_k141_257333 | Not-determined |
| 18-110_k141_97382 | Not-determined |
| 18-110_k141_212494 | Low-quality |
| 18-110_k141_154928 | Not-determined |
| 18-110_k141_108476 | Not-determined |
| 18-110_k141_212495 | Not-determined |
| 18-110_k141_39771 | Not-determined |
| 18-110_k141_143722 | Not-determined |
| 18-110_k141_85241 | Not-determined |
| 18-110_k141_15553 | Not-determined |
| 18-110_k141_97387 | Not-determined |
| 18-110_k141_50890 | Not-determined |
| 18-110_k141_270124 | Not-determined |
| 18-110_k141_154939 | Not-determined |
| 18-110_k141_223629 | Not-determined |
| 18-110_k141_39773 | Not-determined |
| 18-110_k141_97392 | Not-determined |
| 18-110_k141_178754 | Not-determined |
| 18-110_k141_97399 | Not-determined |
| 18-110_k141_189598 | Not-determined |
| 18-110_k141_235993 | Not-determined |
| 18-110_k141_247210 | Not-determined |
| 18-110_k141_63046 | Not-determined |
| 18-110_k141_120054 | Not-determined |
| 18-110_k141_4541 | Not-determined |
| 18-110_k141_28375 | Not-determined |
| 18-110_k141_178762 | Not-determined |
| 18-110_k141_85249 | Not-determined |
| 18-110_k141_143730 | Not-determined |
| 18-110_k141_63058 | Not-determined |
| 18-110_k141_120059 | Not-determined |
| 18-110_k141_178770 | Not-determined |
| 18-110_k141_39791 | Not-determined |
| 18-110_k141_200008 | Not-determined |
| 18-110_k141_247215 | Not-determined |
| 18-110_k141_28387 | Not-determined |
| 18-110_k141_108490 | Not-determined |
| 18-110_k141_28388 | Not-determined |
| 18-110_k141_15564 | Not-determined |
| 18-110_k141_154960 | Not-determined |
| 18-110_k141_236008 | Not-determined |
| 18-110_k141_247222 | Not-determined |
| 18-110_k141_200016 | Low-quality |
| 18-110_k141_257352 | Not-determined |
| 18-110_k141_189622 | Not-determined |
| 18-110_k141_189625 | Not-determined |
| 18-110_k141_270150 | Not-determined |
| 18-110_k141_50905 | Not-determined |
| 18-110_k141_236017 | Not-determined |
| 18-110_k141_257355 | Not-determined |
| 18-110_k141_178788 | Not-determined |
| 18-110_k141_178789 | Not-determined |
| 18-110_k141_131924 | Not-determined |
| 18-110_k141_74512 | Not-determined |
| 18-110_k141_39812 | Not-determined |
| 18-110_k141_178793 | Not-determined |
| 18-110_k141_131929 | Not-determined |
| 18-110_k141_143747 | Not-determined |
| 18-110_k141_63087 | Not-determined |
| 18-110_k141_97413 | Not-determined |
| 18-110_k141_247235 | Not-determined |
| 18-110_k141_247236 | Not-determined |
| 18-110_k141_120090 | Low-quality |
| 18-110_k141_270156 | Not-determined |
| 18-110_k141_200021 | Not-determined |
| 18-110_k141_63095 | Not-determined |
| 18-110_k141_200029 | Not-determined |
| 18-110_k141_15587 | Not-determined |
| 18-110_k141_200042 | Not-determined |
| 18-110_k141_270176 | Not-determined |
| 18-110_k141_74532 | Not-determined |
| 18-110_k141_257377 | Not-determined |
| 18-110_k141_270185 | Not-determined |
| 18-110_k141_28426 | Not-determined |
| 18-110_k141_178826 | Not-determined |
| 18-110_k141_270191 | Not-determined |
| 18-110_k141_154990 | Not-determined |
| 18-110_k141_189652 | Not-determined |
| 18-110_k141_28432 | Not-determined |
| 18-110_k141_189657 | Not-determined |
| 18-110_k141_85284 | Not-determined |
| 18-110_k141_39841 | Not-determined |
| 18-110_k141_108500 | Not-determined |
| 18-110_k141_200059 | Not-determined |
| 18-110_k141_15605 | Not-determined |
| 18-110_k141_270194 | Not-determined |
| 18-110_k141_39845 | Low-quality |
| 18-110_k141_154996 | Not-determined |
| 18-110_k141_39846 | Not-determined |
| 18-110_k141_50923 | Not-determined |
| 18-110_k141_223683 | Not-determined |
| 18-110_k141_74544 | Not-determined |
| 18-110_k141_97444 | Not-determined |
| 18-110_k141_74549 | Not-determined |
| 18-110_k141_15618 | Not-determined |
| 18-110_k141_270203 | Not-determined |
| 18-110_k141_15620 | Not-determined |
| 18-110_k141_270205 | Not-determined |
| 18-110_k141_97452 | Not-determined |
| 18-110_k141_97453 | Not-determined |
| 18-110_k141_15622 | Low-quality |
| 18-110_k141_28468 | Not-determined |
| 18-110_k141_97455 | Low-quality |
| 18-110_k141_108514 | Low-quality |
| 18-110_k141_74556 | Not-determined |
| 18-110_k141_270216 | Not-determined |
| 18-110_k141_39862 | Not-determined |
| 18-110_k141_236072 | Not-determined |
| 18-110_k141_15626 | Not-determined |
| 18-110_k141_108521 | Not-determined |
| 18-110_k141_189672 | Low-quality |
| 18-110_k141_63142 | Not-determined |
| 18-110_k141_50950 | Not-determined |
| 18-110_k141_39869 | Not-determined |
| 18-110_k141_155009 | Not-determined |
| 18-110_k141_39871 | Not-determined |
| 18-110_k141_108525 | Not-determined |
| 18-110_k141_155010 | Not-determined |
| 18-110_k141_28478 | Not-determined |
| 18-110_k141_257389 | Not-determined |
| 18-110_k141_155016 | Not-determined |
| 18-110_k141_257390 | Not-determined |
| 18-110_k141_15634 | Low-quality |
| 18-110_k141_236085 | Not-determined |
| 18-110_k141_236086 | Not-determined |
| 18-110_k141_257395 | Not-determined |
| 18-110_k141_63153 | Not-determined |
| 18-110_k141_28490 | Not-determined |
| 18-110_k141_97484 | Low-quality |
| 18-110_k141_63159 | Not-determined |
| 18-110_k141_15646 | Not-determined |
| 18-110_k141_131989 | Not-determined |
| 18-110_k141_143801 | Not-determined |
| 18-110_k141_131993 | Not-determined |
| 18-110_k141_247302 | Not-determined |
| 18-110_k141_270227 | Not-determined |
| 18-110_k141_131999 | Not-determined |
| 18-110_k141_270231 | Not-determined |
| 18-110_k141_270234 | Not-determined |
| 18-110_k141_143815 | Not-determined |
| 18-110_k141_132013 | Not-determined |
| 18-110_k141_223738 | Not-determined |
| 18-110_k141_247320 | Not-determined |
| 18-110_k141_236099 | Not-determined |
| 18-110_k141_155041 | Not-determined |
| 18-110_k141_166301 | Not-determined |
| 18-110_k141_74618 | Not-determined |
| 18-110_k141_166302 | Not-determined |
| 18-110_k141_85300 | Not-determined |
| 18-110_k141_247329 | Not-determined |
| 18-110_k141_28508 | Not-determined |
| 18-110_k141_15667 | Not-determined |
| 18-110_k141_28511 | Not-determined |
| 18-110_k141_15670 | Not-determined |
| 18-110_k141_223764 | Not-determined |
| 18-110_k141_28514 | Not-determined |
| 18-110_k141_155051 | Not-determined |
| 18-110_k141_85314 | Not-determined |
| 18-110_k141_120134 | Not-determined |
| 18-110_k141_120135 | Not-determined |
| 18-110_k141_15682 | Not-determined |
| 18-110_k141_85322 | Not-determined |
| 18-110_k141_120141 | Not-determined |
| 18-110_k141_236116 | Not-determined |
| 18-110_k141_120142 | Not-determined |
| 18-110_k141_39906 | Not-determined |
| 18-110_k141_236119 | Not-determined |
| 18-110_k141_236121 | Not-determined |
| 18-110_k141_39909 | Not-determined |
| 18-110_k141_28534 | Not-determined |
| 18-110_k141_270255 | Not-determined |
| 18-110_k141_212547 | Not-determined |
| 18-110_k141_236126 | Low-quality |
| 18-110_k141_236128 | Not-determined |
| 18-110_k141_257435 | Not-determined |
| 18-110_k141_39914 | Not-determined |
| 18-110_k141_212551 | Not-determined |
| 18-110_k141_143860 | Not-determined |
| 18-110_k141_155054 | Not-determined |
| 18-110_k141_155057 | Not-determined |
| 18-110_k141_51015 | Not-determined |
| 18-110_k141_257441 | Not-determined |
| 18-110_k141_51017 | Not-determined |
| 18-110_k141_51018 | Not-determined |
| 18-110_k141_247359 | Not-determined |
| 18-110_k141_4594 | Not-determined |
| 18-110_k141_236134 | Low-quality |
| 18-110_k141_4596 | Not-determined |
| 18-110_k141_4599 | Not-determined |
| 18-110_k141_178834 | Not-determined |
| 18-110_k141_85345 | Not-determined |
| 18-110_k141_39921 | Not-determined |
| 18-110_k141_143862 | Not-determined |
| 18-110_k141_247368 | Not-determined |
| 18-110_k141_247375 | Not-determined |
| 18-110_k141_132046 | Low-quality |
| 18-110_k141_28572 | Not-determined |
| 18-110_k141_97535 | Low-quality |
| 18-110_k141_85362 | Not-determined |
| 18-110_k141_85363 | Not-determined |
| 18-110_k141_257458 | Not-determined |
| 18-110_k141_85366 | Not-determined |
| 18-110_k141_74669 | Not-determined |
| 18-110_k141_247381 | Not-determined |
| 18-110_k141_257460 | Not-determined |
| 18-110_k141_51028 | Not-determined |
| 18-110_k141_97541 | Not-determined |
| 18-110_k141_51034 | Not-determined |
| 18-110_k141_4615 | Low-quality |
| 18-110_k141_97544 | Not-determined |
| 18-110_k141_132055 | Not-determined |
| 18-110_k141_51041 | Not-determined |
| 18-110_k141_143900 | Not-determined |
| 18-110_k141_51045 | Not-determined |
| 18-110_k141_270301 | Not-determined |
| 18-110_k141_270306 | Not-determined |
| 18-110_k141_247396 | Not-determined |
| 18-110_k141_247401 | Not-determined |
| 18-110_k141_74682 | Not-determined |
| 18-110_k141_247404 | Not-determined |
| 18-110_k141_63203 | Not-determined |
| 18-110_k141_15710 | Low-quality |
| 18-110_k141_15712 | Not-determined |
| 18-110_k141_178901 | Not-determined |
| 18-110_k141_51063 | Not-determined |
| 18-110_k141_132075 | Low-quality |
| 18-110_k141_97562 | Not-determined |
| 18-110_k141_166327 | Not-determined |
| 18-110_k141_270321 | Not-determined |
| 18-110_k141_15719 | Not-determined |
| 18-110_k141_247414 | Low-quality |
| 18-110_k141_189722 | Not-determined |
| 18-110_k141_15721 | Low-quality |
| 18-110_k141_223832 | Not-determined |
| 18-110_k141_28603 | Low-quality |
| 18-110_k141_15723 | Not-determined |
| 18-110_k141_63216 | Not-determined |
| 18-110_k141_143929 | Not-determined |
| 18-110_k141_143930 | Not-determined |
| 18-110_k141_39969 | Not-determined |
| 18-110_k141_39970 | Low-quality |
| 18-110_k141_74692 | Low-quality |
| 18-110_k141_28610 | Not-determined |
| 18-110_k141_143934 | Not-determined |
| 18-110_k141_97573 | Low-quality |
| 18-110_k141_189735 | Low-quality |
| 18-110_k141_270323 | Not-determined |
| 18-110_k141_132087 | Not-determined |
| 18-110_k141_132088 | Not-determined |
| 18-110_k141_166334 | Not-determined |
| 18-110_k141_39981 | Not-determined |
| 18-110_k141_97577 | Not-determined |
| 18-110_k141_28614 | Low-quality |
| 18-110_k141_74701 | Not-determined |
| 18-110_k141_189741 | Not-determined |
| 18-110_k141_189742 | Not-determined |
| 18-110_k141_166345 | Not-determined |
| 18-110_k141_97590 | Not-determined |
| 18-110_k141_270341 | Low-quality |
| 18-110_k141_178927 | Not-determined |
| 18-110_k141_51095 | Not-determined |
| 18-110_k141_236185 | Not-determined |
| 18-110_k141_28630 | Not-determined |
| 18-110_k141_51097 | Low-quality |
| 18-110_k141_74705 | Not-determined |
| 18-110_k141_51101 | Low-quality |
| 18-110_k141_270356 | Not-determined |
| 18-110_k141_143960 | Not-determined |
| 18-110_k141_143961 | Not-determined |
| 18-110_k141_51108 | Not-determined |
| 18-110_k141_236199 | Not-determined |
| 18-110_k141_132098 | Not-determined |
| 18-110_k141_51115 | Not-determined |
| 18-110_k141_178938 | Not-determined |
| 18-110_k141_178940 | Not-determined |
| 18-110_k141_236206 | Not-determined |
| 18-110_k141_108534 | Not-determined |
| 18-110_k141_51121 | Not-determined |
| 18-110_k141_40011 | Not-determined |
| 18-110_k141_40012 | Not-determined |
| 18-110_k141_236218 | Not-determined |
| 18-110_k141_257490 | Not-determined |
| 18-110_k141_97618 | Not-determined |
| 18-110_k141_51134 | Low-quality |
| 18-110_k141_63250 | Not-determined |
| 18-110_k141_143979 | Not-determined |
| 18-110_k141_200086 | Not-determined |
| 18-110_k141_200087 | Not-determined |
| 18-110_k141_257495 | Not-determined |
| 18-110_k141_97623 | Low-quality |
| 18-110_k141_108557 | Not-determined |
| 18-110_k141_63259 | Not-determined |
| 18-110_k141_143987 | Not-determined |
| 18-110_k141_28644 | Not-determined |
| 18-110_k141_28645 | Not-determined |
| 18-110_k141_28646 | Low-quality |
| 18-110_k141_85415 | Not-determined |
| 18-110_k141_212569 | Not-determined |
| 18-110_k141_85418 | Not-determined |
| 18-110_k141_143992 | Not-determined |
| 18-110_k141_247474 | Not-determined |
| 18-110_k141_143993 | Not-determined |
| 18-110_k141_212572 | Not-determined |
| 18-110_k141_85422 | Not-determined |
| 18-110_k141_212577 | Not-determined |
| 18-110_k141_85427 | Low-quality |
| 18-110_k141_85430 | Not-determined |
| 18-110_k141_247477 | Not-determined |
| 18-110_k141_212584 | Low-quality |
| 18-110_k141_74731 | Not-determined |
| 18-110_k141_166394 | Not-determined |
| 18-110_k141_144005 | Not-determined |
| 18-110_k141_144007 | Not-determined |
| 18-110_k141_200111 | Not-determined |
| 18-110_k141_74733 | Low-quality |
| 18-110_k141_74739 | Not-determined |
| 18-110_k141_51140 | Not-determined |
| 18-110_k141_74741 | Not-determined |
| 18-110_k141_108571 | Not-determined |
| 18-110_k141_144015 | Not-determined |
| 18-110_k141_108580 | Not-determined |
| 18-110_k141_247481 | Not-determined |
| 18-110_k141_108583 | Not-determined |
| 18-110_k141_85443 | Not-determined |
| 18-110_k141_132151 | Not-determined |
| 18-110_k141_63278 | Low-quality |
| 18-110_k141_189780 | Not-determined |
| 18-110_k141_74755 | Low-quality |
| 18-110_k141_155133 | Not-determined |
| 18-110_k141_63281 | Not-determined |
| 18-110_k141_247497 | Not-determined |
| 18-110_k141_63290 | Not-determined |
| 18-110_k141_132164 | Not-determined |
| 18-110_k141_166422 | Not-determined |
| 18-110_k141_257518 | Not-determined |
| 18-110_k141_108599 | Not-determined |
| 18-110_k141_63296 | Not-determined |
| 18-110_k141_155145 | Not-determined |
| 18-110_k141_166431 | Not-determined |
| 18-110_k141_74779 | Not-determined |
| 18-110_k141_108613 | Not-determined |
| 18-110_k141_108614 | Not-determined |
| 18-110_k141_189800 | Not-determined |
| 18-110_k141_189801 | Not-determined |
| 18-110_k141_132181 | Not-determined |
| 18-110_k141_85471 | Low-quality |
| 18-110_k141_51148 | Not-determined |
| 18-110_k141_108621 | Not-determined |
| 18-110_k141_223889 | Low-quality |
| 18-110_k141_74787 | Not-determined |
| 18-110_k141_63321 | Not-determined |
| 18-110_k141_51153 | Not-determined |
| 18-110_k141_247517 | Not-determined |
| 18-110_k141_74790 | Low-quality |
| 18-110_k141_212617 | Low-quality |
| 18-110_k141_120193 | Low-quality |
| 18-110_k141_28689 | Not-determined |
| 18-110_k141_28691 | Not-determined |
| 18-110_k141_28693 | Not-determined |
| 18-110_k141_247519 | Not-determined |
| 18-110_k141_132192 | Not-determined |
| 18-110_k141_189823 | Not-determined |
| 18-110_k141_144058 | Not-determined |
| 18-110_k141_155175 | Not-determined |
| 18-110_k141_212624 | Not-determined |
| 18-110_k141_189825 | Not-determined |
| 18-110_k141_155176 | Not-determined |
| 18-110_k141_257537 | Not-determined |
| 18-110_k141_63337 | Not-determined |
| 18-110_k141_155180 | Not-determined |
| 18-110_k141_212633 | Not-determined |
| 18-110_k141_85500 | Not-determined |
| 18-110_k141_120206 | Not-determined |
| 18-110_k141_120210 | Not-determined |
| 18-110_k141_200160 | Not-determined |
| 18-110_k141_144078 | Not-determined |
| 18-110_k141_120215 | Not-determined |
| 18-110_k141_51170 | Not-determined |
| 18-110_k141_155191 | Low-quality |
| 18-110_k141_4728 | Not-determined |
| 18-110_k141_179001 | Not-determined |
| 18-110_k141_74808 | Not-determined |
| 18-110_k141_144085 | Low-quality |
| 18-110_k141_179012 | Not-determined |
| 18-110_k141_4735 | Not-determined |
| 18-110_k141_212657 | Low-quality |
| 18-110_k141_236245 | Not-determined |
| 18-110_k141_223916 | Not-determined |
| 18-110_k141_28712 | Not-determined |
| 18-110_k141_155201 | Low-quality |
| 18-110_k141_132216 | Not-determined |
| 18-110_k141_155203 | Not-determined |
| 18-110_k141_212662 | Low-quality |
| 18-110_k141_223922 | Not-determined |
| 18-110_k141_144100 | Not-determined |
| 18-110_k141_85510 | Not-determined |
| 18-110_k141_155212 | Low-quality |
| 18-110_k141_85517 | Not-determined |
| 18-110_k141_63364 | Not-determined |
| 18-110_k141_4759 | Not-determined |
| 18-110_k141_108669 | Not-determined |
| 18-110_k141_85522 | Not-determined |
| 18-110_k141_63367 | Not-determined |
| 18-110_k141_85525 | Not-determined |
| 18-110_k141_85526 | Not-determined |
| 18-110_k141_223939 | Not-determined |
| 18-110_k141_63372 | Not-determined |
| 18-110_k141_166454 | Not-determined |
| 18-110_k141_108675 | Not-determined |
| 18-110_k141_179044 | Not-determined |
| 18-110_k141_144122 | Not-determined |
| 18-110_k141_97665 | Not-determined |
| 18-110_k141_28745 | Not-determined |
| 18-110_k141_212679 | Low-quality |
| 18-110_k141_223953 | Not-determined |
| 18-110_k141_85542 | Not-determined |
| 18-110_k141_223954 | Not-determined |
| 18-110_k141_85543 | Low-quality |
| 18-110_k141_108685 | Not-determined |
| 18-110_k141_223955 | Not-determined |
| 18-110_k141_155237 | Not-determined |
| 18-110_k141_189874 | Not-determined |
| 18-110_k141_212686 | Not-determined |
| 18-110_k141_212687 | Not-determined |
| 18-110_k141_51190 | Not-determined |
| 18-110_k141_179051 | Not-determined |
| 18-110_k141_51195 | Not-determined |
| 18-110_k141_257576 | Not-determined |
| 18-110_k141_63398 | Not-determined |
| 18-110_k141_15755 | Not-determined |
| 18-110_k141_120244 | Low-quality |
| 18-110_k141_15758 | Not-determined |
| 18-110_k141_85548 | Not-determined |
| 18-110_k141_74838 | Not-determined |
| 18-110_k141_132234 | Not-determined |
| 18-110_k141_189886 | Not-determined |
| 18-110_k141_120248 | Not-determined |
| 18-110_k141_236278 | Not-determined |
| 18-110_k141_179058 | Not-determined |
| 18-110_k141_132236 | Not-determined |
| 18-110_k141_236279 | Not-determined |
| 18-110_k141_189892 | Not-determined |
| 18-110_k141_155244 | Not-determined |
| 18-110_k141_223962 | Not-determined |
| 18-110_k141_15762 | Not-determined |
| 18-110_k141_63406 | Not-determined |
| 18-110_k141_257589 | Low-quality |
| 18-110_k141_97692 | Not-determined |
| 18-110_k141_236288 | Not-determined |
| 18-110_k141_15770 | Not-determined |
| 18-110_k141_28766 | Not-determined |
| 18-110_k141_97707 | Low-quality |
| 18-110_k141_179065 | Not-determined |
| 18-110_k141_15776 | Not-determined |
| 18-110_k141_144143 | Not-determined |
| 18-110_k141_132242 | Not-determined |
| 18-110_k141_223968 | Not-determined |
| 18-110_k141_236289 | Not-determined |
| 18-110_k141_15777 | Not-determined |
| 18-110_k141_212707 | Not-determined |
| 18-110_k141_247527 | Not-determined |
| 18-110_k141_247528 | Not-determined |
| 18-110_k141_28775 | Not-determined |
| 18-110_k141_270364 | Not-determined |
| 18-110_k141_132246 | Not-determined |
| 18-110_k141_108713 | Not-determined |
| 18-110_k141_247534 | Not-determined |
| 18-110_k141_51224 | Not-determined |
| 18-110_k141_270372 | Not-determined |
| 18-110_k141_85572 | Low-quality |
| 18-110_k141_108727 | Not-determined |
| 18-110_k141_120256 | Not-determined |
| 18-110_k141_247546 | Not-determined |
| 18-110_k141_28781 | Not-determined |
| 18-110_k141_223983 | Not-determined |
| 18-110_k141_166496 | Not-determined |
| 18-110_k141_28784 | Not-determined |
| 18-110_k141_144173 | Not-determined |
| 18-110_k141_236313 | Not-determined |
| 18-110_k141_155270 | Not-determined |
| 18-110_k141_120271 | Low-quality |
| 18-110_k141_144179 | Not-determined |
| 18-110_k141_144181 | Not-determined |
| 18-110_k141_155273 | Not-determined |
| 18-110_k141_247560 | Not-determined |
| 18-110_k141_155277 | Low-quality |
| 18-110_k141_179094 | Not-determined |
| 18-110_k141_132266 | Not-determined |
| 18-110_k141_236330 | Not-determined |
| 18-110_k141_155283 | Not-determined |
| 18-110_k141_108751 | Not-determined |
| 18-110_k141_179103 | Not-determined |
| 18-110_k141_85602 | Not-determined |
| 18-110_k141_51229 | Not-determined |
| 18-110_k141_51238 | Not-determined |
| 18-110_k141_120295 | Not-determined |
| 18-110_k141_40100 | Not-determined |
| 18-110_k141_85610 | Not-determined |
| 18-110_k141_40103 | Low-quality |
| 18-110_k141_212739 | Not-determined |
| 18-110_k141_51242 | Not-determined |
| 18-110_k141_132282 | Not-determined |
| 18-110_k141_224003 | Not-determined |
| 18-110_k141_270400 | Not-determined |
| 18-110_k141_212743 | Not-determined |
| 18-110_k141_4790 | Low-quality |
| 18-110_k141_270404 | Low-quality |
| 18-110_k141_189943 | Not-determined |
| 18-110_k141_40114 | Low-quality |
| 18-110_k141_4793 | Not-determined |
| 18-110_k141_236362 | Not-determined |
| 18-110_k141_28807 | Not-determined |
| 18-110_k141_4796 | Not-determined |
| 18-110_k141_108769 | Not-determined |
| 18-110_k141_120319 | Not-determined |
| 18-110_k141_28814 | Not-determined |
| 18-110_k141_120324 | Not-determined |
| 18-110_k141_40123 | Not-determined |
| 18-110_k141_247577 | Not-determined |
| 18-110_k141_236373 | Not-determined |
| 18-110_k141_132299 | Not-determined |
| 18-110_k141_247578 | Not-determined |
| 18-110_k141_200176 | Not-determined |
| 18-110_k141_28819 | Not-determined |
| 18-110_k141_4804 | Not-determined |
| 18-110_k141_97732 | Not-determined |
| 18-110_k141_270423 | Low-quality |
| 18-110_k141_97733 | Not-determined |
| 18-110_k141_28824 | Not-determined |
| 18-110_k141_270424 | Low-quality |
| 18-110_k141_40134 | Not-determined |
| 18-110_k141_4806 | Not-determined |
| 18-110_k141_166552 | Not-determined |
| 18-110_k141_40136 | Not-determined |
| 18-110_k141_247590 | Not-determined |
| 18-110_k141_51260 | Not-determined |
| 18-110_k141_179133 | Not-determined |
| 18-110_k141_40141 | Not-determined |
| 18-110_k141_247594 | Low-quality |
| 18-110_k141_85648 | Not-determined |
| 18-110_k141_179134 | Not-determined |
| 18-110_k141_28832 | Not-determined |
| 18-110_k141_166561 | Not-determined |
| 18-110_k141_85653 | Not-determined |
| 18-110_k141_51263 | Not-determined |
| 18-110_k141_132312 | Not-determined |
| 18-110_k141_15837 | Not-determined |
| 18-110_k141_212750 | Not-determined |
| 18-110_k141_63424 | Not-determined |
| 18-110_k141_236393 | Not-determined |
| 18-110_k141_212752 | Not-determined |
| 18-110_k141_97754 | Not-determined |
| 18-110_k141_63428 | Not-determined |
| 18-110_k141_132317 | Not-determined |
| 18-110_k141_166572 | Not-determined |

| | |
|---|---|
| 18-110_k141_166576 | Not-determined |
| 18-110_k141_4824 | Not-determined |
| 18-110_k141_74883 | Not-determined |
| 18-110_k141_155339 | Low-quality |
| 18-110_k141_85666 | Not-determined |
| 18-110_k141_236401 | Low-quality |
| 18-110_k141_4828 | Not-determined |
| 18-110_k141_257646 | Low-quality |
| 18-110_k141_166590 | Low-quality |
| 18-110_k141_189972 | Not-determined |
| 18-110_k141_257652 | Not-determined |
| 18-110_k141_166592 | Low-quality |
| 18-110_k141_247607 | Not-determined |
| 18-110_k141_63444 | Not-determined |
| 18-110_k141_166594 | Not-determined |
| 18-110_k141_28865 | Not-determined |
| 18-110_k141_15849 | Low-quality |
| 18-110_k141_166596 | Not-determined |
| 18-110_k141_120349 | Not-determined |
| 18-110_k141_120355 | Not-determined |
| 18-110_k141_85682 | Not-determined |
| 18-110_k141_15857 | Not-determined |
| 18-110_k141_212770 | Not-determined |
| 18-110_k141_63456 | Not-determined |
| 18-110_k141_200211 | Not-determined |
| 18-110_k141_15860 | Not-determined |
| 18-110_k141_51278 | Not-determined |
| 18-110_k141_51282 | Not-determined |
| 18-110_k141_120364 | Not-determined |
| 18-110_k141_200217 | Not-determined |
| 18-110_k141_4836 | Not-determined |
| 18-110_k141_40172 | Low-quality |
| 18-110_k141_74894 | Not-determined |
| 18-110_k141_212777 | Not-determined |
| 18-110_k141_200220 | Not-determined |
| 18-110_k141_247613 | Not-determined |
| 18-110_k141_40176 | Not-determined |
| 18-110_k141_200223 | Not-determined |
| 18-110_k141_247616 | Not-determined |
| 18-110_k141_40182 | Not-determined |
| 18-110_k141_97795 | Not-determined |
| 18-110_k141_97796 | Not-determined |
| 18-110_k141_120374 | Not-determined |
| 18-110_k141_212795 | Not-determined |
| 18-110_k141_74898 | Not-determined |
| 18-110_k141_97801 | Not-determined |
| 18-110_k141_179167 | Not-determined |
| 18-110_k141_179168 | Not-determined |
| 18-110_k141_179170 | Not-determined |
| 18-110_k141_236416 | Not-determined |
| 18-110_k141_247628 | Not-determined |
| 18-110_k141_74907 | Not-determined |
| 18-110_k141_247630 | Low-quality |
| 18-110_k141_4850 | Not-determined |
| 18-110_k141_179176 | Not-determined |
| 18-110_k141_257673 | Not-determined |
| 18-110_k141_63478 | Not-determined |
| 18-110_k141_120383 | Not-determined |
| 18-110_k141_28907 | Not-determined |
| 18-110_k141_224024 | Not-determined |
| 18-110_k141_85698 | Not-determined |
| 18-110_k141_85700 | Not-determined |
| 18-110_k141_74918 | Not-determined |
| 18-110_k141_120388 | Not-determined |
| 18-110_k141_97824 | Not-determined |
| 18-110_k141_200237 | Not-determined |
| 18-110_k141_85704 | Not-determined |
| 18-110_k141_257679 | Not-determined |
| 18-110_k141_97825 | Not-determined |
| 18-110_k141_200239 | Not-determined |
| 18-110_k141_200240 | Not-determined |
| 18-110_k141_247643 | Not-determined |
| 18-110_k141_40204 | Not-determined |
| 18-110_k141_270449 | Low-quality |
| 18-110_k141_85706 | Not-determined |
| 18-110_k141_190008 | Not-determined |
| 18-110_k141_236433 | Not-determined |
| 18-110_k141_74929 | Not-determined |
| 18-110_k141_179183 | Not-determined |
| 18-110_k141_190016 | Not-determined |
| 18-110_k141_120393 | Not-determined |
| 18-110_k141_120395 | Not-determined |
| 18-110_k141_190019 | Not-determined |
| 18-110_k141_247656 | Not-determined |
| 18-110_k141_247658 | Not-determined |
| 18-110_k141_85723 | Not-determined |
| 18-110_k141_224047 | Not-determined |
| 18-110_k141_40225 | Not-determined |
| 18-110_k141_224050 | Not-determined |
| 18-110_k141_179189 | Not-determined |
| 18-110_k141_190028 | Not-determined |
| 18-110_k141_85732 | Low-quality |
| 18-110_k141_85733 | Not-determined |
| 18-110_k141_85734 | Low-quality |
| 18-110_k141_4873 | Not-determined |
| 18-110_k141_190031 | Not-determined |
| 18-110_k141_257696 | Not-determined |
| 18-110_k141_200270 | Not-determined |
| 18-110_k141_190035 | Not-determined |
| 18-110_k141_179194 | Not-determined |
| 18-110_k141_4877 | Not-determined |
| 18-110_k141_4878 | Not-determined |
| 18-110_k141_63512 | Not-determined |
| 18-110_k141_4883 | Not-determined |
| 18-110_k141_51334 | Low-quality |
| 18-110_k141_51336 | Low-quality |
| 18-110_k141_155359 | Not-determined |
| 18-110_k141_179203 | Not-determined |
| 18-110_k141_224063 | Not-determined |
| 18-110_k141_74962 | Not-determined |
| 18-110_k141_4888 | Not-determined |
| 18-110_k141_74966 | Not-determined |
| 18-110_k141_28949 | Not-determined |
| 18-110_k141_51347 | Not-determined |
| 18-110_k141_28952 | Not-determined |
| 18-110_k141_190055 | Not-determined |
| 18-110_k141_51349 | Not-determined |
| 18-110_k141_40247 | Not-determined |
| 18-110_k141_40248 | Not-determined |
| 18-110_k141_200288 | Not-determined |
| 18-110_k141_120439 | Not-determined |
| 18-110_k141_40250 | Not-determined |
| 18-110_k141_74974 | Not-determined |
| 18-110_k141_224072 | Not-determined |
| 18-110_k141_179207 | Not-determined |
| 18-110_k141_155369 | Not-determined |
| 18-110_k141_120442 | Not-determined |
| 18-110_k141_166637 | Not-determined |
| 18-110_k141_51356 | Not-determined |
| 18-110_k141_74977 | Not-determined |
| 18-110_k141_28963 | Not-determined |
| 18-110_k141_179211 | Not-determined |
| 18-110_k141_212847 | Not-determined |
| 18-110_k141_270469 | Not-determined |
| 18-110_k141_212852 | Not-determined |
| 18-110_k141_4896 | Low-quality |
| 18-110_k141_200303 | Low-quality |
| 18-110_k141_85761 | Low-quality |
| 18-110_k141_236449 | Not-determined |
| 18-110_k141_179220 | Not-determined |
| 18-110_k141_63523 | Low-quality |
| 18-110_k141_179221 | Not-determined |
| 18-110_k141_28968 | Not-determined |
| 18-110_k141_270476 | Not-determined |
| 18-110_k141_63530 | Not-determined |
| 18-110_k141_200313 | Not-determined |
| 18-110_k141_224085 | Low-quality |
| 18-110_k141_270480 | Not-determined |
| 18-110_k141_224089 | Not-determined |
| 18-110_k141_85770 | Not-determined |
| 18-110_k141_247677 | Not-determined |
| 18-110_k141_236466 | Not-determined |
| 18-110_k141_28986 | Not-determined |
| 18-110_k141_247683 | Not-determined |
| 18-110_k141_247684 | Not-determined |
| 18-110_k141_212876 | Low-quality |
| 18-110_k141_247685 | Not-determined |
| 18-110_k141_132352 | Not-determined |
| 18-110_k141_132353 | Not-determined |
| 18-110_k141_108813 | Not-determined |
| 18-110_k141_270491 | Not-determined |
| 18-110_k141_132355 | Not-determined |
| 18-110_k141_108821 | Not-determined |
| 18-110_k141_75011 | Not-determined |
| 18-110_k141_247688 | Not-determined |
| 18-110_k141_4917 | Low-quality |
| 18-110_k141_257735 | Not-determined |
| 18-110_k141_63555 | Not-determined |
| 18-110_k141_236482 | Not-determined |
| 18-110_k141_179223 | Not-determined |
| 18-110_k141_212882 | Not-determined |
| 18-110_k141_212883 | Not-determined |
| 18-110_k141_29006 | Not-determined |
| 18-110_k141_179227 | Not-determined |
| 18-110_k141_29007 | Not-determined |
| 18-110_k141_200337 | Not-determined |
| 18-110_k141_247695 | Not-determined |
| 18-110_k141_236488 | Low-quality |
| 18-110_k141_4932 | Not-determined |
| 18-110_k141_29020 | Not-determined |
| 18-110_k141_40274 | Not-determined |
| 18-110_k141_120477 | Not-determined |
| 18-110_k141_166690 | Not-determined |
| 18-110_k141_51410 | Not-determined |
| 18-110_k141_224122 | Low-quality |
| 18-110_k141_51418 | Low-quality |
| 18-110_k141_63572 | Not-determined |
| 18-110_k141_15912 | Low-quality |
| 18-110_k141_212904 | Not-determined |
| 18-110_k141_212913 | Not-determined |
| 18-110_k141_212914 | Not-determined |
| 18-110_k141_270526 | Not-determined |
| 18-110_k141_63582 | Not-determined |
| 18-110_k141_40303 | Not-determined |
| 18-110_k141_200362 | Not-determined |
| 18-110_k141_166712 | Not-determined |
| 18-110_k141_97839 | Not-determined |
| 18-110_k141_200365 | Not-determined |
| 18-110_k141_15928 | Not-determined |
| 18-110_k141_270538 | Not-determined |
| 18-110_k141_166722 | Low-quality |
| 18-110_k141_108831 | Not-determined |
| 18-110_k141_4966 | Not-determined |
| 18-110_k141_29047 | Not-determined |
| 18-110_k141_51437 | Not-determined |
| 18-110_k141_247725 | Not-determined |
| 18-110_k141_179261 | Not-determined |
| 18-110_k141_257778 | Not-determined |
| 18-110_k141_236542 | Not-determined |
| 18-110_k141_132364 | Not-determined |
| 18-110_k141_15943 | Not-determined |
| 18-110_k141_166725 | Not-determined |
| 18-110_k141_270549 | Not-determined |
| 18-110_k141_179271 | Not-determined |
| 18-110_k141_166726 | Not-determined |
| 18-110_k141_51447 | Not-determined |
| 18-110_k141_63590 | Not-determined |
| 18-110_k141_132373 | Not-determined |
| 18-110_k141_257788 | Not-determined |
| 18-110_k141_270565 | Not-determined |
| 18-110_k141_270569 | Not-determined |
| 18-110_k141_155420 | Not-determined |
| 18-110_k141_40319 | Not-determined |
| 18-110_k141_132391 | Low-quality |
| 18-110_k141_51467 | Not-determined |
| 18-110_k141_132396 | Not-determined |
| 18-110_k141_51469 | Not-determined |
| 18-110_k141_108857 | Not-determined |
| 18-110_k141_224155 | Low-quality |
| 18-110_k141_15953 | Not-determined |
| 18-110_k141_132401 | Not-determined |
| 18-110_k141_236555 | Not-determined |
| 18-110_k141_15958 | Not-determined |
| 18-110_k141_179299 | Not-determined |
| 18-110_k141_270595 | Not-determined |
| 18-110_k141_63607 | Not-determined |
| 18-110_k141_200380 | Not-determined |
| 18-110_k141_166767 | Low-quality |
| 18-110_k141_132420 | Low-quality |
| 18-110_k141_236566 | Not-determined |
| 18-110_k141_97849 | Low-quality |
| 18-110_k141_97851 | Not-determined |
| 18-110_k141_40346 | Not-determined |
| 18-110_k141_120503 | Not-determined |
| 18-110_k141_29116 | Not-determined |
| 18-110_k141_97855 | Not-determined |
| 18-110_k141_190126 | Not-determined |
| 18-110_k141_29124 | Not-determined |
| 18-110_k141_179305 | Not-determined |
| 18-110_k141_155448 | Not-determined |
| 18-110_k141_155449 | Not-determined |
| 18-110_k141_212921 | Not-determined |
| 18-110_k141_63621 | Low-quality |
| 18-110_k141_179307 | Not-determined |
| 18-110_k141_63622 | Not-determined |
| 18-110_k141_63624 | Not-determined |
| 18-110_k141_166779 | Not-determined |
| 18-110_k141_247764 | Not-determined |
| 18-110_k141_257801 | Not-determined |
| 18-110_k141_97867 | Not-determined |
| 18-110_k141_247773 | Not-determined |
| 18-110_k141_179322 | Not-determined |
| 18-110_k141_190146 | Not-determined |
| 18-110_k141_40366 | Not-determined |
| 18-110_k141_63639 | Not-determined |
| 18-110_k141_155471 | Not-determined |
| 18-110_k141_166785 | Not-determined |
| 18-110_k141_132442 | Not-determined |
| 18-110_k141_5051 | Not-determined |
| 18-110_k141_179330 | Not-determined |
| 18-110_k141_120527 | Not-determined |
| 18-110_k141_120529 | Not-determined |
| 18-110_k141_179332 | Not-determined |
| 18-110_k141_166791 | Not-determined |
| 18-110_k141_120530 | Not-determined |
| 18-110_k141_29152 | Not-determined |
| 18-110_k141_120531 | Not-determined |
| 18-110_k141_247779 | Not-determined |
| 18-110_k141_120536 | Not-determined |
| 18-110_k141_190167 | Not-determined |
| 18-110_k141_155479 | Not-determined |
| 18-110_k141_270627 | Not-determined |
| 18-110_k141_224168 | Not-determined |
| 18-110_k141_155483 | Not-determined |
| 18-110_k141_257819 | Not-determined |
| 18-110_k141_108880 | Low-quality |
| 18-110_k141_224169 | Not-determined |
| 18-110_k141_190168 | Not-determined |
| 18-110_k141_15986 | Not-determined |
| 18-110_k141_200411 | Not-determined |
| 18-110_k141_40376 | Low-quality |
| 18-110_k141_247788 | Not-determined |
| 18-110_k141_97877 | Not-determined |
| 18-110_k141_270632 | Not-determined |
| 18-110_k141_108886 | Not-determined |
| 18-110_k141_247793 | Not-determined |
| 18-110_k141_270637 | Not-determined |
| 18-110_k141_190177 | Not-determined |
| 18-110_k141_97881 | Not-determined |
| 18-110_k141_120545 | Low-quality |
| 18-110_k141_270642 | Not-determined |
| 18-110_k141_108891 | Not-determined |
| 18-110_k141_270646 | Low-quality |
| 18-110_k141_16005 | Not-determined |
| 18-110_k141_16007 | Not-determined |
| 18-110_k141_166807 | Not-determined |
| 18-110_k141_108901 | Not-determined |
| 18-110_k141_51529 | Not-determined |
| 18-110_k141_40396 | Not-determined |
| 18-110_k141_108904 | Not-determined |
| 18-110_k141_108906 | Not-determined |
| 18-110_k141_29182 | Not-determined |
| 18-110_k141_270653 | Not-determined |
| 18-110_k141_270654 | Not-determined |
| 18-110_k141_247812 | Not-determined |
| 18-110_k141_97900 | Not-determined |
| 18-110_k141_16025 | Not-determined |
| 18-110_k141_212957 | Not-determined |
| 18-110_k141_120549 | Not-determined |
| 18-110_k141_212961 | Not-determined |
| 18-110_k141_155518 | Not-determined |
| 18-110_k141_63650 | Not-determined |
| 18-110_k141_200429 | Not-determined |
| 18-110_k141_29193 | Not-determined |
| 18-110_k141_179340 | Not-determined |
| 18-110_k141_166822 | Not-determined |
| 18-110_k141_166824 | Not-determined |
| 18-110_k141_5066 | Not-determined |
| 18-110_k141_166829 | Not-determined |
| 18-110_k141_29198 | Not-determined |
| 18-110_k141_29201 | Not-determined |
| 18-110_k141_97923 | Not-determined |
| 18-110_k141_270664 | Not-determined |
| 18-110_k141_51538 | Not-determined |
| 18-110_k141_29206 | Not-determined |
| 18-110_k141_224199 | Not-determined |
| 18-110_k141_166837 | Not-determined |
| 18-110_k141_155533 | Not-determined |
| 18-110_k141_97931 | Not-determined |
| 18-110_k141_63665 | Low-quality |
| 18-110_k141_5088 | Not-determined |
| 18-110_k141_120562 | Not-determined |
| 18-110_k141_190190 | Not-determined |
| 18-110_k141_5091 | Not-determined |
| 18-110_k141_51545 | Not-determined |
| 18-110_k141_51549 | Low-quality |
| 18-110_k141_179355 | Not-determined |
| 18-110_k141_155553 | Not-determined |
| 18-110_k141_200450 | Not-determined |
| 18-110_k141_97953 | Not-determined |
| 18-110_k141_270680 | Not-determined |
| 18-110_k141_247819 | Not-determined |
| 18-110_k141_224221 | Low-quality |
| 18-110_k141_97961 | Not-determined |
| 18-110_k141_51566 | Not-determined |
| 18-110_k141_257844 | Not-determined |
| 18-110_k141_51570 | Not-determined |
| 18-110_k141_155572 | Low-quality |
| 18-110_k141_270698 | Not-determined |
| 18-110_k141_155575 | Not-determined |
| 18-110_k141_179377 | Not-determined |
| 18-110_k141_166866 | Not-determined |
| 18-110_k141_16048 | Low-quality |
| 18-110_k141_166874 | Not-determined |
| 18-110_k141_213008 | Low-quality |
| 18-110_k141_97973 | Not-determined |
| 18-110_k141_120593 | Not-determined |
| 18-110_k141_155590 | Not-determined |
| 18-110_k141_213010 | Not-determined |
| 18-110_k141_179389 | Low-quality |
| 18-110_k141_97982 | Not-determined |
| 18-110_k141_247836 | Not-determined |
| 18-110_k141_213015 | Low-quality |
| 18-110_k141_179393 | Low-quality |
| 18-110_k141_166887 | Low-quality |
| 18-110_k141_224234 | Not-determined |
| 18-110_k141_155604 | Not-determined |
| 18-110_k141_224238 | Not-determined |
| 18-110_k141_155607 | Not-determined |
| 18-110_k141_270720 | Not-determined |
| 18-110_k141_236662 | Not-determined |
| 18-110_k141_179399 | Not-determined |
| 18-110_k141_179404 | Not-determined |
| 18-110_k141_270733 | Not-determined |
| 18-110_k141_40452 | Not-determined |
| 18-110_k141_190239 | Not-determined |
| 18-110_k141_257869 | Not-determined |
| 18-110_k141_97990 | Not-determined |
| 18-110_k141_85781 | Not-determined |
| 18-110_k141_270738 | Not-determined |
| 18-110_k141_224253 | Not-determined |
| 18-110_k141_40461 | Low-quality |
| 18-110_k141_224259 | Not-determined |
| 18-110_k141_166932 | Not-determined |
| 18-110_k141_120613 | Not-determined |
| 18-110_k141_40473 | Not-determined |
| 18-110_k141_224262 | Not-determined |
| 18-110_k141_5141 | Low-quality |
| 18-110_k141_224264 | Low-quality |
| 18-110_k141_270743 | Low-quality |
| 18-110_k141_120620 | Not-determined |
| 18-110_k141_155639 | Not-determined |
| 18-110_k141_29268 | Not-determined |
| 18-110_k141_85807 | Not-determined |
| 18-110_k141_200454 | Not-determined |
| 18-110_k141_16102 | Low-quality |
| 18-110_k141_257883 | Not-determined |
| 18-110_k141_247857 | Not-determined |
| 18-110_k141_200472 | Low-quality |
| 18-110_k141_132456 | Not-determined |
| 18-110_k141_257887 | Not-determined |
| 18-110_k141_257889 | Not-determined |
| 18-110_k141_98027 | Not-determined |
| 18-110_k141_98028 | Low-quality |
| 18-110_k141_75041 | Not-determined |
| 18-110_k141_257898 | Not-determined |
| 18-110_k141_5156 | Not-determined |
| 18-110_k141_270777 | Not-determined |
| 18-110_k141_51616 | Low-quality |
| 18-110_k141_224300 | Not-determined |
| 18-110_k141_257908 | Low-quality |
| 18-110_k141_257909 | Low-quality |
| 18-110_k141_120644 | Not-determined |
| 18-110_k141_51622 | Not-determined |
| 18-110_k141_51623 | Not-determined |
| 18-110_k141_85833 | Not-determined |
| 18-110_k141_98050 | Not-determined |
| 18-110_k141_40520 | Not-determined |
| 18-110_k141_179447 | Low-quality |
| 18-110_k141_98057 | Not-determined |
| 18-110_k141_29307 | Not-determined |
| 18-110_k141_224309 | Not-determined |
| 18-110_k141_132494 | Not-determined |
| 18-110_k141_51637 | Not-determined |
| 18-110_k141_270783 | Low-quality |
| 18-110_k141_98065 | Not-determined |
| 18-110_k141_51638 | Not-determined |
| 18-110_k141_5184 | Not-determined |
| 18-110_k141_224315 | Not-determined |
| 18-110_k141_16125 | Low-quality |
| 18-110_k141_224316 | Low-quality |
| 18-110_k141_40533 | Low-quality |
| 18-110_k141_40534 | Not-determined |
| 18-110_k141_16128 | Not-determined |
| 18-110_k141_132506 | Not-determined |
| 18-110_k141_270792 | Not-determined |
| 18-110_k141_132507 | Not-determined |
| 18-110_k141_29324 | Not-determined |
| 18-110_k141_98075 | Not-determined |
| 18-110_k141_85848 | Not-determined |
| 18-110_k141_29329 | Not-determined |
| 18-110_k141_270797 | Not-determined |
| 18-110_k141_257917 | Not-determined |
| 18-110_k141_5194 | Not-determined |
| 18-110_k141_257925 | Not-determined |
| 18-110_k141_85862 | Low-quality |
| 18-110_k141_98080 | Not-determined |
| 18-110_k141_190276 | Not-determined |
| 18-110_k141_85864 | Not-determined |
| 18-110_k141_85866 | Not-determined |
| 18-110_k141_40545 | Not-determined |
| 18-110_k141_85868 | Not-determined |
| 18-110_k141_5199 | Not-determined |
| 18-110_k141_179458 | Not-determined |
| 18-110_k141_200509 | Not-determined |
| 18-110_k141_120665 | Not-determined |
| 18-110_k141_40559 | Low-quality |
| 18-110_k141_224339 | Not-determined |
| 18-110_k141_132528 | Low-quality |
| 18-110_k141_224341 | Not-determined |
| 18-110_k141_200517 | Not-determined |
| 18-110_k141_200519 | Not-determined |
| 18-110_k141_75082 | Not-determined |
| 18-110_k141_29348 | Not-determined |
| 18-110_k141_247888 | Not-determined |
| 18-110_k141_247889 | Not-determined |
| 18-110_k141_247890 | Not-determined |
| 18-110_k141_120675 | Not-determined |
| 18-110_k141_98110 | Not-determined |
| 18-110_k141_5223 | Low-quality |
| 18-110_k141_132545 | Not-determined |
| 18-110_k141_132547 | Not-determined |
| 18-110_k141_270830 | Low-quality |
| 18-110_k141_5228 | Not-determined |
| 18-110_k141_132549 | Not-determined |
| 18-110_k141_98117 | Not-determined |
| 18-110_k141_190295 | Not-determined |
| 18-110_k141_200529 | Not-determined |
| 18-110_k141_29366 | Not-determined |
| 18-110_k141_63704 | Not-determined |
| 18-110_k141_200533 | Not-determined |
| 18-110_k141_236701 | Low-quality |
| 18-110_k141_98126 | Not-determined |
| 18-110_k141_75107 | Not-determined |
| 18-110_k141_63711 | Not-determined |
| 18-110_k141_75109 | Not-determined |
| 18-110_k141_155681 | Not-determined |
| 18-110_k141_190304 | Not-determined |
| 18-110_k141_51693 | Not-determined |
| 18-110_k141_5244 | Not-determined |
| 18-110_k141_85919 | Not-determined |
| 18-110_k141_190308 | Not-determined |
| 18-110_k141_85925 | Not-determined |
| 18-110_k141_155688 | Not-determined |
| 18-110_k141_155689 | Not-determined |
| 18-110_k141_51700 | Not-determined |
| 18-110_k141_16149 | Not-determined |
| 18-110_k141_40591 | Not-determined |
| 18-110_k141_85930 | Not-determined |
| 18-110_k141_247917 | Not-determined |
| 18-110_k141_132580 | Not-determined |
| 18-110_k141_190323 | Not-determined |
| 18-110_k141_75129 | Not-determined |
| 18-110_k141_166943 | Not-determined |
| 18-110_k141_51710 | Not-determined |
| 18-110_k141_29377 | Not-determined |
| 18-110_k141_132583 | Not-determined |
| 18-110_k141_120714 | Not-determined |
| 18-110_k141_98140 | Not-determined |
| 18-110_k141_85939 | Low-quality |
| 18-110_k141_98143 | Not-determined |
| 18-110_k141_108977 | Not-determined |
| 18-110_k141_213066 | Not-determined |
| 18-110_k141_98149 | Not-determined |
| 18-110_k141_29380 | Not-determined |
| 18-110_k141_247930 | Not-determined |
| 18-110_k141_247931 | Not-determined |
| 18-110_k141_132589 | Low-quality |
| 18-110_k141_247932 | Not-determined |
| 18-110_k141_257969 | Not-determined |
| 18-110_k141_16160 | Not-determined |
| 18-110_k141_98155 | Not-determined |
| 18-110_k141_200556 | Low-quality |
| 18-110_k141_200557 | Not-determined |
| 18-110_k141_166950 | Not-determined |
| 18-110_k141_270856 | Not-determined |
| 18-110_k141_16164 | Not-determined |
| 18-110_k141_98160 | Not-determined |
| 18-110_k141_200561 | Not-determined |
| 18-110_k141_108983 | Not-determined |
| 18-110_k141_190338 | Not-determined |
| 18-110_k141_236711 | Not-determined |
| 18-110_k141_190339 | Not-determined |
| 18-110_k141_270866 | Not-determined |
| 18-110_k141_247950 | Not-determined |
| 18-110_k141_270869 | Not-determined |
| 18-110_k141_75138 | Not-determined |
| 18-110_k141_270870 | Not-determined |
| 18-110_k141_224389 | Not-determined |
| 18-110_k141_132612 | Not-determined |
| 18-110_k141_166959 | Not-determined |
| 18-110_k141_132614 | Not-determined |
| 18-110_k141_120720 | Low-quality |
| 18-110_k141_190357 | Not-determined |
| 18-110_k141_98177 | Not-determined |
| 18-110_k141_5249 | Not-determined |
| 18-110_k141_190361 | Not-determined |
| 18-110_k141_224394 | Not-determined |
| 18-110_k141_224395 | Not-determined |
| 18-110_k141_29386 | Not-determined |
| 18-110_k141_16173 | Not-determined |
| 18-110_k141_120733 | Not-determined |
| 18-110_k141_200582 | Not-determined |
| 18-110_k141_98197 | Not-determined |
| 18-110_k141_51752 | Not-determined |
| 18-110_k141_16180 | Not-determined |
| 18-110_k141_190374 | Low-quality |
| 18-110_k141_109009 | Not-determined |
| 18-110_k141_179502 | Not-determined |
| 18-110_k141_16182 | Not-determined |
| 18-110_k141_29404 | Not-determined |
| 18-110_k141_109011 | Low-quality |
| 18-110_k141_29405 | Not-determined |
| 18-110_k141_63730 | Not-determined |
| 18-110_k141_179504 | Not-determined |
| 18-110_k141_85967 | Not-determined |
| 18-110_k141_200598 | Not-determined |
| 18-110_k141_29411 | Not-determined |
| 18-110_k141_224405 | Low-quality |
| 18-110_k141_247968 | Not-determined |
| 18-110_k141_29415 | Not-determined |
| 18-110_k141_120748 | Not-determined |
| 18-110_k141_63739 | Not-determined |
| 18-110_k141_132641 | Not-determined |
| 18-110_k141_85978 | Not-determined |
| 18-110_k141_236724 | Not-determined |
| 18-110_k141_98206 | Not-determined |
| 18-110_k141_236733 | Not-determined |
| 18-110_k141_98211 | Low-quality |

| | |
|---|---|
| 18-110_k141_16198 | Not-determined |
| 18-110_k141_166984 | Not-determined |
| 18-110_k141_155744 | Not-determined |
| 18-110_k141_75192 | Not-determined |
| 18-110_k141_98220 | Not-determined |
| 18-110_k141_132672 | Not-determined |
| 18-110_k141_98229 | Low-quality |
| 18-110_k141_224432 | Low-quality |
| 18-110_k141_270909 | Not-determined |
| 18-110_k141_179531 | Not-determined |
| 18-110_k141_200618 | Not-determined |
| 18-110_k141_98233 | Not-determined |
| 18-110_k141_29444 | Not-determined |
| 18-110_k141_155750 | Not-determined |
| 18-110_k141_29445 | Not-determined |
| 18-110_k141_179533 | Not-determined |
| 18-110_k141_120769 | Low-quality |
| 18-110_k141_63759 | Not-determined |
| 18-110_k141_63760 | Not-determined |
| 18-110_k141_236749 | Not-determined |
| 18-110_k141_248007 | Not-determined |
| 18-110_k141_224445 | Not-determined |
| 18-110_k141_16210 | Not-determined |
| 18-110_k141_179543 | Not-determined |
| 18-110_k141_86012 | Low-quality |
| 18-110_k141_29460 | Low-quality |
| 18-110_k141_236757 | Not-determined |
| 18-110_k141_132688 | Not-determined |
| 18-110_k141_270925 | Not-determined |
| 18-110_k141_179549 | Not-determined |
| 18-110_k141_132692 | Low-quality |
| 18-110_k141_167009 | Not-determined |
| 18-110_k141_5302 | Not-determined |
| 18-110_k141_109056 | Not-determined |
| 18-110_k141_75210 | Not-determined |
| 18-110_k141_132695 | Not-determined |
| 18-110_k141_224453 | Not-determined |
| 18-110_k141_29468 | Not-determined |
| 18-110_k141_224455 | Low-quality |
| 18-110_k141_179559 | Not-determined |
| 18-110_k141_29470 | Not-determined |
| 18-110_k141_144232 | Not-determined |
| 18-110_k141_132702 | Not-determined |
| 18-110_k141_258039 | Not-determined |
| 18-110_k141_40640 | Low-quality |
| 18-110_k141_224462 | Not-determined |
| 18-110_k141_179567 | Not-determined |
| 18-110_k141_51769 | Not-determined |
| 18-110_k141_200654 | Not-determined |
| 18-110_k141_86024 | Not-determined |
| 18-110_k141_155790 | Not-determined |
| 18-110_k141_224474 | Not-determined |
| 18-110_k141_270943 | Not-determined |
| 18-110_k141_270945 | Not-determined |
| 18-110_k141_179581 | Not-determined |
| 18-110_k141_109076 | Not-determined |
| 18-110_k141_248024 | Not-determined |
| 18-110_k141_248028 | Not-determined |
| 18-110_k141_109083 | Not-determined |
| 18-110_k141_258052 | Not-determined |
| 18-110_k141_75215 | Not-determined |
| 18-110_k141_200667 | Not-determined |
| 18-110_k141_224492 | Not-determined |
| 18-110_k141_5323 | Not-determined |
| 18-110_k141_200669 | Not-determined |
| 18-110_k141_144245 | Low-quality |
| 18-110_k141_167031 | Not-determined |
| 18-110_k141_29513 | Not-determined |
| 18-110_k141_224496 | Not-determined |
| 18-110_k141_63803 | Not-determined |
| 18-110_k141_51803 | Not-determined |
| 18-110_k141_132714 | Not-determined |
| 18-110_k141_224499 | Low-quality |
| 18-110_k141_179585 | Not-determined |
| 18-110_k141_120789 | Not-determined |
| 18-110_k141_75230 | Not-determined |
| 18-110_k141_75232 | Not-determined |
| 18-110_k141_132724 | Not-determined |
| 18-110_k141_200679 | Not-determined |
| 18-110_k141_167042 | Not-determined |
| 18-110_k141_29524 | Not-determined |
| 18-110_k141_236807 | Not-determined |
| 18-110_k141_132729 | Not-determined |
| 18-110_k141_5339 | Not-determined |
| 18-110_k141_16235 | Not-determined |
| 18-110_k141_86052 | Not-determined |
| 18-110_k141_144256 | Not-determined |
| 18-110_k141_109095 | Not-determined |
| 18-110_k141_29539 | Not-determined |
| 18-110_k141_86059 | Not-determined |
| 18-110_k141_155816 | Low-quality |
| 18-110_k141_51814 | Not-determined |
| 18-110_k141_5346 | Not-determined |
| 18-110_k141_29544 | Not-determined |
| 18-110_k141_224523 | Not-determined |
| 18-110_k141_179602 | Not-determined |
| 18-110_k141_51819 | Not-determined |
| 18-110_k141_120799 | Not-determined |
| 18-110_k141_144260 | Not-determined |
| 18-110_k141_5348 | Not-determined |
| 18-110_k141_29550 | Not-determined |
| 18-110_k141_144262 | Not-determined |
| 18-110_k141_29552 | Not-determined |
| 18-110_k141_190401 | Low-quality |
| 18-110_k141_144265 | Not-determined |
| 18-110_k141_224530 | Not-determined |
| 18-110_k141_270963 | Not-determined |
| 18-110_k141_51828 | Low-quality |
| 18-110_k141_63829 | Not-determined |
| 18-110_k141_86073 | Not-determined |
| 18-110_k141_132755 | Not-determined |
| 18-110_k141_120806 | Not-determined |
| 18-110_k141_16244 | Not-determined |
| 18-110_k141_224543 | Not-determined |
| 18-110_k141_224548 | Not-determined |
| 18-110_k141_51842 | Low-quality |
| 18-110_k141_120817 | Not-determined |
| 18-110_k141_270982 | Not-determined |
| 18-110_k141_75266 | Not-determined |
| 18-110_k141_63846 | Not-determined |
| 18-110_k141_167075 | Not-determined |
| 18-110_k141_258057 | Low-quality |
| 18-110_k141_155829 | Not-determined |
| 18-110_k141_16255 | Not-determined |
| 18-110_k141_5370 | Not-determined |
| 18-110_k141_63852 | Not-determined |
| 18-110_k141_109115 | Not-determined |
| 18-110_k141_109117 | Not-determined |
| 18-110_k141_132759 | Not-determined |
| 18-110_k141_144271 | Not-determined |
| 18-110_k141_51854 | Not-determined |
| 18-110_k141_16259 | Low-quality |
| 18-110_k141_86077 | Not-determined |
| 18-110_k141_224555 | Not-determined |
| 18-110_k141_29572 | Low-quality |
| 18-110_k141_109124 | Not-determined |
| 18-110_k141_132768 | Not-determined |
| 18-110_k141_16265 | Not-determined |
| 18-110_k141_270991 | Not-determined |
| 18-110_k141_16266 | Not-determined |
| 18-110_k141_270993 | Not-determined |
| 18-110_k141_132773 | Not-determined |
| 18-110_k141_258067 | Not-determined |
| 18-110_k141_190445 | Low-quality |
| 18-110_k141_270997 | Not-determined |
| 18-110_k141_98262 | Not-determined |
| 18-110_k141_120847 | Not-determined |
| 18-110_k141_132778 | Low-quality |
| 18-110_k141_16276 | Not-determined |
| 18-110_k141_248116 | Not-determined |
| 18-110_k141_248117 | Not-determined |
| 18-110_k141_5391 | Low-quality |
| 18-110_k141_5395 | Not-determined |
| 18-110_k141_132782 | Not-determined |
| 18-110_k141_132784 | Not-determined |
| 18-110_k141_5400 | Not-determined |
| 18-110_k141_29601 | Not-determined |
| 18-110_k141_248123 | Not-determined |
| 18-110_k141_258080 | Not-determined |
| 18-110_k141_200719 | Not-determined |
| 18-110_k141_248127 | Not-determined |
| 18-110_k141_224576 | Not-determined |
| 18-110_k141_51886 | Not-determined |
| 18-110_k141_167116 | Low-quality |
| 18-110_k141_155870 | Not-determined |
| 18-110_k141_167121 | Low-quality |
| 18-110_k141_248133 | Not-determined |
| 18-110_k141_155876 | Not-determined |
| 18-110_k141_98276 | Low-quality |
| 18-110_k141_155877 | Not-determined |
| 18-110_k141_16293 | Not-determined |
| 18-110_k141_16296 | Not-determined |
| 18-110_k141_98281 | Not-determined |
| 18-110_k141_155884 | Not-determined |
| 18-110_k141_155885 | Not-determined |
| 18-110_k141_144312 | Not-determined |
| 18-110_k141_5421 | Not-determined |
| 18-110_k141_98284 | Not-determined |
| 18-110_k141_29609 | Not-determined |
| 18-110_k141_155893 | Not-determined |
| 18-110_k141_248148 | Not-determined |
| 18-110_k141_132809 | Not-determined |
| 18-110_k141_236845 | Not-determined |
| 18-110_k141_155902 | Not-determined |
| 18-110_k141_98291 | Not-determined |
| 18-110_k141_109170 | Not-determined |
| 18-110_k141_120886 | Not-determined |
| 18-110_k141_190453 | Not-determined |
| 18-110_k141_109173 | Low-quality |
| 18-110_k141_236858 | Not-determined |
| 18-110_k141_144327 | Not-determined |
| 18-110_k141_190460 | Not-determined |
| 18-110_k141_16318 | Not-determined |
| 18-110_k141_224616 | Not-determined |
| 18-110_k141_75322 | Not-determined |
| 18-110_k141_224620 | Low-quality |
| 18-110_k141_5450 | Not-determined |
| 18-110_k141_29621 | Not-determined |
| 18-110_k141_213117 | Low-quality |
| 18-110_k141_213119 | Not-determined |
| 18-110_k141_109199 | Not-determined |
| 18-110_k141_144344 | Not-determined |
| 18-110_k141_98336 | Low-quality |
| 18-110_k141_109206 | Low-quality |
| 18-110_k141_98337 | Not-determined |
| 18-110_k141_155927 | Not-determined |
| 18-110_k141_51927 | Not-determined |
| 18-110_k141_120915 | Not-determined |
| 18-110_k141_51931 | Not-determined |
| 18-110_k141_155930 | Not-determined |
| 18-110_k141_16360 | Not-determined |
| 18-110_k141_213134 | Not-determined |
| 18-110_k141_98344 | Not-determined |
| 18-110_k141_51940 | Not-determined |
| 18-110_k141_236890 | Not-determined |
| 18-110_k141_248191 | Not-determined |
| 18-110_k141_179700 | Not-determined |
| 18-110_k141_258131 | Not-determined |
| 18-110_k141_155948 | Not-determined |
| 18-110_k141_224649 | Not-determined |
| 18-110_k141_236900 | Not-determined |
| 18-110_k141_51945 | Not-determined |
| 18-110_k141_120922 | Low-quality |
| 18-110_k141_29650 | Not-determined |
| 18-110_k141_29651 | Not-determined |
| 18-110_k141_120929 | Low-quality |
| 18-110_k141_86102 | Low-quality |
| 18-110_k141_258136 | Not-determined |
| 18-110_k141_86103 | Not-determined |
| 18-110_k141_109215 | Not-determined |
| 18-110_k141_236908 | Not-determined |
| 18-110_k141_236909 | Not-determined |
| 18-110_k141_179713 | Not-determined |
| 18-110_k141_236913 | Not-determined |
| 18-110_k141_167200 | Not-determined |
| 18-110_k141_167202 | Low-quality |
| 18-110_k141_86116 | Not-determined |
| 18-110_k141_109223 | Not-determined |
| 18-110_k141_258150 | Not-determined |
| 18-110_k141_63883 | Not-determined |
| 18-110_k141_63886 | Not-determined |
| 18-110_k141_16380 | Not-determined |
| 18-110_k141_144371 | Not-determined |
| 18-110_k141_200758 | Not-determined |
| 18-110_k141_144373 | Low-quality |
| 18-110_k141_63889 | Not-determined |
| 18-110_k141_120942 | Not-determined |
| 18-110_k141_132815 | Not-determined |
| 18-110_k141_40703 | Not-determined |
| 18-110_k141_63894 | Not-determined |
| 18-110_k141_236937 | Not-determined |
| 18-110_k141_271019 | Not-determined |
| 18-110_k141_271020 | Low-quality |
| 18-110_k141_63897 | Not-determined |
| 18-110_k141_167231 | Not-determined |
| 18-110_k141_40712 | Not-determined |
| 18-110_k141_51947 | Not-determined |
| 18-110_k141_40717 | Not-determined |
| 18-110_k141_16393 | Not-determined |
| 18-110_k141_120954 | Not-determined |
| 18-110_k141_40720 | Not-determined |
| 18-110_k141_248234 | Not-determined |
| 18-110_k141_132819 | Not-determined |
| 18-110_k141_40727 | Not-determined |
| 18-110_k141_132824 | Not-determined |
| 18-110_k141_236961 | Not-determined |
| 18-110_k141_86132 | Not-determined |
| 18-110_k141_200781 | Low-quality |
| 18-110_k141_236962 | Not-determined |
| 18-110_k141_213174 | Not-determined |
| 18-110_k141_224676 | Not-determined |
| 18-110_k141_224677 | Not-determined |
| 18-110_k141_248240 | Not-determined |
| 18-110_k141_51958 | Not-determined |
| 18-110_k141_86141 | Not-determined |
| 18-110_k141_200787 | Not-determined |
| 18-110_k141_258169 | Not-determined |
| 18-110_k141_132837 | Not-determined |
| 18-110_k141_5494 | Not-determined |
| 18-110_k141_109238 | Not-determined |
| 18-110_k141_5497 | Not-determined |
| 18-110_k141_86148 | Not-determined |
| 18-110_k141_179736 | Not-determined |
| 18-110_k141_40753 | Not-determined |
| 18-110_k141_29695 | Not-determined |
| 18-110_k141_109244 | Not-determined |
| 18-110_k141_179739 | Not-determined |
| 18-110_k141_258178 | Not-determined |
| 18-110_k141_29698 | Not-determined |
| 18-110_k141_63938 | Not-determined |
| 18-110_k141_40760 | Not-determined |
| 18-110_k141_258184 | Not-determined |
| 18-110_k141_179745 | Low-quality |
| 18-110_k141_224682 | Not-determined |
| 18-110_k141_155964 | Not-determined |
| 18-110_k141_155967 | Not-determined |
| 18-110_k141_86166 | Not-determined |
| 18-110_k141_236973 | Not-determined |
| 18-110_k141_40766 | Not-determined |
| 18-110_k141_213193 | Not-determined |
| 18-110_k141_63947 | Low-quality |
| 18-110_k141_190537 | Low-quality |
| 18-110_k141_5503 | Low-quality |
| 18-110_k141_63950 | Low-quality |
| 18-110_k141_190540 | Not-determined |
| 18-110_k141_224695 | Not-determined |
| 18-110_k141_258190 | Not-determined |
| 18-110_k141_75357 | Not-determined |
| 18-110_k141_258196 | Not-determined |
| 18-110_k141_109265 | Not-determined |
| 18-110_k141_236985 | Not-determined |
| 18-110_k141_190545 | Not-determined |
| 18-110_k141_86184 | Not-determined |
| 18-110_k141_224702 | Not-determined |
| 18-110_k141_224703 | Not-determined |
| 18-110_k141_179760 | Not-determined |
| 18-110_k141_258202 | Not-determined |
| 18-110_k141_86187 | Not-determined |
| 18-110_k141_236997 | Not-determined |
| 18-110_k141_200806 | Not-determined |
| 18-110_k141_120970 | Not-determined |
| 18-110_k141_29709 | Not-determined |
| 18-110_k141_29710 | Not-determined |
| 18-110_k141_155979 | Not-determined |
| 18-110_k141_5515 | Not-determined |
| 18-110_k141_5517 | Not-determined |
| 18-110_k141_179776 | Not-determined |
| 18-110_k141_224713 | Low-quality |
| 18-110_k141_120981 | Not-determined |
| 18-110_k141_29720 | Not-determined |
| 18-110_k141_109280 | Not-determined |
| 18-110_k141_120986 | Not-determined |
| 18-110_k141_179779 | Not-determined |
| 18-110_k141_29722 | Not-determined |
| 18-110_k141_167275 | Not-determined |
| 18-110_k141_132897 | Not-determined |
| 18-110_k141_132898 | Not-determined |
| 18-110_k141_40786 | Not-determined |
| 18-110_k141_29726 | Not-determined |
| 18-110_k141_86197 | Not-determined |
| 18-110_k141_224725 | Not-determined |
| 18-110_k141_40792 | Not-determined |
| 18-110_k141_16442 | Not-determined |
| 18-110_k141_63971 | Not-determined |
| 18-110_k141_63972 | Not-determined |
| 18-110_k141_237024 | Not-determined |
| 18-110_k141_40800 | Low-quality |
| 18-110_k141_179792 | Not-determined |
| 18-110_k141_109295 | Not-determined |
| 18-110_k141_248253 | Not-determined |
| 18-110_k141_190554 | Not-determined |
| 18-110_k141_29732 | Not-determined |
| 18-110_k141_109300 | Not-determined |
| 18-110_k141_179801 | Not-determined |
| 18-110_k141_271050 | Not-determined |
| 18-110_k141_200818 | Not-determined |
| 18-110_k141_167303 | Not-determined |
| 18-110_k141_29740 | Not-determined |
| 18-110_k141_179806 | Not-determined |
| 18-110_k141_144458 | Low-quality |
| 18-110_k141_63988 | Not-determined |
| 18-110_k141_109310 | Not-determined |
| 18-110_k141_179811 | Not-determined |
| 18-110_k141_271062 | Not-determined |
| 18-110_k141_109311 | Not-determined |
| 18-110_k141_237035 | Low-quality |
| 18-110_k141_248272 | Not-determined |
| 18-110_k141_52026 | Not-determined |
| 18-110_k141_155998 | Not-determined |
| 18-110_k141_144470 | Not-determined |
| 18-110_k141_224757 | Not-determined |
| 18-110_k141_167317 | Not-determined |
| 18-110_k141_144482 | Not-determined |
| 18-110_k141_144484 | Not-determined |
| 18-110_k141_98380 | Not-determined |
| 18-110_k141_248287 | Not-determined |
| 18-110_k141_40831 | Not-determined |
| 18-110_k141_64009 | Not-determined |
| 18-110_k141_213236 | Not-determined |
| 18-110_k141_213237 | Not-determined |
| 18-110_k141_271080 | Not-determined |
| 18-110_k141_179820 | Not-determined |
| 18-110_k141_40839 | Not-determined |
| 18-110_k141_132944 | Not-determined |
| 18-110_k141_132946 | Not-determined |
| 18-110_k141_237046 | Not-determined |
| 18-110_k141_213245 | Not-determined |
| 18-110_k141_167341 | Not-determined |
| 18-110_k141_271085 | Not-determined |
| 18-110_k141_190577 | Not-determined |
| 18-110_k141_200852 | Not-determined |
| 18-110_k141_121022 | Not-determined |
| 18-110_k141_179829 | Not-determined |
| 18-110_k141_224780 | Low-quality |
| 18-110_k141_213248 | Not-determined |
| 18-110_k141_144496 | Not-determined |
| 18-110_k141_109342 | Not-determined |
| 18-110_k141_237065 | Low-quality |
| 18-110_k141_213254 | Not-determined |
| 18-110_k141_144506 | Not-determined |
| 18-110_k141_52046 | Not-determined |
| 18-110_k141_86240 | Not-determined |
| 18-110_k141_98385 | Not-determined |
| 18-110_k141_52051 | Not-determined |
| 18-110_k141_109356 | Not-determined |
| 18-110_k141_64036 | Not-determined |
| 18-110_k141_121036 | Not-determined |
| 18-110_k141_86251 | Not-determined |
| 18-110_k141_5569 | Low-quality |
| 18-110_k141_75371 | Not-determined |
| 18-110_k141_167374 | Not-determined |
| 18-110_k141_109363 | Low-quality |
| 18-110_k141_64044 | Not-determined |
| 18-110_k141_75378 | Not-determined |
| 18-110_k141_248317 | Not-determined |
| 18-110_k141_109364 | Not-determined |
| 18-110_k141_167378 | Low-quality |
| 18-110_k141_98388 | Not-determined |
| 18-110_k141_40865 | Not-determined |
| 18-110_k141_86261 | Low-quality |
| 18-110_k141_109367 | Not-determined |
| 18-110_k141_98399 | Not-determined |
| 18-110_k141_86264 | Not-determined |
| 18-110_k141_179859 | Not-determined |
| 18-110_k141_86265 | Not-determined |
| 18-110_k141_109376 | Not-determined |
| 18-110_k141_167392 | Not-determined |
| 18-110_k141_52070 | Not-determined |
| 18-110_k141_179861 | Not-determined |
| 18-110_k141_167393 | Not-determined |
| 18-110_k141_237098 | Not-determined |
| 18-110_k141_52072 | Not-determined |
| 18-110_k141_224813 | Not-determined |
| 18-110_k141_75397 | Not-determined |
| 18-110_k141_190601 | Low-quality |
| 18-110_k141_121063 | Not-determined |
| 18-110_k141_75404 | Not-determined |
| 18-110_k141_167401 | Not-determined |
| 18-110_k141_237107 | Not-determined |
| 18-110_k141_132974 | Not-determined |
| 18-110_k141_98414 | Not-determined |
| 18-110_k141_237110 | Not-determined |
| 18-110_k141_144537 | Low-quality |
| 18-110_k141_132980 | Not-determined |
| 18-110_k141_121079 | Not-determined |
| 18-110_k141_75421 | Not-determined |
| 18-110_k141_132983 | Low-quality |
| 18-110_k141_132984 | Not-determined |
| 18-110_k141_179881 | Not-determined |
| 18-110_k141_109388 | Not-determined |
| 18-110_k141_132988 | Not-determined |
| 18-110_k141_40887 | Not-determined |
| 18-110_k141_109393 | Not-determined |
| 18-110_k141_132993 | Not-determined |
| 18-110_k141_98430 | Not-determined |
| 18-110_k141_237126 | Not-determined |
| 18-110_k141_167420 | Not-determined |
| 18-110_k141_224824 | Not-determined |
| 18-110_k141_224825 | Not-determined |
| 18-110_k141_64079 | Not-determined |
| 18-110_k141_121090 | Not-determined |
| 18-110_k141_213298 | Not-determined |
| 18-110_k141_86291 | Not-determined |
| 18-110_k141_29803 | Not-determined |
| 18-110_k141_224836 | Not-determined |
| 18-110_k141_86297 | Not-determined |
| 18-110_k141_52107 | Not-determined |
| 18-110_k141_86298 | Not-determined |
| 18-110_k141_271131 | Not-determined |
| 18-110_k141_121096 | Low-quality |
| 18-110_k141_86302 | Not-determined |
| 18-110_k141_75444 | Not-determined |
| 18-110_k141_121104 | Not-determined |
| 18-110_k141_237142 | Not-determined |
| 18-110_k141_167430 | Not-determined |
| 18-110_k141_179923 | Not-determined |
| 18-110_k141_64104 | Not-determined |
| 18-110_k141_52117 | Not-determined |
| 18-110_k141_144567 | Not-determined |
| 18-110_k141_52118 | Not-determined |
| 18-110_k141_156023 | Not-determined |
| 18-110_k141_271151 | Not-determined |
| 18-110_k141_190645 | Not-determined |
| 18-110_k141_52121 | Not-determined |
| 18-110_k141_86323 | Not-determined |
| 18-110_k141_52128 | Not-determined |
| 18-110_k141_40913 | Not-determined |
| 18-110_k141_156034 | Not-determined |
| 18-110_k141_213321 | Not-determined |
| 18-110_k141_75471 | Not-determined |
| 18-110_k141_109433 | Not-determined |
| 18-110_k141_109435 | Not-determined |
| 18-110_k141_224883 | Not-determined |
| 18-110_k141_75487 | Not-determined |
| 18-110_k141_190665 | Not-determined |
| 18-110_k141_133010 | Not-determined |
| 18-110_k141_190666 | Not-determined |
| 18-110_k141_213344 | Not-determined |
| 18-110_k141_40935 | Not-determined |
| 18-110_k141_190669 | Not-determined |
| 18-110_k141_75495 | Not-determined |
| 18-110_k141_64144 | Not-determined |
| 18-110_k141_109442 | Not-determined |
| 18-110_k141_75502 | Not-determined |
| 18-110_k141_133015 | Not-determined |
| 18-110_k141_213345 | Not-determined |
| 18-110_k141_75504 | Not-determined |
| 18-110_k141_52156 | Not-determined |
| 18-110_k141_52157 | Not-determined |
| 18-110_k141_213348 | Low-quality |
| 18-110_k141_75509 | Not-determined |
| 18-110_k141_271184 | Not-determined |
| 18-110_k141_133022 | Not-determined |
| 18-110_k141_167458 | Not-determined |
| 18-110_k141_64157 | Not-determined |
| 18-110_k141_40959 | Not-determined |
| 18-110_k141_64160 | Not-determined |
| 18-110_k141_5594 | Not-determined |
| 18-110_k141_64164 | Not-determined |
| 18-110_k141_156051 | Not-determined |
| 18-110_k141_156054 | Not-determined |
| 18-110_k141_271192 | Not-determined |
| 18-110_k141_144603 | Not-determined |
| 18-110_k141_156058 | Low-quality |
| 18-110_k141_248354 | Not-determined |
| 18-110_k141_40970 | Not-determined |
| 18-110_k141_64169 | Not-determined |
| 18-110_k141_248356 | Not-determined |
| 18-110_k141_121143 | Not-determined |
| 18-110_k141_179940 | Not-determined |
| 18-110_k141_190691 | Not-determined |
| 18-110_k141_144613 | Not-determined |
| 18-110_k141_213372 | Not-determined |
| 18-110_k141_213373 | Not-determined |
| 18-110_k141_5606 | Not-determined |
| 18-110_k141_190693 | Not-determined |
| 18-110_k141_121149 | Not-determined |
| 18-110_k141_121151 | Not-determined |
| 18-110_k141_133040 | Not-determined |
| 18-110_k141_156074 | Not-determined |
| 18-110_k141_258235 | Not-determined |
| 18-110_k141_40982 | Not-determined |
| 18-110_k141_40984 | Low-quality |
| 18-110_k141_258237 | Not-determined |
| 18-110_k141_156083 | Not-determined |
| 18-110_k141_248379 | Low-quality |
| 18-110_k141_248380 | Not-determined |
| 18-110_k141_75537 | Not-determined |
| 18-110_k141_121161 | Not-determined |
| 18-110_k141_213396 | Not-determined |
| 18-110_k141_248382 | Not-determined |
| 18-110_k141_133052 | Not-determined |
| 18-110_k141_248384 | Not-determined |
| 18-110_k141_167485 | Not-determined |
| 18-110_k141_248386 | Not-determined |
| 18-110_k141_156097 | Not-determined |
| 18-110_k141_258246 | Not-determined |
| 18-110_k141_224912 | Not-determined |
| 18-110_k141_167490 | Not-determined |
| 18-110_k141_98481 | Not-determined |
| 18-110_k141_98486 | Not-determined |
| 18-110_k141_200924 | Not-determined |
| 18-110_k141_179971 | Not-determined |
| 18-110_k141_179975 | Low-quality |
| 18-110_k141_224930 | Not-determined |
| 18-110_k141_98491 | Not-determined |
| 18-110_k141_179979 | Not-determined |
| 18-110_k141_190712 | Low-quality |
| 18-110_k141_190713 | Low-quality |
| 18-110_k141_41020 | Not-determined |
| 18-110_k141_133070 | Not-determined |
| 18-110_k141_5627 | Not-determined |
| 18-110_k141_29822 | Not-determined |
| 18-110_k141_156120 | Not-determined |
| 18-110_k141_258261 | Not-determined |
| 18-110_k141_213410 | Not-determined |
| 18-110_k141_109514 | Not-determined |
| 18-110_k141_41025 | Not-determined |
| 18-110_k141_109518 | Not-determined |
| 18-110_k141_190724 | Not-determined |
| 18-110_k141_121173 | Not-determined |
| 18-110_k141_156130 | Not-determined |
| 18-110_k141_144647 | Not-determined |
| 18-110_k141_200949 | Not-determined |
| 18-110_k141_86365 | Not-determined |
| 18-110_k141_133096 | Not-determined |
| 18-110_k141_213431 | Not-determined |
| 18-110_k141_16505 | Not-determined |
| 18-110_k141_213433 | Not-determined |

| | |
|---|---|
| 18-110_k141_109539 | Not-determined |
| 18-110_k141_109540 | Not-determined |
| 18-110_k141_98517 | Not-determined |
| 18-110_k141_109541 | Not-determined |
| 18-110_k141_98519 | Not-determined |
| 18-110_k141_213437 | Low-quality |
| 18-110_k141_86373 | Not-determined |
| 18-110_k141_224958 | Not-determined |
| 18-110_k141_86374 | Low-quality |
| 18-110_k141_271250 | Not-determined |
| 18-110_k141_109548 | Not-determined |
| 18-110_k141_213445 | Not-determined |
| 18-110_k141_52216 | Not-determined |
| 18-110_k141_200954 | Not-determined |
| 18-110_k141_86382 | Low-quality |
| 18-110_k141_41031 | Not-determined |
| 18-110_k141_16513 | Not-determined |
| 18-110_k141_200959 | Not-determined |
| 18-110_k141_75583 | Not-determined |
| 18-110_k141_64242 | Not-determined |
| 18-110_k141_271260 | Not-determined |
| 18-110_k141_52225 | Not-determined |
| 18-110_k141_64248 | Not-determined |
| 18-110_k141_190763 | Not-determined |
| 18-110_k141_64249 | Not-determined |
| 18-110_k141_16523 | Not-determined |
| 18-110_k141_224965 | Not-determined |
| 18-110_k141_41051 | Not-determined |
| 18-110_k141_271263 | Not-determined |
| 18-110_k141_224967 | Not-determined |
| 18-110_k141_258277 | Not-determined |
| 18-110_k141_64261 | Not-determined |
| 18-110_k141_271268 | Not-determined |
| 18-110_k141_75594 | Not-determined |
| 18-110_k141_41054 | Low-quality |
| 18-110_k141_75597 | Not-determined |
| 18-110_k141_29841 | Not-determined |
| 18-110_k141_64269 | Not-determined |
| 18-110_k141_133112 | Not-determined |
| 18-110_k141_16527 | Not-determined |
| 18-110_k141_144654 | Not-determined |
| 18-110_k141_213461 | Not-determined |
| 18-110_k141_41059 | Not-determined |
| 18-110_k141_75607 | Not-determined |
| 18-110_k141_86406 | Not-determined |
| 18-110_k141_98537 | Low-quality |
| 18-110_k141_86407 | Not-determined |
| 18-110_k141_248416 | Not-determined |
| 18-110_k141_144664 | Not-determined |
| 18-110_k141_121196 | Not-determined |
| 18-110_k141_200988 | Not-determined |
| 18-110_k141_98546 | Not-determined |
| 18-110_k141_213476 | Not-determined |
| 18-110_k141_121202 | Not-determined |
| 18-110_k141_133134 | Not-determined |
| 18-110_k141_109565 | Not-determined |
| 18-110_k141_41070 | Not-determined |
| 18-110_k141_16544 | Not-determined |
| 18-110_k141_29872 | Not-determined |
| 18-110_k141_29874 | Not-determined |
| 18-110_k141_5663 | Not-determined |
| 18-110_k141_121211 | Not-determined |
| 18-110_k141_109571 | Not-determined |
| 18-110_k141_109572 | Not-determined |
| 18-110_k141_98559 | Not-determined |
| 18-110_k141_109574 | Not-determined |
| 18-110_k141_64295 | Not-determined |
| 18-110_k141_52269 | Not-determined |
| 18-110_k141_98562 | Not-determined |
| 18-110_k141_167563 | Not-determined |
| 18-110_k141_41077 | Not-determined |
| 18-110_k141_156164 | Not-determined |
| 18-110_k141_121228 | Not-determined |
| 18-110_k141_258310 | Low-quality |
| 18-110_k141_213497 | Not-determined |
| 18-110_k141_41087 | Not-determined |
| 18-110_k141_86435 | Not-determined |
| 18-110_k141_64309 | Not-determined |
| 18-110_k141_225004 | Not-determined |
| 18-110_k141_29897 | Not-determined |
| 18-110_k141_258319 | Not-determined |
| 18-110_k141_121237 | Not-determined |
| 18-101_k141_153615 | Medium-quality |
| 18-110_k141_248458 | Not-determined |
| 18-110_k141_156174 | Not-determined |
| 18-110_k141_86445 | Not-determined |
| 18-110_k141_86446 | Not-determined |
| 18-110_k141_190788 | Not-determined |
| 18-110_k141_86447 | Not-determined |
| 18-110_k141_75628 | Not-determined |
| 18-110_k141_86452 | Not-determined |
| 18-110_k141_144704 | Not-determined |
| 18-110_k141_86454 | Low-quality |
| 18-110_k141_156182 | Not-determined |
| 18-110_k141_86459 | Not-determined |
| 18-110_k141_121247 | Low-quality |
| 18-110_k141_109599 | Low-quality |
| 18-110_k141_98595 | Not-determined |
| 18-110_k141_201049 | Not-determined |
| 18-110_k141_133143 | Not-determined |
| 18-110_k141_41120 | Not-determined |
| 18-110_k141_201051 | Low-quality |
| 18-110_k141_86467 | Not-determined |
| 18-110_k141_271278 | Not-determined |
| 18-110_k141_98604 | Not-determined |
| 18-110_k141_213528 | Low-quality |
| 18-110_k141_121258 | Not-determined |
| 18-110_k141_109610 | Not-determined |
| 18-110_k141_41131 | Not-determined |
| 18-110_k141_248473 | Low-quality |
| 18-110_k141_258336 | Not-determined |
| 18-110_k141_248474 | Not-determined |
| 18-110_k141_156215 | Not-determined |
| 18-110_k141_190816 | Not-determined |
| 18-110_k141_75654 | Not-determined |
| 18-110_k141_156218 | Not-determined |
| 18-110_k141_75660 | Low-quality |
| 18-110_k141_133156 | Not-determined |
| 18-110_k141_133159 | Not-determined |
| 18-110_k141_180044 | Not-determined |
| 18-110_k141_16567 | Not-determined |
| 18-110_k141_52323 | Not-determined |
| 18-110_k141_190830 | Not-determined |
| 18-110_k141_29942 | Not-determined |
| 18-110_k141_201055 | Not-determined |
| 18-110_k141_64339 | Not-determined |
| 18-110_k141_16577 | Not-determined |
| 18-110_k141_64346 | Not-determined |
| 18-110_k141_16579 | Low-quality |
| 18-110_k141_258356 | Not-determined |
| 18-110_k141_144737 | Not-determined |
| 18-110_k141_75676 | Not-determined |
| 18-110_k141_225032 | Not-determined |
| 18-110_k141_144738 | Low-quality |
| 18-110_k141_5682 | Not-determined |
| 18-110_k141_201059 | Not-determined |
| 18-110_k141_190839 | Not-determined |
| 18-110_k141_41164 | Not-determined |
| 18-110_k141_64353 | Not-determined |
| 18-110_k141_156238 | Low-quality |
| 18-110_k141_201062 | Not-determined |
| 18-110_k141_41167 | Not-determined |
| 18-110_k141_16591 | Not-determined |
| 18-110_k141_5693 | Not-determined |
| 18-110_k141_41173 | Not-determined |
| 18-110_k141_225042 | Not-determined |
| 18-110_k141_225044 | Not-determined |
| 18-110_k141_29956 | Not-determined |
| 18-110_k141_16601 | Not-determined |
| 18-110_k141_237208 | Not-determined |
| 18-110_k141_271284 | Not-determined |
| 18-110_k141_237219 | Not-determined |
| 18-110_k141_64364 | Not-determined |
| 18-110_k141_64365 | Low-quality |
| 18-110_k141_248487 | Not-determined |
| 18-110_k141_121279 | Not-determined |
| 18-110_k141_271291 | Not-determined |
| 18-110_k141_180070 | Not-determined |
| 18-110_k141_144768 | Not-determined |
| 18-110_k141_52347 | Not-determined |
| 18-110_k141_248501 | Not-determined |
| 18-110_k141_144779 | Not-determined |
| 18-110_k141_271309 | Low-quality |
| 18-110_k141_16619 | Not-determined |
| 18-110_k141_52359 | Not-determined |
| 18-110_k141_180086 | Not-determined |
| 18-110_k141_29966 | Not-determined |
| 18-110_k141_248503 | Not-determined |
| 18-110_k141_16630 | Not-determined |
| 18-110_k141_190849 | Not-determined |
| 18-110_k141_5702 | Not-determined |
| 18-110_k141_52370 | Not-determined |
| 18-110_k141_5704 | Not-determined |
| 18-110_k141_75694 | Not-determined |
| 18-110_k141_75695 | Not-determined |
| 18-110_k141_109641 | Not-determined |
| 18-110_k141_180097 | Not-determined |
| 18-110_k141_52374 | Not-determined |
| 18-110_k141_225068 | Not-determined |
| 18-110_k141_41202 | Not-determined |
| 18-110_k141_29980 | Not-determined |
| 18-110_k141_201079 | Not-determined |
| 18-110_k141_75703 | Not-determined |
| 18-110_k141_121285 | Not-determined |
| 18-110_k141_225080 | Not-determined |
| 18-110_k141_75707 | Not-determined |
| 18-110_k141_52384 | Not-determined |
| 18-110_k141_225081 | Not-determined |
| 18-110_k141_201089 | Not-determined |
| 18-110_k141_121287 | Not-determined |
| 18-110_k141_41205 | Not-determined |
| 18-110_k141_225084 | Not-determined |
| 18-110_k141_5716 | Not-determined |
| 18-110_k141_64407 | Not-determined |
| 18-110_k141_75711 | Not-determined |
| 18-110_k141_41211 | Not-determined |
| 18-110_k141_16640 | Not-determined |
| 18-110_k141_225093 | Not-determined |
| 18-110_k141_167601 | Not-determined |
| 18-110_k141_75714 | Not-determined |
| 18-110_k141_225097 | Not-determined |
| 18-110_k141_213607 | Not-determined |
| 18-110_k141_5725 | Not-determined |
| 18-110_k141_190875 | Not-determined |
| 18-110_k141_271341 | Not-determined |
| 18-110_k141_64416 | Not-determined |
| 18-110_k141_248535 | Not-determined |
| 18-110_k141_167609 | Not-determined |
| 18-110_k141_213612 | Not-determined |
| 18-110_k141_248540 | Not-determined |
| 18-110_k141_52394 | Not-determined |
| 18-110_k141_180119 | Not-determined |
| 18-110_k141_64422 | Not-determined |
| 18-110_k141_133250 | Not-determined |
| 18-110_k141_190882 | Not-determined |
| 18-110_k141_86493 | Not-determined |
| 18-110_k141_167617 | Not-determined |
| 18-110_k141_98616 | Not-determined |
| 18-110_k141_237252 | Not-determined |
| 18-110_k141_64430 | Not-determined |
| 18-110_k141_41231 | Not-determined |
| 18-110_k141_75734 | Not-determined |
| 18-110_k141_109674 | Not-determined |
| 18-110_k141_98628 | Not-determined |
| 18-110_k141_271343 | Low-quality |
| 18-110_k141_75736 | Not-determined |
| 18-110_k141_225112 | Not-determined |
| 18-110_k141_5748 | Not-determined |
| 18-110_k141_271352 | Not-determined |
| 18-110_k141_248556 | Not-determined |
| 18-110_k141_75739 | Not-determined |
| 18-110_k141_225113 | Low-quality |
| 18-110_k141_64441 | Not-determined |
| 18-110_k141_64445 | Not-determined |
| 18-110_k141_248559 | Low-quality |
| 18-110_k141_64446 | Not-determined |
| 18-110_k141_144835 | Not-determined |
| 18-110_k141_30018 | Not-determined |
| 18-110_k141_41244 | Not-determined |
| 18-110_k141_41245 | Not-determined |
| 18-110_k141_64452 | Low-quality |
| 18-110_k141_180143 | Not-determined |
| 18-110_k141_121342 | Not-determined |
| 18-110_k141_237273 | Not-determined |
| 18-110_k141_98652 | Not-determined |
| 18-110_k141_75758 | Not-determined |
| 18-110_k141_201116 | Not-determined |
| 18-110_k141_258390 | Not-determined |
| 18-110_k141_237278 | Not-determined |
| 18-110_k141_144851 | Not-determined |
| 18-110_k141_98662 | Not-determined |
| 18-110_k141_5769 | Not-determined |
| 18-110_k141_5773 | Not-determined |
| 18-110_k141_64471 | Not-determined |
| 18-110_k141_180176 | Not-determined |
| 18-110_k141_64473 | Not-determined |
| 18-110_k141_16684 | Not-determined |
| 18-110_k141_144866 | Not-determined |
| 18-110_k141_180187 | Not-determined |
| 18-110_k141_237304 | Low-quality |
| 18-110_k141_86532 | Low-quality |
| 18-110_k141_201130 | Low-quality |
| 18-110_k141_180192 | Not-determined |
| 18-110_k141_86537 | Not-determined |
| 18-110_k141_75788 | Not-determined |
| 18-110_k141_144874 | Not-determined |
| 18-110_k141_248573 | Not-determined |
| 18-110_k141_144877 | Low-quality |
| 18-110_k141_144879 | Not-determined |
| 18-110_k141_237314 | Not-determined |
| 18-110_k141_121356 | Not-determined |
| 18-110_k141_144882 | Not-determined |
| 18-110_k141_75793 | Low-quality |
| 18-110_k141_144884 | Not-determined |
| 18-110_k141_16692 | Not-determined |
| 18-110_k141_98680 | Not-determined |
| 18-110_k141_201140 | Low-quality |
| 18-110_k141_237322 | Not-determined |
| 18-110_k141_258434 | Not-determined |
| 18-110_k141_30053 | Not-determined |
| 18-110_k141_237325 | Not-determined |
| 18-110_k141_121375 | Not-determined |
| 18-110_k141_144892 | Low-quality |
| 18-110_k141_86556 | Not-determined |
| 18-110_k141_121378 | Not-determined |
| 18-110_k141_52398 | Not-determined |
| 18-110_k141_64500 | Not-determined |
| 18-110_k141_64501 | Not-determined |
| 18-110_k141_180217 | Not-determined |
| 18-110_k141_5791 | Not-determined |
| 18-110_k141_180218 | Not-determined |
| 18-110_k141_201148 | Not-determined |
| 18-110_k141_225115 | Not-determined |
| 18-110_k141_5799 | Not-determined |
| 18-110_k141_156258 | Not-determined |
| 18-110_k141_41306 | Not-determined |
| 18-110_k141_64508 | Not-determined |
| 18-110_k141_41308 | Not-determined |
| 18-101-final-viral-combined_k141_153615\|\|full | Medium-quality |
| 18-110_k141_75802 | Not-determined |
| 18-110_k141_75805 | Not-determined |
| 18-110_k141_225126 | Not-determined |
| 18-110_k141_271405 | Not-determined |
| 18-110_k141_64523 | Not-determined |
| 18-110_k141_109715 | Not-determined |
| 18-110_k141_98700 | Not-determined |
| 18-110_k141_133313 | Not-determined |
| 18-110_k141_75817 | Not-determined |
| 18-110_k141_258445 | Not-determined |
| 18-110_k141_133315 | Not-determined |
| 18-110_k141_121406 | Not-determined |
| 18-110_k141_271414 | Low-quality |
| 18-110_k141_16722 | Low-quality |
| 18-110_k141_225132 | Not-determined |
| 18-110_k141_180235 | Not-determined |
| 18-110_k141_75825 | Not-determined |
| 18-110_k141_75826 | Not-determined |
| 18-110_k141_16724 | Low-quality |
| 18-110_k141_5814 | Not-determined |
| 18-110_k141_271423 | Low-quality |
| 18-110_k141_258457 | Not-determined |
| 18-110_k141_258458 | Not-determined |
| 18-110_k141_271425 | Not-determined |
| 18-110_k141_156274 | Not-determined |
| 18-110_k141_237380 | Low-quality |
| 18-110_k141_225150 | Not-determined |
| 18-110_k141_237381 | Not-determined |
| 18-110_k141_133342 | Not-determined |
| 18-110_k141_133343 | Not-determined |
| 18-110_k141_258467 | Not-determined |
| 18-110_k141_225154 | Not-determined |
| 18-110_k141_64561 | Not-determined |
| 18-110_k141_225158 | Low-quality |
| 18-110_k141_121433 | Not-determined |
| 18-110_k141_52431 | Not-determined |
| 18-110_k141_16730 | Not-determined |
| 18-110_k141_41330 | Not-determined |
| 18-110_k141_41335 | Not-determined |
| 18-110_k141_64573 | Not-determined |
| 18-110_k141_64574 | Not-determined |
| 18-110_k141_41337 | Not-determined |
| 18-110_k141_75836 | Low-quality |
| 18-110_k141_52445 | Not-determined |
| 18-110_k141_156281 | Not-determined |
| 18-110_k141_180252 | Not-determined |
| 18-110_k141_248627 | Not-determined |
| 18-110_k141_248628 | Not-determined |
| 18-110_k141_144918 | Low-quality |
| 18-110_k141_180256 | Not-determined |
| 18-110_k141_156283 | Not-determined |
| 18-110_k141_144924 | Not-determined |
| 18-110_k141_237400 | Not-determined |
| 18-110_k141_271442 | Not-determined |
| 18-110_k141_52453 | Not-determined |
| 18-110_k141_109732 | Not-determined |
| 18-110_k141_271444 | Not-determined |
| 18-110_k141_86601 | Not-determined |
| 18-110_k141_109740 | Not-determined |
| 18-110_k141_144935 | Not-determined |
| 18-110_k141_52462 | Not-determined |
| 18-110_k141_248640 | Not-determined |
| 18-110_k141_86603 | Low-quality |
| 18-110_k141_52463 | Not-determined |
| 18-110_k141_75852 | Not-determined |
| 18-110_k141_86605 | Not-determined |
| 18-110_k141_52465 | Not-determined |
| 18-110_k141_144943 | Not-determined |
| 18-110_k141_258492 | Not-determined |
| 18-110_k141_237411 | Not-determined |
| 18-110_k141_75855 | Not-determined |
| 18-110_k141_144949 | Not-determined |
| 18-110_k141_180280 | Low-quality |
| 18-110_k141_121460 | Not-determined |
| 18-110_k141_258493 | Low-quality |
| 18-110_k141_86613 | Not-determined |
| 18-110_k141_258497 | Low-quality |
| 18-110_k141_41372 | Not-determined |
| 18-110_k141_156309 | Low-quality |
| 18-110_k141_225164 | Not-determined |
| 18-110_k141_190911 | Not-determined |
| 18-110_k141_167758 | Not-determined |
| 18-110_k141_225172 | Low-quality |
| 18-110_k141_180292 | Not-determined |
| 18-110_k141_75876 | Not-determined |
| 18-110_k141_86631 | Not-determined |
| 18-110_k141_75878 | Not-determined |
| 18-110_k141_98732 | Not-determined |
| 18-110_k141_271475 | Not-determined |
| 18-110_k141_180300 | Not-determined |
| 18-110_k141_109776 | Not-determined |
| 18-110_k141_16756 | Low-quality |
| 18-110_k141_52485 | Not-determined |
| 18-110_k141_144972 | Not-determined |
| 18-110_k141_75882 | Not-determined |
| 18-110_k141_144974 | Not-determined |
| 18-110_k141_271484 | Not-determined |
| 18-110_k141_156329 | Not-determined |
| 18-110_k141_75884 | Not-determined |
| 18-110_k141_109784 | Not-determined |
| 18-110_k141_271489 | Low-quality |
| 18-110_k141_213677 | Not-determined |
| 18-110_k141_121488 | Not-determined |
| 18-110_k141_16768 | Not-determined |
| 18-110_k141_5883 | Low-quality |
| 18-110_k141_5885 | Not-determined |
| 18-110_k141_156343 | Not-determined |
| 18-110_k141_5892 | Not-determined |
| 18-110_k141_156347 | Not-determined |
| 18-110_k141_237461 | Not-determined |
| 18-110_k141_16781 | Not-determined |
| 18-110_k141_248656 | Not-determined |
| 18-110_k141_156353 | Low-quality |
| 18-110_k141_5895 | Not-determined |
| 18-110_k141_16787 | Not-determined |
| 18-110_k141_225191 | Not-determined |
| 18-110_k141_156358 | Low-quality |
| 18-110_k141_121502 | Not-determined |
| 18-110_k141_75904 | Not-determined |
| 18-110_k141_52504 | Not-determined |
| 18-110_k141_201273 | Not-determined |
| 18-110_k141_75907 | Not-determined |
| 18-110_k141_201275 | Not-determined |
| 18-110_k141_52507 | Not-determined |
| 18-110_k141_5899 | Not-determined |
| 18-110_k141_109818 | Not-determined |
| 18-110_k141_258520 | Not-determined |
| 18-110_k141_86682 | Not-determined |
| 18-110_k141_86684 | Not-determined |
| 18-110_k141_225197 | Low-quality |
| 18-110_k141_190937 | Not-determined |
| 18-110_k141_167787 | Not-determined |
| 18-110_k141_156373 | Not-determined |
| 18-110_k141_145010 | Not-determined |
| 18-110_k141_156375 | Low-quality |
| 18-110_k141_75918 | Not-determined |
| 18-110_k141_109826 | Low-quality |
| 18-110_k141_201284 | Low-quality |
| 18-110_k141_86690 | Not-determined |
| 18-110_k141_98755 | Not-determined |
| 18-110_k141_133386 | Low-quality |
| 18-110_k141_201290 | Not-determined |
| 18-110_k141_201294 | Not-determined |
| 18-110_k141_109845 | Not-determined |
| 18-110_k141_64595 | Not-determined |
| 18-110_k141_5907 | Not-determined |
| 18-110_k141_167810 | Low-quality |
| 18-110_k141_30086 | Not-determined |
| 18-110_k141_64604 | Not-determined |
| 18-110_k141_133391 | Not-determined |
| 18-110_k141_145026 | Not-determined |
| 18-110_k141_156387 | Not-determined |
| 18-110_k141_109856 | Low-quality |
| 18-110_k141_248684 | Not-determined |
| 18-110_k141_121528 | Not-determined |
| 18-110_k141_5915 | Not-determined |
| 18-110_k141_30092 | Not-determined |
| 18-110_k141_64609 | Not-determined |
| 18-110_k141_133407 | Not-determined |
| 18-110_k141_225219 | Low-quality |
| 18-110_k141_133409 | Not-determined |
| 18-110_k141_156395 | Not-determined |
| 18-110_k141_201309 | Not-determined |
| 18-110_k141_225225 | Not-determined |
| 18-110_k141_225226 | Not-determined |
| 18-110_k141_30102 | Not-determined |
| 18-110_k141_271558 | Not-determined |
| 18-110_k141_41384 | Not-determined |
| 18-110_k141_213710 | Not-determined |
| 18-110_k141_30106 | Not-determined |
| 18-110_k141_201318 | Not-determined |
| 18-110_k141_213713 | Not-determined |
| 18-110_k141_109874 | Not-determined |
| 18-110_k141_41387 | Not-determined |
| 18-110_k141_180357 | Not-determined |
| 18-110_k141_190957 | Not-determined |
| 18-110_k141_180359 | Not-determined |
| 18-110_k141_64625 | Not-determined |
| 18-110_k141_64626 | Not-determined |
| 18-110_k141_190960 | Not-determined |
| 18-110_k141_5938 | Not-determined |
| 18-110_k141_225242 | Not-determined |
| 18-110_k141_133435 | Not-determined |
| 18-110_k141_271580 | Not-determined |
| 18-110_k141_248710 | Not-determined |
| 18-110_k141_180374 | Not-determined |
| 18-110_k141_41401 | Not-determined |
| 18-110_k141_180380 | Not-determined |
| 18-110_k141_190969 | Not-determined |
| 18-110_k141_201340 | Not-determined |
| 18-110_k141_133445 | Not-determined |
| 18-110_k141_190973 | Not-determined |
| 18-110_k141_30135 | Not-determined |
| 18-110_k141_190980 | Not-determined |
| 18-110_k141_213730 | Not-determined |
| 18-110_k141_180393 | Not-determined |
| 18-110_k141_248720 | Not-determined |
| 18-110_k141_145047 | Not-determined |
| 18-110_k141_30141 | Low-quality |
| 18-110_k141_271593 | Not-determined |
| 18-110_k141_180398 | Not-determined |
| 18-110_k141_190989 | Not-determined |
| 18-110_k141_248727 | Not-determined |
| 18-110_k141_121546 | Not-determined |
| 18-110_k141_121547 | Not-determined |
| 18-110_k141_52559 | Not-determined |
| 18-110_k141_248736 | Not-determined |
| 18-110_k141_121552 | Not-determined |
| 18-110_k141_258566 | Not-determined |
| 18-110_k141_41421 | Not-determined |
| 18-110_k141_98814 | Not-determined |
| 18-110_k141_213753 | Not-determined |
| 18-110_k141_52568 | Not-determined |
| 18-110_k141_5972 | Not-determined |
| 18-110_k141_201365 | Not-determined |
| 18-110_k141_225269 | Not-determined |
| 18-110_k141_156454 | Not-determined |
| 18-110_k141_248750 | Not-determined |
| 18-110_k141_16838 | Not-determined |
| 18-110_k141_145076 | Not-determined |
| 18-110_k141_145078 | Low-quality |
| 18-110_k141_225279 | Not-determined |
| 18-110_k141_258577 | Not-determined |
| 18-110_k141_167875 | Not-determined |
| 18-110_k141_52583 | Not-determined |
| 18-110_k141_5980 | Not-determined |
| 18-110_k141_98824 | Not-determined |
| 18-110_k141_30172 | Not-determined |
| 18-110_k141_225293 | Not-determined |
| 18-110_k141_167878 | Not-determined |
| 18-110_k141_16847 | Not-determined |
| 18-110_k141_86695 | Not-determined |
| 18-110_k141_248763 | Low-quality |
| 18-110_k141_156456 | Not-determined |
| 18-110_k141_121575 | Not-determined |
| 18-110_k141_30195 | Not-determined |
| 18-110_k141_225304 | Not-determined |
| 18-110_k141_248775 | Not-determined |
| 18-110_k141_109892 | Not-determined |
| 18-110_k141_225309 | Not-determined |
| 18-110_k141_16864 | Not-determined |
| 18-110_k141_225311 | Low-quality |
| 18-110_k141_98834 | Low-quality |
| 18-110_k141_109898 | Not-determined |
| 18-110_k141_109900 | Not-determined |
| 18-110_k141_213768 | Not-determined |
| 18-110_k141_30206 | Not-determined |
| 18-110_k141_145099 | Not-determined |
| 18-110_k141_271599 | Not-determined |
| 18-110_k141_30208 | Not-determined |
| 18-110_k141_121583 | Not-determined |
| 18-110_k141_109906 | Low-quality |
| 18-110_k141_30212 | Not-determined |
| 18-110_k141_109912 | Not-determined |
| 18-110_k141_121586 | Not-determined |
| 18-110_k141_180449 | Low-quality |
| 18-110_k141_109913 | Not-determined |
| 18-110_k141_225314 | Not-determined |
| 18-110_k141_16877 | Low-quality |
| 18-110_k141_258618 | Not-determined |
| 18-110_k141_75986 | Not-determined |
| 18-110_k141_86727 | Low-quality |
| 18-110_k141_30221 | Not-determined |
| 18-110_k141_213779 | Not-determined |
| 18-110_k141_225320 | Not-determined |
| 18-110_k141_121601 | Not-determined |
| 18-110_k141_52621 | Not-determined |
| 18-110_k141_213780 | Not-determined |
| 18-110_k141_64635 | Not-determined |
| 18-110_k141_225324 | Not-determined |
| 18-110_k141_98851 | Not-determined |
| 18-110_k141_41454 | Not-determined |
| 18-110_k141_98856 | Not-determined |
| 18-110_k141_86741 | Not-determined |
| 18-110_k141_16895 | Not-determined |
| 18-110_k141_156471 | Not-determined |
| 18-110_k141_16898 | Not-determined |
| 18-110_k141_109932 | Low-quality |
| 18-110_k141_213789 | Not-determined |
| 18-110_k141_98862 | Not-determined |
| 18-110_k141_145121 | Not-determined |
| 18-110_k141_30227 | Not-determined |
| 18-110_k141_258639 | Not-determined |
| 18-110_k141_86756 | Not-determined |
| 18-110_k141_271615 | Low-quality |
| 18-110_k141_237521 | Not-determined |
| 18-110_k141_213799 | Not-determined |
| 18-110_k141_133501 | Not-determined |
| 18-110_k141_225355 | Not-determined |
| 18-110_k141_167915 | Not-determined |

| | |
|---|---|
| 18-110_k141_271622 | Not-determined |
| 18-110_k141_98876 | Not-determined |
| 18-110_k141_145141 | Not-determined |
| 18-110_k141_52649 | Not-determined |
| 18-110_k141_133512 | Not-determined |
| 18-110_k141_156480 | Not-determined |
| 18-110_k141_248815 | Not-determined |
| 18-110_k141_6036 | Not-determined |
| 18-110_k141_6037 | Low-quality |
| 18-110_k141_248820 | Not-determined |
| 18-110_k141_237536 | Not-determined |
| 18-110_k141_121616 | Not-determined |
| 18-110_k141_191044 | Not-determined |
| 18-110_k141_225362 | Not-determined |
| 18-110_k141_52663 | Not-determined |
| 18-110_k141_41490 | Not-determined |
| 18-110_k141_41491 | Not-determined |
| 18-110_k141_52666 | Not-determined |
| 18-110_k141_167932 | Not-determined |
| 18-110_k141_109959 | Not-determined |
| 18-110_k141_109961 | Not-determined |
| 18-101-final-viral-combined_k141_153670\|\|full | Medium-quality |
| 18-110_k141_225372 | Low-quality |
| 18-110_k141_41498 | Low-quality |
| 18-110_k141_225378 | Not-determined |
| 18-110_k141_271641 | Not-determined |
| 18-110_k141_121639 | Not-determined |
| 18-110_k141_98882 | Not-determined |
| 18-110_k141_133536 | Low-quality |
| 18-110_k141_271651 | Not-determined |
| 18-110_k141_237565 | Not-determined |
| 18-110_k141_248841 | Not-determined |
| 18-110_k141_86768 | Not-determined |
| 18-110_k141_30240 | Low-quality |
| 18-110_k141_167942 | Not-determined |
| 18-110_k141_213846 | Not-determined |
| 18-110_k141_41514 | Not-determined |
| 18-110_k141_201448 | Not-determined |
| 18-110_k141_41515 | Not-determined |
| 18-110_k141_167946 | Not-determined |
| 18-110_k141_30243 | Not-determined |
| 18-110_k141_201452 | Not-determined |
| 18-110_k141_109979 | Not-determined |
| 18-110_k141_145146 | Not-determined |
| 18-110_k141_145147 | Not-determined |
| 18-110_k141_191063 | Not-determined |
| 18-110_k141_248853 | Not-determined |
| 18-110_k141_76002 | Not-determined |
| 18-110_k141_30249 | Not-determined |
| 18-110_k141_258662 | Not-determined |
| 18-110_k141_76006 | Not-determined |
| 18-110_k141_237581 | Not-determined |
| 18-110_k141_271660 | Low-quality |
| 18-110_k141_271661 | Not-determined |
| 18-110_k141_167960 | Not-determined |
| 18-110_k141_41524 | Low-quality |
| 18-110_k141_213859 | Not-determined |
| 18-110_k141_167963 | Not-determined |
| 18-110_k141_86788 | Not-determined |
| 18-110_k141_109990 | Not-determined |
| 18-110_k141_191070 | Not-determined |
| 18-110_k141_86793 | Not-determined |
| 18-110_k141_145173 | Not-determined |
| 18-110_k141_52702 | Not-determined |
| 18-110_k141_213872 | Not-determined |
| 18-110_k141_121654 | Low-quality |
| 18-110_k141_110002 | Not-determined |
| 18-110_k141_98896 | Not-determined |
| 18-110_k141_191089 | Not-determined |
| 18-110_k141_167984 | Not-determined |
| 18-110_k141_167985 | Not-determined |
| 18-110_k141_191092 | Not-determined |
| 18-110_k141_98901 | Not-determined |
| 18-110_k141_191096 | Not-determined |
| 18-110_k141_213879 | Not-determined |
| 18-110_k141_180492 | Not-determined |
| 18-110_k141_248875 | Low-quality |
| 18-110_k141_180493 | Not-determined |
| 18-110_k141_213881 | Low-quality |
| 18-110_k141_98905 | Not-determined |
| 18-110_k141_76034 | Not-determined |
| 18-110_k141_271682 | Not-determined |
| 18-110_k141_76036 | Not-determined |
| 18-110_k141_225419 | Not-determined |
| 18-110_k141_133545 | Not-determined |
| 18-110_k141_133550 | Not-determined |
| 18-110_k141_98912 | Not-determined |
| 18-110_k141_41538 | Not-determined |
| 18-110_k141_201479 | Not-determined |
| 18-110_k141_271690 | Not-determined |
| 18-110_k141_41541 | Not-determined |
| 18-110_k141_64678 | Not-determined |
| 18-110_k141_258699 | Not-determined |
| 18-110_k141_98920 | Not-determined |
| 18-110_k141_258702 | Not-determined |
| 18-110_k141_180514 | Low-quality |
| 18-110_k141_64681 | Low-quality |
| 18-110_k141_271700 | Not-determined |
| 18-110_k141_191111 | Not-determined |
| 18-110_k141_98928 | Not-determined |
| 18-110_k141_213894 | Not-determined |
| 18-110_k141_213901 | Not-determined |
| 18-110_k141_76050 | Not-determined |
| 18-110_k141_271710 | Not-determined |
| 18-110_k141_121668 | Low-quality |
| 18-110_k141_133584 | Not-determined |
| 18-110_k141_110011 | Low-quality |
| 18-110_k141_248910 | Not-determined |
| 18-110_k141_30288 | Not-determined |
| 18-110_k141_30293 | Not-determined |
| 18-110_k141_76058 | Not-determined |
| 18-110_k141_121676 | Not-determined |
| 18-110_k141_6104 | Not-determined |
| 18-110_k141_121678 | Not-determined |
| 18-110_k141_133597 | Not-determined |
| 18-110_k141_180541 | Not-determined |
| 18-110_k141_213915 | Not-determined |
| 18-110_k141_191129 | Not-determined |
| 18-110_k141_237618 | Not-determined |
| 18-110_k141_98951 | Not-determined |
| 18-110_k141_213916 | Not-determined |
| 18-110_k141_258722 | Low-quality |
| 18-110_k141_213918 | Not-determined |
| 18-110_k141_191139 | Low-quality |
| 18-110_k141_225443 | Not-determined |
| 18-110_k141_225446 | Not-determined |
| 18-110_k141_213924 | Low-quality |
| 18-110_k141_30320 | Not-determined |
| 18-110_k141_30321 | Not-determined |
| 18-110_k141_156564 | Not-determined |
| 18-110_k141_133615 | Not-determined |
| 18-110_k141_191151 | Not-determined |
| 18-110_k141_156571 | Not-determined |
| 18-110_k141_52763 | Not-determined |
| 18-110_k141_248919 | Not-determined |
| 18-110_k141_271722 | Low-quality |
| 18-110_k141_237621 | Not-determined |
| 18-110_k141_248920 | Not-determined |
| 18-110_k141_52769 | Not-determined |
| 18-110_k141_16937 | Not-determined |
| 18-110_k141_16938 | Not-determined |
| 18-110_k141_98978 | Not-determined |
| 18-110_k141_30328 | Not-determined |
| 18-110_k141_86843 | Not-determined |
| 18-110_k141_237631 | Low-quality |
| 18-110_k141_168039 | Not-determined |
| 18-110_k141_156586 | Not-determined |
| 18-110_k141_168042 | Not-determined |
| 18-110_k141_133632 | Not-determined |
| 18-110_k141_248935 | Not-determined |
| 18-110_k141_64699 | Not-determined |
| 18-110_k141_213935 | Low-quality |
| 18-110_k141_191173 | Not-determined |
| 18-110_k141_237636 | Not-determined |
| 18-110_k141_64704 | Not-determined |
| 18-110_k141_145191 | Not-determined |
| 18-110_k141_191180 | Not-determined |
| 18-110_k141_201508 | Not-determined |
| 18-110_k141_248941 | Not-determined |
| 18-110_k141_225473 | Not-determined |
| 18-110_k141_16960 | Low-quality |
| 18-110_k141_191184 | Not-determined |
| 18-110_k141_41566 | Not-determined |
| 18-110_k141_191186 | Not-determined |
| 18-110_k141_30348 | Not-determined |
| 18-110_k141_41570 | Not-determined |
| 18-110_k141_213952 | Not-determined |
| 18-110_k141_156606 | Not-determined |
| 18-110_k141_213953 | Not-determined |
| 18-110_k141_86871 | Not-determined |
| 18-110_k141_6139 | Not-determined |
| 18-110_k141_98999 | Not-determined |
| 18-110_k141_41578 | Low-quality |
| 18-110_k141_99009 | Not-determined |
| 18-110_k141_76079 | Not-determined |
| 18-110_k141_6151 | Not-determined |
| 18-110_k141_86882 | Not-determined |
| 18-110_k141_168059 | Not-determined |
| 18-110_k141_86884 | Not-determined |
| 18-110_k141_237640 | Not-determined |
| 18-110_k141_168065 | Not-determined |
| 18-110_k141_16969 | Not-determined |
| 18-110_k141_271753 | Not-determined |
| 18-110_k141_76094 | Not-determined |
| 18-110_k141_41582 | Low-quality |
| 18-110_k141_41584 | Not-determined |
| 18-110_k141_76098 | Not-determined |
| 18-110_k141_30367 | Low-quality |
| 18-110_k141_76101 | Not-determined |
| 18-110_k141_225489 | Not-determined |
| 18-110_k141_225491 | Not-determined |
| 18-110_k141_156639 | Not-determined |
| 18-110_k141_76104 | Not-determined |
| 18-110_k141_225494 | Not-determined |
| 18-110_k141_191202 | Not-determined |
| 18-110_k141_86894 | Not-determined |
| 18-110_k141_191205 | Not-determined |
| 18-110_k141_258737 | Low-quality |
| 18-110_k141_191210 | Not-determined |
| 18-110_k141_237665 | Not-determined |
| 18-110_k141_258740 | Not-determined |
| 18-110_k141_237666 | Not-determined |
| 18-110_k141_258742 | Not-determined |
| 18-110_k141_133678 | Not-determined |
| 18-110_k141_30390 | Not-determined |
| 18-110_k141_237669 | Not-determined |
| 18-110_k141_225505 | Not-determined |
| 18-110_k141_191219 | Not-determined |
| 18-110_k141_133679 | Low-quality |
| 18-110_k141_191220 | Not-determined |
| 18-110_k141_225509 | Not-determined |
| 18-110_k141_41616 | Not-determined |
| 18-110_k141_156664 | Not-determined |
| 18-110_k141_258752 | Not-determined |
| 18-110_k141_6179 | Not-determined |
| 18-110_k141_156668 | Not-determined |
| 18-110_k141_64743 | Low-quality |
| 18-110_k141_180561 | Not-determined |
| 18-110_k141_237678 | Not-determined |
| 18-110_k141_16999 | Not-determined |
| 18-110_k141_64753 | Not-determined |
| 18-110_k141_64754 | Not-determined |
| 18-110_k141_121720 | Not-determined |
| 18-110_k141_133689 | Not-determined |
| 18-110_k141_191228 | Not-determined |
| 18-110_k141_17004 | Not-determined |
| 18-110_k141_191231 | Not-determined |
| 18-110_k141_30406 | Not-determined |
| 18-110_k141_121725 | Not-determined |
| 18-110_k141_41623 | Not-determined |
| 18-110_k141_145247 | Not-determined |
| 18-110_k141_248959 | Not-determined |
| 18-110_k141_76122 | Not-determined |
| 18-110_k141_248966 | Not-determined |
| 18-110_k141_17014 | Not-determined |
| 18-110_k141_133694 | Low-quality |
| 18-110_k141_191244 | Not-determined |
| 18-110_k141_99062 | Not-determined |
| 18-110_k141_86905 | Not-determined |
| 18-110_k141_225529 | Low-quality |
| 18-110_k141_76128 | Not-determined |
| 18-110_k141_225534 | Not-determined |
| 18-110_k141_30430 | Not-determined |
| 18-110_k141_30431 | Not-determined |
| 18-110_k141_214014 | Not-determined |
| 18-110_k141_225536 | Not-determined |
| 18-110_k141_180583 | Not-determined |
| 18-110_k141_76135 | Not-determined |
| 18-110_k141_6190 | Low-quality |
| 18-110_k141_145252 | Not-determined |
| 18-110_k141_191253 | Low-quality |
| 18-110_k141_180591 | Low-quality |
| 18-110_k141_191257 | Not-determined |
| 18-110_k141_156715 | Not-determined |
| 18-110_k141_225555 | Not-determined |
| 18-110_k141_156726 | Not-determined |
| 18-110_k141_121753 | Not-determined |
| 18-110_k141_121754 | Not-determined |
| 18-110_k141_271801 | Not-determined |
| 18-110_k141_156730 | Not-determined |
| 18-110_k141_271803 | Not-determined |
| 18-110_k141_99099 | Not-determined |
| 18-110_k141_17031 | Not-determined |
| 18-110_k141_180607 | Not-determined |
| 18-110_k141_110103 | Not-determined |
| 18-110_k141_99108 | Not-determined |
| 18-110_k141_201548 | Low-quality |
| 18-110_k141_30471 | Not-determined |
| 18-110_k141_110107 | Not-determined |
| 18-110_k141_168140 | Not-determined |
| 18-110_k141_133729 | Not-determined |
| 18-110_k141_110114 | Low-quality |
| 18-110_k141_64781 | Not-determined |
| 18-110_k141_225589 | Not-determined |
| 18-110_k141_17053 | Not-determined |
| 18-110_k141_145278 | Low-quality |
| 18-110_k141_225592 | Not-determined |
| 18-110_k141_76155 | Not-determined |
| 18-110_k141_76156 | Not-determined |
| 18-110_k141_41666 | Not-determined |
| 18-110_k141_214040 | Not-determined |
| 18-110_k141_6219 | Not-determined |
| 18-110_k141_271814 | Not-determined |
| 18-110_k141_86923 | Not-determined |
| 18-110_k141_41674 | Not-determined |
| 18-110_k141_145293 | Not-determined |
| 18-110_k141_168174 | Not-determined |
| 18-110_k141_17071 | Not-determined |
| 18-110_k141_6228 | Low-quality |
| 18-110_k141_271825 | Not-determined |
| 18-110_k141_168183 | Not-determined |
| 18-110_k141_191274 | Not-determined |
| 18-110_k141_76164 | Not-determined |
| 18-110_k141_76165 | Not-determined |
| 18-110_k141_76169 | Low-quality |
| 18-110_k141_191279 | Not-determined |
| 18-110_k141_191280 | Low-quality |
| 18-110_k141_76171 | Not-determined |
| 18-110_k141_121758 | Low-quality |
| 18-110_k141_86937 | Not-determined |
| 18-110_k141_76181 | Not-determined |
| 18-110_k141_86942 | Not-determined |
| 18-110_k141_168199 | Not-determined |
| 18-110_k141_6239 | Not-determined |
| 18-110_k141_225607 | Not-determined |
| 18-110_k141_201577 | Not-determined |
| 18-110_k141_258800 | Not-determined |
| 18-110_k141_99152 | Not-determined |
| 18-110_k141_168216 | Not-determined |
| 18-110_k141_76199 | Not-determined |
| 18-110_k141_145320 | Not-determined |
| 18-102-final-viral-combined_k141_52006\|\|0_partial | Medium-quality |
| 18-110_k141_271833 | Not-determined |
| 18-110_k141_271835 | Not-determined |
| 18-110_k141_271838 | Not-determined |
| 18-110_k141_76213 | Not-determined |
| 18-110_k141_52795 | Not-determined |
| 18-110_k141_225621 | Low-quality |
| 18-110_k141_145334 | Not-determined |
| 18-110_k141_99161 | Low-quality |
| 18-110_k141_99162 | Not-determined |
| 18-110_k141_191296 | Not-determined |
| 18-110_k141_258821 | Not-determined |
| 18-110_k141_52799 | Not-determined |
| 18-110_k141_99165 | Not-determined |
| 18-110_k141_271847 | Not-determined |
| 18-110_k141_145341 | Low-quality |
| 18-110_k141_225627 | Not-determined |
| 18-110_k141_133788 | Not-determined |
| 18-110_k141_271863 | Not-determined |
| 18-110_k141_201593 | Not-determined |
| 18-110_k141_168228 | Not-determined |
| 18-110_k141_271865 | Low-quality |
| 18-110_k141_168238 | Not-determined |
| 18-110_k141_86967 | Not-determined |
| 18-110_k141_121805 | Not-determined |
| 18-110_k141_145362 | Not-determined |
| 18-110_k141_168240 | Not-determined |
| 18-110_k141_52819 | Not-determined |
| 18-110_k141_64846 | Not-determined |
| 18-110_k141_41734 | Not-determined |
| 18-110_k141_110135 | Low-quality |
| 18-110_k141_52822 | Not-determined |
| 18-110_k141_191310 | Not-determined |
| 18-110_k141_6282 | Low-quality |
| 18-110_k141_180679 | Not-determined |
| 18-110_k141_271874 | Not-determined |
| 18-110_k141_191314 | Not-determined |
| 18-110_k141_271876 | Not-determined |
| 18-110_k141_41740 | Not-determined |
| 18-110_k141_133801 | Not-determined |
| 18-110_k141_30526 | Not-determined |
| 18-110_k141_180682 | Not-determined |
| 18-110_k141_76243 | Not-determined |
| 18-110_k141_110138 | Not-determined |
| 18-110_k141_6287 | Not-determined |
| 18-110_k141_86982 | Low-quality |
| 18-110_k141_86985 | Not-determined |
| 18-110_k141_214093 | Low-quality |
| 18-110_k141_76251 | Not-determined |
| 18-110_k141_41750 | Not-determined |
| 18-110_k141_121832 | Not-determined |
| 18-110_k141_30538 | Not-determined |
| 18-110_k141_214101 | Not-determined |
| 18-110_k141_214104 | Not-determined |
| 18-110_k141_145376 | Not-determined |
| 18-110_k141_271889 | Not-determined |
| 18-110_k141_86994 | Not-determined |
| 18-110_k141_156827 | Not-determined |
| 18-110_k141_133824 | Not-determined |
| 18-110_k141_258858 | Not-determined |
| 18-110_k141_87002 | Low-quality |
| 18-110_k141_156833 | Not-determined |
| 18-110_k141_258863 | Not-determined |
| 18-110_k141_87008 | Not-determined |
| 18-110_k141_180699 | Not-determined |
| 18-110_k141_180701 | Low-quality |
| 18-110_k141_30546 | Not-determined |
| 18-110_k141_133836 | Not-determined |
| 18-110_k141_156841 | Low-quality |
| 18-110_k141_30549 | Not-determined |
| 18-110_k141_225646 | Not-determined |
| 18-110_k141_30551 | Not-determined |
| 18-110_k141_191351 | Not-determined |
| 18-110_k141_168289 | Not-determined |
| 18-110_k141_191353 | Not-determined |
| 18-110_k141_271894 | Not-determined |
| 18-110_k141_271896 | Not-determined |
| 18-110_k141_258878 | Not-determined |
| 18-110_k141_64866 | Not-determined |
| 18-110_k141_271898 | Not-determined |
| 18-110_k141_145402 | Not-determined |
| 18-110_k141_168296 | Not-determined |
| 18-110_k141_214120 | Not-determined |
| 18-110_k141_133847 | Not-determined |
| 18-110_k141_271906 | Not-determined |
| 18-110_k141_214126 | Not-determined |
| 18-110_k141_17109 | Not-determined |
| 18-110_k141_17111 | Low-quality |
| 18-110_k141_214128 | Not-determined |
| 18-110_k141_30577 | Not-determined |
| 18-110_k141_271911 | Not-determined |
| 18-110_k141_225666 | Not-determined |
| 18-110_k141_99202 | Not-determined |
| 18-110_k141_168301 | Low-quality |
| 18-110_k141_6326 | Not-determined |
| 18-110_k141_6328 | Not-determined |
| 18-110_k141_99211 | Not-determined |
| 18-110_k141_6332 | Not-determined |
| 18-110_k141_52867 | Not-determined |
| 18-110_k141_17129 | Not-determined |
| 18-110_k141_17130 | Not-determined |
| 18-110_k141_76297 | Not-determined |
| 18-110_k141_121878 | Low-quality |
| 18-110_k141_76298 | Not-determined |
| 18-110_k141_87041 | Not-determined |
| 18-110_k141_99212 | Not-determined |
| 18-110_k141_30607 | Not-determined |
| 18-110_k141_145433 | Not-determined |
| 18-110_k141_180722 | Not-determined |
| 18-110_k141_258900 | Not-determined |
| 18-110_k141_64871 | Not-determined |
| 18-110_k141_87047 | Not-determined |
| 18-110_k141_87048 | Not-determined |
| 18-110_k141_214141 | Low-quality |
| 18-110_k141_99220 | Not-determined |
| 18-110_k141_249005 | Not-determined |
| 18-110_k141_237774 | Not-determined |
| 18-110_k141_214147 | Not-determined |
| 18-110_k141_133888 | Not-determined |
| 18-110_k141_110181 | Not-determined |
| 18-110_k141_249012 | Not-determined |
| 18-110_k141_249014 | Not-determined |
| 18-110_k141_87059 | Not-determined |
| 18-110_k141_258913 | Not-determined |
| 18-110_k141_30636 | Not-determined |
| 18-110_k141_191397 | Not-determined |
| 18-110_k141_258915 | Not-determined |
| 18-110_k141_180746 | Not-determined |
| 18-110_k141_225703 | Not-determined |
| 18-110_k141_249018 | Not-determined |
| 18-110_k141_30640 | Not-determined |
| 18-110_k141_87067 | Not-determined |
| 18-110_k141_214159 | Not-determined |
| 18-110_k141_258926 | Not-determined |
| 18-110_k141_258933 | Not-determined |
| 18-110_k141_30652 | Not-determined |
| 18-110_k141_214166 | Not-determined |
| 18-110_k141_258938 | Not-determined |
| 18-110_k141_64886 | Low-quality |
| 18-110_k141_168339 | Not-determined |
| 18-110_k141_30654 | Not-determined |
| 18-110_k141_156888 | Not-determined |
| 18-110_k141_145451 | Not-determined |
| 18-110_k141_214176 | Not-determined |
| 18-110_k141_64897 | Not-determined |
| 18-110_k141_249033 | Not-determined |
| 18-110_k141_201604 | Not-determined |
| 18-110_k141_30658 | Not-determined |
| 18-110_k141_201606 | Not-determined |
| 18-110_k141_121904 | Not-determined |
| 18-110_k141_201607 | Not-determined |
| 18-110_k141_76301 | Not-determined |
| 18-110_k141_145458 | Low-quality |
| 18-110_k141_258950 | Not-determined |
| 18-110_k141_6369 | Not-determined |
| 18-110_k141_258955 | Not-determined |
| 18-110_k141_6375 | Not-determined |
| 18-110_k141_191425 | Not-determined |
| 18-110_k141_237799 | Not-determined |
| 18-110_k141_87108 | Not-determined |
| 18-110_k141_76322 | Not-determined |
| 18-110_k141_145483 | Low-quality |
| 18-110_k141_201625 | Not-determined |
| 18-110_k141_133900 | Not-determined |
| 18-110_k141_258960 | Not-determined |
| 18-110_k141_168369 | Low-quality |
| 18-110_k141_76324 | Not-determined |
| 18-110_k141_76325 | Not-determined |
| 18-110_k141_76326 | Not-determined |
| 18-110_k141_168371 | Not-determined |
| 18-110_k141_87129 | Not-determined |
| 18-110_k141_64930 | Not-determined |
| 18-110_k141_133918 | Not-determined |
| 18-110_k141_225745 | Low-quality |
| 18-110_k141_76328 | Low-quality |
| 18-110_k141_249074 | Not-determined |
| 18-110_k141_225750 | Not-determined |
| 18-110_k141_133927 | Not-determined |
| 18-110_k141_191461 | Not-determined |
| 18-110_k141_133929 | Not-determined |
| 18-110_k141_121920 | Not-determined |
| 18-110_k141_99263 | Low-quality |
| 18-110_k141_168379 | Not-determined |
| 18-110_k141_52927 | Not-determined |
| 18-110_k141_237828 | Not-determined |
| 18-110_k141_30704 | Not-determined |
| 18-110_k141_110226 | Not-determined |
| 18-110_k141_121932 | Low-quality |
| 18-110_k141_237834 | Not-determined |
| 18-110_k141_121936 | Not-determined |
| 18-110_k141_110232 | Not-determined |
| 18-110_k141_133941 | Low-quality |
| 18-110_k141_87155 | Not-determined |
| 18-110_k141_64951 | Not-determined |
| 18-110_k141_133947 | Not-determined |
| 18-110_k141_110237 | Not-determined |
| 18-110_k141_191468 | Not-determined |
| 18-110_k141_191469 | Not-determined |
| 18-110_k141_156949 | Not-determined |
| 18-110_k141_52947 | Low-quality |
| 18-110_k141_191471 | Not-determined |
| 18-110_k141_87159 | Not-determined |
| 18-110_k141_156950 | Low-quality |
| 18-110_k141_76344 | Not-determined |
| 18-110_k141_121946 | Not-determined |
| 18-110_k141_258979 | Not-determined |
| 18-110_k141_258981 | Not-determined |
| 18-110_k141_191480 | Not-determined |
| 18-110_k141_168408 | Not-determined |
| 18-110_k141_52956 | Not-determined |
| 18-110_k141_76360 | Not-determined |
| 18-110_k141_121952 | Not-determined |
| 18-110_k141_133963 | Not-determined |
| 18-110_k141_249103 | Not-determined |
| 18-110_k141_76363 | Not-determined |
| 18-110_k141_121953 | Not-determined |
| 18-110_k141_249105 | Not-determined |
| 18-110_k141_272007 | Not-determined |
| 18-110_k141_249106 | Not-determined |
| 18-110_k141_191486 | Low-quality |
| 18-110_k141_191488 | Not-determined |
| 18-110_k141_52963 | Low-quality |
| 18-110_k141_121956 | Low-quality |
| 18-110_k141_168421 | Not-determined |
| 18-110_k141_99298 | Not-determined |
| 18-110_k141_64971 | Not-determined |
| 18-110_k141_99302 | Not-determined |
| 18-110_k141_121962 | Not-determined |
| 18-110_k141_121963 | Not-determined |
| 18-110_k141_110259 | Low-quality |
| 18-110_k141_64974 | Low-quality |
| 18-110_k141_180808 | Not-determined |
| 18-110_k141_110264 | Not-determined |
| 18-110_k141_76382 | Not-determined |
| 18-110_k141_76384 | Not-determined |
| 18-110_k141_225769 | Not-determined |
| 18-110_k141_99309 | Not-determined |
| 18-110_k141_87192 | Not-determined |
| 18-110_k141_121971 | Not-determined |
| 18-110_k141_87193 | Not-determined |
| 18-110_k141_133980 | Not-determined |
| 18-110_k141_156984 | Low-quality |
| 18-110_k141_133984 | Not-determined |
| 18-110_k141_99322 | Not-determined |
| 18-110_k141_87205 | Not-determined |
| 18-110_k141_225781 | Not-determined |
| 18-110_k141_76394 | Not-determined |
| 18-110_k141_76397 | Not-determined |
| 18-110_k141_52987 | Not-determined |
| 18-110_k141_180826 | Not-determined |
| 18-110_k141_180827 | Not-determined |
| 18-110_k141_133993 | Not-determined |
| 18-110_k141_87212 | Not-determined |
| 18-110_k141_17150 | Not-determined |
| 18-110_k141_30728 | Not-determined |
| 18-110_k141_156998 | Not-determined |
| 18-110_k141_225785 | Not-determined |
| 18-110_k141_30731 | Not-determined |
| 18-110_k141_225787 | Not-determined |
| 18-110_k141_87219 | Not-determined |
| 18-110_k141_30736 | Not-determined |
| 18-110_k141_249137 | Not-determined |
| 18-110_k141_6404 | Not-determined |
| 18-110_k141_134001 | Not-determined |
| 18-110_k141_201683 | Not-determined |
| 18-110_k141_201685 | Not-determined |
| 18-110_k141_53004 | Low-quality |
| 18-110_k141_87234 | Not-determined |
| 18-110_k141_225795 | Not-determined |
| 18-110_k141_157005 | Not-determined |
| 18-110_k141_17159 | Not-determined |
| 18-110_k141_99338 | Not-determined |
| 18-110_k141_76412 | Not-determined |
| 18-110_k141_76414 | Not-determined |
| 18-110_k141_76415 | Not-determined |
| 18-110_k141_145529 | Not-determined |
| 18-110_k141_145530 | Not-determined |
| 18-110_k141_65010 | Not-determined |

| | |
|---|---|
| 18-110_k141_225811 | Low-quality |
| 18-110_k141_237911 | Not-determined |
| 18-110_k141_214242 | Not-determined |
| 18-110_k141_201709 | Not-determined |
| 18-110_k141_65016 | Not-determined |
| 18-110_k141_214246 | Low-quality |
| 18-110_k141_134009 | Not-determined |
| 18-110_k141_53027 | Not-determined |
| 18-110_k141_53032 | Not-determined |
| 18-110_k141_30749 | Not-determined |
| 18-110_k141_259043 | Not-determined |
| 18-110_k141_122026 | Not-determined |
| 18-110_k141_225819 | Not-determined |
| 18-110_k141_191524 | Not-determined |
| 18-110_k141_191526 | Not-determined |
| 18-110_k141_6428 | Not-determined |
| 18-110_k141_191527 | Not-determined |
| 18-110_k141_214250 | Not-determined |
| 18-110_k141_157032 | Not-determined |
| 18-110_k141_191533 | Not-determined |
| 18-110_k141_65027 | Not-determined |
| 18-110_k141_30756 | Not-determined |
| 18-110_k141_214262 | Low-quality |
| 18-110_k141_134029 | Not-determined |
| 18-110_k141_6436 | Not-determined |
| 18-110_k141_157049 | Low-quality |
| 18-110_k141_191537 | Low-quality |
| 18-110_k141_237941 | Not-determined |
| 18-110_k141_30768 | Low-quality |
| 18-110_k141_180882 | Not-determined |
| 18-110_k141_157057 | Not-determined |
| 18-110_k141_99376 | Not-determined |
| 18-110_k141_249154 | Not-determined |
| 18-110_k141_157064 | Not-determined |
| 18-110_k141_201720 | Low-quality |
| 18-110_k141_87273 | Not-determined |
| 18-110_k141_157070 | Not-determined |
| 18-110_k141_259057 | Not-determined |
| 18-110_k141_6450 | Not-determined |
| 18-110_k141_134049 | Not-determined |
| 18-110_k141_249163 | Not-determined |
| 18-110_k141_76448 | Not-determined |
| 18-110_k141_65057 | Not-determined |
| 18-110_k141_272095 | Not-determined |
| 18-110_k141_65061 | Not-determined |
| 18-110_k141_134064 | Not-determined |
| 18-110_k141_65065 | Not-determined |
| 18-110_k141_87298 | Not-determined |
| 18-110_k141_65066 | Not-determined |
| 18-110_k141_110344 | Not-determined |
| 18-110_k141_87301 | Not-determined |
| 18-110_k141_87303 | Not-determined |
| 18-110_k141_134073 | Low-quality |
| 18-110_k141_65070 | Not-determined |
| 18-110_k141_17220 | Not-determined |
| 18-110_k141_65073 | Not-determined |
| 18-110_k141_259066 | Not-determined |
| 18-110_k141_180904 | Not-determined |
| 18-110_k141_53051 | Not-determined |
| 18-110_k141_53052 | Not-determined |
| 18-110_k141_17223 | Not-determined |
| 18-110_k141_249180 | Not-determined |
| 18-110_k141_180912 | Not-determined |
| 18-110_k141_249182 | Not-determined |
| 18-110_k141_191568 | Not-determined |
| 18-110_k141_214280 | Not-determined |
| 18-110_k141_237960 | Not-determined |
| 18-110_k141_87325 | Not-determined |
| 18-110_k141_157106 | Not-determined |
| 18-110_k141_65081 | Low-quality |
| 18-110_k141_110359 | Not-determined |
| 18-110_k141_214281 | Not-determined |
| 18-110_k141_17238 | Not-determined |
| 18-110_k141_30809 | Not-determined |
| 18-110_k141_76494 | Not-determined |
| 18-110_k141_134084 | Not-determined |
| 18-110_k141_180919 | Not-determined |
| 18-110_k141_249203 | Not-determined |
| 18-110_k141_157122 | Not-determined |
| 18-110_k141_249205 | Not-determined |
| 18-110_k141_249209 | Not-determined |
| 18-110_k141_259098 | Not-determined |
| 18-110_k141_30821 | Not-determined |
| 18-110_k141_17249 | Not-determined |
| 18-110_k141_87352 | Not-determined |
| 18-110_k141_53078 | Not-determined |
| 18-110_k141_87360 | Not-determined |
| 18-110_k141_201770 | Low-quality |
| 18-110_k141_259105 | Not-determined |
| 18-110_k141_30834 | Not-determined |
| 18-110_k141_180923 | Not-determined |
| 18-110_k141_180925 | Not-determined |
| 18-110_k141_30842 | Not-determined |
| 18-110_k141_87370 | Not-determined |
| 18-110_k141_259113 | Not-determined |
| 18-110_k141_214302 | Not-determined |
| 18-110_k141_225845 | Low-quality |
| 18-110_k141_53083 | Not-determined |
| 18-110_k141_225846 | Low-quality |
| 18-110_k141_214308 | Low-quality |
| 18-110_k141_259133 | Not-determined |
| 18-110_k141_17257 | Not-determined |
| 18-110_k141_214316 | Not-determined |
| 18-110_k141_53090 | Not-determined |
| 18-110_k141_87387 | Not-determined |
| 18-110_k141_168464 | Not-determined |
| 18-110_k141_168466 | Not-determined |
| 18-110_k141_65119 | Not-determined |
| 18-110_k141_191622 | Not-determined |
| 18-110_k141_201797 | Not-determined |
| 18-110_k141_53100 | Not-determined |
| 18-110_k141_201799 | Not-determined |
| 18-110_k141_201801 | Not-determined |
| 18-110_k141_122052 | Not-determined |
| 18-110_k141_17272 | Not-determined |
| 18-110_k141_191631 | Not-determined |
| 18-110_k141_134102 | Not-determined |
| 18-110_k141_122058 | Not-determined |
| 18-110_k141_110386 | Not-determined |
| 18-110_k141_76541 | Not-determined |
| 18-110_k141_41759 | Not-determined |
| 18-110_k141_122060 | Low-quality |
| 18-110_k141_87405 | Low-quality |
| 18-110_k141_122062 | Not-determined |
| 18-110_k141_259149 | Not-determined |
| 18-110_k141_122063 | Low-quality |
| 18-110_k141_168480 | Not-determined |
| 18-110_k141_87411 | Not-determined |
| 18-110_k141_99419 | Not-determined |
| 18-110_k141_122068 | Not-determined |
| 18-110_k141_259154 | Not-determined |
| 18-110_k141_249219 | Not-determined |
| 18-110_k141_272140 | Not-determined |
| 18-110_k141_157150 | Not-determined |
| 18-110_k141_76556 | Not-determined |
| 18-110_k141_134114 | Not-determined |
| 18-110_k141_17294 | Not-determined |
| 18-110_k141_134120 | Not-determined |
| 18-110_k141_41782 | Not-determined |
| 18-110_k141_41783 | Not-determined |
| 18-110_k141_134121 | Low-quality |
| 18-110_k141_41784 | Not-determined |
| 18-110_k141_259168 | Not-determined |
| 18-110_k141_214343 | Not-determined |
| 18-110_k141_87428 | Not-determined |
| 18-110_k141_272149 | Not-determined |
| 18-110_k141_41793 | Not-determined |
| 18-110_k141_272152 | Not-determined |
| 18-110_k141_17302 | Not-determined |
| 18-110_k141_122081 | Not-determined |
| 18-110_k141_99447 | Not-determined |
| 18-110_k141_17310 | Not-determined |
| 18-110_k141_180967 | Not-determined |
| 18-110_k141_17312 | Not-determined |
| 18-110_k141_145655 | Not-determined |
| 18-110_k141_65145 | Not-determined |
| 18-110_k141_259172 | Low-quality |
| 18-110_k141_87440 | Not-determined |
| 18-110_k141_65147 | Not-determined |
| 18-110_k141_180975 | Not-determined |
| 18-110_k141_157166 | Not-determined |
| 18-110_k141_201836 | Not-determined |
| 18-110_k141_168496 | Not-determined |
| 18-110_k141_201839 | Not-determined |
| 18-110_k141_180980 | Not-determined |
| 18-110_k141_122113 | Low-quality |
| 18-110_k141_30890 | Not-determined |
| 18-110_k141_259176 | Not-determined |
| 18-110_k141_76567 | Not-determined |
| 18-110_k141_65153 | Not-determined |
| 18-110_k141_41809 | Not-determined |
| 18-110_k141_157173 | Not-determined |
| 18-110_k141_122123 | Low-quality |
| 18-110_k141_225892 | Low-quality |
| 18-110_k141_272169 | Not-determined |
| 18-110_k141_238063 | Not-determined |
| 18-110_k141_110421 | Not-determined |
| 18-110_k141_238065 | Not-determined |
| 18-110_k141_6472 | Not-determined |
| 18-110_k141_110428 | Not-determined |
| 18-110_k141_30920 | Not-determined |
| 18-110_k141_249227 | Not-determined |
| 18-110_k141_201860 | Low-quality |
| 18-110_k141_145693 | Low-quality |
| 18-110_k141_110432 | Not-determined |
| 18-110_k141_181010 | Not-determined |
| 18-110_k141_87481 | Not-determined |
| 18-110_k141_134159 | Not-determined |
| 18-110_k141_238084 | Not-determined |
| 18-110_k141_181015 | Not-determined |
| 18-110_k141_6493 | Not-determined |
| 18-110_k141_65183 | Not-determined |
| 18-110_k141_65184 | Not-determined |
| 18-110_k141_145708 | Not-determined |
| 18-110_k141_76579 | Not-determined |
| 18-110_k141_134165 | Not-determined |
| 18-110_k141_6500 | Not-determined |
| 18-110_k141_168520 | Not-determined |
| 18-110_k141_122129 | Low-quality |
| 18-110_k141_65191 | Not-determined |
| 18-110_k141_134170 | Not-determined |
| 18-110_k141_157192 | Not-determined |
| 18-110_k141_168526 | Not-determined |
| 18-110_k141_122133 | Not-determined |
| 18-110_k141_87489 | Low-quality |
| 18-110_k141_99470 | Not-determined |
| 18-110_k141_181038 | Not-determined |
| 18-110_k141_157201 | Low-quality |
| 18-110_k141_87492 | Not-determined |
| 18-110_k141_201869 | Not-determined |
| 18-110_k141_191694 | Not-determined |
| 18-110_k141_272192 | Not-determined |
| 18-110_k141_134178 | Not-determined |
| 18-110_k141_134179 | Not-determined |
| 18-110_k141_99485 | Not-determined |
| 18-110_k141_6511 | Not-determined |
| 18-110_k141_110457 | Not-determined |
| 18-110_k141_214367 | Not-determined |
| 18-110_k141_17355 | Not-determined |
| 18-110_k141_87505 | Not-determined |
| 18-110_k141_181050 | Not-determined |
| 18-110_k141_181051 | Not-determined |
| 18-110_k141_145737 | Not-determined |
| 18-110_k141_201883 | Not-determined |
| 18-110_k141_157214 | Not-determined |
| 18-110_k141_53151 | Not-determined |
| 18-110_k141_99494 | Not-determined |
| 18-110_k141_87518 | Not-determined |
| 18-110_k141_99497 | Not-determined |
| 18-110_k141_6523 | Not-determined |
| 18-110_k141_110478 | Not-determined |
| 18-110_k141_145755 | Not-determined |
| 18-110_k141_214378 | Not-determined |
| 18-110_k141_134195 | Not-determined |
| 18-110_k141_214380 | Not-determined |
| 18-110_k141_122152 | Low-quality |
| 18-110_k141_17366 | Not-determined |
| 18-110_k141_168564 | Not-determined |
| 18-110_k141_145762 | Not-determined |
| 18-110_k141_30979 | Not-determined |
| 18-110_k141_110491 | Not-determined |
| 18-110_k141_272218 | Not-determined |
| 18-110_k141_41821 | Low-quality |
| 18-110_k141_249264 | Not-determined |
| 18-110_k141_30988 | Not-determined |
| 18-110_k141_238118 | Not-determined |
| 18-110_k141_134208 | Not-determined |
| 18-110_k141_99506 | Not-determined |
| 18-110_k141_191702 | Not-determined |
| 18-110_k141_134216 | Not-determined |
| 18-110_k141_110493 | Low-quality |
| 18-110_k141_249271 | Not-determined |
| 18-110_k141_30993 | Not-determined |
| 18-110_k141_145781 | Not-determined |
| 18-110_k141_87551 | Not-determined |
| 18-110_k141_168586 | Not-determined |
| 18-110_k141_87552 | Not-determined |
| 18-110_k141_99518 | Not-determined |
| 18-110_k141_201884 | Low-quality |
| 18-110_k141_110501 | Not-determined |
| 18-110_k141_122171 | Low-quality |
| 18-110_k141_145787 | Not-determined |
| 18-110_k141_168595 | Low-quality |
| 18-110_k141_181081 | Not-determined |
| 18-110_k141_122177 | Not-determined |
| 18-110_k141_214399 | Not-determined |
| 18-110_k141_65214 | Not-determined |
| 18-110_k141_53192 | Not-determined |
| 18-110_k141_99527 | Low-quality |
| 18-110_k141_99535 | Not-determined |
| 18-110_k141_134233 | Not-determined |
| 18-110_k141_122182 | Not-determined |
| 18-110_k141_99539 | Not-determined |
| 18-110_k141_41849 | Not-determined |
| 18-110_k141_65233 | Low-quality |
| 18-110_k141_249296 | Not-determined |
| 18-110_k141_225924 | Not-determined |
| 18-110_k141_238142 | Not-determined |
| 18-110_k141_31017 | Not-determined |
| 18-110_k141_134246 | Not-determined |
| 18-110_k141_65240 | Not-determined |
| 18-110_k141_65241 | Not-determined |
| 18-110_k141_87585 | Not-determined |
| 18-110_k141_201903 | Not-determined |
| 18-110_k141_238158 | Low-quality |
| 18-110_k141_214416 | Not-determined |
| 18-110_k141_181122 | Not-determined |
| 18-110_k141_181126 | Low-quality |
| 18-110_k141_214422 | Not-determined |
| 18-110_k141_238168 | Not-determined |
| 18-110_k141_238169 | Not-determined |
| 18-110_k141_249310 | Not-determined |
| 18-110_k141_122210 | Not-determined |
| 18-110_k141_214429 | Not-determined |
| 18-110_k141_76637 | Not-determined |
| 18-110_k141_122212 | Low-quality |
| 18-110_k141_201916 | Not-determined |
| 18-110_k141_87606 | Not-determined |
| 18-110_k141_65258 | Not-determined |
| 18-110_k141_31035 | Not-determined |
| 18-110_k141_31038 | Not-determined |
| 18-110_k141_238181 | Not-determined |
| 18-110_k141_249314 | Not-determined |
| 18-110_k141_134259 | Not-determined |
| 18-110_k141_225950 | Not-determined |
| 18-110_k141_76651 | Not-determined |
| 18-110_k141_201926 | Not-determined |
| 18-110_k141_145825 | Not-determined |
| 18-110_k141_181139 | Not-determined |
| 18-110_k141_157281 | Not-determined |
| 18-110_k141_238199 | Not-determined |
| 18-110_k141_214445 | Not-determined |
| 18-110_k141_145828 | Not-determined |
| 18-110_k141_272256 | Not-determined |
| 18-110_k141_122230 | Low-quality |
| 18-110_k141_214449 | Not-determined |
| 18-110_k141_31044 | Not-determined |
| 18-110_k141_168663 | Not-determined |
| 18-110_k141_238206 | Not-determined |
| 18-110_k141_225962 | Not-determined |
| 18-110_k141_145832 | Not-determined |
| 18-110_k141_76667 | Low-quality |
| 18-110_k141_145833 | Not-determined |
| 18-110_k141_145837 | Not-determined |
| 18-110_k141_249323 | Low-quality |
| 18-110_k141_31055 | Not-determined |
| 18-110_k141_272267 | Not-determined |
| 18-110_k141_249325 | Not-determined |
| 18-110_k141_157295 | Not-determined |
| 18-110_k141_214462 | Not-determined |
| 18-110_k141_99563 | Not-determined |
| 18-110_k141_259205 | Low-quality |
| 18-110_k141_214464 | Low-quality |
| 18-110_k141_145847 | Not-determined |
| 18-110_k141_225979 | Not-determined |
| 18-110_k141_145848 | Low-quality |
| 18-110_k141_122246 | Not-determined |
| 18-110_k141_122247 | Not-determined |
| 18-110_k141_214468 | Not-determined |
| 18-110_k141_238223 | Not-determined |
| 18-110_k141_225981 | Not-determined |
| 18-110_k141_214470 | Not-determined |
| 18-110_k141_99569 | Not-determined |
| 18-110_k141_76674 | Not-determined |
| 18-110_k141_249341 | Not-determined |
| 18-110_k141_157310 | Not-determined |
| 18-110_k141_214478 | Not-determined |
| 18-110_k141_31082 | Low-quality |
| 18-110_k141_225992 | Not-determined |
| 18-110_k141_41893 | Not-determined |
| 18-110_k141_41894 | Not-determined |
| 18-110_k141_214485 | Not-determined |
| 18-110_k141_201947 | Not-determined |
| 18-110_k141_41899 | Not-determined |
| 18-110_k141_214489 | Not-determined |
| 18-110_k141_259217 | Not-determined |
| 18-110_k141_99580 | Not-determined |
| 18-110_k141_238238 | Not-determined |
| 18-110_k141_41904 | Not-determined |
| 18-110_k141_41907 | Not-determined |
| 18-110_k141_238242 | Low-quality |
| 18-110_k141_65302 | Not-determined |
| 18-110_k141_122271 | Not-determined |
| 18-110_k141_226004 | Not-determined |
| 18-110_k141_76685 | Not-determined |
| 18-110_k141_65311 | Not-determined |
| 18-110_k141_226013 | Low-quality |
| 18-110_k141_134288 | Not-determined |
| 18-110_k141_145877 | Not-determined |
| 18-110_k141_201954 | Not-determined |
| 18-110_k141_31103 | Low-quality |
| 18-110_k141_99587 | Not-determined |
| 18-110_k141_99589 | Not-determined |
| 18-110_k141_259228 | Not-determined |
| 18-110_k141_238252 | Not-determined |
| 18-110_k141_41928 | Not-determined |
| 18-110_k141_65326 | Not-determined |
| 18-110_k141_226024 | Low-quality |
| 18-110_k141_214505 | Not-determined |
| 18-110_k141_41934 | Not-determined |
| 18-110_k141_249372 | Low-quality |
| 18-110_k141_99605 | Not-determined |
| 18-110_k141_41939 | Not-determined |
| 18-110_k141_99608 | Not-determined |
| 18-110_k141_249375 | Not-determined |
| 18-110_k141_41940 | Low-quality |
| 18-110_k141_145885 | Not-determined |
| 18-110_k141_249380 | Not-determined |
| 18-110_k141_134305 | Not-determined |
| 18-110_k141_249384 | Not-determined |
| 18-110_k141_122303 | Not-determined |
| 18-110_k141_238275 | Not-determined |
| 18-110_k141_31107 | Low-quality |
| 18-110_k141_134311 | Not-determined |
| 18-110_k141_168704 | Not-determined |
| 18-110_k141_226041 | Not-determined |
| 18-110_k141_249392 | Not-determined |
| 18-110_k141_168705 | Not-determined |
| 18-110_k141_41955 | Not-determined |
| 18-110_k141_122308 | Not-determined |
| 18-110_k141_41959 | Not-determined |
| 18-110_k141_41962 | Not-determined |
| 18-110_k141_201970 | Not-determined |
| 18-110_k141_238289 | Not-determined |
| 18-110_k141_201974 | Not-determined |
| 18-110_k141_41967 | Not-determined |
| 18-110_k141_168715 | Low-quality |
| 18-110_k141_238294 | Not-determined |
| 18-110_k141_168724 | Not-determined |
| 18-110_k141_17384 | Not-determined |
| 18-110_k141_238296 | Low-quality |
| 18-110_k141_157339 | Not-determined |
| 18-110_k141_145896 | Not-determined |
| 18-110_k141_249405 | Not-determined |
| 18-110_k141_17390 | Not-determined |
| 18-110_k141_145899 | Not-determined |
| 18-110_k141_122324 | Not-determined |
| 18-110_k141_214536 | Not-determined |
| 18-110_k141_65354 | Not-determined |
| 18-110_k141_53272 | Not-determined |
| 18-110_k141_31140 | Not-determined |
| 18-110_k141_249413 | Not-determined |
| 18-110_k141_134353 | Not-determined |
| 18-110_k141_76745 | Not-determined |
| 18-110_k141_201993 | Not-determined |
| 18-110_k141_31145 | Not-determined |
| 18-110_k141_41980 | Not-determined |
| 18-110_k141_168738 | Low-quality |
| 18-110_k141_201996 | Not-determined |
| 18-110_k141_201999 | Not-determined |
| 18-110_k141_272279 | Not-determined |
| 18-110_k141_259259 | Not-determined |
| 18-110_k141_238306 | Not-determined |
| 18-110_k141_99613 | Not-determined |
| 18-110_k141_249420 | Not-determined |
| 18-110_k141_249421 | Not-determined |
| 18-110_k141_259262 | Not-determined |
| 18-110_k141_53286 | Not-determined |
| 18-110_k141_53288 | Not-determined |
| 18-110_k141_202001 | Not-determined |
| 18-110_k141_157367 | Low-quality |
| 18-110_k141_65379 | Not-determined |
| 18-110_k141_259268 | Not-determined |
| 18-110_k141_53294 | Low-quality |
| 18-110_k141_214549 | Not-determined |
| 18-110_k141_214550 | Low-quality |
| 18-110_k141_214554 | Low-quality |
| 18-110_k141_145925 | Not-determined |
| 18-110_k141_134373 | Not-determined |
| 18-110_k141_249431 | Not-determined |
| 18-110_k141_238316 | Not-determined |
| 18-110_k141_122350 | Not-determined |
| 18-110_k141_122351 | Not-determined |
| 18-110_k141_42004 | Not-determined |
| 18-110_k141_249435 | Low-quality |
| 18-110_k141_65393 | Not-determined |
| 18-110_k141_238321 | Not-determined |
| 18-110_k141_42009 | Not-determined |
| 18-110_k141_65396 | Not-determined |
| 18-110_k141_122355 | Low-quality |
| 18-110_k141_99639 | Not-determined |
| 18-110_k141_272297 | Not-determined |
| 18-110_k141_214583 | Not-determined |
| 18-110_k141_65409 | Not-determined |
| 18-110_k141_145942 | Not-determined |
| 18-110_k141_168777 | Not-determined |
| 18-110_k141_99644 | Not-determined |
| 18-110_k141_259278 | Not-determined |
| 18-110_k141_214594 | Not-determined |
| 18-110_k141_31180 | Not-determined |
| 18-110_k141_31184 | Low-quality |
| 18-110_k141_272311 | Not-determined |
| 18-110_k141_168787 | Not-determined |
| 18-110_k141_272320 | Not-determined |
| 18-110_k141_99654 | Not-determined |
| 18-110_k141_226057 | Not-determined |
| 18-110_k141_272323 | Not-determined |
| 18-110_k141_31195 | Not-determined |
| 18-110_k141_53330 | Not-determined |
| 18-110_k141_42026 | Not-determined |
| 18-110_k141_31197 | Not-determined |
| 18-110_k141_134397 | Not-determined |
| 18-110_k141_226065 | Not-determined |
| 18-110_k141_87646 | Not-determined |
| 18-110_k141_181170 | Not-determined |
| 18-110_k141_157414 | Not-determined |
| 18-110_k141_168806 | Not-determined |
| 18-110_k141_65440 | Not-determined |
| 18-110_k141_202044 | Not-determined |
| 18-110_k141_87656 | Not-determined |
| 18-110_k141_145974 | Not-determined |
| 18-110_k141_202046 | Not-determined |
| 18-110_k141_134413 | Not-determined |
| 18-110_k141_238371 | Not-determined |
| 18-110_k141_202049 | Not-determined |
| 18-110_k141_238373 | Not-determined |
| 18-110_k141_99676 | Not-determined |
| 18-110_k141_202052 | Not-determined |
| 18-110_k141_202054 | Not-determined |
| 18-110_k141_110524 | Not-determined |
| 18-110_k141_238380 | Not-determined |
| 18-110_k141_110527 | Not-determined |
| 18-110_k141_76751 | Not-determined |
| 18-110_k141_110529 | Not-determined |
| 18-110_k141_6643 | Not-determined |
| 18-110_k141_134419 | Not-determined |
| 18-110_k141_99685 | Low-quality |
| 18-110_k141_31223 | Not-determined |
| 18-110_k141_181186 | Not-determined |
| 18-110_k141_99691 | Not-determined |
| 18-110_k141_6653 | Not-determined |
| 18-110_k141_87666 | Not-determined |
| 18-110_k141_145985 | Not-determined |
| 18-110_k141_110539 | Not-determined |
| 18-110_k141_134437 | Not-determined |
| 18-110_k141_76770 | Not-determined |
| 18-110_k141_31228 | Not-determined |
| 18-110_k141_238395 | Not-determined |
| 18-110_k141_65472 | Low-quality |
| 18-110_k141_134439 | Not-determined |
| 18-110_k141_31233 | Not-determined |
| 18-110_k141_53376 | Not-determined |
| 18-110_k141_181194 | Not-determined |
| 18-110_k141_53377 | Not-determined |
| 18-110_k141_134443 | Not-determined |
| 18-110_k141_145998 | Not-determined |
| 18-110_k141_202083 | Not-determined |
| 18-110_k141_122403 | Not-determined |
| 18-110_k141_181203 | Not-determined |
| 18-110_k141_53380 | Not-determined |
| 18-110_k141_181205 | Not-determined |
| 18-110_k141_42078 | Not-determined |
| 18-110_k141_157432 | Low-quality |
| 18-110_k141_122410 | Not-determined |
| 18-110_k141_146002 | Not-determined |
| 18-110_k141_146004 | Not-determined |
| 18-110_k141_76798 | Not-determined |
| 18-110_k141_272364 | Low-quality |
| 18-110_k141_53394 | Not-determined |
| 18-110_k141_31247 | Not-determined |
| 18-110_k141_146012 | Not-determined |
| 18-110_k141_214620 | Low-quality |
| 18-110_k141_214622 | Low-quality |
| 18-110_k141_42101 | Low-quality |
| 18-110_k141_181223 | Low-quality |
| 18-110_k141_17456 | Not-determined |
| 18-110_k141_157451 | Not-determined |
| 18-110_k141_249455 | Not-determined |
| 18-110_k141_6684 | Low-quality |
| 18-110_k141_76812 | Not-determined |
| 18-110_k141_181233 | Not-determined |
| 18-110_k141_146024 | Not-determined |
| 18-110_k141_134468 | Not-determined |
| 18-110_k141_122420 | Not-determined |
| 18-110_k141_134477 | Not-determined |
| 18-110_k141_181243 | Not-determined |
| 18-110_k141_168862 | Not-determined |
| 18-110_k141_168865 | Not-determined |
| 18-110_k141_134481 | Low-quality |
| 18-105-final-viral-combined_k141_39793\|\|0_partial | High-quality |
| 18-110_k141_110560 | Not-determined |
| 18-110_k141_53413 | Not-determined |
| 18-110_k141_259315 | Not-determined |
| 18-110_k141_122427 | Not-determined |
| 18-110_k141_168871 | Not-determined |
| 18-110_k141_87695 | Not-determined |
| 18-110_k141_259318 | Not-determined |
| 18-110_k141_226081 | Not-determined |
| 18-110_k141_53423 | Not-determined |
| 18-110_k141_31279 | Not-determined |
| 18-110_k141_87701 | Not-determined |
| 18-110_k141_110578 | Not-determined |
| 18-110_k141_238438 | Not-determined |
| 18-110_k141_53432 | Low-quality |
| 18-110_k141_146049 | Not-determined |
| 18-110_k141_53433 | Low-quality |
| 18-110_k141_6710 | Low-quality |
| 18-110_k141_202137 | Not-determined |
| 18-110_k141_122450 | Not-determined |
| 18-110_k141_272383 | Not-determined |
| 18-110_k141_238444 | Low-quality |
| 18-110_k141_87708 | Not-determined |
| 18-110_k141_134501 | Not-determined |
| 18-110_k141_272389 | Not-determined |
| 18-110_k141_31286 | Not-determined |
| 18-110_k141_168894 | Not-determined |
| 18-110_k141_168895 | Not-determined |
| 18-110_k141_238451 | Not-determined |
| 18-110_k141_31289 | Low-quality |

| | |
|---|---|
| 18-110_k141_249494 | Not-determined |
| 18-110_k141_76847 | Not-determined |
| 18-110_k141_76848 | Not-determined |
| 18-110_k141_17485 | Not-determined |
| 18-110_k141_6727 | Not-determined |
| 18-110_k141_259334 | Not-determined |
| 18-110_k141_202150 | Not-determined |
| 18-110_k141_214638 | Not-determined |
| 18-110_k141_202151 | Not-determined |
| 18-110_k141_87718 | Not-determined |
| 18-110_k141_110603 | Low-quality |
| 18-110_k141_272403 | Low-quality |
| 18-110_k141_157480 | Not-determined |
| 18-110_k141_146063 | Not-determined |
| 18-110_k141_214646 | Not-determined |
| 18-110_k141_259340 | Not-determined |
| 18-110_k141_226113 | Not-determined |
| 18-110_k141_17493 | Low-quality |
| 18-110_k141_110618 | Not-determined |
| 18-110_k141_6738 | Not-determined |
| 18-110_k141_76861 | Not-determined |
| 18-110_k141_146077 | Not-determined |
| 18-110_k141_157491 | Not-determined |
| 18-110_k141_31301 | Not-determined |
| 18-110_k141_17505 | Not-determined |
| 18-110_k141_76866 | Not-determined |
| 18-110_k141_87736 | Not-determined |
| 18-110_k141_99736 | Low-quality |
| 18-110_k141_168922 | Not-determined |
| 18-110_k141_146088 | Not-determined |
| 18-110_k141_110635 | Not-determined |
| 18-110_k141_238482 | Not-determined |
| 18-110_k141_17509 | Not-determined |
| 18-110_k141_146090 | Not-determined |
| 18-110_k141_110637 | Not-determined |
| 18-110_k141_259354 | Not-determined |
| 18-110_k141_99740 | Not-determined |
| 18-110_k141_146095 | Not-determined |
| 18-110_k141_238489 | Not-determined |
| 18-110_k141_202182 | Not-determined |
| 18-110_k141_31311 | Low-quality |
| 18-110_k141_226125 | Not-determined |
| 18-110_k141_146105 | Not-determined |
| 18-110_k141_53479 | Not-determined |
| 18-110_k141_168933 | Not-determined |
| 18-110_k141_214654 | Not-determined |
| 18-110_k141_181280 | Not-determined |
| 18-110_k141_168936 | Not-determined |
| 18-110_k141_157520 | Low-quality |
| 18-110_k141_76877 | Not-determined |
| 18-110_k141_259366 | Not-determined |
| 18-106-final-viral-combined_k141_107964\|\|0_partial | High-quality |
| 18-110_k141_110643 | Not-determined |
| 18-110_k141_17521 | Not-determined |
| 18-110_k141_259370 | Not-determined |
| 18-110_k141_202194 | Not-determined |
| 18-110_k141_168944 | Not-determined |
| 18-110_k141_17524 | Not-determined |
| 18-110_k141_226138 | Not-determined |
| 18-110_k141_42161 | Not-determined |
| 18-110_k141_249541 | Not-determined |
| 18-110_k141_110661 | Not-determined |
| 18-110_k141_168949 | Not-determined |
| 18-110_k141_226142 | Not-determined |
| 18-110_k141_238509 | Not-determined |
| 18-110_k141_17530 | Not-determined |
| 18-110_k141_249545 | Not-determined |
| 18-110_k141_146123 | Not-determined |
| 18-110_k141_76884 | Not-determined |
| 18-110_k141_17533 | Not-determined |
| 18-110_k141_259382 | Not-determined |
| 18-110_k141_65526 | Not-determined |
| 18-110_k141_168959 | Not-determined |
| 18-110_k141_226143 | Not-determined |
| 18-110_k141_146129 | Not-determined |
| 18-110_k141_76889 | Not-determined |
| 18-110_k141_214669 | Not-determined |
| 18-110_k141_6762 | Not-determined |
| 18-110_k141_146134 | Low-quality |
| 18-110_k141_65529 | Low-quality |
| 18-110_k141_202213 | Not-determined |
| 18-110_k141_202218 | Not-determined |
| 18-110_k141_31334 | Not-determined |
| 18-110_k141_87751 | Not-determined |
| 18-110_k141_214674 | Not-determined |
| 18-110_k141_249553 | Not-determined |
| 18-110_k141_202221 | Not-determined |
| 18-110_k141_181307 | Not-determined |
| 18-110_k141_76896 | Not-determined |
| 18-110_k141_181308 | Not-determined |
| 18-110_k141_110686 | Low-quality |
| 18-110_k141_214680 | Not-determined |
| 18-110_k141_31338 | Not-determined |
| 18-110_k141_110689 | Not-determined |
| 18-110_k141_42195 | Not-determined |
| 18-110_k141_6772 | Not-determined |
| 18-110_k141_272433 | Not-determined |
| 18-110_k141_65545 | Not-determined |
| 18-110_k141_238548 | Not-determined |
| 18-110_k141_87763 | Not-determined |
| 18-110_k141_110696 | Not-determined |
| 18-110_k141_31345 | Not-determined |
| 18-110_k141_238554 | Not-determined |
| 18-110_k141_87765 | Low-quality |
| 18-110_k141_226161 | Not-determined |
| 18-110_k141_87768 | Not-determined |
| 18-110_k141_76908 | Low-quality |
| 18-110_k141_31349 | Not-determined |
| 18-110_k141_249564 | Not-determined |
| 18-110_k141_134529 | Not-determined |
| 18-110_k141_110711 | Not-determined |
| 18-110_k141_181326 | Not-determined |
| 18-110_k141_181327 | Not-determined |
| 18-110_k141_31355 | Not-determined |
| 18-110_k141_31356 | Not-determined |
| 18-110_k141_99772 | Not-determined |
| 18-110_k141_238568 | Not-determined |
| 18-110_k141_110717 | Not-determined |
| 18-110_k141_146161 | Not-determined |
| 18-110_k141_134538 | Not-determined |
| 18-110_k141_146165 | Not-determined |
| 18-110_k141_42217 | Not-determined |
| 18-110_k141_99778 | Not-determined |
| 18-110_k141_181335 | Low-quality |
| 18-110_k141_76933 | Low-quality |
| 18-110_k141_134545 | Not-determined |
| 18-110_k141_31369 | Not-determined |
| 18-110_k141_99785 | Not-determined |
| 18-110_k141_99786 | Not-determined |
| 18-110_k141_272457 | Not-determined |
| 18-110_k141_53529 | Low-quality |
| 18-110_k141_42225 | Not-determined |
| 18-110_k141_99790 | Not-determined |
| 18-110_k141_169002 | Not-determined |
| 18-110_k141_249575 | Not-determined |
| 18-110_k141_42235 | Not-determined |
| 18-110_k141_272465 | Not-determined |
| 18-110_k141_42237 | Not-determined |
| 18-110_k141_249579 | Not-determined |
| 18-110_k141_238593 | Not-determined |
| 18-110_k141_249581 | Not-determined |
| 18-110_k141_53547 | Not-determined |
| 18-110_k141_259408 | Not-determined |
| 18-110_k141_6803 | Not-determined |
| 18-110_k141_42242 | Not-determined |
| 18-110_k141_157581 | Not-determined |
| 18-110_k141_87790 | Not-determined |
| 18-110_k141_122482 | Not-determined |
| 18-110_k141_181375 | Not-determined |
| 18-110_k141_6805 | Not-determined |
| 18-110_k141_191711 | Not-determined |
| 18-110_k141_249590 | Not-determined |
| 18-110_k141_214710 | Not-determined |
| 18-110_k141_181378 | Low-quality |
| 18-110_k141_122485 | Not-determined |
| 18-110_k141_31388 | Not-determined |
| 18-110_k141_31389 | Low-quality |
| 18-110_k141_249594 | Not-determined |
| 18-110_k141_110746 | Not-determined |
| 18-110_k141_17583 | Not-determined |
| 18-110_k141_181383 | Not-determined |
| 18-110_k141_181386 | Not-determined |
| 18-110_k141_110750 | Not-determined |
| 18-110_k141_17585 | Not-determined |
| 18-110_k141_181390 | Not-determined |
| 18-110_k141_272486 | Not-determined |
| 18-110_k141_53562 | Not-determined |
| 18-110_k141_214717 | Not-determined |
| 18-110_k141_202235 | Low-quality |
| 18-110_k141_6820 | Not-determined |
| 18-110_k141_76981 | Low-quality |
| 18-110_k141_110761 | Not-determined |
| 18-110_k141_65601 | Not-determined |
| 18-110_k141_214724 | Not-determined |
| 18-110_k141_134563 | Not-determined |
| 18-110_k141_17588 | Low-quality |
| 18-110_k141_249605 | Not-determined |
| 18-110_k141_238605 | Not-determined |
| 18-110_k141_6824 | Not-determined |
| 18-110_k141_238607 | Not-determined |
| 18-110_k141_76990 | Not-determined |
| 18-110_k141_76992 | Not-determined |
| 18-110_k141_87803 | Not-determined |
| 18-110_k141_65613 | Not-determined |
| 18-110_k141_31413 | Not-determined |
| 18-110_k141_238620 | Low-quality |
| 18-110_k141_122515 | Not-determined |
| 18-110_k141_87812 | Not-determined |
| 18-110_k141_31415 | Low-quality |
| 18-110_k141_249618 | Not-determined |
| 18-110_k141_110772 | Not-determined |
| 18-110_k141_146200 | Not-determined |
| 18-110_k141_6838 | Not-determined |
| 18-110_k141_31425 | Not-determined |
| 18-110_k141_214746 | Low-quality |
| 18-110_k141_214747 | Not-determined |
| 18-110_k141_214749 | Low-quality |
| 18-110_k141_6842 | Not-determined |
| 18-110_k141_214751 | Not-determined |
| 18-110_k141_181413 | Not-determined |
| 18-110_k141_110792 | Low-quality |
| 18-110_k141_6858 | Not-determined |
| 18-110_k141_202258 | Not-determined |
| 18-110_k141_226186 | Not-determined |
| 18-110_k141_157598 | Not-determined |
| 18-110_k141_77009 | Not-determined |
| 18-110_k141_226194 | Not-determined |
| 18-110_k141_191771 | Low-quality |
| 18-110_k141_226195 | Not-determined |
| 18-110_k141_122546 | Not-determined |
| 18-110_k141_191779 | Low-quality |
| 18-110_k141_77014 | Not-determined |
| 18-110_k141_272493 | Low-quality |
| 18-110_k141_53569 | Not-determined |
| 18-110_k141_110811 | Not-determined |
| 18-110_k141_169026 | Not-determined |
| 18-110_k141_169028 | Low-quality |
| 18-110_k141_146231 | Not-determined |
| 18-110_k141_169029 | Not-determined |
| 18-110_k141_169030 | Not-determined |
| 18-110_k141_53573 | Not-determined |
| 18-110_k141_6870 | Not-determined |
| 18-110_k141_53574 | Not-determined |
| 18-110_k141_146235 | Not-determined |
| 18-110_k141_238638 | Not-determined |
| 18-110_k141_87827 | Not-determined |
| 18-110_k141_77031 | Low-quality |
| 18-110_k141_169045 | Not-determined |
| 18-110_k141_110826 | Low-quality |
| 18-110_k141_134607 | Not-determined |
| 18-110_k141_17638 | Not-determined |
| 18-110_k141_31460 | Not-determined |
| 18-110_k141_17639 | Not-determined |
| 18-110_k141_214791 | Low-quality |
| 18-110_k141_134612 | Not-determined |
| 18-110_k141_226222 | Not-determined |
| 18-110_k141_110832 | Not-determined |
| 18-110_k141_157617 | Not-determined |
| 18-110_k141_157618 | Not-determined |
| 18-110_k141_134618 | Not-determined |
| 18-110_k141_259444 | Not-determined |
| 18-110_k141_226224 | Not-determined |
| 18-110_k141_191813 | Not-determined |
| 18-110_k141_181425 | Not-determined |
| 18-110_k141_31472 | Low-quality |
| 18-110_k141_259447 | Not-determined |
| 18-110_k141_169053 | Not-determined |
| 18-110_k141_226227 | Not-determined |
| 18-110_k141_17648 | Low-quality |
| 18-110_k141_134629 | Not-determined |
| 18-110_k141_99891 | Low-quality |
| 18-110_k141_99892 | Not-determined |
| 18-110_k141_134631 | Not-determined |
| 18-110_k141_6903 | Not-determined |
| 18-110_k141_214794 | Not-determined |
| 18-110_k141_65677 | Not-determined |
| 18-110_k141_31482 | Low-quality |
| 18-110_k141_157635 | Not-determined |
| 18-110_k141_202299 | Not-determined |
| 18-110_k141_122590 | Not-determined |
| 18-110_k141_259461 | Not-determined |
| 18-110_k141_87856 | Not-determined |
| 18-110_k141_17650 | Low-quality |
| 18-110_k141_53602 | Low-quality |
| 18-110_k141_122595 | Low-quality |
| 18-110_k141_6919 | Not-determined |
| 18-110_k141_226246 | Not-determined |
| 18-110_k141_249665 | Low-quality |
| 18-110_k141_249666 | Not-determined |
| 18-110_k141_226249 | Not-determined |
| 18-110_k141_6924 | Not-determined |
| 18-110_k141_65696 | Not-determined |
| 18-110_k141_238654 | Not-determined |
| 18-110_k141_238655 | Not-determined |
| 18-110_k141_99917 | Not-determined |
| 18-110_k141_65701 | Not-determined |
| 18-110_k141_157656 | Not-determined |
| 18-110_k141_122608 | Not-determined |
| 18-110_k141_53624 | Not-determined |
| 18-110_k141_157662 | Not-determined |
| 18-110_k141_99920 | Not-determined |
| 18-110_k141_214819 | Not-determined |
| 18-110_k141_99922 | Not-determined |
| 18-110_k141_259479 | Not-determined |
| 18-110_k141_110858 | Not-determined |
| 18-110_k141_157666 | Not-determined |
| 18-110_k141_146286 | Not-determined |
| 18-110_k141_272520 | Not-determined |
| 18-110_k141_77062 | Not-determined |
| 18-110_k141_42296 | Not-determined |
| 18-110_k141_53638 | Not-determined |
| 18-110_k141_87885 | Not-determined |
| 18-110_k141_181469 | Low-quality |
| 18-110_k141_214835 | Not-determined |
| 18-110_k141_238665 | Not-determined |
| 18-110_k141_157672 | Low-quality |
| 18-110_k141_157673 | Not-determined |
| 18-110_k141_6953 | Low-quality |
| 18-110_k141_122625 | Not-determined |
| 18-110_k141_157675 | Not-determined |
| 18-110_k141_53641 | Not-determined |
| 18-110_k141_122627 | Not-determined |
| 18-110_k141_202332 | Not-determined |
| 18-110_k141_53643 | Not-determined |
| 18-110_k141_226277 | Low-quality |
| 18-110_k141_259492 | Not-determined |
| 18-110_k141_53645 | Low-quality |
| 18-110_k141_181476 | Not-determined |
| 18-110_k141_157679 | Not-determined |
| 18-110_k141_226281 | Not-determined |
| 18-110_k141_259495 | Not-determined |
| 18-110_k141_249683 | Not-determined |
| 18-110_k141_87896 | Not-determined |
| 18-110_k141_65730 | Not-determined |
| 18-110_k141_17686 | Not-determined |
| 18-110_k141_202341 | Not-determined |
| 18-110_k141_249690 | Not-determined |
| 18-110_k141_53649 | Not-determined |
| 18-110_k141_110870 | Not-determined |
| 18-110_k141_87902 | Not-determined |
| 18-110_k141_65736 | Not-determined |
| 18-110_k141_157687 | Not-determined |
| 18-110_k141_191855 | Not-determined |
| 18-110_k141_87905 | Not-determined |
| 18-110_k141_191858 | Not-determined |
| 18-110_k141_6964 | Not-determined |
| 18-110_k141_214844 | Not-determined |
| 18-110_k141_272546 | Not-determined |
| 18-110_k141_146317 | Not-determined |
| 18-110_k141_272548 | Not-determined |
| 18-110_k141_272549 | Not-determined |
| 18-110_k141_87916 | Not-determined |
| 18-110_k141_17701 | Not-determined |
| 18-110_k141_214853 | Not-determined |
| 18-110_k141_191867 | Not-determined |
| 18-110_k141_191868 | Not-determined |
| 18-110_k141_31516 | Low-quality |
| 18-110_k141_272559 | Not-determined |
| 18-110_k141_53658 | Not-determined |
| 18-110_k141_272562 | Not-determined |
| 18-110_k141_157705 | Not-determined |
| 18-110_k141_53663 | Not-determined |
| 18-110_k141_42331 | Not-determined |
| 18-110_k141_65754 | Low-quality |
| 18-110_k141_214861 | Not-determined |
| 18-110_k141_31522 | Not-determined |
| 18-110_k141_53666 | Low-quality |
| 18-110_k141_110884 | Low-quality |
| 18-110_k141_122664 | Not-determined |
| 18-110_k141_191877 | Not-determined |
| 18-110_k141_191882 | Not-determined |
| 18-110_k141_99960 | Not-determined |
| 18-110_k141_191886 | Not-determined |
| 18-110_k141_87929 | Low-quality |
| 18-110_k141_110895 | Not-determined |
| 18-110_k141_53675 | Not-determined |
| 18-110_k141_202352 | Not-determined |
| 18-110_k141_87934 | Not-determined |
| 18-110_k141_249711 | Not-determined |
| 18-110_k141_272564 | Not-determined |
| 18-110_k141_146342 | Not-determined |
| 18-110_k141_31542 | Not-determined |
| 18-110_k141_87943 | Not-determined |
| 18-110_k141_31548 | Not-determined |
| 18-110_k141_214874 | Not-determined |
| 18-110_k141_191912 | Not-determined |
| 18-110_k141_157733 | Not-determined |
| 18-110_k141_87946 | Not-determined |
| 18-110_k141_259503 | Not-determined |
| 18-110_k141_259504 | Not-determined |
| 18-110_k141_17731 | Not-determined |
| 18-110_k141_181543 | Not-determined |
| 18-110_k141_87954 | Low-quality |
| 18-110_k141_214888 | Not-determined |
| 18-110_k141_17734 | Not-determined |
| 18-110_k141_191927 | Not-determined |
| 18-110_k141_226332 | Not-determined |
| 18-110_k141_31560 | Not-determined |
| 18-110_k141_110910 | Not-determined |
| 18-110_k141_17738 | Not-determined |
| 18-110_k141_87959 | Not-determined |
| 18-110_k141_226335 | Low-quality |
| 18-110_k141_17741 | Not-determined |
| 18-110_k141_110912 | Low-quality |
| 18-110_k141_202379 | Not-determined |
| 18-110_k141_146363 | Not-determined |
| 18-110_k141_31567 | Not-determined |
| 18-110_k141_146365 | Not-determined |
| 18-110_k141_7015 | Not-determined |
| 18-110_k141_214896 | Not-determined |
| 18-110_k141_110919 | Not-determined |
| 18-110_k141_259513 | Low-quality |
| 18-110_k141_100004 | Not-determined |
| 18-110_k141_110924 | Not-determined |
| 18-110_k141_17754 | Not-determined |
| 18-110_k141_87968 | Not-determined |
| 18-110_k141_191940 | Not-determined |
| 18-110_k141_191945 | Not-determined |
| 18-110_k141_17764 | Not-determined |
| 18-110_k141_17766 | Not-determined |
| 18-110_k141_17767 | Not-determined |
| 18-110_k141_191950 | Not-determined |
| 18-110_k141_134639 | Not-determined |
| 18-110_k141_146384 | Not-determined |
| 18-110_k141_259527 | Not-determined |
| 18-110_k141_226356 | Not-determined |
| 18-110_k141_31569 | Not-determined |
| 18-110_k141_226359 | Not-determined |
| 18-110_k141_31572 | Not-determined |
| 18-110_k141_157754 | Not-determined |
| 18-110_k141_53721 | Not-determined |
| 18-110_k141_17780 | Not-determined |
| 18-110_k141_259535 | Not-determined |
| 18-110_k141_259536 | Not-determined |
| 18-110_k141_157767 | Not-determined |
| 18-110_k141_146398 | Not-determined |
| 18-110_k141_249733 | Not-determined |
| 18-110_k141_87992 | Not-determined |
| 18-110_k141_146401 | Low-quality |
| 18-110_k141_134654 | Not-determined |
| 18-110_k141_157772 | Not-determined |
| 18-110_k141_202413 | Not-determined |
| 18-110_k141_53738 | Not-determined |
| 18-110_k141_169066 | Not-determined |
| 18-110_k141_272604 | Not-determined |
| 18-110_k141_272609 | Not-determined |
| 18-110_k141_169076 | Not-determined |
| 18-110_k141_259551 | Not-determined |
| 18-110_k141_146414 | Not-determined |
| 18-110_k141_53741 | Not-determined |
| 18-110_k141_53742 | Not-determined |
| 18-110_k141_53744 | Not-determined |
| 18-110_k141_146419 | Not-determined |
| 18-110_k141_17799 | Low-quality |
| 18-110_k141_202425 | Not-determined |
| 18-110_k141_226395 | Not-determined |
| 18-110_k141_214927 | Not-determined |
| 18-110_k141_146423 | Low-quality |
| 18-110_k141_146424 | Low-quality |
| 18-110_k141_169089 | Not-determined |
| 18-110_k141_157788 | Not-determined |
| 18-110_k141_110946 | Low-quality |
| 18-110_k141_134678 | Not-determined |
| 18-110_k141_110948 | Not-determined |
| 18-110_k141_272616 | Not-determined |
| 18-110_k141_226402 | Not-determined |
| 18-110_k141_134679 | Not-determined |
| 18-110_k141_192003 | Not-determined |
| 18-110_k141_192005 | Not-determined |
| 18-110_k141_134684 | Not-determined |
| 18-110_k141_146439 | Not-determined |
| 18-110_k141_214942 | Not-determined |
| 18-110_k141_214944 | Not-determined |
| 18-110_k141_272629 | Not-determined |
| 18-110_k141_214946 | Not-determined |
| 18-110_k141_238793 | Not-determined |
| 18-110_k141_7071 | Not-determined |
| 18-110_k141_134691 | Not-determined |
| 18-110_k141_53763 | Not-determined |
| 18-110_k141_134694 | Not-determined |
| 18-110_k141_157795 | Not-determined |
| 18-110_k141_146443 | Not-determined |
| 18-110_k141_202437 | Not-determined |
| 18-110_k141_181573 | Not-determined |
| 18-110_k141_181574 | Not-determined |
| 18-110_k141_169100 | Not-determined |
| 18-110_k141_122727 | Not-determined |
| 18-110_k141_77091 | Low-quality |
| 18-110_k141_88012 | Not-determined |
| 18-110_k141_122730 | Not-determined |
| 18-110_k141_238804 | Not-determined |
| 18-110_k141_110955 | Not-determined |
| 18-110_k141_272641 | Low-quality |
| 18-110_k141_110959 | Not-determined |
| 18-110_k141_110961 | Not-determined |
| 18-110_k141_181582 | Not-determined |
| 18-110_k141_88023 | Not-determined |
| 18-110_k141_272650 | Low-quality |
| 18-110_k141_249779 | Not-determined |
| 18-110_k141_192010 | Not-determined |
| 18-110_k141_7092 | Not-determined |
| 18-110_k141_249783 | Not-determined |
| 18-110_k141_169116 | Not-determined |
| 18-110_k141_169117 | Not-determined |
| 18-110_k141_249785 | Not-determined |
| 18-110_k141_42382 | Not-determined |
| 18-110_k141_192014 | Not-determined |
| 18-110_k141_249790 | Not-determined |
| 18-110_k141_77108 | Not-determined |
| 18-110_k141_110971 | Not-determined |
| 18-110_k141_31652 | Not-determined |
| 18-110_k141_122736 | Not-determined |
| 18-110_k141_272659 | Not-determined |
| 18-110_k141_181593 | Not-determined |
| 18-110_k141_122738 | Not-determined |
| 18-110_k141_249799 | Not-determined |
| 18-110_k141_181601 | Not-determined |
| 18-110_k141_7102 | Not-determined |
| 18-110_k141_192023 | Not-determined |
| 18-110_k141_238813 | Not-determined |
| 18-110_k141_77122 | Not-determined |
| 18-110_k141_31666 | Not-determined |
| 18-110_k141_122746 | Not-determined |
| 18-110_k141_134726 | Not-determined |
| 18-110_k141_157823 | Not-determined |
| 18-110_k141_146469 | Not-determined |
| 18-110_k141_110995 | Not-determined |
| 18-110_k141_53804 | Not-determined |
| 18-110_k141_17834 | Low-quality |
| 18-110_k141_157834 | Not-determined |
| 18-110_k141_192028 | Not-determined |
| 18-110_k141_77132 | Not-determined |
| 18-110_k141_238825 | Not-determined |
| 18-110_k141_88041 | Not-determined |
| 18-110_k141_122760 | Low-quality |
| 18-110_k141_122761 | Not-determined |
| 18-110_k141_88043 | Not-determined |
| 18-110_k141_181619 | Not-determined |
| 18-110_k141_259607 | Not-determined |
| 18-110_k141_192034 | Not-determined |
| 18-110_k141_157837 | Not-determined |
| 18-110_k141_238835 | Not-determined |
| 18-110_k141_259611 | Not-determined |
| 18-110_k141_249822 | Not-determined |
| 18-110_k141_192037 | Not-determined |
| 18-110_k141_146485 | Not-determined |
| 18-110_k141_122776 | Not-determined |
| 18-110_k141_272673 | Not-determined |
| 18-110_k141_272675 | Not-determined |
| 18-110_k141_259630 | Not-determined |
| 18-110_k141_77154 | Not-determined |
| 18-110_k141_157844 | Not-determined |
| 18-110_k141_88061 | Not-determined |
| 18-110_k141_77155 | Not-determined |
| 18-110_k141_31681 | Not-determined |
| 18-110_k141_181639 | Not-determined |
| 18-110_k141_31687 | Not-determined |
| 18-110_k141_42410 | Not-determined |
| 18-110_k141_259643 | Not-determined |
| 18-110_k141_238861 | Not-determined |
| 18-110_k141_134765 | Not-determined |
| 18-110_k141_77162 | Not-determined |
| 18-110_k141_249836 | Not-determined |
| 18-110_k141_77163 | Not-determined |
| 18-110_k141_134767 | Not-determined |
| 18-110_k141_77164 | Not-determined |
| 18-110_k141_100023 | Not-determined |
| 18-110_k141_146498 | Not-determined |
| 18-110_k141_169142 | Not-determined |
| 18-110_k141_7132 | Not-determined |
| 18-110_k141_111026 | Not-determined |
| 18-110_k141_31698 | Not-determined |
| 18-110_k141_65821 | Not-determined |
| 18-110_k141_88073 | Not-determined |
| 18-110_k141_134773 | Not-determined |
| 18-110_k141_238873 | Not-determined |
| 18-110_k141_17857 | Not-determined |
| 18-110_k141_88079 | Not-determined |
| 18-110_k141_100033 | Not-determined |
| 18-110_k141_53815 | Not-determined |
| 18-110_k141_100034 | Not-determined |
| 18-110_k141_122806 | Not-determined |
| 18-110_k141_42420 | Not-determined |
| 18-110_k141_88083 | Not-determined |
| 18-110_k141_53816 | Not-determined |
| 18-110_k141_65825 | Not-determined |
| 18-110_k141_65827 | Not-determined |
| 18-110_k141_215011 | Not-determined |
| 18-110_k141_88093 | Not-determined |
| 18-110_k141_272680 | Not-determined |
| 18-110_k141_259650 | Not-determined |
| 18-110_k141_226462 | Not-determined |
| 18-110_k141_53830 | Not-determined |
| 18-110_k141_77188 | Not-determined |
| 18-110_k141_169159 | Low-quality |
| 18-110_k141_192068 | Not-determined |
| 18-110_k141_134780 | Not-determined |
| 18-110_k141_157886 | Not-determined |
| 18-110_k141_238887 | Low-quality |
| 18-110_k141_157888 | Not-determined |
| 18-110_k141_192076 | Not-determined |
| 18-110_k141_77196 | Not-determined |
| 18-110_k141_42429 | Not-determined |
| 18-110_k141_42430 | Not-determined |
| 18-110_k141_122829 | Not-determined |
| 18-110_k141_134784 | Low-quality |
| 18-110_k141_134786 | Not-determined |
| 18-110_k141_192085 | Not-determined |
| 18-110_k141_249865 | Not-determined |

| | |
|---|---|
| 18-110_k141_134792 | Low-quality |
| 18-110_k141_111053 | Not-determined |
| 18-110_k141_192094 | Not-determined |
| 18-110_k141_53846 | Not-determined |
| 18-110_k141_77205 | Not-determined |
| 18-110_k141_77207 | Not-determined |
| 18-110_k141_53848 | Not-determined |
| 18-110_k141_53852 | Not-determined |
| 18-110_k141_7146 | Not-determined |
| 18-110_k141_122845 | Not-determined |
| 18-110_k141_226478 | Not-determined |
| 18-110_k141_146538 | Not-determined |
| 18-110_k141_77213 | Not-determined |
| 18-110_k141_134803 | Not-determined |
| 18-110_k141_100059 | Not-determined |
| 18-110_k141_181653 | Not-determined |
| 18-110_k141_272710 | Not-determined |
| 18-110_k141_272711 | Not-determined |
| 18-110_k141_122851 | Low-quality |
| 18-110_k141_146544 | Low-quality |
| 18-110_k141_272714 | Not-determined |
| 18-110_k141_122858 | Low-quality |
| 18-110_k141_169196 | Not-determined |
| 18-110_k141_77219 | Not-determined |
| 18-110_k141_181666 | Not-determined |
| 18-110_k141_169201 | Not-determined |
| 18-110_k141_169204 | Not-determined |
| 18-110_k141_249868 | Not-determined |
| 18-110_k141_134812 | Not-determined |
| 18-110_k141_157905 | Not-determined |
| 18-110_k141_122859 | Low-quality |
| 18-110_k141_146557 | Not-determined |
| 18-110_k141_238897 | Not-determined |
| 18-110_k141_42466 | Not-determined |
| 18-110_k141_100086 | Not-determined |
| 18-110_k141_249875 | Not-determined |
| 18-110_k141_53869 | Not-determined |
| 18-110_k141_134819 | Not-determined |
| 18-110_k141_146561 | Not-determined |
| 18-110_k141_100092 | Not-determined |
| 18-110_k141_134829 | Low-quality |
| 18-110_k141_169217 | Not-determined |
| 18-110_k141_249883 | Not-determined |
| 18-110_k141_238906 | Not-determined |
| 18-110_k141_77242 | Not-determined |
| 18-110_k141_157917 | Low-quality |
| 18-110_k141_249888 | Not-determined |
| 18-110_k141_17913 | Not-determined |
| 18-110_k141_17914 | Not-determined |
| 18-110_k141_226504 | Not-determined |
| 18-110_k141_17918 | Not-determined |
| 18-110_k141_42487 | Not-determined |
| 18-110_k141_272743 | Not-determined |
| 18-110_k141_169224 | Not-determined |
| 18-110_k141_100107 | Not-determined |
| 18-110_k141_169225 | Not-determined |
| 18-110_k141_272744 | Not-determined |
| 18-110_k141_215066 | Not-determined |
| 18-110_k141_226513 | Not-determined |
| 18-110_k141_181697 | Not-determined |
| 18-110_k141_181698 | Not-determined |
| 18-110_k141_77253 | Not-determined |
| 18-110_k141_122882 | Low-quality |
| 18-110_k141_238932 | Not-determined |
| 18-110_k141_238933 | Not-determined |
| 18-110_k141_122890 | Not-determined |
| 18-110_k141_238937 | Not-determined |
| 18-110_k141_181705 | Low-quality |
| 18-110_k141_122896 | Not-determined |
| 18-110_k141_215080 | Not-determined |
| 18-110_k141_249906 | Not-determined |
| 18-110_k141_272759 | Not-determined |
| 18-110_k141_53905 | Not-determined |
| 18-110_k141_88141 | Not-determined |
| 18-110_k141_215083 | Not-determined |
| 18-110_k141_226518 | Not-determined |
| 18-110_k141_226520 | Not-determined |
| 18-110_k141_65899 | Not-determined |
| 18-110_k141_226524 | Not-determined |
| 18-110_k141_42501 | Not-determined |
| 18-110_k141_157950 | Low-quality |
| 18-110_k141_122911 | Not-determined |
| 18-110_k141_17927 | Low-quality |
| 18-110_k141_238957 | Not-determined |
| 18-110_k141_31761 | Not-determined |
| 18-110_k141_272785 | Not-determined |
| 18-110_k141_134851 | Not-determined |
| 18-110_k141_215087 | Not-determined |
| 18-110_k141_134852 | Not-determined |
| 18-110_k141_65903 | Low-quality |
| 18-110_k141_272789 | Not-determined |
| 18-101-final-viral-combined_k141_331896\|\|full | Medium-quality |
| 18-110_k141_238969 | Not-determined |
| 18-110_k141_111059 | Not-determined |
| 18-110_k141_111062 | Not-determined |
| 18-110_k141_181723 | Not-determined |
| 18-110_k141_31770 | Not-determined |
| 18-110_k141_157961 | Not-determined |
| 18-110_k141_272804 | Not-determined |
| 18-110_k141_157963 | Not-determined |
| 18-110_k141_226547 | Not-determined |
| 18-110_k141_53925 | Low-quality |
| 18-110_k141_146576 | Not-determined |
| 18-110_k141_181735 | Not-determined |
| 18-110_k141_53931 | Not-determined |
| 18-110_k141_53932 | Not-determined |
| 18-110_k141_202462 | Not-determined |
| 18-110_k141_202464 | Not-determined |
| 18-110_k141_192135 | Not-determined |
| 18-110_k141_31786 | Not-determined |
| 18-110_k141_157975 | Not-determined |
| 18-110_k141_88153 | Low-quality |
| 18-110_k141_53944 | Not-determined |
| 18-110_k141_238991 | Not-determined |
| 18-110_k141_157977 | Low-quality |
| 18-110_k141_42529 | Not-determined |
| 18-110_k141_226562 | Not-determined |
| 18-110_k141_181743 | Not-determined |
| 18-110_k141_181744 | Not-determined |
| 18-110_k141_111085 | Not-determined |
| 18-110_k141_192151 | Not-determined |
| 18-110_k141_17930 | Not-determined |
| 18-110_k141_239006 | Not-determined |
| 18-110_k141_53954 | Not-determined |
| 18-110_k141_111095 | Not-determined |
| 18-110_k141_181753 | Not-determined |
| 18-110_k141_181754 | Not-determined |
| 18-110_k141_53958 | Not-determined |
| 18-110_k141_239008 | Low-quality |
| 18-110_k141_17939 | Not-determined |
| 18-110_k141_239015 | Low-quality |
| 18-110_k141_192160 | Not-determined |
| 18-110_k141_192161 | Not-determined |
| 18-110_k141_259710 | Not-determined |
| 18-110_k141_31803 | Not-determined |
| 18-110_k141_53970 | Not-determined |
| 18-110_k141_226578 | Not-determined |
| 18-110_k141_122942 | Low-quality |
| 18-110_k141_77292 | Not-determined |
| 18-110_k141_202495 | Not-determined |
| 18-110_k141_202497 | Low-quality |
| 18-110_k141_226589 | Not-determined |
| 18-110_k141_31806 | Not-determined |
| 18-110_k141_17952 | Not-determined |
| 18-110_k141_192177 | Not-determined |
| 18-110_k141_259721 | Not-determined |
| 18-110_k141_239031 | Not-determined |
| 18-110_k141_17960 | Not-determined |
| 18-110_k141_169313 | Low-quality |
| 18-110_k141_169316 | Not-determined |
| 18-110_k141_202507 | Not-determined |
| 18-110_k141_272826 | Not-determined |
| 18-110_k141_53981 | Not-determined |
| 18-110_k141_181778 | Not-determined |
| 18-110_k141_169328 | Not-determined |
| 18-110_k141_169330 | Not-determined |
| 18-110_k141_226612 | Not-determined |
| 18-110_k141_226617 | Low-quality |
| 18-110_k141_65937 | Low-quality |
| 18-110_k141_226621 | Not-determined |
| 18-110_k141_192190 | Not-determined |
| 18-110_k141_259737 | Not-determined |
| 18-110_k141_202519 | Not-determined |
| 18-110_k141_192191 | Not-determined |
| 18-110_k141_65939 | Not-determined |
| 18-110_k141_202520 | Not-determined |
| 18-110_k141_77313 | Low-quality |
| 18-110_k141_100173 | Low-quality |
| 18-110_k141_122955 | Not-determined |
| 18-110_k141_181798 | Not-determined |
| 18-110_k141_158013 | Not-determined |
| 18-110_k141_158014 | Low-quality |
| 18-110_k141_272846 | Not-determined |
| 18-110_k141_259742 | Not-determined |
| 18-110_k141_272848 | Not-determined |
| 18-110_k141_88181 | Low-quality |
| 18-110_k141_272851 | Not-determined |
| 18-110_k141_88185 | Not-determined |
| 18-110_k141_192193 | Not-determined |
| 18-110_k141_111124 | Low-quality |
| 18-110_k141_192199 | Not-determined |
| 18-110_k141_65950 | Not-determined |
| 18-110_k141_169356 | Not-determined |
| 18-110_k141_192204 | Not-determined |
| 18-110_k141_31824 | Not-determined |
| 18-110_k141_181824 | Not-determined |
| 18-110_k141_122978 | Not-determined |
| 18-110_k141_146623 | Not-determined |
| 18-110_k141_111137 | Not-determined |
| 18-110_k141_100192 | Not-determined |
| 18-110_k141_202541 | Not-determined |
| 18-110_k141_77345 | Not-determined |
| 18-110_k141_100194 | Not-determined |
| 18-110_k141_249913 | Not-determined |
| 18-110_k141_202549 | Not-determined |
| 18-110_k141_77353 | Not-determined |
| 18-110_k141_202553 | Not-determined |
| 18-110_k141_42553 | Not-determined |
| 18-110_k141_146630 | Low-quality |
| 18-110_k141_249920 | Not-determined |
| 18-110_k141_7229 | Not-determined |
| 18-110_k141_272878 | Not-determined |
| 18-110_k141_42566 | Not-determined |
| 18-110_k141_249927 | Not-determined |
| 18-110_k141_77376 | Not-determined |
| 18-110_k141_215112 | Not-determined |
| 18-110_k141_111142 | Not-determined |
| 18-110_k141_111143 | Not-determined |
| 18-110_k141_18029 | Not-determined |
| 18-110_k141_111144 | Low-quality |
| 18-110_k141_169392 | Not-determined |
| 18-110_k141_111150 | Not-determined |
| 18-110_k141_77385 | Not-determined |
| 18-110_k141_146656 | Not-determined |
| 18-110_k141_181852 | Not-determined |
| 18-110_k141_146658 | Not-determined |
| 18-110_k141_77388 | Not-determined |
| 18-110_k141_192222 | Low-quality |
| 18-110_k141_123001 | Not-determined |
| 18-110_k141_272905 | Not-determined |
| 18-110_k141_88220 | Not-determined |
| 18-110_k141_31859 | Low-quality |
| 18-110_k141_54045 | Not-determined |
| 18-110_k141_31861 | Not-determined |
| 18-110_k141_202571 | Not-determined |
| 18-110_k141_123017 | Not-determined |
| 18-110_k141_31868 | Not-determined |
| 18-110_k141_239102 | Not-determined |
| 18-110_k141_88236 | Not-determined |
| 18-110_k141_272917 | Not-determined |
| 18-110_k141_134970 | Not-determined |
| 18-110_k141_169416 | Not-determined |
| 18-110_k141_123022 | Not-determined |
| 18-110_k141_249949 | Not-determined |
| 18-110_k141_239117 | Not-determined |
| 18-110_k141_215125 | Not-determined |
| 18-110_k141_272928 | Not-determined |
| 18-110_k141_77412 | Not-determined |
| 18-110_k141_239121 | Not-determined |
| 18-110_k141_169428 | Low-quality |
| 18-110_k141_77418 | Not-determined |
| 18-110_k141_249952 | Not-determined |
| 18-110_k141_123035 | Not-determined |
| 18-110_k141_215126 | Not-determined |
| 18-110_k141_65997 | Not-determined |
| 18-110_k141_134991 | Not-determined |
| 18-110_k141_7279 | Not-determined |
| 18-110_k141_134993 | Not-determined |
| 18-110_k141_215128 | Not-determined |
| 18-110_k141_123039 | Not-determined |
| 18-110_k141_146681 | Low-quality |
| 18-110_k141_181891 | Not-determined |
| 18-110_k141_134994 | Not-determined |
| 18-110_k141_181895 | Not-determined |
| 18-110_k141_66006 | Low-quality |
| 18-110_k141_66007 | Not-determined |
| 18-110_k141_146687 | Not-determined |
| 18-110_k141_272950 | Not-determined |
| 18-110_k141_272951 | Not-determined |
| 18-110_k141_135004 | Not-determined |
| 18-110_k141_111195 | Not-determined |
| 18-110_k141_31897 | Not-determined |
| 18-110_k141_31899 | Not-determined |
| 18-110_k141_169438 | Not-determined |
| 18-110_k141_31900 | Not-determined |
| 18-110_k141_135008 | Not-determined |
| 18-110_k141_7291 | Not-determined |
| 18-110_k141_42630 | Not-determined |
| 18-110_k141_259786 | Not-determined |
| 18-110_k141_226633 | Not-determined |
| 18-110_k141_259789 | Not-determined |
| 18-110_k141_77437 | Not-determined |
| 18-110_k141_259796 | Not-determined |
| 18-110_k141_7297 | Not-determined |
| 18-110_k141_66009 | Not-determined |
| 18-110_k141_54086 | Not-determined |
| 18-110_k141_239141 | Low-quality |
| 18-110_k141_135033 | Not-determined |
| 18-110_k141_259802 | Not-determined |
| 18-110_k141_259803 | Not-determined |
| 18-110_k141_18067 | Not-determined |
| 18-110_k141_146704 | Not-determined |
| 18-110_k141_169454 | Not-determined |
| 18-110_k141_215155 | Not-determined |
| 18-110_k141_123060 | Not-determined |
| 18-110_k141_111225 | Not-determined |
| 18-110_k141_66015 | Not-determined |
| 18-110_k141_54091 | Not-determined |
| 18-110_k141_215160 | Not-determined |
| 18-110_k141_239153 | Not-determined |
| 18-110_k141_31913 | Not-determined |
| 18-110_k141_202620 | Not-determined |
| 18-110_k141_215169 | Not-determined |
| 18-110_k141_169459 | Not-determined |
| 18-110_k141_215175 | Not-determined |
| 18-110_k141_272981 | Not-determined |
| 18-110_k141_31918 | Not-determined |
| 18-110_k141_42649 | Not-determined |
| 18-110_k141_7316 | Not-determined |
| 18-110_k141_54110 | Low-quality |
| 18-110_k141_42656 | Not-determined |
| 18-110_k141_54114 | Not-determined |
| 18-110_k141_202631 | Not-determined |
| 18-110_k141_181935 | Not-determined |
| 18-110_k141_100244 | Not-determined |
| 18-110_k141_77482 | Low-quality |
| 18-110_k141_42662 | Low-quality |
| 18-110_k141_100246 | Not-determined |
| 18-110_k141_239174 | Not-determined |
| 18-110_k141_42669 | Not-determined |
| 18-110_k141_239182 | Not-determined |
| 18-110_k141_135058 | Not-determined |
| 18-110_k141_42671 | Low-quality |
| 18-110_k141_7332 | Not-determined |
| 18-110_k141_169477 | Not-determined |
| 18-110_k141_100254 | Not-determined |
| 18-110_k141_192267 | Low-quality |
| 18-110_k141_273001 | Not-determined |
| 18-110_k141_54126 | Not-determined |
| 18-110_k141_239194 | Not-determined |
| 18-110_k141_77495 | Not-determined |
| 18-110_k141_146731 | Not-determined |
| 18-110_k141_239206 | Not-determined |
| 18-110_k141_215187 | Not-determined |
| 18-110_k141_181952 | Not-determined |
| 18-110_k141_123099 | Not-determined |
| 18-110_k141_42686 | Not-determined |
| 18-110_k141_123100 | Not-determined |
| 18-110_k141_77514 | Not-determined |
| 18-110_k141_192288 | Not-determined |
| 18-110_k141_77521 | Low-quality |
| 18-110_k141_192294 | Not-determined |
| 18-110_k141_192295 | Not-determined |
| 18-110_k141_18113 | Not-determined |
| 18-110_k141_239225 | Low-quality |
| 18-110_k141_123119 | Not-determined |
| 18-110_k141_18118 | Not-determined |
| 18-110_k141_239234 | Low-quality |
| 18-110_k141_123122 | Not-determined |
| 18-110_k141_31944 | Not-determined |
| 18-110_k141_77529 | Not-determined |
| 18-110_k141_226650 | Not-determined |
| 18-110_k141_42718 | Not-determined |
| 18-110_k141_100281 | Not-determined |
| 18-110_k141_239241 | Not-determined |
| 18-110_k141_123131 | Not-determined |
| 18-110_k141_42724 | Not-determined |
| 18-110_k141_192298 | Not-determined |
| 18-110_k141_42734 | Not-determined |
| 18-110_k141_100291 | Not-determined |
| 18-110_k141_239251 | Not-determined |
| 18-110_k141_239252 | Not-determined |
| 18-110_k141_158116 | Not-determined |
| 18-110_k141_273029 | Low-quality |
| 18-110_k141_239253 | Not-determined |
| 18-110_k141_146766 | Not-determined |
| 18-110_k141_239255 | Not-determined |
| 18-110_k141_18134 | Not-determined |
| 18-110_k141_181970 | Not-determined |
| 18-110_k141_135092 | Not-determined |
| 18-110_k141_181973 | Not-determined |
| 18-110_k141_54153 | Not-determined |
| 18-110_k141_77553 | Not-determined |
| 18-110_k141_273049 | Not-determined |
| 18-110_k141_54160 | Not-determined |
| 18-110_k141_42740 | Not-determined |
| 18-110_k141_146778 | Not-determined |
| 18-110_k141_54163 | Not-determined |
| 18-110_k141_249992 | Not-determined |
| 18-110_k141_158126 | Not-determined |
| 18-110_k141_54168 | Not-determined |
| 18-110_k141_100298 | Not-determined |
| 18-110_k141_77562 | Not-determined |
| 18-110_k141_158132 | Not-determined |
| 18-110_k141_146794 | Not-determined |
| 18-110_k141_146795 | Not-determined |
| 18-110_k141_146802 | Not-determined |
| 18-110_k141_54180 | Not-determined |
| 18-110_k141_158138 | Not-determined |
| 18-110_k141_215244 | Not-determined |
| 18-110_k141_182006 | Not-determined |
| 18-110_k141_239297 | Low-quality |
| 18-110_k141_77579 | Not-determined |
| 18-110_k141_146816 | Not-determined |
| 18-110_k141_226675 | Not-determined |
| 18-110_k141_146819 | Low-quality |
| 18-110_k141_239305 | Not-determined |
| 18-110_k141_182016 | Not-determined |
| 18-110_k141_273071 | Not-determined |
| 18-110_k141_202651 | Not-determined |
| 18-110_k141_77591 | Low-quality |
| 18-110_k141_202653 | Low-quality |
| 18-110_k141_215253 | Not-determined |
| 18-110_k141_273075 | Not-determined |
| 18-110_k141_239313 | Not-determined |
| 18-110_k141_158160 | Not-determined |
| 18-110_k141_135124 | Not-determined |
| 18-110_k141_77596 | Not-determined |
| 18-110_k141_250014 | Not-determined |
| 18-110_k141_42776 | Not-determined |
| 18-110_k141_215257 | Not-determined |
| 18-110_k141_66041 | Not-determined |
| 18-110_k141_135128 | Low-quality |
| 18-110_k141_273081 | Not-determined |
| 18-110_k141_169503 | Not-determined |
| 18-110_k141_111269 | Low-quality |
| 18-110_k141_158169 | Not-determined |
| 18-110_k141_169507 | Not-determined |
| 18-110_k141_135133 | Not-determined |
| 18-110_k141_182036 | Not-determined |
| 18-110_k141_135135 | Not-determined |
| 18-110_k141_77605 | Not-determined |
| 18-110_k141_169513 | Not-determined |
| 18-110_k141_239328 | Not-determined |
| 18-110_k141_31996 | Not-determined |
| 18-110_k141_77609 | Not-determined |
| 18-110_k141_158176 | Low-quality |
| 18-110_k141_135143 | Not-determined |
| 18-110_k141_158178 | Not-determined |
| 18-110_k141_66049 | Not-determined |
| 18-110_k141_66050 | Not-determined |
| 18-110_k141_239335 | Low-quality |
| 18-110_k141_182051 | Not-determined |
| 18-110_k141_123208 | Not-determined |
| 18-110_k141_158186 | Low-quality |
| 18-110_k141_135152 | Not-determined |
| 18-110_k141_169532 | Not-determined |
| 18-110_k141_226699 | Not-determined |
| 18-110_k141_135160 | Not-determined |
| 18-110_k141_66063 | Not-determined |
| 18-110_k141_54200 | Not-determined |
| 18-110_k141_100338 | Not-determined |
| 18-110_k141_135162 | Not-determined |
| 18-110_k141_273093 | Not-determined |
| 18-110_k141_100340 | Not-determined |
| 18-110_k141_100343 | Not-determined |
| 18-110_k141_250045 | Not-determined |
| 18-110_k141_66070 | Not-determined |
| 18-110_k141_239352 | Not-determined |
| 18-110_k141_169547 | Not-determined |
| 18-110_k141_111297 | Low-quality |
| 18-110_k141_100348 | Not-determined |
| 18-110_k141_250055 | Low-quality |
| 18-110_k141_111298 | Not-determined |
| 18-110_k141_77645 | Not-determined |
| 18-110_k141_66074 | Not-determined |
| 18-110_k141_215278 | Not-determined |
| 18-110_k141_77650 | Not-determined |
| 18-110_k141_111305 | Not-determined |
| 18-110_k141_66083 | Not-determined |
| 18-110_k141_42824 | Not-determined |
| 18-110_k141_42825 | Not-determined |
| 18-110_k141_250064 | Not-determined |
| 18-110_k141_135179 | Not-determined |
| 18-110_k141_77661 | Not-determined |
| 18-110_k141_146854 | Not-determined |
| 18-110_k141_123243 | Not-determined |
| 18-110_k141_182101 | Not-determined |
| 18-110_k141_146862 | Not-determined |
| 18-110_k141_226726 | Not-determined |
| 18-110_k141_146869 | Not-determined |
| 18-110_k141_42838 | Not-determined |
| 18-110_k141_215293 | Not-determined |
| 18-110_k141_239371 | Not-determined |
| 18-110_k141_77676 | Not-determined |
| 18-110_k141_146874 | Not-determined |
| 18-110_k141_135198 | Not-determined |
| 18-110_k141_77681 | Not-determined |
| 18-110_k141_100381 | Not-determined |
| 18-110_k141_158204 | Not-determined |
| 18-110_k141_66109 | Not-determined |
| 18-110_k141_226735 | Not-determined |
| 18-110_k141_250100 | Not-determined |
| 18-110_k141_192348 | Not-determined |
| 18-110_k141_100384 | Not-determined |
| 18-110_k141_182136 | Not-determined |
| 18-110_k141_66112 | Not-determined |
| 18-110_k141_123275 | Not-determined |
| 18-110_k141_77693 | Not-determined |
| 18-110_k141_215296 | Not-determined |
| 18-110_k141_54255 | Not-determined |
| 18-110_k141_32066 | Not-determined |
| 18-110_k141_215299 | Not-determined |
| 18-110_k141_100391 | Not-determined |
| 18-110_k141_146888 | Not-determined |
| 18-110_k141_77698 | Not-determined |
| 18-110_k141_215306 | Not-determined |
| 18-110_k141_202684 | Not-determined |
| 18-110_k141_182148 | Not-determined |
| 18-110_k141_215308 | Not-determined |
| 18-110_k141_250112 | Low-quality |
| 18-110_k141_250113 | Not-determined |
| 18-110_k141_250114 | Not-determined |
| 18-110_k141_226745 | Not-determined |
| 18-110_k141_32078 | Not-determined |
| 18-110_k141_192356 | Not-determined |
| 18-110_k141_32081 | Not-determined |
| 18-110_k141_192366 | Not-determined |
| 18-110_k141_111346 | Not-determined |
| 18-110_k141_18140 | Not-determined |
| 18-110_k141_77716 | Not-determined |
| 18-110_k141_32095 | Not-determined |
| 18-110_k141_66126 | Not-determined |
| 18-110_k141_169601 | Not-determined |
| 18-110_k141_158248 | Not-determined |
| 18-110_k141_259850 | Not-determined |
| 18-110_k141_215336 | Not-determined |
| 18-110_k141_259852 | Not-determined |
| 18-110_k141_169606 | Not-determined |
| 18-110_k141_42873 | Not-determined |
| 18-110_k141_215339 | Not-determined |
| 18-110_k141_32104 | Not-determined |
| 18-110_k141_77720 | Not-determined |
| 18-110_k141_111356 | Not-determined |
| 18-110_k141_18149 | Not-determined |
| 18-110_k141_192387 | Not-determined |
| 18-110_k141_111366 | Not-determined |
| 18-110_k141_111369 | Not-determined |
| 18-110_k141_32127 | Not-determined |
| 18-110_k141_169629 | Not-determined |
| 18-110_k141_18160 | Not-determined |
| 18-110_k141_135208 | Not-determined |
| 18-110_k141_226765 | Not-determined |
| 18-110_k141_202703 | Not-determined |
| 18-110_k141_42908 | Not-determined |
| 18-110_k141_146949 | Low-quality |
| 18-110_k141_226775 | Not-determined |
| 18-110_k141_146953 | Not-determined |
| 18-110_k141_32140 | Not-determined |
| 18-110_k141_259872 | Not-determined |
| 18-110_k141_146956 | Low-quality |
| 18-110_k141_77754 | Not-determined |
| 18-110_k141_215368 | Not-determined |
| 18-110_k141_42916 | Not-determined |
| 18-110_k141_54274 | Not-determined |
| 18-110_k141_100465 | Not-determined |
| 18-110_k141_202726 | Not-determined |
| 18-110_k141_77763 | Not-determined |
| 18-110_k141_192408 | Not-determined |
| 18-110_k141_192412 | Not-determined |
| 18-110_k141_192413 | Not-determined |
| 18-110_k141_158258 | Not-determined |
| 18-110_k141_192419 | Low-quality |
| 18-110_k141_18177 | Not-determined |
| 18-110_k141_54296 | Not-determined |
| 18-110_k141_77776 | Not-determined |
| 18-110_k141_18179 | Not-determined |
| 18-110_k141_42940 | Low-quality |
| 18-110_k141_54298 | Not-determined |
| 18-110_k141_146970 | Not-determined |
| 18-110_k141_42944 | Not-determined |
| 18-110_k141_111397 | Not-determined |
| 18-110_k141_202741 | Low-quality |
| 18-110_k141_135240 | Not-determined |
| 18-110_k141_169668 | Not-determined |
| 18-110_k141_169669 | Not-determined |
| 18-110_k141_202750 | Not-determined |
| 18-110_k141_111399 | Low-quality |
| 18-110_k141_202752 | Not-determined |
| 18-110_k141_111401 | Not-determined |
| 18-110_k141_42952 | Not-determined |
| 18-110_k141_239402 | Not-determined |
| 18-110_k141_158266 | Not-determined |
| 18-110_k141_146987 | Not-determined |
| 18-110_k141_32160 | Not-determined |
| 18-110_k141_239408 | Not-determined |
| 18-110_k141_215392 | Not-determined |
| 18-110_k141_77787 | Not-determined |
| 18-110_k141_215398 | Not-determined |
| 18-110_k141_202769 | Not-determined |
| 18-110_k141_158281 | Not-determined |
| 18-110_k141_215400 | Not-determined |
| 18-110_k141_202772 | Not-determined |
| 18-110_k141_77791 | Not-determined |
| 18-110_k141_239418 | Not-determined |
| 18-110_k141_226802 | Low-quality |
| 18-110_k141_32175 | Not-determined |
| 18-110_k141_215417 | Not-determined |
| 18-110_k141_169690 | Not-determined |
| 18-110_k141_215419 | Not-determined |
| 18-110_k141_215420 | Not-determined |
| 18-110_k141_202790 | Not-determined |
| 18-110_k141_32178 | Low-quality |
| 18-110_k141_77809 | Not-determined |
| 18-110_k141_77810 | Not-determined |
| 18-110_k141_147020 | Not-determined |
| 18-110_k141_111427 | Not-determined |
| 18-110_k141_147024 | Not-determined |
| 18-110_k141_18222 | Not-determined |

| | |
|---|---|
| 18-110_k141_88285 | Not-determined |
| 18-110_k141_77815 | Not-determined |
| 18-110_k141_147031 | Not-determined |
| 18-110_k141_226806 | Not-determined |
| 18-110_k141_32192 | Low-quality |
| 18-110_k141_100481 | Not-determined |
| 18-110_k141_215429 | Low-quality |
| 18-110_k141_250181 | Not-determined |
| 18-110_k141_147038 | Not-determined |
| 18-110_k141_169697 | Not-determined |
| 18-110_k141_135279 | Not-determined |
| 18-110_k141_135286 | Not-determined |
| 18-110_k141_192485 | Not-determined |
| 18-110_k141_215444 | Low-quality |
| 18-110_k141_169711 | Not-determined |
| 18-110_k141_202817 | Not-determined |
| 18-110_k141_169712 | Not-determined |
| 18-110_k141_192494 | Not-determined |
| 18-110_k141_77832 | Not-determined |
| 18-110_k141_123374 | Not-determined |
| 18-110_k141_158304 | Not-determined |
| 18-110_k141_135298 | Not-determined |
| 18-110_k141_158306 | Not-determined |
| 18-110_k141_123380 | Not-determined |
| 18-110_k141_202828 | Not-determined |
| 18-110_k141_169725 | Not-determined |
| 18-110_k141_239464 | Not-determined |
| 18-110_k141_226840 | Not-determined |
| 18-110_k141_239472 | Not-determined |
| 18-110_k141_42990 | Low-quality |
| 18-110_k141_226843 | Not-determined |
| 18-110_k141_123396 | Not-determined |
| 18-110_k141_226845 | Not-determined |
| 18-110_k141_147068 | Not-determined |
| 18-110_k141_32217 | Low-quality |
| 18-110_k141_250207 | Not-determined |
| 18-110_k141_77841 | Not-determined |
| 18-110_k141_43002 | Not-determined |
| 18-110_k141_250211 | Low-quality |
| 18-110_k141_215473 | Low-quality |
| 18-110_k141_250213 | Not-determined |
| 18-110_k141_250215 | Not-determined |
| 18-110_k141_215478 | Not-determined |
| 18-110_k141_111480 | Not-determined |
| 18-110_k141_111481 | Not-determined |
| 18-110_k141_100490 | Not-determined |
| 18-110_k141_239487 | Not-determined |
| 18-110_k141_226849 | Not-determined |
| 18-110_k141_147081 | Not-determined |
| 18-110_k141_88322 | Not-determined |
| 18-110_k141_226854 | Not-determined |
| 18-110_k141_77852 | Not-determined |
| 18-110_k141_215496 | Not-determined |
| 18-110_k141_100496 | Not-determined |
| 18-110_k141_147088 | Not-determined |
| 18-110_k141_158340 | Not-determined |
| 18-110_k141_147093 | Not-determined |
| 18-110_k141_226858 | Not-determined |
| 18-110_k141_226860 | Not-determined |
| 18-110_k141_32228 | Not-determined |
| 18-110_k141_88343 | Not-determined |
| 18-110_k141_100508 | Not-determined |
| 18-110_k141_158354 | Not-determined |
| 18-110_k141_111496 | Not-determined |
| 18-110_k141_192534 | Not-determined |
| 18-110_k141_192537 | Low-quality |
| 18-110_k141_158359 | Not-determined |
| 18-110_k141_32246 | Not-determined |
| 18-110_k141_135330 | Not-determined |
| 18-110_k141_192545 | Not-determined |
| 18-110_k141_192546 | Not-determined |
| 18-110_k141_77873 | Not-determined |
| 18-110_k141_77874 | Not-determined |
| 18-110_k141_88370 | Not-determined |
| 18-110_k141_7364 | Not-determined |
| 18-110_k141_135336 | Not-determined |
| 18-110_k141_147132 | Not-determined |
| 18-110_k141_32259 | Not-determined |
| 18-110_k141_77882 | Not-determined |
| 18-110_k141_158368 | Not-determined |
| 18-110_k141_169769 | Not-determined |
| 18-110_k141_54366 | Not-determined |
| 18-110_k141_18280 | Low-quality |
| 18-110_k141_7376 | Not-determined |
| 18-110_k141_32266 | Not-determined |
| 18-110_k141_32267 | Low-quality |
| 18-110_k141_192558 | Not-determined |
| 18-110_k141_54378 | Not-determined |
| 18-110_k141_43050 | Not-determined |
| 18-110_k141_54381 | Not-determined |
| 18-110_k141_43051 | Low-quality |
| 18-110_k141_88383 | Not-determined |
| 18-110_k141_250226 | Not-determined |
| 18-110_k141_111510 | Not-determined |
| 18-110_k141_135349 | Not-determined |
| 18-110_k141_226898 | Not-determined |
| 18-110_k141_169787 | Not-determined |
| 18-110_k141_54387 | Not-determined |
| 18-110_k141_100542 | Not-determined |
| 18-110_k141_7382 | Not-determined |
| 18-110_k141_123399 | Not-determined |
| 18-110_k141_226904 | Not-determined |
| 18-110_k141_135356 | Low-quality |
| 18-110_k141_158384 | Not-determined |
| 18-110_k141_43062 | Not-determined |
| 18-110_k141_192575 | Low-quality |
| 18-110_k141_273131 | Not-determined |
| 18-110_k141_32283 | Not-determined |
| 18-110_k141_250234 | Not-determined |
| 18-110_k141_7391 | Not-determined |
| 18-110_k141_32288 | Not-determined |
| 18-110_k141_18306 | Not-determined |
| 18-110_k141_273143 | Low-quality |
| 18-110_k141_111524 | Not-determined |
| 18-110_k141_192585 | Not-determined |
| 18-110_k141_111527 | Not-determined |
| 18-110_k141_135360 | Not-determined |
| 18-110_k141_147170 | Not-determined |
| 18-110_k141_226927 | Not-determined |
| 18-110_k141_135366 | Not-determined |
| 18-110_k141_147177 | Not-determined |
| 18-110_k141_100575 | Not-determined |
| 18-110_k141_111538 | Not-determined |
| 18-110_k141_7403 | Not-determined |
| 18-110_k141_88394 | Not-determined |
| 18-110_k141_169815 | Not-determined |
| 18-110_k141_192592 | Not-determined |
| 18-110_k141_88398 | Not-determined |
| 18-110_k141_169821 | Low-quality |
| 18-110_k141_192599 | Not-determined |
| 18-110_k141_135379 | Not-determined |
| 18-110_k141_100589 | Not-determined |
| 18-110_k141_111547 | Not-determined |
| 18-110_k141_239531 | Not-determined |
| 18-110_k141_88407 | Not-determined |
| 18-110_k141_123443 | Not-determined |
| 18-110_k141_192609 | Not-determined |
| 18-110_k141_192610 | Not-determined |
| 18-110_k141_192611 | Not-determined |
| 18-110_k141_123449 | Not-determined |
| 18-110_k141_54439 | Not-determined |
| 18-110_k141_202880 | Not-determined |
| 18-110_k141_169828 | Not-determined |
| 18-110_k141_135399 | Not-determined |
| 18-110_k141_123453 | Not-determined |
| 18-110_k141_18339 | Not-determined |
| 18-110_k141_239538 | Low-quality |
| 18-110_k141_226949 | Not-determined |
| 18-110_k141_192616 | Not-determined |
| 18-110_k141_192617 | Not-determined |
| 18-110_k141_147187 | Low-quality |
| 18-110_k141_43091 | Not-determined |
| 18-110_k141_226956 | Low-quality |
| 18-110_k141_273162 | Not-determined |
| 18-110_k141_18350 | Not-determined |
| 18-110_k141_147191 | Low-quality |
| 18-110_k141_239544 | Not-determined |
| 18-110_k141_66157 | Not-determined |
| 18-110_k141_273164 | Not-determined |
| 18-110_k141_135415 | Not-determined |
| 18-110_k141_259900 | Low-quality |
| 18-110_k141_54458 | Not-determined |
| 18-110_k141_88409 | Low-quality |
| 18-110_k141_192630 | Not-determined |
| 18-110_k141_239549 | Not-determined |
| 18-110_k141_250288 | Not-determined |
| 18-110_k141_202892 | Not-determined |
| 18-110_k141_250289 | Not-determined |
| 18-110_k141_273167 | Not-determined |
| 18-110_k141_215567 | Not-determined |
| 18-110_k141_123473 | Not-determined |
| 18-110_k141_239554 | Not-determined |
| 18-110_k141_273171 | Low-quality |
| 18-110_k141_66168 | Low-quality |
| 18-110_k141_111569 | Not-determined |
| 18-110_k141_226970 | Not-determined |
| 18-110_k141_54469 | Not-determined |
| 18-110_k141_123481 | Not-determined |
| 18-110_k141_18358 | Low-quality |
| 18-110_k141_273177 | Not-determined |
| 18-110_k141_259903 | Not-determined |
| 18-110_k141_226976 | Not-determined |
| 18-110_k141_18360 | Not-determined |
| 18-110_k141_239567 | Not-determined |
| 18-110_k141_18361 | Not-determined |
| 18-110_k141_250300 | Not-determined |
| 18-110_k141_88426 | Not-determined |
| 18-110_k141_100631 | Not-determined |
| 18-110_k141_192653 | Not-determined |
| 18-110_k141_18371 | Not-determined |
| 18-110_k141_7415 | Not-determined |
| 18-110_k141_111577 | Not-determined |
| 18-110_k141_100635 | Not-determined |
| 18-110_k141_226988 | Not-determined |
| 18-110_k141_88436 | Not-determined |
| 18-110_k141_43124 | Not-determined |
| 18-110_k141_250315 | Not-determined |
| 18-110_k141_226989 | Not-determined |
| 18-110_k141_215590 | Not-determined |
| 18-110_k141_239573 | Low-quality |
| 18-110_k141_54473 | Not-determined |
| 18-110_k141_239579 | Not-determined |
| 18-110_k141_54479 | Not-determined |
| 18-110_k141_192666 | Not-determined |
| 18-110_k141_66186 | Not-determined |
| 18-110_k141_135428 | Not-determined |
| 18-110_k141_77943 | Not-determined |
| 18-110_k141_227004 | Not-determined |
| 18-110_k141_66191 | Not-determined |
| 18-110_k141_273184 | Not-determined |
| 18-110_k141_66192 | Not-determined |
| 18-110_k141_227007 | Low-quality |
| 18-110_k141_32328 | Not-determined |
| 18-110_k141_54493 | Not-determined |
| 18-110_k141_147221 | Not-determined |
| 18-110_k141_77953 | Not-determined |
| 18-110_k141_54498 | Not-determined |
| 18-110_k141_43144 | Not-determined |
| 18-110_k141_147227 | Not-determined |
| 18-110_k141_111605 | Not-determined |
| 18-110_k141_202935 | Low-quality |
| 18-110_k141_111613 | Low-quality |
| 18-110_k141_147234 | Not-determined |
| 18-110_k141_32334 | Not-determined |
| 18-110_k141_250334 | Not-determined |
| 18-110_k141_135455 | Not-determined |
| 18-110_k141_192687 | Not-determined |
| 18-110_k141_43163 | Not-determined |
| 18-110_k141_18413 | Not-determined |
| 18-110_k141_100664 | Not-determined |
| 18-110_k141_135459 | Not-determined |
| 18-110_k141_250341 | Low-quality |
| 18-110_k141_135461 | Not-determined |
| 18-110_k141_147248 | Not-determined |
| 18-110_k141_100666 | Not-determined |
| 18-110_k141_147250 | Not-determined |
| 18-110_k141_239620 | Not-determined |
| 18-110_k141_123515 | Not-determined |
| 18-110_k141_215615 | Low-quality |
| 18-110_k141_169847 | Low-quality |
| 18-110_k141_169848 | Not-determined |
| 18-110_k141_239626 | Not-determined |
| 18-110_k141_202964 | Not-determined |
| 18-110_k141_239628 | Not-determined |
| 18-110_k141_215623 | Not-determined |
| 18-110_k141_273202 | Not-determined |
| 18-110_k141_147266 | Not-determined |
| 18-110_k141_111625 | Low-quality |
| 18-110_k141_43179 | Not-determined |
| 18-110_k141_239641 | Not-determined |
| 18-110_k141_100679 | Not-determined |
| 18-110_k141_169857 | Not-determined |
| 18-110_k141_239645 | Not-determined |
| 18-110_k141_273211 | Not-determined |
| 18-110_k141_227034 | Not-determined |
| 18-110_k141_111636 | Not-determined |
| 18-110_k141_273216 | Not-determined |
| 18-110_k141_147278 | Not-determined |
| 18-110_k141_43190 | Not-determined |
| 18-110_k141_135495 | Low-quality |
| 18-110_k141_18444 | Not-determined |
| 18-110_k141_215642 | Not-determined |
| 18-110_k141_147286 | Not-determined |
| 18-110_k141_7459 | Not-determined |
| 18-110_k141_111648 | Not-determined |
| 18-110_k141_273220 | Low-quality |
| 18-110_k141_7466 | Not-determined |
| 18-110_k141_273221 | Not-determined |
| 18-110_k141_215647 | Not-determined |
| 18-110_k141_215653 | Not-determined |
| 18-110_k141_192691 | Not-determined |
| 18-110_k141_147302 | Not-determined |
| 18-110_k141_135511 | Low-quality |
| 18-110_k141_111662 | Not-determined |
| 18-110_k141_135518 | Not-determined |
| 18-110_k141_239674 | Not-determined |
| 18-110_k141_192708 | Not-determined |
| 18-110_k141_215666 | Not-determined |
| 18-110_k141_239682 | Not-determined |
| 18-110_k141_227049 | Not-determined |
| 18-110_k141_111670 | Not-determined |
| 18-110_k141_239685 | Not-determined |
| 18-110_k141_259955 | Not-determined |
| 18-110_k141_111675 | Not-determined |
| 18-110_k141_215675 | Not-determined |
| 18-110_k141_123565 | Not-determined |
| 18-110_k141_227054 | Low-quality |
| 18-110_k141_135536 | Not-determined |
| 18-110_k141_273242 | Not-determined |
| 18-110_k141_54518 | Low-quality |
| 18-110_k141_100714 | Not-determined |
| 18-110_k141_66221 | Not-determined |
| 18-110_k141_192714 | Not-determined |
| 18-110_k141_273244 | Not-determined |
| 18-110_k141_54523 | Not-determined |
| 18-110_k141_192716 | Not-determined |
| 18-110_k141_123576 | Not-determined |
| 18-110_k141_239702 | Not-determined |
| 18-110_k141_192718 | Not-determined |
| 18-110_k141_147308 | Low-quality |
| 18-110_k141_123579 | Not-determined |
| 18-110_k141_147311 | Not-determined |
| 18-110_k141_111693 | Not-determined |
| 18-110_k141_227060 | Low-quality |
| 18-110_k141_239708 | Not-determined |
| 18-110_k141_100722 | Low-quality |
| 18-110_k141_43229 | Not-determined |
| 18-110_k141_111699 | Not-determined |
| 18-110_k141_227065 | Low-quality |
| 18-110_k141_43236 | Not-determined |
| 18-110_k141_66233 | Not-determined |
| 18-110_k141_100733 | Not-determined |
| 18-110_k141_135552 | Low-quality |
| 18-110_k141_135554 | Not-determined |
| 18-110_k141_123591 | Not-determined |
| 18-110_k141_100739 | Not-determined |
| 18-110_k141_123594 | Not-determined |
| 18-110_k141_111701 | Low-quality |
| 18-110_k141_100742 | Not-determined |
| 18-110_k141_147315 | Low-quality |
| 18-110_k141_239711 | Not-determined |
| 18-110_k141_239714 | Not-determined |
| 18-110_k141_100755 | Not-determined |
| 18-110_k141_227088 | Not-determined |
| 18-110_k141_239718 | Not-determined |
| 18-110_k141_215692 | Not-determined |
| 18-110_k141_239720 | Low-quality |
| 18-110_k141_77998 | Low-quality |
| 18-110_k141_54567 | Not-determined |
| 18-110_k141_147324 | Low-quality |
| 18-110_k141_215698 | Not-determined |
| 18-110_k141_147330 | Not-determined |
| 18-110_k141_215702 | Not-determined |
| 18-110_k141_78010 | Not-determined |
| 18-110_k141_32412 | Not-determined |
| 18-110_k141_7502 | Not-determined |
| 18-110_k141_78016 | Not-determined |
| 18-110_k141_54585 | Not-determined |
| 18-110_k141_78017 | Not-determined |
| 18-110_k141_54586 | Not-determined |
| 18-110_k141_100771 | Not-determined |
| 18-110_k141_182246 | Low-quality |
| 18-110_k141_192768 | Not-determined |
| 18-110_k141_202994 | Not-determined |
| 18-110_k141_147339 | Not-determined |
| 18-110_k141_192775 | Not-determined |
| 18-110_k141_123630 | Not-determined |
| 18-110_k141_273252 | Not-determined |
| 18-110_k141_32424 | Not-determined |
| 18-110_k141_273259 | Not-determined |
| 18-110_k141_203007 | Not-determined |
| 18-110_k141_43270 | Not-determined |
| 18-110_k141_18488 | Not-determined |
| 18-110_k141_18489 | Not-determined |
| 18-110_k141_273272 | Not-determined |
| 18-110_k141_203016 | Not-determined |
| 18-110_k141_273276 | Not-determined |
| 18-110_k141_78036 | Not-determined |
| 18-110_k141_43275 | Not-determined |
| 18-110_k141_147360 | Not-determined |
| 18-110_k141_123646 | Low-quality |
| 18-110_k141_54599 | Not-determined |
| 18-110_k141_273280 | Not-determined |
| 18-110_k141_32430 | Not-determined |
| 18-110_k141_215730 | Not-determined |
| 18-110_k141_273289 | Low-quality |
| 18-110_k141_100778 | Not-determined |
| 18-110_k141_123653 | Not-determined |
| 18-110_k141_88512 | Not-determined |
| 18-110_k141_147373 | Not-determined |
| 18-110_k141_123656 | Not-determined |
| 18-110_k141_78044 | Not-determined |
| 18-110_k141_203029 | Not-determined |
| 18-110_k141_147379 | Not-determined |
| 18-110_k141_147382 | Not-determined |
| 18-110_k141_147386 | Not-determined |
| 18-110_k141_123664 | Not-determined |
| 18-110_k141_54614 | Not-determined |
| 18-110_k141_260003 | Not-determined |
| 18-110_k141_54615 | Low-quality |
| 18-110_k141_100791 | Not-determined |
| 18-110_k141_123670 | Not-determined |
| 18-110_k141_100795 | Not-determined |
| 18-110_k141_54621 | Not-determined |
| 18-110_k141_54622 | Low-quality |
| 18-110_k141_32437 | Not-determined |
| 18-110_k141_66270 | Not-determined |
| 18-110_k141_7536 | Not-determined |
| 18-110_k141_54623 | Not-determined |
| 18-110_k141_260010 | Not-determined |
| 18-110_k141_135592 | Not-determined |
| 18-110_k141_260012 | Not-determined |
| 18-110_k141_18504 | Not-determined |
| 18-110_k141_192813 | Not-determined |
| 18-110_k141_135604 | Not-determined |
| 18-110_k141_147400 | Not-determined |
| 18-110_k141_66280 | Not-determined |
| 18-110_k141_32452 | Not-determined |
| 18-110_k141_169931 | Low-quality |
| 18-110_k141_239726 | Not-determined |
| 18-110_k141_239727 | Not-determined |
| 18-110_k141_32455 | Not-determined |
| 18-110_k141_66285 | Not-determined |
| 18-110_k141_66286 | Not-determined |
| 18-110_k141_273303 | Not-determined |
| 18-110_k141_147405 | Not-determined |
| 18-110_k141_7551 | Low-quality |
| 18-110_k141_18515 | Not-determined |
| 18-110_k141_66290 | Low-quality |
| 18-110_k141_54637 | Not-determined |
| 18-110_k141_100816 | Low-quality |
| 18-110_k141_227129 | Not-determined |
| 18-110_k141_111749 | Not-determined |
| 18-110_k141_260023 | Not-determined |
| 18-110_k141_32470 | Not-determined |
| 18-110_k141_111756 | Not-determined |
| 18-110_k141_123700 | Not-determined |
| 18-110_k141_239738 | Not-determined |
| 18-110_k141_111761 | Not-determined |
| 18-110_k141_7560 | Not-determined |
| 18-110_k141_123702 | Low-quality |
| 18-110_k141_7562 | Not-determined |
| 18-110_k141_147423 | Not-determined |
| 18-110_k141_66300 | Not-determined |
| 18-110_k141_273320 | Not-determined |
| 18-110_k141_158501 | Not-determined |
| 18-110_k141_111769 | Not-determined |
| 18-110_k141_123714 | Not-determined |
| 18-110_k141_182308 | Not-determined |
| 18-110_k141_260037 | Not-determined |
| 18-110_k141_100833 | Low-quality |
| 18-110_k141_192826 | Not-determined |
| 18-110_k141_135640 | Not-determined |
| 18-110_k141_100835 | Not-determined |
| 18-110_k141_239759 | Not-determined |
| 18-110_k141_147440 | Not-determined |
| 18-110_k141_147444 | Not-determined |
| 18-110_k141_123722 | Low-quality |
| 18-110_k141_7576 | Not-determined |
| 18-110_k141_100838 | Not-determined |
| 18-110_k141_135656 | Not-determined |
| 18-110_k141_100845 | Not-determined |
| 18-110_k141_123731 | Not-determined |
| 18-110_k141_192832 | Low-quality |
| 18-110_k141_78072 | Not-determined |
| 18-110_k141_227156 | Not-determined |
| 18-110_k141_7577 | Not-determined |
| 18-110_k141_135663 | Not-determined |
| 18-110_k141_7582 | Low-quality |
| 18-110_k141_227163 | Not-determined |
| 18-110_k141_18536 | Not-determined |
| 18-110_k141_260048 | Not-determined |
| 18-110_k141_135667 | Not-determined |
| 18-110_k141_158525 | Not-determined |
| 18-110_k141_147462 | Not-determined |
| 18-110_k141_18540 | Not-determined |
| 18-110_k141_169951 | Not-determined |
| 18-110_k141_158526 | Not-determined |
| 18-110_k141_147466 | Not-determined |
| 18-110_k141_260054 | Not-determined |
| 18-110_k141_273345 | Low-quality |
| 18-110_k141_78090 | Not-determined |
| 18-110_k141_147471 | Not-determined |
| 18-110_k141_147474 | Not-determined |
| 18-110_k141_66329 | Not-determined |
| 18-110_k141_273351 | Low-quality |
| 18-110_k141_147476 | Not-determined |
| 18-110_k141_135678 | Low-quality |
| 18-110_k141_32522 | Not-determined |
| 18-110_k141_78093 | Low-quality |
| 18-110_k141_78094 | Not-determined |
| 18-110_k141_111796 | Not-determined |
| 18-110_k141_260065 | Low-quality |
| 18-110_k141_78095 | Not-determined |
| 18-110_k141_32526 | Not-determined |
| 18-110_k141_250375 | Not-determined |
| 18-110_k141_18554 | Not-determined |
| 18-110_k141_158541 | Low-quality |
| 18-110_k141_100864 | Not-determined |
| 18-110_k141_111804 | Not-determined |
| 18-110_k141_135689 | Not-determined |
| 18-110_k141_182331 | Not-determined |
| 18-110_k141_66338 | Not-determined |
| 18-110_k141_111808 | Not-determined |
| 18-110_k141_135699 | Not-determined |
| 18-110_k141_111809 | Low-quality |
| 18-110_k141_203106 | Not-determined |
| 18-110_k141_18569 | Not-determined |
| 18-110_k141_260080 | Not-determined |
| 18-110_k141_215778 | Not-determined |
| 18-110_k141_147489 | Not-determined |
| 18-110_k141_54700 | Not-determined |
| 18-110_k141_147492 | Not-determined |
| 18-110_k141_182335 | Low-quality |
| 18-110_k141_135707 | Not-determined |
| 18-110_k141_111817 | Not-determined |
| 18-110_k141_100880 | Not-determined |
| 18-110_k141_135711 | Not-determined |
| 18-110_k141_203121 | Not-determined |
| 18-110_k141_147495 | Not-determined |
| 18-110_k141_111821 | Not-determined |
| 18-110_k141_182340 | Not-determined |
| 18-110_k141_182342 | Not-determined |
| 18-110_k141_66363 | Not-determined |
| 18-110_k141_203125 | Not-determined |
| 18-110_k141_54708 | Not-determined |
| 18-110_k141_54709 | Not-determined |
| 18-110_k141_203130 | Not-determined |
| 18-110_k141_7610 | Not-determined |
| 18-110_k141_7613 | Not-determined |
| 18-110_k141_135728 | Low-quality |
| 18-110_k141_273373 | Not-determined |
| 18-110_k141_7617 | Not-determined |
| 18-110_k141_100892 | Not-determined |
| 18-110_k141_123769 | Low-quality |
| 18-110_k141_215796 | Not-determined |
| 18-110_k141_203136 | Not-determined |
| 18-110_k141_54718 | Not-determined |
| 18-110_k141_111832 | Not-determined |
| 18-110_k141_18580 | Not-determined |
| 18-110_k141_273381 | Not-determined |
| 18-110_k141_135739 | Low-quality |
| 18-110_k141_250411 | Not-determined |
| 18-110_k141_169982 | Not-determined |
| 18-110_k141_215805 | Not-determined |
| 18-110_k141_192883 | Not-determined |
| 18-110_k141_54732 | Not-determined |
| 18-110_k141_203147 | Not-determined |
| 18-110_k141_18590 | Low-quality |
| 18-110_k141_215809 | Low-quality |
| 18-110_k141_215810 | Not-determined |
| 18-110_k141_111834 | Low-quality |
| 18-110_k141_100910 | Not-determined |
| 18-110_k141_111835 | Not-determined |
| 18-110_k141_135758 | Not-determined |
| 18-110_k141_66386 | Not-determined |
| 18-110_k141_192891 | Not-determined |
| 18-110_k141_203153 | Not-determined |
| 18-110_k141_260091 | Not-determined |
| 18-110_k141_66388 | Not-determined |
| 18-110_k141_7629 | Not-determined |
| 18-110_k141_123794 | Not-determined |
| 18-110_k141_54748 | Not-determined |
| 18-110_k141_250429 | Not-determined |
| 18-110_k141_54753 | Not-determined |
| 18-110_k141_123801 | Not-determined |
| 18-110_k141_7637 | Not-determined |
| 18-110_k141_182384 | Low-quality |
| 18-110_k141_227242 | Not-determined |
| 18-110_k141_170001 | Not-determined |
| 18-110_k141_170002 | Not-determined |
| 18-110_k141_182386 | Not-determined |
| 18-110_k141_111842 | Not-determined |
| 18-110_k141_260100 | Not-determined |
| 18-110_k141_158571 | Not-determined |
| 18-110_k141_250435 | Not-determined |
| 18-110_k141_170005 | Low-quality |
| 18-110_k141_260105 | Not-determined |
| 18-110_k141_135769 | Not-determined |
| 18-110_k141_215830 | Not-determined |
| 18-110_k141_182392 | Not-determined |
| 18-110_k141_88589 | Not-determined |
| 18-110_k141_43379 | Not-determined |
| 18-110_k141_78120 | Not-determined |
| 18-110_k141_135772 | Not-determined |
| 18-110_k141_182398 | Not-determined |
| 18-110_k141_18607 | Not-determined |
| 18-110_k141_111860 | Not-determined |
| 18-110_k141_7648 | Not-determined |
| 18-110_k141_7650 | Not-determined |
| 18-110_k141_215847 | Low-quality |
| 18-110_k141_182409 | Not-determined |
| 18-110_k141_135780 | Not-determined |
| 18-110_k141_18622 | Not-determined |
| 18-110_k141_111872 | Not-determined |
| 18-110_k141_147563 | Not-determined |
| 18-110_k141_227259 | Not-determined |
| 18-110_k141_227260 | Not-determined |
| 18-110_k141_170026 | Not-determined |
| 18-110_k141_135793 | Not-determined |
| 18-110_k141_273425 | Not-determined |
| 18-110_k141_54760 | Not-determined |
| 18-110_k141_182433 | Not-determined |
| 18-110_k141_273426 | Not-determined |
| 18-110_k141_273427 | Not-determined |
| 18-110_k141_54764 | Not-determined |
| 18-110_k141_273433 | Not-determined |
| 18-110_k141_192917 | Not-determined |
| 18-110_k141_158591 | Not-determined |

| | |
|---|---|
| 18-110_k141_123811 | Not-determined |
| 18-110_k141_88619 | Low-quality |
| 18-110_k141_239811 | Not-determined |
| 18-110_k141_260144 | Not-determined |
| 18-110_k141_54773 | Not-determined |
| 18-110_k141_203194 | Not-determined |
| 18-110_k141_78155 | Not-determined |
| 18-110_k141_227278 | Not-determined |
| 18-110_k141_227279 | Not-determined |
| 18-110_k141_123829 | Not-determined |
| 18-110_k141_273466 | Not-determined |
| 18-110_k141_273469 | Not-determined |
| 18-110_k141_111902 | Not-determined |
| 18-110_k141_215861 | Low-quality |
| 18-110_k141_135837 | Not-determined |
| 18-110_k141_158622 | Not-determined |
| 18-110_k141_78171 | Not-determined |
| 18-110_k141_215867 | Not-determined |
| 18-110_k141_227298 | Not-determined |
| 18-110_k141_215870 | Not-determined |
| 18-110_k141_54788 | Not-determined |
| 18-110_k141_158628 | Not-determined |
| 18-110_k141_78179 | Low-quality |
| 18-110_k141_250501 | Not-determined |
| 18-110_k141_32610 | Not-determined |
| 18-110_k141_32611 | Not-determined |
| 18-110_k141_250509 | Not-determined |
| 18-110_k141_100922 | Not-determined |
| 18-110_k141_54799 | Not-determined |
| 18-110_k141_7696 | Not-determined |
| 18-110_k141_7697 | Not-determined |
| 18-110_k141_260168 | Not-determined |
| 18-110_k141_273488 | Not-determined |
| 18-110_k141_250519 | Not-determined |
| 18-110_k141_250520 | Not-determined |
| 18-110_k141_260172 | Not-determined |
| 18-110_k141_260173 | Not-determined |
| 18-110_k141_273491 | Low-quality |
| 18-110_k141_100927 | Not-determined |
| 18-110_k141_7702 | Not-determined |
| 18-110_k141_54812 | Not-determined |
| 18-110_k141_123860 | Not-determined |
| 18-110_k141_273499 | Not-determined |
| 18-110_k141_100940 | Not-determined |
| 18-110_k141_273500 | Not-determined |
| 18-110_k141_66471 | Not-determined |
| 18-110_k141_54818 | Not-determined |
| 18-110_k141_215892 | Not-determined |
| 18-110_k141_192927 | Not-determined |
| 18-110_k141_100942 | Not-determined |
| 18-110_k141_7714 | Not-determined |
| 18-110_k141_111908 | Low-quality |
| 18-110_k141_239855 | Not-determined |
| 18-110_k141_135875 | Not-determined |
| 18-110_k141_54821 | Not-determined |
| 18-110_k141_192936 | Not-determined |
| 18-110_k141_100950 | Not-determined |
| 18-110_k141_7720 | Not-determined |
| 18-110_k141_192940 | Not-determined |
| 18-110_k141_43468 | Not-determined |
| 18-110_k141_192941 | Not-determined |
| 18-110_k141_32634 | Not-determined |
| 18-110_k141_182454 | Not-determined |
| 18-110_k141_18652 | Not-determined |
| 18-110_k141_7731 | Not-determined |
| 18-110_k141_192950 | Not-determined |
| 18-110_k141_147582 | Not-determined |
| 18-110_k141_182461 | Not-determined |
| 18-110_k141_7736 | Low-quality |
| 18-110_k141_100968 | Not-determined |
| 18-110_k141_227310 | Not-determined |
| 18-110_k141_260192 | Not-determined |
| 18-110_k141_111942 | Not-determined |
| 18-110_k141_18656 | Not-determined |
| 18-110_k141_54835 | Not-determined |
| 18-110_k141_111945 | Not-determined |
| 18-110_k141_123893 | Low-quality |
| 18-110_k141_32654 | Not-determined |
| 18-110_k141_111950 | Not-determined |
| 18-110_k141_111951 | Not-determined |
| 18-110_k141_54840 | Not-determined |
| 18-110_k141_182478 | Not-determined |
| 18-110_k141_88666 | Not-determined |
| 18-110_k141_54845 | Not-determined |
| 18-110_k141_32659 | Not-determined |
| 18-110_k141_215913 | Low-quality |
| 18-110_k141_18667 | Not-determined |
| 18-110_k141_123900 | Not-determined |
| 18-110_k141_54860 | Not-determined |
| 18-110_k141_32667 | Not-determined |
| 18-110_k141_123904 | Not-determined |
| 18-110_k141_215925 | Not-determined |
| 18-110_k141_43499 | Not-determined |
| 18-110_k141_88683 | Not-determined |
| 18-110_k141_78233 | Not-determined |
| 18-110_k141_18678 | Not-determined |
| 18-110_k141_7757 | Not-determined |
| 18-110_k141_100992 | Not-determined |
| 18-110_k141_78237 | Not-determined |
| 18-110_k141_182488 | Not-determined |
| 18-110_k141_182491 | Low-quality |
| 18-110_k141_182492 | Not-determined |
| 18-110_k141_182493 | Not-determined |
| 18-110_k141_192983 | Low-quality |
| 18-110_k141_158665 | Not-determined |
| 18-110_k141_78249 | Not-determined |
| 18-110_k141_227346 | Not-determined |
| 18-110_k141_123925 | Not-determined |
| 18-110_k141_66483 | Not-determined |
| 18-110_k141_123926 | Not-determined |
| 18-110_k141_182503 | Not-determined |
| 18-110_k141_203241 | Low-quality |
| 18-110_k141_123930 | Not-determined |
| 18-110_k141_66486 | Not-determined |
| 18-110_k141_7774 | Not-determined |
| 18-110_k141_18697 | Not-determined |
| 18-110_k141_135914 | Not-determined |
| 18-110_k141_18698 | Not-determined |
| 18-110_k141_135916 | Not-determined |
| 18-110_k141_43513 | Low-quality |
| 18-110_k141_111956 | Not-determined |
| 18-110_k141_250583 | Not-determined |
| 18-110_k141_147635 | Not-determined |
| 18-110_k141_101009 | Not-determined |
| 18-110_k141_66493 | Not-determined |
| 18-110_k141_123941 | Not-determined |
| 18-110_k141_250587 | Not-determined |
| 18-110_k141_193007 | Not-determined |
| 18-110_k141_66496 | Not-determined |
| 18-110_k141_227360 | Not-determined |
| 18-110_k141_158690 | Not-determined |
| 18-110_k141_215941 | Not-determined |
| 18-110_k141_260222 | Not-determined |
| 18-110_k141_7790 | Not-determined |
| 18-110_k141_215946 | Not-determined |
| 18-110_k141_147646 | Not-determined |
| 18-110_k141_203262 | Not-determined |
| 18-110_k141_54869 | Not-determined |
| 18-110_k141_18719 | Not-determined |
| 18-110_k141_111968 | Not-determined |
| 18-110_k141_260237 | Not-determined |
| 18-110_k141_250606 | Not-determined |
| 18-110_k141_54880 | Low-quality |
| 18-110_k141_18727 | Not-determined |
| 18-110_k141_203267 | Not-determined |
| 18-110_k141_260243 | Not-determined |
| 18-110_k141_101040 | Not-determined |
| 18-110_k141_101043 | Not-determined |
| 18-110_k141_135944 | Not-determined |
| 18-110_k141_215968 | Low-quality |
| 18-110_k141_101046 | Not-determined |
| 18-110_k141_182554 | Not-determined |
| 18-110_k141_250620 | Not-determined |
| 18-110_k141_54893 | Low-quality |
| 18-110_k141_88718 | Not-determined |
| 18-110_k141_88721 | Low-quality |
| 18-110_k141_43545 | Not-determined |
| 18-110_k141_158705 | Low-quality |
| 18-110_k141_18738 | Not-determined |
| 18-110_k141_54898 | Not-determined |
| 18-110_k141_170074 | Not-determined |
| 18-110_k141_54900 | Not-determined |
| 18-110_k141_239888 | Not-determined |
| 18-110_k141_182569 | Not-determined |
| 18-110_k141_147682 | Not-determined |
| 18-110_k141_215984 | Not-determined |
| 18-110_k141_158710 | Low-quality |
| 18-110_k141_182572 | Not-determined |
| 18-110_k141_54905 | Not-determined |
| 18-110_k141_158711 | Not-determined |
| 18-110_k141_135959 | Not-determined |
| 18-110_k141_88734 | Not-determined |
| 18-110_k141_111984 | Low-quality |
| 18-110_k141_239891 | Not-determined |
| 18-110_k141_54908 | Not-determined |
| 18-110_k141_203290 | Not-determined |
| 18-110_k141_78300 | Not-determined |
| 18-110_k141_203293 | Not-determined |
| 18-110_k141_215994 | Not-determined |
| 18-110_k141_7811 | Not-determined |
| 18-110_k141_135963 | Not-determined |
| 18-110_k141_135964 | Not-determined |
| 18-110_k141_182592 | Not-determined |
| 18-110_k141_88752 | Not-determined |
| 18-110_k141_43568 | Not-determined |
| 18-110_k141_147695 | Not-determined |
| 18-110_k141_227399 | Not-determined |
| 18-110_k141_135971 | Not-determined |
| 18-110_k141_273573 | Not-determined |
| 18-110_k141_135973 | Not-determined |
| 18-110_k141_158731 | Not-determined |
| 18-110_k141_54929 | Not-determined |
| 18-110_k141_66546 | Not-determined |
| 18-110_k141_260275 | Low-quality |
| 18-110_k141_43581 | Not-determined |
| 18-110_k141_112002 | Not-determined |
| 18-110_k141_260278 | Not-determined |
| 18-110_k141_170107 | Not-determined |
| 18-110_k141_66553 | Not-determined |
| 18-110_k141_147705 | Not-determined |
| 18-110_k141_227406 | Not-determined |
| 18-110_k141_123983 | Not-determined |
| 18-110_k141_158746 | Not-determined |
| 18-110_k141_216005 | Not-determined |
| 18-110_k141_158753 | Not-determined |
| 18-110_k141_123990 | Not-determined |
| 18-110_k141_7826 | Not-determined |
| 18-110_k141_182607 | Not-determined |
| 18-110_k141_158756 | Not-determined |
| 18-110_k141_170114 | Not-determined |
| 18-110_k141_250626 | Not-determined |
| 18-110_k141_216013 | Not-determined |
| 18-110_k141_43602 | Not-determined |
| 18-110_k141_123999 | Not-determined |
| 18-110_k141_124001 | Not-determined |
| 18-110_k141_135987 | Not-determined |
| 18-110_k141_135988 | Not-determined |
| 18-110_k141_273587 | Low-quality |
| 18-110_k141_147725 | Not-determined |
| 18-110_k141_7838 | Low-quality |
| 18-110_k141_158768 | Not-determined |
| 18-110_k141_66572 | Not-determined |
| 18-110_k141_54941 | Not-determined |
| 18-110_k141_250642 | Not-determined |
| 18-110_k141_216018 | Not-determined |
| 18-110_k141_7840 | Not-determined |
| 18-110_k141_216021 | Not-determined |
| 18-110_k141_273593 | Not-determined |
| 18-110_k141_147733 | Not-determined |
| 18-110_k141_239917 | Not-determined |
| 18-110_k141_32686 | Not-determined |
| 18-110_k141_170134 | Not-determined |
| 18-110_k141_101082 | Not-determined |
| 18-110_k141_66580 | Not-determined |
| 18-110_k141_7848 | Not-determined |
| 18-110_k141_18779 | Not-determined |
| 18-110_k141_43621 | Not-determined |
| 18-110_k141_203315 | Not-determined |
| 18-110_k141_136000 | Not-determined |
| 18-110_k141_170140 | Not-determined |
| 18-110_k141_250654 | Not-determined |
| 18-110_k141_239922 | Not-determined |
| 18-110_k141_54948 | Not-determined |
| 18-110_k141_170144 | Not-determined |
| 18-110_k141_101092 | Not-determined |
| 18-110_k141_18792 | Not-determined |
| 18-110_k141_18793 | Not-determined |
| 18-110_k141_136018 | Not-determined |
| 18-110_k141_216041 | Not-determined |
| 18-110_k141_250660 | Not-determined |
| 18-110_k141_158786 | Low-quality |
| 18-110_k141_239937 | Not-determined |
| 18-110_k141_18798 | Not-determined |
| 18-110_k141_101096 | Not-determined |
| 18-110_k141_239940 | Not-determined |
| 18-110_k141_78324 | Not-determined |
| 18-110_k141_273615 | Low-quality |
| 18-110_k141_227429 | Not-determined |
| 18-110_k141_250669 | Not-determined |
| 18-110_k141_216048 | Not-determined |
| 18-110_k141_193021 | Not-determined |
| 18-110_k141_216053 | Not-determined |
| 18-110_k141_193025 | Not-determined |
| 18-110_k141_216058 | Not-determined |
| 18-110_k141_88804 | Not-determined |
| 18-110_k141_250674 | Not-determined |
| 18-110_k141_170165 | Not-determined |
| 18-110_k141_158806 | Not-determined |
| 18-110_k141_216064 | Not-determined |
| 18-110_k141_112041 | Not-determined |
| 18-110_k141_273635 | Not-determined |
| 18-110_k141_32731 | Not-determined |
| 18-110_k141_54960 | Not-determined |
| 18-110_k141_88834 | Low-quality |
| 18-110_k141_193036 | Not-determined |
| 18-110_k141_158821 | Not-determined |
| 18-110_k141_227441 | Not-determined |
| 18-110_k141_182696 | Not-determined |
| 18-110_k141_170184 | Not-determined |
| 18-110_k141_216083 | Not-determined |
| 18-110_k141_193048 | Not-determined |
| 18-110_k141_43668 | Not-determined |
| 18-110_k141_112055 | Not-determined |
| 18-110_k141_88836 | Not-determined |
| 18-110_k141_182705 | Not-determined |
| 18-110_k141_158830 | Not-determined |
| 18-110_k141_43671 | Low-quality |
| 18-110_k141_43678 | Not-determined |
| 18-110_k141_227457 | Not-determined |
| 18-110_k141_227458 | Not-determined |
| 18-110_k141_250706 | Not-determined |
| 18-110_k141_170204 | Not-determined |
| 18-110_k141_32765 | Not-determined |
| 18-110_k141_170207 | Not-determined |
| 18-110_k141_112073 | Not-determined |
| 18-110_k141_32770 | Not-determined |
| 18-110_k141_32771 | Not-determined |
| 18-110_k141_147741 | Not-determined |
| 18-110_k141_273646 | Not-determined |
| 18-110_k141_147743 | Not-determined |
| 18-110_k141_227472 | Not-determined |
| 18-110_k141_216105 | Not-determined |
| 18-110_k141_136083 | Not-determined |
| 18-110_k141_136084 | Not-determined |
| 18-110_k141_273657 | Not-determined |
| 18-110_k141_193072 | Not-determined |
| 18-110_k141_136091 | Not-determined |
| 18-110_k141_158848 | Low-quality |
| 18-110_k141_147754 | Low-quality |
| 18-110_k141_66627 | Not-determined |
| 18-110_k141_227483 | Not-determined |
| 18-110_k141_136098 | Not-determined |
| 18-110_k141_273667 | Not-determined |
| 18-110_k141_273668 | Not-determined |
| 18-110_k141_250728 | Not-determined |
| 18-110_k141_78357 | Not-determined |
| 18-110_k141_66631 | Not-determined |
| 18-110_k141_273670 | Not-determined |
| 18-110_k141_203365 | Not-determined |
| 18-110_k141_18862 | Not-determined |
| 18-110_k141_203366 | Not-determined |
| 18-110_k141_54988 | Not-determined |
| 18-110_k141_7930 | Not-determined |
| 18-110_k141_203370 | Not-determined |
| 18-110_k141_158862 | Not-determined |
| 18-110_k141_136115 | Not-determined |
| 18-110_k141_136117 | Not-determined |
| 18-110_k141_260311 | Not-determined |
| 18-110_k141_260312 | Not-determined |
| 18-110_k141_158866 | Not-determined |
| 18-110_k141_193087 | Not-determined |
| 18-110_k141_112113 | Not-determined |
| 18-110_k141_136122 | Not-determined |
| 18-110_k141_182760 | Not-determined |
| 18-110_k141_43741 | Not-determined |
| 18-110_k141_55001 | Not-determined |
| 18-110_k141_18875 | Low-quality |
| 18-110_k141_227505 | Not-determined |
| 18-110_k141_88899 | Not-determined |
| 18-110_k141_136132 | Low-quality |
| 18-110_k141_18878 | Not-determined |
| 18-110_k141_112125 | Not-determined |
| 18-110_k141_182768 | Not-determined |
| 18-110_k141_203387 | Not-determined |
| 18-110_k141_182770 | Not-determined |
| 18-110_k141_18880 | Not-determined |
| 18-110_k141_101154 | Not-determined |
| 18-110_k141_182771 | Not-determined |
| 18-110_k141_193107 | Not-determined |
| 18-110_k141_216133 | Not-determined |
| 18-110_k141_182774 | Not-determined |
| 18-110_k141_18884 | Not-determined |
| 18-110_k141_227521 | Not-determined |
| 18-110_k141_182780 | Not-determined |
| 18-110_k141_227523 | Not-determined |
| 18-110_k141_101162 | Not-determined |
| 18-110_k141_250761 | Not-determined |
| 18-110_k141_147789 | Not-determined |
| 18-110_k141_147792 | Low-quality |
| 18-110_k141_216136 | Not-determined |
| 18-110_k141_216137 | Low-quality |
| 18-110_k141_158898 | Not-determined |
| 18-110_k141_170228 | Low-quality |
| 18-110_k141_18895 | Not-determined |
| 18-110_k141_66672 | Not-determined |
| 18-110_k141_170230 | Not-determined |
| 18-110_k141_273702 | Not-determined |
| 18-110_k141_136144 | Not-determined |
| 18-110_k141_78377 | Not-determined |
| 18-110_k141_158908 | Not-determined |
| 18-110_k141_227539 | Not-determined |
| 18-110_k141_170237 | Not-determined |
| 18-110_k141_66679 | Not-determined |
| 18-110_k141_193127 | Not-determined |
| 18-110_k141_101179 | Not-determined |
| 18-110_k141_32847 | Not-determined |
| 18-110_k141_158914 | Not-determined |
| 18-110_k141_158917 | Not-determined |
| 18-110_k141_55029 | Not-determined |
| 18-110_k141_101189 | Not-determined |
| 18-110_k141_250779 | Not-determined |
| 18-110_k141_170242 | Not-determined |
| 18-110_k141_158922 | Not-determined |
| 18-110_k141_55038 | Not-determined |
| 18-110_k141_250782 | Low-quality |
| 18-110_k141_273707 | Low-quality |
| 18-110_k141_88928 | Not-determined |
| 18-110_k141_182809 | Not-determined |
| 18-110_k141_170250 | Not-determined |
| 18-110_k141_66702 | Not-determined |
| 18-110_k141_124008 | Not-determined |
| 18-110_k141_18908 | Low-quality |
| 18-110_k141_55044 | Not-determined |
| 18-110_k141_55045 | Not-determined |
| 18-110_k141_193146 | Not-determined |
| 18-110_k141_88940 | Not-determined |
| 18-110_k141_101207 | Not-determined |
| 18-110_k141_88942 | Low-quality |
| 18-110_k141_260338 | Not-determined |
| 18-110_k141_182827 | Not-determined |
| 18-110_k141_193159 | Not-determined |
| 18-110_k141_203418 | Not-determined |
| 18-110_k141_101211 | Not-determined |
| 18-110_k141_203420 | Not-determined |
| 18-110_k141_32877 | Not-determined |
| 18-110_k141_66712 | Not-determined |
| 18-110_k141_43806 | Not-determined |
| 18-110_k141_227561 | Not-determined |
| 18-110_k141_227562 | Not-determined |
| 18-110_k141_55064 | Low-quality |
| 18-110_k141_66718 | Not-determined |
| 18-110_k141_124020 | Not-determined |
| 18-110_k141_203430 | Not-determined |
| 18-110_k141_182831 | Not-determined |
| 18-110_k141_203432 | Not-determined |
| 18-110_k141_227568 | Not-determined |
| 18-110_k141_216187 | Not-determined |
| 18-110_k141_193176 | Not-determined |
| 18-110_k141_112147 | Not-determined |
| 18-110_k141_203434 | Low-quality |
| 18-110_k141_240034 | Not-determined |
| 18-110_k141_32893 | Not-determined |
| 18-110_k141_203438 | Not-determined |
| 18-110_k141_124031 | Not-determined |
| 18-110_k141_273734 | Not-determined |
| 18-110_k141_136200 | Not-determined |
| 18-110_k141_240041 | Not-determined |
| 18-110_k141_136205 | Not-determined |
| 18-110_k141_66741 | Not-determined |
| 18-110_k141_273744 | Not-determined |
| 18-110_k141_216201 | Not-determined |
| 18-110_k141_273747 | Not-determined |
| 18-110_k141_240046 | Not-determined |
| 18-110_k141_32912 | Low-quality |
| 18-110_k141_170287 | Not-determined |
| 18-110_k141_101236 | Not-determined |
| 18-110_k141_240058 | Low-quality |
| 18-110_k141_216206 | Not-determined |
| 18-110_k141_78410 | Not-determined |
| 18-110_k141_88980 | Not-determined |
| 18-110_k141_193192 | Not-determined |
| 18-110_k141_66760 | Not-determined |
| 18-110_k141_273763 | Not-determined |
| 18-110_k141_240068 | Not-determined |
| 18-110_k141_66765 | Not-determined |
| 18-110_k141_66766 | Not-determined |
| 18-110_k141_147861 | Not-determined |
| 18-110_k141_101248 | Not-determined |
| 18-110_k141_240071 | Low-quality |
| 18-110_k141_193206 | Not-determined |
| 18-110_k141_66778 | Not-determined |
| 18-110_k141_147867 | Not-determined |
| 18-110_k141_240077 | Not-determined |
| 18-110_k141_7963 | Not-determined |
| 18-110_k141_273779 | Not-determined |
| 18-110_k141_55099 | Not-determined |
| 18-110_k141_78435 | Not-determined |
| 18-110_k141_216224 | Not-determined |
| 18-110_k141_89001 | Low-quality |
| 18-110_k141_78441 | Low-quality |
| 18-110_k141_260360 | Not-determined |
| 18-110_k141_78443 | Not-determined |
| 18-110_k141_147876 | Low-quality |
| 18-110_k141_18942 | Not-determined |
| 18-110_k141_89005 | Not-determined |
| 18-110_k141_227609 | Not-determined |
| 18-110_k141_101265 | Not-determined |
| 18-110_k141_260366 | Not-determined |
| 18-110_k141_18948 | Not-determined |
| 18-110_k141_78453 | Low-quality |
| 18-110_k141_216247 | Not-determined |
| 18-110_k141_240093 | Low-quality |
| 18-110_k141_18950 | Not-determined |
| 18-110_k141_7982 | Not-determined |
| 18-110_k141_78457 | Not-determined |
| 18-110_k141_216253 | Not-determined |
| 18-110_k141_273794 | Not-determined |
| 18-110_k141_227626 | Low-quality |
| 18-110_k141_101269 | Not-determined |
| 18-110_k141_78462 | Not-determined |
| 18-110_k141_216256 | Not-determined |
| 18-110_k141_240100 | Not-determined |
| 18-110_k141_89013 | Not-determined |
| 18-110_k141_101274 | Not-determined |
| 18-110_k141_101276 | Not-determined |
| 18-110_k141_7997 | Not-determined |
| 18-110_k141_273804 | Not-determined |
| 18-110_k141_7998 | Not-determined |
| 18-110_k141_136218 | Not-determined |
| 18-110_k141_147894 | Not-determined |
| 18-110_k141_273807 | Not-determined |
| 18-110_k141_147901 | Not-determined |
| 18-110_k141_18965 | Low-quality |
| 18-110_k141_136233 | Not-determined |
| 18-110_k141_147907 | Not-determined |
| 18-110_k141_273814 | Not-determined |
| 18-110_k141_250805 | Not-determined |
| 18-110_k141_8002 | Not-determined |
| 18-110_k141_136237 | Not-determined |
| 18-110_k141_240131 | Not-determined |
| 18-110_k141_78490 | Not-determined |
| 18-110_k141_240136 | Low-quality |
| 18-110_k141_136242 | Not-determined |
| 18-110_k141_260403 | Not-determined |
| 18-110_k141_250816 | Not-determined |
| 18-110_k141_216274 | Not-determined |
| 18-110_k141_260405 | Low-quality |
| 18-110_k141_260409 | Not-determined |
| 18-110_k141_193224 | Not-determined |
| 18-110_k141_66818 | Low-quality |
| 18-110_k141_112199 | Not-determined |
| 18-110_k141_216285 | Not-determined |
| 18-110_k141_147936 | Not-determined |
| 18-110_k141_136258 | Not-determined |
| 18-110_k141_147937 | Not-determined |
| 18-110_k141_136260 | Not-determined |
| 18-110_k141_18989 | Not-determined |
| 18-110_k141_101323 | Not-determined |
| 18-110_k141_101324 | Not-determined |
| 18-110_k141_66834 | Not-determined |
| 18-110_k141_78517 | Low-quality |
| 18-110_k141_216290 | Not-determined |
| 18-110_k141_66839 | Not-determined |
| 18-110_k141_147949 | Not-determined |
| 18-110_k141_78522 | Not-determined |
| 18-110_k141_240161 | Not-determined |
| 18-110_k141_78523 | Not-determined |
| 18-110_k141_273840 | Not-determined |
| 18-110_k141_124086 | Not-determined |
| 18-110_k141_136278 | Not-determined |
| 18-110_k141_203483 | Not-determined |
| 18-110_k141_8028 | Not-determined |
| 18-110_k141_147967 | Not-determined |
| 18-110_k141_250835 | Not-determined |
| 18-110_k141_136281 | Low-quality |
| 18-110_k141_124099 | Not-determined |
| 18-110_k141_136288 | Not-determined |
| 18-110_k141_66849 | Not-determined |
| 18-110_k141_101348 | Low-quality |
| 18-110_k141_112225 | Not-determined |
| 18-110_k141_112226 | Not-determined |
| 18-110_k141_8039 | Not-determined |
| 18-110_k141_216303 | Not-determined |
| 18-110_k141_216305 | Not-determined |
| 18-110_k141_193231 | Not-determined |
| 18-110_k141_193232 | Not-determined |
| 18-110_k141_89072 | Not-determined |
| 18-110_k141_8046 | Not-determined |
| 18-110_k141_273849 | Not-determined |
| 18-110_k141_203492 | Not-determined |
| 18-110_k141_66857 | Not-determined |
| 18-110_k141_43892 | Not-determined |
| 18-110_k141_32967 | Not-determined |
| 18-110_k141_216316 | Not-determined |
| 18-110_k141_43896 | Not-determined |
| 18-110_k141_170372 | Not-determined |
| 18-110_k141_32972 | Not-determined |
| 18-110_k141_170378 | Not-determined |
| 18-110_k141_124123 | Not-determined |
| 18-110_k141_43909 | Not-determined |
| 18-110_k141_78538 | Low-quality |
| 18-110_k141_43914 | Not-determined |
| 18-110_k141_66873 | Low-quality |
| 18-110_k141_193254 | Not-determined |
| 18-110_k141_216326 | Not-determined |
| 18-110_k141_193257 | Not-determined |
| 18-110_k141_193258 | Not-determined |
| 18-110_k141_136309 | Not-determined |
| 18-110_k141_8065 | Low-quality |
| 18-110_k141_203506 | Not-determined |
| 18-110_k141_240191 | Not-determined |
| 18-110_k141_101372 | Not-determined |
| 18-110_k141_240192 | Not-determined |
| 18-110_k141_101373 | Not-determined |
| 18-110_k141_43926 | Not-determined |
| 18-110_k141_216340 | Not-determined |
| 18-110_k141_203514 | Not-determined |
| 18-110_k141_260444 | Not-determined |
| 18-110_k141_101376 | Not-determined |
| 18-110_k141_101377 | Not-determined |
| 18-110_k141_158967 | Not-determined |
| 18-110_k141_19049 | Not-determined |
| 18-110_k141_250847 | Not-determined |
| 18-110_k141_158970 | Not-determined |
| 18-110_k141_136319 | Not-determined |
| 18-110_k141_227689 | Not-determined |
| 18-110_k141_66893 | Not-determined |
| 18-110_k141_78554 | Not-determined |
| 18-110_k141_227690 | Not-determined |

| | |
|---|---|
| 18-110_k141_203525 | Not-determined |
| 18-110_k141_112255 | Not-determined |
| 18-110_k141_124135 | Not-determined |
| 18-110_k141_19053 | Not-determined |
| 18-110_k141_101387 | Not-determined |
| 18-110_k141_66902 | Not-determined |
| 18-110_k141_193280 | Not-determined |
| 18-110_k141_240218 | Not-determined |
| 18-110_k141_147972 | Not-determined |
| 18-110_k141_78568 | Not-determined |
| 18-110_k141_240222 | Not-determined |
| 18-110_k141_124148 | Not-determined |
| 18-110_k141_240224 | Not-determined |
| 18-110_k141_216351 | Not-determined |
| 18-110_k141_101397 | Low-quality |
| 18-110_k141_227712 | Not-determined |
| 18-110_k141_43952 | Not-determined |
| 18-110_k141_158980 | Not-determined |
| 18-110_k141_193293 | Not-determined |
| 18-110_k141_124154 | Not-determined |
| 18-110_k141_193295 | Not-determined |
| 18-110_k141_124155 | Not-determined |
| 18-110_k141_260464 | Not-determined |
| 18-110_k141_227719 | Not-determined |
| 18-110_k141_112262 | Not-determined |
| 18-110_k141_89112 | Not-determined |
| 18-110_k141_66906 | Not-determined |
| 18-110_k141_227722 | Not-determined |
| 18-110_k141_136341 | Not-determined |
| 18-110_k141_66911 | Not-determined |
| 18-110_k141_101410 | Low-quality |
| 18-110_k141_170400 | Not-determined |
| 18-110_k141_33000 | Not-determined |
| 18-110_k141_260473 | Not-determined |
| 18-110_k141_147988 | Not-determined |
| 18-110_k141_19069 | Not-determined |
| 18-110_k141_203560 | Not-determined |
| 18-110_k141_33004 | Not-determined |
| 18-110_k141_240238 | Not-determined |
| 18-110_k141_101418 | Low-quality |
| 18-110_k141_203564 | Not-determined |
| 18-110_k141_227729 | Not-determined |
| 18-110_k141_170402 | Not-determined |
| 18-110_k141_182889 | Not-determined |
| 18-110_k141_89127 | Not-determined |
| 18-110_k141_273878 | Not-determined |
| 18-110_k141_170405 | Low-quality |
| 18-110_k141_250872 | Not-determined |
| 18-110_k141_66922 | Not-determined |
| 18-110_k141_78595 | Not-determined |
| 18-110_k141_124172 | Not-determined |
| 18-110_k141_148006 | Not-determined |
| 18-110_k141_148007 | Not-determined |
| 18-110_k141_78599 | Not-determined |
| 18-110_k141_89136 | Not-determined |
| 18-110_k141_112285 | Not-determined |
| 18-110_k141_158984 | Not-determined |
| 18-110_k141_66928 | Not-determined |
| 18-110_k141_78607 | Not-determined |
| 18-110_k141_33035 | Not-determined |
| 18-110_k141_124182 | Not-determined |
| 18-110_k141_273895 | Not-determined |
| 18-110_k141_66929 | Not-determined |
| 18-110_k141_182912 | Not-determined |
| 18-110_k141_193328 | Not-determined |
| 18-110_k141_260493 | Not-determined |
| 18-110_k141_170428 | Not-determined |
| 18-110_k141_193330 | Not-determined |
| 18-110_k141_19085 | Not-determined |
| 18-110_k141_203577 | Not-determined |
| 18-110_k141_112297 | Not-determined |
| 18-110_k141_260497 | Not-determined |
| 18-110_k141_89152 | Not-determined |
| 18-110_k141_136357 | Not-determined |
| 18-110_k141_158999 | Not-determined |
| 18-110_k141_240273 | Not-determined |
| 18-110_k141_203580 | Not-determined |
| 18-110_k141_66936 | Not-determined |
| 18-110_k141_182927 | Not-determined |
| 18-110_k141_136363 | Not-determined |
| 18-110_k141_193335 | Not-determined |
| 18-110_k141_43993 | Not-determined |
| 18-110_k141_240279 | Not-determined |
| 18-110_k141_112309 | Not-determined |
| 18-110_k141_273909 | Not-determined |
| 18-110_k141_203590 | Not-determined |
| 18-110_k141_240282 | Not-determined |
| 18-110_k141_159017 | Not-determined |
| 18-110_k141_136368 | Not-determined |
| 18-110_k141_19106 | Not-determined |
| 18-110_k141_19110 | Not-determined |
| 18-110_k141_19113 | Not-determined |
| 18-110_k141_227743 | Not-determined |
| 18-110_k141_216375 | Not-determined |
| 18-110_k141_273916 | Low-quality |
| 18-110_k141_112318 | Not-determined |
| 18-110_k141_19114 | Not-determined |
| 18-110_k141_203604 | Not-determined |
| 18-110_k141_78630 | Low-quality |
| 18-110_k141_216377 | Not-determined |
| 18-110_k141_240294 | Not-determined |
| 18-110_k141_203607 | Not-determined |
| 18-110_k141_136374 | Not-determined |
| 18-110_k141_193354 | Not-determined |
| 18-110_k141_250914 | Not-determined |
| 18-110_k141_136378 | Not-determined |
| 18-110_k141_19123 | Not-determined |
| 18-110_k141_112322 | Not-determined |
| 18-110_k141_182970 | Not-determined |
| 18-110_k141_78643 | Not-determined |
| 18-110_k141_240305 | Not-determined |
| 18-110_k141_112327 | Not-determined |
| 18-110_k141_193364 | Not-determined |
| 18-110_k141_78649 | Not-determined |
| 18-110_k141_182974 | Not-determined |
| 18-110_k141_250918 | Not-determined |
| 18-110_k141_193368 | Not-determined |
| 18-110_k141_148034 | Not-determined |
| 18-110_k141_33093 | Not-determined |
| 18-110_k141_19141 | Not-determined |
| 18-110_k141_112348 | Not-determined |
| 18-110_k141_89193 | Not-determined |
| 18-110_k141_216389 | Not-determined |
| 18-110_k141_193380 | Not-determined |
| 18-110_k141_193381 | Not-determined |
| 18-110_k141_148037 | Not-determined |
| 18-110_k141_19148 | Not-determined |
| 18-110_k141_112359 | Not-determined |
| 18-110_k141_159039 | Not-determined |
| 18-110_k141_112361 | Not-determined |
| 18-110_k141_44012 | Not-determined |
| 18-110_k141_112363 | Not-determined |
| 18-110_k141_159043 | Low-quality |
| 18-110_k141_112369 | Not-determined |
| 18-110_k141_250944 | Low-quality |
| 18-110_k141_216408 | Not-determined |
| 18-110_k141_216409 | Not-determined |
| 18-110_k141_250945 | Not-determined |
| 18-110_k141_227765 | Not-determined |
| 18-110_k141_216415 | Not-determined |
| 18-110_k141_182987 | Not-determined |
| 18-110_k141_182990 | Not-determined |
| 18-110_k141_8142 | Not-determined |
| 18-110_k141_170473 | Not-determined |
| 18-110_k141_159053 | Low-quality |
| 18-110_k141_240331 | Not-determined |
| 18-110_k141_227773 | Low-quality |
| 18-110_k141_124260 | Not-determined |
| 18-110_k141_170478 | Not-determined |
| 18-110_k141_124266 | Not-determined |
| 18-110_k141_170479 | Not-determined |
| 18-110_k141_170483 | Not-determined |
| 18-110_k141_240339 | Not-determined |
| 18-110_k141_89217 | Low-quality |
| 18-110_k141_148039 | Low-quality |
| 18-110_k141_240344 | Not-determined |
| 18-110_k141_8149 | Not-determined |
| 18-110_k141_112386 | Not-determined |
| 18-110_k141_19173 | Not-determined |
| 18-110_k141_78689 | Not-determined |
| 18-110_k141_170491 | Not-determined |
| 18-110_k141_44036 | Not-determined |
| 18-110_k141_170493 | Not-determined |
| 18-110_k141_78693 | Not-determined |
| 18-110_k141_159077 | Not-determined |
| 18-110_k141_78695 | Not-determined |
| 18-110_k141_240358 | Not-determined |
| 18-110_k141_89231 | Not-determined |
| 18-110_k141_216433 | Not-determined |
| 18-110_k141_136429 | Not-determined |
| 18-110_k141_216434 | Not-determined |
| 18-110_k141_8163 | Not-determined |
| 18-110_k141_203657 | Low-quality |
| 18-110_k141_8165 | Not-determined |
| 18-110_k141_170505 | Not-determined |
| 18-110_k141_136435 | Not-determined |
| 18-110_k141_136436 | Not-determined |
| 18-110_k141_136437 | Low-quality |
| 18-110_k141_136438 | Not-determined |
| 18-110_k141_8169 | Not-determined |
| 18-110_k141_216445 | Not-determined |
| 18-110_k141_66970 | Not-determined |
| 18-110_k141_78712 | Not-determined |
| 18-110_k141_136446 | Not-determined |
| 18-110_k141_33121 | Not-determined |
| 18-110_k141_33123 | Not-determined |
| 18-110_k141_89243 | Not-determined |
| 18-110_k141_148056 | Not-determined |
| 18-110_k141_183015 | Not-determined |
| 18-110_k141_203664 | Not-determined |
| 18-110_k141_183017 | Not-determined |
| 18-110_k141_89247 | Not-determined |
| 18-110_k141_216453 | Low-quality |
| 18-110_k141_203667 | Not-determined |
| 18-110_k141_203668 | Not-determined |
| 18-110_k141_203669 | Not-determined |
| 18-110_k141_33132 | Not-determined |
| 18-110_k141_101448 | Not-determined |
| 18-110_k141_148063 | Not-determined |
| 18-110_k141_170531 | Not-determined |
| 18-110_k141_227793 | Low-quality |
| 18-110_k141_170532 | Not-determined |
| 18-110_k141_170534 | Not-determined |
| 18-110_k141_78732 | Not-determined |
| 18-110_k141_124282 | Not-determined |
| 18-110_k141_19184 | Not-determined |
| 18-110_k141_19185 | Not-determined |
| 18-110_k141_203683 | Not-determined |
| 18-110_k141_183031 | Not-determined |
| 18-110_k141_148070 | Not-determined |
| 18-110_k141_193415 | Not-determined |
| 18-110_k141_250960 | Not-determined |
| 18-110_k141_227795 | Not-determined |
| 18-110_k141_101461 | Not-determined |
| 18-110_k141_250964 | Not-determined |
| 18-110_k141_183038 | Not-determined |
| 18-110_k141_183039 | Not-determined |
| 18-110_k141_136466 | Not-determined |
| 18-110_k141_33153 | Not-determined |
| 18-110_k141_33154 | Not-determined |
| 18-110_k141_67003 | Not-determined |
| 18-110_k141_101469 | Not-determined |
| 18-110_k141_33157 | Not-determined |
| 18-110_k141_19199 | Not-determined |
| 18-110_k141_33158 | Not-determined |
| 18-110_k141_148087 | Not-determined |
| 18-110_k141_148089 | Not-determined |
| 18-110_k141_124293 | Not-determined |
| 18-110_k141_183055 | Low-quality |
| 18-110_k141_124296 | Not-determined |
| 18-110_k141_183058 | Not-determined |
| 18-110_k141_101488 | Not-determined |
| 18-110_k141_159083 | Not-determined |
| 18-110_k141_250978 | Not-determined |
| 18-110_k141_216482 | Not-determined |
| 18-110_k141_148096 | Not-determined |
| 18-110_k141_101491 | Not-determined |
| 18-110_k141_78758 | Low-quality |
| 18-110_k141_193444 | Not-determined |
| 18-110_k141_101493 | Not-determined |
| 18-110_k141_55122 | Not-determined |
| 18-110_k141_101495 | Not-determined |
| 18-110_k141_78766 | Not-determined |
| 18-110_k141_19226 | Not-determined |
| 18-110_k141_148108 | Not-determined |
| 18-110_k141_274018 | Not-determined |
| 18-110_k141_193458 | Not-determined |
| 18-110_k141_44100 | Not-determined |
| 18-110_k141_8195 | Not-determined |
| 18-110_k141_78776 | Not-determined |
| 18-110_k141_193464 | Not-determined |
| 18-110_k141_193465 | Not-determined |
| 18-110_k141_136503 | Not-determined |
| 18-110_k141_33190 | Not-determined |
| 18-110_k141_148116 | Not-determined |
| 18-110_k141_193467 | Not-determined |
| 18-110_k141_183090 | Not-determined |
| 18-110_k141_203728 | Low-quality |
| 18-110_k141_78787 | Not-determined |
| 18-110_k141_124318 | Not-determined |
| 18-110_k141_193479 | Not-determined |
| 18-110_k141_193481 | Not-determined |
| 18-110_k141_170549 | Not-determined |
| 18-110_k141_19242 | Low-quality |
| 18-110_k141_193488 | Not-determined |
| 18-110_k141_170554 | Low-quality |
| 18-110_k141_78799 | Not-determined |
| 18-110_k141_159121 | Not-determined |
| 18-110_k141_78805 | Not-determined |
| 18-110_k141_227849 | Not-determined |
| 18-110_k141_170563 | Low-quality |
| 18-110_k141_193501 | Not-determined |
| 18-110_k141_193503 | Low-quality |
| 18-110_k141_55153 | Not-determined |
| 18-110_k141_159129 | Low-quality |
| 18-110_k141_203759 | Not-determined |
| 18-110_k141_33220 | Not-determined |
| 18-110_k141_78816 | Not-determined |
| 18-110_k141_33222 | Not-determined |
| 18-110_k141_159136 | Not-determined |
| 18-110_k141_193508 | Not-determined |
| 18-110_k141_112408 | Not-determined |
| 18-110_k141_112412 | Not-determined |
| 18-110_k141_112413 | Not-determined |
| 18-110_k141_89310 | Not-determined |
| 18-110_k141_55167 | Not-determined |
| 18-110_k141_240435 | Not-determined |
| 18-110_k141_227861 | Not-determined |
| 18-110_k141_159148 | Low-quality |
| 18-110_k141_183135 | Not-determined |
| 18-110_k141_251017 | Not-determined |
| 18-110_k141_78833 | Not-determined |
| 18-110_k141_124368 | Not-determined |
| 18-110_k141_203772 | Low-quality |
| 18-110_k141_101557 | Not-determined |
| 18-110_k141_227868 | Low-quality |
| 18-110_k141_274027 | Not-determined |
| 18-110_k141_8221 | Not-determined |
| 18-110_k141_227870 | Not-determined |
| 18-110_k141_101559 | Not-determined |
| 18-110_k141_101563 | Not-determined |
| 18-110_k141_216500 | Not-determined |
| 18-110_k141_216503 | Not-determined |
| 18-110_k141_78845 | Not-determined |
| 18-110_k141_240447 | Not-determined |
| 18-110_k141_101568 | Not-determined |
| 18-110_k141_101569 | Not-determined |
| 18-110_k141_159162 | Not-determined |
| 18-110_k141_78851 | Not-determined |
| 18-110_k141_274035 | Not-determined |
| 18-110_k141_8231 | Not-determined |
| 18-110_k141_44142 | Not-determined |
| 18-110_k141_78854 | Not-determined |
| 18-110_k141_112419 | Not-determined |
| 18-110_k141_170581 | Not-determined |
| 18-110_k141_101577 | Not-determined |
| 18-110_k141_193528 | Not-determined |
| 18-110_k141_148170 | Not-determined |
| 18-110_k141_203790 | Not-determined |
| 18-110_k141_136572 | Not-determined |
| 18-110_k141_55178 | Not-determined |
| 18-110_k141_44145 | Not-determined |
| 18-110_k141_216506 | Not-determined |
| 18-110_k141_183147 | Not-determined |
| 18-110_k141_67011 | Not-determined |
| 18-110_k141_67013 | Not-determined |
| 18-110_k141_89345 | Not-determined |
| 18-110_k141_67014 | Not-determined |
| 18-110_k141_8248 | Not-determined |
| 18-110_k141_170606 | Not-determined |
| 18-110_k141_183161 | Not-determined |
| 18-110_k141_251046 | Low-quality |
| 18-110_k141_124388 | Not-determined |
| 18-110_k141_112434 | Low-quality |
| 18-110_k141_67030 | Not-determined |
| 18-110_k141_274044 | Not-determined |
| 18-110_k141_240471 | Not-determined |
| 18-110_k141_170614 | Not-determined |
| 18-110_k141_159175 | Not-determined |
| 18-110_k141_67039 | Not-determined |
| 18-110_k141_8261 | Not-determined |
| 18-110_k141_33267 | Not-determined |
| 18-110_k141_136590 | Not-determined |
| 18-110_k141_112439 | Not-determined |
| 18-110_k141_240481 | Not-determined |
| 18-110_k141_89357 | Low-quality |
| 18-110_k141_240487 | Not-determined |
| 18-110_k141_183182 | Not-determined |
| 18-110_k141_33277 | Not-determined |
| 18-110_k141_203810 | Not-determined |
| 18-110_k141_78881 | Not-determined |
| 18-110_k141_274052 | Low-quality |
| 18-110_k141_170632 | Not-determined |
| 18-110_k141_183187 | Low-quality |
| 18-110_k141_19307 | Not-determined |
| 18-110_k141_89374 | Not-determined |
| 18-110_k141_89376 | Not-determined |
| 18-110_k141_136608 | Not-determined |
| 18-110_k141_227901 | Not-determined |
| 18-110_k141_240502 | Not-determined |
| 18-110_k141_78887 | Not-determined |
| 18-110_k141_148203 | Low-quality |
| 18-110_k141_148207 | Not-determined |
| 18-110_k141_136615 | Not-determined |
| 18-110_k141_89383 | Not-determined |
| 18-110_k141_159208 | Not-determined |
| 18-110_k141_251063 | Not-determined |
| 18-110_k141_203831 | Not-determined |
| 18-110_k141_274072 | Low-quality |
| 18-110_k141_124416 | Low-quality |
| 18-110_k141_67071 | Low-quality |
| 18-110_k141_19337 | Not-determined |
| 18-110_k141_148225 | Low-quality |
| 18-110_k141_251071 | Not-determined |
| 18-110_k141_55209 | Not-determined |
| 18-110_k141_193565 | Not-determined |
| 18-110_k141_8306 | Not-determined |
| 18-110_k141_193568 | Not-determined |
| 18-110_k141_227912 | Not-determined |
| 18-110_k141_67077 | Not-determined |
| 18-110_k141_55214 | Not-determined |
| 18-110_k141_148233 | Not-determined |
| 18-110_k141_67084 | Not-determined |
| 18-110_k141_251082 | Not-determined |
| 18-110_k141_183206 | Not-determined |
| 18-110_k141_240514 | Low-quality |
| 18-110_k141_33287 | Not-determined |
| 18-110_k141_67096 | Not-determined |
| 18-110_k141_55221 | Not-determined |
| 18-110_k141_78917 | Not-determined |
| 18-110_k141_216576 | Not-determined |
| 18-110_k141_89401 | Not-determined |
| 18-110_k141_67100 | Not-determined |
| 18-110_k141_136647 | Not-determined |
| 18-110_k141_124440 | Not-determined |
| 18-110_k141_136648 | Not-determined |
| 18-110_k141_55231 | Not-determined |
| 18-110_k141_216577 | Low-quality |
| 18-110_k141_55232 | Not-determined |
| 18-110_k141_148258 | Not-determined |
| 18-110_k141_203842 | Low-quality |
| 18-110_k141_89413 | Not-determined |
| 18-110_k141_240520 | Not-determined |
| 18-110_k141_159230 | Not-determined |
| 18-110_k141_148265 | Not-determined |
| 18-110_k141_55240 | Not-determined |
| 18-110_k141_260559 | Not-determined |
| 18-110_k141_170651 | Not-determined |
| 18-110_k141_227931 | Not-determined |
| 18-110_k141_148269 | Low-quality |
| 18-110_k141_216590 | Not-determined |
| 18-110_k141_124464 | Not-determined |
| 18-110_k141_78947 | Not-determined |
| 18-110_k141_8325 | Not-determined |
| 18-110_k141_159246 | Not-determined |
| 18-110_k141_33307 | Not-determined |
| 18-110_k141_33308 | Not-determined |
| 18-110_k141_170676 | Not-determined |
| 18-110_k141_55259 | Not-determined |
| 18-110_k141_112516 | Not-determined |
| 18-110_k141_124474 | Not-determined |
| 18-110_k141_203867 | Low-quality |
| 18-110_k141_193620 | Not-determined |
| 18-110_k141_274096 | Low-quality |
| 18-110_k141_8335 | Not-determined |
| 18-110_k141_227952 | Low-quality |
| 18-110_k141_193622 | Not-determined |
| 18-110_k141_67110 | Not-determined |
| 18-110_k141_89427 | Not-determined |
| 18-110_k141_8339 | Low-quality |
| 18-110_k141_260575 | Low-quality |
| 18-110_k141_193626 | Not-determined |
| 18-110_k141_183257 | Not-determined |
| 18-110_k141_19353 | Not-determined |
| 18-110_k141_8344 | Low-quality |
| 18-110_k141_148293 | Not-determined |
| 18-110_k141_159264 | Not-determined |
| 18-110_k141_193636 | Not-determined |
| 18-110_k141_33327 | Low-quality |
| 18-110_k141_19362 | Not-determined |
| 18-110_k141_159267 | Not-determined |
| 18-110_k141_251109 | Not-determined |
| 18-110_k141_148295 | Not-determined |
| 18-110_k141_19364 | Not-determined |
| 18-110_k141_8347 | Not-determined |
| 18-110_k141_260586 | Not-determined |
| 18-110_k141_136675 | Not-determined |
| 18-110_k141_112528 | Not-determined |
| 18-110_k141_19370 | Not-determined |
| 18-110_k141_274105 | Not-determined |
| 18-110_k141_240554 | Not-determined |
| 18-110_k141_193647 | Not-determined |
| 18-110_k141_112534 | Not-determined |
| 18-110_k141_112538 | Not-determined |
| 18-110_k141_112540 | Low-quality |
| 18-110_k141_227975 | Not-determined |
| 18-110_k141_33337 | Not-determined |
| 18-110_k141_8358 | Not-determined |
| 18-110_k141_78971 | Not-determined |
| 18-110_k141_8362 | Not-determined |
| 18-110_k141_136687 | Not-determined |
| 18-110_k141_101594 | Not-determined |
| 18-110_k141_89460 | Not-determined |
| 18-110_k141_112551 | Not-determined |
| 18-110_k141_170709 | Not-determined |
| 18-110_k141_8366 | Low-quality |
| 18-110_k141_170715 | Not-determined |
| 18-110_k141_159285 | Not-determined |
| 18-110_k141_159286 | Low-quality |
| 18-110_k141_159289 | Not-determined |
| 18-110_k141_216625 | Not-determined |
| 18-110_k141_124509 | Not-determined |
| 18-110_k141_159291 | Not-determined |
| 18-110_k141_78981 | Not-determined |
| 18-110_k141_8377 | Not-determined |
| 18-110_k141_136692 | Low-quality |
| 18-110_k141_78984 | Not-determined |
| 18-110_k141_124512 | Not-determined |
| 18-110_k141_89470 | Not-determined |
| 18-110_k141_78987 | Not-determined |
| 18-110_k141_240585 | Not-determined |
| 18-110_k141_124517 | Not-determined |
| 18-110_k141_8390 | Not-determined |
| 18-110_k141_148318 | Low-quality |
| 18-110_k141_89483 | Not-determined |
| 18-110_k141_78993 | Not-determined |
| 18-110_k141_55295 | Not-determined |
| 18-110_k141_33360 | Not-determined |
| 18-110_k141_159312 | Not-determined |
| 18-110_k141_251152 | Not-determined |
| 18-110_k141_183308 | Not-determined |
| 18-110_k141_124525 | Not-determined |
| 18-110_k141_159320 | Not-determined |
| 18-110_k141_33366 | Not-determined |
| 18-110_k141_274132 | Not-determined |
| 18-110_k141_8402 | Not-determined |
| 18-110_k141_8404 | Not-determined |
| 18-110_k141_79012 | Not-determined |
| 18-110_k141_79014 | Low-quality |
| 18-110_k141_251157 | Not-determined |
| 18-110_k141_124533 | Not-determined |
| 18-110_k141_148338 | Not-determined |
| 18-110_k141_112582 | Not-determined |
| 18-110_k141_112583 | Not-determined |
| 18-110_k141_251161 | Low-quality |
| 18-110_k141_159338 | Not-determined |
| 18-110_k141_79021 | Not-determined |
| 18-110_k141_19385 | Not-determined |
| 18-110_k141_251165 | Not-determined |
| 18-110_k141_19389 | Not-determined |
| 18-110_k141_228007 | Not-determined |
| 18-110_k141_216653 | Not-determined |
| 18-110_k141_228009 | Not-determined |
| 18-110_k141_240626 | Not-determined |
| 18-110_k141_89512 | Not-determined |
| 18-110_k141_148349 | Low-quality |
| 18-110_k141_89516 | Not-determined |
| 18-110_k141_148352 | Not-determined |
| 18-110_k141_183325 | Not-determined |
| 18-110_k141_112603 | Not-determined |
| 18-110_k141_228015 | Not-determined |
| 18-110_k141_44255 | Not-determined |
| 18-110_k141_44256 | Not-determined |
| 18-110_k141_44258 | Not-determined |
| 18-110_k141_216665 | Not-determined |
| 18-110_k141_216667 | Low-quality |
| 18-110_k141_67145 | Low-quality |
| 18-110_k141_274157 | Low-quality |
| 18-110_k141_136713 | Not-determined |
| 18-110_k141_183343 | Not-determined |
| 18-110_k141_228026 | Not-determined |
| 18-110_k141_136715 | Not-determined |
| 18-110_k141_240638 | Not-determined |
| 18-110_k141_8448 | Not-determined |
| 18-110_k141_8449 | Not-determined |
| 18-110_k141_170757 | Not-determined |
| 18-110_k141_55325 | Not-determined |
| 18-110_k141_274171 | Low-quality |
| 18-110_k141_240651 | Not-determined |
| 18-110_k141_136723 | Not-determined |
| 18-110_k141_240658 | Not-determined |
| 18-110_k141_44294 | Not-determined |
| 18-110_k141_112629 | Not-determined |
| 18-110_k141_136725 | Not-determined |
| 18-110_k141_33387 | Not-determined |
| 18-110_k141_183372 | Not-determined |
| 18-110_k141_124562 | Not-determined |
| 18-110_k141_19419 | Not-determined |
| 18-110_k141_124570 | Not-determined |
| 18-110_k141_89556 | Not-determined |
| 18-110_k141_159385 | Not-determined |
| 18-110_k141_8469 | Not-determined |
| 18-110_k141_159387 | Not-determined |
| 18-110_k141_136733 | Not-determined |
| 18-110_k141_19422 | Low-quality |
| 18-110_k141_159390 | Not-determined |
| 18-110_k141_136735 | Not-determined |
| 18-110_k141_203905 | Not-determined |
| 18-110_k141_148396 | Low-quality |
| 18-110_k141_136743 | Not-determined |
| 18-110_k141_203913 | Not-determined |
| 18-110_k141_159400 | Not-determined |
| 18-110_k141_203914 | Not-determined |
| 18-110_k141_55344 | Not-determined |
| 18-110_k141_148397 | Not-determined |
| 18-110_k141_55345 | Not-determined |
| 18-110_k141_136751 | Not-determined |
| 18-110_k141_251211 | Low-quality |
| 18-110_k141_148401 | Not-determined |
| 18-110_k141_274188 | Not-determined |
| 18-110_k141_8477 | Not-determined |
| 18-110_k141_216715 | Not-determined |
| 18-110_k141_33398 | Not-determined |
| 18-110_k141_260663 | Not-determined |
| 18-110_k141_33399 | Not-determined |
| 18-110_k141_33400 | Not-determined |
| 18-110_k141_228057 | Not-determined |
| 18-110_k141_33402 | Not-determined |
| 18-110_k141_8492 | Not-determined |
| 18-110_k141_89587 | Not-determined |
| 18-110_k141_148414 | Not-determined |
| 18-110_k141_203921 | Not-determined |
| 18-110_k141_203926 | Not-determined |
| 18-110_k141_79063 | Not-determined |
| 18-110_k141_203930 | Not-determined |
| 18-110_k141_89594 | Not-determined |
| 18-110_k141_33416 | Not-determined |
| 18-110_k141_183423 | Not-determined |
| 18-110_k141_170778 | Not-determined |

| | |
|---|---|
| 18-110_k141_203935 | Not-determined |
| 18-110_k141_216736 | Low-quality |
| 18-110_k141_216738 | Low-quality |
| 18-110_k141_79074 | Low-quality |
| 18-110_k141_79078 | Not-determined |
| 18-110_k141_79080 | Not-determined |
| 18-110_k141_89608 | Not-determined |
| 18-110_k141_260685 | Low-quality |
| 18-110_k141_216745 | Not-determined |
| 18-110_k141_101642 | Low-quality |
| 18-110_k141_136771 | Not-determined |
| 18-110_k141_101645 | Not-determined |
| 18-110_k141_101648 | Not-determined |
| 18-110_k141_203954 | Not-determined |
| 18-110_k141_136775 | Not-determined |
| 18-110_k141_136777 | Not-determined |
| 18-110_k141_260694 | Not-determined |
| 18-110_k141_216754 | Not-determined |
| 18-110_k141_101655 | Not-determined |
| 18-110_k141_159410 | Not-determined |
| 18-110_k141_240673 | Not-determined |
| 18-110_k141_124614 | Not-determined |
| 18-110_k141_170801 | Not-determined |
| 18-110_k141_193683 | Not-determined |
| 18-110_k141_183450 | Not-determined |
| 18-110_k141_19470 | Not-determined |
| 18-110_k141_79101 | Not-determined |
| 18-110_k141_79102 | Not-determined |
| 18-110_k141_240686 | Not-determined |
| 18-110_k141_79104 | Not-determined |
| 18-110_k141_55353 | Not-determined |
| 18-110_k141_159423 | Not-determined |
| 18-110_k141_124619 | Not-determined |
| 18-110_k141_216774 | Not-determined |
| 18-110_k141_33453 | Not-determined |
| 18-110_k141_67169 | Not-determined |
| 18-110_k141_216775 | Low-quality |
| 18-110_k141_240696 | Low-quality |
| 18-110_k141_79113 | Low-quality |
| 18-110_k141_112687 | Not-determined |
| 18-110_k141_19481 | Not-determined |
| 18-110_k141_216779 | Not-determined |
| 18-110_k141_55366 | Not-determined |
| 18-110_k141_251244 | Not-determined |
| 18-110_k141_55368 | Not-determined |
| 18-110_k141_19491 | Not-determined |
| 18-110_k141_55371 | Not-determined |
| 18-110_k141_136781 | Not-determined |
| 18-110_k141_44372 | Not-determined |
| 18-110_k141_136783 | Not-determined |
| 18-110_k141_216791 | Not-determined |
| 18-110_k141_19495 | Not-determined |
| 18-110_k141_216795 | Low-quality |
| 18-110_k141_228112 | Not-determined |
| 18-110_k141_67181 | Not-determined |
| 18-110_k141_33472 | Not-determined |
| 18-110_k141_228114 | Low-quality |
| 18-110_k141_19507 | Not-determined |
| 18-110_k141_260698 | Not-determined |
| 18-110_k141_216800 | Not-determined |
| 18-110_k141_79143 | Not-determined |
| 18-110_k141_112714 | Not-determined |
| 18-110_k141_251277 | Not-determined |
| 18-110_k141_67198 | Not-determined |
| 18-110_k141_193727 | Not-determined |
| 18-110_k141_136799 | Not-determined |
| 18-110_k141_33478 | Not-determined |
| 18-110_k141_136802 | Not-determined |
| 18-110_k141_55397 | Not-determined |
| 18-110_k141_112726 | Not-determined |
| 18-110_k141_260708 | Not-determined |
| 18-110_k141_79150 | Not-determined |
| 18-110_k141_33488 | Not-determined |
| 18-110_k141_55400 | Not-determined |
| 18-110_k141_33492 | Not-determined |
| 18-110_k141_67207 | Not-determined |
| 18-110_k141_67208 | Not-determined |
| 18-110_k141_89630 | Not-determined |
| 18-110_k141_112731 | Not-determined |
| 18-110_k141_44386 | Not-determined |
| 18-110_k141_228153 | Not-determined |
| 18-110_k141_228154 | Not-determined |
| 18-110_k141_101682 | Not-determined |
| 18-110_k141_101684 | Not-determined |
| 18-110_k141_44390 | Not-determined |
| 18-110_k141_79156 | Not-determined |
| 18-110_k141_159486 | Not-determined |
| 18-110_k141_79158 | Not-determined |
| 18-110_k141_33507 | Not-determined |
| 18-110_k141_79163 | Not-determined |
| 18-110_k141_89639 | Not-determined |
| 18-110_k141_240706 | Not-determined |
| 18-110_k141_216829 | Not-determined |
| 18-110_k141_240707 | Not-determined |
| 18-110_k141_159494 | Not-determined |
| 18-110_k141_112737 | Not-determined |
| 18-110_k141_55417 | Not-determined |
| 18-110_k141_183495 | Not-determined |
| 18-110_k141_183497 | Not-determined |
| 18-110_k141_33511 | Not-determined |
| 18-110_k141_136839 | Not-determined |
| 18-110_k141_148539 | Not-determined |
| 18-110_k141_44395 | Not-determined |
| 18-110_k141_159500 | Not-determined |
| 18-110_k141_19531 | Low-quality |
| 18-110_k141_159505 | Not-determined |
| 18-110_k141_183502 | Not-determined |
| 18-110_k141_19534 | Not-determined |
| 18-110_k141_159509 | Not-determined |
| 18-110_k141_55426 | Not-determined |
| 18-110_k141_183505 | Not-determined |
| 18-110_k141_228181 | Not-determined |
| 18-110_k141_55430 | Not-determined |
| 18-110_k141_44403 | Not-determined |
| 18-110_k141_124677 | Not-determined |
| 18-110_k141_240725 | Not-determined |
| 18-110_k141_112748 | Not-determined |
| 18-110_k141_101708 | Not-determined |
| 18-110_k141_183507 | Not-determined |
| 18-110_k141_228184 | Not-determined |
| 18-110_k141_19539 | Not-determined |
| 18-110_k141_67225 | Not-determined |
| 18-110_k141_55435 | Low-quality |
| 18-110_k141_193753 | Not-determined |
| 18-110_k141_136856 | Not-determined |
| 18-110_k141_19545 | Not-determined |
| 18-110_k141_19548 | Not-determined |
| 18-110_k141_216866 | Not-determined |
| 18-110_k141_159527 | Not-determined |
| 18-110_k141_148570 | Low-quality |
| 18-110_k141_79190 | Not-determined |
| 18-110_k141_170846 | Not-determined |
| 18-110_k141_216871 | Not-determined |
| 18-110_k141_89658 | Not-determined |
| 18-110_k141_101733 | Not-determined |
| 18-110_k141_33541 | Not-determined |
| 18-110_k141_112761 | Not-determined |
| 18-110_k141_159550 | Not-determined |
| 18-110_k141_136884 | Not-determined |
| 18-110_k141_136886 | Not-determined |
| 18-110_k141_183520 | Not-determined |
| 18-110_k141_44431 | Not-determined |
| 18-110_k141_204045 | Not-determined |
| 18-110_k141_89676 | Not-determined |
| 18-110_k141_240768 | Not-determined |
| 18-110_k141_240771 | Not-determined |
| 18-110_k141_240778 | Not-determined |
| 18-110_k141_112778 | Not-determined |
| 18-110_k141_260764 | Not-determined |
| 18-110_k141_101767 | Not-determined |
| 18-110_k141_79213 | Not-determined |
| 18-110_k141_44453 | Not-determined |
| 18-110_k141_79214 | Not-determined |
| 18-110_k141_101771 | Not-determined |
| 18-110_k141_44455 | Not-determined |
| 18-110_k141_204062 | Not-determined |
| 18-110_k141_228204 | Not-determined |
| 18-110_k141_136920 | Not-determined |
| 18-110_k141_112780 | Not-determined |
| 18-110_k141_67270 | Not-determined |
| 18-110_k141_136923 | Not-determined |
| 18-110_k141_67275 | Not-determined |
| 18-110_k141_19593 | Not-determined |
| 18-110_k141_19594 | Not-determined |
| 18-110_k141_228215 | Not-determined |
| 18-110_k141_228216 | Not-determined |
| 18-110_k141_136931 | Not-determined |
| 18-110_k141_67281 | Low-quality |
| 18-110_k141_44470 | Not-determined |
| 18-110_k141_89702 | Not-determined |
| 18-110_k141_79231 | Not-determined |
| 18-110_k141_33549 | Not-determined |
| 18-110_k141_44475 | Not-determined |
| 18-110_k141_204079 | Not-determined |
| 18-110_k141_124722 | Low-quality |
| 18-110_k141_19606 | Not-determined |
| 18-110_k141_19607 | Not-determined |
| 18-110_k141_193800 | Low-quality |
| 18-110_k141_44478 | Not-determined |
| 18-110_k141_67290 | Not-determined |
| 18-110_k141_89710 | Not-determined |
| 18-110_k141_44483 | Not-determined |
| 18-110_k141_193806 | Not-determined |
| 18-110_k141_101791 | Low-quality |
| 18-110_k141_204085 | Low-quality |
| 18-110_k141_79248 | Not-determined |
| 18-110_k141_112805 | Not-determined |
| 18-110_k141_55490 | Not-determined |
| 18-110_k141_112806 | Not-determined |
| 18-110_k141_33564 | Not-determined |
| 18-110_k141_55492 | Not-determined |
| 18-110_k141_204091 | Not-determined |
| 18-110_k141_44491 | Not-determined |
| 18-110_k141_67305 | Not-determined |
| 18-110_k141_79255 | Not-determined |
| 18-110_k141_240822 | Not-determined |
| 18-110_k141_112815 | Not-determined |
| 18-110_k141_44497 | Not-determined |
| 18-110_k141_67312 | Not-determined |
| 18-110_k141_55505 | Not-determined |
| 18-110_k141_89729 | Not-determined |
| 18-110_k141_112817 | Not-determined |
| 18-110_k141_67318 | Not-determined |
| 18-110_k141_159589 | Not-determined |
| 18-110_k141_112824 | Not-determined |
| 18-110_k141_101809 | Not-determined |
| 18-110_k141_251321 | Not-determined |
| 18-110_k141_240834 | Not-determined |
| 18-110_k141_193842 | Not-determined |
| 18-110_k141_251326 | Not-determined |
| 18-110_k141_251328 | Not-determined |
| 18-110_k141_44507 | Not-determined |
| 18-110_k141_124757 | Not-determined |
| 18-110_k141_240844 | Not-determined |
| 18-110_k141_240845 | Not-determined |
| 18-110_k141_33574 | Not-determined |
| 18-110_k141_44513 | Not-determined |
| 18-110_k141_101816 | Not-determined |
| 18-110_k141_159600 | Not-determined |
| 18-110_k141_124765 | Low-quality |
| 18-110_k141_89759 | Not-determined |
| 18-110_k141_89762 | Not-determined |
| 18-110_k141_44521 | Not-determined |
| 18-110_k141_183553 | Not-determined |
| 18-110_k141_19644 | Not-determined |
| 18-110_k141_67343 | Not-determined |
| 18-110_k141_79279 | Not-determined |
| 18-110_k141_44530 | Not-determined |
| 18-110_k141_79280 | Not-determined |
| 18-110_k141_44532 | Not-determined |
| 18-110_k141_193859 | Not-determined |
| 18-110_k141_148630 | Not-determined |
| 18-110_k141_193864 | Not-determined |
| 18-110_k141_44544 | Not-determined |
| 18-110_k141_67354 | Not-determined |
| 18-110_k141_170891 | Not-determined |
| 18-110_k141_240862 | Not-determined |
| 18-110_k141_170895 | Not-determined |
| 18-110_k141_101836 | Not-determined |
| 18-110_k141_228240 | Not-determined |
| 18-110_k141_44553 | Not-determined |
| 18-110_k141_19666 | Not-determined |
| 18-110_k141_183578 | Not-determined |
| 18-110_k141_33601 | Not-determined |
| 18-110_k141_79287 | Not-determined |
| 18-110_k141_240877 | Not-determined |
| 18-110_k141_228246 | Not-determined |
| 18-110_k141_33605 | Not-determined |
| 18-110_k141_183586 | Not-determined |
| 18-110_k141_228250 | Not-determined |
| 18-110_k141_101841 | Not-determined |
| 18-110_k141_170899 | Not-determined |
| 18-110_k141_148651 | Not-determined |
| 18-110_k141_228254 | Not-determined |
| 18-110_k141_89773 | Not-determined |
| 18-110_k141_89774 | Not-determined |
| 18-110_k141_33612 | Not-determined |
| 18-110_k141_260820 | Not-determined |
| 18-110_k141_44572 | Not-determined |
| 18-110_k141_33615 | Not-determined |
| 18-110_k141_251350 | Not-determined |
| 18-110_k141_112860 | Not-determined |
| 18-110_k141_112862 | Not-determined |
| 18-110_k141_193879 | Not-determined |
| 18-110_k141_101846 | Not-determined |
| 18-110_k141_260828 | Not-determined |
| 18-110_k141_19673 | Not-determined |
| 18-110_k141_89780 | Not-determined |
| 18-110_k141_101852 | Not-determined |
| 18-110_k141_44582 | Not-determined |
| 18-110_k141_33626 | Not-determined |
| 18-110_k141_79303 | Not-determined |
| 18-110_k141_148667 | Not-determined |
| 18-110_k141_44585 | Not-determined |
| 18-110_k141_33631 | Not-determined |
| 18-110_k141_204136 | Not-determined |
| 18-110_k141_67399 | Not-determined |
| 18-110_k141_67402 | Not-determined |
| 18-110_k141_183622 | Not-determined |
| 18-110_k141_33637 | Not-determined |
| 18-110_k141_33639 | Not-determined |
| 18-110_k141_136963 | Not-determined |
| 18-110_k141_251364 | Not-determined |
| 18-110_k141_89801 | Low-quality |
| 18-110_k141_159665 | Not-determined |
| 18-110_k141_67405 | Not-determined |
| 18-110_k141_148694 | Not-determined |
| 18-110_k141_251373 | Not-determined |
| 18-110_k141_19700 | Not-determined |
| 18-110_k141_89808 | Low-quality |
| 18-110_k141_33652 | Not-determined |
| 18-110_k141_101873 | Not-determined |
| 18-110_k141_124774 | Not-determined |
| 18-110_k141_33654 | Not-determined |
| 18-110_k141_193899 | Not-determined |
| 18-110_k141_8523 | Not-determined |
| 18-110_k141_260856 | Not-determined |
| 18-110_k141_33658 | Not-determined |
| 18-110_k141_193903 | Not-determined |
| 18-110_k141_193905 | Not-determined |
| 18-110_k141_251384 | Not-determined |
| 18-110_k141_79333 | Not-determined |
| 18-110_k141_44604 | Not-determined |
| 18-110_k141_89817 | Not-determined |
| 18-110_k141_170924 | Not-determined |
| 18-110_k141_112905 | Not-determined |
| 18-110_k141_124790 | Not-determined |
| 18-110_k141_274207 | Not-determined |
| 18-110_k141_170929 | Not-determined |
| 18-110_k141_19718 | Not-determined |
| 18-110_k141_89822 | Not-determined |
| 18-110_k141_159684 | Not-determined |
| 18-110_k141_274209 | Not-determined |
| 18-110_k141_55587 | Not-determined |
| 18-110_k141_124796 | Not-determined |
| 18-110_k141_240891 | Not-determined |
| 18-110_k141_170938 | Not-determined |
| 18-110_k141_89827 | Not-determined |
| 18-110_k141_216925 | Not-determined |
| 18-110_k141_101902 | Not-determined |
| 18-110_k141_240892 | Low-quality |
| 18-110_k141_112916 | Not-determined |
| 18-110_k141_112918 | Not-determined |
| 18-110_k141_124809 | Low-quality |
| 18-110_k141_112920 | Not-determined |
| 18-110_k141_112921 | Not-determined |
| 18-110_k141_274214 | Not-determined |
| 18-110_k141_112922 | Not-determined |
| 18-110_k141_204168 | Not-determined |
| 18-110_k141_19736 | Not-determined |
| 18-110_k141_170948 | Not-determined |
| 18-110_k141_240904 | Not-determined |
| 18-110_k141_193918 | Low-quality |
| 18-110_k141_101918 | Not-determined |
| 18-110_k141_148743 | Not-determined |
| 18-110_k141_183658 | Not-determined |
| 18-110_k141_204174 | Not-determined |
| 18-110_k141_240914 | Low-quality |
| 18-110_k141_148748 | Not-determined |
| 18-110_k141_112933 | Not-determined |
| 18-110_k141_170952 | Not-determined |
| 18-110_k141_216941 | Not-determined |
| 18-110_k141_216942 | Not-determined |
| 18-110_k141_33704 | Not-determined |
| 18-110_k141_19742 | Not-determined |
| 18-110_k141_55608 | Not-determined |
| 18-110_k141_33708 | Not-determined |
| 18-110_k141_124821 | Not-determined |
| 18-110_k141_251396 | Not-determined |
| 18-110_k141_33710 | Not-determined |
| 18-110_k141_274222 | Not-determined |
| 18-110_k141_260862 | Not-determined |
| 18-110_k141_240924 | Not-determined |
| 18-110_k141_101934 | Not-determined |
| 18-110_k141_251400 | Not-determined |
| 18-110_k141_101936 | Not-determined |
| 18-110_k141_251403 | Not-determined |
| 18-110_k141_216949 | Low-quality |
| 18-110_k141_79362 | Not-determined |
| 18-110_k141_183676 | Not-determined |
| 18-110_k141_260868 | Not-determined |
| 18-110_k141_193934 | Not-determined |
| 18-110_k141_148762 | Low-quality |
| 18-110_k141_137011 | Not-determined |
| 18-110_k141_274231 | Not-determined |
| 18-110_k141_260873 | Not-determined |
| 18-110_k141_137013 | Low-quality |
| 18-110_k141_260875 | Not-determined |
| 18-110_k141_137015 | Not-determined |
| 18-110_k141_19757 | Low-quality |
| 18-110_k141_251411 | Not-determined |
| 18-110_k141_137018 | Not-determined |
| 18-110_k141_137020 | Low-quality |
| 18-110_k141_183682 | Not-determined |
| 18-110_k141_251414 | Not-determined |
| 18-110_k141_193942 | Not-determined |
| 18-110_k141_101945 | Not-determined |
| 18-110_k141_170972 | Not-determined |
| 18-110_k141_67466 | Not-determined |
| 18-110_k141_137023 | Not-determined |
| 18-110_k141_148776 | Not-determined |
| 18-110_k141_67471 | Not-determined |
| 18-110_k141_124836 | Not-determined |
| 18-110_k141_240954 | Not-determined |
| 18-110_k141_240956 | Low-quality |
| 18-110_k141_260886 | Not-determined |
| 18-110_k141_124842 | Not-determined |
| 18-110_k141_79380 | Not-determined |
| 18-110_k141_79385 | Not-determined |
| 18-110_k141_260889 | Not-determined |
| 18-110_k141_251433 | Not-determined |
| 18-110_k141_260891 | Not-determined |
| 18-110_k141_79391 | Not-determined |
| 18-110_k141_148782 | Not-determined |
| 18-110_k141_183702 | Not-determined |
| 18-110_k141_170999 | Not-determined |
| 18-110_k141_44638 | Not-determined |
| 18-110_k141_124844 | Low-quality |
| 18-110_k141_44642 | Not-determined |
| 18-110_k141_193958 | Low-quality |
| 18-110_k141_137034 | Not-determined |
| 18-110_k141_67491 | Not-determined |
| 18-110_k141_33731 | Not-determined |
| 18-110_k141_260905 | Not-determined |
| 18-110_k141_137044 | Not-determined |
| 18-110_k141_251449 | Not-determined |
| 18-110_k141_183721 | Not-determined |
| 18-110_k141_8583 | Not-determined |
| 18-110_k141_159758 | Not-determined |
| 18-110_k141_67496 | Not-determined |
| 18-110_k141_137058 | Not-determined |
| 18-110_k141_171028 | Not-determined |
| 18-110_k141_112986 | Not-determined |
| 18-110_k141_137064 | Not-determined |
| 18-110_k141_171031 | Not-determined |
| 18-110_k141_251464 | Not-determined |
| 18-110_k141_101984 | Not-determined |
| 18-110_k141_67501 | Not-determined |
| 18-110_k141_193975 | Not-determined |
| 18-110_k141_67503 | Not-determined |
| 18-110_k141_148803 | Not-determined |
| 18-110_k141_101992 | Not-determined |
| 18-110_k141_148806 | Not-determined |
| 18-110_k141_113003 | Not-determined |
| 18-110_k141_193981 | Not-determined |
| 18-110_k141_113005 | Not-determined |
| 18-110_k141_260914 | Not-determined |
| 18-110_k141_67514 | Not-determined |
| 18-110_k141_102007 | Low-quality |
| 18-110_k141_79414 | Not-determined |
| 18-110_k141_228285 | Not-determined |
| 18-110_k141_33746 | Not-determined |
| 18-110_k141_228288 | Not-determined |
| 18-110_k141_79422 | Not-determined |
| 18-110_k141_33751 | Not-determined |
| 18-110_k141_55677 | Not-determined |
| 18-110_k141_113023 | Not-determined |
| 18-110_k141_148819 | Not-determined |
| 18-110_k141_113026 | Not-determined |
| 18-110_k141_241026 | Not-determined |
| 18-110_k141_55681 | Not-determined |
| 18-110_k141_148824 | Not-determined |
| 18-110_k141_193990 | Not-determined |
| 18-110_k141_44669 | Not-determined |
| 18-110_k141_44670 | Not-determined |
| 18-110_k141_79451 | Not-determined |
| 18-110_k141_113030 | Not-determined |
| 18-110_k141_260948 | Not-determined |
| 18-110_k141_241037 | Not-determined |
| 18-110_k141_113033 | Low-quality |
| 18-110_k141_89869 | Not-determined |
| 18-110_k141_171068 | Low-quality |
| 18-110_k141_171070 | Not-determined |
| 18-110_k141_44687 | Not-determined |
| 18-110_k141_79458 | Not-determined |
| 18-110_k141_228316 | Not-determined |
| 18-110_k141_228317 | Not-determined |
| 18-110_k141_260954 | Not-determined |
| 18-110_k141_260955 | Not-determined |
| 18-110_k141_241055 | Not-determined |
| 18-110_k141_159797 | Not-determined |
| 18-110_k141_194025 | Not-determined |
| 18-110_k141_171075 | Not-determined |
| 18-110_k141_113042 | Not-determined |
| 18-110_k141_241064 | Not-determined |
| 18-110_k141_33763 | Not-determined |
| 18-110_k141_194029 | Not-determined |
| 18-110_k141_124927 | Not-determined |
| 18-110_k141_148849 | Not-determined |
| 18-110_k141_8619 | Not-determined |
| 18-110_k141_124929 | Not-determined |
| 18-110_k141_260964 | Not-determined |
| 18-110_k141_8620 | Not-determined |
| 18-110_k141_33768 | Low-quality |
| 18-110_k141_194032 | Not-determined |
| 18-110_k141_89888 | Not-determined |
| 18-110_k141_33773 | Not-determined |
| 18-110_k141_171080 | Low-quality |
| 18-110_k141_251539 | Not-determined |
| 18-110_k141_89895 | Not-determined |
| 18-110_k141_194037 | Not-determined |
| 18-110_k141_241079 | Not-determined |
| 18-110_k141_228341 | Not-determined |
| 18-110_k141_124948 | Not-determined |
| 18-110_k141_194041 | Not-determined |
| 18-110_k141_194042 | Not-determined |
| 18-110_k141_171098 | Not-determined |
| 18-110_k141_44711 | Not-determined |
| 18-110_k141_204198 | Not-determined |
| 18-110_k141_228354 | Not-determined |
| 18-110_k141_89903 | Low-quality |
| 18-110_k141_79492 | Not-determined |
| 18-110_k141_113071 | Not-determined |
| 18-110_k141_241084 | Not-determined |
| 18-110_k141_113076 | Not-determined |
| 18-110_k141_113077 | Not-determined |
| 18-110_k141_55707 | Low-quality |
| 18-110_k141_44727 | Not-determined |
| 18-110_k141_124965 | Low-quality |
| 18-110_k141_19837 | Low-quality |
| 18-110_k141_183768 | Not-determined |
| 18-110_k141_251570 | Low-quality |
| 18-110_k141_260992 | Not-determined |
| 18-110_k141_79506 | Not-determined |
| 18-110_k141_241106 | Not-determined |
| 18-110_k141_260997 | Not-determined |
| 18-110_k141_19847 | Not-determined |
| 18-110_k141_241110 | Not-determined |
| 18-110_k141_159853 | Low-quality |
| 18-110_k141_241113 | Low-quality |
| 18-110_k141_183790 | Not-determined |
| 18-110_k141_260998 | Low-quality |
| 18-110_k141_261000 | Not-determined |
| 18-110_k141_228392 | Not-determined |
| 18-110_k141_19864 | Not-determined |
| 18-110_k141_19865 | Not-determined |
| 18-110_k141_137143 | Not-determined |
| 18-110_k141_55739 | Not-determined |
| 18-110_k141_19869 | Not-determined |
| 18-110_k141_274271 | Not-determined |
| 18-110_k141_67577 | Not-determined |
| 18-110_k141_194047 | Not-determined |
| 18-110_k141_194050 | Not-determined |
| 18-110_k141_124983 | Not-determined |
| 18-110_k141_194051 | Not-determined |
| 18-110_k141_228405 | Not-determined |
| 18-110_k141_241130 | Not-determined |
| 18-110_k141_274275 | Not-determined |
| 18-110_k141_55753 | Not-determined |
| 18-110_k141_148882 | Not-determined |
| 18-110_k141_194060 | Not-determined |
| 18-110_k141_274276 | Not-determined |
| 18-110_k141_251601 | Low-quality |
| 18-110_k141_261015 | Not-determined |
| 18-110_k141_274278 | Not-determined |
| 18-110_k141_8703 | Not-determined |
| 18-110_k141_148888 | Not-determined |
| 18-110_k141_79555 | Not-determined |
| 18-110_k141_261020 | Not-determined |
| 18-110_k141_137169 | Not-determined |
| 18-110_k141_102093 | Not-determined |
| 18-110_k141_194067 | Not-determined |
| 18-110_k141_44765 | Not-determined |
| 18-110_k141_113129 | Not-determined |
| 18-110_k141_19889 | Low-quality |
| 18-110_k141_102103 | Not-determined |
| 18-110_k141_44772 | Not-determined |
| 18-110_k141_67610 | Low-quality |
| 18-110_k141_55776 | Not-determined |
| 18-110_k141_251625 | Not-determined |
| 18-110_k141_67612 | Not-determined |
| 18-110_k141_44781 | Not-determined |
| 18-110_k141_8722 | Not-determined |
| 18-110_k141_8725 | Not-determined |
| 18-110_k141_33849 | Not-determined |
| 18-110_k141_19902 | Not-determined |
| 18-110_k141_33850 | Not-determined |
| 18-110_k141_241159 | Not-determined |
| 18-110_k141_171130 | Not-determined |
| 18-110_k141_228429 | Not-determined |
| 18-110_k141_102107 | Not-determined |
| 18-110_k141_89959 | Not-determined |
| 18-110_k141_89963 | Not-determined |
| 18-110_k141_19913 | Low-quality |
| 18-110_k141_67632 | Low-quality |
| 18-110_k141_67638 | Not-determined |
| 18-110_k141_89969 | Not-determined |
| 18-110_k141_102115 | Not-determined |
| 18-110_k141_251646 | Not-determined |
| 18-110_k141_251648 | Low-quality |
| 18-110_k141_89970 | Not-determined |
| 18-110_k141_137203 | Not-determined |
| 18-110_k141_8740 | Low-quality |
| 18-110_k141_148912 | Not-determined |
| 18-110_k141_261058 | Not-determined |
| 18-110_k141_261059 | Not-determined |
| 18-110_k141_33886 | Not-determined |
| 18-110_k141_204288 | Not-determined |
| 18-110_k141_19925 | Not-determined |
| 18-110_k141_137216 | Not-determined |
| 18-110_k141_204293 | Not-determined |
| 18-110_k141_137221 | Not-determined |
| 18-110_k141_261063 | Low-quality |
| 18-110_k141_33898 | Not-determined |
| 18-110_k141_33899 | Not-determined |
| 18-110_k141_102132 | Not-determined |
| 18-110_k141_148931 | Not-determined |
| 18-110_k141_102134 | Not-determined |
| 18-110_k141_19939 | Not-determined |

| | |
|---|---|
| 18-110_k141_148936 | Not-determined |
| 18-110_k141_204299 | Not-determined |
| 18-110_k141_148938 | Not-determined |
| 18-110_k141_251677 | Not-determined |
| 18-110_k141_148940 | Not-determined |
| 18-110_k141_89993 | Not-determined |
| 18-110_k141_148943 | Not-determined |
| 18-110_k141_204304 | Not-determined |
| 18-110_k141_194109 | Not-determined |
| 18-110_k141_137233 | Not-determined |
| 18-110_k141_194111 | Not-determined |
| 18-110_k141_137236 | Not-determined |
| 18-110_k141_113166 | Not-determined |
| 18-110_k141_137238 | Not-determined |
| 18-110_k141_194116 | Not-determined |
| 18-110_k141_79572 | Not-determined |
| 18-110_k141_228459 | Not-determined |
| 18-110_k141_274321 | Not-determined |
| 18-110_k141_171155 | Not-determined |
| 18-110_k141_113172 | Not-determined |
| 18-110_k141_228465 | Not-determined |
| 18-110_k141_137248 | Not-determined |
| 18-110_k141_194131 | Not-determined |
| 18-110_k141_113178 | Not-determined |
| 18-110_k141_171162 | Not-determined |
| 18-110_k141_194133 | Not-determined |
| 18-110_k141_44828 | Not-determined |
| 18-110_k141_261073 | Not-determined |
| 18-110_k141_261076 | Not-determined |
| 18-110_k141_113186 | Not-determined |
| 18-110_k141_274340 | Not-determined |
| 18-110_k141_216984 | Not-determined |
| 18-110_k141_204312 | Not-determined |
| 18-110_k141_19964 | Not-determined |
| 18-110_k141_44834 | Not-determined |
| 18-110_k141_79595 | Not-determined |
| 18-110_k141_274344 | Not-determined |
| 18-110_k141_8784 | Not-determined |
| 18-110_k141_137257 | Not-determined |
| 18-110_k141_171173 | Not-determined |
| 18-110_k141_79601 | Not-determined |
| 18-110_k141_194147 | Not-determined |
| 18-110_k141_19980 | Not-determined |
| 18-110_k141_33941 | Not-determined |
| 18-110_k141_194150 | Not-determined |
| 18-110_k141_19985 | Not-determined |
| 18-110_k141_44850 | Not-determined |
| 18-110_k141_204332 | Not-determined |
| 18-110_k141_137266 | Not-determined |
| 18-110_k141_44853 | Not-determined |
| 18-110_k141_79615 | Not-determined |
| 18-110_k141_228498 | Not-determined |
| 18-110_k141_194154 | Not-determined |
| 18-110_k141_90038 | Not-determined |
| 18-110_k141_171183 | Not-determined |
| 18-110_k141_251724 | Not-determined |
| 18-110_k141_90042 | Not-determined |
| 18-110_k141_251727 | Not-determined |
| 18-110_k141_148969 | Not-determined |
| 18-110_k141_79621 | Not-determined |
| 18-110_k141_261092 | Low-quality |
| 18-110_k141_90047 | Not-determined |
| 18-110_k141_137277 | Not-determined |
| 18-110_k141_125008 | Not-determined |
| 18-110_k141_251732 | Not-determined |
| 18-110_k141_148976 | Low-quality |
| 18-110_k141_102160 | Not-determined |
| 18-110_k141_20001 | Not-determined |
| 18-110_k141_228515 | Not-determined |
| 18-110_k141_20004 | Not-determined |
| 18-110_k141_67685 | Not-determined |
| 18-110_k141_90065 | Not-determined |
| 18-110_k141_20005 | Not-determined |
| 18-110_k141_204337 | Not-determined |
| 18-110_k141_171204 | Not-determined |
| 18-110_k141_241198 | Low-quality |
| 18-110_k141_194164 | Not-determined |
| 18-110_k141_79635 | Not-determined |
| 18-110_k141_44870 | Not-determined |
| 18-110_k141_194169 | Not-determined |
| 18-110_k141_137291 | Not-determined |
| 18-110_k141_137294 | Not-determined |
| 18-110_k141_241204 | Not-determined |
| 18-110_k141_55810 | Not-determined |
| 18-110_k141_67695 | Not-determined |
| 18-110_k141_183848 | Not-determined |
| 18-110_k141_274352 | Not-determined |
| 18-110_k141_183850 | Low-quality |
| 18-110_k141_204353 | Not-determined |
| 18-110_k141_79645 | Not-determined |
| 18-110_k141_44875 | Not-determined |
| 18-110_k141_44880 | Not-determined |
| 18-110_k141_204360 | Not-determined |
| 18-110_k141_194176 | Low-quality |
| 18-110_k141_251742 | Not-determined |
| 18-110_k141_274360 | Not-determined |
| 18-110_k141_137308 | Not-determined |
| 18-110_k141_20031 | Not-determined |
| 18-110_k141_44886 | Not-determined |
| 18-110_k141_137315 | Not-determined |
| 18-110_k141_183875 | Not-determined |
| 18-110_k141_67713 | Not-determined |
| 18-110_k141_55830 | Not-determined |
| 18-110_k141_241211 | Not-determined |
| 18-110_k141_183880 | Not-determined |
| 18-110_k141_228529 | Not-determined |
| 18-110_k141_125046 | Not-determined |
| 18-110_k141_125047 | Not-determined |
| 18-110_k141_44894 | Not-determined |
| 18-110_k141_55833 | Not-determined |
| 18-110_k141_274368 | Not-determined |
| 18-110_k141_79664 | Not-determined |
| 18-110_k141_113228 | Not-determined |
| 18-110_k141_228535 | Not-determined |
| 18-110_k141_183889 | Low-quality |
| 18-110_k141_113237 | Not-determined |
| 18-110_k141_33970 | Not-determined |
| 18-110_k141_241227 | Not-determined |
| 18-110_k141_171220 | Not-determined |
| 18-110_k141_67729 | Low-quality |
| 18-110_k141_171221 | Not-determined |
| 18-110_k141_90107 | Not-determined |
| 18-110_k141_274387 | Not-determined |
| 18-110_k141_55857 | Not-determined |
| 18-110_k141_183909 | Not-determined |
| 18-110_k141_8810 | Not-determined |
| 18-110_k141_79690 | Not-determined |
| 18-110_k141_183913 | Not-determined |
| 18-110_k141_183916 | Not-determined |
| 18-110_k141_55859 | Not-determined |
| 18-110_k141_125063 | Not-determined |
| 18-110_k141_171231 | Not-determined |
| 18-110_k141_241242 | Not-determined |
| 18-110_k141_33978 | Not-determined |
| 18-110_k141_55870 | Not-determined |
| 18-110_k141_183920 | Not-determined |
| 18-110_k141_20042 | Low-quality |
| 18-110_k141_171241 | Not-determined |
| 18-110_k141_251775 | Not-determined |
| 18-110_k141_183925 | Low-quality |
| 18-110_k141_251776 | Not-determined |
| 18-110_k141_113258 | Not-determined |
| 18-110_k141_113259 | Not-determined |
| 18-110_k141_274413 | Not-determined |
| 18-110_k141_113263 | Not-determined |
| 18-110_k141_44945 | Not-determined |
| 18-110_k141_204370 | Not-determined |
| 18-110_k141_33985 | Not-determined |
| 18-110_k141_113266 | Not-determined |
| 18-110_k141_55874 | Not-determined |
| 18-110_k141_102180 | Not-determined |
| 18-110_k141_228574 | Not-determined |
| 18-110_k141_79705 | Not-determined |
| 18-110_k141_194203 | Not-determined |
| 18-110_k141_90126 | Not-determined |
| 18-110_k141_261146 | Not-determined |
| 18-110_k141_102188 | Not-determined |
| 18-110_k141_67768 | Low-quality |
| 18-110_k141_149037 | Not-determined |
| 18-110_k141_149041 | Not-determined |
| 18-110_k141_90131 | Not-determined |
| 18-110_k141_90136 | Not-determined |
| 18-110_k141_20057 | Not-determined |
| 18-110_k141_90139 | Not-determined |
| 18-110_k141_34006 | Not-determined |
| 18-110_k141_171275 | Not-determined |
| 18-110_k141_67776 | Low-quality |
| 18-110_k141_204380 | Low-quality |
| 18-110_k141_44961 | Not-determined |
| 18-110_k141_149053 | Low-quality |
| 18-110_k141_149054 | Not-determined |
| 18-110_k141_44962 | Not-determined |
| 18-110_k141_171286 | Low-quality |
| 18-110_k141_34011 | Not-determined |
| 18-110_k141_171290 | Not-determined |
| 18-110_k141_67790 | Not-determined |
| 18-110_k141_125103 | Not-determined |
| 18-110_k141_274436 | Not-determined |
| 18-110_k141_90162 | Low-quality |
| 18-110_k141_102207 | Low-quality |
| 18-110_k141_102210 | Not-determined |
| 18-110_k141_217047 | Not-determined |
| 18-110_k141_183973 | Not-determined |
| 18-110_k141_183974 | Not-determined |
| 18-110_k141_55917 | Not-determined |
| 18-110_k141_44973 | Not-determined |
| 18-110_k141_149069 | Not-determined |
| 18-110_k141_102220 | Not-determined |
| 18-110_k141_20081 | Not-determined |
| 18-110_k141_228609 | Not-determined |
| 18-110_k141_204403 | Not-determined |
| 18-110_k141_113289 | Not-determined |
| 18-110_k141_274454 | Not-determined |
| 18-110_k141_20084 | Not-determined |
| 18-110_k141_79735 | Not-determined |
| 18-110_k141_44983 | Not-determined |
| 18-110_k141_217056 | Not-determined |
| 18-110_k141_44984 | Not-determined |
| 18-110_k141_90181 | Not-determined |
| 18-110_k141_171300 | Not-determined |
| 18-110_k141_171301 | Not-determined |
| 18-110_k141_274462 | Not-determined |
| 18-110_k141_44992 | Not-determined |
| 18-110_k141_171306 | Not-determined |
| 18-110_k141_204414 | Not-determined |
| 18-110_k141_34025 | Low-quality |
| 18-110_k141_44994 | Not-determined |
| 18-110_k141_261150 | Low-quality |
| 18-110_k141_102236 | Not-determined |
| 18-110_k141_90189 | Not-determined |
| 18-110_k141_79745 | Not-determined |
| 18-110_k141_261154 | Not-determined |
| 18-110_k141_113303 | Not-determined |
| 18-110_k141_113304 | Not-determined |
| 18-110_k141_34033 | Not-determined |
| 18-110_k141_79750 | Not-determined |
| 18-110_k141_149103 | Not-determined |
| 18-110_k141_45010 | Not-determined |
| 18-110_k141_113314 | Not-determined |
| 18-110_k141_149108 | Not-determined |
| 18-110_k141_241312 | Not-determined |
| 18-110_k141_159958 | Not-determined |
| 18-110_k141_90194 | Not-determined |
| 18-110_k141_261168 | Not-determined |
| 18-110_k141_45024 | Not-determined |
| 18-110_k141_137400 | Not-determined |
| 18-110_k141_79761 | Not-determined |
| 18-110_k141_79762 | Not-determined |
| 18-110_k141_67838 | Not-determined |
| 18-110_k141_183984 | Not-determined |
| 18-110_k141_34053 | Not-determined |
| 18-110_k141_194268 | Not-determined |
| 18-110_k141_204433 | Not-determined |
| 18-110_k141_102258 | Not-determined |
| 18-110_k141_20097 | Not-determined |
| 18-110_k141_20101 | Not-determined |
| 18-110_k141_274488 | Not-determined |
| 18-110_k141_55945 | Not-determined |
| 18-110_k141_228615 | Not-determined |
| 18-110_k141_217078 | Not-determined |
| 18-110_k141_183997 | Not-determined |
| 18-110_k141_228619 | Not-determined |
| 18-110_k141_194282 | Not-determined |
| 18-110_k141_159983 | Not-determined |
| 18-110_k141_159986 | Not-determined |
| 18-110_k141_102270 | Not-determined |
| 18-110_k141_274506 | Low-quality |
| 18-110_k141_90218 | Not-determined |
| 18-110_k141_217079 | Low-quality |
| 18-110_k141_20117 | Not-determined |
| 18-110_k141_45053 | Not-determined |
| 18-110_k141_34068 | Low-quality |
| 18-110_k141_274519 | Not-determined |
| 18-110_k141_102287 | Not-determined |
| 18-110_k141_274522 | Not-determined |
| 18-110_k141_102289 | Not-determined |
| 18-110_k141_149137 | Not-determined |
| 18-110_k141_55953 | Not-determined |
| 18-110_k141_261172 | Not-determined |
| 18-110_k141_149139 | Not-determined |
| 18-110_k141_194296 | Not-determined |
| 18-110_k141_241334 | Not-determined |
| 18-110_k141_204457 | Not-determined |
| 18-110_k141_194299 | Not-determined |
| 18-110_k141_137419 | Low-quality |
| 18-110_k141_90235 | Not-determined |
| 18-110_k141_171325 | Not-determined |
| 18-110_k141_137423 | Not-determined |
| 18-110_k141_45067 | Not-determined |
| 18-110_k141_137425 | Not-determined |
| 18-110_k141_102305 | Not-determined |
| 18-110_k141_137427 | Not-determined |
| 18-110_k141_251885 | Not-determined |
| 18-110_k141_194307 | Not-determined |
| 18-110_k141_90245 | Not-determined |
| 18-110_k141_90246 | Low-quality |
| 18-110_k141_228647 | Not-determined |
| 18-110_k141_34093 | Not-determined |
| 18-110_k141_137434 | Not-determined |
| 18-110_k141_137436 | Not-determined |
| 18-110_k141_137438 | Not-determined |
| 18-110_k141_90254 | Not-determined |
| 18-110_k141_45091 | Not-determined |
| 18-110_k141_55981 | Not-determined |
| 18-110_k141_149157 | Not-determined |
| 18-110_k141_274555 | Not-determined |
| 18-110_k141_45098 | Not-determined |
| 18-110_k141_274556 | Low-quality |
| 18-110_k141_67906 | Not-determined |
| 18-110_k141_102324 | Not-determined |
| 18-110_k141_113362 | Not-determined |
| 18-110_k141_261199 | Not-determined |
| 18-110_k141_113363 | Not-determined |
| 18-110_k141_261200 | Not-determined |
| 18-110_k141_34106 | Not-determined |
| 18-110_k141_67912 | Not-determined |
| 18-110_k141_261201 | Not-determined |
| 18-110_k141_45102 | Not-determined |
| 18-110_k141_45103 | Low-quality |
| 18-110_k141_160034 | Not-determined |
| 18-110_k141_204476 | Not-determined |
| 18-110_k141_125137 | Not-determined |
| 18-110_k141_171355 | Not-determined |
| 18-110_k141_204481 | Not-determined |
| 18-110_k141_149169 | Not-determined |
| 18-110_k141_125142 | Not-determined |
| 18-110_k141_45112 | Not-determined |
| 18-110_k141_261210 | Not-determined |
| 18-110_k141_8926 | Not-determined |
| 18-110_k141_90278 | Low-quality |
| 18-110_k141_90281 | Not-determined |
| 18-110_k141_20165 | Low-quality |
| 18-110_k141_90286 | Not-determined |
| 18-110_k141_102352 | Not-determined |
| 18-110_k141_67925 | Low-quality |
| 18-110_k141_261215 | Not-determined |
| 18-110_k141_45124 | Not-determined |
| 18-110_k141_149191 | Low-quality |
| 18-110_k141_90290 | Not-determined |
| 18-110_k141_184012 | Not-determined |
| 18-110_k141_55998 | Not-determined |
| 18-110_k141_20171 | Not-determined |
| 18-110_k141_20174 | Not-determined |
| 18-110_k141_261221 | Not-determined |
| 18-110_k141_261224 | Not-determined |
| 18-110_k141_125151 | Not-determined |
| 18-110_k141_90298 | Not-determined |
| 18-110_k141_204497 | Low-quality |
| 18-110_k141_34112 | Not-determined |
| 18-110_k141_171378 | Not-determined |
| 18-110_k141_113399 | Not-determined |
| 18-110_k141_261228 | Not-determined |
| 18-110_k141_171380 | Not-determined |
| 18-110_k141_56005 | Not-determined |
| 18-110_k141_56006 | Not-determined |
| 18-110_k141_20188 | Not-determined |
| 18-110_k141_228681 | Not-determined |
| 18-110_k141_79854 | Low-quality |
| 18-110_k141_45154 | Not-determined |
| 18-110_k141_261239 | Low-quality |
| 18-110_k141_274593 | Not-determined |
| 18-110_k141_8943 | Not-determined |
| 18-110_k141_8944 | Not-determined |
| 18-110_k141_67964 | Not-determined |
| 18-110_k141_56028 | Not-determined |
| 18-110_k141_56031 | Not-determined |
| 18-110_k141_113414 | Low-quality |
| 18-110_k141_194349 | Not-determined |
| 18-110_k141_8955 | Not-determined |
| 18-110_k141_8960 | Not-determined |
| 18-110_k141_251911 | Not-determined |
| 18-110_k141_102358 | Not-determined |
| 18-110_k141_274613 | Not-determined |
| 18-110_k141_34117 | Low-quality |
| 18-110_k141_34118 | Not-determined |
| 18-110_k141_8965 | Not-determined |
| 18-110_k141_45172 | Low-quality |
| 18-110_k141_67980 | Not-determined |
| 18-110_k141_228694 | Not-determined |
| 18-110_k141_241377 | Not-determined |
| 18-110_k141_228695 | Not-determined |
| 18-110_k141_8977 | Not-determined |
| 18-110_k141_20217 | Not-determined |
| 18-110_k141_194370 | Not-determined |
| 18-110_k141_45190 | Not-determined |
| 18-110_k141_79888 | Not-determined |
| 18-110_k141_34123 | Not-determined |
| 18-110_k141_79891 | Not-determined |
| 18-110_k141_20221 | Not-determined |
| 18-110_k141_251923 | Not-determined |
| 18-110_k141_184052 | Not-determined |
| 18-110_k141_79893 | Not-determined |
| 18-110_k141_67988 | Not-determined |
| 18-110_k141_56055 | Not-determined |
| 18-110_k141_149265 | Not-determined |
| 18-110_k141_184064 | Not-determined |
| 18-110_k141_251933 | Not-determined |
| 18-110_k141_149269 | Not-determined |
| 18-110_k141_56060 | Not-determined |
| 18-110_k141_20233 | Not-determined |
| 18-110_k141_217130 | Not-determined |
| 18-110_k141_204527 | Not-determined |
| 18-110_k141_204528 | Not-determined |
| 18-110_k141_217131 | Low-quality |
| 18-110_k141_184076 | Not-determined |
| 18-110_k141_149282 | Not-determined |
| 18-110_k141_149283 | Not-determined |
| 18-110_k141_125215 | Not-determined |
| 18-110_k141_217144 | Not-determined |
| 18-110_k141_228711 | Not-determined |
| 18-110_k141_125217 | Low-quality |
| 18-110_k141_184097 | Not-determined |
| 18-110_k141_204540 | Not-determined |
| 18-110_k141_204543 | Not-determined |
| 18-110_k141_79934 | Not-determined |
| 18-110_k141_113465 | Not-determined |
| 18-110_k141_137536 | Not-determined |
| 18-110_k141_102372 | Not-determined |
| 18-110_k141_79940 | Not-determined |
| 18-110_k141_137541 | Not-determined |
| 18-110_k141_34155 | Low-quality |
| 18-110_k141_194399 | Not-determined |
| 18-110_k141_79946 | Not-determined |
| 18-110_k141_45248 | Not-determined |
| 18-110_k141_56077 | Not-determined |
| 18-110_k141_149307 | Not-determined |
| 18-110_k141_184109 | Low-quality |
| 18-110_k141_149309 | Not-determined |
| 18-110_k141_45256 | Not-determined |
| 18-110_k141_261251 | Not-determined |
| 18-110_k141_102386 | Not-determined |
| 18-110_k141_137553 | Low-quality |
| 18-110_k141_45268 | Not-determined |
| 18-110_k141_34169 | Not-determined |
| 18-110_k141_228728 | Not-determined |
| 18-110_k141_34176 | Not-determined |
| 18-110_k141_184122 | Not-determined |
| 18-110_k141_90414 | Not-determined |
| 18-110_k141_113492 | Not-determined |
| 18-110_k141_45286 | Not-determined |
| 18-110_k141_217171 | Not-determined |
| 18-110_k141_125250 | Not-determined |
| 18-110_k141_45298 | Not-determined |
| 18-110_k141_102396 | Not-determined |
| 18-110_k141_204592 | Low-quality |
| 18-110_k141_90433 | Not-determined |
| 18-110_k141_204597 | Not-determined |
| 18-110_k141_102398 | Low-quality |
| 18-110_k141_204600 | Not-determined |
| 18-110_k141_79982 | Not-determined |
| 18-110_k141_228766 | Not-determined |
| 18-110_k141_171411 | Not-determined |
| 18-110_k141_184162 | Not-determined |
| 18-110_k141_204613 | Not-determined |
| 18-110_k141_228772 | Not-determined |
| 18-110_k141_228774 | Not-determined |
| 18-110_k141_45313 | Not-determined |
| 18-110_k141_56114 | Not-determined |
| 18-110_k141_113513 | Not-determined |
| 18-110_k141_228776 | Not-determined |
| 18-110_k141_217192 | Not-determined |
| 18-110_k141_9036 | Not-determined |
| 18-110_k141_149341 | Not-determined |
| 18-110_k141_204624 | Not-determined |
| 18-110_k141_34190 | Low-quality |
| 18-110_k141_9041 | Not-determined |
| 18-110_k141_160112 | Not-determined |
| 18-110_k141_217197 | Not-determined |
| 18-110_k141_149347 | Not-determined |
| 18-110_k141_80006 | Not-determined |
| 18-110_k141_9045 | Not-determined |
| 18-110_k141_204627 | Not-determined |
| 18-110_k141_113526 | Low-quality |
| 18-110_k141_228787 | Not-determined |
| 18-110_k141_194456 | Not-determined |
| 18-110_k141_9050 | Not-determined |
| 18-110_k141_217206 | Not-determined |
| 18-110_k141_217207 | Low-quality |
| 18-110_k141_194460 | Not-determined |
| 18-110_k141_228794 | Not-determined |
| 18-110_k141_241459 | Not-determined |
| 18-110_k141_68051 | Not-determined |
| 18-110_k141_68053 | Not-determined |
| 18-110_k141_251951 | Not-determined |
| 18-110_k141_241464 | Not-determined |
| 18-110_k141_149368 | Not-determined |
| 18-110_k141_160118 | Not-determined |
| 18-110_k141_102415 | Not-determined |
| 18-110_k141_274674 | Not-determined |
| 18-110_k141_194464 | Low-quality |
| 18-110_k141_56137 | Not-determined |
| 18-110_k141_68058 | Not-determined |
| 18-110_k141_194465 | Low-quality |
| 18-110_k141_102420 | Not-determined |
| 18-110_k141_261319 | Not-determined |
| 18-110_k141_149380 | Not-determined |
| 18-110_k141_68064 | Not-determined |
| 18-110_k141_90443 | Not-determined |
| 18-110_k141_9066 | Not-determined |
| 18-110_k141_90445 | Low-quality |
| 18-110_k141_261328 | Not-determined |
| 18-110_k141_125283 | Low-quality |
| 18-110_k141_251966 | Not-determined |
| 18-110_k141_9075 | Not-determined |
| 18-110_k141_149395 | Low-quality |
| 18-110_k141_137642 | Not-determined |
| 18-110_k141_137643 | Not-determined |
| 18-110_k141_217235 | Not-determined |
| 18-110_k141_194479 | Not-determined |
| 18-110_k141_261331 | Not-determined |
| 18-110_k141_160136 | Not-determined |
| 18-110_k141_34219 | Not-determined |
| 18-110_k141_204638 | Not-determined |
| 18-110_k141_137660 | Low-quality |
| 18-110_k141_160144 | Not-determined |
| 18-110_k141_261349 | Not-determined |
| 18-110_k141_68087 | Not-determined |
| 18-110_k141_149403 | Not-determined |
| 18-110_k141_160148 | Not-determined |
| 18-110_k141_9092 | Not-determined |
| 18-110_k141_113575 | Low-quality |
| 18-110_k141_45356 | Not-determined |
| 18-110_k141_45357 | Not-determined |
| 18-110_k141_228807 | Not-determined |
| 18-110_k141_137675 | Not-determined |
| 18-110_k141_160157 | Not-determined |
| 18-110_k141_160160 | Not-determined |
| 18-110_k141_9097 | Not-determined |
| 18-110_k141_274695 | Not-determined |
| 18-110_k141_228815 | Not-determined |
| 18-110_k141_9099 | Not-determined |
| 18-110_k141_45368 | Not-determined |
| 18-110_k141_149413 | Not-determined |
| 18-110_k141_274698 | Not-determined |
| 18-110_k141_204658 | Not-determined |
| 18-110_k141_160164 | Not-determined |
| 18-110_k141_137691 | Not-determined |
| 18-110_k141_228824 | Not-determined |
| 18-110_k141_125304 | Not-determined |
| 18-110_k141_171482 | Low-quality |
| 18-110_k141_228827 | Not-determined |
| 18-110_k141_68100 | Not-determined |
| 18-110_k141_137693 | Not-determined |
| 18-110_k141_194522 | Not-determined |
| 18-110_k141_160173 | Not-determined |
| 18-110_k141_90481 | Not-determined |
| 18-110_k141_228829 | Not-determined |
| 18-110_k141_137700 | Low-quality |
| 18-110_k141_160178 | Not-determined |
| 18-110_k141_45382 | Not-determined |
| 18-110_k141_160184 | Not-determined |
| 18-110_k141_90489 | Not-determined |
| 18-110_k141_149431 | Not-determined |
| 18-110_k141_228832 | Not-determined |
| 18-110_k141_137706 | Not-determined |
| 18-110_k141_204675 | Not-determined |
| 18-110_k141_194526 | Not-determined |
| 18-110_k141_274705 | Low-quality |
| 18-110_k141_274707 | Not-determined |
| 18-110_k141_204680 | Not-determined |
| 18-110_k141_45389 | Not-determined |
| 18-110_k141_113590 | Not-determined |
| 18-110_k141_9126 | Not-determined |
| 18-110_k141_56194 | Not-determined |
| 18-110_k141_90500 | Not-determined |
| 18-110_k141_34257 | Not-determined |
| 18-110_k141_34260 | Not-determined |
| 18-110_k141_149448 | Not-determined |
| 18-110_k141_45396 | Not-determined |
| 18-110_k141_217270 | Not-determined |
| 18-110_k141_34269 | Not-determined |
| 18-110_k141_171508 | Not-determined |
| 18-110_k141_68126 | Not-determined |
| 18-110_k141_56206 | Not-determined |
| 18-110_k141_113601 | Low-quality |
| 18-110_k141_9134 | Not-determined |
| 18-110_k141_56210 | Not-determined |
| 18-110_k141_56211 | Not-determined |
| 18-110_k141_80041 | Low-quality |
| 18-110_k141_194536 | Not-determined |
| 18-110_k141_9142 | Not-determined |
| 18-110_k141_149457 | Not-determined |
| 18-110_k141_228852 | Not-determined |
| 18-110_k141_45414 | Not-determined |
| 18-110_k141_137719 | Not-determined |
| 18-110_k141_9144 | Not-determined |
| 18-110_k141_137721 | Not-determined |
| 18-110_k141_184243 | Low-quality |
| 18-110_k141_45424 | Not-determined |
| 18-110_k141_274731 | Not-determined |
| 18-110_k141_160208 | Not-determined |
| 18-110_k141_160210 | Not-determined |
| 18-110_k141_274738 | Not-determined |
| 18-110_k141_160217 | Not-determined |
| 18-110_k141_113611 | Not-determined |
| 18-110_k141_160220 | Not-determined |
| 18-110_k141_184250 | Not-determined |
| 18-110_k141_34299 | Not-determined |
| 18-110_k141_274740 | Low-quality |
| 18-110_k141_274741 | Not-determined |
| 18-110_k141_56235 | Not-determined |
| 18-110_k141_125350 | Not-determined |
| 18-110_k141_68167 | Not-determined |
| 18-110_k141_80052 | Not-determined |
| 18-110_k141_68170 | Not-determined |
| 18-110_k141_149488 | Not-determined |
| 18-110_k141_45448 | Not-determined |
| 18-110_k141_34306 | Not-determined |
| 18-110_k141_90505 | Not-determined |

| | |
|---|---|
| 18-110_k141_45452 | Not-determined |
| 18-110_k141_102477 | Not-determined |
| 18-110_k141_80059 | Not-determined |
| 18-110_k141_56242 | Low-quality |
| 18-110_k141_102485 | Not-determined |
| 18-110_k141_20245 | Not-determined |
| 18-110_k141_252015 | Not-determined |
| 18-110_k141_68190 | Not-determined |
| 18-110_k141_90517 | Not-determined |
| 18-110_k141_204708 | Not-determined |
| 18-110_k141_56250 | Not-determined |
| 18-110_k141_137748 | Not-determined |
| 18-110_k141_56255 | Not-determined |
| 18-110_k141_184278 | Low-quality |
| 18-110_k141_113616 | Not-determined |
| 18-110_k141_228902 | Not-determined |
| 18-110_k141_56262 | Not-determined |
| 18-110_k141_184280 | Not-determined |
| 18-110_k141_9161 | Not-determined |
| 18-110_k141_252018 | Not-determined |
| 18-110_k141_56265 | Not-determined |
| 18-110_k141_34342 | Not-determined |
| 18-110_k141_90521 | Not-determined |
| 18-110_k141_252021 | Not-determined |
| 18-110_k141_149498 | Not-determined |
| 18-110_k141_125371 | Not-determined |
| 18-110_k141_68211 | Not-determined |
| 18-110_k141_171584 | Not-determined |
| 18-110_k141_149500 | Not-determined |
| 18-110_k141_217323 | Not-determined |
| 18-110_k141_45479 | Not-determined |
| 18-110_k141_160261 | Not-determined |
| 18-110_k141_204724 | Not-determined |
| 18-110_k141_274778 | Not-determined |
| 18-110_k141_217329 | Not-determined |
| 18-110_k141_125374 | Low-quality |
| 18-110_k141_102522 | Not-determined |
| 18-110_k141_20267 | Not-determined |
| 18-110_k141_171588 | Not-determined |
| 18-110_k141_56283 | Low-quality |
| 18-110_k141_90540 | Not-determined |
| 18-110_k141_204727 | Not-determined |
| 18-110_k141_274786 | Not-determined |
| 18-110_k141_68222 | Low-quality |
| 18-110_k141_171591 | Not-determined |
| 18-110_k141_80068 | Not-determined |
| 18-110_k141_68224 | Not-determined |
| 18-110_k141_274787 | Not-determined |
| 18-110_k141_90546 | Not-determined |
| 18-110_k141_102528 | Not-determined |
| 18-110_k141_252031 | Not-determined |
| 18-110_k141_102530 | Not-determined |
| 18-110_k141_80072 | Not-determined |
| 18-110_k141_160278 | Not-determined |
| 18-110_k141_184296 | Not-determined |
| 18-110_k141_45490 | Not-determined |
| 18-110_k141_274794 | Not-determined |
| 18-110_k141_80078 | Not-determined |
| 18-110_k141_80079 | Not-determined |
| 18-110_k141_217344 | Not-determined |
| 18-110_k141_194575 | Not-determined |
| 18-110_k141_204741 | Not-determined |
| 18-110_k141_90559 | Not-determined |
| 18-110_k141_171607 | Not-determined |
| 18-110_k141_68231 | Not-determined |
| 18-110_k141_160292 | Not-determined |
| 18-110_k141_184304 | Not-determined |
| 18-110_k141_160293 | Not-determined |
| 18-110_k141_20283 | Not-determined |
| 18-110_k141_102540 | Not-determined |
| 18-110_k141_204745 | Not-determined |
| 18-110_k141_125386 | Not-determined |
| 18-110_k141_90564 | Not-determined |
| 18-110_k141_217353 | Not-determined |
| 18-110_k141_45500 | Not-determined |
| 18-110_k141_160297 | Not-determined |
| 18-110_k141_160299 | Not-determined |
| 18-110_k141_274811 | Not-determined |
| 18-110_k141_194591 | Low-quality |
| 18-110_k141_137797 | Not-determined |
| 18-110_k141_252056 | Not-determined |
| 18-110_k141_171618 | Not-determined |
| 18-110_k141_34367 | Not-determined |
| 18-110_k141_194595 | Not-determined |
| 18-110_k141_194596 | Not-determined |
| 18-110_k141_194597 | Not-determined |
| 18-110_k141_68256 | Not-determined |
| 18-110_k141_56316 | Not-determined |
| 18-110_k141_68263 | Not-determined |
| 18-110_k141_194604 | Not-determined |
| 18-110_k141_9196 | Not-determined |
| 18-110_k141_204763 | Not-determined |
| 18-110_k141_204769 | Not-determined |
| 18-110_k141_56333 | Not-determined |
| 18-110_k141_274826 | Not-determined |
| 18-110_k141_34376 | Not-determined |
| 18-110_k141_274831 | Not-determined |
| 18-110_k141_45521 | Low-quality |
| 18-110_k141_261426 | Not-determined |
| 18-110_k141_90591 | Not-determined |
| 18-110_k141_252098 | Not-determined |
| 18-110_k141_34382 | Not-determined |
| 18-110_k141_68276 | Not-determined |
| 18-110_k141_9215 | Not-determined |
| 18-110_k141_274845 | Not-determined |
| [illegible] | [illegible] |

| | |
|---|---|
| 18-110_k141_275216 | Not-determined |
| 18-110_k141_125884 | Not-determined |
| 18-110_k141_113952 | Not-determined |
| 18-110_k141_125886 | Not-determined |
| 18-110_k141_45856 | Not-determined |
| 18-110_k141_171988 | Not-determined |
| 18-110_k141_113956 | Low-quality |
| 18-110_k141_241870 | Not-determined |
| 18-110_k141_261776 | Not-determined |
| 18-110_k141_56554 | Not-determined |
| 18-110_k141_229280 | Not-determined |
| 18-110_k141_205150 | Not-determined |
| 18-110_k141_172000 | Low-quality |
| 18-110_k141_45863 | Not-determined |
| 18-110_k141_125900 | Not-determined |
| 18-110_k141_45867 | Not-determined |
| 18-110_k141_45868 | Not-determined |
| 18-110_k141_149738 | Low-quality |
| 18-110_k141_91132 | Low-quality |
| 18-110_k141_172016 | Not-determined |
| 18-110_k141_91136 | Not-determined |
| 18-110_k141_172018 | Not-determined |
| 18-110_k141_125910 | Not-determined |
| 18-110_k141_9589 | Not-determined |
| 18-110_k141_261783 | Not-determined |
| 18-110_k141_205162 | Not-determined |
| 18-110_k141_172025 | Not-determined |
| 18-110_k141_20639 | Not-determined |
| 18-110_k141_68546 | Not-determined |
| 18-110_k141_261785 | Not-determined |
| 18-110_k141_275232 | Not-determined |
| 18-110_k141_149748 | Not-determined |
| 18-110_k141_172034 | Not-determined |
| 18-110_k141_275234 | Not-determined |
| 18-110_k141_275235 | Not-determined |
| 18-110_k141_205168 | Not-determined |
| 18-110_k141_172043 | Not-determined |
| 18-110_k141_68549 | Not-determined |
| 18-110_k141_20650 | Low-quality |
| 18-110_k141_20651 | Not-determined |
| 18-110_k141_205179 | Not-determined |
| 18-110_k141_205180 | Low-quality |
| 18-110_k141_138197 | Not-determined |
| 18-110_k141_261797 | Not-determined |
| 18-110_k141_9604 | Not-determined |
| 18-110_k141_160686 | Not-determined |
| 18-110_k141_252459 | Low-quality |
| 18-110_k141_172065 | Not-determined |
| 18-110_k141_252463 | Not-determined |
| 18-110_k141_91166 | Not-determined |
| 18-110_k141_252466 | Not-determined |
| 18-110_k141_217857 | Not-determined |
| 18-110_k141_91171 | Not-determined |
| 18-110_k141_20669 | Not-determined |
| 18-110_k141_20670 | Not-determined |
| 18-110_k141_252472 | Not-determined |
| 18-110_k141_45891 | Not-determined |
| 18-110_k141_252475 | Low-quality |
| 18-110_k141_113990 | Low-quality |
| 18-110_k141_252484 | Not-determined |
| 18-110_k141_138207 | Not-determined |
| 18-110_k141_56588 | Not-determined |
| 18-110_k141_275263 | Low-quality |
| 18-110_k141_138211 | Not-determined |
| 18-110_k141_80441 | Not-determined |
| 18-110_k141_113993 | Low-quality |
| 18-110_k141_205204 | Not-determined |
| 18-110_k141_252500 | Not-determined |
| 18-110_k141_113997 | Not-determined |
| 18-110_k141_113998 | Not-determined |
| 18-110_k141_252507 | Not-determined |
| 18-110_k141_56594 | Not-determined |
| 18-110_k141_229342 | Not-determined |
| 18-110_k141_160716 | Not-determined |
| 18-110_k141_125959 | Not-determined |
| 18-110_k141_20691 | Not-determined |
| 18-110_k141_80460 | Not-determined |
| 18-110_k141_275273 | Not-determined |
| 18-110_k141_275274 | Not-determined |
| 18-110_k141_261814 | Not-determined |
| 18-110_k141_261817 | Not-determined |
| 18-110_k141_275281 | Not-determined |
| 18-110_k141_261822 | Not-determined |
| 18-110_k141_275286 | Not-determined |
| 18-110_k141_9612 | Not-determined |
| 18-110_k141_114034 | Not-determined |
| 18-110_k141_114035 | Not-determined |
| 18-110_k141_252541 | Not-determined |
| 18-110_k141_68593 | Low-quality |
| 18-110_k141_252544 | Not-determined |
| 18-110_k141_56637 | Not-determined |
| 18-110_k141_205221 | Not-determined |
| 18-110_k141_20728 | Not-determined |
| 18-110_k141_252560 | Not-determined |
| 18-110_k141_20732 | Low-quality |
| 18-110_k141_68601 | Not-determined |
| 18-110_k141_205227 | Not-determined |
| 18-110_k141_125992 | Not-determined |
| 18-110_k141_125994 | Not-determined |
| 18-110_k141_68617 | Low-quality |
| 18-110_k141_45939 | Not-determined |
| 18-110_k141_252586 | Not-determined |
| 18-110_k141_229382 | Not-determined |
| 18-110_k141_20744 | Not-determined |
| 18-110_k141_252591 | Not-determined |
| 18-110_k141_126007 | Not-determined |
| 18-110_k141_217874 | Not-determined |
| 18-110_k141_45943 | Low-quality |
| 18-110_k141_45944 | Not-determined |
| 18-110_k141_275343 | Not-determined |
| 18-110_k141_56674 | Not-determined |
| 18-110_k141_80489 | Not-determined |
| 18-110_k141_68625 | Not-determined |
| 18-110_k141_20758 | Not-determined |
| 18-110_k141_20759 | Not-determined |
| 18-110_k141_114055 | Low-quality |
| 18-110_k141_45953 | Not-determined |
| 18-110_k141_252607 | Not-determined |
| 18-110_k141_126017 | Not-determined |
| 18-110_k141_172127 | Not-determined |
| 18-110_k141_275363 | Not-determined |
| 18-110_k141_275370 | Not-determined |
| 18-110_k141_172133 | Not-determined |
| 18-110_k141_114059 | Low-quality |
| 18-110_k141_56687 | Not-determined |
| 18-110_k141_172140 | Not-determined |
| 18-110_k141_252615 | Not-determined |
| 18-110_k141_9642 | Not-determined |
| 18-110_k141_114064 | Not-determined |
| 18-110_k141_20779 | Not-determined |
| 18-110_k141_172143 | Not-determined |
| 18-110_k141_20782 | Not-determined |
| 18-110_k141_91218 | Not-determined |
| 18-110_k141_217903 | Not-determined |
| 18-110_k141_45961 | Not-determined |
| 18-110_k141_56702 | Not-determined |
| 18-110_k141_195088 | Not-determined |
| 18-110_k141_20789 | Not-determined |
| 18-110_k141_172156 | Not-determined |
| 18-110_k141_20790 | Low-quality |
| 18-110_k141_160746 | Not-determined |
| 18-110_k141_114080 | Not-determined |
| 18-110_k141_114081 | Not-determined |
| 18-110_k141_229402 | Not-determined |
| 18-110_k141_172164 | Not-determined |
| 18-110_k141_205278 | Not-determined |
| 18-110_k141_195092 | Not-determined |
| 18-110_k141_160751 | Not-determined |
| 18-110_k141_9665 | Not-determined |
| 18-110_k141_114094 | Not-determined |
| 18-110_k141_45977 | Not-determined |
| 18-110_k141_172170 | Not-determined |
| 18-110_k141_45980 | Not-determined |
| 18-110_k141_91242 | Low-quality |
| 18-110_k141_56719 | Not-determined |
| 18-110_k141_160763 | Not-determined |
| 18-110_k141_229416 | Not-determined |
| 18-110_k141_195101 | Not-determined |
| 18-110_k141_229425 | Not-determined |
| 18-110_k141_195106 | Not-determined |
| 18-110_k141_91255 | Not-determined |
| 18-110_k141_275406 | Not-determined |
| 18-110_k141_229430 | Low-quality |
| 18-110_k141_205296 | Not-determined |
| 18-110_k141_160772 | Not-determined |
| 18-110_k141_229433 | Not-determined |
| 18-110_k141_217915 | Not-determined |
| 18-110_k141_160776 | Not-determined |
| 18-110_k141_91263 | Not-determined |
| 18-110_k141_45990 | Not-determined |
| 18-110_k141_205299 | Not-determined |
| 18-110_k141_45997 | Not-determined |
| 18-110_k141_80503 | Not-determined |
| 18-110_k141_229443 | Not-determined |
| 18-110_k141_20816 | Not-determined |
| 18-110_k141_46002 | Not-determined |
| 18-110_k141_275424 | Not-determined |
| 18-110_k141_46005 | Not-determined |
| 18-110_k141_56742 | Not-determined |
| 18-110_k141_275431 | Not-determined |
| 18-110_k141_20826 | Not-determined |
| 18-110_k141_205311 | Not-determined |
| 18-110_k141_275434 | Not-determined |
| 18-110_k141_160797 | Not-determined |
| 18-110_k141_275439 | Not-determined |
| 18-110_k141_275440 | Not-determined |
| 18-110_k141_46016 | Not-determined |
| 18-110_k141_160800 | Not-determined |
| 18-110_k141_195114 | Not-determined |
| 18-110_k141_68638 | Not-determined |
| 18-110_k141_20834 | Not-determined |
| 18-110_k141_80528 | Not-determined |
| 18-110_k141_20837 | Not-determined |
| 18-110_k141_252654 | Not-determined |
| 18-110_k141_205321 | Not-determined |
| 18-110_k141_20841 | Not-determined |
| 18-110_k141_160808 | Not-determined |
| 18-110_k141_252661 | Not-determined |
| 18-110_k141_91289 | Not-determined |
| 18-110_k141_91294 | Not-determined |
| 18-110_k141_217944 | Not-determined |
| 18-110_k141_160811 | Not-determined |
| 18-110_k141_46019 | Not-determined |
| 18-110_k141_46021 | Not-determined |
| 18-110_k141_56776 | Not-determined |
| 18-110_k141_252674 | Low-quality |
| 18-110_k141_126094 | Not-determined |
| 18-110_k141_275465 | Not-determined |
| 18-110_k141_252677 | Not-determined |
| 18-110_k141_68661 | Not-determined |
| 18-110_k141_46023 | Low-quality |
| 18-110_k141_172224 | Not-determined |
| 18-110_k141_217954 | Not-determined |
| 18-110_k141_126097 | Low-quality |
| 18-110_k141_114125 | Not-determined |
| 18-110_k141_20855 | Not-determined |
| 18-110_k141_56783 | Low-quality |
| 18-110_k141_68666 | Not-determined |
| 18-110_k141_252693 | Not-determined |
| 18-110_k141_217960 | Not-determined |
| 18-110_k141_172235 | Not-determined |
| 18-110_k141_20866 | Not-determined |
| 18-110_k141_172239 | Not-determined |
| 18-110_k141_103013 | Not-determined |
| 18-110_k141_252705 | Low-quality |
| 18-110_k141_205349 | Not-determined |
| 18-110_k141_195138 | Not-determined |
| 18-110_k141_275480 | Not-determined |
| 18-110_k141_103018 | Not-determined |
| 18-110_k141_205352 | Not-determined |
| 18-110_k141_172248 | Not-determined |
| 18-110_k141_20875 | Not-determined |
| 18-110_k141_275483 | Not-determined |
| 18-110_k141_80558 | Low-quality |
| 18-110_k141_91316 | Not-determined |
| 18-110_k141_229493 | Not-determined |
| 18-110_k141_114143 | Not-determined |
| 18-110_k141_261868 | Not-determined |
| 18-110_k141_114145 | Not-determined |
| 18-110_k141_80563 | Not-determined |
| 18-110_k141_68687 | Not-determined |
| 18-110_k141_91318 | Not-determined |
| 18-110_k141_261872 | Low-quality |
| 18-110_k141_91320 | Not-determined |
| 18-110_k141_20882 | Low-quality |
| 18-110_k141_217972 | Not-determined |
| 18-110_k141_217973 | Not-determined |
| 18-110_k141_195146 | Not-determined |
| 18-110_k141_205379 | Not-determined |
| 18-110_k141_56811 | Not-determined |
| 18-110_k141_217979 | Low-quality |
| 18-110_k141_217980 | Not-determined |
| 18-110_k141_46058 | Not-determined |
| 18-110_k141_91341 | Not-determined |
| 18-110_k141_91344 | Not-determined |
| 18-110_k141_217989 | Not-determined |
| 18-110_k141_229511 | Not-determined |
| 18-110_k141_56825 | Not-determined |
| 18-110_k141_172262 | Not-determined |
| 18-110_k141_217994 | Not-determined |
| 18-110_k141_229524 | Not-determined |
| 18-110_k141_205390 | Not-determined |
| 18-110_k141_56838 | Low-quality |
| 18-110_k141_229529 | Not-determined |
| 18-110_k141_68729 | Not-determined |
| 18-110_k141_218006 | Low-quality |
| 18-110_k141_80601 | Not-determined |
| 18-110_k141_195182 | Not-determined |
| 18-110_k141_218018 | Not-determined |
| 18-110_k141_103038 | Not-determined |
| 18-110_k141_172287 | Not-determined |
| 18-110_k141_56857 | Not-determined |
| 18-110_k141_172291 | Not-determined |
| 18-110_k141_195195 | Not-determined |
| 18-110_k141_205413 | Not-determined |
| 18-110_k141_205414 | Low-quality |
| 18-110_k141_103043 | Not-determined |
| 18-110_k141_103046 | Not-determined |
| 18-110_k141_195199 | Not-determined |
| 18-110_k141_229553 | Not-determined |
| 18-110_k141_218039 | Not-determined |
| 18-110_k141_126154 | Not-determined |
| 18-110_k141_20911 | Not-determined |
| 18-110_k141_114162 | Not-determined |
| 18-110_k141_252751 | Not-determined |
| 18-110_k141_56865 | Not-determined |
| 18-110_k141_20914 | Not-determined |
| 18-110_k141_160885 | Low-quality |
| 18-110_k141_218046 | Not-determined |
| 18-110_k141_126161 | Low-quality |
| 18-110_k141_56868 | Not-determined |
| 18-110_k141_160893 | Not-determined |
| 18-110_k141_103063 | Not-determined |
| 18-110_k141_114166 | Not-determined |
| 18-110_k141_160895 | Not-determined |
| 18-110_k141_56879 | Not-determined |
| 18-110_k141_275511 | Not-determined |
| 18-110_k141_229576 | Not-determined |
| 18-110_k141_195222 | Not-determined |
| 18-110_k141_229577 | Not-determined |
| 18-110_k141_195223 | Not-determined |
| 18-110_k141_218057 | Not-determined |
| 18-110_k141_9763 | Not-determined |
| 18-110_k141_114180 | Not-determined |
| 18-110_k141_160926 | Not-determined |
| 18-110_k141_275525 | Not-determined |
| 18-110_k141_275526 | Low-quality |
| 18-110_k141_218086 | Not-determined |
| 18-110_k141_172337 | Not-determined |
| 18-110_k141_172338 | Not-determined |
| 18-110_k141_205472 | Not-determined |
| 18-110_k141_20944 | Not-determined |
| 18-110_k141_9779 | Not-determined |
| 18-110_k141_205474 | Not-determined |
| 18-110_k141_275533 | Not-determined |
| 18-110_k141_229603 | Not-determined |
| 18-110_k141_103072 | Not-determined |
| 18-110_k141_20950 | Not-determined |
| 18-110_k141_9790 | Not-determined |
| 18-110_k141_20954 | Not-determined |
| 18-110_k141_205488 | Not-determined |
| 18-110_k141_229610 | Not-determined |
| 18-110_k141_20960 | Not-determined |
| 18-110_k141_9801 | Not-determined |
| 18-110_k141_126168 | Low-quality |
| 18-110_k141_218112 | Not-determined |
| 18-110_k141_160958 | Not-determined |
| 18-110_k141_9809 | Not-determined |
| 18-110_k141_126181 | Not-determined |
| 18-110_k141_56895 | Not-determined |
| 18-110_k141_275539 | Not-determined |
| 18-110_k141_103098 | Not-determined |
| 18-110_k141_9814 | Low-quality |
| 18-110_k141_9815 | Not-determined |
| 18-110_k141_9816 | Not-determined |
| 18-110_k141_172357 | Not-determined |
| 18-110_k141_172358 | Not-determined |
| 18-110_k141_91459 | Not-determined |
| 18-110_k141_56907 | Low-quality |
| 18-110_k141_9823 | Not-determined |
| 18-110_k141_68759 | Not-determined |
| 18-110_k141_9833 | Not-determined |
| 18-110_k141_114216 | Not-determined |
| 18-110_k141_91471 | Low-quality |
| 18-110_k141_68764 | Not-determined |
| 18-110_k141_114219 | Not-determined |
| 18-110_k141_91475 | Low-quality |
| 18-110_k141_172381 | Not-determined |
| 18-110_k141_103116 | Not-determined |
| 18-110_k141_172385 | Not-determined |
| 18-110_k141_275563 | Not-determined |
| 18-110_k141_275565 | Not-determined |
| 18-110_k141_218140 | Low-quality |
| 18-110_k141_261913 | Not-determined |
| 18-110_k141_172402 | Not-determined |
| 18-110_k141_172403 | Not-determined |
| 18-110_k141_218151 | Low-quality |
| 18-110_k141_275580 | Not-determined |
| 18-110_k141_91503 | Not-determined |
| 18-110_k141_229639 | Low-quality |
| 18-110_k141_91505 | Not-determined |
| 18-110_k141_205527 | Not-determined |
| 18-110_k141_103131 | Not-determined |
| 18-110_k141_205528 | Not-determined |
| 18-110_k141_9863 | Not-determined |
| 18-110_k141_275590 | Not-determined |
| 18-110_k141_160992 | Not-determined |
| 18-110_k141_205541 | Not-determined |
| 18-110_k141_91526 | Not-determined |
| 18-110_k141_114240 | Low-quality |
| 18-110_k141_229660 | Not-determined |
| 18-110_k141_229665 | Not-determined |
| 18-110_k141_229666 | Not-determined |
| 18-110_k141_275608 | Low-quality |
| 18-110_k141_229668 | Not-determined |
| 18-110_k141_172450 | Low-quality |
| 18-110_k141_172451 | Not-determined |
| 18-110_k141_229670 | Not-determined |
| 18-110_k141_9880 | Not-determined |
| 18-110_k141_261950 | Not-determined |
| 18-110_k141_126265 | Not-determined |
| 18-110_k141_91557 | Not-determined |
| 18-110_k141_126269 | Low-quality |
| 18-110_k141_20989 | Not-determined |
| 18-110_k141_172475 | Not-determined |
| 18-110_k141_114259 | Not-determined |
| 18-110_k141_161010 | Not-determined |
| 18-110_k141_275619 | Not-determined |
| 18-110_k141_205589 | Not-determined |
| 18-110_k141_161015 | Not-determined |
| 18-110_k141_161016 | Not-determined |
| 18-110_k141_103165 | Not-determined |
| 18-110_k141_114270 | Not-determined |
| 18-110_k141_205591 | Not-determined |
| 18-110_k141_91570 | Not-determined |
| 18-110_k141_126304 | Not-determined |
| 18-110_k141_172490 | Not-determined |
| 18-110_k141_161026 | Not-determined |
| 18-110_k141_229697 | Not-determined |
| 18-110_k141_114286 | Not-determined |
| 18-110_k141_114287 | Not-determined |
| 18-110_k141_126312 | Not-determined |
| 18-110_k141_114289 | Not-determined |
| 18-110_k141_261994 | Not-determined |
| 18-110_k141_114296 | Not-determined |
| 18-110_k141_114298 | Not-determined |
| 18-110_k141_195233 | Not-determined |
| 18-110_k141_114302 | Not-determined |
| 18-110_k141_195235 | Not-determined |
| 18-110_k141_126331 | Not-determined |
| 18-110_k141_161035 | Not-determined |
| 18-110_k141_195243 | Not-determined |
| 18-110_k141_161039 | Not-determined |
| 18-110_k141_56982 | Not-determined |
| 18-110_k141_103201 | Not-determined |
| 18-110_k141_172512 | Not-determined |
| 18-110_k141_205626 | Not-determined |
| 18-110_k141_103207 | Not-determined |
| 18-110_k141_126346 | Not-determined |
| 18-110_k141_126348 | Not-determined |
| 18-110_k141_172523 | Not-determined |
| 18-110_k141_9927 | Not-determined |
| 18-110_k141_114322 | Not-determined |
| 18-110_k141_21026 | Not-determined |
| 18-110_k141_161046 | Not-determined |
| 18-110_k141_161047 | Not-determined |
| 18-110_k141_205638 | Not-determined |
| 18-110_k141_114328 | Not-determined |
| 18-110_k141_262028 | Not-determined |
| 18-110_k141_195266 | Not-determined |
| 18-110_k141_262031 | Not-determined |
| 18-110_k141_21031 | Not-determined |
| 18-110_k141_126359 | Not-determined |
| 18-110_k141_205645 | Not-determined |
| 18-110_k141_262036 | Not-determined |
| 18-110_k141_114340 | Not-determined |
| 18-110_k141_103216 | Low-quality |
| 18-110_k141_195280 | Not-determined |
| 18-110_k141_229732 | Not-determined |
| 18-110_k141_275643 | Not-determined |
| 18-110_k141_114350 | Not-determined |
| 18-110_k141_275648 | Not-determined |
| 18-110_k141_229737 | Not-determined |
| 18-110_k141_114356 | Low-quality |
| 18-110_k141_103233 | Not-determined |
| 18-110_k141_103234 | Not-determined |
| 18-110_k141_114360 | Not-determined |
| 18-110_k141_103235 | Not-determined |
| 18-110_k141_229742 | Not-determined |
| 18-110_k141_21050 | Not-determined |
| 18-110_k141_91619 | Not-determined |
| 18-110_k141_229747 | Not-determined |
| 18-110_k141_56999 | Low-quality |
| 18-110_k141_57000 | Low-quality |
| 18-110_k141_103242 | Not-determined |
| 18-110_k141_172558 | Not-determined |
| 18-110_k141_103245 | Not-determined |
| 18-110_k141_103246 | Not-determined |
| 18-110_k141_195284 | Not-determined |
| 18-110_k141_275664 | Not-determined |
| 18-110_k141_229758 | Not-determined |
| 18-110_k141_275678 | Not-determined |
| 18-110_k141_218271 | Not-determined |
| 18-110_k141_103263 | Not-determined |
| 18-110_k141_275680 | Not-determined |
| 18-110_k141_9986 | Not-determined |
| 18-110_k141_21076 | Not-determined |
| 18-110_k141_172585 | Not-determined |
| 18-110_k141_9987 | Not-determined |
| 18-110_k141_91629 | Not-determined |
| 18-110_k141_21079 | Not-determined |
| 18-110_k141_252782 | Not-determined |
| 18-110_k141_9990 | Not-determined |
| 18-110_k141_205670 | Not-determined |
| 18-110_k141_195307 | Not-determined |
| 18-110_k141_205672 | Not-determined |
| 18-110_k141_195308 | Not-determined |
| 18-110_k141_161066 | Not-determined |
| 18-110_k141_9997 | Not-determined |
| 18-110_k141_205680 | Not-determined |
| 18-110_k141_21091 | Low-quality |
| 18-110_k141_126394 | Not-determined |
| 18-110_k141_252792 | Low-quality |
| 18-110_k141_252795 | Not-determined |
| 18-110_k141_126400 | Low-quality |
| 18-110_k141_10013 | Not-determined |
| 18-110_k141_10014 | Not-determined |
| 18-110_k141_195322 | Not-determined |
| 18-110_k141_57014 | Low-quality |
| 18-110_k141_229789 | Not-determined |
| 18-110_k141_114372 | Not-determined |
| 18-110_k141_126414 | Not-determined |
| 18-110_k141_229794 | Not-determined |
| 18-110_k141_161087 | Not-determined |
| 18-110_k141_126422 | Not-determined |
| 18-110_k141_275710 | Not-determined |
| 18-110_k141_126424 | Low-quality |
| 18-110_k141_262092 | Low-quality |
| 18-110_k141_275714 | Low-quality |
| 18-110_k141_161098 | Not-determined |
| 18-110_k141_57035 | Not-determined |
| 18-110_k141_57036 | Not-determined |
| 18-110_k141_262108 | Not-determined |
| 18-110_k141_205700 | Not-determined |
| 18-110_k141_91654 | Not-determined |
| 18-110_k141_161108 | Not-determined |
| 18-110_k141_195329 | Not-determined |
| 18-110_k141_205708 | Not-determined |
| 18-110_k141_21109 | Not-determined |
| 18-110_k141_114376 | Not-determined |
| 18-110_k141_161117 | Not-determined |
| 18-110_k141_218308 | Not-determined |
| 18-110_k141_262119 | Not-determined |
| 18-110_k141_161121 | Not-determined |
| 18-110_k141_262121 | Not-determined |
| 18-110_k141_91661 | Not-determined |
| 18-110_k141_57054 | Not-determined |
| 18-110_k141_229820 | Not-determined |
| 18-110_k141_252818 | Not-determined |
| 18-110_k141_161126 | Not-determined |
| 18-110_k141_262126 | Not-determined |
| 18-110_k141_57061 | Not-determined |
| 18-110_k141_68850 | Not-determined |
| 18-110_k141_262129 | Not-determined |
| 18-110_k141_205722 | Not-determined |
| 18-110_k141_229833 | Not-determined |
| 18-110_k141_195348 | Not-determined |
| 18-110_k141_229840 | Not-determined |
| 18-110_k141_114401 | Not-determined |
| 18-110_k141_252834 | Not-determined |
| 18-110_k141_275753 | Not-determined |
| 18-110_k141_275763 | Not-determined |
| 18-110_k141_218345 | Not-determined |
| 18-110_k141_252846 | Not-determined |
| 18-110_k141_275766 | Not-determined |
| 18-110_k141_252852 | Low-quality |
| 18-110_k141_205727 | Not-determined |
| 18-110_k141_10027 | Low-quality |
| 18-110_k141_229847 | Not-determined |
| 18-110_k141_68858 | Not-determined |
| 18-110_k141_10031 | Not-determined |
| 18-110_k141_229852 | Low-quality |
| 18-110_k141_91717 | Not-determined |
| 18-110_k141_21139 | Not-determined |
| 18-110_k141_218357 | Not-determined |
| 18-110_k141_126452 | Not-determined |
| 18-110_k141_57076 | Low-quality |
| 18-110_k141_10037 | Not-determined |
| 18-110_k141_161144 | Low-quality |
| 18-110_k141_126461 | Not-determined |
| 18-110_k141_275778 | Not-determined |
| 18-110_k141_57088 | Not-determined |
| 18-110_k141_205750 | Not-determined |
| 18-110_k141_205757 | Not-determined |
| 18-110_k141_91757 | Not-determined |
| 18-110_k141_252879 | Not-determined |
| 18-110_k141_205762 | Not-determined |
| 18-110_k141_10067 | Not-determined |
| 18-110_k141_57100 | Not-determined |
| 18-110_k141_68899 | Not-determined |
| 18-110_k141_252891 | Not-determined |
| 18-110_k141_262140 | Not-determined |
| 18-110_k141_205773 | Not-determined |
| 18-110_k141_275802 | Not-determined |
| 18-110_k141_68907 | Not-determined |
| 18-110_k141_68912 | Low-quality |
| 18-110_k141_68914 | Not-determined |
| 18-110_k141_252900 | Not-determined |
| 18-110_k141_229873 | Not-determined |
| 18-110_k141_161163 | Not-determined |
| 18-110_k141_10081 | Not-determined |
| 18-110_k141_252906 | Not-determined |
| 18-110_k141_205781 | Not-determined |
| 18-110_k141_10084 | Not-determined |
| 18-110_k141_91787 | Not-determined |
| 18-110_k141_57109 | Not-determined |
| 18-110_k141_57114 | Not-determined |
| 18-110_k141_195403 | Not-determined |
| 18-110_k141_218372 | Not-determined |
| 18-110_k141_229878 | Not-determined |
| 18-110_k141_229879 | Not-determined |
| 18-110_k141_68937 | Not-determined |
| 18-110_k141_252915 | Not-determined |
| 18-110_k141_91800 | Not-determined |
| 18-110_k141_205791 | Not-determined |
| 18-110_k141_275825 | Not-determined |
| 18-110_k141_218380 | Not-determined |
| 18-110_k141_103347 | Low-quality |
| 18-110_k141_218384 | Not-determined |

| | |
|---|---|
| 18-110_k141_205794 | Not-determined |
| 18-110_k141_275836 | Not-determined |
| 18-110_k141_205795 | Not-determined |
| 18-110_k141_252920 | Not-determined |
| 18-110_k141_103355 | Not-determined |
| 18-110_k141_205798 | Not-determined |
| 18-110_k141_91806 | Not-determined |
| 18-110_k141_205800 | Not-determined |
| 18-110_k141_275854 | Not-determined |
| 18-110_k141_68950 | Not-determined |
| 18-110_k141_229913 | Not-determined |
| 18-110_k141_195429 | Not-determined |
| 18-110_k141_91818 | Not-determined |
| 18-110_k141_229916 | Not-determined |
| 18-110_k141_68960 | Not-determined |
| 18-110_k141_252933 | Not-determined |
| 18-110_k141_229920 | Not-determined |
| 18-110_k141_172623 | Not-determined |
| 18-110_k141_218404 | Not-determined |
| 18-110_k141_68973 | Not-determined |
| 18-110_k141_114482 | Not-determined |
| 18-110_k141_91830 | Not-determined |
| 18-110_k141_262186 | Not-determined |
| 18-110_k141_205824 | Not-determined |
| 18-110_k141_252954 | Not-determined |
| 18-110_k141_205825 | Not-determined |
| 18-110_k141_114487 | Not-determined |
| 18-110_k141_21201 | Not-determined |
| 18-110_k141_68978 | Not-determined |
| 18-110_k141_262191 | Low-quality |
| 18-110_k141_21205 | Not-determined |
| 18-110_k141_21206 | Not-determined |
| 17-106-final-viral-combined_k141_119640\|\|0_partial | Medium-quality |
| 18-110_k141_91838 | Not-determined |
| 18-110_k141_275865 | Not-determined |
| 18-110_k141_275866 | Not-determined |
| 18-110_k141_57124 | Not-determined |
| 18-110_k141_21213 | Not-determined |
| 18-110_k141_91842 | Not-determined |
| 18-110_k141_252962 | Not-determined |
| 18-110_k141_275871 | Not-determined |
| 18-110_k141_205835 | Not-determined |
| 18-110_k141_275873 | Not-determined |
| 18-110_k141_252967 | Not-determined |
| 18-110_k141_103365 | Not-determined |
| 18-110_k141_229942 | Not-determined |
| 18-110_k141_252972 | Not-determined |
| 18-110_k141_57136 | Not-determined |
| 18-110_k141_10130 | Low-quality |
| 18-110_k141_218411 | Not-determined |
| 18-110_k141_103376 | Not-determined |
| 18-110_k141_91862 | Not-determined |
| 18-110_k141_218420 | Not-determined |
| 18-110_k141_57150 | Not-determined |
| 18-110_k141_114508 | Not-determined |
| 18-110_k141_68991 | Not-determined |
| 18-110_k141_68995 | Not-determined |
| 18-110_k141_68996 | Not-determined |
| 18-110_k141_57158 | Not-determined |
| 18-110_k141_103391 | Not-determined |
| 18-110_k141_91870 | Low-quality |
| 18-110_k141_126522 | Not-determined |
| 18-110_k141_103393 | Not-determined |
| 18-110_k141_91873 | Not-determined |
| 18-110_k141_205863 | Not-determined |
| 18-110_k141_57172 | Not-determined |
| 18-110_k141_103402 | Not-determined |
| 18-110_k141_91876 | Not-determined |
| 18-110_k141_172648 | Low-quality |
| 18-110_k141_172650 | Not-determined |
| 18-110_k141_10155 | Not-determined |
| 18-110_k141_57180 | Not-determined |
| 18-110_k141_252991 | Not-determined |
| 18-110_k141_252996 | Not-determined |
| 18-110_k141_126529 | Not-determined |
| 18-110_k141_126530 | Not-determined |
| 18-110_k141_252999 | Not-determined |
| 18-110_k141_262208 | Low-quality |
| 18-110_k141_229975 | Not-determined |
| 18-110_k141_126536 | Not-determined |
| 18-110_k141_103415 | Not-determined |
| 18-110_k141_126538 | Not-determined |
| 18-110_k141_57192 | Low-quality |
| 18-110_k141_161182 | Not-determined |
| 18-110_k141_69018 | Low-quality |
| 18-110_k141_10179 | Not-determined |
| 18-110_k141_69022 | Not-determined |
| 18-110_k141_195434 | Not-determined |
| 18-110_k141_262218 | Not-determined |
| 18-110_k141_195441 | Not-determined |
| 18-110_k141_21279 | Not-determined |
| 18-110_k141_103444 | Not-determined |
| 18-110_k141_91903 | Not-determined |
| 18-110_k141_172657 | Not-determined |
| 18-110_k141_172658 | Not-determined |
| 18-110_k141_91906 | Not-determined |
| 18-110_k141_103448 | Low-quality |
| 18-110_k141_262224 | Not-determined |
| 18-110_k141_103454 | Not-determined |
| 18-110_k141_230000 | Not-determined |
| 18-110_k141_262226 | Not-determined |
| 18-110_k141_262231 | Not-determined |
| 18-110_k141_195445 | Not-determined |
| 18-110_k141_195446 | Not-determined |
| 18-110_k141_195447 | Not-determined |
| 18-110_k141_21303 | Not-determined |
| 18-110_k141_21304 | Not-determined |
| 18-110_k141_218488 | Not-determined |
| 18-110_k141_195448 | Not-determined |
| 18-110_k141_230014 | Not-determined |
| 18-110_k141_230016 | Not-determined |
| 18-110_k141_275914 | Not-determined |
| 18-110_k141_275916 | Not-determined |
| 18-110_k141_21313 | Not-determined |
| 18-110_k141_172684 | Low-quality |
| 18-110_k141_275927 | Not-determined |
| 18-110_k141_57218 | Not-determined |
| 18-110_k141_161200 | Not-determined |
| 18-110_k141_275929 | Not-determined |
| 18-110_k141_57219 | Not-determined |
| 18-110_k141_218502 | Not-determined |
| 18-110_k141_253009 | Low-quality |
| 18-110_k141_253015 | Not-determined |
| 18-110_k141_262249 | Not-determined |
| 18-110_k141_275940 | Not-determined |
| 18-110_k141_114526 | Low-quality |
| 18-110_k141_69068 | Not-determined |
| 18-110_k141_126579 | Low-quality |
| 18-110_k141_218515 | Not-determined |
| 18-110_k141_205894 | Not-determined |
| 18-110_k141_230058 | Not-determined |
| 18-110_k141_230061 | Not-determined |
| 18-110_k141_205899 | Not-determined |
| 18-110_k141_205901 | Not-determined |
| 18-110_k141_218530 | Not-determined |
| 18-110_k141_195490 | Not-determined |
| 18-110_k141_195491 | Not-determined |
| 18-110_k141_126587 | Not-determined |
| 18-110_k141_126588 | Not-determined |
| 18-110_k141_103475 | Not-determined |
| 18-110_k141_126590 | Not-determined |
| 18-110_k141_57259 | Low-quality |
| 18-110_k141_262276 | Not-determined |
| 18-110_k141_57261 | Not-determined |
| 18-110_k141_253040 | Not-determined |
| 18-110_k141_126596 | Low-quality |
| 18-110_k141_253046 | Not-determined |
| 18-110_k141_253047 | Not-determined |
| 18-110_k141_69090 | Not-determined |
| 18-110_k141_114540 | Not-determined |
| 18-110_k141_262285 | Not-determined |
| 18-110_k141_218551 | Not-determined |
| 18-110_k141_69098 | Not-determined |
| 18-110_k141_253051 | Not-determined |
| 18-110_k141_262295 | Not-determined |
| 18-110_k141_195504 | Not-determined |
| 18-110_k141_57281 | Not-determined |
| 18-110_k141_21346 | Not-determined |
| 18-110_k141_57284 | Not-determined |
| 18-110_k141_262300 | Not-determined |
| 18-110_k141_103511 | Not-determined |
| 18-110_k141_230080 | Not-determined |
| 18-110_k141_230081 | Not-determined |
| 18-110_k141_262306 | Not-determined |
| 18-110_k141_275961 | Not-determined |
| 18-110_k141_21356 | Not-determined |
| 18-110_k141_230088 | Not-determined |
| 18-110_k141_230091 | Not-determined |
| 18-110_k141_114546 | Not-determined |
| 18-110_k141_126605 | Not-determined |
| 18-110_k141_172748 | Low-quality |
| 18-110_k141_262319 | Not-determined |
| 18-110_k141_262320 | Not-determined |
| 18-110_k141_21365 | Not-determined |
| 18-110_k141_172753 | Not-determined |
| 18-110_k141_172754 | Low-quality |
| 18-110_k141_21371 | Not-determined |
| 18-110_k141_172764 | Not-determined |
| 18-110_k141_205942 | Not-determined |
| 18-110_k141_230107 | Not-determined |
| 18-110_k141_253081 | Not-determined |
| 18-110_k141_253083 | Not-determined |
| 18-110_k141_10199 | Not-determined |
| 18-110_k141_205949 | Not-determined |
| 18-110_k141_218582 | Low-quality |
| 18-110_k141_161240 | Not-determined |
| 18-110_k141_195534 | Not-determined |
| 18-110_k141_218587 | Not-determined |
| 18-110_k141_21381 | Not-determined |
| 18-110_k141_57316 | Not-determined |
| 18-110_k141_103538 | Not-determined |
| 18-110_k141_262337 | Not-determined |
| 18-110_k141_161248 | Low-quality |
| 18-110_k141_172780 | Not-determined |
| 18-110_k141_253096 | Not-determined |
| 18-110_k141_262339 | Not-determined |
| 18-110_k141_195541 | Not-determined |
| 18-110_k141_205967 | Not-determined |
| 18-110_k141_205969 | Not-determined |
| 18-110_k141_161256 | Low-quality |
| 18-110_k141_218600 | Low-quality |
| 18-110_k141_218601 | Not-determined |
| 18-110_k141_126641 | Not-determined |
| 18-110_k141_262354 | Not-determined |
| 18-110_k141_161265 | Not-determined |
| 18-110_k141_161266 | Low-quality |
| 18-110_k141_195561 | Not-determined |
| 18-110_k141_10227 | Not-determined |
| 18-110_k141_91975 | Not-determined |
| 18-110_k141_230123 | Not-determined |
| 18-110_k141_114566 | Not-determined |
| 18-110_k141_114567 | Not-determined |
| 18-110_k141_206000 | Low-quality |
| 18-110_k141_230132 | Not-determined |
| 18-110_k141_230139 | Not-determined |
| 18-110_k141_10245 | Low-quality |
| 18-110_k141_57340 | Low-quality |
| 18-110_k141_230141 | Not-determined |
| 18-110_k141_103561 | Not-determined |
| 18-110_k141_57348 | Low-quality |
| 18-110_k141_10256 | Not-determined |
| 18-110_k141_206017 | Not-determined |
| 18-110_k141_57351 | Not-determined |
| 18-110_k141_114586 | Not-determined |
| 18-110_k141_230155 | Not-determined |
| 18-110_k141_21421 | Not-determined |
| 18-110_k141_114591 | Not-determined |
| 18-110_k141_103571 | Not-determined |
| 18-110_k141_21428 | Not-determined |
| 18-110_k141_253164 | Not-determined |
| 18-110_k141_195590 | Not-determined |
| 18-110_k141_195594 | Not-determined |
| 18-110_k141_195596 | Not-determined |
| 18-110_k141_57361 | Not-determined |
| 18-110_k141_126697 | Not-determined |
| 18-110_k141_126704 | Not-determined |
| 18-110_k141_10272 | Not-determined |
| 18-110_k141_195606 | Not-determined |
| 18-110_k141_126712 | Not-determined |
| 18-110_k141_92005 | Not-determined |
| 18-110_k141_114610 | Low-quality |
| 18-110_k141_262391 | Not-determined |
| 18-110_k141_195622 | Not-determined |
| 18-110_k141_57372 | Not-determined |
| 18-110_k141_275988 | Not-determined |
| 18-110_k141_253169 | Not-determined |
| 18-110_k141_10286 | Not-determined |
| 18-110_k141_10289 | Not-determined |
| 18-110_k141_218658 | Not-determined |
| 18-110_k141_195636 | Not-determined |
| 18-110_k141_253181 | Not-determined |
| 18-110_k141_114631 | Not-determined |
| 18-110_k141_57398 | Not-determined |
| 18-110_k141_195640 | Not-determined |
| 18-110_k141_218670 | Low-quality |
| 18-110_k141_230183 | Not-determined |
| 18-110_k141_218680 | Not-determined |
| 18-110_k141_57410 | Not-determined |
| 18-110_k141_253207 | Not-determined |
| 18-110_k141_206036 | Not-determined |
| 18-110_k141_206043 | Low-quality |
| 18-110_k141_262408 | Not-determined |
| 18-110_k141_262409 | Not-determined |
| 18-110_k141_195681 | Low-quality |
| 18-110_k141_21478 | Not-determined |
| 18-110_k141_206052 | Not-determined |
| 18-110_k141_276003 | Not-determined |
| 18-110_k141_57422 | Not-determined |
| 18-110_k141_195690 | Not-determined |
| 18-110_k141_103607 | Not-determined |
| 18-110_k141_276011 | Low-quality |
| 18-110_k141_57427 | Not-determined |
| 18-110_k141_195699 | Not-determined |
| 18-110_k141_253244 | Not-determined |
| 18-110_k141_206067 | Not-determined |
| 18-110_k141_253253 | Not-determined |
| 18-110_k141_195704 | Not-determined |
| 18-110_k141_206072 | Not-determined |
| 18-110_k141_230227 | Not-determined |
| 18-110_k141_218703 | Not-determined |
| 18-110_k141_276030 | Not-determined |
| 18-110_k141_21502 | Not-determined |
| 18-110_k141_195711 | Not-determined |
| 18-110_k141_21507 | Not-determined |
| 18-110_k141_276040 | Low-quality |
| 18-110_k141_195716 | Not-determined |
| 18-110_k141_276044 | Not-determined |
| 18-110_k141_57441 | Not-determined |
| 18-110_k141_253270 | Not-determined |
| 18-110_k141_218718 | Not-determined |
| 18-110_k141_276051 | Not-determined |
| 18-110_k141_276055 | Not-determined |
| 18-110_k141_218723 | Not-determined |
| 18-110_k141_262439 | Not-determined |
| 18-110_k141_57451 | Not-determined |
| 18-110_k141_206084 | Not-determined |
| 18-110_k141_276062 | Not-determined |
| 18-110_k141_218731 | Not-determined |
| 18-110_k141_276069 | Not-determined |
| 18-110_k141_206088 | Not-determined |
| 18-110_k141_206089 | Not-determined |
| 18-110_k141_253287 | Not-determined |
| 18-110_k141_230261 | Not-determined |
| 18-110_k141_206093 | Low-quality |
| 18-110_k141_206094 | Not-determined |
| 18-110_k141_253296 | Not-determined |
| 18-110_k141_218745 | Low-quality |
| 18-110_k141_206108 | Not-determined |
| 18-110_k141_230273 | Not-determined |
| 18-110_k141_276090 | Not-determined |
| 18-110_k141_206139 | Not-determined |
| 18-110_k141_57494 | Low-quality |
| 18-110_k141_57497 | Not-determined |
| 18-110_k141_276098 | Not-determined |
| 18-110_k141_10378 | Not-determined |
| 18-110_k141_10379 | Not-determined |
| 18-110_k141_276103 | Not-determined |
| 18-110_k141_276107 | Not-determined |
| 18-110_k141_92081 | Not-determined |
| 18-110_k141_206153 | Low-quality |
| 18-110_k141_262476 | Not-determined |
| 18-110_k141_21570 | Not-determined |
| 18-110_k141_92089 | Not-determined |
| 18-110_k141_206163 | Not-determined |
| 18-110_k141_103635 | Not-determined |
| 18-110_k141_218753 | Low-quality |
| 18-110_k141_206173 | Not-determined |
| 18-110_k141_218756 | Not-determined |
| 18-110_k141_10401 | Low-quality |
| 18-110_k141_114664 | Not-determined |
| 18-110_k141_103645 | Not-determined |
| 18-110_k141_230299 | Not-determined |
| 18-110_k141_92105 | Not-determined |
| 18-110_k141_10413 | Not-determined |
| 18-110_k141_230306 | Not-determined |
| 18-110_k141_218769 | Not-determined |
| 18-110_k141_276141 | Not-determined |
| 18-110_k141_262506 | Not-determined |
| 18-110_k141_218776 | Not-determined |
| 18-110_k141_10416 | Not-determined |
| 18-110_k141_10417 | Not-determined |
| 18-110_k141_218780 | Not-determined |
| 18-110_k141_21595 | Not-determined |
| 18-110_k141_21596 | Not-determined |
| 18-110_k141_218785 | Not-determined |
| 18-110_k141_230321 | Not-determined |
| 18-110_k141_21603 | Not-determined |
| 18-110_k141_230326 | Not-determined |
| 18-110_k141_218786 | Not-determined |
| 18-110_k141_230328 | Not-determined |
| 18-110_k141_92110 | Low-quality |
| 18-110_k141_21607 | Not-determined |
| 18-110_k141_21608 | Not-determined |
| 18-110_k141_92113 | Not-determined |
| 18-110_k141_57546 | Not-determined |
| 18-110_k141_206205 | Not-determined |
| 18-110_k141_230336 | Low-quality |
| 18-110_k141_276175 | Low-quality |
| 18-110_k141_230342 | Not-determined |
| 18-110_k141_206217 | Not-determined |
| 18-110_k141_276185 | Not-determined |
| 18-110_k141_10422 | Not-determined |
| 18-110_k141_262524 | Not-determined |
| 18-110_k141_10423 | Not-determined |
| 18-110_k141_114716 | Not-determined |
| 18-110_k141_57557 | Not-determined |
| 18-110_k141_21623 | Not-determined |
| 18-110_k141_114726 | Not-determined |
| 18-110_k141_262530 | Not-determined |
| 18-110_k141_21627 | Not-determined |
| 18-110_k141_262533 | Not-determined |
| 18-110_k141_253313 | Not-determined |
| 18-110_k141_10434 | Low-quality |
| 18-110_k141_253317 | Not-determined |
| 18-110_k141_21634 | Not-determined |
| 18-110_k141_230369 | Not-determined |
| 18-110_k141_114735 | Not-determined |
| 18-110_k141_276198 | Not-determined |
| 18-110_k141_218835 | Not-determined |
| 18-110_k141_10445 | Not-determined |
| 18-110_k141_276201 | Not-determined |
| 18-110_k141_21650 | Not-determined |
| 18-110_k141_114738 | Not-determined |
| 18-110_k141_57574 | Low-quality |
| 18-110_k141_218848 | Not-determined |
| 18-110_k141_10457 | Not-determined |
| 18-110_k141_276202 | Not-determined |
| 18-110_k141_10459 | Not-determined |
| 18-110_k141_218859 | Low-quality |
| 18-110_k141_114746 | Low-quality |
| 18-110_k141_57580 | Not-determined |
| 18-110_k141_262557 | Not-determined |
| 18-110_k141_114749 | Not-determined |
| 18-110_k141_262566 | Not-determined |
| 18-110_k141_114770 | Not-determined |
| 18-110_k141_10483 | Not-determined |
| 18-110_k141_10484 | Not-determined |
| 18-110_k141_21669 | Low-quality |
| 18-110_k141_21670 | Not-determined |
| 18-110_k141_195745 | Low-quality |
| 18-110_k141_253328 | Not-determined |
| 18-110_k141_114795 | Low-quality |
| 18-110_k141_21684 | Not-determined |
| 18-110_k141_21685 | Not-determined |
| 18-110_k141_10491 | Not-determined |
| 18-110_k141_10493 | Not-determined |
| 18-110_k141_276206 | Not-determined |
| 18-110_k141_10514 | Not-determined |
| 18-110_k141_195779 | Not-determined |
| 18-110_k141_114815 | Not-determined |
| 18-110_k141_114817 | Not-determined |
| 18-110_k141_10534 | Not-determined |
| 18-110_k141_276214 | Not-determined |
| 18-110_k141_114856 | Not-determined |
| 18-110_k141_10554 | Not-determined |
| 18-110_k141_206317 | Not-determined |
| 18-110_k141_114867 | Not-determined |
| 18-110_k141_276229 | Not-determined |
| 18-110_k141_276230 | Not-determined |
| 18-110_k141_206327 | Not-determined |
| 18-110_k141_276231 | Not-determined |
| 18-110_k141_262634 | Not-determined |
| 18-110_k141_114881 | Not-determined |
| 18-110_k141_276244 | Low-quality |
| 18-110_k141_21761 | Not-determined |
| 18-110_k141_276248 | Not-determined |
| 18-110_k141_276249 | Not-determined |
| 18-110_k141_276252 | Not-determined |
| 18-110_k141_276253 | Not-determined |
| 18-110_k141_114897 | Low-quality |
| 18-110_k141_276255 | Not-determined |
| 18-110_k141_276258 | Not-determined |
| 18-110_k141_262643 | Not-determined |
| 18-110_k141_262644 | Not-determined |
| 18-110_k141_276267 | Not-determined |
| 18-110_k141_10584 | Not-determined |
| 18-110_k141_276282 | Not-determined |
| 18-110_k141_114906 | Not-determined |
| 18-110_k141_10590 | Not-determined |
| 18-110_k141_276286 | Not-determined |
| 18-110_k141_114914 | Not-determined |
| 18-110_k141_114917 | Not-determined |
| 18-110_k141_262655 | Not-determined |
| 18-110_k141_276305 | Not-determined |
| 18-110_k141_276312 | Not-determined |
| 18-110_k141_10605 | Not-determined |
| 18-110_k141_276321 | Not-determined |
| 18-110_k141_276327 | Not-determined |
| 18-110_k141_10616 | Not-determined |
| 18-110_k141_276343 | Not-determined |
| 18-110_k141_195811 | Not-determined |
| 18-110_k141_276348 | Not-determined |
| 18-110_k141_276352 | Not-determined |
| 18-110_k141_276357 | Not-determined |
| 18-110_k141_10622 | Not-determined |
| 18-110_k141_276362 | Not-determined |
| 18-110_k141_276367 | Not-determined |
| 18-110_k141_276373 | Not-determined |
| 18-110_k141_276380 | Not-determined |
| 18-110_k141_276382 | Not-determined |
| 18-110_k141_276383 | Not-determined |
| 18-110_k141_276386 | Not-determined |
| 18-110_k141_276396 | Not-determined |
| 18-110_k141_276398 | Not-determined |
| 18-110_k141_276399 | Not-determined |
| 18-110_k141_276400 | Not-determined |
| 18-110_k141_276406 | Not-determined |
| 18-110_k141_10630 | Low-quality |
| 18-110_k141_276409 | Not-determined |
| 18-110_k141_276411 | Not-determined |
| 18-110_k141_276413 | Not-determined |
| 18-110_k141_276415 | Not-determined |
| 18-110_k141_276418 | Not-determined |
| 18-110_k141_206380 | Not-determined |
| 18-110_k141_276420 | Not-determined |
| 18-110_k141_276423 | Not-determined |
| 18-110_k141_276424 | Not-determined |
| 18-110_k141_276428 | Not-determined |
| 18-110_k141_114944 | Not-determined |
| 18-110_k141_206386 | Not-determined |
| 18-110_k141_276431 | Not-determined |
| 18-110_k141_276432 | Not-determined |
| 18-110_k141_276434 | Not-determined |
| 18-110_k141_276435 | Not-determined |
| 18-110_k141_276436 | Not-determined |
| 18-110_k141_276438 | Not-determined |
| 18-110_k141_276442 | Not-determined |
| 18-110_k141_276444 | Not-determined |
| 18-110_k141_276450 | Not-determined |
| 18-110_k141_276454 | Not-determined |
| 18-110_k141_276456 | Not-determined |
| 18-110_k141_276457 | Not-determined |
| 18-110_k141_114955 | Not-determined |
| 18-110_k141_10659 | Not-determined |
| 18-110_k141_10664 | Not-determined |
| 18-110_k141_206408 | Not-determined |
| 18-110_k141_253371 | Low-quality |
| 18-110_k141_206412 | Not-determined |
| 18-110_k141_206419 | Not-determined |
| 18-110_k141_206424 | Not-determined |
| 18-110_k141_114979 | Not-determined |
| 18-110_k141_10685 | Not-determined |
| 18-110_k141_206434 | Not-determined |
| 18-110_k141_262706 | Not-determined |
| 18-110_k141_10691 | Low-quality |
| 18-110_k141_253382 | Not-determined |
| 18-110_k141_253385 | Not-determined |
| 18-110_k141_253393 | Not-determined |
| 18-110_k141_262722 | Not-determined |
| 18-110_k141_253395 | Low-quality |
| 18-110_k141_262728 | Low-quality |
| 18-110_k141_262734 | Low-quality |
| 18-110_k141_10707 | Not-determined |
| 18-110_k141_262751 | Not-determined |
| 18-110_k141_10726 | Not-determined |
| 18-110_k141_10727 | Not-determined |
| 18-110_k141_115000 | Not-determined |
| 18-110_k141_262764 | Not-determined |
| 18-110_k141_262769 | Low-quality |
| 18-110_k141_262770 | Not-determined |
| 18-110_k141_262772 | Not-determined |
| 18-110_k141_262782 | Not-determined |
| 18-110_k141_10750 | Not-determined |
| 18-110_k141_10758 | Not-determined |
| 18-110_k141_262791 | Not-determined |
| 18-110_k141_115032 | Not-determined |
| 18-110_k141_10766 | Not-determined |
| 18-110_k141_206511 | Not-determined |
| 18-110_k141_10771 | Not-determined |
| 18-110_k141_262803 | Not-determined |
| 18-110_k141_115050 | Not-determined |
| 18-110_k141_10777 | Not-determined |
| 18-110_k141_115051 | Low-quality |
| 18-110_k141_206524 | Not-determined |
| 18-110_k141_21840 | Not-determined |
| 18-110_k141_10787 | Low-quality |
| 18-110_k141_115067 | Not-determined |
| 18-110_k141_10798 | Not-determined |
| 18-110_k141_115079 | Not-determined |
| 18-110_k141_262819 | Low-quality |
| 18-110_k141_115082 | Not-determined |
| 18-110_k141_115084 | Not-determined |
| 18-110_k141_21861 | Not-determined |
| 18-110_k141_21862 | Not-determined |
| 18-110_k141_115092 | Not-determined |
| 18-110_k141_21869 | Not-determined |
| 18-110_k141_206555 | Not-determined |
| 18-110_k141_262833 | Not-determined |
| 18-110_k141_21872 | Not-determined |
| 18-110_k141_21876 | Not-determined |
| 18-110_k141_115103 | Not-determined |
| 18-110_k141_21885 | Not-determined |
| 18-110_k141_262856 | Not-determined |
| 18-110_k141_115136 | Not-determined |
| 18-110_k141_115141 | Not-determined |
| 18-110_k141_262868 | Low-quality |
| 18-110_k141_21919 | Not-determined |
| 18-110_k141_262881 | Not-determined |
| 18-110_k141_206587 | Not-determined |
| 18-110_k141_21933 | Not-determined |
| 18-110_k141_206596 | Not-determined |
| 18-110_k141_262890 | Not-determined |
| 18-110_k141_206601 | Not-determined |
| 18-110_k141_262894 | Not-determined |
| 18-110_k141_262897 | Not-determined |
| 18-110_k141_10822 | Not-determined |
| 18-110_k141_10824 | Not-determined |
| 18-110_k141_262905 | Not-determined |
| 18-110_k141_21950 | Not-determined |
| 18-110_k141_262908 | Low-quality |
| 18-110_k141_262911 | Not-determined |
| 18-110_k141_262915 | Not-determined |
| 18-110_k141_10835 | Not-determined |
| 18-110_k141_262917 | Not-determined |
| 18-110_k141_10840 | Low-quality |
| 18-110_k141_21959 | Not-determined |
| 18-110_k141_115176 | Not-determined |
| 18-110_k141_206644 | Not-determined |
| 18-110_k141_206657 | Not-determined |
| 18-110_k141_206658 | Not-determined |
| 18-110_k141_115184 | Not-determined |
| 18-110_k141_21989 | Not-determined |
| 18-110_k141_262964 | Not-determined |
| 18-110_k141_22000 | Not-determined |
| 18-110_k141_10880 | Low-quality |
| 18-110_k141_262984 | Not-determined |
| 18-110_k141_10886 | Not-determined |
| 18-110_k141_206674 | Not-determined |
| 18-110_k141_206676 | Not-determined |

| | |
|---|---|
| 18-110_k141_10903 | Not-determined |
| 18-110_k141_22018 | Not-determined |
| 18-110_k141_22024 | Not-determined |
| 18-110_k141_263004 | Not-determined |
| 18-110_k141_22039 | Not-determined |
| 18-110_k141_10926 | Not-determined |
| 18-110_k141_22042 | Not-determined |
| 18-110_k141_263018 | Not-determined |
| 18-110_k141_22056 | Not-determined |
| 18-110_k141_206720 | Not-determined |
| 18-110_k141_263024 | Not-determined |
| 18-110_k141_22061 | Low-quality |
| 18-110_k141_206724 | Not-determined |
| 18-110_k141_263043 | Not-determined |
| 18-110_k141_22079 | Not-determined |
| 18-110_k141_206730 | Not-determined |
| 18-110_k141_206731 | Not-determined |
| 18-110_k141_263063 | Not-determined |
| 18-110_k141_10966 | Not-determined |
| 18-110_k141_263071 | Not-determined |
| 18-110_k141_263086 | Not-determined |
| 18-110_k141_22090 | Not-determined |
| 18-110_k141_22095 | Not-determined |
| 18-110_k141_206737 | Not-determined |
| 18-110_k141_263098 | Not-determined |
| 18-110_k141_263110 | Low-quality |
| 18-110_k141_22132 | Not-determined |
| 18-110_k141_206766 | Not-determined |
| 18-110_k141_263118 | Not-determined |
| 18-110_k141_22142 | Not-determined |
| 18-110_k141_263130 | Low-quality |
| 18-110_k141_263134 | Not-determined |
| 18-110_k141_263165 | Low-quality |
| 18-110_k141_263171 | Not-determined |
| 18-110_k141_206800 | Not-determined |
| 18-110_k141_263177 | Not-determined |
| 18-110_k141_10993 | Not-determined |
| 18-110_k141_22213 | Not-determined |
| 18-110_k141_10999 | Low-quality |
| 18-110_k141_22219 | Not-determined |
| 18-110_k141_11005 | Not-determined |
| 18-110_k141_11012 | Not-determined |
| 18-110_k141_263185 | Not-determined |
| 18-110_k141_11019 | Not-determined |
| 18-110_k141_11033 | Not-determined |
| 18-110_k141_206829 | Not-determined |
| 18-110_k141_11061 | Not-determined |
| 18-110_k141_11064 | Low-quality |
| 18-110_k141_263209 | Not-determined |
| 18-110_k141_263213 | Not-determined |
| 18-110_k141_22263 | Low-quality |
| 18-110_k141_263226 | Not-determined |
| 18-110_k141_263227 | Not-determined |
| 18-110_k141_263229 | Not-determined |
| 18-110_k141_11075 | Not-determined |
| 18-110_k141_22272 | Low-quality |
| 18-110_k141_22285 | Not-determined |
| 18-110_k141_22295 | Not-determined |
| 18-110_k141_206836 | Not-determined |
| 18-110_k141_22299 | Low-quality |
| 18-110_k141_263240 | Not-determined |
| 18-110_k141_263244 | Not-determined |
| 18-110_k141_22321 | Not-determined |
| 18-110_k141_263249 | Not-determined |
| 18-110_k141_206862 | Not-determined |
| 18-110_k141_22331 | Not-determined |
| 18-110_k141_22333 | Not-determined |
| 18-110_k141_22354 | Low-quality |
| 18-110_k141_263314 | Not-determined |
| 18-110_k141_263326 | Not-determined |
| 18-110_k141_263329 | Not-determined |
| 18-110_k141_263334 | Not-determined |
| 18-110_k141_22375 | Not-determined |
| 18-110_k141_263336 | Not-determined |
| 18-110_k141_263340 | Not-determined |
| 18-110_k141_263343 | Not-determined |
| 18-110_k141_263344 | Not-determined |
| 18-110_k141_263346 | Not-determined |
| 18-110_k141_206894 | Not-determined |
| 18-110_k141_206895 | Not-determined |
| 18-110_k141_22388 | Not-determined |
| 18-110_k141_22402 | Not-determined |
| 18-110_k141_206909 | Not-determined |
| 18-110_k141_263368 | Low-quality |
| 18-110_k141_263369 | Not-determined |
| 18-110_k141_263372 | Not-determined |
| 18-110_k141_263373 | Not-determined |
| 18-110_k141_263388 | Not-determined |
| 18-110_k141_263389 | Not-determined |
| 18-110_k141_22413 | Not-determined |
| 18-110_k141_22439 | Not-determined |
| 18-110_k141_263441 | Not-determined |
| 18-110_k141_22453 | Not-determined |
| 18-110_k141_206957 | Not-determined |
| 18-110_k141_22460 | Not-determined |
| 18-110_k141_22461 | Not-determined |
| 18-110_k141_22466 | Not-determined |
| 18-110_k141_22474 | Not-determined |
| 18-110_k141_22478 | Not-determined |
| 18-110_k141_263464 | Not-determined |
| 18-110_k141_263476 | Low-quality |
| 18-110_k141_22496 | Not-determined |
| 18-110_k141_22508 | Not-determined |
| 18-110_k141_22517 | Not-determined |
| 18-110_k141_263535 | Not-determined |
| 18-110_k141_263556 | Not-determined |
| 18-110_k141_263568 | Low-quality |
| 18-110_k141_11133 | Not-determined |
| 18-110_k141_207072 | Not-determined |
| 18-110_k141_11149 | Not-determined |
| 18-110_k141_207097 | Not-determined |
| 18-110_k141_263606 | Not-determined |
| 18-110_k141_22556 | Not-determined |
| 18-110_k141_11163 | Not-determined |
| 18-110_k141_11172 | Not-determined |
| 18-110_k141_207131 | Not-determined |
| 18-110_k141_207135 | Not-determined |
| 18-110_k141_263662 | Not-determined |
| 18-110_k141_22586 | Low-quality |
| 18-110_k141_207163 | Not-determined |
| 18-110_k141_207176 | Not-determined |
| 18-110_k141_263694 | Not-determined |
| 18-110_k141_207186 | Not-determined |
| 18-110_k141_11230 | Low-quality |
| 18-110_k141_263699 | Not-determined |
| 18-110_k141_263703 | Not-determined |
| 18-110_k141_11244 | Not-determined |
| 18-110_k141_11250 | Not-determined |
| 18-110_k141_22622 | Not-determined |
| 18-110_k141_207235 | Not-determined |
| 18-110_k141_207236 | Low-quality |
| 18-110_k141_207245 | Not-determined |
| 18-110_k141_22657 | Not-determined |
| 18-110_k141_207251 | Not-determined |
| 18-110_k141_22670 | Not-determined |
| 18-110_k141_263762 | Not-determined |
| 18-110_k141_263769 | Not-determined |
| 18-110_k141_11313 | Not-determined |
| 18-110_k141_263776 | Low-quality |
| 18-110_k141_263783 | Not-determined |
| 18-110_k141_11327 | Not-determined |
| 18-110_k141_207281 | Not-determined |
| 18-110_k141_263838 | Not-determined |
| 18-110_k141_11342 | Not-determined |
| 18-110_k141_263849 | Not-determined |
| 18-110_k141_263863 | Not-determined |
| 18-110_k141_11354 | Not-determined |
| 18-110_k141_263865 | Not-determined |
| 18-110_k141_263867 | Not-determined |
| 18-110_k141_11358 | Not-determined |
| 18-110_k141_207317 | Not-determined |
| 18-110_k141_207325 | Not-determined |
| 18-110_k141_207328 | Not-determined |
| 18-110_k141_11394 | Not-determined |
| 18-110_k141_11401 | Not-determined |
| 18-110_k141_11402 | Not-determined |
| 18-110_k141_207336 | Not-determined |
| 18-110_k141_11408 | Low-quality |
| 18-110_k141_11410 | Low-quality |
| 18-110_k141_11413 | Not-determined |
| 18-110_k141_11421 | Not-determined |
| 18-110_k141_263892 | Not-determined |
| 18-110_k141_22809 | Not-determined |
| 18-110_k141_22814 | Not-determined |
| 18-110_k141_22823 | Not-determined |
| 18-110_k141_263903 | Not-determined |
| 18-110_k141_11447 | Low-quality |
| 18-110_k141_11450 | Not-determined |
| 18-110_k141_11460 | Low-quality |
| 18-110_k141_263927 | Not-determined |
| 18-110_k141_263934 | Not-determined |
| 18-110_k141_22865 | Not-determined |
| 18-110_k141_22878 | Not-determined |
| 18-110_k141_263962 | Not-determined |
| 18-110_k141_263973 | Not-determined |
| 18-110_k141_263982 | Not-determined |
| 18-110_k141_11519 | Not-determined |
| 18-110_k141_264014 | Not-determined |
| 18-110_k141_22923 | Not-determined |
| 18-110_k141_264017 | Not-determined |
| 18-110_k141_264037 | Not-determined |
| 18-110_k141_264044 | Not-determined |
| 18-110_k141_264050 | Not-determined |
| 18-110_k141_264085 | Not-determined |
| 18-110_k141_22995 | Not-determined |
| 18-110_k141_264086 | Not-determined |
| 18-110_k141_23006 | Not-determined |
| 18-110_k141_23009 | Not-determined |
| 18-110_k141_264100 | Not-determined |
| 18-110_k141_264116 | Not-determined |
| 18-110_k141_264179 | Not-determined |
| 18-110_k141_264195 | Not-determined |
| 18-110_k141_264207 | Not-determined |
| 18-110_k141_264225 | Not-determined |
| 18-110_k141_264248 | Not-determined |
| 18-110_k141_264259 | Not-determined |
| 18-110_k141_264265 | Not-determined |
| 18-110_k141_264293 | Not-determined |
| 18-110_k141_264299 | Not-determined |
| 18-110_k141_264302 | Not-determined |
| 18-110_k141_264329 | Not-determined |
| 18-110_k141_264347 | Not-determined |
| 18-110_k141_264408 | Not-determined |
| 18-110_k141_264431 | Not-determined |
| 18-110_k141_264449 | Not-determined |
| 18-110_k141_264461 | Not-determined |
| 18-110_k141_264470 | Not-determined |
| 18-110_k141_264483 | Not-determined |
| 18-110_k141_264484 | Not-determined |
| 18-110_k141_264519 | Not-determined |
| 18-110_k141_264524 | Not-determined |
| 18-110_k141_264539 | Not-determined |
| 18-110_k141_264548 | Not-determined |
| 18-110_k141_264579 | Low-quality |
| 18-110_k141_264604 | Not-determined |
| 18-110_k141_264662 | Not-determined |
| 18-110_k141_264676 | Not-determined |
| 18-110_k141_264707 | Not-determined |
| 18-110_k141_264741 | Not-determined |
| 18-110_k141_264753 | Not-determined |
| 18-110_k141_264811 | Not-determined |
| 18-110_k141_264814 | Not-determined |
| 18-110_k141_264841 | Not-determined |
| 18-110_k141_264891 | Not-determined |
| 18-110_k141_264892 | Not-determined |
| 18-110_k141_264900 | Not-determined |
| 18-110_k141_264901 | Low-quality |
| 18-110_k141_264916 | Not-determined |
| 18-110_k141_264929 | Not-determined |

| | Count | Mean | Median | SD |
|---|---|---|---|---|
| Complete (100% completeness) | 68 | 89327.5 | 83068.5 | 61107.7 |
| High quality (90-100% completeness) | 304 | 48077.1 | 44281 | 28283.9 |
| Medium quality (50-90% completeness) | 589 | 40431.5 | 30929 | 35297.8 |
| Low quality (<50% completeness) | 64500 | 2228 | 1029 | 4674 |
| Undetermined | 196283 | 1029.1 | 466 | 1865.6 |
| Total UViGs | 261744 | 36218.64 | 30929 | 24295.99 |
| Sum of 50-100% genome completeness | 961 | 59278.7 | 44281 | 17285.58 |
